# Comprehensive profiling of the human intestinal DNA virome and prediction of disease-associated bacterial hosts in severe Myalgic Encephalomyelitis/Chronic Fatigue Syndrome (ME/CFS)

**DOI:** 10.1101/2023.06.29.23291738

**Authors:** Shen-Yuan Hsieh, George M. Savva, Andrea Telatin, Sumeet K. Tiwari, Mohammad A. Tariq, Fiona Newberry, Katharine A. Seton, Catherine Booth, Amolak S. Bansal, Thomas Wileman, Evelien M. Adriaenssens, Simon R. Carding

## Abstract

Myalgic encephalomyelitis/chronic fatigue syndrome (ME/CFS) is a disabling disorder of unknown etiology with severely affected patients being house- and/or bedbound. A historical association with chronic virus infection and subsequent recent reports correlating intestinal microbial dysbiosis with disease pathology prompted us to analyze the intestinal virome in a small cohort of severely-affected ME/CFS patients and same household healthy controls (SHHC). Datasets from whole metagenomic sequencing (WMS) and sequencing of virus-like particles (VLP)-enriched metagenomes from the same fecal sample yielded diverse, high-quality vOTUs with high read coverage and high genome completeness. The core intestinal virome was largely composed of tailed phages in the class *Caudoviricetes* with no significant differences in alpha diversity between ME/CFS and SHHC groups. However, the WMS dataset had a higher Shannon measure than the VLP dataset (*p* < 0.0001), with VLP- and WMS-derived sequences indicating differential abundances within several viral families and different viral compositions in beta diversity. This confirms that combining different isolation methodologies identifies a greater diversity of viruses including extracellular phages and integrated prophages. DNA viromes and bacteriomes from ME/CFS and SHHC groups were comparable with no differences in any alpha or beta diversity measures. One vOTU derived from the VLP-derived dataset was assigned to ssDNA human virus smacovirus 1. Using an *in-silico* approach to predict cohort-based bacterial hosts, we identified members of the *Anaerotruncus* genus interacting with unique viruses present in ME/CFS microbiomes; this may contribute to the GI microbial dysbiosis described in ME/CFS patients.

## Introduction

Myalgic Encephalomyelitis/chronic fatigue syndrome (ME/CFS) is a severe disabling and debilitating heterogeneous disorder affecting multiple bodily systems. Symptoms include myalgia, post-exertional malaise (PEM), sleep disturbance, gastrointestinal (GI) dysfunction, neurological and cognitive impairment, and prolonged unexplained fatigue^1,2^. The estimated global prevalence of ME/CFS is between 0.2 and 2.6%, depending on the diagnostic criteria used^3–5^; the majority (70%) of affected individuals are female^4,6^. Approximately 25% of patients are severely affected, i.e., they are housebound or bedbound for prolonged periods of time, requiring in-home assistance, which is often provided by family members^7^. The clinical definition of ME/CFS is controversial due to a lack in distinctive ME/CFS biomarkers resulting in diagnosis being based upon a variety of different clinical criteria (e.g., Fukuda *et al.*^2^; Canadian Consensus^8^; NICE [National Institute for Health and Care Excellence] guideline^9^; the Oxford^10^; International Consensus Criteria [ICC]^1^; and Institute of Medicine (IOM)^11^). Recently, a diagnostic scoring system has also been used to enhance accuracy and distinguish ME/CFS from other diseases with overlapping symptoms such as prolonged depression and fatigue^12^.

The etiology of ME/CFS remains unknown with several hypotheses being proposed including chronic viral infections^13,14^, intestinal microbial dysbiosis^15–18^, metabolic disorders^19,20^, mitochondrial dysfunction^21,22^, and/or autoimmunity^23–25^. Chronic viral infection is supported by clinical- and laboratory-based observations indicating that most ME/CFS cases (50–80%) are associated with or follow prolonged influenza-like symptoms including post-viral fatigue^6,26^. Several human eukaryotic viruses are potential etiological agents of ME/CFS: human herpesviruses 6 (HHV-6) and HHV-7^27^; *Epstein-Barr virus* (EBV) and *Cytomegalovirus* (CMV)^28,29^; *Parvovirus B19*^30–32^; and enteroviruses^33,34^. More recently, numerous clinical cases of coronavirus (18–52%) with overlapping ME/CFS symptomology have been identified including severe acute respiratory syndrome coronavirus (SARS-CoV and SARS-CoV-2) and Middle East respiratory syndrome coronavirus (MERS-CoV) which are both characterized by chronic post-viral fatigue and cognitive deficits^35–37^.

A potential source of pathogenic viruses is the human virome and in particular, the gastrointestinal (GI) virome^38,39^ which comprises eukaryotic viruses, bacteriophages (or phages)^40^, archaeal viruses, endogenous retroviruses, and other rare viruses from diverse environmental sources (e.g., water, animals, and plants^41^). Phages account for the vast majority (>90%) of the intestinal virobiota^42,43^ and have the capability to alter the structural and functional composition of the microbial community (gain or loss of species), thereby contributing to intestinal homeostasis and health of the human host^44^. Persistent alterations in the human intestinal virome have been identified in patients with chronic inflammatory GI disorders such as inflammatory bowel disease (IBD)^38^, suggesting that alterations in intestinal virome may contribute to the pathophysiology of chronic diseases.

The possibility that GI viral infection contributes to the pathology of ME/CFS is supported by clinical- and laboratory-based observations^30,34,45^. Most ME/CFS patients (92%) have persistent or intermittent symptoms of GI dysfunction and suffer from irritable bowel syndrome (IBS)^46^, with associated alterations in the intestinal virome^38,39,47^. In one of the few virome studies in ME/CFS patients to date, alterations in the intestinal (fecal) virome were identified in a pair of identical twins discordant for ME/CFS and living apart from each other for several years; the affected twin displayed a comparative increase in the abundance of phages from the class *Caudoviricetes* (previously the order *Caudovirales*, but reclassified by International Committee on Taxonomy of Viruses [ICTV]^48^), of which the majority were predicted to be siphoviruses and myoviruses^45^.

Here we designed a study to gain a more complete and comprehensive profile of the human GI DNA virome. Specifically, we combined and compared two datasets derived from virus-like particle (VLP)-enriched metagenomes and whole metagenomic sequences (WMS) from stool samples. The stool samples were obtained from a small cohort of severely-affected ME/CFS patients and healthy individuals living in the same environment/ household who cared for these patients (i.e., same household healthy controls; SHHC). To determine whether more high-quality viral genomes could be identified after VLP enrichment, we first compared the VLP-enriched DNA virome with DNA viruses derived from the whole metagenome of the same sample. Secondly, we determined whether there were systematic differences in virome and bacteriome composition of severely-affected ME/CFS patients compared with healthy individuals; the nature of any differences in virome were determined using various viral isolation methodologies. Thirdly, we determined whether it was possible to predict the disease-associated bacterial hosts for the viruses uniquely present in ME/CFS patients.

## Results

### Study cohort and VLP characterization

Nine severely-affected ME/CFS patients who met 2003 Canadian Consensus Criteria and NICE 2007/CG53 guideline were recruited from the CFS Service of the Epsom and St Helier NHS Foundation Trust University Hospital or the East Coast Community Healthcare Centre ME/CFS Service alongside eight SHHC that were parents and/or carers of these patients (**Table 1**). ME/CFS patients were all female with an average age of 33.8 ± 13 years (mean ± SD), consistent with the known higher prevalence of ME/CFS in females than males in the general population. Amongst these four severely affected patients had a self-reported GI disorder. The age at onset and duration of illness varied amongst patients. All patients had received their diagnosis at least 6 months prior to recruitment to the study. SHHC subjects comprised three males and five females of average age 53.6 ± 15.5 years (mean ± SD, *n* = 8). No SHHC reported any GI disorders or symptoms at the time of enrollment and sample collection.

**Table 1.** Cohort characteristics.

|  | ME/CFS | SHHC |
| --- | --- | --- |
| Age; mean ± SD (median) | 33.8 ± 13.0 (37.0) | 53.6 ± 15.5 (58.5) |
| Gender (male:female) | 0:9 | 3:5 |
| Antibiotic use < 4 weeks | 0 | 0 |
| Probiotic capsule<br>supplement use < 4 weeks | 0 | 0 |
| Illness duration > 6 months* | 9 | N/A |
| Self-reported with GI<br>disorder | Four self-reported;<br>Five non-reported | 0 |

\* All severely-affected patients received their confirmed diagnosis at least 6 months prior to sample collection
N/A: not applicable

Transmission electron micrographs (TEM) of VLP samples (*n* = 17) showed a predominance of phages, particularly tailed double-stranded DNA (dsDNA) phages belonging to the class *Caudoviricetes* (including three common morphotypes: siphoviruses, myoviruses, podoviruses) alongside a filamentous virus seen in one sample (**Fig. 1**). SYBR Gold staining and epifluorescence microscopy (EFM) were used to quantify enriched VLPs, which numbered 1.6 × 10^9^ ± 8.1 × 10^8^ VLP/g feces per sample (mean ± SD), with fecal VLPs yielding 2,518.5 ± 1,832.9 ng DNA per sample (mean ± SD) (**Supplementary Table S1**). There was no difference in the number (mean ± SD) of ME/CFS-derived VLPs (1.6 × 10^9^ ± 9.3 × 10^8^ VLP/g feces; *n* = 9) compared with SHHC-derived VLPs (1.5 × 10^9^ ± 7.1 × 10^8^ VLP/g feces; *n* = 8). However, ME/CFS-derived VLPs yielded less DNA (mean ± SD) than SHHC-derived VLPs (1,585.2 ± 1,411.0 ng DNA versus 3,568.4 ± 1738.8 ng DNA) (**Supplementary Table S1**).

**Figure 1.**
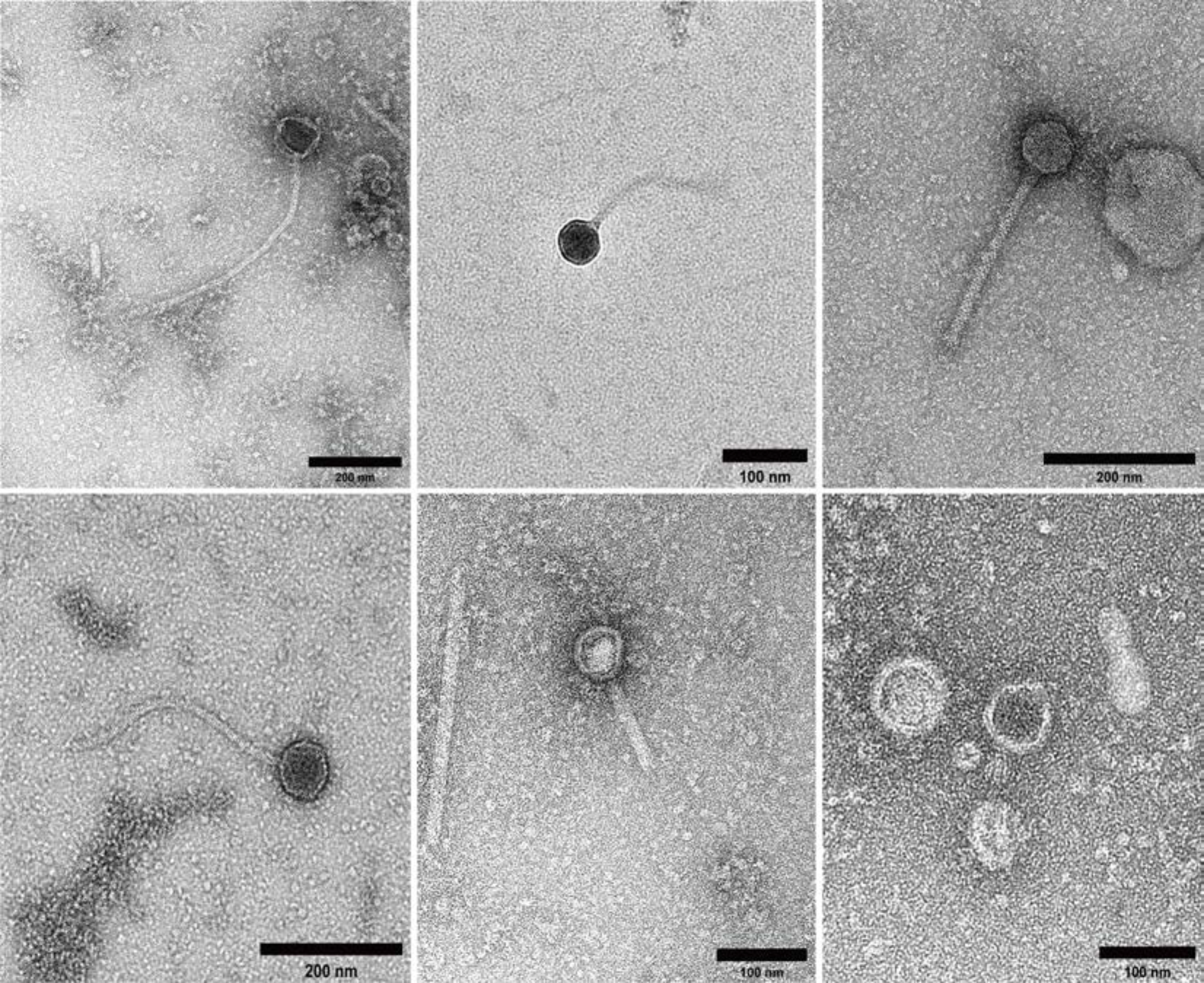
Transmission electron micrographs of fecal VLPs. Representative transmission electron micrographs of VLPs in fecal filtrates derived from one patient and three SHHC subjects. In all samples the majority of intact virions and VLPs detected belonged to the class *Caudoviricetes* which morphologically includes siphoviruses (long tails), myoviruses (long, contractile tails) and podoviruses (short tails); a filamentous virus was seen in one sample (middle bottom). Scale bar = 100–200 nm.

Genomic DNA from bulk fecal samples from seven ME/CFS and five SHHC patients were used to profile non-enriched viral and prokaryotic genomes by whole metagenomic sequencing (WMS). Five fecal samples contained insufficient biomass for parallel WMS and VLP analysis, and were therefore prioritized for VLP enrichment. Three sequence datasets were created: (1) DNA virome derived from VLP-enriched metagenomes, (2) DNA viromes derived from WMS sequences and (3) bacteriomes derived from whole metagenomes (WMS-Bac).

### Quantitative and qualitative assessment of enriched VLP and WMS derived viral genomes

The bioinformatic pipeline used for analyzing the cohort-associated viromes and bacteriomes is depicted in **Fig. 2**. The statistical analyses of both VLP and WMS sequence datasets are detailed in **Supplementary Table S1–S5**.

**Figure 2.**
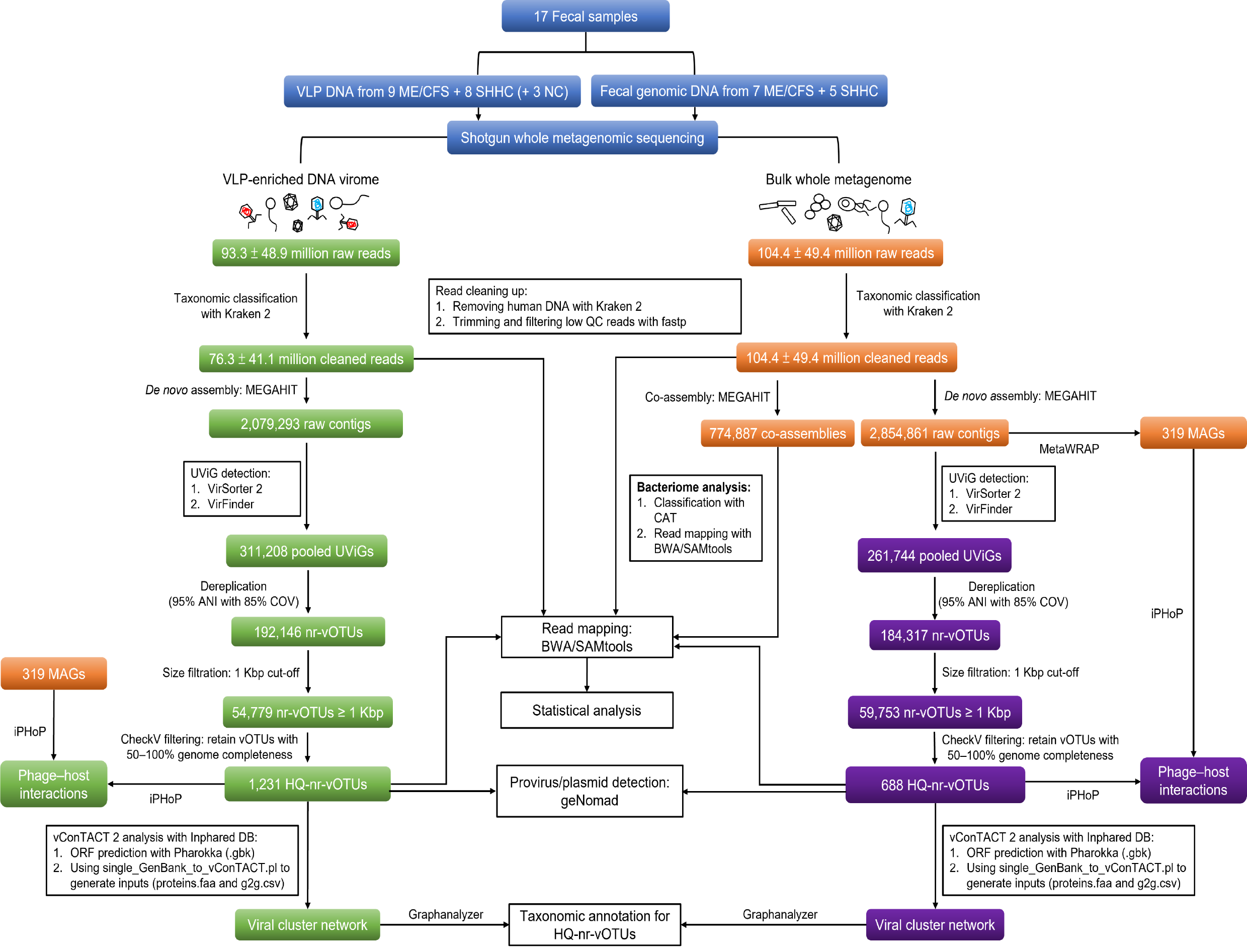
Overview of bioinformatic pipeline for analyzing DNA viromes derived from VLP-enriched metagenomes; and WMS-vOTUs and WMS-Bac derived from whole metagenomes. Blue boxes represent the inputs to the pipeline including enriched isolated VLP DNA and fecal genomic DNA derived from the same samples. Three negative controls were included in the enriched VLP analysis. Green boxes represent the process used to analyze the enriched DNA virome. Orange boxes represent the process used to analyze bacteriomes (WMS-Bac), including prokaryotic metagenome-assembled genome (MAG) binning for host prediction. Purple boxes denote the process used to analyze non-enriched viral genomes derived from whole metagenomes (WMS-vOTUs). The solid lines denote primary analyses connecting with boxes representing further analyses.

From the enriched VLP dataset, 311,208 uncultivated virus genomes (UViGs) were detected (**Supplementary Table S2**). These UViGs were clustered and dereplicated at 95% ANI over 85% of the contig length to generate 192,146 primary non-redundant viral operational taxonomic units (vOTUs). From these 1,231 non-redundant, high-quality (HQ) vOTUs were retained after removal of genome fragments less than 1 Kbp in length, low-quality (<50% genome completeness) genomes, and undetermined genomes; (HQ) vOTUs included 193 complete (62,030.8 ± 40,950.2 bp, mean ± SD), 488 high-quality (>90% completeness) and 550 medium-quality (50–90% completeness) genomes, with an average length of 44,581.9 ± 33,085.6 bp per vOTU (mean ± SD) (**Supplementary Table S3**). In the WMS dataset, 688 non-redundant HQ-vOTUs including 63 complete genomes (90,560.9 ± 63,003.9 bp, mean ± SD) were retained, with an average length of 49,477.2 ± 41,680.2 bp per vOTU (mean ± SD) (**Supplementary Table S4 and S5**). To assess similarity between VLP and WMS sequence datasets, we aligned 688 of WMS-HQ-vOTUs against 1,231 of VLP-HQ-vOTUs resulting in 600 non-redundant sequence alignments with the best hit being obtained. Of these, 193 alignments could be considered as highly likely to be identical between both datasets with high reliability, as determined using several critical thresholds (**Supplementary Table S1**).

Moreover, our data showed that the putative integrated viruses (proviruses or prophages) detected in WMS-HQ-vOTUs outnumbered those detected in VLP-HQ-vOTUs (**Table 2, Supplementary Table S6**). Analysis of plasmid sequences showed that the number of plasmids was reduced in both datasets after removing low quality and undetermined genomes, with very low plasmid contamination being retained in both datasets as indicated by low numbers of plasmid hallmark genes and low marker enrichment scores (**Table 2, Supplementary Table S6**).

**Table 2.** Proviral genomes and plasmid sequences.

|  | Proviruses |  | Plasmids |  |
| --- | --- | --- | --- | --- |
|  | All VLP-nr-vOTUs<br>(n = 192,146) | All WMS-nr-vOTUs<br>(n = 184,317) | All VLP-nr-vOTUs<br>(n = 192,146) | All WMS-nr-vOTUs<br>(n = 184,317) |
| Pre-CheckV filtration | 254 | 537 | 4,119 | 5,359 |
|  | VLP-HQ-vOTUs<br>(n = 1,231) | WMS-HQ-vOTUs<br>(n = 688) | VLP-HQ-vOTUs<br>(n = 1,231) | WMS-HQ-vOTUs<br>(n = 688) |
| Post-CheckV filtration | 123 | 254 | 6 | 6 |

nr: non-redundant; HQ: high quality

Three process control samples (i.e., TBT buffer alone [designated as Negative Controls (NC)] generated very low numbers of non-human sequences (**Supplementary Table S7**). For taxonomic classification, NC-derived reads were primarily assigned to members of the bacterial phyla *Proteobacteria*, *Actinobacteria*, *Bacteroidetes*, and *Firmicutes*. Only 0.48% of reads in NC1 (859/180,120), 0.01% in NC2 (1/9,296) and 1.16% in NC3 (647/55,832), respectively, were assigned to viruses such as *Lactococcus* phages, pahexaviruses, moineauviruses, and human polyomaviruses (**Supplementary Table S7**). In addition, only 6– 15% of NC-derived reads could be mapped to VLP-HQ-vOTUs, with very low coverage seen after VLP enrichment (**Supplementary Table S1**). Using the assembly and quality filtering workflow described above, no HQ-vOTUs were present in the process control samples.

### Cluster analysis and taxonomic assignment of the HQ-vOTUs

To assess the distribution of related groups of viruses, VLP and WMS HQ-vOTUs were grouped into 1,621 and 1,515 viral clusters (VCs), respectively. A total of 1,160 VCs from the VLP dataset that had reference viruses were identified. Of these, 37 VCs were uniquely composed of ME/CFS-associated vOTUs compared with ten VCs uniquely associated with SHHC vOTUs; 147 shared VCs were identified that consisted of communal virus groups present in both samples (**Fig. 3A, Supplementary Table S9**). In parallel, we identified 1,181 VCs from the WMS dataset that had reference viruses. Of these, 17 unique VCs were ME/CFS-associated and ten were SHHC-associated; there were 93 shared VCs composed of communal virus groups (**Fig. 3B, Supplementary Table S10**). Distributions of HQ-vOTUs and VCs in the WMS dataset were different to the VLP dataset, reflecting different viral compositions obtained from both datasets.

**Figure 3.**
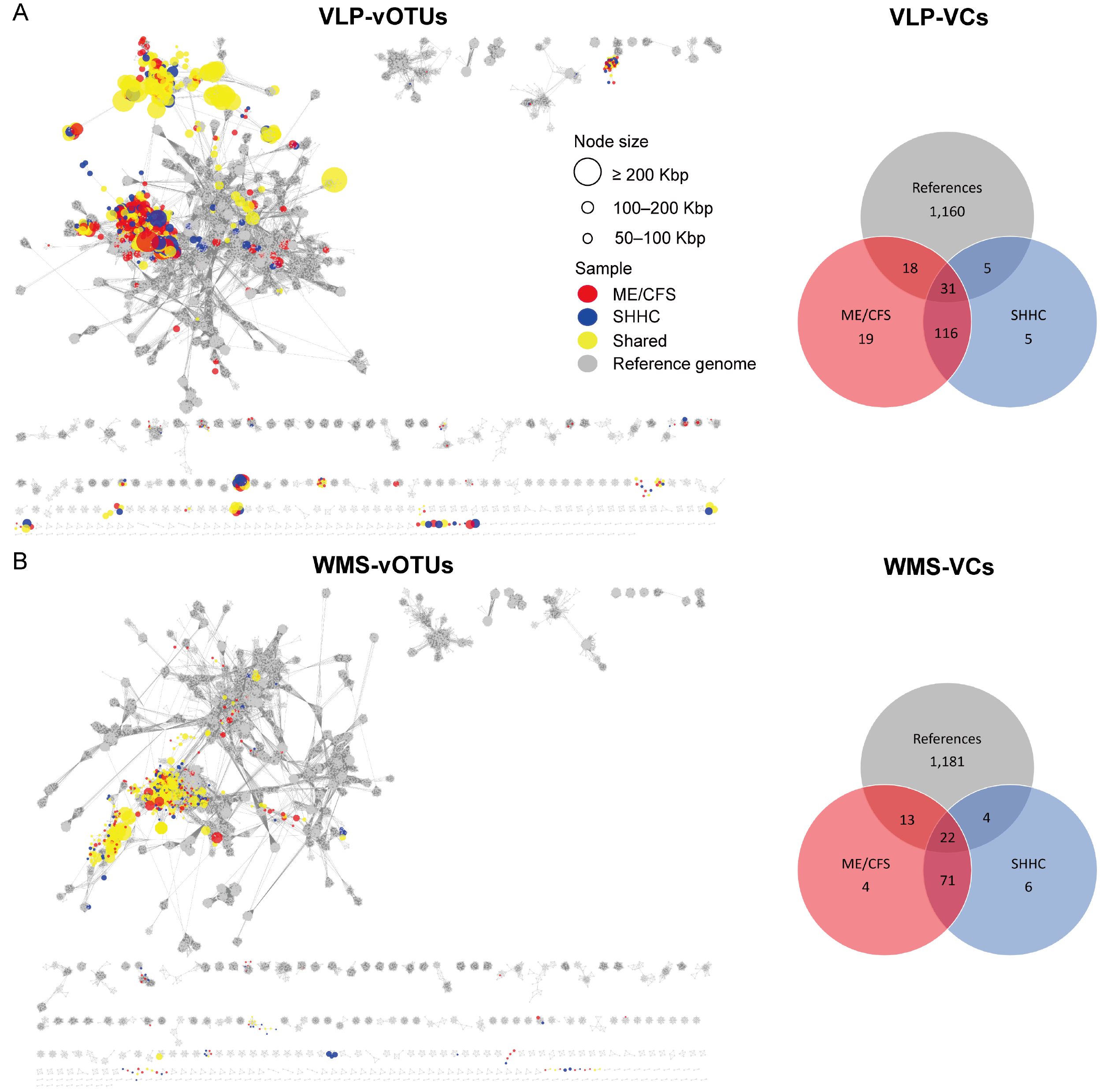
Viral cluster networks showing similarity between high-quality vOTUs and reference viruses. The networks denote subsets of HQ-vOTUs from both (A) VLP-enriched datasets and (B) WMS-derived datasets. HQ-vOTUs were grouped into a viral cluster or subcluster based on genome similarity and amino acid homology against INPHARED-based reference viruses (grey). Based on the mean coverage of mapped reads per vOTU per dataset, each virus (vOTU) was determined based on it being present in at least one dataset. The colored circles (nodes) denote the true presence of viruses (red: uniquely present in ME/CFS-derived samples; blue: uniquely present in SHHC-derived samples; yellow: communal viruses seen in both samples). The edges (grey lines) represent similarities weighted by edge betweenness across the genomes, with the node size reflecting genome length in Kbp. Venn diagrams display the distribution of VCs in samples, based on the mean coverage of mapped reads.

Most VLP-HQ-vOTUs were grouped in the classes *Caudoviricetes*, *Faserviricetes*, and *Malgrandaviricetes*, with the remainder being unknown or unclassified. Most known viruses were tailed dsDNA phages belonging to the class *Caudoviricetes* and non-tailed ssDNA viruses including members of the family *Microviridae*. We also identified members of the family *Inoviridae*, which were filamentous ssDNA phages. Similarly, the WMS-HQ-vOTUs with ‘clustered’ or ‘clustered/singleton’ status were grouped into the classes *Caudoviricetes*, *Faserviricetes*, and *Malgrandaviricetes*, with the remainder being unknown or unclassified. Of note, from the VLP dataset we identified six VCs with ‘clustered’ or ‘clustered/singleton’ vOTUs that contained human-associated *Crassvirales* phages. From the WMS dataset only four VCs composed of ‘clustered’ or ‘clustered/singleton’ vOTUs were associated with *Crassvirales* phages (**Supplementary Table S8**).

### Detection of GI eukaryotic DNA viruses

To identify the presence of eukaryotic DNA viromes in our datasets, mapped viral reads were first retrieved from VLP and WMS HQ-vOTUs. Overall, very few mapped viral reads (<1%) were classified as human-associated eukaryotic viruses; the majority were unclassified or phages (data not shown). Of the classified viral reads, some unique human-associated eukaryotic viruses were seen in the ME/CFS-derived samples including human betaherpesvirus 6A (HHV-6A), papillomaviruses, coronavirus NL63, and adenovirus 54, with others being communal viruses shared in both ME/CFS- and SHHC-derived samples (data not shown). Next, we undertook a more stringent analysis by aligning both VLP and WMS HQ-vOTUs against NCBI viral RefSeq genomes. Of the classified VLP and WMS HQ-vOTUs, only one vOTU derived from the VLP dataset was assigned to human smacovirus 1 (eukaryotic ssDNA virus), with the majority being DNA phages (**Supplementary Table S1**).

### Macrodiversity of intestinal viromes

With respect to the beta diversity of intestinal DNA phages, we found that, within the same sample, there were two distinct clusters represented by VLP and WMS-derived groups, respectively (*p* < 0.001) (**Fig. 4A**). There was no evidence for clustering by ME/CFS status (p = 0.691); the SHHC sample had a similar viral composition to the corresponding ME/CFS sample (PERMANOVA *p* = 0.172) (**Fig 4B**). We also determined mean relative abundances of intestinal viruses across ME/CFS- and SHHC-derived samples (**Fig. 4C**) and measured the relative virus abundances for each individual sample separated by household-matched pairs at family and genus levels (**Supplementary Fig. S2 and S3**). Overall, the most dominant viral families across samples belonged to the class *Caudoviricetes* which is composed of tailed phages (**Fig. 4C**). Although there was no evidence of any differences between ME/CFS- and SHHC-derived samples, there were differences between VLP-enriched datasets and WMS-derived datasets in the relative abundance of viruses from several families (**Fig. 4D**). By comparing VLP-enriched and WMS-derived datasets, we found that enriched VLPs had higher abundances (as measured by centered log-ratio, CLR) of the families *Suoliviridae* (*p* < 0.0001), *Salasmaviridae* (*p* < 0.0001), *Intestiviridae* (*p* < 0.0001), *Steigviridae* (*p* = 0.002), and *Crevaviridae* (*p* < 0.001); and lower abundances of the families *Duneviridae* (*p* < 0.0001), *Winoviridae* (*p* < 0.0001) and other unclassified families (*p* < 0.05) (**Fig. 4D**). Moreover, the Shannon index of alpha diversity was higher in WMS-derived datasets than in VLP-enriched datasets (difference = 2.28, *p* < 0.0001) (**Fig. 4E**).

**Figure 4.**
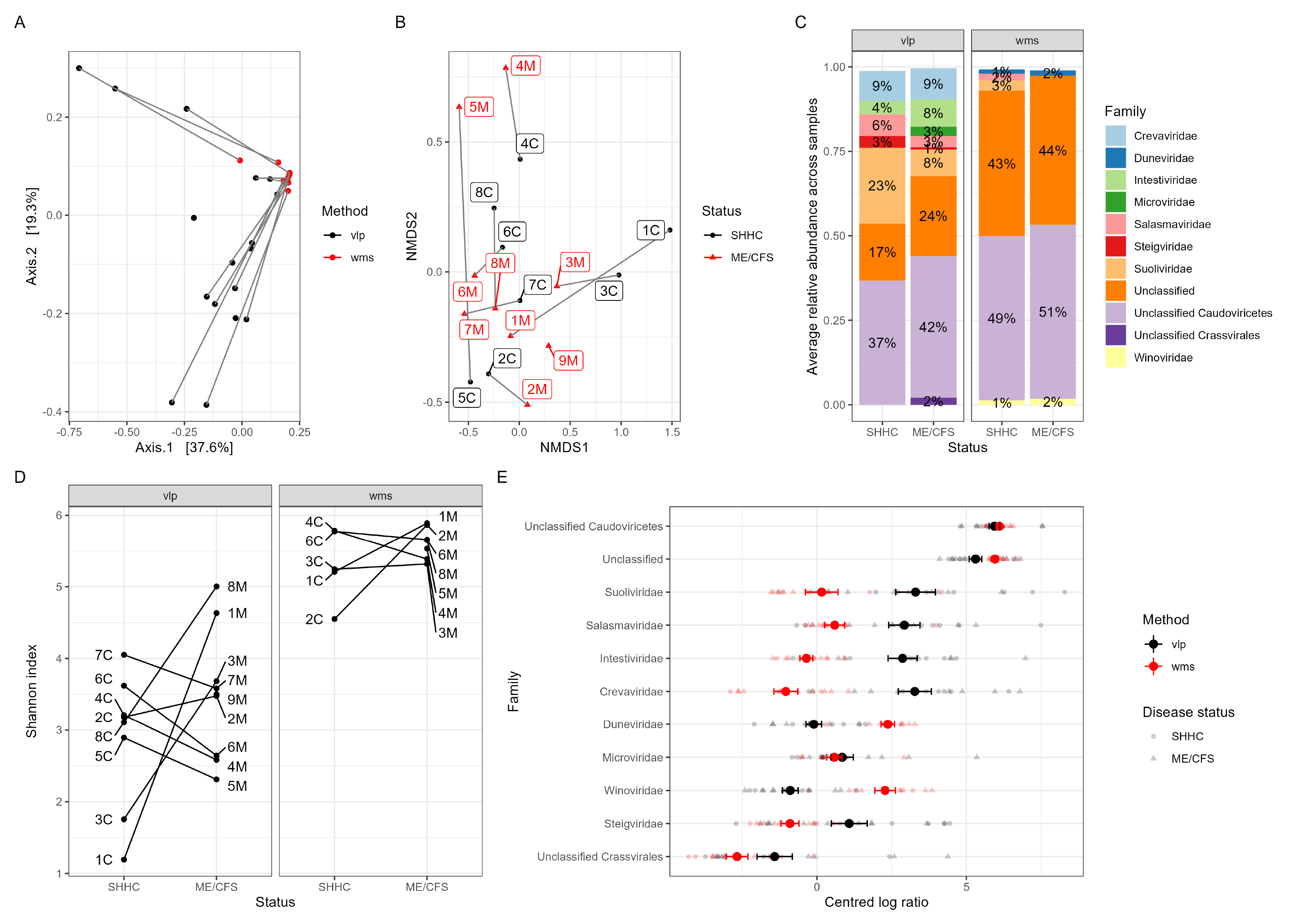
Macrodiversity analyses of intestinal viromes. (A) Analysis of beta diversity of intestinal DNA phages from VLP-enriched (black) and WMS-derived (red) datasets. VLP and WMS isolation methods generated different viral compositions forming two distinct clusters in the ordination analysis (*p* < 0.001). (B) Using NMDS analysis, some household-matched pairs had similar viral compositions while others were more distinct (*p* = 0.172 for overall test of clustering within the household-matched pairs). (C) The 11 most abundant intestinal viruses at the family level across samples separated by VLP- and WMS-derived groups (unclassified categories included). Each assigned family is depicted by different colors, with the average relative abundance per dataset shown as %. (D) Estimation of the alpha diversity of intestinal viromes using Shannon index within household-matched pairs. Diversity was higher for WMS-derived viruses than VLP-enriched viruses (*p* < 0.0001). (E) Analysis of centered log-ratios (CLR) depicting differences in relative abundance estimates of different viral families from VLP-enriched and WMS-derived datasets. The mean CLR of viruses from VLP-enriched datasets are shown as large red circles with the mean CLR of viruses from WMS-derived datasets labeled in black (error bars representing ± 1 standard error) with individual ME/CFS-derived samples depicted as triangles with SHHC-derived samples shown as circles.

### Macrodiversity of intestinal bacteriomes

To assess the impact of ME/CFS on intestinal bacterial communities we first determined beta diversity at the bacterial genus level (**Fig. 5A**). Using Jaccard distance to assess species similarity we found that within each household-matched pair the SHHC subject had similar bacterial compositions to the corresponding ME/CFS patient (*p* = 0.002), with no evidence of any differences between the ME/CFS and SHHC groups (*p* = 0.924) (**Fig. 5A**). Overall, across WMS-derived datasets the three most abundant bacterial phyla were *Firmicutes* (46%), *Bacteroidetes* (25%), and *Actinobacteria* (2.7%) (**Supplementary Fig. S4**). Amongst the 12 most abundant bacteria at the genus level *Alistipes* (8.2%), *Bacteroides* (4.3%) and *Faecalibacterium* (4%) were prominent (unknown genera were excluded; **Fig. 5B**). *Prevotella* dominated the profile of one individual (sample 2C) (**Fig. 5B**). Comparing the centered log-ratios (CLRs) amongst the 12 most abundant bacterial genera showed no significant differences between ME/CFS and SHHC groups (**Supplementary Fig. S5**). ‘Observed richness’, ‘Chao1’ and ‘Shannon’ indices were also used to determine the alpha diversity of intestinal bacteriomes (**Fig. 5C and 5D**). Overall, no significant differences were seen between bateria from ME/CFS- or SHHC-derived samples for any measure of observed species richness (*p* = 0.877), Chao1 (*p* = 0.828) and Shannon index (*p* = 0.975), respectively, although there was a high intra-class correlation within the household-matched pairs for the observed richness and Chao1 measures (ICC = 0.754 and 0.837, respectively).

**Figure 5.**
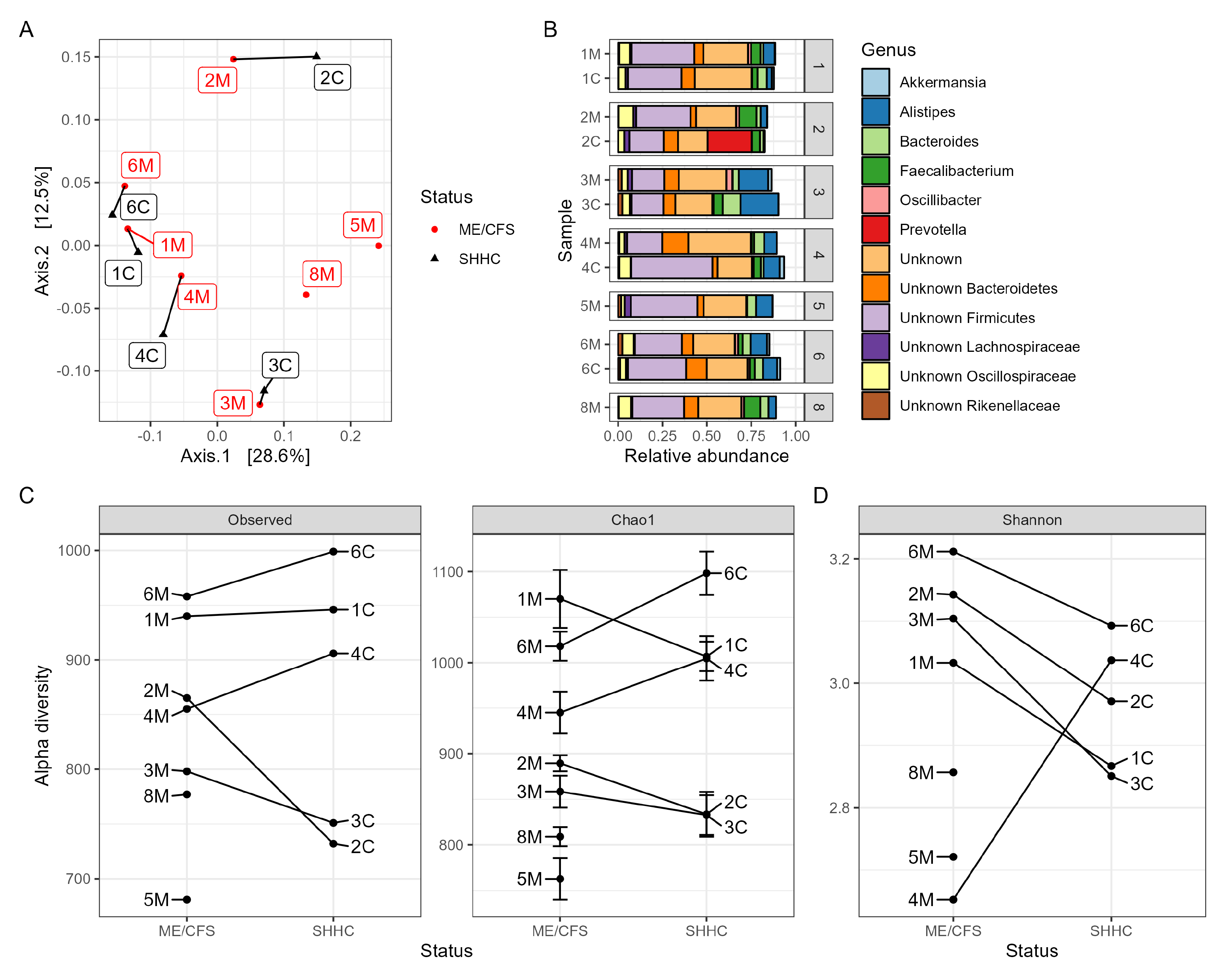
Macrodiversity analysis of intestinal bacteriomes. (A) Beta diversity analysis of the bacteriomes from ME/CFS and SHHC groups displayed in a PCoA plot with Jaccard distance showing similarity within household-matched pairs at the genus level (PERMANOVA *p* = 0.002) (M: ME/CFS samples; C: SHHC samples). (B) Top 12 relative abundances of bacteria at the genus level (without unknown genera) as derived from five household-matched pairs with two unpaired samples. Each assigned genus is depicted in a different color. (C and D) Estimation of the alpha diversity indices. The numbers for average read coverage are observed alongside the estimations of Chao1 richness (C) and Shannon index (D).

Finally, using data on between- and within-household variations in the relative abundance of bacterial genera, we performed a power calculation to estimate the numbers of samples that would be needed in a future study to reliably identify differential abundance of individual genera between groups. The data suggested that at least 15 household-matched pairs would be required to detect ‘true’ differences of the scale of a 10-fold difference in abundances of specific bacterial genera in matched pairs, with a 90% statistical power at a critical threshold of *p* < 0.001, whereas more than 100 pairs would be needed to detect 90% of genera that had a true two-fold change in abundance between groups. (**Supplementary Fig. S6**).

### *Predicting* ME/CFS-*associated bacterial hosts for HQ-vOTUs*

Using a combination of 319 metagenome-assembled genomes (MAGs) from this study and reference bacterial genomes, 81 shared bacterial hosts were identified in the VLP- and WMS-derived datasets; 94 hosts were uniquely detected from the VLP dataset and 25 hosts were uniquely detected from the WMS dataset (**Fig. 6, Supplementary Table S11**). Next, we investigated whether these predicted hosts were ME/CFS- or health-associated by interacting with either ME/CFS- or SHHC-unique viruses, or communal viruses shared in both samples (**Supplementary Table S11**). Of the 81 bacterial hosts shared in both the VLP and WMS-derived datasets, 11 bacteria (labeled in red) including the genera QAMI01, *Erysipelatoclostridium*, *Lacticaseibacillus*, Firm-11, *Christensenella*, *Eubacterium*, *Anaerobutyricum*, *Anaerostipes*, *Acutalibacter*, *Veillonella*, and *Salmonella* potentially associated with ME/CFS-unique viruses (**Supplementary Table S11**). Of the 94 VLP-derived bacterial hosts, 37 bacteria (including *Clostridium*_J and *Clostridium*_AP, and *Anaerotruncus*) had predicted associations with ME/CFS-unique viruses. In addition, only six WMS-derived bacterial genera (including *Bacteroides*_G, *Campylobacter*_D, HGM12998, *Pseudoflavonifractor*, *Oxalobacter* and *Klebsiella*) were associated with ME/CFS-unique viruses (**Supplementary Table S11**).

**Figure 6.**
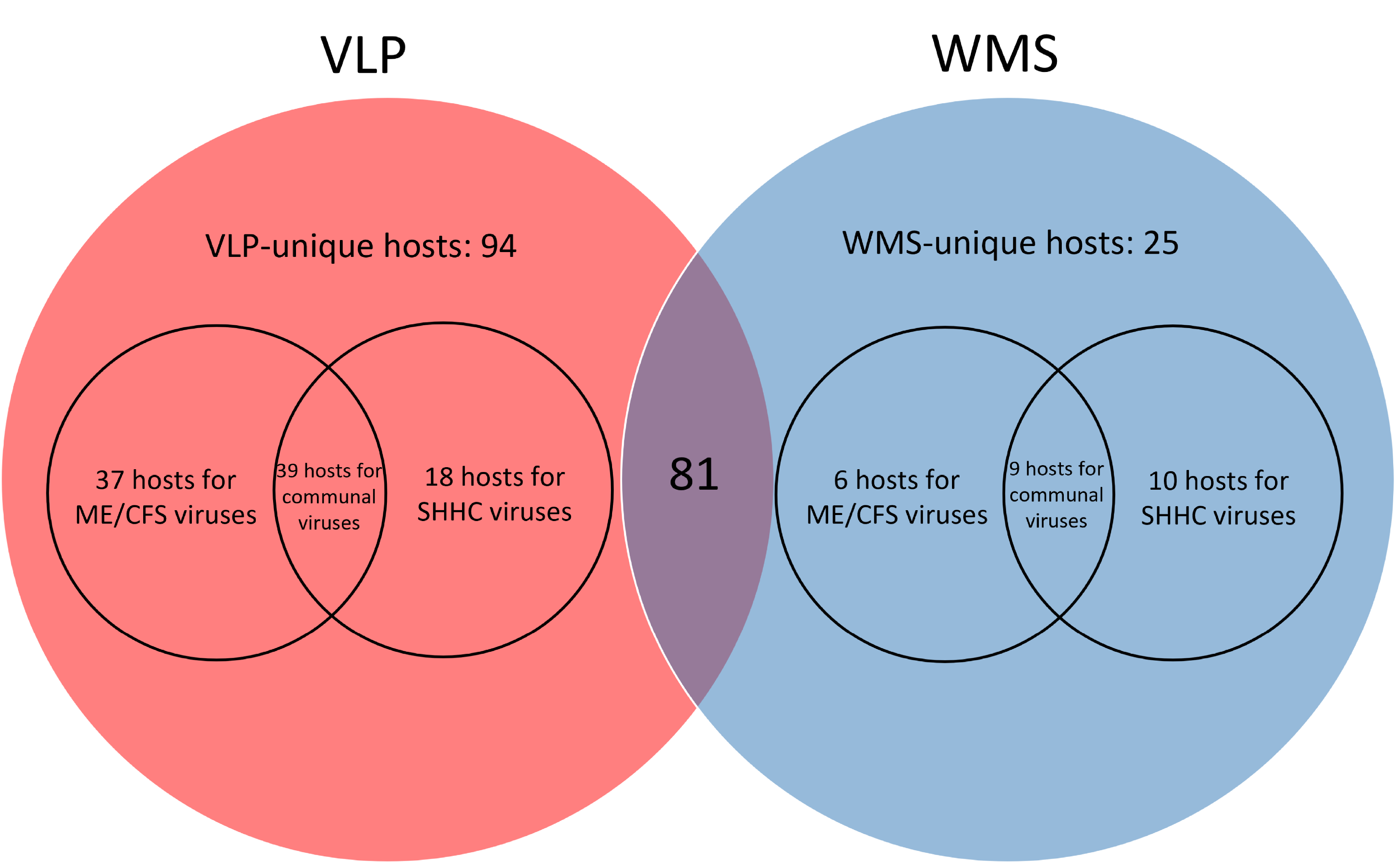
Venn diagrams depicting predicted bacterial hosts from the VLP- and WMS-derived datasets that associate with viruses that are either ME/CFS-unique, SHHC-unique or communal. A total of 81 shared bacterial hosts (middle) were detected in both VLP- and WMS-derived datasets. Of the VLP-derived bacterial hosts (red), 37 bacterial genera were potentially associated with ME/CFS-unique viruses from ME/CFS patient samples with six hosts derived from the WMS dataset (blue) associated with ME/CFS-unique viruses.

Finally, we compared the 20 most abundant bacterial genera from ME/CFS samples with the host predictions to identify possible ME/CFS-associated bacterial hosts (**Table 4**). Within the 20 most abundant genera, most bacterial hosts, such as *Faecalibacterium* and *Roseburia*, associated with communal viruses. Although the genera *Bacteroides*, *Ruminococcus*, *Clostridium* and *Eubacterium* associated with diverse viruses including a mixture of communal viruses and ME/CFS- and SHHC-unique viruses, the genus *Anaerotruncus* from the VLP-derived dataset was the only bacterium associating with ME/CFS-unique viruses with no identifiable interactions with SHHC-derived or communal viruses (one unknown virus and two unclassified *Caudoviricetes* phages) (**Table 4, Supplementary Table S11**).

**Table 3.**
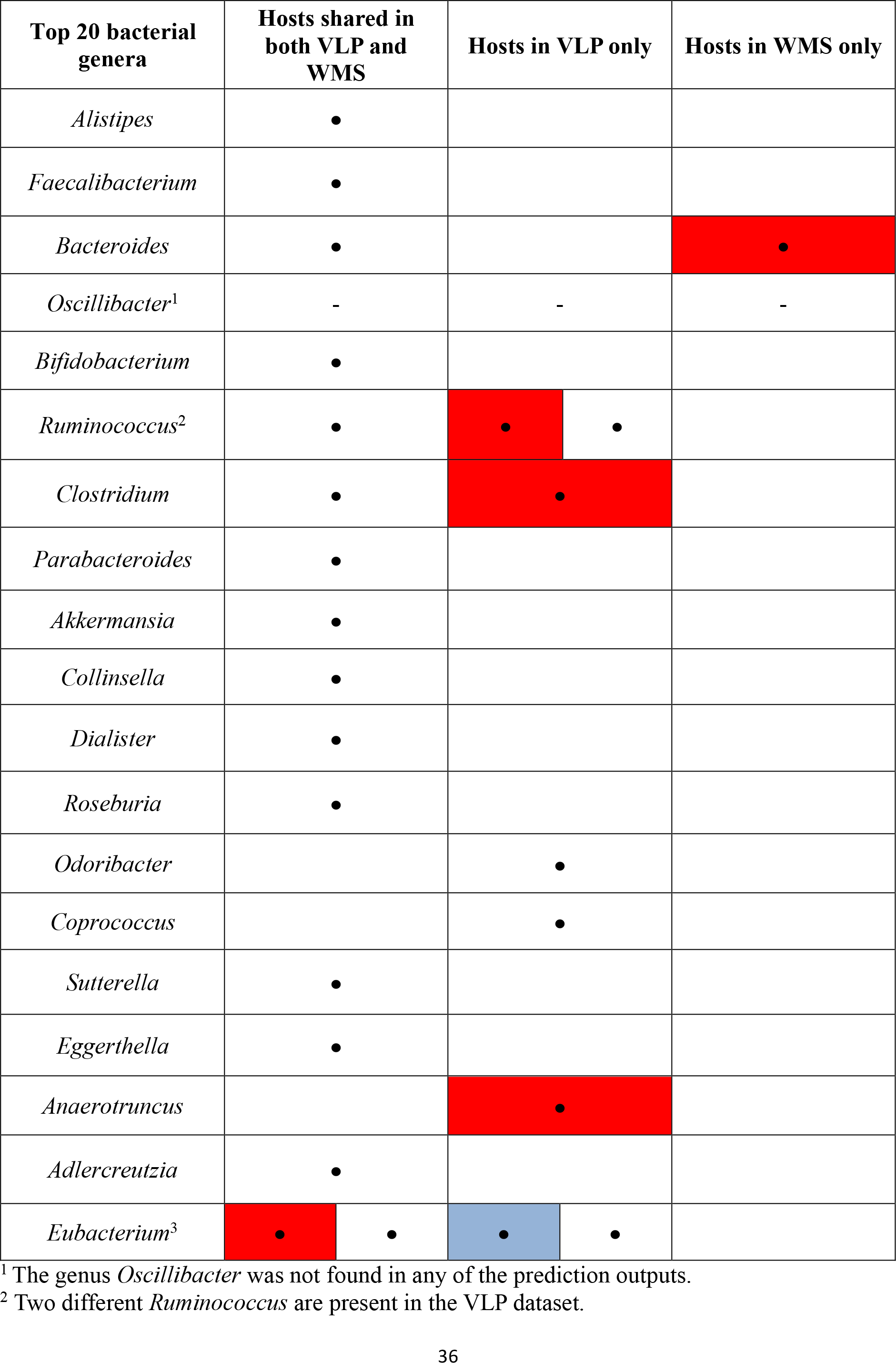
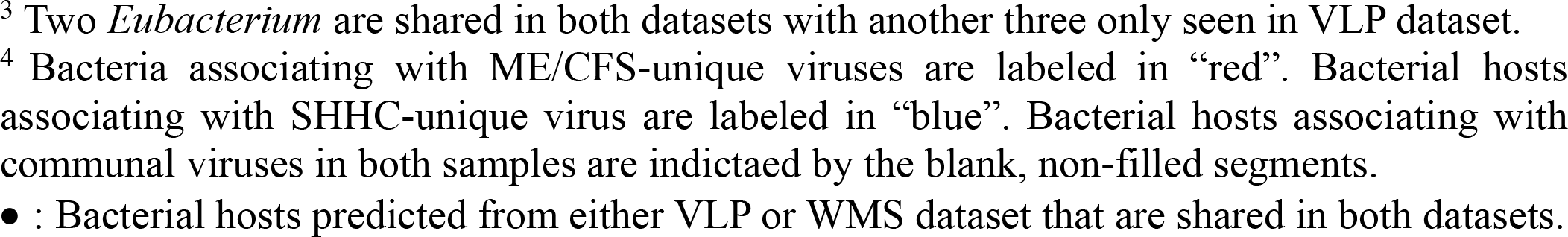
The 20 most abundant ME/CFS-derived bacterial genera associating with unique or communal viruses in VLP and WMS samples.

| Top 20 bacterial genera | Hosts shared in both VLP and WMS |  | Hosts in VLP only |  | Hosts in WMS only |
| --- | --- | --- | --- | --- | --- |
| *Alistipes* | • |  |  |  |  |
| *Faecalibacterium* | • |  |  |  |  |
| *Bacteroides* | • |  |  |  | • |
| *Oscillibacter*<sup>1</sup> | - |  | - |  | - |
| *Bifidobacterium* | • |  |  |  |  |
| *Ruminococcus*<sup>2</sup> | • |  | • | • |  |
| *Clostridium* | • |  | • |  |  |
| *Parabacteroides* | • |  |  |  |  |
| *Akkermansia* | • |  |  |  |  |
| *Collinsella* | • |  |  |  |  |
| *Dialister* | • |  |  |  |  |
| *Roseburia* | • |  |  |  |  |
| *Odoribacter* |  |  | • |  |  |
| *Coprococcus* |  |  | • |  |  |
| *Sutterella* | • |  |  |  |  |
| *Eggerthella* | • |  |  |  |  |
| *Anaerotruncus* |  |  | • |  |  |
| *Adlercreutzia* | • |  |  |  |  |
| *Eubacterium*<sup>3</sup> | • | • | • | • |  |

<sup>1</sup> The genus *Oscillibacter* was not found in any of the prediction outputs.
<sup>2</sup> Two different *Ruminococcus* are present in the VLP dataset.
<sup>3</sup> Two *Eubacterium* are shared in both datasets with another three only seen in VLP dataset. <sup>4</sup> Bacteria associating with ME/CFS-unique viruses are labeled in “red”. Bacterial hosts associating with SHHC-unique virus are labeled in “blue”. Bacterial hosts associating with communal viruses in both samples are indicated by the blank, non-filled segments. • : Bacterial hosts predicted from either VLP or WMS dataset that are shared in both datasets.

## Discussion

By combining VLP and WMS viromes we obtained a more complete and comprehensive profile of the human GI virome in a small cohort of severely-affected ME/CFS patients and non-affected healthy individuals living in the same household as the patient. We also predicted ME/CFS-associated bacterial hosts that interacted with the unique viruses in severely-affected ME/CFS patients; these may provide new insights into GI dysbiosis in ME/CFS.

An important feature of our study design was the inclusion of SHHC subjects to account for and minimize any impacts of environmental factors and the indoor environment on the microbiome (prokaryome)^49^. Individuals sharing the same household have similar microbial compositions compared with those living in different households thereby increasing confidence in attributing differences in microbiomes in ME/CFS patients to the disease itself^49^. Whilst SHHCs co-habit and live in the same environment as the patient, it is important to acknowledge differences in gender (9/9 patients female, 5/8 SHHCs female) and age (average of 33.8 years old for patients and 53.6 years old for SHHCs) that may contribute to variations in individual microbiomes^50,51^.

We recovered ∼10^9^ VLP/g feces per sample after VLP enrichment with the most common viral morphotypes being tailed phages, broadly consistent with other studies^52–55^. Non-human reads derived from VLP enrichment had a higher mapping rate and average read coverage to the sorted high-quality (HQ) vOTUs than those derived from WMS, consistent with most mapped reads derived from viruses after VLP enrichment. Moreover, non-human reads derived from the negative control samples had no or very low mapping rates and coverage to VLP-HQ-vOTUs, suggesting low levels of contamination in our enriched virome dataset. The HQ-vOTU datasets from enriched VLPs and WMS from the same stool sample were overlapping but with distinct components. Most enriched viruses were likely extracellular lytic viruses such as crAss-like phages, depicting typical predator-prey dynamics^56^. However, in general extracellular lytic phages can be sparce and infrequent within the adult GIT, with the exception of lytic *Crassvirales* and *Microviridae* phages that stably reside in the healthy adult GIT^57^. Therefore, many VLP-enriched enteric viruses are likely to be temperate phages that enter the lytic cycle^58,59^. This could explain why numbers of proviruses in the VLP-derived samples are lower than in the WMS-derived samples. In contrast, most viruses detected from WMS are more likely to be prophages dormant within the bacterial genomes with prophage induction being associated with GI inflammation and dysbiosis^60–62^. This suggests that induced or non-induced prophages may also play an important role in modulating GI microbial ecology. Using both VLP- and WMS-based isolation methods for sequencing is advantageous given the dominance of temperate phages or induced prophages interacting with GI microbiota through piggyback-the-winner dynamics^58,61,63^; together VLP- and WMS-based isolation methods provide complementary datasets that include more diverse, high-quality and larger complete viral genomes comprising extracellular viruses and integrated proviruses, which provides a more comprehensive profile of the human intestinal DNA virome, whilst also capturing a small overlap (∼8.5%) in viral populations^43,57^.

Although most VCs were composed of communal viruses shared by both ME/CFS- and SHHC-derived samples, we identified specific VCs comprising unique viruses only present in ME/CFS- or SHHC-derived samples. The VCs and viruses unique to ME/CFS-derived samples are likely associated with the disease. Of the classified VCs, most viruses were *Caudoviricetes* phages including members of human-associated crAss-like phages, with a minority being members of the *Microviridae* and *Inoviridae* families (i.e., ssDNA viruses) and others. Whilst members of the *Microviridae* have been considered predominant members of the human GI virome^57^, they may be overrepresented in virome studies using multiple displacement amplification (MDA) due to biases that preferentially amplify short circular ssDNA^64^. To avoid MDA-based amplification biases and overestimation of ssDNA viruses, we used a conventional PCR method for library preparation prior to sequencing. Moreover, we used a chloroform-free protocol for VLP isolation to avoid depleting enveloped viruses. Members of the order *Crassvirales* are the most abundant tailed dsDNA phages within the human GIT^65,66^. However, although we used vConTACT 2 alongside the recently updated reference viral database, many VCs/vOTUs still remain unknown or unclassified.

To identify the presence of human GI eukaryotic DNA viromes in our samples, we initially retrieved mapped viral reads from VLP and WMS HQ-vOTUs. However, very few viral reads were classified as human-associated eukaryotic viruses. The majority were phages or unclassified, with very few reads identifying unique eukaryotic DNA viruses in the ME/CFS-derived samples; this included HHV-6A (data not shown) which had been associated previously with ME/CFS^27^. Furthermore, using blast-based methodology for HQ-vOTUs against NCBI viral RefSeq database, only one HQ-vOTU derived from the VLP dataset was identified as a eukaryotic virus, and was assigned to human-associated smacovirus 1 isolate (small, circular ssDNA virus); the remainder were unclassified or phages. Although smacoviruses were initially detected in fecal samples from non-human primates and human fecal samples with unexplained diarrhea, their role in human-associated disease remains controversial^67,68^. Of note, no eukaryotic DNA viruses were classified from our cluster analysis due to the current limitation of vConTACT 2^69^. We cannot therefore exclude the possibility that eukaryotic DNA viruses are present in our samples (within the unknown or unclassified VCs) but are undetectable using the tools available. Also, no RNA viruses or phages were detected as the extraction protocols used were not optimized for RNA viruses/phages. Although some human-associated pathogenic GI viruses are RNA viruses (e.g., rotavirus, norovirus and enteroviruses), other enteric RNA phages may originate from plant-based food^70^. For completeness therefore future GI virome studies should attempt to include enteric RNA viruses.

Considering the limited sample size in this study, it was perhaps unsurprising that no significant differences were observed in viral abundance and diversity between the ME/CFS and SHHC groups. Consistent with recent ME/CFS microbiome studies^15,18^, our data indicated that *Firmicutes*, *Bacteroidetes* and *Actinobacteria* were the three most dominant phyla in the intestinal microbiota with our data also showing an intra-class correlation within the household-matched pairs across many bacterial genera. Although no statistical evidence is present for the differences, the intestinal microbiome of severely-affected ME/CFS patients could be characterized by a reduction in *Firmicutes* and an increase in *Bacteroidetes*^15,18,71^. There were no quantitative differences in the alpha and beta diversities of bacteria between the ME/CFS and healthy control samples, which may reflect the limited sample size and, as illustrated by our statistical power calculation, may require more than 100 pairs to detect 90% of genera that had a true two-fold change in abundance between groups.

Phages can modulate the dynamics of microbial communities in the human GIT via direct or indirect trans-kingdom interactions^39,72^. By using an *in silico* predictive approach for host predictions we have potentially identified unique and shared bacterial hosts from the VLP and WMS datasets. Within the 20 most abundant ME/CFS-derived bacteria, the genera *Bacteroides*, *Ruminococcus*, *Clostridium*, *Anaerotruncus*, and *Eubacterium* associated with ME/CFS-unique viruses derived from the patient samples; of these most bacteria were short-chain fatty acid (SCFA) producers. Recent reports identified that reductions in butyrate-producing bacteria such as *Faecalibacterium prausnitzii*, *Roseburia* spp., and *Eubacterium rectale* were associated with butyrate deficiency and disease severity in ME/CFS^16,73^. An increased abundance of the genus *Bacteroides* has also been seen in ME/CFS patients compared with non-affected healthy individuals^15^. Moreover, we noted that most of the abundant bacterial hosts were broadly associated with a variety of communal viruses or ME/CFS- or SHHC-unique viruses, while *Anaerotruncus* derived from VLP-enriched samples was uniquely associated with ME/CFS-unique viruses. This finding supports and extends a previously reported increase in *Anaerotruncus colihominis* in fecal metagenomes from a cohort of ME/CFS patients^74^.

## Conclusions

Using different isolation methodologies and VLP-enriched and whole metagenome-based sequencing methods has enabled us to obtain more diverse and high-quality vOTUs with high read coverage and high genome completeness; this provides a more comprehensive profile of the human GI DNA virome in healthy individuals and severely affected ME/CFS patients. Although no significant differences were seen in viral or bacterial abundance and/or diversity between the ME/CFS and SHHC sample groups there were differential effects in relation to microbes from VLP-enriched samples compared with WMS-derived samples. In addition, we were able to predict a possible ME/CFS-associated bacterial host able to interact with ME/CFS-unique viruses seen in patient samples. This study therefore provides the framework and rationale for studies in larger cohorts of ME/CFS patients to further investigate ME/CFS-associated interactions between the intestinal virome and bacteriome.

## Participants and Methods

### Study participants and sample collection

All participants were registered with the CFS service of the St Helier Hospital, Surrey, UK or the ME/CFS Service of the East Coast Community Healthcare Centre (ECCHC), Norfolk/Suffolk, UK and were recruited to the study between 2017 and 2019. All fulfilled the Canadian^8^ and NICE (National Institute For Health And Care Excellence) 2007/CG53 guideline^9^ diagnostic criteria of CFS which were used alongside clinical history and hospital anxiety depression scale (HADS)^75^ to exclude those with significant clinical depression and anxiety. Participants consuming probiotic capsules or antibiotics within four weeks prior to sample collection were also excluded from the study. Disease status was based on, (1) few or no activities of daily living, (2) severe cognitive difficulties, (3) wheelchair dependency for mobility, (4) unable or rarely able to leave the house or are bed bound requiring assistance with washing, toilet use, feeding and, (5) often displaying significant worsening symptoms with mental or physical exertion and in extreme cases unable to tolerate noise and are light sensitive. The inclusion criteria for SHHC recruitment were: (1) the individual had to be living with or caring for the patient, (2) men or women aged between 18 and 70 years, (3) no current or ongoing medical conditions and (4) able to provide informed consent. SHHC participants having long-term medical conditions (e.g., inflammatory bowel disease, IBS), suffering from autoimmune diseases, significant anxiety, or depression, taking immunomodulatory drugs, statins, beta blocker or steroids and consuming probiotic capsules or antibiotics within four weeks prior to sample collection, were excluded from the study. Ethical approval was obtained from the University of East Anglia Faculty of Medicine and Health Sciences Research Ethics Committee in 2014 (reference FMH20142015–28) and the Health Research Authority NRES Committee London Hampstead (reference 17/LO/1102; IRAS project ID: 218545). Subjects recruited in 2017 also completed a shortened SF-36, the Chalder fatigue questionnaire, a self-efficacy questionnaire, a visual analogue pain rating scale, and the Epworth sleepiness scale. Informed written consent was obtained from all participants.

Fecal samples were collected from participants’ homes, followed by transportation in a chilled and insulated Fecotainer^®^ (Excretas Medical BV, Netherlands) for delivery to the laboratory within 6 h of collection. Upon receipt, samples were aliquoted and stored at −80 °C prior to analysis.

### Fecal VLP and VLP DNA isolation

The fecal VLP and VLP DNA isolation protocol was as described previously^76^. To evaluate the recovery efficiency of VLP isolation, two preliminary spiking-and-recovery assays were done prior to formal experiments (**Supplementary Method S1;** data not shown). For VLP and DNA isolation, frozen fecal aliquots (3–4 g feces per aliquot) were homogenized in sterile TBT buffer (100 mM Tris-HCl, pH 8.0; 100 mM NaCl; 10 mM MgCl_2_·6H_2_O) by vortexing and then incubated on ice for 1 h. The fecal homogenates were then centrifuged at 11,200 × g for 30 min at 10 °C for two rounds. Supernatants were filtered sequentially through 0.8 µm (Sterlitech, USA) and 0.45 µm (Starlab Ltd., UK) polyethersulfone cartridge filters. NaCl (final concentration 6%, w/v) was then added to fecal filtrates and mixed, followed by adding PEG 8000 (final concentration 10%, w/v; Sigma-Aldrich Ltd., UK). After re-suspension samples were incubated at 4 °C for 16 h, PEG-precipitated VLPs harvested by centrifugation at 4500 × g for 60 min at 4 °C, and VLP-containing pellets resuspended in ∼500 μL of TBT buffer. To avoid losing enveloped viruses, no chloroform was included in the protocol. VLP suspensions were then treated with 10 U of TURBO DNase (Invitrogen/Thermo Fisher Scientific, UK) and 20 U of RNase I (Ambion/Thermo Fisher Scientific, UK) at 37 °C for 45 min. EDTA (final concentration 15 mM, pH 8.0) was added to stop the reaction, followed by heat inactivation at 75 °C for 10 min. Proteinase K (100 μg; Ambion/Thermo Fisher Scientific, UK) and 5% (w/v) SDS (final concentration 0.5%, w/v) were added and incubated at 56 °C for 75 min, followed by the addition of lysis buffer (final concentration 133.3 mM Tris-HCl, pH 8.0; 33.3 mM EDTA, pH 8.0; 3.3% SDS, w/v) and incubation at 65 °C for 15 min. The VLP lysate was treated with an equal volume of phenol/chloroform/isoamyl alcohol (25:24:1, v/v/v; Thermo Fisher Scientific, UK) and mixed thoroughly by vortexing for 30 sec, followed by two-round centrifugation at 15,000 × g for 5 min at ambient temperature. The resulting aqueous phase was collected and transferred to a ZR genomic DNA Clean & Concentrator™-25 column (Zymo Research; Cambridge Bioscience Ltd., UK) with purified DNA eluted in low-EDTA elusion buffer. To increase DNA concentrations, 10–12 DNA aliquots isolated from each sample (i.e., average amounts of feces: 30.9 ± 11.8 g per sample, mean ± SD, *n* = 17) were pooled and concentrated to a final volume of 60–100 µL (**Supplementary Fig. S1**). Concentrated DNA was stored at −80 °C. Three negative control (NC) samples (TBT buffer only) were included as process controls to monitor contaminations and false positive signals in high-throughput sequencing. These NC samples (NC1–3) were added at the homogenization step and throughout the process of isolation, library preparation and sequencing. The quantity and quality of recovered VLP DNA was determined using the Nanodrop and Qubit™ 1x dsDNA HS Assay Kit (Thermo Fisher Scientific, UK).

### Epifluorescence microscopy (EFM)-based enumeration of enriched fecal VLPs

The primary Invitrogen™ SYBR™ Gold stock solution (concentration 10,000x; Thermo Fisher Scientific, UK) was diluted to a working concentration of 0.25% (v/v) with sterile Ambion™ nuclease-free water and stored at −20 °C until used. For use, SYBR Gold solution was defrosted at ambient temperature in darkness for 15 min prior to use. 20 µL of PEG-precipitated VLPs were diluted in 900 µL of nuclease-free water to which 100 µL of SYBR Gold working solution (final concentration 0.025%, v/v) was added, mixed gently, and then incubated in the dark for 15 min at ambient temperature. An Omnipore 0.45 µm, 13-mm PTFE backing filter (Millipore/Sigma-Aldrich Ltd., UK) was placed on top of a Swinnex filter holder with a silicone gasket (Millipore/Sigma-Aldrich Ltd., UK). The backing filter was rinsed by nuclease-free water followed by placing a 0.02 µm white Whatman™ Anodisc 13-mm filter membrane (Sigma-Aldrich Ltd., UK) on top of the backing filter. The Anodisc filter membrane was rinsed by nuclease-free water using a low-vacuum pressure (∼20 kPa; Millivac-Maxi vacuum pump, Millipore/Sigma-Aldrich Ltd., UK), as described previously^52,77^. The SYBR Gold-labeled VLP sample (20 µL) was then fixed on the Anodisc membrane using low-vacuum pressure until all liquid had passed through the membrane. The membrane was washed with 1 mL of nuclease-free water to remove excess dye. The filter membrane was then transferred to a Whatman^®^ filter paper disc (Sigma-Aldrich Ltd., UK) and left to dry for 1 min. Prior to adding a coverslip, Fluoromount-G^®^ antifade mounting reagent (SouthernBiotech, USA) was spotted on a microscope slide and the dried filter membrane placed onto the mounted droplet. The slide was left in the dark at ambient temperature for 16 h and slides were subsequently imaged using a Zeiss Axio Imager M2 widefield epifluorescence microscope with Alexa Fluor 488 channel and 100x oil objective lens. For each slide, 20 digital images were captured, and SYBR Gold-labeled viral particles were viewed and counted using ImageJ software with the average number of VLPs per field multiplied by sample dilution factor and microscope conversion factor (i.e., area of 13-mm Anodisc filter / area of field of view) and then divided by sample volume^77^. Additionally, negative control samples were included as process controls to monitor contamination, including non-stained VLP suspension, dye incubated with sterile TBT buffer and dye incubated with nuclease-free water. One VLP sample was used as a positive control for each batch of staining and imaging to monitor experimental consistency.

### Transmission electron microscopy (TEM)

Five microliters of diluted fecal filtrate was applied to carbon-film on copper 400 mesh grids (EM Resolutions, UK) for 1 min and excess liquid removed by wicking the edge of the grid with Whatman filter paper, followed by 2-min incubation with 0.5% (w/v) UA solution. Excess UA was then removed by wicking with filter paper. Each grid was vapor fixed by adding 1 mL of 2.5% (v/v) glutaraldehyde to the dish containing the dried grids for a minimum of 2 h. Imaging was then performed using a Talos F200C TEM microscope at 200 kV with a Gatan One View digital camera.

### Library preparation and whole metagenomic sequencing of enriched VLP DNA

VLP DNA collected from seventeen fecal samples alongside three negative control samples were constructed in a pooled PCR-based barcoding library by the Quadram Institute Bioscience in-house sequencing service using Nextera XT DNA Library Preparation Kit (Illumina Inc., UK). DNA samples were normalized to 0.5 ng/µL with 10 mM Tris-HCl (pH 8.0) prior to library preparation, followed by tagmentation and adapter ligation using 1 ng input DNA template. Input DNA was randomly fragmented with engineered Tn5 transposase generating a mean of ∼300 bp of DNA inserts with adapters, followed by PCR amplification to barcode the adapter-ligated DNA input. The PCR program was 72 °C for 3 min, 95 °C for 1 min, followed by 14 cycles of 95 °C for 10 sec, 55 °C for 20 sec and 72 °C for 3 min. All libraries were then pooled, followed by cleaning with KAPA pure beads (Roche Inc., UK) and quantified using Invitrogen™ Quant-iT dsDNA assay kit (Thermo Fisher Scientific, UK). A pooled library sample was then sequenced using 2 × 150 bp paired-end chemistry (PE150) on the Illumina HiSeq X Ten platform (Novogene Ltd., UK). The raw sequencing data of ME/CFS- and SHHC-derived samples had a Q30 score of >90% generating an average depth of 14 Gb per sample. In total, 1,586,595,530 paired-end raw sequencing reads (93,329,148.8 ± 48,857,207.3 per sample, mean ± SD, *n* = 17) were obtained. Three negative control samples (NC1–3) were included as process controls and had a Q30 score of >88% with around 0.1 Gb of output per sample generating a total of 791,934 paired-end raw sequencing reads (263,978 ± 307,643.7, mean ± SD, *n* = 3). Paired-end sequencing reads were provided as FASTQ format. All raw sequencing reads were pre-processed to trim and remove adapters, low quality (Q-value ≤ 38) and N nucleotides (Novogene Ltd., UK) using readfq^78^ and FxTools (v0.17)^79^. Human genomic DNA detected by Kraken 2 (v2.0.8)^80,81^ against the Genome Reference Consortium Human Build 37 (GRCh37/hg19) database was removed by setting the confidence at 0.5, followed by further cleaning of the reads using fastp (v0.23.1)^82^ with a quality cut-off of 20 prior to genome assembly. Kraken 2 was also used to taxonomically classify the sequences of the negative controls (NC1–3) and the sequences detected were considered as biases or contaminants.

### Fecal genomic DNA isolation, library preparation and whole metagenomic sequencing

As five fecal samples contained insufficient biomass for both whole metagenomic shotgun (WMS) sequencing and VLP analysis, they were prioritized for VLP enrichment. The remaining twelve fecal samples (i.e., seven ME/CFS and five SHHC samples) were processed for WMS sequencing. Briefly, genomic DNA was extracted from ∼250 mg aliquot per sample using FastDNA™ SPIN Kit for Soil (MP Biomedicals, UK) following the manufacturer’s instructions. The quantity and quality of recovered DNA samples were determined using the Nanodrop and Qubit™ 1x dsDNA HS Assay Kit. DNA was stored in DNase/Pyrogen-Free water at 4 °C. A pooled PCR-barcoding sequencing library was prepared by the in-house sequencing service (QIB) using Nextera™ DNA Flex Library Preparation Kit (Illumina Inc., UK) with 14 PCR cycles, followed by sequencing using 2 × 150 bp paired-end chemistry (PE150) on the Illumina NovaSeq 6000 platform (Novogene Ltd., UK). The raw data from ME/CFS- and SHHC-derived samples had a Q30 score of >91% per sample and generated a total of 1,252,729,390 paired-end raw sequencing reads (104,394,115.8 ± 49,361,687 per sample, mean ± SD, *n* = 12). Paired-end sequencing reads were provided as FASTQ format. All raw sequencing reads were pre-processed as described above. No positive and negative control samples were included for WMS analysis.

### Analysis of VLP and WMS HQ-vOTUs

#### Genome assembly and viral mining

Cleaned reads were assembled using MEGAHIT assembler (v1.2.9)^83^ with default parameters. QUAST (v5.0.2)^84^ and SeqFu (v1.8.4)^85^ were used to assess the quality and quantity of assembled genomic contigs. Putative uncultivated virus genomes (UViGs) were then predicted using VirSorter 2 (v2.0)^86^ and VirFinder (v1.1)^87^. All putative viruses and proviruses were sorted and classified into the output of VirSorter 2, and in parallel, all contigs were also run through VirFinder with those having a score of >0.7 and *p* < 0.05 considered viral^43^. All UViGs were pooled and CheckV (v0.7.0)^88^ used to evaluate the quality of both VLP- and WMS-derived UViGs in ‘end-to-end’ mode (with default parameters).

#### Dereplicating and generating non-redundant viral operational taxonomic units (vOTUs)

A rapid genome clustering script computing the pairwise average nucleotide identities (ANI) was used to cluster and dereplicate the UViGs, followed by generating non-redundant viral operational taxonomic units (vOTUs) clustered at 95% ANI over 85% of the length of sequences, as described in Nayfach *et al.* (2021)^88^ (code available at https://bitbucket.org/berkeleylab/checkv/src/master/). Each vOTU was deemed a single viral species and represented in the datasets by the longest sequence. vOTUs <1 Kbp derived from both VLP and WMS datasets were removed. Using CheckV to assess the completeness of vOTUs, low-quality (i.e., <50% genome completeness) and undetermined vOTUs were removed, and the remaining vOTUs including complete genomes, high-quality (i.e., >90% completeness) and medium-quality genomes (i.e., 50–90% completeness) were retained. Finally, high-quality non-redundant vOTUs (HQ-vOTUs) were retrieved. These HQ-vOTUs met certain quality thresholds and were more likely “true” viruses, and were used for read mapping, cluster analysis, taxonomic annotation and virus–host prediction.

#### Similarity identification between VLP and WMS HQ-vOTU datasets

To determine sequence similarity between VLP and WMS HQ-vOTU datasets, BLAST+ (v2.12.0)^89^ was used to align the WMS-HQ-vOTUs against the VLP-HQ-vOTUs. Query sequences with the best alignment hit were obtained, followed by the use of filtering thresholds including e-value (cut-off: 1e-05), bitscore (cut-off: 1000), percentage of identical matches (pident; 95% cut-off) and query coverage per subject sequence (qcovs; 85% cut-off) to retrieve alignments highly likely to be identical between the VLP and WMS datasets.

#### Detecting provirus and plasmids from VLP and WMS vOTU datasets

geNomad (v1.3.3)^90,91^ was used to detect potential proviruses (prophages) and plasmids in both VLP and WMS HQ-vOTU datasets. geNomad alongside its database (v1.2) was run in ‘end-to-end’ mode with --min-score at 0.7 and --cleanup flag.

#### Read mapping, average coverage calculation and count table construction

Cleaned reads from 17 VLP-derived samples and 12 WMS-derived samples were separately mapped to the VLP and WMS HQ-vOTUs, respectively, using BWA (v0.7.17)^92^ with the bwa-mem mode for paired-end manner, followed by use of SAMtools (v1.12)^93^ to sort and index the resulting alignments. To determine the ‘true’ distribution of each virus (i.e., a virus uniquely originating from either ME/CFS- or SHHC-derived samples or commonly present in both samples), the average read coverage (i.e., [read count × average read length] / vOTU length) was calculated using the ‘average-coverage’ module of BamToCov (v2.7.0)^94^ with default parameters. A cut-off threshold of average coverage was set at 10-fold, that is, viruses (vOTUs) were considered ‘truly’ present in a sample when the average coverage was over 10-fold (**Supplementary Table S1**). This analysis was also applied to vOTU cluster analysis and host predictions. In parallel, both VLP and WMS HQ-vOTUs were merged and deduplicated, followed by read mapping for each sample and generating a jointed average coverage-based count table using BamToCov for viral abundance and diversity analyses.

#### vOTU clustering and taxonomic annotation

Cluster analysis to taxonomically classify the VLP-HQ-vOTUs and WMS-HQ-vOTUs was performed using vConTACT 2 (v0.9.19)^69,95^. vConTACT 2 used vOTU genomes and reference viral genomes as the nodes, with its edges being weighted based on their amino acid homology between two genomes (nodes). HQ-vOTUs and bacteriophage isolate reference genomes from the INPHARED-based reference viral genomes^96^ were then clustered to form the networks, based on their shared gene content and sequence similarity^69,95^.

Pharokka (v1.1.0)^97^ was used to predict phage genes within the vOTU genomes in ‘meta’ mode and to generate amino acid files in .gbk format; then both a curated ‘amino acid’ (.faa) and a ‘gene-to-genome’ (.csv) mapping files were generated using a Perl script (code available at https://github.com/RyanCook94/Random-Perl-Scripts^98^). Curated files were then concatenated with INPHARED-based files^96^ (ver. 1Dec2022; available at https://github.com/RyanCook94/inphared^99^). The merged ‘amino acid’ and ‘gene-to-genome’ mapping files were used as inputs for vConTACT 2 with Diamond, MCL and ClusterONE modules (with --db ‘none’). The output (c1.ntw) was then visualized using Cytoscape (v3.9.1)^100^. Some original VCs were divided into subclusters by vConTACT 2, where clusters were deemed to be ICTV (sub)families and subclusters were deemed to be ICTV genera. The clustering status for every vOTU and reference viral genome was also labeled by vConTACT 2, as described previously^95,101^.

To automatically perform taxonomic annotation when using the INPHARED reference viral database^99^, graphanalyzer (v1.5.1)^101^ was used for taxonomic assignments of both VLP and WMS HQ-vOTUs. Graphanalyzer decides an appropriate taxonomy for each vOTU relying on the ‘weight score’ and ‘clustering status’ generated by vConTACT 2. The ‘status’ clustered, “level” Cx and “high weight” are considered as higher reliability, and “level” Nx and “low weight” have less confidence^102^. Taxonomy is inherited until the “subfamily” level if the status is labeled as “Clustered/Singleton” or “Overlap”. It is inherited at the “family” level if vOTUs and reference genomes are not found in the same cluster (i.e., lower levels were therefore omitted and labeled as “O”). “Outlier” is probably at the (sub)family level, but this group is less confident. “Singleton” is unable to be included in a network due to a lack of sufficient information for annotation (i.e., labeled as “n.a.”). For the vOTUs labeling “Clustered”, their taxonomies are inherited until the “genus’ level with high confidence.

#### Detection of human eukaryotic DNA viruses

Briefly, paired-end viral reads mapped to the VLP and WMS HQ-vOTUs were retrieved and sorted using SAMtools, and then BAM were converting to Fastq format using BEDtools (v2.31.0)^103^. Retrieved mapped viral reads were then classified using Kraken 2 against its latest Refeq viral database (available at https://benlangmead.github.io/aws-indexes/k2). In parallel, both VLP and WMS HQ-vOTUs were aligned against NCBI viral RefSeq genomes using BLAST+; query sequences with the best alignment hit were retained.

### Analysis of bacteriome

Briefly, cleaned reads were pooled and then co-assembled across 12 WMS-derived samples using MEGAHIT (v1.2.9). 774,887 contigs were obtained, with an N50 length of 890 bp (**Supplementary Table S1**). Contigs were dereplicated and taxonomically classified using CAT (Contig Annotation Tool)^104^. Reads were then mapped against pooled non-redundant contigs using BWA, sorted and alignments indexed using SAMtools, and a count table generated using BamToCov. Read counts were then normalized by genome lengths per contig per sample for bacterial abundance and diversity analyses.

### Host predictions for VLP and WMS HQ-vOTUs

To predict bacterial hosts for both VLP and WMS HQ-vOTU datasets, iPHoP (v1.2.0)^105^ was used to produce the output of host predictions. First, total contigs from whole metagenomes were binned into metagenome-assembled genomes (MAGs) separately for each sample using metaWRAP (v1.3.2)^106^ which incorporates three binning tools including MetaBAT2 (v2.12.1)^107^, MaxBin2 (v2.2.6)^108^ and CONCOCT (v1.1.0)^109^. CheckM (v1.2.0)^110^ was also integrated into the workflow of the metaWRAP pipeline to assess genome completeness and contamination in the ‘bin_refinement’ module. The final binned MAGs were retained based on the criteria of ≥90% completeness and ≤5% contamination using CheckM, as previously described^111^. 319 non-redundant binned prokaryotic MAGs (including 317 bacterial and two archaeal genomes) were obtained, with 8 - 40 MAGs generated from the WMS-derived samples (26.6 ± 12 MAGs per sample, mean ± SD, *n* = 12).

Next, taxonomic decorate tree files derived from MAGs were generated using the ‘de_novo_wf’ module of GTDB-Tk alongside its database (ver. release 207)^112^, followed by creation of a customized host database by adding MAGs with the default database using the ‘add_to_db’ module of iPHoP with default parameters. Finally, iPHoP was used to predict bacterial hosts for both VLP and WMS HQ-vOTUs against the customized host databases. All bacterial taxa predicted were derived from the GTDB-Tk database and those genera with a suffix name could be considered as different categories. Virus–host interactions were then investigated to determine potential disease-associated hosts that interacted with ME/CFS-unique viruses present in ME/CFS-derived samples, based on the determination of average read coverage, as described above.

### Statistical analysis

Several statistical techniques were used to compare the viromes and bacteriomes of severely-affected ME/CFS patients, SHHC subjects, and processing control samples. All statistical analyses were done in R (v4.2.0), using phyloseq (v1.40.0), microbiome (v1.18.0) and vegan (v2.6-4) packages. Beta diversities were calculated using Bray–Curtis distance to compare abundances at the genus level and Jaccard distance was used to count the number of species in common. Both distance measures were visualized using non-metric multidimensional scaling (NMDS) or Principal Coordinate Analysis (PCoA) with the first and second axes shown.

Statistical significance of similarities in beta diversities within groups was tested using PERMANOVA with appropriate permutation restrictions. Alpha diversity was measured in each dataset using the observed average read coverage of species, the Chao1 richness and the Shannon index. Data were either rarefied to the minimum sample depth (to compare observed species numbers) or unrarefied (for Shannon index and relative abundance calculations) as appropriate, depending on the specific research question. Specific methods are detailed in the main text alongside each analysis. Linear mixed models were used to compare univariate characteristics (i.e., diversity measures, individual taxonomic abundances) between samples, considering the household pairing and individual participant as random effects as appropriate. Centered log-ratio (CLR) transformations were used prior to statistical comparisons of taxon abundances.

## Supporting information

S Table 2 VLP UViGs

S Table 3 VLP OTUs

S Table 4 WMS UViGs

S Table 5 WMS OTUs

S Table 6 proviruses and plasmids

S Table 7 Contaminant monitor

S Table 8 Virla taxonomy

S Table 9 VLP viral cluster analysis

S Table 10 WMS viral cluster analysis

S Table 11 Host predictions

S Table Cohort and sequence stats

## Data Availability

The sequences derived from the VLP-enriched metagenomes have been deposited to ENA (https://www.ebi.ac.uk/ena/browser/home) under the study accession no. PRJEB57952 and the sequences from the whole metagenomes (WMS) have also been deposited to ENA under the project accession no. PRJEB57953.

## Acknowledgements

We are grateful for the assistance provided by the CFS Service team at St Helier Hospital and the ME/CFS Service of the East Coast Community Healthcare Centre in recruiting study participants. We are also grateful to all the patients and their carers who participated in the study. We also thank Mr. David Baker, head of QIB sequencing service, who assisted with sequencing library preparation for Illumina sequencing. We also thank the JIC Bioimaging facility and staff for contributing to the study. The authors gratefully acknowledge the support of the Biotechnology and Biological Sciences Research Council (BBSRC); this research was supported by the BBSRC Institute Strategic Programme Grant Gut Microbes and Health BB/R012490/1 and its constituent projects BBS/E/F/000PR10353, BBS/E/F/000PR10355 and BBS/E/F/000PR10356 (EMA, SRC). GMS was funded by the BBSRC Core Capability Grant BB/CCG1860/1. KAS and FN were supported by PhD studentships jointly funded by Invest in ME Research (UK Charity number 1153730) and the University of East Anglia. FN was funded by a Ramsey Award from SOLVE M.E.

## Disclosure statement

The authors have no competing interests to declare.

## Notes

### Competing Interest Statement

The authors have declared no competing interest.

### Author Declarations

Ethical approval was obtained from the University of East Anglia Faculty of Medicine and Health Sciences Research Ethics Committee in 2014 (reference FMH20142015.28) and the Health Research Authority NRES Committee London Hampstead (reference 17/LO/1102; IRAS project ID: 218545).

