## Supplementary material for "Comprehensive profiling of the human intestinal DNA virome and prediction of disease-associated bacterial hosts in severe Myalgic Encephalomyelitis/Chronic Fatigue Syndrome (ME/CFS)": Methods and Figures

**This file includes:**

Supplementary Method S1

Supplementary Figure S1–S6

**Other supplementary tables (excel spreadsheets) for this manuscript include the following:**

S1. Cohorts and stats of sequences

S2. Quality and quantity of VLP-UViGs

S3. Quality and quantity of VLP-vOTUs

S4. Quality and quantity of WMS-UViGs

S5. Quality and quantity of WMS-vOTUs

S6. Provirus and plasmid detection

S7. NC1–3: contamination monitoring for negative controls

S8. Taxonomic annotation for VLP and WMS HQ-vOTUs

S9. VLP VCs analysis

S10. WMS VCs analysis

S11. VLP/WMS: host predictions

**Supplementary Method S1:**

***Spiking-and-recovery assays for VLP isolation***

To evaluate the recovery efficiency of VLP isolation, two spiking-and-recovery assays were carried out (data not shown). Briefly, 1 mL of a reference *Bacteroides fragilis* phage ΦB124-14 with known titre (~$8.7 \times 10^9$ pfu/ml; PFU: plaque-forming unit) was spiked with three independent fecal homogenates from three different healthy male donors, followed by collecting PEG-VLP suspensions after PEG precipitation for plaque forming assays. Three negative control samples (buffer only) were used as process controls to monitor contamination. Additionally, fecal homogenates without phage spiking were used to monitor aboriginal hosts in the human GI-tract which were potentially infected by ΦB124-14. Technical triplicates for each of three independent samples and negative control samples were performed. In parallel, to corroborate the accuracy of the spiking assay determined by plaque assay, SYBR Gold-prestained spiked phage and epifluorescence microscopy (EFM) were used to visually count fecal VLPs. Briefly, 1 mL of SYBR Gold-prestained phage suspension (ΦB124-14; ~$8.7 \times 10^9$ pfu/ml) was spiked with a fecal homogenate from one healthy male donor, followed by collecting 20 μL of PEG-VLP suspensions after PEG precipitation for observation and enumeration. Three types of negative control samples were included as process controls to monitor contamination, including unstained VLP suspension, dye incubated with sterile TBT buffer and dye incubated with nuclease-free water.

**Supplementary Figure S1.**

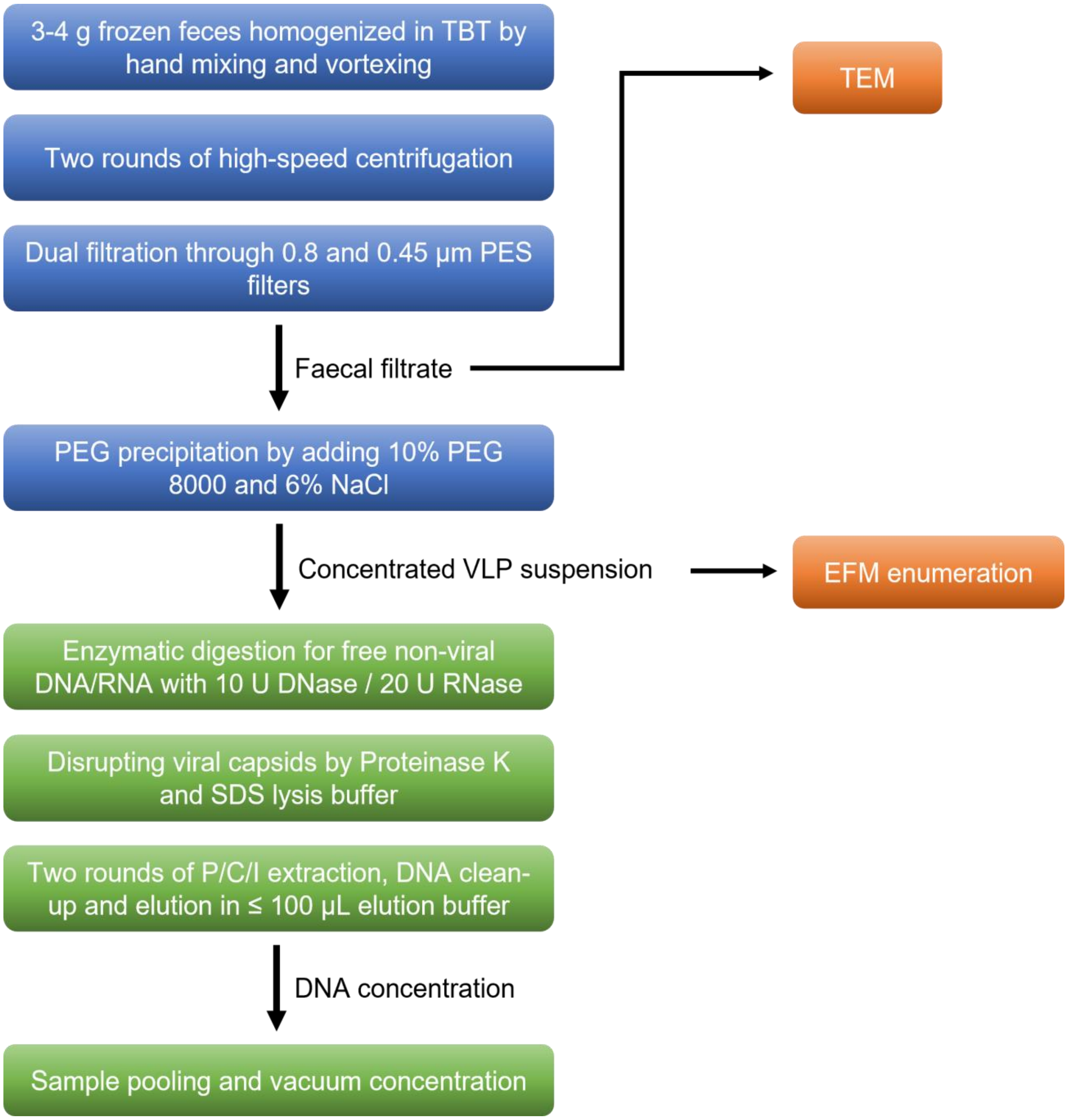


**Fig. S1. An experimental workflow of VLP isolation (blue boxes) and VLP DNA extraction (green boxes) protocol combining epifluorescence (EFM) and electron microscopy (TEM)-based approaches for characterizing the human intestinal virome.** The detailed protocols are described in the section of Methods and Materials.

**Supplementary Figure S2.**

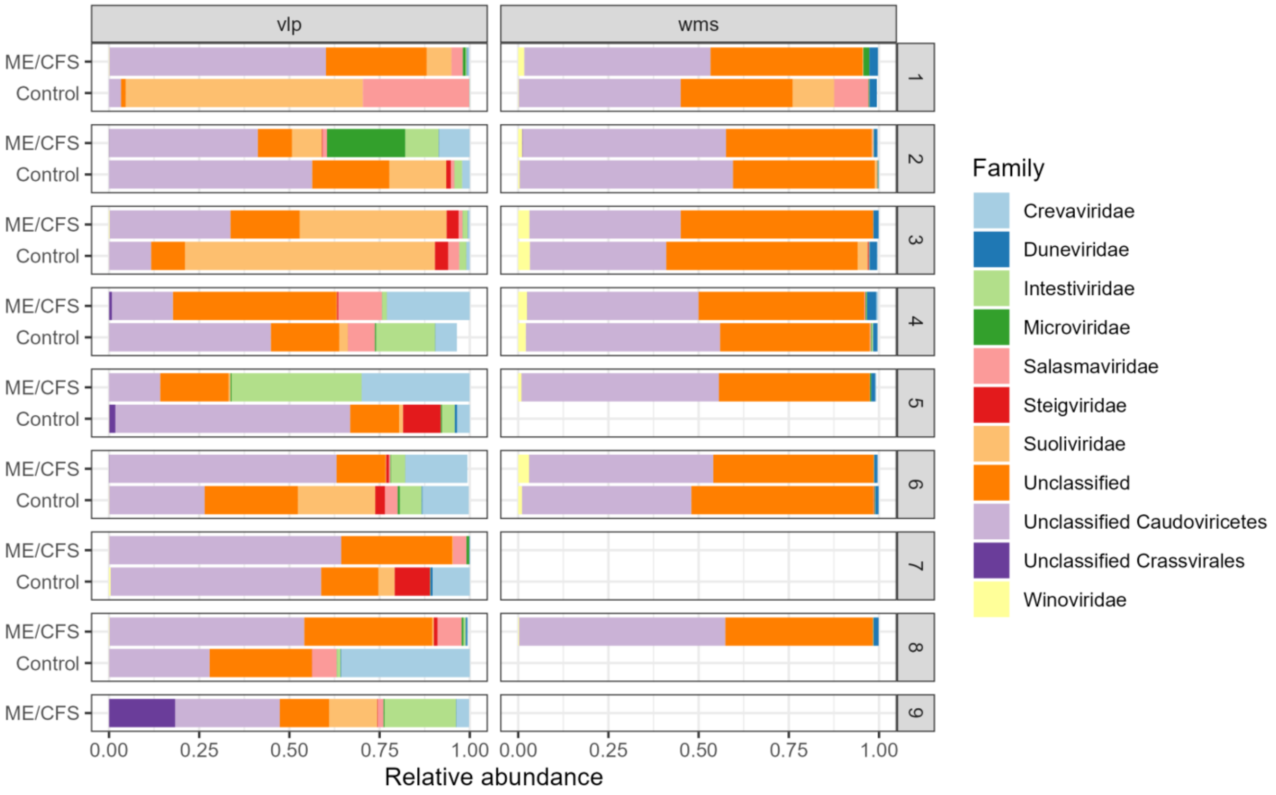


**Figure S2. Distribution of relative abundance of viruses at the family level for individual household-matched pairs.** The 11 most abundant intestinal viruses are shown at the family level separated by individual household-matched pairs (unclassified categories included). Unclassified tailed phages belonging to the *Caudoviricetes* class are the most abundant viruses across all samples. Each assigned family is separate in different colors and the average relative abundance per sample is shown in %.

**Supplementary Figure S3.**

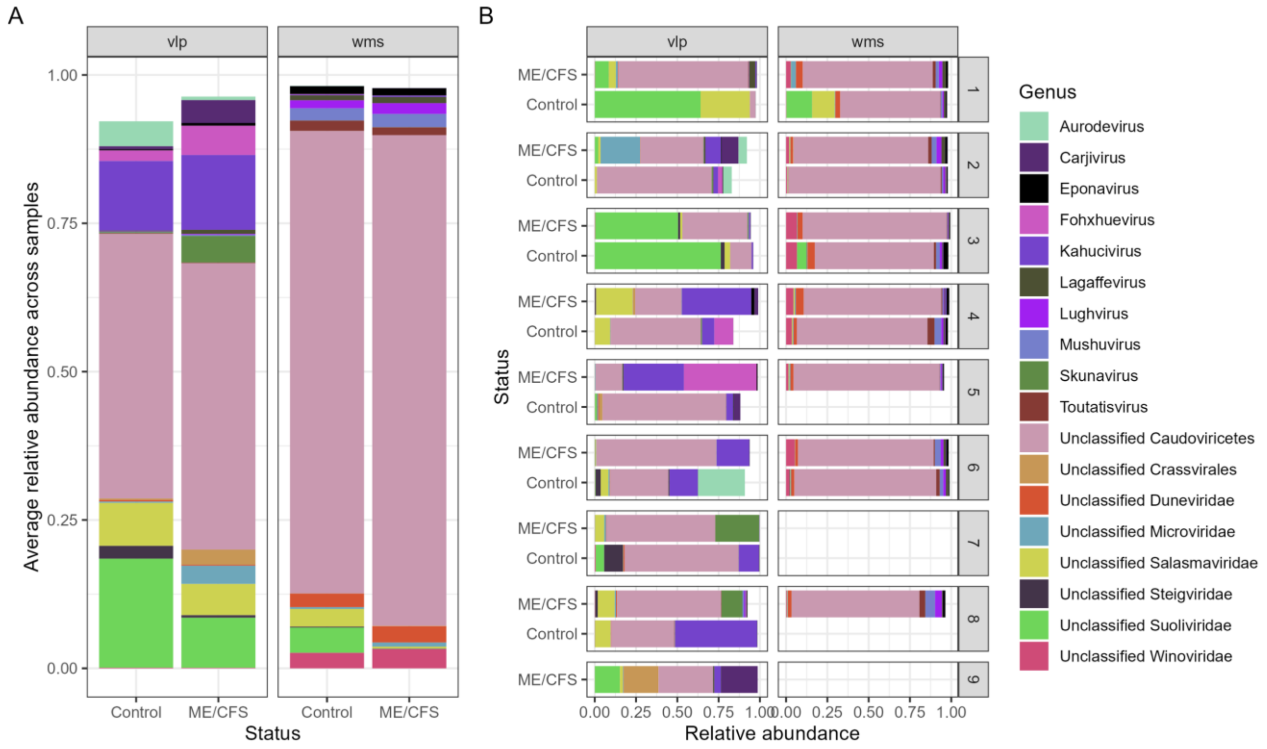


**Figure S3. Distribution of relative abundances of viruses at the genus level separated by disease and control groups or by individual household-matched pairs.** The 18 most abundant intestinal viruses are shown at the genus level separated by individual household-matched pairs (unclassified categories included). Unclassified tailed phages belonging to the *Caudoviricetes* class are the most abundant viruses across all samples. Each assigned genus is separate in different colors and the average relative abundance per sample is shown in %.

**Supplementary Figure S4.**

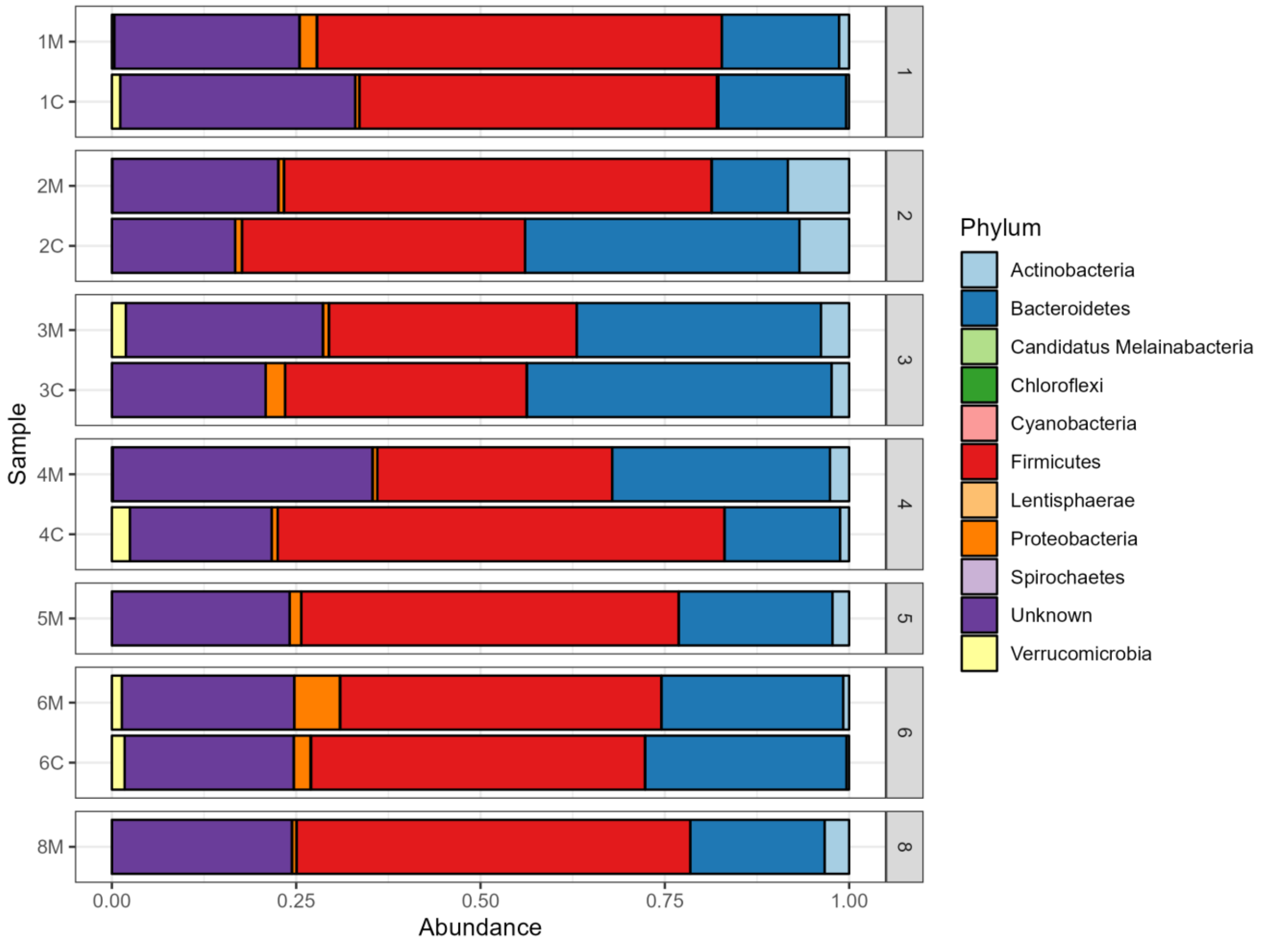


**Figure S4. Top 11 bacterial relative abundances at the phylum rank across ME/CFS- and SHHC-derived samples.** Overall, the three most dominant bacterial phyla derived from five household-matched pairs with two unpaired samples are *Firmicutes* (46%), *Bacteroidetes* (25%), and *Actinobacteria* (2.7%) across the WMS-derived samples. Each assigned phylum is separate in color and the average relative abundance per sample is shown in %.

**Supplementary Figure S5.**

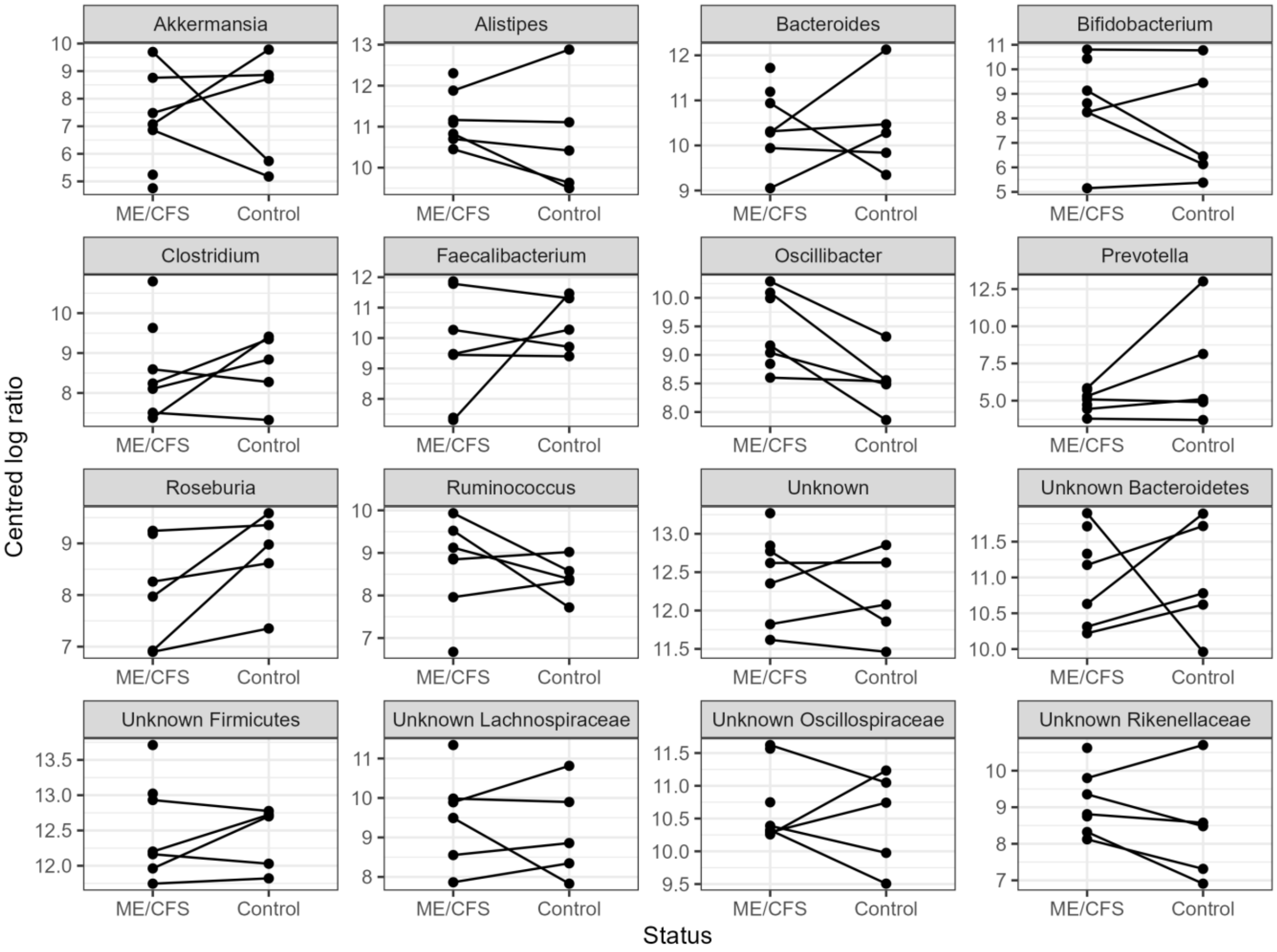


**Figure S5. Analysis of centered log-ratios (CLRs) to determine the similarity/difference of the top 12 bacterial relative abundances at the genus level between the ME/CFS and SHHC derived samples.** There is no significant difference seen between cases and healthy controls, but in many cases, there is some evidence showing a high intra-class correlation within the household-matched pairs, reflecting the statistically significant clustering within household in the beta diversity.

**Supplementary Figure S6.**

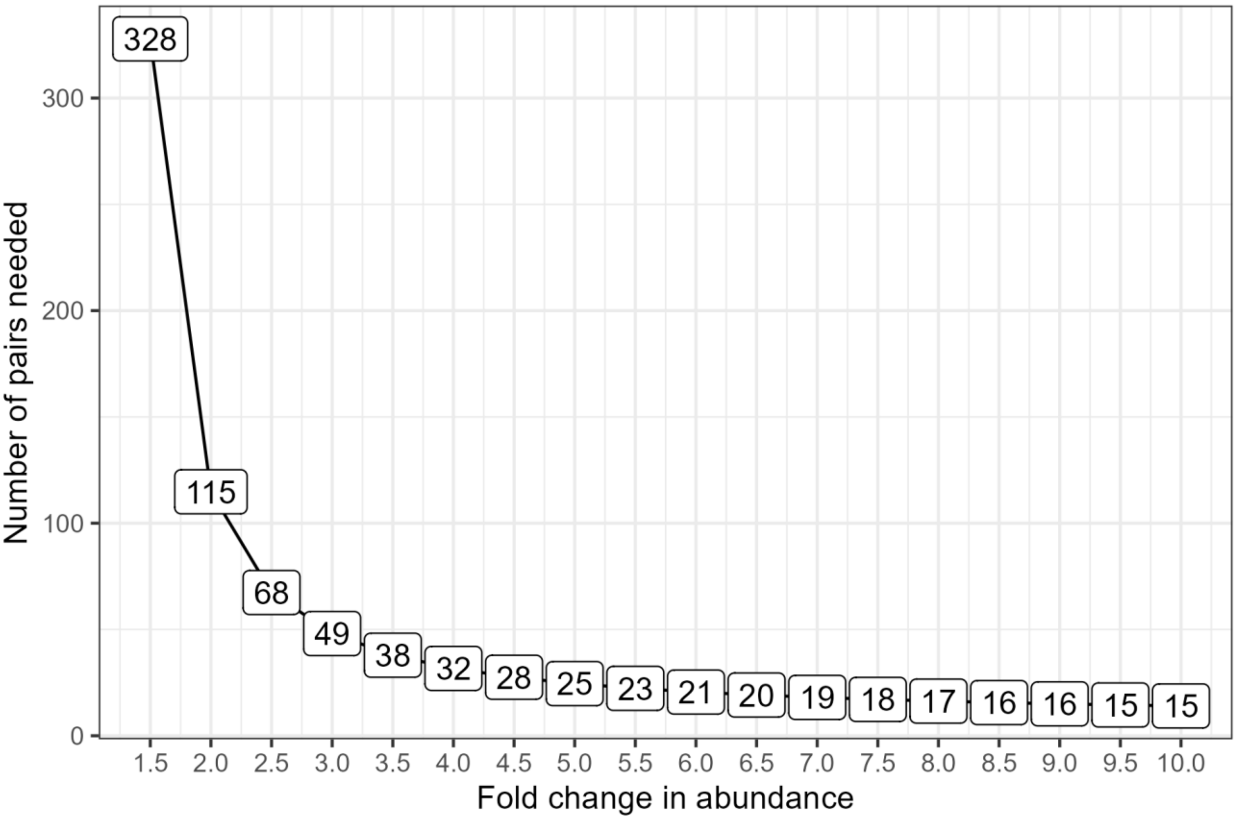


**Figure S6. Power calculation analysis for differential genus abundances of household-matched pairs.** Using data on between- and within-household variations in the relative abundance of bacterial genera, we approximately estimate the numbers of household-matched pairs required to detect "true" differences between the abundance of a specific bacterial genus of the pairwise households, with a 90% statistical power at a critical threshold of $p < 0.001$.
