## Supplementary material for "Comprehensive profiling of the human intestinal DNA virome and prediction of disease-associated bacterial hosts in severe Myalgic Encephalomyelitis/Chronic Fatigue Syndrome (ME/CFS)": S Table 3 VLP OTUs

| contig_id | contig_length | provirus | proviral_length | gene_count | viral_genes | host_genes | checkv_quality | miuvig_quality | completeness | completeness_method | contamination |
|---|---|---|---|---|---|---|---|---|---|---|---|
| S17_extracted_k141_29558 | 1174 | No | NA | 1 | 0 | 1 | Not-determined | Genome-fragment | NA | NA | 0 |
| S9_extracted_k141_55517 | 1000 | No | NA | 2 | 2 | 0 | Low-quality | Genome-fragment | 1.09 | AAI-based (medium-confidence) | 0 |
| S17_extracted_k141_73905 | 4103 | No | NA | 3 | 0 | 2 | Not-determined | Genome-fragment | NA | NA | 0 |
| S17_extracted_k141_44341 | 7337 | No | NA | 5 | 0 | 5 | Not-determined | Genome-fragment | NA | NA | 0 |
| S17_extracted_k141_125625 | 3571 | No | NA | 5 | 0 | 0 | Not-determined | Genome-fragment | NA | NA | 0 |
| S17_extracted_k141_140412 | 1061 | No | NA | 1 | 0 | 0 | Not-determined | Genome-fragment | NA | NA | 0 |
| S17_extracted_k141_88694 | 1059 | No | NA | 2 | 0 | 2 | Not-determined | Genome-fragment | NA | NA | 0 |
| S17_extracted_k141_66541 | 1536 | No | NA | 1 | 0 | 1 | Not-determined | Genome-fragment | NA | NA | 0 |
| S14_extracted_k141_60624 | 1000 | No | NA | 1 | 0 | 1 | Low-quality | Genome-fragment | 2.56 | AAI-based (high-confidence) | 0 |
| S17_extracted_k141_14804 | 1049 | No | NA | 1 | 0 | 0 | Not-determined | Genome-fragment | NA | NA | 0 |
| S14_extracted_k141_11153 | 1000 | No | NA | 1 | 1 | 0 | Low-quality | Genome-fragment | 0.76 | AAI-based (medium-confidence) | 0 |
| S17_extracted_k141_66548 | 1930 | No | NA | 3 | 0 | 1 | Not-determined | Genome-fragment | NA | NA | 0 |
| S1_vs_k141_123264\|\|full | 1000 | No | NA | 4 | 3 | 0 | Low-quality | Genome-fragment | 1.61 | AAI-based (medium-confidence) | 0 |
| S7_extracted_65780 | 1000 | No | NA | 4 | 1 | 0 | Low-quality | Genome-fragment | 2.56 | AAI-based (high-confidence) | 0 |
| S17_extracted_k141_14823 | 1010 | No | NA | 3 | 0 | 0 | Not-determined | Genome-fragment | NA | NA | 0 |
| S17_extracted_k141_36993 | 1031 | No | NA | 1 | 0 | 1 | Not-determined | Genome-fragment | NA | NA | 0 |
| S7_vs_25620\|\|full | 1000 | No | NA | 4 | 2 | 0 | Low-quality | Genome-fragment | 1.99 | AAI-based (high-confidence) | 0 |
| S17_extracted_k141_169985 | 2542 | No | NA | 1 | 0 | 0 | Not-determined | Genome-fragment | NA | NA | 0 |
| S17_extracted_k141_51785 | 6067 | No | NA | 2 | 0 | 2 | Not-determined | Genome-fragment | NA | NA | 0 |
| S17_extracted_k141_96125 | 1301 | No | NA | 3 | 0 | 1 | Not-determined | Genome-fragment | NA | NA | 0 |
| S7_vs_48054\|\|full | 1000 | No | NA | 3 | 0 | 0 | Low-quality | Genome-fragment | 0.65 | AAI-based (high-confidence) | 0 |
| S17_extracted_k141_96132 | 1343 | No | NA | 3 | 0 | 3 | Not-determined | Genome-fragment | NA | NA | 0 |
| S7_vs_16714\|\|lt2gene | 1000 | No | NA | 2 | 0 | 1 | Low-quality | Genome-fragment | 1.12 | AAI-based (medium-confidence) | 0 |
| S10_extracted_25739 | 1000 | No | NA | 2 | 0 | 2 | Low-quality | Genome-fragment | 1.08 | AAI-based (medium-confidence) | 0 |
| S17_extracted_k141_118309 | 1474 | No | NA | 1 | 0 | 1 | Not-determined | Genome-fragment | NA | NA | 0 |
| S17_extracted_k141_96141 | 1262 | No | NA | 2 | 0 | 0 | Not-determined | Genome-fragment | NA | NA | 0 |
| S17_extracted_k141_66582 | 4129 | No | NA | 2 | 0 | 0 | Not-determined | Genome-fragment | NA | NA | 0 |
| S17_extracted_k141_29669 | 3665 | No | NA | 4 | 0 | 2 | Not-determined | Genome-fragment | NA | NA | 0 |
| S17_extracted_k141_29672 | 1006 | No | NA | 1 | 0 | 1 | Not-determined | Genome-fragment | NA | NA | 0 |
| S17_extracted_k141_104 | 4839 | No | NA | 5 | 0 | 1 | Not-determined | Genome-fragment | NA | NA | 0 |
| S17_extracted_k141_133078 | 1049 | No | NA | 2 | 0 | 1 | Not-determined | Genome-fragment | NA | NA | 0 |
| S17_extracted_k141_107 | 7447 | No | NA | 6 | 0 | 5 | Not-determined | Genome-fragment | NA | NA | 0 |
| S17_extracted_k141_162641 | 1132 | No | NA | 2 | 0 | 0 | Not-determined | Genome-fragment | NA | NA | 0 |
| S17_extracted_k141_96154 | 1386 | No | NA | 1 | 0 | 1 | Not-determined | Genome-fragment | NA | NA | 0 |
| S8_extracted_k141_192562 | 1000 | No | NA | 2 | 0 | 0 | Low-quality | Genome-fragment | 1.52 | AAI-based (high-confidence) | 0 |
| S17_extracted_k141_29703 | 2194 | No | NA | 3 | 0 | 1 | Not-determined | Genome-fragment | NA | NA | 0 |
| S17_extracted_k141_140474 | 3147 | No | NA | 4 | 0 | 3 | Not-determined | Genome-fragment | NA | NA | 0 |
| S17_extracted_k141_29713 | 1436 | No | NA | 2 | 0 | 1 | Not-determined | Genome-fragment | NA | NA | 0 |
| S17_extracted_k141_66627 | 1313 | No | NA | 2 | 0 | 0 | Not-determined | Genome-fragment | NA | NA | 0 |
| S17_extracted_k141_74031 | 2593 | No | NA | 2 | 0 | 0 | Not-determined | Genome-fragment | NA | NA | 0 |
| S17_extracted_k141_162664 | 2006 | No | NA | 3 | 0 | 2 | Not-determined | Genome-fragment | NA | NA | 0 |
| S17_extracted_k141_44417 | 1820 | No | NA | 1 | 0 | 1 | Not-determined | Genome-fragment | NA | NA | 0 |
| S17_extracted_k141_22232 | 2815 | No | NA | 3 | 0 | 1 | Not-determined | Genome-fragment | NA | NA | 0 |
| S17_extracted_k141_125697 | 1388 | No | NA | 2 | 0 | 1 | Not-determined | Genome-fragment | NA | NA | 0 |
| S17_extracted_k141_74065 | 1224 | No | NA | 3 | 0 | 0 | Not-determined | Genome-fragment | NA | NA | 0 |
| S17_extracted_k141_74068 | 1372 | No | NA | 2 | 0 | 0 | Not-determined | Genome-fragment | NA | NA | 0 |
| S17_extracted_k141_103593 | 1264 | No | NA | 2 | 0 | 1 | Not-determined | Genome-fragment | NA | NA | 0 |
| S8_extracted_k141_88686 | 1000 | No | NA | 2 | 0 | 0 | Low-quality | Genome-fragment | 1.9 | AAI-based (medium-confidence) | 0 |
| S17_extracted_k141_103596 | 1042 | No | NA | 1 | 0 | 1 | Not-determined | Genome-fragment | NA | NA | 0 |
| S17_extracted_k141_37084 | 1005 | No | NA | 2 | 0 | 1 | Not-determined | Genome-fragment | NA | NA | 0 |
| S8_vs_k141_143174\|\|full | 1000 | No | NA | 3 | 0 | 0 | Low-quality | Genome-fragment | 0.39 | AAI-based (high-confidence) | 0 |
| S17_extracted_k141_88810 | 2336 | No | NA | 4 | 0 | 0 | Not-determined | Genome-fragment | NA | NA | 0 |
| S17_extracted_k141_25378 | 1001 | No | NA | 1 | 1 | 0 | Low-quality | Genome-fragment | 2.24 | AAI-based (high-confidence) | 0 |
| S17_extracted_k141_118382 | 1010 | No | NA | 1 | 0 | 0 | Not-determined | Genome-fragment | NA | NA | 0 |
| S17_extracted_k141_103645 | 1363 | No | NA | 3 | 0 | 0 | Not-determined | Genome-fragment | NA | NA | 0 |
| S17_vs_k141_120886\|\|full | 1001 | No | NA | 3 | 0 | 0 | Low-quality | Genome-fragment | 1.55 | AAI-based (medium-confidence) | 0 |
| S17_extracted_k141_155306 | 1073 | No | NA | 2 | 0 | 0 | Not-determined | Genome-fragment | NA | NA | 0 |
| S17_vs_k141_94174\|\|full | 1001 | No | NA | 2 | 0 | 1 | Low-quality | Genome-fragment | 2.32 | AAI-based (medium-confidence) | 0 |
| S17_extracted_k141_147961 | 4804 | No | NA | 9 | 0 | 4 | Not-determined | Genome-fragment | NA | NA | 0 |
| S17_extracted_k141_51926 | 1543 | No | NA | 1 | 0 | 0 | Not-determined | Genome-fragment | NA | NA | 0 |
| S17_extracted_k141_22300 | 1330 | No | NA | 1 | 0 | 0 | Not-determined | Genome-fragment | NA | NA | 0 |
| S17_extracted_k141_37184 | 1522 | No | NA | 2 | 0 | 1 | Not-determined | Genome-fragment | NA | NA | 0 |
| S17_vs_k141_84320\|\|lt2gene | 1001 | No | NA | 2 | 2 | 0 | Low-quality | Genome-fragment | 1.79 | HMM-based (lower-bound) | 0 |
| S17_extracted_k141_7647 | 1103 | No | NA | 3 | 0 | 0 | Not-determined | Genome-fragment | NA | NA | 0 |
| S17_extracted_k141_37193 | 1111 | No | NA | 1 | 0 | 0 | Not-determined | Genome-fragment | NA | NA | 0 |
| S17_extracted_k141_103677 | 2068 | No | NA | 3 | 0 | 0 | Not-determined | Genome-fragment | NA | NA | 0 |
| S17_extracted_k141_81330 | 4034 | No | NA | 5 | 0 | 4 | Not-determined | Genome-fragment | NA | NA | 0 |
| S17_extracted_k141_140605 | 1322 | No | NA | 2 | 0 | 2 | Not-determined | Genome-fragment | NA | NA | 0 |
| S17_vs_k141_4543\|\|lt2gene | 1001 | No | NA | 1 | 1 | 0 | Low-quality | Genome-fragment | 1.6 | AAI-based (high-confidence) | 0 |
| S17_extracted_k141_118438 | 1737 | No | NA | 4 | 0 | 4 | Not-determined | Genome-fragment | NA | NA | 0 |
| S17_vs_k141_63804\|\|lt2gene | 1001 | No | NA | 2 | 2 | 0 | Low-quality | Genome-fragment | 2.46 | AAI-based (high-confidence) | 0 |
| S17_extracted_k141_96379 | 1323 | No | NA | 1 | 0 | 1 | Not-determined | Genome-fragment | NA | NA | 0 |
| S9_vs_k141_16791\|\|full | 1001 | No | NA | 3 | 1 | 0 | Low-quality | Genome-fragment | 2.2 | AAI-based (high-confidence) | 0 |
| S17_extracted_k141_125784 | 1616 | No | NA | 3 | 0 | 2 | Not-determined | Genome-fragment | NA | NA | 0 |
| S11_vs_k141_51168\|\|full | 1001 | No | NA | 3 | 0 | 0 | Low-quality | Genome-fragment | 2.21 | AAI-based (high-confidence) | 0 |
| S13_extracted_38304 | 1001 | No | NA | 2 | 0 | 1 | Low-quality | Genome-fragment | 2.36 | AAI-based (medium-confidence) | 0 |
| S13_vs_77368\|\|full | 1001 | No | NA | 3 | 0 | 2 | Low-quality | Genome-fragment | 1.16 | AAI-based (medium-confidence) | 0 |
| S17_extracted_k141_103755 | 3384 | No | NA | 5 | 0 | 2 | Not-determined | Genome-fragment | NA | NA | 0 |
| S17_extracted_k141_155401 | 1201 | No | NA | 1 | 0 | 1 | Not-determined | Genome-fragment | NA | NA | 0 |
| S17_extracted_k141_7719 | 8499 | No | NA | 5 | 0 | 2 | Not-determined | Genome-fragment | NA | NA | 0 |
| S17_extracted_k141_96427 | 5567 | No | NA | 4 | 0 | 3 | Not-determined | Genome-fragment | NA | NA | 0 |
| S17_extracted_k141_74143 | 3989 | No | NA | 2 | 0 | 2 | Not-determined | Genome-fragment | NA | NA | 0 |
| S17_extracted_k141_140653 | 29997 | No | NA | 37 | 9 | 1 | High-quality | High-quality | 100 | AAI-based (high-confidence) | 0 |
| S14_vs_k141_103781\|\|full | 1001 | No | NA | 2 | 1 | 0 | Low-quality | Genome-fragment | 2.52 | AAI-based (medium-confidence) | 0 |
| S14_vs_k141_38094\|\|full | 1001 | No | NA | 3 | 1 | 0 | Low-quality | Genome-fragment | 2.34 | AAI-based (high-confidence) | 0 |
| S3_vs_k141_121456\|\|full | 1001 | No | NA | 3 | 0 | 0 | Low-quality | Genome-fragment | 1.42 | AAI-based (high-confidence) | 0 |
| S5_vs_k141_12455\|\|full | 1001 | No | NA | 3 | 0 | 1 | Low-quality | Genome-fragment | 0.84 | AAI-based (medium-confidence) | 0 |
| S17_extracted_k141_125851 | 1538 | No | NA | 2 | 0 | 1 | Not-determined | Genome-fragment | NA | NA | 0 |
| S17_extracted_k141_125863 | 1004 | No | NA | 1 | 0 | 1 | Not-determined | Genome-fragment | NA | NA | 0 |
| S17_extracted_k141_118507 | 4607 | No | NA | 2 | 0 | 1 | Not-determined | Genome-fragment | NA | NA | 0 |
| S17_extracted_k141_148103 | 1806 | No | NA | 2 | 0 | 2 | Not-determined | Genome-fragment | NA | NA | 0 |
| S17_extracted_k141_66746 | 1169 | No | NA | 2 | 0 | 0 | Not-determined | Genome-fragment | NA | NA | 0 |
| S17_extracted_k141_7736 | 10395 | No | NA | 6 | 0 | 4 | Not-determined | Genome-fragment | NA | NA | 0 |
| S7_extracted_42821 | 1001 | No | NA | 2 | 1 | 0 | Low-quality | Genome-fragment | 3.7 | AAI-based (high-confidence) | 0 |
| S17_extracted_k141_29997 | 1014 | No | NA | 1 | 0 | 1 | Not-determined | Genome-fragment | NA | NA | 0 |
| S17_extracted_k141_81413 | 1133 | No | NA | 2 | 0 | 1 | Not-determined | Genome-fragment | NA | NA | 0 |
| S17_extracted_k141_125909 | 2865 | No | NA | 4 | 0 | 4 | Not-determined | Genome-fragment | NA | NA | 0 |
| S7_vs_34835\|\|full | 1001 | No | NA | 3 | 0 | 0 | Low-quality | Genome-fragment | 2.02 | AAI-based (high-confidence) | 0 |
| S17_extracted_k141_140720 | 1189 | No | NA | 2 | 0 | 2 | Not-determined | Genome-fragment | NA | NA | 0 |
| S17_extracted_k141_37341 | 1057 | No | NA | 2 | 0 | 0 | Not-determined | Genome-fragment | NA | NA | 0 |
| S17_extracted_k141_148160 | 1921 | No | NA | 3 | 0 | 1 | Not-determined | Genome-fragment | NA | NA | 0 |
| S17_extracted_k141_170225 | 1194 | No | NA | 1 | 0 | 1 | Not-determined | Genome-fragment | NA | NA | 0 |
| S17_extracted_k141_289 | 1451 | No | NA | 2 | 0 | 2 | Not-determined | Genome-fragment | NA | NA | 0 |
| S17_extracted_k141_125919 | 3054 | No | NA | 2 | 0 | 1 | Not-determined | Genome-fragment | NA | NA | 0 |
| S17_extracted_k141_52092 | 1025 | No | NA | 3 | 0 | 1 | Not-determined | Genome-fragment | NA | NA | 0 |
| S17_extracted_k141_292 | 1320 | No | NA | 1 | 0 | 0 | Not-determined | Genome-fragment | NA | NA | 0 |
| S17_extracted_k141_103850 | 4010 | No | NA | 2 | 0 | 2 | Not-determined | Genome-fragment | NA | NA | 0 |
| S16_vs_54557\|\|full | 1001 | No | NA | 2 | 1 | 0 | Low-quality | Genome-fragment | 2.23 | AAI-based (high-confidence) | 0 |
| S17_extracted_k141_111083 | 1216 | No | NA | 1 | 0 | 0 | Not-determined | Genome-fragment | NA | NA | 0 |
| S17_extracted_k141_140766 | 1060 | No | NA | 2 | 0 | 1 | Not-determined | Genome-fragment | NA | NA | 0 |
| S10_extracted_36455 | 1001 | No | NA | 3 | 0 | 0 | Low-quality | Genome-fragment | 1.67 | AAI-based (medium-confidence) | 0 |
| S17_extracted_k141_308 | 1380 | No | NA | 1 | 0 | 1 | Not-determined | Genome-fragment | NA | NA | 0 |
| S17_extracted_k141_111096 | 1865 | No | NA | 4 | 0 | 1 | Not-determined | Genome-fragment | NA | NA | 0 |
| S10_extracted_121167 | 1001 | No | NA | 2 | 1 | 0 | Low-quality | Genome-fragment | 2.1 | AAI-based (high-confidence) | 0 |
| S15_extracted_35980 | 1001 | No | NA | 1 | 0 | 0 | Low-quality | Genome-fragment | 0.59 | AAI-based (high-confidence) | 0 |
| S17_extracted_k141_59560 | 1097 | No | NA | 1 | 0 | 1 | Not-determined | Genome-fragment | NA | NA | 0 |
| S17_extracted_k141_325 | 1150 | No | NA | 1 | 0 | 1 | Not-determined | Genome-fragment | NA | NA | 0 |
| S17_extracted_k141_103878 | 4149 | No | NA | 2 | 0 | 2 | Not-determined | Genome-fragment | NA | NA | 0 |
| S17_extracted_k141_140802 | 1355 | No | NA | 2 | 0 | 0 | Not-determined | Genome-fragment | NA | NA | 0 |
| S17_extracted_k141_133365 | 1241 | No | NA | 2 | 0 | 0 | Not-determined | Genome-fragment | NA | NA | 0 |
| S17_extracted_k141_103897 | 1916 | No | NA | 2 | 0 | 2 | Not-determined | Genome-fragment | NA | NA | 0 |
| S15_vs_55331\|\|lt2gene | 1001 | No | NA | 1 | 1 | 0 | Low-quality | Genome-fragment | 1.74 | HMM-based (lower-bound) | 0 |
| S17_extracted_k141_15078 | 1148 | No | NA | 1 | 0 | 1 | Not-determined | Genome-fragment | NA | NA | 0 |
| S17_extracted_k141_162900 | 1901 | No | NA | 3 | 0 | 2 | Not-determined | Genome-fragment | NA | NA | 0 |
| S17_extracted_k141_88970 | 1004 | No | NA | 1 | 0 | 0 | Not-determined | Genome-fragment | NA | NA | 0 |
| S17_extracted_k141_66837 | 4239 | No | NA | 6 | 0 | 6 | Not-determined | Genome-fragment | NA | NA | 0 |
| S17_extracted_k141_74318 | 1672 | No | NA | 2 | 0 | 0 | Not-determined | Genome-fragment | NA | NA | 0 |
| S17_extracted_k141_360 | 1192 | No | NA | 1 | 0 | 1 | Not-determined | Genome-fragment | NA | NA | 0 |
| S17_extracted_k141_103928 | 3523 | No | NA | 4 | 0 | 2 | Not-determined | Genome-fragment | NA | NA | 0 |
| S17_extracted_k141_118620 | 7750 | No | NA | 14 | 0 | 3 | Not-determined | Genome-fragment | NA | NA | 0 |
| S8_vs_k141_207951\|\|full | 1001 | No | NA | 3 | 0 | 0 | Low-quality | Genome-fragment | 1.7 | AAI-based (high-confidence) | 0 |
| S17_extracted_k141_162911 | 1228 | No | NA | 2 | 0 | 0 | Not-determined | Genome-fragment | NA | NA | 0 |
| S17_extracted_k141_148243 | 5411 | No | NA | 4 | 0 | 1 | Not-determined | Genome-fragment | NA | NA | 0 |
| S17_extracted_k141_103956 | 1065 | No | NA | 2 | 0 | 0 | Not-determined | Genome-fragment | NA | NA | 0 |
| S17_extracted_k141_19145 | 1002 | No | NA | 3 | 0 | 0 | Low-quality | Genome-fragment | 2.19 | AAI-based (high-confidence) | 0 |
| S17_extracted_k141_148250 | 2661 | No | NA | 3 | 0 | 3 | Not-determined | Genome-fragment | NA | NA | 0 |
| S17_extracted_k141_52179 | 2308 | No | NA | 3 | 0 | 2 | Not-determined | Genome-fragment | NA | NA | 0 |
| S17_extracted_k141_22444 | 1253 | No | NA | 3 | 0 | 0 | Not-determined | Genome-fragment | NA | NA | 0 |
| S5_vs_k141_127737\|\|full | 1002 | No | NA | 3 | 0 | 1 | Low-quality | Genome-fragment | 1.49 | AAI-based (high-confidence) | 0 |
| S17_extracted_k141_375 | 1698 | No | NA | 1 | 0 | 0 | Not-determined | Genome-fragment | NA | NA | 0 |
| S17_extracted_k141_103966 | 3099 | No | NA | 3 | 0 | 1 | Not-determined | Genome-fragment | NA | NA | 0 |
| S17_extracted_k141_52191 | 13853 | No | NA | 12 | 0 | 7 | Not-determined | Genome-fragment | NA | NA | 0 |
| S5_vs_k141_116068\|\|lt2gene | 1002 | No | NA | 2 | 1 | 0 | Low-quality | Genome-fragment | 2.18 | HMM-based (lower-bound) | 0 |
| S17_extracted_k141_30095 | 1520 | No | NA | 2 | 0 | 1 | Not-determined | Genome-fragment | NA | NA | 0 |
| S17_extracted_k141_52200 | 3371 | No | NA | 4 | 0 | 3 | Not-determined | Genome-fragment | NA | NA | 0 |
| S17_extracted_k141_155499 | 2647 | No | NA | 1 | 0 | 1 | Not-determined | Genome-fragment | NA | NA | 0 |
| S17_extracted_k141_155502 | 1385 | No | NA | 1 | 0 | 1 | Not-determined | Genome-fragment | NA | NA | 0 |
| S17_extracted_k141_89020 | 4900 | No | NA | 3 | 0 | 2 | Not-determined | Genome-fragment | NA | NA | 0 |
| S17_extracted_k141_425 | 1082 | No | NA | 1 | 0 | 0 | Not-determined | Genome-fragment | NA | NA | 0 |
| S17_extracted_k141_140917 | 1426 | No | NA | 1 | 0 | 1 | Not-determined | Genome-fragment | NA | NA | 0 |
| S17_extracted_k141_111208 | 1004 | No | NA | 2 | 0 | 0 | Not-determined | Genome-fragment | NA | NA | 0 |
| S17_extracted_k141_22496 | 1039 | No | NA | 1 | 0 | 0 | Not-determined | Genome-fragment | NA | NA | 0 |
| S7_vs_31350\|\|full | 1002 | No | NA | 4 | 0 | 0 | Low-quality | Genome-fragment | 3.11 | AAI-based (high-confidence) | 0 |
| S17_extracted_k141_22498 | 1237 | No | NA | 4 | 0 | 0 | Not-determined | Genome-fragment | NA | NA | 0 |
| S17_extracted_k141_170347 | 2720 | No | NA | 2 | 0 | 1 | Not-determined | Genome-fragment | NA | NA | 0 |
| S16_extracted_19899 | 1002 | No | NA | 3 | 0 | 0 | Low-quality | Genome-fragment | 1.75 | AAI-based (medium-confidence) | 0 |
| S17_extracted_k141_118725 | 1244 | No | NA | 4 | 0 | 0 | Not-determined | Genome-fragment | NA | NA | 0 |
| S16_extracted_143827 | 1002 | No | NA | 1 | 0 | 0 | Low-quality | Genome-fragment | 0.94 | AAI-based (medium-confidence) | 0 |
| S17_extracted_k141_89053 | 5042 | No | NA | 6 | 0 | 4 | Not-determined | Genome-fragment | NA | NA | 0 |
| S17_extracted_k141_148303 | 15913 | No | NA | 20 | 10 | 0 | Medium-quality | Genome-fragment | 70.05 | AAI-based (medium-confidence) | 0 |
| S17_extracted_k141_170364 | 2048 | No | NA | 2 | 0 | 1 | Not-determined | Genome-fragment | NA | NA | 0 |
| S17_extracted_k141_81585 | 1113 | No | NA | 1 | 0 | 1 | Not-determined | Genome-fragment | NA | NA | 0 |
| S17_extracted_k141_74430 | 5357 | No | NA | 4 | 0 | 3 | Not-determined | Genome-fragment | NA | NA | 0 |
| S10_extracted_80128 | 1002 | No | NA | 2 | 1 | 0 | Low-quality | Genome-fragment | 0.87 | AAI-based (medium-confidence) | 0 |
| S12_extracted_k141_105604 | 1002 | No | NA | 2 | 1 | 0 | Low-quality | Genome-fragment | 0.57 | HMM-based (lower-bound) | 0 |
| S12_extracted_k141_37605 | 1002 | No | NA | 1 | 1 | 0 | Low-quality | Genome-fragment | 0.82 | HMM-based (lower-bound) | 0 |
| S6_vs_k141_52326\|\|lt2gene | 1002 | No | NA | 2 | 1 | 0 | Low-quality | Genome-fragment | 2.41 | AAI-based (high-confidence) | 0 |
| S17_extracted_k141_22539 | 1566 | No | NA | 3 | 0 | 1 | Not-determined | Genome-fragment | NA | NA | 0 |
| S17_extracted_k141_507 | 5467 | No | NA | 7 | 0 | 1 | Not-determined | Genome-fragment | NA | NA | 0 |
| S8_extracted_k141_138895 | 1002 | No | NA | 1 | 0 | 0 | Low-quality | Genome-fragment | 1.11 | AAI-based (medium-confidence) | 0 |
| S17_extracted_k141_75586 | 1003 | No | NA | 3 | 1 | 0 | Low-quality | Genome-fragment | 2.83 | AAI-based (high-confidence) | 0 |
| S17_extracted_k141_52317 | 1168 | No | NA | 1 | 0 | 1 | Not-determined | Genome-fragment | NA | NA | 0 |
| S17_extracted_k141_132283 | 1003 | No | NA | 1 | 0 | 1 | Low-quality | Genome-fragment | 0.56 | AAI-based (medium-confidence) | 0 |
| S17_extracted_k141_118801 | 3245 | No | NA | 4 | 0 | 4 | Not-determined | Genome-fragment | NA | NA | 0 |
| S17_extracted_k141_126044 | 1468 | No | NA | 2 | 0 | 1 | Not-determined | Genome-fragment | NA | NA | 0 |
| S17_extracted_k141_111282 | 1013 | No | NA | 1 | 0 | 1 | Not-determined | Genome-fragment | NA | NA | 0 |
| S17_extracted_k141_552 | 5196 | No | NA | 3 | 0 | 2 | Not-determined | Genome-fragment | NA | NA | 0 |
| S17_extracted_k141_8018 | 5489 | No | NA | 6 | 0 | 5 | Not-determined | Genome-fragment | NA | NA | 0 |
| S11_vs_k141_96733\|\|full | 1003 | No | NA | 2 | 0 | 0 | Low-quality | Genome-fragment | 2.23 | AAI-based (high-confidence) | 0 |
| S17_extracted_k141_133531 | 1521 | No | NA | 1 | 0 | 0 | Not-determined | Genome-fragment | NA | NA | 0 |
| S14_extracted_k141_69284 | 1003 | No | NA | 5 | 0 | 0 | Low-quality | Genome-fragment | 7.2 | AAI-based (high-confidence) | 0 |
| S17_extracted_k141_8050 | 2822 | No | NA | 4 | 0 | 3 | Not-determined | Genome-fragment | NA | NA | 0 |
| S17_extracted_k141_584 | 3364 | No | NA | 2 | 0 | 0 | Not-determined | Genome-fragment | NA | NA | 0 |
| S17_extracted_k141_170448 | 1537 | No | NA | 2 | 0 | 0 | Not-determined | Genome-fragment | NA | NA | 0 |
| S1_extracted_k141_81469 | 1003 | No | NA | 2 | 0 | 0 | Low-quality | Genome-fragment | 0.57 | AAI-based (high-confidence) | 0 |
| S17_extracted_k141_66932 | 1547 | No | NA | 1 | 0 | 0 | Not-determined | Genome-fragment | NA | NA | 0 |
| S17_extracted_k141_44926 | 1269 | No | NA | 2 | 0 | 0 | Not-determined | Genome-fragment | NA | NA | 0 |
| S1_extracted_k141_115287 | 1003 | No | NA | 1 | 0 | 1 | Low-quality | Genome-fragment | 2.44 | AAI-based (high-confidence) | 0 |
| S3_vs_k141_32391\|\|full | 1003 | No | NA | 3 | 0 | 0 | Low-quality | Genome-fragment | 1.47 | AAI-based (medium-confidence) | 0 |
| S3_vs_k141_31771\|\|full | 1003 | No | NA | 2 | 0 | 0 | Low-quality | Genome-fragment | 0.47 | AAI-based (medium-confidence) | 0 |
| S17_extracted_k141_30324 | 4069 | No | NA | 11 | 0 | 10 | Not-determined | Genome-fragment | NA | NA | 0 |
| S17_extracted_k141_59748 | 4284 | No | NA | 5 | 0 | 0 | Not-determined | Genome-fragment | NA | NA | 0 |
| S17_extracted_k141_104146 | 9281 | No | NA | 8 | 0 | 4 | Not-determined | Genome-fragment | NA | NA | 0 |
| S17_extracted_k141_37716 | 1127 | No | NA | 1 | 0 | 0 | Not-determined | Genome-fragment | NA | NA | 0 |
| S5_vs_k141_61075\|\|full | 1003 | No | NA | 3 | 0 | 0 | Low-quality | Genome-fragment | 1.87 | AAI-based (medium-confidence) | 0 |
| S5_vs_k141_56289\|\|full | 1003 | No | NA | 4 | 0 | 0 | Low-quality | Genome-fragment | 2.52 | AAI-based (high-confidence) | 0 |
| S17_extracted_k141_30343 | 1381 | No | NA | 2 | 0 | 1 | Not-determined | Genome-fragment | NA | NA | 0 |
| S17_extracted_k141_163112 | 3357 | No | NA | 4 | 0 | 0 | Not-determined | Genome-fragment | NA | NA | 0 |
| S17_extracted_k141_155748 | 1055 | No | NA | 1 | 0 | 0 | Not-determined | Genome-fragment | NA | NA | 0 |
| S5_vs_k141_5104\|\|full | 1003 | No | NA | 2 | 0 | 0 | Low-quality | Genome-fragment | 2.46 | AAI-based (medium-confidence) | 0 |
| S7_extracted_134444 | 1003 | No | NA | 1 | 0 | 0 | Low-quality | Genome-fragment | 1.69 | AAI-based (high-confidence) | 0 |
| S17_extracted_k141_163119 | 3054 | No | NA | 3 | 0 | 0 | Not-determined | Genome-fragment | NA | NA | 0 |
| S17_extracted_k141_81651 | 1298 | No | NA | 2 | 0 | 1 | Not-determined | Genome-fragment | NA | NA | 0 |
| S17_extracted_k141_81661 | 1199 | No | NA | 1 | 0 | 1 | Not-determined | Genome-fragment | NA | NA | 0 |
| S17_extracted_k141_118902 | 1030 | No | NA | 1 | 0 | 1 | Not-determined | Genome-fragment | NA | NA | 0 |
| S17_extracted_k141_104215 | 1034 | No | NA | 2 | 0 | 1 | Not-determined | Genome-fragment | NA | NA | 0 |
| S17_extracted_k141_30360 | 3612 | No | NA | 4 | 0 | 4 | Not-determined | Genome-fragment | NA | NA | 0 |
| S7_vs_98447\|\|full | 1003 | No | NA | 3 | 1 | 0 | Low-quality | Genome-fragment | 2.46 | AAI-based (high-confidence) | 0 |
| S17_extracted_k141_15341 | 1807 | No | NA | 2 | 0 | 0 | Not-determined | Genome-fragment | NA | NA | 0 |
| S17_extracted_k141_74585 | 1420 | No | NA | 2 | 0 | 2 | Not-determined | Genome-fragment | NA | NA | 0 |
| S17_extracted_k141_89166 | 3533 | No | NA | 4 | 0 | 3 | Not-determined | Genome-fragment | NA | NA | 0 |
| S16_extracted_160405 | 1003 | No | NA | 3 | 0 | 0 | Low-quality | Genome-fragment | 1.62 | AAI-based (high-confidence) | 0 |
| S17_extracted_k141_96608 | 5512 | No | NA | 6 | 0 | 2 | Not-determined | Genome-fragment | NA | NA | 0 |
| S17_extracted_k141_148503 | 1613 | No | NA | 1 | 0 | 0 | Not-determined | Genome-fragment | NA | NA | 0 |
| S17_extracted_k141_30375 | 6432 | No | NA | 4 | 0 | 0 | Not-determined | Genome-fragment | NA | NA | 0 |
| S17_extracted_k141_66986 | 4689 | No | NA | 4 | 0 | 3 | Not-determined | Genome-fragment | NA | NA | 0 |
| S17_extracted_k141_104252 | 1602 | No | NA | 1 | 0 | 1 | Not-determined | Genome-fragment | NA | NA | 0 |
| S16_vs_144302\|\|full | 1003 | No | NA | 2 | 0 | 0 | Low-quality | Genome-fragment | 1.65 | AAI-based (high-confidence) | 0 |
| S17_extracted_k141_163185 | 1167 | No | NA | 2 | 0 | 1 | Not-determined | Genome-fragment | NA | NA | 0 |
| S16_vs_65129\|\|lt2gene | 1003 | No | NA | 1 | 1 | 0 | Low-quality | Genome-fragment | 1.69 | HMM-based (lower-bound) | 0 |
| S17_extracted_k141_133619 | 1184 | No | NA | 1 | 0 | 1 | Not-determined | Genome-fragment | NA | NA | 0 |
| S17_extracted_k141_148557 | 1119 | No | NA | 2 | 0 | 2 | Not-determined | Genome-fragment | NA | NA | 0 |
| S10_extracted_29139 | 1003 | No | NA | 1 | 1 | 0 | Low-quality | Genome-fragment | 0.41 | AAI-based (high-confidence) | 0 |
| S17_extracted_k141_104300 | 1884 | No | NA | 2 | 0 | 0 | Not-determined | Genome-fragment | NA | NA | 0 |
| S17_extracted_k141_118943 | 2862 | No | NA | 4 | 0 | 2 | Not-determined | Genome-fragment | NA | NA | 0 |
| S17_extracted_k141_705 | 1588 | No | NA | 1 | 0 | 1 | Not-determined | Genome-fragment | NA | NA | 0 |
| S17_extracted_k141_148586 | 1124 | No | NA | 1 | 0 | 0 | Not-determined | Genome-fragment | NA | NA | 0 |
| S17_extracted_k141_30469 | 1787 | No | NA | 3 | 0 | 3 | Not-determined | Genome-fragment | NA | NA | 0 |
| S17_extracted_k141_111470 | 2103 | No | NA | 2 | 0 | 2 | Not-determined | Genome-fragment | NA | NA | 0 |
| S17_extracted_k141_67042 | 15604 | No | NA | 18 | 7 | 0 | Medium-quality | Genome-fragment | 79.04 | AAI-based (medium-confidence) | 0 |
| S17_extracted_k141_67045 | 1420 | No | NA | 2 | 0 | 2 | Not-determined | Genome-fragment | NA | NA | 0 |
| S17_extracted_k141_30483 | 1021 | No | NA | 1 | 0 | 1 | Not-determined | Genome-fragment | NA | NA | 0 |
| S12_extracted_k141_59921 | 1003 | No | NA | 1 | 1 | 0 | Low-quality | Genome-fragment | 2.48 | AAI-based (high-confidence) | 0 |
| S17_extracted_k141_45033 | 1250 | No | NA | 2 | 0 | 1 | Not-determined | Genome-fragment | NA | NA | 0 |
| S17_extracted_k141_45037 | 3760 | No | NA | 4 | 0 | 4 | Not-determined | Genome-fragment | NA | NA | 0 |
| S17_extracted_k141_89281 | 2613 | No | NA | 2 | 0 | 0 | Not-determined | Genome-fragment | NA | NA | 0 |
| S12_vs_k141_85026\|\|full | 1003 | No | NA | 2 | 0 | 1 | Low-quality | Genome-fragment | 0.59 | AAI-based (medium-confidence) | 0 |
| S8_extracted_k141_191251 | 1003 | No | NA | 2 | 0 | 0 | Low-quality | Genome-fragment | 1.6 | AAI-based (high-confidence) | 0 |
| S17_extracted_k141_74698 | 4576 | No | NA | 6 | 0 | 1 | Not-determined | Genome-fragment | NA | NA | 0 |
| S8_vs_k141_10426\|\|full | 1003 | No | NA | 3 | 1 | 0 | Low-quality | Genome-fragment | 2.87 | AAI-based (high-confidence) | 0 |
| S9_vs_k141_25171\|\|full | 1004 | No | NA | 3 | 2 | 0 | Low-quality | Genome-fragment | 2.84 | AAI-based (high-confidence) | 0 |
| S17_extracted_k141_67107 | 1966 | No | NA | 3 | 0 | 3 | Not-determined | Genome-fragment | NA | NA | 0 |
| S11_extracted_k141_1385 | 1004 | No | NA | 4 | 0 | 0 | Low-quality | Genome-fragment | 1.2 | AAI-based (high-confidence) | 0 |
| S17_extracted_k141_96660 | 5565 | No | NA | 3 | 0 | 3 | Not-determined | Genome-fragment | NA | NA | 0 |
| S17_extracted_k141_52580 | 1925 | No | NA | 2 | 0 | 1 | Not-determined | Genome-fragment | NA | NA | 0 |
| S17_extracted_k141_45090 | 1474 | No | NA | 1 | 0 | 1 | Not-determined | Genome-fragment | NA | NA | 0 |
| S17_extracted_k141_163318 | 35575 | No | NA | 31 | 0 | 13 | Not-determined | Genome-fragment | NA | NA | 0 |
| S17_extracted_k141_723 | 1330 | No | NA | 1 | 0 | 1 | Not-determined | Genome-fragment | NA | NA | 0 |
| S17_extracted_k141_741 | 4944 | No | NA | 4 | 0 | 1 | Not-determined | Genome-fragment | NA | NA | 0 |
| S11_vs_k141_18151\|\|lt2gene | 1004 | No | NA | 1 | 1 | 0 | Low-quality | Genome-fragment | 1.6 | AAI-based (high-confidence) | 0 |
| S1_extracted_k141_90594 | 1004 | No | NA | 2 | 0 | 0 | Low-quality | Genome-fragment | 1.63 | AAI-based (high-confidence) | 0 |
| S17_extracted_k141_156032 | 1388 | No | NA | 2 | 0 | 1 | Not-determined | Genome-fragment | NA | NA | 0 |
| S1_extracted_k141_62774 | 1004 | No | NA | 2 | 0 | 0 | Low-quality | Genome-fragment | 2.23 | AAI-based (high-confidence) | 0 |
| S3_extracted_k141_40043 | 1004 | No | NA | 2 | 0 | 0 | Low-quality | Genome-fragment | 0.53 | AAI-based (medium-confidence) | 0 |
| S17_extracted_k141_52646 | 2055 | No | NA | 2 | 0 | 0 | Not-determined | Genome-fragment | NA | NA | 0 |
| S7_extracted_191 | 1004 | No | NA | 3 | 3 | 0 | Low-quality | Genome-fragment | 3.29 | AAI-based (high-confidence) | 0 |
| S7_vs_118782\|\|full | 1004 | No | NA | 3 | 1 | 0 | Low-quality | Genome-fragment | 2.16 | AAI-based (high-confidence) | 0 |
| S12_extracted_k141_68470 | 1004 | No | NA | 1 | 0 | 0 | Low-quality | Genome-fragment | 2.8 | AAI-based (high-confidence) | 0 |
| S17_extracted_k141_104502 | 1996 | No | NA | 4 | 0 | 4 | Not-determined | Genome-fragment | NA | NA | 0 |
| S15_vs_38927\|\|lt2gene | 1004 | No | NA | 2 | 1 | 0 | Low-quality | Genome-fragment | 0.59 | AAI-based (medium-confidence) | 0 |
| S8_extracted_k141_101213 | 1004 | No | NA | 4 | 0 | 0 | Low-quality | Genome-fragment | 1.65 | AAI-based (high-confidence) | 0 |
| S17_extracted_k141_52666 | 1218 | No | NA | 2 | 0 | 1 | Not-determined | Genome-fragment | NA | NA | 0 |
| S17_extracted_k141_111561 | 2200 | No | NA | 1 | 0 | 1 | Not-determined | Genome-fragment | NA | NA | 0 |
| S17_extracted_k141_30555 | 1218 | No | NA | 1 | 0 | 1 | Not-determined | Genome-fragment | NA | NA | 0 |
| S17_extracted_k141_170798 | 1206 | No | NA | 2 | 0 | 1 | Not-determined | Genome-fragment | NA | NA | 0 |
| S17_extracted_k141_133740 | 1116 | No | NA | 1 | 0 | 1 | Not-determined | Genome-fragment | NA | NA | 0 |
| S8_extracted_k141_23829 | 1004 | No | NA | 1 | 0 | 1 | Low-quality | Genome-fragment | 0.16 | AAI-based (medium-confidence) | 0 |
| S17_extracted_k141_30571 | 2857 | No | NA | 2 | 0 | 2 | Not-determined | Genome-fragment | NA | NA | 0 |
| S17_extracted_k141_813 | 1333 | No | NA | 2 | 0 | 2 | Not-determined | Genome-fragment | NA | NA | 0 |
| S17_extracted_k141_52703 | 1360 | No | NA | 2 | 0 | 2 | Not-determined | Genome-fragment | NA | NA | 0 |
| S17_extracted_k141_22861 | 1299 | No | NA | 4 | 0 | 3 | Not-determined | Genome-fragment | NA | NA | 0 |
| S8_vs_k141_90820\|\|full | 1004 | No | NA | 2 | 1 | 0 | Low-quality | Genome-fragment | 2.04 | AAI-based (medium-confidence) | 0 |
| S8_vs_k141_59297\|\|full | 1004 | No | NA | 4 | 0 | 0 | Low-quality | Genome-fragment | 2.69 | AAI-based (high-confidence) | 0 |
| S9_extracted_k141_57957 | 1005 | No | NA | 2 | 1 | 0 | Low-quality | Genome-fragment | 1.03 | HMM-based (lower-bound) | 0 |
| S17_extracted_k141_15435 | 3278 | No | NA | 1 | 0 | 1 | Not-determined | Genome-fragment | NA | NA | 0 |
| S17_extracted_k141_89416 | 1234 | No | NA | 1 | 0 | 1 | Not-determined | Genome-fragment | NA | NA | 0 |
| S13_vs_47062\|\|full | 1005 | No | NA | 3 | 0 | 0 | Low-quality | Genome-fragment | 1.38 | AAI-based (medium-confidence) | 0 |
| S17_extracted_k141_52735 | 2262 | No | NA | 2 | 0 | 2 | Not-determined | Genome-fragment | NA | NA | 0 |
| S14_vs_k141_12239\|\|lt2gene | 1005 | No | NA | 2 | 1 | 0 | Low-quality | Genome-fragment | 2.53 | AAI-based (high-confidence) | 0 |
| S5_extracted_k141_141743 | 1005 | No | NA | 4 | 0 | 0 | Low-quality | Genome-fragment | 2.27 | AAI-based (medium-confidence) | 0 |
| S5_extracted_k141_82725 | 1005 | No | NA | 1 | 1 | 0 | Low-quality | Genome-fragment | 3.15 | AAI-based (high-confidence) | 0 |
| S7_extracted_94514 | 1005 | No | NA | 2 | 1 | 0 | Low-quality | Genome-fragment | 1.74 | AAI-based (medium-confidence) | 0 |
| S17_extracted_k141_104602 | 6411 | No | NA | 7 | 0 | 3 | Not-determined | Genome-fragment | NA | NA | 0 |
| S17_extracted_k141_163469 | 1318 | No | NA | 2 | 0 | 2 | Not-determined | Genome-fragment | NA | NA | 0 |
| S17_extracted_k141_148795 | 2365 | No | NA | 2 | 0 | 2 | Not-determined | Genome-fragment | NA | NA | 0 |
| S17_extracted_k141_96865 | 1299 | No | NA | 2 | 0 | 2 | Not-determined | Genome-fragment | NA | NA | 0 |
| S17_extracted_k141_52760 | 1200 | No | NA | 2 | 0 | 2 | Not-determined | Genome-fragment | NA | NA | 0 |
| S17_extracted_k141_59974 | 1491 | No | NA | 1 | 0 | 0 | Not-determined | Genome-fragment | NA | NA | 0 |
| S7_extracted_95140 | 1005 | No | NA | 2 | 1 | 0 | Low-quality | Genome-fragment | 2.51 | AAI-based (high-confidence) | 0 |
| S17_extracted_k141_170896 | 1434 | No | NA | 3 | 0 | 3 | Not-determined | Genome-fragment | NA | NA | 0 |
| S7_vs_77460\|\|lt2gene | 1005 | No | NA | 2 | 2 | 0 | Low-quality | Genome-fragment | 1.64 | HMM-based (lower-bound) | 0 |
| S17_extracted_k141_38122 | 2556 | No | NA | 1 | 0 | 0 | Not-determined | Genome-fragment | NA | NA | 0 |
| S7_vs_130378\|\|lt2gene | 1005 | No | NA | 1 | 1 | 0 | Low-quality | Genome-fragment | 2.2 | AAI-based (high-confidence) | 0 |
| S17_extracted_k141_67336 | 1420 | No | NA | 1 | 0 | 1 | Not-determined | Genome-fragment | NA | NA | 0 |
| S17_extracted_k141_156182 | 3292 | No | NA | 4 | 0 | 3 | Not-determined | Genome-fragment | NA | NA | 0 |
| S17_extracted_k141_67348 | 1062 | No | NA | 2 | 0 | 2 | Not-determined | Genome-fragment | NA | NA | 0 |
| S17_extracted_k141_148847 | 1056 | No | NA | 2 | 0 | 0 | Not-determined | Genome-fragment | NA | NA | 0 |
| S17_extracted_k141_38135 | 1366 | No | NA | 1 | 0 | 0 | Not-determined | Genome-fragment | NA | NA | 0 |
| S12_vs_k141_18476\|\|full | 1005 | No | NA | 3 | 1 | 0 | Low-quality | Genome-fragment | 2.58 | AAI-based (high-confidence) | 0 |
| S17_extracted_k141_60005 | 1031 | No | NA | 1 | 0 | 0 | Not-determined | Genome-fragment | NA | NA | 0 |
| S17_extracted_k141_133782 | 1012 | No | NA | 1 | 0 | 0 | Not-determined | Genome-fragment | NA | NA | 0 |
| S17_extracted_k141_917 | 1345 | No | NA | 2 | 0 | 0 | Not-determined | Genome-fragment | NA | NA | 0 |
| S17_extracted_k141_163483 | 13324 | No | NA | 10 | 0 | 8 | Not-determined | Genome-fragment | NA | NA | 0 |
| S12_vs_k141_89895\|\|full | 1005 | No | NA | 3 | 1 | 0 | Low-quality | Genome-fragment | 1.89 | AAI-based (high-confidence) | 0 |
| S17_extracted_k141_15488 | 1385 | No | NA | 2 | 0 | 0 | Not-determined | Genome-fragment | NA | NA | 0 |
| S17_extracted_k141_81895 | 1965 | No | NA | 3 | 0 | 2 | Not-determined | Genome-fragment | NA | NA | 0 |
| S12_vs_k141_72079\|\|lt2gene | 1005 | No | NA | 2 | 1 | 0 | Low-quality | Genome-fragment | 2.67 | AAI-based (high-confidence) | 0 |
| S17_extracted_k141_15500 | 3715 | No | NA | 6 | 0 | 1 | Not-determined | Genome-fragment | NA | NA | 0 |
| S8_extracted_k141_194279 | 1005 | No | NA | 2 | 2 | 0 | Low-quality | Genome-fragment | 1.64 | HMM-based (lower-bound) | 0 |
| S17_extracted_k141_60043 | 3615 | No | NA | 3 | 0 | 3 | Not-determined | Genome-fragment | NA | NA | 0 |
| S17_extracted_k141_126375 | 1153 | No | NA | 2 | 0 | 1 | Not-determined | Genome-fragment | NA | NA | 0 |
| S17_extracted_k141_126377 | 1290 | No | NA | 1 | 0 | 1 | Not-determined | Genome-fragment | NA | NA | 0 |
| S17_extracted_k141_52874 | 1455 | No | NA | 1 | 0 | 1 | Not-determined | Genome-fragment | NA | NA | 0 |
| S17_extracted_k141_8399 | 1895 | No | NA | 1 | 0 | 0 | Not-determined | Genome-fragment | NA | NA | 0 |
| S17_extracted_k141_60070 | 1303 | No | NA | 2 | 0 | 0 | Not-determined | Genome-fragment | NA | NA | 0 |
| S17_extracted_k141_22934 | 1201 | No | NA | 1 | 0 | 1 | Not-determined | Genome-fragment | NA | NA | 0 |
| S17_extracted_k141_60072 | 4027 | No | NA | 10 | 0 | 8 | Not-determined | Genome-fragment | NA | NA | 0 |
| S8_extracted_k141_152570 | 1005 | No | NA | 1 | 1 | 0 | Low-quality | Genome-fragment | 2.51 | AAI-based (medium-confidence) | 0 |
| S17_extracted_k141_170940 | 3949 | No | NA | 2 | 0 | 1 | Not-determined | Genome-fragment | NA | NA | 0 |
| S17_extracted_k141_126401 | 1639 | No | NA | 2 | 0 | 2 | Not-determined | Genome-fragment | NA | NA | 0 |
| S8_vs_k141_151704\|\|full | 1005 | No | NA | 2 | 1 | 0 | Low-quality | Genome-fragment | 0.97 | AAI-based (medium-confidence) | 0 |
| S17_extracted_k141_45310 | 2183 | No | NA | 1 | 0 | 0 | Not-determined | Genome-fragment | NA | NA | 0 |
| S9_vs_k141_21618\|\|lt2gene | 1006 | No | NA | 1 | 1 | 0 | Low-quality | Genome-fragment | 3.25 | HMM-based (lower-bound) | 0 |
| S17_extracted_k141_170950 | 1178 | No | NA | 2 | 0 | 1 | Not-determined | Genome-fragment | NA | NA | 0 |
| S17_extracted_k141_104712 | 1573 | No | NA | 3 | 0 | 3 | Not-determined | Genome-fragment | NA | NA | 0 |
| S17_extracted_k141_133842 | 1459 | No | NA | 3 | 0 | 1 | Not-determined | Genome-fragment | NA | NA | 0 |
| S17_extracted_k141_156302 | 5399 | No | NA | 6 | 0 | 4 | Not-determined | Genome-fragment | NA | NA | 0 |
| S17_extracted_k141_89514 | 4754 | No | NA | 2 | 0 | 2 | Not-determined | Genome-fragment | NA | NA | 0 |
| S17_extracted_k141_60111 | 1054 | No | NA | 1 | 0 | 1 | Not-determined | Genome-fragment | NA | NA | 0 |
| S17_extracted_k141_126410 | 1270 | No | NA | 1 | 0 | 1 | Not-determined | Genome-fragment | NA | NA | 0 |
| S17_extracted_k141_148964 | 1068 | No | NA | 1 | 0 | 0 | Not-determined | Genome-fragment | NA | NA | 0 |
| S17_extracted_k141_156322 | 1261 | No | NA | 1 | 0 | 1 | Not-determined | Genome-fragment | NA | NA | 0 |
| S17_extracted_k141_119240 | 1007 | No | NA | 1 | 0 | 1 | Not-determined | Genome-fragment | NA | NA | 0 |
| S17_extracted_k141_30684 | 1287 | No | NA | 2 | 0 | 0 | Not-determined | Genome-fragment | NA | NA | 0 |
| S11_extracted_k141_73186 | 1006 | No | NA | 3 | 0 | 1 | Low-quality | Genome-fragment | 0.93 | AAI-based (high-confidence) | 0 |
| S17_extracted_k141_15592 | 1389 | No | NA | 3 | 0 | 2 | Not-determined | Genome-fragment | NA | NA | 0 |
| S17_extracted_k141_45353 | 9531 | No | NA | 14 | 0 | 2 | Not-determined | Genome-fragment | NA | NA | 0 |
| S17_extracted_k141_1007 | 4070 | No | NA | 4 | 0 | 2 | Not-determined | Genome-fragment | NA | NA | 0 |
| S17_extracted_k141_67418 | 3510 | No | NA | 4 | 0 | 2 | Not-determined | Genome-fragment | NA | NA | 0 |
| S1_extracted_k141_74959 | 1006 | No | NA | 2 | 1 | 0 | Low-quality | Genome-fragment | 2.96 | AAI-based (high-confidence) | 0 |
| S17_extracted_k141_67427 | 1120 | No | NA | 1 | 0 | 1 | Not-determined | Genome-fragment | NA | NA | 0 |
| S17_extracted_k141_67430 | 1551 | No | NA | 1 | 0 | 1 | Not-determined | Genome-fragment | NA | NA | 0 |
| S17_extracted_k141_104758 | 1573 | No | NA | 3 | 0 | 2 | Not-determined | Genome-fragment | NA | NA | 0 |
| S17_extracted_k141_96972 | 1002 | No | NA | 2 | 0 | 0 | Not-determined | Genome-fragment | NA | NA | 0 |
| S1_vs_k141_153330\|\|full | 1006 | No | NA | 2 | 1 | 0 | Low-quality | Genome-fragment | 1.26 | AAI-based (high-confidence) | 0 |
| S17_extracted_k141_60166 | 2958 | No | NA | 3 | 0 | 2 | Not-determined | Genome-fragment | NA | NA | 0 |
| S17_extracted_k141_96979 | 6240 | No | NA | 7 | 0 | 0 | Not-determined | Genome-fragment | NA | NA | 0 |
| S17_extracted_k141_149025 | 9454 | No | NA | 5 | 0 | 2 | Not-determined | Genome-fragment | NA | NA | 0 |
| S17_extracted_k141_133906 | 2780 | No | NA | 2 | 0 | 0 | Not-determined | Genome-fragment | NA | NA | 0 |
| S17_extracted_k141_133911 | 1132 | No | NA | 2 | 0 | 2 | Not-determined | Genome-fragment | NA | NA | 0 |
| S17_extracted_k141_30735 | 1668 | No | NA | 4 | 0 | 1 | Not-determined | Genome-fragment | NA | NA | 0 |
| S3_extracted_k141_48668 | 1006 | No | NA | 3 | 2 | 0 | Low-quality | Genome-fragment | 2.67 | AAI-based (medium-confidence) | 0 |
| S3_vs_k141_49491\|\|full | 1006 | No | NA | 1 | 0 | 0 | Low-quality | Genome-fragment | 2.08 | AAI-based (high-confidence) | 0 |
| S17_extracted_k141_8537 | 1286 | No | NA | 4 | 0 | 3 | Not-determined | Genome-fragment | NA | NA | 0 |
| S17_extracted_k141_171083 | 3829 | No | NA | 7 | 0 | 0 | Not-determined | Genome-fragment | NA | NA | 0 |
| S3_vs_k141_6461\|\|lt2gene | 1006 | No | NA | 2 | 1 | 0 | Low-quality | Genome-fragment | 1.98 | AAI-based (high-confidence) | 0 |
| S17_extracted_k141_60192 | 2372 | No | NA | 1 | 0 | 1 | Not-determined | Genome-fragment | NA | NA | 0 |
| S17_extracted_k141_133930 | 6713 | No | NA | 6 | 0 | 0 | Not-determined | Genome-fragment | NA | NA | 0 |
| S5_vs_k141_106119\|\|lt2gene | 1006 | No | NA | 2 | 1 | 0 | Low-quality | Genome-fragment | 2.17 | AAI-based (medium-confidence) | 0 |
| S17_extracted_k141_22989 | 1709 | No | NA | 3 | 0 | 2 | Not-determined | Genome-fragment | NA | NA | 0 |
| S7_extracted_15759 | 1006 | No | NA | 1 | 0 | 0 | Low-quality | Genome-fragment | 2.48 | AAI-based (high-confidence) | 0 |
| S17_extracted_k141_119294 | 1922 | No | NA | 2 | 0 | 1 | Not-determined | Genome-fragment | NA | NA | 0 |
| S17_extracted_k141_119298 | 1119 | No | NA | 2 | 0 | 0 | Not-determined | Genome-fragment | NA | NA | 0 |
| S17_extracted_k141_97040 | 1776 | No | NA | 1 | 0 | 1 | Not-determined | Genome-fragment | NA | NA | 0 |
| S7_vs_73256\|\|full | 1006 | No | NA | 2 | 0 | 0 | Low-quality | Genome-fragment | 2.27 | AAI-based (high-confidence) | 0 |
| S17_extracted_k141_156416 | 3077 | No | NA | 2 | 0 | 1 | Not-determined | Genome-fragment | NA | NA | 0 |
| S17_extracted_k141_149089 | 1071 | No | NA | 2 | 0 | 2 | Not-determined | Genome-fragment | NA | NA | 0 |
| S7_vs_92999\|\|full | 1006 | No | NA | 3 | 0 | 0 | Low-quality | Genome-fragment | 2.17 | AAI-based (high-confidence) | 0 |
| S17_extracted_k141_8558 | 1989 | No | NA | 4 | 0 | 2 | Not-determined | Genome-fragment | NA | NA | 0 |
| S17_extracted_k141_81980 | 1155 | No | NA | 2 | 0 | 0 | Not-determined | Genome-fragment | NA | NA | 0 |
| S17_extracted_k141_81983 | 1138 | No | NA | 1 | 0 | 1 | Not-determined | Genome-fragment | NA | NA | 0 |
| S17_extracted_k141_171112 | 1249 | No | NA | 1 | 0 | 0 | Not-determined | Genome-fragment | NA | NA | 0 |
| S17_extracted_k141_119329 | 3081 | No | NA | 4 | 0 | 4 | Not-determined | Genome-fragment | NA | NA | 0 |
| S17_extracted_k141_67505 | 1015 | No | NA | 1 | 0 | 1 | Not-determined | Genome-fragment | NA | NA | 0 |
| S17_extracted_k141_81998 | 1131 | No | NA | 1 | 0 | 1 | Not-determined | Genome-fragment | NA | NA | 0 |
| S17_extracted_k141_38337 | 1755 | No | NA | 1 | 0 | 1 | Not-determined | Genome-fragment | NA | NA | 0 |
| S17_extracted_k141_156478 | 1850 | No | NA | 2 | 0 | 0 | Not-determined | Genome-fragment | NA | NA | 0 |
| S17_extracted_k141_163676 | 1491 | No | NA | 1 | 0 | 1 | Not-determined | Genome-fragment | NA | NA | 0 |
| S17_extracted_k141_8633 | 1273 | No | NA | 1 | 0 | 0 | Not-determined | Genome-fragment | NA | NA | 0 |
| S17_extracted_k141_104907 | 1049 | No | NA | 1 | 0 | 1 | Not-determined | Genome-fragment | NA | NA | 0 |
| S17_extracted_k141_104909 | 1120 | No | NA | 1 | 0 | 1 | Not-determined | Genome-fragment | NA | NA | 0 |
| S17_extracted_k141_60285 | 3748 | No | NA | 4 | 0 | 3 | Not-determined | Genome-fragment | NA | NA | 0 |
| S17_extracted_k141_149148 | 2401 | No | NA | 2 | 0 | 2 | Not-determined | Genome-fragment | NA | NA | 0 |
| S17_extracted_k141_141531 | 2781 | No | NA | 3 | 0 | 2 | Not-determined | Genome-fragment | NA | NA | 0 |
| S17_extracted_k141_23081 | 1380 | No | NA | 1 | 0 | 1 | Not-determined | Genome-fragment | NA | NA | 0 |
| S17_extracted_k141_149160 | 4262 | No | NA | 2 | 0 | 1 | Not-determined | Genome-fragment | NA | NA | 0 |
| S17_extracted_k141_82062 | 6098 | No | NA | 2 | 0 | 1 | Not-determined | Genome-fragment | NA | NA | 0 |
| S17_extracted_k141_141554 | 2085 | No | NA | 3 | 0 | 2 | Not-determined | Genome-fragment | NA | NA | 0 |
| S17_extracted_k141_38390 | 1513 | No | NA | 1 | 0 | 1 | Not-determined | Genome-fragment | NA | NA | 0 |
| S17_extracted_k141_149184 | 6097 | No | NA | 5 | 0 | 4 | Not-determined | Genome-fragment | NA | NA | 0 |
| S17_extracted_k141_53083 | 1464 | No | NA | 3 | 0 | 0 | Not-determined | Genome-fragment | NA | NA | 0 |
| S7_vs_31062\|\|lt2gene | 1006 | No | NA | 2 | 1 | 0 | Low-quality | Genome-fragment | 2.08 | AAI-based (medium-confidence) | 0 |
| S17_extracted_k141_119398 | 1675 | No | NA | 1 | 0 | 0 | Not-determined | Genome-fragment | NA | NA | 0 |
| S17_extracted_k141_60313 | 4923 | No | NA | 4 | 0 | 2 | Not-determined | Genome-fragment | NA | NA | 0 |
| S17_extracted_k141_156536 | 5414 | No | NA | 5 | 0 | 3 | Not-determined | Genome-fragment | NA | NA | 0 |
| S17_extracted_k141_141611 | 1433 | No | NA | 2 | 0 | 0 | Not-determined | Genome-fragment | NA | NA | 0 |
| S17_extracted_k141_111917 | 1460 | No | NA | 1 | 0 | 0 | Not-determined | Genome-fragment | NA | NA | 0 |
| S17_extracted_k141_104992 | 6446 | No | NA | 13 | 0 | 13 | Not-determined | Genome-fragment | NA | NA | 0 |
| S16_vs_70296\|\|full | 1006 | No | NA | 3 | 1 | 0 | Low-quality | Genome-fragment | 0.65 | HMM-based (lower-bound) | 0 |
| S17_extracted_k141_97187 | 1090 | No | NA | 2 | 0 | 2 | Not-determined | Genome-fragment | NA | NA | 0 |
| S17_extracted_k141_97188 | 1222 | No | NA | 1 | 0 | 1 | Not-determined | Genome-fragment | NA | NA | 0 |
| S17_extracted_k141_111931 | 17015 | No | NA | 21 | 9 | 0 | Medium-quality | Genome-fragment | 78.9 | AAI-based (medium-confidence) | 0 |
| S17_extracted_k141_111932 | 1047 | No | NA | 1 | 0 | 0 | Not-determined | Genome-fragment | NA | NA | 0 |
| S17_extracted_k141_67633 | 1376 | No | NA | 1 | 0 | 0 | Not-determined | Genome-fragment | NA | NA | 0 |
| S17_extracted_k141_119454 | 1275 | No | NA | 2 | 0 | 2 | Not-determined | Genome-fragment | NA | NA | 0 |
| S17_extracted_k141_119459 | 1067 | No | NA | 1 | 0 | 0 | Not-determined | Genome-fragment | NA | NA | 0 |
| S17_extracted_k141_1171 | 3021 | No | NA | 3 | 0 | 3 | Not-determined | Genome-fragment | NA | NA | 0 |
| S17_extracted_k141_1173 | 5366 | No | NA | 4 | 0 | 4 | Not-determined | Genome-fragment | NA | NA | 0 |
| S12_extracted_k141_175897 | 1006 | No | NA | 2 | 0 | 0 | Low-quality | Genome-fragment | 2.24 | AAI-based (high-confidence) | 0 |
| S17_extracted_k141_119476 | 2243 | No | NA | 2 | 0 | 1 | Not-determined | Genome-fragment | NA | NA | 0 |
| S17_extracted_k141_15733 | 1091 | No | NA | 2 | 0 | 2 | Not-determined | Genome-fragment | NA | NA | 0 |
| S17_extracted_k141_23181 | 1186 | No | NA | 2 | 0 | 1 | Not-determined | Genome-fragment | NA | NA | 0 |

| | | | | | | | | | | | |
|---|---|---|---|---|---|---|---|---|---|---|---|
| S12_vs_k141_14186\|\|full | 1006 | No | NA | 2 | 0 | 0 | Low-quality | Genome-fragment | 1.65 | AAI-based (medium-confidence) | 0 |
| S17_extracted_k141_134107 | 1604 | No | NA | 2 | 0 | 1 | Not-determined | Genome-fragment | NA | NA | 0 |
| S17_extracted_k141_30960 | 7498 | No | NA | 5 | 0 | 3 | Not-determined | Genome-fragment | NA | NA | 0 |
| S17_extracted_k141_134131 | 1351 | No | NA | 1 | 0 | 0 | Not-determined | Genome-fragment | NA | NA | 0 |
| S17_extracted_k141_1208 | 1847 | No | NA | 1 | 0 | 1 | Not-determined | Genome-fragment | NA | NA | 0 |
| S17_extracted_k141_15750 | 2517 | No | NA | 3 | 0 | 3 | Not-determined | Genome-fragment | NA | NA | 0 |
| S17_extracted_k141_1209 | 1157 | No | NA | 2 | 0 | 1 | Not-determined | Genome-fragment | NA | NA | 0 |
| S4_extracted_15922 | 1006 | No | NA | 3 | 0 | 0 | Low-quality | Genome-fragment | 1.98 | AAI-based (medium-confidence) | 0 |
| S17_extracted_k141_134144 | 1486 | No | NA | 2 | 0 | 1 | Not-determined | Genome-fragment | NA | NA | 0 |
| S17_extracted_k141_1219 | 2097 | No | NA | 4 | 0 | 1 | Not-determined | Genome-fragment | NA | NA | 0 |
| S17_extracted_k141_134152 | 3206 | No | NA | 3 | 0 | 3 | Not-determined | Genome-fragment | NA | NA | 0 |
| S17_extracted_k141_53244 | 2677 | No | NA | 7 | 0 | 7 | Not-determined | Genome-fragment | NA | NA | 0 |
| S17_extracted_k141_67768 | 1228 | No | NA | 3 | 0 | 3 | Not-determined | Genome-fragment | NA | NA | 0 |
| S17_extracted_k141_149358 | 5104 | No | NA | 5 | 0 | 5 | Not-determined | Genome-fragment | NA | NA | 0 |
| S17_extracted_k141_82160 | 1578 | No | NA | 3 | 0 | 0 | Not-determined | Genome-fragment | NA | NA | 0 |
| S17_extracted_k141_97302 | 1877 | No | NA | 4 | 0 | 0 | Not-determined | Genome-fragment | NA | NA | 0 |
| S17_extracted_k141_8811 | 3030 | No | NA | 3 | 0 | 2 | Not-determined | Genome-fragment | NA | NA | 0 |
| S17_extracted_k141_97304 | 1690 | No | NA | 2 | 0 | 0 | Not-determined | Genome-fragment | NA | NA | 0 |
| S17_extracted_k141_45721 | 1391 | No | NA | 1 | 0 | 1 | Not-determined | Genome-fragment | NA | NA | 0 |
| S17_extracted_k141_97313 | 2426 | No | NA | 2 | 0 | 2 | Not-determined | Genome-fragment | NA | NA | 0 |
| S17_extracted_k141_171354 | 1664 | No | NA | 2 | 0 | 1 | Not-determined | Genome-fragment | NA | NA | 0 |
| S17_extracted_k141_31035 | 1170 | No | NA | 1 | 0 | 0 | Not-determined | Genome-fragment | NA | NA | 0 |
| S4_extracted_28750 | 1006 | No | NA | 3 | 0 | 1 | Low-quality | Genome-fragment | 1.24 | AAI-based (high-confidence) | 0 |
| S17_extracted_k141_67808 | 1040 | No | NA | 1 | 0 | 0 | Not-determined | Genome-fragment | NA | NA | 0 |
| S17_extracted_k141_67809 | 3656 | No | NA | 3 | 0 | 2 | Not-determined | Genome-fragment | NA | NA | 0 |
| S17_extracted_k141_45776 | 1110 | No | NA | 1 | 0 | 1 | Not-determined | Genome-fragment | NA | NA | 0 |
| S17_extracted_k141_105174 | 2119 | No | NA | 1 | 0 | 0 | Not-determined | Genome-fragment | NA | NA | 0 |
| S17_extracted_k141_134213 | 1121 | No | NA | 2 | 0 | 0 | Not-determined | Genome-fragment | NA | NA | 0 |
| S17_extracted_k141_23262 | 3488 | No | NA | 2 | 0 | 2 | Not-determined | Genome-fragment | NA | NA | 0 |
| S17_extracted_k141_1305 | 1223 | No | NA | 1 | 0 | 0 | Not-determined | Genome-fragment | NA | NA | 0 |
| S17_extracted_k141_38610 | 1128 | No | NA | 1 | 0 | 1 | Not-determined | Genome-fragment | NA | NA | 0 |
| S11_extracted_k141_13453 | 1007 | No | NA | 2 | 1 | 0 | Low-quality | Genome-fragment | 2.52 | AAI-based (medium-confidence) | 0 |
| S17_extracted_k141_82209 | 1816 | No | NA | 1 | 0 | 0 | Not-determined | Genome-fragment | NA | NA | 0 |
| S14_extracted_k141_64403 | 1007 | No | NA | 2 | 0 | 0 | Low-quality | Genome-fragment | 2.43 | AAI-based (high-confidence) | 0 |
| S17_extracted_k141_1326 | 4515 | No | NA | 3 | 0 | 3 | Not-determined | Genome-fragment | NA | NA | 0 |
| S14_vs_k141_113931\|\|full | 1007 | No | NA | 2 | 0 | 1 | Low-quality | Genome-fragment | 1.75 | AAI-based (medium-confidence) | 0 |
| S14_vs_k141_117104\|\|lt2gene | 1007 | No | NA | 2 | 0 | 0 | Low-quality | Genome-fragment | 3.75 | AAI-based (high-confidence) | 0 |
| S17_extracted_k141_156690 | 1215 | No | NA | 2 | 0 | 0 | Not-determined | Genome-fragment | NA | NA | 0 |
| S1_vs_k141_93075\|\|full | 1007 | No | NA | 2 | 1 | 0 | Low-quality | Genome-fragment | 1.7 | AAI-based (high-confidence) | 0 |
| S3_vs_k141_8400\|\|full | 1007 | No | NA | 2 | 0 | 1 | Low-quality | Genome-fragment | 1.75 | AAI-based (medium-confidence) | 0 |
| S17_extracted_k141_90048 | 1668 | No | NA | 1 | 0 | 1 | Not-determined | Genome-fragment | NA | NA | 0 |
| S17_extracted_k141_67842 | 1020 | No | NA | 2 | 0 | 1 | Not-determined | Genome-fragment | NA | NA | 0 |
| S17_extracted_k141_31113 | 1257 | No | NA | 3 | 0 | 2 | Not-determined | Genome-fragment | NA | NA | 0 |
| S17_extracted_k141_31130 | 1031 | No | NA | 1 | 0 | 0 | Not-determined | Genome-fragment | NA | NA | 0 |
| S17_extracted_k141_45870 | 1058 | No | NA | 2 | 0 | 1 | Not-determined | Genome-fragment | NA | NA | 0 |
| S17_extracted_k141_23322 | 5416 | No | NA | 3 | 0 | 0 | Not-determined | Genome-fragment | NA | NA | 0 |
| S17_extracted_k141_112062 | 9822 | No | NA | 5 | 0 | 4 | Not-determined | Genome-fragment | NA | NA | 0 |
| S17_extracted_k141_141820 | 3748 | No | NA | 2 | 0 | 2 | Not-determined | Genome-fragment | NA | NA | 0 |
| S5_vs_k141_40275\|\|lt2gene | 1007 | No | NA | 2 | 1 | 0 | Low-quality | Genome-fragment | 2.15 | AAI-based (medium-confidence) | 0 |
| S17_extracted_k141_163905 | 1490 | No | NA | 2 | 0 | 1 | Not-determined | Genome-fragment | NA | NA | 0 |
| S17_extracted_k141_38722 | 1330 | No | NA | 1 | 0 | 0 | Not-determined | Genome-fragment | NA | NA | 0 |
| S17_extracted_k141_97482 | 4501 | No | NA | 5 | 0 | 4 | Not-determined | Genome-fragment | NA | NA | 0 |
| S17_extracted_k141_53437 | 1650 | No | NA | 2 | 0 | 2 | Not-determined | Genome-fragment | NA | NA | 0 |
| S17_extracted_k141_156749 | 1027 | No | NA | 1 | 0 | 1 | Not-determined | Genome-fragment | NA | NA | 0 |
| S17_extracted_k141_60570 | 1201 | No | NA | 2 | 0 | 1 | Not-determined | Genome-fragment | NA | NA | 0 |
| S7_extracted_39146 | 1007 | No | NA | 1 | 1 | 0 | Low-quality | Genome-fragment | 17.19 | AAI-based (high-confidence) | 0 |
| S17_extracted_k141_45897 | 1308 | No | NA | 2 | 0 | 2 | Not-determined | Genome-fragment | NA | NA | 0 |
| S17_extracted_k141_97497 | 3585 | No | NA | 3 | 0 | 2 | Not-determined | Genome-fragment | NA | NA | 0 |
| S17_extracted_k141_149552 | 1263 | No | NA | 1 | 0 | 1 | Not-determined | Genome-fragment | NA | NA | 0 |
| S7_vs_83089\|\|full | 1007 | No | NA | 2 | 1 | 0 | Low-quality | Genome-fragment | 2.64 | AAI-based (medium-confidence) | 0 |
| S16_vs_41486\|\|full | 1007 | No | NA | 3 | 0 | 0 | Low-quality | Genome-fragment | 2.75 | AAI-based (medium-confidence) | 0 |
| S17_extracted_k141_67882 | 1492 | No | NA | 1 | 0 | 0 | Not-determined | Genome-fragment | NA | NA | 0 |
| S17_extracted_k141_163947 | 1888 | No | NA | 1 | 0 | 0 | Not-determined | Genome-fragment | NA | NA | 0 |
| S17_extracted_k141_75269 | 2816 | No | NA | 4 | 0 | 0 | Not-determined | Genome-fragment | NA | NA | 0 |
| S17_extracted_k141_171533 | 1139 | No | NA | 4 | 0 | 4 | Not-determined | Genome-fragment | NA | NA | 0 |
| S17_extracted_k141_9035 | 6973 | No | NA | 6 | 0 | 4 | Not-determined | Genome-fragment | NA | NA | 0 |
| S16_vs_101805\|\|full | 1007 | No | NA | 4 | 1 | 0 | Low-quality | Genome-fragment | 1.77 | HMM-based (lower-bound) | 0 |
| S10_extracted_13937 | 1007 | No | NA | 2 | 0 | 1 | Low-quality | Genome-fragment | 2.24 | AAI-based (medium-confidence) | 0 |
| S12_vs_k141_71360\|\|full | 1007 | No | NA | 3 | 1 | 0 | Low-quality | Genome-fragment | 2.12 | HMM-based (lower-bound) | 0 |
| S12_vs_k141_7266\|\|full | 1007 | No | NA | 3 | 0 | 0 | Low-quality | Genome-fragment | 1.58 | AAI-based (medium-confidence) | 0 |
| S12_vs_k141_98405\|\|lt2gene | 1007 | No | NA | 2 | 2 | 0 | Low-quality | Genome-fragment | 2.72 | AAI-based (high-confidence) | 0 |
| S17_extracted_k141_1436 | 1998 | No | NA | 4 | 0 | 4 | Not-determined | Genome-fragment | NA | NA | 0 |
| S4_extracted_30862 | 1007 | No | NA | 1 | 1 | 0 | Low-quality | Genome-fragment | 3.45 | HMM-based (lower-bound) | 0 |
| S17_extracted_k141_119722 | 1020 | No | NA | 2 | 0 | 1 | Not-determined | Genome-fragment | NA | NA | 0 |
| S17_extracted_k141_88673 | 1008 | No | NA | 1 | 0 | 0 | Low-quality | Genome-fragment | 1.13 | AAI-based (medium-confidence) | 0 |
| S17_extracted_k141_90134 | 1431 | No | NA | 4 | 0 | 0 | Not-determined | Genome-fragment | NA | NA | 0 |
| S9_vs_k141_22028\|\|full | 1008 | No | NA | 4 | 1 | 0 | Low-quality | Genome-fragment | 1.7 | HMM-based (lower-bound) | 0 |
| S17_extracted_k141_156859 | 2120 | No | NA | 2 | 0 | 2 | Not-determined | Genome-fragment | NA | NA | 0 |
| S17_extracted_k141_97592 | 2255 | No | NA | 2 | 0 | 0 | Not-determined | Genome-fragment | NA | NA | 0 |
| S17_extracted_k141_105339 | 3885 | No | NA | 4 | 0 | 4 | Not-determined | Genome-fragment | NA | NA | 0 |
| S13_extracted_96258 | 1008 | No | NA | 4 | 0 | 0 | Low-quality | Genome-fragment | 1.65 | AAI-based (high-confidence) | 0 |
| S17_extracted_k141_90152 | 6420 | No | NA | 4 | 0 | 3 | Not-determined | Genome-fragment | NA | NA | 0 |
| S17_extracted_k141_23427 | 3217 | No | NA | 1 | 0 | 0 | Not-determined | Genome-fragment | NA | NA | 0 |
| S1_extracted_k141_85332 | 1008 | No | NA | 3 | 0 | 0 | Low-quality | Genome-fragment | 2.17 | AAI-based (high-confidence) | 0 |
| S1_vs_k141_45610\|\|full | 1008 | No | NA | 3 | 2 | 0 | Low-quality | Genome-fragment | 3.67 | HMM-based (lower-bound) | 0 |
| S17_extracted_k141_126712 | 3710 | No | NA | 1 | 0 | 1 | Not-determined | Genome-fragment | NA | NA | 0 |
| S17_extracted_k141_23438 | 2315 | No | NA | 3 | 0 | 1 | Not-determined | Genome-fragment | NA | NA | 0 |
| S3_vs_k141_22444\|\|lt2gene | 1008 | No | NA | 2 | 2 | 0 | Low-quality | Genome-fragment | 3.52 | AAI-based (high-confidence) | 0 |
| S17_extracted_k141_119783 | 1005 | No | NA | 1 | 0 | 0 | Not-determined | Genome-fragment | NA | NA | 0 |
| S5_extracted_k141_140708 | 1008 | No | NA | 3 | 0 | 0 | Low-quality | Genome-fragment | 2.69 | AAI-based (high-confidence) | 0 |
| S17_extracted_k141_38813 | 3447 | No | NA | 4 | 0 | 3 | Not-determined | Genome-fragment | NA | NA | 0 |
| S17_extracted_k141_60727 | 1024 | No | NA | 1 | 0 | 1 | Not-determined | Genome-fragment | NA | NA | 0 |
| S7_extracted_3121 | 1008 | No | NA | 3 | 1 | 0 | Low-quality | Genome-fragment | 1.92 | AAI-based (medium-confidence) | 0 |
| S17_extracted_k141_68016 | 1886 | No | NA | 3 | 0 | 2 | Not-determined | Genome-fragment | NA | NA | 0 |
| S17_extracted_k141_164110 | 1329 | No | NA | 1 | 0 | 1 | Not-determined | Genome-fragment | NA | NA | 0 |
| S17_extracted_k141_97671 | 19897 | No | NA | 27 | 8 | 0 | Medium-quality | Genome-fragment | 55.59 | AAI-based (medium-confidence) | 0 |
| S17_extracted_k141_164117 | 1197 | No | NA | 1 | 0 | 1 | Not-determined | Genome-fragment | NA | NA | 0 |
| S7_extracted_133101 | 1008 | No | NA | 2 | 2 | 0 | Low-quality | Genome-fragment | 2.18 | AAI-based (medium-confidence) | 0 |
| S7_vs_73002\|\|full | 1008 | No | NA | 2 | 1 | 0 | Low-quality | Genome-fragment | 0.68 | HMM-based (lower-bound) | 0 |
| S17_extracted_k141_149631 | 3122 | No | NA | 2 | 0 | 1 | Not-determined | Genome-fragment | NA | NA | 0 |
| S17_extracted_k141_46061 | 2492 | No | NA | 2 | 0 | 1 | Not-determined | Genome-fragment | NA | NA | 0 |
| S17_extracted_k141_82450 | 1260 | No | NA | 1 | 0 | 0 | Not-determined | Genome-fragment | NA | NA | 0 |
| S17_extracted_k141_31318 | 1092 | No | NA | 2 | 0 | 2 | Not-determined | Genome-fragment | NA | NA | 0 |
| S17_extracted_k141_112209 | 1297 | No | NA | 2 | 0 | 0 | Not-determined | Genome-fragment | NA | NA | 0 |
| S17_extracted_k141_15988 | 1082 | No | NA | 1 | 0 | 1 | Not-determined | Genome-fragment | NA | NA | 0 |
| S7_vs_35481\|\|lt2gene | 1008 | No | NA | 1 | 1 | 0 | Low-quality | Genome-fragment | 2.23 | AAI-based (medium-confidence) | 0 |
| S17_extracted_k141_126784 | 3162 | No | NA | 2 | 0 | 1 | Not-determined | Genome-fragment | NA | NA | 0 |
| S16_extracted_125396 | 1008 | No | NA | 1 | 0 | 0 | Low-quality | Genome-fragment | 1.84 | AAI-based (high-confidence) | 0 |
| S16_vs_106372\|\|full | 1008 | No | NA | 3 | 0 | 2 | Low-quality | Genome-fragment | 1.48 | AAI-based (high-confidence) | 0 |
| S10_extracted_105084 | 1008 | No | NA | 1 | 1 | 0 | Low-quality | Genome-fragment | 0.75 | HMM-based (lower-bound) | 0 |
| S10_vs_50884\|\|full | 1008 | No | NA | 3 | 1 | 0 | Low-quality | Genome-fragment | 2.36 | AAI-based (high-confidence) | 0 |
| S17_extracted_k141_112260 | 7608 | No | NA | 13 | 0 | 3 | Not-determined | Genome-fragment | NA | NA | 0 |
| S17_extracted_k141_112262 | 1617 | No | NA | 2 | 0 | 0 | Not-determined | Genome-fragment | NA | NA | 0 |
| S17_extracted_k141_112263 | 1416 | No | NA | 2 | 0 | 0 | Not-determined | Genome-fragment | NA | NA | 0 |
| S17_extracted_k141_46168 | 1375 | No | NA | 1 | 0 | 1 | Not-determined | Genome-fragment | NA | NA | 0 |
| S12_vs_k141_6004\|\|full | 1008 | No | NA | 2 | 0 | 0 | Low-quality | Genome-fragment | 2.49 | AAI-based (high-confidence) | 0 |
| S17_extracted_k141_119879 | 1140 | No | NA | 1 | 0 | 0 | Not-determined | Genome-fragment | NA | NA | 0 |
| S12_vs_k141_45981\|\|full | 1008 | No | NA | 2 | 1 | 0 | Low-quality | Genome-fragment | 1.49 | HMM-based (lower-bound) | 0 |
| S6_extracted_k141_10626 | 1008 | No | NA | 1 | 0 | 0 | Low-quality | Genome-fragment | 2.94 | AAI-based (medium-confidence) | 0 |
| S17_extracted_k141_31376 | 3477 | No | NA | 3 | 0 | 1 | Not-determined | Genome-fragment | NA | NA | 0 |
| S17_extracted_k141_16048 | 1130 | No | NA | 1 | 0 | 0 | Not-determined | Genome-fragment | NA | NA | 0 |
| S17_extracted_k141_46200 | 1300 | No | NA | 1 | 0 | 1 | Not-determined | Genome-fragment | NA | NA | 0 |
| S8_vs_k141_46080\|\|full | 1008 | No | NA | 3 | 3 | 0 | Low-quality | Genome-fragment | 3.28 | AAI-based (high-confidence) | 0 |
| S8_vs_k141_48494\|\|full | 1008 | No | NA | 3 | 0 | 0 | Low-quality | Genome-fragment | 2.5 | AAI-based (high-confidence) | 0 |
| S17_extracted_k141_173456 | 1009 | No | NA | 3 | 1 | 0 | Low-quality | Genome-fragment | 1.84 | AAI-based (high-confidence) | 0 |
| S17_extracted_k141_23561 | 10746 | No | NA | 10 | 0 | 1 | Not-determined | Genome-fragment | NA | NA | 0 |
| S17_extracted_k141_134536 | 1381 | No | NA | 2 | 0 | 2 | Not-determined | Genome-fragment | NA | NA | 0 |
| S1_vs_k141_65959\|\|full | 1009 | No | NA | 3 | 0 | 0 | Low-quality | Genome-fragment | 1.12 | AAI-based (medium-confidence) | 0 |
| S17_extracted_k141_90199 | 4659 | No | NA | 4 | 0 | 4 | Not-determined | Genome-fragment | NA | NA | 0 |
| S3_vs_k141_115291\|\|lt2gene | 1009 | No | NA | 2 | 1 | 0 | Low-quality | Genome-fragment | 0.83 | HMM-based (lower-bound) | 0 |
| S17_extracted_k141_75386 | 1714 | No | NA | 2 | 0 | 2 | Not-determined | Genome-fragment | NA | NA | 0 |
| S17_extracted_k141_31400 | 1138 | No | NA | 2 | 0 | 2 | Not-determined | Genome-fragment | NA | NA | 0 |
| S17_extracted_k141_126911 | 4870 | No | NA | 11 | 0 | 0 | Not-determined | Genome-fragment | NA | NA | 0 |
| S5_vs_k141_54724\|\|full | 1009 | No | NA | 2 | 2 | 0 | Low-quality | Genome-fragment | 3.34 | AAI-based (medium-confidence) | 0 |
| S7_extracted_92265 | 1009 | No | NA | 1 | 1 | 0 | Low-quality | Genome-fragment | 3.42 | HMM-based (lower-bound) | 0 |
| S17_extracted_k141_53775 | 1687 | No | NA | 1 | 0 | 1 | Not-determined | Genome-fragment | NA | NA | 0 |
| S12_vs_k141_28258\|\|full | 1009 | No | NA | 2 | 1 | 0 | Low-quality | Genome-fragment | 1.47 | AAI-based (medium-confidence) | 0 |
| S17_extracted_k141_126939 | 7777 | No | NA | 5 | 0 | 2 | Not-determined | Genome-fragment | NA | NA | 0 |
| S17_extracted_k141_75428 | 3639 | No | NA | 2 | 0 | 1 | Not-determined | Genome-fragment | NA | NA | 0 |
| S17_extracted_k141_68163 | 2015 | No | NA | 1 | 0 | 0 | Not-determined | Genome-fragment | NA | NA | 0 |
| S17_extracted_k141_9292 | 4260 | No | NA | 1 | 0 | 1 | Not-determined | Genome-fragment | NA | NA | 0 |
| S17_extracted_k141_164171 | 2581 | No | NA | 2 | 0 | 1 | Not-determined | Genome-fragment | NA | NA | 0 |
| S17_extracted_k141_97860 | 1118 | No | NA | 1 | 0 | 0 | Not-determined | Genome-fragment | NA | NA | 0 |
| S12_vs_k141_100528\|\|full | 1009 | No | NA | 3 | 1 | 0 | Low-quality | Genome-fragment | 2.99 | AAI-based (high-confidence) | 0 |
| S15_vs_3058\|\|lt2gene | 1009 | No | NA | 1 | 1 | 0 | Low-quality | Genome-fragment | 2.35 | AAI-based (high-confidence) | 0 |
| S17_extracted_k141_68184 | 2137 | No | NA | 2 | 0 | 0 | Not-determined | Genome-fragment | NA | NA | 0 |
| S17_extracted_k141_126961 | 4475 | No | NA | 8 | 0 | 3 | Not-determined | Genome-fragment | NA | NA | 0 |
| S4_extracted_3078 | 1009 | No | NA | 2 | 1 | 0 | Low-quality | Genome-fragment | 1.87 | HMM-based (lower-bound) | 0 |
| S4_extracted_3732 | 1009 | No | NA | 2 | 1 | 0 | Low-quality | Genome-fragment | 2.46 | AAI-based (high-confidence) | 0 |
| S17_extracted_k141_1715 | 1771 | No | NA | 2 | 0 | 1 | Not-determined | Genome-fragment | NA | NA | 0 |
| S17_extracted_k141_157123 | 1385 | No | NA | 2 | 0 | 1 | Not-determined | Genome-fragment | NA | NA | 0 |
| S17_extracted_k141_1718 | 3785 | No | NA | 3 | 0 | 1 | Not-determined | Genome-fragment | NA | NA | 0 |
| S17_extracted_k141_16139 | 1199 | No | NA | 1 | 0 | 0 | Not-determined | Genome-fragment | NA | NA | 0 |
| S17_extracted_k141_1724 | 1026 | No | NA | 1 | 0 | 1 | Not-determined | Genome-fragment | NA | NA | 0 |
| S17_extracted_k141_75438 | 2354 | No | NA | 2 | 0 | 2 | Not-determined | Genome-fragment | NA | NA | 0 |
| S17_extracted_k141_105667 | 1420 | No | NA | 3 | 0 | 2 | Not-determined | Genome-fragment | NA | NA | 0 |
| S17_extracted_k141_46347 | 1603 | No | NA | 2 | 0 | 1 | Not-determined | Genome-fragment | NA | NA | 0 |
| S17_extracted_k141_16148 | 1008 | No | NA | 1 | 0 | 1 | Not-determined | Genome-fragment | NA | NA | 0 |
| S6_extracted_k141_14830 | 1009 | No | NA | 3 | 1 | 0 | Low-quality | Genome-fragment | 2.21 | AAI-based (high-confidence) | 0 |
| S17_extracted_k141_23738 | 4355 | No | NA | 3 | 0 | 0 | Not-determined | Genome-fragment | NA | NA | 0 |
| S17_extracted_k141_134648 | 1280 | No | NA | 1 | 0 | 0 | Not-determined | Genome-fragment | NA | NA | 0 |
| S6_extracted_k141_7947 | 1009 | No | NA | 1 | 0 | 0 | Low-quality | Genome-fragment | 0.76 | AAI-based (high-confidence) | 0 |
| S17_extracted_k141_6613 | 1010 | No | NA | 1 | 0 | 1 | Low-quality | Genome-fragment | 0.56 | AAI-based (medium-confidence) | 0 |
| S17_extracted_k141_171571 | 1616 | No | NA | 4 | 0 | 4 | Not-determined | Genome-fragment | NA | NA | 0 |
| S17_extracted_k141_127008 | 1838 | No | NA | 1 | 0 | 1 | Not-determined | Genome-fragment | NA | NA | 0 |
| S17_extracted_k141_65150 | 1010 | No | NA | 2 | 0 | 0 | Low-quality | Genome-fragment | 0.58 | AAI-based (medium-confidence) | 0 |
| S17_extracted_k141_53877 | 1017 | No | NA | 2 | 0 | 0 | Not-determined | Genome-fragment | NA | NA | 0 |
| S17_vs_k141_95253\|\|lt2gene | 1010 | No | NA | 1 | 0 | 0 | Low-quality | Genome-fragment | 2.1 | AAI-based (high-confidence) | 0 |
| S17_extracted_k141_171579 | 5947 | No | NA | 6 | 0 | 5 | Not-determined | Genome-fragment | NA | NA | 0 |
| S17_extracted_k141_164225 | 1480 | No | NA | 1 | 0 | 1 | Not-determined | Genome-fragment | NA | NA | 0 |
| S17_extracted_k141_31519 | 1924 | No | NA | 2 | 0 | 2 | Not-determined | Genome-fragment | NA | NA | 0 |
| S11_vs_k141_1814\|\|lt2gene | 1010 | No | NA | 1 | 1 | 0 | Low-quality | Genome-fragment | 2.36 | AAI-based (medium-confidence) | 0 |
| S17_extracted_k141_142297 | 2583 | No | NA | 2 | 0 | 1 | Not-determined | Genome-fragment | NA | NA | 0 |
| S17_extracted_k141_46388 | 23386 | No | NA | 38 | 5 | 0 | Medium-quality | Genome-fragment | 52.03 | AAI-based (high-confidence) | 0 |
| S13_extracted_101293 | 1010 | No | NA | 2 | 0 | 1 | Low-quality | Genome-fragment | 1.46 | AAI-based (medium-confidence) | 0 |
| S17_extracted_k141_157161 | 7596 | No | NA | 5 | 0 | 4 | Not-determined | Genome-fragment | NA | NA | 0 |
| S14_extracted_k141_26471 | 1010 | No | NA | 3 | 3 | 0 | Low-quality | Genome-fragment | 21.17 | AAI-based (high-confidence) | 0 |
| S17_extracted_k141_149941 | 3463 | No | NA | 3 | 0 | 2 | Not-determined | Genome-fragment | NA | NA | 0 |
| S17_extracted_k141_39078 | 1655 | No | NA | 1 | 0 | 1 | Not-determined | Genome-fragment | NA | NA | 0 |
| S17_extracted_k141_134712 | 2690 | No | NA | 2 | 0 | 1 | Not-determined | Genome-fragment | NA | NA | 0 |
| S3_vs_k141_104545\|\|full | 1010 | No | NA | 4 | 0 | 0 | Low-quality | Genome-fragment | 2.39 | AAI-based (high-confidence) | 0 |
| S17_extracted_k141_134726 | 1084 | No | NA | 1 | 0 | 0 | Not-determined | Genome-fragment | NA | NA | 0 |
| S5_extracted_k141_115640 | 1010 | No | NA | 3 | 0 | 0 | Low-quality | Genome-fragment | 1.85 | AAI-based (high-confidence) | 0 |
| S17_extracted_k141_60854 | 1385 | No | NA | 1 | 0 | 0 | Not-determined | Genome-fragment | NA | NA | 0 |
| S17_extracted_k141_82734 | 3208 | No | NA | 3 | 0 | 1 | Not-determined | Genome-fragment | NA | NA | 0 |
| S17_extracted_k141_134742 | 8096 | No | NA | 6 | 0 | 6 | Not-determined | Genome-fragment | NA | NA | 0 |
| S17_extracted_k141_46426 | 1297 | No | NA | 1 | 0 | 0 | Not-determined | Genome-fragment | NA | NA | 0 |
| S17_extracted_k141_134749 | 1792 | No | NA | 2 | 0 | 1 | Not-determined | Genome-fragment | NA | NA | 0 |
| S17_extracted_k141_127121 | 1176 | No | NA | 2 | 0 | 2 | Not-determined | Genome-fragment | NA | NA | 0 |
| S17_extracted_k141_60896 | 1049 | No | NA | 2 | 0 | 1 | Not-determined | Genome-fragment | NA | NA | 0 |
| S17_extracted_k141_150016 | 1186 | No | NA | 3 | 0 | 0 | Not-determined | Genome-fragment | NA | NA | 0 |
| S17_extracted_k141_16303 | 2220 | No | NA | 1 | 0 | 1 | Not-determined | Genome-fragment | NA | NA | 0 |
| S17_extracted_k141_68321 | 2300 | No | NA | 1 | 0 | 0 | Not-determined | Genome-fragment | NA | NA | 0 |
| S17_extracted_k141_90329 | 1021 | No | NA | 2 | 0 | 1 | Not-determined | Genome-fragment | NA | NA | 0 |
| S17_extracted_k141_98059 | 1466 | No | NA | 1 | 0 | 0 | Not-determined | Genome-fragment | NA | NA | 0 |
| S17_extracted_k141_75542 | 3532 | No | NA | 2 | 0 | 2 | Not-determined | Genome-fragment | NA | NA | 0 |
| S17_extracted_k141_98071 | 1190 | No | NA | 2 | 0 | 1 | Not-determined | Genome-fragment | NA | NA | 0 |
| S17_extracted_k141_16327 | 1160 | No | NA | 1 | 0 | 1 | Not-determined | Genome-fragment | NA | NA | 0 |
| S17_extracted_k141_134801 | 1171 | No | NA | 1 | 0 | 0 | Not-determined | Genome-fragment | NA | NA | 0 |
| S17_extracted_k141_112498 | 1078 | No | NA | 1 | 0 | 1 | Not-determined | Genome-fragment | NA | NA | 0 |
| S5_extracted_k141_21571 | 1010 | No | NA | 3 | 0 | 0 | Low-quality | Genome-fragment | 2.14 | AAI-based (medium-confidence) | 0 |
| S17_extracted_k141_157227 | 1099 | No | NA | 1 | 0 | 0 | Not-determined | Genome-fragment | NA | NA | 0 |
| S17_extracted_k141_142431 | 2644 | No | NA | 3 | 0 | 3 | Not-determined | Genome-fragment | NA | NA | 0 |
| S7_extracted_86679 | 1010 | No | NA | 2 | 1 | 0 | Low-quality | Genome-fragment | 2.33 | AAI-based (high-confidence) | 0 |
| S7_vs_65712\|\|full | 1010 | No | NA | 3 | 0 | 0 | Low-quality | Genome-fragment | 2.53 | AAI-based (medium-confidence) | 0 |
| S17_extracted_k141_150078 | 3466 | No | NA | 2 | 0 | 2 | Not-determined | Genome-fragment | NA | NA | 0 |
| S17_extracted_k141_39241 | 1044 | No | NA | 2 | 0 | 1 | Not-determined | Genome-fragment | NA | NA | 0 |
| S7_vs_130232\|\|full | 1010 | No | NA | 3 | 1 | 0 | Low-quality | Genome-fragment | 2.74 | AAI-based (high-confidence) | 0 |
| S17_extracted_k141_134855 | 1869 | No | NA | 1 | 0 | 0 | Not-determined | Genome-fragment | NA | NA | 0 |
| S17_extracted_k141_164399 | 1632 | No | NA | 1 | 0 | 1 | Not-determined | Genome-fragment | NA | NA | 0 |
| S17_extracted_k141_157254 | 1248 | No | NA | 1 | 0 | 1 | Not-determined | Genome-fragment | NA | NA | 0 |
| S17_extracted_k141_31661 | 16460 | No | NA | 11 | 0 | 8 | Not-determined | Genome-fragment | NA | NA | 0 |
| S17_extracted_k141_171729 | 1056 | No | NA | 3 | 0 | 0 | Not-determined | Genome-fragment | NA | NA | 0 |
| S17_extracted_k141_98150 | 1429 | No | NA | 3 | 0 | 1 | Not-determined | Genome-fragment | NA | NA | 0 |
| S17_extracted_k141_98151 | 1051 | No | NA | 1 | 0 | 1 | Not-determined | Genome-fragment | NA | NA | 0 |
| S17_extracted_k141_23923 | 2619 | No | NA | 2 | 0 | 1 | Not-determined | Genome-fragment | NA | NA | 0 |
| S12_extracted_185231 | 1010 | No | NA | 2 | 0 | 0 | Low-quality | Genome-fragment | 1.08 | AAI-based (high-confidence) | 0 |
| S17_extracted_k141_68464 | 1626 | No | NA | 2 | 0 | 1 | Not-determined | Genome-fragment | NA | NA | 0 |
| S17_extracted_k141_164458 | 1129 | No | NA | 1 | 0 | 1 | Not-determined | Genome-fragment | NA | NA | 0 |
| S17_extracted_k141_68471 | 1287 | No | NA | 1 | 0 | 1 | Not-determined | Genome-fragment | NA | NA | 0 |
| S17_extracted_k141_61003 | 6593 | No | NA | 8 | 0 | 1 | Not-determined | Genome-fragment | NA | NA | 0 |
| S17_extracted_k141_23943 | 1300 | No | NA | 2 | 0 | 0 | Not-determined | Genome-fragment | NA | NA | 0 |
| S8_extracted_k141_31236 | 1010 | No | NA | 1 | 1 | 0 | Low-quality | Genome-fragment | 1.56 | HMM-based (lower-bound) | 0 |
| S17_extracted_k141_134904 | 2489 | No | NA | 2 | 0 | 2 | Not-determined | Genome-fragment | NA | NA | 0 |
| S17_extracted_k141_46520 | 1035 | No | NA | 2 | 0 | 2 | Not-determined | Genome-fragment | NA | NA | 0 |
| S8_extracted_k141_169864 | 1010 | No | NA | 2 | 1 | 0 | Low-quality | Genome-fragment | 0.68 | HMM-based (lower-bound) | 0 |
| S17_extracted_k141_16384 | 3560 | No | NA | 2 | 0 | 2 | Not-determined | Genome-fragment | NA | NA | 0 |
| S17_extracted_k141_150177 | 5880 | No | NA | 4 | 0 | 3 | Not-determined | Genome-fragment | NA | NA | 0 |
| S17_extracted_k141_39338 | 1153 | No | NA | 2 | 0 | 1 | Not-determined | Genome-fragment | NA | NA | 0 |
| S17_extracted_k141_68498 | 3540 | No | NA | 4 | 0 | 3 | Not-determined | Genome-fragment | NA | NA | 0 |
| S17_extracted_k141_1963 | 1253 | No | NA | 3 | 0 | 2 | Not-determined | Genome-fragment | NA | NA | 0 |
| S17_extracted_k141_1976 | 1000 | No | NA | 1 | 0 | 1 | Not-determined | Genome-fragment | NA | NA | 0 |
| S17_extracted_k141_75666 | 6339 | No | NA | 3 | 0 | 2 | Not-determined | Genome-fragment | NA | NA | 0 |
| S17_extracted_k141_25648 | 1011 | No | NA | 1 | 1 | 0 | Low-quality | Genome-fragment | 1.77 | HMM-based (lower-bound) | 0 |
| S17_extracted_k141_82878 | 2389 | No | NA | 2 | 0 | 1 | Not-determined | Genome-fragment | NA | NA | 0 |
| S11_vs_k141_4498\|\|full | 1011 | No | NA | 2 | 1 | 0 | Low-quality | Genome-fragment | 2.36 | AAI-based (medium-confidence) | 0 |
| S17_extracted_k141_112668 | 1080 | No | NA | 2 | 0 | 2 | Not-determined | Genome-fragment | NA | NA | 0 |
| S17_extracted_k141_2019 | 7010 | No | NA | 5 | 0 | 5 | Not-determined | Genome-fragment | NA | NA | 0 |
| S17_extracted_k141_112678 | 1222 | No | NA | 1 | 0 | 0 | Not-determined | Genome-fragment | NA | NA | 0 |
| S17_extracted_k141_105981 | 5737 | No | NA | 7 | 0 | 2 | Not-determined | Genome-fragment | NA | NA | 0 |
| S17_extracted_k141_75728 | 1714 | No | NA | 1 | 0 | 1 | Not-determined | Genome-fragment | NA | NA | 0 |
| S17_extracted_k141_31795 | 1059 | No | NA | 1 | 0 | 0 | Not-determined | Genome-fragment | NA | NA | 0 |
| S17_extracted_k141_164530 | 7653 | No | NA | 7 | 0 | 4 | Not-determined | Genome-fragment | NA | NA | 0 |
| S17_extracted_k141_127316 | 7067 | No | NA | 10 | 0 | 0 | Not-determined | Genome-fragment | NA | NA | 0 |
| S11_vs_k141_5958\|\|full | 1011 | No | NA | 3 | 1 | 0 | Low-quality | Genome-fragment | 1.32 | HMM-based (lower-bound) | 0 |
| S17_extracted_k141_106005 | 1539 | No | NA | 2 | 0 | 2 | Not-determined | Genome-fragment | NA | NA | 0 |
| S11_vs_k141_32365\|\|full | 1011 | No | NA | 3 | 0 | 1 | Low-quality | Genome-fragment | 0.76 | AAI-based (medium-confidence) | 0 |
| S14_vs_k141_9507\|\|full | 1011 | No | NA | 2 | 0 | 0 | Low-quality | Genome-fragment | 2.36 | AAI-based (high-confidence) | 0 |
| S17_extracted_k141_24071 | 1625 | No | NA | 2 | 0 | 2 | Not-determined | Genome-fragment | NA | NA | 0 |
| S17_extracted_k141_39472 | 1724 | No | NA | 1 | 0 | 1 | Not-determined | Genome-fragment | NA | NA | 0 |
| S17_extracted_k141_164578 | 2901 | No | NA | 2 | 0 | 2 | Not-determined | Genome-fragment | NA | NA | 0 |
| S17_extracted_k141_164581 | 1059 | No | NA | 1 | 0 | 1 | Not-determined | Genome-fragment | NA | NA | 0 |
| S17_extracted_k141_164594 | 1216 | No | NA | 3 | 0 | 1 | Not-determined | Genome-fragment | NA | NA | 0 |
| S17_extracted_k141_31845 | 1511 | No | NA | 4 | 0 | 4 | Not-determined | Genome-fragment | NA | NA | 0 |
| S17_extracted_k141_120343 | 3440 | No | NA | 1 | 0 | 0 | Not-determined | Genome-fragment | NA | NA | 0 |
| S1_vs_k141_115896\|\|full | 1011 | No | NA | 2 | 2 | 0 | Low-quality | Genome-fragment | 2.44 | AAI-based (medium-confidence) | 0 |
| S17_extracted_k141_135058 | 1399 | No | NA | 1 | 0 | 0 | Not-determined | Genome-fragment | NA | NA | 0 |
| S17_extracted_k141_39487 | 7698 | No | NA | 8 | 0 | 6 | Not-determined | Genome-fragment | NA | NA | 0 |
| S1_vs_k141_119881\|\|full | 1011 | No | NA | 3 | 0 | 0 | Low-quality | Genome-fragment | 1.6 | AAI-based (high-confidence) | 0 |
| S17_extracted_k141_112756 | 1934 | No | NA | 2 | 0 | 0 | Not-determined | Genome-fragment | NA | NA | 0 |
| S17_extracted_k141_39495 | 3196 | No | NA | 4 | 0 | 3 | Not-determined | Genome-fragment | NA | NA | 0 |
| S17_extracted_k141_142688 | 3242 | No | NA | 3 | 0 | 1 | Not-determined | Genome-fragment | NA | NA | 0 |
| S17_extracted_k141_75831 | 1145 | No | NA | 1 | 0 | 1 | Not-determined | Genome-fragment | NA | NA | 0 |
| S17_extracted_k141_171818 | 1090 | No | NA | 1 | 0 | 1 | Not-determined | Genome-fragment | NA | NA | 0 |
| S17_extracted_k141_112769 | 1438 | No | NA | 1 | 0 | 1 | Not-determined | Genome-fragment | NA | NA | 0 |
| S5_extracted_k141_122077 | 1011 | No | NA | 4 | 0 | 0 | Low-quality | Genome-fragment | 2.29 | AAI-based (high-confidence) | 0 |
| S17_extracted_k141_46637 | 1026 | No | NA | 1 | 0 | 1 | Not-determined | Genome-fragment | NA | NA | 0 |
| S17_extracted_k141_16566 | 1005 | No | NA | 2 | 0 | 1 | Not-determined | Genome-fragment | NA | NA | 0 |
| S17_extracted_k141_164656 | 4902 | No | NA | 4 | 0 | 3 | Not-determined | Genome-fragment | NA | NA | 0 |
| S17_extracted_k141_75836 | 7622 | No | NA | 6 | 0 | 6 | Not-determined | Genome-fragment | NA | NA | 0 |
| S17_extracted_k141_16590 | 1143 | No | NA | 2 | 0 | 1 | Not-determined | Genome-fragment | NA | NA | 0 |
| S5_vs_k141_22643\|\|lt2gene | 1011 | No | NA | 1 | 1 | 0 | Low-quality | Genome-fragment | 1.15 | HMM-based (lower-bound) | 0 |
| S17_extracted_k141_90575 | 2804 | No | NA | 4 | 0 | 0 | Not-determined | Genome-fragment | NA | NA | 0 |
| S16_extracted_101286 | 1011 | No | NA | 2 | 1 | 0 | Low-quality | Genome-fragment | 1.69 | AAI-based (medium-confidence) | 0 |
| S17_extracted_k141_106163 | 1382 | No | NA | 2 | 0 | 2 | Not-determined | Genome-fragment | NA | NA | 0 |
| S17_extracted_k141_24139 | 1133 | No | NA | 1 | 0 | 1 | Not-determined | Genome-fragment | NA | NA | 0 |
| S17_extracted_k141_75893 | 1629 | No | NA | 1 | 0 | 1 | Not-determined | Genome-fragment | NA | NA | 0 |
| S17_extracted_k141_112823 | 1975 | No | NA | 3 | 0 | 2 | Not-determined | Genome-fragment | NA | NA | 0 |
| S17_extracted_k141_127417 | 1364 | No | NA | 1 | 0 | 1 | Not-determined | Genome-fragment | NA | NA | 0 |
| S17_extracted_k141_46684 | 3817 | No | NA | 4 | 0 | 0 | Not-determined | Genome-fragment | NA | NA | 0 |
| S17_extracted_k141_68752 | 1547 | No | NA | 2 | 0 | 0 | Not-determined | Genome-fragment | NA | NA | 0 |
| S17_extracted_k141_120441 | 6109 | No | NA | 5 | 0 | 5 | Not-determined | Genome-fragment | NA | NA | 0 |
| S17_extracted_k141_112825 | 10015 | No | NA | 7 | 0 | 5 | Not-determined | Genome-fragment | NA | NA | 0 |
| S17_extracted_k141_90603 | 4504 | No | NA | 2 | 0 | 2 | Not-determined | Genome-fragment | NA | NA | 0 |
| S17_extracted_k141_16642 | 1088 | No | NA | 2 | 0 | 1 | Not-determined | Genome-fragment | NA | NA | 0 |
| S17_extracted_k141_24162 | 1998 | No | NA | 2 | 0 | 2 | Not-determined | Genome-fragment | NA | NA | 0 |
| S16_vs_51425\|\|full | 1011 | No | NA | 2 | 0 | 2 | Low-quality | Genome-fragment | 2.34 | AAI-based (medium-confidence) | 0 |
| S16_vs_103149\|\|full | 1011 | No | NA | 2 | 0 | 0 | Low-quality | Genome-fragment | 0.93 | AAI-based (medium-confidence) | 0 |
| S17_extracted_k141_171935 | 6402 | No | NA | 4 | 0 | 3 | Not-determined | Genome-fragment | NA | NA | 0 |
| S17_extracted_k141_75969 | 1482 | No | NA | 3 | 0 | 2 | Not-determined | Genome-fragment | NA | NA | 0 |
| S17_extracted_k141_75978 | 1323 | No | NA | 2 | 0 | 2 | Not-determined | Genome-fragment | NA | NA | 0 |
| S16_vs_122521\|\|full | 1011 | No | NA | 1 | 1 | 0 | Low-quality | Genome-fragment | 0.55 | HMM-based (lower-bound) | 0 |
| S17_extracted_k141_46758 | 1003 | No | NA | 1 | 0 | 1 | Not-determined | Genome-fragment | NA | NA | 0 |
| S17_extracted_k141_164740 | 1132 | No | NA | 2 | 0 | 0 | Not-determined | Genome-fragment | NA | NA | 0 |
| S16_vs_146223\|\|full | 1011 | No | NA | 2 | 0 | 1 | Low-quality | Genome-fragment | 1.97 | AAI-based (medium-confidence) | 0 |
| S17_extracted_k141_90646 | 1010 | No | NA | 3 | 0 | 0 | Not-determined | Genome-fragment | NA | NA | 0 |
| S12_vs_k141_152159\|\|lt2gene | 1011 | No | NA | 1 | 1 | 0 | Low-quality | Genome-fragment | 2.36 | AAI-based (high-confidence) | 0 |
| S17_extracted_k141_54308 | 1479 | No | NA | 2 | 0 | 2 | Not-determined | Genome-fragment | NA | NA | 0 |
| S17_extracted_k141_75995 | 2091 | No | NA | 2 | 0 | 2 | Not-determined | Genome-fragment | NA | NA | 0 |
| S17_extracted_k141_157646 | 4408 | No | NA | 4 | 0 | 4 | Not-determined | Genome-fragment | NA | NA | 0 |
| S17_extracted_k141_135123 | 1944 | No | NA | 2 | 0 | 1 | Not-determined | Genome-fragment | NA | NA | 0 |
| S17_extracted_k141_16684 | 5628 | No | NA | 4 | 0 | 3 | Not-determined | Genome-fragment | NA | NA | 0 |
| S2_extracted_k141_18439 | 1011 | No | NA | 3 | 2 | 0 | Low-quality | Genome-fragment | 3.35 | AAI-based (high-confidence) | 0 |
| S17_extracted_k141_164794 | 1035 | No | NA | 2 | 0 | 0 | Not-determined | Genome-fragment | NA | NA | 0 |
| S8_extracted_k141_89738 | 1011 | No | NA | 1 | 1 | 0 | Low-quality | Genome-fragment | 0.65 | HMM-based (lower-bound) | 0 |
| S17_extracted_k141_39662 | 2852 | No | NA | 2 | 0 | 2 | Not-determined | Genome-fragment | NA | NA | 0 |
| S11_extracted_k141_9305 | 1012 | No | NA | 2 | 1 | 0 | Low-quality | Genome-fragment | 2.41 | AAI-based (high-confidence) | 0 |
| S17_extracted_k141_157704 | 3316 | No | NA | 2 | 0 | 0 | Not-determined | Genome-fragment | NA | NA | 0 |
| S17_extracted_k141_61145 | 1668 | No | NA | 2 | 0 | 0 | Not-determined | Genome-fragment | NA | NA | 0 |
| S17_extracted_k141_16727 | 1262 | No | NA | 1 | 0 | 1 | Not-determined | Genome-fragment | NA | NA | 0 |
| S17_extracted_k141_61157 | 1615 | No | NA | 2 | 0 | 1 | Not-determined | Genome-fragment | NA | NA | 0 |
| S13_vs_13096\|\|full | 1012 | No | NA | 3 | 0 | 0 | Low-quality | Genome-fragment | 2.34 | AAI-based (high-confidence) | 0 |
| S17_extracted_k141_127523 | 2734 | No | NA | 1 | 0 | 0 | Not-determined | Genome-fragment | NA | NA | 0 |
| S17_extracted_k141_150553 | 5399 | No | NA | 3 | 0 | 3 | Not-determined | Genome-fragment | NA | NA | 0 |
| S17_extracted_k141_142879 | 1431 | No | NA | 2 | 0 | 1 | Not-determined | Genome-fragment | NA | NA | 0 |
| S17_extracted_k141_39705 | 1455 | No | NA | 2 | 0 | 2 | Not-determined | Genome-fragment | NA | NA | 0 |
| S17_extracted_k141_112983 | 1105 | No | NA | 2 | 0 | 0 | Not-determined | Genome-fragment | NA | NA | 0 |
| S17_extracted_k141_142888 | 4620 | No | NA | 3 | 0 | 0 | Not-determined | Genome-fragment | NA | NA | 0 |
| S17_extracted_k141_68904 | 3651 | No | NA | 6 | 0 | 5 | Not-determined | Genome-fragment | NA | NA | 0 |
| S14_vs_k141_29841\|\|full | 1012 | No | NA | 3 | 1 | 0 | Low-quality | Genome-fragment | 3.06 | AAI-based (medium-confidence) | 0 |
| S17_extracted_k141_142906 | 1054 | No | NA | 2 | 0 | 2 | Not-determined | Genome-fragment | NA | NA | 0 |
| S17_extracted_k141_112997 | 3284 | No | NA | 3 | 0 | 2 | Not-determined | Genome-fragment | NA | NA | 0 |
| S17_extracted_k141_120642 | 1289 | No | NA | 3 | 0 | 0 | Not-determined | Genome-fragment | NA | NA | 0 |
| S17_extracted_k141_76093 | 1101 | No | NA | 1 | 0 | 0 | Not-determined | Genome-fragment | NA | NA | 0 |
| S14_vs_k141_100260\|\|full | 1012 | No | NA | 2 | 0 | 1 | Low-quality | Genome-fragment | 0.27 | AAI-based (medium-confidence) | 0 |
| S17_extracted_k141_54395 | 2162 | No | NA | 2 | 0 | 2 | Not-determined | Genome-fragment | NA | NA | 0 |
| S17_extracted_k141_83080 | 3380 | No | NA | 2 | 0 | 1 | Not-determined | Genome-fragment | NA | NA | 0 |
| S17_extracted_k141_142940 | 1161 | No | NA | 2 | 0 | 1 | Not-determined | Genome-fragment | NA | NA | 0 |
| S17_extracted_k141_46832 | 3512 | No | NA | 4 | 0 | 2 | Not-determined | Genome-fragment | NA | NA | 0 |
| S14_vs_k141_56602\|\|full | 1012 | No | NA | 2 | 1 | 0 | Low-quality | Genome-fragment | 2.47 | AAI-based (high-confidence) | 0 |
| S17_extracted_k141_90803 | 1720 | No | NA | 1 | 0 | 1 | Not-determined | Genome-fragment | NA | NA | 0 |
| S17_extracted_k141_32066 | 4099 | No | NA | 4 | 0 | 3 | Not-determined | Genome-fragment | NA | NA | 0 |
| S17_extracted_k141_90804 | 1640 | No | NA | 1 | 0 | 1 | Not-determined | Genome-fragment | NA | NA | 0 |
| S1_vs_k141_50343\|\|full | 1012 | No | NA | 2 | 0 | 1 | Low-quality | Genome-fragment | 1.89 | AAI-based (medium-confidence) | 0 |
| S17_extracted_k141_54411 | 1411 | No | NA | 2 | 0 | 1 | Not-determined | Genome-fragment | NA | NA | 0 |
| S3_extracted_k141_90378 | 1012 | No | NA | 3 | 0 | 0 | Low-quality | Genome-fragment | 1.94 | AAI-based (medium-confidence) | 0 |
| S17_extracted_k141_113064 | 1072 | No | NA | 2 | 0 | 0 | Not-determined | Genome-fragment | NA | NA | 0 |
| S17_extracted_k141_68985 | 1199 | No | NA | 1 | 0 | 1 | Not-determined | Genome-fragment | NA | NA | 0 |
| S17_extracted_k141_16789 | 1185 | No | NA | 1 | 0 | 0 | Not-determined | Genome-fragment | NA | NA | 0 |
| S17_extracted_k141_61271 | 3904 | No | NA | 3 | 0 | 3 | Not-determined | Genome-fragment | NA | NA | 0 |
| S3_extracted_k141_109270 | 1012 | No | NA | 2 | 1 | 0 | Low-quality | Genome-fragment | 2.27 | AAI-based (high-confidence) | 0 |
| S17_extracted_k141_150677 | 1046 | No | NA | 2 | 0 | 2 | Not-determined | Genome-fragment | NA | NA | 0 |
| S17_extracted_k141_113069 | 1020 | No | NA | 2 | 0 | 2 | Not-determined | Genome-fragment | NA | NA | 0 |
| S17_extracted_k141_120721 | 11346 | No | NA | 9 | 0 | 5 | Not-determined | Genome-fragment | NA | NA | 0 |
| S17_extracted_k141_54439 | 1037 | No | NA | 1 | 0 | 0 | Not-determined | Genome-fragment | NA | NA | 0 |
| S17_extracted_k141_32134 | 1378 | No | NA | 2 | 0 | 2 | Not-determined | Genome-fragment | NA | NA | 0 |
| S17_extracted_k141_32137 | 1266 | No | NA | 2 | 0 | 0 | Not-determined | Genome-fragment | NA | NA | 0 |
| S5_vs_k141_100509\|\|lt2gene | 1012 | No | NA | 2 | 1 | 0 | Low-quality | Genome-fragment | 2.58 | AAI-based (high-confidence) | 0 |
| S17_extracted_k141_135243 | 1523 | No | NA | 2 | 0 | 1 | Not-determined | Genome-fragment | NA | NA | 0 |
| S7_vs_133432\|\|lt2gene | 1012 | No | NA | 1 | 1 | 0 | Low-quality | Genome-fragment | 0.58 | HMM-based (lower-bound) | 0 |
| S17_extracted_k141_143068 | 1465 | No | NA | 2 | 0 | 1 | Not-determined | Genome-fragment | NA | NA | 0 |
| S17_extracted_k141_113118 | 5425 | No | NA | 3 | 0 | 3 | Not-determined | Genome-fragment | NA | NA | 0 |
| S17_extracted_k141_9981 | 1510 | No | NA | 1 | 0 | 0 | Not-determined | Genome-fragment | NA | NA | 0 |
| S17_extracted_k141_83209 | 1290 | No | NA | 2 | 0 | 2 | Not-determined | Genome-fragment | NA | NA | 0 |
| S17_extracted_k141_46961 | 1651 | No | NA | 2 | 0 | 0 | Not-determined | Genome-fragment | NA | NA | 0 |
| S17_extracted_k141_83214 | 3812 | No | NA | 2 | 0 | 0 | Not-determined | Genome-fragment | NA | NA | 0 |
| S17_extracted_k141_10000 | 1477 | No | NA | 2 | 0 | 1 | Not-determined | Genome-fragment | NA | NA | 0 |
| S17_extracted_k141_32200 | 2364 | No | NA | 2 | 0 | 2 | Not-determined | Genome-fragment | NA | NA | 0 |
| S17_extracted_k141_46981 | 1092 | No | NA | 1 | 0 | 0 | Not-determined | Genome-fragment | NA | NA | 0 |
| S16_extracted_107579 | 1012 | No | NA | 3 | 0 | 0 | Low-quality | Genome-fragment | 1.43 | AAI-based (high-confidence) | 0 |
| S16_vs_19994\|\|full | 1012 | No | NA | 2 | 2 | 0 | Low-quality | Genome-fragment | 2.89 | AAI-based (medium-confidence) | 0 |
| S17_extracted_k141_69039 | 1324 | No | NA | 2 | 0 | 1 | Not-determined | Genome-fragment | NA | NA | 0 |
| S10_vs_74792\|\|full | 1012 | No | NA | 3 | 1 | 0 | Low-quality | Genome-fragment | 2.14 | AAI-based (high-confidence) | 0 |
| S17_extracted_k141_98675 | 1462 | No | NA | 1 | 0 | 1 | Not-determined | Genome-fragment | NA | NA | 0 |
| S17_extracted_k141_10013 | 2100 | No | NA | 2 | 0 | 1 | Not-determined | Genome-fragment | NA | NA | 0 |
| S12_extracted_k141_71446 | 1012 | No | NA | 1 | 1 | 0 | Low-quality | Genome-fragment | 1.48 | HMM-based (lower-bound) | 0 |
| S17_extracted_k141_143085 | 4077 | No | NA | 1 | 0 | 0 | Not-determined | Genome-fragment | NA | NA | 0 |
| S12_extracted_k141_150701 | 1012 | No | NA | 2 | 1 | 0 | Low-quality | Genome-fragment | 1.54 | HMM-based (lower-bound) | 0 |
| S8_vs_k141_86607\|\|lt2gene | 1012 | No | NA | 2 | 1 | 0 | Low-quality | Genome-fragment | 2.82 | AAI-based (high-confidence) | 0 |
| S17_extracted_k141_157883 | 1250 | No | NA | 2 | 0 | 0 | Not-determined | Genome-fragment | NA | NA | 0 |
| S17_extracted_k141_47026 | 11400 | No | NA | 16 | 0 | 1 | Not-determined | Genome-fragment | NA | NA | 0 |
| S17_extracted_k141_143144 | 3266 | No | NA | 3 | 0 | 0 | Not-determined | Genome-fragment | NA | NA | 0 |
| S17_extracted_k141_172132 | 12235 | No | NA | 19 | 4 | 0 | Complete | High-quality | 100 | ITR (medium-confidence) | 0 |
| S17_vs_k141_140709\|\|full | 1013 | No | NA | 3 | 0 | 0 | Low-quality | Genome-fragment | 0.4 | AAI-based (medium-confidence) | 0 |
| S17_extracted_k141_47033 | 1007 | No | NA | 1 | 0 | 1 | Not-determined | Genome-fragment | NA | NA | 0 |
| S11_vs_k141_66539\|\|full | 1013 | No | NA | 2 | 0 | 0 | Low-quality | Genome-fragment | 0.77 | AAI-based (high-confidence) | 0 |
| S11_vs_k141_28487\|\|full | 1013 | Yes | 863 | 4 | 1 | 1 | Low-quality | Genome-fragment | 1.18 | AAI-based (high-confidence) | 14.81 |
| S17_extracted_k141_150808 | 1882 | No | NA | 1 | 0 | 1 | Not-determined | Genome-fragment | NA | NA | 0 |
| S17_extracted_k141_32304 | 1002 | No | NA | 2 | 0 | 2 | Not-determined | Genome-fragment | NA | NA | 0 |
| S17_extracted_k141_164967 | 4554 | No | NA | 5 | 0 | 4 | Not-determined | Genome-fragment | NA | NA | 0 |
| S11_vs_k141_71619\|\|lt2gene | 1013 | No | NA | 1 | 1 | 0 | Low-quality | Genome-fragment | 2.25 | AAI-based (high-confidence) | 0 |
| S17_extracted_k141_69124 | 3765 | No | NA | 4 | 0 | 4 | Not-determined | Genome-fragment | NA | NA | 0 |
| S17_extracted_k141_83302 | 1240 | No | NA | 2 | 0 | 2 | Not-determined | Genome-fragment | NA | NA | 0 |
| S14_extracted_k141_73544 | 1013 | No | NA | 3 | 3 | 0 | Low-quality | Genome-fragment | 3.31 | AAI-based (high-confidence) | 0 |
| S17_extracted_k141_83315 | 1480 | No | NA | 2 | 0 | 2 | Not-determined | Genome-fragment | NA | NA | 0 |
| S1_extracted_k141_78878 | 1013 | No | NA | 3 | 0 | 0 | Low-quality | Genome-fragment | 2.11 | AAI-based (high-confidence) | 0 |
| S17_extracted_k141_2376 | 1954 | No | NA | 1 | 0 | 1 | Not-determined | Genome-fragment | NA | NA | 0 |
| S17_extracted_k141_98788 | 1906 | No | NA | 1 | 0 | 0 | Not-determined | Genome-fragment | NA | NA | 0 |
| S5_extracted_k141_88667 | 1013 | No | NA | 1 | 0 | 0 | Low-quality | Genome-fragment | 2.6 | AAI-based (high-confidence) | 0 |
| S17_extracted_k141_90996 | 4952 | No | NA | 3 | 0 | 1 | Not-determined | Genome-fragment | NA | NA | 0 |
| S17_extracted_k141_165002 | 1330 | No | NA | 3 | 0 | 0 | Not-determined | Genome-fragment | NA | NA | 0 |
| S17_extracted_k141_98813 | 1495 | No | NA | 3 | 0 | 0 | Not-determined | Genome-fragment | NA | NA | 0 |
| S17_extracted_k141_2395 | 1005 | No | NA | 1 | 0 | 0 | Not-determined | Genome-fragment | NA | NA | 0 |
| S17_extracted_k141_10112 | 1458 | No | NA | 2 | 0 | 0 | Not-determined | Genome-fragment | NA | NA | 0 |
| S5_extracted_k141_60155 | 1013 | No | NA | 1 | 1 | 0 | Low-quality | Genome-fragment | 0.7 | HMM-based (lower-bound) | 0 |
| S5_vs_k141_107997\|\|full | 1013 | No | NA | 3 | 0 | 1 | Low-quality | Genome-fragment | 1.01 | AAI-based (medium-confidence) | 0 |
| S17_extracted_k141_165022 | 9596 | No | NA | 8 | 0 | 3 | Not-determined | Genome-fragment | NA | NA | 0 |
| S17_extracted_k141_16927 | 1061 | No | NA | 1 | 0 | 1 | Not-determined | Genome-fragment | NA | NA | 0 |
| S17_extracted_k141_165026 | 2038 | No | NA | 2 | 0 | 1 | Not-determined | Genome-fragment | NA | NA | 0 |
| S7_vs_66815\|\|full | 1013 | No | NA | 2 | 0 | 0 | Low-quality | Genome-fragment | 2.39 | AAI-based (medium-confidence) | 0 |
| S17_extracted_k141_10128 | 1611 | No | NA | 2 | 0 | 1 | Not-determined | Genome-fragment | NA | NA | 0 |
| S17_extracted_k141_113155 | 1289 | No | NA | 1 | 0 | 1 | Not-determined | Genome-fragment | NA | NA | 0 |
| S17_extracted_k141_165029 | 5378 | No | NA | 7 | 0 | 4 | Not-determined | Genome-fragment | NA | NA | 0 |
| S17_extracted_k141_16942 | 1037 | No | NA | 1 | 0 | 1 | Not-determined | Genome-fragment | NA | NA | 0 |
| S17_extracted_k141_98843 | 3684 | No | NA | 2 | 0 | 1 | Not-determined | Genome-fragment | NA | NA | 0 |
| S17_extracted_k141_135444 | 5253 | No | NA | 4 | 0 | 1 | Not-determined | Genome-fragment | NA | NA | 0 |
| S17_extracted_k141_24488 | 3787 | No | NA | 5 | 0 | 3 | Not-determined | Genome-fragment | NA | NA | 0 |
| S17_extracted_k141_158002 | 1059 | No | NA | 1 | 0 | 0 | Not-determined | Genome-fragment | NA | NA | 0 |
| S17_extracted_k141_113174 | 2910 | No | NA | 2 | 0 | 1 | Not-determined | Genome-fragment | NA | NA | 0 |
| S17_extracted_k141_16972 | 1038 | No | NA | 2 | 0 | 1 | Not-determined | Genome-fragment | NA | NA | 0 |
| S17_extracted_k141_16983 | 1031 | No | NA | 2 | 0 | 0 | Not-determined | Genome-fragment | NA | NA | 0 |
| S16_vs_46507\|\|full | 1013 | No | NA | 2 | 1 | 0 | Low-quality | Genome-fragment | 2.04 | AAI-based (medium-confidence) | 0 |
| S17_extracted_k141_2410 | 1154 | No | NA | 2 | 0 | 1 | Not-determined | Genome-fragment | NA | NA | 0 |

| | | | | | | | | | | | | |
|---|---|---|---|---|---|---|---|---|---|---|---|---|
| S17_extracted_k141_32419 | 1018 | No | NA | 1 | 0 | 0 | Not-determined | Genome-fragment | NA | | NA | 0 |
| S16_vs_111247\|\|full | 1013 | No | NA | 2 | 0 | 1 | Low-quality | Genome-fragment | | 0.99 | AAI-based (medium-confidence) | 0 |
| S12_vs_k141_126925\|\|full | 1013 | No | NA | 2 | 1 | 0 | Low-quality | Genome-fragment | | 19.07 | AAI-based (medium-confidence) | 0 |
| S17_extracted_k141_143283 | 1895 | No | NA | 2 | 0 | 0 | Not-determined | Genome-fragment | NA | | NA | 0 |
| S17_extracted_k141_127876 | 1045 | No | NA | 1 | 0 | 1 | Not-determined | Genome-fragment | NA | | NA | 0 |
| S17_extracted_k141_32453 | 4714 | No | NA | 4 | 0 | 3 | Not-determined | Genome-fragment | NA | | NA | 0 |
| S4_extracted_25533 | 1013 | No | NA | 5 | 0 | 0 | Low-quality | Genome-fragment | | 2.92 | AAI-based (high-confidence) | 0 |
| S17_extracted_k141_135506 | 3610 | No | NA | 4 | 0 | 1 | Not-determined | Genome-fragment | NA | | NA | 0 |
| S17_extracted_k141_32463 | 1153 | No | NA | 2 | 0 | 0 | Not-determined | Genome-fragment | NA | | NA | 0 |
| S17_extracted_k141_2484 | 1037 | No | NA | 2 | 0 | 1 | Not-determined | Genome-fragment | NA | | NA | 0 |
| S8_extracted_k141_45221 | 1013 | No | NA | 2 | 0 | 1 | Low-quality | Genome-fragment | | 0.66 | AAI-based (medium-confidence) | 0 |
| S17_extracted_k141_47224 | 1960 | No | NA | 3 | 0 | 0 | Not-determined | Genome-fragment | NA | | NA | 0 |
| S17_extracted_k141_69260 | 1154 | No | NA | 2 | 0 | 1 | Not-determined | Genome-fragment | NA | | NA | 0 |
| S17_extracted_k141_135538 | 4846 | No | NA | 2 | 0 | 0 | Not-determined | Genome-fragment | NA | | NA | 0 |
| S17_extracted_k141_83367 | 1160 | No | NA | 1 | 0 | 1 | Not-determined | Genome-fragment | NA | | NA | 0 |
| S17_extracted_k141_32516 | 6058 | No | NA | 4 | 0 | 3 | Not-determined | Genome-fragment | NA | | NA | 0 |
| S8_extracted_k141_22478 | 1013 | No | NA | 2 | 1 | 0 | Low-quality | Genome-fragment | | 3.34 | AAI-based (high-confidence) | 0 |
| S17_extracted_k141_91199 | 4540 | No | NA | 5 | 0 | 1 | Not-determined | Genome-fragment | NA | | NA | 0 |
| S17_extracted_k141_172347 | 1148 | No | NA | 1 | 0 | 1 | Not-determined | Genome-fragment | NA | | NA | 0 |
| S17_extracted_k141_113245 | 1447 | No | NA | 3 | 0 | 0 | Not-determined | Genome-fragment | NA | | NA | 0 |
| S17_extracted_k141_61513 | 2746 | No | NA | 6 | 0 | 0 | Not-determined | Genome-fragment | NA | | NA | 0 |
| S17_extracted_k141_61514 | 1402 | No | NA | 2 | 0 | 2 | Not-determined | Genome-fragment | NA | | NA | 0 |
| S17_extracted_k141_113249 | 1358 | No | NA | 1 | 0 | 0 | Not-determined | Genome-fragment | NA | | NA | 0 |
| S17_extracted_k141_17092 | 3545 | No | NA | 2 | 0 | 1 | Not-determined | Genome-fragment | NA | | NA | 0 |
| S17_extracted_k141_165077 | 11958 | No | NA | 8 | 0 | 8 | Not-determined | Genome-fragment | NA | | NA | 0 |
| S17_extracted_k141_120951 | 3892 | No | NA | 6 | 0 | 6 | Not-determined | Genome-fragment | NA | | NA | 0 |
| S17_extracted_k141_172356 | 4512 | No | NA | 6 | 0 | 2 | Not-determined | Genome-fragment | NA | | NA | 0 |
| S17_extracted_k141_10229 | 1273 | No | NA | 1 | 0 | 1 | Not-determined | Genome-fragment | NA | | NA | 0 |
| S8_extracted_k141_137129 | 1013 | No | NA | 2 | 1 | 0 | Low-quality | Genome-fragment | | 2.13 | AAI-based (high-confidence) | 0 |
| S17_extracted_k141_143343 | 1513 | No | NA | 1 | 0 | 1 | Not-determined | Genome-fragment | NA | | NA | 0 |
| S8_extracted_k141_16453 | 1013 | No | NA | 3 | 0 | 0 | Low-quality | Genome-fragment | | 5.18 | AAI-based (high-confidence) | 0 |
| S17_extracted_k141_127961 | 1214 | No | NA | 2 | 0 | 2 | Not-determined | Genome-fragment | NA | | NA | 0 |
| S17_extracted_k141_76534 | 1341 | No | NA | 2 | 0 | 1 | Not-determined | Genome-fragment | NA | | NA | 0 |
| S17_extracted_k141_106687 | 4094 | No | NA | 8 | 0 | 1 | Not-determined | Genome-fragment | NA | | NA | 0 |
| S17_extracted_k141_76540 | 7313 | No | NA | 3 | 0 | 2 | Not-determined | Genome-fragment | NA | | NA | 0 |
| S17_extracted_k141_17133 | 1115 | No | NA | 1 | 0 | 0 | Not-determined | Genome-fragment | NA | | NA | 0 |
| S17_extracted_k141_83414 | 1035 | No | NA | 2 | 0 | 2 | Not-determined | Genome-fragment | NA | | NA | 0 |
| S17_extracted_k141_113298 | 3675 | No | NA | 1 | 0 | 1 | Not-determined | Genome-fragment | NA | | NA | 0 |
| S17_extracted_k141_47309 | 1083 | No | NA | 2 | 0 | 2 | Not-determined | Genome-fragment | NA | | NA | 0 |
| S17_extracted_k141_76550 | 1621 | No | NA | 2 | 0 | 2 | Not-determined | Genome-fragment | NA | | NA | 0 |
| S17_extracted_k141_158159 | 1424 | No | NA | 2 | 0 | 1 | Not-determined | Genome-fragment | NA | | NA | 0 |
| S8_vs_k141_80673\|\|lt2gene | 1013 | No | NA | 1 | 1 | 0 | Low-quality | Genome-fragment | | 1.88 | AAI-based (high-confidence) | 0 |
| S17_extracted_k141_17153 | 1127 | No | NA | 1 | 0 | 0 | Not-determined | Genome-fragment | NA | | NA | 0 |
| S17_extracted_k141_7908 | 1014 | No | NA | 3 | 3 | 0 | Low-quality | Genome-fragment | | 3.35 | AAI-based (high-confidence) | 0 |
| S17_extracted_k141_91274 | 2283 | No | NA | 3 | 0 | 2 | Not-determined | Genome-fragment | NA | | NA | 0 |
| S17_extracted_k141_118052 | 1014 | No | NA | 2 | 0 | 1 | Low-quality | Genome-fragment | | 0.93 | AAI-based (medium-confidence) | 0 |
| S17_extracted_k141_32628 | 1223 | No | NA | 1 | 0 | 1 | Not-determined | Genome-fragment | NA | | NA | 0 |
| S17_extracted_k141_135608 | 1099 | No | NA | 3 | 0 | 2 | Not-determined | Genome-fragment | NA | | NA | 0 |
| S11_vs_k141_23045\|\|full | 1014 | No | NA | 3 | 1 | 0 | Low-quality | Genome-fragment | | 1.32 | AAI-based (high-confidence) | 0 |
| S17_extracted_k141_91289 | 1165 | No | NA | 1 | 0 | 0 | Not-determined | Genome-fragment | NA | | NA | 0 |
| S17_extracted_k141_40252 | 2010 | No | NA | 2 | 0 | 1 | Not-determined | Genome-fragment | NA | | NA | 0 |
| S11_vs_k141_82621\|\|full | 1014 | No | NA | 4 | 0 | 0 | Low-quality | Genome-fragment | | 2.25 | AAI-based (high-confidence) | 0 |
| S17_extracted_k141_24669 | 1202 | No | NA | 1 | 0 | 1 | Not-determined | Genome-fragment | NA | | NA | 0 |
| S13_vs_112381\|\|full | 1014 | No | NA | 2 | 1 | 0 | Low-quality | Genome-fragment | | 2.16 | AAI-based (medium-confidence) | 0 |
| S14_vs_k141_12464\|\|full | 1014 | No | NA | 3 | 0 | 0 | Low-quality | Genome-fragment | | 1.83 | AAI-based (medium-confidence) | 0 |
| S17_extracted_k141_61606 | 1373 | No | NA | 2 | 0 | 2 | Not-determined | Genome-fragment | NA | | NA | 0 |
| S17_extracted_k141_32660 | 2855 | No | NA | 3 | 0 | 1 | Not-determined | Genome-fragment | NA | | NA | 0 |
| S17_extracted_k141_121035 | 1515 | No | NA | 2 | 0 | 2 | Not-determined | Genome-fragment | NA | | NA | 0 |
| S1_extracted_k141_70908 | 1014 | No | NA | 2 | 0 | 0 | Low-quality | Genome-fragment | | 1.99 | AAI-based (high-confidence) | 0 |
| S17_extracted_k141_40279 | 1228 | No | NA | 1 | 0 | 0 | Not-determined | Genome-fragment | NA | | NA | 0 |
| S1_vs_k141_82849\|\|lt2gene | 1014 | No | NA | 1 | 1 | 0 | Low-quality | Genome-fragment | | 17.19 | AAI-based (high-confidence) | 0 |
| S17_extracted_k141_47398 | 3755 | No | NA | 3 | 0 | 3 | Not-determined | Genome-fragment | NA | | NA | 0 |
| S5_vs_k141_98664\|\|full | 1014 | No | NA | 2 | 0 | 1 | Low-quality | Genome-fragment | | 2.48 | AAI-based (high-confidence) | 0 |
| S17_extracted_k141_47402 | 1107 | No | NA | 2 | 0 | 0 | Not-determined | Genome-fragment | NA | | NA | 0 |
| S17_extracted_k141_47404 | 1052 | No | NA | 2 | 0 | 0 | Not-determined | Genome-fragment | NA | | NA | 0 |
| S7_extracted_119169 | 1014 | No | NA | 3 | 0 | 1 | Low-quality | Genome-fragment | | 7.27 | AAI-based (high-confidence) | 0 |
| S17_extracted_k141_10277 | 3441 | No | NA | 6 | 0 | 0 | Not-determined | Genome-fragment | NA | | NA | 0 |
| S16_vs_136002\|\|full | 1014 | No | NA | 4 | 1 | 0 | Low-quality | Genome-fragment | | 1.97 | HMM-based (lower-bound) | 0 |
| S17_extracted_k141_99085 | 2671 | No | NA | 4 | 0 | 0 | Not-determined | Genome-fragment | NA | | NA | 0 |
| S17_extracted_k141_135686 | 1500 | No | NA | 2 | 0 | 1 | Not-determined | Genome-fragment | NA | | NA | 0 |
| S17_extracted_k141_165257 | 1879 | No | NA | 1 | 0 | 1 | Not-determined | Genome-fragment | NA | | NA | 0 |
| S16_vs_138358\|\|lt2gene | 1014 | No | NA | 1 | 1 | 0 | Low-quality | Genome-fragment | | 2.98 | AAI-based (high-confidence) | 0 |
| S17_extracted_k141_69460 | 1043 | No | NA | 1 | 0 | 0 | Not-determined | Genome-fragment | NA | | NA | 0 |
| S17_extracted_k141_17257 | 1242 | No | NA | 2 | 0 | 0 | Not-determined | Genome-fragment | NA | | NA | 0 |
| S17_extracted_k141_10316 | 1125 | No | NA | 2 | 0 | 2 | Not-determined | Genome-fragment | NA | | NA | 0 |
| S10_vs_74415\|\|full | 1014 | No | NA | 3 | 0 | 0 | Low-quality | Genome-fragment | | 1.08 | AAI-based (medium-confidence) | 0 |
| S17_extracted_k141_69478 | 1293 | No | NA | 3 | 0 | 2 | Not-determined | Genome-fragment | NA | | NA | 0 |
| S15_vs_62008\|\|full | 1014 | No | NA | 4 | 0 | 0 | Low-quality | Genome-fragment | | 2 | AAI-based (high-confidence) | 0 |
| S17_extracted_k141_17280 | 4205 | No | NA | 6 | 0 | 0 | Not-determined | Genome-fragment | NA | | NA | 0 |
| S17_extracted_k141_61693 | 4137 | No | NA | 5 | 0 | 2 | Not-determined | Genome-fragment | NA | | NA | 0 |
| S17_extracted_k141_24776 | 7985 | No | NA | 10 | 0 | 1 | Not-determined | Genome-fragment | NA | | NA | 0 |
| S8_extracted_k141_11951 | 1014 | No | NA | 1 | 0 | 0 | Low-quality | Genome-fragment | | 2.02 | AAI-based (high-confidence) | 0 |
| S17_extracted_k141_47492 | 1358 | No | NA | 2 | 0 | 1 | Not-determined | Genome-fragment | NA | | NA | 0 |
| S17_extracted_k141_40343 | 1523 | No | NA | 2 | 0 | 2 | Not-determined | Genome-fragment | NA | | NA | 0 |
| S17_extracted_k141_44548 | 1015 | No | NA | 4 | 0 | 0 | Low-quality | Genome-fragment | | 1.97 | AAI-based (medium-confidence) | 0 |
| S17_extracted_k141_10341 | 1409 | No | NA | 2 | 0 | 2 | Not-determined | Genome-fragment | NA | | NA | 0 |
| S17_extracted_k141_128147 | 7401 | No | NA | 8 | 3 | 0 | High-quality | High-quality | | 100 | AAI-based (high-confidence) | 0 |
| S17_extracted_k141_54729 | 1120 | No | NA | 2 | 0 | 1 | Not-determined | Genome-fragment | NA | | NA | 0 |
| S17_extracted_k141_106936 | 1177 | No | NA | 2 | 0 | 0 | Not-determined | Genome-fragment | NA | | NA | 0 |
| S17_extracted_k141_47545 | 1996 | No | NA | 2 | 0 | 1 | Not-determined | Genome-fragment | NA | | NA | 0 |
| S17_extracted_k141_106942 | 1635 | No | NA | 3 | 0 | 0 | Not-determined | Genome-fragment | NA | | NA | 0 |
| S9_extracted_k141_79590 | 1015 | No | NA | 1 | 0 | 0 | Low-quality | Genome-fragment | | 0.63 | AAI-based (high-confidence) | 0 |
| S17_extracted_k141_76763 | 1268 | No | NA | 1 | 0 | 1 | Not-determined | Genome-fragment | NA | | NA | 0 |
| S17_extracted_k141_69562 | 2039 | No | NA | 3 | 0 | 1 | Not-determined | Genome-fragment | NA | | NA | 0 |
| S9_vs_k141_72672\|\|lt2gene | 1015 | No | NA | 1 | 1 | 0 | Low-quality | Genome-fragment | | 2.78 | AAI-based (high-confidence) | 0 |
| S17_extracted_k141_32701 | 1122 | No | NA | 2 | 0 | 1 | Not-determined | Genome-fragment | NA | | NA | 0 |
| S11_extracted_k141_55902 | 1015 | No | NA | 1 | 1 | 0 | Low-quality | Genome-fragment | | 2.25 | HMM-based (lower-bound) | 0 |
| S14_extracted_k141_59712 | 1015 | No | NA | 1 | 1 | 0 | Low-quality | Genome-fragment | | 0.7 | HMM-based (lower-bound) | 0 |
| S14_extracted_k141_67217 | 1015 | No | NA | 2 | 1 | 0 | Low-quality | Genome-fragment | | 0.65 | HMM-based (lower-bound) | 0 |
| S17_extracted_k141_2831 | 1035 | No | NA | 1 | 0 | 1 | Not-determined | Genome-fragment | NA | | NA | 0 |
| S17_extracted_k141_99239 | 6540 | No | NA | 5 | 0 | 3 | Not-determined | Genome-fragment | NA | | NA | 0 |
| S17_extracted_k141_10394 | 2525 | No | NA | 2 | 0 | 1 | Not-determined | Genome-fragment | NA | | NA | 0 |
| S17_extracted_k141_172666 | 2833 | No | NA | 3 | 0 | 1 | Not-determined | Genome-fragment | NA | | NA | 0 |
| S14_vs_k141_58507\|\|full | 1015 | No | NA | 3 | 3 | 0 | Low-quality | Genome-fragment | | 2.83 | AAI-based (high-confidence) | 0 |
| S17_extracted_k141_143621 | 1108 | No | NA | 1 | 0 | 0 | Not-determined | Genome-fragment | NA | | NA | 0 |
| S1_vs_k141_56077\|\|full | 1015 | No | NA | 3 | 0 | 0 | Low-quality | Genome-fragment | | 1.74 | AAI-based (high-confidence) | 0 |
| S3_extracted_k141_78412 | 1015 | No | NA | 4 | 1 | 0 | Low-quality | Genome-fragment | | 1.52 | AAI-based (high-confidence) | 0 |
| S17_extracted_k141_24859 | 3966 | No | NA | 5 | 0 | 2 | Not-determined | Genome-fragment | NA | | NA | 0 |
| S3_vs_k141_11246\|\|full | 1015 | No | NA | 2 | 0 | 0 | Low-quality | Genome-fragment | | 0.59 | AAI-based (high-confidence) | 0 |
| S17_extracted_k141_113535 | 7478 | No | NA | 3 | 0 | 2 | Not-determined | Genome-fragment | NA | | NA | 0 |
| S17_extracted_k141_135812 | 1045 | No | NA | 1 | 0 | 0 | Not-determined | Genome-fragment | NA | | NA | 0 |
| S17_extracted_k141_69627 | 1153 | No | NA | 1 | 0 | 1 | Not-determined | Genome-fragment | NA | | NA | 0 |
| S17_extracted_k141_69630 | 1234 | No | NA | 2 | 0 | 2 | Not-determined | Genome-fragment | NA | | NA | 0 |
| S5_vs_k141_75564\|\|full | 1015 | No | NA | 3 | 2 | 0 | Low-quality | Genome-fragment | | 2.64 | AAI-based (high-confidence) | 0 |
| S17_extracted_k141_135816 | 3116 | No | NA | 5 | 0 | 0 | Not-determined | Genome-fragment | NA | | NA | 0 |
| S17_extracted_k141_165441 | 1006 | No | NA | 2 | 0 | 2 | Not-determined | Genome-fragment | NA | | NA | 0 |
| S17_extracted_k141_54836 | 2122 | No | NA | 2 | 0 | 0 | Not-determined | Genome-fragment | NA | | NA | 0 |
| S17_extracted_k141_121114 | 3025 | No | NA | 5 | 0 | 5 | Not-determined | Genome-fragment | NA | | NA | 0 |
| S7_extracted_69180 | 1015 | No | NA | 2 | 0 | 2 | Low-quality | Genome-fragment | | 0.66 | AAI-based (medium-confidence) | 0 |
| S17_extracted_k141_47622 | 3169 | No | NA | 3 | 0 | 2 | Not-determined | Genome-fragment | NA | | NA | 0 |
| S17_extracted_k141_91488 | 1507 | No | NA | 2 | 0 | 2 | Not-determined | Genome-fragment | NA | | NA | 0 |
| S10_vs_27874\|\|full | 1015 | No | NA | 3 | 0 | 0 | Low-quality | Genome-fragment | | 2.06 | AAI-based (high-confidence) | 0 |
| S10_vs_60781\|\|full | 1015 | No | NA | 3 | 1 | 0 | Low-quality | Genome-fragment | | 2.87 | AAI-based (high-confidence) | 0 |
| S17_extracted_k141_113568 | 5332 | No | NA | 3 | 0 | 3 | Not-determined | Genome-fragment | NA | | NA | 0 |
| S10_vs_101949\|\|full | 1015 | No | NA | 2 | 0 | 2 | Low-quality | Genome-fragment | | 1.53 | AAI-based (medium-confidence) | 0 |
| S12_vs_k141_80353\|\|full | 1015 | No | NA | 3 | 0 | 0 | Low-quality | Genome-fragment | | 0.72 | AAI-based (medium-confidence) | 0 |
| S17_extracted_k141_135834 | 4929 | No | NA | 7 | 0 | 7 | Not-determined | Genome-fragment | NA | | NA | 0 |
| S17_extracted_k141_91503 | 1152 | No | NA | 2 | 0 | 1 | Not-determined | Genome-fragment | NA | | NA | 0 |
| S12_vs_k141_170246\|\|lt2gene | 1015 | No | NA | 1 | 1 | 0 | Low-quality | Genome-fragment | | 0.7 | HMM-based (lower-bound) | 0 |
| S15_vs_23235\|\|full | 1015 | No | NA | 4 | 0 | 0 | Low-quality | Genome-fragment | | 2.34 | AAI-based (high-confidence) | 0 |
| S17_extracted_k141_121138 | 1016 | No | NA | 2 | 0 | 2 | Not-determined | Genome-fragment | NA | | NA | 0 |
| S17_extracted_k141_2905 | 7170 | No | NA | 4 | 0 | 3 | Not-determined | Genome-fragment | NA | | NA | 0 |
| S17_extracted_k141_121144 | 1793 | No | NA | 2 | 0 | 0 | Not-determined | Genome-fragment | NA | | NA | 0 |
| S17_extracted_k141_143670 | 1060 | No | NA | 2 | 0 | 1 | Not-determined | Genome-fragment | NA | | NA | 0 |
| S8_extracted_k141_78982 | 1015 | No | NA | 2 | 0 | 0 | Low-quality | Genome-fragment | | 2.86 | AAI-based (medium-confidence) | 0 |
| S17_extracted_k141_172729 | 3728 | No | NA | 2 | 0 | 0 | Not-determined | Genome-fragment | NA | | NA | 0 |
| S17_extracted_k141_135861 | 6935 | No | NA | 9 | 0 | 1 | Not-determined | Genome-fragment | NA | | NA | 0 |
| S17_extracted_k141_40500 | 1405 | No | NA | 2 | 0 | 0 | Not-determined | Genome-fragment | NA | | NA | 0 |
| S17_extracted_k141_32767 | 3777 | No | NA | 7 | 0 | 6 | Not-determined | Genome-fragment | NA | | NA | 0 |
| S17_extracted_k141_24929 | 1085 | No | NA | 2 | 0 | 1 | Not-determined | Genome-fragment | NA | | NA | 0 |
| S17_extracted_k141_2939 | 1011 | No | NA | 1 | 0 | 0 | Not-determined | Genome-fragment | NA | | NA | 0 |
| S17_extracted_k141_143697 | 1100 | No | NA | 2 | 0 | 2 | Not-determined | Genome-fragment | NA | | NA | 0 |
| S8_extracted_k141_202132 | 1015 | No | NA | 1 | 0 | 1 | Low-quality | Genome-fragment | | 0.67 | AAI-based (high-confidence) | 0 |
| S17_vs_k141_75192\|\|full | 1016 | No | NA | 3 | 0 | 0 | Low-quality | Genome-fragment | | 1.49 | AAI-based (medium-confidence) | 0 |
| S17_extracted_k141_17492 | 1101 | No | NA | 2 | 0 | 2 | Not-determined | Genome-fragment | NA | | NA | 0 |
| S17_extracted_k141_83608 | 1476 | No | NA | 2 | 0 | 1 | Not-determined | Genome-fragment | NA | | NA | 0 |
| S17_extracted_k141_47714 | 1288 | No | NA | 2 | 0 | 0 | Not-determined | Genome-fragment | NA | | NA | 0 |
| S17_extracted_k141_40555 | 1994 | No | NA | 3 | 0 | 3 | Not-determined | Genome-fragment | NA | | NA | 0 |
| S17_extracted_k141_2962 | 3808 | No | NA | 3 | 0 | 0 | Not-determined | Genome-fragment | NA | | NA | 0 |
| S17_extracted_k141_61932 | 1309 | No | NA | 2 | 0 | 1 | Not-determined | Genome-fragment | NA | | NA | 0 |
| S17_extracted_k141_151366 | 1260 | No | NA | 1 | 0 | 0 | Not-determined | Genome-fragment | NA | | NA | 0 |
| S17_extracted_k141_128373 | 2165 | No | NA | 2 | 0 | 2 | Not-determined | Genome-fragment | NA | | NA | 0 |
| S17_extracted_k141_10597 | 3342 | No | NA | 3 | 0 | 2 | Not-determined | Genome-fragment | NA | | NA | 0 |
| S17_extracted_k141_47768 | 6459 | No | NA | 4 | 0 | 4 | Not-determined | Genome-fragment | NA | | NA | 0 |
| S17_vs_k141_50491\|\|full | 1016 | No | NA | 2 | 0 | 1 | Low-quality | Genome-fragment | | 2.41 | AAI-based (medium-confidence) | 0 |
| S17_extracted_k141_128382 | 3376 | No | NA | 2 | 0 | 2 | Not-determined | Genome-fragment | NA | | NA | 0 |
| S17_extracted_k141_76967 | 1998 | No | NA | 3 | 0 | 0 | Not-determined | Genome-fragment | NA | | NA | 0 |
| S17_extracted_k141_91626 | 1145 | No | NA | 3 | 0 | 2 | Not-determined | Genome-fragment | NA | | NA | 0 |
| S17_extracted_k141_113722 | 4181 | No | NA | 5 | 0 | 2 | Not-determined | Genome-fragment | NA | | NA | 0 |
| S9_vs_k141_16260\|\|full | 1016 | No | NA | 2 | 0 | 0 | Low-quality | Genome-fragment | | 0.72 | AAI-based (medium-confidence) | 0 |
| S17_extracted_k141_3013 | 5275 | No | NA | 3 | 0 | 2 | Not-determined | Genome-fragment | NA | | NA | 0 |
| S17_extracted_k141_99451 | 1607 | No | NA | 3 | 0 | 0 | Not-determined | Genome-fragment | NA | | NA | 0 |
| S17_extracted_k141_158439 | 1314 | No | NA | 2 | 0 | 0 | Not-determined | Genome-fragment | NA | | NA | 0 |
| S17_extracted_k141_128444 | 1122 | No | NA | 2 | 0 | 0 | Not-determined | Genome-fragment | NA | | NA | 0 |
| S14_vs_k141_44112\|\|full | 1016 | No | NA | 3 | 0 | 0 | Low-quality | Genome-fragment | | 1.46 | AAI-based (medium-confidence) | 0 |
| S17_extracted_k141_121302 | 7557 | No | NA | 8 | 0 | 1 | Not-determined | Genome-fragment | NA | | NA | 0 |
| S14_vs_k141_372\|\|full | 1016 | No | NA | 3 | 1 | 0 | Low-quality | Genome-fragment | | 0.54 | HMM-based (lower-bound) | 0 |
| S17_extracted_k141_55010 | 4476 | No | NA | 4 | 0 | 4 | Not-determined | Genome-fragment | NA | | NA | 0 |
| S14_vs_k141_17749\|\|full | 1016 | No | NA | 4 | 0 | 2 | Low-quality | Genome-fragment | | 1.68 | AAI-based (medium-confidence) | 0 |
| S17_extracted_k141_69794 | 2184 | No | NA | 2 | 0 | 2 | Not-determined | Genome-fragment | NA | | NA | 0 |
| S17_extracted_k141_69801 | 1458 | No | NA | 2 | 0 | 2 | Not-determined | Genome-fragment | NA | | NA | 0 |
| S14_vs_k141_46140\|\|lt2gene | 1016 | No | NA | 2 | 1 | 0 | Low-quality | Genome-fragment | | 2.51 | AAI-based (high-confidence) | 0 |
| S17_extracted_k141_77057 | 15807 | No | NA | 21 | 7 | 0 | Medium-quality | Genome-fragment | | 52.24 | HMM-based (lower-bound) | 0 |
| S17_extracted_k141_32939 | 1427 | No | NA | 2 | 0 | 0 | Not-determined | Genome-fragment | NA | | NA | 0 |
| S17_extracted_k141_135978 | 3792 | No | NA | 3 | 0 | 2 | Not-determined | Genome-fragment | NA | | NA | 0 |
| S3_extracted_k141_57908 | 1016 | No | NA | 2 | 0 | 0 | Low-quality | Genome-fragment | | 1.7 | AAI-based (high-confidence) | 0 |
| S17_extracted_k141_128487 | 2506 | No | NA | 3 | 0 | 2 | Not-determined | Genome-fragment | NA | | NA | 0 |
| S3_extracted_k141_55518 | 1016 | No | NA | 2 | 0 | 0 | Low-quality | Genome-fragment | | 2.15 | AAI-based (high-confidence) | 0 |
| S5_extracted_k141_70065 | 1016 | No | NA | 3 | 0 | 0 | Low-quality | Genome-fragment | | 1.98 | AAI-based (medium-confidence) | 0 |
| S17_extracted_k141_107260 | 6587 | No | NA | 5 | 0 | 3 | Not-determined | Genome-fragment | NA | | NA | 0 |
| S17_extracted_k141_165659 | 1064 | No | NA | 2 | 0 | 1 | Not-determined | Genome-fragment | NA | | NA | 0 |
| S17_extracted_k141_47869 | 1987 | No | NA | 3 | 0 | 1 | Not-determined | Genome-fragment | NA | | NA | 0 |
| S5_vs_k141_103797\|\|full | 1016 | No | NA | 2 | 0 | 0 | Low-quality | Genome-fragment | | 1.81 | AAI-based (high-confidence) | 0 |
| S17_extracted_k141_47870 | 1338 | No | NA | 2 | 0 | 2 | Not-determined | Genome-fragment | NA | | NA | 0 |
| S17_extracted_k141_158539 | 3191 | No | NA | 5 | 0 | 0 | Not-determined | Genome-fragment | NA | | NA | 0 |
| S17_extracted_k141_10738 | 1024 | No | NA | 2 | 0 | 0 | Not-determined | Genome-fragment | NA | | NA | 0 |
| S5_vs_k141_134553\|\|full | 1016 | No | NA | 3 | 1 | 0 | Low-quality | Genome-fragment | | 2.31 | AAI-based (medium-confidence) | 0 |
| S17_extracted_k141_17718 | 1135 | No | NA | 1 | 0 | 0 | Not-determined | Genome-fragment | NA | | NA | 0 |
| S17_extracted_k141_55069 | 1018 | No | NA | 1 | 0 | 1 | Not-determined | Genome-fragment | NA | | NA | 0 |
| S17_extracted_k141_32987 | 1249 | No | NA | 2 | 0 | 2 | Not-determined | Genome-fragment | NA | | NA | 0 |
| S17_extracted_k141_17727 | 1594 | No | NA | 2 | 0 | 2 | Not-determined | Genome-fragment | NA | | NA | 0 |
| S17_extracted_k141_33001 | 3096 | No | NA | 3 | 0 | 1 | Not-determined | Genome-fragment | NA | | NA | 0 |
| S17_extracted_k141_10757 | 1285 | No | NA | 2 | 0 | 1 | Not-determined | Genome-fragment | NA | | NA | 0 |
| S17_extracted_k141_121380 | 3699 | No | NA | 4 | 0 | 3 | Not-determined | Genome-fragment | NA | | NA | 0 |
| S5_vs_k141_77059\|\|full | 1016 | No | NA | 2 | 0 | 1 | Low-quality | Genome-fragment | | 0.46 | AAI-based (medium-confidence) | 0 |
| S17_extracted_k141_3164 | 1759 | No | NA | 1 | 0 | 0 | Not-determined | Genome-fragment | NA | | NA | 0 |
| S17_extracted_k141_128556 | 1321 | No | NA | 2 | 0 | 0 | Not-determined | Genome-fragment | NA | | NA | 0 |
| S17_extracted_k141_128558 | 1726 | No | NA | 1 | 0 | 0 | Not-determined | Genome-fragment | NA | | NA | 0 |
| S7_extracted_29856 | 1016 | No | NA | 3 | 0 | 2 | Low-quality | Genome-fragment | | 2.2 | AAI-based (medium-confidence) | 0 |
| S17_extracted_k141_172914 | 1607 | No | NA | 1 | 0 | 1 | Not-determined | Genome-fragment | NA | | NA | 0 |
| S17_extracted_k141_143895 | 2396 | No | NA | 3 | 0 | 1 | Not-determined | Genome-fragment | NA | | NA | 0 |
| S7_extracted_64257 | 1016 | No | NA | 1 | 0 | 0 | Low-quality | Genome-fragment | | 4.61 | AAI-based (high-confidence) | 0 |
| S17_extracted_k141_10794 | 1241 | No | NA | 3 | 0 | 0 | Not-determined | Genome-fragment | NA | | NA | 0 |
| S17_extracted_k141_136061 | 1249 | No | NA | 2 | 0 | 1 | Not-determined | Genome-fragment | NA | | NA | 0 |
| S7_extracted_88438 | 1016 | No | NA | 3 | 0 | 0 | Low-quality | Genome-fragment | | 0.68 | AAI-based (medium-confidence) | 0 |
| S17_extracted_k141_143923 | 1216 | No | NA | 2 | 0 | 0 | Not-determined | Genome-fragment | NA | | NA | 0 |
| S7_vs_32497\|\|lt2gene | 1016 | No | NA | 1 | 1 | 0 | Low-quality | Genome-fragment | | 2.37 | AAI-based (high-confidence) | 0 |
| S16_vs_122382\|\|full | 1016 | No | NA | 2 | 0 | 1 | Low-quality | Genome-fragment | | 1.58 | AAI-based (medium-confidence) | 0 |
| S17_extracted_k141_69937 | 1106 | No | NA | 1 | 0 | 1 | Not-determined | Genome-fragment | NA | | NA | 0 |
| S17_extracted_k141_113927 | 2719 | No | NA | 1 | 0 | 1 | Not-determined | Genome-fragment | NA | | NA | 0 |
| S16_vs_160899\|\|full | 1016 | No | NA | 2 | 0 | 0 | Low-quality | Genome-fragment | | 0.61 | AAI-based (high-confidence) | 0 |
| S16_vs_93496\|\|lt2gene | 1016 | No | NA | 2 | 2 | 0 | Low-quality | Genome-fragment | | 0.54 | HMM-based (lower-bound) | 0 |
| S17_extracted_k141_25170 | 1037 | No | NA | 2 | 0 | 0 | Not-determined | Genome-fragment | NA | | NA | 0 |
| S17_extracted_k141_121476 | 3880 | No | NA | 4 | 0 | 3 | Not-determined | Genome-fragment | NA | | NA | 0 |
| S17_extracted_k141_40811 | 3801 | No | NA | 2 | 0 | 2 | Not-determined | Genome-fragment | NA | | NA | 0 |
| S17_extracted_k141_33135 | 2388 | No | NA | 4 | 0 | 3 | Not-determined | Genome-fragment | NA | | NA | 0 |
| S17_extracted_k141_91849 | 3376 | No | NA | 2 | 0 | 2 | Not-determined | Genome-fragment | NA | | NA | 0 |
| S17_extracted_k141_77220 | 1578 | No | NA | 2 | 0 | 1 | Not-determined | Genome-fragment | NA | | NA | 0 |
| S17_extracted_k141_33161 | 3477 | No | NA | 7 | 0 | 1 | Not-determined | Genome-fragment | NA | | NA | 0 |
| S17_extracted_k141_48012 | 1020 | No | NA | 2 | 0 | 1 | Not-determined | Genome-fragment | NA | | NA | 0 |
| S17_extracted_k141_113987 | 5044 | No | NA | 5 | 0 | 2 | Not-determined | Genome-fragment | NA | | NA | 0 |
| S17_extracted_k141_165831 | 2474 | No | NA | 2 | 0 | 2 | Not-determined | Genome-fragment | NA | | NA | 0 |
| S17_extracted_k141_151557 | 1198 | No | NA | 2 | 0 | 2 | Not-determined | Genome-fragment | NA | | NA | 0 |
| S17_extracted_k141_128640 | 2274 | No | NA | 5 | 0 | 0 | Not-determined | Genome-fragment | NA | | NA | 0 |
| S17_extracted_k141_55267 | 1105 | No | NA | 1 | 0 | 1 | Not-determined | Genome-fragment | NA | | NA | 0 |
| S17_extracted_k141_121530 | 1345 | No | NA | 1 | 0 | 0 | Not-determined | Genome-fragment | NA | | NA | 0 |
| S17_extracted_k141_158699 | 3250 | No | NA | 2 | 0 | 2 | Not-determined | Genome-fragment | NA | | NA | 0 |
| S17_extracted_k141_77273 | 1049 | No | NA | 1 | 0 | 1 | Not-determined | Genome-fragment | NA | | NA | 0 |
| S10_vs_25912\|\|full | 1016 | No | NA | 4 | 0 | 0 | Low-quality | Genome-fragment | | 2.08 | AAI-based (high-confidence) | 0 |
| S17_extracted_k141_136204 | 1779 | No | NA | 2 | 0 | 2 | Not-determined | Genome-fragment | NA | | NA | 0 |
| S17_extracted_k141_70028 | 1043 | No | NA | 1 | 0 | 0 | Not-determined | Genome-fragment | NA | | NA | 0 |
| S17_extracted_k141_33217 | 1928 | No | NA | 3 | 0 | 0 | Not-determined | Genome-fragment | NA | | NA | 0 |
| S17_extracted_k141_3375 | 1432 | No | NA | 2 | 0 | 1 | Not-determined | Genome-fragment | NA | | NA | 0 |
| S17_extracted_k141_121563 | 3364 | No | NA | 4 | 0 | 3 | Not-determined | Genome-fragment | NA | | NA | 0 |
| S17_extracted_k141_10893 | 1486 | No | NA | 1 | 0 | 1 | Not-determined | Genome-fragment | NA | | NA | 0 |
| S17_extracted_k141_3401 | 1451 | No | NA | 2 | 0 | 1 | Not-determined | Genome-fragment | NA | | NA | 0 |
| S17_extracted_k141_70076 | 1142 | No | NA | 2 | 0 | 1 | Not-determined | Genome-fragment | NA | | NA | 0 |
| S12_vs_k141_173318\|\|full | 1016 | No | NA | 2 | 0 | 0 | Low-quality | Genome-fragment | | 1.05 | AAI-based (high-confidence) | 0 |
| S17_extracted_k141_40918 | 5283 | No | NA | 7 | 0 | 1 | Not-determined | Genome-fragment | NA | | NA | 0 |
| S17_extracted_k141_33288 | 1062 | No | NA | 2 | 0 | 2 | Not-determined | Genome-fragment | NA | | NA | 0 |
| S17_extracted_k141_77323 | 1808 | No | NA | 2 | 0 | 0 | Not-determined | Genome-fragment | NA | | NA | 0 |
| S2_vs_k141_11559\|\|full | 1016 | No | NA | 3 | 0 | 0 | Low-quality | Genome-fragment | | 1.06 | AAI-based (high-confidence) | 0 |
| S17_extracted_k141_77328 | 1997 | No | NA | 4 | 0 | 0 | Not-determined | Genome-fragment | NA | | NA | 0 |
| S17_extracted_k141_173117 | 1128 | No | NA | 1 | 0 | 0 | Not-determined | Genome-fragment | NA | | NA | 0 |
| S17_extracted_k141_173126 | 1278 | No | NA | 2 | 0 | 0 | Not-determined | Genome-fragment | NA | | NA | 0 |
| S17_extracted_k141_107508 | 1689 | No | NA | 2 | 0 | 2 | Not-determined | Genome-fragment | NA | | NA | 0 |
| S17_extracted_k141_151625 | 1600 | No | NA | 2 | 0 | 0 | Not-determined | Genome-fragment | NA | | NA | 0 |
| S17_extracted_k141_70139 | 1571 | No | NA | 2 | 0 | 1 | Not-determined | Genome-fragment | NA | | NA | 0 |
| S4_extracted_2119 | 1016 | No | NA | 4 | 3 | 0 | Low-quality | Genome-fragment | | 3.34 | AAI-based (high-confidence) | 0 |
| S17_extracted_k141_84003 | 1570 | No | NA | 1 | 0 | 1 | Not-determined | Genome-fragment | NA | | NA | 0 |
| S17_extracted_k141_55390 | 1780 | No | NA | 1 | 0 | 1 | Not-determined | Genome-fragment | NA | | NA | 0 |
| S4_extracted_6477 | 1016 | No | NA | 3 | 0 | 1 | Low-quality | Genome-fragment | | 0.42 | AAI-based (medium-confidence) | 0 |
| S17_extracted_k141_173166 | 1176 | No | NA | 2 | 0 | 0 | Not-determined | Genome-fragment | NA | | NA | 0 |
| S17_extracted_k141_25328 | 3020 | No | NA | 2 | 0 | 2 | Not-determined | Genome-fragment | NA | | NA | 0 |
| S17_extracted_k141_55396 | 16910 | No | NA | 15 | 0 | 4 | Not-determined | Genome-fragment | NA | | NA | 0 |
| S17_extracted_k141_158886 | 1008 | No | NA | 1 | 0 | 1 | Not-determined | Genome-fragment | NA | | NA | 0 |
| S17_extracted_k141_40991 | 2237 | No | NA | 4 | 0 | 2 | Not-determined | Genome-fragment | NA | | NA | 0 |
| S17_extracted_k141_165929 | 1379 | No | NA | 2 | 0 | 1 | Not-determined | Genome-fragment | NA | | NA | 0 |
| S17_extracted_k141_62315 | 2959 | No | NA | 8 | 0 | 7 | Not-determined | Genome-fragment | NA | | NA | 0 |
| S17_extracted_k141_165933 | 4553 | No | NA | 3 | 0 | 3 | Not-determined | Genome-fragment | NA | | NA | 0 |
| S17_extracted_k141_3523 | 1523 | No | NA | 2 | 0 | 0 | Not-determined | Genome-fragment | NA | | NA | 0 |
| S17_extracted_k141_33368 | 1125 | No | NA | 1 | 0 | 0 | Not-determined | Genome-fragment | NA | | NA | 0 |
| S17_extracted_k141_99845 | 1597 | No | NA | 3 | 0 | 1 | Not-determined | Genome-fragment | NA | | NA | 0 |
| S17_extracted_k141_33381 | 1163 | No | NA | 1 | 0 | 1 | Not-determined | Genome-fragment | NA | | NA | 0 |
| S4_extracted_19463 | 1016 | No | NA | 3 | 0 | 0 | Low-quality | Genome-fragment | | 2.38 | AAI-based (medium-confidence) | 0 |
| S17_extracted_k141_144117 | 1177 | No | NA | 1 | 0 | 0 | Not-determined | Genome-fragment | NA | | NA | 0 |
| S17_extracted_k141_70226 | 1189 | No | NA | 1 | 0 | 1 | Not-determined | Genome-fragment | NA | | NA | 0 |
| S17_extracted_k141_84070 | 3578 | No | NA | 3 | 0 | 1 | Not-determined | Genome-fragment | NA | | NA | 0 |
| S17_extracted_k141_165954 | 1653 | No | NA | 1 | 0 | 1 | Not-determined | Genome-fragment | NA | | NA | 0 |
| S6_extracted_k141_8874 | 1016 | No | NA | 1 | 1 | 0 | Low-quality | Genome-fragment | | 1.72 | AAI-based (high-confidence) | 0 |
| S8_extracted_k141_132247 | 1016 | No | NA | 1 | 0 | 1 | Low-quality | Genome-fragment | | 1.64 | AAI-based (medium-confidence) | 0 |
| S17_extracted_k141_33391 | 1326 | No | NA | 1 | 0 | 1 | Not-determined | Genome-fragment | NA | | NA | 0 |
| S17_extracted_k141_25400 | 4240 | No | NA | 4 | 0 | 3 | Not-determined | Genome-fragment | NA | | NA | 0 |
| S17_extracted_k141_107589 | 2012 | No | NA | 3 | 0 | 1 | Not-determined | Genome-fragment | NA | | NA | 0 |
| S17_extracted_k141_128878 | 10585 | No | NA | 5 | 0 | 4 | Not-determined | Genome-fragment | NA | | NA | 0 |
| S17_extracted_k141_48217 | 2437 | No | NA | 3 | 0 | 2 | Not-determined | Genome-fragment | NA | | NA | 0 |
| S8_vs_k141_142926\|\|full | 1016 | No | NA | 2 | 0 | 0 | Low-quality | Genome-fragment | | 1.61 | AAI-based (medium-confidence) | 0 |
| S17_extracted_k141_84121 | 1087 | No | NA | 1 | 0 | 1 | Not-determined | Genome-fragment | NA | | NA | 0 |
| S17_extracted_k141_84134 | 2528 | No | NA | 3 | 0 | 3 | Not-determined | Genome-fragment | NA | | NA | 0 |
| S17_extracted_k141_70293 | 1099 | No | NA | 1 | 0 | 0 | Not-determined | Genome-fragment | NA | | NA | 0 |
| S17_extracted_k141_121683 | 1161 | No | NA | 3 | 0 | 3 | Not-determined | Genome-fragment | NA | | NA | 0 |
| S17_extracted_k141_152383 | 1017 | No | NA | 1 | 0 | 0 | Low-quality | Genome-fragment | | 0.77 | AAI-based (high-confidence) | 0 |
| S17_extracted_k141_18135 | 1064 | No | NA | 1 | 0 | 0 | Not-determined | Genome-fragment | NA | | NA | 0 |
| S17_extracted_k141_33437 | 2938 | No | NA | 3 | 0 | 2 | Not-determined | Genome-fragment | NA | | NA | 0 |
| S17_extracted_k141_25455 | 1264 | No | NA | 1 | 0 | 0 | Not-determined | Genome-fragment | NA | | NA | 0 |
| S17_extracted_k141_128926 | 3925 | No | NA | 3 | 0 | 1 | Not-determined | Genome-fragment | NA | | NA | 0 |
| S17_extracted_k141_173334 | 1384 | No | NA | 3 | 0 | 2 | Not-determined | Genome-fragment | NA | | NA | 0 |
| S17_extracted_k141_70310 | 1055 | No | NA | 1 | 0 | 0 | Not-determined | Genome-fragment | NA | | NA | 0 |
| S13_vs_146034\|\|full | 1017 | No | NA | 3 | 0 | 0 | Low-quality | Genome-fragment | | 2.57 | AAI-based (medium-confidence) | 0 |
| S17_extracted_k141_77525 | 1604 | No | NA | 3 | 0 | 3 | Not-determined | Genome-fragment | NA | | NA | 0 |
| S17_extracted_k141_77533 | 1114 | No | NA | 2 | 0 | 1 | Not-determined | Genome-fragment | NA | | NA | 0 |
| S14_extracted_k141_78717 | 1017 | No | NA | 1 | 1 | 0 | Low-quality | Genome-fragment | | 3.28 | HMM-based (lower-bound) | 0 |
| S17_extracted_k141_114242 | 1137 | No | NA | 1 | 0 | 1 | Not-determined | Genome-fragment | NA | | NA | 0 |
| S17_extracted_k141_33520 | 1424 | No | NA | 1 | 0 | 1 | Not-determined | Genome-fragment | NA | | NA | 0 |
| S14_vs_k141_110747\|\|lt2gene | 1017 | No | NA | 2 | 2 | 0 | Low-quality | Genome-fragment | | 3.61 | AAI-based (medium-confidence) | 0 |
| S17_extracted_k141_173369 | 1307 | No | NA | 3 | 0 | 0 | Not-determined | Genome-fragment | NA | | NA | 0 |
| S3_extracted_k141_61548 | 1017 | No | NA | 1 | 0 | 0 | Low-quality | Genome-fragment | | 0.16 | AAI-based (high-confidence) | 0 |
| S3_vs_k141_55024\|\|full | 1017 | No | NA | 4 | 0 | 0 | Low-quality | Genome-fragment | | 1.94 | AAI-based (medium-confidence) | 0 |
| S7_extracted_5818 | 1017 | No | NA | 2 | 1 | 0 | Low-quality | Genome-fragment | | 2.16 | AAI-based (medium-confidence) | 0 |
| S7_extracted_30681 | 1017 | No | NA | 2 | 1 | 0 | Low-quality | Genome-fragment | | 3.69 | AAI-based (high-confidence) | 0 |
| S17_extracted_k141_70372 | 1413 | No | NA | 3 | 0 | 2 | Not-determined | Genome-fragment | NA | | NA | 0 |
| S17_extracted_k141_129014 | 1414 | No | NA | 1 | 0 | 0 | Not-determined | Genome-fragment | NA | | NA | 0 |
| S17_extracted_k141_151843 | 5026 | No | NA | 8 | 0 | 1 | Not-determined | Genome-fragment | NA | | NA | 0 |
| S17_extracted_k141_121747 | 3509 | No | NA | 3 | 0 | 2 | Not-determined | Genome-fragment | NA | | NA | 0 |
| S17_extracted_k141_92201 | 1297 | No | NA | 2 | 0 | 0 | Not-determined | Genome-fragment | NA | | NA | 0 |
| S17_extracted_k141_159105 | 1102 | No | NA | 2 | 0 | 1 | Not-determined | Genome-fragment | NA | | NA | 0 |
| S7_vs_114805\|\|full | 1017 | No | NA | 3 | 0 | 0 | Low-quality | Genome-fragment | | 1.63 | AAI-based (medium-confidence) | 0 |
| S17_extracted_k141_70392 | 2292 | No | NA | 3 | 0 | 2 | Not-determined | Genome-fragment | NA | | NA | 0 |
| S17_extracted_k141_33592 | 3767 | No | NA | 6 | 0 | 2 | Not-determined | Genome-fragment | NA | | NA | 0 |
| S17_extracted_k141_92217 | 1387 | No | NA | 1 | 0 | 1 | Not-determined | Genome-fragment | NA | | NA | 0 |
| S16_extracted_129288 | 1017 | No | NA | 4 | 0 | 0 | Low-quality | Genome-fragment | | 0.62 | AAI-based (medium-confidence) | 0 |
| S17_extracted_k141_3670 | 3561 | No | NA | 7 | 0 | 1 | Not-determined | Genome-fragment | NA | | NA | 0 |
| S16_extracted_160631 | 1017 | No | NA | 1 | 0 | 1 | Low-quality | Genome-fragment | | 0.57 | AAI-based (medium-confidence) | 0 |
| S17_extracted_k141_151890 | 2813 | No | NA | 3 | 0 | 1 | Not-determined | Genome-fragment | NA | | NA | 0 |
| S16_vs_132201\|\|full | 1017 | No | NA | 2 | 1 | 0 | Low-quality | Genome-fragment | | 2.48 | AAI-based (high-confidence) | 0 |
| S17_extracted_k141_144308 | 1249 | No | NA | 2 | 0 | 0 | Not-determined | Genome-fragment | NA | | NA | 0 |
| S12_extracted_k141_110294 | 1017 | No | NA | 2 | 0 | 1 | Low-quality | Genome-fragment | | 1.13 | AAI-based (medium-confidence) | 0 |
| S17_extracted_k141_159139 | 2725 | No | NA | 2 | 0 | 2 | Not-determined | Genome-fragment | NA | | NA | 0 |
| S17_extracted_k141_151900 | 1403 | No | NA | 2 | 0 | 2 | Not-determined | Genome-fragment | NA | | NA | 0 |
| S17_extracted_k141_70425 | 3577 | No | NA | 2 | 0 | 2 | Not-determined | Genome-fragment | NA | | NA | 0 |
| S17_extracted_k141_144326 | 2585 | No | NA | 1 | 0 | 1 | Not-determined | Genome-fragment | NA | | NA | 0 |
| S17_extracted_k141_55617 | 3882 | No | NA | 3 | 0 | 1 | Not-determined | Genome-fragment | NA | | NA | 0 |
| S17_extracted_k141_33629 | 3467 | No | NA | 3 | 0 | 3 | Not-determined | Genome-fragment | NA | | NA | 0 |
| S8_vs_k141_174691\|\|full | 1017 | No | NA | 3 | 0 | 0 | Low-quality | Genome-fragment | | 1.69 | AAI-based (high-confidence) | 0 |
| S17_extracted_k141_159152 | 3688 | No | NA | 4 | 0 | 2 | Not-determined | Genome-fragment | NA | | NA | 0 |
| S17_extracted_k141_121777 | 7909 | No | NA | 5 | 0 | 3 | Not-determined | Genome-fragment | NA | | NA | 0 |
| S17_extracted_k141_18303 | 2121 | No | NA | 1 | 0 | 1 | Not-determined | Genome-fragment | NA | | NA | 0 |
| S17_extracted_k141_55628 | 1373 | No | NA | 2 | 0 | 1 | Not-determined | Genome-fragment | NA | | NA | 0 |
| S17_extracted_k141_41187 | 1815 | No | NA | 4 | 0 | 2 | Not-determined | Genome-fragment | NA | | NA | 0 |
| S17_vs_k141_8319\|\|full | 1018 | No | NA | 4 | 0 | 1 | Low-quality | Genome-fragment | | 0.64 | AAI-based (medium-confidence) | 0 |
| S17_extracted_k141_151919 | 5444 | No | NA | 10 | 0 | 1 | Not-determined | Genome-fragment | NA | | NA | 0 |
| S17_extracted_k141_159155 | 1775 | No | NA | 2 | 0 | 1 | Not-determined | Genome-fragment | NA | | NA | 0 |
| S17_extracted_k141_70499 | 1300 | No | NA | 2 | 0 | 1 | Not-determined | Genome-fragment | NA | | NA | 0 |
| S17_extracted_k141_55675 | 1477 | No | NA | 1 | 0 | 1 | Not-determined | Genome-fragment | NA | | NA | 0 |
| S17_extracted_k141_77679 | 4591 | No | NA | 4 | 0 | 3 | Not-determined | Genome-fragment | NA | | NA | 0 |
| S17_extracted_k141_41246 | 1024 | No | NA | 1 | 0 | 1 | Not-determined | Genome-fragment | NA | | NA | 0 |
| S17_extracted_k141_151972 | 1597 | No | NA | 2 | 0 | 2 | Not-determined | Genome-fragment | NA | | NA | 0 |
| S17_extracted_k141_41255 | 1256 | No | NA | 1 | 0 | 1 | Not-determined | Genome-fragment | NA | | NA | 0 |
| S17_extracted_k141_151974 | 3863 | No | NA | 3 | 0 | 0 | Not-determined | Genome-fragment | NA | | NA | 0 |
| S17_extracted_k141_166185 | 2188 | No | NA | 4 | 0 | 0 | Not-determined | Genome-fragment | NA | | NA | 0 |
| S9_vs_k141_17230\|\|full | 1018 | No | NA | 2 | 0 | 1 | Low-quality | Genome-fragment | | 1.79 | AAI-based (medium-confidence) | 0 |
| S9_vs_k141_42901\|\|lt2gene | 1018 | No | NA | 2 | 1 | 0 | Low-quality | Genome-fragment | | 3.05 | AAI-based (high-confidence) | 0 |
| S17_extracted_k141_114499 | 1191 | No | NA | 1 | 0 | 1 | Not-determined | Genome-fragment | NA | | NA | 0 |
| S17_extracted_k141_18388 | 1764 | No | NA | 3 | 0 | 3 | Not-determined | Genome-fragment | NA | | NA | 0 |
| S13_vs_94563\|\|full | 1018 | No | NA | 2 | 1 | 0 | Low-quality | Genome-fragment | | 1.75 | HMM-based (lower-bound) | 0 |
| S17_extracted_k141_70504 | 4613 | No | NA | 6 | 0 | 3 | Not-determined | Genome-fragment | NA | | NA | 0 |
| S17_extracted_k141_41272 | 1562 | No | NA | 1 | 0 | 1 | Not-determined | Genome-fragment | NA | | NA | 0 |
| S14_extracted_k141_22997 | 1018 | No | NA | 1 | 0 | 1 | Low-quality | Genome-fragment | | 0.5 | AAI-based (medium-confidence) | 0 |
| S17_extracted_k141_159180 | 1500 | No | NA | 2 | 0 | 1 | Not-determined | Genome-fragment | NA | | NA | 0 |
| S17_extracted_k141_121844 | 1196 | No | NA | 1 | 0 | 0 | Not-determined | Genome-fragment | NA | | NA | 0 |
| S17_extracted_k141_100049 | 6778 | No | NA | 3 | 0 | 3 | Not-determined | Genome-fragment | NA | | NA | 0 |
| S17_extracted_k141_114505 | 16352 | No | NA | 15 | 0 | 6 | Not-determined | Genome-fragment | NA | | NA | 0 |
| S17_extracted_k141_18413 | 1010 | No | NA | 1 | 0 | 0 | Not-determined | Genome-fragment | NA | | NA | 0 |
| S17_extracted_k141_55722 | 1582 | No | NA | 2 | 0 | 2 | Not-determined | Genome-fragment | NA | | NA | 0 |
| S17_extracted_k141_114515 | 1429 | No | NA | 1 | 0 | 1 | Not-determined | Genome-fragment | NA | | NA | 0 |
| S17_extracted_k141_84286 | 3873 | No | NA | 8 | 0 | 5 | Not-determined | Genome-fragment | NA | | NA | 0 |
| S17_extracted_k141_84294 | 1664 | No | NA | 2 | 0 | 0 | Not-determined | Genome-fragment | NA | | NA | 0 |
| S14_vs_k141_55241\|\|lt2gene | 1018 | No | NA | 2 | 1 | 0 | Low-quality | Genome-fragment | | 3.35 | AAI-based (high-confidence) | 0 |
| S17_extracted_k141_152017 | 2283 | No | NA | 2 | 0 | 2 | Not-determined | Genome-fragment | NA | | NA | 0 |
| S1_vs_k141_56108\|\|full | 1018 | No | NA | 2 | 1 | 0 | Low-quality | Genome-fragment | | 1.35 | HMM-based (lower-bound) | 0 |
| S17_extracted_k141_84311 | 1334 | No | NA | 3 | 0 | 1 | Not-determined | Genome-fragment | NA | | NA | 0 |
| S17_extracted_k141_33780 | 1803 | No | NA | 2 | 0 | 1 | Not-determined | Genome-fragment | NA | | NA | 0 |
| S17_extracted_k141_18479 | 1076 | No | NA | 1 | 0 | 1 | Not-determined | Genome-fragment | NA | | NA | 0 |
| S17_extracted_k141_114570 | 1003 | No | NA | 1 | 0 | 1 | Not-determined | Genome-fragment | NA | | NA | 0 |
| S17_extracted_k141_100097 | 7039 | No | NA | 8 | 0 | 2 | Not-determined | Genome-fragment | NA | | NA | 0 |
| S3_vs_k141_50542\|\|full | 1018 | No | NA | 3 | 0 | 0 | Low-quality | Genome-fragment | | 2.28 | AAI-based (medium-confidence) | 0 |
| S17_extracted_k141_55784 | 3466 | No | NA | 7 | 0 | 7 | Not-determined | Genome-fragment | NA | | NA | 0 |
| S17_extracted_k141_84332 | 3408 | No | NA | 4 | 0 | 1 | Not-determined | Genome-fragment | NA | | NA | 0 |
| S17_extracted_k141_84337 | 1633 | No | NA | 3 | 0 | 2 | Not-determined | Genome-fragment | NA | | NA | 0 |
| S17_extracted_k141_18495 | 1079 | No | NA | 1 | 0 | 1 | Not-determined | Genome-fragment | NA | | NA | 0 |
| S7_vs_133144\|\|full | 1018 | No | NA | 2 | 0 | 0 | Low-quality | Genome-fragment | | 1.96 | AAI-based (medium-confidence) | 0 |
| S17_extracted_k141_33825 | 1127 | No | NA | 2 | 0 | 1 | Not-determined | Genome-fragment | NA | | NA | 0 |
| S17_extracted_k141_173569 | 3479 | No | NA | 5 | 0 | 0 | Not-determined | Genome-fragment | NA | | NA | 0 |
| S17_extracted_k141_55803 | 3557 | No | NA | 2 | 0 | 1 | Not-determined | Genome-fragment | NA | | NA | 0 |
| S7_vs_30888\|\|lt2gene | 1018 | No | NA | 1 | 1 | 0 | Low-quality | Genome-fragment | | 1.93 | HMM-based (lower-bound) | 0 |
| S17_extracted_k141_33845 | 1100 | No | NA | 1 | 0 | 1 | Not-determined | Genome-fragment | NA | | NA | 0 |
| S16_extracted_159849 | 1018 | No | NA | 2 | 0 | 1 | Low-quality | Genome-fragment | | 0.6 | AAI-based (medium-confidence) | 0 |
| S17_extracted_k141_92331 | 1009 | No | NA | 1 | 0 | 1 | Not-determined | Genome-fragment | NA | | NA | 0 |
| S17_extracted_k141_25577 | 1009 | No | NA | 2 | 0 | 0 | Not-determined | Genome-fragment | NA | | NA | 0 |
| S17_extracted_k141_136622 | 1124 | No | NA | 2 | 0 | 2 | Not-determined | Genome-fragment | NA | | NA | 0 |
| S17_extracted_k141_70605 | 1675 | No | NA | 2 | 0 | 2 | Not-determined | Genome-fragment | NA | | NA | 0 |
| S10_vs_93951\|\|full | 1018 | No | NA | 2 | 0 | 1 | Low-quality | Genome-fragment | | 1.82 | AAI-based (medium-confidence) | 0 |
| S6_extracted_k141_37264 | 1018 | No | NA | 2 | 0 | 0 | Low-quality | Genome-fragment | | 1.66 | AAI-based (high-confidence) | 0 |
| S17_extracted_k141_3956 | 1714 | No | NA | 3 | 0 | 3 | Not-determined | Genome-fragment | NA | | NA | 0 |
| S17_extracted_k141_33860 | 7715 | No | NA | 4 | 0 | 1 | Not-determined | Genome-fragment | NA | | NA | 0 |
| S17_extracted_k141_136632 | 2614 | No | NA | 4 | 0 | 2 | Not-determined | Genome-fragment | NA | | NA | 0 |
| S8_vs_k141_70241\|\|full | 1018 | No | NA | 2 | 0 | 0 | Low-quality | Genome-fragment | | 2.28 | AAI-based (high-confidence) | 0 |
| S17_extracted_k141_41360 | 1620 | No | NA | 2 | 0 | 2 | Not-determined | Genome-fragment | NA | | NA | 0 |
| S17_extracted_k141_77883 | 1465 | No | NA | 2 | 0 | 2 | Not-determined | Genome-fragment | NA | | NA | 0 |
| S17_extracted_k141_100206 | 1398 | No | NA | 1 | 0 | 0 | Not-determined | Genome-fragment | NA | | NA | 0 |
| S17_extracted_k141_121975 | 1898 | No | NA | 1 | 0 | 1 | Not-determined | Genome-fragment | NA | | NA | 0 |
| S17_extracted_k141_129375 | 3356 | No | NA | 3 | 0 | 1 | Not-determined | Genome-fragment | NA | | NA | 0 |
| S17_extracted_k141_84413 | 1805 | No | NA | 4 | 0 | 1 | Not-determined | Genome-fragment | NA | | NA | 0 |
| S17_extracted_k141_70616 | 2902 | No | NA | 2 | 0 | 2 | Not-determined | Genome-fragment | NA | | NA | 0 |
| S8_vs_k141_62050\|\|full | 1018 | No | NA | 2 | 0 | 0 | Low-quality | Genome-fragment | | 0.65 | AAI-based (high-confidence) | 0 |
| S17_extracted_k141_70626 | 1014 | No | NA | 2 | 0 | 0 | Not-determined | Genome-fragment | NA | | NA | 0 |
| S17_extracted_k141_11341 | 2920 | No | NA | 2 | 0 | 0 | Not-determined | Genome-fragment | NA | | NA | 0 |
| S17_extracted_k141_121990 | 3078 | No | NA | 4 | 0 | 3 | Not-determined | Genome-fragment | NA | | NA | 0 |
| S17_extracted_k141_121993 | 1674 | No | NA | 2 | 0 | 0 | Not-determined | Genome-fragment | NA | | NA | 0 |
| S17_extracted_k141_107974 | 1214 | No | NA | 2 | 0 | 1 | Not-determined | Genome-fragment | NA | | NA | 0 |
| S17_vs_k141_15486\|\|full | 1019 | No | NA | 3 | 0 | 0 | Low-quality | Genome-fragment | | 2.1 | AAI-based (medium-confidence) | 0 |
| S17_extracted_k141_70670 | 3458 | No | NA | 3 | 0 | 3 | Not-determined | Genome-fragment | NA | | NA | 0 |
| S17_extracted_k141_108003 | 1027 | No | NA | 1 | 0 | 1 | Not-determined | Genome-fragment | NA | | NA | 0 |
| S17_extracted_k141_41478 | 1185 | No | NA | 1 | 0 | 1 | Not-determined | Genome-fragment | NA | | NA | 0 |
| S17_extracted_k141_114748 | 2158 | No | NA | 7 | 0 | 0 | Not-determined | Genome-fragment | NA | | NA | 0 |
| S17_extracted_k141_122046 | 1397 | No | NA | 2 | 0 | 0 | Not-determined | Genome-fragment | NA | | NA | 0 |
| S17_extracted_k141_129448 | 2705 | No | NA | 2 | 0 | 2 | Not-determined | Genome-fragment | NA | | NA | 0 |
| S17_extracted_k141_48660 | 1613 | No | NA | 2 | 0 | 2 | Not-determined | Genome-fragment | NA | | NA | 0 |
| S9_extracted_k141_16829 | 1019 | No | NA | 1 | 1 | 0 | Low-quality | Genome-fragment | | 2.3 | AAI-based (high-confidence) | 0 |
| S17_extracted_k141_84464 | 1017 | No | NA | 1 | 0 | 1 | Not-determined | Genome-fragment | NA | | NA | 0 |
| S17_extracted_k141_108045 | 4890 | No | NA | 6 | 0 | 3 | Not-determined | Genome-fragment | NA | | NA | 0 |
| S17_extracted_k141_166462 | 1002 | No | NA | 2 | 0 | 2 | Not-determined | Genome-fragment | NA | | NA | 0 |
| S17_extracted_k141_33976 | 1508 | No | NA | 1 | 0 | 1 | Not-determined | Genome-fragment | NA | | NA | 0 |

| | | | | | | | | | | | |
|---|---|---|---|---|---|---|---|---|---|---|---|
| S17_extracted_k141_152240 | 1699 | No | NA | 1 | 0 | 0 | Not-determined | Genome-fragment | NA | NA | 0 |
| S17_extracted_k141_159350 | 1825 | No | NA | 2 | 0 | 0 | Not-determined | Genome-fragment | NA | NA | 0 |
| S9_extracted_k141_62960 | 1019 | No | NA | 3 | 0 | 0 | Low-quality | Genome-fragment | 1.8 | AAI-based (medium-confidence) | 0 |
| S17_extracted_k141_84494 | 5641 | No | NA | 5 | 0 | 2 | Not-determined | Genome-fragment | NA | NA | 0 |
| S17_extracted_k141_48696 | 5706 | No | NA | 3 | 0 | 1 | Not-determined | Genome-fragment | NA | NA | 0 |
| S9_vs_k141_60422\|\|full | 1019 | No | NA | 2 | 0 | 0 | Low-quality | Genome-fragment | 0.48 | AAI-based (high-confidence) | 0 |
| S11_vs_k141_3274\|\|full | 1019 | No | NA | 2 | 0 | 0 | Low-quality | Genome-fragment | 1.71 | AAI-based (medium-confidence) | 0 |
| S17_extracted_k141_78031 | 1090 | No | NA | 1 | 0 | 0 | Not-determined | Genome-fragment | NA | NA | 0 |
| S17_extracted_k141_41565 | 8027 | No | NA | 6 | 0 | 4 | Not-determined | Genome-fragment | NA | NA | 0 |
| S17_extracted_k141_55981 | 1136 | No | NA | 2 | 0 | 0 | Not-determined | Genome-fragment | NA | NA | 0 |
| S17_extracted_k141_78033 | 1408 | No | NA | 2 | 0 | 1 | Not-determined | Genome-fragment | NA | NA | 0 |
| S17_extracted_k141_48744 | 1153 | No | NA | 2 | 0 | 0 | Not-determined | Genome-fragment | NA | NA | 0 |
| S11_vs_k141_65707\|\|lt2gene | 1019 | No | NA | 2 | 0 | 0 | Low-quality | Genome-fragment | 2.1 | AAI-based (medium-confidence) | 0 |
| S17_extracted_k141_92577 | 1241 | No | NA | 1 | 0 | 1 | Not-determined | Genome-fragment | NA | NA | 0 |
| S13_vs_92484\|\|full | 1019 | No | NA | 2 | 0 | 2 | Low-quality | Genome-fragment | 0.74 | AAI-based (medium-confidence) | 0 |
| S17_extracted_k141_11431 | 1077 | No | NA | 1 | 0 | 0 | Not-determined | Genome-fragment | NA | NA | 0 |
| S14_extracted_k141_25193 | 1019 | No | NA | 1 | 1 | 0 | Low-quality | Genome-fragment | 2.64 | AAI-based (medium-confidence) | 0 |
| S14_vs_k141_31919\|\|full | 1019 | No | NA | 3 | 1 | 0 | Low-quality | Genome-fragment | 0.63 | HMM-based (lower-bound) | 0 |
| S17_extracted_k141_18702 | 2776 | No | NA | 3 | 0 | 0 | Not-determined | Genome-fragment | NA | NA | 0 |
| S17_extracted_k141_56062 | 1137 | No | NA | 3 | 0 | 1 | Not-determined | Genome-fragment | NA | NA | 0 |
| S1_vs_k141_93429\|\|full | 1019 | No | NA | 3 | 1 | 0 | Low-quality | Genome-fragment | 2.05 | AAI-based (high-confidence) | 0 |
| S17_extracted_k141_114940 | 1190 | No | NA | 2 | 0 | 1 | Not-determined | Genome-fragment | NA | NA | 0 |
| S17_extracted_k141_108191 | 4006 | No | NA | 5 | 0 | 3 | Not-determined | Genome-fragment | NA | NA | 0 |
| S17_extracted_k141_25818 | 2345 | No | NA | 2 | 0 | 0 | Not-determined | Genome-fragment | NA | NA | 0 |
| S17_extracted_k141_152385 | 4905 | No | NA | 11 | 0 | 0 | Not-determined | Genome-fragment | NA | NA | 0 |
| S17_extracted_k141_108205 | 1197 | No | NA | 1 | 0 | 0 | Not-determined | Genome-fragment | NA | NA | 0 |
| S17_extracted_k141_11455 | 1066 | No | NA | 1 | 0 | 0 | Not-determined | Genome-fragment | NA | NA | 0 |
| S17_extracted_k141_41661 | 1053 | No | NA | 1 | 0 | 0 | Not-determined | Genome-fragment | NA | NA | 0 |
| S17_extracted_k141_129614 | 1176 | No | NA | 1 | 0 | 1 | Not-determined | Genome-fragment | NA | NA | 0 |
| S3_extracted_k141_96842 | 1019 | No | NA | 1 | 1 | 0 | Low-quality | Genome-fragment | 2.39 | HMM-based (lower-bound) | 0 |
| S17_extracted_k141_136915 | 1843 | No | NA | 1 | 0 | 0 | Not-determined | Genome-fragment | NA | NA | 0 |
| S17_extracted_k141_78175 | 1066 | No | NA | 1 | 0 | 0 | Not-determined | Genome-fragment | NA | NA | 0 |
| S17_extracted_k141_84590 | 4227 | No | NA | 2 | 0 | 2 | Not-determined | Genome-fragment | NA | NA | 0 |
| S17_extracted_k141_100418 | 7107 | No | NA | 7 | 0 | 2 | Not-determined | Genome-fragment | NA | NA | 0 |
| S17_extracted_k141_84594 | 2347 | No | NA | 2 | 0 | 0 | Not-determined | Genome-fragment | NA | NA | 0 |
| S3_vs_k141_97574\|\|full | 1019 | No | NA | 2 | 1 | 0 | Low-quality | Genome-fragment | 1.89 | AAI-based (medium-confidence) | 0 |
| S17_extracted_k141_56109 | 1564 | No | NA | 2 | 0 | 1 | Not-determined | Genome-fragment | NA | NA | 0 |
| S17_extracted_k141_166613 | 3350 | No | NA | 4 | 0 | 2 | Not-determined | Genome-fragment | NA | NA | 0 |
| S17_extracted_k141_56119 | 3163 | No | NA | 6 | 0 | 3 | Not-determined | Genome-fragment | NA | NA | 0 |
| S17_extracted_k141_100447 | 4151 | No | NA | 3 | 0 | 2 | Not-determined | Genome-fragment | NA | NA | 0 |
| S17_extracted_k141_11506 | 1517 | No | NA | 2 | 0 | 1 | Not-determined | Genome-fragment | NA | NA | 0 |
| S5_vs_k141_136639\|\|full | 1019 | No | NA | 3 | 0 | 0 | Low-quality | Genome-fragment | 1.48 | AAI-based (high-confidence) | 0 |
| S17_extracted_k141_92738 | 1217 | No | NA | 3 | 0 | 2 | Not-determined | Genome-fragment | NA | NA | 0 |
| S17_extracted_k141_115036 | 5012 | No | NA | 2 | 0 | 2 | Not-determined | Genome-fragment | NA | NA | 0 |
| S17_extracted_k141_136954 | 3960 | No | NA | 6 | 0 | 3 | Not-determined | Genome-fragment | NA | NA | 0 |
| S17_extracted_k141_159511 | 5767 | No | NA | 9 | 0 | 1 | Not-determined | Genome-fragment | NA | NA | 0 |
| S17_extracted_k141_11567 | 1008 | No | NA | 2 | 0 | 1 | Not-determined | Genome-fragment | NA | NA | 0 |
| S17_extracted_k141_34173 | 3326 | No | NA | 3 | 0 | 2 | Not-determined | Genome-fragment | NA | NA | 0 |
| S5_vs_k141_65532\|\|full | 1019 | No | NA | 3 | 0 | 2 | Low-quality | Genome-fragment | 0.77 | AAI-based (medium-confidence) | 0 |
| S17_extracted_k141_4110 | 1290 | No | NA | 1 | 0 | 1 | Not-determined | Genome-fragment | NA | NA | 0 |
| S17_extracted_k141_136971 | 1659 | No | NA | 2 | 0 | 1 | Not-determined | Genome-fragment | NA | NA | 0 |
| S17_extracted_k141_115062 | 1140 | No | NA | 3 | 0 | 3 | Not-determined | Genome-fragment | NA | NA | 0 |
| S17_extracted_k141_173835 | 1146 | No | NA | 1 | 0 | 1 | Not-determined | Genome-fragment | NA | NA | 0 |
| S7_extracted_106978 | 1019 | No | NA | 4 | 1 | 0 | Low-quality | Genome-fragment | 4.65 | AAI-based (high-confidence) | 0 |
| S7_extracted_121645 | 1019 | No | NA | 2 | 0 | 0 | Low-quality | Genome-fragment | 2.34 | AAI-based (medium-confidence) | 0 |
| S17_extracted_k141_166693 | 4304 | No | NA | 4 | 0 | 0 | Not-determined | Genome-fragment | NA | NA | 0 |
| S17_extracted_k141_78293 | 4700 | No | NA | 4 | 0 | 3 | Not-determined | Genome-fragment | NA | NA | 0 |
| S17_extracted_k141_129722 | 1301 | No | NA | 1 | 0 | 0 | Not-determined | Genome-fragment | NA | NA | 0 |
| S17_extracted_k141_122195 | 1664 | No | NA | 3 | 0 | 0 | Not-determined | Genome-fragment | NA | NA | 0 |
| S17_extracted_k141_173861 | 1914 | No | NA | 4 | 0 | 3 | Not-determined | Genome-fragment | NA | NA | 0 |
| S17_extracted_k141_137026 | 1078 | No | NA | 1 | 0 | 1 | Not-determined | Genome-fragment | NA | NA | 0 |
| S17_extracted_k141_84714 | 1308 | No | NA | 1 | 0 | 0 | Not-determined | Genome-fragment | NA | NA | 0 |
| S7_vs_98038\|\|full | 1019 | No | NA | 3 | 0 | 0 | Low-quality | Genome-fragment | 1.84 | AAI-based (high-confidence) | 0 |
| S17_extracted_k141_159570 | 3278 | No | NA | 1 | 0 | 1 | Not-determined | Genome-fragment | NA | NA | 0 |
| S17_extracted_k141_108432 | 1642 | No | NA | 2 | 0 | 1 | Not-determined | Genome-fragment | NA | NA | 0 |
| S17_extracted_k141_108435 | 1050 | No | NA | 1 | 0 | 1 | Not-determined | Genome-fragment | NA | NA | 0 |
| S17_extracted_k141_159583 | 1645 | No | NA | 2 | 0 | 1 | Not-determined | Genome-fragment | NA | NA | 0 |
| S7_vs_121638\|\|full | 1019 | No | NA | 4 | 1 | 0 | Low-quality | Genome-fragment | 1.72 | AAI-based (medium-confidence) | 0 |
| S17_extracted_k141_62564 | 1472 | No | NA | 2 | 0 | 1 | Not-determined | Genome-fragment | NA | NA | 0 |
| S16_vs_1853\|\|full | 1019 | No | NA | 2 | 0 | 0 | Low-quality | Genome-fragment | 2.83 | AAI-based (high-confidence) | 0 |
| S17_extracted_k141_159598 | 1222 | No | NA | 3 | 0 | 0 | Not-determined | Genome-fragment | NA | NA | 0 |
| S17_extracted_k141_152617 | 2026 | No | NA | 3 | 0 | 3 | Not-determined | Genome-fragment | NA | NA | 0 |
| S17_extracted_k141_122260 | 1226 | No | NA | 2 | 0 | 1 | Not-determined | Genome-fragment | NA | NA | 0 |
| S17_extracted_k141_144763 | 6846 | No | NA | 9 | 0 | 1 | Not-determined | Genome-fragment | NA | NA | 0 |
| S16_vs_63209\|\|full | 1019 | No | NA | 5 | 0 | 0 | Low-quality | Genome-fragment | 2.25 | AAI-based (high-confidence) | 0 |
| S17_extracted_k141_144770 | 1203 | No | NA | 1 | 0 | 1 | Not-determined | Genome-fragment | NA | NA | 0 |
| S17_extracted_k141_108480 | 5645 | No | NA | 3 | 0 | 3 | Not-determined | Genome-fragment | NA | NA | 0 |
| S17_extracted_k141_56341 | 1041 | No | NA | 1 | 0 | 0 | Not-determined | Genome-fragment | NA | NA | 0 |
| S16_vs_88157\|\|full | 1019 | No | NA | 3 | 1 | 0 | Low-quality | Genome-fragment | 2.71 | AAI-based (medium-confidence) | 0 |
| S17_extracted_k141_173941 | 1861 | No | NA | 2 | 0 | 2 | Not-determined | Genome-fragment | NA | NA | 0 |
| S17_extracted_k141_159635 | 1181 | No | NA | 2 | 0 | 1 | Not-determined | Genome-fragment | NA | NA | 0 |
| S16_vs_130520\|\|lt2gene | 1019 | No | NA | 2 | 2 | 0 | Low-quality | Genome-fragment | 2.94 | AAI-based (high-confidence) | 0 |
| S17_extracted_k141_129831 | 1222 | No | NA | 1 | 0 | 0 | Not-determined | Genome-fragment | NA | NA | 0 |
| S17_extracted_k141_144787 | 4528 | No | NA | 8 | 0 | 0 | Not-determined | Genome-fragment | NA | NA | 0 |
| S17_extracted_k141_84809 | 11189 | No | NA | 11 | 0 | 7 | Not-determined | Genome-fragment | NA | NA | 0 |
| S17_extracted_k141_41892 | 1123 | No | NA | 2 | 0 | 1 | Not-determined | Genome-fragment | NA | NA | 0 |
| S17_extracted_k141_166828 | 1906 | No | NA | 2 | 0 | 2 | Not-determined | Genome-fragment | NA | NA | 0 |
| S17_extracted_k141_100641 | 1167 | No | NA | 1 | 0 | 0 | Not-determined | Genome-fragment | NA | NA | 0 |
| S10_vs_89019\|\|lt2gene | 1019 | No | NA | 2 | 1 | 0 | Low-quality | Genome-fragment | 2.45 | AAI-based (medium-confidence) | 0 |
| S17_extracted_k141_129875 | 1037 | No | NA | 2 | 0 | 0 | Not-determined | Genome-fragment | NA | NA | 0 |
| S17_extracted_k141_26098 | 2415 | No | NA | 4 | 0 | 0 | Not-determined | Genome-fragment | NA | NA | 0 |
| S17_extracted_k141_174012 | 1002 | No | NA | 1 | 0 | 0 | Not-determined | Genome-fragment | NA | NA | 0 |
| S17_extracted_k141_92883 | 4649 | No | NA | 5 | 0 | 4 | Not-determined | Genome-fragment | NA | NA | 0 |
| S17_extracted_k141_100683 | 1081 | No | NA | 1 | 0 | 0 | Not-determined | Genome-fragment | NA | NA | 0 |
| S17_extracted_k141_49151 | 1283 | No | NA | 2 | 0 | 1 | Not-determined | Genome-fragment | NA | NA | 0 |
| S12_vs_k141_79674\|\|full | 1019 | No | NA | 3 | 1 | 0 | Low-quality | Genome-fragment | 0.55 | HMM-based (lower-bound) | 0 |
| S17_extracted_k141_49168 | 1315 | No | NA | 2 | 0 | 1 | Not-determined | Genome-fragment | NA | NA | 0 |
| S17_extracted_k141_41950 | 1423 | No | NA | 2 | 0 | 2 | Not-determined | Genome-fragment | NA | NA | 0 |
| S17_extracted_k141_92916 | 1118 | No | NA | 2 | 0 | 2 | Not-determined | Genome-fragment | NA | NA | 0 |
| S17_extracted_k141_92921 | 1845 | No | NA | 3 | 0 | 1 | Not-determined | Genome-fragment | NA | NA | 0 |
| S17_extracted_k141_144851 | 1082 | No | NA | 1 | 0 | 0 | Not-determined | Genome-fragment | NA | NA | 0 |
| S17_extracted_k141_92929 | 1516 | No | NA | 1 | 0 | 1 | Not-determined | Genome-fragment | NA | NA | 0 |
| S17_extracted_k141_78454 | 1673 | No | NA | 1 | 0 | 0 | Not-determined | Genome-fragment | NA | NA | 0 |
| S17_extracted_k141_102405 | 1020 | No | NA | 4 | 0 | 0 | Low-quality | Genome-fragment | 2.85 | AAI-based (medium-confidence) | 0 |
| S17_extracted_k141_137198 | 3205 | No | NA | 2 | 0 | 2 | Not-determined | Genome-fragment | NA | NA | 0 |
| S17_extracted_k141_36283 | 1020 | No | NA | 1 | 1 | 0 | Low-quality | Genome-fragment | 0.99 | AAI-based (high-confidence) | 0 |
| S17_extracted_k141_78468 | 6906 | No | NA | 9 | 0 | 0 | Not-determined | Genome-fragment | NA | NA | 0 |
| S11_extracted_k141_57199 | 1020 | No | NA | 3 | 0 | 0 | Low-quality | Genome-fragment | 1.41 | AAI-based (high-confidence) | 0 |
| S17_extracted_k141_62763 | 1014 | No | NA | 2 | 0 | 2 | Not-determined | Genome-fragment | NA | NA | 0 |
| S17_extracted_k141_4347 | 10420 | No | NA | 10 | 0 | 8 | Not-determined | Genome-fragment | NA | NA | 0 |
| S11_vs_k141_92472\|\|full | 1020 | No | NA | 3 | 0 | 0 | Low-quality | Genome-fragment | 2.43 | AAI-based (high-confidence) | 0 |
| S17_extracted_k141_84922 | 1028 | No | NA | 1 | 0 | 1 | Not-determined | Genome-fragment | NA | NA | 0 |
| S1_vs_k141_147361\|\|full | 1020 | No | NA | 2 | 1 | 0 | Low-quality | Genome-fragment | 2.64 | AAI-based (high-confidence) | 0 |
| S5_extracted_k141_105375 | 1020 | No | NA | 2 | 0 | 0 | Low-quality | Genome-fragment | 0.92 | AAI-based (medium-confidence) | 0 |
| S17_extracted_k141_84939 | 1083 | No | NA | 1 | 0 | 1 | Not-determined | Genome-fragment | NA | NA | 0 |
| S17_extracted_k141_174033 | 64565 | No | NA | 73 | 16 | 1 | Medium-quality | Genome-fragment | 59.33 | AAI-based (high-confidence) | 0 |
| S17_extracted_k141_84947 | 1227 | No | NA | 1 | 0 | 0 | Not-determined | Genome-fragment | NA | NA | 0 |
| S17_extracted_k141_115377 | 1087 | No | NA | 3 | 0 | 1 | Not-determined | Genome-fragment | NA | NA | 0 |
| S17_extracted_k141_42054 | 2528 | No | NA | 2 | 0 | 2 | Not-determined | Genome-fragment | NA | NA | 0 |
| S17_extracted_k141_84962 | 1055 | No | NA | 1 | 0 | 1 | Not-determined | Genome-fragment | NA | NA | 0 |
| S17_extracted_k141_144952 | 1774 | No | NA | 1 | 0 | 1 | Not-determined | Genome-fragment | NA | NA | 0 |
| S17_extracted_k141_108584 | 1212 | No | NA | 1 | 0 | 1 | Not-determined | Genome-fragment | NA | NA | 0 |
| S17_extracted_k141_144966 | 1261 | No | NA | 1 | 0 | 0 | Not-determined | Genome-fragment | NA | NA | 0 |
| S17_extracted_k141_49335 | 1053 | No | NA | 1 | 0 | 0 | Not-determined | Genome-fragment | NA | NA | 0 |
| S17_extracted_k141_167051 | 1004 | No | NA | 2 | 0 | 1 | Not-determined | Genome-fragment | NA | NA | 0 |
| S17_extracted_k141_49344 | 1033 | No | NA | 2 | 0 | 2 | Not-determined | Genome-fragment | NA | NA | 0 |
| S17_extracted_k141_174097 | 1181 | No | NA | 1 | 0 | 0 | Not-determined | Genome-fragment | NA | NA | 0 |
| S17_extracted_k141_42095 | 1370 | No | NA | 3 | 0 | 0 | Not-determined | Genome-fragment | NA | NA | 0 |
| S5_vs_k141_108586\|\|full | 1020 | No | NA | 3 | 0 | 0 | Low-quality | Genome-fragment | 2.58 | AAI-based (medium-confidence) | 0 |
| S7_extracted_45581 | 1020 | No | NA | 1 | 0 | 0 | Low-quality | Genome-fragment | 1.11 | AAI-based (medium-confidence) | 0 |
| S17_extracted_k141_34488 | 1186 | No | NA | 2 | 0 | 1 | Not-determined | Genome-fragment | NA | NA | 0 |
| S17_extracted_k141_122306 | 1156 | No | NA | 3 | 0 | 0 | Not-determined | Genome-fragment | NA | NA | 0 |
| S17_extracted_k141_93005 | 2471 | No | NA | 1 | 0 | 1 | Not-determined | Genome-fragment | NA | NA | 0 |
| S17_extracted_k141_115467 | 5213 | No | NA | 4 | 0 | 2 | Not-determined | Genome-fragment | NA | NA | 0 |
| S17_extracted_k141_71267 | 8655 | No | NA | 7 | 0 | 6 | Not-determined | Genome-fragment | NA | NA | 0 |
| S7_vs_23436\|\|full | 1020 | No | NA | 3 | 0 | 0 | Low-quality | Genome-fragment | 1.55 | AAI-based (high-confidence) | 0 |
| S17_extracted_k141_122314 | 2184 | No | NA | 4 | 0 | 0 | Not-determined | Genome-fragment | NA | NA | 0 |
| S17_extracted_k141_85013 | 1872 | No | NA | 3 | 0 | 1 | Not-determined | Genome-fragment | NA | NA | 0 |
| S8_extracted_k141_40363 | 1020 | No | NA | 1 | 0 | 0 | Low-quality | Genome-fragment | 1.44 | AAI-based (high-confidence) | 0 |
| S17_extracted_k141_62877 | 2974 | No | NA | 1 | 0 | 1 | Not-determined | Genome-fragment | NA | NA | 0 |
| S13_extracted_132059 | 1021 | No | NA | 2 | 0 | 0 | Low-quality | Genome-fragment | 0.53 | AAI-based (high-confidence) | 0 |
| S17_extracted_k141_122338 | 1084 | No | NA | 2 | 0 | 2 | Not-determined | Genome-fragment | NA | NA | 0 |
| S14_extracted_k141_58377 | 1021 | No | NA | 1 | 1 | 0 | Low-quality | Genome-fragment | 1.56 | AAI-based (medium-confidence) | 0 |
| S17_extracted_k141_152904 | 2360 | No | NA | 2 | 0 | 2 | Not-determined | Genome-fragment | NA | NA | 0 |
| S14_vs_k141_91904\|\|full | 1021 | No | NA | 4 | 1 | 0 | Low-quality | Genome-fragment | 2.42 | AAI-based (medium-confidence) | 0 |
| S17_extracted_k141_78619 | 2795 | No | NA | 4 | 0 | 1 | Not-determined | Genome-fragment | NA | NA | 0 |
| S17_extracted_k141_115507 | 11242 | No | NA | 8 | 0 | 3 | Not-determined | Genome-fragment | NA | NA | 0 |
| S17_extracted_k141_85027 | 8138 | No | NA | 10 | 0 | 2 | Not-determined | Genome-fragment | NA | NA | 0 |
| S17_extracted_k141_56673 | 3416 | No | NA | 5 | 0 | 3 | Not-determined | Genome-fragment | NA | NA | 0 |
| S17_extracted_k141_115510 | 3787 | No | NA | 4 | 0 | 3 | Not-determined | Genome-fragment | NA | NA | 0 |
| S17_extracted_k141_115514 | 1228 | No | NA | 1 | 0 | 0 | Not-determined | Genome-fragment | NA | NA | 0 |
| S17_extracted_k141_4469 | 1108 | No | NA | 2 | 0 | 1 | Not-determined | Genome-fragment | NA | NA | 0 |
| S17_extracted_k141_174179 | 1840 | No | NA | 3 | 0 | 0 | Not-determined | Genome-fragment | NA | NA | 0 |
| S17_extracted_k141_71328 | 1367 | No | NA | 2 | 0 | 0 | Not-determined | Genome-fragment | NA | NA | 0 |
| S1_extracted_k141_17220 | 1021 | No | NA | 1 | 0 | 0 | Low-quality | Genome-fragment | 0.68 | AAI-based (high-confidence) | 0 |
| S17_extracted_k141_78638 | 4491 | No | NA | 6 | 0 | 5 | Not-determined | Genome-fragment | NA | NA | 0 |
| S1_extracted_k141_99067 | 1021 | No | NA | 2 | 2 | 0 | Low-quality | Genome-fragment | 3.37 | HMM-based (lower-bound) | 0 |
| S17_extracted_k141_159953 | 1236 | No | NA | 1 | 0 | 0 | Not-determined | Genome-fragment | NA | NA | 0 |
| S17_extracted_k141_78648 | 1478 | No | NA | 3 | 0 | 1 | Not-determined | Genome-fragment | NA | NA | 0 |
| S17_extracted_k141_145085 | 3503 | No | NA | 7 | 0 | 6 | Not-determined | Genome-fragment | NA | NA | 0 |
| S17_extracted_k141_78653 | 1258 | No | NA | 2 | 0 | 1 | Not-determined | Genome-fragment | NA | NA | 0 |
| S1_vs_k141_100352\|\|full | 1021 | No | NA | 2 | 0 | 0 | Low-quality | Genome-fragment | 1.75 | AAI-based (high-confidence) | 0 |
| S17_extracted_k141_167125 | 20847 | No | NA | 20 | 0 | 6 | Not-determined | Genome-fragment | NA | NA | 0 |
| S3_vs_k141_32295\|\|full | 1021 | No | NA | 1 | 1 | 0 | Low-quality | Genome-fragment | 2.32 | AAI-based (medium-confidence) | 0 |
| S17_extracted_k141_56713 | 1143 | No | NA | 2 | 0 | 1 | Not-determined | Genome-fragment | NA | NA | 0 |
| S17_extracted_k141_167138 | 3494 | No | NA | 2 | 0 | 0 | Not-determined | Genome-fragment | NA | NA | 0 |
| S5_vs_k141_37784\|\|full | 1021 | No | NA | 3 | 1 | 0 | Low-quality | Genome-fragment | 1.32 | AAI-based (medium-confidence) | 0 |
| S17_extracted_k141_93124 | 2907 | No | NA | 3 | 0 | 2 | Not-determined | Genome-fragment | NA | NA | 0 |
| S5_vs_k141_81888\|\|full | 1021 | No | NA | 3 | 0 | 0 | Low-quality | Genome-fragment | 2.44 | AAI-based (high-confidence) | 0 |
| S17_extracted_k141_4530 | 1228 | No | NA | 2 | 0 | 2 | Not-determined | Genome-fragment | NA | NA | 0 |
| S7_extracted_66854 | 1021 | No | NA | 3 | 0 | 0 | Low-quality | Genome-fragment | 0.73 | AAI-based (high-confidence) | 0 |
| S17_extracted_k141_56733 | 4393 | No | NA | 4 | 0 | 3 | Not-determined | Genome-fragment | NA | NA | 0 |
| S17_extracted_k141_115604 | 1214 | No | NA | 2 | 0 | 1 | Not-determined | Genome-fragment | NA | NA | 0 |
| S17_extracted_k141_71384 | 1506 | No | NA | 1 | 0 | 1 | Not-determined | Genome-fragment | NA | NA | 0 |
| S16_extracted_69330 | 1021 | No | NA | 1 | 0 | 1 | Low-quality | Genome-fragment | 0.51 | AAI-based (medium-confidence) | 0 |
| S17_extracted_k141_115634 | 1395 | No | NA | 2 | 0 | 1 | Not-determined | Genome-fragment | NA | NA | 0 |
| S17_extracted_k141_63000 | 2200 | No | NA | 2 | 0 | 2 | Not-determined | Genome-fragment | NA | NA | 0 |
| S17_extracted_k141_49467 | 2073 | No | NA | 4 | 0 | 4 | Not-determined | Genome-fragment | NA | NA | 0 |
| S17_extracted_k141_174203 | 15880 | No | NA | 25 | 7 | 0 | Medium-quality | Genome-fragment | 69.95 | AAI-based (medium-confidence) | 0 |
| S17_extracted_k141_145112 | 1021 | No | NA | 1 | 0 | 1 | Not-determined | Genome-fragment | NA | NA | 0 |
| S17_extracted_k141_153017 | 1014 | No | NA | 1 | 0 | 0 | Not-determined | Genome-fragment | NA | NA | 0 |
| S17_extracted_k141_167206 | 7256 | No | NA | 6 | 0 | 3 | Not-determined | Genome-fragment | NA | NA | 0 |
| S17_extracted_k141_145129 | 2119 | No | NA | 3 | 0 | 1 | Not-determined | Genome-fragment | NA | NA | 0 |
| S17_extracted_k141_167220 | 1062 | No | NA | 2 | 0 | 1 | Not-determined | Genome-fragment | NA | NA | 0 |
| S17_extracted_k141_63035 | 1222 | No | NA | 1 | 0 | 1 | Not-determined | Genome-fragment | NA | NA | 0 |
| S17_extracted_k141_159998 | 1036 | No | NA | 1 | 0 | 1 | Not-determined | Genome-fragment | NA | NA | 0 |
| S17_extracted_k141_93214 | 1054 | No | NA | 1 | 0 | 0 | Not-determined | Genome-fragment | NA | NA | 0 |
| S17_extracted_k141_167232 | 3491 | No | NA | 6 | 0 | 5 | Not-determined | Genome-fragment | NA | NA | 0 |
| S10_vs_87292\|\|full | 1021 | No | NA | 4 | 1 | 0 | Low-quality | Genome-fragment | 1.89 | HMM-based (lower-bound) | 0 |
| S17_extracted_k141_71472 | 1671 | No | NA | 2 | 0 | 1 | Not-determined | Genome-fragment | NA | NA | 0 |
| S17_extracted_k141_153065 | 1018 | No | NA | 2 | 0 | 1 | Not-determined | Genome-fragment | NA | NA | 0 |
| S12_extracted_k141_61873 | 1021 | No | NA | 1 | 1 | 0 | Low-quality | Genome-fragment | 0.92 | HMM-based (lower-bound) | 0 |
| S12_vs_k141_102461\|\|full | 1021 | No | NA | 3 | 0 | 0 | Low-quality | Genome-fragment | 1.44 | AAI-based (medium-confidence) | 0 |
| S17_extracted_k141_12066 | 1224 | No | NA | 1 | 0 | 1 | Not-determined | Genome-fragment | NA | NA | 0 |
| S17_extracted_k141_160051 | 1014 | No | NA | 2 | 0 | 1 | Not-determined | Genome-fragment | NA | NA | 0 |
| S17_extracted_k141_93236 | 1885 | No | NA | 3 | 0 | 2 | Not-determined | Genome-fragment | NA | NA | 0 |
| S17_extracted_k141_71492 | 2668 | No | NA | 4 | 0 | 2 | Not-determined | Genome-fragment | NA | NA | 0 |
| S17_extracted_k141_93237 | 1075 | No | NA | 2 | 0 | 1 | Not-determined | Genome-fragment | NA | NA | 0 |
| S12_vs_k141_40143\|\|full | 1021 | No | NA | 2 | 1 | 0 | Low-quality | Genome-fragment | 1.24 | AAI-based (high-confidence) | 0 |
| S17_extracted_k141_34757 | 1185 | No | NA | 1 | 0 | 0 | Not-determined | Genome-fragment | NA | NA | 0 |
| S17_extracted_k141_71513 | 1041 | No | NA | 2 | 0 | 1 | Not-determined | Genome-fragment | NA | NA | 0 |
| S17_extracted_k141_12086 | 1696 | No | NA | 3 | 0 | 1 | Not-determined | Genome-fragment | NA | NA | 0 |
| S17_extracted_k141_63094 | 1415 | No | NA | 1 | 0 | 0 | Not-determined | Genome-fragment | NA | NA | 0 |
| S17_extracted_k141_101086 | 1216 | No | NA | 2 | 0 | 1 | Not-determined | Genome-fragment | NA | NA | 0 |
| S17_extracted_k141_122498 | 3120 | No | NA | 3 | 0 | 3 | Not-determined | Genome-fragment | NA | NA | 0 |
| S17_extracted_k141_42349 | 9842 | No | NA | 9 | 0 | 1 | Not-determined | Genome-fragment | NA | NA | 0 |
| S17_extracted_k141_56815 | 1049 | No | NA | 1 | 0 | 1 | Not-determined | Genome-fragment | NA | NA | 0 |
| S17_extracted_k141_4688 | 1770 | No | NA | 2 | 0 | 1 | Not-determined | Genome-fragment | NA | NA | 0 |
| S12_vs_k141_68619\|\|full | 1021 | No | NA | 2 | 0 | 1 | Low-quality | Genome-fragment | 0.46 | AAI-based (medium-confidence) | 0 |
| S17_extracted_k141_160077 | 3092 | No | NA | 2 | 0 | 2 | Not-determined | Genome-fragment | NA | NA | 0 |
| S17_extracted_k141_174309 | 1809 | No | NA | 2 | 0 | 0 | Not-determined | Genome-fragment | NA | NA | 0 |
| S15_vs_71291\|\|full | 1021 | No | NA | 3 | 0 | 0 | Low-quality | Genome-fragment | 2.26 | AAI-based (high-confidence) | 0 |
| S17_extracted_k141_115757 | 1197 | No | NA | 1 | 0 | 1 | Not-determined | Genome-fragment | NA | NA | 0 |
| S17_extracted_k141_160092 | 1277 | No | NA | 1 | 0 | 1 | Not-determined | Genome-fragment | NA | NA | 0 |
| S17_extracted_k141_115761 | 3590 | No | NA | 2 | 0 | 2 | Not-determined | Genome-fragment | NA | NA | 0 |
| S8_extracted_k141_127009 | 1021 | No | NA | 1 | 1 | 0 | Low-quality | Genome-fragment | 1.99 | AAI-based (high-confidence) | 0 |
| S17_extracted_k141_145224 | 1206 | No | NA | 2 | 0 | 0 | Not-determined | Genome-fragment | NA | NA | 0 |
| S17_extracted_k141_108849 | 5439 | No | NA | 5 | 0 | 4 | Not-determined | Genome-fragment | NA | NA | 0 |
| S8_extracted_k141_199463 | 1021 | No | NA | 3 | 0 | 0 | Low-quality | Genome-fragment | 1.75 | AAI-based (high-confidence) | 0 |
| S17_extracted_k141_4730 | 4117 | No | NA | 5 | 0 | 0 | Not-determined | Genome-fragment | NA | NA | 0 |
| S17_extracted_k141_108869 | 3207 | No | NA | 3 | 0 | 2 | Not-determined | Genome-fragment | NA | NA | 0 |
| S8_vs_k141_113446\|\|full | 1021 | No | NA | 3 | 0 | 0 | Low-quality | Genome-fragment | 2.24 | AAI-based (medium-confidence) | 0 |
| S9_vs_k141_68576\|\|lt2gene | 1022 | No | NA | 1 | 1 | 0 | Low-quality | Genome-fragment | 2.23 | AAI-based (high-confidence) | 0 |
| S11_extracted_k141_59756 | 1022 | No | NA | 1 | 0 | 1 | Low-quality | Genome-fragment | 1.48 | AAI-based (high-confidence) | 0 |
| S17_extracted_k141_93304 | 2823 | No | NA | 2 | 0 | 2 | Not-determined | Genome-fragment | NA | NA | 0 |
| S17_extracted_k141_115794 | 1160 | No | NA | 3 | 0 | 1 | Not-determined | Genome-fragment | NA | NA | 0 |
| S17_extracted_k141_34874 | 1143 | No | NA | 2 | 0 | 2 | Not-determined | Genome-fragment | NA | NA | 0 |
| S17_extracted_k141_145278 | 1382 | No | NA | 2 | 0 | 0 | Not-determined | Genome-fragment | NA | NA | 0 |
| S17_extracted_k141_63152 | 1037 | No | NA | 3 | 0 | 2 | Not-determined | Genome-fragment | NA | NA | 0 |
| S17_extracted_k141_85313 | 2121 | No | NA | 1 | 0 | 0 | Not-determined | Genome-fragment | NA | NA | 0 |
| S17_extracted_k141_42411 | 1404 | No | NA | 2 | 0 | 1 | Not-determined | Genome-fragment | NA | NA | 0 |
| S17_extracted_k141_160126 | 5830 | No | NA | 4 | 0 | 2 | Not-determined | Genome-fragment | NA | NA | 0 |
| S17_extracted_k141_71607 | 1067 | No | NA | 1 | 0 | 0 | Not-determined | Genome-fragment | NA | NA | 0 |
| S17_extracted_k141_4809 | 1236 | No | NA | 2 | 0 | 1 | Not-determined | Genome-fragment | NA | NA | 0 |
| S11_extracted_k141_41572 | 1022 | No | NA | 2 | 1 | 0 | Low-quality | Genome-fragment | 2.58 | AAI-based (medium-confidence) | 0 |
| S17_extracted_k141_93360 | 1168 | No | NA | 1 | 0 | 0 | Not-determined | Genome-fragment | NA | NA | 0 |
| S17_extracted_k141_4817 | 4757 | No | NA | 4 | 0 | 4 | Not-determined | Genome-fragment | NA | NA | 0 |
| S11_extracted_k141_29503 | 1022 | No | NA | 1 | 0 | 0 | Low-quality | Genome-fragment | 0.81 | AAI-based (high-confidence) | 0 |
| S17_extracted_k141_108948 | 3059 | No | NA | 2 | 0 | 1 | Not-determined | Genome-fragment | NA | NA | 0 |
| S17_extracted_k141_160167 | 2228 | No | NA | 2 | 0 | 1 | Not-determined | Genome-fragment | NA | NA | 0 |
| S11_extracted_k141_36098 | 1022 | No | NA | 2 | 2 | 0 | Low-quality | Genome-fragment | 2.73 | AAI-based (high-confidence) | 0 |
| S17_extracted_k141_78886 | 3179 | No | NA | 2 | 0 | 2 | Not-determined | Genome-fragment | NA | NA | 0 |
| S17_extracted_k141_153269 | 1305 | No | NA | 2 | 0 | 1 | Not-determined | Genome-fragment | NA | NA | 0 |
| S17_extracted_k141_108976 | 2629 | No | NA | 1 | 0 | 1 | Not-determined | Genome-fragment | NA | NA | 0 |
| S17_extracted_k141_130155 | 1992 | No | NA | 2 | 0 | 0 | Not-determined | Genome-fragment | NA | NA | 0 |
| S17_extracted_k141_78914 | 1858 | No | NA | 2 | 0 | 0 | Not-determined | Genome-fragment | NA | NA | 0 |
| S17_extracted_k141_34992 | 6009 | No | NA | 4 | 0 | 2 | Not-determined | Genome-fragment | NA | NA | 0 |
| S17_extracted_k141_160234 | 1538 | No | NA | 1 | 0 | 1 | Not-determined | Genome-fragment | NA | NA | 0 |
| S17_extracted_k141_137848 | 1016 | No | NA | 2 | 0 | 0 | Not-determined | Genome-fragment | NA | NA | 0 |
| S17_extracted_k141_19384 | 1529 | No | NA | 3 | 0 | 0 | Not-determined | Genome-fragment | NA | NA | 0 |
| S17_extracted_k141_78922 | 1067 | No | NA | 1 | 0 | 1 | Not-determined | Genome-fragment | NA | NA | 0 |
| S11_vs_k141_102647\|\|lt2gene | 1022 | No | NA | 2 | 1 | 0 | Low-quality | Genome-fragment | 2.22 | AAI-based (medium-confidence) | 0 |
| S17_extracted_k141_130193 | 1088 | No | NA | 2 | 0 | 2 | Not-determined | Genome-fragment | NA | NA | 0 |
| S13_extracted_133631 | 1022 | No | NA | 2 | 0 | 1 | Low-quality | Genome-fragment | 2.36 | AAI-based (medium-confidence) | 0 |
| S17_extracted_k141_101225 | 3195 | No | NA | 3 | 0 | 2 | Not-determined | Genome-fragment | NA | NA | 0 |
| S17_extracted_k141_85442 | 3639 | No | NA | 3 | 0 | 2 | Not-determined | Genome-fragment | NA | NA | 0 |
| S17_extracted_k141_153303 | 1283 | No | NA | 2 | 0 | 2 | Not-determined | Genome-fragment | NA | NA | 0 |
| S17_extracted_k141_63314 | 2685 | No | NA | 3 | 0 | 1 | Not-determined | Genome-fragment | NA | NA | 0 |
| S17_extracted_k141_122731 | 1137 | No | NA | 3 | 0 | 0 | Not-determined | Genome-fragment | NA | NA | 0 |
| S17_extracted_k141_71693 | 1130 | No | NA | 2 | 0 | 1 | Not-determined | Genome-fragment | NA | NA | 0 |
| S1_extracted_k141_4778 | 1022 | No | NA | 1 | 1 | 0 | Low-quality | Genome-fragment | 2.15 | AAI-based (high-confidence) | 0 |
| S3_extracted_k141_83648 | 1022 | No | NA | 2 | 1 | 0 | Low-quality | Genome-fragment | 3.01 | AAI-based (high-confidence) | 0 |
| S17_extracted_k141_71704 | 1548 | No | NA | 0 | 0 | 0 | Not-determined | Genome-fragment | NA | NA | 0 |
| S17_extracted_k141_35077 | 2807 | No | NA | 1 | 0 | 1 | Not-determined | Genome-fragment | NA | NA | 0 |
| S17_extracted_k141_130222 | 1566 | No | NA | 2 | 0 | 1 | Not-determined | Genome-fragment | NA | NA | 0 |
| S17_extracted_k141_78988 | 2027 | No | NA | 2 | 0 | 2 | Not-determined | Genome-fragment | NA | NA | 0 |
| S17_extracted_k141_71711 | 1325 | No | NA | 2 | 0 | 2 | Not-determined | Genome-fragment | NA | NA | 0 |
| S17_extracted_k141_12207 | 1466 | No | NA | 1 | 0 | 1 | Not-determined | Genome-fragment | NA | NA | 0 |
| S17_extracted_k141_167495 | 1175 | No | NA | 2 | 0 | 1 | Not-determined | Genome-fragment | NA | NA | 0 |
| S17_extracted_k141_174408 | 3448 | No | NA | 2 | 0 | 0 | Not-determined | Genome-fragment | NA | NA | 0 |
| S17_extracted_k141_26560 | 1197 | No | NA | 1 | 0 | 0 | Not-determined | Genome-fragment | NA | NA | 0 |
| S17_extracted_k141_153361 | 1177 | No | NA | 1 | 0 | 1 | Not-determined | Genome-fragment | NA | NA | 0 |
| S17_extracted_k141_4977 | 1502 | No | NA | 1 | 0 | 1 | Not-determined | Genome-fragment | NA | NA | 0 |
| S17_extracted_k141_122793 | 1670 | No | NA | 3 | 0 | 1 | Not-determined | Genome-fragment | NA | NA | 0 |
| S17_extracted_k141_93510 | 2056 | No | NA | 3 | 0 | 2 | Not-determined | Genome-fragment | NA | NA | 0 |
| S17_extracted_k141_19459 | 2664 | No | NA | 3 | 0 | 2 | Not-determined | Genome-fragment | NA | NA | 0 |
| S17_extracted_k141_145430 | 1276 | No | NA | 2 | 0 | 2 | Not-determined | Genome-fragment | NA | NA | 0 |
| S17_extracted_k141_19468 | 3328 | No | NA | 5 | 0 | 1 | Not-determined | Genome-fragment | NA | NA | 0 |
| S17_extracted_k141_26597 | 2445 | No | NA | 2 | 0 | 1 | Not-determined | Genome-fragment | NA | NA | 0 |
| S17_extracted_k141_160321 | 1367 | No | NA | 3 | 0 | 0 | Not-determined | Genome-fragment | NA | NA | 0 |
| S3_vs_k141_54124\|\|full | 1022 | No | NA | 5 | 0 | 0 | Low-quality | Genome-fragment | 2.58 | AAI-based (high-confidence) | 0 |
| S7_vs_55100\|\|lt2gene | 1022 | No | NA | 2 | 1 | 0 | Low-quality | Genome-fragment | 0.78 | HMM-based (lower-bound) | 0 |
| S17_extracted_k141_145513 | 1236 | No | NA | 1 | 0 | 0 | Not-determined | Genome-fragment | NA | NA | 0 |
| S7_vs_127549\|\|lt2gene | 1022 | No | NA | 1 | 1 | 0 | Low-quality | Genome-fragment | 1.35 | HMM-based (lower-bound) | 0 |
| S17_extracted_k141_71821 | 2293 | No | NA | 2 | 0 | 1 | Not-determined | Genome-fragment | NA | NA | 0 |
| S16_extracted_36899 | 1022 | No | NA | 1 | 1 | 0 | Low-quality | Genome-fragment | 3.11 | AAI-based (high-confidence) | 0 |
| S17_extracted_k141_5049 | 1024 | No | NA | 1 | 0 | 1 | Not-determined | Genome-fragment | NA | NA | 0 |
| S17_extracted_k141_35138 | 52045 | No | NA | 76 | 12 | 4 | Medium-quality | Genome-fragment | 73.5 | AAI-based (high-confidence) | 0 |
| S17_extracted_k141_153455 | 1235 | No | NA | 1 | 0 | 0 | Not-determined | Genome-fragment | NA | NA | 0 |
| S17_extracted_k141_109121 | 4422 | No | NA | 4 | 0 | 4 | Not-determined | Genome-fragment | NA | NA | 0 |
| S17_extracted_k141_63391 | 1656 | No | NA | 2 | 0 | 1 | Not-determined | Genome-fragment | NA | NA | 0 |
| S17_extracted_k141_85587 | 1183 | No | NA | 2 | 0 | 1 | Not-determined | Genome-fragment | NA | NA | 0 |
| S17_extracted_k141_42638 | 2759 | No | NA | 2 | 0 | 0 | Not-determined | Genome-fragment | NA | NA | 0 |
| S17_extracted_k141_116099 | 1453 | No | NA | 2 | 0 | 2 | Not-determined | Genome-fragment | NA | NA | 0 |
| S17_extracted_k141_137987 | 1541 | No | NA | 1 | 0 | 1 | Not-determined | Genome-fragment | NA | NA | 0 |
| S17_extracted_k141_116102 | 1777 | No | NA | 1 | 0 | 1 | Not-determined | Genome-fragment | NA | NA | 0 |
| S17_extracted_k141_71823 | 30287 | No | NA | 30 | 0 | 5 | Not-determined | Genome-fragment | NA | NA | 0 |
| S17_extracted_k141_122899 | 5609 | No | NA | 4 | 0 | 4 | Not-determined | Genome-fragment | NA | NA | 0 |
| S17_extracted_k141_49785 | 1498 | No | NA | 3 | 0 | 2 | Not-determined | Genome-fragment | NA | NA | 0 |
| S17_extracted_k141_85621 | 1595 | No | NA | 3 | 0 | 1 | Not-determined | Genome-fragment | NA | NA | 0 |
| S17_extracted_k141_49788 | 2174 | No | NA | 2 | 0 | 1 | Not-determined | Genome-fragment | NA | NA | 0 |
| S16_extracted_124054 | 1022 | No | NA | 4 | 0 | 0 | Low-quality | Genome-fragment | 2.41 | AAI-based (medium-confidence) | 0 |
| S17_extracted_k141_85627 | 1192 | No | NA | 2 | 0 | 0 | Not-determined | Genome-fragment | NA | NA | 0 |
| S16_vs_92380\|\|full | 1022 | No | NA | 2 | 0 | 1 | Low-quality | Genome-fragment | 1.98 | AAI-based (high-confidence) | 0 |
| S16_vs_151457\|\|full | 1022 | No | NA | 3 | 1 | 0 | Low-quality | Genome-fragment | 2.58 | AAI-based (high-confidence) | 0 |
| S17_extracted_k141_12283 | 1799 | No | NA | 1 | 0 | 1 | Not-determined | Genome-fragment | NA | NA | 0 |
| S17_extracted_k141_71848 | 1818 | No | NA | 1 | 0 | 0 | Not-determined | Genome-fragment | NA | NA | 0 |
| S17_extracted_k141_167607 | 1286 | No | NA | 1 | 0 | 1 | Not-determined | Genome-fragment | NA | NA | 0 |
| S12_vs_k141_21427\|\|full | 1022 | No | NA | 3 | 1 | 0 | Low-quality | Genome-fragment | 3.16 | AAI-based (high-confidence) | 0 |
| S17_extracted_k141_160438 | 1291 | No | NA | 3 | 0 | 0 | Not-determined | Genome-fragment | NA | NA | 0 |
| S17_extracted_k141_130426 | 1023 | No | NA | 2 | 0 | 0 | Low-quality | Genome-fragment | 0.61 | AAI-based (medium-confidence) | 0 |
| S17_extracted_k141_160440 | 1422 | No | NA | 1 | 0 | 0 | Not-determined | Genome-fragment | NA | NA | 0 |
| S17_extracted_k141_63436 | 1018 | No | NA | 1 | 0 | 1 | Not-determined | Genome-fragment | NA | NA | 0 |
| S17_extracted_k141_138035 | 2981 | No | NA | 2 | 0 | 2 | Not-determined | Genome-fragment | NA | NA | 0 |
| S17_extracted_k141_35233 | 4097 | No | NA | 3 | 0 | 1 | Not-determined | Genome-fragment | NA | NA | 0 |
| S17_extracted_k141_57240 | 1041 | No | NA | 1 | 0 | 0 | Not-determined | Genome-fragment | NA | NA | 0 |
| S17_extracted_k141_5184 | 1466 | No | NA | 1 | 0 | 1 | Not-determined | Genome-fragment | NA | NA | 0 |
| S17_extracted_k141_42724 | 1492 | No | NA | 2 | 0 | 1 | Not-determined | Genome-fragment | NA | NA | 0 |
| S17_extracted_k141_12340 | 2806 | No | NA | 2 | 0 | 0 | Not-determined | Genome-fragment | NA | NA | 0 |
| S17_extracted_k141_109248 | 1302 | No | NA | 2 | 0 | 2 | Not-determined | Genome-fragment | NA | NA | 0 |
| S17_extracted_k141_79151 | 16088 | No | NA | 20 | 7 | 0 | Medium-quality | Genome-fragment | 74.35 | AAI-based (medium-confidence) | 0 |
| S17_vs_k141_112633\|\|full | 1023 | No | NA | 3 | 0 | 0 | Low-quality | Genome-fragment | 3.08 | AAI-based (high-confidence) | 0 |
| S17_extracted_k141_167640 | 1053 | No | NA | 1 | 0 | 1 | Not-determined | Genome-fragment | NA | NA | 0 |
| S17_extracted_k141_71934 | 1268 | No | NA | 3 | 0 | 1 | Not-determined | Genome-fragment | NA | NA | 0 |
| S17_extracted_k141_116219 | 1645 | No | NA | 1 | 0 | 1 | Not-determined | Genome-fragment | NA | NA | 0 |
| S17_extracted_k141_85721 | 1394 | No | NA | 2 | 0 | 1 | Not-determined | Genome-fragment | NA | NA | 0 |
| S17_vs_k141_32155\|\|full | 1023 | No | NA | 3 | 0 | 0 | Low-quality | Genome-fragment | 2.21 | AAI-based (high-confidence) | 0 |
| S17_vs_k141_65098\|\|full | 1023 | No | NA | 4 | 1 | 0 | Low-quality | Genome-fragment | 0.69 | AAI-based (medium-confidence) | 0 |
| S13_extracted_10615 | 1023 | No | NA | 2 | 0 | 1 | Low-quality | Genome-fragment | 1.04 | AAI-based (medium-confidence) | 0 |
| S17_extracted_k141_49919 | 3868 | No | NA | 2 | 0 | 1 | Not-determined | Genome-fragment | NA | NA | 0 |
| S17_extracted_k141_35372 | 1253 | No | NA | 1 | 0 | 0 | Not-determined | Genome-fragment | NA | NA | 0 |
| S1_extracted_k141_66855 | 1023 | No | NA | 4 | 0 | 0 | Low-quality | Genome-fragment | 2.81 | AAI-based (high-confidence) | 0 |
| S17_extracted_k141_138125 | 4655 | No | NA | 3 | 0 | 2 | Not-determined | Genome-fragment | NA | NA | 0 |
| S17_extracted_k141_145631 | 2065 | No | NA | 1 | 0 | 1 | Not-determined | Genome-fragment | NA | NA | 0 |
| S17_extracted_k141_138133 | 2809 | No | NA | 1 | 0 | 1 | Not-determined | Genome-fragment | NA | NA | 0 |
| S17_extracted_k141_35398 | 1267 | No | NA | 2 | 0 | 1 | Not-determined | Genome-fragment | NA | NA | 0 |
| S17_extracted_k141_145639 | 1451 | No | NA | 1 | 0 | 0 | Not-determined | Genome-fragment | NA | NA | 0 |
| S1_vs_k141_47231\|\|full | 1023 | No | NA | 3 | 1 | 0 | Low-quality | Genome-fragment | 1.96 | AAI-based (high-confidence) | 0 |
| S1_vs_k141_135196\|\|full | 1023 | No | NA | 3 | 1 | 0 | Low-quality | Genome-fragment | 2.59 | AAI-based (high-confidence) | 0 |
| S17_extracted_k141_42808 | 6582 | No | NA | 7 | 0 | 4 | Not-determined | Genome-fragment | NA | NA | 0 |
| S17_extracted_k141_123074 | 1268 | No | NA | 2 | 0 | 1 | Not-determined | Genome-fragment | NA | NA | 0 |
| S17_extracted_k141_5242 | 1908 | No | NA | 1 | 0 | 0 | Not-determined | Genome-fragment | NA | NA | 0 |
| S17_extracted_k141_174610 | 1951 | No | NA | 2 | 0 | 1 | Not-determined | Genome-fragment | NA | NA | 0 |
| S17_extracted_k141_167758 | 6353 | No | NA | 7 | 0 | 0 | Not-determined | Genome-fragment | NA | NA | 0 |
| S17_extracted_k141_72047 | 1139 | No | NA | 1 | 0 | 1 | Not-determined | Genome-fragment | NA | NA | 0 |
| S17_extracted_k141_160567 | 1018 | No | NA | 1 | 0 | 1 | Not-determined | Genome-fragment | NA | NA | 0 |
| S17_extracted_k141_101527 | 1264 | No | NA | 3 | 0 | 0 | Not-determined | Genome-fragment | NA | NA | 0 |
| S17_extracted_k141_19710 | 1957 | No | NA | 1 | 0 | 1 | Not-determined | Genome-fragment | NA | NA | 0 |
| S17_extracted_k141_123096 | 4945 | No | NA | 4 | 0 | 1 | Not-determined | Genome-fragment | NA | NA | 0 |
| S1_vs_k141_57091\|\|lt2gene | 1023 | No | NA | 2 | 2 | 0 | Low-quality | Genome-fragment | 1.89 | HMM-based (lower-bound) | 0 |
| S17_extracted_k141_138181 | 1385 | No | NA | 2 | 0 | 0 | Not-determined | Genome-fragment | NA | NA | 0 |
| S17_extracted_k141_101547 | 1258 | No | NA | 2 | 0 | 2 | Not-determined | Genome-fragment | NA | NA | 0 |
| S3_vs_k141_14819\|\|lt2gene | 1023 | No | NA | 2 | 1 | 0 | Low-quality | Genome-fragment | 2.16 | AAI-based (high-confidence) | 0 |
| S17_extracted_k141_160587 | 3122 | No | NA | 3 | 0 | 3 | Not-determined | Genome-fragment | NA | NA | 0 |
| S17_extracted_k141_167794 | 1695 | No | NA | 1 | 0 | 1 | Not-determined | Genome-fragment | NA | NA | 0 |
| S7_extracted_97000 | 1023 | No | NA | 2 | 0 | 0 | Low-quality | Genome-fragment | 2.32 | AAI-based (medium-confidence) | 0 |
| S17_extracted_k141_79300 | 1417 | No | NA | 3 | 0 | 2 | Not-determined | Genome-fragment | NA | NA | 0 |
| S17_extracted_k141_85874 | 1211 | No | NA | 2 | 0 | 1 | Not-determined | Genome-fragment | NA | NA | 0 |
| S17_extracted_k141_12520 | 1462 | No | NA | 2 | 0 | 1 | Not-determined | Genome-fragment | NA | NA | 0 |
| S17_extracted_k141_116382 | 1032 | No | NA | 1 | 0 | 1 | Not-determined | Genome-fragment | NA | NA | 0 |
| S17_extracted_k141_85887 | 1234 | No | NA | 2 | 0 | 2 | Not-determined | Genome-fragment | NA | NA | 0 |
| S17_extracted_k141_130553 | 1237 | No | NA | 2 | 0 | 2 | Not-determined | Genome-fragment | NA | NA | 0 |
| S7_extracted_132721 | 1023 | No | NA | 3 | 0 | 0 | Low-quality | Genome-fragment | 0.67 | AAI-based (high-confidence) | 0 |
| S17_extracted_k141_26778 | 2884 | No | NA | 3 | 0 | 1 | Not-determined | Genome-fragment | NA | NA | 0 |
| S17_extracted_k141_145709 | 1221 | No | NA | 2 | 0 | 1 | Not-determined | Genome-fragment | NA | NA | 0 |
| S17_extracted_k141_50088 | 1163 | No | NA | 2 | 0 | 2 | Not-determined | Genome-fragment | NA | NA | 0 |
| S16_extracted_10527 | 1023 | No | NA | 2 | 0 | 1 | Low-quality | Genome-fragment | 1.19 | AAI-based (high-confidence) | 0 |
| S16_extracted_93305 | 1023 | No | NA | 2 | 0 | 0 | Low-quality | Genome-fragment | 1.58 | AAI-based (high-confidence) | 0 |
| S17_extracted_k141_109495 | 1059 | No | NA | 1 | 0 | 0 | Not-determined | Genome-fragment | NA | NA | 0 |
| S17_extracted_k141_174696 | 4708 | No | NA | 6 | 0 | 5 | Not-determined | Genome-fragment | NA | NA | 0 |
| S17_extracted_k141_79353 | 2385 | No | NA | 3 | 0 | 2 | Not-determined | Genome-fragment | NA | NA | 0 |
| S17_extracted_k141_93760 | 1281 | No | NA | 2 | 0 | 0 | Not-determined | Genome-fragment | NA | NA | 0 |
| S17_extracted_k141_101626 | 2231 | No | NA | 2 | 0 | 2 | Not-determined | Genome-fragment | NA | NA | 0 |
| S17_extracted_k141_12557 | 4083 | No | NA | 4 | 0 | 4 | Not-determined | Genome-fragment | NA | NA | 0 |
| S17_extracted_k141_79368 | 3369 | No | NA | 2 | 0 | 0 | Not-determined | Genome-fragment | NA | NA | 0 |
| S16_vs_126196\|\|full | 1023 | No | NA | 3 | 2 | 0 | Low-quality | Genome-fragment | 2.55 | AAI-based (high-confidence) | 0 |
| S17_extracted_k141_123173 | 4266 | No | NA | 4 | 0 | 4 | Not-determined | Genome-fragment | NA | NA | 0 |
| S17_extracted_k141_101649 | 2629 | No | NA | 2 | 0 | 1 | Not-determined | Genome-fragment | NA | NA | 0 |
| S17_extracted_k141_174712 | 1184 | No | NA | 1 | 0 | 0 | Not-determined | Genome-fragment | NA | NA | 0 |
| S17_extracted_k141_109530 | 1139 | No | NA | 2 | 0 | 2 | Not-determined | Genome-fragment | NA | NA | 0 |
| S6_vs_k141_26265\|\|lt2gene | 1023 | No | NA | 1 | 1 | 0 | Low-quality | Genome-fragment | 16.77 | AAI-based (high-confidence) | 0 |
| S17_extracted_k141_85940 | 1061 | No | NA | 1 | 0 | 1 | Not-determined | Genome-fragment | NA | NA | 0 |
| S17_extracted_k141_50147 | 1503 | No | NA | 3 | 0 | 2 | Not-determined | Genome-fragment | NA | NA | 0 |
| S17_extracted_k141_93781 | 3917 | No | NA | 6 | 0 | 0 | Not-determined | Genome-fragment | NA | NA | 0 |
| S17_extracted_k141_123210 | 2743 | No | NA | 1 | 0 | 0 | Not-determined | Genome-fragment | NA | NA | 0 |
| S17_extracted_k141_123211 | 3243 | No | NA | 6 | 0 | 1 | Not-determined | Genome-fragment | NA | NA | 0 |
| S8_vs_k141_85083\|\|full | 1023 | No | NA | 3 | 0 | 0 | Low-quality | Genome-fragment | 1.47 | AAI-based (medium-confidence) | 0 |
| S17_extracted_k141_35498 | 1109 | No | NA | 1 | 0 | 1 | Not-determined | Genome-fragment | NA | NA | 0 |
| S17_extracted_k141_145807 | 1046 | No | NA | 1 | 0 | 1 | Not-determined | Genome-fragment | NA | NA | 0 |
| S17_extracted_k141_160715 | 1964 | No | NA | 4 | 0 | 4 | Not-determined | Genome-fragment | NA | NA | 0 |
| S17_extracted_k141_93817 | 1035 | No | NA | 1 | 0 | 0 | Not-determined | Genome-fragment | NA | NA | 0 |
| S17_extracted_k141_123237 | 3814 | No | NA | 2 | 0 | 2 | Not-determined | Genome-fragment | NA | NA | 0 |
| S17_extracted_k141_145831 | 2348 | No | NA | 1 | 0 | 0 | Not-determined | Genome-fragment | NA | NA | 0 |
| S17_extracted_k141_93838 | 1244 | No | NA | 3 | 0 | 1 | Not-determined | Genome-fragment | NA | NA | 0 |
| S17_extracted_k141_93848 | 1525 | No | NA | 1 | 0 | 1 | Not-determined | Genome-fragment | NA | NA | 0 |
| S17_extracted_k141_116509 | 1519 | No | NA | 1 | 0 | 1 | Not-determined | Genome-fragment | NA | NA | 0 |
| S17_extracted_k141_174787 | 1201 | No | NA | 1 | 0 | 1 | Not-determined | Genome-fragment | NA | NA | 0 |
| S17_extracted_k141_167953 | 3701 | No | NA | 3 | 0 | 1 | Not-determined | Genome-fragment | NA | NA | 0 |
| S17_extracted_k141_93869 | 1126 | No | NA | 1 | 0 | 0 | Not-determined | Genome-fragment | NA | NA | 0 |
| S17_extracted_k141_35569 | 1508 | No | NA | 2 | 0 | 1 | Not-determined | Genome-fragment | NA | NA | 0 |
| S17_extracted_k141_50241 | 1260 | No | NA | 3 | 0 | 2 | Not-determined | Genome-fragment | NA | NA | 0 |
| S17_extracted_k141_22641 | 1024 | No | NA | 2 | 0 | 1 | Low-quality | Genome-fragment | 0.18 | AAI-based (medium-confidence) | 0 |
| S17_extracted_k141_72176 | 2648 | No | NA | 3 | 0 | 0 | Not-determined | Genome-fragment | NA | NA | 0 |
| S17_extracted_k141_116566 | 1109 | No | NA | 1 | 0 | 1 | Not-determined | Genome-fragment | NA | NA | 0 |
| S17_extracted_k141_160793 | 1024 | No | NA | 2 | 0 | 1 | Not-determined | Genome-fragment | NA | NA | 0 |
| S17_extracted_k141_123311 | 2840 | No | NA | 1 | 0 | 1 | Not-determined | Genome-fragment | NA | NA | 0 |
| S17_extracted_k141_138322 | 5763 | No | NA | 5 | 0 | 0 | Not-determined | Genome-fragment | NA | NA | 0 |
| S17_extracted_k141_116583 | 4446 | No | NA | 6 | 0 | 0 | Not-determined | Genome-fragment | NA | NA | 0 |
| S17_extracted_k141_5550 | 14093 | No | NA | 12 | 0 | 5 | Not-determined | Genome-fragment | NA | NA | 0 |
| S17_extracted_k141_101824 | 4054 | No | NA | 3 | 0 | 2 | Not-determined | Genome-fragment | NA | NA | 0 |
| S17_extracted_k141_168020 | 5103 | No | NA | 3 | 0 | 3 | Not-determined | Genome-fragment | NA | NA | 0 |
| S17_extracted_k141_35629 | 1272 | No | NA | 1 | 0 | 1 | Not-determined | Genome-fragment | NA | NA | 0 |
| S17_extracted_k141_145924 | 1256 | No | NA | 2 | 0 | 2 | Not-determined | Genome-fragment | NA | NA | 0 |

| | | | | | | | | | | | |
|---|---|---|---|---|---|---|---|---|---|---|---|
| S17_extracted_k141_63845 | 1702 | No | NA | 2 | 0 | 2 | Not-determined | Genome-fragment | NA | NA | 0 |
| S17_vs_k141_79109\|\|lt2gene | 1024 | No | NA | 2 | 1 | 0 | Low-quality | Genome-fragment | 2.2 | HMM-based (lower-bound) | 0 |
| S17_extracted_k141_93932 | 3892 | No | NA | 4 | 0 | 4 | Not-determined | Genome-fragment | NA | NA | 0 |
| S17_extracted_k141_63850 | 3560 | No | NA | 1 | 0 | 1 | Not-determined | Genome-fragment | NA | NA | 0 |
| S17_extracted_k141_109722 | 1606 | No | NA | 3 | 0 | 3 | Not-determined | Genome-fragment | NA | NA | 0 |
| S9_extracted_k141_58529 | 1024 | No | NA | 3 | 0 | 0 | Low-quality | Genome-fragment | 1.86 | AAI-based (medium-confidence) | 0 |
| S11_vs_k141_72045\|\|full | 1024 | No | NA | 2 | 0 | 1 | Low-quality | Genome-fragment | 1.31 | AAI-based (medium-confidence) | 0 |
| S17_extracted_k141_153861 | 1266 | No | NA | 2 | 0 | 0 | Not-determined | Genome-fragment | NA | NA | 0 |
| S17_extracted_k141_109750 | 1174 | No | NA | 2 | 0 | 1 | Not-determined | Genome-fragment | NA | NA | 0 |
| S17_extracted_k141_109758 | 1773 | No | NA | 1 | 0 | 1 | Not-determined | Genome-fragment | NA | NA | 0 |
| S17_extracted_k141_174871 | 1032 | No | NA | 2 | 0 | 1 | Not-determined | Genome-fragment | NA | NA | 0 |
| S17_extracted_k141_50334 | 4996 | No | NA | 7 | 0 | 0 | Not-determined | Genome-fragment | NA | NA | 0 |
| S17_extracted_k141_79551 | 1307 | No | NA | 2 | 0 | 0 | Not-determined | Genome-fragment | NA | NA | 0 |
| S17_extracted_k141_26952 | 1403 | No | NA | 2 | 0 | 0 | Not-determined | Genome-fragment | NA | NA | 0 |
| S17_extracted_k141_19800 | 1312 | No | NA | 2 | 0 | 0 | Not-determined | Genome-fragment | NA | NA | 0 |
| S17_extracted_k141_43116 | 1198 | No | NA | 1 | 0 | 1 | Not-determined | Genome-fragment | NA | NA | 0 |
| S17_extracted_k141_101899 | 5103 | No | NA | 2 | 0 | 1 | Not-determined | Genome-fragment | NA | NA | 0 |
| S17_extracted_k141_109789 | 1626 | No | NA | 1 | 0 | 1 | Not-determined | Genome-fragment | NA | NA | 0 |
| S17_extracted_k141_109809 | 1180 | No | NA | 1 | 0 | 1 | Not-determined | Genome-fragment | NA | NA | 0 |
| S17_extracted_k141_101935 | 1853 | No | NA | 1 | 0 | 0 | Not-determined | Genome-fragment | NA | NA | 0 |
| S17_extracted_k141_109820 | 2827 | No | NA | 2 | 0 | 2 | Not-determined | Genome-fragment | NA | NA | 0 |
| S14_extracted_k141_45701 | 1024 | No | NA | 3 | 1 | 0 | Low-quality | Genome-fragment | 2.65 | AAI-based (high-confidence) | 0 |
| S14_extracted_k141_46220 | 1024 | No | NA | 4 | 4 | 0 | Low-quality | Genome-fragment | 3.33 | AAI-based (high-confidence) | 0 |
| S17_extracted_k141_12742 | 7424 | No | NA | 7 | 0 | 3 | Not-determined | Genome-fragment | NA | NA | 0 |
| S17_extracted_k141_109870 | 1558 | No | NA | 3 | 0 | 2 | Not-determined | Genome-fragment | NA | NA | 0 |
| S1_extracted_k141_150258 | 1024 | No | NA | 2 | 0 | 0 | Low-quality | Genome-fragment | 1.59 | AAI-based (high-confidence) | 0 |
| S17_extracted_k141_5674 | 1513 | No | NA | 2 | 0 | 2 | Not-determined | Genome-fragment | NA | NA | 0 |
| S17_extracted_k141_146046 | 1590 | No | NA | 1 | 0 | 0 | Not-determined | Genome-fragment | NA | NA | 0 |
| S5_extracted_k141_129437 | 1024 | No | NA | 2 | 1 | 0 | Low-quality | Genome-fragment | 2.24 | AAI-based (high-confidence) | 0 |
| S17_extracted_k141_146058 | 2209 | No | NA | 2 | 0 | 1 | Not-determined | Genome-fragment | NA | NA | 0 |
| S17_extracted_k141_50442 | 1438 | No | NA | 2 | 0 | 2 | Not-determined | Genome-fragment | NA | NA | 0 |
| S17_extracted_k141_5700 | 3286 | No | NA | 3 | 0 | 1 | Not-determined | Genome-fragment | NA | NA | 0 |
| S17_extracted_k141_79672 | 1052 | No | NA | 1 | 0 | 0 | Not-determined | Genome-fragment | NA | NA | 0 |
| S17_extracted_k141_5703 | 1013 | No | NA | 2 | 0 | 0 | Not-determined | Genome-fragment | NA | NA | 0 |
| S17_extracted_k141_168215 | 1237 | No | NA | 2 | 0 | 2 | Not-determined | Genome-fragment | NA | NA | 0 |
| S17_extracted_k141_130763 | 1475 | No | NA | 2 | 0 | 1 | Not-determined | Genome-fragment | NA | NA | 0 |
| S17_extracted_k141_109923 | 1050 | No | NA | 2 | 0 | 1 | Not-determined | Genome-fragment | NA | NA | 0 |
| S5_extracted_k141_99822 | 1024 | No | NA | 3 | 3 | 0 | Low-quality | Genome-fragment | 2.51 | AAI-based (high-confidence) | 0 |
| S17_extracted_k141_116791 | 2664 | No | NA | 3 | 0 | 0 | Not-determined | Genome-fragment | NA | NA | 0 |
| S17_extracted_k141_161084 | 1053 | No | NA | 2 | 0 | 0 | Not-determined | Genome-fragment | NA | NA | 0 |
| S5_extracted_k141_96907 | 1024 | No | NA | 2 | 2 | 0 | Low-quality | Genome-fragment | 3.03 | AAI-based (high-confidence) | 0 |
| S17_extracted_k141_27137 | 1560 | No | NA | 2 | 0 | 0 | Not-determined | Genome-fragment | NA | NA | 0 |
| S17_extracted_k141_116812 | 1082 | No | NA | 1 | 0 | 1 | Not-determined | Genome-fragment | NA | NA | 0 |
| S16_extracted_106038 | 1024 | No | NA | 2 | 0 | 2 | Low-quality | Genome-fragment | 0.86 | AAI-based (medium-confidence) | 0 |
| S10_vs_10839\|\|full | 1024 | No | NA | 2 | 0 | 1 | Low-quality | Genome-fragment | 2.73 | AAI-based (high-confidence) | 0 |
| S17_extracted_k141_19933 | 2256 | No | NA | 4 | 0 | 1 | Not-determined | Genome-fragment | NA | NA | 0 |
| S10_vs_43001\|\|full | 1024 | No | NA | 3 | 0 | 0 | Low-quality | Genome-fragment | 2.49 | AAI-based (high-confidence) | 0 |
| S17_extracted_k141_154045 | 1043 | No | NA | 2 | 0 | 0 | Not-determined | Genome-fragment | NA | NA | 0 |
| S17_extracted_k141_146135 | 3232 | No | NA | 1 | 0 | 0 | Not-determined | Genome-fragment | NA | NA | 0 |
| S12_vs_k141_178351\|\|full | 1024 | No | NA | 2 | 0 | 1 | Low-quality | Genome-fragment | 1.79 | AAI-based (medium-confidence) | 0 |
| S17_extracted_k141_64032 | 3053 | No | NA | 2 | 0 | 0 | Not-determined | Genome-fragment | NA | NA | 0 |
| S15_extracted_68588 | 1024 | No | NA | 2 | 0 | 0 | Low-quality | Genome-fragment | 1.71 | AAI-based (high-confidence) | 0 |
| S17_extracted_k141_35899 | 1014 | No | NA | 1 | 0 | 1 | Not-determined | Genome-fragment | NA | NA | 0 |
| S17_extracted_k141_43284 | 1037 | No | NA | 1 | 0 | 0 | Not-determined | Genome-fragment | NA | NA | 0 |
| S17_extracted_k141_12878 | 1825 | No | NA | 3 | 0 | 1 | Not-determined | Genome-fragment | NA | NA | 0 |
| S9_vs_k141_4173\|\|full | 1025 | No | NA | 2 | 1 | 0 | Low-quality | Genome-fragment | 2.07 | AAI-based (high-confidence) | 0 |
| S17_extracted_k141_19964 | 1177 | No | NA | 2 | 0 | 0 | Not-determined | Genome-fragment | NA | NA | 0 |
| S17_extracted_k141_94035 | 1100 | No | NA | 1 | 0 | 0 | Not-determined | Genome-fragment | NA | NA | 0 |
| S17_extracted_k141_5787 | 2546 | No | NA | 1 | 0 | 1 | Not-determined | Genome-fragment | NA | NA | 0 |
| S17_extracted_k141_123589 | 2127 | No | NA | 1 | 0 | 0 | Not-determined | Genome-fragment | NA | NA | 0 |
| S17_extracted_k141_50547 | 4522 | No | NA | 5 | 0 | 5 | Not-determined | Genome-fragment | NA | NA | 0 |
| S17_extracted_k141_138584 | 2142 | No | NA | 1 | 0 | 0 | Not-determined | Genome-fragment | NA | NA | 0 |
| S17_extracted_k141_72472 | 3888 | No | NA | 4 | 0 | 0 | Not-determined | Genome-fragment | NA | NA | 0 |
| S17_extracted_k141_94062 | 1143 | No | NA | 2 | 0 | 0 | Not-determined | Genome-fragment | NA | NA | 0 |
| S17_extracted_k141_94069 | 3995 | No | NA | 3 | 0 | 2 | Not-determined | Genome-fragment | NA | NA | 0 |
| S17_extracted_k141_50562 | 3183 | No | NA | 2 | 0 | 2 | Not-determined | Genome-fragment | NA | NA | 0 |
| S13_extracted_84529 | 1025 | No | NA | 2 | 0 | 0 | Low-quality | Genome-fragment | 1.39 | AAI-based (medium-confidence) | 0 |
| S17_extracted_k141_138623 | 1024 | No | NA | 3 | 0 | 3 | Not-determined | Genome-fragment | NA | NA | 0 |
| S17_extracted_k141_27212 | 1287 | No | NA | 2 | 0 | 2 | Not-determined | Genome-fragment | NA | NA | 0 |
| S17_extracted_k141_138629 | 1034 | No | NA | 1 | 0 | 0 | Not-determined | Genome-fragment | NA | NA | 0 |
| S13_extracted_153322 | 1025 | No | NA | 4 | 0 | 0 | Low-quality | Genome-fragment | 1.67 | AAI-based (high-confidence) | 0 |
| S17_extracted_k141_116926 | 1065 | No | NA | 2 | 0 | 1 | Not-determined | Genome-fragment | NA | NA | 0 |
| S17_extracted_k141_20038 | 3319 | No | NA | 3 | 0 | 2 | Not-determined | Genome-fragment | NA | NA | 0 |
| S17_extracted_k141_79826 | 2349 | No | NA | 1 | 0 | 0 | Not-determined | Genome-fragment | NA | NA | 0 |
| S17_extracted_k141_27243 | 3280 | No | NA | 2 | 0 | 1 | Not-determined | Genome-fragment | NA | NA | 0 |
| S17_extracted_k141_110079 | 1362 | No | NA | 1 | 0 | 0 | Not-determined | Genome-fragment | NA | NA | 0 |
| S17_extracted_k141_27245 | 1114 | No | NA | 2 | 0 | 1 | Not-determined | Genome-fragment | NA | NA | 0 |
| S17_extracted_k141_5881 | 2567 | No | NA | 4 | 0 | 0 | Not-determined | Genome-fragment | NA | NA | 0 |
| S17_extracted_k141_20049 | 2734 | No | NA | 2 | 0 | 0 | Not-determined | Genome-fragment | NA | NA | 0 |
| S17_extracted_k141_102087 | 1343 | No | NA | 2 | 0 | 1 | Not-determined | Genome-fragment | NA | NA | 0 |
| S14_extracted_k141_41194 | 1025 | No | NA | 2 | 1 | 0 | Low-quality | Genome-fragment | 5.3 | AAI-based (medium-confidence) | 0 |
| S14_vs_k141_58635\|\|full | 1025 | No | NA | 3 | 0 | 0 | Low-quality | Genome-fragment | 2.93 | AAI-based (medium-confidence) | 0 |
| S17_extracted_k141_94160 | 1097 | No | NA | 1 | 0 | 0 | Not-determined | Genome-fragment | NA | NA | 0 |
| S17_extracted_k141_64165 | 3167 | No | NA | 3 | 0 | 3 | Not-determined | Genome-fragment | NA | NA | 0 |
| S17_extracted_k141_123697 | 1515 | No | NA | 1 | 0 | 1 | Not-determined | Genome-fragment | NA | NA | 0 |
| S17_extracted_k141_161283 | 2646 | No | NA | 1 | 0 | 0 | Not-determined | Genome-fragment | NA | NA | 0 |
| S14_vs_k141_96915\|\|lt2gene | 1025 | No | NA | 1 | 1 | 0 | Low-quality | Genome-fragment | 3.28 | AAI-based (high-confidence) | 0 |
| S17_extracted_k141_154263 | 1352 | No | NA | 2 | 0 | 2 | Not-determined | Genome-fragment | NA | NA | 0 |
| S17_extracted_k141_175063 | 1431 | No | NA | 1 | 0 | 0 | Not-determined | Genome-fragment | NA | NA | 0 |
| S17_extracted_k141_168275 | 1022 | No | NA | 2 | 0 | 0 | Not-determined | Genome-fragment | NA | NA | 0 |
| S17_extracted_k141_168286 | 1557 | No | NA | 2 | 0 | 0 | Not-determined | Genome-fragment | NA | NA | 0 |
| S17_extracted_k141_175085 | 1030 | No | NA | 1 | 0 | 1 | Not-determined | Genome-fragment | NA | NA | 0 |
| S17_extracted_k141_168287 | 2763 | No | NA | 2 | 0 | 1 | Not-determined | Genome-fragment | NA | NA | 0 |
| S17_extracted_k141_57844 | 1068 | No | NA | 1 | 0 | 0 | Not-determined | Genome-fragment | NA | NA | 0 |
| S14_vs_k141_36386\|\|lt2gene | 1025 | No | NA | 1 | 1 | 0 | Low-quality | Genome-fragment | 1.81 | AAI-based (high-confidence) | 0 |
| S1_vs_k141_41498\|\|full | 1025 | No | NA | 2 | 0 | 0 | Low-quality | Genome-fragment | 1.94 | AAI-based (high-confidence) | 0 |
| S17_extracted_k141_72564 | 1935 | No | NA | 3 | 0 | 3 | Not-determined | Genome-fragment | NA | NA | 0 |
| S17_extracted_k141_36051 | 1411 | No | NA | 2 | 0 | 2 | Not-determined | Genome-fragment | NA | NA | 0 |
| S17_extracted_k141_72586 | 1436 | No | NA | 1 | 0 | 0 | Not-determined | Genome-fragment | NA | NA | 0 |
| S17_extracted_k141_138782 | 1144 | No | NA | 1 | 0 | 0 | Not-determined | Genome-fragment | NA | NA | 0 |
| S17_extracted_k141_154387 | 3189 | No | NA | 5 | 0 | 5 | Not-determined | Genome-fragment | NA | NA | 0 |
| S17_extracted_k141_168332 | 3728 | No | NA | 4 | 0 | 1 | Not-determined | Genome-fragment | NA | NA | 0 |
| S17_extracted_k141_168339 | 1509 | No | NA | 1 | 0 | 1 | Not-determined | Genome-fragment | NA | NA | 0 |
| S1_vs_k141_111277\|\|full | 1025 | No | NA | 3 | 2 | 0 | Low-quality | Genome-fragment | 2.13 | HMM-based (lower-bound) | 0 |
| S3_extracted_k141_68770 | 1025 | No | NA | 3 | 0 | 0 | Low-quality | Genome-fragment | 1.66 | AAI-based (high-confidence) | 0 |
| S3_vs_k141_57446\|\|full | 1025 | No | NA | 3 | 0 | 0 | Low-quality | Genome-fragment | 2.3 | AAI-based (high-confidence) | 0 |
| S3_vs_k141_127211\|\|full | 1025 | No | NA | 4 | 1 | 0 | Low-quality | Genome-fragment | 2.82 | AAI-based (medium-confidence) | 0 |
| S17_extracted_k141_20175 | 3704 | No | NA | 2 | 0 | 1 | Not-determined | Genome-fragment | NA | NA | 0 |
| S17_extracted_k141_168350 | 1055 | No | NA | 3 | 0 | 0 | Not-determined | Genome-fragment | NA | NA | 0 |
| S17_extracted_k141_110238 | 4343 | No | NA | 3 | 0 | 1 | Not-determined | Genome-fragment | NA | NA | 0 |
| S17_extracted_k141_102269 | 1546 | No | NA | 2 | 0 | 1 | Not-determined | Genome-fragment | NA | NA | 0 |
| S17_extracted_k141_94273 | 1228 | No | NA | 4 | 0 | 0 | Not-determined | Genome-fragment | NA | NA | 0 |
| S17_extracted_k141_175125 | 1623 | No | NA | 3 | 0 | 1 | Not-determined | Genome-fragment | NA | NA | 0 |
| S17_extracted_k141_131114 | 1110 | No | NA | 3 | 0 | 1 | Not-determined | Genome-fragment | NA | NA | 0 |
| S5_vs_k141_92007\|\|full | 1025 | No | NA | 3 | 2 | 0 | Low-quality | Genome-fragment | 2.45 | AAI-based (high-confidence) | 0 |
| S17_extracted_k141_57939 | 3337 | No | NA | 2 | 0 | 2 | Not-determined | Genome-fragment | NA | NA | 0 |
| S17_extracted_k141_131125 | 1092 | No | NA | 2 | 0 | 1 | Not-determined | Genome-fragment | NA | NA | 0 |
| S17_extracted_k141_161442 | 1132 | No | NA | 2 | 0 | 2 | Not-determined | Genome-fragment | NA | NA | 0 |
| S17_extracted_k141_168377 | 3402 | No | NA | 4 | 0 | 2 | Not-determined | Genome-fragment | NA | NA | 0 |
| S5_vs_k141_49799\|\|full | 1025 | No | NA | 3 | 1 | 0 | Low-quality | Genome-fragment | 1.68 | AAI-based (high-confidence) | 0 |
| S17_extracted_k141_168389 | 1035 | No | NA | 1 | 0 | 1 | Not-determined | Genome-fragment | NA | NA | 0 |
| S5_vs_k141_111120\|\|full | 1025 | No | NA | 3 | 0 | 1 | Low-quality | Genome-fragment | 0.53 | AAI-based (medium-confidence) | 0 |
| S5_vs_k141_89412\|\|lt2gene | 1025 | No | NA | 1 | 0 | 0 | Low-quality | Genome-fragment | 2.47 | AAI-based (medium-confidence) | 0 |
| S7_extracted_109321 | 1025 | No | NA | 1 | 0 | 0 | Low-quality | Genome-fragment | 2.45 | AAI-based (high-confidence) | 0 |
| S17_extracted_k141_146470 | 1722 | No | NA | 3 | 0 | 2 | Not-determined | Genome-fragment | NA | NA | 0 |
| S17_extracted_k141_64375 | 1241 | No | NA | 3 | 0 | 2 | Not-determined | Genome-fragment | NA | NA | 0 |
| S17_extracted_k141_154472 | 1598 | No | NA | 2 | 0 | 1 | Not-determined | Genome-fragment | NA | NA | 0 |
| S7_vs_102924\|\|full | 1025 | No | NA | 3 | 1 | 0 | Low-quality | Genome-fragment | 2.51 | AAI-based (high-confidence) | 0 |
| S17_extracted_k141_57998 | 2203 | No | NA | 3 | 0 | 2 | Not-determined | Genome-fragment | NA | NA | 0 |
| S17_extracted_k141_58004 | 1710 | No | NA | 1 | 0 | 1 | Not-determined | Genome-fragment | NA | NA | 0 |
| S17_extracted_k141_117222 | 1251 | No | NA | 1 | 0 | 1 | Not-determined | Genome-fragment | NA | NA | 0 |
| S17_extracted_k141_86630 | 3129 | No | NA | 2 | 0 | 0 | Not-determined | Genome-fragment | NA | NA | 0 |
| S17_extracted_k141_146556 | 1435 | No | NA | 3 | 0 | 1 | Not-determined | Genome-fragment | NA | NA | 0 |
| S17_extracted_k141_117234 | 1065 | No | NA | 1 | 0 | 1 | Not-determined | Genome-fragment | NA | NA | 0 |
| S7_vs_112255\|\|lt2gene | 1025 | No | NA | 2 | 1 | 0 | Low-quality | Genome-fragment | 2.91 | AAI-based (high-confidence) | 0 |
| S16_extracted_85231 | 1025 | No | NA | 3 | 2 | 0 | Low-quality | Genome-fragment | 1.71 | AAI-based (high-confidence) | 0 |
| S17_extracted_k141_138964 | 1070 | No | NA | 3 | 0 | 0 | Not-determined | Genome-fragment | NA | NA | 0 |
| S16_extracted_170284 | 1025 | No | NA | 3 | 0 | 0 | Low-quality | Genome-fragment | 1.04 | AAI-based (medium-confidence) | 0 |
| S17_extracted_k141_80015 | 5180 | No | NA | 2 | 0 | 0 | Not-determined | Genome-fragment | NA | NA | 0 |
| S10_extracted_68437 | 1025 | No | NA | 1 | 0 | 0 | Low-quality | Genome-fragment | 2.05 | AAI-based (medium-confidence) | 0 |
| S17_extracted_k141_6209 | 1004 | No | NA | 2 | 0 | 0 | Not-determined | Genome-fragment | NA | NA | 0 |
| S10_vs_19377\|\|lt2gene | 1025 | No | NA | 2 | 2 | 0 | Low-quality | Genome-fragment | 2.74 | AAI-based (high-confidence) | 0 |
| S12_extracted_k141_119354 | 1025 | No | NA | 3 | 0 | 2 | Low-quality | Genome-fragment | 0.86 | AAI-based (medium-confidence) | 0 |
| S12_extracted_k141_108233 | 1025 | No | NA | 1 | 0 | 1 | Low-quality | Genome-fragment | 1.29 | AAI-based (high-confidence) | 0 |
| S17_extracted_k141_64497 | 1035 | No | NA | 1 | 0 | 0 | Not-determined | Genome-fragment | NA | NA | 0 |
| S17_extracted_k141_131207 | 2820 | No | NA | 1 | 0 | 0 | Not-determined | Genome-fragment | NA | NA | 0 |
| S17_extracted_k141_110438 | 1148 | No | NA | 2 | 0 | 2 | Not-determined | Genome-fragment | NA | NA | 0 |
| S12_vs_k141_130491\|\|full | 1025 | No | NA | 4 | 1 | 0 | Low-quality | Genome-fragment | 2.3 | AAI-based (high-confidence) | 0 |
| S17_extracted_k141_36213 | 1301 | No | NA | 2 | 0 | 2 | Not-determined | Genome-fragment | NA | NA | 0 |
| S6_extracted_k141_52909 | 1025 | No | NA | 3 | 1 | 0 | Low-quality | Genome-fragment | 2.08 | AAI-based (high-confidence) | 0 |
| S8_vs_k141_166455\|\|full | 1025 | No | NA | 2 | 0 | 1 | Low-quality | Genome-fragment | 1.75 | AAI-based (medium-confidence) | 0 |
| S8_vs_k141_63960\|\|full | 1025 | No | NA | 2 | 0 | 0 | Low-quality | Genome-fragment | 0.3 | AAI-based (medium-confidence) | 0 |
| S17_extracted_k141_124018 | 12016 | No | NA | 6 | 0 | 5 | Not-determined | Genome-fragment | NA | NA | 0 |
| S17_extracted_k141_175310 | 1016 | No | NA | 1 | 0 | 0 | Not-determined | Genome-fragment | NA | NA | 0 |
| S17_extracted_k141_131244 | 1060 | No | NA | 2 | 0 | 2 | Not-determined | Genome-fragment | NA | NA | 0 |
| S17_extracted_k141_43623 | 1194 | No | NA | 1 | 0 | 0 | Not-determined | Genome-fragment | NA | NA | 0 |
| S9_extracted_k141_21561 | 1026 | No | NA | 1 | 0 | 1 | Low-quality | Genome-fragment | 2.44 | AAI-based (medium-confidence) | 0 |
| S17_extracted_k141_154660 | 2658 | No | NA | 1 | 0 | 1 | Not-determined | Genome-fragment | NA | NA | 0 |
| S9_extracted_k141_78037 | 1026 | No | NA | 4 | 0 | 0 | Low-quality | Genome-fragment | 1.74 | AAI-based (medium-confidence) | 0 |
| S17_extracted_k141_58076 | 3809 | No | NA | 6 | 0 | 0 | Not-determined | Genome-fragment | NA | NA | 0 |
| S17_extracted_k141_94388 | 1398 | No | NA | 4 | 0 | 0 | Not-determined | Genome-fragment | NA | NA | 0 |
| S17_extracted_k141_131286 | 1191 | No | NA | 2 | 0 | 1 | Not-determined | Genome-fragment | NA | NA | 0 |
| S17_extracted_k141_58091 | 1438 | No | NA | 2 | 0 | 1 | Not-determined | Genome-fragment | NA | NA | 0 |
| S17_extracted_k141_72773 | 2132 | No | NA | 4 | 0 | 1 | Not-determined | Genome-fragment | NA | NA | 0 |
| S17_extracted_k141_154670 | 5497 | No | NA | 3 | 0 | 2 | Not-determined | Genome-fragment | NA | NA | 0 |
| S17_extracted_k141_36263 | 1440 | No | NA | 2 | 0 | 1 | Not-determined | Genome-fragment | NA | NA | 0 |
| S13_vs_61038\|\|full | 1026 | No | NA | 2 | 0 | 0 | Low-quality | Genome-fragment | 1.1 | AAI-based (high-confidence) | 0 |
| S17_extracted_k141_80139 | 3585 | No | NA | 2 | 0 | 2 | Not-determined | Genome-fragment | NA | NA | 0 |
| S17_extracted_k141_86783 | 1310 | No | NA | 1 | 0 | 0 | Not-determined | Genome-fragment | NA | NA | 0 |
| S13_vs_103022\|\|full | 1026 | No | NA | 2 | 0 | 0 | Low-quality | Genome-fragment | 0.47 | AAI-based (medium-confidence) | 0 |
| S14_vs_k141_19805\|\|lt2gene | 1026 | No | NA | 2 | 1 | 0 | Low-quality | Genome-fragment | 2.64 | AAI-based (high-confidence) | 0 |
| S1_extracted_k141_122728 | 1026 | No | NA | 1 | 0 | 0 | Low-quality | Genome-fragment | 1.64 | AAI-based (high-confidence) | 0 |
| S17_extracted_k141_131327 | 5392 | No | NA | 7 | 0 | 4 | Not-determined | Genome-fragment | NA | NA | 0 |
| S17_extracted_k141_27691 | 1942 | No | NA | 2 | 0 | 0 | Not-determined | Genome-fragment | NA | NA | 0 |
| S17_extracted_k141_102538 | 1115 | No | NA | 3 | 0 | 1 | Not-determined | Genome-fragment | NA | NA | 0 |
| S17_extracted_k141_175389 | 1837 | No | NA | 1 | 0 | 1 | Not-determined | Genome-fragment | NA | NA | 0 |
| S17_extracted_k141_131345 | 1077 | No | NA | 1 | 0 | 0 | Not-determined | Genome-fragment | NA | NA | 0 |
| S1_vs_k141_16284\|\|full | 1026 | No | NA | 2 | 1 | 0 | Low-quality | Genome-fragment | 1.6 | AAI-based (high-confidence) | 0 |
| S17_extracted_k141_43710 | 1152 | No | NA | 1 | 0 | 0 | Not-determined | Genome-fragment | NA | NA | 0 |
| S1_vs_k141_77944\|\|lt2gene | 1026 | No | NA | 1 | 1 | 0 | Low-quality | Genome-fragment | 1.62 | HMM-based (lower-bound) | 0 |
| S17_extracted_k141_6355 | 1539 | No | NA | 2 | 0 | 1 | Not-determined | Genome-fragment | NA | NA | 0 |
| S17_extracted_k141_13382 | 2338 | No | NA | 3 | 0 | 1 | Not-determined | Genome-fragment | NA | NA | 0 |
| S3_extracted_k141_43795 | 1026 | No | NA | 3 | 0 | 0 | Low-quality | Genome-fragment | 1.93 | AAI-based (high-confidence) | 0 |
| S17_extracted_k141_110556 | 2783 | No | NA | 3 | 0 | 1 | Not-determined | Genome-fragment | NA | NA | 0 |
| S17_extracted_k141_43725 | 2052 | No | NA | 2 | 0 | 0 | Not-determined | Genome-fragment | NA | NA | 0 |
| S17_extracted_k141_86873 | 1609 | No | NA | 2 | 0 | 1 | Not-determined | Genome-fragment | NA | NA | 0 |
| S17_extracted_k141_175434 | 1372 | No | NA | 2 | 0 | 2 | Not-determined | Genome-fragment | NA | NA | 0 |
| S17_extracted_k141_27734 | 1296 | No | NA | 1 | 0 | 1 | Not-determined | Genome-fragment | NA | NA | 0 |
| S3_vs_k141_110104\|\|full | 1026 | No | NA | 3 | 3 | 0 | Low-quality | Genome-fragment | 1.91 | AAI-based (high-confidence) | 0 |
| S17_extracted_k141_175449 | 3423 | No | NA | 1 | 0 | 1 | Not-determined | Genome-fragment | NA | NA | 0 |
| S17_extracted_k141_175451 | 1114 | No | NA | 3 | 0 | 0 | Not-determined | Genome-fragment | NA | NA | 0 |
| S17_extracted_k141_72918 | 1637 | No | NA | 4 | 0 | 3 | Not-determined | Genome-fragment | NA | NA | 0 |
| S5_vs_k141_43122\|\|full | 1026 | No | NA | 3 | 1 | 0 | Low-quality | Genome-fragment | 2.27 | AAI-based (high-confidence) | 0 |
| S5_vs_k141_2888\|\|full | 1026 | No | NA | 4 | 2 | 0 | Low-quality | Genome-fragment | 2.4 | AAI-based (medium-confidence) | 0 |
| S17_extracted_k141_175465 | 1085 | No | NA | 1 | 0 | 1 | Not-determined | Genome-fragment | NA | NA | 0 |
| S17_extracted_k141_154761 | 7711 | No | NA | 1 | 0 | 1 | Not-determined | Genome-fragment | NA | NA | 0 |
| S17_extracted_k141_20525 | 3895 | No | NA | 4 | 0 | 1 | Not-determined | Genome-fragment | NA | NA | 0 |
| S5_vs_k141_127141\|\|full | 1026 | No | NA | 2 | 2 | 0 | Low-quality | Genome-fragment | 2.24 | AAI-based (medium-confidence) | 0 |
| S17_extracted_k141_117503 | 1025 | No | NA | 2 | 0 | 2 | Not-determined | Genome-fragment | NA | NA | 0 |
| S17_extracted_k141_20531 | 1215 | No | NA | 2 | 0 | 2 | Not-determined | Genome-fragment | NA | NA | 0 |
| S17_extracted_k141_36367 | 8162 | No | NA | 5 | 0 | 4 | Not-determined | Genome-fragment | NA | NA | 0 |
| S5_vs_k141_81744\|\|full | 1026 | No | NA | 3 | 0 | 0 | Low-quality | Genome-fragment | 1.73 | AAI-based (medium-confidence) | 0 |
| S17_extracted_k141_58235 | 1074 | No | NA | 3 | 0 | 1 | Not-determined | Genome-fragment | NA | NA | 0 |
| S17_extracted_k141_131451 | 1442 | No | NA | 4 | 0 | 0 | Not-determined | Genome-fragment | NA | NA | 0 |
| S7_extracted_100664 | 1026 | No | NA | 3 | 0 | 0 | Low-quality | Genome-fragment | 2.1 | AAI-based (medium-confidence) | 0 |
| S17_extracted_k141_139165 | 1154 | No | NA | 2 | 0 | 1 | Not-determined | Genome-fragment | NA | NA | 0 |
| S17_extracted_k141_58242 | 1163 | No | NA | 1 | 0 | 1 | Not-determined | Genome-fragment | NA | NA | 0 |
| S17_extracted_k141_94486 | 2038 | No | NA | 2 | 0 | 0 | Not-determined | Genome-fragment | NA | NA | 0 |
| S17_extracted_k141_13500 | 1870 | No | NA | 3 | 0 | 1 | Not-determined | Genome-fragment | NA | NA | 0 |
| S17_extracted_k141_102671 | 1843 | No | NA | 2 | 0 | 1 | Not-determined | Genome-fragment | NA | NA | 0 |
| S17_extracted_k141_51008 | 1144 | No | NA | 4 | 0 | 2 | Not-determined | Genome-fragment | NA | NA | 0 |
| S7_vs_84670\|\|full | 1026 | No | NA | 3 | 0 | 0 | Low-quality | Genome-fragment | 2.06 | AAI-based (medium-confidence) | 0 |
| S17_extracted_k141_110623 | 3281 | No | NA | 3 | 0 | 0 | Not-determined | Genome-fragment | NA | NA | 0 |
| S17_extracted_k141_94502 | 1004 | No | NA | 1 | 0 | 0 | Not-determined | Genome-fragment | NA | NA | 0 |
| S8_vs_k141_123577\|\|full | 1026 | No | NA | 3 | 0 | 1 | Low-quality | Genome-fragment | 1.7 | AAI-based (medium-confidence) | 0 |
| S9_vs_k141_2681\|\|full | 1027 | No | NA | 2 | 1 | 0 | Low-quality | Genome-fragment | 2.31 | AAI-based (high-confidence) | 0 |
| S17_extracted_k141_146926 | 1409 | No | NA | 2 | 0 | 0 | Not-determined | Genome-fragment | NA | NA | 0 |
| S17_extracted_k141_80356 | 1392 | No | NA | 1 | 0 | 0 | Not-determined | Genome-fragment | NA | NA | 0 |
| S14_vs_k141_56805\|\|full | 1027 | No | NA | 3 | 0 | 0 | Low-quality | Genome-fragment | 0.58 | AAI-based (medium-confidence) | 0 |
| S17_extracted_k141_146940 | 1607 | No | NA | 1 | 0 | 1 | Not-determined | Genome-fragment | NA | NA | 0 |
| S17_extracted_k141_51045 | 1331 | No | NA | 1 | 0 | 1 | Not-determined | Genome-fragment | NA | NA | 0 |
| S17_extracted_k141_117540 | 1039 | No | NA | 1 | 0 | 1 | Not-determined | Genome-fragment | NA | NA | 0 |
| S17_extracted_k141_124273 | 1638 | No | NA | 1 | 0 | 1 | Not-determined | Genome-fragment | NA | NA | 0 |
| S17_extracted_k141_87000 | 2873 | No | NA | 4 | 0 | 4 | Not-determined | Genome-fragment | NA | NA | 0 |
| S17_extracted_k141_27845 | 1112 | No | NA | 1 | 0 | 0 | Not-determined | Genome-fragment | NA | NA | 0 |
| S17_extracted_k141_13557 | 3770 | No | NA | 4 | 0 | 3 | Not-determined | Genome-fragment | NA | NA | 0 |
| S17_extracted_k141_161865 | 1794 | No | NA | 2 | 0 | 1 | Not-determined | Genome-fragment | NA | NA | 0 |
| S17_extracted_k141_43877 | 1496 | No | NA | 1 | 0 | 1 | Not-determined | Genome-fragment | NA | NA | 0 |
| S17_extracted_k141_131534 | 1342 | No | NA | 2 | 0 | 1 | Not-determined | Genome-fragment | NA | NA | 0 |
| S1_vs_k141_16329\|\|full | 1027 | No | NA | 2 | 0 | 0 | Low-quality | Genome-fragment | 0.65 | AAI-based (medium-confidence) | 0 |
| S17_extracted_k141_80423 | 1038 | No | NA | 3 | 0 | 1 | Not-determined | Genome-fragment | NA | NA | 0 |
| S17_extracted_k141_168672 | 3573 | No | NA | 3 | 0 | 1 | Not-determined | Genome-fragment | NA | NA | 0 |
| S17_extracted_k141_80450 | 2353 | No | NA | 1 | 0 | 1 | Not-determined | Genome-fragment | NA | NA | 0 |
| S17_extracted_k141_102805 | 1719 | No | NA | 4 | 0 | 0 | Not-determined | Genome-fragment | NA | NA | 0 |
| S17_extracted_k141_124346 | 2132 | No | NA | 2 | 0 | 2 | Not-determined | Genome-fragment | NA | NA | 0 |
| S17_extracted_k141_124349 | 4929 | No | NA | 4 | 0 | 2 | Not-determined | Genome-fragment | NA | NA | 0 |
| S17_extracted_k141_36452 | 16135 | No | NA | 23 | 6 | 0 | Medium-quality | Genome-fragment | 68.35 | AAI-based (high-confidence) | 0 |
| S17_extracted_k141_6569 | 2452 | No | NA | 2 | 0 | 1 | Not-determined | Genome-fragment | NA | NA | 0 |
| S17_extracted_k141_6572 | 3094 | No | NA | 4 | 0 | 2 | Not-determined | Genome-fragment | NA | NA | 0 |
| S1_vs_k141_11615\|\|lt2gene | 1027 | No | NA | 2 | 1 | 0 | Low-quality | Genome-fragment | 2.28 | AAI-based (high-confidence) | 0 |
| S17_extracted_k141_27929 | 1067 | No | NA | 2 | 0 | 1 | Not-determined | Genome-fragment | NA | NA | 0 |
| S16_vs_156030\|\|full | 1027 | No | NA | 2 | 1 | 0 | Low-quality | Genome-fragment | 1.55 | AAI-based (medium-confidence) | 0 |
| S17_extracted_k141_154921 | 4661 | No | NA | 6 | 0 | 0 | Not-determined | Genome-fragment | NA | NA | 0 |
| S17_extracted_k141_154934 | 1792 | No | NA | 2 | 0 | 0 | Not-determined | Genome-fragment | NA | NA | 0 |
| S17_extracted_k141_80488 | 2353 | No | NA | 1 | 0 | 1 | Not-determined | Genome-fragment | NA | NA | 0 |
| S10_vs_14075\|\|full | 1027 | No | NA | 3 | 0 | 0 | Low-quality | Genome-fragment | 1.79 | AAI-based (medium-confidence) | 0 |
| S17_extracted_k141_80497 | 1575 | No | NA | 1 | 0 | 1 | Not-determined | Genome-fragment | NA | NA | 0 |
| S17_extracted_k141_110684 | 4391 | No | NA | 4 | 0 | 3 | Not-determined | Genome-fragment | NA | NA | 0 |
| S17_extracted_k141_147144 | 1004 | No | NA | 1 | 0 | 0 | Not-determined | Genome-fragment | NA | NA | 0 |
| S17_extracted_k141_154972 | 5060 | No | NA | 4 | 0 | 3 | Not-determined | Genome-fragment | NA | NA | 0 |
| S17_extracted_k141_139277 | 9827 | No | NA | 13 | 0 | 2 | Not-determined | Genome-fragment | NA | NA | 0 |
| S17_extracted_k141_6603 | 1068 | No | NA | 1 | 0 | 1 | Not-determined | Genome-fragment | NA | NA | 0 |
| S17_extracted_k141_51206 | 1439 | No | NA | 2 | 0 | 0 | Not-determined | Genome-fragment | NA | NA | 0 |
| S17_extracted_k141_175610 | 4223 | No | NA | 2 | 0 | 2 | Not-determined | Genome-fragment | NA | NA | 0 |
| S10_vs_14796\|\|full | 1027 | No | NA | 3 | 1 | 0 | Low-quality | Genome-fragment | 2.44 | AAI-based (high-confidence) | 0 |
| S12_vs_k141_84258\|\|full | 1027 | No | NA | 3 | 0 | 1 | Low-quality | Genome-fragment | 2.43 | AAI-based (medium-confidence) | 0 |
| S17_extracted_k141_139298 | 1442 | No | NA | 1 | 0 | 0 | Not-determined | Genome-fragment | NA | NA | 0 |
| S17_extracted_k141_175628 | 1863 | No | NA | 3 | 0 | 0 | Not-determined | Genome-fragment | NA | NA | 0 |
| S17_extracted_k141_131642 | 3201 | No | NA | 2 | 0 | 1 | Not-determined | Genome-fragment | NA | NA | 0 |
| S17_extracted_k141_87111 | 20056 | No | NA | 12 | 0 | 4 | Not-determined | Genome-fragment | NA | NA | 0 |
| S17_extracted_k141_44022 | 2379 | No | NA | 2 | 0 | 2 | Not-determined | Genome-fragment | NA | NA | 0 |
| S17_extracted_k141_175638 | 1182 | No | NA | 1 | 0 | 0 | Not-determined | Genome-fragment | NA | NA | 0 |
| S17_extracted_k141_155009 | 1187 | No | NA | 3 | 0 | 1 | Not-determined | Genome-fragment | NA | NA | 0 |
| S17_extracted_k141_175646 | 1020 | No | NA | 1 | 0 | 1 | Not-determined | Genome-fragment | NA | NA | 0 |
| S17_extracted_k141_6631 | 1255 | No | NA | 2 | 0 | 1 | Not-determined | Genome-fragment | NA | NA | 0 |
| S17_extracted_k141_73167 | 1263 | No | NA | 2 | 0 | 0 | Not-determined | Genome-fragment | NA | NA | 0 |
| S17_extracted_k141_162022 | 1536 | No | NA | 3 | 0 | 0 | Not-determined | Genome-fragment | NA | NA | 0 |
| S12_vs_k141_7731\|\|full | 1027 | No | NA | 2 | 1 | 0 | Low-quality | Genome-fragment | 2.37 | AAI-based (high-confidence) | 0 |
| S17_extracted_k141_175670 | 1030 | No | NA | 1 | 0 | 1 | Not-determined | Genome-fragment | NA | NA | 0 |
| S2_extracted_k141_17221 | 1027 | No | NA | 2 | 1 | 0 | Low-quality | Genome-fragment | 2.56 | AAI-based (high-confidence) | 0 |
| S17_extracted_k141_73175 | 1107 | No | NA | 1 | 0 | 0 | Not-determined | Genome-fragment | NA | NA | 0 |
| S17_extracted_k141_80581 | 1280 | No | NA | 1 | 0 | 1 | Not-determined | Genome-fragment | NA | NA | 0 |
| S17_extracted_k141_155047 | 1038 | No | NA | 2 | 0 | 1 | Not-determined | Genome-fragment | NA | NA | 0 |
| S8_vs_k141_92066\|\|full | 1027 | No | NA | 2 | 0 | 1 | Low-quality | Genome-fragment | 0.53 | AAI-based (medium-confidence) | 0 |
| S17_extracted_k141_51284 | 1038 | No | NA | 2 | 0 | 1 | Not-determined | Genome-fragment | NA | NA | 0 |
| S17_extracted_k141_55693 | 1028 | No | NA | 1 | 0 | 1 | Low-quality | Genome-fragment | 3.46 | AAI-based (medium-confidence) | 0 |
| S17_extracted_k141_139373 | 1012 | No | NA | 3 | 0 | 0 | Not-determined | Genome-fragment | NA | NA | 0 |
| S17_extracted_k141_93125 | 1028 | No | NA | 4 | 4 | 0 | Low-quality | Genome-fragment | 3.39 | AAI-based (high-confidence) | 0 |
| S17_extracted_k141_168859 | 1702 | No | NA | 1 | 0 | 0 | Not-determined | Genome-fragment | NA | NA | 0 |
| S17_extracted_k141_80629 | 1125 | No | NA | 1 | 0 | 1 | Not-determined | Genome-fragment | NA | NA | 0 |
| S17_extracted_k141_44060 | 2502 | No | NA | 2 | 0 | 1 | Not-determined | Genome-fragment | NA | NA | 0 |
| S17_extracted_k141_124442 | 1590 | No | NA | 2 | 0 | 0 | Not-determined | Genome-fragment | NA | NA | 0 |
| S17_extracted_k141_94666 | 4014 | No | NA | 4 | 0 | 2 | Not-determined | Genome-fragment | NA | NA | 0 |
| S17_extracted_k141_87208 | 3553 | No | NA | 2 | 0 | 0 | Not-determined | Genome-fragment | NA | NA | 0 |
| S17_extracted_k141_124456 | 1064 | No | NA | 2 | 0 | 1 | Not-determined | Genome-fragment | NA | NA | 0 |
| S17_extracted_k141_13772 | 1177 | No | NA | 2 | 0 | 1 | Not-determined | Genome-fragment | NA | NA | 0 |
| S17_extracted_k141_73250 | 1326 | No | NA | 3 | 0 | 3 | Not-determined | Genome-fragment | NA | NA | 0 |
| S11_vs_k141_71077\|\|full | 1028 | No | NA | 4 | 1 | 0 | Low-quality | Genome-fragment | 2.2 | AAI-based (high-confidence) | 0 |
| S14_vs_k141_115822\|\|full | 1028 | No | NA | 2 | 0 | 2 | Low-quality | Genome-fragment | 2.46 | AAI-based (medium-confidence) | 0 |
| S17_extracted_k141_131755 | 1790 | No | NA | 4 | 0 | 4 | Not-determined | Genome-fragment | NA | NA | 0 |
| S17_extracted_k141_28031 | 1043 | No | NA | 1 | 0 | 0 | Not-determined | Genome-fragment | NA | NA | 0 |
| S17_extracted_k141_6720 | 1323 | No | NA | 2 | 0 | 2 | Not-determined | Genome-fragment | NA | NA | 0 |
| S17_extracted_k141_13792 | 2995 | No | NA | 3 | 0 | 3 | Not-determined | Genome-fragment | NA | NA | 0 |
| S17_extracted_k141_51337 | 2430 | No | NA | 4 | 0 | 0 | Not-determined | Genome-fragment | NA | NA | 0 |
| S17_extracted_k141_44102 | 1160 | No | NA | 1 | 0 | 0 | Not-determined | Genome-fragment | NA | NA | 0 |
| S17_extracted_k141_73279 | 3533 | No | NA | 4 | 0 | 4 | Not-determined | Genome-fragment | NA | NA | 0 |
| S17_extracted_k141_20705 | 1171 | No | NA | 2 | 0 | 1 | Not-determined | Genome-fragment | NA | NA | 0 |
| S17_extracted_k141_147278 | 1052 | No | NA | 1 | 0 | 1 | Not-determined | Genome-fragment | NA | NA | 0 |
| S17_extracted_k141_58523 | 1161 | No | NA | 2 | 0 | 0 | Not-determined | Genome-fragment | NA | NA | 0 |
| S17_extracted_k141_147284 | 1225 | No | NA | 3 | 0 | 3 | Not-determined | Genome-fragment | NA | NA | 0 |
| S1_vs_k141_134818\|\|full | 1028 | No | NA | 3 | 0 | 0 | Low-quality | Genome-fragment | 2.5 | AAI-based (high-confidence) | 0 |
| S17_extracted_k141_28055 | 1854 | No | NA | 3 | 0 | 3 | Not-determined | Genome-fragment | NA | NA | 0 |
| S17_extracted_k141_139438 | 1533 | No | NA | 2 | 0 | 2 | Not-determined | Genome-fragment | NA | NA | 0 |
| S17_extracted_k141_131808 | 1204 | No | NA | 1 | 0 | 1 | Not-determined | Genome-fragment | NA | NA | 0 |
| S1_vs_k141_77523\|\|full | 1028 | No | NA | 3 | 0 | 0 | Low-quality | Genome-fragment | 1.26 | AAI-based (high-confidence) | 0 |
| S1_vs_k141_104936\|\|full | 1028 | No | NA | 4 | 1 | 0 | Low-quality | Genome-fragment | 2.63 | AAI-based (high-confidence) | 0 |
| S1_vs_k141_102248\|\|lt2gene | 1028 | No | NA | 2 | 1 | 0 | Low-quality | Genome-fragment | 2.14 | AAI-based (high-confidence) | 0 |
| S17_extracted_k141_168908 | 1259 | No | NA | 1 | 0 | 1 | Not-determined | Genome-fragment | NA | NA | 0 |
| S17_extracted_k141_139473 | 1057 | No | NA | 2 | 0 | 2 | Not-determined | Genome-fragment | NA | NA | 0 |
| S17_extracted_k141_73322 | 4041 | No | NA | 7 | 0 | 5 | Not-determined | Genome-fragment | NA | NA | 0 |
| S17_extracted_k141_64655 | 5584 | No | NA | 5 | 0 | 2 | Not-determined | Genome-fragment | NA | NA | 0 |
| S17_extracted_k141_6794 | 1289 | No | NA | 3 | 0 | 2 | Not-determined | Genome-fragment | NA | NA | 0 |
| S5_vs_k141_18882\|\|full | 1028 | No | NA | 3 | 0 | 0 | Low-quality | Genome-fragment | 2.64 | AAI-based (medium-confidence) | 0 |
| S17_extracted_k141_51384 | 1174 | No | NA | 1 | 0 | 0 | Not-determined | Genome-fragment | NA | NA | 0 |
| S17_extracted_k141_20776 | 1283 | No | NA | 1 | 0 | 1 | Not-determined | Genome-fragment | NA | NA | 0 |
| S17_extracted_k141_87332 | 1524 | No | NA | 1 | 0 | 0 | Not-determined | Genome-fragment | NA | NA | 0 |
| S17_extracted_k141_44156 | 10711 | No | NA | 6 | 0 | 6 | Not-determined | Genome-fragment | NA | NA | 0 |
| S17_extracted_k141_162120 | 1185 | No | NA | 2 | 0 | 1 | Not-determined | Genome-fragment | NA | NA | 0 |
| S17_extracted_k141_51393 | 2332 | No | NA | 3 | 0 | 1 | Not-determined | Genome-fragment | NA | NA | 0 |
| S17_extracted_k141_20794 | 1065 | No | NA | 1 | 0 | 0 | Not-determined | Genome-fragment | NA | NA | 0 |
| S17_extracted_k141_6807 | 5633 | No | NA | 4 | 0 | 2 | Not-determined | Genome-fragment | NA | NA | 0 |
| S17_extracted_k141_80659 | 1667 | No | NA | 2 | 0 | 1 | Not-determined | Genome-fragment | NA | NA | 0 |
| S17_extracted_k141_168960 | 1921 | No | NA | 1 | 0 | 1 | Not-determined | Genome-fragment | NA | NA | 0 |
| S7_extracted_514 | 1028 | No | NA | 2 | 1 | 0 | Low-quality | Genome-fragment | 4.66 | AAI-based (high-confidence) | 0 |
| S7_vs_18537\|\|full | 1028 | No | NA | 3 | 0 | 0 | Low-quality | Genome-fragment | 2.42 | AAI-based (medium-confidence) | 0 |
| S17_extracted_k141_94823 | 1453 | No | NA | 2 | 0 | 1 | Not-determined | Genome-fragment | NA | NA | 0 |
| S17_extracted_k141_175834 | 2232 | No | NA | 3 | 0 | 1 | Not-determined | Genome-fragment | NA | NA | 0 |
| S17_extracted_k141_64712 | 1438 | No | NA | 4 | 0 | 0 | Not-determined | Genome-fragment | NA | NA | 0 |
| S17_extracted_k141_87397 | 3167 | No | NA | 2 | 0 | 1 | Not-determined | Genome-fragment | NA | NA | 0 |
| S17_extracted_k141_51451 | 1397 | No | NA | 3 | 0 | 1 | Not-determined | Genome-fragment | NA | NA | 0 |
| S17_extracted_k141_162180 | 1141 | No | NA | 1 | 0 | 1 | Not-determined | Genome-fragment | NA | NA | 0 |
| S17_extracted_k141_169034 | 1024 | No | NA | 1 | 0 | 0 | Not-determined | Genome-fragment | NA | NA | 0 |
| S17_extracted_k141_131935 | 1862 | No | NA | 2 | 0 | 2 | Not-determined | Genome-fragment | NA | NA | 0 |
| S17_extracted_k141_13885 | 5568 | No | NA | 4 | 0 | 3 | Not-determined | Genome-fragment | NA | NA | 0 |
| S16_vs_23301\|\|full | 1028 | No | NA | 3 | 0 | 0 | Low-quality | Genome-fragment | 0.63 | AAI-based (medium-confidence) | 0 |
| S17_extracted_k141_162200 | 1175 | No | NA | 1 | 0 | 1 | Not-determined | Genome-fragment | NA | NA | 0 |
| S17_extracted_k141_64767 | 1682 | No | NA | 1 | 0 | 1 | Not-determined | Genome-fragment | NA | NA | 0 |
| S10_vs_119900\|\|full | 1028 | No | NA | 2 | 0 | 0 | Low-quality | Genome-fragment | 1.98 | AAI-based (high-confidence) | 0 |
| S17_extracted_k141_175895 | 1203 | No | NA | 1 | 0 | 0 | Not-determined | Genome-fragment | NA | NA | 0 |
| S17_extracted_k141_80767 | 1617 | No | NA | 3 | 0 | 2 | Not-determined | Genome-fragment | NA | NA | 0 |
| S12_extracted_k141_180636 | 1028 | No | NA | 2 | 0 | 1 | Low-quality | Genome-fragment | 0.66 | AAI-based (medium-confidence) | 0 |
| S17_extracted_k141_175970 | 1389 | No | NA | 1 | 0 | 0 | Not-determined | Genome-fragment | NA | NA | 0 |
| S12_vs_k141_164692\|\|full | 1028 | No | NA | 3 | 0 | 2 | Low-quality | Genome-fragment | 0.73 | AAI-based (medium-confidence) | 0 |
| S17_extracted_k141_13941 | 2223 | No | NA | 3 | 0 | 3 | Not-determined | Genome-fragment | NA | NA | 0 |
| S8_extracted_k141_32456 | 1028 | No | NA | 1 | 0 | 0 | Low-quality | Genome-fragment | 2.3 | AAI-based (high-confidence) | 0 |
| S17_extracted_k141_162287 | 3896 | No | NA | 4 | 0 | 2 | Not-determined | Genome-fragment | NA | NA | 0 |
| S11_vs_k141_88844\|\|full | 1029 | No | NA | 2 | 0 | 1 | Low-quality | Genome-fragment | 0.58 | AAI-based (high-confidence) | 0 |
| S17_extracted_k141_13948 | 1377 | No | NA | 3 | 0 | 2 | Not-determined | Genome-fragment | NA | NA | 0 |
| S17_extracted_k141_51566 | 2063 | No | NA | 2 | 0 | 0 | Not-determined | Genome-fragment | NA | NA | 0 |
| S3_extracted_k141_107231 | 1029 | No | NA | 2 | 0 | 0 | Low-quality | Genome-fragment | 1.02 | AAI-based (medium-confidence) | 0 |
| S17_extracted_k141_139626 | 1352 | No | NA | 2 | 0 | 0 | Not-determined | Genome-fragment | NA | NA | 0 |
| S3_vs_k141_34511\|\|full | 1029 | No | NA | 2 | 0 | 0 | Low-quality | Genome-fragment | 2.06 | AAI-based (medium-confidence) | 0 |
| S3_vs_k141_37161\|\|full | 1029 | No | NA | 2 | 0 | 1 | Low-quality | Genome-fragment | 0.69 | AAI-based (high-confidence) | 0 |
| S5_extracted_k141_137102 | 1029 | No | NA | 2 | 0 | 0 | Low-quality | Genome-fragment | 2.48 | AAI-based (high-confidence) | 0 |
| S17_extracted_k141_64800 | 1114 | No | NA | 1 | 0 | 1 | Not-determined | Genome-fragment | NA | NA | 0 |
| S17_extracted_k141_162326 | 1564 | No | NA | 2 | 0 | 2 | Not-determined | Genome-fragment | NA | NA | 0 |
| S17_extracted_k141_51601 | 7838 | No | NA | 5 | 0 | 3 | Not-determined | Genome-fragment | NA | NA | 0 |
| S17_extracted_k141_80858 | 3602 | No | NA | 4 | 0 | 0 | Not-determined | Genome-fragment | NA | NA | 0 |
| S17_extracted_k141_21028 | 3249 | No | NA | 3 | 0 | 2 | Not-determined | Genome-fragment | NA | NA | 0 |
| S17_extracted_k141_132117 | 6192 | No | NA | 5 | 0 | 1 | Not-determined | Genome-fragment | NA | NA | 0 |
| S17_extracted_k141_132123 | 1067 | No | NA | 1 | 0 | 0 | Not-determined | Genome-fragment | NA | NA | 0 |
| S17_extracted_k141_64847 | 1235 | No | NA | 2 | 0 | 2 | Not-determined | Genome-fragment | NA | NA | 0 |
| S17_extracted_k141_169182 | 1145 | No | NA | 2 | 0 | 1 | Not-determined | Genome-fragment | NA | NA | 0 |
| S17_extracted_k141_176045 | 1006 | No | NA | 1 | 0 | 0 | Not-determined | Genome-fragment | NA | NA | 0 |
| S17_extracted_k141_73623 | 1879 | No | NA | 1 | 0 | 0 | Not-determined | Genome-fragment | NA | NA | 0 |
| S17_extracted_k141_51634 | 1350 | No | NA | 1 | 0 | 1 | Not-determined | Genome-fragment | NA | NA | 0 |
| S17_extracted_k141_147636 | 1818 | No | NA | 2 | 0 | 2 | Not-determined | Genome-fragment | NA | NA | 0 |
| S17_extracted_k141_169208 | 1925 | No | NA | 1 | 0 | 1 | Not-determined | Genome-fragment | NA | NA | 0 |
| S17_extracted_k141_7049 | 1385 | No | NA | 2 | 0 | 2 | Not-determined | Genome-fragment | NA | NA | 0 |
| NC1_vf_k141_1129 | 1029 | No | NA | 1 | 0 | 1 | Low-quality | Genome-fragment | 1.3 | AAI-based (high-confidence) | 0 |
| S17_extracted_k141_28399 | 1119 | No | NA | 2 | 0 | 1 | Not-determined | Genome-fragment | NA | NA | 0 |
| S17_extracted_k141_51658 | 1047 | No | NA | 1 | 0 | 1 | Not-determined | Genome-fragment | NA | NA | 0 |
| S17_extracted_k141_58670 | 1519 | No | NA | 1 | 0 | 1 | Not-determined | Genome-fragment | NA | NA | 0 |
| S17_extracted_k141_7082 | 1090 | No | NA | 1 | 0 | 1 | Not-determined | Genome-fragment | NA | NA | 0 |
| S16_extracted_35204 | 1029 | No | NA | 4 | 1 | 0 | Low-quality | Genome-fragment | 2.05 | AAI-based (high-confidence) | 0 |
| S17_extracted_k141_64909 | 1086 | No | NA | 1 | 0 | 0 | Not-determined | Genome-fragment | NA | NA | 0 |
| S17_extracted_k141_103373 | 3193 | No | NA | 5 | 0 | 4 | Not-determined | Genome-fragment | NA | NA | 0 |
| S17_extracted_k141_95010 | 2119 | No | NA | 1 | 0 | 1 | Not-determined | Genome-fragment | NA | NA | 0 |
| S16_vs_84209\|\|lt2gene | 1029 | No | NA | 2 | 0 | 0 | Low-quality | Genome-fragment | 2.33 | AAI-based (medium-confidence) | 0 |
| S17_extracted_k141_95011 | 1225 | No | NA | 1 | 0 | 0 | Not-determined | Genome-fragment | NA | NA | 0 |
| S17_extracted_k141_28424 | 1147 | No | NA | 1 | 0 | 1 | Not-determined | Genome-fragment | NA | NA | 0 |
| S17_extracted_k141_124776 | 1395 | No | NA | 2 | 0 | 1 | Not-determined | Genome-fragment | NA | NA | 0 |
| S17_extracted_k141_87633 | 1769 | No | NA | 2 | 0 | 1 | Not-determined | Genome-fragment | NA | NA | 0 |
| S17_extracted_k141_87634 | 2007 | No | NA | 1 | 0 | 0 | Not-determined | Genome-fragment | NA | NA | 0 |
| S17_extracted_k141_73684 | 3401 | No | NA | 3 | 0 | 3 | Not-determined | Genome-fragment | NA | NA | 0 |
| S17_extracted_k141_7133 | 1103 | No | NA | 1 | 0 | 1 | Not-determined | Genome-fragment | NA | NA | 0 |
| S12_vs_k141_148654\|\|full | 1029 | No | NA | 3 | 0 | 0 | Low-quality | Genome-fragment | 0.75 | AAI-based (medium-confidence) | 0 |
| S8_vs_k141_103993\|\|full | 1029 | No | NA | 3 | 0 | 1 | Low-quality | Genome-fragment | 0.62 | AAI-based (medium-confidence) | 0 |
| S17_extracted_k141_21170 | 3056 | No | NA | 2 | 0 | 2 | Not-determined | Genome-fragment | NA | NA | 0 |
| S17_extracted_k141_21171 | 1305 | No | NA | 2 | 0 | 1 | Not-determined | Genome-fragment | NA | NA | 0 |
| S17_extracted_k141_162470 | 1801 | No | NA | 3 | 0 | 1 | Not-determined | Genome-fragment | NA | NA | 0 |
| S17_extracted_k141_73695 | 1534 | No | NA | 2 | 0 | 0 | Not-determined | Genome-fragment | NA | NA | 0 |
| S17_extracted_k141_36814 | 3372 | No | NA | 5 | 0 | 2 | Not-determined | Genome-fragment | NA | NA | 0 |
| S17_vs_k141_79878\|\|full | 1030 | No | NA | 2 | 0 | 0 | Low-quality | Genome-fragment | 1.32 | AAI-based (high-confidence) | 0 |
| S17_extracted_k141_169309 | 1092 | No | NA | 2 | 0 | 2 | Not-determined | Genome-fragment | NA | NA | 0 |
| S17_extracted_k141_28487 | 4927 | No | NA | 3 | 0 | 2 | Not-determined | Genome-fragment | NA | NA | 0 |
| S17_extracted_k141_28488 | 1091 | No | NA | 1 | 0 | 1 | Not-determined | Genome-fragment | NA | NA | 0 |
| S9_extracted_k141_51257 | 1030 | No | NA | 2 | 0 | 0 | Low-quality | Genome-fragment | 1.93 | AAI-based (medium-confidence) | 0 |
| S17_extracted_k141_118070 | 3217 | No | NA | 2 | 0 | 0 | Not-determined | Genome-fragment | NA | NA | 0 |
| S17_extracted_k141_169321 | 3501 | No | NA | 2 | 0 | 2 | Not-determined | Genome-fragment | NA | NA | 0 |
| S17_extracted_k141_169344 | 1068 | No | NA | 2 | 0 | 0 | Not-determined | Genome-fragment | NA | NA | 0 |
| S9_vs_k141_2174\|\|lt2gene | 1030 | No | NA | 2 | 1 | 0 | Low-quality | Genome-fragment | 3.05 | AAI-based (medium-confidence) | 0 |
| S17_extracted_k141_81071 | 2062 | No | NA | 2 | 0 | 2 | Not-determined | Genome-fragment | NA | NA | 0 |
| S17_extracted_k141_176200 | 5838 | No | NA | 6 | 0 | 4 | Not-determined | Genome-fragment | NA | NA | 0 |
| S14_extracted_k141_55052 | 1030 | No | NA | 1 | 0 | 0 | Low-quality | Genome-fragment | 3.23 | AAI-based (high-confidence) | 0 |
| S1_extracted_k141_20971 | 1030 | No | NA | 2 | 0 | 0 | Low-quality | Genome-fragment | 3.12 | AAI-based (medium-confidence) | 0 |
| S3_extracted_k141_81932 | 1030 | No | NA | 1 | 0 | 1 | Low-quality | Genome-fragment | 0.45 | AAI-based (medium-confidence) | 0 |
| S17_extracted_k141_169395 | 1029 | No | NA | 2 | 0 | 1 | Not-determined | Genome-fragment | NA | NA | 0 |
| S3_vs_k141_114453\|\|full | 1030 | No | NA | 2 | 0 | 0 | Low-quality | Genome-fragment | 1.13 | AAI-based (high-confidence) | 0 |
| S17_extracted_k141_95131 | 1521 | No | NA | 1 | 0 | 1 | Not-determined | Genome-fragment | NA | NA | 0 |
| S5_extracted_k141_39353 | 1030 | No | NA | 2 | 0 | 0 | Low-quality | Genome-fragment | 1.5 | AAI-based (high-confidence) | 0 |
| S17_extracted_k141_176268 | 1106 | No | NA | 3 | 0 | 2 | Not-determined | Genome-fragment | NA | NA | 0 |
| S17_extracted_k141_176270 | 1171 | No | NA | 1 | 0 | 1 | Not-determined | Genome-fragment | NA | NA | 0 |

| | | | | | | | | | | | |
|---|---|---|---|---|---|---|---|---|---|---|---|
| S17_extracted_k141_73849 | 2792 | No | NA | 2 | 0 | 2 | Not-determined | Genome-fragment | NA | NA | 0 |
| S17_extracted_k141_169445 | 1856 | No | NA | 4 | 0 | 3 | Not-determined | Genome-fragment | NA | NA | 0 |
| S17_extracted_k141_124959 | 1503 | No | NA | 2 | 0 | 0 | Not-determined | Genome-fragment | NA | NA | 0 |
| S17_extracted_k141_124971 | 1126 | No | NA | 1 | 0 | 0 | Not-determined | Genome-fragment | NA | NA | 0 |
| S5_vs_k141_74671\|\|lt2gene | 1030 | No | NA | 1 | 1 | 0 | Low-quality | Genome-fragment | 0.64 | HMM-based (lower-bound) | 0 |
| S17_extracted_k141_36907 | 1642 | No | NA | 3 | 0 | 2 | Not-determined | Genome-fragment | NA | NA | 0 |
| S17_extracted_k141_81160 | 2268 | No | NA | 2 | 0 | 1 | Not-determined | Genome-fragment | NA | NA | 0 |
| S17_extracted_k141_65154 | 10424 | No | NA | 6 | 0 | 3 | Not-determined | Genome-fragment | NA | NA | 0 |
| S16_extracted_9538 | 1030 | No | NA | 1 | 1 | 0 | Low-quality | Genome-fragment | 1.39 | AAI-based (high-confidence) | 0 |
| S17_extracted_k141_7335 | 3164 | No | NA | 6 | 0 | 0 | Not-determined | Genome-fragment | NA | NA | 0 |
| S16_extracted_75305 | 1030 | No | NA | 3 | 0 | 1 | Low-quality | Genome-fragment | 1.84 | AAI-based (medium-confidence) | 0 |
| S17_extracted_k141_169522 | 1532 | No | NA | 4 | 0 | 3 | Not-determined | Genome-fragment | NA | NA | 0 |
| S17_extracted_k141_169529 | 1678 | No | NA | 1 | 0 | 0 | Not-determined | Genome-fragment | NA | NA | 0 |
| S17_extracted_k141_176330 | 4147 | No | NA | 2 | 0 | 1 | Not-determined | Genome-fragment | NA | NA | 0 |
| S17_extracted_k141_21505 | 1108 | No | NA | 2 | 0 | 0 | Not-determined | Genome-fragment | NA | NA | 0 |
| S16_vs_27996\|\|full | 1030 | No | NA | 2 | 2 | 0 | Low-quality | Genome-fragment | 2.2 | AAI-based (medium-confidence) | 0 |
| S17_extracted_k141_169550 | 1006 | No | NA | 1 | 0 | 1 | Not-determined | Genome-fragment | NA | NA | 0 |
| S17_extracted_k141_81214 | 4662 | No | NA | 2 | 0 | 0 | Not-determined | Genome-fragment | NA | NA | 0 |
| S17_extracted_k141_65272 | 1111 | No | NA | 1 | 0 | 1 | Not-determined | Genome-fragment | NA | NA | 0 |
| S17_extracted_k141_14304 | 1215 | No | NA | 2 | 0 | 0 | Not-determined | Genome-fragment | NA | NA | 0 |
| S17_extracted_k141_14331 | 1294 | No | NA | 2 | 0 | 0 | Not-determined | Genome-fragment | NA | NA | 0 |
| S17_extracted_k141_132526 | 1022 | No | NA | 2 | 0 | 1 | Not-determined | Genome-fragment | NA | NA | 0 |
| S17_extracted_k141_81226 | 1338 | No | NA | 3 | 0 | 0 | Not-determined | Genome-fragment | NA | NA | 0 |
| S17_extracted_k141_65321 | 8451 | No | NA | 5 | 0 | 3 | Not-determined | Genome-fragment | NA | NA | 0 |
| S17_extracted_k141_176441 | 3900 | No | NA | 3 | 0 | 2 | Not-determined | Genome-fragment | NA | NA | 0 |
| S17_extracted_k141_95354 | 3319 | No | NA | 4 | 0 | 2 | Not-determined | Genome-fragment | NA | NA | 0 |
| S17_extracted_k141_169722 | 1573 | No | NA | 2 | 0 | 2 | Not-determined | Genome-fragment | NA | NA | 0 |
| S17_extracted_k141_169731 | 7446 | No | NA | 5 | 0 | 4 | Not-determined | Genome-fragment | NA | NA | 0 |
| S17_extracted_k141_169737 | 1043 | No | NA | 1 | 0 | 0 | Not-determined | Genome-fragment | NA | NA | 0 |
| S17_extracted_k141_28883 | 1010 | No | NA | 4 | 0 | 0 | Not-determined | Genome-fragment | NA | NA | 0 |
| S17_extracted_k141_7367 | 5528 | No | NA | 9 | 0 | 1 | Not-determined | Genome-fragment | NA | NA | 0 |
| S17_extracted_k141_132648 | 3640 | No | NA | 4 | 0 | 2 | Not-determined | Genome-fragment | NA | NA | 0 |
| S10_vs_1259\|\|full | 1030 | No | NA | 3 | 0 | 0 | Low-quality | Genome-fragment | 1.75 | AAI-based (high-confidence) | 0 |
| S12_extracted_k141_130268 | 1030 | No | NA | 1 | 1 | 0 | Low-quality | Genome-fragment | 2.34 | AAI-based (high-confidence) | 0 |
| S12_vs_k141_119888\|\|full | 1030 | No | NA | 2 | 0 | 0 | Low-quality | Genome-fragment | 2.14 | AAI-based (high-confidence) | 0 |
| S17_extracted_k141_14436 | 4219 | No | NA | 5 | 0 | 4 | Not-determined | Genome-fragment | NA | NA | 0 |
| S17_extracted_k141_14439 | 1365 | No | NA | 3 | 0 | 3 | Not-determined | Genome-fragment | NA | NA | 0 |
| S17_extracted_k141_59045 | 3456 | No | NA | 3 | 0 | 1 | Not-determined | Genome-fragment | NA | NA | 0 |
| S17_extracted_k141_140087 | 1319 | No | NA | 2 | 0 | 1 | Not-determined | Genome-fragment | NA | NA | 0 |
| S17_extracted_k141_65423 | 1317 | No | NA | 2 | 0 | 1 | Not-determined | Genome-fragment | NA | NA | 0 |
| S17_extracted_k141_65430 | 1468 | No | NA | 2 | 0 | 0 | Not-determined | Genome-fragment | NA | NA | 0 |
| S4_extracted_10627 | 1030 | No | NA | 5 | 5 | 0 | Low-quality | Genome-fragment | 3.38 | AAI-based (high-confidence) | 0 |
| S17_extracted_k141_132696 | 3778 | No | NA | 2 | 0 | 1 | Not-determined | Genome-fragment | NA | NA | 0 |
| S17_extracted_k141_28958 | 1826 | No | NA | 2 | 0 | 1 | Not-determined | Genome-fragment | NA | NA | 0 |
| S17_extracted_k141_14477 | 1040 | No | NA | 1 | 0 | 1 | Not-determined | Genome-fragment | NA | NA | 0 |
| S17_extracted_k141_21718 | 32371 | No | NA | 19 | 0 | 6 | High-quality | High-quality | 93.35 | AAI-based (medium-confidence) | 0 |
| S17_extracted_k141_59082 | 2196 | No | NA | 2 | 0 | 1 | Not-determined | Genome-fragment | NA | NA | 0 |
| S17_extracted_k141_88117 | 1159 | No | NA | 2 | 0 | 1 | Not-determined | Genome-fragment | NA | NA | 0 |
| S17_extracted_k141_81232 | 102314 | No | NA | 103 | 16 | 1 | Complete | High-quality | 100 | DTR (high-confidence) | 0 |
| S17_extracted_k141_132711 | 2332 | No | NA | 3 | 0 | 2 | Not-determined | Genome-fragment | NA | NA | 0 |
| S17_extracted_k141_65455 | 9112 | No | NA | 9 | 0 | 7 | Not-determined | Genome-fragment | NA | NA | 0 |
| S17_extracted_k141_95413 | 6729 | No | NA | 2 | 0 | 2 | Not-determined | Genome-fragment | NA | NA | 0 |
| S17_extracted_k141_140121 | 1089 | No | NA | 1 | 0 | 0 | Not-determined | Genome-fragment | NA | NA | 0 |
| S17_extracted_k141_81250 | 7892 | No | NA | 7 | 0 | 3 | Not-determined | Genome-fragment | NA | NA | 0 |
| S17_extracted_k141_176593 | 2473 | No | NA | 7 | 0 | 6 | Not-determined | Genome-fragment | NA | NA | 0 |
| S17_extracted_k141_81275 | 1629 | No | NA | 1 | 0 | 1 | Not-determined | Genome-fragment | NA | NA | 0 |
| S17_extracted_k141_95462 | 1057 | No | NA | 1 | 0 | 1 | Not-determined | Genome-fragment | NA | NA | 0 |
| S17_extracted_k141_88186 | 3488 | No | NA | 4 | 0 | 1 | Not-determined | Genome-fragment | NA | NA | 0 |
| S8_vs_k141_92992\|\|full | 1030 | No | NA | 2 | 0 | 2 | Low-quality | Genome-fragment | 1.66 | AAI-based (medium-confidence) | 0 |
| S17_extracted_k141_125242 | 2674 | No | NA | 2 | 0 | 1 | Not-determined | Genome-fragment | NA | NA | 0 |
| S17_extracted_k141_125256 | 1575 | No | NA | 1 | 0 | 0 | Not-determined | Genome-fragment | NA | NA | 0 |
| S17_extracted_k141_21787 | 4162 | No | NA | 4 | 0 | 3 | Not-determined | Genome-fragment | NA | NA | 0 |
| S17_extracted_k141_176614 | 1476 | No | NA | 1 | 0 | 1 | Not-determined | Genome-fragment | NA | NA | 0 |
| S17_extracted_k141_132802 | 1353 | No | NA | 3 | 0 | 2 | Not-determined | Genome-fragment | NA | NA | 0 |
| S17_extracted_k141_125285 | 1170 | No | NA | 2 | 0 | 2 | Not-determined | Genome-fragment | NA | NA | 0 |
| S17_extracted_k141_176627 | 1233 | No | NA | 2 | 0 | 1 | Not-determined | Genome-fragment | NA | NA | 0 |
| S17_extracted_k141_65628 | 1324 | No | NA | 1 | 0 | 0 | Not-determined | Genome-fragment | NA | NA | 0 |
| S17_extracted_k141_14643 | 3840 | No | NA | 4 | 0 | 3 | Not-determined | Genome-fragment | NA | NA | 0 |
| S17_extracted_k141_176672 | 4516 | No | NA | 4 | 0 | 3 | Not-determined | Genome-fragment | NA | NA | 0 |
| S17_extracted_k141_95633 | 1150 | No | NA | 1 | 0 | 0 | Not-determined | Genome-fragment | NA | NA | 0 |
| S17_extracted_k141_14666 | 1108 | No | NA | 2 | 0 | 1 | Not-determined | Genome-fragment | NA | NA | 0 |
| S17_extracted_k141_176703 | 2247 | No | NA | 1 | 0 | 1 | Not-determined | Genome-fragment | NA | NA | 0 |
| S17_extracted_k141_14675 | 1279 | No | NA | 3 | 0 | 0 | Not-determined | Genome-fragment | NA | NA | 0 |
| S17_extracted_k141_125411 | 1364 | No | NA | 2 | 0 | 0 | Not-determined | Genome-fragment | NA | NA | 0 |
| S8_vs_k141_14440\|\|lt2gene | 1030 | No | NA | 2 | 1 | 0 | Low-quality | Genome-fragment | 2.11 | AAI-based (medium-confidence) | 0 |
| S17_extracted_k141_65715 | 1055 | No | NA | 1 | 0 | 0 | Not-determined | Genome-fragment | NA | NA | 0 |
| S17_extracted_k141_132991 | 1243 | No | NA | 1 | 0 | 0 | Not-determined | Genome-fragment | NA | NA | 0 |
| S17_extracted_k141_95777 | 2160 | No | NA | 1 | 0 | 1 | Not-determined | Genome-fragment | NA | NA | 0 |
| S17_extracted_k141_29234 | 1089 | No | NA | 2 | 0 | 0 | Not-determined | Genome-fragment | NA | NA | 0 |
| S17_extracted_k141_125509 | 1144 | No | NA | 2 | 0 | 2 | Not-determined | Genome-fragment | NA | NA | 0 |
| S17_extracted_k141_88431 | 1393 | No | NA | 2 | 0 | 0 | Not-determined | Genome-fragment | NA | NA | 0 |
| S17_extracted_k141_14772 | 2905 | No | NA | 2 | 0 | 1 | Not-determined | Genome-fragment | NA | NA | 0 |
| S17_extracted_k141_65791 | 1447 | No | NA | 2 | 0 | 1 | Not-determined | Genome-fragment | NA | NA | 0 |
| S17_extracted_k141_65806 | 1115 | No | NA | 2 | 0 | 2 | Not-determined | Genome-fragment | NA | NA | 0 |
| S17_extracted_k141_125580 | 3895 | No | NA | 4 | 0 | 3 | Not-determined | Genome-fragment | NA | NA | 0 |
| S17_extracted_k141_32900 | 1031 | No | NA | 3 | 2 | 0 | Low-quality | Genome-fragment | 3.28 | AAI-based (high-confidence) | 0 |
| S17_extracted_k141_65846 | 1160 | No | NA | 2 | 0 | 2 | Not-determined | Genome-fragment | NA | NA | 0 |
| S17_extracted_k141_29319 | 1088 | No | NA | 3 | 0 | 3 | Not-determined | Genome-fragment | NA | NA | 0 |
| S17_extracted_k141_29370 | 1967 | No | NA | 1 | 0 | 0 | Not-determined | Genome-fragment | NA | NA | 0 |
| S17_extracted_k141_65935 | 3810 | No | NA | 3 | 0 | 3 | Not-determined | Genome-fragment | NA | NA | 0 |
| S17_vs_k141_2314\|\|full | 1031 | No | NA | 3 | 1 | 0 | Low-quality | Genome-fragment | 1.53 | HMM-based (lower-bound) | 0 |
| S17_vs_k141_71471\|\|full | 1031 | No | NA | 3 | 0 | 0 | Low-quality | Genome-fragment | 0.76 | AAI-based (medium-confidence) | 0 |
| S17_extracted_k141_95856 | 2648 | No | NA | 3 | 0 | 2 | Not-determined | Genome-fragment | NA | NA | 0 |
| S17_extracted_k141_176976 | 2829 | No | NA | 4 | 0 | 2 | Not-determined | Genome-fragment | NA | NA | 0 |
| S17_extracted_k141_29447 | 3467 | No | NA | 4 | 0 | 2 | Not-determined | Genome-fragment | NA | NA | 0 |
| S9_vs_k141_48669\|\|full | 1031 | No | NA | 3 | 0 | 0 | Low-quality | Genome-fragment | 2.29 | AAI-based (high-confidence) | 0 |
| S17_extracted_k141_88604 | 1197 | No | NA | 2 | 0 | 1 | Not-determined | Genome-fragment | NA | NA | 0 |
| S17_extracted_k141_95918 | 3279 | No | NA | 6 | 0 | 4 | Not-determined | Genome-fragment | NA | NA | 0 |
| S17_extracted_k141_177070 | 1599 | No | NA | 1 | 0 | 1 | Not-determined | Genome-fragment | NA | NA | 0 |
| S17_extracted_k141_177088 | 1172 | No | NA | 1 | 0 | 0 | Not-determined | Genome-fragment | NA | NA | 0 |
| S11_extracted_k141_99962 | 1031 | No | NA | 2 | 1 | 0 | Low-quality | Genome-fragment | 0.3 | HMM-based (lower-bound) | 0 |
| S17_extracted_k141_29528 | 10475 | No | NA | 6 | 0 | 1 | Not-determined | Genome-fragment | NA | NA | 0 |
| S17_extracted_k141_29533 | 4147 | No | NA | 3 | 0 | 1 | Not-determined | Genome-fragment | NA | NA | 0 |
| S17_extracted_k141_177147 | 1105 | No | NA | 2 | 0 | 0 | Not-determined | Genome-fragment | NA | NA | 0 |
| S14_vs_k141_34526\|\|lt2gene | 1031 | No | NA | 2 | 0 | 0 | Low-quality | Genome-fragment | 2.44 | AAI-based (medium-confidence) | 0 |
| S1_vs_k141_102446\|\|full | 1031 | No | NA | 2 | 0 | 0 | Low-quality | Genome-fragment | 0.63 | AAI-based (medium-confidence) | 0 |
| S17_extracted_k141_177178 | 3265 | No | NA | 2 | 0 | 1 | Not-determined | Genome-fragment | NA | NA | 0 |
| S17_extracted_k141_177183 | 1046 | No | NA | 2 | 0 | 2 | Not-determined | Genome-fragment | NA | NA | 0 |
| S3_extracted_k141_88430 | 1031 | No | NA | 2 | 1 | 0 | Low-quality | Genome-fragment | 2.79 | AAI-based (high-confidence) | 0 |
| S17_extracted_k141_177197 | 53491 | No | NA | 80 | 12 | 0 | Complete | High-quality | 100 | DTR (high-confidence) | 0 |
| S17_extracted_k141_177199 | 40033 | No | NA | 59 | 44 | 0 | Complete | High-quality | 100 | DTR (high-confidence) | 0 |
| S17_extracted_k141_177200 | 36624 | No | NA | 45 | 10 | 1 | Complete | High-quality | 100 | DTR (high-confidence) | 0 |
| S17_extracted_k141_66356 | 1042 | No | NA | 1 | 0 | 1 | Not-determined | Genome-fragment | NA | NA | 0 |
| S17_extracted_k141_66357 | 1110 | No | NA | 2 | 0 | 0 | Not-determined | Genome-fragment | NA | NA | 0 |
| S17_extracted_k141_177221 | 3089 | No | NA | 5 | 0 | 0 | Not-determined | Genome-fragment | NA | NA | 0 |
| S17_extracted_k141_177254 | 2664 | No | NA | 2 | 0 | 1 | Not-determined | Genome-fragment | NA | NA | 0 |
| S17_vs_k141_15243\|\|full | 5390 | No | NA | 8 | 0 | 0 | Not-determined | Genome-fragment | NA | NA | 0 |
| S17_vs_k141_153490\|\|full | 2132 | No | NA | 2 | 0 | 0 | Not-determined | Genome-fragment | NA | NA | 0 |
| S17_vs_k141_138036\|\|full | 20778 | No | NA | 42 | 5 | 0 | High-quality | High-quality | 98.12 | AAI-based (high-confidence) | 0 |
| S17_vs_k141_57885\|\|full | 2732 | No | NA | 6 | 0 | 2 | Not-determined | Genome-fragment | NA | NA | 0 |
| S17_vs_k141_103009\|\|full | 43347 | No | NA | 59 | 12 | 0 | High-quality | High-quality | 100 | AAI-based (high-confidence) | 0 |
| S17_vs_k141_169681\|\|full | 5594 | No | NA | 9 | 0 | 0 | Not-determined | Genome-fragment | NA | NA | 0 |
| S17_vs_k141_177196\|\|full | 38554 | No | NA | 59 | 8 | 0 | High-quality | High-quality | 96.54 | AAI-based (high-confidence) | 0 |
| S17_vs_k141_177202\|\|full | 39636 | No | NA | 60 | 21 | 3 | High-quality | High-quality | 99.91 | AAI-based (high-confidence) | 0 |
| S17_vs_k141_177203\|\|full | 49241 | No | NA | 74 | 29 | 0 | High-quality | High-quality | 100 | AAI-based (high-confidence) | 0 |
| S17_vs_k141_177210\|\|full | 4760 | No | NA | 9 | 4 | 0 | High-quality | High-quality | 99.66 | AAI-based (high-confidence) | 0 |
| S17_vs_k141_66506\|\|full | 2430 | No | NA | 3 | 0 | 0 | Not-determined | Genome-fragment | NA | NA | 0 |
| S17_vs_k141_133010\|\|full | 4330 | No | NA | 4 | 0 | 1 | Not-determined | Genome-fragment | NA | NA | 0 |
| S17_vs_k141_118249\|\|full | 2249 | No | NA | 3 | 0 | 1 | Not-determined | Genome-fragment | NA | NA | 0 |
| S17_vs_k141_29590\|\|full | 2096 | No | NA | 4 | 0 | 1 | Not-determined | Genome-fragment | NA | NA | 0 |
| S3_extracted_k141_34431 | 1031 | No | NA | 3 | 1 | 0 | Low-quality | Genome-fragment | 4.71 | AAI-based (high-confidence) | 0 |
| S17_vs_k141_125634\|\|full | 1315 | No | NA | 2 | 0 | 0 | Not-determined | Genome-fragment | NA | NA | 0 |
| S3_extracted_k141_14196 | 1031 | No | NA | 2 | 0 | 0 | Low-quality | Genome-fragment | 0.56 | AAI-based (high-confidence) | 0 |
| S17_vs_k141_125657\|\|full | 1266 | No | NA | 3 | 0 | 2 | Not-determined | Genome-fragment | NA | NA | 0 |
| S17_vs_k141_110876\|\|full | 2699 | No | NA | 3 | 0 | 2 | Not-determined | Genome-fragment | NA | NA | 0 |
| S3_vs_k141_9509\|\|lt2gene | 1031 | No | NA | 2 | 2 | 0 | Low-quality | Genome-fragment | 2.78 | AAI-based (medium-confidence) | 0 |
| S7_vs_101812\|\|full | 1031 | No | NA | 2 | 0 | 0 | Low-quality | Genome-fragment | 2.84 | AAI-based (medium-confidence) | 0 |
| S16_vs_45294\|\|full | 1031 | No | NA | 3 | 0 | 0 | Low-quality | Genome-fragment | 2.27 | AAI-based (high-confidence) | 0 |
| S17_vs_k141_44366\|\|full | 1833 | No | NA | 3 | 0 | 0 | Not-determined | Genome-fragment | NA | NA | 0 |
| S17_vs_k141_110905\|\|full | 2324 | No | NA | 3 | 0 | 0 | Not-determined | Genome-fragment | NA | NA | 0 |
| S17_vs_k141_118290\|\|full | 2266 | No | NA | 3 | 0 | 0 | Not-determined | Genome-fragment | NA | NA | 0 |
| S17_vs_k141_44386\|\|full | 1915 | No | NA | 2 | 0 | 2 | Not-determined | Genome-fragment | NA | NA | 0 |
| S17_vs_k141_162600\|\|full | 2860 | No | NA | 3 | 0 | 2 | Not-determined | Genome-fragment | NA | NA | 0 |
| S17_vs_k141_37008\|\|full | 2609 | No | NA | 3 | 0 | 1 | Not-determined | Genome-fragment | NA | NA | 0 |
| S16_vs_93228\|\|full | 1031 | No | NA | 2 | 0 | 0 | Low-quality | Genome-fragment | 1.79 | AAI-based (medium-confidence) | 0 |
| S17_vs_k141_118308\|\|full | 5224 | No | NA | 5 | 0 | 2 | Not-determined | Genome-fragment | NA | NA | 0 |
| S10_vs_103738\|\|full | 1031 | No | NA | 3 | 0 | 0 | Low-quality | Genome-fragment | 1.89 | AAI-based (high-confidence) | 0 |
| S12_extracted_k141_166225 | 1031 | No | NA | 1 | 0 | 0 | Low-quality | Genome-fragment | 0.46 | AAI-based (medium-confidence) | 0 |
| S17_vs_k141_37017\|\|full | 2459 | No | NA | 3 | 0 | 1 | Not-determined | Genome-fragment | NA | NA | 0 |
| S17_vs_k141_59208\|\|full | 2343 | No | NA | 3 | 0 | 2 | Not-determined | Genome-fragment | NA | NA | 0 |
| S17_vs_k141_133074\|\|full | 8311 | No | NA | 8 | 0 | 2 | Not-determined | Genome-fragment | NA | NA | 0 |
| S17_vs_k141_140464\|\|full | 1526 | No | NA | 3 | 0 | 2 | Not-determined | Genome-fragment | NA | NA | 0 |
| S12_extracted_k141_163889 | 1031 | No | NA | 2 | 2 | 0 | Low-quality | Genome-fragment | 3.4 | AAI-based (high-confidence) | 0 |
| S17_vs_k141_162643\|\|full | 1911 | No | NA | 2 | 0 | 1 | Not-determined | Genome-fragment | NA | NA | 0 |
| S8_extracted_k141_177771 | 1031 | No | NA | 1 | 0 | 0 | Low-quality | Genome-fragment | 2.38 | AAI-based (high-confidence) | 0 |
| S17_vs_k141_114\|\|full | 2489 | No | NA | 3 | 0 | 1 | Not-determined | Genome-fragment | NA | NA | 0 |
| S17_vs_k141_59236\|\|full | 1251 | No | NA | 2 | 0 | 1 | Not-determined | Genome-fragment | NA | NA | 0 |
| S17_vs_k141_51825\|\|full | 11791 | No | NA | 13 | 0 | 9 | Not-determined | Genome-fragment | NA | NA | 0 |
| S9_extracted_k141_42856 | 1032 | No | NA | 2 | 0 | 0 | Low-quality | Genome-fragment | 2.08 | AAI-based (high-confidence) | 0 |
| S9_vs_k141_35615\|\|lt2gene | 1032 | No | NA | 2 | 1 | 0 | Low-quality | Genome-fragment | 1.4 | HMM-based (lower-bound) | 0 |
| S13_vs_149911\|\|full | 1032 | No | NA | 4 | 1 | 0 | Low-quality | Genome-fragment | 21.7 | AAI-based (high-confidence) | 0 |
| S14_extracted_k141_77896 | 1032 | No | NA | 1 | 0 | 0 | Low-quality | Genome-fragment | 2.68 | AAI-based (high-confidence) | 0 |
| S17_vs_k141_14886\|\|full | 1083 | No | NA | 3 | 0 | 1 | Not-determined | Genome-fragment | NA | NA | 0 |
| S14_vs_k141_60220\|\|full | 1032 | No | NA | 3 | 2 | 0 | Low-quality | Genome-fragment | 3.6 | AAI-based (high-confidence) | 0 |
| S17_vs_k141_22238\|\|full | 7215 | No | NA | 5 | 0 | 1 | Not-determined | Genome-fragment | NA | NA | 0 |
| S14_vs_k141_17440\|\|full | 1032 | No | NA | 3 | 3 | 0 | Low-quality | Genome-fragment | 3.47 | AAI-based (high-confidence) | 0 |
| S17_vs_k141_96230\|\|full | 4419 | No | NA | 5 | 0 | 1 | Not-determined | Genome-fragment | NA | NA | 0 |
| S1_vs_k141_81813\|\|full | 1032 | No | NA | 3 | 0 | 0 | Low-quality | Genome-fragment | 1.21 | AAI-based (medium-confidence) | 0 |
| S17_vs_k141_66672\|\|full | 2052 | No | NA | 2 | 0 | 2 | Not-determined | Genome-fragment | NA | NA | 0 |
| S17_vs_k141_14905\|\|full | 1795 | No | NA | 3 | 0 | 0 | Not-determined | Genome-fragment | NA | NA | 0 |
| S17_vs_k141_59312\|\|full | 12086 | No | NA | 11 | 0 | 7 | Not-determined | Genome-fragment | NA | NA | 0 |
| S17_vs_k141_118363\|\|full | 1531 | No | NA | 2 | 0 | 1 | Not-determined | Genome-fragment | NA | NA | 0 |
| S17_vs_k141_59322\|\|full | 6478 | No | NA | 6 | 0 | 5 | Not-determined | Genome-fragment | NA | NA | 0 |
| S16_vs_79314\|\|full | 1032 | No | NA | 3 | 1 | 0 | Low-quality | Genome-fragment | 2.46 | AAI-based (high-confidence) | 0 |
| S10_vs_25512\|\|full | 1032 | No | NA | 3 | 0 | 0 | Low-quality | Genome-fragment | 2.68 | AAI-based (medium-confidence) | 0 |
| S10_vs_114072\|\|full | 1032 | No | NA | 2 | 1 | 0 | Low-quality | Genome-fragment | 1.61 | AAI-based (medium-confidence) | 0 |
| S17_vs_k141_133190\|\|full | 4538 | No | NA | 4 | 0 | 4 | Not-determined | Genome-fragment | NA | NA | 0 |
| S15_extracted_66954 | 1032 | No | NA | 2 | 1 | 0 | Low-quality | Genome-fragment | 2.29 | AAI-based (high-confidence) | 0 |
| S17_vs_k141_44495\|\|full | 4223 | No | NA | 5 | 0 | 2 | Not-determined | Genome-fragment | NA | NA | 0 |
| S17_vs_k141_44496\|\|full | 1062 | No | NA | 3 | 0 | 0 | Not-determined | Genome-fragment | NA | NA | 0 |
| S17_vs_k141_118426\|\|full | 3192 | No | NA | 5 | 0 | 1 | Not-determined | Genome-fragment | NA | NA | 0 |
| S15_vs_4966\|\|full | 1032 | No | NA | 3 | 0 | 0 | Low-quality | Genome-fragment | 1.08 | AAI-based (medium-confidence) | 0 |
| S4_extracted_26255 | 1032 | No | NA | 3 | 1 | 0 | Low-quality | Genome-fragment | 1.23 | AAI-based (high-confidence) | 0 |
| S8_extracted_k141_166508 | 1032 | No | NA | 1 | 0 | 0 | Low-quality | Genome-fragment | 1.56 | AAI-based (medium-confidence) | 0 |
| S17_vs_k141_140615\|\|full | 2666 | No | NA | 5 | 0 | 1 | Not-determined | Genome-fragment | NA | NA | 0 |
| S17_vs_k141_7690\|\|full | 2197 | No | NA | 3 | 0 | 1 | Not-determined | Genome-fragment | NA | NA | 0 |
| S8_extracted_k141_64495 | 1032 | No | NA | 1 | 0 | 1 | Low-quality | Genome-fragment | 1.01 | AAI-based (medium-confidence) | 0 |
| S17_vs_k141_148004\|\|full | 1614 | No | NA | 4 | 0 | 2 | Not-determined | Genome-fragment | NA | NA | 0 |
| S17_vs_k141_133231\|\|full | 8959 | No | NA | 11 | 1 | 1 | High-quality | High-quality | 100 | HMM-based (lower-bound) | 0 |
| S17_vs_k141_96376\|\|full | 1431 | No | NA | 4 | 0 | 1 | Not-determined | Genome-fragment | NA | NA | 0 |
| S17_vs_k141_162783\|\|full | 1087 | No | NA | 3 | 0 | 1 | Not-determined | Genome-fragment | NA | NA | 0 |
| S17_vs_k141_44534\|\|full | 4606 | No | NA | 3 | 0 | 2 | Not-determined | Genome-fragment | NA | NA | 0 |
| S8_extracted_k141_178935 | 1032 | No | NA | 1 | 1 | 0 | Low-quality | Genome-fragment | 0.71 | HMM-based (lower-bound) | 0 |
| S17_vs_k141_7712\|\|full | 1074 | No | NA | 2 | 0 | 2 | Not-determined | Genome-fragment | NA | NA | 0 |
| S17_vs_k141_51947\|\|full | 2410 | No | NA | 3 | 0 | 2 | Not-determined | Genome-fragment | NA | NA | 0 |
| S17_vs_k141_155372\|\|full | 3447 | No | NA | 3 | 0 | 0 | Not-determined | Genome-fragment | NA | NA | 0 |
| S17_vs_k141_148060\|\|full | 2757 | No | NA | 5 | 0 | 2 | Not-determined | Genome-fragment | NA | NA | 0 |
| S17_vs_k141_96422\|\|full | 2373 | No | NA | 4 | 0 | 1 | Not-determined | Genome-fragment | NA | NA | 0 |
| S8_extracted_k141_49818 | 1032 | No | NA | 1 | 0 | 0 | Low-quality | Genome-fragment | 2.23 | AAI-based (high-confidence) | 0 |
| S17_vs_k141_155407\|\|full | 1437 | No | NA | 3 | 0 | 0 | Not-determined | Genome-fragment | NA | NA | 0 |
| S8_extracted_k141_188970 | 1032 | No | NA | 1 | 1 | 0 | Low-quality | Genome-fragment | 2.63 | AAI-based (high-confidence) | 0 |
| S8_vs_k141_122003\|\|lt2gene | 1032 | No | NA | 2 | 2 | 0 | Low-quality | Genome-fragment | 4.35 | AAI-based (medium-confidence) | 0 |
| S17_vs_k141_78761\|\|lt2gene | 1033 | No | NA | 2 | 1 | 0 | Low-quality | Genome-fragment | 2.53 | AAI-based (high-confidence) | 0 |
| S17_vs_k141_88906\|\|full | 1671 | No | NA | 2 | 0 | 2 | Not-determined | Genome-fragment | NA | NA | 0 |
| S11_vs_k141_72026\|\|full | 1033 | No | NA | 3 | 0 | 0 | Low-quality | Genome-fragment | 1.84 | AAI-based (high-confidence) | 0 |
| S11_vs_k141_81891\|\|full | 1033 | No | NA | 3 | 2 | 0 | Low-quality | Genome-fragment | 2.06 | AAI-based (medium-confidence) | 0 |
| S17_vs_k141_88918\|\|full | 2262 | No | NA | 2 | 0 | 2 | Not-determined | Genome-fragment | NA | NA | 0 |
| S17_vs_k141_37289\|\|full | 14637 | No | NA | 10 | 0 | 8 | Not-determined | Genome-fragment | NA | NA | 0 |
| S14_vs_k141_111590\|\|full | 1033 | No | NA | 6 | 4 | 0 | Low-quality | Genome-fragment | 3.4 | AAI-based (high-confidence) | 0 |
| S14_vs_k141_54425\|\|full | 1033 | No | NA | 2 | 1 | 0 | Low-quality | Genome-fragment | 1.37 | HMM-based (lower-bound) | 0 |
| S17_vs_k141_125870\|\|full | 2208 | No | NA | 3 | 0 | 1 | Not-determined | Genome-fragment | NA | NA | 0 |
| S7_extracted_70704 | 1033 | No | NA | 3 | 1 | 0 | Low-quality | Genome-fragment | 4.7 | AAI-based (high-confidence) | 0 |
| S7_vs_90197\|\|full | 1033 | No | NA | 2 | 0 | 0 | Low-quality | Genome-fragment | 2.44 | AAI-based (medium-confidence) | 0 |
| S12_vs_k141_184547\|\|lt2gene | 1033 | No | NA | 2 | 0 | 0 | Low-quality | Genome-fragment | 2.51 | AAI-based (high-confidence) | 0 |
| S17_vs_k141_118515\|\|full | 1368 | No | NA | 3 | 0 | 1 | Not-determined | Genome-fragment | NA | NA | 0 |
| S15_vs_42295\|\|full | 1033 | No | NA | 3 | 0 | 0 | Low-quality | Genome-fragment | 1.17 | AAI-based (high-confidence) | 0 |
| S17_vs_k141_96476\|\|full | 1476 | No | NA | 2 | 0 | 2 | Not-determined | Genome-fragment | NA | NA | 0 |
| S17_vs_k141_111014\|\|full | 3315 | No | NA | 2 | 0 | 0 | Not-determined | Genome-fragment | NA | NA | 0 |
| S17_vs_k141_111016\|\|full | 3779 | No | NA | 5 | 0 | 0 | Not-determined | Genome-fragment | NA | NA | 0 |
| S17_vs_k141_162831\|\|full | 1300 | No | NA | 3 | 0 | 0 | Not-determined | Genome-fragment | NA | NA | 0 |
| S15_vs_25639\|\|lt2gene | 1033 | No | NA | 1 | 1 | 0 | Low-quality | Genome-fragment | 2.5 | AAI-based (medium-confidence) | 0 |
| S17_vs_k141_103828\|\|full | 3215 | No | NA | 4 | 0 | 1 | Not-determined | Genome-fragment | NA | NA | 0 |
| S17_vs_k141_74215\|\|full | 4938 | No | NA | 3 | 0 | 1 | Not-determined | Genome-fragment | NA | NA | 0 |
| S4_extracted_18796 | 1033 | No | NA | 3 | 3 | 0 | Low-quality | Genome-fragment | 3.39 | AAI-based (high-confidence) | 0 |
| S17_vs_k141_264\|\|full | 3583 | No | NA | 3 | 0 | 2 | Not-determined | Genome-fragment | NA | NA | 0 |
| S17_vs_k141_44697\|\|full | 4440 | No | NA | 3 | 0 | 2 | Not-determined | Genome-fragment | NA | NA | 0 |
| S17_vs_k141_22356\|\|full | 1661 | No | NA | 3 | 0 | 1 | Not-determined | Genome-fragment | NA | NA | 0 |
| S17_vs_k141_148164\|\|full | 1279 | No | NA | 2 | 0 | 2 | Not-determined | Genome-fragment | NA | NA | 0 |
| S17_vs_k141_30007\|\|full | 15806 | No | NA | 13 | 0 | 9 | Not-determined | Genome-fragment | NA | NA | 0 |
| S17_vs_k141_74255\|\|full | 1745 | No | NA | 3 | 0 | 2 | Not-determined | Genome-fragment | NA | NA | 0 |
| S4_extracted_27105 | 1033 | No | NA | 2 | 0 | 0 | Low-quality | Genome-fragment | 2.23 | AAI-based (high-confidence) | 0 |
| S6_vs_k141_27931\|\|full | 1033 | No | NA | 5 | 1 | 0 | Low-quality | Genome-fragment | 2.03 | AAI-based (medium-confidence) | 0 |
| S6_vs_k141_8807\|\|lt2gene | 1033 | No | NA | 2 | 1 | 0 | Low-quality | Genome-fragment | 2.15 | AAI-based (high-confidence) | 0 |
| S17_vs_k141_66802\|\|full | 2597 | No | NA | 3 | 0 | 3 | Not-determined | Genome-fragment | NA | NA | 0 |
| S17_vs_k141_96529\|\|full | 5117 | No | NA | 5 | 0 | 4 | Not-determined | Genome-fragment | NA | NA | 0 |
| S17_vs_k141_170279\|\|full | 4897 | No | NA | 8 | 0 | 1 | Not-determined | Genome-fragment | NA | NA | 0 |
| S6_vs_k141_11697\|\|lt2gene | 1033 | No | NA | 2 | 0 | 0 | Low-quality | Genome-fragment | 1.54 | AAI-based (medium-confidence) | 0 |
| S8_vs_k141_560\|\|full | 1033 | No | NA | 4 | 0 | 0 | Low-quality | Genome-fragment | 2.9 | AAI-based (medium-confidence) | 0 |
| S17_extracted_k141_13152 | 1034 | No | NA | 2 | 0 | 2 | Low-quality | Genome-fragment | 0.47 | AAI-based (medium-confidence) | 0 |
| S17_vs_k141_170284\|\|full | 3856 | No | NA | 5 | 0 | 3 | Not-determined | Genome-fragment | NA | NA | 0 |
| S17_vs_k141_7807\|\|full | 2117 | No | NA | 3 | 0 | 2 | Not-determined | Genome-fragment | NA | NA | 0 |
| S17_vs_k141_88942\|\|full | 3207 | No | NA | 5 | 0 | 1 | Not-determined | Genome-fragment | NA | NA | 0 |
| S17_vs_k141_137428\|\|full | 1034 | No | NA | 2 | 0 | 0 | Low-quality | Genome-fragment | 0.19 | AAI-based (medium-confidence) | 0 |
| S17_vs_k141_118579\|\|full | 5765 | No | NA | 5 | 0 | 4 | Not-determined | Genome-fragment | NA | NA | 0 |
| S17_vs_k141_139021\|\|full | 1034 | No | NA | 4 | 0 | 0 | Low-quality | Genome-fragment | 2.25 | AAI-based (high-confidence) | 0 |
| S17_vs_k141_162880\|\|full | 13996 | No | NA | 16 | 0 | 1 | Not-determined | Genome-fragment | NA | NA | 0 |
| S17_vs_k141_66827\|\|full | 12355 | No | NA | 11 | 0 | 9 | Not-determined | Genome-fragment | NA | NA | 0 |
| S13_extracted_102950 | 1034 | No | NA | 2 | 1 | 0 | Low-quality | Genome-fragment | 1.65 | AAI-based (high-confidence) | 0 |
| S17_vs_k141_133375\|\|full | 1453 | No | NA | 3 | 0 | 2 | Not-determined | Genome-fragment | NA | NA | 0 |
| S17_vs_k141_111162\|\|full | 1272 | No | NA | 2 | 0 | 1 | Not-determined | Genome-fragment | NA | NA | 0 |
| S5_vs_k141_97065\|\|lt2gene | 1034 | No | NA | 2 | 0 | 0 | Low-quality | Genome-fragment | 2.87 | AAI-based (medium-confidence) | 0 |
| S7_vs_109229\|\|full | 1034 | No | NA | 3 | 1 | 0 | Low-quality | Genome-fragment | 2.66 | AAI-based (high-confidence) | 0 |
| S17_vs_k141_59625\|\|full | 1898 | No | NA | 2 | 0 | 0 | Not-determined | Genome-fragment | NA | NA | 0 |
| S17_vs_k141_88979\|\|full | 5535 | No | NA | 4 | 0 | 3 | Not-determined | Genome-fragment | NA | NA | 0 |
| S17_vs_k141_59631\|\|full | 5677 | No | NA | 5 | 0 | 4 | Not-determined | Genome-fragment | NA | NA | 0 |
| S17_vs_k141_81521\|\|full | 1398 | No | NA | 2 | 0 | 1 | Not-determined | Genome-fragment | NA | NA | 0 |
| S16_extracted_44689 | 1034 | No | NA | 2 | 0 | 0 | Low-quality | Genome-fragment | 1.47 | AAI-based (medium-confidence) | 0 |
| S17_vs_k141_81545\|\|full | 1235 | No | NA | 3 | 0 | 0 | Not-determined | Genome-fragment | NA | NA | 0 |
| S17_vs_k141_37491\|\|full | 1230 | No | NA | 2 | 0 | 2 | Not-determined | Genome-fragment | NA | NA | 0 |
| S17_vs_k141_89008\|\|full | 1083 | No | NA | 3 | 0 | 0 | Not-determined | Genome-fragment | NA | NA | 0 |
| S17_vs_k141_15131\|\|full | 9190 | No | NA | 5 | 0 | 4 | Not-determined | Genome-fragment | NA | NA | 0 |
| S17_vs_k141_388\|\|full | 5427 | No | NA | 6 | 0 | 4 | Not-determined | Genome-fragment | NA | NA | 0 |
| S10_vs_57582\|\|lt2gene | 1034 | No | NA | 2 | 1 | 0 | Low-quality | Genome-fragment | 1.91 | HMM-based (lower-bound) | 0 |
| S17_vs_k141_170328\|\|full | 1587 | No | NA | 3 | 0 | 0 | Not-determined | Genome-fragment | NA | NA | 0 |
| S10_vs_90514\|\|lt2gene | 1034 | No | NA | 1 | 1 | 0 | Low-quality | Genome-fragment | 1.96 | HMM-based (lower-bound) | 0 |
| S17_vs_k141_89026\|\|full | 1591 | No | NA | 3 | 0 | 1 | Not-determined | Genome-fragment | NA | NA | 0 |
| S17_vs_k141_426\|\|full | 1360 | No | NA | 2 | 0 | 2 | Not-determined | Genome-fragment | NA | NA | 0 |
| S17_vs_k141_15165\|\|full | 4870 | No | NA | 3 | 0 | 3 | Not-determined | Genome-fragment | NA | NA | 0 |
| S17_vs_k141_59674\|\|full | 2207 | No | NA | 2 | 0 | 0 | Not-determined | Genome-fragment | NA | NA | 0 |
| S12_vs_k141_68140\|\|full | 1034 | No | NA | 2 | 0 | 1 | Low-quality | Genome-fragment | 1.46 | AAI-based (medium-confidence) | 0 |
| S17_vs_k141_59678\|\|full | 1981 | No | NA | 3 | 0 | 0 | Not-determined | Genome-fragment | NA | NA | 0 |
| S17_vs_k141_170343\|\|full | 2585 | No | NA | 4 | 0 | 2 | Not-determined | Genome-fragment | NA | NA | 0 |
| S2_extracted_k141_4190 | 1034 | No | NA | 2 | 2 | 0 | Low-quality | Genome-fragment | 3.41 | AAI-based (high-confidence) | 0 |
| S8_extracted_k141_75293 | 1034 | No | NA | 2 | 0 | 0 | Low-quality | Genome-fragment | 1.9 | AAI-based (medium-confidence) | 0 |
| S17_vs_k141_30134\|\|full | 3887 | No | NA | 5 | 0 | 2 | Not-determined | Genome-fragment | NA | NA | 0 |
| S17_vs_k141_118719\|\|full | 4658 | No | NA | 3 | 0 | 3 | Not-determined | Genome-fragment | NA | NA | 0 |
| S11_extracted_k141_8711 | 1035 | No | NA | 2 | 0 | 0 | Low-quality | Genome-fragment | 1.82 | AAI-based (medium-confidence) | 0 |
| S17_vs_k141_81581\|\|full | 1738 | No | NA | 2 | 0 | 0 | Not-determined | Genome-fragment | NA | NA | 0 |
| S17_vs_k141_37565\|\|full | 1305 | No | NA | 3 | 0 | 2 | Not-determined | Genome-fragment | NA | NA | 0 |
| S17_vs_k141_111224\|\|full | 1023 | No | NA | 3 | 0 | 1 | Not-determined | Genome-fragment | NA | NA | 0 |
| S11_vs_k141_53263\|\|full | 1035 | No | NA | 2 | 1 | 0 | Low-quality | Genome-fragment | 2.65 | AAI-based (high-confidence) | 0 |
| S13_vs_7485\|\|full | 1035 | No | NA | 3 | 0 | 1 | Low-quality | Genome-fragment | 1.06 | AAI-based (medium-confidence) | 0 |
| S17_vs_k141_22540\|\|full | 3282 | No | NA | 4 | 0 | 4 | Not-determined | Genome-fragment | NA | NA | 0 |
| S13_vs_117100\|\|full | 1035 | No | NA | 2 | 0 | 0 | Low-quality | Genome-fragment | 2.41 | AAI-based (medium-confidence) | 0 |
| S17_vs_k141_37608\|\|full | 1414 | No | NA | 3 | 0 | 1 | Not-determined | Genome-fragment | NA | NA | 0 |
| S14_extracted_k141_88184 | 1035 | No | NA | 3 | 2 | 0 | Low-quality | Genome-fragment | 3.41 | AAI-based (high-confidence) | 0 |
| S17_vs_k141_37615\|\|full | 1415 | No | NA | 4 | 0 | 0 | Not-determined | Genome-fragment | NA | NA | 0 |
| S17_vs_k141_30203\|\|full | 2878 | No | NA | 3 | 0 | 0 | Not-determined | Genome-fragment | NA | NA | 0 |
| S14_extracted_k141_82427 | 1035 | No | NA | 3 | 0 | 0 | Low-quality | Genome-fragment | 1.73 | AAI-based (high-confidence) | 0 |
| S5_extracted_k141_93770 | 1035 | No | NA | 2 | 0 | 0 | Low-quality | Genome-fragment | 1.14 | AAI-based (medium-confidence) | 0 |
| S17_vs_k141_59724\|\|full | 1424 | No | NA | 3 | 0 | 0 | Not-determined | Genome-fragment | NA | NA | 0 |
| S17_vs_k141_22563\|\|full | 1319 | No | NA | 2 | 0 | 1 | Not-determined | Genome-fragment | NA | NA | 0 |
| S17_vs_k141_170402\|\|full | 8713 | No | NA | 4 | 0 | 4 | Not-determined | Genome-fragment | NA | NA | 0 |
| S17_vs_k141_133494\|\|full | 3890 | No | NA | 4 | 0 | 3 | Not-determined | Genome-fragment | NA | NA | 0 |
| S5_vs_k141_113653\|\|full | 1035 | No | NA | 3 | 1 | 0 | Low-quality | Genome-fragment | 2.15 | AAI-based (high-confidence) | 0 |
| S17_vs_k141_74485\|\|full | 1713 | No | NA | 3 | 0 | 0 | Not-determined | Genome-fragment | NA | NA | 0 |
| S7_extracted_82460 | 1035 | No | NA | 3 | 0 | 0 | Low-quality | Genome-fragment | 2.43 | AAI-based (high-confidence) | 0 |
| S17_vs_k141_52328\|\|full | 1512 | No | NA | 3 | 0 | 2 | Not-determined | Genome-fragment | NA | NA | 0 |
| S7_vs_41805\|\|full | 1035 | No | NA | 3 | 2 | 0 | Low-quality | Genome-fragment | 2.4 | AAI-based (high-confidence) | 0 |
| S16_vs_99444\|\|full | 1035 | No | NA | 2 | 0 | 0 | Low-quality | Genome-fragment | 1.69 | AAI-based (high-confidence) | 0 |
| S17_vs_k141_15274\|\|full | 3396 | No | NA | 3 | 0 | 1 | Not-determined | Genome-fragment | NA | NA | 0 |
| S17_vs_k141_118812\|\|full | 1821 | No | NA | 2 | 0 | 2 | Not-determined | Genome-fragment | NA | NA | 0 |
| S17_vs_k141_74490\|\|full | 4592 | No | NA | 6 | 0 | 1 | Not-determined | Genome-fragment | NA | NA | 0 |
| S16_vs_115067\|\|full | 1035 | No | NA | 3 | 2 | 0 | Low-quality | Genome-fragment | 2.2 | AAI-based (high-confidence) | 0 |
| S12_extracted_k141_157587 | 1035 | No | NA | 3 | 3 | 0 | Low-quality | Genome-fragment | 1.73 | AAI-based (high-confidence) | 0 |
| S17_vs_k141_59727\|\|full | 1099 | No | NA | 2 | 0 | 0 | Not-determined | Genome-fragment | NA | NA | 0 |
| S17_vs_k141_126079\|\|full | 3072 | No | NA | 2 | 0 | 0 | Not-determined | Genome-fragment | NA | NA | 0 |
| S12_vs_k141_31762\|\|full | 1035 | No | NA | 3 | 0 | 0 | Low-quality | Genome-fragment | 1.85 | AAI-based (high-confidence) | 0 |
| S12_vs_k141_33397\|\|full | 1035 | No | NA | 3 | 0 | 1 | Low-quality | Genome-fragment | 1.63 | AAI-based (medium-confidence) | 0 |
| S12_vs_k141_36721\|\|full | 1035 | No | NA | 3 | 0 | 2 | Low-quality | Genome-fragment | 0.61 | AAI-based (medium-confidence) | 0 |
| S8_vs_k141_183136\|\|full | 1035 | No | NA | 2 | 0 | 1 | Low-quality | Genome-fragment | 1.35 | AAI-based (high-confidence) | 0 |
| S8_vs_k141_204171\|\|full | 1035 | No | NA | 2 | 0 | 0 | Low-quality | Genome-fragment | 1.21 | AAI-based (medium-confidence) | 0 |
| S17_vs_k141_59733\|\|full | 2405 | No | NA | 3 | 0 | 1 | Not-determined | Genome-fragment | NA | NA | 0 |
| S17_vs_k141_44934\|\|full | 1045 | No | NA | 3 | 0 | 0 | Not-determined | Genome-fragment | NA | NA | 0 |
| S17_vs_k141_37695\|\|full | 1460 | No | NA | 2 | 0 | 1 | Not-determined | Genome-fragment | NA | NA | 0 |
| S17_extracted_k141_146837 | 1036 | No | NA | 2 | 0 | 0 | Low-quality | Genome-fragment | 0.61 | AAI-based (high-confidence) | 0 |
| S17_vs_k141_59854\|\|lt2gene | 1036 | No | NA | 2 | 2 | 0 | Low-quality | Genome-fragment | 1.13 | HMM-based (lower-bound) | 0 |
| S17_vs_k141_126112\|\|full | 15658 | No | NA | 15 | 0 | 6 | Not-determined | Genome-fragment | NA | NA | 0 |
| S17_vs_k141_59749\|\|full | 2986 | No | NA | 3 | 0 | 2 | Not-determined | Genome-fragment | NA | NA | 0 |
| S17_vs_k141_118864\|\|full | 2914 | No | NA | 2 | 0 | 1 | Not-determined | Genome-fragment | NA | NA | 0 |
| S17_vs_k141_111354\|\|full | 16003 | No | NA | 16 | 0 | 11 | Not-determined | Genome-fragment | NA | NA | 0 |
| S17_vs_k141_96574\|\|full | 1246 | No | NA | 2 | 0 | 1 | Not-determined | Genome-fragment | NA | NA | 0 |
| S17_vs_k141_104163\|\|full | 2182 | No | NA | 3 | 0 | 1 | Not-determined | Genome-fragment | NA | NA | 0 |
| S11_extracted_k141_63694 | 1036 | No | NA | 3 | 2 | 0 | Low-quality | Genome-fragment | 2.37 | AAI-based (medium-confidence) | 0 |
| S17_vs_k141_52426\|\|full | 1509 | No | NA | 2 | 0 | 1 | Not-determined | Genome-fragment | NA | NA | 0 |
| S13_extracted_96722 | 1036 | No | NA | 1 | 0 | 1 | Low-quality | Genome-fragment | 0.68 | AAI-based (high-confidence) | 0 |
| S1_vs_k141_118722\|\|full | 1036 | No | NA | 2 | 0 | 1 | Low-quality | Genome-fragment | 0.61 | AAI-based (medium-confidence) | 0 |
| S17_vs_k141_52436\|\|full | 4325 | No | NA | 5 | 0 | 1 | Not-determined | Genome-fragment | NA | NA | 0 |
| S17_vs_k141_111380\|\|full | 1959 | No | NA | 3 | 0 | 0 | Not-determined | Genome-fragment | NA | NA | 0 |
| S1_vs_k141_75675\|\|lt2gene | 1036 | No | NA | 2 | 1 | 0 | Low-quality | Genome-fragment | 2.32 | AAI-based (high-confidence) | 0 |
| S3_vs_k141_111793\|\|full | 1036 | No | NA | 2 | 0 | 0 | Low-quality | Genome-fragment | 1.7 | AAI-based (medium-confidence) | 0 |
| S17_vs_k141_163142\|\|full | 2572 | No | NA | 4 | 0 | 0 | Not-determined | Genome-fragment | NA | NA | 0 |
| S17_vs_k141_74580\|\|full | 2668 | No | NA | 4 | 0 | 1 | Not-determined | Genome-fragment | NA | NA | 0 |
| S17_vs_k141_44971\|\|full | 3168 | No | NA | 4 | 0 | 1 | Not-determined | Genome-fragment | NA | NA | 0 |
| S17_vs_k141_66970\|\|full | 15458 | No | NA | 7 | 0 | 6 | Not-determined | Genome-fragment | NA | NA | 0 |
| S17_vs_k141_96600\|\|full | 2886 | No | NA | 3 | 0 | 3 | Not-determined | Genome-fragment | NA | NA | 0 |
| S5_extracted_k141_112411 | 1036 | No | NA | 4 | 1 | 0 | Low-quality | Genome-fragment | 2.97 | AAI-based (medium-confidence) | 0 |
| S7_vs_97356\|\|full | 1036 | No | NA | 2 | 1 | 0 | Low-quality | Genome-fragment | 2.66 | AAI-based (high-confidence) | 0 |
| S7_vs_110152\|\|full | 1036 | No | NA | 2 | 0 | 0 | Low-quality | Genome-fragment | 2.56 | AAI-based (medium-confidence) | 0 |
| S17_vs_k141_670\|\|full | 4148 | No | NA | 3 | 0 | 3 | Not-determined | Genome-fragment | NA | NA | 0 |
| S16_extracted_13761 | 1036 | No | NA | 2 | 0 | 0 | Low-quality | Genome-fragment | 2.29 | AAI-based (medium-confidence) | 0 |
| S17_vs_k141_89213\|\|full | 5475 | No | NA | 4 | 0 | 1 | Not-determined | Genome-fragment | NA | NA | 0 |
| S17_vs_k141_104257\|\|full | 1318 | No | NA | 3 | 0 | 2 | Not-determined | Genome-fragment | NA | NA | 0 |
| S17_vs_k141_163176\|\|full | 2486 | No | NA | 3 | 0 | 2 | Not-determined | Genome-fragment | NA | NA | 0 |
| S16_extracted_20180 | 1036 | No | NA | 1 | 0 | 1 | Low-quality | Genome-fragment | 0.93 | AAI-based (medium-confidence) | 0 |
| S17_vs_k141_8088\|\|full | 4121 | No | NA | 2 | 0 | 2 | Not-determined | Genome-fragment | NA | NA | 0 |
| S16_extracted_45454 | 1036 | No | NA | 2 | 0 | 2 | Low-quality | Genome-fragment | 0.52 | AAI-based (medium-confidence) | 0 |
| S16_vs_104261\|\|full | 1036 | No | NA | 3 | 1 | 0 | Low-quality | Genome-fragment | 2.33 | AAI-based (high-confidence) | 0 |
| S17_vs_k141_81707\|\|full | 2582 | No | NA | 3 | 0 | 2 | Not-determined | Genome-fragment | NA | NA | 0 |
| S17_vs_k141_111445\|\|full | 4150 | No | NA | 2 | 0 | 2 | Not-determined | Genome-fragment | NA | NA | 0 |
| S17_vs_k141_89241\|\|full | 2214 | No | NA | 3 | 0 | 1 | Not-determined | Genome-fragment | NA | NA | 0 |
| S10_vs_24170\|\|full | 1036 | No | NA | 2 | 1 | 0 | Low-quality | Genome-fragment | 2.47 | AAI-based (high-confidence) | 0 |
| S10_vs_75932\|\|lt2gene | 1036 | No | NA | 1 | 1 | 0 | Low-quality | Genome-fragment | 7.03 | AAI-based (high-confidence) | 0 |
| S17_vs_k141_81728\|\|full | 2419 | No | NA | 3 | 0 | 3 | Not-determined | Genome-fragment | NA | NA | 0 |
| S15_vs_51849\|\|full | 1036 | No | NA | 2 | 0 | 0 | Low-quality | Genome-fragment | 1.82 | AAI-based (medium-confidence) | 0 |
| S17_vs_k141_45041\|\|full | 2580 | No | NA | 4 | 0 | 3 | Not-determined | Genome-fragment | NA | NA | 0 |
| S17_vs_k141_141192\|\|full | 6080 | No | NA | 10 | 0 | 0 | Not-determined | Genome-fragment | NA | NA | 0 |
| S8_vs_k141_191926\|\|full | 1036 | No | NA | 4 | 0 | 0 | Low-quality | Genome-fragment | 1.03 | AAI-based (high-confidence) | 0 |
| S17_vs_k141_52538\|\|full | 2743 | No | NA | 4 | 0 | 2 | Not-determined | Genome-fragment | NA | NA | 0 |
| S11_vs_k141_92349\|\|full | 1037 | No | NA | 2 | 0 | 1 | Low-quality | Genome-fragment | 0.8 | AAI-based (medium-confidence) | 0 |
| S17_vs_k141_126231\|\|full | 7927 | No | NA | 8 | 0 | 2 | Not-determined | Genome-fragment | NA | NA | 0 |
| S13_vs_142655\|\|full | 1037 | No | NA | 3 | 1 | 0 | Low-quality | Genome-fragment | 1.2 | HMM-based (lower-bound) | 0 |
| S17_vs_k141_118977\|\|full | 1482 | No | NA | 2 | 0 | 1 | Not-determined | Genome-fragment | NA | NA | 0 |
| S17_vs_k141_37924\|\|full | 1096 | No | NA | 2 | 0 | 2 | Not-determined | Genome-fragment | NA | NA | 0 |
| S14_extracted_k141_84194 | 1037 | No | NA | 1 | 0 | 0 | Low-quality | Genome-fragment | 2.39 | AAI-based (high-confidence) | 0 |
| S17_vs_k141_74723\|\|full | 2516 | No | NA | 3 | 0 | 0 | Not-determined | Genome-fragment | NA | NA | 0 |
| S17_vs_k141_170699\|\|full | 2957 | No | NA | 4 | 0 | 0 | Not-determined | Genome-fragment | NA | NA | 0 |
| S17_vs_k141_133634\|\|full | 3935 | No | NA | 3 | 0 | 0 | Not-determined | Genome-fragment | NA | NA | 0 |
| S17_vs_k141_96669\|\|full | 2852 | No | NA | 3 | 0 | 2 | Not-determined | Genome-fragment | NA | NA | 0 |
| S17_vs_k141_155977\|\|full | 1844 | No | NA | 3 | 0 | 2 | Not-determined | Genome-fragment | NA | NA | 0 |
| S14_extracted_k141_42593 | 1037 | No | NA | 3 | 0 | 1 | Low-quality | Genome-fragment | 0.77 | AAI-based (medium-confidence) | 0 |
| S1_extracted_k141_94305 | 1037 | No | NA | 1 | 1 | 0 | Low-quality | Genome-fragment | 0.72 | AAI-based (high-confidence) | 0 |
| S17_vs_k141_22742\|\|full | 1070 | No | NA | 3 | 0 | 0 | Not-determined | Genome-fragment | NA | NA | 0 |
| S1_vs_k141_113507\|\|full | 1037 | No | NA | 3 | 0 | 0 | Low-quality | Genome-fragment | 2.43 | AAI-based (high-confidence) | 0 |
| S17_vs_k141_89339\|\|full | 5239 | No | NA | 8 | 0 | 2 | Not-determined | Genome-fragment | NA | NA | 0 |
| S1_vs_k141_141024\|\|full | 1037 | No | NA | 3 | 0 | 0 | Low-quality | Genome-fragment | 2.63 | AAI-based (high-confidence) | 0 |
| S1_vs_k141_51460\|\|full | 1037 | No | NA | 2 | 1 | 0 | Low-quality | Genome-fragment | 2.67 | AAI-based (medium-confidence) | 0 |
| S17_vs_k141_67155\|\|full | 1462 | No | NA | 2 | 0 | 1 | Not-determined | Genome-fragment | NA | NA | 0 |
| S17_vs_k141_111539\|\|full | 2041 | No | NA | 3 | 0 | 1 | Not-determined | Genome-fragment | NA | NA | 0 |
| S17_vs_k141_170747\|\|full | 3684 | No | NA | 3 | 0 | 2 | Not-determined | Genome-fragment | NA | NA | 0 |
| S17_vs_k141_104488\|\|full | 3402 | No | NA | 3 | 0 | 2 | Not-determined | Genome-fragment | NA | NA | 0 |
| S5_vs_k141_15272\|\|lt2gene | 1037 | No | NA | 1 | 1 | 0 | Low-quality | Genome-fragment | 3.02 | AAI-based (high-confidence) | 0 |
| S7_extracted_2254 | 1037 | No | NA | 2 | 0 | 0 | Low-quality | Genome-fragment | 1.98 | AAI-based (medium-confidence) | 0 |
| S17_vs_k141_170767\|\|full | 1966 | No | NA | 2 | 0 | 0 | Not-determined | Genome-fragment | NA | NA | 0 |
| S17_vs_k141_163359\|\|full | 1184 | No | NA | 2 | 0 | 1 | Not-determined | Genome-fragment | NA | NA | 0 |
| S17_vs_k141_45115\|\|full | 4011 | No | NA | 3 | 0 | 3 | Not-determined | Genome-fragment | NA | NA | 0 |
| S7_extracted_19799 | 1037 | No | NA | 2 | 2 | 0 | Low-quality | Genome-fragment | 2.35 | AAI-based (medium-confidence) | 0 |
| S17_vs_k141_148714\|\|full | 2233 | No | NA | 3 | 0 | 2 | Not-determined | Genome-fragment | NA | NA | 0 |
| S17_vs_k141_104552\|\|full | 2893 | No | NA | 3 | 0 | 1 | Not-determined | Genome-fragment | NA | NA | 0 |
| S17_vs_k141_89379\|\|full | 1801 | No | NA | 3 | 0 | 0 | Not-determined | Genome-fragment | NA | NA | 0 |
| S17_vs_k141_89384\|\|full | 1661 | No | NA | 3 | 0 | 1 | Not-determined | Genome-fragment | NA | NA | 0 |
| S17_vs_k141_8288\|\|full | 3779 | No | NA | 4 | 0 | 3 | Not-determined | Genome-fragment | NA | NA | 0 |
| S10_vs_14817\|\|full | 1037 | No | NA | 4 | 1 | 0 | Low-quality | Genome-fragment | 2.39 | AAI-based (high-confidence) | 0 |
| S17_vs_k141_163418\|\|full | 1331 | No | NA | 3 | 0 | 0 | Not-determined | Genome-fragment | NA | NA | 0 |
| S17_vs_k141_170830\|\|full | 1721 | No | NA | 2 | 0 | 1 | Not-determined | Genome-fragment | NA | NA | 0 |
| S12_vs_k141_42432\|\|full | 1037 | No | NA | 3 | 1 | 0 | Low-quality | Genome-fragment | 2.48 | AAI-based (high-confidence) | 0 |
| S12_vs_k141_3566\|\|full | 1037 | No | NA | 3 | 1 | 0 | Low-quality | Genome-fragment | 2.2 | AAI-based (high-confidence) | 0 |
| S17_vs_k141_170840\|\|full | 1732 | No | NA | 3 | 0 | 1 | Not-determined | Genome-fragment | NA | NA | 0 |
| S17_vs_k141_38033\|\|full | 2229 | No | NA | 3 | 0 | 2 | Not-determined | Genome-fragment | NA | NA | 0 |
| S17_vs_k141_96831\|\|full | 4298 | No | NA | 5 | 0 | 4 | Not-determined | Genome-fragment | NA | NA | 0 |
| S17_vs_k141_104572\|\|full | 2681 | No | NA | 2 | 0 | 1 | Not-determined | Genome-fragment | NA | NA | 0 |
| S17_vs_k141_126346\|\|full | 1681 | No | NA | 3 | 0 | 1 | Not-determined | Genome-fragment | NA | NA | 0 |
| S2_vs_k141_11866\|\|full | 1037 | No | NA | 2 | 1 | 0 | Low-quality | Genome-fragment | 2.53 | AAI-based (high-confidence) | 0 |
| S17_vs_k141_45179\|\|full | 4107 | No | NA | 5 | 0 | 3 | Not-determined | Genome-fragment | NA | NA | 0 |
| S6_extracted_k141_17310 | 1037 | No | NA | 2 | 1 | 0 | Low-quality | Genome-fragment | 2.69 | AAI-based (high-confidence) | 0 |
| S8_extracted_k141_61347 | 1037 | No | NA | 2 | 0 | 1 | Low-quality | Genome-fragment | 0.65 | AAI-based (medium-confidence) | 0 |
| S8_extracted_k141_28656 | 1037 | No | NA | 3 | 0 | 0 | Low-quality | Genome-fragment | 2.21 | AAI-based (medium-confidence) | 0 |
| S8_extracted_k141_155679 | 1037 | No | NA | 3 | 1 | 0 | Low-quality | Genome-fragment | 2.04 | AAI-based (high-confidence) | 0 |
| S8_vs_k141_38085\|\|full | 1037 | No | NA | 3 | 1 | 0 | Low-quality | Genome-fragment | 2.74 | AAI-based (high-confidence) | 0 |
| S9_vs_k141_68053\|\|full | 1038 | No | NA | 3 | 0 | 0 | Low-quality | Genome-fragment | 2.74 | AAI-based (high-confidence) | 0 |
| S9_vs_k141_68274\|\|lt2gene | 1038 | No | NA | 2 | 1 | 0 | Low-quality | Genome-fragment | 2.37 | AAI-based (high-confidence) | 0 |
| S17_vs_k141_52789\|\|full | 2778 | No | NA | 5 | 0 | 0 | Not-determined | Genome-fragment | NA | NA | 0 |
| S17_vs_k141_148834\|\|full | 1784 | No | NA | 3 | 0 | 0 | Not-determined | Genome-fragment | NA | NA | 0 |

| | | | | | | | | | | | |
|---|---|---|---|---|---|---|---|---|---|---|---|
| S17_vs_k141_52806\|\|full | 2949 | No | NA | 4 | 0 | 1 | Not-determined | Genome-fragment | NA | NA | 0 |
| S17_vs_k141_111618\|\|full | 3060 | No | NA | 4 | 0 | 0 | Not-determined | Genome-fragment | NA | NA | 0 |
| S17_vs_k141_133785\|\|full | 3328 | No | NA | 5 | 0 | 4 | Not-determined | Genome-fragment | NA | NA | 0 |
| S17_vs_k141_67359\|\|full | 1056 | No | NA | 2 | 0 | 0 | Not-determined | Genome-fragment | NA | NA | 0 |
| S17_vs_k141_96922\|\|full | 4978 | No | NA | 7 | 0 | 6 | Not-determined | Genome-fragment | NA | NA | 0 |
| S11_vs_k141_68451\|\|full | 1038 | No | NA | 3 | 1 | 0 | Low-quality | Genome-fragment | 2.28 | AAI-based (high-confidence) | 0 |
| S17_vs_k141_67361\|\|full | 3174 | No | NA | 4 | 0 | 3 | Not-determined | Genome-fragment | NA | NA | 0 |
| S17_vs_k141_52838\|\|full | 1603 | No | NA | 3 | 0 | 0 | Not-determined | Genome-fragment | NA | NA | 0 |
| S11_vs_k141_95781\|\|full | 1038 | No | NA | 2 | 0 | 0 | Low-quality | Genome-fragment | 1.47 | AAI-based (medium-confidence) | 0 |
| S17_vs_k141_81899\|\|full | 1971 | No | NA | 4 | 0 | 3 | Not-determined | Genome-fragment | NA | NA | 0 |
| S17_vs_k141_22912\|\|full | 2795 | No | NA | 5 | 0 | 0 | Not-determined | Genome-fragment | NA | NA | 0 |
| S17_vs_k141_111663\|\|full | 1316 | No | NA | 2 | 0 | 1 | Not-determined | Genome-fragment | NA | NA | 0 |
| S13_vs_110212\|\|full | 1038 | No | NA | 3 | 3 | 0 | Low-quality | Genome-fragment | 2.7 | AAI-based (high-confidence) | 0 |
| S14_extracted_k141_31515 | 1038 | No | NA | 4 | 4 | 0 | Low-quality | Genome-fragment | 3.43 | AAI-based (high-confidence) | 0 |
| S17_vs_k141_30648\|\|full | 2061 | No | NA | 2 | 0 | 0 | Not-determined | Genome-fragment | NA | NA | 0 |
| S17_vs_k141_8424\|\|full | 4529 | No | NA | 4 | 0 | 2 | Not-determined | Genome-fragment | NA | NA | 0 |
| S1_extracted_k141_119134 | 1038 | No | NA | 2 | 2 | 0 | Low-quality | Genome-fragment | 1.52 | AAI-based (medium-confidence) | 0 |
| S1_vs_k141_155040\|\|lt2gene | 1038 | No | NA | 1 | 1 | 0 | Low-quality | Genome-fragment | 2.22 | HMM-based (lower-bound) | 0 |
| S3_extracted_k141_65325 | 1038 | No | NA | 1 | 0 | 1 | Low-quality | Genome-fragment | 1.06 | AAI-based (high-confidence) | 0 |
| S17_vs_k141_45336\|\|full | 1927 | No | NA | 3 | 0 | 2 | Not-determined | Genome-fragment | NA | NA | 0 |
| S16_vs_92419\|\|full | 1038 | No | NA | 1 | 1 | 0 | Low-quality | Genome-fragment | 2.64 | AAI-based (medium-confidence) | 0 |
| S17_vs_k141_984\|\|full | 18858 | No | NA | 19 | 0 | 11 | Not-determined | Genome-fragment | NA | NA | 0 |
| S12_vs_k141_155264\|\|full | 1038 | No | NA | 4 | 1 | 0 | Low-quality | Genome-fragment | 2.28 | AAI-based (medium-confidence) | 0 |
| S17_vs_k141_141371\|\|full | 1398 | No | NA | 3 | 0 | 0 | Not-determined | Genome-fragment | NA | NA | 0 |
| S15_extracted_24635 | 1038 | No | NA | 1 | 0 | 1 | Low-quality | Genome-fragment | 0.45 | AAI-based (medium-confidence) | 0 |
| S8_vs_k141_203659\|\|full | 1038 | No | NA | 2 | 0 | 0 | Low-quality | Genome-fragment | 2.19 | AAI-based (medium-confidence) | 0 |
| S17_vs_k141_52890\|\|full | 60210 | No | NA | 53 | 0 | 14 | Medium-quality | Genome-fragment | 63.54 | AAI-based (medium-confidence) | 0 |
| S17_vs_k141_81929\|\|full | 7371 | No | NA | 12 | 1 | 2 | Not-determined | Genome-fragment | NA | NA | 0 |
| S8_vs_k141_25231\|\|full | 1038 | No | NA | 3 | 0 | 0 | Low-quality | Genome-fragment | 2.19 | AAI-based (medium-confidence) | 0 |
| S17_vs_k141_119269\|\|full | 5736 | No | NA | 6 | 0 | 1 | Not-determined | Genome-fragment | NA | NA | 0 |
| S17_vs_k141_156356\|\|full | 6580 | No | NA | 11 | 0 | 1 | Not-determined | Genome-fragment | NA | NA | 0 |
| S17_vs_k141_8497\|\|full | 2292 | No | NA | 3 | 0 | 0 | Not-determined | Genome-fragment | NA | NA | 0 |
| S17_vs_k141_111739\|\|full | 1176 | No | NA | 2 | 0 | 0 | Not-determined | Genome-fragment | NA | NA | 0 |
| S17_vs_k141_81943\|\|full | 2051 | No | NA | 2 | 0 | 1 | Not-determined | Genome-fragment | NA | NA | 0 |
| S17_vs_k141_96991\|\|full | 2727 | No | NA | 4 | 0 | 2 | Not-determined | Genome-fragment | NA | NA | 0 |
| S14_extracted_k141_77244 | 1039 | No | NA | 2 | 2 | 0 | Low-quality | Genome-fragment | 3.11 | AAI-based (high-confidence) | 0 |
| S14_extracted_k141_21771 | 1039 | No | NA | 2 | 0 | 1 | Low-quality | Genome-fragment | 0.29 | AAI-based (high-confidence) | 0 |
| S14_vs_k141_100590\|\|full | 1039 | No | NA | 3 | 1 | 0 | Low-quality | Genome-fragment | 2.69 | AAI-based (medium-confidence) | 0 |
| S3_extracted_k141_30189 | 1039 | No | NA | 2 | 0 | 0 | Low-quality | Genome-fragment | 1.78 | AAI-based (medium-confidence) | 0 |
| S17_vs_k141_38278\|\|full | 1134 | No | NA | 3 | 0 | 0 | Not-determined | Genome-fragment | NA | NA | 0 |
| S17_vs_k141_104812\|\|full | 1030 | No | NA | 3 | 0 | 1 | Not-determined | Genome-fragment | NA | NA | 0 |
| S17_vs_k141_81960\|\|full | 4779 | No | NA | 4 | 0 | 2 | Not-determined | Genome-fragment | NA | NA | 0 |
| S17_vs_k141_156425\|\|full | 1620 | No | NA | 4 | 0 | 2 | Not-determined | Genome-fragment | NA | NA | 0 |
| S17_vs_k141_104840\|\|full | 2046 | No | NA | 3 | 0 | 0 | Not-determined | Genome-fragment | NA | NA | 0 |
| S17_vs_k141_97048\|\|full | 1478 | No | NA | 2 | 0 | 0 | Not-determined | Genome-fragment | NA | NA | 0 |
| S17_vs_k141_1050\|\|full | 3656 | No | NA | 3 | 0 | 1 | Not-determined | Genome-fragment | NA | NA | 0 |
| S17_vs_k141_104847\|\|full | 1258 | No | NA | 3 | 0 | 0 | Not-determined | Genome-fragment | NA | NA | 0 |
| S17_vs_k141_111843\|\|full | 1471 | No | NA | 3 | 0 | 0 | Not-determined | Genome-fragment | NA | NA | 0 |
| S17_vs_k141_163659\|\|full | 1883 | No | NA | 3 | 0 | 1 | Not-determined | Genome-fragment | NA | NA | 0 |
| S5_extracted_k141_68264 | 1039 | No | NA | 3 | 2 | 0 | Low-quality | Genome-fragment | 3.01 | AAI-based (high-confidence) | 0 |
| S17_vs_k141_111855\|\|full | 3234 | No | NA | 3 | 0 | 0 | Not-determined | Genome-fragment | NA | NA | 0 |
| S17_vs_k141_119353\|\|full | 1123 | No | NA | 3 | 0 | 2 | Not-determined | Genome-fragment | NA | NA | 0 |
| S5_vs_k141_69955\|\|lt2gene | 1039 | No | NA | 1 | 1 | 0 | Low-quality | Genome-fragment | 0.93 | HMM-based (lower-bound) | 0 |
| S17_vs_k141_119367\|\|full | 1485 | No | NA | 3 | 0 | 0 | Not-determined | Genome-fragment | NA | NA | 0 |
| S16_extracted_19515 | 1039 | No | NA | 2 | 0 | 0 | Low-quality | Genome-fragment | 2.45 | AAI-based (high-confidence) | 0 |
| S16_vs_2499\|\|full | 1039 | No | NA | 3 | 0 | 0 | Low-quality | Genome-fragment | 0.68 | AAI-based (high-confidence) | 0 |
| S17_vs_k141_163680\|\|full | 1254 | No | NA | 2 | 0 | 0 | Not-determined | Genome-fragment | NA | NA | 0 |
| S17_vs_k141_1087\|\|full | 9719 | No | NA | 7 | 0 | 4 | Not-determined | Genome-fragment | NA | NA | 0 |
| S17_vs_k141_111875\|\|full | 2270 | No | NA | 4 | 0 | 2 | Not-determined | Genome-fragment | NA | NA | 0 |
| S17_vs_k141_8631\|\|full | 3209 | No | NA | 4 | 0 | 0 | Not-determined | Genome-fragment | NA | NA | 0 |
| S17_vs_k141_149144\|\|full | 4107 | No | NA | 6 | 0 | 2 | Not-determined | Genome-fragment | NA | NA | 0 |
| S17_vs_k141_111889\|\|full | 2311 | No | NA | 2 | 0 | 0 | Not-determined | Genome-fragment | NA | NA | 0 |
| S17_vs_k141_67579\|\|full | 1260 | No | NA | 2 | 0 | 0 | Not-determined | Genome-fragment | NA | NA | 0 |
| S16_vs_93314\|\|full | 1039 | No | NA | 3 | 0 | 0 | Low-quality | Genome-fragment | 2.52 | AAI-based (high-confidence) | 0 |
| S16_vs_54357\|\|lt2gene | 1039 | No | NA | 1 | 1 | 0 | Low-quality | Genome-fragment | 2.37 | AAI-based (high-confidence) | 0 |
| S17_vs_k141_171198\|\|full | 6378 | No | NA | 7 | 0 | 5 | Not-determined | Genome-fragment | NA | NA | 0 |
| S17_vs_k141_60305\|\|full | 1505 | No | NA | 3 | 0 | 1 | Not-determined | Genome-fragment | NA | NA | 0 |
| S10_vs_42914\|\|full | 1039 | No | NA | 3 | 0 | 0 | Low-quality | Genome-fragment | 2.17 | AAI-based (high-confidence) | 0 |
| S17_vs_k141_171212\|\|full | 1111 | No | NA | 2 | 0 | 2 | Not-determined | Genome-fragment | NA | NA | 0 |
| S17_vs_k141_53092\|\|full | 4196 | No | NA | 4 | 0 | 4 | Not-determined | Genome-fragment | NA | NA | 0 |
| S17_vs_k141_38429\|\|full | 2968 | No | NA | 4 | 0 | 0 | Not-determined | Genome-fragment | NA | NA | 0 |
| S10_vs_98456\|\|full | 1039 | No | NA | 3 | 0 | 0 | Low-quality | Genome-fragment | 2.53 | AAI-based (medium-confidence) | 0 |
| S10_vs_42217\|\|lt2gene | 1039 | No | NA | 2 | 2 | 0 | Low-quality | Genome-fragment | 2.24 | AAI-based (high-confidence) | 0 |
| S17_vs_k141_163733\|\|full | 2210 | No | NA | 3 | 0 | 2 | Not-determined | Genome-fragment | NA | NA | 0 |
| S17_vs_k141_74975\|\|full | 2686 | No | NA | 4 | 0 | 0 | Not-determined | Genome-fragment | NA | NA | 0 |
| S12_vs_k141_37844\|\|full | 1039 | No | NA | 3 | 0 | 0 | Low-quality | Genome-fragment | 2.99 | AAI-based (high-confidence) | 0 |
| S12_vs_k141_62762\|\|full | 1039 | No | NA | 3 | 0 | 0 | Low-quality | Genome-fragment | 2.15 | AAI-based (medium-confidence) | 0 |
| S17_vs_k141_119426\|\|full | 3378 | No | NA | 10 | 0 | 0 | Not-determined | Genome-fragment | NA | NA | 0 |
| S6_extracted_k141_9328 | 1039 | No | NA | 2 | 0 | 2 | Low-quality | Genome-fragment | 0.47 | AAI-based (medium-confidence) | 0 |
| S17_vs_k141_163760\|\|full | 1833 | No | NA | 2 | 0 | 1 | Not-determined | Genome-fragment | NA | NA | 0 |
| S17_vs_k141_119449\|\|full | 1087 | No | NA | 3 | 0 | 0 | Not-determined | Genome-fragment | NA | NA | 0 |
| S8_extracted_k141_143743 | 1039 | No | NA | 3 | 0 | 0 | Low-quality | Genome-fragment | 1.67 | AAI-based (high-confidence) | 0 |
| S17_vs_k141_1160\|\|full | 1261 | No | NA | 3 | 0 | 0 | Not-determined | Genome-fragment | NA | NA | 0 |
| S8_extracted_k141_49111 | 1039 | No | NA | 1 | 0 | 1 | Low-quality | Genome-fragment | 1.73 | AAI-based (medium-confidence) | 0 |
| S17_vs_k141_23145\|\|full | 1616 | No | NA | 2 | 0 | 0 | Not-determined | Genome-fragment | NA | NA | 0 |
| S8_vs_k141_87414\|\|full | 1039 | No | NA | 2 | 0 | 1 | Low-quality | Genome-fragment | 3.32 | AAI-based (medium-confidence) | 0 |
| S8_vs_k141_141295\|\|lt2gene | 1039 | No | NA | 1 | 1 | 0 | Low-quality | Genome-fragment | 3.34 | AAI-based (high-confidence) | 0 |
| S17_vs_k141_45678\|\|full | 1410 | No | NA | 3 | 0 | 2 | Not-determined | Genome-fragment | NA | NA | 0 |
| S17_vs_k141_67686\|\|full | 1381 | No | NA | 2 | 0 | 1 | Not-determined | Genome-fragment | NA | NA | 0 |
| S17_vs_k141_141684\|\|full | 2505 | No | NA | 3 | 0 | 2 | Not-determined | Genome-fragment | NA | NA | 0 |
| S17_vs_k141_89829\|\|full | 1547 | No | NA | 4 | 0 | 0 | Not-determined | Genome-fragment | NA | NA | 0 |
| S17_vs_k141_141701\|\|full | 4755 | No | NA | 6 | 0 | 2 | Not-determined | Genome-fragment | NA | NA | 0 |
| S8_vs_k141_158478\|\|lt2gene | 1039 | No | NA | 1 | 1 | 0 | Low-quality | Genome-fragment | 1.73 | AAI-based (high-confidence) | 0 |
| S9_vs_k141_66290\|\|full | 1040 | No | NA | 2 | 0 | 0 | Low-quality | Genome-fragment | 1.68 | AAI-based (high-confidence) | 0 |
| S17_vs_k141_134110\|\|full | 4589 | No | NA | 4 | 0 | 3 | Not-determined | Genome-fragment | NA | NA | 0 |
| S17_vs_k141_171294\|\|full | 3024 | No | NA | 6 | 0 | 1 | Not-determined | Genome-fragment | NA | NA | 0 |
| S11_extracted_k141_37886 | 1040 | No | NA | 1 | 1 | 0 | Low-quality | Genome-fragment | 0.74 | HMM-based (lower-bound) | 0 |
| S11_vs_k141_57753\|\|lt2gene | 1040 | No | NA | 2 | 1 | 0 | Low-quality | Genome-fragment | 2.75 | AAI-based (medium-confidence) | 0 |
| S17_vs_k141_111982\|\|full | 1405 | No | NA | 3 | 0 | 0 | Not-determined | Genome-fragment | NA | NA | 0 |
| S17_vs_k141_1206\|\|full | 1418 | No | NA | 4 | 0 | 0 | Not-determined | Genome-fragment | NA | NA | 0 |
| S17_vs_k141_8760\|\|full | 1371 | No | NA | 2 | 0 | 0 | Not-determined | Genome-fragment | NA | NA | 0 |
| S17_vs_k141_53227\|\|full | 3000 | No | NA | 3 | 0 | 1 | Not-determined | Genome-fragment | NA | NA | 0 |
| S5_extracted_k141_16013 | 1040 | No | NA | 2 | 0 | 0 | Low-quality | Genome-fragment | 1.7 | AAI-based (high-confidence) | 0 |
| S17_vs_k141_97282\|\|full | 1357 | No | NA | 3 | 0 | 0 | Not-determined | Genome-fragment | NA | NA | 0 |
| S17_vs_k141_8774\|\|full | 3827 | No | NA | 12 | 0 | 1 | Not-determined | Genome-fragment | NA | NA | 0 |
| S17_vs_k141_171307\|\|full | 4725 | No | NA | 6 | 0 | 1 | Not-determined | Genome-fragment | NA | NA | 0 |
| S17_vs_k141_23201\|\|full | 2501 | No | NA | 4 | 0 | 0 | Not-determined | Genome-fragment | NA | NA | 0 |
| S5_extracted_k141_59429 | 1040 | No | NA | 2 | 2 | 0 | Low-quality | Genome-fragment | 2.16 | AAI-based (high-confidence) | 0 |
| S17_vs_k141_1240\|\|full | 1474 | No | NA | 4 | 0 | 0 | Not-determined | Genome-fragment | NA | NA | 0 |
| S5_vs_k141_37589\|\|full | 1040 | No | NA | 3 | 0 | 0 | Low-quality | Genome-fragment | 2.83 | AAI-based (high-confidence) | 0 |
| S7_vs_71164\|\|lt2gene | 1040 | No | NA | 1 | 0 | 0 | Low-quality | Genome-fragment | 1.78 | AAI-based (medium-confidence) | 0 |
| S15_vs_4097\|\|full | 1040 | No | NA | 2 | 1 | 0 | Low-quality | Genome-fragment | 3.52 | HMM-based (lower-bound) | 0 |
| S17_vs_k141_75118\|\|full | 8203 | No | NA | 6 | 0 | 6 | Not-determined | Genome-fragment | NA | NA | 0 |
| S8_vs_k141_135352\|\|lt2gene | 1040 | No | NA | 1 | 1 | 0 | Low-quality | Genome-fragment | 2.35 | AAI-based (medium-confidence) | 0 |
| S17_vs_k141_134176\|\|full | 4021 | No | NA | 3 | 0 | 2 | Not-determined | Genome-fragment | NA | NA | 0 |
| S17_vs_k141_31041\|\|full | 1965 | No | NA | 3 | 0 | 0 | Not-determined | Genome-fragment | NA | NA | 0 |
| S9_vs_k141_2527\|\|full | 1041 | No | NA | 3 | 2 | 0 | Low-quality | Genome-fragment | 2.53 | AAI-based (high-confidence) | 0 |
| S17_vs_k141_1276\|\|full | 1265 | No | NA | 2 | 0 | 0 | Not-determined | Genome-fragment | NA | NA | 0 |
| S11_vs_k141_86434\|\|lt2gene | 1041 | No | NA | 2 | 0 | 0 | Low-quality | Genome-fragment | 2.47 | AAI-based (high-confidence) | 0 |
| S17_vs_k141_89992\|\|full | 3705 | No | NA | 5 | 0 | 3 | Not-determined | Genome-fragment | NA | NA | 0 |
| S17_vs_k141_53310\|\|full | 1747 | No | NA | 4 | 0 | 0 | Not-determined | Genome-fragment | NA | NA | 0 |
| S17_vs_k141_171396\|\|full | 2818 | No | NA | 3 | 0 | 2 | Not-determined | Genome-fragment | NA | NA | 0 |
| S11_vs_k141_51727\|\|lt2gene | 1041 | No | NA | 2 | 1 | 0 | Low-quality | Genome-fragment | 2.37 | AAI-based (medium-confidence) | 0 |
| S17_vs_k141_156670\|\|full | 2928 | No | NA | 3 | 0 | 1 | Not-determined | Genome-fragment | NA | NA | 0 |
| S17_vs_k141_134221\|\|full | 1238 | No | NA | 3 | 0 | 1 | Not-determined | Genome-fragment | NA | NA | 0 |
| S17_vs_k141_141732\|\|full | 33997 | No | NA | 31 | 0 | 6 | Not-determined | Genome-fragment | NA | NA | 0 |
| S17_vs_k141_97406\|\|full | 2281 | No | NA | 3 | 0 | 2 | Not-determined | Genome-fragment | NA | NA | 0 |
| S1_extracted_k141_66750 | 1041 | No | NA | 3 | 0 | 0 | Low-quality | Genome-fragment | 2.68 | AAI-based (high-confidence) | 0 |
| S17_vs_k141_90033\|\|full | 2987 | No | NA | 4 | 0 | 1 | Not-determined | Genome-fragment | NA | NA | 0 |
| S17_vs_k141_31099\|\|full | 1649 | No | NA | 4 | 0 | 0 | Not-determined | Genome-fragment | NA | NA | 0 |
| S1_vs_k141_38562\|\|lt2gene | 1041 | No | NA | 1 | 1 | 0 | Low-quality | Genome-fragment | 3.57 | HMM-based (lower-bound) | 0 |
| S17_vs_k141_97431\|\|full | 3513 | No | NA | 2 | 0 | 2 | Not-determined | Genome-fragment | NA | NA | 0 |
| S3_extracted_k141_60602 | 1041 | No | NA | 2 | 0 | 0 | Low-quality | Genome-fragment | 0.37 | AAI-based (medium-confidence) | 0 |
| S3_vs_k141_105337\|\|full | 1041 | No | NA | 2 | 0 | 1 | Low-quality | Genome-fragment | 1.54 | AAI-based (medium-confidence) | 0 |
| S5_vs_k141_102865\|\|lt2gene | 1041 | No | NA | 2 | 1 | 0 | Low-quality | Genome-fragment | 2.53 | AAI-based (high-confidence) | 0 |
| S17_vs_k141_171449\|\|full | 3180 | No | NA | 4 | 0 | 1 | Not-determined | Genome-fragment | NA | NA | 0 |
| S16_vs_101034\|\|full | 1041 | No | NA | 3 | 1 | 0 | Low-quality | Genome-fragment | 1.54 | HMM-based (lower-bound) | 0 |
| S17_vs_k141_75187\|\|full | 1609 | No | NA | 3 | 0 | 2 | Not-determined | Genome-fragment | NA | NA | 0 |
| S10_extracted_16727 | 1041 | No | NA | 2 | 0 | 0 | Low-quality | Genome-fragment | 1.27 | AAI-based (medium-confidence) | 0 |
| S17_vs_k141_126592\|\|full | 1863 | No | NA | 3 | 0 | 1 | Not-determined | Genome-fragment | NA | NA | 0 |
| S12_vs_k141_146545\|\|full | 1041 | No | NA | 3 | 1 | 0 | Low-quality | Genome-fragment | 2.35 | AAI-based (high-confidence) | 0 |
| S12_vs_k141_164809\|\|full | 1041 | No | NA | 3 | 0 | 0 | Low-quality | Genome-fragment | 0.97 | AAI-based (medium-confidence) | 0 |
| S17_vs_k141_38710\|\|full | 2166 | No | NA | 2 | 0 | 1 | Not-determined | Genome-fragment | NA | NA | 0 |
| S17_vs_k141_75205\|\|full | 6192 | No | NA | 8 | 0 | 4 | Not-determined | Genome-fragment | NA | NA | 0 |
| S17_vs_k141_23341\|\|full | 3963 | No | NA | 4 | 0 | 4 | Not-determined | Genome-fragment | NA | NA | 0 |
| S15_vs_2427\|\|lt2gene | 1041 | No | NA | 1 | 1 | 0 | Low-quality | Genome-fragment | 2.87 | AAI-based (high-confidence) | 0 |
| S15_vs_46359\|\|lt2gene | 1041 | No | NA | 2 | 1 | 0 | Low-quality | Genome-fragment | 19.19 | AAI-based (high-confidence) | 0 |
| S6_vs_k141_38021\|\|lt2gene | 1041 | No | NA | 2 | 1 | 0 | Low-quality | Genome-fragment | 2.42 | AAI-based (high-confidence) | 0 |
| S8_extracted_k141_16237 | 1041 | No | NA | 1 | 0 | 0 | Low-quality | Genome-fragment | 1.85 | AAI-based (medium-confidence) | 0 |
| S17_vs_k141_38734\|\|full | 1589 | No | NA | 1 | 0 | 0 | Not-determined | Genome-fragment | NA | NA | 0 |
| S11_vs_k141_69969\|\|lt2gene | 1042 | No | NA | 2 | 1 | 0 | Low-quality | Genome-fragment | 3.14 | AAI-based (medium-confidence) | 0 |
| S14_extracted_k141_93236 | 1042 | No | NA | 2 | 1 | 0 | Low-quality | Genome-fragment | 0.55 | HMM-based (lower-bound) | 0 |
| S17_vs_k141_53461\|\|full | 1071 | No | NA | 1 | 0 | 0 | Not-determined | Genome-fragment | NA | NA | 0 |
| S17_vs_k141_75256\|\|full | 1617 | No | NA | 2 | 0 | 0 | Not-determined | Genome-fragment | NA | NA | 0 |
| S17_vs_k141_141847\|\|full | 2225 | No | NA | 2 | 0 | 2 | Not-determined | Genome-fragment | NA | NA | 0 |
| S4_vs_k141_13263\|\|full | 1042 | No | NA | 3 | 2 | 0 | Low-quality | Genome-fragment | 3.17 | AAI-based (high-confidence) | 0 |
| S5_vs_k141_24818\|\|lt2gene | 1042 | No | NA | 2 | 1 | 0 | Low-quality | Genome-fragment | 2.3 | AAI-based (high-confidence) | 0 |
| S17_vs_k141_9036\|\|full | 1202 | No | NA | 2 | 0 | 1 | Not-determined | Genome-fragment | NA | NA | 0 |
| S17_vs_k141_105280\|\|full | 4616 | No | NA | 4 | 0 | 2 | Not-determined | Genome-fragment | NA | NA | 0 |
| S17_vs_k141_67887\|\|full | 2009 | No | NA | 5 | 0 | 2 | Not-determined | Genome-fragment | NA | NA | 0 |
| S17_vs_k141_156808\|\|full | 18506 | No | NA | 13 | 0 | 10 | Not-determined | Genome-fragment | NA | NA | 0 |
| S17_vs_k141_163977\|\|full | 2884 | No | NA | 4 | 0 | 1 | Not-determined | Genome-fragment | NA | NA | 0 |
| S17_vs_k141_163987\|\|full | 2373 | No | NA | 3 | 0 | 1 | Not-determined | Genome-fragment | NA | NA | 0 |
| S16_extracted_79229 | 1042 | No | NA | 2 | 0 | 2 | Low-quality | Genome-fragment | 1.87 | AAI-based (medium-confidence) | 0 |
| S16_extracted_129768 | 1042 | No | NA | 3 | 0 | 1 | Low-quality | Genome-fragment | 1 | AAI-based (medium-confidence) | 0 |
| S16_vs_22423\|\|full | 1042 | No | NA | 2 | 1 | 0 | Low-quality | Genome-fragment | 0.67 | HMM-based (lower-bound) | 0 |
| S17_vs_k141_97596\|\|full | 2313 | No | NA | 2 | 0 | 1 | Not-determined | Genome-fragment | NA | NA | 0 |
| S17_vs_k141_105343\|\|full | 1543 | No | NA | 3 | 0 | 1 | Not-determined | Genome-fragment | NA | NA | 0 |
| S12_extracted_k141_131550 | 1042 | No | NA | 2 | 0 | 0 | Low-quality | Genome-fragment | 2.85 | AAI-based (medium-confidence) | 0 |
| S8_extracted_k141_54561 | 1042 | No | NA | 3 | 0 | 2 | Low-quality | Genome-fragment | 1.08 | AAI-based (medium-confidence) | 0 |
| S17_vs_k141_15928\|\|full | 2623 | No | NA | 4 | 0 | 2 | Not-determined | Genome-fragment | NA | NA | 0 |
| S17_vs_k141_53561\|\|full | 1203 | No | NA | 3 | 0 | 1 | Not-determined | Genome-fragment | NA | NA | 0 |
| S8_vs_k141_194644\|\|full | 1042 | No | NA | 2 | 0 | 0 | Low-quality | Genome-fragment | 2.48 | AAI-based (medium-confidence) | 0 |
| S17_vs_k141_37906\|\|lt2gene | 1043 | No | NA | 2 | 2 | 0 | Low-quality | Genome-fragment | 2.18 | AAI-based (high-confidence) | 0 |
| S13_vs_61342\|\|full | 1043 | No | NA | 2 | 0 | 0 | Low-quality | Genome-fragment | 2.28 | AAI-based (high-confidence) | 0 |
| S13_vs_98618\|\|full | 1043 | No | NA | 3 | 0 | 0 | Low-quality | Genome-fragment | 0.63 | AAI-based (medium-confidence) | 0 |
| S17_vs_k141_82367\|\|full | 3224 | No | NA | 7 | 0 | 2 | Not-determined | Genome-fragment | NA | NA | 0 |
| S17_vs_k141_90192\|\|full | 4884 | No | NA | 6 | 0 | 4 | Not-determined | Genome-fragment | NA | NA | 0 |
| S17_vs_k141_68010\|\|full | 1396 | No | NA | 3 | 0 | 2 | Not-determined | Genome-fragment | NA | NA | 0 |
| S17_vs_k141_142025\|\|full | 1908 | No | NA | 3 | 0 | 1 | Not-determined | Genome-fragment | NA | NA | 0 |
| S17_vs_k141_105412\|\|full | 1441 | No | NA | 2 | 0 | 2 | Not-determined | Genome-fragment | NA | NA | 0 |
| S17_vs_k141_82403\|\|full | 1940 | No | NA | 2 | 0 | 1 | Not-determined | Genome-fragment | NA | NA | 0 |
| S14_vs_k141_74444\|\|full | 1043 | No | NA | 3 | 0 | 1 | Low-quality | Genome-fragment | 0.76 | AAI-based (medium-confidence) | 0 |
| S17_vs_k141_53601\|\|full | 3089 | No | NA | 7 | 0 | 1 | Not-determined | Genome-fragment | NA | NA | 0 |
| S14_vs_k141_62284\|\|lt2gene | 1043 | No | NA | 1 | 1 | 0 | Low-quality | Genome-fragment | 2.54 | AAI-based (high-confidence) | 0 |
| S17_vs_k141_68037\|\|full | 3109 | No | NA | 4 | 0 | 2 | Not-determined | Genome-fragment | NA | NA | 0 |
| S1_vs_k141_44502\|\|full | 1043 | No | NA | 3 | 1 | 0 | Low-quality | Genome-fragment | 0.63 | HMM-based (lower-bound) | 0 |
| S17_vs_k141_53605\|\|full | 2061 | No | NA | 3 | 0 | 0 | Not-determined | Genome-fragment | NA | NA | 0 |
| S17_vs_k141_53617\|\|full | 2393 | No | NA | 4 | 0 | 3 | Not-determined | Genome-fragment | NA | NA | 0 |
| S17_vs_k141_82435\|\|full | 3118 | No | NA | 6 | 0 | 2 | Not-determined | Genome-fragment | NA | NA | 0 |
| S17_vs_k141_156938\|\|full | 1098 | No | NA | 2 | 0 | 1 | Not-determined | Genome-fragment | NA | NA | 0 |
| S3_extracted_k141_49070 | 1043 | No | NA | 2 | 0 | 0 | Low-quality | Genome-fragment | 1.83 | AAI-based (high-confidence) | 0 |
| S3_extracted_k141_97202 | 1043 | No | NA | 2 | 0 | 0 | Low-quality | Genome-fragment | 1.59 | AAI-based (medium-confidence) | 0 |
| S17_vs_k141_105470\|\|full | 2182 | No | NA | 2 | 0 | 2 | Not-determined | Genome-fragment | NA | NA | 0 |
| S17_vs_k141_23496\|\|full | 2285 | No | NA | 3 | 0 | 2 | Not-determined | Genome-fragment | NA | NA | 0 |
| S17_vs_k141_38878\|\|full | 2096 | No | NA | 3 | 0 | 0 | Not-determined | Genome-fragment | NA | NA | 0 |
| S5_vs_k141_121435\|\|full | 1043 | No | NA | 3 | 0 | 0 | Low-quality | Genome-fragment | 1.61 | AAI-based (medium-confidence) | 0 |
| S17_vs_k141_9210\|\|full | 2151 | No | NA | 3 | 0 | 1 | Not-determined | Genome-fragment | NA | NA | 0 |
| S17_vs_k141_97755\|\|full | 2629 | No | NA | 3 | 0 | 1 | Not-determined | Genome-fragment | NA | NA | 0 |
| S5_vs_k141_33466\|\|full | 1043 | No | NA | 4 | 0 | 0 | Low-quality | Genome-fragment | 3.03 | AAI-based (high-confidence) | 0 |
| S7_extracted_86450 | 1043 | No | NA | 3 | 3 | 0 | Low-quality | Genome-fragment | 3.37 | AAI-based (high-confidence) | 0 |
| S17_vs_k141_82499\|\|full | 5107 | No | NA | 3 | 0 | 2 | Not-determined | Genome-fragment | NA | NA | 0 |
| S7_extracted_108755 | 1043 | No | NA | 2 | 2 | 0 | Low-quality | Genome-fragment | 1.73 | HMM-based (lower-bound) | 0 |
| S17_vs_k141_60786\|\|full | 2383 | No | NA | 3 | 0 | 3 | Not-determined | Genome-fragment | NA | NA | 0 |
| S17_vs_k141_134513\|\|full | 1727 | No | NA | 2 | 0 | 1 | Not-determined | Genome-fragment | NA | NA | 0 |
| S17_vs_k141_46188\|\|full | 1203 | No | NA | 3 | 0 | 1 | Not-determined | Genome-fragment | NA | NA | 0 |
| S7_vs_45525\|\|full | 1043 | No | NA | 4 | 1 | 0 | Low-quality | Genome-fragment | 2.73 | AAI-based (high-confidence) | 0 |
| S17_vs_k141_126869\|\|full | 2904 | No | NA | 2 | 0 | 1 | Not-determined | Genome-fragment | NA | NA | 0 |
| S17_vs_k141_1621\|\|full | 1167 | No | NA | 4 | 0 | 0 | Not-determined | Genome-fragment | NA | NA | 0 |
| S10_extracted_106271 | 1043 | No | NA | 1 | 0 | 0 | Low-quality | Genome-fragment | 2.4 | AAI-based (high-confidence) | 0 |
| S17_vs_k141_16054\|\|full | 1799 | No | NA | 2 | 0 | 0 | Not-determined | Genome-fragment | NA | NA | 0 |
| S10_extracted_121228 | 1043 | No | NA | 2 | 2 | 0 | Low-quality | Genome-fragment | 3.44 | HMM-based (lower-bound) | 0 |
| S17_vs_k141_75384\|\|full | 1620 | No | NA | 3 | 0 | 0 | Not-determined | Genome-fragment | NA | NA | 0 |
| S12_extracted_k141_133254 | 1043 | No | NA | 2 | 0 | 0 | Low-quality | Genome-fragment | 1.66 | AAI-based (medium-confidence) | 0 |
| S15_vs_616\|\|full | 1043 | No | NA | 2 | 0 | 1 | Low-quality | Genome-fragment | 2.26 | AAI-based (high-confidence) | 0 |
| S6_vs_k141_6492\|\|full | 1043 | No | NA | 3 | 1 | 0 | Low-quality | Genome-fragment | 1.96 | AAI-based (medium-confidence) | 0 |
| S17_vs_k141_31409\|\|full | 1746 | No | NA | 3 | 0 | 2 | Not-determined | Genome-fragment | NA | NA | 0 |
| S8_vs_k141_147154\|\|full | 1043 | No | NA | 2 | 0 | 0 | Low-quality | Genome-fragment | 2.14 | AAI-based (high-confidence) | 0 |
| S8_vs_k141_121783\|\|lt2gene | 1043 | No | NA | 2 | 1 | 0 | Low-quality | Genome-fragment | 2.69 | AAI-based (high-confidence) | 0 |
| S17_vs_k141_75406\|\|full | 2421 | No | NA | 2 | 0 | 1 | Not-determined | Genome-fragment | NA | NA | 0 |
| S14_extracted_k141_89399 | 1044 | No | NA | 2 | 1 | 0 | Low-quality | Genome-fragment | 3.44 | AAI-based (high-confidence) | 0 |
| S17_vs_k141_31426\|\|full | 2076 | No | NA | 3 | 0 | 1 | Not-determined | Genome-fragment | NA | NA | 0 |
| S17_vs_k141_38955\|\|full | 2053 | No | NA | 3 | 0 | 0 | Not-determined | Genome-fragment | NA | NA | 0 |
| S17_vs_k141_142202\|\|full | 1727 | No | NA | 3 | 0 | 0 | Not-determined | Genome-fragment | NA | NA | 0 |
| S17_vs_k141_164175\|\|full | 9143 | No | NA | 5 | 0 | 2 | Not-determined | Genome-fragment | NA | NA | 0 |
| S17_vs_k141_68201\|\|full | 1509 | No | NA | 3 | 0 | 0 | Not-determined | Genome-fragment | NA | NA | 0 |
| S1_extracted_k141_27060 | 1044 | No | NA | 1 | 0 | 0 | Low-quality | Genome-fragment | 0.59 | AAI-based (high-confidence) | 0 |
| S17_vs_k141_97887\|\|full | 1075 | No | NA | 2 | 0 | 1 | Not-determined | Genome-fragment | NA | NA | 0 |
| S17_vs_k141_82601\|\|full | 10815 | No | NA | 11 | 0 | 3 | Not-determined | Genome-fragment | NA | NA | 0 |
| S17_vs_k141_126974\|\|full | 1613 | No | NA | 3 | 0 | 0 | Not-determined | Genome-fragment | NA | NA | 0 |
| S17_vs_k141_75441\|\|full | 1211 | No | NA | 2 | 0 | 0 | Not-determined | Genome-fragment | NA | NA | 0 |
| S1_extracted_k141_2421 | 1044 | No | NA | 1 | 0 | 0 | Low-quality | Genome-fragment | 1.06 | AAI-based (high-confidence) | 0 |
| S1_vs_k141_123222\|\|full | 1044 | No | NA | 3 | 0 | 0 | Low-quality | Genome-fragment | 1.3 | AAI-based (high-confidence) | 0 |
| S1_vs_k141_63395\|\|lt2gene | 1044 | No | NA | 2 | 1 | 0 | Low-quality | Genome-fragment | 2.77 | AAI-based (medium-confidence) | 0 |
| S5_vs_k141_5563\|\|full | 1044 | No | NA | 2 | 0 | 1 | Low-quality | Genome-fragment | 0.89 | AAI-based (medium-confidence) | 0 |
| S17_vs_k141_120044\|\|full | 2432 | No | NA | 7 | 0 | 0 | Not-determined | Genome-fragment | NA | NA | 0 |
| S7_vs_25695\|\|full | 1044 | No | NA | 3 | 0 | 0 | Low-quality | Genome-fragment | 2.18 | AAI-based (high-confidence) | 0 |
| S16_extracted_95025 | 1044 | No | NA | 1 | 0 | 0 | Low-quality | Genome-fragment | 1.78 | AAI-based (high-confidence) | 0 |
| S10_extracted_114375 | 1044 | No | NA | 2 | 1 | 0 | Low-quality | Genome-fragment | 0.45 | AAI-based (medium-confidence) | 0 |
| S17_vs_k141_60805\|\|full | 3558 | No | NA | 3 | 0 | 1 | Not-determined | Genome-fragment | NA | NA | 0 |
| S10_vs_54546\|\|full | 1044 | No | NA | 5 | 1 | 0 | Low-quality | Genome-fragment | 1.78 | AAI-based (medium-confidence) | 0 |
| S12_extracted_k141_84821 | 1044 | No | NA | 2 | 0 | 1 | Low-quality | Genome-fragment | 2.97 | AAI-based (medium-confidence) | 0 |
| S17_vs_k141_68264\|\|full | 1509 | No | NA | 3 | 0 | 0 | Not-determined | Genome-fragment | NA | NA | 0 |
| S15_vs_26781\|\|full | 1044 | No | NA | 2 | 0 | 0 | Low-quality | Genome-fragment | 2.17 | AAI-based (medium-confidence) | 0 |
| S17_vs_k141_164233\|\|full | 2012 | No | NA | 3 | 0 | 0 | Not-determined | Genome-fragment | NA | NA | 0 |
| S6_vs_k141_16440\|\|full | 1044 | No | NA | 3 | 0 | 0 | Low-quality | Genome-fragment | 2.28 | AAI-based (medium-confidence) | 0 |
| S17_vs_k141_82692\|\|full | 2752 | No | NA | 3 | 0 | 2 | Not-determined | Genome-fragment | NA | NA | 0 |
| S17_vs_k141_16228\|\|full | 1589 | No | NA | 2 | 0 | 0 | Not-determined | Genome-fragment | NA | NA | 0 |
| S17_vs_k141_164253\|\|full | 1661 | No | NA | 2 | 0 | 2 | Not-determined | Genome-fragment | NA | NA | 0 |
| S8_extracted_k141_15708 | 1044 | No | NA | 2 | 0 | 1 | Low-quality | Genome-fragment | 3.33 | AAI-based (medium-confidence) | 0 |
| S17_vs_k141_82714\|\|full | 1389 | No | NA | 3 | 0 | 1 | Not-determined | Genome-fragment | NA | NA | 0 |
| S17_extracted_k141_90452 | 1045 | No | NA | 3 | 1 | 0 | Low-quality | Genome-fragment | 1.35 | AAI-based (high-confidence) | 0 |
| S17_vs_k141_23832\|\|full | 1497 | No | NA | 2 | 0 | 1 | Not-determined | Genome-fragment | NA | NA | 0 |
| S17_vs_k141_60847\|\|full | 9520 | No | NA | 11 | 0 | 5 | Not-determined | Genome-fragment | NA | NA | 0 |
| S17_vs_k141_164262\|\|full | 2816 | No | NA | 3 | 0 | 0 | Not-determined | Genome-fragment | NA | NA | 0 |
| S17_vs_k141_60861\|\|full | 2435 | No | NA | 4 | 0 | 0 | Not-determined | Genome-fragment | NA | NA | 0 |
| S9_extracted_k141_24091 | 1045 | No | NA | 2 | 0 | 1 | Low-quality | Genome-fragment | 0.07 | AAI-based (medium-confidence) | 0 |
| S17_vs_k141_23842\|\|full | 5450 | No | NA | 5 | 0 | 2 | Not-determined | Genome-fragment | NA | NA | 0 |
| S9_vs_k141_76100\|\|lt2gene | 1045 | No | NA | 2 | 1 | 0 | Low-quality | Genome-fragment | 2.39 | AAI-based (high-confidence) | 0 |
| S17_vs_k141_23845\|\|full | 2983 | No | NA | 4 | 0 | 0 | Not-determined | Genome-fragment | NA | NA | 0 |
| S17_vs_k141_75506\|\|full | 2179 | No | NA | 4 | 0 | 1 | Not-determined | Genome-fragment | NA | NA | 0 |
| S14_vs_k141_19064\|\|full | 1045 | No | NA | 3 | 3 | 0 | Low-quality | Genome-fragment | 3.14 | AAI-based (high-confidence) | 0 |
| S3_extracted_k141_53225 | 1045 | No | NA | 4 | 0 | 0 | Low-quality | Genome-fragment | 3.35 | AAI-based (high-confidence) | 0 |
| S17_vs_k141_120199\|\|full | 1512 | No | NA | 4 | 0 | 0 | Not-determined | Genome-fragment | NA | NA | 0 |
| S17_vs_k141_127139\|\|full | 3352 | No | NA | 3 | 0 | 1 | Not-determined | Genome-fragment | NA | NA | 0 |
| S5_extracted_k141_136375 | 1045 | No | NA | 3 | 0 | 0 | Low-quality | Genome-fragment | 1.38 | AAI-based (high-confidence) | 0 |
| S17_vs_k141_120216\|\|full | 3257 | No | NA | 3 | 0 | 1 | Not-determined | Genome-fragment | NA | NA | 0 |
| S5_extracted_k141_127117 | 1045 | No | NA | 3 | 0 | 0 | Low-quality | Genome-fragment | 2.91 | AAI-based (high-confidence) | 0 |
| S17_vs_k141_157209\|\|full | 43342 | No | NA | 70 | 13 | 0 | High-quality | High-quality | 95.84 | AAI-based (high-confidence) | 0 |
| S17_vs_k141_1851\|\|full | 3456 | No | NA | 3 | 0 | 0 | Not-determined | Genome-fragment | NA | NA | 0 |
| S17_vs_k141_16330\|\|full | 3470 | No | NA | 7 | 0 | 0 | Not-determined | Genome-fragment | NA | NA | 0 |
| S17_vs_k141_23889\|\|full | 2613 | No | NA | 2 | 0 | 1 | Not-determined | Genome-fragment | NA | NA | 0 |
| S17_vs_k141_60940\|\|full | 1761 | No | NA | 3 | 0 | 2 | Not-determined | Genome-fragment | NA | NA | 0 |
| S5_extracted_k141_76939 | 1045 | No | NA | 1 | 1 | 0 | Low-quality | Genome-fragment | 1.38 | HMM-based (lower-bound) | 0 |
| S7_extracted_16454 | 1045 | No | NA | 2 | 0 | 1 | Low-quality | Genome-fragment | 1.75 | AAI-based (high-confidence) | 0 |
| S17_vs_k141_112524\|\|full | 2434 | No | NA | 3 | 0 | 1 | Not-determined | Genome-fragment | NA | NA | 0 |
| S17_vs_k141_31654\|\|full | 24962 | No | NA | 33 | 18 | 0 | Medium-quality | Genome-fragment | 66.18 | AAI-based (high-confidence) | 0 |
| S17_vs_k141_54041\|\|full | 2856 | No | NA | 7 | 0 | 0 | Not-determined | Genome-fragment | NA | NA | 0 |
| S17_vs_k141_82770\|\|full | 3259 | No | NA | 4 | 0 | 0 | Not-determined | Genome-fragment | NA | NA | 0 |
| S7_extracted_129735 | 1045 | No | NA | 2 | 1 | 0 | Low-quality | Genome-fragment | 2.65 | AAI-based (medium-confidence) | 0 |
| S17_vs_k141_82775\|\|full | 1519 | No | NA | 2 | 0 | 1 | Not-determined | Genome-fragment | NA | NA | 0 |
| S17_vs_k141_112544\|\|full | 1514 | No | NA | 3 | 0 | 1 | Not-determined | Genome-fragment | NA | NA | 0 |
| S17_vs_k141_171722\|\|full | 3411 | No | NA | 4 | 0 | 3 | Not-determined | Genome-fragment | NA | NA | 0 |
| S17_vs_k141_127200\|\|full | 2057 | No | NA | 2 | 0 | 0 | Not-determined | Genome-fragment | NA | NA | 0 |
| S7_vs_93782\|\|full | 1045 | No | NA | 2 | 2 | 0 | Low-quality | Genome-fragment | 2.52 | AAI-based (high-confidence) | 0 |
| S16_extracted_109140 | 1045 | No | NA | 4 | 0 | 0 | Low-quality | Genome-fragment | 2.27 | AAI-based (medium-confidence) | 0 |
| S16_vs_9175\|\|lt2gene | 1045 | No | NA | 2 | 1 | 0 | Low-quality | Genome-fragment | 3.54 | HMM-based (lower-bound) | 0 |
| S17_vs_k141_150136\|\|full | 2043 | No | NA | 3 | 0 | 0 | Not-determined | Genome-fragment | NA | NA | 0 |
| S10_vs_33835\|\|full | 1045 | No | NA | 3 | 0 | 0 | Low-quality | Genome-fragment | 2.77 | AAI-based (high-confidence) | 0 |
| S17_vs_k141_171756\|\|full | 3545 | No | NA | 3 | 0 | 0 | Not-determined | Genome-fragment | NA | NA | 0 |
| S12_extracted_k141_187883 | 1045 | No | NA | 3 | 0 | 1 | Low-quality | Genome-fragment | 3.34 | AAI-based (medium-confidence) | 0 |
| S17_vs_k141_134911\|\|full | 4231 | No | NA | 4 | 0 | 3 | Not-determined | Genome-fragment | NA | NA | 0 |
| S12_vs_k141_90388\|\|lt2gene | 1045 | No | NA | 1 | 1 | 0 | Low-quality | Genome-fragment | 1.84 | AAI-based (high-confidence) | 0 |
| S17_vs_k141_112617\|\|full | 1094 | No | NA | 3 | 0 | 0 | Not-determined | Genome-fragment | NA | NA | 0 |
| S17_vs_k141_9459\|\|full | 2150 | No | NA | 5 | 0 | 0 | Not-determined | Genome-fragment | NA | NA | 0 |
| S17_vs_k141_150181\|\|full | 2655 | No | NA | 6 | 0 | 2 | Not-determined | Genome-fragment | NA | NA | 0 |
| S6_extracted_k141_30897 | 1045 | No | NA | 1 | 0 | 0 | Low-quality | Genome-fragment | 2.73 | AAI-based (medium-confidence) | 0 |
| S6_extracted_k141_8320 | 1045 | No | NA | 1 | 0 | 1 | Low-quality | Genome-fragment | 3.51 | AAI-based (medium-confidence) | 0 |
| S17_vs_k141_98183\|\|full | 1557 | No | NA | 3 | 0 | 1 | Not-determined | Genome-fragment | NA | NA | 0 |
| S17_vs_k141_134954\|\|full | 1666 | No | NA | 2 | 0 | 1 | Not-determined | Genome-fragment | NA | NA | 0 |
| S6_extracted_k141_27937 | 1045 | No | NA | 2 | 0 | 0 | Low-quality | Genome-fragment | 1.11 | AAI-based (high-confidence) | 0 |
| S17_vs_k141_31721\|\|full | 1253 | No | NA | 2 | 0 | 1 | Not-determined | Genome-fragment | NA | NA | 0 |
| S17_vs_k141_98200\|\|full | 1647 | No | NA | 3 | 0 | 0 | Not-determined | Genome-fragment | NA | NA | 0 |
| S8_extracted_k141_192876 | 1045 | No | NA | 2 | 0 | 1 | Low-quality | Genome-fragment | 1.21 | AAI-based (high-confidence) | 0 |
| S8_vs_k141_108783\|\|full | 1045 | No | NA | 3 | 1 | 0 | Low-quality | Genome-fragment | 1.99 | AAI-based (high-confidence) | 0 |
| S17_extracted_k141_124887 | 1046 | No | NA | 2 | 0 | 1 | Low-quality | Genome-fragment | 0.51 | AAI-based (medium-confidence) | 0 |
| S9_vs_k141_77381\|\|full | 1046 | No | NA | 3 | 0 | 0 | Low-quality | Genome-fragment | 2.41 | AAI-based (high-confidence) | 0 |
| S14_extracted_k141_105691 | 1046 | No | NA | 2 | 0 | 0 | Low-quality | Genome-fragment | 1.77 | AAI-based (high-confidence) | 0 |
| S17_vs_k141_150216\|\|full | 2771 | No | NA | 3 | 0 | 1 | Not-determined | Genome-fragment | NA | NA | 0 |
| S14_vs_k141_12538\|\|full | 1046 | No | NA | 3 | 0 | 1 | Low-quality | Genome-fragment | 0.94 | AAI-based (medium-confidence) | 0 |
| S1_extracted_k141_156687 | 1046 | No | NA | 2 | 0 | 0 | Low-quality | Genome-fragment | 2.46 | AAI-based (medium-confidence) | 0 |
| S17_vs_k141_75711\|\|full | 7915 | No | NA | 4 | 0 | 1 | Not-determined | Genome-fragment | NA | NA | 0 |
| S3_extracted_k141_10146 | 1046 | No | NA | 2 | 0 | 0 | Low-quality | Genome-fragment | 2.27 | AAI-based (medium-confidence) | 0 |
| S5_vs_k141_121607\|\|lt2gene | 1046 | No | NA | 2 | 1 | 0 | Low-quality | Genome-fragment | 2.61 | AAI-based (high-confidence) | 0 |
| S7_vs_13668\|\|lt2gene | 1046 | No | NA | 1 | 1 | 0 | Low-quality | Genome-fragment | 2.59 | AAI-based (high-confidence) | 0 |
| S17_vs_k141_134992\|\|full | 6478 | No | NA | 5 | 0 | 4 | Not-determined | Genome-fragment | NA | NA | 0 |
| S16_vs_35640\|\|full | 1046 | No | NA | 3 | 0 | 0 | Low-quality | Genome-fragment | 2.67 | AAI-based (medium-confidence) | 0 |
| S12_extracted_k141_96362 | 1046 | No | NA | 3 | 0 | 0 | Low-quality | Genome-fragment | 2.25 | AAI-based (high-confidence) | 0 |
| S12_extracted_k141_13937 | 1046 | No | NA | 4 | 0 | 0 | Low-quality | Genome-fragment | 2.29 | AAI-based (high-confidence) | 0 |
| S17_vs_k141_150251\|\|full | 5468 | No | NA | 8 | 0 | 0 | Not-determined | Genome-fragment | NA | NA | 0 |
| S15_extracted_22433 | 1046 | No | NA | 1 | 1 | 0 | Low-quality | Genome-fragment | 2.32 | AAI-based (high-confidence) | 0 |
| S17_vs_k141_39421\|\|full | 2606 | No | NA | 3 | 0 | 0 | Not-determined | Genome-fragment | NA | NA | 0 |
| S17_vs_k141_68589\|\|full | 6833 | No | NA | 4 | 0 | 4 | Not-determined | Genome-fragment | NA | NA | 0 |
| S17_vs_k141_2044\|\|full | 1051 | No | NA | 2 | 0 | 0 | Not-determined | Genome-fragment | NA | NA | 0 |
| S15_vs_11101\|\|full | 1046 | No | NA | 3 | 2 | 0 | Low-quality | Genome-fragment | 2.46 | AAI-based (high-confidence) | 0 |
| S17_vs_k141_127333\|\|full | 3969 | No | NA | 4 | 0 | 2 | Not-determined | Genome-fragment | NA | NA | 0 |
| S17_vs_k141_75757\|\|full | 2423 | No | NA | 3 | 0 | 2 | Not-determined | Genome-fragment | NA | NA | 0 |
| S6_vs_k141_14134\|\|full | 1046 | No | NA | 2 | 0 | 0 | Low-quality | Genome-fragment | 1.71 | AAI-based (high-confidence) | 0 |
| S17_vs_k141_90499\|\|full | 2176 | No | NA | 4 | 0 | 0 | Not-determined | Genome-fragment | NA | NA | 0 |
| S6_vs_k141_49090\|\|full | 1046 | No | NA | 2 | 0 | 0 | Low-quality | Genome-fragment | 1.47 | AAI-based (medium-confidence) | 0 |
| S17_vs_k141_9602\|\|full | 1252 | No | NA | 3 | 0 | 1 | Not-determined | Genome-fragment | NA | NA | 0 |
| S17_vs_k141_39473\|\|full | 1412 | No | NA | 3 | 0 | 1 | Not-determined | Genome-fragment | NA | NA | 0 |
| S17_vs_k141_68628\|\|full | 1904 | No | NA | 2 | 0 | 1 | Not-determined | Genome-fragment | NA | NA | 0 |
| S17_vs_k141_164589\|\|full | 3344 | No | NA | 5 | 0 | 1 | Not-determined | Genome-fragment | NA | NA | 0 |
| S8_vs_k141_18937\|\|full | 1046 | No | NA | 3 | 1 | 0 | Low-quality | Genome-fragment | 1.8 | AAI-based (high-confidence) | 0 |
| S17_vs_k141_24083\|\|full | 1032 | No | NA | 3 | 0 | 0 | Not-determined | Genome-fragment | NA | NA | 0 |
| S17_vs_k141_54212\|\|full | 2860 | No | NA | 3 | 0 | 2 | Not-determined | Genome-fragment | NA | NA | 0 |
| S17_vs_k141_68641\|\|full | 2059 | No | NA | 3 | 0 | 1 | Not-determined | Genome-fragment | NA | NA | 0 |
| S17_vs_k141_2079\|\|full | 25077 | No | NA | 32 | 2 | 2 | Medium-quality | Genome-fragment | 58.55 | AAI-based (high-confidence) | 0 |
| S17_vs_k141_135064\|\|full | 1484 | No | NA | 3 | 0 | 2 | Not-determined | Genome-fragment | NA | NA | 0 |
| S8_vs_k141_74921\|\|full | 1046 | No | NA | 3 | 0 | 0 | Low-quality | Genome-fragment | 1.97 | AAI-based (medium-confidence) | 0 |
| S17_vs_k141_150372\|\|full | 1126 | No | NA | 2 | 0 | 0 | Not-determined | Genome-fragment | NA | NA | 0 |
| S17_vs_k141_164625\|\|full | 4132 | No | NA | 6 | 0 | 0 | Not-determined | Genome-fragment | NA | NA | 0 |
| S17_vs_k141_16552\|\|full | 2146 | No | NA | 2 | 0 | 0 | Not-determined | Genome-fragment | NA | NA | 0 |
| S17_vs_k141_39496\|\|full | 12869 | No | NA | 20 | 1 | 2 | Not-determined | Genome-fragment | NA | NA | 0 |
| S17_extracted_k141_9003 | 1047 | No | NA | 1 | 1 | 0 | Low-quality | Genome-fragment | 1.65 | HMM-based (lower-bound) | 0 |
| S17_extracted_k141_34746 | 1047 | No | NA | 2 | 1 | 0 | Low-quality | Genome-fragment | 2.48 | AAI-based (medium-confidence) | 0 |
| S9_extracted_k141_36682 | 1047 | No | NA | 2 | 0 | 0 | Low-quality | Genome-fragment | 0.6 | AAI-based (high-confidence) | 0 |
| S17_vs_k141_68706\|\|full | 1097 | No | NA | 1 | 0 | 1 | Not-determined | Genome-fragment | NA | NA | 0 |
| S17_vs_k141_157510\|\|full | 1668 | No | NA | 2 | 0 | 1 | Not-determined | Genome-fragment | NA | NA | 0 |
| S9_vs_k141_87941\|\|full | 1047 | No | NA | 3 | 0 | 1 | Low-quality | Genome-fragment | 49.34 | AAI-based (medium-confidence) | 0 |
| S17_vs_k141_150412\|\|full | 1903 | No | NA | 2 | 0 | 2 | Not-determined | Genome-fragment | NA | NA | 0 |
| S11_extracted_k141_25118 | 1047 | No | NA | 2 | 0 | 0 | Low-quality | Genome-fragment | 2.48 | AAI-based (medium-confidence) | 0 |
| S17_vs_k141_157531\|\|full | 4333 | No | NA | 7 | 0 | 0 | Not-determined | Genome-fragment | NA | NA | 0 |
| S17_vs_k141_171894\|\|full | 2904 | No | NA | 4 | 0 | 1 | Not-determined | Genome-fragment | NA | NA | 0 |
| S17_vs_k141_39515\|\|full | 36323 | No | NA | 45 | 4 | 1 | Medium-quality | Genome-fragment | 58.53 | AAI-based (high-confidence) | 0 |
| S17_vs_k141_171906\|\|full | 5839 | No | NA | 7 | 0 | 4 | Not-determined | Genome-fragment | NA | NA | 0 |
| S17_vs_k141_150440\|\|full | 1873 | No | NA | 4 | 0 | 0 | Not-determined | Genome-fragment | NA | NA | 0 |
| S17_vs_k141_171912\|\|full | 1604 | No | NA | 3 | 0 | 1 | Not-determined | Genome-fragment | NA | NA | 0 |
| S17_vs_k141_68775\|\|full | 2942 | No | NA | 4 | 0 | 2 | Not-determined | Genome-fragment | NA | NA | 0 |
| S13_extracted_48175 | 1047 | No | NA | 2 | 0 | 1 | Low-quality | Genome-fragment | 0.48 | AAI-based (medium-confidence) | 0 |
| S14_vs_k141_2639\|\|full | 1047 | No | NA | 3 | 0 | 0 | Low-quality | Genome-fragment | 2.63 | AAI-based (high-confidence) | 0 |
| S17_vs_k141_24165\|\|full | 1715 | No | NA | 2 | 0 | 1 | Not-determined | Genome-fragment | NA | NA | 0 |
| S17_vs_k141_112863\|\|full | 1042 | No | NA | 2 | 0 | 1 | Not-determined | Genome-fragment | NA | NA | 0 |
| S3_extracted_k141_19230 | 1047 | No | NA | 1 | 0 | 0 | Low-quality | Genome-fragment | 2.22 | AAI-based (high-confidence) | 0 |
| S17_vs_k141_61049\|\|full | 2524 | No | NA | 3 | 0 | 2 | Not-determined | Genome-fragment | NA | NA | 0 |
| S17_vs_k141_171944\|\|full | 2454 | No | NA | 3 | 0 | 1 | Not-determined | Genome-fragment | NA | NA | 0 |
| S17_vs_k141_90642\|\|full | 2686 | No | NA | 5 | 0 | 0 | Not-determined | Genome-fragment | NA | NA | 0 |
| S3_vs_k141_40918\|\|lt2gene | 1047 | No | NA | 2 | 2 | 0 | Low-quality | Genome-fragment | 1.71 | HMM-based (lower-bound) | 0 |
| S17_vs_k141_61079\|\|full | 1048 | No | NA | 3 | 0 | 1 | Not-determined | Genome-fragment | NA | NA | 0 |
| S7_vs_57199\|\|full | 1047 | No | NA | 4 | 0 | 0 | Low-quality | Genome-fragment | 2.32 | AAI-based (medium-confidence) | 0 |
| S17_vs_k141_24204\|\|full | 3060 | No | NA | 3 | 0 | 1 | Not-determined | Genome-fragment | NA | NA | 0 |
| S17_vs_k141_82943\|\|full | 3063 | No | NA | 4 | 0 | 3 | Not-determined | Genome-fragment | NA | NA | 0 |
| S16_extracted_143699 | 1047 | No | NA | 2 | 0 | 1 | Low-quality | Genome-fragment | 1.87 | AAI-based (medium-confidence) | 0 |
| S17_vs_k141_120527\|\|full | 1479 | No | NA | 1 | 0 | 1 | Not-determined | Genome-fragment | NA | NA | 0 |
| S17_vs_k141_61104\|\|full | 1741 | No | NA | 2 | 0 | 0 | Not-determined | Genome-fragment | NA | NA | 0 |
| S10_vs_38074\|\|full | 1047 | No | NA | 2 | 1 | 0 | Low-quality | Genome-fragment | 2.02 | AAI-based (medium-confidence) | 0 |
| S12_extracted_k141_98567 | 1047 | No | NA | 2 | 1 | 0 | Low-quality | Genome-fragment | 3.08 | AAI-based (high-confidence) | 0 |
| S17_vs_k141_39651\|\|full | 1222 | No | NA | 3 | 0 | 1 | Not-determined | Genome-fragment | NA | NA | 0 |
| S17_vs_k141_68824\|\|full | 4822 | No | NA | 5 | 0 | 1 | Not-determined | Genome-fragment | NA | NA | 0 |
| S12_vs_k141_165443\|\|full | 1047 | No | NA | 2 | 0 | 1 | Low-quality | Genome-fragment | 0.5 | AAI-based (high-confidence) | 0 |
| S17_vs_k141_142838\|\|full | 2281 | No | NA | 4 | 0 | 3 | Not-determined | Genome-fragment | NA | NA | 0 |
| S4_extracted_25211 | 1047 | No | NA | 2 | 0 | 1 | Low-quality | Genome-fragment | 3.43 | AAI-based (high-confidence) | 0 |
| S17_vs_k141_90714\|\|full | 4701 | No | NA | 8 | 0 | 7 | Not-determined | Genome-fragment | NA | NA | 0 |
| S6_vs_k141_43922\|\|full | 1047 | No | NA | 2 | 0 | 0 | Low-quality | Genome-fragment | 1.74 | AAI-based (high-confidence) | 0 |
| S17_vs_k141_32000\|\|full | 1399 | No | NA | 3 | 0 | 0 | Not-determined | Genome-fragment | NA | NA | 0 |
| S8_extracted_k141_70115 | 1047 | No | NA | 3 | 2 | 0 | Low-quality | Genome-fragment | 1.6 | AAI-based (medium-confidence) | 0 |
| S8_extracted_k141_49668 | 1047 | No | NA | 2 | 0 | 1 | Low-quality | Genome-fragment | 0.59 | AAI-based (medium-confidence) | 0 |
| S17_vs_k141_54340\|\|full | 1404 | No | NA | 2 | 0 | 0 | Not-determined | Genome-fragment | NA | NA | 0 |
| S8_vs_k141_105928\|\|full | 1047 | No | NA | 2 | 0 | 0 | Low-quality | Genome-fragment | 1.05 | AAI-based (medium-confidence) | 0 |
| S8_vs_k141_147124\|\|full | 1047 | No | NA | 4 | 2 | 0 | Low-quality | Genome-fragment | 2.72 | AAI-based (high-confidence) | 0 |
| S17_vs_k141_76048\|\|full | 13221 | No | NA | 16 | 0 | 4 | Not-determined | Genome-fragment | NA | NA | 0 |
| S17_vs_k141_127519\|\|full | 31736 | No | NA | 37 | 7 | 0 | Medium-quality | Genome-fragment | 72.13 | AAI-based (high-confidence) | 0 |
| S8_vs_k141_179216\|\|lt2gene | 1047 | No | NA | 1 | 1 | 0 | Low-quality | Genome-fragment | 0.6 | HMM-based (lower-bound) | 0 |
| S17_vs_k141_127533\|\|full | 1164 | No | NA | 3 | 0 | 1 | Not-determined | Genome-fragment | NA | NA | 0 |
| S17_vs_k141_24273\|\|full | 1048 | No | NA | 2 | 0 | 1 | Low-quality | Genome-fragment | 2.11 | AAI-based (high-confidence) | 0 |
| S11_extracted_k141_65743 | 1048 | No | NA | 2 | 0 | 0 | Low-quality | Genome-fragment | 1.33 | AAI-based (high-confidence) | 0 |
| S17_vs_k141_157729\|\|full | 3582 | No | NA | 3 | 0 | 1 | Not-determined | Genome-fragment | NA | NA | 0 |
| S11_vs_k141_98032\|\|full | 1048 | No | NA | 3 | 0 | 1 | Low-quality | Genome-fragment | 0.47 | AAI-based (medium-confidence) | 0 |
| S11_vs_k141_8141\|\|full | 1048 | No | NA | 2 | 0 | 2 | Low-quality | Genome-fragment | 1.52 | AAI-based (high-confidence) | 0 |
| S17_vs_k141_157755\|\|full | 1895 | No | NA | 2 | 0 | 2 | Not-determined | Genome-fragment | NA | NA | 0 |
| S17_vs_k141_24311\|\|full | 2588 | No | NA | 6 | 0 | 0 | Not-determined | Genome-fragment | NA | NA | 0 |
| S13_extracted_45611 | 1048 | No | NA | 2 | 1 | 0 | Low-quality | Genome-fragment | 2.17 | AAI-based (medium-confidence) | 0 |
| S14_extracted_k141_105021 | 1048 | No | NA | 2 | 1 | 0 | Low-quality | Genome-fragment | 3.81 | AAI-based (high-confidence) | 0 |
| S3_extracted_k141_19782 | 1048 | No | NA | 1 | 1 | 0 | Low-quality | Genome-fragment | 2.84 | AAI-based (high-confidence) | 0 |
| S3_vs_k141_123138\|\|full | 1048 | No | NA | 2 | 1 | 0 | Low-quality | Genome-fragment | 1.92 | AAI-based (medium-confidence) | 0 |
| S3_vs_k141_69775\|\|lt2gene | 1048 | No | NA | 1 | 1 | 0 | Low-quality | Genome-fragment | 0.72 | HMM-based (lower-bound) | 0 |
| S5_extracted_k141_134057 | 1048 | No | NA | 3 | 2 | 0 | Low-quality | Genome-fragment | 1.41 | AAI-based (medium-confidence) | 0 |
| S5_vs_k141_27260\|\|lt2gene | 1048 | No | NA | 2 | 1 | 0 | Low-quality | Genome-fragment | 1.5 | AAI-based (medium-confidence) | 0 |
| S7_vs_39072\|\|full | 1048 | No | NA | 2 | 1 | 0 | Low-quality | Genome-fragment | 1.98 | HMM-based (lower-bound) | 0 |
| S16_extracted_83952 | 1048 | No | NA | 3 | 0 | 0 | Low-quality | Genome-fragment | 1.97 | AAI-based (high-confidence) | 0 |
| S17_vs_k141_16767\|\|full | 2779 | No | NA | 3 | 0 | 0 | Not-determined | Genome-fragment | NA | NA | 0 |
| S17_vs_k141_46857\|\|full | 2195 | No | NA | 2 | 0 | 1 | Not-determined | Genome-fragment | NA | NA | 0 |

| | | | | | | | | | | | | |
|---|---|---|---|---|---|---|---|---|---|---|---|---|
| S16_vs_173119\|\|full | 1048 | No | NA | 2 | 1 | 0 | Low-quality | Genome-fragment | | 0.93 | HMM-based (lower-bound) | 0 |
| S16_vs_65962\|\|full | 1048 | No | NA | 3 | 0 | 1 | Low-quality | Genome-fragment | | 1.83 | AAI-based (medium-confidence) | 0 |
| S16_vs_141177\|\|full | 1048 | No | NA | 2 | 0 | 1 | Low-quality | Genome-fragment | | 2.06 | AAI-based (medium-confidence) | 0 |
| S10_vs_94607\|\|full | 1048 | No | NA | 3 | 0 | 1 | Low-quality | Genome-fragment | | 2.26 | AAI-based (medium-confidence) | 0 |
| S17_vs_k141_113061\|\|full | 3152 | No | NA | 5 | 0 | 3 | Not-determined | Genome-fragment | NA | | NA | 0 |
| S10_vs_115859\|\|full | 1048 | No | NA | 3 | 0 | 1 | Low-quality | Genome-fragment | | 3.98 | AAI-based (medium-confidence) | 0 |
| S10_vs_40352\|\|lt2gene | 1048 | No | NA | 1 | 1 | 0 | Low-quality | Genome-fragment | | 2.14 | AAI-based (high-confidence) | 0 |
| S17_vs_k141_46917\|\|full | 1773 | No | NA | 3 | 0 | 1 | Not-determined | Genome-fragment | NA | | NA | 0 |
| S17_vs_k141_2270\|\|full | 1181 | No | NA | 2 | 0 | 0 | Not-determined | Genome-fragment | NA | | NA | 0 |
| S15_vs_42177\|\|lt2gene | 1048 | No | NA | 2 | 1 | 0 | Low-quality | Genome-fragment | | 19.33 | AAI-based (high-confidence) | 0 |
| S17_vs_k141_127663\|\|full | 1358 | No | NA | 3 | 0 | 1 | Not-determined | Genome-fragment | NA | | NA | 0 |
| S17_vs_k141_135251\|\|full | 1088 | No | NA | 2 | 0 | 2 | Not-determined | Genome-fragment | NA | | NA | 0 |
| S8_vs_k141_46095\|\|full | 1048 | No | NA | 3 | 0 | 0 | Low-quality | Genome-fragment | | 2.48 | AAI-based (high-confidence) | 0 |
| S9_extracted_k141_75804 | 1049 | No | NA | 3 | 1 | 0 | Low-quality | Genome-fragment | | 2.24 | AAI-based (high-confidence) | 0 |
| S17_vs_k141_54483\|\|full | 1063 | No | NA | 4 | 0 | 1 | Not-determined | Genome-fragment | NA | | NA | 0 |
| S17_vs_k141_98661\|\|full | 3663 | No | NA | 5 | 0 | 3 | Not-determined | Genome-fragment | NA | | NA | 0 |
| S9_vs_k141_88850\|\|full | 1049 | No | NA | 2 | 0 | 0 | Low-quality | Genome-fragment | | 2.36 | AAI-based (medium-confidence) | 0 |
| S17_vs_k141_76223\|\|full | 2293 | No | NA | 3 | 0 | 2 | Not-determined | Genome-fragment | NA | | NA | 0 |
| S17_vs_k141_90885\|\|full | 16374 | No | NA | 9 | 0 | 7 | Not-determined | Genome-fragment | NA | | NA | 0 |
| S1_extracted_k141_7128 | 1049 | No | NA | 1 | 0 | 1 | Low-quality | Genome-fragment | | 2.05 | AAI-based (high-confidence) | 0 |
| S3_vs_k141_120317\|\|full | 1049 | No | NA | 4 | 0 | 0 | Low-quality | Genome-fragment | | 0.6 | AAI-based (high-confidence) | 0 |
| S3_vs_k141_123189\|\|lt2gene | 1049 | No | NA | 2 | 1 | 0 | Low-quality | Genome-fragment | | 1.67 | HMM-based (lower-bound) | 0 |
| S5_vs_k141_129802\|\|full | 1049 | No | NA | 2 | 1 | 0 | Low-quality | Genome-fragment | | 2.55 | AAI-based (high-confidence) | 0 |
| S7_extracted_72280 | 1049 | No | NA | 3 | 0 | 0 | Low-quality | Genome-fragment | | 1.94 | AAI-based (medium-confidence) | 0 |
| S7_extracted_98467 | 1049 | No | NA | 3 | 1 | 0 | Low-quality | Genome-fragment | | 16.58 | AAI-based (high-confidence) | 0 |
| S17_vs_k141_90924\|\|full | 3914 | No | NA | 4 | 0 | 3 | Not-determined | Genome-fragment | NA | | NA | 0 |
| S17_vs_k141_10067\|\|full | 1313 | No | NA | 2 | 0 | 2 | Not-determined | Genome-fragment | NA | | NA | 0 |
| S16_vs_152872\|\|full | 1049 | No | NA | 2 | 0 | 1 | Low-quality | Genome-fragment | | 1.41 | AAI-based (high-confidence) | 0 |
| S17_vs_k141_24394\|\|full | 1376 | No | NA | 3 | 0 | 1 | Not-determined | Genome-fragment | NA | | NA | 0 |
| S12_vs_k141_173976\|\|full | 1049 | No | NA | 2 | 0 | 0 | Low-quality | Genome-fragment | | 2.11 | AAI-based (high-confidence) | 0 |
| S15_extracted_67508 | 1049 | No | NA | 2 | 0 | 1 | Low-quality | Genome-fragment | | 0.99 | AAI-based (medium-confidence) | 0 |
| S17_vs_k141_54578\|\|full | 1640 | No | NA | 4 | 0 | 0 | Not-determined | Genome-fragment | NA | | NA | 0 |
| S17_vs_k141_157905\|\|full | 1856 | No | NA | 5 | 0 | 0 | Not-determined | Genome-fragment | NA | | NA | 0 |
| S17_vs_k141_90970\|\|full | 1703 | No | NA | 3 | 0 | 1 | Not-determined | Genome-fragment | NA | | NA | 0 |
| S17_vs_k141_47043\|\|full | 2708 | No | NA | 4 | 0 | 0 | Not-determined | Genome-fragment | NA | | NA | 0 |
| S17_extracted_k141_70719 | 1050 | No | NA | 2 | 0 | 0 | Low-quality | Genome-fragment | | 0.72 | AAI-based (medium-confidence) | 0 |
| S17_vs_k141_2369\|\|full | 2195 | No | NA | 4 | 0 | 0 | Not-determined | Genome-fragment | NA | | NA | 0 |
| S17_extracted_k141_34391 | 1050 | No | NA | 3 | 0 | 0 | Low-quality | Genome-fragment | | 2.48 | AAI-based (medium-confidence) | 0 |
| S17_vs_k141_106209\|\|full | 1050 | No | NA | 3 | 0 | 0 | Low-quality | Genome-fragment | | 0.44 | AAI-based (medium-confidence) | 0 |
| S17_vs_k141_157957\|\|full | 2826 | No | NA | 9 | 0 | 0 | Not-determined | Genome-fragment | NA | | NA | 0 |
| S17_vs_k141_120805\|\|full | 3394 | No | NA | 6 | 0 | 0 | Not-determined | Genome-fragment | NA | | NA | 0 |
| S17_vs_k141_47991\|\|full | 1050 | No | NA | 3 | 0 | 0 | Low-quality | Genome-fragment | | 1.42 | AAI-based (high-confidence) | 0 |
| S17_vs_k141_10139\|\|full | 1110 | No | NA | 2 | 0 | 1 | Not-determined | Genome-fragment | NA | | NA | 0 |
| S17_vs_k141_165035\|\|full | 4736 | No | NA | 3 | 0 | 3 | Not-determined | Genome-fragment | NA | | NA | 0 |
| S9_vs_k141_960\|\|lt2gene | 1050 | No | NA | 2 | 1 | 0 | Low-quality | Genome-fragment | | 2.86 | AAI-based (high-confidence) | 0 |
| S17_vs_k141_113165\|\|full | 1623 | No | NA | 2 | 0 | 0 | Not-determined | Genome-fragment | NA | | NA | 0 |
| S17_vs_k141_47126\|\|full | 8421 | No | NA | 9 | 0 | 2 | Not-determined | Genome-fragment | NA | | NA | 0 |
| S11_vs_k141_87074\|\|full | 1050 | No | NA | 2 | 0 | 0 | Low-quality | Genome-fragment | | 2.05 | AAI-based (high-confidence) | 0 |
| S1_extracted_k141_27617 | 1050 | No | NA | 2 | 0 | 0 | Low-quality | Genome-fragment | | 1.99 | AAI-based (high-confidence) | 0 |
| S3_vs_k141_39819\|\|full | 1050 | No | NA | 2 | 0 | 0 | Low-quality | Genome-fragment | | 2.13 | AAI-based (medium-confidence) | 0 |
| S17_vs_k141_127837\|\|full | 2039 | No | NA | 3 | 0 | 1 | Not-determined | Genome-fragment | NA | | NA | 0 |
| S17_vs_k141_32414\|\|full | 1992 | No | NA | 3 | 0 | 2 | Not-determined | Genome-fragment | NA | | NA | 0 |
| S5_extracted_k141_105048 | 1050 | No | NA | 1 | 0 | 0 | Low-quality | Genome-fragment | | 2.62 | AAI-based (high-confidence) | 0 |
| S5_extracted_k141_120259 | 1050 | No | NA | 2 | 1 | 0 | Low-quality | Genome-fragment | | 1.82 | AAI-based (medium-confidence) | 0 |
| S7_vs_121810\|\|lt2gene | 1050 | No | NA | 1 | 1 | 0 | Low-quality | Genome-fragment | | 1.21 | HMM-based (lower-bound) | 0 |
| S7_vs_134063\|\|lt2gene | 1050 | No | NA | 2 | 1 | 0 | Low-quality | Genome-fragment | | 2.31 | AAI-based (high-confidence) | 0 |
| S16_vs_84572\|\|full | 1050 | No | NA | 5 | 0 | 0 | Low-quality | Genome-fragment | | 1.31 | AAI-based (high-confidence) | 0 |
| S17_vs_k141_91135\|\|full | 2476 | No | NA | 3 | 0 | 0 | Not-determined | Genome-fragment | NA | | NA | 0 |
| S10_extracted_71736 | 1050 | No | NA | 3 | 0 | 0 | Low-quality | Genome-fragment | | 2.09 | AAI-based (high-confidence) | 0 |
| S12_vs_k141_80313\|\|lt2gene | 1050 | No | NA | 1 | 1 | 0 | Low-quality | Genome-fragment | | 3.12 | AAI-based (high-confidence) | 0 |
| S2_extracted_k141_18125 | 1050 | No | NA | 1 | 0 | 0 | Low-quality | Genome-fragment | | 2.4 | AAI-based (medium-confidence) | 0 |
| S8_extracted_k141_146915 | 1050 | No | NA | 2 | 0 | 1 | Low-quality | Genome-fragment | | 0.61 | AAI-based (medium-confidence) | 0 |
| S17_vs_k141_32472\|\|full | 5255 | No | NA | 3 | 0 | 3 | Not-determined | Genome-fragment | NA | | NA | 0 |
| S17_vs_k141_150947\|\|full | 2708 | No | NA | 3 | 0 | 0 | Not-determined | Genome-fragment | NA | | NA | 0 |
| S11_extracted_k141_37806 | 1051 | No | NA | 3 | 0 | 0 | Low-quality | Genome-fragment | | 1.56 | AAI-based (high-confidence) | 0 |
| S13_extracted_131425 | 1051 | No | NA | 2 | 0 | 0 | Low-quality | Genome-fragment | | 0.78 | AAI-based (medium-confidence) | 0 |
| S17_vs_k141_98933\|\|full | 6340 | No | NA | 7 | 0 | 3 | Not-determined | Genome-fragment | NA | | NA | 0 |
| S17_vs_k141_143306\|\|full | 1648 | No | NA | 4 | 0 | 0 | Not-determined | Genome-fragment | NA | | NA | 0 |
| S17_vs_k141_24563\|\|full | 2125 | No | NA | 4 | 0 | 2 | Not-determined | Genome-fragment | NA | | NA | 0 |
| S17_vs_k141_40130\|\|full | 2891 | No | NA | 3 | 0 | 1 | Not-determined | Genome-fragment | NA | | NA | 0 |
| S13_extracted_137445 | 1051 | No | NA | 1 | 0 | 1 | Low-quality | Genome-fragment | | 0.11 | AAI-based (medium-confidence) | 0 |
| S1_extracted_k141_99973 | 1051 | No | NA | 2 | 0 | 1 | Low-quality | Genome-fragment | | 0.94 | AAI-based (high-confidence) | 0 |
| S1_vs_k141_91764\|\|full | 1051 | No | NA | 3 | 0 | 0 | Low-quality | Genome-fragment | | 0.35 | AAI-based (medium-confidence) | 0 |
| S17_vs_k141_69269\|\|full | 1848 | No | NA | 2 | 0 | 1 | Not-determined | Genome-fragment | NA | | NA | 0 |
| S17_vs_k141_83373\|\|full | 4616 | No | NA | 6 | 0 | 2 | Not-determined | Genome-fragment | NA | | NA | 0 |
| S3_vs_k141_14890\|\|full | 1051 | No | NA | 3 | 0 | 1 | Low-quality | Genome-fragment | | 1.3 | AAI-based (medium-confidence) | 0 |
| S16_vs_133838\|\|full | 1051 | No | NA | 3 | 0 | 0 | Low-quality | Genome-fragment | | 2.94 | AAI-based (medium-confidence) | 0 |
| S10_extracted_104661 | 1051 | No | NA | 3 | 0 | 0 | Low-quality | Genome-fragment | | 2.36 | AAI-based (high-confidence) | 0 |
| S17_vs_k141_61543\|\|full | 1451 | No | NA | 3 | 0 | 0 | Not-determined | Genome-fragment | NA | | NA | 0 |
| S17_vs_k141_32568\|\|full | 1025 | No | NA | 2 | 0 | 0 | Not-determined | Genome-fragment | NA | | NA | 0 |
| S17_vs_k141_17137\|\|full | 2634 | No | NA | 5 | 0 | 1 | Not-determined | Genome-fragment | NA | | NA | 0 |
| S17_vs_k141_47316\|\|full | 2542 | No | NA | 2 | 0 | 2 | Not-determined | Genome-fragment | NA | | NA | 0 |
| S10_vs_84954\|\|lt2gene | 1051 | No | NA | 2 | 1 | 0 | Low-quality | Genome-fragment | | 2.31 | AAI-based (high-confidence) | 0 |
| S17_vs_k141_32599\|\|full | 1896 | No | NA | 2 | 0 | 1 | Not-determined | Genome-fragment | NA | | NA | 0 |
| S8_extracted_k141_64148 | 1051 | No | NA | 2 | 0 | 2 | Low-quality | Genome-fragment | | 0.8 | AAI-based (medium-confidence) | 0 |
| S17_vs_k141_2625\|\|full | 2967 | No | NA | 3 | 0 | 1 | Not-determined | Genome-fragment | NA | | NA | 0 |
| S8_vs_k141_78705\|\|full | 1051 | No | NA | 3 | 0 | 0 | Low-quality | Genome-fragment | | 0.91 | AAI-based (medium-confidence) | 0 |
| S8_vs_k141_98624\|\|lt2gene | 1051 | No | NA | 1 | 1 | 0 | Low-quality | Genome-fragment | | 2.31 | AAI-based (high-confidence) | 0 |
| S17_vs_k141_83471\|\|full | 1144 | No | NA | 3 | 0 | 0 | Not-determined | Genome-fragment | NA | | NA | 0 |
| S17_vs_k141_32647\|\|full | 1601 | No | NA | 2 | 0 | 0 | Not-determined | Genome-fragment | NA | | NA | 0 |
| S17_vs_k141_99040\|\|full | 2676 | No | NA | 3 | 0 | 3 | Not-determined | Genome-fragment | NA | | NA | 0 |
| S17_vs_k141_106787\|\|full | 1594 | No | NA | 2 | 0 | 0 | Not-determined | Genome-fragment | NA | | NA | 0 |
| S9_extracted_k141_53740 | 1052 | No | NA | 1 | 0 | 1 | Low-quality | Genome-fragment | | 0.29 | AAI-based (medium-confidence) | 0 |
| S17_vs_k141_76609\|\|full | 1941 | No | NA | 3 | 0 | 2 | Not-determined | Genome-fragment | NA | | NA | 0 |
| S17_vs_k141_113337\|\|full | 1090 | No | NA | 2 | 0 | 0 | Not-determined | Genome-fragment | NA | | NA | 0 |
| S13_vs_33660\|\|lt2gene | 1052 | No | NA | 1 | 1 | 0 | Low-quality | Genome-fragment | | 2.21 | HMM-based (lower-bound) | 0 |
| S17_vs_k141_61609\|\|full | 3164 | No | NA | 5 | 0 | 0 | Not-determined | Genome-fragment | NA | | NA | 0 |
| S17_vs_k141_40268\|\|full | 4471 | No | NA | 5 | 0 | 0 | Not-determined | Genome-fragment | NA | | NA | 0 |
| S5_vs_k141_119230\|\|full | 1052 | No | NA | 3 | 0 | 0 | Low-quality | Genome-fragment | | 3.01 | AAI-based (medium-confidence) | 0 |
| S17_vs_k141_69420\|\|full | 3055 | No | NA | 3 | 0 | 3 | Not-determined | Genome-fragment | NA | | NA | 0 |
| S17_vs_k141_106822\|\|full | 1326 | No | NA | 3 | 0 | 1 | Not-determined | Genome-fragment | NA | | NA | 0 |
| S17_vs_k141_113378\|\|full | 3339 | No | NA | 6 | 0 | 0 | Not-determined | Genome-fragment | NA | | NA | 0 |
| S7_extracted_21812 | 1052 | No | NA | 4 | 0 | 0 | Low-quality | Genome-fragment | | 2.21 | AAI-based (high-confidence) | 0 |
| S7_vs_66685\|\|full | 1052 | No | NA | 2 | 0 | 1 | Low-quality | Genome-fragment | | 0.98 | AAI-based (medium-confidence) | 0 |
| S7_vs_130592\|\|full | 1052 | No | NA | 3 | 0 | 0 | Low-quality | Genome-fragment | | 2.41 | AAI-based (high-confidence) | 0 |
| S17_vs_k141_54682\|\|full | 4092 | No | NA | 5 | 0 | 5 | Not-determined | Genome-fragment | NA | | NA | 0 |
| S17_vs_k141_113389\|\|full | 11255 | No | NA | 8 | 0 | 3 | Not-determined | Genome-fragment | NA | | NA | 0 |
| S17_vs_k141_128103\|\|full | 12626 | No | NA | 11 | 0 | 1 | Not-determined | Genome-fragment | NA | | NA | 0 |
| S17_vs_k141_10304\|\|full | 2538 | No | NA | 2 | 0 | 1 | Not-determined | Genome-fragment | NA | | NA | 0 |
| S17_vs_k141_121041\|\|full | 34329 | No | NA | 65 | 19 | 0 | Medium-quality | Genome-fragment | | 72.74 | AAI-based (high-confidence) | 0 |
| S16_extracted_41482 | 1052 | No | NA | 3 | 1 | 0 | Low-quality | Genome-fragment | | 0.96 | HMM-based (lower-bound) | 0 |
| S12_extracted_k141_16034 | 1052 | No | NA | 1 | 0 | 1 | Low-quality | Genome-fragment | | 0.47 | AAI-based (medium-confidence) | 0 |
| S17_vs_k141_135708\|\|full | 3275 | No | NA | 3 | 0 | 2 | Not-determined | Genome-fragment | NA | | NA | 0 |
| S17_vs_k141_10324\|\|full | 2223 | No | NA | 4 | 0 | 2 | Not-determined | Genome-fragment | NA | | NA | 0 |
| S12_extracted_k141_137914 | 1052 | No | NA | 2 | 1 | 0 | Low-quality | Genome-fragment | | 1.06 | AAI-based (medium-confidence) | 0 |
| S15_extracted_12472 | 1052 | No | NA | 3 | 0 | 0 | Low-quality | Genome-fragment | | 0.61 | AAI-based (high-confidence) | 0 |
| S17_vs_k141_97347\|\|full | 1053 | No | NA | 2 | 0 | 2 | Low-quality | Genome-fragment | | 1.77 | AAI-based (medium-confidence) | 0 |
| S17_vs_k141_83550\|\|full | 1152 | No | NA | 3 | 0 | 1 | Not-determined | Genome-fragment | NA | | NA | 0 |
| S17_vs_k141_61720\|\|full | 2718 | No | NA | 3 | 0 | 0 | Not-determined | Genome-fragment | NA | | NA | 0 |
| S17_vs_k141_17307\|\|full | 1374 | No | NA | 2 | 0 | 2 | Not-determined | Genome-fragment | NA | | NA | 0 |
| S11_vs_k141_34808\|\|full | 1053 | No | NA | 3 | 2 | 0 | Low-quality | Genome-fragment | | 1.93 | AAI-based (high-confidence) | 0 |
| S17_vs_k141_61726\|\|full | 2777 | No | NA | 3 | 0 | 2 | Not-determined | Genome-fragment | NA | | NA | 0 |
| S11_vs_k141_12685\|\|full | 1053 | No | NA | 3 | 0 | 0 | Low-quality | Genome-fragment | | 2.92 | AAI-based (medium-confidence) | 0 |
| S17_vs_k141_113436\|\|full | 1861 | No | NA | 2 | 0 | 1 | Not-determined | Genome-fragment | NA | | NA | 0 |
| S14_extracted_k141_29162 | 1053 | No | NA | 2 | 2 | 0 | Low-quality | Genome-fragment | | 2.37 | AAI-based (high-confidence) | 0 |
| S17_vs_k141_61752\|\|full | 3289 | No | NA | 5 | 0 | 1 | Not-determined | Genome-fragment | NA | | NA | 0 |
| S17_vs_k141_165348\|\|full | 3266 | No | NA | 3 | 0 | 3 | Not-determined | Genome-fragment | NA | | NA | 0 |
| S17_vs_k141_61765\|\|full | 1157 | No | NA | 2 | 0 | 1 | Not-determined | Genome-fragment | NA | | NA | 0 |
| S17_vs_k141_47560\|\|full | 1961 | No | NA | 7 | 0 | 0 | Not-determined | Genome-fragment | NA | | NA | 0 |
| S17_vs_k141_135783\|\|full | 1733 | No | NA | 3 | 0 | 0 | Not-determined | Genome-fragment | NA | | NA | 0 |
| S3_vs_k141_35594\|\|lt2gene | 1053 | No | NA | 2 | 0 | 0 | Low-quality | Genome-fragment | | 1.78 | AAI-based (high-confidence) | 0 |
| S3_vs_k141_4879\|\|lt2gene | 1053 | No | NA | 1 | 1 | 0 | Low-quality | Genome-fragment | | 2.39 | AAI-based (high-confidence) | 0 |
| S17_vs_k141_32731\|\|full | 1295 | No | NA | 3 | 0 | 1 | Not-determined | Genome-fragment | NA | | NA | 0 |
| S17_vs_k141_99249\|\|full | 1729 | No | NA | 2 | 0 | 2 | Not-determined | Genome-fragment | NA | | NA | 0 |
| S5_vs_k141_88980\|\|full | 1053 | No | NA | 2 | 0 | 0 | Low-quality | Genome-fragment | | 1.77 | AAI-based (high-confidence) | 0 |
| S7_vs_39023\|\|full | 1053 | No | NA | 3 | 1 | 0 | Low-quality | Genome-fragment | | 2.72 | AAI-based (medium-confidence) | 0 |
| S16_vs_74218\|\|full | 1053 | No | NA | 3 | 1 | 0 | Low-quality | Genome-fragment | | 1.59 | HMM-based (lower-bound) | 0 |
| S10_extracted_21761 | 1053 | No | NA | 2 | 0 | 0 | Low-quality | Genome-fragment | | 0.97 | AAI-based (high-confidence) | 0 |
| S17_vs_k141_121107\|\|full | 2013 | No | NA | 3 | 0 | 2 | Not-determined | Genome-fragment | NA | | NA | 0 |
| S17_vs_k141_135822\|\|full | 2265 | No | NA | 5 | 0 | 1 | Not-determined | Genome-fragment | NA | | NA | 0 |
| S17_vs_k141_143643\|\|full | 18147 | No | NA | 25 | 0 | 9 | Not-determined | Genome-fragment | NA | | NA | 0 |
| S17_vs_k141_121119\|\|full | 7296 | No | NA | 11 | 0 | 1 | Not-determined | Genome-fragment | NA | | NA | 0 |
| S17_vs_k141_40477\|\|full | 1189 | No | NA | 2 | 0 | 2 | Not-determined | Genome-fragment | NA | | NA | 0 |
| S12_vs_k141_175421\|\|full | 1053 | No | NA | 3 | 0 | 0 | Low-quality | Genome-fragment | | 0.75 | AAI-based (medium-confidence) | 0 |
| S17_vs_k141_143657\|\|full | 2675 | No | NA | 3 | 0 | 3 | Not-determined | Genome-fragment | NA | | NA | 0 |
| S15_vs_60020\|\|lt2gene | 1053 | No | NA | 1 | 0 | 0 | Low-quality | Genome-fragment | | 2.39 | AAI-based (high-confidence) | 0 |
| S2_extracted_k141_597 | 1053 | No | NA | 5 | 0 | 0 | Low-quality | Genome-fragment | | 16.92 | AAI-based (high-confidence) | 0 |
| S17_vs_k141_143665\|\|full | 2460 | No | NA | 5 | 0 | 2 | Not-determined | Genome-fragment | NA | | NA | 0 |
| S17_vs_k141_99346\|\|full | 1104 | No | NA | 2 | 0 | 2 | Not-determined | Genome-fragment | NA | | NA | 0 |
| S17_vs_k141_107017\|\|full | 5514 | No | NA | 7 | 0 | 5 | Not-determined | Genome-fragment | NA | | NA | 0 |
| S17_vs_k141_91533\|\|full | 1938 | No | NA | 5 | 0 | 1 | Not-determined | Genome-fragment | NA | | NA | 0 |
| S17_vs_k141_128287\|\|full | 1063 | No | NA | 2 | 0 | 1 | Not-determined | Genome-fragment | NA | | NA | 0 |
| S8_vs_k141_60654\|\|full | 1053 | No | NA | 3 | 0 | 0 | Low-quality | Genome-fragment | | 2.36 | AAI-based (high-confidence) | 0 |
| S17_vs_k141_7831\|\|full | 1054 | No | NA | 4 | 0 | 0 | Low-quality | Genome-fragment | | 2.44 | AAI-based (medium-confidence) | 0 |
| S17_vs_k141_26285\|\|full | 1054 | No | NA | 2 | 0 | 0 | Low-quality | Genome-fragment | | 2 | AAI-based (high-confidence) | 0 |
| S14_extracted_k141_110487 | 1054 | No | NA | 5 | 2 | 0 | Low-quality | Genome-fragment | | 2.11 | AAI-based (high-confidence) | 0 |
| S17_vs_k141_24943\|\|full | 4386 | No | NA | 9 | 0 | 3 | Not-determined | Genome-fragment | NA | | NA | 0 |
| S17_vs_k141_158356\|\|full | 1323 | No | NA | 2 | 0 | 1 | Not-determined | Genome-fragment | NA | | NA | 0 |
| S14_extracted_k141_97949 | 1054 | No | NA | 2 | 2 | 0 | Low-quality | Genome-fragment | | 3.12 | AAI-based (high-confidence) | 0 |
| S17_vs_k141_17493\|\|full | 1585 | No | NA | 2 | 0 | 0 | Not-determined | Genome-fragment | NA | | NA | 0 |
| S17_vs_k141_113670\|\|full | 1844 | No | NA | 2 | 0 | 2 | Not-determined | Genome-fragment | NA | | NA | 0 |
| S14_extracted_k141_71120 | 1054 | No | NA | 2 | 1 | 0 | Low-quality | Genome-fragment | | 1.89 | HMM-based (lower-bound) | 0 |
| S17_vs_k141_24989\|\|full | 3139 | No | NA | 4 | 0 | 3 | Not-determined | Genome-fragment | NA | | NA | 0 |
| S1_extracted_k141_85452 | 1054 | No | NA | 1 | 0 | 0 | Low-quality | Genome-fragment | | 1.09 | AAI-based (medium-confidence) | 0 |
| S1_extracted_k141_119607 | 1054 | No | NA | 4 | 0 | 0 | Low-quality | Genome-fragment | | 1.05 | AAI-based (high-confidence) | 0 |
| S1_vs_k141_92661\|\|full | 1054 | No | NA | 2 | 0 | 1 | Low-quality | Genome-fragment | | 1.65 | AAI-based (medium-confidence) | 0 |
| S3_vs_k141_63101\|\|full | 1054 | No | NA | 4 | 1 | 0 | Low-quality | Genome-fragment | | 1.96 | AAI-based (high-confidence) | 0 |
| S17_vs_k141_113746\|\|full | 1142 | No | NA | 2 | 0 | 2 | Not-determined | Genome-fragment | NA | | NA | 0 |
| S17_vs_k141_3053\|\|full | 1501 | No | NA | 3 | 0 | 0 | Not-determined | Genome-fragment | NA | | NA | 0 |
| S16_extracted_89665 | 1054 | No | NA | 3 | 1 | 0 | Low-quality | Genome-fragment | | 1.41 | AAI-based (high-confidence) | 0 |
| S16_vs_24263\|\|full | 1054 | No | NA | 2 | 0 | 0 | Low-quality | Genome-fragment | | 1.59 | AAI-based (medium-confidence) | 0 |
| S17_vs_k141_158480\|\|full | 1625 | No | NA | 2 | 0 | 0 | Not-determined | Genome-fragment | NA | | NA | 0 |
| S17_vs_k141_107231\|\|full | 2096 | No | NA | 2 | 0 | 2 | Not-determined | Genome-fragment | NA | | NA | 0 |
| S10_extracted_14471 | 1054 | No | NA | 2 | 0 | 0 | Low-quality | Genome-fragment | | 1.04 | AAI-based (medium-confidence) | 0 |
| S10_vs_106229\|\|full | 1054 | No | NA | 3 | 0 | 0 | Low-quality | Genome-fragment | | 2.25 | AAI-based (medium-confidence) | 0 |
| S12_extracted_k141_126369 | 1054 | No | NA | 1 | 0 | 1 | Low-quality | Genome-fragment | | 2.97 | AAI-based (medium-confidence) | 0 |
| S12_extracted_k141_27289 | 1054 | No | NA | 1 | 0 | 0 | Low-quality | Genome-fragment | | 1.27 | AAI-based (high-confidence) | 0 |
| S17_vs_k141_62032\|\|full | 2884 | No | NA | 3 | 0 | 2 | Not-determined | Genome-fragment | NA | | NA | 0 |
| S6_extracted_k141_40989 | 1054 | No | NA | 2 | 1 | 0 | Low-quality | Genome-fragment | | 2.26 | AAI-based (high-confidence) | 0 |
| S17_vs_k141_3109\|\|full | 3171 | No | NA | 2 | 0 | 2 | Not-determined | Genome-fragment | NA | | NA | 0 |
| S8_extracted_k141_133252 | 1054 | No | NA | 3 | 0 | 1 | Low-quality | Genome-fragment | | 0.67 | AAI-based (medium-confidence) | 0 |
| S17_vs_k141_121345\|\|full | 5794 | No | NA | 5 | 0 | 4 | Not-determined | Genome-fragment | NA | | NA | 0 |
| S17_vs_k141_10723\|\|full | 4730 | No | NA | 4 | 0 | 1 | Not-determined | Genome-fragment | NA | | NA | 0 |
| S8_vs_k141_202469\|\|full | 1054 | No | NA | 2 | 0 | 0 | Low-quality | Genome-fragment | | 2.34 | AAI-based (high-confidence) | 0 |
| S17_extracted_k141_10115 | 1055 | No | NA | 3 | 0 | 0 | Low-quality | Genome-fragment | | 1.73 | AAI-based (high-confidence) | 0 |
| S9_extracted_k141_30789 | 1055 | No | NA | 2 | 0 | 2 | Low-quality | Genome-fragment | | 0.58 | AAI-based (medium-confidence) | 0 |
| S17_vs_k141_69850\|\|full | 5944 | No | NA | 6 | 0 | 0 | Not-determined | Genome-fragment | NA | | NA | 0 |
| S11_extracted_k141_18389 | 1055 | No | NA | 1 | 0 | 0 | Low-quality | Genome-fragment | | 2.49 | AAI-based (high-confidence) | 0 |
| S17_vs_k141_25100\|\|full | 4648 | No | NA | 6 | 0 | 0 | Not-determined | Genome-fragment | NA | | NA | 0 |
| S17_vs_k141_128520\|\|full | 3502 | No | NA | 4 | 0 | 3 | Not-determined | Genome-fragment | NA | | NA | 0 |
| S13_extracted_153455 | 1055 | No | NA | 2 | 0 | 0 | Low-quality | Genome-fragment | | 1.6 | AAI-based (high-confidence) | 0 |
| S17_vs_k141_151453\|\|full | 3291 | No | NA | 3 | 0 | 2 | Not-determined | Genome-fragment | NA | | NA | 0 |
| S1_vs_k141_87697\|\|full | 1055 | No | NA | 3 | 0 | 0 | Low-quality | Genome-fragment | | 1.79 | AAI-based (high-confidence) | 0 |
| S17_vs_k141_107298\|\|full | 1176 | No | NA | 3 | 0 | 1 | Not-determined | Genome-fragment | NA | | NA | 0 |
| S17_vs_k141_25117\|\|full | 1214 | No | NA | 2 | 0 | 2 | Not-determined | Genome-fragment | NA | | NA | 0 |
| S17_vs_k141_107307\|\|full | 1198 | No | NA | 2 | 0 | 1 | Not-determined | Genome-fragment | NA | | NA | 0 |
| S17_vs_k141_25121\|\|full | 1609 | No | NA | 2 | 0 | 0 | Not-determined | Genome-fragment | NA | | NA | 0 |
| S17_vs_k141_128574\|\|full | 1377 | No | NA | 3 | 0 | 1 | Not-determined | Genome-fragment | NA | | NA | 0 |
| S3_vs_k141_58041\|\|full | 1055 | No | NA | 2 | 0 | 1 | Low-quality | Genome-fragment | | 1.74 | AAI-based (medium-confidence) | 0 |
| S5_vs_k141_50144\|\|full | 1055 | No | NA | 3 | 1 | 0 | Low-quality | Genome-fragment | | 1.75 | AAI-based (high-confidence) | 0 |
| S17_vs_k141_172925\|\|full | 4853 | No | NA | 4 | 0 | 3 | Not-determined | Genome-fragment | NA | | NA | 0 |
| S17_vs_k141_136057\|\|full | 8893 | No | NA | 12 | 0 | 2 | Not-determined | Genome-fragment | NA | | NA | 0 |
| S17_vs_k141_10796\|\|full | 2327 | No | NA | 3 | 0 | 2 | Not-determined | Genome-fragment | NA | | NA | 0 |
| S17_vs_k141_3183\|\|full | 1127 | No | NA | 3 | 0 | 0 | Not-determined | Genome-fragment | NA | | NA | 0 |
| S5_vs_k141_69264\|\|full | 1055 | No | NA | 3 | 1 | 0 | Low-quality | Genome-fragment | | 1.58 | HMM-based (lower-bound) | 0 |
| S17_vs_k141_33113\|\|full | 1262 | No | NA | 3 | 0 | 2 | Not-determined | Genome-fragment | NA | | NA | 0 |
| S17_vs_k141_91805\|\|full | 2989 | No | NA | 3 | 0 | 1 | Not-determined | Genome-fragment | NA | | NA | 0 |
| S17_vs_k141_17787\|\|full | 6708 | No | NA | 6 | 0 | 5 | Not-determined | Genome-fragment | NA | | NA | 0 |
| S7_vs_75734\|\|lt2gene | 1055 | No | NA | 2 | 0 | 0 | Low-quality | Genome-fragment | | 2.84 | AAI-based (medium-confidence) | 0 |
| S16_vs_2073\|\|full | 1055 | No | NA | 2 | 0 | 0 | Low-quality | Genome-fragment | | 2.06 | AAI-based (high-confidence) | 0 |
| S16_vs_40286\|\|full | 1055 | No | NA | 4 | 0 | 0 | Low-quality | Genome-fragment | | 2.21 | AAI-based (high-confidence) | 0 |
| S12_extracted_k141_135897 | 1055 | No | NA | 1 | 0 | 1 | Low-quality | Genome-fragment | | 0.25 | AAI-based (medium-confidence) | 0 |
| S12_vs_k141_155319\|\|full | 1055 | No | NA | 2 | 0 | 0 | Low-quality | Genome-fragment | | 1.86 | AAI-based (medium-confidence) | 0 |
| S17_vs_k141_48007\|\|full | 1359 | No | NA | 3 | 0 | 2 | Not-determined | Genome-fragment | NA | | NA | 0 |
| S8_extracted_k141_110601 | 1055 | No | NA | 3 | 0 | 0 | Low-quality | Genome-fragment | | 1.92 | AAI-based (medium-confidence) | 0 |
| S17_vs_k141_113983\|\|full | 2211 | No | NA | 4 | 0 | 0 | Not-determined | Genome-fragment | NA | | NA | 0 |
| S17_vs_k141_128661\|\|full | 1133 | No | NA | 2 | 0 | 1 | Not-determined | Genome-fragment | NA | | NA | 0 |
| S17_vs_k141_55292\|\|full | 4166 | No | NA | 3 | 0 | 3 | Not-determined | Genome-fragment | NA | | NA | 0 |
| S17_vs_k141_10860\|\|full | 1082 | No | NA | 2 | 0 | 2 | Not-determined | Genome-fragment | NA | | NA | 0 |
| S17_vs_k141_144023\|\|full | 1517 | No | NA | 3 | 0 | 1 | Not-determined | Genome-fragment | NA | | NA | 0 |
| S8_vs_k141_150814\|\|full | 1055 | No | NA | 2 | 0 | 1 | Low-quality | Genome-fragment | | 1.53 | AAI-based (medium-confidence) | 0 |
| S8_vs_k141_118838\|\|full | 1055 | No | NA | 3 | 0 | 1 | Low-quality | Genome-fragment | | 1.02 | AAI-based (medium-confidence) | 0 |
| S17_vs_k141_121542\|\|full | 1823 | No | NA | 2 | 0 | 1 | Not-determined | Genome-fragment | NA | | NA | 0 |
| S8_vs_k141_180044\|\|full | 1055 | No | NA | 2 | 0 | 2 | Low-quality | Genome-fragment | | 2.38 | AAI-based (medium-confidence) | 0 |
| S8_vs_k141_14823\|\|full | 1055 | No | NA | 2 | 0 | 0 | Low-quality | Genome-fragment | | 2.35 | AAI-based (high-confidence) | 0 |
| S17_vs_k141_77305\|\|full | 3723 | No | NA | 4 | 0 | 1 | Not-determined | Genome-fragment | NA | | NA | 0 |
| S17_extracted_k141_172645 | 1056 | No | NA | 1 | 0 | 0 | Low-quality | Genome-fragment | | 2.7 | AAI-based (high-confidence) | 0 |
| S17_vs_k141_48093\|\|full | 1082 | No | NA | 2 | 0 | 0 | Not-determined | Genome-fragment | NA | | NA | 0 |
| S17_vs_k141_99736\|\|full | 1215 | No | NA | 3 | 0 | 1 | Not-determined | Genome-fragment | NA | | NA | 0 |
| S17_vs_k141_3415\|\|full | 2339 | No | NA | 3 | 0 | 3 | Not-determined | Genome-fragment | NA | | NA | 0 |
| S17_vs_k141_151576\|\|full | 3618 | No | NA | 4 | 0 | 2 | Not-determined | Genome-fragment | NA | | NA | 0 |
| S17_vs_k141_114072\|\|full | 2054 | No | NA | 2 | 0 | 0 | Not-determined | Genome-fragment | NA | | NA | 0 |
| S17_extracted_k141_48131 | 1056 | No | NA | 1 | 0 | 1 | Low-quality | Genome-fragment | | 0.28 | AAI-based (high-confidence) | 0 |
| S17_extracted_k141_64033 | 1056 | No | NA | 2 | 1 | 0 | Low-quality | Genome-fragment | | 1.94 | AAI-based (high-confidence) | 0 |
| S17_vs_k141_128784\|\|full | 1423 | No | NA | 2 | 0 | 0 | Not-determined | Genome-fragment | NA | | NA | 0 |
| S9_extracted_k141_42461 | 1056 | No | NA | 1 | 0 | 1 | Low-quality | Genome-fragment | | 0.37 | AAI-based (medium-confidence) | 0 |
| S17_vs_k141_70145\|\|full | 2275 | No | NA | 2 | 0 | 2 | Not-determined | Genome-fragment | NA | | NA | 0 |
| S9_extracted_k141_76907 | 1056 | No | NA | 2 | 0 | 2 | Low-quality | Genome-fragment | | 0.89 | AAI-based (medium-confidence) | 0 |
| S9_vs_k141_11803\|\|lt2gene | 1056 | No | NA | 2 | 2 | 0 | Low-quality | Genome-fragment | | 2.75 | AAI-based (medium-confidence) | 0 |
| S11_vs_k141_89126\|\|lt2gene | 1056 | No | NA | 1 | 1 | 0 | Low-quality | Genome-fragment | | 2.28 | AAI-based (high-confidence) | 0 |
| S13_vs_126828\|\|full | 1056 | No | NA | 3 | 0 | 0 | Low-quality | Genome-fragment | | 0.62 | AAI-based (high-confidence) | 0 |
| S17_vs_k141_99815\|\|full | 1613 | No | NA | 1 | 0 | 1 | Not-determined | Genome-fragment | NA | | NA | 0 |
| S17_vs_k141_77403\|\|full | 1033 | No | NA | 3 | 0 | 2 | Not-determined | Genome-fragment | NA | | NA | 0 |
| S17_vs_k141_25322\|\|full | 1392 | No | NA | 4 | 0 | 0 | Not-determined | Genome-fragment | NA | | NA | 0 |
| S17_vs_k141_33342\|\|full | 1516 | No | NA | 2 | 0 | 1 | Not-determined | Genome-fragment | NA | | NA | 0 |
| S17_vs_k141_40965\|\|full | 2693 | No | NA | 3 | 0 | 1 | Not-determined | Genome-fragment | NA | | NA | 0 |
| S17_vs_k141_18017\|\|full | 2038 | No | NA | 3 | 0 | 1 | Not-determined | Genome-fragment | NA | | NA | 0 |
| S17_vs_k141_158876\|\|full | 2418 | No | NA | 4 | 0 | 0 | Not-determined | Genome-fragment | NA | | NA | 0 |
| S17_vs_k141_128820\|\|full | 2482 | No | NA | 3 | 0 | 0 | Not-determined | Genome-fragment | NA | | NA | 0 |
| S17_vs_k141_121617\|\|full | 1423 | No | NA | 2 | 0 | 0 | Not-determined | Genome-fragment | NA | | NA | 0 |
| S14_extracted_k141_76193 | 1056 | No | NA | 1 | 0 | 1 | Low-quality | Genome-fragment | | 0.71 | AAI-based (high-confidence) | 0 |
| S17_vs_k141_158911\|\|full | 1660 | No | NA | 2 | 0 | 0 | Not-determined | Genome-fragment | NA | | NA | 0 |
| S17_vs_k141_25377\|\|full | 2754 | No | NA | 2 | 0 | 1 | Not-determined | Genome-fragment | NA | | NA | 0 |
| S14_vs_k141_43337\|\|full | 1056 | No | NA | 3 | 0 | 2 | Low-quality | Genome-fragment | | 0.62 | AAI-based (medium-confidence) | 0 |
| S14_vs_k141_36175\|\|lt2gene | 1056 | No | NA | 1 | 1 | 0 | Low-quality | Genome-fragment | | 1.7 | HMM-based (lower-bound) | 0 |
| S17_vs_k141_25387\|\|full | 1054 | No | NA | 3 | 0 | 1 | Not-determined | Genome-fragment | NA | | NA | 0 |
| S17_vs_k141_114181\|\|full | 1436 | No | NA | 3 | 0 | 1 | Not-determined | Genome-fragment | NA | | NA | 0 |
| S1_extracted_k141_102343 | 1056 | No | NA | 3 | 0 | 0 | Low-quality | Genome-fragment | | 0.69 | AAI-based (high-confidence) | 0 |
| S17_vs_k141_48200\|\|full | 1320 | No | NA | 1 | 0 | 0 | Not-determined | Genome-fragment | NA | | NA | 0 |
| S3_extracted_k141_117065 | 1056 | No | NA | 3 | 3 | 0 | Low-quality | Genome-fragment | | 3.48 | AAI-based (high-confidence) | 0 |
| S3_vs_k141_969\|\|full | 1056 | No | NA | 3 | 1 | 0 | Low-quality | Genome-fragment | | 1.73 | AAI-based (high-confidence) | 0 |
| S3_vs_k141_87651\|\|lt2gene | 1056 | No | NA | 1 | 1 | 0 | Low-quality | Genome-fragment | | 0.95 | HMM-based (lower-bound) | 0 |
| S7_vs_117612\|\|full | 1056 | No | NA | 3 | 0 | 0 | Low-quality | Genome-fragment | | 3.01 | AAI-based (medium-confidence) | 0 |
| S17_vs_k141_62378\|\|full | 1672 | No | NA | 3 | 0 | 0 | Not-determined | Genome-fragment | NA | | NA | 0 |
| S17_vs_k141_62385\|\|full | 1104 | No | NA | 2 | 0 | 1 | Not-determined | Genome-fragment | NA | | NA | 0 |
| S7_vs_71477\|\|lt2gene | 1056 | No | NA | 2 | 0 | 1 | Low-quality | Genome-fragment | | 1.4 | AAI-based (medium-confidence) | 0 |
| S17_vs_k141_173285\|\|full | 1092 | No | NA | 2 | 0 | 1 | Not-determined | Genome-fragment | NA | | NA | 0 |
| S10_vs_108870\|\|full | 1056 | No | NA | 2 | 0 | 1 | Low-quality | Genome-fragment | | 0.66 | AAI-based (medium-confidence) | 0 |
| S17_vs_k141_62399\|\|full | 4343 | No | NA | 2 | 0 | 1 | Not-determined | Genome-fragment | NA | | NA | 0 |
| S17_vs_k141_55472\|\|full | 1039 | No | NA | 2 | 0 | 0 | Not-determined | Genome-fragment | NA | | NA | 0 |
| S17_vs_k141_173300\|\|full | 4591 | No | NA | 4 | 0 | 3 | Not-determined | Genome-fragment | NA | | NA | 0 |
| S17_vs_k141_62406\|\|full | 3543 | No | NA | 5 | 0 | 2 | Not-determined | Genome-fragment | NA | | NA | 0 |
| S17_vs_k141_33450\|\|full | 1879 | No | NA | 3 | 0 | 2 | Not-determined | Genome-fragment | NA | | NA | 0 |
| S17_vs_k141_48255\|\|full | 2180 | No | NA | 3 | 0 | 0 | Not-determined | Genome-fragment | NA | | NA | 0 |
| S17_vs_k141_25462\|\|full | 3423 | No | NA | 9 | 0 | 0 | Not-determined | Genome-fragment | NA | | NA | 0 |
| S17_vs_k141_62429\|\|full | 1545 | No | NA | 2 | 0 | 1 | Not-determined | Genome-fragment | NA | | NA | 0 |
| S17_vs_k141_136436\|\|full | 7964 | No | NA | 5 | 0 | 1 | Not-determined | Genome-fragment | NA | | NA | 0 |
| S12_vs_k141_88342\|\|full | 1056 | No | NA | 2 | 1 | 0 | Low-quality | Genome-fragment | | 1.9 | AAI-based (medium-confidence) | 0 |
| S17_vs_k141_165992\|\|full | 1114 | No | NA | 3 | 0 | 1 | Not-determined | Genome-fragment | NA | | NA | 0 |
| S17_vs_k141_165993\|\|full | 1729 | No | NA | 2 | 0 | 1 | Not-determined | Genome-fragment | NA | | NA | 0 |
| S12_vs_k141_46813\|\|full | 1056 | No | NA | 3 | 0 | 1 | Low-quality | Genome-fragment | | 1.92 | AAI-based (medium-confidence) | 0 |
| S17_vs_k141_84197\|\|full | 2189 | No | NA | 3 | 0 | 0 | Not-determined | Genome-fragment | NA | | NA | 0 |
| S15_extracted_66338 | 1056 | No | NA | 1 | 0 | 1 | Low-quality | Genome-fragment | | 0.74 | AAI-based (medium-confidence) | 0 |
| S17_vs_k141_92127\|\|full | 1269 | No | NA | 2 | 0 | 0 | Not-determined | Genome-fragment | NA | | NA | 0 |
| S17_vs_k141_114220\|\|full | 21659 | No | NA | 24 | 0 | 17 | Not-determined | Genome-fragment | NA | | NA | 0 |
| S15_vs_25502\|\|full | 1056 | No | NA | 2 | 0 | 0 | Low-quality | Genome-fragment | | 0.83 | AAI-based (medium-confidence) | 0 |
| S17_vs_k141_166017\|\|full | 3894 | No | NA | 2 | 0 | 1 | Not-determined | Genome-fragment | NA | | NA | 0 |
| S17_vs_k141_92130\|\|full | 3412 | No | NA | 4 | 0 | 3 | Not-determined | Genome-fragment | NA | | NA | 0 |
| S17_vs_k141_48290\|\|full | 1751 | No | NA | 3 | 0 | 0 | Not-determined | Genome-fragment | NA | | NA | 0 |
| S17_vs_k141_3566\|\|full | 2839 | No | NA | 3 | 0 | 1 | Not-determined | Genome-fragment | NA | | NA | 0 |
| S8_vs_k141_81267\|\|full | 1056 | No | NA | 3 | 1 | 0 | Low-quality | Genome-fragment | | 2.56 | AAI-based (medium-confidence) | 0 |
| S17_extracted_k141_82324 | 1057 | No | NA | 2 | 2 | 0 | Low-quality | Genome-fragment | | 3.47 | AAI-based (high-confidence) | 0 |
| S17_vs_k141_84214\|\|full | 6444 | No | NA | 14 | 0 | 3 | Not-determined | Genome-fragment | NA | | NA | 0 |
| S17_vs_k141_9983\|\|full | 1057 | No | NA | 2 | 1 | 0 | Low-quality | Genome-fragment | | 2.03 | AAI-based (medium-confidence) | 0 |
| S17_vs_k141_11011\|\|full | 15698 | No | NA | 12 | 0 | 11 | Not-determined | Genome-fragment | NA | | NA | 0 |
| S17_vs_k141_26407\|\|full | 1057 | No | NA | 2 | 0 | 0 | Low-quality | Genome-fragment | | 0.82 | AAI-based (high-confidence) | 0 |
| S17_vs_k141_55538\|\|full | 1415 | No | NA | 2 | 0 | 0 | Not-determined | Genome-fragment | NA | | NA | 0 |
| S17_vs_k141_41099\|\|full | 1045 | No | NA | 3 | 0 | 1 | Not-determined | Genome-fragment | NA | | NA | 0 |
| S17_vs_k141_128985\|\|full | 2353 | No | NA | 3 | 0 | 0 | Not-determined | Genome-fragment | NA | | NA | 0 |
| S17_vs_k141_128992\|\|full | 2016 | No | NA | 3 | 0 | 1 | Not-determined | Genome-fragment | NA | | NA | 0 |
| S9_extracted_k141_78433 | 1057 | No | NA | 4 | 3 | 0 | Low-quality | Genome-fragment | | 1.7 | HMM-based (lower-bound) | 0 |
| S17_vs_k141_3609\|\|full | 2404 | No | NA | 4 | 0 | 0 | Not-determined | Genome-fragment | NA | | NA | 0 |
| S17_vs_k141_166050\|\|full | 3224 | No | NA | 3 | 0 | 0 | Not-determined | Genome-fragment | NA | | NA | 0 |
| S17_vs_k141_114311\|\|full | 1986 | No | NA | 4 | 0 | 0 | Not-determined | Genome-fragment | NA | | NA | 0 |
| S17_vs_k141_151849\|\|full | 1615 | No | NA | 2 | 0 | 1 | Not-determined | Genome-fragment | NA | | NA | 0 |
| S11_vs_k141_33160\|\|full | 1057 | No | NA | 3 | 0 | 0 | Low-quality | Genome-fragment | | 2.1 | AAI-based (medium-confidence) | 0 |
| S11_vs_k141_51279\|\|lt2gene | 1057 | No | NA | 2 | 1 | 0 | Low-quality | Genome-fragment | | 2.59 | AAI-based (high-confidence) | 0 |
| S13_extracted_27958 | 1057 | No | NA | 2 | 0 | 1 | Low-quality | Genome-fragment | | 1.22 | AAI-based (medium-confidence) | 0 |
| S13_vs_29156\|\|full | 1057 | No | NA | 2 | 0 | 1 | Low-quality | Genome-fragment | | 1.88 | AAI-based (medium-confidence) | 0 |
| S17_vs_k141_121763\|\|full | 1941 | No | NA | 3 | 0 | 1 | Not-determined | Genome-fragment | NA | | NA | 0 |
| S17_vs_k141_114365\|\|full | 1255 | No | NA | 3 | 0 | 1 | Not-determined | Genome-fragment | NA | | NA | 0 |
| S17_vs_k141_114367\|\|full | 1604 | No | NA | 2 | 0 | 1 | Not-determined | Genome-fragment | NA | | NA | 0 |
| S13_vs_49000\|\|full | 1057 | No | NA | 2 | 0 | 0 | Low-quality | Genome-fragment | | 0.94 | AAI-based (medium-confidence) | 0 |
| S17_vs_k141_62501\|\|full | 4229 | No | NA | 5 | 0 | 3 | Not-determined | Genome-fragment | NA | | NA | 0 |
| S1_vs_k141_82686\|\|full | 1057 | No | NA | 2 | 1 | 0 | Low-quality | Genome-fragment | | 0.68 | HMM-based (lower-bound) | 0 |
| S17_vs_k141_18300\|\|full | 3334 | No | NA | 3 | 0 | 0 | Not-determined | Genome-fragment | NA | | NA | 0 |
| S17_vs_k141_3728\|\|full | 1385 | No | NA | 2 | 0 | 1 | Not-determined | Genome-fragment | NA | | NA | 0 |
| S3_extracted_k141_116160 | 1057 | No | NA | 4 | 0 | 0 | Low-quality | Genome-fragment | | 1.81 | AAI-based (high-confidence) | 0 |
| S17_vs_k141_99979\|\|full | 1155 | No | NA | 3 | 0 | 0 | Not-determined | Genome-fragment | NA | | NA | 0 |
| S17_vs_k141_11165\|\|full | 1257 | No | NA | 3 | 0 | 0 | Not-determined | Genome-fragment | NA | | NA | 0 |
| S3_vs_k141_78643\|\|full | 1057 | No | NA | 2 | 0 | 0 | Low-quality | Genome-fragment | | 1.32 | AAI-based (medium-confidence) | 0 |
| S17_vs_k141_166157\|\|full | 2140 | No | NA | 2 | 0 | 1 | Not-determined | Genome-fragment | NA | | NA | 0 |
| S17_vs_k141_33677\|\|full | 3377 | No | NA | 3 | 0 | 3 | Not-determined | Genome-fragment | NA | | NA | 0 |
| S5_vs_k141_130524\|\|full | 1057 | No | NA | 4 | 0 | 0 | Low-quality | Genome-fragment | | 2.46 | AAI-based (high-confidence) | 0 |
| S5_vs_k141_5887\|\|full | 1057 | No | NA | 2 | 1 | 0 | Low-quality | Genome-fragment | | 0.61 | HMM-based (lower-bound) | 0 |
| S7_vs_110557\|\|lt2gene | 1057 | No | NA | 2 | 1 | 0 | Low-quality | Genome-fragment | | 2.46 | AAI-based (high-confidence) | 0 |
| S17_vs_k141_11187\|\|full | 1751 | No | NA | 4 | 0 | 0 | Not-determined | Genome-fragment | NA | | NA | 0 |
| S17_vs_k141_3771\|\|full | 1151 | No | NA | 2 | 0 | 0 | Not-determined | Genome-fragment | NA | | NA | 0 |
| S16_extracted_90187 | 1057 | No | NA | 3 | 0 | 0 | Low-quality | Genome-fragment | | 2.76 | AAI-based (medium-confidence) | 0 |
| S16_extracted_105478 | 1057 | No | NA | 1 | 0 | 0 | Low-quality | Genome-fragment | | 1.68 | AAI-based (high-confidence) | 0 |
| S16_vs_66644\|\|full | 1057 | No | NA | 3 | 1 | 0 | Low-quality | Genome-fragment | | 1.95 | AAI-based (high-confidence) | 0 |
| S17_vs_k141_3780\|\|full | 2161 | No | NA | 3 | 0 | 1 | Not-determined | Genome-fragment | NA | | NA | 0 |
| S17_vs_k141_77718\|\|full | 1437 | No | NA | 2 | 0 | 0 | Not-determined | Genome-fragment | NA | | NA | 0 |
| S16_vs_106899\|\|full | 1057 | No | NA | 4 | 0 | 0 | Low-quality | Genome-fragment | | 1.88 | AAI-based (high-confidence) | 0 |
| S16_vs_115489\|\|full | 1057 | No | NA | 5 | 1 | 0 | Low-quality | Genome-fragment | | 1.37 | AAI-based (high-confidence) | 0 |
| S17_vs_k141_41300\|\|full | 2037 | No | NA | 3 | 0 | 1 | Not-determined | Genome-fragment | NA | | NA | 0 |
| S16_vs_143020\|\|full | 1057 | No | NA | 3 | 0 | 0 | Low-quality | Genome-fragment | | 1.34 | AAI-based (medium-confidence) | 0 |
| S10_vs_20372\|\|full | 1057 | No | NA | 2 | 1 | 0 | Low-quality | Genome-fragment | | 1.55 | AAI-based (medium-confidence) | 0 |
| S17_vs_k141_143965\|\|full | 1058 | No | NA | 2 | 0 | 1 | Low-quality | Genome-fragment | | 2.28 | AAI-based (medium-confidence) | 0 |
| S17_vs_k141_136524\|\|full | 1246 | No | NA | 2 | 0 | 0 | Not-determined | Genome-fragment | NA | | NA | 0 |
| S9_extracted_k141_52850 | 1058 | No | NA | 2 | 2 | 0 | Low-quality | Genome-fragment | | 3.49 | AAI-based (high-confidence) | 0 |
| S17_vs_k141_25550\|\|full | 2296 | No | NA | 5 | 0 | 2 | Not-determined | Genome-fragment | NA | | NA | 0 |
| S17_vs_k141_77779\|\|full | 1269 | No | NA | 3 | 0 | 1 | Not-determined | Genome-fragment | NA | | NA | 0 |
| S17_vs_k141_33791\|\|full | 1739 | No | NA | 3 | 0 | 0 | Not-determined | Genome-fragment | NA | | NA | 0 |
| S9_vs_k141_80611\|\|full | 1058 | No | NA | 2 | 1 | 0 | Low-quality | Genome-fragment | | 1.87 | AAI-based (high-confidence) | 0 |
| S17_vs_k141_107832\|\|full | 1344 | No | NA | 2 | 0 | 0 | Not-determined | Genome-fragment | NA | | NA | 0 |
| S9_vs_k141_83938\|\|lt2gene | 1058 | No | NA | 1 | 1 | 0 | Low-quality | Genome-fragment | | 25.42 | HMM-based (lower-bound) | 0 |
| S17_vs_k141_77818\|\|full | 4201 | No | NA | 4 | 0 | 3 | Not-determined | Genome-fragment | NA | | NA | 0 |
| S14_extracted_k141_50271 | 1058 | No | NA | 3 | 0 | 1 | Low-quality | Genome-fragment | | 2.33 | AAI-based (medium-confidence) | 0 |
| S14_vs_k141_55203\|\|lt2gene | 1058 | No | NA | 1 | 1 | 0 | Low-quality | Genome-fragment | | 1.55 | HMM-based (lower-bound) | 0 |
| S17_vs_k141_152063\|\|full | 1205 | No | NA | 2 | 0 | 1 | Not-determined | Genome-fragment | NA | | NA | 0 |
| S14_vs_k141_33399\|\|lt2gene | 1058 | No | NA | 2 | 1 | 0 | Low-quality | Genome-fragment | | 2.46 | AAI-based (high-confidence) | 0 |
| S17_vs_k141_18502\|\|full | 1370 | No | NA | 3 | 0 | 2 | Not-determined | Genome-fragment | NA | | NA | 0 |
| S16_extracted_113958 | 1058 | No | NA | 1 | 0 | 0 | Low-quality | Genome-fragment | | 1.79 | AAI-based (high-confidence) | 0 |
| S16_extracted_162756 | 1058 | No | NA | 1 | 0 | 1 | Low-quality | Genome-fragment | | 0.61 | AAI-based (medium-confidence) | 0 |
| S17_vs_k141_152065\|\|full | 7120 | No | NA | 7 | 0 | 4 | Not-determined | Genome-fragment | NA | | NA | 0 |
| S16_vs_123264\|\|full | 1058 | No | NA | 3 | 1 | 0 | Low-quality | Genome-fragment | | 3.1 | AAI-based (high-confidence) | 0 |
| S17_vs_k141_166296\|\|full | 2673 | No | NA | 4 | 0 | 4 | Not-determined | Genome-fragment | NA | | NA | 0 |
| S17_vs_k141_77836\|\|full | 2510 | No | NA | 2 | 0 | 1 | Not-determined | Genome-fragment | NA | | NA | 0 |
| S10_extracted_85023 | 1058 | No | NA | 3 | 0 | 0 | Low-quality | Genome-fragment | | 2.58 | AAI-based (high-confidence) | 0 |
| S12_extracted_k141_28338 | 1058 | No | NA | 3 | 2 | 0 | Low-quality | Genome-fragment | | 3.02 | AAI-based (high-confidence) | 0 |
| S17_vs_k141_55828\|\|full | 1914 | No | NA | 3 | 0 | 1 | Not-determined | Genome-fragment | NA | | NA | 0 |
| S12_extracted_k141_123574 | 1058 | No | NA | 1 | 0 | 0 | Low-quality | Genome-fragment | | 0.4 | AAI-based (medium-confidence) | 0 |
| S15_extracted_41870 | 1058 | No | NA | 2 | 1 | 0 | Low-quality | Genome-fragment | | 1.71 | AAI-based (high-confidence) | 0 |
| S17_vs_k141_18509\|\|full | 26539 | No | NA | 35 | 0 | 7 | Not-determined | Genome-fragment | NA | | NA | 0 |
| S17_vs_k141_121950\|\|full | 2302 | No | NA | 6 | 0 | 0 | Not-determined | Genome-fragment | NA | | NA | 0 |
| S17_vs_k141_33879\|\|full | 4685 | No | NA | 3 | 0 | 2 | Not-determined | Genome-fragment | NA | | NA | 0 |
| S17_vs_k141_114668\|\|full | 2270 | No | NA | 4 | 0 | 0 | Not-determined | Genome-fragment | NA | | NA | 0 |
| S15_vs_71951\|\|full | 1058 | No | NA | 2 | 0 | 1 | Low-quality | Genome-fragment | | 1.59 | AAI-based (medium-confidence) | 0 |
| S17_vs_k141_48618\|\|full | 3009 | No | NA | 9 | 0 | 0 | Not-determined | Genome-fragment | NA | | NA | 0 |
| S8_vs_k141_27511\|\|full | 1058 | No | NA | 3 | 1 | 0 | Low-quality | Genome-fragment | | 2.79 | AAI-based (high-confidence) | 0 |
| S17_vs_k141_11352\|\|full | 2840 | No | NA | 4 | 0 | 1 | Not-determined | Genome-fragment | NA | | NA | 0 |
| S17_vs_k141_100230\|\|full | 1213 | No | NA | 3 | 0 | 0 | Not-determined | Genome-fragment | NA | | NA | 0 |
| S17_extracted_k141_16716 | 1059 | No | NA | 1 | 0 | 0 | Low-quality | Genome-fragment | | 0.75 | AAI-based (medium-confidence) | 0 |
| S17_vs_k141_100238\|\|full | 1523 | No | NA | 2 | 0 | 1 | Not-determined | Genome-fragment | NA | | NA | 0 |
| S17_vs_k141_70672\|\|full | 3505 | No | NA | 4 | 0 | 0 | Not-determined | Genome-fragment | NA | | NA | 0 |
| S17_extracted_k141_47771 | 1059 | No | NA | 2 | 0 | 0 | Low-quality | Genome-fragment | | 1.67 | AAI-based (medium-confidence) | 0 |
| S9_extracted_k141_31862 | 1059 | No | NA | 1 | 1 | 0 | Low-quality | Genome-fragment | | 2.26 | AAI-based (high-confidence) | 0 |
| S17_vs_k141_41492\|\|full | 1012 | No | NA | 2 | 0 | 1 | Not-determined | Genome-fragment | NA | | NA | 0 |
| S17_vs_k141_77993\|\|full | 1730 | No | NA | 3 | 0 | 1 | Not-determined | Genome-fragment | NA | | NA | 0 |
| S17_vs_k141_136760\|\|full | 5057 | No | NA | 3 | 0 | 1 | Not-determined | Genome-fragment | NA | | NA | 0 |
| S17_vs_k141_41534\|\|full | 1789 | No | NA | 3 | 0 | 2 | Not-determined | Genome-fragment | NA | | NA | 0 |
| S11_extracted_k141_16131 | 1059 | No | NA | 1 | 0 | 0 | Low-quality | Genome-fragment | | 1.42 | AAI-based (medium-confidence) | 0 |
| S14_extracted_k141_58365 | 1059 | No | NA | 2 | 1 | 0 | Low-quality | Genome-fragment | | 3.51 | AAI-based (high-confidence) | 0 |
| S17_vs_k141_129488\|\|full | 2481 | No | NA | 3 | 0 | 2 | Not-determined | Genome-fragment | NA | | NA | 0 |
| S17_vs_k141_122091\|\|full | 2210 | No | NA | 3 | 0 | 2 | Not-determined | Genome-fragment | NA | | NA | 0 |
| S1_vs_k141_19827\|\|full | 1059 | No | NA | 2 | 0 | 0 | Low-quality | Genome-fragment | | 1.61 | AAI-based (high-confidence) | 0 |
| S3_extracted_k141_47994 | 1059 | No | NA | 3 | 0 | 0 | Low-quality | Genome-fragment | | 0.49 | AAI-based (high-confidence) | 0 |
| S17_vs_k141_41597\|\|full | 2911 | No | NA | 3 | 0 | 0 | Not-determined | Genome-fragment | NA | | NA | 0 |
| S17_vs_k141_56009\|\|full | 2064 | No | NA | 3 | 0 | 1 | Not-determined | Genome-fragment | NA | | NA | 0 |
| S17_vs_k141_56012\|\|full | 2054 | No | NA | 3 | 0 | 0 | Not-determined | Genome-fragment | NA | | NA | 0 |
| S3_extracted_k141_101747 | 1059 | No | NA | 4 | 0 | 0 | Low-quality | Genome-fragment | | 0.79 | AAI-based (high-confidence) | 0 |
| S17_vs_k141_11420\|\|full | 1115 | No | NA | 3 | 0 | 0 | Not-determined | Genome-fragment | NA | | NA | 0 |
| S17_vs_k141_114902\|\|full | 1026 | No | NA | 2 | 0 | 1 | Not-determined | Genome-fragment | NA | | NA | 0 |
| S17_vs_k141_84558\|\|full | 9526 | No | NA | 6 | 0 | 4 | Not-determined | Genome-fragment | NA | | NA | 0 |
| S17_vs_k141_166574\|\|full | 1214 | No | NA | 2 | 0 | 1 | Not-determined | Genome-fragment | NA | | NA | 0 |
| S7_vs_87497\|\|full | 1059 | No | NA | 3 | 0 | 1 | Low-quality | Genome-fragment | | 0.37 | AAI-based (medium-confidence) | 0 |
| S17_vs_k141_122165\|\|full | 1761 | No | NA | 3 | 0 | 2 | Not-determined | Genome-fragment | NA | | NA | 0 |
| S7_vs_99160\|\|full | 1059 | No | NA | 3 | 0 | 1 | Low-quality | Genome-fragment | | 0.59 | AAI-based (medium-confidence) | 0 |
| S17_vs_k141_56074\|\|full | 2540 | No | NA | 3 | 0 | 3 | Not-determined | Genome-fragment | NA | | NA | 0 |
| S7_vs_100394\|\|full | 1059 | No | NA | 3 | 0 | 0 | Low-quality | Genome-fragment | | 2.59 | AAI-based (medium-confidence) | 0 |
| S16_vs_24146\|\|full | 1059 | No | NA | 2 | 0 | 2 | Low-quality | Genome-fragment | | 1.61 | AAI-based (medium-confidence) | 0 |
| S17_vs_k141_166588\|\|full | 1099 | No | NA | 3 | 0 | 1 | Not-determined | Genome-fragment | NA | | NA | 0 |
| S17_vs_k141_78154\|\|full | 1459 | No | NA | 3 | 0 | 1 | Not-determined | Genome-fragment | NA | | NA | 0 |
| S17_vs_k141_41689\|\|full | 1334 | No | NA | 3 | 0 | 0 | Not-determined | Genome-fragment | NA | | NA | 0 |
| S17_vs_k141_70836\|\|full | 2928 | No | NA | 3 | 0 | 2 | Not-determined | Genome-fragment | NA | | NA | 0 |
| S16_vs_50583\|\|full | 1059 | No | NA | 3 | 0 | 0 | Low-quality | Genome-fragment | | 1.78 | AAI-based (medium-confidence) | 0 |
| S17_vs_k141_144583\|\|full | 2042 | No | NA | 2 | 0 | 0 | Not-determined | Genome-fragment | NA | | NA | 0 |
| S10_vs_46279\|\|full | 1059 | No | NA | 3 | 0 | 0 | Low-quality | Genome-fragment | | 1.62 | AAI-based (medium-confidence) | 0 |
| S17_vs_k141_78194\|\|full | 1259 | No | NA | 2 | 0 | 0 | Not-determined | Genome-fragment | NA | | NA | 0 |
| S17_vs_k141_18793\|\|full | 2728 | No | NA | 2 | 1 | 0 | Not-determined | Genome-fragment | NA | | NA | 0 |
| S17_vs_k141_132360\|\|full | 1060 | No | NA | 2 | 0 | 0 | Low-quality | Genome-fragment | | 1.96 | AAI-based (high-confidence) | 0 |
| S17_vs_k141_144593\|\|full | 3236 | No | NA | 3 | 0 | 0 | Not-determined | Genome-fragment | NA | | NA | 0 |

| | | | | | | | | | | | | |
|---|---|---|---|---|---|---|---|---|---|---|---|---|
| S9_extracted_k141_31318 | 1060 | No | NA | 3 | 0 | 0 | Low-quality | Genome-fragment | | 1.92 | AAI-based (high-confidence) | 0 |
| S9_vs_k141_40976||full | 1060 | No | NA | 5 | 2 | 0 | Low-quality | Genome-fragment | | 2.91 | AAI-based (high-confidence) | 0 |
| S17_vs_k141_152455||full | 2020 | No | NA | 3 | 0 | 0 | Not-determined | Genome-fragment | NA | | NA | 0 |
| S17_vs_k141_166626||full | 1687 | No | NA | 3 | 0 | 0 | Not-determined | Genome-fragment | NA | | NA | 0 |
| S17_vs_k141_173767||full | 1935 | No | NA | 4 | 0 | 0 | Not-determined | Genome-fragment | NA | | NA | 0 |
| S13_extracted_70542 | 1060 | No | NA | 2 | 1 | 0 | Low-quality | Genome-fragment | | 2.8 | AAI-based (high-confidence) | 0 |
| S1_extracted_k141_101237 | 1060 | No | NA | 1 | 0 | 0 | Low-quality | Genome-fragment | | 0.5 | AAI-based (high-confidence) | 0 |
| S1_vs_k141_102432||full | 1060 | No | NA | 3 | 0 | 0 | Low-quality | Genome-fragment | | 1.22 | AAI-based (medium-confidence) | 0 |
| S17_vs_k141_70893||full | 2090 | No | NA | 1 | 0 | 1 | Not-determined | Genome-fragment | NA | | NA | 0 |
| S1_vs_k141_114456||lt2gene | 1060 | No | NA | 2 | 1 | 0 | Low-quality | Genome-fragment | | 1.49 | HMM-based (lower-bound) | 0 |
| S17_vs_k141_152479||full | 2283 | No | NA | 5 | 0 | 0 | Not-determined | Genome-fragment | NA | | NA | 0 |
| S17_vs_k141_11547||full | 1685 | No | NA | 4 | 0 | 1 | Not-determined | Genome-fragment | NA | | NA | 0 |
| S17_vs_k141_100478||full | 12154 | No | NA | 9 | 0 | 7 | Not-determined | Genome-fragment | NA | | NA | 0 |
| S17_vs_k141_129667||full | 2348 | No | NA | 2 | 0 | 0 | Not-determined | Genome-fragment | NA | | NA | 0 |
| S3_vs_k141_60206||full | 1060 | No | NA | 4 | 3 | 0 | Low-quality | Genome-fragment | | 1.74 | AAI-based (high-confidence) | 0 |
| S17_vs_k141_100503||full | 1569 | No | NA | 2 | 0 | 2 | Not-determined | Genome-fragment | NA | | NA | 0 |
| S17_vs_k141_56185||full | 1224 | No | NA | 3 | 0 | 0 | Not-determined | Genome-fragment | NA | | NA | 0 |
| S17_vs_k141_129687||full | 7736 | No | NA | 9 | 0 | 8 | Not-determined | Genome-fragment | NA | | NA | 0 |
| S17_vs_k141_136968||full | 2828 | No | NA | 5 | 0 | 0 | Not-determined | Genome-fragment | NA | | NA | 0 |
| S3_vs_k141_110495||full | 1060 | No | NA | 3 | 1 | 0 | Low-quality | Genome-fragment | | 1.53 | AAI-based (medium-confidence) | 0 |
| S17_vs_k141_92770||full | 1169 | No | NA | 1 | 0 | 0 | Not-determined | Genome-fragment | NA | | NA | 0 |
| S17_vs_k141_25940||full | 5251 | No | NA | 7 | 0 | 0 | Not-determined | Genome-fragment | NA | | NA | 0 |
| S17_vs_k141_25947||full | 2172 | No | NA | 3 | 0 | 2 | Not-determined | Genome-fragment | NA | | NA | 0 |
| S5_extracted_k141_93184 | 1060 | No | NA | 3 | 1 | 0 | Low-quality | Genome-fragment | | 2.86 | AAI-based (medium-confidence) | 0 |
| S17_vs_k141_108372||full | 8279 | No | NA | 8 | 0 | 2 | Not-determined | Genome-fragment | NA | | NA | 0 |
| S17_vs_k141_129715||full | 2864 | No | NA | 5 | 0 | 2 | Not-determined | Genome-fragment | NA | | NA | 0 |
| S5_vs_k141_129934||full | 1060 | No | NA | 2 | 0 | 1 | Low-quality | Genome-fragment | | 0.78 | AAI-based (medium-confidence) | 0 |
| S17_vs_k141_56227||full | 1672 | No | NA | 4 | 0 | 1 | Not-determined | Genome-fragment | NA | | NA | 0 |
| S17_vs_k141_11577||full | 1758 | No | NA | 3 | 0 | 1 | Not-determined | Genome-fragment | NA | | NA | 0 |
| S17_vs_k141_122194||full | 5526 | No | NA | 7 | 0 | 4 | Not-determined | Genome-fragment | NA | | NA | 0 |
| S5_vs_k141_96637||full | 1060 | No | NA | 2 | 1 | 0 | Low-quality | Genome-fragment | | 0.67 | HMM-based (lower-bound) | 0 |
| S7_vs_39076||full | 1060 | No | NA | 4 | 1 | 0 | Low-quality | Genome-fragment | | 2.55 | AAI-based (medium-confidence) | 0 |
| S7_vs_59447||full | 1060 | No | NA | 2 | 0 | 1 | Low-quality | Genome-fragment | | 1.71 | AAI-based (medium-confidence) | 0 |
| S17_vs_k141_144720||full | 2616 | No | NA | 5 | 0 | 2 | Not-determined | Genome-fragment | NA | | NA | 0 |
| S17_vs_k141_137039||full | 1086 | No | NA | 3 | 0 | 1 | Not-determined | Genome-fragment | NA | | NA | 0 |
| S17_vs_k141_84733||full | 5566 | No | NA | 4 | 0 | 2 | Not-determined | Genome-fragment | NA | | NA | 0 |
| S7_vs_70066||lt2gene | 1060 | No | NA | 2 | 1 | 0 | Low-quality | Genome-fragment | | 2.65 | AAI-based (high-confidence) | 0 |
| S7_vs_82530||lt2gene | 1060 | No | NA | 2 | 1 | 0 | Low-quality | Genome-fragment | | 2.61 | AAI-based (high-confidence) | 0 |
| S17_vs_k141_166752||full | 1515 | No | NA | 2 | 0 | 1 | Not-determined | Genome-fragment | NA | | NA | 0 |
| S17_vs_k141_173917||full | 2462 | No | NA | 2 | 0 | 2 | Not-determined | Genome-fragment | NA | | NA | 0 |
| S17_vs_k141_71032||full | 1199 | No | NA | 2 | 0 | 1 | Not-determined | Genome-fragment | NA | | NA | 0 |
| S16_extracted_33684 | 1060 | No | NA | 1 | 0 | 1 | Low-quality | Genome-fragment | | 0.49 | AAI-based (medium-confidence) | 0 |
| S17_vs_k141_34293||full | 2097 | No | NA | 2 | 0 | 2 | Not-determined | Genome-fragment | NA | | NA | 0 |
| S17_vs_k141_41845||full | 37248 | No | NA | 54 | 18 | 2 | High-quality | High-quality | | 100 | AAI-based (high-confidence) | 0 |
| S17_vs_k141_115136||full | 1429 | No | NA | 3 | 0 | 2 | Not-determined | Genome-fragment | NA | | NA | 0 |
| S17_vs_k141_137088||full | 4342 | No | NA | 7 | 0 | 0 | Not-determined | Genome-fragment | NA | | NA | 0 |
| S16_vs_109843||full | 1060 | No | NA | 2 | 2 | 0 | Low-quality | Genome-fragment | | 3.13 | AAI-based (medium-confidence) | 0 |
| S16_vs_168338||lt2gene | 1060 | No | NA | 1 | 1 | 0 | Low-quality | Genome-fragment | | 2.5 | HMM-based (lower-bound) | 0 |
| S17_vs_k141_152663||full | 2239 | No | NA | 3 | 0 | 2 | Not-determined | Genome-fragment | NA | | NA | 0 |
| S10_vs_68327||lt2gene | 1060 | No | NA | 1 | 1 | 0 | Low-quality | Genome-fragment | | 1.96 | HMM-based (lower-bound) | 0 |
| S17_vs_k141_62635||full | 2053 | No | NA | 3 | 0 | 1 | Not-determined | Genome-fragment | NA | | NA | 0 |
| S12_extracted_k141_2159 | 1060 | No | NA | 2 | 2 | 0 | Low-quality | Genome-fragment | | 4.88 | AAI-based (medium-confidence) | 0 |
| S9_extracted_k141_68277 | 1061 | No | NA | 2 | 0 | 0 | Low-quality | Genome-fragment | | 0.64 | AAI-based (high-confidence) | 0 |
| S17_vs_k141_115200||full | 2063 | No | NA | 2 | 0 | 2 | Not-determined | Genome-fragment | NA | | NA | 0 |
| S11_vs_k141_86309||full | 1061 | No | NA | 2 | 0 | 1 | Low-quality | Genome-fragment | | 1.21 | AAI-based (medium-confidence) | 0 |
| S11_vs_k141_57563||full | 1061 | No | NA | 3 | 1 | 0 | Low-quality | Genome-fragment | | 1.26 | AAI-based (high-confidence) | 0 |
| S1_extracted_k141_59913 | 1061 | No | NA | 1 | 0 | 0 | Low-quality | Genome-fragment | | 2.87 | AAI-based (medium-confidence) | 0 |
| S17_vs_k141_129862||full | 3228 | No | NA | 5 | 0 | 1 | Not-determined | Genome-fragment | NA | | NA | 0 |
| S5_vs_k141_28825||full | 1061 | No | NA | 3 | 1 | 0 | Low-quality | Genome-fragment | | 2.38 | AAI-based (medium-confidence) | 0 |
| S7_extracted_15686 | 1061 | No | NA | 1 | 1 | 0 | Low-quality | Genome-fragment | | 2.58 | AAI-based (high-confidence) | 0 |
| S7_vs_20871||full | 1061 | No | NA | 2 | 1 | 0 | Low-quality | Genome-fragment | | 2.19 | AAI-based (high-confidence) | 0 |
| S7_vs_72855||full | 1061 | No | NA | 5 | 1 | 0 | Low-quality | Genome-fragment | | 2.08 | AAI-based (high-confidence) | 0 |
| S17_vs_k141_56377||full | 3619 | No | NA | 4 | 0 | 3 | Not-determined | Genome-fragment | NA | | NA | 0 |
| S7_vs_45559||lt2gene | 1061 | No | NA | 1 | 1 | 0 | Low-quality | Genome-fragment | | 0.73 | HMM-based (lower-bound) | 0 |
| S17_vs_k141_144822||full | 3614 | No | NA | 4 | 0 | 0 | Not-determined | Genome-fragment | NA | | NA | 0 |
| S17_vs_k141_115262||full | 1327 | No | NA | 3 | 0 | 1 | Not-determined | Genome-fragment | NA | | NA | 0 |
| S17_vs_k141_108523||full | 5831 | No | NA | 6 | 0 | 1 | Not-determined | Genome-fragment | NA | | NA | 0 |
| S16_extracted_43610 | 1061 | No | NA | 1 | 0 | 1 | Low-quality | Genome-fragment | | 0.4 | AAI-based (high-confidence) | 0 |
| S17_vs_k141_56458||full | 1803 | No | NA | 3 | 0 | 0 | Not-determined | Genome-fragment | NA | | NA | 0 |
| S17_vs_k141_92955||full | 2738 | No | NA | 4 | 0 | 1 | Not-determined | Genome-fragment | NA | | NA | 0 |
| S17_vs_k141_159801||full | 1111 | No | NA | 3 | 0 | 0 | Not-determined | Genome-fragment | NA | | NA | 0 |
| S16_extracted_83869 | 1061 | No | NA | 1 | 0 | 0 | Low-quality | Genome-fragment | | 2.66 | AAI-based (high-confidence) | 0 |
| S17_vs_k141_26183||full | 1364 | No | NA | 3 | 0 | 2 | Not-determined | Genome-fragment | NA | | NA | 0 |
| S16_extracted_107625 | 1061 | No | NA | 1 | 0 | 0 | Low-quality | Genome-fragment | | 0.2 | AAI-based (medium-confidence) | 0 |
| S17_vs_k141_159812||full | 4734 | No | NA | 6 | 0 | 4 | Not-determined | Genome-fragment | NA | | NA | 0 |
| S17_vs_k141_49247||full | 1208 | No | NA | 3 | 0 | 0 | Not-determined | Genome-fragment | NA | | NA | 0 |
| S17_vs_k141_115327||full | 2522 | No | NA | 3 | 0 | 2 | Not-determined | Genome-fragment | NA | | NA | 0 |
| S17_vs_k141_71191||full | 2031 | No | NA | 3 | 0 | 1 | Not-determined | Genome-fragment | NA | | NA | 0 |
| S16_vs_75436||full | 1061 | No | NA | 2 | 0 | 1 | Low-quality | Genome-fragment | | 0.55 | AAI-based (medium-confidence) | 0 |
| S10_vs_802||full | 1061 | No | NA | 3 | 0 | 0 | Low-quality | Genome-fragment | | 2.88 | AAI-based (high-confidence) | 0 |
| S17_vs_k141_62814||full | 1771 | No | NA | 2 | 0 | 0 | Not-determined | Genome-fragment | NA | | NA | 0 |
| S10_vs_110892||full | 1061 | No | NA | 2 | 0 | 1 | Low-quality | Genome-fragment | | 1.33 | AAI-based (medium-confidence) | 0 |
| S17_vs_k141_166992||full | 2328 | No | NA | 3 | 0 | 3 | Not-determined | Genome-fragment | NA | | NA | 0 |
| S17_vs_k141_137270||full | 4843 | No | NA | 4 | 0 | 4 | Not-determined | Genome-fragment | NA | | NA | 0 |
| S12_extracted_k141_57161 | 1061 | No | NA | 2 | 0 | 0 | Low-quality | Genome-fragment | | 11.67 | AAI-based (medium-confidence) | 0 |
| S17_vs_k141_34431||full | 2138 | No | NA | 4 | 0 | 3 | Not-determined | Genome-fragment | NA | | NA | 0 |
| S17_vs_k141_11809||full | 2491 | No | NA | 3 | 0 | 0 | Not-determined | Genome-fragment | NA | | NA | 0 |
| S8_vs_k141_48105||full | 1061 | No | NA | 4 | 1 | 0 | Low-quality | Genome-fragment | | 2.42 | AAI-based (high-confidence) | 0 |
| S17_extracted_k141_112107 | 1062 | No | NA | 3 | 0 | 1 | Low-quality | Genome-fragment | | 0.6 | AAI-based (medium-confidence) | 0 |
| S17_vs_k141_159857||full | 2442 | No | NA | 3 | 0 | 1 | Not-determined | Genome-fragment | NA | | NA | 0 |
| S17_vs_k141_55170||lt2gene | 1062 | No | NA | 2 | 2 | 0 | Low-quality | Genome-fragment | | 3.12 | AAI-based (high-confidence) | 0 |
| S9_vs_k141_87514||lt2gene | 1062 | No | NA | 2 | 1 | 0 | Low-quality | Genome-fragment | | 2.32 | AAI-based (high-confidence) | 0 |
| S17_vs_k141_26239||full | 3663 | No | NA | 4 | 0 | 1 | Not-determined | Genome-fragment | NA | | NA | 0 |
| S17_vs_k141_26244||full | 2700 | No | NA | 5 | 0 | 2 | Not-determined | Genome-fragment | NA | | NA | 0 |
| S17_vs_k141_122281||full | 4063 | No | NA | 3 | 0 | 2 | Not-determined | Genome-fragment | NA | | NA | 0 |
| S17_vs_k141_174098||full | 1684 | No | NA | 2 | 0 | 1 | Not-determined | Genome-fragment | NA | | NA | 0 |
| S13_vs_90249||full | 1062 | No | NA | 3 | 0 | 0 | Low-quality | Genome-fragment | | 2.01 | AAI-based (medium-confidence) | 0 |
| S1_vs_k141_48476||full | 1062 | No | NA | 2 | 1 | 0 | Low-quality | Genome-fragment | | 2.6 | AAI-based (medium-confidence) | 0 |
| S17_vs_k141_56626||full | 2387 | No | NA | 3 | 0 | 2 | Not-determined | Genome-fragment | NA | | NA | 0 |
| S17_vs_k141_137360||full | 2525 | No | NA | 3 | 0 | 2 | Not-determined | Genome-fragment | NA | | NA | 0 |
| S5_vs_k141_116341||full | 1062 | No | NA | 3 | 1 | 0 | Low-quality | Genome-fragment | | 1.76 | HMM-based (lower-bound) | 0 |
| S7_extracted_108397 | 1062 | No | NA | 2 | 0 | 0 | Low-quality | Genome-fragment | | 2.53 | AAI-based (medium-confidence) | 0 |
| S17_vs_k141_19135||full | 3363 | No | NA | 4 | 0 | 2 | Not-determined | Genome-fragment | NA | | NA | 0 |
| S7_vs_66491||full | 1062 | No | NA | 3 | 0 | 0 | Low-quality | Genome-fragment | | 2.53 | AAI-based (medium-confidence) | 0 |
| S17_vs_k141_108639||full | 9342 | No | NA | 5 | 0 | 3 | Not-determined | Genome-fragment | NA | | NA | 0 |
| S17_vs_k141_174134||full | 6085 | No | NA | 10 | 0 | 1 | Not-determined | Genome-fragment | NA | | NA | 0 |
| S16_vs_105984||full | 1062 | No | NA | 4 | 0 | 1 | Low-quality | Genome-fragment | | 0.83 | AAI-based (medium-confidence) | 0 |
| S17_vs_k141_167094||full | 2063 | No | NA | 2 | 0 | 0 | Not-determined | Genome-fragment | NA | | NA | 0 |
| S10_vs_78267||full | 1062 | No | NA | 3 | 0 | 0 | Low-quality | Genome-fragment | | 2.42 | AAI-based (medium-confidence) | 0 |
| S12_extracted_k141_34227 | 1062 | No | NA | 2 | 1 | 0 | Low-quality | Genome-fragment | | 0.59 | HMM-based (lower-bound) | 0 |
| S17_vs_k141_19181||full | 1126 | No | NA | 2 | 0 | 1 | Not-determined | Genome-fragment | NA | | NA | 0 |
| S17_vs_k141_34565||full | 1023 | No | NA | 2 | 0 | 1 | Not-determined | Genome-fragment | NA | | NA | 0 |
| S17_vs_k141_159949||full | 1216 | No | NA | 3 | 0 | 0 | Not-determined | Genome-fragment | NA | | NA | 0 |
| S8_extracted_k141_140561 | 1062 | No | NA | 4 | 0 | 0 | Low-quality | Genome-fragment | | 2.28 | AAI-based (medium-confidence) | 0 |
| S17_vs_k141_26315||full | 1521 | No | NA | 2 | 0 | 2 | Not-determined | Genome-fragment | NA | | NA | 0 |
| S17_vs_k141_71342||full | 3702 | No | NA | 5 | 0 | 2 | Not-determined | Genome-fragment | NA | | NA | 0 |
| S17_vs_k141_26316||full | 3004 | No | NA | 3 | 0 | 2 | Not-determined | Genome-fragment | NA | | NA | 0 |
| S17_vs_k141_42214||full | 2026 | No | NA | 3 | 0 | 2 | Not-determined | Genome-fragment | NA | | NA | 0 |
| S17_vs_k141_121365||full | 1063 | No | NA | 3 | 0 | 0 | Low-quality | Genome-fragment | | 1.67 | AAI-based (high-confidence) | 0 |
| S11_extracted_k141_57362 | 1063 | No | NA | 1 | 0 | 0 | Low-quality | Genome-fragment | | 1.87 | AAI-based (high-confidence) | 0 |
| S14_vs_k141_39877||full | 1063 | No | NA | 5 | 1 | 0 | Low-quality | Genome-fragment | | 2.21 | AAI-based (medium-confidence) | 0 |
| S17_vs_k141_167139||full | 1557 | No | NA | 2 | 0 | 1 | Not-determined | Genome-fragment | NA | | NA | 0 |
| S1_extracted_k141_89111 | 1063 | No | NA | 4 | 0 | 0 | Low-quality | Genome-fragment | | 1.74 | AAI-based (high-confidence) | 0 |
| S17_vs_k141_62957||full | 4368 | No | NA | 3 | 0 | 2 | Not-determined | Genome-fragment | NA | | NA | 0 |
| S1_vs_k141_42880||full | 1063 | No | NA | 4 | 0 | 0 | Low-quality | Genome-fragment | | 2.48 | AAI-based (high-confidence) | 0 |
| S17_vs_k141_71375||full | 2068 | No | NA | 3 | 0 | 2 | Not-determined | Genome-fragment | NA | | NA | 0 |
| S17_vs_k141_49424||full | 1129 | No | NA | 2 | 0 | 1 | Not-determined | Genome-fragment | NA | | NA | 0 |
| S17_vs_k141_137479||full | 1690 | No | NA | 3 | 0 | 2 | Not-determined | Genome-fragment | NA | | NA | 0 |
| S17_vs_k141_93151||full | 1160 | No | NA | 3 | 0 | 2 | Not-determined | Genome-fragment | NA | | NA | 0 |
| S1_vs_k141_112167||lt2gene | 1063 | No | NA | 2 | 2 | 0 | Low-quality | Genome-fragment | | 1.35 | HMM-based (lower-bound) | 0 |
| S3_extracted_k141_9341 | 1063 | No | NA | 1 | 0 | 0 | Low-quality | Genome-fragment | | 0.85 | AAI-based (high-confidence) | 0 |
| S17_vs_k141_4590||full | 4596 | No | NA | 5 | 0 | 0 | Not-determined | Genome-fragment | NA | | NA | 0 |
| S3_vs_k141_98704||full | 1063 | No | NA | 3 | 0 | 0 | Low-quality | Genome-fragment | | 1.59 | AAI-based (high-confidence) | 0 |
| S5_extracted_k141_40126 | 1063 | No | NA | 2 | 0 | 0 | Low-quality | Genome-fragment | | 1.46 | AAI-based (high-confidence) | 0 |
| S17_vs_k141_85133||full | 3137 | No | NA | 3 | 0 | 3 | Not-determined | Genome-fragment | NA | | NA | 0 |
| S5_vs_k141_124638||full | 1063 | No | NA | 2 | 0 | 0 | Low-quality | Genome-fragment | | 1.93 | AAI-based (medium-confidence) | 0 |
| S7_vs_119250||full | 1063 | No | NA | 5 | 0 | 2 | Low-quality | Genome-fragment | | 0.28 | AAI-based (medium-confidence) | 0 |
| S17_vs_k141_71467||full | 3446 | No | NA | 4 | 0 | 1 | Not-determined | Genome-fragment | NA | | NA | 0 |
| S17_vs_k141_137587||full | 1465 | No | NA | 2 | 0 | 0 | Not-determined | Genome-fragment | NA | | NA | 0 |
| S17_vs_k141_160037||full | 1951 | No | NA | 2 | 0 | 2 | Not-determined | Genome-fragment | NA | | NA | 0 |
| S16_vs_86635||lt2gene | 1063 | No | NA | 1 | 1 | 0 | Low-quality | Genome-fragment | | 2.81 | AAI-based (medium-confidence) | 0 |
| S17_vs_k141_101066||full | 2238 | No | NA | 3 | 0 | 2 | Not-determined | Genome-fragment | NA | | NA | 0 |
| S17_vs_k141_160045||full | 1814 | No | NA | 3 | 0 | 0 | Not-determined | Genome-fragment | NA | | NA | 0 |
| S10_extracted_6449 | 1063 | No | NA | 1 | 0 | 0 | Low-quality | Genome-fragment | | 0.47 | AAI-based (high-confidence) | 0 |
| S15_extracted_24958 | 1063 | No | NA | 1 | 1 | 0 | Low-quality | Genome-fragment | | 2.47 | AAI-based (high-confidence) | 0 |
| S17_vs_k141_167258||full | 1438 | No | NA | 2 | 0 | 1 | Not-determined | Genome-fragment | NA | | NA | 0 |
| S17_vs_k141_4674||full | 1816 | No | NA | 3 | 0 | 2 | Not-determined | Genome-fragment | NA | | NA | 0 |
| S17_vs_k141_85178||full | 7811 | No | NA | 9 | 0 | 6 | Not-determined | Genome-fragment | NA | | NA | 0 |
| S17_vs_k141_4687||full | 1374 | No | NA | 2 | 0 | 0 | Not-determined | Genome-fragment | NA | | NA | 0 |
| S8_vs_k141_153443||lt2gene | 1063 | No | NA | 2 | 1 | 0 | Low-quality | Genome-fragment | | 2.49 | HMM-based (lower-bound) | 0 |
| S17_vs_k141_12091||full | 6399 | No | NA | 18 | 0 | 1 | Not-determined | Genome-fragment | NA | | NA | 0 |
| S9_vs_k141_25597||full | 1064 | No | NA | 3 | 1 | 0 | Low-quality | Genome-fragment | | 3.48 | AAI-based (medium-confidence) | 0 |
| S17_vs_k141_42371||full | 1821 | No | NA | 3 | 0 | 0 | Not-determined | Genome-fragment | NA | | NA | 0 |
| S17_vs_k141_42384||full | 2308 | No | NA | 3 | 0 | 2 | Not-determined | Genome-fragment | NA | | NA | 0 |
| S17_vs_k141_137690||full | 2143 | No | NA | 3 | 0 | 2 | Not-determined | Genome-fragment | NA | | NA | 0 |
| S17_vs_k141_78836||full | 3282 | No | NA | 4 | 0 | 1 | Not-determined | Genome-fragment | NA | | NA | 0 |
| S17_vs_k141_160115||full | 3771 | No | NA | 5 | 0 | 0 | Not-determined | Genome-fragment | NA | | NA | 0 |
| S17_vs_k141_160117||full | 1162 | No | NA | 3 | 0 | 2 | Not-determined | Genome-fragment | NA | | NA | 0 |
| S9_vs_k141_82559||full | 1064 | No | NA | 3 | 0 | 0 | Low-quality | Genome-fragment | | 2.46 | AAI-based (high-confidence) | 0 |
| S17_vs_k141_115796||full | 2215 | No | NA | 2 | 0 | 1 | Not-determined | Genome-fragment | NA | | NA | 0 |
| S13_vs_7876||full | 1064 | No | NA | 3 | 1 | 0 | Low-quality | Genome-fragment | | 3.02 | AAI-based (high-confidence) | 0 |
| S17_vs_k141_71587||full | 2310 | No | NA | 3 | 0 | 1 | Not-determined | Genome-fragment | NA | | NA | 0 |
| S14_extracted_k141_105208 | 1064 | No | NA | 3 | 2 | 0 | Low-quality | Genome-fragment | | 2.06 | AAI-based (high-confidence) | 0 |
| S14_vs_k141_96618||full | 1064 | No | NA | 2 | 1 | 0 | Low-quality | Genome-fragment | | 2.65 | AAI-based (high-confidence) | 0 |
| S17_vs_k141_56916||full | 1292 | No | NA | 2 | 0 | 0 | Not-determined | Genome-fragment | NA | | NA | 0 |
| S1_vs_k141_73850||lt2gene | 1064 | No | NA | 1 | 1 | 0 | Low-quality | Genome-fragment | | 2.53 | AAI-based (high-confidence) | 0 |
| S17_vs_k141_101146||full | 1872 | No | NA | 2 | 0 | 1 | Not-determined | Genome-fragment | NA | | NA | 0 |
| S3_extracted_k141_75647 | 1064 | No | NA | 2 | 0 | 0 | Low-quality | Genome-fragment | | 2.29 | AAI-based (high-confidence) | 0 |
| S3_vs_k141_52985||full | 1064 | No | NA | 3 | 0 | 0 | Low-quality | Genome-fragment | | 2.55 | AAI-based (high-confidence) | 0 |
| S17_vs_k141_93338||full | 5920 | No | NA | 4 | 0 | 4 | Not-determined | Genome-fragment | NA | | NA | 0 |
| S17_vs_k141_49634||full | 4770 | No | NA | 3 | 0 | 1 | Not-determined | Genome-fragment | NA | | NA | 0 |
| S5_vs_k141_73100||full | 1064 | No | NA | 2 | 1 | 0 | Low-quality | Genome-fragment | | 1.78 | AAI-based (high-confidence) | 0 |
| S17_vs_k141_130110||full | 1861 | No | NA | 3 | 0 | 1 | Not-determined | Genome-fragment | NA | | NA | 0 |
| S17_vs_k141_115847||full | 2568 | No | NA | 3 | 0 | 1 | Not-determined | Genome-fragment | NA | | NA | 0 |
| S10_vs_95893||full | 1064 | No | NA | 3 | 0 | 0 | Low-quality | Genome-fragment | | 0.87 | AAI-based (medium-confidence) | 0 |
| S17_vs_k141_78864||full | 2355 | No | NA | 4 | 0 | 3 | Not-determined | Genome-fragment | NA | | NA | 0 |
| S10_vs_119001||full | 1064 | No | NA | 2 | 0 | 1 | Low-quality | Genome-fragment | | 2.17 | AAI-based (high-confidence) | 0 |
| S17_vs_k141_78870||full | 3318 | No | NA | 9 | 0 | 0 | Not-determined | Genome-fragment | NA | | NA | 0 |
| S17_vs_k141_56953||full | 1748 | No | NA | 3 | 0 | 1 | Not-determined | Genome-fragment | NA | | NA | 0 |
| S17_vs_k141_56964||full | 2597 | No | NA | 3 | 0 | 2 | Not-determined | Genome-fragment | NA | | NA | 0 |
| S17_vs_k141_4851||full | 3066 | No | NA | 4 | 0 | 1 | Not-determined | Genome-fragment | NA | | NA | 0 |
| S17_vs_k141_85402||full | 2977 | No | NA | 6 | 0 | 0 | Not-determined | Genome-fragment | NA | | NA | 0 |
| S17_vs_k141_26500||full | 2234 | No | NA | 3 | 0 | 2 | Not-determined | Genome-fragment | NA | | NA | 0 |
| S17_vs_k141_93411||full | 1319 | No | NA | 2 | 0 | 1 | Not-determined | Genome-fragment | NA | | NA | 0 |
| S17_vs_k141_49675||full | 1868 | No | NA | 3 | 0 | 2 | Not-determined | Genome-fragment | NA | | NA | 0 |
| S17_vs_k141_130189||full | 1974 | No | NA | 3 | 0 | 1 | Not-determined | Genome-fragment | NA | | NA | 0 |
| S2_vs_k141_19992||full | 1064 | No | NA | 3 | 2 | 0 | Low-quality | Genome-fragment | | 3.01 | AAI-based (high-confidence) | 0 |
| S17_vs_k141_42503||full | 1738 | No | NA | 2 | 0 | 1 | Not-determined | Genome-fragment | NA | | NA | 0 |
| S17_vs_k141_35008||full | 1842 | No | NA | 3 | 0 | 2 | Not-determined | Genome-fragment | NA | | NA | 0 |
| S17_vs_k141_42515||full | 1107 | No | NA | 3 | 0 | 1 | Not-determined | Genome-fragment | NA | | NA | 0 |
| S17_vs_k141_101233||full | 3555 | No | NA | 4 | 0 | 0 | Not-determined | Genome-fragment | NA | | NA | 0 |
| S17_vs_k141_153305||full | 3910 | No | NA | 4 | 0 | 2 | Not-determined | Genome-fragment | NA | | NA | 0 |
| S17_vs_k141_78957||full | 1589 | No | NA | 3 | 0 | 2 | Not-determined | Genome-fragment | NA | | NA | 0 |
| S17_vs_k141_35065||full | 2941 | No | NA | 5 | 0 | 2 | Not-determined | Genome-fragment | NA | | NA | 0 |
| S8_vs_k141_174498||full | 1064 | No | NA | 3 | 0 | 0 | Low-quality | Genome-fragment | | 2.47 | AAI-based (high-confidence) | 0 |
| S17_vs_k141_78973||full | 1260 | No | NA | 2 | 0 | 0 | Not-determined | Genome-fragment | NA | | NA | 0 |
| S17_vs_k141_115976||full | 1370 | No | NA | 3 | 0 | 0 | Not-determined | Genome-fragment | NA | | NA | 0 |
| S17_extracted_k141_160862 | 1065 | No | NA | 3 | 0 | 0 | Low-quality | Genome-fragment | | 1.49 | AAI-based (medium-confidence) | 0 |
| S17_vs_k141_45223||full | 1065 | No | NA | 3 | 0 | 1 | Low-quality | Genome-fragment | | 0.75 | AAI-based (medium-confidence) | 0 |
| S17_vs_k141_167510||full | 1486 | No | NA | 2 | 0 | 1 | Not-determined | Genome-fragment | NA | | NA | 0 |
| S9_extracted_k141_49929 | 1065 | No | NA | 1 | 0 | 0 | Low-quality | Genome-fragment | | 0.62 | AAI-based (high-confidence) | 0 |
| S17_vs_k141_109097||full | 3015 | No | NA | 3 | 0 | 3 | Not-determined | Genome-fragment | NA | | NA | 0 |
| S17_vs_k141_137910||full | 1052 | No | NA | 2 | 0 | 1 | Not-determined | Genome-fragment | NA | | NA | 0 |
| S11_extracted_k141_56870 | 1065 | No | NA | 3 | 0 | 2 | Low-quality | Genome-fragment | | 0.63 | AAI-based (medium-confidence) | 0 |
| S17_vs_k141_63375||full | 2084 | No | NA | 3 | 0 | 2 | Not-determined | Genome-fragment | NA | | NA | 0 |
| S17_vs_k141_71772||full | 2322 | No | NA | 2 | 0 | 0 | Not-determined | Genome-fragment | NA | | NA | 0 |
| S17_vs_k141_4982||full | 4719 | No | NA | 4 | 0 | 1 | Not-determined | Genome-fragment | NA | | NA | 0 |
| S14_extracted_k141_73676 | 1065 | No | NA | 1 | 0 | 0 | Low-quality | Genome-fragment | | 1.63 | AAI-based (high-confidence) | 0 |
| S17_vs_k141_145479||full | 2766 | No | NA | 4 | 0 | 1 | Not-determined | Genome-fragment | NA | | NA | 0 |
| S17_vs_k141_19499||full | 2051 | No | NA | 2 | 0 | 0 | Not-determined | Genome-fragment | NA | | NA | 0 |
| S17_vs_k141_145497||full | 1172 | No | NA | 3 | 0 | 1 | Not-determined | Genome-fragment | NA | | NA | 0 |
| S17_vs_k141_19527||full | 1944 | No | NA | 3 | 0 | 0 | Not-determined | Genome-fragment | NA | | NA | 0 |
| S17_vs_k141_49723||full | 2250 | No | NA | 3 | 0 | 2 | Not-determined | Genome-fragment | NA | | NA | 0 |
| S17_vs_k141_122864||full | 1361 | No | NA | 3 | 0 | 3 | Not-determined | Genome-fragment | NA | | NA | 0 |
| S1_extracted_k141_64402 | 1065 | No | NA | 3 | 0 | 0 | Low-quality | Genome-fragment | | 1.53 | AAI-based (medium-confidence) | 0 |
| S1_extracted_k141_71491 | 1065 | No | NA | 1 | 1 | 0 | Low-quality | Genome-fragment | | 2.22 | AAI-based (high-confidence) | 0 |
| S17_vs_k141_42661||full | 1258 | No | NA | 3 | 0 | 1 | Not-determined | Genome-fragment | NA | | NA | 0 |
| S17_vs_k141_138008||full | 1831 | No | NA | 1 | 0 | 1 | Not-determined | Genome-fragment | NA | | NA | 0 |
| S17_vs_k141_5130||full | 2653 | No | NA | 2 | 0 | 2 | Not-determined | Genome-fragment | NA | | NA | 0 |
| S17_vs_k141_174479||full | 6977 | No | NA | 5 | 0 | 4 | Not-determined | Genome-fragment | NA | | NA | 0 |
| S17_vs_k141_138013||full | 1629 | No | NA | 3 | 0 | 0 | Not-determined | Genome-fragment | NA | | NA | 0 |
| S17_vs_k141_167601||full | 1289 | No | NA | 2 | 0 | 2 | Not-determined | Genome-fragment | NA | | NA | 0 |
| S17_vs_k141_116129||full | 1519 | No | NA | 2 | 0 | 0 | Not-determined | Genome-fragment | NA | | NA | 0 |
| S17_vs_k141_167606||full | 2900 | No | NA | 4 | 0 | 2 | Not-determined | Genome-fragment | NA | | NA | 0 |
| S17_vs_k141_138021||full | 2427 | No | NA | 3 | 0 | 2 | Not-determined | Genome-fragment | NA | | NA | 0 |
| S17_vs_k141_71864||full | 2659 | No | NA | 5 | 0 | 0 | Not-determined | Genome-fragment | NA | | NA | 0 |
| S1_vs_k141_94459||lt2gene | 1065 | No | NA | 2 | 1 | 0 | Low-quality | Genome-fragment | | 3.12 | AAI-based (medium-confidence) | 0 |
| S17_vs_k141_57234||full | 4523 | No | NA | 4 | 0 | 1 | Not-determined | Genome-fragment | NA | | NA | 0 |
| S17_vs_k141_42715||full | 1310 | No | NA | 2 | 0 | 0 | Not-determined | Genome-fragment | NA | | NA | 0 |
| S17_vs_k141_49836||full | 1996 | No | NA | 2 | 0 | 0 | Not-determined | Genome-fragment | NA | | NA | 0 |
| S3_extracted_k141_51822 | 1065 | No | NA | 3 | 0 | 0 | Low-quality | Genome-fragment | | 0.7 | AAI-based (high-confidence) | 0 |
| S17_vs_k141_85673||full | 1094 | No | NA | 3 | 0 | 0 | Not-determined | Genome-fragment | NA | | NA | 0 |
| S7_extracted_35615 | 1065 | No | NA | 1 | 0 | 0 | Low-quality | Genome-fragment | | 1.93 | AAI-based (high-confidence) | 0 |
| S7_vs_52987||full | 1065 | No | NA | 3 | 0 | 1 | Low-quality | Genome-fragment | | 1.82 | AAI-based (high-confidence) | 0 |
| S7_vs_61176||full | 1065 | No | NA | 3 | 0 | 0 | Low-quality | Genome-fragment | | 1.75 | AAI-based (medium-confidence) | 0 |
| S7_vs_124975||full | 1065 | No | NA | 2 | 0 | 0 | Low-quality | Genome-fragment | | 1.29 | AAI-based (high-confidence) | 0 |
| S16_extracted_142915 | 1065 | No | NA | 2 | 0 | 0 | Low-quality | Genome-fragment | | 1.92 | AAI-based (medium-confidence) | 0 |
| S16_vs_28960||full | 1065 | No | NA | 2 | 0 | 0 | Low-quality | Genome-fragment | | 0.56 | AAI-based (medium-confidence) | 0 |
| S17_vs_k141_153564||full | 1415 | No | NA | 3 | 0 | 2 | Not-determined | Genome-fragment | NA | | NA | 0 |
| S17_vs_k141_42746||full | 4467 | No | NA | 4 | 0 | 4 | Not-determined | Genome-fragment | NA | | NA | 0 |
| S16_vs_121844||full | 1065 | No | NA | 2 | 0 | 0 | Low-quality | Genome-fragment | | 2.55 | AAI-based (high-confidence) | 0 |
| S16_vs_171006||full | 1065 | No | NA | 3 | 1 | 0 | Low-quality | Genome-fragment | | 2.09 | AAI-based (high-confidence) | 0 |
| S12_vs_k141_83854||lt2gene | 1065 | No | NA | 1 | 1 | 0 | Low-quality | Genome-fragment | | 2.11 | AAI-based (medium-confidence) | 0 |
| S17_vs_k141_35299||full | 1482 | No | NA | 3 | 0 | 1 | Not-determined | Genome-fragment | NA | | NA | 0 |
| S17_extracted_k141_14995 | 1066 | No | NA | 2 | 1 | 0 | Low-quality | Genome-fragment | | 2.98 | AAI-based (high-confidence) | 0 |
| S17_vs_k141_63511||full | 3493 | No | NA | 4 | 0 | 1 | Not-determined | Genome-fragment | NA | | NA | 0 |
| S17_vs_k141_116247||full | 1066 | No | NA | 3 | 0 | 0 | Not-determined | Genome-fragment | NA | | NA | 0 |
| S17_extracted_k141_51481 | 1066 | No | NA | 2 | 0 | 1 | Low-quality | Genome-fragment | | 0.32 | AAI-based (medium-confidence) | 0 |
| S17_vs_k141_85735||full | 1376 | No | NA | 3 | 0 | 0 | Not-determined | Genome-fragment | NA | | NA | 0 |
| S17_vs_k141_31268||lt2gene | 1066 | No | NA | 2 | 0 | 0 | Low-quality | Genome-fragment | | 1.93 | AAI-based (medium-confidence) | 0 |
| S17_vs_k141_71972||full | 3832 | No | NA | 2 | 0 | 1 | Not-determined | Genome-fragment | NA | | NA | 0 |
| S17_vs_k141_160502||full | 37084 | No | NA | 47 | 6 | 1 | Medium-quality | Genome-fragment | | 63.41 | AAI-based (high-confidence) | 0 |
| S11_extracted_k141_42082 | 1066 | No | NA | 2 | 0 | 0 | Low-quality | Genome-fragment | | 0.96 | AAI-based (medium-confidence) | 0 |
| S13_extracted_154021 | 1066 | No | NA | 2 | 0 | 0 | Low-quality | Genome-fragment | | 2.06 | AAI-based (medium-confidence) | 0 |
| S14_extracted_k141_111522 | 1066 | No | NA | 3 | 0 | 0 | Low-quality | Genome-fragment | | 1.95 | AAI-based (medium-confidence) | 0 |
| S14_vs_k141_39961||full | 1066 | No | NA | 2 | 2 | 0 | Low-quality | Genome-fragment | | 2.95 | AAI-based (medium-confidence) | 0 |
| S17_vs_k141_123050||full | 1605 | No | NA | 1 | 0 | 1 | Not-determined | Genome-fragment | NA | | NA | 0 |
| S17_vs_k141_63567||full | 1566 | No | NA | 3 | 0 | 0 | Not-determined | Genome-fragment | NA | | NA | 0 |
| S1_vs_k141_12121||full | 1066 | No | NA | 3 | 1 | 0 | Low-quality | Genome-fragment | | 1.34 | AAI-based (high-confidence) | 0 |
| S17_vs_k141_123077||full | 1135 | No | NA | 2 | 0 | 1 | Not-determined | Genome-fragment | NA | | NA | 0 |
| S17_vs_k141_130470||full | 1370 | No | NA | 2 | 0 | 2 | Not-determined | Genome-fragment | NA | | NA | 0 |
| S1_vs_k141_106049||full | 1066 | No | NA | 3 | 2 | 0 | Low-quality | Genome-fragment | | 2.47 | AAI-based (high-confidence) | 0 |
| S17_vs_k141_174617||full | 4836 | No | NA | 5 | 0 | 2 | Not-determined | Genome-fragment | NA | | NA | 0 |
| S5_extracted_k141_78054 | 1066 | No | NA | 1 | 0 | 1 | Low-quality | Genome-fragment | | 1.18 | AAI-based (medium-confidence) | 0 |
| S17_vs_k141_26758||full | 2873 | No | NA | 5 | 0 | 2 | Not-determined | Genome-fragment | NA | | NA | 0 |
| S17_vs_k141_79284||full | 5362 | No | NA | 4 | 0 | 4 | Not-determined | Genome-fragment | NA | | NA | 0 |
| S5_vs_k141_50748||full | 1066 | No | NA | 3 | 1 | 0 | Low-quality | Genome-fragment | | 3.04 | AAI-based (high-confidence) | 0 |
| S5_vs_k141_9380||full | 1066 | No | NA | 2 | 1 | 0 | Low-quality | Genome-fragment | | 1.76 | AAI-based (medium-confidence) | 0 |
| S17_vs_k141_160590||full | 1167 | No | NA | 2 | 0 | 1 | Not-determined | Genome-fragment | NA | | NA | 0 |
| S7_vs_57064||full | 1066 | No | NA | 3 | 1 | 0 | Low-quality | Genome-fragment | | 2.64 | AAI-based (high-confidence) | 0 |
| S17_vs_k141_145688||full | 2180 | No | NA | 3 | 0 | 3 | Not-determined | Genome-fragment | NA | | NA | 0 |
| S17_vs_k141_42860||full | 3909 | No | NA | 7 | 0 | 0 | Not-determined | Genome-fragment | NA | | NA | 0 |
| S7_vs_96792||full | 1066 | No | NA | 4 | 0 | 0 | Low-quality | Genome-fragment | | 2.37 | AAI-based (medium-confidence) | 0 |
| S17_vs_k141_26774||full | 5536 | No | NA | 6 | 0 | 2 | Not-determined | Genome-fragment | NA | | NA | 0 |
| S17_vs_k141_12521||full | 8202 | No | NA | 8 | 0 | 3 | Not-determined | Genome-fragment | NA | | NA | 0 |
| S17_vs_k141_12528||full | 1368 | No | NA | 3 | 0 | 2 | Not-determined | Genome-fragment | NA | | NA | 0 |
| S17_vs_k141_153613||full | 2518 | No | NA | 3 | 0 | 3 | Not-determined | Genome-fragment | NA | | NA | 0 |
| S17_vs_k141_12536||full | 2088 | No | NA | 4 | 0 | 0 | Not-determined | Genome-fragment | NA | | NA | 0 |
| S17_vs_k141_85914||full | 1963 | No | NA | 2 | 0 | 2 | Not-determined | Genome-fragment | NA | | NA | 0 |
| S7_vs_20194||lt2gene | 1066 | No | NA | 1 | 1 | 0 | Low-quality | Genome-fragment | | 2.45 | AAI-based (high-confidence) | 0 |
| S16_extracted_15520 | 1066 | No | NA | 2 | 0 | 0 | Low-quality | Genome-fragment | | 1.92 | AAI-based (medium-confidence) | 0 |
| S17_vs_k141_93770||full | 4223 | No | NA | 2 | 0 | 2 | Not-determined | Genome-fragment | NA | | NA | 0 |
| S10_extracted_40093 | 1066 | No | NA | 2 | 0 | 2 | Low-quality | Genome-fragment | | 0.57 | AAI-based (medium-confidence) | 0 |
| S17_vs_k141_5401||full | 2562 | No | NA | 6 | 0 | 1 | Not-determined | Genome-fragment | NA | | NA | 0 |
| S10_extracted_57163 | 1066 | No | NA | 2 | 2 | 0 | Low-quality | Genome-fragment | | 0.62 | HMM-based (lower-bound) | 0 |
| S17_vs_k141_160676||full | 3039 | No | NA | 4 | 0 | 2 | Not-determined | Genome-fragment | NA | | NA | 0 |
| S12_extracted_k141_299 | 1066 | No | NA | 3 | 0 | 0 | Low-quality | Genome-fragment | | 2.45 | AAI-based (high-confidence) | 0 |
| S17_vs_k141_123195||full | 1709 | No | NA | 3 | 0 | 3 | Not-determined | Genome-fragment | NA | | NA | 0 |
| S12_vs_k141_169357||full | 1066 | No | NA | 3 | 2 | 0 | Low-quality | Genome-fragment | | 2.52 | AAI-based (medium-confidence) | 0 |
| S17_vs_k141_109558||full | 3109 | No | NA | 3 | 0 | 1 | Not-determined | Genome-fragment | NA | | NA | 0 |
| S6_extracted_k141_42682 | 1066 | No | NA | 1 | 0 | 0 | Low-quality | Genome-fragment | | 1.45 | AAI-based (medium-confidence) | 0 |
| S17_vs_k141_145811||full | 2683 | No | NA | 5 | 0 | 0 | Not-determined | Genome-fragment | NA | | NA | 0 |
| S8_vs_k141_21776||lt2gene | 1066 | No | NA | 2 | 1 | 0 | Low-quality | Genome-fragment | | 0.57 | HMM-based (lower-bound) | 0 |
| S17_vs_k141_72104||full | 3372 | No | NA | 4 | 0 | 2 | Not-determined | Genome-fragment | NA | | NA | 0 |
| S17_extracted_k141_52075 | 1067 | No | NA | 1 | 0 | 0 | Low-quality | Genome-fragment | | 1.72 | AAI-based (high-confidence) | 0 |
| S17_vs_k141_85962||full | 11079 | No | NA | 8 | 0 | 5 | Not-determined | Genome-fragment | NA | | NA | 0 |
| S17_extracted_k141_76021 | 1067 | No | NA | 1 | 0 | 0 | Low-quality | Genome-fragment | | 0.81 | AAI-based (high-confidence) | 0 |
| S17_vs_k141_160741||full | 1537 | No | NA | 3 | 0 | 2 | Not-determined | Genome-fragment | NA | | NA | 0 |
| S17_vs_k141_155014||full | 1067 | No | NA | 2 | 0 | 0 | Low-quality | Genome-fragment | | 2.79 | AAI-based (high-confidence) | 0 |
| S17_vs_k141_174752||full | 8637 | No | NA | 5 | 0 | 2 | Not-determined | Genome-fragment | NA | | NA | 0 |
| S17_vs_k141_174758||full | 1602 | No | NA | 3 | 0 | 0 | Not-determined | Genome-fragment | NA | | NA | 0 |
| S9_vs_k141_14419||full | 1067 | No | NA | 3 | 1 | 0 | Low-quality | Genome-fragment | | 0.69 | HMM-based (lower-bound) | 0 |
| S17_vs_k141_109611||full | 5287 | No | NA | 5 | 0 | 3 | Not-determined | Genome-fragment | NA | | NA | 0 |
| S17_vs_k141_12665||full | 2292 | No | NA | 3 | 0 | 2 | Not-determined | Genome-fragment | NA | | NA | 0 |
| S17_vs_k141_79436||full | 1143 | No | NA | 3 | 0 | 1 | Not-determined | Genome-fragment | NA | | NA | 0 |
| S17_vs_k141_93855||full | 4428 | No | NA | 5 | 0 | 1 | Not-determined | Genome-fragment | NA | | NA | 0 |
| S14_vs_k141_13086||full | 1067 | No | NA | 3 | 1 | 0 | Low-quality | Genome-fragment | | 1.86 | AAI-based (high-confidence) | 0 |
| S1_vs_k141_108436||full | 1067 | No | NA | 2 | 0 | 0 | Low-quality | Genome-fragment | | 0.89 | AAI-based (medium-confidence) | 0 |
| S17_vs_k141_174798||full | 1919 | No | NA | 3 | 0 | 1 | Not-determined | Genome-fragment | NA | | NA | 0 |
| S1_vs_k141_152021||lt2gene | 1067 | No | NA | 1 | 0 | 0 | Low-quality | Genome-fragment | | 2.64 | AAI-based (high-confidence) | 0 |
| S3_vs_k141_33587||full | 1067 | No | NA | 2 | 0 | 0 | Low-quality | Genome-fragment | | 2.67 | AAI-based (high-confidence) | 0 |
| S3_vs_k141_28622||full | 1067 | No | NA | 3 | 0 | 0 | Low-quality | Genome-fragment | | 1.36 | AAI-based (high-confidence) | 0 |
| S17_vs_k141_138306||full | 3504 | No | NA | 3 | 0 | 2 | Not-determined | Genome-fragment | NA | | NA | 0 |
| S5_vs_k141_13926||full | 1067 | No | NA | 3 | 0 | 0 | Low-quality | Genome-fragment | | 2.14 | AAI-based (high-confidence) | 0 |
| S5_vs_k141_18839||lt2gene | 1067 | No | NA | 2 | 1 | 0 | Low-quality | Genome-fragment | | 2.48 | AAI-based (medium-confidence) | 0 |
| S17_vs_k141_138320||full | 3006 | No | NA | 2 | 0 | 2 | Not-determined | Genome-fragment | NA | | NA | 0 |
| S17_vs_k141_35594||full | 3539 | No | NA | 4 | 0 | 4 | Not-determined | Genome-fragment | NA | | NA | 0 |
| S7_extracted_71002 | 1067 | No | NA | 4 | 1 | 0 | Low-quality | Genome-fragment | | 1.96 | AAI-based (high-confidence) | 0 |
| S7_vs_103637||full | 1067 | No | NA | 3 | 0 | 0 | Low-quality | Genome-fragment | | 2.58 | AAI-based (medium-confidence) | 0 |
| S17_vs_k141_160844||full | 2077 | No | NA | 2 | 0 | 1 | Not-determined | Genome-fragment | NA | | NA | 0 |
| S7_vs_117667||full | 1067 | No | NA | 3 | 0 | 0 | Low-quality | Genome-fragment | | 2.57 | AAI-based (high-confidence) | 0 |
| S16_extracted_5952 | 1067 | No | NA | 2 | 0 | 0 | Low-quality | Genome-fragment | | 2.01 | AAI-based (medium-confidence) | 0 |
| S17_vs_k141_116634||full | 1597 | No | NA | 3 | 0 | 1 | Not-determined | Genome-fragment | NA | | NA | 0 |
| S17_vs_k141_168049||full | 2405 | No | NA | 4 | 0 | 2 | Not-determined | Genome-fragment | NA | | NA | 0 |
| S17_vs_k141_57559||full | 2144 | No | NA | 4 | 0 | 0 | Not-determined | Genome-fragment | NA | | NA | 0 |
| S10_extracted_6298 | 1067 | No | NA | 2 | 0 | 1 | Low-quality | Genome-fragment | | 1.43 | AAI-based (medium-confidence) | 0 |
| S17_vs_k141_26945||full | 1026 | No | NA | 2 | 0 | 1 | Not-determined | Genome-fragment | NA | | NA | 0 |
| S12_vs_k141_157599||full | 1067 | No | NA | 3 | 1 | 0 | Low-quality | Genome-fragment | | 1.06 | AAI-based (high-confidence) | 0 |
| S17_vs_k141_116640||full | 20807 | No | NA | 28 | 10 | 0 | Medium-quality | Genome-fragment | | 53.37 | AAI-based (medium-confidence) | 0 |
| S17_vs_k141_153891||full | 1420 | No | NA | 2 | 0 | 2 | Not-determined | Genome-fragment | NA | | NA | 0 |
| S12_vs_k141_32262||lt2gene | 1067 | No | NA | 1 | 1 | 0 | Low-quality | Genome-fragment | | 2.33 | HMM-based (lower-bound) | 0 |
| S8_extracted_k141_24192 | 1067 | No | NA | 2 | 0 | 0 | Low-quality | Genome-fragment | | 1.62 | AAI-based (high-confidence) | 0 |
| S17_vs_k141_12692||full | 2752 | No | NA | 3 | 0 | 2 | Not-determined | Genome-fragment | NA | | NA | 0 |
| S9_extracted_k141_10150 | 1068 | No | NA | 2 | 1 | 0 | Low-quality | Genome-fragment | | 1.7 | AAI-based (medium-confidence) | 0 |
| S17_vs_k141_19843||full | 2329 | No | NA | 3 | 0 | 2 | Not-determined | Genome-fragment | NA | | NA | 0 |
| S17_vs_k141_79589||full | 3051 | No | NA | 3 | 0 | 2 | Not-determined | Genome-fragment | NA | | NA | 0 |
| S11_vs_k141_70769||full | 1068 | No | NA | 3 | 1 | 0 | Low-quality | Genome-fragment | | 2.41 | AAI-based (medium-confidence) | 0 |
| S17_vs_k141_130684||full | 2502 | No | NA | 3 | 0 | 0 | Not-determined | Genome-fragment | NA | | NA | 0 |
| S17_vs_k141_43165||full | 1485 | No | NA | 3 | 0 | 0 | Not-determined | Genome-fragment | NA | | NA | 0 |
| S17_vs_k141_79599||full | 2260 | No | NA | 2 | 0 | 2 | Not-determined | Genome-fragment | NA | | NA | 0 |
| S17_vs_k141_35766||full | 3011 | No | NA | 3 | 0 | 3 | Not-determined | Genome-fragment | NA | | NA | 0 |
| S17_vs_k141_168159||full | 1861 | No | NA | 3 | 0 | 0 | Not-determined | Genome-fragment | NA | | NA | 0 |
| S13_extracted_97585 | 1068 | No | NA | 1 | 0 | 0 | Low-quality | Genome-fragment | | 2.43 | AAI-based (medium-confidence) | 0 |
| S13_vs_77||full | 1068 | No | NA | 4 | 0 | 1 | Low-quality | Genome-fragment | | 0.67 | AAI-based (medium-confidence) | 0 |
| S17_vs_k141_35783||full | 2658 | No | NA | 2 | 0 | 1 | Not-determined | Genome-fragment | NA | | NA | 0 |
| S17_vs_k141_161002||full | 1008 | No | NA | 2 | 0 | 1 | Not-determined | Genome-fragment | NA | | NA | 0 |
| S17_vs_k141_50403||full | 2220 | No | NA | 4 | 0 | 0 | Not-determined | Genome-fragment | NA | | NA | 0 |
| S17_vs_k141_79628||full | 3350 | No | NA | 3 | 0 | 1 | Not-determined | Genome-fragment | NA | | NA | 0 |
| S13_vs_23914||full | 1068 | No | NA | 4 | 0 | 0 | Low-quality | Genome-fragment | | 1.39 | AAI-based (high-confidence) | 0 |
| S17_vs_k141_130721||full | 2637 | No | NA | 4 | 0 | 2 | Not-determined | Genome-fragment | NA | | NA | 0 |
| S17_vs_k141_174936||full | 3030 | No | NA | 3 | 0 | 1 | Not-determined | Genome-fragment | NA | | NA | 0 |
| S13_vs_28431||full | 1068 | No | NA | 4 | 0 | 0 | Low-quality | Genome-fragment | | 1.99 | AAI-based (medium-confidence) | 0 |
| S14_vs_k141_85495||full | 1068 | No | NA | 3 | 0 | 0 | Low-quality | Genome-fragment | | 6.24 | AAI-based (high-confidence) | 0 |
| S17_vs_k141_93957||full | 2029 | No | NA | 3 | 0 | 2 | Not-determined | Genome-fragment | NA | | NA | 0 |
| S14_vs_k141_63984||full | 1068 | No | NA | 2 | 1 | 0 | Low-quality | Genome-fragment | | 2.18 | AAI-based (medium-confidence) | 0 |
| S17_vs_k141_161038||full | 1714 | No | NA | 3 | 0 | 3 | Not-determined | Genome-fragment | NA | | NA | 0 |
| S17_vs_k141_57650||full | 2168 | No | NA | 4 | 0 | 0 | Not-determined | Genome-fragment | NA | | NA | 0 |
| S14_vs_k141_90761||lt2gene | 1068 | No | NA | 1 | 1 | 0 | Low-quality | Genome-fragment | | 2.77 | AAI-based (high-confidence) | 0 |
| S1_vs_k141_55346||full | 1068 | No | NA | 2 | 0 | 1 | Low-quality | Genome-fragment | | 2.08 | AAI-based (medium-confidence) | 0 |
| S1_vs_k141_134739||lt2gene | 1068 | No | NA | 1 | 1 | 0 | Low-quality | Genome-fragment | | 2.23 | AAI-based (high-confidence) | 0 |
| S17_vs_k141_27108||full | 1309 | No | NA | 3 | 0 | 0 | Not-determined | Genome-fragment | NA | | NA | 0 |
| S3_extracted_k141_45575 | 1068 | No | NA | 2 | 0 | 0 | Low-quality | Genome-fragment | | 1.58 | AAI-based (high-confidence) | 0 |
| S3_vs_k141_124683||full | 1068 | No | NA | 3 | 2 | 0 | Low-quality | Genome-fragment | | 1.73 | AAI-based (high-confidence) | 0 |
| S5_vs_k141_133756||full | 1068 | No | NA | 4 | 1 | 0 | Low-quality | Genome-fragment | | 1.77 | AAI-based (medium-confidence) | 0 |
| S17_vs_k141_5725||full | 1733 | No | NA | 3 | 0 | 0 | Not-determined | Genome-fragment | NA | | NA | 0 |
| S17_vs_k141_63999||full | 1478 | No | NA | 4 | 0 | 2 | Not-determined | Genome-fragment | NA | | NA | 0 |
| S5_vs_k141_117809||full | 1068 | No | NA | 3 | 0 | 0 | Low-quality | Genome-fragment | | 2.68 | AAI-based (high-confidence) | 0 |
| S5_vs_k141_43255||lt2gene | 1068 | No | NA | 2 | 2 | 0 | Low-quality | Genome-fragment | | 3.29 | AAI-based (high-confidence) | 0 |
| S17_vs_k141_161090||full | 1804 | No | NA | 3 | 0 | 0 | Not-determined | Genome-fragment | NA | | NA | 0 |
| S7_vs_30328||full | 1068 | No | NA | 3 | 2 | 0 | Low-quality | Genome-fragment | | 2.56 | AAI-based (medium-confidence) | 0 |
| S7_vs_136428||full | 1068 | No | NA | 3 | 0 | 0 | Low-quality | Genome-fragment | | 2.27 | AAI-based (high-confidence) | 0 |
| S16_vs_9347||full | 1068 | No | NA | 3 | 0 | 0 | Low-quality | Genome-fragment | | 1.93 | AAI-based (medium-confidence) | 0 |
| S16_vs_90939||full | 1068 | No | NA | 3 | 1 | 0 | Low-quality | Genome-fragment | | 2.48 | AAI-based (medium-confidence) | 0 |
| S12_vs_k141_95253||full | 1068 | No | NA | 3 | 0 | 0 | Low-quality | Genome-fragment | | 1.99 | AAI-based (high-confidence) | 0 |
| S12_vs_k141_125410||full | 1068 | No | NA | 2 | 0 | 0 | Low-quality | Genome-fragment | | 2.94 | AAI-based (medium-confidence) | 0 |
| S17_vs_k141_5753||full | 2907 | No | NA | 5 | 0 | 1 | Not-determined | Genome-fragment | NA | | NA | 0 |
| S6_extracted_k141_37223 | 1068 | No | NA | 1 | 0 | 1 | Low-quality | Genome-fragment | | 0.34 | AAI-based (medium-confidence) | 0 |
| S6_extracted_k141_51098 | 1068 | No | NA | 2 | 0 | 0 | Low-quality | Genome-fragment | | 2.3 | AAI-based (medium-confidence) | 0 |
| S17_vs_k141_123573||full | 1088 | No | NA | 2 | 0 | 2 | Not-determined | Genome-fragment | NA | | NA | 0 |
| S6_vs_k141_56155||full | 1068 | No | NA | 2 | 0 | 0 | Low-quality | Genome-fragment | | 3.05 | AAI-based (high-confidence) | 0 |
| S17_vs_k141_123582||full | 4011 | No | NA | 5 | 0 | 2 | Not-determined | Genome-fragment | NA | | NA | 0 |
| S17_vs_k141_12881||full | 3280 | No | NA | 4 | 0 | 0 | Not-determined | Genome-fragment | NA | | NA | 0 |
| S17_vs_k141_138575||full | 1560 | No | NA | 4 | 0 | 1 | Not-determined | Genome-fragment | NA | | NA | 0 |
| S17_vs_k141_43300||full | 1255 | No | NA | 3 | 0 | 0 | Not-determined | Genome-fragment | NA | | NA | 0 |
| S17_vs_k141_19974||full | 1570 | No | NA | 4 | 0 | 0 | Not-determined | Genome-fragment | NA | | NA | 0 |
| S11_vs_k141_95490||full | 1069 | No | NA | 4 | 0 | 0 | Low-quality | Genome-fragment | | 1.88 | AAI-based (medium-confidence) | 0 |
| S11_vs_k141_36482||lt2gene | 1069 | No | NA | 2 | 2 | 0 | Low-quality | Genome-fragment | | 1.3 | HMM-based (lower-bound) | 0 |
| S17_vs_k141_168235||full | 57169 | No | NA | 50 | 0 | 10 | Not-determined | Genome-fragment | NA | | NA | 0 |
| S17_vs_k141_27183||full | 1632 | No | NA | 3 | 0 | 0 | Not-determined | Genome-fragment | NA | | NA | 0 |
| S13_vs_153118||full | 1069 | No | NA | 3 | 0 | 3 | Low-quality | Genome-fragment | | 0.33 | AAI-based (medium-confidence) | 0 |
| S14_extracted_k141_94958 | 1069 | No | NA | 3 | 1 | 0 | Low-quality | Genome-fragment | | 2.56 | AAI-based (high-confidence) | 0 |
| S17_vs_k141_50578||full | 1234 | No | NA | 3 | 0 | 0 | Not-determined | Genome-fragment | NA | | NA | 0 |
| S14_vs_k141_53370||full | 1069 | No | NA | 4 | 0 | 0 | Low-quality | Genome-fragment | | 2.77 | AAI-based (medium-confidence) | 0 |
| S17_vs_k141_174992||full | 1414 | No | NA | 3 | 0 | 0 | Not-determined | Genome-fragment | NA | | NA | 0 |
| S17_vs_k141_130920||full | 1370 | No | NA | 3 | 0 | 2 | Not-determined | Genome-fragment | NA | | NA | 0 |
| S17_vs_k141_138634||full | 1695 | No | NA | 3 | 0 | 2 | Not-determined | Genome-fragment | NA | | NA | 0 |
| S17_vs_k141_102068||full | 16475 | No | NA | 19 | 0 | 2 | Not-determined | Genome-fragment | NA | | NA | 0 |
| S17_vs_k141_110068||full | 4540 | No | NA | 4 | 0 | 2 | Not-determined | Genome-fragment | NA | | NA | 0 |
| S17_vs_k141_86370||full | 3666 | No | NA | 5 | 0 | 3 | Not-determined | Genome-fragment | NA | | NA | 0 |
| S1_vs_k141_42858||full | 1069 | No | NA | 3 | 0 | 2 | Low-quality | Genome-fragment | | 0.68 | AAI-based (medium-confidence) | 0 |
| S17_vs_k141_86377||full | 1716 | No | NA | 3 | 0 | 0 | Not-determined | Genome-fragment | NA | | NA | 0 |
| S17_vs_k141_43307||full | 42499 | No | NA | 34 | 5 | 1 | Medium-quality | Genome-fragment | | 66.94 | AAI-based (high-confidence) | 0 |
| S5_vs_k141_67019||full | 1069 | No | NA | 3 | 2 | 0 | Low-quality | Genome-fragment | | 2.44 | AAI-based (medium-confidence) | 0 |
| S16_extracted_125168 | 1069 | No | NA | 2 | 0 | 0 | Low-quality | Genome-fragment | | 1.23 | AAI-based (medium-confidence) | 0 |
| S17_vs_k141_35957||full | 3203 | No | NA | 2 | 0 | 0 | Not-determined | Genome-fragment | NA | | NA | 0 |
| S17_vs_k141_154186||full | 4674 | No | NA | 2 | 0 | 1 | Not-determined | Genome-fragment | NA | | NA | 0 |
| S17_vs_k141_57762||full | 5559 | No | NA | 3 | 0 | 2 | Not-determined | Genome-fragment | NA | | NA | 0 |
| S17_vs_k141_110100||full | 1024 | No | NA | 3 | 0 | 0 | Not-determined | Genome-fragment | NA | | NA | 0 |
| S16_vs_14820||lt2gene | 1069 | No | NA | 2 | 2 | 0 | Low-quality | Genome-fragment | | 2.24 | AAI-based (medium-confidence) | 0 |
| S17_vs_k141_27285||full | 3403 | No | NA | 4 | 0 | 2 | Not-determined | Genome-fragment | NA | | NA | 0 |
| S17_vs_k141_35986||full | 3275 | No | NA | 3 | 0 | 1 | Not-determined | Genome-fragment | NA | | NA | 0 |
| S17_vs_k141_123703||full | 1880 | No | NA | 3 | 0 | 2 | Not-determined | Genome-fragment | NA | | NA | 0 |
| S17_vs_k141_79861||full | 3497 | No | NA | 2 | 0 | 1 | Not-determined | Genome-fragment | NA | | NA | 0 |

| | | | | | | | | | | | |
|---|---|---|---|---|---|---|---|---|---|---|---|
| S17_vs_k141_64184\|\|full | 1015 | No | NA | 3 | 0 | 0 | Not-determined | Genome-fragment | NA | NA | 0 |
| S17_vs_k141_146279\|\|full | 4454 | No | NA | 5 | 0 | 3 | Not-determined | Genome-fragment | NA | NA | 0 |
| S17_vs_k141_161306\|\|full | 1357 | No | NA | 3 | 0 | 2 | Not-determined | Genome-fragment | NA | NA | 0 |
| S17_vs_k141_146291\|\|full | 3160 | No | NA | 4 | 0 | 2 | Not-determined | Genome-fragment | NA | NA | 0 |
| S17_vs_k141_79875\|\|full | 5986 | No | NA | 7 | 0 | 2 | Not-determined | Genome-fragment | NA | NA | 0 |
| S10_vs_10905\|\|lt2gene | 1069 | No | NA | 2 | 1 | 0 | Low-quality | Genome-fragment | 1.42 | HMM-based (lower-bound) | 0 |
| S17_vs_k141_146308\|\|full | 2362 | No | NA | 2 | 0 | 0 | Not-determined | Genome-fragment | NA | NA | 0 |
| S12_extracted_k141_122422 | 1069 | No | NA | 3 | 0 | 0 | Low-quality | Genome-fragment | 1.02 | AAI-based (medium-confidence) | 0 |
| S17_vs_k141_102193\|\|full | 2735 | No | NA | 5 | 0 | 1 | Not-determined | Genome-fragment | NA | NA | 0 |
| S17_vs_k141_131017\|\|full | 1644 | No | NA | 2 | 0 | 2 | Not-determined | Genome-fragment | NA | NA | 0 |
| S17_vs_k141_161338\|\|full | 2199 | No | NA | 2 | 0 | 1 | Not-determined | Genome-fragment | NA | NA | 0 |
| S17_vs_k141_146333\|\|full | 2058 | No | NA | 2 | 0 | 1 | Not-determined | Genome-fragment | NA | NA | 0 |
| S17_vs_k141_50691\|\|full | 1413 | No | NA | 2 | 0 | 2 | Not-determined | Genome-fragment | NA | NA | 0 |
| S12_vs_k141_141597\|\|full | 1069 | No | NA | 3 | 1 | 0 | Low-quality | Genome-fragment | 2.36 | AAI-based (medium-confidence) | 0 |
| S17_vs_k141_146356\|\|full | 2160 | No | NA | 3 | 0 | 3 | Not-determined | Genome-fragment | NA | NA | 0 |
| S15_vs_47120\|\|lt2gene | 1069 | No | NA | 1 | 0 | 0 | Low-quality | Genome-fragment | 1.67 | AAI-based (high-confidence) | 0 |
| S17_vs_k141_146384\|\|full | 1048 | No | NA | 3 | 0 | 1 | Not-determined | Genome-fragment | NA | NA | 0 |
| S17_vs_k141_72599\|\|full | 2644 | No | NA | 2 | 0 | 1 | Not-determined | Genome-fragment | NA | NA | 0 |
| S17_vs_k141_8258\|\|lt2gene | 1070 | No | NA | 2 | 2 | 0 | Low-quality | Genome-fragment | 2.07 | AAI-based (high-confidence) | 0 |
| S17_vs_k141_86500\|\|full | 2110 | No | NA | 3 | 0 | 2 | Not-determined | Genome-fragment | NA | NA | 0 |
| S17_vs_k141_161388\|\|full | 1838 | No | NA | 3 | 0 | 0 | Not-determined | Genome-fragment | NA | NA | 0 |
| S9_vs_k141_5041\|\|full | 1070 | No | NA | 3 | 0 | 0 | Low-quality | Genome-fragment | 1.25 | AAI-based (medium-confidence) | 0 |
| S17_vs_k141_43451\|\|full | 1906 | No | NA | 3 | 0 | 1 | Not-determined | Genome-fragment | NA | NA | 0 |
| S11_extracted_k141_97615 | 1070 | No | NA | 1 | 0 | 1 | Low-quality | Genome-fragment | 0.34 | AAI-based (medium-confidence) | 0 |
| S13_extracted_5611 | 1070 | No | NA | 1 | 0 | 1 | Low-quality | Genome-fragment | 0.29 | AAI-based (medium-confidence) | 0 |
| S17_vs_k141_102301\|\|full | 5022 | No | NA | 4 | 0 | 3 | Not-determined | Genome-fragment | NA | NA | 0 |
| S14_extracted_k141_54336 | 1070 | No | NA | 1 | 0 | 0 | Low-quality | Genome-fragment | 2.06 | AAI-based (medium-confidence) | 0 |
| S17_vs_k141_102315\|\|full | 2003 | No | NA | 3 | 0 | 0 | Not-determined | Genome-fragment | NA | NA | 0 |
| S17_vs_k141_64350\|\|full | 1708 | No | NA | 3 | 0 | 1 | Not-determined | Genome-fragment | NA | NA | 0 |
| S17_vs_k141_110303\|\|full | 1796 | No | NA | 2 | 0 | 2 | Not-determined | Genome-fragment | NA | NA | 0 |
| S17_vs_k141_13155\|\|full | 1065 | No | NA | 2 | 0 | 0 | Not-determined | Genome-fragment | NA | NA | 0 |
| S14_extracted_k141_83943 | 1070 | No | NA | 2 | 0 | 0 | Low-quality | Genome-fragment | 5.61 | AAI-based (high-confidence) | 0 |
| S17_vs_k141_138876\|\|full | 4152 | No | NA | 5 | 0 | 3 | Not-determined | Genome-fragment | NA | NA | 0 |
| S14_vs_k141_112058\|\|lt2gene | 1070 | No | NA | 2 | 1 | 0 | Low-quality | Genome-fragment | 1.8 | AAI-based (medium-confidence) | 0 |
| S5_vs_k141_84901\|\|full | 1070 | No | NA | 3 | 0 | 1 | Low-quality | Genome-fragment | 2.77 | AAI-based (high-confidence) | 0 |
| S17_vs_k141_161481\|\|full | 1262 | No | NA | 3 | 0 | 0 | Not-determined | Genome-fragment | NA | NA | 0 |
| S17_vs_k141_57995\|\|full | 5499 | No | NA | 3 | 0 | 1 | Not-determined | Genome-fragment | NA | NA | 0 |
| S7_extracted_61998 | 1070 | No | NA | 3 | 1 | 0 | Low-quality | Genome-fragment | 2 | AAI-based (medium-confidence) | 0 |
| S17_vs_k141_131168\|\|full | 2367 | No | NA | 5 | 0 | 1 | Not-determined | Genome-fragment | NA | NA | 0 |
| S17_vs_k141_168442\|\|full | 7584 | No | NA | 7 | 0 | 4 | Not-determined | Genome-fragment | NA | NA | 0 |
| S17_vs_k141_86622\|\|full | 1213 | No | NA | 2 | 0 | 1 | Not-determined | Genome-fragment | NA | NA | 0 |
| S17_vs_k141_20302\|\|full | 1403 | No | NA | 3 | 0 | 0 | Not-determined | Genome-fragment | NA | NA | 0 |
| S17_vs_k141_154524\|\|full | 2078 | No | NA | 5 | 0 | 0 | Not-determined | Genome-fragment | NA | NA | 0 |
| S17_vs_k141_146564\|\|full | 1235 | No | NA | 2 | 0 | 0 | Not-determined | Genome-fragment | NA | NA | 0 |
| S17_vs_k141_131194\|\|full | 1888 | No | NA | 3 | 0 | 3 | Not-determined | Genome-fragment | NA | NA | 0 |
| S17_vs_k141_64447\|\|full | 3164 | No | NA | 4 | 0 | 3 | Not-determined | Genome-fragment | NA | NA | 0 |
| S7_vs_106088\|\|full | 1070 | No | NA | 3 | 1 | 0 | Low-quality | Genome-fragment | 1.99 | AAI-based (medium-confidence) | 0 |
| S16_extracted_101339 | 1070 | No | NA | 2 | 0 | 0 | Low-quality | Genome-fragment | 1.77 | AAI-based (high-confidence) | 0 |
| S17_vs_k141_102370\|\|full | 1139 | No | NA | 2 | 0 | 1 | Not-determined | Genome-fragment | NA | NA | 0 |
| S17_vs_k141_175234\|\|full | 2654 | No | NA | 5 | 0 | 1 | Not-determined | Genome-fragment | NA | NA | 0 |
| S17_vs_k141_20317\|\|full | 15455 | No | NA | 14 | 0 | 11 | Not-determined | Genome-fragment | NA | NA | 0 |
| S16_extracted_104121 | 1070 | No | NA | 2 | 1 | 0 | Low-quality | Genome-fragment | 2.53 | AAI-based (medium-confidence) | 0 |
| S17_vs_k141_110426\|\|full | 1985 | No | NA | 3 | 0 | 2 | Not-determined | Genome-fragment | NA | NA | 0 |
| S17_vs_k141_58025\|\|full | 12945 | No | NA | 16 | 5 | 0 | High-quality | High-quality | 94.49 | AAI-based (medium-confidence) | 0 |
| S17_vs_k141_102412\|\|full | 1321 | No | NA | 2 | 0 | 0 | Not-determined | Genome-fragment | NA | NA | 0 |
| S16_vs_98242\|\|full | 1070 | No | NA | 2 | 0 | 0 | Low-quality | Genome-fragment | 0.67 | AAI-based (medium-confidence) | 0 |
| S10_extracted_71334 | 1070 | No | NA | 3 | 0 | 0 | Low-quality | Genome-fragment | 1.33 | AAI-based (high-confidence) | 0 |
| S17_vs_k141_36188\|\|full | 1817 | No | NA | 3 | 0 | 0 | Not-determined | Genome-fragment | NA | NA | 0 |
| S17_vs_k141_27588\|\|full | 2801 | No | NA | 3 | 0 | 3 | Not-determined | Genome-fragment | NA | NA | 0 |
| S17_vs_k141_110435\|\|full | 1042 | No | NA | 4 | 0 | 2 | Not-determined | Genome-fragment | NA | NA | 0 |
| S10_extracted_91113 | 1070 | No | NA | 2 | 0 | 2 | Low-quality | Genome-fragment | 1.47 | AAI-based (high-confidence) | 0 |
| S17_vs_k141_64516\|\|full | 1113 | No | NA | 3 | 0 | 1 | Not-determined | Genome-fragment | NA | NA | 0 |
| S17_vs_k141_161600\|\|full | 1471 | No | NA | 2 | 0 | 1 | Not-determined | Genome-fragment | NA | NA | 0 |
| S17_vs_k141_175302\|\|full | 1342 | No | NA | 2 | 0 | 1 | Not-determined | Genome-fragment | NA | NA | 0 |
| S17_vs_k141_117355\|\|full | 2667 | No | NA | 7 | 0 | 0 | Not-determined | Genome-fragment | NA | NA | 0 |
| S17_vs_k141_64543\|\|full | 9122 | No | NA | 4 | 0 | 4 | Not-determined | Genome-fragment | NA | NA | 0 |
| S10_vs_61861\|\|full | 1070 | No | NA | 3 | 0 | 0 | Low-quality | Genome-fragment | 1.66 | AAI-based (high-confidence) | 0 |
| S17_vs_k141_6263\|\|full | 5531 | No | NA | 5 | 0 | 1 | Not-determined | Genome-fragment | NA | NA | 0 |
| S12_vs_k141_19710\|\|full | 1070 | No | NA | 2 | 0 | 0 | Low-quality | Genome-fragment | 2.49 | AAI-based (high-confidence) | 0 |
| S17_vs_k141_94401\|\|full | 1616 | No | NA | 2 | 0 | 1 | Not-determined | Genome-fragment | NA | NA | 0 |
| S12_vs_k141_94784\|\|lt2gene | 1070 | No | NA | 1 | 1 | 0 | Low-quality | Genome-fragment | 2.41 | AAI-based (medium-confidence) | 0 |
| S17_vs_k141_64602\|\|full | 1934 | No | NA | 2 | 0 | 2 | Not-determined | Genome-fragment | NA | NA | 0 |
| S17_vs_k141_64611\|\|full | 1679 | No | NA | 2 | 0 | 1 | Not-determined | Genome-fragment | NA | NA | 0 |
| S4_extracted_31063 | 1070 | No | NA | 2 | 0 | 0 | Low-quality | Genome-fragment | 1.18 | AAI-based (medium-confidence) | 0 |
| S17_vs_k141_50896\|\|full | 2502 | No | NA | 2 | 0 | 2 | Not-determined | Genome-fragment | NA | NA | 0 |
| S17_vs_k141_175417\|\|full | 1183 | No | NA | 3 | 0 | 0 | Not-determined | Genome-fragment | NA | NA | 0 |
| S17_vs_k141_154719\|\|full | 1631 | No | NA | 3 | 0 | 2 | Not-determined | Genome-fragment | NA | NA | 0 |
| S17_vs_k141_72874\|\|full | 1060 | No | NA | 2 | 0 | 0 | Not-determined | Genome-fragment | NA | NA | 0 |
| S8_extracted_k141_36618 | 1070 | No | NA | 1 | 0 | 1 | Low-quality | Genome-fragment | 3.42 | AAI-based (high-confidence) | 0 |
| S17_vs_k141_20493\|\|full | 1320 | No | NA | 2 | 0 | 2 | Not-determined | Genome-fragment | NA | NA | 0 |
| S8_vs_k141_37439\|\|full | 1070 | No | NA | 3 | 0 | 0 | Low-quality | Genome-fragment | 0.66 | AAI-based (high-confidence) | 0 |
| S17_vs_k141_154736\|\|full | 3675 | No | NA | 3 | 0 | 3 | Not-determined | Genome-fragment | NA | NA | 0 |
| S17_vs_k141_85751\|\|full | 1071 | No | NA | 2 | 0 | 1 | Low-quality | Genome-fragment | 1.29 | AAI-based (medium-confidence) | 0 |
| S17_vs_k141_168494\|\|full | 1946 | No | NA | 3 | 0 | 1 | Not-determined | Genome-fragment | NA | NA | 0 |
| S13_vs_31337\|\|full | 1071 | No | NA | 3 | 0 | 0 | Low-quality | Genome-fragment | 1.81 | AAI-based (medium-confidence) | 0 |
| S1_extracted_k141_97017 | 1071 | No | NA | 3 | 0 | 0 | Low-quality | Genome-fragment | 0.72 | AAI-based (high-confidence) | 0 |
| S17_vs_k141_110565\|\|full | 2561 | No | NA | 4 | 0 | 4 | Not-determined | Genome-fragment | NA | NA | 0 |
| S17_vs_k141_154742\|\|full | 2553 | No | NA | 3 | 0 | 2 | Not-determined | Genome-fragment | NA | NA | 0 |
| S1_extracted_k141_79737 | 1071 | No | NA | 1 | 0 | 1 | Low-quality | Genome-fragment | 0.97 | AAI-based (high-confidence) | 0 |
| S17_vs_k141_168507\|\|full | 1617 | No | NA | 2 | 0 | 2 | Not-determined | Genome-fragment | NA | NA | 0 |
| S7_extracted_75779 | 1071 | No | NA | 2 | 0 | 0 | Low-quality | Genome-fragment | 2.67 | AAI-based (high-confidence) | 0 |
| S17_vs_k141_86903\|\|full | 2562 | No | NA | 3 | 0 | 3 | Not-determined | Genome-fragment | NA | NA | 0 |
| S7_vs_29590\|\|full | 1071 | No | NA | 3 | 1 | 0 | Low-quality | Genome-fragment | 2.44 | AAI-based (medium-confidence) | 0 |
| S16_extracted_142393 | 1071 | No | NA | 4 | 0 | 0 | Low-quality | Genome-fragment | 1.79 | AAI-based (high-confidence) | 0 |
| S17_vs_k141_161749\|\|full | 1437 | No | NA | 2 | 0 | 1 | Not-determined | Genome-fragment | NA | NA | 0 |
| S17_vs_k141_131428\|\|full | 2094 | No | NA | 3 | 0 | 2 | Not-determined | Genome-fragment | NA | NA | 0 |
| S16_vs_124728\|\|lt2gene | 1071 | No | NA | 2 | 2 | 0 | Low-quality | Genome-fragment | 2.68 | AAI-based (high-confidence) | 0 |
| S10_vs_55809\|\|full | 1071 | No | NA | 4 | 0 | 0 | Low-quality | Genome-fragment | 2.1 | AAI-based (high-confidence) | 0 |
| S17_vs_k141_175502\|\|full | 1337 | No | NA | 2 | 0 | 0 | Not-determined | Genome-fragment | NA | NA | 0 |
| S17_vs_k141_139161\|\|full | 1819 | No | NA | 3 | 0 | 1 | Not-determined | Genome-fragment | NA | NA | 0 |
| S17_vs_k141_20542\|\|full | 2070 | No | NA | 4 | 0 | 1 | Not-determined | Genome-fragment | NA | NA | 0 |
| S17_vs_k141_168565\|\|full | 1354 | No | NA | 2 | 0 | 2 | Not-determined | Genome-fragment | NA | NA | 0 |
| S17_vs_k141_154785\|\|full | 7721 | No | NA | 4 | 0 | 2 | Not-determined | Genome-fragment | NA | NA | 0 |
| S17_vs_k141_110614\|\|full | 1986 | No | NA | 3 | 0 | 0 | Not-determined | Genome-fragment | NA | NA | 0 |
| S17_vs_k141_117507\|\|full | 25659 | No | NA | 25 | 1 | 1 | Medium-quality | Genome-fragment | 58.11 | AAI-based (medium-confidence) | 0 |
| S10_vs_92079\|\|full | 1071 | No | NA | 3 | 0 | 0 | Low-quality | Genome-fragment | 2.16 | AAI-based (medium-confidence) | 0 |
| S17_vs_k141_80348\|\|full | 1578 | No | NA | 4 | 0 | 1 | Not-determined | Genome-fragment | NA | NA | 0 |
| S10_vs_53381\|\|lt2gene | 1071 | No | NA | 2 | 0 | 0 | Low-quality | Genome-fragment | 2.57 | AAI-based (high-confidence) | 0 |
| S17_vs_k141_20580\|\|full | 3576 | No | NA | 3 | 0 | 2 | Not-determined | Genome-fragment | NA | NA | 0 |
| S12_extracted_k141_136650 | 1071 | No | NA | 2 | 0 | 0 | Low-quality | Genome-fragment | 0.47 | AAI-based (medium-confidence) | 0 |
| S17_vs_k141_73008\|\|full | 1541 | No | NA | 3 | 0 | 1 | Not-determined | Genome-fragment | NA | NA | 0 |
| S17_vs_k141_36418\|\|full | 1089 | No | NA | 2 | 0 | 1 | Not-determined | Genome-fragment | NA | NA | 0 |
| S15_extracted_76136 | 1071 | No | NA | 2 | 0 | 1 | Low-quality | Genome-fragment | 3.63 | AAI-based (medium-confidence) | 0 |
| S17_vs_k141_36437\|\|full | 5135 | No | NA | 2 | 0 | 0 | Not-determined | Genome-fragment | NA | NA | 0 |
| S17_vs_k141_62546\|\|full | 1072 | No | NA | 3 | 0 | 0 | Low-quality | Genome-fragment | 2.5 | AAI-based (high-confidence) | 0 |
| S17_vs_k141_58303\|\|full | 1788 | No | NA | 2 | 0 | 0 | Not-determined | Genome-fragment | NA | NA | 0 |
| S17_vs_k141_161889\|\|full | 1088 | No | NA | 2 | 0 | 1 | Not-determined | Genome-fragment | NA | NA | 0 |
| S17_vs_k141_58306\|\|full | 1324 | No | NA | 3 | 0 | 0 | Not-determined | Genome-fragment | NA | NA | 0 |
| S17_vs_k141_124322\|\|full | 1515 | No | NA | 3 | 0 | 1 | Not-determined | Genome-fragment | NA | NA | 0 |
| S11_extracted_k141_60911 | 1072 | No | NA | 1 | 1 | 0 | Low-quality | Genome-fragment | 2.26 | AAI-based (high-confidence) | 0 |
| S17_vs_k141_58318\|\|full | 2399 | No | NA | 2 | 0 | 2 | Not-determined | Genome-fragment | NA | NA | 0 |
| S14_extracted_k141_25949 | 1072 | No | NA | 3 | 2 | 0 | Low-quality | Genome-fragment | 2.52 | AAI-based (medium-confidence) | 0 |
| S1_vs_k141_43311\|\|full | 1072 | No | NA | 2 | 1 | 0 | Low-quality | Genome-fragment | 2.8 | AAI-based (high-confidence) | 0 |
| S1_vs_k141_158460\|\|full | 1072 | No | NA | 3 | 0 | 0 | Low-quality | Genome-fragment | 2.54 | AAI-based (high-confidence) | 0 |
| S3_extracted_k141_112047 | 1072 | No | NA | 1 | 0 | 0 | Low-quality | Genome-fragment | 1.36 | AAI-based (high-confidence) | 0 |
| S17_vs_k141_168702\|\|full | 1441 | No | NA | 3 | 0 | 1 | Not-determined | Genome-fragment | NA | NA | 0 |
| S17_vs_k141_147095\|\|full | 1533 | No | NA | 3 | 0 | 1 | Not-determined | Genome-fragment | NA | NA | 0 |
| S17_vs_k141_110666\|\|full | 70348 | Yes | 66114 | 105 | 29 | 4 | High-quality | High-quality | 100 | AAI-based (high-confidence) | 6.02 |
| S17_vs_k141_51172\|\|full | 2156 | No | NA | 3 | 0 | 1 | Not-determined | Genome-fragment | NA | NA | 0 |
| S3_extracted_k141_63290 | 1072 | No | NA | 1 | 0 | 0 | Low-quality | Genome-fragment | 0.81 | AAI-based (high-confidence) | 0 |
| S3_vs_k141_33429\|\|full | 1072 | No | NA | 3 | 0 | 0 | Low-quality | Genome-fragment | 0.77 | AAI-based (high-confidence) | 0 |
| S17_vs_k141_43968\|\|full | 1150 | No | NA | 2 | 0 | 1 | Not-determined | Genome-fragment | NA | NA | 0 |
| S3_vs_k141_78199\|\|full | 1072 | No | NA | 2 | 1 | 0 | Low-quality | Genome-fragment | 2.44 | AAI-based (high-confidence) | 0 |
| S17_vs_k141_175574\|\|full | 1460 | No | NA | 2 | 0 | 1 | Not-determined | Genome-fragment | NA | NA | 0 |
| S17_vs_k141_94612\|\|full | 2214 | No | NA | 3 | 0 | 0 | Not-determined | Genome-fragment | NA | NA | 0 |
| S17_vs_k141_80501\|\|full | 2034 | No | NA | 3 | 0 | 1 | Not-determined | Genome-fragment | NA | NA | 0 |
| S3_vs_k141_102351\|\|full | 1072 | No | NA | 4 | 0 | 0 | Low-quality | Genome-fragment | 2.44 | AAI-based (high-confidence) | 0 |
| S17_vs_k141_175591\|\|full | 2267 | No | NA | 4 | 0 | 1 | Not-determined | Genome-fragment | NA | NA | 0 |
| S17_vs_k141_73104\|\|full | 1848 | No | NA | 2 | 0 | 2 | Not-determined | Genome-fragment | NA | NA | 0 |
| S17_vs_k141_124387\|\|full | 1432 | No | NA | 3 | 0 | 0 | Not-determined | Genome-fragment | NA | NA | 0 |
| S5_extracted_k141_9123 | 1072 | No | NA | 2 | 1 | 0 | Low-quality | Genome-fragment | 2.86 | AAI-based (high-confidence) | 0 |
| S17_vs_k141_168769\|\|full | 2432 | No | NA | 2 | 0 | 1 | Not-determined | Genome-fragment | NA | NA | 0 |
| S5_extracted_k141_136936 | 1072 | No | NA | 2 | 0 | 0 | Low-quality | Genome-fragment | 2.73 | AAI-based (medium-confidence) | 0 |
| S5_vs_k141_20889\|\|full | 1072 | No | NA | 2 | 0 | 0 | Low-quality | Genome-fragment | 1.3 | AAI-based (high-confidence) | 0 |
| S17_vs_k141_44024\|\|full | 1803 | No | NA | 2 | 0 | 1 | Not-determined | Genome-fragment | NA | NA | 0 |
| S5_vs_k141_35187\|\|lt2gene | 1072 | No | NA | 2 | 1 | 0 | Low-quality | Genome-fragment | 2.64 | AAI-based (high-confidence) | 0 |
| S17_vs_k141_6635\|\|full | 1330 | No | NA | 3 | 0 | 0 | Not-determined | Genome-fragment | NA | NA | 0 |
| S17_vs_k141_147198\|\|full | 3174 | No | NA | 5 | 0 | 3 | Not-determined | Genome-fragment | NA | NA | 0 |
| S7_vs_14679\|\|full | 1072 | No | NA | 1 | 0 | 0 | Low-quality | Genome-fragment | 2.36 | AAI-based (high-confidence) | 0 |
| S17_vs_k141_147203\|\|full | 1722 | No | NA | 5 | 0 | 0 | Not-determined | Genome-fragment | NA | NA | 0 |
| S17_vs_k141_73174\|\|full | 2806 | No | NA | 2 | 0 | 2 | Not-determined | Genome-fragment | NA | NA | 0 |
| S17_vs_k141_131679\|\|full | 1049 | No | NA | 3 | 0 | 2 | Not-determined | Genome-fragment | NA | NA | 0 |
| S17_vs_k141_139339\|\|full | 2499 | No | NA | 2 | 0 | 1 | Not-determined | Genome-fragment | NA | NA | 0 |
| S7_vs_73401\|\|full | 1072 | No | NA | 2 | 0 | 0 | Low-quality | Genome-fragment | 2.15 | AAI-based (high-confidence) | 0 |
| S17_vs_k141_51265\|\|full | 1680 | No | NA | 3 | 0 | 1 | Not-determined | Genome-fragment | NA | NA | 0 |
| S17_vs_k141_168824\|\|full | 1848 | No | NA | 3 | 0 | 2 | Not-determined | Genome-fragment | NA | NA | 0 |
| S16_extracted_22507 | 1072 | No | NA | 1 | 0 | 0 | Low-quality | Genome-fragment | 0.7 | AAI-based (high-confidence) | 0 |
| S17_vs_k141_80606\|\|full | 1026 | No | NA | 2 | 0 | 1 | Not-determined | Genome-fragment | NA | NA | 0 |
| S17_vs_k141_51291\|\|full | 7510 | No | NA | 9 | 0 | 7 | Not-determined | Genome-fragment | NA | NA | 0 |
| S17_vs_k141_131733\|\|full | 2932 | No | NA | 4 | 0 | 3 | Not-determined | Genome-fragment | NA | NA | 0 |
| S17_vs_k141_87206\|\|full | 1915 | No | NA | 3 | 0 | 2 | Not-determined | Genome-fragment | NA | NA | 0 |
| S17_vs_k141_6697\|\|full | 3115 | No | NA | 3 | 0 | 2 | Not-determined | Genome-fragment | NA | NA | 0 |
| S16_vs_93993\|\|full | 1072 | No | NA | 5 | 0 | 0 | Low-quality | Genome-fragment | 3.2 | AAI-based (high-confidence) | 0 |
| S17_vs_k141_73257\|\|full | 1007 | No | NA | 3 | 0 | 0 | Not-determined | Genome-fragment | NA | NA | 0 |
| S17_vs_k141_80638\|\|full | 2092 | No | NA | 3 | 0 | 2 | Not-determined | Genome-fragment | NA | NA | 0 |
| S17_vs_k141_117707\|\|full | 1958 | No | NA | 3 | 0 | 1 | Not-determined | Genome-fragment | NA | NA | 0 |
| S17_vs_k141_44076\|\|full | 3167 | No | NA | 6 | 0 | 0 | Not-determined | Genome-fragment | NA | NA | 0 |
| S17_vs_k141_51330\|\|full | 1547 | No | NA | 3 | 0 | 0 | Not-determined | Genome-fragment | NA | NA | 0 |
| S10_vs_23091\|\|lt2gene | 1072 | No | NA | 2 | 1 | 0 | Low-quality | Genome-fragment | 2.29 | AAI-based (medium-confidence) | 0 |
| S12_vs_k141_175954\|\|full | 1072 | No | NA | 2 | 0 | 1 | Low-quality | Genome-fragment | 1.82 | AAI-based (medium-confidence) | 0 |
| S17_vs_k141_139423\|\|full | 1624 | No | NA | 3 | 0 | 0 | Not-determined | Genome-fragment | NA | NA | 0 |
| S17_vs_k141_73288\|\|full | 1067 | No | NA | 2 | 0 | 0 | Not-determined | Genome-fragment | NA | NA | 0 |
| S15_extracted_58187 | 1072 | No | NA | 3 | 1 | 0 | Low-quality | Genome-fragment | 1.21 | AAI-based (high-confidence) | 0 |
| S17_vs_k141_44103\|\|full | 8521 | No | NA | 4 | 0 | 3 | Not-determined | Genome-fragment | NA | NA | 0 |
| S6_vs_k141_6681\|\|full | 1072 | No | NA | 3 | 0 | 0 | Low-quality | Genome-fragment | 2.05 | AAI-based (high-confidence) | 0 |
| S8_vs_k141_8904\|\|full | 1072 | No | NA | 2 | 1 | 0 | Low-quality | Genome-fragment | 0.67 | AAI-based (high-confidence) | 0 |
| S8_vs_k141_104000\|\|full | 1072 | No | NA | 2 | 0 | 1 | Low-quality | Genome-fragment | 0.3 | AAI-based (medium-confidence) | 0 |
| S17_vs_k141_168898\|\|full | 7403 | No | NA | 6 | 0 | 2 | Not-determined | Genome-fragment | NA | NA | 0 |
| S17_vs_k141_13823\|\|full | 1715 | No | NA | 2 | 0 | 2 | Not-determined | Genome-fragment | NA | NA | 0 |
| S17_vs_k141_139463\|\|full | 1506 | No | NA | 3 | 0 | 1 | Not-determined | Genome-fragment | NA | NA | 0 |
| S17_vs_k141_110805\|\|full | 1880 | No | NA | 3 | 0 | 1 | Not-determined | Genome-fragment | NA | NA | 0 |
| S17_vs_k141_73330\|\|full | 3568 | No | NA | 5 | 0 | 3 | Not-determined | Genome-fragment | NA | NA | 0 |
| S17_vs_k141_20768\|\|full | 2161 | No | NA | 4 | 0 | 1 | Not-determined | Genome-fragment | NA | NA | 0 |
| S17_extracted_k141_138867 | 1073 | No | NA | 1 | 0 | 1 | Low-quality | Genome-fragment | 0.4 | AAI-based (medium-confidence) | 0 |
| S17_vs_k141_64236\|\|lt2gene | 1073 | No | NA | 2 | 0 | 0 | Low-quality | Genome-fragment | 2.6 | AAI-based (medium-confidence) | 0 |
| S9_extracted_k141_49587 | 1073 | No | NA | 2 | 1 | 0 | Low-quality | Genome-fragment | 2.91 | AAI-based (high-confidence) | 0 |
| S17_vs_k141_64684\|\|full | 1741 | No | NA | 4 | 0 | 0 | Not-determined | Genome-fragment | NA | NA | 0 |
| S9_vs_k141_26177\|\|lt2gene | 1073 | No | NA | 1 | 1 | 0 | Low-quality | Genome-fragment | 1.57 | HMM-based (lower-bound) | 0 |
| S13_vs_15121\|\|full | 1073 | No | NA | 3 | 0 | 2 | Low-quality | Genome-fragment | 1.56 | AAI-based (high-confidence) | 0 |
| S17_vs_k141_117802\|\|full | 2878 | No | NA | 2 | 0 | 1 | Not-determined | Genome-fragment | NA | NA | 0 |
| S17_vs_k141_169010\|\|full | 1509 | No | NA | 2 | 0 | 1 | Not-determined | Genome-fragment | NA | NA | 0 |
| S17_vs_k141_117830\|\|full | 3089 | No | NA | 3 | 0 | 2 | Not-determined | Genome-fragment | NA | NA | 0 |
| S17_vs_k141_131917\|\|full | 1628 | No | NA | 2 | 0 | 2 | Not-determined | Genome-fragment | NA | NA | 0 |
| S14_vs_k141_62476\|\|full | 1073 | No | NA | 3 | 0 | 0 | Low-quality | Genome-fragment | 2.75 | AAI-based (medium-confidence) | 0 |
| S17_vs_k141_80727\|\|full | 1386 | No | NA | 2 | 0 | 1 | Not-determined | Genome-fragment | NA | NA | 0 |
| S17_vs_k141_51471\|\|full | 1623 | No | NA | 3 | 0 | 1 | Not-determined | Genome-fragment | NA | NA | 0 |
| S17_vs_k141_103150\|\|full | 4415 | No | NA | 5 | 0 | 1 | Not-determined | Genome-fragment | NA | NA | 0 |
| S1_vs_k141_15803\|\|full | 1073 | No | NA | 2 | 0 | 1 | Low-quality | Genome-fragment | 0.45 | AAI-based (medium-confidence) | 0 |
| S17_vs_k141_117862\|\|full | 1685 | No | NA | 2 | 0 | 1 | Not-determined | Genome-fragment | NA | NA | 0 |
| S17_vs_k141_131974\|\|full | 1371 | No | NA | 3 | 0 | 2 | Not-determined | Genome-fragment | NA | NA | 0 |
| S1_vs_k141_16026\|\|lt2gene | 1073 | No | NA | 2 | 0 | 0 | Low-quality | Genome-fragment | 2.13 | AAI-based (high-confidence) | 0 |
| S3_extracted_k141_90201 | 1073 | No | NA | 3 | 0 | 0 | Low-quality | Genome-fragment | 0.92 | AAI-based (medium-confidence) | 0 |
| S3_vs_k141_101249\|\|full | 1073 | No | NA | 3 | 0 | 0 | Low-quality | Genome-fragment | 2.69 | AAI-based (high-confidence) | 0 |
| S5_extracted_k141_47797 | 1073 | No | NA | 2 | 2 | 0 | Low-quality | Genome-fragment | 1.67 | HMM-based (lower-bound) | 0 |
| S5_vs_k141_17854\|\|full | 1073 | No | NA | 3 | 1 | 0 | Low-quality | Genome-fragment | 2.94 | AAI-based (high-confidence) | 0 |
| S17_vs_k141_169079\|\|full | 1043 | No | NA | 2 | 0 | 1 | Not-determined | Genome-fragment | NA | NA | 0 |
| S17_vs_k141_117885\|\|full | 4158 | No | NA | 4 | 0 | 3 | Not-determined | Genome-fragment | NA | NA | 0 |
| S17_vs_k141_13929\|\|full | 2290 | No | NA | 2 | 0 | 0 | Not-determined | Genome-fragment | NA | NA | 0 |
| S17_vs_k141_132049\|\|full | 2229 | No | NA | 3 | 0 | 3 | Not-determined | Genome-fragment | NA | NA | 0 |
| S17_vs_k141_13953\|\|full | 2247 | No | NA | 3 | 0 | 2 | Not-determined | Genome-fragment | NA | NA | 0 |
| S17_vs_k141_94924\|\|full | 1608 | No | NA | 2 | 0 | 1 | Not-determined | Genome-fragment | NA | NA | 0 |
| S17_vs_k141_162305\|\|full | 1272 | No | NA | 2 | 0 | 1 | Not-determined | Genome-fragment | NA | NA | 0 |
| S17_vs_k141_162310\|\|full | 2392 | No | NA | 4 | 0 | 2 | Not-determined | Genome-fragment | NA | NA | 0 |
| S17_vs_k141_28327\|\|full | 3221 | No | NA | 4 | 0 | 1 | Not-determined | Genome-fragment | NA | NA | 0 |
| S5_vs_k141_6797\|\|lt2gene | 1073 | No | NA | 2 | 1 | 0 | Low-quality | Genome-fragment | 1.65 | HMM-based (lower-bound) | 0 |
| S17_vs_k141_162319\|\|full | 1554 | No | NA | 3 | 0 | 2 | Not-determined | Genome-fragment | NA | NA | 0 |
| S7_extracted_15893 | 1073 | No | NA | 2 | 0 | 0 | Low-quality | Genome-fragment | 1.34 | AAI-based (medium-confidence) | 0 |
| S17_vs_k141_87548\|\|full | 14884 | No | NA | 20 | 0 | 2 | Not-determined | Genome-fragment | NA | NA | 0 |
| S17_vs_k141_103311\|\|full | 1314 | No | NA | 2 | 0 | 1 | Not-determined | Genome-fragment | NA | NA | 0 |
| S17_vs_k141_73585\|\|full | 7545 | No | NA | 5 | 0 | 3 | Not-determined | Genome-fragment | NA | NA | 0 |
| S7_extracted_63375 | 1073 | No | NA | 2 | 1 | 0 | Low-quality | Genome-fragment | 1.61 | HMM-based (lower-bound) | 0 |
| S17_vs_k141_64834\|\|full | 2213 | No | NA | 4 | 0 | 1 | Not-determined | Genome-fragment | NA | NA | 0 |
| S17_vs_k141_176033\|\|full | 2894 | No | NA | 5 | 0 | 1 | Not-determined | Genome-fragment | NA | NA | 0 |
| S17_vs_k141_6999\|\|full | 1106 | No | NA | 2 | 0 | 2 | Not-determined | Genome-fragment | NA | NA | 0 |
| S7_extracted_113821 | 1073 | No | NA | 2 | 0 | 1 | Low-quality | Genome-fragment | 0.36 | AAI-based (high-confidence) | 0 |
| S17_vs_k141_7008\|\|full | 1644 | No | NA | 2 | 0 | 2 | Not-determined | Genome-fragment | NA | NA | 0 |
| S16_extracted_155746 | 1073 | No | NA | 2 | 0 | 0 | Low-quality | Genome-fragment | 1.73 | AAI-based (high-confidence) | 0 |
| S17_vs_k141_117991\|\|full | 15700 | No | NA | 14 | 0 | 7 | Not-determined | Genome-fragment | NA | NA | 0 |
| S16_vs_139847\|\|lt2gene | 1073 | No | NA | 1 | 1 | 0 | Low-quality | Genome-fragment | 1.57 | HMM-based (lower-bound) | 0 |
| S12_vs_k141_70947\|\|full | 1073 | No | NA | 3 | 0 | 1 | Low-quality | Genome-fragment | 0.24 | AAI-based (medium-confidence) | 0 |
| S17_vs_k141_139660\|\|full | 1319 | No | NA | 2 | 0 | 1 | Not-determined | Genome-fragment | NA | NA | 0 |
| S17_vs_k141_169230\|\|full | 2341 | No | NA | 4 | 0 | 1 | Not-determined | Genome-fragment | NA | NA | 0 |
| S17_vs_k141_124738\|\|full | 1058 | No | NA | 3 | 0 | 0 | Not-determined | Genome-fragment | NA | NA | 0 |
| S6_vs_k141_1511\|\|full | 1073 | No | NA | 3 | 0 | 0 | Low-quality | Genome-fragment | 2.63 | AAI-based (high-confidence) | 0 |
| S17_vs_k141_21113\|\|full | 3178 | No | NA | 3 | 0 | 1 | Not-determined | Genome-fragment | NA | NA | 0 |
| S8_extracted_k141_31700 | 1073 | No | NA | 2 | 0 | 2 | Low-quality | Genome-fragment | 0.56 | AAI-based (medium-confidence) | 0 |
| S17_vs_k141_36792\|\|full | 3641 | No | NA | 7 | 0 | 4 | Not-determined | Genome-fragment | NA | NA | 0 |
| S17_vs_k141_103398\|\|full | 3245 | No | NA | 3 | 0 | 2 | Not-determined | Genome-fragment | NA | NA | 0 |
| S17_vs_k141_7140\|\|full | 3504 | No | NA | 5 | 0 | 4 | Not-determined | Genome-fragment | NA | NA | 0 |
| S8_vs_k141_143907\|\|full | 1073 | No | NA | 2 | 0 | 2 | Low-quality | Genome-fragment | 0.34 | AAI-based (medium-confidence) | 0 |
| S17_vs_k141_64955\|\|full | 3060 | No | NA | 3 | 0 | 2 | Not-determined | Genome-fragment | NA | NA | 0 |
| S9_extracted_k141_49826 | 1074 | No | NA | 1 | 0 | 0 | Low-quality | Genome-fragment | 0.45 | AAI-based (medium-confidence) | 0 |
| S11_vs_k141_38106\|\|lt2gene | 1074 | No | NA | 2 | 1 | 0 | Low-quality | Genome-fragment | 0.65 | HMM-based (lower-bound) | 0 |
| S17_vs_k141_7189\|\|full | 2483 | No | NA | 2 | 0 | 2 | Not-determined | Genome-fragment | NA | NA | 0 |
| S17_vs_k141_58752\|\|full | 1467 | No | NA | 1 | 0 | 0 | Not-determined | Genome-fragment | NA | NA | 0 |
| S17_vs_k141_21232\|\|full | 3069 | No | NA | 3 | 0 | 0 | Not-determined | Genome-fragment | NA | NA | 0 |
| S14_extracted_k141_79415 | 1074 | No | NA | 2 | 0 | 1 | Low-quality | Genome-fragment | 0.7 | AAI-based (medium-confidence) | 0 |
| S17_vs_k141_64984\|\|full | 1444 | No | NA | 3 | 0 | 0 | Not-determined | Genome-fragment | NA | NA | 0 |
| S14_vs_k141_3519\|\|full | 1074 | No | NA | 3 | 0 | 0 | Low-quality | Genome-fragment | 2.66 | AAI-based (high-confidence) | 0 |
| S1_extracted_k141_127786 | 1074 | No | NA | 4 | 0 | 0 | Low-quality | Genome-fragment | 1.99 | AAI-based (medium-confidence) | 0 |
| S5_vs_k141_17337\|\|full | 1074 | No | NA | 4 | 0 | 0 | Low-quality | Genome-fragment | 2.29 | AAI-based (high-confidence) | 0 |
| S5_vs_k141_111665\|\|lt2gene | 1074 | No | NA | 2 | 0 | 2 | Low-quality | Genome-fragment | 1.92 | AAI-based (medium-confidence) | 0 |
| S7_vs_25223\|\|full | 1074 | No | NA | 3 | 0 | 0 | Low-quality | Genome-fragment | 1.09 | AAI-based (medium-confidence) | 0 |
| S7_vs_116076\|\|lt2gene | 1074 | No | NA | 2 | 1 | 0 | Low-quality | Genome-fragment | 1.59 | HMM-based (lower-bound) | 0 |
| S10_vs_114234\|\|full | 1074 | No | NA | 4 | 0 | 1 | Low-quality | Genome-fragment | 3.15 | AAI-based (high-confidence) | 0 |
| S17_vs_k141_14197\|\|full | 2462 | No | NA | 3 | 0 | 2 | Not-determined | Genome-fragment | NA | NA | 0 |
| S17_vs_k141_124906\|\|full | 3165 | No | NA | 3 | 0 | 0 | Not-determined | Genome-fragment | NA | NA | 0 |
| S17_vs_k141_176255\|\|full | 1776 | No | NA | 2 | 0 | 1 | Not-determined | Genome-fragment | NA | NA | 0 |
| S10_vs_23815\|\|lt2gene | 1074 | Yes | 476 | 2 | 1 | 1 | Low-quality | Genome-fragment | 0.84 | AAI-based (medium-confidence) | 55.68 |
| S12_extracted_k141_123376 | 1074 | No | NA | 2 | 1 | 0 | Low-quality | Genome-fragment | 0.74 | HMM-based (lower-bound) | 0 |
| S17_vs_k141_73830\|\|full | 32605 | No | NA | 56 | 11 | 0 | High-quality | High-quality | 94.89 | AAI-based (high-confidence) | 0 |
| S8_extracted_k141_68565 | 1074 | No | NA | 2 | 1 | 0 | Low-quality | Genome-fragment | 1.85 | AAI-based (high-confidence) | 0 |
| S17_extracted_k141_37667 | 1075 | No | NA | 2 | 0 | 1 | Low-quality | Genome-fragment | 0.66 | AAI-based (medium-confidence) | 0 |
| S13_vs_53444\|\|lt2gene | 1075 | No | NA | 1 | 1 | 0 | Low-quality | Genome-fragment | 0.87 | HMM-based (lower-bound) | 0 |
| S17_vs_k141_169440\|\|full | 3772 | No | NA | 3 | 0 | 3 | Not-determined | Genome-fragment | NA | NA | 0 |
| S17_vs_k141_124939\|\|full | 1109 | No | NA | 3 | 0 | 1 | Not-determined | Genome-fragment | NA | NA | 0 |
| S17_vs_k141_81137\|\|full | 3786 | No | NA | 3 | 0 | 2 | Not-determined | Genome-fragment | NA | NA | 0 |
| S14_extracted_k141_13289 | 1075 | No | NA | 4 | 4 | 0 | Low-quality | Genome-fragment | 3.55 | AAI-based (high-confidence) | 0 |
| S14_vs_k141_39284\|\|lt2gene | 1075 | No | NA | 2 | 1 | 0 | Low-quality | Genome-fragment | 2.85 | AAI-based (high-confidence) | 0 |
| S14_vs_k141_10185\|\|lt2gene | 1075 | No | NA | 2 | 1 | 0 | Low-quality | Genome-fragment | 2.29 | AAI-based (high-confidence) | 0 |
| S5_extracted_k141_94815 | 1075 | No | NA | 1 | 0 | 0 | Low-quality | Genome-fragment | 2.7 | AAI-based (high-confidence) | 0 |
| S5_vs_k141_127394\|\|full | 1075 | No | NA | 3 | 0 | 0 | Low-quality | Genome-fragment | 49.27 | AAI-based (medium-confidence) | 0 |
| S17_vs_k141_95200\|\|full | 4727 | No | NA | 4 | 0 | 3 | Not-determined | Genome-fragment | NA | NA | 0 |
| S5_vs_k141_12913\|\|lt2gene | 1075 | No | NA | 3 | 2 | 0 | Low-quality | Genome-fragment | 2.06 | AAI-based (high-confidence) | 0 |
| S17_vs_k141_81202\|\|full | 7113 | No | NA | 6 | 0 | 1 | Not-determined | Genome-fragment | NA | NA | 0 |
| S17_vs_k141_36924\|\|full | 1478 | No | NA | 4 | 0 | 1 | Not-determined | Genome-fragment | NA | NA | 0 |
| S17_vs_k141_58920\|\|full | 1520 | No | NA | 4 | 0 | 2 | Not-determined | Genome-fragment | NA | NA | 0 |
| S5_vs_k141_136154\|\|lt2gene | 1075 | No | NA | 1 | 1 | 0 | Low-quality | Genome-fragment | 0.65 | HMM-based (lower-bound) | 0 |
| S17_vs_k141_21523\|\|full | 2431 | No | NA | 3 | 0 | 2 | Not-determined | Genome-fragment | NA | NA | 0 |
| S17_vs_k141_124990\|\|full | 2515 | No | NA | 3 | 0 | 1 | Not-determined | Genome-fragment | NA | NA | 0 |
| S17_vs_k141_176388\|\|full | 3007 | No | NA | 5 | 0 | 1 | Not-determined | Genome-fragment | NA | NA | 0 |
| S17_vs_k141_125004\|\|full | 1602 | No | NA | 4 | 0 | 0 | Not-determined | Genome-fragment | NA | NA | 0 |
| S17_vs_k141_14290\|\|full | 1158 | No | NA | 2 | 0 | 1 | Not-determined | Genome-fragment | NA | NA | 0 |
| S17_vs_k141_21577\|\|full | 1830 | No | NA | 4 | 0 | 0 | Not-determined | Genome-fragment | NA | NA | 0 |
| S16_vs_507\|\|full | 1075 | No | NA | 2 | 0 | 0 | Low-quality | Genome-fragment | 0.66 | AAI-based (medium-confidence) | 0 |
| S17_vs_k141_65286\|\|full | 1871 | No | NA | 3 | 0 | 2 | Not-determined | Genome-fragment | NA | NA | 0 |
| S17_vs_k141_58946\|\|full | 6748 | No | NA | 8 | 0 | 7 | Not-determined | Genome-fragment | NA | NA | 0 |
| S17_vs_k141_14365\|\|full | 2194 | No | NA | 4 | 0 | 0 | Not-determined | Genome-fragment | NA | NA | 0 |
| S10_vs_29910\|\|full | 1075 | No | NA | 2 | 1 | 0 | Low-quality | Genome-fragment | 0.95 | HMM-based (lower-bound) | 0 |
| S17_vs_k141_7360\|\|full | 6605 | No | NA | 10 | 0 | 4 | Not-determined | Genome-fragment | NA | NA | 0 |
| S17_vs_k141_14373\|\|full | 2105 | No | NA | 3 | 0 | 3 | Not-determined | Genome-fragment | NA | NA | 0 |
| S17_vs_k141_21652\|\|full | 2817 | No | NA | 3 | 0 | 2 | Not-determined | Genome-fragment | NA | NA | 0 |
| S17_vs_k141_14405\|\|full | 1414 | No | NA | 4 | 0 | 0 | Not-determined | Genome-fragment | NA | NA | 0 |
| S17_vs_k141_58999\|\|full | 3428 | No | NA | 4 | 0 | 3 | Not-determined | Genome-fragment | NA | NA | 0 |
| S12_extracted_k141_114046 | 1075 | No | NA | 2 | 1 | 0 | Low-quality | Genome-fragment | 0.92 | AAI-based (medium-confidence) | 0 |
| S17_vs_k141_169786\|\|full | 1560 | No | NA | 3 | 0 | 0 | Not-determined | Genome-fragment | NA | NA | 0 |
| S17_vs_k141_59022\|\|full | 2525 | No | NA | 4 | 0 | 0 | Not-determined | Genome-fragment | NA | NA | 0 |
| S12_vs_k141_29223\|\|full | 1075 | No | NA | 2 | 0 | 0 | Low-quality | Genome-fragment | 2.62 | AAI-based (high-confidence) | 0 |
| S17_vs_k141_28919\|\|full | 2779 | No | NA | 3 | 0 | 2 | Not-determined | Genome-fragment | NA | NA | 0 |
| S12_vs_k141_85119\|\|full | 1075 | No | NA | 2 | 2 | 0 | Low-quality | Genome-fragment | 2.58 | AAI-based (medium-confidence) | 0 |
| S17_vs_k141_95372\|\|full | 3364 | No | NA | 2 | 0 | 1 | Not-determined | Genome-fragment | NA | NA | 0 |
| S8_extracted_k141_176416 | 1075 | No | NA | 1 | 0 | 1 | Low-quality | Genome-fragment | 0.58 | AAI-based (medium-confidence) | 0 |
| S17_vs_k141_14510\|\|full | 3092 | No | NA | 4 | 0 | 1 | Not-determined | Genome-fragment | NA | NA | 0 |
| S17_vs_k141_169875\|\|full | 1757 | No | NA | 3 | 0 | 3 | Not-determined | Genome-fragment | NA | NA | 0 |
| S17_vs_k141_81255\|\|full | 1693 | No | NA | 3 | 0 | 1 | Not-determined | Genome-fragment | NA | NA | 0 |
| S17_vs_k141_65485\|\|full | 10873 | No | NA | 18 | 0 | 2 | Not-determined | Genome-fragment | NA | NA | 0 |
| S17_vs_k141_65502\|\|full | 1067 | No | NA | 2 | 0 | 0 | Not-determined | Genome-fragment | NA | NA | 0 |
| S17_vs_k141_169921\|\|full | 2614 | No | NA | 4 | 0 | 1 | Not-determined | Genome-fragment | NA | NA | 0 |
| S17_vs_k141_88218\|\|full | 1047 | No | NA | 2 | 0 | 1 | Not-determined | Genome-fragment | NA | NA | 0 |
| S17_vs_k141_14569\|\|full | 2864 | No | NA | 4 | 0 | 1 | Not-determined | Genome-fragment | NA | NA | 0 |
| S11_vs_k141_13482\|\|full | 1076 | No | NA | 3 | 0 | 1 | Low-quality | Genome-fragment | 0.36 | AAI-based (medium-confidence) | 0 |
| S17_vs_k141_21790\|\|full | 4254 | No | NA | 3 | 0 | 2 | Not-determined | Genome-fragment | NA | NA | 0 |
| S13_extracted_91984 | 1076 | No | NA | 3 | 0 | 2 | Low-quality | Genome-fragment | 2.4 | AAI-based (medium-confidence) | 0 |
| S13_vs_96206\|\|full | 1076 | No | NA | 3 | 0 | 0 | Low-quality | Genome-fragment | 3.38 | AAI-based (medium-confidence) | 0 |
| S1_extracted_k141_131374 | 1076 | No | NA | 2 | 1 | 0 | Low-quality | Genome-fragment | 0.7 | AAI-based (medium-confidence) | 0 |
| S17_vs_k141_125312\|\|full | 1275 | No | NA | 2 | 0 | 0 | Not-determined | Genome-fragment | NA | NA | 0 |
| S3_vs_k141_21642\|\|full | 1076 | No | NA | 3 | 1 | 0 | Low-quality | Genome-fragment | 3.13 | AAI-based (high-confidence) | 0 |
| S17_vs_k141_176686\|\|full | 1672 | No | NA | 2 | 0 | 0 | Not-determined | Genome-fragment | NA | NA | 0 |
| S5_extracted_k141_45881 | 1076 | No | NA | 5 | 1 | 0 | Low-quality | Genome-fragment | 3.13 | AAI-based (medium-confidence) | 0 |
| S17_vs_k141_132895\|\|full | 1282 | No | NA | 3 | 0 | 1 | Not-determined | Genome-fragment | NA | NA | 0 |
| S7_extracted_15476 | 1076 | No | NA | 2 | 0 | 0 | Low-quality | Genome-fragment | 0.91 | AAI-based (medium-confidence) | 0 |
| S17_vs_k141_125458\|\|full | 5928 | No | NA | 6 | 0 | 5 | Not-determined | Genome-fragment | NA | NA | 0 |
| S17_vs_k141_65746\|\|full | 1409 | No | NA | 3 | 0 | 3 | Not-determined | Genome-fragment | NA | NA | 0 |
| S7_extracted_34758 | 1076 | No | NA | 1 | 1 | 0 | Low-quality | Genome-fragment | 0.82 | HMM-based (lower-bound) | 0 |
| S17_vs_k141_125556\|\|full | 1602 | No | NA | 3 | 0 | 2 | Not-determined | Genome-fragment | NA | NA | 0 |
| S17_vs_k141_21993\|\|full | 5187 | No | NA | 6 | 0 | 1 | Not-determined | Genome-fragment | NA | NA | 0 |
| S7_vs_105065\|\|full | 1076 | No | NA | 3 | 1 | 0 | Low-quality | Genome-fragment | 3.13 | AAI-based (medium-confidence) | 0 |
| S17_vs_k141_65882\|\|full | 1940 | No | NA | 2 | 0 | 2 | Not-determined | Genome-fragment | NA | NA | 0 |
| S17_vs_k141_88522\|\|full | 1850 | No | NA | 3 | 0 | 1 | Not-determined | Genome-fragment | NA | NA | 0 |
| S12_extracted_k141_63888 | 1076 | No | NA | 2 | 1 | 0 | Low-quality | Genome-fragment | 0.8 | HMM-based (lower-bound) | 0 |
| S17_vs_k141_95855\|\|full | 1922 | No | NA | 3 | 0 | 1 | Not-determined | Genome-fragment | NA | NA | 0 |
| S12_extracted_k141_125692 | 1076 | No | NA | 2 | 0 | 1 | Low-quality | Genome-fragment | 0.33 | AAI-based (medium-confidence) | 0 |
| S17_vs_k141_95866\|\|full | 3044 | No | NA | 1 | 0 | 1 | Not-determined | Genome-fragment | NA | NA | 0 |
| S12_vs_k141_51695\|\|full | 1076 | No | NA | 2 | 0 | 0 | Low-quality | Genome-fragment | 2.4 | AAI-based (medium-confidence) | 0 |
| S12_vs_k141_119048\|\|lt2gene | 1076 | No | NA | 1 | 1 | 0 | Low-quality | Genome-fragment | 2.15 | AAI-based (high-confidence) | 0 |
| S15_vs_12954\|\|lt2gene | 1076 | No | NA | 1 | 1 | 0 | Low-quality | Genome-fragment | 2.82 | AAI-based (high-confidence) | 0 |
| S17_vs_k141_88632\|\|full | 2378 | No | NA | 3 | 0 | 3 | Not-determined | Genome-fragment | NA | NA | 0 |
| S17_vs_k141_22071\|\|full | 1071 | No | NA | 1 | 0 | 1 | Not-determined | Genome-fragment | NA | NA | 0 |
| S8_extracted_k141_3812 | 1076 | No | NA | 3 | 1 | 0 | Low-quality | Genome-fragment | 3.34 | AAI-based (high-confidence) | 0 |
| S8_extracted_k141_151538 | 1076 | No | NA | 1 | 0 | 0 | Low-quality | Genome-fragment | 2.47 | AAI-based (high-confidence) | 0 |
| S8_extracted_k141_169697 | 1076 | No | NA | 4 | 0 | 0 | Low-quality | Genome-fragment | 1.64 | AAI-based (medium-confidence) | 0 |
| S17_vs_k141_95960\|\|full | 10489 | No | NA | 9 | 0 | 0 | Not-determined | Genome-fragment | NA | NA | 0 |
| S17_vs_k141_170180\|\|full | 1077 | No | NA | 2 | 2 | 0 | Low-quality | Genome-fragment | 2.57 | AAI-based (high-confidence) | 0 |
| S17_vs_k141_177118\|\|full | 2563 | No | NA | 4 | 0 | 2 | Not-determined | Genome-fragment | NA | NA | 0 |
| S9_extracted_k141_67479 | 1077 | No | NA | 2 | 0 | 0 | Low-quality | Genome-fragment | 2.6 | AAI-based (high-confidence) | 0 |
| S17_vs_k141_177134\|\|full | 2939 | No | NA | 6 | 0 | 2 | Not-determined | Genome-fragment | NA | NA | 0 |
| S9_extracted_k141_52138 | 1077 | No | NA | 2 | 0 | 1 | Low-quality | Genome-fragment | 1.97 | AAI-based (high-confidence) | 0 |
| S14_vs_k141_45479\|\|full | 1077 | No | NA | 3 | 0 | 0 | Low-quality | Genome-fragment | 6.12 | AAI-based (medium-confidence) | 0 |
| S17_vs_k141_66121\|\|full | 1567 | No | NA | 3 | 0 | 0 | Not-determined | Genome-fragment | NA | NA | 0 |
| S17_vs_k141_66298\|\|full | 2012 | No | NA | 3 | 0 | 0 | Not-determined | Genome-fragment | NA | NA | 0 |
| S17_vs_k141_66330\|\|full | 3080 | No | NA | 4 | 0 | 2 | Not-determined | Genome-fragment | NA | NA | 0 |
| S17_vs_k141_66388\|\|full | 5961 | No | NA | 4 | 0 | 2 | Not-determined | Genome-fragment | NA | NA | 0 |
| S17_vs_k141_147886\|\|0_partial | 13305 | No | NA | 14 | 0 | 2 | Not-determined | Genome-fragment | NA | NA | 0 |
| S17_vs_k141_141233\|\|0_partial | 15752 | No | NA | 12 | 0 | 3 | Not-determined | Genome-fragment | NA | NA | 0 |
| S17_vs_k141_45389\|\|0_partial | 13282 | No | NA | 9 | 0 | 7 | Not-determined | Genome-fragment | NA | NA | 0 |
| S17_vs_k141_8690\|\|0_partial | 17288 | No | NA | 26 | 1 | 6 | High-quality | High-quality | 92.42 | HMM-based (lower-bound) | 0 |
| S17_vs_k141_82185\|\|0_partial | 12363 | No | NA | 15 | 0 | 6 | Not-determined | Genome-fragment | NA | NA | 0 |
| S14_vs_k141_57566\|\|full | 1077 | No | NA | 3 | 0 | 0 | Low-quality | Genome-fragment | 2.56 | AAI-based (high-confidence) | 0 |
| S1_extracted_k141_138855 | 1077 | No | NA | 1 | 0 | 0 | Low-quality | Genome-fragment | 0.81 | AAI-based (high-confidence) | 0 |
| S17_vs_k141_91924\|\|0_partial | 11124 | No | NA | 9 | 0 | 2 | Not-determined | Genome-fragment | NA | NA | 0 |
| S17_vs_k141_11405\|\|0_partial | 17913 | No | NA | 15 | 0 | 7 | Not-determined | Genome-fragment | NA | NA | 0 |
| S17_vs_k141_62552\|\|0_partial | 2111 | No | NA | 2 | 0 | 0 | Not-determined | Genome-fragment | NA | NA | 0 |
| S3_vs_k141_96475\|\|full | 1077 | No | NA | 2 | 0 | 0 | Low-quality | Genome-fragment | 2.16 | AAI-based (medium-confidence) | 0 |
| S17_vs_k141_139583\|\|0_partial | 11782 | No | NA | 11 | 0 | 4 | Not-determined | Genome-fragment | NA | NA | 0 |
| S3_vs_k141_123845\|\|full | 1077 | No | NA | 3 | 2 | 0 | Low-quality | Genome-fragment | 0.68 | HMM-based (lower-bound) | 0 |
| S17_vs_k141_88824\|\|lt2gene | 1808 | No | NA | 1 | 0 | 0 | Not-determined | Genome-fragment | NA | NA | 0 |
| S5_vs_k141_134347\|\|full | 1077 | No | NA | 2 | 0 | 0 | Low-quality | Genome-fragment | 2.72 | AAI-based (medium-confidence) | 0 |
| S5_vs_k141_118117\|\|full | 1077 | No | NA | 3 | 0 | 0 | Low-quality | Genome-fragment | 2.15 | AAI-based (medium-confidence) | 0 |
| S17_vs_k141_81505\|\|lt2gene | 1189 | No | NA | 1 | 0 | 0 | Not-determined | Genome-fragment | NA | NA | 0 |
| S7_extracted_34772 | 1077 | No | NA | 2 | 0 | 0 | Low-quality | Genome-fragment | 2.44 | AAI-based (high-confidence) | 0 |
| S17_vs_k141_155556\|\|lt2gene | 1068 | No | NA | 2 | 0 | 0 | Not-determined | Genome-fragment | NA | NA | 0 |
| S12_vs_k141_152342\|\|full | 1077 | No | NA | 4 | 1 | 0 | Low-quality | Genome-fragment | 2.33 | AAI-based (medium-confidence) | 0 |
| S17_vs_k141_133585\|\|lt2gene | 1338 | No | NA | 1 | 0 | 1 | Not-determined | Genome-fragment | NA | NA | 0 |
| S12_vs_k141_47872\|\|lt2gene | 1077 | No | NA | 2 | 0 | 0 | Low-quality | Genome-fragment | 2.56 | AAI-based (high-confidence) | 0 |
| S15_extracted_28290 | 1077 | No | NA | 2 | 0 | 0 | Low-quality | Genome-fragment | 2.5 | AAI-based (medium-confidence) | 0 |
| S8_extracted_k141_153948 | 1077 | No | NA | 4 | 0 | 0 | Low-quality | Genome-fragment | 2.6 | AAI-based (medium-confidence) | 0 |
| S17_vs_k141_45087\|\|lt2gene | 1325 | No | NA | 1 | 0 | 0 | Not-determined | Genome-fragment | NA | NA | 0 |
| S8_vs_k141_130323\|\|full | 1077 | No | NA | 2 | 0 | 0 | Low-quality | Genome-fragment | 1.13 | AAI-based (medium-confidence) | 0 |
| S8_vs_k141_48748\|\|full | 1077 | No | NA | 1 | 1 | 0 | Low-quality | Genome-fragment | 2.62 | AAI-based (medium-confidence) | 0 |
| S17_vs_k141_141291\|\|lt2gene | 1622 | No | NA | 1 | 0 | 0 | Not-determined | Genome-fragment | NA | NA | 0 |
| S17_vs_k141_96898\|\|lt2gene | 2521 | No | NA | 1 | 0 | 0 | Not-determined | Genome-fragment | NA | NA | 0 |
| S17_vs_k141_89542\|\|lt2gene | 1435 | No | NA | 2 | 0 | 0 | Not-determined | Genome-fragment | NA | NA | 0 |
| S17_vs_k141_119364\|\|lt2gene | 1558 | No | NA | 1 | 0 | 0 | Not-determined | Genome-fragment | NA | NA | 0 |
| S17_extracted_k141_10740 | 1078 | No | NA | 3 | 0 | 0 | Low-quality | Genome-fragment | 1.07 | AAI-based (high-confidence) | 0 |
| S17_vs_k141_23394\|\|lt2gene | 1702 | No | NA | 2 | 0 | 0 | Not-determined | Genome-fragment | NA | NA | 0 |
| S9_vs_k141_25616\|\|full | 1078 | No | NA | 4 | 2 | 0 | Low-quality | Genome-fragment | 1.67 | AAI-based (medium-confidence) | 0 |
| S17_vs_k141_157074\|\|lt2gene | 1475 | No | NA | 2 | 0 | 0 | Not-determined | Genome-fragment | NA | NA | 0 |
| S11_extracted_k141_47459 | 1078 | No | NA | 3 | 1 | 0 | Low-quality | Genome-fragment | 2.43 | AAI-based (medium-confidence) | 0 |
| S3_extracted_k141_24052 | 1078 | No | NA | 1 | 0 | 1 | Low-quality | Genome-fragment | 0.31 | AAI-based (high-confidence) | 0 |
| S3_vs_k141_102208\|\|full | 1078 | No | NA | 2 | 0 | 0 | Low-quality | Genome-fragment | 1.58 | AAI-based (medium-confidence) | 0 |
| S17_vs_k141_134997\|\|lt2gene | 1221 | No | NA | 1 | 0 | 0 | Not-determined | Genome-fragment | NA | NA | 0 |
| S5_extracted_k141_51723 | 1078 | No | NA | 2 | 0 | 0 | Low-quality | Genome-fragment | 2.38 | AAI-based (medium-confidence) | 0 |
| S5_extracted_k141_107656 | 1078 | No | NA | 1 | 0 | 0 | Low-quality | Genome-fragment | 1.84 | AAI-based (high-confidence) | 0 |
| S5_vs_k141_134055\|\|full | 1078 | No | NA | 2 | 0 | 0 | Low-quality | Genome-fragment | 0.63 | AAI-based (medium-confidence) | 0 |
| S5_vs_k141_107266\|\|lt2gene | 1078 | No | NA | 2 | 2 | 0 | Low-quality | Genome-fragment | 1.48 | AAI-based (high-confidence) | 0 |
| S7_vs_71434\|\|full | 1078 | No | NA | 3 | 2 | 0 | Low-quality | Genome-fragment | 1.94 | AAI-based (medium-confidence) | 0 |
| S16_vs_102575\|\|full | 1078 | No | NA | 3 | 0 | 1 | Low-quality | Genome-fragment | 0.77 | AAI-based (medium-confidence) | 0 |
| S10_vs_26620\|\|full | 1078 | No | NA | 3 | 0 | 0 | Low-quality | Genome-fragment | 2.62 | AAI-based (high-confidence) | 0 |
| S12_vs_k141_140503\|\|full | 1078 | No | NA | 2 | 0 | 0 | Low-quality | Genome-fragment | 1.53 | AAI-based (medium-confidence) | 0 |
| S17_vs_k141_54925\|\|lt2gene | 1471 | No | NA | 1 | 0 | 1 | Not-determined | Genome-fragment | NA | NA | 0 |
| S6_vs_k141_27339\|\|full | 1078 | No | NA | 3 | 1 | 0 | Low-quality | Genome-fragment | 2.39 | AAI-based (high-confidence) | 0 |
| S6_vs_k141_54547\|\|lt2gene | 1078 | No | NA | 2 | 1 | 0 | Low-quality | Genome-fragment | 2.69 | AAI-based (high-confidence) | 0 |
| S8_extracted_k141_23517 | 1078 | No | NA | 2 | 1 | 0 | Low-quality | Genome-fragment | 0.68 | HMM-based (lower-bound) | 0 |
| S8_vs_k141_202906\|\|full | 1078 | No | NA | 3 | 0 | 2 | Low-quality | Genome-fragment | 0.28 | AAI-based (high-confidence) | 0 |
| S8_vs_k141_143483\|\|full | 1078 | No | NA | 3 | 0 | 1 | Low-quality | Genome-fragment | 1.97 | AAI-based (medium-confidence) | 0 |
| S17_vs_k141_33057\|\|lt2gene | 1078 | No | NA | 1 | 0 | 0 | Not-determined | Genome-fragment | NA | NA | 0 |
| S17_extracted_k141_104258 | 1079 | No | NA | 3 | 3 | 0 | Low-quality | Genome-fragment | 3.56 | AAI-based (high-confidence) | 0 |
| S17_vs_k141_176443\|\|full | 1079 | No | NA | 4 | 2 | 0 | Low-quality | Genome-fragment | 1.77 | AAI-based (high-confidence) | 0 |
| S1_vs_k141_140546\|\|lt2gene | 1079 | No | NA | 1 | 1 | 0 | Low-quality | Genome-fragment | 0.75 | HMM-based (lower-bound) | 0 |
| S5_vs_k141_102881\|\|full | 1079 | No | NA | 4 | 0 | 0 | Low-quality | Genome-fragment | 2.62 | AAI-based (high-confidence) | 0 |
| S17_vs_k141_128746\|\|lt2gene | 2097 | No | NA | 2 | 0 | 0 | Not-determined | Genome-fragment | NA | NA | 0 |
| S7_extracted_76500 | 1079 | No | NA | 3 | 1 | 0 | Low-quality | Genome-fragment | 2.75 | AAI-based (high-confidence) | 0 |
| S17_vs_k141_62516\|\|lt2gene | 1161 | No | NA | 1 | 0 | 0 | Not-determined | Genome-fragment | NA | NA | 0 |

| | | | | | | | | | | | |
|---|---|---|---|---|---|---|---|---|---|---|---|
| S7_extracted_127658 | 1079 | No | NA | 2 | 0 | 0 | Low-quality | Genome-fragment | 1.17 | AAI-based (medium-confidence) | 0 |
| S7_vs_61050\|\|full | 1079 | No | NA | 3 | 0 | 0 | Low-quality | Genome-fragment | 2.86 | AAI-based (medium-confidence) | 0 |
| S16_vs_158565\|\|full | 1079 | No | NA | 3 | 2 | 0 | Low-quality | Genome-fragment | 1.32 | HMM-based (lower-bound) | 0 |
| S17_vs_k141_77956\|\|lt2gene | 1295 | No | NA | 2 | 0 | 0 | Not-determined | Genome-fragment | NA | NA | 0 |
| S10_extracted_17814 | 1079 | No | NA | 2 | 0 | 1 | Low-quality | Genome-fragment | 2.53 | AAI-based (medium-confidence) | 0 |
| S10_extracted_61604 | 1079 | No | NA | 2 | 1 | 0 | Low-quality | Genome-fragment | 3.85 | HMM-based (lower-bound) | 0 |
| S10_vs_53249\|\|full | 1079 | No | NA | 4 | 0 | 0 | Low-quality | Genome-fragment | 2.39 | AAI-based (medium-confidence) | 0 |
| S12_vs_k141_159175\|\|full | 1079 | No | NA | 2 | 0 | 1 | Low-quality | Genome-fragment | 1.24 | AAI-based (high-confidence) | 0 |
| S15_vs_54942\|\|full | 1079 | No | NA | 4 | 0 | 0 | Low-quality | Genome-fragment | 2.32 | AAI-based (medium-confidence) | 0 |
| S8_extracted_k141_142835 | 1079 | No | NA | 1 | 1 | 0 | Low-quality | Genome-fragment | 1.87 | AAI-based (medium-confidence) | 0 |
| S17_vs_k141_27129\|\|lt2gene | 1080 | No | NA | 2 | 2 | 0 | Low-quality | Genome-fragment | 0.88 | HMM-based (lower-bound) | 0 |
| S17_vs_k141_153143\|\|lt2gene | 1025 | No | NA | 2 | 0 | 0 | Not-determined | Genome-fragment | NA | NA | 0 |
| S11_extracted_k141_22553 | 1080 | No | NA | 3 | 0 | 0 | Low-quality | Genome-fragment | 2.52 | AAI-based (high-confidence) | 0 |
| S11_vs_k141_32582\|\|full | 1080 | No | NA | 3 | 0 | 0 | Low-quality | Genome-fragment | 1.06 | AAI-based (medium-confidence) | 0 |
| S11_vs_k141_43895\|\|full | 1080 | No | NA | 4 | 1 | 0 | Low-quality | Genome-fragment | 2.72 | AAI-based (high-confidence) | 0 |
| S17_vs_k141_160591\|\|lt2gene | 2291 | No | NA | 2 | 0 | 2 | Not-determined | Genome-fragment | NA | NA | 0 |
| S17_vs_k141_5455\|\|lt2gene | 3605 | No | NA | 2 | 0 | 1 | Not-determined | Genome-fragment | NA | NA | 0 |
| S13_extracted_95417 | 1080 | No | NA | 2 | 0 | 0 | Low-quality | Genome-fragment | 1.12 | AAI-based (high-confidence) | 0 |
| S1_extracted_k141_32200 | 1080 | No | NA | 2 | 1 | 0 | Low-quality | Genome-fragment | 2.98 | AAI-based (high-confidence) | 0 |
| S1_vs_k141_101898\|\|full | 1080 | No | NA | 2 | 0 | 0 | Low-quality | Genome-fragment | 2.25 | AAI-based (medium-confidence) | 0 |
| S3_extracted_k141_38625 | 1080 | No | NA | 2 | 1 | 0 | Low-quality | Genome-fragment | 0.66 | AAI-based (high-confidence) | 0 |
| S17_vs_k141_168156\|\|lt2gene | 1693 | No | NA | 2 | 0 | 0 | Not-determined | Genome-fragment | NA | NA | 0 |
| S3_vs_k141_4095\|\|full | 1080 | No | NA | 4 | 0 | 0 | Low-quality | Genome-fragment | 2.19 | AAI-based (medium-confidence) | 0 |
| S5_extracted_k141_93304 | 1080 | No | NA | 3 | 2 | 0 | Low-quality | Genome-fragment | 1.91 | AAI-based (high-confidence) | 0 |
| S16_vs_71457\|\|full | 1080 | No | NA | 3 | 0 | 0 | Low-quality | Genome-fragment | 2.04 | AAI-based (medium-confidence) | 0 |
| S16_vs_129382\|\|full | 1080 | No | NA | 4 | 0 | 1 | Low-quality | Genome-fragment | 0.99 | AAI-based (medium-confidence) | 0 |
| S12_vs_k141_24915\|\|full | 1080 | No | NA | 3 | 1 | 0 | Low-quality | Genome-fragment | 2.46 | AAI-based (medium-confidence) | 0 |
| S17_vs_k141_138810\|\|lt2gene | 1558 | No | NA | 2 | 0 | 1 | Not-determined | Genome-fragment | NA | NA | 0 |
| S15_extracted_31831 | 1080 | No | NA | 2 | 0 | 0 | Low-quality | Genome-fragment | 0.65 | AAI-based (medium-confidence) | 0 |
| S17_vs_k141_57967\|\|lt2gene | 1634 | No | NA | 2 | 0 | 1 | Not-determined | Genome-fragment | NA | NA | 0 |
| S8_extracted_k141_58987 | 1080 | No | NA | 2 | 2 | 0 | Low-quality | Genome-fragment | 1.79 | AAI-based (medium-confidence) | 0 |
| S8_extracted_k141_171887 | 1080 | No | NA | 3 | 1 | 0 | Low-quality | Genome-fragment | 1.79 | AAI-based (high-confidence) | 0 |
| S17_vs_k141_51073\|\|lt2gene | 1165 | No | NA | 2 | 0 | 1 | Not-determined | Genome-fragment | NA | NA | 0 |
| S17_vs_k141_96710\|\|full | 1081 | No | NA | 5 | 0 | 0 | Low-quality | Genome-fragment | 2.72 | AAI-based (high-confidence) | 0 |
| S17_vs_k141_13703\|\|lt2gene | 1514 | No | NA | 1 | 0 | 0 | Not-determined | Genome-fragment | NA | NA | 0 |
| S17_vs_k141_152734\|\|full | 1081 | No | NA | 2 | 0 | 0 | Low-quality | Genome-fragment | 2.39 | AAI-based (medium-confidence) | 0 |
| S13_vs_45543\|\|full | 1081 | No | NA | 3 | 1 | 0 | Low-quality | Genome-fragment | 2.89 | AAI-based (high-confidence) | 0 |
| S17_vs_k141_80879\|\|lt2gene | 1004 | No | NA | 1 | 0 | 0 | Not-determined | Genome-fragment | NA | NA | 0 |
| S1_extracted_k141_158236 | 1081 | No | NA | 2 | 1 | 0 | Low-quality | Genome-fragment | 0.71 | AAI-based (high-confidence) | 0 |
| S3_extracted_k141_46748 | 1081 | No | NA | 3 | 1 | 0 | Low-quality | Genome-fragment | 2.65 | AAI-based (high-confidence) | 0 |
| S5_vs_k141_113577\|\|lt2gene | 1081 | No | NA | 1 | 1 | 0 | Low-quality | Genome-fragment | 2.83 | AAI-based (high-confidence) | 0 |
| S9_extracted_k141_15162 | 1230 | No | NA | 2 | 0 | 1 | Not-determined | Genome-fragment | NA | NA | 0 |
| S9_extracted_k141_45474 | 8137 | No | NA | 4 | 0 | 3 | Not-determined | Genome-fragment | NA | NA | 0 |
| S9_extracted_k141_79586 | 1738 | No | NA | 1 | 0 | 0 | Not-determined | Genome-fragment | NA | NA | 0 |
| S9_extracted_k141_87166 | 1231 | No | NA | 1 | 0 | 1 | Not-determined | Genome-fragment | NA | NA | 0 |
| S7_extracted_93724 | 1081 | No | NA | 2 | 0 | 0 | Low-quality | Genome-fragment | 0.96 | AAI-based (medium-confidence) | 0 |
| S9_extracted_k141_11404 | 1253 | No | NA | 1 | 0 | 0 | Not-determined | Genome-fragment | NA | NA | 0 |
| S9_extracted_k141_83380 | 4599 | No | NA | 2 | 0 | 0 | Not-determined | Genome-fragment | NA | NA | 0 |
| S7_vs_91304\|\|full | 1081 | No | NA | 2 | 2 | 0 | Low-quality | Genome-fragment | 2.25 | AAI-based (high-confidence) | 0 |
| S12_vs_k141_43679\|\|full | 1081 | No | NA | 2 | 0 | 0 | Low-quality | Genome-fragment | 1.57 | AAI-based (medium-confidence) | 0 |
| S9_extracted_k141_34148 | 1119 | No | NA | 5 | 0 | 0 | Not-determined | Genome-fragment | NA | NA | 0 |
| S12_vs_k141_37254\|\|full | 1081 | No | NA | 3 | 0 | 0 | Low-quality | Genome-fragment | 0.61 | AAI-based (medium-confidence) | 0 |
| S9_extracted_k141_75808 | 1444 | No | NA | 2 | 0 | 1 | Not-determined | Genome-fragment | NA | NA | 0 |
| S9_extracted_k141_34159 | 1607 | No | NA | 1 | 0 | 0 | Not-determined | Genome-fragment | NA | NA | 0 |
| S8_extracted_k141_29239 | 1081 | No | NA | 2 | 0 | 1 | Low-quality | Genome-fragment | 0.27 | AAI-based (high-confidence) | 0 |
| S9_extracted_k141_87232 | 1512 | No | NA | 2 | 0 | 1 | Not-determined | Genome-fragment | NA | NA | 0 |
| S8_extracted_k141_127447 | 1081 | No | NA | 2 | 0 | 1 | Low-quality | Genome-fragment | 2.02 | AAI-based (medium-confidence) | 0 |
| S8_vs_k141_4137\|\|full | 1081 | No | NA | 3 | 0 | 0 | Low-quality | Genome-fragment | 2.54 | AAI-based (high-confidence) | 0 |
| S9_extracted_k141_22831 | 1266 | No | NA | 1 | 0 | 0 | Not-determined | Genome-fragment | NA | NA | 0 |
| S9_extracted_k141_37942 | 7031 | No | NA | 14 | 0 | 13 | Not-determined | Genome-fragment | NA | NA | 0 |
| S17_extracted_k141_147931 | 1082 | No | NA | 3 | 0 | 0 | Low-quality | Genome-fragment | 1.86 | AAI-based (high-confidence) | 0 |
| S9_extracted_k141_37951 | 1447 | No | NA | 2 | 0 | 0 | Not-determined | Genome-fragment | NA | NA | 0 |
| S9_extracted_k141_75883 | 1008 | No | NA | 1 | 0 | 1 | Not-determined | Genome-fragment | NA | NA | 0 |
| S9_extracted_k141_83471 | 1494 | No | NA | 1 | 0 | 0 | Not-determined | Genome-fragment | NA | NA | 0 |
| S9_extracted_k141_53150 | 1751 | No | NA | 2 | 0 | 1 | Not-determined | Genome-fragment | NA | NA | 0 |
| S9_extracted_k141_53172 | 1119 | No | NA | 1 | 0 | 0 | Not-determined | Genome-fragment | NA | NA | 0 |
| S17_extracted_k141_4694 | 1082 | No | NA | 2 | 0 | 0 | Low-quality | Genome-fragment | 2.11 | AAI-based (high-confidence) | 0 |
| S17_vs_k141_4142\|\|full | 1082 | No | NA | 2 | 0 | 0 | Low-quality | Genome-fragment | 0.75 | AAI-based (medium-confidence) | 0 |
| S9_extracted_k141_75905 | 1120 | No | NA | 3 | 0 | 3 | Not-determined | Genome-fragment | NA | NA | 0 |
| S9_extracted_k141_75906 | 1872 | No | NA | 4 | 0 | 0 | Not-determined | Genome-fragment | NA | NA | 0 |
| S9_extracted_k141_22903 | 1896 | No | NA | 2 | 0 | 2 | Not-determined | Genome-fragment | NA | NA | 0 |
| S9_extracted_k141_83536 | 1607 | No | NA | 2 | 0 | 0 | Not-determined | Genome-fragment | NA | NA | 0 |
| S14_extracted_k141_58947 | 1082 | No | NA | 3 | 3 | 0 | Low-quality | Genome-fragment | 3.33 | AAI-based (high-confidence) | 0 |
| S14_extracted_k141_72534 | 1082 | No | NA | 2 | 1 | 0 | Low-quality | Genome-fragment | 0.68 | HMM-based (lower-bound) | 0 |
| S1_vs_k141_125924\|\|full | 1082 | No | NA | 2 | 0 | 2 | Low-quality | Genome-fragment | 0.43 | AAI-based (medium-confidence) | 0 |
| S3_vs_k141_77715\|\|full | 1082 | No | NA | 2 | 1 | 0 | Low-quality | Genome-fragment | 2.85 | AAI-based (medium-confidence) | 0 |
| S5_extracted_k141_49161 | 1082 | No | NA | 3 | 0 | 0 | Low-quality | Genome-fragment | 1.92 | AAI-based (medium-confidence) | 0 |
| S9_extracted_k141_79752 | 4531 | No | NA | 5 | 0 | 1 | Not-determined | Genome-fragment | NA | NA | 0 |
| S9_extracted_k141_45612 | 1161 | No | NA | 1 | 0 | 1 | Not-determined | Genome-fragment | NA | NA | 0 |
| S9_extracted_k141_34362 | 1050 | No | NA | 2 | 0 | 1 | Not-determined | Genome-fragment | NA | NA | 0 |
| S9_extracted_k141_19155 | 2404 | No | NA | 1 | 0 | 1 | Not-determined | Genome-fragment | NA | NA | 0 |
| S5_vs_k141_84736\|\|full | 1082 | No | NA | 2 | 0 | 2 | Low-quality | Genome-fragment | 1.23 | AAI-based (medium-confidence) | 0 |
| S7_extracted_113858 | 1082 | No | NA | 1 | 0 | 1 | Low-quality | Genome-fragment | 1.66 | AAI-based (high-confidence) | 0 |
| S9_extracted_k141_7714 | 1769 | No | NA | 1 | 0 | 1 | Not-determined | Genome-fragment | NA | NA | 0 |
| S9_extracted_k141_34421 | 1339 | No | NA | 2 | 0 | 2 | Not-determined | Genome-fragment | NA | NA | 0 |
| S9_extracted_k141_64618 | 1987 | No | NA | 2 | 0 | 2 | Not-determined | Genome-fragment | NA | NA | 0 |
| S9_extracted_k141_11648 | 4594 | No | NA | 7 | 0 | 2 | Not-determined | Genome-fragment | NA | NA | 0 |
| S9_extracted_k141_68384 | 3758 | No | NA | 4 | 0 | 0 | Not-determined | Genome-fragment | NA | NA | 0 |
| S16_extracted_10876 | 1082 | No | NA | 3 | 0 | 1 | Low-quality | Genome-fragment | 1.87 | AAI-based (medium-confidence) | 0 |
| S9_extracted_k141_64631 | 1008 | No | NA | 2 | 0 | 2 | Not-determined | Genome-fragment | NA | NA | 0 |
| S9_extracted_k141_53360 | 1042 | No | NA | 1 | 0 | 0 | Not-determined | Genome-fragment | NA | NA | 0 |
| S9_extracted_k141_49425 | 1162 | No | NA | 2 | 0 | 1 | Not-determined | Genome-fragment | NA | NA | 0 |
| S16_vs_151113\|\|lt2gene | 1082 | No | NA | 2 | 1 | 0 | Low-quality | Genome-fragment | 3.34 | AAI-based (high-confidence) | 0 |
| S10_vs_20268\|\|full | 1082 | No | NA | 4 | 0 | 0 | Low-quality | Genome-fragment | 2.4 | AAI-based (high-confidence) | 0 |
| S9_extracted_k141_19272 | 1051 | No | NA | 2 | 0 | 1 | Not-determined | Genome-fragment | NA | NA | 0 |
| S12_extracted_k141_90926 | 1082 | No | NA | 2 | 0 | 0 | Low-quality | Genome-fragment | 2.03 | AAI-based (high-confidence) | 0 |
| S9_extracted_k141_68444 | 1032 | No | NA | 1 | 0 | 1 | Not-determined | Genome-fragment | NA | NA | 0 |
| S9_extracted_k141_26772 | 1290 | No | NA | 5 | 0 | 0 | Not-determined | Genome-fragment | NA | NA | 0 |
| S12_vs_k141_163893\|\|full | 1082 | No | NA | 5 | 0 | 0 | Low-quality | Genome-fragment | 2.8 | AAI-based (high-confidence) | 0 |
| S9_extracted_k141_57068 | 1118 | No | NA | 1 | 0 | 1 | Not-determined | Genome-fragment | NA | NA | 0 |
| S9_extracted_k141_4115 | 33438 | No | NA | 41 | 33 | 0 | Medium-quality | Genome-fragment | 86.27 | AAI-based (high-confidence) | 0 |
| S9_extracted_k141_38261 | 11379 | No | NA | 12 | 0 | 3 | Not-determined | Genome-fragment | NA | NA | 0 |
| S9_extracted_k141_26805 | 6114 | No | NA | 7 | 3 | 0 | Complete | High-quality | 100 | DTR (high-confidence) | 0 |
| S9_extracted_k141_38286 | 1280 | No | NA | 1 | 0 | 1 | Not-determined | Genome-fragment | NA | NA | 0 |
| S9_extracted_k141_15519 | 1111 | No | NA | 2 | 0 | 1 | Not-determined | Genome-fragment | NA | NA | 0 |
| S9_extracted_k141_4137 | 1085 | No | NA | 2 | 0 | 0 | Not-determined | Genome-fragment | NA | NA | 0 |
| S9_extracted_k141_87556 | 2982 | No | NA | 1 | 0 | 0 | Not-determined | Genome-fragment | NA | NA | 0 |
| S9_extracted_k141_42048 | 1848 | No | NA | 2 | 0 | 1 | Not-determined | Genome-fragment | NA | NA | 0 |
| S9_extracted_k141_11720 | 1105 | No | NA | 2 | 0 | 1 | Not-determined | Genome-fragment | NA | NA | 0 |
| S9_extracted_k141_87566 | 1379 | No | NA | 2 | 0 | 2 | Not-determined | Genome-fragment | NA | NA | 0 |
| S9_extracted_k141_15545 | 3035 | No | NA | 3 | 0 | 1 | Not-determined | Genome-fragment | NA | NA | 0 |
| S9_extracted_k141_19284 | 41326 | No | NA | 69 | 28 | 1 | Complete | High-quality | 100 | DTR (high-confidence) | 0 |
| S9_extracted_k141_79919 | 34073 | No | NA | 60 | 23 | 0 | Complete | High-quality | 100 | DTR (high-confidence) | 0 |
| S8_extracted_k141_4008 | 1082 | No | NA | 2 | 0 | 1 | Low-quality | Genome-fragment | 0.67 | AAI-based (medium-confidence) | 0 |
| S9_extracted_k141_68558 | 1138 | No | NA | 3 | 0 | 0 | Not-determined | Genome-fragment | NA | NA | 0 |
| S9_extracted_k141_72340 | 3672 | No | NA | 4 | 0 | 3 | Not-determined | Genome-fragment | NA | NA | 0 |
| S17_vs_k141_66907\|\|full | 1083 | Yes | 750 | 3 | 1 | 1 | Low-quality | Genome-fragment | 0.47 | HMM-based (lower-bound) | 30.75 |
| S9_extracted_k141_68561 | 4316 | No | NA | 4 | 0 | 2 | Not-determined | Genome-fragment | NA | NA | 0 |
| S9_extracted_k141_30776 | 1030 | No | NA | 1 | 0 | 1 | Not-determined | Genome-fragment | NA | NA | 0 |
| S9_extracted_k141_465 | 1076 | No | NA | 1 | 0 | 0 | Not-determined | Genome-fragment | NA | NA | 0 |
| S13_vs_92212\|\|full | 1083 | No | NA | 3 | 0 | 0 | Low-quality | Genome-fragment | 2.79 | AAI-based (medium-confidence) | 0 |
| S14_vs_k141_43092\|\|full | 1083 | No | NA | 3 | 0 | 0 | Low-quality | Genome-fragment | 2.78 | AAI-based (medium-confidence) | 0 |
| S3_extracted_k141_55244 | 1083 | No | NA | 5 | 0 | 0 | Low-quality | Genome-fragment | 2.07 | AAI-based (medium-confidence) | 0 |
| S3_vs_k141_114930\|\|full | 1083 | No | NA | 2 | 0 | 0 | Low-quality | Genome-fragment | 2.13 | AAI-based (high-confidence) | 0 |
| S3_vs_k141_52678\|\|full | 1083 | No | NA | 3 | 0 | 2 | Low-quality | Genome-fragment | 0.29 | AAI-based (medium-confidence) | 0 |
| S9_extracted_k141_49616 | 1796 | No | NA | 3 | 0 | 3 | Not-determined | Genome-fragment | NA | NA | 0 |
| S9_extracted_k141_7886 | 1062 | No | NA | 1 | 0 | 1 | Not-determined | Genome-fragment | NA | NA | 0 |
| S9_extracted_k141_61027 | 5332 | No | NA | 10 | 0 | 1 | Not-determined | Genome-fragment | NA | NA | 0 |
| S3_vs_k141_55184\|\|full | 1083 | No | NA | 4 | 0 | 0 | Low-quality | Genome-fragment | 2.78 | AAI-based (high-confidence) | 0 |
| S16_extracted_115106 | 1083 | No | NA | 1 | 0 | 0 | Low-quality | Genome-fragment | 1.19 | AAI-based (medium-confidence) | 0 |
| S9_extracted_k141_79979 | 1494 | No | NA | 1 | 0 | 1 | Not-determined | Genome-fragment | NA | NA | 0 |
| S10_extracted_117308 | 1083 | No | NA | 2 | 0 | 1 | Low-quality | Genome-fragment | 2.17 | AAI-based (medium-confidence) | 0 |
| S2_vs_k141_13749\|\|full | 1083 | No | NA | 5 | 0 | 0 | Low-quality | Genome-fragment | 2.99 | AAI-based (high-confidence) | 0 |
| S9_extracted_k141_42166 | 5560 | No | NA | 1 | 0 | 1 | Not-determined | Genome-fragment | NA | NA | 0 |
| S2_vs_k141_7008\|\|lt2gene | 1083 | No | NA | 1 | 1 | 0 | Low-quality | Genome-fragment | 2.88 | AAI-based (medium-confidence) | 0 |
| S9_extracted_k141_79995 | 1043 | No | NA | 1 | 0 | 0 | Not-determined | Genome-fragment | NA | NA | 0 |
| S9_extracted_k141_546 | 1374 | No | NA | 1 | 0 | 1 | Not-determined | Genome-fragment | NA | NA | 0 |
| S9_extracted_k141_26970 | 1062 | No | NA | 1 | 0 | 0 | Not-determined | Genome-fragment | NA | NA | 0 |
| S9_extracted_k141_15693 | 4140 | No | NA | 2 | 0 | 1 | Not-determined | Genome-fragment | NA | NA | 0 |
| S8_extracted_k141_166420 | 1083 | No | NA | 3 | 0 | 0 | Low-quality | Genome-fragment | 1.75 | AAI-based (medium-confidence) | 0 |
| S8_vs_k141_104812\|\|full | 1083 | No | NA | 3 | 0 | 1 | Low-quality | Genome-fragment | 0.15 | AAI-based (high-confidence) | 0 |
| S9_extracted_k141_76134 | 2323 | No | NA | 1 | 0 | 1 | Not-determined | Genome-fragment | NA | NA | 0 |
| S17_vs_k141_164243\|\|full | 1084 | No | NA | 3 | 0 | 0 | Low-quality | Genome-fragment | 2.78 | AAI-based (medium-confidence) | 0 |
| S9_extracted_k141_61094 | 1036 | No | NA | 1 | 0 | 1 | Not-determined | Genome-fragment | NA | NA | 0 |
| S17_vs_k141_135548\|\|full | 1084 | No | NA | 2 | 0 | 0 | Low-quality | Genome-fragment | 0.85 | AAI-based (medium-confidence) | 0 |
| S9_vs_k141_9220\|\|full | 1084 | No | NA | 3 | 1 | 0 | Low-quality | Genome-fragment | 2.61 | AAI-based (medium-confidence) | 0 |
| S11_extracted_k141_49316 | 1084 | No | NA | 1 | 0 | 0 | Low-quality | Genome-fragment | 2.51 | AAI-based (high-confidence) | 0 |
| S9_extracted_k141_80043 | 1126 | No | NA | 2 | 0 | 2 | Not-determined | Genome-fragment | NA | NA | 0 |
| S9_extracted_k141_72451 | 1737 | No | NA | 2 | 0 | 1 | Not-determined | Genome-fragment | NA | NA | 0 |
| S9_extracted_k141_83857 | 1662 | No | NA | 2 | 0 | 0 | Not-determined | Genome-fragment | NA | NA | 0 |
| S9_extracted_k141_27023 | 1346 | No | NA | 1 | 0 | 1 | Not-determined | Genome-fragment | NA | NA | 0 |
| S9_extracted_k141_87720 | 1279 | No | NA | 1 | 0 | 0 | Not-determined | Genome-fragment | NA | NA | 0 |
| S9_extracted_k141_8008 | 1339 | No | NA | 2 | 0 | 0 | Not-determined | Genome-fragment | NA | NA | 0 |
| S9_extracted_k141_42252 | 1586 | No | NA | 1 | 0 | 0 | Not-determined | Genome-fragment | NA | NA | 0 |
| S1_vs_k141_81303\|\|full | 1084 | No | NA | 4 | 0 | 0 | Low-quality | Genome-fragment | 2.64 | AAI-based (high-confidence) | 0 |
| S1_vs_k141_88472\|\|full | 1084 | No | NA | 4 | 0 | 0 | Low-quality | Genome-fragment | 1.79 | AAI-based (high-confidence) | 0 |
| S9_extracted_k141_618 | 1994 | No | NA | 2 | 0 | 1 | Not-determined | Genome-fragment | NA | NA | 0 |
| S3_extracted_k141_92112 | 1084 | No | NA | 3 | 0 | 0 | Low-quality | Genome-fragment | 3.03 | AAI-based (high-confidence) | 0 |
| S9_extracted_k141_80081 | 1028 | No | NA | 2 | 0 | 0 | Not-determined | Genome-fragment | NA | NA | 0 |
| S3_vs_k141_48797\|\|full | 1084 | No | NA | 2 | 1 | 0 | Low-quality | Genome-fragment | 1.38 | AAI-based (high-confidence) | 0 |
| S9_extracted_k141_87778 | 3280 | No | NA | 3 | 0 | 2 | Not-determined | Genome-fragment | NA | NA | 0 |
| S9_extracted_k141_53606 | 1038 | No | NA | 1 | 0 | 0 | Not-determined | Genome-fragment | NA | NA | 0 |
| S3_vs_k141_105027\|\|full | 1084 | No | NA | 3 | 1 | 0 | Low-quality | Genome-fragment | 2.31 | AAI-based (high-confidence) | 0 |
| S9_extracted_k141_34660 | 1263 | No | NA | 2 | 0 | 0 | Not-determined | Genome-fragment | NA | NA | 0 |
| S9_extracted_k141_68624 | 1163 | No | NA | 2 | 0 | 1 | Not-determined | Genome-fragment | NA | NA | 0 |
| S5_extracted_k141_20412 | 1084 | No | NA | 3 | 1 | 0 | Low-quality | Genome-fragment | 2.65 | AAI-based (medium-confidence) | 0 |
| S9_extracted_k141_53641 | 1160 | No | NA | 3 | 0 | 0 | Not-determined | Genome-fragment | NA | NA | 0 |
| S9_extracted_k141_80145 | 1047 | No | NA | 1 | 0 | 1 | Not-determined | Genome-fragment | NA | NA | 0 |
| S9_extracted_k141_64844 | 93233 | No | NA | 143 | 42 | 1 | High-quality | High-quality | 100 | AAI-based (high-confidence) | 0 |
| S9_extracted_k141_8094 | 1279 | No | NA | 1 | 0 | 0 | Not-determined | Genome-fragment | NA | NA | 0 |
| S9_extracted_k141_45930 | 1535 | No | NA | 2 | 0 | 0 | Not-determined | Genome-fragment | NA | NA | 0 |
| S5_vs_k141_36770\|\|full | 1084 | No | NA | 3 | 1 | 0 | Low-quality | Genome-fragment | 1.97 | AAI-based (medium-confidence) | 0 |
| S9_extracted_k141_57277 | 56189 | No | NA | 96 | 21 | 0 | Complete | High-quality | 100 | DTR (high-confidence) | 0 |
| S9_extracted_k141_87867 | 1339 | No | NA | 2 | 0 | 0 | Not-determined | Genome-fragment | NA | NA | 0 |
| S9_extracted_k141_12085 | 1104 | No | NA | 2 | 0 | 0 | Not-determined | Genome-fragment | NA | NA | 0 |
| S9_extracted_k141_15874 | 1579 | No | NA | 2 | 0 | 0 | Not-determined | Genome-fragment | NA | NA | 0 |
| S5_vs_k141_56588\|\|full | 1084 | No | NA | 3 | 1 | 0 | Low-quality | Genome-fragment | 2.46 | AAI-based (high-confidence) | 0 |
| S7_vs_1280\|\|full | 1084 | No | NA | 4 | 0 | 0 | Low-quality | Genome-fragment | 2.27 | AAI-based (medium-confidence) | 0 |
| S9_extracted_k141_64939 | 5390 | No | NA | 6 | 0 | 1 | Not-determined | Genome-fragment | NA | NA | 0 |
| S9_extracted_k141_34736 | 13774 | No | NA | 17 | 8 | 0 | Medium-quality | Genome-fragment | 62.9 | AAI-based (medium-confidence) | 0 |
| S7_vs_128244\|\|lt2gene | 1084 | No | NA | 1 | 1 | 0 | Low-quality | Genome-fragment | 1.29 | HMM-based (lower-bound) | 0 |
| S16_extracted_147087 | 1084 | No | NA | 2 | 2 | 0 | Low-quality | Genome-fragment | 1.98 | AAI-based (high-confidence) | 0 |
| S9_extracted_k141_53786 | 1510 | No | NA | 1 | 0 | 0 | Not-determined | Genome-fragment | NA | NA | 0 |
| S9_extracted_k141_64976 | 1596 | No | NA | 4 | 0 | 0 | Not-determined | Genome-fragment | NA | NA | 0 |
| S9_extracted_k141_15949 | 2000 | No | NA | 5 | 0 | 0 | Not-determined | Genome-fragment | NA | NA | 0 |
| S9_extracted_k141_68770 | 2433 | No | NA | 3 | 0 | 3 | Not-determined | Genome-fragment | NA | NA | 0 |
| S9_extracted_k141_23394 | 1283 | No | NA | 1 | 0 | 1 | Not-determined | Genome-fragment | NA | NA | 0 |
| S16_vs_114605\|\|full | 1084 | No | NA | 2 | 1 | 0 | Low-quality | Genome-fragment | 2.98 | AAI-based (high-confidence) | 0 |
| S9_extracted_k141_57339 | 2674 | No | NA | 1 | 0 | 1 | Not-determined | Genome-fragment | NA | NA | 0 |
| S16_vs_111946\|\|lt2gene | 1084 | No | NA | 2 | 2 | 0 | Low-quality | Genome-fragment | 1.52 | HMM-based (lower-bound) | 0 |
| S12_vs_k141_50836\|\|full | 1084 | No | NA | 3 | 1 | 0 | Low-quality | Genome-fragment | 2.53 | AAI-based (medium-confidence) | 0 |
| S9_extracted_k141_27298 | 2254 | No | NA | 2 | 0 | 1 | Not-determined | Genome-fragment | NA | NA | 0 |
| S9_extracted_k141_27304 | 1015 | No | NA | 1 | 0 | 1 | Not-determined | Genome-fragment | NA | NA | 0 |
| S9_extracted_k141_53821 | 1663 | No | NA | 1 | 0 | 0 | Not-determined | Genome-fragment | NA | NA | 0 |
| S9_extracted_k141_49765 | 3353 | No | NA | 3 | 0 | 3 | Not-determined | Genome-fragment | NA | NA | 0 |
| S15_extracted_72892 | 1084 | No | NA | 4 | 1 | 0 | Low-quality | Genome-fragment | 3.11 | AAI-based (high-confidence) | 0 |
| S8_extracted_k141_66518 | 1084 | No | NA | 2 | 0 | 1 | Low-quality | Genome-fragment | 0.66 | AAI-based (medium-confidence) | 0 |
| S9_extracted_k141_737 | 1500 | No | NA | 2 | 0 | 0 | Not-determined | Genome-fragment | NA | NA | 0 |
| S9_extracted_k141_84074 | 1049 | No | NA | 1 | 0 | 0 | Not-determined | Genome-fragment | NA | NA | 0 |
| S9_extracted_k141_80362 | 1232 | No | NA | 2 | 0 | 1 | Not-determined | Genome-fragment | NA | NA | 0 |
| S9_extracted_k141_76394 | 1226 | No | NA | 1 | 0 | 1 | Not-determined | Genome-fragment | NA | NA | 0 |
| S9_extracted_k141_34831 | 1682 | No | NA | 3 | 0 | 1 | Not-determined | Genome-fragment | NA | NA | 0 |
| S14_extracted_k141_4674 | 1085 | No | NA | 3 | 1 | 0 | Low-quality | Genome-fragment | 0.5 | HMM-based (lower-bound) | 0 |
| S9_extracted_k141_46144 | 1593 | No | NA | 1 | 0 | 1 | Not-determined | Genome-fragment | NA | NA | 0 |
| S1_vs_k141_26151\|\|lt2gene | 1085 | No | NA | 2 | 2 | 0 | Low-quality | Genome-fragment | 3.81 | HMM-based (lower-bound) | 0 |
| S5_extracted_k141_15954 | 1085 | No | NA | 4 | 0 | 0 | Low-quality | Genome-fragment | 0.39 | AAI-based (medium-confidence) | 0 |
| S7_vs_106147\|\|lt2gene | 1085 | No | NA | 2 | 1 | 0 | Low-quality | Genome-fragment | 1.38 | AAI-based (medium-confidence) | 0 |
| S9_extracted_k141_23500 | 1103 | No | NA | 1 | 0 | 0 | Not-determined | Genome-fragment | NA | NA | 0 |
| S9_extracted_k141_4544 | 2738 | No | NA | 3 | 0 | 0 | Not-determined | Genome-fragment | NA | NA | 0 |
| S16_vs_57192\|\|full | 1085 | No | NA | 3 | 1 | 0 | Low-quality | Genome-fragment | 1.6 | HMM-based (lower-bound) | 0 |
| S12_vs_k141_185004\|\|full | 1085 | No | NA | 3 | 0 | 0 | Low-quality | Genome-fragment | 1.75 | AAI-based (medium-confidence) | 0 |
| S4_extracted_8626 | 1085 | No | NA | 1 | 0 | 0 | Low-quality | Genome-fragment | 2.26 | AAI-based (high-confidence) | 0 |
| S9_extracted_k141_53940 | 1178 | No | NA | 2 | 0 | 1 | Not-determined | Genome-fragment | NA | NA | 0 |
| S9_extracted_k141_65093 | 1222 | No | NA | 2 | 0 | 1 | Not-determined | Genome-fragment | NA | NA | 0 |
| S9_extracted_k141_813 | 3334 | No | NA | 2 | 0 | 2 | Not-determined | Genome-fragment | NA | NA | 0 |
| S8_extracted_k141_159645 | 1085 | No | NA | 1 | 0 | 1 | Low-quality | Genome-fragment | 0.77 | AAI-based (high-confidence) | 0 |
| S8_vs_k141_200203\|\|full | 1085 | No | NA | 2 | 1 | 0 | Low-quality | Genome-fragment | 1.87 | AAI-based (medium-confidence) | 0 |
| S8_vs_k141_29377\|\|full | 1085 | No | NA | 3 | 0 | 0 | Low-quality | Genome-fragment | 1.06 | AAI-based (high-confidence) | 0 |
| S17_extracted_k141_173337 | 1086 | No | NA | 2 | 0 | 0 | Low-quality | Genome-fragment | 3.45 | AAI-based (high-confidence) | 0 |
| S9_extracted_k141_80448 | 3552 | No | NA | 5 | 0 | 0 | Not-determined | Genome-fragment | NA | NA | 0 |
| S17_vs_k141_12842\|\|lt2gene | 1086 | No | NA | 1 | 1 | 0 | Low-quality | Genome-fragment | 1.78 | HMM-based (lower-bound) | 0 |
| S9_extracted_k141_12317 | 1213 | No | NA | 2 | 0 | 0 | Not-determined | Genome-fragment | NA | NA | 0 |
| S9_extracted_k141_61530 | 1135 | No | NA | 1 | 0 | 1 | Not-determined | Genome-fragment | NA | NA | 0 |
| S9_extracted_k141_57492 | 3971 | No | NA | 4 | 0 | 1 | Not-determined | Genome-fragment | NA | NA | 0 |
| S9_extracted_k141_79709 | 1086 | No | NA | 4 | 1 | 0 | Low-quality | Genome-fragment | 18.23 | AAI-based (high-confidence) | 0 |
| S9_extracted_k141_23571 | 18101 | No | NA | 13 | 0 | 4 | Not-determined | Genome-fragment | NA | NA | 0 |
| S11_vs_k141_40461\|\|full | 1086 | No | NA | 2 | 0 | 0 | Low-quality | Genome-fragment | 2.03 | AAI-based (medium-confidence) | 0 |
| S9_extracted_k141_16167 | 1003 | No | NA | 1 | 0 | 1 | Not-determined | Genome-fragment | NA | NA | 0 |
| S13_extracted_80374 | 1086 | No | NA | 1 | 0 | 1 | Low-quality | Genome-fragment | 0.19 | AAI-based (high-confidence) | 0 |
| S13_extracted_116309 | 1086 | No | NA | 5 | 0 | 0 | Low-quality | Genome-fragment | 17.09 | AAI-based (high-confidence) | 0 |
| S13_extracted_148066 | 1086 | No | NA | 1 | 0 | 0 | Low-quality | Genome-fragment | 1.92 | AAI-based (medium-confidence) | 0 |
| S9_extracted_k141_72894 | 3500 | No | NA | 7 | 0 | 1 | Not-determined | Genome-fragment | NA | NA | 0 |
| S9_extracted_k141_4600 | 1366 | No | NA | 1 | 0 | 0 | Not-determined | Genome-fragment | NA | NA | 0 |
| S9_extracted_k141_38992 | 1726 | No | NA | 1 | 0 | 1 | Not-determined | Genome-fragment | NA | NA | 0 |
| S9_extracted_k141_34996 | 1397 | No | NA | 1 | 0 | 0 | Not-determined | Genome-fragment | NA | NA | 0 |
| S9_extracted_k141_57529 | 3813 | No | NA | 4 | 0 | 4 | Not-determined | Genome-fragment | NA | NA | 0 |
| S9_extracted_k141_35001 | 1138 | No | NA | 2 | 0 | 0 | Not-determined | Genome-fragment | NA | NA | 0 |
| S9_extracted_k141_72908 | 1091 | No | NA | 1 | 0 | 0 | Not-determined | Genome-fragment | NA | NA | 0 |
| S14_vs_k141_19955\|\|full | 1086 | No | NA | 3 | 0 | 0 | Low-quality | Genome-fragment | 3.13 | AAI-based (high-confidence) | 0 |
| S9_extracted_k141_39012 | 1024 | No | NA | 1 | 0 | 1 | Not-determined | Genome-fragment | NA | NA | 0 |
| S9_extracted_k141_934 | 9848 | No | NA | 12 | 8 | 0 | Medium-quality | Genome-fragment | 62.07 | AAI-based (medium-confidence) | 0 |
| S14_vs_k141_63281\|\|full | 1086 | No | NA | 3 | 0 | 0 | Low-quality | Genome-fragment | 2.56 | AAI-based (high-confidence) | 0 |
| S9_extracted_k141_31391 | 1081 | No | NA | 1 | 0 | 1 | Not-determined | Genome-fragment | NA | NA | 0 |
| S9_extracted_k141_954 | 1653 | No | NA | 2 | 0 | 0 | Not-determined | Genome-fragment | NA | NA | 0 |
| S7_vs_84049\|\|lt2gene | 1086 | No | NA | 2 | 2 | 0 | Low-quality | Genome-fragment | 2.37 | AAI-based (high-confidence) | 0 |
| S16_vs_71180\|\|full | 1086 | No | NA | 2 | 0 | 1 | Low-quality | Genome-fragment | 0.43 | AAI-based (medium-confidence) | 0 |
| S9_extracted_k141_57599 | 1130 | No | NA | 2 | 0 | 0 | Not-determined | Genome-fragment | NA | NA | 0 |
| S9_extracted_k141_72985 | 1232 | No | NA | 2 | 0 | 1 | Not-determined | Genome-fragment | NA | NA | 0 |
| S12_extracted_k141_48450 | 1086 | No | NA | 1 | 1 | 0 | Low-quality | Genome-fragment | 17.71 | AAI-based (medium-confidence) | 0 |
| S9_extracted_k141_8251 | 1523 | No | NA | 1 | 0 | 1 | Not-determined | Genome-fragment | NA | NA | 0 |
| S15_vs_17886\|\|full | 1086 | No | NA | 3 | 0 | 0 | Low-quality | Genome-fragment | 2.25 | AAI-based (medium-confidence) | 0 |
| S6_extracted_k141_32873 | 1086 | No | NA | 1 | 1 | 0 | Low-quality | Genome-fragment | 1.54 | AAI-based (high-confidence) | 0 |
| S9_extracted_k141_19850 | 1469 | No | NA | 1 | 0 | 0 | Not-determined | Genome-fragment | NA | NA | 0 |
| S9_extracted_k141_8262 | 3644 | No | NA | 5 | 0 | 1 | Not-determined | Genome-fragment | NA | NA | 0 |
| S9_extracted_k141_65289 | 1316 | No | NA | 1 | 0 | 0 | Not-determined | Genome-fragment | NA | NA | 0 |
| S9_extracted_k141_76513 | 4827 | No | NA | 5 | 0 | 4 | Not-determined | Genome-fragment | NA | NA | 0 |
| S17_vs_k141_84815\|\|full | 1087 | No | NA | 3 | 1 | 0 | Low-quality | Genome-fragment | 3.04 | AAI-based (high-confidence) | 0 |
| S9_extracted_k141_35133 | 1139 | No | NA | 1 | 0 | 1 | Not-determined | Genome-fragment | NA | NA | 0 |
| S13_extracted_91904 | 1087 | No | NA | 1 | 1 | 0 | Low-quality | Genome-fragment | 3.72 | HMM-based (lower-bound) | 0 |
| S9_extracted_k141_12527 | 4188 | No | NA | 1 | 0 | 1 | Not-determined | Genome-fragment | NA | NA | 0 |
| S14_extracted_k141_24179 | 1087 | No | NA | 2 | 0 | 0 | Low-quality | Genome-fragment | 1.5 | AAI-based (medium-confidence) | 0 |
| S9_extracted_k141_16432 | 1024 | No | NA | 3 | 0 | 0 | Not-determined | Genome-fragment | NA | NA | 0 |
| S9_extracted_k141_54257 | 1410 | No | NA | 3 | 0 | 1 | Not-determined | Genome-fragment | NA | NA | 0 |
| S9_extracted_k141_1071 | 4979 | No | NA | 6 | 0 | 0 | Not-determined | Genome-fragment | NA | NA | 0 |
| S14_vs_k141_5340\|\|full | 1087 | No | NA | 4 | 0 | 0 | Low-quality | Genome-fragment | 2.34 | AAI-based (high-confidence) | 0 |
| S9_extracted_k141_42841 | 1165 | No | NA | 2 | 0 | 0 | Not-determined | Genome-fragment | NA | NA | 0 |
| S14_vs_k141_59037\|\|lt2gene | 1087 | No | NA | 1 | 1 | 0 | Low-quality | Genome-fragment | 2.51 | AAI-based (high-confidence) | 0 |
| S9_extracted_k141_19957 | 1155 | No | NA | 1 | 0 | 1 | Not-determined | Genome-fragment | NA | NA | 0 |
| S3_vs_k141_102413\|\|lt2gene | 1087 | No | NA | 2 | 1 | 0 | Low-quality | Genome-fragment | 1.67 | AAI-based (high-confidence) | 0 |
| S9_extracted_k141_8366 | 1940 | No | NA | 3 | 0 | 1 | Not-determined | Genome-fragment | NA | NA | 0 |
| S9_extracted_k141_42865 | 1233 | No | NA | 2 | 0 | 1 | Not-determined | Genome-fragment | NA | NA | 0 |
| S5_extracted_k141_13214 | 1087 | No | NA | 2 | 1 | 0 | Low-quality | Genome-fragment | 4.96 | AAI-based (high-confidence) | 0 |
| S9_extracted_k141_1131 | 1065 | No | NA | 2 | 0 | 0 | Not-determined | Genome-fragment | NA | NA | 0 |
| S9_extracted_k141_84250 | 3064 | No | NA | 3 | 0 | 3 | Not-determined | Genome-fragment | NA | NA | 0 |
| S5_vs_k141_88287\|\|lt2gene | 1087 | No | NA | 1 | 1 | 0 | Low-quality | Genome-fragment | 2.69 | AAI-based (high-confidence) | 0 |
| S9_extracted_k141_39221 | 1909 | No | NA | 2 | 0 | 2 | Not-determined | Genome-fragment | NA | NA | 0 |
| S9_extracted_k141_61670 | 5794 | No | NA | 4 | 0 | 1 | Not-determined | Genome-fragment | NA | NA | 0 |
| S7_extracted_26937 | 1087 | No | NA | 1 | 1 | 0 | Low-quality | Genome-fragment | 17.06 | AAI-based (high-confidence) | 0 |
| S16_extracted_63489 | 1087 | No | NA | 2 | 0 | 0 | Low-quality | Genome-fragment | 2.64 | AAI-based (medium-confidence) | 0 |
| S9_extracted_k141_57740 | 1107 | No | NA | 1 | 0 | 0 | Not-determined | Genome-fragment | NA | NA | 0 |
| S9_extracted_k141_12647 | 1990 | No | NA | 2 | 0 | 0 | Not-determined | Genome-fragment | NA | NA | 0 |
| S9_extracted_k141_27660 | 1149 | No | NA | 1 | 0 | 1 | Not-determined | Genome-fragment | NA | NA | 0 |
| S16_vs_158279\|\|full | 1087 | No | NA | 2 | 0 | 0 | Low-quality | Genome-fragment | 2.22 | AAI-based (high-confidence) | 0 |
| S9_extracted_k141_4863 | 1032 | No | NA | 2 | 0 | 0 | Not-determined | Genome-fragment | NA | NA | 0 |
| S9_extracted_k141_80519 | 1118 | No | NA | 2 | 0 | 0 | Not-determined | Genome-fragment | NA | NA | 0 |
| S9_extracted_k141_35300 | 3671 | No | NA | 1 | 0 | 1 | Not-determined | Genome-fragment | NA | NA | 0 |
| S10_vs_22617\|\|lt2gene | 1087 | No | NA | 1 | 1 | 0 | Low-quality | Genome-fragment | 2.18 | AAI-based (high-confidence) | 0 |
| S9_extracted_k141_12677 | 3406 | No | NA | 8 | 0 | 8 | Not-determined | Genome-fragment | NA | NA | 0 |
| S12_extracted_k141_59832 | 1087 | No | NA | 3 | 0 | 0 | Low-quality | Genome-fragment | 1.72 | AAI-based (high-confidence) | 0 |
| S9_extracted_k141_35345 | 1412 | No | NA | 2 | 0 | 2 | Not-determined | Genome-fragment | NA | NA | 0 |
| S4_extracted_18450 | 1087 | No | NA | 2 | 1 | 0 | Low-quality | Genome-fragment | 1.29 | HMM-based (lower-bound) | 0 |
| S9_extracted_k141_1247 | 3784 | No | NA | 5 | 0 | 3 | Not-determined | Genome-fragment | NA | NA | 0 |
| S9_extracted_k141_20115 | 1118 | No | NA | 1 | 0 | 0 | Not-determined | Genome-fragment | NA | NA | 0 |
| S9_extracted_k141_61772 | 1589 | No | NA | 2 | 0 | 1 | Not-determined | Genome-fragment | NA | NA | 0 |
| S8_vs_k141_99936\|\|full | 1087 | No | NA | 2 | 2 | 0 | Low-quality | Genome-fragment | 1.13 | HMM-based (lower-bound) | 0 |
| S17_vs_k141_68634\|\|full | 1088 | No | NA | 2 | 1 | 0 | Low-quality | Genome-fragment | 2.32 | AAI-based (medium-confidence) | 0 |
| S9_extracted_k141_27161 | 1088 | No | NA | 4 | 0 | 0 | Low-quality | Genome-fragment | 1.38 | AAI-based (high-confidence) | 0 |
| S9_vs_k141_23134\|\|lt2gene | 1088 | No | NA | 2 | 1 | 0 | Low-quality | Genome-fragment | 3.03 | AAI-based (high-confidence) | 0 |
| S9_extracted_k141_46626 | 1083 | No | NA | 2 | 0 | 0 | Not-determined | Genome-fragment | NA | NA | 0 |
| S9_extracted_k141_57833 | 1245 | No | NA | 2 | 0 | 0 | Not-determined | Genome-fragment | NA | NA | 0 |
| S9_extracted_k141_39417 | 1310 | No | NA | 2 | 0 | 0 | Not-determined | Genome-fragment | NA | NA | 0 |
| S9_extracted_k141_31740 | 1324 | No | NA | 2 | 0 | 0 | Not-determined | Genome-fragment | NA | NA | 0 |
| S11_vs_k141_2603\|\|lt2gene | 1088 | No | NA | 1 | 1 | 0 | Low-quality | Genome-fragment | 2.45 | AAI-based (medium-confidence) | 0 |
| S14_extracted_k141_82213 | 1088 | No | NA | 2 | 1 | 0 | Low-quality | Genome-fragment | 0.6 | AAI-based (medium-confidence) | 0 |
| S9_extracted_k141_31753 | 1987 | No | NA | 3 | 0 | 1 | Not-determined | Genome-fragment | NA | NA | 0 |
| S14_extracted_k141_91780 | 1088 | No | NA | 2 | 0 | 1 | Low-quality | Genome-fragment | 0.26 | AAI-based (high-confidence) | 0 |
| S9_extracted_k141_88564 | 1185 | No | NA | 2 | 0 | 1 | Not-determined | Genome-fragment | NA | NA | 0 |
| S9_extracted_k141_73311 | 38973 | No | NA | 54 | 26 | 0 | Medium-quality | Genome-fragment | 70.19 | AAI-based (medium-confidence) | 0 |
| S9_extracted_k141_31788 | 1152 | No | NA | 1 | 0 | 0 | Not-determined | Genome-fragment | NA | NA | 0 |
| S9_extracted_k141_23893 | 1602 | No | NA | 1 | 0 | 1 | Not-determined | Genome-fragment | NA | NA | 0 |
| S1_extracted_k141_41842 | 1088 | No | NA | 2 | 0 | 0 | Low-quality | Genome-fragment | 1.8 | AAI-based (medium-confidence) | 0 |
| S9_extracted_k141_31819 | 1213 | No | NA | 1 | 0 | 1 | Not-determined | Genome-fragment | NA | NA | 0 |
| S1_extracted_k141_25626 | 1088 | No | NA | 2 | 0 | 0 | Low-quality | Genome-fragment | 2.62 | AAI-based (high-confidence) | 0 |
| S9_extracted_k141_23901 | 5984 | No | NA | 11 | 3 | 0 | Complete | High-quality | 100 | DTR (high-confidence) | 0 |
| S1_extracted_k141_52827 | 1088 | No | NA | 1 | 0 | 1 | Low-quality | Genome-fragment | 0.66 | AAI-based (medium-confidence) | 0 |
| S9_extracted_k141_5131 | 1796 | No | NA | 2 | 0 | 2 | Not-determined | Genome-fragment | NA | NA | 0 |
| S1_vs_k141_53713\|\|full | 1088 | No | NA | 3 | 0 | 1 | Low-quality | Genome-fragment | 1.02 | AAI-based (medium-confidence) | 0 |
| S1_vs_k141_33705\|\|full | 1088 | No | NA | 2 | 1 | 0 | Low-quality | Genome-fragment | 2.73 | AAI-based (medium-confidence) | 0 |
| S9_extracted_k141_20288 | 1212 | No | NA | 2 | 0 | 1 | Not-determined | Genome-fragment | NA | NA | 0 |
| S3_extracted_k141_66984 | 1088 | No | NA | 4 | 1 | 0 | Low-quality | Genome-fragment | 2 | AAI-based (high-confidence) | 0 |
| S9_extracted_k141_16668 | 1376 | No | NA | 1 | 0 | 1 | Not-determined | Genome-fragment | NA | NA | 0 |
| S3_vs_k141_73430\|\|full | 1088 | No | NA | 3 | 2 | 0 | Low-quality | Genome-fragment | 2.25 | AAI-based (medium-confidence) | 0 |
| S3_vs_k141_74728\|\|full | 1088 | No | NA | 3 | 1 | 0 | Low-quality | Genome-fragment | 2.45 | AAI-based (medium-confidence) | 0 |
| S9_extracted_k141_54426 | 1088 | No | NA | 1 | 0 | 0 | Not-determined | Genome-fragment | NA | NA | 0 |
| S5_vs_k141_6335\|\|full | 1088 | No | NA | 4 | 0 | 0 | Low-quality | Genome-fragment | 1.26 | AAI-based (medium-confidence) | 0 |
| S9_extracted_k141_69324 | 1056 | No | NA | 1 | 0 | 0 | Not-determined | Genome-fragment | NA | NA | 0 |
| S5_vs_k141_37682\|\|full | 1088 | No | NA | 3 | 0 | 1 | Low-quality | Genome-fragment | 0.75 | AAI-based (medium-confidence) | 0 |
| S9_extracted_k141_58032 | 1108 | No | NA | 2 | 0 | 0 | Not-determined | Genome-fragment | NA | NA | 0 |
| S9_extracted_k141_46788 | 1152 | No | NA | 2 | 0 | 1 | Not-determined | Genome-fragment | NA | NA | 0 |
| S9_extracted_k141_80851 | 1252 | No | NA | 2 | 0 | 0 | Not-determined | Genome-fragment | NA | NA | 0 |
| S9_extracted_k141_76769 | 98250 | No | NA | 97 | 14 | 2 | High-quality | High-quality | 97.31 | AAI-based (high-confidence) | 0 |
| S5_vs_k141_35018\|\|lt2gene | 1088 | No | NA | 2 | 2 | 0 | Low-quality | Genome-fragment | 1.72 | HMM-based (lower-bound) | 0 |
| S7_extracted_84334 | 1088 | No | NA | 4 | 0 | 0 | Low-quality | Genome-fragment | 0.59 | AAI-based (high-confidence) | 0 |
| S9_extracted_k141_69385 | 1019 | No | NA | 2 | 0 | 1 | Not-determined | Genome-fragment | NA | NA | 0 |
| S9_extracted_k141_76779 | 2860 | No | NA | 6 | 0 | 0 | Not-determined | Genome-fragment | NA | NA | 0 |
| S9_extracted_k141_76783 | 1312 | No | NA | 2 | 0 | 1 | Not-determined | Genome-fragment | NA | NA | 0 |
| S7_vs_14857\|\|full | 1088 | No | NA | 2 | 0 | 0 | Low-quality | Genome-fragment | 0.74 | AAI-based (high-confidence) | 0 |
| S7_vs_47775\|\|full | 1088 | No | NA | 2 | 2 | 0 | Low-quality | Genome-fragment | 2.81 | AAI-based (high-confidence) | 0 |
| S9_extracted_k141_5316 | 1047 | No | NA | 2 | 0 | 1 | Not-determined | Genome-fragment | NA | NA | 0 |
| S9_extracted_k141_65780 | 1086 | No | NA | 2 | 0 | 1 | Not-determined | Genome-fragment | NA | NA | 0 |
| S9_extracted_k141_39537 | 1349 | No | NA | 1 | 0 | 0 | Not-determined | Genome-fragment | NA | NA | 0 |
| S10_extracted_18947 | 1088 | No | NA | 3 | 0 | 0 | Low-quality | Genome-fragment | 0.64 | AAI-based (medium-confidence) | 0 |
| S9_extracted_k141_20447 | 1121 | No | NA | 2 | 0 | 1 | Not-determined | Genome-fragment | NA | NA | 0 |
| S2_extracted_k141_11048 | 1088 | No | NA | 2 | 0 | 0 | Low-quality | Genome-fragment | 1.13 | AAI-based (high-confidence) | 0 |
| S8_extracted_k141_63358 | 1088 | No | NA | 3 | 0 | 0 | Low-quality | Genome-fragment | 1.87 | AAI-based (high-confidence) | 0 |
| S8_extracted_k141_111334 | 1088 | No | NA | 2 | 1 | 0 | Low-quality | Genome-fragment | 2.58 | AAI-based (high-confidence) | 0 |
| S9_vs_k141_83371\|\|full | 1089 | No | NA | 3 | 2 | 0 | Low-quality | Genome-fragment | 2.69 | AAI-based (high-confidence) | 0 |
| S9_extracted_k141_32126 | 2334 | No | NA | 2 | 0 | 1 | Not-determined | Genome-fragment | NA | NA | 0 |
| S9_extracted_k141_20491 | 1409 | No | NA | 2 | 0 | 1 | Not-determined | Genome-fragment | NA | NA | 0 |
| S9_extracted_k141_28093 | 1178 | No | NA | 1 | 0 | 1 | Not-determined | Genome-fragment | NA | NA | 0 |
| S9_extracted_k141_73520 | 1346 | No | NA | 3 | 0 | 3 | Not-determined | Genome-fragment | NA | NA | 0 |
| S9_extracted_k141_35749 | 1357 | No | NA | 2 | 0 | 2 | Not-determined | Genome-fragment | NA | NA | 0 |
| S9_extracted_k141_28105 | 1143 | No | NA | 2 | 0 | 1 | Not-determined | Genome-fragment | NA | NA | 0 |
| S11_extracted_k141_22454 | 1089 | No | NA | 2 | 0 | 0 | Low-quality | Genome-fragment | 0.83 | AAI-based (high-confidence) | 0 |
| S11_vs_k141_86436\|\|lt2gene | 1089 | No | NA | 1 | 1 | 0 | Low-quality | Genome-fragment | 2.52 | AAI-based (high-confidence) | 0 |
| S14_vs_k141_81224\|\|full | 1089 | No | NA | 4 | 3 | 0 | Low-quality | Genome-fragment | 3.59 | AAI-based (high-confidence) | 0 |
| S9_extracted_k141_32180 | 1605 | No | NA | 2 | 0 | 2 | Not-determined | Genome-fragment | NA | NA | 0 |
| S14_vs_k141_44606\|\|full | 1089 | No | NA | 2 | 0 | 0 | Low-quality | Genome-fragment | 1.51 | AAI-based (medium-confidence) | 0 |
| S9_extracted_k141_32195 | 1839 | No | NA | 2 | 0 | 1 | Not-determined | Genome-fragment | NA | NA | 0 |
| S9_extracted_k141_16931 | 1551 | No | NA | 2 | 0 | 2 | Not-determined | Genome-fragment | NA | NA | 0 |
| S9_extracted_k141_46824 | 1071 | No | NA | 2 | 0 | 2 | Not-determined | Genome-fragment | NA | NA | 0 |
| S9_extracted_k141_5431 | 4359 | No | NA | 1 | 0 | 1 | Not-determined | Genome-fragment | NA | NA | 0 |
| S14_vs_k141_18923\|\|lt2gene | 1089 | No | NA | 1 | 1 | 0 | Low-quality | Genome-fragment | 1.59 | HMM-based (lower-bound) | 0 |
| S1_vs_k141_106704\|\|full | 1089 | No | NA | 3 | 1 | 0 | Low-quality | Genome-fragment | 2.09 | AAI-based (medium-confidence) | 0 |
| S3_vs_k141_39815\|\|full | 1089 | No | NA | 3 | 0 | 0 | Low-quality | Genome-fragment | 1.93 | AAI-based (high-confidence) | 0 |
| S5_extracted_k141_139293 | 1089 | No | NA | 4 | 0 | 0 | Low-quality | Genome-fragment | 1.93 | AAI-based (medium-confidence) | 0 |
| S5_vs_k141_117021\|\|lt2gene | 1089 | No | NA | 2 | 2 | 0 | Low-quality | Genome-fragment | 3.15 | AAI-based (high-confidence) | 0 |
| S9_extracted_k141_12952 | 3697 | No | NA | 1 | 0 | 1 | Not-determined | Genome-fragment | NA | NA | 0 |
| S9_extracted_k141_32250 | 1856 | No | NA | 1 | 0 | 1 | Not-determined | Genome-fragment | NA | NA | 0 |
| S7_vs_21835\|\|full | 1089 | No | NA | 3 | 0 | 1 | Low-quality | Genome-fragment | 2.54 | AAI-based (medium-confidence) | 0 |
| S9_extracted_k141_54703 | 1857 | No | NA | 2 | 0 | 0 | Not-determined | Genome-fragment | NA | NA | 0 |
| S9_extracted_k141_12961 | 1347 | No | NA | 2 | 0 | 1 | Not-determined | Genome-fragment | NA | NA | 0 |
| S7_vs_91359\|\|full | 1089 | No | NA | 3 | 1 | 0 | Low-quality | Genome-fragment | 1.59 | AAI-based (medium-confidence) | 0 |
| S9_extracted_k141_39684 | 1198 | No | NA | 2 | 0 | 0 | Not-determined | Genome-fragment | NA | NA | 0 |
| S9_extracted_k141_69601 | 2134 | No | NA | 5 | 0 | 5 | Not-determined | Genome-fragment | NA | NA | 0 |
| S9_extracted_k141_17030 | 1425 | No | NA | 1 | 0 | 0 | Not-determined | Genome-fragment | NA | NA | 0 |
| S7_vs_131933\|\|full | 1089 | No | NA | 2 | 1 | 0 | Low-quality | Genome-fragment | 2.38 | AAI-based (medium-confidence) | 0 |
| S9_extracted_k141_28205 | 1353 | No | NA | 1 | 0 | 0 | Not-determined | Genome-fragment | NA | NA | 0 |
| S9_extracted_k141_76974 | 1317 | No | NA | 1 | 0 | 0 | Not-determined | Genome-fragment | NA | NA | 0 |
| S9_extracted_k141_35860 | 1587 | No | NA | 3 | 0 | 3 | Not-determined | Genome-fragment | NA | NA | 0 |
| S10_vs_68920\|\|full | 1089 | No | NA | 4 | 0 | 0 | Low-quality | Genome-fragment | 1.95 | AAI-based (medium-confidence) | 0 |
| S9_extracted_k141_5516 | 1055 | No | NA | 2 | 0 | 1 | Not-determined | Genome-fragment | NA | NA | 0 |
| S8_extracted_k141_84226 | 1089 | No | NA | 2 | 0 | 1 | Low-quality | Genome-fragment | 2.43 | AAI-based (medium-confidence) | 0 |
| S8_vs_k141_86922\|\|full | 1089 | No | NA | 2 | 0 | 0 | Low-quality | Genome-fragment | 1.86 | AAI-based (medium-confidence) | 0 |
| S8_vs_k141_94645\|\|full | 1089 | No | NA | 3 | 0 | 0 | Low-quality | Genome-fragment | 1.17 | AAI-based (medium-confidence) | 0 |
| S9_extracted_k141_69697 | 1415 | No | NA | 1 | 0 | 0 | Not-determined | Genome-fragment | NA | NA | 0 |
| S1_extracted_k141_66744 | 1090 | No | NA | 1 | 0 | 0 | Low-quality | Genome-fragment | 0.62 | AAI-based (high-confidence) | 0 |
| S1_vs_k141_154793\|\|full | 1090 | No | NA | 3 | 1 | 0 | Low-quality | Genome-fragment | 1.81 | AAI-based (high-confidence) | 0 |
| S9_extracted_k141_28299 | 6409 | No | NA | 8 | 0 | 2 | Not-determined | Genome-fragment | NA | NA | 0 |
| S9_extracted_k141_32322 | 3137 | No | NA | 2 | 0 | 2 | Not-determined | Genome-fragment | NA | NA | 0 |
| S9_extracted_k141_28300 | 1138 | No | NA | 2 | 0 | 1 | Not-determined | Genome-fragment | NA | NA | 0 |
| S1_vs_k141_98940\|\|full | 1090 | No | NA | 3 | 1 | 0 | Low-quality | Genome-fragment | 2.9 | AAI-based (medium-confidence) | 0 |
| S9_extracted_k141_58259 | 2127 | No | NA | 3 | 0 | 1 | Not-determined | Genome-fragment | NA | NA | 0 |
| S9_extracted_k141_9136 | 1429 | No | NA | 3 | 0 | 1 | Not-determined | Genome-fragment | NA | NA | 0 |
| S9_extracted_k141_81186 | 1262 | No | NA | 1 | 0 | 1 | Not-determined | Genome-fragment | NA | NA | 0 |
| S9_extracted_k141_28347 | 1001 | No | NA | 1 | 0 | 1 | Not-determined | Genome-fragment | NA | NA | 0 |
| S3_extracted_k141_104533 | 1090 | No | NA | 2 | 0 | 1 | Low-quality | Genome-fragment | 1.98 | AAI-based (high-confidence) | 0 |
| S9_extracted_k141_17185 | 3341 | No | NA | 3 | 0 | 0 | Not-determined | Genome-fragment | NA | NA | 0 |
| S9_extracted_k141_81215 | 3209 | No | NA | 2 | 0 | 2 | Not-determined | Genome-fragment | NA | NA | 0 |
| S9_extracted_k141_1891 | 1675 | No | NA | 1 | 0 | 0 | Not-determined | Genome-fragment | NA | NA | 0 |
| S9_extracted_k141_5661 | 1421 | No | NA | 1 | 0 | 0 | Not-determined | Genome-fragment | NA | NA | 0 |
| S9_extracted_k141_20707 | 1316 | No | NA | 1 | 0 | 0 | Not-determined | Genome-fragment | NA | NA | 0 |
| S5_vs_k141_7647\|\|lt2gene | 1090 | No | NA | 3 | 1 | 0 | Low-quality | Genome-fragment | 1.83 | HMM-based (lower-bound) | 0 |
| S7_vs_56769\|\|full | 1090 | No | NA | 3 | 1 | 0 | Low-quality | Genome-fragment | 2.71 | AAI-based (high-confidence) | 0 |
| S7_vs_130938\|\|full | 1090 | No | NA | 3 | 0 | 0 | Low-quality | Genome-fragment | 1.87 | AAI-based (medium-confidence) | 0 |
| S16_extracted_12819 | 1090 | No | NA | 1 | 1 | 0 | Low-quality | Genome-fragment | 22.84 | AAI-based (high-confidence) | 0 |
| S6_vs_k141_51975\|\|full | 1090 | No | NA | 3 | 1 | 0 | Low-quality | Genome-fragment | 2.84 | AAI-based (medium-confidence) | 0 |
| S8_vs_k141_26094\|\|full | 1090 | No | NA | 3 | 0 | 0 | Low-quality | Genome-fragment | 2.58 | AAI-based (medium-confidence) | 0 |
| S9_extracted_k141_43672 | 1195 | No | NA | 1 | 0 | 1 | Not-determined | Genome-fragment | NA | NA | 0 |
| S9_extracted_k141_58362 | 1473 | No | NA | 2 | 0 | 1 | Not-determined | Genome-fragment | NA | NA | 0 |
| S8_vs_k141_207170\|\|full | 1090 | No | NA | 4 | 0 | 0 | Low-quality | Genome-fragment | 2.45 | AAI-based (high-confidence) | 0 |
| S9_extracted_k141_65917 | 5221 | No | NA | 7 | 0 | 2 | Not-determined | Genome-fragment | NA | NA | 0 |
| S9_extracted_k141_43686 | 3599 | No | NA | 1 | 0 | 0 | Not-determined | Genome-fragment | NA | NA | 0 |

| | | | | | | | | | | | | |
|---|---|---|---|---|---|---|---|---|---|---|---|---|
| S9_extracted_k141_24523 | 1936 | No | NA | | 2 | 0 | 2 | Not-determined | Genome-fragment | NA | NA | 0 |
| S9_extracted_k141_73675 | 1711 | No | NA | | 3 | 0 | 0 | Not-determined | Genome-fragment | NA | NA | 0 |
| S9_extracted_k141_43710 | 1328 | No | NA | | 1 | 0 | 0 | Not-determined | Genome-fragment | NA | NA | 0 |
| S9_vs_k141_88793\|\|full | 1091 | No | NA | | 3 | 0 | 0 | Low-quality | Genome-fragment | 2.58 | AAI-based (medium-confidence) | 0 |
| S9_extracted_k141_1965 | 1319 | No | NA | | 2 | 0 | 0 | Not-determined | Genome-fragment | NA | NA | 0 |
| S9_extracted_k141_20781 | 1010 | No | NA | | 1 | 0 | 0 | Not-determined | Genome-fragment | NA | NA | 0 |
| S1_extracted_k141_81236 | 1091 | No | NA | | 1 | 0 | 1 | Low-quality | Genome-fragment | 0.52 | AAI-based (medium-confidence) | 0 |
| S9_extracted_k141_13186 | 1026 | No | NA | | 2 | 0 | 0 | Not-determined | Genome-fragment | NA | NA | 0 |
| S1_vs_k141_121073\|\|full | 1091 | No | NA | | 2 | 0 | 0 | Low-quality | Genome-fragment | 2.31 | AAI-based (medium-confidence) | 0 |
| S9_extracted_k141_9190 | 1123 | No | NA | | 2 | 0 | 0 | Not-determined | Genome-fragment | NA | NA | 0 |
| S1_vs_k141_53319\|\|full | 1091 | No | NA | | 3 | 1 | 0 | Low-quality | Genome-fragment | 2.73 | AAI-based (high-confidence) | 0 |
| S9_extracted_k141_32388 | 1336 | No | NA | | 2 | 0 | 0 | Not-determined | Genome-fragment | NA | NA | 0 |
| S9_extracted_k141_39992 | 1085 | No | NA | | 2 | 0 | 2 | Not-determined | Genome-fragment | NA | NA | 0 |
| S9_extracted_k141_73708 | 1206 | No | NA | | 1 | 0 | 1 | Not-determined | Genome-fragment | NA | NA | 0 |
| S5_extracted_k141_22457 | 1091 | No | NA | | 3 | 2 | 0 | Low-quality | Genome-fragment | 2.46 | AAI-based (high-confidence) | 0 |
| S9_extracted_k141_5709 | 1769 | No | NA | | 2 | 0 | 1 | Not-determined | Genome-fragment | NA | NA | 0 |
| S9_extracted_k141_81333 | 3956 | No | NA | | 2 | 0 | 0 | Not-determined | Genome-fragment | NA | NA | 0 |
| S5_vs_k141_138270\|\|full | 1091 | No | NA | | 2 | 0 | 1 | Low-quality | Genome-fragment | 1.48 | AAI-based (medium-confidence) | 0 |
| S9_extracted_k141_88842 | 1211 | No | NA | | 2 | 0 | 1 | Not-determined | Genome-fragment | NA | NA | 0 |
| S9_extracted_k141_69945 | 3312 | No | NA | | 3 | 0 | 2 | Not-determined | Genome-fragment | NA | NA | 0 |
| S9_extracted_k141_47102 | 1964 | No | NA | | 2 | 0 | 0 | Not-determined | Genome-fragment | NA | NA | 0 |
| S9_extracted_k141_20844 | 1130 | No | NA | | 1 | 0 | 0 | Not-determined | Genome-fragment | NA | NA | 0 |
| S9_extracted_k141_81345 | 3301 | No | NA | | 6 | 0 | 0 | Not-determined | Genome-fragment | NA | NA | 0 |
| S7_vs_9310\|\|full | 1091 | No | NA | | 3 | 0 | 0 | Low-quality | Genome-fragment | 2.16 | AAI-based (medium-confidence) | 0 |
| S9_extracted_k141_58446 | 5423 | No | NA | | 9 | 0 | 0 | Not-determined | Genome-fragment | NA | NA | 0 |
| S9_extracted_k141_77294 | 1358 | No | NA | | 1 | 0 | 1 | Not-determined | Genome-fragment | NA | NA | 0 |
| S9_extracted_k141_17375 | 1179 | No | NA | | 2 | 0 | 0 | Not-determined | Genome-fragment | NA | NA | 0 |
| S9_extracted_k141_55106 | 1365 | No | NA | | 1 | 0 | 0 | Not-determined | Genome-fragment | NA | NA | 0 |
| S9_extracted_k141_24658 | 1393 | No | NA | | 1 | 0 | 1 | Not-determined | Genome-fragment | NA | NA | 0 |
| S9_extracted_k141_20880 | 1664 | No | NA | | 2 | 0 | 0 | Not-determined | Genome-fragment | NA | NA | 0 |
| S16_extracted_39772 | 1091 | No | NA | | 3 | 0 | 0 | Low-quality | Genome-fragment | 2.32 | AAI-based (medium-confidence) | 0 |
| S9_extracted_k141_32477 | 1281 | No | NA | | 2 | 0 | 0 | Not-determined | Genome-fragment | NA | NA | 0 |
| S10_vs_95651\|\|full | 1091 | No | NA | | 3 | 0 | 0 | Low-quality | Genome-fragment | 3.22 | AAI-based (medium-confidence) | 0 |
| S9_extracted_k141_77343 | 1117 | No | NA | | 2 | 0 | 0 | Not-determined | Genome-fragment | NA | NA | 0 |
| S9_extracted_k141_24691 | 1155 | No | NA | | 1 | 0 | 1 | Not-determined | Genome-fragment | NA | NA | 0 |
| S12_vs_k141_28671\|\|full | 1091 | No | NA | | 4 | 0 | 0 | Low-quality | Genome-fragment | 2.33 | AAI-based (medium-confidence) | 0 |
| S12_vs_k141_154828\|\|full | 1091 | No | NA | | 2 | 0 | 2 | Low-quality | Genome-fragment | 1.47 | AAI-based (medium-confidence) | 0 |
| S9_extracted_k141_13319 | 1419 | No | NA | | 3 | 0 | 0 | Not-determined | Genome-fragment | NA | NA | 0 |
| S9_extracted_k141_32500 | 2362 | No | NA | | 1 | 0 | 0 | Not-determined | Genome-fragment | NA | NA | 0 |
| S9_extracted_k141_55146 | 1491 | No | NA | | 1 | 0 | 0 | Not-determined | Genome-fragment | NA | NA | 0 |
| S8_vs_k141_53768\|\|lt2gene | 1091 | No | NA | | 1 | 1 | 0 | Low-quality | Genome-fragment | 1.82 | HMM-based (lower-bound) | 0 |
| S17_extracted_k141_19736 | 1092 | No | NA | | 3 | 0 | 1 | Low-quality | Genome-fragment | 0.47 | AAI-based (medium-confidence) | 0 |
| S9_extracted_k141_88874 | 1602 | No | NA | | 2 | 0 | 0 | Not-determined | Genome-fragment | NA | NA | 0 |
| S9_extracted_k141_24707 | 1280 | No | NA | | 1 | 0 | 1 | Not-determined | Genome-fragment | NA | NA | 0 |
| S9_extracted_k141_66045 | 1228 | No | NA | | 2 | 0 | 1 | Not-determined | Genome-fragment | NA | NA | 0 |
| S9_extracted_k141_5826 | 1143 | No | NA | | 2 | 0 | 2 | Not-determined | Genome-fragment | NA | NA | 0 |
| S17_vs_k141_42995\|\|lt2gene | 1092 | No | NA | | 1 | 0 | 0 | Low-quality | Genome-fragment | 2.54 | AAI-based (medium-confidence) | 0 |
| S11_vs_k141_78395\|\|full | 1092 | No | NA | | 2 | 1 | 0 | Low-quality | Genome-fragment | 0.58 | HMM-based (lower-bound) | 0 |
| S9_extracted_k141_47228 | 1779 | No | NA | | 1 | 0 | 1 | Not-determined | Genome-fragment | NA | NA | 0 |
| S9_extracted_k141_17441 | 1874 | No | NA | | 2 | 0 | 0 | Not-determined | Genome-fragment | NA | NA | 0 |
| S11_vs_k141_87340\|\|full | 1092 | No | NA | | 4 | 0 | 0 | Low-quality | Genome-fragment | 2.24 | AAI-based (high-confidence) | 0 |
| S9_extracted_k141_24749 | 3918 | No | NA | | 9 | 0 | 0 | Not-determined | Genome-fragment | NA | NA | 0 |
| S11_vs_k141_30414\|\|full | 1092 | No | NA | | 2 | 0 | 0 | Low-quality | Genome-fragment | 2.77 | AAI-based (high-confidence) | 0 |
| S9_extracted_k141_43907 | 1720 | No | NA | | 2 | 0 | 1 | Not-determined | Genome-fragment | NA | NA | 0 |
| S14_vs_k141_110103\|\|full | 1092 | No | NA | | 3 | 2 | 0 | Low-quality | Genome-fragment | 3.53 | AAI-based (high-confidence) | 0 |
| S5_extracted_k141_110159 | 1092 | No | NA | | 3 | 3 | 0 | Low-quality | Genome-fragment | 3.26 | AAI-based (medium-confidence) | 0 |
| S9_extracted_k141_28639 | 1107 | No | NA | | 3 | 0 | 2 | Not-determined | Genome-fragment | NA | NA | 0 |
| S9_extracted_k141_36220 | 1134 | No | NA | | 2 | 0 | 2 | Not-determined | Genome-fragment | NA | NA | 0 |
| S16_vs_70414\|\|full | 1092 | No | NA | | 3 | 1 | 0 | Low-quality | Genome-fragment | 1.83 | AAI-based (high-confidence) | 0 |
| S9_extracted_k141_47276 | 1116 | No | NA | | 1 | 0 | 1 | Not-determined | Genome-fragment | NA | NA | 0 |
| S9_extracted_k141_55187 | 4387 | No | NA | | 4 | 0 | 1 | Not-determined | Genome-fragment | NA | NA | 0 |
| S16_vs_97670\|\|full | 1092 | No | NA | | 3 | 1 | 0 | Low-quality | Genome-fragment | 2.2 | AAI-based (medium-confidence) | 0 |
| S16_vs_156329\|\|lt2gene | 1092 | No | NA | | 1 | 1 | 0 | Low-quality | Genome-fragment | 1.95 | HMM-based (lower-bound) | 0 |
| S10_extracted_8952 | 1092 | No | NA | | 4 | 0 | 0 | Low-quality | Genome-fragment | 2.29 | AAI-based (high-confidence) | 0 |
| S12_vs_k141_166976\|\|full | 1092 | No | NA | | 3 | 1 | 0 | Low-quality | Genome-fragment | 2.65 | AAI-based (medium-confidence) | 0 |
| S12_vs_k141_21664\|\|full | 1092 | No | NA | | 4 | 1 | 0 | Low-quality | Genome-fragment | 1.76 | AAI-based (high-confidence) | 0 |
| S15_extracted_13793 | 1092 | No | NA | | 2 | 0 | 0 | Low-quality | Genome-fragment | 0.75 | AAI-based (high-confidence) | 0 |
| S9_extracted_k141_2108 | 1667 | No | NA | | 2 | 0 | 1 | Not-determined | Genome-fragment | NA | NA | 0 |
| S2_extracted_k141_14227 | 1092 | No | NA | | 1 | 1 | 0 | Low-quality | Genome-fragment | 1.77 | AAI-based (high-confidence) | 0 |
| S11_vs_k141_75722\|\|full | 1093 | No | NA | | 3 | 1 | 0 | Low-quality | Genome-fragment | 1.64 | AAI-based (high-confidence) | 0 |
| S9_extracted_k141_66139 | 1152 | No | NA | | 2 | 0 | 0 | Not-determined | Genome-fragment | NA | NA | 0 |
| S9_extracted_k141_66159 | 1780 | No | NA | | 2 | 0 | 1 | Not-determined | Genome-fragment | NA | NA | 0 |
| S9_extracted_k141_89028 | 1195 | No | NA | | 1 | 0 | 1 | Not-determined | Genome-fragment | NA | NA | 0 |
| S9_extracted_k141_13504 | 1489 | No | NA | | 2 | 0 | 1 | Not-determined | Genome-fragment | NA | NA | 0 |
| S9_extracted_k141_40283 | 1253 | No | NA | | 1 | 0 | 1 | Not-determined | Genome-fragment | NA | NA | 0 |
| S9_extracted_k141_5957 | 3815 | No | NA | | 4 | 0 | 0 | Not-determined | Genome-fragment | NA | NA | 0 |
| S13_extracted_82067 | 1093 | No | NA | | 2 | 0 | 0 | Low-quality | Genome-fragment | 1.69 | AAI-based (medium-confidence) | 0 |
| S9_extracted_k141_5963 | 2157 | No | NA | | 2 | 0 | 1 | Not-determined | Genome-fragment | NA | NA | 0 |
| S9_extracted_k141_24881 | 1492 | No | NA | | 2 | 0 | 1 | Not-determined | Genome-fragment | NA | NA | 0 |
| S9_extracted_k141_28771 | 2714 | No | NA | | 3 | 0 | 0 | Not-determined | Genome-fragment | NA | NA | 0 |
| S9_extracted_k141_81547 | 1188 | No | NA | | 2 | 0 | 1 | Not-determined | Genome-fragment | NA | NA | 0 |
| S14_vs_k141_104029\|\|full | 1093 | No | NA | | 4 | 0 | 0 | Low-quality | Genome-fragment | 1.77 | AAI-based (high-confidence) | 0 |
| S14_vs_k141_31367\|\|full | 1093 | No | NA | | 3 | 1 | 0 | Low-quality | Genome-fragment | 2.6 | AAI-based (high-confidence) | 0 |
| S9_extracted_k141_28794 | 1055 | No | NA | | 1 | 0 | 0 | Not-determined | Genome-fragment | NA | NA | 0 |
| S1_extracted_k141_86381 | 1093 | No | NA | | 1 | 0 | 1 | Low-quality | Genome-fragment | 2.6 | AAI-based (high-confidence) | 0 |
| S9_extracted_k141_36375 | 3496 | No | NA | | 3 | 0 | 3 | Not-determined | Genome-fragment | NA | NA | 0 |
| S9_extracted_k141_44101 | 2428 | No | NA | | 3 | 0 | 0 | Not-determined | Genome-fragment | NA | NA | 0 |
| S9_extracted_k141_2221 | 2081 | No | NA | | 2 | 0 | 1 | Not-determined | Genome-fragment | NA | NA | 0 |
| S9_extracted_k141_85234 | 1514 | No | NA | | 3 | 0 | 0 | Not-determined | Genome-fragment | NA | NA | 0 |
| S9_extracted_k141_62736 | 1353 | No | NA | | 4 | 0 | 0 | Not-determined | Genome-fragment | NA | NA | 0 |
| S9_extracted_k141_81631 | 1180 | No | NA | | 3 | 0 | 2 | Not-determined | Genome-fragment | NA | NA | 0 |
| S9_extracted_k141_89107 | 3673 | No | NA | | 1 | 0 | 0 | Not-determined | Genome-fragment | NA | NA | 0 |
| S1_vs_k141_85869\|\|lt2gene | 1093 | No | NA | | 2 | 2 | 0 | Low-quality | Genome-fragment | 2.26 | AAI-based (high-confidence) | 0 |
| S9_extracted_k141_66213 | 1204 | No | NA | | 1 | 0 | 0 | Not-determined | Genome-fragment | NA | NA | 0 |
| S9_extracted_k141_36433 | 1069 | No | NA | | 2 | 0 | 0 | Not-determined | Genome-fragment | NA | NA | 0 |
| S9_extracted_k141_9600 | 1771 | No | NA | | 1 | 0 | 1 | Not-determined | Genome-fragment | NA | NA | 0 |
| S3_extracted_k141_25462 | 1093 | No | NA | | 2 | 1 | 0 | Low-quality | Genome-fragment | 1.26 | HMM-based (lower-bound) | 0 |
| S9_extracted_k141_36461 | 1451 | No | NA | | 1 | 0 | 0 | Not-determined | Genome-fragment | NA | NA | 0 |
| S9_extracted_k141_70236 | 15768 | No | NA | | 16 | 10 | 0 | Medium-quality | Genome-fragment | 67.84 | AAI-based (medium-confidence) | 0 |
| S9_extracted_k141_74038 | 2447 | No | NA | | 1 | 0 | 1 | Not-determined | Genome-fragment | NA | NA | 0 |
| S9_extracted_k141_28923 | 1049 | No | NA | | 3 | 0 | 1 | Not-determined | Genome-fragment | NA | NA | 0 |
| S3_vs_k141_27693\|\|full | 1093 | No | NA | | 3 | 0 | 0 | Low-quality | Genome-fragment | 1.37 | AAI-based (medium-confidence) | 0 |
| S16_extracted_168173 | 1093 | No | NA | | 2 | 1 | 0 | Low-quality | Genome-fragment | 1 | AAI-based (high-confidence) | 0 |
| S16_vs_44425\|\|full | 1093 | No | NA | | 3 | 1 | 0 | Low-quality | Genome-fragment | 2.26 | AAI-based (high-confidence) | 0 |
| S9_extracted_k141_81682 | 1003 | No | NA | | 1 | 0 | 0 | Not-determined | Genome-fragment | NA | NA | 0 |
| S16_vs_108796\|\|full | 1093 | No | NA | | 3 | 1 | 0 | Low-quality | Genome-fragment | 1.76 | AAI-based (high-confidence) | 0 |
| S9_extracted_k141_55400 | 1430 | No | NA | | 1 | 0 | 0 | Not-determined | Genome-fragment | NA | NA | 0 |
| S9_extracted_k141_9674 | 1550 | No | NA | | 1 | 0 | 1 | Not-determined | Genome-fragment | NA | NA | 0 |
| S9_extracted_k141_66331 | 1145 | No | NA | | 2 | 0 | 0 | Not-determined | Genome-fragment | NA | NA | 0 |
| S16_vs_129364\|\|full | 1093 | No | NA | | 2 | 0 | 0 | Low-quality | Genome-fragment | 1.48 | AAI-based (medium-confidence) | 0 |
| S9_extracted_k141_50888 | 13484 | No | NA | | 5 | 0 | 4 | Not-determined | Genome-fragment | NA | NA | 0 |
| S9_extracted_k141_17723 | 1078 | No | NA | | 2 | 0 | 1 | Not-determined | Genome-fragment | NA | NA | 0 |
| S9_extracted_k141_74117 | 2399 | No | NA | | 4 | 0 | 4 | Not-determined | Genome-fragment | NA | NA | 0 |
| S9_extracted_k141_36576 | 1313 | No | NA | | 4 | 0 | 0 | Not-determined | Genome-fragment | NA | NA | 0 |
| S9_extracted_k141_9718 | 1058 | No | NA | | 2 | 0 | 1 | Not-determined | Genome-fragment | NA | NA | 0 |
| S6_vs_k141_9436\|\|full | 1093 | No | NA | | 2 | 0 | 0 | Low-quality | Genome-fragment | 2.79 | AAI-based (high-confidence) | 0 |
| S9_extracted_k141_58822 | 3940 | No | NA | | 3 | 0 | 1 | Not-determined | Genome-fragment | NA | NA | 0 |
| S8_vs_k141_199717\|\|full | 1093 | No | NA | | 3 | 0 | 1 | Low-quality | Genome-fragment | 0.48 | AAI-based (medium-confidence) | 0 |
| S9_extracted_k141_9741 | 1061 | No | NA | | 1 | 0 | 1 | Not-determined | Genome-fragment | NA | NA | 0 |
| S9_extracted_k141_58845 | 1357 | No | NA | | 3 | 0 | 2 | Not-determined | Genome-fragment | NA | NA | 0 |
| S9_extracted_k141_21337 | 1338 | No | NA | | 2 | 0 | 1 | Not-determined | Genome-fragment | NA | NA | 0 |
| S9_extracted_k141_81753 | 1096 | No | NA | | 1 | 0 | 0 | Not-determined | Genome-fragment | NA | NA | 0 |
| S9_extracted_k141_36623 | 2760 | No | NA | | 1 | 0 | 0 | Not-determined | Genome-fragment | NA | NA | 0 |
| S8_vs_k141_20591\|\|full | 1093 | No | NA | | 4 | 0 | 0 | Low-quality | Genome-fragment | 0.63 | AAI-based (high-confidence) | 0 |
| S9_extracted_k141_55474 | 4966 | No | NA | | 3 | 0 | 1 | Not-determined | Genome-fragment | NA | NA | 0 |
| S9_vs_k141_84055\|\|lt2gene | 1094 | No | NA | | 2 | 2 | 0 | Low-quality | Genome-fragment | 1.63 | AAI-based (medium-confidence) | 0 |
| S13_extracted_940 | 1094 | No | NA | | 2 | 0 | 0 | Low-quality | Genome-fragment | 0.66 | AAI-based (medium-confidence) | 0 |
| S13_extracted_144109 | 1094 | No | NA | | 2 | 0 | 0 | Low-quality | Genome-fragment | 0.51 | AAI-based (medium-confidence) | 0 |
| S9_extracted_k141_13507 | 231917 | Yes | | 109327 | 242 | 18 | 73 | Complete | High-quality | 100 | Provirus (high-confidence) | 52.86 |
| S14_extracted_k141_70514 | 1094 | No | NA | | 1 | 1 | 0 | Low-quality | Genome-fragment | 2.5 | AAI-based (high-confidence) | 0 |
| S1_vs_k141_29331\|\|full | 1094 | No | NA | | 3 | 0 | 1 | Low-quality | Genome-fragment | 0.84 | AAI-based (high-confidence) | 0 |
| S1_vs_k141_57328\|\|full | 1094 | No | NA | | 3 | 0 | 2 | Low-quality | Genome-fragment | 0.39 | AAI-based (medium-confidence) | 0 |
| S1_vs_k141_106644\|\|full | 1094 | No | NA | | 4 | 1 | 0 | Low-quality | Genome-fragment | 1.78 | AAI-based (medium-confidence) | 0 |
| S9_extracted_k141_36665 | 1460 | No | NA | | 2 | 0 | 2 | Not-determined | Genome-fragment | NA | NA | 0 |
| S9_extracted_k141_9790 | 1867 | No | NA | | 3 | 0 | 0 | Not-determined | Genome-fragment | NA | NA | 0 |
| S3_vs_k141_106322\|\|full | 1094 | No | NA | | 4 | 3 | 0 | Low-quality | Genome-fragment | 1.81 | AAI-based (high-confidence) | 0 |
| S9_extracted_k141_21444 | 2754 | No | NA | | 3 | 0 | 2 | Not-determined | Genome-fragment | NA | NA | 0 |
| S9_extracted_k141_50939 | 1055 | No | NA | | 2 | 0 | 1 | Not-determined | Genome-fragment | NA | NA | 0 |
| S9_extracted_k141_6287 | 6831 | No | NA | | 9 | 0 | 4 | Not-determined | Genome-fragment | NA | NA | 0 |
| S9_extracted_k141_25217 | 1127 | No | NA | | 1 | 0 | 1 | Not-determined | Genome-fragment | NA | NA | 0 |
| S5_extracted_k141_71637 | 1094 | No | NA | | 3 | 2 | 0 | Low-quality | Genome-fragment | 2.63 | AAI-based (medium-confidence) | 0 |
| S9_extracted_k141_47539 | 1078 | No | NA | | 1 | 0 | 1 | Not-determined | Genome-fragment | NA | NA | 0 |
| S9_extracted_k141_58958 | 1108 | No | NA | | 1 | 0 | 1 | Not-determined | Genome-fragment | NA | NA | 0 |
| S7_extracted_129409 | 1094 | No | NA | | 5 | 0 | 0 | Low-quality | Genome-fragment | 0.65 | AAI-based (high-confidence) | 0 |
| S16_extracted_34442 | 1094 | No | NA | | 2 | 0 | 1 | Low-quality | Genome-fragment | 1.5 | AAI-based (medium-confidence) | 0 |
| S9_extracted_k141_36717 | 1516 | No | NA | | 2 | 0 | 1 | Not-determined | Genome-fragment | NA | NA | 0 |
| S9_extracted_k141_6319 | 3370 | No | NA | | 2 | 0 | 2 | Not-determined | Genome-fragment | NA | NA | 0 |
| S9_extracted_k141_25234 | 25733 | No | NA | | 18 | 0 | 11 | Not-determined | Genome-fragment | NA | NA | 0 |
| S9_extracted_k141_47602 | 1959 | No | NA | | 1 | 0 | 0 | Not-determined | Genome-fragment | NA | NA | 0 |
| S9_extracted_k141_36771 | 1366 | No | NA | | 1 | 0 | 0 | Not-determined | Genome-fragment | NA | NA | 0 |
| S9_extracted_k141_2574 | 1220 | No | NA | | 4 | 0 | 3 | Not-determined | Genome-fragment | NA | NA | 0 |
| S16_vs_153524\|\|full | 1094 | No | NA | | 2 | 1 | 0 | Low-quality | Genome-fragment | 2.95 | AAI-based (high-confidence) | 0 |
| S9_extracted_k141_32827 | 1033 | No | NA | | 2 | 0 | 0 | Not-determined | Genome-fragment | NA | NA | 0 |
| S9_extracted_k141_29205 | 2373 | No | NA | | 2 | 0 | 1 | Not-determined | Genome-fragment | NA | NA | 0 |
| S9_extracted_k141_66576 | 1168 | No | NA | | 1 | 0 | 0 | Not-determined | Genome-fragment | NA | NA | 0 |
| S9_extracted_k141_29215 | 3433 | No | NA | | 4 | 0 | 2 | Not-determined | Genome-fragment | NA | NA | 0 |
| S10_extracted_78510 | 1094 | No | NA | | 4 | 1 | 0 | Low-quality | Genome-fragment | 1.74 | AAI-based (high-confidence) | 0 |
| S9_extracted_k141_59039 | 5248 | No | NA | | 5 | 0 | 5 | Not-determined | Genome-fragment | NA | NA | 0 |
| S9_extracted_k141_9947 | 1097 | No | NA | | 1 | 0 | 1 | Not-determined | Genome-fragment | NA | NA | 0 |
| S10_vs_77518\|\|lt2gene | 1094 | No | NA | | 2 | 1 | 0 | Low-quality | Genome-fragment | 1.14 | HMM-based (lower-bound) | 0 |
| S12_extracted_k141_101704 | 1094 | No | NA | | 2 | 0 | 1 | Low-quality | Genome-fragment | 0.5 | AAI-based (medium-confidence) | 0 |
| S9_extracted_k141_66585 | 1352 | No | NA | | 2 | 0 | 2 | Not-determined | Genome-fragment | NA | NA | 0 |
| S9_extracted_k141_62967 | 3466 | No | NA | | 6 | 0 | 5 | Not-determined | Genome-fragment | NA | NA | 0 |
| S9_extracted_k141_6410 | 1435 | No | NA | | 1 | 0 | 1 | Not-determined | Genome-fragment | NA | NA | 0 |
| S9_extracted_k141_74265 | 4099 | No | NA | | 7 | 0 | 4 | Not-determined | Genome-fragment | NA | NA | 0 |
| S9_extracted_k141_89246 | 1182 | No | NA | | 2 | 0 | 2 | Not-determined | Genome-fragment | NA | NA | 0 |
| S9_extracted_k141_59098 | 1030 | No | NA | | 1 | 0 | 0 | Not-determined | Genome-fragment | NA | NA | 0 |
| S9_extracted_k141_13711 | 3319 | No | NA | | 1 | 0 | 1 | Not-determined | Genome-fragment | NA | NA | 0 |
| S9_extracted_k141_89291 | 1323 | No | NA | | 2 | 0 | 1 | Not-determined | Genome-fragment | NA | NA | 0 |
| S9_extracted_k141_2641 | 1480 | No | NA | | 1 | 0 | 0 | Not-determined | Genome-fragment | NA | NA | 0 |
| S12_vs_k141_50154\|\|full | 1094 | No | NA | | 2 | 2 | 0 | Low-quality | Genome-fragment | 2.8 | AAI-based (high-confidence) | 0 |
| S9_extracted_k141_32859 | 1187 | No | NA | | 1 | 0 | 0 | Not-determined | Genome-fragment | NA | NA | 0 |
| S9_extracted_k141_47706 | 1327 | No | NA | | 1 | 0 | 0 | Not-determined | Genome-fragment | NA | NA | 0 |
| S9_extracted_k141_85602 | 2301 | No | NA | | 1 | 0 | 1 | Not-determined | Genome-fragment | NA | NA | 0 |
| S8_extracted_k141_55590 | 1094 | No | NA | | 4 | 0 | 0 | Low-quality | Genome-fragment | 2.38 | AAI-based (medium-confidence) | 0 |
| S9_extracted_k141_13735 | 1554 | No | NA | | 2 | 0 | 1 | Not-determined | Genome-fragment | NA | NA | 0 |
| S8_vs_k141_4029\|\|full | 1094 | No | NA | | 2 | 0 | 0 | Low-quality | Genome-fragment | 3.04 | AAI-based (medium-confidence) | 0 |
| S8_vs_k141_180306\|\|full | 1094 | No | NA | | 3 | 0 | 1 | Low-quality | Genome-fragment | 1.99 | AAI-based (medium-confidence) | 0 |
| S17_extracted_k141_94913 | 1095 | No | NA | | 3 | 0 | 0 | Low-quality | Genome-fragment | 0.69 | AAI-based (medium-confidence) | 0 |
| S17_vs_k141_91736\|\|lt2gene | 1095 | No | NA | | 1 | 1 | 0 | Low-quality | Genome-fragment | 1.85 | AAI-based (high-confidence) | 0 |
| S9_vs_k141_56398\|\|full | 1095 | No | NA | | 3 | 0 | 0 | Low-quality | Genome-fragment | 1.99 | AAI-based (medium-confidence) | 0 |
| S9_extracted_k141_25316 | 1406 | No | NA | | 1 | 0 | 1 | Not-determined | Genome-fragment | NA | NA | 0 |
| S9_extracted_k141_32924 | 1528 | No | NA | | 1 | 0 | 0 | Not-determined | Genome-fragment | NA | NA | 0 |
| S14_vs_k141_51100\|\|full | 1095 | No | NA | | 3 | 0 | 0 | Low-quality | Genome-fragment | 2.02 | AAI-based (high-confidence) | 0 |
| S9_extracted_k141_10085 | 2405 | No | NA | | 2 | 0 | 1 | Not-determined | Genome-fragment | NA | NA | 0 |
| S9_extracted_k141_85681 | 1045 | No | NA | | 1 | 0 | 1 | Not-determined | Genome-fragment | NA | NA | 0 |
| S9_extracted_k141_10116 | 1232 | No | NA | | 4 | 0 | 3 | Not-determined | Genome-fragment | NA | NA | 0 |
| S9_extracted_k141_74407 | 3092 | No | NA | | 5 | 0 | 4 | Not-determined | Genome-fragment | NA | NA | 0 |
| S9_extracted_k141_29344 | 1160 | No | NA | | 1 | 0 | 0 | Not-determined | Genome-fragment | NA | NA | 0 |
| S3_extracted_k141_48901 | 1095 | No | NA | | 2 | 0 | 0 | Low-quality | Genome-fragment | 3.12 | AAI-based (medium-confidence) | 0 |
| S3_vs_k141_75860\|\|lt2gene | 1095 | No | NA | | 2 | 1 | 0 | Low-quality | Genome-fragment | 2.78 | AAI-based (medium-confidence) | 0 |
| S9_extracted_k141_55756 | 1507 | No | NA | | 2 | 0 | 2 | Not-determined | Genome-fragment | NA | NA | 0 |
| S5_extracted_k141_39607 | 1095 | No | NA | | 1 | 0 | 0 | Low-quality | Genome-fragment | 1.83 | AAI-based (medium-confidence) | 0 |
| S9_extracted_k141_44594 | 1641 | No | NA | | 4 | 0 | 0 | Not-determined | Genome-fragment | NA | NA | 0 |
| S9_extracted_k141_47766 | 1108 | No | NA | | 2 | 0 | 1 | Not-determined | Genome-fragment | NA | NA | 0 |
| S9_extracted_k141_29415 | 1261 | No | NA | | 3 | 0 | 0 | Not-determined | Genome-fragment | NA | NA | 0 |
| S9_extracted_k141_36980 | 2754 | No | NA | | 1 | 0 | 1 | Not-determined | Genome-fragment | NA | NA | 0 |
| S5_vs_k141_63688\|\|full | 1095 | No | NA | | 3 | 0 | 0 | Low-quality | Genome-fragment | 2.85 | AAI-based (medium-confidence) | 0 |
| S7_vs_38680\|\|full | 1095 | No | NA | | 2 | 2 | 0 | Low-quality | Genome-fragment | 2.46 | AAI-based (medium-confidence) | 0 |
| S9_extracted_k141_33051 | 1473 | No | NA | | 4 | 0 | 0 | Not-determined | Genome-fragment | NA | NA | 0 |
| S9_extracted_k141_51061 | 2998 | No | NA | | 1 | 0 | 0 | Not-determined | Genome-fragment | NA | NA | 0 |
| S16_extracted_120938 | 1095 | No | NA | | 3 | 2 | 0 | Low-quality | Genome-fragment | 2.49 | AAI-based (high-confidence) | 0 |
| S9_extracted_k141_6640 | 1420 | No | NA | | 2 | 0 | 1 | Not-determined | Genome-fragment | NA | NA | 0 |
| S9_extracted_k141_70503 | 1561 | No | NA | | 2 | 0 | 0 | Not-determined | Genome-fragment | NA | NA | 0 |
| S12_vs_k141_180863\|\|full | 1095 | No | NA | | 3 | 1 | 0 | Low-quality | Genome-fragment | 2.04 | AAI-based (medium-confidence) | 0 |
| S9_extracted_k141_47843 | 2390 | No | NA | | 3 | 0 | 0 | Not-determined | Genome-fragment | NA | NA | 0 |
| S9_extracted_k141_66782 | 1131 | No | NA | | 2 | 0 | 0 | Not-determined | Genome-fragment | NA | NA | 0 |
| S9_extracted_k141_55835 | 1529 | No | NA | | 1 | 0 | 1 | Not-determined | Genome-fragment | NA | NA | 0 |
| S9_extracted_k141_82136 | 2293 | No | NA | | 3 | 0 | 0 | Not-determined | Genome-fragment | NA | NA | 0 |
| S15_vs_5851\|\|full | 1095 | No | NA | | 2 | 0 | 1 | Low-quality | Genome-fragment | 0.4 | AAI-based (medium-confidence) | 0 |
| S6_vs_k141_31308\|\|full | 1095 | No | NA | | 3 | 0 | 0 | Low-quality | Genome-fragment | 2.66 | AAI-based (high-confidence) | 0 |
| S8_extracted_k141_109502 | 1095 | No | NA | | 1 | 0 | 0 | Low-quality | Genome-fragment | 0.61 | AAI-based (high-confidence) | 0 |
| S17_extracted_k141_82822 | 1096 | No | NA | | 2 | 0 | 0 | Low-quality | Genome-fragment | 2.1 | AAI-based (medium-confidence) | 0 |
| S9_extracted_k141_89440 | 1524 | No | NA | | 1 | 0 | 0 | Not-determined | Genome-fragment | NA | NA | 0 |
| S11_extracted_k141_78443 | 1096 | No | NA | | 3 | 2 | 0 | Low-quality | Genome-fragment | 3.61 | AAI-based (high-confidence) | 0 |
| S9_extracted_k141_85864 | 4282 | No | NA | | 3 | 0 | 0 | Not-determined | Genome-fragment | NA | NA | 0 |
| S11_vs_k141_19836\|\|full | 1096 | No | NA | | 2 | 0 | 0 | Low-quality | Genome-fragment | 1.84 | AAI-based (high-confidence) | 0 |
| S9_extracted_k141_14006 | 1045 | No | NA | | 1 | 0 | 1 | Not-determined | Genome-fragment | NA | NA | 0 |
| S9_extracted_k141_89460 | 1507 | No | NA | | 3 | 0 | 0 | Not-determined | Genome-fragment | NA | NA | 0 |
| S9_extracted_k141_25537 | 1408 | No | NA | | 1 | 0 | 1 | Not-determined | Genome-fragment | NA | NA | 0 |
| S13_extracted_105258 | 1096 | No | NA | | 2 | 0 | 0 | Low-quality | Genome-fragment | 0.69 | AAI-based (high-confidence) | 0 |
| S9_extracted_k141_66867 | 1053 | No | NA | | 1 | 0 | 0 | Not-determined | Genome-fragment | NA | NA | 0 |
| S9_extracted_k141_59321 | 2315 | No | NA | | 1 | 0 | 0 | Not-determined | Genome-fragment | NA | NA | 0 |
| S9_extracted_k141_2837 | 1124 | No | NA | | 2 | 0 | 0 | Not-determined | Genome-fragment | NA | NA | 0 |
| S9_extracted_k141_89493 | 1836 | No | NA | | 2 | 0 | 1 | Not-determined | Genome-fragment | NA | NA | 0 |
| S1_vs_k141_60808\|\|lt2gene | 1096 | No | NA | | 1 | 1 | 0 | Low-quality | Genome-fragment | 3.62 | AAI-based (medium-confidence) | 0 |
| S9_extracted_k141_10323 | 8828 | No | NA | | 4 | 0 | 1 | Not-determined | Genome-fragment | NA | NA | 0 |
| S5_vs_k141_41401\|\|full | 1096 | No | NA | | 2 | 0 | 0 | Low-quality | Genome-fragment | 0.57 | AAI-based (medium-confidence) | 0 |
| S7_extracted_26600 | 1096 | No | NA | | 2 | 1 | 0 | Low-quality | Genome-fragment | 0.47 | HMM-based (lower-bound) | 0 |
| S9_extracted_k141_59331 | 2093 | No | NA | | 2 | 0 | 2 | Not-determined | Genome-fragment | NA | NA | 0 |
| S9_extracted_k141_37116 | 1134 | No | NA | | 3 | 0 | 1 | Not-determined | Genome-fragment | NA | NA | 0 |
| S7_vs_129330\|\|full | 1096 | No | NA | | 4 | 0 | 0 | Low-quality | Genome-fragment | 2.68 | AAI-based (high-confidence) | 0 |
| S9_extracted_k141_89560 | 1148 | No | NA | | 1 | 0 | 1 | Not-determined | Genome-fragment | NA | NA | 0 |
| S9_extracted_k141_6729 | 1076 | No | NA | | 1 | 0 | 1 | Not-determined | Genome-fragment | NA | NA | 0 |
| S9_extracted_k141_59366 | 1628 | No | NA | | 1 | 0 | 1 | Not-determined | Genome-fragment | NA | NA | 0 |
| S9_extracted_k141_40903 | 1896 | No | NA | | 2 | 0 | 2 | Not-determined | Genome-fragment | NA | NA | 0 |
| S7_vs_77724\|\|lt2gene | 1096 | No | NA | | 1 | 1 | 0 | Low-quality | Genome-fragment | 2.92 | AAI-based (high-confidence) | 0 |
| S9_extracted_k141_18227 | 11201 | No | NA | | 15 | 0 | 5 | Not-determined | Genome-fragment | NA | NA | 0 |
| S9_extracted_k141_55939 | 1380 | No | NA | | 3 | 0 | 0 | Not-determined | Genome-fragment | NA | NA | 0 |
| S16_extracted_10465 | 1096 | No | NA | | 1 | 0 | 1 | Low-quality | Genome-fragment | 0.82 | AAI-based (high-confidence) | 0 |
| S9_extracted_k141_85975 | 1604 | No | NA | | 3 | 0 | 0 | Not-determined | Genome-fragment | NA | NA | 0 |
| S9_extracted_k141_55955 | 1229 | No | NA | | 2 | 0 | 0 | Not-determined | Genome-fragment | NA | NA | 0 |
| S10_extracted_80304 | 1096 | No | NA | | 2 | 0 | 0 | Low-quality | Genome-fragment | 0.57 | AAI-based (medium-confidence) | 0 |
| S9_extracted_k141_89596 | 4963 | No | NA | | 5 | 0 | 2 | Not-determined | Genome-fragment | NA | NA | 0 |
| S9_extracted_k141_2932 | 1163 | No | NA | | 1 | 0 | 1 | Not-determined | Genome-fragment | NA | NA | 0 |
| S9_extracted_k141_74514 | 4153 | No | NA | | 5 | 0 | 2 | Not-determined | Genome-fragment | NA | NA | 0 |
| S10_vs_4693\|\|full | 1096 | No | NA | | 4 | 0 | 0 | Low-quality | Genome-fragment | 2.48 | AAI-based (high-confidence) | 0 |
| S9_extracted_k141_25637 | 1166 | No | NA | | 2 | 0 | 2 | Not-determined | Genome-fragment | NA | NA | 0 |
| S9_extracted_k141_44783 | 1613 | No | NA | | 4 | 0 | 3 | Not-determined | Genome-fragment | NA | NA | 0 |
| S12_extracted_k141_80715 | 1096 | No | NA | | 1 | 0 | 0 | Low-quality | Genome-fragment | 1.91 | AAI-based (high-confidence) | 0 |
| S9_extracted_k141_33323 | 7178 | No | NA | | 10 | 0 | 3 | Not-determined | Genome-fragment | NA | NA | 0 |
| S9_extracted_k141_59423 | 1141 | No | NA | | 1 | 0 | 0 | Not-determined | Genome-fragment | NA | NA | 0 |
| S9_extracted_k141_10446 | 1321 | No | NA | | 2 | 0 | 1 | Not-determined | Genome-fragment | NA | NA | 0 |
| S12_extracted_k141_11943 | 1096 | No | NA | | 1 | 1 | 0 | Low-quality | Genome-fragment | 1.54 | HMM-based (lower-bound) | 0 |
| S12_extracted_k141_68087 | 1096 | No | NA | | 1 | 0 | 0 | Low-quality | Genome-fragment | 3.43 | AAI-based (high-confidence) | 0 |
| S9_extracted_k141_18346 | 2750 | No | NA | | 9 | 0 | 0 | Not-determined | Genome-fragment | NA | NA | 0 |
| S9_extracted_k141_86070 | 1129 | No | NA | | 1 | 0 | 1 | Not-determined | Genome-fragment | NA | NA | 0 |
| S9_extracted_k141_51350 | 2103 | No | NA | | 1 | 0 | 0 | Not-determined | Genome-fragment | NA | NA | 0 |
| S9_extracted_k141_82302 | 1008 | No | NA | | 1 | 0 | 1 | Not-determined | Genome-fragment | NA | NA | 0 |
| S9_extracted_k141_66986 | 1477 | No | NA | | 1 | 0 | 1 | Not-determined | Genome-fragment | NA | NA | 0 |
| S9_extracted_k141_82314 | 1602 | No | NA | | 3 | 0 | 0 | Not-determined | Genome-fragment | NA | NA | 0 |
| S9_extracted_k141_82326 | 1000 | No | NA | | 1 | 0 | 0 | Not-determined | Genome-fragment | NA | NA | 0 |
| S15_extracted_63749 | 1096 | No | NA | | 2 | 2 | 0 | Low-quality | Genome-fragment | 3.62 | HMM-based (lower-bound) | 0 |
| S6_vs_k141_44388\|\|full | 1096 | No | NA | | 3 | 0 | 0 | Low-quality | Genome-fragment | 1.73 | AAI-based (high-confidence) | 0 |
| S9_extracted_k141_56109 | 1038 | No | NA | | 3 | 0 | 0 | Not-determined | Genome-fragment | NA | NA | 0 |
| S9_extracted_k141_67031 | 1050 | No | NA | | 3 | 0 | 0 | Not-determined | Genome-fragment | NA | NA | 0 |
| S8_extracted_k141_168335 | 1096 | No | NA | | 3 | 0 | 0 | Low-quality | Genome-fragment | 1.67 | AAI-based (high-confidence) | 0 |
| S9_extracted_k141_70768 | 1091 | No | NA | | 2 | 0 | 0 | Not-determined | Genome-fragment | NA | NA | 0 |
| S8_vs_k141_27888\|\|lt2gene | 1096 | No | NA | | 2 | 1 | 0 | Low-quality | Genome-fragment | 2.89 | AAI-based (medium-confidence) | 0 |
| S9_extracted_k141_44893 | 1038 | No | NA | | 2 | 0 | 1 | Not-determined | Genome-fragment | NA | NA | 0 |
| S9_extracted_k141_74678 | 1237 | No | NA | | 2 | 0 | 1 | Not-determined | Genome-fragment | NA | NA | 0 |
| S9_vs_k141_31572\|\|full | 1097 | No | NA | | 4 | 2 | 0 | Low-quality | Genome-fragment | 2.34 | AAI-based (high-confidence) | 0 |
| S9_extracted_k141_48204 | 1048 | No | NA | | 2 | 0 | 0 | Not-determined | Genome-fragment | NA | NA | 0 |
| S9_extracted_k141_25827 | 2303 | No | NA | | 1 | 0 | 1 | Not-determined | Genome-fragment | NA | NA | 0 |
| S9_vs_k141_63319\|\|full | 1097 | No | NA | | 3 | 1 | 0 | Low-quality | Genome-fragment | 3 | AAI-based (medium-confidence) | 0 |
| S9_extracted_k141_10606 | 1531 | No | NA | | 3 | 0 | 1 | Not-determined | Genome-fragment | NA | NA | 0 |
| S9_extracted_k141_67104 | 2332 | No | NA | | 1 | 0 | 0 | Not-determined | Genome-fragment | NA | NA | 0 |
| S9_extracted_k141_74734 | 1311 | No | NA | | 2 | 0 | 2 | Not-determined | Genome-fragment | NA | NA | 0 |
| S9_extracted_k141_18510 | 1223 | No | NA | | 2 | 0 | 1 | Not-determined | Genome-fragment | NA | NA | 0 |
| S9_extracted_k141_78266 | 1660 | No | NA | | 1 | 0 | 1 | Not-determined | Genome-fragment | NA | NA | 0 |
| S9_extracted_k141_37419 | 1079 | No | NA | | 1 | 0 | 0 | Not-determined | Genome-fragment | NA | NA | 0 |
| S13_extracted_108627 | 1097 | No | NA | | 2 | 0 | 2 | Low-quality | Genome-fragment | 0.11 | AAI-based (medium-confidence) | 0 |
| S9_extracted_k141_25902 | 1332 | No | NA | | 1 | 0 | 0 | Not-determined | Genome-fragment | NA | NA | 0 |
| S9_extracted_k141_14135 | 1011 | No | NA | | 2 | 0 | 0 | Not-determined | Genome-fragment | NA | NA | 0 |
| S9_extracted_k141_48248 | 1441 | No | NA | | 2 | 0 | 2 | Not-determined | Genome-fragment | NA | NA | 0 |
| S1_extracted_k141_123605 | 1097 | No | NA | | 3 | 0 | 3 | Low-quality | Genome-fragment | 2.05 | AAI-based (medium-confidence) | 0 |
| S1_vs_k141_128139\|\|full | 1097 | No | NA | | 3 | 0 | 3 | Low-quality | Genome-fragment | 0.67 | AAI-based (medium-confidence) | 0 |
| S9_extracted_k141_89883 | 1250 | No | NA | | 2 | 0 | 2 | Not-determined | Genome-fragment | NA | NA | 0 |
| S1_vs_k141_61111\|\|full | 1097 | No | NA | | 3 | 0 | 1 | Low-quality | Genome-fragment | 1.37 | AAI-based (high-confidence) | 0 |
| S9_extracted_k141_56230 | 3090 | No | NA | | 4 | 0 | 2 | Not-determined | Genome-fragment | NA | NA | 0 |
| S1_vs_k141_62162\|\|full | 1097 | No | NA | | 3 | 0 | 1 | Low-quality | Genome-fragment | 1.98 | AAI-based (high-confidence) | 0 |
| S9_extracted_k141_45037 | 1857 | No | NA | | 2 | 0 | 0 | Not-determined | Genome-fragment | NA | NA | 0 |
| S9_extracted_k141_29952 | 1036 | No | NA | | 1 | 0 | 0 | Not-determined | Genome-fragment | NA | NA | 0 |
| S1_vs_k141_111676\|\|full | 1097 | No | NA | | 2 | 0 | 0 | Low-quality | Genome-fragment | 0.47 | AAI-based (medium-confidence) | 0 |
| S3_extracted_k141_44113 | 1097 | No | NA | | 2 | 1 | 0 | Low-quality | Genome-fragment | 2.72 | AAI-based (high-confidence) | 0 |
| S9_extracted_k141_70957 | 1486 | No | NA | | 3 | 0 | 1 | Not-determined | Genome-fragment | NA | NA | 0 |
| S9_extracted_k141_25973 | 1010 | No | NA | | 1 | 0 | 0 | Not-determined | Genome-fragment | NA | NA | 0 |
| S9_extracted_k141_41244 | 1260 | No | NA | | 3 | 0 | 1 | Not-determined | Genome-fragment | NA | NA | 0 |
| S9_extracted_k141_25985 | 3145 | No | NA | | 8 | 0 | 7 | Not-determined | Genome-fragment | NA | NA | 0 |
| S3_vs_k141_47838\|\|full | 1097 | No | NA | | 3 | 0 | 3 | Low-quality | Genome-fragment | 0.46 | AAI-based (medium-confidence) | 0 |
| S9_extracted_k141_22208 | 2890 | No | NA | | 5 | 0 | 0 | Not-determined | Genome-fragment | NA | NA | 0 |
| S5_extracted_k141_110548 | 1097 | No | NA | | 3 | 1 | 0 | Low-quality | Genome-fragment | 1.73 | AAI-based (medium-confidence) | 0 |
| S9_extracted_k141_30069 | 1269 | No | NA | | 2 | 0 | 0 | Not-determined | Genome-fragment | NA | NA | 0 |
| S5_vs_k141_4044\|\|full | 1097 | No | NA | | 3 | 0 | 0 | Low-quality | Genome-fragment | 3.2 | AAI-based (medium-confidence) | 0 |
| S5_vs_k141_96767\|\|full | 1097 | No | NA | | 2 | 0 | 1 | Low-quality | Genome-fragment | 1.42 | AAI-based (medium-confidence) | 0 |
| S9_extracted_k141_59802 | 1572 | No | NA | | 3 | 0 | 2 | Not-determined | Genome-fragment | NA | NA | 0 |
| S9_extracted_k141_51517 | 2672 | No | NA | | 3 | 0 | 0 | Not-determined | Genome-fragment | NA | NA | 0 |
| S9_extracted_k141_22292 | 3271 | No | NA | | 5 | 0 | 1 | Not-determined | Genome-fragment | NA | NA | 0 |
| S7_extracted_66657 | 1097 | No | NA | | 1 | 0 | 0 | Low-quality | Genome-fragment | 1.88 | AAI-based (high-confidence) | 0 |
| S16_vs_88094\|\|full | 1097 | No | NA | | 4 | 0 | 0 | Low-quality | Genome-fragment | 1.39 | AAI-based (high-confidence) | 0 |
| S9_extracted_k141_78467 | 1280 | No | NA | | 2 | 0 | 1 | Not-determined | Genome-fragment | NA | NA | 0 |
| S10_vs_56023\|\|full | 1097 | No | NA | | 2 | 0 | 1 | Low-quality | Genome-fragment | 0.44 | AAI-based (medium-confidence) | 0 |
| S9_extracted_k141_89994 | 3696 | No | NA | | 5 | 0 | 4 | Not-determined | Genome-fragment | NA | NA | 0 |
| S9_extracted_k141_41351 | 1516 | No | NA | | 2 | 0 | 0 | Not-determined | Genome-fragment | NA | NA | 0 |
| S10_vs_81581\|\|full | 1097 | No | NA | | 2 | 0 | 0 | Low-quality | Genome-fragment | 2.68 | AAI-based (medium-confidence) | 0 |
| S12_extracted_k141_156880 | 1097 | No | NA | | 2 | 0 | 2 | Low-quality | Genome-fragment | 0.95 | AAI-based (medium-confidence) | 0 |
| S9_extracted_k141_26086 | 4294 | No | NA | | 1 | 0 | 0 | Not-determined | Genome-fragment | NA | NA | 0 |
| S9_extracted_k141_14270 | 4183 | No | NA | | 1 | 0 | 1 | Not-determined | Genome-fragment | NA | NA | 0 |
| S15_vs_53867\|\|full | 1097 | No | NA | | 3 | 0 | 0 | Low-quality | Genome-fragment | 2.04 | AAI-based (high-confidence) | 0 |
| S9_extracted_k141_90019 | 3373 | No | NA | | 4 | 0 | 2 | Not-determined | Genome-fragment | NA | NA | 0 |
| S4_extracted_2088 | 1097 | No | NA | | 4 | 4 | 0 | Low-quality | Genome-fragment | 3.62 | AAI-based (high-confidence) | 0 |
| S9_extracted_k141_26118 | 1335 | No | NA | | 1 | 0 | 0 | Not-determined | Genome-fragment | NA | NA | 0 |
| S9_extracted_k141_26125 | 1232 | No | NA | | 1 | 0 | 0 | Not-determined | Genome-fragment | NA | NA | 0 |
| S9_extracted_k141_67316 | 1519 | No | NA | | 2 | 0 | 0 | Not-determined | Genome-fragment | NA | NA | 0 |
| S8_vs_k141_183208\|\|full | 1097 | No | NA | | 3 | 0 | 0 | Low-quality | Genome-fragment | 1.79 | AAI-based (medium-confidence) | 0 |
| S9_extracted_k141_67326 | 1259 | No | NA | | 1 | 0 | 0 | Not-determined | Genome-fragment | NA | NA | 0 |
| S9_extracted_k141_51641 | 1004 | No | NA | | 2 | 0 | 2 | Not-determined | Genome-fragment | NA | NA | 0 |
| S9_extracted_k141_75056 | 1150 | No | NA | | 2 | 0 | 2 | Not-determined | Genome-fragment | NA | NA | 0 |
| S9_extracted_k141_14345 | 4486 | No | NA | | 4 | 0 | 4 | Not-determined | Genome-fragment | NA | NA | 0 |
| S9_extracted_k141_30226 | 4981 | No | NA | | 8 | 0 | 7 | Not-determined | Genome-fragment | NA | NA | 0 |
| S8_vs_k141_23594\|\|full | 1097 | No | NA | | 3 | 0 | 0 | Low-quality | Genome-fragment | 0.7 | AAI-based (medium-confidence) | 0 |
| S9_extracted_k141_30228 | 1207 | No | NA | | 3 | 0 | 1 | Not-determined | Genome-fragment | NA | NA | 0 |
| S8_vs_k141_50893\|\|full | 1097 | No | NA | | 3 | 1 | 0 | Low-quality | Genome-fragment | 3.72 | HMM-based (lower-bound) | 0 |
| S17_extracted_k141_84151 | 1098 | No | NA | | 2 | 0 | 1 | Low-quality | Genome-fragment | 1.3 | AAI-based (high-confidence) | 0 |
| S9_extracted_k141_3412 | 1411 | No | NA | | 3 | 0 | 1 | Not-determined | Genome-fragment | NA | NA | 0 |
| S9_extracted_k141_78642 | 1126 | No | NA | | 1 | 0 | 0 | Not-determined | Genome-fragment | NA | NA | 0 |
| S9_extracted_k141_33829 | 1031 | No | NA | | 3 | 0 | 0 | Not-determined | Genome-fragment | NA | NA | 0 |
| S9_extracted_k141_56487 | 1288 | No | NA | | 2 | 0 | 2 | Not-determined | Genome-fragment | NA | NA | 0 |
| S9_extracted_k141_56492 | 1284 | No | NA | | 1 | 0 | 1 | Not-determined | Genome-fragment | NA | NA | 0 |
| S9_extracted_k141_45370 | 2473 | No | NA | | 3 | 0 | 2 | Not-determined | Genome-fragment | NA | NA | 0 |
| S17_extracted_k141_102256 | 1098 | No | NA | | 1 | 0 | 0 | Low-quality | Genome-fragment | 1.17 | AAI-based (high-confidence) | 0 |
| S9_extracted_k141_67438 | 2000 | No | NA | | 1 | 0 | 1 | Not-determined | Genome-fragment | NA | NA | 0 |
| S9_extracted_k141_3435 | 1002 | No | NA | | 1 | 0 | 1 | Not-determined | Genome-fragment | NA | NA | 0 |
| S9_extracted_k141_67454 | 1903 | No | NA | | 2 | 0 | 0 | Not-determined | Genome-fragment | NA | NA | 0 |
| S9_extracted_k141_22480 | 1092 | No | NA | | 2 | 0 | 1 | Not-determined | Genome-fragment | NA | NA | 0 |
| S9_extracted_k141_86490 | 1150 | No | NA | | 1 | 0 | 1 | Not-determined | Genome-fragment | NA | NA | 0 |
| S9_extracted_k141_90195 | 1259 | No | NA | | 3 | 0 | 0 | Not-determined | Genome-fragment | NA | NA | 0 |
| S9_extracted_k141_67476 | 1634 | No | NA | | 1 | 0 | 0 | Not-determined | Genome-fragment | NA | NA | 0 |
| S14_extracted_k141_60170 | 1098 | No | NA | | 3 | 0 | 0 | Low-quality | Genome-fragment | 2.3 | AAI-based (medium-confidence) | 0 |
| S14_vs_k141_105888\|\|lt2gene | 1098 | No | NA | | 2 | 1 | 0 | Low-quality | Genome-fragment | 1.2 | HMM-based (lower-bound) | 0 |
| S9_extracted_k141_86527 | 1035 | No | NA | | 1 | 0 | 0 | Not-determined | Genome-fragment | NA | NA | 0 |
| S1_vs_k141_28261\|\|full | 1098 | No | NA | | 2 | 0 | 0 | Low-quality | Genome-fragment | 3.28 | AAI-based (high-confidence) | 0 |
| S1_vs_k141_145089\|\|full | 1098 | No | NA | | 3 | 1 | 0 | Low-quality | Genome-fragment | 2.54 | AAI-based (medium-confidence) | 0 |
| S3_vs_k141_39268\|\|full | 1098 | No | NA | | 3 | 1 | 0 | Low-quality | Genome-fragment | 1.75 | AAI-based (high-confidence) | 0 |
| S9_extracted_k141_67530 | 11239 | No | NA | | 2 | 0 | 1 | Not-determined | Genome-fragment | NA | NA | 0 |
| S9_extracted_k141_51656 | 128272 | No | NA | | 138 | 12 | 0 | Complete | High-quality | 100 | DTR (high-confidence) | 0 |
| S9_extracted_k141_75235 | 1699 | No | NA | | 3 | 0 | 1 | Not-determined | Genome-fragment | NA | NA | 0 |
| S9_extracted_k141_56596 | 1883 | No | NA | | 2 | 0 | 1 | Not-determined | Genome-fragment | NA | NA | 0 |
| S5_extracted_k141_121134 | 1098 | No | NA | | 1 | 1 | 0 | Low-quality | Genome-fragment | 2.94 | AAI-based (high-confidence) | 0 |
| S9_extracted_k141_56605 | 1353 | No | NA | | 1 | 0 | 0 | Not-determined | Genome-fragment | NA | NA | 0 |
| S9_extracted_k141_86572 | 3424 | No | NA | | 2 | 0 | 2 | Not-determined | Genome-fragment | NA | NA | 0 |
| S5_extracted_k141_137369 | 1098 | No | NA | | 2 | 0 | 0 | Low-quality | Genome-fragment | 0.36 | AAI-based (medium-confidence) | 0 |
| S7_extracted_127051 | 1098 | No | NA | | 3 | 1 | 0 | Low-quality | Genome-fragment | 2.39 | AAI-based (medium-confidence) | 0 |
| S9_extracted_k141_22636 | 2174 | No | NA | | 1 | 0 | 0 | Not-determined | Genome-fragment | NA | NA | 0 |
| S9_extracted_k141_3532 | 1285 | No | NA | | 1 | 0 | 0 | Not-determined | Genome-fragment | NA | NA | 0 |
| S9_extracted_k141_56633 | 3319 | No | NA | | 6 | 0 | 1 | Not-determined | Genome-fragment | NA | NA | 0 |
| S9_extracted_k141_60099 | 1110 | No | NA | | 1 | 0 | 0 | Not-determined | Genome-fragment | NA | NA | 0 |
| S16_extracted_136437 | 1098 | No | NA | | 3 | 0 | 1 | Low-quality | Genome-fragment | 2.75 | AAI-based (medium-confidence) | 0 |
| S9_extracted_k141_48578 | 1502 | No | NA | | 1 | 0 | 0 | Not-determined | Genome-fragment | NA | NA | 0 |
| S9_extracted_k141_56645 | 1484 | No | NA | | 1 | 0 | 0 | Not-determined | Genome-fragment | NA | NA | 0 |
| S9_extracted_k141_11138 | 3105 | No | NA | | 3 | 0 | 2 | Not-determined | Genome-fragment | NA | NA | 0 |
| S12_vs_k141_150035\|\|full | 1098 | No | NA | | 5 | 0 | 0 | Low-quality | Genome-fragment | 4.41 | AAI-based (high-confidence) | 0 |
| S12_vs_k141_97001\|\|full | 1098 | No | NA | | 2 | 0 | 0 | Low-quality | Genome-fragment | 3.06 | AAI-based (medium-confidence) | 0 |
| S9_extracted_k141_60119 | 1375 | No | NA | | 2 | 0 | 2 | Not-determined | Genome-fragment | NA | NA | 0 |
| S9_extracted_k141_56671 | 1006 | No | NA | | 1 | 0 | 1 | Not-determined | Genome-fragment | NA | NA | 0 |
| S9_extracted_k141_3575 | 4004 | No | NA | | 2 | 0 | 2 | Not-determined | Genome-fragment | NA | NA | 0 |
| S12_vs_k141_141236\|\|lt2gene | 1098 | No | NA | | 1 | 1 | 0 | Low-quality | Genome-fragment | 2.63 | AAI-based (high-confidence) | 0 |
| S8_extracted_k141_27949 | 1098 | No | NA | | 2 | 0 | 0 | Low-quality | Genome-fragment | 1.09 | AAI-based (high-confidence) | 0 |
| S8_vs_k141_84928\|\|full | 1098 | No | NA | | 3 | 0 | 0 | Low-quality | Genome-fragment | 1.96 | AAI-based (medium-confidence) | 0 |
| S9_extracted_k141_48607 | 1272 | No | NA | | 3 | 0 | 0 | Not-determined | Genome-fragment | NA | NA | 0 |
| S9_extracted_k141_78974 | 1142 | No | NA | | 3 | 0 | 0 | Not-determined | Genome-fragment | NA | NA | 0 |
| S9_extracted_k141_71609 | 1107 | No | NA | | 1 | 0 | 0 | Not-determined | Genome-fragment | NA | NA | 0 |
| S9_extracted_k141_75363 | 12595 | No | NA | | 14 | 7 | 1 | Medium-quality | Genome-fragment | 53.41 | AAI-based (medium-confidence) | 0 |
| S17_vs_k141_36429\|\|lt2gene | 1099 | No | NA | | 1 | 1 | 0 | Low-quality | Genome-fragment | 19.85 | AAI-based (high-confidence) | 0 |
| S9_vs_k141_20745\|\|full | 1099 | No | NA | | 2 | 0 | 0 | Low-quality | Genome-fragment | 1.03 | AAI-based (medium-confidence) | 0 |
| S11_vs_k141_102790\|\|full | 1099 | No | NA | | 3 | 0 | 0 | Low-quality | Genome-fragment | 1.33 | AAI-based (medium-confidence) | 0 |
| S9_extracted_k141_56791 | 1364 | No | NA | | 2 | 0 | 1 | Not-determined | Genome-fragment | NA | NA | 0 |
| S9_extracted_k141_82808 | 1046 | No | NA | | 2 | 0 | 0 | Not-determined | Genome-fragment | NA | NA | 0 |
| S9_extracted_k141_11316 | 1025 | No | NA | | 1 | 0 | 0 | Not-determined | Genome-fragment | NA | NA | 0 |
| S9_extracted_k141_3716 | 1591 | No | NA | | 1 | 0 | 1 | Not-determined | Genome-fragment | NA | NA | 0 |
| S3_extracted_k141_60395 | 1099 | No | NA | | 2 | 0 | 0 | Low-quality | Genome-fragment | 2.17 | AAI-based (medium-confidence) | 0 |
| S5_vs_k141_30480\|\|full | 1099 | No | NA | | 3 | 1 | 0 | Low-quality | Genome-fragment | 2.42 | AAI-based (medium-confidence) | 0 |
| S5_vs_k141_18084\|\|full | 1099 | No | NA | | 2 | 0 | 0 | Low-quality | Genome-fragment | 1.47 | AAI-based (high-confidence) | 0 |
| S9_extracted_k141_86860 | 2634 | No | NA | | 7 | 0 | 7 | Not-determined | Genome-fragment | NA | NA | 0 |
| S7_vs_1637\|\|full | 1099 | No | NA | | 4 | 1 | 0 | Low-quality | Genome-fragment | 1.22 | AAI-based (high-confidence) | 0 |
| S9_extracted_k141_60357 | 1184 | No | NA | | 2 | 0 | 2 | Not-determined | Genome-fragment | NA | NA | 0 |
| S9_extracted_k141_75457 | 1058 | No | NA | | 2 | 0 | 1 | Not-determined | Genome-fragment | NA | NA | 0 |
| S9_extracted_k141_79101 | 1043 | No | NA | | 1 | 0 | 0 | Not-determined | Genome-fragment | NA | NA | 0 |
| S7_vs_112865\|\|full | 1099 | No | NA | | 3 | 1 | 0 | Low-quality | Genome-fragment | 2.8 | AAI-based (high-confidence) | 0 |
| S9_extracted_k141_82928 | 1209 | No | NA | | 2 | 0 | 2 | Not-determined | Genome-fragment | NA | NA | 0 |
| S16_extracted_122732 | 1099 | No | NA | | 3 | 1 | 0 | Low-quality | Genome-fragment | 1.73 | AAI-based (high-confidence) | 0 |
| S16_extracted_129711 | 1099 | No | NA | | 1 | 0 | 0 | Low-quality | Genome-fragment | 4.31 | AAI-based (high-confidence) | 0 |
| S9_extracted_k141_48824 | 3182 | No | NA | | 2 | 0 | 0 | Not-determined | Genome-fragment | NA | NA | 0 |
| S9_extracted_k141_71753 | 1169 | No | NA | | 2 | 0 | 1 | Not-determined | Genome-fragment | NA | NA | 0 |
| S12_vs_k141_65584\|\|full | 1099 | No | NA | | 3 | 0 | 0 | Low-quality | Genome-fragment | 0.95 | AAI-based (medium-confidence) | 0 |
| S8_extracted_k141_19081 | 1099 | No | NA | | 3 | 0 | 1 | Low-quality | Genome-fragment | 2.12 | AAI-based (medium-confidence) | 0 |
| S9_extracted_k141_14958 | 1097 | No | NA | | 2 | 0 | 1 | Not-determined | Genome-fragment | NA | NA | 0 |
| S9_extracted_k141_83016 | 2056 | No | NA | | 2 | 0 | 0 | Not-determined | Genome-fragment | NA | NA | 0 |
| S8_vs_k141_147359\|\|full | 1099 | No | NA | | 3 | 0 | 0 | Low-quality | Genome-fragment | 0.6 | AAI-based (medium-confidence) | 0 |
| S9_extracted_k141_75608 | 1733 | No | NA | | 2 | 0 | 2 | Not-determined | Genome-fragment | NA | NA | 0 |
| S9_extracted_k141_90548 | 3857 | No | NA | | 3 | 0 | 2 | Not-determined | Genome-fragment | NA | NA | 0 |
| S9_extracted_k141_67910 | 1406 | No | NA | | 3 | 0 | 0 | Not-determined | Genome-fragment | NA | NA | 0 |
| S14_vs_k141_99173\|\|full | 1100 | No | NA | | 2 | 1 | 0 | Low-quality | Genome-fragment | 0.67 | AAI-based (high-confidence) | 0 |

| | | | | | | | | | | | |
|---|---|---|---|---|---|---|---|---|---|---|---|
| S9_extracted_k141_83062 | 1135 | No | NA | 1 | 0 | 1 | Not-determined | Genome-fragment | NA | NA | 0 |
| S1_extracted_k141_156811 | 1100 | No | NA | 1 | 1 | 0 | Low-quality | Genome-fragment | 2.36 | HMM-based (lower-bound) | 0 |
| S9_extracted_k141_79269 | 3026 | No | NA | 4 | 0 | 2 | Not-determined | Genome-fragment | NA | NA | 0 |
| S1_extracted_k141_39224 | 1100 | No | NA | 3 | 0 | 0 | Low-quality | Genome-fragment | 1.81 | AAI-based (high-confidence) | 0 |
| S3_extracted_k141_76931 | 1100 | No | NA | 4 | 0 | 0 | Low-quality | Genome-fragment | 1.24 | AAI-based (high-confidence) | 0 |
| S9_extracted_k141_15012 | 32097 | No | NA | 49 | 30 | 0 | High-quality | High-quality | 99.61 | AAI-based (high-confidence) | 0 |
| S3_vs_k141_98659||full | 1100 | No | NA | 2 | 0 | 1 | Low-quality | Genome-fragment | 0.77 | AAI-based (high-confidence) | 0 |
| S5_vs_k141_24718||full | 1100 | No | NA | 4 | 1 | 0 | Low-quality | Genome-fragment | 0.67 | HMM-based (lower-bound) | 0 |
| S9_extracted_k141_64018 | 1220 | No | NA | 2 | 0 | 0 | Not-determined | Genome-fragment | NA | NA | 0 |
| S5_vs_k141_13116||full | 1100 | No | NA | 3 | 0 | 0 | Low-quality | Genome-fragment | 1.7 | AAI-based (medium-confidence) | 0 |
| S9_extracted_k141_75753 | 1126 | No | NA | 1 | 0 | 0 | Not-determined | Genome-fragment | NA | NA | 0 |
| S5_vs_k141_32349||full | 1100 | No | NA | 4 | 2 | 0 | Low-quality | Genome-fragment | 2.56 | AAI-based (medium-confidence) | 0 |
| S7_vs_98994||full | 1100 | No | NA | 3 | 1 | 0 | Low-quality | Genome-fragment | 2.7 | AAI-based (medium-confidence) | 0 |
| S9_extracted_k141_68095 | 2518 | No | NA | 2 | 0 | 2 | Not-determined | Genome-fragment | NA | NA | 0 |
| S9_extracted_k141_68100 | 7276 | No | NA | 9 | 0 | 5 | Not-determined | Genome-fragment | NA | NA | 0 |
| S9_extracted_k141_83140 | 1466 | No | NA | 1 | 0 | 0 | Not-determined | Genome-fragment | NA | NA | 0 |
| S7_vs_99918||full | 1100 | No | NA | 3 | 0 | 1 | Low-quality | Genome-fragment | 2.11 | AAI-based (medium-confidence) | 0 |
| S16_extracted_19198 | 1100 | No | NA | 3 | 0 | 0 | Low-quality | Genome-fragment | 2.54 | AAI-based (medium-confidence) | 0 |
| S16_extracted_121252 | 1100 | No | NA | 1 | 0 | 1 | Low-quality | Genome-fragment | 0.3 | AAI-based (medium-confidence) | 0 |
| S10_vs_48301||full | 1100 | No | NA | 3 | 1 | 0 | Low-quality | Genome-fragment | 1.37 | AAI-based (medium-confidence) | 0 |
| S9_extracted_k141_64183 | 1032 | No | NA | 2 | 0 | 0 | Not-determined | Genome-fragment | NA | NA | 0 |
| S9_extracted_k141_83236 | 1073 | No | NA | 1 | 0 | 0 | Not-determined | Genome-fragment | NA | NA | 0 |
| S9_extracted_k141_79553 | 1274 | No | NA | 1 | 0 | 1 | Not-determined | Genome-fragment | NA | NA | 0 |
| S8_extracted_k141_35395 | 1100 | No | NA | 1 | 0 | 1 | Low-quality | Genome-fragment | 2.35 | AAI-based (medium-confidence) | 0 |
| S9_extracted_k141_64246 | 1689 | No | NA | 1 | 0 | 1 | Not-determined | Genome-fragment | NA | NA | 0 |
| S9_extracted_k141_83314 | 1133 | No | NA | 2 | 0 | 1 | Not-determined | Genome-fragment | NA | NA | 0 |
| S9_extracted_k141_64257 | 1130 | No | NA | 1 | 0 | 1 | Not-determined | Genome-fragment | NA | NA | 0 |
| S9_extracted_k141_83353 | 1501 | No | NA | 1 | 0 | 0 | Not-determined | Genome-fragment | NA | NA | 0 |
| S9_extracted_k141_49239 | 1499 | No | NA | 2 | 0 | 1 | Not-determined | Genome-fragment | NA | NA | 0 |
| S8_vs_k141_202435||lt2gene | 1100 | No | NA | 2 | 1 | 0 | Low-quality | Genome-fragment | 0.71 | HMM-based (lower-bound) | 0 |
| S9_extracted_k141_90808 | 1399 | No | NA | 1 | 0 | 1 | Not-determined | Genome-fragment | NA | NA | 0 |
| S9_extracted_k141_52611 | 1424 | No | NA | 2 | 0 | 1 | Not-determined | Genome-fragment | NA | NA | 0 |
| S9_extracted_k141_90839 | 1011 | No | NA | 1 | 0 | 0 | Not-determined | Genome-fragment | NA | NA | 0 |
| S9_extracted_k141_52629 | 2197 | No | NA | 3 | 0 | 1 | Not-determined | Genome-fragment | NA | NA | 0 |
| S17_extracted_k141_50502 | 1101 | No | NA | 2 | 2 | 0 | Low-quality | Genome-fragment | 2.87 | AAI-based (high-confidence) | 0 |
| S9_extracted_k141_52713 | 1192 | No | NA | 2 | 0 | 1 | Not-determined | Genome-fragment | NA | NA | 0 |
| S9_extracted_k141_52828 | 2215 | No | NA | 2 | 0 | 0 | Not-determined | Genome-fragment | NA | NA | 0 |
| S17_vs_k141_177075||full | 1101 | No | NA | 3 | 2 | 0 | Low-quality | Genome-fragment | 2.8 | AAI-based (medium-confidence) | 0 |
| S9_extracted_k141_52880 | 1131 | No | NA | 2 | 0 | 1 | Not-determined | Genome-fragment | NA | NA | 0 |
| S9_extracted_k141_52938 | 1147 | No | NA | 2 | 0 | 1 | Not-determined | Genome-fragment | NA | NA | 0 |
| S9_extracted_k141_90847 | 148940 | No | NA | 173 | 39 | 5 | Complete | High-quality | 100 | DTR (medium-confidence) | 0 |
| S9_extracted_k141_53030 | 1299 | No | NA | 2 | 0 | 0 | Not-determined | Genome-fragment | NA | NA | 0 |
| S9_extracted_k141_90852 | 5449 | No | NA | 11 | 0 | 0 | Complete | High-quality | 100 | DTR (high-confidence) | 0 |
| S14_extracted_k141_49984 | 1101 | No | NA | 1 | 0 | 0 | Low-quality | Genome-fragment | 0.12 | AAI-based (medium-confidence) | 0 |
| S9_extracted_k141_90856 | 38132 | No | NA | 49 | 36 | 0 | Complete | High-quality | 100 | DTR (high-confidence) | 0 |
| S9_extracted_k141_90866 | 9389 | No | NA | 15 | 6 | 0 | Complete | High-quality | 100 | DTR (high-confidence) | 0 |
| S9_extracted_k141_90871 | 36948 | No | NA | 59 | 26 | 0 | Complete | High-quality | 100 | DTR (high-confidence) | 0 |
| S9_vs_k141_45812||full | 36359 | No | NA | 56 | 13 | 2 | Medium-quality | Genome-fragment | 69.64 | HMM-based (lower-bound) | 0 |
| S9_vs_k141_49943||full | 6495 | No | NA | 5 | 0 | 0 | Not-determined | Genome-fragment | NA | NA | 0 |
| S9_vs_k141_88286||full | 46223 | No | NA | 61 | 12 | 1 | High-quality | High-quality | 100 | AAI-based (high-confidence) | 0 |
| S9_vs_k141_50439||full | 56327 | No | NA | 95 | 21 | 1 | High-quality | High-quality | 100 | AAI-based (high-confidence) | 0 |
| S1_vs_k141_143883||full | 1101 | No | NA | 2 | 1 | 0 | Low-quality | Genome-fragment | 1.77 | AAI-based (high-confidence) | 0 |
| S9_vs_k141_5676||full | 34189 | No | NA | 55 | 17 | 0 | Complete | High-quality | 100 | DTR (medium-confidence) | 0 |
| S9_vs_k141_66877||full | 36296 | No | NA | 48 | 14 | 0 | High-quality | High-quality | 94.24 | AAI-based (high-confidence) | 0 |
| S9_vs_k141_75050||full | 32777 | No | NA | 43 | 14 | 2 | High-quality | High-quality | 100 | AAI-based (high-confidence) | 0 |
| S9_vs_k141_48424||full | 54788 | No | NA | 86 | 15 | 4 | High-quality | High-quality | 97.96 | AAI-based (high-confidence) | 0 |
| S9_vs_k141_90846||full | 38274 | No | NA | 57 | 21 | 1 | High-quality | High-quality | 93.75 | AAI-based (high-confidence) | 0 |
| S9_vs_k141_90848||full | 42668 | No | NA | 73 | 19 | 1 | Medium-quality | Genome-fragment | 79.27 | AAI-based (high-confidence) | 0 |
| S9_vs_k141_90849||full | 40590 | No | NA | 57 | 10 | 0 | High-quality | High-quality | 100 | AAI-based (high-confidence) | 0 |
| S9_vs_k141_90850||full | 38814 | No | NA | 62 | 21 | 0 | High-quality | High-quality | 98.71 | AAI-based (high-confidence) | 0 |
| S9_vs_k141_90851||full | 45923 | No | NA | 63 | 30 | 0 | High-quality | High-quality | 100 | AAI-based (high-confidence) | 0 |
| S9_vs_k141_90857||full | 37664 | No | NA | 51 | 8 | 0 | High-quality | High-quality | 96.18 | AAI-based (high-confidence) | 0 |
| S9_vs_k141_90859||full | 44116 | No | NA | 69 | 30 | 4 | High-quality | High-quality | 100 | AAI-based (high-confidence) | 0 |
| S9_vs_k141_90860||full | 28818 | No | NA | 50 | 20 | 1 | High-quality | High-quality | 95.78 | AAI-based (medium-confidence) | 0 |
| S9_vs_k141_90862||full | 2114 | No | NA | 3 | 0 | 0 | Not-determined | Genome-fragment | NA | NA | 0 |
| S9_vs_k141_90863||full | 5218 | No | NA | 6 | 1 | 0 | High-quality | High-quality | 94.97 | AAI-based (high-confidence) | 0 |
| S3_vs_k141_104758||full | 1101 | No | NA | 3 | 1 | 0 | Low-quality | Genome-fragment | 2.09 | AAI-based (high-confidence) | 0 |
| S5_vs_k141_53809||full | 1101 | No | NA | 3 | 1 | 0 | Low-quality | Genome-fragment | 2.06 | AAI-based (medium-confidence) | 0 |
| S5_vs_k141_132815||full | 1101 | No | NA | 3 | 1 | 0 | Low-quality | Genome-fragment | 1.78 | AAI-based (medium-confidence) | 0 |
| S5_vs_k141_58172||full | 1101 | No | NA | 2 | 1 | 0 | Low-quality | Genome-fragment | 2 | AAI-based (high-confidence) | 0 |
| S9_vs_k141_90894||full | 2663 | No | NA | 5 | 0 | 0 | Not-determined | Genome-fragment | NA | NA | 0 |
| S16_extracted_148976 | 1101 | No | NA | 2 | 0 | 2 | Low-quality | Genome-fragment | 0.84 | AAI-based (medium-confidence) | 0 |
| S12_extracted_k141_105606 | 1101 | No | NA | 2 | 1 | 0 | Low-quality | Genome-fragment | 0.76 | HMM-based (lower-bound) | 0 |
| S12_vs_k141_173460||full | 1101 | No | NA | 3 | 1 | 0 | Low-quality | Genome-fragment | 2.45 | AAI-based (medium-confidence) | 0 |
| S12_vs_k141_186590||full | 1101 | No | NA | 2 | 0 | 1 | Low-quality | Genome-fragment | 1.39 | AAI-based (medium-confidence) | 0 |
| S9_vs_k141_3810||full | 1527 | No | NA | 2 | 0 | 1 | Not-determined | Genome-fragment | NA | NA | 0 |
| S15_vs_17990||full | 1101 | No | NA | 3 | 0 | 0 | Low-quality | Genome-fragment | 2.62 | AAI-based (high-confidence) | 0 |
| S9_vs_k141_53086||full | 3341 | No | NA | 6 | 0 | 0 | Not-determined | Genome-fragment | NA | NA | 0 |
| S9_vs_k141_87184||full | 2429 | No | NA | 2 | 0 | 2 | Not-determined | Genome-fragment | NA | NA | 0 |
| S9_vs_k141_22773||full | 6884 | No | NA | 7 | 0 | 2 | Not-determined | Genome-fragment | NA | NA | 0 |
| S9_vs_k141_34149||full | 1372 | No | NA | 3 | 0 | 0 | Not-determined | Genome-fragment | NA | NA | 0 |
| S17_vs_k141_71306||full | 1102 | No | NA | 2 | 0 | 1 | Low-quality | Genome-fragment | 1.71 | AAI-based (high-confidence) | 0 |
| S11_vs_k141_76168||full | 1102 | No | NA | 3 | 2 | 0 | Low-quality | Genome-fragment | 1.66 | HMM-based (lower-bound) | 0 |
| S9_vs_k141_83413||full | 1331 | No | NA | 2 | 0 | 0 | Not-determined | Genome-fragment | NA | NA | 0 |
| S11_vs_k141_84279||full | 1102 | No | NA | 2 | 0 | 0 | Low-quality | Genome-fragment | 2.65 | AAI-based (medium-confidence) | 0 |
| S13_extracted_8580 | 1102 | No | NA | 1 | 0 | 0 | Low-quality | Genome-fragment | 2.15 | AAI-based (high-confidence) | 0 |
| S9_vs_k141_53092||full | 3603 | No | NA | 2 | 0 | 1 | Not-determined | Genome-fragment | NA | NA | 0 |
| S9_vs_k141_30347||full | 2258 | No | NA | 3 | 0 | 1 | Not-determined | Genome-fragment | NA | NA | 0 |
| S14_vs_k141_33050||full | 1102 | No | NA | 3 | 0 | 0 | Low-quality | Genome-fragment | 3.13 | AAI-based (medium-confidence) | 0 |
| S1_vs_k141_137903||full | 1102 | No | NA | 4 | 2 | 0 | Low-quality | Genome-fragment | 1.82 | AAI-based (high-confidence) | 0 |
| S9_vs_k141_34191||full | 2271 | No | NA | 1 | 0 | 0 | Not-determined | Genome-fragment | NA | NA | 0 |
| S1_vs_k141_125730||lt2gene | 1102 | No | NA | 1 | 1 | 0 | Low-quality | Genome-fragment | 2.47 | AAI-based (high-confidence) | 0 |
| S9_vs_k141_22811||full | 2443 | No | NA | 3 | 0 | 1 | Not-determined | Genome-fragment | NA | NA | 0 |
| S7_extracted_61465 | 1102 | No | NA | 1 | 0 | 0 | Low-quality | Genome-fragment | 2.04 | AAI-based (high-confidence) | 0 |
| S9_vs_k141_15247||full | 4948 | No | NA | 4 | 0 | 3 | Not-determined | Genome-fragment | NA | NA | 0 |
| S9_vs_k141_18991||full | 25513 | No | NA | 26 | 0 | 16 | Not-determined | Genome-fragment | NA | NA | 0 |
| S9_vs_k141_7655||full | 1540 | No | NA | 2 | 0 | 0 | Not-determined | Genome-fragment | NA | NA | 0 |
| S9_vs_k141_22823||full | 1298 | No | NA | 2 | 0 | 0 | Not-determined | Genome-fragment | NA | NA | 0 |
| S9_vs_k141_83457||full | 1517 | No | NA | 2 | 0 | 0 | Not-determined | Genome-fragment | NA | NA | 0 |
| S9_vs_k141_64510||full | 1350 | No | NA | 3 | 0 | 0 | Not-determined | Genome-fragment | NA | NA | 0 |
| S9_vs_k141_22825||full | 2802 | No | NA | 3 | 0 | 0 | Not-determined | Genome-fragment | NA | NA | 0 |
| S9_vs_k141_68270||full | 1498 | No | NA | 3 | 0 | 1 | Not-determined | Genome-fragment | NA | NA | 0 |
| S9_vs_k141_60696||full | 2402 | No | NA | 4 | 0 | 1 | Not-determined | Genome-fragment | NA | NA | 0 |
| S9_vs_k141_83469||full | 3457 | No | NA | 5 | 0 | 3 | Not-determined | Genome-fragment | NA | NA | 0 |
| S16_extracted_66528 | 1102 | No | NA | 4 | 1 | 0 | Low-quality | Genome-fragment | 3.52 | AAI-based (medium-confidence) | 0 |
| S10_vs_48234||full | 1102 | No | NA | 2 | 0 | 1 | Low-quality | Genome-fragment | 0.74 | AAI-based (high-confidence) | 0 |
| S4_extracted_22682 | 1102 | No | NA | 2 | 2 | 0 | Low-quality | Genome-fragment | 2.99 | HMM-based (lower-bound) | 0 |
| S9_vs_k141_97||full | 2246 | No | NA | 2 | 0 | 0 | Not-determined | Genome-fragment | NA | NA | 0 |
| S8_extracted_k141_93720 | 1102 | No | NA | 2 | 0 | 2 | Low-quality | Genome-fragment | 3.63 | AAI-based (high-confidence) | 0 |
| S17_extracted_k141_134380 | 1103 | No | NA | 3 | 0 | 1 | Low-quality | Genome-fragment | 0.49 | AAI-based (medium-confidence) | 0 |
| S11_vs_k141_80990||full | 1103 | No | NA | 3 | 0 | 0 | Low-quality | Genome-fragment | 1.51 | AAI-based (high-confidence) | 0 |
| S9_vs_k141_19064||full | 4826 | No | NA | 6 | 0 | 3 | Not-determined | Genome-fragment | NA | NA | 0 |
| S9_vs_k141_79697||full | 1411 | No | NA | 2 | 0 | 0 | Not-determined | Genome-fragment | NA | NA | 0 |
| S9_vs_k141_11522||full | 2557 | No | NA | 3 | 0 | 3 | Not-determined | Genome-fragment | NA | NA | 0 |
| S11_vs_k141_4988||lt2gene | 1103 | No | NA | 2 | 2 | 0 | Low-quality | Genome-fragment | 2.45 | AAI-based (high-confidence) | 0 |
| S13_vs_21737||full | 1103 | No | NA | 2 | 0 | 1 | Low-quality | Genome-fragment | 2.51 | AAI-based (medium-confidence) | 0 |
| S1_extracted_k141_20215 | 1103 | No | NA | 3 | 1 | 0 | Low-quality | Genome-fragment | 2.17 | AAI-based (medium-confidence) | 0 |
| S1_vs_k141_83346||lt2gene | 1103 | No | NA | 1 | 1 | 0 | Low-quality | Genome-fragment | 2.19 | AAI-based (high-confidence) | 0 |
| S9_vs_k141_68298||full | 1833 | No | NA | 4 | 0 | 2 | Not-determined | Genome-fragment | NA | NA | 0 |
| S3_vs_k141_82950||lt2gene | 1103 | No | NA | 1 | 1 | 0 | Low-quality | Genome-fragment | 0.93 | AAI-based (high-confidence) | 0 |
| S9_vs_k141_45576||full | 2493 | No | NA | 3 | 0 | 2 | Not-determined | Genome-fragment | NA | NA | 0 |
| S5_vs_k141_128832||full | 1103 | No | NA | 2 | 2 | 0 | Low-quality | Genome-fragment | 1.34 | HMM-based (lower-bound) | 0 |
| S9_vs_k141_34333||full | 1059 | No | NA | 3 | 0 | 1 | Not-determined | Genome-fragment | NA | NA | 0 |
| S5_vs_k141_93374||lt2gene | 1103 | No | NA | 1 | 1 | 0 | Low-quality | Genome-fragment | 2.09 | HMM-based (lower-bound) | 0 |
| S7_extracted_122271 | 1103 | No | NA | 2 | 0 | 0 | Low-quality | Genome-fragment | 2.91 | AAI-based (high-confidence) | 0 |
| S9_vs_k141_30556||full | 3672 | No | NA | 4 | 0 | 3 | Not-determined | Genome-fragment | NA | NA | 0 |
| S16_vs_39452||full | 1103 | No | NA | 2 | 0 | 0 | Low-quality | Genome-fragment | 1.89 | AAI-based (high-confidence) | 0 |
| S9_vs_k141_11606||full | 1342 | No | NA | 3 | 0 | 0 | Not-determined | Genome-fragment | NA | NA | 0 |
| S10_extracted_34872 | 1103 | No | NA | 3 | 0 | 1 | Low-quality | Genome-fragment | 0.7 | AAI-based (high-confidence) | 0 |
| S9_vs_k141_34388||full | 1016 | No | NA | 2 | 0 | 0 | Not-determined | Genome-fragment | NA | NA | 0 |
| S10_vs_51142||full | 1103 | No | NA | 2 | 0 | 0 | Low-quality | Genome-fragment | 1.58 | AAI-based (high-confidence) | 0 |
| S9_vs_k141_45651||full | 1568 | No | NA | 3 | 0 | 0 | Not-determined | Genome-fragment | NA | NA | 0 |
| S12_vs_k141_58178||full | 1103 | No | NA | 2 | 0 | 0 | Low-quality | Genome-fragment | 0.64 | AAI-based (high-confidence) | 0 |
| S9_vs_k141_57003||full | 3482 | No | NA | 2 | 0 | 2 | Not-determined | Genome-fragment | NA | NA | 0 |
| S9_vs_k141_4043||full | 4732 | No | NA | 5 | 0 | 2 | Not-determined | Genome-fragment | NA | NA | 0 |
| S9_vs_k141_49380||full | 2405 | No | NA | 4 | 0 | 0 | Not-determined | Genome-fragment | NA | NA | 0 |
| S9_vs_k141_68371||full | 1104 | No | NA | 3 | 0 | 1 | Not-determined | Genome-fragment | NA | NA | 0 |
| S9_vs_k141_83574||full | 3376 | No | NA | 3 | 0 | 1 | Not-determined | Genome-fragment | NA | NA | 0 |
| S2_vs_k141_2873||lt2gene | 1103 | No | NA | 2 | 1 | 0 | Low-quality | Genome-fragment | 2.65 | AAI-based (high-confidence) | 0 |
| S6_extracted_k141_38690 | 1103 | No | NA | 3 | 0 | 3 | Low-quality | Genome-fragment | 0.31 | AAI-based (medium-confidence) | 0 |
| S6_vs_k141_49137||full | 1103 | No | NA | 3 | 0 | 1 | Low-quality | Genome-fragment | 0.58 | AAI-based (medium-confidence) | 0 |
| S9_vs_k141_34461||full | 1172 | No | NA | 2 | 0 | 1 | Not-determined | Genome-fragment | NA | NA | 0 |
| S9_vs_k141_322||full | 2056 | No | NA | 5 | 0 | 0 | Not-determined | Genome-fragment | NA | NA | 0 |
| S8_vs_k141_45946||full | 1103 | No | NA | 3 | 0 | 0 | Low-quality | Genome-fragment | 1.64 | AAI-based (high-confidence) | 0 |
| S8_vs_k141_3882||full | 1103 | No | NA | 2 | 0 | 0 | Low-quality | Genome-fragment | 2.04 | AAI-based (high-confidence) | 0 |
| S17_extracted_k141_62101 | 1104 | No | NA | 1 | 0 | 0 | Low-quality | Genome-fragment | 0.19 | AAI-based (high-confidence) | 0 |
| S9_vs_k141_38192||full | 2521 | No | NA | 3 | 0 | 1 | Not-determined | Genome-fragment | NA | NA | 0 |
| S9_vs_k141_41933||full | 1628 | No | NA | 3 | 0 | 0 | Not-determined | Genome-fragment | NA | NA | 0 |
| S9_vs_k141_68415||full | 1387 | No | NA | 2 | 0 | 0 | Not-determined | Genome-fragment | NA | NA | 0 |
| S17_extracted_k141_87196 | 1104 | No | NA | 1 | 0 | 0 | Low-quality | Genome-fragment | 0.18 | AAI-based (medium-confidence) | 0 |
| S9_vs_k141_38198||full | 1854 | No | NA | 4 | 0 | 1 | Not-determined | Genome-fragment | NA | NA | 0 |
| S9_vs_k141_87452||full | 9268 | No | NA | 8 | 0 | 3 | Not-determined | Genome-fragment | NA | NA | 0 |
| S9_vs_k141_64671||full | 1260 | No | NA | 3 | 0 | 2 | Not-determined | Genome-fragment | NA | NA | 0 |
| S9_extracted_k141_76468 | 1104 | No | NA | 3 | 0 | 1 | Low-quality | Genome-fragment | 2.02 | AAI-based (medium-confidence) | 0 |
| S9_vs_k141_57044||full | 4309 | No | NA | 6 | 0 | 0 | Not-determined | Genome-fragment | NA | NA | 0 |
| S9_extracted_k141_37590 | 1104 | No | NA | 2 | 0 | 0 | Low-quality | Genome-fragment | 1.1 | AAI-based (high-confidence) | 0 |
| S9_vs_k141_3373||full | 1104 | No | NA | 4 | 0 | 0 | Low-quality | Genome-fragment | 2.61 | AAI-based (high-confidence) | 0 |
| S9_vs_k141_47797||lt2gene | 1104 | No | NA | 2 | 2 | 0 | Low-quality | Genome-fragment | 2.54 | AAI-based (high-confidence) | 0 |
| S9_vs_k141_57053||full | 1275 | No | NA | 3 | 0 | 0 | Not-determined | Genome-fragment | NA | NA | 0 |
| S11_extracted_k141_64746 | 1104 | No | NA | 2 | 1 | 0 | Low-quality | Genome-fragment | 2.66 | AAI-based (high-confidence) | 0 |
| S13_vs_140372||full | 1104 | No | NA | 3 | 0 | 0 | Low-quality | Genome-fragment | 2.46 | AAI-based (medium-confidence) | 0 |
| S14_extracted_k141_52132 | 1104 | No | NA | 1 | 0 | 0 | Low-quality | Genome-fragment | 3.54 | AAI-based (medium-confidence) | 0 |
| S9_vs_k141_49443||full | 1031 | No | NA | 3 | 0 | 1 | Not-determined | Genome-fragment | NA | NA | 0 |
| S1_vs_k141_136180||full | 1104 | No | NA | 5 | 2 | 0 | Low-quality | Genome-fragment | 2.08 | AAI-based (high-confidence) | 0 |
| S7_extracted_42177 | 1104 | No | NA | 3 | 0 | 0 | Low-quality | Genome-fragment | 1.56 | AAI-based (high-confidence) | 0 |
| S9_vs_k141_38250||full | 2884 | No | NA | 7 | 0 | 1 | Not-determined | Genome-fragment | NA | NA | 0 |
| S7_vs_115857||lt2gene | 1104 | No | NA | 2 | 2 | 0 | Low-quality | Genome-fragment | 2.73 | AAI-based (high-confidence) | 0 |
| S9_vs_k141_60867||full | 2934 | No | NA | 4 | 0 | 3 | Not-determined | Genome-fragment | NA | NA | 0 |
| S12_extracted_k141_117961 | 1104 | No | NA | 1 | 0 | 0 | Low-quality | Genome-fragment | 1.1 | AAI-based (high-confidence) | 0 |
| S9_vs_k141_23088||full | 1717 | No | NA | 4 | 0 | 1 | Not-determined | Genome-fragment | NA | NA | 0 |
| S12_vs_k141_154112||full | 1104 | No | NA | 4 | 0 | 0 | Low-quality | Genome-fragment | 1.97 | AAI-based (high-confidence) | 0 |
| S2_extracted_k141_14184 | 1104 | No | NA | 1 | 0 | 1 | Low-quality | Genome-fragment | 0.49 | AAI-based (medium-confidence) | 0 |
| S6_vs_k141_862||full | 1104 | No | NA | 2 | 0 | 0 | Low-quality | Genome-fragment | 3.08 | AAI-based (medium-confidence) | 0 |
| S8_vs_k141_4171||full | 1104 | No | NA | 3 | 0 | 0 | Low-quality | Genome-fragment | 0.68 | AAI-based (high-confidence) | 0 |
| S17_extracted_k141_119617 | 1105 | No | NA | 2 | 1 | 0 | Low-quality | Genome-fragment | 5.04 | AAI-based (high-confidence) | 0 |
| S9_vs_k141_15509||full | 2005 | No | NA | 3 | 0 | 1 | Not-determined | Genome-fragment | NA | NA | 0 |
| S9_vs_k141_7834||full | 1920 | No | NA | 2 | 0 | 2 | Not-determined | Genome-fragment | NA | NA | 0 |
| S17_vs_k141_93970||full | 1105 | No | NA | 2 | 0 | 1 | Low-quality | Genome-fragment | 0.86 | AAI-based (medium-confidence) | 0 |
| S9_vs_k141_64753||full | 1328 | No | NA | 2 | 0 | 2 | Not-determined | Genome-fragment | NA | NA | 0 |
| S9_extracted_k141_28637 | 1105 | No | NA | 2 | 1 | 0 | Low-quality | Genome-fragment | 2.83 | AAI-based (high-confidence) | 0 |
| S9_vs_k141_83685||full | 1071 | No | NA | 2 | 0 | 1 | Not-determined | Genome-fragment | NA | NA | 0 |
| S9_vs_k141_55364||full | 1105 | No | NA | 4 | 1 | 0 | Low-quality | Genome-fragment | 1.49 | HMM-based (lower-bound) | 0 |
| S11_extracted_k141_58544 | 1105 | No | NA | 1 | 1 | 0 | Low-quality | Genome-fragment | 2.01 | AAI-based (medium-confidence) | 0 |
| S13_extracted_2292 | 1105 | No | NA | 3 | 1 | 0 | Low-quality | Genome-fragment | 1.84 | AAI-based (high-confidence) | 0 |
| S14_vs_k141_68324||full | 1105 | No | NA | 3 | 0 | 1 | Low-quality | Genome-fragment | 1.7 | AAI-based (high-confidence) | 0 |
| S3_extracted_k141_74441 | 1105 | No | NA | 1 | 1 | 0 | Low-quality | Genome-fragment | 1.87 | AAI-based (high-confidence) | 0 |
| S9_vs_k141_64797||full | 4492 | No | NA | 7 | 0 | 3 | Not-determined | Genome-fragment | NA | NA | 0 |
| S9_vs_k141_30768||full | 3203 | No | NA | 6 | 0 | 0 | Not-determined | Genome-fragment | NA | NA | 0 |
| S5_vs_k141_114618||full | 1105 | No | NA | 3 | 2 | 0 | Low-quality | Genome-fragment | 2.93 | AAI-based (high-confidence) | 0 |
| S7_extracted_2837 | 1105 | No | NA | 4 | 4 | 0 | Low-quality | Genome-fragment | 3.64 | AAI-based (high-confidence) | 0 |
| S9_vs_k141_463||full | 1196 | No | NA | 2 | 0 | 0 | Not-determined | Genome-fragment | NA | NA | 0 |
| S9_vs_k141_76015||full | 1502 | No | NA | 2 | 0 | 1 | Not-determined | Genome-fragment | NA | NA | 0 |
| S7_vs_83481||lt2gene | 1105 | No | NA | 1 | 1 | 0 | Low-quality | Genome-fragment | 3.19 | AAI-based (high-confidence) | 0 |
| S16_extracted_127802 | 1105 | No | NA | 3 | 1 | 0 | Low-quality | Genome-fragment | 1.97 | AAI-based (medium-confidence) | 0 |
| S16_vs_72966||full | 1105 | No | NA | 2 | 0 | 0 | Low-quality | Genome-fragment | 0.64 | AAI-based (high-confidence) | 0 |
| S16_vs_95188||full | 1105 | No | NA | 4 | 0 | 0 | Low-quality | Genome-fragment | 0.67 | AAI-based (high-confidence) | 0 |
| S9_vs_k141_76029||full | 3635 | No | NA | 4 | 0 | 4 | Not-determined | Genome-fragment | NA | NA | 0 |
| S9_vs_k141_492||full | 3802 | No | NA | 5 | 0 | 0 | Not-determined | Genome-fragment | NA | NA | 0 |
| S12_extracted_k141_115834 | 1105 | No | NA | 2 | 1 | 0 | Low-quality | Genome-fragment | 1.53 | AAI-based (medium-confidence) | 0 |
| S12_extracted_k141_21917 | 1105 | No | NA | 1 | 1 | 0 | Low-quality | Genome-fragment | 2.07 | HMM-based (lower-bound) | 0 |
| S9_vs_k141_7881||full | 28302 | No | NA | 40 | 23 | 0 | Medium-quality | Genome-fragment | 61.82 | AAI-based (medium-confidence) | 0 |
| S9_vs_k141_61025||full | 1701 | No | NA | 3 | 0 | 2 | Not-determined | Genome-fragment | NA | NA | 0 |
| S6_extracted_k141_39793 | 1105 | No | NA | 2 | 0 | 0 | Low-quality | Genome-fragment | 0.41 | AAI-based (medium-confidence) | 0 |
| S8_vs_k141_112073||full | 1105 | No | NA | 3 | 0 | 1 | Low-quality | Genome-fragment | 2.01 | AAI-based (medium-confidence) | 0 |
| S8_vs_k141_107188||lt2gene | 1105 | No | NA | 1 | 1 | 0 | Low-quality | Genome-fragment | 1.61 | HMM-based (lower-bound) | 0 |
| S9_vs_k141_53442||full | 1190 | No | NA | 3 | 0 | 0 | Not-determined | Genome-fragment | NA | NA | 0 |
| S9_vs_k141_72385||full | 2757 | No | NA | 4 | 0 | 0 | Not-determined | Genome-fragment | NA | NA | 0 |
| S9_vs_k141_23184||full | 1027 | No | NA | 2 | 0 | 0 | Not-determined | Genome-fragment | NA | NA | 0 |
| S9_vs_k141_15647||full | 1381 | No | NA | 2 | 0 | 0 | Not-determined | Genome-fragment | NA | NA | 0 |
| S17_vs_k141_106261||full | 1106 | No | NA | 5 | 5 | 0 | Low-quality | Genome-fragment | 3.65 | AAI-based (high-confidence) | 0 |
| S11_extracted_k141_32274 | 1106 | No | NA | 2 | 0 | 0 | Low-quality | Genome-fragment | 1.71 | AAI-based (medium-confidence) | 0 |
| S9_vs_k141_19367||full | 1103 | No | NA | 2 | 0 | 2 | Not-determined | Genome-fragment | NA | NA | 0 |
| S9_vs_k141_26934||full | 1804 | No | NA | 2 | 0 | 0 | Not-determined | Genome-fragment | NA | NA | 0 |
| S14_extracted_k141_7451 | 1106 | No | NA | 2 | 2 | 0 | Low-quality | Genome-fragment | 3.64 | AAI-based (high-confidence) | 0 |
| S9_vs_k141_34601||full | 1093 | No | NA | 2 | 0 | 1 | Not-determined | Genome-fragment | NA | NA | 0 |
| S1_extracted_k141_157185 | 1106 | No | NA | 2 | 1 | 0 | Low-quality | Genome-fragment | 2.06 | HMM-based (lower-bound) | 0 |
| S1_vs_k141_1116||full | 1106 | No | NA | 2 | 0 | 0 | Low-quality | Genome-fragment | 0.92 | AAI-based (medium-confidence) | 0 |
| S9_vs_k141_11881||full | 1047 | No | NA | 1 | 0 | 0 | Not-determined | Genome-fragment | NA | NA | 0 |
| S1_vs_k141_72928||full | 1106 | No | NA | 2 | 0 | 0 | Low-quality | Genome-fragment | 1.95 | AAI-based (high-confidence) | 0 |
| S9_vs_k141_72431||full | 3372 | No | NA | 4 | 0 | 1 | Not-determined | Genome-fragment | NA | NA | 0 |
| S9_vs_k141_61077||full | 1639 | No | NA | 2 | 0 | 0 | Not-determined | Genome-fragment | NA | NA | 0 |
| S9_vs_k141_80026||full | 1601 | No | NA | 4 | 0 | 0 | Not-determined | Genome-fragment | NA | NA | 0 |
| S9_vs_k141_30850||full | 1620 | No | NA | 4 | 0 | 2 | Not-determined | Genome-fragment | NA | NA | 0 |
| S5_vs_k141_67181||lt2gene | 1106 | No | NA | 1 | 1 | 0 | Low-quality | Genome-fragment | 2.46 | AAI-based (high-confidence) | 0 |
| S7_extracted_28574 | 1106 | No | NA | 2 | 0 | 0 | Low-quality | Genome-fragment | 1.27 | AAI-based (high-confidence) | 0 |
| S7_vs_49187||full | 1106 | No | NA | 4 | 2 | 0 | Low-quality | Genome-fragment | 2.77 | AAI-based (high-confidence) | 0 |
| S7_vs_83840||full | 1106 | No | NA | 3 | 0 | 1 | Low-quality | Genome-fragment | 0.98 | AAI-based (medium-confidence) | 0 |
| S9_vs_k141_7993||full | 1170 | No | NA | 2 | 0 | 1 | Not-determined | Genome-fragment | NA | NA | 0 |
| S9_vs_k141_30888||full | 1675 | No | NA | 4 | 0 | 0 | Not-determined | Genome-fragment | NA | NA | 0 |
| S7_vs_85287||full | 1106 | No | NA | 3 | 0 | 0 | Low-quality | Genome-fragment | 1.09 | AAI-based (medium-confidence) | 0 |
| S16_extracted_25375 | 1106 | No | NA | 3 | 0 | 0 | Low-quality | Genome-fragment | 0.64 | AAI-based (high-confidence) | 0 |
| S9_vs_k141_38472||full | 10302 | No | NA | 5 | 0 | 5 | Not-determined | Genome-fragment | NA | NA | 0 |
| S16_extracted_80254 | 1106 | No | NA | 1 | 0 | 0 | Low-quality | Genome-fragment | 2.58 | AAI-based (medium-confidence) | 0 |
| S16_vs_104245||lt2gene | 1106 | No | NA | 2 | 1 | 0 | Low-quality | Genome-fragment | 2.8 | AAI-based (high-confidence) | 0 |
| S10_extracted_86464 | 1106 | No | NA | 3 | 0 | 2 | Low-quality | Genome-fragment | 2.89 | AAI-based (medium-confidence) | 0 |
| S17_extracted_k141_29393 | 1107 | No | NA | 1 | 0 | 1 | Low-quality | Genome-fragment | 0.37 | AAI-based (medium-confidence) | 0 |
| S17_vs_k141_99620||full | 1107 | No | NA | 4 | 0 | 0 | Low-quality | Genome-fragment | 1.33 | AAI-based (high-confidence) | 0 |
| S17_vs_k141_17958||full | 1107 | No | NA | 2 | 0 | 0 | Low-quality | Genome-fragment | 2.13 | AAI-based (medium-confidence) | 0 |
| S14_extracted_k141_45144 | 1107 | No | NA | 4 | 4 | 0 | Low-quality | Genome-fragment | 3.65 | AAI-based (high-confidence) | 0 |
| S9_vs_k141_8062||full | 2135 | No | NA | 3 | 0 | 1 | Not-determined | Genome-fragment | NA | NA | 0 |
| S14_extracted_k141_46476 | 1107 | No | NA | 3 | 0 | 0 | Low-quality | Genome-fragment | 8.72 | AAI-based (high-confidence) | 0 |
| S5_extracted_k141_120159 | 1107 | No | NA | 3 | 3 | 0 | Low-quality | Genome-fragment | 3.65 | AAI-based (high-confidence) | 0 |
| S9_vs_k141_30963||full | 1579 | No | NA | 3 | 0 | 1 | Not-determined | Genome-fragment | NA | NA | 0 |
| S9_vs_k141_61223||full | 1110 | No | NA | 3 | 0 | 0 | Not-determined | Genome-fragment | NA | NA | 0 |
| S7_vs_78050||full | 1107 | No | NA | 3 | 0 | 0 | Low-quality | Genome-fragment | 2.86 | AAI-based (medium-confidence) | 0 |
| S9_vs_k141_45865||full | 2976 | No | NA | 9 | 0 | 0 | Not-determined | Genome-fragment | NA | NA | 0 |
| S16_extracted_72062 | 1107 | No | NA | 3 | 0 | 0 | Low-quality | Genome-fragment | 2.28 | AAI-based (high-confidence) | 0 |
| S9_vs_k141_4343||full | 37691 | No | NA | 48 | 8 | 0 | High-quality | High-quality | 95.71 | AAI-based (high-confidence) | 0 |
| S16_extracted_90695 | 1107 | No | NA | 4 | 0 | 0 | Low-quality | Genome-fragment | 0.81 | AAI-based (medium-confidence) | 0 |
| S16_vs_20451||full | 1107 | No | NA | 2 | 0 | 1 | Low-quality | Genome-fragment | 1.86 | AAI-based (medium-confidence) | 0 |
| S9_vs_k141_72581||full | 3001 | No | NA | 4 | 0 | 1 | Not-determined | Genome-fragment | NA | NA | 0 |
| S9_vs_k141_34682||full | 2453 | No | NA | 4 | 0 | 0 | Not-determined | Genome-fragment | NA | NA | 0 |
| S10_extracted_11953 | 1107 | No | NA | 2 | 0 | 0 | Low-quality | Genome-fragment | 2.4 | AAI-based (high-confidence) | 0 |
| S10_vs_82978||full | 1107 | No | NA | 2 | 0 | 1 | Low-quality | Genome-fragment | 0.2 | AAI-based (medium-confidence) | 0 |
| S9_vs_k141_15835||full | 1184 | No | NA | 2 | 0 | 2 | Not-determined | Genome-fragment | NA | NA | 0 |
| S9_vs_k141_68658||full | 1481 | No | NA | 3 | 0 | 1 | Not-determined | Genome-fragment | NA | NA | 0 |
| S12_extracted_k141_99775 | 1107 | No | NA | 4 | 1 | 0 | Low-quality | Genome-fragment | 2.52 | AAI-based (medium-confidence) | 0 |
| S12_extracted_k141_131011 | 1107 | No | NA | 2 | 0 | 1 | Low-quality | Genome-fragment | 0.72 | AAI-based (high-confidence) | 0 |
| S8_extracted_k141_62215 | 1107 | No | NA | 2 | 0 | 1 | Low-quality | Genome-fragment | 0.2 | AAI-based (medium-confidence) | 0 |
| S9_vs_k141_61287||full | 5030 | No | NA | 5 | 0 | 3 | Not-determined | Genome-fragment | NA | NA | 0 |
| S9_vs_k141_4360||full | 1267 | No | NA | 2 | 0 | 0 | Not-determined | Genome-fragment | NA | NA | 0 |
| S17_extracted_k141_90930 | 1108 | No | NA | 2 | 0 | 0 | Low-quality | Genome-fragment | 2.17 | AAI-based (high-confidence) | 0 |
| S17_vs_k141_132777||full | 1108 | No | NA | 3 | 1 | 0 | Low-quality | Genome-fragment | 2.03 | HMM-based (lower-bound) | 0 |
| S17_vs_k141_128416||lt2gene | 1108 | No | NA | 2 | 1 | 0 | Low-quality | Genome-fragment | 2.64 | AAI-based (high-confidence) | 0 |
| S9_vs_k141_23349||full | 1781 | No | NA | 4 | 0 | 1 | Not-determined | Genome-fragment | NA | NA | 0 |
| S9_vs_k141_80211||full | 12108 | No | NA | 7 | 0 | 0 | Not-determined | Genome-fragment | NA | NA | 0 |
| S11_extracted_k141_86790 | 1108 | No | NA | 2 | 0 | 1 | Low-quality | Genome-fragment | 0.99 | AAI-based (medium-confidence) | 0 |
| S9_vs_k141_61326||full | 1810 | No | NA | 4 | 0 | 0 | Not-determined | Genome-fragment | NA | NA | 0 |
| S13_extracted_38147 | 1108 | No | NA | 2 | 0 | 1 | Low-quality | Genome-fragment | 1.1 | AAI-based (high-confidence) | 0 |
| S14_extracted_k141_101954 | 1108 | No | NA | 3 | 2 | 0 | Low-quality | Genome-fragment | 3.66 | AAI-based (high-confidence) | 0 |
| S9_vs_k141_87923||full | 3500 | No | NA | 5 | 0 | 3 | Not-determined | Genome-fragment | NA | NA | 0 |
| S1_vs_k141_1371||full | 1108 | No | NA | 3 | 3 | 0 | Low-quality | Genome-fragment | 2.44 | AAI-based (high-confidence) | 0 |
| S1_vs_k141_6030||full | 1108 | No | NA | 3 | 1 | 0 | Low-quality | Genome-fragment | 2.28 | AAI-based (high-confidence) | 0 |
| S9_vs_k141_84011||full | 2228 | No | NA | 4 | 0 | 1 | Not-determined | Genome-fragment | NA | NA | 0 |
| S3_vs_k141_17758||full | 1108 | No | NA | 2 | 0 | 0 | Low-quality | Genome-fragment | 2.02 | AAI-based (medium-confidence) | 0 |
| S3_vs_k141_106606||full | 1108 | No | NA | 3 | 1 | 0 | Low-quality | Genome-fragment | 1.99 | AAI-based (medium-confidence) | 0 |
| S5_extracted_k141_62752 | 1108 | No | NA | 5 | 1 | 0 | Low-quality | Genome-fragment | 2.06 | AAI-based (medium-confidence) | 0 |
| S5_vs_k141_55093||full | 1108 | No | NA | 2 | 1 | 0 | Low-quality | Genome-fragment | 1.64 | HMM-based (lower-bound) | 0 |
| S9_vs_k141_27274||full | 7496 | No | NA | 8 | 0 | 3 | Not-determined | Genome-fragment | NA | NA | 0 |
| S5_vs_k141_138218||full | 1108 | No | NA | 4 | 1 | 0 | Low-quality | Genome-fragment | 2.15 | AAI-based (high-confidence) | 0 |
| S9_vs_k141_4422||full | 1031 | No | NA | 2 | 0 | 0 | Not-determined | Genome-fragment | NA | NA | 0 |
| S7_vs_25343||full | 1108 | No | NA | 3 | 0 | 0 | Low-quality | Genome-fragment | 2.7 | AAI-based (medium-confidence) | 0 |
| S9_vs_k141_53808||full | 4644 | No | NA | 4 | 0 | 2 | Not-determined | Genome-fragment | NA | NA | 0 |
| S16_extracted_71420 | 1108 | No | NA | 5 | 0 | 0 | Low-quality | Genome-fragment | 2.21 | AAI-based (high-confidence) | 0 |
| S10_extracted_95679 | 1108 | No | NA | 2 | 0 | 0 | Low-quality | Genome-fragment | 2.07 | AAI-based (medium-confidence) | 0 |
| S9_vs_k141_80322||full | 1564 | No | NA | 5 | 0 | 0 | Not-determined | Genome-fragment | NA | NA | 0 |
| S10_vs_78830||full | 1108 | No | NA | 3 | 2 | 0 | Low-quality | Genome-fragment | 1.38 | AAI-based (high-confidence) | 0 |
| S12_vs_k141_87824||full | 1108 | No | NA | 2 | 0 | 0 | Low-quality | Genome-fragment | 1.92 | AAI-based (high-confidence) | 0 |
| S9_vs_k141_57345||full | 1385 | No | NA | 2 | 0 | 0 | Not-determined | Genome-fragment | NA | NA | 0 |
| S11_extracted_k141_65833 | 1109 | No | NA | 3 | 0 | 0 | Low-quality | Genome-fragment | 0.72 | AAI-based (high-confidence) | 0 |
| S9_vs_k141_61358||full | 1048 | No | NA | 2 | 0 | 0 | Not-determined | Genome-fragment | NA | NA | 0 |
| S9_vs_k141_72712||full | 1214 | No | NA | 2 | 0 | 1 | Not-determined | Genome-fragment | NA | NA | 0 |
| S9_vs_k141_68803||full | 2539 | No | NA | 3 | 0 | 1 | Not-determined | Genome-fragment | NA | NA | 0 |
| S11_extracted_k141_44061 | 1109 | No | NA | 2 | 2 | 0 | Low-quality | Genome-fragment | 3.66 | AAI-based (high-confidence) | 0 |
| S11_vs_k141_8963||full | 1109 | No | NA | 2 | 0 | 0 | Low-quality | Genome-fragment | 1 | AAI-based (high-confidence) | 0 |
| S14_vs_k141_1212||full | 1109 | No | NA | 3 | 1 | 0 | Low-quality | Genome-fragment | 3.01 | AAI-based (high-confidence) | 0 |
| S1_extracted_k141_149014 | 1109 | No | NA | 1 | 1 | 0 | Low-quality | Genome-fragment | 1.84 | AAI-based (high-confidence) | 0 |
| S1_vs_k141_127040||full | 1109 | No | NA | 4 | 1 | 0 | Low-quality | Genome-fragment | 1.76 | AAI-based (high-confidence) | 0 |
| S9_vs_k141_27315||full | 1300 | No | NA | 3 | 0 | 1 | Not-determined | Genome-fragment | NA | NA | 0 |
| S9_vs_k141_84046||full | 29842 | No | NA | 46 | 14 | 1 | High-quality | High-quality | 93.21 | AAI-based (medium-confidence) | 0 |
| S7_extracted_7212 | 1109 | No | NA | 3 | 0 | 0 | Low-quality | Genome-fragment | 0.71 | AAI-based (high-confidence) | 0 |
| S9_vs_k141_49763||full | 1493 | No | NA | 3 | 0 | 1 | Not-determined | Genome-fragment | NA | NA | 0 |
| S9_vs_k141_76374||full | 4193 | No | NA | 4 | 0 | 2 | Not-determined | Genome-fragment | NA | NA | 0 |
| S9_vs_k141_57388||full | 2324 | No | NA | 2 | 0 | 2 | Not-determined | Genome-fragment | NA | NA | 0 |
| S16_extracted_86065 | 1109 | No | NA | 3 | 1 | 0 | Low-quality | Genome-fragment | 2.87 | AAI-based (high-confidence) | 0 |
| S9_vs_k141_65082||full | 1340 | No | NA | 3 | 0 | 2 | Not-determined | Genome-fragment | NA | NA | 0 |
| S16_extracted_163305 | 1109 | No | NA | 5 | 0 | 0 | Low-quality | Genome-fragment | 0.7 | AAI-based (high-confidence) | 0 |
| S9_vs_k141_23450||full | 10166 | No | NA | 4 | 0 | 4 | Not-determined | Genome-fragment | NA | NA | 0 |
| S16_vs_113778||full | 1109 | No | NA | 3 | 1 | 0 | Low-quality | Genome-fragment | 2.32 | AAI-based (high-confidence) | 0 |
| S16_vs_143661||lt2gene | 1109 | No | NA | 1 | 1 | 0 | Low-quality | Genome-fragment | 1.89 | AAI-based (high-confidence) | 0 |
| S9_vs_k141_87990||full | 16416 | No | NA | 15 | 0 | 12 | Not-determined | Genome-fragment | NA | NA | 0 |
| S9_vs_k141_23454||full | 1044 | No | NA | 2 | 0 | 2 | Not-determined | Genome-fragment | NA | NA | 0 |
| S12_vs_k141_110403||full | 1109 | No | NA | 3 | 1 | 0 | Low-quality | Genome-fragment | 1.98 | AAI-based (high-confidence) | 0 |
| S9_vs_k141_38799||full | 1961 | No | NA | 2 | 0 | 2 | Not-determined | Genome-fragment | NA | NA | 0 |
| S9_vs_k141_19649||full | 1458 | No | NA | 1 | 0 | 0 | Not-determined | Genome-fragment | NA | NA | 0 |
| S9_vs_k141_34830||full | 1695 | No | NA | 2 | 0 | 1 | Not-determined | Genome-fragment | NA | NA | 0 |
| S9_vs_k141_46141||full | 1265 | No | NA | 3 | 0 | 0 | Not-determined | Genome-fragment | NA | NA | 0 |
| S9_vs_k141_49807||full | 1073 | No | NA | 3 | 0 | 0 | Not-determined | Genome-fragment | NA | NA | 0 |
| S9_vs_k141_57424||full | 2742 | No | NA | 4 | 0 | 0 | Not-determined | Genome-fragment | NA | NA | 0 |
| S8_vs_k141_93135||full | 1109 | Yes | 656 | 3 | 1 | 1 | Low-quality | Genome-fragment | 0.42 | HMM-based (lower-bound) | 40.85 |
| S9_extracted_k141_23839 | 1110 | No | NA | 3 | 0 | 0 | Low-quality | Genome-fragment | 1.85 | AAI-based (high-confidence) | 0 |
| S9_vs_k141_4441||full | 1110 | No | NA | 3 | 0 | 0 | Low-quality | Genome-fragment | 2.19 | AAI-based (medium-confidence) | 0 |
| S13_vs_98422||full | 1110 | No | NA | 3 | 1 | 0 | Low-quality | Genome-fragment | 1.92 | AAI-based (medium-confidence) | 0 |
| S9_vs_k141_16064||full | 1021 | No | NA | 2 | 0 | 1 | Not-determined | Genome-fragment | NA | NA | 0 |
| S9_vs_k141_46171||full | 2740 | No | NA | 3 | 0 | 2 | Not-determined | Genome-fragment | NA | NA | 0 |
| S9_vs_k141_38853||full | 2292 | No | NA | 3 | 0 | 0 | Not-determined | Genome-fragment | NA | NA | 0 |
| S9_vs_k141_23518||full | 1352 | No | NA | 2 | 0 | 1 | Not-determined | Genome-fragment | NA | NA | 0 |
| S14_extracted_k141_10015 | 1110 | No | NA | 2 | 0 | 0 | Low-quality | Genome-fragment | 1.59 | AAI-based (high-confidence) | 0 |
| S1_vs_k141_15744||lt2gene | 1110 | No | NA | 1 | 1 | 0 | Low-quality | Genome-fragment | 2.43 | AAI-based (high-confidence) | 0 |
| S5_extracted_k141_61372 | 1110 | No | NA | 1 | 0 | 1 | Low-quality | Genome-fragment | 0.74 | AAI-based (medium-confidence) | 0 |
| S7_extracted_121283 | 1110 | No | NA | 2 | 0 | 0 | Low-quality | Genome-fragment | 0.7 | AAI-based (medium-confidence) | 0 |
| S7_extracted_126006 | 1110 | No | NA | 2 | 0 | 0 | Low-quality | Genome-fragment | 1.71 | AAI-based (high-confidence) | 0 |
| S7_vs_47980||full | 1110 | No | NA | 3 | 0 | 0 | Low-quality | Genome-fragment | 2.33 | AAI-based (high-confidence) | 0 |
| S7_vs_89428||full | 1110 | No | NA | 3 | 0 | 1 | Low-quality | Genome-fragment | 0.3 | AAI-based (medium-confidence) | 0 |
| S9_vs_k141_19689||full | 28742 | No | NA | 44 | 11 | 1 | Medium-quality | Genome-fragment | 62.32 | AAI-based (high-confidence) | 0 |
| S7_vs_118831||full | 1110 | No | NA | 5 | 0 | 0 | Low-quality | Genome-fragment | 1.45 | AAI-based (medium-confidence) | 0 |
| S10_extracted_32826 | 1110 | No | NA | 1 | 0 | 1 | Low-quality | Genome-fragment | 0.33 | AAI-based (medium-confidence) | 0 |
| S9_vs_k141_61504||full | 2310 | No | NA | 2 | 0 | 2 | Not-determined | Genome-fragment | NA | NA | 0 |
| S9_vs_k141_65132||full | 1662 | No | NA | 2 | 0 | 1 | Not-determined | Genome-fragment | NA | NA | 0 |
| S9_vs_k141_27348||full | 1187 | No | NA | 2 | 0 | 0 | Not-determined | Genome-fragment | NA | NA | 0 |
| S9_vs_k141_42619||full | 1494 | No | NA | 3 | 0 | 1 | Not-determined | Genome-fragment | NA | NA | 0 |
| S12_vs_k141_118277||full | 1110 | No | NA | 3 | 1 | 0 | Low-quality | Genome-fragment | 1.76 | AAI-based (high-confidence) | 0 |
| S9_vs_k141_12307||full | 3099 | No | NA | 4 | 0 | 4 | Not-determined | Genome-fragment | NA | NA | 0 |
| S9_vs_k141_34898||full | 31642 | No | NA | 54 | 25 | 0 | Medium-quality | Genome-fragment | 78.72 | AAI-based (medium-confidence) | 0 |
| S9_vs_k141_72853||full | 1486 | No | NA | 2 | 0 | 0 | Not-determined | Genome-fragment | NA | NA | 0 |
| S9_vs_k141_65155||full | 1349 | No | NA | 2 | 0 | 0 | Not-determined | Genome-fragment | NA | NA | 0 |
| S9_vs_k141_61525||full | 6375 | No | NA | 6 | 0 | 6 | Not-determined | Genome-fragment | NA | NA | 0 |
| S12_vs_k141_53502||full | 1110 | No | NA | 4 | 1 | 0 | Low-quality | Genome-fragment | 2.17 | AAI-based (medium-confidence) | 0 |
| S9_vs_k141_38942||full | 1617 | No | NA | 2 | 0 | 1 | Not-determined | Genome-fragment | NA | NA | 0 |
| S6_extracted_k141_7331 | 1110 | No | NA | 1 | 0 | 0 | Low-quality | Genome-fragment | 1.15 | AAI-based (medium-confidence) | 0 |
| S9_vs_k141_4558||full | 61044 | No | NA | 90 | 32 | 0 | High-quality | High-quality | 96.55 | AAI-based (high-confidence) | 0 |
| S8_extracted_k141_36514 | 1110 | No | NA | 1 | 0 | 0 | Low-quality | Genome-fragment | 0.55 | AAI-based (medium-confidence) | 0 |
| S9_vs_k141_31299||full | 2184 | No | NA | 4 | 0 | 0 | Not-determined | Genome-fragment | NA | NA | 0 |
| S8_vs_k141_57242||full | 1110 | No | NA | 3 | 1 | 0 | Low-quality | Genome-fragment | 1.84 | AAI-based (high-confidence) | 0 |
| S14_extracted_k141_1057 | 1111 | No | NA | 1 | 1 | 0 | Low-quality | Genome-fragment | 2.9 | AAI-based (high-confidence) | 0 |
| S9_vs_k141_68847||full | 1551 | No | NA | 3 | 0 | 0 | Not-determined | Genome-fragment | NA | NA | 0 |
| S3_extracted_k141_84807 | 1111 | No | NA | 3 | 0 | 0 | Low-quality | Genome-fragment | 1.8 | AAI-based (high-confidence) | 0 |
| S3_vs_k141_24177||full | 1111 | No | NA | 3 | 0 | 0 | Low-quality | Genome-fragment | 1.06 | AAI-based (high-confidence) | 0 |
| S3_vs_k141_15830||full | 1111 | No | NA | 2 | 0 | 0 | Low-quality | Genome-fragment | 0.91 | AAI-based (high-confidence) | 0 |
| S5_extracted_k141_25424 | 1111 | No | NA | 2 | 1 | 0 | Low-quality | Genome-fragment | 2.25 | AAI-based (high-confidence) | 0 |
| S9_vs_k141_65203||full | 1661 | No | NA | 3 | 0 | 0 | Not-determined | Genome-fragment | NA | NA | 0 |
| S9_vs_k141_46337||full | 3419 | No | NA | 6 | 0 | 0 | Not-determined | Genome-fragment | NA | NA | 0 |
| S7_extracted_130720 | 1111 | No | NA | 2 | 0 | 0 | Low-quality | Genome-fragment | 1.98 | AAI-based (high-confidence) | 0 |
| S9_vs_k141_42715||full | 3388 | No | NA | 3 | 0 | 2 | Not-determined | Genome-fragment | NA | NA | 0 |
| S7_vs_84606||full | 1111 | No | NA | 2 | 0 | 0 | Low-quality | Genome-fragment | 2.46 | AAI-based (medium-confidence) | 0 |
| S7_vs_86235||full | 1111 | No | NA | 2 | 0 | 0 | Low-quality | Genome-fragment | 1.52 | AAI-based (medium-confidence) | 0 |
| S16_extracted_82636 | 1111 | No | NA | 1 | 1 | 0 | Low-quality | Genome-fragment | 23.29 | AAI-based (high-confidence) | 0 |
| S16_extracted_109468 | 1111 | No | NA | 2 | 1 | 0 | Low-quality | Genome-fragment | 1.39 | AAI-based (medium-confidence) | 0 |
| S16_vs_12247||full | 1111 | No | NA | 2 | 1 | 0 | Low-quality | Genome-fragment | 0.77 | HMM-based (lower-bound) | 0 |
| S16_vs_105559||full | 1111 | No | NA | 3 | 1 | 0 | Low-quality | Genome-fragment | 1.95 | AAI-based (high-confidence) | 0 |
| S10_extracted_26796 | 1111 | No | NA | 1 | 1 | 0 | Low-quality | Genome-fragment | 2.79 | AAI-based (high-confidence) | 0 |
| S10_vs_50717||full | 1111 | No | NA | 1 | 0 | 0 | Low-quality | Genome-fragment | 2.4 | AAI-based (medium-confidence) | 0 |
| S9_vs_k141_35043||full | 1014 | No | NA | 3 | 0 | 2 | Not-determined | Genome-fragment | NA | NA | 0 |
| S12_vs_k141_153035||full | 1111 | No | NA | 3 | 1 | 0 | Low-quality | Genome-fragment | 0.56 | HMM-based (lower-bound) | 0 |
| S9_vs_k141_16270||full | 3386 | No | NA | 3 | 0 | 1 | Not-determined | Genome-fragment | NA | NA | 0 |
| S12_vs_k141_69652||full | 1111 | No | NA | 3 | 2 | 0 | Low-quality | Genome-fragment | 3.11 | AAI-based (high-confidence) | 0 |
| S15_vs_59162||full | 1111 | No | NA | 3 | 3 | 0 | Low-quality | Genome-fragment | 2.91 | AAI-based (medium-confidence) | 0 |
| S9_vs_k141_54131||full | 2687 | No | NA | 4 | 0 | 0 | Not-determined | Genome-fragment | NA | NA | 0 |
| S9_vs_k141_27444||full | 5218 | No | NA | 7 | 0 | 5 | Not-determined | Genome-fragment | NA | NA | 0 |
| S8_extracted_k141_130510 | 1111 | No | NA | 2 | 2 | 0 | Low-quality | Genome-fragment | 0.6 | HMM-based (lower-bound) | 0 |
| S8_extracted_k141_129608 | 1111 | No | NA | 3 | 0 | 1 | Low-quality | Genome-fragment | 0.88 | AAI-based (medium-confidence) | 0 |
| S17_vs_k141_167005||full | 1112 | No | NA | 3 | 1 | 0 | Low-quality | Genome-fragment | 2.17 | AAI-based (medium-confidence) | 0 |
| S9_vs_k141_19846||full | 1199 | No | NA | 2 | 0 | 1 | Not-determined | Genome-fragment | NA | NA | 0 |
| S9_vs_k141_41002||lt2gene | 1112 | No | NA | 2 | 1 | 0 | Low-quality | Genome-fragment | 2.57 | AAI-based (high-confidence) | 0 |
| S11_vs_k141_35350||full | 1112 | No | NA | 4 | 1 | 0 | Low-quality | Genome-fragment | 1.68 | AAI-based (high-confidence) | 0 |
| S13_vs_149361||full | 1112 | No | NA | 2 | 0 | 0 | Low-quality | Genome-fragment | 0.97 | AAI-based (medium-confidence) | 0 |
| S1_extracted_k141_138468 | 1112 | No | NA | 1 | 0 | 0 | Low-quality | Genome-fragment | 1.94 | AAI-based (high-confidence) | 0 |
| S1_vs_k141_25360||full | 1112 | No | NA | 2 | 2 | 0 | Low-quality | Genome-fragment | 2.56 | HMM-based (lower-bound) | 0 |
| S5_extracted_k141_145507 | 1112 | No | NA | 3 | 2 | 0 | Low-quality | Genome-fragment | 2.89 | AAI-based (medium-confidence) | 0 |
| S10_vs_7700||full | 1112 | No | NA | 3 | 0 | 0 | Low-quality | Genome-fragment | 2.05 | AAI-based (high-confidence) | 0 |
| S12_extracted_k141_151512 | 1112 | No | NA | 2 | 0 | 0 | Low-quality | Genome-fragment | 1.96 | AAI-based (high-confidence) | 0 |
| S9_vs_k141_23643||full | 1785 | No | NA | 2 | 0 | 1 | Not-determined | Genome-fragment | NA | NA | 0 |
| S12_extracted_k141_42555 | 1112 | No | NA | 2 | 2 | 0 | Low-quality | Genome-fragment | 1.18 | HMM-based (lower-bound) | 0 |
| S12_extracted_k141_132308 | 1112 | No | NA | 1 | 1 | 0 | Low-quality | Genome-fragment | 2.45 | AAI-based (high-confidence) | 0 |
| S15_extracted_70105 | 1112 | No | NA | 2 | 2 | 0 | Low-quality | Genome-fragment | 1.89 | AAI-based (medium-confidence) | 0 |
| S9_vs_k141_57660||full | 1696 | No | NA | 3 | 0 | 1 | Not-determined | Genome-fragment | NA | NA | 0 |
| S9_vs_k141_19872||full | 1604 | No | NA | 2 | 0 | 0 | Not-determined | Genome-fragment | NA | NA | 0 |
| S9_vs_k141_27506||full | 3442 | No | NA | 5 | 0 | 3 | Not-determined | Genome-fragment | NA | NA | 0 |
| S8_extracted_k141_72612 | 1112 | No | NA | 1 | 0 | 1 | Low-quality | Genome-fragment | 0.6 | AAI-based (medium-confidence) | 0 |
| S8_vs_k141_151134||full | 1112 | No | NA | 2 | 2 | 0 | Low-quality | Genome-fragment | 2.68 | AAI-based (high-confidence) | 0 |
| S9_vs_k141_50152||full | 1630 | No | NA | 5 | 0 | 0 | Not-determined | Genome-fragment | NA | NA | 0 |
| S9_vs_k141_39152||full | 1316 | No | NA | 2 | 0 | 1 | Not-determined | Genome-fragment | NA | NA | 0 |
| S9_vs_k141_54219||full | 2213 | No | NA | 3 | 0 | 0 | Not-determined | Genome-fragment | NA | NA | 0 |
| S9_vs_k141_27562||full | 1058 | No | NA | 2 | 0 | 0 | Not-determined | Genome-fragment | NA | NA | 0 |
| S9_vs_k141_50157||full | 2113 | No | NA | 3 | 0 | 2 | Not-determined | Genome-fragment | NA | NA | 0 |
| S17_extracted_k141_16517 | 1113 | No | NA | 2 | 2 | 0 | Low-quality | Genome-fragment | 2.51 | AAI-based (high-confidence) | 0 |
| S9_vs_k141_68947||full | 1674 | No | NA | 3 | 0 | 0 | Not-determined | Genome-fragment | NA | NA | 0 |
| S17_vs_k141_126985||full | 1113 | No | NA | 2 | 0 | 0 | Low-quality | Genome-fragment | 0.67 | AAI-based (medium-confidence) | 0 |
| S14_vs_k141_19021||lt2gene | 1113 | No | NA | 1 | 1 | 0 | Low-quality | Genome-fragment | 1.79 | HMM-based (lower-bound) | 0 |
| S9_vs_k141_68949||full | 2898 | No | NA | 2 | 0 | 0 | Not-determined | Genome-fragment | NA | NA | 0 |
| S9_vs_k141_57702||full | 1022 | No | NA | 2 | 0 | 0 | Not-determined | Genome-fragment | NA | NA | 0 |
| S3_vs_k141_56466||full | 1113 | No | NA | 4 | 1 | 0 | Low-quality | Genome-fragment | 2.13 | AAI-based (medium-confidence) | 0 |
| S3_vs_k141_120696||full | 1113 | No | NA | 3 | 1 | 0 | Low-quality | Genome-fragment | 0.72 | AAI-based (high-confidence) | 0 |

| | | | | | | | | | | | |
|---|---|---|---|---|---|---|---|---|---|---|---|
| S9_vs_k141_84214||full | 1903 | No | NA | 3 | 0 | 1 | Not-determined | Genome-fragment | NA | NA | 0 |
| S9_vs_k141_1092||full | 1625 | No | NA | 2 | 0 | 0 | Not-determined | Genome-fragment | NA | NA | 0 |
| S9_vs_k141_88318||full | 1454 | No | NA | 2 | 0 | 1 | Not-determined | Genome-fragment | NA | NA | 0 |
| S3_vs_k141_13077||lt2gene | 1113 | No | NA | 2 | 1 | 0 | Low-quality | Genome-fragment | 1.81 | AAI-based (medium-confidence) | 0 |
| S5_vs_k141_66159||full | 1113 | No | NA | 3 | 0 | 0 | Low-quality | Genome-fragment | 2.84 | AAI-based (high-confidence) | 0 |
| S7_vs_34176||full | 1113 | No | NA | 2 | 0 | 1 | Low-quality | Genome-fragment | 0.87 | AAI-based (medium-confidence) | 0 |
| S9_vs_k141_46498||full | 20897 | Yes | 17942 | 34 | 12 | 4 | Medium-quality | Genome-fragment | 50.53 | AAI-based (medium-confidence) | 14.14 |
| S12_extracted_k141_48581 | 1113 | No | NA | 2 | 1 | 0 | Low-quality | Genome-fragment | 1.43 | AAI-based (high-confidence) | 0 |
| S12_extracted_k141_180519 | 1113 | No | NA | 3 | 0 | 0 | Low-quality | Genome-fragment | 1.71 | AAI-based (high-confidence) | 0 |
| S9_vs_k141_4824||full | 10095 | No | NA | 8 | 0 | 1 | Not-determined | Genome-fragment | NA | NA | 0 |
| S9_vs_k141_46511||full | 1930 | No | NA | 2 | 0 | 2 | Not-determined | Genome-fragment | NA | NA | 0 |
| S8_extracted_k141_145937 | 1113 | No | NA | 3 | 0 | 0 | Low-quality | Genome-fragment | 1.52 | AAI-based (medium-confidence) | 0 |
| S9_vs_k141_1123||full | 1430 | No | NA | 2 | 0 | 1 | Not-determined | Genome-fragment | NA | NA | 0 |
| S1_vs_k141_154006||full | 1114 | No | NA | 3 | 0 | 0 | Low-quality | Genome-fragment | 2.21 | AAI-based (high-confidence) | 0 |
| S3_vs_k141_88247||lt2gene | 1114 | No | NA | 1 | 1 | 0 | Low-quality | Genome-fragment | 3.1 | AAI-based (high-confidence) | 0 |
| S9_vs_k141_88353||full | 2480 | No | NA | 2 | 0 | 2 | Not-determined | Genome-fragment | NA | NA | 0 |
| S9_vs_k141_50232||full | 1520 | No | NA | 3 | 0 | 1 | Not-determined | Genome-fragment | NA | NA | 0 |
| S5_extracted_k141_123651 | 1114 | No | NA | 2 | 2 | 0 | Low-quality | Genome-fragment | 3.34 | AAI-based (high-confidence) | 0 |
| S9_vs_k141_16516||full | 1291 | No | NA | 2 | 0 | 1 | Not-determined | Genome-fragment | NA | NA | 0 |
| S5_vs_k141_32662||full | 1114 | No | NA | 3 | 2 | 0 | Low-quality | Genome-fragment | 2.78 | AAI-based (high-confidence) | 0 |
| S5_vs_k141_130955||full | 1114 | No | NA | 2 | 0 | 1 | Low-quality | Genome-fragment | 0.37 | AAI-based (medium-confidence) | 0 |
| S9_vs_k141_1163||full | 3827 | No | NA | 3 | 0 | 1 | Not-determined | Genome-fragment | NA | NA | 0 |
| S5_vs_k141_40078||full | 1114 | No | NA | 3 | 1 | 0 | Low-quality | Genome-fragment | 1.99 | AAI-based (medium-confidence) | 0 |
| S16_vs_27176||full | 1114 | No | NA | 3 | 1 | 0 | Low-quality | Genome-fragment | 3.07 | AAI-based (medium-confidence) | 0 |
| S12_vs_k141_145808||full | 1114 | No | NA | 2 | 0 | 0 | Low-quality | Genome-fragment | 2.17 | AAI-based (high-confidence) | 0 |
| S9_vs_k141_57763||full | 1000 | No | NA | 2 | 0 | 1 | Not-determined | Genome-fragment | NA | NA | 0 |
| S9_vs_k141_88382||full | 2419 | No | NA | 3 | 0 | 0 | Not-determined | Genome-fragment | NA | NA | 0 |
| S6_vs_k141_49362||lt2gene | 1114 | No | NA | 2 | 1 | 0 | Low-quality | Genome-fragment | 2.47 | AAI-based (high-confidence) | 0 |
| S8_extracted_k141_123306 | 1114 | No | NA | 1 | 0 | 1 | Low-quality | Genome-fragment | 0.62 | AAI-based (medium-confidence) | 0 |
| S9_vs_k141_12660||full | 2932 | No | NA | 5 | 0 | 1 | Not-determined | Genome-fragment | NA | NA | 0 |
| S8_vs_k141_177801||lt2gene | 1114 | No | NA | 1 | 1 | 0 | Low-quality | Genome-fragment | 2.02 | AAI-based (high-confidence) | 0 |
| S9_vs_k141_61710||full | 2013 | No | NA | 2 | 0 | 0 | Not-determined | Genome-fragment | NA | NA | 0 |
| S9_vs_k141_20047||full | 1466 | No | NA | 3 | 0 | 0 | Not-determined | Genome-fragment | NA | NA | 0 |
| S9_vs_k141_57781||full | 1721 | No | NA | 4 | 0 | 2 | Not-determined | Genome-fragment | NA | NA | 0 |
| S17_vs_k141_111351||full | 1115 | No | NA | 2 | 0 | 0 | Low-quality | Genome-fragment | 2.83 | AAI-based (medium-confidence) | 0 |
| S9_vs_k141_16562||full | 3295 | No | NA | 3 | 0 | 1 | Not-determined | Genome-fragment | NA | NA | 0 |
| S9_extracted_k141_82932 | 1115 | No | NA | 1 | 0 | 1 | Low-quality | Genome-fragment | 0.48 | AAI-based (high-confidence) | 0 |
| S1_extracted_k141_155062 | 1115 | No | NA | 2 | 0 | 0 | Low-quality | Genome-fragment | 0.63 | AAI-based (medium-confidence) | 0 |
| S1_extracted_k141_65053 | 1115 | No | NA | 3 | 0 | 2 | Low-quality | Genome-fragment | 2.13 | AAI-based (high-confidence) | 0 |
| S9_vs_k141_61739||full | 1068 | No | NA | 2 | 0 | 1 | Not-determined | Genome-fragment | NA | NA | 0 |
| S3_vs_k141_48341||full | 1115 | No | NA | 3 | 0 | 0 | Low-quality | Genome-fragment | 1.62 | AAI-based (medium-confidence) | 0 |
| S9_vs_k141_54410||full | 3249 | No | NA | 5 | 0 | 1 | Not-determined | Genome-fragment | NA | NA | 0 |
| S5_vs_k141_24728||full | 1115 | No | NA | 3 | 0 | 0 | Low-quality | Genome-fragment | 2.81 | AAI-based (medium-confidence) | 0 |
| S9_vs_k141_27764||full | 1154 | No | NA | 3 | 0 | 1 | Not-determined | Genome-fragment | NA | NA | 0 |
| S7_vs_96531||full | 1115 | No | NA | 3 | 0 | 0 | Low-quality | Genome-fragment | 2.03 | AAI-based (high-confidence) | 0 |
| S9_vs_k141_35366||full | 1361 | No | NA | 2 | 0 | 0 | Not-determined | Genome-fragment | NA | NA | 0 |
| S9_vs_k141_80599||full | 2506 | No | NA | 5 | 0 | 2 | Not-determined | Genome-fragment | NA | NA | 0 |
| S9_vs_k141_65448||full | 2858 | No | NA | 9 | 0 | 0 | Not-determined | Genome-fragment | NA | NA | 0 |
| S16_extracted_66421 | 1115 | No | NA | 4 | 0 | 0 | Low-quality | Genome-fragment | 2.23 | AAI-based (high-confidence) | 0 |
| S12_extracted_k141_137235 | 1115 | No | NA | 1 | 0 | 1 | Low-quality | Genome-fragment | 0.27 | AAI-based (medium-confidence) | 0 |
| S12_extracted_k141_98660 | 1115 | No | NA | 4 | 0 | 0 | Low-quality | Genome-fragment | 1.8 | AAI-based (high-confidence) | 0 |
| S8_vs_k141_187193||full | 1115 | No | NA | 2 | 0 | 0 | Low-quality | Genome-fragment | 2.3 | AAI-based (high-confidence) | 0 |
| S8_vs_k141_49241||full | 1115 | No | NA | 3 | 1 | 0 | Low-quality | Genome-fragment | 2.62 | AAI-based (high-confidence) | 0 |
| S9_vs_k141_69154||full | 4501 | No | NA | 4 | 0 | 1 | Not-determined | Genome-fragment | NA | NA | 0 |
| S17_vs_k141_161343||lt2gene | 1116 | No | NA | 2 | 2 | 0 | Low-quality | Genome-fragment | 2.57 | AAI-based (high-confidence) | 0 |
| S9_vs_k141_21629||full | 1116 | No | NA | 3 | 2 | 0 | Low-quality | Genome-fragment | 2.78 | AAI-based (high-confidence) | 0 |
| S1_vs_k141_41443||full | 1116 | No | NA | 2 | 0 | 0 | Low-quality | Genome-fragment | 1.96 | AAI-based (high-confidence) | 0 |
| S9_vs_k141_35410||full | 1059 | No | NA | 2 | 0 | 1 | Not-determined | Genome-fragment | NA | NA | 0 |
| S9_vs_k141_23801||full | 1451 | No | NA | 3 | 0 | 0 | Not-determined | Genome-fragment | NA | NA | 0 |
| S9_vs_k141_39399||full | 1102 | No | NA | 4 | 0 | 0 | Not-determined | Genome-fragment | NA | NA | 0 |
| S3_extracted_k141_80886 | 1116 | No | NA | 2 | 1 | 0 | Low-quality | Genome-fragment | 0.83 | AAI-based (high-confidence) | 0 |
| S3_extracted_k141_74088 | 1116 | No | NA | 2 | 1 | 0 | Low-quality | Genome-fragment | 1.96 | AAI-based (high-confidence) | 0 |
| S9_vs_k141_43089||full | 2699 | No | NA | 2 | 0 | 1 | Not-determined | Genome-fragment | NA | NA | 0 |
| S9_vs_k141_69207||full | 1103 | No | NA | 2 | 0 | 0 | Not-determined | Genome-fragment | NA | NA | 0 |
| S10_vs_10326||full | 1116 | No | NA | 2 | 0 | 0 | Low-quality | Genome-fragment | 0.98 | AAI-based (high-confidence) | 0 |
| S10_vs_103190||full | 1116 | No | NA | 3 | 3 | 0 | Low-quality | Genome-fragment | 7.55 | AAI-based (high-confidence) | 0 |
| S9_vs_k141_27813||full | 43436 | No | NA | 69 | 17 | 1 | High-quality | High-quality | 96.78 | AAI-based (high-confidence) | 0 |
| S12_extracted_k141_100684 | 1116 | No | NA | 1 | 0 | 1 | Low-quality | Genome-fragment | 1.35 | AAI-based (medium-confidence) | 0 |
| S9_vs_k141_88561||full | 1717 | No | NA | 5 | 0 | 0 | Not-determined | Genome-fragment | NA | NA | 0 |
| S9_vs_k141_57881||full | 2890 | No | NA | 1 | 0 | 0 | Not-determined | Genome-fragment | NA | NA | 0 |
| S9_vs_k141_76712||full | 1296 | No | NA | 3 | 0 | 1 | Not-determined | Genome-fragment | NA | NA | 0 |
| S9_vs_k141_5055||full | 1699 | No | NA | 3 | 0 | 0 | Not-determined | Genome-fragment | NA | NA | 0 |
| S12_vs_k141_183287||full | 1116 | No | NA | 3 | 0 | 0 | Low-quality | Genome-fragment | 1.64 | AAI-based (high-confidence) | 0 |
| S9_vs_k141_88571||full | 3357 | No | NA | 3 | 0 | 3 | Not-determined | Genome-fragment | NA | NA | 0 |
| S9_vs_k141_5070||full | 2620 | No | NA | 3 | 0 | 3 | Not-determined | Genome-fragment | NA | NA | 0 |
| S8_extracted_k141_31065 | 1116 | No | NA | 2 | 0 | 0 | Low-quality | Genome-fragment | 2.56 | AAI-based (medium-confidence) | 0 |
| S8_extracted_k141_137145 | 1116 | No | NA | 3 | 0 | 1 | Low-quality | Genome-fragment | 1.73 | AAI-based (medium-confidence) | 0 |
| S8_vs_k141_49103||lt2gene | 1116 | No | NA | 1 | 1 | 0 | Low-quality | Genome-fragment | 1.97 | AAI-based (medium-confidence) | 0 |
| S9_vs_k141_1400||full | 1399 | No | NA | 2 | 0 | 0 | Not-determined | Genome-fragment | NA | NA | 0 |
| S17_extracted_k141_169811 | 1117 | No | NA | 3 | 1 | 0 | Low-quality | Genome-fragment | 1.89 | AAI-based (medium-confidence) | 0 |
| S9_vs_k141_23914||full | 1498 | No | NA | 5 | 0 | 0 | Not-determined | Genome-fragment | NA | NA | 0 |
| S9_vs_k141_1407||full | 2816 | No | NA | 2 | 0 | 1 | Not-determined | Genome-fragment | NA | NA | 0 |
| S9_vs_k141_39459||full | 1632 | No | NA | 3 | 0 | 1 | Not-determined | Genome-fragment | NA | NA | 0 |
| S11_vs_k141_65106||lt2gene | 1117 | No | NA | 2 | 1 | 0 | Low-quality | Genome-fragment | 2.71 | AAI-based (high-confidence) | 0 |
| S1_extracted_k141_94498 | 1117 | No | NA | 2 | 0 | 0 | Low-quality | Genome-fragment | 2.72 | AAI-based (high-confidence) | 0 |
| S5_vs_k141_7309||full | 1117 | No | NA | 3 | 0 | 0 | Low-quality | Genome-fragment | 1.07 | AAI-based (high-confidence) | 0 |
| S9_vs_k141_23945||full | 2418 | No | NA | 3 | 0 | 1 | Not-determined | Genome-fragment | NA | NA | 0 |
| S9_vs_k141_20284||full | 1192 | No | NA | 3 | 0 | 1 | Not-determined | Genome-fragment | NA | NA | 0 |
| S5_vs_k141_71386||full | 1117 | No | NA | 3 | 1 | 0 | Low-quality | Genome-fragment | 3.04 | AAI-based (medium-confidence) | 0 |
| S5_vs_k141_94334||lt2gene | 1117 | No | NA | 2 | 1 | 0 | Low-quality | Genome-fragment | 2.85 | AAI-based (medium-confidence) | 0 |
| S9_vs_k141_43200||full | 1301 | No | NA | 3 | 0 | 1 | Not-determined | Genome-fragment | NA | NA | 0 |
| S9_vs_k141_23985||full | 2717 | No | NA | 9 | 0 | 0 | Not-determined | Genome-fragment | NA | NA | 0 |
| S7_vs_79136||full | 1117 | No | NA | 3 | 1 | 0 | Low-quality | Genome-fragment | 2.52 | AAI-based (high-confidence) | 0 |
| S16_extracted_143432 | 1117 | No | NA | 2 | 0 | 2 | Low-quality | Genome-fragment | 0.72 | AAI-based (medium-confidence) | 0 |
| S16_vs_29224||full | 1117 | No | NA | 2 | 1 | 0 | Low-quality | Genome-fragment | 0.71 | AAI-based (high-confidence) | 0 |
| S12_extracted_k141_31809 | 1117 | No | NA | 2 | 0 | 0 | Low-quality | Genome-fragment | 1.53 | AAI-based (medium-confidence) | 0 |
| S9_vs_k141_69322||full | 27451 | No | NA | 30 | 3 | 0 | Medium-quality | Genome-fragment | 68.73 | AAI-based (medium-confidence) | 0 |
| S12_extracted_k141_39912 | 1117 | No | NA | 3 | 0 | 0 | Low-quality | Genome-fragment | 2.37 | AAI-based (medium-confidence) | 0 |
| S9_vs_k141_84512||full | 2150 | No | NA | 2 | 0 | 0 | Not-determined | Genome-fragment | NA | NA | 0 |
| S9_vs_k141_43255||full | 1148 | No | NA | 3 | 0 | 0 | Not-determined | Genome-fragment | NA | NA | 0 |
| S12_extracted_k141_151507 | 1117 | No | NA | 3 | 0 | 1 | Low-quality | Genome-fragment | 1.14 | AAI-based (high-confidence) | 0 |
| S17_vs_k141_74821||full | 1118 | No | NA | 2 | 0 | 0 | Low-quality | Genome-fragment | 2.04 | AAI-based (medium-confidence) | 0 |
| S1_vs_k141_131912||full | 1118 | No | NA | 2 | 0 | 0 | Low-quality | Genome-fragment | 1.47 | AAI-based (medium-confidence) | 0 |
| S9_vs_k141_1531||full | 3372 | No | NA | 4 | 0 | 2 | Not-determined | Genome-fragment | NA | NA | 0 |
| S3_vs_k141_37891||full | 1118 | No | NA | 4 | 1 | 0 | Low-quality | Genome-fragment | 3.44 | AAI-based (high-confidence) | 0 |
| S7_extracted_84298 | 1118 | No | NA | 3 | 0 | 0 | Low-quality | Genome-fragment | 1.43 | AAI-based (medium-confidence) | 0 |
| S7_vs_43857||full | 1118 | No | NA | 2 | 0 | 0 | Low-quality | Genome-fragment | 0.87 | AAI-based (medium-confidence) | 0 |
| S9_vs_k141_54512||full | 3125 | No | NA | 5 | 0 | 0 | Not-determined | Genome-fragment | NA | NA | 0 |
| S7_vs_131256||lt2gene | 1118 | No | NA | 1 | 1 | 0 | Low-quality | Genome-fragment | 2.5 | AAI-based (medium-confidence) | 0 |
| S16_vs_128966||full | 1118 | No | NA | 2 | 1 | 0 | Low-quality | Genome-fragment | 1.57 | AAI-based (medium-confidence) | 0 |
| S9_vs_k141_65752||full | 2477 | No | NA | 4 | 0 | 0 | Not-determined | Genome-fragment | NA | NA | 0 |
| S6_vs_k141_20143||full | 1118 | No | NA | 4 | 0 | 0 | Low-quality | Genome-fragment | 3.12 | AAI-based (medium-confidence) | 0 |
| S9_vs_k141_8782||full | 1515 | No | NA | 2 | 0 | 0 | Not-determined | Genome-fragment | NA | NA | 0 |
| S9_vs_k141_65771||full | 2824 | No | NA | 5 | 0 | 1 | Not-determined | Genome-fragment | NA | NA | 0 |
| S9_vs_k141_76801||full | 1703 | No | NA | 2 | 0 | 1 | Not-determined | Genome-fragment | NA | NA | 0 |
| S9_vs_k141_8811||full | 1040 | No | NA | 2 | 0 | 1 | Not-determined | Genome-fragment | NA | NA | 0 |
| S9_vs_k141_28017||full | 1910 | No | NA | 3 | 0 | 0 | Not-determined | Genome-fragment | NA | NA | 0 |
| S8_vs_k141_151875||full | 1118 | No | NA | 3 | 1 | 0 | Low-quality | Genome-fragment | 0.8 | HMM-based (lower-bound) | 0 |
| S11_extracted_k141_53197 | 1119 | No | NA | 2 | 0 | 0 | Low-quality | Genome-fragment | 2.33 | AAI-based (high-confidence) | 0 |
| S9_vs_k141_80939||full | 2812 | No | NA | 3 | 0 | 2 | Not-determined | Genome-fragment | NA | NA | 0 |
| S9_vs_k141_28050||full | 1485 | No | NA | 3 | 0 | 0 | Not-determined | Genome-fragment | NA | NA | 0 |
| S9_vs_k141_69466||full | 3517 | No | NA | 5 | 0 | 2 | Not-determined | Genome-fragment | NA | NA | 0 |
| S11_vs_k141_1953||full | 1119 | No | NA | 3 | 0 | 0 | Low-quality | Genome-fragment | 2.53 | AAI-based (medium-confidence) | 0 |
| S9_vs_k141_62110||full | 2179 | No | NA | 4 | 0 | 1 | Not-determined | Genome-fragment | NA | NA | 0 |
| S14_extracted_k141_64504 | 1119 | No | NA | 1 | 0 | 1 | Low-quality | Genome-fragment | 0.54 | AAI-based (medium-confidence) | 0 |
| S9_vs_k141_80965||full | 1423 | No | NA | 3 | 0 | 0 | Not-determined | Genome-fragment | NA | NA | 0 |
| S3_vs_k141_28434||full | 1119 | No | NA | 3 | 0 | 0 | Low-quality | Genome-fragment | 2.67 | AAI-based (medium-confidence) | 0 |
| S3_vs_k141_86726||full | 1119 | No | NA | 2 | 1 | 0 | Low-quality | Genome-fragment | 0.45 | HMM-based (lower-bound) | 0 |
| S3_vs_k141_86985||lt2gene | 1119 | No | NA | 1 | 1 | 0 | Low-quality | Genome-fragment | 2.93 | AAI-based (high-confidence) | 0 |
| S9_vs_k141_73523||full | 2790 | No | NA | 3 | 0 | 1 | Not-determined | Genome-fragment | NA | NA | 0 |
| S5_vs_k141_6038||full | 1119 | No | NA | 3 | 2 | 0 | Low-quality | Genome-fragment | 2.47 | AAI-based (medium-confidence) | 0 |
| S7_vs_102789||full | 1119 | No | NA | 4 | 1 | 0 | Low-quality | Genome-fragment | 3.07 | AAI-based (high-confidence) | 0 |
| S16_vs_4203||full | 1119 | No | NA | 3 | 0 | 1 | Low-quality | Genome-fragment | 0.58 | AAI-based (medium-confidence) | 0 |
| S16_vs_150929||full | 1119 | No | NA | 3 | 1 | 0 | Low-quality | Genome-fragment | 2.86 | AAI-based (high-confidence) | 0 |
| S10_extracted_40388 | 1119 | No | NA | 3 | 0 | 0 | Low-quality | Genome-fragment | 1.78 | AAI-based (high-confidence) | 0 |
| S9_vs_k141_54656||full | 1558 | No | NA | 3 | 0 | 0 | Not-determined | Genome-fragment | NA | NA | 0 |
| S12_extracted_k141_5959 | 1119 | No | NA | 2 | 0 | 1 | Low-quality | Genome-fragment | 1.49 | AAI-based (medium-confidence) | 0 |
| S9_vs_k141_54660||full | 1831 | No | NA | 3 | 0 | 0 | Not-determined | Genome-fragment | NA | NA | 0 |
| S12_vs_k141_165004||full | 1119 | No | NA | 3 | 1 | 0 | Low-quality | Genome-fragment | 2.02 | AAI-based (medium-confidence) | 0 |
| S8_extracted_k141_86993 | 1119 | No | NA | 2 | 0 | 1 | Low-quality | Genome-fragment | 2.88 | AAI-based (medium-confidence) | 0 |
| S17_extracted_k141_73525 | 1120 | No | NA | 1 | 0 | 0 | Low-quality | Genome-fragment | 1.19 | AAI-based (medium-confidence) | 0 |
| S9_vs_k141_16933||full | 4431 | No | NA | 6 | 0 | 2 | Not-determined | Genome-fragment | NA | NA | 0 |
| S9_vs_k141_58086||full | 27326 | No | NA | 51 | 12 | 2 | Medium-quality | Genome-fragment | 77.03 | AAI-based (high-confidence) | 0 |
| S9_vs_k141_8553||full | 1120 | No | NA | 2 | 2 | 0 | Low-quality | Genome-fragment | 2.5 | AAI-based (medium-confidence) | 0 |
| S13_vs_8166||full | 1120 | No | NA | 2 | 0 | 1 | Low-quality | Genome-fragment | 0.44 | AAI-based (medium-confidence) | 0 |
| S14_vs_k141_39309||lt2gene | 1120 | No | NA | 1 | 1 | 0 | Low-quality | Genome-fragment | 1.64 | HMM-based (lower-bound) | 0 |
| S14_vs_k141_19519||lt2gene | 1120 | No | NA | 2 | 1 | 0 | Low-quality | Genome-fragment | 2.29 | AAI-based (high-confidence) | 0 |
| S9_vs_k141_39674||full | 1290 | No | NA | 3 | 0 | 1 | Not-determined | Genome-fragment | NA | NA | 0 |
| S9_vs_k141_35808||full | 3513 | No | NA | 3 | 0 | 1 | Not-determined | Genome-fragment | NA | NA | 0 |
| S3_extracted_k141_124092 | 1120 | No | NA | 3 | 0 | 0 | Low-quality | Genome-fragment | 1.19 | AAI-based (high-confidence) | 0 |
| S9_vs_k141_84678||full | 1991 | No | NA | 3 | 0 | 2 | Not-determined | Genome-fragment | NA | NA | 0 |
| S10_extracted_21573 | 1120 | No | NA | 1 | 0 | 0 | Low-quality | Genome-fragment | 0.5 | AAI-based (high-confidence) | 0 |
| S12_extracted_k141_124236 | 1120 | No | NA | 2 | 0 | 1 | Low-quality | Genome-fragment | 3.04 | AAI-based (medium-confidence) | 0 |
| S9_vs_k141_35820||full | 4800 | No | NA | 5 | 0 | 2 | Not-determined | Genome-fragment | NA | NA | 0 |
| S15_vs_45881||full | 1120 | No | NA | 2 | 0 | 0 | Low-quality | Genome-fragment | 0.45 | AAI-based (high-confidence) | 0 |
| S9_vs_k141_35830||full | 1271 | No | NA | 2 | 0 | 1 | Not-determined | Genome-fragment | NA | NA | 0 |
| S9_vs_k141_28197||full | 27879 | No | NA | 41 | 10 | 0 | Medium-quality | Genome-fragment | 74.59 | AAI-based (medium-confidence) | 0 |
| S15_vs_70494||full | 1120 | No | NA | 3 | 0 | 3 | Low-quality | Genome-fragment | 1.85 | AAI-based (medium-confidence) | 0 |
| S9_vs_k141_1748||full | 1704 | No | NA | 3 | 0 | 1 | Not-determined | Genome-fragment | NA | NA | 0 |
| S9_vs_k141_20616||full | 1812 | No | NA | 2 | 0 | 1 | Not-determined | Genome-fragment | NA | NA | 0 |
| S9_vs_k141_35857||full | 1403 | No | NA | 3 | 0 | 1 | Not-determined | Genome-fragment | NA | NA | 0 |
| S8_extracted_k141_198848 | 1120 | No | NA | 2 | 0 | 0 | Low-quality | Genome-fragment | 2.8 | AAI-based (high-confidence) | 0 |
| S9_vs_k141_54753||full | 1321 | No | NA | 2 | 0 | 0 | Not-determined | Genome-fragment | NA | NA | 0 |
| S9_vs_k141_65837||full | 1297 | No | NA | 2 | 0 | 0 | Not-determined | Genome-fragment | NA | NA | 0 |
| S9_vs_k141_43488||full | 2065 | No | NA | 4 | 0 | 1 | Not-determined | Genome-fragment | NA | NA | 0 |
| S17_extracted_k141_76555 | 1121 | No | NA | 2 | 0 | 0 | Low-quality | Genome-fragment | 2.12 | AAI-based (high-confidence) | 0 |
| S9_vs_k141_39742||full | 2342 | No | NA | 4 | 0 | 1 | Not-determined | Genome-fragment | NA | NA | 0 |
| S9_vs_k141_81100||full | 4207 | No | NA | 3 | 0 | 2 | Not-determined | Genome-fragment | NA | NA | 0 |
| S9_vs_k141_46866||full | 2827 | No | NA | 2 | 0 | 1 | Not-determined | Genome-fragment | NA | NA | 0 |
| S17_vs_k141_146081||full | 1121 | No | NA | 2 | 0 | 2 | Low-quality | Genome-fragment | 0.88 | AAI-based (medium-confidence) | 0 |
| S9_vs_k141_58197||full | 1946 | No | NA | 3 | 0 | 1 | Not-determined | Genome-fragment | NA | NA | 0 |
| S9_extracted_k141_17367 | 1121 | No | NA | 1 | 0 | 0 | Low-quality | Genome-fragment | 2.32 | AAI-based (high-confidence) | 0 |
| S9_vs_k141_46875||full | 2309 | No | NA | 3 | 0 | 1 | Not-determined | Genome-fragment | NA | NA | 0 |
| S11_vs_k141_87086||full | 1121 | No | NA | 3 | 0 | 1 | Low-quality | Genome-fragment | 0.64 | AAI-based (high-confidence) | 0 |
| S9_vs_k141_77037||full | 1693 | No | NA | 5 | 0 | 0 | Not-determined | Genome-fragment | NA | NA | 0 |
| S11_vs_k141_95559||full | 1121 | No | NA | 3 | 1 | 0 | Low-quality | Genome-fragment | 1.97 | AAI-based (medium-confidence) | 0 |
| S13_vs_61906||full | 1121 | No | NA | 4 | 0 | 0 | Low-quality | Genome-fragment | 2.56 | AAI-based (medium-confidence) | 0 |
| S14_vs_k141_46494||full | 1121 | No | NA | 3 | 1 | 0 | Low-quality | Genome-fragment | 2.69 | AAI-based (high-confidence) | 0 |
| S14_vs_k141_60212||lt2gene | 1121 | No | NA | 1 | 1 | 0 | Low-quality | Genome-fragment | 1.64 | HMM-based (lower-bound) | 0 |
| S9_vs_k141_46908||full | 1193 | No | NA | 3 | 0 | 0 | Not-determined | Genome-fragment | NA | NA | 0 |
| S9_vs_k141_62311||full | 3021 | No | NA | 3 | 0 | 3 | Not-determined | Genome-fragment | NA | NA | 0 |
| S1_extracted_k141_158632 | 1121 | No | NA | 1 | 1 | 0 | Low-quality | Genome-fragment | 1.87 | HMM-based (lower-bound) | 0 |
| S3_extracted_k141_41132 | 1121 | No | NA | 3 | 1 | 0 | Low-quality | Genome-fragment | 1.44 | AAI-based (high-confidence) | 0 |
| S9_vs_k141_17140||full | 2130 | No | NA | 4 | 0 | 2 | Not-determined | Genome-fragment | NA | NA | 0 |
| S3_vs_k141_122188||full | 1121 | No | NA | 2 | 0 | 0 | Low-quality | Genome-fragment | 1.61 | AAI-based (high-confidence) | 0 |
| S7_extracted_5108 | 1121 | No | NA | 2 | 2 | 0 | Low-quality | Genome-fragment | 3.7 | AAI-based (high-confidence) | 0 |
| S7_vs_102738||full | 1121 | No | NA | 2 | 0 | 0 | Low-quality | Genome-fragment | 1.94 | AAI-based (high-confidence) | 0 |
| S7_vs_54853||lt2gene | 1121 | No | NA | 1 | 1 | 0 | Low-quality | Genome-fragment | 1.64 | HMM-based (lower-bound) | 0 |
| S9_vs_k141_5600||full | 2302 | No | NA | 4 | 0 | 0 | Not-determined | Genome-fragment | NA | NA | 0 |
| S9_vs_k141_62334||full | 1009 | No | NA | 4 | 0 | 0 | Not-determined | Genome-fragment | NA | NA | 0 |
| S16_extracted_15380 | 1121 | No | NA | 2 | 0 | 0 | Low-quality | Genome-fragment | 1.9 | AAI-based (high-confidence) | 0 |
| S9_vs_k141_54878||full | 2947 | No | NA | 4 | 0 | 2 | Not-determined | Genome-fragment | NA | NA | 0 |
| S16_extracted_86079 | 1121 | No | NA | 3 | 1 | 0 | Low-quality | Genome-fragment | 3.48 | AAI-based (high-confidence) | 0 |
| S9_vs_k141_24425||full | 1804 | No | NA | 3 | 0 | 1 | Not-determined | Genome-fragment | NA | NA | 0 |
| S10_vs_88078||full | 1121 | No | NA | 2 | 0 | 0 | Low-quality | Genome-fragment | 2.23 | AAI-based (medium-confidence) | 0 |
| S12_vs_k141_46009||full | 1121 | No | NA | 3 | 0 | 1 | Low-quality | Genome-fragment | 0.36 | AAI-based (medium-confidence) | 0 |
| S9_vs_k141_62353||full | 2087 | No | NA | 3 | 0 | 0 | Not-determined | Genome-fragment | NA | NA | 0 |
| S9_vs_k141_17175||full | 2248 | No | NA | 3 | 0 | 0 | Not-determined | Genome-fragment | NA | NA | 0 |
| S4_extracted_13057 | 1121 | No | NA | 2 | 0 | 0 | Low-quality | Genome-fragment | 1.06 | AAI-based (high-confidence) | 0 |
| S8_extracted_k141_152045 | 1121 | No | NA | 2 | 0 | 0 | Low-quality | Genome-fragment | 0.66 | AAI-based (high-confidence) | 0 |
| S17_vs_k141_149950||full | 1122 | No | NA | 3 | 0 | 0 | Low-quality | Genome-fragment | 1.67 | AAI-based (high-confidence) | 0 |
| S9_vs_k141_81209||full | 1203 | No | NA | 4 | 0 | 2 | Not-determined | Genome-fragment | NA | NA | 0 |
| S1_vs_k141_102255||full | 1122 | No | NA | 3 | 0 | 0 | Low-quality | Genome-fragment | 2.06 | AAI-based (medium-confidence) | 0 |
| S9_vs_k141_46927||full | 2539 | No | NA | 4 | 0 | 1 | Not-determined | Genome-fragment | NA | NA | 0 |
| S1_vs_k141_32263||full | 1122 | No | NA | 3 | 1 | 0 | Low-quality | Genome-fragment | 1.83 | AAI-based (high-confidence) | 0 |
| S3_extracted_k141_106953 | 1122 | No | NA | 3 | 0 | 0 | Low-quality | Genome-fragment | 1.87 | AAI-based (high-confidence) | 0 |
| S5_extracted_k141_144238 | 1122 | No | NA | 2 | 1 | 0 | Low-quality | Genome-fragment | 2.32 | AAI-based (high-confidence) | 0 |
| S9_vs_k141_24472||full | 4571 | No | NA | 4 | 0 | 3 | Not-determined | Genome-fragment | NA | NA | 0 |
| S7_extracted_10877 | 1122 | No | NA | 1 | 1 | 0 | Low-quality | Genome-fragment | 3.62 | AAI-based (high-confidence) | 0 |
| S9_vs_k141_1894||full | 3001 | No | NA | 4 | 0 | 0 | Not-determined | Genome-fragment | NA | NA | 0 |
| S7_vs_21364||full | 1122 | No | NA | 2 | 1 | 0 | Low-quality | Genome-fragment | 2.23 | AAI-based (medium-confidence) | 0 |
| S16_vs_56427||lt2gene | 1122 | No | NA | 2 | 1 | 0 | Low-quality | Genome-fragment | 3.29 | AAI-based (high-confidence) | 0 |
| S9_vs_k141_62410||full | 3130 | No | NA | 6 | 0 | 1 | Not-determined | Genome-fragment | NA | NA | 0 |
| S10_vs_61189||full | 1122 | No | NA | 2 | 0 | 1 | Low-quality | Genome-fragment | 2.4 | AAI-based (medium-confidence) | 0 |
| S9_vs_k141_65910||full | 1709 | No | NA | 3 | 0 | 2 | Not-determined | Genome-fragment | NA | NA | 0 |
| S10_vs_79667||full | 1122 | No | NA | 3 | 0 | 0 | Low-quality | Genome-fragment | 3.04 | AAI-based (high-confidence) | 0 |
| S9_vs_k141_28450||full | 1747 | No | NA | 3 | 0 | 0 | Not-determined | Genome-fragment | NA | NA | 0 |
| S9_vs_k141_1930||full | 1187 | No | NA | 3 | 0 | 0 | Not-determined | Genome-fragment | NA | NA | 0 |
| S12_extracted_k141_53204 | 1122 | No | NA | 1 | 1 | 0 | Low-quality | Genome-fragment | 2.51 | AAI-based (high-confidence) | 0 |
| S9_vs_k141_81246||full | 4676 | No | NA | 5 | 0 | 4 | Not-determined | Genome-fragment | NA | NA | 0 |
| S8_extracted_k141_12309 | 1122 | No | NA | 1 | 1 | 0 | Low-quality | Genome-fragment | 2.15 | AAI-based (high-confidence) | 0 |
| S9_vs_k141_77195||full | 2418 | No | NA | 3 | 0 | 0 | Not-determined | Genome-fragment | NA | NA | 0 |
| S9_vs_k141_65934||full | 1137 | No | NA | 1 | 0 | 0 | Not-determined | Genome-fragment | NA | NA | 0 |
| S8_vs_k141_155728||full | 1122 | No | NA | 4 | 1 | 0 | Low-quality | Genome-fragment | 1.14 | AAI-based (high-confidence) | 0 |
| S17_vs_k141_116428||full | 1123 | No | NA | 2 | 0 | 0 | Low-quality | Genome-fragment | 2.97 | AAI-based (high-confidence) | 0 |
| S9_vs_k141_24546||full | 2567 | No | NA | 2 | 0 | 2 | Not-determined | Genome-fragment | NA | NA | 0 |
| S17_vs_k141_109864||full | 1123 | No | NA | 3 | 0 | 0 | Low-quality | Genome-fragment | 1.3 | AAI-based (medium-confidence) | 0 |
| S11_vs_k141_22162||full | 1123 | No | NA | 4 | 0 | 0 | Low-quality | Genome-fragment | 2.2 | AAI-based (medium-confidence) | 0 |
| S9_vs_k141_9176||full | 1601 | No | NA | 3 | 0 | 0 | Not-determined | Genome-fragment | NA | NA | 0 |
| S9_vs_k141_17303||full | 2811 | No | NA | 4 | 0 | 2 | Not-determined | Genome-fragment | NA | NA | 0 |
| S14_extracted_k141_73218 | 1123 | No | NA | 1 | 1 | 0 | Low-quality | Genome-fragment | 1.99 | AAI-based (medium-confidence) | 0 |
| S9_vs_k141_88789||full | 3032 | No | NA | 4 | 0 | 1 | Not-determined | Genome-fragment | NA | NA | 0 |
| S14_vs_k141_70484||full | 1123 | No | NA | 3 | 0 | 0 | Low-quality | Genome-fragment | 2.58 | AAI-based (high-confidence) | 0 |
| S5_vs_k141_92942||full | 1123 | No | NA | 3 | 2 | 0 | Low-quality | Genome-fragment | 2.81 | AAI-based (high-confidence) | 0 |
| S7_extracted_92695 | 1123 | No | NA | 3 | 0 | 0 | Low-quality | Genome-fragment | 1.44 | AAI-based (high-confidence) | 0 |
| S15_vs_62041||full | 1123 | No | NA | 2 | 1 | 0 | Low-quality | Genome-fragment | 2.16 | AAI-based (medium-confidence) | 0 |
| S6_vs_k141_8983||full | 1123 | No | NA | 2 | 0 | 1 | Low-quality | Genome-fragment | 1.64 | AAI-based (medium-confidence) | 0 |
| S17_vs_k141_112893||full | 1124 | No | NA | 3 | 0 | 2 | Low-quality | Genome-fragment | 0.38 | AAI-based (medium-confidence) | 0 |
| S9_vs_k141_1966||full | 26960 | No | NA | 47 | 17 | 1 | Medium-quality | Genome-fragment | 75.45 | AAI-based (high-confidence) | 0 |
| S9_vs_k141_58402||full | 22074 | No | NA | 29 | 8 | 0 | Medium-quality | Genome-fragment | 56.74 | AAI-based (high-confidence) | 0 |
| S9_vs_k141_90785||full | 1124 | No | NA | 3 | 2 | 0 | Low-quality | Genome-fragment | 1.86 | AAI-based (high-confidence) | 0 |
| S9_vs_k141_55072||full | 1894 | No | NA | 2 | 0 | 2 | Not-determined | Genome-fragment | NA | NA | 0 |
| S9_vs_k141_9250||full | 2508 | No | NA | 3 | 0 | 2 | Not-determined | Genome-fragment | NA | NA | 0 |
| S11_vs_k141_50521||lt2gene | 1124 | No | NA | 1 | 1 | 0 | Low-quality | Genome-fragment | 1.7 | AAI-based (medium-confidence) | 0 |
| S9_vs_k141_36121||full | 4057 | No | NA | 6 | 0 | 0 | Not-determined | Genome-fragment | NA | NA | 0 |
| S14_vs_k141_24499||full | 1124 | No | NA | 3 | 0 | 0 | Low-quality | Genome-fragment | 2.39 | AAI-based (high-confidence) | 0 |
| S9_vs_k141_20869||full | 6576 | No | NA | 8 | 0 | 6 | Not-determined | Genome-fragment | NA | NA | 0 |
| S1_extracted_k141_48823 | 1124 | No | NA | 2 | 0 | 0 | Low-quality | Genome-fragment | 1.08 | AAI-based (high-confidence) | 0 |
| S9_vs_k141_81370||full | 2392 | No | NA | 2 | 0 | 2 | Not-determined | Genome-fragment | NA | NA | 0 |
| S3_extracted_k141_124043 | 1124 | No | NA | 1 | 1 | 0 | Low-quality | Genome-fragment | 0.32 | HMM-based (lower-bound) | 0 |
| S9_vs_k141_36142||full | 2166 | No | NA | 4 | 0 | 0 | Not-determined | Genome-fragment | NA | NA | 0 |
| S5_extracted_k141_113719 | 1124 | No | NA | 4 | 0 | 0 | Low-quality | Genome-fragment | 3.36 | AAI-based (medium-confidence) | 0 |
| S9_vs_k141_73764||full | 25774 | No | NA | 41 | 18 | 0 | Medium-quality | Genome-fragment | 78.08 | AAI-based (high-confidence) | 0 |
| S16_vs_170524||full | 1124 | No | NA | 2 | 0 | 0 | Low-quality | Genome-fragment | 2.3 | AAI-based (high-confidence) | 0 |
| S9_vs_k141_88847||full | 33348 | No | NA | 37 | 9 | 0 | Medium-quality | Genome-fragment | 88.26 | AAI-based (medium-confidence) | 0 |
| S16_vs_139341||lt2gene | 1124 | No | NA | 2 | 1 | 0 | Low-quality | Genome-fragment | 3.08 | AAI-based (medium-confidence) | 0 |
| S10_extracted_112729 | 1124 | No | NA | 3 | 1 | 0 | Low-quality | Genome-fragment | 1.04 | AAI-based (high-confidence) | 0 |
| S12_vs_k141_21250||full | 1124 | No | NA | 2 | 0 | 0 | Low-quality | Genome-fragment | 1 | AAI-based (medium-confidence) | 0 |
| S9_vs_k141_43844||full | 1574 | No | NA | 3 | 0 | 0 | Not-determined | Genome-fragment | NA | NA | 0 |
| S9_vs_k141_36162||full | 4153 | No | NA | 6 | 0 | 3 | Not-determined | Genome-fragment | NA | NA | 0 |
| S12_vs_k141_61261||lt2gene | 1124 | No | NA | 2 | 2 | 0 | Low-quality | Genome-fragment | 2.76 | AAI-based (high-confidence) | 0 |
| S6_vs_k141_48732||full | 1124 | No | NA | 2 | 0 | 0 | Low-quality | Genome-fragment | 2.37 | AAI-based (medium-confidence) | 0 |
| S17_extracted_k141_51822 | 1125 | No | NA | 3 | 1 | 0 | Low-quality | Genome-fragment | 1.69 | AAI-based (high-confidence) | 0 |
| S9_vs_k141_62557||full | 1146 | No | NA | 3 | 0 | 2 | Not-determined | Genome-fragment | NA | NA | 0 |
| S9_vs_k141_40115||full | 1737 | No | NA | 3 | 0 | 1 | Not-determined | Genome-fragment | NA | NA | 0 |
| S17_vs_k141_122952||full | 1125 | No | NA | 3 | 0 | 1 | Low-quality | Genome-fragment | 2.11 | AAI-based (high-confidence) | 0 |
| S9_vs_k141_62572||full | 2174 | No | NA | 4 | 0 | 0 | Not-determined | Genome-fragment | NA | NA | 0 |
| S9_vs_k141_34976||full | 1125 | No | NA | 3 | 1 | 0 | Low-quality | Genome-fragment | 2.04 | AAI-based (high-confidence) | 0 |
| S9_vs_k141_13377||full | 2188 | No | NA | 2 | 0 | 1 | Not-determined | Genome-fragment | NA | NA | 0 |
| S9_vs_k141_13380||full | 1265 | No | NA | 2 | 0 | 0 | Not-determined | Genome-fragment | NA | NA | 0 |
| S11_vs_k141_71841||lt2gene | 1125 | No | NA | 1 | 1 | 0 | Low-quality | Genome-fragment | 2.76 | AAI-based (high-confidence) | 0 |
| S14_vs_k141_24264||full | 1125 | No | NA | 2 | 0 | 0 | Low-quality | Genome-fragment | 2.14 | AAI-based (medium-confidence) | 0 |
| S14_vs_k141_29137||full | 1125 | No | NA | 4 | 0 | 0 | Low-quality | Genome-fragment | 2.63 | AAI-based (medium-confidence) | 0 |
| S3_extracted_k141_94321 | 1125 | No | NA | 2 | 0 | 0 | Low-quality | Genome-fragment | 2.68 | AAI-based (high-confidence) | 0 |
| S5_extracted_k141_109354 | 1125 | No | NA | 1 | 0 | 1 | Low-quality | Genome-fragment | 0.35 | AAI-based (medium-confidence) | 0 |
| S5_vs_k141_73531||full | 1125 | No | NA | 3 | 1 | 0 | Low-quality | Genome-fragment | 2.46 | AAI-based (medium-confidence) | 0 |
| S7_vs_46212||full | 1125 | No | NA | 3 | 1 | 0 | Low-quality | Genome-fragment | 3.21 | AAI-based (high-confidence) | 0 |
| S16_vs_16529||full | 1125 | No | NA | 2 | 0 | 0 | Low-quality | Genome-fragment | 2.25 | AAI-based (medium-confidence) | 0 |
| S9_vs_k141_9442||full | 1218 | No | NA | 2 | 0 | 1 | Not-determined | Genome-fragment | NA | NA | 0 |
| S16_vs_158820||full | 1125 | No | NA | 3 | 0 | 0 | Low-quality | Genome-fragment | 2.68 | AAI-based (high-confidence) | 0 |
| S9_vs_k141_58596||full | 1859 | No | NA | 2 | 0 | 1 | Not-determined | Genome-fragment | NA | NA | 0 |
| S10_extracted_41297 | 1125 | No | NA | 2 | 0 | 0 | Low-quality | Genome-fragment | 2.51 | AAI-based (medium-confidence) | 0 |
| S10_vs_1314||lt2gene | 1125 | No | NA | 2 | 1 | 0 | Low-quality | Genome-fragment | 2.46 | AAI-based (medium-confidence) | 0 |
| S6_vs_k141_13941||full | 1125 | No | NA | 4 | 2 | 0 | Low-quality | Genome-fragment | 2.61 | AAI-based (medium-confidence) | 0 |
| S17_vs_k141_111698||full | 1126 | No | NA | 2 | 0 | 1 | Low-quality | Genome-fragment | 0.87 | AAI-based (medium-confidence) | 0 |
| S9_vs_k141_88997||full | 2085 | No | NA | 3 | 0 | 2 | Not-determined | Genome-fragment | NA | NA | 0 |
| S17_vs_k141_151045||lt2gene | 1126 | No | NA | 1 | 1 | 0 | Low-quality | Genome-fragment | 2.97 | AAI-based (medium-confidence) | 0 |
| S11_vs_k141_95814||full | 1126 | No | NA | 3 | 1 | 0 | Low-quality | Genome-fragment | 2.71 | AAI-based (medium-confidence) | 0 |
| S9_vs_k141_66152||full | 3204 | No | NA | 3 | 0 | 1 | Not-determined | Genome-fragment | NA | NA | 0 |
| S9_vs_k141_17578||full | 1803 | No | NA | 4 | 0 | 0 | Not-determined | Genome-fragment | NA | NA | 0 |
| S13_vs_138363||full | 1126 | No | NA | 2 | 0 | 0 | Low-quality | Genome-fragment | 1.33 | AAI-based (medium-confidence) | 0 |
| S9_vs_k141_13487||full | 1165 | No | NA | 2 | 0 | 0 | Not-determined | Genome-fragment | NA | NA | 0 |
| S9_vs_k141_36290||full | 1556 | No | NA | 3 | 0 | 0 | Not-determined | Genome-fragment | NA | NA | 0 |
| S14_vs_k141_30519||full | 1126 | No | NA | 2 | 1 | 0 | Low-quality | Genome-fragment | 1.8 | HMM-based (lower-bound) | 0 |
| S9_vs_k141_28749||full | 2895 | No | NA | 4 | 0 | 3 | Not-determined | Genome-fragment | NA | NA | 0 |
| S9_vs_k141_9508||full | 1892 | No | NA | 3 | 0 | 2 | Not-determined | Genome-fragment | NA | NA | 0 |
| S1_vs_k141_12021||full | 1126 | No | NA | 3 | 0 | 0 | Low-quality | Genome-fragment | 2.94 | AAI-based (high-confidence) | 0 |
| S9_vs_k141_21058||full | 1246 | No | NA | 3 | 0 | 1 | Not-determined | Genome-fragment | NA | NA | 0 |
| S3_vs_k141_82536||full | 1126 | No | NA | 3 | 2 | 0 | Low-quality | Genome-fragment | 0.86 | AAI-based (high-confidence) | 0 |
| S3_vs_k141_7149||full | 1126 | No | NA | 4 | 0 | 0 | Low-quality | Genome-fragment | 2.56 | AAI-based (medium-confidence) | 0 |
| S5_extracted_k141_12803 | 1126 | No | NA | 3 | 0 | 0 | Low-quality | Genome-fragment | 2.95 | AAI-based (high-confidence) | 0 |
| S9_vs_k141_32588||full | 54215 | No | NA | 67 | 27 | 1 | Medium-quality | Genome-fragment | 84.63 | AAI-based (high-confidence) | 0 |
| S5_vs_k141_138904||full | 1126 | No | NA | 2 | 0 | 0 | Low-quality | Genome-fragment | 1.95 | AAI-based (high-confidence) | 0 |
| S16_vs_141421||lt2gene | 1126 | No | NA | 2 | 1 | 0 | Low-quality | Genome-fragment | 3.23 | AAI-based (medium-confidence) | 0 |
| S9_vs_k141_81572||full | 1019 | No | NA | 2 | 0 | 1 | Not-determined | Genome-fragment | NA | NA | 0 |
| S12_vs_k141_142803||full | 1126 | No | NA | 2 | 0 | 0 | Low-quality | Genome-fragment | 1.65 | AAI-based (medium-confidence) | 0 |
| S9_vs_k141_44103||full | 2280 | No | NA | 6 | 0 | 0 | Not-determined | Genome-fragment | NA | NA | 0 |
| S9_vs_k141_89083||full | 7414 | No | NA | 6 | 0 | 4 | Not-determined | Genome-fragment | NA | NA | 0 |
| S9_vs_k141_36392||full | 5703 | No | NA | 8 | 0 | 2 | Not-determined | Genome-fragment | NA | NA | 0 |
| S9_vs_k141_2231||full | 1421 | No | NA | 3 | 0 | 1 | Not-determined | Genome-fragment | NA | NA | 0 |
| S12_vs_k141_168638||full | 1126 | No | NA | 3 | 0 | 0 | Low-quality | Genome-fragment | 3.07 | AAI-based (medium-confidence) | 0 |
| S9_vs_k141_2235||full | 1239 | No | NA | 3 | 0 | 2 | Not-determined | Genome-fragment | NA | NA | 0 |
| S9_vs_k141_62746||full | 4484 | No | NA | 2 | 0 | 2 | Not-determined | Genome-fragment | NA | NA | 0 |
| S4_extracted_8746 | 1126 | No | NA | 2 | 0 | 0 | Low-quality | Genome-fragment | 2.9 | AAI-based (medium-confidence) | 0 |
| S9_vs_k141_81633||full | 1762 | No | NA | 2 | 0 | 2 | Not-determined | Genome-fragment | NA | NA | 0 |
| S8_vs_k141_201210||full | 1126 | No | NA | 3 | 2 | 0 | Low-quality | Genome-fragment | 3.28 | AAI-based (medium-confidence) | 0 |
| S17_extracted_k141_52658 | 1127 | No | NA | 1 | 1 | 0 | Low-quality | Genome-fragment | 3.61 | AAI-based (high-confidence) | 0 |
| S17_vs_k141_13519||full | 1127 | No | NA | 4 | 0 | 2 | Low-quality | Genome-fragment | 2.52 | AAI-based (high-confidence) | 0 |
| S9_vs_k141_8020||full | 1127 | No | NA | 3 | 0 | 0 | Low-quality | Genome-fragment | 1.34 | AAI-based (high-confidence) | 0 |
| S9_vs_k141_73990||full | 2245 | No | NA | 3 | 0 | 2 | Not-determined | Genome-fragment | NA | NA | 0 |
| S11_extracted_k141_87614 | 1127 | No | NA | 3 | 0 | 0 | Low-quality | Genome-fragment | 1.86 | AAI-based (medium-confidence) | 0 |
| S9_vs_k141_85272||full | 1760 | No | NA | 3 | 0 | 0 | Not-determined | Genome-fragment | NA | NA | 0 |
| S11_vs_k141_9702||full | 1127 | No | NA | 3 | 1 | 0 | Low-quality | Genome-fragment | 3.3 | AAI-based (high-confidence) | 0 |
| S1_extracted_k141_113854 | 1127 | No | NA | 2 | 1 | 0 | Low-quality | Genome-fragment | 1.75 | AAI-based (high-confidence) | 0 |
| S9_vs_k141_9612||full | 1504 | No | NA | 2 | 0 | 0 | Not-determined | Genome-fragment | NA | NA | 0 |
| S1_extracted_k141_75703 | 1127 | No | NA | 1 | 0 | 0 | Low-quality | Genome-fragment | 0.56 | AAI-based (high-confidence) | 0 |
| S9_vs_k141_40397||full | 2863 | No | NA | 3 | 0 | 0 | Not-determined | Genome-fragment | NA | NA | 0 |
| S1_extracted_k141_30075 | 1127 | No | NA | 3 | 0 | 0 | Low-quality | Genome-fragment | 1.91 | AAI-based (high-confidence) | 0 |
| S1_extracted_k141_106666 | 1127 | No | NA | 5 | 0 | 0 | Low-quality | Genome-fragment | 2.38 | AAI-based (high-confidence) | 0 |
| S3_extracted_k141_102509 | 1127 | No | NA | 1 | 0 | 1 | Low-quality | Genome-fragment | 0.44 | AAI-based (high-confidence) | 0 |
| S5_vs_k141_142054||full | 1127 | No | NA | 3 | 2 | 0 | Low-quality | Genome-fragment | 3.71 | AAI-based (high-confidence) | 0 |
| S5_vs_k141_32237||lt2gene | 1127 | No | NA | 2 | 2 | 0 | Low-quality | Genome-fragment | 1.92 | AAI-based (medium-confidence) | 0 |
| S7_vs_8367||full | 1127 | No | NA | 3 | 0 | 1 | Low-quality | Genome-fragment | 0.37 | AAI-based (high-confidence) | 0 |
| S16_vs_50226||lt2gene | 1127 | No | NA | 1 | 1 | 0 | Low-quality | Genome-fragment | 0.7 | HMM-based (lower-bound) | 0 |
| S12_vs_k141_116123||full | 1127 | No | NA | 2 | 0 | 0 | Low-quality | Genome-fragment | 2.47 | AAI-based (high-confidence) | 0 |
| S12_vs_k141_155768||full | 1127 | No | NA | 3 | 1 | 0 | Low-quality | Genome-fragment | 2.45 | AAI-based (high-confidence) | 0 |
| S9_vs_k141_21276||full | 6754 | No | NA | 8 | 0 | 0 | Not-determined | Genome-fragment | NA | NA | 0 |
| S9_vs_k141_2346||full | 1720 | No | NA | 2 | 0 | 0 | Not-determined | Genome-fragment | NA | NA | 0 |
| S12_vs_k141_15250||full | 1127 | No | NA | 3 | 2 | 0 | Low-quality | Genome-fragment | 2.43 | AAI-based (medium-confidence) | 0 |
| S9_vs_k141_28986||full | 5065 | No | NA | 5 | 0 | 0 | Not-determined | Genome-fragment | NA | NA | 0 |
| S17_vs_k141_174782||full | 1128 | No | NA | 3 | 0 | 0 | Low-quality | Genome-fragment | 2.67 | AAI-based (high-confidence) | 0 |
| S9_vs_k141_40475||full | 3413 | No | NA | 5 | 0 | 0 | Not-determined | Genome-fragment | NA | NA | 0 |
| S17_vs_k141_14222||full | 1128 | No | NA | 4 | 1 | 0 | Low-quality | Genome-fragment | 1.81 | HMM-based (lower-bound) | 0 |
| S9_vs_k141_55416||full | 1091 | No | NA | 3 | 0 | 0 | Not-determined | Genome-fragment | NA | NA | 0 |
| S14_extracted_k141_15526 | 1128 | No | NA | 2 | 0 | 1 | Low-quality | Genome-fragment | 2.56 | AAI-based (medium-confidence) | 0 |
| S9_vs_k141_55422||full | 2145 | No | NA | 3 | 0 | 3 | Not-determined | Genome-fragment | NA | NA | 0 |
| S9_vs_k141_58805||full | 7465 | No | NA | 9 | 0 | 0 | Not-determined | Genome-fragment | NA | NA | 0 |
| S3_extracted_k141_88101 | 1128 | No | NA | 2 | 0 | 0 | Low-quality | Genome-fragment | 2.28 | AAI-based (medium-confidence) | 0 |
| S5_extracted_k141_35633 | 1128 | No | NA | 2 | 2 | 0 | Low-quality | Genome-fragment | 3.03 | AAI-based (high-confidence) | 0 |
| S9_vs_k141_47405||full | 1555 | No | NA | 4 | 0 | 0 | Not-determined | Genome-fragment | NA | NA | 0 |
| S5_vs_k141_24027||lt2gene | 1128 | No | NA | 2 | 2 | 0 | Low-quality | Genome-fragment | 2.33 | AAI-based (medium-confidence) | 0 |
| S7_extracted_137933 | 1128 | No | NA | 1 | 1 | 0 | Low-quality | Genome-fragment | 4.25 | AAI-based (high-confidence) | 0 |
| S9_vs_k141_81734||full | 1253 | No | NA | 2 | 0 | 2 | Not-determined | Genome-fragment | NA | NA | 0 |
| S9_vs_k141_55437||full | 6960 | No | NA | 12 | 0 | 2 | Not-determined | Genome-fragment | NA | NA | 0 |
| S16_extracted_91343 | 1128 | No | NA | 3 | 1 | 0 | Low-quality | Genome-fragment | 0.94 | AAI-based (high-confidence) | 0 |
| S9_vs_k141_21334||full | 2389 | No | NA | 3 | 0 | 1 | Not-determined | Genome-fragment | NA | NA | 0 |
| S16_vs_8061||full | 1128 | No | NA | 4 | 2 | 0 | Low-quality | Genome-fragment | 2.65 | AAI-based (medium-confidence) | 0 |
| S9_vs_k141_6187||full | 3967 | No | NA | 2 | 0 | 1 | Not-determined | Genome-fragment | NA | NA | 0 |
| S16_vs_169690||full | 1128 | No | NA | 4 | 0 | 0 | Low-quality | Genome-fragment | 3 | AAI-based (medium-confidence) | 0 |
| S9_vs_k141_62803||full | 35651 | No | NA | 53 | 21 | 0 | High-quality | High-quality | 92.44 | AAI-based (high-confidence) | 0 |
| S9_vs_k141_55481||full | 1772 | No | NA | 4 | 0 | 2 | Not-determined | Genome-fragment | NA | NA | 0 |
| S16_vs_138041||lt2gene | 1128 | No | NA | 2 | 2 | 0 | Low-quality | Genome-fragment | 2.89 | AAI-based (high-confidence) | 0 |
| S10_vs_6587||full | 1128 | No | NA | 3 | 0 | 0 | Low-quality | Genome-fragment | 2.88 | AAI-based (medium-confidence) | 0 |
| S9_vs_k141_36646||full | 1075 | No | NA | 2 | 0 | 1 | Not-determined | Genome-fragment | NA | NA | 0 |
| S12_extracted_k141_96113 | 1128 | No | NA | 3 | 2 | 0 | Low-quality | Genome-fragment | 1.02 | AAI-based (high-confidence) | 0 |
| S9_vs_k141_62810||full | 8469 | No | NA | 11 | 0 | 1 | Not-determined | Genome-fragment | NA | NA | 0 |
| S8_vs_k141_33646||full | 1128 | No | NA | 2 | 0 | 0 | Low-quality | Genome-fragment | 3.16 | AAI-based (medium-confidence) | 0 |
| S17_vs_k141_155591||lt2gene | 1129 | No | NA | 1 | 1 | 0 | Low-quality | Genome-fragment | 3.3 | AAI-based (high-confidence) | 0 |
| S9_extracted_k141_67775 | 1129 | No | NA | 2 | 0 | 1 | Low-quality | Genome-fragment | 0.43 | AAI-based (medium-confidence) | 0 |
| S9_vs_k141_62818||full | 1541 | No | NA | 4 | 0 | 0 | Not-determined | Genome-fragment | NA | NA | 0 |
| S9_vs_k141_74184||full | 2382 | No | NA | 3 | 0 | 2 | Not-determined | Genome-fragment | NA | NA | 0 |
| S9_vs_k141_87652||lt2gene | 1129 | No | NA | 1 | 1 | 0 | Low-quality | Genome-fragment | 2.41 | AAI-based (high-confidence) | 0 |
| S11_vs_k141_33782||lt2gene | 1129 | No | NA | 1 | 1 | 0 | Low-quality | Genome-fragment | 3.49 | AAI-based (medium-confidence) | 0 |
| S14_extracted_k141_34036 | 1129 | No | NA | 3 | 0 | 0 | Low-quality | Genome-fragment | 1.71 | AAI-based (medium-confidence) | 0 |
| S1_extracted_k141_28410 | 1129 | No | NA | 2 | 0 | 0 | Low-quality | Genome-fragment | 2.03 | AAI-based (medium-confidence) | 0 |
| S9_vs_k141_55518||full | 4802 | No | NA | 4 | 0 | 4 | Not-determined | Genome-fragment | NA | NA | 0 |
| S9_vs_k141_66463||full | 1371 | No | NA | 2 | 0 | 1 | Not-determined | Genome-fragment | NA | NA | 0 |
| S9_vs_k141_77579||full | 1799 | No | NA | 2 | 0 | 2 | Not-determined | Genome-fragment | NA | NA | 0 |
| S9_vs_k141_85472||full | 1704 | No | NA | 2 | 0 | 2 | Not-determined | Genome-fragment | NA | NA | 0 |
| S1_vs_k141_157935||lt2gene | 1129 | No | NA | 1 | 1 | 0 | Low-quality | Genome-fragment | 2.84 | AAI-based (high-confidence) | 0 |
| S9_vs_k141_55540||full | 2285 | No | NA | 6 | 0 | 2 | Not-determined | Genome-fragment | NA | NA | 0 |
| S9_vs_k141_74227||full | 6647 | No | NA | 8 | 0 | 3 | Not-determined | Genome-fragment | NA | NA | 0 |
| S3_extracted_k141_97477 | 1129 | No | NA | 1 | 1 | 0 | Low-quality | Genome-fragment | 1.87 | AAI-based (high-confidence) | 0 |
| S5_extracted_k141_122939 | 1129 | No | NA | 2 | 0 | 0 | Low-quality | Genome-fragment | 2.3 | AAI-based (high-confidence) | 0 |
| S5_vs_k141_19878||full | 1129 | No | NA | 2 | 0 | 1 | Low-quality | Genome-fragment | 1.72 | AAI-based (high-confidence) | 0 |
| S9_vs_k141_32768||full | 2633 | No | NA | 3 | 0 | 0 | Not-determined | Genome-fragment | NA | NA | 0 |
| S9_vs_k141_44331||full | 29758 | No | NA | 48 | 18 | 1 | Medium-quality | Genome-fragment | 71.77 | AAI-based (high-confidence) | 0 |
| S5_vs_k141_9143||full | 1129 | No | NA | 3 | 0 | 0 | Low-quality | Genome-fragment | 2.37 | AAI-based (medium-confidence) | 0 |
| S16_vs_131016||lt2gene | 1129 | No | NA | 1 | 0 | 0 | Low-quality | Genome-fragment | 2.67 | AAI-based (high-confidence) | 0 |
| S10_vs_65988||full | 1129 | No | NA | 3 | 0 | 1 | Low-quality | Genome-fragment | 2.76 | AAI-based (medium-confidence) | 0 |
| S9_vs_k141_77604||full | 1279 | No | NA | 2 | 0 | 0 | Not-determined | Genome-fragment | NA | NA | 0 |
| S10_vs_633||lt2gene | 1129 | No | NA | 2 | 1 | 0 | Low-quality | Genome-fragment | 2.04 | AAI-based (medium-confidence) | 0 |
| S10_vs_45215||lt2gene | 1129 | No | NA | 1 | 1 | 0 | Low-quality | Genome-fragment | 2.62 | AAI-based (high-confidence) | 0 |
| S12_vs_k141_88623||full | 1129 | No | NA | 4 | 0 | 0 | Low-quality | Genome-fragment | 2.62 | AAI-based (high-confidence) | 0 |
| S12_vs_k141_94397||lt2gene | 1129 | No | NA | 2 | 1 | 0 | Low-quality | Genome-fragment | 1.23 | HMM-based (lower-bound) | 0 |
| S15_vs_75206||full | 1129 | No | NA | 3 | 1 | 0 | Low-quality | Genome-fragment | 3.87 | HMM-based (lower-bound) | 0 |
| S8_vs_k141_104888||full | 1129 | No | NA | 3 | 1 | 0 | Low-quality | Genome-fragment | 1.39 | HMM-based (lower-bound) | 0 |
| S8_vs_k141_93129||lt2gene | 1129 | No | NA | 2 | 1 | 0 | Low-quality | Genome-fragment | 2.93 | AAI-based (medium-confidence) | 0 |
| S9_extracted_k141_86591 | 1130 | No | NA | 3 | 2 | 0 | Low-quality | Genome-fragment | 3.73 | AAI-based (high-confidence) | 0 |
| S13_extracted_32821 | 1130 | No | NA | 1 | 0 | 1 | Low-quality | Genome-fragment | 1.91 | AAI-based (high-confidence) | 0 |
| S9_vs_k141_50979||full | 1788 | No | NA | 4 | 0 | 2 | Not-determined | Genome-fragment | NA | NA | 0 |
| S9_vs_k141_40606||full | 1481 | No | NA | 6 | 0 | 4 | Not-determined | Genome-fragment | NA | NA | 0 |
| S9_vs_k141_17865||full | 8606 | No | NA | 8 | 0 | 6 | Not-determined | Genome-fragment | NA | NA | 0 |
| S9_vs_k141_77639||full | 3496 | No | NA | 6 | 0 | 4 | Not-determined | Genome-fragment | NA | NA | 0 |
| S9_vs_k141_59008||full | 1502 | No | NA | 5 | 0 | 2 | Not-determined | Genome-fragment | NA | NA | 0 |
| S9_vs_k141_29176||full | 2685 | No | NA | 4 | 0 | 3 | Not-determined | Genome-fragment | NA | NA | 0 |
| S13_vs_123526||full | 1130 | No | NA | 3 | 2 | 0 | Low-quality | Genome-fragment | 2.4 | AAI-based (medium-confidence) | 0 |
| S9_vs_k141_89205||full | 2853 | No | NA | 3 | 0 | 2 | Not-determined | Genome-fragment | NA | NA | 0 |
| S9_vs_k141_77658||full | 9388 | No | NA | 6 | 0 | 1 | Not-determined | Genome-fragment | NA | NA | 0 |
| S9_vs_k141_47631||full | 1754 | No | NA | 3 | 0 | 0 | Not-determined | Genome-fragment | NA | NA | 0 |
| S5_vs_k141_5904||full | 1130 | No | NA | 4 | 1 | 0 | Low-quality | Genome-fragment | 2.55 | AAI-based (medium-confidence) | 0 |
| S9_vs_k141_9939||full | 1994 | No | NA | 3 | 0 | 1 | Not-determined | Genome-fragment | NA | NA | 0 |
| S9_vs_k141_74250||full | 1012 | No | NA | 3 | 0 | 1 | Not-determined | Genome-fragment | NA | NA | 0 |

| | | | | | | | | | | | |
|---|---|---|---|---|---|---|---|---|---|---|---|
| S7_vs_25330\|\|full | 1130 | No | NA | 4 | 0 | 1 | Low-quality | Genome-fragment | 2.33 | AAI-based (medium-confidence) | 0 |
| S9_vs_k141_47634\|\|full | 1683 | No | NA | 3 | 0 | 0 | Not-determined | Genome-fragment | NA | NA | 0 |
| S2_vs_k141_18296\|\|full | 1130 | No | NA | 3 | 1 | 0 | Low-quality | Genome-fragment | 1.11 | AAI-based (high-confidence) | 0 |
| S9_vs_k141_2600\|\|full | 3220 | No | NA | 2 | 0 | 1 | Not-determined | Genome-fragment | NA | NA | 0 |
| S2_vs_k141_17325\|\|full | 1130 | No | NA | 3 | 0 | 0 | Low-quality | Genome-fragment | 2.11 | AAI-based (medium-confidence) | 0 |
| S9_vs_k141_74262\|\|full | 1019 | No | NA | 2 | 0 | 0 | Not-determined | Genome-fragment | NA | NA | 0 |
| S9_vs_k141_85541\|\|full | 9159 | No | NA | 12 | 0 | 9 | Not-determined | Genome-fragment | NA | NA | 0 |
| S8_extracted_k141_47816 | 1130 | No | NA | 1 | 0 | 1 | Low-quality | Genome-fragment | 3.61 | AAI-based (medium-confidence) | 0 |
| S8_vs_k141_96104\|\|full | 1130 | No | NA | 3 | 0 | 1 | Low-quality | Genome-fragment | 1.48 | AAI-based (medium-confidence) | 0 |
| S9_vs_k141_44434\|\|full | 1117 | No | NA | 2 | 0 | 0 | Not-determined | Genome-fragment | NA | NA | 0 |
| S9_vs_k141_70504\|\|lt2gene | 1131 | No | NA | 1 | 1 | 0 | Low-quality | Genome-fragment | 2.6 | AAI-based (high-confidence) | 0 |
| S9_vs_k141_81934\|\|full | 3894 | No | NA | 5 | 0 | 4 | Not-determined | Genome-fragment | NA | NA | 0 |
| S9_vs_k141_67723\|\|lt2gene | 1131 | No | NA | 1 | 1 | 0 | Low-quality | Genome-fragment | 1.78 | HMM-based (lower-bound) | 0 |
| S11_vs_k141_93520\|\|full | 1131 | No | NA | 4 | 1 | 0 | Low-quality | Genome-fragment | 2.01 | AAI-based (high-confidence) | 0 |
| S9_vs_k141_17948\|\|full | 1377 | No | NA | 3 | 0 | 1 | Not-determined | Genome-fragment | NA | NA | 0 |
| S9_vs_k141_10015\|\|full | 1215 | No | NA | 2 | 0 | 0 | Not-determined | Genome-fragment | NA | NA | 0 |
| S9_vs_k141_59105\|\|full | 3312 | No | NA | 3 | 0 | 2 | Not-determined | Genome-fragment | NA | NA | 0 |
| S11_vs_k141_58881\|\|lt2gene | 1131 | No | NA | 1 | 1 | 0 | Low-quality | Genome-fragment | 1.99 | AAI-based (medium-confidence) | 0 |
| S9_vs_k141_13727\|\|full | 1944 | No | NA | 2 | 0 | 2 | Not-determined | Genome-fragment | NA | NA | 0 |
| S9_vs_k141_74336\|\|full | 1776 | No | NA | 4 | 0 | 1 | Not-determined | Genome-fragment | NA | NA | 0 |
| S13_extracted_69943 | 1131 | No | NA | 2 | 0 | 2 | Low-quality | Genome-fragment | 0.54 | AAI-based (medium-confidence) | 0 |
| S3_vs_k141_90406\|\|full | 1131 | No | NA | 4 | 0 | 0 | Low-quality | Genome-fragment | 2.56 | AAI-based (high-confidence) | 0 |
| S5_extracted_k141_117369 | 1131 | No | NA | 3 | 2 | 0 | Low-quality | Genome-fragment | 5.16 | AAI-based (high-confidence) | 0 |
| S5_vs_k141_141546\|\|full | 1131 | No | NA | 3 | 0 | 0 | Low-quality | Genome-fragment | 0.65 | AAI-based (medium-confidence) | 0 |
| S9_vs_k141_32888\|\|full | 4622 | No | NA | 4 | 0 | 2 | Not-determined | Genome-fragment | NA | NA | 0 |
| S9_vs_k141_10064\|\|full | 1191 | No | NA | 2 | 0 | 1 | Not-determined | Genome-fragment | NA | NA | 0 |
| S9_vs_k141_17983\|\|full | 1848 | No | NA | 5 | 0 | 0 | Not-determined | Genome-fragment | NA | NA | 0 |
| S5_vs_k141_3366\|\|full | 1131 | No | NA | 3 | 3 | 0 | Low-quality | Genome-fragment | 2.57 | AAI-based (high-confidence) | 0 |
| S5_vs_k141_22754\|\|lt2gene | 1131 | No | NA | 2 | 1 | 0 | Low-quality | Genome-fragment | 0.65 | HMM-based (lower-bound) | 0 |
| S16_extracted_51965 | 1131 | No | NA | 4 | 0 | 0 | Low-quality | Genome-fragment | 1.04 | AAI-based (high-confidence) | 0 |
| S12_vs_k141_21127\|\|lt2gene | 1131 | No | NA | 2 | 2 | 0 | Low-quality | Genome-fragment | 1.37 | HMM-based (lower-bound) | 0 |
| S6_extracted_k141_45552 | 1131 | No | NA | 3 | 1 | 0 | Low-quality | Genome-fragment | 2.51 | AAI-based (medium-confidence) | 0 |
| S8_extracted_k141_18075 | 1131 | No | NA | 3 | 0 | 0 | Low-quality | Genome-fragment | 2.98 | AAI-based (medium-confidence) | 0 |
| S8_extracted_k141_54877 | 1131 | No | NA | 2 | 0 | 0 | Low-quality | Genome-fragment | 2.13 | AAI-based (medium-confidence) | 0 |
| S8_extracted_k141_136583 | 1131 | No | NA | 2 | 0 | 0 | Low-quality | Genome-fragment | 1.76 | AAI-based (medium-confidence) | 0 |
| S9_vs_k141_32919\|\|full | 1773 | No | NA | 3 | 0 | 1 | Not-determined | Genome-fragment | NA | NA | 0 |
| S9_vs_k141_70348\|\|full | 2120 | No | NA | 3 | 0 | 0 | Not-determined | Genome-fragment | NA | NA | 0 |
| S9_vs_k141_63089\|\|full | 1811 | No | NA | 2 | 0 | 0 | Not-determined | Genome-fragment | NA | NA | 0 |
| S8_extracted_k141_68324 | 1131 | No | NA | 2 | 0 | 0 | Low-quality | Genome-fragment | 2.33 | AAI-based (medium-confidence) | 0 |
| S9_vs_k141_66635\|\|full | 1560 | No | NA | 3 | 0 | 1 | Not-determined | Genome-fragment | NA | NA | 0 |
| S8_vs_k141_253\|\|full | 1131 | No | NA | 3 | 0 | 0 | Low-quality | Genome-fragment | 1.78 | AAI-based (high-confidence) | 0 |
| S9_vs_k141_18020\|\|full | 1251 | No | NA | 2 | 0 | 0 | Not-determined | Genome-fragment | NA | NA | 0 |
| S9_vs_k141_29319\|\|full | 1397 | No | NA | 2 | 0 | 0 | Not-determined | Genome-fragment | NA | NA | 0 |
| S17_extracted_k141_16774 | 1132 | No | NA | 4 | 1 | 0 | Low-quality | Genome-fragment | 2.7 | AAI-based (high-confidence) | 0 |
| S17_extracted_k141_83384 | 1132 | No | NA | 2 | 0 | 0 | Low-quality | Genome-fragment | 0.92 | AAI-based (medium-confidence) | 0 |
| S9_vs_k141_89328\|\|full | 1543 | No | NA | 2 | 0 | 0 | Not-determined | Genome-fragment | NA | NA | 0 |
| S17_extracted_k141_113570 | 1132 | No | NA | 1 | 0 | 1 | Low-quality | Genome-fragment | 0.5 | AAI-based (medium-confidence) | 0 |
| S17_vs_k141_125948\|\|lt2gene | 1132 | No | NA | 2 | 1 | 0 | Low-quality | Genome-fragment | 3.08 | AAI-based (high-confidence) | 0 |
| S9_vs_k141_10117\|\|full | 1303 | No | NA | 3 | 0 | 0 | Not-determined | Genome-fragment | NA | NA | 0 |
| S9_vs_k141_2713\|\|full | 12132 | No | NA | 13 | 0 | 5 | Not-determined | Genome-fragment | NA | NA | 0 |
| S9_vs_k141_10119\|\|full | 1021 | No | NA | 3 | 0 | 1 | Not-determined | Genome-fragment | NA | NA | 0 |
| S9_vs_k141_21713\|\|full | 2116 | No | NA | 2 | 0 | 2 | Not-determined | Genome-fragment | NA | NA | 0 |
| S9_vs_k141_13824\|\|full | 1070 | No | NA | 2 | 0 | 0 | Not-determined | Genome-fragment | NA | NA | 0 |
| S9_vs_k141_63127\|\|full | 5787 | No | NA | 7 | 0 | 3 | Not-determined | Genome-fragment | NA | NA | 0 |
| S11_vs_k141_69975\|\|lt2gene | 1132 | No | NA | 1 | 1 | 0 | Low-quality | Genome-fragment | 1.98 | HMM-based (lower-bound) | 0 |
| S9_vs_k141_77830\|\|full | 8617 | No | NA | 13 | 0 | 2 | Not-determined | Genome-fragment | NA | NA | 0 |
| S14_extracted_k141_103280 | 1132 | No | NA | 3 | 3 | 0 | Low-quality | Genome-fragment | 2.83 | AAI-based (high-confidence) | 0 |
| S9_vs_k141_55738\|\|full | 43096 | No | NA | 64 | 18 | 1 | Medium-quality | Genome-fragment | 80.9 | AAI-based (high-confidence) | 0 |
| S9_vs_k141_6555\|\|full | 1165 | No | NA | 4 | 0 | 0 | Not-determined | Genome-fragment | NA | NA | 0 |
| S9_vs_k141_77844\|\|full | 1912 | No | NA | 2 | 0 | 1 | Not-determined | Genome-fragment | NA | NA | 0 |
| S14_vs_k141_1043\|\|full | 1132 | No | NA | 3 | 0 | 0 | Low-quality | Genome-fragment | 2.19 | AAI-based (high-confidence) | 0 |
| S1_vs_k141_77820\|\|full | 1132 | No | NA | 3 | 0 | 0 | Low-quality | Genome-fragment | 4.5 | AAI-based (high-confidence) | 0 |
| S1_vs_k141_25683\|\|full | 1132 | No | NA | 4 | 0 | 0 | Low-quality | Genome-fragment | 0.78 | AAI-based (high-confidence) | 0 |
| S7_vs_126972\|\|lt2gene | 1132 | No | NA | 1 | 1 | 0 | Low-quality | Genome-fragment | 3.3 | AAI-based (high-confidence) | 0 |
| S16_vs_58698\|\|full | 1132 | No | NA | 2 | 0 | 0 | Low-quality | Genome-fragment | 1.21 | AAI-based (medium-confidence) | 0 |
| S16_vs_153118\|\|full | 1132 | No | NA | 3 | 1 | 0 | Low-quality | Genome-fragment | 2.45 | AAI-based (medium-confidence) | 0 |
| S9_extracted_k141_75258 | 1133 | No | NA | 2 | 0 | 1 | Low-quality | Genome-fragment | 1.55 | AAI-based (medium-confidence) | 0 |
| S9_vs_k141_18113\|\|full | 1934 | No | NA | 3 | 0 | 3 | Not-determined | Genome-fragment | NA | NA | 0 |
| S9_vs_k141_88190\|\|lt2gene | 1133 | No | NA | 2 | 1 | 0 | Low-quality | Genome-fragment | 3.15 | AAI-based (medium-confidence) | 0 |
| S11_extracted_k141_5403 | 1133 | No | NA | 4 | 0 | 0 | Low-quality | Genome-fragment | 3.89 | AAI-based (medium-confidence) | 0 |
| S9_vs_k141_13880\|\|full | 2243 | No | NA | 2 | 0 | 2 | Not-determined | Genome-fragment | NA | NA | 0 |
| S13_vs_81330\|\|full | 1133 | No | NA | 3 | 1 | 0 | Low-quality | Genome-fragment | 1.94 | AAI-based (high-confidence) | 0 |
| S9_vs_k141_37010\|\|full | 1330 | No | NA | 3 | 0 | 0 | Not-determined | Genome-fragment | NA | NA | 0 |
| S9_vs_k141_47813\|\|full | 3961 | No | NA | 4 | 0 | 2 | Not-determined | Genome-fragment | NA | NA | 0 |
| S3_extracted_k141_104139 | 1133 | No | NA | 2 | 1 | 0 | Low-quality | Genome-fragment | 3.35 | AAI-based (high-confidence) | 0 |
| S3_vs_k141_110177\|\|full | 1133 | No | NA | 5 | 0 | 0 | Low-quality | Genome-fragment | 3.22 | AAI-based (medium-confidence) | 0 |
| S5_extracted_k141_20921 | 1133 | No | NA | 1 | 0 | 0 | Low-quality | Genome-fragment | 0.91 | AAI-based (high-confidence) | 0 |
| S5_vs_k141_117424\|\|full | 1133 | No | NA | 3 | 0 | 1 | Low-quality | Genome-fragment | 2.51 | AAI-based (high-confidence) | 0 |
| S9_vs_k141_6633\|\|full | 1443 | No | NA | 3 | 0 | 0 | Not-determined | Genome-fragment | NA | NA | 0 |
| S5_vs_k141_84609\|\|full | 1133 | No | NA | 2 | 1 | 0 | Low-quality | Genome-fragment | 2.22 | AAI-based (high-confidence) | 0 |
| S12_vs_k141_6706\|\|full | 1133 | No | NA | 4 | 0 | 0 | Low-quality | Genome-fragment | 3.04 | AAI-based (high-confidence) | 0 |
| S9_vs_k141_21806\|\|full | 1579 | No | NA | 4 | 0 | 1 | Not-determined | Genome-fragment | NA | NA | 0 |
| S12_vs_k141_23078\|\|full | 1133 | No | NA | 1 | 0 | 0 | Low-quality | Genome-fragment | 1.78 | AAI-based (medium-confidence) | 0 |
| S15_vs_69194\|\|full | 1133 | No | NA | 2 | 0 | 1 | Low-quality | Genome-fragment | 3.62 | AAI-based (high-confidence) | 0 |
| S8_vs_k141_187155\|\|full | 1133 | No | NA | 4 | 1 | 0 | Low-quality | Genome-fragment | 2.28 | AAI-based (medium-confidence) | 0 |
| S9_vs_k141_10243\|\|full | 2339 | No | NA | 3 | 0 | 0 | Not-determined | Genome-fragment | NA | NA | 0 |
| S9_vs_k141_10246\|\|full | 2410 | No | NA | 5 | 0 | 3 | Not-determined | Genome-fragment | NA | NA | 0 |
| S17_extracted_k141_91804 | 1134 | No | NA | 2 | 0 | 2 | Low-quality | Genome-fragment | 2.02 | AAI-based (medium-confidence) | 0 |
| S9_extracted_k141_61570 | 1134 | No | NA | 3 | 3 | 0 | Low-quality | Genome-fragment | 3.74 | AAI-based (high-confidence) | 0 |
| S9_vs_k141_31151\|\|full | 1134 | No | NA | 3 | 1 | 0 | Low-quality | Genome-fragment | 2.18 | AAI-based (high-confidence) | 0 |
| S9_vs_k141_40767\|\|full | 2151 | No | NA | 4 | 0 | 1 | Not-determined | Genome-fragment | NA | NA | 0 |
| S9_vs_k141_21848\|\|full | 1596 | No | NA | 3 | 0 | 0 | Not-determined | Genome-fragment | NA | NA | 0 |
| S9_vs_k141_44251\|\|full | 1134 | No | NA | 3 | 0 | 0 | Low-quality | Genome-fragment | 2.87 | AAI-based (medium-confidence) | 0 |
| S9_vs_k141_33105\|\|full | 1163 | No | NA | 2 | 0 | 0 | Not-determined | Genome-fragment | NA | NA | 0 |
| S13_vs_34092\|\|full | 1134 | No | NA | 3 | 2 | 0 | Low-quality | Genome-fragment | 2.78 | AAI-based (medium-confidence) | 0 |
| S9_vs_k141_40778\|\|full | 1502 | No | NA | 3 | 0 | 0 | Not-determined | Genome-fragment | NA | NA | 0 |
| S9_vs_k141_33115\|\|full | 1258 | No | NA | 3 | 0 | 2 | Not-determined | Genome-fragment | NA | NA | 0 |
| S9_vs_k141_63264\|\|full | 1475 | No | NA | 3 | 0 | 1 | Not-determined | Genome-fragment | NA | NA | 0 |
| S1_vs_k141_121762\|\|full | 1134 | No | NA | 5 | 0 | 0 | Low-quality | Genome-fragment | 2.48 | AAI-based (high-confidence) | 0 |
| S1_vs_k141_154910\|\|lt2gene | 1134 | No | NA | 2 | 1 | 0 | Low-quality | Genome-fragment | 1.76 | AAI-based (high-confidence) | 0 |
| S3_vs_k141_19074\|\|full | 1134 | No | NA | 2 | 0 | 0 | Low-quality | Genome-fragment | 2.7 | AAI-based (high-confidence) | 0 |
| S9_vs_k141_2799\|\|full | 60546 | No | NA | 57 | 3 | 33 | High-quality | High-quality | 90 | HMM-based (lower-bound) | 0 |
| S9_vs_k141_44663\|\|full | 1854 | No | NA | 3 | 0 | 1 | Not-determined | Genome-fragment | NA | NA | 0 |
| S5_extracted_k141_40093 | 1134 | No | NA | 3 | 1 | 0 | Low-quality | Genome-fragment | 2.61 | AAI-based (medium-confidence) | 0 |
| S9_vs_k141_21850\|\|full | 37209 | No | NA | 46 | 9 | 2 | Medium-quality | Genome-fragment | 77.19 | AAI-based (medium-confidence) | 0 |
| S10_vs_97719\|\|full | 1134 | No | NA | 4 | 1 | 0 | Low-quality | Genome-fragment | 2.06 | AAI-based (high-confidence) | 0 |
| S9_vs_k141_63304\|\|full | 1015 | No | NA | 3 | 0 | 2 | Not-determined | Genome-fragment | NA | NA | 0 |
| S15_vs_20366\|\|full | 1134 | No | NA | 4 | 1 | 0 | Low-quality | Genome-fragment | 2.22 | AAI-based (high-confidence) | 0 |
| S6_vs_k141_5703\|\|full | 1134 | No | NA | 3 | 0 | 0 | Low-quality | Genome-fragment | 1.83 | AAI-based (medium-confidence) | 0 |
| S8_extracted_k141_112622 | 1134 | No | NA | 4 | 3 | 0 | Low-quality | Genome-fragment | 3.53 | AAI-based (high-confidence) | 0 |
| S17_vs_k141_120018\|\|lt2gene | 1135 | No | NA | 2 | 0 | 0 | Low-quality | Genome-fragment | 3.01 | AAI-based (medium-confidence) | 0 |
| S9_extracted_k141_17626 | 1135 | No | NA | 2 | 1 | 0 | Low-quality | Genome-fragment | 2.78 | AAI-based (high-confidence) | 0 |
| S9_extracted_k141_60322 | 1135 | No | NA | 4 | 2 | 0 | Low-quality | Genome-fragment | 2.08 | AAI-based (high-confidence) | 0 |
| S11_extracted_k141_58025 | 1135 | No | NA | 3 | 0 | 3 | Low-quality | Genome-fragment | 0.33 | AAI-based (high-confidence) | 0 |
| S11_vs_k141_34497\|\|full | 1135 | No | NA | 3 | 0 | 0 | Low-quality | Genome-fragment | 2.21 | AAI-based (medium-confidence) | 0 |
| S3_extracted_k141_79745 | 1135 | No | NA | 1 | 0 | 1 | Low-quality | Genome-fragment | 0.57 | AAI-based (medium-confidence) | 0 |
| S3_vs_k141_55016\|\|full | 1135 | No | NA | 3 | 0 | 0 | Low-quality | Genome-fragment | 2 | AAI-based (medium-confidence) | 0 |
| S9_vs_k141_6726\|\|full | 1601 | No | NA | 4 | 0 | 2 | Not-determined | Genome-fragment | NA | NA | 0 |
| S5_vs_k141_34717\|\|full | 1135 | No | NA | 4 | 1 | 0 | Low-quality | Genome-fragment | 0.58 | AAI-based (medium-confidence) | 0 |
| S7_vs_128173\|\|full | 1135 | No | NA | 3 | 0 | 0 | Low-quality | Genome-fragment | 3.01 | AAI-based (high-confidence) | 0 |
| S9_vs_k141_48006\|\|full | 1970 | No | NA | 3 | 0 | 1 | Not-determined | Genome-fragment | NA | NA | 0 |
| S9_vs_k141_18228\|\|full | 1808 | No | NA | 3 | 0 | 3 | Not-determined | Genome-fragment | NA | NA | 0 |
| S9_vs_k141_70641\|\|full | 1927 | No | NA | 3 | 0 | 1 | Not-determined | Genome-fragment | NA | NA | 0 |
| S16_vs_8265\|\|full | 1135 | No | NA | 1 | 0 | 0 | Low-quality | Genome-fragment | 2.43 | AAI-based (medium-confidence) | 0 |
| S16_vs_63762\|\|full | 1135 | No | NA | 5 | 0 | 0 | Low-quality | Genome-fragment | 2.62 | AAI-based (high-confidence) | 0 |
| S9_vs_k141_85961\|\|full | 2724 | No | NA | 4 | 0 | 2 | Not-determined | Genome-fragment | NA | NA | 0 |
| S16_vs_139405\|\|full | 1135 | No | NA | 3 | 2 | 0 | Low-quality | Genome-fragment | 2.5 | AAI-based (high-confidence) | 0 |
| S9_vs_k141_70662\|\|full | 2372 | No | NA | 4 | 0 | 0 | Not-determined | Genome-fragment | NA | NA | 0 |
| S9_vs_k141_25607\|\|full | 1426 | No | NA | 3 | 0 | 2 | Not-determined | Genome-fragment | NA | NA | 0 |
| S12_extracted_k141_152017 | 1135 | No | NA | 2 | 1 | 0 | Low-quality | Genome-fragment | 2.55 | AAI-based (high-confidence) | 0 |
| S8_extracted_k141_3883 | 1135 | No | NA | 3 | 0 | 2 | Low-quality | Genome-fragment | 0.76 | AAI-based (medium-confidence) | 0 |
| S17_extracted_k141_26566 | 1136 | No | NA | 4 | 3 | 0 | Low-quality | Genome-fragment | 3.74 | AAI-based (high-confidence) | 0 |
| S17_vs_k141_74407\|\|full | 1136 | No | NA | 3 | 0 | 0 | Low-quality | Genome-fragment | 3.01 | AAI-based (high-confidence) | 0 |
| S17_vs_k141_100148\|\|full | 1136 | No | NA | 5 | 2 | 0 | Low-quality | Genome-fragment | 2.74 | AAI-based (high-confidence) | 0 |
| S17_vs_k141_20204\|\|full | 1136 | No | NA | 3 | 0 | 0 | Low-quality | Genome-fragment | 1.16 | AAI-based (high-confidence) | 0 |
| S11_vs_k141_23751\|\|full | 1136 | No | NA | 3 | 0 | 0 | Low-quality | Genome-fragment | 3.3 | AAI-based (high-confidence) | 0 |
| S9_vs_k141_44770\|\|full | 1446 | No | NA | 3 | 0 | 1 | Not-determined | Genome-fragment | NA | NA | 0 |
| S9_vs_k141_48067\|\|full | 1514 | No | NA | 2 | 0 | 2 | Not-determined | Genome-fragment | NA | NA | 0 |
| S9_vs_k141_37191\|\|full | 1543 | No | NA | 3 | 0 | 2 | Not-determined | Genome-fragment | NA | NA | 0 |
| S9_vs_k141_29653\|\|full | 2771 | No | NA | 4 | 0 | 1 | Not-determined | Genome-fragment | NA | NA | 0 |
| S9_vs_k141_59411\|\|full | 2227 | No | NA | 3 | 0 | 3 | Not-determined | Genome-fragment | NA | NA | 0 |
| S13_extracted_149942 | 1136 | No | NA | 2 | 0 | 1 | Low-quality | Genome-fragment | 2.57 | AAI-based (medium-confidence) | 0 |
| S9_vs_k141_10424\|\|full | 23985 | No | NA | 50 | 5 | 3 | Medium-quality | Genome-fragment | 56.13 | AAI-based (high-confidence) | 0 |
| S9_vs_k141_48082\|\|full | 1592 | No | NA | 2 | 0 | 2 | Not-determined | Genome-fragment | NA | NA | 0 |
| S9_vs_k141_37200\|\|full | 7178 | No | NA | 6 | 0 | 4 | Not-determined | Genome-fragment | NA | NA | 0 |
| S9_vs_k141_40959\|\|full | 1827 | No | NA | 4 | 0 | 0 | Not-determined | Genome-fragment | NA | NA | 0 |
| S9_vs_k141_2970\|\|full | 2323 | No | NA | 2 | 0 | 2 | Not-determined | Genome-fragment | NA | NA | 0 |
| S13_vs_136969\|\|full | 1136 | No | NA | 3 | 0 | 1 | Low-quality | Genome-fragment | 0.87 | AAI-based (medium-confidence) | 0 |
| S1_vs_k141_106864\|\|full | 1136 | No | NA | 3 | 1 | 0 | Low-quality | Genome-fragment | 2.08 | HMM-based (lower-bound) | 0 |
| S3_extracted_k141_13677 | 1136 | No | NA | 4 | 0 | 0 | Low-quality | Genome-fragment | 1.9 | AAI-based (medium-confidence) | 0 |
| S5_vs_k141_92314\|\|full | 1136 | No | NA | 2 | 0 | 0 | Low-quality | Genome-fragment | 2.25 | AAI-based (high-confidence) | 0 |
| S5_vs_k141_25687\|\|full | 1136 | No | NA | 2 | 0 | 0 | Low-quality | Genome-fragment | 2.11 | AAI-based (high-confidence) | 0 |
| S7_vs_118689\|\|full | 1136 | No | NA | 2 | 0 | 0 | Low-quality | Genome-fragment | 2.65 | AAI-based (medium-confidence) | 0 |
| S9_vs_k141_25699\|\|full | 4342 | No | NA | 5 | 0 | 1 | Not-determined | Genome-fragment | NA | NA | 0 |
| S7_vs_122559\|\|full | 1136 | No | NA | 2 | 0 | 1 | Low-quality | Genome-fragment | 2.44 | AAI-based (high-confidence) | 0 |
| S9_vs_k141_40992\|\|full | 1427 | No | NA | 3 | 0 | 0 | Not-determined | Genome-fragment | NA | NA | 0 |
| S9_vs_k141_33375\|\|full | 2634 | No | NA | 3 | 0 | 0 | Not-determined | Genome-fragment | NA | NA | 0 |
| S9_vs_k141_56047\|\|full | 1638 | No | NA | 4 | 0 | 3 | Not-determined | Genome-fragment | NA | NA | 0 |
| S9_vs_k141_51348\|\|full | 1318 | No | NA | 2 | 0 | 1 | Not-determined | Genome-fragment | NA | NA | 0 |
| S16_extracted_60206 | 1136 | No | NA | 2 | 1 | 0 | Low-quality | Genome-fragment | 2.95 | AAI-based (high-confidence) | 0 |
| S10_extracted_44082 | 1136 | No | NA | 1 | 0 | 1 | Low-quality | Genome-fragment | 0.73 | AAI-based (high-confidence) | 0 |
| S10_vs_116684\|\|full | 1136 | No | NA | 2 | 1 | 0 | Low-quality | Genome-fragment | 2.09 | AAI-based (high-confidence) | 0 |
| S9_vs_k141_22020\|\|full | 1592 | No | NA | 2 | 0 | 1 | Not-determined | Genome-fragment | NA | NA | 0 |
| S17_extracted_k141_173374 | 1137 | No | NA | 2 | 0 | 1 | Low-quality | Genome-fragment | 0.49 | AAI-based (medium-confidence) | 0 |
| S9_vs_k141_79949\|\|lt2gene | 1137 | No | NA | 2 | 2 | 0 | Low-quality | Genome-fragment | 2.64 | AAI-based (high-confidence) | 0 |
| S9_vs_k141_89698\|\|full | 1683 | No | NA | 3 | 0 | 2 | Not-determined | Genome-fragment | NA | NA | 0 |
| S9_vs_k141_41007\|\|full | 2345 | No | NA | 3 | 1 | 0 | Not-determined | Genome-fragment | NA | NA | 0 |
| S1_extracted_k141_92214 | 1137 | No | NA | 2 | 0 | 1 | Low-quality | Genome-fragment | 6.77 | AAI-based (medium-confidence) | 0 |
| S9_vs_k141_41009\|\|full | 5828 | No | NA | 5 | 0 | 3 | Not-determined | Genome-fragment | NA | NA | 0 |
| S9_vs_k141_56089\|\|full | 2615 | No | NA | 2 | 0 | 2 | Not-determined | Genome-fragment | NA | NA | 0 |
| S3_extracted_k141_106306 | 1137 | No | NA | 1 | 0 | 1 | Low-quality | Genome-fragment | 0.5 | AAI-based (medium-confidence) | 0 |
| S9_vs_k141_59549\|\|full | 1510 | No | NA | 2 | 0 | 1 | Not-determined | Genome-fragment | NA | NA | 0 |
| S9_vs_k141_18424\|\|full | 1805 | No | NA | 3 | 0 | 1 | Not-determined | Genome-fragment | NA | NA | 0 |
| S3_extracted_k141_71125 | 1137 | No | NA | 1 | 0 | 1 | Low-quality | Genome-fragment | 0.65 | AAI-based (high-confidence) | 0 |
| S9_vs_k141_74653\|\|full | 1985 | No | NA | 3 | 0 | 2 | Not-determined | Genome-fragment | NA | NA | 0 |
| S9_vs_k141_63485\|\|full | 37897 | No | NA | 61 | 22 | 2 | High-quality | High-quality | 93.66 | AAI-based (high-confidence) | 0 |
| S3_vs_k141_3351\|\|lt2gene | 1137 | No | NA | 1 | 1 | 0 | Low-quality | Genome-fragment | 2.66 | AAI-based (high-confidence) | 0 |
| S3_vs_k141_44483\|\|lt2gene | 1137 | No | NA | 2 | 1 | 0 | Low-quality | Genome-fragment | 1.79 | HMM-based (lower-bound) | 0 |
| S9_vs_k141_37315\|\|full | 3807 | No | NA | 3 | 0 | 2 | Not-determined | Genome-fragment | NA | NA | 0 |
| S9_vs_k141_22070\|\|full | 3659 | No | NA | 5 | 0 | 1 | Not-determined | Genome-fragment | NA | NA | 0 |
| S9_vs_k141_67052\|\|full | 1923 | No | NA | 2 | 0 | 0 | Not-determined | Genome-fragment | NA | NA | 0 |
| S9_vs_k141_25816\|\|full | 6412 | No | NA | 10 | 0 | 0 | Not-determined | Genome-fragment | NA | NA | 0 |
| S9_vs_k141_14089\|\|full | 2051 | No | NA | 3 | 0 | 0 | Not-determined | Genome-fragment | NA | NA | 0 |
| S9_vs_k141_33457\|\|full | 1798 | No | NA | 3 | 0 | 1 | Not-determined | Genome-fragment | NA | NA | 0 |
| S9_vs_k141_67059\|\|full | 2275 | No | NA | 2 | 0 | 2 | Not-determined | Genome-fragment | NA | NA | 0 |
| S5_extracted_k141_33956 | 1137 | No | NA | 4 | 1 | 0 | Low-quality | Genome-fragment | 3.53 | AAI-based (high-confidence) | 0 |
| S9_vs_k141_56154\|\|full | 1360 | No | NA | 2 | 0 | 1 | Not-determined | Genome-fragment | NA | NA | 0 |
| S9_vs_k141_14098\|\|full | 3743 | No | NA | 4 | 0 | 1 | Not-determined | Genome-fragment | NA | NA | 0 |
| S9_vs_k141_74705\|\|full | 1095 | No | NA | 3 | 0 | 0 | Not-determined | Genome-fragment | NA | NA | 0 |
| S9_vs_k141_89776\|\|full | 4216 | No | NA | 5 | 0 | 1 | Not-determined | Genome-fragment | NA | NA | 0 |
| S5_vs_k141_93278\|\|full | 1137 | No | NA | 4 | 0 | 0 | Low-quality | Genome-fragment | 2.43 | AAI-based (high-confidence) | 0 |
| S7_extracted_125208 | 1137 | No | NA | 3 | 0 | 0 | Low-quality | Genome-fragment | 3.11 | AAI-based (medium-confidence) | 0 |
| S7_vs_53135\|\|full | 1137 | No | NA | 5 | 0 | 0 | Low-quality | Genome-fragment | 1.91 | AAI-based (high-confidence) | 0 |
| S10_vs_52426\|\|full | 1137 | No | NA | 3 | 0 | 0 | Low-quality | Genome-fragment | 2.86 | AAI-based (high-confidence) | 0 |
| S12_extracted_k141_122333 | 1137 | No | NA | 1 | 0 | 1 | Low-quality | Genome-fragment | 1.23 | AAI-based (high-confidence) | 0 |
| S9_vs_k141_25853\|\|full | 2102 | No | NA | 3 | 0 | 1 | Not-determined | Genome-fragment | NA | NA | 0 |
| S9_vs_k141_70838\|\|full | 4529 | No | NA | 6 | 0 | 2 | Not-determined | Genome-fragment | NA | NA | 0 |
| S9_vs_k141_63560\|\|full | 2973 | No | NA | 3 | 0 | 2 | Not-determined | Genome-fragment | NA | NA | 0 |
| S9_vs_k141_56170\|\|full | 2794 | No | NA | 4 | 0 | 3 | Not-determined | Genome-fragment | NA | NA | 0 |
| S9_vs_k141_78246\|\|full | 2400 | No | NA | 2 | 0 | 1 | Not-determined | Genome-fragment | NA | NA | 0 |
| S9_vs_k141_59640\|\|full | 1908 | No | NA | 3 | 0 | 0 | Not-determined | Genome-fragment | NA | NA | 0 |
| S12_vs_k141_52758\|\|full | 1137 | No | NA | 3 | 2 | 0 | Low-quality | Genome-fragment | 1.29 | HMM-based (lower-bound) | 0 |
| S9_vs_k141_86218\|\|full | 4083 | No | NA | 7 | 0 | 7 | Not-determined | Genome-fragment | NA | NA | 0 |
| S9_vs_k141_22123\|\|full | 1458 | No | NA | 2 | 0 | 1 | Not-determined | Genome-fragment | NA | NA | 0 |
| S8_vs_k141_49040\|\|full | 1137 | No | NA | 2 | 0 | 2 | Low-quality | Genome-fragment | 2.39 | AAI-based (medium-confidence) | 0 |
| S9_vs_k141_62172\|\|full | 1138 | No | NA | 3 | 3 | 0 | Low-quality | Genome-fragment | 1.91 | AAI-based (high-confidence) | 0 |
| S11_vs_k141_48803\|\|full | 1138 | No | NA | 3 | 1 | 0 | Low-quality | Genome-fragment | 2.62 | AAI-based (high-confidence) | 0 |
| S11_vs_k141_71244\|\|full | 1138 | No | NA | 2 | 0 | 0 | Low-quality | Genome-fragment | 0.67 | AAI-based (high-confidence) | 0 |
| S9_vs_k141_48240\|\|full | 33693 | No | NA | 52 | 12 | 0 | Medium-quality | Genome-fragment | 81.63 | AAI-based (high-confidence) | 0 |
| S9_vs_k141_22136\|\|full | 2939 | No | NA | 4 | 0 | 1 | Not-determined | Genome-fragment | NA | NA | 0 |
| S9_vs_k141_10676\|\|full | 1937 | No | NA | 2 | 0 | 2 | Not-determined | Genome-fragment | NA | NA | 0 |
| S9_vs_k141_41157\|\|full | 2817 | No | NA | 3 | 0 | 1 | Not-determined | Genome-fragment | NA | NA | 0 |
| S9_vs_k141_7060\|\|full | 1613 | No | NA | 2 | 0 | 1 | Not-determined | Genome-fragment | NA | NA | 0 |
| S9_vs_k141_18577\|\|full | 4323 | No | NA | 7 | 0 | 0 | Not-determined | Genome-fragment | NA | NA | 0 |
| S9_vs_k141_56234\|\|full | 6567 | No | NA | 8 | 0 | 0 | Not-determined | Genome-fragment | NA | NA | 0 |
| S9_vs_k141_14166\|\|full | 3122 | No | NA | 6 | 0 | 0 | Not-determined | Genome-fragment | NA | NA | 0 |
| S9_vs_k141_59709\|\|full | 3039 | No | NA | 3 | 0 | 2 | Not-determined | Genome-fragment | NA | NA | 0 |
| S13_extracted_81669 | 1138 | No | NA | 2 | 0 | 0 | Low-quality | Genome-fragment | 1.86 | AAI-based (medium-confidence) | 0 |
| S9_vs_k141_70952\|\|full | 2393 | No | NA | 3 | 0 | 1 | Not-determined | Genome-fragment | NA | NA | 0 |
| S14_vs_k141_64558\|\|full | 1138 | No | NA | 4 | 1 | 0 | Low-quality | Genome-fragment | 2.61 | AAI-based (medium-confidence) | 0 |
| S3_extracted_k141_32085 | 1138 | No | NA | 1 | 0 | 1 | Low-quality | Genome-fragment | 0.64 | AAI-based (medium-confidence) | 0 |
| S9_vs_k141_10703\|\|full | 3368 | No | NA | 4 | 0 | 1 | Not-determined | Genome-fragment | NA | NA | 0 |
| S5_vs_k141_64903\|\|full | 1138 | No | NA | 3 | 0 | 0 | Low-quality | Genome-fragment | 2.18 | AAI-based (medium-confidence) | 0 |
| S9_vs_k141_89926\|\|full | 1815 | No | NA | 2 | 0 | 2 | Not-determined | Genome-fragment | NA | NA | 0 |
| S5_vs_k141_76730\|\|full | 1138 | No | NA | 2 | 0 | 1 | Low-quality | Genome-fragment | 0.35 | AAI-based (medium-confidence) | 0 |
| S7_extracted_65908 | 1138 | No | NA | 3 | 1 | 0 | Low-quality | Genome-fragment | 2.63 | AAI-based (high-confidence) | 0 |
| S7_vs_115295\|\|full | 1138 | No | NA | 5 | 2 | 0 | Low-quality | Genome-fragment | 1.78 | AAI-based (high-confidence) | 0 |
| S9_vs_k141_48309\|\|full | 1039 | No | NA | 2 | 0 | 0 | Not-determined | Genome-fragment | NA | NA | 0 |
| S7_vs_130930\|\|full | 1138 | No | NA | 3 | 2 | 0 | Low-quality | Genome-fragment | 3.13 | AAI-based (high-confidence) | 0 |
| S9_vs_k141_33625\|\|full | 1839 | No | NA | 3 | 0 | 1 | Not-determined | Genome-fragment | NA | NA | 0 |
| S9_vs_k141_3173\|\|full | 1320 | No | NA | 2 | 0 | 2 | Not-determined | Genome-fragment | NA | NA | 0 |
| S4_extracted_19944 | 1138 | No | NA | 3 | 0 | 0 | Low-quality | Genome-fragment | 2.27 | AAI-based (high-confidence) | 0 |
| S4_extracted_30757 | 1138 | No | NA | 4 | 3 | 0 | Low-quality | Genome-fragment | 3.76 | AAI-based (high-confidence) | 0 |
| S9_vs_k141_30030\|\|full | 3073 | No | NA | 3 | 0 | 3 | Not-determined | Genome-fragment | NA | NA | 0 |
| S9_vs_k141_48325\|\|full | 8794 | No | NA | 13 | 0 | 2 | Not-determined | Genome-fragment | NA | NA | 0 |
| S9_vs_k141_59760\|\|full | 1549 | No | NA | 2 | 0 | 1 | Not-determined | Genome-fragment | NA | NA | 0 |
| S9_vs_k141_90037\|\|full | 1139 | No | NA | 3 | 0 | 0 | Low-quality | Genome-fragment | 1.85 | AAI-based (high-confidence) | 0 |
| S9_vs_k141_37437\|\|full | 41219 | No | NA | 68 | 20 | 1 | High-quality | High-quality | 98.78 | AAI-based (high-confidence) | 0 |
| S9_vs_k141_30075\|\|full | 2621 | No | NA | 3 | 0 | 1 | Not-determined | Genome-fragment | NA | NA | 0 |
| S9_vs_k141_3241\|\|full | 1192 | No | NA | 3 | 0 | 0 | Not-determined | Genome-fragment | NA | NA | 0 |
| S11_extracted_k141_3884 | 1139 | No | NA | 3 | 0 | 0 | Low-quality | Genome-fragment | 2.38 | AAI-based (high-confidence) | 0 |
| S9_vs_k141_71037\|\|full | 2512 | No | NA | 3 | 0 | 2 | Not-determined | Genome-fragment | NA | NA | 0 |
| S9_vs_k141_37449\|\|full | 1798 | No | NA | 3 | 0 | 1 | Not-determined | Genome-fragment | NA | NA | 0 |
| S5_extracted_k141_32384 | 1139 | No | NA | 2 | 1 | 0 | Low-quality | Genome-fragment | 2.05 | AAI-based (high-confidence) | 0 |
| S16_vs_44927\|\|full | 1139 | No | NA | 3 | 1 | 0 | Low-quality | Genome-fragment | 3.14 | AAI-based (high-confidence) | 0 |
| S9_vs_k141_89957\|\|full | 1976 | No | NA | 4 | 0 | 4 | Not-determined | Genome-fragment | NA | NA | 0 |
| S12_extracted_k141_5387 | 1139 | No | NA | 2 | 0 | 0 | Low-quality | Genome-fragment | 2.21 | AAI-based (high-confidence) | 0 |
| S12_vs_k141_101040\|\|full | 1139 | No | NA | 2 | 0 | 0 | Low-quality | Genome-fragment | 0.61 | AAI-based (high-confidence) | 0 |
| S9_vs_k141_48347\|\|full | 35042 | No | NA | 64 | 21 | 0 | High-quality | High-quality | 100 | AAI-based (high-confidence) | 0 |
| S12_vs_k141_125694\|\|full | 1139 | No | NA | 3 | 1 | 0 | Low-quality | Genome-fragment | 2.86 | AAI-based (high-confidence) | 0 |
| S9_vs_k141_18744\|\|full | 3213 | No | NA | 3 | 0 | 2 | Not-determined | Genome-fragment | NA | NA | 0 |
| S9_vs_k141_14235\|\|full | 21474 | No | NA | 32 | 5 | 2 | Medium-quality | Genome-fragment | 57.29 | AAI-based (medium-confidence) | 0 |
| S9_vs_k141_51532\|\|full | 1569 | No | NA | 3 | 0 | 2 | Not-determined | Genome-fragment | NA | NA | 0 |
| S12_vs_k141_15562\|\|full | 1139 | No | NA | 2 | 0 | 0 | Low-quality | Genome-fragment | 2.01 | AAI-based (high-confidence) | 0 |
| S11_vs_k141_74885\|\|full | 1140 | No | NA | 3 | 0 | 0 | Low-quality | Genome-fragment | 0.4 | AAI-based (medium-confidence) | 0 |
| S9_vs_k141_51543\|\|full | 2126 | No | NA | 3 | 0 | 3 | Not-determined | Genome-fragment | NA | NA | 0 |
| S9_vs_k141_86383\|\|full | 1326 | No | NA | 3 | 0 | 0 | Not-determined | Genome-fragment | NA | NA | 0 |
| S11_vs_k141_88753\|\|full | 1140 | No | NA | 3 | 2 | 0 | Low-quality | Genome-fragment | 3.9 | AAI-based (medium-confidence) | 0 |
| S9_vs_k141_10867\|\|full | 1479 | No | NA | 3 | 0 | 0 | Not-determined | Genome-fragment | NA | NA | 0 |
| S9_vs_k141_41361\|\|full | 2627 | No | NA | 4 | 0 | 0 | Not-determined | Genome-fragment | NA | NA | 0 |
| S9_vs_k141_56354\|\|full | 44039 | No | NA | 66 | 17 | 2 | High-quality | High-quality | 98.12 | AAI-based (high-confidence) | 0 |
| S11_vs_k141_64489\|\|lt2gene | 1140 | No | NA | 2 | 2 | 0 | Low-quality | Genome-fragment | 3.28 | AAI-based (high-confidence) | 0 |
| S14_extracted_k141_110710 | 1140 | No | NA | 1 | 0 | 0 | Low-quality | Genome-fragment | 1.97 | AAI-based (high-confidence) | 0 |
| S14_extracted_k141_7073 | 1140 | No | NA | 1 | 0 | 1 | Low-quality | Genome-fragment | 1.21 | AAI-based (medium-confidence) | 0 |
| S9_vs_k141_67291\|\|full | 1594 | No | NA | 2 | 0 | 2 | Not-determined | Genome-fragment | NA | NA | 0 |
| S14_vs_k141_40567\|\|full | 1140 | No | NA | 4 | 2 | 0 | Low-quality | Genome-fragment | 3.28 | AAI-based (high-confidence) | 0 |
| S1_extracted_k141_119646 | 1140 | No | NA | 2 | 0 | 0 | Low-quality | Genome-fragment | 1.79 | AAI-based (medium-confidence) | 0 |
| S1_vs_k141_157295\|\|lt2gene | 1140 | No | NA | 1 | 1 | 0 | Low-quality | Genome-fragment | 0.66 | AAI-based (high-confidence) | 0 |
| S3_vs_k141_48255\|\|full | 1140 | No | NA | 3 | 1 | 0 | Low-quality | Genome-fragment | 0.5 | HMM-based (lower-bound) | 0 |
| S5_extracted_k141_122802 | 1140 | No | NA | 1 | 1 | 0 | Low-quality | Genome-fragment | 3.86 | HMM-based (lower-bound) | 0 |
| S9_vs_k141_41371\|\|full | 25415 | No | NA | 35 | 21 | 0 | Medium-quality | Genome-fragment | 58.18 | AAI-based (high-confidence) | 0 |
| S5_extracted_k141_104979 | 1140 | No | NA | 3 | 2 | 0 | Low-quality | Genome-fragment | 1.64 | HMM-based (lower-bound) | 0 |
| S5_vs_k141_115528\|\|full | 1140 | No | NA | 3 | 2 | 0 | Low-quality | Genome-fragment | 2.24 | AAI-based (high-confidence) | 0 |
| S9_vs_k141_59919\|\|full | 1641 | No | NA | 2 | 0 | 1 | Not-determined | Genome-fragment | NA | NA | 0 |
| S5_vs_k141_128205\|\|lt2gene | 1140 | No | NA | 2 | 2 | 0 | Low-quality | Genome-fragment | 2.45 | AAI-based (high-confidence) | 0 |
| S9_vs_k141_41397\|\|full | 4342 | No | NA | 5 | 0 | 0 | Not-determined | Genome-fragment | NA | NA | 0 |
| S9_vs_k141_37560\|\|full | 4684 | No | NA | 5 | 0 | 2 | Not-determined | Genome-fragment | NA | NA | 0 |
| S7_vs_45577\|\|full | 1140 | No | NA | 3 | 0 | 1 | Low-quality | Genome-fragment | 0.44 | AAI-based (medium-confidence) | 0 |
| S9_vs_k141_71174\|\|full | 1108 | No | NA | 4 | 0 | 0 | Not-determined | Genome-fragment | NA | NA | 0 |
| S9_vs_k141_78576\|\|full | 1205 | No | NA | 4 | 0 | 0 | Not-determined | Genome-fragment | NA | NA | 0 |
| S7_vs_85470\|\|full | 1140 | No | NA | 2 | 0 | 0 | Low-quality | Genome-fragment | 2.22 | AAI-based (medium-confidence) | 0 |
| S10_vs_7250\|\|full | 1140 | No | NA | 3 | 0 | 0 | Low-quality | Genome-fragment | 0.62 | AAI-based (medium-confidence) | 0 |
| S4_extracted_21185 | 1140 | No | NA | 3 | 2 | 0 | Low-quality | Genome-fragment | 2.3 | AAI-based (high-confidence) | 0 |
| S9_vs_k141_56427\|\|full | 3567 | No | NA | 4 | 0 | 0 | Not-determined | Genome-fragment | NA | NA | 0 |
| S8_vs_k141_122922\|\|full | 1140 | No | NA | 3 | 0 | 3 | Low-quality | Genome-fragment | 1.59 | AAI-based (medium-confidence) | 0 |
| S9_vs_k141_14352\|\|full | 1618 | No | NA | 3 | 0 | 1 | Not-determined | Genome-fragment | NA | NA | 0 |
| S9_vs_k141_78619\|\|full | 1717 | No | NA | 2 | 0 | 1 | Not-determined | Genome-fragment | NA | NA | 0 |
| S11_vs_k141_506\|\|full | 1141 | No | NA | 5 | 0 | 0 | Low-quality | Genome-fragment | 1.89 | AAI-based (high-confidence) | 0 |
| S9_vs_k141_30243\|\|full | 1921 | No | NA | 3 | 0 | 1 | Not-determined | Genome-fragment | NA | NA | 0 |
| S11_vs_k141_46607\|\|full | 1141 | No | NA | 4 | 1 | 0 | Low-quality | Genome-fragment | 1.98 | AAI-based (high-confidence) | 0 |
| S9_vs_k141_75109\|\|full | 1744 | No | NA | 3 | 0 | 0 | Not-determined | Genome-fragment | NA | NA | 0 |
| S11_vs_k141_88227\|\|full | 1141 | No | NA | 5 | 1 | 0 | Low-quality | Genome-fragment | 2.67 | AAI-based (medium-confidence) | 0 |
| S9_vs_k141_26179\|\|full | 23328 | No | NA | 48 | 16 | 1 | Medium-quality | Genome-fragment | 68.67 | AAI-based (high-confidence) | 0 |
| S13_extracted_27313 | 1141 | No | NA | 1 | 0 | 0 | Low-quality | Genome-fragment | 2.06 | AAI-based (medium-confidence) | 0 |
| S3_vs_k141_21646\|\|full | 1141 | No | NA | 4 | 0 | 0 | Low-quality | Genome-fragment | 1.95 | AAI-based (medium-confidence) | 0 |
| S3_vs_k141_6333\|\|lt2gene | 1141 | No | NA | 2 | 2 | 0 | Low-quality | Genome-fragment | 2.77 | AAI-based (high-confidence) | 0 |
| S9_vs_k141_3417\|\|full | 1347 | No | NA | 2 | 0 | 1 | Not-determined | Genome-fragment | NA | NA | 0 |
| S5_vs_k141_100771\|\|full | 1141 | No | NA | 2 | 1 | 0 | Low-quality | Genome-fragment | 2.66 | AAI-based (high-confidence) | 0 |
| S9_vs_k141_67414\|\|full | 1011 | No | NA | 2 | 0 | 1 | Not-determined | Genome-fragment | NA | NA | 0 |
| S7_vs_14867\|\|full | 1141 | No | NA | 2 | 0 | 1 | Low-quality | Genome-fragment | 0.7 | AAI-based (medium-confidence) | 0 |
| S16_extracted_85965 | 1141 | No | NA | 4 | 1 | 0 | Low-quality | Genome-fragment | 2.43 | AAI-based (high-confidence) | 0 |
| S9_vs_k141_41462\|\|full | 1525 | No | NA | 5 | 0 | 0 | Not-determined | Genome-fragment | NA | NA | 0 |
| S9_vs_k141_67419\|\|full | 3255 | No | NA | 4 | 0 | 2 | Not-determined | Genome-fragment | NA | NA | 0 |
| S16_extracted_119077 | 1141 | No | NA | 3 | 3 | 0 | Low-quality | Genome-fragment | 2.42 | AAI-based (medium-confidence) | 0 |
| S16_vs_131369\|\|full | 1141 | No | NA | 4 | 1 | 0 | Low-quality | Genome-fragment | 2.57 | AAI-based (high-confidence) | 0 |
| S9_vs_k141_41467\|\|full | 1921 | No | NA | 3 | 0 | 0 | Not-determined | Genome-fragment | NA | NA | 0 |
| S12_extracted_k141_143058 | 1141 | No | NA | 2 | 0 | 0 | Low-quality | Genome-fragment | 2.41 | AAI-based (medium-confidence) | 0 |
| S12_vs_k141_112448\|\|full | 1141 | No | NA | 2 | 0 | 0 | Low-quality | Genome-fragment | 0.84 | AAI-based (medium-confidence) | 0 |
| S15_vs_17424\|\|full | 1141 | No | NA | 2 | 0 | 0 | Low-quality | Genome-fragment | 1.34 | AAI-based (high-confidence) | 0 |
| S9_vs_k141_14450\|\|full | 2476 | No | NA | 3 | 0 | 2 | Not-determined | Genome-fragment | NA | NA | 0 |
| S6_vs_k141_13956\|\|full | 1141 | No | NA | 3 | 2 | 0 | Low-quality | Genome-fragment | 2.9 | AAI-based (high-confidence) | 0 |
| S8_vs_k141_26228\|\|full | 1141 | No | NA | 2 | 0 | 0 | Low-quality | Genome-fragment | 2.59 | AAI-based (medium-confidence) | 0 |
| S9_vs_k141_90185\|\|full | 1085 | No | NA | 2 | 0 | 0 | Not-determined | Genome-fragment | NA | NA | 0 |
| S17_extracted_k141_139099 | 1142 | No | NA | 2 | 0 | 0 | Low-quality | Genome-fragment | 0.57 | AAI-based (medium-confidence) | 0 |
| S9_vs_k141_11044\|\|full | 1063 | No | NA | 3 | 0 | 0 | Not-determined | Genome-fragment | NA | NA | 0 |
| S9_vs_k141_75168\|\|full | 1467 | No | NA | 3 | 0 | 0 | Not-determined | Genome-fragment | NA | NA | 0 |
| S17_extracted_k141_95097 | 1142 | No | NA | 2 | 0 | 1 | Low-quality | Genome-fragment | 2.26 | AAI-based (high-confidence) | 0 |
| S9_vs_k141_75175\|\|full | 1093 | No | NA | 2 | 0 | 2 | Not-determined | Genome-fragment | NA | NA | 0 |
| S9_vs_k141_3467\|\|full | 2177 | No | NA | 3 | 0 | 1 | Not-determined | Genome-fragment | NA | NA | 0 |
| S9_vs_k141_78689\|\|full | 1098 | No | NA | 2 | 0 | 0 | Not-determined | Genome-fragment | NA | NA | 0 |
| S9_vs_k141_7440\|\|full | 1321 | No | NA | 3 | 0 | 1 | Not-determined | Genome-fragment | NA | NA | 0 |
| S9_vs_k141_75195\|\|full | 2839 | No | NA | 2 | 0 | 2 | Not-determined | Genome-fragment | NA | NA | 0 |
| S13_extracted_9908 | 1142 | No | NA | 3 | 0 | 0 | Low-quality | Genome-fragment | 2.05 | AAI-based (medium-confidence) | 0 |
| S9_vs_k141_56556\|\|full | 1197 | No | NA | 2 | 0 | 2 | Not-determined | Genome-fragment | NA | NA | 0 |
| S13_extracted_145962 | 1142 | No | NA | 2 | 0 | 0 | Low-quality | Genome-fragment | 2.47 | AAI-based (medium-confidence) | 0 |
| S13_vs_133156\|\|lt2gene | 1142 | No | NA | 1 | 1 | 0 | Low-quality | Genome-fragment | 2.19 | AAI-based (medium-confidence) | 0 |
| S9_vs_k141_26324\|\|full | 1969 | No | NA | 3 | 0 | 1 | Not-determined | Genome-fragment | NA | NA | 0 |
| S14_extracted_k141_10151 | 1142 | No | NA | 4 | 2 | 0 | Low-quality | Genome-fragment | 3.75 | AAI-based (high-confidence) | 0 |
| S9_vs_k141_67538\|\|full | 2639 | No | NA | 4 | 0 | 1 | Not-determined | Genome-fragment | NA | NA | 0 |
| S1_vs_k141_158659\|\|full | 1142 | No | NA | 3 | 0 | 0 | Low-quality | Genome-fragment | 0.74 | AAI-based (high-confidence) | 0 |
| S3_vs_k141_58133\|\|full | 1142 | No | NA | 3 | 0 | 0 | Low-quality | Genome-fragment | 2.36 | AAI-based (medium-confidence) | 0 |
| S9_vs_k141_51676\|\|full | 2849 | No | NA | 5 | 0 | 1 | Not-determined | Genome-fragment | NA | NA | 0 |
| S5_extracted_k141_137757 | 1142 | No | NA | 1 | 0 | 1 | Low-quality | Genome-fragment | 0.43 | AAI-based (high-confidence) | 0 |
| S9_vs_k141_78779\|\|full | 1306 | No | NA | 2 | 0 | 2 | Not-determined | Genome-fragment | NA | NA | 0 |
| S9_vs_k141_37768\|\|full | 5737 | No | NA | 4 | 0 | 0 | Not-determined | Genome-fragment | NA | NA | 0 |
| S9_vs_k141_48538\|\|full | 1402 | No | NA | 3 | 0 | 3 | Not-determined | Genome-fragment | NA | NA | 0 |
| S7_extracted_674 | 1142 | No | NA | 1 | 1 | 0 | Low-quality | Genome-fragment | 2.72 | HMM-based (lower-bound) | 0 |
| S9_vs_k141_78796\|\|full | 3577 | No | NA | 6 | 0 | 3 | Not-determined | Genome-fragment | NA | NA | 0 |
| S7_vs_13867\|\|full | 1142 | No | NA | 2 | 2 | 0 | Low-quality | Genome-fragment | 2.5 | AAI-based (high-confidence) | 0 |
| S7_vs_42483\|\|full | 1142 | No | NA | 2 | 1 | 0 | Low-quality | Genome-fragment | 2.35 | AAI-based (medium-confidence) | 0 |
| S7_vs_74338\|\|lt2gene | 1142 | No | NA | 2 | 1 | 0 | Low-quality | Genome-fragment | 0.44 | HMM-based (lower-bound) | 0 |
| S9_vs_k141_3557\|\|full | 2130 | No | NA | 3 | 0 | 0 | Not-determined | Genome-fragment | NA | NA | 0 |
| S9_vs_k141_75290\|\|full | 12648 | No | NA | 16 | 6 | 0 | High-quality | High-quality | 92.6 | AAI-based (medium-confidence) | 0 |
| S9_vs_k141_14662\|\|full | 2418 | No | NA | 3 | 0 | 1 | Not-determined | Genome-fragment | NA | NA | 0 |
| S16_extracted_58727 | 1142 | No | NA | 3 | 0 | 1 | Low-quality | Genome-fragment | 1.94 | AAI-based (medium-confidence) | 0 |
| S9_vs_k141_90304\|\|full | 3180 | No | NA | 6 | 0 | 3 | Not-determined | Genome-fragment | NA | NA | 0 |
| S16_vs_14664\|\|full | 1142 | No | NA | 3 | 0 | 1 | Low-quality | Genome-fragment | 0.82 | AAI-based (high-confidence) | 0 |
| S10_extracted_17657 | 1142 | No | NA | 2 | 0 | 0 | Low-quality | Genome-fragment | 2.11 | AAI-based (high-confidence) | 0 |
| S9_vs_k141_51796\|\|full | 2447 | No | NA | 4 | 0 | 2 | Not-determined | Genome-fragment | NA | NA | 0 |
| S10_extracted_104192 | 1142 | No | NA | 2 | 0 | 1 | Low-quality | Genome-fragment | 1.57 | AAI-based (medium-confidence) | 0 |
| S12_vs_k141_49689\|\|full | 1142 | No | NA | 3 | 0 | 0 | Low-quality | Genome-fragment | 2.7 | AAI-based (medium-confidence) | 0 |
| S9_vs_k141_67662\|\|full | 1693 | No | NA | 3 | 0 | 0 | Not-determined | Genome-fragment | NA | NA | 0 |
| S9_vs_k141_14698\|\|full | 3810 | No | NA | 4 | 0 | 3 | Not-determined | Genome-fragment | NA | NA | 0 |
| S12_vs_k141_28971\|\|full | 1142 | No | NA | 2 | 0 | 0 | Low-quality | Genome-fragment | 1.43 | AAI-based (medium-confidence) | 0 |
| S9_vs_k141_51847\|\|full | 3948 | No | NA | 4 | 0 | 1 | Not-determined | Genome-fragment | NA | NA | 0 |
| S9_vs_k141_67686\|\|full | 1662 | No | NA | 3 | 0 | 0 | Not-determined | Genome-fragment | NA | NA | 0 |
| S12_vs_k141_101059\|\|full | 1142 | No | NA | 4 | 0 | 0 | Low-quality | Genome-fragment | 2.46 | AAI-based (medium-confidence) | 0 |
| S17_vs_k141_10063\|\|full | 1143 | No | NA | 3 | 1 | 0 | Low-quality | Genome-fragment | 1.32 | HMM-based (lower-bound) | 0 |
| S9_vs_k141_46315\|\|full | 1143 | No | NA | 4 | 3 | 0 | Low-quality | Genome-fragment | 2.05 | HMM-based (lower-bound) | 0 |
| S9_vs_k141_67696\|\|full | 1223 | No | NA | 2 | 0 | 0 | Not-determined | Genome-fragment | NA | NA | 0 |
| S9_vs_k141_14734\|\|full | 3251 | No | NA | 6 | 0 | 2 | Not-determined | Genome-fragment | NA | NA | 0 |
| S9_vs_k141_56747\|\|full | 2542 | No | NA | 7 | 0 | 0 | Not-determined | Genome-fragment | NA | NA | 0 |
| S11_vs_k141_54093\|\|full | 1143 | No | NA | 2 | 0 | 0 | Low-quality | Genome-fragment | 2.23 | AAI-based (high-confidence) | 0 |
| S9_vs_k141_60256\|\|full | 1825 | No | NA | 5 | 0 | 0 | Not-determined | Genome-fragment | NA | NA | 0 |
| S11_vs_k141_74297\|\|full | 1143 | No | NA | 4 | 0 | 0 | Low-quality | Genome-fragment | 1.12 | AAI-based (medium-confidence) | 0 |
| S9_vs_k141_3678\|\|full | 1004 | No | NA | 3 | 0 | 1 | Not-determined | Genome-fragment | NA | NA | 0 |
| S1_extracted_k141_81508 | 1143 | No | NA | 3 | 1 | 0 | Low-quality | Genome-fragment | 1.42 | AAI-based (high-confidence) | 0 |
| S1_extracted_k141_159237 | 1143 | No | NA | 1 | 0 | 0 | Low-quality | Genome-fragment | 1.43 | AAI-based (medium-confidence) | 0 |
| S9_vs_k141_48692\|\|full | 1272 | No | NA | 2 | 0 | 0 | Not-determined | Genome-fragment | NA | NA | 0 |
| S9_vs_k141_78993\|\|full | 3810 | No | NA | 4 | 0 | 4 | Not-determined | Genome-fragment | NA | NA | 0 |
| S9_vs_k141_63732\|\|full | 2535 | No | NA | 4 | 0 | 2 | Not-determined | Genome-fragment | NA | NA | 0 |
| S9_vs_k141_75406\|\|full | 7706 | No | NA | 8 | 0 | 8 | Not-determined | Genome-fragment | NA | NA | 0 |
| S5_extracted_k141_107443 | 1143 | No | NA | 1 | 1 | 0 | Low-quality | Genome-fragment | 2.94 | AAI-based (medium-confidence) | 0 |
| S9_vs_k141_11339\|\|full | 1242 | No | NA | 3 | 0 | 0 | Not-determined | Genome-fragment | NA | NA | 0 |
| S9_vs_k141_71681\|\|full | 3964 | No | NA | 5 | 0 | 1 | Not-determined | Genome-fragment | NA | NA | 0 |
| S5_vs_k141_51455\|\|full | 1143 | No | NA | 3 | 0 | 0 | Low-quality | Genome-fragment | 0.39 | AAI-based (medium-confidence) | 0 |
| S7_vs_55124\|\|full | 1143 | No | NA | 3 | 0 | 1 | Low-quality | Genome-fragment | 0.95 | AAI-based (medium-confidence) | 0 |
| S9_vs_k141_52069\|\|full | 2367 | No | NA | 3 | 0 | 0 | Not-determined | Genome-fragment | NA | NA | 0 |
| S9_vs_k141_63810\|\|full | 2147 | No | NA | 3 | 0 | 2 | Not-determined | Genome-fragment | NA | NA | 0 |
| S9_vs_k141_52089\|\|full | 1576 | No | NA | 4 | 0 | 1 | Not-determined | Genome-fragment | NA | NA | 0 |
| S9_vs_k141_86892\|\|full | 2008 | No | NA | 3 | 0 | 0 | Not-determined | Genome-fragment | NA | NA | 0 |
| S7_vs_133452\|\|lt2gene | 1143 | No | NA | 2 | 2 | 0 | Low-quality | Genome-fragment | 1.63 | HMM-based (lower-bound) | 0 |
| S9_vs_k141_67792\|\|full | 72275 | Yes | 39401 | 78 | 26 | 18 | Complete | High-quality | 100 | Provirus (high-confidence) | 45.48 |
| S9_vs_k141_90476\|\|full | 1030 | No | NA | 3 | 0 | 0 | Not-determined | Genome-fragment | NA | NA | 0 |
| S9_vs_k141_71754\|\|full | 1222 | No | NA | 3 | 0 | 1 | Not-determined | Genome-fragment | NA | NA | 0 |
| S10_extracted_13057 | 1143 | No | NA | 1 | 0 | 1 | Low-quality | Genome-fragment | 0.31 | AAI-based (high-confidence) | 0 |
| S9_vs_k141_71756\|\|full | 1667 | No | NA | 3 | 0 | 1 | Not-determined | Genome-fragment | NA | NA | 0 |
| S9_vs_k141_82948\|\|full | 2515 | No | NA | 2 | 0 | 0 | Not-determined | Genome-fragment | NA | NA | 0 |
| S9_vs_k141_86943\|\|full | 4274 | No | NA | 7 | 0 | 1 | Not-determined | Genome-fragment | NA | NA | 0 |
| S9_vs_k141_71761\|\|full | 1652 | No | NA | 2 | 0 | 1 | Not-determined | Genome-fragment | NA | NA | 0 |
| S9_vs_k141_82962\|\|full | 1878 | No | NA | 3 | 0 | 1 | Not-determined | Genome-fragment | NA | NA | 0 |
| S10_extracted_48853 | 1143 | No | NA | 1 | 0 | 0 | Low-quality | Genome-fragment | 1.42 | AAI-based (high-confidence) | 0 |
| S9_vs_k141_67844\|\|full | 3582 | No | NA | 3 | 0 | 2 | Not-determined | Genome-fragment | NA | NA | 0 |
| S10_vs_82184\|\|lt2gene | 1143 | No | NA | 2 | 2 | 0 | Low-quality | Genome-fragment | 3.02 | AAI-based (high-confidence) | 0 |
| S9_vs_k141_63859\|\|full | 1979 | No | NA | 4 | 0 | 0 | Not-determined | Genome-fragment | NA | NA | 0 |
| S9_vs_k141_48874\|\|full | 2126 | No | NA | 6 | 0 | 0 | Not-determined | Genome-fragment | NA | NA | 0 |
| S12_vs_k141_176760\|\|lt2gene | 1143 | No | NA | 1 | 1 | 0 | Low-quality | Genome-fragment | 2.16 | HMM-based (lower-bound) | 0 |
| S15_extracted_41530 | 1143 | No | NA | 2 | 0 | 0 | Low-quality | Genome-fragment | 1.23 | AAI-based (medium-confidence) | 0 |
| S9_vs_k141_71837\|\|full | 1358 | No | NA | 3 | 0 | 1 | Not-determined | Genome-fragment | NA | NA | 0 |
| S2_vs_k141_255\|\|full | 1143 | No | NA | 3 | 0 | 1 | Low-quality | Genome-fragment | 0.49 | AAI-based (medium-confidence) | 0 |
| S9_vs_k141_14967\|\|full | 3863 | No | NA | 3 | 0 | 3 | Not-determined | Genome-fragment | NA | NA | 0 |
| S9_vs_k141_79217\|\|full | 1208 | No | NA | 3 | 0 | 2 | Not-determined | Genome-fragment | NA | NA | 0 |
| S17_extracted_k141_117086 | 1144 | No | NA | 3 | 0 | 0 | Low-quality | Genome-fragment | 2.99 | AAI-based (high-confidence) | 0 |
| S9_vs_k141_9712\|\|full | 1144 | No | NA | 3 | 2 | 0 | Low-quality | Genome-fragment | 2.63 | AAI-based (high-confidence) | 0 |
| S9_vs_k141_34092\|\|full | 1647 | No | NA | 3 | 0 | 0 | Not-determined | Genome-fragment | NA | NA | 0 |
| S9_vs_k141_52222\|\|full | 3291 | No | NA | 3 | 0 | 0 | Not-determined | Genome-fragment | NA | NA | 0 |
| S9_vs_k141_15014\|\|full | 2014 | No | NA | 2 | 0 | 2 | Not-determined | Genome-fragment | NA | NA | 0 |
| S11_extracted_k141_38071 | 1144 | No | NA | 1 | 0 | 0 | Low-quality | Genome-fragment | 3.05 | AAI-based (medium-confidence) | 0 |
| S13_vs_22969\|\|full | 1144 | No | NA | 3 | 0 | 0 | Low-quality | Genome-fragment | 2.85 | AAI-based (high-confidence) | 0 |
| S9_vs_k141_68005\|\|full | 2933 | No | NA | 3 | 0 | 1 | Not-determined | Genome-fragment | NA | NA | 0 |
| S13_vs_68772\|\|full | 1144 | No | NA | 2 | 0 | 0 | Low-quality | Genome-fragment | 2 | AAI-based (medium-confidence) | 0 |
| S9_vs_k141_64004\|\|full | 1613 | No | NA | 3 | 0 | 2 | Not-determined | Genome-fragment | NA | NA | 0 |
| S9_vs_k141_60430\|\|full | 1505 | No | NA | 2 | 0 | 1 | Not-determined | Genome-fragment | NA | NA | 0 |

| | | | | | | | | | | | | |
|---|---|---|---|---|---|---|---|---|---|---|---|---|
| S13_vs_104546\|\|full | 1144 | No | NA | 3 | 0 | 0 | Low-quality | Genome-fragment | | 2.79 | AAI-based (high-confidence) | 0 |
| S1_vs_k141_85486\|\|lt2gene | 1144 | No | NA | 2 | 2 | 0 | Low-quality | Genome-fragment | | 2.81 | AAI-based (high-confidence) | 0 |
| S3_vs_k141_95685\|\|full | 1144 | No | NA | 3 | 3 | 0 | Low-quality | Genome-fragment | | 3.75 | AAI-based (high-confidence) | 0 |
| S5_extracted_k141_93878 | 1144 | No | NA | 2 | 1 | 0 | Low-quality | Genome-fragment | | 1.81 | AAI-based (high-confidence) | 0 |
| S7_vs_33958\|\|lt2gene | 1144 | No | NA | 2 | 1 | 0 | Low-quality | Genome-fragment | | 2.94 | AAI-based (high-confidence) | 0 |
| S9_vs_k141_49063\|\|full | 1896 | No | NA | 3 | 0 | 1 | Not-determined | Genome-fragment | NA | | NA | 0 |
| S10_vs_58081\|\|full | 1144 | No | NA | 3 | 0 | 0 | Low-quality | Genome-fragment | | 1.98 | AAI-based (high-confidence) | 0 |
| S10_vs_75924\|\|lt2gene | 1144 | No | NA | 2 | 1 | 0 | Low-quality | Genome-fragment | | 3.07 | AAI-based (medium-confidence) | 0 |
| S9_vs_k141_83108\|\|full | 88175 | No | NA | 132 | 16 | 1 | High-quality | High-quality | | 100 | AAI-based (high-confidence) | 0 |
| S9_vs_k141_83137\|\|full | 1010 | No | NA | 3 | 0 | 0 | Not-determined | Genome-fragment | NA | | NA | 0 |
| S9_vs_k141_52280\|\|full | 5521 | No | NA | 6 | 0 | 3 | Not-determined | Genome-fragment | NA | | NA | 0 |
| S12_extracted_k141_46902 | 1144 | No | NA | 2 | 1 | 0 | Low-quality | Genome-fragment | | 2.75 | AAI-based (high-confidence) | 0 |
| S8_vs_k141_111140\|\|full | 1144 | No | NA | 2 | 0 | 1 | Low-quality | Genome-fragment | | 2.46 | AAI-based (medium-confidence) | 0 |
| S17_extracted_k141_156129 | 1145 | No | NA | 3 | 1 | 0 | Low-quality | Genome-fragment | | 2.07 | AAI-based (high-confidence) | 0 |
| S17_extracted_k141_62048 | 1145 | No | NA | 2 | 2 | 0 | Low-quality | Genome-fragment | | 2.69 | AAI-based (high-confidence) | 0 |
| S11_vs_k141_83167\|\|full | 1145 | No | NA | 4 | 0 | 0 | Low-quality | Genome-fragment | | 3.15 | AAI-based (high-confidence) | 0 |
| S14_extracted_k141_7749 | 1145 | No | NA | 2 | 1 | 0 | Low-quality | Genome-fragment | | 3.78 | AAI-based (high-confidence) | 0 |
| S14_extracted_k141_9687 | 1145 | No | NA | 3 | 3 | 0 | Low-quality | Genome-fragment | | 3.74 | AAI-based (high-confidence) | 0 |
| S9_vs_k141_49152\|\|full | 2871 | No | NA | 4 | 0 | 1 | Not-determined | Genome-fragment | NA | | NA | 0 |
| S14_extracted_k141_76355 | 1145 | No | NA | 2 | 0 | 0 | Low-quality | Genome-fragment | | 2.43 | AAI-based (medium-confidence) | 0 |
| S1_extracted_k141_17172 | 1145 | No | NA | 1 | 0 | 1 | Low-quality | Genome-fragment | | 1.95 | AAI-based (high-confidence) | 0 |
| S9_vs_k141_83290\|\|full | 2163 | No | NA | 4 | 0 | 0 | Not-determined | Genome-fragment | NA | | NA | 0 |
| S3_vs_k141_2162\|\|full | 1145 | No | NA | 3 | 0 | 0 | Low-quality | Genome-fragment | | 2.16 | AAI-based (medium-confidence) | 0 |
| S9_vs_k141_49246\|\|full | 1053 | No | NA | 2 | 0 | 2 | Not-determined | Genome-fragment | NA | | NA | 0 |
| S9_vs_k141_90727\|\|full | 173711 | Yes | 66238 | 166 | 26 | 68 | High-quality | High-quality | | 97.83 | AAI-based (high-confidence) | 61.87 |
| S9_vs_k141_52543\|\|full | 1797 | No | NA | 2 | 0 | 0 | Not-determined | Genome-fragment | NA | | NA | 0 |
| S5_extracted_k141_103184 | 1145 | No | NA | 3 | 0 | 1 | Low-quality | Genome-fragment | | 2.68 | AAI-based (high-confidence) | 0 |
| S16_extracted_58709 | 1145 | No | NA | 2 | 1 | 0 | Low-quality | Genome-fragment | | 2.5 | AAI-based (high-confidence) | 0 |
| S9_vs_k141_52757\|\|full | 1526 | No | NA | 2 | 0 | 0 | Not-determined | Genome-fragment | NA | | NA | 0 |
| S16_vs_41073\|\|full | 1145 | No | NA | 3 | 0 | 0 | Low-quality | Genome-fragment | | 2.86 | AAI-based (high-confidence) | 0 |
| S12_extracted_k141_181715 | 1145 | No | NA | 2 | 0 | 1 | Low-quality | Genome-fragment | | 1.48 | AAI-based (medium-confidence) | 0 |
| S12_extracted_k141_72947 | 1145 | No | NA | 3 | 3 | 0 | Low-quality | Genome-fragment | | 3.78 | AAI-based (high-confidence) | 0 |
| S9_vs_k141_16723\|\|full | 1146 | No | NA | 2 | 0 | 1 | Low-quality | Genome-fragment | | 0.59 | AAI-based (medium-confidence) | 0 |
| S9_vs_k141_23649\|\|0_partial | 18591 | No | NA | 12 | 0 | 1 | Not-determined | Genome-fragment | NA | | NA | 0 |
| S9_vs_k141_55642\|\|lt2gene | 1146 | No | NA | 1 | 1 | 0 | Low-quality | Genome-fragment | | 2.83 | AAI-based (medium-confidence) | 0 |
| S1_extracted_k141_119796 | 1146 | No | NA | 2 | 1 | 0 | Low-quality | Genome-fragment | | 1.92 | AAI-based (high-confidence) | 0 |
| S1_extracted_k141_106755 | 1146 | No | NA | 2 | 0 | 0 | Low-quality | Genome-fragment | | 2.17 | AAI-based (high-confidence) | 0 |
| S1_vs_k141_48609\|\|full | 1146 | No | NA | 2 | 0 | 0 | Low-quality | Genome-fragment | | 1.87 | AAI-based (medium-confidence) | 0 |
| S5_extracted_k141_53674 | 1146 | No | NA | 3 | 0 | 0 | Low-quality | Genome-fragment | | 1.41 | AAI-based (medium-confidence) | 0 |
| S9_vs_k141_38095\|\|lt2gene | 1878 | No | NA | 2 | 0 | 0 | Not-determined | Genome-fragment | NA | | NA | 0 |
| S5_vs_k141_68374\|\|full | 1146 | No | NA | 2 | 1 | 0 | Low-quality | Genome-fragment | | 3.06 | AAI-based (high-confidence) | 0 |
| S5_vs_k141_73428\|\|lt2gene | 1146 | No | NA | 1 | 1 | 0 | Low-quality | Genome-fragment | | 2.68 | HMM-based (lower-bound) | 0 |
| S16_extracted_29876 | 1146 | No | NA | 2 | 1 | 0 | Low-quality | Genome-fragment | | 2.9 | AAI-based (medium-confidence) | 0 |
| S16_extracted_122610 | 1146 | No | NA | 2 | 2 | 0 | Low-quality | Genome-fragment | | 2.54 | AAI-based (high-confidence) | 0 |
| S16_extracted_152200 | 1146 | No | NA | 3 | 0 | 0 | Low-quality | Genome-fragment | | 1.81 | AAI-based (high-confidence) | 0 |
| S16_vs_30547\|\|full | 1146 | No | NA | 2 | 0 | 0 | Low-quality | Genome-fragment | | 0.55 | AAI-based (medium-confidence) | 0 |
| S15_extracted_45120 | 1146 | No | NA | 3 | 0 | 1 | Low-quality | Genome-fragment | | 1.58 | AAI-based (medium-confidence) | 0 |
| S8_extracted_k141_73088 | 1146 | No | NA | 3 | 1 | 0 | Low-quality | Genome-fragment | | 0.62 | HMM-based (lower-bound) | 0 |
| S8_extracted_k141_127981 | 1146 | No | NA | 3 | 0 | 0 | Low-quality | Genome-fragment | | 1.68 | AAI-based (medium-confidence) | 0 |
| S8_vs_k141_105702\|\|full | 1146 | No | NA | 4 | 0 | 0 | Low-quality | Genome-fragment | | 2.8 | AAI-based (high-confidence) | 0 |
| S17_vs_k141_24566\|\|full | 1147 | No | NA | 3 | 0 | 1 | Low-quality | Genome-fragment | | 0.37 | AAI-based (medium-confidence) | 0 |
| S11_extracted_k141_12492 | 1147 | No | NA | 2 | 1 | 0 | Low-quality | Genome-fragment | | 2.56 | AAI-based (high-confidence) | 0 |
| S11_vs_k141_898\|\|full | 1147 | No | NA | 3 | 1 | 0 | Low-quality | Genome-fragment | | 2.68 | AAI-based (medium-confidence) | 0 |
| S11_vs_k141_54717\|\|full | 1147 | No | NA | 4 | 1 | 0 | Low-quality | Genome-fragment | | 2.65 | AAI-based (high-confidence) | 0 |
| S9_vs_k141_42609\|\|lt2gene | 2243 | No | NA | 1 | 1 | 0 | Medium-quality | Genome-fragment | | 53.9 | HMM-based (lower-bound) | 0 |
| S14_vs_k141_106703\|\|full | 1147 | No | NA | 4 | 4 | 0 | Low-quality | Genome-fragment | | 2.99 | AAI-based (high-confidence) | 0 |
| S9_vs_k141_31261\|\|lt2gene | 1197 | No | NA | 2 | 0 | 0 | Not-determined | Genome-fragment | NA | | NA | 0 |
| S14_vs_k141_13631\|\|lt2gene | 1147 | No | NA | 2 | 0 | 0 | Low-quality | Genome-fragment | | 1.88 | AAI-based (high-confidence) | 0 |
| S3_extracted_k141_33312 | 1147 | No | NA | 3 | 1 | 0 | Low-quality | Genome-fragment | | 2.64 | AAI-based (high-confidence) | 0 |
| S7_vs_50932\|\|full | 1147 | No | NA | 3 | 0 | 0 | Low-quality | Genome-fragment | | 2.81 | AAI-based (high-confidence) | 0 |
| S7_vs_90137\|\|full | 1147 | No | NA | 2 | 2 | 0 | Low-quality | Genome-fragment | | 2.47 | HMM-based (lower-bound) | 0 |
| S9_vs_k141_27450\|\|lt2gene | 1002 | No | NA | 1 | 0 | 0 | Not-determined | Genome-fragment | NA | | NA | 0 |
| S9_vs_k141_35128\|\|lt2gene | 1910 | No | NA | 1 | 0 | 0 | Not-determined | Genome-fragment | NA | | NA | 0 |
| S16_extracted_89805 | 1147 | No | NA | 4 | 0 | 0 | Low-quality | Genome-fragment | | 1.81 | AAI-based (high-confidence) | 0 |
| S9_vs_k141_46489\|\|lt2gene | 1824 | No | NA | 1 | 0 | 0 | Not-determined | Genome-fragment | NA | | NA | 0 |
| S16_extracted_132968 | 1147 | No | NA | 2 | 0 | 1 | Low-quality | Genome-fragment | | 2.21 | AAI-based (medium-confidence) | 0 |
| S16_vs_23055\|\|lt2gene | 1147 | No | NA | 2 | 0 | 0 | Low-quality | Genome-fragment | | 2.77 | AAI-based (medium-confidence) | 0 |
| S10_extracted_48292 | 1147 | No | NA | 2 | 0 | 1 | Low-quality | Genome-fragment | | 2.73 | AAI-based (high-confidence) | 0 |
| S9_vs_k141_16515\|\|lt2gene | 1254 | No | NA | 1 | 0 | 0 | Not-determined | Genome-fragment | NA | | NA | 0 |
| S10_vs_52161\|\|full | 1147 | No | NA | 3 | 1 | 0 | Low-quality | Genome-fragment | | 1.67 | AAI-based (high-confidence) | 0 |
| S8_vs_k141_143740\|\|full | 1147 | No | NA | 3 | 1 | 0 | Low-quality | Genome-fragment | | 3.05 | AAI-based (high-confidence) | 0 |
| S8_vs_k141_154279\|\|full | 1147 | No | NA | 4 | 1 | 0 | Low-quality | Genome-fragment | | 2.12 | AAI-based (high-confidence) | 0 |
| S17_vs_k141_27246\|\|full | 1148 | No | NA | 3 | 2 | 0 | Low-quality | Genome-fragment | | 2.11 | AAI-based (high-confidence) | 0 |
| S9_vs_k141_30531\|\|full | 1148 | No | NA | 3 | 2 | 0 | Low-quality | Genome-fragment | | 0.91 | HMM-based (lower-bound) | 0 |
| S3_vs_k141_9681\|\|full | 1148 | No | NA | 4 | 1 | 0 | Low-quality | Genome-fragment | | 1.88 | AAI-based (high-confidence) | 0 |
| S5_extracted_k141_38231 | 1148 | No | NA | 3 | 3 | 0 | Low-quality | Genome-fragment | | 3.77 | AAI-based (high-confidence) | 0 |
| S7_extracted_111877 | 1148 | No | NA | 5 | 0 | 0 | Low-quality | Genome-fragment | | 2.47 | AAI-based (medium-confidence) | 0 |
| S9_vs_k141_28254\|\|lt2gene | 1863 | No | NA | 1 | 0 | 0 | Not-determined | Genome-fragment | NA | | NA | 0 |
| S16_vs_35774\|\|lt2gene | 1148 | No | NA | 2 | 1 | 0 | Low-quality | Genome-fragment | | 2.76 | AAI-based (high-confidence) | 0 |
| S9_vs_k141_35931\|\|lt2gene | 2363 | No | NA | 1 | 0 | 1 | Not-determined | Genome-fragment | NA | | NA | 0 |
| S12_vs_k141_124732\|\|full | 1148 | No | NA | 4 | 1 | 0 | Low-quality | Genome-fragment | | 1.89 | AAI-based (medium-confidence) | 0 |
| S2_vs_k141_15593\|\|full | 1148 | No | NA | 3 | 0 | 0 | Low-quality | Genome-fragment | | 2.01 | AAI-based (medium-confidence) | 0 |
| S9_vs_k141_62430\|\|lt2gene | 1184 | No | NA | 1 | 0 | 0 | Not-determined | Genome-fragment | NA | | NA | 0 |
| S8_vs_k141_189104\|\|full | 1148 | No | NA | 2 | 0 | 0 | Low-quality | Genome-fragment | | 0.55 | AAI-based (medium-confidence) | 0 |
| S11_vs_k141_56219\|\|full | 1149 | No | NA | 3 | 1 | 0 | Low-quality | Genome-fragment | | 8.51 | AAI-based (high-confidence) | 0 |
| S13_extracted_47576 | 1149 | No | NA | 3 | 1 | 0 | Low-quality | Genome-fragment | | 3.18 | AAI-based (high-confidence) | 0 |
| S14_extracted_k141_114742 | 1149 | No | NA | 1 | 0 | 1 | Low-quality | Genome-fragment | | 0.33 | AAI-based (medium-confidence) | 0 |
| S1_extracted_k141_21632 | 1149 | No | NA | 1 | 0 | 0 | Low-quality | Genome-fragment | | 0.72 | AAI-based (high-confidence) | 0 |
| S1_vs_k141_110286\|\|full | 1149 | No | NA | 4 | 0 | 0 | Low-quality | Genome-fragment | | 1.75 | AAI-based (high-confidence) | 0 |
| S3_extracted_k141_10466 | 1149 | No | NA | 3 | 2 | 0 | Low-quality | Genome-fragment | | 2.01 | AAI-based (high-confidence) | 0 |
| S7_extracted_109515 | 1149 | No | NA | 1 | 0 | 0 | Low-quality | Genome-fragment | | 1.76 | AAI-based (medium-confidence) | 0 |
| S7_vs_84701\|\|full | 1149 | No | NA | 2 | 0 | 0 | Low-quality | Genome-fragment | | 2.08 | AAI-based (medium-confidence) | 0 |
| S16_extracted_75336 | 1149 | No | NA | 2 | 1 | 0 | Low-quality | Genome-fragment | | 2.74 | AAI-based (high-confidence) | 0 |
| S16_vs_144209\|\|full | 1149 | No | NA | 1 | 1 | 0 | Low-quality | Genome-fragment | | 0.66 | AAI-based (high-confidence) | 0 |
| S16_vs_164770\|\|full | 1149 | No | NA | 2 | 0 | 0 | Low-quality | Genome-fragment | | 2.43 | AAI-based (high-confidence) | 0 |
| S10_extracted_72212 | 1149 | No | NA | 2 | 1 | 0 | Low-quality | Genome-fragment | | 2.01 | AAI-based (medium-confidence) | 0 |
| S4_extracted_2012 | 1149 | No | NA | 3 | 1 | 0 | Low-quality | Genome-fragment | | 2.1 | AAI-based (high-confidence) | 0 |
| S8_extracted_k141_110426 | 1149 | No | NA | 3 | 0 | 3 | Low-quality | Genome-fragment | | 1.02 | AAI-based (medium-confidence) | 0 |
| S8_vs_k141_60392\|\|full | 1149 | No | NA | 3 | 0 | 1 | Low-quality | Genome-fragment | | 0.77 | AAI-based (high-confidence) | 0 |
| S17_vs_k141_80458\|\|full | 1150 | No | NA | 3 | 0 | 0 | Low-quality | Genome-fragment | | 0.82 | AAI-based (medium-confidence) | 0 |
| S17_vs_k141_28362\|\|full | 1150 | No | NA | 3 | 0 | 0 | Low-quality | Genome-fragment | | 0.85 | AAI-based (medium-confidence) | 0 |
| S11_extracted_k141_99225 | 1150 | No | NA | 2 | 1 | 0 | Low-quality | Genome-fragment | | 1.86 | AAI-based (high-confidence) | 0 |
| S13_vs_87198\|\|full | 1150 | No | NA | 4 | 0 | 0 | Low-quality | Genome-fragment | | 0.97 | AAI-based (medium-confidence) | 0 |
| S14_vs_k141_88714\|\|full | 1150 | No | NA | 3 | 0 | 0 | Low-quality | Genome-fragment | | 0.37 | AAI-based (medium-confidence) | 0 |
| S5_vs_k141_24321\|\|lt2gene | 1150 | No | NA | 2 | 2 | 0 | Low-quality | Genome-fragment | | 3.38 | AAI-based (high-confidence) | 0 |
| S7_extracted_27797 | 1150 | No | NA | 2 | 0 | 2 | Low-quality | Genome-fragment | | 0.69 | AAI-based (high-confidence) | 0 |
| S7_vs_37168\|\|full | 1150 | No | NA | 4 | 1 | 0 | Low-quality | Genome-fragment | | 3.34 | AAI-based (medium-confidence) | 0 |
| S16_vs_68\|\|full | 1150 | No | NA | 3 | 1 | 0 | Low-quality | Genome-fragment | | 2.28 | AAI-based (high-confidence) | 0 |
| S16_vs_152325\|\|full | 1150 | No | NA | 2 | 0 | 0 | Low-quality | Genome-fragment | | 1.07 | AAI-based (high-confidence) | 0 |
| S10_vs_104152\|\|full | 1150 | No | NA | 4 | 0 | 0 | Low-quality | Genome-fragment | | 1.88 | AAI-based (high-confidence) | 0 |
| S10_vs_87428\|\|lt2gene | 1150 | No | NA | 2 | 1 | 0 | Low-quality | Genome-fragment | | 2.66 | AAI-based (medium-confidence) | 0 |
| S8_extracted_k141_29890 | 1150 | No | NA | 3 | 0 | 0 | Low-quality | Genome-fragment | | 2.99 | AAI-based (high-confidence) | 0 |
| S8_vs_k141_167776\|\|full | 1150 | No | NA | 3 | 2 | 0 | Low-quality | Genome-fragment | | 1.9 | AAI-based (high-confidence) | 0 |
| S9_extracted_k141_16518 | 1151 | No | NA | 4 | 2 | 0 | Low-quality | Genome-fragment | | 2.42 | AAI-based (medium-confidence) | 0 |
| S9_vs_k141_20273\|\|full | 1151 | No | NA | 2 | 0 | 0 | Low-quality | Genome-fragment | | 1.52 | AAI-based (medium-confidence) | 0 |
| S9_vs_k141_85064\|\|full | 1151 | No | NA | 4 | 2 | 0 | Low-quality | Genome-fragment | | 1.71 | AAI-based (medium-confidence) | 0 |
| S11_extracted_k141_71708 | 1151 | No | NA | 1 | 0 | 0 | Low-quality | Genome-fragment | | 1.96 | AAI-based (high-confidence) | 0 |
| S11_extracted_k141_65509 | 1151 | No | NA | 4 | 2 | 0 | Low-quality | Genome-fragment | | 2.83 | AAI-based (medium-confidence) | 0 |
| S9_vs_k141_52163\|\|lt2gene | 1272 | No | NA | 2 | 0 | 0 | Not-determined | Genome-fragment | NA | | NA | 0 |
| S9_vs_k141_75650\|\|lt2gene | 1377 | No | NA | 1 | 0 | 0 | Not-determined | Genome-fragment | NA | | NA | 0 |
| S11_vs_k141_89411\|\|full | 1151 | No | NA | 3 | 1 | 0 | Low-quality | Genome-fragment | | 0.79 | HMM-based (lower-bound) | 0 |
| S9_vs_k141_15103\|\|lt2gene | 1056 | No | NA | 1 | 0 | 0 | Not-determined | Genome-fragment | NA | | NA | 0 |
| S13_vs_137798\|\|full | 1151 | No | NA | 5 | 0 | 0 | Low-quality | Genome-fragment | | 0.72 | AAI-based (high-confidence) | 0 |
| S14_extracted_k141_46672 | 1151 | No | NA | 3 | 2 | 0 | Low-quality | Genome-fragment | | 2.95 | AAI-based (high-confidence) | 0 |
| S1_vs_k141_7927\|\|full | 1151 | No | NA | 3 | 1 | 0 | Low-quality | Genome-fragment | | 1.51 | HMM-based (lower-bound) | 0 |
| S11_extracted_k141_91116 | 2262 | No | NA | 2 | 0 | 0 | Not-determined | Genome-fragment | NA | | NA | 0 |
| S5_vs_k141_54589\|\|full | 1151 | No | NA | 2 | 0 | 0 | Low-quality | Genome-fragment | | 1.86 | AAI-based (medium-confidence) | 0 |
| S11_extracted_k141_71598 | 1001 | No | NA | 2 | 0 | 0 | Not-determined | Genome-fragment | NA | | NA | 0 |
| S11_extracted_k141_65124 | 1440 | No | NA | 2 | 0 | 1 | Not-determined | Genome-fragment | NA | | NA | 0 |
| S11_extracted_k141_19546 | 5780 | No | NA | 4 | 0 | 3 | Not-determined | Genome-fragment | NA | | NA | 0 |
| S11_extracted_k141_58587 | 16434 | No | NA | 21 | 6 | 0 | Medium-quality | Genome-fragment | | 74.1 | AAI-based (medium-confidence) | 0 |
| S11_extracted_k141_45605 | 4665 | No | NA | 6 | 0 | 1 | Not-determined | Genome-fragment | NA | | NA | 0 |
| S11_extracted_k141_97663 | 1041 | No | NA | 2 | 0 | 1 | Not-determined | Genome-fragment | NA | | NA | 0 |
| S11_extracted_k141_52127 | 1056 | No | NA | 2 | 0 | 1 | Not-determined | Genome-fragment | NA | | NA | 0 |
| S11_extracted_k141_71663 | 1488 | No | NA | 1 | 0 | 1 | Not-determined | Genome-fragment | NA | | NA | 0 |
| S11_extracted_k141_32596 | 1899 | No | NA | 1 | 0 | 0 | Not-determined | Genome-fragment | NA | | NA | 0 |
| S11_extracted_k141_97 | 2740 | No | NA | 3 | 0 | 2 | Not-determined | Genome-fragment | NA | | NA | 0 |
| S11_extracted_k141_13033 | 60444 | No | NA | 92 | 35 | 1 | High-quality | High-quality | | 100 | AAI-based (high-confidence) | 0 |
| S11_extracted_k141_58676 | 1049 | No | NA | 1 | 0 | 1 | Not-determined | Genome-fragment | NA | | NA | 0 |
| S7_extracted_26222 | 1151 | No | NA | 2 | 1 | 0 | Low-quality | Genome-fragment | | 2.92 | AAI-based (high-confidence) | 0 |
| S16_extracted_140387 | 1151 | No | NA | 2 | 1 | 0 | Low-quality | Genome-fragment | | 0.97 | AAI-based (high-confidence) | 0 |
| S16_vs_160053\|\|full | 1151 | No | NA | 5 | 1 | 0 | Low-quality | Genome-fragment | | 2.3 | AAI-based (high-confidence) | 0 |
| S12_extracted_k141_16621 | 1151 | No | NA | 1 | 0 | 0 | Low-quality | Genome-fragment | | 1.85 | AAI-based (high-confidence) | 0 |
| S11_extracted_k141_52218 | 1456 | No | NA | 2 | 0 | 1 | Not-determined | Genome-fragment | NA | | NA | 0 |
| S11_extracted_k141_32645 | 4659 | No | NA | 9 | 0 | 2 | Not-determined | Genome-fragment | NA | | NA | 0 |
| S12_vs_k141_166537\|\|full | 1151 | No | NA | 4 | 0 | 0 | Low-quality | Genome-fragment | | 2.29 | AAI-based (medium-confidence) | 0 |
| S11_extracted_k141_65285 | 1010 | No | NA | 3 | 0 | 0 | Not-determined | Genome-fragment | NA | | NA | 0 |
| S12_vs_k141_104741\|\|full | 1151 | No | NA | 2 | 0 | 0 | Low-quality | Genome-fragment | | 1.57 | AAI-based (high-confidence) | 0 |
| S17_extracted_k141_108623 | 1152 | No | NA | 2 | 0 | 1 | Low-quality | Genome-fragment | | 2.52 | AAI-based (medium-confidence) | 0 |
| S17_vs_k141_163292\|\|lt2gene | 1152 | No | NA | 1 | 1 | 0 | Low-quality | Genome-fragment | | 2.09 | HMM-based (lower-bound) | 0 |
| S11_extracted_k141_91332 | 1343 | No | NA | 1 | 0 | 1 | Not-determined | Genome-fragment | NA | | NA | 0 |
| S9_extracted_k141_86863 | 1152 | No | NA | 2 | 2 | 0 | Low-quality | Genome-fragment | | 2.22 | AAI-based (high-confidence) | 0 |
| S1_extracted_k141_61101 | 1152 | No | NA | 1 | 0 | 1 | Low-quality | Genome-fragment | | 1.14 | AAI-based (medium-confidence) | 0 |
| S3_extracted_k141_11592 | 1152 | No | NA | 2 | 0 | 0 | Low-quality | Genome-fragment | | 2.76 | AAI-based (high-confidence) | 0 |
| S5_vs_k141_100476\|\|full | 1152 | No | NA | 3 | 0 | 0 | Low-quality | Genome-fragment | | 1.99 | AAI-based (medium-confidence) | 0 |
| S11_extracted_k141_78244 | 1414 | No | NA | 2 | 0 | 2 | Not-determined | Genome-fragment | NA | | NA | 0 |
| S11_extracted_k141_32744 | 3750 | No | NA | 5 | 0 | 1 | Not-determined | Genome-fragment | NA | | NA | 0 |
| S11_extracted_k141_39225 | 1409 | No | NA | 3 | 0 | 1 | Not-determined | Genome-fragment | NA | | NA | 0 |
| S7_vs_32211\|\|full | 1152 | No | NA | 3 | 0 | 0 | Low-quality | Genome-fragment | | 2.58 | AAI-based (high-confidence) | 0 |
| S10_vs_76504\|\|full | 1152 | No | NA | 2 | 0 | 1 | Low-quality | Genome-fragment | | 1.35 | AAI-based (medium-confidence) | 0 |
| S12_vs_k141_137101\|\|full | 1152 | No | NA | 2 | 0 | 2 | Low-quality | Genome-fragment | | 0.62 | AAI-based (high-confidence) | 0 |
| S11_extracted_k141_58780 | 1026 | No | NA | 3 | 0 | 0 | Not-determined | Genome-fragment | NA | | NA | 0 |
| S11_extracted_k141_52371 | 3167 | No | NA | 1 | 0 | 1 | Not-determined | Genome-fragment | NA | | NA | 0 |
| S11_extracted_k141_32786 | 7443 | No | NA | 6 | 0 | 3 | Not-determined | Genome-fragment | NA | | NA | 0 |
| S11_extracted_k141_407 | 1472 | No | NA | 2 | 0 | 0 | Not-determined | Genome-fragment | NA | | NA | 0 |
| S6_extracted_k141_41535 | 1152 | No | NA | 2 | 0 | 1 | Low-quality | Genome-fragment | | 0.27 | AAI-based (high-confidence) | 0 |
| S11_extracted_k141_417 | 1761 | No | NA | 2 | 0 | 1 | Not-determined | Genome-fragment | NA | | NA | 0 |
| S11_extracted_k141_45673 | 1052 | No | NA | 2 | 0 | 0 | Not-determined | Genome-fragment | NA | | NA | 0 |
| S11_extracted_k141_78350 | 1012 | No | NA | 1 | 0 | 0 | Not-determined | Genome-fragment | NA | | NA | 0 |
| S11_extracted_k141_39277 | 1658 | No | NA | 4 | 0 | 0 | Not-determined | Genome-fragment | NA | | NA | 0 |
| S11_extracted_k141_65366 | 1302 | No | NA | 2 | 0 | 2 | Not-determined | Genome-fragment | NA | | NA | 0 |
| S11_extracted_k141_45707 | 7039 | No | NA | 9 | 0 | 0 | Not-determined | Genome-fragment | NA | | NA | 0 |
| S11_extracted_k141_58799 | 1722 | No | NA | 3 | 0 | 1 | Not-determined | Genome-fragment | NA | | NA | 0 |
| S11_extracted_k141_91455 | 1020 | No | NA | 2 | 0 | 2 | Not-determined | Genome-fragment | NA | | NA | 0 |
| S11_extracted_k141_6724 | 2253 | No | NA | 1 | 0 | 1 | Not-determined | Genome-fragment | NA | | NA | 0 |
| S11_extracted_k141_58810 | 2027 | No | NA | 1 | 0 | 0 | Not-determined | Genome-fragment | NA | | NA | 0 |
| S17_vs_k141_68913\|\|full | 1153 | No | NA | 4 | 0 | 1 | Low-quality | Genome-fragment | | 2.69 | AAI-based (medium-confidence) | 0 |
| S9_vs_k141_23189\|\|full | 1153 | No | NA | 3 | 2 | 0 | Low-quality | Genome-fragment | | 2.51 | AAI-based (medium-confidence) | 0 |
| S11_extracted_k141_78408 | 1305 | No | NA | 2 | 0 | 2 | Not-determined | Genome-fragment | NA | | NA | 0 |
| S11_extracted_k141_39325 | 2033 | No | NA | 4 | 0 | 3 | Not-determined | Genome-fragment | NA | | NA | 0 |
| S14_extracted_k141_92527 | 1153 | No | NA | 1 | 0 | 0 | Low-quality | Genome-fragment | | 0.92 | AAI-based (high-confidence) | 0 |
| S1_vs_k141_100373\|\|full | 1153 | No | NA | 2 | 1 | 0 | Low-quality | Genome-fragment | | 1.58 | AAI-based (high-confidence) | 0 |
| S11_extracted_k141_19840 | 1194 | No | NA | 1 | 0 | 1 | Not-determined | Genome-fragment | NA | | NA | 0 |
| S11_extracted_k141_84982 | 1589 | No | NA | 2 | 0 | 0 | Not-determined | Genome-fragment | NA | | NA | 0 |
| S3_extracted_k141_9318 | 1153 | No | NA | 3 | 0 | 0 | Low-quality | Genome-fragment | | 1.75 | AAI-based (high-confidence) | 0 |
| S11_extracted_k141_91514 | 1247 | No | NA | 2 | 0 | 1 | Not-determined | Genome-fragment | NA | | NA | 0 |
| S11_extracted_k141_26374 | 1612 | No | NA | 2 | 0 | 0 | Not-determined | Genome-fragment | NA | | NA | 0 |
| S11_extracted_k141_45751 | 1271 | No | NA | 2 | 0 | 0 | Not-determined | Genome-fragment | NA | | NA | 0 |
| S3_extracted_k141_35499 | 1153 | No | NA | 2 | 0 | 0 | Low-quality | Genome-fragment | | 0.65 | AAI-based (medium-confidence) | 0 |
| S11_extracted_k141_91528 | 1647 | No | NA | 2 | 0 | 0 | Not-determined | Genome-fragment | NA | | NA | 0 |
| S3_vs_k141_78702\|\|lt2gene | 1153 | No | NA | 1 | 1 | 0 | Low-quality | Genome-fragment | | 0.74 | HMM-based (lower-bound) | 0 |
| S11_extracted_k141_19884 | 3116 | No | NA | 2 | 0 | 2 | Not-determined | Genome-fragment | NA | | NA | 0 |
| S11_extracted_k141_26403 | 1610 | No | NA | 2 | 0 | 0 | Not-determined | Genome-fragment | NA | | NA | 0 |
| S5_extracted_k141_287 | 1153 | No | NA | 2 | 1 | 0 | Low-quality | Genome-fragment | | 3.35 | AAI-based (high-confidence) | 0 |
| S11_extracted_k141_39400 | 2668 | No | NA | 3 | 0 | 2 | Not-determined | Genome-fragment | NA | | NA | 0 |
| S11_extracted_k141_85051 | 1304 | No | NA | 2 | 0 | 0 | Not-determined | Genome-fragment | NA | | NA | 0 |
| S11_extracted_k141_78504 | 3191 | No | NA | 3 | 0 | 2 | Not-determined | Genome-fragment | NA | | NA | 0 |
| S11_extracted_k141_6824 | 4654 | No | NA | 8 | 0 | 0 | Not-determined | Genome-fragment | NA | | NA | 0 |
| S11_extracted_k141_45810 | 1336 | No | NA | 1 | 0 | 0 | Not-determined | Genome-fragment | NA | | NA | 0 |
| S7_vs_90963\|\|full | 1153 | No | NA | 3 | 1 | 0 | Low-quality | Genome-fragment | | 2.12 | AAI-based (high-confidence) | 0 |
| S11_extracted_k141_6846 | 1202 | No | NA | 2 | 0 | 1 | Not-determined | Genome-fragment | NA | | NA | 0 |
| S16_extracted_168110 | 1153 | No | NA | 3 | 0 | 0 | Low-quality | Genome-fragment | | 3.04 | AAI-based (high-confidence) | 0 |
| S16_vs_54099\|\|full | 1153 | No | NA | 3 | 1 | 0 | Low-quality | Genome-fragment | | 2.36 | AAI-based (medium-confidence) | 0 |
| S16_vs_130652\|\|lt2gene | 1153 | No | NA | 1 | 1 | 0 | Low-quality | Genome-fragment | | 0.8 | HMM-based (lower-bound) | 0 |
| S11_extracted_k141_633 | 1329 | No | NA | 4 | 0 | 0 | Not-determined | Genome-fragment | NA | | NA | 0 |
| S11_extracted_k141_78593 | 1461 | No | NA | 2 | 0 | 2 | Not-determined | Genome-fragment | NA | | NA | 0 |
| S11_extracted_k141_71993 | 1219 | No | NA | 3 | 0 | 3 | Not-determined | Genome-fragment | NA | | NA | 0 |
| S10_extracted_67249 | 1153 | No | NA | 2 | 0 | 0 | Low-quality | Genome-fragment | | 1.45 | AAI-based (high-confidence) | 0 |
| S10_vs_66379\|\|full | 1153 | No | NA | 3 | 1 | 0 | Low-quality | Genome-fragment | | 2.36 | AAI-based (high-confidence) | 0 |
| S11_extracted_k141_13236 | 7845 | No | NA | 7 | 0 | 5 | Not-determined | Genome-fragment | NA | | NA | 0 |
| S11_extracted_k141_45926 | 2077 | No | NA | 2 | 0 | 0 | Not-determined | Genome-fragment | NA | | NA | 0 |
| S12_vs_k141_112426\|\|full | 1153 | No | NA | 3 | 0 | 1 | Low-quality | Genome-fragment | | 0.67 | AAI-based (medium-confidence) | 0 |
| S6_extracted_k141_38142 | 1153 | No | NA | 3 | 0 | 1 | Low-quality | Genome-fragment | | 2.54 | AAI-based (high-confidence) | 0 |
| S17_vs_k141_156772\|\|full | 1154 | No | NA | 2 | 0 | 0 | Low-quality | Genome-fragment | | 2.32 | AAI-based (medium-confidence) | 0 |
| S11_extracted_k141_45939 | 3349 | No | NA | 2 | 0 | 2 | Not-determined | Genome-fragment | NA | | NA | 0 |
| S11_extracted_k141_39557 | 1289 | No | NA | 4 | 0 | 0 | Not-determined | Genome-fragment | NA | | NA | 0 |
| S11_extracted_k141_66113 | 1154 | No | NA | 2 | 2 | 0 | Low-quality | Genome-fragment | | 2.03 | AAI-based (high-confidence) | 0 |
| S11_vs_k141_15787\|\|full | 1154 | No | NA | 3 | 0 | 0 | Low-quality | Genome-fragment | | 2.7 | AAI-based (medium-confidence) | 0 |
| S13_extracted_37506 | 1154 | No | NA | 2 | 0 | 2 | Low-quality | Genome-fragment | | 0.6 | AAI-based (medium-confidence) | 0 |
| S11_extracted_k141_703 | 1047 | No | NA | 2 | 0 | 0 | Not-determined | Genome-fragment | NA | | NA | 0 |
| S5_vs_k141_33345\|\|full | 1154 | No | NA | 2 | 0 | 0 | Low-quality | Genome-fragment | | 1.25 | AAI-based (medium-confidence) | 0 |
| S11_extracted_k141_85156 | 4011 | No | NA | 2 | 0 | 0 | Not-determined | Genome-fragment | NA | | NA | 0 |
| S11_extracted_k141_85163 | 1792 | No | NA | 2 | 0 | 1 | Not-determined | Genome-fragment | NA | | NA | 0 |
| S16_extracted_121096 | 1154 | No | NA | 2 | 0 | 2 | Low-quality | Genome-fragment | | 0.35 | AAI-based (high-confidence) | 0 |
| S10_vs_85836\|\|full | 1154 | No | NA | 4 | 0 | 0 | Low-quality | Genome-fragment | | 2.51 | AAI-based (high-confidence) | 0 |
| S11_extracted_k141_52609 | 1017 | No | NA | 3 | 0 | 1 | Not-determined | Genome-fragment | NA | | NA | 0 |
| S11_extracted_k141_78714 | 1192 | No | NA | 2 | 0 | 2 | Not-determined | Genome-fragment | NA | | NA | 0 |
| S11_extracted_k141_78717 | 1030 | No | NA | 2 | 0 | 0 | Not-determined | Genome-fragment | NA | | NA | 0 |
| S11_extracted_k141_98202 | 1137 | No | NA | 2 | 0 | 0 | Not-determined | Genome-fragment | NA | | NA | 0 |
| S2_extracted_k141_16985 | 1154 | No | NA | 2 | 1 | 0 | Low-quality | Genome-fragment | | 0.72 | HMM-based (lower-bound) | 0 |
| S8_vs_k141_91626\|\|full | 1154 | No | NA | 4 | 0 | 0 | Low-quality | Genome-fragment | | 2.83 | AAI-based (medium-confidence) | 0 |
| S8_vs_k141_40278\|\|full | 1154 | No | NA | 3 | 1 | 0 | Low-quality | Genome-fragment | | 0.7 | HMM-based (lower-bound) | 0 |
| S11_extracted_k141_78761 | 3234 | No | NA | 3 | 0 | 3 | Not-determined | Genome-fragment | NA | | NA | 0 |
| S17_extracted_k141_77325 | 1155 | No | NA | 3 | 3 | 0 | Low-quality | Genome-fragment | | 3.81 | AAI-based (high-confidence) | 0 |
| S11_extracted_k141_13413 | 3231 | No | NA | 7 | 0 | 0 | Not-determined | Genome-fragment | NA | | NA | 0 |
| S11_extracted_k141_13418 | 2000 | No | NA | 2 | 0 | 1 | Not-determined | Genome-fragment | NA | | NA | 0 |
| S17_vs_k141_68082\|\|full | 1155 | No | NA | 3 | 0 | 0 | Low-quality | Genome-fragment | | 2.64 | AAI-based (medium-confidence) | 0 |
| S11_extracted_k141_39676 | 1432 | No | NA | 1 | 0 | 1 | Not-determined | Genome-fragment | NA | | NA | 0 |
| S11_extracted_k141_58999 | 1094 | No | NA | 1 | 0 | 1 | Not-determined | Genome-fragment | NA | | NA | 0 |
| S11_extracted_k141_33117 | 52675 | No | NA | 77 | 16 | 2 | Complete | High-quality | | 100 | DTR (high-confidence) | 0 |
| S9_vs_k141_86426\|\|lt2gene | 1155 | No | NA | 2 | 0 | 0 | Low-quality | Genome-fragment | | 2.02 | AAI-based (high-confidence) | 0 |
| S14_vs_k141_93273\|\|full | 1155 | No | NA | 1 | 0 | 0 | Low-quality | Genome-fragment | | 2.84 | AAI-based (medium-confidence) | 0 |
| S11_extracted_k141_842 | 1929 | No | NA | 2 | 0 | 2 | Not-determined | Genome-fragment | NA | | NA | 0 |
| S1_vs_k141_39887\|\|full | 1155 | No | NA | 3 | 0 | 1 | Low-quality | Genome-fragment | | 1.75 | AAI-based (medium-confidence) | 0 |
| S3_vs_k141_91146\|\|full | 1155 | No | NA | 3 | 2 | 0 | Low-quality | Genome-fragment | | 1.45 | AAI-based (high-confidence) | 0 |
| S11_extracted_k141_65695 | 2244 | No | NA | 2 | 0 | 0 | Not-determined | Genome-fragment | NA | | NA | 0 |
| S5_vs_k141_133720\|\|lt2gene | 1155 | No | NA | 1 | 0 | 0 | Low-quality | Genome-fragment | | 3.22 | AAI-based (medium-confidence) | 0 |
| S11_extracted_k141_59071 | 2869 | No | NA | 2 | 0 | 1 | Not-determined | Genome-fragment | NA | | NA | 0 |
| S7_extracted_12927 | 1155 | No | NA | 1 | 0 | 1 | Low-quality | Genome-fragment | | 2.07 | AAI-based (medium-confidence) | 0 |
| S11_extracted_k141_13559 | 3022 | No | NA | 1 | 0 | 0 | Not-determined | Genome-fragment | NA | | NA | 0 |
| S7_vs_47519\|\|full | 1155 | No | NA | 3 | 0 | 0 | Low-quality | Genome-fragment | | 1.12 | AAI-based (medium-confidence) | 0 |
| S11_extracted_k141_33253 | 10572 | No | NA | 10 | 0 | 2 | Not-determined | Genome-fragment | NA | | NA | 0 |
| S11_extracted_k141_46264 | 1206 | No | NA | 3 | 0 | 1 | Not-determined | Genome-fragment | NA | | NA | 0 |
| S11_extracted_k141_947 | 4657 | No | NA | 5 | 0 | 0 | Not-determined | Genome-fragment | NA | | NA | 0 |
| S16_extracted_169289 | 1155 | No | NA | 2 | 0 | 2 | Low-quality | Genome-fragment | | 2.06 | AAI-based (medium-confidence) | 0 |
| S11_extracted_k141_7050 | 4088 | No | NA | 2 | 0 | 2 | Not-determined | Genome-fragment | NA | | NA | 0 |
| S11_extracted_k141_85456 | 1042 | No | NA | 1 | 0 | 1 | Not-determined | Genome-fragment | NA | | NA | 0 |
| S16_vs_124138\|\|full | 1155 | No | NA | 3 | 0 | 0 | Low-quality | Genome-fragment | | 1.31 | AAI-based (medium-confidence) | 0 |
| S10_vs_103769\|\|full | 1155 | No | NA | 2 | 1 | 0 | Low-quality | Genome-fragment | | 2.28 | AAI-based (high-confidence) | 0 |
| S12_extracted_k141_53375 | 1155 | No | NA | 5 | 0 | 0 | Low-quality | Genome-fragment | | 3.12 | AAI-based (high-confidence) | 0 |
| S11_extracted_k141_39837 | 2372 | No | NA | 2 | 0 | 1 | Not-determined | Genome-fragment | NA | | NA | 0 |
| S11_extracted_k141_46334 | 1142 | No | NA | 1 | 0 | 0 | Not-determined | Genome-fragment | NA | | NA | 0 |
| S11_extracted_k141_46338 | 1720 | No | NA | 2 | 0 | 2 | Not-determined | Genome-fragment | NA | | NA | 0 |
| S11_extracted_k141_65804 | 2192 | No | NA | 5 | 0 | 0 | Not-determined | Genome-fragment | NA | | NA | 0 |
| S11_extracted_k141_65814 | 1078 | No | NA | 2 | 0 | 0 | Not-determined | Genome-fragment | NA | | NA | 0 |
| S12_vs_k141_18455\|\|full | 1155 | No | NA | 2 | 0 | 1 | Low-quality | Genome-fragment | | 2.59 | AAI-based (medium-confidence) | 0 |
| S11_extracted_k141_46370 | 1132 | No | NA | 2 | 0 | 2 | Not-determined | Genome-fragment | NA | | NA | 0 |
| S12_vs_k141_75208\|\|full | 1155 | No | NA | 2 | 0 | 0 | Low-quality | Genome-fragment | | 2.35 | AAI-based (medium-confidence) | 0 |
| S11_extracted_k141_1059 | 2401 | No | NA | 2 | 0 | 0 | Not-determined | Genome-fragment | NA | | NA | 0 |
| S11_extracted_k141_92052 | 1020 | No | NA | 1 | 0 | 1 | Not-determined | Genome-fragment | NA | | NA | 0 |
| S11_extracted_k141_52862 | 3191 | No | NA | 3 | 0 | 1 | Not-determined | Genome-fragment | NA | | NA | 0 |
| S11_extracted_k141_1069 | 7194 | No | NA | 11 | 0 | 5 | Not-determined | Genome-fragment | NA | | NA | 0 |
| S11_extracted_k141_33410 | 3095 | No | NA | 1 | 0 | 1 | Not-determined | Genome-fragment | NA | | NA | 0 |
| S12_vs_k141_52642\|\|lt2gene | 1155 | No | NA | 1 | 1 | 0 | Low-quality | Genome-fragment | | 0.63 | HMM-based (lower-bound) | 0 |
| S11_extracted_k141_98610 | 1354 | No | NA | 1 | 0 | 1 | Not-determined | Genome-fragment | NA | | NA | 0 |
| S8_extracted_k141_84220 | 1155 | No | NA | 2 | 0 | 1 | Low-quality | Genome-fragment | | 2.58 | AAI-based (medium-confidence) | 0 |
| S8_vs_k141_62168\|\|full | 1155 | No | NA | 3 | 0 | 1 | Low-quality | Genome-fragment | | 1.98 | AAI-based (medium-confidence) | 0 |
| S11_extracted_k141_98636 | 1002 | No | NA | 1 | 0 | 0 | Not-determined | Genome-fragment | NA | | NA | 0 |
| S11_extracted_k141_85620 | 1309 | No | NA | 2 | 0 | 2 | Not-determined | Genome-fragment | NA | | NA | 0 |
| S8_vs_k141_142732\|\|full | 1155 | No | NA | 3 | 0 | 2 | Low-quality | Genome-fragment | | 0.77 | AAI-based (medium-confidence) | 0 |
| S11_extracted_k141_46429 | 1319 | No | NA | 2 | 0 | 2 | Not-determined | Genome-fragment | NA | | NA | 0 |
| S11_extracted_k141_85629 | 2803 | No | NA | 2 | 0 | 0 | Not-determined | Genome-fragment | NA | | NA | 0 |
| S11_extracted_k141_52958 | 4896 | No | NA | 4 | 0 | 2 | Not-determined | Genome-fragment | NA | | NA | 0 |
| S11_extracted_k141_7239 | 1171 | No | NA | 2 | 0 | 1 | Not-determined | Genome-fragment | NA | | NA | 0 |
| S8_vs_k141_127699\|\|full | 1155 | No | NA | 2 | 0 | 0 | Low-quality | Genome-fragment | | 1.84 | AAI-based (medium-confidence) | 0 |
| S11_extracted_k141_7248 | 1145 | No | NA | 3 | 0 | 0 | Not-determined | Genome-fragment | NA | | NA | 0 |
| S11_extracted_k141_65889 | 1223 | No | NA | 2 | 0 | 0 | Not-determined | Genome-fragment | NA | | NA | 0 |
| S11_extracted_k141_52977 | 1258 | No | NA | 1 | 0 | 1 | Not-determined | Genome-fragment | NA | | NA | 0 |
| S8_vs_k141_120135\|\|full | 1155 | No | NA | 3 | 1 | 0 | Low-quality | Genome-fragment | | 2.94 | AAI-based (high-confidence) | 0 |
| S11_extracted_k141_79209 | 1733 | No | NA | 3 | 0 | 3 | Not-determined | Genome-fragment | NA | | NA | 0 |
| S11_extracted_k141_33433 | 77933 | No | NA | 88 | 16 | 1 | Complete | High-quality | | 100 | DTR (high-confidence) | 0 |
| S11_extracted_k141_46482 | 1006 | No | NA | 1 | 0 | 1 | Not-determined | Genome-fragment | NA | | NA | 0 |
| S11_extracted_k141_59366 | 3910 | No | NA | 2 | 0 | 1 | Not-determined | Genome-fragment | NA | | NA | 0 |
| S17_extracted_k141_69129 | 1156 | No | NA | 3 | 0 | 3 | Low-quality | Genome-fragment | | 2.61 | AAI-based (medium-confidence) | 0 |
| S11_extracted_k141_7335 | 3375 | No | NA | 2 | 0 | 2 | Not-determined | Genome-fragment | NA | | NA | 0 |
| S11_extracted_k141_40007 | 1338 | No | NA | 1 | 0 | 0 | Not-determined | Genome-fragment | NA | | NA | 0 |
| S17_vs_k141_19595\|\|full | 1156 | No | NA | 2 | 0 | 0 | Low-quality | Genome-fragment | | 1.15 | AAI-based (high-confidence) | 0 |
| S11_extracted_k141_66001 | 5907 | No | NA | 6 | 0 | 5 | Not-determined | Genome-fragment | NA | | NA | 0 |
| S11_extracted_k141_72691 | 1070 | No | NA | 1 | 0 | 1 | Not-determined | Genome-fragment | NA | | NA | 0 |
| S11_extracted_k141_92196 | 2226 | No | NA | 3 | 0 | 3 | Not-determined | Genome-fragment | NA | | NA | 0 |
| S11_extracted_k141_79267 | 1219 | No | NA | 2 | 0 | 2 | Not-determined | Genome-fragment | NA | | NA | 0 |
| S11_extracted_k141_7400 | 1653 | No | NA | 2 | 0 | 0 | Not-determined | Genome-fragment | NA | | NA | 0 |
| S11_extracted_k141_53099 | 2880 | No | NA | 3 | 0 | 1 | Not-determined | Genome-fragment | NA | | NA | 0 |
| S11_extracted_k141_53110 | 3815 | No | NA | 5 | 0 | 4 | Not-determined | Genome-fragment | NA | | NA | 0 |
| S11_extracted_k141_27020 | 1222 | No | NA | 4 | 0 | 1 | Not-determined | Genome-fragment | NA | | NA | 0 |
| S11_extracted_k141_7442 | 12033 | No | NA | 9 | 0 | 4 | Not-determined | Genome-fragment | NA | | NA | 0 |
| S11_extracted_k141_53165 | 1758 | No | NA | 2 | 0 | 2 | Not-determined | Genome-fragment | NA | | NA | 0 |
| S11_extracted_k141_53186 | 4689 | No | NA | 2 | 0 | 1 | Not-determined | Genome-fragment | NA | | NA | 0 |
| S17_vs_k141_50957\|\|full | 1156 | No | NA | 3 | 0 | 0 | Low-quality | Genome-fragment | | 2.47 | AAI-based (medium-confidence) | 0 |
| S11_extracted_k141_79342 | 1259 | No | NA | 1 | 0 | 1 | Not-determined | Genome-fragment | NA | | NA | 0 |
| S11_extracted_k141_40129 | 2977 | No | NA | 3 | 0 | 1 | Not-determined | Genome-fragment | NA | | NA | 0 |
| S9_vs_k141_38100\|\|full | 1156 | No | NA | 4 | 0 | 0 | Low-quality | Genome-fragment | | 0.67 | AAI-based (medium-confidence) | 0 |
| S11_extracted_k141_33661 | 1113 | No | NA | 2 | 0 | 1 | Not-determined | Genome-fragment | NA | | NA | 0 |
| S11_extracted_k141_66148 | 1299 | No | NA | 2 | 0 | 1 | Not-determined | Genome-fragment | NA | | NA | 0 |
| S9_vs_k141_89126\|\|full | 1156 | No | NA | 4 | 1 | 0 | Low-quality | Genome-fragment | | 3.05 | AAI-based (high-confidence) | 0 |
| S11_extracted_k141_7534 | 4233 | No | NA | 2 | 0 | 2 | Not-determined | Genome-fragment | NA | | NA | 0 |
| S11_extracted_k141_7536 | 1716 | No | NA | 1 | 0 | 1 | Not-determined | Genome-fragment | NA | | NA | 0 |
| S9_vs_k141_72840\|\|lt2gene | 1156 | No | NA | 1 | 1 | 0 | Low-quality | Genome-fragment | | 2.66 | AAI-based (high-confidence) | 0 |
| S11_extracted_k141_1317 | 1151 | No | NA | 3 | 0 | 3 | Not-determined | Genome-fragment | NA | | NA | 0 |
| S11_extracted_k141_55358 | 1156 | No | NA | 1 | 1 | 0 | Low-quality | Genome-fragment | | 0.79 | HMM-based (lower-bound) | 0 |
| S11_extracted_k141_53287 | 1191 | No | NA | 2 | 0 | 0 | Not-determined | Genome-fragment | NA | | NA | 0 |
| S13_vs_141710\|\|full | 1156 | No | NA | 2 | 0 | 1 | Low-quality | Genome-fragment | | 2 | AAI-based (medium-confidence) | 0 |
| S14_extracted_k141_104017 | 1156 | No | NA | 1 | 1 | 0 | Low-quality | Genome-fragment | | 2.71 | HMM-based (lower-bound) | 0 |
| S11_extracted_k141_66160 | 56970 | No | NA | 87 | 13 | 1 | High-quality | High-quality | | 100 | AAI-based (high-confidence) | 0 |
| S1_vs_k141_144315\|\|full | 1156 | No | NA | 3 | 0 | 0 | Low-quality | Genome-fragment | | 2.94 | AAI-based (high-confidence) | 0 |
| S11_extracted_k141_20813 | 1902 | No | NA | 2 | 0 | 2 | Not-determined | Genome-fragment | NA | | NA | 0 |
| S11_extracted_k141_92319 | 2152 | No | NA | 2 | 0 | 1 | Not-determined | Genome-fragment | NA | | NA | 0 |
| S11_extracted_k141_72985 | 1054 | No | NA | 1 | 0 | 0 | Not-determined | Genome-fragment | NA | | NA | 0 |
| S11_extracted_k141_59708 | 1080 | No | NA | 1 | 0 | 0 | Not-determined | Genome-fragment | NA | | NA | 0 |
| S3_vs_k141_88852\|\|full | 1156 | No | NA | 2 | 0 | 1 | Low-quality | Genome-fragment | | 1.52 | AAI-based (medium-confidence) | 0 |
| S3_vs_k141_31033\|\|full | 1156 | No | NA | 2 | 0 | 1 | Low-quality | Genome-fragment | | 0.68 | AAI-based (medium-confidence) | 0 |
| S5_vs_k141_458\|\|lt2gene | 1156 | No | NA | 2 | 1 | 0 | Low-quality | Genome-fragment | | 1.96 | AAI-based (medium-confidence) | 0 |
| S7_extracted_8118 | 1156 | No | NA | 4 | 0 | 0 | Low-quality | Genome-fragment | | 1.13 | AAI-based (medium-confidence) | 0 |
| S11_extracted_k141_20878 | 1001 | No | NA | 2 | 0 | 2 | Not-determined | Genome-fragment | NA | | NA | 0 |
| S11_extracted_k141_27236 | 1002 | No | NA | 1 | 0 | 1 | Not-determined | Genome-fragment | NA | | NA | 0 |
| S11_extracted_k141_40221 | 1468 | No | NA | 2 | 0 | 1 | Not-determined | Genome-fragment | NA | | NA | 0 |
| S11_extracted_k141_27241 | 1866 | No | NA | 4 | 0 | 1 | Not-determined | Genome-fragment | NA | | NA | 0 |
| S16_vs_24111\|\|full | 1156 | No | NA | 2 | 0 | 0 | Low-quality | Genome-fragment | | 1.25 | AAI-based (high-confidence) | 0 |
| S11_extracted_k141_1494 | 1064 | No | NA | 1 | 0 | 1 | Not-determined | Genome-fragment | NA | | NA | 0 |
| S11_extracted_k141_86087 | 4605 | No | NA | 3 | 0 | 2 | Not-determined | Genome-fragment | NA | | NA | 0 |
| S11_extracted_k141_33774 | 2045 | No | NA | 1 | 0 | 0 | Not-determined | Genome-fragment | NA | | NA | 0 |
| S11_extracted_k141_92391 | 15893 | No | NA | 22 | 7 | 0 | Medium-quality | Genome-fragment | | 52.52 | HMM-based (lower-bound) | 0 |
| S11_extracted_k141_20907 | 1597 | No | NA | 2 | 0 | 0 | Not-determined | Genome-fragment | NA | | NA | 0 |
| S11_extracted_k141_92396 | 1328 | No | NA | 1 | 0 | 0 | Not-determined | Genome-fragment | NA | | NA | 0 |
| S12_vs_k141_1138\|\|full | 1156 | No | NA | 2 | 0 | 0 | Low-quality | Genome-fragment | | 1.97 | AAI-based (high-confidence) | 0 |
| S11_extracted_k141_79449 | 1032 | No | NA | 1 | 0 | 1 | Not-determined | Genome-fragment | NA | | NA | 0 |
| S11_extracted_k141_46742 | 5120 | No | NA | 4 | 0 | 0 | Not-determined | Genome-fragment | NA | | NA | 0 |
| S12_vs_k141_1548\|\|full | 1156 | No | NA | 4 | 0 | 0 | Low-quality | Genome-fragment | | 1.38 | AAI-based (high-confidence) | 0 |
| S11_extracted_k141_59774 | 1165 | No | NA | 1 | 0 | 1 | Not-determined | Genome-fragment | NA | | NA | 0 |
| S15_extracted_53618 | 1156 | No | NA | 1 | 0 | 0 | Low-quality | Genome-fragment | | 0.82 | AAI-based (high-confidence) | 0 |
| S4_extracted_9057 | 1156 | No | NA | 3 | 0 | 0 | Low-quality | Genome-fragment | | 1.22 | AAI-based (high-confidence) | 0 |
| S11_extracted_k141_27297 | 1425 | No | NA | 2 | 0 | 2 | Not-determined | Genome-fragment | NA | | NA | 0 |
| S8_vs_k141_9370\|\|lt2gene | 1156 | No | NA | 2 | 1 | 0 | Low-quality | Genome-fragment | | 1.95 | AAI-based (high-confidence) | 0 |
| S11_extracted_k141_46758 | 1182 | No | NA | 1 | 0 | 0 | Not-determined | Genome-fragment | NA | | NA | 0 |
| S11_extracted_k141_99014 | 1989 | No | NA | 5 | 0 | 1 | Not-determined | Genome-fragment | NA | | NA | 0 |
| S11_extracted_k141_73154 | 4914 | No | NA | 4 | 0 | 3 | Not-determined | Genome-fragment | NA | | NA | 0 |
| S17_vs_k141_170932\|\|full | 1157 | No | NA | 2 | 1 | 0 | Low-quality | Genome-fragment | | 0.82 | HMM-based (lower-bound) | 0 |
| S11_extracted_k141_100340 | 1157 | No | NA | 2 | 1 | 0 | Low-quality | Genome-fragment | | 0.81 | AAI-based (high-confidence) | 0 |
| S11_extracted_k141_1605 | 1038 | No | NA | 1 | 0 | 0 | Not-determined | Genome-fragment | NA | | NA | 0 |
| S11_extracted_k141_27335 | 1249 | No | NA | 1 | 0 | 0 | Not-determined | Genome-fragment | NA | | NA | 0 |
| S11_vs_k141_774\|\|full | 1157 | No | NA | 4 | 1 | 0 | Low-quality | Genome-fragment | | 7.36 | AAI-based (high-confidence) | 0 |
| S11_vs_k141_74087\|\|full | 1157 | No | NA | 2 | 0 | 0 | Low-quality | Genome-fragment | | 2.29 | AAI-based (high-confidence) | 0 |
| S11_extracted_k141_99045 | 3480 | No | NA | 3 | 0 | 1 | Not-determined | Genome-fragment | NA | | NA | 0 |
| S11_extracted_k141_14040 | 3195 | No | NA | 3 | 0 | 2 | Not-determined | Genome-fragment | NA | | NA | 0 |
| S11_extracted_k141_86211 | 4231 | No | NA | 4 | 0 | 1 | Not-determined | Genome-fragment | NA | | NA | 0 |
| S13_extracted_115423 | 1157 | No | NA | 1 | 0 | 1 | Low-quality | Genome-fragment | | 0.51 | AAI-based (high-confidence) | 0 |
| S11_extracted_k141_14072 | 1455 | No | NA | 2 | 0 | 2 | Not-determined | Genome-fragment | NA | | NA | 0 |
| S11_extracted_k141_27391 | 2140 | No | NA | 1 | 0 | 0 | Not-determined | Genome-fragment | NA | | NA | 0 |
| S11_extracted_k141_27395 | 1026 | No | NA | 1 | 0 | 0 | Not-determined | Genome-fragment | NA | | NA | 0 |
| S3_vs_k141_20908\|\|full | 1157 | No | NA | 2 | 0 | 1 | Low-quality | Genome-fragment | | 2.21 | AAI-based (medium-confidence) | 0 |
| S11_extracted_k141_21034 | 2249 | No | NA | 3 | 0 | 2 | Not-determined | Genome-fragment | NA | | NA | 0 |
| S11_extracted_k141_53524 | 1237 | No | NA | 2 | 0 | 1 | Not-determined | Genome-fragment | NA | | NA | 0 |
| S11_extracted_k141_21059 | 1014 | No | NA | 1 | 0 | 0 | Not-determined | Genome-fragment | NA | | NA | 0 |
| S11_extracted_k141_86314 | 1739 | No | NA | 2 | 0 | 1 | Not-determined | Genome-fragment | NA | | NA | 0 |
| S11_extracted_k141_99068 | 3554 | No | NA | 3 | 0 | 0 | Not-determined | Genome-fragment | NA | | NA | 0 |
| S7_extracted_63068 | 1157 | No | NA | 2 | 0 | 1 | Low-quality | Genome-fragment | | 2.34 | AAI-based (medium-confidence) | 0 |
| S11_extracted_k141_33868 | 1996 | No | NA | 2 | 0 | 1 | Not-determined | Genome-fragment | NA | | NA | 0 |
| S16_vs_102148\|\|full | 1157 | No | NA | 3 | 0 | 1 | Low-quality | Genome-fragment | | 1.73 | AAI-based (medium-confidence) | 0 |
| S11_extracted_k141_53586 | 1723 | No | NA | 3 | 0 | 0 | Not-determined | Genome-fragment | NA | | NA | 0 |
| S11_extracted_k141_53591 | 6063 | No | NA | 6 | 0 | 2 | Not-determined | Genome-fragment | NA | | NA | 0 |
| S11_extracted_k141_46871 | 4490 | No | NA | 3 | 0 | 1 | Not-determined | Genome-fragment | NA | | NA | 0 |
| S11_extracted_k141_14176 | 1669 | No | NA | 1 | 0 | 1 | Not-determined | Genome-fragment | NA | | NA | 0 |
| S11_extracted_k141_66409 | 1005 | No | NA | 2 | 0 | 0 | Not-determined | Genome-fragment | NA | | NA | 0 |
| S11_extracted_k141_40437 | 1008 | No | NA | 3 | 0 | 0 | Not-determined | Genome-fragment | NA | | NA | 0 |
| S11_extracted_k141_7978 | 1084 | No | NA | 2 | 0 | 1 | Not-determined | Genome-fragment | NA | | NA | 0 |
| S16_vs_149086\|\|full | 1157 | No | NA | 2 | 0 | 1 | Low-quality | Genome-fragment | | 1.29 | AAI-based (medium-confidence) | 0 |
| S11_extracted_k141_7990 | 1291 | No | NA | 2 | 0 | 0 | Not-determined | Genome-fragment | NA | | NA | 0 |
| S11_extracted_k141_21136 | 1342 | No | NA | 1 | 0 | 1 | Not-determined | Genome-fragment | NA | | NA | 0 |
| S12_vs_k141_25892\|\|full | 1157 | No | NA | 4 | 0 | 0 | Low-quality | Genome-fragment | | 2.68 | AAI-based (medium-confidence) | 0 |
| S15_extracted_58179 | 1157 | No | NA | 3 | 1 | 0 | Low-quality | Genome-fragment | | 2.67 | AAI-based (high-confidence) | 0 |
| S11_extracted_k141_66456 | 1122 | No | NA | 1 | 0 | 1 | Not-determined | Genome-fragment | NA | | NA | 0 |
| S11_extracted_k141_21148 | 1433 | No | NA | 3 | 0 | 0 | Not-determined | Genome-fragment | NA | | NA | 0 |
| S8_extracted_k141_158982 | 1157 | No | NA | 3 | 0 | 1 | Low-quality | Genome-fragment | | 0.9 | AAI-based (medium-confidence) | 0 |
| S11_extracted_k141_27614 | 1621 | No | NA | 2 | 0 | 2 | Not-determined | Genome-fragment | NA | | NA | 0 |
| S11_extracted_k141_99134 | 3626 | No | NA | 7 | 0 | 5 | Not-determined | Genome-fragment | NA | | NA | 0 |
| S11_extracted_k141_8030 | 1367 | No | NA | 2 | 0 | 2 | Not-determined | Genome-fragment | NA | | NA | 0 |
| S11_extracted_k141_46975 | 4628 | No | NA | 3 | 0 | 3 | Not-determined | Genome-fragment | NA | | NA | 0 |
| S11_extracted_k141_46978 | 2729 | No | NA | 1 | 0 | 1 | Not-determined | Genome-fragment | NA | | NA | 0 |
| S8_extracted_k141_15754 | 1157 | No | NA | 5 | 1 | 0 | Low-quality | Genome-fragment | | 1.06 | AAI-based (high-confidence) | 0 |
| S11_extracted_k141_8076 | 1204 | No | NA | 2 | 0 | 1 | Not-determined | Genome-fragment | NA | | NA | 0 |
| S17_vs_k141_107758\|\|full | 1158 | No | NA | 3 | 2 | 0 | Low-quality | Genome-fragment | | 2.09 | HMM-based (lower-bound) | 0 |
| S11_vs_k141_46942\|\|lt2gene | 1158 | No | NA | 1 | 1 | 0 | Low-quality | Genome-fragment | | 2.91 | AAI-based (medium-confidence) | 0 |
| S11_vs_k141_2540\|\|lt2gene | 1158 | No | NA | 2 | 1 | 0 | Low-quality | Genome-fragment | | 1.38 | HMM-based (lower-bound) | 0 |
| S11_extracted_k141_99201 | 1147 | No | NA | 2 | 0 | 0 | Not-determined | Genome-fragment | NA | | NA | 0 |
| S11_extracted_k141_53689 | 2158 | No | NA | 2 | 0 | 0 | Not-determined | Genome-fragment | NA | | NA | 0 |
| S11_extracted_k141_66522 | 27814 | No | NA | 38 | 20 | 0 | High-quality | High-quality | | 92.74 | AAI-based (high-confidence) | 0 |
| S11_extracted_k141_8167 | 1061 | No | NA | 2 | 0 | 1 | Not-determined | Genome-fragment | NA | | NA | 0 |
| S1_extracted_k141_151569 | 1158 | No | NA | 2 | 0 | 2 | Low-quality | Genome-fragment | | 0.87 | AAI-based (medium-confidence) | 0 |
| S11_extracted_k141_92575 | 1046 | No | NA | 3 | 0 | 1 | Not-determined | Genome-fragment | NA | | NA | 0 |
| S11_extracted_k141_53719 | 2053 | No | NA | 3 | 0 | 0 | Not-determined | Genome-fragment | NA | | NA | 0 |
| S11_extracted_k141_27751 | 4200 | No | NA | 6 | 0 | 0 | Not-determined | Genome-fragment | NA | | NA | 0 |
| S11_extracted_k141_40471 | 1793 | No | NA | 3 | 0 | 1 | Not-determined | Genome-fragment | NA | | NA | 0 |
| S11_extracted_k141_14426 | 1611 | No | NA | 3 | 0 | 1 | Not-determined | Genome-fragment | NA | | NA | 0 |

| | | | | | | | | | | | |
|---|---|---|---|---|---|---|---|---|---|---|---|
| S3_extracted_k141_123490 | 1158 | No | NA | 2 | 0 | 0 | Low-quality | Genome-fragment | 2.39 | AAI-based (high-confidence) | 0 |
| S7_vs_32454\|\|full | 1158 | No | NA | 4 | 0 | 0 | Low-quality | Genome-fragment | 2.77 | AAI-based (medium-confidence) | 0 |
| S16_extracted_79347 | 1158 | No | NA | 4 | 0 | 0 | Low-quality | Genome-fragment | 2.14 | AAI-based (high-confidence) | 0 |
| S16_extracted_107975 | 1158 | No | NA | 2 | 0 | 0 | Low-quality | Genome-fragment | 0.56 | AAI-based (medium-confidence) | 0 |
| S11_extracted_k141_53758 | 1024 | No | NA | 1 | 0 | 1 | Not-determined | Genome-fragment | NA | NA | 0 |
| S11_extracted_k141_47124 | 3784 | No | NA | 4 | 0 | 3 | Not-determined | Genome-fragment | NA | NA | 0 |
| S11_extracted_k141_40509 | 1419 | No | NA | 1 | 0 | 1 | Not-determined | Genome-fragment | NA | NA | 0 |
| S11_extracted_k141_99255 | 1210 | No | NA | 3 | 0 | 1 | Not-determined | Genome-fragment | NA | NA | 0 |
| S11_extracted_k141_60072 | 13530 | No | NA | 7 | 0 | 3 | Not-determined | Genome-fragment | NA | NA | 0 |
| S11_extracted_k141_27813 | 1190 | No | NA | 1 | 0 | 1 | Not-determined | Genome-fragment | NA | NA | 0 |
| S11_extracted_k141_40546 | 1513 | No | NA | 2 | 0 | 0 | Not-determined | Genome-fragment | NA | NA | 0 |
| S12_vs_k141_40531\|\|full | 1158 | No | NA | 4 | 0 | 0 | Low-quality | Genome-fragment | 2.87 | AAI-based (high-confidence) | 0 |
| S12_vs_k141_81249\|\|full | 1158 | No | NA | 2 | 0 | 1 | Low-quality | Genome-fragment | 0.3 | AAI-based (medium-confidence) | 0 |
| S11_extracted_k141_21396 | 4666 | No | NA | 6 | 0 | 0 | Not-determined | Genome-fragment | NA | NA | 0 |
| S2_extracted_k141_14126 | 1158 | No | NA | 2 | 0 | 0 | Low-quality | Genome-fragment | 1.21 | AAI-based (high-confidence) | 0 |
| S11_extracted_k141_66655 | 1439 | No | NA | 1 | 0 | 0 | Not-determined | Genome-fragment | NA | NA | 0 |
| S11_extracted_k141_53833 | 1290 | No | NA | 1 | 0 | 1 | Not-determined | Genome-fragment | NA | NA | 0 |
| S11_extracted_k141_47144 | 1378 | No | NA | 2 | 0 | 1 | Not-determined | Genome-fragment | NA | NA | 0 |
| S2_vs_k141_18637\|\|full | 1158 | No | NA | 2 | 0 | 0 | Low-quality | Genome-fragment | 1.7 | AAI-based (high-confidence) | 0 |
| S4_extracted_31092 | 1158 | No | NA | 3 | 1 | 0 | Low-quality | Genome-fragment | 2.06 | AAI-based (medium-confidence) | 0 |
| S11_extracted_k141_47177 | 1300 | No | NA | 2 | 0 | 1 | Not-determined | Genome-fragment | NA | NA | 0 |
| S11_extracted_k141_99342 | 1035 | No | NA | 2 | 0 | 2 | Not-determined | Genome-fragment | NA | NA | 0 |
| S11_extracted_k141_33919 | 2717 | No | NA | 3 | 0 | 3 | Not-determined | Genome-fragment | NA | NA | 0 |
| S11_extracted_k141_53889 | 4381 | No | NA | 5 | 0 | 2 | Not-determined | Genome-fragment | NA | NA | 0 |
| S11_extracted_k141_27845 | 60409 | No | NA | 74 | 8 | 0 | High-quality | High-quality | 95.09 | AAI-based (high-confidence) | 0 |
| S8_vs_k141_180324\|\|full | 1158 | No | NA | 5 | 0 | 0 | Low-quality | Genome-fragment | 1.58 | AAI-based (medium-confidence) | 0 |
| S11_extracted_k141_47215 | 2802 | No | NA | 3 | 0 | 0 | Not-determined | Genome-fragment | NA | NA | 0 |
| S11_extracted_k141_92779 | 1129 | No | NA | 2 | 0 | 0 | Not-determined | Genome-fragment | NA | NA | 0 |
| S17_extracted_k141_60079 | 1159 | No | NA | 2 | 0 | 2 | Low-quality | Genome-fragment | 1.43 | AAI-based (medium-confidence) | 0 |
| S11_extracted_k141_86572 | 1317 | No | NA | 2 | 0 | 1 | Not-determined | Genome-fragment | NA | NA | 0 |
| S9_vs_k141_84680\|\|full | 1159 | No | NA | 5 | 0 | 0 | Low-quality | Genome-fragment | 0.67 | AAI-based (high-confidence) | 0 |
| S9_vs_k141_65710\|\|lt2gene | 1159 | No | NA | 2 | 1 | 0 | Low-quality | Genome-fragment | 3.01 | AAI-based (medium-confidence) | 0 |
| S11_extracted_k141_8373 | 1135 | No | NA | 2 | 0 | 0 | Not-determined | Genome-fragment | NA | NA | 0 |
| S11_extracted_k141_73606 | 7868 | No | NA | 3 | 0 | 2 | Not-determined | Genome-fragment | NA | NA | 0 |
| S11_extracted_k141_21555 | 1239 | No | NA | 2 | 0 | 0 | Not-determined | Genome-fragment | NA | NA | 0 |
| S14_extracted_k141_48525 | 1159 | No | NA | 2 | 0 | 1 | Low-quality | Genome-fragment | 0.39 | AAI-based (high-confidence) | 0 |
| S11_extracted_k141_2087 | 3565 | No | NA | 4 | 0 | 0 | Not-determined | Genome-fragment | NA | NA | 0 |
| S1_vs_k141_72493\|\|full | 1159 | No | NA | 2 | 0 | 1 | Low-quality | Genome-fragment | 1.1 | AAI-based (medium-confidence) | 0 |
| S11_extracted_k141_40691 | 1159 | No | NA | 1 | 0 | 1 | Not-determined | Genome-fragment | NA | NA | 0 |
| S11_extracted_k141_53988 | 1857 | No | NA | 1 | 0 | 1 | Not-determined | Genome-fragment | NA | NA | 0 |
| S5_vs_k141_68194\|\|full | 1159 | No | NA | 3 | 0 | 0 | Low-quality | Genome-fragment | 2.44 | AAI-based (medium-confidence) | 0 |
| S11_extracted_k141_92796 | 1983 | No | NA | 1 | 0 | 1 | Not-determined | Genome-fragment | NA | NA | 0 |
| S11_extracted_k141_54012 | 1680 | No | NA | 2 | 0 | 2 | Not-determined | Genome-fragment | NA | NA | 0 |
| S11_extracted_k141_54014 | 1650 | No | NA | 1 | 0 | 1 | Not-determined | Genome-fragment | NA | NA | 0 |
| S11_extracted_k141_8430 | 1454 | No | NA | 1 | 0 | 1 | Not-determined | Genome-fragment | NA | NA | 0 |
| S11_extracted_k141_40715 | 1597 | No | NA | 1 | 0 | 1 | Not-determined | Genome-fragment | NA | NA | 0 |
| S11_extracted_k141_34008 | 27571 | No | NA | 43 | 12 | 0 | Medium-quality | Genome-fragment | 57.97 | AAI-based (high-confidence) | 0 |
| S11_extracted_k141_86695 | 5588 | No | NA | 5 | 0 | 1 | Not-determined | Genome-fragment | NA | NA | 0 |
| S11_extracted_k141_86707 | 1466 | No | NA | 2 | 0 | 0 | Not-determined | Genome-fragment | NA | NA | 0 |
| S11_extracted_k141_14595 | 1068 | No | NA | 1 | 0 | 0 | Not-determined | Genome-fragment | NA | NA | 0 |
| S7_extracted_13529 | 1159 | No | NA | 2 | 1 | 0 | Low-quality | Genome-fragment | 2.67 | AAI-based (high-confidence) | 0 |
| S11_extracted_k141_40783 | 1140 | No | NA | 1 | 0 | 1 | Not-determined | Genome-fragment | NA | NA | 0 |
| S11_extracted_k141_27953 | 5256 | No | NA | 8 | 0 | 3 | Not-determined | Genome-fragment | NA | NA | 0 |
| S7_vs_39541\|\|lt2gene | 1159 | No | NA | 1 | 1 | 0 | Low-quality | Genome-fragment | 2.92 | AAI-based (medium-confidence) | 0 |
| S11_extracted_k141_73693 | 1120 | No | NA | 2 | 0 | 1 | Not-determined | Genome-fragment | NA | NA | 0 |
| S11_extracted_k141_86736 | 3513 | No | NA | 7 | 0 | 0 | Not-determined | Genome-fragment | NA | NA | 0 |
| S12_extracted_k141_145385 | 1159 | No | NA | 2 | 1 | 0 | Low-quality | Genome-fragment | 2.73 | AAI-based (high-confidence) | 0 |
| S11_extracted_k141_99562 | 1789 | No | NA | 2 | 0 | 2 | Not-determined | Genome-fragment | NA | NA | 0 |
| S11_extracted_k141_92887 | 5261 | No | NA | 5 | 0 | 5 | Not-determined | Genome-fragment | NA | NA | 0 |
| S11_extracted_k141_99565 | 1028 | No | NA | 1 | 0 | 1 | Not-determined | Genome-fragment | NA | NA | 0 |
| S11_extracted_k141_99578 | 1047 | No | NA | 3 | 0 | 2 | Not-determined | Genome-fragment | NA | NA | 0 |
| S11_extracted_k141_34082 | 1218 | No | NA | 7 | 0 | 0 | Not-determined | Genome-fragment | NA | NA | 0 |
| S11_extracted_k141_8512 | 1378 | No | NA | 2 | 0 | 0 | Not-determined | Genome-fragment | NA | NA | 0 |
| S11_extracted_k141_73735 | 1311 | No | NA | 2 | 0 | 1 | Not-determined | Genome-fragment | NA | NA | 0 |
| S11_extracted_k141_21656 | 1209 | No | NA | 2 | 0 | 1 | Not-determined | Genome-fragment | NA | NA | 0 |
| S12_vs_k141_15638\|\|full | 1159 | No | NA | 2 | 1 | 0 | Low-quality | Genome-fragment | 2.08 | AAI-based (medium-confidence) | 0 |
| S12_vs_k141_150129\|\|lt2gene | 1159 | No | NA | 2 | 2 | 0 | Low-quality | Genome-fragment | 1.43 | HMM-based (lower-bound) | 0 |
| S11_extracted_k141_34112 | 3707 | No | NA | 4 | 0 | 2 | Not-determined | Genome-fragment | NA | NA | 0 |
| S11_extracted_k141_54156 | 1725 | No | NA | 2 | 0 | 1 | Not-determined | Genome-fragment | NA | NA | 0 |
| S11_extracted_k141_73794 | 1004 | No | NA | 2 | 0 | 0 | Not-determined | Genome-fragment | NA | NA | 0 |
| S15_extracted_36576 | 1159 | No | NA | 2 | 0 | 2 | Low-quality | Genome-fragment | 0.88 | AAI-based (medium-confidence) | 0 |
| S11_extracted_k141_99619 | 1403 | No | NA | 3 | 0 | 0 | Not-determined | Genome-fragment | NA | NA | 0 |
| S11_extracted_k141_21727 | 1151 | No | NA | 1 | 0 | 1 | Not-determined | Genome-fragment | NA | NA | 0 |
| S11_extracted_k141_99652 | 1659 | No | NA | 2 | 0 | 1 | Not-determined | Genome-fragment | NA | NA | 0 |
| S6_vs_k141_32329\|\|full | 1159 | No | NA | 2 | 0 | 0 | Low-quality | Genome-fragment | 2.54 | AAI-based (high-confidence) | 0 |
| S8_extracted_k141_45249 | 1159 | No | NA | 2 | 0 | 0 | Low-quality | Genome-fragment | 0.57 | AAI-based (medium-confidence) | 0 |
| S8_extracted_k141_68741 | 1159 | No | NA | 2 | 1 | 0 | Low-quality | Genome-fragment | 2.63 | AAI-based (high-confidence) | 0 |
| S11_extracted_k141_66801 | 3201 | No | NA | 4 | 0 | 1 | Not-determined | Genome-fragment | NA | NA | 0 |
| S11_extracted_k141_99734 | 1475 | No | NA | 1 | 0 | 1 | Not-determined | Genome-fragment | NA | NA | 0 |
| S11_extracted_k141_8654 | 1063 | No | NA | 1 | 0 | 1 | Not-determined | Genome-fragment | NA | NA | 0 |
| S11_extracted_k141_34224 | 35904 | No | NA | 59 | 26 | 0 | Complete | High-quality | 100 | DTR (high-confidence) | 0 |
| S8_vs_k141_146534\|\|full | 1159 | No | NA | 3 | 0 | 0 | Low-quality | Genome-fragment | 2.77 | AAI-based (high-confidence) | 0 |
| S11_extracted_k141_80275 | 3120 | No | NA | 2 | 0 | 2 | Not-determined | Genome-fragment | NA | NA | 0 |
| S17_extracted_k141_140779 | 1160 | No | NA | 2 | 0 | 2 | Low-quality | Genome-fragment | 2.07 | AAI-based (medium-confidence) | 0 |
| S11_extracted_k141_54299 | 1392 | No | NA | 1 | 0 | 0 | Not-determined | Genome-fragment | NA | NA | 0 |
| S11_extracted_k141_73937 | 19632 | No | NA | 14 | 0 | 8 | Not-determined | Genome-fragment | NA | NA | 0 |
| S11_extracted_k141_8699 | 1143 | No | NA | 2 | 0 | 1 | Not-determined | Genome-fragment | NA | NA | 0 |
| S11_extracted_k141_60438 | 1538 | No | NA | 3 | 0 | 0 | Not-determined | Genome-fragment | NA | NA | 0 |
| S17_extracted_k141_33599 | 1160 | No | NA | 1 | 1 | 0 | Low-quality | Genome-fragment | 3.44 | AAI-based (medium-confidence) | 0 |
| S14_extracted_k141_9779 | 1160 | No | NA | 2 | 2 | 0 | Low-quality | Genome-fragment | 3.83 | AAI-based (high-confidence) | 0 |
| S11_extracted_k141_2441 | 18509 | No | NA | 21 | 0 | 5 | Not-determined | Genome-fragment | NA | NA | 0 |
| S11_extracted_k141_54327 | 1344 | No | NA | 1 | 0 | 0 | Not-determined | Genome-fragment | NA | NA | 0 |
| S11_extracted_k141_28186 | 1238 | No | NA | 3 | 0 | 0 | Not-determined | Genome-fragment | NA | NA | 0 |
| S1_vs_k141_132280\|\|full | 1160 | No | NA | 3 | 1 | 0 | Low-quality | Genome-fragment | 1.98 | AAI-based (high-confidence) | 0 |
| S11_extracted_k141_99751 | 72288 | No | NA | 100 | 19 | 1 | Medium-quality | Genome-fragment | 65.25 | AAI-based (medium-confidence) | 0 |
| S11_extracted_k141_66851 | 1209 | No | NA | 1 | 0 | 0 | Not-determined | Genome-fragment | NA | NA | 0 |
| S11_extracted_k141_28195 | 2999 | No | NA | 5 | 0 | 4 | Not-determined | Genome-fragment | NA | NA | 0 |
| S11_extracted_k141_87121 | 5231 | No | NA | 5 | 0 | 4 | Not-determined | Genome-fragment | NA | NA | 0 |
| S11_extracted_k141_54381 | 1326 | No | NA | 3 | 0 | 0 | Not-determined | Genome-fragment | NA | NA | 0 |
| S11_extracted_k141_87138 | 1160 | No | NA | 1 | 0 | 0 | Not-determined | Genome-fragment | NA | NA | 0 |
| S11_extracted_k141_66887 | 1519 | No | NA | 3 | 0 | 1 | Not-determined | Genome-fragment | NA | NA | 0 |
| S11_extracted_k141_34329 | 1487 | No | NA | 2 | 0 | 1 | Not-determined | Genome-fragment | NA | NA | 0 |
| S11_extracted_k141_14916 | 2889 | No | NA | 5 | 0 | 0 | Not-determined | Genome-fragment | NA | NA | 0 |
| S3_vs_k141_96306\|\|full | 1160 | No | NA | 3 | 1 | 0 | Low-quality | Genome-fragment | 2.64 | AAI-based (high-confidence) | 0 |
| S3_vs_k141_111023\|\|full | 1160 | No | NA | 4 | 0 | 0 | Low-quality | Genome-fragment | 2.39 | AAI-based (medium-confidence) | 0 |
| S11_extracted_k141_54393 | 17935 | No | NA | 18 | 0 | 2 | Not-determined | Genome-fragment | NA | NA | 0 |
| S5_vs_k141_41793\|\|lt2gene | 1160 | No | NA | 1 | 1 | 0 | Low-quality | Genome-fragment | 2.82 | AAI-based (high-confidence) | 0 |
| S7_extracted_72734 | 1160 | No | NA | 2 | 2 | 0 | Low-quality | Genome-fragment | 2.47 | AAI-based (high-confidence) | 0 |
| S11_extracted_k141_8810 | 1111 | No | NA | 2 | 0 | 0 | Not-determined | Genome-fragment | NA | NA | 0 |
| S11_extracted_k141_54442 | 1332 | No | NA | 2 | 0 | 1 | Not-determined | Genome-fragment | NA | NA | 0 |
| S11_extracted_k141_21967 | 11502 | No | NA | 14 | 0 | 0 | Not-determined | Genome-fragment | NA | NA | 0 |
| S7_vs_100695\|\|full | 1160 | No | NA | 3 | 1 | 0 | Low-quality | Genome-fragment | 0.65 | HMM-based (lower-bound) | 0 |
| S11_extracted_k141_34408 | 1367 | No | NA | 1 | 0 | 0 | Not-determined | Genome-fragment | NA | NA | 0 |
| S12_vs_k141_160188\|\|full | 1160 | No | NA | 2 | 0 | 0 | Low-quality | Genome-fragment | 2.24 | AAI-based (medium-confidence) | 0 |
| S11_extracted_k141_41251 | 1359 | No | NA | 2 | 0 | 1 | Not-determined | Genome-fragment | NA | NA | 0 |
| S11_extracted_k141_54483 | 1132 | No | NA | 1 | 0 | 0 | Not-determined | Genome-fragment | NA | NA | 0 |
| S11_extracted_k141_2582 | 1417 | No | NA | 2 | 0 | 0 | Not-determined | Genome-fragment | NA | NA | 0 |
| S11_extracted_k141_87231 | 1358 | No | NA | 2 | 0 | 1 | Not-determined | Genome-fragment | NA | NA | 0 |
| S11_extracted_k141_54504 | 1833 | No | NA | 2 | 0 | 0 | Not-determined | Genome-fragment | NA | NA | 0 |
| S12_vs_k141_61885\|\|full | 1160 | No | NA | 3 | 0 | 0 | Low-quality | Genome-fragment | 2.92 | AAI-based (high-confidence) | 0 |
| S11_extracted_k141_15068 | 1812 | No | NA | 3 | 0 | 2 | Not-determined | Genome-fragment | NA | NA | 0 |
| S11_extracted_k141_67017 | 1571 | No | NA | 1 | 0 | 0 | Not-determined | Genome-fragment | NA | NA | 0 |
| S8_extracted_k141_69731 | 1160 | No | NA | 2 | 0 | 0 | Low-quality | Genome-fragment | 1.41 | AAI-based (medium-confidence) | 0 |
| S11_extracted_k141_99943 | 4382 | No | NA | 5 | 0 | 3 | Not-determined | Genome-fragment | NA | NA | 0 |
| S11_extracted_k141_67030 | 6158 | No | NA | 4 | 0 | 3 | Not-determined | Genome-fragment | NA | NA | 0 |
| S8_extracted_k141_197521 | 1160 | No | NA | 1 | 0 | 1 | Low-quality | Genome-fragment | 0.34 | AAI-based (medium-confidence) | 0 |
| S17_extracted_k141_122098 | 1161 | No | NA | 2 | 0 | 1 | Low-quality | Genome-fragment | 2.47 | AAI-based (medium-confidence) | 0 |
| S9_vs_k141_76359\|\|full | 1161 | No | NA | 3 | 1 | 0 | Low-quality | Genome-fragment | 2.01 | AAI-based (high-confidence) | 0 |
| S11_extracted_k141_67055 | 1404 | No | NA | 1 | 0 | 0 | Not-determined | Genome-fragment | NA | NA | 0 |
| S13_vs_144237\|\|full | 1161 | No | NA | 3 | 1 | 0 | Low-quality | Genome-fragment | 1.13 | AAI-based (medium-confidence) | 0 |
| S11_extracted_k141_93354 | 2089 | No | NA | 4 | 0 | 4 | Not-determined | Genome-fragment | NA | NA | 0 |
| S11_extracted_k141_93355 | 1019 | No | NA | 1 | 0 | 0 | Not-determined | Genome-fragment | NA | NA | 0 |
| S11_extracted_k141_34501 | 1936 | No | NA | 4 | 0 | 0 | Not-determined | Genome-fragment | NA | NA | 0 |
| S14_vs_k141_71621\|\|full | 1161 | No | NA | 3 | 0 | 0 | Low-quality | Genome-fragment | 2.02 | AAI-based (medium-confidence) | 0 |
| S11_extracted_k141_74200 | 1479 | No | NA | 2 | 0 | 0 | Not-determined | Genome-fragment | NA | NA | 0 |
| S1_extracted_k141_44115 | 1161 | No | NA | 2 | 0 | 1 | Low-quality | Genome-fragment | 2 | AAI-based (medium-confidence) | 0 |
| S11_extracted_k141_8946 | 5632 | No | NA | 4 | 0 | 3 | Not-determined | Genome-fragment | NA | NA | 0 |
| S5_extracted_k141_124763 | 1161 | No | NA | 1 | 0 | 0 | Low-quality | Genome-fragment | 1.44 | AAI-based (high-confidence) | 0 |
| S11_extracted_k141_15133 | 1099 | No | NA | 1 | 0 | 0 | Not-determined | Genome-fragment | NA | NA | 0 |
| S11_extracted_k141_54611 | 1128 | No | NA | 2 | 0 | 1 | Not-determined | Genome-fragment | NA | NA | 0 |
| S5_extracted_k141_22198 | 1161 | No | NA | 3 | 2 | 0 | Low-quality | Genome-fragment | 5.39 | AAI-based (medium-confidence) | 0 |
| S11_extracted_k141_15178 | 1176 | No | NA | 2 | 0 | 1 | Not-determined | Genome-fragment | NA | NA | 0 |
| S5_extracted_k141_22212 | 1161 | No | NA | 3 | 2 | 0 | Low-quality | Genome-fragment | 5.39 | AAI-based (medium-confidence) | 0 |
| S11_extracted_k141_93425 | 3437 | No | NA | 3 | 0 | 2 | Not-determined | Genome-fragment | NA | NA | 0 |
| S5_vs_k141_99473\|\|full | 1161 | No | NA | 3 | 0 | 0 | Low-quality | Genome-fragment | 1.86 | AAI-based (medium-confidence) | 0 |
| S11_extracted_k141_47632 | 32534 | No | NA | 48 | 17 | 1 | Complete | High-quality | 100 | DTR (high-confidence) | 0 |
| S11_extracted_k141_47634 | 1114 | No | NA | 3 | 0 | 0 | Not-determined | Genome-fragment | NA | NA | 0 |
| S5_vs_k141_39675\|\|full | 1161 | No | NA | 3 | 1 | 0 | Low-quality | Genome-fragment | 2.85 | AAI-based (medium-confidence) | 0 |
| S5_vs_k141_18210\|\|full | 1161 | No | NA | 2 | 0 | 0 | Low-quality | Genome-fragment | 2.23 | AAI-based (medium-confidence) | 0 |
| S11_extracted_k141_80531 | 3414 | No | NA | 4 | 0 | 2 | Not-determined | Genome-fragment | NA | NA | 0 |
| S7_extracted_33864 | 1161 | No | NA | 1 | 1 | 0 | Low-quality | Genome-fragment | 3.93 | HMM-based (lower-bound) | 0 |
| S7_vs_9317\|\|full | 1161 | No | NA | 3 | 0 | 0 | Low-quality | Genome-fragment | 1.09 | AAI-based (high-confidence) | 0 |
| S11_extracted_k141_22099 | 1751 | No | NA | 3 | 0 | 2 | Not-determined | Genome-fragment | NA | NA | 0 |
| S7_vs_15860\|\|full | 1161 | No | NA | 4 | 0 | 3 | Low-quality | Genome-fragment | 1.8 | AAI-based (medium-confidence) | 0 |
| S11_extracted_k141_100052 | 3506 | No | NA | 2 | 0 | 2 | Not-determined | Genome-fragment | NA | NA | 0 |
| S11_extracted_k141_15271 | 1055 | No | NA | 3 | 0 | 0 | Not-determined | Genome-fragment | NA | NA | 0 |
| S11_extracted_k141_2744 | 1585 | No | NA | 3 | 0 | 0 | Not-determined | Genome-fragment | NA | NA | 0 |
| S11_extracted_k141_9068 | 1003 | No | NA | 1 | 0 | 0 | Not-determined | Genome-fragment | NA | NA | 0 |
| S11_extracted_k141_87488 | 2110 | No | NA | 3 | 0 | 1 | Not-determined | Genome-fragment | NA | NA | 0 |
| S11_extracted_k141_100104 | 1110 | No | NA | 2 | 0 | 0 | Not-determined | Genome-fragment | NA | NA | 0 |
| S12_extracted_k141_125042 | 1161 | No | NA | 2 | 2 | 0 | Low-quality | Genome-fragment | 2.78 | AAI-based (high-confidence) | 0 |
| S12_extracted_k141_70755 | 1161 | No | NA | 1 | 0 | 1 | Low-quality | Genome-fragment | 0.28 | AAI-based (medium-confidence) | 0 |
| S11_extracted_k141_9110 | 2243 | No | NA | 3 | 0 | 0 | Not-determined | Genome-fragment | NA | NA | 0 |
| S11_extracted_k141_80571 | 1025 | No | NA | 1 | 0 | 1 | Not-determined | Genome-fragment | NA | NA | 0 |
| S12_vs_k141_136873\|\|full | 1161 | No | NA | 4 | 1 | 0 | Low-quality | Genome-fragment | 2.79 | AAI-based (high-confidence) | 0 |
| S11_extracted_k141_41493 | 1085 | No | NA | 2 | 0 | 1 | Not-determined | Genome-fragment | NA | NA | 0 |
| S8_extracted_k141_28176 | 1161 | No | NA | 3 | 1 | 0 | Low-quality | Genome-fragment | 2.67 | AAI-based (high-confidence) | 0 |
| S11_extracted_k141_15359 | 1924 | No | NA | 1 | 0 | 1 | Not-determined | Genome-fragment | NA | NA | 0 |
| S11_extracted_k141_22215 | 3327 | No | NA | 4 | 0 | 4 | Not-determined | Genome-fragment | NA | NA | 0 |
| S11_extracted_k141_93559 | 2791 | No | NA | 3 | 0 | 3 | Not-determined | Genome-fragment | NA | NA | 0 |
| S11_extracted_k141_80669 | 3257 | No | NA | 3 | 0 | 0 | Not-determined | Genome-fragment | NA | NA | 0 |
| S17_vs_k141_74577\|\|full | 1162 | No | NA | 2 | 0 | 0 | Low-quality | Genome-fragment | 2.2 | AAI-based (high-confidence) | 0 |
| S17_vs_k141_128358\|\|full | 1162 | No | NA | 3 | 3 | 0 | Low-quality | Genome-fragment | 1.76 | HMM-based (lower-bound) | 0 |
| S11_extracted_k141_67169 | 1002 | No | NA | 2 | 0 | 1 | Not-determined | Genome-fragment | NA | NA | 0 |
| S11_extracted_k141_9180 | 1558 | No | NA | 3 | 0 | 3 | Not-determined | Genome-fragment | NA | NA | 0 |
| S11_extracted_k141_28515 | 98716 | No | NA | 96 | 26 | 0 | Medium-quality | Genome-fragment | 61.32 | AAI-based (high-confidence) | 0 |
| S11_extracted_k141_15367 | 1572 | No | NA | 2 | 0 | 2 | Not-determined | Genome-fragment | NA | NA | 0 |
| S13_extracted_7335 | 1162 | No | NA | 3 | 1 | 0 | Low-quality | Genome-fragment | 0.73 | AAI-based (high-confidence) | 0 |
| S11_extracted_k141_47793 | 1019 | No | NA | 1 | 0 | 0 | Not-determined | Genome-fragment | NA | NA | 0 |
| S14_extracted_k141_95700 | 1162 | No | NA | 2 | 0 | 1 | Low-quality | Genome-fragment | 0.31 | AAI-based (medium-confidence) | 0 |
| S14_extracted_k141_65315 | 1162 | No | NA | 4 | 0 | 0 | Low-quality | Genome-fragment | 1.19 | AAI-based (high-confidence) | 0 |
| S14_vs_k141_47975\|\|lt2gene | 1162 | No | NA | 1 | 1 | 0 | Low-quality | Genome-fragment | 2.66 | AAI-based (high-confidence) | 0 |
| S11_extracted_k141_2943 | 1054 | No | NA | 2 | 0 | 1 | Not-determined | Genome-fragment | NA | NA | 0 |
| S3_extracted_k141_83847 | 1162 | No | NA | 1 | 1 | 0 | Low-quality | Genome-fragment | 0.8 | HMM-based (lower-bound) | 0 |
| S3_vs_k141_56186\|\|lt2gene | 1162 | No | NA | 1 | 1 | 0 | Low-quality | Genome-fragment | 2.26 | AAI-based (high-confidence) | 0 |
| S11_extracted_k141_67213 | 1319 | No | NA | 2 | 0 | 0 | Not-determined | Genome-fragment | NA | NA | 0 |
| S11_extracted_k141_93644 | 1002 | No | NA | 3 | 0 | 1 | Not-determined | Genome-fragment | NA | NA | 0 |
| S11_extracted_k141_41654 | 1076 | No | NA | 1 | 0 | 1 | Not-determined | Genome-fragment | NA | NA | 0 |
| S5_extracted_k141_124961 | 1162 | No | NA | 1 | 1 | 0 | Low-quality | Genome-fragment | 1.32 | HMM-based (lower-bound) | 0 |
| S5_vs_k141_9582\|\|lt2gene | 1162 | No | NA | 1 | 1 | 0 | Low-quality | Genome-fragment | 0.96 | HMM-based (lower-bound) | 0 |
| S7_vs_92253\|\|full | 1162 | No | NA | 2 | 0 | 1 | Low-quality | Genome-fragment | 2.31 | AAI-based (medium-confidence) | 0 |
| S11_extracted_k141_93708 | 4288 | No | NA | 2 | 0 | 1 | Not-determined | Genome-fragment | NA | NA | 0 |
| S11_extracted_k141_9286 | 1295 | No | NA | 2 | 0 | 1 | Not-determined | Genome-fragment | NA | NA | 0 |
| S11_extracted_k141_15442 | 4063 | No | NA | 2 | 0 | 2 | Not-determined | Genome-fragment | NA | NA | 0 |
| S7_vs_132828\|\|full | 1162 | No | NA | 2 | 0 | 0 | Low-quality | Genome-fragment | 1.66 | AAI-based (high-confidence) | 0 |
| S16_vs_162622\|\|full | 1162 | No | NA | 3 | 1 | 0 | Low-quality | Genome-fragment | 2.8 | AAI-based (high-confidence) | 0 |
| S11_extracted_k141_22355 | 1246 | No | NA | 1 | 0 | 1 | Not-determined | Genome-fragment | NA | NA | 0 |
| S11_extracted_k141_74598 | 1052 | No | NA | 1 | 0 | 1 | Not-determined | Genome-fragment | NA | NA | 0 |
| S10_extracted_61977 | 1162 | No | NA | 2 | 0 | 1 | Low-quality | Genome-fragment | 0.7 | AAI-based (medium-confidence) | 0 |
| S10_vs_69703\|\|full | 1162 | No | NA | 4 | 0 | 0 | Low-quality | Genome-fragment | 1.88 | AAI-based (medium-confidence) | 0 |
| S11_extracted_k141_2982 | 1984 | No | NA | 2 | 0 | 1 | Not-determined | Genome-fragment | NA | NA | 0 |
| S11_extracted_k141_60896 | 9398 | No | NA | 6 | 0 | 1 | Not-determined | Genome-fragment | NA | NA | 0 |
| S10_vs_57753\|\|lt2gene | 1162 | No | NA | 2 | 2 | 0 | Low-quality | Genome-fragment | 2.79 | AAI-based (high-confidence) | 0 |
| S11_extracted_k141_80743 | 1557 | No | NA | 1 | 0 | 1 | Not-determined | Genome-fragment | NA | NA | 0 |
| S15_vs_77043\|\|lt2gene | 1162 | No | NA | 1 | 1 | 0 | Low-quality | Genome-fragment | 2.28 | AAI-based (high-confidence) | 0 |
| S8_vs_k141_192440\|\|lt2gene | 1162 | No | NA | 2 | 2 | 0 | Low-quality | Genome-fragment | 3.4 | AAI-based (medium-confidence) | 0 |
| S11_extracted_k141_39502 | 1163 | No | NA | 3 | 0 | 0 | Low-quality | Genome-fragment | 2.95 | AAI-based (medium-confidence) | 0 |
| S11_extracted_k141_34754 | 1022 | No | NA | 3 | 0 | 0 | Not-determined | Genome-fragment | NA | NA | 0 |
| S11_extracted_k141_67326 | 1505 | No | NA | 2 | 0 | 2 | Not-determined | Genome-fragment | NA | NA | 0 |
| S11_extracted_k141_74663 | 2597 | No | NA | 4 | 0 | 1 | Not-determined | Genome-fragment | NA | NA | 0 |
| S1_extracted_k141_939 | 1163 | No | NA | 1 | 1 | 0 | Low-quality | Genome-fragment | 1.8 | AAI-based (high-confidence) | 0 |
| S1_vs_k141_47516\|\|full | 1163 | No | NA | 3 | 0 | 0 | Low-quality | Genome-fragment | 2.17 | AAI-based (medium-confidence) | 0 |
| S11_extracted_k141_60983 | 4343 | No | NA | 4 | 0 | 1 | Not-determined | Genome-fragment | NA | NA | 0 |
| S11_extracted_k141_9391 | 3538 | No | NA | 4 | 0 | 0 | Not-determined | Genome-fragment | NA | NA | 0 |
| S1_vs_k141_94420\|\|full | 1163 | Yes | 639 | 3 | 1 | 1 | Low-quality | Genome-fragment | 0.97 | HMM-based (lower-bound) | 45.06 |
| S3_extracted_k141_119931 | 1163 | No | NA | 3 | 3 | 0 | Low-quality | Genome-fragment | 3.84 | AAI-based (high-confidence) | 0 |
| S11_extracted_k141_28570 | 1451 | No | NA | 2 | 0 | 2 | Not-determined | Genome-fragment | NA | NA | 0 |
| S11_extracted_k141_54768 | 1688 | No | NA | 2 | 0 | 2 | Not-determined | Genome-fragment | NA | NA | 0 |
| S11_extracted_k141_34776 | 1050 | No | NA | 1 | 0 | 1 | Not-determined | Genome-fragment | NA | NA | 0 |
| S11_extracted_k141_48017 | 1441 | No | NA | 2 | 0 | 2 | Not-determined | Genome-fragment | NA | NA | 0 |
| S11_extracted_k141_34783 | 3491 | No | NA | 2 | 0 | 2 | Not-determined | Genome-fragment | NA | NA | 0 |
| S3_extracted_k141_37377 | 1163 | No | NA | 1 | 1 | 0 | Low-quality | Genome-fragment | 1.7 | HMM-based (lower-bound) | 0 |
| S3_vs_k141_45294\|\|full | 1163 | No | NA | 3 | 0 | 0 | Low-quality | Genome-fragment | 2.76 | AAI-based (high-confidence) | 0 |
| S3_vs_k141_45418\|\|full | 1163 | No | NA | 3 | 0 | 0 | Low-quality | Genome-fragment | 3.16 | AAI-based (high-confidence) | 0 |
| S7_extracted_129296 | 1163 | No | NA | 3 | 0 | 0 | Low-quality | Genome-fragment | 2.03 | AAI-based (high-confidence) | 0 |
| S16_extracted_64690 | 1163 | No | NA | 2 | 0 | 1 | Low-quality | Genome-fragment | 1.15 | AAI-based (medium-confidence) | 0 |
| S11_extracted_k141_28645 | 11934 | No | NA | 18 | 0 | 1 | Complete | High-quality | 100 | DTR (high-confidence) | 0 |
| S11_extracted_k141_61113 | 3404 | No | NA | 4 | 0 | 0 | Not-determined | Genome-fragment | NA | NA | 0 |
| S11_extracted_k141_67506 | 1577 | No | NA | 1 | 0 | 0 | Not-determined | Genome-fragment | NA | NA | 0 |
| S16_extracted_142390 | 1163 | No | NA | 2 | 0 | 0 | Low-quality | Genome-fragment | 2.74 | AAI-based (high-confidence) | 0 |
| S11_extracted_k141_3165 | 1201 | No | NA | 1 | 0 | 0 | Not-determined | Genome-fragment | NA | NA | 0 |
| S10_extracted_102502 | 1163 | No | NA | 3 | 0 | 0 | Low-quality | Genome-fragment | 0.89 | AAI-based (high-confidence) | 0 |
| S11_extracted_k141_15628 | 1024 | No | NA | 1 | 0 | 0 | Not-determined | Genome-fragment | NA | NA | 0 |
| S11_extracted_k141_22620 | 1484 | No | NA | 1 | 0 | 0 | Not-determined | Genome-fragment | NA | NA | 0 |
| S11_extracted_k141_61128 | 1115 | No | NA | 2 | 0 | 0 | Not-determined | Genome-fragment | NA | NA | 0 |
| S11_extracted_k141_48078 | 2214 | No | NA | 2 | 0 | 2 | Not-determined | Genome-fragment | NA | NA | 0 |
| S10_vs_52934\|\|lt2gene | 1163 | No | NA | 2 | 1 | 0 | Low-quality | Genome-fragment | 2.85 | AAI-based (medium-confidence) | 0 |
| S11_extracted_k141_41885 | 1144 | No | NA | 1 | 0 | 0 | Not-determined | Genome-fragment | NA | NA | 0 |
| S11_extracted_k141_22638 | 1250 | No | NA | 1 | 0 | 1 | Not-determined | Genome-fragment | NA | NA | 0 |
| S11_extracted_k141_100498 | 1293 | No | NA | 1 | 0 | 1 | Not-determined | Genome-fragment | NA | NA | 0 |
| S8_vs_k141_166516\|\|full | 1163 | No | NA | 2 | 0 | 0 | Low-quality | Genome-fragment | 1.41 | AAI-based (high-confidence) | 0 |
| S11_extracted_k141_9502 | 1063 | No | NA | 1 | 0 | 1 | Not-determined | Genome-fragment | NA | NA | 0 |
| S17_vs_k141_76264\|\|full | 1164 | No | NA | 2 | 0 | 1 | Low-quality | Genome-fragment | 0.72 | AAI-based (medium-confidence) | 0 |
| S11_extracted_k141_61175 | 1159 | No | NA | 2 | 0 | 1 | Not-determined | Genome-fragment | NA | NA | 0 |
| S11_vs_k141_7825\|\|full | 1164 | No | NA | 3 | 2 | 0 | Low-quality | Genome-fragment | 2.87 | AAI-based (high-confidence) | 0 |
| S11_extracted_k141_28701 | 4323 | No | NA | 4 | 0 | 0 | Not-determined | Genome-fragment | NA | NA | 0 |
| S11_extracted_k141_61176 | 3129 | No | NA | 3 | 0 | 0 | Not-determined | Genome-fragment | NA | NA | 0 |
| S11_vs_k141_70856\|\|full | 1164 | No | NA | 4 | 0 | 0 | Low-quality | Genome-fragment | 2.12 | AAI-based (high-confidence) | 0 |
| S11_extracted_k141_100558 | 3356 | No | NA | 2 | 0 | 1 | Not-determined | Genome-fragment | NA | NA | 0 |
| S11_extracted_k141_9540 | 3839 | No | NA | 7 | 0 | 4 | Not-determined | Genome-fragment | NA | NA | 0 |
| S14_extracted_k141_89493 | 1164 | No | NA | 3 | 2 | 0 | Low-quality | Genome-fragment | 3.79 | AAI-based (high-confidence) | 0 |
| S11_extracted_k141_41935 | 1137 | No | NA | 1 | 0 | 1 | Not-determined | Genome-fragment | NA | NA | 0 |
| S14_vs_k141_44322\|\|full | 1164 | No | NA | 4 | 1 | 0 | Low-quality | Genome-fragment | 1.29 | HMM-based (lower-bound) | 0 |
| S11_extracted_k141_74806 | 5375 | No | NA | 4 | 0 | 2 | Not-determined | Genome-fragment | NA | NA | 0 |
| S5_vs_k141_62654\|\|full | 1164 | No | NA | 3 | 1 | 0 | Low-quality | Genome-fragment | 2.61 | AAI-based (high-confidence) | 0 |
| S11_extracted_k141_34949 | 1220 | No | NA | 2 | 0 | 2 | Not-determined | Genome-fragment | NA | NA | 0 |
| S7_vs_76151\|\|full | 1164 | No | NA | 4 | 1 | 0 | Low-quality | Genome-fragment | 3.76 | HMM-based (lower-bound) | 0 |
| S12_extracted_k141_6339 | 1164 | No | NA | 1 | 0 | 1 | Low-quality | Genome-fragment | 3.72 | AAI-based (medium-confidence) | 0 |
| S11_extracted_k141_81064 | 1188 | No | NA | 2 | 0 | 2 | Not-determined | Genome-fragment | NA | NA | 0 |
| S11_extracted_k141_22740 | 5571 | No | NA | 4 | 0 | 3 | Not-determined | Genome-fragment | NA | NA | 0 |
| S11_extracted_k141_67616 | 1163 | No | NA | 1 | 0 | 0 | Not-determined | Genome-fragment | NA | NA | 0 |
| S6_vs_k141_18142\|\|full | 1164 | No | NA | 3 | 2 | 0 | Low-quality | Genome-fragment | 1.72 | AAI-based (high-confidence) | 0 |
| S6_vs_k141_11455\|\|full | 1164 | No | NA | 4 | 1 | 0 | Low-quality | Genome-fragment | 2.62 | AAI-based (high-confidence) | 0 |
| S11_extracted_k141_35028 | 3567 | No | NA | 2 | 0 | 2 | Not-determined | Genome-fragment | NA | NA | 0 |
| S11_extracted_k141_80303 | 1165 | No | NA | 2 | 2 | 0 | Low-quality | Genome-fragment | 0.88 | AAI-based (high-confidence) | 0 |
| S11_extracted_k141_94162 | 1066 | No | NA | 3 | 0 | 0 | Not-determined | Genome-fragment | NA | NA | 0 |
| S11_extracted_k141_60868 | 1165 | No | NA | 1 | 1 | 0 | Low-quality | Genome-fragment | 2.88 | AAI-based (medium-confidence) | 0 |
| S11_extracted_k141_22844 | 1832 | No | NA | 2 | 0 | 2 | Not-determined | Genome-fragment | NA | NA | 0 |
| S11_vs_k141_67640\|\|lt2gene | 1165 | No | NA | 1 | 1 | 0 | Low-quality | Genome-fragment | 2.86 | AAI-based (high-confidence) | 0 |
| S11_extracted_k141_48199 | 6111 | No | NA | 10 | 0 | 0 | Not-determined | Genome-fragment | NA | NA | 0 |
| S14_vs_k141_26906\|\|full | 1165 | Yes | 724 | 3 | 1 | 1 | Low-quality | Genome-fragment | 0.44 | HMM-based (lower-bound) | 37.85 |
| S11_extracted_k141_75038 | 1033 | No | NA | 1 | 0 | 0 | Not-determined | Genome-fragment | NA | NA | 0 |
| S11_extracted_k141_35140 | 2562 | No | NA | 2 | 0 | 1 | Not-determined | Genome-fragment | NA | NA | 0 |
| S1_extracted_k141_64061 | 1165 | No | NA | 3 | 0 | 0 | Low-quality | Genome-fragment | 2.61 | AAI-based (medium-confidence) | 0 |
| S11_extracted_k141_15990 | 1117 | No | NA | 3 | 0 | 0 | Not-determined | Genome-fragment | NA | NA | 0 |
| S1_vs_k141_38478\|\|full | 1165 | No | NA | 3 | 2 | 0 | Low-quality | Genome-fragment | 4.24 | HMM-based (lower-bound) | 0 |
| S3_extracted_k141_24682 | 1165 | No | NA | 2 | 0 | 0 | Low-quality | Genome-fragment | 4.04 | AAI-based (medium-confidence) | 0 |
| S11_extracted_k141_42165 | 1488 | No | NA | 3 | 0 | 3 | Not-determined | Genome-fragment | NA | NA | 0 |
| S11_extracted_k141_28957 | 1020 | No | NA | 1 | 0 | 0 | Not-determined | Genome-fragment | NA | NA | 0 |
| S11_extracted_k141_3379 | 3481 | No | NA | 3 | 0 | 0 | Not-determined | Genome-fragment | NA | NA | 0 |
| S11_extracted_k141_94319 | 1407 | No | NA | 2 | 0 | 2 | Not-determined | Genome-fragment | NA | NA | 0 |
| S5_extracted_k141_17309 | 1165 | No | NA | 3 | 3 | 0 | Low-quality | Genome-fragment | 2.57 | AAI-based (medium-confidence) | 0 |
| S11_extracted_k141_100864 | 1045 | No | NA | 1 | 0 | 1 | Not-determined | Genome-fragment | NA | NA | 0 |
| S11_extracted_k141_48327 | 1068 | No | NA | 3 | 0 | 0 | Not-determined | Genome-fragment | NA | NA | 0 |
| S11_extracted_k141_61393 | 1981 | No | NA | 4 | 0 | 0 | Not-determined | Genome-fragment | NA | NA | 0 |
| S11_extracted_k141_61405 | 1830 | No | NA | 2 | 0 | 0 | Not-determined | Genome-fragment | NA | NA | 0 |
| S11_extracted_k141_94343 | 3252 | No | NA | 3 | 0 | 0 | Not-determined | Genome-fragment | NA | NA | 0 |
| S11_extracted_k141_94351 | 1342 | No | NA | 1 | 0 | 1 | Not-determined | Genome-fragment | NA | NA | 0 |
| S11_extracted_k141_48355 | 1798 | No | NA | 1 | 0 | 1 | Not-determined | Genome-fragment | NA | NA | 0 |
| S11_extracted_k141_94357 | 2454 | No | NA | 3 | 0 | 3 | Not-determined | Genome-fragment | NA | NA | 0 |
| S7_vs_129641\|\|full | 1165 | No | NA | 3 | 1 | 0 | Low-quality | Genome-fragment | 2.82 | AAI-based (medium-confidence) | 0 |
| S10_vs_116827\|\|lt2gene | 1165 | No | NA | 2 | 0 | 0 | Low-quality | Genome-fragment | 2.43 | AAI-based (high-confidence) | 0 |
| S11_extracted_k141_81190 | 1503 | No | NA | 2 | 0 | 1 | Not-determined | Genome-fragment | NA | NA | 0 |
| S11_extracted_k141_61439 | 1179 | No | NA | 2 | 0 | 2 | Not-determined | Genome-fragment | NA | NA | 0 |
| S15_extracted_24945 | 1165 | No | NA | 1 | 0 | 1 | Low-quality | Genome-fragment | 0.43 | AAI-based (medium-confidence) | 0 |
| S8_extracted_k141_102429 | 1165 | No | NA | 2 | 0 | 2 | Low-quality | Genome-fragment | 2.24 | AAI-based (medium-confidence) | 0 |
| S11_extracted_k141_3469 | 1088 | No | NA | 1 | 0 | 0 | Not-determined | Genome-fragment | NA | NA | 0 |
| S11_extracted_k141_16111 | 1257 | No | NA | 2 | 0 | 0 | Not-determined | Genome-fragment | NA | NA | 0 |
| S11_extracted_k141_67826 | 1436 | No | NA | 3 | 0 | 0 | Not-determined | Genome-fragment | NA | NA | 0 |
| S11_extracted_k141_75172 | 3634 | No | NA | 3 | 0 | 3 | Not-determined | Genome-fragment | NA | NA | 0 |
| S11_extracted_k141_87976 | 1325 | No | NA | 2 | 0 | 1 | Not-determined | Genome-fragment | NA | NA | 0 |
| S11_extracted_k141_43257 | 1166 | No | NA | 3 | 1 | 0 | Low-quality | Genome-fragment | 2.77 | AAI-based (high-confidence) | 0 |
| S13_extracted_124335 | 1166 | No | NA | 2 | 0 | 1 | Low-quality | Genome-fragment | 4.34 | AAI-based (medium-confidence) | 0 |
| S11_extracted_k141_87985 | 1080 | No | NA | 2 | 0 | 0 | Not-determined | Genome-fragment | NA | NA | 0 |
| S11_extracted_k141_81229 | 1781 | No | NA | 1 | 0 | 1 | Not-determined | Genome-fragment | NA | NA | 0 |
| S14_vs_k141_8785\|\|full | 1166 | No | NA | 1 | 0 | 1 | Low-quality | Genome-fragment | 0.52 | AAI-based (medium-confidence) | 0 |
| S5_vs_k141_140910\|\|full | 1166 | No | NA | 3 | 1 | 0 | Low-quality | Genome-fragment | 2.92 | AAI-based (high-confidence) | 0 |
| S5_vs_k141_118882\|\|full | 1166 | No | NA | 3 | 0 | 0 | Low-quality | Genome-fragment | 2.28 | AAI-based (high-confidence) | 0 |
| S11_extracted_k141_75208 | 1008 | No | NA | 1 | 0 | 1 | Not-determined | Genome-fragment | NA | NA | 0 |
| S5_vs_k141_142700\|\|full | 1166 | No | NA | 5 | 1 | 0 | Low-quality | Genome-fragment | 2.69 | AAI-based (high-confidence) | 0 |
| S11_extracted_k141_3531 | 1574 | No | NA | 3 | 0 | 0 | Not-determined | Genome-fragment | NA | NA | 0 |
| S5_vs_k141_18230\|\|lt2gene | 1166 | No | NA | 1 | 1 | 0 | Low-quality | Genome-fragment | 1.76 | AAI-based (high-confidence) | 0 |
| S11_extracted_k141_75229 | 1263 | No | NA | 1 | 0 | 1 | Not-determined | Genome-fragment | NA | NA | 0 |
| S11_extracted_k141_35418 | 1548 | No | NA | 2 | 0 | 2 | Not-determined | Genome-fragment | NA | NA | 0 |
| S7_extracted_64967 | 1166 | No | NA | 1 | 1 | 0 | Low-quality | Genome-fragment | 2.46 | AAI-based (high-confidence) | 0 |
| S11_extracted_k141_55302 | 5347 | No | NA | 8 | 0 | 4 | Not-determined | Genome-fragment | NA | NA | 0 |
| S11_extracted_k141_55320 | 2533 | No | NA | 2 | 0 | 0 | Not-determined | Genome-fragment | NA | NA | 0 |
| S7_vs_15522\|\|full | 1166 | No | NA | 3 | 2 | 0 | Low-quality | Genome-fragment | 2.31 | AAI-based (medium-confidence) | 0 |
| S11_extracted_k141_61544 | 1130 | No | NA | 2 | 0 | 1 | Not-determined | Genome-fragment | NA | NA | 0 |
| S16_extracted_136147 | 1166 | No | NA | 3 | 0 | 0 | Low-quality | Genome-fragment | 2.65 | AAI-based (high-confidence) | 0 |
| S16_vs_91033\|\|full | 1166 | No | NA | 5 | 2 | 0 | Low-quality | Genome-fragment | 2.5 | AAI-based (high-confidence) | 0 |
| S11_extracted_k141_67898 | 1096 | No | NA | 2 | 0 | 0 | Not-determined | Genome-fragment | NA | NA | 0 |
| S11_extracted_k141_67911 | 3271 | No | NA | 3 | 0 | 2 | Not-determined | Genome-fragment | NA | NA | 0 |
| S11_extracted_k141_75295 | 1069 | No | NA | 3 | 0 | 3 | Not-determined | Genome-fragment | NA | NA | 0 |
| S16_vs_77365\|\|lt2gene | 1166 | No | NA | 1 | 1 | 0 | Low-quality | Genome-fragment | 2.65 | AAI-based (high-confidence) | 0 |
| S11_extracted_k141_29237 | 1063 | No | NA | 2 | 0 | 1 | Not-determined | Genome-fragment | NA | NA | 0 |
| S11_extracted_k141_42430 | 1893 | No | NA | 3 | 0 | 2 | Not-determined | Genome-fragment | NA | NA | 0 |
| S11_extracted_k141_23217 | 6302 | No | NA | 8 | 0 | 5 | Not-determined | Genome-fragment | NA | NA | 0 |
| S10_extracted_50937 | 1166 | No | NA | 3 | 3 | 0 | Low-quality | Genome-fragment | 3.85 | AAI-based (high-confidence) | 0 |
| S11_extracted_k141_75326 | 1067 | No | NA | 1 | 0 | 0 | Not-determined | Genome-fragment | NA | NA | 0 |
| S11_extracted_k141_101025 | 1950 | No | NA | 2 | 0 | 1 | Not-determined | Genome-fragment | NA | NA | 0 |
| S10_vs_37673\|\|lt2gene | 1166 | No | NA | 1 | 1 | 0 | Low-quality | Genome-fragment | 2.27 | AAI-based (high-confidence) | 0 |
| S15_vs_69876\|\|full | 1166 | No | NA | 4 | 0 | 0 | Low-quality | Genome-fragment | 0.62 | AAI-based (high-confidence) | 0 |
| S11_extracted_k141_101042 | 1757 | No | NA | 4 | 0 | 0 | Not-determined | Genome-fragment | NA | NA | 0 |
| S11_extracted_k141_61631 | 1010 | No | NA | 1 | 0 | 0 | Not-determined | Genome-fragment | NA | NA | 0 |
| S11_extracted_k141_23250 | 7789 | No | NA | 9 | 0 | 8 | Not-determined | Genome-fragment | NA | NA | 0 |
| S11_extracted_k141_35598 | 1004 | No | NA | 2 | 0 | 2 | Not-determined | Genome-fragment | NA | NA | 0 |
| S11_extracted_k141_16321 | 4331 | No | NA | 6 | 0 | 3 | Not-determined | Genome-fragment | NA | NA | 0 |
| S6_vs_k141_14507\|\|lt2gene | 1166 | No | NA | 1 | 1 | 0 | Low-quality | Genome-fragment | 2.48 | AAI-based (high-confidence) | 0 |
| S11_extracted_k141_10111 | 4023 | No | NA | 5 | 0 | 1 | Not-determined | Genome-fragment | NA | NA | 0 |
| S11_extracted_k141_29282 | 1977 | No | NA | 1 | 0 | 1 | Not-determined | Genome-fragment | NA | NA | 0 |
| S8_extracted_k141_71933 | 1166 | No | NA | 2 | 0 | 1 | Low-quality | Genome-fragment | 0.83 | AAI-based (medium-confidence) | 0 |
| S3_vs_k141_76013\|\|full | 1167 | No | NA | 3 | 1 | 0 | Low-quality | Genome-fragment | 0.73 | HMM-based (lower-bound) | 0 |
| S11_extracted_k141_23318 | 1008 | No | NA | 1 | 0 | 1 | Not-determined | Genome-fragment | NA | NA | 0 |
| S11_extracted_k141_29301 | 1444 | No | NA | 1 | 0 | 0 | Not-determined | Genome-fragment | NA | NA | 0 |
| S16_extracted_51039 | 1167 | No | NA | 3 | 0 | 1 | Low-quality | Genome-fragment | 1.45 | AAI-based (high-confidence) | 0 |
| S11_extracted_k141_68072 | 1454 | No | NA | 2 | 0 | 2 | Not-determined | Genome-fragment | NA | NA | 0 |
| S11_extracted_k141_10152 | 2776 | No | NA | 2 | 0 | 2 | Not-determined | Genome-fragment | NA | NA | 0 |
| S11_extracted_k141_68083 | 1046 | No | NA | 2 | 0 | 1 | Not-determined | Genome-fragment | NA | NA | 0 |
| S16_vs_30852\|\|lt2gene | 1167 | No | NA | 1 | 1 | 0 | Low-quality | Genome-fragment | 2.48 | AAI-based (medium-confidence) | 0 |
| S11_extracted_k141_42585 | 6244 | No | NA | 6 | 0 | 0 | Not-determined | Genome-fragment | NA | NA | 0 |
| S10_vs_21459\|\|full | 1167 | No | NA | 3 | 1 | 0 | Low-quality | Genome-fragment | 3.15 | AAI-based (high-confidence) | 0 |
| S11_extracted_k141_42598 | 1367 | No | NA | 2 | 0 | 0 | Not-determined | Genome-fragment | NA | NA | 0 |
| S11_extracted_k141_75543 | 1372 | No | NA | 2 | 0 | 0 | Not-determined | Genome-fragment | NA | NA | 0 |
| S11_extracted_k141_75547 | 1273 | No | NA | 3 | 0 | 0 | Not-determined | Genome-fragment | NA | NA | 0 |
| S10_vs_75992\|\|full | 1167 | No | NA | 3 | 0 | 0 | Low-quality | Genome-fragment | 2.75 | AAI-based (high-confidence) | 0 |
| S12_extracted_k141_22308 | 1167 | No | NA | 4 | 2 | 0 | Low-quality | Genome-fragment | 2.49 | AAI-based (high-confidence) | 0 |
| S11_extracted_k141_55601 | 1059 | No | NA | 2 | 0 | 0 | Not-determined | Genome-fragment | NA | NA | 0 |
| S11_extracted_k141_23377 | 1182 | No | NA | 2 | 0 | 0 | Not-determined | Genome-fragment | NA | NA | 0 |
| S11_extracted_k141_61748 | 3808 | No | NA | 5 | 0 | 0 | Not-determined | Genome-fragment | NA | NA | 0 |
| S11_extracted_k141_61755 | 1036 | No | NA | 1 | 0 | 1 | Not-determined | Genome-fragment | NA | NA | 0 |
| S11_extracted_k141_75559 | 1986 | No | NA | 5 | 0 | 0 | Not-determined | Genome-fragment | NA | NA | 0 |
| S12_vs_k141_144705\|\|full | 1167 | No | NA | 4 | 0 | 0 | Low-quality | Genome-fragment | 3.13 | AAI-based (high-confidence) | 0 |
| S11_extracted_k141_75583 | 1013 | No | NA | 1 | 0 | 1 | Not-determined | Genome-fragment | NA | NA | 0 |
| S11_extracted_k141_88266 | 1416 | No | NA | 1 | 0 | 0 | Not-determined | Genome-fragment | NA | NA | 0 |
| S11_extracted_k141_23415 | 1069 | No | NA | 1 | 0 | 1 | Not-determined | Genome-fragment | NA | NA | 0 |
| S8_vs_k141_67392\|\|full | 1167 | No | NA | 2 | 0 | 0 | Low-quality | Genome-fragment | 1.58 | AAI-based (medium-confidence) | 0 |
| S11_extracted_k141_88299 | 3732 | No | NA | 3 | 0 | 1 | Not-determined | Genome-fragment | NA | NA | 0 |
| S11_extracted_k141_81557 | 1042 | No | NA | 1 | 0 | 0 | Not-determined | Genome-fragment | NA | NA | 0 |
| S11_extracted_k141_101316 | 1061 | No | NA | 2 | 0 | 1 | Not-determined | Genome-fragment | NA | NA | 0 |
| S11_extracted_k141_88349 | 1326 | No | NA | 1 | 0 | 0 | Not-determined | Genome-fragment | NA | NA | 0 |
| S17_vs_k141_3810\|\|full | 1168 | No | NA | 3 | 0 | 0 | Low-quality | Genome-fragment | 2.8 | AAI-based (medium-confidence) | 0 |
| S11_extracted_k141_55736 | 1087 | No | NA | 1 | 0 | 1 | Not-determined | Genome-fragment | NA | NA | 0 |
| S11_extracted_k141_94853 | 1213 | No | NA | 2 | 0 | 0 | Not-determined | Genome-fragment | NA | NA | 0 |
| S17_vs_k141_94801\|\|full | 1168 | No | NA | 3 | 1 | 0 | Low-quality | Genome-fragment | 0.82 | AAI-based (high-confidence) | 0 |
| S11_extracted_k141_55780 | 3055 | No | NA | 3 | 0 | 2 | Not-determined | Genome-fragment | NA | NA | 0 |
| S11_extracted_k141_61920 | 1007 | No | NA | 1 | 0 | 1 | Not-determined | Genome-fragment | NA | NA | 0 |
| S11_extracted_k141_55786 | 1106 | No | NA | 2 | 0 | 1 | Not-determined | Genome-fragment | NA | NA | 0 |
| S11_extracted_k141_101360 | 1032 | No | NA | 2 | 0 | 0 | Not-determined | Genome-fragment | NA | NA | 0 |
| S11_extracted_k141_23503 | 3158 | No | NA | 6 | 0 | 5 | Not-determined | Genome-fragment | NA | NA | 0 |
| S11_extracted_k141_61974 | 3132 | No | NA | 3 | 0 | 2 | Not-determined | Genome-fragment | NA | NA | 0 |
| S11_extracted_k141_55840 | 1022 | No | NA | 2 | 0 | 1 | Not-determined | Genome-fragment | NA | NA | 0 |
| S11_extracted_k141_91862 | 1168 | No | NA | 1 | 1 | 0 | Low-quality | Genome-fragment | 2.98 | AAI-based (high-confidence) | 0 |
| S11_extracted_k141_94948 | 1174 | No | NA | 2 | 0 | 2 | Not-determined | Genome-fragment | NA | NA | 0 |
| S1_vs_k141_154695\|\|full | 1168 | No | NA | 4 | 0 | 0 | Low-quality | Genome-fragment | 5.96 | AAI-based (high-confidence) | 0 |
| S3_extracted_k141_71523 | 1168 | No | NA | 1 | 0 | 0 | Low-quality | Genome-fragment | 2.22 | AAI-based (high-confidence) | 0 |
| S11_extracted_k141_81653 | 1419 | No | NA | 2 | 0 | 0 | Not-determined | Genome-fragment | NA | NA | 0 |
| S11_extracted_k141_42847 | 1025 | No | NA | 1 | 0 | 0 | Not-determined | Genome-fragment | NA | NA | 0 |
| S3_vs_k141_33513\|\|full | 1168 | No | NA | 3 | 0 | 0 | Low-quality | Genome-fragment | 2.57 | AAI-based (high-confidence) | 0 |
| S3_vs_k141_17945\|\|full | 1168 | No | NA | 2 | 0 | 0 | Low-quality | Genome-fragment | 2.32 | AAI-based (medium-confidence) | 0 |
| S11_extracted_k141_36030 | 1387 | No | NA | 1 | 0 | 1 | Not-determined | Genome-fragment | NA | NA | 0 |
| S11_extracted_k141_29549 | 3102 | No | NA | 3 | 0 | 2 | Not-determined | Genome-fragment | NA | NA | 0 |
| S11_extracted_k141_81702 | 1004 | No | NA | 2 | 0 | 0 | Not-determined | Genome-fragment | NA | NA | 0 |
| S11_extracted_k141_88513 | 2190 | No | NA | 1 | 0 | 1 | Not-determined | Genome-fragment | NA | NA | 0 |
| S11_extracted_k141_3790 | 65685 | No | NA | 72 | 16 | 2 | Medium-quality | Genome-fragment | 69.9 | AAI-based (high-confidence) | 0 |
| S16_vs_27899\|\|full | 1168 | No | NA | 2 | 0 | 0 | Low-quality | Genome-fragment | 2.59 | AAI-based (medium-confidence) | 0 |
| S16_vs_169854\|\|full | 1168 | No | NA | 3 | 0 | 1 | Low-quality | Genome-fragment | 0.97 | AAI-based (medium-confidence) | 0 |
| S16_vs_31322\|\|lt2gene | 1168 | No | NA | 2 | 1 | 0 | Low-quality | Genome-fragment | 3.46 | AAI-based (high-confidence) | 0 |
| S11_extracted_k141_3822 | 1462 | No | NA | 1 | 0 | 0 | Not-determined | Genome-fragment | NA | NA | 0 |
| S10_extracted_53462 | 1168 | No | NA | 3 | 0 | 0 | Low-quality | Genome-fragment | 2.07 | AAI-based (medium-confidence) | 0 |
| S10_vs_76289\|\|full | 1168 | No | NA | 3 | 0 | 0 | Low-quality | Genome-fragment | 1.47 | AAI-based (high-confidence) | 0 |
| S11_extracted_k141_88588 | 1155 | No | NA | 2 | 0 | 1 | Not-determined | Genome-fragment | NA | NA | 0 |
| S11_extracted_k141_62133 | 1472 | No | NA | 2 | 0 | 1 | Not-determined | Genome-fragment | NA | NA | 0 |
| S17_vs_k141_88760\|\|full | 1169 | No | NA | 2 | 0 | 0 | Low-quality | Genome-fragment | 4.11 | AAI-based (medium-confidence) | 0 |
| S17_vs_k141_172922\|\|lt2gene | 1169 | No | NA | 2 | 0 | 0 | Low-quality | Genome-fragment | 2.66 | AAI-based (high-confidence) | 0 |
| S11_extracted_k141_16657 | 3140 | No | NA | 2 | 0 | 1 | Not-determined | Genome-fragment | NA | NA | 0 |
| S11_extracted_k141_88618 | 1336 | No | NA | 1 | 0 | 1 | Not-determined | Genome-fragment | NA | NA | 0 |
| S9_extracted_k141_44920 | 1169 | No | NA | 4 | 0 | 0 | Low-quality | Genome-fragment | 1.95 | AAI-based (high-confidence) | 0 |
| S11_extracted_k141_103070 | 1169 | No | NA | 4 | 4 | 0 | Low-quality | Genome-fragment | 3.86 | AAI-based (high-confidence) | 0 |
| S11_extracted_k141_56049 | 1070 | No | NA | 1 | 0 | 0 | Not-determined | Genome-fragment | NA | NA | 0 |
| S11_extracted_k141_36148 | 5666 | No | NA | 4 | 0 | 1 | Not-determined | Genome-fragment | NA | NA | 0 |
| S11_extracted_k141_29631 | 3215 | No | NA | 3 | 0 | 3 | Not-determined | Genome-fragment | NA | NA | 0 |
| S11_extracted_k141_29642 | 1685 | No | NA | 3 | 0 | 1 | Not-determined | Genome-fragment | NA | NA | 0 |

| | | | | | | | | | | | |
|---|---|---|---|---|---|---|---|---|---|---|---|
| S11_extracted_k141_62195 | 1016 | No | NA | 2 | 0 | 1 | Not-determined | Genome-fragment | NA | NA | 0 |
| S11_extracted_k141_36182 | 1189 | No | NA | 1 | 0 | 1 | Not-determined | Genome-fragment | NA | NA | 0 |
| S11_extracted_k141_42929 | 1221 | No | NA | 2 | 0 | 0 | Not-determined | Genome-fragment | NA | NA | 0 |
| S11_extracted_k141_23731 | 1095 | No | NA | 1 | 0 | 1 | Not-determined | Genome-fragment | NA | NA | 0 |
| S11_vs_k141_100876\|\|full | 1169 | No | NA | 3 | 1 | 0 | Low-quality | Genome-fragment | 3.07 | AAI-based (high-confidence) | 0 |
| S11_extracted_k141_36218 | 3377 | No | NA | 4 | 0 | 2 | Not-determined | Genome-fragment | NA | NA | 0 |
| S11_extracted_k141_36240 | 1244 | No | NA | 1 | 0 | 0 | Not-determined | Genome-fragment | NA | NA | 0 |
| S11_extracted_k141_75911 | 3629 | No | NA | 4 | 0 | 4 | Not-determined | Genome-fragment | NA | NA | 0 |
| S11_extracted_k141_95067 | 46485 | No | NA | 69 | 12 | 0 | Complete | High-quality | 100 | DTR (high-confidence) | 0 |
| S13_vs_115006\|\|full | 1169 | No | NA | 2 | 0 | 0 | Low-quality | Genome-fragment | 2.59 | AAI-based (medium-confidence) | 0 |
| S11_extracted_k141_23838 | 1474 | No | NA | 1 | 0 | 1 | Not-determined | Genome-fragment | NA | NA | 0 |
| S14_vs_k141_55512\|\|full | 1169 | No | NA | 3 | 0 | 0 | Low-quality | Genome-fragment | 2.2 | AAI-based (medium-confidence) | 0 |
| S14_vs_k141_72419\|\|full | 1169 | Yes | 657 | 3 | 1 | 2 | Low-quality | Genome-fragment | 0.93 | AAI-based (medium-confidence) | 43.8 |
| S11_extracted_k141_43097 | 1746 | No | NA | 3 | 0 | 2 | Not-determined | Genome-fragment | NA | NA | 0 |
| S3_extracted_k141_121262 | 1169 | No | NA | 3 | 0 | 1 | Low-quality | Genome-fragment | 1.17 | AAI-based (medium-confidence) | 0 |
| S11_extracted_k141_56262 | 3992 | No | NA | 2 | 0 | 2 | Not-determined | Genome-fragment | NA | NA | 0 |
| S11_extracted_k141_36352 | 1573 | No | NA | 2 | 0 | 1 | Not-determined | Genome-fragment | NA | NA | 0 |
| S3_extracted_k141_62064 | 1169 | No | NA | 2 | 2 | 0 | Low-quality | Genome-fragment | 0.69 | AAI-based (high-confidence) | 0 |
| S11_extracted_k141_81971 | 1261 | No | NA | 2 | 0 | 2 | Not-determined | Genome-fragment | NA | NA | 0 |
| S11_extracted_k141_75936 | 1458 | No | NA | 1 | 0 | 0 | Not-determined | Genome-fragment | NA | NA | 0 |
| S11_extracted_k141_4121 | 1171 | No | NA | 1 | 0 | 1 | Not-determined | Genome-fragment | NA | NA | 0 |
| S5_vs_k141_22647\|\|full | 1169 | No | NA | 2 | 0 | 0 | Low-quality | Genome-fragment | 0.5 | AAI-based (medium-confidence) | 0 |
| S11_extracted_k141_29745 | 1380 | No | NA | 1 | 0 | 0 | Not-determined | Genome-fragment | NA | NA | 0 |
| S11_extracted_k141_43191 | 1206 | No | NA | 1 | 0 | 1 | Not-determined | Genome-fragment | NA | NA | 0 |
| S11_extracted_k141_49085 | 3490 | No | NA | 5 | 0 | 4 | Not-determined | Genome-fragment | NA | NA | 0 |
| S11_extracted_k141_36434 | 1514 | No | NA | 1 | 0 | 1 | Not-determined | Genome-fragment | NA | NA | 0 |
| S11_extracted_k141_24027 | 1184 | No | NA | 1 | 0 | 1 | Not-determined | Genome-fragment | NA | NA | 0 |
| S11_extracted_k141_29763 | 3116 | No | NA | 3 | 0 | 3 | Not-determined | Genome-fragment | NA | NA | 0 |
| S7_vs_87313\|\|full | 1169 | No | NA | 3 | 1 | 0 | Low-quality | Genome-fragment | 2.16 | AAI-based (high-confidence) | 0 |
| S7_vs_71643\|\|lt2gene | 1169 | No | NA | 1 | 1 | 0 | Low-quality | Genome-fragment | 1.31 | HMM-based (lower-bound) | 0 |
| S11_extracted_k141_36446 | 2100 | No | NA | 1 | 0 | 0 | Not-determined | Genome-fragment | NA | NA | 0 |
| S16_extracted_41996 | 1169 | No | NA | 1 | 0 | 1 | Low-quality | Genome-fragment | 0.77 | AAI-based (medium-confidence) | 0 |
| S11_extracted_k141_56401 | 1831 | No | NA | 2 | 0 | 2 | Not-determined | Genome-fragment | NA | NA | 0 |
| S11_extracted_k141_49107 | 3077 | No | NA | 3 | 0 | 0 | Not-determined | Genome-fragment | NA | NA | 0 |
| S11_extracted_k141_62498 | 1860 | No | NA | 1 | 0 | 0 | Not-determined | Genome-fragment | NA | NA | 0 |
| S11_extracted_k141_56426 | 3100 | No | NA | 3 | 0 | 3 | Not-determined | Genome-fragment | NA | NA | 0 |
| S11_extracted_k141_43229 | 1808 | No | NA | 2 | 0 | 0 | Not-determined | Genome-fragment | NA | NA | 0 |
| S11_extracted_k141_95288 | 1414 | No | NA | 2 | 0 | 2 | Not-determined | Genome-fragment | NA | NA | 0 |
| S16_vs_23825\|\|full | 1169 | No | NA | 2 | 0 | 1 | Low-quality | Genome-fragment | 2.2 | AAI-based (high-confidence) | 0 |
| S10_extracted_9431 | 1169 | No | NA | 2 | 2 | 0 | Low-quality | Genome-fragment | 3.35 | AAI-based (high-confidence) | 0 |
| S11_extracted_k141_95313 | 1994 | No | NA | 3 | 0 | 2 | Not-determined | Genome-fragment | NA | NA | 0 |
| S11_extracted_k141_101831 | 4961 | No | NA | 6 | 0 | 0 | Not-determined | Genome-fragment | NA | NA | 0 |
| S10_vs_6013\|\|full | 1169 | No | NA | 5 | 1 | 0 | Low-quality | Genome-fragment | 1.96 | HMM-based (lower-bound) | 0 |
| S12_vs_k141_146033\|\|full | 1169 | No | NA | 5 | 0 | 0 | Low-quality | Genome-fragment | 0.64 | AAI-based (high-confidence) | 0 |
| S11_extracted_k141_76016 | 1151 | No | NA | 1 | 0 | 0 | Not-determined | Genome-fragment | NA | NA | 0 |
| S15_vs_67512\|\|lt2gene | 1169 | No | NA | 1 | 0 | 0 | Low-quality | Genome-fragment | 2.88 | AAI-based (high-confidence) | 0 |
| S11_extracted_k141_88939 | 1720 | No | NA | 1 | 0 | 1 | Not-determined | Genome-fragment | NA | NA | 0 |
| S11_extracted_k141_43300 | 1969 | No | NA | 3 | 0 | 0 | Not-determined | Genome-fragment | NA | NA | 0 |
| S11_extracted_k141_17017 | 1452 | No | NA | 1 | 0 | 1 | Not-determined | Genome-fragment | NA | NA | 0 |
| S11_extracted_k141_88980 | 2110 | No | NA | 2 | 0 | 2 | Not-determined | Genome-fragment | NA | NA | 0 |
| S8_vs_k141_121670\|\|full | 1169 | No | NA | 2 | 0 | 0 | Low-quality | Genome-fragment | 2.46 | AAI-based (medium-confidence) | 0 |
| S11_extracted_k141_36596 | 1172 | No | NA | 2 | 0 | 0 | Not-determined | Genome-fragment | NA | NA | 0 |
| S8_vs_k141_167797\|\|full | 1169 | No | NA | 3 | 2 | 0 | Low-quality | Genome-fragment | 1.92 | AAI-based (high-confidence) | 0 |
| S11_extracted_k141_101916 | 3198 | No | NA | 2 | 0 | 2 | Not-determined | Genome-fragment | NA | NA | 0 |
| S11_extracted_k141_68693 | 40422 | No | NA | 53 | 10 | 0 | Medium-quality | Genome-fragment | 67.28 | AAI-based (high-confidence) | 0 |
| S11_extracted_k141_68697 | 1391 | No | NA | 3 | 0 | 1 | Not-determined | Genome-fragment | NA | NA | 0 |
| S11_extracted_k141_82163 | 4572 | No | NA | 4 | 0 | 0 | Not-determined | Genome-fragment | NA | NA | 0 |
| S11_extracted_k141_101974 | 2218 | No | NA | 4 | 0 | 0 | Not-determined | Genome-fragment | NA | NA | 0 |
| S11_extracted_k141_24295 | 3213 | No | NA | 4 | 0 | 1 | Not-determined | Genome-fragment | NA | NA | 0 |
| S11_extracted_k141_17133 | 1044 | No | NA | 1 | 0 | 0 | Not-determined | Genome-fragment | NA | NA | 0 |
| S11_extracted_k141_29970 | 2353 | No | NA | 3 | 0 | 2 | Not-determined | Genome-fragment | NA | NA | 0 |
| S8_vs_k141_173292\|\|full | 1169 | No | NA | 3 | 3 | 0 | Low-quality | Genome-fragment | 3.16 | AAI-based (high-confidence) | 0 |
| S11_extracted_k141_62662 | 1065 | No | NA | 1 | 0 | 1 | Not-determined | Genome-fragment | NA | NA | 0 |
| S11_extracted_k141_17162 | 1047 | No | NA | 2 | 0 | 0 | Not-determined | Genome-fragment | NA | NA | 0 |
| S11_extracted_k141_36741 | 1117 | No | NA | 3 | 0 | 1 | Not-determined | Genome-fragment | NA | NA | 0 |
| S11_extracted_k141_10907 | 37668 | No | NA | 50 | 24 | 0 | Complete | High-quality | 100 | DTR (medium-confidence) | 0 |
| S17_vs_k141_160863\|\|full | 1170 | No | NA | 3 | 0 | 0 | Low-quality | Genome-fragment | 2.04 | AAI-based (high-confidence) | 0 |
| S9_extracted_k141_4929 | 1170 | No | NA | 2 | 0 | 0 | Low-quality | Genome-fragment | 2.37 | AAI-based (medium-confidence) | 0 |
| S11_extracted_k141_62703 | 1582 | No | NA | 2 | 0 | 2 | Not-determined | Genome-fragment | NA | NA | 0 |
| S11_extracted_k141_62754 | 1378 | No | NA | 2 | 0 | 1 | Not-determined | Genome-fragment | NA | NA | 0 |
| S11_vs_k141_9562\|\|full | 1170 | Yes | 276 | 4 | 1 | 1 | Low-quality | Genome-fragment | 0.71 | AAI-based (medium-confidence) | 76.41 |
| S11_extracted_k141_24442 | 2377 | No | NA | 4 | 0 | 0 | Not-determined | Genome-fragment | NA | NA | 0 |
| S11_extracted_k141_10977 | 1176 | No | NA | 1 | 0 | 1 | Not-determined | Genome-fragment | NA | NA | 0 |
| S11_extracted_k141_82286 | 1747 | No | NA | 2 | 0 | 0 | Not-determined | Genome-fragment | NA | NA | 0 |
| S13_vs_51444\|\|full | 1170 | No | NA | 6 | 0 | 0 | Low-quality | Genome-fragment | 3.13 | AAI-based (high-confidence) | 0 |
| S11_extracted_k141_24522 | 1004 | No | NA | 1 | 0 | 0 | Not-determined | Genome-fragment | NA | NA | 0 |
| S11_extracted_k141_49370 | 1036 | No | NA | 2 | 0 | 1 | Not-determined | Genome-fragment | NA | NA | 0 |
| S11_extracted_k141_95700 | 2538 | No | NA | 2 | 0 | 2 | Not-determined | Genome-fragment | NA | NA | 0 |
| S11_extracted_k141_82301 | 3138 | No | NA | 2 | 0 | 0 | Not-determined | Genome-fragment | NA | NA | 0 |
| S11_extracted_k141_24568 | 1024 | No | NA | 1 | 0 | 0 | Not-determined | Genome-fragment | NA | NA | 0 |
| S13_vs_105436\|\|full | 1170 | No | NA | 2 | 0 | 0 | Low-quality | Genome-fragment | 1.32 | AAI-based (medium-confidence) | 0 |
| S11_extracted_k141_95795 | 1759 | No | NA | 2 | 0 | 0 | Not-determined | Genome-fragment | NA | NA | 0 |
| S11_extracted_k141_24655 | 1017 | No | NA | 1 | 0 | 1 | Not-determined | Genome-fragment | NA | NA | 0 |
| S11_extracted_k141_49474 | 1123 | No | NA | 2 | 0 | 1 | Not-determined | Genome-fragment | NA | NA | 0 |
| S11_extracted_k141_76214 | 2256 | No | NA | 1 | 0 | 0 | Not-determined | Genome-fragment | NA | NA | 0 |
| S11_extracted_k141_49488 | 3298 | No | NA | 3 | 0 | 2 | Not-determined | Genome-fragment | NA | NA | 0 |
| S1_vs_k141_52643\|\|lt2gene | 1170 | No | NA | 1 | 1 | 0 | Low-quality | Genome-fragment | 3.2 | AAI-based (high-confidence) | 0 |
| S11_extracted_k141_68972 | 1666 | No | NA | 5 | 0 | 0 | Not-determined | Genome-fragment | NA | NA | 0 |
| S11_extracted_k141_17399 | 1485 | No | NA | 1 | 0 | 0 | Not-determined | Genome-fragment | NA | NA | 0 |
| S11_extracted_k141_24701 | 1005 | No | NA | 1 | 0 | 0 | Not-determined | Genome-fragment | NA | NA | 0 |
| S11_extracted_k141_56775 | 1215 | No | NA | 2 | 0 | 1 | Not-determined | Genome-fragment | NA | NA | 0 |
| S11_extracted_k141_102325 | 1043 | No | NA | 1 | 0 | 0 | Not-determined | Genome-fragment | NA | NA | 0 |
| S7_vs_76401\|\|full | 1170 | No | NA | 2 | 0 | 1 | Low-quality | Genome-fragment | 0.8 | AAI-based (medium-confidence) | 0 |
| S7_vs_97361\|\|full | 1170 | No | NA | 4 | 0 | 0 | Low-quality | Genome-fragment | 3.02 | AAI-based (high-confidence) | 0 |
| S11_extracted_k141_36986 | 1000 | No | NA | 3 | 0 | 2 | Not-determined | Genome-fragment | NA | NA | 0 |
| S11_extracted_k141_76315 | 1072 | No | NA | 2 | 0 | 1 | Not-determined | Genome-fragment | NA | NA | 0 |
| S6_vs_k141_19391\|\|full | 1170 | No | NA | 3 | 0 | 0 | Low-quality | Genome-fragment | 2.76 | AAI-based (high-confidence) | 0 |
| S11_extracted_k141_82359 | 4084 | No | NA | 3 | 0 | 3 | Not-determined | Genome-fragment | NA | NA | 0 |
| S11_extracted_k141_49566 | 3229 | No | NA | 2 | 0 | 1 | Not-determined | Genome-fragment | NA | NA | 0 |
| S11_extracted_k141_95962 | 3031 | No | NA | 3 | 0 | 0 | Not-determined | Genome-fragment | NA | NA | 0 |
| S11_extracted_k141_49593 | 3023 | No | NA | 3 | 0 | 2 | Not-determined | Genome-fragment | NA | NA | 0 |
| S11_extracted_k141_56861 | 5414 | No | NA | 4 | 0 | 1 | Not-determined | Genome-fragment | NA | NA | 0 |
| S8_vs_k141_166468\|\|full | 1170 | No | NA | 3 | 1 | 0 | Low-quality | Genome-fragment | 0.09 | HMM-based (lower-bound) | 0 |
| S11_extracted_k141_30255 | 1075 | No | NA | 1 | 0 | 1 | Not-determined | Genome-fragment | NA | NA | 0 |
| S8_vs_k141_180878\|\|full | 1170 | No | NA | 3 | 0 | 0 | Low-quality | Genome-fragment | 2.24 | AAI-based (medium-confidence) | 0 |
| S11_extracted_k141_37096 | 11660 | No | NA | 6 | 0 | 3 | Not-determined | Genome-fragment | NA | NA | 0 |
| S11_extracted_k141_63060 | 1517 | No | NA | 5 | 0 | 0 | Not-determined | Genome-fragment | NA | NA | 0 |
| S11_extracted_k141_96002 | 1367 | No | NA | 2 | 0 | 0 | Not-determined | Genome-fragment | NA | NA | 0 |
| S11_extracted_k141_89309 | 46753 | No | NA | 42 | 1 | 9 | Medium-quality | Genome-fragment | 52.48 | HMM-based (lower-bound) | 0 |
| S14_extracted_k141_759 | 1171 | No | NA | 4 | 0 | 0 | Low-quality | Genome-fragment | 1.69 | AAI-based (high-confidence) | 0 |
| S1_extracted_k141_133667 | 1171 | No | NA | 1 | 0 | 0 | Low-quality | Genome-fragment | 0.67 | AAI-based (medium-confidence) | 0 |
| S1_vs_k141_146673\|\|full | 1171 | No | NA | 3 | 1 | 0 | Low-quality | Genome-fragment | 2.27 | AAI-based (high-confidence) | 0 |
| S11_extracted_k141_82422 | 1239 | No | NA | 1 | 0 | 0 | Not-determined | Genome-fragment | NA | NA | 0 |
| S1_vs_k141_69091\|\|lt2gene | 1171 | No | NA | 2 | 1 | 0 | Low-quality | Genome-fragment | 0.89 | HMM-based (lower-bound) | 0 |
| S3_extracted_k141_111730 | 1171 | No | NA | 2 | 0 | 2 | Low-quality | Genome-fragment | 0.72 | AAI-based (medium-confidence) | 0 |
| S11_extracted_k141_102467 | 1369 | No | NA | 2 | 0 | 2 | Not-determined | Genome-fragment | NA | NA | 0 |
| S11_extracted_k141_56972 | 3541 | No | NA | 4 | 0 | 2 | Not-determined | Genome-fragment | NA | NA | 0 |
| S11_extracted_k141_63109 | 2306 | No | NA | 4 | 0 | 0 | Not-determined | Genome-fragment | NA | NA | 0 |
| S3_extracted_k141_10428 | 1171 | No | NA | 1 | 1 | 0 | Low-quality | Genome-fragment | 2.85 | AAI-based (high-confidence) | 0 |
| S11_extracted_k141_63112 | 3493 | No | NA | 2 | 0 | 1 | Not-determined | Genome-fragment | NA | NA | 0 |
| S11_extracted_k141_102494 | 1035 | No | NA | 1 | 0 | 1 | Not-determined | Genome-fragment | NA | NA | 0 |
| S3_vs_k141_40935\|\|lt2gene | 1171 | No | NA | 2 | 0 | 0 | Low-quality | Genome-fragment | 3.06 | AAI-based (medium-confidence) | 0 |
| S11_extracted_k141_43784 | 1566 | No | NA | 1 | 0 | 1 | Not-determined | Genome-fragment | NA | NA | 0 |
| S11_extracted_k141_30406 | 1054 | No | NA | 1 | 0 | 1 | Not-determined | Genome-fragment | NA | NA | 0 |
| S11_extracted_k141_43815 | 3915 | No | NA | 5 | 0 | 0 | Not-determined | Genome-fragment | NA | NA | 0 |
| S16_vs_28247\|\|full | 1171 | No | NA | 4 | 0 | 0 | Low-quality | Genome-fragment | 3.27 | AAI-based (high-confidence) | 0 |
| S11_extracted_k141_4900 | 1353 | No | NA | 1 | 0 | 0 | Not-determined | Genome-fragment | NA | NA | 0 |
| S11_extracted_k141_57079 | 1101 | No | NA | 1 | 0 | 1 | Not-determined | Genome-fragment | NA | NA | 0 |
| S11_extracted_k141_30434 | 1064 | No | NA | 1 | 0 | 1 | Not-determined | Genome-fragment | NA | NA | 0 |
| S11_extracted_k141_17679 | 2114 | No | NA | 3 | 0 | 1 | Not-determined | Genome-fragment | NA | NA | 0 |
| S12_extracted_k141_126875 | 1171 | No | NA | 1 | 0 | 1 | Low-quality | Genome-fragment | 0.33 | AAI-based (medium-confidence) | 0 |
| S11_extracted_k141_4957 | 3205 | No | NA | 2 | 0 | 2 | Not-determined | Genome-fragment | NA | NA | 0 |
| S12_vs_k141_94302\|\|full | 1171 | Yes | 442 | 2 | 1 | 1 | Low-quality | Genome-fragment | 0.69 | HMM-based (lower-bound) | 62.25 |
| S11_extracted_k141_25034 | 1024 | No | NA | 1 | 0 | 1 | Not-determined | Genome-fragment | NA | NA | 0 |
| S12_vs_k141_14935\|\|full | 1171 | No | NA | 2 | 0 | 0 | Low-quality | Genome-fragment | 1.86 | AAI-based (high-confidence) | 0 |
| S11_extracted_k141_37313 | 2299 | No | NA | 4 | 0 | 3 | Not-determined | Genome-fragment | NA | NA | 0 |
| S3_extracted_k141_94968 | 1172 | No | NA | 3 | 0 | 0 | Low-quality | Genome-fragment | 3.46 | AAI-based (high-confidence) | 0 |
| S5_vs_k141_140154\|\|full | 1172 | No | NA | 2 | 0 | 0 | Low-quality | Genome-fragment | 2.7 | AAI-based (medium-confidence) | 0 |
| S5_vs_k141_137565\|\|full | 1172 | No | NA | 3 | 1 | 0 | Low-quality | Genome-fragment | 2.05 | AAI-based (medium-confidence) | 0 |
| S11_extracted_k141_37339 | 1991 | No | NA | 2 | 0 | 0 | Not-determined | Genome-fragment | NA | NA | 0 |
| S5_vs_k141_110443\|\|lt2gene | 1172 | No | NA | 2 | 1 | 0 | Low-quality | Genome-fragment | 1.33 | HMM-based (lower-bound) | 0 |
| S11_extracted_k141_49943 | 1035 | No | NA | 1 | 0 | 0 | Not-determined | Genome-fragment | NA | NA | 0 |
| S11_extracted_k141_82626 | 1398 | No | NA | 2 | 0 | 2 | Not-determined | Genome-fragment | NA | NA | 0 |
| S16_vs_163537\|\|full | 1172 | No | NA | 4 | 4 | 0 | Low-quality | Genome-fragment | 2.91 | AAI-based (high-confidence) | 0 |
| S11_extracted_k141_11431 | 1696 | No | NA | 2 | 0 | 1 | Not-determined | Genome-fragment | NA | NA | 0 |
| S11_extracted_k141_57263 | 2874 | No | NA | 5 | 0 | 0 | Not-determined | Genome-fragment | NA | NA | 0 |
| S10_vs_80342\|\|full | 1172 | No | NA | 3 | 1 | 0 | Low-quality | Genome-fragment | 0.88 | AAI-based (high-confidence) | 0 |
| S11_extracted_k141_49977 | 5269 | No | NA | 9 | 0 | 3 | Not-determined | Genome-fragment | NA | NA | 0 |
| S15_vs_24092\|\|full | 1172 | No | NA | 3 | 2 | 0 | Low-quality | Genome-fragment | 3.44 | AAI-based (high-confidence) | 0 |
| S11_extracted_k141_17813 | 1868 | No | NA | 1 | 0 | 1 | Not-determined | Genome-fragment | NA | NA | 0 |
| S8_extracted_k141_45495 | 1172 | No | NA | 2 | 0 | 0 | Low-quality | Genome-fragment | 0.74 | AAI-based (high-confidence) | 0 |
| S11_extracted_k141_69409 | 1381 | No | NA | 3 | 0 | 0 | Not-determined | Genome-fragment | NA | NA | 0 |
| S8_extracted_k141_199134 | 1172 | No | NA | 4 | 1 | 0 | Low-quality | Genome-fragment | 2.61 | AAI-based (high-confidence) | 0 |
| S8_vs_k141_36357\|\|full | 1172 | No | NA | 3 | 0 | 1 | Low-quality | Genome-fragment | 0.47 | AAI-based (medium-confidence) | 0 |
| S8_vs_k141_75737\|\|full | 1172 | No | NA | 3 | 0 | 0 | Low-quality | Genome-fragment | 0.74 | AAI-based (high-confidence) | 0 |
| S17_extracted_k141_69656 | 1173 | No | NA | 1 | 0 | 0 | Low-quality | Genome-fragment | 2.71 | AAI-based (medium-confidence) | 0 |
| S11_extracted_k141_11503 | 4617 | No | NA | 5 | 0 | 4 | Not-determined | Genome-fragment | NA | NA | 0 |
| S17_extracted_k141_55024 | 1173 | No | NA | 3 | 0 | 1 | Low-quality | Genome-fragment | 0.24 | AAI-based (medium-confidence) | 0 |
| S11_extracted_k141_43997 | 59939 | No | NA | 76 | 5 | 0 | Medium-quality | Genome-fragment | 65.91 | AAI-based (high-confidence) | 0 |
| S11_extracted_k141_44020 | 1095 | No | NA | 2 | 0 | 0 | Not-determined | Genome-fragment | NA | NA | 0 |
| S17_vs_k141_141043\|\|full | 1173 | No | NA | 3 | 1 | 0 | Low-quality | Genome-fragment | 1.95 | AAI-based (high-confidence) | 0 |
| S17_vs_k141_71525\|\|lt2gene | 1173 | No | NA | 2 | 2 | 0 | Low-quality | Genome-fragment | 2.99 | AAI-based (high-confidence) | 0 |
| S9_vs_k141_63331\|\|full | 1173 | No | NA | 4 | 0 | 3 | Low-quality | Genome-fragment | 0.34 | AAI-based (medium-confidence) | 0 |
| S11_extracted_k141_89587 | 1438 | No | NA | 3 | 0 | 3 | Not-determined | Genome-fragment | NA | NA | 0 |
| S1_vs_k141_149876\|\|full | 1173 | No | NA | 3 | 1 | 0 | Low-quality | Genome-fragment | 3 | AAI-based (high-confidence) | 0 |
| S11_extracted_k141_11565 | 1354 | No | NA | 1 | 0 | 1 | Not-determined | Genome-fragment | NA | NA | 0 |
| S3_extracted_k141_119425 | 1173 | No | NA | 1 | 0 | 0 | Low-quality | Genome-fragment | 2.42 | AAI-based (high-confidence) | 0 |
| S5_extracted_k141_136215 | 1173 | No | NA | 3 | 1 | 0 | Low-quality | Genome-fragment | 1.83 | AAI-based (medium-confidence) | 0 |
| S11_extracted_k141_17961 | 1465 | No | NA | 3 | 0 | 2 | Not-determined | Genome-fragment | NA | NA | 0 |
| S11_extracted_k141_96576 | 1442 | No | NA | 2 | 0 | 1 | Not-determined | Genome-fragment | NA | NA | 0 |
| S5_extracted_k141_82676 | 1173 | No | NA | 3 | 1 | 0 | Low-quality | Genome-fragment | 2.67 | HMM-based (lower-bound) | 0 |
| S5_vs_k141_2165\|\|full | 1173 | No | NA | 3 | 0 | 0 | Low-quality | Genome-fragment | 3.46 | AAI-based (high-confidence) | 0 |
| S11_extracted_k141_57448 | 5582 | No | NA | 5 | 0 | 4 | Not-determined | Genome-fragment | NA | NA | 0 |
| S5_vs_k141_61906\|\|lt2gene | 1173 | No | NA | 2 | 0 | 0 | Low-quality | Genome-fragment | 2.09 | AAI-based (medium-confidence) | 0 |
| S11_extracted_k141_96619 | 1790 | No | NA | 2 | 0 | 1 | Not-determined | Genome-fragment | NA | NA | 0 |
| S11_extracted_k141_76823 | 1242 | No | NA | 1 | 0 | 1 | Not-determined | Genome-fragment | NA | NA | 0 |
| S7_vs_72508\|\|full | 1173 | No | NA | 3 | 2 | 0 | Low-quality | Genome-fragment | 4.42 | AAI-based (medium-confidence) | 0 |
| S11_extracted_k141_5139 | 5470 | No | NA | 5 | 0 | 3 | Not-determined | Genome-fragment | NA | NA | 0 |
| S16_vs_30253\|\|full | 1173 | No | NA | 3 | 0 | 0 | Low-quality | Genome-fragment | 1.81 | AAI-based (medium-confidence) | 0 |
| S11_extracted_k141_69584 | 5002 | No | NA | 3 | 0 | 1 | Not-determined | Genome-fragment | NA | NA | 0 |
| S11_extracted_k141_44177 | 20533 | No | NA | 22 | 0 | 1 | Not-determined | Genome-fragment | NA | NA | 0 |
| S16_vs_159757\|\|full | 1173 | No | NA | 3 | 3 | 0 | Low-quality | Genome-fragment | 1.94 | AAI-based (high-confidence) | 0 |
| S10_vs_110434\|\|lt2gene | 1173 | No | NA | 2 | 0 | 0 | Low-quality | Genome-fragment | 2.79 | AAI-based (medium-confidence) | 0 |
| S11_extracted_k141_25366 | 1001 | No | NA | 3 | 0 | 2 | Not-determined | Genome-fragment | NA | NA | 0 |
| S11_extracted_k141_63427 | 2791 | No | NA | 3 | 0 | 2 | Not-determined | Genome-fragment | NA | NA | 0 |
| S11_extracted_k141_37722 | 4253 | No | NA | 4 | 0 | 4 | Not-determined | Genome-fragment | NA | NA | 0 |
| S11_extracted_k141_63439 | 1041 | No | NA | 1 | 0 | 0 | Not-determined | Genome-fragment | NA | NA | 0 |
| S12_vs_k141_113261\|\|full | 1173 | No | NA | 2 | 0 | 0 | Low-quality | Genome-fragment | 3.44 | AAI-based (medium-confidence) | 0 |
| S11_extracted_k141_44266 | 1320 | No | NA | 2 | 0 | 2 | Not-determined | Genome-fragment | NA | NA | 0 |
| S4_extracted_25281 | 1173 | No | NA | 2 | 2 | 0 | Low-quality | Genome-fragment | 1.94 | AAI-based (high-confidence) | 0 |
| S11_extracted_k141_44276 | 2507 | No | NA | 2 | 0 | 2 | Not-determined | Genome-fragment | NA | NA | 0 |
| S11_extracted_k141_76969 | 1272 | No | NA | 2 | 0 | 0 | Not-determined | Genome-fragment | NA | NA | 0 |
| S8_vs_k141_1021\|\|full | 1173 | No | NA | 3 | 0 | 2 | Low-quality | Genome-fragment | 0.67 | AAI-based (medium-confidence) | 0 |
| S11_extracted_k141_37758 | 3043 | No | NA | 5 | 0 | 0 | Not-determined | Genome-fragment | NA | NA | 0 |
| S11_extracted_k141_69710 | 1094 | No | NA | 1 | 0 | 1 | Not-determined | Genome-fragment | NA | NA | 0 |
| S11_extracted_k141_89716 | 1037 | No | NA | 1 | 0 | 1 | Not-determined | Genome-fragment | NA | NA | 0 |
| S11_extracted_k141_96735 | 1289 | No | NA | 2 | 0 | 1 | Not-determined | Genome-fragment | NA | NA | 0 |
| S11_extracted_k141_83018 | 1668 | No | NA | 3 | 0 | 1 | Not-determined | Genome-fragment | NA | NA | 0 |
| S8_vs_k141_197901\|\|full | 1173 | No | NA | 3 | 1 | 0 | Low-quality | Genome-fragment | 2.79 | HMM-based (lower-bound) | 0 |
| S11_extracted_k141_63524 | 1399 | No | NA | 1 | 0 | 0 | Not-determined | Genome-fragment | NA | NA | 0 |
| S11_extracted_k141_69748 | 2236 | No | NA | 1 | 0 | 0 | Not-determined | Genome-fragment | NA | NA | 0 |
| S11_extracted_k141_50116 | 1298 | No | NA | 2 | 0 | 2 | Not-determined | Genome-fragment | NA | NA | 0 |
| S8_vs_k141_155736\|\|full | 1173 | No | NA | 2 | 0 | 1 | Low-quality | Genome-fragment | 1.47 | AAI-based (medium-confidence) | 0 |
| S11_extracted_k141_25448 | 1808 | No | NA | 3 | 0 | 0 | Not-determined | Genome-fragment | NA | NA | 0 |
| S11_extracted_k141_96761 | 1542 | No | NA | 1 | 0 | 1 | Not-determined | Genome-fragment | NA | NA | 0 |
| S9_extracted_k141_83057 | 1174 | No | NA | 3 | 1 | 0 | Low-quality | Genome-fragment | 2.37 | AAI-based (medium-confidence) | 0 |
| S11_vs_k141_60470\|\|full | 1174 | No | NA | 4 | 0 | 0 | Low-quality | Genome-fragment | 2.48 | AAI-based (high-confidence) | 0 |
| S11_extracted_k141_31004 | 1001 | No | NA | 2 | 0 | 1 | Not-determined | Genome-fragment | NA | NA | 0 |
| S11_extracted_k141_18183 | 1708 | No | NA | 2 | 0 | 1 | Not-determined | Genome-fragment | NA | NA | 0 |
| S14_extracted_k141_47656 | 1174 | No | NA | 2 | 0 | 0 | Low-quality | Genome-fragment | 2.36 | AAI-based (high-confidence) | 0 |
| S11_extracted_k141_5328 | 1394 | No | NA | 1 | 0 | 0 | Not-determined | Genome-fragment | NA | NA | 0 |
| S11_extracted_k141_96786 | 4251 | No | NA | 6 | 0 | 1 | Not-determined | Genome-fragment | NA | NA | 0 |
| S11_extracted_k141_96795 | 1921 | No | NA | 3 | 0 | 1 | Not-determined | Genome-fragment | NA | NA | 0 |
| S11_extracted_k141_89785 | 5915 | No | NA | 5 | 0 | 4 | Not-determined | Genome-fragment | NA | NA | 0 |
| S3_extracted_k141_119239 | 1174 | No | NA | 2 | 0 | 1 | Low-quality | Genome-fragment | 0.38 | AAI-based (medium-confidence) | 0 |
| S3_vs_k141_112316\|\|full | 1174 | No | NA | 3 | 0 | 0 | Low-quality | Genome-fragment | 2.03 | AAI-based (high-confidence) | 0 |
| S11_extracted_k141_37887 | 2052 | No | NA | 2 | 0 | 2 | Not-determined | Genome-fragment | NA | NA | 0 |
| S11_extracted_k141_18207 | 3188 | No | NA | 4 | 0 | 3 | Not-determined | Genome-fragment | NA | NA | 0 |
| S11_extracted_k141_5345 | 1077 | No | NA | 1 | 0 | 1 | Not-determined | Genome-fragment | NA | NA | 0 |
| S11_extracted_k141_11726 | 1745 | No | NA | 2 | 0 | 1 | Not-determined | Genome-fragment | NA | NA | 0 |
| S11_extracted_k141_57619 | 1581 | No | NA | 2 | 0 | 0 | Not-determined | Genome-fragment | NA | NA | 0 |
| S11_extracted_k141_63649 | 2324 | No | NA | 2 | 0 | 1 | Not-determined | Genome-fragment | NA | NA | 0 |
| S11_extracted_k141_69850 | 1299 | No | NA | 4 | 0 | 0 | Not-determined | Genome-fragment | NA | NA | 0 |
| S11_extracted_k141_5388 | 1283 | No | NA | 1 | 0 | 1 | Not-determined | Genome-fragment | NA | NA | 0 |
| S11_extracted_k141_89818 | 2175 | No | NA | 1 | 0 | 1 | Not-determined | Genome-fragment | NA | NA | 0 |
| S5_extracted_k141_117229 | 1174 | No | NA | 2 | 0 | 0 | Low-quality | Genome-fragment | 1.71 | AAI-based (medium-confidence) | 0 |
| S11_extracted_k141_25547 | 10782 | No | NA | 10 | 0 | 0 | Not-determined | Genome-fragment | NA | NA | 0 |
| S11_extracted_k141_5400 | 2890 | No | NA | 4 | 0 | 3 | Not-determined | Genome-fragment | NA | NA | 0 |
| S7_extracted_86406 | 1174 | No | NA | 1 | 1 | 0 | Low-quality | Genome-fragment | 2.94 | AAI-based (high-confidence) | 0 |
| S11_extracted_k141_96871 | 1110 | No | NA | 1 | 0 | 1 | Not-determined | Genome-fragment | NA | NA | 0 |
| S11_extracted_k141_57691 | 1754 | No | NA | 2 | 0 | 2 | Not-determined | Genome-fragment | NA | NA | 0 |
| S7_vs_12235\|\|full | 1174 | No | NA | 3 | 0 | 0 | Low-quality | Genome-fragment | 2.29 | AAI-based (high-confidence) | 0 |
| S7_vs_60872\|\|full | 1174 | No | NA | 3 | 1 | 0 | Low-quality | Genome-fragment | 2.51 | AAI-based (medium-confidence) | 0 |
| S11_extracted_k141_83187 | 1476 | No | NA | 1 | 0 | 0 | Not-determined | Genome-fragment | NA | NA | 0 |
| S11_extracted_k141_63734 | 3939 | No | NA | 3 | 0 | 3 | Not-determined | Genome-fragment | NA | NA | 0 |
| S11_extracted_k141_50275 | 1377 | No | NA | 2 | 0 | 1 | Not-determined | Genome-fragment | NA | NA | 0 |
| S11_extracted_k141_37938 | 1041 | No | NA | 2 | 0 | 0 | Not-determined | Genome-fragment | NA | NA | 0 |
| S16_vs_61736\|\|lt2gene | 1174 | No | NA | 2 | 1 | 0 | Low-quality | Genome-fragment | 2.18 | HMM-based (lower-bound) | 0 |
| S11_extracted_k141_25609 | 1043 | No | NA | 2 | 0 | 0 | Not-determined | Genome-fragment | NA | NA | 0 |
| S11_extracted_k141_31176 | 1161 | No | NA | 1 | 0 | 1 | Not-determined | Genome-fragment | NA | NA | 0 |
| S11_extracted_k141_77180 | 1381 | No | NA | 2 | 0 | 1 | Not-determined | Genome-fragment | NA | NA | 0 |
| S11_extracted_k141_57741 | 1168 | No | NA | 3 | 0 | 2 | Not-determined | Genome-fragment | NA | NA | 0 |
| S12_extracted_k141_135775 | 1174 | No | NA | 2 | 1 | 0 | Low-quality | Genome-fragment | 2.9 | AAI-based (high-confidence) | 0 |
| S12_extracted_k141_153379 | 1174 | No | NA | 1 | 1 | 0 | Low-quality | Genome-fragment | 2.71 | AAI-based (high-confidence) | 0 |
| S15_extracted_63938 | 1174 | No | NA | 1 | 0 | 1 | Low-quality | Genome-fragment | 0.43 | AAI-based (high-confidence) | 0 |
| S11_extracted_k141_44508 | 1173 | No | NA | 2 | 0 | 1 | Not-determined | Genome-fragment | NA | NA | 0 |
| S4_extracted_24138 | 1174 | No | NA | 3 | 0 | 0 | Low-quality | Genome-fragment | 2.84 | AAI-based (high-confidence) | 0 |
| S11_extracted_k141_44510 | 3984 | No | NA | 4 | 0 | 3 | Not-determined | Genome-fragment | NA | NA | 0 |
| S11_extracted_k141_102921 | 1056 | No | NA | 1 | 0 | 0 | Not-determined | Genome-fragment | NA | NA | 0 |
| S11_extracted_k141_18344 | 1251 | No | NA | 2 | 0 | 0 | Not-determined | Genome-fragment | NA | NA | 0 |
| S8_extracted_k141_203326 | 1174 | No | NA | 1 | 0 | 0 | Low-quality | Genome-fragment | 2.74 | AAI-based (high-confidence) | 0 |
| S11_extracted_k141_44541 | 2340 | No | NA | 3 | 0 | 1 | Not-determined | Genome-fragment | NA | NA | 0 |
| S11_extracted_k141_11852 | 1148 | No | NA | 1 | 0 | 0 | Not-determined | Genome-fragment | NA | NA | 0 |
| S8_vs_k141_59170\|\|full | 1174 | No | NA | 3 | 1 | 0 | Low-quality | Genome-fragment | 2.01 | AAI-based (medium-confidence) | 0 |
| S9_vs_k141_85361\|\|full | 1175 | No | NA | 3 | 0 | 0 | Low-quality | Genome-fragment | 2.95 | AAI-based (medium-confidence) | 0 |
| S11_vs_k141_15163\|\|full | 1175 | No | NA | 3 | 0 | 0 | Low-quality | Genome-fragment | 2.73 | AAI-based (medium-confidence) | 0 |
| S11_extracted_k141_11872 | 4519 | No | NA | 6 | 0 | 3 | Not-determined | Genome-fragment | NA | NA | 0 |
| S11_extracted_k141_44573 | 1469 | No | NA | 2 | 0 | 2 | Not-determined | Genome-fragment | NA | NA | 0 |
| S14_extracted_k141_84342 | 1175 | No | NA | 3 | 3 | 0 | Low-quality | Genome-fragment | 1.96 | AAI-based (medium-confidence) | 0 |
| S11_extracted_k141_97010 | 1225 | No | NA | 2 | 0 | 2 | Not-determined | Genome-fragment | NA | NA | 0 |
| S11_extracted_k141_97016 | 4653 | No | NA | 3 | 0 | 0 | Not-determined | Genome-fragment | NA | NA | 0 |
| S1_vs_k141_133659\|\|full | 1175 | No | NA | 3 | 1 | 0 | Low-quality | Genome-fragment | 3.38 | AAI-based (high-confidence) | 0 |
| S11_extracted_k141_38086 | 1830 | No | NA | 2 | 0 | 1 | Not-determined | Genome-fragment | NA | NA | 0 |
| S11_extracted_k141_5617 | 1856 | No | NA | 2 | 0 | 2 | Not-determined | Genome-fragment | NA | NA | 0 |
| S11_extracted_k141_5618 | 1186 | No | NA | 2 | 0 | 1 | Not-determined | Genome-fragment | NA | NA | 0 |
| S11_extracted_k141_18431 | 3727 | No | NA | 4 | 0 | 1 | Not-determined | Genome-fragment | NA | NA | 0 |
| S11_extracted_k141_31312 | 1881 | No | NA | 1 | 0 | 1 | Not-determined | Genome-fragment | NA | NA | 0 |
| S3_extracted_k141_120358 | 1175 | No | NA | 5 | 0 | 0 | Low-quality | Genome-fragment | 0.68 | AAI-based (high-confidence) | 0 |
| S11_extracted_k141_31315 | 1372 | No | NA | 2 | 0 | 2 | Not-determined | Genome-fragment | NA | NA | 0 |
| S3_vs_k141_37046\|\|lt2gene | 1175 | No | NA | 2 | 1 | 0 | Low-quality | Genome-fragment | 0.81 | AAI-based (medium-confidence) | 0 |
| S11_extracted_k141_97064 | 3434 | No | NA | 1 | 0 | 1 | Not-determined | Genome-fragment | NA | NA | 0 |
| S11_extracted_k141_83379 | 1036 | No | NA | 1 | 0 | 0 | Not-determined | Genome-fragment | NA | NA | 0 |
| S11_extracted_k141_50465 | 1503 | No | NA | 2 | 0 | 1 | Not-determined | Genome-fragment | NA | NA | 0 |
| S11_extracted_k141_5673 | 1077 | No | NA | 1 | 0 | 1 | Not-determined | Genome-fragment | NA | NA | 0 |
| S5_extracted_k141_19576 | 1175 | No | NA | 1 | 1 | 0 | Low-quality | Genome-fragment | 3.26 | AAI-based (medium-confidence) | 0 |
| S11_extracted_k141_44712 | 2398 | No | NA | 2 | 0 | 2 | Not-determined | Genome-fragment | NA | NA | 0 |
| S11_extracted_k141_70015 | 53828 | No | NA | 83 | 14 | 3 | High-quality | High-quality | 92.95 | AAI-based (medium-confidence) | 0 |
| S11_extracted_k141_50487 | 12577 | No | NA | 14 | 0 | 2 | Not-determined | Genome-fragment | NA | NA | 0 |
| S11_extracted_k141_25779 | 1383 | No | NA | 1 | 0 | 1 | Not-determined | Genome-fragment | NA | NA | 0 |
| S11_extracted_k141_18539 | 3172 | No | NA | 3 | 0 | 2 | Not-determined | Genome-fragment | NA | NA | 0 |
| S11_extracted_k141_31409 | 1546 | No | NA | 3 | 0 | 2 | Not-determined | Genome-fragment | NA | NA | 0 |
| S11_extracted_k141_63984 | 3479 | No | NA | 3 | 0 | 3 | Not-determined | Genome-fragment | NA | NA | 0 |
| S11_extracted_k141_90071 | 1073 | No | NA | 1 | 0 | 1 | Not-determined | Genome-fragment | NA | NA | 0 |
| S7_extracted_69815 | 1175 | No | NA | 2 | 0 | 0 | Low-quality | Genome-fragment | 7.1 | AAI-based (high-confidence) | 0 |
| S11_extracted_k141_38233 | 2270 | No | NA | 1 | 0 | 0 | Not-determined | Genome-fragment | NA | NA | 0 |
| S11_extracted_k141_12032 | 3322 | No | NA | 1 | 0 | 1 | Not-determined | Genome-fragment | NA | NA | 0 |
| S11_extracted_k141_57792 | 1115 | No | NA | 3 | 0 | 3 | Not-determined | Genome-fragment | NA | NA | 0 |
| S11_extracted_k141_44795 | 6103 | No | NA | 3 | 0 | 3 | Not-determined | Genome-fragment | NA | NA | 0 |
| S11_extracted_k141_5778 | 4623 | No | NA | 4 | 0 | 1 | Not-determined | Genome-fragment | NA | NA | 0 |
| S11_extracted_k141_57834 | 5233 | No | NA | 4 | 0 | 2 | Not-determined | Genome-fragment | NA | NA | 0 |
| S11_extracted_k141_12093 | 1002 | No | NA | 1 | 0 | 0 | Not-determined | Genome-fragment | NA | NA | 0 |
| S11_extracted_k141_83570 | 1174 | No | NA | 1 | 0 | 0 | Not-determined | Genome-fragment | NA | NA | 0 |
| S11_extracted_k141_57848 | 1040 | No | NA | 4 | 0 | 0 | Not-determined | Genome-fragment | NA | NA | 0 |
| S16_vs_13649\|\|full | 1175 | No | NA | 3 | 0 | 0 | Low-quality | Genome-fragment | 1.81 | AAI-based (high-confidence) | 0 |
| S11_extracted_k141_50584 | 1266 | No | NA | 2 | 0 | 1 | Not-determined | Genome-fragment | NA | NA | 0 |
| S11_extracted_k141_83579 | 1037 | No | NA | 2 | 0 | 0 | Not-determined | Genome-fragment | NA | NA | 0 |
| S11_extracted_k141_25896 | 1729 | No | NA | 2 | 0 | 2 | Not-determined | Genome-fragment | NA | NA | 0 |
| S11_extracted_k141_57853 | 5314 | No | NA | 8 | 0 | 3 | Not-determined | Genome-fragment | NA | NA | 0 |
| S11_extracted_k141_50602 | 1339 | No | NA | 1 | 0 | 0 | Not-determined | Genome-fragment | NA | NA | 0 |
| S16_vs_154245\|\|lt2gene | 1175 | No | NA | 1 | 1 | 0 | Low-quality | Genome-fragment | 1.72 | HMM-based (lower-bound) | 0 |
| S11_extracted_k141_70164 | 1310 | No | NA | 2 | 0 | 2 | Not-determined | Genome-fragment | NA | NA | 0 |
| S12_extracted_k141_93756 | 1175 | No | NA | 2 | 0 | 0 | Low-quality | Genome-fragment | 2.79 | AAI-based (medium-confidence) | 0 |
| S12_vs_k141_40229\|\|lt2gene | 1175 | No | NA | 2 | 2 | 0 | Low-quality | Genome-fragment | 1.65 | AAI-based (medium-confidence) | 0 |
| S11_extracted_k141_18719 | 2230 | No | NA | 3 | 0 | 1 | Not-determined | Genome-fragment | NA | NA | 0 |
| S11_extracted_k141_57892 | 1469 | No | NA | 2 | 0 | 1 | Not-determined | Genome-fragment | NA | NA | 0 |
| S11_extracted_k141_90228 | 1336 | No | NA | 2 | 0 | 1 | Not-determined | Genome-fragment | NA | NA | 0 |
| S11_extracted_k141_70198 | 1480 | No | NA | 2 | 0 | 0 | Not-determined | Genome-fragment | NA | NA | 0 |
| S11_extracted_k141_12156 | 1573 | No | NA | 3 | 0 | 0 | Not-determined | Genome-fragment | NA | NA | 0 |
| S8_extracted_k141_150071 | 1175 | No | NA | 2 | 0 | 0 | Low-quality | Genome-fragment | 4.08 | AAI-based (medium-confidence) | 0 |
| S11_extracted_k141_50660 | 1083 | No | NA | 1 | 0 | 0 | Not-determined | Genome-fragment | NA | NA | 0 |
| S8_extracted_k141_124435 | 1175 | No | NA | 2 | 0 | 1 | Low-quality | Genome-fragment | 4.38 | AAI-based (medium-confidence) | 0 |
| S8_vs_k141_13439\|\|full | 1175 | No | NA | 3 | 1 | 0 | Low-quality | Genome-fragment | 2.56 | AAI-based (medium-confidence) | 0 |
| S11_extracted_k141_64141 | 1021 | No | NA | 1 | 0 | 0 | Not-determined | Genome-fragment | NA | NA | 0 |
| S11_extracted_k141_97222 | 1425 | No | NA | 1 | 0 | 0 | Not-determined | Genome-fragment | NA | NA | 0 |
| S11_extracted_k141_38370 | 1765 | No | NA | 2 | 0 | 0 | Not-determined | Genome-fragment | NA | NA | 0 |
| S11_extracted_k141_97227 | 1500 | No | NA | 1 | 0 | 1 | Not-determined | Genome-fragment | NA | NA | 0 |
| S11_extracted_k141_97239 | 1157 | No | NA | 3 | 0 | 0 | Not-determined | Genome-fragment | NA | NA | 0 |
| S11_extracted_k141_70231 | 1521 | No | NA | 1 | 0 | 1 | Not-determined | Genome-fragment | NA | NA | 0 |
| S9_vs_k141_9709\|\|full | 1176 | No | NA | 3 | 0 | 1 | Low-quality | Genome-fragment | 3.76 | AAI-based (medium-confidence) | 0 |
| S5_extracted_k141_129726 | 1176 | No | NA | 1 | 0 | 0 | Low-quality | Genome-fragment | 3.2 | AAI-based (high-confidence) | 0 |
| S5_extracted_k141_96865 | 1176 | No | NA | 3 | 2 | 0 | Low-quality | Genome-fragment | 3.41 | AAI-based (medium-confidence) | 0 |
| S11_extracted_k141_18852 | 1221 | No | NA | 2 | 0 | 2 | Not-determined | Genome-fragment | NA | NA | 0 |
| S11_extracted_k141_103213 | 1958 | No | NA | 2 | 0 | 0 | Not-determined | Genome-fragment | NA | NA | 0 |
| S5_vs_k141_39654\|\|full | 1176 | No | NA | 3 | 1 | 0 | Low-quality | Genome-fragment | 2.2 | AAI-based (high-confidence) | 0 |
| S5_vs_k141_61214\|\|lt2gene | 1176 | No | NA | 2 | 1 | 0 | Low-quality | Genome-fragment | 3.37 | AAI-based (high-confidence) | 0 |
| S7_extracted_95070 | 1176 | No | NA | 2 | 1 | 0 | Low-quality | Genome-fragment | 2.5 | AAI-based (high-confidence) | 0 |
| S7_vs_106314\|\|full | 1176 | No | NA | 2 | 1 | 0 | Low-quality | Genome-fragment | 1.08 | HMM-based (lower-bound) | 0 |
| S7_vs_136216\|\|full | 1176 | No | NA | 4 | 0 | 0 | Low-quality | Genome-fragment | 2.24 | AAI-based (medium-confidence) | 0 |
| S16_vs_55382\|\|full | 1176 | No | NA | 2 | 0 | 1 | Low-quality | Genome-fragment | 2.68 | AAI-based (medium-confidence) | 0 |
| S11_extracted_k141_12334 | 1041 | No | NA | 2 | 0 | 1 | Not-determined | Genome-fragment | NA | NA | 0 |
| S10_extracted_98687 | 1176 | No | NA | 2 | 0 | 1 | Low-quality | Genome-fragment | 0.38 | AAI-based (medium-confidence) | 0 |
| S11_extracted_k141_90359 | 1332 | No | NA | 1 | 0 | 0 | Not-determined | Genome-fragment | NA | NA | 0 |
| S11_extracted_k141_12339 | 1026 | No | NA | 2 | 0 | 1 | Not-determined | Genome-fragment | NA | NA | 0 |
| S11_extracted_k141_64290 | 1582 | No | NA | 1 | 0 | 1 | Not-determined | Genome-fragment | NA | NA | 0 |
| S11_extracted_k141_5987 | 3464 | No | NA | 3 | 0 | 0 | Not-determined | Genome-fragment | NA | NA | 0 |
| S11_extracted_k141_5997 | 5679 | No | NA | 7 | 0 | 7 | Not-determined | Genome-fragment | NA | NA | 0 |
| S12_extracted_k141_21515 | 1176 | No | NA | 2 | 0 | 0 | Low-quality | Genome-fragment | 2.27 | AAI-based (high-confidence) | 0 |
| S12_vs_k141_897\|\|lt2gene | 1176 | No | NA | 1 | 1 | 0 | Low-quality | Genome-fragment | 3.12 | AAI-based (high-confidence) | 0 |
| S6_vs_k141_33822\|\|full | 1176 | No | NA | 4 | 0 | 0 | Low-quality | Genome-fragment | 2.25 | AAI-based (medium-confidence) | 0 |
| S8_extracted_k141_95934 | 1176 | No | NA | 2 | 0 | 0 | Low-quality | Genome-fragment | 1.97 | AAI-based (medium-confidence) | 0 |
| S17_vs_k141_9374\|\|full | 1177 | No | NA | 4 | 1 | 0 | Low-quality | Genome-fragment | 2.58 | AAI-based (medium-confidence) | 0 |
| S11_extracted_k141_77763 | 1034 | No | NA | 1 | 0 | 0 | Not-determined | Genome-fragment | NA | NA | 0 |
| S11_extracted_k141_6030 | 1459 | No | NA | 1 | 0 | 0 | Not-determined | Genome-fragment | NA | NA | 0 |
| S11_extracted_k141_97386 | 1015 | No | NA | 1 | 0 | 1 | Not-determined | Genome-fragment | NA | NA | 0 |
| S11_extracted_k141_45012 | 1848 | No | NA | 2 | 0 | 0 | Not-determined | Genome-fragment | NA | NA | 0 |
| S11_extracted_k141_103284 | 2526 | No | NA | 2 | 0 | 2 | Not-determined | Genome-fragment | NA | NA | 0 |
| S11_extracted_k141_90457 | 1429 | No | NA | 2 | 0 | 0 | Not-determined | Genome-fragment | NA | NA | 0 |
| S11_extracted_k141_14023 | 1177 | No | NA | 2 | 0 | 2 | Low-quality | Genome-fragment | 0.47 | AAI-based (medium-confidence) | 0 |
| S11_extracted_k141_12430 | 1005 | No | NA | 2 | 0 | 0 | Not-determined | Genome-fragment | NA | NA | 0 |
| S11_extracted_k141_83856 | 2747 | No | NA | 3 | 0 | 2 | Not-determined | Genome-fragment | NA | NA | 0 |
| S11_extracted_k141_64342 | 1093 | No | NA | 2 | 0 | 0 | Not-determined | Genome-fragment | NA | NA | 0 |
| S11_extracted_k141_50840 | 1006 | No | NA | 1 | 0 | 0 | Not-determined | Genome-fragment | NA | NA | 0 |
| S11_extracted_k141_103299 | 1334 | No | NA | 1 | 0 | 0 | Not-determined | Genome-fragment | NA | NA | 0 |
| S11_extracted_k141_6088 | 5241 | No | NA | 4 | 0 | 3 | Not-determined | Genome-fragment | NA | NA | 0 |
| S14_extracted_k141_89723 | 1177 | No | NA | 2 | 2 | 0 | Low-quality | Genome-fragment | 3.88 | AAI-based (high-confidence) | 0 |
| S11_extracted_k141_31776 | 4997 | No | NA | 5 | 0 | 4 | Not-determined | Genome-fragment | NA | NA | 0 |
| S11_extracted_k141_83896 | 1305 | No | NA | 3 | 0 | 0 | Not-determined | Genome-fragment | NA | NA | 0 |
| S3_vs_k141_105526\|\|full | 1177 | No | NA | 2 | 0 | 0 | Low-quality | Genome-fragment | 3.01 | AAI-based (high-confidence) | 0 |
| S11_extracted_k141_103346 | 1159 | No | NA | 3 | 0 | 2 | Not-determined | Genome-fragment | NA | NA | 0 |
| S11_extracted_k141_31806 | 1930 | No | NA | 1 | 0 | 0 | Not-determined | Genome-fragment | NA | NA | 0 |
| S11_extracted_k141_31810 | 3188 | No | NA | 2 | 0 | 0 | Not-determined | Genome-fragment | NA | NA | 0 |
| S5_extracted_k141_99842 | 1177 | No | NA | 2 | 1 | 0 | Low-quality | Genome-fragment | 3.12 | AAI-based (high-confidence) | 0 |
| S11_extracted_k141_12534 | 1380 | No | NA | 2 | 0 | 1 | Not-determined | Genome-fragment | NA | NA | 0 |
| S5_vs_k141_47536\|\|full | 1177 | No | NA | 2 | 0 | 0 | Low-quality | Genome-fragment | 1.63 | AAI-based (medium-confidence) | 0 |
| S7_vs_36556\|\|full | 1177 | No | NA | 5 | 1 | 0 | Low-quality | Genome-fragment | 2.72 | AAI-based (medium-confidence) | 0 |
| S7_vs_125564\|\|full | 1177 | No | NA | 3 | 1 | 0 | Low-quality | Genome-fragment | 2.97 | AAI-based (medium-confidence) | 0 |
| S11_extracted_k141_12586 | 1312 | No | NA | 2 | 0 | 1 | Not-determined | Genome-fragment | NA | NA | 0 |
| S11_extracted_k141_64478 | 1174 | No | NA | 2 | 0 | 1 | Not-determined | Genome-fragment | NA | NA | 0 |
| S16_vs_54462\|\|full | 1177 | No | NA | 3 | 2 | 0 | Low-quality | Genome-fragment | 2.74 | AAI-based (high-confidence) | 0 |
| S16_vs_95575\|\|full | 1177 | No | NA | 3 | 2 | 0 | Low-quality | Genome-fragment | 2.65 | AAI-based (medium-confidence) | 0 |
| S11_extracted_k141_51062 | 7054 | No | NA | 4 | 0 | 1 | Not-determined | Genome-fragment | NA | NA | 0 |
| S11_extracted_k141_103387 | 1218 | No | NA | 1 | 0 | 0 | Not-determined | Genome-fragment | NA | NA | 0 |
| S10_extracted_31884 | 1177 | No | NA | 2 | 2 | 0 | Low-quality | Genome-fragment | 3.7 | AAI-based (high-confidence) | 0 |
| S6_vs_k141_36677\|\|full | 1177 | No | NA | 3 | 2 | 0 | Low-quality | Genome-fragment | 2.57 | AAI-based (high-confidence) | 0 |
| S8_vs_k141_1243\|\|full | 1177 | No | NA | 4 | 0 | 0 | Low-quality | Genome-fragment | 2.28 | AAI-based (high-confidence) | 0 |
| S11_extracted_k141_84088 | 1044 | No | NA | 1 | 0 | 1 | Not-determined | Genome-fragment | NA | NA | 0 |
| S11_extracted_k141_84092 | 1210 | No | NA | 1 | 0 | 0 | Not-determined | Genome-fragment | NA | NA | 0 |
| S11_extracted_k141_64548 | 1053 | No | NA | 1 | 0 | 1 | Not-determined | Genome-fragment | NA | NA | 0 |
| S11_extracted_k141_84107 | 1261 | No | NA | 2 | 0 | 2 | Not-determined | Genome-fragment | NA | NA | 0 |
| S11_extracted_k141_38632 | 1021 | No | NA | 1 | 0 | 0 | Not-determined | Genome-fragment | NA | NA | 0 |
| S8_vs_k141_70093\|\|lt2gene | 1177 | No | NA | 1 | 1 | 0 | Low-quality | Genome-fragment | 3.2 | AAI-based (medium-confidence) | 0 |
| S17_vs_k141_85955\|\|full | 1178 | No | NA | 2 | 0 | 0 | Low-quality | Genome-fragment | 2.9 | AAI-based (high-confidence) | 0 |
| S11_vs_k141_57710\|\|full | 1178 | No | NA | 3 | 0 | 3 | Low-quality | Genome-fragment | 0.75 | AAI-based (medium-confidence) | 0 |
| S3_vs_k141_42597\|\|full | 1178 | No | NA | 3 | 1 | 0 | Low-quality | Genome-fragment | 1.98 | AAI-based (high-confidence) | 0 |
| S11_extracted_k141_6217 | 1181 | No | NA | 1 | 0 | 1 | Not-determined | Genome-fragment | NA | NA | 0 |
| S11_extracted_k141_31956 | 1869 | No | NA | 3 | 0 | 0 | Not-determined | Genome-fragment | NA | NA | 0 |
| S11_extracted_k141_45384 | 1234 | No | NA | 1 | 0 | 1 | Not-determined | Genome-fragment | NA | NA | 0 |
| S11_extracted_k141_45411 | 1508 | No | NA | 4 | 0 | 1 | Not-determined | Genome-fragment | NA | NA | 0 |
| S3_vs_k141_119446\|\|full | 1178 | No | NA | 2 | 1 | 0 | Low-quality | Genome-fragment | 0.76 | HMM-based (lower-bound) | 0 |
| S11_extracted_k141_84172 | 2976 | No | NA | 3 | 0 | 2 | Not-determined | Genome-fragment | NA | NA | 0 |
| S5_extracted_k141_81525 | 1178 | No | NA | 2 | 1 | 0 | Low-quality | Genome-fragment | 2.56 | AAI-based (high-confidence) | 0 |
| S11_extracted_k141_84225 | 1090 | No | NA | 1 | 0 | 1 | Not-determined | Genome-fragment | NA | NA | 0 |
| S5_vs_k141_136774\|\|full | 1178 | No | NA | 3 | 1 | 0 | Low-quality | Genome-fragment | 2.06 | HMM-based (lower-bound) | 0 |
| S11_extracted_k141_64662 | 1129 | No | NA | 2 | 0 | 1 | Not-determined | Genome-fragment | NA | NA | 0 |
| S11_extracted_k141_38795 | 3286 | No | NA | 3 | 0 | 3 | Not-determined | Genome-fragment | NA | NA | 0 |
| S5_vs_k141_11353\|\|full | 1178 | No | NA | 5 | 1 | 0 | Low-quality | Genome-fragment | 2.68 | AAI-based (high-confidence) | 0 |
| S7_extracted_97172 | 1178 | No | NA | 2 | 1 | 0 | Low-quality | Genome-fragment | 1.4 | HMM-based (lower-bound) | 0 |
| S11_extracted_k141_64684 | 3253 | No | NA | 4 | 0 | 2 | Not-determined | Genome-fragment | NA | NA | 0 |
| S7_vs_52502\|\|full | 1178 | No | NA | 3 | 0 | 0 | Low-quality | Genome-fragment | 2.94 | AAI-based (high-confidence) | 0 |
| S7_vs_63781\|\|full | 1178 | No | NA | 3 | 0 | 0 | Low-quality | Genome-fragment | 1.3 | AAI-based (medium-confidence) | 0 |
| S11_extracted_k141_70640 | 2242 | No | NA | 2 | 0 | 1 | Not-determined | Genome-fragment | NA | NA | 0 |
| S11_extracted_k141_64711 | 1164 | No | NA | 1 | 0 | 1 | Not-determined | Genome-fragment | NA | NA | 0 |
| S11_extracted_k141_70645 | 1082 | No | NA | 2 | 0 | 1 | Not-determined | Genome-fragment | NA | NA | 0 |
| S11_extracted_k141_58452 | 1062 | No | NA | 2 | 0 | 1 | Not-determined | Genome-fragment | NA | NA | 0 |
| S10_vs_2091\|\|full | 1178 | No | NA | 2 | 0 | 0 | Low-quality | Genome-fragment | 2.27 | AAI-based (medium-confidence) | 0 |
| S11_extracted_k141_58473 | 1268 | No | NA | 2 | 0 | 0 | Not-determined | Genome-fragment | NA | NA | 0 |
| S10_vs_108072\|\|full | 1178 | No | NA | 3 | 0 | 0 | Low-quality | Genome-fragment | 2.8 | AAI-based (high-confidence) | 0 |
| S10_vs_18541\|\|lt2gene | 1178 | No | NA | 2 | 0 | 0 | Low-quality | Genome-fragment | 2.2 | AAI-based (high-confidence) | 0 |
| S11_extracted_k141_6372 | 2542 | No | NA | 6 | 0 | 0 | Not-determined | Genome-fragment | NA | NA | 0 |
| S11_extracted_k141_58502 | 1157 | No | NA | 1 | 0 | 1 | Not-determined | Genome-fragment | NA | NA | 0 |
| S11_extracted_k141_103619 | 1018 | No | NA | 1 | 0 | 0 | Not-determined | Genome-fragment | NA | NA | 0 |
| S11_extracted_k141_90866 | 1666 | No | NA | 2 | 0 | 1 | Not-determined | Genome-fragment | NA | NA | 0 |
| S11_extracted_k141_6409 | 2941 | No | NA | 3 | 0 | 2 | Not-determined | Genome-fragment | NA | NA | 0 |

| | | | | | | | | | | | |
|---|---|---|---|---|---|---|---|---|---|---|---|
| S12_vs_k141_166114\|\|full | 1178 | No | NA | 4 | 2 | 0 | Low-quality | Genome-fragment | 3.05 | AAI-based (high-confidence) | 0 |
| S11_extracted_k141_84303 | 45395 | No | NA | 63 | 15 | 0 | Complete | High-quality | 100 | DTR (high-confidence) | 0 |
| S12_vs_k141_112914\|\|full | 1178 | No | NA | 4 | 1 | 0 | Low-quality | Genome-fragment | 2.71 | AAI-based (medium-confidence) | 0 |
| S11_extracted_k141_51350 | 1941 | No | NA | 1 | 0 | 1 | Not-determined | Genome-fragment | NA | NA | 0 |
| S12_vs_k141_137656\|\|full | 1178 | No | NA | 2 | 0 | 0 | Low-quality | Genome-fragment | 3.63 | AAI-based (high-confidence) | 0 |
| S15_extracted_41729 | 1178 | No | NA | 1 | 0 | 1 | Low-quality | Genome-fragment | 1.16 | AAI-based (high-confidence) | 0 |
| S11_extracted_k141_84373 | 2609 | No | NA | 1 | 0 | 1 | Not-determined | Genome-fragment | NA | NA | 0 |
| S11_extracted_k141_45513 | 1229 | No | NA | 2 | 0 | 2 | Not-determined | Genome-fragment | NA | NA | 0 |
| S11_extracted_k141_38929 | 1479 | No | NA | 1 | 0 | 0 | Not-determined | Genome-fragment | NA | NA | 0 |
| S8_extracted_k141_138842 | 1178 | No | NA | 1 | 1 | 0 | Low-quality | Genome-fragment | 1.96 | HMM-based (lower-bound) | 0 |
| S11_extracted_k141_103696 | 5294 | No | NA | 6 | 0 | 3 | Not-determined | Genome-fragment | NA | NA | 0 |
| S11_extracted_k141_70820 | 1391 | No | NA | 2 | 0 | 0 | Not-determined | Genome-fragment | NA | NA | 0 |
| S11_extracted_k141_103699 | 4593 | No | NA | 5 | 0 | 4 | Not-determined | Genome-fragment | NA | NA | 0 |
| S11_extracted_k141_38998 | 1205 | No | NA | 2 | 0 | 1 | Not-determined | Genome-fragment | NA | NA | 0 |
| S11_extracted_k141_19520 | 1825 | No | NA | 4 | 0 | 3 | Not-determined | Genome-fragment | NA | NA | 0 |
| S11_extracted_k141_51476 | 5408 | No | NA | 5 | 0 | 4 | Not-determined | Genome-fragment | NA | NA | 0 |
| S11_extracted_k141_64913 | 1116 | No | NA | 2 | 0 | 0 | Not-determined | Genome-fragment | NA | NA | 0 |
| S11_extracted_k141_64947 | 1196 | No | NA | 2 | 0 | 0 | Not-determined | Genome-fragment | NA | NA | 0 |
| S11_extracted_k141_51561 | 1192 | No | NA | 1 | 0 | 1 | Not-determined | Genome-fragment | NA | NA | 0 |
| S11_extracted_k141_71006 | 3753 | No | NA | 4 | 0 | 1 | Not-determined | Genome-fragment | NA | NA | 0 |
| S11_extracted_k141_51616 | 1428 | No | NA | 2 | 0 | 2 | Not-determined | Genome-fragment | NA | NA | 0 |
| S11_extracted_k141_65041 | 1664 | No | NA | 1 | 0 | 1 | Not-determined | Genome-fragment | NA | NA | 0 |
| S11_extracted_k141_103935 | 1056 | No | NA | 1 | 0 | 0 | Not-determined | Genome-fragment | NA | NA | 0 |
| S11_extracted_k141_103938 | 3772 | No | NA | 2 | 0 | 1 | Not-determined | Genome-fragment | NA | NA | 0 |
| S8_vs_k141_44125\|\|full | 1178 | No | NA | 2 | 0 | 0 | Low-quality | Genome-fragment | 2.01 | AAI-based (high-confidence) | 0 |
| S11_extracted_k141_103954 | 1054 | No | NA | 1 | 0 | 1 | Not-determined | Genome-fragment | NA | NA | 0 |
| S8_vs_k141_13179\|\|full | 1178 | No | NA | 2 | 0 | 0 | Low-quality | Genome-fragment | 2.56 | AAI-based (medium-confidence) | 0 |
| S8_vs_k141_153066\|\|lt2gene | 1178 | No | NA | 2 | 1 | 0 | Low-quality | Genome-fragment | 2.82 | AAI-based (medium-confidence) | 0 |
| S9_extracted_k141_54963 | 1179 | No | NA | 2 | 1 | 0 | Low-quality | Genome-fragment | 2.52 | AAI-based (medium-confidence) | 0 |
| S11_extracted_k141_71168 | 1061 | No | NA | 1 | 0 | 1 | Not-determined | Genome-fragment | NA | NA | 0 |
| S14_extracted_k141_41150 | 1179 | No | NA | 1 | 0 | 0 | Low-quality | Genome-fragment | 2.09 | AAI-based (high-confidence) | 0 |
| S14_vs_k141_49072\|\|full | 1179 | No | NA | 3 | 0 | 0 | Low-quality | Genome-fragment | 2.2 | AAI-based (medium-confidence) | 0 |
| S11_extracted_k141_51793 | 2814 | No | NA | 5 | 0 | 0 | Not-determined | Genome-fragment | NA | NA | 0 |
| S11_extracted_k141_32332 | 1100 | No | NA | 1 | 0 | 0 | Not-determined | Genome-fragment | NA | NA | 0 |
| S14_vs_k141_65877\|\|lt2gene | 1179 | No | NA | 1 | 1 | 0 | Low-quality | Genome-fragment | 1.98 | HMM-based (lower-bound) | 0 |
| S11_extracted_k141_71383 | 5505 | No | NA | 5 | 0 | 3 | Not-determined | Genome-fragment | NA | NA | 0 |
| S11_extracted_k141_32476 | 1166 | No | NA | 1 | 0 | 1 | Not-determined | Genome-fragment | NA | NA | 0 |
| S11_extracted_k141_104033 | 106247 | No | NA | 133 | 18 | 4 | Complete | High-quality | 100 | DTR (high-confidence) | 0 |
| S11_extracted_k141_32502 | 1131 | No | NA | 1 | 0 | 0 | Not-determined | Genome-fragment | NA | NA | 0 |
| S11_extracted_k141_104034 | 40797 | No | NA | 76 | 18 | 3 | Complete | High-quality | 100 | DTR (high-confidence) | 0 |
| S11_extracted_k141_71463 | 1721 | No | NA | 2 | 0 | 0 | Not-determined | Genome-fragment | NA | NA | 0 |
| S11_extracted_k141_104035 | 99625 | No | NA | 102 | 15 | 2 | Complete | High-quality | 100 | DTR (high-confidence) | 0 |
| S11_extracted_k141_104044 | 100812 | No | NA | 113 | 22 | 0 | Complete | High-quality | 100 | DTR (high-confidence) | 0 |
| S11_extracted_k141_104046 | 164379 | No | NA | 181 | 17 | 4 | Complete | High-quality | 100 | DTR (high-confidence) | 0 |
| S11_extracted_k141_104048 | 37508 | No | NA | 45 | 9 | 0 | Complete | High-quality | 100 | DTR (high-confidence) | 0 |
| S11_extracted_k141_104052 | 40792 | No | NA | 55 | 22 | 0 | Complete | High-quality | 100 | DTR (high-confidence) | 0 |
| S11_extracted_k141_104059 | 91418 | No | NA | 91 | 8 | 1 | Complete | High-quality | 100 | DTR (high-confidence) | 0 |
| S11_extracted_k141_104062 | 34888 | No | NA | 50 | 12 | 0 | Complete | High-quality | 100 | DTR (high-confidence) | 0 |
| S11_extracted_k141_104070 | 36162 | No | NA | 58 | 27 | 0 | Complete | High-quality | 100 | DTR (high-confidence) | 0 |
| S11_extracted_k141_104071 | 36582 | No | NA | 42 | 8 | 0 | Complete | High-quality | 100 | DTR (medium-confidence) | 0 |
| S11_vs_k141_84627\|\|full | 39772 | No | NA | 67 | 10 | 0 | High-quality | High-quality | 100 | AAI-based (medium-confidence) | 0 |
| S1_extracted_k141_40235 | 1179 | No | NA | 1 | 0 | 1 | Low-quality | Genome-fragment | 0.31 | AAI-based (medium-confidence) | 0 |
| S11_vs_k141_20912\|\|full | 2051 | No | NA | 3 | 0 | 0 | Not-determined | Genome-fragment | NA | NA | 0 |
| S11_vs_k141_40945\|\|full | 44575 | No | NA | 67 | 17 | 0 | High-quality | High-quality | 99.31 | AAI-based (high-confidence) | 0 |
| S11_vs_k141_93101\|\|full | 23434 | No | NA | 53 | 7 | 0 | High-quality | High-quality | 99.44 | AAI-based (high-confidence) | 0 |
| S11_vs_k141_3006\|\|full | 32415 | No | NA | 47 | 17 | 0 | Medium-quality | Genome-fragment | 88.09 | AAI-based (medium-confidence) | 0 |
| S11_vs_k141_75554\|\|full | 16322 | No | NA | 24 | 7 | 0 | Medium-quality | Genome-fragment | 83.95 | AAI-based (medium-confidence) | 0 |
| S11_vs_k141_49071\|\|full | 89156 | No | NA | 156 | 29 | 0 | High-quality | High-quality | 100 | AAI-based (high-confidence) | 0 |
| S11_vs_k141_36748\|\|full | 46177 | No | NA | 74 | 14 | 1 | High-quality | High-quality | 100 | AAI-based (high-confidence) | 0 |
| S11_vs_k141_82368\|\|full | 45487 | No | NA | 66 | 22 | 0 | High-quality | High-quality | 100 | AAI-based (high-confidence) | 0 |
| S11_vs_k141_104032\|\|full | 44004 | No | NA | 74 | 23 | 2 | High-quality | High-quality | 100 | AAI-based (high-confidence) | 0 |
| S11_vs_k141_104036\|\|full | 48912 | No | NA | 60 | 15 | 1 | High-quality | High-quality | 96.17 | AAI-based (high-confidence) | 0 |
| S11_vs_k141_104037\|\|full | 54218 | No | NA | 79 | 15 | 0 | High-quality | High-quality | 100 | AAI-based (medium-confidence) | 0 |
| S11_vs_k141_104038\|\|full | 34262 | No | NA | 53 | 22 | 0 | Medium-quality | Genome-fragment | 89.66 | AAI-based (high-confidence) | 0 |
| S11_vs_k141_104041\|\|full | 36201 | No | NA | 59 | 16 | 2 | High-quality | High-quality | 100 | AAI-based (high-confidence) | 0 |
| S11_vs_k141_104042\|\|full | 37290 | Yes | 35553 | 59 | 18 | 2 | Medium-quality | Genome-fragment | 89.78 | AAI-based (high-confidence) | 4.66 |
| S11_vs_k141_104047\|\|full | 33359 | Yes | 30866 | 54 | 19 | 2 | Medium-quality | Genome-fragment | 85.83 | AAI-based (high-confidence) | 7.47 |
| S11_vs_k141_104049\|\|full | 38479 | No | NA | 56 | 13 | 0 | High-quality | High-quality | 100 | AAI-based (high-confidence) | 0 |
| S3_extracted_k141_49619 | 1179 | No | NA | 4 | 0 | 0 | Low-quality | Genome-fragment | 0.77 | AAI-based (high-confidence) | 0 |
| S3_extracted_k141_84800 | 1179 | No | NA | 3 | 0 | 0 | Low-quality | Genome-fragment | 2.04 | AAI-based (high-confidence) | 0 |
| S11_vs_k141_104053\|\|full | 13498 | No | NA | 20 | 6 | 0 | High-quality | High-quality | 99.05 | AAI-based (high-confidence) | 0 |
| S11_vs_k141_104054\|\|full | 29886 | No | NA | 45 | 14 | 0 | Medium-quality | Genome-fragment | 74.75 | AAI-based (medium-confidence) | 0 |
| S11_vs_k141_104061\|\|full | 15739 | No | NA | 20 | 0 | 3 | Not-determined | Genome-fragment | NA | NA | 0 |
| S11_vs_k141_104064\|\|full | 58041 | No | NA | 86 | 35 | 0 | High-quality | High-quality | 96.25 | AAI-based (high-confidence) | 0 |
| S11_vs_k141_104069\|\|full | 44378 | No | NA | 64 | 7 | 0 | High-quality | High-quality | 99.41 | AAI-based (high-confidence) | 0 |
| S3_vs_k141_43810\|\|full | 1179 | No | NA | 3 | 0 | 1 | Low-quality | Genome-fragment | 1.46 | AAI-based (medium-confidence) | 0 |
| S5_extracted_k141_62607 | 1179 | No | NA | 2 | 1 | 0 | Low-quality | Genome-fragment | 1.76 | HMM-based (lower-bound) | 0 |
| S5_extracted_k141_27629 | 1179 | No | NA | 2 | 1 | 0 | Low-quality | Genome-fragment | 2.91 | AAI-based (high-confidence) | 0 |
| S11_vs_k141_97632\|\|full | 3846 | No | NA | 6 | 0 | 0 | Not-determined | Genome-fragment | NA | NA | 0 |
| S7_vs_106547\|\|lt2gene | 1179 | No | NA | 2 | 1 | 0 | Low-quality | Genome-fragment | 3.44 | AAI-based (high-confidence) | 0 |
| S11_vs_k141_52091\|\|full | 1570 | No | NA | 4 | 0 | 0 | Not-determined | Genome-fragment | NA | NA | 0 |
| S11_vs_k141_38\|\|full | 1170 | No | NA | 3 | 0 | 0 | Not-determined | Genome-fragment | NA | NA | 0 |
| S11_vs_k141_26054\|\|full | 2834 | No | NA | 3 | 0 | 0 | Not-determined | Genome-fragment | NA | NA | 0 |
| S7_vs_110702\|\|lt2gene | 1179 | No | NA | 2 | 2 | 0 | Low-quality | Genome-fragment | 3.94 | AAI-based (high-confidence) | 0 |
| S16_extracted_67340 | 1179 | No | NA | 2 | 1 | 0 | Low-quality | Genome-fragment | 3.14 | AAI-based (high-confidence) | 0 |
| S16_vs_68739\|\|full | 1179 | No | NA | 3 | 0 | 0 | Low-quality | Genome-fragment | 2.84 | AAI-based (medium-confidence) | 0 |
| S10_vs_3709\|\|full | 1179 | No | NA | 4 | 0 | 0 | Low-quality | Genome-fragment | 2.7 | AAI-based (medium-confidence) | 0 |
| S11_vs_k141_19558\|\|full | 1174 | No | NA | 2 | 0 | 1 | Not-determined | Genome-fragment | NA | NA | 0 |
| S11_vs_k141_78148\|\|full | 6383 | No | NA | 7 | 0 | 3 | Not-determined | Genome-fragment | NA | NA | 0 |
| S10_vs_108096\|\|full | 1179 | No | NA | 3 | 0 | 0 | Low-quality | Genome-fragment | 2.07 | AAI-based (medium-confidence) | 0 |
| S11_vs_k141_39092\|\|full | 1911 | No | NA | 4 | 0 | 2 | Not-determined | Genome-fragment | NA | NA | 0 |
| S11_vs_k141_89\|\|full | 1511 | No | NA | 3 | 0 | 0 | Not-determined | Genome-fragment | NA | NA | 0 |
| S11_vs_k141_58652\|\|full | 1387 | No | NA | 2 | 0 | 1 | Not-determined | Genome-fragment | NA | NA | 0 |
| S11_vs_k141_58656\|\|full | 2995 | No | NA | 2 | 0 | 2 | Not-determined | Genome-fragment | NA | NA | 0 |
| S12_vs_k141_13669\|\|full | 1179 | No | NA | 3 | 0 | 0 | Low-quality | Genome-fragment | 2.43 | AAI-based (high-confidence) | 0 |
| S4_extracted_3543 | 1179 | No | NA | 2 | 2 | 0 | Low-quality | Genome-fragment | 3.88 | AAI-based (high-confidence) | 0 |
| S11_vs_k141_97678\|\|full | 1483 | No | NA | 2 | 0 | 0 | Not-determined | Genome-fragment | NA | NA | 0 |
| S11_vs_k141_97680\|\|full | 4205 | No | NA | 7 | 0 | 2 | Not-determined | Genome-fragment | NA | NA | 0 |
| S11_vs_k141_97686\|\|full | 5948 | No | NA | 6 | 0 | 5 | Not-determined | Genome-fragment | NA | NA | 0 |
| S11_vs_k141_32630\|\|full | 1482 | No | NA | 3 | 0 | 0 | Not-determined | Genome-fragment | NA | NA | 0 |
| S11_vs_k141_141\|\|full | 2228 | No | NA | 2 | 0 | 0 | Not-determined | Genome-fragment | NA | NA | 0 |
| S4_extracted_15274 | 1179 | No | NA | 2 | 2 | 0 | Low-quality | Genome-fragment | 3.75 | AAI-based (high-confidence) | 0 |
| S11_vs_k141_164\|\|full | 1960 | No | NA | 2 | 0 | 0 | Not-determined | Genome-fragment | NA | NA | 0 |
| S11_vs_k141_52268\|\|full | 1298 | No | NA | 3 | 0 | 2 | Not-determined | Genome-fragment | NA | NA | 0 |
| S8_extracted_k141_195776 | 1179 | No | NA | 3 | 0 | 0 | Low-quality | Genome-fragment | 2.97 | AAI-based (high-confidence) | 0 |
| S11_vs_k141_198\|\|full | 1435 | No | NA | 3 | 0 | 1 | Not-determined | Genome-fragment | NA | NA | 0 |
| S8_vs_k141_77053\|\|full | 1179 | No | NA | 3 | 0 | 0 | Low-quality | Genome-fragment | 1.62 | AAI-based (high-confidence) | 0 |
| S9_extracted_k141_76326 | 1180 | No | NA | 3 | 2 | 0 | Low-quality | Genome-fragment | 1.96 | AAI-based (high-confidence) | 0 |
| S11_vs_k141_78168\|\|full | 1789 | No | NA | 3 | 0 | 0 | Not-determined | Genome-fragment | NA | NA | 0 |
| S11_vs_k141_241\|\|full | 2577 | No | NA | 3 | 0 | 2 | Not-determined | Genome-fragment | NA | NA | 0 |
| S9_vs_k141_16353\|\|full | 1180 | No | NA | 3 | 1 | 0 | Low-quality | Genome-fragment | 1.94 | AAI-based (high-confidence) | 0 |
| S9_vs_k141_89494\|\|full | 1180 | No | NA | 3 | 1 | 0 | Low-quality | Genome-fragment | 1.69 | AAI-based (high-confidence) | 0 |
| S11_vs_k141_91307\|\|full | 1605 | No | NA | 4 | 0 | 1 | Not-determined | Genome-fragment | NA | NA | 0 |
| S13_vs_19606\|\|full | 1180 | No | NA | 2 | 0 | 0 | Low-quality | Genome-fragment | 2.26 | AAI-based (medium-confidence) | 0 |
| S14_vs_k141_71229\|\|full | 1180 | No | NA | 3 | 0 | 0 | Low-quality | Genome-fragment | 2.6 | AAI-based (medium-confidence) | 0 |
| S11_vs_k141_78182\|\|full | 1379 | No | NA | 2 | 0 | 0 | Not-determined | Genome-fragment | NA | NA | 0 |
| S11_vs_k141_58702\|\|full | 22627 | No | NA | 27 | 22 | 0 | Medium-quality | Genome-fragment | 50.16 | AAI-based (high-confidence) | 0 |
| S5_vs_k141_99160\|\|full | 1180 | No | NA | 3 | 0 | 0 | Low-quality | Genome-fragment | 2.98 | AAI-based (medium-confidence) | 0 |
| S7_vs_38683\|\|full | 1180 | No | NA | 2 | 0 | 1 | Low-quality | Genome-fragment | 3.79 | AAI-based (medium-confidence) | 0 |
| S16_vs_50070\|\|full | 1180 | No | NA | 4 | 1 | 0 | Low-quality | Genome-fragment | 3.43 | AAI-based (high-confidence) | 0 |
| S11_vs_k141_91331\|\|full | 3980 | No | NA | 3 | 0 | 2 | Not-determined | Genome-fragment | NA | NA | 0 |
| S16_vs_74182\|\|full | 1180 | No | NA | 4 | 4 | 0 | Low-quality | Genome-fragment | 2.74 | AAI-based (high-confidence) | 0 |
| S12_vs_k141_57041\|\|full | 1180 | No | NA | 2 | 0 | 0 | Low-quality | Genome-fragment | 0.62 | AAI-based (medium-confidence) | 0 |
| S12_vs_k141_105454\|\|full | 1180 | No | NA | 2 | 2 | 0 | Low-quality | Genome-fragment | 3.37 | AAI-based (medium-confidence) | 0 |
| S11_vs_k141_71754\|\|full | 1501 | No | NA | 2 | 0 | 2 | Not-determined | Genome-fragment | NA | NA | 0 |
| S11_vs_k141_39217\|\|full | 8026 | No | NA | 7 | 0 | 4 | Not-determined | Genome-fragment | NA | NA | 0 |
| S12_vs_k141_77407\|\|full | 1180 | No | NA | 3 | 0 | 0 | Low-quality | Genome-fragment | 0.92 | AAI-based (medium-confidence) | 0 |
| S11_vs_k141_84824\|\|full | 1517 | No | NA | 3 | 0 | 1 | Not-determined | Genome-fragment | NA | NA | 0 |
| S11_vs_k141_6632\|\|full | 2719 | No | NA | 3 | 0 | 2 | Not-determined | Genome-fragment | NA | NA | 0 |
| S11_vs_k141_65304\|\|full | 1578 | No | NA | 3 | 0 | 2 | Not-determined | Genome-fragment | NA | NA | 0 |
| S11_vs_k141_78305\|\|full | 2042 | No | NA | 5 | 0 | 0 | Not-determined | Genome-fragment | NA | NA | 0 |
| S15_extracted_46552 | 1180 | No | NA | 1 | 0 | 1 | Low-quality | Genome-fragment | 0.51 | AAI-based (medium-confidence) | 0 |
| S11_vs_k141_65307\|\|full | 3715 | No | NA | 4 | 0 | 2 | Not-determined | Genome-fragment | NA | NA | 0 |
| S4_extracted_30904 | 1180 | No | NA | 1 | 1 | 0 | Low-quality | Genome-fragment | 3.62 | AAI-based (high-confidence) | 0 |
| S11_vs_k141_58776\|\|full | 6054 | No | NA | 10 | 0 | 0 | Not-determined | Genome-fragment | NA | NA | 0 |
| S11_vs_k141_45658\|\|full | 173195 | Yes | 44246 | 183 | 12 | 97 | Medium-quality | Genome-fragment | 68.29 | AAI-based (high-confidence) | 74.45 |
| S8_extracted_k141_117554 | 1180 | No | NA | 6 | 1 | 0 | Low-quality | Genome-fragment | 0.94 | AAI-based (high-confidence) | 0 |
| S11_vs_k141_84848\|\|full | 41635 | No | NA | 45 | 7 | 4 | High-quality | High-quality | 97.21 | AAI-based (high-confidence) | 0 |
| S8_extracted_k141_189188 | 1180 | No | NA | 2 | 0 | 0 | Low-quality | Genome-fragment | 0.73 | AAI-based (medium-confidence) | 0 |
| S9_extracted_k141_56314 | 1181 | No | NA | 1 | 0 | 0 | Low-quality | Genome-fragment | 1.33 | AAI-based (high-confidence) | 0 |
| S11_vs_k141_75246\|\|full | 1181 | No | NA | 4 | 0 | 0 | Low-quality | Genome-fragment | 2.01 | AAI-based (medium-confidence) | 0 |
| S13_vs_149550\|\|full | 1181 | No | NA | 3 | 0 | 0 | Low-quality | Genome-fragment | 2.28 | AAI-based (medium-confidence) | 0 |
| S11_vs_k141_32832\|\|full | 9578 | No | NA | 6 | 0 | 1 | Not-determined | Genome-fragment | NA | NA | 0 |
| S11_vs_k141_45708\|\|full | 3235 | No | NA | 3 | 0 | 2 | Not-determined | Genome-fragment | NA | NA | 0 |
| S14_vs_k141_84436\|\|full | 1181 | No | NA | 3 | 1 | 0 | Low-quality | Genome-fragment | 1.37 | HMM-based (lower-bound) | 0 |
| S11_vs_k141_39279\|\|full | 11329 | No | NA | 8 | 0 | 6 | Not-determined | Genome-fragment | NA | NA | 0 |
| S11_vs_k141_84917\|\|full | 1039 | No | NA | 2 | 0 | 1 | Not-determined | Genome-fragment | NA | NA | 0 |
| S14_vs_k141_55408\|\|lt2gene | 1181 | No | NA | 2 | 1 | 0 | Low-quality | Genome-fragment | 3.03 | AAI-based (high-confidence) | 0 |
| S3_vs_k141_48729\|\|lt2gene | 1181 | No | NA | 2 | 1 | 0 | Low-quality | Genome-fragment | 2.75 | AAI-based (high-confidence) | 0 |
| S3_vs_k141_18921\|\|lt2gene | 1181 | No | NA | 1 | 1 | 0 | Low-quality | Genome-fragment | 2.72 | AAI-based (high-confidence) | 0 |
| S5_vs_k141_127576\|\|full | 1181 | No | NA | 2 | 0 | 0 | Low-quality | Genome-fragment | 3.31 | AAI-based (medium-confidence) | 0 |
| S5_vs_k141_144078\|\|full | 1181 | No | NA | 2 | 1 | 0 | Low-quality | Genome-fragment | 1.48 | HMM-based (lower-bound) | 0 |
| S11_vs_k141_97971\|\|full | 1274 | No | NA | 2 | 0 | 2 | Not-determined | Genome-fragment | NA | NA | 0 |
| S7_vs_80234\|\|full | 1181 | No | NA | 2 | 1 | 0 | Low-quality | Genome-fragment | 2.67 | AAI-based (high-confidence) | 0 |
| S16_vs_130283\|\|full | 1181 | No | NA | 2 | 0 | 0 | Low-quality | Genome-fragment | 2.54 | AAI-based (medium-confidence) | 0 |
| S11_vs_k141_6776\|\|full | 1283 | No | NA | 2 | 0 | 0 | Not-determined | Genome-fragment | NA | NA | 0 |
| S11_vs_k141_65387\|\|full | 1118 | No | NA | 3 | 0 | 1 | Not-determined | Genome-fragment | NA | NA | 0 |
| S11_vs_k141_522\|\|full | 1458 | No | NA | 4 | 0 | 0 | Not-determined | Genome-fragment | NA | NA | 0 |
| S10_extracted_27010 | 1181 | No | NA | 1 | 0 | 0 | Low-quality | Genome-fragment | 0.48 | AAI-based (high-confidence) | 0 |
| S11_vs_k141_71899\|\|full | 1384 | No | NA | 2 | 0 | 1 | Not-determined | Genome-fragment | NA | NA | 0 |
| S11_vs_k141_52430\|\|full | 1413 | No | NA | 5 | 0 | 0 | Not-determined | Genome-fragment | NA | NA | 0 |
| S11_vs_k141_26383\|\|full | 4496 | No | NA | 8 | 0 | 0 | Not-determined | Genome-fragment | NA | NA | 0 |
| S10_extracted_29788 | 1181 | No | NA | 2 | 0 | 0 | Low-quality | Genome-fragment | 1.09 | AAI-based (high-confidence) | 0 |
| S10_vs_3849\|\|full | 1181 | No | NA | 3 | 0 | 1 | Low-quality | Genome-fragment | 2.88 | AAI-based (medium-confidence) | 0 |
| S12_extracted_k141_37863 | 1181 | No | NA | 2 | 0 | 0 | Low-quality | Genome-fragment | 3.07 | AAI-based (medium-confidence) | 0 |
| S12_vs_k141_21771\|\|lt2gene | 1181 | No | NA | 1 | 1 | 0 | Low-quality | Genome-fragment | 2.79 | AAI-based (high-confidence) | 0 |
| S6_vs_k141_47337\|\|full | 1181 | No | NA | 3 | 0 | 0 | Low-quality | Genome-fragment | 2.71 | AAI-based (high-confidence) | 0 |
| S9_extracted_k141_39952 | 1182 | No | NA | 1 | 1 | 0 | Low-quality | Genome-fragment | 2.77 | AAI-based (medium-confidence) | 0 |
| S9_extracted_k141_90161 | 1182 | No | NA | 3 | 0 | 0 | Low-quality | Genome-fragment | 2.96 | AAI-based (high-confidence) | 0 |
| S11_vs_k141_19892\|\|full | 31610 | No | NA | 41 | 14 | 1 | Medium-quality | Genome-fragment | 70.43 | AAI-based (high-confidence) | 0 |
| S11_vs_k141_78527\|\|full | 1440 | No | NA | 2 | 0 | 1 | Not-determined | Genome-fragment | NA | NA | 0 |
| S13_extracted_147053 | 1182 | No | NA | 1 | 1 | 0 | Low-quality | Genome-fragment | 0.82 | HMM-based (lower-bound) | 0 |
| S11_vs_k141_78534\|\|full | 2945 | No | NA | 5 | 0 | 1 | Not-determined | Genome-fragment | NA | NA | 0 |
| S14_extracted_k141_30302 | 1182 | No | NA | 3 | 2 | 0 | Low-quality | Genome-fragment | 3.9 | AAI-based (high-confidence) | 0 |
| S11_vs_k141_98028\|\|full | 4940 | No | NA | 7 | 0 | 1 | Not-determined | Genome-fragment | NA | NA | 0 |
| S14_extracted_k141_56841 | 1182 | No | NA | 2 | 2 | 0 | Low-quality | Genome-fragment | 1.97 | AAI-based (medium-confidence) | 0 |
| S11_vs_k141_32947\|\|full | 3095 | No | NA | 4 | 0 | 1 | Not-determined | Genome-fragment | NA | NA | 0 |
| S1_extracted_k141_2888 | 1182 | No | NA | 2 | 2 | 0 | Low-quality | Genome-fragment | 2.12 | HMM-based (lower-bound) | 0 |
| S11_vs_k141_32954\|\|full | 6115 | No | NA | 4 | 0 | 2 | Not-determined | Genome-fragment | NA | NA | 0 |
| S3_extracted_k141_89126 | 1182 | No | NA | 2 | 0 | 0 | Low-quality | Genome-fragment | 0.67 | AAI-based (medium-confidence) | 0 |
| S3_vs_k141_91791\|\|full | 1182 | No | NA | 3 | 0 | 0 | Low-quality | Genome-fragment | 2.15 | AAI-based (high-confidence) | 0 |
| S5_vs_k141_43053\|\|full | 1182 | No | NA | 3 | 1 | 0 | Low-quality | Genome-fragment | 2.72 | AAI-based (high-confidence) | 0 |
| S5_vs_k141_73724\|\|full | 1182 | No | NA | 4 | 1 | 0 | Low-quality | Genome-fragment | 3.41 | AAI-based (high-confidence) | 0 |
| S5_vs_k141_78804\|\|full | 1182 | No | NA | 4 | 0 | 0 | Low-quality | Genome-fragment | 2.7 | AAI-based (medium-confidence) | 0 |
| S11_vs_k141_52494\|\|full | 10446 | No | NA | 10 | 0 | 3 | Not-determined | Genome-fragment | NA | NA | 0 |
| S5_vs_k141_45282\|\|lt2gene | 1182 | No | NA | 1 | 1 | 0 | Low-quality | Genome-fragment | 2.69 | AAI-based (high-confidence) | 0 |
| S16_vs_40552\|\|lt2gene | 1182 | No | NA | 2 | 1 | 0 | Low-quality | Genome-fragment | 1.92 | HMM-based (lower-bound) | 0 |
| S11_vs_k141_6878\|\|full | 42579 | No | NA | 72 | 23 | 2 | High-quality | High-quality | 97.62 | AAI-based (high-confidence) | 0 |
| S11_vs_k141_58930\|\|full | 23499 | No | NA | 33 | 6 | 3 | Medium-quality | Genome-fragment | 55.51 | AAI-based (high-confidence) | 0 |
| S11_vs_k141_98098\|\|full | 3879 | No | NA | 7 | 0 | 2 | Not-determined | Genome-fragment | NA | NA | 0 |
| S16_vs_113871\|\|lt2gene | 1182 | No | NA | 2 | 2 | 0 | Low-quality | Genome-fragment | 2.42 | AAI-based (high-confidence) | 0 |
| S11_vs_k141_13239\|\|full | 1246 | No | NA | 2 | 0 | 0 | Not-determined | Genome-fragment | NA | NA | 0 |
| S16_vs_120355\|\|lt2gene | 1182 | No | NA | 2 | 1 | 0 | Low-quality | Genome-fragment | 0.81 | HMM-based (lower-bound) | 0 |
| S10_vs_8751\|\|full | 1182 | No | NA | 2 | 0 | 0 | Low-quality | Genome-fragment | 2.02 | AAI-based (medium-confidence) | 0 |
| S4_extracted_26909 | 1182 | No | NA | 4 | 3 | 0 | Low-quality | Genome-fragment | 3.83 | AAI-based (high-confidence) | 0 |
| S8_vs_k141_68947\|\|lt2gene | 1182 | No | NA | 2 | 1 | 0 | Low-quality | Genome-fragment | 2.05 | HMM-based (lower-bound) | 0 |
| S9_extracted_k141_24390 | 1183 | No | NA | 2 | 1 | 0 | Low-quality | Genome-fragment | 2.89 | AAI-based (high-confidence) | 0 |
| S11_vs_k141_78682\|\|full | 1764 | No | NA | 3 | 0 | 2 | Not-determined | Genome-fragment | NA | NA | 0 |
| S9_extracted_k141_40858 | 1183 | No | NA | 1 | 0 | 1 | Low-quality | Genome-fragment | 0.32 | AAI-based (high-confidence) | 0 |
| S11_vs_k141_85116\|\|full | 1784 | No | NA | 4 | 0 | 3 | Not-determined | Genome-fragment | NA | NA | 0 |
| S11_vs_k141_5224\|\|full | 1183 | No | NA | 3 | 0 | 0 | Low-quality | Genome-fragment | 0.61 | AAI-based (medium-confidence) | 0 |
| S13_extracted_152511 | 1183 | No | NA | 1 | 0 | 1 | Low-quality | Genome-fragment | 0.52 | AAI-based (high-confidence) | 0 |
| S14_extracted_k141_58953 | 1183 | No | NA | 3 | 0 | 0 | Low-quality | Genome-fragment | 1.4 | AAI-based (high-confidence) | 0 |
| S11_vs_k141_72109\|\|full | 4146 | No | NA | 6 | 0 | 3 | Not-determined | Genome-fragment | NA | NA | 0 |
| S1_extracted_k141_104424 | 1183 | No | NA | 1 | 0 | 1 | Low-quality | Genome-fragment | 1.42 | AAI-based (high-confidence) | 0 |
| S11_vs_k141_52577\|\|full | 31692 | No | NA | 45 | 15 | 1 | Medium-quality | Genome-fragment | 88.54 | AAI-based (medium-confidence) | 0 |
| S3_extracted_k141_75140 | 1183 | No | NA | 3 | 0 | 0 | Low-quality | Genome-fragment | 2.81 | AAI-based (high-confidence) | 0 |
| S11_vs_k141_20079\|\|full | 6325 | No | NA | 8 | 0 | 0 | Not-determined | Genome-fragment | NA | NA | 0 |
| S11_vs_k141_72161\|\|full | 2047 | No | NA | 3 | 0 | 1 | Not-determined | Genome-fragment | NA | NA | 0 |
| S5_vs_k141_11909\|\|full | 1183 | No | NA | 3 | 0 | 0 | Low-quality | Genome-fragment | 2.57 | AAI-based (high-confidence) | 0 |
| S11_vs_k141_91702\|\|full | 1569 | No | NA | 3 | 0 | 1 | Not-determined | Genome-fragment | NA | NA | 0 |
| S5_vs_k141_68699\|\|lt2gene | 1183 | No | NA | 2 | 2 | 0 | Low-quality | Genome-fragment | 3.3 | AAI-based (medium-confidence) | 0 |
| S7_vs_4944\|\|full | 1183 | No | NA | 2 | 2 | 0 | Low-quality | Genome-fragment | 2.15 | AAI-based (high-confidence) | 0 |
| S7_vs_45447\|\|full | 1183 | No | NA | 2 | 1 | 0 | Low-quality | Genome-fragment | 2.46 | AAI-based (high-confidence) | 0 |
| S11_vs_k141_98193\|\|full | 34086 | No | NA | 41 | 8 | 2 | Medium-quality | Genome-fragment | 80.53 | AAI-based (medium-confidence) | 0 |
| S11_vs_k141_98194\|\|full | 2531 | No | NA | 3 | 0 | 2 | Not-determined | Genome-fragment | NA | NA | 0 |
| S16_vs_24630\|\|full | 1183 | No | NA | 3 | 0 | 0 | Low-quality | Genome-fragment | 2.24 | AAI-based (medium-confidence) | 0 |
| S11_vs_k141_78726\|\|full | 4258 | No | NA | 6 | 0 | 0 | Not-determined | Genome-fragment | NA | NA | 0 |
| S10_vs_81126\|\|full | 1183 | No | NA | 3 | 0 | 2 | Low-quality | Genome-fragment | 3.78 | AAI-based (medium-confidence) | 0 |
| S17_extracted_k141_78152 | 1184 | No | NA | 5 | 1 | 0 | Low-quality | Genome-fragment | 1.9 | HMM-based (lower-bound) | 0 |
| S17_extracted_k141_93659 | 1184 | No | NA | 3 | 1 | 0 | Low-quality | Genome-fragment | 3.01 | AAI-based (medium-confidence) | 0 |
| S11_extracted_k141_103628 | 1184 | No | NA | 2 | 0 | 0 | Low-quality | Genome-fragment | 2.59 | AAI-based (medium-confidence) | 0 |
| S11_vs_k141_7012\|\|full | 1020 | No | NA | 3 | 0 | 0 | Not-determined | Genome-fragment | NA | NA | 0 |
| S13_extracted_21061 | 1184 | No | NA | 2 | 0 | 0 | Low-quality | Genome-fragment | 1.18 | AAI-based (high-confidence) | 0 |
| S14_vs_k141_12224\|\|full | 1184 | No | NA | 3 | 2 | 0 | Low-quality | Genome-fragment | 2.76 | AAI-based (high-confidence) | 0 |
| S11_vs_k141_39684\|\|full | 1446 | No | NA | 2 | 0 | 2 | Not-determined | Genome-fragment | NA | NA | 0 |
| S3_vs_k141_81839\|\|full | 1184 | No | NA | 3 | 0 | 0 | Low-quality | Genome-fragment | 0.89 | AAI-based (high-confidence) | 0 |
| S5_extracted_k141_137803 | 1184 | No | NA | 1 | 0 | 0 | Low-quality | Genome-fragment | 0.56 | AAI-based (high-confidence) | 0 |
| S5_vs_k141_37301\|\|lt2gene | 1184 | No | NA | 2 | 1 | 0 | Low-quality | Genome-fragment | 3.01 | AAI-based (high-confidence) | 0 |
| S7_extracted_50670 | 1184 | No | NA | 1 | 0 | 1 | Low-quality | Genome-fragment | 0.32 | AAI-based (high-confidence) | 0 |
| S7_vs_24685\|\|full | 1184 | No | NA | 3 | 1 | 0 | Low-quality | Genome-fragment | 3.37 | AAI-based (high-confidence) | 0 |
| S16_vs_61831\|\|full | 1184 | No | NA | 3 | 0 | 0 | Low-quality | Genome-fragment | 1.62 | AAI-based (high-confidence) | 0 |
| S16_vs_70465\|\|full | 1184 | No | NA | 3 | 0 | 0 | Low-quality | Genome-fragment | 2.13 | AAI-based (medium-confidence) | 0 |
| S10_extracted_114582 | 1184 | No | NA | 1 | 0 | 1 | Low-quality | Genome-fragment | 2.38 | AAI-based (high-confidence) | 0 |
| S10_vs_101445\|\|full | 1184 | No | NA | 3 | 2 | 0 | Low-quality | Genome-fragment | 3.65 | AAI-based (high-confidence) | 0 |
| S8_extracted_k141_185375 | 1184 | No | NA | 2 | 1 | 0 | Low-quality | Genome-fragment | 0.65 | HMM-based (lower-bound) | 0 |
| S11_vs_k141_13488\|\|full | 4117 | No | NA | 4 | 0 | 3 | Not-determined | Genome-fragment | NA | NA | 0 |
| S8_vs_k141_104154\|\|full | 1184 | No | NA | 3 | 1 | 0 | Low-quality | Genome-fragment | 0.45 | HMM-based (lower-bound) | 0 |
| S17_vs_k141_138528\|\|full | 1185 | No | NA | 4 | 3 | 0 | Low-quality | Genome-fragment | 5.4 | AAI-based (high-confidence) | 0 |
| S11_vs_k141_20225\|\|full | 36396 | No | NA | 38 | 20 | 0 | High-quality | High-quality | 94.18 | AAI-based (high-confidence) | 0 |
| S11_vs_k141_46148\|\|full | 3225 | No | NA | 5 | 0 | 1 | Not-determined | Genome-fragment | NA | NA | 0 |
| S14_vs_k141_84329\|\|full | 1185 | No | NA | 2 | 0 | 0 | Low-quality | Genome-fragment | 3.03 | AAI-based (high-confidence) | 0 |
| S11_vs_k141_65688\|\|full | 1028 | No | NA | 2 | 0 | 0 | Not-determined | Genome-fragment | NA | NA | 0 |
| S1_vs_k141_96189\|\|full | 1185 | No | NA | 4 | 1 | 0 | Low-quality | Genome-fragment | 2.95 | AAI-based (high-confidence) | 0 |
| S5_vs_k141_68210\|\|full | 1185 | No | NA | 4 | 1 | 0 | Low-quality | Genome-fragment | 3.22 | AAI-based (high-confidence) | 0 |
| S11_vs_k141_91868\|\|full | 4655 | No | NA | 9 | 0 | 5 | Not-determined | Genome-fragment | NA | NA | 0 |
| S5_vs_k141_134998\|\|full | 1185 | No | NA | 3 | 0 | 0 | Low-quality | Genome-fragment | 2 | AAI-based (medium-confidence) | 0 |
| S11_vs_k141_78885\|\|full | 2249 | No | NA | 3 | 0 | 3 | Not-determined | Genome-fragment | NA | NA | 0 |
| S7_extracted_74842 | 1185 | No | NA | 1 | 0 | 1 | Low-quality | Genome-fragment | 1.3 | AAI-based (high-confidence) | 0 |
| S11_vs_k141_78888\|\|full | 6125 | No | NA | 5 | 0 | 5 | Not-determined | Genome-fragment | NA | NA | 0 |
| S7_vs_58799\|\|lt2gene | 1185 | No | NA | 1 | 1 | 0 | Low-quality | Genome-fragment | 4.01 | HMM-based (lower-bound) | 0 |
| S10_extracted_1440 | 1185 | No | NA | 2 | 1 | 0 | Low-quality | Genome-fragment | 2.9 | AAI-based (medium-confidence) | 0 |
| S10_extracted_71399 | 1185 | No | NA | 2 | 0 | 0 | Low-quality | Genome-fragment | 1.22 | AAI-based (medium-confidence) | 0 |
| S12_extracted_k141_20737 | 1185 | No | NA | 1 | 1 | 0 | Low-quality | Genome-fragment | 1.93 | AAI-based (high-confidence) | 0 |
| S12_extracted_k141_22664 | 1185 | No | NA | 2 | 0 | 0 | Low-quality | Genome-fragment | 2.15 | AAI-based (medium-confidence) | 0 |
| S11_vs_k141_26767\|\|full | 24781 | No | NA | 35 | 22 | 0 | Medium-quality | Genome-fragment | 56.42 | AAI-based (high-confidence) | 0 |
| S11_vs_k141_85405\|\|full | 1500 | No | NA | 2 | 0 | 1 | Not-determined | Genome-fragment | NA | NA | 0 |
| S11_vs_k141_20321\|\|full | 6360 | No | NA | 8 | 0 | 0 | Not-determined | Genome-fragment | NA | NA | 0 |
| S11_vs_k141_85411\|\|full | 1250 | No | NA | 2 | 0 | 1 | Not-determined | Genome-fragment | NA | NA | 0 |
| S11_vs_k141_46258\|\|full | 6617 | No | NA | 9 | 0 | 8 | Not-determined | Genome-fragment | NA | NA | 0 |
| S12_extracted_k141_38280 | 1185 | No | NA | 2 | 1 | 0 | Low-quality | Genome-fragment | 1.41 | HMM-based (lower-bound) | 0 |
| S12_vs_k141_65050\|\|full | 1185 | No | NA | 3 | 2 | 0 | Low-quality | Genome-fragment | 2.68 | AAI-based (high-confidence) | 0 |
| S15_vs_208\|\|full | 1185 | No | NA | 4 | 1 | 0 | Low-quality | Genome-fragment | 2.43 | AAI-based (high-confidence) | 0 |
| S11_vs_k141_39758\|\|full | 6229 | No | NA | 12 | 0 | 4 | Not-determined | Genome-fragment | NA | NA | 0 |
| S11_vs_k141_965\|\|full | 1768 | No | NA | 3 | 0 | 0 | Not-determined | Genome-fragment | NA | NA | 0 |
| S4_extracted_4201 | 1185 | No | NA | 2 | 2 | 0 | Low-quality | Genome-fragment | 3.22 | HMM-based (lower-bound) | 0 |
| S11_vs_k141_26824\|\|full | 2034 | No | NA | 2 | 0 | 1 | Not-determined | Genome-fragment | NA | NA | 0 |
| S8_extracted_k141_79984 | 1185 | No | NA | 3 | 1 | 0 | Low-quality | Genome-fragment | 2.86 | AAI-based (high-confidence) | 0 |
| S8_extracted_k141_144545 | 1185 | No | NA | 1 | 0 | 0 | Low-quality | Genome-fragment | 0.33 | AAI-based (medium-confidence) | 0 |
| S11_vs_k141_52777\|\|full | 6559 | No | NA | 4 | 0 | 3 | Not-determined | Genome-fragment | NA | NA | 0 |
| S8_extracted_k141_127308 | 1185 | No | NA | 1 | 0 | 0 | Low-quality | Genome-fragment | 2.27 | AAI-based (medium-confidence) | 0 |
| S11_vs_k141_65767\|\|full | 4200 | No | NA | 7 | 0 | 0 | Not-determined | Genome-fragment | NA | NA | 0 |
| S8_vs_k141_69494\|\|full | 1185 | No | NA | 2 | 0 | 2 | Low-quality | Genome-fragment | 0.31 | AAI-based (high-confidence) | 0 |
| S17_vs_k141_134599\|\|full | 1186 | No | NA | 3 | 0 | 0 | Low-quality | Genome-fragment | 0.69 | AAI-based (high-confidence) | 0 |
| S14_extracted_k141_64368 | 1186 | No | NA | 3 | 3 | 0 | Low-quality | Genome-fragment | 3.92 | AAI-based (high-confidence) | 0 |
| S1_vs_k141_85948\|\|full | 1186 | No | NA | 3 | 0 | 0 | Low-quality | Genome-fragment | 2.8 | AAI-based (medium-confidence) | 0 |
| S11_vs_k141_85463\|\|full | 1083 | No | NA | 2 | 0 | 2 | Not-determined | Genome-fragment | NA | NA | 0 |
| S11_vs_k141_79013\|\|full | 2418 | No | NA | 5 | 0 | 0 | Not-determined | Genome-fragment | NA | NA | 0 |
| S3_vs_k141_10946\|\|full | 1186 | No | NA | 5 | 0 | 0 | Low-quality | Genome-fragment | 2.74 | AAI-based (high-confidence) | 0 |
| S3_vs_k141_60165\|\|full | 1186 | No | NA | 3 | 2 | 0 | Low-quality | Genome-fragment | 2.6 | AAI-based (medium-confidence) | 0 |
| S5_extracted_k141_23080 | 1186 | No | NA | 3 | 1 | 0 | Low-quality | Genome-fragment | 1.95 | AAI-based (high-confidence) | 0 |
| S5_vs_k141_74629\|\|full | 1186 | No | NA | 3 | 0 | 0 | Low-quality | Genome-fragment | 2.78 | AAI-based (high-confidence) | 0 |
| S7_extracted_9509 | 1186 | No | NA | 1 | 0 | 1 | Low-quality | Genome-fragment | 0.23 | AAI-based (high-confidence) | 0 |
| S11_vs_k141_85502\|\|full | 1487 | No | NA | 3 | 0 | 0 | Not-determined | Genome-fragment | NA | NA | 0 |
| S16_vs_17937\|\|full | 1186 | No | NA | 3 | 0 | 0 | Low-quality | Genome-fragment | 0.87 | AAI-based (high-confidence) | 0 |
| S11_vs_k141_98521\|\|full | 2633 | No | NA | 5 | 0 | 0 | Not-determined | Genome-fragment | NA | NA | 0 |
| S12_extracted_k141_88724 | 1186 | No | NA | 1 | 0 | 0 | Low-quality | Genome-fragment | 0.8 | AAI-based (high-confidence) | 0 |
| S11_vs_k141_65830\|\|full | 3231 | No | NA | 3 | 0 | 2 | Not-determined | Genome-fragment | NA | NA | 0 |
| S2_extracted_k141_2522 | 1186 | No | NA | 2 | 0 | 1 | Low-quality | Genome-fragment | 1.53 | AAI-based (medium-confidence) | 0 |
| S11_vs_k141_46373\|\|full | 2650 | No | NA | 3 | 0 | 0 | Not-determined | Genome-fragment | NA | NA | 0 |
| S8_extracted_k141_81977 | 1186 | No | NA | 2 | 1 | 0 | Low-quality | Genome-fragment | 3.83 | HMM-based (lower-bound) | 0 |
| S8_extracted_k141_51630 | 1186 | No | NA | 2 | 0 | 1 | Low-quality | Genome-fragment | 0.39 | AAI-based (medium-confidence) | 0 |
| S11_vs_k141_79090\|\|full | 1197 | No | NA | 3 | 0 | 2 | Not-determined | Genome-fragment | NA | NA | 0 |
| S11_vs_k141_59235\|\|full | 3991 | No | NA | 4 | 0 | 3 | Not-determined | Genome-fragment | NA | NA | 0 |
| S11_vs_k141_20449\|\|full | 6871 | No | NA | 10 | 0 | 3 | Not-determined | Genome-fragment | NA | NA | 0 |
| S8_vs_k141_154981\|\|full | 1186 | No | NA | 2 | 1 | 0 | Low-quality | Genome-fragment | 1.55 | AAI-based (medium-confidence) | 0 |
| S8_vs_k141_172236\|\|lt2gene | 1186 | No | NA | 1 | 1 | 0 | Low-quality | Genome-fragment | 2.39 | HMM-based (lower-bound) | 0 |
| S11_vs_k141_59245\|\|full | 1158 | No | NA | 2 | 0 | 2 | Not-determined | Genome-fragment | NA | NA | 0 |
| S11_vs_k141_52907\|\|full | 2121 | No | NA | 3 | 0 | 0 | Not-determined | Genome-fragment | NA | NA | 0 |
| S11_vs_k141_72554\|\|full | 4244 | No | NA | 9 | 0 | 3 | Not-determined | Genome-fragment | NA | NA | 0 |
| S11_vs_k141_7188\|\|full | 1579 | No | NA | 6 | 0 | 0 | Not-determined | Genome-fragment | NA | NA | 0 |
| S17_extracted_k141_111789 | 1187 | No | NA | 4 | 4 | 0 | Low-quality | Genome-fragment | 5.42 | AAI-based (high-confidence) | 0 |
| S11_vs_k141_52920\|\|full | 1762 | No | NA | 5 | 0 | 0 | Not-determined | Genome-fragment | NA | NA | 0 |
| S17_extracted_k141_166294 | 1187 | No | NA | 4 | 0 | 0 | Low-quality | Genome-fragment | 0.69 | AAI-based (high-confidence) | 0 |
| S17_vs_k141_31399\|\|full | 1187 | No | NA | 4 | 0 | 0 | Low-quality | Genome-fragment | 1.07 | AAI-based (medium-confidence) | 0 |
| S17_vs_k141_151357\|\|lt2gene | 1187 | No | NA | 2 | 1 | 0 | Low-quality | Genome-fragment | 1.55 | HMM-based (lower-bound) | 0 |
| S11_vs_k141_92124\|\|full | 1227 | No | NA | 3 | 0 | 1 | Not-determined | Genome-fragment | NA | NA | 0 |
| S11_vs_k141_46452\|\|full | 1006 | No | NA | 2 | 0 | 1 | Not-determined | Genome-fragment | NA | NA | 0 |
| S14_vs_k141_53726\|\|full | 1187 | No | NA | 2 | 0 | 0 | Low-quality | Genome-fragment | 3.08 | AAI-based (medium-confidence) | 0 |
| S1_vs_k141_67432\|\|full | 1187 | No | NA | 3 | 1 | 0 | Low-quality | Genome-fragment | 2.05 | AAI-based (high-confidence) | 0 |
| S3_vs_k141_63788\|\|full | 1187 | No | NA | 4 | 4 | 0 | Low-quality | Genome-fragment | 3.91 | AAI-based (high-confidence) | 0 |
| S5_vs_k141_99637\|\|full | 1187 | No | NA | 4 | 0 | 1 | Low-quality | Genome-fragment | 2.38 | AAI-based (medium-confidence) | 0 |
| S7_vs_38451\|\|full | 1187 | No | NA | 2 | 1 | 0 | Low-quality | Genome-fragment | 0.28 | HMM-based (lower-bound) | 0 |
| S11_vs_k141_39961\|\|full | 1839 | No | NA | 3 | 0 | 0 | Not-determined | Genome-fragment | NA | NA | 0 |
| S11_vs_k141_72617\|\|full | 4667 | No | NA | 4 | 0 | 4 | Not-determined | Genome-fragment | NA | NA | 0 |
| S11_vs_k141_72636\|\|full | 1804 | No | NA | 5 | 0 | 0 | Not-determined | Genome-fragment | NA | NA | 0 |
| S11_vs_k141_72639\|\|full | 1176 | No | NA | 2 | 0 | 2 | Not-determined | Genome-fragment | NA | NA | 0 |
| S7_vs_86635\|\|full | 1187 | No | NA | 3 | 1 | 0 | Low-quality | Genome-fragment | 2.75 | AAI-based (medium-confidence) | 0 |
| S11_vs_k141_33476\|\|full | 1406 | No | NA | 2 | 0 | 1 | Not-determined | Genome-fragment | NA | NA | 0 |
| S7_vs_96974\|\|lt2gene | 1187 | No | NA | 2 | 1 | 0 | Low-quality | Genome-fragment | 1.94 | HMM-based (lower-bound) | 0 |
| S12_extracted_k141_182681 | 1187 | No | NA | 1 | 0 | 1 | Low-quality | Genome-fragment | 3.03 | AAI-based (medium-confidence) | 0 |
| S12_extracted_k141_156797 | 1187 | No | NA | 2 | 0 | 0 | Low-quality | Genome-fragment | 0.84 | AAI-based (high-confidence) | 0 |
| S8_extracted_k141_101923 | 1187 | No | NA | 3 | 0 | 0 | Low-quality | Genome-fragment | 1.81 | AAI-based (high-confidence) | 0 |
| S8_extracted_k141_51254 | 1187 | No | NA | 2 | 2 | 0 | Low-quality | Genome-fragment | 1.58 | HMM-based (lower-bound) | 0 |
| S11_vs_k141_92188\|\|full | 2499 | No | NA | 4 | 0 | 1 | Not-determined | Genome-fragment | NA | NA | 0 |
| S11_vs_k141_79242\|\|full | 1442 | No | NA | 2 | 0 | 0 | Not-determined | Genome-fragment | NA | NA | 0 |
| S11_vs_k141_13608\|\|full | 1520 | No | NA | 3 | 0 | 0 | Not-determined | Genome-fragment | NA | NA | 0 |
| S17_extracted_k141_94680 | 1188 | No | NA | 2 | 0 | 1 | Low-quality | Genome-fragment | 0.91 | AAI-based (medium-confidence) | 0 |
| S9_extracted_k141_34801 | 1188 | No | NA | 3 | 1 | 0 | Low-quality | Genome-fragment | 2.51 | AAI-based (high-confidence) | 0 |
| S11_vs_k141_72718\|\|full | 2084 | No | NA | 3 | 0 | 1 | Not-determined | Genome-fragment | NA | NA | 0 |
| S9_extracted_k141_43750 | 1188 | No | NA | 3 | 0 | 0 | Low-quality | Genome-fragment | 0.68 | AAI-based (high-confidence) | 0 |
| S14_extracted_k141_113768 | 1188 | No | NA | 2 | 0 | 0 | Low-quality | Genome-fragment | 3.16 | AAI-based (medium-confidence) | 0 |
| S11_vs_k141_66035\|\|full | 1472 | No | NA | 2 | 0 | 1 | Not-determined | Genome-fragment | NA | NA | 0 |
| S14_vs_k141_21522\|\|lt2gene | 1188 | No | NA | 1 | 1 | 0 | Low-quality | Genome-fragment | 2.25 | HMM-based (lower-bound) | 0 |
| S3_extracted_k141_77597 | 1188 | No | NA | 2 | 2 | 0 | Low-quality | Genome-fragment | 3.42 | AAI-based (high-confidence) | 0 |
| S11_vs_k141_66046\|\|full | 3435 | No | NA | 5 | 0 | 1 | Not-determined | Genome-fragment | NA | NA | 0 |
| S3_vs_k141_9337\|\|lt2gene | 1188 | No | NA | 2 | 2 | 0 | Low-quality | Genome-fragment | 1.98 | AAI-based (high-confidence) | 0 |
| S11_vs_k141_72738\|\|full | 7722 | No | NA | 6 | 0 | 3 | Not-determined | Genome-fragment | NA | NA | 0 |
| S5_vs_k141_108212\|\|full | 1188 | No | NA | 3 | 0 | 1 | Low-quality | Genome-fragment | 0.71 | AAI-based (medium-confidence) | 0 |
| S7_extracted_82107 | 1188 | No | NA | 1 | 0 | 0 | Low-quality | Genome-fragment | 2.86 | AAI-based (high-confidence) | 0 |
| S11_vs_k141_79275\|\|full | 35525 | No | NA | 47 | 8 | 3 | High-quality | High-quality | 91.26 | AAI-based (high-confidence) | 0 |
| S11_vs_k141_13652\|\|full | 23079 | No | NA | 37 | 18 | 1 | Medium-quality | Genome-fragment | 63.85 | AAI-based (high-confidence) | 0 |
| S7_vs_23210\|\|full | 1188 | No | NA | 4 | 0 | 0 | Low-quality | Genome-fragment | 2.85 | AAI-based (high-confidence) | 0 |
| S16_extracted_80512 | 1188 | No | NA | 2 | 0 | 0 | Low-quality | Genome-fragment | 2.6 | AAI-based (high-confidence) | 0 |
| S16_extracted_94149 | 1188 | No | NA | 1 | 0 | 0 | Low-quality | Genome-fragment | 1.21 | AAI-based (high-confidence) | 0 |
| S11_vs_k141_27052\|\|full | 1404 | No | NA | 2 | 0 | 1 | Not-determined | Genome-fragment | NA | NA | 0 |
| S11_vs_k141_1270\|\|full | 3222 | No | NA | 3 | 0 | 2 | Not-determined | Genome-fragment | NA | NA | 0 |
| S16_vs_58525\|\|full | 1188 | No | NA | 3 | 0 | 0 | Low-quality | Genome-fragment | 2.87 | AAI-based (medium-confidence) | 0 |
| S12_extracted_k141_140314 | 1188 | No | NA | 1 | 0 | 1 | Low-quality | Genome-fragment | 1.06 | AAI-based (medium-confidence) | 0 |
| S11_vs_k141_20687\|\|full | 3996 | No | NA | 5 | 0 | 2 | Not-determined | Genome-fragment | NA | NA | 0 |
| S12_vs_k141_69120\|\|full | 1188 | No | NA | 1 | 0 | 0 | Low-quality | Genome-fragment | 3.79 | AAI-based (medium-confidence) | 0 |
| S4_extracted_5677 | 1188 | No | NA | 2 | 2 | 0 | Low-quality | Genome-fragment | 3.92 | AAI-based (high-confidence) | 0 |
| S6_extracted_k141_30838 | 1188 | No | NA | 1 | 0 | 1 | Low-quality | Genome-fragment | 0.73 | AAI-based (medium-confidence) | 0 |
| S8_extracted_k141_106538 | 1188 | No | NA | 3 | 0 | 1 | Low-quality | Genome-fragment | 1.86 | AAI-based (medium-confidence) | 0 |
| S11_vs_k141_40119\|\|full | 5231 | No | NA | 4 | 0 | 3 | Not-determined | Genome-fragment | NA | NA | 0 |
| S8_extracted_k141_71536 | 1188 | No | NA | 2 | 0 | 0 | Low-quality | Genome-fragment | 2.82 | AAI-based (medium-confidence) | 0 |
| S8_extracted_k141_4533 | 1188 | No | NA | 3 | 2 | 0 | Low-quality | Genome-fragment | 2.11 | AAI-based (medium-confidence) | 0 |
| S11_vs_k141_13693\|\|full | 6355 | No | NA | 7 | 0 | 1 | Not-determined | Genome-fragment | NA | NA | 0 |
| S11_vs_k141_98793\|\|full | 1600 | No | NA | 3 | 0 | 0 | Not-determined | Genome-fragment | NA | NA | 0 |
| S17_extracted_k141_127838 | 1189 | No | NA | 1 | 0 | 1 | Low-quality | Genome-fragment | 0.83 | AAI-based (high-confidence) | 0 |
| S11_vs_k141_91879\|\|lt2gene | 1189 | No | NA | 1 | 1 | 0 | Low-quality | Genome-fragment | 18.32 | AAI-based (high-confidence) | 0 |
| S14_extracted_k141_67432 | 1189 | No | NA | 2 | 0 | 1 | Low-quality | Genome-fragment | 1.25 | AAI-based (medium-confidence) | 0 |
| S11_vs_k141_7511\|\|full | 1510 | No | NA | 2 | 0 | 0 | Not-determined | Genome-fragment | NA | NA | 0 |
| S16_vs_48620\|\|full | 1189 | No | NA | 2 | 0 | 1 | Low-quality | Genome-fragment | 2.38 | AAI-based (medium-confidence) | 0 |
| S16_vs_74430\|\|full | 1189 | No | NA | 2 | 1 | 0 | Low-quality | Genome-fragment | 0.8 | HMM-based (lower-bound) | 0 |
| S12_extracted_k141_144315 | 1189 | No | NA | 3 | 0 | 0 | Low-quality | Genome-fragment | 0.87 | AAI-based (medium-confidence) | 0 |
| S6_vs_k141_4871\|\|full | 1189 | No | NA | 2 | 0 | 0 | Low-quality | Genome-fragment | 0.71 | AAI-based (high-confidence) | 0 |
| S9_vs_k141_31082\|\|full | 1190 | No | NA | 4 | 0 | 0 | Low-quality | Genome-fragment | 2.92 | AAI-based (high-confidence) | 0 |
| S11_vs_k141_13767\|\|full | 1914 | No | NA | 2 | 0 | 1 | Not-determined | Genome-fragment | NA | NA | 0 |
| S13_extracted_66471 | 1190 | No | NA | 1 | 0 | 0 | Low-quality | Genome-fragment | 2.49 | AAI-based (medium-confidence) | 0 |
| S14_extracted_k141_30633 | 1190 | No | NA | 3 | 0 | 0 | Low-quality | Genome-fragment | 1.8 | AAI-based (high-confidence) | 0 |
| S1_extracted_k141_131566 | 1190 | No | NA | 4 | 0 | 0 | Low-quality | Genome-fragment | 2.56 | AAI-based (medium-confidence) | 0 |
| S1_extracted_k141_139543 | 1190 | No | NA | 2 | 0 | 0 | Low-quality | Genome-fragment | 2.86 | AAI-based (high-confidence) | 0 |
| S3_extracted_k141_20024 | 1190 | No | NA | 2 | 2 | 0 | Low-quality | Genome-fragment | 3.19 | AAI-based (high-confidence) | 0 |
| S3_vs_k141_94117\|\|full | 1190 | No | NA | 4 | 0 | 0 | Low-quality | Genome-fragment | 2.6 | AAI-based (high-confidence) | 0 |
| S11_vs_k141_1393\|\|full | 1236 | No | NA | 3 | 0 | 0 | Not-determined | Genome-fragment | NA | NA | 0 |
| S5_extracted_k141_3517 | 1190 | No | NA | 2 | 2 | 0 | Low-quality | Genome-fragment | 3.16 | AAI-based (high-confidence) | 0 |
| S7_extracted_99074 | 1190 | No | NA | 1 | 0 | 0 | Low-quality | Genome-fragment | 0.53 | AAI-based (high-confidence) | 0 |
| S11_vs_k141_59667\|\|full | 2662 | No | NA | 4 | 0 | 0 | Not-determined | Genome-fragment | NA | NA | 0 |
| S16_extracted_32260 | 1190 | No | NA | 2 | 0 | 0 | Low-quality | Genome-fragment | 1.64 | AAI-based (medium-confidence) | 0 |
| S11_vs_k141_59693\|\|full | 2632 | No | NA | 2 | 0 | 0 | Not-determined | Genome-fragment | NA | NA | 0 |
| S16_vs_133152\|\|full | 1190 | No | NA | 3 | 1 | 0 | Low-quality | Genome-fragment | 1.48 | AAI-based (high-confidence) | 0 |
| S10_vs_34362\|\|full | 1190 | No | NA | 4 | 0 | 0 | Low-quality | Genome-fragment | 1.93 | AAI-based (high-confidence) | 0 |
| S10_vs_66847\|\|full | 1190 | No | NA | 3 | 0 | 0 | Low-quality | Genome-fragment | 1.51 | AAI-based (medium-confidence) | 0 |
| S10_vs_77325\|\|full | 1190 | No | NA | 2 | 0 | 0 | Low-quality | Genome-fragment | 2.2 | AAI-based (medium-confidence) | 0 |
| S8_extracted_k141_78162 | 1190 | No | NA | 3 | 0 | 2 | Low-quality | Genome-fragment | 0.59 | AAI-based (medium-confidence) | 0 |
| S8_vs_k141_196615\|\|full | 1190 | No | NA | 2 | 0 | 2 | Low-quality | Genome-fragment | 0.35 | AAI-based (medium-confidence) | 0 |
| S9_extracted_k141_83091 | 1191 | No | NA | 1 | 0 | 0 | Low-quality | Genome-fragment | 1.71 | AAI-based (medium-confidence) | 0 |
| S9_vs_k141_2321\|\|lt2gene | 1191 | No | NA | 1 | 1 | 0 | Low-quality | Genome-fragment | 3.04 | AAI-based (medium-confidence) | 0 |
| S11_vs_k141_1493\|\|full | 2438 | No | NA | 3 | 0 | 1 | Not-determined | Genome-fragment | NA | NA | 0 |
| S11_vs_k141_66225\|\|full | 2448 | No | NA | 4 | 0 | 0 | Not-determined | Genome-fragment | NA | NA | 0 |
| S11_vs_k141_66230\|\|full | 1175 | No | NA | 3 | 0 | 0 | Not-determined | Genome-fragment | NA | NA | 0 |
| S11_vs_k141_1511\|\|full | 2739 | No | NA | 6 | 0 | 0 | Not-determined | Genome-fragment | NA | NA | 0 |
| S13_vs_97140\|\|full | 1191 | Yes | 378 | 3 | 1 | 1 | Low-quality | Genome-fragment | 0.91 | AAI-based (high-confidence) | 68.26 |
| S14_extracted_k141_81119 | 1191 | No | NA | 5 | 5 | 0 | Low-quality | Genome-fragment | 3.92 | AAI-based (high-confidence) | 0 |
| S1_extracted_k141_965 | 1191 | No | NA | 3 | 0 | 0 | Low-quality | Genome-fragment | 2.14 | AAI-based (medium-confidence) | 0 |
| S1_vs_k141_99502\|\|full | 1191 | No | NA | 3 | 0 | 0 | Low-quality | Genome-fragment | 2.23 | AAI-based (medium-confidence) | 0 |
| S3_vs_k141_104097\|\|full | 1191 | No | NA | 3 | 0 | 0 | Low-quality | Genome-fragment | 3.12 | AAI-based (high-confidence) | 0 |
| S11_vs_k141_86123\|\|full | 3390 | No | NA | 3 | 0 | 1 | Not-determined | Genome-fragment | NA | NA | 0 |
| S11_vs_k141_46770\|\|full | 2725 | No | NA | 6 | 0 | 1 | Not-determined | Genome-fragment | NA | NA | 0 |
| S5_extracted_k141_122928 | 1191 | No | NA | 1 | 0 | 1 | Low-quality | Genome-fragment | 0.4 | AAI-based (medium-confidence) | 0 |
| S11_vs_k141_13996\|\|full | 1462 | No | NA | 1 | 0 | 1 | Not-determined | Genome-fragment | NA | NA | 0 |
| S11_vs_k141_59813\|\|full | 2836 | No | NA | 3 | 0 | 1 | Not-determined | Genome-fragment | NA | NA | 0 |
| S11_vs_k141_73172\|\|full | 6391 | No | NA | 12 | 0 | 0 | Not-determined | Genome-fragment | NA | NA | 0 |
| S11_vs_k141_66326\|\|full | 1626 | No | NA | 3 | 0 | 3 | Not-determined | Genome-fragment | NA | NA | 0 |
| S5_extracted_k141_78386 | 1191 | No | NA | 4 | 0 | 0 | Low-quality | Genome-fragment | 2.78 | AAI-based (high-confidence) | 0 |
| S16_extracted_123553 | 1191 | No | NA | 3 | 0 | 0 | Low-quality | Genome-fragment | 1.57 | AAI-based (high-confidence) | 0 |
| S16_vs_89394\|\|full | 1191 | No | NA | 2 | 0 | 0 | Low-quality | Genome-fragment | 3.05 | AAI-based (medium-confidence) | 0 |
| S11_vs_k141_86204\|\|full | 4969 | No | NA | 7 | 0 | 2 | Not-determined | Genome-fragment | NA | NA | 0 |
| S16_vs_154841\|\|full | 1191 | No | NA | 2 | 0 | 0 | Low-quality | Genome-fragment | 2.4 | AAI-based (high-confidence) | 0 |
| S11_vs_k141_79545\|\|full | 5080 | No | NA | 3 | 0 | 3 | Not-determined | Genome-fragment | NA | NA | 0 |
| S12_vs_k141_20764\|\|full | 1191 | No | NA | 3 | 0 | 0 | Low-quality | Genome-fragment | 2.27 | AAI-based (medium-confidence) | 0 |
| S11_vs_k141_27397\|\|full | 1767 | No | NA | 3 | 0 | 1 | Not-determined | Genome-fragment | NA | NA | 0 |

| | | | | | | | | | | | |
|---|---|---|---|---|---|---|---|---|---|---|---|
| S8_extracted_k141_154703 | 1191 | No | NA | 2 | 2 | 0 | Low-quality | Genome-fragment | 6.09 | AAI-based (high-confidence) | 0 |
| S8_vs_k141_18408\|\|full | 1191 | No | NA | 4 | 2 | 0 | Low-quality | Genome-fragment | 2.32 | AAI-based (medium-confidence) | 0 |
| S11_vs_k141_27404\|\|full | 2695 | No | NA | 3 | 0 | 0 | Not-determined | Genome-fragment | NA | NA | 0 |
| S17_extracted_k141_171577 | 1192 | No | NA | 1 | 1 | 0 | Low-quality | Genome-fragment | 3.2 | AAI-based (high-confidence) | 0 |
| S17_extracted_k141_64912 | 1192 | No | NA | 4 | 0 | 0 | Low-quality | Genome-fragment | 2.64 | AAI-based (high-confidence) | 0 |
| S11_vs_k141_51872\|\|full | 1192 | No | NA | 3 | 1 | 0 | Low-quality | Genome-fragment | 2.8 | AAI-based (high-confidence) | 0 |
| S14_extracted_k141_95824 | 1192 | No | NA | 2 | 0 | 0 | Low-quality | Genome-fragment | 3.68 | AAI-based (high-confidence) | 0 |
| S11_vs_k141_86317\|\|full | 6858 | No | NA | 7 | 0 | 1 | Not-determined | Genome-fragment | NA | NA | 0 |
| S14_extracted_k141_50131 | 1192 | No | NA | 1 | 0 | 0 | Low-quality | Genome-fragment | 2.78 | AAI-based (medium-confidence) | 0 |
| S5_vs_k141_101905\|\|full | 1192 | No | NA | 4 | 0 | 0 | Low-quality | Genome-fragment | 3.18 | AAI-based (high-confidence) | 0 |
| S11_vs_k141_73247\|\|full | 2646 | No | NA | 4 | 0 | 2 | Not-determined | Genome-fragment | NA | NA | 0 |
| S5_vs_k141_131114\|\|lt2gene | 1192 | No | NA | 1 | 1 | 0 | Low-quality | Genome-fragment | 2.61 | AAI-based (medium-confidence) | 0 |
| S7_vs_62320\|\|full | 1192 | No | NA | 2 | 1 | 0 | Low-quality | Genome-fragment | 1.99 | AAI-based (high-confidence) | 0 |
| S11_vs_k141_40397\|\|full | 1978 | No | NA | 2 | 0 | 2 | Not-determined | Genome-fragment | NA | NA | 0 |
| S11_vs_k141_1731\|\|full | 1203 | No | NA | 3 | 0 | 2 | Not-determined | Genome-fragment | NA | NA | 0 |
| S10_vs_62678\|\|full | 1192 | No | NA | 3 | 0 | 1 | Low-quality | Genome-fragment | 1.55 | AAI-based (medium-confidence) | 0 |
| S11_vs_k141_99088\|\|full | 1254 | No | NA | 3 | 0 | 0 | Not-determined | Genome-fragment | NA | NA | 0 |
| S6_vs_k141_54231\|\|full | 1192 | No | NA | 3 | 0 | 0 | Low-quality | Genome-fragment | 0.78 | AAI-based (high-confidence) | 0 |
| S11_vs_k141_40421\|\|full | 2754 | No | NA | 4 | 0 | 1 | Not-determined | Genome-fragment | NA | NA | 0 |
| S11_vs_k141_66411\|\|full | 1880 | No | NA | 3 | 0 | 0 | Not-determined | Genome-fragment | NA | NA | 0 |
| S11_vs_k141_14202\|\|full | 1121 | No | NA | 2 | 0 | 0 | Not-determined | Genome-fragment | NA | NA | 0 |
| S8_extracted_k141_61476 | 1192 | No | NA | 2 | 0 | 0 | Low-quality | Genome-fragment | 3.09 | AAI-based (medium-confidence) | 0 |
| S11_vs_k141_21120\|\|full | 2540 | No | NA | 3 | 0 | 1 | Not-determined | Genome-fragment | NA | NA | 0 |
| S8_vs_k141_132707\|\|full | 1192 | No | NA | 3 | 0 | 0 | Low-quality | Genome-fragment | 2.62 | AAI-based (high-confidence) | 0 |
| S11_vs_k141_46940\|\|full | 3866 | No | NA | 5 | 0 | 0 | Not-determined | Genome-fragment | NA | NA | 0 |
| S11_vs_k141_73316\|\|full | 2989 | No | NA | 3 | 0 | 2 | Not-determined | Genome-fragment | NA | NA | 0 |
| S11_vs_k141_93090\|\|lt2gene | 1193 | No | NA | 1 | 1 | 0 | Low-quality | Genome-fragment | 2.12 | HMM-based (lower-bound) | 0 |
| S11_vs_k141_79712\|\|full | 1603 | No | NA | 3 | 0 | 1 | Not-determined | Genome-fragment | NA | NA | 0 |
| S11_vs_k141_21189\|\|full | 1599 | No | NA | 3 | 0 | 0 | Not-determined | Genome-fragment | NA | NA | 0 |
| S11_vs_k141_99152\|\|full | 1242 | No | NA | 3 | 0 | 0 | Not-determined | Genome-fragment | NA | NA | 0 |
| S14_extracted_k141_97583 | 1193 | No | NA | 2 | 1 | 0 | Low-quality | Genome-fragment | 4.11 | HMM-based (lower-bound) | 0 |
| S11_vs_k141_79756\|\|full | 1939 | No | NA | 2 | 0 | 2 | Not-determined | Genome-fragment | NA | NA | 0 |
| S14_vs_k141_19691\|\|full | 1193 | No | NA | 2 | 0 | 0 | Low-quality | Genome-fragment | 3.01 | AAI-based (high-confidence) | 0 |
| S3_extracted_k141_25389 | 1193 | No | NA | 2 | 2 | 0 | Low-quality | Genome-fragment | 3.8 | AAI-based (high-confidence) | 0 |
| S11_vs_k141_14357\|\|full | 2431 | No | NA | 2 | 0 | 1 | Not-determined | Genome-fragment | NA | NA | 0 |
| S11_vs_k141_1899\|\|full | 5275 | No | NA | 4 | 0 | 4 | Not-determined | Genome-fragment | NA | NA | 0 |
| S3_extracted_k141_103569 | 1193 | No | NA | 2 | 1 | 0 | Low-quality | Genome-fragment | 0.78 | AAI-based (medium-confidence) | 0 |
| S3_vs_k141_118817\|\|full | 1193 | No | NA | 5 | 1 | 0 | Low-quality | Genome-fragment | 2.67 | AAI-based (high-confidence) | 0 |
| S11_vs_k141_53675\|\|full | 20682 | No | NA | 31 | 9 | 2 | Medium-quality | Genome-fragment | 57.33 | AAI-based (medium-confidence) | 0 |
| S11_vs_k141_27717\|\|full | 1172 | No | NA | 3 | 0 | 0 | Not-determined | Genome-fragment | NA | NA | 0 |
| S11_vs_k141_14383\|\|full | 1752 | No | NA | 3 | 0 | 1 | Not-determined | Genome-fragment | NA | NA | 0 |
| S11_vs_k141_21281\|\|full | 1985 | No | NA | 3 | 0 | 1 | Not-determined | Genome-fragment | NA | NA | 0 |
| S11_vs_k141_53702\|\|full | 1891 | No | NA | 2 | 0 | 1 | Not-determined | Genome-fragment | NA | NA | 0 |
| S5_extracted_k141_4128 | 1193 | No | NA | 2 | 0 | 0 | Low-quality | Genome-fragment | 2.39 | AAI-based (medium-confidence) | 0 |
| S5_extracted_k141_64239 | 1193 | No | NA | 3 | 2 | 0 | Low-quality | Genome-fragment | 0.62 | HMM-based (lower-bound) | 0 |
| S5_vs_k141_93698\|\|full | 1193 | No | NA | 3 | 0 | 0 | Low-quality | Genome-fragment | 2.24 | AAI-based (medium-confidence) | 0 |
| S7_vs_109186\|\|full | 1193 | No | NA | 3 | 0 | 0 | Low-quality | Genome-fragment | 2.85 | AAI-based (high-confidence) | 0 |
| S7_vs_137050\|\|lt2gene | 1193 | No | NA | 1 | 1 | 0 | Low-quality | Genome-fragment | 2.13 | AAI-based (medium-confidence) | 0 |
| S16_extracted_91382 | 1193 | No | NA | 3 | 0 | 0 | Low-quality | Genome-fragment | 0.87 | AAI-based (medium-confidence) | 0 |
| S11_vs_k141_60051\|\|full | 2152 | No | NA | 4 | 0 | 1 | Not-determined | Genome-fragment | NA | NA | 0 |
| S10_extracted_50806 | 1193 | No | NA | 1 | 1 | 0 | Low-quality | Genome-fragment | 2.58 | AAI-based (high-confidence) | 0 |
| S12_extracted_k141_159636 | 1193 | No | NA | 3 | 0 | 0 | Low-quality | Genome-fragment | 2.73 | AAI-based (medium-confidence) | 0 |
| S15_vs_7036\|\|lt2gene | 1193 | No | NA | 2 | 1 | 0 | Low-quality | Genome-fragment | 1.9 | AAI-based (high-confidence) | 0 |
| S11_vs_k141_47131\|\|full | 1286 | No | NA | 2 | 0 | 0 | Not-determined | Genome-fragment | NA | NA | 0 |
| S11_vs_k141_79831\|\|full | 40938 | No | NA | 72 | 6 | 3 | Medium-quality | Genome-fragment | 72.66 | AAI-based (high-confidence) | 0 |
| S8_extracted_k141_46933 | 1193 | No | NA | 4 | 1 | 0 | Low-quality | Genome-fragment | 1.21 | AAI-based (high-confidence) | 0 |
| S11_vs_k141_1937\|\|full | 35578 | Yes | 32094 | 46 | 5 | 3 | Medium-quality | Genome-fragment | 55.12 | AAI-based (high-confidence) | 9.79 |
| S8_vs_k141_147582\|\|full | 1193 | No | NA | 3 | 0 | 1 | Low-quality | Genome-fragment | 1.06 | AAI-based (medium-confidence) | 0 |
| S17_vs_k141_153854\|\|lt2gene | 1194 | No | NA | 2 | 0 | 0 | Low-quality | Genome-fragment | 2.78 | AAI-based (high-confidence) | 0 |
| S17_vs_k141_176021\|\|lt2gene | 1194 | No | NA | 1 | 1 | 0 | Low-quality | Genome-fragment | 3.75 | AAI-based (medium-confidence) | 0 |
| S11_vs_k141_21394\|\|full | 8478 | No | NA | 10 | 0 | 5 | Not-determined | Genome-fragment | NA | NA | 0 |
| S11_vs_k141_64419\|\|full | 1194 | No | NA | 3 | 1 | 0 | Low-quality | Genome-fragment | 2.97 | HMM-based (lower-bound) | 0 |
| S14_extracted_k141_83317 | 1194 | No | NA | 3 | 1 | 0 | Low-quality | Genome-fragment | 2.18 | AAI-based (medium-confidence) | 0 |
| S11_vs_k141_53818\|\|full | 2374 | No | NA | 3 | 0 | 0 | Not-determined | Genome-fragment | NA | NA | 0 |
| S11_vs_k141_66652\|\|full | 3285 | No | NA | 4 | 0 | 1 | Not-determined | Genome-fragment | NA | NA | 0 |
| S11_vs_k141_60106\|\|full | 1680 | No | NA | 2 | 0 | 0 | Not-determined | Genome-fragment | NA | NA | 0 |
| S3_extracted_k141_110651 | 1194 | No | NA | 1 | 1 | 0 | Low-quality | Genome-fragment | 0.85 | AAI-based (high-confidence) | 0 |
| S11_vs_k141_1976\|\|full | 3017 | No | NA | 3 | 0 | 2 | Not-determined | Genome-fragment | NA | NA | 0 |
| S5_vs_k141_44105\|\|lt2gene | 1194 | No | NA | 2 | 2 | 0 | Low-quality | Genome-fragment | 1.74 | HMM-based (lower-bound) | 0 |
| S11_vs_k141_73489\|\|full | 2445 | No | NA | 3 | 0 | 3 | Not-determined | Genome-fragment | NA | NA | 0 |
| S11_vs_k141_66673\|\|full | 5522 | No | NA | 3 | 0 | 3 | Not-determined | Genome-fragment | NA | NA | 0 |
| S11_vs_k141_86484\|\|full | 3508 | No | NA | 3 | 0 | 2 | Not-determined | Genome-fragment | NA | NA | 0 |
| S16_extracted_52725 | 1194 | No | NA | 3 | 0 | 0 | Low-quality | Genome-fragment | 2.91 | AAI-based (high-confidence) | 0 |
| S11_vs_k141_99330\|\|full | 2707 | No | NA | 4 | 0 | 1 | Not-determined | Genome-fragment | NA | NA | 0 |
| S11_vs_k141_53878\|\|full | 1038 | No | NA | 2 | 0 | 0 | Not-determined | Genome-fragment | NA | NA | 0 |
| S11_vs_k141_92722\|\|full | 4325 | No | NA | 5 | 0 | 2 | Not-determined | Genome-fragment | NA | NA | 0 |
| S11_vs_k141_47188\|\|full | 2317 | No | NA | 5 | 0 | 1 | Not-determined | Genome-fragment | NA | NA | 0 |
| S16_extracted_57774 | 1194 | No | NA | 4 | 0 | 0 | Low-quality | Genome-fragment | 0.59 | AAI-based (high-confidence) | 0 |
| S11_vs_k141_53886\|\|full | 4505 | No | NA | 6 | 0 | 0 | Not-determined | Genome-fragment | NA | NA | 0 |
| S11_vs_k141_21461\|\|full | 4894 | No | NA | 9 | 0 | 0 | Not-determined | Genome-fragment | NA | NA | 0 |
| S10_extracted_102235 | 1194 | No | NA | 2 | 0 | 0 | Low-quality | Genome-fragment | 2.02 | AAI-based (high-confidence) | 0 |
| S11_vs_k141_2034\|\|full | 1493 | No | NA | 3 | 0 | 1 | Not-determined | Genome-fragment | NA | NA | 0 |
| S12_extracted_k141_114951 | 1194 | No | NA | 1 | 0 | 0 | Low-quality | Genome-fragment | 1.07 | AAI-based (high-confidence) | 0 |
| S11_vs_k141_33950\|\|full | 2392 | No | NA | 2 | 0 | 2 | Not-determined | Genome-fragment | NA | NA | 0 |
| S11_vs_k141_79921\|\|full | 1300 | No | NA | 2 | 0 | 0 | Not-determined | Genome-fragment | NA | NA | 0 |
| S11_vs_k141_92769\|\|full | 1220 | No | NA | 2 | 0 | 0 | Not-determined | Genome-fragment | NA | NA | 0 |
| S12_vs_k141_64858\|\|lt2gene | 1194 | No | NA | 2 | 1 | 0 | Low-quality | Genome-fragment | 2.29 | AAI-based (medium-confidence) | 0 |
| S11_vs_k141_73576\|\|full | 1293 | No | NA | 2 | 0 | 1 | Not-determined | Genome-fragment | NA | NA | 0 |
| S15_extracted_37891 | 1194 | No | NA | 3 | 1 | 0 | Low-quality | Genome-fragment | 1.49 | HMM-based (lower-bound) | 0 |
| S11_vs_k141_86583\|\|full | 1848 | No | NA | 3 | 0 | 0 | Not-determined | Genome-fragment | NA | NA | 0 |
| S8_extracted_k141_177800 | 1194 | No | NA | 4 | 0 | 2 | Low-quality | Genome-fragment | 0.69 | AAI-based (medium-confidence) | 0 |
| S17_vs_k141_156381\|\|full | 1195 | No | NA | 3 | 0 | 0 | Low-quality | Genome-fragment | 1.7 | AAI-based (high-confidence) | 0 |
| S11_vs_k141_21552\|\|full | 4212 | No | NA | 4 | 0 | 2 | Not-determined | Genome-fragment | NA | NA | 0 |
| S11_vs_k141_86617\|\|full | 4112 | No | NA | 3 | 0 | 2 | Not-determined | Genome-fragment | NA | NA | 0 |
| S17_vs_k141_127297\|\|full | 1195 | No | NA | 3 | 1 | 0 | Low-quality | Genome-fragment | 1.38 | HMM-based (lower-bound) | 0 |
| S11_vs_k141_86626\|\|full | 1996 | No | NA | 4 | 0 | 2 | Not-determined | Genome-fragment | NA | NA | 0 |
| S11_vs_k141_54007\|\|full | 1790 | No | NA | 4 | 0 | 0 | Not-determined | Genome-fragment | NA | NA | 0 |
| S9_extracted_k141_88072 | 1195 | No | NA | 4 | 0 | 0 | Low-quality | Genome-fragment | 2.29 | AAI-based (medium-confidence) | 0 |
| S11_vs_k141_23719\|\|full | 1195 | No | NA | 3 | 2 | 0 | Low-quality | Genome-fragment | 2.37 | AAI-based (high-confidence) | 0 |
| S13_extracted_122087 | 1195 | No | NA | 2 | 0 | 1 | Low-quality | Genome-fragment | 0.29 | AAI-based (medium-confidence) | 0 |
| S13_vs_16317\|\|full | 1195 | No | NA | 2 | 0 | 0 | Low-quality | Genome-fragment | 1.99 | AAI-based (medium-confidence) | 0 |
| S13_vs_16798\|\|full | 1195 | No | NA | 3 | 0 | 1 | Low-quality | Genome-fragment | 3.1 | AAI-based (medium-confidence) | 0 |
| S14_vs_k141_55971\|\|full | 1195 | No | NA | 5 | 1 | 0 | Low-quality | Genome-fragment | 0.89 | AAI-based (medium-confidence) | 0 |
| S3_vs_k141_59012\|\|full | 1195 | No | NA | 3 | 2 | 0 | Low-quality | Genome-fragment | 3.4 | AAI-based (high-confidence) | 0 |
| S11_vs_k141_73621\|\|full | 1524 | No | NA | 2 | 0 | 1 | Not-determined | Genome-fragment | NA | NA | 0 |
| S5_vs_k141_97034\|\|full | 1195 | No | NA | 3 | 0 | 0 | Low-quality | Genome-fragment | 0.77 | AAI-based (medium-confidence) | 0 |
| S5_vs_k141_45148\|\|lt2gene | 1195 | No | NA | 2 | 1 | 0 | Low-quality | Genome-fragment | 2.6 | AAI-based (medium-confidence) | 0 |
| S11_vs_k141_86682\|\|full | 1098 | No | NA | 2 | 0 | 0 | Not-determined | Genome-fragment | NA | NA | 0 |
| S11_vs_k141_80057\|\|full | 6788 | No | NA | 12 | 0 | 2 | Not-determined | Genome-fragment | NA | NA | 0 |
| S11_vs_k141_99505\|\|full | 3263 | No | NA | 4 | 0 | 4 | Not-determined | Genome-fragment | NA | NA | 0 |
| S11_vs_k141_40762\|\|full | 2864 | No | NA | 3 | 0 | 3 | Not-determined | Genome-fragment | NA | NA | 0 |
| S7_extracted_76225 | 1195 | No | NA | 2 | 0 | 0 | Low-quality | Genome-fragment | 1.8 | AAI-based (high-confidence) | 0 |
| S11_vs_k141_92845\|\|full | 1288 | No | NA | 3 | 0 | 0 | Not-determined | Genome-fragment | NA | NA | 0 |
| S7_vs_46246\|\|full | 1195 | No | NA | 2 | 0 | 1 | Low-quality | Genome-fragment | 0.4 | AAI-based (medium-confidence) | 0 |
| S10_vs_37263\|\|lt2gene | 1195 | No | NA | 2 | 1 | 0 | Low-quality | Genome-fragment | 1.75 | HMM-based (lower-bound) | 0 |
| S11_vs_k141_8456\|\|full | 36244 | No | NA | 55 | 19 | 0 | High-quality | High-quality | 98.52 | AAI-based (medium-confidence) | 0 |
| S11_vs_k141_14610\|\|full | 2102 | No | NA | 3 | 0 | 3 | Not-determined | Genome-fragment | NA | NA | 0 |
| S12_extracted_k141_67938 | 1195 | No | NA | 2 | 0 | 1 | Low-quality | Genome-fragment | 2.7 | AAI-based (high-confidence) | 0 |
| S12_vs_k141_39535\|\|full | 1195 | No | NA | 3 | 0 | 2 | Low-quality | Genome-fragment | 1.1 | AAI-based (medium-confidence) | 0 |
| S12_vs_k141_51935\|\|full | 1195 | No | NA | 3 | 2 | 0 | Low-quality | Genome-fragment | 3.37 | AAI-based (high-confidence) | 0 |
| S11_vs_k141_2226\|\|full | 1458 | No | NA | 3 | 0 | 0 | Not-determined | Genome-fragment | NA | NA | 0 |
| S11_vs_k141_14655\|\|full | 1957 | No | NA | 3 | 0 | 0 | Not-determined | Genome-fragment | NA | NA | 0 |
| S11_vs_k141_8498\|\|full | 2540 | No | NA | 2 | 0 | 0 | Not-determined | Genome-fragment | NA | NA | 0 |
| S11_vs_k141_92917\|\|full | 2788 | No | NA | 3 | 0 | 1 | Not-determined | Genome-fragment | NA | NA | 0 |
| S11_vs_k141_40869\|\|full | 11467 | No | NA | 15 | 0 | 3 | Not-determined | Genome-fragment | NA | NA | 0 |
| S11_vs_k141_34100\|\|full | 3751 | No | NA | 5 | 0 | 0 | Not-determined | Genome-fragment | NA | NA | 0 |
| S11_vs_k141_86785\|\|full | 1612 | No | NA | 3 | 0 | 1 | Not-determined | Genome-fragment | NA | NA | 0 |
| S8_vs_k141_184279\|\|full | 1195 | No | NA | 3 | 0 | 2 | Low-quality | Genome-fragment | 0.35 | AAI-based (medium-confidence) | 0 |
| S11_vs_k141_86792\|\|full | 1085 | No | NA | 1 | 0 | 1 | Not-determined | Genome-fragment | NA | NA | 0 |
| S17_vs_k141_159026\|\|full | 1196 | No | NA | 4 | 0 | 1 | Low-quality | Genome-fragment | 2.21 | AAI-based (medium-confidence) | 0 |
| S11_extracted_k141_8986 | 1196 | No | NA | 2 | 2 | 0 | Low-quality | Genome-fragment | 1.93 | AAI-based (medium-confidence) | 0 |
| S11_vs_k141_14723\|\|full | 1439 | No | NA | 3 | 0 | 0 | Not-determined | Genome-fragment | NA | NA | 0 |
| S11_vs_k141_40939\|\|full | 4008 | No | NA | 4 | 0 | 3 | Not-determined | Genome-fragment | NA | NA | 0 |
| S13_vs_16730\|\|full | 1196 | No | NA | 2 | 0 | 0 | Low-quality | Genome-fragment | 2.33 | AAI-based (medium-confidence) | 0 |
| S11_vs_k141_86849\|\|full | 1507 | No | NA | 3 | 0 | 0 | Not-determined | Genome-fragment | NA | NA | 0 |
| S3_extracted_k141_79465 | 1196 | No | NA | 4 | 4 | 0 | Low-quality | Genome-fragment | 3.27 | AAI-based (high-confidence) | 0 |
| S3_vs_k141_59728\|\|full | 1196 | No | NA | 4 | 0 | 0 | Low-quality | Genome-fragment | 1.28 | AAI-based (medium-confidence) | 0 |
| S5_extracted_k141_67576 | 1196 | No | NA | 3 | 1 | 0 | Low-quality | Genome-fragment | 2.3 | AAI-based (medium-confidence) | 0 |
| S5_vs_k141_109565\|\|full | 1196 | No | NA | 2 | 0 | 0 | Low-quality | Genome-fragment | 2.51 | AAI-based (high-confidence) | 0 |
| S11_vs_k141_34137\|\|full | 35941 | No | NA | 54 | 18 | 0 | High-quality | High-quality | 100 | AAI-based (high-confidence) | 0 |
| S16_extracted_123987 | 1196 | No | NA | 2 | 0 | 1 | Low-quality | Genome-fragment | 0.75 | AAI-based (high-confidence) | 0 |
| S16_vs_20735\|\|full | 1196 | No | NA | 3 | 2 | 0 | Low-quality | Genome-fragment | 1.9 | AAI-based (medium-confidence) | 0 |
| S11_vs_k141_86885\|\|full | 1638 | No | NA | 3 | 0 | 0 | Not-determined | Genome-fragment | NA | NA | 0 |
| S16_vs_81322\|\|full | 1196 | No | NA | 2 | 0 | 0 | Low-quality | Genome-fragment | 1.46 | AAI-based (high-confidence) | 0 |
| S16_vs_153413\|\|full | 1196 | No | NA | 4 | 0 | 0 | Low-quality | Genome-fragment | 2.79 | AAI-based (medium-confidence) | 0 |
| S12_extracted_k141_48502 | 1196 | No | NA | 5 | 0 | 0 | Low-quality | Genome-fragment | 1.87 | AAI-based (high-confidence) | 0 |
| S11_vs_k141_60347\|\|full | 12444 | No | NA | 14 | 0 | 6 | Not-determined | Genome-fragment | NA | NA | 0 |
| S15_vs_31640\|\|full | 1196 | No | NA | 5 | 0 | 0 | Low-quality | Genome-fragment | 1.96 | AAI-based (high-confidence) | 0 |
| S11_vs_k141_21765\|\|full | 6186 | No | NA | 8 | 0 | 1 | Not-determined | Genome-fragment | NA | NA | 0 |
| S11_vs_k141_86938\|\|full | 1002 | No | NA | 2 | 0 | 0 | Not-determined | Genome-fragment | NA | NA | 0 |
| S11_vs_k141_73858\|\|full | 2205 | No | NA | 5 | 0 | 2 | Not-determined | Genome-fragment | NA | NA | 0 |
| S11_vs_k141_21782\|\|full | 1073 | No | NA | 2 | 0 | 0 | Not-determined | Genome-fragment | NA | NA | 0 |
| S6_vs_k141_17099\|\|full | 1196 | No | NA | 2 | 2 | 0 | Low-quality | Genome-fragment | 2.74 | AAI-based (high-confidence) | 0 |
| S8_extracted_k141_181481 | 1196 | No | NA | 1 | 1 | 0 | Low-quality | Genome-fragment | 1.08 | HMM-based (lower-bound) | 0 |
| S8_extracted_k141_34286 | 1196 | No | NA | 3 | 0 | 0 | Low-quality | Genome-fragment | 2.82 | AAI-based (medium-confidence) | 0 |
| S11_vs_k141_28075\|\|full | 4381 | No | NA | 4 | 0 | 2 | Not-determined | Genome-fragment | NA | NA | 0 |
| S8_vs_k141_89209\|\|full | 1196 | No | NA | 3 | 0 | 0 | Low-quality | Genome-fragment | 1.47 | AAI-based (medium-confidence) | 0 |
| S9_vs_k141_37532\|\|full | 1197 | No | NA | 3 | 0 | 0 | Low-quality | Genome-fragment | 0.76 | AAI-based (medium-confidence) | 0 |
| S11_vs_k141_73902\|\|full | 1286 | No | NA | 2 | 0 | 0 | Not-determined | Genome-fragment | NA | NA | 0 |
| S13_vs_27135\|\|full | 1197 | No | NA | 3 | 1 | 0 | Low-quality | Genome-fragment | 2.4 | AAI-based (high-confidence) | 0 |
| S14_extracted_k141_752 | 1197 | No | NA | 5 | 5 | 0 | Low-quality | Genome-fragment | 3.94 | AAI-based (high-confidence) | 0 |
| S11_vs_k141_2427\|\|full | 2363 | No | NA | 3 | 0 | 1 | Not-determined | Genome-fragment | NA | NA | 0 |
| S14_extracted_k141_22560 | 1197 | No | NA | 2 | 2 | 0 | Low-quality | Genome-fragment | 3.95 | AAI-based (high-confidence) | 0 |
| S14_extracted_k141_9403 | 1197 | No | NA | 1 | 1 | 0 | Low-quality | Genome-fragment | 3.83 | AAI-based (high-confidence) | 0 |
| S5_extracted_k141_22260 | 1197 | No | NA | 3 | 0 | 0 | Low-quality | Genome-fragment | 1.56 | AAI-based (medium-confidence) | 0 |
| S11_vs_k141_21871\|\|full | 2329 | No | NA | 2 | 0 | 1 | Not-determined | Genome-fragment | NA | NA | 0 |
| S7_extracted_8646 | 1197 | No | NA | 4 | 1 | 0 | Low-quality | Genome-fragment | 3.65 | AAI-based (high-confidence) | 0 |
| S7_extracted_25302 | 1197 | No | NA | 1 | 0 | 0 | Low-quality | Genome-fragment | 1.87 | AAI-based (high-confidence) | 0 |
| S11_vs_k141_93139\|\|full | 1470 | No | NA | 2 | 0 | 0 | Not-determined | Genome-fragment | NA | NA | 0 |
| S7_vs_108610\|\|full | 1197 | No | NA | 1 | 0 | 1 | Low-quality | Genome-fragment | 0.69 | AAI-based (medium-confidence) | 0 |
| S7_vs_119473\|\|full | 1197 | No | NA | 3 | 1 | 0 | Low-quality | Genome-fragment | 3.07 | AAI-based (high-confidence) | 0 |
| S16_vs_159890\|\|full | 1197 | No | NA | 3 | 0 | 0 | Low-quality | Genome-fragment | 0.59 | AAI-based (medium-confidence) | 0 |
| S11_vs_k141_14882\|\|full | 38803 | No | NA | 49 | 9 | 0 | Medium-quality | Genome-fragment | 64.08 | HMM-based (lower-bound) | 0 |
| S12_extracted_k141_42999 | 1197 | No | NA | 1 | 0 | 1 | Low-quality | Genome-fragment | 0.49 | AAI-based (high-confidence) | 0 |
| S11_vs_k141_34301\|\|full | 1855 | No | NA | 3 | 0 | 2 | Not-determined | Genome-fragment | NA | NA | 0 |
| S11_vs_k141_41107\|\|full | 37369 | No | NA | 49 | 31 | 0 | Medium-quality | Genome-fragment | 81.54 | AAI-based (high-confidence) | 0 |
| S11_vs_k141_8752\|\|full | 3137 | No | NA | 3 | 0 | 2 | Not-determined | Genome-fragment | NA | NA | 0 |
| S11_vs_k141_41131\|\|full | 3159 | No | NA | 6 | 0 | 0 | Not-determined | Genome-fragment | NA | NA | 0 |
| S11_vs_k141_80399\|\|full | 1938 | No | NA | 2 | 0 | 0 | Not-determined | Genome-fragment | NA | NA | 0 |
| S11_vs_k141_93204\|\|full | 2320 | No | NA | 4 | 0 | 2 | Not-determined | Genome-fragment | NA | NA | 0 |
| S6_vs_k141_45493\|\|full | 1197 | No | NA | 3 | 0 | 1 | Low-quality | Genome-fragment | 1.29 | AAI-based (medium-confidence) | 0 |
| S8_vs_k141_98928\|\|full | 1197 | No | NA | 3 | 0 | 1 | Low-quality | Genome-fragment | 1.24 | AAI-based (high-confidence) | 0 |
| S8_vs_k141_203715\|\|lt2gene | 1197 | No | NA | 1 | 1 | 0 | Low-quality | Genome-fragment | 1.75 | HMM-based (lower-bound) | 0 |
| S11_vs_k141_47490\|\|full | 1179 | No | NA | 3 | 0 | 2 | Not-determined | Genome-fragment | NA | NA | 0 |
| S17_vs_k141_93394\|\|lt2gene | 1198 | No | NA | 2 | 1 | 0 | Low-quality | Genome-fragment | 3.23 | AAI-based (high-confidence) | 0 |
| S11_vs_k141_80417\|\|full | 3607 | No | NA | 4 | 0 | 1 | Not-determined | Genome-fragment | NA | NA | 0 |
| S9_extracted_k141_36525 | 1198 | No | NA | 3 | 0 | 1 | Low-quality | Genome-fragment | 0.95 | AAI-based (high-confidence) | 0 |
| S11_vs_k141_97757\|\|full | 1198 | No | NA | 3 | 0 | 0 | Low-quality | Genome-fragment | 0.73 | AAI-based (high-confidence) | 0 |
| S11_vs_k141_93269\|\|full | 1542 | No | NA | 2 | 0 | 0 | Not-determined | Genome-fragment | NA | NA | 0 |
| S5_vs_k141_86223\|\|full | 1198 | No | NA | 3 | 3 | 0 | Low-quality | Genome-fragment | 2.94 | AAI-based (medium-confidence) | 0 |
| S11_vs_k141_15052\|\|full | 1590 | No | NA | 3 | 0 | 2 | Not-determined | Genome-fragment | NA | NA | 0 |
| S11_vs_k141_87244\|\|full | 1549 | No | NA | 3 | 0 | 3 | Not-determined | Genome-fragment | NA | NA | 0 |
| S11_vs_k141_47561\|\|full | 3902 | No | NA | 4 | 0 | 0 | Not-determined | Genome-fragment | NA | NA | 0 |
| S7_extracted_15649 | 1198 | No | NA | 2 | 1 | 0 | Low-quality | Genome-fragment | 2.27 | HMM-based (lower-bound) | 0 |
| S11_vs_k141_8884\|\|full | 4177 | No | NA | 7 | 0 | 4 | Not-determined | Genome-fragment | NA | NA | 0 |
| S16_extracted_131919 | 1198 | No | NA | 2 | 0 | 0 | Low-quality | Genome-fragment | 0.93 | AAI-based (high-confidence) | 0 |
| S16_extracted_149231 | 1198 | No | NA | 2 | 1 | 0 | Low-quality | Genome-fragment | 2.03 | AAI-based (high-confidence) | 0 |
| S11_vs_k141_60540\|\|full | 1163 | No | NA | 2 | 0 | 0 | Not-determined | Genome-fragment | NA | NA | 0 |
| S16_vs_48369\|\|full | 1198 | No | NA | 3 | 3 | 0 | Low-quality | Genome-fragment | 2.28 | AAI-based (medium-confidence) | 0 |
| S16_vs_68179\|\|full | 1198 | No | NA | 2 | 1 | 0 | Low-quality | Genome-fragment | 1.99 | AAI-based (high-confidence) | 0 |
| S16_vs_99345\|\|lt2gene | 1198 | No | NA | 1 | 1 | 0 | Low-quality | Genome-fragment | 0.64 | HMM-based (lower-bound) | 0 |
| S11_vs_k141_67052\|\|full | 1187 | No | NA | 2 | 0 | 1 | Not-determined | Genome-fragment | NA | NA | 0 |
| S10_extracted_90086 | 1198 | No | NA | 3 | 0 | 0 | Low-quality | Genome-fragment | 0.9 | AAI-based (high-confidence) | 0 |
| S12_extracted_k141_106885 | 1198 | No | NA | 1 | 0 | 0 | Low-quality | Genome-fragment | 1.87 | AAI-based (medium-confidence) | 0 |
| S12_vs_k141_113484\|\|full | 1198 | No | NA | 3 | 1 | 0 | Low-quality | Genome-fragment | 2.09 | AAI-based (medium-confidence) | 0 |
| S15_vs_4774\|\|full | 1198 | No | NA | 4 | 1 | 0 | Low-quality | Genome-fragment | 2.05 | AAI-based (medium-confidence) | 0 |
| S11_vs_k141_22001\|\|full | 3752 | No | NA | 3 | 0 | 2 | Not-determined | Genome-fragment | NA | NA | 0 |
| S8_vs_k141_72213\|\|full | 1198 | No | NA | 2 | 0 | 1 | Low-quality | Genome-fragment | 1.9 | AAI-based (medium-confidence) | 0 |
| S8_vs_k141_16012\|\|full | 1198 | No | NA | 4 | 1 | 0 | Low-quality | Genome-fragment | 2.19 | AAI-based (high-confidence) | 0 |
| S11_extracted_k141_95392 | 1199 | No | NA | 1 | 1 | 0 | Low-quality | Genome-fragment | 27.31 | AAI-based (medium-confidence) | 0 |
| S13_vs_36497\|\|full | 1199 | No | NA | 2 | 0 | 1 | Low-quality | Genome-fragment | 2.96 | AAI-based (high-confidence) | 0 |
| S1_extracted_k141_30281 | 1199 | No | NA | 2 | 1 | 0 | Low-quality | Genome-fragment | 2.87 | AAI-based (medium-confidence) | 0 |
| S1_extracted_k141_111936 | 1199 | No | NA | 2 | 0 | 0 | Low-quality | Genome-fragment | 1.33 | AAI-based (medium-confidence) | 0 |
| S1_vs_k141_81723\|\|full | 1199 | No | NA | 3 | 0 | 0 | Low-quality | Genome-fragment | 0.93 | AAI-based (medium-confidence) | 0 |
| S3_vs_k141_46577\|\|lt2gene | 1199 | No | NA | 2 | 1 | 0 | Low-quality | Genome-fragment | 3.55 | AAI-based (high-confidence) | 0 |
| S11_vs_k141_54639\|\|full | 1771 | No | NA | 4 | 0 | 3 | Not-determined | Genome-fragment | NA | NA | 0 |
| S5_vs_k141_101688\|\|full | 1199 | No | NA | 3 | 2 | 0 | Low-quality | Genome-fragment | 3.2 | AAI-based (medium-confidence) | 0 |
| S7_vs_22199\|\|full | 1199 | No | NA | 3 | 2 | 0 | Low-quality | Genome-fragment | 3.02 | AAI-based (high-confidence) | 0 |
| S16_extracted_86975 | 1199 | No | NA | 2 | 1 | 0 | Low-quality | Genome-fragment | 1.52 | AAI-based (high-confidence) | 0 |
| S11_vs_k141_15209\|\|full | 2716 | No | NA | 5 | 0 | 0 | Not-determined | Genome-fragment | NA | NA | 0 |
| S6_vs_k141_33835\|\|full | 1199 | No | NA | 2 | 0 | 0 | Low-quality | Genome-fragment | 2.29 | AAI-based (medium-confidence) | 0 |
| S8_vs_k141_167435\|\|full | 1199 | No | NA | 2 | 0 | 1 | Low-quality | Genome-fragment | 1.48 | AAI-based (medium-confidence) | 0 |
| S8_vs_k141_181366\|\|full | 1199 | No | NA | 2 | 0 | 0 | Low-quality | Genome-fragment | 2.83 | AAI-based (high-confidence) | 0 |
| S17_extracted_k141_34567 | 1200 | No | NA | 4 | 0 | 0 | Low-quality | Genome-fragment | 21.71 | AAI-based (high-confidence) | 0 |
| S9_extracted_k141_40631 | 1200 | No | NA | 3 | 0 | 1 | Low-quality | Genome-fragment | 2.22 | AAI-based (high-confidence) | 0 |
| S11_vs_k141_47673\|\|full | 1708 | No | NA | 3 | 0 | 2 | Not-determined | Genome-fragment | NA | NA | 0 |
| S11_vs_k141_41441\|\|full | 3712 | No | NA | 4 | 0 | 1 | Not-determined | Genome-fragment | NA | NA | 0 |
| S11_vs_k141_80539\|\|full | 1102 | No | NA | 3 | 0 | 0 | Not-determined | Genome-fragment | NA | NA | 0 |
| S11_vs_k141_28479\|\|full | 1767 | No | NA | 4 | 0 | 0 | Not-determined | Genome-fragment | NA | NA | 0 |
| S9_extracted_k141_79492 | 1200 | No | NA | 3 | 1 | 0 | Low-quality | Genome-fragment | 2.69 | AAI-based (high-confidence) | 0 |
| S11_vs_k141_27118\|\|lt2gene | 1200 | No | NA | 2 | 0 | 0 | Low-quality | Genome-fragment | 2.66 | AAI-based (high-confidence) | 0 |
| S3_vs_k141_83057\|\|full | 1200 | No | NA | 1 | 0 | 0 | Low-quality | Genome-fragment | 2.39 | AAI-based (medium-confidence) | 0 |
| S11_vs_k141_34539\|\|full | 1116 | No | NA | 3 | 0 | 0 | Not-determined | Genome-fragment | NA | NA | 0 |
| S11_vs_k141_34548\|\|full | 1053 | No | NA | 2 | 0 | 2 | Not-determined | Genome-fragment | NA | NA | 0 |
| S3_vs_k141_86703\|\|full | 1200 | No | NA | 3 | 0 | 0 | Low-quality | Genome-fragment | 1.37 | AAI-based (high-confidence) | 0 |
| S11_vs_k141_22135\|\|full | 1131 | No | NA | 2 | 0 | 0 | Not-determined | Genome-fragment | NA | NA | 0 |
| S5_vs_k141_71295\|\|full | 1200 | No | NA | 4 | 1 | 0 | Low-quality | Genome-fragment | 2.79 | AAI-based (medium-confidence) | 0 |
| S11_vs_k141_47723\|\|full | 1432 | No | NA | 3 | 0 | 0 | Not-determined | Genome-fragment | NA | NA | 0 |
| S7_extracted_32817 | 1200 | No | NA | 4 | 0 | 0 | Low-quality | Genome-fragment | 2.04 | AAI-based (medium-confidence) | 0 |
| S7_vs_82705\|\|full | 1200 | No | NA | 3 | 1 | 0 | Low-quality | Genome-fragment | 1.8 | HMM-based (lower-bound) | 0 |
| S7_vs_117526\|\|lt2gene | 1200 | No | NA | 2 | 1 | 0 | Low-quality | Genome-fragment | 1.95 | HMM-based (lower-bound) | 0 |
| S2_extracted_k141_14342 | 1200 | No | NA | 2 | 0 | 0 | Low-quality | Genome-fragment | 1.05 | AAI-based (medium-confidence) | 0 |
| S11_vs_k141_67157\|\|full | 6496 | No | NA | 5 | 0 | 2 | Not-determined | Genome-fragment | NA | NA | 0 |
| S6_extracted_k141_27333 | 1200 | No | NA | 4 | 2 | 0 | Low-quality | Genome-fragment | 1.7 | AAI-based (high-confidence) | 0 |
| S11_vs_k141_15311\|\|full | 3279 | No | NA | 8 | 0 | 0 | Not-determined | Genome-fragment | NA | NA | 0 |
| S11_vs_k141_87524\|\|full | 1990 | No | NA | 2 | 0 | 0 | Not-determined | Genome-fragment | NA | NA | 0 |
| S11_vs_k141_34610\|\|full | 3769 | No | NA | 8 | 0 | 0 | Not-determined | Genome-fragment | NA | NA | 0 |
| S8_extracted_k141_202276 | 1200 | No | NA | 2 | 0 | 2 | Low-quality | Genome-fragment | 0.2 | AAI-based (medium-confidence) | 0 |
| S8_extracted_k141_39293 | 1200 | No | NA | 2 | 0 | 1 | Low-quality | Genome-fragment | 0.56 | AAI-based (high-confidence) | 0 |
| S8_vs_k141_34708\|\|full | 1200 | No | NA | 4 | 1 | 0 | Low-quality | Genome-fragment | 2.79 | AAI-based (high-confidence) | 0 |
| S11_vs_k141_15330\|\|full | 1208 | No | NA | 2 | 0 | 1 | Not-determined | Genome-fragment | NA | NA | 0 |
| S11_vs_k141_2839\|\|full | 3916 | No | NA | 7 | 0 | 1 | Not-determined | Genome-fragment | NA | NA | 0 |
| S9_extracted_k141_58746 | 1201 | No | NA | 1 | 1 | 0 | Low-quality | Genome-fragment | 1.8 | HMM-based (lower-bound) | 0 |
| S11_vs_k141_87570\|\|full | 2305 | No | NA | 3 | 0 | 1 | Not-determined | Genome-fragment | NA | NA | 0 |
| S11_vs_k141_60756\|\|full | 2003 | No | NA | 3 | 0 | 0 | Not-determined | Genome-fragment | NA | NA | 0 |
| S11_vs_k141_34659\|\|full | 2829 | No | NA | 4 | 0 | 0 | Not-determined | Genome-fragment | NA | NA | 0 |
| S13_extracted_65857 | 1201 | No | NA | 2 | 0 | 1 | Low-quality | Genome-fragment | 1.23 | AAI-based (medium-confidence) | 0 |
| S11_vs_k141_74430\|\|full | 2198 | No | NA | 5 | 0 | 0 | Not-determined | Genome-fragment | NA | NA | 0 |
| S13_vs_76971\|\|full | 1201 | No | NA | 4 | 0 | 0 | Low-quality | Genome-fragment | 2.35 | AAI-based (high-confidence) | 0 |
| S1_extracted_k141_37481 | 1201 | No | NA | 2 | 0 | 0 | Low-quality | Genome-fragment | 2.97 | AAI-based (high-confidence) | 0 |
| S1_vs_k141_142003\|\|full | 1201 | No | NA | 4 | 0 | 0 | Low-quality | Genome-fragment | 2.46 | AAI-based (high-confidence) | 0 |
| S1_vs_k141_72705\|\|full | 1201 | No | NA | 3 | 2 | 0 | Low-quality | Genome-fragment | 1.97 | AAI-based (high-confidence) | 0 |
| S3_vs_k141_59969\|\|full | 1201 | No | NA | 3 | 0 | 0 | Low-quality | Genome-fragment | 1.52 | AAI-based (high-confidence) | 0 |
| S11_vs_k141_74510\|\|full | 1419 | No | NA | 4 | 0 | 1 | Not-determined | Genome-fragment | NA | NA | 0 |
| S7_extracted_34197 | 1201 | No | NA | 2 | 1 | 0 | Low-quality | Genome-fragment | 0.93 | AAI-based (high-confidence) | 0 |
| S11_vs_k141_93671\|\|full | 4595 | No | NA | 4 | 0 | 4 | Not-determined | Genome-fragment | NA | NA | 0 |
| S7_extracted_81790 | 1201 | No | NA | 2 | 0 | 0 | Low-quality | Genome-fragment | 2.62 | AAI-based (high-confidence) | 0 |
| S16_vs_52685\|\|full | 1201 | No | NA | 2 | 0 | 2 | Low-quality | Genome-fragment | 0.53 | AAI-based (medium-confidence) | 0 |
| S11_vs_k141_9270\|\|full | 1662 | No | NA | 3 | 0 | 2 | Not-determined | Genome-fragment | NA | NA | 0 |
| S12_extracted_k141_150820 | 1201 | No | NA | 1 | 0 | 0 | Low-quality | Genome-fragment | 2.3 | AAI-based (medium-confidence) | 0 |
| S11_vs_k141_47874\|\|full | 3713 | No | NA | 4 | 0 | 0 | Not-determined | Genome-fragment | NA | NA | 0 |
| S12_extracted_k141_104477 | 1201 | No | NA | 3 | 0 | 0 | Low-quality | Genome-fragment | 1.56 | AAI-based (medium-confidence) | 0 |
| S11_vs_k141_60867\|\|full | 3384 | No | NA | 5 | 0 | 0 | Not-determined | Genome-fragment | NA | NA | 0 |
| S8_extracted_k141_150419 | 1201 | No | NA | 1 | 0 | 1 | Low-quality | Genome-fragment | 0.32 | AAI-based (high-confidence) | 0 |
| S11_vs_k141_54666\|\|full | 1050 | No | NA | 2 | 0 | 0 | Not-determined | Genome-fragment | NA | NA | 0 |
| S17_vs_k141_110316\|\|full | 1202 | No | NA | 2 | 0 | 0 | Low-quality | Genome-fragment | 2.41 | AAI-based (medium-confidence) | 0 |
| S13_extracted_122232 | 1202 | No | NA | 1 | 0 | 0 | Low-quality | Genome-fragment | 0.67 | AAI-based (medium-confidence) | 0 |
| S13_extracted_124805 | 1202 | No | NA | 1 | 0 | 1 | Low-quality | Genome-fragment | 1.94 | AAI-based (high-confidence) | 0 |
| S13_vs_86639\|\|full | 1202 | No | NA | 3 | 0 | 0 | Low-quality | Genome-fragment | 0.89 | AAI-based (medium-confidence) | 0 |
| S1_vs_k141_117465\|\|full | 1202 | No | NA | 5 | 0 | 0 | Low-quality | Genome-fragment | 1.94 | AAI-based (high-confidence) | 0 |
| S5_extracted_k141_38525 | 1202 | No | NA | 1 | 1 | 0 | Low-quality | Genome-fragment | 3.43 | AAI-based (high-confidence) | 0 |
| S7_vs_49512\|\|full | 1202 | No | NA | 3 | 0 | 0 | Low-quality | Genome-fragment | 1.36 | AAI-based (medium-confidence) | 0 |
| S11_vs_k141_34729\|\|full | 2123 | No | NA | 2 | 0 | 1 | Not-determined | Genome-fragment | NA | NA | 0 |
| S11_vs_k141_74619\|\|full | 2587 | No | NA | 5 | 0 | 4 | Not-determined | Genome-fragment | NA | NA | 0 |
| S11_vs_k141_54710\|\|full | 2150 | No | NA | 2 | 0 | 0 | Not-determined | Genome-fragment | NA | NA | 0 |
| S16_extracted_150035 | 1202 | No | NA | 5 | 4 | 0 | Low-quality | Genome-fragment | 3.95 | AAI-based (high-confidence) | 0 |
| S16_vs_67684\|\|full | 1202 | No | NA | 4 | 0 | 2 | Low-quality | Genome-fragment | 2.55 | AAI-based (medium-confidence) | 0 |
| S10_vs_103901\|\|full | 1202 | No | NA | 3 | 0 | 0 | Low-quality | Genome-fragment | 2.91 | AAI-based (high-confidence) | 0 |
| S12_vs_k141_184717\|\|full | 1202 | No | NA | 3 | 0 | 0 | Low-quality | Genome-fragment | 2.62 | AAI-based (high-confidence) | 0 |
| S6_extracted_k141_54222 | 1202 | No | NA | 2 | 0 | 0 | Low-quality | Genome-fragment | 3.27 | AAI-based (medium-confidence) | 0 |
| S17_vs_k141_109594\|\|full | 1203 | No | NA | 2 | 0 | 0 | Low-quality | Genome-fragment | 1.46 | AAI-based (high-confidence) | 0 |
| S11_vs_k141_22418\|\|full | 1030 | No | NA | 2 | 0 | 1 | Not-determined | Genome-fragment | NA | NA | 0 |
| S11_vs_k141_22422\|\|full | 5427 | No | NA | 5 | 0 | 1 | Not-determined | Genome-fragment | NA | NA | 0 |
| S9_extracted_k141_74800 | 1203 | No | NA | 3 | 0 | 0 | Low-quality | Genome-fragment | 2.92 | AAI-based (high-confidence) | 0 |
| S11_vs_k141_93825\|\|full | 1865 | No | NA | 4 | 0 | 0 | Not-determined | Genome-fragment | NA | NA | 0 |
| S9_vs_k141_34821\|\|full | 1203 | No | NA | 4 | 1 | 0 | Low-quality | Genome-fragment | 2.8 | AAI-based (high-confidence) | 0 |
| S11_vs_k141_67390\|\|full | 1584 | No | NA | 2 | 0 | 2 | Not-determined | Genome-fragment | NA | NA | 0 |
| S5_vs_k141_36823\|\|lt2gene | 1203 | No | NA | 2 | 2 | 0 | Low-quality | Genome-fragment | 2.99 | AAI-based (high-confidence) | 0 |
| S7_vs_39647\|\|full | 1203 | No | NA | 3 | 1 | 0 | Low-quality | Genome-fragment | 2.91 | AAI-based (medium-confidence) | 0 |
| S7_vs_66305\|\|full | 1203 | No | NA | 3 | 1 | 0 | Low-quality | Genome-fragment | 2.8 | AAI-based (high-confidence) | 0 |
| S11_vs_k141_15548\|\|full | 9412 | No | NA | 8 | 0 | 5 | Not-determined | Genome-fragment | NA | NA | 0 |
| S11_vs_k141_80866\|\|full | 1272 | No | NA | 2 | 0 | 1 | Not-determined | Genome-fragment | NA | NA | 0 |
| S16_vs_680\|\|full | 1203 | No | NA | 5 | 1 | 0 | Low-quality | Genome-fragment | 2.06 | AAI-based (high-confidence) | 0 |
| S11_vs_k141_54773\|\|full | 2932 | No | NA | 4 | 0 | 1 | Not-determined | Genome-fragment | NA | NA | 0 |
| S10_extracted_35230 | 1203 | No | NA | 2 | 0 | 0 | Low-quality | Genome-fragment | 2.91 | AAI-based (high-confidence) | 0 |
| S15_extracted_1224 | 1203 | No | NA | 4 | 0 | 0 | Low-quality | Genome-fragment | 2.24 | AAI-based (medium-confidence) | 0 |
| S11_vs_k141_41802\|\|full | 1759 | No | NA | 3 | 0 | 1 | Not-determined | Genome-fragment | NA | NA | 0 |
| S11_vs_k141_34803\|\|full | 2090 | No | NA | 3 | 0 | 1 | Not-determined | Genome-fragment | NA | NA | 0 |
| S8_vs_k141_150718\|\|lt2gene | 1203 | No | NA | 1 | 1 | 0 | Low-quality | Genome-fragment | 6.16 | AAI-based (high-confidence) | 0 |
| S11_vs_k141_28617\|\|full | 3051 | No | NA | 5 | 0 | 3 | Not-determined | Genome-fragment | NA | NA | 0 |
| S9_vs_k141_76079\|\|full | 1204 | No | NA | 3 | 1 | 0 | Low-quality | Genome-fragment | 2.28 | HMM-based (lower-bound) | 0 |
| S13_extracted_58191 | 1204 | No | NA | 1 | 0 | 1 | Low-quality | Genome-fragment | 0.73 | AAI-based (medium-confidence) | 0 |
| S14_vs_k141_6236\|\|full | 1204 | No | NA | 3 | 1 | 0 | Low-quality | Genome-fragment | 1.72 | AAI-based (high-confidence) | 0 |
| S1_extracted_k141_132168 | 1204 | No | NA | 2 | 1 | 0 | Low-quality | Genome-fragment | 3.31 | AAI-based (medium-confidence) | 0 |
| S11_vs_k141_9442\|\|full | 2004 | No | NA | 2 | 0 | 1 | Not-determined | Genome-fragment | NA | NA | 0 |
| S11_vs_k141_34852\|\|full | 2827 | No | NA | 3 | 0 | 3 | Not-determined | Genome-fragment | NA | NA | 0 |
| S5_vs_k141_117266\|\|full | 1204 | No | NA | 3 | 0 | 2 | Low-quality | Genome-fragment | 0.81 | AAI-based (medium-confidence) | 0 |
| S11_vs_k141_9471\|\|full | 1245 | No | NA | 3 | 0 | 0 | Not-determined | Genome-fragment | NA | NA | 0 |
| S7_extracted_16758 | 1204 | No | NA | 3 | 1 | 0 | Low-quality | Genome-fragment | 2.82 | AAI-based (high-confidence) | 0 |
| S11_vs_k141_28678\|\|full | 2260 | No | NA | 3 | 0 | 2 | Not-determined | Genome-fragment | NA | NA | 0 |
| S7_vs_82854\|\|full | 1204 | No | NA | 4 | 0 | 0 | Low-quality | Genome-fragment | 2.78 | AAI-based (high-confidence) | 0 |
| S16_extracted_163916 | 1204 | No | NA | 3 | 0 | 1 | Low-quality | Genome-fragment | 2.84 | AAI-based (medium-confidence) | 0 |
| S11_vs_k141_61162\|\|full | 1922 | No | NA | 2 | 0 | 0 | Not-determined | Genome-fragment | NA | NA | 0 |
| S11_vs_k141_93995\|\|full | 3366 | No | NA | 3 | 0 | 0 | Not-determined | Genome-fragment | NA | NA | 0 |
| S16_vs_167354\|\|full | 1204 | No | NA | 4 | 0 | 0 | Low-quality | Genome-fragment | 2.83 | AAI-based (medium-confidence) | 0 |
| S6_extracted_k141_51171 | 1204 | No | NA | 3 | 0 | 0 | Low-quality | Genome-fragment | 1.09 | AAI-based (high-confidence) | 0 |
| S8_extracted_k141_113654 | 1204 | No | NA | 2 | 0 | 1 | Low-quality | Genome-fragment | 0.91 | AAI-based (medium-confidence) | 0 |
| S11_vs_k141_34927\|\|full | 6373 | No | NA | 6 | 0 | 1 | Not-determined | Genome-fragment | NA | NA | 0 |
| S8_vs_k141_178110\|\|full | 1204 | No | NA | 2 | 0 | 0 | Low-quality | Genome-fragment | 2.76 | AAI-based (high-confidence) | 0 |
| S11_vs_k141_34931\|\|full | 3374 | No | NA | 3 | 0 | 1 | Not-determined | Genome-fragment | NA | NA | 0 |
| S11_vs_k141_41934\|\|full | 2364 | No | NA | 2 | 0 | 0 | Not-determined | Genome-fragment | NA | NA | 0 |
| S11_vs_k141_67573\|\|full | 27945 | No | NA | 35 | 3 | 1 | Medium-quality | Genome-fragment | 72.1 | AAI-based (high-confidence) | 0 |
| S17_vs_k141_132155\|\|full | 1205 | No | NA | 2 | 0 | 0 | Low-quality | Genome-fragment | 1.92 | AAI-based (high-confidence) | 0 |
| S9_extracted_k141_75802 | 1205 | No | NA | 3 | 1 | 0 | Low-quality | Genome-fragment | 1.72 | AAI-based (high-confidence) | 0 |
| S14_extracted_k141_47615 | 1205 | No | NA | 4 | 3 | 0 | Low-quality | Genome-fragment | 3.97 | AAI-based (high-confidence) | 0 |
| S14_extracted_k141_35945 | 1205 | No | NA | 3 | 2 | 0 | Low-quality | Genome-fragment | 3.97 | AAI-based (high-confidence) | 0 |
| S7_vs_111176\|\|full | 1205 | No | NA | 4 | 1 | 0 | Low-quality | Genome-fragment | 3.12 | AAI-based (high-confidence) | 0 |
| S11_vs_k141_54914\|\|full | 2054 | No | NA | 3 | 0 | 0 | Not-determined | Genome-fragment | NA | NA | 0 |
| S16_vs_161606\|\|full | 1205 | No | NA | 2 | 0 | 0 | Low-quality | Genome-fragment | 2.44 | AAI-based (medium-confidence) | 0 |
| S10_extracted_56940 | 1205 | No | NA | 3 | 0 | 0 | Low-quality | Genome-fragment | 1.09 | AAI-based (high-confidence) | 0 |
| S11_vs_k141_34954\|\|full | 3579 | No | NA | 4 | 0 | 3 | Not-determined | Genome-fragment | NA | NA | 0 |
| S12_extracted_k141_93397 | 1205 | No | NA | 2 | 0 | 0 | Low-quality | Genome-fragment | 0.97 | AAI-based (high-confidence) | 0 |
| S12_vs_k141_167480\|\|full | 1205 | No | NA | 3 | 1 | 0 | Low-quality | Genome-fragment | 1.98 | AAI-based (high-confidence) | 0 |
| S11_vs_k141_34964\|\|full | 4930 | No | NA | 3 | 0 | 0 | Not-determined | Genome-fragment | NA | NA | 0 |
| S11_vs_k141_48109\|\|full | 65738 | Yes | 32360 | 77 | 17 | 25 | Medium-quality | Genome-fragment | 65.76 | AAI-based (high-confidence) | 50.77 |
| S12_vs_k141_51985\|\|full | 1205 | No | NA | 4 | 1 | 0 | Low-quality | Genome-fragment | 2.2 | AAI-based (medium-confidence) | 0 |
| S11_vs_k141_54965\|\|full | 1661 | No | NA | 3 | 0 | 1 | Not-determined | Genome-fragment | NA | NA | 0 |
| S2_extracted_k141_7975 | 1205 | No | NA | 3 | 2 | 0 | Low-quality | Genome-fragment | 2.74 | AAI-based (high-confidence) | 0 |
| S11_vs_k141_3277\|\|full | 37231 | Yes | 29695 | 41 | 7 | 5 | Medium-quality | Genome-fragment | 69.33 | AAI-based (medium-confidence) | 20.24 |
| S8_vs_k141_72217\|\|full | 1205 | No | NA | 3 | 1 | 0 | Low-quality | Genome-fragment | 2.33 | AAI-based (high-confidence) | 0 |
| S8_vs_k141_103742\|\|full | 1205 | No | NA | 2 | 0 | 0 | Low-quality | Genome-fragment | 2.04 | AAI-based (medium-confidence) | 0 |
| S11_vs_k141_3292\|\|full | 3162 | No | NA | 5 | 0 | 3 | Not-determined | Genome-fragment | NA | NA | 0 |
| S17_extracted_k141_76287 | 1206 | No | NA | 3 | 1 | 0 | Low-quality | Genome-fragment | 2.9 | AAI-based (medium-confidence) | 0 |
| S17_extracted_k141_55556 | 1206 | No | NA | 1 | 0 | 1 | Low-quality | Genome-fragment | 0.67 | AAI-based (high-confidence) | 0 |
| S11_vs_k141_81101\|\|full | 4926 | No | NA | 3 | 0 | 2 | Not-determined | Genome-fragment | NA | NA | 0 |
| S9_vs_k141_58556\|\|full | 1206 | No | NA | 2 | 0 | 0 | Low-quality | Genome-fragment | 1.28 | AAI-based (high-confidence) | 0 |
| S14_extracted_k141_65722 | 1206 | No | NA | 1 | 0 | 1 | Low-quality | Genome-fragment | 0.34 | AAI-based (medium-confidence) | 0 |
| S11_vs_k141_55019\|\|full | 5611 | No | NA | 7 | 0 | 0 | Not-determined | Genome-fragment | NA | NA | 0 |
| S1_vs_k141_140529\|\|full | 1206 | No | NA | 5 | 3 | 0 | Low-quality | Genome-fragment | 2.8 | AAI-based (high-confidence) | 0 |
| S11_vs_k141_67662\|\|full | 2458 | No | NA | 3 | 0 | 1 | Not-determined | Genome-fragment | NA | NA | 0 |
| S3_vs_k141_122779\|\|full | 1206 | No | NA | 1 | 0 | 0 | Low-quality | Genome-fragment | 0.75 | AAI-based (high-confidence) | 0 |
| S11_vs_k141_42031\|\|full | 1719 | No | NA | 3 | 0 | 1 | Not-determined | Genome-fragment | NA | NA | 0 |
| S5_vs_k141_38520\|\|full | 1206 | No | NA | 2 | 0 | 0 | Low-quality | Genome-fragment | 3.14 | AAI-based (high-confidence) | 0 |
| S10_extracted_71855 | 1206 | No | NA | 1 | 0 | 1 | Low-quality | Genome-fragment | 0.29 | AAI-based (high-confidence) | 0 |
| S10_extracted_118980 | 1206 | No | NA | 3 | 0 | 0 | Low-quality | Genome-fragment | 2.6 | AAI-based (medium-confidence) | 0 |
| S11_vs_k141_48187\|\|full | 2683 | No | NA | 3 | 0 | 2 | Not-determined | Genome-fragment | NA | NA | 0 |
| S4_extracted_31423 | 1206 | No | NA | 1 | 1 | 0 | Low-quality | Genome-fragment | 2.82 | HMM-based (lower-bound) | 0 |
| S8_extracted_k141_112826 | 1206 | Yes | 319 | 3 | 1 | 1 | Low-quality | Genome-fragment | 0.34 | HMM-based (lower-bound) | 73.55 |
| S9_vs_k141_45412\|\|full | 1207 | No | NA | 4 | 2 | 0 | Low-quality | Genome-fragment | 2.52 | AAI-based (medium-confidence) | 0 |
| S14_vs_k141_48569\|\|full | 1207 | No | NA | 2 | 1 | 0 | Low-quality | Genome-fragment | 1.86 | HMM-based (lower-bound) | 0 |
| S14_vs_k141_78858\|\|full | 1207 | No | NA | 4 | 0 | 0 | Low-quality | Genome-fragment | 3.18 | AAI-based (high-confidence) | 0 |
| S5_extracted_k141_93723 | 1207 | No | NA | 4 | 1 | 0 | Low-quality | Genome-fragment | 3.51 | AAI-based (medium-confidence) | 0 |
| S11_vs_k141_9725\|\|full | 11703 | No | NA | 6 | 0 | 6 | Not-determined | Genome-fragment | NA | NA | 0 |
| S11_vs_k141_100710\|\|full | 8113 | No | NA | 7 | 0 | 4 | Not-determined | Genome-fragment | NA | NA | 0 |
| S11_vs_k141_22912\|\|full | 2086 | No | NA | 3 | 0 | 1 | Not-determined | Genome-fragment | NA | NA | 0 |
| S7_extracted_123646 | 1207 | No | NA | 4 | 1 | 0 | Low-quality | Genome-fragment | 2.68 | AAI-based (high-confidence) | 0 |
| S11_vs_k141_42137\|\|full | 4849 | No | NA | 6 | 0 | 3 | Not-determined | Genome-fragment | NA | NA | 0 |
| S11_vs_k141_15985\|\|full | 2236 | No | NA | 3 | 0 | 1 | Not-determined | Genome-fragment | NA | NA | 0 |
| S16_extracted_109158 | 1207 | No | NA | 2 | 0 | 1 | Low-quality | Genome-fragment | 1.48 | AAI-based (medium-confidence) | 0 |
| S16_vs_3734\|\|full | 1207 | No | NA | 2 | 1 | 0 | Low-quality | Genome-fragment | 0.81 | HMM-based (lower-bound) | 0 |
| S12_vs_k141_16234\|\|full | 1207 | No | NA | 2 | 1 | 0 | Low-quality | Genome-fragment | 2.87 | AAI-based (high-confidence) | 0 |
| S6_vs_k141_18467\|\|full | 1207 | No | NA | 4 | 0 | 0 | Low-quality | Genome-fragment | 3.51 | AAI-based (high-confidence) | 0 |
| S8_extracted_k141_61698 | 1207 | No | NA | 1 | 1 | 0 | Low-quality | Genome-fragment | 1.79 | HMM-based (lower-bound) | 0 |
| S8_vs_k141_5222\|\|full | 1207 | No | NA | 3 | 1 | 0 | Low-quality | Genome-fragment | 1.92 | AAI-based (medium-confidence) | 0 |
| S11_vs_k141_16017\|\|full | 1835 | No | NA | 2 | 0 | 2 | Not-determined | Genome-fragment | NA | NA | 0 |
| S11_vs_k141_55144\|\|full | 39745 | No | NA | 56 | 25 | 0 | High-quality | High-quality | 99.69 | AAI-based (high-confidence) | 0 |
| S11_vs_k141_28962\|\|full | 2589 | No | NA | 3 | 0 | 2 | Not-determined | Genome-fragment | NA | NA | 0 |
| S11_vs_k141_61383\|\|full | 1757 | No | NA | 4 | 0 | 0 | Not-determined | Genome-fragment | NA | NA | 0 |
| S11_vs_k141_3377\|\|full | 1976 | No | NA | 3 | 0 | 1 | Not-determined | Genome-fragment | NA | NA | 0 |
| S11_vs_k141_16034\|\|full | 2220 | No | NA | 4 | 0 | 0 | Not-determined | Genome-fragment | NA | NA | 0 |
| S17_extracted_k141_165749 | 1208 | No | NA | 1 | 0 | 0 | Low-quality | Genome-fragment | 1.16 | AAI-based (high-confidence) | 0 |
| S11_vs_k141_48312\|\|full | 2966 | No | NA | 3 | 0 | 3 | Not-determined | Genome-fragment | NA | NA | 0 |
| S11_vs_k141_81150\|\|full | 7979 | No | NA | 13 | 0 | 0 | Not-determined | Genome-fragment | NA | NA | 0 |
| S9_extracted_k141_8890 | 1208 | No | NA | 2 | 1 | 0 | Low-quality | Genome-fragment | 1.17 | AAI-based (high-confidence) | 0 |
| S14_vs_k141_77836\|\|full | 1208 | No | NA | 3 | 0 | 0 | Low-quality | Genome-fragment | 3.47 | AAI-based (high-confidence) | 0 |
| S11_vs_k141_48339\|\|full | 2997 | No | NA | 4 | 0 | 1 | Not-determined | Genome-fragment | NA | NA | 0 |
| S1_extracted_k141_47787 | 1208 | No | NA | 2 | 0 | 0 | Low-quality | Genome-fragment | 0.23 | AAI-based (medium-confidence) | 0 |
| S3_vs_k141_50966\|\|lt2gene | 1208 | No | NA | 1 | 1 | 0 | Low-quality | Genome-fragment | 2.56 | AAI-based (high-confidence) | 0 |
| S11_vs_k141_81185\|\|full | 1255 | No | NA | 3 | 0 | 0 | Not-determined | Genome-fragment | NA | NA | 0 |
| S7_vs_62483\|\|full | 1208 | No | NA | 3 | 1 | 0 | Low-quality | Genome-fragment | 2.93 | AAI-based (high-confidence) | 0 |
| S16_extracted_35649 | 1208 | No | NA | 2 | 1 | 0 | Low-quality | Genome-fragment | 2.18 | AAI-based (high-confidence) | 0 |
| S11_vs_k141_3444\|\|full | 1493 | No | NA | 3 | 0 | 0 | Not-determined | Genome-fragment | NA | NA | 0 |
| S11_vs_k141_81192\|\|full | 7035 | No | NA | 11 | 0 | 1 | Not-determined | Genome-fragment | NA | NA | 0 |
| S11_vs_k141_29066\|\|full | 2269 | No | NA | 3 | 0 | 2 | Not-determined | Genome-fragment | NA | NA | 0 |
| S16_vs_116486\|\|full | 1208 | No | NA | 2 | 0 | 1 | Low-quality | Genome-fragment | 2.35 | AAI-based (medium-confidence) | 0 |
| S10_extracted_67002 | 1208 | No | NA | 1 | 0 | 1 | Low-quality | Genome-fragment | 2.88 | AAI-based (high-confidence) | 0 |
| S12_extracted_k141_138258 | 1208 | No | NA | 2 | 1 | 0 | Low-quality | Genome-fragment | 3.01 | AAI-based (high-confidence) | 0 |
| S11_vs_k141_29080\|\|full | 4324 | No | NA | 4 | 0 | 4 | Not-determined | Genome-fragment | NA | NA | 0 |
| S2_vs_k141_13401\|\|full | 1208 | No | NA | 2 | 0 | 0 | Low-quality | Genome-fragment | 1.76 | AAI-based (high-confidence) | 0 |
| S8_vs_k141_27776\|\|full | 1208 | No | NA | 3 | 1 | 0 | Low-quality | Genome-fragment | 1.46 | HMM-based (lower-bound) | 0 |
| S17_vs_k141_12513\|\|full | 1209 | No | NA | 3 | 0 | 1 | Low-quality | Genome-fragment | 3.34 | AAI-based (high-confidence) | 0 |
| S11_vs_k141_3484\|\|full | 1658 | No | NA | 3 | 0 | 3 | Not-determined | Genome-fragment | NA | NA | 0 |
| S14_vs_k141_111915\|\|full | 1209 | No | NA | 4 | 0 | 0 | Low-quality | Genome-fragment | 3.33 | AAI-based (high-confidence) | 0 |

| | | | | | | | | | | | |
|---|---|---|---|---|---|---|---|---|---|---|---|
| S11_vs_k141_100886\|\|full | 2998 | No | NA | 6 | 0 | 1 | Not-determined | Genome-fragment | NA | NA | 0 |
| S11_vs_k141_55246\|\|full | 2740 | No | NA | 5 | 0 | 1 | Not-determined | Genome-fragment | NA | NA | 0 |
| S14_vs_k141_107471\|\|full | 1209 | No | NA | 3 | 1 | 0 | Low-quality | Genome-fragment | 2.84 | AAI-based (medium-confidence) | 0 |
| S1_extracted_k141_69012 | 1209 | No | NA | 2 | 0 | 2 | Low-quality | Genome-fragment | 0.88 | AAI-based (high-confidence) | 0 |
| S11_vs_k141_100894\|\|full | 1819 | No | NA | 4 | 0 | 0 | Not-determined | Genome-fragment | NA | NA | 0 |
| S11_vs_k141_87998\|\|full | 1530 | No | NA | 2 | 0 | 0 | Not-determined | Genome-fragment | NA | NA | 0 |
| S11_vs_k141_67858\|\|full | 3259 | No | NA | 4 | 0 | 2 | Not-determined | Genome-fragment | NA | NA | 0 |
| S11_vs_k141_23153\|\|full | 1492 | No | NA | 3 | 0 | 0 | Not-determined | Genome-fragment | NA | NA | 0 |
| S1_extracted_k141_42763 | 1209 | No | NA | 1 | 0 | 1 | Low-quality | Genome-fragment | 0.67 | AAI-based (high-confidence) | 0 |
| S5_vs_k141_78070\|\|full | 1209 | No | NA | 3 | 3 | 0 | Low-quality | Genome-fragment | 2.65 | AAI-based (medium-confidence) | 0 |
| S11_vs_k141_61510\|\|full | 2350 | No | NA | 3 | 0 | 1 | Not-determined | Genome-fragment | NA | NA | 0 |
| S16_vs_30111\|\|full | 1209 | No | NA | 3 | 0 | 0 | Low-quality | Genome-fragment | 0.7 | AAI-based (high-confidence) | 0 |
| S17_extracted_k141_4554 | 1210 | No | NA | 2 | 0 | 0 | Low-quality | Genome-fragment | 1.96 | AAI-based (high-confidence) | 0 |
| S17_vs_k141_72261\|\|lt2gene | 1210 | No | NA | 2 | 1 | 0 | Low-quality | Genome-fragment | 1.36 | HMM-based (lower-bound) | 0 |
| S11_vs_k141_30436\|\|full | 1210 | No | NA | 2 | 0 | 0 | Low-quality | Genome-fragment | 2.6 | AAI-based (high-confidence) | 0 |
| S11_vs_k141_75241\|\|full | 1073 | No | NA | 2 | 0 | 1 | Not-determined | Genome-fragment | NA | NA | 0 |
| S13_vs_19701\|\|full | 1210 | No | NA | 3 | 0 | 1 | Low-quality | Genome-fragment | 2.09 | AAI-based (medium-confidence) | 0 |
| S1_extracted_k141_103466 | 1210 | No | NA | 2 | 1 | 0 | Low-quality | Genome-fragment | 1.23 | AAI-based (high-confidence) | 0 |
| S11_vs_k141_61540\|\|full | 1819 | No | NA | 3 | 0 | 1 | Not-determined | Genome-fragment | NA | NA | 0 |
| S3_vs_k141_60629\|\|full | 1210 | No | NA | 3 | 1 | 0 | Low-quality | Genome-fragment | 2.64 | AAI-based (high-confidence) | 0 |
| S11_vs_k141_55331\|\|full | 2157 | No | NA | 3 | 0 | 2 | Not-determined | Genome-fragment | NA | NA | 0 |
| S11_vs_k141_61541\|\|full | 5499 | No | NA | 5 | 0 | 2 | Not-determined | Genome-fragment | NA | NA | 0 |
| S5_vs_k141_55446\|\|full | 1210 | No | NA | 3 | 1 | 0 | Low-quality | Genome-fragment | 1.95 | AAI-based (high-confidence) | 0 |
| S11_vs_k141_61552\|\|full | 1071 | No | NA | 3 | 0 | 1 | Not-determined | Genome-fragment | NA | NA | 0 |
| S5_vs_k141_55117\|\|lt2gene | 1210 | No | NA | 1 | 1 | 0 | Low-quality | Genome-fragment | 2.63 | AAI-based (high-confidence) | 0 |
| S7_vs_21167\|\|full | 1210 | No | NA | 4 | 1 | 0 | Low-quality | Genome-fragment | 2.66 | AAI-based (medium-confidence) | 0 |
| S11_vs_k141_81327\|\|full | 3131 | No | NA | 3 | 0 | 1 | Not-determined | Genome-fragment | NA | NA | 0 |
| S11_vs_k141_23188\|\|full | 35761 | No | NA | 50 | 12 | 3 | Medium-quality | Genome-fragment | 60.13 | AAI-based (high-confidence) | 0 |
| S11_vs_k141_94430\|\|full | 1372 | No | NA | 3 | 0 | 0 | Not-determined | Genome-fragment | NA | NA | 0 |
| S11_vs_k141_29249\|\|full | 1306 | No | NA | 2 | 0 | 0 | Not-determined | Genome-fragment | NA | NA | 0 |
| S7_vs_27795\|\|full | 1210 | No | NA | 4 | 0 | 0 | Low-quality | Genome-fragment | 2.77 | AAI-based (high-confidence) | 0 |
| S7_vs_101993\|\|full | 1210 | No | NA | 4 | 1 | 0 | Low-quality | Genome-fragment | 2.43 | AAI-based (high-confidence) | 0 |
| S11_vs_k141_88095\|\|full | 2792 | No | NA | 5 | 0 | 2 | Not-determined | Genome-fragment | NA | NA | 0 |
| S11_vs_k141_88097\|\|full | 1063 | No | NA | 2 | 0 | 1 | Not-determined | Genome-fragment | NA | NA | 0 |
| S16_extracted_104099 | 1210 | No | NA | 1 | 1 | 0 | Low-quality | Genome-fragment | 2.83 | HMM-based (lower-bound) | 0 |
| S16_vs_45027\|\|full | 1210 | No | NA | 5 | 0 | 0 | Low-quality | Genome-fragment | 2.38 | AAI-based (high-confidence) | 0 |
| S11_vs_k141_35575\|\|full | 5637 | No | NA | 4 | 0 | 1 | Not-determined | Genome-fragment | NA | NA | 0 |
| S10_vs_29888\|\|full | 1210 | No | NA | 4 | 1 | 0 | Low-quality | Genome-fragment | 2.41 | AAI-based (high-confidence) | 0 |
| S12_extracted_k141_40616 | 1210 | No | NA | 4 | 1 | 0 | Low-quality | Genome-fragment | 1.9 | AAI-based (high-confidence) | 0 |
| S11_vs_k141_10103\|\|full | 1419 | No | NA | 4 | 0 | 0 | Not-determined | Genome-fragment | NA | NA | 0 |
| S12_vs_k141_112154\|\|full | 1210 | No | NA | 2 | 0 | 0 | Low-quality | Genome-fragment | 3.4 | AAI-based (medium-confidence) | 0 |
| S12_vs_k141_112170\|\|full | 1210 | No | NA | 2 | 0 | 0 | Low-quality | Genome-fragment | 3.51 | AAI-based (medium-confidence) | 0 |
| S15_extracted_19045 | 1210 | No | NA | 2 | 0 | 0 | Low-quality | Genome-fragment | 0.7 | AAI-based (high-confidence) | 0 |
| S11_vs_k141_61652\|\|full | 3331 | No | NA | 4 | 0 | 3 | Not-determined | Genome-fragment | NA | NA | 0 |
| S8_extracted_k141_161671 | 1210 | No | NA | 3 | 0 | 0 | Low-quality | Genome-fragment | 2.57 | AAI-based (high-confidence) | 0 |
| S8_vs_k141_116185\|\|full | 1210 | No | NA | 3 | 1 | 0 | Low-quality | Genome-fragment | 2.84 | AAI-based (medium-confidence) | 0 |
| S8_vs_k141_134442\|\|full | 1210 | No | NA | 4 | 0 | 0 | Low-quality | Genome-fragment | 2.68 | AAI-based (high-confidence) | 0 |
| S11_vs_k141_68056\|\|full | 4842 | No | NA | 3 | 0 | 1 | Not-determined | Genome-fragment | NA | NA | 0 |
| S8_vs_k141_190236\|\|full | 1210 | No | NA | 2 | 0 | 1 | Low-quality | Genome-fragment | 0.98 | AAI-based (medium-confidence) | 0 |
| S13_vs_12876\|\|full | 1211 | No | NA | 2 | 0 | 2 | Low-quality | Genome-fragment | 0.45 | AAI-based (medium-confidence) | 0 |
| S14_extracted_k141_66794 | 1211 | No | NA | 2 | 0 | 0 | Low-quality | Genome-fragment | 1.98 | AAI-based (high-confidence) | 0 |
| S14_vs_k141_59387\|\|full | 1211 | No | NA | 3 | 1 | 0 | Low-quality | Genome-fragment | 1.52 | AAI-based (medium-confidence) | 0 |
| S14_vs_k141_15197\|\|lt2gene | 1211 | No | NA | 2 | 1 | 0 | Low-quality | Genome-fragment | 3.27 | AAI-based (high-confidence) | 0 |
| S14_vs_k141_81713\|\|lt2gene | 1211 | No | NA | 1 | 1 | 0 | Low-quality | Genome-fragment | 4.05 | AAI-based (high-confidence) | 0 |
| S1_extracted_k141_116521 | 1211 | No | NA | 2 | 0 | 0 | Low-quality | Genome-fragment | 1.05 | AAI-based (medium-confidence) | 0 |
| S11_vs_k141_35706\|\|full | 1694 | No | NA | 4 | 0 | 0 | Not-determined | Genome-fragment | NA | NA | 0 |
| S11_vs_k141_68099\|\|full | 2656 | No | NA | 2 | 0 | 0 | Not-determined | Genome-fragment | NA | NA | 0 |
| S11_vs_k141_75542\|\|full | 4094 | No | NA | 6 | 0 | 1 | Not-determined | Genome-fragment | NA | NA | 0 |
| S11_vs_k141_42616\|\|full | 2352 | No | NA | 5 | 0 | 1 | Not-determined | Genome-fragment | NA | NA | 0 |
| S11_vs_k141_23357\|\|full | 43390 | No | NA | 67 | 17 | 1 | High-quality | High-quality | 96.67 | AAI-based (high-confidence) | 0 |
| S11_vs_k141_10172\|\|full | 43295 | No | NA | 66 | 16 | 1 | High-quality | High-quality | 96.46 | AAI-based (high-confidence) | 0 |
| S11_vs_k141_10181\|\|full | 1394 | No | NA | 2 | 0 | 1 | Not-determined | Genome-fragment | NA | NA | 0 |
| S3_extracted_k141_98052 | 1211 | No | NA | 2 | 1 | 0 | Low-quality | Genome-fragment | 3.06 | AAI-based (medium-confidence) | 0 |
| S11_vs_k141_23381\|\|full | 2152 | No | NA | 6 | 0 | 0 | Not-determined | Genome-fragment | NA | NA | 0 |
| S3_vs_k141_1468\|\|full | 1211 | No | NA | 2 | 0 | 0 | Low-quality | Genome-fragment | 3.31 | AAI-based (high-confidence) | 0 |
| S16_vs_129304\|\|lt2gene | 1211 | No | NA | 2 | 1 | 0 | Low-quality | Genome-fragment | 1.8 | HMM-based (lower-bound) | 0 |
| S11_vs_k141_29335\|\|full | 2880 | No | NA | 7 | 0 | 0 | Not-determined | Genome-fragment | NA | NA | 0 |
| S2_vs_k141_15364\|\|full | 1211 | No | NA | 4 | 1 | 0 | Low-quality | Genome-fragment | 2.59 | AAI-based (high-confidence) | 0 |
| S8_extracted_k141_69284 | 1211 | No | NA | 3 | 3 | 0 | Low-quality | Genome-fragment | 2.08 | AAI-based (high-confidence) | 0 |
| S11_vs_k141_75576\|\|full | 5089 | No | NA | 8 | 0 | 4 | Not-determined | Genome-fragment | NA | NA | 0 |
| S11_vs_k141_3569\|\|full | 7783 | No | NA | 7 | 0 | 6 | Not-determined | Genome-fragment | NA | NA | 0 |
| S8_vs_k141_10279\|\|full | 1211 | No | NA | 2 | 0 | 1 | Low-quality | Genome-fragment | 2.25 | AAI-based (medium-confidence) | 0 |
| S17_extracted_k141_45967 | 1212 | No | NA | 1 | 1 | 0 | Low-quality | Genome-fragment | 5.52 | AAI-based (high-confidence) | 0 |
| S11_vs_k141_94765\|\|full | 1877 | No | NA | 3 | 0 | 1 | Not-determined | Genome-fragment | NA | NA | 0 |
| S17_extracted_k141_18984 | 1212 | No | NA | 2 | 0 | 2 | Low-quality | Genome-fragment | 0.33 | AAI-based (medium-confidence) | 0 |
| S9_vs_k141_15310\|\|full | 1212 | No | NA | 3 | 0 | 0 | Low-quality | Genome-fragment | 2.8 | AAI-based (medium-confidence) | 0 |
| S11_vs_k141_81513\|\|full | 2094 | No | NA | 3 | 0 | 1 | Not-determined | Genome-fragment | NA | NA | 0 |
| S9_vs_k141_87322\|\|full | 1212 | No | NA | 2 | 2 | 0 | Low-quality | Genome-fragment | 2.56 | AAI-based (high-confidence) | 0 |
| S9_vs_k141_34909\|\|full | 1212 | No | NA | 2 | 0 | 0 | Low-quality | Genome-fragment | 2.76 | AAI-based (medium-confidence) | 0 |
| S11_vs_k141_10276\|\|full | 1117 | No | NA | 2 | 0 | 2 | Not-determined | Genome-fragment | NA | NA | 0 |
| S11_vs_k141_35855\|\|full | 8706 | No | NA | 6 | 0 | 0 | Not-determined | Genome-fragment | NA | NA | 0 |
| S11_extracted_k141_46908 | 1212 | No | NA | 1 | 0 | 1 | Low-quality | Genome-fragment | 0.22 | AAI-based (high-confidence) | 0 |
| S11_vs_k141_94808\|\|full | 1612 | No | NA | 2 | 0 | 0 | Not-determined | Genome-fragment | NA | NA | 0 |
| S11_vs_k141_88318\|\|full | 1275 | No | NA | 1 | 0 | 1 | Not-determined | Genome-fragment | NA | NA | 0 |
| S14_vs_k141_24184\|\|full | 1212 | No | NA | 3 | 1 | 0 | Low-quality | Genome-fragment | 2.59 | AAI-based (high-confidence) | 0 |
| S14_vs_k141_15268\|\|lt2gene | 1212 | No | NA | 2 | 1 | 0 | Low-quality | Genome-fragment | 19.86 | AAI-based (high-confidence) | 0 |
| S3_extracted_k141_3370 | 1212 | No | NA | 4 | 0 | 0 | Low-quality | Genome-fragment | 3.47 | AAI-based (medium-confidence) | 0 |
| S5_extracted_k141_115210 | 1212 | No | NA | 4 | 1 | 0 | Low-quality | Genome-fragment | 1.45 | HMM-based (lower-bound) | 0 |
| S11_vs_k141_42677\|\|full | 1491 | No | NA | 2 | 0 | 2 | Not-determined | Genome-fragment | NA | NA | 0 |
| S16_extracted_102535 | 1212 | No | NA | 3 | 1 | 0 | Low-quality | Genome-fragment | 3.08 | AAI-based (high-confidence) | 0 |
| S16_vs_19092\|\|full | 1212 | No | NA | 3 | 1 | 0 | Low-quality | Genome-fragment | 2 | AAI-based (medium-confidence) | 0 |
| S11_vs_k141_55768\|\|full | 2974 | No | NA | 5 | 0 | 3 | Not-determined | Genome-fragment | NA | NA | 0 |
| S11_vs_k141_94868\|\|full | 4952 | No | NA | 7 | 0 | 3 | Not-determined | Genome-fragment | NA | NA | 0 |
| S11_vs_k141_68151\|\|full | 61398 | No | NA | 91 | 32 | 1 | High-quality | High-quality | 100 | AAI-based (high-confidence) | 0 |
| S10_vs_116625\|\|full | 1212 | No | NA | 3 | 0 | 2 | Low-quality | Genome-fragment | 0.83 | AAI-based (medium-confidence) | 0 |
| S11_vs_k141_35952\|\|full | 1840 | No | NA | 5 | 0 | 0 | Not-determined | Genome-fragment | NA | NA | 0 |
| S10_vs_96162\|\|lt2gene | 1212 | No | NA | 1 | 1 | 0 | Low-quality | Genome-fragment | 3.11 | AAI-based (high-confidence) | 0 |
| S12_vs_k141_183141\|\|full | 1212 | No | NA | 2 | 1 | 0 | Low-quality | Genome-fragment | 0.82 | AAI-based (medium-confidence) | 0 |
| S15_vs_11321\|\|full | 1212 | No | NA | 2 | 0 | 0 | Low-quality | Genome-fragment | 2.7 | AAI-based (medium-confidence) | 0 |
| S6_vs_k141_7563\|\|full | 1212 | No | NA | 3 | 1 | 0 | Low-quality | Genome-fragment | 2.73 | AAI-based (medium-confidence) | 0 |
| S11_vs_k141_101401\|\|full | 1526 | No | NA | 2 | 0 | 0 | Not-determined | Genome-fragment | NA | NA | 0 |
| S8_vs_k141_101777\|\|lt2gene | 1212 | No | NA | 2 | 0 | 0 | Low-quality | Genome-fragment | 2.01 | AAI-based (high-confidence) | 0 |
| S17_extracted_k141_86508 | 1213 | No | NA | 2 | 2 | 0 | Low-quality | Genome-fragment | 4 | AAI-based (high-confidence) | 0 |
| S13_extracted_27511 | 1213 | No | NA | 5 | 1 | 0 | Low-quality | Genome-fragment | 2.05 | AAI-based (high-confidence) | 0 |
| S11_vs_k141_3771\|\|full | 1729 | No | NA | 2 | 0 | 0 | Not-determined | Genome-fragment | NA | NA | 0 |
| S11_vs_k141_94925\|\|full | 2984 | No | NA | 3 | 0 | 3 | Not-determined | Genome-fragment | NA | NA | 0 |
| S14_vs_k141_10132\|\|full | 1213 | No | NA | 2 | 0 | 0 | Low-quality | Genome-fragment | 3.04 | AAI-based (high-confidence) | 0 |
| S1_vs_k141_33401\|\|full | 1213 | No | NA | 3 | 1 | 0 | Low-quality | Genome-fragment | 2.41 | AAI-based (high-confidence) | 0 |
| S5_extracted_k141_84257 | 1213 | No | NA | 1 | 0 | 0 | Low-quality | Genome-fragment | 1.64 | AAI-based (high-confidence) | 0 |
| S7_vs_5511\|\|full | 1213 | No | NA | 2 | 0 | 0 | Low-quality | Genome-fragment | 2.94 | AAI-based (medium-confidence) | 0 |
| S7_vs_68390\|\|full | 1213 | No | NA | 4 | 0 | 0 | Low-quality | Genome-fragment | 2.43 | AAI-based (high-confidence) | 0 |
| S11_vs_k141_55875\|\|full | 2427 | No | NA | 2 | 0 | 2 | Not-determined | Genome-fragment | NA | NA | 0 |
| S7_vs_105602\|\|lt2gene | 1213 | No | NA | 1 | 1 | 0 | Low-quality | Genome-fragment | 2.31 | AAI-based (high-confidence) | 0 |
| S16_vs_97470\|\|full | 1213 | No | NA | 3 | 1 | 0 | Low-quality | Genome-fragment | 2.41 | AAI-based (high-confidence) | 0 |
| S16_vs_152168\|\|full | 1213 | No | NA | 3 | 2 | 0 | Low-quality | Genome-fragment | 3.39 | AAI-based (medium-confidence) | 0 |
| S11_vs_k141_101466\|\|full | 1232 | No | NA | 3 | 0 | 0 | Not-determined | Genome-fragment | NA | NA | 0 |
| S11_vs_k141_29529\|\|full | 1614 | No | NA | 3 | 0 | 1 | Not-determined | Genome-fragment | NA | NA | 0 |
| S10_vs_7558\|\|full | 1213 | No | NA | 4 | 1 | 0 | Low-quality | Genome-fragment | 2.83 | AAI-based (high-confidence) | 0 |
| S11_vs_k141_36043\|\|full | 1901 | No | NA | 2 | 0 | 0 | Not-determined | Genome-fragment | NA | NA | 0 |
| S10_vs_31861\|\|full | 1213 | No | NA | 2 | 0 | 0 | Low-quality | Genome-fragment | 1.16 | AAI-based (medium-confidence) | 0 |
| S10_vs_44453\|\|full | 1213 | No | NA | 4 | 2 | 0 | Low-quality | Genome-fragment | 2.97 | AAI-based (medium-confidence) | 0 |
| S11_vs_k141_101501\|\|full | 3316 | No | NA | 5 | 0 | 1 | Not-determined | Genome-fragment | NA | NA | 0 |
| S12_vs_k141_58472\|\|full | 1213 | No | NA | 3 | 1 | 0 | Low-quality | Genome-fragment | 2.43 | HMM-based (lower-bound) | 0 |
| S11_vs_k141_101525\|\|full | 1051 | No | NA | 3 | 0 | 1 | Not-determined | Genome-fragment | NA | NA | 0 |
| S6_vs_k141_24983\|\|full | 1213 | No | NA | 3 | 0 | 1 | Low-quality | Genome-fragment | 0.24 | AAI-based (medium-confidence) | 0 |
| S8_extracted_k141_61520 | 1213 | No | NA | 3 | 3 | 0 | Low-quality | Genome-fragment | 3.91 | AAI-based (high-confidence) | 0 |
| S17_vs_k141_175888\|\|full | 1214 | No | NA | 2 | 0 | 1 | Low-quality | Genome-fragment | 3.41 | AAI-based (medium-confidence) | 0 |
| S9_vs_k141_60276\|\|full | 1214 | No | NA | 3 | 0 | 2 | Low-quality | Genome-fragment | 1.03 | AAI-based (medium-confidence) | 0 |
| S11_extracted_k141_101587 | 1214 | No | NA | 2 | 0 | 1 | Low-quality | Genome-fragment | 0.61 | AAI-based (medium-confidence) | 0 |
| S11_vs_k141_75850\|\|full | 1254 | No | NA | 2 | 0 | 1 | Not-determined | Genome-fragment | NA | NA | 0 |
| S11_vs_k141_8098\|\|full | 1214 | No | NA | 3 | 1 | 0 | Low-quality | Genome-fragment | 3.05 | AAI-based (medium-confidence) | 0 |
| S11_vs_k141_3836\|\|full | 3311 | No | NA | 4 | 0 | 1 | Not-determined | Genome-fragment | NA | NA | 0 |
| S11_vs_k141_62127\|\|full | 1933 | No | NA | 3 | 0 | 1 | Not-determined | Genome-fragment | NA | NA | 0 |
| S11_vs_k141_48807\|\|full | 1214 | No | NA | 3 | 1 | 0 | Low-quality | Genome-fragment | 3.04 | AAI-based (high-confidence) | 0 |
| S11_vs_k141_36105\|\|full | 1840 | No | NA | 3 | 0 | 0 | Not-determined | Genome-fragment | NA | NA | 0 |
| S13_extracted_91847 | 1214 | No | NA | 1 | 1 | 0 | Low-quality | Genome-fragment | 2.89 | HMM-based (lower-bound) | 0 |
| S11_vs_k141_42921\|\|full | 4413 | No | NA | 6 | 0 | 2 | Not-determined | Genome-fragment | NA | NA | 0 |
| S11_vs_k141_56002\|\|full | 1297 | No | NA | 3 | 0 | 0 | Not-determined | Genome-fragment | NA | NA | 0 |
| S11_vs_k141_16667\|\|full | 2976 | No | NA | 4 | 0 | 0 | Not-determined | Genome-fragment | NA | NA | 0 |
| S13_vs_57190\|\|full | 1214 | No | NA | 3 | 0 | 1 | Low-quality | Genome-fragment | 0.39 | AAI-based (medium-confidence) | 0 |
| S11_vs_k141_88633\|\|full | 1875 | No | NA | 2 | 0 | 1 | Not-determined | Genome-fragment | NA | NA | 0 |
| S11_vs_k141_56021\|\|full | 1243 | No | NA | 2 | 0 | 1 | Not-determined | Genome-fragment | NA | NA | 0 |
| S13_vs_65421\|\|full | 1214 | No | NA | 3 | 0 | 0 | Low-quality | Genome-fragment | 2.33 | AAI-based (medium-confidence) | 0 |
| S13_vs_76182\|\|full | 1214 | No | NA | 2 | 0 | 0 | Low-quality | Genome-fragment | 1.47 | AAI-based (medium-confidence) | 0 |
| S1_vs_k141_64238\|\|full | 1214 | No | NA | 3 | 1 | 0 | Low-quality | Genome-fragment | 21.96 | AAI-based (high-confidence) | 0 |
| S11_vs_k141_88667\|\|full | 1708 | No | NA | 3 | 0 | 1 | Not-determined | Genome-fragment | NA | NA | 0 |
| S5_vs_k141_30472\|\|full | 1214 | No | NA | 3 | 2 | 0 | Low-quality | Genome-fragment | 2.92 | AAI-based (medium-confidence) | 0 |
| S7_extracted_122436 | 1214 | No | NA | 3 | 0 | 3 | Low-quality | Genome-fragment | 0.32 | AAI-based (medium-confidence) | 0 |
| S7_vs_58149\|\|full | 1214 | No | NA | 3 | 1 | 0 | Low-quality | Genome-fragment | 0.95 | AAI-based (high-confidence) | 0 |
| S11_vs_k141_3914\|\|full | 1311 | No | NA | 3 | 0 | 1 | Not-determined | Genome-fragment | NA | NA | 0 |
| S11_vs_k141_48936\|\|full | 1139 | No | NA | 2 | 0 | 1 | Not-determined | Genome-fragment | NA | NA | 0 |
| S11_vs_k141_88692\|\|full | 4608 | No | NA | 6 | 0 | 4 | Not-determined | Genome-fragment | NA | NA | 0 |
| S11_vs_k141_81775\|\|full | 36215 | No | NA | 56 | 17 | 2 | High-quality | High-quality | 93 | AAI-based (high-confidence) | 0 |
| S7_vs_87422\|\|full | 1214 | No | NA | 3 | 2 | 0 | Low-quality | Genome-fragment | 2.64 | AAI-based (high-confidence) | 0 |
| S11_vs_k141_3934\|\|full | 4465 | No | NA | 6 | 0 | 0 | Not-determined | Genome-fragment | NA | NA | 0 |
| S11_vs_k141_68348\|\|full | 41562 | No | NA | 56 | 26 | 2 | High-quality | High-quality | 100 | AAI-based (high-confidence) | 0 |
| S11_vs_k141_42927\|\|full | 56290 | Yes | 48437 | 60 | 23 | 6 | Medium-quality | Genome-fragment | 87.04 | AAI-based (high-confidence) | 13.95 |
| S11_vs_k141_23724\|\|full | 1307 | No | NA | 1 | 0 | 1 | Not-determined | Genome-fragment | NA | NA | 0 |
| S11_vs_k141_68355\|\|full | 3816 | No | NA | 4 | 0 | 0 | Not-determined | Genome-fragment | NA | NA | 0 |
| S7_vs_94461\|\|full | 1214 | No | NA | 3 | 0 | 0 | Low-quality | Genome-fragment | 2.58 | AAI-based (medium-confidence) | 0 |
| S16_extracted_20791 | 1214 | No | NA | 1 | 1 | 0 | Low-quality | Genome-fragment | 0.74 | AAI-based (high-confidence) | 0 |
| S11_vs_k141_42950\|\|full | 3259 | No | NA | 8 | 0 | 0 | Not-determined | Genome-fragment | NA | NA | 0 |
| S11_vs_k141_42953\|\|full | 4654 | No | NA | 3 | 0 | 2 | Not-determined | Genome-fragment | NA | NA | 0 |
| S16_extracted_129151 | 1214 | No | NA | 1 | 0 | 0 | Low-quality | Genome-fragment | 2.46 | AAI-based (high-confidence) | 0 |
| S10_extracted_86686 | 1214 | No | NA | 1 | 1 | 0 | Low-quality | Genome-fragment | 0.64 | HMM-based (lower-bound) | 0 |
| S12_extracted_k141_1337 | 1214 | No | NA | 2 | 0 | 0 | Low-quality | Genome-fragment | 2.59 | AAI-based (high-confidence) | 0 |
| S15_vs_68187\|\|lt2gene | 1214 | No | NA | 2 | 1 | 0 | Low-quality | Genome-fragment | 3.98 | AAI-based (high-confidence) | 0 |
| S11_vs_k141_49002\|\|full | 1761 | No | NA | 4 | 0 | 1 | Not-determined | Genome-fragment | NA | NA | 0 |
| S11_vs_k141_62260\|\|full | 1272 | No | NA | 3 | 0 | 1 | Not-determined | Genome-fragment | NA | NA | 0 |
| S8_extracted_k141_23257 | 1214 | No | NA | 1 | 0 | 0 | Low-quality | Genome-fragment | 1.54 | AAI-based (medium-confidence) | 0 |
| S11_vs_k141_62262\|\|full | 1091 | No | NA | 3 | 0 | 0 | Not-determined | Genome-fragment | NA | NA | 0 |
| S8_vs_k141_74976\|\|full | 1214 | No | NA | 2 | 0 | 2 | Low-quality | Genome-fragment | 1.03 | AAI-based (medium-confidence) | 0 |
| S11_extracted_k141_80965 | 1215 | No | NA | 3 | 1 | 0 | Low-quality | Genome-fragment | 2.96 | AAI-based (high-confidence) | 0 |
| S14_vs_k141_60258\|\|full | 1215 | No | NA | 2 | 0 | 0 | Low-quality | Genome-fragment | 0.8 | AAI-based (medium-confidence) | 0 |
| S14_vs_k141_27046\|\|full | 1215 | No | NA | 4 | 3 | 0 | Low-quality | Genome-fragment | 2.18 | AAI-based (medium-confidence) | 0 |
| S3_extracted_k141_89808 | 1215 | No | NA | 3 | 0 | 0 | Low-quality | Genome-fragment | 0.87 | AAI-based (medium-confidence) | 0 |
| S3_extracted_k141_94477 | 1215 | No | NA | 2 | 0 | 1 | Low-quality | Genome-fragment | 0.3 | AAI-based (medium-confidence) | 0 |
| S11_vs_k141_68455\|\|full | 1021 | No | NA | 3 | 0 | 2 | Not-determined | Genome-fragment | NA | NA | 0 |
| S5_vs_k141_35022\|\|full | 1215 | No | NA | 4 | 0 | 0 | Low-quality | Genome-fragment | 3.15 | AAI-based (medium-confidence) | 0 |
| S5_vs_k141_5739\|\|full | 1215 | No | NA | 2 | 0 | 1 | Low-quality | Genome-fragment | 0.52 | AAI-based (medium-confidence) | 0 |
| S7_vs_107802\|\|lt2gene | 1215 | No | NA | 1 | 1 | 0 | Low-quality | Genome-fragment | 2.6 | AAI-based (medium-confidence) | 0 |
| S10_vs_99694\|\|full | 1215 | No | NA | 3 | 0 | 0 | Low-quality | Genome-fragment | 2.66 | AAI-based (medium-confidence) | 0 |
| S12_vs_k141_2524\|\|full | 1215 | No | NA | 3 | 2 | 0 | Low-quality | Genome-fragment | 2.89 | AAI-based (medium-confidence) | 0 |
| S12_vs_k141_11139\|\|full | 1215 | No | NA | 3 | 0 | 0 | Low-quality | Genome-fragment | 2.53 | AAI-based (medium-confidence) | 0 |
| S11_vs_k141_36288\|\|full | 1104 | No | NA | 2 | 0 | 1 | Not-determined | Genome-fragment | NA | NA | 0 |
| S4_extracted_7510 | 1215 | No | NA | 4 | 1 | 0 | Low-quality | Genome-fragment | 2.22 | AAI-based (high-confidence) | 0 |
| S11_vs_k141_56210\|\|full | 5955 | No | NA | 4 | 0 | 2 | Not-determined | Genome-fragment | NA | NA | 0 |
| S11_vs_k141_4040\|\|full | 1593 | No | NA | 5 | 0 | 0 | Not-determined | Genome-fragment | NA | NA | 0 |
| S6_extracted_k141_17562 | 1215 | No | NA | 3 | 0 | 0 | Low-quality | Genome-fragment | 0.85 | AAI-based (medium-confidence) | 0 |
| S8_extracted_k141_162271 | 1215 | No | NA | 2 | 0 | 1 | Low-quality | Genome-fragment | 0.74 | AAI-based (medium-confidence) | 0 |
| S11_vs_k141_23923\|\|full | 1334 | No | NA | 3 | 0 | 1 | Not-determined | Genome-fragment | NA | NA | 0 |
| S11_vs_k141_23924\|\|full | 4713 | No | NA | 5 | 0 | 2 | Not-determined | Genome-fragment | NA | NA | 0 |
| S11_vs_k141_62365\|\|full | 2238 | No | NA | 4 | 0 | 2 | Not-determined | Genome-fragment | NA | NA | 0 |
| S11_vs_k141_81966\|\|full | 1403 | No | NA | 3 | 0 | 1 | Not-determined | Genome-fragment | NA | NA | 0 |
| S8_extracted_k141_76440 | 1215 | No | NA | 1 | 0 | 0 | Low-quality | Genome-fragment | 1.42 | AAI-based (high-confidence) | 0 |
| S9_vs_k141_9652\|\|lt2gene | 1216 | No | NA | 2 | 1 | 0 | Low-quality | Genome-fragment | 2.22 | HMM-based (lower-bound) | 0 |
| S11_extracted_k141_23760 | 1216 | No | NA | 1 | 0 | 0 | Low-quality | Genome-fragment | 1.96 | AAI-based (high-confidence) | 0 |
| S11_vs_k141_29683\|\|full | 2642 | No | NA | 4 | 0 | 2 | Not-determined | Genome-fragment | NA | NA | 0 |
| S11_vs_k141_23943\|\|full | 1978 | No | NA | 3 | 0 | 0 | Not-determined | Genome-fragment | NA | NA | 0 |
| S11_vs_k141_4104\|\|full | 11761 | No | NA | 11 | 0 | 1 | Not-determined | Genome-fragment | NA | NA | 0 |
| S1_extracted_k141_9364 | 1216 | No | NA | 2 | 1 | 0 | Low-quality | Genome-fragment | 0.6 | HMM-based (lower-bound) | 0 |
| S3_vs_k141_8361\|\|full | 1216 | No | NA | 4 | 0 | 0 | Low-quality | Genome-fragment | 3.05 | AAI-based (high-confidence) | 0 |
| S5_vs_k141_73890\|\|full | 1216 | No | NA | 3 | 1 | 0 | Low-quality | Genome-fragment | 2.77 | AAI-based (medium-confidence) | 0 |
| S7_extracted_4228 | 1216 | No | NA | 2 | 0 | 0 | Low-quality | Genome-fragment | 0.77 | AAI-based (high-confidence) | 0 |
| S7_vs_19896\|\|full | 1216 | No | NA | 3 | 1 | 0 | Low-quality | Genome-fragment | 3 | AAI-based (medium-confidence) | 0 |
| S11_vs_k141_10613\|\|full | 3376 | No | NA | 5 | 0 | 3 | Not-determined | Genome-fragment | NA | NA | 0 |
| S16_vs_11222\|\|full | 1216 | Yes | 682 | 3 | 1 | 1 | Low-quality | Genome-fragment | 1.4 | AAI-based (medium-confidence) | 43.91 |
| S16_vs_102206\|\|full | 1216 | No | NA | 2 | 0 | 0 | Low-quality | Genome-fragment | 1.34 | AAI-based (high-confidence) | 0 |
| S11_vs_k141_49082\|\|full | 2753 | No | NA | 5 | 0 | 1 | Not-determined | Genome-fragment | NA | NA | 0 |
| S16_vs_90509\|\|lt2gene | 1216 | No | NA | 1 | 1 | 0 | Low-quality | Genome-fragment | 2.95 | AAI-based (high-confidence) | 0 |
| S8_vs_k141_32227\|\|full | 1216 | No | NA | 3 | 1 | 0 | Low-quality | Genome-fragment | 2.66 | AAI-based (high-confidence) | 0 |
| S8_vs_k141_101750\|\|full | 1216 | No | NA | 4 | 0 | 0 | Low-quality | Genome-fragment | 0.83 | AAI-based (high-confidence) | 0 |
| S11_vs_k141_29765\|\|full | 1207 | No | NA | 3 | 0 | 0 | Not-determined | Genome-fragment | NA | NA | 0 |
| S11_vs_k141_62486\|\|full | 2490 | No | NA | 3 | 0 | 1 | Not-determined | Genome-fragment | NA | NA | 0 |
| S17_vs_k141_35265\|\|full | 1217 | No | NA | 2 | 0 | 1 | Low-quality | Genome-fragment | 0.73 | AAI-based (high-confidence) | 0 |
| S14_extracted_k141_33479 | 1217 | No | NA | 1 | 0 | 0 | Low-quality | Genome-fragment | 2.38 | AAI-based (medium-confidence) | 0 |
| S1_vs_k141_19729\|\|lt2gene | 1217 | No | NA | 2 | 1 | 0 | Low-quality | Genome-fragment | 2.56 | AAI-based (high-confidence) | 0 |
| S11_vs_k141_88890\|\|full | 2360 | No | NA | 2 | 0 | 1 | Not-determined | Genome-fragment | NA | NA | 0 |
| S3_vs_k141_124544\|\|lt2gene | 1217 | No | NA | 2 | 2 | 0 | Low-quality | Genome-fragment | 2.52 | AAI-based (high-confidence) | 0 |
| S7_vs_9156\|\|full | 1217 | No | NA | 3 | 2 | 0 | Low-quality | Genome-fragment | 3.62 | AAI-based (high-confidence) | 0 |
| S11_vs_k141_24067\|\|full | 1103 | No | NA | 2 | 0 | 0 | Not-determined | Genome-fragment | NA | NA | 0 |
| S16_extracted_122863 | 1217 | No | NA | 4 | 0 | 0 | Low-quality | Genome-fragment | 2.45 | AAI-based (high-confidence) | 0 |
| S11_vs_k141_36484\|\|full | 3995 | No | NA | 3 | 0 | 2 | Not-determined | Genome-fragment | NA | NA | 0 |
| S11_vs_k141_24082\|\|full | 3699 | No | NA | 3 | 0 | 2 | Not-determined | Genome-fragment | NA | NA | 0 |
| S16_vs_41545\|\|full | 1217 | No | NA | 2 | 0 | 0 | Low-quality | Genome-fragment | 2.32 | AAI-based (high-confidence) | 0 |
| S16_vs_122421\|\|full | 1217 | No | NA | 4 | 0 | 1 | Low-quality | Genome-fragment | 3.17 | AAI-based (medium-confidence) | 0 |
| S12_vs_k141_162161\|\|lt2gene | 1217 | No | NA | 1 | 1 | 0 | Low-quality | Genome-fragment | 1.87 | HMM-based (lower-bound) | 0 |
| S11_vs_k141_29831\|\|full | 6592 | No | NA | 9 | 0 | 2 | Not-determined | Genome-fragment | NA | NA | 0 |
| S6_vs_k141_13824\|\|full | 1217 | No | NA | 3 | 0 | 0 | Low-quality | Genome-fragment | 2.68 | AAI-based (high-confidence) | 0 |
| S11_vs_k141_56481\|\|full | 2106 | No | NA | 3 | 0 | 0 | Not-determined | Genome-fragment | NA | NA | 0 |
| S11_vs_k141_4195\|\|full | 14081 | No | NA | 14 | 0 | 2 | Not-determined | Genome-fragment | NA | NA | 0 |
| S17_vs_k141_50768\|\|full | 1218 | No | NA | 3 | 0 | 0 | Low-quality | Genome-fragment | 1.88 | AAI-based (medium-confidence) | 0 |
| S9_vs_k141_3054\|\|lt2gene | 1218 | No | NA | 1 | 1 | 0 | Low-quality | Genome-fragment | 3.19 | AAI-based (high-confidence) | 0 |
| S11_vs_k141_36577\|\|full | 1871 | No | NA | 3 | 0 | 1 | Not-determined | Genome-fragment | NA | NA | 0 |
| S11_vs_k141_36578\|\|full | 1590 | No | NA | 3 | 0 | 1 | Not-determined | Genome-fragment | NA | NA | 0 |
| S11_vs_k141_1445\|\|full | 1218 | No | NA | 3 | 1 | 0 | Low-quality | Genome-fragment | 2.57 | AAI-based (high-confidence) | 0 |
| S13_extracted_41207 | 1218 | No | NA | 3 | 0 | 0 | Low-quality | Genome-fragment | 1.26 | AAI-based (high-confidence) | 0 |
| S11_vs_k141_101884\|\|full | 1152 | No | NA | 2 | 0 | 1 | Not-determined | Genome-fragment | NA | NA | 0 |
| S14_vs_k141_68794\|\|full | 1218 | No | NA | 2 | 0 | 1 | Low-quality | Genome-fragment | 2.01 | AAI-based (medium-confidence) | 0 |
| S14_vs_k141_80373\|\|full | 1218 | No | NA | 2 | 0 | 1 | Low-quality | Genome-fragment | 0.63 | AAI-based (medium-confidence) | 0 |
| S11_vs_k141_62573\|\|full | 54058 | No | NA | 80 | 13 | 2 | High-quality | High-quality | 99.29 | AAI-based (high-confidence) | 0 |
| S14_vs_k141_9377\|\|lt2gene | 1218 | No | NA | 2 | 0 | 0 | Low-quality | Genome-fragment | 2.08 | AAI-based (high-confidence) | 0 |
| S1_vs_k141_78568\|\|full | 1218 | No | NA | 4 | 0 | 0 | Low-quality | Genome-fragment | 2.72 | AAI-based (high-confidence) | 0 |
| S11_vs_k141_101924\|\|full | 1029 | No | NA | 2 | 0 | 0 | Not-determined | Genome-fragment | NA | NA | 0 |
| S5_vs_k141_143327\|\|full | 1218 | No | NA | 1 | 0 | 0 | Low-quality | Genome-fragment | 2.84 | AAI-based (medium-confidence) | 0 |
| S11_vs_k141_10842\|\|full | 1475 | No | NA | 2 | 0 | 1 | Not-determined | Genome-fragment | NA | NA | 0 |
| S5_vs_k141_11043\|\|full | 1218 | No | NA | 2 | 0 | 1 | Low-quality | Genome-fragment | 1.41 | AAI-based (medium-confidence) | 0 |
| S5_vs_k141_101845\|\|full | 1218 | No | NA | 3 | 2 | 0 | Low-quality | Genome-fragment | 2.81 | AAI-based (medium-confidence) | 0 |
| S16_vs_32133\|\|full | 1218 | No | NA | 3 | 1 | 0 | Low-quality | Genome-fragment | 2.85 | AAI-based (high-confidence) | 0 |
| S11_vs_k141_82176\|\|full | 2924 | No | NA | 2 | 0 | 0 | Not-determined | Genome-fragment | NA | NA | 0 |
| S16_vs_51655\|\|full | 1218 | No | NA | 3 | 0 | 0 | Low-quality | Genome-fragment | 3.59 | AAI-based (high-confidence) | 0 |
| S16_vs_91369\|\|full | 1218 | No | NA | 3 | 1 | 0 | Low-quality | Genome-fragment | 2.01 | AAI-based (high-confidence) | 0 |
| S16_vs_122429\|\|full | 1218 | No | NA | 2 | 0 | 2 | Low-quality | Genome-fragment | 1.27 | AAI-based (medium-confidence) | 0 |
| S11_vs_k141_43389\|\|full | 1431 | No | NA | 3 | 0 | 3 | Not-determined | Genome-fragment | NA | NA | 0 |
| S12_vs_k141_90339\|\|full | 1218 | No | NA | 3 | 1 | 0 | Low-quality | Genome-fragment | 2.47 | AAI-based (medium-confidence) | 0 |
| S6_vs_k141_537\|\|full | 1218 | No | NA | 3 | 0 | 1 | Low-quality | Genome-fragment | 0.75 | AAI-based (medium-confidence) | 0 |
| S11_vs_k141_10901\|\|full | 1428 | No | NA | 3 | 0 | 0 | Not-determined | Genome-fragment | NA | NA | 0 |
| S11_vs_k141_43404\|\|full | 1935 | No | NA | 3 | 0 | 1 | Not-determined | Genome-fragment | NA | NA | 0 |
| S9_vs_k141_69175\|\|lt2gene | 1219 | No | NA | 1 | 0 | 0 | Low-quality | Genome-fragment | 2.33 | AAI-based (high-confidence) | 0 |
| S11_vs_k141_17154\|\|full | 1847 | No | NA | 3 | 0 | 1 | Not-determined | Genome-fragment | NA | NA | 0 |
| S11_vs_k141_4371\|\|full | 1019 | No | NA | 3 | 0 | 0 | Not-determined | Genome-fragment | NA | NA | 0 |
| S11_extracted_k141_102695 | 1219 | No | NA | 3 | 2 | 0 | Low-quality | Genome-fragment | 2.69 | HMM-based (lower-bound) | 0 |
| S11_vs_k141_62673\|\|full | 4574 | No | NA | 6 | 0 | 1 | Not-determined | Genome-fragment | NA | NA | 0 |
| S11_vs_k141_17187\|\|full | 1844 | No | NA | 4 | 0 | 2 | Not-determined | Genome-fragment | NA | NA | 0 |
| S14_vs_k141_61880\|\|lt2gene | 1219 | No | NA | 1 | 1 | 0 | Low-quality | Genome-fragment | 2.83 | AAI-based (high-confidence) | 0 |
| S11_vs_k141_43449\|\|full | 8786 | No | NA | 10 | 0 | 5 | Not-determined | Genome-fragment | NA | NA | 0 |
| S1_extracted_k141_10420 | 1219 | No | NA | 5 | 1 | 0 | Low-quality | Genome-fragment | 3.3 | AAI-based (high-confidence) | 0 |
| S3_vs_k141_109021\|\|full | 1219 | No | NA | 3 | 0 | 0 | Low-quality | Genome-fragment | 2.5 | AAI-based (high-confidence) | 0 |
| S7_vs_129180\|\|lt2gene | 1219 | No | NA | 1 | 1 | 0 | Low-quality | Genome-fragment | 0.38 | HMM-based (lower-bound) | 0 |
| S11_vs_k141_49275\|\|full | 2126 | No | NA | 4 | 0 | 1 | Not-determined | Genome-fragment | NA | NA | 0 |
| S11_vs_k141_10924\|\|full | 4257 | No | NA | 6 | 0 | 0 | Not-determined | Genome-fragment | NA | NA | 0 |
| S16_vs_140160\|\|full | 1219 | Yes | 532 | 2 | 1 | 1 | Low-quality | Genome-fragment | 0.6 | HMM-based (lower-bound) | 56.36 |
| S10_extracted_61490 | 1219 | Yes | 786 | 4 | 3 | 1 | Low-quality | Genome-fragment | 2.4 | AAI-based (medium-confidence) | 35.52 |
| S10_vs_84224\|\|full | 1219 | No | NA | 2 | 1 | 0 | Low-quality | Genome-fragment | 1.47 | AAI-based (high-confidence) | 0 |
| S12_vs_k141_10097\|\|full | 1219 | No | NA | 4 | 3 | 0 | Low-quality | Genome-fragment | 1.99 | AAI-based (high-confidence) | 0 |
| S8_vs_k141_112174\|\|full | 1219 | No | NA | 3 | 0 | 0 | Low-quality | Genome-fragment | 2.27 | AAI-based (high-confidence) | 0 |
| S11_vs_k141_62773\|\|full | 1351 | No | NA | 3 | 0 | 0 | Not-determined | Genome-fragment | NA | NA | 0 |
| S17_extracted_k141_117251 | 1220 | No | NA | 1 | 0 | 0 | Low-quality | Genome-fragment | 1.61 | AAI-based (medium-confidence) | 0 |
| S11_vs_k141_68842\|\|full | 1948 | No | NA | 2 | 0 | 1 | Not-determined | Genome-fragment | NA | NA | 0 |
| S11_vs_k141_43482\|\|full | 1135 | No | NA | 3 | 0 | 0 | Not-determined | Genome-fragment | NA | NA | 0 |
| S11_vs_k141_49362\|\|full | 8365 | No | NA | 6 | 0 | 6 | Not-determined | Genome-fragment | NA | NA | 0 |
| S17_vs_k141_7868\|\|full | 1220 | No | NA | 4 | 1 | 0 | Low-quality | Genome-fragment | 2.86 | AAI-based (high-confidence) | 0 |
| S17_vs_k141_3436\|\|lt2gene | 1220 | No | NA | 2 | 2 | 0 | Low-quality | Genome-fragment | 3.11 | AAI-based (high-confidence) | 0 |
| S9_vs_k141_16329\|\|full | 1220 | No | NA | 2 | 0 | 0 | Low-quality | Genome-fragment | 3.63 | AAI-based (high-confidence) | 0 |
| S11_vs_k141_82863\|\|full | 1220 | No | NA | 3 | 0 | 0 | Low-quality | Genome-fragment | 3.07 | AAI-based (high-confidence) | 0 |
| S13_vs_112026\|\|full | 1220 | No | NA | 2 | 0 | 0 | Low-quality | Genome-fragment | 0.8 | AAI-based (high-confidence) | 0 |
| S14_vs_k141_23295\|\|full | 1220 | No | NA | 3 | 1 | 0 | Low-quality | Genome-fragment | 2.88 | AAI-based (medium-confidence) | 0 |
| S5_vs_k141_9459\|\|lt2gene | 1220 | No | NA | 2 | 1 | 0 | Low-quality | Genome-fragment | 0.84 | HMM-based (lower-bound) | 0 |
| S11_vs_k141_43533\|\|full | 1191 | No | NA | 2 | 0 | 0 | Not-determined | Genome-fragment | NA | NA | 0 |
| S11_vs_k141_68913\|\|full | 5864 | No | NA | 5 | 0 | 2 | Not-determined | Genome-fragment | NA | NA | 0 |
| S5_vs_k141_88384\|\|lt2gene | 1220 | No | NA | 1 | 1 | 0 | Low-quality | Genome-fragment | 2.53 | AAI-based (high-confidence) | 0 |
| S7_extracted_36307 | 1220 | No | NA | 2 | 0 | 0 | Low-quality | Genome-fragment | 10.67 | AAI-based (medium-confidence) | 0 |
| S11_vs_k141_43557\|\|full | 1689 | No | NA | 3 | 0 | 1 | Not-determined | Genome-fragment | NA | NA | 0 |
| S7_vs_22225\|\|lt2gene | 1220 | No | NA | 2 | 1 | 0 | Low-quality | Genome-fragment | 2.83 | AAI-based (medium-confidence) | 0 |
| S15_extracted_952 | 1220 | No | NA | 5 | 0 | 0 | Low-quality | Genome-fragment | 0.71 | AAI-based (medium-confidence) | 0 |
| S4_extracted_4499 | 1220 | No | NA | 3 | 3 | 0 | Low-quality | Genome-fragment | 4.04 | AAI-based (high-confidence) | 0 |
| S6_vs_k141_25592\|\|full | 1220 | No | NA | 2 | 0 | 0 | Low-quality | Genome-fragment | 0.77 | AAI-based (medium-confidence) | 0 |
| S11_vs_k141_43567\|\|full | 1422 | No | NA | 2 | 0 | 0 | Not-determined | Genome-fragment | NA | NA | 0 |
| S8_extracted_k141_69420 | 1220 | No | NA | 2 | 1 | 0 | Low-quality | Genome-fragment | 0.74 | AAI-based (high-confidence) | 0 |
| S11_vs_k141_49455\|\|full | 2150 | No | NA | 3 | 0 | 2 | Not-determined | Genome-fragment | NA | NA | 0 |
| S11_vs_k141_36895\|\|full | 4309 | No | NA | 6 | 0 | 3 | Not-determined | Genome-fragment | NA | NA | 0 |
| S8_vs_k141_154938\|\|full | 1220 | No | NA | 3 | 0 | 0 | Low-quality | Genome-fragment | 3.44 | AAI-based (medium-confidence) | 0 |
| S9_vs_k141_59855\|\|full | 1221 | No | NA | 3 | 0 | 0 | Low-quality | Genome-fragment | 2.53 | AAI-based (high-confidence) | 0 |
| S7_vs_39984\|\|full | 1221 | No | NA | 3 | 0 | 0 | Low-quality | Genome-fragment | 2.97 | AAI-based (medium-confidence) | 0 |
| S7_vs_78585\|\|full | 1221 | No | NA | 3 | 2 | 0 | Low-quality | Genome-fragment | 3.1 | AAI-based (medium-confidence) | 0 |
| S11_vs_k141_62884\|\|full | 4096 | No | NA | 10 | 0 | 0 | Not-determined | Genome-fragment | NA | NA | 0 |
| S7_vs_133793\|\|full | 1221 | No | NA | 2 | 0 | 0 | Low-quality | Genome-fragment | 2.96 | AAI-based (high-confidence) | 0 |
| S11_vs_k141_95849\|\|full | 2455 | No | NA | 2 | 0 | 0 | Not-determined | Genome-fragment | NA | NA | 0 |
| S16_vs_114919\|\|lt2gene | 1221 | No | NA | 2 | 2 | 0 | Low-quality | Genome-fragment | 2.74 | AAI-based (high-confidence) | 0 |
| S12_vs_k141_125345\|\|lt2gene | 1221 | No | NA | 2 | 1 | 0 | Low-quality | Genome-fragment | 3.71 | AAI-based (medium-confidence) | 0 |
| S17_vs_k141_107860\|\|full | 1222 | No | NA | 2 | 0 | 1 | Low-quality | Genome-fragment | 0.53 | AAI-based (high-confidence) | 0 |
| S11_vs_k141_84565\|\|full | 1222 | No | NA | 2 | 0 | 0 | Low-quality | Genome-fragment | 2.42 | AAI-based (high-confidence) | 0 |
| S11_vs_k141_56762\|\|full | 27623 | No | NA | 51 | 6 | 2 | Medium-quality | Genome-fragment | 70.15 | AAI-based (medium-confidence) | 0 |
| S14_extracted_k141_22810 | 1222 | No | NA | 2 | 1 | 0 | Low-quality | Genome-fragment | 2.71 | AAI-based (high-confidence) | 0 |
| S14_vs_k141_85743\|\|full | 1222 | No | NA | 4 | 0 | 0 | Low-quality | Genome-fragment | 2.83 | AAI-based (high-confidence) | 0 |
| S11_vs_k141_95908\|\|full | 1515 | No | NA | 3 | 0 | 2 | Not-determined | Genome-fragment | NA | NA | 0 |
| S11_vs_k141_36967\|\|full | 4853 | No | NA | 7 | 0 | 2 | Not-determined | Genome-fragment | NA | NA | 0 |
| S11_vs_k141_82331\|\|full | 97795 | Yes | 37251 | 103 | 16 | 31 | Complete | High-quality | 100 | Provirus (high-confidence) | 61.91 |
| S14_vs_k141_100565\|\|full | 1222 | No | NA | 2 | 2 | 0 | Low-quality | Genome-fragment | 3.28 | AAI-based (high-confidence) | 0 |
| S3_extracted_k141_40058 | 1222 | No | NA | 1 | 0 | 1 | Low-quality | Genome-fragment | 0.86 | AAI-based (medium-confidence) | 0 |
| S3_vs_k141_70453\|\|full | 1222 | No | NA | 4 | 0 | 0 | Low-quality | Genome-fragment | 3.8 | AAI-based (high-confidence) | 0 |
| S3_vs_k141_68252\|\|lt2gene | 1222 | No | NA | 2 | 0 | 0 | Low-quality | Genome-fragment | 2.13 | AAI-based (high-confidence) | 0 |
| S5_vs_k141_7298\|\|full | 1222 | No | NA | 3 | 0 | 0 | Low-quality | Genome-fragment | 2.47 | AAI-based (medium-confidence) | 0 |
| S7_vs_86065\|\|lt2gene | 1222 | No | NA | 2 | 1 | 0 | Low-quality | Genome-fragment | 19.34 | AAI-based (high-confidence) | 0 |
| S16_extracted_2449 | 1222 | No | NA | 3 | 2 | 0 | Low-quality | Genome-fragment | 2.91 | AAI-based (high-confidence) | 0 |
| S11_vs_k141_95982\|\|full | 1542 | No | NA | 3 | 0 | 1 | Not-determined | Genome-fragment | NA | NA | 0 |
| S10_vs_51300\|\|full | 1222 | No | NA | 2 | 0 | 1 | Low-quality | Genome-fragment | 2.17 | AAI-based (medium-confidence) | 0 |
| S12_extracted_k141_158287 | 1222 | No | NA | 3 | 0 | 0 | Low-quality | Genome-fragment | 3.15 | AAI-based (medium-confidence) | 0 |
| S11_vs_k141_30253\|\|full | 2281 | No | NA | 3 | 0 | 0 | Not-determined | Genome-fragment | NA | NA | 0 |
| S12_vs_k141_33280\|\|full | 1222 | No | NA | 4 | 0 | 2 | Low-quality | Genome-fragment | 2.23 | AAI-based (medium-confidence) | 0 |
| S11_vs_k141_24817\|\|full | 8558 | No | NA | 7 | 0 | 3 | Not-determined | Genome-fragment | NA | NA | 0 |
| S8_extracted_k141_127452 | 1222 | No | NA | 1 | 1 | 0 | Low-quality | Genome-fragment | 2.97 | HMM-based (lower-bound) | 0 |
| S11_vs_k141_4776\|\|full | 2359 | No | NA | 3 | 0 | 3 | Not-determined | Genome-fragment | NA | NA | 0 |
| S9_vs_k141_90716\|\|full | 1223 | No | NA | 3 | 0 | 0 | Low-quality | Genome-fragment | 0.75 | AAI-based (medium-confidence) | 0 |
| S11_vs_k141_102435\|\|full | 1731 | No | NA | 3 | 0 | 0 | Not-determined | Genome-fragment | NA | NA | 0 |
| S5_extracted_k141_6992 | 1223 | No | NA | 1 | 0 | 1 | Low-quality | Genome-fragment | 0.3 | AAI-based (medium-confidence) | 0 |
| S5_extracted_k141_39032 | 1223 | No | NA | 2 | 1 | 0 | Low-quality | Genome-fragment | 2.03 | AAI-based (high-confidence) | 0 |
| S11_vs_k141_89320\|\|full | 2657 | No | NA | 3 | 0 | 1 | Not-determined | Genome-fragment | NA | NA | 0 |
| S5_vs_k141_141280\|\|full | 1223 | No | NA | 3 | 0 | 0 | Low-quality | Genome-fragment | 2.86 | AAI-based (high-confidence) | 0 |
| S11_vs_k141_96031\|\|full | 2428 | No | NA | 4 | 0 | 2 | Not-determined | Genome-fragment | NA | NA | 0 |
| S7_vs_97560\|\|full | 1223 | No | NA | 4 | 0 | 0 | Low-quality | Genome-fragment | 2.86 | AAI-based (medium-confidence) | 0 |
| S16_extracted_21084 | 1223 | No | NA | 2 | 0 | 0 | Low-quality | Genome-fragment | 2.59 | AAI-based (high-confidence) | 0 |
| S11_vs_k141_96048\|\|full | 1962 | No | NA | 3 | 0 | 0 | Not-determined | Genome-fragment | NA | NA | 0 |
| S10_extracted_70242 | 1223 | No | NA | 3 | 0 | 0 | Low-quality | Genome-fragment | 2.13 | AAI-based (high-confidence) | 0 |
| S11_vs_k141_96059\|\|full | 4942 | No | NA | 7 | 0 | 5 | Not-determined | Genome-fragment | NA | NA | 0 |
| S10_vs_87408\|\|lt2gene | 1223 | No | NA | 2 | 0 | 0 | Low-quality | Genome-fragment | 2.92 | AAI-based (high-confidence) | 0 |
| S12_vs_k141_166154\|\|lt2gene | 1223 | No | NA | 2 | 0 | 1 | Low-quality | Genome-fragment | 1.39 | AAI-based (high-confidence) | 0 |
| S11_vs_k141_37171\|\|full | 2068 | No | NA | 4 | 0 | 0 | Not-determined | Genome-fragment | NA | NA | 0 |
| S11_vs_k141_4842\|\|full | 5747 | No | NA | 3 | 0 | 2 | Not-determined | Genome-fragment | NA | NA | 0 |
| S9_extracted_k141_22052 | 1224 | No | NA | 1 | 0 | 1 | Low-quality | Genome-fragment | 2.09 | AAI-based (high-confidence) | 0 |
| S11_vs_k141_56994\|\|full | 1346 | No | NA | 2 | 0 | 1 | Not-determined | Genome-fragment | NA | NA | 0 |
| S11_vs_k141_56995\|\|full | 2553 | No | NA | 4 | 0 | 0 | Not-determined | Genome-fragment | NA | NA | 0 |
| S11_vs_k141_3840\|\|full | 1224 | No | NA | 3 | 1 | 0 | Low-quality | Genome-fragment | 2.38 | AAI-based (high-confidence) | 0 |
| S11_vs_k141_4868\|\|full | 1571 | No | NA | 3 | 0 | 0 | Not-determined | Genome-fragment | NA | NA | 0 |
| S11_vs_k141_23369\|\|lt2gene | 1224 | No | NA | 2 | 2 | 0 | Low-quality | Genome-fragment | 3.16 | AAI-based (high-confidence) | 0 |
| S11_vs_k141_37193\|\|full | 1203 | No | NA | 2 | 0 | 2 | Not-determined | Genome-fragment | NA | NA | 0 |
| S11_vs_k141_69191\|\|full | 2372 | No | NA | 3 | 0 | 1 | Not-determined | Genome-fragment | NA | NA | 0 |
| S11_vs_k141_82477\|\|full | 1424 | No | NA | 1 | 0 | 0 | Not-determined | Genome-fragment | NA | NA | 0 |
| S13_extracted_33151 | 1224 | No | NA | 1 | 0 | 1 | Low-quality | Genome-fragment | 0.8 | AAI-based (high-confidence) | 0 |
| S14_extracted_k141_13264 | 1224 | No | NA | 1 | 0 | 0 | Low-quality | Genome-fragment | 2.12 | AAI-based (high-confidence) | 0 |
| S11_vs_k141_17611\|\|full | 2345 | No | NA | 4 | 0 | 0 | Not-determined | Genome-fragment | NA | NA | 0 |
| S14_vs_k141_94290\|\|full | 1224 | No | NA | 4 | 0 | 0 | Low-quality | Genome-fragment | 2.84 | AAI-based (high-confidence) | 0 |
| S14_vs_k141_30837\|\|lt2gene | 1224 | No | NA | 2 | 1 | 0 | Low-quality | Genome-fragment | 3.14 | AAI-based (high-confidence) | 0 |
| S11_vs_k141_89403\|\|full | 1617 | No | NA | 3 | 0 | 0 | Not-determined | Genome-fragment | NA | NA | 0 |
| S14_vs_k141_76474\|\|lt2gene | 1224 | No | NA | 2 | 2 | 0 | Low-quality | Genome-fragment | 2.89 | AAI-based (high-confidence) | 0 |
| S1_extracted_k141_109621 | 1224 | No | NA | 1 | 1 | 0 | Low-quality | Genome-fragment | 0.66 | HMM-based (lower-bound) | 0 |
| S11_vs_k141_17628\|\|full | 1499 | No | NA | 2 | 0 | 0 | Not-determined | Genome-fragment | NA | NA | 0 |
| S5_extracted_k141_33384 | 1224 | No | NA | 2 | 2 | 0 | Low-quality | Genome-fragment | 3.59 | AAI-based (high-confidence) | 0 |
| S5_vs_k141_132720\|\|full | 1224 | No | NA | 2 | 0 | 0 | Low-quality | Genome-fragment | 4.3 | AAI-based (high-confidence) | 0 |
| S11_vs_k141_11267\|\|full | 18789 | No | NA | 28 | 13 | 2 | Medium-quality | Genome-fragment | 53.56 | AAI-based (medium-confidence) | 0 |
| S11_vs_k141_17649\|\|full | 1971 | No | NA | 4 | 0 | 0 | Not-determined | Genome-fragment | NA | NA | 0 |
| S16_extracted_16127 | 1224 | No | NA | 2 | 2 | 0 | Low-quality | Genome-fragment | 3.29 | AAI-based (high-confidence) | 0 |
| S16_vs_37872\|\|full | 1224 | No | NA | 3 | 3 | 0 | Low-quality | Genome-fragment | 3.49 | AAI-based (high-confidence) | 0 |
| S10_vs_110997\|\|full | 1224 | No | NA | 3 | 0 | 0 | Low-quality | Genome-fragment | 2.9 | AAI-based (medium-confidence) | 0 |
| S11_vs_k141_49813\|\|full | 1709 | No | NA | 2 | 0 | 2 | Not-determined | Genome-fragment | NA | NA | 0 |
| S12_vs_k141_142966\|\|full | 1224 | No | NA | 3 | 0 | 3 | Low-quality | Genome-fragment | 0.31 | AAI-based (medium-confidence) | 0 |
| S12_vs_k141_19188\|\|full | 1224 | No | NA | 3 | 0 | 0 | Low-quality | Genome-fragment | 3.47 | AAI-based (high-confidence) | 0 |
| S12_vs_k141_51157\|\|full | 1224 | No | NA | 2 | 1 | 0 | Low-quality | Genome-fragment | 2.27 | HMM-based (lower-bound) | 0 |
| S6_extracted_k141_17865 | 1224 | No | NA | 1 | 0 | 1 | Low-quality | Genome-fragment | 0.81 | AAI-based (high-confidence) | 0 |
| S6_extracted_k141_22681 | 1224 | No | NA | 2 | 0 | 1 | Low-quality | Genome-fragment | 2.66 | AAI-based (medium-confidence) | 0 |
| S8_extracted_k141_57635 | 1224 | No | NA | 3 | 0 | 1 | Low-quality | Genome-fragment | 1.09 | AAI-based (high-confidence) | 0 |
| S11_vs_k141_37293\|\|full | 1747 | No | NA | 4 | 0 | 0 | Not-determined | Genome-fragment | NA | NA | 0 |
| S11_vs_k141_11205\|\|full | 1225 | No | NA | 2 | 2 | 0 | Low-quality | Genome-fragment | 2.85 | AAI-based (medium-confidence) | 0 |
| S13_vs_26799\|\|full | 1225 | No | NA | 2 | 0 | 1 | Low-quality | Genome-fragment | 2.29 | AAI-based (medium-confidence) | 0 |
| S13_vs_91521\|\|full | 1225 | No | NA | 3 | 0 | 0 | Low-quality | Genome-fragment | 1.28 | AAI-based (high-confidence) | 0 |
| S14_extracted_k141_52289 | 1225 | No | NA | 2 | 1 | 0 | Low-quality | Genome-fragment | 3.2 | AAI-based (high-confidence) | 0 |
| S14_extracted_k141_34433 | 1225 | No | NA | 2 | 2 | 0 | Low-quality | Genome-fragment | 2.72 | AAI-based (high-confidence) | 0 |
| S11_vs_k141_96269\|\|full | 1044 | No | NA | 3 | 0 | 0 | Not-determined | Genome-fragment | NA | NA | 0 |
| S14_vs_k141_86365\|\|full | 1225 | No | NA | 4 | 0 | 0 | Low-quality | Genome-fragment | 2.29 | AAI-based (medium-confidence) | 0 |
| S5_extracted_k141_108995 | 1225 | No | NA | 2 | 0 | 0 | Low-quality | Genome-fragment | 0.9 | AAI-based (medium-confidence) | 0 |
| S11_vs_k141_37345\|\|full | 1269 | No | NA | 3 | 0 | 0 | Not-determined | Genome-fragment | NA | NA | 0 |
| S5_vs_k141_113291\|\|full | 1225 | No | NA | 4 | 0 | 1 | Low-quality | Genome-fragment | 2.08 | AAI-based (medium-confidence) | 0 |
| S11_vs_k141_82619\|\|full | 1827 | No | NA | 2 | 0 | 1 | Not-determined | Genome-fragment | NA | NA | 0 |
| S7_extracted_11633 | 1225 | No | NA | 3 | 0 | 0 | Low-quality | Genome-fragment | 1.94 | AAI-based (medium-confidence) | 0 |
| S7_vs_22404\|\|full | 1225 | No | NA | 3 | 3 | 0 | Low-quality | Genome-fragment | 2.77 | AAI-based (high-confidence) | 0 |
| S7_vs_31669\|\|full | 1225 | No | NA | 3 | 0 | 0 | Low-quality | Genome-fragment | 2.57 | AAI-based (high-confidence) | 0 |
| S11_vs_k141_76655\|\|full | 8533 | No | NA | 7 | 0 | 6 | Not-determined | Genome-fragment | NA | NA | 0 |
| S7_vs_118160\|\|lt2gene | 1225 | No | NA | 2 | 1 | 0 | Low-quality | Genome-fragment | 2.84 | AAI-based (high-confidence) | 0 |
| S16_vs_52841\|\|full | 1225 | No | NA | 3 | 0 | 1 | Low-quality | Genome-fragment | 0.96 | AAI-based (medium-confidence) | 0 |
| S10_vs_82234\|\|full | 1225 | No | NA | 4 | 2 | 0 | Low-quality | Genome-fragment | 2.66 | AAI-based (high-confidence) | 0 |
| S11_vs_k141_11437\|\|full | 1055 | No | NA | 3 | 0 | 0 | Not-determined | Genome-fragment | NA | NA | 0 |
| S11_vs_k141_17808\|\|full | 1776 | No | NA | 4 | 0 | 0 | Not-determined | Genome-fragment | NA | NA | 0 |
| S11_vs_k141_25132\|\|full | 8110 | No | NA | 7 | 0 | 3 | Not-determined | Genome-fragment | NA | NA | 0 |
| S11_vs_k141_30549\|\|full | 1473 | No | NA | 3 | 0 | 0 | Not-determined | Genome-fragment | NA | NA | 0 |
| S8_extracted_k141_50638 | 1225 | No | NA | 1 | 1 | 0 | Low-quality | Genome-fragment | 3.71 | AAI-based (high-confidence) | 0 |
| S11_vs_k141_57283\|\|full | 2179 | No | NA | 3 | 0 | 2 | Not-determined | Genome-fragment | NA | NA | 0 |
| S11_vs_k141_63229\|\|full | 1615 | No | NA | 3 | 0 | 1 | Not-determined | Genome-fragment | NA | NA | 0 |
| S11_vs_k141_82705\|\|full | 2661 | No | NA | 4 | 0 | 0 | Not-determined | Genome-fragment | NA | NA | 0 |
| S8_vs_k141_17338\|\|full | 1225 | No | NA | 3 | 1 | 0 | Low-quality | Genome-fragment | 2.43 | AAI-based (high-confidence) | 0 |
| S11_vs_k141_69425\|\|full | 2356 | No | NA | 2 | 0 | 0 | Not-determined | Genome-fragment | NA | NA | 0 |
| S17_vs_k141_155691\|\|full | 1226 | No | NA | 3 | 0 | 0 | Low-quality | Genome-fragment | 2.59 | AAI-based (high-confidence) | 0 |
| S11_extracted_k141_73351 | 1226 | No | NA | 3 | 1 | 0 | Low-quality | Genome-fragment | 2.23 | AAI-based (medium-confidence) | 0 |
| S11_extracted_k141_29494 | 1226 | No | NA | 2 | 0 | 1 | Low-quality | Genome-fragment | 0.56 | AAI-based (medium-confidence) | 0 |

| | | | | | | | | | | | |
|---|---|---|---|---|---|---|---|---|---|---|---|
| S11_vs_k141_5037\|\|full | 1847 | No | NA | 2 | 0 | 2 | Not-determined | Genome-fragment | NA | NA | 0 |
| S11_vs_k141_11512\|\|full | 3908 | No | NA | 3 | 0 | 0 | Not-determined | Genome-fragment | NA | NA | 0 |
| S11_vs_k141_76745\|\|full | 1724 | No | NA | 3 | 0 | 1 | Not-determined | Genome-fragment | NA | NA | 0 |
| S13_vs_38685\|\|full | 1226 | No | NA | 2 | 0 | 2 | Low-quality | Genome-fragment | 0.7 | AAI-based (medium-confidence) | 0 |
| S11_vs_k141_63264\|\|full | 1935 | No | NA | 3 | 0 | 2 | Not-determined | Genome-fragment | NA | NA | 0 |
| S14_extracted_k141_22460 | 1226 | No | NA | 4 | 4 | 0 | Low-quality | Genome-fragment | 1.98 | AAI-based (medium-confidence) | 0 |
| S14_extracted_k141_47784 | 1226 | No | NA | 2 | 1 | 0 | Low-quality | Genome-fragment | 2.72 | AAI-based (high-confidence) | 0 |
| S11_vs_k141_30652\|\|full | 2531 | No | NA | 3 | 0 | 2 | Not-determined | Genome-fragment | NA | NA | 0 |
| S14_vs_k141_16732\|\|full | 1226 | No | NA | 3 | 1 | 0 | Low-quality | Genome-fragment | 1.99 | HMM-based (lower-bound) | 0 |
| S1_extracted_k141_35181 | 1226 | No | NA | 2 | 0 | 1 | Low-quality | Genome-fragment | 0.14 | AAI-based (medium-confidence) | 0 |
| S1_vs_k141_3778\|\|full | 1226 | No | NA | 2 | 0 | 1 | Low-quality | Genome-fragment | 1.11 | AAI-based (medium-confidence) | 0 |
| S11_vs_k141_76784\|\|full | 34647 | No | NA | 55 | 20 | 2 | High-quality | High-quality | 99.4 | AAI-based (high-confidence) | 0 |
| S11_vs_k141_17934\|\|full | 3338 | No | NA | 4 | 0 | 0 | Not-determined | Genome-fragment | NA | NA | 0 |
| S11_vs_k141_11563\|\|full | 3943 | No | NA | 5 | 0 | 2 | Not-determined | Genome-fragment | NA | NA | 0 |
| S11_vs_k141_17935\|\|full | 4841 | No | NA | 7 | 0 | 5 | Not-determined | Genome-fragment | NA | NA | 0 |
| S11_vs_k141_37561\|\|full | 2630 | No | NA | 2 | 0 | 2 | Not-determined | Genome-fragment | NA | NA | 0 |
| S5_vs_k141_135346\|\|full | 1226 | No | NA | 2 | 1 | 0 | Low-quality | Genome-fragment | 2.43 | AAI-based (high-confidence) | 0 |
| S11_vs_k141_57410\|\|full | 2300 | No | NA | 4 | 0 | 0 | Not-determined | Genome-fragment | NA | NA | 0 |
| S11_vs_k141_25206\|\|full | 2010 | No | NA | 2 | 0 | 1 | Not-determined | Genome-fragment | NA | NA | 0 |
| S11_vs_k141_37581\|\|full | 3140 | No | NA | 5 | 0 | 0 | Not-determined | Genome-fragment | NA | NA | 0 |
| S11_vs_k141_5077\|\|full | 31125 | No | NA | 40 | 24 | 0 | High-quality | High-quality | 100 | AAI-based (high-confidence) | 0 |
| S7_vs_72170\|\|full | 1226 | No | NA | 2 | 0 | 0 | Low-quality | Genome-fragment | 3.61 | AAI-based (high-confidence) | 0 |
| S11_vs_k141_82825\|\|full | 2412 | No | NA | 4 | 0 | 4 | Not-determined | Genome-fragment | NA | NA | 0 |
| S7_vs_113329\|\|full | 1226 | No | NA | 2 | 0 | 1 | Low-quality | Genome-fragment | 1.93 | AAI-based (medium-confidence) | 0 |
| S11_vs_k141_30739\|\|full | 21433 | No | NA | 37 | 7 | 1 | Medium-quality | Genome-fragment | 66.44 | AAI-based (high-confidence) | 0 |
| S12_extracted_k141_107482 | 1226 | No | NA | 1 | 1 | 0 | Low-quality | Genome-fragment | 0.7 | HMM-based (lower-bound) | 0 |
| S12_vs_k141_174156\|\|full | 1226 | No | NA | 3 | 1 | 0 | Low-quality | Genome-fragment | 2.3 | AAI-based (medium-confidence) | 0 |
| S12_vs_k141_136286\|\|full | 1226 | No | NA | 3 | 0 | 0 | Low-quality | Genome-fragment | 3.83 | AAI-based (high-confidence) | 0 |
| S12_vs_k141_58496\|\|full | 1226 | No | NA | 3 | 0 | 0 | Low-quality | Genome-fragment | 3.04 | AAI-based (medium-confidence) | 0 |
| S11_vs_k141_30764\|\|full | 1235 | No | NA | 3 | 0 | 0 | Not-determined | Genome-fragment | NA | NA | 0 |
| S12_vs_k141_125306\|\|full | 1226 | No | NA | 3 | 0 | 0 | Low-quality | Genome-fragment | 1.98 | AAI-based (medium-confidence) | 0 |
| S11_vs_k141_69566\|\|full | 1407 | No | NA | 4 | 0 | 0 | Not-determined | Genome-fragment | NA | NA | 0 |
| S6_vs_k141_9901\|\|full | 1226 | No | NA | 3 | 0 | 1 | Low-quality | Genome-fragment | 0.96 | AAI-based (high-confidence) | 0 |
| S6_vs_k141_31519\|\|full | 1226 | No | NA | 3 | 0 | 0 | Low-quality | Genome-fragment | 0.81 | AAI-based (medium-confidence) | 0 |
| S8_extracted_k141_139112 | 1226 | No | NA | 3 | 0 | 0 | Low-quality | Genome-fragment | 2.48 | AAI-based (high-confidence) | 0 |
| S11_vs_k141_5140\|\|full | 1113 | No | NA | 3 | 0 | 0 | Not-determined | Genome-fragment | NA | NA | 0 |
| S8_vs_k141_207254\|\|full | 1226 | No | NA | 2 | 0 | 0 | Low-quality | Genome-fragment | 2.63 | AAI-based (medium-confidence) | 0 |
| S13_vs_115283\|\|full | 1227 | No | NA | 2 | 0 | 1 | Low-quality | Genome-fragment | 1.3 | AAI-based (medium-confidence) | 0 |
| S11_vs_k141_25347\|\|full | 1140 | No | NA | 2 | 0 | 1 | Not-determined | Genome-fragment | NA | NA | 0 |
| S11_vs_k141_82944\|\|full | 1390 | No | NA | 3 | 0 | 2 | Not-determined | Genome-fragment | NA | NA | 0 |
| S11_vs_k141_37666\|\|full | 13735 | No | NA | 17 | 7 | 0 | Medium-quality | Genome-fragment | 60.5 | AAI-based (medium-confidence) | 0 |
| S3_extracted_k141_69732 | 1227 | No | NA | 1 | 0 | 0 | Low-quality | Genome-fragment | 1.55 | AAI-based (high-confidence) | 0 |
| S3_extracted_k141_95557 | 1227 | No | NA | 2 | 0 | 0 | Low-quality | Genome-fragment | 1.64 | AAI-based (high-confidence) | 0 |
| S11_vs_k141_76900\|\|full | 2217 | No | NA | 3 | 0 | 2 | Not-determined | Genome-fragment | NA | NA | 0 |
| S11_vs_k141_44221\|\|full | 3661 | No | NA | 5 | 0 | 0 | Not-determined | Genome-fragment | NA | NA | 0 |
| S11_vs_k141_69626\|\|full | 1297 | No | NA | 4 | 0 | 1 | Not-determined | Genome-fragment | NA | NA | 0 |
| S5_extracted_k141_45716 | 1227 | No | NA | 2 | 1 | 0 | Low-quality | Genome-fragment | 0.82 | HMM-based (lower-bound) | 0 |
| S16_vs_61187\|\|full | 1227 | No | NA | 3 | 0 | 0 | Low-quality | Genome-fragment | 2.78 | AAI-based (high-confidence) | 0 |
| S2_extracted_k141_13623 | 1227 | No | NA | 2 | 2 | 0 | Low-quality | Genome-fragment | 4.05 | AAI-based (high-confidence) | 0 |
| S4_extracted_7205 | 1227 | No | NA | 2 | 0 | 2 | Low-quality | Genome-fragment | 0.42 | AAI-based (medium-confidence) | 0 |
| S11_vs_k141_44255\|\|full | 1780 | No | NA | 2 | 0 | 1 | Not-determined | Genome-fragment | NA | NA | 0 |
| S6_extracted_k141_51808 | 1227 | No | NA | 2 | 0 | 1 | Low-quality | Genome-fragment | 0.41 | AAI-based (high-confidence) | 0 |
| S8_extracted_k141_115441 | 1227 | No | NA | 2 | 0 | 0 | Low-quality | Genome-fragment | 0.95 | AAI-based (medium-confidence) | 0 |
| S17_extracted_k141_94303 | 1228 | No | NA | 5 | 0 | 0 | Low-quality | Genome-fragment | 0.32 | AAI-based (medium-confidence) | 0 |
| S9_extracted_k141_37832 | 1228 | No | NA | 3 | 1 | 0 | Low-quality | Genome-fragment | 2.24 | AAI-based (high-confidence) | 0 |
| S9_vs_k141_89331\|\|full | 1228 | No | NA | 2 | 0 | 1 | Low-quality | Genome-fragment | 2.18 | AAI-based (medium-confidence) | 0 |
| S9_vs_k141_47758\|\|full | 1228 | No | NA | 3 | 0 | 2 | Low-quality | Genome-fragment | 0.69 | AAI-based (medium-confidence) | 0 |
| S11_extracted_k141_53942 | 1228 | No | NA | 2 | 1 | 0 | Low-quality | Genome-fragment | 2.7 | AAI-based (high-confidence) | 0 |
| S14_extracted_k141_66112 | 1228 | No | NA | 3 | 1 | 0 | Low-quality | Genome-fragment | 9.11 | AAI-based (medium-confidence) | 0 |
| S11_vs_k141_69740\|\|full | 1310 | No | NA | 2 | 0 | 1 | Not-determined | Genome-fragment | NA | NA | 0 |
| S1_vs_k141_133999\|\|full | 1228 | No | NA | 3 | 1 | 0 | Low-quality | Genome-fragment | 2.91 | AAI-based (high-confidence) | 0 |
| S1_vs_k141_72470\|\|full | 1228 | No | NA | 2 | 0 | 1 | Low-quality | Genome-fragment | 1.25 | AAI-based (medium-confidence) | 0 |
| S11_vs_k141_50108\|\|full | 8747 | No | NA | 9 | 0 | 7 | Not-determined | Genome-fragment | NA | NA | 0 |
| S3_vs_k141_73375\|\|full | 1228 | No | NA | 4 | 3 | 0 | Low-quality | Genome-fragment | 2.02 | AAI-based (high-confidence) | 0 |
| S3_vs_k141_104851\|\|full | 1228 | No | NA | 3 | 0 | 0 | Low-quality | Genome-fragment | 2.15 | AAI-based (high-confidence) | 0 |
| S5_vs_k141_58948\|\|full | 1228 | No | NA | 3 | 1 | 0 | Low-quality | Genome-fragment | 3.1 | AAI-based (high-confidence) | 0 |
| S11_vs_k141_25447\|\|full | 1107 | No | NA | 2 | 0 | 0 | Not-determined | Genome-fragment | NA | NA | 0 |
| S7_extracted_123190 | 1228 | No | NA | 3 | 0 | 0 | Low-quality | Genome-fragment | 1.65 | AAI-based (high-confidence) | 0 |
| S7_vs_75108\|\|full | 1228 | No | NA | 2 | 1 | 0 | Low-quality | Genome-fragment | 2.54 | AAI-based (medium-confidence) | 0 |
| S7_vs_86528\|\|full | 1228 | No | NA | 3 | 0 | 1 | Low-quality | Genome-fragment | 0.86 | AAI-based (medium-confidence) | 0 |
| S16_extracted_5441 | 1228 | No | NA | 5 | 0 | 1 | Low-quality | Genome-fragment | 3.8 | AAI-based (high-confidence) | 0 |
| S11_vs_k141_63563\|\|full | 1104 | No | NA | 2 | 0 | 0 | Not-determined | Genome-fragment | NA | NA | 0 |
| S16_extracted_32823 | 1228 | No | NA | 4 | 4 | 0 | Low-quality | Genome-fragment | 3.31 | AAI-based (high-confidence) | 0 |
| S12_extracted_k141_130123 | 1228 | No | NA | 3 | 0 | 3 | Low-quality | Genome-fragment | 0.32 | AAI-based (medium-confidence) | 0 |
| S12_extracted_k141_170907 | 1228 | No | NA | 2 | 0 | 1 | Low-quality | Genome-fragment | 0.76 | AAI-based (medium-confidence) | 0 |
| S12_vs_k141_147023\|\|full | 1228 | No | NA | 5 | 0 | 0 | Low-quality | Genome-fragment | 2.93 | AAI-based (medium-confidence) | 0 |
| S11_vs_k141_69816\|\|full | 3190 | No | NA | 8 | 0 | 0 | Not-determined | Genome-fragment | NA | NA | 0 |
| S11_vs_k141_25485\|\|full | 1338 | No | NA | 3 | 0 | 0 | Not-determined | Genome-fragment | NA | NA | 0 |
| S11_vs_k141_77087\|\|full | 2359 | No | NA | 3 | 0 | 0 | Not-determined | Genome-fragment | NA | NA | 0 |
| S15_extracted_70730 | 1228 | No | NA | 2 | 0 | 0 | Low-quality | Genome-fragment | 2.73 | AAI-based (medium-confidence) | 0 |
| S6_extracted_k141_40405 | 1228 | No | NA | 3 | 1 | 0 | Low-quality | Genome-fragment | 3.46 | AAI-based (high-confidence) | 0 |
| S11_vs_k141_18191\|\|full | 7289 | No | NA | 10 | 0 | 4 | Not-determined | Genome-fragment | NA | NA | 0 |
| S11_vs_k141_77101\|\|full | 3383 | No | NA | 3 | 0 | 0 | Not-determined | Genome-fragment | NA | NA | 0 |
| S11_vs_k141_44383\|\|full | 5070 | No | NA | 5 | 0 | 1 | Not-determined | Genome-fragment | NA | NA | 0 |
| S11_vs_k141_37881\|\|full | 2675 | No | NA | 4 | 0 | 0 | Not-determined | Genome-fragment | NA | NA | 0 |
| S8_vs_k141_53007\|\|full | 1228 | No | NA | 4 | 0 | 0 | Low-quality | Genome-fragment | 2.03 | AAI-based (high-confidence) | 0 |
| S11_vs_k141_77117\|\|full | 9045 | No | NA | 11 | 0 | 7 | Not-determined | Genome-fragment | NA | NA | 0 |
| S11_vs_k141_77067\|\|full | 1229 | No | NA | 4 | 0 | 2 | Low-quality | Genome-fragment | 0.37 | AAI-based (medium-confidence) | 0 |
| S11_vs_k141_50200\|\|full | 4454 | No | NA | 6 | 0 | 0 | Not-determined | Genome-fragment | NA | NA | 0 |
| S13_vs_130289\|\|full | 1229 | No | NA | 2 | 0 | 0 | Low-quality | Genome-fragment | 2.36 | AAI-based (medium-confidence) | 0 |
| S11_vs_k141_102841\|\|full | 1649 | No | NA | 5 | 0 | 0 | Not-determined | Genome-fragment | NA | NA | 0 |
| S11_vs_k141_31087\|\|full | 2171 | No | NA | 3 | 0 | 1 | Not-determined | Genome-fragment | NA | NA | 0 |
| S11_vs_k141_57664\|\|full | 1652 | No | NA | 3 | 0 | 2 | Not-determined | Genome-fragment | NA | NA | 0 |
| S1_extracted_k141_90407 | 1229 | No | NA | 2 | 0 | 1 | Low-quality | Genome-fragment | 0.86 | AAI-based (medium-confidence) | 0 |
| S1_vs_k141_104488\|\|full | 1229 | No | NA | 2 | 1 | 0 | Low-quality | Genome-fragment | 0.53 | HMM-based (lower-bound) | 0 |
| S5_extracted_k141_68693 | 1229 | No | NA | 3 | 0 | 0 | Low-quality | Genome-fragment | 3.27 | AAI-based (medium-confidence) | 0 |
| S5_vs_k141_8004\|\|full | 1229 | No | NA | 3 | 0 | 0 | Low-quality | Genome-fragment | 2.5 | AAI-based (high-confidence) | 0 |
| S7_vs_86975\|\|lt2gene | 1229 | No | NA | 1 | 1 | 0 | Low-quality | Genome-fragment | 3.29 | AAI-based (high-confidence) | 0 |
| S11_vs_k141_50227\|\|full | 1013 | No | NA | 4 | 0 | 0 | Not-determined | Genome-fragment | NA | NA | 0 |
| S16_extracted_171065 | 1229 | No | NA | 2 | 0 | 0 | Low-quality | Genome-fragment | 0.76 | AAI-based (high-confidence) | 0 |
| S16_vs_167261\|\|full | 1229 | No | NA | 5 | 0 | 0 | Low-quality | Genome-fragment | 0.71 | AAI-based (high-confidence) | 0 |
| S11_vs_k141_96864\|\|full | 1137 | No | NA | 3 | 0 | 1 | Not-determined | Genome-fragment | NA | NA | 0 |
| S10_extracted_26754 | 1229 | No | NA | 2 | 0 | 0 | Low-quality | Genome-fragment | 0.94 | AAI-based (high-confidence) | 0 |
| S11_vs_k141_96876\|\|full | 2907 | No | NA | 4 | 0 | 1 | Not-determined | Genome-fragment | NA | NA | 0 |
| S11_vs_k141_18256\|\|full | 1229 | No | NA | 2 | 0 | 1 | Not-determined | Genome-fragment | NA | NA | 0 |
| S10_vs_117586\|\|full | 1229 | No | NA | 3 | 0 | 0 | Low-quality | Genome-fragment | 2.74 | AAI-based (high-confidence) | 0 |
| S11_vs_k141_69897\|\|full | 6612 | No | NA | 5 | 0 | 1 | Not-determined | Genome-fragment | NA | NA | 0 |
| S12_vs_k141_137900\|\|full | 1229 | No | NA | 4 | 0 | 0 | Low-quality | Genome-fragment | 1.53 | AAI-based (high-confidence) | 0 |
| S12_vs_k141_139452\|\|full | 1229 | No | NA | 2 | 1 | 0 | Low-quality | Genome-fragment | 2.9 | AAI-based (medium-confidence) | 0 |
| S11_vs_k141_31140\|\|full | 7306 | No | NA | 10 | 0 | 1 | Not-determined | Genome-fragment | NA | NA | 0 |
| S6_extracted_k141_46334 | 1229 | No | NA | 1 | 0 | 1 | Low-quality | Genome-fragment | 1.35 | AAI-based (medium-confidence) | 0 |
| S8_extracted_k141_99578 | 1229 | No | NA | 2 | 0 | 1 | Low-quality | Genome-fragment | 1.19 | AAI-based (high-confidence) | 0 |
| S11_vs_k141_37929\|\|full | 3496 | No | NA | 4 | 0 | 3 | Not-determined | Genome-fragment | NA | NA | 0 |
| S11_vs_k141_57720\|\|full | 2092 | No | NA | 3 | 0 | 1 | Not-determined | Genome-fragment | NA | NA | 0 |
| S11_vs_k141_69925\|\|full | 1313 | No | NA | 2 | 0 | 0 | Not-determined | Genome-fragment | NA | NA | 0 |
| S8_vs_k141_70713\|\|full | 1229 | No | NA | 2 | 0 | 0 | Low-quality | Genome-fragment | 2.35 | AAI-based (medium-confidence) | 0 |
| S11_vs_k141_63756\|\|full | 2659 | No | NA | 3 | 0 | 3 | Not-determined | Genome-fragment | NA | NA | 0 |
| S9_extracted_k141_58587 | 1230 | No | NA | 4 | 1 | 0 | Low-quality | Genome-fragment | 0.72 | AAI-based (high-confidence) | 0 |
| S11_vs_k141_44489\|\|full | 2230 | No | NA | 4 | 0 | 0 | Not-determined | Genome-fragment | NA | NA | 0 |
| S9_vs_k141_45602\|\|full | 1230 | No | NA | 3 | 2 | 0 | Low-quality | Genome-fragment | 2.88 | AAI-based (medium-confidence) | 0 |
| S11_extracted_k141_60766 | 1230 | No | NA | 1 | 1 | 0 | Low-quality | Genome-fragment | 1.8 | HMM-based (lower-bound) | 0 |
| S11_vs_k141_44506\|\|full | 4736 | No | NA | 3 | 0 | 3 | Not-determined | Genome-fragment | NA | NA | 0 |
| S11_vs_k141_31201\|\|full | 8735 | No | NA | 6 | 0 | 3 | Not-determined | Genome-fragment | NA | NA | 0 |
| S14_vs_k141_6521\|\|full | 1230 | No | NA | 2 | 0 | 1 | Low-quality | Genome-fragment | 2.14 | AAI-based (medium-confidence) | 0 |
| S1_extracted_k141_130935 | 1230 | No | NA | 1 | 0 | 1 | Low-quality | Genome-fragment | 0.65 | AAI-based (high-confidence) | 0 |
| S3_extracted_k141_55063 | 1230 | No | NA | 1 | 0 | 0 | Low-quality | Genome-fragment | 2.51 | AAI-based (high-confidence) | 0 |
| S11_vs_k141_63825\|\|full | 1365 | No | NA | 3 | 0 | 0 | Not-determined | Genome-fragment | NA | NA | 0 |
| S11_vs_k141_50337\|\|full | 5860 | No | NA | 6 | 0 | 4 | Not-determined | Genome-fragment | NA | NA | 0 |
| S11_vs_k141_11850\|\|full | 2031 | No | NA | 2 | 0 | 0 | Not-determined | Genome-fragment | NA | NA | 0 |
| S7_extracted_137464 | 1230 | No | NA | 2 | 0 | 1 | Low-quality | Genome-fragment | 2.81 | AAI-based (medium-confidence) | 0 |
| S7_vs_5041\|\|full | 1230 | No | NA | 4 | 1 | 0 | Low-quality | Genome-fragment | 0.6 | AAI-based (high-confidence) | 0 |
| S11_vs_k141_38049\|\|full | 1508 | No | NA | 2 | 0 | 2 | Not-determined | Genome-fragment | NA | NA | 0 |
| S11_vs_k141_38054\|\|full | 4709 | No | NA | 5 | 0 | 1 | Not-determined | Genome-fragment | NA | NA | 0 |
| S12_extracted_k141_47410 | 1230 | No | NA | 2 | 0 | 1 | Low-quality | Genome-fragment | 0.8 | AAI-based (medium-confidence) | 0 |
| S11_vs_k141_69953\|\|full | 1017 | No | NA | 2 | 0 | 1 | Not-determined | Genome-fragment | NA | NA | 0 |
| S6_extracted_k141_14751 | 1230 | No | NA | 2 | 0 | 2 | Low-quality | Genome-fragment | 0.5 | AAI-based (medium-confidence) | 0 |
| S6_vs_k141_19489\|\|full | 1230 | No | NA | 3 | 0 | 0 | Low-quality | Genome-fragment | 4.23 | AAI-based (high-confidence) | 0 |
| S6_vs_k141_32703\|\|full | 1230 | No | NA | 5 | 1 | 0 | Low-quality | Genome-fragment | 2.37 | AAI-based (medium-confidence) | 0 |
| S11_vs_k141_83298\|\|full | 1402 | No | NA | 3 | 0 | 0 | Not-determined | Genome-fragment | NA | NA | 0 |
| S11_vs_k141_89911\|\|full | 22236 | No | NA | 31 | 13 | 0 | Medium-quality | Genome-fragment | 52.7 | AAI-based (medium-confidence) | 0 |
| S8_extracted_k141_124191 | 1230 | No | NA | 4 | 1 | 0 | Low-quality | Genome-fragment | 1.79 | HMM-based (lower-bound) | 0 |
| S11_vs_k141_83299\|\|full | 7677 | No | NA | 5 | 0 | 2 | Not-determined | Genome-fragment | NA | NA | 0 |
| S11_vs_k141_11918\|\|full | 5619 | No | NA | 4 | 0 | 4 | Not-determined | Genome-fragment | NA | NA | 0 |
| S14_vs_k141_111631\|\|full | 1231 | No | NA | 3 | 0 | 0 | Low-quality | Genome-fragment | 0.83 | AAI-based (medium-confidence) | 0 |
| S11_vs_k141_97035\|\|full | 2454 | No | NA | 2 | 0 | 1 | Not-determined | Genome-fragment | NA | NA | 0 |
| S1_extracted_k141_66689 | 1231 | No | NA | 3 | 0 | 0 | Low-quality | Genome-fragment | 1.24 | AAI-based (high-confidence) | 0 |
| S11_vs_k141_83349\|\|full | 1278 | No | NA | 2 | 0 | 2 | Not-determined | Genome-fragment | NA | NA | 0 |
| S3_extracted_k141_43974 | 1231 | No | NA | 2 | 0 | 0 | Low-quality | Genome-fragment | 2.83 | AAI-based (high-confidence) | 0 |
| S5_extracted_k141_121720 | 1231 | No | NA | 2 | 1 | 0 | Low-quality | Genome-fragment | 3.56 | AAI-based (high-confidence) | 0 |
| S11_vs_k141_77381\|\|full | 1467 | No | NA | 3 | 0 | 2 | Not-determined | Genome-fragment | NA | NA | 0 |
| S11_vs_k141_90010\|\|full | 1668 | No | NA | 3 | 0 | 0 | Not-determined | Genome-fragment | NA | NA | 0 |
| S5_vs_k141_144667\|\|full | 1231 | No | NA | 2 | 0 | 2 | Low-quality | Genome-fragment | 1.38 | AAI-based (medium-confidence) | 0 |
| S11_vs_k141_83417\|\|full | 2904 | No | NA | 3 | 0 | 1 | Not-determined | Genome-fragment | NA | NA | 0 |
| S11_vs_k141_31372\|\|full | 2596 | No | NA | 3 | 0 | 0 | Not-determined | Genome-fragment | NA | NA | 0 |
| S11_vs_k141_97071\|\|full | 19839 | No | NA | 13 | 0 | 8 | Not-determined | Genome-fragment | NA | NA | 0 |
| S11_vs_k141_25790\|\|full | 1684 | No | NA | 3 | 0 | 2 | Not-determined | Genome-fragment | NA | NA | 0 |
| S11_vs_k141_83455\|\|full | 1386 | No | NA | 3 | 0 | 0 | Not-determined | Genome-fragment | NA | NA | 0 |
| S11_vs_k141_25793\|\|full | 1932 | No | NA | 7 | 0 | 0 | Not-determined | Genome-fragment | NA | NA | 0 |
| S7_vs_87621\|\|full | 1231 | No | NA | 3 | 1 | 0 | Low-quality | Genome-fragment | 3.06 | AAI-based (medium-confidence) | 0 |
| S11_vs_k141_11994\|\|full | 54490 | No | NA | 85 | 23 | 1 | High-quality | High-quality | 98.71 | AAI-based (high-confidence) | 0 |
| S11_vs_k141_50530\|\|full | 1647 | No | NA | 3 | 0 | 0 | Not-determined | Genome-fragment | NA | NA | 0 |
| S11_vs_k141_90076\|\|full | 3761 | No | NA | 3 | 0 | 1 | Not-determined | Genome-fragment | NA | NA | 0 |
| S11_vs_k141_57778\|\|full | 2256 | No | NA | 3 | 0 | 2 | Not-determined | Genome-fragment | NA | NA | 0 |
| S7_vs_109694\|\|full | 1231 | No | NA | 3 | 0 | 1 | Low-quality | Genome-fragment | 2.7 | AAI-based (high-confidence) | 0 |
| S16_extracted_20661 | 1231 | No | NA | 3 | 0 | 0 | Low-quality | Genome-fragment | 3.43 | AAI-based (medium-confidence) | 0 |
| S16_extracted_144695 | 1231 | No | NA | 2 | 0 | 2 | Low-quality | Genome-fragment | 2.38 | AAI-based (medium-confidence) | 0 |
| S11_vs_k141_38239\|\|full | 2043 | No | NA | 3 | 0 | 3 | Not-determined | Genome-fragment | NA | NA | 0 |
| S11_vs_k141_18596\|\|full | 1347 | No | NA | 3 | 0 | 2 | Not-determined | Genome-fragment | NA | NA | 0 |
| S16_vs_71434\|\|full | 1231 | No | NA | 3 | 0 | 0 | Low-quality | Genome-fragment | 0.75 | AAI-based (high-confidence) | 0 |
| S10_vs_25691\|\|lt2gene | 1231 | No | NA | 1 | 1 | 0 | Low-quality | Genome-fragment | 0.5 | AAI-based (high-confidence) | 0 |
| S11_vs_k141_57809\|\|full | 1354 | No | NA | 2 | 0 | 0 | Not-determined | Genome-fragment | NA | NA | 0 |
| S12_extracted_k141_106748 | 1231 | No | NA | 2 | 0 | 1 | Low-quality | Genome-fragment | 2.21 | AAI-based (high-confidence) | 0 |
| S12_vs_k141_69372\|\|full | 1231 | No | NA | 2 | 0 | 1 | Low-quality | Genome-fragment | 2.64 | AAI-based (medium-confidence) | 0 |
| S11_vs_k141_25858\|\|full | 1371 | No | NA | 2 | 0 | 1 | Not-determined | Genome-fragment | NA | NA | 0 |
| S15_vs_5425\|\|full | 1231 | No | NA | 3 | 0 | 1 | Low-quality | Genome-fragment | 2.05 | AAI-based (high-confidence) | 0 |
| S8_extracted_k141_79399 | 1231 | No | NA | 3 | 2 | 0 | Low-quality | Genome-fragment | 2.22 | AAI-based (medium-confidence) | 0 |
| S8_extracted_k141_53834 | 1231 | No | NA | 3 | 3 | 0 | Low-quality | Genome-fragment | 4.06 | AAI-based (high-confidence) | 0 |
| S11_vs_k141_5792\|\|full | 1547 | No | NA | 3 | 0 | 2 | Not-determined | Genome-fragment | NA | NA | 0 |
| S13_extracted_68291 | 1232 | No | NA | 5 | 1 | 0 | Low-quality | Genome-fragment | 3.31 | AAI-based (high-confidence) | 0 |
| S1_vs_k141_130107\|\|full | 1232 | Yes | 630 | 3 | 1 | 1 | Low-quality | Genome-fragment | 0.78 | HMM-based (lower-bound) | 48.86 |
| S11_vs_k141_57847\|\|full | 2032 | No | NA | 4 | 0 | 0 | Not-determined | Genome-fragment | NA | NA | 0 |
| S11_vs_k141_5809\|\|full | 1452 | No | NA | 4 | 0 | 0 | Not-determined | Genome-fragment | NA | NA | 0 |
| S11_vs_k141_77528\|\|full | 1665 | No | NA | 4 | 0 | 0 | Not-determined | Genome-fragment | NA | NA | 0 |
| S5_vs_k141_137467\|\|full | 1232 | No | NA | 4 | 0 | 0 | Low-quality | Genome-fragment | 2.02 | AAI-based (medium-confidence) | 0 |
| S11_vs_k141_31477\|\|full | 4484 | No | NA | 6 | 0 | 3 | Not-determined | Genome-fragment | NA | NA | 0 |
| S11_vs_k141_31479\|\|full | 1137 | No | NA | 3 | 0 | 0 | Not-determined | Genome-fragment | NA | NA | 0 |
| S11_vs_k141_64093\|\|full | 2811 | No | NA | 3 | 0 | 2 | Not-determined | Genome-fragment | NA | NA | 0 |
| S11_vs_k141_64109\|\|full | 2007 | No | NA | 3 | 0 | 2 | Not-determined | Genome-fragment | NA | NA | 0 |
| S5_vs_k141_144420\|\|full | 1232 | No | NA | 4 | 1 | 0 | Low-quality | Genome-fragment | 2.32 | AAI-based (medium-confidence) | 0 |
| S7_vs_131367\|\|full | 1232 | No | NA | 2 | 0 | 0 | Low-quality | Genome-fragment | 3.5 | AAI-based (medium-confidence) | 0 |
| S16_vs_52643\|\|full | 1232 | No | NA | 2 | 0 | 0 | Low-quality | Genome-fragment | 0.81 | AAI-based (medium-confidence) | 0 |
| S11_vs_k141_18728\|\|full | 1420 | No | NA | 3 | 0 | 0 | Not-determined | Genome-fragment | NA | NA | 0 |
| S11_vs_k141_44822\|\|full | 1269 | No | NA | 3 | 0 | 2 | Not-determined | Genome-fragment | NA | NA | 0 |
| S11_vs_k141_70211\|\|full | 1808 | No | NA | 3 | 0 | 0 | Not-determined | Genome-fragment | NA | NA | 0 |
| S8_vs_k141_66351\|\|full | 1232 | No | NA | 2 | 0 | 0 | Low-quality | Genome-fragment | 1.91 | AAI-based (high-confidence) | 0 |
| S17_vs_k141_172720\|\|lt2gene | 1233 | No | NA | 1 | 1 | 0 | Low-quality | Genome-fragment | 2.33 | HMM-based (lower-bound) | 0 |
| S9_vs_k141_70216\|\|full | 1233 | No | NA | 2 | 1 | 0 | Low-quality | Genome-fragment | 0.46 | HMM-based (lower-bound) | 0 |
| S11_vs_k141_26012\|\|full | 1580 | No | NA | 4 | 0 | 1 | Not-determined | Genome-fragment | NA | NA | 0 |
| S14_vs_k141_55410\|\|full | 1233 | No | NA | 2 | 2 | 0 | Low-quality | Genome-fragment | 2.39 | AAI-based (high-confidence) | 0 |
| S11_vs_k141_70227\|\|full | 1241 | No | NA | 3 | 0 | 0 | Not-determined | Genome-fragment | NA | NA | 0 |
| S11_vs_k141_12207\|\|full | 2734 | No | NA | 4 | 0 | 4 | Not-determined | Genome-fragment | NA | NA | 0 |
| S11_vs_k141_31557\|\|full | 2341 | No | NA | 4 | 0 | 2 | Not-determined | Genome-fragment | NA | NA | 0 |
| S11_vs_k141_103170\|\|full | 3988 | No | NA | 2 | 0 | 1 | Not-determined | Genome-fragment | NA | NA | 0 |
| S1_vs_k141_54295\|\|full | 1233 | No | NA | 3 | 1 | 0 | Low-quality | Genome-fragment | 2.73 | AAI-based (high-confidence) | 0 |
| S1_vs_k141_57817\|\|full | 1233 | No | NA | 4 | 0 | 0 | Low-quality | Genome-fragment | 3.05 | AAI-based (high-confidence) | 0 |
| S11_vs_k141_83666\|\|full | 24312 | No | NA | 39 | 18 | 0 | Medium-quality | Genome-fragment | 60.55 | AAI-based (high-confidence) | 0 |
| S3_extracted_k141_17374 | 1233 | No | NA | 1 | 1 | 0 | Low-quality | Genome-fragment | 0.85 | HMM-based (lower-bound) | 0 |
| S11_vs_k141_64232\|\|full | 1107 | No | NA | 2 | 0 | 1 | Not-determined | Genome-fragment | NA | NA | 0 |
| S11_vs_k141_70263\|\|full | 4163 | No | NA | 6 | 0 | 0 | Not-determined | Genome-fragment | NA | NA | 0 |
| S11_vs_k141_90335\|\|full | 9662 | No | NA | 14 | 0 | 10 | Not-determined | Genome-fragment | NA | NA | 0 |
| S16_extracted_24279 | 1233 | No | NA | 3 | 0 | 0 | Low-quality | Genome-fragment | 1.89 | AAI-based (medium-confidence) | 0 |
| S11_vs_k141_77704\|\|full | 1516 | No | NA | 3 | 0 | 0 | Not-determined | Genome-fragment | NA | NA | 0 |
| S16_extracted_46016 | 1233 | No | NA | 1 | 1 | 0 | Low-quality | Genome-fragment | 2.19 | AAI-based (medium-confidence) | 0 |
| S11_vs_k141_103252\|\|full | 1200 | No | NA | 2 | 0 | 1 | Not-determined | Genome-fragment | NA | NA | 0 |
| S11_vs_k141_77734\|\|full | 1915 | No | NA | 2 | 0 | 2 | Not-determined | Genome-fragment | NA | NA | 0 |
| S16_extracted_139978 | 1233 | No | NA | 2 | 1 | 0 | Low-quality | Genome-fragment | 2.1 | AAI-based (high-confidence) | 0 |
| S12_vs_k141_176805\|\|full | 1233 | No | NA | 3 | 1 | 0 | Low-quality | Genome-fragment | 3.05 | AAI-based (high-confidence) | 0 |
| S12_vs_k141_160554\|\|lt2gene | 1233 | No | NA | 1 | 1 | 0 | Low-quality | Genome-fragment | 2.71 | AAI-based (medium-confidence) | 0 |
| S17_extracted_k141_16240 | 1234 | No | NA | 3 | 0 | 2 | Low-quality | Genome-fragment | 1.73 | AAI-based (medium-confidence) | 0 |
| S11_vs_k141_103273\|\|full | 1743 | No | NA | 3 | 0 | 1 | Not-determined | Genome-fragment | NA | NA | 0 |
| S9_vs_k141_80834\|\|full | 1234 | No | NA | 2 | 0 | 2 | Low-quality | Genome-fragment | 0.74 | AAI-based (high-confidence) | 0 |
| S11_vs_k141_33683\|\|full | 1234 | No | NA | 3 | 0 | 0 | Low-quality | Genome-fragment | 3.01 | AAI-based (medium-confidence) | 0 |
| S11_vs_k141_64329\|\|full | 2790 | No | NA | 3 | 0 | 2 | Not-determined | Genome-fragment | NA | NA | 0 |
| S13_vs_28539\|\|full | 1234 | No | NA | 3 | 0 | 0 | Low-quality | Genome-fragment | 3.04 | AAI-based (medium-confidence) | 0 |
| S14_extracted_k141_97064 | 1234 | No | NA | 2 | 0 | 1 | Low-quality | Genome-fragment | 2.7 | AAI-based (medium-confidence) | 0 |
| S11_vs_k141_97429\|\|full | 1487 | No | NA | 3 | 0 | 1 | Not-determined | Genome-fragment | NA | NA | 0 |
| S14_extracted_k141_84338 | 1234 | No | NA | 4 | 1 | 0 | Low-quality | Genome-fragment | 2.44 | HMM-based (lower-bound) | 0 |
| S1_extracted_k141_79606 | 1234 | No | NA | 2 | 0 | 2 | Low-quality | Genome-fragment | 0.48 | AAI-based (high-confidence) | 0 |
| S3_vs_k141_53025\|\|full | 1234 | No | NA | 3 | 0 | 0 | Low-quality | Genome-fragment | 1.9 | AAI-based (medium-confidence) | 0 |
| S11_vs_k141_70434\|\|full | 2303 | No | NA | 3 | 0 | 2 | Not-determined | Genome-fragment | NA | NA | 0 |
| S11_vs_k141_90510\|\|full | 3345 | No | NA | 7 | 0 | 0 | Not-determined | Genome-fragment | NA | NA | 0 |
| S11_vs_k141_12483\|\|full | 8822 | No | NA | 8 | 0 | 8 | Not-determined | Genome-fragment | NA | NA | 0 |
| S5_vs_k141_11282\|\|full | 1234 | No | NA | 4 | 1 | 0 | Low-quality | Genome-fragment | 2.27 | AAI-based (medium-confidence) | 0 |
| S7_extracted_689 | 1234 | No | NA | 1 | 0 | 1 | Low-quality | Genome-fragment | 0.13 | AAI-based (medium-confidence) | 0 |
| S7_vs_45576\|\|full | 1234 | No | NA | 3 | 0 | 0 | Low-quality | Genome-fragment | 2.97 | AAI-based (high-confidence) | 0 |
| S16_extracted_22487 | 1234 | No | NA | 2 | 0 | 0 | Low-quality | Genome-fragment | 2.61 | AAI-based (medium-confidence) | 0 |
| S16_vs_134475\|\|full | 1234 | No | NA | 3 | 1 | 0 | Low-quality | Genome-fragment | 2.18 | AAI-based (high-confidence) | 0 |
| S11_vs_k141_45120\|\|full | 3148 | No | NA | 3 | 0 | 1 | Not-determined | Genome-fragment | NA | NA | 0 |
| S12_extracted_k141_30055 | 1234 | No | NA | 3 | 2 | 0 | Low-quality | Genome-fragment | 2.1 | AAI-based (high-confidence) | 0 |
| S15_vs_28877\|\|full | 1234 | No | NA | 3 | 0 | 1 | Low-quality | Genome-fragment | 2.17 | AAI-based (medium-confidence) | 0 |
| S6_vs_k141_41379\|\|lt2gene | 1234 | No | NA | 2 | 0 | 0 | Low-quality | Genome-fragment | 2.89 | AAI-based (high-confidence) | 0 |
| S8_extracted_k141_3486 | 1234 | No | NA | 2 | 0 | 0 | Low-quality | Genome-fragment | 1.67 | AAI-based (high-confidence) | 0 |
| S11_vs_k141_83997\|\|full | 1706 | No | NA | 5 | 0 | 0 | Not-determined | Genome-fragment | NA | NA | 0 |
| S17_vs_k141_128268\|\|full | 1235 | No | NA | 3 | 0 | 0 | Low-quality | Genome-fragment | 2.39 | AAI-based (medium-confidence) | 0 |
| S11_vs_k141_97591\|\|full | 2207 | No | NA | 4 | 0 | 3 | Not-determined | Genome-fragment | NA | NA | 0 |
| S9_extracted_k141_50310 | 1235 | No | NA | 2 | 0 | 0 | Low-quality | Genome-fragment | 3.29 | AAI-based (medium-confidence) | 0 |
| S11_vs_k141_45223\|\|full | 1705 | No | NA | 3 | 0 | 2 | Not-determined | Genome-fragment | NA | NA | 0 |
| S11_vs_k141_51044\|\|full | 1453 | No | NA | 3 | 0 | 2 | Not-determined | Genome-fragment | NA | NA | 0 |
| S11_vs_k141_51057\|\|full | 5975 | No | NA | 5 | 0 | 4 | Not-determined | Genome-fragment | NA | NA | 0 |
| S14_extracted_k141_96922 | 1235 | No | NA | 4 | 4 | 0 | Low-quality | Genome-fragment | 4.06 | AAI-based (high-confidence) | 0 |
| S11_vs_k141_103384\|\|full | 2468 | No | NA | 2 | 0 | 0 | Not-determined | Genome-fragment | NA | NA | 0 |
| S11_vs_k141_58291\|\|full | 3276 | No | NA | 4 | 0 | 0 | Not-determined | Genome-fragment | NA | NA | 0 |
| S1_vs_k141_21973\|\|full | 1235 | No | NA | 3 | 1 | 0 | Low-quality | Genome-fragment | 2.58 | AAI-based (medium-confidence) | 0 |
| S7_extracted_134409 | 1235 | No | NA | 2 | 0 | 0 | Low-quality | Genome-fragment | 1.29 | AAI-based (high-confidence) | 0 |
| S7_vs_125290\|\|full | 1235 | No | NA | 2 | 0 | 0 | Low-quality | Genome-fragment | 2.06 | AAI-based (medium-confidence) | 0 |
| S10_vs_74752\|\|full | 1235 | No | NA | 4 | 1 | 0 | Low-quality | Genome-fragment | 3.33 | AAI-based (high-confidence) | 0 |
| S12_extracted_k141_49937 | 1235 | No | NA | 1 | 1 | 0 | Low-quality | Genome-fragment | 0.67 | HMM-based (lower-bound) | 0 |
| S11_vs_k141_90650\|\|full | 3140 | No | NA | 6 | 0 | 1 | Not-determined | Genome-fragment | NA | NA | 0 |
| S12_extracted_k141_61254 | 1235 | No | NA | 1 | 0 | 0 | Low-quality | Genome-fragment | 1.43 | AAI-based (medium-confidence) | 0 |
| S12_vs_k141_91977\|\|full | 1235 | No | NA | 2 | 0 | 0 | Low-quality | Genome-fragment | 2.82 | AAI-based (medium-confidence) | 0 |
| S12_vs_k141_93061\|\|full | 1235 | No | NA | 2 | 0 | 1 | Low-quality | Genome-fragment | 3.4 | AAI-based (high-confidence) | 0 |
| S12_vs_k141_82246\|\|lt2gene | 1235 | No | NA | 2 | 1 | 0 | Low-quality | Genome-fragment | 3.21 | AAI-based (high-confidence) | 0 |
| S11_vs_k141_38700\|\|full | 2384 | No | NA | 3 | 0 | 2 | Not-determined | Genome-fragment | NA | NA | 0 |
| S8_vs_k141_17906\|\|full | 1235 | No | NA | 2 | 0 | 1 | Low-quality | Genome-fragment | 2.75 | AAI-based (high-confidence) | 0 |
| S11_vs_k141_6246\|\|full | 1946 | No | NA | 3 | 0 | 0 | Not-determined | Genome-fragment | NA | NA | 0 |
| S11_vs_k141_51161\|\|full | 9355 | No | NA | 6 | 0 | 4 | Not-determined | Genome-fragment | NA | NA | 0 |
| S8_vs_k141_207072\|\|full | 1235 | No | NA | 3 | 0 | 0 | Low-quality | Genome-fragment | 8.55 | AAI-based (medium-confidence) | 0 |
| S11_vs_k141_70533\|\|full | 1218 | No | NA | 2 | 0 | 1 | Not-determined | Genome-fragment | NA | NA | 0 |
| S11_extracted_k141_48429 | 1236 | No | NA | 3 | 3 | 0 | Low-quality | Genome-fragment | 3.31 | AAI-based (high-confidence) | 0 |
| S11_vs_k141_86521\|\|lt2gene | 1236 | No | NA | 2 | 2 | 0 | Low-quality | Genome-fragment | 2.82 | AAI-based (high-confidence) | 0 |
| S11_vs_k141_38773\|\|full | 3397 | No | NA | 4 | 0 | 2 | Not-determined | Genome-fragment | NA | NA | 0 |
| S11_vs_k141_64642\|\|full | 1604 | No | NA | 3 | 0 | 0 | Not-determined | Genome-fragment | NA | NA | 0 |
| S13_extracted_2627 | 1236 | No | NA | 1 | 0 | 0 | Low-quality | Genome-fragment | 0.77 | AAI-based (high-confidence) | 0 |
| S11_vs_k141_64657\|\|full | 2196 | No | NA | 2 | 0 | 1 | Not-determined | Genome-fragment | NA | NA | 0 |
| S13_extracted_109394 | 1236 | No | NA | 2 | 0 | 0 | Low-quality | Genome-fragment | 2.52 | AAI-based (high-confidence) | 0 |
| S1_extracted_k141_67804 | 1236 | No | NA | 1 | 0 | 1 | Low-quality | Genome-fragment | 1.52 | AAI-based (high-confidence) | 0 |
| S11_vs_k141_103546\|\|full | 1957 | No | NA | 2 | 0 | 0 | Not-determined | Genome-fragment | NA | NA | 0 |
| S11_vs_k141_6328\|\|full | 10738 | No | NA | 11 | 0 | 5 | Not-determined | Genome-fragment | NA | NA | 0 |
| S5_vs_k141_122165\|\|full | 1236 | No | NA | 4 | 1 | 0 | Low-quality | Genome-fragment | 3.61 | AAI-based (medium-confidence) | 0 |
| S11_vs_k141_84269\|\|full | 7459 | No | NA | 12 | 0 | 0 | Not-determined | Genome-fragment | NA | NA | 0 |
| S11_vs_k141_64715\|\|full | 1673 | No | NA | 4 | 0 | 0 | Not-determined | Genome-fragment | NA | NA | 0 |
| S7_vs_34056\|\|full | 1236 | No | NA | 4 | 0 | 0 | Low-quality | Genome-fragment | 2.13 | AAI-based (high-confidence) | 0 |
| S16_extracted_94711 | 1236 | No | NA | 1 | 0 | 0 | Low-quality | Genome-fragment | 0.24 | AAI-based (medium-confidence) | 0 |
| S10_extracted_3306 | 1236 | No | NA | 2 | 0 | 0 | Low-quality | Genome-fragment | 2.68 | AAI-based (high-confidence) | 0 |
| S11_vs_k141_19361\|\|full | 2505 | No | NA | 5 | 0 | 1 | Not-determined | Genome-fragment | NA | NA | 0 |
| S11_vs_k141_90835\|\|full | 1818 | No | NA | 2 | 0 | 1 | Not-determined | Genome-fragment | NA | NA | 0 |
| S10_extracted_50801 | 1236 | No | NA | 5 | 0 | 0 | Low-quality | Genome-fragment | 2.71 | AAI-based (high-confidence) | 0 |
| S11_vs_k141_38860\|\|full | 39451 | No | NA | 58 | 21 | 0 | High-quality | High-quality | 100 | AAI-based (high-confidence) | 0 |
| S12_extracted_k141_47018 | 1236 | No | NA | 2 | 0 | 0 | Low-quality | Genome-fragment | 2.29 | AAI-based (high-confidence) | 0 |
| S12_vs_k141_156013\|\|full | 1236 | No | NA | 3 | 3 | 0 | Low-quality | Genome-fragment | 2.96 | AAI-based (medium-confidence) | 0 |
| S11_vs_k141_32067\|\|full | 1541 | No | NA | 2 | 0 | 2 | Not-determined | Genome-fragment | NA | NA | 0 |
| S11_vs_k141_51342\|\|full | 2431 | No | NA | 2 | 0 | 2 | Not-determined | Genome-fragment | NA | NA | 0 |
| S12_vs_k141_86325\|\|full | 1236 | No | NA | 2 | 0 | 1 | Low-quality | Genome-fragment | 1.96 | AAI-based (medium-confidence) | 0 |
| S15_extracted_14150 | 1236 | No | NA | 4 | 0 | 1 | Low-quality | Genome-fragment | 0.82 | AAI-based (medium-confidence) | 0 |
| S8_extracted_k141_141653 | 1236 | No | NA | 2 | 0 | 1 | Low-quality | Genome-fragment | 1.4 | AAI-based (medium-confidence) | 0 |
| S8_vs_k141_124572\|\|full | 1236 | No | NA | 3 | 1 | 0 | Low-quality | Genome-fragment | 1.06 | HMM-based (lower-bound) | 0 |
| S8_vs_k141_47209\|\|lt2gene | 1236 | No | NA | 2 | 1 | 0 | Low-quality | Genome-fragment | 2.86 | AAI-based (high-confidence) | 0 |
| S11_vs_k141_6458\|\|full | 42936 | Yes | 37546 | 48 | 12 | 2 | Medium-quality | Genome-fragment | 81.42 | AAI-based (high-confidence) | 12.55 |
| S17_vs_k141_119243\|\|full | 1237 | No | NA | 2 | 0 | 0 | Low-quality | Genome-fragment | 3.07 | AAI-based (high-confidence) | 0 |
| S11_vs_k141_103698\|\|full | 2943 | No | NA | 2 | 0 | 1 | Not-determined | Genome-fragment | NA | NA | 0 |
| S17_vs_k141_41027\|\|full | 1237 | No | NA | 3 | 2 | 0 | Low-quality | Genome-fragment | 2.21 | AAI-based (medium-confidence) | 0 |
| S11_vs_k141_51428\|\|full | 1929 | No | NA | 3 | 0 | 1 | Not-determined | Genome-fragment | NA | NA | 0 |
| S11_vs_k141_246\|\|lt2gene | 1237 | No | NA | 2 | 2 | 0 | Low-quality | Genome-fragment | 1.72 | HMM-based (lower-bound) | 0 |
| S11_vs_k141_39010\|\|full | 1750 | No | NA | 3 | 0 | 0 | Not-determined | Genome-fragment | NA | NA | 0 |
| S14_vs_k141_92469\|\|full | 1237 | No | NA | 4 | 0 | 0 | Low-quality | Genome-fragment | 2.68 | AAI-based (medium-confidence) | 0 |
| S11_vs_k141_70860\|\|full | 1261 | No | NA | 2 | 0 | 0 | Not-determined | Genome-fragment | NA | NA | 0 |
| S3_vs_k141_37261\|\|full | 1237 | No | NA | 3 | 2 | 0 | Low-quality | Genome-fragment | 3.1 | AAI-based (high-confidence) | 0 |
| S11_vs_k141_39036\|\|full | 5425 | No | NA | 3 | 0 | 2 | Not-determined | Genome-fragment | NA | NA | 0 |
| S11_vs_k141_70895\|\|full | 3632 | No | NA | 2 | 0 | 1 | Not-determined | Genome-fragment | NA | NA | 0 |
| S3_vs_k141_21529\|\|full | 1237 | No | NA | 4 | 1 | 0 | Low-quality | Genome-fragment | 2.41 | AAI-based (high-confidence) | 0 |
| S11_vs_k141_91047\|\|full | 1917 | No | NA | 3 | 0 | 2 | Not-determined | Genome-fragment | NA | NA | 0 |
| S7_vs_8737\|\|lt2gene | 1237 | No | NA | 2 | 2 | 0 | Low-quality | Genome-fragment | 3.11 | AAI-based (high-confidence) | 0 |
| S11_vs_k141_91058\|\|full | 1866 | No | NA | 2 | 0 | 2 | Not-determined | Genome-fragment | NA | NA | 0 |
| S11_vs_k141_103809\|\|full | 2276 | No | NA | 4 | 0 | 1 | Not-determined | Genome-fragment | NA | NA | 0 |
| S11_vs_k141_70991\|\|full | 3237 | No | NA | 6 | 0 | 1 | Not-determined | Genome-fragment | NA | NA | 0 |
| S7_vs_75987\|\|lt2gene | 1237 | No | NA | 2 | 1 | 0 | Low-quality | Genome-fragment | 2.94 | AAI-based (medium-confidence) | 0 |
| S16_extracted_70387 | 1237 | No | NA | 2 | 0 | 0 | Low-quality | Genome-fragment | 1.18 | AAI-based (medium-confidence) | 0 |
| S11_vs_k141_91106\|\|full | 1940 | No | NA | 3 | 0 | 2 | Not-determined | Genome-fragment | NA | NA | 0 |
| S4_extracted_23361 | 1237 | No | NA | 2 | 2 | 0 | Low-quality | Genome-fragment | 4.07 | AAI-based (high-confidence) | 0 |
| S11_vs_k141_32189\|\|full | 2949 | No | NA | 2 | 0 | 2 | Not-determined | Genome-fragment | NA | NA | 0 |
| S14_vs_k141_45506\|\|full | 1238 | No | NA | 3 | 2 | 0 | Low-quality | Genome-fragment | 2.66 | AAI-based (high-confidence) | 0 |
| S11_vs_k141_84480\|\|full | 1541 | No | NA | 3 | 0 | 0 | Not-determined | Genome-fragment | NA | NA | 0 |
| S11_vs_k141_51614\|\|full | 1430 | No | NA | 2 | 0 | 1 | Not-determined | Genome-fragment | NA | NA | 0 |
| S11_vs_k141_103885\|\|full | 1800 | No | NA | 3 | 0 | 1 | Not-determined | Genome-fragment | NA | NA | 0 |
| S1_extracted_k141_135120 | 1238 | No | NA | 2 | 1 | 0 | Low-quality | Genome-fragment | 0.83 | AAI-based (medium-confidence) | 0 |
| S11_vs_k141_51642\|\|full | 2465 | No | NA | 3 | 0 | 1 | Not-determined | Genome-fragment | NA | NA | 0 |
| S1_vs_k141_49297\|\|full | 1238 | No | NA | 1 | 0 | 1 | Low-quality | Genome-fragment | 2.4 | AAI-based (high-confidence) | 0 |
| S1_vs_k141_148305\|\|full | 1238 | No | NA | 2 | 0 | 0 | Low-quality | Genome-fragment | 0.86 | AAI-based (medium-confidence) | 0 |
| S11_vs_k141_71085\|\|full | 1687 | No | NA | 4 | 0 | 2 | Not-determined | Genome-fragment | NA | NA | 0 |
| S1_vs_k141_144137\|\|lt2gene | 1238 | No | NA | 1 | 1 | 0 | Low-quality | Genome-fragment | 0.85 | AAI-based (high-confidence) | 0 |
| S11_vs_k141_71090\|\|full | 1326 | No | NA | 2 | 0 | 1 | Not-determined | Genome-fragment | NA | NA | 0 |
| S3_extracted_k141_101749 | 1238 | No | NA | 3 | 0 | 0 | Low-quality | Genome-fragment | 0.76 | AAI-based (high-confidence) | 0 |
| S5_extracted_k141_141965 | 1238 | No | NA | 3 | 2 | 0 | Low-quality | Genome-fragment | 2.71 | AAI-based (high-confidence) | 0 |
| S7_vs_100120\|\|full | 1238 | No | NA | 2 | 0 | 0 | Low-quality | Genome-fragment | 2.28 | AAI-based (medium-confidence) | 0 |
| S7_vs_105515\|\|full | 1238 | No | NA | 3 | 1 | 0 | Low-quality | Genome-fragment | 2.79 | AAI-based (high-confidence) | 0 |
| S11_vs_k141_71208\|\|full | 2112 | No | NA | 3 | 0 | 1 | Not-determined | Genome-fragment | NA | NA | 0 |
| S16_extracted_43350 | 1238 | No | NA | 4 | 1 | 0 | Low-quality | Genome-fragment | 3.84 | AAI-based (high-confidence) | 0 |
| S11_vs_k141_71262\|\|full | 2913 | No | NA | 4 | 0 | 1 | Not-determined | Genome-fragment | NA | NA | 0 |
| S16_vs_33377\|\|full | 1238 | No | NA | 3 | 0 | 0 | Low-quality | Genome-fragment | 2.6 | AAI-based (medium-confidence) | 0 |
| S10_extracted_18727 | 1238 | No | NA | 3 | 0 | 0 | Low-quality | Genome-fragment | 1.65 | AAI-based (medium-confidence) | 0 |
| S11_vs_k141_32438\|\|full | 1900 | No | NA | 3 | 0 | 0 | Not-determined | Genome-fragment | NA | NA | 0 |
| S12_vs_k141_178716\|\|full | 1238 | No | NA | 2 | 2 | 0 | Low-quality | Genome-fragment | 1.99 | HMM-based (lower-bound) | 0 |
| S11_vs_k141_71365\|\|full | 1373 | No | NA | 2 | 0 | 1 | Not-determined | Genome-fragment | NA | NA | 0 |
| S15_vs_12115\|\|full | 1238 | No | NA | 4 | 1 | 0 | Low-quality | Genome-fragment | 2.72 | AAI-based (high-confidence) | 0 |
| S11_vs_k141_92442\|\|0_partial | 15242 | No | NA | 12 | 0 | 7 | Not-determined | Genome-fragment | NA | NA | 0 |
| S11_vs_k141_2527\|\|0_partial | 10385 | No | NA | 8 | 0 | 8 | Not-determined | Genome-fragment | NA | NA | 0 |
| S8_vs_k141_61814\|\|full | 1238 | No | NA | 4 | 1 | 0 | Low-quality | Genome-fragment | 2.97 | AAI-based (high-confidence) | 0 |
| S8_vs_k141_8216\|\|full | 1238 | No | NA | 3 | 0 | 0 | Low-quality | Genome-fragment | 3.81 | AAI-based (high-confidence) | 0 |
| S17_extracted_k141_105503 | 1239 | No | NA | 3 | 0 | 0 | Low-quality | Genome-fragment | 2.33 | AAI-based (medium-confidence) | 0 |
| S9_extracted_k141_64137 | 1239 | No | NA | 3 | 0 | 1 | Low-quality | Genome-fragment | 0.48 | AAI-based (medium-confidence) | 0 |
| S11_vs_k141_64544\|\|0_partial | 12817 | No | NA | 12 | 0 | 6 | Not-determined | Genome-fragment | NA | NA | 0 |
| S13_vs_31328\|\|full | 1239 | No | NA | 3 | 0 | 0 | Low-quality | Genome-fragment | 0.76 | AAI-based (medium-confidence) | 0 |
| S13_vs_12091\|\|lt2gene | 1239 | No | NA | 1 | 1 | 0 | Low-quality | Genome-fragment | 3.55 | AAI-based (high-confidence) | 0 |
| S3_vs_k141_58424\|\|lt2gene | 1239 | No | NA | 2 | 1 | 0 | Low-quality | Genome-fragment | 2.63 | AAI-based (high-confidence) | 0 |
| S5_vs_k141_66264\|\|full | 1239 | No | NA | 2 | 0 | 0 | Low-quality | Genome-fragment | 2.49 | AAI-based (medium-confidence) | 0 |
| S5_vs_k141_41331\|\|lt2gene | 1239 | No | NA | 1 | 1 | 0 | Low-quality | Genome-fragment | 2.04 | AAI-based (high-confidence) | 0 |
| S15_vs_42528\|\|full | 1239 | No | NA | 4 | 0 | 0 | Low-quality | Genome-fragment | 2.02 | AAI-based (high-confidence) | 0 |
| S11_vs_k141_32963\|\|lt2gene | 1001 | No | NA | 1 | 0 | 0 | Not-determined | Genome-fragment | NA | NA | 0 |
| S6_vs_k141_53387\|\|full | 1239 | No | NA | 3 | 0 | 0 | Low-quality | Genome-fragment | 3.02 | AAI-based (high-confidence) | 0 |
| S8_extracted_k141_167048 | 1239 | No | NA | 1 | 0 | 1 | Low-quality | Genome-fragment | 0.66 | AAI-based (medium-confidence) | 0 |
| S8_extracted_k141_32148 | 1239 | No | NA | 1 | 0 | 0 | Low-quality | Genome-fragment | 2.89 | AAI-based (medium-confidence) | 0 |
| S17_extracted_k141_90748 | 1240 | No | NA | 2 | 0 | 0 | Low-quality | Genome-fragment | 3 | AAI-based (high-confidence) | 0 |
| S17_vs_k141_125277\|\|lt2gene | 1240 | No | NA | 2 | 2 | 0 | Low-quality | Genome-fragment | 3.11 | AAI-based (medium-confidence) | 0 |
| S11_extracted_k141_42162 | 1240 | No | NA | 4 | 0 | 0 | Low-quality | Genome-fragment | 0.81 | AAI-based (high-confidence) | 0 |
| S11_vs_k141_86737\|\|full | 1240 | No | NA | 2 | 1 | 0 | Low-quality | Genome-fragment | 0.67 | HMM-based (lower-bound) | 0 |
| S11_vs_k141_34107\|\|full | 1240 | No | NA | 4 | 0 | 0 | Low-quality | Genome-fragment | 2.22 | AAI-based (high-confidence) | 0 |
| S11_vs_k141_33431\|\|lt2gene | 2048 | No | NA | 1 | 0 | 1 | Not-determined | Genome-fragment | NA | NA | 0 |
| S13_extracted_30379 | 1240 | No | NA | 2 | 0 | 0 | Low-quality | Genome-fragment | 3.7 | AAI-based (medium-confidence) | 0 |
| S13_extracted_46447 | 1240 | No | NA | 2 | 0 | 1 | Low-quality | Genome-fragment | 0.78 | AAI-based (high-confidence) | 0 |
| S14_vs_k141_99770\|\|lt2gene | 1240 | No | NA | 2 | 1 | 0 | Low-quality | Genome-fragment | 2.85 | AAI-based (medium-confidence) | 0 |
| S1_vs_k141_84960\|\|full | 1240 | No | NA | 3 | 1 | 0 | Low-quality | Genome-fragment | 0.35 | HMM-based (lower-bound) | 0 |
| S11_vs_k141_59570\|\|lt2gene | 1352 | No | NA | 1 | 0 | 0 | Not-determined | Genome-fragment | NA | NA | 0 |
| S1_vs_k141_19116\|\|full | 1240 | No | NA | 2 | 0 | 0 | Low-quality | Genome-fragment | 2.74 | AAI-based (medium-confidence) | 0 |
| S3_extracted_k141_16330 | 1240 | No | NA | 2 | 0 | 0 | Low-quality | Genome-fragment | 1.94 | AAI-based (medium-confidence) | 0 |
| S5_extracted_k141_89611 | 1240 | No | NA | 1 | 0 | 1 | Low-quality | Genome-fragment | 3.17 | AAI-based (high-confidence) | 0 |
| S12_extracted_k141_49159 | 1240 | No | NA | 4 | 3 | 0 | Low-quality | Genome-fragment | 2.75 | AAI-based (high-confidence) | 0 |
| S15_extracted_72823 | 1240 | No | NA | 1 | 0 | 1 | Low-quality | Genome-fragment | 0.7 | AAI-based (high-confidence) | 0 |
| S2_extracted_k141_16713 | 1240 | No | NA | 2 | 2 | 0 | Low-quality | Genome-fragment | 4.09 | AAI-based (high-confidence) | 0 |
| S8_extracted_k141_70924 | 1240 | No | NA | 2 | 0 | 2 | Low-quality | Genome-fragment | 3.96 | AAI-based (medium-confidence) | 0 |
| S13_vs_130515\|\|full | 1241 | No | NA | 2 | 1 | 0 | Low-quality | Genome-fragment | 0.8 | HMM-based (lower-bound) | 0 |
| S11_vs_k141_79633\|\|lt2gene | 2239 | No | NA | 1 | 0 | 0 | Not-determined | Genome-fragment | NA | NA | 0 |
| S14_extracted_k141_86540 | 1241 | No | NA | 3 | 1 | 0 | Low-quality | Genome-fragment | 1.56 | AAI-based (medium-confidence) | 0 |
| S16_extracted_28551 | 1241 | No | NA | 4 | 1 | 0 | Low-quality | Genome-fragment | 2.79 | AAI-based (high-confidence) | 0 |
| S16_vs_18703\|\|full | 1241 | No | NA | 2 | 0 | 1 | Low-quality | Genome-fragment | 2.01 | AAI-based (medium-confidence) | 0 |
| S10_extracted_5422 | 1241 | No | NA | 4 | 2 | 0 | Low-quality | Genome-fragment | 3.18 | AAI-based (high-confidence) | 0 |
| S12_vs_k141_42601\|\|full | 1241 | No | NA | 3 | 1 | 0 | Low-quality | Genome-fragment | 3.05 | AAI-based (high-confidence) | 0 |
| S8_vs_k141_165190\|\|full | 1241 | No | NA | 5 | 1 | 0 | Low-quality | Genome-fragment | 0.8 | AAI-based (high-confidence) | 0 |
| S11_extracted_k141_79420 | 1242 | No | NA | 3 | 0 | 2 | Low-quality | Genome-fragment | 2.77 | AAI-based (medium-confidence) | 0 |
| S11_vs_k141_58174\|\|full | 1242 | No | NA | 2 | 0 | 0 | Low-quality | Genome-fragment | 3.08 | AAI-based (high-confidence) | 0 |
| S11_vs_k141_54000\|\|lt2gene | 1170 | No | NA | 2 | 0 | 0 | Not-determined | Genome-fragment | NA | NA | 0 |
| S11_vs_k141_8425\|\|lt2gene | 1104 | No | NA | 1 | 0 | 0 | Not-determined | Genome-fragment | NA | NA | 0 |
| S14_vs_k141_23890\|\|full | 1242 | No | NA | 3 | 0 | 0 | Low-quality | Genome-fragment | 2.36 | AAI-based (high-confidence) | 0 |
| S14_vs_k141_18459\|\|full | 1242 | No | NA | 3 | 0 | 0 | Low-quality | Genome-fragment | 1.48 | AAI-based (medium-confidence) | 0 |
| S14_vs_k141_55003\|\|full | 1242 | No | NA | 3 | 0 | 0 | Low-quality | Genome-fragment | 3.18 | AAI-based (medium-confidence) | 0 |
| S14_vs_k141_55608\|\|lt2gene | 1242 | No | NA | 2 | 2 | 0 | Low-quality | Genome-fragment | 3.68 | AAI-based (high-confidence) | 0 |
| S3_extracted_k141_70218 | 1242 | No | NA | 5 | 1 | 0 | Low-quality | Genome-fragment | 3.14 | AAI-based (high-confidence) | 0 |
| S5_vs_k141_126040\|\|full | 1242 | No | NA | 3 | 0 | 0 | Low-quality | Genome-fragment | 2.07 | AAI-based (high-confidence) | 0 |
| S5_vs_k141_53769\|\|full | 1242 | No | NA | 4 | 1 | 0 | Low-quality | Genome-fragment | 2.25 | HMM-based (lower-bound) | 0 |
| S11_vs_k141_87368\|\|lt2gene | 2836 | No | NA | 2 | 0 | 0 | Not-determined | Genome-fragment | NA | NA | 0 |
| S11_vs_k141_80521\|\|lt2gene | 1323 | No | NA | 1 | 0 | 0 | Not-determined | Genome-fragment | NA | NA | 0 |
| S5_vs_k141_50774\|\|lt2gene | 1242 | No | NA | 1 | 1 | 0 | Low-quality | Genome-fragment | 1.81 | HMM-based (lower-bound) | 0 |
| S7_extracted_119757 | 1242 | No | NA | 2 | 0 | 1 | Low-quality | Genome-fragment | 0.48 | AAI-based (medium-confidence) | 0 |
| S7_vs_23029\|\|full | 1242 | No | NA | 4 | 1 | 0 | Low-quality | Genome-fragment | 2.93 | HMM-based (lower-bound) | 0 |
| S16_extracted_84914 | 1242 | No | NA | 1 | 0 | 0 | Low-quality | Genome-fragment | 5.1 | AAI-based (medium-confidence) | 0 |
| S12_vs_k141_131199\|\|full | 1242 | No | NA | 2 | 1 | 0 | Low-quality | Genome-fragment | 0.67 | HMM-based (lower-bound) | 0 |
| S8_vs_k141_116731\|\|full | 1242 | No | NA | 3 | 3 | 0 | Low-quality | Genome-fragment | 2.08 | AAI-based (high-confidence) | 0 |
| S8_vs_k141_50089\|\|full | 1242 | No | NA | 3 | 3 | 0 | Low-quality | Genome-fragment | 3.12 | AAI-based (high-confidence) | 0 |
| S11_vs_k141_28987\|\|lt2gene | 2732 | No | NA | 2 | 0 | 1 | Not-determined | Genome-fragment | NA | NA | 0 |
| S17_vs_k141_87837\|\|lt2gene | 1243 | No | NA | 2 | 0 | 0 | Low-quality | Genome-fragment | 2.4 | AAI-based (high-confidence) | 0 |
| S11_vs_k141_65342\|\|full | 1243 | No | NA | 3 | 0 | 0 | Low-quality | Genome-fragment | 3.71 | AAI-based (medium-confidence) | 0 |
| S13_vs_8351\|\|full | 1243 | No | NA | 4 | 0 | 0 | Low-quality | Genome-fragment | 2.95 | AAI-based (medium-confidence) | 0 |
| S14_extracted_k141_5373 | 1243 | No | NA | 2 | 1 | 0 | Low-quality | Genome-fragment | 7.35 | AAI-based (high-confidence) | 0 |
| S14_vs_k141_86814\|\|full | 1243 | No | NA | 4 | 1 | 0 | Low-quality | Genome-fragment | 0.8 | AAI-based (medium-confidence) | 0 |

| | | | | | | | | | | | |
|---|---|---|---|---|---|---|---|---|---|---|---|
| S5_vs_k141_120462\|\|full | 1243 | No | NA | 3 | 1 | 0 | Low-quality | Genome-fragment | 0.53 | AAI-based (medium-confidence) | 0 |
| S16_extracted_105999 | 1243 | No | NA | 1 | 0 | 0 | Low-quality | Genome-fragment | 0.9 | AAI-based (high-confidence) | 0 |
| S16_vs_113140\|\|lt2gene | 1243 | No | NA | 1 | 1 | 0 | Low-quality | Genome-fragment | 1.68 | AAI-based (high-confidence) | 0 |
| S15_vs_35113\|\|full | 1243 | No | NA | 3 | 0 | 0 | Low-quality | Genome-fragment | 2.24 | AAI-based (high-confidence) | 0 |
| S11_vs_k141_29797\|\|lt2gene | 2426 | No | NA | 2 | 0 | 1 | Not-determined | Genome-fragment | NA | NA | 0 |
| S15_vs_29426\|\|lt2gene | 1243 | No | NA | 2 | 1 | 0 | Low-quality | Genome-fragment | 0.62 | HMM-based (lower-bound) | 0 |
| S11_vs_k141_49195\|\|lt2gene | 1383 | No | NA | 2 | 0 | 2 | Not-determined | Genome-fragment | NA | NA | 0 |
| S11_vs_k141_49235\|\|lt2gene | 1583 | No | NA | 2 | 0 | 1 | Not-determined | Genome-fragment | NA | NA | 0 |
| S11_vs_k141_49323\|\|lt2gene | 1202 | No | NA | 1 | 0 | 0 | Not-determined | Genome-fragment | NA | NA | 0 |
| S2_vs_k141_10117\|\|full | 1243 | No | NA | 2 | 1 | 0 | Low-quality | Genome-fragment | 2.99 | AAI-based (high-confidence) | 0 |
| S8_extracted_k141_3969 | 1243 | No | NA | 3 | 0 | 0 | Low-quality | Genome-fragment | 2.89 | AAI-based (medium-confidence) | 0 |
| S8_extracted_k141_111184 | 1243 | No | NA | 2 | 1 | 0 | Low-quality | Genome-fragment | 2.99 | AAI-based (medium-confidence) | 0 |
| S8_vs_k141_188628\|\|lt2gene | 1243 | No | NA | 1 | 1 | 0 | Low-quality | Genome-fragment | 2.27 | HMM-based (lower-bound) | 0 |
| S17_extracted_k141_123545 | 1244 | No | NA | 2 | 0 | 1 | Low-quality | Genome-fragment | 2.17 | AAI-based (medium-confidence) | 0 |
| S9_extracted_k141_44224 | 1244 | No | NA | 2 | 0 | 0 | Low-quality | Genome-fragment | 1.9 | AAI-based (medium-confidence) | 0 |
| S11_extracted_k141_84135 | 1244 | No | NA | 2 | 2 | 0 | Low-quality | Genome-fragment | 3.07 | AAI-based (high-confidence) | 0 |
| S11_vs_k141_68876\|\|full | 1244 | No | NA | 2 | 1 | 0 | Low-quality | Genome-fragment | 1.11 | HMM-based (lower-bound) | 0 |
| S11_vs_k141_30788\|\|full | 1244 | No | NA | 5 | 0 | 0 | Low-quality | Genome-fragment | 2.71 | AAI-based (high-confidence) | 0 |
| S11_vs_k141_49946\|\|lt2gene | 1267 | No | NA | 1 | 0 | 0 | Not-determined | Genome-fragment | NA | NA | 0 |
| S11_vs_k141_51127\|\|full | 1244 | No | NA | 3 | 0 | 0 | Low-quality | Genome-fragment | 0.63 | AAI-based (medium-confidence) | 0 |
| S13_extracted_86527 | 1244 | No | NA | 2 | 0 | 1 | Low-quality | Genome-fragment | 0.12 | AAI-based (medium-confidence) | 0 |
| S1_extracted_k141_60162 | 1244 | No | NA | 1 | 1 | 0 | Low-quality | Genome-fragment | 2.83 | AAI-based (medium-confidence) | 0 |
| S1_extracted_k141_39340 | 1244 | No | NA | 1 | 0 | 1 | Low-quality | Genome-fragment | 0.81 | AAI-based (high-confidence) | 0 |
| S11_vs_k141_37902\|\|lt2gene | 1084 | No | NA | 1 | 0 | 0 | Not-determined | Genome-fragment | NA | NA | 0 |
| S3_extracted_k141_55295 | 1244 | No | NA | 1 | 0 | 0 | Low-quality | Genome-fragment | 1.51 | AAI-based (high-confidence) | 0 |
| S5_vs_k141_29198\|\|lt2gene | 1244 | No | NA | 1 | 1 | 0 | Low-quality | Genome-fragment | 1.84 | HMM-based (lower-bound) | 0 |
| S7_vs_70898\|\|full | 1244 | No | NA | 3 | 1 | 0 | Low-quality | Genome-fragment | 2.65 | AAI-based (high-confidence) | 0 |
| S7_vs_6651\|\|lt2gene | 1244 | No | NA | 2 | 1 | 0 | Low-quality | Genome-fragment | 3.62 | AAI-based (high-confidence) | 0 |
| S16_vs_148696\|\|lt2gene | 1244 | No | NA | 1 | 1 | 0 | Low-quality | Genome-fragment | 3.27 | AAI-based (high-confidence) | 0 |
| S17_extracted_k141_115320 | 1245 | No | NA | 5 | 2 | 0 | Low-quality | Genome-fragment | 5.68 | AAI-based (high-confidence) | 0 |
| S17_vs_k141_164283\|\|full | 1245 | No | NA | 3 | 2 | 0 | Low-quality | Genome-fragment | 2.8 | AAI-based (medium-confidence) | 0 |
| S17_vs_k141_124979\|\|full | 1245 | No | NA | 3 | 0 | 3 | Low-quality | Genome-fragment | 0.51 | AAI-based (medium-confidence) | 0 |
| S11_extracted_k141_65788 | 1245 | No | NA | 3 | 0 | 0 | Low-quality | Genome-fragment | 0.48 | AAI-based (high-confidence) | 0 |
| S11_vs_k141_103736\|\|full | 1245 | No | NA | 3 | 0 | 0 | Low-quality | Genome-fragment | 3.13 | AAI-based (high-confidence) | 0 |
| S14_vs_k141_88706\|\|full | 1245 | No | NA | 2 | 0 | 0 | Low-quality | Genome-fragment | 2.51 | AAI-based (high-confidence) | 0 |
| S14_vs_k141_14394\|\|full | 1245 | No | NA | 2 | 0 | 0 | Low-quality | Genome-fragment | 2.38 | AAI-based (high-confidence) | 0 |
| S1_vs_k141_81243\|\|full | 1245 | No | NA | 3 | 0 | 2 | Low-quality | Genome-fragment | 1.69 | AAI-based (medium-confidence) | 0 |
| S11_vs_k141_38829\|\|lt2gene | 1137 | No | NA | 1 | 0 | 0 | Not-determined | Genome-fragment | NA | NA | 0 |
| S3_vs_k141_11953\|\|full | 1245 | No | NA | 2 | 2 | 0 | Low-quality | Genome-fragment | 2.68 | AAI-based (high-confidence) | 0 |
| S3_vs_k141_54117\|\|full | 1245 | No | NA | 2 | 2 | 0 | Low-quality | Genome-fragment | 2.58 | AAI-based (high-confidence) | 0 |
| S5_extracted_k141_115611 | 1245 | No | NA | 2 | 1 | 0 | Low-quality | Genome-fragment | 2.3 | AAI-based (medium-confidence) | 0 |
| S5_extracted_k141_123730 | 1245 | No | NA | 5 | 1 | 0 | Low-quality | Genome-fragment | 0.95 | AAI-based (medium-confidence) | 0 |
| S11_vs_k141_51732\|\|lt2gene | 1672 | No | NA | 1 | 0 | 0 | Not-determined | Genome-fragment | NA | NA | 0 |
| S11_vs_k141_32329\|\|lt2gene | 1050 | No | NA | 1 | 0 | 0 | Not-determined | Genome-fragment | NA | NA | 0 |
| S5_vs_k141_135196\|\|full | 1245 | No | NA | 3 | 0 | 1 | Low-quality | Genome-fragment | 2.34 | AAI-based (medium-confidence) | 0 |
| S5_vs_k141_45135\|\|lt2gene | 1245 | No | NA | 1 | 1 | 0 | Low-quality | Genome-fragment | 2.18 | HMM-based (lower-bound) | 0 |
| S7_extracted_7544 | 1245 | No | NA | 1 | 1 | 0 | Low-quality | Genome-fragment | 2.34 | AAI-based (high-confidence) | 0 |
| S13_extracted_601 | 1023 | No | NA | 3 | 0 | 2 | Not-determined | Genome-fragment | NA | NA | 0 |
| S13_extracted_712 | 1156 | No | NA | 1 | 0 | 0 | Not-determined | Genome-fragment | NA | NA | 0 |
| S13_extracted_758 | 1342 | No | NA | 2 | 0 | 1 | Not-determined | Genome-fragment | NA | NA | 0 |
| S7_extracted_9316 | 1245 | No | NA | 3 | 0 | 0 | Low-quality | Genome-fragment | 0.28 | AAI-based (medium-confidence) | 0 |
| S7_extracted_81635 | 1245 | No | NA | 1 | 0 | 1 | Low-quality | Genome-fragment | 4.15 | AAI-based (high-confidence) | 0 |
| S13_extracted_966 | 1345 | No | NA | 2 | 0 | 0 | Not-determined | Genome-fragment | NA | NA | 0 |
| S13_extracted_1137 | 1045 | No | NA | 1 | 0 | 0 | Not-determined | Genome-fragment | NA | NA | 0 |
| S6_vs_k141_52459\|\|full | 1245 | No | NA | 2 | 0 | 2 | Low-quality | Genome-fragment | 2.77 | AAI-based (medium-confidence) | 0 |
| S13_extracted_1546 | 1311 | No | NA | 1 | 0 | 1 | Not-determined | Genome-fragment | NA | NA | 0 |
| S13_extracted_1750 | 1789 | No | NA | 2 | 0 | 2 | Not-determined | Genome-fragment | NA | NA | 0 |
| S13_extracted_1770 | 12617 | No | NA | 16 | 4 | 0 | High-quality | High-quality | 100 | AAI-based (medium-confidence) | 0 |
| S13_extracted_1820 | 1432 | No | NA | 2 | 0 | 0 | Not-determined | Genome-fragment | NA | NA | 0 |
| S13_extracted_2054 | 1291 | No | NA | 2 | 0 | 0 | Not-determined | Genome-fragment | NA | NA | 0 |
| S13_extracted_2135 | 1182 | No | NA | 2 | 0 | 1 | Not-determined | Genome-fragment | NA | NA | 0 |
| S13_extracted_2158 | 1144 | No | NA | 1 | 0 | 0 | Not-determined | Genome-fragment | NA | NA | 0 |
| S13_extracted_2229 | 1337 | No | NA | 1 | 0 | 0 | Not-determined | Genome-fragment | NA | NA | 0 |
| S17_extracted_k141_176089 | 1246 | No | NA | 4 | 1 | 0 | Low-quality | Genome-fragment | 2.28 | AAI-based (medium-confidence) | 0 |
| S13_extracted_2348 | 2157 | No | NA | 1 | 0 | 0 | Not-determined | Genome-fragment | NA | NA | 0 |
| S13_extracted_2436 | 2073 | No | NA | 2 | 0 | 1 | Not-determined | Genome-fragment | NA | NA | 0 |
| S13_extracted_2463 | 2074 | No | NA | 1 | 0 | 0 | Not-determined | Genome-fragment | NA | NA | 0 |
| S13_extracted_2522 | 3804 | No | NA | 4 | 0 | 1 | Not-determined | Genome-fragment | NA | NA | 0 |
| S13_extracted_2524 | 15055 | No | NA | 20 | 0 | 10 | Not-determined | Genome-fragment | NA | NA | 0 |
| S17_vs_k141_102707\|\|full | 1246 | No | NA | 2 | 0 | 0 | Low-quality | Genome-fragment | 0.86 | AAI-based (medium-confidence) | 0 |
| S13_extracted_2772 | 2651 | No | NA | 6 | 0 | 6 | Not-determined | Genome-fragment | NA | NA | 0 |
| S13_extracted_3092 | 2021 | No | NA | 2 | 0 | 2 | Not-determined | Genome-fragment | NA | NA | 0 |
| S13_extracted_3140 | 1386 | No | NA | 2 | 0 | 1 | Not-determined | Genome-fragment | NA | NA | 0 |
| S13_extracted_3645 | 3970 | No | NA | 7 | 0 | 5 | Not-determined | Genome-fragment | NA | NA | 0 |
| S13_extracted_3728 | 93098 | No | NA | 82 | 6 | 0 | Complete | High-quality | 100 | DTR (high-confidence) | 0 |
| S13_extracted_3819 | 1139 | No | NA | 2 | 0 | 1 | Not-determined | Genome-fragment | NA | NA | 0 |
| S13_extracted_3860 | 1467 | No | NA | 2 | 0 | 1 | Not-determined | Genome-fragment | NA | NA | 0 |
| S13_extracted_4181 | 3410 | No | NA | 5 | 0 | 1 | Not-determined | Genome-fragment | NA | NA | 0 |
| S14_vs_k141_16362\|\|lt2gene | 1246 | No | NA | 1 | 1 | 0 | Low-quality | Genome-fragment | 1.86 | HMM-based (lower-bound) | 0 |
| S13_extracted_4314 | 3653 | No | NA | 2 | 0 | 1 | Not-determined | Genome-fragment | NA | NA | 0 |
| S13_extracted_4416 | 3047 | No | NA | 1 | 0 | 1 | Not-determined | Genome-fragment | NA | NA | 0 |
| S13_extracted_4553 | 1049 | No | NA | 1 | 0 | 1 | Not-determined | Genome-fragment | NA | NA | 0 |
| S13_extracted_4598 | 4754 | No | NA | 1 | 0 | 1 | Not-determined | Genome-fragment | NA | NA | 0 |
| S13_extracted_4610 | 9974 | No | NA | 13 | 0 | 6 | Not-determined | Genome-fragment | NA | NA | 0 |
| S13_extracted_4717 | 1117 | No | NA | 1 | 0 | 1 | Not-determined | Genome-fragment | NA | NA | 0 |
| S13_extracted_4752 | 14343 | No | NA | 8 | 0 | 4 | Not-determined | Genome-fragment | NA | NA | 0 |
| S13_extracted_4771 | 1189 | No | NA | 2 | 0 | 0 | Not-determined | Genome-fragment | NA | NA | 0 |
| S13_extracted_4798 | 1117 | No | NA | 1 | 0 | 1 | Not-determined | Genome-fragment | NA | NA | 0 |
| S13_extracted_4831 | 1116 | No | NA | 2 | 0 | 0 | Not-determined | Genome-fragment | NA | NA | 0 |
| S13_extracted_4901 | 95651 | No | NA | 146 | 20 | 3 | Medium-quality | Genome-fragment | 70.74 | AAI-based (high-confidence) | 0 |
| S3_extracted_k141_19519 | 1246 | No | NA | 4 | 2 | 0 | Low-quality | Genome-fragment | 2.24 | AAI-based (medium-confidence) | 0 |
| S13_extracted_5117 | 1707 | No | NA | 2 | 0 | 2 | Not-determined | Genome-fragment | NA | NA | 0 |
| S13_extracted_5203 | 1143 | No | NA | 1 | 0 | 0 | Not-determined | Genome-fragment | NA | NA | 0 |
| S13_extracted_5273 | 1032 | No | NA | 1 | 0 | 0 | Not-determined | Genome-fragment | NA | NA | 0 |
| S13_extracted_5409 | 1020 | No | NA | 1 | 0 | 1 | Not-determined | Genome-fragment | NA | NA | 0 |
| S5_vs_k141_142076\|\|full | 1246 | No | NA | 3 | 1 | 0 | Low-quality | Genome-fragment | 2.69 | AAI-based (high-confidence) | 0 |
| S13_extracted_5726 | 1048 | No | NA | 2 | 0 | 0 | Not-determined | Genome-fragment | NA | NA | 0 |
| S13_extracted_5768 | 1337 | No | NA | 2 | 0 | 0 | Not-determined | Genome-fragment | NA | NA | 0 |
| S13_extracted_5905 | 5593 | No | NA | 7 | 0 | 1 | Not-determined | Genome-fragment | NA | NA | 0 |
| S13_extracted_5977 | 2307 | No | NA | 1 | 0 | 1 | Not-determined | Genome-fragment | NA | NA | 0 |
| S13_extracted_6036 | 1061 | No | NA | 2 | 0 | 1 | Not-determined | Genome-fragment | NA | NA | 0 |
| S13_extracted_6085 | 1826 | No | NA | 2 | 0 | 1 | Not-determined | Genome-fragment | NA | NA | 0 |
| S13_extracted_6387 | 1041 | No | NA | 1 | 0 | 1 | Not-determined | Genome-fragment | NA | NA | 0 |
| S7_vs_12884\|\|full | 1246 | No | NA | 3 | 0 | 0 | Low-quality | Genome-fragment | 2.4 | AAI-based (medium-confidence) | 0 |
| S13_extracted_6782 | 1179 | No | NA | 2 | 0 | 2 | Not-determined | Genome-fragment | NA | NA | 0 |
| S13_extracted_6940 | 2133 | No | NA | 1 | 0 | 0 | Not-determined | Genome-fragment | NA | NA | 0 |
| S13_extracted_6962 | 4397 | No | NA | 5 | 0 | 3 | Not-determined | Genome-fragment | NA | NA | 0 |
| S13_extracted_7323 | 3586 | No | NA | 6 | 0 | 2 | Not-determined | Genome-fragment | NA | NA | 0 |
| S7_vs_117406\|\|full | 1246 | No | NA | 2 | 0 | 0 | Low-quality | Genome-fragment | 2.9 | AAI-based (high-confidence) | 0 |
| S13_extracted_7588 | 1201 | No | NA | 1 | 0 | 0 | Not-determined | Genome-fragment | NA | NA | 0 |
| S13_extracted_7604 | 1293 | No | NA | 2 | 0 | 1 | Not-determined | Genome-fragment | NA | NA | 0 |
| S13_extracted_7636 | 1307 | No | NA | 2 | 0 | 1 | Not-determined | Genome-fragment | NA | NA | 0 |
| S13_extracted_7670 | 67724 | No | NA | 75 | 8 | 0 | High-quality | High-quality | 96.7 | AAI-based (high-confidence) | 0 |
| S13_extracted_7728 | 1122 | No | NA | 2 | 0 | 1 | Not-determined | Genome-fragment | NA | NA | 0 |
| S13_extracted_7780 | 1170 | No | NA | 2 | 0 | 2 | Not-determined | Genome-fragment | NA | NA | 0 |
| S13_extracted_7884 | 1078 | No | NA | 2 | 0 | 2 | Not-determined | Genome-fragment | NA | NA | 0 |
| S13_extracted_7952 | 1753 | No | NA | 3 | 0 | 0 | Not-determined | Genome-fragment | NA | NA | 0 |
| S13_extracted_8039 | 1146 | No | NA | 2 | 0 | 1 | Not-determined | Genome-fragment | NA | NA | 0 |
| S13_extracted_8102 | 1177 | No | NA | 1 | 0 | 1 | Not-determined | Genome-fragment | NA | NA | 0 |
| S13_extracted_8239 | 1345 | No | NA | 2 | 0 | 2 | Not-determined | Genome-fragment | NA | NA | 0 |
| S13_extracted_8285 | 3496 | No | NA | 3 | 0 | 0 | Not-determined | Genome-fragment | NA | NA | 0 |
| S10_vs_12494\|\|lt2gene | 1246 | No | NA | 2 | 1 | 0 | Low-quality | Genome-fragment | 2.87 | AAI-based (high-confidence) | 0 |
| S12_extracted_k141_182053 | 1246 | No | NA | 2 | 1 | 0 | Low-quality | Genome-fragment | 2.55 | AAI-based (high-confidence) | 0 |
| S13_extracted_8496 | 2524 | No | NA | 2 | 0 | 1 | Not-determined | Genome-fragment | NA | NA | 0 |
| S13_extracted_8551 | 1235 | No | NA | 1 | 0 | 1 | Not-determined | Genome-fragment | NA | NA | 0 |
| S12_extracted_k141_145945 | 1246 | No | NA | 2 | 0 | 2 | Low-quality | Genome-fragment | 0.79 | AAI-based (high-confidence) | 0 |
| S13_extracted_8583 | 2182 | No | NA | 2 | 0 | 0 | Not-determined | Genome-fragment | NA | NA | 0 |
| S13_extracted_8646 | 1170 | No | NA | 2 | 0 | 1 | Not-determined | Genome-fragment | NA | NA | 0 |
| S13_extracted_8731 | 1003 | No | NA | 2 | 0 | 1 | Not-determined | Genome-fragment | NA | NA | 0 |
| S13_extracted_8754 | 5140 | No | NA | 5 | 0 | 3 | Not-determined | Genome-fragment | NA | NA | 0 |
| S13_extracted_8761 | 2942 | No | NA | 1 | 0 | 1 | Not-determined | Genome-fragment | NA | NA | 0 |
| S13_extracted_8857 | 1319 | No | NA | 4 | 0 | 3 | Not-determined | Genome-fragment | NA | NA | 0 |
| S13_extracted_8859 | 3135 | No | NA | 4 | 0 | 1 | Not-determined | Genome-fragment | NA | NA | 0 |
| S13_extracted_9024 | 5188 | No | NA | 9 | 0 | 1 | Not-determined | Genome-fragment | NA | NA | 0 |
| S13_extracted_9094 | 1145 | No | NA | 1 | 0 | 1 | Not-determined | Genome-fragment | NA | NA | 0 |
| S13_extracted_9170 | 1383 | No | NA | 1 | 0 | 0 | Not-determined | Genome-fragment | NA | NA | 0 |
| S13_extracted_9294 | 1245 | No | NA | 3 | 0 | 3 | Not-determined | Genome-fragment | NA | NA | 0 |
| S13_extracted_9296 | 2045 | No | NA | 2 | 0 | 0 | Not-determined | Genome-fragment | NA | NA | 0 |
| S13_extracted_9324 | 1023 | No | NA | 1 | 0 | 1 | Not-determined | Genome-fragment | NA | NA | 0 |
| S13_extracted_9372 | 1081 | No | NA | 2 | 0 | 0 | Not-determined | Genome-fragment | NA | NA | 0 |
| S12_vs_k141_56737\|\|full | 1246 | No | NA | 2 | 0 | 2 | Low-quality | Genome-fragment | 1.18 | AAI-based (medium-confidence) | 0 |
| S13_extracted_9429 | 1767 | No | NA | 2 | 0 | 0 | Not-determined | Genome-fragment | NA | NA | 0 |
| S13_extracted_9537 | 1150 | No | NA | 2 | 0 | 1 | Not-determined | Genome-fragment | NA | NA | 0 |
| S13_extracted_9545 | 1184 | No | NA | 2 | 0 | 1 | Not-determined | Genome-fragment | NA | NA | 0 |
| S13_extracted_9648 | 1730 | No | NA | 3 | 0 | 1 | Not-determined | Genome-fragment | NA | NA | 0 |
| S13_extracted_9811 | 1154 | No | NA | 2 | 0 | 0 | Not-determined | Genome-fragment | NA | NA | 0 |
| S13_extracted_9884 | 1566 | No | NA | 2 | 0 | 1 | Not-determined | Genome-fragment | NA | NA | 0 |
| S15_vs_76957\|\|full | 1246 | No | NA | 3 | 0 | 0 | Low-quality | Genome-fragment | 3.23 | AAI-based (high-confidence) | 0 |
| S13_extracted_9993 | 1156 | No | NA | 2 | 0 | 2 | Not-determined | Genome-fragment | NA | NA | 0 |
| S13_extracted_10119 | 1010 | No | NA | 2 | 0 | 2 | Not-determined | Genome-fragment | NA | NA | 0 |
| S13_extracted_10127 | 17386 | No | NA | 24 | 0 | 17 | Not-determined | Genome-fragment | NA | NA | 0 |
| S6_vs_k141_38699\|\|full | 1246 | No | NA | 4 | 1 | 0 | Low-quality | Genome-fragment | 2.05 | AAI-based (high-confidence) | 0 |
| S13_extracted_10480 | 1994 | No | NA | 2 | 0 | 1 | Not-determined | Genome-fragment | NA | NA | 0 |
| S8_extracted_k141_122770 | 1246 | No | NA | 2 | 0 | 1 | Low-quality | Genome-fragment | 0.38 | AAI-based (medium-confidence) | 0 |
| S13_extracted_10557 | 1568 | No | NA | 1 | 0 | 0 | Not-determined | Genome-fragment | NA | NA | 0 |
| S8_vs_k141_173478\|\|full | 1246 | No | NA | 3 | 1 | 0 | Low-quality | Genome-fragment | 2.87 | AAI-based (medium-confidence) | 0 |
| S9_vs_k141_56843\|\|full | 1247 | No | NA | 2 | 2 | 0 | Low-quality | Genome-fragment | 2.95 | AAI-based (high-confidence) | 0 |
| S13_extracted_10620 | 1190 | No | NA | 2 | 0 | 2 | Not-determined | Genome-fragment | NA | NA | 0 |
| S13_extracted_10715 | 1349 | No | NA | 2 | 0 | 1 | Not-determined | Genome-fragment | NA | NA | 0 |
| S13_extracted_10859 | 1907 | No | NA | 3 | 0 | 2 | Not-determined | Genome-fragment | NA | NA | 0 |
| S9_vs_k141_44260\|\|full | 1247 | No | NA | 2 | 0 | 0 | Low-quality | Genome-fragment | 2.45 | AAI-based (high-confidence) | 0 |
| S13_extracted_11193 | 1715 | No | NA | 2 | 0 | 2 | Not-determined | Genome-fragment | NA | NA | 0 |
| S13_extracted_11246 | 97342 | No | NA | 97 | 15 | 2 | Complete | High-quality | 100 | DTR (high-confidence) | 0 |
| S13_extracted_11316 | 1263 | No | NA | 1 | 0 | 1 | Not-determined | Genome-fragment | NA | NA | 0 |
| S9_vs_k141_21080\|\|lt2gene | 1247 | No | NA | 2 | 1 | 0 | Low-quality | Genome-fragment | 3.36 | AAI-based (high-confidence) | 0 |
| S13_extracted_11639 | 1329 | No | NA | 2 | 0 | 1 | Not-determined | Genome-fragment | NA | NA | 0 |
| S14_extracted_k141_55813 | 1247 | No | NA | 1 | 0 | 0 | Low-quality | Genome-fragment | 3.12 | AAI-based (high-confidence) | 0 |
| S13_extracted_11763 | 2640 | No | NA | 3 | 0 | 3 | Not-determined | Genome-fragment | NA | NA | 0 |
| S13_extracted_11877 | 3266 | No | NA | 2 | 0 | 1 | Not-determined | Genome-fragment | NA | NA | 0 |
| S14_vs_k141_33797\|\|full | 1247 | No | NA | 3 | 1 | 0 | Low-quality | Genome-fragment | 2.22 | AAI-based (medium-confidence) | 0 |
| S14_vs_k141_13391\|\|lt2gene | 1247 | No | NA | 2 | 2 | 0 | Low-quality | Genome-fragment | 2.92 | HMM-based (lower-bound) | 0 |
| S13_extracted_12007 | 1036 | No | NA | 1 | 0 | 0 | Not-determined | Genome-fragment | NA | NA | 0 |
| S13_extracted_12080 | 1299 | No | NA | 2 | 0 | 2 | Not-determined | Genome-fragment | NA | NA | 0 |
| S13_extracted_12088 | 2331 | No | NA | 3 | 0 | 0 | Not-determined | Genome-fragment | NA | NA | 0 |
| S5_vs_k141_9564\|\|full | 1247 | No | NA | 5 | 0 | 0 | Low-quality | Genome-fragment | 2.94 | AAI-based (medium-confidence) | 0 |
| S13_extracted_12447 | 1468 | No | NA | 1 | 0 | 0 | Not-determined | Genome-fragment | NA | NA | 0 |
| S13_extracted_12589 | 1046 | No | NA | 2 | 0 | 1 | Not-determined | Genome-fragment | NA | NA | 0 |
| S13_extracted_12623 | 1017 | No | NA | 1 | 0 | 1 | Not-determined | Genome-fragment | NA | NA | 0 |
| S13_extracted_12796 | 1198 | No | NA | 2 | 0 | 2 | Not-determined | Genome-fragment | NA | NA | 0 |
| S13_extracted_12862 | 1241 | No | NA | 3 | 0 | 2 | Not-determined | Genome-fragment | NA | NA | 0 |
| S13_extracted_13010 | 1425 | No | NA | 1 | 0 | 1 | Not-determined | Genome-fragment | NA | NA | 0 |
| S5_vs_k141_65089\|\|full | 1247 | No | NA | 4 | 0 | 0 | Low-quality | Genome-fragment | 2.17 | AAI-based (medium-confidence) | 0 |
| S13_extracted_13219 | 1050 | No | NA | 2 | 0 | 0 | Not-determined | Genome-fragment | NA | NA | 0 |
| S13_extracted_13245 | 1947 | No | NA | 1 | 0 | 0 | Not-determined | Genome-fragment | NA | NA | 0 |
| S13_extracted_13537 | 1309 | No | NA | 2 | 0 | 2 | Not-determined | Genome-fragment | NA | NA | 0 |
| S13_extracted_13607 | 1565 | No | NA | 2 | 0 | 0 | Not-determined | Genome-fragment | NA | NA | 0 |
| S13_extracted_13729 | 2211 | No | NA | 2 | 0 | 1 | Not-determined | Genome-fragment | NA | NA | 0 |
| S13_extracted_13949 | 2626 | No | NA | 1 | 0 | 1 | Not-determined | Genome-fragment | NA | NA | 0 |
| S13_extracted_14074 | 1873 | No | NA | 2 | 0 | 0 | Not-determined | Genome-fragment | NA | NA | 0 |
| S5_vs_k141_60458\|\|full | 1247 | No | NA | 3 | 2 | 0 | Low-quality | Genome-fragment | 2.78 | AAI-based (high-confidence) | 0 |
| S13_extracted_14328 | 1189 | No | NA | 1 | 0 | 0 | Not-determined | Genome-fragment | NA | NA | 0 |
| S7_extracted_103359 | 1247 | No | NA | 2 | 0 | 0 | Low-quality | Genome-fragment | 1.82 | AAI-based (medium-confidence) | 0 |
| S7_vs_88133\|\|lt2gene | 1247 | No | NA | 1 | 1 | 0 | Low-quality | Genome-fragment | 0.71 | HMM-based (lower-bound) | 0 |
| S16_vs_25467\|\|full | 1247 | No | NA | 2 | 0 | 2 | Low-quality | Genome-fragment | 0.67 | AAI-based (medium-confidence) | 0 |
| S13_extracted_14765 | 2005 | No | NA | 3 | 0 | 1 | Not-determined | Genome-fragment | NA | NA | 0 |
| S16_vs_77497\|\|full | 1247 | No | NA | 3 | 0 | 0 | Low-quality | Genome-fragment | 1.48 | AAI-based (high-confidence) | 0 |
| S13_extracted_14929 | 1076 | No | NA | 2 | 0 | 0 | Not-determined | Genome-fragment | NA | NA | 0 |
| S12_vs_k141_183487\|\|lt2gene | 1247 | No | NA | 1 | 1 | 0 | Low-quality | Genome-fragment | 2.91 | AAI-based (high-confidence) | 0 |
| S13_extracted_15054 | 2783 | No | NA | 6 | 0 | 0 | Not-determined | Genome-fragment | NA | NA | 0 |
| S13_extracted_15194 | 3033 | No | NA | 6 | 0 | 6 | Not-determined | Genome-fragment | NA | NA | 0 |
| S6_vs_k141_52358\|\|full | 1247 | No | NA | 4 | 0 | 0 | Low-quality | Genome-fragment | 3.28 | AAI-based (medium-confidence) | 0 |
| S13_extracted_15300 | 1773 | No | NA | 1 | 0 | 1 | Not-determined | Genome-fragment | NA | NA | 0 |
| S13_extracted_15353 | 1314 | No | NA | 2 | 0 | 2 | Not-determined | Genome-fragment | NA | NA | 0 |
| S13_extracted_15426 | 1501 | No | NA | 1 | 0 | 0 | Not-determined | Genome-fragment | NA | NA | 0 |
| S13_extracted_15463 | 1776 | No | NA | 3 | 0 | 2 | Not-determined | Genome-fragment | NA | NA | 0 |
| S8_extracted_k141_18612 | 1247 | No | NA | 3 | 0 | 1 | Low-quality | Genome-fragment | 3.98 | AAI-based (medium-confidence) | 0 |
| S13_extracted_15684 | 1439 | No | NA | 2 | 0 | 1 | Not-determined | Genome-fragment | NA | NA | 0 |
| S13_extracted_15729 | 1058 | No | NA | 1 | 0 | 1 | Not-determined | Genome-fragment | NA | NA | 0 |
| S13_extracted_15817 | 2390 | No | NA | 2 | 0 | 0 | Not-determined | Genome-fragment | NA | NA | 0 |
| S13_extracted_15910 | 1203 | No | NA | 1 | 0 | 1 | Not-determined | Genome-fragment | NA | NA | 0 |
| S13_extracted_16030 | 1324 | No | NA | 3 | 0 | 1 | Not-determined | Genome-fragment | NA | NA | 0 |
| S13_extracted_16073 | 1239 | No | NA | 2 | 0 | 1 | Not-determined | Genome-fragment | NA | NA | 0 |
| S13_extracted_16144 | 1175 | No | NA | 2 | 0 | 1 | Not-determined | Genome-fragment | NA | NA | 0 |
| S13_extracted_16281 | 1719 | No | NA | 1 | 0 | 0 | Not-determined | Genome-fragment | NA | NA | 0 |
| S13_extracted_16327 | 1255 | No | NA | 3 | 0 | 0 | Not-determined | Genome-fragment | NA | NA | 0 |
| S13_extracted_16419 | 1063 | No | NA | 1 | 0 | 0 | Not-determined | Genome-fragment | NA | NA | 0 |
| S13_extracted_16719 | 1754 | No | NA | 1 | 0 | 1 | Not-determined | Genome-fragment | NA | NA | 0 |
| S13_extracted_16913 | 1375 | No | NA | 1 | 0 | 1 | Not-determined | Genome-fragment | NA | NA | 0 |
| S13_extracted_17085 | 1096 | No | NA | 1 | 0 | 1 | Not-determined | Genome-fragment | NA | NA | 0 |
| S13_extracted_17099 | 1021 | No | NA | 1 | 0 | 1 | Not-determined | Genome-fragment | NA | NA | 0 |
| S8_extracted_k141_197950 | 1247 | No | NA | 3 | 0 | 0 | Low-quality | Genome-fragment | 3.84 | AAI-based (high-confidence) | 0 |
| S13_extracted_17191 | 2180 | No | NA | 3 | 0 | 2 | Not-determined | Genome-fragment | NA | NA | 0 |
| S13_extracted_17355 | 2178 | No | NA | 3 | 0 | 0 | Not-determined | Genome-fragment | NA | NA | 0 |
| S13_extracted_17422 | 5395 | No | NA | 6 | 0 | 2 | Not-determined | Genome-fragment | NA | NA | 0 |
| S17_vs_k141_133632\|\|full | 1248 | No | NA | 2 | 1 | 0 | Low-quality | Genome-fragment | 1.91 | AAI-based (medium-confidence) | 0 |
| S13_extracted_17699 | 1058 | No | NA | 2 | 0 | 1 | Not-determined | Genome-fragment | NA | NA | 0 |
| S13_extracted_17733 | 2216 | No | NA | 3 | 0 | 1 | Not-determined | Genome-fragment | NA | NA | 0 |
| S13_extracted_17833 | 4738 | No | NA | 5 | 0 | 4 | Not-determined | Genome-fragment | NA | NA | 0 |
| S13_extracted_17878 | 1123 | No | NA | 3 | 0 | 1 | Not-determined | Genome-fragment | NA | NA | 0 |
| S13_extracted_17981 | 1061 | No | NA | 1 | 0 | 0 | Not-determined | Genome-fragment | NA | NA | 0 |
| S9_vs_k141_21094\|\|full | 1248 | No | NA | 5 | 0 | 0 | Low-quality | Genome-fragment | 2.41 | AAI-based (high-confidence) | 0 |
| S13_extracted_18325 | 1525 | No | NA | 2 | 0 | 2 | Not-determined | Genome-fragment | NA | NA | 0 |
| S13_extracted_18384 | 1255 | No | NA | 2 | 0 | 1 | Not-determined | Genome-fragment | NA | NA | 0 |
| S13_extracted_18461 | 1187 | No | NA | 1 | 0 | 1 | Not-determined | Genome-fragment | NA | NA | 0 |
| S13_extracted_18564 | 1230 | No | NA | 4 | 0 | 1 | Not-determined | Genome-fragment | NA | NA | 0 |
| S13_extracted_18606 | 1240 | No | NA | 1 | 0 | 1 | Not-determined | Genome-fragment | NA | NA | 0 |
| S9_vs_k141_6112\|\|lt2gene | 1248 | No | NA | 1 | 1 | 0 | Low-quality | Genome-fragment | 2.81 | AAI-based (medium-confidence) | 0 |
| S13_extracted_18724 | 3333 | No | NA | 1 | 0 | 1 | Not-determined | Genome-fragment | NA | NA | 0 |
| S13_extracted_18811 | 1607 | No | NA | 2 | 0 | 2 | Not-determined | Genome-fragment | NA | NA | 0 |
| S13_extracted_18863 | 1418 | No | NA | 1 | 0 | 1 | Not-determined | Genome-fragment | NA | NA | 0 |
| S14_vs_k141_31350\|\|full | 1248 | No | NA | 2 | 0 | 0 | Low-quality | Genome-fragment | 2.58 | AAI-based (high-confidence) | 0 |
| S13_extracted_19021 | 1130 | No | NA | 1 | 0 | 1 | Not-determined | Genome-fragment | NA | NA | 0 |
| S13_extracted_19051 | 1677 | No | NA | 2 | 0 | 2 | Not-determined | Genome-fragment | NA | NA | 0 |
| S13_extracted_19114 | 4320 | No | NA | 4 | 0 | 3 | Not-determined | Genome-fragment | NA | NA | 0 |
| S13_extracted_19187 | 2371 | No | NA | 2 | 0 | 0 | Not-determined | Genome-fragment | NA | NA | 0 |
| S13_extracted_19236 | 1243 | No | NA | 2 | 0 | 2 | Not-determined | Genome-fragment | NA | NA | 0 |
| S13_extracted_19243 | 4864 | No | NA | 7 | 0 | 3 | Not-determined | Genome-fragment | NA | NA | 0 |
| S13_extracted_19343 | 4820 | No | NA | 1 | 0 | 0 | Not-determined | Genome-fragment | NA | NA | 0 |
| S13_extracted_19414 | 1460 | No | NA | 1 | 0 | 0 | Not-determined | Genome-fragment | NA | NA | 0 |
| S14_vs_k141_10823\|\|full | 1248 | No | NA | 3 | 0 | 1 | Low-quality | Genome-fragment | 1.61 | AAI-based (medium-confidence) | 0 |
| S13_extracted_19586 | 1573 | No | NA | 1 | 0 | 0 | Not-determined | Genome-fragment | NA | NA | 0 |
| S13_extracted_19630 | 1595 | No | NA | 2 | 0 | 2 | Not-determined | Genome-fragment | NA | NA | 0 |
| S13_extracted_19825 | 1031 | No | NA | 2 | 0 | 1 | Not-determined | Genome-fragment | NA | NA | 0 |
| S13_extracted_20386 | 3735 | No | NA | 4 | 0 | 3 | Not-determined | Genome-fragment | NA | NA | 0 |
| S1_vs_k141_32986\|\|full | 1248 | No | NA | 5 | 2 | 0 | Low-quality | Genome-fragment | 2.22 | AAI-based (high-confidence) | 0 |
| S13_extracted_20450 | 1061 | No | NA | 1 | 0 | 1 | Not-determined | Genome-fragment | NA | NA | 0 |
| S13_extracted_20573 | 1157 | No | NA | 1 | 0 | 0 | Not-determined | Genome-fragment | NA | NA | 0 |
| S13_extracted_20633 | 6616 | No | NA | 9 | 0 | 5 | Not-determined | Genome-fragment | NA | NA | 0 |
| S13_extracted_20669 | 1463 | No | NA | 1 | 0 | 0 | Not-determined | Genome-fragment | NA | NA | 0 |
| S13_extracted_20741 | 3969 | No | NA | 4 | 0 | 2 | Not-determined | Genome-fragment | NA | NA | 0 |
| S13_extracted_20840 | 1488 | No | NA | 2 | 0 | 0 | Not-determined | Genome-fragment | NA | NA | 0 |
| S13_extracted_20954 | 1086 | No | NA | 2 | 0 | 2 | Not-determined | Genome-fragment | NA | NA | 0 |
| S13_extracted_21012 | 1595 | No | NA | 4 | 0 | 2 | Not-determined | Genome-fragment | NA | NA | 0 |
| S5_vs_k141_72943\|\|full | 1248 | No | NA | 3 | 0 | 1 | Low-quality | Genome-fragment | 1.84 | AAI-based (medium-confidence) | 0 |
| S13_extracted_21128 | 2712 | No | NA | 3 | 0 | 2 | Not-determined | Genome-fragment | NA | NA | 0 |
| S13_extracted_21174 | 1627 | No | NA | 2 | 0 | 0 | Not-determined | Genome-fragment | NA | NA | 0 |
| S5_vs_k141_19175\|\|full | 1248 | No | NA | 2 | 0 | 1 | Low-quality | Genome-fragment | 1.82 | AAI-based (medium-confidence) | 0 |
| S13_extracted_21343 | 1053 | No | NA | 2 | 0 | 0 | Not-determined | Genome-fragment | NA | NA | 0 |
| S5_vs_k141_76162\|\|full | 1248 | No | NA | 3 | 1 | 0 | Low-quality | Genome-fragment | 2.99 | AAI-based (high-confidence) | 0 |
| S5_vs_k141_52073\|\|lt2gene | 1248 | No | NA | 2 | 1 | 0 | Low-quality | Genome-fragment | 0.76 | HMM-based (lower-bound) | 0 |
| S13_extracted_21692 | 1060 | No | NA | 1 | 0 | 1 | Not-determined | Genome-fragment | NA | NA | 0 |
| S13_extracted_21694 | 1666 | No | NA | 2 | 0 | 0 | Not-determined | Genome-fragment | NA | NA | 0 |
| S13_extracted_21772 | 1352 | No | NA | 5 | 0 | 5 | Not-determined | Genome-fragment | NA | NA | 0 |
| S13_extracted_21779 | 1185 | No | NA | 2 | 0 | 0 | Not-determined | Genome-fragment | NA | NA | 0 |
| S7_vs_5767\|\|full | 1248 | No | NA | 3 | 0 | 1 | Low-quality | Genome-fragment | 2.96 | AAI-based (medium-confidence) | 0 |
| S13_extracted_21949 | 13143 | No | NA | 6 | 0 | 0 | Not-determined | Genome-fragment | NA | NA | 0 |
| S13_extracted_21997 | 1376 | No | NA | 1 | 0 | 0 | Not-determined | Genome-fragment | NA | NA | 0 |
| S13_extracted_22175 | 36043 | No | NA | 56 | 17 | 0 | High-quality | High-quality | 100 | AAI-based (high-confidence) | 0 |
| S13_extracted_22186 | 7085 | No | NA | 9 | 0 | 5 | Not-determined | Genome-fragment | NA | NA | 0 |
| S7_vs_108664\|\|full | 1248 | No | NA | 2 | 0 | 1 | Low-quality | Genome-fragment | 1 | AAI-based (medium-confidence) | 0 |
| S13_extracted_22368 | 1273 | No | NA | 2 | 0 | 1 | Not-determined | Genome-fragment | NA | NA | 0 |
| S13_extracted_22384 | 2330 | No | NA | 2 | 0 | 2 | Not-determined | Genome-fragment | NA | NA | 0 |
| S7_vs_111735\|\|full | 1248 | No | NA | 3 | 0 | 0 | Low-quality | Genome-fragment | 2.93 | AAI-based (medium-confidence) | 0 |
| S7_vs_89927\|\|lt2gene | 1248 | No | NA | 1 | 0 | 0 | Low-quality | Genome-fragment | 2.48 | AAI-based (medium-confidence) | 0 |
| S13_extracted_23268 | 1222 | No | NA | 1 | 0 | 0 | Not-determined | Genome-fragment | NA | NA | 0 |
| S13_extracted_23350 | 3802 | No | NA | 5 | 0 | 1 | Not-determined | Genome-fragment | NA | NA | 0 |
| S13_extracted_23357 | 3039 | No | NA | 3 | 0 | 2 | Not-determined | Genome-fragment | NA | NA | 0 |
| S13_extracted_23452 | 2219 | No | NA | 1 | 0 | 0 | Not-determined | Genome-fragment | NA | NA | 0 |
| S13_extracted_23474 | 1060 | No | NA | 3 | 0 | 0 | Not-determined | Genome-fragment | NA | NA | 0 |
| S13_extracted_23488 | 5970 | No | NA | 12 | 0 | 12 | Not-determined | Genome-fragment | NA | NA | 0 |
| S6_vs_k141_12491\|\|full | 1248 | No | NA | 4 | 0 | 0 | Low-quality | Genome-fragment | 2.93 | AAI-based (high-confidence) | 0 |
| S13_extracted_23533 | 1026 | No | NA | 1 | 0 | 1 | Not-determined | Genome-fragment | NA | NA | 0 |
| S13_extracted_23579 | 1165 | No | NA | 2 | 0 | 1 | Not-determined | Genome-fragment | NA | NA | 0 |
| S13_extracted_23593 | 1067 | No | NA | 1 | 0 | 0 | Not-determined | Genome-fragment | NA | NA | 0 |
| S13_extracted_23641 | 3120 | No | NA | 2 | 0 | 0 | Not-determined | Genome-fragment | NA | NA | 0 |
| S8_vs_k141_150679\|\|full | 1248 | No | NA | 3 | 0 | 0 | Low-quality | Genome-fragment | 2.86 | AAI-based (high-confidence) | 0 |
| S13_extracted_23724 | 1326 | No | NA | 1 | 0 | 0 | Not-determined | Genome-fragment | NA | NA | 0 |
| S13_extracted_23744 | 1129 | No | NA | 1 | 0 | 1 | Not-determined | Genome-fragment | NA | NA | 0 |
| S13_extracted_23803 | 1856 | No | NA | 1 | 0 | 1 | Not-determined | Genome-fragment | NA | NA | 0 |
| S13_extracted_24071 | 1039 | No | NA | 2 | 0 | 0 | Not-determined | Genome-fragment | NA | NA | 0 |
| S13_extracted_24236 | 3946 | No | NA | 3 | 0 | 3 | Not-determined | Genome-fragment | NA | NA | 0 |
| S13_extracted_24273 | 1452 | No | NA | 1 | 0 | 1 | Not-determined | Genome-fragment | NA | NA | 0 |
| S13_extracted_24337 | 1179 | No | NA | 1 | 0 | 0 | Not-determined | Genome-fragment | NA | NA | 0 |
| S13_extracted_24418 | 1714 | No | NA | 1 | 0 | 1 | Not-determined | Genome-fragment | NA | NA | 0 |
| S13_extracted_24524 | 1215 | No | NA | 1 | 0 | 1 | Not-determined | Genome-fragment | NA | NA | 0 |
| S13_extracted_24765 | 1207 | No | NA | 3 | 0 | 1 | Not-determined | Genome-fragment | NA | NA | 0 |
| S13_extracted_24881 | 1125 | No | NA | 1 | 0 | 1 | Not-determined | Genome-fragment | NA | NA | 0 |
| S13_extracted_24995 | 11236 | No | NA | 7 | 0 | 1 | Not-determined | Genome-fragment | NA | NA | 0 |
| S13_extracted_25047 | 3179 | No | NA | 5 | 0 | 3 | Not-determined | Genome-fragment | NA | NA | 0 |
| S13_extracted_25103 | 1034 | No | NA | 2 | 0 | 1 | Not-determined | Genome-fragment | NA | NA | 0 |
| S13_extracted_25244 | 1239 | No | NA | 1 | 0 | 1 | Not-determined | Genome-fragment | NA | NA | 0 |
| S13_extracted_25483 | 1106 | No | NA | 2 | 0 | 0 | Not-determined | Genome-fragment | NA | NA | 0 |
| S13_extracted_25506 | 3382 | No | NA | 3 | 0 | 1 | Not-determined | Genome-fragment | NA | NA | 0 |
| S13_extracted_25553 | 2329 | No | NA | 3 | 0 | 0 | Not-determined | Genome-fragment | NA | NA | 0 |
| S13_extracted_25693 | 1018 | No | NA | 2 | 0 | 2 | Not-determined | Genome-fragment | NA | NA | 0 |
| S13_extracted_26011 | 1109 | No | NA | 1 | 0 | 0 | Not-determined | Genome-fragment | NA | NA | 0 |
| S13_extracted_26115 | 4882 | No | NA | 8 | 0 | 2 | Not-determined | Genome-fragment | NA | NA | 0 |
| S8_vs_k141_74180\|\|full | 1248 | No | NA | 3 | 1 | 0 | Low-quality | Genome-fragment | 2.85 | AAI-based (high-confidence) | 0 |
| S13_extracted_26492 | 3932 | No | NA | 1 | 0 | 1 | Not-determined | Genome-fragment | NA | NA | 0 |
| S13_extracted_26536 | 1750 | No | NA | 1 | 0 | 1 | Not-determined | Genome-fragment | NA | NA | 0 |
| S13_extracted_26634 | 1031 | No | NA | 1 | 0 | 1 | Not-determined | Genome-fragment | NA | NA | 0 |
| S17_vs_k141_18271\|\|full | 1249 | No | NA | 3 | 0 | 0 | Low-quality | Genome-fragment | 2.11 | AAI-based (high-confidence) | 0 |
| S13_extracted_26805 | 1043 | No | NA | 1 | 0 | 0 | Not-determined | Genome-fragment | NA | NA | 0 |
| S13_extracted_26904 | 1406 | No | NA | 1 | 0 | 1 | Not-determined | Genome-fragment | NA | NA | 0 |
| S13_extracted_27027 | 1775 | No | NA | 2 | 0 | 0 | Not-determined | Genome-fragment | NA | NA | 0 |
| S13_extracted_27042 | 1448 | No | NA | 1 | 0 | 1 | Not-determined | Genome-fragment | NA | NA | 0 |
| S13_extracted_70987 | 1249 | No | NA | 2 | 0 | 1 | Low-quality | Genome-fragment | 3.03 | AAI-based (high-confidence) | 0 |
| S13_extracted_27161 | 3156 | No | NA | 3 | 0 | 1 | Not-determined | Genome-fragment | NA | NA | 0 |
| S13_extracted_27184 | 1091 | No | NA | 1 | 0 | 0 | Not-determined | Genome-fragment | NA | NA | 0 |
| S13_vs_145058\|\|full | 1249 | No | NA | 3 | 0 | 0 | Low-quality | Genome-fragment | 2.69 | AAI-based (medium-confidence) | 0 |
| S13_extracted_27347 | 2637 | No | NA | 3 | 0 | 2 | Not-determined | Genome-fragment | NA | NA | 0 |
| S13_extracted_27390 | 1228 | No | NA | 1 | 0 | 0 | Not-determined | Genome-fragment | NA | NA | 0 |
| S13_vs_51561\|\|lt2gene | 1249 | No | NA | 2 | 2 | 0 | Low-quality | Genome-fragment | 2.49 | AAI-based (high-confidence) | 0 |
| S14_vs_k141_109225\|\|lt2gene | 1249 | No | NA | 2 | 1 | 0 | Low-quality | Genome-fragment | 2.92 | AAI-based (high-confidence) | 0 |
| S13_extracted_27542 | 1070 | No | NA | 2 | 0 | 2 | Not-determined | Genome-fragment | NA | NA | 0 |
| S13_extracted_27648 | 6378 | No | NA | 7 | 0 | 2 | Not-determined | Genome-fragment | NA | NA | 0 |
| S3_extracted_k141_63420 | 1249 | No | NA | 2 | 0 | 0 | Low-quality | Genome-fragment | 2.08 | AAI-based (high-confidence) | 0 |
| S3_vs_k141_41718\|\|lt2gene | 1249 | No | NA | 1 | 1 | 0 | Low-quality | Genome-fragment | 3.02 | AAI-based (high-confidence) | 0 |
| S13_extracted_28309 | 15508 | No | NA | 18 | 7 | 0 | Medium-quality | Genome-fragment | 66.4 | AAI-based (medium-confidence) | 0 |
| S7_vs_34948\|\|full | 1249 | No | NA | 3 | 2 | 0 | Low-quality | Genome-fragment | 2.82 | AAI-based (high-confidence) | 0 |
| S7_vs_113193\|\|full | 1249 | No | NA | 4 | 0 | 0 | Low-quality | Genome-fragment | 3.17 | AAI-based (medium-confidence) | 0 |
| S13_extracted_28773 | 1021 | No | NA | 2 | 0 | 0 | Not-determined | Genome-fragment | NA | NA | 0 |
| S13_extracted_28790 | 2692 | No | NA | 2 | 0 | 2 | Not-determined | Genome-fragment | NA | NA | 0 |
| S13_extracted_29085 | 1131 | No | NA | 2 | 0 | 0 | Not-determined | Genome-fragment | NA | NA | 0 |
| S13_extracted_29503 | 5147 | No | NA | 5 | 0 | 2 | Not-determined | Genome-fragment | NA | NA | 0 |
| S13_extracted_29559 | 1175 | No | NA | 3 | 0 | 3 | Not-determined | Genome-fragment | NA | NA | 0 |
| S10_extracted_66597 | 1249 | No | NA | 1 | 1 | 0 | Low-quality | Genome-fragment | 1.19 | HMM-based (lower-bound) | 0 |
| S13_extracted_29706 | 1132 | No | NA | 1 | 0 | 1 | Not-determined | Genome-fragment | NA | NA | 0 |
| S10_vs_49323\|\|full | 1249 | No | NA | 2 | 0 | 0 | Low-quality | Genome-fragment | 2.41 | AAI-based (high-confidence) | 0 |
| S13_extracted_30218 | 1463 | No | NA | 2 | 0 | 0 | Not-determined | Genome-fragment | NA | NA | 0 |
| S13_extracted_30219 | 1012 | No | NA | 1 | 0 | 0 | Not-determined | Genome-fragment | NA | NA | 0 |
| S13_extracted_30353 | 5700 | No | NA | 6 | 0 | 5 | Not-determined | Genome-fragment | NA | NA | 0 |
| S12_extracted_k141_173554 | 1249 | No | NA | 1 | 1 | 0 | Low-quality | Genome-fragment | 3.4 | AAI-based (high-confidence) | 0 |
| S13_extracted_30394 | 1105 | No | NA | 1 | 0 | 1 | Not-determined | Genome-fragment | NA | NA | 0 |
| S6_vs_k141_980\|\|full | 1249 | No | NA | 4 | 3 | 0 | Low-quality | Genome-fragment | 3.59 | AAI-based (high-confidence) | 0 |
| S13_extracted_30465 | 1879 | No | NA | 3 | 0 | 0 | Not-determined | Genome-fragment | NA | NA | 0 |
| S13_extracted_30474 | 1529 | No | NA | 2 | 0 | 2 | Not-determined | Genome-fragment | NA | NA | 0 |
| S13_extracted_30500 | 1230 | No | NA | 2 | 0 | 1 | Not-determined | Genome-fragment | NA | NA | 0 |
| S13_extracted_30599 | 1190 | No | NA | 1 | 0 | 1 | Not-determined | Genome-fragment | NA | NA | 0 |
| S17_vs_k141_112565\|\|full | 1250 | No | NA | 2 | 0 | 0 | Low-quality | Genome-fragment | 2.11 | AAI-based (medium-confidence) | 0 |
| S13_extracted_31218 | 10922 | No | NA | 7 | 0 | 3 | Not-determined | Genome-fragment | NA | NA | 0 |
| S9_vs_k141_88324\|\|full | 1250 | No | NA | 3 | 1 | 0 | Low-quality | Genome-fragment | 2 | AAI-based (high-confidence) | 0 |
| S11_vs_k141_51995\|\|lt2gene | 1250 | No | NA | 1 | 1 | 0 | Low-quality | Genome-fragment | 1.83 | HMM-based (lower-bound) | 0 |
| S13_extracted_31537 | 1395 | No | NA | 1 | 0 | 0 | Not-determined | Genome-fragment | NA | NA | 0 |
| S13_extracted_31848 | 2339 | No | NA | 3 | 0 | 1 | Not-determined | Genome-fragment | NA | NA | 0 |
| S13_extracted_31967 | 1090 | No | NA | 2 | 0 | 1 | Not-determined | Genome-fragment | NA | NA | 0 |
| S13_extracted_32040 | 2150 | No | NA | 2 | 0 | 1 | Not-determined | Genome-fragment | NA | NA | 0 |
| S13_extracted_32063 | 1624 | No | NA | 2 | 0 | 0 | Not-determined | Genome-fragment | NA | NA | 0 |
| S13_extracted_32097 | 1064 | No | NA | 2 | 0 | 1 | Not-determined | Genome-fragment | NA | NA | 0 |
| S13_extracted_32198 | 1722 | No | NA | 1 | 0 | 0 | Not-determined | Genome-fragment | NA | NA | 0 |
| S13_extracted_32207 | 2109 | No | NA | 3 | 0 | 0 | Not-determined | Genome-fragment | NA | NA | 0 |
| S13_extracted_110816 | 1250 | No | NA | 2 | 0 | 1 | Low-quality | Genome-fragment | 1.69 | AAI-based (medium-confidence) | 0 |
| S13_extracted_32344 | 1184 | No | NA | 1 | 0 | 1 | Not-determined | Genome-fragment | NA | NA | 0 |
| S14_extracted_k141_109332 | 1250 | No | NA | 1 | 1 | 0 | Low-quality | Genome-fragment | 1.83 | HMM-based (lower-bound) | 0 |
| S13_extracted_32475 | 1029 | No | NA | 1 | 0 | 1 | Not-determined | Genome-fragment | NA | NA | 0 |
| S13_extracted_32523 | 1553 | No | NA | 1 | 0 | 0 | Not-determined | Genome-fragment | NA | NA | 0 |
| S13_extracted_32554 | 1152 | No | NA | 1 | 0 | 1 | Not-determined | Genome-fragment | NA | NA | 0 |
| S13_extracted_32708 | 3516 | No | NA | 4 | 0 | 1 | Not-determined | Genome-fragment | NA | NA | 0 |
| S1_vs_k141_62792\|\|full | 1250 | No | NA | 3 | 1 | 0 | Low-quality | Genome-fragment | 2.08 | AAI-based (high-confidence) | 0 |
| S13_extracted_32838 | 1131 | No | NA | 4 | 0 | 2 | Not-determined | Genome-fragment | NA | NA | 0 |
| S13_extracted_32847 | 11215 | No | NA | 7 | 0 | 7 | Not-determined | Genome-fragment | NA | NA | 0 |
| S1_vs_k141_155973\|\|full | 1250 | No | NA | 2 | 0 | 0 | Low-quality | Genome-fragment | 5.24 | AAI-based (medium-confidence) | 0 |
| S3_extracted_k141_73121 | 1250 | No | NA | 2 | 0 | 0 | Low-quality | Genome-fragment | 1.32 | AAI-based (medium-confidence) | 0 |
| S7_vs_58436\|\|lt2gene | 1250 | No | NA | 1 | 1 | 0 | Low-quality | Genome-fragment | 1.66 | AAI-based (medium-confidence) | 0 |
| S13_extracted_33317 | 1487 | No | NA | 3 | 0 | 2 | Not-determined | Genome-fragment | NA | NA | 0 |
| S13_extracted_33420 | 1609 | No | NA | 2 | 0 | 2 | Not-determined | Genome-fragment | NA | NA | 0 |
| S15_extracted_51952 | 1250 | No | NA | 3 | 0 | 0 | Low-quality | Genome-fragment | 1.5 | AAI-based (high-confidence) | 0 |
| S13_extracted_33794 | 1144 | No | NA | 1 | 0 | 1 | Not-determined | Genome-fragment | NA | NA | 0 |
| S13_extracted_33914 | 1612 | No | NA | 3 | 0 | 2 | Not-determined | Genome-fragment | NA | NA | 0 |
| S13_extracted_34105 | 1775 | No | NA | 2 | 0 | 1 | Not-determined | Genome-fragment | NA | NA | 0 |
| S13_extracted_34215 | 1077 | No | NA | 2 | 0 | 1 | Not-determined | Genome-fragment | NA | NA | 0 |
| S13_extracted_34235 | 1225 | No | NA | 2 | 0 | 1 | Not-determined | Genome-fragment | NA | NA | 0 |
| S13_extracted_34315 | 2375 | No | NA | 1 | 0 | 1 | Not-determined | Genome-fragment | NA | NA | 0 |
| S13_extracted_34514 | 2057 | No | NA | 1 | 0 | 1 | Not-determined | Genome-fragment | NA | NA | 0 |
| S13_extracted_34683 | 4310 | No | NA | 6 | 0 | 1 | Not-determined | Genome-fragment | NA | NA | 0 |
| S13_extracted_34837 | 1168 | No | NA | 2 | 0 | 2 | Not-determined | Genome-fragment | NA | NA | 0 |
| S13_extracted_34912 | 1126 | No | NA | 1 | 0 | 0 | Not-determined | Genome-fragment | NA | NA | 0 |
| S13_extracted_35054 | 1045 | No | NA | 2 | 0 | 1 | Not-determined | Genome-fragment | NA | NA | 0 |
| S13_extracted_35308 | 1268 | No | NA | 2 | 0 | 0 | Not-determined | Genome-fragment | NA | NA | 0 |
| S13_extracted_35315 | 1821 | No | NA | 2 | 0 | 0 | Not-determined | Genome-fragment | NA | NA | 0 |
| S13_extracted_35402 | 1236 | No | NA | 2 | 0 | 1 | Not-determined | Genome-fragment | NA | NA | 0 |
| S13_extracted_35686 | 3014 | No | NA | 2 | 0 | 2 | Not-determined | Genome-fragment | NA | NA | 0 |
| S13_extracted_35800 | 1386 | No | NA | 2 | 0 | 0 | Not-determined | Genome-fragment | NA | NA | 0 |
| S13_extracted_35844 | 1285 | No | NA | 1 | 0 | 1 | Not-determined | Genome-fragment | NA | NA | 0 |
| S13_extracted_36096 | 10981 | No | NA | 10 | 0 | 2 | Not-determined | Genome-fragment | NA | NA | 0 |
| S13_extracted_36156 | 1113 | No | NA | 2 | 0 | 1 | Not-determined | Genome-fragment | NA | NA | 0 |
| S13_extracted_36200 | 1056 | No | NA | 2 | 0 | 2 | Not-determined | Genome-fragment | NA | NA | 0 |
| S13_extracted_36452 | 4534 | No | NA | 5 | 0 | 5 | Not-determined | Genome-fragment | NA | NA | 0 |
| S13_extracted_36508 | 2731 | No | NA | 2 | 0 | 1 | Not-determined | Genome-fragment | NA | NA | 0 |
| S13_extracted_37036 | 1013 | No | NA | 2 | 0 | 2 | Not-determined | Genome-fragment | NA | NA | 0 |
| S9_vs_k141_54104\|\|full | 1251 | No | NA | 3 | 0 | 0 | Low-quality | Genome-fragment | 2.75 | AAI-based (high-confidence) | 0 |
| S13_extracted_37164 | 2685 | No | NA | 1 | 0 | 1 | Not-determined | Genome-fragment | NA | NA | 0 |
| S13_extracted_37293 | 1423 | No | NA | 2 | 0 | 1 | Not-determined | Genome-fragment | NA | NA | 0 |

| | | | | | | | | | | | | | |
|---|---|---|---|---|---|---|---|---|---|---|---|---|---|
| S11_vs_k141_27891\|\|full | 1251 | No | NA | | 4 | 0 | 0 | Low-quality | Genome-fragment | | 3.3 | AAI-based (medium-confidence) | 0 |
| S13_extracted_37722 | 1185 | No | NA | | 1 | 0 | 1 | Not-determined | Genome-fragment | NA | | NA | 0 |
| S3_vs_k141_115593\|\|full | 1251 | No | NA | | 2 | 0 | 0 | Low-quality | Genome-fragment | | 1.48 | AAI-based (medium-confidence) | 0 |
| S13_extracted_38143 | 1199 | No | NA | | 2 | 0 | 1 | Not-determined | Genome-fragment | NA | | NA | 0 |
| S3_vs_k141_6607\|\|full | 1251 | No | NA | | 3 | 1 | 0 | Low-quality | Genome-fragment | | 3.75 | HMM-based (lower-bound) | 0 |
| S5_extracted_k141_76625 | 1251 | No | NA | | 1 | 0 | 0 | Low-quality | Genome-fragment | | 2.5 | AAI-based (high-confidence) | 0 |
| S7_vs_28158\|\|full | 1251 | No | NA | | 4 | 0 | 0 | Low-quality | Genome-fragment | | 0.91 | AAI-based (medium-confidence) | 0 |
| S7_vs_80674\|\|full | 1251 | No | NA | | 4 | 2 | 0 | Low-quality | Genome-fragment | | 2.84 | AAI-based (high-confidence) | 0 |
| S16_extracted_91732 | 1251 | No | NA | | 2 | 2 | 0 | Low-quality | Genome-fragment | | 2.85 | AAI-based (medium-confidence) | 0 |
| S13_extracted_38708 | 29135 | No | NA | | 42 | 16 | 1 | Medium-quality | Genome-fragment | | 53.17 | AAI-based (high-confidence) | 0 |
| S13_extracted_38984 | 1075 | No | NA | | 1 | 0 | 0 | Not-determined | Genome-fragment | NA | | NA | 0 |
| S13_extracted_39069 | 2066 | No | NA | | 2 | 0 | 1 | Not-determined | Genome-fragment | NA | | NA | 0 |
| S10_extracted_108721 | 1251 | No | NA | | 2 | 0 | 1 | Low-quality | Genome-fragment | | 0.22 | AAI-based (medium-confidence) | 0 |
| S13_extracted_39196 | 1478 | No | NA | | 2 | 0 | 1 | Not-determined | Genome-fragment | NA | | NA | 0 |
| S8_vs_k141_166224\|\|full | 1251 | No | NA | | 2 | 0 | 1 | Low-quality | Genome-fragment | | 1.41 | AAI-based (medium-confidence) | 0 |
| S13_extracted_39531 | 1490 | No | NA | | 2 | 0 | 0 | Not-determined | Genome-fragment | NA | | NA | 0 |
| S13_extracted_39533 | 1416 | No | NA | | 2 | 0 | 0 | Not-determined | Genome-fragment | NA | | NA | 0 |
| S13_extracted_39622 | 1153 | No | NA | | 2 | 0 | 1 | Not-determined | Genome-fragment | NA | | NA | 0 |
| S13_extracted_39908 | 2135 | No | NA | | 2 | 0 | 0 | Not-determined | Genome-fragment | NA | | NA | 0 |
| S8_vs_k141_49063\|\|full | 1251 | No | NA | | 3 | 0 | 1 | Low-quality | Genome-fragment | | 0.89 | AAI-based (medium-confidence) | 0 |
| S13_extracted_40220 | 1432 | No | NA | | 1 | 0 | 1 | Not-determined | Genome-fragment | NA | | NA | 0 |
| S17_extracted_k141_174688 | 1252 | No | NA | | 2 | 0 | 1 | Low-quality | Genome-fragment | | 1.12 | AAI-based (medium-confidence) | 0 |
| S13_extracted_40432 | 1647 | No | NA | | 2 | 0 | 2 | Not-determined | Genome-fragment | NA | | NA | 0 |
| S13_extracted_40443 | 1132 | No | NA | | 3 | 0 | 0 | Not-determined | Genome-fragment | NA | | NA | 0 |
| S13_extracted_40791 | 1478 | No | NA | | 3 | 0 | 0 | Not-determined | Genome-fragment | NA | | NA | 0 |
| S13_extracted_41076 | 1379 | No | NA | | 1 | 0 | 1 | Not-determined | Genome-fragment | NA | | NA | 0 |
| S17_vs_k141_82881\|\|full | 1252 | No | NA | | 3 | 0 | 0 | Low-quality | Genome-fragment | | 0.86 | AAI-based (medium-confidence) | 0 |
| S9_vs_k141_68296\|\|full | 1252 | No | NA | | 3 | 1 | 0 | Low-quality | Genome-fragment | | 3.49 | AAI-based (medium-confidence) | 0 |
| S13_extracted_41273 | 1579 | No | NA | | 2 | 0 | 0 | Not-determined | Genome-fragment | NA | | NA | 0 |
| S13_extracted_41311 | 1216 | No | NA | | 2 | 0 | 0 | Not-determined | Genome-fragment | NA | | NA | 0 |
| S13_extracted_41404 | 1084 | No | NA | | 2 | 0 | 1 | Not-determined | Genome-fragment | NA | | NA | 0 |
| S13_extracted_41514 | 2207 | No | NA | | 5 | 0 | 0 | Not-determined | Genome-fragment | NA | | NA | 0 |
| S13_extracted_41842 | 3240 | No | NA | | 3 | 0 | 2 | Not-determined | Genome-fragment | NA | | NA | 0 |
| S9_vs_k141_65499\|\|full | 1252 | No | NA | | 3 | 0 | 1 | Low-quality | Genome-fragment | | 2.53 | AAI-based (high-confidence) | 0 |
| S13_extracted_41980 | 4238 | No | NA | | 7 | 0 | 3 | Not-determined | Genome-fragment | NA | | NA | 0 |
| S13_extracted_42250 | 2041 | No | NA | | 3 | 0 | 1 | Not-determined | Genome-fragment | NA | | NA | 0 |
| S13_extracted_42434 | 2831 | No | NA | | 2 | 0 | 1 | Not-determined | Genome-fragment | NA | | NA | 0 |
| S13_extracted_42450 | 1322 | No | NA | | 1 | 0 | 1 | Not-determined | Genome-fragment | NA | | NA | 0 |
| S13_extracted_42655 | 1037 | No | NA | | 1 | 0 | 1 | Not-determined | Genome-fragment | NA | | NA | 0 |
| S11_vs_k141_89027\|\|full | 1252 | No | NA | | 4 | 0 | 0 | Low-quality | Genome-fragment | | 3.65 | AAI-based (high-confidence) | 0 |
| S13_extracted_43171 | 1480 | No | NA | | 1 | 0 | 1 | Not-determined | Genome-fragment | NA | | NA | 0 |
| S11_vs_k141_49270\|\|full | 1252 | No | NA | | 2 | 0 | 2 | Low-quality | Genome-fragment | | 0.56 | AAI-based (medium-confidence) | 0 |
| S7_extracted_73218 | 1252 | No | NA | | 5 | 0 | 0 | Low-quality | Genome-fragment | | 2.6 | AAI-based (high-confidence) | 0 |
| S13_extracted_43858 | 1405 | No | NA | | 3 | 0 | 1 | Not-determined | Genome-fragment | NA | | NA | 0 |
| S13_extracted_43871 | 1852 | No | NA | | 1 | 0 | 0 | Not-determined | Genome-fragment | NA | | NA | 0 |
| S13_extracted_43889 | 3568 | No | NA | | 2 | 0 | 2 | Not-determined | Genome-fragment | NA | | NA | 0 |
| S13_extracted_44034 | 1428 | No | NA | | 1 | 0 | 1 | Not-determined | Genome-fragment | NA | | NA | 0 |
| S13_extracted_44078 | 2275 | No | NA | | 2 | 0 | 0 | Not-determined | Genome-fragment | NA | | NA | 0 |
| S7_vs_14671\|\|full | 1252 | No | NA | | 3 | 1 | 0 | Low-quality | Genome-fragment | | 3.26 | AAI-based (medium-confidence) | 0 |
| S13_extracted_44381 | 3548 | No | NA | | 5 | 0 | 3 | Not-determined | Genome-fragment | NA | | NA | 0 |
| S13_extracted_44394 | 3855 | No | NA | | 4 | 0 | 1 | Not-determined | Genome-fragment | NA | | NA | 0 |
| S16_extracted_104425 | 1252 | No | NA | | 1 | 0 | 0 | Low-quality | Genome-fragment | | 3.28 | AAI-based (medium-confidence) | 0 |
| S13_extracted_44585 | 1065 | No | NA | | 3 | 0 | 0 | Not-determined | Genome-fragment | NA | | NA | 0 |
| S13_extracted_44822 | 1270 | No | NA | | 2 | 0 | 1 | Not-determined | Genome-fragment | NA | | NA | 0 |
| S13_extracted_44879 | 1058 | No | NA | | 1 | 0 | 0 | Not-determined | Genome-fragment | NA | | NA | 0 |
| S10_extracted_19554 | 1252 | No | NA | | 3 | 1 | 0 | Low-quality | Genome-fragment | | 2.53 | AAI-based (high-confidence) | 0 |
| S13_extracted_44944 | 1776 | No | NA | | 4 | 0 | 2 | Not-determined | Genome-fragment | NA | | NA | 0 |
| S13_extracted_45035 | 1185 | No | NA | | 1 | 0 | 1 | Not-determined | Genome-fragment | NA | | NA | 0 |
| S13_extracted_45039 | 1645 | No | NA | | 3 | 0 | 2 | Not-determined | Genome-fragment | NA | | NA | 0 |
| S13_extracted_45087 | 1595 | No | NA | | 2 | 0 | 2 | Not-determined | Genome-fragment | NA | | NA | 0 |
| S13_extracted_45330 | 1918 | No | NA | | 2 | 0 | 1 | Not-determined | Genome-fragment | NA | | NA | 0 |
| S13_extracted_45386 | 1703 | No | NA | | 3 | 0 | 2 | Not-determined | Genome-fragment | NA | | NA | 0 |
| S13_extracted_45424 | 1399 | No | NA | | 1 | 0 | 1 | Not-determined | Genome-fragment | NA | | NA | 0 |
| S12_extracted_k141_41441 | 1252 | Yes | | 506 | 3 | 1 | 1 | Low-quality | Genome-fragment | | 1.55 | AAI-based (high-confidence) | 59.58 |
| S12_extracted_k141_105976 | 1252 | No | NA | | 2 | 1 | 0 | Low-quality | Genome-fragment | | 2.66 | AAI-based (high-confidence) | 0 |
| S13_extracted_45646 | 1111 | No | NA | | 2 | 0 | 2 | Not-determined | Genome-fragment | NA | | NA | 0 |
| S8_extracted_k141_35181 | 1252 | No | NA | | 2 | 0 | 0 | Low-quality | Genome-fragment | | 2.98 | AAI-based (medium-confidence) | 0 |
| S13_extracted_45790 | 1350 | No | NA | | 3 | 0 | 0 | Not-determined | Genome-fragment | NA | | NA | 0 |
| S13_extracted_45811 | 1723 | No | NA | | 3 | 0 | 3 | Not-determined | Genome-fragment | NA | | NA | 0 |
| S13_extracted_45898 | 1288 | No | NA | | 1 | 0 | 1 | Not-determined | Genome-fragment | NA | | NA | 0 |
| S13_extracted_45986 | 1298 | No | NA | | 2 | 0 | 1 | Not-determined | Genome-fragment | NA | | NA | 0 |
| S13_extracted_45999 | 1032 | No | NA | | 1 | 0 | 1 | Not-determined | Genome-fragment | NA | | NA | 0 |
| S8_extracted_k141_123126 | 1252 | No | NA | | 3 | 1 | 0 | Low-quality | Genome-fragment | | 0.97 | AAI-based (medium-confidence) | 0 |
| S13_extracted_46568 | 1462 | No | NA | | 1 | 0 | 0 | Not-determined | Genome-fragment | NA | | NA | 0 |
| S13_extracted_46617 | 1133 | No | NA | | 2 | 0 | 2 | Not-determined | Genome-fragment | NA | | NA | 0 |
| S13_extracted_46767 | 5986 | No | NA | | 4 | 0 | 4 | Not-determined | Genome-fragment | NA | | NA | 0 |
| S13_extracted_46865 | 3211 | No | NA | | 3 | 0 | 2 | Not-determined | Genome-fragment | NA | | NA | 0 |
| S13_extracted_46957 | 1601 | No | NA | | 2 | 0 | 1 | Not-determined | Genome-fragment | NA | | NA | 0 |
| S13_extracted_46985 | 1457 | No | NA | | 5 | 0 | 0 | Not-determined | Genome-fragment | NA | | NA | 0 |
| S13_extracted_47343 | 4457 | No | NA | | 3 | 0 | 1 | Not-determined | Genome-fragment | NA | | NA | 0 |
| S9_vs_k141_47816\|\|full | 1253 | No | NA | | 3 | 0 | 0 | Low-quality | Genome-fragment | | 1.94 | AAI-based (medium-confidence) | 0 |
| S13_extracted_47662 | 1814 | No | NA | | 2 | 0 | 2 | Not-determined | Genome-fragment | NA | | NA | 0 |
| S13_extracted_47937 | 2752 | No | NA | | 4 | 0 | 4 | Not-determined | Genome-fragment | NA | | NA | 0 |
| S14_extracted_k141_53246 | 1253 | No | NA | | 4 | 0 | 1 | Low-quality | Genome-fragment | | 1.81 | AAI-based (medium-confidence) | 0 |
| S13_extracted_47994 | 4534 | No | NA | | 3 | 0 | 2 | Not-determined | Genome-fragment | NA | | NA | 0 |
| S13_extracted_48067 | 3220 | No | NA | | 1 | 0 | 1 | Not-determined | Genome-fragment | NA | | NA | 0 |
| S13_extracted_48100 | 1064 | No | NA | | 2 | 0 | 2 | Not-determined | Genome-fragment | NA | | NA | 0 |
| S13_extracted_48122 | 1468 | No | NA | | 2 | 0 | 1 | Not-determined | Genome-fragment | NA | | NA | 0 |
| S5_extracted_k141_138377 | 1253 | No | NA | | 2 | 2 | 0 | Low-quality | Genome-fragment | | 3.04 | AAI-based (medium-confidence) | 0 |
| S13_extracted_48359 | 3225 | No | NA | | 4 | 0 | 1 | Not-determined | Genome-fragment | NA | | NA | 0 |
| S13_extracted_48416 | 1281 | No | NA | | 1 | 0 | 0 | Not-determined | Genome-fragment | NA | | NA | 0 |
| S13_extracted_48447 | 1035 | No | NA | | 1 | 0 | 0 | Not-determined | Genome-fragment | NA | | NA | 0 |
| S7_vs_60415\|\|full | 1253 | No | NA | | 2 | 1 | 0 | Low-quality | Genome-fragment | | 1.93 | HMM-based (lower-bound) | 0 |
| S13_extracted_48635 | 3757 | No | NA | | 4 | 0 | 2 | Not-determined | Genome-fragment | NA | | NA | 0 |
| S16_vs_16200\|\|full | 1253 | No | NA | | 2 | 0 | 0 | Low-quality | Genome-fragment | | 2.66 | AAI-based (medium-confidence) | 0 |
| S13_extracted_49501 | 1212 | No | NA | | 2 | 0 | 1 | Not-determined | Genome-fragment | NA | | NA | 0 |
| S16_vs_58752\|\|lt2gene | 1253 | No | NA | | 1 | 1 | 0 | Low-quality | Genome-fragment | | 2.04 | HMM-based (lower-bound) | 0 |
| S10_extracted_94592 | 1253 | No | NA | | 4 | 1 | 0 | Low-quality | Genome-fragment | | 1.43 | AAI-based (high-confidence) | 0 |
| S13_extracted_50140 | 1231 | No | NA | | 1 | 0 | 1 | Not-determined | Genome-fragment | NA | | NA | 0 |
| S13_extracted_50510 | 1074 | No | NA | | 2 | 0 | 0 | Not-determined | Genome-fragment | NA | | NA | 0 |
| S13_extracted_50625 | 1153 | No | NA | | 2 | 0 | 1 | Not-determined | Genome-fragment | NA | | NA | 0 |
| S12_vs_k141_143702\|\|full | 1253 | No | NA | | 2 | 0 | 2 | Low-quality | Genome-fragment | | 0.42 | AAI-based (medium-confidence) | 0 |
| S13_extracted_50864 | 3774 | No | NA | | 3 | 0 | 1 | Not-determined | Genome-fragment | NA | | NA | 0 |
| S13_extracted_50892 | 1348 | No | NA | | 2 | 0 | 1 | Not-determined | Genome-fragment | NA | | NA | 0 |
| S13_extracted_51115 | 1580 | No | NA | | 1 | 0 | 1 | Not-determined | Genome-fragment | NA | | NA | 0 |
| S13_extracted_51149 | 1082 | No | NA | | 1 | 0 | 1 | Not-determined | Genome-fragment | NA | | NA | 0 |
| S13_extracted_51168 | 1330 | No | NA | | 3 | 0 | 0 | Not-determined | Genome-fragment | NA | | NA | 0 |
| S13_extracted_51365 | 1825 | No | NA | | 2 | 0 | 0 | Not-determined | Genome-fragment | NA | | NA | 0 |
| S13_extracted_51417 | 2711 | No | NA | | 3 | 0 | 0 | Not-determined | Genome-fragment | NA | | NA | 0 |
| S13_extracted_51592 | 1410 | No | NA | | 2 | 0 | 2 | Not-determined | Genome-fragment | NA | | NA | 0 |
| S13_extracted_51902 | 4871 | No | NA | | 9 | 0 | 4 | Not-determined | Genome-fragment | NA | | NA | 0 |
| S13_extracted_52128 | 1556 | No | NA | | 1 | 0 | 1 | Not-determined | Genome-fragment | NA | | NA | 0 |
| S13_extracted_52421 | 3062 | No | NA | | 3 | 0 | 3 | Not-determined | Genome-fragment | NA | | NA | 0 |
| S13_extracted_52450 | 1376 | No | NA | | 1 | 0 | 0 | Not-determined | Genome-fragment | NA | | NA | 0 |
| S13_extracted_52546 | 3435 | No | NA | | 5 | 0 | 5 | Not-determined | Genome-fragment | NA | | NA | 0 |
| S13_extracted_52597 | 1168 | No | NA | | 1 | 0 | 1 | Not-determined | Genome-fragment | NA | | NA | 0 |
| S13_extracted_52694 | 1031 | No | NA | | 1 | 0 | 0 | Not-determined | Genome-fragment | NA | | NA | 0 |
| S13_extracted_52697 | 3591 | No | NA | | 3 | 0 | 0 | Not-determined | Genome-fragment | NA | | NA | 0 |
| S13_extracted_52722 | 1744 | No | NA | | 4 | 0 | 0 | Not-determined | Genome-fragment | NA | | NA | 0 |
| S13_extracted_52798 | 1071 | No | NA | | 1 | 0 | 1 | Not-determined | Genome-fragment | NA | | NA | 0 |
| S13_extracted_52824 | 1393 | No | NA | | 1 | 0 | 0 | Not-determined | Genome-fragment | NA | | NA | 0 |
| S13_extracted_53023 | 1647 | No | NA | | 2 | 0 | 1 | Not-determined | Genome-fragment | NA | | NA | 0 |
| S13_extracted_53177 | 1403 | No | NA | | 1 | 0 | 1 | Not-determined | Genome-fragment | NA | | NA | 0 |
| S13_extracted_53284 | 3273 | No | NA | | 1 | 0 | 1 | Not-determined | Genome-fragment | NA | | NA | 0 |
| S13_extracted_53341 | 9067 | No | NA | | 3 | 0 | 2 | Not-determined | Genome-fragment | NA | | NA | 0 |
| S13_extracted_53383 | 1222 | No | NA | | 2 | 0 | 2 | Not-determined | Genome-fragment | NA | | NA | 0 |
| S13_extracted_53405 | 1110 | No | NA | | 2 | 0 | 2 | Not-determined | Genome-fragment | NA | | NA | 0 |
| S13_extracted_53407 | 3218 | No | NA | | 1 | 0 | 0 | Not-determined | Genome-fragment | NA | | NA | 0 |
| S13_extracted_53527 | 1020 | No | NA | | 2 | 0 | 2 | Not-determined | Genome-fragment | NA | | NA | 0 |
| S13_extracted_53633 | 5163 | No | NA | | 3 | 0 | 2 | Not-determined | Genome-fragment | NA | | NA | 0 |
| S2_extracted_k141_14500 | 1253 | No | NA | | 2 | 0 | 0 | Low-quality | Genome-fragment | | 0.59 | AAI-based (medium-confidence) | 0 |
| S13_extracted_53649 | 1052 | No | NA | | 3 | 0 | 0 | Not-determined | Genome-fragment | NA | | NA | 0 |
| S13_extracted_53729 | 1082 | No | NA | | 2 | 0 | 0 | Not-determined | Genome-fragment | NA | | NA | 0 |
| S13_extracted_53842 | 2803 | No | NA | | 3 | 0 | 2 | Not-determined | Genome-fragment | NA | | NA | 0 |
| S13_extracted_53926 | 1792 | No | NA | | 2 | 0 | 2 | Not-determined | Genome-fragment | NA | | NA | 0 |
| S13_extracted_53959 | 2407 | No | NA | | 3 | 0 | 0 | Not-determined | Genome-fragment | NA | | NA | 0 |
| S13_extracted_54201 | 37189 | No | NA | | 53 | 13 | 1 | Medium-quality | Genome-fragment | | 64.56 | AAI-based (high-confidence) | 0 |
| S13_extracted_54371 | 1318 | No | NA | | 1 | 0 | 1 | Not-determined | Genome-fragment | NA | | NA | 0 |
| S8_vs_k141_159407\|\|full | 1253 | No | NA | | 2 | 0 | 0 | Low-quality | Genome-fragment | | 1.08 | AAI-based (medium-confidence) | 0 |
| S13_extracted_54541 | 3266 | No | NA | | 2 | 0 | 2 | Not-determined | Genome-fragment | NA | | NA | 0 |
| S13_extracted_54653 | 1847 | No | NA | | 4 | 0 | 4 | Not-determined | Genome-fragment | NA | | NA | 0 |
| S8_vs_k141_188622\|\|full | 1253 | No | NA | | 2 | 0 | 1 | Low-quality | Genome-fragment | | 2.12 | AAI-based (high-confidence) | 0 |
| S13_extracted_54882 | 2652 | No | NA | | 4 | 0 | 3 | Not-determined | Genome-fragment | NA | | NA | 0 |
| S13_extracted_55148 | 1299 | No | NA | | 2 | 0 | 1 | Not-determined | Genome-fragment | NA | | NA | 0 |
| S13_extracted_55592 | 1564 | No | NA | | 2 | 0 | 0 | Not-determined | Genome-fragment | NA | | NA | 0 |
| S13_extracted_55632 | 1152 | No | NA | | 1 | 0 | 0 | Not-determined | Genome-fragment | NA | | NA | 0 |
| S13_extracted_55637 | 1275 | No | NA | | 2 | 0 | 1 | Not-determined | Genome-fragment | NA | | NA | 0 |
| S13_extracted_55653 | 5110 | No | NA | | 8 | 0 | 3 | Not-determined | Genome-fragment | NA | | NA | 0 |
| S13_extracted_55711 | 3024 | No | NA | | 3 | 0 | 0 | Not-determined | Genome-fragment | NA | | NA | 0 |
| S13_extracted_55802 | 3058 | No | NA | | 4 | 0 | 3 | Not-determined | Genome-fragment | NA | | NA | 0 |
| S13_extracted_55962 | 1192 | No | NA | | 2 | 0 | 2 | Not-determined | Genome-fragment | NA | | NA | 0 |
| S13_extracted_56074 | 77817 | No | NA | | 109 | 16 | 0 | Complete | High-quality | | 100 | DTR (high-confidence) | 0 |
| S13_extracted_56100 | 1722 | No | NA | | 1 | 0 | 1 | Not-determined | Genome-fragment | NA | | NA | 0 |
| S13_extracted_56229 | 1036 | No | NA | | 1 | 0 | 1 | Not-determined | Genome-fragment | NA | | NA | 0 |
| S13_extracted_56790 | 1399 | No | NA | | 2 | 0 | 0 | Not-determined | Genome-fragment | NA | | NA | 0 |
| S13_extracted_56873 | 4377 | No | NA | | 1 | 0 | 1 | Not-determined | Genome-fragment | NA | | NA | 0 |
| S13_vs_138714\|\|full | 1254 | No | NA | | 3 | 0 | 1 | Low-quality | Genome-fragment | | 2.97 | AAI-based (medium-confidence) | 0 |
| S13_extracted_57051 | 1588 | No | NA | | 3 | 0 | 1 | Not-determined | Genome-fragment | NA | | NA | 0 |
| S13_extracted_57234 | 1107 | No | NA | | 2 | 0 | 0 | Not-determined | Genome-fragment | NA | | NA | 0 |
| S13_extracted_57276 | 1277 | No | NA | | 2 | 0 | 0 | Not-determined | Genome-fragment | NA | | NA | 0 |
| S13_extracted_57330 | 1169 | No | NA | | 1 | 0 | 1 | Not-determined | Genome-fragment | NA | | NA | 0 |
| S13_extracted_57766 | 3631 | No | NA | | 5 | 0 | 3 | Not-determined | Genome-fragment | NA | | NA | 0 |
| S13_extracted_57881 | 1753 | No | NA | | 1 | 0 | 1 | Not-determined | Genome-fragment | NA | | NA | 0 |
| S13_extracted_57910 | 1509 | No | NA | | 1 | 0 | 0 | Not-determined | Genome-fragment | NA | | NA | 0 |
| S13_extracted_57965 | 1348 | No | NA | | 1 | 0 | 1 | Not-determined | Genome-fragment | NA | | NA | 0 |
| S14_extracted_k141_88861 | 1254 | No | NA | | 2 | 0 | 0 | Low-quality | Genome-fragment | | 1.62 | AAI-based (high-confidence) | 0 |
| S1_extracted_k141_89491 | 1254 | No | NA | | 2 | 0 | 0 | Low-quality | Genome-fragment | | 1.25 | AAI-based (high-confidence) | 0 |
| S1_extracted_k141_136946 | 1254 | No | NA | | 1 | 0 | 1 | Low-quality | Genome-fragment | | 0.69 | AAI-based (medium-confidence) | 0 |
| S13_extracted_58207 | 1612 | No | NA | | 3 | 0 | 2 | Not-determined | Genome-fragment | NA | | NA | 0 |
| S13_extracted_58274 | 1173 | No | NA | | 2 | 0 | 1 | Not-determined | Genome-fragment | NA | | NA | 0 |
| S13_extracted_58303 | 1202 | No | NA | | 2 | 0 | 1 | Not-determined | Genome-fragment | NA | | NA | 0 |
| S13_extracted_58351 | 1578 | No | NA | | 2 | 0 | 1 | Not-determined | Genome-fragment | NA | | NA | 0 |
| S13_extracted_58449 | 1562 | No | NA | | 4 | 0 | 2 | Not-determined | Genome-fragment | NA | | NA | 0 |
| S13_extracted_58466 | 1417 | No | NA | | 4 | 0 | 1 | Not-determined | Genome-fragment | NA | | NA | 0 |
| S13_extracted_58581 | 1502 | No | NA | | 2 | 0 | 1 | Not-determined | Genome-fragment | NA | | NA | 0 |
| S3_extracted_k141_86816 | 1254 | No | NA | | 1 | 1 | 0 | Low-quality | Genome-fragment | | 0.27 | HMM-based (lower-bound) | 0 |
| S13_extracted_58862 | 3312 | No | NA | | 5 | 0 | 2 | Not-determined | Genome-fragment | NA | | NA | 0 |
| S13_extracted_58878 | 1097 | No | NA | | 3 | 0 | 1 | Not-determined | Genome-fragment | NA | | NA | 0 |
| S13_extracted_58986 | 1614 | No | NA | | 3 | 0 | 2 | Not-determined | Genome-fragment | NA | | NA | 0 |
| S13_extracted_59075 | 1223 | No | NA | | 3 | 0 | 0 | Not-determined | Genome-fragment | NA | | NA | 0 |
| S5_extracted_k141_64679 | 1254 | No | NA | | 5 | 2 | 0 | Low-quality | Genome-fragment | | 3.53 | AAI-based (medium-confidence) | 0 |
| S13_extracted_59450 | 1292 | No | NA | | 1 | 0 | 0 | Not-determined | Genome-fragment | NA | | NA | 0 |
| S13_extracted_59570 | 1010 | No | NA | | 2 | 0 | 1 | Not-determined | Genome-fragment | NA | | NA | 0 |
| S5_vs_k141_94871\|\|full | 1254 | No | NA | | 2 | 1 | 0 | Low-quality | Genome-fragment | | 3.24 | AAI-based (high-confidence) | 0 |
| S13_extracted_59894 | 3378 | No | NA | | 3 | 0 | 1 | Not-determined | Genome-fragment | NA | | NA | 0 |
| S13_extracted_59944 | 1131 | No | NA | | 1 | 0 | 1 | Not-determined | Genome-fragment | NA | | NA | 0 |
| S13_extracted_60194 | 3130 | No | NA | | 2 | 0 | 1 | Not-determined | Genome-fragment | NA | | NA | 0 |
| S13_extracted_60250 | 1402 | No | NA | | 1 | 0 | 0 | Not-determined | Genome-fragment | NA | | NA | 0 |
| S13_extracted_60487 | 5241 | No | NA | | 8 | 0 | 4 | Not-determined | Genome-fragment | NA | | NA | 0 |
| S13_extracted_60600 | 3724 | No | NA | | 3 | 0 | 3 | Not-determined | Genome-fragment | NA | | NA | 0 |
| S13_extracted_60925 | 5318 | No | NA | | 5 | 0 | 5 | Not-determined | Genome-fragment | NA | | NA | 0 |
| S5_vs_k141_102115\|\|full | 1254 | No | NA | | 3 | 1 | 0 | Low-quality | Genome-fragment | | 2.21 | AAI-based (high-confidence) | 0 |
| S13_extracted_61109 | 1604 | No | NA | | 2 | 0 | 2 | Not-determined | Genome-fragment | NA | | NA | 0 |
| S13_extracted_61328 | 1046 | No | NA | | 2 | 0 | 1 | Not-determined | Genome-fragment | NA | | NA | 0 |
| S13_extracted_61378 | 1693 | No | NA | | 2 | 0 | 2 | Not-determined | Genome-fragment | NA | | NA | 0 |
| S7_extracted_80519 | 1254 | No | NA | | 2 | 2 | 0 | Low-quality | Genome-fragment | | 0.48 | HMM-based (lower-bound) | 0 |
| S13_extracted_61475 | 1511 | No | NA | | 1 | 0 | 1 | Not-determined | Genome-fragment | NA | | NA | 0 |
| S13_extracted_61504 | 1324 | No | NA | | 2 | 0 | 1 | Not-determined | Genome-fragment | NA | | NA | 0 |
| S13_extracted_61518 | 1447 | No | NA | | 2 | 0 | 0 | Not-determined | Genome-fragment | NA | | NA | 0 |
| S13_extracted_61613 | 1263 | No | NA | | 1 | 0 | 1 | Not-determined | Genome-fragment | NA | | NA | 0 |
| S16_extracted_123255 | 1254 | No | NA | | 3 | 0 | 0 | Low-quality | Genome-fragment | | 2.16 | AAI-based (high-confidence) | 0 |
| S13_extracted_61719 | 1943 | No | NA | | 3 | 0 | 2 | Not-determined | Genome-fragment | NA | | NA | 0 |
| S13_extracted_61886 | 1473 | No | NA | | 4 | 0 | 0 | Not-determined | Genome-fragment | NA | | NA | 0 |
| S13_extracted_61904 | 1026 | No | NA | | 1 | 0 | 1 | Not-determined | Genome-fragment | NA | | NA | 0 |
| S13_extracted_62001 | 1678 | No | NA | | 1 | 0 | 1 | Not-determined | Genome-fragment | NA | | NA | 0 |
| S13_extracted_62006 | 1256 | No | NA | | 1 | 0 | 1 | Not-determined | Genome-fragment | NA | | NA | 0 |
| S13_extracted_62048 | 1338 | No | NA | | 1 | 0 | 1 | Not-determined | Genome-fragment | NA | | NA | 0 |
| S13_extracted_62189 | 1311 | No | NA | | 4 | 0 | 0 | Not-determined | Genome-fragment | NA | | NA | 0 |
| S13_extracted_62262 | 1201 | No | NA | | 3 | 0 | 0 | Not-determined | Genome-fragment | NA | | NA | 0 |
| S13_extracted_62321 | 1693 | No | NA | | 1 | 0 | 1 | Not-determined | Genome-fragment | NA | | NA | 0 |
| S13_extracted_62411 | 1858 | No | NA | | 3 | 0 | 1 | Not-determined | Genome-fragment | NA | | NA | 0 |
| S13_extracted_62432 | 3547 | No | NA | | 1 | 0 | 0 | Not-determined | Genome-fragment | NA | | NA | 0 |
| S16_extracted_162747 | 1254 | No | NA | | 2 | 1 | 0 | Low-quality | Genome-fragment | | 2.31 | AAI-based (high-confidence) | 0 |
| S10_vs_71029\|\|full | 1254 | No | NA | | 2 | 0 | 2 | Low-quality | Genome-fragment | | 0.75 | AAI-based (medium-confidence) | 0 |
| S13_extracted_62500 | 1042 | No | NA | | 2 | 0 | 0 | Not-determined | Genome-fragment | NA | | NA | 0 |
| S13_extracted_62746 | 1392 | No | NA | | 1 | 0 | 0 | Not-determined | Genome-fragment | NA | | NA | 0 |
| S13_extracted_62752 | 1223 | No | NA | | 2 | 0 | 1 | Not-determined | Genome-fragment | NA | | NA | 0 |
| S13_extracted_62783 | 1641 | No | NA | | 1 | 0 | 1 | Not-determined | Genome-fragment | NA | | NA | 0 |
| S13_extracted_62858 | 1997 | No | NA | | 3 | 0 | 2 | Not-determined | Genome-fragment | NA | | NA | 0 |
| S13_extracted_62907 | 2312 | No | NA | | 2 | 0 | 1 | Not-determined | Genome-fragment | NA | | NA | 0 |
| S13_extracted_63300 | 1243 | No | NA | | 2 | 0 | 1 | Not-determined | Genome-fragment | NA | | NA | 0 |
| S13_extracted_63395 | 4027 | No | NA | | 4 | 0 | 2 | Not-determined | Genome-fragment | NA | | NA | 0 |
| S13_extracted_63587 | 1837 | No | NA | | 3 | 0 | 0 | Not-determined | Genome-fragment | NA | | NA | 0 |
| S13_extracted_63723 | 1271 | No | NA | | 2 | 0 | 0 | Not-determined | Genome-fragment | NA | | NA | 0 |
| S12_extracted_k141_138942 | 1254 | No | NA | | 3 | 0 | 1 | Low-quality | Genome-fragment | | 0.68 | AAI-based (medium-confidence) | 0 |
| S13_extracted_64182 | 1292 | No | NA | | 2 | 0 | 1 | Not-determined | Genome-fragment | NA | | NA | 0 |
| S13_extracted_64298 | 3333 | No | NA | | 3 | 0 | 3 | Not-determined | Genome-fragment | NA | | NA | 0 |
| S13_extracted_64323 | 1255 | No | NA | | 1 | 0 | 0 | Not-determined | Genome-fragment | NA | | NA | 0 |
| S13_extracted_64360 | 1255 | No | NA | | 2 | 0 | 1 | Not-determined | Genome-fragment | NA | | NA | 0 |
| S12_extracted_k141_85877 | 1254 | No | NA | | 3 | 0 | 0 | Low-quality | Genome-fragment | | 2.96 | AAI-based (high-confidence) | 0 |
| S13_extracted_64596 | 1954 | No | NA | | 1 | 0 | 1 | Not-determined | Genome-fragment | NA | | NA | 0 |
| S13_extracted_64664 | 2467 | No | NA | | 2 | 0 | 1 | Not-determined | Genome-fragment | NA | | NA | 0 |
| S13_extracted_64872 | 1211 | No | NA | | 1 | 0 | 1 | Not-determined | Genome-fragment | NA | | NA | 0 |
| S13_extracted_64877 | 1119 | No | NA | | 1 | 0 | 1 | Not-determined | Genome-fragment | NA | | NA | 0 |
| S13_extracted_65108 | 1320 | No | NA | | 2 | 0 | 0 | Not-determined | Genome-fragment | NA | | NA | 0 |
| S13_extracted_65151 | 1415 | No | NA | | 1 | 0 | 1 | Not-determined | Genome-fragment | NA | | NA | 0 |
| S13_extracted_65300 | 1325 | No | NA | | 2 | 0 | 0 | Not-determined | Genome-fragment | NA | | NA | 0 |
| S13_extracted_65333 | 1322 | No | NA | | 3 | 0 | 1 | Not-determined | Genome-fragment | NA | | NA | 0 |
| S13_extracted_65400 | 1338 | No | NA | | 2 | 0 | 0 | Not-determined | Genome-fragment | NA | | NA | 0 |
| S13_extracted_65465 | 1202 | No | NA | | 1 | 0 | 1 | Not-determined | Genome-fragment | NA | | NA | 0 |
| S12_vs_k141_67797\|\|full | 1254 | No | NA | | 3 | 0 | 1 | Low-quality | Genome-fragment | | 2.82 | AAI-based (medium-confidence) | 0 |
| S6_extracted_k141_3017 | 1254 | No | NA | | 4 | 2 | 0 | Low-quality | Genome-fragment | | 2.39 | AAI-based (high-confidence) | 0 |
| S9_extracted_k141_1383 | 1255 | No | NA | | 2 | 0 | 1 | Low-quality | Genome-fragment | | 0.58 | AAI-based (medium-confidence) | 0 |
| S13_extracted_66039 | 2070 | No | NA | | 1 | 0 | 1 | Not-determined | Genome-fragment | NA | | NA | 0 |
| S13_extracted_66170 | 1172 | No | NA | | 2 | 0 | 0 | Not-determined | Genome-fragment | NA | | NA | 0 |
| S13_vs_38192\|\|full | 1255 | No | NA | | 3 | 0 | 0 | Low-quality | Genome-fragment | | 2.22 | AAI-based (medium-confidence) | 0 |
| S13_extracted_66348 | 3470 | No | NA | | 2 | 0 | 2 | Not-determined | Genome-fragment | NA | | NA | 0 |
| S13_extracted_66365 | 7357 | No | NA | | 5 | 0 | 3 | Not-determined | Genome-fragment | NA | | NA | 0 |
| S14_extracted_k141_83046 | 1255 | No | NA | | 3 | 2 | 0 | Low-quality | Genome-fragment | | 2.85 | AAI-based (high-confidence) | 0 |
| S13_extracted_66508 | 38005 | No | NA | | 61 | 5 | 0 | High-quality | High-quality | | 100 | AAI-based (high-confidence) | 0 |
| S13_extracted_66738 | 1091 | No | NA | | 2 | 0 | 2 | Not-determined | Genome-fragment | NA | | NA | 0 |
| S13_extracted_66791 | 1410 | No | NA | | 3 | 0 | 0 | Not-determined | Genome-fragment | NA | | NA | 0 |
| S13_extracted_66808 | 3321 | No | NA | | 4 | 0 | 1 | Not-determined | Genome-fragment | NA | | NA | 0 |
| S13_extracted_67092 | 3525 | No | NA | | 6 | 0 | 0 | Not-determined | Genome-fragment | NA | | NA | 0 |
| S13_extracted_67095 | 1044 | No | NA | | 1 | 0 | 1 | Not-determined | Genome-fragment | NA | | NA | 0 |
| S13_extracted_67785 | 1490 | No | NA | | 2 | 0 | 1 | Not-determined | Genome-fragment | NA | | NA | 0 |
| S13_extracted_68135 | 1244 | No | NA | | 1 | 0 | 1 | Not-determined | Genome-fragment | NA | | NA | 0 |
| S13_extracted_68234 | 1054 | No | NA | | 1 | 0 | 0 | Not-determined | Genome-fragment | NA | | NA | 0 |
| S1_extracted_k141_76531 | 1255 | No | NA | | 2 | 0 | 1 | Low-quality | Genome-fragment | | 0.34 | AAI-based (medium-confidence) | 0 |
| S5_extracted_k141_130708 | 1255 | No | NA | | 1 | 1 | 0 | Low-quality | Genome-fragment | | 2.71 | AAI-based (high-confidence) | 0 |
| S13_extracted_68715 | 1846 | No | NA | | 3 | 0 | 0 | Not-determined | Genome-fragment | NA | | NA | 0 |
| S13_extracted_69063 | 3885 | No | NA | | 1 | 0 | 1 | Not-determined | Genome-fragment | NA | | NA | 0 |
| S13_extracted_69155 | 1748 | No | NA | | 3 | 0 | 0 | Not-determined | Genome-fragment | NA | | NA | 0 |
| S13_extracted_69250 | 6052 | No | NA | | 6 | 0 | 1 | Not-determined | Genome-fragment | NA | | NA | 0 |
| S5_vs_k141_10744\|\|full | 1255 | No | NA | | 3 | 0 | 0 | Low-quality | Genome-fragment | | 3.22 | AAI-based (medium-confidence) | 0 |
| S13_extracted_69467 | 3270 | No | NA | | 5 | 0 | 4 | Not-determined | Genome-fragment | NA | | NA | 0 |
| S13_extracted_69784 | 1145 | No | NA | | 1 | 0 | 1 | Not-determined | Genome-fragment | NA | | NA | 0 |
| S13_extracted_69789 | 1020 | No | NA | | 1 | 0 | 0 | Not-determined | Genome-fragment | NA | | NA | 0 |
| S13_extracted_69796 | 1153 | No | NA | | 1 | 0 | 0 | Not-determined | Genome-fragment | NA | | NA | 0 |
| S13_extracted_69822 | 2053 | No | NA | | 5 | 0 | 0 | Not-determined | Genome-fragment | NA | | NA | 0 |
| S13_extracted_69886 | 1795 | No | NA | | 2 | 0 | 0 | Not-determined | Genome-fragment | NA | | NA | 0 |
| S7_vs_86323\|\|full | 1255 | No | NA | | 4 | 1 | 0 | Low-quality | Genome-fragment | | 3.03 | AAI-based (medium-confidence) | 0 |
| S13_extracted_69978 | 1971 | No | NA | | 2 | 0 | 2 | Not-determined | Genome-fragment | NA | | NA | 0 |
| S13_extracted_70081 | 1005 | No | NA | | 2 | 0 | 0 | Not-determined | Genome-fragment | NA | | NA | 0 |
| S13_extracted_70190 | 1329 | No | NA | | 2 | 0 | 2 | Not-determined | Genome-fragment | NA | | NA | 0 |
| S13_extracted_70223 | 1837 | No | NA | | 3 | 0 | 1 | Not-determined | Genome-fragment | NA | | NA | 0 |
| S13_extracted_70237 | 1080 | No | NA | | 1 | 0 | 0 | Not-determined | Genome-fragment | NA | | NA | 0 |
| S13_extracted_70293 | 1063 | No | NA | | 1 | 0 | 0 | Not-determined | Genome-fragment | NA | | NA | 0 |
| S13_extracted_70399 | 1690 | No | NA | | 3 | 0 | 0 | Not-determined | Genome-fragment | NA | | NA | 0 |
| S13_extracted_70486 | 1177 | No | NA | | 3 | 0 | 3 | Not-determined | Genome-fragment | NA | | NA | 0 |
| S16_vs_153164\|\|lt2gene | 1255 | No | NA | | 2 | 2 | 0 | Low-quality | Genome-fragment | | 2.86 | AAI-based (high-confidence) | 0 |
| S13_extracted_70630 | 1091 | No | NA | | 2 | 0 | 0 | Not-determined | Genome-fragment | NA | | NA | 0 |
| S13_extracted_70825 | 1334 | No | NA | | 1 | 0 | 1 | Not-determined | Genome-fragment | NA | | NA | 0 |
| S12_vs_k141_119282\|\|full | 1255 | Yes | | 980 | 3 | 1 | 1 | Low-quality | Genome-fragment | | 2.15 | AAI-based (high-confidence) | 21.91 |
| S13_extracted_71043 | 2094 | No | NA | | 2 | 0 | 0 | Not-determined | Genome-fragment | NA | | NA | 0 |
| S13_extracted_71066 | 1359 | No | NA | | 1 | 0 | 1 | Not-determined | Genome-fragment | NA | | NA | 0 |
| S13_extracted_71130 | 1049 | No | NA | | 1 | 0 | 1 | Not-determined | Genome-fragment | NA | | NA | 0 |
| S13_extracted_71544 | 1666 | No | NA | | 2 | 0 | 1 | Not-determined | Genome-fragment | NA | | NA | 0 |
| S13_extracted_71753 | 1440 | No | NA | | 1 | 0 | 0 | Not-determined | Genome-fragment | NA | | NA | 0 |
| S13_extracted_71802 | 3467 | No | NA | | 3 | 0 | 3 | Not-determined | Genome-fragment | NA | | NA | 0 |
| S13_extracted_71960 | 1603 | No | NA | | 4 | 0 | 0 | Not-determined | Genome-fragment | NA | | NA | 0 |
| S13_extracted_71990 | 5240 | No | NA | | 6 | 0 | 4 | Not-determined | Genome-fragment | NA | | NA | 0 |
| S8_vs_k141_73332\|\|full | 1255 | No | NA | | 2 | 0 | 1 | Low-quality | Genome-fragment | | 3.13 | AAI-based (medium-confidence) | 0 |
| S13_extracted_72193 | 3930 | No | NA | | 7 | 0 | 7 | Not-determined | Genome-fragment | NA | | NA | 0 |
| S13_extracted_72233 | 1865 | No | NA | | 2 | 0 | 0 | Not-determined | Genome-fragment | NA | | NA | 0 |
| S13_extracted_72390 | 1347 | No | NA | | 4 | 0 | 4 | Not-determined | Genome-fragment | NA | | NA | 0 |
| S13_extracted_72772 | 1153 | No | NA | | 1 | 0 | 1 | Not-determined | Genome-fragment | NA | | NA | 0 |
| S13_extracted_72858 | 1372 | No | NA | | 2 | 0 | 2 | Not-determined | Genome-fragment | NA | | NA | 0 |
| S13_extracted_72894 | 1689 | No | NA | | 3 | 0 | 2 | Not-determined | Genome-fragment | NA | | NA | 0 |
| S13_extracted_73357 | 1647 | No | NA | | 1 | 0 | 0 | Not-determined | Genome-fragment | NA | | NA | 0 |
| S17_extracted_k141_98381 | 1256 | No | NA | | 2 | 0 | 0 | Low-quality | Genome-fragment | | 0.77 | AAI-based (medium-confidence) | 0 |
| S13_extracted_73777 | 1107 | No | NA | | 1 | 0 | 0 | Not-determined | Genome-fragment | NA | | NA | 0 |
| S13_extracted_73779 | 2479 | No | NA | | 3 | 0 | 1 | Not-determined | Genome-fragment | NA | | NA | 0 |
| S13_extracted_73788 | 1321 | No | NA | | 1 | 0 | 0 | Not-determined | Genome-fragment | NA | | NA | 0 |
| S13_extracted_73971 | 5982 | No | NA | | 2 | 0 | 2 | Not-determined | Genome-fragment | NA | | NA | 0 |
| S13_extracted_73973 | 1926 | No | NA | | 3 | 0 | 0 | Not-determined | Genome-fragment | NA | | NA | 0 |
| S14_extracted_k141_67265 | 1256 | No | NA | | 1 | 0 | 0 | Low-quality | Genome-fragment | | 1.53 | AAI-based (high-confidence) | 0 |
| S13_extracted_74648 | 1302 | No | NA | | 1 | 0 | 0 | Not-determined | Genome-fragment | NA | | NA | 0 |
| S13_extracted_74771 | 1002 | No | NA | | 1 | 0 | 0 | Not-determined | Genome-fragment | NA | | NA | 0 |
| S13_extracted_75153 | 1262 | No | NA | | 2 | 0 | 2 | Not-determined | Genome-fragment | NA | | NA | 0 |
| S13_extracted_75230 | 2133 | No | NA | | 4 | 0 | 2 | Not-determined | Genome-fragment | NA | | NA | 0 |
| S13_extracted_75400 | 1099 | No | NA | | 1 | 0 | 1 | Not-determined | Genome-fragment | NA | | NA | 0 |
| S13_extracted_75499 | 1021 | No | NA | | 2 | 0 | 0 | Not-determined | Genome-fragment | NA | | NA | 0 |
| S13_extracted_75593 | 3184 | No | NA | | 1 | 0 | 0 | Not-determined | Genome-fragment | NA | | NA | 0 |
| S13_extracted_75606 | 1172 | No | NA | | 1 | 0 | 1 | Not-determined | Genome-fragment | NA | | NA | 0 |
| S13_extracted_75610 | 1009 | No | NA | | 1 | 0 | 0 | Not-determined | Genome-fragment | NA | | NA | 0 |
| S13_extracted_75783 | 3144 | No | NA | | 3 | 0 | 1 | Not-determined | Genome-fragment | NA | | NA | 0 |
| S13_extracted_76319 | 1133 | No | NA | | 1 | 0 | 1 | Not-determined | Genome-fragment | NA | | NA | 0 |
| S13_extracted_76364 | 1536 | No | NA | | 3 | 0 | 0 | Not-determined | Genome-fragment | NA | | NA | 0 |
| S13_extracted_76418 | 1782 | No | NA | | 2 | 0 | 2 | Not-determined | Genome-fragment | NA | | NA | 0 |
| S13_extracted_76436 | 1002 | No | NA | | 1 | 0 | 1 | Not-determined | Genome-fragment | NA | | NA | 0 |
| S13_extracted_76684 | 2420 | No | NA | | 3 | 0 | 1 | Not-determined | Genome-fragment | NA | | NA | 0 |
| S13_extracted_77042 | 1567 | No | NA | | 1 | 0 | 0 | Not-determined | Genome-fragment | NA | | NA | 0 |
| S14_extracted_k141_57446 | 1256 | No | NA | | 4 | 1 | 0 | Low-quality | Genome-fragment | | 0.71 | HMM-based (lower-bound) | 0 |
| S14_vs_k141_15907\|\|full | 1256 | No | NA | | 3 | 1 | 0 | Low-quality | Genome-fragment | | 2.08 | AAI-based (medium-confidence) | 0 |
| S13_extracted_77819 | 4150 | No | NA | | 3 | 0 | 1 | Not-determined | Genome-fragment | NA | | NA | 0 |
| S13_extracted_77923 | 1396 | No | NA | | 1 | 0 | 0 | Not-determined | Genome-fragment | NA | | NA | 0 |
| S13_extracted_77947 | 1612 | No | NA | | 2 | 0 | 1 | Not-determined | Genome-fragment | NA | | NA | 0 |
| S13_extracted_78009 | 4094 | No | NA | | 5 | 0 | 3 | Not-determined | Genome-fragment | NA | | NA | 0 |
| S13_extracted_78277 | 1030 | No | NA | | 2 | 0 | 2 | Not-determined | Genome-fragment | NA | | NA | 0 |
| S14_vs_k141_70375\|\|full | 1256 | No | NA | | 3 | 1 | 0 | Low-quality | Genome-fragment | | 2.8 | AAI-based (high-confidence) | 0 |
| S13_extracted_78386 | 8729 | No | NA | | 9 | 0 | 7 | Not-determined | Genome-fragment | NA | | NA | 0 |
| S13_extracted_78436 | 1079 | No | NA | | 2 | 0 | 1 | Not-determined | Genome-fragment | NA | | NA | 0 |
| S13_extracted_78675 | 2265 | No | NA | | 3 | 0 | 2 | Not-determined | Genome-fragment | NA | | NA | 0 |
| S13_extracted_79092 | 1368 | No | NA | | 1 | 0 | 1 | Not-determined | Genome-fragment | NA | | NA | 0 |
| S13_extracted_79107 | 2094 | No | NA | | 3 | 0 | 3 | Not-determined | Genome-fragment | NA | | NA | 0 |
| S13_extracted_79215 | 1290 | No | NA | | 1 | 0 | 1 | Not-determined | Genome-fragment | NA | | NA | 0 |
| S1_extracted_k141_32196 | 1256 | No | NA | | 1 | 0 | 0 | Low-quality | Genome-fragment | | 0.93 | AAI-based (high-confidence) | 0 |
| S13_extracted_79450 | 14684 | No | NA | | 19 | 0 | 10 | Not-determined | Genome-fragment | NA | | NA | 0 |
| S13_extracted_79483 | 1133 | No | NA | | 2 | 0 | 0 | Not-determined | Genome-fragment | NA | | NA | 0 |
| S1_vs_k141_95489\|\|full | 1256 | No | NA | | 2 | 0 | 1 | Low-quality | Genome-fragment | | 0.39 | AAI-based (medium-confidence) | 0 |
| S13_extracted_79527 | 2353 | No | NA | | 1 | 0 | 0 | Not-determined | Genome-fragment | NA | | NA | 0 |
| S13_extracted_79555 | 1188 | No | NA | | 3 | 0 | 0 | Not-determined | Genome-fragment | NA | | NA | 0 |
| S13_extracted_79633 | 1108 | No | NA | | 2 | 0 | 0 | Not-determined | Genome-fragment | NA | | NA | 0 |
| S13_extracted_79689 | 3765 | No | NA | | 1 | 0 | 1 | Not-determined | Genome-fragment | NA | | NA | 0 |
| S13_extracted_79713 | 1118 | No | NA | | 3 | 0 | 1 | Not-determined | Genome-fragment | NA | | NA | 0 |
| S3_vs_k141_111667\|\|full | 1256 | No | NA | | 3 | 2 | 0 | Low-quality | Genome-fragment | | 2.87 | AAI-based (high-confidence) | 0 |
| S13_extracted_79898 | 1227 | No | NA | | 1 | 0 | 1 | Not-determined | Genome-fragment | NA | | NA | 0 |
| S13_extracted_80065 | 1370 | No | NA | | 2 | 0 | 1 | Not-determined | Genome-fragment | NA | | NA | 0 |
| S13_extracted_80238 | 3478 | No | NA | | 4 | 0 | 0 | Not-determined | Genome-fragment | NA | | NA | 0 |
| S5_vs_k141_75054\|\|full | 1256 | No | NA | | 4 | 0 | 0 | Low-quality | Genome-fragment | | 2.91 | AAI-based (high-confidence) | 0 |
| S13_extracted_80378 | 1127 | No | NA | | 1 | 0 | 0 | Not-determined | Genome-fragment | NA | | NA | 0 |
| S13_extracted_80626 | 2278 | No | NA | | 2 | 0 | 0 | Not-determined | Genome-fragment | NA | | NA | 0 |
| S13_extracted_80794 | 9918 | No | NA | | 15 | 0 | 3 | Not-determined | Genome-fragment | NA | | NA | 0 |
| S13_extracted_80933 | 1742 | No | NA | | 3 | 0 | 0 | Not-determined | Genome-fragment | NA | | NA | 0 |
| S13_extracted_81124 | 1155 | No | NA | | 1 | 0 | 0 | Not-determined | Genome-fragment | NA | | NA | 0 |
| S13_extracted_81283 | 1831 | No | NA | | 2 | 0 | 1 | Not-determined | Genome-fragment | NA | | NA | 0 |
| S13_extracted_81406 | 4038 | No | NA | | 1 | 0 | 1 | Not-determined | Genome-fragment | NA | | NA | 0 |
| S13_extracted_81471 | 1257 | No | NA | | 1 | 0 | 1 | Not-determined | Genome-fragment | NA | | NA | 0 |
| S13_extracted_81557 | 1176 | No | NA | | 1 | 0 | 1 | Not-determined | Genome-fragment | NA | | NA | 0 |
| S13_extracted_81658 | 1117 | No | NA | | 1 | 0 | 0 | Not-determined | Genome-fragment | NA | | NA | 0 |
| S7_vs_102374\|\|full | 1256 | No | NA | | 5 | 0 | 0 | Low-quality | Genome-fragment | | 2.12 | AAI-based (high-confidence) | 0 |
| S13_extracted_81703 | 1008 | No | NA | | 1 | 0 | 1 | Not-determined | Genome-fragment | NA | | NA | 0 |
| S13_extracted_81707 | 1327 | No | NA | | 3 | 0 | 1 | Not-determined | Genome-fragment | NA | | NA | 0 |
| S13_extracted_81765 | 2046 | No | NA | | 4 | 0 | 0 | Not-determined | Genome-fragment | NA | | NA | 0 |
| S13_extracted_81891 | 2797 | No | NA | | 2 | 0 | 1 | Not-determined | Genome-fragment | NA | | NA | 0 |
| S16_extracted_125527 | 1256 | No | NA | | 1 | 0 | 0 | Low-quality | Genome-fragment | | 1.07 | AAI-based (high-confidence) | 0 |
| S13_extracted_82089 | 1443 | No | NA | | 2 | 0 | 2 | Not-determined | Genome-fragment | NA | | NA | 0 |
| S13_extracted_82254 | 1147 | No | NA | | 1 | 0 | 0 | Not-determined | Genome-fragment | NA | | NA | 0 |
| S13_extracted_82326 | 1387 | No | NA | | 1 | 0 | 0 | Not-determined | Genome-fragment | NA | | NA | 0 |
| S13_extracted_82348 | 1417 | No | NA | | 2 | 0 | 1 | Not-determined | Genome-fragment | NA | | NA | 0 |
| S13_extracted_82355 | 1202 | No | NA | | 2 | 0 | 1 | Not-determined | Genome-fragment | NA | | NA | 0 |
| S13_extracted_82368 | 1117 | No | NA | | 2 | 0 | 1 | Not-determined | Genome-fragment | NA | | NA | 0 |
| S16_vs_10984\|\|full | 1256 | No | NA | | 3 | 0 | 1 | Low-quality | Genome-fragment | | 0.47 | AAI-based (medium-confidence) | 0 |
| S13_extracted_82762 | 1795 | No | NA | | 1 | 0 | 1 | Not-determined | Genome-fragment | NA | | NA | 0 |
| S13_extracted_83038 | 1140 | No | NA | | 2 | 0 | 0 | Not-determined | Genome-fragment | NA | | NA | 0 |
| S13_extracted_83303 | 1066 | No | NA | | 2 | 0 | 0 | Not-determined | Genome-fragment | NA | | NA | 0 |
| S13_extracted_83540 | 1248 | No | NA | | 2 | 0 | 1 | Not-determined | Genome-fragment | NA | | NA | 0 |
| S13_extracted_83747 | 1231 | No | NA | | 3 | 0 | 0 | Not-determined | Genome-fragment | NA | | NA | 0 |
| S13_extracted_83803 | 1064 | No | NA | | 2 | 0 | 0 | Not-determined | Genome-fragment | NA | | NA | 0 |
| S13_extracted_83808 | 1687 | No | NA | | 3 | 0 | 0 | Not-determined | Genome-fragment | NA | | NA | 0 |
| S13_extracted_83847 | 1535 | No | NA | | 1 | 0 | 0 | Not-determined | Genome-fragment | NA | | NA | 0 |
| S13_extracted_83872 | 1164 | No | NA | | 2 | 0 | 1 | Not-determined | Genome-fragment | NA | | NA | 0 |
| S13_extracted_83932 | 1131 | No | NA | | 3 | 0 | 0 | Not-determined | Genome-fragment | NA | | NA | 0 |
| S13_extracted_83959 | 1244 | No | NA | | 1 | 0 | 1 | Not-determined | Genome-fragment | NA | | NA | 0 |
| S16_vs_55970\|\|full | 1256 | No | NA | | 4 | 0 | 0 | Low-quality | Genome-fragment | | 0.9 | AAI-based (medium-confidence) | 0 |
| S13_extracted_84102 | 1378 | No | NA | | 3 | 0 | 0 | Not-determined | Genome-fragment | NA | | NA | 0 |
| S16_vs_156640\|\|lt2gene | 1256 | No | NA | | 1 | 1 | 0 | Low-quality | Genome-fragment | | 3.65 | AAI-based (high-confidence) | 0 |
| S13_extracted_84223 | 6089 | No | NA | | 4 | 0 | 2 | Not-determined | Genome-fragment | NA | | NA | 0 |
| S13_extracted_84372 | 4350 | No | NA | | 1 | 0 | 1 | Not-determined | Genome-fragment | NA | | NA | 0 |
| S13_extracted_84409 | 1085 | No | NA | | 2 | 0 | 1 | Not-determined | Genome-fragment | NA | | NA | 0 |
| S13_extracted_84419 | 1020 | No | NA | | 1 | 0 | 1 | Not-determined | Genome-fragment | NA | | NA | 0 |
| S12_extracted_k141_44254 | 1256 | No | NA | | 1 | 0 | 0 | Low-quality | Genome-fragment | | 1.03 | AAI-based (high-confidence) | 0 |
| S13_extracted_84544 | 1032 | No | NA | | 2 | 0 | 0 | Not-determined | Genome-fragment | NA | | NA | 0 |
| S13_extracted_84688 | 1075 | No | NA | | 1 | 0 | 0 | Not-determined | Genome-fragment | NA | | NA | 0 |
| S13_extracted_84721 | 1650 | No | NA | | 2 | 0 | 2 | Not-determined | Genome-fragment | NA | | NA | 0 |
| S13_extracted_84829 | 1003 | No | NA | | 1 | 0 | 0 | Not-determined | Genome-fragment | NA | | NA | 0 |
| S12_vs_k141_38565\|\|full | 1256 | No | NA | | 4 | 0 | 0 | Low-quality | Genome-fragment | | 2.24 | AAI-based (high-confidence) | 0 |
| S13_extracted_84866 | 1501 | No | NA | | 2 | 0 | 0 | Not-determined | Genome-fragment | NA | | NA | 0 |
| S8_extracted_k141_98228 | 1256 | No | NA | | 2 | 0 | 0 | Low-quality | Genome-fragment | | 0.98 | AAI-based (high-confidence) | 0 |
| S13_extracted_85171 | 1032 | No | NA | | 2 | 0 | 0 | Not-determined | Genome-fragment | NA | | NA | 0 |
| S13_extracted_85281 | 1713 | No | NA | | 1 | 0 | 1 | Not-determined | Genome-fragment | NA | | NA | 0 |
| S13_extracted_85475 | 1448 | No | NA | | 2 | 0 | 1 | Not-determined | Genome-fragment | NA | | NA | 0 |
| S13_extracted_85525 | 2055 | No | NA | | 4 | 0 | 3 | Not-determined | Genome-fragment | NA | | NA | 0 |
| S13_extracted_85624 | 1352 | No | NA | | 1 | 0 | 0 | Not-determined | Genome-fragment | NA | | NA | 0 |
| S13_extracted_85626 | 1082 | No | NA | | 4 | 0 | 0 | Not-determined | Genome-fragment | NA | | NA | 0 |
| S13_extracted_85946 | 1924 | No | NA | | 1 | 0 | 0 | Not-determined | Genome-fragment | NA | | NA | 0 |
| S13_extracted_86064 | 1012 | No | NA | | 2 | 0 | 0 | Not-determined | Genome-fragment | NA | | NA | 0 |
| S13_extracted_86088 | 1092 | No | NA | | 1 | 0 | 1 | Not-determined | Genome-fragment | NA | | NA | 0 |
| S13_extracted_86178 | 3244 | No | NA | | 1 | 0 | 0 | Not-determined | Genome-fragment | NA | | NA | 0 |
| S13_extracted_86475 | 1452 | No | NA | | 2 | 0 | 1 | Not-determined | Genome-fragment | NA | | NA | 0 |
| S8_extracted_k141_31011 | 1256 | No | NA | | 3 | 2 | 0 | Low-quality | Genome-fragment | | 2.16 | AAI-based (high-confidence) | 0 |
| S13_extracted_86916 | 1102 | No | NA | | 2 | 0 | 1 | Not-determined | Genome-fragment | NA | | NA | 0 |

| | | | | | | | | | | | |
|---|---|---|---|---|---|---|---|---|---|---|---|
| S13_extracted_86955 | 1408 | No | NA | 1 | 0 | 0 | Not-determined | Genome-fragment | NA | NA | 0 |
| S13_extracted_87022 | 1131 | No | NA | 2 | 0 | 2 | Not-determined | Genome-fragment | NA | NA | 0 |
| S13_extracted_87437 | 1079 | No | NA | 1 | 0 | 1 | Not-determined | Genome-fragment | NA | NA | 0 |
| S8_vs_k141_47687||full | 1256 | No | NA | 2 | 0 | 1 | Low-quality | Genome-fragment | 1.71 | AAI-based (high-confidence) | 0 |
| S8_vs_k141_7155||lt2gene | 1256 | No | NA | 2 | 1 | 0 | Low-quality | Genome-fragment | 2.75 | AAI-based (high-confidence) | 0 |
| S13_extracted_87601 | 1735 | No | NA | 2 | 0 | 1 | Not-determined | Genome-fragment | NA | NA | 0 |
| S13_extracted_87873 | 38348 | No | NA | 47 | 10 | 1 | Medium-quality | Genome-fragment | 84.63 | AAI-based (high-confidence) | 0 |
| S13_extracted_87952 | 1120 | No | NA | 3 | 0 | 1 | Not-determined | Genome-fragment | NA | NA | 0 |
| S9_vs_k141_4192||full | 1257 | No | NA | 3 | 2 | 0 | Low-quality | Genome-fragment | 2.97 | AAI-based (high-confidence) | 0 |
| S13_extracted_88021 | 1249 | No | NA | 1 | 0 | 1 | Not-determined | Genome-fragment | NA | NA | 0 |
| S14_vs_k141_73757||full | 1257 | No | NA | 4 | 1 | 0 | Low-quality | Genome-fragment | 2.17 | AAI-based (high-confidence) | 0 |
| S13_extracted_88235 | 1015 | No | NA | 1 | 0 | 0 | Not-determined | Genome-fragment | NA | NA | 0 |
| S14_vs_k141_95921||full | 1257 | No | NA | 2 | 0 | 0 | Low-quality | Genome-fragment | 1.18 | AAI-based (medium-confidence) | 0 |
| S13_extracted_88479 | 1582 | No | NA | 1 | 0 | 1 | Not-determined | Genome-fragment | NA | NA | 0 |
| S13_extracted_88871 | 2213 | No | NA | 1 | 0 | 0 | Not-determined | Genome-fragment | NA | NA | 0 |
| S1_vs_k141_98270||full | 1257 | No | NA | 3 | 1 | 0 | Low-quality | Genome-fragment | 2.91 | AAI-based (high-confidence) | 0 |
| S13_extracted_89182 | 1633 | No | NA | 2 | 0 | 0 | Not-determined | Genome-fragment | NA | NA | 0 |
| S13_extracted_89247 | 3819 | No | NA | 3 | 0 | 1 | Not-determined | Genome-fragment | NA | NA | 0 |
| S13_extracted_89262 | 4236 | No | NA | 2 | 0 | 1 | Not-determined | Genome-fragment | NA | NA | 0 |
| S13_extracted_89409 | 2277 | No | NA | 5 | 0 | 1 | Not-determined | Genome-fragment | NA | NA | 0 |
| S13_extracted_89412 | 1133 | No | NA | 2 | 0 | 1 | Not-determined | Genome-fragment | NA | NA | 0 |
| S13_extracted_89432 | 3569 | No | NA | 4 | 0 | 3 | Not-determined | Genome-fragment | NA | NA | 0 |
| S13_extracted_89476 | 1034 | No | NA | 3 | 0 | 0 | Not-determined | Genome-fragment | NA | NA | 0 |
| S5_vs_k141_71156||full | 1257 | No | NA | 4 | 0 | 0 | Low-quality | Genome-fragment | 2.64 | AAI-based (medium-confidence) | 0 |
| S5_vs_k141_102939||full | 1257 | No | NA | 3 | 0 | 0 | Low-quality | Genome-fragment | 1.53 | AAI-based (high-confidence) | 0 |
| S13_extracted_90239 | 1502 | No | NA | 3 | 0 | 2 | Not-determined | Genome-fragment | NA | NA | 0 |
| S5_vs_k141_50056||lt2gene | 1257 | No | NA | 1 | 1 | 0 | Low-quality | Genome-fragment | 2.83 | AAI-based (high-confidence) | 0 |
| S13_extracted_90364 | 2349 | No | NA | 3 | 0 | 2 | Not-determined | Genome-fragment | NA | NA | 0 |
| S13_extracted_90794 | 1062 | No | NA | 2 | 0 | 1 | Not-determined | Genome-fragment | NA | NA | 0 |
| S13_extracted_90904 | 4356 | No | NA | 5 | 0 | 1 | Not-determined | Genome-fragment | NA | NA | 0 |
| S13_extracted_91133 | 1037 | No | NA | 2 | 0 | 2 | Not-determined | Genome-fragment | NA | NA | 0 |
| S13_extracted_91189 | 13057 | No | NA | 18 | 4 | 0 | High-quality | High-quality | 95.48 | AAI-based (medium-confidence) | 0 |
| S13_extracted_91229 | 1327 | No | NA | 3 | 0 | 1 | Not-determined | Genome-fragment | NA | NA | 0 |
| S13_extracted_91252 | 1014 | No | NA | 2 | 0 | 1 | Not-determined | Genome-fragment | NA | NA | 0 |
| S13_extracted_91576 | 1175 | No | NA | 1 | 0 | 1 | Not-determined | Genome-fragment | NA | NA | 0 |
| S13_extracted_91625 | 1351 | No | NA | 2 | 0 | 1 | Not-determined | Genome-fragment | NA | NA | 0 |
| S13_extracted_91653 | 3776 | No | NA | 5 | 0 | 2 | Not-determined | Genome-fragment | NA | NA | 0 |
| S13_extracted_91710 | 1089 | No | NA | 2 | 0 | 1 | Not-determined | Genome-fragment | NA | NA | 0 |
| S13_extracted_91789 | 1044 | No | NA | 1 | 0 | 1 | Not-determined | Genome-fragment | NA | NA | 0 |
| S7_extracted_40879 | 1257 | No | NA | 2 | 1 | 0 | Low-quality | Genome-fragment | 0.87 | HMM-based (lower-bound) | 0 |
| S7_vs_10926||full | 1257 | No | NA | 3 | 0 | 0 | Low-quality | Genome-fragment | 3.25 | AAI-based (high-confidence) | 0 |
| S13_extracted_91939 | 3015 | No | NA | 2 | 0 | 2 | Not-determined | Genome-fragment | NA | NA | 0 |
| S10_extracted_20110 | 1257 | No | NA | 2 | 0 | 0 | Low-quality | Genome-fragment | 2.58 | AAI-based (high-confidence) | 0 |
| S13_extracted_91986 | 1381 | No | NA | 1 | 0 | 1 | Not-determined | Genome-fragment | NA | NA | 0 |
| S13_extracted_92008 | 1292 | No | NA | 1 | 0 | 0 | Not-determined | Genome-fragment | NA | NA | 0 |
| S13_extracted_92143 | 8774 | No | NA | 8 | 0 | 2 | Not-determined | Genome-fragment | NA | NA | 0 |
| S13_extracted_92159 | 1194 | No | NA | 1 | 0 | 0 | Not-determined | Genome-fragment | NA | NA | 0 |
| S10_extracted_54661 | 1257 | No | NA | 2 | 0 | 1 | Low-quality | Genome-fragment | 0.44 | AAI-based (medium-confidence) | 0 |
| S13_extracted_92401 | 1993 | No | NA | 2 | 0 | 1 | Not-determined | Genome-fragment | NA | NA | 0 |
| S13_extracted_92448 | 1371 | No | NA | 2 | 0 | 2 | Not-determined | Genome-fragment | NA | NA | 0 |
| S13_extracted_92452 | 1415 | No | NA | 2 | 0 | 1 | Not-determined | Genome-fragment | NA | NA | 0 |
| S13_extracted_92525 | 5294 | No | NA | 9 | 0 | 0 | Not-determined | Genome-fragment | NA | NA | 0 |
| S13_extracted_92555 | 1098 | No | NA | 1 | 0 | 1 | Not-determined | Genome-fragment | NA | NA | 0 |
| S12_extracted_k141_168808 | 1257 | No | NA | 2 | 1 | 0 | Low-quality | Genome-fragment | 3.23 | AAI-based (high-confidence) | 0 |
| S13_extracted_92813 | 1223 | No | NA | 1 | 0 | 0 | Not-determined | Genome-fragment | NA | NA | 0 |
| S13_extracted_92924 | 1347 | No | NA | 1 | 0 | 1 | Not-determined | Genome-fragment | NA | NA | 0 |
| S13_extracted_93038 | 1246 | No | NA | 3 | 0 | 0 | Not-determined | Genome-fragment | NA | NA | 0 |
| S13_extracted_93060 | 1735 | No | NA | 6 | 0 | 6 | Not-determined | Genome-fragment | NA | NA | 0 |
| S13_extracted_93220 | 1029 | No | NA | 3 | 0 | 0 | Not-determined | Genome-fragment | NA | NA | 0 |
| S12_vs_k141_166606||full | 1257 | No | NA | 4 | 1 | 0 | Low-quality | Genome-fragment | 2.43 | AAI-based (high-confidence) | 0 |
| S13_extracted_93329 | 1082 | No | NA | 1 | 0 | 0 | Not-determined | Genome-fragment | NA | NA | 0 |
| S12_vs_k141_82904||lt2gene | 1257 | No | NA | 1 | 1 | 0 | Low-quality | Genome-fragment | 2.09 | HMM-based (lower-bound) | 0 |
| S15_vs_57503||full | 1257 | No | NA | 3 | 3 | 0 | Low-quality | Genome-fragment | 3.15 | AAI-based (high-confidence) | 0 |
| S13_extracted_93976 | 1453 | No | NA | 2 | 0 | 1 | Not-determined | Genome-fragment | NA | NA | 0 |
| S13_extracted_94041 | 2304 | No | NA | 5 | 0 | 0 | Not-determined | Genome-fragment | NA | NA | 0 |
| S13_extracted_94050 | 1054 | No | NA | 1 | 0 | 1 | Not-determined | Genome-fragment | NA | NA | 0 |
| S6_vs_k141_35194||full | 1257 | No | NA | 4 | 0 | 0 | Low-quality | Genome-fragment | 1.76 | AAI-based (medium-confidence) | 0 |
| S13_extracted_94221 | 1640 | No | NA | 1 | 0 | 1 | Not-determined | Genome-fragment | NA | NA | 0 |
| S13_extracted_94349 | 1168 | No | NA | 1 | 0 | 0 | Not-determined | Genome-fragment | NA | NA | 0 |
| S13_extracted_94403 | 2386 | No | NA | 2 | 0 | 1 | Not-determined | Genome-fragment | NA | NA | 0 |
| S8_vs_k141_177881||full | 1257 | No | NA | 2 | 0 | 1 | Low-quality | Genome-fragment | 0.92 | AAI-based (medium-confidence) | 0 |
| S13_extracted_94529 | 1028 | No | NA | 2 | 0 | 1 | Not-determined | Genome-fragment | NA | NA | 0 |
| S13_extracted_94588 | 1123 | No | NA | 2 | 0 | 0 | Not-determined | Genome-fragment | NA | NA | 0 |
| S13_extracted_94618 | 5304 | No | NA | 3 | 0 | 3 | Not-determined | Genome-fragment | NA | NA | 0 |
| S13_extracted_94854 | 1439 | No | NA | 3 | 0 | 3 | Not-determined | Genome-fragment | NA | NA | 0 |
| S13_extracted_94945 | 7259 | No | NA | 8 | 0 | 1 | Not-determined | Genome-fragment | NA | NA | 0 |
| S8_vs_k141_68909||full | 1257 | No | NA | 2 | 0 | 0 | Low-quality | Genome-fragment | 1.85 | AAI-based (medium-confidence) | 0 |
| S13_extracted_95036 | 1011 | No | NA | 3 | 0 | 2 | Not-determined | Genome-fragment | NA | NA | 0 |
| S13_extracted_95107 | 1056 | No | NA | 2 | 0 | 0 | Not-determined | Genome-fragment | NA | NA | 0 |
| S14_extracted_k141_9601 | 1258 | No | NA | 2 | 0 | 0 | Low-quality | Genome-fragment | 3.67 | AAI-based (high-confidence) | 0 |
| S14_vs_k141_102235||full | 1258 | No | NA | 3 | 0 | 0 | Low-quality | Genome-fragment | 3.2 | AAI-based (medium-confidence) | 0 |
| S13_extracted_95548 | 1401 | No | NA | 3 | 0 | 2 | Not-determined | Genome-fragment | NA | NA | 0 |
| S1_vs_k141_150050||full | 1258 | No | NA | 3 | 0 | 0 | Low-quality | Genome-fragment | 2.81 | AAI-based (medium-confidence) | 0 |
| S13_extracted_95868 | 1108 | No | NA | 2 | 0 | 2 | Not-determined | Genome-fragment | NA | NA | 0 |
| S13_extracted_96205 | 1640 | No | NA | 2 | 0 | 2 | Not-determined | Genome-fragment | NA | NA | 0 |
| S3_extracted_k141_7336 | 1258 | No | NA | 2 | 0 | 0 | Low-quality | Genome-fragment | 2.05 | AAI-based (high-confidence) | 0 |
| S13_extracted_96326 | 6011 | No | NA | 3 | 0 | 2 | Not-determined | Genome-fragment | NA | NA | 0 |
| S13_extracted_96646 | 1012 | No | NA | 1 | 0 | 0 | Not-determined | Genome-fragment | NA | NA | 0 |
| S3_vs_k141_96451||full | 1258 | No | NA | 3 | 0 | 0 | Low-quality | Genome-fragment | 3.37 | AAI-based (high-confidence) | 0 |
| S13_extracted_96889 | 1009 | No | NA | 1 | 0 | 0 | Not-determined | Genome-fragment | NA | NA | 0 |
| S13_extracted_97175 | 4831 | No | NA | 4 | 0 | 3 | Not-determined | Genome-fragment | NA | NA | 0 |
| S13_extracted_97386 | 1080 | No | NA | 2 | 0 | 0 | Not-determined | Genome-fragment | NA | NA | 0 |
| S13_extracted_97520 | 1095 | No | NA | 2 | 0 | 1 | Not-determined | Genome-fragment | NA | NA | 0 |
| S5_extracted_k141_110972 | 1258 | No | NA | 3 | 0 | 0 | Low-quality | Genome-fragment | 1.98 | AAI-based (medium-confidence) | 0 |
| S13_extracted_97612 | 1324 | No | NA | 3 | 0 | 0 | Not-determined | Genome-fragment | NA | NA | 0 |
| S5_extracted_k141_136558 | 1258 | No | NA | 3 | 0 | 0 | Low-quality | Genome-fragment | 0.62 | AAI-based (high-confidence) | 0 |
| S13_extracted_97678 | 1017 | No | NA | 2 | 0 | 0 | Not-determined | Genome-fragment | NA | NA | 0 |
| S13_extracted_97702 | 16409 | No | NA | 19 | 7 | 0 | Complete | High-quality | 100 | ITR (high-confidence) | 0 |
| S13_extracted_97950 | 2297 | No | NA | 3 | 0 | 2 | Not-determined | Genome-fragment | NA | NA | 0 |
| S13_extracted_98168 | 1023 | No | NA | 1 | 0 | 1 | Not-determined | Genome-fragment | NA | NA | 0 |
| S13_extracted_98187 | 4579 | No | NA | 4 | 0 | 4 | Not-determined | Genome-fragment | NA | NA | 0 |
| S13_extracted_98335 | 1343 | No | NA | 2 | 0 | 1 | Not-determined | Genome-fragment | NA | NA | 0 |
| S5_vs_k141_118645||full | 1258 | No | NA | 3 | 0 | 0 | Low-quality | Genome-fragment | 2.36 | AAI-based (high-confidence) | 0 |
| S13_extracted_98478 | 1079 | No | NA | 2 | 0 | 1 | Not-determined | Genome-fragment | NA | NA | 0 |
| S13_extracted_98499 | 1392 | No | NA | 1 | 0 | 1 | Not-determined | Genome-fragment | NA | NA | 0 |
| S13_extracted_99191 | 1186 | No | NA | 1 | 0 | 1 | Not-determined | Genome-fragment | NA | NA | 0 |
| S13_extracted_99397 | 4582 | No | NA | 5 | 0 | 3 | Not-determined | Genome-fragment | NA | NA | 0 |
| S13_extracted_99469 | 3884 | No | NA | 5 | 0 | 2 | Not-determined | Genome-fragment | NA | NA | 0 |
| S13_extracted_99590 | 4465 | No | NA | 1 | 0 | 1 | Not-determined | Genome-fragment | NA | NA | 0 |
| S13_extracted_99745 | 1562 | No | NA | 4 | 0 | 1 | Not-determined | Genome-fragment | NA | NA | 0 |
| S13_extracted_99858 | 3420 | No | NA | 2 | 0 | 2 | Not-determined | Genome-fragment | NA | NA | 0 |
| S13_extracted_99965 | 1594 | No | NA | 2 | 0 | 2 | Not-determined | Genome-fragment | NA | NA | 0 |
| S13_extracted_100155 | 1743 | No | NA | 3 | 0 | 3 | Not-determined | Genome-fragment | NA | NA | 0 |
| S13_extracted_100161 | 1126 | No | NA | 1 | 0 | 0 | Not-determined | Genome-fragment | NA | NA | 0 |
| S13_extracted_100335 | 2389 | No | NA | 3 | 0 | 1 | Not-determined | Genome-fragment | NA | NA | 0 |
| S13_extracted_100565 | 1669 | No | NA | 2 | 0 | 1 | Not-determined | Genome-fragment | NA | NA | 0 |
| S13_extracted_100584 | 2490 | No | NA | 4 | 0 | 1 | Not-determined | Genome-fragment | NA | NA | 0 |
| S7_extracted_53064 | 1258 | No | NA | 3 | 1 | 0 | Low-quality | Genome-fragment | 4.15 | AAI-based (high-confidence) | 0 |
| S13_extracted_101065 | 4529 | No | NA | 2 | 0 | 2 | Not-determined | Genome-fragment | NA | NA | 0 |
| S13_extracted_101153 | 3006 | No | NA | 3 | 0 | 2 | Not-determined | Genome-fragment | NA | NA | 0 |
| S16_vs_50425||full | 1258 | No | NA | 3 | 1 | 0 | Low-quality | Genome-fragment | 2.15 | AAI-based (medium-confidence) | 0 |
| S16_vs_126823||full | 1258 | No | NA | 3 | 0 | 1 | Low-quality | Genome-fragment | 0.76 | AAI-based (high-confidence) | 0 |
| S13_extracted_101568 | 1221 | No | NA | 1 | 0 | 1 | Not-determined | Genome-fragment | NA | NA | 0 |
| S13_extracted_101593 | 1077 | No | NA | 1 | 0 | 1 | Not-determined | Genome-fragment | NA | NA | 0 |
| S13_extracted_101654 | 1162 | No | NA | 3 | 0 | 0 | Not-determined | Genome-fragment | NA | NA | 0 |
| S13_extracted_101739 | 2211 | No | NA | 4 | 0 | 2 | Not-determined | Genome-fragment | NA | NA | 0 |
| S13_extracted_101961 | 5952 | No | NA | 9 | 0 | 5 | Not-determined | Genome-fragment | NA | NA | 0 |
| S13_extracted_102007 | 1034 | No | NA | 1 | 0 | 0 | Not-determined | Genome-fragment | NA | NA | 0 |
| S13_extracted_102027 | 3246 | No | NA | 1 | 0 | 1 | Not-determined | Genome-fragment | NA | NA | 0 |
| S13_extracted_102212 | 1072 | No | NA | 1 | 0 | 0 | Not-determined | Genome-fragment | NA | NA | 0 |
| S13_extracted_102321 | 1136 | No | NA | 2 | 0 | 2 | Not-determined | Genome-fragment | NA | NA | 0 |
| S13_extracted_102363 | 1159 | No | NA | 3 | 0 | 0 | Not-determined | Genome-fragment | NA | NA | 0 |
| S13_extracted_102587 | 1393 | No | NA | 2 | 0 | 0 | Not-determined | Genome-fragment | NA | NA | 0 |
| S16_vs_164949||lt2gene | 1258 | No | NA | 2 | 1 | 0 | Low-quality | Genome-fragment | 2.55 | AAI-based (high-confidence) | 0 |
| S13_extracted_102752 | 1163 | No | NA | 1 | 0 | 1 | Not-determined | Genome-fragment | NA | NA | 0 |
| S10_vs_92896||full | 1258 | No | NA | 2 | 0 | 2 | Low-quality | Genome-fragment | 0.67 | AAI-based (medium-confidence) | 0 |
| S13_extracted_103174 | 1007 | No | NA | 1 | 0 | 0 | Not-determined | Genome-fragment | NA | NA | 0 |
| S13_extracted_103185 | 3318 | No | NA | 5 | 0 | 0 | Not-determined | Genome-fragment | NA | NA | 0 |
| S13_extracted_103332 | 1116 | No | NA | 2 | 0 | 1 | Not-determined | Genome-fragment | NA | NA | 0 |
| S13_extracted_103635 | 5862 | No | NA | 8 | 0 | 0 | Not-determined | Genome-fragment | NA | NA | 0 |
| S13_extracted_103867 | 1288 | No | NA | 1 | 0 | 0 | Not-determined | Genome-fragment | NA | NA | 0 |
| S13_extracted_104032 | 1196 | No | NA | 1 | 0 | 1 | Not-determined | Genome-fragment | NA | NA | 0 |
| S12_extracted_k141_17430 | 1258 | No | NA | 1 | 0 | 1 | Low-quality | Genome-fragment | 0.3 | AAI-based (medium-confidence) | 0 |
| S15_extracted_70581 | 1258 | No | NA | 2 | 0 | 2 | Low-quality | Genome-fragment | 3.3 | AAI-based (medium-confidence) | 0 |
| S13_extracted_104352 | 1775 | No | NA | 4 | 0 | 1 | Not-determined | Genome-fragment | NA | NA | 0 |
| S13_extracted_104525 | 1177 | No | NA | 2 | 0 | 0 | Not-determined | Genome-fragment | NA | NA | 0 |
| S13_extracted_104575 | 1228 | No | NA | 1 | 0 | 1 | Not-determined | Genome-fragment | NA | NA | 0 |
| S9_vs_k141_42419||full | 1259 | No | NA | 1 | 0 | 0 | Low-quality | Genome-fragment | 2.45 | AAI-based (medium-confidence) | 0 |
| S13_extracted_104907 | 1160 | No | NA | 1 | 0 | 1 | Not-determined | Genome-fragment | NA | NA | 0 |
| S13_extracted_105049 | 1054 | No | NA | 1 | 0 | 1 | Not-determined | Genome-fragment | NA | NA | 0 |
| S13_extracted_105223 | 1037 | No | NA | 4 | 0 | 1 | Not-determined | Genome-fragment | NA | NA | 0 |
| S11_extracted_k141_41554 | 1259 | No | NA | 2 | 0 | 0 | Low-quality | Genome-fragment | 3.09 | AAI-based (high-confidence) | 0 |
| S13_extracted_105328 | 1759 | No | NA | 1 | 0 | 1 | Not-determined | Genome-fragment | NA | NA | 0 |
| S13_extracted_105449 | 7609 | No | NA | 5 | 0 | 4 | Not-determined | Genome-fragment | NA | NA | 0 |
| S14_vs_k141_46413||full | 1259 | No | NA | 3 | 2 | 0 | Low-quality | Genome-fragment | 2.16 | HMM-based (lower-bound) | 0 |
| S13_extracted_105480 | 1180 | No | NA | 1 | 0 | 1 | Not-determined | Genome-fragment | NA | NA | 0 |
| S13_extracted_105611 | 1205 | No | NA | 2 | 0 | 0 | Not-determined | Genome-fragment | NA | NA | 0 |
| S13_extracted_105726 | 1318 | No | NA | 1 | 0 | 1 | Not-determined | Genome-fragment | NA | NA | 0 |
| S13_extracted_105790 | 1056 | No | NA | 2 | 0 | 0 | Not-determined | Genome-fragment | NA | NA | 0 |
| S13_extracted_105992 | 1353 | No | NA | 2 | 0 | 0 | Not-determined | Genome-fragment | NA | NA | 0 |
| S13_extracted_106111 | 1466 | No | NA | 1 | 0 | 0 | Not-determined | Genome-fragment | NA | NA | 0 |
| S14_vs_k141_63340||full | 1259 | No | NA | 1 | 0 | 0 | Low-quality | Genome-fragment | 3.4 | AAI-based (high-confidence) | 0 |
| S13_extracted_106340 | 1593 | No | NA | 1 | 0 | 1 | Not-determined | Genome-fragment | NA | NA | 0 |
| S13_extracted_106464 | 1383 | No | NA | 1 | 0 | 1 | Not-determined | Genome-fragment | NA | NA | 0 |
| S13_extracted_106617 | 1019 | No | NA | 2 | 0 | 1 | Not-determined | Genome-fragment | NA | NA | 0 |
| S13_extracted_106814 | 1437 | No | NA | 1 | 0 | 0 | Not-determined | Genome-fragment | NA | NA | 0 |
| S13_extracted_106856 | 1496 | No | NA | 1 | 0 | 0 | Not-determined | Genome-fragment | NA | NA | 0 |
| S13_extracted_107014 | 1501 | No | NA | 2 | 0 | 1 | Not-determined | Genome-fragment | NA | NA | 0 |
| S13_extracted_107182 | 1904 | No | NA | 3 | 0 | 1 | Not-determined | Genome-fragment | NA | NA | 0 |
| S13_extracted_107471 | 1083 | No | NA | 1 | 0 | 0 | Not-determined | Genome-fragment | NA | NA | 0 |
| S13_extracted_107618 | 1383 | No | NA | 2 | 0 | 2 | Not-determined | Genome-fragment | NA | NA | 0 |
| S1_extracted_k141_123403 | 1259 | No | NA | 3 | 0 | 0 | Low-quality | Genome-fragment | 1.88 | AAI-based (medium-confidence) | 0 |
| S1_vs_k141_5403||full | 1259 | No | NA | 2 | 0 | 0 | Low-quality | Genome-fragment | 1.96 | AAI-based (medium-confidence) | 0 |
| S13_extracted_108098 | 1072 | No | NA | 1 | 0 | 1 | Not-determined | Genome-fragment | NA | NA | 0 |
| S13_extracted_108177 | 1167 | No | NA | 2 | 0 | 0 | Not-determined | Genome-fragment | NA | NA | 0 |
| S13_extracted_108231 | 1364 | No | NA | 1 | 0 | 1 | Not-determined | Genome-fragment | NA | NA | 0 |
| S13_extracted_108272 | 1012 | No | NA | 2 | 0 | 0 | Not-determined | Genome-fragment | NA | NA | 0 |
| S13_extracted_108288 | 1746 | No | NA | 3 | 0 | 1 | Not-determined | Genome-fragment | NA | NA | 0 |
| S7_vs_99556||full | 1259 | No | NA | 4 | 2 | 0 | Low-quality | Genome-fragment | 3.13 | AAI-based (high-confidence) | 0 |
| S16_vs_51659||full | 1259 | No | NA | 3 | 1 | 0 | Low-quality | Genome-fragment | 2.32 | AAI-based (medium-confidence) | 0 |
| S13_extracted_108473 | 3128 | No | NA | 2 | 0 | 0 | Not-determined | Genome-fragment | NA | NA | 0 |
| S13_extracted_108559 | 1009 | No | NA | 1 | 0 | 0 | Not-determined | Genome-fragment | NA | NA | 0 |
| S10_vs_68985||full | 1259 | No | NA | 3 | 0 | 0 | Low-quality | Genome-fragment | 2.36 | AAI-based (medium-confidence) | 0 |
| S13_extracted_108714 | 1650 | No | NA | 3 | 0 | 3 | Not-determined | Genome-fragment | NA | NA | 0 |
| S12_extracted_k141_136256 | 1259 | No | NA | 2 | 0 | 0 | Low-quality | Genome-fragment | 2.18 | AAI-based (medium-confidence) | 0 |
| S12_vs_k141_167761||lt2gene | 1259 | No | NA | 2 | 1 | 0 | Low-quality | Genome-fragment | 2.06 | AAI-based (medium-confidence) | 0 |
| S13_extracted_108857 | 1105 | No | NA | 1 | 0 | 1 | Not-determined | Genome-fragment | NA | NA | 0 |
| S13_extracted_108877 | 1477 | No | NA | 2 | 0 | 1 | Not-determined | Genome-fragment | NA | NA | 0 |
| S13_extracted_108898 | 1154 | No | NA | 2 | 0 | 1 | Not-determined | Genome-fragment | NA | NA | 0 |
| S13_extracted_108970 | 1426 | No | NA | 2 | 0 | 1 | Not-determined | Genome-fragment | NA | NA | 0 |
| S13_extracted_109133 | 1458 | No | NA | 2 | 0 | 2 | Not-determined | Genome-fragment | NA | NA | 0 |
| S13_extracted_109235 | 45453 | No | NA | 66 | 14 | 0 | Complete | High-quality | 100 | DTR (high-confidence) | 0 |
| S8_extracted_k141_131161 | 1259 | No | NA | 3 | 0 | 1 | Low-quality | Genome-fragment | 0.77 | AAI-based (medium-confidence) | 0 |
| S8_extracted_k141_207108 | 1259 | No | NA | 2 | 2 | 0 | Low-quality | Genome-fragment | 4.15 | AAI-based (high-confidence) | 0 |
| S13_extracted_109446 | 1296 | No | NA | 2 | 0 | 1 | Not-determined | Genome-fragment | NA | NA | 0 |
| S13_extracted_109458 | 1335 | No | NA | 2 | 0 | 0 | Not-determined | Genome-fragment | NA | NA | 0 |
| S13_extracted_109577 | 1369 | No | NA | 2 | 0 | 0 | Not-determined | Genome-fragment | NA | NA | 0 |
| S11_vs_k141_93602||full | 1260 | No | NA | 3 | 0 | 0 | Low-quality | Genome-fragment | 2 | AAI-based (high-confidence) | 0 |
| S13_extracted_109825 | 1069 | No | NA | 1 | 0 | 0 | Not-determined | Genome-fragment | NA | NA | 0 |
| S13_extracted_109869 | 1065 | No | NA | 2 | 0 | 1 | Not-determined | Genome-fragment | NA | NA | 0 |
| S11_vs_k141_83183||lt2gene | 1260 | No | NA | 2 | 1 | 0 | Low-quality | Genome-fragment | 1.67 | HMM-based (lower-bound) | 0 |
| S13_extracted_110219 | 1202 | No | NA | 2 | 0 | 1 | Not-determined | Genome-fragment | NA | NA | 0 |
| S13_extracted_110313 | 1442 | No | NA | 1 | 0 | 1 | Not-determined | Genome-fragment | NA | NA | 0 |
| S13_extracted_110345 | 1562 | No | NA | 1 | 0 | 1 | Not-determined | Genome-fragment | NA | NA | 0 |
| S13_extracted_110462 | 3453 | No | NA | 4 | 0 | 4 | Not-determined | Genome-fragment | NA | NA | 0 |
| S13_extracted_110593 | 1133 | No | NA | 2 | 0 | 1 | Not-determined | Genome-fragment | NA | NA | 0 |
| S13_extracted_110670 | 2840 | No | NA | 4 | 0 | 3 | Not-determined | Genome-fragment | NA | NA | 0 |
| S13_extracted_110701 | 1341 | No | NA | 3 | 0 | 1 | Not-determined | Genome-fragment | NA | NA | 0 |
| S13_extracted_110710 | 1688 | No | NA | 1 | 0 | 1 | Not-determined | Genome-fragment | NA | NA | 0 |
| S13_extracted_110747 | 1218 | No | NA | 1 | 0 | 1 | Not-determined | Genome-fragment | NA | NA | 0 |
| S14_extracted_k141_47197 | 1260 | No | NA | 6 | 6 | 0 | Low-quality | Genome-fragment | 4.17 | AAI-based (high-confidence) | 0 |
| S13_extracted_110833 | 1072 | No | NA | 1 | 0 | 1 | Not-determined | Genome-fragment | NA | NA | 0 |
| S13_extracted_110855 | 1010 | No | NA | 1 | 0 | 0 | Not-determined | Genome-fragment | NA | NA | 0 |
| S13_extracted_110911 | 2311 | No | NA | 1 | 0 | 1 | Not-determined | Genome-fragment | NA | NA | 0 |
| S13_extracted_111050 | 1077 | No | NA | 2 | 0 | 2 | Not-determined | Genome-fragment | NA | NA | 0 |
| S14_vs_k141_92583||full | 1260 | No | NA | 2 | 0 | 0 | Low-quality | Genome-fragment | 1.71 | AAI-based (medium-confidence) | 0 |
| S13_extracted_111207 | 1363 | No | NA | 2 | 0 | 2 | Not-determined | Genome-fragment | NA | NA | 0 |
| S13_extracted_111234 | 3561 | No | NA | 7 | 0 | 1 | Not-determined | Genome-fragment | NA | NA | 0 |
| S13_extracted_111334 | 1693 | No | NA | 1 | 0 | 0 | Not-determined | Genome-fragment | NA | NA | 0 |
| S1_extracted_k141_23259 | 1260 | No | NA | 4 | 0 | 0 | Low-quality | Genome-fragment | 2.2 | AAI-based (high-confidence) | 0 |
| S13_extracted_111443 | 1367 | No | NA | 4 | 0 | 0 | Not-determined | Genome-fragment | NA | NA | 0 |
| S3_extracted_k141_42441 | 1260 | No | NA | 1 | 0 | 0 | Low-quality | Genome-fragment | 0.69 | AAI-based (medium-confidence) | 0 |
| S13_extracted_111860 | 1489 | No | NA | 1 | 0 | 1 | Not-determined | Genome-fragment | NA | NA | 0 |
| S13_extracted_111876 | 3461 | No | NA | 2 | 0 | 1 | Not-determined | Genome-fragment | NA | NA | 0 |
| S13_extracted_111914 | 14845 | No | NA | 30 | 0 | 28 | Not-determined | Genome-fragment | NA | NA | 0 |
| S13_extracted_112004 | 1150 | No | NA | 1 | 0 | 1 | Not-determined | Genome-fragment | NA | NA | 0 |
| S13_extracted_112155 | 1295 | No | NA | 1 | 0 | 0 | Not-determined | Genome-fragment | NA | NA | 0 |
| S13_extracted_112178 | 1383 | No | NA | 1 | 0 | 1 | Not-determined | Genome-fragment | NA | NA | 0 |
| S13_extracted_112423 | 1927 | No | NA | 2 | 0 | 1 | Not-determined | Genome-fragment | NA | NA | 0 |
| S13_extracted_112484 | 2391 | No | NA | 4 | 0 | 0 | Not-determined | Genome-fragment | NA | NA | 0 |
| S13_extracted_112561 | 1101 | No | NA | 1 | 0 | 1 | Not-determined | Genome-fragment | NA | NA | 0 |
| S13_extracted_112625 | 1121 | No | NA | 2 | 0 | 2 | Not-determined | Genome-fragment | NA | NA | 0 |
| S13_extracted_112953 | 1132 | No | NA | 1 | 0 | 1 | Not-determined | Genome-fragment | NA | NA | 0 |
| S13_extracted_113063 | 1805 | No | NA | 1 | 0 | 1 | Not-determined | Genome-fragment | NA | NA | 0 |
| S13_extracted_113237 | 1598 | No | NA | 1 | 0 | 0 | Not-determined | Genome-fragment | NA | NA | 0 |
| S13_extracted_113577 | 1239 | No | NA | 2 | 0 | 2 | Not-determined | Genome-fragment | NA | NA | 0 |
| S13_extracted_114078 | 1024 | No | NA | 1 | 0 | 0 | Not-determined | Genome-fragment | NA | NA | 0 |
| S13_extracted_114193 | 1192 | No | NA | 2 | 0 | 1 | Not-determined | Genome-fragment | NA | NA | 0 |
| S13_extracted_114421 | 1906 | No | NA | 1 | 0 | 1 | Not-determined | Genome-fragment | NA | NA | 0 |
| S13_extracted_114496 | 1535 | No | NA | 1 | 0 | 1 | Not-determined | Genome-fragment | NA | NA | 0 |
| S13_extracted_114766 | 3934 | No | NA | 2 | 0 | 1 | Not-determined | Genome-fragment | NA | NA | 0 |
| S13_extracted_114935 | 1405 | No | NA | 2 | 0 | 0 | Not-determined | Genome-fragment | NA | NA | 0 |
| S13_extracted_115021 | 7179 | No | NA | 8 | 0 | 5 | Not-determined | Genome-fragment | NA | NA | 0 |
| S13_extracted_115047 | 1016 | No | NA | 1 | 0 | 0 | Not-determined | Genome-fragment | NA | NA | 0 |
| S13_extracted_115081 | 1057 | No | NA | 2 | 0 | 0 | Not-determined | Genome-fragment | NA | NA | 0 |
| S13_extracted_115173 | 1414 | No | NA | 2 | 0 | 1 | Not-determined | Genome-fragment | NA | NA | 0 |
| S13_extracted_115373 | 4139 | No | NA | 5 | 0 | 0 | Not-determined | Genome-fragment | NA | NA | 0 |
| S13_extracted_115391 | 4833 | No | NA | 1 | 0 | 0 | Not-determined | Genome-fragment | NA | NA | 0 |
| S3_vs_k141_104569||lt2gene | 1260 | No | NA | 2 | 2 | 0 | Low-quality | Genome-fragment | 3.07 | AAI-based (medium-confidence) | 0 |
| S13_extracted_115743 | 2103 | No | NA | 2 | 0 | 0 | Not-determined | Genome-fragment | NA | NA | 0 |
| S13_extracted_115818 | 3692 | No | NA | 3 | 0 | 2 | Not-determined | Genome-fragment | NA | NA | 0 |
| S5_extracted_k141_100800 | 1260 | No | NA | 3 | 0 | 0 | Low-quality | Genome-fragment | 3.54 | AAI-based (high-confidence) | 0 |
| S13_extracted_116190 | 1491 | No | NA | 1 | 0 | 0 | Not-determined | Genome-fragment | NA | NA | 0 |
| S5_vs_k141_126304||full | 1260 | No | NA | 2 | 1 | 0 | Low-quality | Genome-fragment | 2.92 | AAI-based (medium-confidence) | 0 |
| S13_extracted_116383 | 2144 | No | NA | 3 | 0 | 3 | Not-determined | Genome-fragment | NA | NA | 0 |
| S13_extracted_116533 | 1102 | No | NA | 1 | 0 | 0 | Not-determined | Genome-fragment | NA | NA | 0 |
| S13_extracted_116538 | 7022 | No | NA | 9 | 0 | 5 | Not-determined | Genome-fragment | NA | NA | 0 |
| S13_extracted_116830 | 1668 | No | NA | 1 | 0 | 0 | Not-determined | Genome-fragment | NA | NA | 0 |
| S13_extracted_116840 | 2494 | No | NA | 3 | 0 | 2 | Not-determined | Genome-fragment | NA | NA | 0 |
| S13_extracted_116935 | 1563 | No | NA | 1 | 0 | 1 | Not-determined | Genome-fragment | NA | NA | 0 |
| S13_extracted_117035 | 2114 | No | NA | 2 | 0 | 1 | Not-determined | Genome-fragment | NA | NA | 0 |
| S5_vs_k141_46839||full | 1260 | No | NA | 2 | 0 | 0 | Low-quality | Genome-fragment | 3.17 | AAI-based (medium-confidence) | 0 |
| S13_extracted_117093 | 1293 | No | NA | 1 | 0 | 1 | Not-determined | Genome-fragment | NA | NA | 0 |
| S13_extracted_117109 | 3553 | No | NA | 5 | 0 | 3 | Not-determined | Genome-fragment | NA | NA | 0 |
| S13_extracted_117175 | 1535 | No | NA | 2 | 0 | 2 | Not-determined | Genome-fragment | NA | NA | 0 |
| S13_extracted_117237 | 7084 | No | NA | 5 | 0 | 2 | Not-determined | Genome-fragment | NA | NA | 0 |
| S13_extracted_117292 | 3362 | No | NA | 1 | 0 | 1 | Not-determined | Genome-fragment | NA | NA | 0 |
| S13_extracted_117327 | 9600 | No | NA | 9 | 0 | 6 | Not-determined | Genome-fragment | NA | NA | 0 |
| S13_extracted_117364 | 3460 | No | NA | 2 | 0 | 2 | Not-determined | Genome-fragment | NA | NA | 0 |
| S13_extracted_117530 | 1351 | No | NA | 3 | 0 | 2 | Not-determined | Genome-fragment | NA | NA | 0 |
| S13_extracted_117580 | 1035 | No | NA | 3 | 0 | 1 | Not-determined | Genome-fragment | NA | NA | 0 |
| S7_vs_74294||full | 1260 | No | NA | 3 | 1 | 0 | Low-quality | Genome-fragment | 0.88 | AAI-based (high-confidence) | 0 |
| S13_extracted_118092 | 1001 | No | NA | 2 | 0 | 0 | Not-determined | Genome-fragment | NA | NA | 0 |
| S13_extracted_118458 | 3076 | No | NA | 3 | 0 | 2 | Not-determined | Genome-fragment | NA | NA | 0 |
| S13_extracted_118613 | 2072 | No | NA | 1 | 0 | 0 | Not-determined | Genome-fragment | NA | NA | 0 |
| S13_extracted_118713 | 1757 | No | NA | 2 | 0 | 0 | Not-determined | Genome-fragment | NA | NA | 0 |
| S13_extracted_118920 | 1023 | No | NA | 2 | 0 | 1 | Not-determined | Genome-fragment | NA | NA | 0 |
| S13_extracted_119018 | 3380 | No | NA | 2 | 0 | 0 | Not-determined | Genome-fragment | NA | NA | 0 |
| S13_extracted_119101 | 1179 | No | NA | 2 | 0 | 1 | Not-determined | Genome-fragment | NA | NA | 0 |
| S13_extracted_119154 | 1232 | No | NA | 2 | 0 | 1 | Not-determined | Genome-fragment | NA | NA | 0 |
| S13_extracted_119240 | 1196 | No | NA | 2 | 0 | 1 | Not-determined | Genome-fragment | NA | NA | 0 |
| S13_extracted_119295 | 1061 | No | NA | 2 | 0 | 0 | Not-determined | Genome-fragment | NA | NA | 0 |
| S13_extracted_119320 | 1271 | No | NA | 1 | 0 | 0 | Not-determined | Genome-fragment | NA | NA | 0 |
| S13_extracted_119397 | 4766 | No | NA | 5 | 0 | 3 | Not-determined | Genome-fragment | NA | NA | 0 |
| S13_extracted_119588 | 6426 | No | NA | 7 | 0 | 5 | Not-determined | Genome-fragment | NA | NA | 0 |
| S13_extracted_119595 | 1246 | No | NA | 1 | 0 | 1 | Not-determined | Genome-fragment | NA | NA | 0 |
| S13_extracted_119750 | 1195 | No | NA | 1 | 0 | 1 | Not-determined | Genome-fragment | NA | NA | 0 |
| S7_vs_79226||full | 1260 | No | NA | 3 | 1 | 0 | Low-quality | Genome-fragment | 2.17 | AAI-based (medium-confidence) | 0 |
| S13_extracted_119949 | 1295 | No | NA | 1 | 0 | 0 | Not-determined | Genome-fragment | NA | NA | 0 |
| S13_extracted_119994 | 8479 | No | NA | 8 | 0 | 2 | Not-determined | Genome-fragment | NA | NA | 0 |
| S13_extracted_120223 | 1309 | No | NA | 2 | 0 | 1 | Not-determined | Genome-fragment | NA | NA | 0 |
| S13_extracted_120241 | 2435 | No | NA | 4 | 0 | 3 | Not-determined | Genome-fragment | NA | NA | 0 |
| S13_extracted_120273 | 1214 | No | NA | 1 | 0 | 1 | Not-determined | Genome-fragment | NA | NA | 0 |
| S13_extracted_120391 | 1243 | No | NA | 2 | 0 | 1 | Not-determined | Genome-fragment | NA | NA | 0 |
| S13_extracted_120399 | 1002 | No | NA | 2 | 0 | 2 | Not-determined | Genome-fragment | NA | NA | 0 |
| S13_extracted_120426 | 1157 | No | NA | 1 | 0 | 1 | Not-determined | Genome-fragment | NA | NA | 0 |
| S13_extracted_120429 | 1676 | No | NA | 1 | 0 | 1 | Not-determined | Genome-fragment | NA | NA | 0 |
| S13_extracted_120732 | 1774 | No | NA | 2 | 0 | 1 | Not-determined | Genome-fragment | NA | NA | 0 |
| S13_extracted_120984 | 1588 | No | NA | 2 | 0 | 1 | Not-determined | Genome-fragment | NA | NA | 0 |
| S16_extracted_26284 | 1260 | No | NA | 2 | 0 | 1 | Low-quality | Genome-fragment | 2.54 | AAI-based (medium-confidence) | 0 |
| S13_extracted_121167 | 1101 | No | NA | 1 | 0 | 1 | Not-determined | Genome-fragment | NA | NA | 0 |
| S16_extracted_111925 | 1260 | No | NA | 5 | 1 | 0 | Low-quality | Genome-fragment | 3.29 | AAI-based (high-confidence) | 0 |
| S13_extracted_121265 | 1000 | No | NA | 1 | 0 | 1 | Not-determined | Genome-fragment | NA | NA | 0 |
| S13_extracted_121388 | 1049 | No | NA | 1 | 0 | 0 | Not-determined | Genome-fragment | NA | NA | 0 |
| S13_extracted_121431 | 1330 | No | NA | 1 | 0 | 0 | Not-determined | Genome-fragment | NA | NA | 0 |
| S10_extracted_43997 | 1260 | No | NA | 1 | 0 | 0 | Low-quality | Genome-fragment | 1.49 | AAI-based (high-confidence) | 0 |
| S10_extracted_98594 | 1260 | No | NA | 1 | 0 | 0 | Low-quality | Genome-fragment | 0.62 | AAI-based (medium-confidence) | 0 |
| S10_vs_52359||full | 1260 | No | NA | 2 | 0 | 0 | Low-quality | Genome-fragment | 2.6 | AAI-based (medium-confidence) | 0 |
| S13_extracted_122210 | 7410 | No | NA | 8 | 0 | 7 | Not-determined | Genome-fragment | NA | NA | 0 |
| S6_vs_k141_25105||full | 1260 | No | NA | 2 | 0 | 0 | Low-quality | Genome-fragment | 2.18 | AAI-based (high-confidence) | 0 |
| S13_extracted_122392 | 5301 | No | NA | 3 | 0 | 0 | Not-determined | Genome-fragment | NA | NA | 0 |
| S6_vs_k141_4465||full | 1260 | No | NA | 3 | 0 | 0 | Low-quality | Genome-fragment | 2.09 | AAI-based (high-confidence) | 0 |
| S13_extracted_122580 | 3684 | No | NA | 3 | 0 | 0 | Not-determined | Genome-fragment | NA | NA | 0 |
| S13_extracted_122585 | 1490 | No | NA | 2 | 0 | 1 | Not-determined | Genome-fragment | NA | NA | 0 |
| S13_extracted_122617 | 2298 | No | NA | 2 | 0 | 1 | Not-determined | Genome-fragment | NA | NA | 0 |
| S13_extracted_122673 | 1277 | No | NA | 1 | 0 | 0 | Not-determined | Genome-fragment | NA | NA | 0 |
| S13_extracted_122804 | 1285 | No | NA | 1 | 0 | 0 | Not-determined | Genome-fragment | NA | NA | 0 |
| S13_extracted_122837 | 6382 | No | NA | 8 | 0 | 5 | Not-determined | Genome-fragment | NA | NA | 0 |
| S13_extracted_123025 | 2544 | No | NA | 2 | 0 | 2 | Not-determined | Genome-fragment | NA | NA | 0 |
| S13_extracted_123203 | 1444 | No | NA | 2 | 0 | 0 | Not-determined | Genome-fragment | NA | NA | 0 |
| S6_vs_k141_33116||lt2gene | 1260 | No | NA | 2 | 1 | 0 | Low-quality | Genome-fragment | 2.95 | HMM-based (lower-bound) | 0 |
| S13_extracted_123302 | 1019 | No | NA | 1 | 0 | 0 | Not-determined | Genome-fragment | NA | NA | 0 |
| S13_extracted_123367 | 4054 | No | NA | 3 | 0 | 2 | Not-determined | Genome-fragment | NA | NA | 0 |
| S13_extracted_123375 | 2221 | No | NA | 3 | 0 | 0 | Not-determined | Genome-fragment | NA | NA | 0 |
| S13_extracted_123462 | 1186 | No | NA | 1 | 0 | 1 | Not-determined | Genome-fragment | NA | NA | 0 |
| S13_extracted_123490 | 1267 | No | NA | 1 | 0 | 1 | Not-determined | Genome-fragment | NA | NA | 0 |
| S13_extracted_123496 | 1253 | No | NA | 3 | 0 | 2 | Not-determined | Genome-fragment | NA | NA | 0 |
| S11_vs_k141_20007||full | 1261 | No | NA | 3 | 0 | 1 | Low-quality | Genome-fragment | 0.6 | AAI-based (high-confidence) | 0 |
| S13_extracted_123554 | 2306 | No | NA | 1 | 0 | 0 | Not-determined | Genome-fragment | NA | NA | 0 |
| S13_extracted_123567 | 1487 | No | NA | 2 | 0 | 0 | Not-determined | Genome-fragment | NA | NA | 0 |
| S13_extracted_123865 | 1405 | No | NA | 1 | 0 | 0 | Not-determined | Genome-fragment | NA | NA | 0 |
| S13_extracted_124304 | 1408 | No | NA | 4 | 0 | 2 | Not-determined | Genome-fragment | NA | NA | 0 |
| S11_vs_k141_92655||full | 1261 | No | NA | 2 | 0 | 0 | Low-quality | Genome-fragment | 0.71 | AAI-based (high-confidence) | 0 |
| S13_extracted_124357 | 1102 | No | NA | 1 | 0 | 0 | Not-determined | Genome-fragment | NA | NA | 0 |
| S13_extracted_124500 | 1455 | No | NA | 1 | 0 | 1 | Not-determined | Genome-fragment | NA | NA | 0 |
| S13_extracted_124706 | 1325 | No | NA | 2 | 0 | 0 | Not-determined | Genome-fragment | NA | NA | 0 |
| S13_extracted_124759 | 1267 | No | NA | 2 | 0 | 2 | Not-determined | Genome-fragment | NA | NA | 0 |
| S14_vs_k141_67520||full | 1261 | No | NA | 4 | 1 | 0 | Low-quality | Genome-fragment | 2.75 | AAI-based (high-confidence) | 0 |
| S13_extracted_124842 | 2079 | No | NA | 4 | 0 | 0 | Not-determined | Genome-fragment | NA | NA | 0 |
| S14_vs_k141_99477||full | 1261 | No | NA | 2 | 0 | 0 | Low-quality | Genome-fragment | 1.66 | AAI-based (medium-confidence) | 0 |
| S1_vs_k141_122886||full | 1261 | No | NA | 3 | 0 | 0 | Low-quality | Genome-fragment | 3.22 | AAI-based (high-confidence) | 0 |
| S13_extracted_125105 | 1016 | No | NA | 2 | 0 | 1 | Not-determined | Genome-fragment | NA | NA | 0 |
| S13_extracted_125190 | 1573 | No | NA | 2 | 0 | 1 | Not-determined | Genome-fragment | NA | NA | 0 |
| S13_extracted_125265 | 1145 | No | NA | 2 | 0 | 2 | Not-determined | Genome-fragment | NA | NA | 0 |
| S13_extracted_125326 | 3593 | No | NA | 4 | 0 | 4 | Not-determined | Genome-fragment | NA | NA | 0 |
| S13_extracted_125352 | 1104 | No | NA | 2 | 0 | 1 | Not-determined | Genome-fragment | NA | NA | 0 |
| S13_extracted_125372 | 1824 | No | NA | 3 | 0 | 1 | Not-determined | Genome-fragment | NA | NA | 0 |
| S13_extracted_125449 | 1815 | No | NA | 1 | 0 | 0 | Not-determined | Genome-fragment | NA | NA | 0 |
| S13_extracted_125461 | 1091 | No | NA | 2 | 0 | 2 | Not-determined | Genome-fragment | NA | NA | 0 |
| S13_extracted_125516 | 1987 | No | NA | 3 | 0 | 1 | Not-determined | Genome-fragment | NA | NA | 0 |
| S13_extracted_125519 | 1048 | No | NA | 2 | 0 | 0 | Not-determined | Genome-fragment | NA | NA | 0 |
| S13_extracted_125527 | 1028 | No | NA | 2 | 0 | 2 | Not-determined | Genome-fragment | NA | NA | 0 |
| S13_extracted_125695 | 1361 | No | NA | 2 | 0 | 0 | Not-determined | Genome-fragment | NA | NA | 0 |
| S13_extracted_125726 | 1671 | No | NA | 1 | 0 | 0 | Not-determined | Genome-fragment | NA | NA | 0 |
| S1_vs_k141_12843||full | 1261 | No | NA | 4 | 2 | 0 | Low-quality | Genome-fragment | 2.18 | AAI-based (medium-confidence) | 0 |
| S3_vs_k141_41611||full | 1261 | No | NA | 4 | 0 | 1 | Low-quality | Genome-fragment | 0.72 | AAI-based (high-confidence) | 0 |
| S13_extracted_125908 | 1009 | No | NA | 2 | 0 | 1 | Not-determined | Genome-fragment | NA | NA | 0 |
| S13_extracted_125952 | 1184 | No | NA | 2 | 0 | 1 | Not-determined | Genome-fragment | NA | NA | 0 |
| S13_extracted_126032 | 1196 | No | NA | 1 | 0 | 1 | Not-determined | Genome-fragment | NA | NA | 0 |
| S13_extracted_126042 | 1269 | No | NA | 3 | 0 | 1 | Not-determined | Genome-fragment | NA | NA | 0 |
| S13_extracted_126106 | 4377 | No | NA | 4 | 0 | 2 | Not-determined | Genome-fragment | NA | NA | 0 |
| S13_extracted_126274 | 1932 | No | NA | 2 | 0 | 1 | Not-determined | Genome-fragment | NA | NA | 0 |
| S13_extracted_126350 | 1536 | No | NA | 1 | 0 | 1 | Not-determined | Genome-fragment | NA | NA | 0 |
| S13_extracted_126429 | 1028 | No | NA | 1 | 0 | 0 | Not-determined | Genome-fragment | NA | NA | 0 |
| S13_extracted_126444 | 2384 | No | NA | 4 | 0 | 1 | Not-determined | Genome-fragment | NA | NA | 0 |
| S13_extracted_126505 | 1381 | No | NA | 3 | 0 | 2 | Not-determined | Genome-fragment | NA | NA | 0 |
| S13_extracted_126603 | 1404 | No | NA | 1 | 0 | 1 | Not-determined | Genome-fragment | NA | NA | 0 |
| S13_extracted_126620 | 1403 | No | NA | 2 | 0 | 0 | Not-determined | Genome-fragment | NA | NA | 0 |
| S13_extracted_126693 | 1036 | No | NA | 1 | 0 | 1 | Not-determined | Genome-fragment | NA | NA | 0 |
| S13_extracted_126766 | 2095 | No | NA | 4 | 0 | 0 | Not-determined | Genome-fragment | NA | NA | 0 |
| S13_extracted_126801 | 1493 | No | NA | 2 | 0 | 2 | Not-determined | Genome-fragment | NA | NA | 0 |
| S3_vs_k141_27410||full | 1261 | No | NA | 3 | 0 | 0 | Low-quality | Genome-fragment | 2.19 | AAI-based (medium-confidence) | 0 |
| S5_vs_k141_58438||lt2gene | 1261 | No | NA | 1 | 1 | 0 | Low-quality | Genome-fragment | 3.33 | AAI-based (high-confidence) | 0 |
| S13_extracted_127020 | 2210 | No | NA | 1 | 0 | 1 | Not-determined | Genome-fragment | NA | NA | 0 |
| S13_extracted_127028 | 3392 | No | NA | 5 | 0 | 3 | Not-determined | Genome-fragment | NA | NA | 0 |
| S13_extracted_127078 | 1183 | No | NA | 1 | 0 | 1 | Not-determined | Genome-fragment | NA | NA | 0 |
| S13_extracted_127286 | 1853 | No | NA | 1 | 0 | 0 | Not-determined | Genome-fragment | NA | NA | 0 |
| S13_extracted_127376 | 1770 | No | NA | 2 | 0 | 2 | Not-determined | Genome-fragment | NA | NA | 0 |
| S13_extracted_127402 | 1044 | No | NA | 1 | 0 | 0 | Not-determined | Genome-fragment | NA | NA | 0 |
| S13_extracted_127556 | 1042 | No | NA | 3 | 0 | 1 | Not-determined | Genome-fragment | NA | NA | 0 |
| S13_extracted_127744 | 2245 | No | NA | 3 | 0 | 2 | Not-determined | Genome-fragment | NA | NA | 0 |
| S13_extracted_128030 | 1485 | No | NA | 2 | 0 | 2 | Not-determined | Genome-fragment | NA | NA | 0 |
| S13_extracted_128194 | 1920 | No | NA | 4 | 0 | 1 | Not-determined | Genome-fragment | NA | NA | 0 |
| S13_extracted_128222 | 3446 | No | NA | 2 | 0 | 2 | Not-determined | Genome-fragment | NA | NA | 0 |
| S13_extracted_128254 | 1348 | No | NA | 4 | 0 | 4 | Not-determined | Genome-fragment | NA | NA | 0 |
| S7_vs_16087||full | 1261 | No | NA | 2 | 0 | 0 | Low-quality | Genome-fragment | 1.85 | AAI-based (medium-confidence) | 0 |
| S13_extracted_128572 | 6755 | No | NA | 6 | 0 | 5 | Not-determined | Genome-fragment | NA | NA | 0 |
| S13_extracted_128736 | 6841 | No | NA | 7 | 0 | 5 | Not-determined | Genome-fragment | NA | NA | 0 |
| S13_extracted_128812 | 2280 | No | NA | 2 | 0 | 2 | Not-determined | Genome-fragment | NA | NA | 0 |
| S13_extracted_128983 | 2570 | No | NA | 4 | 0 | 2 | Not-determined | Genome-fragment | NA | NA | 0 |
| S7_vs_54071||full | 1261 | No | NA | 3 | 0 | 0 | Low-quality | Genome-fragment | 3.01 | AAI-based (high-confidence) | 0 |
| S13_extracted_129213 | 1043 | No | NA | 2 | 0 | 1 | Not-determined | Genome-fragment | NA | NA | 0 |
| S13_extracted_129313 | 1008 | No | NA | 2 | 0 | 0 | Not-determined | Genome-fragment | NA | NA | 0 |
| S13_extracted_129534 | 6670 | No | NA | 7 | 0 | 4 | Not-determined | Genome-fragment | NA | NA | 0 |
| S13_extracted_129666 | 4721 | No | NA | 10 | 0 | 9 | Not-determined | Genome-fragment | NA | NA | 0 |
| S13_extracted_129729 | 1606 | No | NA | 3 | 0 | 3 | Not-determined | Genome-fragment | NA | NA | 0 |
| S13_extracted_129941 | 2449 | No | NA | 5 | 0 | 5 | Not-determined | Genome-fragment | NA | NA | 0 |
| S13_extracted_130013 | 3785 | No | NA | 4 | 0 | 1 | Not-determined | Genome-fragment | NA | NA | 0 |
| S13_extracted_130272 | 1127 | No | NA | 1 | 0 | 0 | Not-determined | Genome-fragment | NA | NA | 0 |
| S13_extracted_130299 | 3852 | No | NA | 10 | 0 | 1 | Not-determined | Genome-fragment | NA | NA | 0 |
| S13_extracted_130331 | 9236 | No | NA | 13 | 0 | 6 | Not-determined | Genome-fragment | NA | NA | 0 |
| S13_extracted_130594 | 1182 | No | NA | 1 | 0 | 1 | Not-determined | Genome-fragment | NA | NA | 0 |
| S13_extracted_130640 | 1124 | No | NA | 3 | 0 | 0 | Not-determined | Genome-fragment | NA | NA | 0 |
| S13_extracted_130670 | 1745 | No | NA | 2 | 0 | 1 | Not-determined | Genome-fragment | NA | NA | 0 |
| S13_extracted_130688 | 1072 | No | NA | 2 | 0 | 1 | Not-determined | Genome-fragment | NA | NA | 0 |
| S13_extracted_130760 | 2001 | No | NA | 1 | 0 | 0 | Not-determined | Genome-fragment | NA | NA | 0 |
| S13_extracted_130968 | 1041 | No | NA | 2 | 0 | 1 | Not-determined | Genome-fragment | NA | NA | 0 |
| S13_extracted_131208 | 13280 | No | NA | 13 | 8 | 0 | Medium-quality | Genome-fragment | 64.98 | AAI-based (medium-confidence) | 0 |
| S13_extracted_131280 | 1180 | No | NA | 1 | 0 | 0 | Not-determined | Genome-fragment | NA | NA | 0 |
| S13_extracted_131313 | 1016 | No | NA | 1 | 0 | 1 | Not-determined | Genome-fragment | NA | NA | 0 |
| S12_vs_k141_21577||full | 1261 | No | NA | 2 | 0 | 0 | Low-quality | Genome-fragment | 1.81 | AAI-based (medium-confidence) | 0 |
| S12_vs_k141_47559||lt2gene | 1261 | No | NA | 1 | 1 | 0 | Low-quality | Genome-fragment | 2.12 | HMM-based (lower-bound) | 0 |
| S6_vs_k141_45266||full | 1261 | No | NA | 4 | 0 | 0 | Low-quality | Genome-fragment | 3.23 | AAI-based (high-confidence) | 0 |

| | | | | | | | | | | | |
|---|---|---|---|---|---|---|---|---|---|---|---|
| S13_extracted_131479 | 1423 | No | NA | 1 | 0 | 1 | Not-determined | Genome-fragment | NA | NA | 0 |
| S6_vs_k141_54481\|\|full | 1261 | No | NA | 2 | 0 | 1 | Low-quality | Genome-fragment | 1.57 | AAI-based (high-confidence) | 0 |
| S13_extracted_131633 | 3054 | No | NA | 4 | 0 | 2 | Not-determined | Genome-fragment | NA | NA | 0 |
| S13_extracted_131650 | 1076 | No | NA | 2 | 0 | 1 | Not-determined | Genome-fragment | NA | NA | 0 |
| S13_extracted_131686 | 1741 | No | NA | 3 | 0 | 2 | Not-determined | Genome-fragment | NA | NA | 0 |
| S13_extracted_131708 | 3127 | No | NA | 3 | 0 | 1 | Not-determined | Genome-fragment | NA | NA | 0 |
| S13_extracted_131854 | 3813 | No | NA | 7 | 0 | 4 | Not-determined | Genome-fragment | NA | NA | 0 |
| S13_extracted_131861 | 21298 | No | NA | 21 | 0 | 4 | Not-determined | Genome-fragment | NA | NA | 0 |
| S13_extracted_131889 | 1720 | No | NA | 2 | 0 | 1 | Not-determined | Genome-fragment | NA | NA | 0 |
| S13_extracted_132020 | 1163 | No | NA | 1 | 0 | 1 | Not-determined | Genome-fragment | NA | NA | 0 |
| S13_extracted_132022 | 1143 | No | NA | 2 | 0 | 1 | Not-determined | Genome-fragment | NA | NA | 0 |
| S8_extracted_k141_155948 | 1261 | No | NA | 2 | 0 | 1 | Low-quality | Genome-fragment | 2.5 | AAI-based (medium-confidence) | 0 |
| S8_vs_k141_5668\|\|full | 1261 | No | NA | 3 | 0 | 0 | Low-quality | Genome-fragment | 3.41 | AAI-based (medium-confidence) | 0 |
| S17_vs_k141_78851\|\|full | 1262 | No | NA | 3 | 0 | 1 | Low-quality | Genome-fragment | 0.59 | AAI-based (medium-confidence) | 0 |
| S13_extracted_132198 | 1021 | No | NA | 2 | 0 | 0 | Not-determined | Genome-fragment | NA | NA | 0 |
| S13_extracted_132372 | 1220 | No | NA | 1 | 0 | 1 | Not-determined | Genome-fragment | NA | NA | 0 |
| S13_extracted_132407 | 1199 | No | NA | 2 | 0 | 0 | Not-determined | Genome-fragment | NA | NA | 0 |
| S17_vs_k141_177009\|\|full | 1262 | No | NA | 2 | 0 | 2 | Low-quality | Genome-fragment | 0.59 | AAI-based (high-confidence) | 0 |
| S13_extracted_132643 | 1514 | No | NA | 2 | 0 | 2 | Not-determined | Genome-fragment | NA | NA | 0 |
| S13_extracted_133001 | 1298 | No | NA | 1 | 0 | 1 | Not-determined | Genome-fragment | NA | NA | 0 |
| S13_extracted_133136 | 5641 | No | NA | 1 | 0 | 0 | Not-determined | Genome-fragment | NA | NA | 0 |
| S9_vs_k141_47657\|\|full | 1262 | No | NA | 3 | 0 | 1 | Low-quality | Genome-fragment | 1.63 | AAI-based (medium-confidence) | 0 |
| S13_extracted_133211 | 1009 | No | NA | 1 | 0 | 0 | Not-determined | Genome-fragment | NA | NA | 0 |
| S11_extracted_k141_58714 | 1262 | No | NA | 2 | 0 | 0 | Low-quality | Genome-fragment | 2.05 | AAI-based (high-confidence) | 0 |
| S13_extracted_133621 | 1305 | No | NA | 1 | 0 | 1 | Not-determined | Genome-fragment | NA | NA | 0 |
| S13_extracted_122158 | 1262 | No | NA | 1 | 0 | 0 | Low-quality | Genome-fragment | 0.96 | AAI-based (high-confidence) | 0 |
| S14_extracted_k141_5033 | 1262 | No | NA | 2 | 1 | 0 | Low-quality | Genome-fragment | 2.82 | AAI-based (high-confidence) | 0 |
| S13_extracted_134065 | 1982 | No | NA | 2 | 0 | 1 | Not-determined | Genome-fragment | NA | NA | 0 |
| S3_vs_k141_111350\|\|full | 1262 | No | NA | 3 | 0 | 0 | Low-quality | Genome-fragment | 2.55 | AAI-based (medium-confidence) | 0 |
| S3_vs_k141_52350\|\|lt2gene | 1262 | No | NA | 1 | 1 | 0 | Low-quality | Genome-fragment | 2.45 | AAI-based (high-confidence) | 0 |
| S13_extracted_134641 | 2928 | No | NA | 2 | 0 | 0 | Not-determined | Genome-fragment | NA | NA | 0 |
| S13_extracted_134691 | 3308 | No | NA | 2 | 0 | 1 | Not-determined | Genome-fragment | NA | NA | 0 |
| S13_extracted_134726 | 1227 | No | NA | 1 | 0 | 1 | Not-determined | Genome-fragment | NA | NA | 0 |
| S13_extracted_134794 | 1742 | No | NA | 2 | 0 | 0 | Not-determined | Genome-fragment | NA | NA | 0 |
| S13_extracted_134844 | 1562 | No | NA | 2 | 0 | 2 | Not-determined | Genome-fragment | NA | NA | 0 |
| S13_extracted_135000 | 1414 | No | NA | 2 | 0 | 0 | Not-determined | Genome-fragment | NA | NA | 0 |
| S13_extracted_135009 | 1051 | No | NA | 2 | 0 | 2 | Not-determined | Genome-fragment | NA | NA | 0 |
| S13_extracted_135426 | 1963 | No | NA | 4 | 0 | 1 | Not-determined | Genome-fragment | NA | NA | 0 |
| S5_vs_k141_74521\|\|lt2gene | 1262 | No | NA | 1 | 1 | 0 | Low-quality | Genome-fragment | 2.6 | AAI-based (high-confidence) | 0 |
| S13_extracted_135562 | 1036 | No | NA | 2 | 0 | 1 | Not-determined | Genome-fragment | NA | NA | 0 |
| S13_extracted_135658 | 1214 | No | NA | 2 | 0 | 2 | Not-determined | Genome-fragment | NA | NA | 0 |
| S13_extracted_135823 | 1146 | No | NA | 2 | 0 | 0 | Not-determined | Genome-fragment | NA | NA | 0 |
| S13_extracted_135824 | 1374 | No | NA | 4 | 0 | 2 | Not-determined | Genome-fragment | NA | NA | 0 |
| S13_extracted_135899 | 1788 | No | NA | 1 | 0 | 1 | Not-determined | Genome-fragment | NA | NA | 0 |
| S13_extracted_135900 | 1302 | No | NA | 3 | 0 | 0 | Not-determined | Genome-fragment | NA | NA | 0 |
| S13_extracted_135901 | 1125 | No | NA | 2 | 0 | 0 | Not-determined | Genome-fragment | NA | NA | 0 |
| S13_extracted_135938 | 1089 | No | NA | 2 | 0 | 0 | Not-determined | Genome-fragment | NA | NA | 0 |
| S13_extracted_136077 | 1424 | No | NA | 3 | 0 | 0 | Not-determined | Genome-fragment | NA | NA | 0 |
| S13_extracted_136142 | 1090 | No | NA | 1 | 0 | 0 | Not-determined | Genome-fragment | NA | NA | 0 |
| S13_extracted_136171 | 1764 | No | NA | 2 | 0 | 0 | Not-determined | Genome-fragment | NA | NA | 0 |
| S13_extracted_136186 | 1203 | No | NA | 1 | 0 | 1 | Not-determined | Genome-fragment | NA | NA | 0 |
| S7_vs_60829\|\|full | 1262 | No | NA | 4 | 0 | 0 | Low-quality | Genome-fragment | 3.55 | AAI-based (medium-confidence) | 0 |
| S13_extracted_136508 | 1334 | No | NA | 1 | 0 | 1 | Not-determined | Genome-fragment | NA | NA | 0 |
| S13_extracted_136806 | 1561 | No | NA | 3 | 0 | 1 | Not-determined | Genome-fragment | NA | NA | 0 |
| S8_vs_k141_158171\|\|full | 1262 | No | NA | 3 | 0 | 0 | Low-quality | Genome-fragment | 0.81 | AAI-based (high-confidence) | 0 |
| S8_vs_k141_66903\|\|full | 1262 | No | NA | 4 | 0 | 0 | Low-quality | Genome-fragment | 1.09 | AAI-based (medium-confidence) | 0 |
| S13_extracted_137194 | 1252 | No | NA | 2 | 0 | 2 | Not-determined | Genome-fragment | NA | NA | 0 |
| S13_extracted_137426 | 1371 | No | NA | 1 | 0 | 1 | Not-determined | Genome-fragment | NA | NA | 0 |
| S14_vs_k141_89202\|\|full | 1263 | No | NA | 2 | 0 | 0 | Low-quality | Genome-fragment | 2.86 | AAI-based (high-confidence) | 0 |
| S13_extracted_137634 | 1121 | No | NA | 2 | 0 | 1 | Not-determined | Genome-fragment | NA | NA | 0 |
| S14_vs_k141_68930\|\|full | 1263 | No | NA | 4 | 0 | 0 | Low-quality | Genome-fragment | 2.28 | AAI-based (medium-confidence) | 0 |
| S13_extracted_138116 | 1779 | No | NA | 3 | 0 | 0 | Not-determined | Genome-fragment | NA | NA | 0 |
| S1_vs_k141_21038\|\|full | 1263 | No | NA | 3 | 0 | 0 | Low-quality | Genome-fragment | 2.8 | AAI-based (high-confidence) | 0 |
| S13_extracted_138157 | 1327 | No | NA | 1 | 0 | 0 | Not-determined | Genome-fragment | NA | NA | 0 |
| S13_extracted_138200 | 3163 | No | NA | 5 | 0 | 1 | Not-determined | Genome-fragment | NA | NA | 0 |
| S13_extracted_138272 | 1155 | No | NA | 1 | 0 | 0 | Not-determined | Genome-fragment | NA | NA | 0 |
| S13_extracted_138457 | 4326 | No | NA | 2 | 0 | 2 | Not-determined | Genome-fragment | NA | NA | 0 |
| S13_extracted_138583 | 1521 | No | NA | 1 | 0 | 0 | Not-determined | Genome-fragment | NA | NA | 0 |
| S13_extracted_138701 | 1028 | No | NA | 2 | 0 | 2 | Not-determined | Genome-fragment | NA | NA | 0 |
| S13_extracted_139314 | 1027 | No | NA | 4 | 0 | 3 | Not-determined | Genome-fragment | NA | NA | 0 |
| S13_extracted_139332 | 1567 | No | NA | 3 | 0 | 1 | Not-determined | Genome-fragment | NA | NA | 0 |
| S13_extracted_139453 | 1241 | No | NA | 2 | 0 | 0 | Not-determined | Genome-fragment | NA | NA | 0 |
| S13_extracted_139550 | 1497 | No | NA | 2 | 0 | 2 | Not-determined | Genome-fragment | NA | NA | 0 |
| S3_vs_k141_122910\|\|full | 1263 | No | NA | 2 | 0 | 0 | Low-quality | Genome-fragment | 1.7 | AAI-based (high-confidence) | 0 |
| S5_extracted_k141_143162 | 1263 | No | NA | 2 | 0 | 0 | Low-quality | Genome-fragment | 3.16 | AAI-based (high-confidence) | 0 |
| S13_extracted_139924 | 1856 | No | NA | 2 | 0 | 1 | Not-determined | Genome-fragment | NA | NA | 0 |
| S13_extracted_140062 | 1399 | No | NA | 1 | 0 | 1 | Not-determined | Genome-fragment | NA | NA | 0 |
| S13_extracted_140136 | 9760 | No | NA | 10 | 0 | 2 | Not-determined | Genome-fragment | NA | NA | 0 |
| S13_extracted_140199 | 1105 | No | NA | 2 | 0 | 1 | Not-determined | Genome-fragment | NA | NA | 0 |
| S13_extracted_140461 | 1144 | No | NA | 2 | 0 | 0 | Not-determined | Genome-fragment | NA | NA | 0 |
| S13_extracted_140611 | 1087 | No | NA | 2 | 0 | 1 | Not-determined | Genome-fragment | NA | NA | 0 |
| S13_extracted_140647 | 1075 | No | NA | 1 | 0 | 0 | Not-determined | Genome-fragment | NA | NA | 0 |
| S13_extracted_140697 | 4199 | No | NA | 4 | 0 | 3 | Not-determined | Genome-fragment | NA | NA | 0 |
| S5_extracted_k141_66409 | 1263 | No | NA | 2 | 1 | 0 | Low-quality | Genome-fragment | 2.1 | AAI-based (high-confidence) | 0 |
| S13_extracted_140954 | 4667 | No | NA | 1 | 0 | 1 | Not-determined | Genome-fragment | NA | NA | 0 |
| S5_vs_k141_80850\|\|full | 1263 | No | NA | 3 | 0 | 0 | Low-quality | Genome-fragment | 2.36 | AAI-based (medium-confidence) | 0 |
| S13_extracted_141115 | 1397 | No | NA | 1 | 0 | 1 | Not-determined | Genome-fragment | NA | NA | 0 |
| S7_extracted_119109 | 1263 | No | NA | 1 | 0 | 1 | Low-quality | Genome-fragment | 4.21 | AAI-based (high-confidence) | 0 |
| S13_extracted_141133 | 1287 | No | NA | 2 | 0 | 0 | Not-determined | Genome-fragment | NA | NA | 0 |
| S13_extracted_141184 | 1149 | No | NA | 1 | 0 | 0 | Not-determined | Genome-fragment | NA | NA | 0 |
| S16_extracted_142073 | 1263 | No | NA | 2 | 0 | 0 | Low-quality | Genome-fragment | 2.29 | AAI-based (medium-confidence) | 0 |
| S17_extracted_k141_76082 | 1264 | No | NA | 2 | 0 | 1 | Low-quality | Genome-fragment | 0.59 | AAI-based (medium-confidence) | 0 |
| S13_extracted_141355 | 1387 | No | NA | 2 | 0 | 2 | Not-determined | Genome-fragment | NA | NA | 0 |
| S13_extracted_141394 | 1164 | No | NA | 4 | 0 | 0 | Not-determined | Genome-fragment | NA | NA | 0 |
| S13_extracted_141397 | 1184 | No | NA | 1 | 0 | 1 | Not-determined | Genome-fragment | NA | NA | 0 |
| S13_extracted_141444 | 1107 | No | NA | 3 | 0 | 0 | Not-determined | Genome-fragment | NA | NA | 0 |
| S9_vs_k141_12870\|\|full | 1264 | No | NA | 2 | 2 | 0 | Low-quality | Genome-fragment | 3.59 | AAI-based (high-confidence) | 0 |
| S13_extracted_141793 | 1273 | No | NA | 1 | 0 | 1 | Not-determined | Genome-fragment | NA | NA | 0 |
| S11_vs_k141_42027\|\|full | 1264 | No | NA | 3 | 1 | 0 | Low-quality | Genome-fragment | 3.33 | AAI-based (high-confidence) | 0 |
| S13_extracted_142191 | 1055 | No | NA | 1 | 0 | 1 | Not-determined | Genome-fragment | NA | NA | 0 |
| S13_extracted_142563 | 1445 | No | NA | 2 | 0 | 1 | Not-determined | Genome-fragment | NA | NA | 0 |
| S13_extracted_142922 | 1269 | No | NA | 3 | 0 | 1 | Not-determined | Genome-fragment | NA | NA | 0 |
| S13_extracted_143029 | 1029 | No | NA | 2 | 0 | 2 | Not-determined | Genome-fragment | NA | NA | 0 |
| S13_extracted_143121 | 1446 | No | NA | 2 | 0 | 0 | Not-determined | Genome-fragment | NA | NA | 0 |
| S13_extracted_143129 | 1643 | No | NA | 2 | 0 | 1 | Not-determined | Genome-fragment | NA | NA | 0 |
| S13_extracted_143335 | 1694 | No | NA | 2 | 0 | 1 | Not-determined | Genome-fragment | NA | NA | 0 |
| S13_extracted_143377 | 1305 | Yes | 670 | 2 | 1 | 1 | Not-determined | Genome-fragment | NA | NA | 48.66 |
| S13_extracted_143402 | 1079 | No | NA | 3 | 0 | 1 | Not-determined | Genome-fragment | NA | NA | 0 |
| S13_extracted_143422 | 54390 | No | NA | 42 | 8 | 0 | Medium-quality | Genome-fragment | 54.31 | AAI-based (high-confidence) | 0 |
| S13_extracted_143435 | 1198 | No | NA | 1 | 0 | 1 | Not-determined | Genome-fragment | NA | NA | 0 |
| S13_extracted_143536 | 1018 | No | NA | 2 | 0 | 1 | Not-determined | Genome-fragment | NA | NA | 0 |
| S13_extracted_143599 | 1673 | No | NA | 6 | 0 | 6 | Not-determined | Genome-fragment | NA | NA | 0 |
| S13_extracted_143633 | 4901 | No | NA | 3 | 0 | 2 | Not-determined | Genome-fragment | NA | NA | 0 |
| S13_extracted_143838 | 2295 | No | NA | 3 | 0 | 2 | Not-determined | Genome-fragment | NA | NA | 0 |
| S13_extracted_143946 | 1121 | No | NA | 3 | 0 | 1 | Not-determined | Genome-fragment | NA | NA | 0 |
| S13_extracted_144048 | 1692 | No | NA | 4 | 0 | 0 | Not-determined | Genome-fragment | NA | NA | 0 |
| S13_vs_9701\|\|full | 1264 | No | NA | 2 | 0 | 0 | Low-quality | Genome-fragment | 1.08 | AAI-based (medium-confidence) | 0 |
| S13_extracted_144132 | 3181 | No | NA | 5 | 0 | 4 | Not-determined | Genome-fragment | NA | NA | 0 |
| S1_vs_k141_130024\|\|full | 1264 | No | NA | 3 | 0 | 0 | Low-quality | Genome-fragment | 3.47 | AAI-based (high-confidence) | 0 |
| S13_extracted_144367 | 3494 | No | NA | 6 | 0 | 4 | Not-determined | Genome-fragment | NA | NA | 0 |
| S13_extracted_144578 | 1770 | No | NA | 2 | 0 | 0 | Not-determined | Genome-fragment | NA | NA | 0 |
| S13_extracted_144777 | 1583 | No | NA | 1 | 0 | 1 | Not-determined | Genome-fragment | NA | NA | 0 |
| S13_extracted_144902 | 1480 | No | NA | 3 | 0 | 2 | Not-determined | Genome-fragment | NA | NA | 0 |
| S13_extracted_144920 | 9511 | No | NA | 11 | 0 | 3 | Not-determined | Genome-fragment | NA | NA | 0 |
| S1_vs_k141_50975\|\|full | 1264 | No | NA | 5 | 1 | 0 | Low-quality | Genome-fragment | 2.36 | AAI-based (high-confidence) | 0 |
| S13_extracted_145254 | 2069 | No | NA | 3 | 0 | 0 | Not-determined | Genome-fragment | NA | NA | 0 |
| S13_extracted_145421 | 1959 | No | NA | 3 | 0 | 2 | Not-determined | Genome-fragment | NA | NA | 0 |
| S13_extracted_145573 | 2452 | No | NA | 2 | 0 | 1 | Not-determined | Genome-fragment | NA | NA | 0 |
| S13_extracted_145746 | 1119 | No | NA | 2 | 0 | 1 | Not-determined | Genome-fragment | NA | NA | 0 |
| S13_extracted_145859 | 1002 | No | NA | 1 | 0 | 1 | Not-determined | Genome-fragment | NA | NA | 0 |
| S13_extracted_145870 | 1452 | No | NA | 1 | 0 | 1 | Not-determined | Genome-fragment | NA | NA | 0 |
| S13_extracted_145932 | 1380 | No | NA | 1 | 0 | 1 | Not-determined | Genome-fragment | NA | NA | 0 |
| S3_extracted_k141_78558 | 1264 | No | NA | 3 | 1 | 0 | Low-quality | Genome-fragment | 3.16 | AAI-based (medium-confidence) | 0 |
| S13_extracted_146229 | 1708 | No | NA | 2 | 0 | 1 | Not-determined | Genome-fragment | NA | NA | 0 |
| S13_extracted_146385 | 2155 | No | NA | 2 | 0 | 0 | Not-determined | Genome-fragment | NA | NA | 0 |
| S13_extracted_146437 | 1026 | No | NA | 1 | 0 | 1 | Not-determined | Genome-fragment | NA | NA | 0 |
| S13_extracted_146449 | 1003 | No | NA | 1 | 0 | 0 | Not-determined | Genome-fragment | NA | NA | 0 |
| S13_extracted_146453 | 2273 | No | NA | 3 | 0 | 2 | Not-determined | Genome-fragment | NA | NA | 0 |
| S13_extracted_146550 | 1138 | No | NA | 2 | 0 | 0 | Not-determined | Genome-fragment | NA | NA | 0 |
| S13_extracted_146731 | 1505 | No | NA | 2 | 0 | 2 | Not-determined | Genome-fragment | NA | NA | 0 |
| S13_extracted_146803 | 3541 | No | NA | 4 | 0 | 0 | Not-determined | Genome-fragment | NA | NA | 0 |
| S13_extracted_146808 | 1054 | No | NA | 2 | 0 | 0 | Not-determined | Genome-fragment | NA | NA | 0 |
| S13_extracted_146971 | 1005 | No | NA | 2 | 0 | 0 | Not-determined | Genome-fragment | NA | NA | 0 |
| S5_extracted_k141_136897 | 1264 | No | NA | 3 | 0 | 0 | Low-quality | Genome-fragment | 2.96 | AAI-based (medium-confidence) | 0 |
| S13_extracted_147094 | 1268 | No | NA | 1 | 0 | 0 | Not-determined | Genome-fragment | NA | NA | 0 |
| S7_vs_68218\|\|full | 1264 | No | NA | 3 | 0 | 0 | Low-quality | Genome-fragment | 3.26 | AAI-based (medium-confidence) | 0 |
| S7_vs_63225\|\|lt2gene | 1264 | No | NA | 1 | 1 | 0 | Low-quality | Genome-fragment | 2.11 | HMM-based (lower-bound) | 0 |
| S13_extracted_147343 | 1182 | No | NA | 1 | 0 | 1 | Not-determined | Genome-fragment | NA | NA | 0 |
| S13_extracted_147385 | 3535 | No | NA | 7 | 0 | 0 | Not-determined | Genome-fragment | NA | NA | 0 |
| S13_extracted_147396 | 35175 | No | NA | 41 | 13 | 0 | Complete | High-quality | 100 | DTR (high-confidence) | 0 |
| S13_extracted_147469 | 1249 | No | NA | 1 | 0 | 1 | Not-determined | Genome-fragment | NA | NA | 0 |
| S16_extracted_144183 | 1264 | No | NA | 2 | 1 | 0 | Low-quality | Genome-fragment | 3.1 | AAI-based (high-confidence) | 0 |
| S16_vs_91731\|\|full | 1264 | No | NA | 3 | 0 | 0 | Low-quality | Genome-fragment | 2.09 | AAI-based (medium-confidence) | 0 |
| S13_extracted_147589 | 11817 | No | NA | 17 | 2 | 1 | Complete | High-quality | 100 | DTR (high-confidence) | 0 |
| S13_extracted_147593 | 1755 | No | NA | 4 | 0 | 0 | Not-determined | Genome-fragment | NA | NA | 0 |
| S13_extracted_147649 | 2010 | No | NA | 1 | 0 | 1 | Not-determined | Genome-fragment | NA | NA | 0 |
| S13_extracted_147660 | 1048 | No | NA | 2 | 0 | 1 | Not-determined | Genome-fragment | NA | NA | 0 |
| S13_extracted_147900 | 3661 | No | NA | 1 | 0 | 0 | Not-determined | Genome-fragment | NA | NA | 0 |
| S13_extracted_148054 | 3081 | No | NA | 2 | 0 | 0 | Not-determined | Genome-fragment | NA | NA | 0 |
| S16_vs_139864\|\|full | 1264 | No | NA | 3 | 0 | 0 | Low-quality | Genome-fragment | 0.32 | AAI-based (medium-confidence) | 0 |
| S13_extracted_148190 | 1009 | No | NA | 1 | 0 | 1 | Not-determined | Genome-fragment | NA | NA | 0 |
| S16_vs_45769\|\|lt2gene | 1264 | No | NA | 1 | 1 | 0 | Low-quality | Genome-fragment | 0.87 | HMM-based (lower-bound) | 0 |
| S13_extracted_148387 | 1006 | No | NA | 1 | 0 | 0 | Not-determined | Genome-fragment | NA | NA | 0 |
| S13_extracted_148631 | 1138 | No | NA | 1 | 0 | 1 | Not-determined | Genome-fragment | NA | NA | 0 |
| S13_extracted_148898 | 1061 | No | NA | 2 | 0 | 2 | Not-determined | Genome-fragment | NA | NA | 0 |
| S13_extracted_148949 | 1074 | No | NA | 2 | 0 | 0 | Not-determined | Genome-fragment | NA | NA | 0 |
| S13_extracted_148978 | 1349 | No | NA | 2 | 0 | 1 | Not-determined | Genome-fragment | NA | NA | 0 |
| S13_extracted_148979 | 1082 | No | NA | 2 | 0 | 0 | Not-determined | Genome-fragment | NA | NA | 0 |
| S12_vs_k141_118784\|\|full | 1264 | No | NA | 4 | 0 | 0 | Low-quality | Genome-fragment | 2.78 | AAI-based (high-confidence) | 0 |
| S13_extracted_149267 | 1437 | No | NA | 1 | 0 | 0 | Not-determined | Genome-fragment | NA | NA | 0 |
| S17_extracted_k141_135066 | 1265 | No | NA | 1 | 0 | 0 | Low-quality | Genome-fragment | 1.89 | AAI-based (medium-confidence) | 0 |
| S13_extracted_149436 | 1027 | No | NA | 2 | 0 | 0 | Not-determined | Genome-fragment | NA | NA | 0 |
| S9_vs_k141_79703\|\|full | 1265 | No | NA | 1 | 0 | 0 | Low-quality | Genome-fragment | 2.53 | AAI-based (high-confidence) | 0 |
| S13_extracted_11748 | 1265 | No | NA | 3 | 1 | 0 | Low-quality | Genome-fragment | 3 | AAI-based (high-confidence) | 0 |
| S13_extracted_149888 | 1192 | No | NA | 2 | 0 | 1 | Not-determined | Genome-fragment | NA | NA | 0 |
| S13_extracted_149903 | 1155 | No | NA | 1 | 0 | 1 | Not-determined | Genome-fragment | NA | NA | 0 |
| S14_extracted_k141_54825 | 1265 | No | NA | 1 | 1 | 0 | Low-quality | Genome-fragment | 0.87 | HMM-based (lower-bound) | 0 |
| S13_extracted_150246 | 1380 | No | NA | 2 | 0 | 1 | Not-determined | Genome-fragment | NA | NA | 0 |
| S14_extracted_k141_76885 | 1265 | No | NA | 2 | 2 | 0 | Low-quality | Genome-fragment | 4.23 | AAI-based (high-confidence) | 0 |
| S13_extracted_150317 | 3751 | No | NA | 2 | 0 | 0 | Not-determined | Genome-fragment | NA | NA | 0 |
| S13_extracted_150407 | 5403 | No | NA | 9 | 0 | 0 | Complete | High-quality | 100 | DTR (high-confidence) | 0 |
| S13_extracted_150506 | 1176 | No | NA | 2 | 0 | 1 | Not-determined | Genome-fragment | NA | NA | 0 |
| S13_extracted_150691 | 1090 | No | NA | 1 | 0 | 1 | Not-determined | Genome-fragment | NA | NA | 0 |
| S13_extracted_150779 | 1615 | No | NA | 2 | 0 | 1 | Not-determined | Genome-fragment | NA | NA | 0 |
| S1_vs_k141_140360\|\|full | 1265 | No | NA | 3 | 0 | 0 | Low-quality | Genome-fragment | 2.14 | AAI-based (high-confidence) | 0 |
| S13_extracted_151045 | 86139 | No | NA | 110 | 17 | 2 | Complete | High-quality | 100 | DTR (high-confidence) | 0 |
| S13_extracted_151086 | 1639 | No | NA | 2 | 0 | 1 | Not-determined | Genome-fragment | NA | NA | 0 |
| S13_extracted_151312 | 1060 | No | NA | 2 | 0 | 0 | Not-determined | Genome-fragment | NA | NA | 0 |
| S13_extracted_151344 | 1368 | No | NA | 1 | 0 | 0 | Not-determined | Genome-fragment | NA | NA | 0 |
| S13_extracted_151589 | 1298 | No | NA | 1 | 0 | 1 | Not-determined | Genome-fragment | NA | NA | 0 |
| S13_extracted_151697 | 5076 | No | NA | 1 | 0 | 0 | Not-determined | Genome-fragment | NA | NA | 0 |
| S13_extracted_151755 | 43394 | No | NA | 50 | 28 | 0 | Complete | High-quality | 100 | DTR (high-confidence) | 0 |
| S13_extracted_151847 | 6800 | No | NA | 4 | 0 | 3 | Not-determined | Genome-fragment | NA | NA | 0 |
| S13_extracted_151901 | 1286 | No | NA | 1 | 0 | 0 | Not-determined | Genome-fragment | NA | NA | 0 |
| S13_extracted_152065 | 1094 | No | NA | 1 | 0 | 1 | Not-determined | Genome-fragment | NA | NA | 0 |
| S3_extracted_k141_121667 | 1265 | No | NA | 3 | 0 | 1 | Low-quality | Genome-fragment | 2.16 | AAI-based (high-confidence) | 0 |
| S3_extracted_k141_2484 | 1265 | No | NA | 1 | 1 | 0 | Low-quality | Genome-fragment | 0.87 | HMM-based (lower-bound) | 0 |
| S13_extracted_152250 | 1056 | No | NA | 2 | 0 | 1 | Not-determined | Genome-fragment | NA | NA | 0 |
| S13_extracted_152340 | 1834 | No | NA | 1 | 0 | 1 | Not-determined | Genome-fragment | NA | NA | 0 |
| S13_extracted_152369 | 1114 | No | NA | 1 | 0 | 1 | Not-determined | Genome-fragment | NA | NA | 0 |
| S13_extracted_152435 | 1256 | No | NA | 1 | 0 | 0 | Not-determined | Genome-fragment | NA | NA | 0 |
| S3_extracted_k141_109451 | 1265 | No | NA | 3 | 0 | 0 | Low-quality | Genome-fragment | 1.02 | AAI-based (medium-confidence) | 0 |
| S13_extracted_152536 | 1752 | No | NA | 2 | 0 | 0 | Not-determined | Genome-fragment | NA | NA | 0 |
| S13_extracted_152747 | 1057 | No | NA | 1 | 0 | 1 | Not-determined | Genome-fragment | NA | NA | 0 |
| S3_extracted_k141_39040 | 1265 | No | NA | 4 | 0 | 0 | Low-quality | Genome-fragment | 1.63 | AAI-based (high-confidence) | 0 |
| S13_extracted_153057 | 3638 | No | NA | 5 | 0 | 2 | Not-determined | Genome-fragment | NA | NA | 0 |
| S13_extracted_153076 | 1003 | No | NA | 1 | 0 | 0 | Not-determined | Genome-fragment | NA | NA | 0 |
| S5_vs_k141_132565\|\|full | 1265 | No | NA | 4 | 0 | 0 | Low-quality | Genome-fragment | 2.58 | AAI-based (medium-confidence) | 0 |
| S13_extracted_153288 | 1190 | No | NA | 2 | 0 | 1 | Not-determined | Genome-fragment | NA | NA | 0 |
| S7_vs_4807\|\|full | 1265 | No | NA | 2 | 0 | 1 | Low-quality | Genome-fragment | 0.71 | AAI-based (medium-confidence) | 0 |
| S13_extracted_153390 | 1145 | No | NA | 2 | 0 | 2 | Not-determined | Genome-fragment | NA | NA | 0 |
| S16_vs_126000\|\|full | 1265 | No | NA | 3 | 0 | 0 | Low-quality | Genome-fragment | 2.27 | AAI-based (high-confidence) | 0 |
| S13_extracted_153526 | 1151 | No | NA | 1 | 0 | 1 | Not-determined | Genome-fragment | NA | NA | 0 |
| S13_extracted_153577 | 1260 | No | NA | 1 | 0 | 0 | Not-determined | Genome-fragment | NA | NA | 0 |
| S13_extracted_153612 | 1133 | No | NA | 2 | 0 | 2 | Not-determined | Genome-fragment | NA | NA | 0 |
| S13_extracted_153622 | 1280 | No | NA | 1 | 0 | 1 | Not-determined | Genome-fragment | NA | NA | 0 |
| S13_extracted_153657 | 1833 | No | NA | 1 | 0 | 1 | Not-determined | Genome-fragment | NA | NA | 0 |
| S10_extracted_113182 | 1265 | No | NA | 3 | 1 | 0 | Low-quality | Genome-fragment | 3.12 | AAI-based (high-confidence) | 0 |
| S13_extracted_153743 | 1174 | No | NA | 3 | 0 | 2 | Not-determined | Genome-fragment | NA | NA | 0 |
| S10_vs_16269\|\|full | 1265 | No | NA | 2 | 0 | 0 | Low-quality | Genome-fragment | 2.74 | AAI-based (medium-confidence) | 0 |
| S13_extracted_154047 | 1362 | No | NA | 2 | 0 | 0 | Not-determined | Genome-fragment | NA | NA | 0 |
| S13_extracted_154130 | 1192 | No | NA | 1 | 0 | 0 | Not-determined | Genome-fragment | NA | NA | 0 |
| S13_extracted_154245 | 1186 | No | NA | 2 | 0 | 0 | Not-determined | Genome-fragment | NA | NA | 0 |
| S13_vs_9029\|\|full | 6727 | No | NA | 12 | 0 | 3 | Not-determined | Genome-fragment | NA | NA | 0 |
| S13_vs_12957\|\|full | 15580 | No | NA | 18 | 8 | 0 | Medium-quality | Genome-fragment | 51 | HMM-based (lower-bound) | 0 |
| S13_vs_15645\|\|full | 153823 | No | NA | 203 | 18 | 4 | High-quality | High-quality | 100 | AAI-based (high-confidence) | 0 |
| S13_vs_27462\|\|full | 65150 | No | NA | 98 | 27 | 4 | Medium-quality | Genome-fragment | 82.87 | AAI-based (high-confidence) | 0 |
| S13_vs_28316\|\|full | 3119 | No | NA | 3 | 0 | 3 | Not-determined | Genome-fragment | NA | NA | 0 |
| S13_vs_148294\|\|full | 43488 | No | NA | 68 | 26 | 2 | High-quality | High-quality | 100 | AAI-based (high-confidence) | 0 |
| S13_vs_149860\|\|full | 19576 | No | NA | 25 | 8 | 0 | High-quality | High-quality | 100 | AAI-based (medium-confidence) | 0 |
| S13_vs_150477\|\|full | 8781 | No | NA | 9 | 0 | 3 | Not-determined | Genome-fragment | NA | NA | 0 |
| S13_vs_151510\|\|full | 97768 | No | NA | 110 | 22 | 0 | High-quality | High-quality | 98.51 | AAI-based (high-confidence) | 0 |
| S6_vs_k141_31212\|\|full | 1265 | No | NA | 3 | 1 | 0 | Low-quality | Genome-fragment | 2.06 | HMM-based (lower-bound) | 0 |
| S13_vs_152\|\|full | 1242 | No | NA | 2 | 0 | 2 | Not-determined | Genome-fragment | NA | NA | 0 |
| S17_extracted_k141_34243 | 1266 | No | NA | 4 | 4 | 0 | Low-quality | Genome-fragment | 4.16 | AAI-based (high-confidence) | 0 |
| S13_vs_502\|\|full | 1557 | No | NA | 4 | 0 | 0 | Not-determined | Genome-fragment | NA | NA | 0 |
| S13_vs_587\|\|full | 1337 | No | NA | 3 | 0 | 1 | Not-determined | Genome-fragment | NA | NA | 0 |
| S13_vs_702\|\|full | 2780 | No | NA | 6 | 0 | 0 | Not-determined | Genome-fragment | NA | NA | 0 |
| S13_vs_763\|\|full | 2724 | No | NA | 4 | 0 | 0 | Not-determined | Genome-fragment | NA | NA | 0 |
| S13_vs_837\|\|full | 2915 | No | NA | 2 | 0 | 1 | Not-determined | Genome-fragment | NA | NA | 0 |
| S13_vs_845\|\|full | 3137 | No | NA | 4 | 0 | 4 | Not-determined | Genome-fragment | NA | NA | 0 |
| S13_vs_852\|\|full | 1813 | No | NA | 4 | 0 | 1 | Not-determined | Genome-fragment | NA | NA | 0 |
| S13_vs_1092\|\|full | 3269 | No | NA | 3 | 0 | 1 | Not-determined | Genome-fragment | NA | NA | 0 |
| S13_vs_1200\|\|full | 4539 | No | NA | 5 | 0 | 0 | Not-determined | Genome-fragment | NA | NA | 0 |
| S13_vs_1676\|\|full | 6275 | No | NA | 6 | 0 | 4 | Not-determined | Genome-fragment | NA | NA | 0 |
| S17_vs_k141_252\|\|full | 1266 | No | NA | 3 | 0 | 0 | Low-quality | Genome-fragment | 2.79 | AAI-based (medium-confidence) | 0 |
| S13_vs_2215\|\|full | 1171 | No | NA | 3 | 0 | 0 | Not-determined | Genome-fragment | NA | NA | 0 |
| S13_vs_2276\|\|full | 1216 | No | NA | 3 | 0 | 0 | Not-determined | Genome-fragment | NA | NA | 0 |
| S13_vs_2380\|\|full | 1505 | No | NA | 3 | 0 | 1 | Not-determined | Genome-fragment | NA | NA | 0 |
| S13_vs_2416\|\|full | 2071 | No | NA | 2 | 0 | 1 | Not-determined | Genome-fragment | NA | NA | 0 |
| S9_vs_k141_32504\|\|full | 1266 | No | NA | 5 | 0 | 0 | Low-quality | Genome-fragment | 2.79 | AAI-based (high-confidence) | 0 |
| S11_vs_k141_1000\|\|full | 1266 | No | NA | 3 | 0 | 0 | Low-quality | Genome-fragment | 2.49 | AAI-based (high-confidence) | 0 |
| S3_extracted_k141_109899 | 1266 | No | NA | 2 | 1 | 0 | Low-quality | Genome-fragment | 1.5 | AAI-based (high-confidence) | 0 |
| S13_vs_2841\|\|full | 2224 | No | NA | 3 | 0 | 0 | Not-determined | Genome-fragment | NA | NA | 0 |
| S3_extracted_k141_38782 | 1266 | No | NA | 2 | 0 | 0 | Low-quality | Genome-fragment | 2.11 | AAI-based (high-confidence) | 0 |
| S13_vs_3423\|\|full | 2061 | No | NA | 3 | 0 | 0 | Not-determined | Genome-fragment | NA | NA | 0 |
| S13_vs_3488\|\|full | 1676 | No | NA | 3 | 0 | 1 | Not-determined | Genome-fragment | NA | NA | 0 |
| S5_extracted_k141_132812 | 1266 | No | NA | 3 | 0 | 1 | Low-quality | Genome-fragment | 2.75 | AAI-based (high-confidence) | 0 |
| S13_vs_3666\|\|full | 1569 | No | NA | 3 | 0 | 1 | Not-determined | Genome-fragment | NA | NA | 0 |
| S13_vs_3823\|\|full | 1310 | No | NA | 2 | 0 | 1 | Not-determined | Genome-fragment | NA | NA | 0 |
| S13_vs_3899\|\|full | 1676 | No | NA | 4 | 0 | 2 | Not-determined | Genome-fragment | NA | NA | 0 |
| S13_vs_3910\|\|full | 71167 | No | NA | 76 | 0 | 18 | Not-determined | Genome-fragment | NA | NA | 0 |
| S7_vs_8492\|\|full | 1266 | No | NA | 3 | 2 | 0 | Low-quality | Genome-fragment | 1.73 | HMM-based (lower-bound) | 0 |
| S7_vs_40647\|\|lt2gene | 1266 | Yes | 660 | 2 | 1 | 1 | Low-quality | Genome-fragment | 0.31 | AAI-based (medium-confidence) | 47.87 |
| S10_extracted_44117 | 1266 | No | NA | 1 | 0 | 0 | Low-quality | Genome-fragment | 1.51 | AAI-based (medium-confidence) | 0 |
| S13_vs_4534\|\|full | 2588 | No | NA | 3 | 0 | 1 | Not-determined | Genome-fragment | NA | NA | 0 |
| S12_extracted_k141_3019 | 1266 | No | NA | 2 | 1 | 0 | Low-quality | Genome-fragment | 2.83 | AAI-based (medium-confidence) | 0 |
| S13_vs_4729\|\|full | 2577 | No | NA | 4 | 0 | 1 | Not-determined | Genome-fragment | NA | NA | 0 |
| S13_vs_4837\|\|full | 5053 | No | NA | 9 | 0 | 2 | Not-determined | Genome-fragment | NA | NA | 0 |
| S13_vs_4910\|\|full | 2336 | No | NA | 4 | 0 | 2 | Not-determined | Genome-fragment | NA | NA | 0 |
| S13_vs_4968\|\|full | 1120 | No | NA | 2 | 0 | 1 | Not-determined | Genome-fragment | NA | NA | 0 |
| S2_extracted_k141_6638 | 1266 | No | NA | 2 | 0 | 0 | Low-quality | Genome-fragment | 1.25 | AAI-based (high-confidence) | 0 |
| S13_vs_5336\|\|full | 2313 | No | NA | 4 | 0 | 2 | Not-determined | Genome-fragment | NA | NA | 0 |
| S13_vs_5350\|\|full | 2473 | No | NA | 3 | 0 | 1 | Not-determined | Genome-fragment | NA | NA | 0 |
| S13_vs_5352\|\|full | 1290 | No | NA | 3 | 0 | 0 | Not-determined | Genome-fragment | NA | NA | 0 |
| S13_vs_5404\|\|full | 1243 | No | NA | 2 | 0 | 2 | Not-determined | Genome-fragment | NA | NA | 0 |
| S8_extracted_k141_1829 | 1266 | No | NA | 2 | 0 | 1 | Low-quality | Genome-fragment | 2.1 | AAI-based (medium-confidence) | 0 |
| S8_extracted_k141_182051 | 1266 | No | NA | 1 | 1 | 0 | Low-quality | Genome-fragment | 0.87 | HMM-based (lower-bound) | 0 |
| S17_vs_k141_160787\|\|full | 1267 | No | NA | 3 | 0 | 0 | Low-quality | Genome-fragment | 2.53 | AAI-based (medium-confidence) | 0 |
| S13_vs_5898\|\|full | 6773 | No | NA | 12 | 0 | 2 | Not-determined | Genome-fragment | NA | NA | 0 |
| S9_extracted_k141_53222 | 1267 | No | NA | 3 | 3 | 0 | Low-quality | Genome-fragment | 4.18 | AAI-based (high-confidence) | 0 |
| S11_extracted_k141_8934 | 1267 | No | NA | 2 | 1 | 0 | Low-quality | Genome-fragment | 2.51 | AAI-based (high-confidence) | 0 |
| S13_vs_6287\|\|full | 2182 | No | NA | 4 | 0 | 2 | Not-determined | Genome-fragment | NA | NA | 0 |
| S1_extracted_k141_154107 | 1267 | No | NA | 2 | 0 | 0 | Low-quality | Genome-fragment | 2.52 | AAI-based (high-confidence) | 0 |
| S13_vs_6366\|\|full | 1734 | No | NA | 3 | 0 | 3 | Not-determined | Genome-fragment | NA | NA | 0 |
| S13_vs_6385\|\|full | 1870 | No | NA | 2 | 0 | 1 | Not-determined | Genome-fragment | NA | NA | 0 |
| S5_vs_k141_55972\|\|full | 1267 | No | NA | 3 | 0 | 1 | Low-quality | Genome-fragment | 1.89 | AAI-based (medium-confidence) | 0 |
| S13_vs_6558\|\|full | 1757 | No | NA | 3 | 0 | 1 | Not-determined | Genome-fragment | NA | NA | 0 |
| S13_vs_6769\|\|full | 1674 | No | NA | 2 | 0 | 1 | Not-determined | Genome-fragment | NA | NA | 0 |
| S13_vs_6794\|\|full | 6292 | No | NA | 9 | 0 | 5 | Not-determined | Genome-fragment | NA | NA | 0 |
| S7_vs_13386\|\|full | 1267 | No | NA | 2 | 1 | 0 | Low-quality | Genome-fragment | 2.36 | AAI-based (medium-confidence) | 0 |
| S10_extracted_1275 | 1267 | No | NA | 1 | 1 | 0 | Low-quality | Genome-fragment | 7.05 | AAI-based (high-confidence) | 0 |
| S13_vs_7574\|\|full | 4813 | No | NA | 7 | 0 | 2 | Not-determined | Genome-fragment | NA | NA | 0 |
| S13_vs_7581\|\|full | 1471 | No | NA | 2 | 0 | 0 | Not-determined | Genome-fragment | NA | NA | 0 |
| S13_vs_7659\|\|full | 2718 | No | NA | 3 | 0 | 0 | Not-determined | Genome-fragment | NA | NA | 0 |
| S10_vs_29865\|\|full | 1267 | No | NA | 3 | 1 | 0 | Low-quality | Genome-fragment | 1.82 | HMM-based (lower-bound) | 0 |
| S13_vs_7762\|\|full | 1117 | No | NA | 2 | 0 | 1 | Not-determined | Genome-fragment | NA | NA | 0 |
| S13_vs_7863\|\|full | 1658 | No | NA | 2 | 0 | 0 | Not-determined | Genome-fragment | NA | NA | 0 |
| S15_vs_14821\|\|lt2gene | 1267 | No | NA | 2 | 2 | 0 | Low-quality | Genome-fragment | 3.36 | AAI-based (high-confidence) | 0 |
| S13_vs_7911\|\|full | 1196 | No | NA | 4 | 0 | 0 | Not-determined | Genome-fragment | NA | NA | 0 |
| S13_vs_7963\|\|full | 3195 | No | NA | 4 | 0 | 1 | Not-determined | Genome-fragment | NA | NA | 0 |
| S13_vs_7970\|\|full | 1100 | No | NA | 2 | 0 | 0 | Not-determined | Genome-fragment | NA | NA | 0 |
| S13_vs_8070\|\|full | 3385 | No | NA | 4 | 0 | 1 | Not-determined | Genome-fragment | NA | NA | 0 |
| S8_vs_k141_48567\|\|full | 1267 | No | NA | 3 | 1 | 0 | Low-quality | Genome-fragment | 4.34 | HMM-based (lower-bound) | 0 |
| S13_vs_8259\|\|full | 1671 | No | NA | 4 | 0 | 1 | Not-determined | Genome-fragment | NA | NA | 0 |
| S13_vs_8262\|\|full | 5396 | No | NA | 3 | 0 | 1 | Not-determined | Genome-fragment | NA | NA | 0 |
| S17_vs_k141_84245\|\|full | 1268 | No | NA | 5 | 2 | 0 | Low-quality | Genome-fragment | 2.26 | AAI-based (medium-confidence) | 0 |
| S13_extracted_94955 | 1268 | No | NA | 2 | 0 | 0 | Low-quality | Genome-fragment | 2.38 | AAI-based (medium-confidence) | 0 |
| S14_extracted_k141_66485 | 1268 | No | NA | 4 | 4 | 0 | Low-quality | Genome-fragment | 4.18 | AAI-based (high-confidence) | 0 |
| S13_vs_8785\|\|full | 1604 | No | NA | 2 | 0 | 0 | Not-determined | Genome-fragment | NA | NA | 0 |
| S13_vs_8788\|\|full | 1256 | No | NA | 2 | 0 | 2 | Not-determined | Genome-fragment | NA | NA | 0 |
| S13_vs_8817\|\|full | 1618 | No | NA | 2 | 0 | 1 | Not-determined | Genome-fragment | NA | NA | 0 |
| S14_vs_k141_15080\|\|full | 1268 | No | NA | 2 | 0 | 0 | Low-quality | Genome-fragment | 0.75 | AAI-based (medium-confidence) | 0 |
| S1_vs_k141_18524\|\|full | 1268 | No | NA | 2 | 0 | 0 | Low-quality | Genome-fragment | 0.72 | AAI-based (medium-confidence) | 0 |
| S13_vs_9055\|\|full | 4684 | No | NA | 4 | 0 | 0 | Not-determined | Genome-fragment | NA | NA | 0 |
| S13_vs_9066\|\|full | 2726 | No | NA | 4 | 0 | 0 | Not-determined | Genome-fragment | NA | NA | 0 |
| S13_vs_9438\|\|full | 32269 | Yes | 27620 | 38 | 8 | 3 | Medium-quality | Genome-fragment | 68.61 | AAI-based (high-confidence) | 14.41 |
| S13_vs_9631\|\|full | 2059 | No | NA | 3 | 0 | 2 | Not-determined | Genome-fragment | NA | NA | 0 |
| S13_vs_9653\|\|full | 2171 | No | NA | 3 | 0 | 2 | Not-determined | Genome-fragment | NA | NA | 0 |
| S3_extracted_k141_60767 | 1268 | No | NA | 4 | 0 | 0 | Low-quality | Genome-fragment | 2.91 | AAI-based (high-confidence) | 0 |
| S13_vs_9751\|\|full | 2512 | No | NA | 3 | 0 | 0 | Not-determined | Genome-fragment | NA | NA | 0 |
| S13_vs_9791\|\|full | 2136 | No | NA | 5 | 0 | 2 | Not-determined | Genome-fragment | NA | NA | 0 |
| S5_vs_k141_34111\|\|full | 1268 | No | NA | 3 | 0 | 1 | Low-quality | Genome-fragment | 2.19 | AAI-based (medium-confidence) | 0 |
| S13_vs_9896\|\|full | 2767 | No | NA | 3 | 0 | 3 | Not-determined | Genome-fragment | NA | NA | 0 |
| S13_vs_10001\|\|full | 1313 | No | NA | 4 | 0 | 1 | Not-determined | Genome-fragment | NA | NA | 0 |
| S13_vs_10334\|\|full | 1585 | No | NA | 2 | 0 | 1 | Not-determined | Genome-fragment | NA | NA | 0 |
| S7_vs_101554\|\|full | 1268 | No | NA | 3 | 0 | 0 | Low-quality | Genome-fragment | 2.61 | AAI-based (medium-confidence) | 0 |
| S13_vs_10518\|\|full | 1370 | No | NA | 3 | 0 | 0 | Not-determined | Genome-fragment | NA | NA | 0 |
| S13_vs_10995\|\|full | 1638 | No | NA | 3 | 0 | 1 | Not-determined | Genome-fragment | NA | NA | 0 |
| S13_vs_11099\|\|full | 1683 | No | NA | 3 | 0 | 1 | Not-determined | Genome-fragment | NA | NA | 0 |
| S16_extracted_109420 | 1268 | No | NA | 5 | 0 | 0 | Low-quality | Genome-fragment | 0.61 | AAI-based (medium-confidence) | 0 |
| S13_vs_11653\|\|full | 1935 | No | NA | 2 | 0 | 1 | Not-determined | Genome-fragment | NA | NA | 0 |
| S16_vs_97611\|\|full | 1268 | No | NA | 2 | 0 | 1 | Low-quality | Genome-fragment | 3.5 | AAI-based (high-confidence) | 0 |
| S13_vs_11809\|\|full | 1258 | No | NA | 3 | 0 | 0 | Not-determined | Genome-fragment | NA | NA | 0 |
| S13_vs_11917\|\|full | 1280 | No | NA | 2 | 0 | 1 | Not-determined | Genome-fragment | NA | NA | 0 |
| S13_vs_12015\|\|full | 3839 | No | NA | 3 | 0 | 3 | Not-determined | Genome-fragment | NA | NA | 0 |
| S13_vs_12037\|\|full | 1234 | No | NA | 3 | 0 | 2 | Not-determined | Genome-fragment | NA | NA | 0 |
| S13_vs_12198\|\|full | 2353 | No | NA | 4 | 0 | 2 | Not-determined | Genome-fragment | NA | NA | 0 |
| S13_vs_12215\|\|full | 1148 | No | NA | 2 | 0 | 0 | Not-determined | Genome-fragment | NA | NA | 0 |
| S12_extracted_k141_92007 | 1268 | No | NA | 1 | 0 | 1 | Low-quality | Genome-fragment | 0.91 | AAI-based (high-confidence) | 0 |
| S12_vs_k141_5711\|\|full | 1268 | No | NA | 3 | 1 | 0 | Low-quality | Genome-fragment | 3.21 | AAI-based (medium-confidence) | 0 |
| S13_vs_12406\|\|full | 1870 | No | NA | 3 | 0 | 0 | Not-determined | Genome-fragment | NA | NA | 0 |
| S13_vs_12724\|\|full | 3972 | No | NA | 6 | 0 | 1 | Not-determined | Genome-fragment | NA | NA | 0 |
| S12_vs_k141_163011\|\|full | 1268 | No | NA | 4 | 0 | 0 | Low-quality | Genome-fragment | 2.35 | AAI-based (high-confidence) | 0 |
| S13_vs_12867\|\|full | 1608 | No | NA | 4 | 0 | 1 | Not-determined | Genome-fragment | NA | NA | 0 |
| S6_vs_k141_24085\|\|lt2gene | 1268 | No | NA | 1 | 1 | 0 | Low-quality | Genome-fragment | 3.25 | AAI-based (high-confidence) | 0 |
| S8_extracted_k141_124153 | 1268 | No | NA | 3 | 0 | 0 | Low-quality | Genome-fragment | 1.92 | AAI-based (high-confidence) | 0 |
| S13_vs_13129\|\|full | 34076 | No | NA | 51 | 13 | 1 | High-quality | High-quality | 99.76 | AAI-based (high-confidence) | 0 |
| S13_vs_13498\|\|full | 1021 | No | NA | 2 | 0 | 2 | Not-determined | Genome-fragment | NA | NA | 0 |
| S11_vs_k141_19919\|\|full | 1269 | No | NA | 4 | 1 | 0 | Low-quality | Genome-fragment | 1.07 | AAI-based (medium-confidence) | 0 |
| S13_vs_13710\|\|full | 1196 | No | NA | 4 | 0 | 1 | Not-determined | Genome-fragment | NA | NA | 0 |
| S13_vs_13825\|\|full | 4706 | No | NA | 5 | 0 | 1 | Not-determined | Genome-fragment | NA | NA | 0 |
| S11_vs_k141_26580\|\|full | 1269 | No | NA | 3 | 0 | 0 | Low-quality | Genome-fragment | 3.35 | AAI-based (high-confidence) | 0 |
| S13_vs_14086\|\|full | 1467 | No | NA | 2 | 0 | 1 | Not-determined | Genome-fragment | NA | NA | 0 |
| S13_vs_14117\|\|full | 3311 | No | NA | 3 | 0 | 0 | Not-determined | Genome-fragment | NA | NA | 0 |
| S13_vs_14207\|\|full | 1665 | No | NA | 2 | 0 | 0 | Not-determined | Genome-fragment | NA | NA | 0 |
| S13_vs_88285\|\|lt2gene | 1269 | No | NA | 2 | 2 | 0 | Low-quality | Genome-fragment | 27.17 | AAI-based (high-confidence) | 0 |
| S13_vs_14701\|\|full | 2400 | No | NA | 5 | 0 | 0 | Not-determined | Genome-fragment | NA | NA | 0 |
| S13_vs_14774\|\|full | 8676 | No | NA | 11 | 0 | 1 | Not-determined | Genome-fragment | NA | NA | 0 |
| S13_vs_14794\|\|full | 36336 | No | NA | 34 | 1 | 0 | Medium-quality | Genome-fragment | 88.55 | AAI-based (high-confidence) | 0 |
| S7_vs_7907\|\|lt2gene | 1269 | No | NA | 2 | 1 | 0 | Low-quality | Genome-fragment | 2.65 | AAI-based (high-confidence) | 0 |
| S13_vs_14981\|\|full | 3969 | No | NA | 4 | 0 | 2 | Not-determined | Genome-fragment | NA | NA | 0 |
| S16_extracted_6210 | 1269 | No | NA | 2 | 0 | 0 | Low-quality | Genome-fragment | 1.54 | AAI-based (medium-confidence) | 0 |
| S12_vs_k141_130005\|\|full | 1269 | No | NA | 2 | 0 | 0 | Low-quality | Genome-fragment | 0.83 | AAI-based (medium-confidence) | 0 |
| S13_vs_15162\|\|full | 1392 | No | NA | 3 | 0 | 0 | Not-determined | Genome-fragment | NA | NA | 0 |
| S6_vs_k141_22443\|\|full | 1269 | No | NA | 2 | 0 | 0 | Low-quality | Genome-fragment | 0.81 | AAI-based (medium-confidence) | 0 |
| S13_vs_15459\|\|full | 1287 | No | NA | 2 | 0 | 0 | Not-determined | Genome-fragment | NA | NA | 0 |
| S13_vs_15502\|\|full | 4554 | No | NA | 7 | 0 | 4 | Not-determined | Genome-fragment | NA | NA | 0 |
| S13_vs_15781\|\|full | 1119 | No | NA | 2 | 0 | 2 | Not-determined | Genome-fragment | NA | NA | 0 |
| S13_vs_15802\|\|full | 2613 | No | NA | 2 | 0 | 2 | Not-determined | Genome-fragment | NA | NA | 0 |
| S13_vs_16063\|\|full | 1371 | No | NA | 2 | 0 | 2 | Not-determined | Genome-fragment | NA | NA | 0 |
| S13_vs_16168\|\|full | 8163 | No | NA | 11 | 0 | 1 | Not-determined | Genome-fragment | NA | NA | 0 |
| S17_vs_k141_44916\|\|full | 1270 | No | NA | 3 | 1 | 0 | Low-quality | Genome-fragment | 1.9 | AAI-based (high-confidence) | 0 |
| S13_vs_16282\|\|full | 1653 | No | NA | 3 | 0 | 1 | Not-determined | Genome-fragment | NA | NA | 0 |
| S17_vs_k141_42127\|\|full | 1270 | No | NA | 4 | 0 | 0 | Low-quality | Genome-fragment | 1.84 | AAI-based (high-confidence) | 0 |
| S9_extracted_k141_41203 | 1270 | No | NA | 3 | 2 | 0 | Low-quality | Genome-fragment | 4.17 | AAI-based (high-confidence) | 0 |
| S13_vs_16572\|\|full | 1519 | No | NA | 3 | 0 | 0 | Not-determined | Genome-fragment | NA | NA | 0 |
| S11_vs_k141_72248\|\|full | 1270 | No | NA | 3 | 0 | 0 | Low-quality | Genome-fragment | 1.52 | AAI-based (high-confidence) | 0 |
| S14_extracted_k141_108111 | 1270 | No | NA | 1 | 1 | 0 | Low-quality | Genome-fragment | 2.78 | AAI-based (high-confidence) | 0 |
| S3_extracted_k141_33605 | 1270 | No | NA | 4 | 0 | 0 | Low-quality | Genome-fragment | 1.22 | AAI-based (high-confidence) | 0 |
| S3_extracted_k141_105613 | 1270 | No | NA | 5 | 2 | 0 | Low-quality | Genome-fragment | 2.87 | AAI-based (medium-confidence) | 0 |
| S3_vs_k141_101610\|\|full | 1270 | No | NA | 3 | 1 | 0 | Low-quality | Genome-fragment | 1.84 | AAI-based (medium-confidence) | 0 |
| S13_vs_17289\|\|full | 4552 | No | NA | 9 | 0 | 0 | Not-determined | Genome-fragment | NA | NA | 0 |
| S3_vs_k141_110559\|\|lt2gene | 1270 | No | NA | 2 | 0 | 1 | Low-quality | Genome-fragment | 3.62 | AAI-based (high-confidence) | 0 |
| S7_vs_91455\|\|full | 1270 | No | NA | 2 | 0 | 0 | Low-quality | Genome-fragment | 2.44 | AAI-based (high-confidence) | 0 |
| S13_vs_17713\|\|full | 1909 | No | NA | 4 | 0 | 0 | Not-determined | Genome-fragment | NA | NA | 0 |
| S16_extracted_63446 | 1270 | No | NA | 3 | 1 | 0 | Low-quality | Genome-fragment | 2.89 | AAI-based (medium-confidence) | 0 |
| S13_vs_18143\|\|full | 1296 | No | NA | 2 | 0 | 1 | Not-determined | Genome-fragment | NA | NA | 0 |
| S13_vs_18161\|\|full | 1236 | No | NA | 2 | 0 | 0 | Not-determined | Genome-fragment | NA | NA | 0 |
| S16_extracted_113782 | 1270 | No | NA | 2 | 0 | 2 | Low-quality | Genome-fragment | 3.32 | AAI-based (medium-confidence) | 0 |
| S13_vs_18217\|\|full | 1330 | No | NA | 2 | 0 | 0 | Not-determined | Genome-fragment | NA | NA | 0 |
| S13_vs_18318\|\|full | 1932 | No | NA | 3 | 0 | 0 | Not-determined | Genome-fragment | NA | NA | 0 |
| S13_vs_18429\|\|full | 5542 | No | NA | 6 | 0 | 1 | Not-determined | Genome-fragment | NA | NA | 0 |
| S13_vs_18535\|\|full | 1503 | No | NA | 4 | 0 | 2 | Not-determined | Genome-fragment | NA | NA | 0 |
| S10_vs_68477\|\|full | 1270 | No | NA | 2 | 0 | 1 | Low-quality | Genome-fragment | 0.94 | AAI-based (high-confidence) | 0 |
| S13_vs_18613\|\|full | 4301 | No | NA | 6 | 0 | 2 | Not-determined | Genome-fragment | NA | NA | 0 |
| S13_vs_18693\|\|full | 3367 | No | NA | 6 | 0 | 0 | Not-determined | Genome-fragment | NA | NA | 0 |
| S13_vs_18744\|\|full | 1399 | No | NA | 4 | 0 | 0 | Not-determined | Genome-fragment | NA | NA | 0 |
| S6_vs_k141_54578\|\|full | 1270 | No | NA | 2 | 0 | 0 | Low-quality | Genome-fragment | 1.71 | AAI-based (high-confidence) | 0 |
| S6_vs_k141_53968\|\|lt2gene | 1270 | No | NA | 2 | 2 | 0 | Low-quality | Genome-fragment | 1.82 | HMM-based (lower-bound) | 0 |
| S13_vs_19256\|\|full | 1004 | No | NA | 2 | 0 | 1 | Not-determined | Genome-fragment | NA | NA | 0 |
| S9_extracted_k141_57744 | 1271 | No | NA | 2 | 0 | 0 | Low-quality | Genome-fragment | 4.32 | AAI-based (high-confidence) | 0 |
| S9_vs_k141_45526\|\|full | 1271 | No | NA | 4 | 4 | 0 | Low-quality | Genome-fragment | 3.09 | AAI-based (medium-confidence) | 0 |
| S11_vs_k141_5456\|\|full | 1271 | No | NA | 2 | 0 | 1 | Low-quality | Genome-fragment | 3.49 | AAI-based (high-confidence) | 0 |
| S13_vs_19692\|\|full | 3107 | No | NA | 4 | 0 | 3 | Not-determined | Genome-fragment | NA | NA | 0 |
| S14_extracted_k141_114032 | 1271 | No | NA | 1 | 0 | 1 | Low-quality | Genome-fragment | 0.69 | AAI-based (medium-confidence) | 0 |
| S13_vs_19784\|\|full | 1529 | No | NA | 2 | 0 | 2 | Not-determined | Genome-fragment | NA | NA | 0 |
| S13_vs_19808\|\|full | 35526 | No | NA | 31 | 0 | 22 | Not-determined | Genome-fragment | NA | NA | 0 |
| S13_vs_19886\|\|full | 1874 | No | NA | 3 | 0 | 1 | Not-determined | Genome-fragment | NA | NA | 0 |
| S13_vs_19953\|\|full | 1895 | No | NA | 3 | 0 | 2 | Not-determined | Genome-fragment | NA | NA | 0 |
| S13_vs_20101\|\|full | 2539 | No | NA | 4 | 0 | 3 | Not-determined | Genome-fragment | NA | NA | 0 |
| S14_vs_k141_2009\|\|full | 1271 | No | NA | 3 | 0 | 0 | Low-quality | Genome-fragment | 2.94 | AAI-based (medium-confidence) | 0 |
| S13_vs_20167\|\|full | 4851 | No | NA | 7 | 0 | 4 | Not-determined | Genome-fragment | NA | NA | 0 |

| | | | | | | | | | | | |
|---|---|---|---|---|---|---|---|---|---|---|---|
| S13_vs_20171\|\|full | 1095 | No | NA | 3 | 0 | 0 | Not-determined | Genome-fragment | NA | NA | 0 |
| S1_extracted_k141_8547 | 1271 | No | NA | 1 | 0 | 1 | Low-quality | Genome-fragment | 0.84 | AAI-based (high-confidence) | 0 |
| S13_vs_20301\|\|full | 3358 | No | NA | 3 | 0 | 2 | Not-determined | Genome-fragment | NA | NA | 0 |
| S5_extracted_k141_19161 | 1271 | No | NA | 3 | 0 | 0 | Low-quality | Genome-fragment | 1.61 | AAI-based (high-confidence) | 0 |
| S13_vs_20790\|\|full | 1686 | No | NA | 3 | 0 | 2 | Not-determined | Genome-fragment | NA | NA | 0 |
| S13_vs_21614\|\|full | 1892 | No | NA | 3 | 0 | 1 | Not-determined | Genome-fragment | NA | NA | 0 |
| S13_vs_21657\|\|full | 2544 | No | NA | 4 | 0 | 0 | Not-determined | Genome-fragment | NA | NA | 0 |
| S5_vs_k141_44843\|\|full | 1271 | No | NA | 2 | 0 | 0 | Low-quality | Genome-fragment | 0.38 | AAI-based (medium-confidence) | 0 |
| S13_vs_21844\|\|full | 1085 | No | NA | 4 | 0 | 0 | Not-determined | Genome-fragment | NA | NA | 0 |
| S13_vs_21924\|\|full | 4702 | No | NA | 5 | 0 | 3 | Not-determined | Genome-fragment | NA | NA | 0 |
| S13_vs_22455\|\|full | 1831 | No | NA | 3 | 0 | 0 | Not-determined | Genome-fragment | NA | NA | 0 |
| S13_vs_22476\|\|full | 2178 | No | NA | 3 | 0 | 0 | Not-determined | Genome-fragment | NA | NA | 0 |
| S5_vs_k141_9038\|\|full | 1271 | No | NA | 2 | 0 | 0 | Low-quality | Genome-fragment | 0.99 | AAI-based (high-confidence) | 0 |
| S13_vs_22748\|\|full | 1645 | No | NA | 2 | 0 | 1 | Not-determined | Genome-fragment | NA | NA | 0 |
| S7_extracted_17219 | 1271 | No | NA | 2 | 2 | 0 | Low-quality | Genome-fragment | 2.86 | AAI-based (medium-confidence) | 0 |
| S13_vs_22995\|\|full | 1520 | No | NA | 3 | 0 | 2 | Not-determined | Genome-fragment | NA | NA | 0 |
| S13_vs_23014\|\|full | 2064 | No | NA | 2 | 0 | 2 | Not-determined | Genome-fragment | NA | NA | 0 |
| S13_vs_23211\|\|full | 1416 | No | NA | 3 | 0 | 1 | Not-determined | Genome-fragment | NA | NA | 0 |
| S13_vs_23314\|\|full | 52988 | Yes | 45151 | 63 | 25 | 7 | High-quality | High-quality | 95.28 | AAI-based (high-confidence) | 14.79 |
| S7_extracted_42931 | 1271 | No | NA | 2 | 0 | 0 | Low-quality | Genome-fragment | 3.04 | AAI-based (high-confidence) | 0 |
| S7_vs_88727\|\|full | 1271 | No | NA | 5 | 2 | 0 | Low-quality | Genome-fragment | 2.59 | AAI-based (medium-confidence) | 0 |
| S13_vs_23467\|\|full | 1353 | No | NA | 2 | 0 | 0 | Not-determined | Genome-fragment | NA | NA | 0 |
| S7_vs_98924\|\|full | 1271 | No | NA | 2 | 0 | 0 | Low-quality | Genome-fragment | 2.89 | AAI-based (medium-confidence) | 0 |
| S7_vs_106000\|\|lt2gene | 1271 | No | NA | 2 | 2 | 0 | Low-quality | Genome-fragment | 2.61 | AAI-based (medium-confidence) | 0 |
| S13_vs_24002\|\|full | 2185 | No | NA | 3 | 0 | 1 | Not-determined | Genome-fragment | NA | NA | 0 |
| S16_extracted_79390 | 1271 | No | NA | 5 | 4 | 0 | Low-quality | Genome-fragment | 4.19 | AAI-based (high-confidence) | 0 |
| S10_extracted_51127 | 1271 | No | NA | 4 | 0 | 1 | Low-quality | Genome-fragment | 2.28 | AAI-based (medium-confidence) | 0 |
| S10_extracted_53454 | 1271 | No | NA | 2 | 1 | 0 | Low-quality | Genome-fragment | 2.18 | HMM-based (lower-bound) | 0 |
| S10_extracted_89129 | 1271 | No | NA | 2 | 1 | 0 | Low-quality | Genome-fragment | 2.14 | AAI-based (medium-confidence) | 0 |
| S12_extracted_k141_117343 | 1271 | No | NA | 2 | 0 | 0 | Low-quality | Genome-fragment | 2.38 | AAI-based (high-confidence) | 0 |
| S13_vs_24914\|\|full | 1471 | No | NA | 2 | 0 | 0 | Not-determined | Genome-fragment | NA | NA | 0 |
| S13_vs_24994\|\|full | 1445 | No | NA | 2 | 0 | 0 | Not-determined | Genome-fragment | NA | NA | 0 |
| S13_vs_25004\|\|full | 2759 | No | NA | 2 | 0 | 1 | Not-determined | Genome-fragment | NA | NA | 0 |
| S13_vs_25035\|\|full | 1841 | No | NA | 3 | 0 | 0 | Not-determined | Genome-fragment | NA | NA | 0 |
| S15_extracted_6432 | 1271 | No | NA | 3 | 0 | 0 | Low-quality | Genome-fragment | 2.81 | AAI-based (high-confidence) | 0 |
| S13_vs_25266\|\|full | 2297 | No | NA | 5 | 0 | 0 | Not-determined | Genome-fragment | NA | NA | 0 |
| S13_vs_25271\|\|full | 1400 | No | NA | 3 | 0 | 0 | Not-determined | Genome-fragment | NA | NA | 0 |
| S13_vs_25595\|\|full | 1751 | No | NA | 3 | 0 | 0 | Not-determined | Genome-fragment | NA | NA | 0 |
| S13_vs_25599\|\|full | 2907 | No | NA | 2 | 0 | 2 | Not-determined | Genome-fragment | NA | NA | 0 |
| S13_vs_25675\|\|full | 1438 | No | NA | 2 | 0 | 1 | Not-determined | Genome-fragment | NA | NA | 0 |
| S13_vs_25687\|\|full | 3831 | No | NA | 3 | 0 | 3 | Not-determined | Genome-fragment | NA | NA | 0 |
| S13_vs_25708\|\|full | 3030 | No | NA | 4 | 0 | 1 | Not-determined | Genome-fragment | NA | NA | 0 |
| S13_vs_25862\|\|full | 7705 | No | NA | 7 | 0 | 2 | Not-determined | Genome-fragment | NA | NA | 0 |
| S13_vs_25894\|\|full | 2449 | No | NA | 3 | 0 | 2 | Not-determined | Genome-fragment | NA | NA | 0 |
| S9_vs_k141_7134\|\|full | 1272 | No | NA | 3 | 0 | 0 | Low-quality | Genome-fragment | 2.37 | AAI-based (medium-confidence) | 0 |
| S13_vs_26057\|\|full | 1450 | No | NA | 2 | 0 | 1 | Not-determined | Genome-fragment | NA | NA | 0 |
| S13_vs_26127\|\|full | 1802 | No | NA | 5 | 0 | 0 | Not-determined | Genome-fragment | NA | NA | 0 |
| S14_extracted_k141_77215 | 1272 | No | NA | 2 | 0 | 1 | Low-quality | Genome-fragment | 1.39 | AAI-based (medium-confidence) | 0 |
| S13_vs_26756\|\|full | 48783 | Yes | 36490 | 77 | 15 | 8 | High-quality | High-quality | 99.69 | AAI-based (high-confidence) | 25.2 |
| S14_vs_k141_41343\|\|lt2gene | 1272 | No | NA | 1 | 1 | 0 | Low-quality | Genome-fragment | 30.57 | HMM-based (lower-bound) | 0 |
| S13_vs_26900\|\|full | 2817 | No | NA | 4 | 0 | 2 | Not-determined | Genome-fragment | NA | NA | 0 |
| S13_vs_26909\|\|full | 2375 | No | NA | 4 | 0 | 2 | Not-determined | Genome-fragment | NA | NA | 0 |
| S13_vs_27107\|\|full | 3604 | No | NA | 6 | 0 | 0 | Not-determined | Genome-fragment | NA | NA | 0 |
| S13_vs_27115\|\|full | 1120 | No | NA | 1 | 0 | 1 | Not-determined | Genome-fragment | NA | NA | 0 |
| S3_extracted_k141_105316 | 1272 | No | NA | 3 | 0 | 0 | Low-quality | Genome-fragment | 2.9 | AAI-based (medium-confidence) | 0 |
| S3_extracted_k141_106628 | 1272 | No | NA | 3 | 0 | 0 | Low-quality | Genome-fragment | 3.62 | AAI-based (medium-confidence) | 0 |
| S13_vs_27427\|\|full | 4793 | No | NA | 3 | 0 | 0 | Not-determined | Genome-fragment | NA | NA | 0 |
| S13_vs_27534\|\|full | 1131 | No | NA | 3 | 0 | 0 | Not-determined | Genome-fragment | NA | NA | 0 |
| S13_vs_27539\|\|full | 3445 | No | NA | 4 | 0 | 2 | Not-determined | Genome-fragment | NA | NA | 0 |
| S13_vs_27659\|\|full | 1448 | No | NA | 3 | 0 | 0 | Not-determined | Genome-fragment | NA | NA | 0 |
| S13_vs_27698\|\|full | 7904 | No | NA | 10 | 0 | 4 | Not-determined | Genome-fragment | NA | NA | 0 |
| S7_extracted_97666 | 1272 | No | NA | 2 | 0 | 1 | Low-quality | Genome-fragment | 2.53 | AAI-based (medium-confidence) | 0 |
| S7_vs_68816\|\|lt2gene | 1272 | No | NA | 1 | 1 | 0 | Low-quality | Genome-fragment | 5.68 | AAI-based (high-confidence) | 0 |
| S16_extracted_41443 | 1272 | No | NA | 1 | 0 | 1 | Low-quality | Genome-fragment | 0.2 | AAI-based (high-confidence) | 0 |
| S16_vs_18269\|\|full | 1272 | No | NA | 3 | 1 | 0 | Low-quality | Genome-fragment | 2.41 | AAI-based (high-confidence) | 0 |
| S13_vs_29089\|\|full | 2228 | No | NA | 3 | 0 | 2 | Not-determined | Genome-fragment | NA | NA | 0 |
| S10_vs_56444\|\|full | 1272 | No | NA | 3 | 0 | 0 | Low-quality | Genome-fragment | 0.93 | AAI-based (medium-confidence) | 0 |
| S6_vs_k141_25731\|\|lt2gene | 1272 | No | NA | 1 | 1 | 0 | Low-quality | Genome-fragment | 3.65 | AAI-based (medium-confidence) | 0 |
| S13_vs_29425\|\|full | 1430 | No | NA | 1 | 0 | 0 | Not-determined | Genome-fragment | NA | NA | 0 |
| S8_extracted_k141_5867 | 1272 | No | NA | 3 | 2 | 0 | Low-quality | Genome-fragment | 3.5 | AAI-based (high-confidence) | 0 |
| S13_vs_29974\|\|full | 1729 | No | NA | 3 | 0 | 0 | Not-determined | Genome-fragment | NA | NA | 0 |
| S13_vs_29985\|\|full | 8122 | No | NA | 5 | 0 | 5 | Not-determined | Genome-fragment | NA | NA | 0 |
| S13_vs_30080\|\|full | 1967 | No | NA | 3 | 0 | 1 | Not-determined | Genome-fragment | NA | NA | 0 |
| S13_vs_30101\|\|full | 113152 | Yes | 36858 | 92 | 23 | 27 | Medium-quality | Genome-fragment | 77.39 | AAI-based (high-confidence) | 67.43 |
| S13_vs_30167\|\|full | 62048 | Yes | 54104 | 68 | 25 | 9 | High-quality | High-quality | 100 | AAI-based (high-confidence) | 12.8 |
| S13_vs_30241\|\|full | 6511 | No | NA | 5 | 0 | 1 | Not-determined | Genome-fragment | NA | NA | 0 |
| S13_vs_30515\|\|full | 2537 | No | NA | 3 | 0 | 2 | Not-determined | Genome-fragment | NA | NA | 0 |
| S17_extracted_k141_36613 | 1273 | No | NA | 1 | 1 | 0 | Low-quality | Genome-fragment | 2.33 | HMM-based (lower-bound) | 0 |
| S13_vs_30709\|\|full | 2376 | No | NA | 4 | 0 | 1 | Not-determined | Genome-fragment | NA | NA | 0 |
| S13_vs_31033\|\|full | 1197 | No | NA | 3 | 0 | 0 | Not-determined | Genome-fragment | NA | NA | 0 |
| S13_vs_31065\|\|full | 1808 | No | NA | 3 | 0 | 1 | Not-determined | Genome-fragment | NA | NA | 0 |
| S9_extracted_k141_10271 | 1273 | No | NA | 3 | 2 | 0 | Low-quality | Genome-fragment | 1.8 | HMM-based (lower-bound) | 0 |
| S14_extracted_k141_112565 | 1273 | No | NA | 2 | 0 | 1 | Low-quality | Genome-fragment | 2.83 | AAI-based (medium-confidence) | 0 |
| S13_vs_31463\|\|full | 5566 | No | NA | 7 | 0 | 4 | Not-determined | Genome-fragment | NA | NA | 0 |
| S1_vs_k141_10607\|\|full | 1273 | No | NA | 4 | 1 | 0 | Low-quality | Genome-fragment | 3.01 | AAI-based (high-confidence) | 0 |
| S13_vs_31824\|\|full | 3787 | No | NA | 6 | 0 | 1 | Not-determined | Genome-fragment | NA | NA | 0 |
| S13_vs_32037\|\|full | 2702 | No | NA | 4 | 0 | 1 | Not-determined | Genome-fragment | NA | NA | 0 |
| S3_extracted_k141_19562 | 1273 | No | NA | 2 | 0 | 1 | Low-quality | Genome-fragment | 0.95 | AAI-based (high-confidence) | 0 |
| S13_vs_32150\|\|full | 2088 | No | NA | 4 | 0 | 3 | Not-determined | Genome-fragment | NA | NA | 0 |
| S13_vs_32488\|\|full | 1283 | No | NA | 2 | 0 | 1 | Not-determined | Genome-fragment | NA | NA | 0 |
| S3_vs_k141_87595\|\|full | 1273 | No | NA | 4 | 0 | 0 | Low-quality | Genome-fragment | 1.61 | AAI-based (high-confidence) | 0 |
| S5_vs_k141_118041\|\|full | 1273 | No | NA | 3 | 2 | 0 | Low-quality | Genome-fragment | 2.71 | AAI-based (high-confidence) | 0 |
| S13_vs_33145\|\|full | 6257 | No | NA | 9 | 0 | 1 | Not-determined | Genome-fragment | NA | NA | 0 |
| S13_vs_33417\|\|full | 1719 | No | NA | 2 | 0 | 1 | Not-determined | Genome-fragment | NA | NA | 0 |
| S13_vs_33558\|\|full | 2852 | No | NA | 9 | 0 | 0 | Not-determined | Genome-fragment | NA | NA | 0 |
| S13_vs_33645\|\|full | 1705 | No | NA | 3 | 0 | 1 | Not-determined | Genome-fragment | NA | NA | 0 |
| S5_vs_k141_77180\|\|full | 1273 | No | NA | 2 | 0 | 1 | Low-quality | Genome-fragment | 0.55 | AAI-based (medium-confidence) | 0 |
| S7_extracted_112181 | 1273 | No | NA | 2 | 1 | 0 | Low-quality | Genome-fragment | 2.71 | AAI-based (high-confidence) | 0 |
| S7_vs_47861\|\|full | 1273 | No | NA | 2 | 0 | 0 | Low-quality | Genome-fragment | 0.88 | AAI-based (high-confidence) | 0 |
| S13_vs_33947\|\|full | 86062 | Yes | 28387 | 77 | 5 | 43 | Medium-quality | Genome-fragment | 70.13 | AAI-based (medium-confidence) | 67.02 |
| S7_vs_99015\|\|full | 1273 | No | NA | 3 | 0 | 2 | Low-quality | Genome-fragment | 0.68 | AAI-based (medium-confidence) | 0 |
| S16_vs_21212\|\|full | 1273 | No | NA | 3 | 0 | 0 | Low-quality | Genome-fragment | 0.81 | AAI-based (medium-confidence) | 0 |
| S13_vs_34111\|\|full | 2130 | No | NA | 3 | 0 | 2 | Not-determined | Genome-fragment | NA | NA | 0 |
| S16_vs_115477\|\|full | 1273 | No | NA | 3 | 0 | 0 | Low-quality | Genome-fragment | 2.46 | AAI-based (high-confidence) | 0 |
| S13_vs_34344\|\|full | 1650 | No | NA | 3 | 0 | 0 | Not-determined | Genome-fragment | NA | NA | 0 |
| S12_vs_k141_171020\|\|full | 1273 | No | NA | 2 | 0 | 0 | Low-quality | Genome-fragment | 3.51 | AAI-based (high-confidence) | 0 |
| S13_vs_34512\|\|full | 2406 | No | NA | 3 | 0 | 1 | Not-determined | Genome-fragment | NA | NA | 0 |
| S13_vs_34627\|\|full | 1243 | No | NA | 2 | 0 | 0 | Not-determined | Genome-fragment | NA | NA | 0 |
| S12_vs_k141_56846\|\|lt2gene | 1273 | No | NA | 1 | 0 | 0 | Low-quality | Genome-fragment | 2.28 | AAI-based (high-confidence) | 0 |
| S2_vs_k141_13092\|\|full | 1273 | No | NA | 3 | 1 | 0 | Low-quality | Genome-fragment | 3.24 | AAI-based (high-confidence) | 0 |
| S13_vs_35130\|\|full | 1807 | No | NA | 1 | 0 | 0 | Not-determined | Genome-fragment | NA | NA | 0 |
| S6_vs_k141_39180\|\|full | 1273 | No | NA | 2 | 0 | 0 | Low-quality | Genome-fragment | 1.92 | AAI-based (medium-confidence) | 0 |
| S13_vs_35347\|\|full | 1027 | No | NA | 2 | 0 | 0 | Not-determined | Genome-fragment | NA | NA | 0 |
| S13_vs_35464\|\|full | 1080 | No | NA | 3 | 0 | 3 | Not-determined | Genome-fragment | NA | NA | 0 |
| S8_extracted_k141_139681 | 1273 | No | NA | 1 | 0 | 0 | Low-quality | Genome-fragment | 4.05 | AAI-based (high-confidence) | 0 |
| S8_extracted_k141_186517 | 1273 | No | NA | 3 | 0 | 1 | Low-quality | Genome-fragment | 0.73 | AAI-based (high-confidence) | 0 |
| S13_vs_35816\|\|full | 57224 | Yes | 42012 | 78 | 22 | 6 | Complete | High-quality | 100 | Provirus (medium-confidence) | 26.58 |
| S13_vs_36251\|\|full | 1342 | No | NA | 2 | 0 | 1 | Not-determined | Genome-fragment | NA | NA | 0 |
| S9_vs_k141_13557\|\|full | 1274 | No | NA | 3 | 2 | 0 | Low-quality | Genome-fragment | 2.68 | AAI-based (high-confidence) | 0 |
| S11_vs_k141_31199\|\|full | 1274 | No | NA | 3 | 0 | 0 | Low-quality | Genome-fragment | 0.86 | AAI-based (high-confidence) | 0 |
| S13_vs_36460\|\|full | 1409 | No | NA | 3 | 0 | 1 | Not-determined | Genome-fragment | NA | NA | 0 |
| S11_vs_k141_89650\|\|lt2gene | 1274 | No | NA | 1 | 1 | 0 | Low-quality | Genome-fragment | 1.13 | HMM-based (lower-bound) | 0 |
| S14_vs_k141_41042\|\|lt2gene | 1274 | No | NA | 2 | 2 | 0 | Low-quality | Genome-fragment | 2.9 | AAI-based (high-confidence) | 0 |
| S5_extracted_k141_67469 | 1274 | No | NA | 2 | 0 | 0 | Low-quality | Genome-fragment | 0.75 | AAI-based (high-confidence) | 0 |
| S13_vs_36932\|\|full | 1837 | No | NA | 5 | 0 | 1 | Not-determined | Genome-fragment | NA | NA | 0 |
| S13_vs_37340\|\|full | 1639 | No | NA | 4 | 0 | 0 | Not-determined | Genome-fragment | NA | NA | 0 |
| S16_extracted_48525 | 1274 | No | NA | 2 | 2 | 0 | Low-quality | Genome-fragment | 2.04 | HMM-based (lower-bound) | 0 |
| S16_vs_82032\|\|full | 1274 | No | NA | 3 | 0 | 0 | Low-quality | Genome-fragment | 1.84 | AAI-based (medium-confidence) | 0 |
| S16_vs_142672\|\|full | 1274 | No | NA | 3 | 0 | 0 | Low-quality | Genome-fragment | 3.03 | AAI-based (medium-confidence) | 0 |
| S13_vs_37853\|\|full | 2622 | No | NA | 4 | 0 | 0 | Not-determined | Genome-fragment | NA | NA | 0 |
| S13_vs_38096\|\|full | 1629 | No | NA | 4 | 0 | 1 | Not-determined | Genome-fragment | NA | NA | 0 |
| S13_vs_38153\|\|full | 6669 | No | NA | 8 | 0 | 5 | Not-determined | Genome-fragment | NA | NA | 0 |
| S13_vs_38187\|\|full | 2343 | No | NA | 5 | 0 | 1 | Not-determined | Genome-fragment | NA | NA | 0 |
| S12_vs_k141_66813\|\|full | 1274 | No | NA | 2 | 0 | 0 | Low-quality | Genome-fragment | 0.87 | AAI-based (high-confidence) | 0 |
| S12_vs_k141_170128\|\|full | 1274 | No | NA | 3 | 0 | 2 | Low-quality | Genome-fragment | 1.22 | AAI-based (high-confidence) | 0 |
| S13_vs_38579\|\|full | 2034 | No | NA | 3 | 0 | 1 | Not-determined | Genome-fragment | NA | NA | 0 |
| S6_extracted_k141_8670 | 1274 | No | NA | 2 | 0 | 0 | Low-quality | Genome-fragment | 1.54 | AAI-based (medium-confidence) | 0 |
| S13_vs_38664\|\|full | 4569 | No | NA | 5 | 0 | 3 | Not-determined | Genome-fragment | NA | NA | 0 |
| S8_vs_k141_125699\|\|full | 1274 | No | NA | 4 | 2 | 0 | Low-quality | Genome-fragment | 2.1 | AAI-based (high-confidence) | 0 |
| S8_vs_k141_179418\|\|full | 1274 | No | NA | 2 | 0 | 0 | Low-quality | Genome-fragment | 2.47 | AAI-based (high-confidence) | 0 |
| S13_vs_38724\|\|full | 1820 | No | NA | 4 | 0 | 0 | Not-determined | Genome-fragment | NA | NA | 0 |
| S13_vs_39245\|\|full | 1204 | No | NA | 3 | 0 | 1 | Not-determined | Genome-fragment | NA | NA | 0 |
| S17_extracted_k141_134651 | 1275 | No | NA | 3 | 0 | 1 | Low-quality | Genome-fragment | 3.22 | AAI-based (medium-confidence) | 0 |
| S13_vs_39536\|\|full | 2782 | No | NA | 2 | 0 | 1 | Not-determined | Genome-fragment | NA | NA | 0 |
| S14_vs_k141_18853\|\|full | 1275 | No | NA | 3 | 1 | 0 | Low-quality | Genome-fragment | 3 | AAI-based (high-confidence) | 0 |
| S13_vs_39702\|\|full | 2408 | No | NA | 3 | 0 | 1 | Not-determined | Genome-fragment | NA | NA | 0 |
| S13_vs_39800\|\|full | 1161 | No | NA | 3 | 0 | 1 | Not-determined | Genome-fragment | NA | NA | 0 |
| S13_vs_39884\|\|full | 1523 | No | NA | 4 | 0 | 0 | Not-determined | Genome-fragment | NA | NA | 0 |
| S13_vs_39954\|\|full | 2284 | No | NA | 4 | 0 | 1 | Not-determined | Genome-fragment | NA | NA | 0 |
| S13_vs_40008\|\|full | 3203 | No | NA | 3 | 0 | 1 | Not-determined | Genome-fragment | NA | NA | 0 |
| S13_vs_40043\|\|full | 1300 | No | NA | 2 | 0 | 2 | Not-determined | Genome-fragment | NA | NA | 0 |
| S1_extracted_k141_154648 | 1275 | No | NA | 3 | 1 | 0 | Low-quality | Genome-fragment | 0.82 | AAI-based (high-confidence) | 0 |
| S1_extracted_k141_56493 | 1275 | No | NA | 2 | 0 | 0 | Low-quality | Genome-fragment | 2.99 | AAI-based (high-confidence) | 0 |
| S13_vs_40526\|\|full | 1707 | No | NA | 3 | 0 | 1 | Not-determined | Genome-fragment | NA | NA | 0 |
| S3_vs_k141_90167\|\|full | 1275 | No | NA | 3 | 0 | 0 | Low-quality | Genome-fragment | 1.43 | AAI-based (high-confidence) | 0 |
| S13_vs_40645\|\|full | 3012 | No | NA | 3 | 0 | 2 | Not-determined | Genome-fragment | NA | NA | 0 |
| S5_vs_k141_30851\|\|full | 1275 | No | NA | 3 | 0 | 0 | Low-quality | Genome-fragment | 3.51 | AAI-based (high-confidence) | 0 |
| S5_vs_k141_96935\|\|full | 1275 | No | NA | 3 | 0 | 0 | Low-quality | Genome-fragment | 2.19 | AAI-based (high-confidence) | 0 |
| S10_extracted_8195 | 1275 | No | NA | 2 | 0 | 0 | Low-quality | Genome-fragment | 3.09 | AAI-based (high-confidence) | 0 |
| S12_extracted_k141_10514 | 1275 | No | NA | 1 | 0 | 0 | Low-quality | Genome-fragment | 2.71 | AAI-based (high-confidence) | 0 |
| S15_vs_10023\|\|full | 1275 | No | NA | 3 | 1 | 0 | Low-quality | Genome-fragment | 3.14 | AAI-based (high-confidence) | 0 |
| S13_vs_41418\|\|full | 1717 | No | NA | 2 | 0 | 2 | Not-determined | Genome-fragment | NA | NA | 0 |
| S13_vs_42028\|\|full | 1761 | No | NA | 2 | 0 | 2 | Not-determined | Genome-fragment | NA | NA | 0 |
| S13_vs_42165\|\|full | 1523 | No | NA | 3 | 0 | 0 | Not-determined | Genome-fragment | NA | NA | 0 |
| S6_vs_k141_48769\|\|full | 1275 | No | NA | 3 | 0 | 0 | Low-quality | Genome-fragment | 0.81 | AAI-based (high-confidence) | 0 |
| S8_extracted_k141_111337 | 1275 | No | NA | 5 | 2 | 0 | Low-quality | Genome-fragment | 3.13 | AAI-based (medium-confidence) | 0 |
| S13_vs_42345\|\|full | 2140 | No | NA | 2 | 0 | 0 | Not-determined | Genome-fragment | NA | NA | 0 |
| S8_vs_k141_52595\|\|full | 1275 | No | NA | 4 | 0 | 4 | Low-quality | Genome-fragment | 0.4 | AAI-based (medium-confidence) | 0 |
| S13_vs_42582\|\|full | 1524 | No | NA | 3 | 0 | 2 | Not-determined | Genome-fragment | NA | NA | 0 |
| S13_vs_43414\|\|full | 1721 | No | NA | 3 | 0 | 2 | Not-determined | Genome-fragment | NA | NA | 0 |
| S13_vs_43648\|\|full | 1269 | No | NA | 2 | 0 | 1 | Not-determined | Genome-fragment | NA | NA | 0 |
| S13_vs_43828\|\|full | 1312 | No | NA | 3 | 0 | 1 | Not-determined | Genome-fragment | NA | NA | 0 |
| S17_extracted_k141_116055 | 1276 | No | NA | 2 | 0 | 0 | Low-quality | Genome-fragment | 1.72 | AAI-based (medium-confidence) | 0 |
| S13_vs_43920\|\|full | 1413 | No | NA | 3 | 0 | 2 | Not-determined | Genome-fragment | NA | NA | 0 |
| S17_vs_k141_131420\|\|lt2gene | 1276 | No | NA | 2 | 1 | 0 | Low-quality | Genome-fragment | 2.59 | AAI-based (high-confidence) | 0 |
| S11_vs_k141_84220\|\|lt2gene | 1276 | No | NA | 2 | 1 | 0 | Low-quality | Genome-fragment | 3.25 | AAI-based (high-confidence) | 0 |
| S14_vs_k141_59883\|\|full | 1276 | No | NA | 4 | 0 | 0 | Low-quality | Genome-fragment | 2.36 | AAI-based (high-confidence) | 0 |
| S13_vs_44629\|\|full | 2155 | No | NA | 3 | 0 | 2 | Not-determined | Genome-fragment | NA | NA | 0 |
| S1_vs_k141_68252\|\|full | 1276 | No | NA | 3 | 0 | 0 | Low-quality | Genome-fragment | 0.73 | AAI-based (high-confidence) | 0 |
| S13_vs_44939\|\|full | 1392 | No | NA | 3 | 0 | 0 | Not-determined | Genome-fragment | NA | NA | 0 |
| S5_extracted_k141_82592 | 1276 | No | NA | 2 | 2 | 0 | Low-quality | Genome-fragment | 1.83 | HMM-based (lower-bound) | 0 |
| S13_vs_45268\|\|full | 2449 | No | NA | 3 | 0 | 0 | Not-determined | Genome-fragment | NA | NA | 0 |
| S13_vs_45282\|\|full | 2098 | No | NA | 2 | 0 | 1 | Not-determined | Genome-fragment | NA | NA | 0 |
| S13_vs_45320\|\|full | 1116 | No | NA | 3 | 0 | 0 | Not-determined | Genome-fragment | NA | NA | 0 |
| S13_vs_45335\|\|full | 3068 | No | NA | 3 | 0 | 0 | Not-determined | Genome-fragment | NA | NA | 0 |
| S13_vs_45460\|\|full | 1747 | No | NA | 2 | 0 | 2 | Not-determined | Genome-fragment | NA | NA | 0 |
| S13_vs_45488\|\|full | 124456 | Yes | 35117 | 134 | 26 | 44 | Medium-quality | Genome-fragment | 75.61 | AAI-based (high-confidence) | 71.78 |
| S5_extracted_k141_29300 | 1276 | No | NA | 4 | 0 | 0 | Low-quality | Genome-fragment | 3.53 | AAI-based (high-confidence) | 0 |
| S7_vs_13001\|\|full | 1276 | No | NA | 3 | 0 | 1 | Low-quality | Genome-fragment | 1.96 | AAI-based (medium-confidence) | 0 |
| S13_vs_45600\|\|full | 2417 | No | NA | 3 | 0 | 1 | Not-determined | Genome-fragment | NA | NA | 0 |
| S13_vs_45609\|\|full | 3544 | No | NA | 3 | 0 | 1 | Not-determined | Genome-fragment | NA | NA | 0 |
| S13_vs_45671\|\|full | 1477 | No | NA | 3 | 0 | 1 | Not-determined | Genome-fragment | NA | NA | 0 |
| S12_vs_k141_175093\|\|full | 1276 | No | NA | 3 | 0 | 0 | Low-quality | Genome-fragment | 1.94 | AAI-based (medium-confidence) | 0 |
| S13_vs_45868\|\|full | 1425 | No | NA | 3 | 0 | 2 | Not-determined | Genome-fragment | NA | NA | 0 |
| S12_vs_k141_115965\|\|full | 1276 | No | NA | 2 | 0 | 1 | Low-quality | Genome-fragment | 2.75 | AAI-based (high-confidence) | 0 |
| S12_vs_k141_53847\|\|full | 1276 | No | NA | 2 | 0 | 1 | Low-quality | Genome-fragment | 1.88 | AAI-based (medium-confidence) | 0 |
| S8_vs_k141_74946\|\|full | 1276 | No | NA | 3 | 0 | 2 | Low-quality | Genome-fragment | 0.66 | AAI-based (medium-confidence) | 0 |
| S17_vs_k141_52998\|\|full | 1277 | No | NA | 5 | 1 | 0 | Low-quality | Genome-fragment | 2.76 | AAI-based (high-confidence) | 0 |
| S13_vs_46348\|\|full | 12354 | No | NA | 6 | 0 | 3 | Not-determined | Genome-fragment | NA | NA | 0 |
| S13_vs_46551\|\|full | 6163 | No | NA | 7 | 0 | 4 | Not-determined | Genome-fragment | NA | NA | 0 |
| S11_vs_k141_68188\|\|lt2gene | 1277 | No | NA | 1 | 1 | 0 | Low-quality | Genome-fragment | 1.67 | HMM-based (lower-bound) | 0 |
| S13_vs_46672\|\|full | 1695 | No | NA | 2 | 0 | 0 | Not-determined | Genome-fragment | NA | NA | 0 |
| S14_vs_k141_65229\|\|full | 1277 | No | NA | 5 | 0 | 0 | Low-quality | Genome-fragment | 2.71 | AAI-based (high-confidence) | 0 |
| S1_vs_k141_17207\|\|full | 1277 | No | NA | 4 | 0 | 0 | Low-quality | Genome-fragment | 2.52 | AAI-based (high-confidence) | 0 |
| S5_vs_k141_10793\|\|full | 1277 | No | NA | 2 | 1 | 0 | Low-quality | Genome-fragment | 2.58 | AAI-based (high-confidence) | 0 |
| S13_vs_47612\|\|full | 2840 | No | NA | 4 | 0 | 3 | Not-determined | Genome-fragment | NA | NA | 0 |
| S13_vs_47615\|\|full | 2539 | No | NA | 4 | 0 | 2 | Not-determined | Genome-fragment | NA | NA | 0 |
| S13_vs_47999\|\|full | 1691 | No | NA | 3 | 0 | 1 | Not-determined | Genome-fragment | NA | NA | 0 |
| S13_vs_48224\|\|full | 1011 | No | NA | 2 | 0 | 1 | Not-determined | Genome-fragment | NA | NA | 0 |
| S13_vs_48345\|\|full | 1749 | No | NA | 2 | 0 | 0 | Not-determined | Genome-fragment | NA | NA | 0 |
| S13_vs_48471\|\|full | 4607 | No | NA | 3 | 0 | 1 | Not-determined | Genome-fragment | NA | NA | 0 |
| S7_extracted_57944 | 1277 | No | NA | 4 | 2 | 0 | Low-quality | Genome-fragment | 5.83 | AAI-based (high-confidence) | 0 |
| S13_vs_48790\|\|full | 1626 | No | NA | 3 | 0 | 2 | Not-determined | Genome-fragment | NA | NA | 0 |
| S13_vs_48932\|\|full | 1558 | No | NA | 2 | 0 | 2 | Not-determined | Genome-fragment | NA | NA | 0 |
| S7_vs_55379\|\|full | 1277 | No | NA | 2 | 0 | 0 | Low-quality | Genome-fragment | 2.78 | AAI-based (high-confidence) | 0 |
| S13_vs_49513\|\|full | 3808 | No | NA | 4 | 0 | 0 | Not-determined | Genome-fragment | NA | NA | 0 |
| S16_extracted_122042 | 1277 | No | NA | 1 | 0 | 1 | Low-quality | Genome-fragment | 3.6 | AAI-based (medium-confidence) | 0 |
| S13_vs_49675\|\|full | 2956 | No | NA | 3 | 0 | 3 | Not-determined | Genome-fragment | NA | NA | 0 |
| S13_vs_49823\|\|full | 1056 | No | NA | 2 | 0 | 0 | Not-determined | Genome-fragment | NA | NA | 0 |
| S13_vs_49889\|\|full | 31718 | No | NA | 41 | 12 | 0 | Medium-quality | Genome-fragment | 72.11 | AAI-based (medium-confidence) | 0 |
| S16_vs_116913\|\|full | 1277 | No | NA | 4 | 1 | 0 | Low-quality | Genome-fragment | 3.74 | AAI-based (high-confidence) | 0 |
| S13_vs_50451\|\|full | 6090 | No | NA | 3 | 0 | 1 | Not-determined | Genome-fragment | NA | NA | 0 |
| S13_vs_50693\|\|full | 2330 | No | NA | 4 | 0 | 0 | Not-determined | Genome-fragment | NA | NA | 0 |
| S10_vs_34060\|\|full | 1277 | No | NA | 2 | 0 | 0 | Low-quality | Genome-fragment | 10.64 | AAI-based (high-confidence) | 0 |
| S12_vs_k141_640\|\|full | 1277 | No | NA | 3 | 0 | 0 | Low-quality | Genome-fragment | 3.17 | AAI-based (high-confidence) | 0 |
| S4_extracted_757 | 1277 | No | NA | 4 | 4 | 0 | Low-quality | Genome-fragment | 4.23 | AAI-based (high-confidence) | 0 |
| S6_vs_k141_6942\|\|lt2gene | 1277 | No | NA | 2 | 1 | 0 | Low-quality | Genome-fragment | 2.86 | AAI-based (high-confidence) | 0 |
| S13_vs_51939\|\|full | 2594 | No | NA | 3 | 0 | 3 | Not-determined | Genome-fragment | NA | NA | 0 |
| S8_vs_k141_86756\|\|full | 1277 | No | NA | 1 | 0 | 0 | Low-quality | Genome-fragment | 1.61 | AAI-based (high-confidence) | 0 |
| S9_vs_k141_68651\|\|full | 1278 | No | NA | 4 | 0 | 0 | Low-quality | Genome-fragment | 1.71 | AAI-based (high-confidence) | 0 |
| S11_extracted_k141_95412 | 1278 | No | NA | 2 | 0 | 0 | Low-quality | Genome-fragment | 2.02 | AAI-based (high-confidence) | 0 |
| S11_vs_k141_30184\|\|full | 1278 | No | NA | 2 | 0 | 1 | Low-quality | Genome-fragment | 2.19 | AAI-based (medium-confidence) | 0 |
| S14_vs_k141_4504\|\|full | 1278 | No | NA | 2 | 0 | 1 | Low-quality | Genome-fragment | 2.17 | AAI-based (medium-confidence) | 0 |
| S3_extracted_k141_111379 | 1278 | No | NA | 3 | 0 | 0 | Low-quality | Genome-fragment | 0.9 | AAI-based (medium-confidence) | 0 |
| S13_vs_52652\|\|full | 1933 | No | NA | 3 | 0 | 0 | Not-determined | Genome-fragment | NA | NA | 0 |
| S13_vs_52666\|\|full | 3156 | No | NA | 3 | 0 | 1 | Not-determined | Genome-fragment | NA | NA | 0 |
| S13_vs_52912\|\|full | 6979 | No | NA | 7 | 0 | 5 | Not-determined | Genome-fragment | NA | NA | 0 |
| S13_vs_52924\|\|full | 1822 | No | NA | 3 | 0 | 2 | Not-determined | Genome-fragment | NA | NA | 0 |
| S5_vs_k141_56350\|\|full | 1278 | No | NA | 3 | 0 | 0 | Low-quality | Genome-fragment | 2.85 | AAI-based (medium-confidence) | 0 |
| S10_extracted_86478 | 1278 | No | NA | 2 | 0 | 2 | Low-quality | Genome-fragment | 0.59 | AAI-based (medium-confidence) | 0 |
| S13_vs_53676\|\|full | 2043 | No | NA | 7 | 0 | 0 | Not-determined | Genome-fragment | NA | NA | 0 |
| S10_vs_38500\|\|full | 1278 | No | NA | 3 | 0 | 2 | Low-quality | Genome-fragment | 2.18 | AAI-based (medium-confidence) | 0 |
| S13_vs_53857\|\|full | 1058 | No | NA | 2 | 0 | 1 | Not-determined | Genome-fragment | NA | NA | 0 |
| S13_vs_54076\|\|full | 6304 | No | NA | 9 | 0 | 0 | Not-determined | Genome-fragment | NA | NA | 0 |
| S13_vs_54183\|\|full | 2025 | No | NA | 4 | 0 | 0 | Not-determined | Genome-fragment | NA | NA | 0 |
| S12_extracted_k141_126683 | 1278 | No | NA | 5 | 1 | 0 | Low-quality | Genome-fragment | 2.9 | AAI-based (medium-confidence) | 0 |
| S13_vs_54602\|\|full | 1656 | No | NA | 2 | 0 | 0 | Not-determined | Genome-fragment | NA | NA | 0 |
| S13_vs_54826\|\|full | 1288 | No | NA | 3 | 0 | 1 | Not-determined | Genome-fragment | NA | NA | 0 |
| S13_vs_54920\|\|full | 1507 | No | NA | 3 | 0 | 3 | Not-determined | Genome-fragment | NA | NA | 0 |
| S13_vs_55038\|\|full | 1012 | No | NA | 3 | 0 | 0 | Not-determined | Genome-fragment | NA | NA | 0 |
| S12_vs_k141_119160\|\|full | 1278 | No | NA | 4 | 1 | 0 | Low-quality | Genome-fragment | 1.3 | AAI-based (medium-confidence) | 0 |
| S13_vs_55658\|\|full | 1059 | No | NA | 2 | 0 | 0 | Not-determined | Genome-fragment | NA | NA | 0 |
| S13_vs_55660\|\|full | 2230 | No | NA | 2 | 0 | 0 | Not-determined | Genome-fragment | NA | NA | 0 |
| S6_vs_k141_48708\|\|full | 1278 | No | NA | 3 | 2 | 0 | Low-quality | Genome-fragment | 2.14 | AAI-based (high-confidence) | 0 |
| S13_vs_27729\|\|lt2gene | 1279 | No | NA | 2 | 1 | 0 | Low-quality | Genome-fragment | 9.37 | AAI-based (medium-confidence) | 0 |
| S14_extracted_k141_23002 | 1279 | No | NA | 2 | 1 | 0 | Low-quality | Genome-fragment | 0.49 | HMM-based (lower-bound) | 0 |
| S13_vs_55906\|\|full | 1038 | No | NA | 3 | 0 | 1 | Not-determined | Genome-fragment | NA | NA | 0 |
| S1_vs_k141_103529\|\|full | 1279 | No | NA | 4 | 0 | 0 | Low-quality | Genome-fragment | 2.64 | AAI-based (high-confidence) | 0 |
| S13_vs_55997\|\|full | 7716 | No | NA | 7 | 0 | 6 | Not-determined | Genome-fragment | NA | NA | 0 |
| S10_extracted_93894 | 1279 | No | NA | 3 | 0 | 0 | Low-quality | Genome-fragment | 3.01 | AAI-based (medium-confidence) | 0 |
| S13_vs_56260\|\|full | 3422 | No | NA | 4 | 0 | 2 | Not-determined | Genome-fragment | NA | NA | 0 |
| S13_vs_56327\|\|full | 1089 | No | NA | 2 | 0 | 1 | Not-determined | Genome-fragment | NA | NA | 0 |
| S10_vs_41166\|\|full | 1279 | No | NA | 2 | 0 | 0 | Low-quality | Genome-fragment | 0.56 | AAI-based (medium-confidence) | 0 |
| S12_vs_k141_123229\|\|full | 1279 | No | NA | 2 | 0 | 0 | Low-quality | Genome-fragment | 2.42 | AAI-based (high-confidence) | 0 |
| S13_vs_56606\|\|full | 1028 | No | NA | 2 | 0 | 0 | Not-determined | Genome-fragment | NA | NA | 0 |
| S8_vs_k141_73029\|\|full | 1279 | No | NA | 3 | 0 | 0 | Low-quality | Genome-fragment | 2.83 | AAI-based (high-confidence) | 0 |
| S8_vs_k141_91532\|\|full | 1279 | No | NA | 2 | 0 | 0 | Low-quality | Genome-fragment | 2.71 | AAI-based (medium-confidence) | 0 |
| S9_vs_k141_19154\|\|full | 1280 | No | NA | 2 | 0 | 0 | Low-quality | Genome-fragment | 2 | AAI-based (high-confidence) | 0 |
| S9_vs_k141_82472\|\|lt2gene | 1280 | No | NA | 2 | 1 | 0 | Low-quality | Genome-fragment | 3.15 | AAI-based (high-confidence) | 0 |
| S13_vs_57079\|\|full | 2091 | No | NA | 3 | 0 | 2 | Not-determined | Genome-fragment | NA | NA | 0 |
| S13_vs_57133\|\|full | 5135 | No | NA | 4 | 0 | 3 | Not-determined | Genome-fragment | NA | NA | 0 |
| S13_vs_57144\|\|full | 2467 | No | NA | 3 | 0 | 2 | Not-determined | Genome-fragment | NA | NA | 0 |
| S14_vs_k141_94380\|\|full | 1280 | No | NA | 4 | 4 | 0 | Low-quality | Genome-fragment | 2.64 | AAI-based (high-confidence) | 0 |
| S13_vs_57251\|\|full | 1178 | No | NA | 2 | 0 | 0 | Not-determined | Genome-fragment | NA | NA | 0 |
| S13_vs_57267\|\|full | 289184 | No | NA | 380 | 24 | 9 | Medium-quality | Genome-fragment | 72.12 | AAI-based (high-confidence) | 0 |
| S13_vs_57417\|\|full | 1880 | No | NA | 3 | 0 | 0 | Not-determined | Genome-fragment | NA | NA | 0 |
| S14_vs_k141_13370\|\|full | 1280 | No | NA | 3 | 0 | 1 | Low-quality | Genome-fragment | 2.91 | AAI-based (medium-confidence) | 0 |
| S13_vs_57889\|\|full | 1107 | No | NA | 3 | 0 | 0 | Not-determined | Genome-fragment | NA | NA | 0 |
| S13_vs_58129\|\|full | 1686 | No | NA | 2 | 0 | 0 | Not-determined | Genome-fragment | NA | NA | 0 |
| S13_vs_58230\|\|full | 3969 | No | NA | 5 | 0 | 1 | Not-determined | Genome-fragment | NA | NA | 0 |
| S13_vs_58609\|\|full | 1051 | No | NA | 3 | 0 | 0 | Not-determined | Genome-fragment | NA | NA | 0 |
| S13_vs_58699\|\|full | 2595 | No | NA | 2 | 0 | 1 | Not-determined | Genome-fragment | NA | NA | 0 |
| S13_vs_58864\|\|full | 1296 | No | NA | 3 | 0 | 1 | Not-determined | Genome-fragment | NA | NA | 0 |
| S1_vs_k141_22927\|\|full | 1280 | No | NA | 3 | 0 | 0 | Low-quality | Genome-fragment | 1.42 | AAI-based (medium-confidence) | 0 |
| S13_vs_58904\|\|full | 10659 | No | NA | 10 | 0 | 7 | Not-determined | Genome-fragment | NA | NA | 0 |
| S1_vs_k141_25391\|\|full | 1280 | No | NA | 2 | 0 | 0 | Low-quality | Genome-fragment | 2.89 | AAI-based (medium-confidence) | 0 |
| S13_vs_59103\|\|full | 1615 | No | NA | 3 | 0 | 1 | Not-determined | Genome-fragment | NA | NA | 0 |
| S13_vs_59225\|\|full | 3306 | No | NA | 4 | 0 | 2 | Not-determined | Genome-fragment | NA | NA | 0 |
| S3_extracted_k141_68139 | 1280 | No | NA | 1 | 0 | 0 | Low-quality | Genome-fragment | 3.07 | AAI-based (high-confidence) | 0 |
| S13_vs_59490\|\|full | 8533 | No | NA | 14 | 0 | 0 | Not-determined | Genome-fragment | NA | NA | 0 |
| S13_vs_59547\|\|full | 1549 | No | NA | 4 | 0 | 1 | Not-determined | Genome-fragment | NA | NA | 0 |
| S13_vs_59961\|\|full | 2007 | No | NA | 3 | 0 | 1 | Not-determined | Genome-fragment | NA | NA | 0 |
| S13_vs_59975\|\|full | 1570 | No | NA | 2 | 0 | 0 | Not-determined | Genome-fragment | NA | NA | 0 |
| S13_vs_60281\|\|full | 1249 | No | NA | 2 | 0 | 2 | Not-determined | Genome-fragment | NA | NA | 0 |
| S13_vs_60368\|\|full | 2153 | No | NA | 4 | 0 | 2 | Not-determined | Genome-fragment | NA | NA | 0 |
| S13_vs_60467\|\|full | 1121 | No | NA | 2 | 0 | 1 | Not-determined | Genome-fragment | NA | NA | 0 |
| S13_vs_60534\|\|full | 4786 | No | NA | 3 | 0 | 0 | Not-determined | Genome-fragment | NA | NA | 0 |
| S13_vs_60535\|\|full | 1589 | No | NA | 3 | 0 | 0 | Not-determined | Genome-fragment | NA | NA | 0 |
| S3_vs_k141_4460\|\|full | 1280 | No | NA | 4 | 0 | 0 | Low-quality | Genome-fragment | 3.45 | AAI-based (high-confidence) | 0 |
| S5_vs_k141_68558\|\|full | 1280 | No | NA | 3 | 0 | 0 | Low-quality | Genome-fragment | 1.97 | AAI-based (medium-confidence) | 0 |
| S13_vs_60997\|\|full | 1281 | No | NA | 2 | 0 | 1 | Not-determined | Genome-fragment | NA | NA | 0 |
| S7_extracted_110715 | 1280 | No | NA | 3 | 0 | 0 | Low-quality | Genome-fragment | 3.03 | AAI-based (medium-confidence) | 0 |
| S7_vs_23211\|\|full | 1280 | No | NA | 3 | 0 | 0 | Low-quality | Genome-fragment | 0.89 | AAI-based (medium-confidence) | 0 |
| S13_vs_61225\|\|full | 3354 | No | NA | 3 | 0 | 3 | Not-determined | Genome-fragment | NA | NA | 0 |
| S13_vs_61262\|\|full | 5859 | No | NA | 6 | 0 | 2 | Not-determined | Genome-fragment | NA | NA | 0 |
| S7_vs_134278\|\|lt2gene | 1280 | No | NA | 2 | 1 | 0 | Low-quality | Genome-fragment | 2.09 | HMM-based (lower-bound) | 0 |
| S16_vs_5266\|\|full | 1280 | No | NA | 2 | 0 | 0 | Low-quality | Genome-fragment | 0.71 | AAI-based (high-confidence) | 0 |
| S13_vs_61677\|\|full | 2595 | No | NA | 3 | 0 | 1 | Not-determined | Genome-fragment | NA | NA | 0 |
| S16_vs_53255\|\|full | 1280 | No | NA | 2 | 0 | 0 | Low-quality | Genome-fragment | 2.35 | AAI-based (high-confidence) | 0 |
| S13_vs_62031\|\|full | 1518 | No | NA | 3 | 0 | 1 | Not-determined | Genome-fragment | NA | NA | 0 |
| S12_extracted_k141_6775 | 1280 | No | NA | 1 | 0 | 0 | Low-quality | Genome-fragment | 1.31 | AAI-based (high-confidence) | 0 |
| S13_vs_62142\|\|full | 32085 | No | NA | 39 | 0 | 8 | Not-determined | Genome-fragment | NA | NA | 0 |
| S13_vs_62243\|\|full | 1976 | No | NA | 3 | 0 | 0 | Not-determined | Genome-fragment | NA | NA | 0 |
| S13_vs_62489\|\|full | 1832 | No | NA | 3 | 0 | 0 | Not-determined | Genome-fragment | NA | NA | 0 |
| S13_vs_62728\|\|full | 2197 | No | NA | 4 | 0 | 1 | Not-determined | Genome-fragment | NA | NA | 0 |
| S12_extracted_k141_21779 | 1280 | No | NA | 2 | 0 | 1 | Low-quality | Genome-fragment | 0.84 | AAI-based (high-confidence) | 0 |
| S12_vs_k141_142876\|\|full | 1280 | No | NA | 3 | 1 | 0 | Low-quality | Genome-fragment | 3.06 | AAI-based (medium-confidence) | 0 |
| S13_vs_63080\|\|full | 2295 | No | NA | 2 | 0 | 2 | Not-determined | Genome-fragment | NA | NA | 0 |
| S6_vs_k141_24207\|\|full | 1280 | No | NA | 3 | 0 | 1 | Low-quality | Genome-fragment | 2.08 | AAI-based (medium-confidence) | 0 |
| S13_vs_63425\|\|full | 2104 | No | NA | 2 | 0 | 0 | Not-determined | Genome-fragment | NA | NA | 0 |
| S13_vs_63455\|\|full | 4154 | No | NA | 4 | 0 | 2 | Not-determined | Genome-fragment | NA | NA | 0 |
| S13_vs_63794\|\|full | 2820 | No | NA | 5 | 0 | 4 | Not-determined | Genome-fragment | NA | NA | 0 |
| S13_vs_63833\|\|full | 1280 | No | NA | 2 | 0 | 1 | Not-determined | Genome-fragment | NA | NA | 0 |
| S8_vs_k141_11646\|\|lt2gene | 1280 | No | NA | 2 | 2 | 0 | Low-quality | Genome-fragment | 3.16 | AAI-based (medium-confidence) | 0 |
| S13_vs_63949\|\|full | 1893 | No | NA | 2 | 0 | 1 | Not-determined | Genome-fragment | NA | NA | 0 |
| S13_vs_64129\|\|full | 1127 | No | NA | 2 | 0 | 0 | Not-determined | Genome-fragment | NA | NA | 0 |
| S17_vs_k141_162569\|\|full | 1281 | No | NA | 3 | 0 | 0 | Low-quality | Genome-fragment | 3.07 | AAI-based (medium-confidence) | 0 |
| S13_vs_64380\|\|full | 1357 | No | NA | 3 | 0 | 0 | Not-determined | Genome-fragment | NA | NA | 0 |
| S13_vs_64381\|\|full | 1173 | No | NA | 3 | 0 | 1 | Not-determined | Genome-fragment | NA | NA | 0 |
| S13_vs_64514\|\|full | 1255 | No | NA | 3 | 0 | 1 | Not-determined | Genome-fragment | NA | NA | 0 |
| S13_vs_64701\|\|full | 1746 | No | NA | 2 | 0 | 0 | Not-determined | Genome-fragment | NA | NA | 0 |
| S13_vs_64745\|\|full | 11958 | No | NA | 13 | 0 | 6 | Not-determined | Genome-fragment | NA | NA | 0 |
| S17_vs_k141_153266\|\|lt2gene | 1281 | No | NA | 2 | 1 | 0 | Low-quality | Genome-fragment | 3.05 | AAI-based (high-confidence) | 0 |
| S13_extracted_66306 | 1281 | No | NA | 2 | 2 | 0 | Low-quality | Genome-fragment | 3.36 | AAI-based (high-confidence) | 0 |
| S13_vs_65215\|\|full | 3526 | No | NA | 6 | 0 | 3 | Not-determined | Genome-fragment | NA | NA | 0 |
| S14_extracted_k141_15501 | 1281 | No | NA | 4 | 2 | 0 | Low-quality | Genome-fragment | 4.23 | AAI-based (high-confidence) | 0 |
| S14_extracted_k141_82287 | 1281 | No | NA | 1 | 1 | 0 | Low-quality | Genome-fragment | 4.22 | AAI-based (high-confidence) | 0 |
| S13_vs_65782\|\|full | 1128 | No | NA | 3 | 0 | 1 | Not-determined | Genome-fragment | NA | NA | 0 |
| S13_vs_65800\|\|full | 3219 | No | NA | 2 | 0 | 0 | Not-determined | Genome-fragment | NA | NA | 0 |
| S13_vs_65904\|\|full | 5998 | No | NA | 8 | 0 | 0 | Not-determined | Genome-fragment | NA | NA | 0 |
| S1_vs_k141_155635\|\|full | 1281 | No | NA | 3 | 0 | 1 | Low-quality | Genome-fragment | 0.75 | AAI-based (medium-confidence) | 0 |
| S3_extracted_k141_94221 | 1281 | No | NA | 3 | 0 | 0 | Low-quality | Genome-fragment | 1.23 | AAI-based (high-confidence) | 0 |
| S5_extracted_k141_59290 | 1281 | No | NA | 3 | 0 | 0 | Low-quality | Genome-fragment | 5.85 | AAI-based (high-confidence) | 0 |
| S13_vs_66485\|\|full | 1368 | No | NA | 2 | 0 | 1 | Not-determined | Genome-fragment | NA | NA | 0 |
| S5_vs_k141_38715\|\|full | 1281 | No | NA | 4 | 0 | 0 | Low-quality | Genome-fragment | 3.39 | AAI-based (high-confidence) | 0 |
| S7_extracted_42135 | 1281 | No | NA | 3 | 0 | 0 | Low-quality | Genome-fragment | 2.83 | AAI-based (high-confidence) | 0 |
| S7_extracted_129772 | 1281 | No | NA | 3 | 1 | 0 | Low-quality | Genome-fragment | 0.93 | HMM-based (lower-bound) | 0 |
| S13_vs_67201\|\|full | 1663 | No | NA | 2 | 0 | 0 | Not-determined | Genome-fragment | NA | NA | 0 |
| S7_vs_79045\|\|full | 1281 | No | NA | 3 | 0 | 0 | Low-quality | Genome-fragment | 1.75 | AAI-based (high-confidence) | 0 |
| S13_vs_67277\|\|full | 1275 | No | NA | 4 | 0 | 0 | Not-determined | Genome-fragment | NA | NA | 0 |
| S13_vs_67429\|\|full | 91295 | Yes | 41498 | 116 | 17 | 24 | High-quality | High-quality | 100 | AAI-based (high-confidence) | 54.55 |
| S13_vs_67502\|\|full | 6834 | No | NA | 7 | 0 | 5 | Not-determined | Genome-fragment | NA | NA | 0 |
| S13_vs_67604\|\|full | 1527 | No | NA | 3 | 0 | 2 | Not-determined | Genome-fragment | NA | NA | 0 |
| S16_vs_40708\|\|full | 1281 | No | NA | 3 | 1 | 0 | Low-quality | Genome-fragment | 2.73 | AAI-based (high-confidence) | 0 |
| S13_vs_67787\|\|full | 1424 | No | NA | 3 | 0 | 1 | Not-determined | Genome-fragment | NA | NA | 0 |
| S13_vs_67875\|\|full | 1039 | No | NA | 3 | 0 | 0 | Not-determined | Genome-fragment | NA | NA | 0 |
| S13_vs_68102\|\|full | 15847 | No | NA | 22 | 8 | 0 | Medium-quality | Genome-fragment | 73.21 | AAI-based (medium-confidence) | 0 |
| S13_vs_68142\|\|full | 2602 | No | NA | 3 | 0 | 2 | Not-determined | Genome-fragment | NA | NA | 0 |
| S13_vs_68179\|\|full | 1865 | No | NA | 4 | 0 | 3 | Not-determined | Genome-fragment | NA | NA | 0 |
| S16_vs_27331\|\|lt2gene | 1281 | Yes | 670 | 2 | 1 | 1 | Low-quality | Genome-fragment | 1.11 | AAI-based (high-confidence) | 47.7 |
| S12_vs_k141_64935\|\|full | 1281 | No | NA | 3 | 0 | 0 | Low-quality | Genome-fragment | 2.47 | AAI-based (medium-confidence) | 0 |
| S13_vs_68492\|\|full | 1821 | No | NA | 3 | 0 | 3 | Not-determined | Genome-fragment | NA | NA | 0 |
| S13_vs_68615\|\|full | 1126 | No | NA | 3 | 0 | 1 | Not-determined | Genome-fragment | NA | NA | 0 |
| S13_vs_68630\|\|full | 1761 | No | NA | 3 | 0 | 1 | Not-determined | Genome-fragment | NA | NA | 0 |
| S13_vs_68703\|\|full | 1857 | No | NA | 2 | 0 | 0 | Not-determined | Genome-fragment | NA | NA | 0 |
| S13_vs_68752\|\|full | 2679 | No | NA | 3 | 0 | 1 | Not-determined | Genome-fragment | NA | NA | 0 |
| S6_vs_k141_36418\|\|full | 1281 | No | NA | 3 | 1 | 0 | Low-quality | Genome-fragment | 2.86 | AAI-based (high-confidence) | 0 |
| S17_extracted_k141_141142 | 1282 | No | NA | 5 | 0 | 0 | Low-quality | Genome-fragment | 2.03 | AAI-based (high-confidence) | 0 |
| S13_vs_68813\|\|full | 2305 | No | NA | 3 | 0 | 0 | Not-determined | Genome-fragment | NA | NA | 0 |
| S13_vs_68986\|\|full | 4011 | No | NA | 3 | 0 | 1 | Not-determined | Genome-fragment | NA | NA | 0 |
| S13_vs_69027\|\|full | 2527 | No | NA | 5 | 0 | 4 | Not-determined | Genome-fragment | NA | NA | 0 |
| S9_vs_k141_82496\|\|full | 1282 | No | NA | 2 | 0 | 0 | Low-quality | Genome-fragment | 2.32 | AAI-based (high-confidence) | 0 |
| S13_vs_69186\|\|full | 2670 | No | NA | 3 | 0 | 2 | Not-determined | Genome-fragment | NA | NA | 0 |
| S11_vs_k141_35949\|\|full | 1282 | No | NA | 3 | 0 | 1 | Low-quality | Genome-fragment | 0.39 | AAI-based (high-confidence) | 0 |
| S13_vs_69722\|\|full | 1121 | No | NA | 2 | 0 | 0 | Not-determined | Genome-fragment | NA | NA | 0 |
| S1_vs_k141_64244\|\|full | 1282 | No | NA | 3 | 1 | 0 | Low-quality | Genome-fragment | 0.94 | AAI-based (high-confidence) | 0 |
| S13_vs_69959\|\|full | 1767 | No | NA | 2 | 0 | 1 | Not-determined | Genome-fragment | NA | NA | 0 |
| S13_vs_69998\|\|full | 1203 | No | NA | 2 | 0 | 1 | Not-determined | Genome-fragment | NA | NA | 0 |
| S13_vs_70063\|\|full | 1103 | No | NA | 3 | 0 | 1 | Not-determined | Genome-fragment | NA | NA | 0 |
| S5_extracted_k141_69147 | 1282 | No | NA | 3 | 0 | 0 | Low-quality | Genome-fragment | 3.17 | AAI-based (medium-confidence) | 0 |
| S13_vs_70347\|\|full | 4307 | No | NA | 7 | 0 | 1 | Not-determined | Genome-fragment | NA | NA | 0 |
| S5_vs_k141_104527\|\|lt2gene | 1282 | No | NA | 2 | 1 | 0 | Low-quality | Genome-fragment | 3.52 | HMM-based (lower-bound) | 0 |
| S16_extracted_132659 | 1282 | No | NA | 1 | 0 | 1 | Low-quality | Genome-fragment | 1.65 | AAI-based (high-confidence) | 0 |
| S13_vs_71060\|\|full | 1388 | No | NA | 3 | 0 | 1 | Not-determined | Genome-fragment | NA | NA | 0 |
| S13_vs_71086\|\|full | 1311 | No | NA | 3 | 0 | 2 | Not-determined | Genome-fragment | NA | NA | 0 |
| S16_vs_38174\|\|full | 1282 | No | NA | 3 | 0 | 2 | Low-quality | Genome-fragment | 0.37 | AAI-based (medium-confidence) | 0 |
| S13_vs_71319\|\|full | 1296 | No | NA | 3 | 0 | 2 | Not-determined | Genome-fragment | NA | NA | 0 |
| S13_vs_71448\|\|full | 1226 | No | NA | 3 | 0 | 0 | Not-determined | Genome-fragment | NA | NA | 0 |
| S13_vs_71450\|\|full | 2701 | No | NA | 4 | 0 | 2 | Not-determined | Genome-fragment | NA | NA | 0 |
| S13_vs_71476\|\|full | 1263 | No | NA | 3 | 0 | 1 | Not-determined | Genome-fragment | NA | NA | 0 |
| S13_vs_71522\|\|full | 1917 | No | NA | 4 | 0 | 0 | Not-determined | Genome-fragment | NA | NA | 0 |
| S13_vs_71851\|\|full | 1745 | No | NA | 3 | 0 | 3 | Not-determined | Genome-fragment | NA | NA | 0 |
| S15_extracted_52124 | 1282 | No | NA | 5 | 0 | 0 | Low-quality | Genome-fragment | 0.88 | AAI-based (high-confidence) | 0 |
| S15_vs_5582\|\|full | 1282 | No | NA | 2 | 0 | 0 | Low-quality | Genome-fragment | 3.15 | AAI-based (medium-confidence) | 0 |
| S13_vs_72212\|\|full | 3714 | No | NA | 3 | 0 | 3 | Not-determined | Genome-fragment | NA | NA | 0 |
| S13_vs_72276\|\|full | 2562 | No | NA | 2 | 0 | 0 | Not-determined | Genome-fragment | NA | NA | 0 |
| S13_vs_72637\|\|full | 2218 | No | NA | 3 | 0 | 2 | Not-determined | Genome-fragment | NA | NA | 0 |
| S13_vs_72654\|\|full | 2035 | No | NA | 3 | 0 | 2 | Not-determined | Genome-fragment | NA | NA | 0 |
| S13_vs_72668\|\|full | 1954 | No | NA | 3 | 0 | 1 | Not-determined | Genome-fragment | NA | NA | 0 |
| S13_vs_72848\|\|full | 1109 | No | NA | 2 | 0 | 1 | Not-determined | Genome-fragment | NA | NA | 0 |
| S13_vs_72996\|\|full | 1123 | No | NA | 3 | 0 | 0 | Not-determined | Genome-fragment | NA | NA | 0 |
| S13_vs_73159\|\|full | 3773 | No | NA | 4 | 0 | 2 | Not-determined | Genome-fragment | NA | NA | 0 |
| S4_extracted_14061 | 1282 | No | NA | 2 | 1 | 0 | Low-quality | Genome-fragment | 4.23 | AAI-based (high-confidence) | 0 |
| S13_vs_73333\|\|full | 1654 | No | NA | 2 | 0 | 0 | Not-determined | Genome-fragment | NA | NA | 0 |
| S8_vs_k141_70989\|\|full | 1282 | No | NA | 3 | 2 | 0 | Low-quality | Genome-fragment | 2.86 | AAI-based (high-confidence) | 0 |
| S13_vs_73418\|\|full | 12113 | No | NA | 11 | 0 | 6 | Not-determined | Genome-fragment | NA | NA | 0 |
| S13_vs_73517\|\|full | 1238 | No | NA | 3 | 0 | 0 | Not-determined | Genome-fragment | NA | NA | 0 |
| S13_vs_73658\|\|full | 2266 | No | NA | 3 | 0 | 2 | Not-determined | Genome-fragment | NA | NA | 0 |
| S13_vs_73837\|\|full | 1701 | No | NA | 4 | 0 | 0 | Not-determined | Genome-fragment | NA | NA | 0 |
| S13_vs_73855\|\|full | 1794 | No | NA | 4 | 0 | 0 | Not-determined | Genome-fragment | NA | NA | 0 |
| S13_vs_73864\|\|full | 2323 | No | NA | 3 | 0 | 3 | Not-determined | Genome-fragment | NA | NA | 0 |
| S13_vs_73911\|\|full | 1870 | No | NA | 2 | 0 | 2 | Not-determined | Genome-fragment | NA | NA | 0 |
| S17_vs_k141_95044\|\|full | 1283 | No | NA | 3 | 0 | 0 | Low-quality | Genome-fragment | 1.86 | AAI-based (high-confidence) | 0 |

| | | | | | | | | | | | |
|---|---|---|---|---|---|---|---|---|---|---|---|
| S13_vs_74518\|\|full | 1246 | No | NA | 3 | 0 | 3 | Not-determined | Genome-fragment | NA | NA | 0 |
| S13_vs_74542\|\|full | 1222 | No | NA | 5 | 0 | 0 | Not-determined | Genome-fragment | NA | NA | 0 |
| S13_vs_74580\|\|full | 1755 | No | NA | 4 | 0 | 0 | Not-determined | Genome-fragment | NA | NA | 0 |
| S9_vs_k141_72767\|\|full | 1283 | No | NA | 3 | 0 | 0 | Low-quality | Genome-fragment | 2.63 | AAI-based (medium-confidence) | 0 |
| S13_vs_74849\|\|full | 1180 | No | NA | 2 | 0 | 1 | Not-determined | Genome-fragment | NA | NA | 0 |
| S11_extracted_k141_37737 | 1283 | No | NA | 3 | 1 | 0 | Low-quality | Genome-fragment | 3.2 | AAI-based (high-confidence) | 0 |
| S13_vs_74971\|\|full | 1256 | No | NA | 2 | 0 | 1 | Not-determined | Genome-fragment | NA | NA | 0 |
| S13_vs_75012\|\|full | 1617 | No | NA | 2 | 0 | 0 | Not-determined | Genome-fragment | NA | NA | 0 |
| S14_vs_k141_10958\|\|full | 1283 | No | NA | 3 | 0 | 0 | Low-quality | Genome-fragment | 2.11 | AAI-based (high-confidence) | 0 |
| S13_vs_75369\|\|full | 1534 | No | NA | 3 | 0 | 1 | Not-determined | Genome-fragment | NA | NA | 0 |
| S7_vs_79482\|\|full | 1283 | No | NA | 3 | 1 | 0 | Low-quality | Genome-fragment | 3.49 | AAI-based (medium-confidence) | 0 |
| S13_vs_75681\|\|full | 2457 | No | NA | 4 | 0 | 0 | Not-determined | Genome-fragment | NA | NA | 0 |
| S13_vs_75695\|\|full | 2239 | No | NA | 3 | 0 | 3 | Not-determined | Genome-fragment | NA | NA | 0 |
| S16_extracted_50777 | 1283 | No | NA | 2 | 0 | 1 | Low-quality | Genome-fragment | 1.56 | AAI-based (high-confidence) | 0 |
| S13_vs_75867\|\|full | 2155 | No | NA | 2 | 0 | 1 | Not-determined | Genome-fragment | NA | NA | 0 |
| S13_vs_76004\|\|full | 1613 | No | NA | 3 | 0 | 0 | Not-determined | Genome-fragment | NA | NA | 0 |
| S13_vs_76115\|\|full | 2506 | No | NA | 2 | 0 | 0 | Not-determined | Genome-fragment | NA | NA | 0 |
| S6_vs_k141_42820\|\|full | 1283 | No | NA | 3 | 1 | 0 | Low-quality | Genome-fragment | 3.21 | AAI-based (high-confidence) | 0 |
| S13_vs_76193\|\|full | 3550 | No | NA | 4 | 0 | 1 | Not-determined | Genome-fragment | NA | NA | 0 |
| S8_extracted_k141_51330 | 1283 | No | NA | 2 | 2 | 0 | Low-quality | Genome-fragment | 2.83 | AAI-based (high-confidence) | 0 |
| S17_vs_k141_71998\|\|full | 1284 | No | NA | 3 | 0 | 2 | Low-quality | Genome-fragment | 0.45 | AAI-based (medium-confidence) | 0 |
| S13_vs_76573\|\|full | 1219 | No | NA | 2 | 0 | 1 | Not-determined | Genome-fragment | NA | NA | 0 |
| S9_vs_k141_84577\|\|full | 1284 | No | NA | 3 | 2 | 0 | Low-quality | Genome-fragment | 3.28 | AAI-based (high-confidence) | 0 |
| S13_vs_76934\|\|full | 1842 | No | NA | 3 | 0 | 3 | Not-determined | Genome-fragment | NA | NA | 0 |
| S13_vs_76950\|\|full | 2058 | No | NA | 3 | 0 | 3 | Not-determined | Genome-fragment | NA | NA | 0 |
| S11_vs_k141_101017\|\|full | 1284 | No | NA | 3 | 1 | 0 | Low-quality | Genome-fragment | 3.37 | AAI-based (high-confidence) | 0 |
| S13_vs_76980\|\|full | 3115 | No | NA | 3 | 0 | 3 | Not-determined | Genome-fragment | NA | NA | 0 |
| S13_vs_77128\|\|full | 1561 | No | NA | 4 | 0 | 0 | Not-determined | Genome-fragment | NA | NA | 0 |
| S14_extracted_k141_102871 | 1284 | No | NA | 4 | 0 | 0 | Low-quality | Genome-fragment | 1.54 | AAI-based (high-confidence) | 0 |
| S14_extracted_k141_52055 | 1284 | No | NA | 1 | 1 | 0 | Low-quality | Genome-fragment | 1.88 | HMM-based (lower-bound) | 0 |
| S13_vs_77370\|\|full | 1424 | No | NA | 3 | 0 | 2 | Not-determined | Genome-fragment | NA | NA | 0 |
| S13_vs_77981\|\|full | 20756 | No | NA | 26 | 0 | 13 | Not-determined | Genome-fragment | NA | NA | 0 |
| S13_vs_78017\|\|full | 2517 | No | NA | 5 | 0 | 0 | Not-determined | Genome-fragment | NA | NA | 0 |
| S13_vs_78620\|\|full | 1780 | No | NA | 2 | 0 | 2 | Not-determined | Genome-fragment | NA | NA | 0 |
| S13_vs_78659\|\|full | 1301 | No | NA | 2 | 0 | 1 | Not-determined | Genome-fragment | NA | NA | 0 |
| S13_vs_78895\|\|full | 1172 | No | NA | 3 | 0 | 0 | Not-determined | Genome-fragment | NA | NA | 0 |
| S13_vs_79793\|\|full | 1219 | No | NA | 3 | 0 | 2 | Not-determined | Genome-fragment | NA | NA | 0 |
| S14_extracted_k141_11983 | 1284 | No | NA | 2 | 0 | 0 | Low-quality | Genome-fragment | 1.29 | AAI-based (high-confidence) | 0 |
| S13_vs_79966\|\|full | 5152 | No | NA | 8 | 0 | 1 | Not-determined | Genome-fragment | NA | NA | 0 |
| S13_vs_80078\|\|full | 1659 | No | NA | 2 | 0 | 1 | Not-determined | Genome-fragment | NA | NA | 0 |
| S13_vs_80446\|\|full | 1971 | No | NA | 3 | 0 | 0 | Not-determined | Genome-fragment | NA | NA | 0 |
| S13_vs_80456\|\|full | 1646 | No | NA | 2 | 0 | 0 | Not-determined | Genome-fragment | NA | NA | 0 |
| S13_vs_80570\|\|full | 2050 | No | NA | 3 | 0 | 2 | Not-determined | Genome-fragment | NA | NA | 0 |
| S13_vs_80579\|\|full | 3982 | No | NA | 7 | 0 | 3 | Not-determined | Genome-fragment | NA | NA | 0 |
| S13_vs_80645\|\|full | 2428 | No | NA | 4 | 0 | 0 | Not-determined | Genome-fragment | NA | NA | 0 |
| S14_extracted_k141_56808 | 1284 | No | NA | 3 | 0 | 0 | Low-quality | Genome-fragment | 2.87 | AAI-based (medium-confidence) | 0 |
| S13_vs_80839\|\|full | 1368 | No | NA | 1 | 0 | 0 | Not-determined | Genome-fragment | NA | NA | 0 |
| S14_vs_k141_107498\|\|full | 1284 | No | NA | 2 | 0 | 2 | Low-quality | Genome-fragment | 1.59 | AAI-based (medium-confidence) | 0 |
| S13_vs_81140\|\|full | 10596 | No | NA | 8 | 0 | 1 | Not-determined | Genome-fragment | NA | NA | 0 |
| S13_vs_81222\|\|full | 3992 | No | NA | 9 | 0 | 0 | Not-determined | Genome-fragment | NA | NA | 0 |
| S1_extracted_k141_56901 | 1284 | No | NA | 3 | 2 | 0 | Low-quality | Genome-fragment | 2.88 | AAI-based (high-confidence) | 0 |
| S13_vs_81364\|\|full | 2131 | No | NA | 1 | 0 | 1 | Not-determined | Genome-fragment | NA | NA | 0 |
| S13_vs_81494\|\|full | 2502 | No | NA | 5 | 0 | 1 | Not-determined | Genome-fragment | NA | NA | 0 |
| S7_vs_95122\|\|full | 1284 | No | NA | 3 | 0 | 0 | Low-quality | Genome-fragment | 2.86 | AAI-based (medium-confidence) | 0 |
| S13_vs_81789\|\|full | 2508 | No | NA | 4 | 0 | 1 | Not-determined | Genome-fragment | NA | NA | 0 |
| S16_extracted_14634 | 1284 | No | NA | 4 | 0 | 0 | Low-quality | Genome-fragment | 2.14 | AAI-based (high-confidence) | 0 |
| S13_vs_82039\|\|full | 2238 | No | NA | 3 | 0 | 0 | Not-determined | Genome-fragment | NA | NA | 0 |
| S13_vs_82277\|\|full | 1259 | No | NA | 2 | 0 | 2 | Not-determined | Genome-fragment | NA | NA | 0 |
| S13_vs_82498\|\|full | 4264 | No | NA | 3 | 0 | 2 | Not-determined | Genome-fragment | NA | NA | 0 |
| S13_vs_82520\|\|full | 2505 | No | NA | 3 | 0 | 1 | Not-determined | Genome-fragment | NA | NA | 0 |
| S13_vs_83104\|\|full | 1136 | No | NA | 2 | 0 | 1 | Not-determined | Genome-fragment | NA | NA | 0 |
| S13_vs_83228\|\|full | 2576 | No | NA | 3 | 0 | 2 | Not-determined | Genome-fragment | NA | NA | 0 |
| S10_vs_20402\|\|full | 1284 | No | NA | 3 | 0 | 1 | Low-quality | Genome-fragment | 2.84 | AAI-based (high-confidence) | 0 |
| S13_vs_83326\|\|full | 2083 | No | NA | 4 | 0 | 1 | Not-determined | Genome-fragment | NA | NA | 0 |
| S10_vs_27043\|\|full | 1284 | No | NA | 3 | 1 | 0 | Low-quality | Genome-fragment | 1.9 | HMM-based (lower-bound) | 0 |
| S13_vs_83404\|\|full | 1719 | No | NA | 3 | 0 | 0 | Not-determined | Genome-fragment | NA | NA | 0 |
| S13_vs_83447\|\|full | 56650 | No | NA | 77 | 16 | 4 | High-quality | High-quality | 100 | AAI-based (high-confidence) | 0 |
| S12_vs_k141_187991\|\|full | 1284 | No | NA | 4 | 1 | 0 | Low-quality | Genome-fragment | 3 | AAI-based (high-confidence) | 0 |
| S13_vs_83622\|\|full | 1779 | No | NA | 3 | 0 | 0 | Not-determined | Genome-fragment | NA | NA | 0 |
| S13_vs_83809\|\|full | 1711 | No | NA | 2 | 0 | 0 | Not-determined | Genome-fragment | NA | NA | 0 |
| S13_vs_84187\|\|full | 1969 | No | NA | 2 | 0 | 0 | Not-determined | Genome-fragment | NA | NA | 0 |
| S13_vs_84458\|\|full | 1386 | No | NA | 3 | 0 | 0 | Not-determined | Genome-fragment | NA | NA | 0 |
| S13_vs_84539\|\|full | 2149 | No | NA | 4 | 0 | 3 | Not-determined | Genome-fragment | NA | NA | 0 |
| S4_extracted_6170 | 1284 | No | NA | 4 | 0 | 1 | Low-quality | Genome-fragment | 1.62 | AAI-based (medium-confidence) | 0 |
| S8_extracted_k141_54823 | 1284 | No | NA | 3 | 0 | 1 | Low-quality | Genome-fragment | 0.96 | AAI-based (medium-confidence) | 0 |
| S8_extracted_k141_49838 | 1284 | No | NA | 2 | 0 | 0 | Low-quality | Genome-fragment | 1.19 | AAI-based (high-confidence) | 0 |
| S13_vs_85204\|\|full | 1840 | No | NA | 3 | 0 | 0 | Not-determined | Genome-fragment | NA | NA | 0 |
| S13_vs_85252\|\|full | 1262 | No | NA | 3 | 0 | 0 | Not-determined | Genome-fragment | NA | NA | 0 |
| S13_vs_85254\|\|full | 2127 | No | NA | 6 | 0 | 2 | Not-determined | Genome-fragment | NA | NA | 0 |
| S8_vs_k141_105554\|\|full | 1284 | No | NA | 3 | 0 | 0 | Low-quality | Genome-fragment | 3.08 | AAI-based (high-confidence) | 0 |
| S17_vs_k141_11988\|\|full | 1285 | No | NA | 3 | 1 | 0 | Low-quality | Genome-fragment | 2.2 | HMM-based (lower-bound) | 0 |
| S13_vs_85535\|\|full | 19277 | No | NA | 23 | 0 | 11 | Not-determined | Genome-fragment | NA | NA | 0 |
| S13_vs_85647\|\|full | 1206 | No | NA | 4 | 0 | 0 | Not-determined | Genome-fragment | NA | NA | 0 |
| S9_vs_k141_75327\|\|lt2gene | 1285 | No | NA | 2 | 1 | 0 | Low-quality | Genome-fragment | 2.86 | AAI-based (high-confidence) | 0 |
| S13_vs_85853\|\|full | 1588 | No | NA | 4 | 0 | 2 | Not-determined | Genome-fragment | NA | NA | 0 |
| S13_vs_85895\|\|full | 2221 | No | NA | 3 | 0 | 0 | Not-determined | Genome-fragment | NA | NA | 0 |
| S11_extracted_k141_12025 | 1285 | No | NA | 2 | 2 | 0 | Low-quality | Genome-fragment | 3.17 | AAI-based (high-confidence) | 0 |
| S13_vs_86100\|\|full | 1320 | No | NA | 4 | 0 | 0 | Not-determined | Genome-fragment | NA | NA | 0 |
| S11_vs_k141_88175\|\|full | 1285 | No | NA | 4 | 1 | 0 | Low-quality | Genome-fragment | 2.21 | AAI-based (high-confidence) | 0 |
| S13_vs_86212\|\|full | 1859 | No | NA | 4 | 0 | 0 | Not-determined | Genome-fragment | NA | NA | 0 |
| S13_vs_86222\|\|full | 1736 | No | NA | 2 | 0 | 1 | Not-determined | Genome-fragment | NA | NA | 0 |
| S13_vs_86251\|\|full | 1452 | No | NA | 5 | 0 | 0 | Not-determined | Genome-fragment | NA | NA | 0 |
| S11_vs_k141_79894\|\|lt2gene | 1285 | No | NA | 1 | 1 | 0 | Low-quality | Genome-fragment | 1.18 | HMM-based (lower-bound) | 0 |
| S13_extracted_108812 | 1285 | No | NA | 2 | 1 | 0 | Low-quality | Genome-fragment | 0.82 | HMM-based (lower-bound) | 0 |
| S13_vs_86796\|\|full | 2442 | No | NA | 4 | 0 | 1 | Not-determined | Genome-fragment | NA | NA | 0 |
| S1_vs_k141_138039\|\|full | 1285 | No | NA | 3 | 0 | 1 | Low-quality | Genome-fragment | 0.34 | AAI-based (medium-confidence) | 0 |
| S13_vs_86872\|\|full | 1389 | No | NA | 3 | 0 | 2 | Not-determined | Genome-fragment | NA | NA | 0 |
| S13_vs_87084\|\|full | 1231 | No | NA | 2 | 0 | 1 | Not-determined | Genome-fragment | NA | NA | 0 |
| S13_vs_87108\|\|full | 10612 | No | NA | 16 | 3 | 0 | Medium-quality | Genome-fragment | 77.8 | AAI-based (medium-confidence) | 0 |
| S3_vs_k141_76574\|\|full | 1285 | No | NA | 3 | 0 | 0 | Low-quality | Genome-fragment | 2.46 | AAI-based (medium-confidence) | 0 |
| S13_vs_87197\|\|full | 2147 | No | NA | 2 | 0 | 0 | Not-determined | Genome-fragment | NA | NA | 0 |
| S3_vs_k141_117145\|\|full | 1285 | No | NA | 5 | 0 | 0 | Low-quality | Genome-fragment | 0.87 | AAI-based (medium-confidence) | 0 |
| S13_vs_87204\|\|full | 5785 | No | NA | 3 | 0 | 1 | Not-determined | Genome-fragment | NA | NA | 0 |
| S5_extracted_k141_122911 | 1285 | No | NA | 1 | 0 | 1 | Low-quality | Genome-fragment | 0.84 | AAI-based (medium-confidence) | 0 |
| S13_vs_87782\|\|full | 1064 | No | NA | 3 | 0 | 1 | Not-determined | Genome-fragment | NA | NA | 0 |
| S5_vs_k141_141907\|\|full | 1285 | No | NA | 3 | 0 | 1 | Low-quality | Genome-fragment | 2.25 | AAI-based (medium-confidence) | 0 |
| S16_extracted_27364 | 1285 | No | NA | 4 | 1 | 0 | Low-quality | Genome-fragment | 2.38 | AAI-based (medium-confidence) | 0 |
| S13_vs_87981\|\|full | 1128 | No | NA | 2 | 0 | 0 | Not-determined | Genome-fragment | NA | NA | 0 |
| S13_vs_88012\|\|full | 1296 | No | NA | 3 | 0 | 0 | Not-determined | Genome-fragment | NA | NA | 0 |
| S13_vs_88059\|\|full | 2900 | No | NA | 3 | 0 | 1 | Not-determined | Genome-fragment | NA | NA | 0 |
| S13_vs_88097\|\|full | 1737 | No | NA | 3 | 0 | 3 | Not-determined | Genome-fragment | NA | NA | 0 |
| S13_vs_88101\|\|full | 1253 | No | NA | 2 | 0 | 1 | Not-determined | Genome-fragment | NA | NA | 0 |
| S13_vs_88103\|\|full | 3441 | No | NA | 4 | 0 | 0 | Not-determined | Genome-fragment | NA | NA | 0 |
| S13_vs_88234\|\|full | 1011 | No | NA | 3 | 0 | 0 | Not-determined | Genome-fragment | NA | NA | 0 |
| S16_extracted_101202 | 1285 | No | NA | 2 | 0 | 0 | Low-quality | Genome-fragment | 1.14 | AAI-based (medium-confidence) | 0 |
| S10_extracted_37968 | 1285 | No | NA | 3 | 0 | 0 | Low-quality | Genome-fragment | 0.75 | AAI-based (high-confidence) | 0 |
| S13_vs_88573\|\|full | 1529 | No | NA | 3 | 0 | 2 | Not-determined | Genome-fragment | NA | NA | 0 |
| S13_vs_88674\|\|full | 3292 | No | NA | 5 | 0 | 1 | Not-determined | Genome-fragment | NA | NA | 0 |
| S13_vs_88907\|\|full | 1169 | No | NA | 2 | 0 | 1 | Not-determined | Genome-fragment | NA | NA | 0 |
| S13_vs_88917\|\|full | 1484 | No | NA | 2 | 0 | 0 | Not-determined | Genome-fragment | NA | NA | 0 |
| S13_vs_89299\|\|full | 2154 | No | NA | 3 | 0 | 1 | Not-determined | Genome-fragment | NA | NA | 0 |
| S13_vs_89656\|\|full | 6197 | No | NA | 7 | 0 | 3 | Not-determined | Genome-fragment | NA | NA | 0 |
| S13_vs_89871\|\|full | 4373 | No | NA | 4 | 0 | 2 | Not-determined | Genome-fragment | NA | NA | 0 |
| S13_vs_90223\|\|full | 1074 | No | NA | 3 | 0 | 1 | Not-determined | Genome-fragment | NA | NA | 0 |
| S10_vs_118236\|\|full | 1285 | No | NA | 4 | 1 | 0 | Low-quality | Genome-fragment | 3.3 | AAI-based (medium-confidence) | 0 |
| S2_extracted_k141_5390 | 1285 | No | NA | 3 | 0 | 2 | Low-quality | Genome-fragment | 1.09 | AAI-based (high-confidence) | 0 |
| S13_vs_90280\|\|full | 1610 | No | NA | 4 | 0 | 0 | Not-determined | Genome-fragment | NA | NA | 0 |
| S13_vs_90430\|\|full | 2930 | No | NA | 3 | 0 | 2 | Not-determined | Genome-fragment | NA | NA | 0 |
| S13_vs_90441\|\|full | 1457 | No | NA | 2 | 0 | 0 | Not-determined | Genome-fragment | NA | NA | 0 |
| S13_vs_90489\|\|full | 1751 | No | NA | 2 | 0 | 0 | Not-determined | Genome-fragment | NA | NA | 0 |
| S13_vs_90500\|\|full | 3490 | No | NA | 4 | 0 | 3 | Not-determined | Genome-fragment | NA | NA | 0 |
| S8_vs_k141_138678\|\|full | 1285 | No | NA | 3 | 1 | 0 | Low-quality | Genome-fragment | 2.97 | AAI-based (high-confidence) | 0 |
| S9_vs_k141_61351\|\|lt2gene | 1286 | No | NA | 2 | 1 | 0 | Low-quality | Genome-fragment | 2.44 | AAI-based (medium-confidence) | 0 |
| S13_vs_90967\|\|full | 4316 | No | NA | 6 | 1 | 0 | Medium-quality | Genome-fragment | 78.07 | AAI-based (high-confidence) | 0 |
| S13_vs_91158\|\|full | 1446 | No | NA | 3 | 0 | 1 | Not-determined | Genome-fragment | NA | NA | 0 |
| S11_extracted_k141_45261 | 1286 | No | NA | 1 | 0 | 0 | Low-quality | Genome-fragment | 1.63 | AAI-based (medium-confidence) | 0 |
| S13_vs_91417\|\|full | 6246 | No | NA | 5 | 0 | 2 | Not-determined | Genome-fragment | NA | NA | 0 |
| S11_vs_k141_22246\|\|full | 1286 | No | NA | 3 | 2 | 0 | Low-quality | Genome-fragment | 2.98 | AAI-based (medium-confidence) | 0 |
| S13_vs_91633\|\|full | 2211 | No | NA | 3 | 0 | 1 | Not-determined | Genome-fragment | NA | NA | 0 |
| S13_vs_92136\|\|full | 1399 | No | NA | 3 | 0 | 0 | Not-determined | Genome-fragment | NA | NA | 0 |
| S13_vs_152907\|\|full | 1286 | No | NA | 2 | 0 | 1 | Low-quality | Genome-fragment | 0.83 | AAI-based (medium-confidence) | 0 |
| S13_vs_92264\|\|full | 1952 | No | NA | 2 | 0 | 1 | Not-determined | Genome-fragment | NA | NA | 0 |
| S1_vs_k141_114212\|\|full | 1286 | No | NA | 3 | 0 | 0 | Low-quality | Genome-fragment | 2.82 | AAI-based (high-confidence) | 0 |
| S13_vs_92307\|\|full | 1348 | No | NA | 2 | 0 | 1 | Not-determined | Genome-fragment | NA | NA | 0 |
| S3_vs_k141_35958\|\|lt2gene | 1286 | No | NA | 1 | 1 | 0 | Low-quality | Genome-fragment | 2.16 | HMM-based (lower-bound) | 0 |
| S5_vs_k141_51254\|\|lt2gene | 1286 | No | NA | 2 | 1 | 0 | Low-quality | Genome-fragment | 1.53 | HMM-based (lower-bound) | 0 |
| S7_extracted_115588 | 1286 | No | NA | 2 | 1 | 0 | Low-quality | Genome-fragment | 4.59 | HMM-based (lower-bound) | 0 |
| S13_vs_92693\|\|full | 1634 | No | NA | 2 | 0 | 2 | Not-determined | Genome-fragment | NA | NA | 0 |
| S7_vs_46396\|\|lt2gene | 1286 | No | NA | 2 | 2 | 0 | Low-quality | Genome-fragment | 2.97 | AAI-based (high-confidence) | 0 |
| S13_vs_92846\|\|full | 2098 | No | NA | 5 | 0 | 0 | Not-determined | Genome-fragment | NA | NA | 0 |
| S7_vs_81246\|\|lt2gene | 1286 | No | NA | 2 | 0 | 0 | Low-quality | Genome-fragment | 3.27 | AAI-based (medium-confidence) | 0 |
| S13_vs_93081\|\|full | 1228 | No | NA | 2 | 0 | 1 | Not-determined | Genome-fragment | NA | NA | 0 |
| S13_vs_93182\|\|full | 1368 | No | NA | 2 | 0 | 1 | Not-determined | Genome-fragment | NA | NA | 0 |
| S13_vs_93326\|\|full | 1589 | No | NA | 3 | 0 | 0 | Not-determined | Genome-fragment | NA | NA | 0 |
| S16_vs_117654\|\|full | 1286 | No | NA | 2 | 0 | 0 | Low-quality | Genome-fragment | 1.24 | AAI-based (medium-confidence) | 0 |
| S13_vs_93466\|\|full | 2981 | No | NA | 2 | 0 | 0 | Not-determined | Genome-fragment | NA | NA | 0 |
| S12_extracted_k141_96472 | 1286 | No | NA | 2 | 0 | 0 | Low-quality | Genome-fragment | 2.05 | AAI-based (high-confidence) | 0 |
| S12_extracted_k141_136339 | 1286 | No | NA | 3 | 2 | 0 | Low-quality | Genome-fragment | 2.1 | AAI-based (high-confidence) | 0 |
| S13_vs_93782\|\|full | 2343 | No | NA | 3 | 0 | 3 | Not-determined | Genome-fragment | NA | NA | 0 |
| S8_extracted_k141_183915 | 1286 | No | NA | 2 | 0 | 0 | Low-quality | Genome-fragment | 2.63 | AAI-based (high-confidence) | 0 |
| S8_extracted_k141_178362 | 1286 | No | NA | 5 | 0 | 0 | Low-quality | Genome-fragment | 1.74 | AAI-based (medium-confidence) | 0 |
| S13_vs_94233\|\|full | 2290 | No | NA | 4 | 0 | 3 | Not-determined | Genome-fragment | NA | NA | 0 |
| S17_extracted_k141_74432 | 1287 | No | NA | 2 | 0 | 1 | Low-quality | Genome-fragment | 0.84 | AAI-based (high-confidence) | 0 |
| S13_vs_94522\|\|full | 1744 | No | NA | 2 | 0 | 0 | Not-determined | Genome-fragment | NA | NA | 0 |
| S11_extracted_k141_89684 | 1287 | No | NA | 4 | 0 | 0 | Low-quality | Genome-fragment | 2.31 | AAI-based (high-confidence) | 0 |
| S13_extracted_108364 | 1287 | No | NA | 2 | 0 | 1 | Low-quality | Genome-fragment | 0.95 | AAI-based (medium-confidence) | 0 |
| S14_vs_k141_76106\|\|full | 1287 | No | NA | 4 | 1 | 0 | Low-quality | Genome-fragment | 3.56 | AAI-based (high-confidence) | 0 |
| S13_vs_94899\|\|full | 1743 | No | NA | 3 | 0 | 1 | Not-determined | Genome-fragment | NA | NA | 0 |
| S13_vs_94981\|\|full | 5188 | No | NA | 5 | 0 | 3 | Not-determined | Genome-fragment | NA | NA | 0 |
| S3_vs_k141_118553\|\|full | 1287 | No | NA | 3 | 0 | 0 | Low-quality | Genome-fragment | 3.34 | AAI-based (high-confidence) | 0 |
| S13_vs_95217\|\|full | 1558 | No | NA | 3 | 0 | 2 | Not-determined | Genome-fragment | NA | NA | 0 |
| S13_vs_95293\|\|full | 2221 | No | NA | 3 | 0 | 1 | Not-determined | Genome-fragment | NA | NA | 0 |
| S13_vs_95520\|\|full | 1201 | No | NA | 4 | 0 | 0 | Not-determined | Genome-fragment | NA | NA | 0 |
| S5_extracted_k141_69756 | 1287 | No | NA | 2 | 1 | 0 | Low-quality | Genome-fragment | 2.33 | AAI-based (high-confidence) | 0 |
| S7_vs_6672\|\|full | 1287 | No | NA | 2 | 2 | 0 | Low-quality | Genome-fragment | 2.88 | AAI-based (high-confidence) | 0 |
| S16_extracted_28204 | 1287 | No | NA | 4 | 1 | 0 | Low-quality | Genome-fragment | 3.16 | AAI-based (high-confidence) | 0 |
| S16_extracted_75737 | 1287 | No | NA | 2 | 1 | 0 | Low-quality | Genome-fragment | 3.21 | AAI-based (medium-confidence) | 0 |
| S13_vs_96242\|\|full | 2606 | No | NA | 4 | 0 | 2 | Not-determined | Genome-fragment | NA | NA | 0 |
| S16_vs_44219\|\|full | 1287 | No | NA | 3 | 0 | 0 | Low-quality | Genome-fragment | 1.74 | AAI-based (high-confidence) | 0 |
| S10_extracted_64265 | 1287 | No | NA | 3 | 0 | 0 | Low-quality | Genome-fragment | 3.19 | AAI-based (medium-confidence) | 0 |
| S10_vs_54100\|\|full | 1287 | No | NA | 3 | 1 | 0 | Low-quality | Genome-fragment | 2.72 | AAI-based (high-confidence) | 0 |
| S13_vs_97142\|\|full | 4751 | No | NA | 6 | 0 | 1 | Not-determined | Genome-fragment | NA | NA | 0 |
| S4_extracted_15117 | 1287 | No | NA | 1 | 1 | 0 | Low-quality | Genome-fragment | 2.13 | AAI-based (high-confidence) | 0 |
| S13_vs_97247\|\|full | 6062 | No | NA | 7 | 0 | 5 | Not-determined | Genome-fragment | NA | NA | 0 |
| S13_vs_97313\|\|full | 1310 | No | NA | 2 | 0 | 1 | Not-determined | Genome-fragment | NA | NA | 0 |
| S8_extracted_k141_191341 | 1287 | No | NA | 2 | 0 | 0 | Low-quality | Genome-fragment | 1.38 | AAI-based (medium-confidence) | 0 |
| S17_extracted_k141_15290 | 1288 | No | NA | 4 | 0 | 0 | Low-quality | Genome-fragment | 2.31 | AAI-based (high-confidence) | 0 |
| S9_vs_k141_1288\|\|full | 1288 | No | NA | 2 | 0 | 1 | Low-quality | Genome-fragment | 3.01 | AAI-based (medium-confidence) | 0 |
| S13_vs_97832\|\|full | 1372 | No | NA | 2 | 0 | 1 | Not-determined | Genome-fragment | NA | NA | 0 |
| S11_extracted_k141_30481 | 1288 | No | NA | 4 | 1 | 0 | Low-quality | Genome-fragment | 3.24 | AAI-based (medium-confidence) | 0 |
| S13_vs_98160\|\|full | 1125 | No | NA | 2 | 0 | 0 | Not-determined | Genome-fragment | NA | NA | 0 |
| S13_vs_67734\|\|full | 1288 | No | NA | 2 | 0 | 1 | Low-quality | Genome-fragment | 0.91 | AAI-based (high-confidence) | 0 |
| S1_vs_k141_159562\|\|full | 1288 | No | NA | 2 | 0 | 0 | Low-quality | Genome-fragment | 0.86 | AAI-based (high-confidence) | 0 |
| S5_vs_k141_322\|\|full | 1288 | No | NA | 3 | 0 | 1 | Low-quality | Genome-fragment | 1.1 | AAI-based (medium-confidence) | 0 |
| S13_vs_98591\|\|full | 36816 | No | NA | 54 | 21 | 0 | Medium-quality | Genome-fragment | 85.63 | AAI-based (high-confidence) | 0 |
| S16_vs_44766\|\|full | 1288 | No | NA | 4 | 2 | 0 | Low-quality | Genome-fragment | 2.52 | AAI-based (high-confidence) | 0 |
| S6_extracted_k141_36680 | 1288 | No | NA | 3 | 1 | 0 | Low-quality | Genome-fragment | 2.2 | AAI-based (medium-confidence) | 0 |
| S6_vs_k141_708\|\|full | 1288 | No | NA | 2 | 0 | 0 | Low-quality | Genome-fragment | 0.99 | AAI-based (high-confidence) | 0 |
| S17_vs_k141_47807\|\|full | 1289 | No | NA | 2 | 0 | 0 | Low-quality | Genome-fragment | 3.13 | AAI-based (medium-confidence) | 0 |
| S13_vs_98880\|\|full | 1254 | No | NA | 3 | 0 | 0 | Not-determined | Genome-fragment | NA | NA | 0 |
| S13_vs_98886\|\|full | 1136 | No | NA | 4 | 0 | 0 | Not-determined | Genome-fragment | NA | NA | 0 |
| S9_vs_k141_66802\|\|full | 1289 | No | NA | 3 | 0 | 0 | Low-quality | Genome-fragment | 1.72 | AAI-based (high-confidence) | 0 |
| S13_vs_98986\|\|full | 5752 | No | NA | 5 | 0 | 4 | Not-determined | Genome-fragment | NA | NA | 0 |
| S9_vs_k141_51475\|\|lt2gene | 1289 | No | NA | 1 | 1 | 0 | Low-quality | Genome-fragment | 2.17 | HMM-based (lower-bound) | 0 |
| S11_vs_k141_32783\|\|full | 1289 | No | NA | 3 | 0 | 0 | Low-quality | Genome-fragment | 3.47 | AAI-based (medium-confidence) | 0 |
| S13_vs_17846\|\|full | 1289 | No | NA | 4 | 1 | 0 | Low-quality | Genome-fragment | 2.05 | AAI-based (high-confidence) | 0 |
| S13_vs_99539\|\|full | 1370 | No | NA | 4 | 0 | 0 | Not-determined | Genome-fragment | NA | NA | 0 |
| S1_vs_k141_104821\|\|full | 1289 | No | NA | 2 | 0 | 0 | Low-quality | Genome-fragment | 1.82 | AAI-based (high-confidence) | 0 |
| S1_vs_k141_16554\|\|lt2gene | 1289 | No | NA | 2 | 1 | 0 | Low-quality | Genome-fragment | 0.89 | HMM-based (lower-bound) | 0 |
| S7_extracted_7051 | 1289 | No | NA | 4 | 0 | 0 | Low-quality | Genome-fragment | 3.13 | AAI-based (medium-confidence) | 0 |
| S10_extracted_90424 | 1289 | No | NA | 3 | 0 | 1 | Low-quality | Genome-fragment | 1.63 | AAI-based (medium-confidence) | 0 |
| S13_vs_100013\|\|full | 2521 | No | NA | 2 | 0 | 1 | Not-determined | Genome-fragment | NA | NA | 0 |
| S13_vs_100077\|\|full | 16410 | No | NA | 16 | 0 | 7 | Not-determined | Genome-fragment | NA | NA | 0 |
| S10_vs_4849\|\|lt2gene | 1289 | No | NA | 2 | 1 | 0 | Low-quality | Genome-fragment | 1.71 | HMM-based (lower-bound) | 0 |
| S13_vs_100367\|\|full | 2131 | No | NA | 2 | 0 | 2 | Not-determined | Genome-fragment | NA | NA | 0 |
| S13_vs_100371\|\|full | 2696 | No | NA | 3 | 0 | 0 | Not-determined | Genome-fragment | NA | NA | 0 |
| S13_vs_100468\|\|full | 2141 | No | NA | 3 | 0 | 0 | Not-determined | Genome-fragment | NA | NA | 0 |
| S13_vs_100559\|\|full | 3871 | No | NA | 5 | 0 | 1 | Not-determined | Genome-fragment | NA | NA | 0 |
| S13_vs_100631\|\|full | 3266 | No | NA | 3 | 0 | 0 | Not-determined | Genome-fragment | NA | NA | 0 |
| S13_vs_100690\|\|full | 3484 | No | NA | 5 | 0 | 2 | Not-determined | Genome-fragment | NA | NA | 0 |
| S12_vs_k141_116559\|\|full | 1289 | No | NA | 2 | 0 | 0 | Low-quality | Genome-fragment | 2.33 | AAI-based (medium-confidence) | 0 |
| S8_extracted_k141_123850 | 1289 | No | NA | 2 | 0 | 0 | Low-quality | Genome-fragment | 2.34 | AAI-based (medium-confidence) | 0 |
| S13_vs_101541\|\|full | 1097 | No | NA | 2 | 0 | 0 | Not-determined | Genome-fragment | NA | NA | 0 |
| S8_extracted_k141_11415 | 1289 | No | NA | 2 | 0 | 1 | Low-quality | Genome-fragment | 0.85 | AAI-based (high-confidence) | 0 |
| S13_vs_102146\|\|full | 1036 | No | NA | 2 | 0 | 0 | Not-determined | Genome-fragment | NA | NA | 0 |
| S13_vs_102287\|\|full | 8212 | No | NA | 10 | 0 | 8 | Not-determined | Genome-fragment | NA | NA | 0 |
| S13_vs_102356\|\|full | 3628 | No | NA | 3 | 0 | 0 | Not-determined | Genome-fragment | NA | NA | 0 |
| S13_vs_102884\|\|full | 4274 | No | NA | 5 | 0 | 0 | Not-determined | Genome-fragment | NA | NA | 0 |
| S13_vs_102899\|\|full | 1378 | No | NA | 3 | 0 | 0 | Not-determined | Genome-fragment | NA | NA | 0 |
| S13_vs_103000\|\|full | 1147 | No | NA | 2 | 0 | 0 | Not-determined | Genome-fragment | NA | NA | 0 |
| S17_extracted_k141_118112 | 1290 | No | NA | 2 | 0 | 1 | Low-quality | Genome-fragment | 2.21 | AAI-based (high-confidence) | 0 |
| S13_vs_103161\|\|full | 2251 | No | NA | 3 | 0 | 0 | Not-determined | Genome-fragment | NA | NA | 0 |
| S13_vs_103278\|\|full | 1165 | No | NA | 2 | 0 | 0 | Not-determined | Genome-fragment | NA | NA | 0 |
| S13_vs_103322\|\|full | 1014 | No | NA | 3 | 0 | 2 | Not-determined | Genome-fragment | NA | NA | 0 |
| S13_vs_103595\|\|full | 1842 | No | NA | 3 | 0 | 2 | Not-determined | Genome-fragment | NA | NA | 0 |
| S13_vs_103751\|\|full | 1992 | No | NA | 2 | 0 | 0 | Not-determined | Genome-fragment | NA | NA | 0 |
| S9_extracted_k141_29218 | 1290 | No | NA | 1 | 0 | 0 | Low-quality | Genome-fragment | 0.77 | AAI-based (high-confidence) | 0 |
| S9_vs_k141_2355\|\|lt2gene | 1290 | No | NA | 2 | 1 | 0 | Low-quality | Genome-fragment | 0.8 | HMM-based (lower-bound) | 0 |
| S13_vs_104198\|\|full | 1369 | No | NA | 2 | 0 | 0 | Not-determined | Genome-fragment | NA | NA | 0 |
| S13_vs_104367\|\|full | 5223 | No | NA | 12 | 0 | 0 | Not-determined | Genome-fragment | NA | NA | 0 |
| S13_vs_104383\|\|full | 6492 | No | NA | 6 | 0 | 3 | Not-determined | Genome-fragment | NA | NA | 0 |
| S14_extracted_k141_72646 | 1290 | No | NA | 1 | 0 | 0 | Low-quality | Genome-fragment | 0.21 | AAI-based (high-confidence) | 0 |
| S13_vs_104418\|\|full | 1132 | No | NA | 2 | 0 | 1 | Not-determined | Genome-fragment | NA | NA | 0 |
| S13_vs_104517\|\|full | 1065 | No | NA | 2 | 0 | 1 | Not-determined | Genome-fragment | NA | NA | 0 |
| S14_vs_k141_52739\|\|full | 1290 | No | NA | 4 | 3 | 0 | Low-quality | Genome-fragment | 3.55 | AAI-based (high-confidence) | 0 |
| S13_vs_104606\|\|full | 1419 | No | NA | 3 | 0 | 2 | Not-determined | Genome-fragment | NA | NA | 0 |
| S14_vs_k141_13520\|\|full | 1290 | No | NA | 3 | 0 | 0 | Low-quality | Genome-fragment | 2.64 | AAI-based (medium-confidence) | 0 |
| S13_vs_104940\|\|full | 2344 | No | NA | 3 | 0 | 1 | Not-determined | Genome-fragment | NA | NA | 0 |
| S13_vs_104946\|\|full | 1006 | No | NA | 3 | 0 | 0 | Not-determined | Genome-fragment | NA | NA | 0 |
| S13_vs_105097\|\|full | 2626 | No | NA | 3 | 0 | 2 | Not-determined | Genome-fragment | NA | NA | 0 |
| S13_vs_105337\|\|full | 2850 | No | NA | 3 | 0 | 2 | Not-determined | Genome-fragment | NA | NA | 0 |
| S5_extracted_k141_88410 | 1290 | No | NA | 1 | 0 | 0 | Low-quality | Genome-fragment | 2.22 | AAI-based (medium-confidence) | 0 |
| S5_vs_k141_17308\|\|full | 1290 | No | NA | 3 | 0 | 0 | Low-quality | Genome-fragment | 0.92 | AAI-based (medium-confidence) | 0 |
| S7_extracted_29858 | 1290 | No | NA | 1 | 1 | 0 | Low-quality | Genome-fragment | 1.73 | HMM-based (lower-bound) | 0 |
| S13_vs_105491\|\|full | 4169 | No | NA | 4 | 0 | 2 | Not-determined | Genome-fragment | NA | NA | 0 |
| S13_vs_105758\|\|full | 2284 | No | NA | 3 | 0 | 1 | Not-determined | Genome-fragment | NA | NA | 0 |
| S13_vs_105886\|\|full | 4455 | No | NA | 5 | 0 | 3 | Not-determined | Genome-fragment | NA | NA | 0 |
| S7_extracted_126646 | 1290 | No | NA | 3 | 2 | 0 | Low-quality | Genome-fragment | 9.56 | AAI-based (high-confidence) | 0 |
| S13_vs_105974\|\|full | 1148 | No | NA | 2 | 0 | 0 | Not-determined | Genome-fragment | NA | NA | 0 |
| S13_vs_106129\|\|full | 1878 | No | NA | 3 | 0 | 2 | Not-determined | Genome-fragment | NA | NA | 0 |
| S7_vs_5428\|\|full | 1290 | No | NA | 3 | 1 | 0 | Low-quality | Genome-fragment | 2.3 | AAI-based (high-confidence) | 0 |
| S13_vs_106474\|\|full | 1262 | No | NA | 1 | 0 | 0 | Not-determined | Genome-fragment | NA | NA | 0 |
| S13_vs_106504\|\|full | 3345 | No | NA | 4 | 0 | 3 | Not-determined | Genome-fragment | NA | NA | 0 |
| S13_vs_106667\|\|full | 2997 | No | NA | 5 | 0 | 1 | Not-determined | Genome-fragment | NA | NA | 0 |
| S13_vs_107105\|\|full | 1192 | No | NA | 2 | 0 | 1 | Not-determined | Genome-fragment | NA | NA | 0 |
| S13_vs_107223\|\|full | 1733 | No | NA | 2 | 0 | 2 | Not-determined | Genome-fragment | NA | NA | 0 |
| S13_vs_107405\|\|full | 2804 | No | NA | 3 | 0 | 2 | Not-determined | Genome-fragment | NA | NA | 0 |
| S13_vs_107465\|\|full | 1375 | No | NA | 2 | 0 | 0 | Not-determined | Genome-fragment | NA | NA | 0 |
| S7_vs_52309\|\|full | 1290 | No | NA | 4 | 0 | 0 | Low-quality | Genome-fragment | 2.21 | AAI-based (high-confidence) | 0 |
| S7_vs_40830\|\|lt2gene | 1290 | No | NA | 1 | 1 | 0 | Low-quality | Genome-fragment | 3.03 | AAI-based (high-confidence) | 0 |
| S13_vs_107719\|\|full | 1437 | No | NA | 4 | 0 | 1 | Not-determined | Genome-fragment | NA | NA | 0 |
| S13_vs_107727\|\|full | 1423 | No | NA | 3 | 0 | 0 | Not-determined | Genome-fragment | NA | NA | 0 |
| S13_vs_107963\|\|full | 5570 | No | NA | 5 | 0 | 1 | Not-determined | Genome-fragment | NA | NA | 0 |
| S13_vs_107990\|\|full | 1577 | No | NA | 2 | 0 | 2 | Not-determined | Genome-fragment | NA | NA | 0 |
| S13_vs_108031\|\|full | 2148 | No | NA | 3 | 0 | 0 | Not-determined | Genome-fragment | NA | NA | 0 |
| S13_vs_108047\|\|full | 1096 | No | NA | 2 | 0 | 0 | Not-determined | Genome-fragment | NA | NA | 0 |
| S13_vs_108069\|\|full | 4513 | No | NA | 5 | 0 | 5 | Not-determined | Genome-fragment | NA | NA | 0 |
| S13_vs_108120\|\|full | 1447 | No | NA | 3 | 0 | 2 | Not-determined | Genome-fragment | NA | NA | 0 |
| S13_vs_108252\|\|full | 7266 | No | NA | 6 | 0 | 4 | Not-determined | Genome-fragment | NA | NA | 0 |
| S10_extracted_79366 | 1290 | No | NA | 4 | 0 | 0 | Low-quality | Genome-fragment | 3.26 | AAI-based (medium-confidence) | 0 |
| S13_vs_108381\|\|full | 2328 | No | NA | 5 | 0 | 1 | Not-determined | Genome-fragment | NA | NA | 0 |
| S13_vs_108445\|\|full | 1389 | No | NA | 2 | 0 | 0 | Not-determined | Genome-fragment | NA | NA | 0 |
| S13_vs_108463\|\|full | 1954 | No | NA | 3 | 0 | 1 | Not-determined | Genome-fragment | NA | NA | 0 |
| S8_vs_k141_199772\|\|full | 1290 | No | NA | 3 | 2 | 0 | Low-quality | Genome-fragment | 3.01 | AAI-based (high-confidence) | 0 |
| S8_vs_k141_188276\|\|full | 1290 | No | NA | 2 | 0 | 0 | Low-quality | Genome-fragment | 2.93 | AAI-based (high-confidence) | 0 |
| S13_vs_109160\|\|full | 2588 | No | NA | 3 | 0 | 3 | Not-determined | Genome-fragment | NA | NA | 0 |
| S17_vs_k141_150732\|\|full | 1291 | No | NA | 3 | 2 | 0 | Low-quality | Genome-fragment | 2.76 | AAI-based (high-confidence) | 0 |
| S13_vs_109515\|\|full | 1863 | No | NA | 3 | 0 | 1 | Not-determined | Genome-fragment | NA | NA | 0 |
| S13_vs_109661\|\|full | 3021 | No | NA | 3 | 0 | 0 | Not-determined | Genome-fragment | NA | NA | 0 |
| S13_vs_110059\|\|full | 2899 | No | NA | 4 | 0 | 3 | Not-determined | Genome-fragment | NA | NA | 0 |
| S9_vs_k141_22626\|\|full | 1291 | No | NA | 2 | 0 | 0 | Low-quality | Genome-fragment | 2.05 | AAI-based (high-confidence) | 0 |
| S13_vs_110572\|\|full | 1798 | No | NA | 2 | 0 | 1 | Not-determined | Genome-fragment | NA | NA | 0 |
| S13_vs_110622\|\|full | 1771 | No | NA | 2 | 0 | 2 | Not-determined | Genome-fragment | NA | NA | 0 |
| S13_vs_110673\|\|full | 6112 | No | NA | 6 | 0 | 3 | Not-determined | Genome-fragment | NA | NA | 0 |
| S13_vs_110741\|\|full | 3263 | No | NA | 3 | 0 | 1 | Not-determined | Genome-fragment | NA | NA | 0 |
| S13_vs_110818\|\|full | 1634 | No | NA | 3 | 0 | 1 | Not-determined | Genome-fragment | NA | NA | 0 |
| S13_extracted_45787 | 1291 | No | NA | 2 | 0 | 1 | Low-quality | Genome-fragment | 0.4 | AAI-based (medium-confidence) | 0 |
| S13_vs_111037\|\|full | 2328 | No | NA | 2 | 0 | 2 | Not-determined | Genome-fragment | NA | NA | 0 |
| S13_vs_111098\|\|full | 3576 | No | NA | 4 | 0 | 0 | Not-determined | Genome-fragment | NA | NA | 0 |
| S13_vs_111128\|\|full | 1931 | No | NA | 2 | 0 | 2 | Not-determined | Genome-fragment | NA | NA | 0 |
| S13_extracted_141214 | 1291 | No | NA | 3 | 1 | 0 | Low-quality | Genome-fragment | 2.15 | AAI-based (high-confidence) | 0 |
| S13_vs_35761\|\|full | 1291 | No | NA | 3 | 0 | 1 | Low-quality | Genome-fragment | 2.25 | AAI-based (medium-confidence) | 0 |
| S13_vs_111398\|\|full | 1271 | No | NA | 3 | 0 | 1 | Not-determined | Genome-fragment | NA | NA | 0 |
| S13_vs_111863\|\|full | 1183 | No | NA | 3 | 0 | 2 | Not-determined | Genome-fragment | NA | NA | 0 |
| S13_vs_133097\|\|full | 1291 | No | NA | 2 | 0 | 0 | Low-quality | Genome-fragment | 2.71 | AAI-based (high-confidence) | 0 |
| S1_extracted_k141_100640 | 1291 | No | NA | 4 | 2 | 0 | Low-quality | Genome-fragment | 3.81 | AAI-based (high-confidence) | 0 |
| S3_extracted_k141_38349 | 1291 | No | NA | 5 | 1 | 0 | Low-quality | Genome-fragment | 0.45 | HMM-based (lower-bound) | 0 |
| S5_vs_k141_32344\|\|full | 1291 | No | NA | 4 | 0 | 0 | Low-quality | Genome-fragment | 2.92 | AAI-based (high-confidence) | 0 |
| S5_vs_k141_30110\|\|full | 1291 | No | NA | 2 | 0 | 1 | Low-quality | Genome-fragment | 0.95 | AAI-based (medium-confidence) | 0 |
| S13_vs_112463\|\|full | 2925 | No | NA | 5 | 0 | 1 | Not-determined | Genome-fragment | NA | NA | 0 |
| S13_vs_112470\|\|full | 1103 | No | NA | 3 | 0 | 0 | Not-determined | Genome-fragment | NA | NA | 0 |
| S13_vs_112701\|\|full | 1685 | No | NA | 2 | 0 | 1 | Not-determined | Genome-fragment | NA | NA | 0 |
| S13_vs_112817\|\|full | 2759 | No | NA | 3 | 0 | 1 | Not-determined | Genome-fragment | NA | NA | 0 |
| S13_vs_112820\|\|full | 2031 | No | NA | 3 | 0 | 1 | Not-determined | Genome-fragment | NA | NA | 0 |
| S7_extracted_125837 | 1291 | No | NA | 3 | 2 | 0 | Low-quality | Genome-fragment | 4.48 | AAI-based (high-confidence) | 0 |
| S13_vs_113085\|\|full | 1789 | No | NA | 3 | 0 | 2 | Not-determined | Genome-fragment | NA | NA | 0 |
| S13_vs_113185\|\|full | 1179 | No | NA | 3 | 0 | 0 | Not-determined | Genome-fragment | NA | NA | 0 |
| S7_vs_29759\|\|full | 1291 | No | NA | 4 | 0 | 0 | Low-quality | Genome-fragment | 2.74 | AAI-based (high-confidence) | 0 |
| S7_vs_37556\|\|full | 1291 | No | NA | 3 | 0 | 1 | Low-quality | Genome-fragment | 3.03 | AAI-based (medium-confidence) | 0 |
| S7_vs_80048\|\|full | 1291 | No | NA | 3 | 1 | 0 | Low-quality | Genome-fragment | 0.73 | HMM-based (lower-bound) | 0 |
| S13_vs_113536\|\|full | 5702 | No | NA | 9 | 0 | 0 | Not-determined | Genome-fragment | NA | NA | 0 |
| S13_vs_113593\|\|full | 56024 | No | NA | 74 | 34 | 1 | High-quality | High-quality | 98.71 | AAI-based (high-confidence) | 0 |
| S10_vs_48135\|\|full | 1291 | No | NA | 3 | 0 | 0 | Low-quality | Genome-fragment | 2.68 | AAI-based (high-confidence) | 0 |
| S13_vs_114070\|\|full | 1215 | No | NA | 3 | 0 | 2 | Not-determined | Genome-fragment | NA | NA | 0 |
| S8_extracted_k141_201120 | 1291 | No | NA | 2 | 0 | 1 | Low-quality | Genome-fragment | 1.37 | AAI-based (high-confidence) | 0 |
| S13_vs_114213\|\|full | 34608 | Yes | 28952 | 43 | 11 | 6 | Medium-quality | Genome-fragment | 58.7 | HMM-based (lower-bound) | 16.34 |
| S13_vs_114471\|\|full | 2625 | No | NA | 3 | 0 | 0 | Not-determined | Genome-fragment | NA | NA | 0 |
| S13_vs_114648\|\|full | 2630 | No | NA | 3 | 0 | 2 | Not-determined | Genome-fragment | NA | NA | 0 |
| S8_extracted_k141_126903 | 1291 | No | NA | 2 | 0 | 1 | Low-quality | Genome-fragment | 1.74 | AAI-based (medium-confidence) | 0 |
| S13_vs_114874\|\|full | 1279 | No | NA | 3 | 0 | 1 | Not-determined | Genome-fragment | NA | NA | 0 |
| S13_vs_114899\|\|full | 1384 | No | NA | 2 | 0 | 1 | Not-determined | Genome-fragment | NA | NA | 0 |
| S13_vs_114958\|\|full | 6369 | No | NA | 6 | 0 | 4 | Not-determined | Genome-fragment | NA | NA | 0 |
| S8_extracted_k141_155630 | 1291 | No | NA | 1 | 0 | 0 | Low-quality | Genome-fragment | 2.12 | AAI-based (high-confidence) | 0 |
| S13_vs_115136\|\|full | 4632 | No | NA | 5 | 0 | 4 | Not-determined | Genome-fragment | NA | NA | 0 |
| S8_vs_k141_177024\|\|full | 1291 | No | NA | 3 | 0 | 0 | Low-quality | Genome-fragment | 0.97 | AAI-based (medium-confidence) | 0 |
| S13_vs_115285\|\|full | 1073 | No | NA | 1 | 0 | 1 | Not-determined | Genome-fragment | NA | NA | 0 |
| S13_vs_115310\|\|full | 1895 | No | NA | 4 | 0 | 0 | Not-determined | Genome-fragment | NA | NA | 0 |
| S13_vs_115584\|\|full | 2170 | No | NA | 3 | 0 | 0 | Not-determined | Genome-fragment | NA | NA | 0 |
| S13_vs_115675\|\|full | 2048 | No | NA | 2 | 0 | 1 | Not-determined | Genome-fragment | NA | NA | 0 |
| S13_vs_115730\|\|full | 1766 | No | NA | 2 | 0 | 1 | Not-determined | Genome-fragment | NA | NA | 0 |
| S11_vs_k141_64128\|\|full | 1292 | No | NA | 3 | 0 | 0 | Low-quality | Genome-fragment | 0.63 | AAI-based (medium-confidence) | 0 |
| S13_vs_116049\|\|full | 3425 | No | NA | 4 | 0 | 3 | Not-determined | Genome-fragment | NA | NA | 0 |
| S13_vs_116102\|\|full | 1401 | No | NA | 3 | 0 | 0 | Not-determined | Genome-fragment | NA | NA | 0 |
| S14_vs_k141_68605\|\|full | 1292 | No | NA | 3 | 1 | 0 | Low-quality | Genome-fragment | 0.74 | HMM-based (lower-bound) | 0 |
| S13_vs_116294\|\|full | 1699 | No | NA | 3 | 0 | 0 | Not-determined | Genome-fragment | NA | NA | 0 |
| S14_vs_k141_56199\|\|full | 1292 | No | NA | 2 | 0 | 0 | Low-quality | Genome-fragment | 3.91 | AAI-based (medium-confidence) | 0 |
| S14_vs_k141_64889\|\|full | 1292 | No | NA | 3 | 1 | 0 | Low-quality | Genome-fragment | 2.77 | AAI-based (medium-confidence) | 0 |
| S13_vs_117422\|\|full | 1512 | No | NA | 2 | 0 | 1 | Not-determined | Genome-fragment | NA | NA | 0 |
| S13_vs_117683\|\|full | 2387 | No | NA | 4 | 0 | 2 | Not-determined | Genome-fragment | NA | NA | 0 |
| S1_vs_k141_78912\|\|full | 1292 | No | NA | 3 | 1 | 0 | Low-quality | Genome-fragment | 2.54 | AAI-based (high-confidence) | 0 |
| S5_vs_k141_4462\|\|full | 1292 | No | NA | 3 | 0 | 0 | Low-quality | Genome-fragment | 3.67 | AAI-based (high-confidence) | 0 |
| S5_vs_k141_53234\|\|lt2gene | 1292 | No | NA | 2 | 1 | 0 | Low-quality | Genome-fragment | 2.97 | AAI-based (medium-confidence) | 0 |
| S13_vs_118442\|\|full | 5123 | No | NA | 6 | 0 | 4 | Not-determined | Genome-fragment | NA | NA | 0 |
| S13_vs_118461\|\|full | 3154 | No | NA | 4 | 0 | 3 | Not-determined | Genome-fragment | NA | NA | 0 |
| S13_vs_118462\|\|full | 1551 | No | NA | 2 | 0 | 2 | Not-determined | Genome-fragment | NA | NA | 0 |
| S7_extracted_59357 | 1292 | No | NA | 2 | 1 | 0 | Low-quality | Genome-fragment | 2.26 | HMM-based (lower-bound) | 0 |
| S7_extracted_79538 | 1292 | No | NA | 1 | 0 | 0 | Low-quality | Genome-fragment | 2.33 | AAI-based (high-confidence) | 0 |
| S13_vs_119066\|\|full | 1615 | No | NA | 2 | 0 | 1 | Not-determined | Genome-fragment | NA | NA | 0 |
| S13_vs_119122\|\|full | 4839 | No | NA | 6 | 0 | 3 | Not-determined | Genome-fragment | NA | NA | 0 |
| S13_vs_119144\|\|full | 1050 | No | NA | 3 | 0 | 2 | Not-determined | Genome-fragment | NA | NA | 0 |
| S13_vs_119401\|\|full | 1538 | No | NA | 3 | 0 | 1 | Not-determined | Genome-fragment | NA | NA | 0 |
| S7_vs_35639\|\|full | 1292 | No | NA | 3 | 1 | 0 | Low-quality | Genome-fragment | 2.73 | AAI-based (medium-confidence) | 0 |
| S7_vs_106077\|\|full | 1292 | No | NA | 2 | 0 | 0 | Low-quality | Genome-fragment | 3.34 | AAI-based (medium-confidence) | 0 |
| S16_vs_166188\|\|full | 1292 | No | NA | 2 | 0 | 0 | Low-quality | Genome-fragment | 2.01 | AAI-based (medium-confidence) | 0 |
| S12_extracted_k141_126570 | 1292 | No | NA | 3 | 0 | 1 | Low-quality | Genome-fragment | 1.21 | AAI-based (medium-confidence) | 0 |
| S13_vs_120386\|\|full | 1763 | No | NA | 2 | 0 | 0 | Not-determined | Genome-fragment | NA | NA | 0 |
| S13_vs_120521\|\|full | 2684 | No | NA | 4 | 0 | 1 | Not-determined | Genome-fragment | NA | NA | 0 |
| S13_vs_120725\|\|full | 2144 | No | NA | 4 | 0 | 2 | Not-determined | Genome-fragment | NA | NA | 0 |
| S13_vs_120787\|\|full | 1346 | No | NA | 3 | 0 | 2 | Not-determined | Genome-fragment | NA | NA | 0 |
| S13_vs_120793\|\|full | 2757 | No | NA | 5 | 0 | 3 | Not-determined | Genome-fragment | NA | NA | 0 |
| S8_vs_k141_246\|\|full | 1292 | No | NA | 2 | 0 | 1 | Low-quality | Genome-fragment | 2.25 | AAI-based (medium-confidence) | 0 |
| S13_vs_120975\|\|full | 1928 | No | NA | 3 | 0 | 1 | Not-determined | Genome-fragment | NA | NA | 0 |
| S8_vs_k141_31266\|\|full | 1292 | No | NA | 2 | 1 | 0 | Low-quality | Genome-fragment | 3.17 | AAI-based (medium-confidence) | 0 |
| S13_extracted_137915 | 1293 | No | NA | 1 | 0 | 0 | Low-quality | Genome-fragment | 2.14 | AAI-based (high-confidence) | 0 |
| S13_vs_121307\|\|full | 20430 | No | NA | 29 | 3 | 1 | Medium-quality | Genome-fragment | 53.16 | AAI-based (high-confidence) | 0 |
| S13_vs_121392\|\|full | 1571 | No | NA | 3 | 0 | 1 | Not-determined | Genome-fragment | NA | NA | 0 |
| S13_vs_121608\|\|full | 3029 | No | NA | 6 | 0 | 1 | Not-determined | Genome-fragment | NA | NA | 0 |
| S13_extracted_152131 | 1293 | No | NA | 3 | 0 | 1 | Low-quality | Genome-fragment | 2.59 | AAI-based (medium-confidence) | 0 |
| S1_vs_k141_6465\|\|full | 1293 | No | NA | 3 | 0 | 0 | Low-quality | Genome-fragment | 2.86 | AAI-based (high-confidence) | 0 |
| S13_vs_122124\|\|full | 3545 | No | NA | 3 | 0 | 1 | Not-determined | Genome-fragment | NA | NA | 0 |
| S13_vs_122581\|\|full | 2752 | No | NA | 3 | 0 | 2 | Not-determined | Genome-fragment | NA | NA | 0 |
| S13_vs_122600\|\|full | 2803 | No | NA | 5 | 0 | 3 | Not-determined | Genome-fragment | NA | NA | 0 |
| S13_vs_122910\|\|full | 2212 | No | NA | 3 | 0 | 0 | Not-determined | Genome-fragment | NA | NA | 0 |
| S3_extracted_k141_10869 | 1293 | No | NA | 3 | 0 | 1 | Low-quality | Genome-fragment | 1.09 | AAI-based (high-confidence) | 0 |
| S13_vs_123058\|\|full | 2879 | No | NA | 3 | 0 | 3 | Not-determined | Genome-fragment | NA | NA | 0 |
| S5_vs_k141_96618\|\|full | 1293 | No | NA | 4 | 0 | 0 | Low-quality | Genome-fragment | 3.4 | AAI-based (medium-confidence) | 0 |
| S13_vs_123251\|\|full | 1026 | No | NA | 3 | 0 | 2 | Not-determined | Genome-fragment | NA | NA | 0 |
| S13_vs_123253\|\|full | 1819 | No | NA | 3 | 0 | 1 | Not-determined | Genome-fragment | NA | NA | 0 |
| S13_vs_123304\|\|full | 1973 | No | NA | 2 | 0 | 0 | Not-determined | Genome-fragment | NA | NA | 0 |
| S13_vs_123486\|\|full | 1724 | No | NA | 2 | 0 | 1 | Not-determined | Genome-fragment | NA | NA | 0 |
| S13_vs_123495\|\|full | 1868 | No | NA | 3 | 0 | 0 | Not-determined | Genome-fragment | NA | NA | 0 |
| S7_extracted_14239 | 1293 | No | NA | 3 | 2 | 0 | Low-quality | Genome-fragment | 2.31 | AAI-based (high-confidence) | 0 |
| S13_vs_123651\|\|full | 5432 | No | NA | 6 | 0 | 1 | Not-determined | Genome-fragment | NA | NA | 0 |
| S7_vs_135405\|\|full | 1293 | No | NA | 3 | 1 | 0 | Low-quality | Genome-fragment | 3.76 | AAI-based (high-confidence) | 0 |
| S13_vs_123987\|\|full | 2301 | No | NA | 3 | 0 | 3 | Not-determined | Genome-fragment | NA | NA | 0 |
| S13_vs_124264\|\|full | 1420 | No | NA | 3 | 0 | 0 | Not-determined | Genome-fragment | NA | NA | 0 |
| S16_extracted_114416 | 1293 | No | NA | 2 | 0 | 1 | Low-quality | Genome-fragment | 2.77 | AAI-based (medium-confidence) | 0 |
| S8_extracted_k141_26516 | 1293 | No | NA | 1 | 0 | 0 | Low-quality | Genome-fragment | 0.97 | AAI-based (high-confidence) | 0 |
| S13_vs_124622\|\|full | 1614 | No | NA | 2 | 0 | 0 | Not-determined | Genome-fragment | NA | NA | 0 |
| S13_vs_124830\|\|full | 4025 | No | NA | 4 | 0 | 3 | Not-determined | Genome-fragment | NA | NA | 0 |
| S13_vs_124929\|\|full | 1697 | No | NA | 4 | 0 | 0 | Not-determined | Genome-fragment | NA | NA | 0 |
| S13_vs_125459\|\|full | 1356 | No | NA | 3 | 0 | 1 | Not-determined | Genome-fragment | NA | NA | 0 |
| S8_vs_k141_94479\|\|full | 1293 | No | NA | 4 | 2 | 0 | Low-quality | Genome-fragment | 2.3 | AAI-based (high-confidence) | 0 |
| S13_vs_126016\|\|full | 1340 | No | NA | 3 | 0 | 0 | Not-determined | Genome-fragment | NA | NA | 0 |
| S13_vs_126059\|\|full | 3127 | No | NA | 5 | 0 | 3 | Not-determined | Genome-fragment | NA | NA | 0 |
| S13_vs_126245\|\|full | 2569 | No | NA | 3 | 0 | 2 | Not-determined | Genome-fragment | NA | NA | 0 |
| S13_vs_126340\|\|full | 1261 | No | NA | 2 | 0 | 2 | Not-determined | Genome-fragment | NA | NA | 0 |
| S11_extracted_k141_96330 | 1294 | No | NA | 3 | 0 | 0 | Low-quality | Genome-fragment | 2.68 | AAI-based (high-confidence) | 0 |
| S13_vs_91288\|\|full | 1294 | No | NA | 3 | 0 | 0 | Low-quality | Genome-fragment | 0.92 | AAI-based (medium-confidence) | 0 |
| S13_vs_126858\|\|full | 1379 | No | NA | 3 | 0 | 2 | Not-determined | Genome-fragment | NA | NA | 0 |
| S13_vs_126870\|\|full | 2884 | No | NA | 3 | 0 | 1 | Not-determined | Genome-fragment | NA | NA | 0 |
| S14_extracted_k141_93334 | 1294 | No | NA | 4 | 3 | 0 | Low-quality | Genome-fragment | 2.15 | AAI-based (medium-confidence) | 0 |
| S13_vs_127009\|\|full | 1088 | No | NA | 3 | 0 | 0 | Not-determined | Genome-fragment | NA | NA | 0 |

| | | | | | | | | | | | |
|---|---|---|---|---|---|---|---|---|---|---|---|
| S13_vs_127088\|\|full | 4864 | No | NA | 5 | 0 | 0 | Not-determined | Genome-fragment | NA | NA | 0 |
| S13_vs_127196\|\|full | 2079 | No | NA | 6 | 0 | 0 | Not-determined | Genome-fragment | NA | NA | 0 |
| S13_vs_127354\|\|full | 1124 | No | NA | 2 | 0 | 0 | Not-determined | Genome-fragment | NA | NA | 0 |
| S13_vs_127603\|\|full | 6598 | No | NA | 7 | 0 | 4 | Not-determined | Genome-fragment | NA | NA | 0 |
| S14_vs_k141_12821\|\|full | 1294 | No | NA | 3 | 1 | 0 | Low-quality | Genome-fragment | 4.16 | AAI-based (medium-confidence) | 0 |
| S14_vs_k141_56878\|\|full | 1294 | No | NA | 4 | 0 | 0 | Low-quality | Genome-fragment | 1.37 | AAI-based (medium-confidence) | 0 |
| S5_vs_k141_87135\|\|full | 1294 | No | NA | 2 | 0 | 2 | Low-quality | Genome-fragment | 0.43 | AAI-based (medium-confidence) | 0 |
| S13_vs_127862\|\|full | 2861 | No | NA | 2 | 0 | 0 | Not-determined | Genome-fragment | NA | NA | 0 |
| S5_vs_k141_137066\|\|full | 1294 | No | NA | 3 | 0 | 1 | Low-quality | Genome-fragment | 0.81 | AAI-based (medium-confidence) | 0 |
| S7_vs_63696\|\|full | 1294 | No | NA | 3 | 0 | 0 | Low-quality | Genome-fragment | 1.08 | AAI-based (medium-confidence) | 0 |
| S7_vs_122511\|\|full | 1294 | No | NA | 3 | 0 | 0 | Low-quality | Genome-fragment | 2.66 | AAI-based (medium-confidence) | 0 |
| S13_vs_128358\|\|full | 2319 | No | NA | 3 | 0 | 1 | Not-determined | Genome-fragment | NA | NA | 0 |
| S16_extracted_158911 | 1294 | No | NA | 1 | 1 | 0 | Low-quality | Genome-fragment | 3.08 | HMM-based (lower-bound) | 0 |
| S13_vs_128552\|\|full | 1433 | No | NA | 3 | 0 | 1 | Not-determined | Genome-fragment | NA | NA | 0 |
| S13_vs_128637\|\|full | 3131 | No | NA | 4 | 0 | 1 | Not-determined | Genome-fragment | NA | NA | 0 |
| S13_vs_128781\|\|full | 3637 | No | NA | 3 | 0 | 1 | Not-determined | Genome-fragment | NA | NA | 0 |
| S13_vs_129132\|\|full | 1509 | No | NA | 5 | 0 | 0 | Not-determined | Genome-fragment | NA | NA | 0 |
| S16_vs_130471\|\|full | 1294 | No | NA | 3 | 0 | 0 | Low-quality | Genome-fragment | 2.09 | AAI-based (medium-confidence) | 0 |
| S8_vs_k141_107910\|\|full | 1294 | No | NA | 3 | 2 | 0 | Low-quality | Genome-fragment | 2.73 | AAI-based (high-confidence) | 0 |
| S13_vs_129367\|\|full | 2078 | No | NA | 4 | 0 | 3 | Not-determined | Genome-fragment | NA | NA | 0 |
| S13_vs_129410\|\|full | 1471 | No | NA | 3 | 0 | 1 | Not-determined | Genome-fragment | NA | NA | 0 |
| S13_vs_129498\|\|full | 1829 | No | NA | 2 | 0 | 1 | Not-determined | Genome-fragment | NA | NA | 0 |
| S13_vs_129602\|\|full | 2446 | No | NA | 5 | 0 | 1 | Not-determined | Genome-fragment | NA | NA | 0 |
| S13_vs_129705\|\|full | 1110 | No | NA | 3 | 0 | 1 | Not-determined | Genome-fragment | NA | NA | 0 |
| S17_vs_k141_138596\|\|full | 1295 | No | NA | 3 | 0 | 0 | Low-quality | Genome-fragment | 0.88 | AAI-based (high-confidence) | 0 |
| S13_vs_130179\|\|full | 1167 | No | NA | 3 | 0 | 1 | Not-determined | Genome-fragment | NA | NA | 0 |
| S9_vs_k141_69232\|\|full | 1295 | No | NA | 4 | 1 | 0 | Low-quality | Genome-fragment | 3.18 | AAI-based (high-confidence) | 0 |
| S13_vs_130445\|\|full | 1163 | No | NA | 2 | 0 | 1 | Not-determined | Genome-fragment | NA | NA | 0 |
| S11_extracted_k141_11699 | 1295 | No | NA | 1 | 0 | 1 | Low-quality | Genome-fragment | 0.68 | AAI-based (medium-confidence) | 0 |
| S13_vs_130713\|\|full | 2238 | No | NA | 3 | 0 | 0 | Not-determined | Genome-fragment | NA | NA | 0 |
| S13_vs_130925\|\|full | 6001 | No | NA | 6 | 0 | 4 | Not-determined | Genome-fragment | NA | NA | 0 |
| S13_vs_130939\|\|full | 1559 | No | NA | 2 | 0 | 0 | Not-determined | Genome-fragment | NA | NA | 0 |
| S11_vs_k141_103680\|\|lt2gene | 1295 | No | NA | 2 | 1 | 0 | Low-quality | Genome-fragment | 2.45 | HMM-based (lower-bound) | 0 |
| S13_vs_131191\|\|full | 2851 | No | NA | 4 | 0 | 2 | Not-determined | Genome-fragment | NA | NA | 0 |
| S14_extracted_k141_95604 | 1295 | No | NA | 2 | 2 | 0 | Low-quality | Genome-fragment | 4.55 | AAI-based (high-confidence) | 0 |
| S13_vs_131227\|\|full | 1535 | No | NA | 2 | 0 | 1 | Not-determined | Genome-fragment | NA | NA | 0 |
| S5_extracted_k141_830 | 1295 | No | NA | 3 | 1 | 0 | Low-quality | Genome-fragment | 3.12 | AAI-based (medium-confidence) | 0 |
| S5_extracted_k141_138969 | 1295 | No | NA | 1 | 0 | 0 | Low-quality | Genome-fragment | 2.37 | AAI-based (high-confidence) | 0 |
| S13_vs_131689\|\|full | 6448 | No | NA | 10 | 0 | 2 | Not-determined | Genome-fragment | NA | NA | 0 |
| S13_vs_131724\|\|full | 1281 | No | NA | 2 | 0 | 2 | Not-determined | Genome-fragment | NA | NA | 0 |
| S5_vs_k141_26013\|\|full | 1295 | No | NA | 4 | 1 | 0 | Low-quality | Genome-fragment | 3.56 | HMM-based (lower-bound) | 0 |
| S13_vs_132105\|\|full | 3775 | No | NA | 5 | 0 | 1 | Not-determined | Genome-fragment | NA | NA | 0 |
| S13_vs_132182\|\|full | 1740 | No | NA | 3 | 0 | 3 | Not-determined | Genome-fragment | NA | NA | 0 |
| S5_vs_k141_59388\|\|full | 1295 | No | NA | 3 | 0 | 1 | Low-quality | Genome-fragment | 3.74 | AAI-based (medium-confidence) | 0 |
| S12_vs_k141_41682\|\|full | 1295 | No | NA | 2 | 0 | 0 | Low-quality | Genome-fragment | 2.07 | AAI-based (medium-confidence) | 0 |
| S15_extracted_38900 | 1295 | No | NA | 2 | 0 | 1 | Low-quality | Genome-fragment | 0.81 | AAI-based (high-confidence) | 0 |
| S6_extracted_k141_20615 | 1295 | No | NA | 2 | 1 | 0 | Low-quality | Genome-fragment | 2.18 | HMM-based (lower-bound) | 0 |
| S9_vs_k141_73603\|\|full | 1296 | No | NA | 3 | 1 | 0 | Low-quality | Genome-fragment | 2.04 | AAI-based (high-confidence) | 0 |
| S13_vs_133300\|\|full | 5077 | No | NA | 5 | 0 | 2 | Not-determined | Genome-fragment | NA | NA | 0 |
| S13_vs_133375\|\|full | 1159 | No | NA | 2 | 0 | 0 | Not-determined | Genome-fragment | NA | NA | 0 |
| S9_vs_k141_30565\|\|lt2gene | 1296 | No | NA | 2 | 2 | 0 | Low-quality | Genome-fragment | 3.16 | AAI-based (high-confidence) | 0 |
| S9_vs_k141_82546\|\|lt2gene | 1296 | No | NA | 2 | 2 | 0 | Low-quality | Genome-fragment | 2.16 | AAI-based (high-confidence) | 0 |
| S13_vs_133917\|\|full | 1423 | No | NA | 3 | 0 | 1 | Not-determined | Genome-fragment | NA | NA | 0 |
| S13_vs_134004\|\|full | 1281 | No | NA | 2 | 0 | 0 | Not-determined | Genome-fragment | NA | NA | 0 |
| S13_vs_134115\|\|full | 1931 | No | NA | 3 | 0 | 3 | Not-determined | Genome-fragment | NA | NA | 0 |
| S11_extracted_k141_92098 | 1296 | No | NA | 2 | 2 | 0 | Low-quality | Genome-fragment | 2.91 | AAI-based (medium-confidence) | 0 |
| S13_vs_54555\|\|full | 1296 | No | NA | 4 | 2 | 0 | Low-quality | Genome-fragment | 2.31 | AAI-based (high-confidence) | 0 |
| S13_vs_135018\|\|full | 1917 | No | NA | 2 | 0 | 0 | Not-determined | Genome-fragment | NA | NA | 0 |
| S13_vs_64160\|\|full | 1296 | No | NA | 2 | 0 | 2 | Low-quality | Genome-fragment | 0.49 | AAI-based (medium-confidence) | 0 |
| S13_vs_135119\|\|full | 1119 | No | NA | 4 | 0 | 0 | Not-determined | Genome-fragment | NA | NA | 0 |
| S1_extracted_k141_8143 | 1296 | No | NA | 3 | 0 | 0 | Low-quality | Genome-fragment | 1.32 | AAI-based (high-confidence) | 0 |
| S13_vs_135200\|\|full | 2217 | No | NA | 2 | 0 | 1 | Not-determined | Genome-fragment | NA | NA | 0 |
| S13_vs_135507\|\|full | 2766 | No | NA | 3 | 0 | 1 | Not-determined | Genome-fragment | NA | NA | 0 |
| S13_vs_135750\|\|full | 1962 | No | NA | 4 | 0 | 0 | Not-determined | Genome-fragment | NA | NA | 0 |
| S13_vs_136187\|\|full | 1837 | No | NA | 4 | 0 | 0 | Not-determined | Genome-fragment | NA | NA | 0 |
| S13_vs_136271\|\|full | 2450 | No | NA | 4 | 0 | 0 | Not-determined | Genome-fragment | NA | NA | 0 |
| S13_vs_136290\|\|full | 1007 | No | NA | 2 | 0 | 1 | Not-determined | Genome-fragment | NA | NA | 0 |
| S13_vs_136502\|\|full | 9223 | No | NA | 13 | 0 | 1 | Not-determined | Genome-fragment | NA | NA | 0 |
| S13_vs_136561\|\|full | 2403 | No | NA | 4 | 0 | 1 | Not-determined | Genome-fragment | NA | NA | 0 |
| S13_vs_136583\|\|full | 1517 | No | NA | 2 | 0 | 2 | Not-determined | Genome-fragment | NA | NA | 0 |
| S13_vs_136824\|\|full | 1341 | No | NA | 2 | 0 | 0 | Not-determined | Genome-fragment | NA | NA | 0 |
| S13_vs_136914\|\|full | 1152 | No | NA | 3 | 0 | 1 | Not-determined | Genome-fragment | NA | NA | 0 |
| S7_vs_72006\|\|lt2gene | 1296 | No | NA | 1 | 0 | 0 | Low-quality | Genome-fragment | 3.19 | AAI-based (high-confidence) | 0 |
| S13_vs_137086\|\|full | 2919 | No | NA | 2 | 0 | 1 | Not-determined | Genome-fragment | NA | NA | 0 |
| S7_vs_104014\|\|lt2gene | 1296 | No | NA | 1 | 1 | 0 | Low-quality | Genome-fragment | 3.24 | AAI-based (high-confidence) | 0 |
| S13_vs_137288\|\|full | 2043 | No | NA | 3 | 0 | 0 | Not-determined | Genome-fragment | NA | NA | 0 |
| S13_vs_137365\|\|full | 1741 | No | NA | 3 | 0 | 1 | Not-determined | Genome-fragment | NA | NA | 0 |
| S16_extracted_165043 | 1296 | No | NA | 1 | 0 | 0 | Low-quality | Genome-fragment | 2.04 | AAI-based (high-confidence) | 0 |
| S13_vs_137452\|\|full | 1103 | No | NA | 3 | 0 | 1 | Not-determined | Genome-fragment | NA | NA | 0 |
| S13_vs_137587\|\|full | 2297 | No | NA | 3 | 0 | 2 | Not-determined | Genome-fragment | NA | NA | 0 |
| S16_vs_640\|\|full | 1296 | No | NA | 4 | 1 | 0 | Low-quality | Genome-fragment | 2.06 | AAI-based (medium-confidence) | 0 |
| S13_vs_137913\|\|full | 2266 | No | NA | 3 | 0 | 1 | Not-determined | Genome-fragment | NA | NA | 0 |
| S13_vs_138152\|\|full | 1009 | No | NA | 2 | 0 | 0 | Not-determined | Genome-fragment | NA | NA | 0 |
| S13_vs_138161\|\|full | 2903 | No | NA | 4 | 0 | 4 | Not-determined | Genome-fragment | NA | NA | 0 |
| S10_vs_79821\|\|full | 1296 | No | NA | 3 | 0 | 0 | Low-quality | Genome-fragment | 2.45 | AAI-based (high-confidence) | 0 |
| S12_extracted_k141_54397 | 1296 | No | NA | 3 | 0 | 0 | Low-quality | Genome-fragment | 3.86 | AAI-based (high-confidence) | 0 |
| S13_vs_138801\|\|full | 2015 | No | NA | 4 | 0 | 3 | Not-determined | Genome-fragment | NA | NA | 0 |
| S13_vs_138810\|\|full | 2994 | No | NA | 3 | 0 | 0 | Not-determined | Genome-fragment | NA | NA | 0 |
| S13_vs_138825\|\|full | 7682 | No | NA | 12 | 0 | 2 | Not-determined | Genome-fragment | NA | NA | 0 |
| S13_vs_138862\|\|full | 1916 | No | NA | 3 | 0 | 1 | Not-determined | Genome-fragment | NA | NA | 0 |
| S8_extracted_k141_53368 | 1296 | No | NA | 2 | 0 | 0 | Low-quality | Genome-fragment | 1.97 | AAI-based (high-confidence) | 0 |
| S13_vs_139091\|\|full | 6761 | No | NA | 6 | 0 | 1 | Not-determined | Genome-fragment | NA | NA | 0 |
| S13_vs_139181\|\|full | 1397 | No | NA | 2 | 0 | 0 | Not-determined | Genome-fragment | NA | NA | 0 |
| S8_vs_k141_48294\|\|full | 1296 | No | NA | 2 | 0 | 1 | Low-quality | Genome-fragment | 2.9 | AAI-based (high-confidence) | 0 |
| S13_vs_139693\|\|full | 3276 | No | NA | 5 | 0 | 3 | Not-determined | Genome-fragment | NA | NA | 0 |
| S13_vs_139750\|\|full | 1306 | No | NA | 2 | 0 | 0 | Not-determined | Genome-fragment | NA | NA | 0 |
| S9_extracted_k141_18881 | 1297 | No | NA | 1 | 0 | 1 | Low-quality | Genome-fragment | 0.91 | AAI-based (high-confidence) | 0 |
| S13_vs_140391\|\|full | 2551 | No | NA | 3 | 0 | 1 | Not-determined | Genome-fragment | NA | NA | 0 |
| S13_vs_140564\|\|full | 3593 | No | NA | 5 | 0 | 1 | Not-determined | Genome-fragment | NA | NA | 0 |
| S3_extracted_k141_36240 | 1297 | No | NA | 2 | 0 | 0 | Low-quality | Genome-fragment | 0.8 | AAI-based (high-confidence) | 0 |
| S13_vs_141050\|\|full | 1811 | No | NA | 2 | 0 | 1 | Not-determined | Genome-fragment | NA | NA | 0 |
| S13_vs_141153\|\|full | 1057 | No | NA | 2 | 0 | 1 | Not-determined | Genome-fragment | NA | NA | 0 |
| S3_extracted_k141_83902 | 1297 | No | NA | 4 | 0 | 0 | Low-quality | Genome-fragment | 2.17 | AAI-based (medium-confidence) | 0 |
| S13_vs_141329\|\|full | 15942 | No | NA | 12 | 0 | 3 | Not-determined | Genome-fragment | NA | NA | 0 |
| S10_vs_106645\|\|full | 1297 | No | NA | 4 | 0 | 0 | Low-quality | Genome-fragment | 0.36 | AAI-based (medium-confidence) | 0 |
| S13_vs_141580\|\|full | 2873 | No | NA | 3 | 0 | 2 | Not-determined | Genome-fragment | NA | NA | 0 |
| S12_extracted_k141_47014 | 1297 | No | NA | 3 | 1 | 0 | Low-quality | Genome-fragment | 2.34 | AAI-based (medium-confidence) | 0 |
| S13_vs_141772\|\|full | 1636 | No | NA | 3 | 0 | 2 | Not-determined | Genome-fragment | NA | NA | 0 |
| S13_vs_141825\|\|full | 1282 | No | NA | 2 | 0 | 0 | Not-determined | Genome-fragment | NA | NA | 0 |
| S13_vs_141933\|\|full | 5929 | No | NA | 4 | 0 | 2 | Not-determined | Genome-fragment | NA | NA | 0 |
| S13_vs_142082\|\|full | 1376 | No | NA | 2 | 0 | 0 | Not-determined | Genome-fragment | NA | NA | 0 |
| S13_vs_142214\|\|full | 1067 | No | NA | 2 | 0 | 1 | Not-determined | Genome-fragment | NA | NA | 0 |
| S13_vs_142359\|\|full | 2493 | No | NA | 3 | 0 | 2 | Not-determined | Genome-fragment | NA | NA | 0 |
| S13_vs_142485\|\|full | 1039 | No | NA | 3 | 0 | 1 | Not-determined | Genome-fragment | NA | NA | 0 |
| S4_extracted_30620 | 1297 | No | NA | 3 | 2 | 0 | Low-quality | Genome-fragment | 2.3 | AAI-based (medium-confidence) | 0 |
| S6_vs_k141_25903\|\|full | 1297 | No | NA | 4 | 0 | 0 | Low-quality | Genome-fragment | 3.72 | AAI-based (medium-confidence) | 0 |
| S13_vs_142740\|\|full | 1422 | No | NA | 3 | 0 | 0 | Not-determined | Genome-fragment | NA | NA | 0 |
| S13_vs_142808\|\|full | 1888 | No | NA | 4 | 0 | 0 | Not-determined | Genome-fragment | NA | NA | 0 |
| S13_vs_142865\|\|full | 11419 | No | NA | 7 | 0 | 4 | Not-determined | Genome-fragment | NA | NA | 0 |
| S13_vs_142913\|\|full | 1331 | No | NA | 3 | 0 | 0 | Not-determined | Genome-fragment | NA | NA | 0 |
| S13_vs_143030\|\|full | 5790 | No | NA | 7 | 0 | 1 | Not-determined | Genome-fragment | NA | NA | 0 |
| S13_vs_143068\|\|full | 1367 | No | NA | 3 | 0 | 1 | Not-determined | Genome-fragment | NA | NA | 0 |
| S8_extracted_k141_14442 | 1297 | No | NA | 3 | 0 | 0 | Low-quality | Genome-fragment | 2.47 | AAI-based (high-confidence) | 0 |
| S13_vs_143304\|\|full | 2356 | No | NA | 4 | 0 | 2 | Not-determined | Genome-fragment | NA | NA | 0 |
| S13_vs_143385\|\|full | 2261 | No | NA | 4 | 0 | 2 | Not-determined | Genome-fragment | NA | NA | 0 |
| S17_extracted_k141_76243 | 1298 | No | NA | 2 | 0 | 0 | Low-quality | Genome-fragment | 0.81 | AAI-based (high-confidence) | 0 |
| S13_vs_143739\|\|full | 1131 | No | NA | 2 | 0 | 2 | Not-determined | Genome-fragment | NA | NA | 0 |
| S17_extracted_k141_41339 | 1298 | No | NA | 4 | 3 | 0 | Low-quality | Genome-fragment | 4.26 | AAI-based (high-confidence) | 0 |
| S9_vs_k141_14415\|\|full | 1298 | No | NA | 3 | 1 | 0 | Low-quality | Genome-fragment | 2.4 | AAI-based (high-confidence) | 0 |
| S13_vs_144384\|\|full | 1268 | No | NA | 3 | 0 | 1 | Not-determined | Genome-fragment | NA | NA | 0 |
| S11_vs_k141_30313\|\|full | 1298 | No | NA | 3 | 1 | 0 | Low-quality | Genome-fragment | 2.18 | HMM-based (lower-bound) | 0 |
| S13_vs_144572\|\|full | 1290 | No | NA | 4 | 0 | 0 | Not-determined | Genome-fragment | NA | NA | 0 |
| S13_extracted_8430 | 1298 | No | NA | 2 | 0 | 0 | Low-quality | Genome-fragment | 1.44 | AAI-based (medium-confidence) | 0 |
| S13_vs_144645\|\|full | 1897 | No | NA | 3 | 0 | 1 | Not-determined | Genome-fragment | NA | NA | 0 |
| S13_vs_144853\|\|full | 2097 | No | NA | 5 | 0 | 0 | Not-determined | Genome-fragment | NA | NA | 0 |
| S13_vs_145001\|\|full | 1051 | No | NA | 3 | 0 | 0 | Not-determined | Genome-fragment | NA | NA | 0 |
| S3_extracted_k141_35828 | 1298 | No | NA | 3 | 3 | 0 | Low-quality | Genome-fragment | 2.13 | AAI-based (high-confidence) | 0 |
| S13_vs_145278\|\|full | 2773 | No | NA | 7 | 0 | 6 | Not-determined | Genome-fragment | NA | NA | 0 |
| S13_vs_145288\|\|full | 1285 | No | NA | 3 | 0 | 2 | Not-determined | Genome-fragment | NA | NA | 0 |
| S13_vs_145546\|\|full | 1772 | No | NA | 2 | 0 | 2 | Not-determined | Genome-fragment | NA | NA | 0 |
| S13_vs_145594\|\|full | 4475 | No | NA | 10 | 0 | 0 | Not-determined | Genome-fragment | NA | NA | 0 |
| S3_vs_k141_119885\|\|full | 1298 | No | NA | 3 | 0 | 1 | Low-quality | Genome-fragment | 2.69 | AAI-based (medium-confidence) | 0 |
| S7_extracted_40420 | 1298 | No | NA | 1 | 1 | 0 | Low-quality | Genome-fragment | 3.25 | AAI-based (high-confidence) | 0 |
| S7_vs_40293\|\|lt2gene | 1298 | No | NA | 1 | 1 | 0 | Low-quality | Genome-fragment | 1.72 | HMM-based (lower-bound) | 0 |
| S13_vs_146187\|\|full | 3181 | No | NA | 2 | 0 | 2 | Not-determined | Genome-fragment | NA | NA | 0 |
| S12_vs_k141_66844\|\|full | 1298 | No | NA | 2 | 1 | 0 | Low-quality | Genome-fragment | 2.78 | AAI-based (medium-confidence) | 0 |
| S13_vs_146579\|\|full | 1767 | No | NA | 3 | 0 | 2 | Not-determined | Genome-fragment | NA | NA | 0 |
| S13_vs_146666\|\|full | 1983 | No | NA | 3 | 0 | 1 | Not-determined | Genome-fragment | NA | NA | 0 |
| S13_vs_146761\|\|full | 1275 | No | NA | 2 | 0 | 1 | Not-determined | Genome-fragment | NA | NA | 0 |
| S12_vs_k141_44116\|\|lt2gene | 1298 | No | NA | 2 | 1 | 0 | Low-quality | Genome-fragment | 3.47 | AAI-based (medium-confidence) | 0 |
| S13_vs_147159\|\|full | 1158 | No | NA | 3 | 0 | 3 | Not-determined | Genome-fragment | NA | NA | 0 |
| S13_vs_147712\|\|full | 1412 | No | NA | 3 | 0 | 1 | Not-determined | Genome-fragment | NA | NA | 0 |
| S8_vs_k141_177812\|\|full | 1298 | Yes | 539 | 3 | 1 | 1 | Low-quality | Genome-fragment | 0.23 | HMM-based (lower-bound) | 58.47 |
| S13_vs_148446\|\|full | 30296 | No | NA | 26 | 0 | 10 | Not-determined | Genome-fragment | NA | NA | 0 |
| S13_vs_148495\|\|full | 2561 | No | NA | 3 | 0 | 3 | Not-determined | Genome-fragment | NA | NA | 0 |
| S13_vs_148622\|\|full | 3284 | No | NA | 3 | 0 | 2 | Not-determined | Genome-fragment | NA | NA | 0 |
| S13_vs_148648\|\|full | 3188 | No | NA | 2 | 0 | 2 | Not-determined | Genome-fragment | NA | NA | 0 |
| S13_vs_148952\|\|full | 1391 | No | NA | 3 | 0 | 0 | Not-determined | Genome-fragment | NA | NA | 0 |
| S13_vs_149085\|\|full | 3786 | No | NA | 4 | 0 | 4 | Not-determined | Genome-fragment | NA | NA | 0 |
| S13_vs_149345\|\|full | 1010 | No | NA | 2 | 0 | 1 | Not-determined | Genome-fragment | NA | NA | 0 |
| S8_vs_k141_15244\|\|full | 1298 | No | NA | 2 | 0 | 1 | Low-quality | Genome-fragment | 1.3 | AAI-based (medium-confidence) | 0 |
| S17_vs_k141_57247\|\|full | 1299 | No | NA | 3 | 3 | 0 | Low-quality | Genome-fragment | 3.12 | AAI-based (high-confidence) | 0 |
| S13_vs_149596\|\|full | 1630 | No | NA | 3 | 0 | 3 | Not-determined | Genome-fragment | NA | NA | 0 |
| S13_vs_149597\|\|full | 1013 | No | NA | 2 | 0 | 0 | Not-determined | Genome-fragment | NA | NA | 0 |
| S11_extracted_k141_55583 | 1299 | No | NA | 4 | 0 | 0 | Low-quality | Genome-fragment | 1.82 | AAI-based (high-confidence) | 0 |
| S11_vs_k141_93299\|\|full | 1299 | No | NA | 2 | 0 | 1 | Low-quality | Genome-fragment | 2.62 | AAI-based (medium-confidence) | 0 |
| S13_vs_150073\|\|full | 1649 | No | NA | 3 | 0 | 1 | Not-determined | Genome-fragment | NA | NA | 0 |
| S13_vs_150216\|\|full | 1067 | No | NA | 3 | 0 | 1 | Not-determined | Genome-fragment | NA | NA | 0 |
| S13_vs_150252\|\|full | 1403 | No | NA | 2 | 0 | 1 | Not-determined | Genome-fragment | NA | NA | 0 |
| S13_vs_150379\|\|full | 4654 | No | NA | 6 | 0 | 0 | Not-determined | Genome-fragment | NA | NA | 0 |
| S14_extracted_k141_81689 | 1299 | No | NA | 3 | 3 | 0 | Low-quality | Genome-fragment | 4.38 | AAI-based (high-confidence) | 0 |
| S13_vs_150977\|\|full | 2355 | No | NA | 3 | 0 | 0 | Not-determined | Genome-fragment | NA | NA | 0 |
| S13_vs_151150\|\|full | 1841 | No | NA | 2 | 0 | 2 | Not-determined | Genome-fragment | NA | NA | 0 |
| S13_vs_151217\|\|full | 1415 | No | NA | 4 | 0 | 0 | Not-determined | Genome-fragment | NA | NA | 0 |
| S1_extracted_k141_33147 | 1299 | No | NA | 1 | 0 | 0 | Low-quality | Genome-fragment | 0.83 | AAI-based (high-confidence) | 0 |
| S13_vs_151601\|\|full | 2576 | No | NA | 3 | 0 | 0 | Not-determined | Genome-fragment | NA | NA | 0 |
| S5_extracted_k141_139114 | 1299 | No | NA | 2 | 2 | 0 | Low-quality | Genome-fragment | 3 | AAI-based (high-confidence) | 0 |
| S13_vs_152214\|\|full | 1370 | No | NA | 3 | 0 | 1 | Not-determined | Genome-fragment | NA | NA | 0 |
| S13_vs_152223\|\|full | 60897 | No | NA | 99 | 33 | 0 | High-quality | High-quality | 100 | AAI-based (high-confidence) | 0 |
| S13_vs_152287\|\|full | 2421 | No | NA | 3 | 0 | 3 | Not-determined | Genome-fragment | NA | NA | 0 |
| S13_vs_152414\|\|full | 3507 | No | NA | 4 | 0 | 0 | Not-determined | Genome-fragment | NA | NA | 0 |
| S13_vs_152436\|\|full | 1509 | No | NA | 2 | 0 | 2 | Not-determined | Genome-fragment | NA | NA | 0 |
| S13_vs_152697\|\|full | 1946 | No | NA | 3 | 0 | 1 | Not-determined | Genome-fragment | NA | NA | 0 |
| S13_vs_152718\|\|full | 1459 | No | NA | 3 | 0 | 0 | Not-determined | Genome-fragment | NA | NA | 0 |
| S13_vs_152750\|\|full | 6397 | No | NA | 8 | 0 | 4 | Not-determined | Genome-fragment | NA | NA | 0 |
| S7_vs_100659\|\|full | 1299 | No | NA | 3 | 3 | 0 | Low-quality | Genome-fragment | 2.95 | AAI-based (high-confidence) | 0 |
| S7_vs_119853\|\|full | 1299 | No | NA | 5 | 1 | 0 | Low-quality | Genome-fragment | 2.81 | AAI-based (high-confidence) | 0 |
| S16_vs_55817\|\|full | 1299 | No | NA | 3 | 0 | 0 | Low-quality | Genome-fragment | 0.81 | AAI-based (high-confidence) | 0 |
| S13_vs_153321\|\|full | 2323 | No | NA | 2 | 0 | 2 | Not-determined | Genome-fragment | NA | NA | 0 |
| S13_vs_153332\|\|full | 2938 | No | NA | 3 | 0 | 1 | Not-determined | Genome-fragment | NA | NA | 0 |
| S13_vs_153584\|\|full | 5104 | No | NA | 5 | 0 | 5 | Not-determined | Genome-fragment | NA | NA | 0 |
| S10_vs_100998\|\|full | 1299 | No | NA | 3 | 0 | 0 | Low-quality | Genome-fragment | 1.39 | AAI-based (medium-confidence) | 0 |
| S13_vs_153906\|\|full | 1972 | No | NA | 2 | 0 | 2 | Not-determined | Genome-fragment | NA | NA | 0 |
| S13_vs_153952\|\|full | 3090 | No | NA | 3 | 0 | 2 | Not-determined | Genome-fragment | NA | NA | 0 |
| S13_vs_154019\|\|full | 1741 | No | NA | 3 | 0 | 2 | Not-determined | Genome-fragment | NA | NA | 0 |
| S8_extracted_k141_97465 | 1299 | No | NA | 2 | 0 | 0 | Low-quality | Genome-fragment | 1.39 | AAI-based (high-confidence) | 0 |
| S13_vs_154229\|\|full | 1227 | No | NA | 2 | 0 | 0 | Not-determined | Genome-fragment | NA | NA | 0 |
| S8_extracted_k141_198187 | 1299 | No | NA | 2 | 0 | 1 | Low-quality | Genome-fragment | 1.99 | AAI-based (medium-confidence) | 0 |
| S13_vs_22523\|\|0_partial | 12564 | No | NA | 7 | 0 | 4 | Not-determined | Genome-fragment | NA | NA | 0 |
| S8_extracted_k141_164548 | 1299 | No | NA | 2 | 1 | 0 | Low-quality | Genome-fragment | 0.89 | HMM-based (lower-bound) | 0 |
| S8_vs_k141_169169\|\|full | 1299 | No | NA | 3 | 2 | 0 | Low-quality | Genome-fragment | 3.39 | AAI-based (medium-confidence) | 0 |
| S14_vs_k141_17462\|\|lt2gene | 1300 | No | NA | 2 | 0 | 0 | Low-quality | Genome-fragment | 2.45 | AAI-based (medium-confidence) | 0 |
| S1_extracted_k141_140219 | 1300 | No | NA | 4 | 2 | 0 | Low-quality | Genome-fragment | 2.93 | AAI-based (high-confidence) | 0 |
| S1_extracted_k141_116946 | 1300 | No | NA | 3 | 1 | 0 | Low-quality | Genome-fragment | 3.03 | AAI-based (high-confidence) | 0 |
| S3_extracted_k141_73210 | 1300 | No | NA | 3 | 3 | 0 | Low-quality | Genome-fragment | 4.27 | AAI-based (high-confidence) | 0 |
| S13_vs_123344\|\|0_partial | 11899 | No | NA | 11 | 0 | 9 | Not-determined | Genome-fragment | NA | NA | 0 |
| S5_vs_k141_53705\|\|full | 1300 | No | NA | 3 | 0 | 0 | Low-quality | Genome-fragment | 2.73 | AAI-based (medium-confidence) | 0 |
| S16_vs_51807\|\|full | 1300 | No | NA | 2 | 0 | 0 | Low-quality | Genome-fragment | 0.75 | AAI-based (medium-confidence) | 0 |
| S13_vs_149741\|\|0_partial | 14767 | No | NA | 23 | 0 | 5 | Not-determined | Genome-fragment | NA | NA | 0 |
| S16_vs_141221\|\|lt2gene | 1300 | No | NA | 2 | 1 | 0 | Low-quality | Genome-fragment | 2.18 | AAI-based (high-confidence) | 0 |
| S15_vs_17827\|\|full | 1300 | No | NA | 3 | 0 | 0 | Low-quality | Genome-fragment | 0.8 | AAI-based (high-confidence) | 0 |
| S8_vs_k141_40527\|\|full | 1300 | No | NA | 2 | 0 | 0 | Low-quality | Genome-fragment | 2.96 | AAI-based (medium-confidence) | 0 |
| S13_vs_11769\|\|lt2gene | 1002 | No | NA | 2 | 0 | 0 | Not-determined | Genome-fragment | NA | NA | 0 |
| S9_extracted_k141_54530 | 1301 | No | NA | 3 | 0 | 3 | Low-quality | Genome-fragment | 1.11 | AAI-based (medium-confidence) | 0 |
| S13_vs_92628\|\|full | 1301 | No | NA | 2 | 1 | 0 | Low-quality | Genome-fragment | 9.68 | AAI-based (high-confidence) | 0 |
| S13_vs_21278\|\|lt2gene | 1071 | No | NA | 2 | 0 | 0 | Not-determined | Genome-fragment | NA | NA | 0 |
| S3_extracted_k141_59911 | 1301 | No | NA | 3 | 1 | 0 | Low-quality | Genome-fragment | 0.66 | AAI-based (medium-confidence) | 0 |
| S5_vs_k141_86782\|\|full | 1301 | No | NA | 3 | 0 | 1 | Low-quality | Genome-fragment | 3.18 | AAI-based (high-confidence) | 0 |
| S7_extracted_43073 | 1301 | No | NA | 3 | 1 | 0 | Low-quality | Genome-fragment | 4.42 | AAI-based (medium-confidence) | 0 |
| S13_vs_31411\|\|lt2gene | 2111 | No | NA | 1 | 0 | 0 | Not-determined | Genome-fragment | NA | NA | 0 |
| S16_vs_27638\|\|full | 1301 | No | NA | 3 | 0 | 0 | Low-quality | Genome-fragment | 2.36 | AAI-based (medium-confidence) | 0 |
| S12_vs_k141_107193\|\|full | 1301 | No | NA | 3 | 2 | 0 | Low-quality | Genome-fragment | 3.71 | AAI-based (medium-confidence) | 0 |
| S13_vs_36246\|\|lt2gene | 1475 | No | NA | 1 | 0 | 0 | Not-determined | Genome-fragment | NA | NA | 0 |
| S13_vs_48407\|\|lt2gene | 1067 | No | NA | 2 | 0 | 1 | Not-determined | Genome-fragment | NA | NA | 0 |
| S13_vs_49985\|\|lt2gene | 1211 | No | NA | 1 | 0 | 0 | Not-determined | Genome-fragment | NA | NA | 0 |
| S15_extracted_17672 | 1301 | No | NA | 2 | 0 | 2 | Low-quality | Genome-fragment | 1.6 | AAI-based (medium-confidence) | 0 |
| S15_extracted_25053 | 1301 | No | NA | 4 | 0 | 0 | Low-quality | Genome-fragment | 1.62 | AAI-based (high-confidence) | 0 |
| S13_vs_57461\|\|lt2gene | 1208 | No | NA | 1 | 0 | 0 | Not-determined | Genome-fragment | NA | NA | 0 |
| S13_vs_59104\|\|lt2gene | 1504 | No | NA | 1 | 0 | 0 | Not-determined | Genome-fragment | NA | NA | 0 |
| S13_vs_60714\|\|lt2gene | 1303 | No | NA | 1 | 0 | 0 | Not-determined | Genome-fragment | NA | NA | 0 |
| S13_vs_64050\|\|lt2gene | 2430 | No | NA | 1 | 0 | 0 | Not-determined | Genome-fragment | NA | NA | 0 |
| S13_vs_64338\|\|lt2gene | 1013 | No | NA | 1 | 0 | 0 | Not-determined | Genome-fragment | NA | NA | 0 |
| S15_vs_21198\|\|full | 1301 | No | NA | 4 | 0 | 1 | Low-quality | Genome-fragment | 3 | AAI-based (high-confidence) | 0 |
| S13_vs_71256\|\|lt2gene | 1958 | No | NA | 2 | 0 | 0 | Not-determined | Genome-fragment | NA | NA | 0 |
| S6_vs_k141_46151\|\|full | 1301 | No | NA | 4 | 0 | 0 | Low-quality | Genome-fragment | 4.06 | AAI-based (high-confidence) | 0 |
| S8_vs_k141_105795\|\|full | 1301 | No | NA | 2 | 0 | 0 | Low-quality | Genome-fragment | 2.02 | AAI-based (medium-confidence) | 0 |
| S8_vs_k141_40743\|\|full | 1301 | No | NA | 3 | 1 | 0 | Low-quality | Genome-fragment | 1.63 | AAI-based (medium-confidence) | 0 |
| S13_vs_94682\|\|lt2gene | 1120 | No | NA | 2 | 0 | 0 | Not-determined | Genome-fragment | NA | NA | 0 |
| S11_vs_k141_79319\|\|lt2gene | 1302 | No | NA | 1 | 1 | 0 | Low-quality | Genome-fragment | 0.9 | HMM-based (lower-bound) | 0 |
| S13_extracted_40051 | 1302 | No | NA | 1 | 0 | 1 | Low-quality | Genome-fragment | 0.35 | AAI-based (high-confidence) | 0 |
| S16_extracted_155875 | 1302 | No | NA | 2 | 1 | 0 | Low-quality | Genome-fragment | 2.19 | AAI-based (high-confidence) | 0 |
| S13_vs_112434\|\|lt2gene | 1189 | No | NA | 1 | 0 | 0 | Not-determined | Genome-fragment | NA | NA | 0 |
| S15_extracted_51745 | 1302 | No | NA | 1 | 1 | 0 | Low-quality | Genome-fragment | 2.82 | AAI-based (high-confidence) | 0 |
| S6_extracted_k141_45904 | 1302 | No | NA | 3 | 0 | 0 | Low-quality | Genome-fragment | 2.4 | AAI-based (medium-confidence) | 0 |
| S13_vs_124194\|\|lt2gene | 1133 | No | NA | 1 | 0 | 0 | Not-determined | Genome-fragment | NA | NA | 0 |
| S6_vs_k141_54648\|\|full | 1302 | No | NA | 3 | 0 | 0 | Low-quality | Genome-fragment | 2.88 | AAI-based (medium-confidence) | 0 |
| S13_vs_124659\|\|lt2gene | 1383 | No | NA | 2 | 0 | 0 | Not-determined | Genome-fragment | NA | NA | 0 |
| S8_vs_k141_5836\|\|full | 1302 | No | NA | 3 | 0 | 1 | Low-quality | Genome-fragment | 1.84 | AAI-based (medium-confidence) | 0 |
| S13_vs_135988\|\|lt2gene | 1465 | No | NA | 1 | 0 | 0 | Not-determined | Genome-fragment | NA | NA | 0 |
| S13_vs_138220\|\|lt2gene | 1021 | No | NA | 1 | 0 | 0 | Not-determined | Genome-fragment | NA | NA | 0 |
| S13_vs_145476\|\|lt2gene | 1158 | No | NA | 1 | 0 | 0 | Not-determined | Genome-fragment | NA | NA | 0 |
| S9_vs_k141_11129\|\|full | 1303 | No | NA | 4 | 0 | 0 | Low-quality | Genome-fragment | 2.1 | AAI-based (medium-confidence) | 0 |
| S14_extracted_k141_36657 | 3239 | No | NA | 3 | 0 | 2 | Not-determined | Genome-fragment | NA | NA | 0 |
| S1_vs_k141_57969\|\|full | 1303 | No | NA | 4 | 1 | 0 | Low-quality | Genome-fragment | 3.87 | AAI-based (high-confidence) | 0 |
| S5_vs_k141_86000\|\|full | 1303 | No | NA | 5 | 2 | 0 | Low-quality | Genome-fragment | 3.55 | AAI-based (high-confidence) | 0 |
| S14_extracted_k141_36727 | 1212 | No | NA | 1 | 0 | 1 | Not-determined | Genome-fragment | NA | NA | 0 |
| S5_vs_k141_41702\|\|lt2gene | 1303 | No | NA | 1 | 1 | 0 | Low-quality | Genome-fragment | 3.8 | AAI-based (high-confidence) | 0 |
| S16_extracted_84472 | 1303 | No | NA | 2 | 0 | 0 | Low-quality | Genome-fragment | 1.39 | AAI-based (high-confidence) | 0 |
| S10_vs_57416\|\|full | 1303 | No | NA | 4 | 0 | 0 | Low-quality | Genome-fragment | 2.99 | AAI-based (high-confidence) | 0 |
| S14_extracted_k141_22061 | 1788 | No | NA | 2 | 0 | 1 | Not-determined | Genome-fragment | NA | NA | 0 |
| S14_extracted_k141_120 | 1809 | No | NA | 2 | 0 | 2 | Not-determined | Genome-fragment | NA | NA | 0 |
| S6_extracted_k141_8420 | 1303 | No | NA | 3 | 0 | 0 | Low-quality | Genome-fragment | 3.23 | AAI-based (medium-confidence) | 0 |
| S6_vs_k141_52354\|\|full | 1303 | No | NA | 2 | 0 | 0 | Low-quality | Genome-fragment | 3 | AAI-based (high-confidence) | 0 |
| S11_extracted_k141_63477 | 1304 | No | NA | 5 | 0 | 0 | Low-quality | Genome-fragment | 3.2 | AAI-based (high-confidence) | 0 |
| S14_vs_k141_16003\|\|lt2gene | 1304 | No | NA | 1 | 1 | 0 | Low-quality | Genome-fragment | 2.1 | HMM-based (lower-bound) | 0 |
| S14_vs_k141_91573\|\|lt2gene | 1304 | No | NA | 1 | 1 | 0 | Low-quality | Genome-fragment | 2.33 | AAI-based (high-confidence) | 0 |
| S1_vs_k141_13388\|\|full | 1304 | No | NA | 2 | 0 | 0 | Low-quality | Genome-fragment | 2.98 | AAI-based (high-confidence) | 0 |
| S14_extracted_k141_36795 | 1039 | No | NA | 2 | 0 | 0 | Not-determined | Genome-fragment | NA | NA | 0 |
| S7_vs_28972\|\|full | 1304 | No | NA | 3 | 0 | 0 | Low-quality | Genome-fragment | 3.06 | AAI-based (high-confidence) | 0 |
| S7_vs_31206\|\|full | 1304 | No | NA | 4 | 1 | 0 | Low-quality | Genome-fragment | 2.93 | AAI-based (high-confidence) | 0 |
| S7_vs_74001\|\|full | 1304 | No | NA | 3 | 1 | 0 | Low-quality | Genome-fragment | 1.57 | HMM-based (lower-bound) | 0 |
| S12_vs_k141_17957\|\|full | 1304 | Yes | 414 | 3 | 1 | 1 | Low-quality | Genome-fragment | 0.96 | AAI-based (medium-confidence) | 68.25 |
| S14_extracted_k141_102690 | 1069 | No | NA | 2 | 0 | 1 | Not-determined | Genome-fragment | NA | NA | 0 |
| S15_extracted_44290 | 1304 | No | NA | 3 | 1 | 0 | Low-quality | Genome-fragment | 1.37 | AAI-based (high-confidence) | 0 |
| S14_extracted_k141_44139 | 1329 | No | NA | 1 | 0 | 1 | Not-determined | Genome-fragment | NA | NA | 0 |
| S14_extracted_k141_185 | 2056 | No | NA | 2 | 0 | 0 | Not-determined | Genome-fragment | NA | NA | 0 |
| S15_extracted_45304 | 1304 | No | NA | 3 | 0 | 1 | Low-quality | Genome-fragment | 1.47 | AAI-based (high-confidence) | 0 |
| S8_extracted_k141_196969 | 1304 | No | NA | 2 | 0 | 1 | Low-quality | Genome-fragment | 0.81 | AAI-based (high-confidence) | 0 |
| S8_vs_k141_30867\|\|lt2gene | 1304 | No | NA | 1 | 1 | 0 | Low-quality | Genome-fragment | 1.91 | HMM-based (lower-bound) | 0 |
| S14_extracted_k141_22148 | 3080 | No | NA | 5 | 0 | 1 | Not-determined | Genome-fragment | NA | NA | 0 |
| S14_extracted_k141_110024 | 1225 | No | NA | 2 | 0 | 2 | Not-determined | Genome-fragment | NA | NA | 0 |
| S14_extracted_k141_51465 | 1863 | No | NA | 3 | 0 | 2 | Not-determined | Genome-fragment | NA | NA | 0 |
| S17_extracted_k141_102259 | 1305 | No | NA | 5 | 0 | 0 | Low-quality | Genome-fragment | 1.96 | AAI-based (high-confidence) | 0 |
| S17_vs_k141_137101\|\|full | 1305 | No | NA | 4 | 0 | 0 | Low-quality | Genome-fragment | 2.44 | AAI-based (high-confidence) | 0 |
| S9_vs_k141_2425\|\|full | 1305 | No | NA | 3 | 0 | 0 | Low-quality | Genome-fragment | 1.59 | AAI-based (high-confidence) | 0 |
| S14_extracted_k141_7526 | 3042 | No | NA | 2 | 0 | 0 | Not-determined | Genome-fragment | NA | NA | 0 |
| S14_extracted_k141_14815 | 5011 | No | NA | 2 | 0 | 2 | Not-determined | Genome-fragment | NA | NA | 0 |
| S14_extracted_k141_29530 | 7915 | No | NA | 6 | 0 | 4 | Not-determined | Genome-fragment | NA | NA | 0 |
| S14_extracted_k141_95430 | 1128 | No | NA | 2 | 0 | 1 | Not-determined | Genome-fragment | NA | NA | 0 |
| S13_vs_14802\|\|full | 1305 | No | NA | 3 | 0 | 1 | Low-quality | Genome-fragment | 1.33 | AAI-based (medium-confidence) | 0 |
| S14_extracted_k141_26238 | 1305 | No | NA | 4 | 4 | 0 | Low-quality | Genome-fragment | 4.3 | AAI-based (high-confidence) | 0 |
| S14_extracted_k141_36905 | 4092 | No | NA | 3 | 0 | 2 | Not-determined | Genome-fragment | NA | NA | 0 |
| S14_extracted_k141_58879 | 1049 | No | NA | 2 | 0 | 1 | Not-determined | Genome-fragment | NA | NA | 0 |
| S14_extracted_k141_49714 | 1305 | No | NA | 2 | 2 | 0 | Low-quality | Genome-fragment | 2.03 | HMM-based (lower-bound) | 0 |
| S14_extracted_k141_29537 | 1021 | No | NA | 1 | 0 | 1 | Not-determined | Genome-fragment | NA | NA | 0 |
| S14_extracted_k141_51552 | 1471 | No | NA | 1 | 0 | 1 | Not-determined | Genome-fragment | NA | NA | 0 |
| S14_extracted_k141_7612 | 15544 | No | NA | 11 | 0 | 9 | Not-determined | Genome-fragment | NA | NA | 0 |
| S3_extracted_k141_120184 | 1305 | No | NA | 2 | 0 | 1 | Low-quality | Genome-fragment | 2.61 | AAI-based (medium-confidence) | 0 |
| S14_extracted_k141_29569 | 1239 | No | NA | 2 | 0 | 1 | Not-determined | Genome-fragment | NA | NA | 0 |
| S5_vs_k141_22359\|\|full | 1305 | No | NA | 2 | 0 | 1 | Low-quality | Genome-fragment | 1.43 | AAI-based (medium-confidence) | 0 |
| S14_extracted_k141_95520 | 4407 | No | NA | 4 | 0 | 1 | Not-determined | Genome-fragment | NA | NA | 0 |
| S5_vs_k141_103035\|\|full | 1305 | No | NA | 3 | 0 | 0 | Low-quality | Genome-fragment | 1.9 | AAI-based (high-confidence) | 0 |
| S14_extracted_k141_102768 | 1621 | No | NA | 1 | 0 | 1 | Not-determined | Genome-fragment | NA | NA | 0 |
| S14_extracted_k141_58919 | 36704 | No | NA | 54 | 17 | 0 | High-quality | High-quality | 100 | AAI-based (high-confidence) | 0 |
| S14_extracted_k141_359 | 3192 | No | NA | 5 | 0 | 3 | Not-determined | Genome-fragment | NA | NA | 0 |
| S14_extracted_k141_22259 | 1723 | No | NA | 3 | 0 | 0 | Not-determined | Genome-fragment | NA | NA | 0 |
| S5_vs_k141_135674\|\|lt2gene | 1305 | No | NA | 2 | 0 | 1 | Low-quality | Genome-fragment | 2.51 | AAI-based (medium-confidence) | 0 |
| S7_extracted_25897 | 1305 | No | NA | 3 | 2 | 0 | Low-quality | Genome-fragment | 4.53 | AAI-based (high-confidence) | 0 |
| S7_extracted_114900 | 1305 | No | NA | 3 | 0 | 0 | Low-quality | Genome-fragment | 3.26 | AAI-based (high-confidence) | 0 |
| S6_vs_k141_46149\|\|full | 1305 | No | NA | 2 | 2 | 0 | Low-quality | Genome-fragment | 0.99 | HMM-based (lower-bound) | 0 |
| S8_extracted_k141_95775 | 1305 | No | NA | 4 | 1 | 0 | Low-quality | Genome-fragment | 2.01 | HMM-based (lower-bound) | 0 |
| S11_extracted_k141_59769 | 1306 | No | NA | 5 | 0 | 0 | Low-quality | Genome-fragment | 3.21 | AAI-based (medium-confidence) | 0 |
| S11_vs_k141_6594\|\|full | 1306 | No | NA | 2 | 0 | 0 | Low-quality | Genome-fragment | 1.23 | AAI-based (high-confidence) | 0 |
| S1_vs_k141_132692\|\|full | 1306 | No | NA | 3 | 0 | 0 | Low-quality | Genome-fragment | 3.32 | AAI-based (medium-confidence) | 0 |
| S14_extracted_k141_66179 | 1577 | No | NA | 1 | 0 | 1 | Not-determined | Genome-fragment | NA | NA | 0 |
| S14_extracted_k141_59002 | 3793 | No | NA | 3 | 0 | 3 | Not-determined | Genome-fragment | NA | NA | 0 |
| S1_vs_k141_125994\|\|full | 1306 | No | NA | 2 | 0 | 1 | Low-quality | Genome-fragment | 2.43 | AAI-based (high-confidence) | 0 |
| S3_vs_k141_55947\|\|lt2gene | 1306 | No | NA | 2 | 1 | 0 | Low-quality | Genome-fragment | 2.37 | AAI-based (medium-confidence) | 0 |
| S5_extracted_k141_47500 | 1306 | No | NA | 2 | 0 | 0 | Low-quality | Genome-fragment | 2.67 | AAI-based (medium-confidence) | 0 |
| S14_extracted_k141_95598 | 1663 | No | NA | 2 | 0 | 2 | Not-determined | Genome-fragment | NA | NA | 0 |
| S5_vs_k141_98396\|\|full | 1306 | No | NA | 2 | 0 | 1 | Low-quality | Genome-fragment | 2.26 | AAI-based (medium-confidence) | 0 |
| S14_extracted_k141_88250 | 1009 | No | NA | 1 | 0 | 1 | Not-determined | Genome-fragment | NA | NA | 0 |
| S14_extracted_k141_102882 | 2026 | No | NA | 4 | 0 | 2 | Not-determined | Genome-fragment | NA | NA | 0 |
| S14_extracted_k141_59018 | 1377 | No | NA | 1 | 0 | 1 | Not-determined | Genome-fragment | NA | NA | 0 |
| S14_extracted_k141_14989 | 1515 | No | NA | 3 | 0 | 2 | Not-determined | Genome-fragment | NA | NA | 0 |
| S14_extracted_k141_80935 | 2433 | No | NA | 4 | 0 | 2 | Not-determined | Genome-fragment | NA | NA | 0 |
| S14_extracted_k141_88269 | 1016 | No | NA | 1 | 0 | 0 | Not-determined | Genome-fragment | NA | NA | 0 |
| S14_extracted_k141_110231 | 1177 | No | NA | 2 | 0 | 0 | Not-determined | Genome-fragment | NA | NA | 0 |
| S7_extracted_20297 | 1306 | No | NA | 2 | 0 | 1 | Low-quality | Genome-fragment | 0.56 | AAI-based (high-confidence) | 0 |
| S14_extracted_k141_88291 | 1037 | No | NA | 1 | 0 | 1 | Not-determined | Genome-fragment | NA | NA | 0 |
| S14_extracted_k141_44255 | 1392 | No | NA | 4 | 0 | 0 | Not-determined | Genome-fragment | NA | NA | 0 |
| S14_extracted_k141_44263 | 1174 | No | NA | 2 | 0 | 0 | Not-determined | Genome-fragment | NA | NA | 0 |
| S7_vs_6860\|\|full | 1306 | No | NA | 2 | 0 | 1 | Low-quality | Genome-fragment | 0.96 | AAI-based (medium-confidence) | 0 |
| S14_extracted_k141_545 | 1074 | No | NA | 3 | 0 | 0 | Not-determined | Genome-fragment | NA | NA | 0 |
| S14_extracted_k141_59081 | 1552 | No | NA | 2 | 0 | 2 | Not-determined | Genome-fragment | NA | NA | 0 |
| S10_vs_77029\|\|full | 1306 | No | NA | 4 | 4 | 0 | Low-quality | Genome-fragment | 2.43 | AAI-based (high-confidence) | 0 |
| S12_extracted_k141_161737 | 1306 | No | NA | 1 | 0 | 1 | Low-quality | Genome-fragment | 3.07 | AAI-based (medium-confidence) | 0 |
| S14_extracted_k141_59083 | 5551 | No | NA | 5 | 0 | 5 | Not-determined | Genome-fragment | NA | NA | 0 |
| S14_extracted_k141_102954 | 4906 | No | NA | 2 | 0 | 2 | Not-determined | Genome-fragment | NA | NA | 0 |
| S12_vs_k141_170049\|\|full | 1306 | No | NA | 3 | 0 | 0 | Low-quality | Genome-fragment | 2.2 | AAI-based (high-confidence) | 0 |
| S9_extracted_k141_13318 | 1307 | No | NA | 3 | 1 | 0 | Low-quality | Genome-fragment | 2.63 | AAI-based (medium-confidence) | 0 |
| S3_vs_k141_1497\|\|full | 1307 | No | NA | 3 | 0 | 0 | Low-quality | Genome-fragment | 1.65 | AAI-based (high-confidence) | 0 |
| S14_extracted_k141_102984 | 1263 | No | NA | 1 | 0 | 1 | Not-determined | Genome-fragment | NA | NA | 0 |
| S14_extracted_k141_612 | 2510 | No | NA | 7 | 0 | 7 | Not-determined | Genome-fragment | NA | NA | 0 |
| S5_vs_k141_136056\|\|full | 1307 | No | NA | 2 | 0 | 1 | Low-quality | Genome-fragment | 1.78 | AAI-based (medium-confidence) | 0 |
| S14_extracted_k141_73540 | 1278 | No | NA | 2 | 0 | 0 | Not-determined | Genome-fragment | NA | NA | 0 |
| S5_vs_k141_21625\|\|full | 1307 | No | NA | 3 | 0 | 0 | Low-quality | Genome-fragment | 2.35 | AAI-based (high-confidence) | 0 |
| S14_extracted_k141_44343 | 3324 | No | NA | 7 | 0 | 6 | Not-determined | Genome-fragment | NA | NA | 0 |
| S5_vs_k141_22886\|\|full | 1307 | No | NA | 3 | 1 | 0 | Low-quality | Genome-fragment | 3.18 | AAI-based (medium-confidence) | 0 |
| S5_vs_k141_71528\|\|full | 1307 | No | NA | 4 | 1 | 0 | Low-quality | Genome-fragment | 2.39 | HMM-based (lower-bound) | 0 |
| S14_extracted_k141_110287 | 1312 | No | NA | 2 | 0 | 2 | Not-determined | Genome-fragment | NA | NA | 0 |
| S14_extracted_k141_15074 | 1069 | No | NA | 1 | 0 | 0 | Not-determined | Genome-fragment | NA | NA | 0 |
| S7_vs_44914\|\|full | 1307 | No | NA | 4 | 1 | 0 | Low-quality | Genome-fragment | 2.08 | HMM-based (lower-bound) | 0 |
| S14_extracted_k141_103085 | 3143 | No | NA | 6 | 0 | 0 | Not-determined | Genome-fragment | NA | NA | 0 |
| S12_vs_k141_99338\|\|full | 1307 | No | NA | 4 | 0 | 0 | Low-quality | Genome-fragment | 0.95 | AAI-based (medium-confidence) | 0 |
| S6_vs_k141_18819\|\|full | 1307 | No | NA | 4 | 3 | 0 | Low-quality | Genome-fragment | 3.69 | AAI-based (medium-confidence) | 0 |
| S14_extracted_k141_44364 | 1323 | No | NA | 2 | 0 | 0 | Not-determined | Genome-fragment | NA | NA | 0 |
| S8_vs_k141_18419\|\|full | 1307 | No | NA | 3 | 0 | 0 | Low-quality | Genome-fragment | 2.07 | AAI-based (high-confidence) | 0 |
| S8_vs_k141_124229\|\|full | 1307 | No | NA | 4 | 1 | 0 | Low-quality | Genome-fragment | 2.08 | AAI-based (medium-confidence) | 0 |
| S17_vs_k141_108507\|\|full | 1308 | No | NA | 2 | 0 | 1 | Low-quality | Genome-fragment | 0.94 | AAI-based (medium-confidence) | 0 |
| S14_extracted_k141_37228 | 2297 | No | NA | 1 | 0 | 0 | Not-determined | Genome-fragment | NA | NA | 0 |
| S14_extracted_k141_22398 | 3993 | No | NA | 4 | 0 | 0 | Not-determined | Genome-fragment | NA | NA | 0 |
| S14_extracted_k141_95844 | 2922 | No | NA | 2 | 0 | 2 | Not-determined | Genome-fragment | NA | NA | 0 |
| S17_vs_k141_28907\|\|full | 1308 | No | NA | 2 | 0 | 0 | Low-quality | Genome-fragment | 3.79 | AAI-based (medium-confidence) | 0 |
| S14_extracted_k141_22405 | 1632 | No | NA | 1 | 0 | 0 | Not-determined | Genome-fragment | NA | NA | 0 |
| S11_vs_k141_104103\|\|full | 1308 | No | NA | 3 | 1 | 0 | Low-quality | Genome-fragment | 0.43 | HMM-based (lower-bound) | 0 |
| S3_vs_k141_95167\|\|full | 1308 | No | NA | 4 | 0 | 0 | Low-quality | Genome-fragment | 2.92 | AAI-based (medium-confidence) | 0 |
| S14_extracted_k141_37243 | 4105 | No | NA | 4 | 0 | 4 | Not-determined | Genome-fragment | NA | NA | 0 |
| S14_extracted_k141_44388 | 1995 | No | NA | 1 | 0 | 1 | Not-determined | Genome-fragment | NA | NA | 0 |
| S14_extracted_k141_29876 | 2183 | No | NA | 2 | 0 | 2 | Not-determined | Genome-fragment | NA | NA | 0 |
| S7_vs_21781\|\|full | 1308 | No | NA | 5 | 1 | 0 | Low-quality | Genome-fragment | 3.35 | AAI-based (high-confidence) | 0 |
| S16_extracted_163112 | 1308 | No | NA | 3 | 1 | 0 | Low-quality | Genome-fragment | 2 | AAI-based (high-confidence) | 0 |
| S10_vs_51888\|\|full | 1308 | No | NA | 3 | 0 | 0 | Low-quality | Genome-fragment | 1.04 | AAI-based (high-confidence) | 0 |
| S14_extracted_k141_81128 | 1269 | No | NA | 1 | 0 | 1 | Not-determined | Genome-fragment | NA | NA | 0 |
| S14_extracted_k141_95871 | 1075 | No | NA | 1 | 0 | 1 | Not-determined | Genome-fragment | NA | NA | 0 |
| S12_vs_k141_55696\|\|full | 1308 | No | NA | 5 | 0 | 0 | Low-quality | Genome-fragment | 2.12 | AAI-based (high-confidence) | 0 |
| S14_extracted_k141_73643 | 3685 | No | NA | 2 | 0 | 0 | Not-determined | Genome-fragment | NA | NA | 0 |
| S14_extracted_k141_88499 | 5417 | No | NA | 6 | 0 | 6 | Not-determined | Genome-fragment | NA | NA | 0 |
| S12_vs_k141_82084\|\|full | 1308 | No | NA | 3 | 0 | 0 | Low-quality | Genome-fragment | 1.11 | AAI-based (medium-confidence) | 0 |
| S14_extracted_k141_103154 | 5450 | No | NA | 4 | 0 | 0 | Not-determined | Genome-fragment | NA | NA | 0 |
| S12_vs_k141_60776\|\|full | 1308 | No | NA | 2 | 0 | 0 | Low-quality | Genome-fragment | 1.18 | AAI-based (medium-confidence) | 0 |
| S12_vs_k141_37196\|\|full | 1308 | No | NA | 2 | 1 | 0 | Low-quality | Genome-fragment | 3.17 | AAI-based (high-confidence) | 0 |
| S15_vs_1706\|\|full | 1308 | No | NA | 4 | 0 | 0 | Low-quality | Genome-fragment | 3.46 | AAI-based (high-confidence) | 0 |
| S14_extracted_k141_7878 | 2152 | No | NA | 1 | 0 | 1 | Not-determined | Genome-fragment | NA | NA | 0 |
| S8_extracted_k141_195952 | 1308 | No | NA | 1 | 0 | 0 | Low-quality | Genome-fragment | 3.13 | AAI-based (high-confidence) | 0 |
| S14_extracted_k141_59210 | 1995 | No | NA | 3 | 0 | 3 | Not-determined | Genome-fragment | NA | NA | 0 |
| S14_extracted_k141_59214 | 4267 | No | NA | 5 | 0 | 3 | Not-determined | Genome-fragment | NA | NA | 0 |
| S14_extracted_k141_44452 | 31164 | No | NA | 43 | 16 | 0 | High-quality | High-quality | 99.12 | AAI-based (high-confidence) | 0 |
| S17_extracted_k141_138048 | 1309 | No | NA | 2 | 0 | 0 | Low-quality | Genome-fragment | 5.86 | AAI-based (medium-confidence) | 0 |
| S9_vs_k141_63617\|\|full | 1309 | No | NA | 3 | 2 | 0 | Low-quality | Genome-fragment | 1.93 | HMM-based (lower-bound) | 0 |
| S3_vs_k141_40548\|\|full | 1309 | No | NA | 2 | 0 | 1 | Low-quality | Genome-fragment | 3.67 | AAI-based (medium-confidence) | 0 |
| S5_vs_k141_129684\|\|full | 1309 | No | NA | 3 | 0 | 0 | Low-quality | Genome-fragment | 2.26 | AAI-based (high-confidence) | 0 |
| S14_extracted_k141_770 | 1252 | No | NA | 2 | 0 | 1 | Not-determined | Genome-fragment | NA | NA | 0 |
| S16_extracted_142589 | 1309 | No | NA | 2 | 1 | 0 | Low-quality | Genome-fragment | 2.48 | HMM-based (lower-bound) | 0 |
| S14_extracted_k141_110439 | 3250 | No | NA | 2 | 0 | 1 | Not-determined | Genome-fragment | NA | NA | 0 |
| S14_extracted_k141_66399 | 15983 | No | NA | 20 | 0 | 2 | Not-determined | Genome-fragment | NA | NA | 0 |
| S16_vs_115313\|\|full | 1309 | No | NA | 2 | 0 | 0 | Low-quality | Genome-fragment | 1.99 | AAI-based (medium-confidence) | 0 |
| S14_extracted_k141_51983 | 1938 | No | NA | 4 | 0 | 3 | Not-determined | Genome-fragment | NA | NA | 0 |
| S12_extracted_k141_97987 | 1309 | No | NA | 2 | 0 | 0 | Low-quality | Genome-fragment | 1.66 | AAI-based (medium-confidence) | 0 |
| S12_vs_k141_120502\|\|full | 1309 | No | NA | 3 | 0 | 1 | Low-quality | Genome-fragment | 1.92 | AAI-based (medium-confidence) | 0 |
| S14_extracted_k141_110464 | 1891 | No | NA | 2 | 0 | 2 | Not-determined | Genome-fragment | NA | NA | 0 |
| S14_extracted_k141_110468 | 1341 | No | NA | 2 | 0 | 2 | Not-determined | Genome-fragment | NA | NA | 0 |
| S14_extracted_k141_59294 | 16909 | No | NA | 15 | 0 | 4 | Not-determined | Genome-fragment | NA | NA | 0 |
| S12_vs_k141_149370\|\|full | 1309 | No | NA | 3 | 0 | 0 | Low-quality | Genome-fragment | 1.06 | AAI-based (medium-confidence) | 0 |
| S8_extracted_k141_167303 | 1309 | No | NA | 1 | 1 | 0 | Low-quality | Genome-fragment | 2.75 | AAI-based (high-confidence) | 0 |
| S14_extracted_k141_52011 | 19473 | No | NA | 19 | 0 | 7 | Not-determined | Genome-fragment | NA | NA | 0 |
| S8_extracted_k141_89444 | 1309 | No | NA | 2 | 2 | 0 | Low-quality | Genome-fragment | 1.78 | AAI-based (medium-confidence) | 0 |
| S8_extracted_k141_142212 | 1309 | No | NA | 2 | 0 | 2 | Low-quality | Genome-fragment | 0.87 | AAI-based (high-confidence) | 0 |
| S14_extracted_k141_44513 | 1374 | No | NA | 2 | 0 | 1 | Not-determined | Genome-fragment | NA | NA | 0 |
| S8_extracted_k141_121126 | 1309 | No | NA | 2 | 0 | 0 | Low-quality | Genome-fragment | 4.04 | AAI-based (high-confidence) | 0 |
| S14_extracted_k141_22541 | 1272 | No | NA | 1 | 0 | 1 | Not-determined | Genome-fragment | NA | NA | 0 |
| S14_extracted_k141_110513 | 4889 | No | NA | 7 | 0 | 2 | Not-determined | Genome-fragment | NA | NA | 0 |
| S17_vs_k141_49114\|\|full | 1310 | No | NA | 4 | 2 | 0 | Low-quality | Genome-fragment | 3.06 | AAI-based (medium-confidence) | 0 |

| | | | | | | | | | | | |
|---|---|---|---|---|---|---|---|---|---|---|---|
| S14_extracted_k141_831 | 1268 | No | NA | 1 | 0 | 1 | Not-determined | Genome-fragment | NA | NA | 0 |
| S14_extracted_k141_30031 | 3647 | No | NA | 2 | 0 | 2 | Not-determined | Genome-fragment | NA | NA | 0 |
| S14_extracted_k141_52056 | 4360 | No | NA | 2 | 0 | 1 | Not-determined | Genome-fragment | NA | NA | 0 |
| S14_extracted_k141_52114 | 4614 | No | NA | 9 | 0 | 0 | Not-determined | Genome-fragment | NA | NA | 0 |
| S14_extracted_k141_37467 | 2627 | No | NA | 2 | 0 | 2 | Not-determined | Genome-fragment | NA | NA | 0 |
| S14_extracted_k141_73803 | 3754 | No | NA | 5 | 0 | 3 | Not-determined | Genome-fragment | NA | NA | 0 |
| S14_extracted_k141_30074 | 1478 | No | NA | 2 | 0 | 0 | Not-determined | Genome-fragment | NA | NA | 0 |
| S9_vs_k141_35544\|\|full | 1310 | No | NA | 3 | 2 | 0 | Low-quality | Genome-fragment | 3.2 | AAI-based (high-confidence) | 0 |
| S14_extracted_k141_15293 | 1373 | No | NA | 1 | 0 | 1 | Not-determined | Genome-fragment | NA | NA | 0 |
| S14_vs_k141_84162\|\|full | 1310 | No | NA | 5 | 2 | 0 | Low-quality | Genome-fragment | 2.37 | AAI-based (high-confidence) | 0 |
| S14_extracted_k141_37500 | 5961 | No | NA | 8 | 2 | 0 | High-quality | High-quality | 97.65 | AAI-based (high-confidence) | 0 |
| S1_extracted_k141_119212 | 1310 | No | NA | 1 | 0 | 0 | Low-quality | Genome-fragment | 1.33 | AAI-based (high-confidence) | 0 |
| S14_extracted_k141_869 | 1038 | No | NA | 2 | 0 | 1 | Not-determined | Genome-fragment | NA | NA | 0 |
| S14_extracted_k141_37513 | 3440 | No | NA | 6 | 0 | 0 | Not-determined | Genome-fragment | NA | NA | 0 |
| S5_vs_k141_129240\|\|full | 1310 | No | NA | 3 | 2 | 0 | Low-quality | Genome-fragment | 2.76 | AAI-based (medium-confidence) | 0 |
| S5_vs_k141_50476\|\|lt2gene | 1310 | No | NA | 1 | 1 | 0 | Low-quality | Genome-fragment | 1.84 | HMM-based (lower-bound) | 0 |
| S7_extracted_75574 | 1310 | No | NA | 2 | 0 | 0 | Low-quality | Genome-fragment | 2.72 | AAI-based (high-confidence) | 0 |
| S16_vs_15401\|\|full | 1310 | No | NA | 2 | 1 | 0 | Low-quality | Genome-fragment | 2.89 | AAI-based (high-confidence) | 0 |
| S14_extracted_k141_103285 | 5607 | No | NA | 6 | 0 | 3 | Not-determined | Genome-fragment | NA | NA | 0 |
| S14_extracted_k141_103308 | 1541 | No | NA | 1 | 0 | 1 | Not-determined | Genome-fragment | NA | NA | 0 |
| S16_vs_117110\|\|full | 1310 | No | NA | 3 | 0 | 1 | Low-quality | Genome-fragment | 3.56 | AAI-based (medium-confidence) | 0 |
| S14_extracted_k141_920 | 1095 | No | NA | 1 | 0 | 1 | Not-determined | Genome-fragment | NA | NA | 0 |
| S12_vs_k141_72718\|\|full | 1310 | No | NA | 3 | 1 | 0 | Low-quality | Genome-fragment | 3.18 | AAI-based (high-confidence) | 0 |
| S15_extracted_7794 | 1310 | No | NA | 4 | 1 | 0 | Low-quality | Genome-fragment | 3.84 | AAI-based (high-confidence) | 0 |
| S6_vs_k141_18032\|\|full | 1310 | No | NA | 2 | 0 | 2 | Low-quality | Genome-fragment | 2.11 | AAI-based (medium-confidence) | 0 |
| S6_vs_k141_34549\|\|full | 1310 | No | NA | 2 | 0 | 0 | Low-quality | Genome-fragment | 2.51 | AAI-based (medium-confidence) | 0 |
| S8_extracted_k141_147986 | 1310 | No | NA | 1 | 0 | 0 | Low-quality | Genome-fragment | 4.68 | AAI-based (medium-confidence) | 0 |
| S8_vs_k141_50616\|\|full | 1310 | No | NA | 3 | 0 | 2 | Low-quality | Genome-fragment | 3 | AAI-based (medium-confidence) | 0 |
| S17_extracted_k141_19202 | 1311 | No | NA | 2 | 2 | 0 | Low-quality | Genome-fragment | 4.16 | AAI-based (high-confidence) | 0 |
| S17_vs_k141_58513\|\|full | 1311 | No | NA | 2 | 0 | 1 | Low-quality | Genome-fragment | 1.14 | AAI-based (medium-confidence) | 0 |
| S11_vs_k141_10661\|\|full | 1311 | No | NA | 3 | 0 | 1 | Low-quality | Genome-fragment | 0.83 | AAI-based (medium-confidence) | 0 |
| S14_extracted_k141_52297 | 1474 | No | NA | 1 | 0 | 0 | Not-determined | Genome-fragment | NA | NA | 0 |
| S13_extracted_28634 | 1311 | No | NA | 2 | 2 | 0 | Low-quality | Genome-fragment | 2.49 | HMM-based (lower-bound) | 0 |
| S14_extracted_k141_968 | 7070 | No | NA | 3 | 0 | 2 | Not-determined | Genome-fragment | NA | NA | 0 |
| S14_extracted_k141_53794 | 1311 | No | NA | 4 | 3 | 0 | Low-quality | Genome-fragment | 4.33 | AAI-based (high-confidence) | 0 |
| S3_extracted_k141_92703 | 1311 | No | NA | 1 | 0 | 1 | Low-quality | Genome-fragment | 0.34 | AAI-based (high-confidence) | 0 |
| S16_vs_115322\|\|full | 1311 | No | NA | 2 | 1 | 0 | Low-quality | Genome-fragment | 3.83 | AAI-based (medium-confidence) | 0 |
| S14_extracted_k141_59503 | 3918 | No | NA | 2 | 0 | 0 | Not-determined | Genome-fragment | NA | NA | 0 |
| S12_extracted_k141_33500 | 1311 | No | NA | 3 | 0 | 0 | Low-quality | Genome-fragment | 3.05 | AAI-based (high-confidence) | 0 |
| S14_extracted_k141_22685 | 1305 | No | NA | 1 | 0 | 0 | Not-determined | Genome-fragment | NA | NA | 0 |
| S14_extracted_k141_44726 | 2309 | No | NA | 4 | 0 | 0 | Not-determined | Genome-fragment | NA | NA | 0 |
| S15_vs_50440\|\|full | 1311 | No | NA | 2 | 0 | 0 | Low-quality | Genome-fragment | 0.89 | AAI-based (high-confidence) | 0 |
| S8_vs_k141_3951\|\|full | 1311 | No | NA | 3 | 0 | 0 | Low-quality | Genome-fragment | 0.4 | AAI-based (medium-confidence) | 0 |
| S14_extracted_k141_110722 | 5514 | No | NA | 8 | 0 | 2 | Not-determined | Genome-fragment | NA | NA | 0 |
| S14_extracted_k141_74063 | 1125 | No | NA | 1 | 0 | 0 | Not-determined | Genome-fragment | NA | NA | 0 |
| S14_extracted_k141_103457 | 1499 | No | NA | 1 | 0 | 1 | Not-determined | Genome-fragment | NA | NA | 0 |
| S14_vs_k141_88892\|\|full | 1312 | No | NA | 4 | 3 | 0 | Low-quality | Genome-fragment | 4.33 | AAI-based (high-confidence) | 0 |
| S14_extracted_k141_95984 | 1300 | No | NA | 2 | 0 | 0 | Not-determined | Genome-fragment | NA | NA | 0 |
| S1_extracted_k141_67911 | 1312 | No | NA | 2 | 0 | 1 | Low-quality | Genome-fragment | 2.57 | AAI-based (medium-confidence) | 0 |
| S3_vs_k141_99797\|\|full | 1312 | No | NA | 3 | 0 | 0 | Low-quality | Genome-fragment | 2.81 | AAI-based (medium-confidence) | 0 |
| S14_extracted_k141_88879 | 3493 | No | NA | 5 | 0 | 2 | Not-determined | Genome-fragment | NA | NA | 0 |
| S14_extracted_k141_30256 | 1004 | No | NA | 1 | 0 | 0 | Not-determined | Genome-fragment | NA | NA | 0 |
| S16_extracted_72925 | 1312 | No | NA | 3 | 0 | 0 | Low-quality | Genome-fragment | 2.23 | AAI-based (medium-confidence) | 0 |
| S14_extracted_k141_44777 | 1144 | No | NA | 1 | 0 | 0 | Not-determined | Genome-fragment | NA | NA | 0 |
| S16_vs_4218\|\|full | 1312 | No | NA | 4 | 0 | 0 | Low-quality | Genome-fragment | 1.01 | AAI-based (medium-confidence) | 0 |
| S10_extracted_47430 | 1312 | No | NA | 3 | 1 | 0 | Low-quality | Genome-fragment | 3.06 | AAI-based (high-confidence) | 0 |
| S14_extracted_k141_110756 | 3692 | No | NA | 8 | 0 | 2 | Not-determined | Genome-fragment | NA | NA | 0 |
| S10_extracted_82493 | 1312 | No | NA | 1 | 0 | 0 | Low-quality | Genome-fragment | 1.88 | AAI-based (medium-confidence) | 0 |
| S12_extracted_k141_163068 | 1312 | No | NA | 1 | 0 | 1 | Low-quality | Genome-fragment | 3.35 | AAI-based (medium-confidence) | 0 |
| S12_vs_k141_126376\|\|full | 1312 | No | NA | 3 | 0 | 3 | Low-quality | Genome-fragment | 2.02 | AAI-based (medium-confidence) | 0 |
| S12_vs_k141_142138\|\|full | 1312 | No | NA | 3 | 0 | 0 | Low-quality | Genome-fragment | 3.27 | AAI-based (medium-confidence) | 0 |
| S4_extracted_6192 | 1312 | No | NA | 4 | 3 | 0 | Low-quality | Genome-fragment | 4.26 | AAI-based (high-confidence) | 0 |
| S14_extracted_k141_44805 | 1097 | No | NA | 3 | 0 | 0 | Not-determined | Genome-fragment | NA | NA | 0 |
| S14_extracted_k141_66584 | 3254 | No | NA | 3 | 0 | 0 | Not-determined | Genome-fragment | NA | NA | 0 |
| S14_extracted_k141_110832 | 8404 | No | NA | 9 | 0 | 4 | Not-determined | Genome-fragment | NA | NA | 0 |
| S14_extracted_k141_103602 | 1185 | No | NA | 3 | 0 | 0 | Not-determined | Genome-fragment | NA | NA | 0 |
| S6_vs_k141_32332\|\|full | 1312 | No | NA | 3 | 0 | 0 | Low-quality | Genome-fragment | 0.93 | AAI-based (medium-confidence) | 0 |
| S8_vs_k141_33139\|\|lt2gene | 1312 | No | NA | 1 | 1 | 0 | Low-quality | Genome-fragment | 2.8 | HMM-based (lower-bound) | 0 |
| S17_extracted_k141_171820 | 1313 | No | NA | 5 | 0 | 0 | Low-quality | Genome-fragment | 2.31 | AAI-based (high-confidence) | 0 |
| S1_extracted_k141_138870 | 1313 | No | NA | 4 | 0 | 0 | Low-quality | Genome-fragment | 2.02 | AAI-based (medium-confidence) | 0 |
| S14_extracted_k141_96087 | 2968 | No | NA | 1 | 0 | 1 | Not-determined | Genome-fragment | NA | NA | 0 |
| S1_vs_k141_119558\|\|full | 1313 | No | NA | 2 | 0 | 0 | Low-quality | Genome-fragment | 1.7 | AAI-based (high-confidence) | 0 |
| S3_vs_k141_122730\|\|full | 1313 | No | NA | 5 | 0 | 5 | Low-quality | Genome-fragment | 0.39 | AAI-based (medium-confidence) | 0 |
| S14_extracted_k141_1147 | 5130 | No | NA | 10 | 0 | 1 | Not-determined | Genome-fragment | NA | NA | 0 |
| S14_extracted_k141_103671 | 1030 | No | NA | 2 | 0 | 0 | Not-determined | Genome-fragment | NA | NA | 0 |
| S5_extracted_k141_101653 | 1313 | No | NA | 2 | 2 | 0 | Low-quality | Genome-fragment | 2.24 | HMM-based (lower-bound) | 0 |
| S5_vs_k141_57043\|\|full | 1313 | No | NA | 3 | 1 | 0 | Low-quality | Genome-fragment | 3.49 | AAI-based (high-confidence) | 0 |
| S14_extracted_k141_89006 | 4108 | No | NA | 4 | 0 | 2 | Not-determined | Genome-fragment | NA | NA | 0 |
| S5_vs_k141_43343\|\|lt2gene | 1313 | No | NA | 1 | 1 | 0 | Low-quality | Genome-fragment | 1.92 | HMM-based (lower-bound) | 0 |
| S7_extracted_85081 | 1313 | No | NA | 3 | 0 | 0 | Low-quality | Genome-fragment | 2.47 | AAI-based (high-confidence) | 0 |
| S14_extracted_k141_96112 | 1032 | No | NA | 2 | 0 | 0 | Not-determined | Genome-fragment | NA | NA | 0 |
| S7_vs_34954\|\|full | 1313 | No | NA | 3 | 2 | 0 | Low-quality | Genome-fragment | 0.76 | AAI-based (medium-confidence) | 0 |
| S7_vs_117717\|\|full | 1313 | No | NA | 3 | 0 | 0 | Low-quality | Genome-fragment | 3.09 | AAI-based (high-confidence) | 0 |
| S14_extracted_k141_22844 | 4594 | No | NA | 4 | 0 | 2 | Not-determined | Genome-fragment | NA | NA | 0 |
| S7_vs_130315\|\|full | 1313 | No | NA | 2 | 0 | 0 | Low-quality | Genome-fragment | 0.89 | AAI-based (medium-confidence) | 0 |
| S10_vs_114248\|\|full | 1313 | No | NA | 4 | 0 | 0 | Low-quality | Genome-fragment | 3.43 | AAI-based (high-confidence) | 0 |
| S12_vs_k141_1601\|\|full | 1313 | No | NA | 4 | 0 | 0 | Low-quality | Genome-fragment | 1.52 | AAI-based (medium-confidence) | 0 |
| S14_extracted_k141_59699 | 3882 | No | NA | 4 | 0 | 1 | Not-determined | Genome-fragment | NA | NA | 0 |
| S2_extracted_k141_5288 | 1313 | No | NA | 3 | 1 | 0 | Low-quality | Genome-fragment | 3.36 | AAI-based (high-confidence) | 0 |
| S6_vs_k141_22807\|\|full | 1313 | No | NA | 3 | 1 | 0 | Low-quality | Genome-fragment | 3.29 | AAI-based (medium-confidence) | 0 |
| S14_extracted_k141_1223 | 1010 | No | NA | 4 | 0 | 0 | Not-determined | Genome-fragment | NA | NA | 0 |
| S14_extracted_k141_44932 | 2774 | No | NA | 4 | 0 | 2 | Not-determined | Genome-fragment | NA | NA | 0 |
| S8_vs_k141_152270\|\|full | 1313 | No | NA | 3 | 0 | 0 | Low-quality | Genome-fragment | 3.36 | AAI-based (medium-confidence) | 0 |
| S13_vs_84613\|\|full | 1314 | No | NA | 3 | 0 | 0 | Low-quality | Genome-fragment | 1.74 | AAI-based (high-confidence) | 0 |
| S14_vs_k141_90494\|\|full | 1314 | No | NA | 3 | 1 | 0 | Low-quality | Genome-fragment | 3.55 | AAI-based (high-confidence) | 0 |
| S3_extracted_k141_50768 | 1314 | No | NA | 2 | 2 | 0 | Low-quality | Genome-fragment | 3.26 | AAI-based (high-confidence) | 0 |
| S3_vs_k141_97692\|\|full | 1314 | No | NA | 5 | 0 | 0 | Low-quality | Genome-fragment | 0.9 | AAI-based (high-confidence) | 0 |
| S5_vs_k141_139041\|\|full | 1314 | No | NA | 3 | 0 | 0 | Low-quality | Genome-fragment | 3.6 | AAI-based (high-confidence) | 0 |
| S14_extracted_k141_15803 | 1113 | No | NA | 1 | 0 | 1 | Not-determined | Genome-fragment | NA | NA | 0 |
| S14_extracted_k141_66689 | 1322 | No | NA | 2 | 0 | 1 | Not-determined | Genome-fragment | NA | NA | 0 |
| S5_vs_k141_37093\|\|lt2gene | 1314 | No | NA | 2 | 2 | 0 | Low-quality | Genome-fragment | 3.44 | AAI-based (high-confidence) | 0 |
| S16_vs_119293\|\|full | 1314 | No | NA | 3 | 0 | 1 | Low-quality | Genome-fragment | 1.39 | AAI-based (medium-confidence) | 0 |
| S14_extracted_k141_30537 | 6067 | No | NA | 6 | 0 | 5 | Not-determined | Genome-fragment | NA | NA | 0 |
| S14_extracted_k141_74213 | 1021 | No | NA | 1 | 0 | 0 | Not-determined | Genome-fragment | NA | NA | 0 |
| S14_extracted_k141_89129 | 5241 | No | NA | 7 | 0 | 5 | Not-determined | Genome-fragment | NA | NA | 0 |
| S15_extracted_54875 | 1314 | No | NA | 2 | 1 | 0 | Low-quality | Genome-fragment | 2.91 | AAI-based (high-confidence) | 0 |
| S15_vs_21341\|\|full | 1314 | No | NA | 5 | 0 | 0 | Low-quality | Genome-fragment | 2.86 | AAI-based (medium-confidence) | 0 |
| S2_extracted_k141_12410 | 1314 | No | NA | 5 | 0 | 0 | Low-quality | Genome-fragment | 1.37 | AAI-based (high-confidence) | 0 |
| S14_extracted_k141_81646 | 2214 | No | NA | 1 | 0 | 1 | Not-determined | Genome-fragment | NA | NA | 0 |
| S14_extracted_k141_66702 | 2616 | No | NA | 3 | 0 | 0 | Not-determined | Genome-fragment | NA | NA | 0 |
| S8_vs_k141_182519\|\|lt2gene | 1314 | No | NA | 2 | 1 | 0 | Low-quality | Genome-fragment | 2.29 | AAI-based (high-confidence) | 0 |
| S14_extracted_k141_66713 | 1093 | No | NA | 1 | 0 | 0 | Not-determined | Genome-fragment | NA | NA | 0 |
| S14_extracted_k141_8436 | 1771 | No | NA | 2 | 0 | 0 | Not-determined | Genome-fragment | NA | NA | 0 |
| S14_extracted_k141_74225 | 3044 | No | NA | 5 | 0 | 4 | Not-determined | Genome-fragment | NA | NA | 0 |
| S14_extracted_k141_81680 | 1352 | No | NA | 2 | 0 | 2 | Not-determined | Genome-fragment | NA | NA | 0 |
| S8_vs_k141_5131\|\|lt2gene | 1314 | No | NA | 1 | 1 | 0 | Low-quality | Genome-fragment | 0.51 | HMM-based (lower-bound) | 0 |
| S17_extracted_k141_61894 | 1315 | No | NA | 3 | 2 | 0 | Low-quality | Genome-fragment | 6 | AAI-based (high-confidence) | 0 |
| S14_extracted_k141_89176 | 1130 | No | NA | 1 | 0 | 0 | Not-determined | Genome-fragment | NA | NA | 0 |
| S14_extracted_k141_22988 | 1262 | No | NA | 2 | 0 | 1 | Not-determined | Genome-fragment | NA | NA | 0 |
| S14_extracted_k141_74270 | 1002 | No | NA | 1 | 0 | 0 | Not-determined | Genome-fragment | NA | NA | 0 |
| S9_vs_k141_82145\|\|lt2gene | 1315 | No | NA | 1 | 1 | 0 | Low-quality | Genome-fragment | 1.85 | HMM-based (lower-bound) | 0 |
| S11_extracted_k141_55613 | 1315 | No | NA | 3 | 0 | 0 | Low-quality | Genome-fragment | 2.77 | AAI-based (high-confidence) | 0 |
| S14_extracted_k141_81729 | 4095 | No | NA | 3 | 0 | 3 | Not-determined | Genome-fragment | NA | NA | 0 |
| S14_vs_k141_99593\|\|full | 1315 | No | NA | 3 | 2 | 0 | Low-quality | Genome-fragment | 2.82 | AAI-based (medium-confidence) | 0 |
| S14_vs_k141_79868\|\|full | 1315 | No | NA | 4 | 2 | 0 | Low-quality | Genome-fragment | 3.16 | AAI-based (medium-confidence) | 0 |
| S14_extracted_k141_8467 | 6853 | No | NA | 7 | 0 | 1 | Not-determined | Genome-fragment | NA | NA | 0 |
| S3_vs_k141_60616\|\|full | 1315 | No | NA | 4 | 1 | 0 | Low-quality | Genome-fragment | 2.82 | AAI-based (high-confidence) | 0 |
| S14_extracted_k141_111238 | 1674 | No | NA | 2 | 0 | 2 | Not-determined | Genome-fragment | NA | NA | 0 |
| S14_extracted_k141_59845 | 6195 | No | NA | 10 | 0 | 2 | Not-determined | Genome-fragment | NA | NA | 0 |
| S14_extracted_k141_15976 | 1735 | No | NA | 2 | 0 | 2 | Not-determined | Genome-fragment | NA | NA | 0 |
| S14_extracted_k141_23057 | 1299 | No | NA | 2 | 0 | 0 | Not-determined | Genome-fragment | NA | NA | 0 |
| S14_extracted_k141_81814 | 1304 | No | NA | 2 | 0 | 0 | Not-determined | Genome-fragment | NA | NA | 0 |
| S14_extracted_k141_52548 | 1063 | No | NA | 1 | 0 | 1 | Not-determined | Genome-fragment | NA | NA | 0 |
| S3_vs_k141_45925\|\|full | 1315 | No | NA | 3 | 1 | 0 | Low-quality | Genome-fragment | 2.15 | HMM-based (lower-bound) | 0 |
| S14_extracted_k141_66796 | 9524 | No | NA | 8 | 0 | 7 | Not-determined | Genome-fragment | NA | NA | 0 |
| S14_extracted_k141_111271 | 2245 | No | NA | 3 | 0 | 0 | Not-determined | Genome-fragment | NA | NA | 0 |
| S7_vs_119932\|\|full | 1315 | No | NA | 3 | 1 | 0 | Low-quality | Genome-fragment | 0.51 | HMM-based (lower-bound) | 0 |
| S16_extracted_41013 | 1315 | No | NA | 1 | 1 | 0 | Low-quality | Genome-fragment | 2.2 | AAI-based (high-confidence) | 0 |
| S16_vs_46186\|\|full | 1315 | No | NA | 3 | 1 | 0 | Low-quality | Genome-fragment | 2.2 | AAI-based (high-confidence) | 0 |
| S16_vs_134788\|\|full | 1315 | No | NA | 2 | 0 | 1 | Low-quality | Genome-fragment | 1.79 | AAI-based (high-confidence) | 0 |
| S10_vs_40126\|\|full | 1315 | No | NA | 2 | 1 | 0 | Low-quality | Genome-fragment | 0.69 | HMM-based (lower-bound) | 0 |
| S12_vs_k141_181388\|\|full | 1315 | No | NA | 2 | 0 | 2 | Low-quality | Genome-fragment | 0.71 | AAI-based (medium-confidence) | 0 |
| S15_vs_35563\|\|full | 1315 | No | NA | 2 | 2 | 0 | Low-quality | Genome-fragment | 3.08 | AAI-based (high-confidence) | 0 |
| S14_extracted_k141_96265 | 1128 | No | NA | 3 | 0 | 0 | Not-determined | Genome-fragment | NA | NA | 0 |
| S14_extracted_k141_104059 | 3118 | No | NA | 5 | 0 | 0 | Not-determined | Genome-fragment | NA | NA | 0 |
| S6_vs_k141_56015\|\|lt2gene | 1315 | No | NA | 2 | 1 | 0 | Low-quality | Genome-fragment | 3.1 | AAI-based (high-confidence) | 0 |
| S8_vs_k141_58818\|\|lt2gene | 1315 | No | NA | 2 | 1 | 0 | Low-quality | Genome-fragment | 4.81 | AAI-based (medium-confidence) | 0 |
| S14_extracted_k141_30721 | 10765 | No | NA | 12 | 0 | 0 | Not-determined | Genome-fragment | NA | NA | 0 |
| S14_extracted_k141_1265 | 3459 | No | NA | 3 | 0 | 3 | Not-determined | Genome-fragment | NA | NA | 0 |
| S14_extracted_k141_8598 | 1008 | No | NA | 2 | 0 | 1 | Not-determined | Genome-fragment | NA | NA | 0 |
| S9_vs_k141_90036\|\|full | 1316 | No | NA | 3 | 0 | 0 | Low-quality | Genome-fragment | 1.98 | AAI-based (medium-confidence) | 0 |
| S14_extracted_k141_81943 | 5207 | No | NA | 4 | 0 | 2 | Not-determined | Genome-fragment | NA | NA | 0 |
| S14_extracted_k141_8630 | 1076 | No | NA | 2 | 0 | 1 | Not-determined | Genome-fragment | NA | NA | 0 |
| S11_vs_k141_28463\|\|full | 1316 | No | NA | 2 | 0 | 0 | Low-quality | Genome-fragment | 2.46 | AAI-based (high-confidence) | 0 |
| S14_extracted_k141_16123 | 1726 | No | NA | 4 | 0 | 2 | Not-determined | Genome-fragment | NA | NA | 0 |
| S14_extracted_k141_111391 | 3001 | No | NA | 5 | 0 | 1 | Not-determined | Genome-fragment | NA | NA | 0 |
| S11_vs_k141_56916\|\|full | 1316 | No | NA | 3 | 0 | 1 | Low-quality | Genome-fragment | 3.55 | AAI-based (medium-confidence) | 0 |
| S14_extracted_k141_82210 | 1316 | No | NA | 4 | 4 | 0 | Low-quality | Genome-fragment | 4.34 | AAI-based (high-confidence) | 0 |
| S14_extracted_k141_96349 | 1036 | No | NA | 2 | 0 | 0 | Not-determined | Genome-fragment | NA | NA | 0 |
| S14_extracted_k141_23250 | 1591 | No | NA | 1 | 0 | 1 | Not-determined | Genome-fragment | NA | NA | 0 |
| S14_vs_k141_90418\|\|full | 1316 | No | NA | 5 | 4 | 0 | Low-quality | Genome-fragment | 2.16 | HMM-based (lower-bound) | 0 |
| S1_vs_k141_56918\|\|full | 1316 | No | NA | 3 | 2 | 0 | Low-quality | Genome-fragment | 3.63 | AAI-based (high-confidence) | 0 |
| S14_extracted_k141_66956 | 1434 | No | NA | 2 | 0 | 2 | Not-determined | Genome-fragment | NA | NA | 0 |
| S14_extracted_k141_60073 | 3245 | No | NA | 3 | 0 | 3 | Not-determined | Genome-fragment | NA | NA | 0 |
| S1_vs_k141_84319\|\|lt2gene | 1316 | No | NA | 1 | 1 | 0 | Low-quality | Genome-fragment | 1.74 | HMM-based (lower-bound) | 0 |
| S5_extracted_k141_55843 | 1316 | No | NA | 2 | 0 | 0 | Low-quality | Genome-fragment | 2.46 | AAI-based (high-confidence) | 0 |
| S5_vs_k141_60873\|\|full | 1316 | No | NA | 3 | 2 | 0 | Low-quality | Genome-fragment | 1.73 | HMM-based (lower-bound) | 0 |
| S5_vs_k141_18792\|\|full | 1316 | No | NA | 6 | 0 | 0 | Low-quality | Genome-fragment | 2.25 | AAI-based (high-confidence) | 0 |
| S14_extracted_k141_66996 | 3672 | No | NA | 2 | 0 | 2 | Not-determined | Genome-fragment | NA | NA | 0 |
| S5_vs_k141_19134\|\|lt2gene | 1316 | No | NA | 2 | 2 | 0 | Low-quality | Genome-fragment | 2.57 | AAI-based (medium-confidence) | 0 |
| S16_vs_79629\|\|full | 1316 | No | NA | 4 | 1 | 0 | Low-quality | Genome-fragment | 2.63 | AAI-based (high-confidence) | 0 |
| S14_extracted_k141_52790 | 1315 | No | NA | 2 | 0 | 1 | Not-determined | Genome-fragment | NA | NA | 0 |
| S14_extracted_k141_52799 | 1239 | No | NA | 1 | 0 | 1 | Not-determined | Genome-fragment | NA | NA | 0 |
| S12_vs_k141_112074\|\|full | 1316 | No | NA | 4 | 0 | 0 | Low-quality | Genome-fragment | 3.2 | AAI-based (high-confidence) | 0 |
| S12_vs_k141_43773\|\|lt2gene | 1316 | No | NA | 2 | 1 | 0 | Low-quality | Genome-fragment | 2.12 | HMM-based (lower-bound) | 0 |
| S6_vs_k141_50666\|\|lt2gene | 1316 | No | NA | 2 | 2 | 0 | Low-quality | Genome-fragment | 2.88 | AAI-based (high-confidence) | 0 |
| S8_extracted_k141_46864 | 1316 | No | NA | 1 | 0 | 1 | Low-quality | Genome-fragment | 3.83 | AAI-based (high-confidence) | 0 |
| S17_extracted_k141_95487 | 1317 | No | NA | 1 | 1 | 0 | Low-quality | Genome-fragment | 0.77 | HMM-based (lower-bound) | 0 |
| S14_extracted_k141_38277 | 1137 | No | NA | 2 | 0 | 1 | Not-determined | Genome-fragment | NA | NA | 0 |
| S14_extracted_k141_96446 | 1307 | No | NA | 2 | 0 | 2 | Not-determined | Genome-fragment | NA | NA | 0 |
| S14_extracted_k141_52830 | 3044 | No | NA | 6 | 0 | 1 | Not-determined | Genome-fragment | NA | NA | 0 |
| S17_vs_k141_127535\|\|full | 1317 | No | NA | 3 | 0 | 0 | Low-quality | Genome-fragment | 2.08 | AAI-based (high-confidence) | 0 |
| S14_extracted_k141_38309 | 2187 | No | NA | 1 | 0 | 1 | Not-determined | Genome-fragment | NA | NA | 0 |
| S3_vs_k141_28779\|\|full | 1317 | No | NA | 4 | 0 | 0 | Low-quality | Genome-fragment | 2.13 | AAI-based (high-confidence) | 0 |
| S14_extracted_k141_60222 | 1087 | No | NA | 1 | 0 | 1 | Not-determined | Genome-fragment | NA | NA | 0 |
| S14_extracted_k141_89472 | 1286 | No | NA | 2 | 0 | 2 | Not-determined | Genome-fragment | NA | NA | 0 |
| S16_vs_124981\|\|full | 1317 | No | NA | 5 | 0 | 0 | Low-quality | Genome-fragment | 2.34 | AAI-based (medium-confidence) | 0 |
| S16_vs_74591\|\|lt2gene | 1317 | No | NA | 2 | 0 | 0 | Low-quality | Genome-fragment | 2.89 | AAI-based (high-confidence) | 0 |
| S10_extracted_76897 | 1317 | No | NA | 2 | 0 | 0 | Low-quality | Genome-fragment | 9.73 | AAI-based (medium-confidence) | 0 |
| S14_extracted_k141_60283 | 2226 | No | NA | 2 | 0 | 1 | Not-determined | Genome-fragment | NA | NA | 0 |
| S14_extracted_k141_8901 | 1658 | No | NA | 2 | 0 | 1 | Not-determined | Genome-fragment | NA | NA | 0 |
| S12_extracted_k141_166559 | 1317 | No | NA | 3 | 0 | 2 | Low-quality | Genome-fragment | 3.07 | AAI-based (high-confidence) | 0 |
| S8_extracted_k141_79229 | 1317 | No | NA | 2 | 0 | 0 | Low-quality | Genome-fragment | 0.24 | AAI-based (medium-confidence) | 0 |
| S14_extracted_k141_89511 | 1991 | No | NA | 2 | 0 | 2 | Not-determined | Genome-fragment | NA | NA | 0 |
| S14_extracted_k141_104417 | 3813 | No | NA | 5 | 0 | 4 | Not-determined | Genome-fragment | NA | NA | 0 |
| S8_vs_k141_64176\|\|full | 1317 | No | NA | 3 | 2 | 0 | Low-quality | Genome-fragment | 2.14 | HMM-based (lower-bound) | 0 |
| S17_vs_k141_111001\|\|full | 1318 | No | NA | 3 | 1 | 0 | Low-quality | Genome-fragment | 2.67 | AAI-based (high-confidence) | 0 |
| S14_extracted_k141_16392 | 1036 | No | NA | 1 | 0 | 0 | Not-determined | Genome-fragment | NA | NA | 0 |
| S14_extracted_k141_8927 | 27261 | No | NA | 20 | 0 | 5 | Not-determined | Genome-fragment | NA | NA | 0 |
| S13_vs_105438\|\|full | 1318 | No | NA | 3 | 0 | 1 | Low-quality | Genome-fragment | 1.63 | AAI-based (medium-confidence) | 0 |
| S14_extracted_k141_111792 | 6647 | No | NA | 8 | 0 | 3 | Not-determined | Genome-fragment | NA | NA | 0 |
| S14_extracted_k141_14866 | 1318 | No | NA | 4 | 4 | 0 | Low-quality | Genome-fragment | 4.35 | AAI-based (high-confidence) | 0 |
| S14_extracted_k141_74724 | 3216 | No | NA | 2 | 0 | 2 | Not-determined | Genome-fragment | NA | NA | 0 |
| S14_vs_k141_86850\|\|full | 1318 | No | NA | 2 | 0 | 1 | Low-quality | Genome-fragment | 0.95 | AAI-based (medium-confidence) | 0 |
| S3_extracted_k141_104992 | 1318 | No | NA | 4 | 0 | 0 | Low-quality | Genome-fragment | 0.86 | AAI-based (high-confidence) | 0 |
| S3_vs_k141_64324\|\|full | 1318 | No | NA | 3 | 1 | 0 | Low-quality | Genome-fragment | 1.88 | AAI-based (medium-confidence) | 0 |
| S14_extracted_k141_1515 | 1279 | No | NA | 1 | 0 | 1 | Not-determined | Genome-fragment | NA | NA | 0 |
| S3_vs_k141_108476\|\|lt2gene | 1318 | No | NA | 1 | 1 | 0 | Low-quality | Genome-fragment | 1.68 | AAI-based (high-confidence) | 0 |
| S5_vs_k141_95556\|\|full | 1318 | No | NA | 2 | 0 | 1 | Low-quality | Genome-fragment | 2.2 | AAI-based (high-confidence) | 0 |
| S7_vs_46134\|\|full | 1318 | No | NA | 3 | 1 | 0 | Low-quality | Genome-fragment | 2.02 | HMM-based (lower-bound) | 0 |
| S14_extracted_k141_38470 | 1629 | No | NA | 4 | 0 | 0 | Not-determined | Genome-fragment | NA | NA | 0 |
| S14_extracted_k141_74768 | 1690 | No | NA | 3 | 0 | 0 | Not-determined | Genome-fragment | NA | NA | 0 |
| S14_extracted_k141_104530 | 1082 | No | NA | 2 | 0 | 0 | Not-determined | Genome-fragment | NA | NA | 0 |
| S14_extracted_k141_74780 | 1087 | No | NA | 1 | 0 | 1 | Not-determined | Genome-fragment | NA | NA | 0 |
| S14_extracted_k141_31099 | 1019 | No | NA | 1 | 0 | 1 | Not-determined | Genome-fragment | NA | NA | 0 |
| S7_vs_81343\|\|full | 1318 | No | NA | 4 | 1 | 0 | Low-quality | Genome-fragment | 2.25 | AAI-based (medium-confidence) | 0 |
| S7_vs_74253\|\|lt2gene | 1318 | No | NA | 1 | 1 | 0 | Low-quality | Genome-fragment | 3.73 | AAI-based (medium-confidence) | 0 |
| S14_extracted_k141_1539 | 1091 | No | NA | 3 | 0 | 0 | Not-determined | Genome-fragment | NA | NA | 0 |
| S14_extracted_k141_38501 | 1146 | No | NA | 1 | 0 | 1 | Not-determined | Genome-fragment | NA | NA | 0 |
| S14_extracted_k141_82271 | 1503 | No | NA | 2 | 0 | 1 | Not-determined | Genome-fragment | NA | NA | 0 |
| S16_vs_39803\|\|full | 1318 | No | NA | 4 | 0 | 1 | Low-quality | Genome-fragment | 2.08 | AAI-based (medium-confidence) | 0 |
| S16_vs_123704\|\|full | 1318 | No | NA | 2 | 0 | 1 | Low-quality | Genome-fragment | 2.32 | AAI-based (medium-confidence) | 0 |
| S14_extracted_k141_111875 | 1004 | No | NA | 2 | 0 | 0 | Not-determined | Genome-fragment | NA | NA | 0 |
| S13_extracted_63849 | 1319 | No | NA | 2 | 0 | 1 | Low-quality | Genome-fragment | 1.25 | AAI-based (high-confidence) | 0 |
| S14_extracted_k141_53022 | 1778 | No | NA | 3 | 0 | 2 | Not-determined | Genome-fragment | NA | NA | 0 |
| S14_extracted_k141_67575 | 1319 | No | NA | 3 | 2 | 0 | Low-quality | Genome-fragment | 4.34 | AAI-based (high-confidence) | 0 |
| S14_vs_k141_111951\|\|full | 1319 | No | NA | 4 | 4 | 0 | Low-quality | Genome-fragment | 4.33 | AAI-based (high-confidence) | 0 |
| S1_extracted_k141_125844 | 1319 | No | NA | 1 | 0 | 1 | Low-quality | Genome-fragment | 0.93 | AAI-based (high-confidence) | 0 |
| S15_extracted_14624 | 1319 | No | NA | 2 | 2 | 0 | Low-quality | Genome-fragment | 2.94 | AAI-based (high-confidence) | 0 |
| S14_extracted_k141_31158 | 2002 | No | NA | 5 | 0 | 2 | Not-determined | Genome-fragment | NA | NA | 0 |
| S8_vs_k141_66979\|\|full | 1319 | No | NA | 3 | 1 | 0 | Low-quality | Genome-fragment | 2.14 | AAI-based (medium-confidence) | 0 |
| S14_extracted_k141_74961 | 1118 | No | NA | 2 | 0 | 0 | Not-determined | Genome-fragment | NA | NA | 0 |
| S14_extracted_k141_1622 | 1035 | No | NA | 2 | 0 | 1 | Not-determined | Genome-fragment | NA | NA | 0 |
| S14_extracted_k141_104679 | 1312 | No | NA | 1 | 0 | 0 | Not-determined | Genome-fragment | NA | NA | 0 |
| S14_vs_k141_81134\|\|lt2gene | 1320 | No | NA | 2 | 1 | 0 | Low-quality | Genome-fragment | 4.04 | AAI-based (high-confidence) | 0 |
| S14_extracted_k141_1648 | 1077 | No | NA | 2 | 0 | 1 | Not-determined | Genome-fragment | NA | NA | 0 |
| S1_extracted_k141_155679 | 1320 | No | NA | 2 | 1 | 0 | Low-quality | Genome-fragment | 1.32 | AAI-based (high-confidence) | 0 |
| S14_extracted_k141_104691 | 1062 | No | NA | 1 | 0 | 0 | Not-determined | Genome-fragment | NA | NA | 0 |
| S1_extracted_k141_59699 | 1320 | No | NA | 1 | 1 | 0 | Low-quality | Genome-fragment | 2.22 | HMM-based (lower-bound) | 0 |
| S14_extracted_k141_8940 | 1836 | No | NA | 2 | 0 | 1 | Not-determined | Genome-fragment | NA | NA | 0 |
| S14_extracted_k141_38611 | 1374 | No | NA | 4 | 0 | 0 | Not-determined | Genome-fragment | NA | NA | 0 |
| S1_extracted_k141_60061 | 1320 | No | NA | 1 | 0 | 1 | Low-quality | Genome-fragment | 3.72 | AAI-based (medium-confidence) | 0 |
| S14_extracted_k141_96781 | 1201 | No | NA | 2 | 0 | 1 | Not-determined | Genome-fragment | NA | NA | 0 |
| S1_vs_k141_107954\|\|full | 1320 | No | NA | 2 | 1 | 0 | Low-quality | Genome-fragment | 2.34 | AAI-based (high-confidence) | 0 |
| S14_extracted_k141_104708 | 3584 | No | NA | 7 | 0 | 1 | Not-determined | Genome-fragment | NA | NA | 0 |
| S14_extracted_k141_96802 | 3107 | No | NA | 2 | 0 | 2 | Not-determined | Genome-fragment | NA | NA | 0 |
| S3_extracted_k141_78924 | 1320 | No | NA | 2 | 0 | 1 | Low-quality | Genome-fragment | 2.61 | AAI-based (medium-confidence) | 0 |
| S14_extracted_k141_82400 | 2898 | No | NA | 2 | 0 | 1 | Not-determined | Genome-fragment | NA | NA | 0 |
| S14_extracted_k141_8957 | 1163 | No | NA | 1 | 0 | 1 | Not-determined | Genome-fragment | NA | NA | 0 |
| S3_vs_k141_117904\|\|full | 1320 | No | NA | 3 | 0 | 0 | Low-quality | Genome-fragment | 0.58 | AAI-based (medium-confidence) | 0 |
| S14_extracted_k141_31233 | 1778 | No | NA | 1 | 0 | 1 | Not-determined | Genome-fragment | NA | NA | 0 |
| S14_extracted_k141_45366 | 1647 | No | NA | 3 | 0 | 0 | Not-determined | Genome-fragment | NA | NA | 0 |
| S10_extracted_54035 | 1320 | No | NA | 2 | 1 | 0 | Low-quality | Genome-fragment | 0.54 | AAI-based (high-confidence) | 0 |
| S8_extracted_k141_81983 | 1320 | No | NA | 3 | 0 | 2 | Low-quality | Genome-fragment | 1.88 | AAI-based (high-confidence) | 0 |
| S14_extracted_k141_53183 | 1317 | No | NA | 3 | 0 | 3 | Not-determined | Genome-fragment | NA | NA | 0 |
| S17_vs_k141_158721\|\|lt2gene | 1321 | No | NA | 1 | 1 | 0 | Low-quality | Genome-fragment | 1.93 | HMM-based (lower-bound) | 0 |
| S11_extracted_k141_65649 | 1321 | No | NA | 3 | 0 | 2 | Low-quality | Genome-fragment | 0.83 | AAI-based (high-confidence) | 0 |
| S14_extracted_k141_75076 | 1108 | No | NA | 1 | 0 | 1 | Not-determined | Genome-fragment | NA | NA | 0 |
| S14_extracted_k141_45391 | 8902 | No | NA | 6 | 0 | 4 | Not-determined | Genome-fragment | NA | NA | 0 |
| S14_extracted_k141_60691 | 3424 | No | NA | 5 | 0 | 2 | Not-determined | Genome-fragment | NA | NA | 0 |
| S14_extracted_k141_82455 | 1026 | No | NA | 2 | 0 | 1 | Not-determined | Genome-fragment | NA | NA | 0 |
| S14_extracted_k141_31311 | 1398 | No | NA | 3 | 0 | 3 | Not-determined | Genome-fragment | NA | NA | 0 |
| S11_vs_k141_44550\|\|full | 1321 | No | NA | 4 | 3 | 0 | Low-quality | Genome-fragment | 2.18 | AAI-based (medium-confidence) | 0 |
| S14_extracted_k141_82467 | 1084 | No | NA | 3 | 0 | 1 | Not-determined | Genome-fragment | NA | NA | 0 |
| S14_extracted_k141_38726 | 1305 | No | NA | 1 | 0 | 1 | Not-determined | Genome-fragment | NA | NA | 0 |
| S14_extracted_k141_22200 | 1321 | No | NA | 3 | 1 | 0 | Low-quality | Genome-fragment | 0.69 | HMM-based (lower-bound) | 0 |
| S14_extracted_k141_45447 | 1534 | No | NA | 1 | 0 | 0 | Not-determined | Genome-fragment | NA | NA | 0 |
| S14_extracted_k141_31343 | 2128 | No | NA | 4 | 0 | 1 | Not-determined | Genome-fragment | NA | NA | 0 |
| S1_vs_k141_44768\|\|full | 1321 | No | NA | 2 | 0 | 1 | Low-quality | Genome-fragment | 1.1 | AAI-based (medium-confidence) | 0 |
| S5_vs_k141_68623\|\|full | 1321 | No | NA | 2 | 0 | 1 | Low-quality | Genome-fragment | 1.63 | AAI-based (medium-confidence) | 0 |
| S14_extracted_k141_60763 | 6623 | No | NA | 6 | 0 | 6 | Not-determined | Genome-fragment | NA | NA | 0 |
| S14_extracted_k141_23748 | 1188 | No | NA | 2 | 0 | 0 | Not-determined | Genome-fragment | NA | NA | 0 |
| S5_vs_k141_15854\|\|full | 1321 | No | NA | 3 | 0 | 0 | Low-quality | Genome-fragment | 2.62 | AAI-based (medium-confidence) | 0 |
| S14_extracted_k141_38851 | 1491 | No | NA | 1 | 0 | 1 | Not-determined | Genome-fragment | NA | NA | 0 |
| S5_vs_k141_36342\|\|full | 1321 | No | NA | 3 | 1 | 0 | Low-quality | Genome-fragment | 0.89 | HMM-based (lower-bound) | 0 |
| S16_extracted_36088 | 1321 | No | NA | 4 | 0 | 0 | Low-quality | Genome-fragment | 3.17 | AAI-based (high-confidence) | 0 |
| S16_vs_33023\|\|full | 1321 | No | NA | 2 | 0 | 0 | Low-quality | Genome-fragment | 0.32 | AAI-based (medium-confidence) | 0 |
| S14_extracted_k141_53416 | 1213 | No | NA | 1 | 0 | 0 | Not-determined | Genome-fragment | NA | NA | 0 |
| S14_extracted_k141_53417 | 1036 | No | NA | 2 | 0 | 0 | Not-determined | Genome-fragment | NA | NA | 0 |
| S14_extracted_k141_104954 | 1227 | No | NA | 2 | 0 | 1 | Not-determined | Genome-fragment | NA | NA | 0 |
| S14_extracted_k141_67690 | 3398 | No | NA | 4 | 0 | 0 | Not-determined | Genome-fragment | NA | NA | 0 |
| S12_vs_k141_126208\|\|full | 1321 | No | NA | 2 | 0 | 2 | Low-quality | Genome-fragment | 1.71 | AAI-based (medium-confidence) | 0 |
| S14_extracted_k141_38960 | 1167 | No | NA | 1 | 0 | 1 | Not-determined | Genome-fragment | NA | NA | 0 |
| S14_extracted_k141_67732 | 4357 | No | NA | 6 | 0 | 2 | Not-determined | Genome-fragment | NA | NA | 0 |
| S14_extracted_k141_112173 | 1189 | No | NA | 2 | 0 | 2 | Not-determined | Genome-fragment | NA | NA | 0 |
| S15_vs_16156\|\|full | 1321 | No | NA | 5 | 0 | 0 | Low-quality | Genome-fragment | 2.37 | AAI-based (medium-confidence) | 0 |
| S15_vs_65294\|\|full | 1321 | No | NA | 4 | 0 | 0 | Low-quality | Genome-fragment | 1.5 | AAI-based (high-confidence) | 0 |
| S14_extracted_k141_105002 | 1249 | No | NA | 2 | 0 | 1 | Not-determined | Genome-fragment | NA | NA | 0 |
| S14_extracted_k141_82736 | 5689 | No | NA | 4 | 0 | 2 | Not-determined | Genome-fragment | NA | NA | 0 |
| S8_vs_k141_155412\|\|lt2gene | 1321 | No | NA | 2 | 1 | 0 | Low-quality | Genome-fragment | 0.71 | HMM-based (lower-bound) | 0 |
| S17_vs_k141_164906\|\|full | 1322 | No | NA | 2 | 1 | 0 | Low-quality | Genome-fragment | 1.12 | AAI-based (medium-confidence) | 0 |
| S17_vs_k141_51377\|\|full | 1322 | No | NA | 3 | 0 | 1 | Low-quality | Genome-fragment | 2.77 | AAI-based (medium-confidence) | 0 |
| S9_extracted_k141_31622 | 1322 | No | NA | 2 | 1 | 0 | Low-quality | Genome-fragment | 2.95 | AAI-based (high-confidence) | 0 |
| S14_extracted_k141_105058 | 3059 | No | NA | 2 | 0 | 0 | Not-determined | Genome-fragment | NA | NA | 0 |
| S9_vs_k141_6541\|\|full | 1322 | No | NA | 3 | 0 | 0 | Low-quality | Genome-fragment | 2.73 | AAI-based (high-confidence) | 0 |
| S14_extracted_k141_75300 | 1174 | No | NA | 1 | 0 | 1 | Not-determined | Genome-fragment | NA | NA | 0 |
| S14_extracted_k141_75303 | 1164 | No | NA | 3 | 0 | 3 | Not-determined | Genome-fragment | NA | NA | 0 |
| S14_extracted_k141_31437 | 1076 | No | NA | 2 | 0 | 0 | Not-determined | Genome-fragment | NA | NA | 0 |
| S13_vs_68786\|\|full | 1322 | No | NA | 2 | 0 | 0 | Low-quality | Genome-fragment | 2.35 | AAI-based (high-confidence) | 0 |
| S14_extracted_k141_82770 | 1135 | No | NA | 1 | 0 | 1 | Not-determined | Genome-fragment | NA | NA | 0 |
| S14_extracted_k141_31441 | 1954 | No | NA | 3 | 0 | 1 | Not-determined | Genome-fragment | NA | NA | 0 |
| S14_vs_k141_82046\|\|full | 1322 | No | NA | 3 | 0 | 0 | Low-quality | Genome-fragment | 2.81 | AAI-based (medium-confidence) | 0 |
| S14_vs_k141_35185\|\|full | 1322 | No | NA | 3 | 0 | 0 | Low-quality | Genome-fragment | 1.84 | AAI-based (medium-confidence) | 0 |
| S1_extracted_k141_140797 | 1322 | No | NA | 2 | 0 | 2 | Low-quality | Genome-fragment | 0.46 | AAI-based (medium-confidence) | 0 |
| S5_vs_k141_25542\|\|lt2gene | 1322 | No | NA | 1 | 1 | 0 | Low-quality | Genome-fragment | 2.68 | AAI-based (high-confidence) | 0 |
| S14_extracted_k141_1927 | 6101 | No | NA | 4 | 0 | 3 | Not-determined | Genome-fragment | NA | NA | 0 |
| S14_extracted_k141_31444 | 13377 | No | NA | 18 | 0 | 5 | Not-determined | Genome-fragment | NA | NA | 0 |
| S5_vs_k141_58617\|\|lt2gene | 1322 | No | NA | 2 | 1 | 0 | Low-quality | Genome-fragment | 0.82 | HMM-based (lower-bound) | 0 |
| S14_extracted_k141_90110 | 3828 | No | NA | 6 | 0 | 1 | Not-determined | Genome-fragment | NA | NA | 0 |
| S12_vs_k141_63426\|\|full | 1322 | No | NA | 3 | 0 | 1 | Low-quality | Genome-fragment | 0.83 | AAI-based (medium-confidence) | 0 |
| S12_vs_k141_87776\|\|full | 1322 | No | NA | 3 | 1 | 0 | Low-quality | Genome-fragment | 3.09 | AAI-based (medium-confidence) | 0 |
| S8_vs_k141_157830\|\|full | 1322 | No | NA | 3 | 2 | 0 | Low-quality | Genome-fragment | 3.22 | HMM-based (lower-bound) | 0 |
| S14_extracted_k141_75365 | 1530 | No | NA | 3 | 0 | 3 | Not-determined | Genome-fragment | NA | NA | 0 |
| S8_vs_k141_5716\|\|full | 1322 | No | NA | 2 | 0 | 2 | Low-quality | Genome-fragment | 1.55 | AAI-based (high-confidence) | 0 |
| S14_extracted_k141_53605 | 2061 | No | NA | 2 | 0 | 1 | Not-determined | Genome-fragment | NA | NA | 0 |
| S14_extracted_k141_112258 | 1778 | No | NA | 2 | 0 | 2 | Not-determined | Genome-fragment | NA | NA | 0 |
| S14_extracted_k141_67918 | 3793 | No | NA | 6 | 0 | 1 | Not-determined | Genome-fragment | NA | NA | 0 |
| S17_extracted_k141_41175 | 1323 | No | NA | 2 | 0 | 0 | Low-quality | Genome-fragment | 1.32 | AAI-based (high-confidence) | 0 |
| S11_vs_k141_72422\|\|full | 1323 | No | NA | 3 | 0 | 0 | Low-quality | Genome-fragment | 2.9 | AAI-based (high-confidence) | 0 |
| S14_extracted_k141_24079 | 3164 | No | NA | 2 | 0 | 1 | Not-determined | Genome-fragment | NA | NA | 0 |
| S14_extracted_k141_97067 | 1126 | No | NA | 1 | 0 | 0 | Not-determined | Genome-fragment | NA | NA | 0 |
| S14_extracted_k141_16969 | 2222 | No | NA | 2 | 0 | 2 | Not-determined | Genome-fragment | NA | NA | 0 |
| S13_vs_33032\|\|full | 1323 | No | NA | 4 | 1 | 0 | Low-quality | Genome-fragment | 1.02 | AAI-based (high-confidence) | 0 |
| S14_extracted_k141_7832 | 1323 | No | NA | 4 | 3 | 0 | Low-quality | Genome-fragment | 4.34 | AAI-based (high-confidence) | 0 |
| S14_extracted_k141_53687 | 3574 | No | NA | 8 | 0 | 8 | Not-determined | Genome-fragment | NA | NA | 0 |
| S14_extracted_k141_82897 | 1206 | No | NA | 1 | 0 | 1 | Not-determined | Genome-fragment | NA | NA | 0 |
| S14_extracted_k141_82904 | 1253 | No | NA | 1 | 0 | 1 | Not-determined | Genome-fragment | NA | NA | 0 |
| S14_vs_k141_47996\|\|full | 1323 | No | NA | 6 | 1 | 0 | Low-quality | Genome-fragment | 2.09 | AAI-based (medium-confidence) | 0 |
| S14_extracted_k141_17002 | 1517 | No | NA | 1 | 0 | 1 | Not-determined | Genome-fragment | NA | NA | 0 |
| S14_extracted_k141_24114 | 3461 | No | NA | 3 | 0 | 2 | Not-determined | Genome-fragment | NA | NA | 0 |
| S14_vs_k141_7973\|\|lt2gene | 1323 | No | NA | 2 | 2 | 0 | Low-quality | Genome-fragment | 0.94 | AAI-based (medium-confidence) | 0 |
| S5_vs_k141_2004\|\|full | 1323 | No | NA | 3 | 2 | 0 | Low-quality | Genome-fragment | 3.43 | AAI-based (medium-confidence) | 0 |
| S14_extracted_k141_97184 | 3694 | No | NA | 5 | 0 | 2 | Not-determined | Genome-fragment | NA | NA | 0 |
| S14_extracted_k141_82953 | 3044 | No | NA | 6 | 0 | 0 | Not-determined | Genome-fragment | NA | NA | 0 |
| S14_extracted_k141_60910 | 1086 | No | NA | 1 | 0 | 0 | Not-determined | Genome-fragment | NA | NA | 0 |
| S5_vs_k141_60502\|\|full | 1323 | No | NA | 4 | 0 | 0 | Low-quality | Genome-fragment | 3.02 | AAI-based (medium-confidence) | 0 |
| S14_extracted_k141_97193 | 1206 | No | NA | 1 | 0 | 0 | Not-determined | Genome-fragment | NA | NA | 0 |
| S7_extracted_115796 | 1323 | No | NA | 6 | 0 | 0 | Low-quality | Genome-fragment | 2.95 | AAI-based (medium-confidence) | 0 |
| S14_extracted_k141_90260 | 3109 | No | NA | 4 | 0 | 2 | Not-determined | Genome-fragment | NA | NA | 0 |
| S16_extracted_162941 | 1323 | No | NA | 4 | 0 | 0 | Low-quality | Genome-fragment | 3.28 | AAI-based (medium-confidence) | 0 |
| S8_extracted_k141_184555 | 1323 | No | NA | 2 | 0 | 0 | Low-quality | Genome-fragment | 4.2 | AAI-based (high-confidence) | 0 |
| S14_extracted_k141_17041 | 3953 | No | NA | 4 | 0 | 2 | Not-determined | Genome-fragment | NA | NA | 0 |
| S14_extracted_k141_83010 | 2188 | No | NA | 3 | 0 | 3 | Not-determined | Genome-fragment | NA | NA | 0 |
| S14_extracted_k141_17083 | 1095 | No | NA | 1 | 0 | 0 | Not-determined | Genome-fragment | NA | NA | 0 |
| S8_vs_k141_52145\|\|full | 1323 | No | NA | 2 | 0 | 1 | Low-quality | Genome-fragment | 1.49 | AAI-based (medium-confidence) | 0 |
| S14_extracted_k141_112398 | 1142 | No | NA | 1 | 0 | 1 | Not-determined | Genome-fragment | NA | NA | 0 |
| S17_extracted_k141_117296 | 1324 | No | NA | 2 | 1 | 0 | Low-quality | Genome-fragment | 0.11 | HMM-based (lower-bound) | 0 |
| S14_extracted_k141_90268 | 1152 | No | NA | 2 | 0 | 0 | Not-determined | Genome-fragment | NA | NA | 0 |
| S9_vs_k141_66609\|\|full | 1324 | No | NA | 2 | 1 | 0 | Low-quality | Genome-fragment | 3.81 | AAI-based (medium-confidence) | 0 |
| S9_vs_k141_74947\|\|full | 1324 | No | NA | 5 | 0 | 0 | Low-quality | Genome-fragment | 2.42 | AAI-based (high-confidence) | 0 |
| S11_vs_k141_69717\|\|full | 1324 | No | NA | 5 | 1 | 0 | Low-quality | Genome-fragment | 0.78 | HMM-based (lower-bound) | 0 |
| S14_extracted_k141_90305 | 1716 | No | NA | 2 | 0 | 1 | Not-determined | Genome-fragment | NA | NA | 0 |
| S14_extracted_k141_9557 | 1140 | No | NA | 2 | 0 | 2 | Not-determined | Genome-fragment | NA | NA | 0 |
| S11_vs_k141_34404\|\|lt2gene | 1324 | No | NA | 2 | 2 | 0 | Low-quality | Genome-fragment | 3.35 | AAI-based (high-confidence) | 0 |
| S13_vs_133143\|\|full | 1324 | No | NA | 5 | 2 | 0 | Low-quality | Genome-fragment | 3.41 | AAI-based (high-confidence) | 0 |
| S14_vs_k141_84218\|\|full | 1324 | No | NA | 4 | 0 | 0 | Low-quality | Genome-fragment | 2.79 | AAI-based (high-confidence) | 0 |
| S1_vs_k141_7354\|\|full | 1324 | No | NA | 3 | 0 | 0 | Low-quality | Genome-fragment | 0.8 | AAI-based (medium-confidence) | 0 |
| S14_extracted_k141_31717 | 1099 | No | NA | 2 | 0 | 2 | Not-determined | Genome-fragment | NA | NA | 0 |
| S5_vs_k141_65718\|\|full | 1324 | No | NA | 4 | 1 | 0 | Low-quality | Genome-fragment | 2.12 | HMM-based (lower-bound) | 0 |
| S14_extracted_k141_97357 | 1057 | No | NA | 2 | 0 | 1 | Not-determined | Genome-fragment | NA | NA | 0 |
| S14_extracted_k141_61147 | 1017 | No | NA | 2 | 0 | 0 | Not-determined | Genome-fragment | NA | NA | 0 |
| S14_extracted_k141_105461 | 4381 | No | NA | 7 | 0 | 0 | Not-determined | Genome-fragment | NA | NA | 0 |
| S7_vs_5077\|\|full | 1324 | No | NA | 3 | 1 | 0 | Low-quality | Genome-fragment | 2.52 | AAI-based (medium-confidence) | 0 |
| S14_extracted_k141_68215 | 1920 | No | NA | 1 | 0 | 0 | Not-determined | Genome-fragment | NA | NA | 0 |
| S16_vs_169807\|\|full | 1324 | No | NA | 4 | 2 | 0 | Low-quality | Genome-fragment | 3.09 | AAI-based (high-confidence) | 0 |
| S14_extracted_k141_61182 | 1089 | No | NA | 1 | 0 | 1 | Not-determined | Genome-fragment | NA | NA | 0 |
| S14_extracted_k141_68229 | 1136 | No | NA | 2 | 0 | 0 | Not-determined | Genome-fragment | NA | NA | 0 |
| S10_extracted_58799 | 1324 | No | NA | 3 | 0 | 0 | Low-quality | Genome-fragment | 1.86 | AAI-based (high-confidence) | 0 |
| S12_vs_k141_28552\|\|full | 1324 | No | NA | 3 | 0 | 0 | Low-quality | Genome-fragment | 4.63 | AAI-based (medium-confidence) | 0 |
| S14_extracted_k141_17277 | 1169 | No | NA | 2 | 0 | 1 | Not-determined | Genome-fragment | NA | NA | 0 |
| S14_extracted_k141_112586 | 1811 | No | NA | 3 | 0 | 1 | Not-determined | Genome-fragment | NA | NA | 0 |
| S14_extracted_k141_75894 | 1103 | No | NA | 2 | 0 | 0 | Not-determined | Genome-fragment | NA | NA | 0 |
| S14_extracted_k141_24254 | 12666 | No | NA | 13 | 0 | 1 | Not-determined | Genome-fragment | NA | NA | 0 |
| S14_extracted_k141_31817 | 3613 | No | NA | 5 | 0 | 4 | Not-determined | Genome-fragment | NA | NA | 0 |
| S15_extracted_41353 | 1324 | No | NA | 5 | 0 | 0 | Low-quality | Genome-fragment | 2.83 | AAI-based (high-confidence) | 0 |
| S14_extracted_k141_31824 | 1265 | No | NA | 1 | 0 | 0 | Not-determined | Genome-fragment | NA | NA | 0 |
| S2_extracted_k141_2535 | 1324 | No | NA | 3 | 2 | 0 | Low-quality | Genome-fragment | 4.82 | HMM-based (lower-bound) | 0 |
| S17_vs_k141_60781\|\|full | 1325 | No | NA | 3 | 0 | 0 | Low-quality | Genome-fragment | 3.16 | AAI-based (medium-confidence) | 0 |
| S14_extracted_k141_24315 | 1176 | No | NA | 1 | 0 | 0 | Not-determined | Genome-fragment | NA | NA | 0 |
| S14_extracted_k141_112623 | 1439 | No | NA | 1 | 0 | 0 | Not-determined | Genome-fragment | NA | NA | 0 |
| S14_extracted_k141_97461 | 5020 | No | NA | 2 | 0 | 0 | Not-determined | Genome-fragment | NA | NA | 0 |
| S14_extracted_k141_105582 | 1189 | No | NA | 2 | 0 | 0 | Not-determined | Genome-fragment | NA | NA | 0 |
| S17_vs_k141_135069\|\|full | 1325 | No | NA | 3 | 0 | 0 | Low-quality | Genome-fragment | 2.79 | AAI-based (medium-confidence) | 0 |
| S14_extracted_k141_24349 | 1861 | No | NA | 1 | 0 | 1 | Not-determined | Genome-fragment | NA | NA | 0 |
| S14_extracted_k141_31851 | 13966 | No | NA | 8 | 0 | 5 | Not-determined | Genome-fragment | NA | NA | 0 |
| S14_extracted_k141_76012 | 2222 | No | NA | 2 | 0 | 1 | Not-determined | Genome-fragment | NA | NA | 0 |
| S17_vs_k141_139750\|\|full | 1325 | No | NA | 4 | 0 | 0 | Low-quality | Genome-fragment | 3.51 | AAI-based (high-confidence) | 0 |
| S17_vs_k141_38488\|\|lt2gene | 1325 | No | NA | 2 | 1 | 0 | Low-quality | Genome-fragment | 2.42 | AAI-based (high-confidence) | 0 |
| S14_extracted_k141_83263 | 3852 | No | NA | 3 | 0 | 3 | Not-determined | Genome-fragment | NA | NA | 0 |
| S14_extracted_k141_31916 | 1167 | No | NA | 2 | 0 | 2 | Not-determined | Genome-fragment | NA | NA | 0 |
| S11_vs_k141_30155\|\|full | 1325 | No | NA | 2 | 0 | 0 | Low-quality | Genome-fragment | 2.06 | AAI-based (high-confidence) | 0 |
| S14_extracted_k141_68491 | 1605 | No | NA | 3 | 0 | 1 | Not-determined | Genome-fragment | NA | NA | 0 |
| S13_vs_4371\|\|full | 1325 | No | NA | 3 | 0 | 1 | Low-quality | Genome-fragment | 0.79 | AAI-based (medium-confidence) | 0 |
| S14_vs_k141_76814\|\|full | 1325 | No | NA | 3 | 0 | 0 | Low-quality | Genome-fragment | 2.64 | AAI-based (high-confidence) | 0 |
| S1_vs_k141_81277\|\|lt2gene | 1325 | No | NA | 1 | 0 | 0 | Low-quality | Genome-fragment | 3.61 | AAI-based (high-confidence) | 0 |
| S14_extracted_k141_39524 | 4455 | No | NA | 3 | 0 | 0 | Not-determined | Genome-fragment | NA | NA | 0 |
| S5_extracted_k141_123004 | 1325 | No | NA | 1 | 0 | 1 | Low-quality | Genome-fragment | 0.63 | AAI-based (medium-confidence) | 0 |
| S14_extracted_k141_112686 | 1572 | No | NA | 1 | 0 | 0 | Not-determined | Genome-fragment | NA | NA | 0 |
| S7_extracted_76682 | 1325 | No | NA | 2 | 0 | 0 | Low-quality | Genome-fragment | 1.91 | AAI-based (medium-confidence) | 0 |
| S14_extracted_k141_76109 | 1366 | No | NA | 1 | 0 | 1 | Not-determined | Genome-fragment | NA | NA | 0 |
| S14_extracted_k141_2471 | 6585 | No | NA | 3 | 0 | 2 | Not-determined | Genome-fragment | NA | NA | 0 |
| S7_vs_52331\|\|full | 1325 | No | NA | 5 | 1 | 0 | Low-quality | Genome-fragment | 1.98 | AAI-based (high-confidence) | 0 |
| S7_vs_92878\|\|lt2gene | 1325 | No | NA | 2 | 1 | 0 | Low-quality | Genome-fragment | 2.93 | AAI-based (high-confidence) | 0 |
| S14_extracted_k141_54118 | 1328 | No | NA | 1 | 0 | 1 | Not-determined | Genome-fragment | NA | NA | 0 |
| S16_vs_161848\|\|lt2gene | 1325 | No | NA | 1 | 1 | 0 | Low-quality | Genome-fragment | 0.51 | HMM-based (lower-bound) | 0 |
| S14_extracted_k141_97635 | 2199 | No | NA | 3 | 0 | 0 | Not-determined | Genome-fragment | NA | NA | 0 |
| S14_extracted_k141_54171 | 4154 | No | NA | 5 | 0 | 1 | Not-determined | Genome-fragment | NA | NA | 0 |
| S14_extracted_k141_68566 | 7027 | No | NA | 5 | 0 | 4 | Not-determined | Genome-fragment | NA | NA | 0 |
| S10_extracted_98339 | 1325 | No | NA | 2 | 0 | 0 | Low-quality | Genome-fragment | 2.42 | AAI-based (high-confidence) | 0 |
| S14_extracted_k141_97654 | 3624 | No | NA | 3 | 0 | 2 | Not-determined | Genome-fragment | NA | NA | 0 |
| S6_extracted_k141_13275 | 1325 | No | NA | 3 | 0 | 0 | Low-quality | Genome-fragment | 2.47 | AAI-based (medium-confidence) | 0 |
| S8_extracted_k141_112733 | 1325 | No | NA | 1 | 0 | 1 | Low-quality | Genome-fragment | 0.38 | AAI-based (high-confidence) | 0 |
| S14_extracted_k141_9897 | 1312 | No | NA | 3 | 0 | 1 | Not-determined | Genome-fragment | NA | NA | 0 |

| | | | | | | | | | | | |
|---|---|---|---|---|---|---|---|---|---|---|---|
| S8_vs_k141_196761\|\|full | 1325 | No | NA | 4 | 1 | 0 | Low-quality | Genome-fragment | 3.19 | AAI-based (medium-confidence) | 0 |
| S14_extracted_k141_90781 | 1220 | No | NA | 2 | 0 | 0 | Not-determined | Genome-fragment | NA | NA | 0 |
| S8_vs_k141_100981\|\|full | 1325 | No | NA | 3 | 0 | 0 | Low-quality | Genome-fragment | 3.07 | AAI-based (high-confidence) | 0 |
| S8_vs_k141_205417\|\|full | 1325 | No | NA | 2 | 2 | 0 | Low-quality | Genome-fragment | 3.18 | AAI-based (medium-confidence) | 0 |
| S9_extracted_k141_5634 | 1326 | No | NA | 4 | 2 | 0 | Low-quality | Genome-fragment | 2.02 | AAI-based (high-confidence) | 0 |
| S9_vs_k141_69304\|\|full | 1326 | No | NA | 1 | 0 | 1 | Low-quality | Genome-fragment | 0.63 | AAI-based (medium-confidence) | 0 |
| S14_extracted_k141_2591 | 1075 | No | NA | 1 | 0 | 1 | Not-determined | Genome-fragment | NA | NA | 0 |
| S14_extracted_k141_81252 | 1326 | No | NA | 3 | 2 | 0 | Low-quality | Genome-fragment | 4.36 | AAI-based (high-confidence) | 0 |
| S14_vs_k141_62872\|\|full | 1326 | No | NA | 3 | 3 | 0 | Low-quality | Genome-fragment | 3.41 | AAI-based (high-confidence) | 0 |
| S14_extracted_k141_83439 | 3399 | No | NA | 5 | 0 | 3 | Not-determined | Genome-fragment | NA | NA | 0 |
| S14_extracted_k141_9988 | 4885 | No | NA | 4 | 0 | 2 | Not-determined | Genome-fragment | NA | NA | 0 |
| S14_vs_k141_12141\|\|lt2gene | 1326 | No | NA | 2 | 1 | 0 | Low-quality | Genome-fragment | 2.17 | AAI-based (medium-confidence) | 0 |
| S14_extracted_k141_32110 | 1164 | No | NA | 4 | 0 | 0 | Not-determined | Genome-fragment | NA | NA | 0 |
| S1_vs_k141_72545\|\|full | 1326 | No | NA | 3 | 0 | 0 | Low-quality | Genome-fragment | 3.29 | AAI-based (high-confidence) | 0 |
| S7_extracted_35119 | 1326 | No | NA | 2 | 0 | 1 | Low-quality | Genome-fragment | 1 | AAI-based (high-confidence) | 0 |
| S14_extracted_k141_54344 | 1041 | No | NA | 2 | 0 | 1 | Not-determined | Genome-fragment | NA | NA | 0 |
| S14_extracted_k141_32142 | 3086 | No | NA | 2 | 0 | 0 | Not-determined | Genome-fragment | NA | NA | 0 |
| S14_extracted_k141_61371 | 1369 | No | NA | 2 | 0 | 1 | Not-determined | Genome-fragment | NA | NA | 0 |
| S14_extracted_k141_54357 | 1678 | No | NA | 2 | 0 | 0 | Not-determined | Genome-fragment | NA | NA | 0 |
| S14_extracted_k141_68679 | 1180 | No | NA | 2 | 0 | 1 | Not-determined | Genome-fragment | NA | NA | 0 |
| S7_vs_134704\|\|full | 1326 | No | NA | 2 | 0 | 1 | Low-quality | Genome-fragment | 0.93 | AAI-based (high-confidence) | 0 |
| S14_extracted_k141_24621 | 1020 | No | NA | 1 | 0 | 0 | Not-determined | Genome-fragment | NA | NA | 0 |
| S14_extracted_k141_68699 | 1392 | No | NA | 3 | 0 | 1 | Not-determined | Genome-fragment | NA | NA | 0 |
| S14_extracted_k141_39753 | 1147 | No | NA | 1 | 0 | 0 | Not-determined | Genome-fragment | NA | NA | 0 |
| S14_extracted_k141_10028 | 1872 | No | NA | 1 | 0 | 0 | Not-determined | Genome-fragment | NA | NA | 0 |
| S7_vs_135584\|\|lt2gene | 1326 | No | NA | 2 | 2 | 0 | Low-quality | Genome-fragment | 2.66 | AAI-based (high-confidence) | 0 |
| S16_vs_65755\|\|full | 1326 | No | NA | 2 | 0 | 1 | Low-quality | Genome-fragment | 2.79 | AAI-based (high-confidence) | 0 |
| S14_extracted_k141_76366 | 1217 | No | NA | 2 | 0 | 1 | Not-determined | Genome-fragment | NA | NA | 0 |
| S14_extracted_k141_61448 | 1097 | No | NA | 1 | 0 | 1 | Not-determined | Genome-fragment | NA | NA | 0 |
| S16_vs_3909\|\|lt2gene | 1326 | No | NA | 2 | 0 | 0 | Low-quality | Genome-fragment | 3.67 | AAI-based (medium-confidence) | 0 |
| S14_extracted_k141_46380 | 1369 | No | NA | 1 | 0 | 1 | Not-determined | Genome-fragment | NA | NA | 0 |
| S14_extracted_k141_2640 | 1206 | No | NA | 2 | 0 | 1 | Not-determined | Genome-fragment | NA | NA | 0 |
| S14_extracted_k141_83574 | 3061 | No | NA | 4 | 0 | 0 | Not-determined | Genome-fragment | NA | NA | 0 |
| S16_vs_6204\|\|lt2gene | 1326 | No | NA | 1 | 1 | 0 | Low-quality | Genome-fragment | 0.92 | HMM-based (lower-bound) | 0 |
| S10_extracted_17256 | 1326 | No | NA | 1 | 1 | 0 | Low-quality | Genome-fragment | 0.91 | HMM-based (lower-bound) | 0 |
| S14_extracted_k141_83593 | 3568 | No | NA | 3 | 0 | 0 | Not-determined | Genome-fragment | NA | NA | 0 |
| S14_extracted_k141_68748 | 3107 | No | NA | 3 | 0 | 3 | Not-determined | Genome-fragment | NA | NA | 0 |
| S14_extracted_k141_105895 | 1070 | No | NA | 1 | 0 | 0 | Not-determined | Genome-fragment | NA | NA | 0 |
| S14_extracted_k141_39817 | 1318 | No | NA | 3 | 0 | 0 | Not-determined | Genome-fragment | NA | NA | 0 |
| S12_vs_k141_150592\|\|full | 1326 | No | NA | 3 | 3 | 0 | Low-quality | Genome-fragment | 3.54 | AAI-based (medium-confidence) | 0 |
| S14_extracted_k141_76464 | 2303 | No | NA | 2 | 0 | 2 | Not-determined | Genome-fragment | NA | NA | 0 |
| S17_extracted_k141_4720 | 1327 | No | NA | 3 | 1 | 0 | Low-quality | Genome-fragment | 2.7 | AAI-based (high-confidence) | 0 |
| S14_extracted_k141_32295 | 2490 | No | NA | 1 | 0 | 0 | Not-determined | Genome-fragment | NA | NA | 0 |
| S14_extracted_k141_76480 | 1420 | No | NA | 1 | 0 | 1 | Not-determined | Genome-fragment | NA | NA | 0 |
| S14_extracted_k141_54508 | 6837 | No | NA | 6 | 0 | 5 | Not-determined | Genome-fragment | NA | NA | 0 |
| S17_vs_k141_129691\|\|full | 1327 | No | NA | 3 | 0 | 0 | Low-quality | Genome-fragment | 2.57 | AAI-based (medium-confidence) | 0 |
| S14_extracted_k141_76491 | 4131 | No | NA | 4 | 0 | 4 | Not-determined | Genome-fragment | NA | NA | 0 |
| S9_extracted_k141_20317 | 1327 | No | NA | 4 | 4 | 0 | Low-quality | Genome-fragment | 4.35 | AAI-based (high-confidence) | 0 |
| S14_extracted_k141_68855 | 1238 | No | NA | 1 | 0 | 1 | Not-determined | Genome-fragment | NA | NA | 0 |
| S13_vs_98603\|\|full | 1327 | No | NA | 4 | 0 | 0 | Low-quality | Genome-fragment | 1.86 | AAI-based (medium-confidence) | 0 |
| S14_extracted_k141_2758 | 1387 | No | NA | 2 | 0 | 0 | Not-determined | Genome-fragment | NA | NA | 0 |
| S14_extracted_k141_68862 | 1730 | No | NA | 2 | 0 | 1 | Not-determined | Genome-fragment | NA | NA | 0 |
| S14_vs_k141_44677\|\|full | 1327 | No | NA | 2 | 2 | 0 | Low-quality | Genome-fragment | 2.94 | AAI-based (medium-confidence) | 0 |
| S14_extracted_k141_68882 | 2765 | No | NA | 1 | 0 | 1 | Not-determined | Genome-fragment | NA | NA | 0 |
| S14_extracted_k141_54590 | 1126 | No | NA | 2 | 0 | 1 | Not-determined | Genome-fragment | NA | NA | 0 |
| S14_extracted_k141_2792 | 3545 | No | NA | 5 | 0 | 2 | Not-determined | Genome-fragment | NA | NA | 0 |
| S14_extracted_k141_105991 | 1511 | No | NA | 1 | 0 | 0 | Not-determined | Genome-fragment | NA | NA | 0 |
| S14_extracted_k141_39919 | 1044 | No | NA | 1 | 0 | 1 | Not-determined | Genome-fragment | NA | NA | 0 |
| S1_vs_k141_44939\|\|full | 1327 | No | NA | 3 | 2 | 0 | Low-quality | Genome-fragment | 2.48 | HMM-based (lower-bound) | 0 |
| S14_extracted_k141_68913 | 4025 | No | NA | 6 | 0 | 1 | Not-determined | Genome-fragment | NA | NA | 0 |
| S14_extracted_k141_83735 | 1395 | No | NA | 2 | 0 | 1 | Not-determined | Genome-fragment | NA | NA | 0 |
| S14_extracted_k141_97887 | 1238 | No | NA | 2 | 0 | 1 | Not-determined | Genome-fragment | NA | NA | 0 |
| S1_vs_k141_25685\|\|lt2gene | 1327 | No | NA | 1 | 1 | 0 | Low-quality | Genome-fragment | 2.86 | AAI-based (high-confidence) | 0 |
| S3_extracted_k141_90402 | 1327 | No | NA | 4 | 1 | 0 | Low-quality | Genome-fragment | 2.43 | AAI-based (medium-confidence) | 0 |
| S14_extracted_k141_91032 | 2114 | No | NA | 2 | 0 | 2 | Not-determined | Genome-fragment | NA | NA | 0 |
| S14_extracted_k141_106055 | 1156 | No | NA | 2 | 0 | 1 | Not-determined | Genome-fragment | NA | NA | 0 |
| S14_extracted_k141_83790 | 1995 | No | NA | 2 | 0 | 0 | Not-determined | Genome-fragment | NA | NA | 0 |
| S14_extracted_k141_46606 | 1393 | No | NA | 4 | 0 | 0 | Not-determined | Genome-fragment | NA | NA | 0 |
| S3_extracted_k141_90524 | 1327 | No | NA | 1 | 0 | 0 | Low-quality | Genome-fragment | 2.22 | AAI-based (high-confidence) | 0 |
| S14_extracted_k141_76587 | 3525 | No | NA | 3 | 0 | 2 | Not-determined | Genome-fragment | NA | NA | 0 |
| S16_vs_61095\|\|full | 1327 | No | NA | 2 | 0 | 2 | Low-quality | Genome-fragment | 1.13 | AAI-based (medium-confidence) | 0 |
| S14_extracted_k141_10311 | 10317 | No | NA | 7 | 0 | 6 | Not-determined | Genome-fragment | NA | NA | 0 |
| S14_extracted_k141_76606 | 6166 | No | NA | 13 | 0 | 1 | Not-determined | Genome-fragment | NA | NA | 0 |
| S10_extracted_114219 | 1327 | No | NA | 3 | 1 | 0 | Low-quality | Genome-fragment | 2.52 | AAI-based (high-confidence) | 0 |
| S12_vs_k141_85514\|\|full | 1327 | No | NA | 5 | 1 | 0 | Low-quality | Genome-fragment | 2.1 | AAI-based (high-confidence) | 0 |
| S14_extracted_k141_24893 | 1188 | No | NA | 2 | 0 | 0 | Not-determined | Genome-fragment | NA | NA | 0 |
| S14_extracted_k141_68996 | 1491 | No | NA | 1 | 0 | 1 | Not-determined | Genome-fragment | NA | NA | 0 |
| S8_vs_k141_150677\|\|full | 1327 | No | NA | 2 | 0 | 0 | Low-quality | Genome-fragment | 0.55 | AAI-based (high-confidence) | 0 |
| S14_extracted_k141_76699 | 1044 | No | NA | 2 | 0 | 1 | Not-determined | Genome-fragment | NA | NA | 0 |
| S17_vs_k141_54314\|\|full | 1328 | No | NA | 2 | 0 | 0 | Low-quality | Genome-fragment | 1.19 | AAI-based (medium-confidence) | 0 |
| S14_extracted_k141_98032 | 3079 | No | NA | 6 | 0 | 0 | Not-determined | Genome-fragment | NA | NA | 0 |
| S13_vs_132887\|\|full | 1328 | No | NA | 2 | 0 | 1 | Low-quality | Genome-fragment | 2.17 | AAI-based (medium-confidence) | 0 |
| S14_extracted_k141_76707 | 1517 | No | NA | 1 | 0 | 0 | Not-determined | Genome-fragment | NA | NA | 0 |
| S13_vs_99840\|\|lt2gene | 1328 | No | NA | 2 | 2 | 0 | Low-quality | Genome-fragment | 2.18 | HMM-based (lower-bound) | 0 |
| S1_extracted_k141_21219 | 1328 | No | NA | 5 | 0 | 0 | Low-quality | Genome-fragment | 2.19 | AAI-based (high-confidence) | 0 |
| S14_extracted_k141_91205 | 1098 | No | NA | 1 | 0 | 1 | Not-determined | Genome-fragment | NA | NA | 0 |
| S14_extracted_k141_2883 | 1235 | No | NA | 1 | 0 | 0 | Not-determined | Genome-fragment | NA | NA | 0 |
| S14_extracted_k141_113149 | 1084 | No | NA | 1 | 0 | 1 | Not-determined | Genome-fragment | NA | NA | 0 |
| S3_extracted_k141_42206 | 1328 | No | NA | 2 | 1 | 0 | Low-quality | Genome-fragment | 0.47 | HMM-based (lower-bound) | 0 |
| S14_extracted_k141_106156 | 31621 | No | NA | 49 | 26 | 0 | Medium-quality | Genome-fragment | 86.11 | AAI-based (high-confidence) | 0 |
| S14_extracted_k141_106157 | 1224 | No | NA | 2 | 0 | 2 | Not-determined | Genome-fragment | NA | NA | 0 |
| S14_extracted_k141_98082 | 1145 | No | NA | 1 | 0 | 1 | Not-determined | Genome-fragment | NA | NA | 0 |
| S14_extracted_k141_69104 | 1119 | No | NA | 1 | 0 | 1 | Not-determined | Genome-fragment | NA | NA | 0 |
| S14_extracted_k141_32520 | 1061 | No | NA | 2 | 0 | 2 | Not-determined | Genome-fragment | NA | NA | 0 |
| S5_vs_k141_9591\|\|full | 1328 | No | NA | 4 | 1 | 0 | Low-quality | Genome-fragment | 3.06 | AAI-based (high-confidence) | 0 |
| S14_extracted_k141_98105 | 11803 | No | NA | 3 | 0 | 2 | Not-determined | Genome-fragment | NA | NA | 0 |
| S7_extracted_108211 | 1328 | No | NA | 2 | 0 | 0 | Low-quality | Genome-fragment | 2.4 | AAI-based (high-confidence) | 0 |
| S14_extracted_k141_24983 | 1077 | No | NA | 1 | 0 | 0 | Not-determined | Genome-fragment | NA | NA | 0 |
| S16_vs_7893\|\|full | 1328 | No | NA | 3 | 0 | 1 | Low-quality | Genome-fragment | 1.01 | AAI-based (medium-confidence) | 0 |
| S12_extracted_k141_89421 | 1328 | No | NA | 1 | 0 | 1 | Low-quality | Genome-fragment | 0.88 | AAI-based (high-confidence) | 0 |
| S14_extracted_k141_106205 | 2557 | No | NA | 2 | 0 | 2 | Not-determined | Genome-fragment | NA | NA | 0 |
| S14_extracted_k141_69156 | 1021 | No | NA | 2 | 0 | 1 | Not-determined | Genome-fragment | NA | NA | 0 |
| S14_extracted_k141_25012 | 1655 | No | NA | 1 | 0 | 1 | Not-determined | Genome-fragment | NA | NA | 0 |
| S14_extracted_k141_106220 | 1718 | No | NA | 1 | 0 | 1 | Not-determined | Genome-fragment | NA | NA | 0 |
| S12_vs_k141_166283\|\|full | 1328 | No | NA | 3 | 0 | 0 | Low-quality | Genome-fragment | 1.96 | AAI-based (medium-confidence) | 0 |
| S12_vs_k141_98207\|\|full | 1328 | No | NA | 3 | 2 | 0 | Low-quality | Genome-fragment | 0.71 | AAI-based (high-confidence) | 0 |
| S14_extracted_k141_40243 | 1245 | No | NA | 2 | 0 | 2 | Not-determined | Genome-fragment | NA | NA | 0 |
| S14_extracted_k141_91334 | 3214 | No | NA | 8 | 0 | 0 | Not-determined | Genome-fragment | NA | NA | 0 |
| S12_vs_k141_8819\|\|lt2gene | 1328 | No | NA | 1 | 1 | 0 | Low-quality | Genome-fragment | 0.92 | HMM-based (lower-bound) | 0 |
| S9_extracted_k141_13237 | 1329 | No | NA | 3 | 0 | 0 | Low-quality | Genome-fragment | 1.9 | AAI-based (high-confidence) | 0 |
| S14_extracted_k141_69191 | 3685 | No | NA | 3 | 0 | 2 | Not-determined | Genome-fragment | NA | NA | 0 |
| S5_vs_k141_33957\|\|full | 1329 | No | NA | 4 | 3 | 0 | Low-quality | Genome-fragment | 3.51 | AAI-based (medium-confidence) | 0 |
| S5_vs_k141_83158\|\|full | 1329 | No | NA | 3 | 0 | 0 | Low-quality | Genome-fragment | 3.66 | AAI-based (medium-confidence) | 0 |
| S16_vs_43249\|\|full | 1329 | No | NA | 3 | 2 | 0 | Low-quality | Genome-fragment | 3.44 | AAI-based (high-confidence) | 0 |
| S14_extracted_k141_113325 | 4853 | No | NA | 4 | 0 | 2 | Not-determined | Genome-fragment | NA | NA | 0 |
| S10_vs_523\|\|full | 1329 | No | NA | 3 | 0 | 0 | Low-quality | Genome-fragment | 3.18 | AAI-based (medium-confidence) | 0 |
| S12_vs_k141_6379\|\|full | 1329 | No | NA | 2 | 0 | 2 | Low-quality | Genome-fragment | 1.84 | AAI-based (medium-confidence) | 0 |
| S4_extracted_11230 | 1329 | No | NA | 3 | 3 | 0 | Low-quality | Genome-fragment | 4.38 | AAI-based (high-confidence) | 0 |
| S14_extracted_k141_25101 | 1061 | No | NA | 1 | 0 | 1 | Not-determined | Genome-fragment | NA | NA | 0 |
| S14_extracted_k141_25108 | 1486 | No | NA | 1 | 0 | 1 | Not-determined | Genome-fragment | NA | NA | 0 |
| S6_extracted_k141_21738 | 1329 | No | NA | 6 | 4 | 0 | Low-quality | Genome-fragment | 4.37 | AAI-based (high-confidence) | 0 |
| S14_extracted_k141_18146 | 2067 | No | NA | 2 | 0 | 2 | Not-determined | Genome-fragment | NA | NA | 0 |
| S6_vs_k141_43398\|\|full | 1329 | No | NA | 3 | 0 | 3 | Low-quality | Genome-fragment | 0.4 | AAI-based (medium-confidence) | 0 |
| S14_extracted_k141_46921 | 1783 | No | NA | 3 | 0 | 1 | Not-determined | Genome-fragment | NA | NA | 0 |
| S14_extracted_k141_54871 | 1080 | No | NA | 2 | 0 | 0 | Not-determined | Genome-fragment | NA | NA | 0 |
| S14_extracted_k141_69319 | 3078 | No | NA | 3 | 0 | 0 | Not-determined | Genome-fragment | NA | NA | 0 |
| S14_extracted_k141_2152 | 1098 | No | NA | 1 | 0 | 1 | Not-determined | Genome-fragment | NA | NA | 0 |
| S6_vs_k141_49402\|\|full | 1329 | No | NA | 2 | 1 | 0 | Low-quality | Genome-fragment | 2.17 | HMM-based (lower-bound) | 0 |
| S17_vs_k141_113869\|\|full | 1330 | No | NA | 2 | 0 | 1 | Low-quality | Genome-fragment | 0.95 | AAI-based (high-confidence) | 0 |
| S14_extracted_k141_10581 | 1238 | No | NA | 1 | 0 | 1 | Not-determined | Genome-fragment | NA | NA | 0 |
| S9_vs_k141_15823\|\|full | 1330 | No | NA | 3 | 0 | 2 | Low-quality | Genome-fragment | 0.67 | AAI-based (medium-confidence) | 0 |
| S14_extracted_k141_106328 | 1372 | No | NA | 2 | 0 | 0 | Not-determined | Genome-fragment | NA | NA | 0 |
| S9_vs_k141_20494\|\|full | 1330 | No | NA | 2 | 0 | 0 | Low-quality | Genome-fragment | 2.89 | AAI-based (medium-confidence) | 0 |
| S14_extracted_k141_98350 | 1050 | No | NA | 1 | 0 | 1 | Not-determined | Genome-fragment | NA | NA | 0 |
| S9_vs_k141_62200\|\|full | 1330 | No | NA | 2 | 0 | 1 | Low-quality | Genome-fragment | 0.91 | AAI-based (high-confidence) | 0 |
| S11_extracted_k141_5957 | 1330 | No | NA | 2 | 0 | 1 | Low-quality | Genome-fragment | 3.5 | AAI-based (medium-confidence) | 0 |
| S14_extracted_k141_69419 | 7561 | No | NA | 11 | 0 | 4 | Not-determined | Genome-fragment | NA | NA | 0 |
| S11_vs_k141_85368\|\|full | 1330 | No | NA | 3 | 3 | 0 | Low-quality | Genome-fragment | 3.05 | AAI-based (high-confidence) | 0 |
| S14_extracted_k141_69420 | 5878 | No | NA | 6 | 0 | 5 | Not-determined | Genome-fragment | NA | NA | 0 |
| S14_extracted_k141_18245 | 2048 | No | NA | 3 | 0 | 1 | Not-determined | Genome-fragment | NA | NA | 0 |
| S11_vs_k141_70606\|\|full | 1330 | No | NA | 4 | 1 | 0 | Low-quality | Genome-fragment | 2.06 | AAI-based (medium-confidence) | 0 |
| S14_extracted_k141_84240 | 1888 | No | NA | 3 | 0 | 2 | Not-determined | Genome-fragment | NA | NA | 0 |
| S1_vs_k141_145672\|\|full | 1330 | No | NA | 4 | 0 | 1 | Low-quality | Genome-fragment | 2.83 | AAI-based (medium-confidence) | 0 |
| S14_extracted_k141_3246 | 15693 | No | NA | 13 | 0 | 7 | Not-determined | Genome-fragment | NA | NA | 0 |
| S5_vs_k141_67965\|\|full | 1330 | No | NA | 3 | 0 | 0 | Low-quality | Genome-fragment | 1.01 | AAI-based (medium-confidence) | 0 |
| S14_extracted_k141_69446 | 1108 | No | NA | 2 | 0 | 0 | Not-determined | Genome-fragment | NA | NA | 0 |
| S14_extracted_k141_10662 | 1926 | No | NA | 2 | 0 | 0 | Not-determined | Genome-fragment | NA | NA | 0 |
| S14_extracted_k141_98415 | 1090 | No | NA | 2 | 0 | 0 | Not-determined | Genome-fragment | NA | NA | 0 |
| S14_extracted_k141_47035 | 6427 | No | NA | 5 | 0 | 4 | Not-determined | Genome-fragment | NA | NA | 0 |
| S14_extracted_k141_25250 | 1733 | No | NA | 2 | 0 | 1 | Not-determined | Genome-fragment | NA | NA | 0 |
| S7_vs_82191\|\|full | 1330 | No | NA | 2 | 0 | 0 | Low-quality | Genome-fragment | 2.83 | AAI-based (medium-confidence) | 0 |
| S14_extracted_k141_47059 | 1238 | No | NA | 2 | 0 | 1 | Not-determined | Genome-fragment | NA | NA | 0 |
| S14_extracted_k141_32805 | 1054 | No | NA | 2 | 0 | 0 | Not-determined | Genome-fragment | NA | NA | 0 |
| S14_extracted_k141_32808 | 1859 | No | NA | 4 | 0 | 1 | Not-determined | Genome-fragment | NA | NA | 0 |
| S14_extracted_k141_3252 | 3478 | No | NA | 5 | 0 | 2 | Not-determined | Genome-fragment | NA | NA | 0 |
| S14_extracted_k141_40424 | 1497 | No | NA | 2 | 0 | 1 | Not-determined | Genome-fragment | NA | NA | 0 |
| S10_extracted_78265 | 1330 | No | NA | 1 | 0 | 1 | Low-quality | Genome-fragment | 0.63 | AAI-based (medium-confidence) | 0 |
| S10_vs_95461\|\|lt2gene | 1330 | No | NA | 1 | 0 | 0 | Low-quality | Genome-fragment | 3.09 | AAI-based (high-confidence) | 0 |
| S14_extracted_k141_3277 | 1089 | No | NA | 1 | 0 | 1 | Not-determined | Genome-fragment | NA | NA | 0 |
| S14_extracted_k141_47141 | 1251 | No | NA | 2 | 0 | 1 | Not-determined | Genome-fragment | NA | NA | 0 |
| S14_extracted_k141_69503 | 2668 | No | NA | 4 | 0 | 1 | Not-determined | Genome-fragment | NA | NA | 0 |
| S14_extracted_k141_18371 | 2275 | No | NA | 2 | 0 | 1 | Not-determined | Genome-fragment | NA | NA | 0 |
| S14_extracted_k141_61913 | 1320 | No | NA | 1 | 0 | 1 | Not-determined | Genome-fragment | NA | NA | 0 |
| S14_extracted_k141_113596 | 1231 | No | NA | 3 | 0 | 3 | Not-determined | Genome-fragment | NA | NA | 0 |
| S12_vs_k141_153269\|\|lt2gene | 1330 | No | NA | 1 | 1 | 0 | Low-quality | Genome-fragment | 2.95 | AAI-based (high-confidence) | 0 |
| S8_vs_k141_71950\|\|full | 1330 | No | NA | 2 | 0 | 1 | Low-quality | Genome-fragment | 2.24 | AAI-based (medium-confidence) | 0 |
| S14_extracted_k141_18392 | 1106 | No | NA | 1 | 0 | 0 | Not-determined | Genome-fragment | NA | NA | 0 |
| S14_extracted_k141_98502 | 1895 | No | NA | 2 | 0 | 1 | Not-determined | Genome-fragment | NA | NA | 0 |
| S14_extracted_k141_10776 | 5036 | No | NA | 4 | 0 | 4 | Not-determined | Genome-fragment | NA | NA | 0 |
| S17_extracted_k141_19858 | 1331 | No | NA | 4 | 2 | 0 | Low-quality | Genome-fragment | 2.9 | AAI-based (medium-confidence) | 0 |
| S17_vs_k141_124452\|\|full | 1331 | No | NA | 2 | 0 | 0 | Low-quality | Genome-fragment | 1.36 | AAI-based (high-confidence) | 0 |
| S9_vs_k141_30094\|\|full | 1331 | No | NA | 3 | 0 | 0 | Low-quality | Genome-fragment | 1.58 | AAI-based (medium-confidence) | 0 |
| S14_extracted_k141_10796 | 1050 | No | NA | 1 | 0 | 1 | Not-determined | Genome-fragment | NA | NA | 0 |
| S11_vs_k141_96321\|\|full | 1331 | No | NA | 3 | 1 | 0 | Low-quality | Genome-fragment | 3.27 | AAI-based (medium-confidence) | 0 |
| S14_extracted_k141_53017 | 1331 | No | NA | 2 | 1 | 0 | Low-quality | Genome-fragment | 3.57 | AAI-based (high-confidence) | 0 |
| S14_extracted_k141_91701 | 1007 | No | NA | 2 | 0 | 2 | Not-determined | Genome-fragment | NA | NA | 0 |
| S14_extracted_k141_10819 | 1003 | No | NA | 2 | 0 | 1 | Not-determined | Genome-fragment | NA | NA | 0 |
| S14_extracted_k141_61962 | 1013 | No | NA | 2 | 0 | 0 | Not-determined | Genome-fragment | NA | NA | 0 |
| S14_extracted_k141_47248 | 1549 | No | NA | 1 | 0 | 1 | Not-determined | Genome-fragment | NA | NA | 0 |
| S14_extracted_k141_32918 | 1655 | No | NA | 2 | 0 | 2 | Not-determined | Genome-fragment | NA | NA | 0 |
| S14_extracted_k141_69583 | 2247 | No | NA | 3 | 0 | 1 | Not-determined | Genome-fragment | NA | NA | 0 |
| S14_vs_k141_25710\|\|full | 1331 | No | NA | 2 | 0 | 0 | Low-quality | Genome-fragment | 2.31 | AAI-based (high-confidence) | 0 |
| S14_extracted_k141_18461 | 2065 | No | NA | 2 | 0 | 2 | Not-determined | Genome-fragment | NA | NA | 0 |
| S14_extracted_k141_18465 | 1129 | No | NA | 1 | 0 | 1 | Not-determined | Genome-fragment | NA | NA | 0 |
| S14_extracted_k141_113680 | 1906 | No | NA | 3 | 0 | 0 | Not-determined | Genome-fragment | NA | NA | 0 |
| S7_vs_82078\|\|full | 1331 | No | NA | 3 | 2 | 0 | Low-quality | Genome-fragment | 3.39 | AAI-based (high-confidence) | 0 |
| S14_extracted_k141_55018 | 1843 | No | NA | 3 | 0 | 2 | Not-determined | Genome-fragment | NA | NA | 0 |
| S14_extracted_k141_10903 | 1048 | No | NA | 2 | 0 | 2 | Not-determined | Genome-fragment | NA | NA | 0 |
| S7_vs_90049\|\|full | 1331 | No | NA | 3 | 0 | 0 | Low-quality | Genome-fragment | 2.29 | AAI-based (medium-confidence) | 0 |
| S10_extracted_94973 | 1331 | No | NA | 2 | 0 | 0 | Low-quality | Genome-fragment | 2.85 | AAI-based (high-confidence) | 0 |
| S14_extracted_k141_84472 | 1631 | No | NA | 1 | 0 | 0 | Not-determined | Genome-fragment | NA | NA | 0 |
| S12_extracted_k141_143394 | 1331 | No | NA | 2 | 0 | 0 | Low-quality | Genome-fragment | 1.44 | AAI-based (medium-confidence) | 0 |
| S14_extracted_k141_10946 | 1046 | No | NA | 2 | 0 | 1 | Not-determined | Genome-fragment | NA | NA | 0 |
| S12_vs_k141_15086\|\|full | 1331 | No | NA | 3 | 1 | 0 | Low-quality | Genome-fragment | 2.56 | AAI-based (medium-confidence) | 0 |
| S8_vs_k141_124259\|\|full | 1331 | No | NA | 3 | 2 | 0 | Low-quality | Genome-fragment | 2.27 | AAI-based (high-confidence) | 0 |
| S14_extracted_k141_113802 | 1132 | No | NA | 1 | 0 | 0 | Not-determined | Genome-fragment | NA | NA | 0 |
| S9_vs_k141_82359\|\|full | 1332 | No | NA | 5 | 0 | 0 | Low-quality | Genome-fragment | 1.89 | AAI-based (high-confidence) | 0 |
| S14_extracted_k141_113808 | 1113 | No | NA | 1 | 0 | 0 | Not-determined | Genome-fragment | NA | NA | 0 |
| S14_extracted_k141_106620 | 1635 | No | NA | 1 | 0 | 0 | Not-determined | Genome-fragment | NA | NA | 0 |
| S1_extracted_k141_94667 | 1332 | No | NA | 2 | 0 | 2 | Low-quality | Genome-fragment | 2.26 | AAI-based (high-confidence) | 0 |
| S14_extracted_k141_33021 | 1728 | No | NA | 2 | 0 | 1 | Not-determined | Genome-fragment | NA | NA | 0 |
| S14_extracted_k141_91868 | 1210 | No | NA | 2 | 0 | 0 | Not-determined | Genome-fragment | NA | NA | 0 |
| S14_extracted_k141_40735 | 1973 | No | NA | 3 | 0 | 3 | Not-determined | Genome-fragment | NA | NA | 0 |
| S14_extracted_k141_47394 | 1011 | No | NA | 2 | 0 | 2 | Not-determined | Genome-fragment | NA | NA | 0 |
| S14_extracted_k141_33032 | 1049 | No | NA | 1 | 0 | 1 | Not-determined | Genome-fragment | NA | NA | 0 |
| S1_vs_k141_123503\|\|full | 1332 | No | NA | 2 | 2 | 0 | Low-quality | Genome-fragment | 2.27 | AAI-based (high-confidence) | 0 |
| S7_vs_70070\|\|full | 1332 | No | NA | 4 | 0 | 0 | Low-quality | Genome-fragment | 0.76 | AAI-based (high-confidence) | 0 |
| S12_vs_k141_95934\|\|full | 1332 | No | NA | 3 | 0 | 0 | Low-quality | Genome-fragment | 0.79 | AAI-based (medium-confidence) | 0 |
| S14_extracted_k141_47440 | 1241 | No | NA | 2 | 0 | 0 | Not-determined | Genome-fragment | NA | NA | 0 |
| S8_vs_k141_193229\|\|full | 1332 | No | NA | 4 | 0 | 0 | Low-quality | Genome-fragment | 2.86 | AAI-based (high-confidence) | 0 |
| S14_extracted_k141_106687 | 1390 | No | NA | 1 | 0 | 1 | Not-determined | Genome-fragment | NA | NA | 0 |
| S14_extracted_k141_77123 | 1533 | No | NA | 2 | 0 | 2 | Not-determined | Genome-fragment | NA | NA | 0 |
| S8_vs_k141_53754\|\|lt2gene | 1332 | No | NA | 2 | 1 | 0 | Low-quality | Genome-fragment | 3.07 | AAI-based (high-confidence) | 0 |
| S17_vs_k141_143922\|\|full | 1333 | No | NA | 5 | 1 | 0 | Low-quality | Genome-fragment | 2.91 | AAI-based (high-confidence) | 0 |
| S14_extracted_k141_47474 | 6913 | No | NA | 6 | 0 | 5 | Not-determined | Genome-fragment | NA | NA | 0 |
| S9_extracted_k141_90844 | 1333 | No | NA | 5 | 0 | 0 | Low-quality | Genome-fragment | 3.46 | AAI-based (medium-confidence) | 0 |
| S14_extracted_k141_113926 | 1035 | No | NA | 1 | 0 | 0 | Not-determined | Genome-fragment | NA | NA | 0 |
| S11_extracted_k141_30320 | 1333 | No | NA | 3 | 0 | 0 | Low-quality | Genome-fragment | 2.5 | AAI-based (high-confidence) | 0 |
| S14_extracted_k141_106751 | 1054 | No | NA | 2 | 0 | 2 | Not-determined | Genome-fragment | NA | NA | 0 |
| S14_extracted_k141_47510 | 1484 | No | NA | 2 | 0 | 1 | Not-determined | Genome-fragment | NA | NA | 0 |
| S14_extracted_k141_62219 | 3024 | No | NA | 3 | 0 | 3 | Not-determined | Genome-fragment | NA | NA | 0 |
| S14_extracted_k141_47523 | 2522 | No | NA | 4 | 0 | 2 | Not-determined | Genome-fragment | NA | NA | 0 |
| S14_extracted_k141_3572 | 3647 | No | NA | 5 | 0 | 2 | Not-determined | Genome-fragment | NA | NA | 0 |
| S11_vs_k141_63050\|\|full | 1333 | No | NA | 3 | 0 | 0 | Low-quality | Genome-fragment | 2.35 | AAI-based (high-confidence) | 0 |
| S11_vs_k141_56622\|\|lt2gene | 1333 | No | NA | 2 | 1 | 0 | Low-quality | Genome-fragment | 3.05 | AAI-based (high-confidence) | 0 |
| S14_extracted_k141_113960 | 1013 | No | NA | 2 | 0 | 0 | Not-determined | Genome-fragment | NA | NA | 0 |
| S14_extracted_k141_59472 | 1333 | No | NA | 2 | 2 | 0 | Low-quality | Genome-fragment | 4.4 | AAI-based (high-confidence) | 0 |
| S14_extracted_k141_92004 | 3841 | No | NA | 8 | 0 | 8 | Not-determined | Genome-fragment | NA | NA | 0 |
| S1_extracted_k141_127828 | 1333 | No | NA | 1 | 1 | 0 | Low-quality | Genome-fragment | 2.96 | AAI-based (high-confidence) | 0 |
| S14_extracted_k141_11223 | 5104 | No | NA | 6 | 0 | 0 | Not-determined | Genome-fragment | NA | NA | 0 |
| S1_vs_k141_154342\|\|full | 1333 | No | NA | 5 | 0 | 0 | Low-quality | Genome-fragment | 2.9 | AAI-based (high-confidence) | 0 |
| S14_extracted_k141_77256 | 1235 | No | NA | 1 | 0 | 1 | Not-determined | Genome-fragment | NA | NA | 0 |
| S14_extracted_k141_18746 | 1461 | No | NA | 2 | 0 | 0 | Not-determined | Genome-fragment | NA | NA | 0 |
| S14_extracted_k141_25680 | 3738 | No | NA | 3 | 0 | 3 | Not-determined | Genome-fragment | NA | NA | 0 |
| S1_vs_k141_66033\|\|full | 1333 | No | NA | 3 | 2 | 0 | Low-quality | Genome-fragment | 3.27 | AAI-based (high-confidence) | 0 |
| S3_vs_k141_101199\|\|full | 1333 | No | NA | 2 | 0 | 0 | Low-quality | Genome-fragment | 2.45 | AAI-based (high-confidence) | 0 |
| S14_extracted_k141_18794 | 1086 | No | NA | 2 | 0 | 0 | Not-determined | Genome-fragment | NA | NA | 0 |
| S5_extracted_k141_21896 | 1333 | No | NA | 1 | 1 | 0 | Low-quality | Genome-fragment | 3.48 | AAI-based (high-confidence) | 0 |
| S5_vs_k141_41513\|\|full | 1333 | No | NA | 3 | 2 | 0 | Low-quality | Genome-fragment | 3.07 | AAI-based (medium-confidence) | 0 |
| S14_extracted_k141_92092 | 5347 | No | NA | 7 | 0 | 5 | Not-determined | Genome-fragment | NA | NA | 0 |
| S7_vs_41724\|\|full | 1333 | No | NA | 3 | 0 | 1 | Low-quality | Genome-fragment | 2.71 | AAI-based (medium-confidence) | 0 |
| S14_extracted_k141_98914 | 1734 | No | NA | 3 | 0 | 1 | Not-determined | Genome-fragment | NA | NA | 0 |
| S14_extracted_k141_11298 | 1688 | No | NA | 1 | 0 | 1 | Not-determined | Genome-fragment | NA | NA | 0 |
| S10_extracted_20902 | 1333 | No | NA | 3 | 0 | 0 | Low-quality | Genome-fragment | 0.62 | AAI-based (medium-confidence) | 0 |
| S10_vs_8936\|\|full | 1333 | No | NA | 4 | 0 | 0 | Low-quality | Genome-fragment | 3.62 | AAI-based (medium-confidence) | 0 |
| S14_extracted_k141_25736 | 1195 | No | NA | 2 | 0 | 1 | Not-determined | Genome-fragment | NA | NA | 0 |
| S10_vs_111703\|\|full | 1333 | No | NA | 2 | 1 | 0 | Low-quality | Genome-fragment | 3.28 | AAI-based (medium-confidence) | 0 |
| S12_vs_k141_45331\|\|full | 1333 | No | NA | 3 | 0 | 0 | Low-quality | Genome-fragment | 1.52 | AAI-based (medium-confidence) | 0 |
| S14_extracted_k141_18951 | 1088 | No | NA | 2 | 0 | 1 | Not-determined | Genome-fragment | NA | NA | 0 |
| S12_vs_k141_109600\|\|full | 1333 | No | NA | 2 | 0 | 1 | Low-quality | Genome-fragment | 0.79 | AAI-based (high-confidence) | 0 |
| S14_extracted_k141_98981 | 1565 | No | NA | 1 | 0 | 0 | Not-determined | Genome-fragment | NA | NA | 0 |
| S14_extracted_k141_11409 | 3564 | No | NA | 4 | 0 | 2 | Not-determined | Genome-fragment | NA | NA | 0 |
| S14_extracted_k141_77487 | 4141 | No | NA | 6 | 0 | 0 | Not-determined | Genome-fragment | NA | NA | 0 |
| S15_extracted_29627 | 1333 | No | NA | 4 | 1 | 0 | Low-quality | Genome-fragment | 1.07 | AAI-based (high-confidence) | 0 |
| S14_extracted_k141_47592 | 5359 | No | NA | 3 | 0 | 3 | Not-determined | Genome-fragment | NA | NA | 0 |
| S14_extracted_k141_92240 | 34251 | No | NA | 52 | 15 | 2 | High-quality | High-quality | 98.36 | AAI-based (high-confidence) | 0 |
| S14_extracted_k141_41141 | 1232 | No | NA | 2 | 0 | 1 | Not-determined | Genome-fragment | NA | NA | 0 |
| S14_extracted_k141_3905 | 1147 | No | NA | 1 | 0 | 0 | Not-determined | Genome-fragment | NA | NA | 0 |
| S11_extracted_k141_41325 | 1334 | No | NA | 4 | 1 | 0 | Low-quality | Genome-fragment | 2.06 | HMM-based (lower-bound) | 0 |
| S14_extracted_k141_40659 | 1334 | No | NA | 4 | 1 | 0 | Low-quality | Genome-fragment | 3.44 | AAI-based (medium-confidence) | 0 |
| S14_extracted_k141_93973 | 1334 | No | NA | 2 | 0 | 0 | Low-quality | Genome-fragment | 2.11 | AAI-based (medium-confidence) | 0 |
| S14_extracted_k141_62370 | 1084 | No | NA | 1 | 0 | 1 | Not-determined | Genome-fragment | NA | NA | 0 |
| S14_extracted_k141_19075 | 1411 | No | NA | 2 | 0 | 1 | Not-determined | Genome-fragment | NA | NA | 0 |
| S14_vs_k141_109833\|\|full | 1334 | No | NA | 2 | 0 | 0 | Low-quality | Genome-fragment | 2.11 | AAI-based (medium-confidence) | 0 |
| S1_vs_k141_43802\|\|full | 1334 | No | NA | 3 | 0 | 0 | Low-quality | Genome-fragment | 1.62 | AAI-based (medium-confidence) | 0 |
| S14_extracted_k141_41188 | 1461 | No | NA | 2 | 0 | 0 | Not-determined | Genome-fragment | NA | NA | 0 |
| S14_extracted_k141_11491 | 2946 | No | NA | 4 | 0 | 1 | Not-determined | Genome-fragment | NA | NA | 0 |
| S1_vs_k141_65375\|\|full | 1334 | No | NA | 3 | 0 | 0 | Low-quality | Genome-fragment | 0.95 | AAI-based (high-confidence) | 0 |
| S14_extracted_k141_11495 | 1368 | No | NA | 1 | 0 | 1 | Not-determined | Genome-fragment | NA | NA | 0 |
| S14_extracted_k141_11496 | 1522 | No | NA | 1 | 0 | 1 | Not-determined | Genome-fragment | NA | NA | 0 |
| S14_extracted_k141_33433 | 1167 | No | NA | 1 | 0 | 1 | Not-determined | Genome-fragment | NA | NA | 0 |
| S3_extracted_k141_44617 | 1334 | No | NA | 2 | 0 | 0 | Low-quality | Genome-fragment | 0.92 | AAI-based (medium-confidence) | 0 |
| S14_extracted_k141_99109 | 6380 | No | NA | 4 | 0 | 3 | Not-determined | Genome-fragment | NA | NA | 0 |
| S14_extracted_k141_62475 | 1002 | No | NA | 2 | 0 | 1 | Not-determined | Genome-fragment | NA | NA | 0 |
| S14_extracted_k141_85020 | 7582 | No | NA | 9 | 0 | 0 | Not-determined | Genome-fragment | NA | NA | 0 |
| S3_vs_k141_28149\|\|full | 1334 | No | NA | 2 | 0 | 1 | Low-quality | Genome-fragment | 0.71 | AAI-based (medium-confidence) | 0 |
| S5_extracted_k141_59529 | 1334 | No | NA | 4 | 1 | 0 | Low-quality | Genome-fragment | 4.06 | AAI-based (medium-confidence) | 0 |
| S14_extracted_k141_99127 | 1028 | No | NA | 1 | 0 | 1 | Not-determined | Genome-fragment | NA | NA | 0 |
| S5_vs_k141_47804\|\|full | 1334 | No | NA | 3 | 1 | 0 | Low-quality | Genome-fragment | 0.86 | HMM-based (lower-bound) | 0 |
| S14_extracted_k141_4031 | 1118 | No | NA | 2 | 0 | 2 | Not-determined | Genome-fragment | NA | NA | 0 |
| S7_vs_27154\|\|full | 1334 | No | NA | 3 | 0 | 0 | Low-quality | Genome-fragment | 3.15 | AAI-based (high-confidence) | 0 |
| S14_extracted_k141_33501 | 1686 | No | NA | 3 | 0 | 2 | Not-determined | Genome-fragment | NA | NA | 0 |
| S14_extracted_k141_99138 | 1133 | No | NA | 1 | 0 | 1 | Not-determined | Genome-fragment | NA | NA | 0 |
| S14_extracted_k141_114328 | 4159 | No | NA | 4 | 0 | 2 | Not-determined | Genome-fragment | NA | NA | 0 |
| S14_extracted_k141_4043 | 2663 | No | NA | 2 | 0 | 0 | Not-determined | Genome-fragment | NA | NA | 0 |
| S14_extracted_k141_92354 | 1604 | No | NA | 1 | 0 | 0 | Not-determined | Genome-fragment | NA | NA | 0 |
| S8_vs_k141_97665\|\|full | 1334 | No | NA | 3 | 0 | 1 | Low-quality | Genome-fragment | 0.88 | AAI-based (medium-confidence) | 0 |
| S9_extracted_k141_43205 | 1335 | No | NA | 3 | 0 | 0 | Low-quality | Genome-fragment | 3.59 | AAI-based (medium-confidence) | 0 |
| S9_vs_k141_14726\|\|lt2gene | 1335 | No | NA | 2 | 0 | 0 | Low-quality | Genome-fragment | 1.7 | AAI-based (high-confidence) | 0 |
| S14_extracted_k141_33556 | 1870 | No | NA | 1 | 0 | 0 | Not-determined | Genome-fragment | NA | NA | 0 |
| S11_extracted_k141_97061 | 1335 | No | NA | 2 | 0 | 1 | Low-quality | Genome-fragment | 1.07 | AAI-based (high-confidence) | 0 |
| S14_extracted_k141_70201 | 2125 | No | NA | 1 | 0 | 1 | Not-determined | Genome-fragment | NA | NA | 0 |
| S14_extracted_k141_62589 | 1100 | No | NA | 1 | 0 | 0 | Not-determined | Genome-fragment | NA | NA | 0 |
| S14_extracted_k141_81207 | 1335 | No | NA | 2 | 2 | 0 | Low-quality | Genome-fragment | 4.4 | AAI-based (high-confidence) | 0 |
| S14_extracted_k141_70216 | 1024 | No | NA | 1 | 0 | 0 | Not-determined | Genome-fragment | NA | NA | 0 |
| S14_extracted_k141_114373 | 3886 | No | NA | 4 | 0 | 2 | Not-determined | Genome-fragment | NA | NA | 0 |
| S14_extracted_k141_77711 | 1929 | No | NA | 1 | 0 | 0 | Not-determined | Genome-fragment | NA | NA | 0 |
| S3_extracted_k141_6103 | 1335 | No | NA | 3 | 0 | 1 | Low-quality | Genome-fragment | 2.29 | AAI-based (medium-confidence) | 0 |
| S14_extracted_k141_25939 | 1967 | No | NA | 4 | 0 | 0 | Not-determined | Genome-fragment | NA | NA | 0 |
| S14_extracted_k141_11631 | 1814 | No | NA | 2 | 0 | 2 | Not-determined | Genome-fragment | NA | NA | 0 |
| S3_vs_k141_82265\|\|full | 1335 | No | NA | 3 | 0 | 1 | Low-quality | Genome-fragment | 0.94 | AAI-based (medium-confidence) | 0 |
| S14_extracted_k141_19383 | 1753 | No | NA | 2 | 0 | 0 | Not-determined | Genome-fragment | NA | NA | 0 |
| S14_extracted_k141_85122 | 32993 | No | NA | 47 | 19 | 0 | Medium-quality | Genome-fragment | 79.58 | AAI-based (high-confidence) | 0 |
| S14_extracted_k141_62647 | 1196 | No | NA | 2 | 0 | 1 | Not-determined | Genome-fragment | NA | NA | 0 |
| S14_extracted_k141_107177 | 1031 | No | NA | 1 | 0 | 1 | Not-determined | Genome-fragment | NA | NA | 0 |
| S14_extracted_k141_62679 | 1334 | No | NA | 1 | 0 | 0 | Not-determined | Genome-fragment | NA | NA | 0 |
| S14_extracted_k141_92498 | 1342 | No | NA | 1 | 0 | 0 | Not-determined | Genome-fragment | NA | NA | 0 |
| S14_extracted_k141_41423 | 1123 | No | NA | 1 | 0 | 1 | Not-determined | Genome-fragment | NA | NA | 0 |
| S14_extracted_k141_19444 | 1035 | No | NA | 3 | 0 | 1 | Not-determined | Genome-fragment | NA | NA | 0 |
| S14_extracted_k141_41455 | 1314 | No | NA | 2 | 0 | 0 | Not-determined | Genome-fragment | NA | NA | 0 |
| S5_vs_k141_34246\|\|lt2gene | 1335 | No | NA | 2 | 2 | 0 | Low-quality | Genome-fragment | 5.78 | AAI-based (medium-confidence) | 0 |
| S14_extracted_k141_26000 | 11615 | No | NA | 8 | 0 | 3 | Not-determined | Genome-fragment | NA | NA | 0 |
| S14_extracted_k141_33647 | 1180 | No | NA | 1 | 0 | 0 | Not-determined | Genome-fragment | NA | NA | 0 |
| S12_extracted_k141_13845 | 1335 | No | NA | 3 | 2 | 0 | Low-quality | Genome-fragment | 3.02 | AAI-based (high-confidence) | 0 |
| S14_extracted_k141_99344 | 15862 | No | NA | 21 | 8 | 0 | Medium-quality | Genome-fragment | 66.98 | AAI-based (medium-confidence) | 0 |
| S12_extracted_k141_155969 | 1335 | No | NA | 2 | 0 | 1 | Low-quality | Genome-fragment | 0.31 | AAI-based (medium-confidence) | 0 |
| S14_extracted_k141_99366 | 4930 | No | NA | 4 | 0 | 4 | Not-determined | Genome-fragment | NA | NA | 0 |
| S12_vs_k141_124699\|\|full | 1335 | No | NA | 3 | 1 | 0 | Low-quality | Genome-fragment | 3.67 | AAI-based (high-confidence) | 0 |
| S14_extracted_k141_114491 | 15913 | No | NA | 19 | 7 | 0 | Medium-quality | Genome-fragment | 68.55 | AAI-based (medium-confidence) | 0 |
| S8_extracted_k141_90167 | 1335 | No | NA | 4 | 0 | 0 | Low-quality | Genome-fragment | 2.03 | AAI-based (high-confidence) | 0 |
| S8_extracted_k141_66557 | 1335 | No | NA | 2 | 1 | 0 | Low-quality | Genome-fragment | 2.38 | HMM-based (lower-bound) | 0 |
| S14_extracted_k141_55819 | 1183 | No | NA | 3 | 0 | 2 | Not-determined | Genome-fragment | NA | NA | 0 |
| S14_extracted_k141_99411 | 3646 | No | NA | 4 | 0 | 0 | Not-determined | Genome-fragment | NA | NA | 0 |
| S17_vs_k141_16748\|\|full | 1336 | No | NA | 3 | 1 | 0 | Low-quality | Genome-fragment | 3.31 | AAI-based (medium-confidence) | 0 |
| S14_extracted_k141_92593 | 2587 | No | NA | 5 | 0 | 0 | Not-determined | Genome-fragment | NA | NA | 0 |
| S9_extracted_k141_71797 | 1336 | No | NA | 4 | 0 | 0 | Low-quality | Genome-fragment | 0.81 | AAI-based (high-confidence) | 0 |
| S14_extracted_k141_4242 | 1389 | No | NA | 2 | 0 | 1 | Not-determined | Genome-fragment | NA | NA | 0 |
| S14_extracted_k141_70368 | 3037 | No | NA | 3 | 0 | 3 | Not-determined | Genome-fragment | NA | NA | 0 |
| S14_extracted_k141_19580 | 1236 | No | NA | 3 | 0 | 2 | Not-determined | Genome-fragment | NA | NA | 0 |
| S14_extracted_k141_55885 | 2981 | No | NA | 4 | 0 | 1 | Not-determined | Genome-fragment | NA | NA | 0 |
| S9_vs_k141_75216\|\|full | 1336 | No | NA | 3 | 0 | 1 | Low-quality | Genome-fragment | 3.39 | AAI-based (medium-confidence) | 0 |
| S11_vs_k141_70130\|\|full | 1336 | No | NA | 5 | 0 | 0 | Low-quality | Genome-fragment | 2.4 | AAI-based (medium-confidence) | 0 |
| S14_extracted_k141_4266 | 5455 | No | NA | 13 | 0 | 1 | Not-determined | Genome-fragment | NA | NA | 0 |
| S14_extracted_k141_47883 | 1115 | No | NA | 1 | 0 | 1 | Not-determined | Genome-fragment | NA | NA | 0 |
| S14_extracted_k141_70435 | 1082 | No | NA | 2 | 0 | 0 | Not-determined | Genome-fragment | NA | NA | 0 |
| S14_extracted_k141_107398 | 1430 | No | NA | 3 | 0 | 1 | Not-determined | Genome-fragment | NA | NA | 0 |
| S11_vs_k141_365\|\|lt2gene | 1336 | No | NA | 2 | 1 | 0 | Low-quality | Genome-fragment | 3.89 | AAI-based (high-confidence) | 0 |
| S14_extracted_k141_47942 | 1283 | No | NA | 1 | 0 | 0 | Not-determined | Genome-fragment | NA | NA | 0 |
| S14_extracted_k141_4306 | 1105 | No | NA | 2 | 0 | 0 | Not-determined | Genome-fragment | NA | NA | 0 |
| S14_extracted_k141_86188 | 1336 | No | NA | 6 | 5 | 0 | Low-quality | Genome-fragment | 4.41 | AAI-based (high-confidence) | 0 |
| S1_extracted_k141_152750 | 1336 | No | NA | 3 | 0 | 0 | Low-quality | Genome-fragment | 2.69 | AAI-based (high-confidence) | 0 |
| S5_extracted_k141_143719 | 1336 | No | NA | 4 | 2 | 0 | Low-quality | Genome-fragment | 3.88 | AAI-based (medium-confidence) | 0 |
| S14_extracted_k141_47961 | 1332 | No | NA | 1 | 0 | 1 | Not-determined | Genome-fragment | NA | NA | 0 |
| S14_extracted_k141_19642 | 1296 | No | NA | 3 | 0 | 3 | Not-determined | Genome-fragment | NA | NA | 0 |
| S14_extracted_k141_114686 | 1005 | No | NA | 1 | 0 | 1 | Not-determined | Genome-fragment | NA | NA | 0 |
| S7_vs_87919\|\|full | 1336 | No | NA | 2 | 1 | 0 | Low-quality | Genome-fragment | 3.1 | AAI-based (high-confidence) | 0 |
| S10_extracted_23266 | 1336 | No | NA | 2 | 0 | 1 | Low-quality | Genome-fragment | 0.47 | AAI-based (medium-confidence) | 0 |
| S4_extracted_31525 | 1336 | No | NA | 6 | 5 | 0 | Low-quality | Genome-fragment | 4.41 | AAI-based (high-confidence) | 0 |
| S8_vs_k141_55239\|\|full | 1336 | No | NA | 3 | 0 | 2 | Low-quality | Genome-fragment | 0.5 | AAI-based (medium-confidence) | 0 |
| S17_vs_k141_160861\|\|full | 1337 | No | NA | 3 | 0 | 3 | Low-quality | Genome-fragment | 0.82 | AAI-based (medium-confidence) | 0 |
| S9_extracted_k141_54700 | 1337 | No | NA | 4 | 0 | 0 | Low-quality | Genome-fragment | 0.77 | AAI-based (high-confidence) | 0 |
| S11_vs_k141_2151\|\|lt2gene | 1337 | No | NA | 2 | 1 | 0 | Low-quality | Genome-fragment | 0.92 | HMM-based (lower-bound) | 0 |
| S14_extracted_k141_19753 | 2474 | No | NA | 1 | 0 | 1 | Not-determined | Genome-fragment | NA | NA | 0 |
| S14_extracted_k141_63009 | 1016 | No | NA | 2 | 0 | 1 | Not-determined | Genome-fragment | NA | NA | 0 |
| S11_vs_k141_32115\|\|lt2gene | 1337 | No | NA | 2 | 1 | 0 | Low-quality | Genome-fragment | 2.65 | AAI-based (high-confidence) | 0 |
| S14_vs_k141_85880\|\|full | 1337 | No | NA | 2 | 0 | 0 | Low-quality | Genome-fragment | 0.77 | AAI-based (high-confidence) | 0 |
| S14_vs_k141_100233\|\|full | 1337 | No | NA | 3 | 0 | 1 | Low-quality | Genome-fragment | 2.7 | AAI-based (high-confidence) | 0 |
| S3_vs_k141_98361\|\|full | 1337 | No | NA | 3 | 2 | 0 | Low-quality | Genome-fragment | 3.3 | AAI-based (high-confidence) | 0 |
| S14_extracted_k141_78138 | 6833 | No | NA | 4 | 0 | 3 | Not-determined | Genome-fragment | NA | NA | 0 |
| S14_extracted_k141_99647 | 1903 | No | NA | 2 | 0 | 2 | Not-determined | Genome-fragment | NA | NA | 0 |
| S7_extracted_78555 | 1337 | No | NA | 2 | 0 | 0 | Low-quality | Genome-fragment | 4.37 | AAI-based (high-confidence) | 0 |
| S14_extracted_k141_70715 | 1256 | No | NA | 2 | 0 | 1 | Not-determined | Genome-fragment | NA | NA | 0 |
| S14_extracted_k141_85442 | 6385 | No | NA | 7 | 0 | 1 | Not-determined | Genome-fragment | NA | NA | 0 |
| S7_vs_62064\|\|full | 1337 | No | NA | 2 | 0 | 2 | Low-quality | Genome-fragment | 0.5 | AAI-based (medium-confidence) | 0 |
| S14_extracted_k141_107595 | 1089 | No | NA | 2 | 0 | 0 | Not-determined | Genome-fragment | NA | NA | 0 |
| S14_extracted_k141_33937 | 2098 | No | NA | 1 | 0 | 0 | Not-determined | Genome-fragment | NA | NA | 0 |
| S14_extracted_k141_78139 | 53948 | Yes | 50221 | 63 | 30 | 2 | High-quality | High-quality | 93.75 | AAI-based (high-confidence) | 6.91 |
| S7_vs_131218\|\|full | 1337 | No | NA | 3 | 1 | 0 | Low-quality | Genome-fragment | 3.1 | AAI-based (high-confidence) | 0 |
| S14_extracted_k141_33974 | 1620 | No | NA | 1 | 0 | 1 | Not-determined | Genome-fragment | NA | NA | 0 |
| S16_vs_18403\|\|full | 1337 | No | NA | 3 | 0 | 3 | Low-quality | Genome-fragment | 0.63 | AAI-based (medium-confidence) | 0 |
| S16_vs_96575\|\|full | 1337 | No | NA | 3 | 0 | 1 | Low-quality | Genome-fragment | 0.92 | AAI-based (high-confidence) | 0 |
| S10_vs_38501\|\|full | 1337 | No | NA | 2 | 0 | 0 | Low-quality | Genome-fragment | 1.51 | AAI-based (medium-confidence) | 0 |
| S12_extracted_k141_54063 | 1337 | No | NA | 3 | 1 | 0 | Low-quality | Genome-fragment | 2.68 | AAI-based (high-confidence) | 0 |
| S6_extracted_k141_48550 | 1337 | No | NA | 2 | 1 | 0 | Low-quality | Genome-fragment | 3.4 | AAI-based (high-confidence) | 0 |
| S14_extracted_k141_19914 | 1145 | No | NA | 1 | 0 | 1 | Not-determined | Genome-fragment | NA | NA | 0 |
| S14_extracted_k141_107712 | 3196 | No | NA | 2 | 0 | 2 | Not-determined | Genome-fragment | NA | NA | 0 |
| S14_extracted_k141_19928 | 1194 | No | NA | 1 | 0 | 0 | Not-determined | Genome-fragment | NA | NA | 0 |
| S14_extracted_k141_107723 | 1689 | No | NA | 2 | 0 | 2 | Not-determined | Genome-fragment | NA | NA | 0 |
| S14_extracted_k141_63195 | 1321 | No | NA | 2 | 0 | 1 | Not-determined | Genome-fragment | NA | NA | 0 |
| S8_vs_k141_134666\|\|full | 1337 | No | NA | 3 | 0 | 0 | Low-quality | Genome-fragment | 2.69 | AAI-based (medium-confidence) | 0 |
| S8_vs_k141_57369\|\|full | 1337 | No | NA | 2 | 0 | 0 | Low-quality | Genome-fragment | 3.53 | AAI-based (medium-confidence) | 0 |
| S14_extracted_k141_56130 | 46683 | No | NA | 68 | 31 | 0 | High-quality | High-quality | 100 | AAI-based (high-confidence) | 0 |
| S9_extracted_k141_68583 | 1338 | No | NA | 1 | 0 | 0 | Low-quality | Genome-fragment | 2.05 | AAI-based (high-confidence) | 0 |
| S11_vs_k141_93031\|\|full | 1338 | No | NA | 3 | 0 | 0 | Low-quality | Genome-fragment | 3.81 | AAI-based (medium-confidence) | 0 |
| S14_extracted_k141_41930 | 1121 | No | NA | 1 | 0 | 1 | Not-determined | Genome-fragment | NA | NA | 0 |
| S13_vs_8881\|\|full | 1338 | No | NA | 3 | 0 | 2 | Low-quality | Genome-fragment | 0.47 | AAI-based (medium-confidence) | 0 |
| S14_extracted_k141_93078 | 7888 | No | NA | 5 | 0 | 4 | Not-determined | Genome-fragment | NA | NA | 0 |
| S13_vs_8915\|\|full | 1338 | No | NA | 3 | 0 | 2 | Low-quality | Genome-fragment | 2.09 | AAI-based (medium-confidence) | 0 |
| S14_extracted_k141_41984 | 3037 | No | NA | 6 | 0 | 1 | Not-determined | Genome-fragment | NA | NA | 0 |
| S14_extracted_k141_93134 | 15540 | No | NA | 21 | 6 | 0 | Medium-quality | Genome-fragment | 65.86 | AAI-based (high-confidence) | 0 |
| S14_extracted_k141_107789 | 1050 | No | NA | 2 | 0 | 1 | Not-determined | Genome-fragment | NA | NA | 0 |
| S3_vs_k141_23258\|\|full | 1338 | No | NA | 3 | 0 | 0 | Low-quality | Genome-fragment | 2.63 | AAI-based (medium-confidence) | 0 |
| S14_extracted_k141_56170 | 1858 | No | NA | 1 | 0 | 1 | Not-determined | Genome-fragment | NA | NA | 0 |
| S14_extracted_k141_114964 | 4653 | No | NA | 5 | 0 | 3 | Not-determined | Genome-fragment | NA | NA | 0 |
| S14_extracted_k141_99815 | 1208 | No | NA | 2 | 0 | 1 | Not-determined | Genome-fragment | NA | NA | 0 |
| S7_vs_32732\|\|full | 1338 | No | NA | 3 | 1 | 0 | Low-quality | Genome-fragment | 2.33 | AAI-based (high-confidence) | 0 |
| S7_vs_69113\|\|full | 1338 | No | NA | 2 | 1 | 0 | Low-quality | Genome-fragment | 3.55 | AAI-based (medium-confidence) | 0 |
| S14_extracted_k141_85720 | 1145 | No | NA | 2 | 0 | 2 | Not-determined | Genome-fragment | NA | NA | 0 |
| S14_extracted_k141_114981 | 1084 | No | NA | 1 | 0 | 0 | Not-determined | Genome-fragment | NA | NA | 0 |
| S7_vs_109696\|\|full | 1338 | No | NA | 3 | 1 | 0 | Low-quality | Genome-fragment | 3.32 | AAI-based (high-confidence) | 0 |
| S14_extracted_k141_42063 | 1068 | No | NA | 2 | 0 | 2 | Not-determined | Genome-fragment | NA | NA | 0 |
| S7_vs_92612\|\|lt2gene | 1338 | No | NA | 2 | 0 | 1 | Low-quality | Genome-fragment | 1.57 | AAI-based (medium-confidence) | 0 |
| S14_extracted_k141_99886 | 1949 | No | NA | 7 | 0 | 0 | Not-determined | Genome-fragment | NA | NA | 0 |
| S14_extracted_k141_48325 | 1005 | No | NA | 1 | 0 | 0 | Not-determined | Genome-fragment | NA | NA | 0 |
| S14_extracted_k141_4834 | 1303 | No | NA | 1 | 0 | 0 | Not-determined | Genome-fragment | NA | NA | 0 |
| S14_extracted_k141_34150 | 1752 | No | NA | 1 | 0 | 0 | Not-determined | Genome-fragment | NA | NA | 0 |
| S16_extracted_118634 | 1338 | No | NA | 3 | 0 | 0 | Low-quality | Genome-fragment | 2.19 | AAI-based (medium-confidence) | 0 |
| S8_extracted_k141_63621 | 1338 | No | NA | 3 | 0 | 0 | Low-quality | Genome-fragment | 2.09 | AAI-based (high-confidence) | 0 |
| S14_extracted_k141_78302 | 1100 | No | NA | 1 | 0 | 1 | Not-determined | Genome-fragment | NA | NA | 0 |
| S14_extracted_k141_26450 | 1067 | No | NA | 2 | 0 | 1 | Not-determined | Genome-fragment | NA | NA | 0 |
| S8_vs_k141_192648\|\|full | 1338 | No | NA | 4 | 1 | 0 | Low-quality | Genome-fragment | 3.08 | AAI-based (high-confidence) | 0 |
| S14_extracted_k141_93264 | 1062 | No | NA | 1 | 0 | 1 | Not-determined | Genome-fragment | NA | NA | 0 |
| S17_vs_k141_141040\|\|full | 1339 | No | NA | 2 | 0 | 0 | Low-quality | Genome-fragment | 1.12 | AAI-based (high-confidence) | 0 |

| | | | | | | | | | | | |
|---|---|---|---|---|---|---|---|---|---|---|---|
| S14_extracted_k141_93279 | 4544 | No | NA | 3 | 0 | 3 | Not-determined | Genome-fragment | NA | NA | 0 |
| S14_extracted_k141_115093 | 1428 | No | NA | 2 | 0 | 0 | Not-determined | Genome-fragment | NA | NA | 0 |
| S14_extracted_k141_107921 | 1127 | No | NA | 1 | 0 | 1 | Not-determined | Genome-fragment | NA | NA | 0 |
| S9_vs_k141_25426\|\|full | 1339 | No | NA | 2 | 0 | 0 | Low-quality | Genome-fragment | 1.79 | AAI-based (high-confidence) | 0 |
| S14_extracted_k141_107934 | 1571 | No | NA | 2 | 0 | 2 | Not-determined | Genome-fragment | NA | NA | 0 |
| S11_extracted_k141_30599 | 1339 | No | NA | 2 | 1 | 0 | Low-quality | Genome-fragment | 2.85 | AAI-based (medium-confidence) | 0 |
| S1_vs_k141_21666\|\|full | 1339 | No | NA | 3 | 1 | 0 | Low-quality | Genome-fragment | 2.31 | AAI-based (high-confidence) | 0 |
| S1_vs_k141_149452\|\|lt2gene | 1339 | No | NA | 2 | 1 | 0 | Low-quality | Genome-fragment | 1.75 | HMM-based (lower-bound) | 0 |
| S5_vs_k141_94404\|\|full | 1339 | No | NA | 5 | 0 | 0 | Low-quality | Genome-fragment | 3.09 | AAI-based (high-confidence) | 0 |
| S14_extracted_k141_115149 | 3692 | No | NA | 3 | 0 | 1 | Not-determined | Genome-fragment | NA | NA | 0 |
| S14_extracted_k141_78441 | 1053 | No | NA | 1 | 0 | 1 | Not-determined | Genome-fragment | NA | NA | 0 |
| S14_extracted_k141_4942 | 2893 | No | NA | 2 | 0 | 0 | Not-determined | Genome-fragment | NA | NA | 0 |
| S7_vs_128301\|\|full | 1339 | No | NA | 2 | 0 | 0 | Low-quality | Genome-fragment | 3.33 | AAI-based (medium-confidence) | 0 |
| S14_extracted_k141_34309 | 1042 | No | NA | 2 | 0 | 2 | Not-determined | Genome-fragment | NA | NA | 0 |
| S14_extracted_k141_78486 | 1234 | No | NA | 2 | 0 | 2 | Not-determined | Genome-fragment | NA | NA | 0 |
| S14_extracted_k141_48453 | 1837 | No | NA | 3 | 0 | 1 | Not-determined | Genome-fragment | NA | NA | 0 |
| S10_extracted_32872 | 1339 | No | NA | 1 | 0 | 1 | Low-quality | Genome-fragment | 1.33 | AAI-based (high-confidence) | 0 |
| S10_extracted_85362 | 1339 | No | NA | 4 | 0 | 0 | Low-quality | Genome-fragment | 3.06 | AAI-based (high-confidence) | 0 |
| S15_extracted_43484 | 1339 | No | NA | 2 | 1 | 0 | Low-quality | Genome-fragment | 3.38 | AAI-based (medium-confidence) | 0 |
| S17_extracted_k141_130326 | 1340 | No | NA | 2 | 0 | 1 | Low-quality | Genome-fragment | 9.57 | AAI-based (medium-confidence) | 0 |
| S17_vs_k141_47490\|\|full | 1340 | No | NA | 3 | 1 | 0 | Low-quality | Genome-fragment | 1.2 | HMM-based (lower-bound) | 0 |
| S14_extracted_k141_56439 | 4099 | No | NA | 5 | 0 | 1 | Not-determined | Genome-fragment | NA | NA | 0 |
| S14_extracted_k141_71097 | 1201 | No | NA | 2 | 0 | 2 | Not-determined | Genome-fragment | NA | NA | 0 |
| S14_extracted_k141_71106 | 1150 | No | NA | 2 | 0 | 1 | Not-determined | Genome-fragment | NA | NA | 0 |
| S14_extracted_k141_63590 | 1190 | No | NA | 3 | 0 | 3 | Not-determined | Genome-fragment | NA | NA | 0 |
| S9_vs_k141_17873\|\|full | 1340 | No | NA | 4 | 1 | 0 | Low-quality | Genome-fragment | 3.65 | AAI-based (high-confidence) | 0 |
| S11_vs_k141_91805\|\|lt2gene | 1340 | No | NA | 2 | 2 | 0 | Low-quality | Genome-fragment | 3.29 | AAI-based (high-confidence) | 0 |
| S14_extracted_k141_115269 | 2062 | No | NA | 2 | 0 | 1 | Not-determined | Genome-fragment | NA | NA | 0 |
| S14_extracted_k141_20023 | 1091 | No | NA | 1 | 0 | 1 | Not-determined | Genome-fragment | NA | NA | 0 |
| S14_extracted_k141_26627 | 1319 | No | NA | 3 | 0 | 3 | Not-determined | Genome-fragment | NA | NA | 0 |
| S14_extracted_k141_31947 | 1340 | No | NA | 4 | 4 | 0 | Low-quality | Genome-fragment | 4.42 | AAI-based (high-confidence) | 0 |
| S14_extracted_k141_63633 | 1090 | No | NA | 2 | 0 | 1 | Not-determined | Genome-fragment | NA | NA | 0 |
| S14_extracted_k141_78634 | 1219 | No | NA | 1 | 0 | 1 | Not-determined | Genome-fragment | NA | NA | 0 |
| S14_extracted_k141_48582 | 4530 | No | NA | 5 | 0 | 1 | Not-determined | Genome-fragment | NA | NA | 0 |
| S14_extracted_k141_85902 | 2237 | No | NA | 7 | 0 | 0 | Not-determined | Genome-fragment | NA | NA | 0 |
| S14_vs_k141_82119\|\|full | 1340 | No | NA | 1 | 0 | 0 | Low-quality | Genome-fragment | 1.56 | AAI-based (high-confidence) | 0 |
| S14_extracted_k141_100232 | 6510 | No | NA | 2 | 0 | 1 | Not-determined | Genome-fragment | NA | NA | 0 |
| S14_vs_k141_49925\|\|lt2gene | 1340 | No | NA | 2 | 1 | 0 | Low-quality | Genome-fragment | 2.93 | AAI-based (high-confidence) | 0 |
| S14_extracted_k141_63692 | 1171 | No | NA | 2 | 0 | 1 | Not-determined | Genome-fragment | NA | NA | 0 |
| S14_extracted_k141_63693 | 1236 | No | NA | 2 | 0 | 1 | Not-determined | Genome-fragment | NA | NA | 0 |
| S14_extracted_k141_108145 | 1184 | No | NA | 4 | 0 | 0 | Not-determined | Genome-fragment | NA | NA | 0 |
| S1_vs_k141_92702\|\|full | 1340 | No | NA | 3 | 0 | 0 | Low-quality | Genome-fragment | 4.56 | AAI-based (high-confidence) | 0 |
| S14_extracted_k141_63726 | 2676 | No | NA | 3 | 0 | 2 | Not-determined | Genome-fragment | NA | NA | 0 |
| S3_extracted_k141_89675 | 1340 | No | NA | 1 | 0 | 0 | Low-quality | Genome-fragment | 0.79 | AAI-based (high-confidence) | 0 |
| S14_extracted_k141_115379 | 4325 | No | NA | 2 | 0 | 2 | Not-determined | Genome-fragment | NA | NA | 0 |
| S3_vs_k141_74446\|\|full | 1340 | No | NA | 4 | 4 | 0 | Low-quality | Genome-fragment | 3.09 | AAI-based (high-confidence) | 0 |
| S14_extracted_k141_48696 | 1223 | No | NA | 3 | 0 | 2 | Not-determined | Genome-fragment | NA | NA | 0 |
| S14_extracted_k141_86022 | 1351 | No | NA | 2 | 0 | 1 | Not-determined | Genome-fragment | NA | NA | 0 |
| S14_extracted_k141_5123 | 1651 | No | NA | 3 | 0 | 1 | Not-determined | Genome-fragment | NA | NA | 0 |
| S14_extracted_k141_100331 | 4305 | No | NA | 6 | 0 | 6 | Not-determined | Genome-fragment | NA | NA | 0 |
| S14_extracted_k141_100339 | 2318 | No | NA | 2 | 0 | 2 | Not-determined | Genome-fragment | NA | NA | 0 |
| S7_vs_134898\|\|full | 1340 | No | NA | 3 | 1 | 0 | Low-quality | Genome-fragment | 2 | HMM-based (lower-bound) | 0 |
| S7_vs_117919\|\|lt2gene | 1340 | No | NA | 1 | 1 | 0 | Low-quality | Genome-fragment | 2.62 | AAI-based (high-confidence) | 0 |
| S14_extracted_k141_42605 | 1118 | No | NA | 1 | 0 | 1 | Not-determined | Genome-fragment | NA | NA | 0 |
| S14_extracted_k141_42614 | 1139 | No | NA | 1 | 0 | 1 | Not-determined | Genome-fragment | NA | NA | 0 |
| S16_extracted_28900 | 1340 | No | NA | 3 | 2 | 0 | Low-quality | Genome-fragment | 2.22 | AAI-based (high-confidence) | 0 |
| S14_extracted_k141_34693 | 2179 | No | NA | 1 | 0 | 0 | Not-determined | Genome-fragment | NA | NA | 0 |
| S16_vs_58431\|\|full | 1340 | No | NA | 3 | 0 | 0 | Low-quality | Genome-fragment | 2.48 | AAI-based (medium-confidence) | 0 |
| S14_extracted_k141_100431 | 1134 | No | NA | 1 | 0 | 0 | Not-determined | Genome-fragment | NA | NA | 0 |
| S14_extracted_k141_34715 | 1533 | No | NA | 2 | 0 | 2 | Not-determined | Genome-fragment | NA | NA | 0 |
| S14_extracted_k141_12570 | 1054 | No | NA | 1 | 0 | 0 | Not-determined | Genome-fragment | NA | NA | 0 |
| S14_extracted_k141_26904 | 1001 | No | NA | 1 | 0 | 1 | Not-determined | Genome-fragment | NA | NA | 0 |
| S14_extracted_k141_20278 | 1069 | No | NA | 3 | 0 | 0 | Not-determined | Genome-fragment | NA | NA | 0 |
| S14_extracted_k141_26931 | 3111 | No | NA | 4 | 0 | 1 | Not-determined | Genome-fragment | NA | NA | 0 |
| S14_extracted_k141_56681 | 1532 | No | NA | 1 | 0 | 1 | Not-determined | Genome-fragment | NA | NA | 0 |
| S10_vs_87486\|\|lt2gene | 1340 | No | NA | 2 | 2 | 0 | Low-quality | Genome-fragment | 2.38 | HMM-based (lower-bound) | 0 |
| S14_extracted_k141_48775 | 36317 | No | NA | 42 | 9 | 0 | Medium-quality | Genome-fragment | 60.96 | AAI-based (high-confidence) | 0 |
| S14_extracted_k141_86189 | 7440 | No | NA | 5 | 0 | 4 | Not-determined | Genome-fragment | NA | NA | 0 |
| S14_extracted_k141_20320 | 1130 | No | NA | 1 | 0 | 0 | Not-determined | Genome-fragment | NA | NA | 0 |
| S12_vs_k141_21369\|\|lt2gene | 1340 | No | NA | 1 | 1 | 0 | Low-quality | Genome-fragment | 3.04 | AAI-based (high-confidence) | 0 |
| S2_vs_k141_10032\|\|full | 1340 | No | NA | 3 | 0 | 1 | Low-quality | Genome-fragment | 3.13 | AAI-based (medium-confidence) | 0 |
| S8_vs_k141_81846\|\|full | 1340 | No | NA | 4 | 1 | 0 | Low-quality | Genome-fragment | 2.8 | AAI-based (high-confidence) | 0 |
| S11_vs_k141_42928\|\|lt2gene | 1341 | No | NA | 2 | 1 | 0 | Low-quality | Genome-fragment | 3.26 | AAI-based (high-confidence) | 0 |
| S3_extracted_k141_58222 | 1341 | No | NA | 4 | 2 | 0 | Low-quality | Genome-fragment | 3.88 | AAI-based (high-confidence) | 0 |
| S14_extracted_k141_115516 | 1430 | No | NA | 1 | 0 | 1 | Not-determined | Genome-fragment | NA | NA | 0 |
| S14_extracted_k141_71465 | 1069 | No | NA | 1 | 0 | 1 | Not-determined | Genome-fragment | NA | NA | 0 |
| S14_extracted_k141_34850 | 3283 | No | NA | 3 | 0 | 1 | Not-determined | Genome-fragment | NA | NA | 0 |
| S5_vs_k141_51996\|\|full | 1341 | No | NA | 4 | 1 | 0 | Low-quality | Genome-fragment | 2.39 | HMM-based (lower-bound) | 0 |
| S7_extracted_87481 | 1341 | No | NA | 3 | 3 | 0 | Low-quality | Genome-fragment | 6.12 | AAI-based (high-confidence) | 0 |
| S14_extracted_k141_100571 | 1365 | No | NA | 1 | 0 | 1 | Not-determined | Genome-fragment | NA | NA | 0 |
| S7_vs_23402\|\|full | 1341 | No | NA | 2 | 0 | 1 | Low-quality | Genome-fragment | 1.35 | AAI-based (medium-confidence) | 0 |
| S16_vs_29714\|\|full | 1341 | No | NA | 3 | 0 | 0 | Low-quality | Genome-fragment | 3.19 | AAI-based (medium-confidence) | 0 |
| S14_extracted_k141_48853 | 1161 | No | NA | 1 | 0 | 0 | Not-determined | Genome-fragment | NA | NA | 0 |
| S14_extracted_k141_12710 | 1195 | No | NA | 2 | 0 | 1 | Not-determined | Genome-fragment | NA | NA | 0 |
| S14_extracted_k141_48872 | 1260 | No | NA | 1 | 0 | 0 | Not-determined | Genome-fragment | NA | NA | 0 |
| S16_vs_158116\|\|lt2gene | 1341 | No | NA | 1 | 1 | 0 | Low-quality | Genome-fragment | 3.09 | AAI-based (high-confidence) | 0 |
| S10_vs_51114\|\|full | 1341 | No | NA | 4 | 0 | 0 | Low-quality | Genome-fragment | 1.01 | AAI-based (medium-confidence) | 0 |
| S14_extracted_k141_108397 | 1944 | No | NA | 2 | 0 | 2 | Not-determined | Genome-fragment | NA | NA | 0 |
| S10_vs_69176\|\|full | 1341 | No | NA | 3 | 0 | 0 | Low-quality | Genome-fragment | 3.1 | AAI-based (high-confidence) | 0 |
| S14_extracted_k141_34967 | 5414 | No | NA | 6 | 0 | 5 | Not-determined | Genome-fragment | NA | NA | 0 |
| S12_extracted_k141_103393 | 1341 | No | NA | 3 | 0 | 2 | Low-quality | Genome-fragment | 0.43 | AAI-based (medium-confidence) | 0 |
| S12_extracted_k141_5094 | 1341 | No | NA | 2 | 0 | 0 | Low-quality | Genome-fragment | 3.15 | AAI-based (medium-confidence) | 0 |
| S12_vs_k141_63255\|\|full | 1341 | No | NA | 3 | 0 | 2 | Low-quality | Genome-fragment | 1.11 | AAI-based (medium-confidence) | 0 |
| S14_extracted_k141_20496 | 1028 | No | NA | 1 | 0 | 0 | Not-determined | Genome-fragment | NA | NA | 0 |
| S14_extracted_k141_78836 | 1257 | No | NA | 2 | 0 | 1 | Not-determined | Genome-fragment | NA | NA | 0 |
| S14_extracted_k141_71669 | 1535 | No | NA | 2 | 0 | 2 | Not-determined | Genome-fragment | NA | NA | 0 |
| S14_extracted_k141_12895 | 1098 | No | NA | 3 | 0 | 0 | Not-determined | Genome-fragment | NA | NA | 0 |
| S14_extracted_k141_49049 | 11402 | No | NA | 5 | 0 | 5 | Not-determined | Genome-fragment | NA | NA | 0 |
| S14_extracted_k141_71686 | 1284 | No | NA | 1 | 0 | 0 | Not-determined | Genome-fragment | NA | NA | 0 |
| S14_extracted_k141_78877 | 1267 | No | NA | 1 | 0 | 1 | Not-determined | Genome-fragment | NA | NA | 0 |
| S14_extracted_k141_43040 | 1651 | No | NA | 4 | 0 | 4 | Not-determined | Genome-fragment | NA | NA | 0 |
| S14_extracted_k141_64096 | 1046 | No | NA | 2 | 0 | 2 | Not-determined | Genome-fragment | NA | NA | 0 |
| S12_vs_k141_171775\|\|full | 1341 | No | NA | 3 | 0 | 0 | Low-quality | Genome-fragment | 2.52 | AAI-based (medium-confidence) | 0 |
| S15_extracted_55764 | 1341 | No | NA | 3 | 0 | 0 | Low-quality | Genome-fragment | 2.18 | AAI-based (high-confidence) | 0 |
| S14_extracted_k141_108556 | 3793 | No | NA | 2 | 0 | 2 | Not-determined | Genome-fragment | NA | NA | 0 |
| S14_extracted_k141_12989 | 1655 | No | NA | 3 | 0 | 0 | Not-determined | Genome-fragment | NA | NA | 0 |
| S17_extracted_k141_58485 | 1342 | No | NA | 2 | 0 | 0 | Low-quality | Genome-fragment | 0.88 | AAI-based (medium-confidence) | 0 |
| S14_extracted_k141_86483 | 5439 | No | NA | 1 | 0 | 1 | Not-determined | Genome-fragment | NA | NA | 0 |
| S14_extracted_k141_115736 | 42416 | No | NA | 76 | 19 | 0 | Complete | High-quality | 100 | DTR (high-confidence) | 0 |
| S14_extracted_k141_27277 | 2083 | No | NA | 4 | 0 | 3 | Not-determined | Genome-fragment | NA | NA | 0 |
| S14_extracted_k141_5570 | 1063 | No | NA | 1 | 0 | 1 | Not-determined | Genome-fragment | NA | NA | 0 |
| S14_extracted_k141_13035 | 4885 | No | NA | 4 | 0 | 3 | Not-determined | Genome-fragment | NA | NA | 0 |
| S9_vs_k141_32350\|\|full | 1342 | No | NA | 3 | 1 | 0 | Low-quality | Genome-fragment | 2.76 | AAI-based (medium-confidence) | 0 |
| S14_extracted_k141_43182 | 1458 | No | NA | 2 | 0 | 0 | Not-determined | Genome-fragment | NA | NA | 0 |
| S14_extracted_k141_108632 | 1202 | No | NA | 1 | 0 | 1 | Not-determined | Genome-fragment | NA | NA | 0 |
| S11_vs_k141_7711\|\|full | 1342 | No | NA | 3 | 1 | 0 | Low-quality | Genome-fragment | 3.25 | HMM-based (lower-bound) | 0 |
| S14_extracted_k141_43184 | 1096 | No | NA | 2 | 0 | 0 | Not-determined | Genome-fragment | NA | NA | 0 |
| S14_extracted_k141_108648 | 1630 | No | NA | 2 | 0 | 0 | Not-determined | Genome-fragment | NA | NA | 0 |
| S14_extracted_k141_78958 | 2157 | No | NA | 1 | 0 | 1 | Not-determined | Genome-fragment | NA | NA | 0 |
| S14_extracted_k141_20684 | 6711 | No | NA | 5 | 0 | 2 | Not-determined | Genome-fragment | NA | NA | 0 |
| S14_extracted_k141_20694 | 4549 | No | NA | 1 | 0 | 1 | Not-determined | Genome-fragment | NA | NA | 0 |
| S14_extracted_k141_108686 | 3267 | No | NA | 3 | 0 | 3 | Not-determined | Genome-fragment | NA | NA | 0 |
| S14_extracted_k141_71887 | 3669 | No | NA | 3 | 0 | 2 | Not-determined | Genome-fragment | NA | NA | 0 |
| S13_vs_142648\|\|full | 1342 | No | NA | 1 | 0 | 1 | Low-quality | Genome-fragment | 1.67 | AAI-based (high-confidence) | 0 |
| S14_extracted_k141_43235 | 1092 | No | NA | 1 | 0 | 1 | Not-determined | Genome-fragment | NA | NA | 0 |
| S14_extracted_k141_49103 | 1793 | No | NA | 1 | 0 | 0 | Not-determined | Genome-fragment | NA | NA | 0 |
| S14_extracted_k141_78989 | 1241 | No | NA | 1 | 0 | 1 | Not-determined | Genome-fragment | NA | NA | 0 |
| S14_extracted_k141_57011 | 1211 | No | NA | 2 | 0 | 0 | Not-determined | Genome-fragment | NA | NA | 0 |
| S14_extracted_k141_100082 | 1342 | No | NA | 2 | 2 | 0 | Low-quality | Genome-fragment | 0.73 | HMM-based (lower-bound) | 0 |
| S14_extracted_k141_64230 | 3991 | No | NA | 2 | 0 | 2 | Not-determined | Genome-fragment | NA | NA | 0 |
| S14_extracted_k141_49195 | 1359 | No | NA | 2 | 0 | 1 | Not-determined | Genome-fragment | NA | NA | 0 |
| S4_extracted_7231 | 1342 | No | NA | 3 | 0 | 0 | Low-quality | Genome-fragment | 3.44 | AAI-based (medium-confidence) | 0 |
| S14_extracted_k141_108837 | 1081 | No | NA | 2 | 0 | 0 | Not-determined | Genome-fragment | NA | NA | 0 |
| S14_extracted_k141_5774 | 1041 | No | NA | 1 | 0 | 0 | Not-determined | Genome-fragment | NA | NA | 0 |
| S14_extracted_k141_108851 | 1141 | No | NA | 2 | 0 | 1 | Not-determined | Genome-fragment | NA | NA | 0 |
| S8_vs_k141_185532\|\|full | 1342 | No | NA | 2 | 0 | 0 | Low-quality | Genome-fragment | 1.4 | AAI-based (medium-confidence) | 0 |
| S14_extracted_k141_5788 | 1163 | No | NA | 2 | 0 | 1 | Not-determined | Genome-fragment | NA | NA | 0 |
| S9_vs_k141_18480\|\|lt2gene | 1343 | No | NA | 2 | 2 | 0 | Low-quality | Genome-fragment | 3.77 | AAI-based (high-confidence) | 0 |
| S13_vs_139321\|\|full | 1343 | No | NA | 3 | 0 | 1 | Low-quality | Genome-fragment | 3.03 | AAI-based (medium-confidence) | 0 |
| S1_vs_k141_27995\|\|full | 1343 | No | NA | 3 | 0 | 0 | Low-quality | Genome-fragment | 2.7 | AAI-based (medium-confidence) | 0 |
| S14_extracted_k141_43340 | 6047 | No | NA | 9 | 3 | 0 | High-quality | High-quality | 97.55 | AAI-based (high-confidence) | 0 |
| S14_extracted_k141_101065 | 1069 | No | NA | 2 | 0 | 2 | Not-determined | Genome-fragment | NA | NA | 0 |
| S14_extracted_k141_64288 | 1654 | No | NA | 1 | 0 | 1 | Not-determined | Genome-fragment | NA | NA | 0 |
| S14_extracted_k141_79107 | 1583 | No | NA | 1 | 0 | 1 | Not-determined | Genome-fragment | NA | NA | 0 |
| S3_extracted_k141_112626 | 1343 | No | NA | 3 | 1 | 0 | Low-quality | Genome-fragment | 0.96 | AAI-based (high-confidence) | 0 |
| S3_vs_k141_110122\|\|full | 1343 | No | NA | 4 | 3 | 0 | Low-quality | Genome-fragment | 3.24 | AAI-based (medium-confidence) | 0 |
| S14_extracted_k141_35347 | 1355 | No | NA | 2 | 0 | 1 | Not-determined | Genome-fragment | NA | NA | 0 |
| S14_extracted_k141_57142 | 1397 | No | NA | 3 | 0 | 0 | Not-determined | Genome-fragment | NA | NA | 0 |
| S14_extracted_k141_79124 | 5693 | No | NA | 6 | 0 | 4 | Not-determined | Genome-fragment | NA | NA | 0 |
| S14_extracted_k141_13197 | 1234 | No | NA | 1 | 0 | 1 | Not-determined | Genome-fragment | NA | NA | 0 |
| S14_extracted_k141_5843 | 1182 | No | NA | 1 | 0 | 1 | Not-determined | Genome-fragment | NA | NA | 0 |
| S14_extracted_k141_108936 | 1454 | No | NA | 1 | 0 | 1 | Not-determined | Genome-fragment | NA | NA | 0 |
| S14_extracted_k141_35366 | 1363 | No | NA | 2 | 0 | 2 | Not-determined | Genome-fragment | NA | NA | 0 |
| S5_extracted_k141_49886 | 1343 | No | NA | 2 | 1 | 0 | Low-quality | Genome-fragment | 3.35 | AAI-based (high-confidence) | 0 |
| S7_vs_73531\|\|full | 1343 | No | NA | 6 | 1 | 0 | Low-quality | Genome-fragment | 2.75 | AAI-based (medium-confidence) | 0 |
| S14_extracted_k141_5869 | 1100 | No | NA | 2 | 0 | 2 | Not-determined | Genome-fragment | NA | NA | 0 |
| S14_extracted_k141_27638 | 1131 | No | NA | 1 | 0 | 0 | Not-determined | Genome-fragment | NA | NA | 0 |
| S14_extracted_k141_79177 | 1524 | No | NA | 4 | 0 | 1 | Not-determined | Genome-fragment | NA | NA | 0 |
| S7_vs_130683\|\|lt2gene | 1343 | No | NA | 2 | 2 | 0 | Low-quality | Genome-fragment | 3.61 | AAI-based (high-confidence) | 0 |
| S16_extracted_120016 | 1343 | No | NA | 2 | 1 | 0 | Low-quality | Genome-fragment | 0.54 | HMM-based (lower-bound) | 0 |
| S14_extracted_k141_86870 | 7486 | No | NA | 6 | 0 | 4 | Not-determined | Genome-fragment | NA | NA | 0 |
| S14_extracted_k141_57240 | 2280 | No | NA | 2 | 0 | 0 | Not-determined | Genome-fragment | NA | NA | 0 |
| S16_vs_111582\|\|full | 1343 | No | NA | 2 | 0 | 0 | Low-quality | Genome-fragment | 3.06 | AAI-based (high-confidence) | 0 |
| S12_vs_k141_152211\|\|full | 1343 | No | NA | 4 | 0 | 0 | Low-quality | Genome-fragment | 19.54 | AAI-based (medium-confidence) | 0 |
| S8_vs_k141_161771\|\|full | 1343 | No | NA | 3 | 0 | 0 | Low-quality | Genome-fragment | 3.34 | AAI-based (medium-confidence) | 0 |
| S14_extracted_k141_86942 | 1043 | No | NA | 1 | 0 | 0 | Not-determined | Genome-fragment | NA | NA | 0 |
| S14_extracted_k141_21002 | 1712 | No | NA | 2 | 0 | 0 | Not-determined | Genome-fragment | NA | NA | 0 |
| S17_vs_k141_81020\|\|full | 1344 | No | NA | 2 | 0 | 0 | Low-quality | Genome-fragment | 2.51 | AAI-based (medium-confidence) | 0 |
| S14_extracted_k141_101201 | 1023 | No | NA | 1 | 0 | 1 | Not-determined | Genome-fragment | NA | NA | 0 |
| S9_vs_k141_20008\|\|full | 1344 | No | NA | 3 | 0 | 3 | Low-quality | Genome-fragment | 0.73 | AAI-based (medium-confidence) | 0 |
| S14_extracted_k141_79275 | 3188 | No | NA | 1 | 0 | 1 | Not-determined | Genome-fragment | NA | NA | 0 |
| S9_vs_k141_83160\|\|lt2gene | 1344 | No | NA | 2 | 1 | 0 | Low-quality | Genome-fragment | 2.93 | AAI-based (high-confidence) | 0 |
| S14_extracted_k141_79278 | 1497 | No | NA | 3 | 0 | 0 | Not-determined | Genome-fragment | NA | NA | 0 |
| S14_extracted_k141_27715 | 3368 | No | NA | 4 | 0 | 2 | Not-determined | Genome-fragment | NA | NA | 0 |
| S14_extracted_k141_101223 | 3346 | No | NA | 2 | 0 | 2 | Not-determined | Genome-fragment | NA | NA | 0 |
| S11_vs_k141_65792\|\|full | 1344 | No | NA | 1 | 0 | 0 | Low-quality | Genome-fragment | 1.92 | AAI-based (high-confidence) | 0 |
| S14_extracted_k141_109060 | 1217 | No | NA | 1 | 0 | 0 | Not-determined | Genome-fragment | NA | NA | 0 |
| S14_extracted_k141_87043 | 3019 | No | NA | 3 | 0 | 2 | Not-determined | Genome-fragment | NA | NA | 0 |
| S14_extracted_k141_79335 | 2684 | No | NA | 2 | 0 | 0 | Not-determined | Genome-fragment | NA | NA | 0 |
| S14_extracted_k141_64390 | 1172 | No | NA | 1 | 0 | 0 | Not-determined | Genome-fragment | NA | NA | 0 |
| S14_extracted_k141_72101 | 2953 | No | NA | 3 | 0 | 1 | Not-determined | Genome-fragment | NA | NA | 0 |
| S14_extracted_k141_94537 | 1609 | No | NA | 4 | 0 | 4 | Not-determined | Genome-fragment | NA | NA | 0 |
| S14_extracted_k141_94547 | 2351 | No | NA | 2 | 0 | 0 | Not-determined | Genome-fragment | NA | NA | 0 |
| S14_extracted_k141_35619 | 1138 | No | NA | 1 | 0 | 1 | Not-determined | Genome-fragment | NA | NA | 0 |
| S13_vs_129291\|\|full | 1344 | No | NA | 3 | 0 | 0 | Low-quality | Genome-fragment | 2.16 | AAI-based (high-confidence) | 0 |
| S14_extracted_k141_94568 | 1483 | No | NA | 2 | 0 | 1 | Not-determined | Genome-fragment | NA | NA | 0 |
| S1_extracted_k141_150898 | 1344 | No | NA | 1 | 1 | 0 | Low-quality | Genome-fragment | 1.97 | AAI-based (high-confidence) | 0 |
| S3_extracted_k141_46539 | 1344 | No | NA | 2 | 2 | 0 | Low-quality | Genome-fragment | 26.13 | AAI-based (high-confidence) | 0 |
| S14_extracted_k141_64419 | 1364 | No | NA | 2 | 0 | 1 | Not-determined | Genome-fragment | NA | NA | 0 |
| S3_vs_k141_23265\|\|full | 1344 | No | NA | 4 | 0 | 0 | Low-quality | Genome-fragment | 1.99 | AAI-based (high-confidence) | 0 |
| S14_extracted_k141_64425 | 1142 | No | NA | 2 | 0 | 2 | Not-determined | Genome-fragment | NA | NA | 0 |
| S5_extracted_k141_111068 | 1344 | No | NA | 1 | 0 | 0 | Low-quality | Genome-fragment | 1.02 | AAI-based (high-confidence) | 0 |
| S14_extracted_k141_94602 | 4564 | No | NA | 4 | 0 | 3 | Not-determined | Genome-fragment | NA | NA | 0 |
| S14_extracted_k141_6137 | 1697 | No | NA | 1 | 0 | 1 | Not-determined | Genome-fragment | NA | NA | 0 |
| S5_vs_k141_70073\|\|full | 1344 | No | NA | 4 | 0 | 0 | Low-quality | Genome-fragment | 2.1 | AAI-based (high-confidence) | 0 |
| S14_extracted_k141_101376 | 1167 | No | NA | 3 | 0 | 0 | Not-determined | Genome-fragment | NA | NA | 0 |
| S14_extracted_k141_57452 | 1045 | No | NA | 3 | 0 | 1 | Not-determined | Genome-fragment | NA | NA | 0 |
| S7_vs_18238\|\|lt2gene | 1344 | No | NA | 2 | 2 | 0 | Low-quality | Genome-fragment | 3.08 | AAI-based (high-confidence) | 0 |
| S14_extracted_k141_27933 | 3495 | No | NA | 3 | 0 | 2 | Not-determined | Genome-fragment | NA | NA | 0 |
| S14_extracted_k141_49689 | 2605 | No | NA | 3 | 0 | 0 | Not-determined | Genome-fragment | NA | NA | 0 |
| S12_vs_k141_166953\|\|full | 1344 | No | NA | 3 | 0 | 0 | Low-quality | Genome-fragment | 0.9 | AAI-based (medium-confidence) | 0 |
| S17_extracted_k141_38287 | 1345 | No | NA | 1 | 0 | 1 | Low-quality | Genome-fragment | 1.2 | AAI-based (medium-confidence) | 0 |
| S11_extracted_k141_73665 | 1345 | No | NA | 1 | 0 | 0 | Low-quality | Genome-fragment | 2.36 | AAI-based (high-confidence) | 0 |
| S11_extracted_k141_69904 | 1345 | No | NA | 5 | 0 | 0 | Low-quality | Genome-fragment | 1.29 | AAI-based (medium-confidence) | 0 |
| S14_extracted_k141_94719 | 1179 | No | NA | 1 | 0 | 1 | Not-determined | Genome-fragment | NA | NA | 0 |
| S14_extracted_k141_13495 | 1471 | No | NA | 1 | 0 | 1 | Not-determined | Genome-fragment | NA | NA | 0 |
| S14_extracted_k141_72253 | 3055 | No | NA | 2 | 0 | 2 | Not-determined | Genome-fragment | NA | NA | 0 |
| S14_extracted_k141_87276 | 1015 | No | NA | 2 | 0 | 1 | Not-determined | Genome-fragment | NA | NA | 0 |
| S14_extracted_k141_94750 | 1396 | No | NA | 2 | 0 | 0 | Not-determined | Genome-fragment | NA | NA | 0 |
| S14_extracted_k141_43787 | 1242 | No | NA | 2 | 0 | 2 | Not-determined | Genome-fragment | NA | NA | 0 |
| S14_extracted_k141_21265 | 1028 | No | NA | 1 | 0 | 0 | Not-determined | Genome-fragment | NA | NA | 0 |
| S14_extracted_k141_35829 | 20156 | No | NA | 17 | 0 | 4 | Not-determined | Genome-fragment | NA | NA | 0 |
| S7_vs_134439\|\|full | 1345 | No | NA | 3 | 0 | 0 | Low-quality | Genome-fragment | 3.72 | AAI-based (high-confidence) | 0 |
| S12_extracted_k141_150016 | 1345 | No | NA | 2 | 0 | 1 | Low-quality | Genome-fragment | 3.05 | AAI-based (medium-confidence) | 0 |
| S14_extracted_k141_57629 | 1299 | No | NA | 3 | 0 | 0 | Not-determined | Genome-fragment | NA | NA | 0 |
| S12_extracted_k141_159644 | 1345 | No | NA | 3 | 0 | 2 | Low-quality | Genome-fragment | 0.88 | AAI-based (medium-confidence) | 0 |
| S12_vs_k141_24636\|\|full | 1345 | No | NA | 4 | 0 | 0 | Low-quality | Genome-fragment | 0.92 | AAI-based (high-confidence) | 0 |
| S14_extracted_k141_94800 | 1212 | No | NA | 2 | 0 | 0 | Not-determined | Genome-fragment | NA | NA | 0 |
| S14_extracted_k141_94815 | 1090 | No | NA | 2 | 0 | 0 | Not-determined | Genome-fragment | NA | NA | 0 |
| S12_vs_k141_35422\|\|full | 1345 | No | NA | 2 | 0 | 1 | Low-quality | Genome-fragment | 0.6 | AAI-based (medium-confidence) | 0 |
| S15_vs_10652\|\|full | 1345 | No | NA | 4 | 0 | 1 | Low-quality | Genome-fragment | 0.58 | AAI-based (medium-confidence) | 0 |
| S14_extracted_k141_72309 | 1219 | No | NA | 4 | 0 | 0 | Not-determined | Genome-fragment | NA | NA | 0 |
| S14_extracted_k141_116541 | 1012 | No | NA | 1 | 0 | 1 | Not-determined | Genome-fragment | NA | NA | 0 |
| S14_extracted_k141_64639 | 1014 | No | NA | 2 | 0 | 0 | Not-determined | Genome-fragment | NA | NA | 0 |
| S8_vs_k141_176045\|\|full | 1345 | No | NA | 5 | 0 | 0 | Low-quality | Genome-fragment | 1.43 | AAI-based (high-confidence) | 0 |
| S8_vs_k141_21875\|\|full | 1345 | No | NA | 3 | 0 | 0 | Low-quality | Genome-fragment | 2.17 | AAI-based (high-confidence) | 0 |
| S9_vs_k141_54286\|\|full | 1346 | No | NA | 4 | 1 | 0 | Low-quality | Genome-fragment | 2.65 | AAI-based (medium-confidence) | 0 |
| S14_extracted_k141_109431 | 1046 | No | NA | 4 | 0 | 0 | Not-determined | Genome-fragment | NA | NA | 0 |
| S14_extracted_k141_6450 | 1979 | No | NA | 3 | 0 | 0 | Not-determined | Genome-fragment | NA | NA | 0 |
| S14_extracted_k141_57689 | 34453 | No | NA | 30 | 12 | 0 | Medium-quality | Genome-fragment | 60.77 | AAI-based (high-confidence) | 0 |
| S9_vs_k141_69014\|\|full | 1346 | No | NA | 1 | 0 | 0 | Low-quality | Genome-fragment | 2.97 | AAI-based (medium-confidence) | 0 |
| S14_extracted_k141_109467 | 1110 | No | NA | 3 | 0 | 0 | Not-determined | Genome-fragment | NA | NA | 0 |
| S14_extracted_k141_109480 | 3843 | No | NA | 3 | 0 | 2 | Not-determined | Genome-fragment | NA | NA | 0 |
| S11_vs_k141_37137\|\|full | 1346 | No | NA | 2 | 1 | 0 | Low-quality | Genome-fragment | 3.18 | AAI-based (high-confidence) | 0 |
| S14_extracted_k141_36016 | 1698 | No | NA | 1 | 0 | 0 | Not-determined | Genome-fragment | NA | NA | 0 |
| S14_extracted_k141_6532 | 2348 | No | NA | 3 | 0 | 0 | Not-determined | Genome-fragment | NA | NA | 0 |
| S14_extracted_k141_57830 | 8089 | No | NA | 14 | 0 | 0 | Not-determined | Genome-fragment | NA | NA | 0 |
| S14_extracted_k141_109545 | 26025 | No | NA | 41 | 5 | 1 | Medium-quality | Genome-fragment | 60.65 | AAI-based (high-confidence) | 0 |
| S14_extracted_k141_101686 | 2436 | No | NA | 2 | 0 | 1 | Not-determined | Genome-fragment | NA | NA | 0 |
| S14_vs_k141_73558\|\|lt2gene | 1346 | No | NA | 2 | 1 | 0 | Low-quality | Genome-fragment | 3.96 | AAI-based (high-confidence) | 0 |
| S3_extracted_k141_23050 | 1346 | No | NA | 3 | 1 | 0 | Low-quality | Genome-fragment | 2.95 | AAI-based (medium-confidence) | 0 |
| S3_vs_k141_71340\|\|lt2gene | 1346 | No | NA | 1 | 1 | 0 | Low-quality | Genome-fragment | 3.55 | AAI-based (high-confidence) | 0 |
| S14_extracted_k141_72335 | 43571 | No | NA | 62 | 19 | 0 | High-quality | High-quality | 100 | AAI-based (medium-confidence) | 0 |
| S5_vs_k141_23703\|\|lt2gene | 1346 | No | NA | 1 | 1 | 0 | Low-quality | Genome-fragment | 1.97 | HMM-based (lower-bound) | 0 |
| S14_extracted_k141_36118 | 5322 | No | NA | 5 | 0 | 3 | Not-determined | Genome-fragment | NA | NA | 0 |
| S14_extracted_k141_116747 | 1043 | No | NA | 0 | 0 | 0 | Not-determined | Genome-fragment | NA | NA | 0 |
| S14_extracted_k141_6638 | 1109 | No | NA | 2 | 0 | 2 | Not-determined | Genome-fragment | NA | NA | 0 |
| S7_extracted_30359 | 1346 | No | NA | 2 | 1 | 0 | Low-quality | Genome-fragment | 1.97 | HMM-based (lower-bound) | 0 |
| S14_extracted_k141_109721 | 5728 | No | NA | 5 | 0 | 4 | Not-determined | Genome-fragment | NA | NA | 0 |
| S7_extracted_57893 | 1346 | No | NA | 3 | 0 | 0 | Low-quality | Genome-fragment | 3.64 | AAI-based (high-confidence) | 0 |
| S14_extracted_k141_109753 | 1146 | No | NA | 2 | 0 | 0 | Not-determined | Genome-fragment | NA | NA | 0 |
| S7_vs_82712\|\|full | 1346 | No | NA | 3 | 1 | 0 | Low-quality | Genome-fragment | 3.14 | AAI-based (medium-confidence) | 0 |
| S14_extracted_k141_57993 | 1356 | No | NA | 3 | 0 | 0 | Not-determined | Genome-fragment | NA | NA | 0 |
| S16_extracted_6990 | 1346 | No | NA | 4 | 0 | 0 | Low-quality | Genome-fragment | 2.78 | AAI-based (high-confidence) | 0 |
| S14_extracted_k141_80055 | 1535 | No | NA | 1 | 0 | 0 | Not-determined | Genome-fragment | NA | NA | 0 |
| S14_extracted_k141_6730 | 1117 | No | NA | 2 | 0 | 1 | Not-determined | Genome-fragment | NA | NA | 0 |
| S16_vs_91880\|\|lt2gene | 1346 | No | NA | 1 | 1 | 0 | Low-quality | Genome-fragment | 1.78 | HMM-based (lower-bound) | 0 |
| S14_extracted_k141_101865 | 1972 | No | NA | 2 | 0 | 0 | Not-determined | Genome-fragment | NA | NA | 0 |
| S14_extracted_k141_6741 | 1156 | No | NA | 1 | 0 | 0 | Not-determined | Genome-fragment | NA | NA | 0 |
| S10_extracted_46005 | 1346 | No | NA | 3 | 0 | 3 | Low-quality | Genome-fragment | 0.96 | AAI-based (medium-confidence) | 0 |
| S14_extracted_k141_109814 | 1065 | No | NA | 1 | 0 | 1 | Not-determined | Genome-fragment | NA | NA | 0 |
| S14_extracted_k141_109817 | 1648 | No | NA | 2 | 0 | 2 | Not-determined | Genome-fragment | NA | NA | 0 |
| S14_extracted_k141_80088 | 3610 | No | NA | 4 | 0 | 3 | Not-determined | Genome-fragment | NA | NA | 0 |
| S10_vs_103886\|\|full | 1346 | No | NA | 3 | 0 | 0 | Low-quality | Genome-fragment | 2.54 | AAI-based (medium-confidence) | 0 |
| S12_vs_k141_83422\|\|full | 1346 | No | NA | 3 | 0 | 0 | Low-quality | Genome-fragment | 2.6 | AAI-based (medium-confidence) | 0 |
| S14_extracted_k141_101880 | 1950 | No | NA | 1 | 0 | 1 | Not-determined | Genome-fragment | NA | NA | 0 |
| S6_extracted_k141_27256 | 1346 | No | NA | 3 | 0 | 2 | Low-quality | Genome-fragment | 1.14 | AAI-based (medium-confidence) | 0 |
| S17_vs_k141_62665\|\|full | 1347 | No | NA | 2 | 0 | 0 | Low-quality | Genome-fragment | 1.86 | AAI-based (medium-confidence) | 0 |
| S11_vs_k141_32282\|\|full | 1347 | No | NA | 3 | 0 | 0 | Low-quality | Genome-fragment | 3.09 | AAI-based (medium-confidence) | 0 |
| S14_extracted_k141_116900 | 1114 | No | NA | 1 | 0 | 0 | Not-determined | Genome-fragment | NA | NA | 0 |
| S14_extracted_k141_32599 | 1347 | No | NA | 3 | 1 | 0 | Low-quality | Genome-fragment | 0.91 | HMM-based (lower-bound) | 0 |
| S7_vs_45990\|\|full | 1347 | No | NA | 3 | 0 | 0 | Low-quality | Genome-fragment | 3.06 | AAI-based (medium-confidence) | 0 |
| S14_extracted_k141_116951 | 1672 | No | NA | 2 | 0 | 0 | Not-determined | Genome-fragment | NA | NA | 0 |
| S14_extracted_k141_36332 | 1858 | No | NA | 1 | 0 | 1 | Not-determined | Genome-fragment | NA | NA | 0 |
| S14_extracted_k141_21833 | 1896 | No | NA | 1 | 0 | 0 | Not-determined | Genome-fragment | NA | NA | 0 |
| S16_extracted_22217 | 1347 | No | NA | 2 | 0 | 0 | Low-quality | Genome-fragment | 4.2 | AAI-based (high-confidence) | 0 |
| S14_extracted_k141_50256 | 1864 | No | NA | 2 | 0 | 1 | Not-determined | Genome-fragment | NA | NA | 0 |
| S10_vs_90902\|\|lt2gene | 1347 | No | NA | 1 | 1 | 0 | Low-quality | Genome-fragment | 3.32 | AAI-based (high-confidence) | 0 |
| S15_extracted_9761 | 1347 | No | NA | 1 | 1 | 0 | Low-quality | Genome-fragment | 0.81 | AAI-based (high-confidence) | 0 |
| S2_vs_k141_879\|\|lt2gene | 1347 | No | NA | 2 | 1 | 0 | Low-quality | Genome-fragment | 3.13 | AAI-based (medium-confidence) | 0 |
| S8_extracted_k141_174696 | 1347 | No | NA | 2 | 0 | 0 | Low-quality | Genome-fragment | 1.07 | AAI-based (medium-confidence) | 0 |
| S14_extracted_k141_58246 | 2107 | No | NA | 3 | 0 | 3 | Not-determined | Genome-fragment | NA | NA | 0 |
| S14_extracted_k141_6879 | 1603 | No | NA | 2 | 0 | 0 | Not-determined | Genome-fragment | NA | NA | 0 |
| S14_extracted_k141_21932 | 3016 | No | NA | 1 | 0 | 1 | Not-determined | Genome-fragment | NA | NA | 0 |
| S14_extracted_k141_80214 | 3820 | No | NA | 5 | 0 | 2 | Not-determined | Genome-fragment | NA | NA | 0 |
| S14_extracted_k141_80216 | 1172 | No | NA | 2 | 0 | 1 | Not-determined | Genome-fragment | NA | NA | 0 |
| S14_extracted_k141_65153 | 3547 | No | NA | 4 | 0 | 1 | Not-determined | Genome-fragment | NA | NA | 0 |
| S17_extracted_k141_164718 | 1348 | No | NA | 3 | 0 | 0 | Low-quality | Genome-fragment | 2.76 | AAI-based (medium-confidence) | 0 |
| S14_extracted_k141_65173 | 1079 | No | NA | 2 | 0 | 0 | Not-determined | Genome-fragment | NA | NA | 0 |
| S14_extracted_k141_102028 | 1078 | No | NA | 2 | 0 | 0 | Not-determined | Genome-fragment | NA | NA | 0 |
| S17_vs_k141_78481\|\|full | 1348 | No | NA | 4 | 1 | 0 | Low-quality | Genome-fragment | 3.27 | AAI-based (high-confidence) | 0 |
| S14_extracted_k141_117135 | 1300 | No | NA | 1 | 0 | 0 | Not-determined | Genome-fragment | NA | NA | 0 |
| S14_extracted_k141_14179 | 5264 | No | NA | 3 | 0 | 2 | Not-determined | Genome-fragment | NA | NA | 0 |
| S14_extracted_k141_58353 | 2968 | No | NA | 2 | 0 | 1 | Not-determined | Genome-fragment | NA | NA | 0 |
| S9_vs_k141_8420\|\|full | 1348 | No | NA | 2 | 2 | 0 | Low-quality | Genome-fragment | 2.39 | AAI-based (high-confidence) | 0 |
| S14_extracted_k141_28518 | 1041 | No | NA | 2 | 0 | 2 | Not-determined | Genome-fragment | NA | NA | 0 |
| S11_vs_k141_79995\|\|lt2gene | 1348 | No | NA | 2 | 2 | 0 | Low-quality | Genome-fragment | 3.51 | AAI-based (high-confidence) | 0 |
| S1_vs_k141_48655\|\|lt2gene | 1348 | No | NA | 2 | 1 | 0 | Low-quality | Genome-fragment | 3.32 | AAI-based (medium-confidence) | 0 |
| S14_extracted_k141_117147 | 35338 | No | NA | 55 | 16 | 0 | Complete | High-quality | 100 | DTR (high-confidence) | 0 |
| S7_vs_127316\|\|full | 1348 | No | NA | 2 | 0 | 0 | Low-quality | Genome-fragment | 0.77 | AAI-based (medium-confidence) | 0 |
| S16_extracted_45096 | 1348 | No | NA | 2 | 1 | 0 | Low-quality | Genome-fragment | 3.35 | AAI-based (high-confidence) | 0 |
| S14_extracted_k141_72909 | 2652 | No | NA | 1 | 0 | 0 | Not-determined | Genome-fragment | NA | NA | 0 |
| S16_vs_37694\|\|full | 1348 | No | NA | 2 | 1 | 0 | Low-quality | Genome-fragment | 0.8 | AAI-based (high-confidence) | 0 |
| S6_extracted_k141_27934 | 1348 | No | NA | 3 | 0 | 0 | Low-quality | Genome-fragment | 1.74 | AAI-based (high-confidence) | 0 |
| S14_extracted_k141_28591 | 1494 | No | NA | 2 | 0 | 1 | Not-determined | Genome-fragment | NA | NA | 0 |
| S14_extracted_k141_102190 | 1204 | No | NA | 2 | 0 | 1 | Not-determined | Genome-fragment | NA | NA | 0 |
| S14_extracted_k141_7100 | 1141 | No | NA | 1 | 0 | 1 | Not-determined | Genome-fragment | NA | NA | 0 |
| S14_extracted_k141_117150 | 39147 | No | NA | 56 | 17 | 1 | Complete | High-quality | 100 | DTR (high-confidence) | 0 |
| S17_vs_k141_3366\|\|full | 1349 | No | NA | 3 | 0 | 0 | Low-quality | Genome-fragment | 0.97 | AAI-based (medium-confidence) | 0 |
| S14_extracted_k141_72953 | 38044 | No | NA | 53 | 26 | 1 | High-quality | High-quality | 98.41 | AAI-based (high-confidence) | 0 |
| S14_extracted_k141_50595 | 1169 | No | NA | 2 | 0 | 0 | Not-determined | Genome-fragment | NA | NA | 0 |
| S14_extracted_k141_72990 | 1011 | No | NA | 3 | 0 | 0 | Not-determined | Genome-fragment | NA | NA | 0 |
| S14_extracted_k141_58520 | 1186 | No | NA | 1 | 0 | 0 | Not-determined | Genome-fragment | NA | NA | 0 |
| S14_extracted_k141_65431 | 3366 | No | NA | 5 | 0 | 0 | Not-determined | Genome-fragment | NA | NA | 0 |
| S14_extracted_k141_28683 | 1962 | No | NA | 3 | 0 | 2 | Not-determined | Genome-fragment | NA | NA | 0 |
| S14_extracted_k141_73050 | 1158 | No | NA | 2 | 0 | 0 | Not-determined | Genome-fragment | NA | NA | 0 |
| S14_extracted_k141_80473 | 1865 | No | NA | 1 | 0 | 1 | Not-determined | Genome-fragment | NA | NA | 0 |
| S14_extracted_k141_7262 | 5959 | No | NA | 7 | 0 | 3 | Not-determined | Genome-fragment | NA | NA | 0 |
| S14_extracted_k141_7265 | 1478 | No | NA | 2 | 0 | 0 | Not-determined | Genome-fragment | NA | NA | 0 |
| S14_extracted_k141_28740 | 1182 | No | NA | 1 | 0 | 0 | Not-determined | Genome-fragment | NA | NA | 0 |
| S14_extracted_k141_65495 | 1331 | No | NA | 1 | 0 | 0 | Not-determined | Genome-fragment | NA | NA | 0 |
| S14_extracted_k141_17329 | 1349 | No | NA | 1 | 0 | 0 | Low-quality | Genome-fragment | 3.15 | AAI-based (medium-confidence) | 0 |
| S14_extracted_k141_80532 | 2600 | No | NA | 2 | 0 | 2 | Not-determined | Genome-fragment | NA | NA | 0 |
| S14_extracted_k141_107562 | 1349 | No | NA | 3 | 1 | 0 | Low-quality | Genome-fragment | 3.1 | AAI-based (medium-confidence) | 0 |
| S14_vs_k141_104005\|\|full | 1349 | No | NA | 3 | 2 | 0 | Low-quality | Genome-fragment | 3.47 | AAI-based (medium-confidence) | 0 |
| S14_extracted_k141_102427 | 1282 | No | NA | 3 | 0 | 0 | Not-determined | Genome-fragment | NA | NA | 0 |
| S14_extracted_k141_117155 | 38997 | No | NA | 52 | 27 | 0 | Medium-quality | Genome-fragment | 65.14 | AAI-based (medium-confidence) | 0 |
| S14_extracted_k141_50778 | 1051 | No | NA | 2 | 0 | 2 | Not-determined | Genome-fragment | NA | NA | 0 |
| S14_extracted_k141_73196 | 1454 | No | NA | 3 | 0 | 3 | Not-determined | Genome-fragment | NA | NA | 0 |
| S3_vs_k141_120725\|\|full | 1349 | No | NA | 3 | 0 | 0 | Low-quality | Genome-fragment | 2.06 | AAI-based (medium-confidence) | 0 |
| S14_extracted_k141_28910 | 1126 | No | NA | 2 | 0 | 0 | Not-determined | Genome-fragment | NA | NA | 0 |
| S14_extracted_k141_117157 | 59976 | No | NA | 80 | 9 | 0 | Complete | High-quality | 100 | DTR (high-confidence) | 0 |
| S14_extracted_k141_73262 | 3969 | No | NA | 5 | 0 | 3 | Not-determined | Genome-fragment | NA | NA | 0 |
| S14_extracted_k141_28966 | 1397 | No | NA | 2 | 0 | 2 | Not-determined | Genome-fragment | NA | NA | 0 |
| S5_vs_k141_38199\|\|full | 1349 | No | NA | 2 | 1 | 0 | Low-quality | Genome-fragment | 2.88 | AAI-based (high-confidence) | 0 |
| S14_extracted_k141_65676 | 1233 | No | NA | 2 | 0 | 1 | Not-determined | Genome-fragment | NA | NA | 0 |
| S5_vs_k141_145346\|\|full | 1349 | No | NA | 2 | 1 | 0 | Low-quality | Genome-fragment | 0.4 | HMM-based (lower-bound) | 0 |
| S14_extracted_k141_117162 | 36948 | No | NA | 45 | 11 | 1 | Complete | High-quality | 100 | DTR (high-confidence) | 0 |
| S7_vs_69467\|\|lt2gene | 1349 | No | NA | 2 | 2 | 0 | Low-quality | Genome-fragment | 4.34 | AAI-based (high-confidence) | 0 |
| S14_extracted_k141_117164 | 14258 | No | NA | 17 | 6 | 0 | Medium-quality | Genome-fragment | 65.51 | AAI-based (medium-confidence) | 0 |
| S14_extracted_k141_51134 | 33359 | No | NA | 47 | 24 | 1 | Complete | High-quality | 100 | DTR (high-confidence) | 0 |
| S14_extracted_k141_65861 | 3397 | No | NA | 2 | 0 | 2 | Not-determined | Genome-fragment | NA | NA | 0 |
| S16_extracted_10659 | 1349 | No | NA | 3 | 0 | 0 | Low-quality | Genome-fragment | 1.4 | AAI-based (high-confidence) | 0 |
| S14_extracted_k141_117194 | 1393 | No | NA | 2 | 0 | 0 | Not-determined | Genome-fragment | NA | NA | 0 |
| S14_vs_k141_15228\|\|full | 150458 | No | NA | 195 | 13 | 5 | High-quality | High-quality | 97.82 | AAI-based (high-confidence) | 0 |
| S14_vs_k141_88666\|\|full | 36753 | No | NA | 55 | 17 | 1 | High-quality | High-quality | 90.08 | AAI-based (high-confidence) | 0 |
| S14_vs_k141_30439\|\|full | 34044 | No | NA | 53 | 16 | 0 | High-quality | High-quality | 99.24 | AAI-based (medium-confidence) | 0 |
| S14_vs_k141_96286\|\|full | 30986 | No | NA | 46 | 16 | 0 | High-quality | High-quality | 91.93 | AAI-based (high-confidence) | 0 |
| S14_vs_k141_9676\|\|full | 42662 | No | NA | 52 | 12 | 0 | High-quality | High-quality | 100 | AAI-based (high-confidence) | 0 |
| S14_vs_k141_69166\|\|full | 4148 | No | NA | 9 | 0 | 2 | Not-determined | Genome-fragment | NA | NA | 0 |
| S14_vs_k141_3251\|\|full | 34146 | No | NA | 58 | 14 | 2 | High-quality | High-quality | 100 | AAI-based (high-confidence) | 0 |
| S14_vs_k141_26320\|\|full | 172200 | No | NA | 229 | 33 | 6 | High-quality | High-quality | 100 | AAI-based (high-confidence) | 0 |
| S14_vs_k141_4887\|\|full | 40549 | No | NA | 49 | 7 | 2 | High-quality | High-quality | 97.58 | AAI-based (high-confidence) | 0 |
| S14_vs_k141_42401\|\|full | 2574 | No | NA | 6 | 0 | 0 | Not-determined | Genome-fragment | NA | NA | 0 |
| S14_vs_k141_94397\|\|full | 5962 | No | NA | 9 | 0 | 0 | Not-determined | Genome-fragment | NA | NA | 0 |
| S16_vs_129987\|\|lt2gene | 1349 | No | NA | 2 | 1 | 0 | Low-quality | Genome-fragment | 2.52 | AAI-based (high-confidence) | 0 |
| S14_vs_k141_117153\|\|full | 37231 | No | NA | 50 | 13 | 0 | High-quality | High-quality | 100 | AAI-based (high-confidence) | 0 |
| S10_extracted_2042 | 1349 | No | NA | 4 | 0 | 0 | Low-quality | Genome-fragment | 4.09 | AAI-based (high-confidence) | 0 |
| S14_vs_k141_117158\|\|full | 40845 | No | NA | 70 | 23 | 2 | High-quality | High-quality | 93.32 | AAI-based (high-confidence) | 0 |
| S14_vs_k141_117160\|\|full | 34792 | No | NA | 64 | 24 | 1 | Medium-quality | Genome-fragment | 89.27 | AAI-based (medium-confidence) | 0 |
| S14_vs_k141_117161\|\|full | 35177 | No | NA | 50 | 18 | 0 | High-quality | High-quality | 99.84 | AAI-based (high-confidence) | 0 |
| S14_vs_k141_117163\|\|full | 14322 | No | NA | 26 | 0 | 0 | Not-determined | Genome-fragment | NA | NA | 0 |
| S14_vs_k141_117165\|\|full | 40247 | No | NA | 65 | 8 | 0 | High-quality | High-quality | 100 | AAI-based (high-confidence) | 0 |
| S14_vs_k141_117166\|\|full | 38784 | No | NA | 68 | 18 | 2 | High-quality | High-quality | 94.06 | AAI-based (high-confidence) | 0 |
| S14_vs_k141_117171\|\|full | 2414 | No | NA | 3 | 0 | 0 | Not-determined | Genome-fragment | NA | NA | 0 |
| S14_vs_k141_7331\|\|full | 2679 | No | NA | 2 | 0 | 1 | Not-determined | Genome-fragment | NA | NA | 0 |
| S10_extracted_9812 | 1349 | No | NA | 3 | 0 | 0 | Low-quality | Genome-fragment | 1.82 | AAI-based (high-confidence) | 0 |
| S14_vs_k141_29315\|\|full | 1729 | No | NA | 3 | 0 | 2 | Not-determined | Genome-fragment | NA | NA | 0 |
| S14_vs_k141_22006\|\|full | 1509 | No | NA | 2 | 0 | 2 | Not-determined | Genome-fragment | NA | NA | 0 |
| S14_vs_k141_43992\|\|full | 3614 | No | NA | 5 | 0 | 1 | Not-determined | Genome-fragment | NA | NA | 0 |
| S14_vs_k141_65985\|\|full | 2717 | No | NA | 2 | 0 | 2 | Not-determined | Genome-fragment | NA | NA | 0 |
| S14_vs_k141_44020\|\|full | 2139 | No | NA | 3 | 0 | 1 | Not-determined | Genome-fragment | NA | NA | 0 |
| S14_vs_k141_95278\|\|full | 1316 | No | NA | 3 | 0 | 0 | Not-determined | Genome-fragment | NA | NA | 0 |
| S12_extracted_k141_35360 | 1349 | No | NA | 1 | 0 | 0 | Low-quality | Genome-fragment | 2.55 | AAI-based (high-confidence) | 0 |
| S8_extracted_k141_14313 | 1349 | No | NA | 1 | 1 | 0 | Low-quality | Genome-fragment | 1.97 | HMM-based (lower-bound) | 0 |
| S14_vs_k141_29356\|\|full | 2981 | No | NA | 3 | 0 | 2 | Not-determined | Genome-fragment | NA | NA | 0 |
| S14_vs_k141_7403\|\|full | 1459 | No | NA | 3 | 0 | 1 | Not-determined | Genome-fragment | NA | NA | 0 |
| S14_vs_k141_87956\|\|full | 2397 | No | NA | 2 | 0 | 1 | Not-determined | Genome-fragment | NA | NA | 0 |
| S14_vs_k141_73301\|\|full | 29973 | No | NA | 46 | 17 | 0 | Medium-quality | Genome-fragment | 71.49 | AAI-based (high-confidence) | 0 |
| S14_vs_k141_36754\|\|full | 1784 | No | NA | 3 | 0 | 1 | Not-determined | Genome-fragment | NA | NA | 0 |
| S14_vs_k141_29398\|\|full | 1185 | No | NA | 2 | 0 | 1 | Not-determined | Genome-fragment | NA | NA | 0 |
| S11_extracted_k141_52622 | 1350 | No | NA | 3 | 0 | 1 | Low-quality | Genome-fragment | 0.41 | AAI-based (medium-confidence) | 0 |
| S14_vs_k141_14737\|\|full | 35673 | No | NA | 51 | 16 | 1 | High-quality | High-quality | 98.83 | AAI-based (high-confidence) | 0 |
| S11_vs_k141_43808\|\|full | 1350 | No | NA | 5 | 2 | 0 | Low-quality | Genome-fragment | 3.64 | AAI-based (medium-confidence) | 0 |
| S7_vs_99743\|\|full | 1350 | No | NA | 3 | 0 | 0 | Low-quality | Genome-fragment | 2.75 | AAI-based (high-confidence) | 0 |
| S16_vs_119454\|\|full | 1350 | No | NA | 2 | 0 | 0 | Low-quality | Genome-fragment | 2.73 | AAI-based (medium-confidence) | 0 |
| S16_vs_121129\|\|full | 1350 | No | NA | 2 | 0 | 2 | Low-quality | Genome-fragment | 0.38 | AAI-based (medium-confidence) | 0 |
| S14_vs_k141_36807\|\|full | 3398 | No | NA | 5 | 0 | 5 | Not-determined | Genome-fragment | NA | NA | 0 |
| S10_vs_11356\|\|full | 1350 | No | NA | 3 | 0 | 1 | Low-quality | Genome-fragment | 1.83 | AAI-based (medium-confidence) | 0 |
| S14_vs_k141_7465\|\|full | 2883 | No | NA | 3 | 0 | 0 | Not-determined | Genome-fragment | NA | NA | 0 |
| S12_vs_k141_77072\|\|full | 1350 | No | NA | 3 | 0 | 0 | Low-quality | Genome-fragment | 2.64 | AAI-based (medium-confidence) | 0 |
| S4_extracted_7875 | 1350 | No | NA | 2 | 1 | 0 | Low-quality | Genome-fragment | 4.45 | AAI-based (high-confidence) | 0 |
| S11_vs_k141_81040\|\|lt2gene | 1351 | No | NA | 1 | 1 | 0 | Low-quality | Genome-fragment | 3.56 | AAI-based (high-confidence) | 0 |
| S14_vs_k141_183\|\|full | 11611 | No | NA | 15 | 0 | 4 | Not-determined | Genome-fragment | NA | NA | 0 |
| S14_vs_k141_95415\|\|full | 1125 | No | NA | 3 | 0 | 0 | Not-determined | Genome-fragment | NA | NA | 0 |
| S14_vs_k141_102698\|\|full | 22313 | No | NA | 36 | 15 | 0 | Medium-quality | Genome-fragment | 66.53 | AAI-based (high-confidence) | 0 |
| S13_vs_69345\|\|full | 1351 | No | NA | 3 | 1 | 0 | Low-quality | Genome-fragment | 2.75 | AAI-based (medium-confidence) | 0 |
| S14_vs_k141_51463\|\|full | 3038 | No | NA | 3 | 0 | 2 | Not-determined | Genome-fragment | NA | NA | 0 |
| S14_extracted_k141_113114 | 1351 | No | NA | 1 | 0 | 1 | Low-quality | Genome-fragment | 0.23 | AAI-based (high-confidence) | 0 |
| S14_vs_k141_58815\|\|full | 11138 | No | NA | 7 | 0 | 3 | Not-determined | Genome-fragment | NA | NA | 0 |
| S14_vs_k141_102726\|\|full | 5385 | No | NA | 7 | 0 | 3 | Not-determined | Genome-fragment | NA | NA | 0 |
| S14_vs_k141_246\|\|full | 1072 | No | NA | 3 | 0 | 0 | Not-determined | Genome-fragment | NA | NA | 0 |
| S14_vs_k141_47251\|\|full | 1351 | No | NA | 2 | 0 | 1 | Low-quality | Genome-fragment | 3.5 | AAI-based (medium-confidence) | 0 |
| S3_vs_k141_49962\|\|full | 1351 | No | NA | 2 | 1 | 0 | Low-quality | Genome-fragment | 3.37 | AAI-based (high-confidence) | 0 |
| S7_vs_132893\|\|full | 1351 | No | NA | 3 | 1 | 0 | Low-quality | Genome-fragment | 3.33 | AAI-based (medium-confidence) | 0 |
| S14_vs_k141_58870\|\|full | 4165 | No | NA | 5 | 0 | 1 | Not-determined | Genome-fragment | NA | NA | 0 |
| S12_extracted_k141_38943 | 1351 | No | NA | 2 | 0 | 1 | Low-quality | Genome-fragment | 3.25 | AAI-based (medium-confidence) | 0 |
| S14_vs_k141_80799\|\|full | 2854 | No | NA | 4 | 0 | 2 | Not-determined | Genome-fragment | NA | NA | 0 |
| S14_vs_k141_51515\|\|full | 1891 | No | NA | 3 | 0 | 1 | Not-determined | Genome-fragment | NA | NA | 0 |
| S12_vs_k141_138448\|\|full | 1351 | No | NA | 3 | 1 | 0 | Low-quality | Genome-fragment | 3.24 | AAI-based (high-confidence) | 0 |
| S14_vs_k141_80815\|\|full | 2192 | No | NA | 4 | 0 | 1 | Not-determined | Genome-fragment | NA | NA | 0 |
| S14_vs_k141_7605\|\|full | 3759 | No | NA | 5 | 0 | 1 | Not-determined | Genome-fragment | NA | NA | 0 |
| S2_extracted_k141_8635 | 1351 | No | NA | 1 | 1 | 0 | Low-quality | Genome-fragment | 3.16 | HMM-based (lower-bound) | 0 |

| | | | | | | | | | | | | |
|---|---|---|---|---|---|---|---|---|---|---|---|---|
| S17_vs_k141_32974\|\|full | 1352 | No | NA | 3 | 0 | 0 | Low-quality | Genome-fragment | | 2.29 | AAI-based (high-confidence) | 0 |
| S14_vs_k141_102744\|\|full | 3274 | No | NA | 4 | 0 | 0 | Not-determined | Genome-fragment | NA | | NA | 0 |
| S14_extracted_k141_2227 | 1352 | No | NA | 6 | 5 | 0 | Low-quality | Genome-fragment | | 4.47 | AAI-based (high-confidence) | 0 |
| S14_extracted_k141_71607 | 1352 | No | NA | 3 | 0 | 0 | Low-quality | Genome-fragment | | 3.14 | AAI-based (medium-confidence) | 0 |
| S14_vs_k141_80831\|\|full | 9744 | No | NA | 11 | 0 | 8 | Not-determined | Genome-fragment | NA | | NA | 0 |
| S14_vs_k141_10901\|\|full | 1352 | No | NA | 4 | 0 | 0 | Low-quality | Genome-fragment | | 2.16 | AAI-based (medium-confidence) | 0 |
| S14_vs_k141_26487\|\|full | 1352 | No | NA | 3 | 0 | 0 | Low-quality | Genome-fragment | | 0.77 | AAI-based (high-confidence) | 0 |
| S14_vs_k141_93590\|\|full | 1352 | No | NA | 3 | 0 | 0 | Low-quality | Genome-fragment | | 1.74 | AAI-based (medium-confidence) | 0 |
| S14_vs_k141_88156\|\|full | 1778 | No | NA | 3 | 0 | 0 | Not-determined | Genome-fragment | NA | | NA | 0 |
| S14_vs_k141_29595\|\|full | 3392 | No | NA | 7 | 0 | 4 | Not-determined | Genome-fragment | NA | | NA | 0 |
| S14_vs_k141_7635\|\|full | 7248 | No | NA | 7 | 0 | 1 | Not-determined | Genome-fragment | NA | | NA | 0 |
| S14_vs_k141_102804\|\|full | 1730 | No | NA | 3 | 0 | 0 | Not-determined | Genome-fragment | NA | | NA | 0 |
| S3_extracted_k141_108191 | 1352 | No | NA | 2 | 2 | 0 | Low-quality | Genome-fragment | | 2.63 | AAI-based (high-confidence) | 0 |
| S3_vs_k141_115556\|\|full | 1352 | No | NA | 4 | 1 | 0 | Low-quality | Genome-fragment | | 2.52 | AAI-based (high-confidence) | 0 |
| S3_vs_k141_103361\|\|full | 1352 | No | NA | 3 | 1 | 0 | Low-quality | Genome-fragment | | 0.75 | HMM-based (lower-bound) | 0 |
| S7_vs_36722\|\|full | 1352 | No | NA | 3 | 0 | 2 | Low-quality | Genome-fragment | | 1.87 | AAI-based (medium-confidence) | 0 |
| S14_vs_k141_66139\|\|full | 198450 | Yes | 56174 | 202 | 26 | 89 | Medium-quality | Genome-fragment | | 82.23 | HMM-based (lower-bound) | 71.69 |
| S14_vs_k141_22271\|\|full | 4976 | No | NA | 4 | 0 | 3 | Not-determined | Genome-fragment | NA | | NA | 0 |
| S7_vs_83683\|\|full | 1352 | No | NA | 4 | 1 | 0 | Low-quality | Genome-fragment | | 2.16 | HMM-based (lower-bound) | 0 |
| S14_vs_k141_7683\|\|full | 6337 | No | NA | 5 | 0 | 0 | Not-determined | Genome-fragment | NA | | NA | 0 |
| S8_extracted_k141_183956 | 1352 | No | NA | 1 | 0 | 0 | Low-quality | Genome-fragment | | 1.6 | AAI-based (high-confidence) | 0 |
| S9_extracted_k141_41983 | 1353 | No | NA | 1 | 1 | 0 | Low-quality | Genome-fragment | | 1.54 | HMM-based (lower-bound) | 0 |
| S11_vs_k141_61010\|\|full | 1353 | No | NA | 4 | 0 | 0 | Low-quality | Genome-fragment | | 1.79 | AAI-based (medium-confidence) | 0 |
| S14_vs_k141_51657\|\|full | 1692 | No | NA | 3 | 0 | 2 | Not-determined | Genome-fragment | NA | | NA | 0 |
| S14_vs_k141_51676\|\|full | 3457 | No | NA | 2 | 0 | 1 | Not-determined | Genome-fragment | NA | | NA | 0 |
| S14_vs_k141_80917\|\|full | 1866 | No | NA | 3 | 0 | 2 | Not-determined | Genome-fragment | NA | | NA | 0 |
| S14_vs_k141_14985\|\|full | 7969 | No | NA | 5 | 0 | 5 | Not-determined | Genome-fragment | NA | | NA | 0 |
| S14_vs_k141_491\|\|full | 1685 | No | NA | 6 | 0 | 0 | Not-determined | Genome-fragment | NA | | NA | 0 |
| S14_vs_k141_44218\|\|full | 1578 | No | NA | 2 | 0 | 1 | Not-determined | Genome-fragment | NA | | NA | 0 |
| S14_vs_k141_102917\|\|full | 5647 | No | NA | 4 | 0 | 2 | Not-determined | Genome-fragment | NA | | NA | 0 |
| S11_vs_k141_15578\|\|full | 1353 | No | NA | 5 | 1 | 0 | Low-quality | Genome-fragment | | 3.38 | AAI-based (high-confidence) | 0 |
| S14_vs_k141_88282\|\|full | 1914 | No | NA | 2 | 0 | 0 | Not-determined | Genome-fragment | NA | | NA | 0 |
| S14_vs_k141_547\|\|full | 1476 | No | NA | 2 | 0 | 0 | Not-determined | Genome-fragment | NA | | NA | 0 |
| S14_vs_k141_66208\|\|full | 67174 | Yes | 48001 | 80 | 14 | 10 | Complete | High-quality | | 100 | Provirus (high-confidence) | 28.54 |
| S13_extracted_28117 | 1353 | No | NA | 1 | 0 | 1 | Low-quality | Genome-fragment | | 2.26 | AAI-based (high-confidence) | 0 |
| S13_extracted_39161 | 1353 | No | NA | 2 | 0 | 0 | Low-quality | Genome-fragment | | 1.4 | AAI-based (high-confidence) | 0 |
| S1_vs_k141_126895\|\|full | 1353 | No | NA | 3 | 0 | 0 | Low-quality | Genome-fragment | | 2.59 | AAI-based (medium-confidence) | 0 |
| S14_vs_k141_29751\|\|full | 1144 | No | NA | 2 | 0 | 2 | Not-determined | Genome-fragment | NA | | NA | 0 |
| S1_vs_k141_152456\|\|full | 1353 | No | NA | 2 | 1 | 0 | Low-quality | Genome-fragment | | 2.26 | AAI-based (high-confidence) | 0 |
| S1_vs_k141_41883\|\|lt2gene | 1353 | No | NA | 1 | 0 | 0 | Low-quality | Genome-fragment | | 2.31 | AAI-based (medium-confidence) | 0 |
| S3_extracted_k141_96516 | 1353 | No | NA | 2 | 0 | 1 | Low-quality | Genome-fragment | | 2.66 | AAI-based (medium-confidence) | 0 |
| S3_extracted_k141_82331 | 1353 | No | NA | 5 | 0 | 0 | Low-quality | Genome-fragment | | 2.02 | AAI-based (high-confidence) | 0 |
| S14_vs_k141_44323\|\|full | 3074 | No | NA | 3 | 0 | 1 | Not-determined | Genome-fragment | NA | | NA | 0 |
| S3_vs_k141_111919\|\|full | 1353 | No | NA | 7 | 0 | 0 | Low-quality | Genome-fragment | | 2.25 | AAI-based (high-confidence) | 0 |
| S14_vs_k141_103003\|\|full | 8979 | No | NA | 8 | 0 | 6 | Not-determined | Genome-fragment | NA | | NA | 0 |
| S5_extracted_k141_35285 | 1353 | No | NA | 3 | 0 | 2 | Low-quality | Genome-fragment | | 0.65 | AAI-based (medium-confidence) | 0 |
| S14_vs_k141_29805\|\|full | 1440 | No | NA | 3 | 0 | 0 | Not-determined | Genome-fragment | NA | | NA | 0 |
| S5_vs_k141_78886\|\|full | 1353 | No | NA | 5 | 0 | 0 | Low-quality | Genome-fragment | | 3.08 | AAI-based (medium-confidence) | 0 |
| S10_extracted_7419 | 1353 | No | NA | 2 | 1 | 0 | Low-quality | Genome-fragment | | 1.25 | HMM-based (lower-bound) | 0 |
| S10_vs_1307\|\|full | 1353 | No | NA | 4 | 0 | 0 | Low-quality | Genome-fragment | | 2.49 | AAI-based (high-confidence) | 0 |
| S10_vs_23387\|\|full | 1353 | No | NA | 4 | 0 | 0 | Low-quality | Genome-fragment | | 2.68 | AAI-based (high-confidence) | 0 |
| S14_vs_k141_29833\|\|full | 12367 | No | NA | 8 | 0 | 3 | Not-determined | Genome-fragment | NA | | NA | 0 |
| S14_vs_k141_37199\|\|full | 1114 | No | NA | 3 | 0 | 0 | Not-determined | Genome-fragment | NA | | NA | 0 |
| S12_vs_k141_122661\|\|full | 1353 | No | NA | 3 | 0 | 2 | Low-quality | Genome-fragment | | 3.13 | AAI-based (high-confidence) | 0 |
| S8_extracted_k141_111370 | 1353 | No | NA | 3 | 1 | 0 | Low-quality | Genome-fragment | | 2.74 | AAI-based (high-confidence) | 0 |
| S14_vs_k141_632\|\|full | 37447 | No | NA | 54 | 33 | 1 | Medium-quality | Genome-fragment | | 83.64 | AAI-based (high-confidence) | 0 |
| S14_vs_k141_110290\|\|full | 33612 | No | NA | 57 | 22 | 1 | High-quality | High-quality | | 99.36 | AAI-based (high-confidence) | 0 |
| S14_vs_k141_7771\|\|full | 1350 | No | NA | 4 | 0 | 0 | Not-determined | Genome-fragment | NA | | NA | 0 |
| S17_vs_k141_54149\|\|full | 1354 | No | NA | 3 | 1 | 0 | Low-quality | Genome-fragment | | 3.17 | AAI-based (high-confidence) | 0 |
| S14_vs_k141_95835\|\|full | 3057 | No | NA | 3 | 0 | 2 | Not-determined | Genome-fragment | NA | | NA | 0 |
| S14_extracted_k141_92119 | 1354 | No | NA | 1 | 1 | 0 | Low-quality | Genome-fragment | | 4.37 | AAI-based (high-confidence) | 0 |
| S16_extracted_163388 | 1354 | No | NA | 2 | 0 | 0 | Low-quality | Genome-fragment | | 2.23 | AAI-based (high-confidence) | 0 |
| S14_vs_k141_88409\|\|full | 6371 | No | NA | 4 | 0 | 2 | Not-determined | Genome-fragment | NA | | NA | 0 |
| S8_vs_k141_171538\|\|full | 1354 | No | NA | 4 | 3 | 0 | Low-quality | Genome-fragment | | 2.43 | HMM-based (lower-bound) | 0 |
| S14_vs_k141_7808\|\|full | 1252 | No | NA | 2 | 0 | 1 | Not-determined | Genome-fragment | NA | | NA | 0 |
| S17_extracted_k141_75034 | 1355 | No | NA | 2 | 0 | 1 | Low-quality | Genome-fragment | | 0.88 | AAI-based (high-confidence) | 0 |
| S14_vs_k141_44383\|\|full | 15296 | No | NA | 9 | 0 | 5 | Not-determined | Genome-fragment | NA | | NA | 0 |
| S17_extracted_k141_27867 | 1355 | No | NA | 2 | 0 | 0 | Low-quality | Genome-fragment | | 4.19 | AAI-based (high-confidence) | 0 |
| S9_vs_k141_53017\|\|full | 1355 | No | NA | 3 | 0 | 0 | Low-quality | Genome-fragment | | 3.38 | AAI-based (high-confidence) | 0 |
| S11_vs_k141_77053\|\|full | 1355 | No | NA | 5 | 1 | 0 | Low-quality | Genome-fragment | | 3.99 | AAI-based (medium-confidence) | 0 |
| S14_vs_k141_44424\|\|full | 1599 | No | NA | 3 | 0 | 2 | Not-determined | Genome-fragment | NA | | NA | 0 |
| S14_extracted_k141_81858 | 1355 | No | NA | 4 | 0 | 0 | Low-quality | Genome-fragment | | 3.01 | AAI-based (high-confidence) | 0 |
| S14_vs_k141_88484\|\|full | 2285 | No | NA | 4 | 0 | 1 | Not-determined | Genome-fragment | NA | | NA | 0 |
| S14_extracted_k141_77655 | 1355 | No | NA | 4 | 4 | 0 | Low-quality | Genome-fragment | | 4.47 | AAI-based (high-confidence) | 0 |
| S3_vs_k141_57587\|\|full | 1355 | No | NA | 3 | 0 | 0 | Low-quality | Genome-fragment | | 3.84 | AAI-based (medium-confidence) | 0 |
| S5_vs_k141_138674\|\|lt2gene | 1355 | No | NA | 1 | 1 | 0 | Low-quality | Genome-fragment | | 2.35 | HMM-based (lower-bound) | 0 |
| S7_vs_70866\|\|lt2gene | 1355 | No | NA | 2 | 2 | 0 | Low-quality | Genome-fragment | | 3.68 | AAI-based (high-confidence) | 0 |
| S16_vs_54005\|\|full | 1355 | No | NA | 4 | 0 | 0 | Low-quality | Genome-fragment | | 1.87 | AAI-based (high-confidence) | 0 |
| S14_vs_k141_51883\|\|full | 2205 | No | NA | 4 | 0 | 0 | Not-determined | Genome-fragment | NA | | NA | 0 |
| S12_extracted_k141_178625 | 1355 | No | NA | 3 | 0 | 0 | Low-quality | Genome-fragment | | 3.29 | AAI-based (high-confidence) | 0 |
| S12_vs_k141_167253\|\|full | 1355 | No | NA | 5 | 0 | 0 | Low-quality | Genome-fragment | | 2.67 | AAI-based (high-confidence) | 0 |
| S4_extracted_22209 | 1355 | No | NA | 2 | 1 | 0 | Low-quality | Genome-fragment | | 2.98 | AAI-based (high-confidence) | 0 |
| S8_vs_k141_144063\|\|full | 1355 | No | NA | 2 | 0 | 0 | Low-quality | Genome-fragment | | 0.75 | AAI-based (medium-confidence) | 0 |
| S8_vs_k141_50777\|\|full | 1355 | No | NA | 2 | 0 | 0 | Low-quality | Genome-fragment | | 1.93 | AAI-based (medium-confidence) | 0 |
| S17_vs_k141_90330\|\|full | 1356 | No | NA | 4 | 0 | 0 | Low-quality | Genome-fragment | | 3.02 | AAI-based (high-confidence) | 0 |
| S17_vs_k141_6827\|\|full | 1356 | No | NA | 3 | 0 | 0 | Low-quality | Genome-fragment | | 2.21 | AAI-based (medium-confidence) | 0 |
| S14_vs_k141_110424\|\|full | 2404 | No | NA | 3 | 0 | 1 | Not-determined | Genome-fragment | NA | | NA | 0 |
| S11_extracted_k141_67110 | 1356 | No | NA | 3 | 0 | 0 | Low-quality | Genome-fragment | | 2.14 | AAI-based (medium-confidence) | 0 |
| S11_vs_k141_93905\|\|full | 1356 | No | NA | 3 | 0 | 0 | Low-quality | Genome-fragment | | 2.26 | AAI-based (high-confidence) | 0 |
| S14_vs_k141_59292\|\|full | 1957 | No | NA | 3 | 0 | 3 | Not-determined | Genome-fragment | NA | | NA | 0 |
| S11_vs_k141_97178\|\|full | 1356 | No | NA | 4 | 0 | 0 | Low-quality | Genome-fragment | | 2.37 | AAI-based (high-confidence) | 0 |
| S14_vs_k141_66412\|\|full | 1301 | No | NA | 3 | 0 | 1 | Not-determined | Genome-fragment | NA | | NA | 0 |
| S3_vs_k141_93773\|\|full | 1356 | No | NA | 2 | 1 | 0 | Low-quality | Genome-fragment | | 3.1 | AAI-based (medium-confidence) | 0 |
| S14_vs_k141_59300\|\|full | 1107 | No | NA | 3 | 0 | 0 | Not-determined | Genome-fragment | NA | | NA | 0 |
| S16_vs_123110\|\|lt2gene | 1356 | No | NA | 1 | 1 | 0 | Low-quality | Genome-fragment | | 3.8 | AAI-based (high-confidence) | 0 |
| S14_vs_k141_818\|\|full | 1657 | No | NA | 3 | 0 | 2 | Not-determined | Genome-fragment | NA | | NA | 0 |
| S14_vs_k141_110493\|\|full | 3090 | No | NA | 4 | 0 | 0 | Not-determined | Genome-fragment | NA | | NA | 0 |
| S10_extracted_5550 | 1356 | No | NA | 2 | 0 | 0 | Low-quality | Genome-fragment | | 1.26 | AAI-based (high-confidence) | 0 |
| S14_vs_k141_88639\|\|full | 1549 | No | NA | 3 | 0 | 2 | Not-determined | Genome-fragment | NA | | NA | 0 |
| S14_vs_k141_73784\|\|full | 2348 | No | NA | 3 | 0 | 2 | Not-determined | Genome-fragment | NA | | NA | 0 |
| S14_vs_k141_110522\|\|full | 2727 | No | NA | 5 | 0 | 2 | Not-determined | Genome-fragment | NA | | NA | 0 |
| S10_vs_24272\|\|full | 1356 | No | NA | 5 | 1 | 0 | Low-quality | Genome-fragment | | 2.46 | AAI-based (high-confidence) | 0 |
| S14_vs_k141_30032\|\|full | 1717 | No | NA | 3 | 0 | 0 | Not-determined | Genome-fragment | NA | | NA | 0 |
| S14_vs_k141_52058\|\|full | 1146 | No | NA | 2 | 0 | 1 | Not-determined | Genome-fragment | NA | | NA | 0 |
| S14_vs_k141_15237\|\|full | 3935 | No | NA | 5 | 0 | 2 | Not-determined | Genome-fragment | NA | | NA | 0 |
| S14_vs_k141_44531\|\|full | 18080 | No | NA | 15 | 0 | 12 | Not-determined | Genome-fragment | NA | | NA | 0 |
| S12_extracted_k141_50389 | 1356 | No | NA | 3 | 0 | 2 | Low-quality | Genome-fragment | | 0.88 | AAI-based (medium-confidence) | 0 |
| S8_vs_k141_109433\|\|full | 1356 | No | NA | 3 | 0 | 0 | Low-quality | Genome-fragment | | 3.65 | AAI-based (medium-confidence) | 0 |
| S9_vs_k141_6602\|\|full | 1357 | No | NA | 5 | 1 | 0 | Low-quality | Genome-fragment | | 0.98 | AAI-based (medium-confidence) | 0 |
| S11_extracted_k141_3220 | 1357 | No | NA | 3 | 2 | 0 | Low-quality | Genome-fragment | | 1.14 | AAI-based (high-confidence) | 0 |
| S14_vs_k141_8024\|\|full | 4821 | No | NA | 2 | 0 | 0 | Not-determined | Genome-fragment | NA | | NA | 0 |
| S14_vs_k141_52133\|\|full | 2018 | No | NA | 3 | 0 | 1 | Not-determined | Genome-fragment | NA | | NA | 0 |
| S11_vs_k141_28871\|\|lt2gene | 1357 | No | NA | 1 | 1 | 0 | Low-quality | Genome-fragment | | 2.28 | HMM-based (lower-bound) | 0 |
| S14_vs_k141_88695\|\|full | 2964 | No | NA | 4 | 0 | 3 | Not-determined | Genome-fragment | NA | | NA | 0 |
| S14_vs_k141_59347\|\|full | 36660 | No | NA | 56 | 15 | 1 | High-quality | High-quality | | 95.27 | AAI-based (medium-confidence) | 0 |
| S14_extracted_k141_22878 | 1357 | No | NA | 2 | 2 | 0 | Low-quality | Genome-fragment | | 4.48 | AAI-based (high-confidence) | 0 |
| S14_vs_k141_89450\|\|full | 1357 | No | NA | 4 | 2 | 0 | Low-quality | Genome-fragment | | 3.22 | AAI-based (medium-confidence) | 0 |
| S14_vs_k141_73857\|\|full | 1511 | No | NA | 2 | 0 | 0 | Not-determined | Genome-fragment | NA | | NA | 0 |
| S14_vs_k141_73862\|\|full | 1171 | No | NA | 3 | 0 | 0 | Not-determined | Genome-fragment | NA | | NA | 0 |
| S14_vs_k141_37522\|\|full | 3825 | No | NA | 4 | 0 | 1 | Not-determined | Genome-fragment | NA | | NA | 0 |
| S14_vs_k141_30108\|\|full | 28701 | No | NA | 68 | 9 | 1 | Medium-quality | Genome-fragment | | 66.39 | AAI-based (high-confidence) | 0 |
| S1_extracted_k141_55607 | 1357 | No | NA | 2 | 1 | 0 | Low-quality | Genome-fragment | | 3.43 | AAI-based (high-confidence) | 0 |
| S3_vs_k141_76999\|\|lt2gene | 1357 | No | NA | 2 | 1 | 0 | Low-quality | Genome-fragment | | 3.41 | AAI-based (high-confidence) | 0 |
| S14_vs_k141_15354\|\|full | 2887 | No | NA | 3 | 0 | 0 | Not-determined | Genome-fragment | NA | | NA | 0 |
| S14_vs_k141_52160\|\|full | 28351 | No | NA | 19 | 0 | 5 | Not-determined | Genome-fragment | NA | | NA | 0 |
| S14_vs_k141_52177\|\|full | 6451 | No | NA | 9 | 0 | 2 | Not-determined | Genome-fragment | NA | | NA | 0 |
| S16_vs_924\|\|full | 1357 | No | NA | 4 | 0 | 0 | Low-quality | Genome-fragment | | 2.16 | AAI-based (high-confidence) | 0 |
| S12_vs_k141_4461\|\|full | 1357 | No | NA | 3 | 0 | 1 | Low-quality | Genome-fragment | | 0.6 | AAI-based (medium-confidence) | 0 |
| S12_vs_k141_172835\|\|full | 1357 | No | NA | 2 | 1 | 0 | Low-quality | Genome-fragment | | 2 | HMM-based (lower-bound) | 0 |
| S15_extracted_57319 | 1357 | No | NA | 1 | 0 | 0 | Low-quality | Genome-fragment | | 3.27 | AAI-based (medium-confidence) | 0 |
| S8_extracted_k141_165629 | 1357 | No | NA | 2 | 1 | 0 | Low-quality | Genome-fragment | | 2.55 | AAI-based (high-confidence) | 0 |
| S14_vs_k141_8104\|\|full | 1164 | No | NA | 2 | 0 | 1 | Not-determined | Genome-fragment | NA | | NA | 0 |
| S14_vs_k141_110634\|\|full | 4559 | No | NA | 5 | 0 | 3 | Not-determined | Genome-fragment | NA | | NA | 0 |
| S8_vs_k141_144151\|\|full | 1357 | No | NA | 3 | 0 | 2 | Low-quality | Genome-fragment | | 0.72 | AAI-based (medium-confidence) | 0 |
| S14_vs_k141_88767\|\|full | 1752 | No | NA | 2 | 0 | 2 | Not-determined | Genome-fragment | NA | | NA | 0 |
| S14_vs_k141_30152\|\|full | 12244 | No | NA | 14 | 0 | 1 | Not-determined | Genome-fragment | NA | | NA | 0 |
| S17_vs_k141_93825\|\|full | 1358 | No | NA | 5 | 0 | 0 | Low-quality | Genome-fragment | | 2.98 | AAI-based (medium-confidence) | 0 |
| S9_extracted_k141_87125 | 1358 | No | NA | 4 | 1 | 0 | Low-quality | Genome-fragment | | 1.46 | AAI-based (high-confidence) | 0 |
| S11_vs_k141_11234\|\|full | 1358 | No | NA | 2 | 1 | 0 | Low-quality | Genome-fragment | | 2.01 | HMM-based (lower-bound) | 0 |
| S14_vs_k141_103381\|\|full | 1246 | No | NA | 3 | 0 | 0 | Not-determined | Genome-fragment | NA | | NA | 0 |
| S14_vs_k141_37607\|\|full | 14720 | No | NA | 16 | 0 | 5 | Not-determined | Genome-fragment | NA | | NA | 0 |
| S3_extracted_k141_112240 | 1358 | No | NA | 2 | 0 | 1 | Low-quality | Genome-fragment | | 2.14 | AAI-based (medium-confidence) | 0 |
| S14_vs_k141_15427\|\|full | 15353 | No | NA | 24 | 0 | 0 | Not-determined | Genome-fragment | NA | | NA | 0 |
| S5_vs_k141_122580\|\|full | 1358 | No | NA | 4 | 0 | 0 | Low-quality | Genome-fragment | | 3.07 | AAI-based (medium-confidence) | 0 |
| S14_vs_k141_44661\|\|full | 5538 | No | NA | 4 | 0 | 2 | Not-determined | Genome-fragment | NA | | NA | 0 |
| S5_vs_k141_44964\|\|full | 1358 | No | NA | 3 | 2 | 0 | Low-quality | Genome-fragment | | 1.89 | HMM-based (lower-bound) | 0 |
| S5_vs_k141_107597\|\|full | 1358 | No | NA | 3 | 0 | 2 | Low-quality | Genome-fragment | | 1.84 | AAI-based (high-confidence) | 0 |
| S5_vs_k141_71837\|\|lt2gene | 1358 | No | NA | 2 | 1 | 0 | Low-quality | Genome-fragment | | 3.7 | AAI-based (high-confidence) | 0 |
| S14_vs_k141_15457\|\|full | 7828 | No | NA | 7 | 0 | 6 | Not-determined | Genome-fragment | NA | | NA | 0 |
| S14_vs_k141_1001\|\|full | 2856 | No | NA | 5 | 0 | 1 | Not-determined | Genome-fragment | NA | | NA | 0 |
| S16_vs_120819\|\|lt2gene | 1358 | No | NA | 2 | 1 | 0 | Low-quality | Genome-fragment | | 2.67 | AAI-based (high-confidence) | 0 |
| S14_vs_k141_110714\|\|full | 3424 | No | NA | 5 | 0 | 0 | Not-determined | Genome-fragment | NA | | NA | 0 |
| S12_vs_k141_40558\|\|full | 1358 | No | NA | 3 | 0 | 0 | Low-quality | Genome-fragment | | 2.12 | AAI-based (medium-confidence) | 0 |
| S14_vs_k141_103450\|\|full | 1959 | No | NA | 3 | 0 | 1 | Not-determined | Genome-fragment | NA | | NA | 0 |
| S4_extracted_20267 | 1358 | No | NA | 3 | 0 | 0 | Low-quality | Genome-fragment | | 3.31 | AAI-based (medium-confidence) | 0 |
| S6_vs_k141_46420\|\|full | 1358 | No | NA | 2 | 0 | 0 | Low-quality | Genome-fragment | | 2.12 | AAI-based (high-confidence) | 0 |
| S9_extracted_k141_60163 | 1359 | No | NA | 3 | 0 | 1 | Low-quality | Genome-fragment | | 0.78 | AAI-based (high-confidence) | 0 |
| S13_vs_16671\|\|full | 1359 | No | NA | 4 | 1 | 0 | Low-quality | Genome-fragment | | 3.5 | AAI-based (high-confidence) | 0 |
| S14_vs_k141_52314\|\|full | 2335 | No | NA | 2 | 0 | 2 | Not-determined | Genome-fragment | NA | | NA | 0 |
| S14_vs_k141_1037\|\|full | 1969 | No | NA | 2 | 0 | 2 | Not-determined | Genome-fragment | NA | | NA | 0 |
| S14_vs_k141_52317\|\|full | 1742 | No | NA | 3 | 0 | 3 | Not-determined | Genome-fragment | NA | | NA | 0 |
| S14_extracted_k141_86741 | 1359 | No | NA | 2 | 2 | 0 | Low-quality | Genome-fragment | | 3.1 | AAI-based (high-confidence) | 0 |
| S7_extracted_66914 | 1359 | No | NA | 3 | 0 | 3 | Low-quality | Genome-fragment | | 0.36 | AAI-based (medium-confidence) | 0 |
| S14_vs_k141_59556\|\|full | 1289 | No | NA | 4 | 0 | 0 | Not-determined | Genome-fragment | NA | | NA | 0 |
| S7_vs_95006\|\|full | 1359 | No | NA | 3 | 1 | 0 | Low-quality | Genome-fragment | | 3.36 | AAI-based (medium-confidence) | 0 |
| S14_vs_k141_52352\|\|full | 3814 | No | NA | 4 | 0 | 3 | Not-determined | Genome-fragment | NA | | NA | 0 |
| S12_vs_k141_17503\|\|full | 1359 | No | NA | 3 | 0 | 1 | Low-quality | Genome-fragment | | 1.87 | AAI-based (medium-confidence) | 0 |
| S14_vs_k141_15541\|\|full | 1642 | No | NA | 3 | 0 | 2 | Not-determined | Genome-fragment | NA | | NA | 0 |
| S12_vs_k141_28176\|\|full | 1359 | No | NA | 4 | 0 | 0 | Low-quality | Genome-fragment | | 3.75 | AAI-based (high-confidence) | 0 |
| S14_vs_k141_52360\|\|full | 1084 | No | NA | 2 | 0 | 0 | Not-determined | Genome-fragment | NA | | NA | 0 |
| S14_vs_k141_44798\|\|full | 1522 | No | NA | 2 | 0 | 1 | Not-determined | Genome-fragment | NA | | NA | 0 |
| S2_vs_k141_6123\|\|full | 1359 | No | NA | 7 | 0 | 0 | Low-quality | Genome-fragment | | 2.2 | AAI-based (high-confidence) | 0 |
| S14_vs_k141_1071\|\|full | 3191 | No | NA | 3 | 0 | 1 | Not-determined | Genome-fragment | NA | | NA | 0 |
| S14_vs_k141_37733\|\|full | 2075 | No | NA | 4 | 0 | 1 | Not-determined | Genome-fragment | NA | | NA | 0 |
| S14_vs_k141_81430\|\|full | 1371 | No | NA | 3 | 0 | 0 | Not-determined | Genome-fragment | NA | | NA | 0 |
| S14_vs_k141_103549\|\|full | 1247 | No | NA | 2 | 0 | 0 | Not-determined | Genome-fragment | NA | | NA | 0 |
| S14_vs_k141_1083\|\|full | 4929 | No | NA | 3 | 0 | 2 | Not-determined | Genome-fragment | NA | | NA | 0 |
| S14_vs_k141_66590\|\|full | 1266 | No | NA | 3 | 0 | 1 | Not-determined | Genome-fragment | NA | | NA | 0 |
| S14_vs_k141_30336\|\|full | 20033 | No | NA | 18 | 0 | 10 | Not-determined | Genome-fragment | NA | | NA | 0 |
| S8_extracted_k141_35610 | 1359 | No | NA | 2 | 0 | 2 | Low-quality | Genome-fragment | | 0.41 | AAI-based (medium-confidence) | 0 |
| S8_vs_k141_72768\|\|full | 1359 | No | NA | 3 | 1 | 0 | Low-quality | Genome-fragment | | 3.1 | AAI-based (medium-confidence) | 0 |
| S8_vs_k141_146296\|\|full | 1359 | No | NA | 2 | 0 | 0 | Low-quality | Genome-fragment | | 1.45 | AAI-based (medium-confidence) | 0 |
| S9_vs_k141_4798\|\|lt2gene | 1360 | No | NA | 1 | 0 | 0 | Low-quality | Genome-fragment | | 1.55 | AAI-based (medium-confidence) | 0 |
| S13_extracted_94146 | 1360 | No | NA | 4 | 0 | 0 | Low-quality | Genome-fragment | | 7.36 | AAI-based (high-confidence) | 0 |
| S14_vs_k141_103625\|\|full | 1891 | No | NA | 4 | 0 | 2 | Not-determined | Genome-fragment | NA | | NA | 0 |
| S13_vs_85916\|\|full | 1360 | No | NA | 3 | 0 | 0 | Low-quality | Genome-fragment | | 0.75 | AAI-based (medium-confidence) | 0 |
| S7_vs_92108\|\|full | 1360 | No | NA | 3 | 0 | 0 | Low-quality | Genome-fragment | | 3.49 | AAI-based (high-confidence) | 0 |
| S7_vs_60639\|\|lt2gene | 1360 | No | NA | 1 | 1 | 0 | Low-quality | Genome-fragment | | 1.25 | HMM-based (lower-bound) | 0 |
| S15_extracted_32737 | 1360 | No | NA | 2 | 1 | 0 | Low-quality | Genome-fragment | | 1.29 | AAI-based (medium-confidence) | 0 |
| S17_vs_k141_135125\|\|lt2gene | 1361 | No | NA | 1 | 1 | 0 | Low-quality | Genome-fragment | | 2.87 | AAI-based (medium-confidence) | 0 |
| S14_vs_k141_8212\|\|full | 1156 | No | NA | 3 | 0 | 1 | Not-determined | Genome-fragment | NA | | NA | 0 |
| S14_vs_k141_52429\|\|full | 1836 | No | NA | 4 | 0 | 3 | Not-determined | Genome-fragment | NA | | NA | 0 |
| S9_vs_k141_835\|\|full | 1361 | No | NA | 2 | 0 | 1 | Low-quality | Genome-fragment | | 1.77 | AAI-based (medium-confidence) | 0 |
| S14_vs_k141_103700\|\|full | 1403 | No | NA | 4 | 0 | 0 | Not-determined | Genome-fragment | NA | | NA | 0 |
| S14_vs_k141_96111\|\|full | 29345 | Yes | 26132 | 35 | 19 | 5 | Medium-quality | Genome-fragment | | 56.04 | AAI-based (high-confidence) | 10.95 |
| S14_vs_k141_103805\|\|full | 1361 | No | NA | 4 | 1 | 0 | Low-quality | Genome-fragment | | 3.23 | AAI-based (high-confidence) | 0 |
| S1_extracted_k141_150655 | 1361 | No | NA | 5 | 2 | 0 | Low-quality | Genome-fragment | | 3.03 | AAI-based (high-confidence) | 0 |
| S14_vs_k141_59667\|\|full | 2508 | No | NA | 3 | 0 | 0 | Not-determined | Genome-fragment | NA | | NA | 0 |
| S1_vs_k141_40202\|\|full | 1361 | No | NA | 2 | 1 | 0 | Low-quality | Genome-fragment | | 2.27 | AAI-based (medium-confidence) | 0 |
| S5_extracted_k141_106398 | 1361 | No | NA | 4 | 0 | 0 | Low-quality | Genome-fragment | | 0.89 | AAI-based (medium-confidence) | 0 |
| S14_vs_k141_59695\|\|full | 3871 | No | NA | 4 | 0 | 3 | Not-determined | Genome-fragment | NA | | NA | 0 |
| S7_extracted_77819 | 1361 | No | NA | 4 | 3 | 0 | Low-quality | Genome-fragment | | 6.21 | AAI-based (high-confidence) | 0 |
| S14_vs_k141_89023\|\|full | 1475 | No | NA | 3 | 0 | 0 | Not-determined | Genome-fragment | NA | | NA | 0 |
| S7_vs_8555\|\|full | 1361 | No | NA | 4 | 1 | 0 | Low-quality | Genome-fragment | | 3.05 | AAI-based (high-confidence) | 0 |
| S16_extracted_172999 | 1361 | No | NA | 1 | 0 | 1 | Low-quality | Genome-fragment | | 1.63 | AAI-based (high-confidence) | 0 |
| S10_extracted_69050 | 1361 | No | NA | 3 | 0 | 0 | Low-quality | Genome-fragment | | 3.5 | AAI-based (high-confidence) | 0 |
| S14_vs_k141_74067\|\|full | 252822 | Yes | 59702 | 234 | 13 | 94 | Medium-quality | Genome-fragment | | 51.55 | HMM-based (lower-bound) | 76.39 |
| S14_vs_k141_111017\|\|full | 2698 | No | NA | 3 | 0 | 2 | Not-determined | Genome-fragment | NA | | NA | 0 |
| S14_vs_k141_1221\|\|full | 1249 | No | NA | 2 | 0 | 2 | Not-determined | Genome-fragment | NA | | NA | 0 |
| S14_vs_k141_1231\|\|full | 1373 | No | NA | 3 | 0 | 0 | Not-determined | Genome-fragment | NA | | NA | 0 |
| S10_extracted_105209 | 1361 | No | NA | 1 | 0 | 1 | Low-quality | Genome-fragment | | 0.63 | AAI-based (medium-confidence) | 0 |
| S12_extracted_k141_113589 | 1361 | No | NA | 3 | 0 | 1 | Low-quality | Genome-fragment | | 0.47 | AAI-based (medium-confidence) | 0 |
| S14_vs_k141_66662\|\|full | 1152 | No | NA | 2 | 0 | 0 | Not-determined | Genome-fragment | NA | | NA | 0 |
| S14_vs_k141_37896\|\|full | 1013 | No | NA | 2 | 0 | 0 | Not-determined | Genome-fragment | NA | | NA | 0 |
| S14_vs_k141_59756\|\|full | 2239 | No | NA | 3 | 0 | 0 | Not-determined | Genome-fragment | NA | | NA | 0 |
| S8_extracted_k141_178962 | 1361 | No | NA | 2 | 0 | 0 | Low-quality | Genome-fragment | | 2.62 | AAI-based (medium-confidence) | 0 |
| S14_vs_k141_15757\|\|full | 2033 | No | NA | 2 | 0 | 1 | Not-determined | Genome-fragment | NA | | NA | 0 |
| S8_extracted_k141_128463 | 1361 | No | NA | 1 | 1 | 0 | Low-quality | Genome-fragment | | 3.8 | AAI-based (high-confidence) | 0 |
| S14_vs_k141_111050\|\|full | 1066 | No | NA | 3 | 0 | 0 | Not-determined | Genome-fragment | NA | | NA | 0 |
| S17_extracted_k141_94173 | 1362 | No | NA | 2 | 0 | 1 | Low-quality | Genome-fragment | | 0.16 | AAI-based (medium-confidence) | 0 |
| S14_vs_k141_37935\|\|full | 4930 | No | NA | 3 | 0 | 2 | Not-determined | Genome-fragment | NA | | NA | 0 |
| S11_extracted_k141_96207 | 1362 | No | NA | 5 | 4 | 0 | Low-quality | Genome-fragment | | 3.66 | AAI-based (high-confidence) | 0 |
| S13_vs_36254\|\|full | 1362 | No | NA | 3 | 1 | 0 | Low-quality | Genome-fragment | | 1.86 | AAI-based (high-confidence) | 0 |
| S14_extracted_k141_30313 | 1362 | No | NA | 3 | 2 | 0 | Low-quality | Genome-fragment | | 2.27 | AAI-based (medium-confidence) | 0 |
| S14_vs_k141_52461\|\|full | 4562 | No | NA | 3 | 0 | 3 | Not-determined | Genome-fragment | NA | | NA | 0 |
| S14_vs_k141_56985\|\|full | 1362 | No | NA | 3 | 1 | 0 | Low-quality | Genome-fragment | | 2.49 | HMM-based (lower-bound) | 0 |
| S1_vs_k141_6753\|\|full | 1362 | No | NA | 3 | 0 | 0 | Low-quality | Genome-fragment | | 3.65 | AAI-based (high-confidence) | 0 |
| S14_vs_k141_30566\|\|full | 34961 | No | NA | 65 | 27 | 0 | Medium-quality | Genome-fragment | | 87.84 | AAI-based (medium-confidence) | 0 |
| S14_vs_k141_59802\|\|full | 4600 | No | NA | 3 | 0 | 1 | Not-determined | Genome-fragment | NA | | NA | 0 |
| S14_vs_k141_15896\|\|full | 3422 | No | NA | 4 | 0 | 1 | Not-determined | Genome-fragment | NA | | NA | 0 |
| S3_extracted_k141_14732 | 1362 | No | NA | 2 | 0 | 0 | Low-quality | Genome-fragment | | 1.99 | AAI-based (medium-confidence) | 0 |
| S5_extracted_k141_90290 | 1362 | No | NA | 1 | 0 | 1 | Low-quality | Genome-fragment | | 2.11 | AAI-based (medium-confidence) | 0 |
| S5_vs_k141_133407\|\|full | 1362 | No | NA | 5 | 0 | 5 | Low-quality | Genome-fragment | | 0.4 | AAI-based (medium-confidence) | 0 |
| S14_vs_k141_45054\|\|full | 3586 | No | NA | 3 | 0 | 2 | Not-determined | Genome-fragment | NA | | NA | 0 |
| S14_vs_k141_8459\|\|full | 1120 | No | NA | 3 | 0 | 2 | Not-determined | Genome-fragment | NA | | NA | 0 |
| S7_vs_128067\|\|full | 1362 | No | NA | 3 | 0 | 0 | Low-quality | Genome-fragment | | 3.12 | AAI-based (high-confidence) | 0 |
| S14_vs_k141_45073\|\|full | 1523 | No | NA | 2 | 0 | 1 | Not-determined | Genome-fragment | NA | | NA | 0 |
| S16_vs_32918\|\|full | 1362 | No | NA | 3 | 0 | 0 | Low-quality | Genome-fragment | | 2.21 | AAI-based (high-confidence) | 0 |
| S14_vs_k141_89208\|\|full | 1752 | No | NA | 2 | 0 | 1 | Not-determined | Genome-fragment | NA | | NA | 0 |
| S14_vs_k141_15965\|\|full | 1809 | No | NA | 4 | 0 | 0 | Not-determined | Genome-fragment | NA | | NA | 0 |
| S14_vs_k141_66757\|\|full | 2330 | No | NA | 2 | 0 | 2 | Not-determined | Genome-fragment | NA | | NA | 0 |
| S14_vs_k141_74328\|\|full | 1897 | No | NA | 3 | 0 | 0 | Not-determined | Genome-fragment | NA | | NA | 0 |
| S14_vs_k141_66768\|\|full | 5433 | No | NA | 6 | 0 | 4 | Not-determined | Genome-fragment | NA | | NA | 0 |
| S10_vs_39248\|\|lt2gene | 1362 | No | NA | 2 | 1 | 0 | Low-quality | Genome-fragment | | 3.61 | AAI-based (medium-confidence) | 0 |
| S12_extracted_k141_86511 | 1362 | No | NA | 2 | 0 | 0 | Low-quality | Genome-fragment | | 0.89 | AAI-based (medium-confidence) | 0 |
| S6_extracted_k141_5407 | 1362 | No | NA | 3 | 1 | 0 | Low-quality | Genome-fragment | | 4.61 | HMM-based (lower-bound) | 0 |
| S9_extracted_k141_76465 | 1363 | No | NA | 4 | 4 | 0 | Low-quality | Genome-fragment | | 3.92 | AAI-based (medium-confidence) | 0 |
| S14_vs_k141_96194\|\|full | 1472 | No | NA | 3 | 0 | 1 | Not-determined | Genome-fragment | NA | | NA | 0 |
| S14_vs_k141_89271\|\|full | 1485 | No | NA | 5 | 0 | 2 | Not-determined | Genome-fragment | NA | | NA | 0 |
| S9_extracted_k141_51293 | 1363 | No | NA | 1 | 1 | 0 | Low-quality | Genome-fragment | | 3.58 | AAI-based (high-confidence) | 0 |
| S1_vs_k141_60681\|\|full | 1363 | No | NA | 3 | 0 | 0 | Low-quality | Genome-fragment | | 4.02 | AAI-based (high-confidence) | 0 |
| S3_vs_k141_74111\|\|full | 1363 | No | NA | 3 | 1 | 0 | Low-quality | Genome-fragment | | 1.73 | AAI-based (high-confidence) | 0 |
| S14_vs_k141_38083\|\|full | 1370 | No | NA | 4 | 0 | 0 | Not-determined | Genome-fragment | NA | | NA | 0 |
| S14_vs_k141_23115\|\|full | 3392 | No | NA | 4 | 0 | 2 | Not-determined | Genome-fragment | NA | | NA | 0 |
| S3_vs_k141_13350\|\|full | 1363 | No | NA | 3 | 2 | 0 | Low-quality | Genome-fragment | | 3.6 | AAI-based (medium-confidence) | 0 |
| S16_vs_125137\|\|full | 1363 | No | NA | 3 | 0 | 0 | Low-quality | Genome-fragment | | 2.58 | AAI-based (medium-confidence) | 0 |
| S10_vs_109406\|\|full | 1363 | No | NA | 3 | 0 | 0 | Low-quality | Genome-fragment | | 1.52 | AAI-based (medium-confidence) | 0 |
| S8_extracted_k141_104269 | 1363 | No | NA | 2 | 0 | 0 | Low-quality | Genome-fragment | | 1.37 | AAI-based (high-confidence) | 0 |
| S9_vs_k141_8467\|\|lt2gene | 1364 | No | NA | 2 | 2 | 0 | Low-quality | Genome-fragment | | 2.95 | AAI-based (medium-confidence) | 0 |
| S13_vs_153832\|\|full | 1364 | No | NA | 2 | 0 | 1 | Low-quality | Genome-fragment | | 0.97 | AAI-based (high-confidence) | 0 |
| S14_vs_k141_89302\|\|full | 1020 | No | NA | 2 | 0 | 0 | Not-determined | Genome-fragment | NA | | NA | 0 |
| S14_vs_k141_73517\|\|lt2gene | 1364 | No | NA | 2 | 1 | 0 | Low-quality | Genome-fragment | | 3.05 | AAI-based (medium-confidence) | 0 |
| S1_extracted_k141_116595 | 1364 | No | NA | 2 | 2 | 0 | Low-quality | Genome-fragment | | 3.42 | AAI-based (high-confidence) | 0 |
| S14_vs_k141_104053\|\|full | 3454 | No | NA | 2 | 0 | 2 | Not-determined | Genome-fragment | NA | | NA | 0 |
| S14_vs_k141_66841\|\|full | 19282 | No | NA | 26 | 13 | 0 | Medium-quality | Genome-fragment | | 55 | AAI-based (high-confidence) | 0 |
| S14_vs_k141_23188\|\|full | 5216 | No | NA | 4 | 0 | 1 | Not-determined | Genome-fragment | NA | | NA | 0 |
| S14_vs_k141_38162\|\|full | 1044 | No | NA | 2 | 0 | 0 | Not-determined | Genome-fragment | NA | | NA | 0 |
| S14_vs_k141_23208\|\|full | 1302 | No | NA | 2 | 0 | 1 | Not-determined | Genome-fragment | NA | | NA | 0 |
| S1_vs_k141_115769\|\|full | 1364 | No | NA | 1 | 0 | 1 | Low-quality | Genome-fragment | | 2.08 | AAI-based (high-confidence) | 0 |
| S14_vs_k141_96317\|\|full | 2505 | No | NA | 3 | 0 | 0 | Not-determined | Genome-fragment | NA | | NA | 0 |
| S14_vs_k141_38182\|\|full | 3429 | No | NA | 3 | 0 | 2 | Not-determined | Genome-fragment | NA | | NA | 0 |
| S14_vs_k141_111399\|\|full | 3680 | No | NA | 6 | 0 | 0 | Not-determined | Genome-fragment | NA | | NA | 0 |
| S3_extracted_k141_94417 | 1364 | No | NA | 2 | 0 | 1 | Low-quality | Genome-fragment | | 2.26 | AAI-based (high-confidence) | 0 |
| S5_vs_k141_87119\|\|full | 1364 | No | NA | 3 | 0 | 1 | Low-quality | Genome-fragment | | 0.87 | AAI-based (medium-confidence) | 0 |
| S14_vs_k141_23227\|\|full | 1100 | No | NA | 3 | 0 | 0 | Not-determined | Genome-fragment | NA | | NA | 0 |
| S16_vs_68659\|\|full | 1364 | No | NA | 4 | 0 | 0 | Low-quality | Genome-fragment | | 2.39 | AAI-based (high-confidence) | 0 |
| S12_vs_k141_82128\|\|full | 1364 | No | NA | 4 | 0 | 0 | Low-quality | Genome-fragment | | 3.63 | AAI-based (medium-confidence) | 0 |
| S14_vs_k141_60036\|\|full | 14901 | No | NA | 13 | 0 | 10 | Not-determined | Genome-fragment | NA | | NA | 0 |
| S4_extracted_13994 | 1364 | No | NA | 4 | 1 | 0 | Low-quality | Genome-fragment | | 5.02 | HMM-based (lower-bound) | 0 |
| S17_extracted_k141_167426 | 1365 | No | NA | 2 | 0 | 1 | Low-quality | Genome-fragment | | 0.94 | AAI-based (high-confidence) | 0 |
| S9_vs_k141_58080\|\|full | 1365 | No | NA | 4 | 0 | 0 | Low-quality | Genome-fragment | | 0.79 | AAI-based (high-confidence) | 0 |
| S11_vs_k141_63115\|\|lt2gene | 1365 | No | NA | 1 | 1 | 0 | Low-quality | Genome-fragment | | 3.18 | AAI-based (high-confidence) | 0 |
| S14_vs_k141_66951\|\|full | 1516 | No | NA | 3 | 0 | 0 | Not-determined | Genome-fragment | NA | | NA | 0 |
| S14_vs_k141_52679\|\|full | 1075 | No | NA | 4 | 0 | 1 | Not-determined | Genome-fragment | NA | | NA | 0 |
| S14_extracted_k141_30507 | 1365 | No | NA | 2 | 2 | 0 | Low-quality | Genome-fragment | | 3.59 | AAI-based (high-confidence) | 0 |
| S14_vs_k141_60066\|\|full | 1870 | No | NA | 2 | 0 | 1 | Not-determined | Genome-fragment | NA | | NA | 0 |
| S14_vs_k141_96358\|\|full | 2894 | No | NA | 4 | 0 | 3 | Not-determined | Genome-fragment | NA | | NA | 0 |
| S14_extracted_k141_18318 | 1365 | No | NA | 3 | 3 | 0 | Low-quality | Genome-fragment | | 4.51 | AAI-based (high-confidence) | 0 |
| S14_vs_k141_96364\|\|full | 2604 | No | NA | 3 | 0 | 2 | Not-determined | Genome-fragment | NA | | NA | 0 |
| S3_extracted_k141_6734 | 1365 | No | NA | 2 | 1 | 0 | Low-quality | Genome-fragment | | 2.21 | AAI-based (high-confidence) | 0 |
| S7_extracted_80286 | 1365 | No | NA | 5 | 1 | 0 | Low-quality | Genome-fragment | | 0.79 | HMM-based (lower-bound) | 0 |
| S7_vs_5751\|\|lt2gene | 1365 | No | NA | 2 | 2 | 0 | Low-quality | Genome-fragment | | 3.22 | AAI-based (high-confidence) | 0 |
| S16_vs_66753\|\|full | 1365 | No | NA | 3 | 1 | 0 | Low-quality | Genome-fragment | | 3.82 | AAI-based (high-confidence) | 0 |
| S10_extracted_2684 | 1365 | No | NA | 3 | 3 | 0 | Low-quality | Genome-fragment | | 4.82 | HMM-based (lower-bound) | 0 |
| S14_vs_k141_8783\|\|full | 1326 | No | NA | 2 | 0 | 2 | Not-determined | Genome-fragment | NA | | NA | 0 |
| S14_vs_k141_52800\|\|full | 1674 | No | NA | 3 | 0 | 1 | Not-determined | Genome-fragment | NA | | NA | 0 |
| S12_extracted_k141_148408 | 1365 | No | NA | 4 | 2 | 0 | Low-quality | Genome-fragment | | 4.21 | HMM-based (lower-bound) | 0 |
| S14_vs_k141_16241\|\|full | 6920 | No | NA | 6 | 0 | 4 | Not-determined | Genome-fragment | NA | | NA | 0 |
| S3_vs_k141_8908\|\|full | 1366 | No | NA | 2 | 2 | 0 | Low-quality | Genome-fragment | | 3.61 | AAI-based (high-confidence) | 0 |
| S5_extracted_k141_140681 | 1366 | No | NA | 4 | 1 | 0 | Low-quality | Genome-fragment | | 0.92 | HMM-based (lower-bound) | 0 |
| S14_vs_k141_52814\|\|full | 2134 | No | NA | 2 | 0 | 2 | Not-determined | Genome-fragment | NA | | NA | 0 |
| S14_vs_k141_30852\|\|full | 1841 | No | NA | 2 | 0 | 0 | Not-determined | Genome-fragment | NA | | NA | 0 |
| S7_extracted_97012 | 1366 | No | NA | 5 | 0 | 0 | Low-quality | Genome-fragment | | 6.23 | AAI-based (high-confidence) | 0 |
| S16_vs_10205\|\|full | 1366 | No | NA | 3 | 1 | 0 | Low-quality | Genome-fragment | | 3.57 | AAI-based (high-confidence) | 0 |
| S16_vs_150937\|\|full | 1366 | No | NA | 2 | 0 | 2 | Low-quality | Genome-fragment | | 0.5 | AAI-based (medium-confidence) | 0 |
| S14_vs_k141_8818\|\|full | 2981 | No | NA | 4 | 0 | 1 | Not-determined | Genome-fragment | NA | | NA | 0 |
| S10_vs_98692\|\|full | 1366 | No | NA | 2 | 1 | 0 | Low-quality | Genome-fragment | | 2.94 | AAI-based (medium-confidence) | 0 |
| S12_vs_k141_157172\|\|full | 1366 | No | NA | 4 | 1 | 0 | Low-quality | Genome-fragment | | 10.28 | AAI-based (high-confidence) | 0 |
| S8_extracted_k141_186486 | 1366 | No | NA | 2 | 2 | 0 | Low-quality | Genome-fragment | | 3.71 | HMM-based (lower-bound) | 0 |
| S17_vs_k141_15652\|\|full | 1367 | No | NA | 3 | 0 | 0 | Low-quality | Genome-fragment | | 3.27 | AAI-based (medium-confidence) | 0 |
| S14_vs_k141_74599\|\|full | 1384 | No | NA | 2 | 0 | 0 | Not-determined | Genome-fragment | NA | | NA | 0 |
| S14_vs_k141_1419\|\|full | 2561 | No | NA | 3 | 0 | 0 | Not-determined | Genome-fragment | NA | | NA | 0 |
| S14_vs_k141_30904\|\|full | 4240 | No | NA | 3 | 0 | 2 | Not-determined | Genome-fragment | NA | | NA | 0 |
| S17_vs_k141_97077\|\|full | 1367 | No | NA | 3 | 0 | 0 | Low-quality | Genome-fragment | | 2.94 | AAI-based (high-confidence) | 0 |
| S14_vs_k141_111617\|\|full | 1770 | No | NA | 3 | 0 | 1 | Not-determined | Genome-fragment | NA | | NA | 0 |
| S14_vs_k141_104343\|\|full | 2671 | No | NA | 3 | 0 | 1 | Not-determined | Genome-fragment | NA | | NA | 0 |
| S11_vs_k141_80040\|\|full | 1367 | No | NA | 5 | 2 | 0 | Low-quality | Genome-fragment | | 2.69 | AAI-based (high-confidence) | 0 |
| S13_extracted_44086 | 1367 | No | NA | 2 | 0 | 0 | Low-quality | Genome-fragment | | 3.06 | AAI-based (high-confidence) | 0 |
| S14_vs_k141_96492\|\|full | 1910 | No | NA | 3 | 0 | 3 | Not-determined | Genome-fragment | NA | | NA | 0 |
| S14_vs_k141_38328\|\|full | 1982 | No | NA | 4 | 0 | 0 | Not-determined | Genome-fragment | NA | | NA | 0 |
| S14_extracted_k141_106805 | 1367 | No | NA | 1 | 0 | 1 | Low-quality | Genome-fragment | | 0.38 | AAI-based (high-confidence) | 0 |
| S14_vs_k141_1441\|\|full | 6365 | No | NA | 5 | 0 | 3 | Not-determined | Genome-fragment | NA | | NA | 0 |
| S14_vs_k141_61906\|\|full | 1367 | No | NA | 2 | 0 | 0 | Low-quality | Genome-fragment | | 2.13 | AAI-based (medium-confidence) | 0 |
| S14_vs_k141_23457\|\|full | 3017 | No | NA | 3 | 0 | 2 | Not-determined | Genome-fragment | NA | | NA | 0 |
| S1_vs_k141_138013\|\|full | 1367 | No | NA | 2 | 0 | 1 | Low-quality | Genome-fragment | | 1.86 | AAI-based (high-confidence) | 0 |
| S14_vs_k141_104402\|\|full | 1784 | No | NA | 2 | 0 | 2 | Not-determined | Genome-fragment | NA | | NA | 0 |
| S1_vs_k141_38942\|\|full | 1367 | No | NA | 4 | 1 | 0 | Low-quality | Genome-fragment | | 2.83 | AAI-based (high-confidence) | 0 |
| S14_vs_k141_96557\|\|full | 2897 | No | NA | 4 | 0 | 1 | Not-determined | Genome-fragment | NA | | NA | 0 |
| S14_vs_k141_52947\|\|full | 2733 | No | NA | 2 | 0 | 1 | Not-determined | Genome-fragment | NA | | NA | 0 |
| S5_vs_k141_135898\|\|full | 1367 | No | NA | 3 | 0 | 0 | Low-quality | Genome-fragment | | 2.9 | AAI-based (high-confidence) | 0 |
| S5_vs_k141_28040\|\|full | 1367 | No | NA | 2 | 0 | 0 | Low-quality | Genome-fragment | | 3.17 | AAI-based (medium-confidence) | 0 |
| S14_vs_k141_74643\|\|full | 9713 | No | NA | 5 | 0 | 5 | Not-determined | Genome-fragment | NA | | NA | 0 |
| S14_vs_k141_82148\|\|full | 2850 | No | NA | 7 | 0 | 2 | Not-determined | Genome-fragment | NA | | NA | 0 |
| S7_extracted_3752 | 1367 | No | NA | 2 | 2 | 0 | Low-quality | Genome-fragment | | 4.5 | AAI-based (high-confidence) | 0 |
| S7_vs_34735\|\|full | 1367 | No | NA | 2 | 0 | 0 | Low-quality | Genome-fragment | | 3.03 | AAI-based (medium-confidence) | 0 |
| S14_vs_k141_74671\|\|full | 1453 | No | NA | 3 | 0 | 0 | Not-determined | Genome-fragment | NA | | NA | 0 |
| S7_vs_39428\|\|lt2gene | 1367 | No | NA | 2 | 2 | 0 | Low-quality | Genome-fragment | | 3.59 | AAI-based (high-confidence) | 0 |
| S16_extracted_10036 | 1367 | No | NA | 4 | 0 | 0 | Low-quality | Genome-fragment | | 2.4 | AAI-based (medium-confidence) | 0 |
| S14_vs_k141_111766\|\|full | 2192 | No | NA | 2 | 0 | 0 | Not-determined | Genome-fragment | NA | | NA | 0 |
| S16_vs_107837\|\|full | 1367 | No | NA | 3 | 1 | 0 | Low-quality | Genome-fragment | | 2.19 | HMM-based (lower-bound) | 0 |
| S16_vs_128627\|\|full | 1367 | No | NA | 4 | 2 | 0 | Low-quality | Genome-fragment | | 2.2 | AAI-based (high-confidence) | 0 |
| S17_extracted_k141_56342 | 1368 | No | NA | 2 | 0 | 0 | Low-quality | Genome-fragment | | 2.28 | AAI-based (medium-confidence) | 0 |
| S14_vs_k141_8929\|\|full | 1124 | No | NA | 2 | 0 | 1 | Not-determined | Genome-fragment | NA | | NA | 0 |
| S17_vs_k141_134548\|\|full | 1368 | No | NA | 4 | 1 | 0 | Low-quality | Genome-fragment | | 2.91 | AAI-based (medium-confidence) | 0 |
| S9_extracted_k141_89500 | 1368 | No | NA | 4 | 0 | 4 | Low-quality | Genome-fragment | | 0.39 | AAI-based (medium-confidence) | 0 |
| S14_vs_k141_82187\|\|full | 2753 | No | NA | 4 | 0 | 4 | Not-determined | Genome-fragment | NA | | NA | 0 |
| S9_vs_k141_26568\|\|full | 1368 | No | NA | 6 | 0 | 0 | Low-quality | Genome-fragment | | 2.65 | AAI-based (high-confidence) | 0 |
| S9_vs_k141_4541\|\|full | 1368 | No | NA | 3 | 2 | 0 | Low-quality | Genome-fragment | | 2.99 | AAI-based (high-confidence) | 0 |
| S14_vs_k141_82211\|\|full | 3090 | No | NA | 2 | 0 | 1 | Not-determined | Genome-fragment | NA | | NA | 0 |
| S14_vs_k141_23497\|\|full | 3535 | No | NA | 2 | 0 | 0 | Not-determined | Genome-fragment | NA | | NA | 0 |
| S14_vs_k141_96674\|\|full | 5446 | No | NA | 8 | 0 | 2 | Not-determined | Genome-fragment | NA | | NA | 0 |
| S11_vs_k141_37237\|\|full | 1368 | No | NA | 4 | 2 | 0 | Low-quality | Genome-fragment | | 3.36 | AAI-based (high-confidence) | 0 |
| S13_extracted_109353 | 1368 | No | NA | 2 | 0 | 2 | Low-quality | Genome-fragment | | 0.24 | AAI-based (medium-confidence) | 0 |
| S14_vs_k141_89620\|\|full | 5152 | No | NA | 7 | 0 | 0 | Not-determined | Genome-fragment | NA | | NA | 0 |
| S14_vs_k141_67200\|\|full | 1097 | No | NA | 2 | 0 | 0 | Not-determined | Genome-fragment | NA | | NA | 0 |
| S14_vs_k141_89631\|\|full | 2205 | No | NA | 3 | 0 | 3 | Not-determined | Genome-fragment | NA | | NA | 0 |
| S14_vs_k141_38480\|\|full | 8999 | No | NA | 10 | 0 | 5 | Not-determined | Genome-fragment | NA | | NA | 0 |
| S13_vs_6013\|\|full | 1368 | No | NA | 3 | 1 | 0 | Low-quality | Genome-fragment | | 1.59 | AAI-based (medium-confidence) | 0 |
| S14_vs_k141_49820\|\|full | 1368 | No | NA | 4 | 0 | 0 | Low-quality | Genome-fragment | | 0.7 | AAI-based (medium-confidence) | 0 |
| S14_vs_k141_16523\|\|full | 1420 | No | NA | 3 | 0 | 1 | Not-determined | Genome-fragment | NA | | NA | 0 |
| S1_extracted_k141_145077 | 1368 | No | NA | 1 | 1 | 0 | Low-quality | Genome-fragment | | 1.38 | AAI-based (high-confidence) | 0 |
| S5_vs_k141_54348\|\|full | 1368 | No | NA | 3 | 0 | 0 | Low-quality | Genome-fragment | | 3.56 | AAI-based (high-confidence) | 0 |
| S5_vs_k141_11145\|\|full | 1368 | No | NA | 4 | 1 | 0 | Low-quality | Genome-fragment | | 1.53 | HMM-based (lower-bound) | 0 |
| S5_vs_k141_77176\|\|full | 1368 | No | NA | 3 | 1 | 0 | Low-quality | Genome-fragment | | 3.2 | AAI-based (high-confidence) | 0 |
| S14_vs_k141_53034\|\|full | 3242 | No | NA | 4 | 0 | 2 | Not-determined | Genome-fragment | NA | | NA | 0 |
| S16_vs_1124\|\|full | 1368 | No | NA | 2 | 0 | 0 | Low-quality | Genome-fragment | | 3.76 | AAI-based (high-confidence) | 0 |
| S16_vs_43334\|\|full | 1368 | No | NA | 3 | 0 | 2 | Low-quality | Genome-fragment | | 2.18 | AAI-based (medium-confidence) | 0 |
| S14_vs_k141_111927\|\|full | 1361 | No | NA | 2 | 0 | 0 | Not-determined | Genome-fragment | NA | | NA | 0 |
| S14_vs_k141_16562\|\|full | 5592 | No | NA | 4 | 0 | 1 | Not-determined | Genome-fragment | NA | | NA | 0 |
| S14_vs_k141_67324\|\|full | 1290 | No | NA | 2 | 0 | 1 | Not-determined | Genome-fragment | NA | | NA | 0 |
| S14_vs_k141_31134\|\|full | 1625 | No | NA | 3 | 0 | 1 | Not-determined | Genome-fragment | NA | | NA | 0 |
| S10_extracted_48765 | 1368 | No | NA | 2 | 1 | 0 | Low-quality | Genome-fragment | | 2.98 | AAI-based (high-confidence) | 0 |
| S14_vs_k141_74925\|\|full | 4250 | No | NA | 4 | 0 | 1 | Not-determined | Genome-fragment | NA | | NA | 0 |
| S14_vs_k141_31145\|\|full | 2043 | No | NA | 2 | 0 | 1 | Not-determined | Genome-fragment | NA | | NA | 0 |
| S14_vs_k141_60507\|\|full | 1221 | No | NA | 2 | 0 | 2 | Not-determined | Genome-fragment | NA | | NA | 0 |
| S10_vs_23883\|\|full | 1368 | No | NA | 3 | 0 | 1 | Low-quality | Genome-fragment | | 0.71 | AAI-based (medium-confidence) | 0 |
| S14_vs_k141_53096\|\|full | 1354 | No | NA | 2 | 0 | 0 | Not-determined | Genome-fragment | NA | | NA | 0 |
| S14_vs_k141_82321\|\|full | 1187 | No | NA | 2 | 0 | 1 | Not-determined | Genome-fragment | NA | | NA | 0 |
| S14_vs_k141_38581\|\|full | 4784 | No | NA | 5 | 0 | 0 | Not-determined | Genome-fragment | NA | | NA | 0 |
| S14_vs_k141_67402\|\|full | 3438 | No | NA | 4 | 0 | 1 | Not-determined | Genome-fragment | NA | | NA | 0 |
| S8_extracted_k141_53806 | 1368 | No | NA | 2 | 0 | 1 | Low-quality | Genome-fragment | | 2.5 | AAI-based (medium-confidence) | 0 |
| S11_extracted_k141_98114 | 1369 | No | NA | 3 | 0 | 1 | Low-quality | Genome-fragment | | 3.56 | AAI-based (medium-confidence) | 0 |
| S14_vs_k141_38607\|\|full | 1196 | No | NA | 3 | 0 | 1 | Not-determined | Genome-fragment | NA | | NA | 0 |
| S14_vs_k141_46088\|\|full | 1369 | No | NA | 3 | 0 | 0 | Low-quality | Genome-fragment | | 2.23 | AAI-based (medium-confidence) | 0 |
| S14_vs_k141_31199\|\|full | 8760 | No | NA | 6 | 0 | 6 | Not-determined | Genome-fragment | NA | | NA | 0 |
| S14_vs_k141_89794\|\|full | 1059 | No | NA | 2 | 0 | 0 | Not-determined | Genome-fragment | NA | | NA | 0 |
| S14_vs_k141_5845\|\|lt2gene | 1369 | No | NA | 2 | 2 | 0 | Low-quality | Genome-fragment | | 3.31 | AAI-based (high-confidence) | 0 |
| S14_vs_k141_75004\|\|full | 1291 | No | NA | 3 | 0 | 1 | Not-determined | Genome-fragment | NA | | NA | 0 |
| S14_vs_k141_67439\|\|full | 1640 | No | NA | 2 | 0 | 0 | Not-determined | Genome-fragment | NA | | NA | 0 |
| S3_vs_k141_72110\|\|full | 1369 | No | NA | 5 | 0 | 1 | Low-quality | Genome-fragment | | 1.39 | AAI-based (medium-confidence) | 0 |
| S14_vs_k141_104703\|\|full | 1010 | No | NA | 2 | 0 | 1 | Not-determined | Genome-fragment | NA | | NA | 0 |
| S3_vs_k141_37197\|\|full | 1369 | No | NA | 3 | 0 | 1 | Low-quality | Genome-fragment | | 1.86 | AAI-based (medium-confidence) | 0 |
| S7_vs_86461\|\|full | 1369 | No | NA | 2 | 0 | 1 | Low-quality | Genome-fragment | | 0.63 | AAI-based (medium-confidence) | 0 |
| S14_vs_k141_31221\|\|full | 1946 | No | NA | 2 | 0 | 1 | Not-determined | Genome-fragment | NA | | NA | 0 |
| S16_extracted_38579 | 1369 | No | NA | 3 | 0 | 2 | Low-quality | Genome-fragment | | 1.08 | AAI-based (medium-confidence) | 0 |
| S14_vs_k141_75014\|\|full | 20775 | No | NA | 15 | 0 | 10 | Not-determined | Genome-fragment | NA | | NA | 0 |
| S16_vs_95076\|\|full | 1369 | No | NA | 3 | 0 | 0 | Low-quality | Genome-fragment | | 3.19 | AAI-based (high-confidence) | 0 |
| S14_vs_k141_82401\|\|full | 1330 | No | NA | 3 | 0 | 2 | Not-determined | Genome-fragment | NA | | NA | 0 |
| S14_vs_k141_31231\|\|full | 2687 | No | NA | 3 | 0 | 3 | Not-determined | Genome-fragment | NA | | NA | 0 |
| S10_vs_84953\|\|full | 1369 | No | NA | 3 | 0 | 0 | Low-quality | Genome-fragment | | 1.88 | AAI-based (medium-confidence) | 0 |
| S14_vs_k141_45387\|\|full | 2232 | No | NA | 3 | 0 | 3 | Not-determined | Genome-fragment | NA | | NA | 0 |
| S14_vs_k141_16694\|\|full | 2897 | No | NA | 4 | 0 | 1 | Not-determined | Genome-fragment | NA | | NA | 0 |
| S12_vs_k141_146677\|\|lt2gene | 1369 | No | NA | 2 | 1 | 0 | Low-quality | Genome-fragment | | 1.7 | AAI-based (high-confidence) | 0 |
| S8_vs_k141_139715\|\|full | 1369 | No | NA | 4 | 1 | 0 | Low-quality | Genome-fragment | | 1.64 | AAI-based (medium-confidence) | 0 |
| S14_vs_k141_31314\|\|full | 1682 | No | NA | 3 | 0 | 0 | Not-determined | Genome-fragment | NA | | NA | 0 |
| S14_vs_k141_23710\|\|full | 2952 | No | NA | 4 | 0 | 2 | Not-determined | Genome-fragment | NA | | NA | 0 |
| S14_vs_k141_112068\|\|full | 44297 | No | NA | 81 | 16 | 0 | High-quality | High-quality | | 99.28 | AAI-based (high-confidence) | 0 |
| S17_vs_k141_97177\|\|full | 1370 | No | NA | 3 | 1 | 0 | Low-quality | Genome-fragment | | 2.23 | HMM-based (lower-bound) | 0 |
| S14_vs_k141_16759\|\|full | 1007 | No | NA | 3 | 0 | 0 | Not-determined | Genome-fragment | NA | | NA | 0 |
| S14_vs_k141_16763\|\|full | 1605 | No | NA | 3 | 0 | 0 | Not-determined | Genome-fragment | NA | | NA | 0 |
| S9_extracted_k141_60412 | 1370 | No | NA | 2 | 2 | 0 | Low-quality | Genome-fragment | | 2.28 | AAI-based (high-confidence) | 0 |
| S14_vs_k141_53298\|\|full | 2001 | No | NA | 4 | 0 | 3 | Not-determined | Genome-fragment | NA | | NA | 0 |
| S14_vs_k141_67541\|\|full | 3281 | No | NA | 3 | 0 | 1 | Not-determined | Genome-fragment | NA | | NA | 0 |

| | | | | | | | | | | | |
|---|---|---|---|---|---|---|---|---|---|---|---|
| S9_vs_k141_774\|\|full | 1370 | No | NA | 3 | 0 | 0 | Low-quality | Genome-fragment | 2.44 | AAI-based (high-confidence) | 0 |
| S11_vs_k141_94531\|\|full | 1370 | No | NA | 3 | 0 | 1 | Low-quality | Genome-fragment | 1.98 | AAI-based (medium-confidence) | 0 |
| S1_vs_k141_113503\|\|full | 1370 | No | NA | 3 | 1 | 0 | Low-quality | Genome-fragment | 1.84 | AAI-based (high-confidence) | 0 |
| S3_extracted_k141_45311 | 1370 | No | NA | 2 | 0 | 0 | Low-quality | Genome-fragment | 1.95 | AAI-based (high-confidence) | 0 |
| S5_vs_k141_1297\|\|full | 1370 | No | NA | 2 | 0 | 0 | Low-quality | Genome-fragment | 2.21 | AAI-based (medium-confidence) | 0 |
| S14_vs_k141_104835\|\|full | 1006 | No | NA | 2 | 0 | 0 | Not-determined | Genome-fragment | NA | NA | 0 |
| S14_vs_k141_53311\|\|full | 2002 | No | NA | 4 | 0 | 3 | Not-determined | Genome-fragment | NA | NA | 0 |
| S14_vs_k141_104841\|\|full | 1965 | No | NA | 2 | 0 | 0 | Not-determined | Genome-fragment | NA | NA | 0 |
| S7_extracted_100935 | 1370 | No | NA | 3 | 0 | 0 | Low-quality | Genome-fragment | 2.22 | AAI-based (high-confidence) | 0 |
| S7_vs_88748\|\|lt2gene | 1370 | No | NA | 2 | 1 | 0 | Low-quality | Genome-fragment | 3.54 | AAI-based (high-confidence) | 0 |
| S16_vs_108365\|\|full | 1370 | No | NA | 3 | 0 | 0 | Low-quality | Genome-fragment | 3.33 | AAI-based (high-confidence) | 0 |
| S12_vs_k141_31787\|\|full | 1370 | No | NA | 3 | 1 | 0 | Low-quality | Genome-fragment | 0.86 | AAI-based (high-confidence) | 0 |
| S6_extracted_k141_52258 | 1370 | No | NA | 1 | 0 | 0 | Low-quality | Genome-fragment | 0.52 | AAI-based (medium-confidence) | 0 |
| S14_vs_k141_67601\|\|full | 3822 | No | NA | 3 | 0 | 2 | Not-determined | Genome-fragment | NA | NA | 0 |
| S8_extracted_k141_122766 | 1370 | No | NA | 1 | 1 | 0 | Low-quality | Genome-fragment | 2 | HMM-based (lower-bound) | 0 |
| S8_extracted_k141_149318 | 1370 | No | NA | 3 | 0 | 0 | Low-quality | Genome-fragment | 0.87 | AAI-based (medium-confidence) | 0 |
| S17_vs_k141_144567\|\|full | 1371 | No | NA | 3 | 1 | 0 | Low-quality | Genome-fragment | 2.96 | AAI-based (medium-confidence) | 0 |
| S14_vs_k141_45534\|\|full | 2778 | No | NA | 4 | 0 | 1 | Not-determined | Genome-fragment | NA | NA | 0 |
| S14_vs_k141_38905\|\|full | 29314 | No | NA | 43 | 12 | 1 | Medium-quality | Genome-fragment | 72.3 | AAI-based (medium-confidence) | 0 |
| S11_vs_k141_52317\|\|full | 1371 | No | NA | 3 | 0 | 0 | Low-quality | Genome-fragment | 3.61 | AAI-based (high-confidence) | 0 |
| S14_vs_k141_23805\|\|full | 1706 | No | NA | 2 | 0 | 2 | Not-determined | Genome-fragment | NA | NA | 0 |
| S14_vs_k141_53419\|\|full | 2993 | No | NA | 4 | 0 | 4 | Not-determined | Genome-fragment | NA | NA | 0 |
| S14_vs_k141_1828\|\|full | 1316 | No | NA | 3 | 0 | 0 | Not-determined | Genome-fragment | NA | NA | 0 |
| S14_extracted_k141_21403 | 1371 | No | NA | 3 | 2 | 0 | Low-quality | Genome-fragment | 4.54 | AAI-based (high-confidence) | 0 |
| S1_extracted_k141_102536 | 1371 | No | NA | 2 | 0 | 0 | Low-quality | Genome-fragment | 1.39 | AAI-based (high-confidence) | 0 |
| S5_vs_k141_97188\|\|full | 1371 | No | NA | 5 | 0 | 0 | Low-quality | Genome-fragment | 2.7 | AAI-based (medium-confidence) | 0 |
| S7_extracted_67454 | 1371 | No | NA | 2 | 2 | 0 | Low-quality | Genome-fragment | 3.12 | AAI-based (high-confidence) | 0 |
| S14_vs_k141_67670\|\|full | 2697 | No | NA | 3 | 0 | 0 | Not-determined | Genome-fragment | NA | NA | 0 |
| S7_vs_12292\|\|full | 1371 | No | NA | 3 | 0 | 0 | Low-quality | Genome-fragment | 3.2 | AAI-based (high-confidence) | 0 |
| S14_vs_k141_45610\|\|full | 2650 | No | NA | 3 | 0 | 0 | Not-determined | Genome-fragment | NA | NA | 0 |
| S7_vs_32131\|\|lt2gene | 1371 | No | NA | 1 | 1 | 0 | Low-quality | Genome-fragment | 3.09 | AAI-based (high-confidence) | 0 |
| S12_extracted_k141_58716 | 1371 | No | NA | 3 | 0 | 0 | Low-quality | Genome-fragment | 2.2 | AAI-based (high-confidence) | 0 |
| S14_vs_k141_9162\|\|full | 102634 | Yes | 20846 | 106 | 16 | 57 | Medium-quality | Genome-fragment | 60.28 | AAI-based (high-confidence) | 79.69 |
| S14_vs_k141_67727\|\|full | 1070 | No | NA | 2 | 0 | 1 | Not-determined | Genome-fragment | NA | NA | 0 |
| S6_vs_k141_38247\|\|full | 1371 | No | NA | 3 | 0 | 0 | Low-quality | Genome-fragment | 2.26 | AAI-based (high-confidence) | 0 |
| S8_vs_k141_156209\|\|full | 1371 | No | NA | 3 | 0 | 2 | Low-quality | Genome-fragment | 2.75 | AAI-based (high-confidence) | 0 |
| S14_vs_k141_112176\|\|full | 1077 | No | NA | 2 | 0 | 1 | Not-determined | Genome-fragment | NA | NA | 0 |
| S14_vs_k141_53490\|\|full | 7848 | No | NA | 5 | 0 | 4 | Not-determined | Genome-fragment | NA | NA | 0 |
| S8_vs_k141_141534\|\|full | 1371 | No | NA | 4 | 0 | 0 | Low-quality | Genome-fragment | 3.09 | AAI-based (high-confidence) | 0 |
| S9_vs_k141_81303\|\|full | 1372 | No | NA | 3 | 2 | 0 | Low-quality | Genome-fragment | 3.36 | AAI-based (medium-confidence) | 0 |
| S14_extracted_k141_116897 | 1372 | No | NA | 3 | 0 | 0 | Low-quality | Genome-fragment | 2.9 | AAI-based (high-confidence) | 0 |
| S14_vs_k141_11785\|\|lt2gene | 1372 | No | NA | 1 | 0 | 0 | Low-quality | Genome-fragment | 3.49 | AAI-based (medium-confidence) | 0 |
| S1_vs_k141_85411\|\|full | 1372 | No | NA | 3 | 0 | 2 | Low-quality | Genome-fragment | 1.83 | AAI-based (medium-confidence) | 0 |
| S5_vs_k141_124024\|\|full | 1372 | No | NA | 2 | 0 | 0 | Low-quality | Genome-fragment | 2.91 | AAI-based (high-confidence) | 0 |
| S5_vs_k141_11792\|\|full | 1372 | No | NA | 3 | 0 | 0 | Low-quality | Genome-fragment | 2.29 | AAI-based (medium-confidence) | 0 |
| S14_vs_k141_96959\|\|full | 5171 | No | NA | 7 | 0 | 0 | Not-determined | Genome-fragment | NA | NA | 0 |
| S14_vs_k141_105116\|\|full | 2820 | No | NA | 3 | 0 | 2 | Not-determined | Genome-fragment | NA | NA | 0 |
| S14_vs_k141_53567\|\|full | 5575 | No | NA | 4 | 0 | 3 | Not-determined | Genome-fragment | NA | NA | 0 |
| S16_extracted_66212 | 1372 | No | NA | 1 | 1 | 0 | Low-quality | Genome-fragment | 2 | HMM-based (lower-bound) | 0 |
| S14_vs_k141_90123\|\|full | 6521 | No | NA | 7 | 0 | 1 | Not-determined | Genome-fragment | NA | NA | 0 |
| S14_vs_k141_9286\|\|full | 3885 | No | NA | 5 | 0 | 5 | Not-determined | Genome-fragment | NA | NA | 0 |
| S16_extracted_113444 | 1372 | No | NA | 3 | 0 | 0 | Low-quality | Genome-fragment | 2.6 | AAI-based (high-confidence) | 0 |
| S14_vs_k141_112252\|\|full | 22565 | No | NA | 45 | 11 | 0 | Medium-quality | Genome-fragment | 67.86 | AAI-based (high-confidence) | 0 |
| S16_extracted_153405 | 1372 | No | NA | 1 | 0 | 0 | Low-quality | Genome-fragment | 2.15 | AAI-based (high-confidence) | 0 |
| S16_vs_47675\|\|full | 1372 | No | NA | 2 | 0 | 0 | Low-quality | Genome-fragment | 2.13 | AAI-based (medium-confidence) | 0 |
| S16_vs_55725\|\|full | 1372 | No | NA | 3 | 2 | 0 | Low-quality | Genome-fragment | 2.73 | AAI-based (high-confidence) | 0 |
| S16_vs_94098\|\|full | 1372 | No | NA | 3 | 0 | 0 | Low-quality | Genome-fragment | 1.95 | AAI-based (medium-confidence) | 0 |
| S14_vs_k141_60862\|\|full | 1411 | No | NA | 2 | 0 | 2 | Not-determined | Genome-fragment | NA | NA | 0 |
| S14_vs_k141_31464\|\|full | 2955 | No | NA | 3 | 0 | 1 | Not-determined | Genome-fragment | NA | NA | 0 |
| S14_vs_k141_53611\|\|full | 3930 | No | NA | 3 | 0 | 1 | Not-determined | Genome-fragment | NA | NA | 0 |
| S10_extracted_57122 | 1372 | No | NA | 1 | 0 | 1 | Low-quality | Genome-fragment | 2.31 | AAI-based (high-confidence) | 0 |
| S14_vs_k141_75419\|\|full | 2364 | No | NA | 3 | 0 | 2 | Not-determined | Genome-fragment | NA | NA | 0 |
| S10_vs_41650\|\|full | 1372 | No | NA | 2 | 1 | 0 | Low-quality | Genome-fragment | 1.79 | AAI-based (high-confidence) | 0 |
| S10_vs_78305\|\|lt2gene | 1372 | No | NA | 1 | 1 | 0 | Low-quality | Genome-fragment | 2 | HMM-based (lower-bound) | 0 |
| S14_vs_k141_60904\|\|full | 1796 | No | NA | 5 | 0 | 0 | Not-determined | Genome-fragment | NA | NA | 0 |
| S15_vs_16308\|\|full | 1372 | No | NA | 3 | 0 | 0 | Low-quality | Genome-fragment | 2.49 | AAI-based (medium-confidence) | 0 |
| S6_extracted_k141_25186 | 1372 | No | NA | 2 | 0 | 0 | Low-quality | Genome-fragment | 0.48 | AAI-based (medium-confidence) | 0 |
| S8_extracted_k141_7244 | 1372 | No | NA | 2 | 1 | 0 | Low-quality | Genome-fragment | 0.89 | HMM-based (lower-bound) | 0 |
| S14_vs_k141_75475\|\|full | 1739 | No | NA | 3 | 0 | 0 | Not-determined | Genome-fragment | NA | NA | 0 |
| S14_vs_k141_9413\|\|full | 2307 | No | NA | 2 | 0 | 1 | Not-determined | Genome-fragment | NA | NA | 0 |
| S14_vs_k141_53688\|\|full | 11920 | No | NA | 18 | 0 | 1 | Not-determined | Genome-fragment | NA | NA | 0 |
| S17_extracted_k141_109331 | 1373 | No | NA | 1 | 1 | 0 | Low-quality | Genome-fragment | 0.79 | HMM-based (lower-bound) | 0 |
| S17_vs_k141_109540\|\|full | 1373 | No | NA | 4 | 0 | 1 | Low-quality | Genome-fragment | 4.68 | AAI-based (medium-confidence) | 0 |
| S9_extracted_k141_24236 | 1373 | No | NA | 4 | 2 | 0 | Low-quality | Genome-fragment | 3.3 | AAI-based (medium-confidence) | 0 |
| S9_vs_k141_83412\|\|full | 1373 | No | NA | 3 | 0 | 0 | Low-quality | Genome-fragment | 2.82 | AAI-based (high-confidence) | 0 |
| S14_vs_k141_24123\|\|full | 1185 | No | NA | 2 | 0 | 0 | Not-determined | Genome-fragment | NA | NA | 0 |
| S14_vs_k141_53731\|\|full | 1606 | No | NA | 3 | 0 | 0 | Not-determined | Genome-fragment | NA | NA | 0 |
| S14_vs_k141_2016\|\|full | 9685 | No | NA | 10 | 0 | 5 | Not-determined | Genome-fragment | NA | NA | 0 |
| S11_extracted_k141_83742 | 1373 | No | NA | 5 | 0 | 0 | Low-quality | Genome-fragment | 3.45 | AAI-based (high-confidence) | 0 |
| S14_vs_k141_60930\|\|full | 2090 | No | NA | 4 | 0 | 1 | Not-determined | Genome-fragment | NA | NA | 0 |
| S13_extracted_141093 | 1373 | No | NA | 3 | 0 | 0 | Low-quality | Genome-fragment | 1.67 | AAI-based (medium-confidence) | 0 |
| S14_vs_k141_32426\|\|lt2gene | 1373 | No | NA | 1 | 1 | 0 | Low-quality | Genome-fragment | 1.93 | HMM-based (lower-bound) | 0 |
| S14_vs_k141_2037\|\|full | 3989 | No | NA | 3 | 0 | 3 | Not-determined | Genome-fragment | NA | NA | 0 |
| S3_vs_k141_103988\|\|full | 1373 | No | NA | 3 | 1 | 0 | Low-quality | Genome-fragment | 0.92 | HMM-based (lower-bound) | 0 |
| S14_vs_k141_90264\|\|full | 3293 | No | NA | 4 | 0 | 1 | Not-determined | Genome-fragment | NA | NA | 0 |
| S3_vs_k141_98974\|\|full | 1373 | No | NA | 2 | 0 | 0 | Low-quality | Genome-fragment | 2.97 | AAI-based (high-confidence) | 0 |
| S14_vs_k141_2069\|\|full | 2524 | No | NA | 2 | 0 | 1 | Not-determined | Genome-fragment | NA | NA | 0 |
| S14_vs_k141_9453\|\|full | 6415 | No | NA | 6 | 0 | 4 | Not-determined | Genome-fragment | NA | NA | 0 |
| S3_vs_k141_116147\|\|full | 1373 | No | NA | 3 | 0 | 0 | Low-quality | Genome-fragment | 2.51 | AAI-based (high-confidence) | 0 |
| S14_vs_k141_24191\|\|full | 1410 | No | NA | 3 | 0 | 1 | Not-determined | Genome-fragment | NA | NA | 0 |
| S14_vs_k141_17049\|\|full | 4522 | No | NA | 5 | 0 | 3 | Not-determined | Genome-fragment | NA | NA | 0 |
| S14_vs_k141_112345\|\|full | 38263 | No | NA | 26 | 1 | 17 | Medium-quality | Genome-fragment | 71.14 | AAI-based (high-confidence) | 0 |
| S14_vs_k141_105287\|\|full | 1628 | No | NA | 3 | 0 | 1 | Not-determined | Genome-fragment | NA | NA | 0 |
| S14_vs_k141_39266\|\|full | 37426 | Yes | 26099 | 49 | 12 | 6 | Medium-quality | Genome-fragment | 62.52 | AAI-based (medium-confidence) | 30.27 |
| S14_vs_k141_9464\|\|full | 2304 | No | NA | 2 | 0 | 2 | Not-determined | Genome-fragment | NA | NA | 0 |
| S14_vs_k141_2115\|\|full | 2479 | No | NA | 5 | 0 | 0 | Not-determined | Genome-fragment | NA | NA | 0 |
| S7_extracted_75915 | 1373 | No | NA | 3 | 3 | 0 | Low-quality | Genome-fragment | 3.03 | AAI-based (high-confidence) | 0 |
| S16_extracted_10843 | 1373 | No | NA | 2 | 0 | 0 | Low-quality | Genome-fragment | 4.21 | AAI-based (high-confidence) | 0 |
| S16_vs_54432\|\|full | 1373 | No | NA | 2 | 0 | 1 | Low-quality | Genome-fragment | 1.13 | AAI-based (high-confidence) | 0 |
| S14_vs_k141_31623\|\|full | 1488 | No | NA | 4 | 0 | 0 | Not-determined | Genome-fragment | NA | NA | 0 |
| S12_extracted_k141_158381 | 1373 | No | NA | 2 | 0 | 0 | Low-quality | Genome-fragment | 3.36 | AAI-based (medium-confidence) | 0 |
| S14_vs_k141_97256\|\|full | 2134 | No | NA | 3 | 0 | 0 | Not-determined | Genome-fragment | NA | NA | 0 |
| S14_vs_k141_17121\|\|full | 2752 | No | NA | 6 | 0 | 0 | Not-determined | Genome-fragment | NA | NA | 0 |
| S6_extracted_k141_36604 | 1373 | No | NA | 2 | 0 | 1 | Low-quality | Genome-fragment | 0.45 | AAI-based (high-confidence) | 0 |
| S6_vs_k141_14364\|\|full | 1373 | No | NA | 4 | 1 | 0 | Low-quality | Genome-fragment | 0.85 | HMM-based (lower-bound) | 0 |
| S14_vs_k141_2191\|\|full | 1952 | No | NA | 5 | 0 | 0 | Not-determined | Genome-fragment | NA | NA | 0 |
| S8_vs_k141_105291\|\|full | 1373 | No | NA | 3 | 0 | 0 | Low-quality | Genome-fragment | 3.1 | AAI-based (medium-confidence) | 0 |
| S14_vs_k141_75686\|\|full | 13410 | No | NA | 13 | 0 | 2 | Not-determined | Genome-fragment | NA | NA | 0 |
| S14_vs_k141_9574\|\|full | 2241 | No | NA | 6 | 0 | 0 | Not-determined | Genome-fragment | NA | NA | 0 |
| S17_extracted_k141_119139 | 1374 | No | NA | 3 | 3 | 0 | Low-quality | Genome-fragment | 2.3 | AAI-based (medium-confidence) | 0 |
| S17_vs_k141_167981\|\|full | 1374 | No | NA | 3 | 0 | 0 | Low-quality | Genome-fragment | 1.63 | AAI-based (high-confidence) | 0 |
| S9_extracted_k141_65481 | 1374 | No | NA | 2 | 2 | 0 | Low-quality | Genome-fragment | 4.09 | AAI-based (high-confidence) | 0 |
| S9_vs_k141_79846\|\|full | 1374 | No | NA | 3 | 1 | 0 | Low-quality | Genome-fragment | 3.56 | AAI-based (medium-confidence) | 0 |
| S9_vs_k141_81230\|\|full | 1374 | No | NA | 3 | 0 | 0 | Low-quality | Genome-fragment | 26.63 | AAI-based (medium-confidence) | 0 |
| S14_vs_k141_83117\|\|full | 2419 | No | NA | 3 | 0 | 0 | Not-determined | Genome-fragment | NA | NA | 0 |
| S14_extracted_k141_45456 | 1374 | No | NA | 2 | 2 | 0 | Low-quality | Genome-fragment | 4.42 | AAI-based (high-confidence) | 0 |
| S14_vs_k141_97324\|\|full | 1882 | No | NA | 3 | 0 | 0 | Not-determined | Genome-fragment | NA | NA | 0 |
| S14_vs_k141_111011\|\|full | 1374 | No | NA | 2 | 0 | 0 | Low-quality | Genome-fragment | 3 | AAI-based (high-confidence) | 0 |
| S14_vs_k141_112522\|\|full | 2080 | No | NA | 3 | 0 | 1 | Not-determined | Genome-fragment | NA | NA | 0 |
| S14_vs_k141_61114\|\|full | 1126 | No | NA | 2 | 0 | 1 | Not-determined | Genome-fragment | NA | NA | 0 |
| S14_vs_k141_31725\|\|full | 4332 | No | NA | 8 | 0 | 2 | Not-determined | Genome-fragment | NA | NA | 0 |
| S14_vs_k141_97349\|\|full | 5142 | No | NA | 5 | 0 | 1 | Not-determined | Genome-fragment | NA | NA | 0 |
| S14_vs_k141_70246\|\|full | 1374 | No | NA | 4 | 1 | 0 | Low-quality | Genome-fragment | 2.75 | AAI-based (high-confidence) | 0 |
| S3_extracted_k141_73281 | 1374 | No | NA | 1 | 1 | 0 | Low-quality | Genome-fragment | 2.21 | HMM-based (lower-bound) | 0 |
| S14_vs_k141_97352\|\|full | 2232 | No | NA | 3 | 0 | 0 | Not-determined | Genome-fragment | NA | NA | 0 |
| S14_vs_k141_31732\|\|full | 1951 | No | NA | 3 | 0 | 2 | Not-determined | Genome-fragment | NA | NA | 0 |
| S16_vs_170074\|\|full | 1374 | No | NA | 4 | 1 | 0 | Low-quality | Genome-fragment | 0.8 | AAI-based (high-confidence) | 0 |
| S10_vs_24438\|\|lt2gene | 1374 | No | NA | 2 | 1 | 0 | Low-quality | Genome-fragment | 2.6 | HMM-based (lower-bound) | 0 |
| S14_vs_k141_75808\|\|full | 5698 | No | NA | 5 | 0 | 3 | Not-determined | Genome-fragment | NA | NA | 0 |
| S2_extracted_k141_11527 | 1374 | No | NA | 2 | 1 | 0 | Low-quality | Genome-fragment | 4.53 | AAI-based (high-confidence) | 0 |
| S14_vs_k141_97371\|\|full | 2114 | No | NA | 3 | 0 | 2 | Not-determined | Genome-fragment | NA | NA | 0 |
| S8_vs_k141_176661\|\|full | 1374 | No | NA | 3 | 0 | 0 | Low-quality | Genome-fragment | 3.03 | AAI-based (medium-confidence) | 0 |
| S14_vs_k141_53854\|\|full | 9579 | No | NA | 10 | 0 | 6 | Not-determined | Genome-fragment | NA | NA | 0 |
| S14_vs_k141_53857\|\|full | 2562 | No | NA | 3 | 0 | 3 | Not-determined | Genome-fragment | NA | NA | 0 |
| S14_vs_k141_31768\|\|full | 1004 | No | NA | 3 | 0 | 0 | Not-determined | Genome-fragment | NA | NA | 0 |
| S1_vs_k141_32753\|\|full | 1375 | No | NA | 2 | 0 | 1 | Low-quality | Genome-fragment | 0.86 | AAI-based (medium-confidence) | 0 |
| S1_vs_k141_5446\|\|full | 1375 | No | NA | 3 | 0 | 1 | Low-quality | Genome-fragment | 1.35 | AAI-based (high-confidence) | 0 |
| S14_vs_k141_17287\|\|full | 4161 | No | NA | 7 | 0 | 3 | Not-determined | Genome-fragment | NA | NA | 0 |
| S3_vs_k141_97863\|\|full | 1375 | No | NA | 4 | 2 | 0 | Low-quality | Genome-fragment | 2.78 | AAI-based (high-confidence) | 0 |
| S5_vs_k141_72878\|\|full | 1375 | No | NA | 3 | 0 | 0 | Low-quality | Genome-fragment | 1.17 | AAI-based (medium-confidence) | 0 |
| S5_vs_k141_109895\|\|full | 1375 | No | NA | 3 | 2 | 0 | Low-quality | Genome-fragment | 2.87 | AAI-based (medium-confidence) | 0 |
| S5_vs_k141_81338\|\|lt2gene | 1375 | No | NA | 2 | 1 | 0 | Low-quality | Genome-fragment | 2.76 | AAI-based (medium-confidence) | 0 |
| S16_vs_43957\|\|full | 1375 | No | NA | 3 | 0 | 0 | Low-quality | Genome-fragment | 2.87 | AAI-based (high-confidence) | 0 |
| S14_vs_k141_31811\|\|full | 5106 | No | NA | 6 | 0 | 1 | Not-determined | Genome-fragment | NA | NA | 0 |
| S16_vs_110218\|\|full | 1375 | No | NA | 4 | 3 | 0 | Low-quality | Genome-fragment | 2.24 | AAI-based (high-confidence) | 0 |
| S14_vs_k141_2294\|\|full | 1221 | No | NA | 2 | 0 | 0 | Not-determined | Genome-fragment | NA | NA | 0 |
| S10_vs_54538\|\|full | 1375 | No | NA | 1 | 0 | 1 | Low-quality | Genome-fragment | 2.33 | AAI-based (high-confidence) | 0 |
| S10_vs_114138\|\|lt2gene | 1375 | No | NA | 2 | 1 | 0 | Low-quality | Genome-fragment | 1.81 | HMM-based (lower-bound) | 0 |
| S14_vs_k141_97429\|\|full | 3446 | No | NA | 4 | 0 | 1 | Not-determined | Genome-fragment | NA | NA | 0 |
| S14_vs_k141_9702\|\|full | 3884 | No | NA | 4 | 0 | 0 | Not-determined | Genome-fragment | NA | NA | 0 |
| S14_vs_k141_9704\|\|full | 1377 | No | NA | 3 | 0 | 0 | Not-determined | Genome-fragment | NA | NA | 0 |
| S14_vs_k141_105536\|\|full | 1101 | No | NA | 3 | 0 | 1 | Not-determined | Genome-fragment | NA | NA | 0 |
| S14_vs_k141_68318\|\|full | 2322 | No | NA | 3 | 0 | 1 | Not-determined | Genome-fragment | NA | NA | 0 |
| S14_vs_k141_105549\|\|full | 1169 | No | NA | 2 | 0 | 1 | Not-determined | Genome-fragment | NA | NA | 0 |
| S12_vs_k141_17673\|\|full | 1375 | No | NA | 4 | 1 | 0 | Low-quality | Genome-fragment | 3.26 | AAI-based (high-confidence) | 0 |
| S14_vs_k141_75965\|\|full | 2251 | No | NA | 3 | 0 | 1 | Not-determined | Genome-fragment | NA | NA | 0 |
| S14_vs_k141_75967\|\|full | 1242 | No | NA | 3 | 0 | 1 | Not-determined | Genome-fragment | NA | NA | 0 |
| S2_vs_k141_5055\|\|lt2gene | 1375 | No | NA | 1 | 1 | 0 | Low-quality | Genome-fragment | 2.6 | HMM-based (lower-bound) | 0 |
| S14_vs_k141_31846\|\|full | 34007 | No | NA | 21 | 0 | 16 | Not-determined | Genome-fragment | NA | NA | 0 |
| S4_extracted_6710 | 1375 | No | NA | 3 | 0 | 1 | Low-quality | Genome-fragment | 3.25 | AAI-based (high-confidence) | 0 |
| S14_vs_k141_31848\|\|full | 2062 | No | NA | 3 | 0 | 2 | Not-determined | Genome-fragment | NA | NA | 0 |
| S8_vs_k141_19850\|\|full | 1375 | No | NA | 4 | 0 | 0 | Low-quality | Genome-fragment | 3.19 | AAI-based (medium-confidence) | 0 |
| S8_vs_k141_72513\|\|full | 1375 | No | NA | 3 | 0 | 1 | Low-quality | Genome-fragment | 2.02 | AAI-based (medium-confidence) | 0 |
| S9_vs_k141_16185\|\|lt2gene | 1376 | No | NA | 1 | 1 | 0 | Low-quality | Genome-fragment | 1.26 | HMM-based (lower-bound) | 0 |
| S13_extracted_11453 | 1376 | No | NA | 5 | 2 | 0 | Low-quality | Genome-fragment | 2.31 | AAI-based (high-confidence) | 0 |
| S14_vs_k141_68407\|\|full | 1705 | No | NA | 2 | 0 | 0 | Not-determined | Genome-fragment | NA | NA | 0 |
| S14_vs_k141_24356\|\|full | 2308 | No | NA | 4 | 0 | 1 | Not-determined | Genome-fragment | NA | NA | 0 |
| S14_vs_k141_54000\|\|full | 1450 | No | NA | 2 | 0 | 2 | Not-determined | Genome-fragment | NA | NA | 0 |
| S14_vs_k141_60518\|\|full | 1376 | No | NA | 3 | 0 | 0 | Low-quality | Genome-fragment | 3.47 | AAI-based (medium-confidence) | 0 |
| S14_vs_k141_49403\|\|full | 1376 | No | NA | 3 | 1 | 0 | Low-quality | Genome-fragment | 2.09 | HMM-based (lower-bound) | 0 |
| S1_vs_k141_146922\|\|full | 1376 | No | NA | 5 | 0 | 0 | Low-quality | Genome-fragment | 2.81 | AAI-based (high-confidence) | 0 |
| S14_vs_k141_24380\|\|full | 1635 | No | NA | 2 | 0 | 1 | Not-determined | Genome-fragment | NA | NA | 0 |
| S1_vs_k141_119898\|\|full | 1376 | No | NA | 2 | 1 | 0 | Low-quality | Genome-fragment | 0.87 | HMM-based (lower-bound) | 0 |
| S14_vs_k141_2383\|\|full | 2456 | No | NA | 3 | 0 | 1 | Not-determined | Genome-fragment | NA | NA | 0 |
| S14_vs_k141_68453\|\|full | 1130 | No | NA | 3 | 0 | 0 | Not-determined | Genome-fragment | NA | NA | 0 |
| S5_vs_k141_53758\|\|full | 1376 | No | NA | 3 | 0 | 0 | Low-quality | Genome-fragment | 1.81 | AAI-based (high-confidence) | 0 |
| S14_vs_k141_31917\|\|full | 7512 | No | NA | 8 | 0 | 6 | Not-determined | Genome-fragment | NA | NA | 0 |
| S5_vs_k141_89901\|\|full | 1376 | No | NA | 2 | 0 | 0 | Low-quality | Genome-fragment | 2.98 | AAI-based (medium-confidence) | 0 |
| S5_vs_k141_66618\|\|full | 1376 | No | NA | 2 | 1 | 0 | Low-quality | Genome-fragment | 3.1 | AAI-based (high-confidence) | 0 |
| S14_vs_k141_68470\|\|full | 6116 | No | NA | 9 | 0 | 7 | Not-determined | Genome-fragment | NA | NA | 0 |
| S16_vs_107426\|\|full | 1376 | No | NA | 3 | 0 | 1 | Low-quality | Genome-fragment | 1.93 | AAI-based (medium-confidence) | 0 |
| S16_vs_107916\|\|full | 1376 | No | NA | 3 | 3 | 0 | Low-quality | Genome-fragment | 3.22 | AAI-based (medium-confidence) | 0 |
| S10_vs_20087\|\|full | 1376 | No | NA | 2 | 0 | 0 | Low-quality | Genome-fragment | 3.84 | AAI-based (high-confidence) | 0 |
| S12_extracted_k141_98132 | 1376 | No | NA | 1 | 0 | 1 | Low-quality | Genome-fragment | 1.42 | AAI-based (medium-confidence) | 0 |
| S12_vs_k141_71122\|\|full | 1376 | No | NA | 4 | 0 | 0 | Low-quality | Genome-fragment | 2.34 | AAI-based (high-confidence) | 0 |
| S14_vs_k141_105634\|\|full | 2820 | No | NA | 6 | 0 | 0 | Not-determined | Genome-fragment | NA | NA | 0 |
| S11_extracted_k141_16151 | 1377 | No | NA | 1 | 0 | 0 | Low-quality | Genome-fragment | 1.74 | AAI-based (high-confidence) | 0 |
| S11_extracted_k141_90601 | 1377 | No | NA | 3 | 0 | 0 | Low-quality | Genome-fragment | 1.54 | AAI-based (high-confidence) | 0 |
| S14_vs_k141_83347\|\|full | 3204 | No | NA | 3 | 0 | 2 | Not-determined | Genome-fragment | NA | NA | 0 |
| S14_extracted_k141_70783 | 1377 | No | NA | 4 | 0 | 0 | Low-quality | Genome-fragment | 3.58 | AAI-based (high-confidence) | 0 |
| S14_vs_k141_105672\|\|full | 1755 | No | NA | 3 | 0 | 0 | Not-determined | Genome-fragment | NA | NA | 0 |
| S14_vs_k141_61302\|\|full | 1366 | No | NA | 2 | 0 | 1 | Not-determined | Genome-fragment | NA | NA | 0 |
| S14_vs_k141_2523\|\|full | 2604 | No | NA | 3 | 0 | 2 | Not-determined | Genome-fragment | NA | NA | 0 |
| S14_vs_k141_83374\|\|full | 1167 | No | NA | 2 | 0 | 0 | Not-determined | Genome-fragment | NA | NA | 0 |
| S14_vs_k141_97664\|\|full | 1798 | No | NA | 3 | 0 | 0 | Not-determined | Genome-fragment | NA | NA | 0 |
| S14_vs_k141_39602\|\|full | 1517 | No | NA | 2 | 0 | 2 | Not-determined | Genome-fragment | NA | NA | 0 |
| S14_vs_k141_32497\|\|full | 1377 | No | NA | 3 | 0 | 0 | Low-quality | Genome-fragment | 2.4 | AAI-based (medium-confidence) | 0 |
| S1_extracted_k141_137327 | 1377 | No | NA | 1 | 1 | 0 | Low-quality | Genome-fragment | 2.15 | AAI-based (high-confidence) | 0 |
| S3_vs_k141_114478\|\|full | 1377 | No | NA | 3 | 1 | 0 | Low-quality | Genome-fragment | 2.57 | AAI-based (medium-confidence) | 0 |
| S5_extracted_k141_11534 | 1377 | No | NA | 1 | 0 | 0 | Low-quality | Genome-fragment | 0.52 | AAI-based (medium-confidence) | 0 |
| S5_vs_k141_18763\|\|lt2gene | 1377 | No | NA | 2 | 2 | 0 | Low-quality | Genome-fragment | 3.19 | AAI-based (high-confidence) | 0 |
| S5_vs_k141_143057\|\|lt2gene | 1377 | No | NA | 2 | 2 | 0 | Low-quality | Genome-fragment | 2.96 | AAI-based (high-confidence) | 0 |
| S7_vs_28955\|\|full | 1377 | No | NA | 2 | 0 | 0 | Low-quality | Genome-fragment | 0.75 | AAI-based (high-confidence) | 0 |
| S14_vs_k141_32051\|\|full | 1199 | No | NA | 3 | 0 | 0 | Not-determined | Genome-fragment | NA | NA | 0 |
| S16_vs_170351\|\|full | 1377 | No | NA | 3 | 1 | 0 | Low-quality | Genome-fragment | 2.88 | AAI-based (high-confidence) | 0 |
| S12_extracted_k141_154730 | 1377 | No | NA | 2 | 0 | 0 | Low-quality | Genome-fragment | 1.96 | AAI-based (medium-confidence) | 0 |
| S14_vs_k141_112800\|\|full | 1117 | No | NA | 3 | 0 | 2 | Not-determined | Genome-fragment | NA | NA | 0 |
| S14_vs_k141_105739\|\|full | 1273 | No | NA | 3 | 0 | 0 | Not-determined | Genome-fragment | NA | NA | 0 |
| S14_vs_k141_17574\|\|full | 1959 | No | NA | 2 | 0 | 2 | Not-determined | Genome-fragment | NA | NA | 0 |
| S12_vs_k141_81336\|\|full | 1377 | No | NA | 3 | 2 | 0 | Low-quality | Genome-fragment | 3.02 | AAI-based (high-confidence) | 0 |
| S12_vs_k141_184260\|\|full | 1377 | No | NA | 3 | 0 | 3 | Low-quality | Genome-fragment | 1.83 | AAI-based (medium-confidence) | 0 |
| S12_vs_k141_90736\|\|full | 1377 | No | NA | 4 | 0 | 0 | Low-quality | Genome-fragment | 3.45 | AAI-based (high-confidence) | 0 |
| S15_vs_63955\|\|full | 1377 | No | NA | 4 | 0 | 0 | Low-quality | Genome-fragment | 3.03 | AAI-based (high-confidence) | 0 |
| S8_extracted_k141_18023 | 1377 | No | NA | 3 | 2 | 0 | Low-quality | Genome-fragment | 21.6 | AAI-based (high-confidence) | 0 |
| S8_extracted_k141_177074 | 1377 | No | NA | 2 | 1 | 0 | Low-quality | Genome-fragment | 1.99 | HMM-based (lower-bound) | 0 |
| S17_vs_k141_50495\|\|full | 1378 | No | NA | 2 | 0 | 1 | Low-quality | Genome-fragment | 0.84 | AAI-based (high-confidence) | 0 |
| S14_vs_k141_83488\|\|full | 3065 | No | NA | 4 | 0 | 2 | Not-determined | Genome-fragment | NA | NA | 0 |
| S9_extracted_k141_37803 | 1378 | No | NA | 1 | 0 | 1 | Low-quality | Genome-fragment | 2.42 | AAI-based (medium-confidence) | 0 |
| S14_vs_k141_68670\|\|full | 1498 | No | NA | 3 | 0 | 1 | Not-determined | Genome-fragment | NA | NA | 0 |
| S14_vs_k141_54361\|\|full | 5517 | No | NA | 5 | 0 | 4 | Not-determined | Genome-fragment | NA | NA | 0 |
| S11_vs_k141_43115\|\|full | 1378 | No | NA | 3 | 1 | 0 | Low-quality | Genome-fragment | 2.75 | AAI-based (medium-confidence) | 0 |
| S14_vs_k141_17652\|\|full | 1193 | No | NA | 3 | 0 | 0 | Not-determined | Genome-fragment | NA | NA | 0 |
| S13_vs_100308\|\|full | 1378 | No | NA | 4 | 2 | 0 | Low-quality | Genome-fragment | 2.3 | AAI-based (high-confidence) | 0 |
| S14_vs_k141_90910\|\|full | 2032 | No | NA | 4 | 0 | 0 | Not-determined | Genome-fragment | NA | NA | 0 |
| S14_vs_k141_61400\|\|full | 2394 | No | NA | 3 | 0 | 2 | Not-determined | Genome-fragment | NA | NA | 0 |
| S14_vs_k141_61420\|\|full | 1443 | No | NA | 2 | 0 | 1 | Not-determined | Genome-fragment | NA | NA | 0 |
| S14_vs_k141_76342\|\|full | 3893 | No | NA | 3 | 0 | 2 | Not-determined | Genome-fragment | NA | NA | 0 |
| S14_vs_k141_17668\|\|full | 5466 | No | NA | 3 | 0 | 0 | Not-determined | Genome-fragment | NA | NA | 0 |
| S14_vs_k141_39784\|\|full | 5300 | No | NA | 3 | 0 | 3 | Not-determined | Genome-fragment | NA | NA | 0 |
| S14_vs_k141_61445\|\|full | 1783 | No | NA | 2 | 0 | 1 | Not-determined | Genome-fragment | NA | NA | 0 |
| S1_vs_k141_105741\|\|full | 1378 | No | NA | 2 | 0 | 2 | Low-quality | Genome-fragment | 0.67 | AAI-based (medium-confidence) | 0 |
| S14_vs_k141_17695\|\|full | 1995 | No | NA | 3 | 0 | 3 | Not-determined | Genome-fragment | NA | NA | 0 |
| S1_vs_k141_25700\|\|lt2gene | 1378 | No | NA | 1 | 1 | 0 | Low-quality | Genome-fragment | 2.26 | AAI-based (high-confidence) | 0 |
| S3_vs_k141_19591\|\|full | 1378 | No | NA | 4 | 0 | 0 | Low-quality | Genome-fragment | 2.15 | AAI-based (medium-confidence) | 0 |
| S14_vs_k141_90934\|\|full | 37471 | No | NA | 46 | 5 | 1 | High-quality | High-quality | 98.98 | AAI-based (high-confidence) | 0 |
| S5_extracted_k141_145298 | 1378 | No | NA | 4 | 0 | 0 | Low-quality | Genome-fragment | 2.86 | AAI-based (high-confidence) | 0 |
| S14_vs_k141_105866\|\|full | 3237 | No | NA | 6 | 0 | 4 | Not-determined | Genome-fragment | NA | NA | 0 |
| S5_vs_k141_97230\|\|full | 1378 | No | NA | 3 | 1 | 0 | Low-quality | Genome-fragment | 3.44 | AAI-based (medium-confidence) | 0 |
| S5_vs_k141_67333\|\|full | 1378 | No | NA | 5 | 1 | 0 | Low-quality | Genome-fragment | 3.67 | AAI-based (medium-confidence) | 0 |
| S5_vs_k141_94272\|\|lt2gene | 1378 | No | NA | 1 | 1 | 0 | Low-quality | Genome-fragment | 2.01 | HMM-based (lower-bound) | 0 |
| S14_vs_k141_112964\|\|full | 2506 | No | NA | 3 | 0 | 2 | Not-determined | Genome-fragment | NA | NA | 0 |
| S14_vs_k141_61487\|\|full | 2173 | No | NA | 5 | 0 | 1 | Not-determined | Genome-fragment | NA | NA | 0 |
| S14_vs_k141_39797\|\|full | 41367 | No | NA | 54 | 20 | 1 | High-quality | High-quality | 95.9 | AAI-based (high-confidence) | 0 |
| S16_vs_54472\|\|full | 1378 | No | NA | 2 | 0 | 1 | Low-quality | Genome-fragment | 1.87 | AAI-based (high-confidence) | 0 |
| S16_vs_77919\|\|full | 1378 | No | NA | 2 | 1 | 0 | Low-quality | Genome-fragment | 3.26 | AAI-based (high-confidence) | 0 |
| S14_vs_k141_68777\|\|full | 1174 | No | NA | 3 | 0 | 1 | Not-determined | Genome-fragment | NA | NA | 0 |
| S16_vs_116915\|\|full | 1378 | No | NA | 4 | 1 | 0 | Low-quality | Genome-fragment | 1.24 | HMM-based (lower-bound) | 0 |
| S12_vs_k141_178102\|\|full | 1378 | Yes | 1069 | 4 | 1 | 1 | Low-quality | Genome-fragment | 1.77 | AAI-based (medium-confidence) | 22.42 |
| S15_vs_40212\|\|lt2gene | 1378 | No | NA | 2 | 1 | 0 | Low-quality | Genome-fragment | 3.18 | AAI-based (high-confidence) | 0 |
| S14_vs_k141_112976\|\|full | 11171 | No | NA | 11 | 0 | 9 | Not-determined | Genome-fragment | NA | NA | 0 |
| S4_extracted_8487 | 1378 | No | NA | 2 | 1 | 0 | Low-quality | Genome-fragment | 3.47 | AAI-based (high-confidence) | 0 |
| S14_vs_k141_105918\|\|full | 1250 | No | NA | 2 | 0 | 0 | Not-determined | Genome-fragment | NA | NA | 0 |
| S14_vs_k141_68824\|\|full | 1225 | No | NA | 2 | 0 | 0 | Not-determined | Genome-fragment | NA | NA | 0 |
| S14_vs_k141_54526\|\|full | 8733 | No | NA | 9 | 0 | 8 | Not-determined | Genome-fragment | NA | NA | 0 |
| S14_vs_k141_39857\|\|full | 12636 | No | NA | 9 | 0 | 6 | Not-determined | Genome-fragment | NA | NA | 0 |
| S9_extracted_k141_10196 | 1379 | No | NA | 2 | 0 | 0 | Low-quality | Genome-fragment | 1.81 | AAI-based (high-confidence) | 0 |
| S11_extracted_k141_84920 | 1379 | No | NA | 3 | 2 | 0 | Low-quality | Genome-fragment | 21.85 | AAI-based (high-confidence) | 0 |
| S13_extracted_132163 | 1379 | No | NA | 3 | 1 | 0 | Low-quality | Genome-fragment | 3.73 | AAI-based (medium-confidence) | 0 |
| S14_vs_k141_68858\|\|full | 3209 | No | NA | 3 | 0 | 1 | Not-determined | Genome-fragment | NA | NA | 0 |
| S3_vs_k141_109339\|\|full | 1379 | No | NA | 3 | 1 | 0 | Low-quality | Genome-fragment | 2.82 | AAI-based (medium-confidence) | 0 |
| S14_vs_k141_54569\|\|full | 2671 | No | NA | 3 | 0 | 2 | Not-determined | Genome-fragment | NA | NA | 0 |
| S14_vs_k141_91001\|\|full | 1185 | No | NA | 3 | 0 | 0 | Not-determined | Genome-fragment | NA | NA | 0 |
| S16_vs_4414\|\|lt2gene | 1379 | No | NA | 2 | 2 | 0 | Low-quality | Genome-fragment | 3.46 | AAI-based (medium-confidence) | 0 |
| S14_vs_k141_97835\|\|full | 2070 | No | NA | 4 | 1 | 0 | Medium-quality | Genome-fragment | 60.76 | AAI-based (high-confidence) | 0 |
| S10_extracted_8693 | 1379 | No | NA | 1 | 0 | 1 | Low-quality | Genome-fragment | 2.45 | AAI-based (medium-confidence) | 0 |
| S6_extracted_k141_22610 | 1379 | No | NA | 2 | 0 | 1 | Low-quality | Genome-fragment | 0.46 | AAI-based (medium-confidence) | 0 |
| S14_vs_k141_24772\|\|full | 1042 | No | NA | 3 | 0 | 0 | Not-determined | Genome-fragment | NA | NA | 0 |
| S8_extracted_k141_32252 | 1379 | No | NA | 2 | 0 | 1 | Low-quality | Genome-fragment | 0.86 | AAI-based (medium-confidence) | 0 |
| S17_vs_k141_144224\|\|full | 1380 | No | NA | 5 | 5 | 0 | Low-quality | Genome-fragment | 3.17 | AAI-based (medium-confidence) | 0 |
| S9_vs_k141_49330\|\|full | 1380 | No | NA | 3 | 0 | 1 | Low-quality | Genome-fragment | 2.37 | AAI-based (medium-confidence) | 0 |
| S1_extracted_k141_23277 | 1380 | No | NA | 1 | 0 | 0 | Low-quality | Genome-fragment | 0.79 | AAI-based (high-confidence) | 0 |
| S1_vs_k141_95975\|\|full | 1380 | No | NA | 1 | 0 | 0 | Low-quality | Genome-fragment | 3.44 | AAI-based (high-confidence) | 0 |
| S14_vs_k141_24800\|\|full | 1099 | No | NA | 2 | 0 | 2 | Not-determined | Genome-fragment | NA | NA | 0 |
| S14_vs_k141_46577\|\|full | 2198 | No | NA | 4 | 0 | 1 | Not-determined | Genome-fragment | NA | NA | 0 |
| S3_extracted_k141_108605 | 1380 | No | NA | 3 | 0 | 0 | Low-quality | Genome-fragment | 2.23 | AAI-based (high-confidence) | 0 |
| S5_vs_k141_135332\|\|full | 1380 | No | NA | 3 | 0 | 0 | Low-quality | Genome-fragment | 2.47 | AAI-based (medium-confidence) | 0 |
| S5_vs_k141_88284\|\|full | 1380 | No | NA | 2 | 1 | 0 | Low-quality | Genome-fragment | 2.67 | AAI-based (high-confidence) | 0 |
| S5_vs_k141_34900\|\|full | 1380 | Yes | 711 | 3 | 1 | 1 | Low-quality | Genome-fragment | 0.42 | HMM-based (lower-bound) | 48.48 |
| S14_vs_k141_46599\|\|full | 1732 | No | NA | 3 | 0 | 2 | Not-determined | Genome-fragment | NA | NA | 0 |
| S16_extracted_71116 | 1380 | No | NA | 4 | 0 | 0 | Low-quality | Genome-fragment | 2.27 | AAI-based (high-confidence) | 0 |
| S16_vs_2248\|\|full | 1380 | No | NA | 4 | 0 | 0 | Low-quality | Genome-fragment | 1.26 | AAI-based (high-confidence) | 0 |
| S10_extracted_30512 | 1380 | No | NA | 2 | 0 | 0 | Low-quality | Genome-fragment | 3 | AAI-based (high-confidence) | 0 |
| S8_vs_k141_145874\|\|full | 1380 | No | NA | 3 | 0 | 0 | Low-quality | Genome-fragment | 1.39 | AAI-based (high-confidence) | 0 |
| S8_vs_k141_156036\|\|full | 1380 | No | NA | 3 | 0 | 2 | Low-quality | Genome-fragment | 1.79 | AAI-based (medium-confidence) | 0 |
| S14_vs_k141_17836\|\|full | 6466 | No | NA | 7 | 0 | 3 | Not-determined | Genome-fragment | NA | NA | 0 |
| S17_vs_k141_52467\|\|full | 1381 | No | NA | 3 | 2 | 0 | Low-quality | Genome-fragment | 4.06 | AAI-based (high-confidence) | 0 |
| S14_vs_k141_91116\|\|full | 2208 | No | NA | 5 | 0 | 4 | Not-determined | Genome-fragment | NA | NA | 0 |
| S14_vs_k141_10330\|\|full | 2034 | No | NA | 3 | 0 | 3 | Not-determined | Genome-fragment | NA | NA | 0 |
| S14_vs_k141_83863\|\|full | 6807 | No | NA | 10 | 0 | 0 | Not-determined | Genome-fragment | NA | NA | 0 |
| S17_vs_k141_153054\|\|full | 1381 | No | NA | 2 | 1 | 0 | Low-quality | Genome-fragment | 1.63 | AAI-based (medium-confidence) | 0 |
| S14_vs_k141_61529\|\|full | 1407 | No | NA | 3 | 0 | 2 | Not-determined | Genome-fragment | NA | NA | 0 |
| S14_vs_k141_83875\|\|full | 5690 | No | NA | 4 | 0 | 4 | Not-determined | Genome-fragment | NA | NA | 0 |
| S14_vs_k141_68974\|\|full | 2394 | No | NA | 4 | 0 | 1 | Not-determined | Genome-fragment | NA | NA | 0 |
| S14_extracted_k141_116320 | 1381 | No | NA | 3 | 0 | 0 | Low-quality | Genome-fragment | 2.35 | AAI-based (high-confidence) | 0 |
| S14_vs_k141_113085\|\|full | 12500 | No | NA | 9 | 0 | 5 | Not-determined | Genome-fragment | NA | NA | 0 |
| S14_vs_k141_46693\|\|full | 3578 | No | NA | 7 | 0 | 1 | Not-determined | Genome-fragment | NA | NA | 0 |
| S14_vs_k141_61530\|\|full | 30159 | No | NA | 49 | 15 | 0 | Medium-quality | Genome-fragment | 89.33 | AAI-based (medium-confidence) | 0 |
| S14_vs_k141_2870\|\|full | 88314 | Yes | 46524 | 94 | 30 | 19 | Medium-quality | Genome-fragment | 86.01 | AAI-based (high-confidence) | 47.32 |
| S14_vs_k141_24922\|\|full | 1973 | No | NA | 3 | 0 | 0 | Not-determined | Genome-fragment | NA | NA | 0 |
| S14_vs_k141_106136\|\|full | 1920 | No | NA | 2 | 0 | 1 | Not-determined | Genome-fragment | NA | NA | 0 |
| S14_vs_k141_17906\|\|full | 2916 | No | NA | 2 | 0 | 1 | Not-determined | Genome-fragment | NA | NA | 0 |
| S14_vs_k141_107252\|\|full | 1381 | No | NA | 4 | 1 | 0 | Low-quality | Genome-fragment | 2.95 | AAI-based (medium-confidence) | 0 |
| S14_vs_k141_11792\|\|full | 1381 | No | NA | 3 | 0 | 0 | Low-quality | Genome-fragment | 3.16 | AAI-based (medium-confidence) | 0 |
| S14_vs_k141_76700\|\|full | 3812 | No | NA | 7 | 0 | 5 | Not-determined | Genome-fragment | NA | NA | 0 |
| S1_extracted_k141_108885 | 1381 | No | NA | 2 | 0 | 0 | Low-quality | Genome-fragment | 2.58 | AAI-based (high-confidence) | 0 |
| S14_vs_k141_32499\|\|full | 78663 | Yes | 27007 | 95 | 11 | 38 | Complete | High-quality | 100 | Provirus (medium-confidence) | 65.67 |
| S14_vs_k141_24969\|\|full | 1705 | No | NA | 3 | 0 | 0 | Not-determined | Genome-fragment | NA | NA | 0 |
| S14_vs_k141_61629\|\|full | 1599 | No | NA | 3 | 0 | 0 | Not-determined | Genome-fragment | NA | NA | 0 |
| S14_vs_k141_32522\|\|full | 1856 | No | NA | 3 | 0 | 3 | Not-determined | Genome-fragment | NA | NA | 0 |
| S14_vs_k141_113169\|\|full | 1691 | No | NA | 7 | 0 | 0 | Not-determined | Genome-fragment | NA | NA | 0 |
| S1_vs_k141_98109\|\|full | 1381 | No | NA | 3 | 1 | 0 | Low-quality | Genome-fragment | 3.92 | AAI-based (high-confidence) | 0 |
| S14_vs_k141_61662\|\|full | 1136 | No | NA | 2 | 0 | 1 | Not-determined | Genome-fragment | NA | NA | 0 |
| S14_vs_k141_61668\|\|full | 1581 | No | NA | 3 | 0 | 1 | Not-determined | Genome-fragment | NA | NA | 0 |
| S3_vs_k141_1664\|\|full | 1381 | No | NA | 2 | 0 | 0 | Low-quality | Genome-fragment | 2.68 | AAI-based (medium-confidence) | 0 |
| S7_vs_47615\|\|full | 1381 | No | NA | 4 | 3 | 0 | Low-quality | Genome-fragment | 3.82 | AAI-based (high-confidence) | 0 |
| S7_vs_69477\|\|full | 1381 | No | NA | 4 | 1 | 0 | Low-quality | Genome-fragment | 2.72 | AAI-based (medium-confidence) | 0 |
| S14_vs_k141_113226\|\|full | 1637 | No | NA | 2 | 0 | 2 | Not-determined | Genome-fragment | NA | NA | 0 |
| S14_vs_k141_113227\|\|full | 1621 | No | NA | 2 | 0 | 2 | Not-determined | Genome-fragment | NA | NA | 0 |
| S7_vs_70385\|\|full | 1381 | No | NA | 3 | 0 | 0 | Low-quality | Genome-fragment | 0.76 | AAI-based (medium-confidence) | 0 |
| S7_vs_87594\|\|full | 1381 | No | NA | 2 | 0 | 0 | Low-quality | Genome-fragment | 0.87 | AAI-based (medium-confidence) | 0 |
| S14_vs_k141_91308\|\|full | 1840 | No | NA | 3 | 0 | 1 | Not-determined | Genome-fragment | NA | NA | 0 |
| S14_vs_k141_83976\|\|full | 1404 | No | NA | 4 | 0 | 0 | Not-determined | Genome-fragment | NA | NA | 0 |
| S14_vs_k141_98150\|\|full | 1205 | No | NA | 2 | 0 | 0 | Not-determined | Genome-fragment | NA | NA | 0 |
| S14_vs_k141_113258\|\|full | 2176 | No | NA | 2 | 0 | 2 | Not-determined | Genome-fragment | NA | NA | 0 |
| S7_vs_105916\|\|full | 1381 | No | NA | 3 | 0 | 0 | Low-quality | Genome-fragment | 3.62 | AAI-based (medium-confidence) | 0 |
| S14_vs_k141_83996\|\|full | 1589 | No | NA | 2 | 0 | 1 | Not-determined | Genome-fragment | NA | NA | 0 |
| S9_vs_k141_14951\|\|full | 1382 | No | NA | 3 | 2 | 0 | Low-quality | Genome-fragment | 3.11 | AAI-based (high-confidence) | 0 |
| S14_vs_k141_69183\|\|full | 6075 | No | NA | 4 | 0 | 2 | Not-determined | Genome-fragment | NA | NA | 0 |
| S14_vs_k141_69184\|\|full | 3022 | No | NA | 4 | 0 | 3 | Not-determined | Genome-fragment | NA | NA | 0 |
| S14_vs_k141_98169\|\|full | 3743 | No | NA | 5 | 0 | 2 | Not-determined | Genome-fragment | NA | NA | 0 |
| S14_extracted_k141_78974 | 1382 | No | NA | 3 | 0 | 2 | Low-quality | Genome-fragment | 1.02 | AAI-based (medium-confidence) | 0 |
| S14_vs_k141_18066\|\|full | 6339 | No | NA | 7 | 0 | 4 | Not-determined | Genome-fragment | NA | NA | 0 |
| S1_extracted_k141_132069 | 1382 | No | NA | 2 | 1 | 0 | Low-quality | Genome-fragment | 3.48 | AAI-based (high-confidence) | 0 |
| S14_vs_k141_106240\|\|full | 3450 | No | NA | 4 | 0 | 2 | Not-determined | Genome-fragment | NA | NA | 0 |
| S14_vs_k141_46812\|\|full | 1449 | No | NA | 3 | 0 | 1 | Not-determined | Genome-fragment | NA | NA | 0 |
| S14_vs_k141_106248\|\|full | 1253 | No | NA | 3 | 0 | 0 | Not-determined | Genome-fragment | NA | NA | 0 |
| S1_vs_k141_42951\|\|full | 1382 | No | NA | 3 | 0 | 0 | Low-quality | Genome-fragment | 3.05 | AAI-based (medium-confidence) | 0 |
| S14_vs_k141_91404\|\|full | 1322 | No | NA | 3 | 0 | 1 | Not-determined | Genome-fragment | NA | NA | 0 |
| S14_vs_k141_76874\|\|full | 2340 | No | NA | 4 | 0 | 1 | Not-determined | Genome-fragment | NA | NA | 0 |
| S14_vs_k141_69262\|\|full | 1839 | No | NA | 3 | 0 | 2 | Not-determined | Genome-fragment | NA | NA | 0 |
| S3_extracted_k141_84137 | 1382 | No | NA | 3 | 0 | 0 | Low-quality | Genome-fragment | 0.71 | AAI-based (high-confidence) | 0 |
| S3_vs_k141_51278\|\|full | 1382 | No | NA | 3 | 1 | 0 | Low-quality | Genome-fragment | 2.25 | HMM-based (lower-bound) | 0 |
| S7_extracted_19795 | 1382 | No | NA | 3 | 0 | 0 | Low-quality | Genome-fragment | 2.17 | AAI-based (high-confidence) | 0 |
| S14_vs_k141_25088\|\|full | 1510 | No | NA | 4 | 0 | 1 | Not-determined | Genome-fragment | NA | NA | 0 |
| S14_vs_k141_10527\|\|full | 17551 | No | NA | 12 | 0 | 8 | Not-determined | Genome-fragment | NA | NA | 0 |
| S14_vs_k141_76938\|\|full | 2518 | No | NA | 3 | 0 | 1 | Not-determined | Genome-fragment | NA | NA | 0 |
| S14_vs_k141_84126\|\|full | 1471 | No | NA | 2 | 0 | 0 | Not-determined | Genome-fragment | NA | NA | 0 |
| S14_vs_k141_113384\|\|full | 4977 | No | NA | 7 | 0 | 0 | Not-determined | Genome-fragment | NA | NA | 0 |
| S14_vs_k141_54846\|\|full | 8729 | No | NA | 6 | 0 | 4 | Not-determined | Genome-fragment | NA | NA | 0 |
| S14_vs_k141_98261\|\|full | 3306 | No | NA | 2 | 0 | 2 | Not-determined | Genome-fragment | NA | NA | 0 |
| S14_vs_k141_3122\|\|full | 2469 | No | NA | 3 | 0 | 3 | Not-determined | Genome-fragment | NA | NA | 0 |
| S14_vs_k141_76981\|\|full | 1449 | No | NA | 2 | 0 | 1 | Not-determined | Genome-fragment | NA | NA | 0 |
| S14_vs_k141_76994\|\|full | 1802 | No | NA | 5 | 0 | 0 | Not-determined | Genome-fragment | NA | NA | 0 |
| S14_vs_k141_46941\|\|full | 2061 | No | NA | 3 | 0 | 1 | Not-determined | Genome-fragment | NA | NA | 0 |
| S7_vs_7281\|\|full | 1382 | No | NA | 3 | 1 | 0 | Low-quality | Genome-fragment | 2.08 | HMM-based (lower-bound) | 0 |
| S16_extracted_75195 | 1382 | No | NA | 3 | 0 | 3 | Low-quality | Genome-fragment | 0.52 | AAI-based (medium-confidence) | 0 |
| S10_vs_73191\|\|full | 1382 | No | NA | 3 | 0 | 1 | Low-quality | Genome-fragment | 1.83 | AAI-based (high-confidence) | 0 |
| S14_vs_k141_10582\|\|full | 2481 | No | NA | 5 | 0 | 2 | Not-determined | Genome-fragment | NA | NA | 0 |
| S12_vs_k141_148868\|\|lt2gene | 1382 | No | NA | 2 | 2 | 0 | Low-quality | Genome-fragment | 2.23 | AAI-based (medium-confidence) | 0 |
| S17_extracted_k141_13938 | 1383 | No | NA | 3 | 3 | 0 | Low-quality | Genome-fragment | 3.71 | AAI-based (high-confidence) | 0 |
| S9_vs_k141_5696\|\|full | 1383 | No | NA | 3 | 0 | 0 | Low-quality | Genome-fragment | 3.12 | AAI-based (high-confidence) | 0 |
| S11_vs_k141_101500\|\|full | 1383 | No | NA | 2 | 0 | 0 | Low-quality | Genome-fragment | 3.31 | AAI-based (medium-confidence) | 0 |
| S11_vs_k141_24944\|\|full | 1383 | No | NA | 4 | 1 | 0 | Low-quality | Genome-fragment | 2.96 | AAI-based (high-confidence) | 0 |
| S14_vs_k141_113456\|\|full | 2705 | No | NA | 4 | 0 | 1 | Not-determined | Genome-fragment | NA | NA | 0 |
| S13_vs_81796\|\|full | 1383 | No | NA | 3 | 0 | 0 | Low-quality | Genome-fragment | 1.61 | AAI-based (medium-confidence) | 0 |
| S14_vs_k141_91543\|\|full | 1765 | No | NA | 3 | 0 | 3 | Not-determined | Genome-fragment | NA | NA | 0 |
| S5_vs_k141_53533\|\|full | 1383 | No | NA | 5 | 0 | 0 | Low-quality | Genome-fragment | 3.13 | AAI-based (high-confidence) | 0 |
| S7_vs_12553\|\|full | 1383 | No | NA | 5 | 0 | 0 | Low-quality | Genome-fragment | 2.24 | AAI-based (medium-confidence) | 0 |
| S7_vs_46650\|\|full | 1383 | No | NA | 2 | 0 | 2 | Low-quality | Genome-fragment | 1.68 | AAI-based (medium-confidence) | 0 |
| S14_vs_k141_3209\|\|full | 1530 | No | NA | 2 | 0 | 1 | Not-determined | Genome-fragment | NA | NA | 0 |
| S14_vs_k141_25213\|\|full | 2637 | No | NA | 7 | 0 | 0 | Not-determined | Genome-fragment | NA | NA | 0 |
| S7_vs_62906\|\|full | 1383 | No | NA | 3 | 0 | 0 | Low-quality | Genome-fragment | 2.53 | AAI-based (high-confidence) | 0 |
| S14_vs_k141_32748\|\|full | 1064 | No | NA | 2 | 0 | 0 | Not-determined | Genome-fragment | NA | NA | 0 |
| S14_vs_k141_47026\|\|full | 1189 | No | NA | 3 | 0 | 1 | Not-determined | Genome-fragment | NA | NA | 0 |
| S10_extracted_82233 | 1383 | No | NA | 2 | 0 | 0 | Low-quality | Genome-fragment | 0.85 | AAI-based (high-confidence) | 0 |
| S14_vs_k141_25239\|\|full | 1687 | No | NA | 2 | 0 | 2 | Not-determined | Genome-fragment | NA | NA | 0 |
| S10_vs_5252\|\|full | 1383 | No | NA | 4 | 1 | 0 | Low-quality | Genome-fragment | 2.26 | HMM-based (lower-bound) | 0 |
| S6_vs_k141_7338\|\|full | 1383 | No | NA | 4 | 1 | 0 | Low-quality | Genome-fragment | 3.59 | AAI-based (medium-confidence) | 0 |
| S6_vs_k141_3053\|\|full | 1383 | No | NA | 2 | 0 | 0 | Low-quality | Genome-fragment | 2.27 | AAI-based (medium-confidence) | 0 |
| S8_vs_k141_206885\|\|full | 1383 | No | NA | 3 | 0 | 1 | Low-quality | Genome-fragment | 2.41 | AAI-based (medium-confidence) | 0 |
| S14_vs_k141_84289\|\|full | 2031 | No | NA | 2 | 0 | 0 | Not-determined | Genome-fragment | NA | NA | 0 |
| S14_vs_k141_18333\|\|full | 1463 | No | NA | 3 | 0 | 1 | Not-determined | Genome-fragment | NA | NA | 0 |
| S17_vs_k141_37189\|\|full | 1384 | No | NA | 3 | 0 | 0 | Low-quality | Genome-fragment | 0.8 | AAI-based (high-confidence) | 0 |
| S14_vs_k141_32799\|\|full | 5606 | No | NA | 7 | 0 | 4 | Not-determined | Genome-fragment | NA | NA | 0 |
| S14_vs_k141_25269\|\|full | 4579 | No | NA | 5 | 0 | 4 | Not-determined | Genome-fragment | NA | NA | 0 |
| S14_vs_k141_54951\|\|full | 1405 | No | NA | 2 | 0 | 0 | Not-determined | Genome-fragment | NA | NA | 0 |
| S9_vs_k141_71291\|\|full | 1384 | No | NA | 4 | 1 | 0 | Low-quality | Genome-fragment | 2.92 | AAI-based (medium-confidence) | 0 |
| S14_vs_k141_10693\|\|full | 1831 | No | NA | 4 | 0 | 1 | Not-determined | Genome-fragment | NA | NA | 0 |
| S11_extracted_k141_29156 | 1384 | No | NA | 2 | 0 | 0 | Low-quality | Genome-fragment | 1.16 | AAI-based (high-confidence) | 0 |
| S14_vs_k141_10729\|\|full | 7461 | No | NA | 11 | 0 | 8 | Not-determined | Genome-fragment | NA | NA | 0 |
| S13_vs_36916\|\|full | 1384 | No | NA | 2 | 0 | 0 | Low-quality | Genome-fragment | 1.74 | AAI-based (high-confidence) | 0 |
| S14_vs_k141_40471\|\|full | 1940 | No | NA | 2 | 0 | 2 | Not-determined | Genome-fragment | NA | NA | 0 |
| S13_vs_70694\|\|full | 1384 | No | NA | 4 | 1 | 0 | Low-quality | Genome-fragment | 2.66 | AAI-based (high-confidence) | 0 |
| S3_extracted_k141_28889 | 1384 | No | NA | 2 | 0 | 2 | Low-quality | Genome-fragment | 0.96 | AAI-based (high-confidence) | 0 |
| S14_vs_k141_106466\|\|full | 4249 | No | NA | 6 | 0 | 2 | Not-determined | Genome-fragment | NA | NA | 0 |
| S14_vs_k141_113588\|\|full | 1728 | No | NA | 4 | 0 | 0 | Not-determined | Genome-fragment | NA | NA | 0 |
| S14_vs_k141_69519\|\|full | 2019 | No | NA | 4 | 0 | 3 | Not-determined | Genome-fragment | NA | NA | 0 |
| S3_vs_k141_68959\|\|full | 1384 | No | NA | 3 | 0 | 1 | Low-quality | Genome-fragment | 1.11 | AAI-based (medium-confidence) | 0 |
| S5_vs_k141_49702\|\|full | 1384 | No | NA | 3 | 0 | 0 | Low-quality | Genome-fragment | 0.87 | AAI-based (medium-confidence) | 0 |
| S5_vs_k141_129266\|\|full | 1384 | No | NA | 3 | 1 | 0 | Low-quality | Genome-fragment | 4.25 | AAI-based (high-confidence) | 0 |
| S14_vs_k141_98500\|\|full | 5742 | No | NA | 4 | 0 | 1 | Not-determined | Genome-fragment | NA | NA | 0 |
| S14_vs_k141_84332\|\|full | 5215 | No | NA | 4 | 0 | 0 | Not-determined | Genome-fragment | NA | NA | 0 |
| S7_extracted_137936 | 1384 | No | NA | 3 | 2 | 0 | Low-quality | Genome-fragment | 2.86 | AAI-based (high-confidence) | 0 |
| S16_vs_172299\|\|full | 1384 | No | NA | 2 | 0 | 0 | Low-quality | Genome-fragment | 3.53 | AAI-based (high-confidence) | 0 |
| S10_vs_37395\|\|full | 1384 | No | NA | 2 | 1 | 0 | Low-quality | Genome-fragment | 2.55 | AAI-based (high-confidence) | 0 |
| S12_vs_k141_66690\|\|full | 1384 | No | NA | 3 | 0 | 1 | Low-quality | Genome-fragment | 0.33 | AAI-based (medium-confidence) | 0 |
| S12_vs_k141_171019\|\|full | 1384 | No | NA | 3 | 1 | 0 | Low-quality | Genome-fragment | 3.04 | AAI-based (high-confidence) | 0 |
| S2_vs_k141_1248\|\|full | 1384 | No | NA | 4 | 0 | 0 | Low-quality | Genome-fragment | 2.39 | AAI-based (high-confidence) | 0 |
| S6_vs_k141_4356\|\|full | 1384 | No | NA | 3 | 0 | 2 | Low-quality | Genome-fragment | 0.41 | AAI-based (medium-confidence) | 0 |
| S17_extracted_k141_98763 | 1385 | No | NA | 1 | 0 | 1 | Low-quality | Genome-fragment | 0.75 | AAI-based (medium-confidence) | 0 |
| S13_vs_146182\|\|full | 1385 | No | NA | 4 | 0 | 4 | Low-quality | Genome-fragment | 0.41 | AAI-based (medium-confidence) | 0 |
| S14_vs_k141_47250\|\|full | 6014 | No | NA | 6 | 0 | 4 | Not-determined | Genome-fragment | NA | NA | 0 |
| S7_extracted_73242 | 1385 | No | NA | 1 | 0 | 1 | Low-quality | Genome-fragment | 0.68 | AAI-based (medium-confidence) | 0 |

| | | | | | | | | | | | |
|---|---|---|---|---|---|---|---|---|---|---|---|
| S14_vs_k141_98559\|\|full | 2459 | No | NA | 3 | 0 | 2 | Not-determined | Genome-fragment | NA | NA | 0 |
| S8_vs_k141_61696\|\|full | 1385 | No | NA | 3 | 1 | 0 | Low-quality | Genome-fragment | 3.87 | AAI-based (high-confidence) | 0 |
| S14_vs_k141_18451\|\|full | 2063 | No | NA | 3 | 0 | 0 | Not-determined | Genome-fragment | NA | NA | 0 |
| S8_vs_k141_111418\|\|full | 1385 | No | NA | 6 | 2 | 0 | Low-quality | Genome-fragment | 2.53 | AAI-based (high-confidence) | 0 |
| S17_vs_k141_35792\|\|lt2gene | 1386 | No | NA | 2 | 1 | 0 | Low-quality | Genome-fragment | 2.43 | HMM-based (lower-bound) | 0 |
| S9_vs_k141_30841\|\|full | 1386 | No | NA | 5 | 0 | 0 | Low-quality | Genome-fragment | 2.87 | AAI-based (high-confidence) | 0 |
| S14_vs_k141_32922\|\|full | 1890 | No | NA | 3 | 0 | 1 | Not-determined | Genome-fragment | NA | NA | 0 |
| S14_vs_k141_47259\|\|full | 1261 | No | NA | 4 | 0 | 0 | Not-determined | Genome-fragment | NA | NA | 0 |
| S11_vs_k141_65836\|\|full | 1386 | No | NA | 2 | 0 | 2 | Low-quality | Genome-fragment | 0.56 | AAI-based (medium-confidence) | 0 |
| S14_vs_k141_47260\|\|full | 4701 | No | NA | 7 | 0 | 2 | Not-determined | Genome-fragment | NA | NA | 0 |
| S11_vs_k141_33677\|\|full | 1386 | No | NA | 4 | 1 | 0 | Low-quality | Genome-fragment | 4.15 | HMM-based (lower-bound) | 0 |
| S14_extracted_k141_65050 | 1386 | No | NA | 2 | 2 | 0 | Low-quality | Genome-fragment | 4.57 | AAI-based (high-confidence) | 0 |
| S5_vs_k141_76806\|\|full | 1386 | No | NA | 3 | 1 | 0 | Low-quality | Genome-fragment | 2.07 | HMM-based (lower-bound) | 0 |
| S7_extracted_134303 | 1386 | No | NA | 4 | 0 | 0 | Low-quality | Genome-fragment | 4.2 | AAI-based (medium-confidence) | 0 |
| S14_vs_k141_10866\|\|full | 2313 | No | NA | 4 | 0 | 3 | Not-determined | Genome-fragment | NA | NA | 0 |
| S7_vs_59453\|\|full | 1386 | No | NA | 3 | 1 | 0 | Low-quality | Genome-fragment | 2.73 | AAI-based (medium-confidence) | 0 |
| S7_vs_79004\|\|full | 1386 | No | NA | 3 | 1 | 0 | Low-quality | Genome-fragment | 0.85 | HMM-based (lower-bound) | 0 |
| S7_vs_88274\|\|full | 1386 | No | NA | 2 | 1 | 0 | Low-quality | Genome-fragment | 2.26 | AAI-based (high-confidence) | 0 |
| S14_vs_k141_32970\|\|full | 4939 | No | NA | 5 | 0 | 1 | Not-determined | Genome-fragment | NA | NA | 0 |
| S7_vs_41681\|\|lt2gene | 1386 | No | NA | 1 | 1 | 0 | Low-quality | Genome-fragment | 1.28 | HMM-based (lower-bound) | 0 |
| S16_extracted_30145 | 1386 | No | NA | 4 | 0 | 0 | Low-quality | Genome-fragment | 3.23 | AAI-based (medium-confidence) | 0 |
| S8_vs_k141_26077\|\|full | 1386 | No | NA | 2 | 1 | 0 | Low-quality | Genome-fragment | 1.59 | HMM-based (lower-bound) | 0 |
| S11_vs_k141_91648\|\|lt2gene | 1387 | No | NA | 1 | 1 | 0 | Low-quality | Genome-fragment | 1.3 | HMM-based (lower-bound) | 0 |
| S1_vs_k141_58506\|\|full | 1387 | No | NA | 2 | 0 | 0 | Low-quality | Genome-fragment | 2.13 | AAI-based (medium-confidence) | 0 |
| S1_vs_k141_71541\|\|full | 1387 | No | NA | 3 | 1 | 0 | Low-quality | Genome-fragment | 4.54 | AAI-based (high-confidence) | 0 |
| S5_vs_k141_52984\|\|full | 1387 | No | NA | 3 | 3 | 0 | Low-quality | Genome-fragment | 3.17 | AAI-based (high-confidence) | 0 |
| S16_extracted_125584 | 1387 | No | NA | 4 | 0 | 0 | Low-quality | Genome-fragment | 2.59 | AAI-based (medium-confidence) | 0 |
| S14_vs_k141_69659\|\|full | 2454 | No | NA | 5 | 0 | 1 | Not-determined | Genome-fragment | NA | NA | 0 |
| S16_vs_45824\|\|full | 1387 | No | NA | 3 | 0 | 0 | Low-quality | Genome-fragment | 0.91 | AAI-based (high-confidence) | 0 |
| S16_vs_107866\|\|full | 1387 | No | NA | 2 | 0 | 0 | Low-quality | Genome-fragment | 2.33 | AAI-based (medium-confidence) | 0 |
| S12_extracted_k141_155104 | 1387 | No | NA | 6 | 0 | 0 | Low-quality | Genome-fragment | 2.5 | AAI-based (medium-confidence) | 0 |
| S14_vs_k141_10942\|\|full | 2020 | No | NA | 3 | 0 | 2 | Not-determined | Genome-fragment | NA | NA | 0 |
| S14_vs_k141_69695\|\|full | 2972 | No | NA | 8 | 0 | 0 | Not-determined | Genome-fragment | NA | NA | 0 |
| S14_vs_k141_106576\|\|full | 3111 | No | NA | 4 | 0 | 0 | Not-determined | Genome-fragment | NA | NA | 0 |
| S11_extracted_k141_61902 | 1388 | No | NA | 4 | 0 | 0 | Low-quality | Genome-fragment | 2.04 | AAI-based (high-confidence) | 0 |
| S14_vs_k141_98666\|\|full | 3074 | No | NA | 2 | 0 | 1 | Not-determined | Genome-fragment | NA | NA | 0 |
| S14_vs_k141_84519\|\|full | 2180 | No | NA | 4 | 0 | 1 | Not-determined | Genome-fragment | NA | NA | 0 |
| S11_vs_k141_55272\|\|full | 1388 | No | NA | 3 | 0 | 2 | Low-quality | Genome-fragment | 0.42 | AAI-based (medium-confidence) | 0 |
| S11_vs_k141_89019\|\|full | 1388 | No | NA | 4 | 3 | 0 | Low-quality | Genome-fragment | 2.3 | AAI-based (medium-confidence) | 0 |
| S14_vs_k141_98685\|\|full | 6174 | No | NA | 6 | 0 | 5 | Not-determined | Genome-fragment | NA | NA | 0 |
| S14_vs_k141_106635\|\|full | 3014 | No | NA | 4 | 0 | 0 | Not-determined | Genome-fragment | NA | NA | 0 |
| S13_vs_87859\|\|full | 1388 | No | NA | 3 | 1 | 0 | Low-quality | Genome-fragment | 3.81 | AAI-based (medium-confidence) | 0 |
| S14_extracted_k141_92591 | 1388 | No | NA | 2 | 1 | 0 | Low-quality | Genome-fragment | 0.86 | HMM-based (lower-bound) | 0 |
| S14_vs_k141_7991\|\|full | 1388 | No | NA | 2 | 2 | 0 | Low-quality | Genome-fragment | 3.29 | AAI-based (high-confidence) | 0 |
| S14_vs_k141_47392\|\|full | 2444 | No | NA | 3 | 0 | 3 | Not-determined | Genome-fragment | NA | NA | 0 |
| S14_vs_k141_34827\|\|lt2gene | 1388 | No | NA | 2 | 1 | 0 | Low-quality | Genome-fragment | 3.29 | AAI-based (high-confidence) | 0 |
| S3_extracted_k141_26197 | 1388 | No | NA | 2 | 0 | 1 | Low-quality | Genome-fragment | 1.76 | AAI-based (high-confidence) | 0 |
| S14_vs_k141_3453\|\|full | 2470 | No | NA | 3 | 0 | 1 | Not-determined | Genome-fragment | NA | NA | 0 |
| S14_vs_k141_69758\|\|full | 4571 | No | NA | 6 | 0 | 2 | Not-determined | Genome-fragment | NA | NA | 0 |
| S14_vs_k141_33079\|\|full | 3922 | No | NA | 3 | 0 | 2 | Not-determined | Genome-fragment | NA | NA | 0 |
| S14_vs_k141_55119\|\|full | 1554 | No | NA | 3 | 0 | 1 | Not-determined | Genome-fragment | NA | NA | 0 |
| S14_vs_k141_40812\|\|full | 2053 | No | NA | 3 | 0 | 2 | Not-determined | Genome-fragment | NA | NA | 0 |
| S14_vs_k141_18636\|\|full | 1562 | No | NA | 3 | 0 | 1 | Not-determined | Genome-fragment | NA | NA | 0 |
| S5_vs_k141_116016\|\|full | 1388 | No | NA | 4 | 0 | 0 | Low-quality | Genome-fragment | 4.03 | AAI-based (medium-confidence) | 0 |
| S14_vs_k141_106697\|\|full | 1636 | No | NA | 4 | 0 | 0 | Not-determined | Genome-fragment | NA | NA | 0 |
| S5_vs_k141_117133\|\|full | 1388 | No | NA | 4 | 2 | 0 | Low-quality | Genome-fragment | 4.16 | AAI-based (medium-confidence) | 0 |
| S16_extracted_103824 | 1388 | No | NA | 3 | 1 | 0 | Low-quality | Genome-fragment | 3.83 | AAI-based (high-confidence) | 0 |
| S14_vs_k141_106711\|\|full | 1255 | No | NA | 5 | 0 | 0 | Not-determined | Genome-fragment | NA | NA | 0 |
| S14_vs_k141_69847\|\|full | 1356 | No | NA | 3 | 0 | 3 | Not-determined | Genome-fragment | NA | NA | 0 |
| S16_vs_125892\|\|full | 1388 | No | NA | 2 | 1 | 0 | Low-quality | Genome-fragment | 0.91 | AAI-based (medium-confidence) | 0 |
| S16_vs_130550\|\|full | 1388 | No | NA | 4 | 2 | 0 | Low-quality | Genome-fragment | 3.95 | AAI-based (high-confidence) | 0 |
| S10_extracted_36677 | 1388 | No | NA | 3 | 1 | 0 | Low-quality | Genome-fragment | 1.23 | AAI-based (high-confidence) | 0 |
| S14_vs_k141_69864\|\|full | 2237 | No | NA | 5 | 0 | 3 | Not-determined | Genome-fragment | NA | NA | 0 |
| S2_vs_k141_20480\|\|full | 1388 | No | NA | 2 | 0 | 2 | Low-quality | Genome-fragment | 2.32 | AAI-based (medium-confidence) | 0 |
| S14_vs_k141_77148\|\|full | 2324 | No | NA | 3 | 0 | 3 | Not-determined | Genome-fragment | NA | NA | 0 |
| S14_vs_k141_62178\|\|full | 32459 | No | NA | 51 | 20 | 2 | High-quality | High-quality | 96.78 | AAI-based (high-confidence) | 0 |
| S8_extracted_k141_31199 | 1388 | No | NA | 2 | 0 | 0 | Low-quality | Genome-fragment | 0.75 | AAI-based (medium-confidence) | 0 |
| S14_vs_k141_69870\|\|full | 3892 | No | NA | 4 | 0 | 3 | Not-determined | Genome-fragment | NA | NA | 0 |
| S9_vs_k141_44698\|\|full | 1389 | No | NA | 3 | 0 | 0 | Low-quality | Genome-fragment | 3.35 | AAI-based (high-confidence) | 0 |
| S14_vs_k141_47488\|\|full | 1058 | No | NA | 2 | 0 | 0 | Not-determined | Genome-fragment | NA | NA | 0 |
| S14_vs_k141_33133\|\|full | 1087 | No | NA | 3 | 0 | 1 | Not-determined | Genome-fragment | NA | NA | 0 |
| S14_vs_k141_40918\|\|full | 1059 | No | NA | 1 | 0 | 0 | Not-determined | Genome-fragment | NA | NA | 0 |
| S14_extracted_k141_95333 | 1389 | No | NA | 4 | 0 | 0 | Low-quality | Genome-fragment | 2.37 | AAI-based (high-confidence) | 0 |
| S14_vs_k141_30990\|\|full | 1389 | No | NA | 3 | 1 | 0 | Low-quality | Genome-fragment | 3.12 | AAI-based (high-confidence) | 0 |
| S14_vs_k141_84654\|\|full | 7602 | No | NA | 4 | 0 | 2 | Not-determined | Genome-fragment | NA | NA | 0 |
| S3_vs_k141_88962\|\|full | 1389 | No | NA | 3 | 1 | 0 | Low-quality | Genome-fragment | 3.03 | HMM-based (lower-bound) | 0 |
| S3_vs_k141_127030\|\|full | 1389 | No | NA | 3 | 0 | 0 | Low-quality | Genome-fragment | 1.14 | AAI-based (medium-confidence) | 0 |
| S5_vs_k141_91956\|\|full | 1389 | No | NA | 3 | 0 | 0 | Low-quality | Genome-fragment | 1.48 | AAI-based (high-confidence) | 0 |
| S14_vs_k141_55267\|\|full | 10555 | No | NA | 8 | 0 | 6 | Not-determined | Genome-fragment | NA | NA | 0 |
| S5_vs_k141_99062\|\|full | 1389 | No | NA | 3 | 1 | 0 | Low-quality | Genome-fragment | 1.33 | AAI-based (high-confidence) | 0 |
| S14_vs_k141_18734\|\|full | 1782 | No | NA | 3 | 0 | 0 | Not-determined | Genome-fragment | NA | NA | 0 |
| S14_vs_k141_3620\|\|full | 2231 | No | NA | 3 | 0 | 1 | Not-determined | Genome-fragment | NA | NA | 0 |
| S16_vs_14716\|\|full | 1389 | No | NA | 3 | 2 | 0 | Low-quality | Genome-fragment | 3.24 | AAI-based (medium-confidence) | 0 |
| S16_vs_76194\|\|full | 1389 | No | NA | 2 | 0 | 0 | Low-quality | Genome-fragment | 2.33 | AAI-based (high-confidence) | 0 |
| S14_vs_k141_113984\|\|full | 2105 | No | NA | 3 | 0 | 2 | Not-determined | Genome-fragment | NA | NA | 0 |
| S8_extracted_k141_201328 | 1389 | No | NA | 2 | 0 | 2 | Low-quality | Genome-fragment | 0.32 | AAI-based (high-confidence) | 0 |
| S17_vs_k141_149532\|\|full | 1390 | No | NA | 2 | 1 | 0 | Low-quality | Genome-fragment | 3.39 | AAI-based (medium-confidence) | 0 |
| S14_vs_k141_77304\|\|full | 2322 | No | NA | 4 | 0 | 1 | Not-determined | Genome-fragment | NA | NA | 0 |
| S14_vs_k141_114022\|\|full | 1080 | No | NA | 3 | 0 | 1 | Not-determined | Genome-fragment | NA | NA | 0 |
| S17_vs_k141_24527\|\|full | 1390 | No | NA | 4 | 0 | 0 | Low-quality | Genome-fragment | 3.53 | AAI-based (medium-confidence) | 0 |
| S14_vs_k141_92076\|\|full | 1165 | No | NA | 2 | 0 | 0 | Not-determined | Genome-fragment | NA | NA | 0 |
| S14_vs_k141_98909\|\|full | 1749 | No | NA | 2 | 0 | 2 | Not-determined | Genome-fragment | NA | NA | 0 |
| S9_vs_k141_64690\|\|full | 1390 | No | NA | 2 | 1 | 0 | Low-quality | Genome-fragment | 3.53 | AAI-based (high-confidence) | 0 |
| S7_extracted_74414 | 1390 | No | NA | 2 | 2 | 0 | Low-quality | Genome-fragment | 4.88 | HMM-based (lower-bound) | 0 |
| S14_vs_k141_18845\|\|full | 2772 | No | NA | 4 | 0 | 3 | Not-determined | Genome-fragment | NA | NA | 0 |
| S7_vs_25008\|\|full | 1390 | No | NA | 4 | 0 | 0 | Low-quality | Genome-fragment | 2.63 | AAI-based (medium-confidence) | 0 |
| S10_vs_4214\|\|full | 1390 | No | NA | 4 | 0 | 0 | Low-quality | Genome-fragment | 2.56 | AAI-based (high-confidence) | 0 |
| S12_extracted_k141_156391 | 1390 | No | NA | 3 | 0 | 1 | Low-quality | Genome-fragment | 0.4 | AAI-based (medium-confidence) | 0 |
| S14_vs_k141_25741\|\|full | 1456 | No | NA | 2 | 0 | 0 | Not-determined | Genome-fragment | NA | NA | 0 |
| S12_vs_k141_100679\|\|full | 1390 | No | NA | 2 | 1 | 0 | Low-quality | Genome-fragment | 3.18 | AAI-based (high-confidence) | 0 |
| S14_vs_k141_33280\|\|full | 1445 | No | NA | 3 | 0 | 0 | Not-determined | Genome-fragment | NA | NA | 0 |
| S4_extracted_7057 | 1390 | No | NA | 5 | 0 | 1 | Low-quality | Genome-fragment | 1.18 | AAI-based (medium-confidence) | 0 |
| S14_vs_k141_55376\|\|full | 1101 | No | NA | 2 | 0 | 1 | Not-determined | Genome-fragment | NA | NA | 0 |
| S14_vs_k141_18962\|\|full | 1969 | No | NA | 3 | 0 | 1 | Not-determined | Genome-fragment | NA | NA | 0 |
| S14_vs_k141_98965\|\|full | 3251 | No | NA | 6 | 0 | 0 | Not-determined | Genome-fragment | NA | NA | 0 |
| S14_vs_k141_55407\|\|full | 6005 | No | NA | 7 | 0 | 1 | Not-determined | Genome-fragment | NA | NA | 0 |
| S8_vs_k141_32596\|\|full | 1390 | No | NA | 3 | 1 | 0 | Low-quality | Genome-fragment | 2.57 | HMM-based (lower-bound) | 0 |
| S14_vs_k141_92222\|\|full | 2602 | No | NA | 3 | 0 | 2 | Not-determined | Genome-fragment | NA | NA | 0 |
| S17_extracted_k141_91998 | 1391 | No | NA | 1 | 0 | 0 | Low-quality | Genome-fragment | 0.97 | AAI-based (high-confidence) | 0 |
| S14_vs_k141_106971\|\|full | 1839 | No | NA | 5 | 0 | 0 | Not-determined | Genome-fragment | NA | NA | 0 |
| S14_vs_k141_99003\|\|full | 6352 | No | NA | 6 | 0 | 3 | Not-determined | Genome-fragment | NA | NA | 0 |
| S17_extracted_k141_85169 | 1391 | No | NA | 2 | 0 | 1 | Low-quality | Genome-fragment | 1.33 | AAI-based (medium-confidence) | 0 |
| S14_vs_k141_99022\|\|full | 3870 | No | NA | 4 | 0 | 0 | Not-determined | Genome-fragment | NA | NA | 0 |
| S17_vs_k141_49192\|\|lt2gene | 1391 | No | NA | 2 | 2 | 0 | Low-quality | Genome-fragment | 3.24 | AAI-based (high-confidence) | 0 |
| S11_vs_k141_32019\|\|full | 1391 | No | NA | 3 | 0 | 0 | Low-quality | Genome-fragment | 3 | AAI-based (high-confidence) | 0 |
| S14_vs_k141_69959\|\|full | 1334 | No | NA | 2 | 0 | 2 | Not-determined | Genome-fragment | NA | NA | 0 |
| S13_vs_42233\|\|full | 1391 | No | NA | 3 | 0 | 0 | Low-quality | Genome-fragment | 2.16 | AAI-based (medium-confidence) | 0 |
| S14_vs_k141_55498\|\|full | 5183 | No | NA | 4 | 0 | 3 | Not-determined | Genome-fragment | NA | NA | 0 |
| S14_vs_k141_99040\|\|full | 2478 | No | NA | 3 | 0 | 0 | Not-determined | Genome-fragment | NA | NA | 0 |
| S14_vs_k141_55502\|\|full | 2524 | No | NA | 4 | 0 | 2 | Not-determined | Genome-fragment | NA | NA | 0 |
| S14_vs_k141_55503\|\|full | 1302 | No | NA | 2 | 0 | 2 | Not-determined | Genome-fragment | NA | NA | 0 |
| S14_vs_k141_42462\|\|full | 1391 | No | NA | 3 | 1 | 0 | Low-quality | Genome-fragment | 3.12 | AAI-based (high-confidence) | 0 |
| S14_vs_k141_59523\|\|lt2gene | 1391 | No | NA | 2 | 1 | 0 | Low-quality | Genome-fragment | 2.63 | HMM-based (lower-bound) | 0 |
| S14_vs_k141_99048\|\|full | 3089 | No | NA | 4 | 0 | 2 | Not-determined | Genome-fragment | NA | NA | 0 |
| S14_vs_k141_54815\|\|lt2gene | 1391 | No | NA | 2 | 1 | 0 | Low-quality | Genome-fragment | 3.31 | AAI-based (high-confidence) | 0 |
| S3_vs_k141_81564\|\|full | 1391 | No | NA | 4 | 2 | 0 | Low-quality | Genome-fragment | 3.88 | AAI-based (high-confidence) | 0 |
| S5_vs_k141_61841\|\|full | 1391 | Yes | 667 | 4 | 1 | 1 | Low-quality | Genome-fragment | 1.16 | AAI-based (medium-confidence) | 52.05 |
| S5_vs_k141_106711\|\|full | 1391 | No | NA | 4 | 0 | 0 | Low-quality | Genome-fragment | 1.15 | AAI-based (medium-confidence) | 0 |
| S7_vs_85762\|\|full | 1391 | No | NA | 3 | 0 | 0 | Low-quality | Genome-fragment | 3.01 | AAI-based (medium-confidence) | 0 |
| S14_vs_k141_41178\|\|full | 5267 | No | NA | 7 | 0 | 2 | Not-determined | Genome-fragment | NA | NA | 0 |
| S10_vs_41304\|\|full | 1391 | No | NA | 2 | 0 | 0 | Low-quality | Genome-fragment | 2.7 | AAI-based (medium-confidence) | 0 |
| S14_vs_k141_3973\|\|full | 3779 | No | NA | 5 | 0 | 3 | Not-determined | Genome-fragment | NA | NA | 0 |
| S14_vs_k141_70031\|\|full | 1742 | No | NA | 4 | 0 | 0 | Not-determined | Genome-fragment | NA | NA | 0 |
| S10_vs_39275\|\|lt2gene | 1391 | No | NA | 2 | 0 | 0 | Low-quality | Genome-fragment | 1.9 | AAI-based (medium-confidence) | 0 |
| S14_vs_k141_19163\|\|full | 2168 | No | NA | 1 | 0 | 1 | Not-determined | Genome-fragment | NA | NA | 0 |
| S8_extracted_k141_161109 | 1391 | No | NA | 3 | 1 | 0 | Low-quality | Genome-fragment | 2.8 | AAI-based (high-confidence) | 0 |
| S14_vs_k141_85005\|\|full | 13517 | No | NA | 13 | 0 | 4 | Not-determined | Genome-fragment | NA | NA | 0 |
| S14_vs_k141_19175\|\|full | 2075 | No | NA | 3 | 0 | 1 | Not-determined | Genome-fragment | NA | NA | 0 |
| S8_vs_k141_164079\|\|full | 1391 | No | NA | 4 | 0 | 0 | Low-quality | Genome-fragment | 3.32 | AAI-based (medium-confidence) | 0 |
| S14_vs_k141_25852\|\|full | 107963 | No | NA | 113 | 2 | 29 | Medium-quality | Genome-fragment | 69.62 | HMM-based (lower-bound) | 0 |
| S13_extracted_136965 | 1392 | No | NA | 4 | 2 | 0 | Low-quality | Genome-fragment | 3.46 | AAI-based (high-confidence) | 0 |
| S14_vs_k141_36772\|\|full | 1392 | No | NA | 4 | 0 | 0 | Low-quality | Genome-fragment | 3.39 | AAI-based (high-confidence) | 0 |
| S14_vs_k141_57506\|\|full | 1392 | No | NA | 2 | 0 | 0 | Low-quality | Genome-fragment | 3.47 | AAI-based (medium-confidence) | 0 |
| S14_vs_k141_92342\|\|full | 2953 | No | NA | 6 | 0 | 0 | Not-determined | Genome-fragment | NA | NA | 0 |
| S1_vs_k141_58112\|\|full | 1392 | No | NA | 3 | 1 | 0 | Low-quality | Genome-fragment | 29.78 | AAI-based (high-confidence) | 0 |
| S14_vs_k141_25877\|\|full | 2507 | No | NA | 2 | 0 | 0 | Not-determined | Genome-fragment | NA | NA | 0 |
| S14_vs_k141_19223\|\|full | 1561 | No | NA | 2 | 0 | 0 | Not-determined | Genome-fragment | NA | NA | 0 |
| S1_vs_k141_34393\|\|lt2gene | 1392 | No | NA | 2 | 1 | 0 | Low-quality | Genome-fragment | 3.88 | AAI-based (medium-confidence) | 0 |
| S14_vs_k141_11510\|\|full | 66098 | No | NA | 66 | 1 | 19 | Medium-quality | Genome-fragment | 58.68 | HMM-based (lower-bound) | 0 |
| S3_extracted_k141_79286 | 1392 | No | NA | 5 | 0 | 0 | Low-quality | Genome-fragment | 1.47 | AAI-based (high-confidence) | 0 |
| S14_vs_k141_4037\|\|full | 4235 | No | NA | 5 | 0 | 1 | Not-determined | Genome-fragment | NA | NA | 0 |
| S14_vs_k141_11522\|\|full | 1867 | No | NA | 4 | 0 | 2 | Not-determined | Genome-fragment | NA | NA | 0 |
| S14_vs_k141_85078\|\|full | 2209 | No | NA | 3 | 0 | 2 | Not-determined | Genome-fragment | NA | NA | 0 |
| S5_extracted_k141_87538 | 1392 | No | NA | 4 | 3 | 0 | Low-quality | Genome-fragment | 4.13 | AAI-based (high-confidence) | 0 |
| S5_extracted_k141_130972 | 1392 | No | NA | 3 | 1 | 0 | Low-quality | Genome-fragment | 3.82 | AAI-based (medium-confidence) | 0 |
| S14_vs_k141_33541\|\|full | 5493 | No | NA | 9 | 0 | 4 | Not-determined | Genome-fragment | NA | NA | 0 |
| S7_vs_84737\|\|lt2gene | 1392 | No | NA | 1 | 1 | 0 | Low-quality | Genome-fragment | 3.16 | AAI-based (high-confidence) | 0 |
| S14_vs_k141_4097\|\|full | 1472 | No | NA | 2 | 0 | 1 | Not-determined | Genome-fragment | NA | NA | 0 |
| S10_vs_89160\|\|full | 1392 | No | NA | 3 | 1 | 0 | Low-quality | Genome-fragment | 3.73 | AAI-based (medium-confidence) | 0 |
| S14_vs_k141_62577\|\|full | 1294 | No | NA | 4 | 0 | 0 | Not-determined | Genome-fragment | NA | NA | 0 |
| S14_vs_k141_19329\|\|full | 5498 | No | NA | 9 | 0 | 1 | Not-determined | Genome-fragment | NA | NA | 0 |
| S8_extracted_k141_86084 | 1392 | No | NA | 2 | 0 | 1 | Low-quality | Genome-fragment | 1.1 | AAI-based (medium-confidence) | 0 |
| S14_vs_k141_41354\|\|full | 2674 | No | NA | 4 | 0 | 2 | Not-determined | Genome-fragment | NA | NA | 0 |
| S9_vs_k141_55411\|\|lt2gene | 1393 | No | NA | 2 | 1 | 0 | Low-quality | Genome-fragment | 2.24 | HMM-based (lower-bound) | 0 |
| S14_vs_k141_25933\|\|full | 5352 | No | NA | 7 | 0 | 2 | Not-determined | Genome-fragment | NA | NA | 0 |
| S1_vs_k141_89819\|\|full | 1393 | No | NA | 4 | 0 | 2 | Low-quality | Genome-fragment | 1.89 | AAI-based (medium-confidence) | 0 |
| S14_vs_k141_92465\|\|full | 1666 | No | NA | 3 | 0 | 0 | Not-determined | Genome-fragment | NA | NA | 0 |
| S5_extracted_k141_120953 | 1393 | No | NA | 2 | 1 | 0 | Low-quality | Genome-fragment | 0.41 | HMM-based (lower-bound) | 0 |
| S7_extracted_71777 | 1393 | No | NA | 2 | 0 | 1 | Low-quality | Genome-fragment | 0.98 | AAI-based (medium-confidence) | 0 |
| S7_vs_47193\|\|lt2gene | 1393 | No | NA | 2 | 1 | 0 | Low-quality | Genome-fragment | 4.5 | HMM-based (lower-bound) | 0 |
| S16_extracted_107656 | 1393 | No | NA | 2 | 0 | 0 | Low-quality | Genome-fragment | 1.17 | AAI-based (high-confidence) | 0 |
| S14_vs_k141_41420\|\|full | 1560 | No | NA | 2 | 0 | 0 | Not-determined | Genome-fragment | NA | NA | 0 |
| S14_vs_k141_114422\|\|full | 3118 | No | NA | 5 | 0 | 1 | Not-determined | Genome-fragment | NA | NA | 0 |
| S14_vs_k141_41422\|\|full | 5436 | No | NA | 4 | 0 | 4 | Not-determined | Genome-fragment | NA | NA | 0 |
| S16_vs_24186\|\|full | 1393 | No | NA | 4 | 1 | 0 | Low-quality | Genome-fragment | 3.16 | AAI-based (medium-confidence) | 0 |
| S14_vs_k141_19445\|\|full | 1634 | No | NA | 3 | 0 | 2 | Not-determined | Genome-fragment | NA | NA | 0 |
| S14_vs_k141_41437\|\|full | 3743 | No | NA | 3 | 0 | 3 | Not-determined | Genome-fragment | NA | NA | 0 |
| S10_vs_51363\|\|full | 1393 | No | NA | 5 | 1 | 0 | Low-quality | Genome-fragment | 1.2 | HMM-based (lower-bound) | 0 |
| S14_vs_k141_92517\|\|full | 2876 | No | NA | 3 | 0 | 2 | Not-determined | Genome-fragment | NA | NA | 0 |
| S6_vs_k141_40758\|\|full | 1393 | No | NA | 4 | 0 | 0 | Low-quality | Genome-fragment | 2.36 | AAI-based (high-confidence) | 0 |
| S8_extracted_k141_72173 | 1393 | No | NA | 2 | 0 | 0 | Low-quality | Genome-fragment | 1.26 | AAI-based (medium-confidence) | 0 |
| S8_vs_k141_3475\|\|full | 1393 | No | NA | 2 | 0 | 1 | Low-quality | Genome-fragment | 0.92 | AAI-based (medium-confidence) | 0 |
| S9_vs_k141_12174\|\|full | 1394 | No | NA | 2 | 0 | 0 | Low-quality | Genome-fragment | 1.01 | AAI-based (medium-confidence) | 0 |
| S1_extracted_k141_52848 | 1394 | No | NA | 2 | 1 | 0 | Low-quality | Genome-fragment | 2.99 | AAI-based (high-confidence) | 0 |
| S1_vs_k141_107800\|\|full | 1394 | No | NA | 2 | 0 | 1 | Low-quality | Genome-fragment | 2.57 | AAI-based (medium-confidence) | 0 |
| S3_vs_k141_100339\|\|full | 1394 | No | NA | 3 | 0 | 0 | Low-quality | Genome-fragment | 2.28 | AAI-based (high-confidence) | 0 |
| S14_vs_k141_62734\|\|full | 1270 | No | NA | 2 | 0 | 0 | Not-determined | Genome-fragment | NA | NA | 0 |
| S5_extracted_k141_107956 | 1394 | No | NA | 2 | 0 | 0 | Low-quality | Genome-fragment | 0.95 | AAI-based (high-confidence) | 0 |
| S14_vs_k141_19517\|\|full | 1113 | No | NA | 2 | 0 | 0 | Not-determined | Genome-fragment | NA | NA | 0 |
| S5_vs_k141_98819\|\|full | 1394 | No | NA | 5 | 0 | 0 | Low-quality | Genome-fragment | 2.41 | AAI-based (medium-confidence) | 0 |
| S14_vs_k141_62743\|\|full | 5683 | No | NA | 5 | 0 | 4 | Not-determined | Genome-fragment | NA | NA | 0 |
| S7_vs_83540\|\|full | 1394 | No | NA | 6 | 0 | 0 | Low-quality | Genome-fragment | 3.12 | AAI-based (high-confidence) | 0 |
| S14_vs_k141_114499\|\|full | 1098 | No | NA | 2 | 0 | 1 | Not-determined | Genome-fragment | NA | NA | 0 |
| S14_vs_k141_4191\|\|full | 2115 | No | NA | 5 | 0 | 0 | Not-determined | Genome-fragment | NA | NA | 0 |
| S14_vs_k141_62771\|\|full | 1042 | No | NA | 3 | 0 | 0 | Not-determined | Genome-fragment | NA | NA | 0 |
| S7_vs_132921\|\|full | 1394 | No | NA | 5 | 1 | 0 | Low-quality | Genome-fragment | 3.01 | HMM-based (lower-bound) | 0 |
| S10_extracted_14017 | 1394 | No | NA | 1 | 0 | 0 | Low-quality | Genome-fragment | 3.11 | AAI-based (medium-confidence) | 0 |
| S12_extracted_k141_104870 | 1394 | No | NA | 3 | 0 | 0 | Low-quality | Genome-fragment | 2.25 | AAI-based (high-confidence) | 0 |
| S8_extracted_k141_118988 | 1394 | No | NA | 3 | 1 | 0 | Low-quality | Genome-fragment | 2.45 | AAI-based (medium-confidence) | 0 |
| S14_vs_k141_85269\|\|full | 1040 | No | NA | 2 | 0 | 1 | Not-determined | Genome-fragment | NA | NA | 0 |
| S14_vs_k141_4253\|\|full | 1855 | No | NA | 2 | 0 | 2 | Not-determined | Genome-fragment | NA | NA | 0 |
| S8_vs_k141_46417\|\|full | 1394 | No | NA | 2 | 0 | 0 | Low-quality | Genome-fragment | 3.11 | AAI-based (medium-confidence) | 0 |
| S17_vs_k141_148982\|\|full | 1395 | No | NA | 2 | 0 | 2 | Low-quality | Genome-fragment | 0.98 | AAI-based (medium-confidence) | 0 |
| S17_vs_k141_154910\|\|full | 1395 | No | NA | 2 | 2 | 0 | Low-quality | Genome-fragment | 2.15 | HMM-based (lower-bound) | 0 |
| S14_vs_k141_55884\|\|full | 2308 | No | NA | 3 | 0 | 2 | Not-determined | Genome-fragment | NA | NA | 0 |
| S11_vs_k141_22903\|\|full | 1395 | No | NA | 2 | 0 | 2 | Low-quality | Genome-fragment | 0.63 | AAI-based (medium-confidence) | 0 |
| S14_vs_k141_19598\|\|full | 1829 | No | NA | 3 | 0 | 0 | Not-determined | Genome-fragment | NA | NA | 0 |
| S14_vs_k141_4291\|\|full | 1672 | No | NA | 3 | 0 | 0 | Not-determined | Genome-fragment | NA | NA | 0 |
| S14_extracted_k141_44872 | 1395 | No | NA | 4 | 4 | 0 | Low-quality | Genome-fragment | 4.51 | AAI-based (high-confidence) | 0 |
| S14_vs_k141_114690\|\|lt2gene | 1395 | No | NA | 2 | 2 | 0 | Low-quality | Genome-fragment | 3.37 | AAI-based (high-confidence) | 0 |
| S1_vs_k141_38014\|\|full | 1395 | No | NA | 3 | 1 | 0 | Low-quality | Genome-fragment | 3.24 | AAI-based (high-confidence) | 0 |
| S3_extracted_k141_59311 | 1395 | No | NA | 6 | 0 | 0 | Low-quality | Genome-fragment | 5.19 | AAI-based (high-confidence) | 0 |
| S14_vs_k141_33682\|\|full | 2448 | No | NA | 4 | 0 | 1 | Not-determined | Genome-fragment | NA | NA | 0 |
| S14_vs_k141_11660\|\|full | 1885 | No | NA | 2 | 0 | 1 | Not-determined | Genome-fragment | NA | NA | 0 |
| S3_vs_k141_83117\|\|full | 1395 | No | NA | 3 | 1 | 0 | Low-quality | Genome-fragment | 2.73 | AAI-based (high-confidence) | 0 |
| S7_extracted_59193 | 1395 | No | NA | 1 | 0 | 1 | Low-quality | Genome-fragment | 2.39 | AAI-based (high-confidence) | 0 |
| S14_vs_k141_70455\|\|full | 2464 | No | NA | 6 | 0 | 0 | Not-determined | Genome-fragment | NA | NA | 0 |
| S7_extracted_108526 | 1395 | Yes | 326 | 3 | 1 | 1 | Low-quality | Genome-fragment | 0.7 | AAI-based (medium-confidence) | 76.63 |
| S8_vs_k141_14240\|\|lt2gene | 1395 | No | NA | 1 | 1 | 0 | Low-quality | Genome-fragment | 1.57 | HMM-based (lower-bound) | 0 |
| S14_vs_k141_114648\|\|full | 3604 | No | NA | 4 | 0 | 3 | Not-determined | Genome-fragment | NA | NA | 0 |
| S14_vs_k141_64335\|\|full | 1396 | No | NA | 2 | 0 | 0 | Low-quality | Genome-fragment | 2.98 | AAI-based (high-confidence) | 0 |
| S14_vs_k141_33707\|\|full | 1013 | No | NA | 2 | 0 | 0 | Not-determined | Genome-fragment | NA | NA | 0 |
| S14_vs_k141_4342\|\|full | 1943 | No | NA | 4 | 0 | 0 | Not-determined | Genome-fragment | NA | NA | 0 |
| S3_vs_k141_82510\|\|full | 1396 | No | NA | 4 | 1 | 0 | Low-quality | Genome-fragment | 3.39 | AAI-based (high-confidence) | 0 |
| S14_vs_k141_114657\|\|full | 4493 | No | NA | 3 | 0 | 1 | Not-determined | Genome-fragment | NA | NA | 0 |
| S16_extracted_154831 | 1396 | No | NA | 3 | 0 | 2 | Low-quality | Genome-fragment | 3.94 | AAI-based (medium-confidence) | 0 |
| S14_vs_k141_33719\|\|full | 2099 | No | NA | 2 | 0 | 1 | Not-determined | Genome-fragment | NA | NA | 0 |
| S10_vs_101297\|\|lt2gene | 1396 | No | NA | 2 | 2 | 0 | Low-quality | Genome-fragment | 2.4 | HMM-based (lower-bound) | 0 |
| S14_vs_k141_26380\|\|full | 1397 | No | NA | 3 | 0 | 0 | Low-quality | Genome-fragment | 4.05 | AAI-based (high-confidence) | 0 |
| S14_vs_k141_107472\|\|full | 4976 | No | NA | 6 | 0 | 4 | Not-determined | Genome-fragment | NA | NA | 0 |
| S14_vs_k141_85373\|\|full | 1468 | No | NA | 3 | 0 | 0 | Not-determined | Genome-fragment | NA | NA | 0 |
| S14_vs_k141_92797\|\|full | 1356 | No | NA | 2 | 0 | 0 | Not-determined | Genome-fragment | NA | NA | 0 |
| S1_vs_k141_155456\|\|lt2gene | 1397 | No | NA | 2 | 1 | 0 | Low-quality | Genome-fragment | 3.06 | AAI-based (high-confidence) | 0 |
| S3_extracted_k141_91242 | 1397 | No | NA | 4 | 2 | 0 | Low-quality | Genome-fragment | 2.29 | AAI-based (high-confidence) | 0 |
| S5_extracted_k141_114417 | 1397 | No | NA | 3 | 0 | 0 | Low-quality | Genome-fragment | 3.38 | AAI-based (medium-confidence) | 0 |
| S14_vs_k141_92821\|\|full | 12227 | No | NA | 17 | 0 | 2 | Not-determined | Genome-fragment | NA | NA | 0 |
| S14_vs_k141_26180\|\|full | 3982 | No | NA | 3 | 0 | 2 | Not-determined | Genome-fragment | NA | NA | 0 |
| S7_extracted_137347 | 1397 | No | NA | 3 | 1 | 0 | Low-quality | Genome-fragment | 2.43 | AAI-based (medium-confidence) | 0 |
| S14_vs_k141_70579\|\|full | 1289 | No | NA | 3 | 0 | 0 | Not-determined | Genome-fragment | NA | NA | 0 |
| S16_vs_75404\|\|full | 1397 | No | NA | 2 | 2 | 0 | Low-quality | Genome-fragment | 2.99 | AAI-based (high-confidence) | 0 |
| S14_vs_k141_78078\|\|full | 5167 | No | NA | 7 | 0 | 3 | Not-determined | Genome-fragment | NA | NA | 0 |
| S12_vs_k141_63513\|\|full | 1397 | No | NA | 2 | 0 | 0 | Low-quality | Genome-fragment | 2.45 | AAI-based (medium-confidence) | 0 |
| S4_extracted_25293 | 1397 | No | NA | 3 | 0 | 2 | Low-quality | Genome-fragment | 0.78 | AAI-based (medium-confidence) | 0 |
| S14_vs_k141_26208\|\|full | 2870 | No | NA | 5 | 0 | 2 | Not-determined | Genome-fragment | NA | NA | 0 |
| S8_vs_k141_888\|\|full | 1397 | No | NA | 2 | 0 | 1 | Low-quality | Genome-fragment | 3.23 | AAI-based (medium-confidence) | 0 |
| S14_vs_k141_19724\|\|full | 1917 | No | NA | 3 | 0 | 1 | Not-determined | Genome-fragment | NA | NA | 0 |
| S17_vs_k141_96326\|\|full | 1398 | No | NA | 3 | 0 | 0 | Low-quality | Genome-fragment | 2.11 | AAI-based (high-confidence) | 0 |
| S14_extracted_k141_79927 | 1398 | No | NA | 4 | 1 | 0 | Low-quality | Genome-fragment | 2.13 | HMM-based (lower-bound) | 0 |
| S1_vs_k141_120906\|\|full | 1398 | No | NA | 3 | 1 | 0 | Low-quality | Genome-fragment | 3.97 | AAI-based (high-confidence) | 0 |
| S14_vs_k141_85424\|\|full | 1455 | No | NA | 2 | 0 | 0 | Not-determined | Genome-fragment | NA | NA | 0 |
| S1_vs_k141_141061\|\|full | 1398 | No | NA | 4 | 0 | 0 | Low-quality | Genome-fragment | 3.46 | AAI-based (high-confidence) | 0 |
| S14_vs_k141_107540\|\|full | 1834 | No | NA | 2 | 0 | 0 | Not-determined | Genome-fragment | NA | NA | 0 |
| S14_vs_k141_41743\|\|full | 1184 | No | NA | 2 | 0 | 1 | Not-determined | Genome-fragment | NA | NA | 0 |
| S1_vs_k141_2797\|\|full | 1398 | No | NA | 3 | 0 | 0 | Low-quality | Genome-fragment | 2.96 | AAI-based (high-confidence) | 0 |
| S14_vs_k141_107544\|\|full | 5718 | No | NA | 6 | 0 | 4 | Not-determined | Genome-fragment | NA | NA | 0 |
| S3_extracted_k141_115575 | 1398 | No | NA | 2 | 2 | 0 | Low-quality | Genome-fragment | 4.61 | AAI-based (high-confidence) | 0 |
| S3_vs_k141_35725\|\|full | 1398 | No | NA | 2 | 1 | 0 | Low-quality | Genome-fragment | 0.46 | HMM-based (lower-bound) | 0 |
| S14_vs_k141_19787\|\|full | 1806 | No | NA | 4 | 0 | 0 | Not-determined | Genome-fragment | NA | NA | 0 |
| S14_vs_k141_26283\|\|full | 2262 | No | NA | 4 | 0 | 2 | Not-determined | Genome-fragment | NA | NA | 0 |
| S14_vs_k141_107564\|\|full | 2240 | No | NA | 3 | 0 | 2 | Not-determined | Genome-fragment | NA | NA | 0 |
| S7_vs_92674\|\|full | 1398 | No | NA | 4 | 0 | 0 | Low-quality | Genome-fragment | 3.8 | AAI-based (high-confidence) | 0 |
| S16_extracted_123900 | 1398 | No | NA | 2 | 0 | 1 | Low-quality | Genome-fragment | 2.64 | AAI-based (high-confidence) | 0 |
| S14_vs_k141_107573\|\|full | 1168 | No | NA | 4 | 0 | 0 | Not-determined | Genome-fragment | NA | NA | 0 |
| S6_vs_k141_11680\|\|full | 1398 | No | NA | 2 | 0 | 2 | Low-quality | Genome-fragment | 0.44 | AAI-based (medium-confidence) | 0 |
| S8_vs_k141_97942\|\|lt2gene | 1398 | No | NA | 2 | 1 | 0 | Low-quality | Genome-fragment | 3.27 | AAI-based (high-confidence) | 0 |
| S17_extracted_k141_98213 | 1399 | No | NA | 2 | 1 | 0 | Low-quality | Genome-fragment | 2.34 | AAI-based (high-confidence) | 0 |
| S17_extracted_k141_110641 | 1399 | No | NA | 3 | 0 | 0 | Low-quality | Genome-fragment | 3.52 | AAI-based (medium-confidence) | 0 |
| S14_vs_k141_107611\|\|full | 1403 | No | NA | 2 | 0 | 1 | Not-determined | Genome-fragment | NA | NA | 0 |
| S14_vs_k141_41831\|\|full | 1573 | No | NA | 3 | 0 | 2 | Not-determined | Genome-fragment | NA | NA | 0 |
| S9_vs_k141_3349\|\|full | 1399 | No | NA | 2 | 0 | 1 | Low-quality | Genome-fragment | 3.32 | AAI-based (high-confidence) | 0 |
| S14_vs_k141_85509\|\|full | 1445 | No | NA | 4 | 0 | 0 | Not-determined | Genome-fragment | NA | NA | 0 |
| S11_extracted_k141_19923 | 1399 | No | NA | 2 | 0 | 0 | Low-quality | Genome-fragment | 4.09 | AAI-based (high-confidence) | 0 |
| S14_vs_k141_112796\|\|full | 1399 | No | NA | 3 | 1 | 0 | Low-quality | Genome-fragment | 3.57 | AAI-based (high-confidence) | 0 |
| S14_vs_k141_70801\|\|full | 2350 | No | NA | 3 | 0 | 2 | Not-determined | Genome-fragment | NA | NA | 0 |
| S1_extracted_k141_54785 | 1399 | No | NA | 3 | 1 | 0 | Low-quality | Genome-fragment | 2.84 | AAI-based (high-confidence) | 0 |
| S14_vs_k141_107669\|\|full | 4491 | No | NA | 5 | 0 | 4 | Not-determined | Genome-fragment | NA | NA | 0 |
| S14_vs_k141_70810\|\|full | 2275 | No | NA | 5 | 0 | 1 | Not-determined | Genome-fragment | NA | NA | 0 |
| S14_vs_k141_41869\|\|full | 1392 | No | NA | 3 | 0 | 0 | Not-determined | Genome-fragment | NA | NA | 0 |
| S14_vs_k141_114886\|\|full | 1230 | No | NA | 3 | 0 | 0 | Not-determined | Genome-fragment | NA | NA | 0 |
| S3_extracted_k141_73391 | 1399 | No | NA | 1 | 0 | 0 | Low-quality | Genome-fragment | 2.31 | AAI-based (high-confidence) | 0 |
| S14_vs_k141_78159\|\|full | 1525 | No | NA | 2 | 0 | 1 | Not-determined | Genome-fragment | NA | NA | 0 |
| S5_vs_k141_25872\|\|full | 1399 | No | NA | 3 | 0 | 1 | Low-quality | Genome-fragment | 1.91 | AAI-based (medium-confidence) | 0 |
| S14_vs_k141_85558\|\|full | 2362 | No | NA | 3 | 0 | 2 | Not-determined | Genome-fragment | NA | NA | 0 |
| S14_vs_k141_34001\|\|full | 3147 | No | NA | 4 | 0 | 2 | Not-determined | Genome-fragment | NA | NA | 0 |
| S14_vs_k141_41888\|\|full | 1174 | No | NA | 3 | 0 | 1 | Not-determined | Genome-fragment | NA | NA | 0 |
| S14_vs_k141_4604\|\|full | 1300 | No | NA | 2 | 0 | 1 | Not-determined | Genome-fragment | NA | NA | 0 |
| S14_vs_k141_4607\|\|full | 3109 | No | NA | 3 | 0 | 3 | Not-determined | Genome-fragment | NA | NA | 0 |
| S14_vs_k141_85617\|\|full | 6246 | No | NA | 12 | 0 | 0 | Not-determined | Genome-fragment | NA | NA | 0 |
| S5_vs_k141_15035\|\|full | 1399 | No | NA | 5 | 1 | 0 | Low-quality | Genome-fragment | 3.67 | AAI-based (medium-confidence) | 0 |
| S14_vs_k141_78164\|\|full | 38093 | Yes | 33442 | 55 | 8 | 6 | Medium-quality | Genome-fragment | 78.08 | AAI-based (medium-confidence) | 12.21 |
| S14_vs_k141_19952\|\|full | 2360 | No | NA | 3 | 0 | 1 | Not-determined | Genome-fragment | NA | NA | 0 |
| S12_extracted_k141_37641 | 1399 | No | NA | 3 | 1 | 0 | Low-quality | Genome-fragment | 5.15 | HMM-based (lower-bound) | 0 |
| S12_vs_k141_180556\|\|full | 1399 | No | NA | 4 | 1 | 0 | Low-quality | Genome-fragment | 2.45 | AAI-based (high-confidence) | 0 |
| S14_vs_k141_89424\|\|full | 1400 | No | NA | 3 | 1 | 0 | Low-quality | Genome-fragment | 2.59 | AAI-based (medium-confidence) | 0 |
| S5_extracted_k141_51130 | 1400 | No | NA | 3 | 0 | 0 | Low-quality | Genome-fragment | 2.47 | AAI-based (medium-confidence) | 0 |
| S14_vs_k141_48196\|\|full | 2244 | No | NA | 4 | 0 | 0 | Not-determined | Genome-fragment | NA | NA | 0 |
| S5_vs_k141_18831\|\|full | 1400 | No | NA | 2 | 0 | 1 | Low-quality | Genome-fragment | 1.07 | AAI-based (high-confidence) | 0 |
| S14_vs_k141_114951\|\|full | 1129 | No | NA | 2 | 0 | 0 | Not-determined | Genome-fragment | NA | NA | 0 |
| S5_vs_k141_71824\|\|lt2gene | 1400 | No | NA | 2 | 2 | 0 | Low-quality | Genome-fragment | 3.81 | AAI-based (high-confidence) | 0 |
| S14_vs_k141_78218\|\|full | 3306 | No | NA | 5 | 0 | 1 | Not-determined | Genome-fragment | NA | NA | 0 |
| S14_vs_k141_26348\|\|full | 1470 | No | NA | 3 | 0 | 1 | Not-determined | Genome-fragment | NA | NA | 0 |
| S14_vs_k141_26351\|\|full | 3103 | No | NA | 4 | 0 | 3 | Not-determined | Genome-fragment | NA | NA | 0 |
| S14_vs_k141_4745\|\|full | 3236 | No | NA | 6 | 0 | 0 | Not-determined | Genome-fragment | NA | NA | 0 |
| S16_vs_583\|\|full | 1400 | No | NA | 2 | 0 | 0 | Low-quality | Genome-fragment | 1.01 | AAI-based (medium-confidence) | 0 |
| S16_vs_87424\|\|full | 1400 | No | NA | 4 | 1 | 0 | Low-quality | Genome-fragment | 2.7 | HMM-based (lower-bound) | 0 |
| S16_vs_150908\|\|full | 1400 | No | NA | 5 | 1 | 0 | Low-quality | Genome-fragment | 2.47 | AAI-based (medium-confidence) | 0 |
| S14_vs_k141_99849\|\|full | 2765 | No | NA | 4 | 0 | 1 | Not-determined | Genome-fragment | NA | NA | 0 |
| S14_vs_k141_12004\|\|full | 5072 | No | NA | 9 | 0 | 0 | Not-determined | Genome-fragment | NA | NA | 0 |
| S12_vs_k141_33631\|\|full | 1400 | No | NA | 3 | 2 | 0 | Low-quality | Genome-fragment | 2.6 | AAI-based (medium-confidence) | 0 |
| S4_extracted_15963 | 1400 | No | NA | 5 | 0 | 0 | Low-quality | Genome-fragment | 3.51 | AAI-based (high-confidence) | 0 |
| S6_vs_k141_24720\|\|full | 1400 | No | NA | 2 | 1 | 0 | Low-quality | Genome-fragment | 1.7 | HMM-based (lower-bound) | 0 |
| S14_vs_k141_42052\|\|full | 1004 | No | NA | 3 | 0 | 1 | Not-determined | Genome-fragment | NA | NA | 0 |
| S14_vs_k141_4799\|\|full | 3945 | No | NA | 4 | 0 | 2 | Not-determined | Genome-fragment | NA | NA | 0 |
| S8_vs_k141_137626\|\|full | 1400 | No | NA | 3 | 1 | 0 | Low-quality | Genome-fragment | 2.76 | AAI-based (high-confidence) | 0 |
| S14_vs_k141_85759\|\|full | 5913 | No | NA | 7 | 0 | 4 | Not-determined | Genome-fragment | NA | NA | 0 |
| S14_vs_k141_93211\|\|full | 1920 | No | NA | 5 | 0 | 0 | Not-determined | Genome-fragment | NA | NA | 0 |
| S14_vs_k141_99908\|\|full | 1210 | No | NA | 3 | 0 | 3 | Not-determined | Genome-fragment | NA | NA | 0 |
| S14_vs_k141_107829\|\|full | 2104 | No | NA | 3 | 0 | 1 | Not-determined | Genome-fragment | NA | NA | 0 |
| S14_vs_k141_4847\|\|full | 2572 | No | NA | 5 | 0 | 2 | Not-determined | Genome-fragment | NA | NA | 0 |
| S14_vs_k141_78282\|\|full | 2080 | No | NA | 2 | 0 | 0 | Not-determined | Genome-fragment | NA | NA | 0 |
| S14_vs_k141_78296\|\|full | 1384 | No | NA | 2 | 0 | 0 | Not-determined | Genome-fragment | NA | NA | 0 |
| S14_vs_k141_93237\|\|full | 1219 | No | NA | 2 | 0 | 0 | Not-determined | Genome-fragment | NA | NA | 0 |
| S17_vs_k141_173398\|\|full | 1401 | No | NA | 4 | 1 | 0 | Low-quality | Genome-fragment | 1.57 | HMM-based (lower-bound) | 0 |
| S9_vs_k141_47537\|\|full | 1401 | No | NA | 4 | 0 | 1 | Low-quality | Genome-fragment | 2.9 | AAI-based (medium-confidence) | 0 |
| S14_vs_k141_12087\|\|full | 1496 | No | NA | 5 | 0 | 1 | Not-determined | Genome-fragment | NA | NA | 0 |
| S13_vs_4321\|\|full | 1401 | No | NA | 2 | 1 | 0 | Low-quality | Genome-fragment | 0.63 | HMM-based (lower-bound) | 0 |
| S14_vs_k141_99951\|\|full | 1343 | No | NA | 3 | 0 | 3 | Not-determined | Genome-fragment | NA | NA | 0 |
| S14_vs_k141_53659\|\|full | 1401 | No | NA | 3 | 0 | 1 | Low-quality | Genome-fragment | 3.01 | AAI-based (medium-confidence) | 0 |
| S3_extracted_k141_113642 | 1401 | No | NA | 3 | 2 | 0 | Low-quality | Genome-fragment | 2.22 | AAI-based (high-confidence) | 0 |
| S5_vs_k141_11718\|\|full | 1401 | No | NA | 2 | 0 | 1 | Low-quality | Genome-fragment | 0.43 | AAI-based (medium-confidence) | 0 |
| S14_vs_k141_56251\|\|full | 31100 | No | NA | 45 | 18 | 1 | Medium-quality | Genome-fragment | 75.74 | AAI-based (medium-confidence) | 0 |
| S14_vs_k141_115062\|\|full | 1735 | No | NA | 3 | 0 | 0 | Not-determined | Genome-fragment | NA | NA | 0 |
| S14_vs_k141_56261\|\|full | 2466 | No | NA | 3 | 0 | 1 | Not-determined | Genome-fragment | NA | NA | 0 |
| S14_vs_k141_115075\|\|full | 1248 | No | NA | 2 | 0 | 0 | Not-determined | Genome-fragment | NA | NA | 0 |
| S14_vs_k141_70985\|\|full | 158000 | No | NA | 153 | 4 | 46 | Medium-quality | Genome-fragment | 77.59 | HMM-based (lower-bound) | 0 |
| S7_vs_854\|\|full | 1401 | No | NA | 4 | 0 | 1 | Low-quality | Genome-fragment | 2.45 | AAI-based (medium-confidence) | 0 |
| S7_vs_44556\|\|full | 1401 | No | NA | 3 | 0 | 0 | Low-quality | Genome-fragment | 2.42 | AAI-based (high-confidence) | 0 |
| S14_vs_k141_93284\|\|full | 1281 | No | NA | 3 | 0 | 1 | Not-determined | Genome-fragment | NA | NA | 0 |
| S14_vs_k141_63402\|\|full | 1196 | No | NA | 2 | 0 | 1 | Not-determined | Genome-fragment | NA | NA | 0 |
| S10_vs_25876\|\|lt2gene | 1401 | No | NA | 1 | 1 | 0 | Low-quality | Genome-fragment | 3.12 | AAI-based (high-confidence) | 0 |
| S15_extracted_59383 | 1401 | No | NA | 3 | 0 | 1 | Low-quality | Genome-fragment | 0.54 | AAI-based (medium-confidence) | 0 |
| S8_extracted_k141_206020 | 1401 | No | NA | 3 | 0 | 2 | Low-quality | Genome-fragment | 3.24 | AAI-based (medium-confidence) | 0 |
| S14_vs_k141_42195\|\|full | 2371 | No | NA | 3 | 0 | 1 | Not-determined | Genome-fragment | NA | NA | 0 |
| S8_vs_k141_43913\|\|lt2gene | 1401 | No | NA | 1 | 1 | 0 | Low-quality | Genome-fragment | 3.47 | AAI-based (high-confidence) | 0 |
| S14_vs_k141_93354\|\|full | 2137 | No | NA | 4 | 0 | 2 | Not-determined | Genome-fragment | NA | NA | 0 |
| S14_vs_k141_48416\|\|full | 5289 | No | NA | 5 | 0 | 3 | Not-determined | Genome-fragment | NA | NA | 0 |
| S14_vs_k141_12167\|\|full | 1275 | No | NA | 2 | 0 | 0 | Not-determined | Genome-fragment | NA | NA | 0 |
| S14_vs_k141_71051\|\|full | 1241 | No | NA | 2 | 0 | 2 | Not-determined | Genome-fragment | NA | NA | 0 |
| S3_extracted_k141_64860 | 1402 | No | NA | 5 | 3 | 0 | Low-quality | Genome-fragment | 6.4 | AAI-based (high-confidence) | 0 |
| S14_vs_k141_34300\|\|full | 4816 | No | NA | 7 | 0 | 3 | Not-determined | Genome-fragment | NA | NA | 0 |
| S12_vs_k141_140580\|\|full | 1402 | No | NA | 2 | 0 | 0 | Low-quality | Genome-fragment | 3.6 | AAI-based (medium-confidence) | 0 |
| S12_vs_k141_59957\|\|lt2gene | 1402 | No | NA | 1 | 1 | 0 | Low-quality | Genome-fragment | 3.35 | AAI-based (medium-confidence) | 0 |
| S15_vs_70553\|\|full | 1402 | No | NA | 4 | 0 | 0 | Low-quality | Genome-fragment | 2.21 | AAI-based (medium-confidence) | 0 |
| S17_extracted_k141_135821 | 1403 | No | NA | 4 | 4 | 0 | Low-quality | Genome-fragment | 4.6 | AAI-based (high-confidence) | 0 |
| S3_vs_k141_48910\|\|full | 1403 | No | NA | 3 | 0 | 0 | Low-quality | Genome-fragment | 1.75 | AAI-based (high-confidence) | 0 |
| S3_vs_k141_22838\|\|full | 1403 | No | NA | 1 | 0 | 1 | Low-quality | Genome-fragment | 2.06 | AAI-based (medium-confidence) | 0 |
| S5_vs_k141_72607\|\|full | 1403 | No | NA | 3 | 0 | 1 | Low-quality | Genome-fragment | 1.91 | AAI-based (medium-confidence) | 0 |
| S14_vs_k141_115202\|\|full | 1132 | No | NA | 2 | 0 | 1 | Not-determined | Genome-fragment | NA | NA | 0 |
| S16_vs_117760\|\|full | 1403 | No | NA | 2 | 0 | 2 | Low-quality | Genome-fragment | 1.16 | AAI-based (medium-confidence) | 0 |
| S14_vs_k141_26522\|\|full | 6819 | No | NA | 6 | 0 | 4 | Not-determined | Genome-fragment | NA | NA | 0 |
| S16_vs_146828\|\|full | 1403 | No | NA | 3 | 0 | 0 | Low-quality | Genome-fragment | 3.39 | AAI-based (medium-confidence) | 0 |
| S14_vs_k141_63514\|\|full | 1201 | No | NA | 2 | 0 | 0 | Not-determined | Genome-fragment | NA | NA | 0 |
| S10_extracted_12866 | 1403 | No | NA | 4 | 1 | 0 | Low-quality | Genome-fragment | 1.12 | HMM-based (lower-bound) | 0 |
| S14_vs_k141_56410\|\|full | 19996 | No | NA | 28 | 7 | 0 | Medium-quality | Genome-fragment | 50.21 | AAI-based (medium-confidence) | 0 |
| S14_vs_k141_78500\|\|full | 1635 | No | NA | 3 | 0 | 0 | Not-determined | Genome-fragment | NA | NA | 0 |
| S12_vs_k141_176841\|\|full | 1403 | No | NA | 3 | 0 | 1 | Low-quality | Genome-fragment | 0.99 | AAI-based (medium-confidence) | 0 |
| S14_extracted_k141_11450 | 1404 | No | NA | 2 | 2 | 0 | Low-quality | Genome-fragment | 2.34 | AAI-based (medium-confidence) | 0 |
| S14_vs_k141_108029\|\|full | 3548 | No | NA | 4 | 0 | 4 | Not-determined | Genome-fragment | NA | NA | 0 |
| S14_vs_k141_12216\|\|full | 6583 | No | NA | 5 | 0 | 5 | Not-determined | Genome-fragment | NA | NA | 0 |
| S1_extracted_k141_60373 | 1404 | No | NA | 3 | 0 | 0 | Low-quality | Genome-fragment | 1.02 | AAI-based (high-confidence) | 0 |
| S1_extracted_k141_42742 | 1404 | No | NA | 2 | 0 | 2 | Low-quality | Genome-fragment | 0.57 | AAI-based (medium-confidence) | 0 |
| S14_vs_k141_108054\|\|full | 7289 | No | NA | 4 | 0 | 4 | Not-determined | Genome-fragment | NA | NA | 0 |
| S14_vs_k141_63578\|\|full | 1325 | No | NA | 3 | 0 | 1 | Not-determined | Genome-fragment | NA | NA | 0 |
| S1_vs_k141_63380\|\|lt2gene | 1404 | No | NA | 2 | 2 | 0 | Low-quality | Genome-fragment | 3.44 | AAI-based (medium-confidence) | 0 |
| S3_vs_k141_87816\|\|full | 1404 | No | NA | 2 | 1 | 0 | Low-quality | Genome-fragment | 6.41 | AAI-based (high-confidence) | 0 |
| S14_vs_k141_42386\|\|full | 2102 | No | NA | 3 | 0 | 0 | Not-determined | Genome-fragment | NA | NA | 0 |
| S14_vs_k141_5085\|\|full | 1142 | No | NA | 2 | 0 | 2 | Not-determined | Genome-fragment | NA | NA | 0 |
| S3_vs_k141_25561\|\|lt2gene | 1404 | No | NA | 1 | 1 | 0 | Low-quality | Genome-fragment | 2.66 | HMM-based (lower-bound) | 0 |
| S5_extracted_k141_85074 | 1404 | No | NA | 2 | 2 | 0 | Low-quality | Genome-fragment | 4.48 | AAI-based (high-confidence) | 0 |
| S14_vs_k141_78589\|\|full | 1220 | No | NA | 2 | 0 | 0 | Not-determined | Genome-fragment | NA | NA | 0 |
| S5_vs_k141_99881\|\|full | 1404 | No | NA | 2 | 1 | 0 | Low-quality | Genome-fragment | 1.74 | HMM-based (lower-bound) | 0 |
| S14_vs_k141_26585\|\|full | 1106 | No | NA | 3 | 0 | 1 | Not-determined | Genome-fragment | NA | NA | 0 |
| S5_vs_k141_106625\|\|lt2gene | 1404 | No | NA | 2 | 2 | 0 | Low-quality | Genome-fragment | 2.59 | AAI-based (medium-confidence) | 0 |
| S7_vs_48363\|\|lt2gene | 1404 | No | NA | 2 | 1 | 0 | Low-quality | Genome-fragment | 0.87 | HMM-based (lower-bound) | 0 |
| NC3_vf_k141_707 | 1404 | No | NA | 3 | 0 | 0 | Low-quality | Genome-fragment | 2.76 | AAI-based (medium-confidence) | 0 |
| S16_vs_129366\|\|full | 1404 | No | NA | 3 | 0 | 0 | Low-quality | Genome-fragment | 1.2 | AAI-based (medium-confidence) | 0 |
| S16_vs_164497\|\|full | 1404 | No | NA | 4 | 1 | 0 | Low-quality | Genome-fragment | 3.24 | AAI-based (medium-confidence) | 0 |
| S14_vs_k141_100227\|\|full | 24160 | No | NA | 52 | 19 | 1 | Medium-quality | Genome-fragment | 56.88 | AAI-based (high-confidence) | 0 |
| S12_extracted_k141_52512 | 1404 | No | NA | 2 | 2 | 0 | Low-quality | Genome-fragment | 2.34 | AAI-based (high-confidence) | 0 |
| S12_vs_k141_130395\|\|full | 1404 | No | NA | 3 | 0 | 0 | Low-quality | Genome-fragment | 3.33 | AAI-based (medium-confidence) | 0 |
| S4_extracted_10611 | 1404 | No | NA | 3 | 0 | 0 | Low-quality | Genome-fragment | 1.34 | AAI-based (medium-confidence) | 0 |
| S14_vs_k141_115370\|\|full | 2045 | No | NA | 5 | 0 | 1 | Not-determined | Genome-fragment | NA | NA | 0 |
| S14_vs_k141_42466\|\|full | 1978 | No | NA | 5 | 0 | 0 | Not-determined | Genome-fragment | NA | NA | 0 |
| S8_vs_k141_49092\|\|full | 1404 | No | NA | 3 | 1 | 0 | Low-quality | Genome-fragment | 3.19 | AAI-based (high-confidence) | 0 |
| S14_vs_k141_85948\|\|full | 1467 | No | NA | 2 | 0 | 1 | Not-determined | Genome-fragment | NA | NA | 0 |
| S14_vs_k141_85950\|\|full | 1369 | No | NA | 3 | 0 | 0 | Not-determined | Genome-fragment | NA | NA | 0 |
| S17_vs_k141_44782\|\|full | 1405 | No | NA | 3 | 0 | 0 | Low-quality | Genome-fragment | 3.09 | AAI-based (high-confidence) | 0 |
| S14_vs_k141_42489\|\|full | 4768 | No | NA | 6 | 0 | 5 | Not-determined | Genome-fragment | NA | NA | 0 |
| S11_vs_k141_90718\|\|full | 1405 | No | NA | 2 | 0 | 0 | Low-quality | Genome-fragment | 3.72 | AAI-based (high-confidence) | 0 |
| S13_vs_149929\|\|full | 1405 | No | NA | 3 | 0 | 0 | Low-quality | Genome-fragment | 3.55 | AAI-based (medium-confidence) | 0 |
| S14_vs_k141_100017\|\|full | 1405 | No | NA | 4 | 1 | 0 | Low-quality | Genome-fragment | 3.11 | AAI-based (high-confidence) | 0 |
| S14_vs_k141_108159\|\|full | 1381 | No | NA | 3 | 0 | 0 | Not-determined | Genome-fragment | NA | NA | 0 |
| S14_vs_k141_115377\|\|full | 24316 | No | NA | 28 | 10 | 0 | Medium-quality | Genome-fragment | 53.35 | AAI-based (medium-confidence) | 0 |
| S1_extracted_k141_126854 | 1405 | No | NA | 2 | 0 | 0 | Low-quality | Genome-fragment | 2.45 | AAI-based (high-confidence) | 0 |
| S14_vs_k141_34548\|\|full | 4626 | No | NA | 4 | 0 | 4 | Not-determined | Genome-fragment | NA | NA | 0 |
| S1_extracted_k141_119293 | 1405 | No | NA | 1 | 0 | 0 | Low-quality | Genome-fragment | 0.96 | AAI-based (high-confidence) | 0 |
| S3_vs_k141_104964\|\|full | 1405 | No | NA | 6 | 0 | 0 | Low-quality | Genome-fragment | 3.53 | AAI-based (high-confidence) | 0 |
| S14_vs_k141_63741\|\|full | 2056 | No | NA | 3 | 0 | 2 | Not-determined | Genome-fragment | NA | NA | 0 |
| S3_vs_k141_91808\|\|full | 1405 | No | NA | 2 | 2 | 0 | Low-quality | Genome-fragment | 0.71 | AAI-based (high-confidence) | 0 |
| S3_vs_k141_92178\|\|lt2gene | 1405 | No | NA | 1 | 1 | 0 | Low-quality | Genome-fragment | 2.36 | HMM-based (lower-bound) | 0 |
| S14_vs_k141_34568\|\|full | 1452 | No | NA | 3 | 0 | 0 | Not-determined | Genome-fragment | NA | NA | 0 |
| S7_extracted_57687 | 1405 | No | NA | 1 | 0 | 0 | Low-quality | Genome-fragment | 2.35 | AAI-based (high-confidence) | 0 |
| S14_vs_k141_34583\|\|full | 2716 | No | NA | 2 | 0 | 2 | Not-determined | Genome-fragment | NA | NA | 0 |

| | | | | | | | | | | | |
|---|---|---|---|---|---|---|---|---|---|---|---|
| S14_vs_k141_26768\|\|full | 8432 | No | NA | 5 | 0 | 5 | Not-determined | Genome-fragment | NA | NA | 0 |
| S14_vs_k141_108193\|\|full | 2805 | No | NA | 4 | 0 | 0 | Not-determined | Genome-fragment | NA | NA | 0 |
| S16_vs_171681\|\|full | 1405 | No | NA | 3 | 0 | 1 | Low-quality | Genome-fragment | 1.06 | AAI-based (medium-confidence) | 0 |
| S10_vs_87897\|\|full | 1405 | No | NA | 3 | 1 | 0 | Low-quality | Genome-fragment | 1.77 | AAI-based (high-confidence) | 0 |
| S14_vs_k141_42577\|\|full | 12726 | No | NA | 12 | 0 | 11 | Not-determined | Genome-fragment | NA | NA | 0 |
| S4_extracted_28323 | 1405 | No | NA | 4 | 0 | 0 | Low-quality | Genome-fragment | 2.12 | AAI-based (high-confidence) | 0 |
| S14_vs_k141_108218\|\|full | 1290 | No | NA | 3 | 0 | 0 | Not-determined | Genome-fragment | NA | NA | 0 |
| S8_vs_k141_155891\|\|full | 1405 | No | NA | 3 | 0 | 0 | Low-quality | Genome-fragment | 4.17 | AAI-based (high-confidence) | 0 |
| S14_vs_k141_93565\|\|full | 16641 | No | NA | 22 | 8 | 0 | Medium-quality | Genome-fragment | 70.27 | AAI-based (high-confidence) | 0 |
| S17_vs_k141_78281\|\|full | 1406 | No | NA | 3 | 1 | 0 | Low-quality | Genome-fragment | 0.11 | HMM-based (lower-bound) | 0 |
| S14_vs_k141_71397\|\|full | 3157 | No | NA | 5 | 0 | 1 | Not-determined | Genome-fragment | NA | NA | 0 |
| S14_extracted_k141_44157 | 1406 | No | NA | 3 | 2 | 0 | Low-quality | Genome-fragment | 1.51 | AAI-based (medium-confidence) | 0 |
| S14_vs_k141_56642\|\|full | 1310 | No | NA | 3 | 0 | 0 | Not-determined | Genome-fragment | NA | NA | 0 |
| S1_vs_k141_151255\|\|full | 1406 | No | NA | 2 | 0 | 1 | Low-quality | Genome-fragment | 0.82 | AAI-based (medium-confidence) | 0 |
| S14_vs_k141_56643\|\|full | 1542 | No | NA | 3 | 0 | 0 | Not-determined | Genome-fragment | NA | NA | 0 |
| S14_vs_k141_12542\|\|full | 2130 | No | NA | 3 | 0 | 0 | Not-determined | Genome-fragment | NA | NA | 0 |
| S14_vs_k141_20193\|\|full | 48326 | No | NA | 79 | 22 | 1 | High-quality | High-quality | 100 | AAI-based (high-confidence) | 0 |
| S14_vs_k141_26872\|\|full | 2316 | No | NA | 6 | 0 | 0 | Not-determined | Genome-fragment | NA | NA | 0 |
| S3_vs_k141_42453\|\|full | 1406 | No | NA | 4 | 3 | 0 | Low-quality | Genome-fragment | 4.65 | AAI-based (high-confidence) | 0 |
| S14_vs_k141_34725\|\|full | 1219 | No | NA | 2 | 0 | 0 | Not-determined | Genome-fragment | NA | NA | 0 |
| S14_vs_k141_42705\|\|full | 7484 | No | NA | 5 | 0 | 5 | Not-determined | Genome-fragment | NA | NA | 0 |
| S5_extracted_k141_38634 | 1406 | No | NA | 2 | 0 | 0 | Low-quality | Genome-fragment | 4.14 | AAI-based (high-confidence) | 0 |
| S5_vs_k141_85486\|\|lt2gene | 1406 | No | NA | 2 | 2 | 0 | Low-quality | Genome-fragment | 2.49 | HMM-based (lower-bound) | 0 |
| S7_vs_55740\|\|full | 1406 | No | NA | 2 | 0 | 1 | Low-quality | Genome-fragment | 1.64 | AAI-based (medium-confidence) | 0 |
| S14_vs_k141_42718\|\|full | 1167 | No | NA | 2 | 0 | 0 | Not-determined | Genome-fragment | NA | NA | 0 |
| S14_vs_k141_100470\|\|full | 1226 | No | NA | 3 | 0 | 0 | Not-determined | Genome-fragment | NA | NA | 0 |
| S14_vs_k141_34768\|\|full | 2455 | No | NA | 3 | 0 | 2 | Not-determined | Genome-fragment | NA | NA | 0 |
| S14_vs_k141_93661\|\|full | 5972 | No | NA | 4 | 0 | 3 | Not-determined | Genome-fragment | NA | NA | 0 |
| S7_vs_137273\|\|full | 1406 | No | NA | 2 | 0 | 1 | Low-quality | Genome-fragment | 0.44 | AAI-based (medium-confidence) | 0 |
| S12_extracted_k141_161611 | 1406 | No | NA | 3 | 0 | 0 | Low-quality | Genome-fragment | 3.54 | AAI-based (medium-confidence) | 0 |
| S12_vs_k141_143949\|\|full | 1406 | No | NA | 3 | 0 | 3 | Low-quality | Genome-fragment | 2.05 | AAI-based (medium-confidence) | 0 |
| S14_vs_k141_115455\|\|full | 40864 | No | NA | 62 | 15 | 1 | Medium-quality | Genome-fragment | 67.27 | HMM-based (lower-bound) | 0 |
| S14_vs_k141_100488\|\|full | 2151 | No | NA | 4 | 0 | 2 | Not-determined | Genome-fragment | NA | NA | 0 |
| S8_extracted_k141_69444 | 1406 | No | NA | 2 | 0 | 2 | Low-quality | Genome-fragment | 0.91 | AAI-based (medium-confidence) | 0 |
| S8_vs_k141_109649\|\|full | 1406 | No | NA | 3 | 0 | 0 | Low-quality | Genome-fragment | 3.09 | AAI-based (medium-confidence) | 0 |
| S14_vs_k141_71438\|\|full | 1169 | No | NA | 3 | 0 | 1 | Not-determined | Genome-fragment | NA | NA | 0 |
| S14_vs_k141_5339\|\|full | 2460 | No | NA | 3 | 0 | 3 | Not-determined | Genome-fragment | NA | NA | 0 |
| S11_extracted_k141_30556 | 1407 | No | NA | 1 | 1 | 0 | Low-quality | Genome-fragment | 3.45 | AAI-based (high-confidence) | 0 |
| S14_vs_k141_26993\|\|full | 1270 | No | NA | 3 | 0 | 1 | Not-determined | Genome-fragment | NA | NA | 0 |
| S14_vs_k141_56733\|\|full | 2777 | No | NA | 3 | 0 | 3 | Not-determined | Genome-fragment | NA | NA | 0 |
| S14_extracted_k141_6679 | 1407 | No | NA | 3 | 1 | 0 | Low-quality | Genome-fragment | 1.52 | AAI-based (high-confidence) | 0 |
| S1_extracted_k141_157233 | 1407 | No | NA | 3 | 1 | 0 | Low-quality | Genome-fragment | 2.67 | AAI-based (high-confidence) | 0 |
| S14_vs_k141_108337\|\|full | 1043 | No | NA | 3 | 0 | 0 | Not-determined | Genome-fragment | NA | NA | 0 |
| S5_extracted_k141_111514 | 1407 | No | NA | 3 | 0 | 0 | Low-quality | Genome-fragment | 2.42 | AAI-based (high-confidence) | 0 |
| S14_vs_k141_12662\|\|full | 2278 | No | NA | 5 | 0 | 0 | Not-determined | Genome-fragment | NA | NA | 0 |
| S7_vs_119069\|\|lt2gene | 1407 | No | NA | 1 | 1 | 0 | Low-quality | Genome-fragment | 0.97 | HMM-based (lower-bound) | 0 |
| S16_extracted_136490 | 1407 | No | NA | 3 | 0 | 0 | Low-quality | Genome-fragment | 2.96 | AAI-based (medium-confidence) | 0 |
| S10_vs_35382\|\|full | 1407 | No | NA | 4 | 0 | 1 | Low-quality | Genome-fragment | 1.17 | AAI-based (high-confidence) | 0 |
| S12_vs_k141_40346\|\|full | 1407 | No | NA | 3 | 0 | 0 | Low-quality | Genome-fragment | 2.53 | AAI-based (high-confidence) | 0 |
| S8_vs_k141_95861\|\|full | 1407 | No | NA | 3 | 0 | 0 | Low-quality | Genome-fragment | 1.74 | AAI-based (medium-confidence) | 0 |
| S13_vs_52568\|\|full | 1408 | No | NA | 5 | 0 | 0 | Low-quality | Genome-fragment | 2.8 | AAI-based (medium-confidence) | 0 |
| S14_vs_k141_583\|\|full | 1408 | No | NA | 3 | 0 | 2 | Low-quality | Genome-fragment | 1.52 | AAI-based (high-confidence) | 0 |
| S14_vs_k141_34899\|\|full | 1878 | No | NA | 2 | 0 | 1 | Not-determined | Genome-fragment | NA | NA | 0 |
| S1_vs_k141_103446\|\|full | 1408 | No | NA | 4 | 1 | 0 | Low-quality | Genome-fragment | 2.79 | AAI-based (high-confidence) | 0 |
| S14_vs_k141_27049\|\|full | 6269 | No | NA | 5 | 0 | 3 | Not-determined | Genome-fragment | NA | NA | 0 |
| S14_vs_k141_108376\|\|full | 19905 | No | NA | 35 | 9 | 1 | Medium-quality | Genome-fragment | 55 | AAI-based (medium-confidence) | 0 |
| S7_vs_99448\|\|full | 1408 | No | NA | 4 | 1 | 0 | Low-quality | Genome-fragment | 4.32 | AAI-based (high-confidence) | 0 |
| S7_vs_131214\|\|lt2gene | 1408 | No | NA | 1 | 1 | 0 | Low-quality | Genome-fragment | 4.65 | AAI-based (high-confidence) | 0 |
| S16_vs_130658\|\|lt2gene | 1408 | No | NA | 2 | 1 | 0 | Low-quality | Genome-fragment | 2.45 | AAI-based (medium-confidence) | 0 |
| S14_vs_k141_78735\|\|full | 5404 | No | NA | 3 | 0 | 2 | Not-determined | Genome-fragment | NA | NA | 0 |
| S14_vs_k141_27062\|\|full | 1623 | No | NA | 3 | 0 | 3 | Not-determined | Genome-fragment | NA | NA | 0 |
| S10_vs_75252\|\|full | 1408 | No | NA | 2 | 0 | 2 | Low-quality | Genome-fragment | 0.69 | AAI-based (medium-confidence) | 0 |
| S14_vs_k141_108396\|\|full | 1311 | No | NA | 2 | 0 | 1 | Not-determined | Genome-fragment | NA | NA | 0 |
| S14_vs_k141_48894\|\|full | 1024 | No | NA | 3 | 0 | 0 | Not-determined | Genome-fragment | NA | NA | 0 |
| S12_vs_k141_105313\|\|lt2gene | 1408 | No | NA | 2 | 1 | 0 | Low-quality | Genome-fragment | 2.93 | AAI-based (high-confidence) | 0 |
| S14_vs_k141_34934\|\|full | 1159 | No | NA | 2 | 0 | 0 | Not-determined | Genome-fragment | NA | NA | 0 |
| S17_vs_k141_89510\|\|full | 1409 | No | NA | 3 | 0 | 0 | Low-quality | Genome-fragment | 2 | AAI-based (high-confidence) | 0 |
| S9_extracted_k141_76644 | 1409 | No | NA | 2 | 0 | 2 | Low-quality | Genome-fragment | 0.71 | AAI-based (medium-confidence) | 0 |
| S14_vs_k141_5418\|\|full | 1797 | No | NA | 3 | 0 | 3 | Not-determined | Genome-fragment | NA | NA | 0 |
| S14_vs_k141_63981\|\|full | 1651 | No | NA | 4 | 0 | 1 | Not-determined | Genome-fragment | NA | NA | 0 |
| S11_vs_k141_93428\|\|full | 1409 | No | NA | 4 | 0 | 0 | Low-quality | Genome-fragment | 3.22 | AAI-based (high-confidence) | 0 |
| S14_vs_k141_65827\|\|full | 1409 | No | NA | 3 | 2 | 0 | Low-quality | Genome-fragment | 4.41 | AAI-based (high-confidence) | 0 |
| S14_vs_k141_56834\|\|full | 1466 | No | NA | 3 | 0 | 2 | Not-determined | Genome-fragment | NA | NA | 0 |
| S14_vs_k141_78789\|\|full | 1389 | No | NA | 2 | 0 | 1 | Not-determined | Genome-fragment | NA | NA | 0 |
| S3_vs_k141_84863\|\|full | 1409 | No | NA | 3 | 0 | 0 | Low-quality | Genome-fragment | 0.83 | AAI-based (high-confidence) | 0 |
| S14_vs_k141_12788\|\|full | 1568 | No | NA | 4 | 0 | 2 | Not-determined | Genome-fragment | NA | NA | 0 |
| S3_vs_k141_20480\|\|lt2gene | 1409 | No | NA | 2 | 1 | 0 | Low-quality | Genome-fragment | 3.35 | AAI-based (high-confidence) | 0 |
| S14_vs_k141_20437\|\|full | 1511 | No | NA | 2 | 0 | 1 | Not-determined | Genome-fragment | NA | NA | 0 |
| S14_vs_k141_86335\|\|full | 5924 | No | NA | 8 | 0 | 4 | Not-determined | Genome-fragment | NA | NA | 0 |
| S14_vs_k141_27153\|\|full | 1640 | No | NA | 2 | 0 | 1 | Not-determined | Genome-fragment | NA | NA | 0 |
| S5_vs_k141_19190\|\|lt2gene | 1409 | No | NA | 1 | 1 | 0 | Low-quality | Genome-fragment | 2.06 | HMM-based (lower-bound) | 0 |
| S16_extracted_1005 | 1409 | No | NA | 2 | 1 | 0 | Low-quality | Genome-fragment | 4.07 | AAI-based (high-confidence) | 0 |
| S16_extracted_80526 | 1409 | No | NA | 2 | 2 | 0 | Low-quality | Genome-fragment | 4.39 | AAI-based (high-confidence) | 0 |
| S14_vs_k141_42940\|\|full | 2953 | No | NA | 2 | 0 | 2 | Not-determined | Genome-fragment | NA | NA | 0 |
| S16_vs_98547\|\|full | 1409 | No | NA | 4 | 1 | 0 | Low-quality | Genome-fragment | 0.87 | HMM-based (lower-bound) | 0 |
| S12_vs_k141_17400\|\|lt2gene | 1409 | No | NA | 1 | 1 | 0 | Low-quality | Genome-fragment | 0.97 | HMM-based (lower-bound) | 0 |
| S14_vs_k141_71631\|\|full | 2207 | No | NA | 5 | 0 | 1 | Not-determined | Genome-fragment | NA | NA | 0 |
| S8_extracted_k141_196984 | 1409 | No | NA | 1 | 1 | 0 | Low-quality | Genome-fragment | 22.1 | AAI-based (high-confidence) | 0 |
| S9_vs_k141_6056\|\|full | 1410 | No | NA | 4 | 0 | 1 | Low-quality | Genome-fragment | 3.07 | AAI-based (high-confidence) | 0 |
| S14_vs_k141_12855\|\|full | 1006 | No | NA | 2 | 0 | 0 | Not-determined | Genome-fragment | NA | NA | 0 |
| S13_extracted_95667 | 1410 | No | NA | 3 | 2 | 0 | Low-quality | Genome-fragment | 2.31 | AAI-based (medium-confidence) | 0 |
| S14_vs_k141_12862\|\|full | 2615 | No | NA | 4 | 0 | 1 | Not-determined | Genome-fragment | NA | NA | 0 |
| S14_vs_k141_12868\|\|full | 3275 | No | NA | 4 | 0 | 3 | Not-determined | Genome-fragment | NA | NA | 0 |
| S14_vs_k141_78833\|\|full | 2621 | No | NA | 2 | 0 | 2 | Not-determined | Genome-fragment | NA | NA | 0 |
| S14_vs_k141_71653\|\|full | 2524 | No | NA | 4 | 0 | 0 | Not-determined | Genome-fragment | NA | NA | 0 |
| S5_extracted_k141_61455 | 1410 | No | NA | 2 | 1 | 0 | Low-quality | Genome-fragment | 1.59 | AAI-based (high-confidence) | 0 |
| S7_vs_118181\|\|full | 1410 | No | NA | 2 | 0 | 1 | Low-quality | Genome-fragment | 3.43 | AAI-based (medium-confidence) | 0 |
| S14_vs_k141_49023\|\|full | 10696 | No | NA | 10 | 0 | 7 | Not-determined | Genome-fragment | NA | NA | 0 |
| S14_vs_k141_86421\|\|full | 2387 | No | NA | 3 | 0 | 2 | Not-determined | Genome-fragment | NA | NA | 0 |
| S14_vs_k141_27178\|\|full | 2103 | No | NA | 4 | 0 | 4 | Not-determined | Genome-fragment | NA | NA | 0 |
| S7_vs_120441\|\|full | 1410 | No | NA | 3 | 0 | 0 | Low-quality | Genome-fragment | 3.07 | AAI-based (high-confidence) | 0 |
| S14_vs_k141_27179\|\|full | 3105 | No | NA | 3 | 0 | 1 | Not-determined | Genome-fragment | NA | NA | 0 |
| S14_vs_k141_71678\|\|full | 2819 | No | NA | 4 | 0 | 4 | Not-determined | Genome-fragment | NA | NA | 0 |
| S14_vs_k141_27195\|\|full | 1277 | No | NA | 3 | 0 | 1 | Not-determined | Genome-fragment | NA | NA | 0 |
| S8_vs_k141_176717\|\|full | 1410 | No | NA | 3 | 0 | 0 | Low-quality | Genome-fragment | 4.5 | AAI-based (high-confidence) | 0 |
| S14_vs_k141_12914\|\|full | 1472 | No | NA | 3 | 0 | 0 | Not-determined | Genome-fragment | NA | NA | 0 |
| S14_vs_k141_93937\|\|full | 2891 | No | NA | 3 | 0 | 1 | Not-determined | Genome-fragment | NA | NA | 0 |
| S14_vs_k141_78884\|\|full | 1133 | No | NA | 3 | 0 | 0 | Not-determined | Genome-fragment | NA | NA | 0 |
| S9_vs_k141_17991\|\|full | 1411 | No | NA | 4 | 0 | 0 | Low-quality | Genome-fragment | 4.5 | AAI-based (high-confidence) | 0 |
| S11_extracted_k141_62570 | 1411 | No | NA | 1 | 0 | 1 | Low-quality | Genome-fragment | 2.82 | AAI-based (medium-confidence) | 0 |
| S14_vs_k141_13626\|\|full | 1411 | No | NA | 4 | 2 | 0 | Low-quality | Genome-fragment | 1.3 | HMM-based (lower-bound) | 0 |
| S1_vs_k141_43629\|\|full | 1411 | No | NA | 4 | 3 | 0 | Low-quality | Genome-fragment | 3.66 | AAI-based (high-confidence) | 0 |
| S14_vs_k141_27241\|\|full | 2940 | No | NA | 3 | 0 | 3 | Not-determined | Genome-fragment | NA | NA | 0 |
| S3_vs_k141_100542\|\|lt2gene | 1411 | No | NA | 2 | 1 | 0 | Low-quality | Genome-fragment | 3.21 | AAI-based (high-confidence) | 0 |
| S14_vs_k141_108553\|\|full | 1194 | No | NA | 2 | 0 | 0 | Not-determined | Genome-fragment | NA | NA | 0 |
| S5_vs_k141_136814\|\|full | 1411 | No | NA | 3 | 2 | 0 | Low-quality | Genome-fragment | 3.17 | AAI-based (medium-confidence) | 0 |
| S16_extracted_103142 | 1411 | No | NA | 3 | 0 | 0 | Low-quality | Genome-fragment | 1.95 | AAI-based (medium-confidence) | 0 |
| S16_vs_134070\|\|full | 1411 | No | NA | 2 | 0 | 1 | Low-quality | Genome-fragment | 1 | AAI-based (medium-confidence) | 0 |
| S14_vs_k141_43112\|\|full | 1521 | No | NA | 2 | 0 | 1 | Not-determined | Genome-fragment | NA | NA | 0 |
| S14_vs_k141_108570\|\|full | 4484 | No | NA | 6 | 0 | 6 | Not-determined | Genome-fragment | NA | NA | 0 |
| S10_vs_21324\|\|full | 1411 | No | NA | 3 | 2 | 0 | Low-quality | Genome-fragment | 2.26 | AAI-based (medium-confidence) | 0 |
| S14_vs_k141_27279\|\|full | 7875 | No | NA | 9 | 0 | 5 | Not-determined | Genome-fragment | NA | NA | 0 |
| S10_vs_28166\|\|full | 1411 | No | NA | 2 | 0 | 0 | Low-quality | Genome-fragment | 2.94 | AAI-based (medium-confidence) | 0 |
| S14_vs_k141_35144\|\|full | 1243 | No | NA | 2 | 0 | 1 | Not-determined | Genome-fragment | NA | NA | 0 |
| S10_vs_77663\|\|full | 1411 | No | NA | 3 | 1 | 0 | Low-quality | Genome-fragment | 2.19 | AAI-based (high-confidence) | 0 |
| S6_vs_k141_52038\|\|full | 1411 | No | NA | 3 | 0 | 2 | Low-quality | Genome-fragment | 0.83 | AAI-based (medium-confidence) | 0 |
| S14_vs_k141_56989\|\|full | 1458 | No | NA | 2 | 0 | 0 | Not-determined | Genome-fragment | NA | NA | 0 |
| S6_vs_k141_26403\|\|full | 1411 | No | NA | 3 | 0 | 1 | Low-quality | Genome-fragment | 0.6 | AAI-based (medium-confidence) | 0 |
| S11_vs_k141_52479\|\|full | 1412 | No | NA | 3 | 0 | 1 | Low-quality | Genome-fragment | 1.08 | AAI-based (medium-confidence) | 0 |
| S11_vs_k141_70817\|\|full | 1412 | No | NA | 3 | 0 | 0 | Low-quality | Genome-fragment | 1.08 | AAI-based (medium-confidence) | 0 |
| S14_vs_k141_20673\|\|full | 3315 | No | NA | 4 | 0 | 1 | Not-determined | Genome-fragment | NA | NA | 0 |
| S1_extracted_k141_68132 | 1412 | No | NA | 2 | 0 | 0 | Low-quality | Genome-fragment | 1.45 | AAI-based (high-confidence) | 0 |
| S14_vs_k141_43189\|\|full | 1556 | No | NA | 3 | 0 | 1 | Not-determined | Genome-fragment | NA | NA | 0 |
| S14_vs_k141_35204\|\|full | 2783 | No | NA | 4 | 0 | 4 | Not-determined | Genome-fragment | NA | NA | 0 |
| S1_extracted_k141_18605 | 1412 | No | NA | 3 | 0 | 1 | Low-quality | Genome-fragment | 3.09 | AAI-based (high-confidence) | 0 |
| S14_vs_k141_5616\|\|full | 2681 | No | NA | 3 | 0 | 2 | Not-determined | Genome-fragment | NA | NA | 0 |
| S1_vs_k141_16591\|\|full | 1412 | No | NA | 4 | 0 | 1 | Low-quality | Genome-fragment | 0.81 | AAI-based (high-confidence) | 0 |
| S7_vs_92812\|\|full | 1412 | No | NA | 4 | 1 | 0 | Low-quality | Genome-fragment | 3.58 | AAI-based (high-confidence) | 0 |
| S7_vs_94296\|\|full | 1412 | No | NA | 4 | 1 | 0 | Low-quality | Genome-fragment | 2.84 | AAI-based (high-confidence) | 0 |
| S14_vs_k141_64161\|\|full | 1362 | No | NA | 3 | 0 | 0 | Not-determined | Genome-fragment | NA | NA | 0 |
| S7_vs_109090\|\|full | 1412 | No | NA | 3 | 0 | 1 | Low-quality | Genome-fragment | 3.39 | AAI-based (medium-confidence) | 0 |
| S14_vs_k141_43225\|\|full | 2028 | No | NA | 3 | 0 | 2 | Not-determined | Genome-fragment | NA | NA | 0 |
| S14_vs_k141_78979\|\|full | 2092 | No | NA | 3 | 0 | 1 | Not-determined | Genome-fragment | NA | NA | 0 |
| S16_vs_109211\|\|full | 1412 | No | NA | 2 | 1 | 0 | Low-quality | Genome-fragment | 1 | HMM-based (lower-bound) | 0 |
| S10_extracted_103947 | 1412 | No | NA | 1 | 0 | 1 | Low-quality | Genome-fragment | 3.98 | AAI-based (medium-confidence) | 0 |
| S14_vs_k141_78984\|\|full | 12703 | No | NA | 14 | 0 | 8 | Not-determined | Genome-fragment | NA | NA | 0 |
| S14_vs_k141_100965\|\|full | 1539 | No | NA | 3 | 0 | 0 | Not-determined | Genome-fragment | NA | NA | 0 |
| S12_vs_k141_13851\|\|full | 1412 | No | NA | 2 | 0 | 2 | Low-quality | Genome-fragment | 0.45 | AAI-based (medium-confidence) | 0 |
| S3_vs_k141_3225\|\|lt2gene | 1413 | No | NA | 2 | 2 | 0 | Low-quality | Genome-fragment | 3.13 | AAI-based (high-confidence) | 0 |
| S14_vs_k141_64199\|\|full | 1278 | No | NA | 4 | 0 | 1 | Not-determined | Genome-fragment | NA | NA | 0 |
| S5_vs_k141_87956\|\|lt2gene | 1413 | No | NA | 2 | 2 | 0 | Low-quality | Genome-fragment | 3.34 | AAI-based (medium-confidence) | 0 |
| S14_vs_k141_27446\|\|full | 2485 | No | NA | 4 | 0 | 2 | Not-determined | Genome-fragment | NA | NA | 0 |
| S7_vs_3284\|\|full | 1413 | No | NA | 3 | 1 | 0 | Low-quality | Genome-fragment | 2.58 | HMM-based (lower-bound) | 0 |
| S7_vs_75071\|\|full | 1413 | No | NA | 2 | 0 | 0 | Low-quality | Genome-fragment | 3.32 | AAI-based (medium-confidence) | 0 |
| S14_vs_k141_20836\|\|full | 1414 | No | NA | 3 | 0 | 2 | Not-determined | Genome-fragment | NA | NA | 0 |
| S2_extracted_k141_3536 | 1413 | No | NA | 1 | 0 | 1 | Low-quality | Genome-fragment | 0.79 | AAI-based (medium-confidence) | 0 |
| S14_vs_k141_94273\|\|full | 1871 | No | NA | 3 | 0 | 2 | Not-determined | Genome-fragment | NA | NA | 0 |
| S6_vs_k141_43211\|\|full | 1413 | No | NA | 3 | 2 | 0 | Low-quality | Genome-fragment | 2.01 | AAI-based (medium-confidence) | 0 |
| S14_vs_k141_20849\|\|full | 1418 | No | NA | 3 | 0 | 0 | Not-determined | Genome-fragment | NA | NA | 0 |
| S8_extracted_k141_101026 | 1413 | No | NA | 1 | 0 | 1 | Low-quality | Genome-fragment | 0.87 | AAI-based (high-confidence) | 0 |
| S8_vs_k141_112230\|\|full | 1413 | No | NA | 5 | 1 | 0 | Low-quality | Genome-fragment | 3.22 | AAI-based (medium-confidence) | 0 |
| S11_extracted_k141_6979 | 1414 | No | NA | 4 | 0 | 0 | Low-quality | Genome-fragment | 3.64 | AAI-based (high-confidence) | 0 |
| S1_vs_k141_980\|\|lt2gene | 1414 | No | NA | 2 | 2 | 0 | Low-quality | Genome-fragment | 2.6 | AAI-based (medium-confidence) | 0 |
| S10_vs_20219\|\|full | 1414 | No | NA | 3 | 0 | 0 | Low-quality | Genome-fragment | 3.36 | AAI-based (high-confidence) | 0 |
| S14_vs_k141_94320\|\|full | 3415 | No | NA | 3 | 0 | 2 | Not-determined | Genome-fragment | NA | NA | 0 |
| S14_vs_k141_35348\|\|full | 3970 | No | NA | 6 | 0 | 4 | Not-determined | Genome-fragment | NA | NA | 0 |
| S12_extracted_k141_959 | 1414 | No | NA | 2 | 0 | 0 | Low-quality | Genome-fragment | 2.79 | AAI-based (high-confidence) | 0 |
| S12_extracted_k141_187394 | 1414 | No | NA | 2 | 0 | 0 | Low-quality | Genome-fragment | 2.23 | AAI-based (high-confidence) | 0 |
| S15_extracted_23267 | 1414 | No | NA | 3 | 0 | 0 | Low-quality | Genome-fragment | 0.84 | AAI-based (high-confidence) | 0 |
| S14_vs_k141_101105\|\|full | 2766 | No | NA | 3 | 0 | 2 | Not-determined | Genome-fragment | NA | NA | 0 |
| S15_vs_5155\|\|full | 1414 | No | NA | 3 | 1 | 0 | Low-quality | Genome-fragment | 2.76 | AAI-based (high-confidence) | 0 |
| S8_vs_k141_122027\|\|full | 1414 | No | NA | 3 | 0 | 0 | Low-quality | Genome-fragment | 3.3 | AAI-based (high-confidence) | 0 |
| S8_vs_k141_151320\|\|full | 1414 | No | NA | 3 | 1 | 0 | Low-quality | Genome-fragment | 2.47 | AAI-based (medium-confidence) | 0 |
| S11_vs_k141_16282\|\|full | 1415 | No | NA | 2 | 0 | 0 | Low-quality | Genome-fragment | 0.8 | AAI-based (medium-confidence) | 0 |
| S1_extracted_k141_36038 | 1415 | No | NA | 1 | 0 | 1 | Low-quality | Genome-fragment | 0.24 | AAI-based (high-confidence) | 0 |
| S5_extracted_k141_8403 | 1415 | No | NA | 1 | 0 | 1 | Low-quality | Genome-fragment | 2.98 | AAI-based (high-confidence) | 0 |
| S14_vs_k141_35410\|\|full | 1147 | No | NA | 2 | 0 | 1 | Not-determined | Genome-fragment | NA | NA | 0 |
| S14_vs_k141_5887\|\|full | 3883 | No | NA | 2 | 0 | 2 | Not-determined | Genome-fragment | NA | NA | 0 |
| S5_vs_k141_61261\|\|lt2gene | 1415 | No | NA | 1 | 1 | 0 | Low-quality | Genome-fragment | 2.77 | AAI-based (medium-confidence) | 0 |
| S16_extracted_160726 | 1415 | No | NA | 3 | 0 | 0 | Low-quality | Genome-fragment | 2.42 | AAI-based (high-confidence) | 0 |
| S14_vs_k141_49309\|\|full | 1234 | No | NA | 3 | 0 | 1 | Not-determined | Genome-fragment | NA | NA | 0 |
| S16_vs_13456\|\|full | 1415 | No | NA | 2 | 0 | 0 | Low-quality | Genome-fragment | 1.84 | AAI-based (high-confidence) | 0 |
| S16_vs_159084\|\|full | 1415 | No | NA | 3 | 0 | 1 | Low-quality | Genome-fragment | 0.87 | AAI-based (medium-confidence) | 0 |
| S10_vs_34981\|\|full | 1415 | No | NA | 2 | 0 | 0 | Low-quality | Genome-fragment | 3.11 | AAI-based (high-confidence) | 0 |
| S14_vs_k141_13248\|\|full | 2233 | No | NA | 3 | 0 | 2 | Not-determined | Genome-fragment | NA | NA | 0 |
| S14_vs_k141_94393\|\|full | 1553 | No | NA | 4 | 0 | 0 | Not-determined | Genome-fragment | NA | NA | 0 |
| S14_vs_k141_35463\|\|full | 2960 | No | NA | 2 | 0 | 2 | Not-determined | Genome-fragment | NA | NA | 0 |
| S17_vs_k141_142064\|\|full | 1416 | No | NA | 2 | 0 | 1 | Low-quality | Genome-fragment | 1.81 | AAI-based (medium-confidence) | 0 |
| S14_vs_k141_101172\|\|full | 2607 | No | NA | 4 | 0 | 1 | Not-determined | Genome-fragment | NA | NA | 0 |
| S14_vs_k141_94409\|\|full | 2076 | No | NA | 6 | 0 | 0 | Not-determined | Genome-fragment | NA | NA | 0 |
| S14_vs_k141_57266\|\|full | 1911 | No | NA | 2 | 0 | 0 | Not-determined | Genome-fragment | NA | NA | 0 |
| S14_vs_k141_13306\|\|full | 4614 | No | NA | 6 | 0 | 1 | Not-determined | Genome-fragment | NA | NA | 0 |
| S9_vs_k141_79683\|\|lt2gene | 1416 | No | NA | 2 | 1 | 0 | Low-quality | Genome-fragment | 3.26 | AAI-based (high-confidence) | 0 |
| S14_vs_k141_13310\|\|full | 6460 | No | NA | 8 | 0 | 1 | Not-determined | Genome-fragment | NA | NA | 0 |
| S14_extracted_k141_96924 | 1416 | No | NA | 4 | 0 | 0 | Low-quality | Genome-fragment | 1.98 | AAI-based (medium-confidence) | 0 |
| S14_vs_k141_21021\|\|full | 1139 | No | NA | 2 | 0 | 1 | Not-determined | Genome-fragment | NA | NA | 0 |
| S14_vs_k141_101211\|\|full | 1118 | No | NA | 4 | 0 | 0 | Not-determined | Genome-fragment | NA | NA | 0 |
| S3_vs_k141_70307\|\|full | 1416 | No | NA | 3 | 0 | 2 | Low-quality | Genome-fragment | 0.91 | AAI-based (medium-confidence) | 0 |
| S5_vs_k141_33952\|\|full | 1416 | No | NA | 4 | 1 | 0 | Low-quality | Genome-fragment | 3.59 | AAI-based (high-confidence) | 0 |
| S14_vs_k141_94458\|\|full | 2680 | No | NA | 6 | 0 | 1 | Not-determined | Genome-fragment | NA | NA | 0 |
| S12_extracted_k141_65880 | 1416 | No | NA | 2 | 1 | 0 | Low-quality | Genome-fragment | 0.87 | HMM-based (lower-bound) | 0 |
| S12_vs_k141_7898\|\|full | 1416 | No | NA | 2 | 0 | 1 | Low-quality | Genome-fragment | 2.92 | AAI-based (high-confidence) | 0 |
| S14_vs_k141_57342\|\|full | 1175 | No | NA | 3 | 0 | 0 | Not-determined | Genome-fragment | NA | NA | 0 |
| S12_vs_k141_37086\|\|full | 1416 | No | NA | 2 | 0 | 1 | Low-quality | Genome-fragment | 1.66 | AAI-based (medium-confidence) | 0 |
| S14_vs_k141_5990\|\|full | 1389 | No | NA | 2 | 0 | 1 | Not-determined | Genome-fragment | NA | NA | 0 |
| S14_vs_k141_94475\|\|full | 2316 | No | NA | 4 | 0 | 2 | Not-determined | Genome-fragment | NA | NA | 0 |
| S8_extracted_k141_112695 | 1416 | No | NA | 3 | 0 | 1 | Low-quality | Genome-fragment | 0.32 | AAI-based (high-confidence) | 0 |
| S8_vs_k141_107404\|\|lt2gene | 1416 | No | NA | 2 | 2 | 0 | Low-quality | Genome-fragment | 4.33 | AAI-based (medium-confidence) | 0 |
| S14_vs_k141_21040\|\|full | 1712 | No | NA | 3 | 0 | 1 | Not-determined | Genome-fragment | NA | NA | 0 |
| S17_extracted_k141_163096 | 1417 | No | NA | 4 | 0 | 1 | Low-quality | Genome-fragment | 2.59 | AAI-based (medium-confidence) | 0 |
| S14_vs_k141_101256\|\|full | 1364 | No | NA | 3 | 0 | 0 | Not-determined | Genome-fragment | NA | NA | 0 |
| S9_vs_k141_70418\|\|full | 1417 | No | NA | 4 | 1 | 0 | Low-quality | Genome-fragment | 3.24 | AAI-based (high-confidence) | 0 |
| S11_vs_k141_46549\|\|full | 1417 | No | NA | 3 | 0 | 0 | Low-quality | Genome-fragment | 1.28 | AAI-based (medium-confidence) | 0 |
| S14_vs_k141_8711\|\|full | 1417 | No | NA | 3 | 0 | 0 | Low-quality | Genome-fragment | 1.99 | AAI-based (medium-confidence) | 0 |
| S14_vs_k141_72072\|\|full | 2635 | No | NA | 3 | 0 | 0 | Not-determined | Genome-fragment | NA | NA | 0 |
| S5_extracted_k141_133017 | 1417 | No | NA | 5 | 4 | 0 | Low-quality | Genome-fragment | 3.84 | AAI-based (high-confidence) | 0 |
| S14_vs_k141_64389\|\|full | 13951 | No | NA | 14 | 0 | 4 | Not-determined | Genome-fragment | NA | NA | 0 |
| S14_vs_k141_94519\|\|full | 1490 | No | NA | 3 | 0 | 0 | Not-determined | Genome-fragment | NA | NA | 0 |
| S14_vs_k141_57396\|\|full | 1746 | No | NA | 4 | 0 | 1 | Not-determined | Genome-fragment | NA | NA | 0 |
| S5_vs_k141_4647\|\|full | 1417 | No | NA | 3 | 2 | 0 | Low-quality | Genome-fragment | 2.44 | AAI-based (high-confidence) | 0 |
| S16_vs_69945\|\|full | 1417 | No | NA | 3 | 0 | 0 | Low-quality | Genome-fragment | 2.32 | AAI-based (medium-confidence) | 0 |
| S10_vs_14888\|\|full | 1417 | No | NA | 3 | 0 | 0 | Low-quality | Genome-fragment | 2.78 | AAI-based (high-confidence) | 0 |
| S14_vs_k141_87100\|\|full | 4035 | No | NA | 5 | 0 | 4 | Not-determined | Genome-fragment | NA | NA | 0 |
| S12_extracted_k141_186779 | 1417 | No | NA | 2 | 0 | 0 | Low-quality | Genome-fragment | 2.92 | AAI-based (high-confidence) | 0 |
| S4_extracted_27892 | 1417 | No | NA | 2 | 0 | 0 | Low-quality | Genome-fragment | 3.26 | AAI-based (high-confidence) | 0 |
| S14_vs_k141_79397\|\|full | 3669 | No | NA | 5 | 0 | 1 | Not-determined | Genome-fragment | NA | NA | 0 |
| S14_vs_k141_116265\|\|full | 1196 | No | NA | 3 | 0 | 1 | Not-determined | Genome-fragment | NA | NA | 0 |
| S14_vs_k141_101324\|\|full | 1009 | No | NA | 2 | 0 | 0 | Not-determined | Genome-fragment | NA | NA | 0 |
| S8_extracted_k141_113378 | 1417 | No | NA | 2 | 0 | 2 | Low-quality | Genome-fragment | 0.39 | AAI-based (medium-confidence) | 0 |
| S14_vs_k141_94577\|\|full | 1677 | No | NA | 2 | 0 | 2 | Not-determined | Genome-fragment | NA | NA | 0 |
| S14_vs_k141_6095\|\|full | 1538 | No | NA | 3 | 0 | 0 | Not-determined | Genome-fragment | NA | NA | 0 |
| S8_extracted_k141_51683 | 1417 | No | NA | 2 | 0 | 0 | Low-quality | Genome-fragment | 3.28 | AAI-based (high-confidence) | 0 |
| S14_vs_k141_27862\|\|full | 1188 | No | NA | 3 | 0 | 0 | Not-determined | Genome-fragment | NA | NA | 0 |
| S14_vs_k141_27867\|\|full | 2101 | No | NA | 3 | 0 | 3 | Not-determined | Genome-fragment | NA | NA | 0 |
| S14_vs_k141_109181\|\|full | 1136 | No | NA | 2 | 0 | 0 | Not-determined | Genome-fragment | NA | NA | 0 |
| S8_vs_k141_80750\|\|full | 1417 | No | NA | 2 | 2 | 0 | Low-quality | Genome-fragment | 3.23 | AAI-based (high-confidence) | 0 |
| S14_vs_k141_116327\|\|full | 1323 | No | NA | 4 | 0 | 1 | Not-determined | Genome-fragment | NA | NA | 0 |
| S9_extracted_k141_12981 | 1418 | No | NA | 1 | 1 | 0 | Low-quality | Genome-fragment | 3.5 | AAI-based (high-confidence) | 0 |
| S14_vs_k141_49637\|\|full | 1821 | No | NA | 3 | 0 | 0 | Not-determined | Genome-fragment | NA | NA | 0 |
| S14_vs_k141_116336\|\|full | 1553 | No | NA | 3 | 0 | 0 | Not-determined | Genome-fragment | NA | NA | 0 |
| S9_vs_k141_62021\|\|full | 1418 | No | NA | 2 | 0 | 0 | Low-quality | Genome-fragment | 2.48 | AAI-based (high-confidence) | 0 |
| S14_vs_k141_57454\|\|full | 2204 | No | NA | 3 | 0 | 3 | Not-determined | Genome-fragment | NA | NA | 0 |
| S13_vs_35207\|\|full | 1418 | No | NA | 3 | 1 | 0 | Low-quality | Genome-fragment | 2.83 | AAI-based (high-confidence) | 0 |
| S14_vs_k141_57458\|\|full | 1308 | No | NA | 3 | 0 | 1 | Not-determined | Genome-fragment | NA | NA | 0 |
| S14_vs_k141_72204\|\|full | 2908 | No | NA | 4 | 0 | 1 | Not-determined | Genome-fragment | NA | NA | 0 |
| S13_vs_86462\|\|full | 1418 | No | NA | 2 | 0 | 0 | Low-quality | Genome-fragment | 3.12 | AAI-based (high-confidence) | 0 |
| S14_vs_k141_94660\|\|full | 1009 | No | NA | 2 | 0 | 0 | Not-determined | Genome-fragment | NA | NA | 0 |
| S14_vs_k141_116376\|\|full | 1353 | No | NA | 2 | 0 | 2 | Not-determined | Genome-fragment | NA | NA | 0 |
| S14_vs_k141_13430\|\|full | 2591 | No | NA | 4 | 0 | 1 | Not-determined | Genome-fragment | NA | NA | 0 |
| S14_vs_k141_35029\|\|full | 1418 | No | NA | 4 | 3 | 0 | Low-quality | Genome-fragment | 3.7 | AAI-based (high-confidence) | 0 |
| S14_vs_k141_27920\|\|full | 3203 | No | NA | 5 | 0 | 2 | Not-determined | Genome-fragment | NA | NA | 0 |
| S14_vs_k141_87221\|\|full | 3754 | No | NA | 5 | 0 | 5 | Not-determined | Genome-fragment | NA | NA | 0 |
| S14_vs_k141_87627\|\|full | 1418 | No | NA | 3 | 1 | 0 | Low-quality | Genome-fragment | 3.05 | AAI-based (high-confidence) | 0 |
| S14_vs_k141_27925\|\|full | 1477 | No | NA | 2 | 0 | 0 | Not-determined | Genome-fragment | NA | NA | 0 |
| S14_vs_k141_64472\|\|full | 2223 | No | NA | 2 | 0 | 2 | Not-determined | Genome-fragment | NA | NA | 0 |
| S14_vs_k141_13436\|\|full | 1184 | No | NA | 3 | 0 | 0 | Not-determined | Genome-fragment | NA | NA | 0 |
| S14_vs_k141_94673\|\|full | 2357 | No | NA | 5 | 0 | 2 | Not-determined | Genome-fragment | NA | NA | 0 |
| S14_vs_k141_79505\|\|full | 40116 | No | NA | 48 | 7 | 3 | Medium-quality | Genome-fragment | 89.03 | AAI-based (medium-confidence) | 0 |
| S14_vs_k141_14658\|\|lt2gene | 1418 | No | NA | 2 | 2 | 0 | Low-quality | Genome-fragment | 3.01 | AAI-based (high-confidence) | 0 |
| S14_vs_k141_13451\|\|full | 2692 | No | NA | 3 | 0 | 0 | Not-determined | Genome-fragment | NA | NA | 0 |
| S14_vs_k141_87250\|\|full | 2419 | No | NA | 4 | 0 | 3 | Not-determined | Genome-fragment | NA | NA | 0 |
| S3_extracted_k141_59130 | 1418 | No | NA | 2 | 0 | 2 | Low-quality | Genome-fragment | 0.96 | AAI-based (medium-confidence) | 0 |
| S14_vs_k141_72219\|\|full | 1426 | No | NA | 3 | 0 | 0 | Not-determined | Genome-fragment | NA | NA | 0 |
| S14_vs_k141_49704\|\|full | 1683 | No | NA | 3 | 0 | 1 | Not-determined | Genome-fragment | NA | NA | 0 |
| S7_extracted_66072 | 1418 | No | NA | 2 | 0 | 0 | Low-quality | Genome-fragment | 2.94 | AAI-based (medium-confidence) | 0 |
| S14_vs_k141_21224\|\|full | 6635 | No | NA | 5 | 0 | 4 | Not-determined | Genome-fragment | NA | NA | 0 |
| S14_vs_k141_57540\|\|full | 2759 | No | NA | 3 | 0 | 2 | Not-determined | Genome-fragment | NA | NA | 0 |
| S7_vs_73472\|\|full | 1418 | No | NA | 4 | 1 | 0 | Low-quality | Genome-fragment | 2.57 | AAI-based (high-confidence) | 0 |
| S14_vs_k141_27951\|\|full | 16144 | No | NA | 11 | 0 | 7 | Not-determined | Genome-fragment | NA | NA | 0 |
| S7_vs_98220\|\|full | 1418 | No | NA | 3 | 0 | 0 | Low-quality | Genome-fragment | 1.08 | AAI-based (medium-confidence) | 0 |
| S12_vs_k141_112996\|\|full | 1418 | No | NA | 4 | 0 | 0 | Low-quality | Genome-fragment | 1.76 | AAI-based (medium-confidence) | 0 |
| S14_vs_k141_87280\|\|full | 1427 | No | NA | 3 | 0 | 0 | Not-determined | Genome-fragment | NA | NA | 0 |
| S14_vs_k141_35822\|\|full | 1493 | No | NA | 3 | 0 | 0 | Not-determined | Genome-fragment | NA | NA | 0 |
| S15_vs_16090\|\|lt2gene | 1418 | No | NA | 1 | 1 | 0 | Low-quality | Genome-fragment | 2.28 | HMM-based (lower-bound) | 0 |
| S6_vs_k141_10674\|\|full | 1418 | No | NA | 3 | 1 | 0 | Low-quality | Genome-fragment | 2.4 | AAI-based (medium-confidence) | 0 |
| S14_vs_k141_109316\|\|full | 14598 | No | NA | 7 | 0 | 5 | Not-determined | Genome-fragment | NA | NA | 0 |
| S14_vs_k141_21278\|\|full | 4498 | No | NA | 7 | 0 | 2 | Not-determined | Genome-fragment | NA | NA | 0 |
| S14_vs_k141_13533\|\|full | 2306 | No | NA | 5 | 0 | 0 | Not-determined | Genome-fragment | NA | NA | 0 |
| S14_vs_k141_79558\|\|full | 1870 | No | NA | 3 | 0 | 0 | Not-determined | Genome-fragment | NA | NA | 0 |
| S14_vs_k141_101482\|\|full | 20333 | No | NA | 25 | 3 | 1 | Medium-quality | Genome-fragment | 54.32 | AAI-based (high-confidence) | 0 |
| S14_vs_k141_94798\|\|full | 1992 | No | NA | 6 | 0 | 1 | Not-determined | Genome-fragment | NA | NA | 0 |
| S14_vs_k141_94804\|\|full | 2031 | No | NA | 3 | 0 | 2 | Not-determined | Genome-fragment | NA | NA | 0 |
| S8_vs_k141_128199\|\|full | 1418 | No | NA | 4 | 2 | 0 | Low-quality | Genome-fragment | 2.39 | AAI-based (medium-confidence) | 0 |
| S17_vs_k141_165235\|\|full | 1419 | No | NA | 3 | 0 | 3 | Low-quality | Genome-fragment | 0.44 | AAI-based (medium-confidence) | 0 |
| S14_vs_k141_28016\|\|full | 5302 | No | NA | 4 | 0 | 3 | Not-determined | Genome-fragment | NA | NA | 0 |
| S14_vs_k141_72292\|\|full | 1989 | No | NA | 3 | 0 | 1 | Not-determined | Genome-fragment | NA | NA | 0 |
| S14_vs_k141_79637\|\|full | 1758 | No | NA | 3 | 0 | 1 | Not-determined | Genome-fragment | NA | NA | 0 |
| S9_extracted_k141_61511 | 1419 | No | NA | 1 | 0 | 1 | Low-quality | Genome-fragment | 0.93 | AAI-based (high-confidence) | 0 |
| S14_vs_k141_79638\|\|full | 2363 | No | NA | 3 | 0 | 1 | Not-determined | Genome-fragment | NA | NA | 0 |
| S9_vs_k141_30624\|\|full | 1419 | No | NA | 5 | 1 | 0 | Low-quality | Genome-fragment | 3.56 | AAI-based (high-confidence) | 0 |
| S11_vs_k141_92066\|\|lt2gene | 1419 | No | NA | 2 | 2 | 0 | Low-quality | Genome-fragment | 3.23 | AAI-based (high-confidence) | 0 |
| S14_vs_k141_43890\|\|full | 1927 | No | NA | 2 | 0 | 2 | Not-determined | Genome-fragment | NA | NA | 0 |
| S14_vs_k141_94829\|\|full | 1682 | No | NA | 3 | 0 | 1 | Not-determined | Genome-fragment | NA | NA | 0 |
| S14_vs_k141_6417\|\|full | 2930 | No | NA | 3 | 0 | 3 | Not-determined | Genome-fragment | NA | NA | 0 |
| S11_vs_k141_9767\|\|lt2gene | 1419 | No | NA | 2 | 1 | 0 | Low-quality | Genome-fragment | 3.22 | AAI-based (medium-confidence) | 0 |
| S14_vs_k141_43908\|\|full | 2231 | No | NA | 3 | 0 | 3 | Not-determined | Genome-fragment | NA | NA | 0 |
| S5_vs_k141_53577\|\|full | 1419 | No | NA | 2 | 0 | 0 | Low-quality | Genome-fragment | 2.27 | AAI-based (medium-confidence) | 0 |
| S7_vs_1468\|\|full | 1419 | No | NA | 3 | 0 | 1 | Low-quality | Genome-fragment | 2.02 | AAI-based (medium-confidence) | 0 |
| S12_extracted_k141_70529 | 1419 | No | NA | 1 | 1 | 0 | Low-quality | Genome-fragment | 4.13 | AAI-based (high-confidence) | 0 |
| S2_extracted_k141_17149 | 1419 | No | NA | 3 | 1 | 0 | Low-quality | Genome-fragment | 1.48 | AAI-based (high-confidence) | 0 |
| S4_extracted_28836 | 1419 | No | NA | 3 | 2 | 0 | Low-quality | Genome-fragment | 2.24 | AAI-based (high-confidence) | 0 |
| S14_vs_k141_87448\|\|full | 7482 | No | NA | 6 | 0 | 3 | Not-determined | Genome-fragment | NA | NA | 0 |
| S14_vs_k141_28118\|\|full | 4479 | No | NA | 5 | 0 | 1 | Not-determined | Genome-fragment | NA | NA | 0 |
| S9_vs_k141_88781\|\|full | 1420 | No | NA | 4 | 3 | 0 | Low-quality | Genome-fragment | 3.25 | AAI-based (medium-confidence) | 0 |
| S5_vs_k141_119001\|\|full | 1420 | No | NA | 5 | 2 | 0 | Low-quality | Genome-fragment | 3.64 | AAI-based (high-confidence) | 0 |
| S7_vs_93072\|\|full | 1420 | No | NA | 3 | 1 | 0 | Low-quality | Genome-fragment | 2.56 | AAI-based (high-confidence) | 0 |
| S16_vs_39531\|\|full | 1420 | No | NA | 5 | 0 | 0 | Low-quality | Genome-fragment | 1.05 | AAI-based (medium-confidence) | 0 |
| S14_vs_k141_21476\|\|full | 1577 | No | NA | 3 | 0 | 0 | Not-determined | Genome-fragment | NA | NA | 0 |
| S10_extracted_55869 | 1420 | No | NA | 2 | 1 | 0 | Low-quality | Genome-fragment | 3.15 | AAI-based (medium-confidence) | 0 |
| S14_vs_k141_21484\|\|full | 2366 | No | NA | 4 | 0 | 1 | Not-determined | Genome-fragment | NA | NA | 0 |
| S14_vs_k141_109503\|\|full | 4521 | No | NA | 5 | 0 | 2 | Not-determined | Genome-fragment | NA | NA | 0 |
| S14_vs_k141_28145\|\|full | 2842 | No | NA | 5 | 0 | 0 | Not-determined | Genome-fragment | NA | NA | 0 |
| S10_vs_8778\|\|full | 1420 | No | NA | 3 | 1 | 0 | Low-quality | Genome-fragment | 4.06 | AAI-based (high-confidence) | 0 |
| S12_extracted_k141_17655 | 1420 | No | NA | 4 | 0 | 2 | Low-quality | Genome-fragment | 3.26 | AAI-based (medium-confidence) | 0 |
| S12_extracted_k141_147964 | 1420 | No | NA | 2 | 0 | 0 | Low-quality | Genome-fragment | 2.27 | AAI-based (high-confidence) | 0 |
| S14_vs_k141_13706\|\|full | 1800 | No | NA | 4 | 0 | 0 | Not-determined | Genome-fragment | NA | NA | 0 |
| S14_vs_k141_49927\|\|full | 1234 | No | NA | 3 | 0 | 0 | Not-determined | Genome-fragment | NA | NA | 0 |
| S12_vs_k141_27326\|\|full | 1420 | No | NA | 4 | 0 | 0 | Low-quality | Genome-fragment | 2.65 | AAI-based (medium-confidence) | 0 |
| S12_vs_k141_116501\|\|lt2gene | 1420 | No | NA | 1 | 1 | 0 | Low-quality | Genome-fragment | 2.07 | HMM-based (lower-bound) | 0 |
| S14_vs_k141_116693\|\|full | 1978 | No | NA | 3 | 0 | 2 | Not-determined | Genome-fragment | NA | NA | 0 |
| S15_extracted_71598 | 1420 | No | NA | 4 | 0 | 0 | Low-quality | Genome-fragment | 3.46 | AAI-based (medium-confidence) | 0 |
| S14_vs_k141_57853\|\|full | 2028 | No | NA | 3 | 0 | 1 | Not-determined | Genome-fragment | NA | NA | 0 |
| S14_vs_k141_79875\|\|full | 1818 | No | NA | 2 | 0 | 0 | Not-determined | Genome-fragment | NA | NA | 0 |
| S14_vs_k141_109568\|\|full | 2434 | No | NA | 2 | 0 | 0 | Not-determined | Genome-fragment | NA | NA | 0 |
| S14_vs_k141_109571\|\|full | 1991 | No | NA | 4 | 0 | 2 | Not-determined | Genome-fragment | NA | NA | 0 |
| S14_vs_k141_36087\|\|full | 1433 | No | NA | 3 | 0 | 0 | Not-determined | Genome-fragment | NA | NA | 0 |
| S14_vs_k141_116718\|\|full | 13281 | No | NA | 13 | 0 | 13 | Not-determined | Genome-fragment | NA | NA | 0 |
| S14_vs_k141_64836\|\|full | 1644 | No | NA | 3 | 0 | 0 | Not-determined | Genome-fragment | NA | NA | 0 |
| S14_vs_k141_57926\|\|full | 2036 | No | NA | 6 | 0 | 1 | Not-determined | Genome-fragment | NA | NA | 0 |
| S15_vs_73875\|\|lt2gene | 1420 | No | NA | 2 | 1 | 0 | Low-quality | Genome-fragment | 3.16 | AAI-based (high-confidence) | 0 |
| S14_vs_k141_109674\|\|full | 2282 | No | NA | 3 | 0 | 1 | Not-determined | Genome-fragment | NA | NA | 0 |
| S8_extracted_k141_202403 | 1420 | No | NA | 2 | 0 | 0 | Low-quality | Genome-fragment | 0.75 | AAI-based (medium-confidence) | 0 |
| S8_vs_k141_147736\|\|full | 1420 | No | NA | 3 | 2 | 0 | Low-quality | Genome-fragment | 3.17 | AAI-based (high-confidence) | 0 |
| S14_vs_k141_13852\|\|full | 1470 | No | NA | 2 | 0 | 0 | Not-determined | Genome-fragment | NA | NA | 0 |
| S14_vs_k141_116805\|\|full | 1806 | No | NA | 3 | 0 | 0 | Not-determined | Genome-fragment | NA | NA | 0 |
| S8_vs_k141_31178\|\|full | 1420 | No | NA | 5 | 0 | 0 | Low-quality | Genome-fragment | 4.27 | AAI-based (high-confidence) | 0 |
| S14_vs_k141_101829\|\|full | 1501 | No | NA | 2 | 0 | 0 | Not-determined | Genome-fragment | NA | NA | 0 |
| S14_vs_k141_13877\|\|full | 1748 | No | NA | 3 | 0 | 3 | Not-determined | Genome-fragment | NA | NA | 0 |
| S14_vs_k141_21677\|\|full | 3026 | No | NA | 3 | 0 | 1 | Not-determined | Genome-fragment | NA | NA | 0 |
| S8_vs_k141_196674\|\|full | 1420 | No | NA | 3 | 0 | 0 | Low-quality | Genome-fragment | 1.95 | AAI-based (medium-confidence) | 0 |
| S14_vs_k141_80027\|\|full | 2750 | No | NA | 3 | 0 | 0 | Not-determined | Genome-fragment | NA | NA | 0 |
| S14_vs_k141_101843\|\|full | 3513 | No | NA | 2 | 0 | 1 | Not-determined | Genome-fragment | NA | NA | 0 |
| S14_vs_k141_80033\|\|full | 1014 | No | NA | 1 | 0 | 1 | Not-determined | Genome-fragment | NA | NA | 0 |
| S14_vs_k141_109770\|\|full | 2208 | No | NA | 3 | 0 | 1 | Not-determined | Genome-fragment | NA | NA | 0 |
| S14_vs_k141_95159\|\|full | 1160 | No | NA | 3 | 0 | 0 | Not-determined | Genome-fragment | NA | NA | 0 |
| S14_vs_k141_58026\|\|full | 1037 | No | NA | 3 | 0 | 1 | Not-determined | Genome-fragment | NA | NA | 0 |
| S14_vs_k141_87517\|\|full | 1507 | No | NA | 3 | 0 | 0 | Not-determined | Genome-fragment | NA | NA | 0 |
| S13_vs_4655\|\|full | 1421 | No | NA | 4 | 2 | 0 | Low-quality | Genome-fragment | 3.81 | AAI-based (high-confidence) | 0 |
| S14_vs_k141_21729\|\|full | 2295 | No | NA | 2 | 0 | 1 | Not-determined | Genome-fragment | NA | NA | 0 |
| S14_vs_k141_64981\|\|full | 2193 | No | NA | 6 | 0 | 0 | Not-determined | Genome-fragment | NA | NA | 0 |
| S14_vs_k141_72511\|\|full | 1765 | No | NA | 3 | 0 | 0 | Not-determined | Genome-fragment | NA | NA | 0 |
| S7_vs_123439\|\|full | 1421 | No | NA | 3 | 0 | 0 | Low-quality | Genome-fragment | 2.54 | AAI-based (high-confidence) | 0 |
| S14_vs_k141_109830\|\|full | 4423 | No | NA | 8 | 0 | 0 | Not-determined | Genome-fragment | NA | NA | 0 |
| S12_vs_k141_42674\|\|full | 1421 | No | NA | 2 | 0 | 0 | Low-quality | Genome-fragment | 3.13 | AAI-based (high-confidence) | 0 |
| S17_vs_k141_143883\|\|lt2gene | 1422 | No | NA | 2 | 0 | 0 | Low-quality | Genome-fragment | 2.95 | AAI-based (high-confidence) | 0 |
| S14_vs_k141_21774\|\|full | 2981 | No | NA | 3 | 0 | 0 | Not-determined | Genome-fragment | NA | NA | 0 |
| S14_vs_k141_6767\|\|full | 6666 | No | NA | 3 | 0 | 3 | Not-determined | Genome-fragment | NA | NA | 0 |
| S14_vs_k141_21796\|\|full | 2075 | No | NA | 3 | 0 | 0 | Not-determined | Genome-fragment | NA | NA | 0 |
| S14_vs_k141_65030\|\|full | 2336 | No | NA | 4 | 0 | 0 | Not-determined | Genome-fragment | NA | NA | 0 |
| S14_vs_k141_116910\|\|full | 1192 | No | NA | 2 | 0 | 0 | Not-determined | Genome-fragment | NA | NA | 0 |
| S9_vs_k141_18750\|\|full | 1422 | No | NA | 5 | 1 | 0 | Low-quality | Genome-fragment | 2.45 | AAI-based (high-confidence) | 0 |
| S11_vs_k141_52063\|\|lt2gene | 1422 | No | NA | 1 | 1 | 0 | Low-quality | Genome-fragment | 4.14 | AAI-based (high-confidence) | 0 |
| S14_vs_k141_83076\|\|full | 1422 | No | NA | 3 | 0 | 0 | Low-quality | Genome-fragment | 1.89 | AAI-based (medium-confidence) | 0 |
| S14_vs_k141_58114\|\|full | 1547 | No | NA | 2 | 0 | 2 | Not-determined | Genome-fragment | NA | NA | 0 |
| S14_vs_k141_21811\|\|full | 1999 | No | NA | 8 | 0 | 0 | Not-determined | Genome-fragment | NA | NA | 0 |
| S3_vs_k141_108196\|\|full | 1422 | No | NA | 2 | 0 | 0 | Low-quality | Genome-fragment | 2.58 | AAI-based (high-confidence) | 0 |
| S14_vs_k141_21824\|\|full | 1000 | No | NA | 3 | 0 | 1 | Not-determined | Genome-fragment | NA | NA | 0 |
| S14_vs_k141_65072\|\|full | 14656 | No | NA | 11 | 0 | 7 | Not-determined | Genome-fragment | NA | NA | 0 |
| S7_vs_34597\|\|full | 1422 | No | NA | 4 | 0 | 0 | Low-quality | Genome-fragment | 4.29 | AAI-based (high-confidence) | 0 |
| S14_vs_k141_36336\|\|full | 1760 | No | NA | 5 | 0 | 1 | Not-determined | Genome-fragment | NA | NA | 0 |
| S14_vs_k141_21843\|\|full | 3139 | No | NA | 4 | 0 | 1 | Not-determined | Genome-fragment | NA | NA | 0 |
| S7_vs_79578\|\|full | 1422 | No | NA | 3 | 0 | 2 | Low-quality | Genome-fragment | 1.09 | AAI-based (medium-confidence) | 0 |
| S14_vs_k141_87657\|\|full | 1018 | No | NA | 2 | 0 | 0 | Not-determined | Genome-fragment | NA | NA | 0 |
| S16_extracted_18907 | 1422 | No | NA | 3 | 0 | 0 | Low-quality | Genome-fragment | 0.92 | AAI-based (medium-confidence) | 0 |
| S14_vs_k141_87661\|\|full | 3308 | No | NA | 4 | 0 | 3 | Not-determined | Genome-fragment | NA | NA | 0 |
| S16_extracted_32536 | 1422 | No | NA | 3 | 0 | 0 | Low-quality | Genome-fragment | 2.66 | AAI-based (high-confidence) | 0 |
| S16_vs_59042\|\|full | 1422 | No | NA | 3 | 0 | 0 | Low-quality | Genome-fragment | 1.14 | AAI-based (high-confidence) | 0 |
| S14_vs_k141_58206\|\|full | 2088 | No | NA | 2 | 0 | 1 | Not-determined | Genome-fragment | NA | NA | 0 |
| S16_vs_129772\|\|full | 1422 | No | NA | 3 | 1 | 0 | Low-quality | Genome-fragment | 3.46 | AAI-based (medium-confidence) | 0 |
| S14_vs_k141_21892\|\|full | 1873 | No | NA | 3 | 0 | 2 | Not-determined | Genome-fragment | NA | NA | 0 |
| S14_vs_k141_36382\|\|full | 4637 | No | NA | 6 | 0 | 4 | Not-determined | Genome-fragment | NA | NA | 0 |
| S10_vs_91843\|\|full | 1422 | No | NA | 3 | 0 | 0 | Low-quality | Genome-fragment | 1.73 | AAI-based (medium-confidence) | 0 |
| S14_vs_k141_14074\|\|full | 1465 | No | NA | 1 | 0 | 0 | Not-determined | Genome-fragment | NA | NA | 0 |
| S14_vs_k141_87711\|\|full | 2312 | No | NA | 3 | 0 | 2 | Not-determined | Genome-fragment | NA | NA | 0 |
| S12_extracted_k141_169593 | 1422 | No | NA | 2 | 1 | 0 | Low-quality | Genome-fragment | 2.9 | AAI-based (high-confidence) | 0 |
| S12_vs_k141_51315\|\|full | 1422 | No | NA | 5 | 1 | 0 | Low-quality | Genome-fragment | 3.09 | AAI-based (medium-confidence) | 0 |
| S14_vs_k141_21935\|\|full | 2402 | No | NA | 4 | 0 | 2 | Not-determined | Genome-fragment | NA | NA | 0 |
| S2_extracted_k141_10568 | 1422 | No | NA | 2 | 2 | 0 | Low-quality | Genome-fragment | 4.67 | AAI-based (high-confidence) | 0 |
| S14_extracted_k141_88333 | 1423 | No | NA | 3 | 3 | 0 | Low-quality | Genome-fragment | 3.87 | HMM-based (lower-bound) | 0 |
| S14_vs_k141_36482\|\|full | 1533 | No | NA | 2 | 0 | 2 | Not-determined | Genome-fragment | NA | NA | 0 |
| S14_vs_k141_75667\|\|full | 1423 | No | NA | 3 | 1 | 0 | Low-quality | Genome-fragment | 2.38 | AAI-based (high-confidence) | 0 |
| S14_vs_k141_21978\|\|full | 1408 | No | NA | 5 | 0 | 0 | Not-determined | Genome-fragment | NA | NA | 0 |
| S14_vs_k141_11455\|\|full | 1423 | No | NA | 6 | 0 | 0 | Low-quality | Genome-fragment | 3.39 | AAI-based (medium-confidence) | 0 |
| S14_vs_k141_117106\|\|full | 1089 | No | NA | 3 | 0 | 1 | Not-determined | Genome-fragment | NA | NA | 0 |
| S1_extracted_k141_22986 | 1423 | No | NA | 3 | 1 | 0 | Low-quality | Genome-fragment | 1.47 | AAI-based (high-confidence) | 0 |
| S5_extracted_k141_92035 | 1423 | No | NA | 2 | 1 | 0 | Low-quality | Genome-fragment | 3.13 | AAI-based (high-confidence) | 0 |
| S5_vs_k141_81489\|\|lt2gene | 1423 | No | NA | 2 | 1 | 0 | Low-quality | Genome-fragment | 0.88 | HMM-based (lower-bound) | 0 |
| S7_extracted_62238 | 1423 | No | NA | 4 | 3 | 0 | Low-quality | Genome-fragment | 6.49 | AAI-based (high-confidence) | 0 |
| S7_vs_42284\|\|full | 1423 | No | NA | 4 | 0 | 0 | Low-quality | Genome-fragment | 3.81 | AAI-based (high-confidence) | 0 |
| S14_vs_k141_6981\|\|full | 9629 | No | NA | 8 | 0 | 4 | Not-determined | Genome-fragment | NA | NA | 0 |
| S16_vs_5293\|\|full | 1423 | No | NA | 3 | 0 | 0 | Low-quality | Genome-fragment | 3.79 | AAI-based (medium-confidence) | 0 |
| S16_vs_115968\|\|full | 1423 | No | NA | 4 | 0 | 0 | Low-quality | Genome-fragment | 3.07 | AAI-based (medium-confidence) | 0 |
| S14_vs_k141_65232\|\|full | 1570 | No | NA | 3 | 0 | 2 | Not-determined | Genome-fragment | NA | NA | 0 |
| S14_vs_k141_65248\|\|full | 2434 | No | NA | 3 | 0 | 0 | Not-determined | Genome-fragment | NA | NA | 0 |
| S14_vs_k141_58376\|\|full | 1425 | No | NA | 3 | 0 | 2 | Not-determined | Genome-fragment | NA | NA | 0 |
| S14_vs_k141_87827\|\|full | 1072 | No | NA | 2 | 0 | 1 | Not-determined | Genome-fragment | NA | NA | 0 |
| S14_vs_k141_80342\|\|full | 2389 | No | NA | 4 | 0 | 2 | Not-determined | Genome-fragment | NA | NA | 0 |
| S8_vs_k141_180752\|\|full | 1423 | No | NA | 5 | 0 | 0 | Low-quality | Genome-fragment | 4.93 | AAI-based (high-confidence) | 0 |
| S14_vs_k141_80362\|\|full | 2241 | No | NA | 3 | 0 | 2 | Not-determined | Genome-fragment | NA | NA | 0 |
| S14_vs_k141_58392\|\|full | 1151 | No | NA | 3 | 0 | 0 | Not-determined | Genome-fragment | NA | NA | 0 |
| S8_vs_k141_195028\|\|lt2gene | 1423 | No | NA | 2 | 1 | 0 | Low-quality | Genome-fragment | 3.14 | AAI-based (high-confidence) | 0 |
| S17_vs_k141_155672\|\|full | 1424 | No | NA | 2 | 0 | 0 | Low-quality | Genome-fragment | 1.52 | AAI-based (medium-confidence) | 0 |
| S17_vs_k141_113735\|\|full | 1424 | No | NA | 2 | 2 | 0 | Low-quality | Genome-fragment | 3.88 | AAI-based (high-confidence) | 0 |

| | | | | | | | | | | | | | |
|---|---|---|---|---|---|---|---|---|---|---|---|---|---|
| S13_vs_87153\|\|full | 1424 | No | NA | | 2 | 0 | 0 | Low-quality | Genome-fragment | | 1.51 | AAI-based (medium-confidence) | 0 |
| S14_vs_k141_36632\|\|full | 4294 | No | NA | | 3 | 0 | 1 | Not-determined | Genome-fragment | NA | | NA | 0 |
| S14_vs_k141_50531\|\|full | 1023 | No | NA | | 2 | 0 | 1 | Not-determined | Genome-fragment | NA | | NA | 0 |
| S14_vs_k141_102181\|\|full | 1086 | No | NA | | 3 | 0 | 0 | Not-determined | Genome-fragment | NA | | NA | 0 |
| S14_vs_k141_21980\|\|full | 1424 | No | NA | | 4 | 0 | 0 | Low-quality | Genome-fragment | | 3.03 | AAI-based (high-confidence) | 0 |
| S14_vs_k141_14355\|\|full | 1911 | No | NA | | 3 | 0 | 1 | Not-determined | Genome-fragment | NA | | NA | 0 |
| S14_vs_k141_7109\|\|full | 3087 | No | NA | | 4 | 0 | 2 | Not-determined | Genome-fragment | NA | | NA | 0 |
| S1_extracted_k141_4241 | 1424 | No | NA | | 3 | 1 | 0 | Low-quality | Genome-fragment | | 1.98 | AAI-based (high-confidence) | 0 |
| S14_vs_k141_50587\|\|full | 1196 | No | NA | | 2 | 0 | 0 | Not-determined | Genome-fragment | NA | | NA | 0 |
| S14_vs_k141_80415\|\|full | 2354 | No | NA | | 4 | 0 | 1 | Not-determined | Genome-fragment | NA | | NA | 0 |
| S1_vs_k141_68338\|\|full | 1424 | No | NA | | 3 | 2 | 0 | Low-quality | Genome-fragment | | 1.79 | HMM-based (lower-bound) | 0 |
| S1_vs_k141_96483\|\|full | 1424 | No | NA | | 4 | 1 | 0 | Low-quality | Genome-fragment | | 3.37 | AAI-based (high-confidence) | 0 |
| S14_vs_k141_58459\|\|full | 7263 | No | NA | | 7 | 0 | 2 | Not-determined | Genome-fragment | NA | | NA | 0 |
| S14_vs_k141_80431\|\|full | 6489 | No | NA | | 12 | 0 | 3 | Not-determined | Genome-fragment | NA | | NA | 0 |
| S14_vs_k141_14403\|\|full | 1488 | No | NA | | 3 | 0 | 0 | Not-determined | Genome-fragment | NA | | NA | 0 |
| S1_vs_k141_151943\|\|full | 1424 | No | NA | | 2 | 0 | 1 | Low-quality | Genome-fragment | | 1.02 | AAI-based (medium-confidence) | 0 |
| S14_vs_k141_7200\|\|full | 1013 | No | NA | | 3 | 0 | 1 | Not-determined | Genome-fragment | NA | | NA | 0 |
| S3_vs_k141_19344\|\|full | 1424 | No | NA | | 4 | 1 | 0 | Low-quality | Genome-fragment | | 2.53 | AAI-based (high-confidence) | 0 |
| S14_vs_k141_102300\|\|full | 1063 | No | NA | | 2 | 0 | 1 | Not-determined | Genome-fragment | NA | | NA | 0 |
| S3_vs_k141_20785\|\|full | 1424 | No | NA | | 3 | 1 | 0 | Low-quality | Genome-fragment | | 3.21 | AAI-based (medium-confidence) | 0 |
| S14_vs_k141_73014\|\|full | 4861 | No | NA | | 3 | 0 | 2 | Not-determined | Genome-fragment | NA | | NA | 0 |
| S14_vs_k141_80468\|\|full | 1627 | No | NA | | 3 | 0 | 0 | Not-determined | Genome-fragment | NA | | NA | 0 |
| S14_vs_k141_73040\|\|full | 2739 | No | NA | | 2 | 0 | 2 | Not-determined | Genome-fragment | NA | | NA | 0 |
| S14_vs_k141_7229\|\|full | 8865 | No | NA | | 11 | 0 | 3 | Not-determined | Genome-fragment | NA | | NA | 0 |
| S14_vs_k141_7242\|\|full | 3085 | No | NA | | 3 | 0 | 1 | Not-determined | Genome-fragment | NA | | NA | 0 |
| S7_extracted_76059 | 1424 | No | NA | | 4 | 0 | 0 | Low-quality | Genome-fragment | | 3.06 | AAI-based (high-confidence) | 0 |
| S14_vs_k141_65452\|\|full | 5696 | No | NA | | 5 | 0 | 4 | Not-determined | Genome-fragment | NA | | NA | 0 |
| S7_vs_117621\|\|full | 1424 | No | NA | | 3 | 2 | 0 | Low-quality | Genome-fragment | | 4.57 | AAI-based (medium-confidence) | 0 |
| S16_vs_60511\|\|full | 1424 | No | NA | | 4 | 1 | 0 | Low-quality | Genome-fragment | | 2.84 | AAI-based (medium-confidence) | 0 |
| S14_vs_k141_28727\|\|full | 1591 | No | NA | | 3 | 0 | 3 | Not-determined | Genome-fragment | NA | | NA | 0 |
| S14_vs_k141_73106\|\|full | 1466 | No | NA | | 4 | 0 | 0 | Not-determined | Genome-fragment | NA | | NA | 0 |
| S14_vs_k141_65483\|\|full | 1939 | No | NA | | 3 | 0 | 0 | Not-determined | Genome-fragment | NA | | NA | 0 |
| S14_vs_k141_14468\|\|full | 1884 | No | NA | | 3 | 0 | 0 | Not-determined | Genome-fragment | NA | | NA | 0 |
| S14_vs_k141_73126\|\|full | 6496 | No | NA | | 11 | 0 | 5 | Not-determined | Genome-fragment | NA | | NA | 0 |
| S14_vs_k141_80588\|\|full | 1012 | No | NA | | 2 | 0 | 2 | Not-determined | Genome-fragment | NA | | NA | 0 |
| S14_vs_k141_73214\|\|full | 2863 | No | NA | | 7 | 0 | 0 | Not-determined | Genome-fragment | NA | | NA | 0 |

[illegible]

| | | | | | | | | | | | | |
|---|---|---|---|---|---|---|---|---|---|---|---|---|
| S1_extracted_k141_101268 | 1027 | No | NA | 1 | 0 | 1 | Not-determined | Genome-fragment | NA | | NA | 0 |
| S1_extracted_k141_953 | 1808 | No | NA | 2 | 0 | 1 | Not-determined | Genome-fragment | NA | | NA | 0 |
| S5_vs_k141_71530\|\|full | 1444 | No | NA | 2 | 0 | 0 | Low-quality | Genome-fragment | | 3.73 | AAI-based (high-confidence) | 0 |
| S5_vs_k141_58314\|\|full | 1444 | No | NA | 3 | 0 | 0 | Low-quality | Genome-fragment | | 2.66 | AAI-based (medium-confidence) | 0 |
| S1_extracted_k141_41260 | 3465 | No | NA | 1 | 0 | 1 | Not-determined | Genome-fragment | NA | | NA | 0 |
| S7_vs_118432\|\|full | 1444 | No | NA | 4 | 2 | 0 | Low-quality | Genome-fragment | | 1.75 | HMM-based (lower-bound) | 0 |
| S10_vs_37280\|\|full | 1444 | No | NA | 4 | 1 | 0 | Low-quality | Genome-fragment | | 2.49 | AAI-based (medium-confidence) | 0 |
| S1_extracted_k141_27748 | 3990 | No | NA | 2 | 0 | 2 | Not-determined | Genome-fragment | NA | | NA | 0 |
| S1_extracted_k141_966 | 1018 | No | NA | 1 | 0 | 0 | Not-determined | Genome-fragment | NA | | NA | 0 |
| S1_extracted_k141_141293 | 1037 | No | NA | 2 | 0 | 2 | Not-determined | Genome-fragment | NA | | NA | 0 |
| S1_extracted_k141_94656 | 1004 | No | NA | 2 | 0 | 1 | Not-determined | Genome-fragment | NA | | NA | 0 |
| S2_vs_k141_5131\|\|lt2gene | 1444 | No | NA | 2 | 1 | 0 | Low-quality | Genome-fragment | | 3.53 | AAI-based (high-confidence) | 0 |
| S6_vs_k141_24342\|\|full | 1444 | No | NA | 3 | 0 | 0 | Low-quality | Genome-fragment | | 0.84 | AAI-based (medium-confidence) | 0 |
| S6_vs_k141_3378\|\|full | 1444 | No | NA | 5 | 4 | 0 | Low-quality | Genome-fragment | | 2.95 | AAI-based (medium-confidence) | 0 |
| S1_extracted_k141_54574 | 1015 | No | NA | 1 | 0 | 1 | Not-determined | Genome-fragment | NA | | NA | 0 |
| S1_extracted_k141_54579 | 1411 | No | NA | 1 | 0 | 1 | Not-determined | Genome-fragment | NA | | NA | 0 |
| S1_extracted_k141_81320 | 4631 | No | NA | 2 | 0 | 2 | Not-determined | Genome-fragment | NA | | NA | 0 |
| S1_extracted_k141_141315 | 1102 | No | NA | 2 | 0 | 1 | Not-determined | Genome-fragment | NA | | NA | 0 |
| S1_extracted_k141_134524 | 3159 | No | NA | 1 | 0 | 1 | Not-determined | Genome-fragment | NA | | NA | 0 |
| S1_extracted_k141_47875 | 1388 | No | NA | 1 | 0 | 0 | Not-determined | Genome-fragment | NA | | NA | 0 |
| S1_extracted_k141_41313 | 3619 | No | NA | 4 | 0 | 3 | Not-determined | Genome-fragment | NA | | NA | 0 |
| S1_extracted_k141_94697 | 3328 | No | NA | 3 | 0 | 0 | Not-determined | Genome-fragment | NA | | NA | 0 |
| S1_extracted_k141_21189 | 1281 | No | NA | 2 | 0 | 1 | Not-determined | Genome-fragment | NA | | NA | 0 |
| S1_extracted_k141_154202 | 3124 | No | NA | 2 | 0 | 2 | Not-determined | Genome-fragment | NA | | NA | 0 |
| S1_extracted_k141_101369 | 1662 | No | NA | 2 | 0 | 0 | Not-determined | Genome-fragment | NA | | NA | 0 |
| S1_extracted_k141_94715 | 3042 | No | NA | 2 | 0 | 2 | Not-determined | Genome-fragment | NA | | NA | 0 |
| S1_extracted_k141_101376 | 2034 | No | NA | 1 | 0 | 1 | Not-determined | Genome-fragment | NA | | NA | 0 |
| S8_vs_k141_166915\|\|full | 1444 | No | NA | 3 | 0 | 0 | Low-quality | Genome-fragment | | 3.5 | AAI-based (high-confidence) | 0 |
| S17_extracted_k141_94363 | 1445 | No | NA | 2 | 0 | 2 | Low-quality | Genome-fragment | | 0.41 | AAI-based (medium-confidence) | 0 |
| S1_extracted_k141_128035 | 1220 | No | NA | 1 | 0 | 1 | Not-determined | Genome-fragment | NA | | NA | 0 |
| S1_extracted_k141_94726 | 1067 | No | NA | 2 | 0 | 1 | Not-determined | Genome-fragment | NA | | NA | 0 |
| S1_extracted_k141_67892 | 1502 | No | NA | 2 | 0 | 2 | Not-determined | Genome-fragment | NA | | NA | 0 |
| S1_extracted_k141_94735 | 1190 | No | NA | 1 | 0 | 1 | Not-determined | Genome-fragment | NA | | NA | 0 |
| S17_vs_k141_145686\|\|full | 1445 | No | NA | 2 | 0 | 0 | Low-quality | Genome-fragment | | 1.02 | AAI-based (high-confidence) | 0 |
| S9_extracted_k141_82418 | 1445 | No | NA | 4 | 4 | 0 | Low-quality | Genome-fragment | | 4.77 | AAI-based (high-confidence) | 0 |
| S1_extracted_k141_107924 | 1207 | No | NA | 1 | 0 | 0 | Not-determined | Genome-fragment | NA | | NA | 0 |
| S1_extracted_k141_21250 | 1012 | No | NA | 1 | 0 | 0 | Not-determined | Genome-fragment | NA | | NA | 0 |
| S1_extracted_k141_101426 | 1010 | No | NA | 1 | 0 | 1 | Not-determined | Genome-fragment | NA | | NA | 0 |
| S13_extracted_59751 | 1445 | No | NA | 2 | 0 | 1 | Low-quality | Genome-fragment | | 0.8 | AAI-based (high-confidence) | 0 |
| S14_extracted_k141_95559 | 1445 | No | NA | 4 | 3 | 0 | Low-quality | Genome-fragment | | 4.76 | AAI-based (high-confidence) | 0 |
| S14_extracted_k141_106355 | 1445 | No | NA | 4 | 0 | 0 | Low-quality | Genome-fragment | | 2.64 | AAI-based (high-confidence) | 0 |
| S14_vs_k141_3196\|\|full | 1445 | No | NA | 3 | 0 | 0 | Low-quality | Genome-fragment | | 1.58 | AAI-based (high-confidence) | 0 |
| S1_extracted_k141_67948 | 1193 | No | NA | 1 | 0 | 1 | Not-determined | Genome-fragment | NA | | NA | 0 |
| S1_vs_k141_48454\|\|full | 1445 | No | NA | 3 | 1 | 0 | Low-quality | Genome-fragment | | 0.94 | AAI-based (high-confidence) | 0 |
| S1_extracted_k141_114627 | 1032 | No | NA | 1 | 0 | 1 | Not-determined | Genome-fragment | NA | | NA | 0 |
| S1_extracted_k141_107958 | 2640 | No | NA | 3 | 0 | 2 | Not-determined | Genome-fragment | NA | | NA | 0 |
| S1_extracted_k141_81431 | 4981 | No | NA | 7 | 0 | 0 | Not-determined | Genome-fragment | NA | | NA | 0 |
| S5_vs_k141_114577\|\|full | 1445 | No | NA | 3 | 0 | 0 | Low-quality | Genome-fragment | | 3.07 | AAI-based (medium-confidence) | 0 |
| S1_extracted_k141_67966 | 1166 | No | NA | 1 | 0 | 1 | Not-determined | Genome-fragment | NA | | NA | 0 |
| S1_extracted_k141_14940 | 1111 | No | NA | 1 | 0 | 1 | Not-determined | Genome-fragment | NA | | NA | 0 |
| S1_extracted_k141_60828 | 4008 | No | NA | 3 | 0 | 2 | Not-determined | Genome-fragment | NA | | NA | 0 |
| S16_extracted_146369 | 1445 | No | NA | 2 | 0 | 0 | Low-quality | Genome-fragment | | 3.64 | AAI-based (medium-confidence) | 0 |
| S1_extracted_k141_88092 | 1319 | No | NA | 1 | 0 | 1 | Not-determined | Genome-fragment | NA | | NA | 0 |
| S1_extracted_k141_121238 | 1900 | No | NA | 2 | 0 | 0 | Not-determined | Genome-fragment | NA | | NA | 0 |
| S1_extracted_k141_41463 | 1095 | No | NA | 1 | 0 | 1 | Not-determined | Genome-fragment | NA | | NA | 0 |
| S10_vs_81305\|\|full | 1445 | No | NA | 2 | 0 | 0 | Low-quality | Genome-fragment | | 2.48 | AAI-based (medium-confidence) | 0 |
| S1_extracted_k141_8103 | 1132 | No | NA | 3 | 0 | 3 | Not-determined | Genome-fragment | NA | | NA | 0 |
| S1_extracted_k141_147441 | 1644 | No | NA | 1 | 0 | 1 | Not-determined | Genome-fragment | NA | | NA | 0 |
| S15_extracted_56155 | 1445 | No | NA | 3 | 1 | 0 | Low-quality | Genome-fragment | | 3.15 | AAI-based (high-confidence) | 0 |
| S8_extracted_k141_131413 | 1445 | No | NA | 3 | 3 | 0 | Low-quality | Genome-fragment | | 4.76 | AAI-based (high-confidence) | 0 |
| S9_vs_k141_11271\|\|full | 1446 | No | NA | 3 | 2 | 0 | Low-quality | Genome-fragment | | 3.85 | AAI-based (high-confidence) | 0 |
| S1_extracted_k141_154283 | 1589 | No | NA | 1 | 0 | 0 | Not-determined | Genome-fragment | NA | | NA | 0 |
| S1_extracted_k141_108033 | 1919 | No | NA | 2 | 0 | 2 | Not-determined | Genome-fragment | NA | | NA | 0 |
| S1_vs_k141_5688\|\|full | 1446 | No | NA | 3 | 0 | 0 | Low-quality | Genome-fragment | | 1.14 | AAI-based (medium-confidence) | 0 |
| S1_extracted_k141_121270 | 1457 | No | NA | 1 | 0 | 0 | Not-determined | Genome-fragment | NA | | NA | 0 |
| S3_extracted_k141_97385 | 1446 | No | NA | 4 | 0 | 0 | Low-quality | Genome-fragment | | 1.13 | AAI-based (medium-confidence) | 0 |
| S3_vs_k141_103772\|\|full | 1446 | No | NA | 3 | 0 | 2 | Low-quality | Genome-fragment | | 0.81 | AAI-based (medium-confidence) | 0 |
| S1_extracted_k141_94880 | 1423 | No | NA | 3 | 0 | 0 | Not-determined | Genome-fragment | NA | | NA | 0 |
| S3_vs_k141_8308\|\|full | 1446 | No | NA | 3 | 0 | 2 | Low-quality | Genome-fragment | | 3.34 | AAI-based (medium-confidence) | 0 |
| S1_extracted_k141_68084 | 1122 | No | NA | 2 | 0 | 2 | Not-determined | Genome-fragment | NA | | NA | 0 |
| S7_vs_19803\|\|full | 1446 | No | NA | 2 | 0 | 2 | Low-quality | Genome-fragment | | 1.14 | AAI-based (medium-confidence) | 0 |
| S16_extracted_11764 | 1446 | No | NA | 3 | 0 | 3 | Low-quality | Genome-fragment | | 0.47 | AAI-based (medium-confidence) | 0 |
| S12_vs_k141_171152\|\|full | 1446 | No | NA | 5 | 1 | 0 | Low-quality | Genome-fragment | | 3.73 | AAI-based (medium-confidence) | 0 |
| S1_extracted_k141_141498 | 1078 | No | NA | 1 | 0 | 1 | Not-determined | Genome-fragment | NA | | NA | 0 |
| S11_vs_k141_24625\|\|full | 1447 | No | NA | 3 | 0 | 1 | Low-quality | Genome-fragment | | 2.22 | AAI-based (medium-confidence) | 0 |
| S14_extracted_k141_17134 | 1447 | No | NA | 1 | 1 | 0 | Low-quality | Genome-fragment | | 4.6 | AAI-based (high-confidence) | 0 |
| S1_extracted_k141_28075 | 1285 | No | NA | 3 | 0 | 0 | Not-determined | Genome-fragment | NA | | NA | 0 |
| S1_extracted_k141_141510 | 4322 | No | NA | 4 | 0 | 4 | Not-determined | Genome-fragment | NA | | NA | 0 |
| S14_extracted_k141_40953 | 1447 | No | NA | 1 | 0 | 1 | Low-quality | Genome-fragment | | 1.66 | AAI-based (medium-confidence) | 0 |
| S1_extracted_k141_114828 | 7077 | No | NA | 3 | 0 | 2 | Not-determined | Genome-fragment | NA | | NA | 0 |
| S1_extracted_k141_147489 | 1621 | No | NA | 1 | 0 | 0 | Not-determined | Genome-fragment | NA | | NA | 0 |
| S1_extracted_k141_1188 | 1029 | No | NA | 1 | 0 | 0 | Not-determined | Genome-fragment | NA | | NA | 0 |
| S1_extracted_k141_74844 | 1515 | No | NA | 2 | 0 | 2 | Not-determined | Genome-fragment | NA | | NA | 0 |
| S14_vs_k141_6642\|\|lt2gene | 1447 | No | NA | 2 | 2 | 0 | Low-quality | Genome-fragment | | 3.04 | AAI-based (high-confidence) | 0 |
| S1_extracted_k141_74858 | 1002 | No | NA | 1 | 0 | 0 | Not-determined | Genome-fragment | NA | | NA | 0 |
| S5_vs_k141_117816\|\|full | 1447 | No | NA | 4 | 2 | 0 | Low-quality | Genome-fragment | | 3.32 | AAI-based (medium-confidence) | 0 |
| S16_extracted_133238 | 1447 | No | NA | 4 | 4 | 0 | Low-quality | Genome-fragment | | 4.75 | AAI-based (high-confidence) | 0 |
| S1_extracted_k141_95017 | 1231 | No | NA | 2 | 0 | 2 | Not-determined | Genome-fragment | NA | | NA | 0 |
| S16_vs_31282\|\|full | 1447 | No | NA | 3 | 2 | 0 | Low-quality | Genome-fragment | | 3.79 | AAI-based (high-confidence) | 0 |
| S1_extracted_k141_147516 | 1124 | No | NA | 1 | 0 | 1 | Not-determined | Genome-fragment | NA | | NA | 0 |
| S1_extracted_k141_54886 | 1008 | No | NA | 1 | 0 | 1 | Not-determined | Genome-fragment | NA | | NA | 0 |
| S10_vs_92493\|\|full | 1447 | No | NA | 3 | 0 | 2 | Low-quality | Genome-fragment | | 1.08 | AAI-based (medium-confidence) | 0 |
| S1_extracted_k141_1259 | 3368 | No | NA | 1 | 0 | 0 | Not-determined | Genome-fragment | NA | | NA | 0 |
| S1_extracted_k141_95036 | 1064 | No | NA | 1 | 0 | 1 | Not-determined | Genome-fragment | NA | | NA | 0 |
| S10_vs_119860\|\|lt2gene | 1447 | No | NA | 1 | 1 | 0 | Low-quality | Genome-fragment | | 2.43 | HMM-based (lower-bound) | 0 |
| S1_extracted_k141_147526 | 1170 | No | NA | 1 | 0 | 1 | Not-determined | Genome-fragment | NA | | NA | 0 |
| S6_vs_k141_19919\|\|full | 1447 | No | NA | 3 | 0 | 0 | Low-quality | Genome-fragment | | 2.8 | AAI-based (high-confidence) | 0 |
| S1_extracted_k141_81649 | 1236 | No | NA | 1 | 0 | 0 | Not-determined | Genome-fragment | NA | | NA | 0 |
| S1_extracted_k141_101740 | 1050 | No | NA | 1 | 0 | 0 | Not-determined | Genome-fragment | NA | | NA | 0 |
| S1_extracted_k141_8333 | 3161 | No | NA | 1 | 0 | 1 | Not-determined | Genome-fragment | NA | | NA | 0 |
| S8_extracted_k141_44652 | 1447 | No | NA | 3 | 0 | 0 | Low-quality | Genome-fragment | | 3.19 | AAI-based (high-confidence) | 0 |
| S8_vs_k141_138125\|\|full | 1447 | No | NA | 2 | 0 | 0 | Low-quality | Genome-fragment | | 2.99 | AAI-based (high-confidence) | 0 |
| S1_extracted_k141_48301 | 1153 | No | NA | 1 | 0 | 1 | Not-determined | Genome-fragment | NA | | NA | 0 |
| S1_extracted_k141_28222 | 1731 | No | NA | 1 | 0 | 1 | Not-determined | Genome-fragment | NA | | NA | 0 |
| S1_extracted_k141_41677 | 1199 | No | NA | 1 | 0 | 1 | Not-determined | Genome-fragment | NA | | NA | 0 |
| S8_vs_k141_207778\|\|full | 1447 | No | NA | 2 | 0 | 1 | Low-quality | Genome-fragment | | 1.72 | AAI-based (high-confidence) | 0 |
| S1_extracted_k141_88319 | 1142 | No | NA | 2 | 0 | 0 | Not-determined | Genome-fragment | NA | | NA | 0 |
| S1_extracted_k141_81711 | 1066 | No | NA | 1 | 0 | 0 | Not-determined | Genome-fragment | NA | | NA | 0 |
| S1_extracted_k141_15273 | 1139 | No | NA | 2 | 0 | 0 | Not-determined | Genome-fragment | NA | | NA | 0 |
| S1_extracted_k141_55012 | 1054 | No | NA | 1 | 0 | 1 | Not-determined | Genome-fragment | NA | | NA | 0 |
| S1_extracted_k141_68301 | 1591 | No | NA | 1 | 0 | 1 | Not-determined | Genome-fragment | NA | | NA | 0 |
| S1_extracted_k141_21607 | 1264 | No | NA | 1 | 0 | 0 | Not-determined | Genome-fragment | NA | | NA | 0 |
| S1_extracted_k141_147574 | 2406 | No | NA | 1 | 0 | 1 | Not-determined | Genome-fragment | NA | | NA | 0 |
| S17_extracted_k141_61364 | 1448 | No | NA | 2 | 0 | 0 | Low-quality | Genome-fragment | | 1.47 | AAI-based (high-confidence) | 0 |
| S1_extracted_k141_75103 | 1177 | No | NA | 1 | 0 | 1 | Not-determined | Genome-fragment | NA | | NA | 0 |
| S1_extracted_k141_68343 | 1152 | No | NA | 1 | 0 | 1 | Not-determined | Genome-fragment | NA | | NA | 0 |
| S1_extracted_k141_147578 | 5782 | No | NA | 9 | 0 | 1 | Not-determined | Genome-fragment | NA | | NA | 0 |
| S9_vs_k141_45889\|\|full | 1448 | No | NA | 5 | 2 | 0 | Low-quality | Genome-fragment | | 2.26 | AAI-based (medium-confidence) | 0 |
| S1_extracted_k141_95168 | 1320 | No | NA | 2 | 0 | 1 | Not-determined | Genome-fragment | NA | | NA | 0 |
| S1_extracted_k141_66394 | 1448 | No | NA | 1 | 0 | 0 | Low-quality | Genome-fragment | | 1.47 | AAI-based (high-confidence) | 0 |
| S1_extracted_k141_34779 | 2133 | No | NA | 1 | 0 | 1 | Not-determined | Genome-fragment | NA | | NA | 0 |
| S1_extracted_k141_28375 | 1135 | No | NA | 1 | 0 | 1 | Not-determined | Genome-fragment | NA | | NA | 0 |
| S1_extracted_k141_95205 | 1126 | No | NA | 2 | 0 | 2 | Not-determined | Genome-fragment | NA | | NA | 0 |
| S1_extracted_k141_88413 | 1225 | No | NA | 2 | 0 | 2 | Not-determined | Genome-fragment | NA | | NA | 0 |
| S1_extracted_k141_101877 | 1000 | No | NA | 1 | 0 | 0 | Not-determined | Genome-fragment | NA | | NA | 0 |
| S1_extracted_k141_135081 | 1613 | No | NA | 1 | 0 | 1 | Not-determined | Genome-fragment | NA | | NA | 0 |
| S1_extracted_k141_21695 | 1552 | No | NA | 2 | 0 | 2 | Not-determined | Genome-fragment | NA | | NA | 0 |
| S1_extracted_k141_154570 | 1837 | No | NA | 3 | 0 | 1 | Not-determined | Genome-fragment | NA | | NA | 0 |
| S1_vs_k141_105720\|\|full | 1448 | No | NA | 2 | 0 | 1 | Low-quality | Genome-fragment | | 1.02 | AAI-based (high-confidence) | 0 |
| S5_vs_k141_13759\|\|full | 1448 | No | NA | 5 | 3 | 0 | Low-quality | Genome-fragment | | 4.53 | AAI-based (high-confidence) | 0 |
| S16_vs_32137\|\|full | 1448 | No | NA | 2 | 2 | 0 | Low-quality | Genome-fragment | | 3.09 | AAI-based (medium-confidence) | 0 |
| S1_extracted_k141_8521 | 1053 | No | NA | 2 | 0 | 0 | Not-determined | Genome-fragment | NA | | NA | 0 |
| S1_extracted_k141_95225 | 3571 | No | NA | 4 | 0 | 4 | Not-determined | Genome-fragment | NA | | NA | 0 |
| S1_extracted_k141_28398 | 28234 | No | NA | 44 | 10 | 0 | Medium-quality | Genome-fragment | | 61.16 | AAI-based (high-confidence) | 0 |
| S16_vs_38779\|\|full | 1448 | No | NA | 2 | 1 | 0 | Low-quality | Genome-fragment | | 3.52 | AAI-based (high-confidence) | 0 |
| S8_vs_k141_193282\|\|full | 1448 | No | NA | 2 | 0 | 1 | Low-quality | Genome-fragment | | 1.68 | AAI-based (medium-confidence) | 0 |
| S8_vs_k141_153029\|\|full | 1448 | No | NA | 5 | 1 | 0 | Low-quality | Genome-fragment | | 1 | HMM-based (lower-bound) | 0 |
| S8_vs_k141_205520\|\|full | 1448 | No | NA | 5 | 2 | 0 | Low-quality | Genome-fragment | | 3 | AAI-based (high-confidence) | 0 |
| S8_vs_k141_111510\|\|full | 1448 | No | NA | 3 | 0 | 0 | Low-quality | Genome-fragment | | 2.01 | AAI-based (medium-confidence) | 0 |
| S8_vs_k141_103165\|\|lt2gene | 1448 | No | NA | 1 | 1 | 0 | Low-quality | Genome-fragment | | 2.15 | HMM-based (lower-bound) | 0 |
| S1_extracted_k141_21719 | 1083 | No | NA | 1 | 0 | 1 | Not-determined | Genome-fragment | NA | | NA | 0 |
| S1_extracted_k141_68461 | 1175 | No | NA | 2 | 0 | 0 | Not-determined | Genome-fragment | NA | | NA | 0 |
| S11_vs_k141_81053\|\|full | 1449 | No | NA | 5 | 0 | 0 | Low-quality | Genome-fragment | | 3.6 | AAI-based (high-confidence) | 0 |
| S1_extracted_k141_108445 | 2027 | No | NA | 3 | 0 | 1 | Not-determined | Genome-fragment | NA | | NA | 0 |
| S1_extracted_k141_81934 | 4695 | No | NA | 4 | 0 | 1 | Not-determined | Genome-fragment | NA | | NA | 0 |
| S1_extracted_k141_48565 | 2287 | No | NA | 3 | 0 | 0 | Not-determined | Genome-fragment | NA | | NA | 0 |
| S1_extracted_k141_147632 | 4887 | No | NA | 3 | 0 | 2 | Not-determined | Genome-fragment | NA | | NA | 0 |
| S1_extracted_k141_88510 | 3252 | No | NA | 4 | 0 | 3 | Not-determined | Genome-fragment | NA | | NA | 0 |
| S1_extracted_k141_15508 | 1154 | No | NA | 1 | 0 | 0 | Not-determined | Genome-fragment | NA | | NA | 0 |
| S13_vs_128473\|\|full | 1449 | No | NA | 6 | 0 | 0 | Low-quality | Genome-fragment | | 2.4 | AAI-based (high-confidence) | 0 |
| S1_extracted_k141_154623 | 6139 | No | NA | 4 | 0 | 4 | Not-determined | Genome-fragment | NA | | NA | 0 |
| S13_vs_139016\|\|full | 1449 | No | NA | 4 | 0 | 0 | Low-quality | Genome-fragment | | 3.39 | AAI-based (medium-confidence) | 0 |
| S1_extracted_k141_28489 | 1396 | No | NA | 4 | 0 | 0 | Not-determined | Genome-fragment | NA | | NA | 0 |
| S1_extracted_k141_75237 | 1294 | No | NA | 1 | 0 | 1 | Not-determined | Genome-fragment | NA | | NA | 0 |
| S1_extracted_k141_41958 | 1098 | No | NA | 1 | 0 | 1 | Not-determined | Genome-fragment | NA | | NA | 0 |
| S14_extracted_k141_63361 | 1449 | No | NA | 4 | 3 | 0 | Low-quality | Genome-fragment | | 2.96 | AAI-based (medium-confidence) | 0 |
| S1_extracted_k141_81992 | 1872 | No | NA | 1 | 0 | 0 | Not-determined | Genome-fragment | NA | | NA | 0 |
| S1_extracted_k141_141955 | 1529 | No | NA | 3 | 0 | 3 | Not-determined | Genome-fragment | NA | | NA | 0 |
| S1_extracted_k141_102022 | 3835 | No | NA | 3 | 0 | 1 | Not-determined | Genome-fragment | NA | | NA | 0 |
| S1_extracted_k141_68564 | 1025 | No | NA | 2 | 0 | 1 | Not-determined | Genome-fragment | NA | | NA | 0 |
| S14_vs_k141_85100\|\|full | 1449 | No | NA | 3 | 0 | 3 | Low-quality | Genome-fragment | | 0.65 | AAI-based (medium-confidence) | 0 |
| S1_extracted_k141_115281 | 1208 | No | NA | 2 | 0 | 0 | Not-determined | Genome-fragment | NA | | NA | 0 |
| S1_extracted_k141_34916 | 1370 | No | NA | 1 | 0 | 1 | Not-determined | Genome-fragment | NA | | NA | 0 |
| S1_vs_k141_43354\|\|full | 1449 | No | NA | 3 | 1 | 0 | Low-quality | Genome-fragment | | 3.77 | AAI-based (high-confidence) | 0 |
| S1_extracted_k141_48647 | 1043 | No | NA | 1 | 0 | 1 | Not-determined | Genome-fragment | NA | | NA | 0 |
| S1_extracted_k141_15611 | 4104 | No | NA | 1 | 0 | 1 | Not-determined | Genome-fragment | NA | | NA | 0 |
| S1_extracted_k141_28540 | 1010 | No | NA | 2 | 0 | 0 | Not-determined | Genome-fragment | NA | | NA | 0 |
| S1_extracted_k141_128717 | 1871 | No | NA | 1 | 0 | 1 | Not-determined | Genome-fragment | NA | | NA | 0 |
| S1_extracted_k141_1564 | 1335 | No | NA | 3 | 0 | 0 | Not-determined | Genome-fragment | NA | | NA | 0 |
| S1_extracted_k141_34932 | 1355 | No | NA | 1 | 0 | 0 | Not-determined | Genome-fragment | NA | | NA | 0 |
| S1_extracted_k141_21844 | 1174 | No | NA | 1 | 0 | 0 | Not-determined | Genome-fragment | NA | | NA | 0 |
| S1_extracted_k141_135283 | 1303 | No | NA | 2 | 0 | 0 | Not-determined | Genome-fragment | NA | | NA | 0 |
| S1_extracted_k141_61093 | 1279 | No | NA | 2 | 0 | 2 | Not-determined | Genome-fragment | NA | | NA | 0 |
| S1_extracted_k141_1582 | 4432 | No | NA | 4 | 0 | 3 | Not-determined | Genome-fragment | NA | | NA | 0 |
| S1_extracted_k141_95423 | 1337 | No | NA | 1 | 0 | 1 | Not-determined | Genome-fragment | NA | | NA | 0 |
| S1_extracted_k141_34969 | 1404 | No | NA | 1 | 0 | 0 | Not-determined | Genome-fragment | NA | | NA | 0 |
| S5_vs_k141_110760\|\|lt2gene | 1449 | No | NA | 1 | 1 | 0 | Low-quality | Genome-fragment | | 3.9 | AAI-based (medium-confidence) | 0 |
| S1_extracted_k141_121822 | 2917 | No | NA | 1 | 0 | 1 | Not-determined | Genome-fragment | NA | | NA | 0 |
| S1_extracted_k141_142055 | 1260 | No | NA | 1 | 0 | 0 | Not-determined | Genome-fragment | NA | | NA | 0 |
| S1_extracted_k141_82098 | 2055 | No | NA | 2 | 0 | 2 | Not-determined | Genome-fragment | NA | | NA | 0 |
| S1_extracted_k141_42065 | 1248 | No | NA | 1 | 0 | 1 | Not-determined | Genome-fragment | NA | | NA | 0 |
| S16_vs_75800\|\|full | 1449 | No | NA | 2 | 0 | 0 | Low-quality | Genome-fragment | | 3.29 | AAI-based (high-confidence) | 0 |
| S1_extracted_k141_88704 | 3182 | No | NA | 4 | 0 | 1 | Not-determined | Genome-fragment | NA | | NA | 0 |
| S1_extracted_k141_68680 | 1144 | No | NA | 1 | 0 | 1 | Not-determined | Genome-fragment | NA | | NA | 0 |
| S1_extracted_k141_61136 | 1391 | No | NA | 2 | 0 | 1 | Not-determined | Genome-fragment | NA | | NA | 0 |
| S1_extracted_k141_68699 | 1225 | No | NA | 2 | 0 | 1 | Not-determined | Genome-fragment | NA | | NA | 0 |
| S1_extracted_k141_135391 | 8825 | No | NA | 4 | 0 | 3 | Not-determined | Genome-fragment | NA | | NA | 0 |
| S10_vs_69281\|\|full | 1449 | No | NA | 3 | 0 | 1 | Low-quality | Genome-fragment | | 0.89 | AAI-based (medium-confidence) | 0 |
| S1_extracted_k141_21932 | 3638 | No | NA | 1 | 0 | 1 | Not-determined | Genome-fragment | NA | | NA | 0 |
| S12_vs_k141_16455\|\|full | 1449 | No | NA | 3 | 0 | 1 | Low-quality | Genome-fragment | | 2.02 | AAI-based (high-confidence) | 0 |
| S1_extracted_k141_75451 | 1607 | No | NA | 2 | 0 | 1 | Not-determined | Genome-fragment | NA | | NA | 0 |
| S8_extracted_k141_24435 | 1449 | No | NA | 2 | 1 | 0 | Low-quality | Genome-fragment | | 2.46 | AAI-based (high-confidence) | 0 |
| S1_extracted_k141_55475 | 3175 | No | NA | 3 | 0 | 2 | Not-determined | Genome-fragment | NA | | NA | 0 |
| S8_vs_k141_144187\|\|full | 1449 | No | NA | 6 | 1 | 0 | Low-quality | Genome-fragment | | 3.22 | AAI-based (high-confidence) | 0 |
| S9_extracted_k141_67795 | 1450 | No | NA | 2 | 2 | 0 | Low-quality | Genome-fragment | | 3.86 | AAI-based (high-confidence) | 0 |
| S1_extracted_k141_82207 | 2774 | No | NA | 4 | 0 | 1 | Not-determined | Genome-fragment | NA | | NA | 0 |
| S1_extracted_k141_8819 | 3185 | No | NA | 1 | 0 | 1 | Not-determined | Genome-fragment | NA | | NA | 0 |
| S1_extracted_k141_135466 | 1290 | No | NA | 1 | 0 | 1 | Not-determined | Genome-fragment | NA | | NA | 0 |
| S1_extracted_k141_48855 | 1002 | No | NA | 1 | 0 | 1 | Not-determined | Genome-fragment | NA | | NA | 0 |
| S1_extracted_k141_88804 | 1045 | No | NA | 1 | 0 | 1 | Not-determined | Genome-fragment | NA | | NA | 0 |
| S1_extracted_k141_128913 | 1002 | No | NA | 1 | 0 | 1 | Not-determined | Genome-fragment | NA | | NA | 0 |
| S1_extracted_k141_42196 | 1742 | No | NA | 2 | 0 | 1 | Not-determined | Genome-fragment | NA | | NA | 0 |
| S1_extracted_k141_22047 | 1021 | No | NA | 2 | 0 | 0 | Not-determined | Genome-fragment | NA | | NA | 0 |
| S11_vs_k141_59745\|\|full | 1450 | No | NA | 3 | 1 | 0 | Low-quality | Genome-fragment | | 3.42 | AAI-based (high-confidence) | 0 |
| S1_extracted_k141_42214 | 2194 | No | NA | 3 | 0 | 0 | Not-determined | Genome-fragment | NA | | NA | 0 |
| S1_extracted_k141_61257 | 1310 | No | NA | 2 | 0 | 0 | Not-determined | Genome-fragment | NA | | NA | 0 |
| S1_extracted_k141_8884 | 1135 | No | NA | 1 | 0 | 0 | Not-determined | Genome-fragment | NA | | NA | 0 |
| S3_vs_k141_2522\|\|full | 1450 | No | NA | 3 | 2 | 0 | Low-quality | Genome-fragment | | 2.52 | HMM-based (lower-bound) | 0 |
| S1_extracted_k141_108820 | 1148 | No | NA | 2 | 0 | 0 | Not-determined | Genome-fragment | NA | | NA | 0 |
| S5_extracted_k141_89722 | 1450 | No | NA | 5 | 3 | 0 | Low-quality | Genome-fragment | | 4.75 | AAI-based (high-confidence) | 0 |
| S1_extracted_k141_128998 | 1045 | No | NA | 1 | 0 | 1 | Not-determined | Genome-fragment | NA | | NA | 0 |
| S7_vs_109915\|\|full | 1450 | No | NA | 4 | 1 | 0 | Low-quality | Genome-fragment | | 2.54 | HMM-based (lower-bound) | 0 |
| S12_vs_k141_135131\|\|lt2gene | 1450 | No | NA | 2 | 1 | 0 | Low-quality | Genome-fragment | | 3.11 | AAI-based (medium-confidence) | 0 |
| S1_extracted_k141_95707 | 2500 | No | NA | 2 | 0 | 2 | Not-determined | Genome-fragment | NA | | NA | 0 |
| S15_vs_15419\|\|full | 1450 | No | NA | 3 | 0 | 1 | Low-quality | Genome-fragment | | 2.85 | AAI-based (medium-confidence) | 0 |
| S1_extracted_k141_135557 | 2844 | No | NA | 3 | 0 | 2 | Not-determined | Genome-fragment | NA | | NA | 0 |
| S1_extracted_k141_75630 | 2568 | No | NA | 2 | 0 | 0 | Not-determined | Genome-fragment | NA | | NA | 0 |
| S1_extracted_k141_135573 | 1168 | No | NA | 1 | 0 | 1 | Not-determined | Genome-fragment | NA | | NA | 0 |
| S1_extracted_k141_108884 | 1323 | No | NA | 2 | 0 | 1 | Not-determined | Genome-fragment | NA | | NA | 0 |
| S4_extracted_10849 | 1450 | No | NA | 2 | 2 | 0 | Low-quality | Genome-fragment | | 4.77 | AAI-based (high-confidence) | 0 |
| S4_extracted_12542 | 1450 | No | NA | 2 | 2 | 0 | Low-quality | Genome-fragment | | 4.78 | AAI-based (high-confidence) | 0 |
| S1_extracted_k141_61296 | 1461 | No | NA | 1 | 0 | 0 | Not-determined | Genome-fragment | NA | | NA | 0 |
| S1_extracted_k141_115648 | 1224 | No | NA | 1 | 0 | 1 | Not-determined | Genome-fragment | NA | | NA | 0 |
| S17_extracted_k141_158510 | 1451 | No | NA | 2 | 0 | 0 | Low-quality | Genome-fragment | | 2.87 | AAI-based (high-confidence) | 0 |
| S1_extracted_k141_16044 | 3449 | No | NA | 6 | 0 | 1 | Not-determined | Genome-fragment | NA | | NA | 0 |
| S1_extracted_k141_95759 | 1220 | No | NA | 1 | 0 | 1 | Not-determined | Genome-fragment | NA | | NA | 0 |
| S1_extracted_k141_122178 | 1080 | No | NA | 1 | 0 | 1 | Not-determined | Genome-fragment | NA | | NA | 0 |
| S1_extracted_k141_16052 | 1062 | No | NA | 1 | 0 | 1 | Not-determined | Genome-fragment | NA | | NA | 0 |
| S1_extracted_k141_16056 | 1213 | No | NA | 1 | 0 | 0 | Not-determined | Genome-fragment | NA | | NA | 0 |
| S17_extracted_k141_48468 | 1451 | No | NA | 5 | 0 | 0 | Low-quality | Genome-fragment | | 1.96 | AAI-based (high-confidence) | 0 |
| S1_extracted_k141_55657 | 1413 | No | NA | 1 | 0 | 1 | Not-determined | Genome-fragment | NA | | NA | 0 |
| S1_extracted_k141_115694 | 1016 | No | NA | 1 | 0 | 1 | Not-determined | Genome-fragment | NA | | NA | 0 |
| S1_extracted_k141_108930 | 1100 | No | NA | 2 | 0 | 1 | Not-determined | Genome-fragment | NA | | NA | 0 |
| S9_extracted_k141_81429 | 1451 | No | NA | 1 | 0 | 0 | Low-quality | Genome-fragment | | 1.66 | AAI-based (medium-confidence) | 0 |
| S1_extracted_k141_75689 | 1140 | No | NA | 1 | 0 | 1 | Not-determined | Genome-fragment | NA | | NA | 0 |
| S1_extracted_k141_22149 | 42111 | No | NA | 63 | 9 | 0 | High-quality | High-quality | | 91.64 | AAI-based (high-confidence) | 0 |
| S9_vs_k141_49470\|\|full | 1451 | No | NA | 2 | 0 | 1 | Low-quality | Genome-fragment | | 2.23 | AAI-based (high-confidence) | 0 |
| S1_extracted_k141_154951 | 1193 | No | NA | 3 | 0 | 0 | Not-determined | Genome-fragment | NA | | NA | 0 |
| S1_extracted_k141_108949 | 3616 | No | NA | 4 | 0 | 2 | Not-determined | Genome-fragment | NA | | NA | 0 |
| S1_extracted_k141_68949 | 8689 | No | NA | 22 | 0 | 18 | Not-determined | Genome-fragment | NA | | NA | 0 |
| S14_vs_k141_114582\|\|full | 1451 | No | NA | 3 | 0 | 0 | Low-quality | Genome-fragment | | 3.4 | AAI-based (medium-confidence) | 0 |
| S1_extracted_k141_82457 | 1004 | No | NA | 1 | 0 | 0 | Not-determined | Genome-fragment | NA | | NA | 0 |
| S1_extracted_k141_75735 | 4761 | No | NA | 5 | 0 | 1 | Not-determined | Genome-fragment | NA | | NA | 0 |
| S1_extracted_k141_75746 | 1454 | No | NA | 1 | 0 | 1 | Not-determined | Genome-fragment | NA | | NA | 0 |
| S1_extracted_k141_115759 | 1194 | No | NA | 1 | 0 | 1 | Not-determined | Genome-fragment | NA | | NA | 0 |
| S5_extracted_k141_45159 | 1451 | No | NA | 2 | 0 | 0 | Low-quality | Genome-fragment | | 5.17 | AAI-based (medium-confidence) | 0 |
| S1_extracted_k141_122279 | 3177 | No | NA | 3 | 0 | 1 | Not-determined | Genome-fragment | NA | | NA | 0 |
| S1_extracted_k141_69000 | 1358 | No | NA | 2 | 0 | 0 | Not-determined | Genome-fragment | NA | | NA | 0 |
| S12_vs_k141_163464\|\|lt2gene | 1451 | No | NA | 2 | 2 | 0 | Low-quality | Genome-fragment | | 4.23 | AAI-based (high-confidence) | 0 |
| S2_extracted_k141_5097 | 1451 | No | NA | 2 | 1 | 0 | Low-quality | Genome-fragment | | 1.43 | AAI-based (high-confidence) | 0 |
| S8_vs_k141_160091\|\|full | 1451 | No | NA | 2 | 0 | 1 | Low-quality | Genome-fragment | | 1.14 | AAI-based (high-confidence) | 0 |
| S17_extracted_k141_134432 | 1452 | No | NA | 3 | 1 | 0 | Low-quality | Genome-fragment | | 2.4 | AAI-based (medium-confidence) | 0 |
| S11_vs_k141_45741\|\|full | 1452 | No | NA | 3 | 0 | 0 | Low-quality | Genome-fragment | | 3.08 | AAI-based (medium-confidence) | 0 |
| S1_extracted_k141_29055 | 1317 | No | NA | 3 | 0 | 0 | Not-determined | Genome-fragment | NA | | NA | 0 |
| S1_vs_k141_125595\|\|full | 1452 | No | NA | 2 | 0 | 1 | Low-quality | Genome-fragment | | 3.1 | AAI-based (high-confidence) | 0 |
| S5_extracted_k141_19858 | 1452 | No | NA | 2 | 1 | 0 | Low-quality | Genome-fragment | | 2.79 | HMM-based (lower-bound) | 0 |
| S1_extracted_k141_115845 | 1012 | No | NA | 1 | 0 | 1 | Not-determined | Genome-fragment | NA | | NA | 0 |
| S1_extracted_k141_29064 | 1081 | No | NA | 1 | 0 | 1 | Not-determined | Genome-fragment | NA | | NA | 0 |
| S1_extracted_k141_42513 | 2938 | No | NA | 2 | 0 | 2 | Not-determined | Genome-fragment | NA | | NA | 0 |
| S1_extracted_k141_16230 | 1248 | No | NA | 1 | 0 | 1 | Not-determined | Genome-fragment | NA | | NA | 0 |
| S1_extracted_k141_95960 | 1221 | No | NA | 1 | 0 | 1 | Not-determined | Genome-fragment | NA | | NA | 0 |
| S7_vs_26865\|\|full | 1452 | No | NA | 2 | 0 | 1 | Low-quality | Genome-fragment | | 1.81 | AAI-based (high-confidence) | 0 |
| S1_extracted_k141_142484 | 1120 | No | NA | 1 | 0 | 0 | Not-determined | Genome-fragment | NA | | NA | 0 |
| S16_extracted_170077 | 1452 | No | NA | 3 | 0 | 0 | Low-quality | Genome-fragment | | 2.43 | AAI-based (medium-confidence) | 0 |
| S1_extracted_k141_42533 | 1102 | No | NA | 3 | 0 | 1 | Not-determined | Genome-fragment | NA | | NA | 0 |
| S10_vs_115054\|\|full | 1452 | No | NA | 2 | 0 | 1 | Low-quality | Genome-fragment | | 0.52 | AAI-based (medium-confidence) | 0 |
| S1_extracted_k141_9195 | 1340 | No | NA | 2 | 0 | 2 | Not-determined | Genome-fragment | NA | | NA | 0 |
| S1_extracted_k141_16245 | 1468 | No | NA | 1 | 0 | 0 | Not-determined | Genome-fragment | NA | | NA | 0 |
| S1_extracted_k141_29103 | 1273 | No | NA | 2 | 0 | 0 | Not-determined | Genome-fragment | NA | | NA | 0 |
| S1_extracted_k141_42555 | 4632 | No | NA | 5 | 0 | 5 | Not-determined | Genome-fragment | NA | | NA | 0 |
| S1_extracted_k141_9224 | 1302 | No | NA | 1 | 0 | 1 | Not-determined | Genome-fragment | NA | | NA | 0 |
| S1_extracted_k141_147938 | 2391 | No | NA | 2 | 0 | 1 | Not-determined | Genome-fragment | NA | | NA | 0 |
| S1_extracted_k141_42576 | 1165 | No | NA | 1 | 0 | 1 | Not-determined | Genome-fragment | NA | | NA | 0 |
| S12_extracted_k141_123490 | 1452 | No | NA | 4 | 0 | 0 | Low-quality | Genome-fragment | | 2.66 | AAI-based (high-confidence) | 0 |
| S1_extracted_k141_155049 | 1616 | No | NA | 1 | 0 | 1 | Not-determined | Genome-fragment | NA | | NA | 0 |
| S1_extracted_k141_75888 | 1058 | No | NA | 1 | 0 | 0 | Not-determined | Genome-fragment | NA | | NA | 0 |
| S4_extracted_8680 | 1452 | No | NA | 1 | 0 | 0 | Low-quality | Genome-fragment | | 2.44 | AAI-based (medium-confidence) | 0 |
| S17_vs_k141_50321\|\|full | 1453 | No | NA | 2 | 0 | 1 | Low-quality | Genome-fragment | | 1.97 | AAI-based (medium-confidence) | 0 |
| S1_extracted_k141_16353 | 1476 | No | NA | 2 | 0 | 2 | Not-determined | Genome-fragment | NA | | NA | 0 |
| S1_extracted_k141_96071 | 1491 | No | NA | 1 | 0 | 1 | Not-determined | Genome-fragment | NA | | NA | 0 |
| S17_vs_k141_142206\|\|lt2gene | 1453 | No | NA | 1 | 1 | 0 | Low-quality | Genome-fragment | | 3.78 | AAI-based (high-confidence) | 0 |
| S14_extracted_k141_54153 | 1453 | No | NA | 4 | 1 | 0 | Low-quality | Genome-fragment | | 2.24 | HMM-based (lower-bound) | 0 |
| S1_extracted_k141_35409 | 1071 | No | NA | 2 | 0 | 1 | Not-determined | Genome-fragment | NA | | NA | 0 |
| S3_vs_k141_45230\|\|full | 1453 | No | NA | 4 | 1 | 0 | Low-quality | Genome-fragment | | 4.02 | AAI-based (high-confidence) | 0 |
| S7_vs_78210\|\|full | 1453 | No | NA | 3 | 0 | 0 | Low-quality | Genome-fragment | | 3.73 | AAI-based (high-confidence) | 0 |
| S1_extracted_k141_142633 | 1053 | No | NA | 1 | 0 | 1 | Not-determined | Genome-fragment | NA | | NA | 0 |
| S1_extracted_k141_82718 | 1078 | No | NA | 1 | 0 | 0 | Not-determined | Genome-fragment | NA | | NA | 0 |
| S16_vs_125296\|\|full | 1453 | No | NA | 3 | 1 | 0 | Low-quality | Genome-fragment | | 0.94 | HMM-based (lower-bound) | 0 |
| S16_vs_144420\|\|lt2gene | 1453 | No | NA | 2 | 2 | 0 | Low-quality | Genome-fragment | | 4.52 | AAI-based (high-confidence) | 0 |
| S1_extracted_k141_61560 | 1350 | No | NA | 1 | 0 | 0 | Not-determined | Genome-fragment | NA | | NA | 0 |
| S1_extracted_k141_61563 | 1051 | No | NA | 4 | 0 | 0 | Not-determined | Genome-fragment | NA | | NA | 0 |
| S1_extracted_k141_142647 | 3583 | No | NA | 3 | 0 | 0 | Not-determined | Genome-fragment | NA | | NA | 0 |
| S1_extracted_k141_96125 | 1062 | No | NA | 2 | 0 | 0 | Not-determined | Genome-fragment | NA | | NA | 0 |
| S1_extracted_k141_49391 | 1164 | No | NA | 1 | 0 | 1 | Not-determined | Genome-fragment | NA | | NA | 0 |
| S1_extracted_k141_82765 | 1135 | No | NA | 2 | 0 | 2 | Not-determined | Genome-fragment | NA | | NA | 0 |
| S1_extracted_k141_135963 | 1358 | No | NA | 3 | 0 | 2 | Not-determined | Genome-fragment | NA | | NA | 0 |
| S6_vs_k141_42379\|\|full | 1453 | No | NA | 3 | 0 | 2 | Low-quality | Genome-fragment | | 1.35 | AAI-based (medium-confidence) | 0 |
| S1_extracted_k141_89262 | 1117 | No | NA | 1 | 0 | 1 | Not-determined | Genome-fragment | NA | | NA | 0 |
| S8_vs_k141_4557\|\|full | 1453 | No | NA | 2 | 0 | 0 | Low-quality | Genome-fragment | | 2.92 | AAI-based (high-confidence) | 0 |
| S9_vs_k141_73088\|\|full | 1454 | No | NA | 2 | 0 | 0 | Low-quality | Genome-fragment | | 1.47 | AAI-based (medium-confidence) | 0 |
| S7_extracted_21155 | 1454 | No | NA | 3 | 0 | 0 | Low-quality | Genome-fragment | | 2.71 | AAI-based (high-confidence) | 0 |
| S7_vs_114936\|\|full | 1454 | No | NA | 3 | 0 | 0 | Low-quality | Genome-fragment | | 3.31 | AAI-based (medium-confidence) | 0 |
| S1_extracted_k141_135985 | 2078 | No | NA | 2 | 0 | 0 | Not-determined | Genome-fragment | NA | | NA | 0 |
| S1_extracted_k141_116098 | 1204 | No | NA | 1 | 0 | 1 | Not-determined | Genome-fragment | NA | | NA | 0 |
| S16_vs_95190\|\|full | 1454 | No | NA | 2 | 0 | 0 | Low-quality | Genome-fragment | | 1.85 | AAI-based (medium-confidence) | 0 |
| S1_extracted_k141_129462 | 3149 | No | NA | 1 | 0 | 1 | Not-determined | Genome-fragment | NA | | NA | 0 |
| S10_vs_38948\|\|full | 1454 | No | NA | 2 | 0 | 0 | Low-quality | Genome-fragment | | 2.55 | AAI-based (medium-confidence) | 0 |
| S10_vs_42752\|\|full | 1454 | No | NA | 3 | 0 | 0 | Low-quality | Genome-fragment | | 1.81 | AAI-based (high-confidence) | 0 |
| S1_extracted_k141_22491 | 1652 | No | NA | 2 | 0 | 1 | Not-determined | Genome-fragment | NA | | NA | 0 |
| S8_extracted_k141_176338 | 1454 | No | NA | 2 | 0 | 0 | Low-quality | Genome-fragment | | 0.95 | AAI-based (medium-confidence) | 0 |
| S1_extracted_k141_122563 | 2311 | No | NA | 2 | 0 | 1 | Not-determined | Genome-fragment | NA | | NA | 0 |
| S1_extracted_k141_82852 | 5260 | No | NA | 3 | 0 | 3 | Not-determined | Genome-fragment | NA | | NA | 0 |
| S1_extracted_k141_16536 | 1266 | No | NA | 1 | 0 | 0 | Not-determined | Genome-fragment | NA | | NA | 0 |
| S8_vs_k141_1278\|\|full | 1454 | No | NA | 2 | 1 | 0 | Low-quality | Genome-fragment | | 2.87 | AAI-based (medium-confidence) | 0 |
| S1_extracted_k141_116191 | 3158 | No | NA | 4 | 0 | 3 | Not-determined | Genome-fragment | NA | | NA | 0 |
| S1_extracted_k141_96240 | 3214 | No | NA | 2 | 0 | 1 | Not-determined | Genome-fragment | NA | | NA | 0 |
| S1_extracted_k141_16567 | 7933 | No | NA | 4 | 0 | 3 | Not-determined | Genome-fragment | NA | | NA | 0 |
| S1_extracted_k141_29446 | 1095 | No | NA | 1 | 0 | 1 | Not-determined | Genome-fragment | NA | | NA | 0 |
| S1_extracted_k141_148035 | 1163 | No | NA | 1 | 0 | 1 | Not-determined | Genome-fragment | NA | | NA | 0 |
| S1_extracted_k141_29459 | 1316 | No | NA | 3 | 0 | 2 | Not-determined | Genome-fragment | NA | | NA | 0 |
| S1_extracted_k141_35579 | 1254 | No | NA | 1 | 0 | 0 | Not-determined | Genome-fragment | NA | | NA | 0 |
| S17_extracted_k141_81467 | 1455 | No | NA | 3 | 0 | 1 | Low-quality | Genome-fragment | | 0.24 | AAI-based (medium-confidence) | 0 |
| S1_extracted_k141_89384 | 1030 | No | NA | 2 | 0 | 1 | Not-determined | Genome-fragment | NA | | NA | 0 |
| S1_extracted_k141_129574 | 3585 | No | NA | 3 | 0 | 2 | Not-determined | Genome-fragment | NA | | NA | 0 |
| S1_extracted_k141_136113 | 10754 | No | NA | 11 | 0 | 1 | Not-determined | Genome-fragment | NA | | NA | 0 |
| S1_extracted_k141_22608 | 1128 | No | NA | 2 | 0 | 1 | Not-determined | Genome-fragment | NA | | NA | 0 |
| S1_extracted_k141_96286 | 2133 | No | NA | 5 | 0 | 5 | Not-determined | Genome-fragment | NA | | NA | 0 |
| S1_extracted_k141_76174 | 1097 | No | NA | 2 | 0 | 2 | Not-determined | Genome-fragment | NA | | NA | 0 |
| S1_extracted_k141_142833 | 10106 | No | NA | 9 | 0 | 2 | Not-determined | Genome-fragment | NA | | NA | 0 |
| S1_extracted_k141_9562 | 1001 | No | NA | 2 | 0 | 1 | Not-determined | Genome-fragment | NA | | NA | 0 |
| S1_extracted_k141_69400 | 3726 | No | NA | 6 | 0 | 0 | Not-determined | Genome-fragment | NA | | NA | 0 |
| S1_extracted_k141_82994 | 2897 | No | NA | 1 | 0 | 1 | Not-determined | Genome-fragment | NA | | NA | 0 |
| S1_extracted_k141_42929 | 1535 | No | NA | 1 | 0 | 0 | Not-determined | Genome-fragment | NA | | NA | 0 |
| S1_extracted_k141_102997 | 1042 | No | NA | 2 | 0 | 0 | Not-determined | Genome-fragment | NA | | NA | 0 |
| S1_extracted_k141_116313 | 6847 | No | NA | 4 | 0 | 2 | Not-determined | Genome-fragment | NA | | NA | 0 |
| S17_vs_k141_71883\|\|full | 1455 | No | NA | 3 | 0 | 2 | Low-quality | Genome-fragment | | 3.36 | AAI-based (medium-confidence) | 0 |
| S17_vs_k141_83829\|\|lt2gene | 1455 | No | NA | 2 | 2 | 0 | Low-quality | Genome-fragment | | 3.78 | AAI-based (high-confidence) | 0 |
| S3_extracted_k141_22767 | 1455 | No | NA | 5 | 0 | 0 | Low-quality | Genome-fragment | | 1.39 | AAI-based (high-confidence) | 0 |
| S10_extracted_8897 | 1455 | No | NA | 2 | 1 | 0 | Low-quality | Genome-fragment | | 2.77 | HMM-based (lower-bound) | 0 |
| S1_extracted_k141_109471 | 3326 | No | NA | 1 | 0 | 1 | Not-determined | Genome-fragment | NA | | NA | 0 |
| S12_extracted_k141_28710 | 1455 | No | NA | 3 | 0 | 0 | Low-quality | Genome-fragment | | 1.6 | AAI-based (high-confidence) | 0 |
| S1_extracted_k141_116358 | 1128 | No | NA | 2 | 0 | 2 | Not-determined | Genome-fragment | NA | | NA | 0 |
| S12_extracted_k141_86373 | 1455 | No | NA | 6 | 1 | 0 | Low-quality | Genome-fragment | | 2.59 | AAI-based (high-confidence) | 0 |
| S6_vs_k141_41848\|\|full | 1455 | No | NA | 4 | 2 | 0 | Low-quality | Genome-fragment | | 1.96 | AAI-based (high-confidence) | 0 |
| S8_vs_k141_14467\|\|full | 1455 | No | NA | 3 | 1 | 0 | Low-quality | Genome-fragment | | 3.85 | AAI-based (high-confidence) | 0 |
| S17_vs_k141_126480\|\|full | 1456 | No | NA | 2 | 1 | 0 | Low-quality | Genome-fragment | | 2.48 | AAI-based (medium-confidence) | 0 |
| S1_extracted_k141_69520 | 1693 | No | NA | 1 | 0 | 0 | Not-determined | Genome-fragment | NA | | NA | 0 |
| S1_extracted_k141_89557 | 1094 | No | NA | 1 | 0 | 1 | Not-determined | Genome-fragment | NA | | NA | 0 |
| S1_extracted_k141_22729 | 1492 | No | NA | 1 | 0 | 1 | Not-determined | Genome-fragment | NA | | NA | 0 |
| S1_extracted_k141_16789 | 2245 | No | NA | 2 | 0 | 1 | Not-determined | Genome-fragment | NA | | NA | 0 |
| S1_extracted_k141_2434 | 1248 | No | NA | 1 | 0 | 1 | Not-determined | Genome-fragment | NA | | NA | 0 |
| S1_extracted_k141_142898 | 6791 | No | NA | 4 | 0 | 4 | Not-determined | Genome-fragment | NA | | NA | 0 |
| S1_extracted_k141_96440 | 1638 | No | NA | 2 | 0 | 1 | Not-determined | Genome-fragment | NA | | NA | 0 |
| S1_extracted_k141_109567 | 1466 | No | NA | 2 | 0 | 1 | Not-determined | Genome-fragment | NA | | NA | 0 |
| S17_vs_k141_86785\|\|lt2gene | 1456 | No | NA | 2 | 1 | 0 | Low-quality | Genome-fragment | | 2.45 | AAI-based (medium-confidence) | 0 |
| S1_extracted_k141_22753 | 4084 | No | NA | 2 | 0 | 1 | Not-determined | Genome-fragment | NA | | NA | 0 |
| S9_vs_k141_73181\|\|lt2gene | 1456 | No | NA | 1 | 1 | 0 | Low-quality | Genome-fragment | | 3.63 | AAI-based (medium-confidence) | 0 |
| S1_extracted_k141_68197 | 1456 | No | NA | 2 | 0 | 0 | Low-quality | Genome-fragment | | 2.84 | AAI-based (medium-confidence) | 0 |
| S1_extracted_k141_16839 | 1449 | No | NA | 2 | 0 | 0 | Not-determined | Genome-fragment | NA | | NA | 0 |
| S1_extracted_k141_136336 | 1018 | No | NA | 1 | 0 | 1 | Not-determined | Genome-fragment | NA | | NA | 0 |
| S3_extracted_k141_24725 | 1456 | No | NA | 1 | 0 | 0 | Low-quality | Genome-fragment | | 0.71 | AAI-based (medium-confidence) | 0 |
| S1_extracted_k141_9763 | 1226 | No | NA | 1 | 0 | 1 | Not-determined | Genome-fragment | NA | | NA | 0 |
| S1_extracted_k141_116484 | 1715 | No | NA | 1 | 0 | 1 | Not-determined | Genome-fragment | NA | | NA | 0 |
| S5_vs_k141_2878\|\|full | 1456 | No | NA | 4 | 1 | 0 | Low-quality | Genome-fragment | | 2.68 | AAI-based (high-confidence) | 0 |
| S1_extracted_k141_89646 | 2992 | No | NA | 1 | 0 | 1 | Not-determined | Genome-fragment | NA | | NA | 0 |
| S7_vs_104590\|\|full | 1456 | No | NA | 4 | 1 | 0 | Low-quality | Genome-fragment | | 3.53 | AAI-based (high-confidence) | 0 |
| S16_vs_18749\|\|full | 1456 | No | NA | 3 | 0 | 0 | Low-quality | Genome-fragment | | 4.17 | AAI-based (high-confidence) | 0 |
| S1_extracted_k141_103225 | 1243 | No | NA | 1 | 0 | 1 | Not-determined | Genome-fragment | NA | | NA | 0 |
| S1_extracted_k141_56380 | 3848 | No | NA | 6 | 0 | 0 | Not-determined | Genome-fragment | NA | | NA | 0 |
| S1_extracted_k141_9800 | 2539 | No | NA | 2 | 0 | 2 | Not-determined | Genome-fragment | NA | | NA | 0 |
| S16_vs_113652\|\|full | 1456 | No | NA | 3 | 0 | 0 | Low-quality | Genome-fragment | | 2.98 | AAI-based (medium-confidence) | 0 |
| S1_extracted_k141_69626 | 2807 | No | NA | 3 | 0 | 2 | Not-determined | Genome-fragment | NA | | NA | 0 |
| S1_extracted_k141_89674 | 1542 | No | NA | 2 | 0 | 2 | Not-determined | Genome-fragment | NA | | NA | 0 |
| S1_extracted_k141_109673 | 1324 | No | NA | 1 | 0 | 1 | Not-determined | Genome-fragment | NA | | NA | 0 |
| S1_extracted_k141_122911 | 1072 | No | NA | 1 | 0 | 0 | Not-determined | Genome-fragment | NA | | NA | 0 |
| S1_extracted_k141_22869 | 1038 | No | NA | 1 | 0 | 1 | Not-determined | Genome-fragment | NA | | NA | 0 |
| S1_extracted_k141_9828 | 1696 | No | NA | 3 | 0 | 0 | Not-determined | Genome-fragment | NA | | NA | 0 |
| S1_extracted_k141_89686 | 1240 | No | NA | 1 | 0 | 1 | Not-determined | Genome-fragment | NA | | NA | 0 |
| S1_extracted_k141_16933 | 1890 | No | NA | 1 | 0 | 1 | Not-determined | Genome-fragment | NA | | NA | 0 |
| S13_extracted_14545 | 1457 | No | NA | 2 | 0 | 1 | Low-quality | Genome-fragment | | 2.82 | AAI-based (medium-confidence) | 0 |
| S1_extracted_k141_107387 | 1457 | No | NA | 2 | 0 | 0 | Low-quality | Genome-fragment | | 3.94 | AAI-based (high-confidence) | 0 |
| S1_extracted_k141_76457 | 2165 | No | NA | 2 | 0 | 1 | Not-determined | Genome-fragment | NA | | NA | 0 |
| S1_extracted_k141_49928 | 2295 | No | NA | 3 | 0 | 3 | Not-determined | Genome-fragment | NA | | NA | 0 |
| S1_extracted_k141_148170 | 1797 | No | NA | 1 | 0 | 1 | Not-determined | Genome-fragment | NA | | NA | 0 |
| S1_extracted_k141_136427 | 1058 | No | NA | 1 | 0 | 0 | Not-determined | Genome-fragment | NA | | NA | 0 |
| S1_extracted_k141_49939 | 1129 | No | NA | 1 | 0 | 1 | Not-determined | Genome-fragment | NA | | NA | 0 |
| S1_extracted_k141_143049 | 3404 | No | NA | 4 | 0 | 3 | Not-determined | Genome-fragment | NA | | NA | 0 |
| S1_extracted_k141_158229 | 1457 | No | NA | 3 | 0 | 0 | Low-quality | Genome-fragment | | 2.54 | AAI-based (high-confidence) | 0 |
| S3_vs_k141_116586\|\|full | 1457 | No | NA | 4 | 0 | 0 | Low-quality | Genome-fragment | | 1.32 | AAI-based (high-confidence) | 0 |
| S1_extracted_k141_83249 | 1682 | No | NA | 4 | 0 | 0 | Not-determined | Genome-fragment | NA | | NA | 0 |
| S1_extracted_k141_129928 | 1078 | No | NA | 2 | 0 | 0 | Not-determined | Genome-fragment | NA | | NA | 0 |
| S5_vs_k141_1043\|\|full | 1457 | No | NA | 3 | 1 | 0 | Low-quality | Genome-fragment | | 2.16 | HMM-based (lower-bound) | 0 |
| S16_extracted_210 | 1457 | No | NA | 3 | 0 | 2 | Low-quality | Genome-fragment | | 0.46 | AAI-based (medium-confidence) | 0 |
| S16_extracted_125389 | 1457 | No | NA | 2 | 0 | 1 | Low-quality | Genome-fragment | | 0.61 | AAI-based (medium-confidence) | 0 |
| S1_extracted_k141_155378 | 4100 | No | NA | 1 | 0 | 1 | Not-determined | Genome-fragment | NA | | NA | 0 |
| S10_vs_110152\|\|full | 1457 | No | NA | 5 | 1 | 0 | Low-quality | Genome-fragment | | 3.29 | AAI-based (medium-confidence) | 0 |
| S1_extracted_k141_103361 | 1266 | No | NA | 2 | 0 | 2 | Not-determined | Genome-fragment | NA | | NA | 0 |
| S15_extracted_13212 | 1457 | No | NA | 2 | 0 | 0 | Low-quality | Genome-fragment | | 3.52 | AAI-based (medium-confidence) | 0 |
| S1_extracted_k141_29861 | 1061 | No | NA | 1 | 0 | 1 | Not-determined | Genome-fragment | NA | | NA | 0 |
| S8_extracted_k141_165040 | 1457 | No | NA | 3 | 0 | 0 | Low-quality | Genome-fragment | | 4.15 | AAI-based (medium-confidence) | 0 |
| S8_extracted_k141_127845 | 1457 | No | NA | 2 | 0 | 0 | Low-quality | Genome-fragment | | 1.37 | AAI-based (high-confidence) | 0 |
| S1_extracted_k141_148262 | 3046 | No | NA | 1 | 0 | 1 | Not-determined | Genome-fragment | NA | | NA | 0 |
| S1_extracted_k141_29877 | 3715 | No | NA | 6 | 0 | 2 | Not-determined | Genome-fragment | NA | | NA | 0 |
| S1_extracted_k141_69755 | 1815 | No | NA | 2 | 0 | 1 | Not-determined | Genome-fragment | NA | | NA | 0 |
| S1_extracted_k141_103436 | 2678 | No | NA | 2 | 0 | 2 | Not-determined | Genome-fragment | NA | | NA | 0 |
| S1_extracted_k141_130079 | 1017 | No | NA | 1 | 0 | 0 | Not-determined | Genome-fragment | NA | | NA | 0 |
| S1_extracted_k141_83406 | 1046 | No | NA | 1 | 0 | 0 | Not-determined | Genome-fragment | NA | | NA | 0 |
| S8_vs_k141_121910\|\|full | 1457 | No | NA | 2 | 0 | 0 | Low-quality | Genome-fragment | | 2.72 | AAI-based (medium-confidence) | 0 |
| S17_vs_k141_22275\|\|full | 1458 | No | NA | 2 | 0 | 1 | Low-quality | Genome-fragment | | 1.17 | AAI-based (medium-confidence) | 0 |
| S9_extracted_k141_87916 | 1458 | No | NA | 2 | 0 | 2 | Low-quality | Genome-fragment | | 0.95 | AAI-based (high-confidence) | 0 |
| S1_extracted_k141_96814 | 1225 | No | NA | 2 | 0 | 0 | Not-determined | Genome-fragment | NA | | NA | 0 |
| S1_extracted_k141_136526 | 31147 | No | NA | 53 | 12 | 0 | Medium-quality | Genome-fragment | | 73.92 | AAI-based (high-confidence) | 0 |
| S1_extracted_k141_56618 | 1843 | No | NA | 2 | 0 | 2 | Not-determined | Genome-fragment | NA | | NA | 0 |

| | | | | | | | | | | | |
|---|---|---|---|---|---|---|---|---|---|---|---|
| S3_extracted_k141_89338 | 1458 | No | NA | 1 | 0 | 1 | Low-quality | Genome-fragment | 1.85 | AAI-based (high-confidence) | 0 |
| S1_extracted_k141_83421 | 1721 | No | NA | 4 | 0 | 1 | Not-determined | Genome-fragment | NA | NA | 0 |
| S1_extracted_k141_23016 | 1129 | No | NA | 1 | 0 | 1 | Not-determined | Genome-fragment | NA | NA | 0 |
| S1_extracted_k141_96829 | 4581 | No | NA | 2 | 0 | 2 | Not-determined | Genome-fragment | NA | NA | 0 |
| S1_extracted_k141_56635 | 1333 | No | NA | 2 | 0 | 1 | Not-determined | Genome-fragment | NA | NA | 0 |
| S1_extracted_k141_143225 | 1119 | No | NA | 1 | 0 | 1 | Not-determined | Genome-fragment | NA | NA | 0 |
| S1_extracted_k141_23029 | 1189 | No | NA | 1 | 0 | 0 | Not-determined | Genome-fragment | NA | NA | 0 |
| S1_extracted_k141_143228 | 1400 | No | NA | 2 | 0 | 0 | Not-determined | Genome-fragment | NA | NA | 0 |
| S1_extracted_k141_83448 | 1198 | No | NA | 1 | 0 | 1 | Not-determined | Genome-fragment | NA | NA | 0 |
| S1_extracted_k141_10087 | 1293 | No | NA | 2 | 0 | 0 | Not-determined | Genome-fragment | NA | NA | 0 |
| S5_extracted_k141_80449 | 1458 | No | NA | 3 | 0 | 0 | Low-quality | Genome-fragment | 2.88 | AAI-based (medium-confidence) | 0 |
| S1_extracted_k141_69812 | 1503 | No | NA | 1 | 0 | 0 | Not-determined | Genome-fragment | NA | NA | 0 |
| NC3_vf_k141_2069 | 1458 | No | NA | 1 | 0 | 0 | Low-quality | Genome-fragment | 2.27 | AAI-based (medium-confidence) | 0 |
| S1_extracted_k141_56686 | 1162 | No | NA | 1 | 0 | 0 | Not-determined | Genome-fragment | NA | NA | 0 |
| S10_vs_39050\|\|full | 1458 | No | NA | 4 | 0 | 0 | Low-quality | Genome-fragment | 2.86 | AAI-based (high-confidence) | 0 |
| S12_extracted_k141_28707 | 1458 | No | NA | 3 | 0 | 0 | Low-quality | Genome-fragment | 3.32 | AAI-based (medium-confidence) | 0 |
| S12_extracted_k141_116611 | 1458 | No | NA | 2 | 0 | 0 | Low-quality | Genome-fragment | 1.5 | AAI-based (medium-confidence) | 0 |
| S8_extracted_k141_133117 | 1458 | No | NA | 2 | 1 | 0 | Low-quality | Genome-fragment | 3.89 | AAI-based (high-confidence) | 0 |
| S8_vs_k141_178635\|\|full | 1458 | No | NA | 3 | 0 | 0 | Low-quality | Genome-fragment | 3.66 | AAI-based (medium-confidence) | 0 |
| S1_extracted_k141_103548 | 1066 | No | NA | 1 | 0 | 0 | Not-determined | Genome-fragment | NA | NA | 0 |
| S17_vs_k141_155589\|\|full | 1459 | No | NA | 3 | 2 | 0 | Low-quality | Genome-fragment | 3.57 | AAI-based (high-confidence) | 0 |
| S1_extracted_k141_110052 | 1049 | No | NA | 1 | 0 | 1 | Not-determined | Genome-fragment | NA | NA | 0 |
| S1_extracted_k141_69838 | 2215 | No | NA | 1 | 0 | 1 | Not-determined | Genome-fragment | NA | NA | 0 |
| S1_extracted_k141_106818 | 1459 | No | NA | 1 | 0 | 0 | Low-quality | Genome-fragment | 1.47 | AAI-based (high-confidence) | 0 |
| S1_extracted_k141_69839 | 1060 | No | NA | 2 | 0 | 1 | Not-determined | Genome-fragment | NA | NA | 0 |
| S5_vs_k141_17501\|\|full | 1459 | No | NA | 3 | 0 | 1 | Low-quality | Genome-fragment | 1.15 | AAI-based (medium-confidence) | 0 |
| S1_extracted_k141_90008 | 1201 | No | NA | 1 | 0 | 1 | Not-determined | Genome-fragment | NA | NA | 0 |
| S1_extracted_k141_50288 | 1767 | No | NA | 1 | 0 | 1 | Not-determined | Genome-fragment | NA | NA | 0 |
| S16_vs_37247\|\|full | 1459 | No | NA | 4 | 0 | 0 | Low-quality | Genome-fragment | 2.57 | AAI-based (medium-confidence) | 0 |
| S10_extracted_54021 | 1459 | No | NA | 2 | 0 | 0 | Low-quality | Genome-fragment | 4.39 | AAI-based (medium-confidence) | 0 |
| S1_extracted_k141_17258 | 1182 | No | NA | 1 | 0 | 0 | Not-determined | Genome-fragment | NA | NA | 0 |
| S1_extracted_k141_103612 | 2326 | No | NA | 2 | 0 | 1 | Not-determined | Genome-fragment | NA | NA | 0 |
| S6_vs_k141_17079\|\|full | 1459 | No | NA | 2 | 0 | 0 | Low-quality | Genome-fragment | 3.73 | AAI-based (high-confidence) | 0 |
| S1_extracted_k141_43548 | 1228 | No | NA | 4 | 0 | 3 | Not-determined | Genome-fragment | NA | NA | 0 |
| S8_extracted_k141_123889 | 1459 | No | NA | 2 | 0 | 1 | Low-quality | Genome-fragment | 1.44 | AAI-based (medium-confidence) | 0 |
| S1_extracted_k141_155550 | 1282 | No | NA | 1 | 0 | 1 | Not-determined | Genome-fragment | NA | NA | 0 |
| S8_vs_k141_180109\|\|full | 1459 | No | NA | 2 | 2 | 0 | Low-quality | Genome-fragment | 4.64 | AAI-based (high-confidence) | 0 |
| S11_vs_k141_53711\|\|full | 1460 | No | NA | 3 | 0 | 0 | Low-quality | Genome-fragment | 2.59 | AAI-based (medium-confidence) | 0 |
| S1_extracted_k141_62237 | 1727 | No | NA | 1 | 0 | 0 | Not-determined | Genome-fragment | NA | NA | 0 |
| S1_extracted_k141_17302 | 1559 | No | NA | 1 | 0 | 1 | Not-determined | Genome-fragment | NA | NA | 0 |
| S1_extracted_k141_2838 | 1528 | No | NA | 2 | 0 | 1 | Not-determined | Genome-fragment | NA | NA | 0 |
| S1_extracted_k141_123326 | 4263 | No | NA | 5 | 0 | 2 | Not-determined | Genome-fragment | NA | NA | 0 |
| S1_extracted_k141_90062 | 1041 | No | NA | 4 | 0 | 3 | Not-determined | Genome-fragment | NA | NA | 0 |
| S1_extracted_k141_116985 | 1042 | No | NA | 2 | 0 | 1 | Not-determined | Genome-fragment | NA | NA | 0 |
| S1_extracted_k141_36130 | 7262 | No | NA | 7 | 0 | 1 | Not-determined | Genome-fragment | NA | NA | 0 |
| S13_extracted_835 | 1460 | No | NA | 2 | 0 | 1 | Low-quality | Genome-fragment | 0.27 | AAI-based (medium-confidence) | 0 |
| S1_extracted_k141_143400 | 1410 | No | NA | 2 | 0 | 2 | Not-determined | Genome-fragment | NA | NA | 0 |
| S13_extracted_29615 | 1460 | No | NA | 2 | 0 | 0 | Low-quality | Genome-fragment | 1.51 | AAI-based (high-confidence) | 0 |
| S13_extracted_32269 | 1460 | No | NA | 4 | 0 | 0 | Low-quality | Genome-fragment | 0.78 | AAI-based (medium-confidence) | 0 |
| S1_extracted_k141_130252 | 3536 | No | NA | 4 | 0 | 0 | Not-determined | Genome-fragment | NA | NA | 0 |
| S1_extracted_k141_123368 | 1190 | No | NA | 1 | 0 | 0 | Not-determined | Genome-fragment | NA | NA | 0 |
| S13_vs_79868\|\|full | 1460 | No | NA | 3 | 0 | 0 | Low-quality | Genome-fragment | 2.46 | AAI-based (medium-confidence) | 0 |
| S13_vs_81708\|\|full | 1460 | No | NA | 2 | 0 | 0 | Low-quality | Genome-fragment | 0.79 | AAI-based (medium-confidence) | 0 |
| S1_extracted_k141_136793 | 3930 | No | NA | 3 | 0 | 2 | Not-determined | Genome-fragment | NA | NA | 0 |
| S1_extracted_k141_117040 | 1065 | No | NA | 1 | 0 | 1 | Not-determined | Genome-fragment | NA | NA | 0 |
| S1_vs_k141_37656\|\|full | 1460 | No | NA | 2 | 1 | 0 | Low-quality | Genome-fragment | 3.44 | AAI-based (high-confidence) | 0 |
| S1_extracted_k141_69917 | 1404 | No | NA | 3 | 0 | 2 | Not-determined | Genome-fragment | NA | NA | 0 |
| S5_vs_k141_94120\|\|full | 1460 | No | NA | 3 | 0 | 2 | Low-quality | Genome-fragment | 1.41 | AAI-based (medium-confidence) | 0 |
| S1_extracted_k141_110192 | 1386 | No | NA | 1 | 0 | 1 | Not-determined | Genome-fragment | NA | NA | 0 |
| S1_extracted_k141_148424 | 1009 | No | NA | 2 | 0 | 0 | Not-determined | Genome-fragment | NA | NA | 0 |
| S5_vs_k141_21134\|\|full | 1460 | No | NA | 3 | 0 | 0 | Low-quality | Genome-fragment | 1.81 | AAI-based (medium-confidence) | 0 |
| S1_extracted_k141_50380 | 1010 | No | NA | 3 | 0 | 3 | Not-determined | Genome-fragment | NA | NA | 0 |
| S1_extracted_k141_36172 | 1684 | No | NA | 2 | 0 | 2 | Not-determined | Genome-fragment | NA | NA | 0 |
| S1_extracted_k141_83763 | 1342 | No | NA | 2 | 0 | 1 | Not-determined | Genome-fragment | NA | NA | 0 |
| S5_vs_k141_104079\|\|lt2gene | 1460 | No | NA | 2 | 1 | 0 | Low-quality | Genome-fragment | 4.07 | AAI-based (high-confidence) | 0 |
| S1_extracted_k141_136867 | 3269 | No | NA | 2 | 0 | 1 | Not-determined | Genome-fragment | NA | NA | 0 |
| S1_extracted_k141_62357 | 3259 | No | NA | 1 | 0 | 1 | Not-determined | Genome-fragment | NA | NA | 0 |
| S5_vs_k141_113275\|\|lt2gene | 1460 | No | NA | 1 | 1 | 0 | Low-quality | Genome-fragment | 1.01 | HMM-based (lower-bound) | 0 |
| S1_extracted_k141_148488 | 1183 | No | NA | 1 | 0 | 0 | Not-determined | Genome-fragment | NA | NA | 0 |
| S1_extracted_k141_57012 | 1058 | No | NA | 1 | 0 | 1 | Not-determined | Genome-fragment | NA | NA | 0 |
| S16_vs_62086\|\|full | 1460 | No | NA | 2 | 0 | 1 | Low-quality | Genome-fragment | 0.88 | AAI-based (medium-confidence) | 0 |
| S1_extracted_k141_103761 | 1542 | No | NA | 1 | 0 | 1 | Not-determined | Genome-fragment | NA | NA | 0 |
| S16_vs_143001\|\|full | 1460 | No | NA | 3 | 0 | 1 | Low-quality | Genome-fragment | 2.99 | AAI-based (medium-confidence) | 0 |
| S10_vs_27376\|\|lt2gene | 1460 | No | NA | 2 | 2 | 0 | Low-quality | Genome-fragment | 3.65 | AAI-based (medium-confidence) | 0 |
| S1_extracted_k141_43732 | 1607 | No | NA | 2 | 0 | 2 | Not-determined | Genome-fragment | NA | NA | 0 |
| S1_extracted_k141_97169 | 4458 | No | NA | 3 | 0 | 0 | Not-determined | Genome-fragment | NA | NA | 0 |
| S12_extracted_k141_47931 | 1460 | No | NA | 4 | 0 | 3 | Low-quality | Genome-fragment | 0.43 | AAI-based (medium-confidence) | 0 |
| S12_vs_k141_917\|\|full | 1460 | No | NA | 4 | 0 | 0 | Low-quality | Genome-fragment | 3.69 | AAI-based (high-confidence) | 0 |
| S1_extracted_k141_43750 | 1158 | No | NA | 1 | 0 | 1 | Not-determined | Genome-fragment | NA | NA | 0 |
| S12_vs_k141_146903\|\|full | 1460 | No | NA | 3 | 0 | 0 | Low-quality | Genome-fragment | 3.83 | AAI-based (medium-confidence) | 0 |
| S12_vs_k141_86205\|\|full | 1460 | No | NA | 2 | 0 | 1 | Low-quality | Genome-fragment | 2.28 | AAI-based (medium-confidence) | 0 |
| S1_extracted_k141_43755 | 1150 | No | NA | 1 | 0 | 1 | Not-determined | Genome-fragment | NA | NA | 0 |
| S1_extracted_k141_90242 | 1243 | No | NA | 1 | 0 | 1 | Not-determined | Genome-fragment | NA | NA | 0 |
| S8_vs_k141_151754\|\|full | 1460 | No | NA | 2 | 0 | 0 | Low-quality | Genome-fragment | 1.74 | AAI-based (medium-confidence) | 0 |
| S17_extracted_k141_76622 | 1461 | No | NA | 2 | 0 | 0 | Low-quality | Genome-fragment | 1.52 | AAI-based (high-confidence) | 0 |
| S14_vs_k141_71691\|\|full | 1461 | No | NA | 3 | 0 | 1 | Low-quality | Genome-fragment | 0.39 | AAI-based (medium-confidence) | 0 |
| S3_extracted_k141_80188 | 1461 | No | NA | 3 | 0 | 0 | Low-quality | Genome-fragment | 1 | AAI-based (high-confidence) | 0 |
| S3_extracted_k141_912 | 1461 | No | NA | 1 | 1 | 0 | Low-quality | Genome-fragment | 3.35 | AAI-based (high-confidence) | 0 |
| S1_extracted_k141_97255 | 1022 | No | NA | 1 | 0 | 1 | Not-determined | Genome-fragment | NA | NA | 0 |
| S1_extracted_k141_148545 | 1523 | No | NA | 1 | 0 | 1 | Not-determined | Genome-fragment | NA | NA | 0 |
| S1_extracted_k141_110384 | 2237 | No | NA | 2 | 0 | 2 | Not-determined | Genome-fragment | NA | NA | 0 |
| S1_extracted_k141_77132 | 4630 | No | NA | 1 | 0 | 1 | Not-determined | Genome-fragment | NA | NA | 0 |
| S16_extracted_121181 | 1461 | No | NA | 2 | 0 | 0 | Low-quality | Genome-fragment | 2.42 | AAI-based (medium-confidence) | 0 |
| S1_extracted_k141_30421 | 1215 | No | NA | 2 | 0 | 0 | Not-determined | Genome-fragment | NA | NA | 0 |
| S1_extracted_k141_155721 | 1001 | No | NA | 1 | 0 | 0 | Not-determined | Genome-fragment | NA | NA | 0 |
| S1_extracted_k141_77142 | 1012 | No | NA | 2 | 0 | 2 | Not-determined | Genome-fragment | NA | NA | 0 |
| S1_extracted_k141_103879 | 1181 | No | NA | 1 | 0 | 1 | Not-determined | Genome-fragment | NA | NA | 0 |
| S1_extracted_k141_123597 | 1402 | No | NA | 1 | 0 | 0 | Not-determined | Genome-fragment | NA | NA | 0 |
| S1_extracted_k141_10553 | 3606 | No | NA | 2 | 0 | 2 | Not-determined | Genome-fragment | NA | NA | 0 |
| S1_extracted_k141_155727 | 3198 | No | NA | 1 | 0 | 1 | Not-determined | Genome-fragment | NA | NA | 0 |
| S1_extracted_k141_23533 | 1046 | No | NA | 2 | 0 | 1 | Not-determined | Genome-fragment | NA | NA | 0 |
| S1_extracted_k141_50591 | 21026 | No | NA | 19 | 13 | 0 | Medium-quality | Genome-fragment | 54.77 | AAI-based (high-confidence) | 0 |
| S1_extracted_k141_83910 | 1017 | No | NA | 1 | 0 | 1 | Not-determined | Genome-fragment | NA | NA | 0 |
| S16_vs_19706\|\|lt2gene | 1461 | No | NA | 2 | 1 | 0 | Low-quality | Genome-fragment | 3.8 | AAI-based (high-confidence) | 0 |
| S1_extracted_k141_143598 | 3049 | No | NA | 3 | 0 | 2 | Not-determined | Genome-fragment | NA | NA | 0 |
| S1_extracted_k141_110433 | 1182 | No | NA | 2 | 0 | 1 | Not-determined | Genome-fragment | NA | NA | 0 |
| S1_extracted_k141_57143 | 2641 | No | NA | 2 | 0 | 2 | Not-determined | Genome-fragment | NA | NA | 0 |
| S1_extracted_k141_10578 | 1168 | No | NA | 2 | 0 | 2 | Not-determined | Genome-fragment | NA | NA | 0 |
| S1_extracted_k141_83927 | 1077 | No | NA | 2 | 0 | 0 | Not-determined | Genome-fragment | NA | NA | 0 |
| S10_vs_27420\|\|lt2gene | 1461 | No | NA | 2 | 0 | 0 | Low-quality | Genome-fragment | 3.84 | AAI-based (high-confidence) | 0 |
| S12_vs_k141_24348\|\|full | 1461 | No | NA | 4 | 0 | 0 | Low-quality | Genome-fragment | 2.55 | AAI-based (medium-confidence) | 0 |
| S8_extracted_k141_21967 | 1461 | No | NA | 2 | 0 | 0 | Low-quality | Genome-fragment | 0.98 | AAI-based (high-confidence) | 0 |
| S1_extracted_k141_43878 | 1229 | No | NA | 1 | 0 | 1 | Not-determined | Genome-fragment | NA | NA | 0 |
| S1_extracted_k141_3007 | 6093 | No | NA | 3 | 0 | 2 | Not-determined | Genome-fragment | NA | NA | 0 |
| S8_vs_k141_41771\|\|full | 1461 | No | NA | 3 | 1 | 0 | Low-quality | Genome-fragment | 28.46 | AAI-based (high-confidence) | 0 |
| S17_vs_k141_18398\|\|full | 1462 | No | NA | 2 | 1 | 0 | Low-quality | Genome-fragment | 3.92 | AAI-based (high-confidence) | 0 |
| S1_extracted_k141_148585 | 3370 | No | NA | 1 | 0 | 1 | Not-determined | Genome-fragment | NA | NA | 0 |
| S1_extracted_k141_143642 | 1156 | No | NA | 1 | 0 | 1 | Not-determined | Genome-fragment | NA | NA | 0 |
| S1_extracted_k141_43913 | 1014 | No | NA | 1 | 0 | 1 | Not-determined | Genome-fragment | NA | NA | 0 |
| S1_extracted_k141_30523 | 1281 | No | NA | 1 | 0 | 0 | Not-determined | Genome-fragment | NA | NA | 0 |
| S1_extracted_k141_70073 | 3036 | No | NA | 1 | 0 | 1 | Not-determined | Genome-fragment | NA | NA | 0 |
| S1_extracted_k141_57208 | 1304 | No | NA | 2 | 0 | 2 | Not-determined | Genome-fragment | NA | NA | 0 |
| S1_extracted_k141_137145 | 1120 | No | NA | 4 | 0 | 3 | Not-determined | Genome-fragment | NA | NA | 0 |
| S1_extracted_k141_97392 | 1083 | No | NA | 1 | 0 | 0 | Not-determined | Genome-fragment | NA | NA | 0 |
| S1_extracted_k141_97399 | 1473 | No | NA | 2 | 0 | 1 | Not-determined | Genome-fragment | NA | NA | 0 |
| S9_extracted_k141_24225 | 1462 | No | NA | 4 | 0 | 0 | Low-quality | Genome-fragment | 2.44 | AAI-based (high-confidence) | 0 |
| S1_extracted_k141_50698 | 3528 | No | NA | 1 | 0 | 1 | Not-determined | Genome-fragment | NA | NA | 0 |
| S1_extracted_k141_50699 | 1396 | No | NA | 2 | 0 | 0 | Not-determined | Genome-fragment | NA | NA | 0 |
| S11_vs_k141_73889\|\|full | 1462 | No | NA | 3 | 0 | 0 | Low-quality | Genome-fragment | 1.13 | AAI-based (medium-confidence) | 0 |
| S1_extracted_k141_117322 | 2463 | No | NA | 2 | 0 | 1 | Not-determined | Genome-fragment | NA | NA | 0 |
| S1_extracted_k141_84012 | 1124 | No | NA | 1 | 0 | 1 | Not-determined | Genome-fragment | NA | NA | 0 |
| S1_extracted_k141_50719 | 2488 | No | NA | 7 | 0 | 6 | Not-determined | Genome-fragment | NA | NA | 0 |
| S14_vs_k141_28905\|\|full | 1462 | No | NA | 2 | 2 | 0 | Low-quality | Genome-fragment | 4.35 | AAI-based (high-confidence) | 0 |
| S1_extracted_k141_155788 | 1737 | No | NA | 4 | 0 | 3 | Not-determined | Genome-fragment | NA | NA | 0 |
| S1_extracted_k141_123695 | 1033 | No | NA | 1 | 0 | 1 | Not-determined | Genome-fragment | NA | NA | 0 |
| S3_extracted_k141_34872 | 1462 | No | NA | 3 | 0 | 1 | Low-quality | Genome-fragment | 4 | AAI-based (medium-confidence) | 0 |
| S5_extracted_k141_25102 | 1462 | No | NA | 2 | 1 | 0 | Low-quality | Genome-fragment | 6.67 | AAI-based (high-confidence) | 0 |
| S1_extracted_k141_155791 | 1472 | No | NA | 1 | 0 | 0 | Not-determined | Genome-fragment | NA | NA | 0 |
| S1_extracted_k141_84068 | 1879 | No | NA | 4 | 0 | 4 | Not-determined | Genome-fragment | NA | NA | 0 |
| S7_vs_421\|\|lt2gene | 1462 | No | NA | 2 | 2 | 0 | Low-quality | Genome-fragment | 3.5 | AAI-based (medium-confidence) | 0 |
| S16_vs_81252\|\|full | 1462 | No | NA | 5 | 1 | 0 | Low-quality | Genome-fragment | 2.52 | AAI-based (high-confidence) | 0 |
| S1_extracted_k141_17738 | 1089 | No | NA | 1 | 0 | 1 | Not-determined | Genome-fragment | NA | NA | 0 |
| S1_extracted_k141_123727 | 1190 | No | NA | 2 | 0 | 2 | Not-determined | Genome-fragment | NA | NA | 0 |
| S2_extracted_k141_834 | 1462 | No | NA | 3 | 3 | 0 | Low-quality | Genome-fragment | 4.82 | AAI-based (high-confidence) | 0 |
| S1_extracted_k141_97528 | 2819 | No | NA | 1 | 0 | 1 | Not-determined | Genome-fragment | NA | NA | 0 |
| S8_extracted_k141_5234 | 1462 | No | NA | 2 | 0 | 1 | Low-quality | Genome-fragment | 0.73 | AAI-based (medium-confidence) | 0 |
| S1_extracted_k141_30690 | 1262 | No | NA | 1 | 0 | 1 | Not-determined | Genome-fragment | NA | NA | 0 |
| S1_extracted_k141_137267 | 3726 | No | NA | 2 | 0 | 0 | Not-determined | Genome-fragment | NA | NA | 0 |
| S1_extracted_k141_110651 | 1309 | No | NA | 2 | 0 | 1 | Not-determined | Genome-fragment | NA | NA | 0 |
| S1_extracted_k141_110652 | 1023 | No | NA | 1 | 0 | 1 | Not-determined | Genome-fragment | NA | NA | 0 |
| S1_extracted_k141_137269 | 1115 | No | NA | 1 | 0 | 0 | Not-determined | Genome-fragment | NA | NA | 0 |
| S8_extracted_k141_121484 | 1462 | No | NA | 2 | 0 | 0 | Low-quality | Genome-fragment | 3.48 | AAI-based (medium-confidence) | 0 |
| S8_vs_k141_57316\|\|full | 1462 | No | NA | 2 | 0 | 2 | Low-quality | Genome-fragment | 0.39 | AAI-based (medium-confidence) | 0 |
| S17_vs_k141_107707\|\|full | 1463 | No | NA | 3 | 0 | 0 | Low-quality | Genome-fragment | 1.88 | AAI-based (medium-confidence) | 0 |
| S9_vs_k141_27356\|\|full | 1463 | No | NA | 2 | 0 | 0 | Low-quality | Genome-fragment | 0.7 | AAI-based (high-confidence) | 0 |
| S1_extracted_k141_57367 | 1357 | No | NA | 2 | 0 | 1 | Not-determined | Genome-fragment | NA | NA | 0 |
| S1_extracted_k141_90621 | 1224 | No | NA | 2 | 0 | 2 | Not-determined | Genome-fragment | NA | NA | 0 |
| S14_vs_k141_46095\|\|full | 1463 | No | NA | 2 | 0 | 0 | Low-quality | Genome-fragment | 2.45 | AAI-based (medium-confidence) | 0 |
| S1_extracted_k141_62742 | 1270 | No | NA | 1 | 0 | 1 | Not-determined | Genome-fragment | NA | NA | 0 |
| S14_vs_k141_11819\|\|full | 1463 | No | NA | 5 | 0 | 1 | Low-quality | Genome-fragment | 1.91 | AAI-based (high-confidence) | 0 |
| S1_extracted_k141_116718 | 1463 | No | NA | 2 | 0 | 0 | Low-quality | Genome-fragment | 2.37 | AAI-based (high-confidence) | 0 |
| S3_vs_k141_76750\|\|full | 1463 | No | NA | 3 | 2 | 0 | Low-quality | Genome-fragment | 3.23 | AAI-based (high-confidence) | 0 |
| S3_vs_k141_93734\|\|full | 1463 | No | NA | 4 | 1 | 0 | Low-quality | Genome-fragment | 1.52 | AAI-based (high-confidence) | 0 |
| S1_extracted_k141_143862 | 2154 | No | NA | 3 | 0 | 3 | Not-determined | Genome-fragment | NA | NA | 0 |
| S5_vs_k141_112100\|\|full | 1463 | No | NA | 2 | 0 | 1 | Low-quality | Genome-fragment | 0.98 | AAI-based (high-confidence) | 0 |
| S1_extracted_k141_17871 | 1164 | No | NA | 3 | 0 | 0 | Not-determined | Genome-fragment | NA | NA | 0 |
| S1_extracted_k141_57459 | 1823 | No | NA | 2 | 0 | 2 | Not-determined | Genome-fragment | NA | NA | 0 |
| S1_extracted_k141_44184 | 1128 | No | NA | 1 | 0 | 1 | Not-determined | Genome-fragment | NA | NA | 0 |
| S1_extracted_k141_97644 | 1097 | No | NA | 1 | 0 | 1 | Not-determined | Genome-fragment | NA | NA | 0 |
| S1_extracted_k141_84213 | 1022 | No | NA | 1 | 0 | 1 | Not-determined | Genome-fragment | NA | NA | 0 |
| S5_vs_k141_29351\|\|full | 1463 | No | NA | 3 | 0 | 0 | Low-quality | Genome-fragment | 2.83 | AAI-based (medium-confidence) | 0 |
| S1_extracted_k141_137412 | 1337 | No | NA | 2 | 0 | 2 | Not-determined | Genome-fragment | NA | NA | 0 |
| S1_extracted_k141_57488 | 1135 | No | NA | 1 | 0 | 0 | Not-determined | Genome-fragment | NA | NA | 0 |
| S1_extracted_k141_130847 | 1836 | No | NA | 2 | 0 | 2 | Not-determined | Genome-fragment | NA | NA | 0 |
| S1_extracted_k141_30848 | 1442 | No | NA | 2 | 0 | 1 | Not-determined | Genome-fragment | NA | NA | 0 |
| S1_extracted_k141_62808 | 1089 | No | NA | 1 | 0 | 1 | Not-determined | Genome-fragment | NA | NA | 0 |
| S1_extracted_k141_143927 | 1929 | No | NA | 3 | 0 | 1 | Not-determined | Genome-fragment | NA | NA | 0 |
| S1_extracted_k141_104236 | 1517 | No | NA | 1 | 0 | 1 | Not-determined | Genome-fragment | NA | NA | 0 |
| S16_vs_140370\|\|full | 1463 | No | NA | 4 | 0 | 0 | Low-quality | Genome-fragment | 3.49 | AAI-based (high-confidence) | 0 |
| S12_vs_k141_116997\|\|full | 1463 | No | NA | 3 | 1 | 0 | Low-quality | Genome-fragment | 3.29 | AAI-based (medium-confidence) | 0 |
| S1_extracted_k141_110833 | 3494 | No | NA | 4 | 0 | 1 | Not-determined | Genome-fragment | NA | NA | 0 |
| S1_extracted_k141_117635 | 1302 | No | NA | 1 | 0 | 1 | Not-determined | Genome-fragment | NA | NA | 0 |
| S1_extracted_k141_117647 | 1003 | No | NA | 2 | 0 | 2 | Not-determined | Genome-fragment | NA | NA | 0 |
| S6_vs_k141_25409\|\|full | 1463 | No | NA | 3 | 0 | 0 | Low-quality | Genome-fragment | 3.17 | AAI-based (medium-confidence) | 0 |
| S1_extracted_k141_90788 | 1299 | No | NA | 3 | 0 | 0 | Not-determined | Genome-fragment | NA | NA | 0 |
| S17_vs_k141_46016\|\|full | 1464 | No | NA | 2 | 0 | 0 | Low-quality | Genome-fragment | 2.85 | AAI-based (medium-confidence) | 0 |
| S1_extracted_k141_117689 | 1085 | No | NA | 1 | 0 | 1 | Not-determined | Genome-fragment | NA | NA | 0 |
| S9_extracted_k141_81991 | 1464 | No | NA | 4 | 0 | 0 | Low-quality | Genome-fragment | 3.6 | AAI-based (medium-confidence) | 0 |
| S9_vs_k141_61809\|\|full | 1464 | No | NA | 3 | 2 | 0 | Low-quality | Genome-fragment | 3.52 | AAI-based (high-confidence) | 0 |
| S1_extracted_k141_62923 | 2083 | No | NA | 1 | 0 | 1 | Not-determined | Genome-fragment | NA | NA | 0 |
| S9_vs_k141_70343\|\|full | 1464 | No | NA | 4 | 4 | 0 | Low-quality | Genome-fragment | 4.18 | AAI-based (medium-confidence) | 0 |
| S1_extracted_k141_51122 | 1038 | No | NA | 2 | 0 | 2 | Not-determined | Genome-fragment | NA | NA | 0 |
| S1_extracted_k141_18041 | 1391 | No | NA | 1 | 0 | 0 | Not-determined | Genome-fragment | NA | NA | 0 |
| S1_extracted_k141_130965 | 2947 | No | NA | 1 | 0 | 1 | Not-determined | Genome-fragment | NA | NA | 0 |
| S1_extracted_k141_90852 | 1229 | No | NA | 1 | 0 | 1 | Not-determined | Genome-fragment | NA | NA | 0 |
| S1_extracted_k141_18056 | 1030 | No | NA | 1 | 0 | 1 | Not-determined | Genome-fragment | NA | NA | 0 |
| S1_extracted_k141_90858 | 1915 | No | NA | 1 | 0 | 1 | Not-determined | Genome-fragment | NA | NA | 0 |
| S1_extracted_k141_62954 | 1052 | No | NA | 1 | 0 | 0 | Not-determined | Genome-fragment | NA | NA | 0 |
| S1_extracted_k141_130991 | 1524 | No | NA | 4 | 0 | 4 | Not-determined | Genome-fragment | NA | NA | 0 |
| S1_extracted_k141_51145 | 1040 | No | NA | 2 | 0 | 1 | Not-determined | Genome-fragment | NA | NA | 0 |
| S1_extracted_k141_131012 | 1943 | No | NA | 2 | 0 | 1 | Not-determined | Genome-fragment | NA | NA | 0 |
| S1_extracted_k141_3369 | 1440 | No | NA | 1 | 0 | 1 | Not-determined | Genome-fragment | NA | NA | 0 |
| S1_extracted_k141_155941 | 1872 | No | NA | 1 | 0 | 1 | Not-determined | Genome-fragment | NA | NA | 0 |
| S13_vs_68533\|\|lt2gene | 1464 | No | NA | 1 | 0 | 0 | Low-quality | Genome-fragment | 3.79 | AAI-based (high-confidence) | 0 |
| S14_vs_k141_15307\|\|full | 1464 | No | NA | 4 | 1 | 0 | Low-quality | Genome-fragment | 3.7 | AAI-based (medium-confidence) | 0 |
| S1_extracted_k141_117780 | 1699 | No | NA | 2 | 0 | 2 | Not-determined | Genome-fragment | NA | NA | 0 |
| S14_vs_k141_78100\|\|full | 1464 | No | NA | 3 | 1 | 0 | Low-quality | Genome-fragment | 2.65 | AAI-based (high-confidence) | 0 |
| S1_extracted_k141_57591 | 3064 | No | NA | 3 | 0 | 0 | Not-determined | Genome-fragment | NA | NA | 0 |
| S1_extracted_k141_31058 | 1016 | No | NA | 3 | 0 | 2 | Not-determined | Genome-fragment | NA | NA | 0 |
| S5_extracted_k141_61990 | 1464 | No | NA | 3 | 0 | 0 | Low-quality | Genome-fragment | 3.13 | AAI-based (high-confidence) | 0 |
| S5_vs_k141_84600\|\|full | 1464 | No | NA | 4 | 0 | 0 | Low-quality | Genome-fragment | 1.99 | AAI-based (high-confidence) | 0 |
| S7_vs_77082\|\|full | 1464 | No | NA | 2 | 0 | 2 | Low-quality | Genome-fragment | 0.79 | AAI-based (medium-confidence) | 0 |
| S16_vs_156482\|\|full | 1464 | No | NA | 2 | 1 | 0 | Low-quality | Genome-fragment | 3.1 | AAI-based (high-confidence) | 0 |
| S1_extracted_k141_97869 | 1250 | No | NA | 1 | 0 | 1 | Not-determined | Genome-fragment | NA | NA | 0 |
| S1_extracted_k141_36772 | 3064 | No | NA | 2 | 0 | 2 | Not-determined | Genome-fragment | NA | NA | 0 |
| S12_vs_k141_73916\|\|full | 1464 | No | NA | 3 | 2 | 0 | Low-quality | Genome-fragment | 3.46 | AAI-based (high-confidence) | 0 |
| S1_extracted_k141_90967 | 1619 | No | NA | 2 | 0 | 1 | Not-determined | Genome-fragment | NA | NA | 0 |
| S8_extracted_k141_145824 | 1464 | No | NA | 2 | 1 | 0 | Low-quality | Genome-fragment | 2.35 | AAI-based (high-confidence) | 0 |
| S1_extracted_k141_36794 | 1005 | No | NA | 1 | 0 | 0 | Not-determined | Genome-fragment | NA | NA | 0 |
| S1_extracted_k141_104449 | 4635 | No | NA | 13 | 0 | 13 | Not-determined | Genome-fragment | NA | NA | 0 |
| S9_vs_k141_7102\|\|full | 1465 | No | NA | 3 | 2 | 0 | Low-quality | Genome-fragment | 3.32 | AAI-based (medium-confidence) | 0 |
| S1_extracted_k141_44479 | 1052 | No | NA | 2 | 0 | 1 | Not-determined | Genome-fragment | NA | NA | 0 |
| S1_extracted_k141_111091 | 1204 | No | NA | 1 | 0 | 1 | Not-determined | Genome-fragment | NA | NA | 0 |
| S1_extracted_k141_36840 | 1078 | No | NA | 2 | 0 | 2 | Not-determined | Genome-fragment | NA | NA | 0 |
| S1_extracted_k141_57667 | 2232 | No | NA | 1 | 0 | 1 | Not-determined | Genome-fragment | NA | NA | 0 |
| S1_extracted_k141_137696 | 1257 | No | NA | 2 | 0 | 2 | Not-determined | Genome-fragment | NA | NA | 0 |
| S1_extracted_k141_31125 | 1041 | No | NA | 2 | 0 | 2 | Not-determined | Genome-fragment | NA | NA | 0 |
| S1_extracted_k141_44512 | 2608 | No | NA | 5 | 0 | 5 | Not-determined | Genome-fragment | NA | NA | 0 |
| S1_extracted_k141_31142 | 3297 | No | NA | 1 | 0 | 1 | Not-determined | Genome-fragment | NA | NA | 0 |
| S1_extracted_k141_77822 | 1114 | No | NA | 1 | 0 | 0 | Not-determined | Genome-fragment | NA | NA | 0 |
| S1_extracted_k141_51325 | 1143 | No | NA | 3 | 0 | 0 | Not-determined | Genome-fragment | NA | NA | 0 |
| S1_extracted_k141_117889 | 1162 | No | NA | 1 | 0 | 1 | Not-determined | Genome-fragment | NA | NA | 0 |
| S1_extracted_k141_104531 | 1974 | No | NA | 2 | 0 | 0 | Not-determined | Genome-fragment | NA | NA | 0 |
| S1_extracted_k141_51368 | 1185 | No | NA | 1 | 0 | 1 | Not-determined | Genome-fragment | NA | NA | 0 |
| S1_extracted_k141_131203 | 1000 | No | NA | 2 | 0 | 2 | Not-determined | Genome-fragment | NA | NA | 0 |
| S1_extracted_k141_91099 | 1320 | No | NA | 1 | 0 | 0 | Not-determined | Genome-fragment | NA | NA | 0 |
| S1_extracted_k141_131220 | 1912 | No | NA | 3 | 0 | 3 | Not-determined | Genome-fragment | NA | NA | 0 |
| S11_vs_k141_76120\|\|full | 1465 | No | NA | 4 | 0 | 0 | Low-quality | Genome-fragment | 3.68 | AAI-based (medium-confidence) | 0 |
| S1_extracted_k141_97996 | 1365 | No | NA | 1 | 0 | 1 | Not-determined | Genome-fragment | NA | NA | 0 |
| S1_extracted_k141_70551 | 1803 | No | NA | 1 | 0 | 0 | Not-determined | Genome-fragment | NA | NA | 0 |
| S1_extracted_k141_70556 | 1273 | No | NA | 1 | 0 | 1 | Not-determined | Genome-fragment | NA | NA | 0 |
| S1_extracted_k141_18319 | 1045 | No | NA | 3 | 0 | 0 | Not-determined | Genome-fragment | NA | NA | 0 |
| S1_extracted_k141_36973 | 1016 | No | NA | 3 | 0 | 0 | Not-determined | Genome-fragment | NA | NA | 0 |
| S1_extracted_k141_18346 | 1098 | No | NA | 2 | 0 | 1 | Not-determined | Genome-fragment | NA | NA | 0 |
| S1_extracted_k141_63229 | 1806 | No | NA | 2 | 0 | 1 | Not-determined | Genome-fragment | NA | NA | 0 |
| S1_extracted_k141_70620 | 2137 | No | NA | 2 | 0 | 2 | Not-determined | Genome-fragment | NA | NA | 0 |
| S3_extracted_k141_91295 | 1465 | No | NA | 1 | 0 | 1 | Low-quality | Genome-fragment | 0.25 | AAI-based (high-confidence) | 0 |
| S1_extracted_k141_44663 | 1338 | No | NA | 1 | 0 | 1 | Not-determined | Genome-fragment | NA | NA | 0 |
| S1_extracted_k141_137886 | 1106 | No | NA | 1 | 0 | 0 | Not-determined | Genome-fragment | NA | NA | 0 |
| S1_extracted_k141_31334 | 2089 | No | NA | 1 | 0 | 1 | Not-determined | Genome-fragment | NA | NA | 0 |
| S1_extracted_k141_84663 | 1473 | No | NA | 3 | 0 | 1 | Not-determined | Genome-fragment | NA | NA | 0 |
| S1_extracted_k141_63260 | 2783 | No | NA | 1 | 0 | 1 | Not-determined | Genome-fragment | NA | NA | 0 |
| S1_extracted_k141_104674 | 1239 | No | NA | 4 | 0 | 1 | Not-determined | Genome-fragment | NA | NA | 0 |
| S3_extracted_k141_47223 | 1465 | No | NA | 1 | 0 | 0 | Low-quality | Genome-fragment | 0.25 | AAI-based (high-confidence) | 0 |
| S5_extracted_k141_45943 | 1465 | No | NA | 3 | 2 | 0 | Low-quality | Genome-fragment | 3.41 | AAI-based (medium-confidence) | 0 |
| S1_extracted_k141_51544 | 1038 | No | NA | 2 | 0 | 0 | Not-determined | Genome-fragment | NA | NA | 0 |
| S1_extracted_k141_118045 | 3160 | No | NA | 1 | 0 | 1 | Not-determined | Genome-fragment | NA | NA | 0 |
| S1_extracted_k141_24400 | 1031 | No | NA | 1 | 0 | 0 | Not-determined | Genome-fragment | NA | NA | 0 |
| S5_vs_k141_34232\|\|lt2gene | 1465 | No | NA | 2 | 2 | 0 | Low-quality | Genome-fragment | 3.1 | AAI-based (high-confidence) | 0 |
| S1_extracted_k141_3648 | 1244 | No | NA | 2 | 0 | 1 | Not-determined | Genome-fragment | NA | NA | 0 |
| S1_extracted_k141_137981 | 1493 | No | NA | 1 | 0 | 1 | Not-determined | Genome-fragment | NA | NA | 0 |
| S1_extracted_k141_124386 | 1550 | No | NA | 1 | 0 | 1 | Not-determined | Genome-fragment | NA | NA | 0 |
| S1_extracted_k141_137997 | 1842 | No | NA | 2 | 0 | 1 | Not-determined | Genome-fragment | NA | NA | 0 |
| S1_extracted_k141_111367 | 1210 | No | NA | 1 | 0 | 1 | Not-determined | Genome-fragment | NA | NA | 0 |
| S1_extracted_k141_78107 | 1087 | No | NA | 1 | 0 | 1 | Not-determined | Genome-fragment | NA | NA | 0 |
| S7_vs_105216\|\|full | 1465 | No | NA | 5 | 0 | 1 | Low-quality | Genome-fragment | 2.1 | AAI-based (medium-confidence) | 0 |
| S1_extracted_k141_148981 | 1468 | No | NA | 1 | 0 | 0 | Not-determined | Genome-fragment | NA | NA | 0 |
| S16_vs_49373\|\|full | 1465 | No | NA | 4 | 0 | 0 | Low-quality | Genome-fragment | 3.02 | AAI-based (medium-confidence) | 0 |
| S1_extracted_k141_138042 | 3254 | No | NA | 4 | 0 | 1 | Not-determined | Genome-fragment | NA | NA | 0 |
| S1_extracted_k141_91354 | 1385 | No | NA | 2 | 0 | 0 | Not-determined | Genome-fragment | NA | NA | 0 |
| S1_extracted_k141_118183 | 2201 | No | NA | 1 | 0 | 1 | Not-determined | Genome-fragment | NA | NA | 0 |
| S10_vs_52443\|\|full | 1465 | No | NA | 2 | 2 | 0 | Low-quality | Genome-fragment | 3.45 | AAI-based (high-confidence) | 0 |
| S1_extracted_k141_58053 | 1428 | No | NA | 1 | 0 | 0 | Not-determined | Genome-fragment | NA | NA | 0 |
| S1_extracted_k141_111439 | 3365 | No | NA | 1 | 0 | 0 | Not-determined | Genome-fragment | NA | NA | 0 |
| S1_extracted_k141_51683 | 2151 | No | NA | 3 | 0 | 3 | Not-determined | Genome-fragment | NA | NA | 0 |
| S1_extracted_k141_78185 | 1262 | No | NA | 3 | 0 | 1 | Not-determined | Genome-fragment | NA | NA | 0 |
| S10_vs_68299\|\|full | 1465 | No | NA | 5 | 1 | 0 | Low-quality | Genome-fragment | 4.67 | AAI-based (high-confidence) | 0 |
| S1_extracted_k141_84840 | 7118 | No | NA | 4 | 0 | 3 | Not-determined | Genome-fragment | NA | NA | 0 |
| S1_extracted_k141_24596 | 1332 | No | NA | 3 | 0 | 2 | Not-determined | Genome-fragment | NA | NA | 0 |
| S12_vs_k141_19554\|\|full | 1465 | No | NA | 2 | 0 | 0 | Low-quality | Genome-fragment | 4.08 | AAI-based (high-confidence) | 0 |
| S1_extracted_k141_149006 | 1193 | No | NA | 1 | 0 | 0 | Not-determined | Genome-fragment | NA | NA | 0 |
| S15_vs_33688\|\|lt2gene | 1465 | No | NA | 2 | 2 | 0 | Low-quality | Genome-fragment | 3.32 | AAI-based (high-confidence) | 0 |
| S8_extracted_k141_53232 | 1465 | No | NA | 3 | 0 | 1 | Low-quality | Genome-fragment | 0.37 | AAI-based (medium-confidence) | 0 |
| S1_extracted_k141_98311 | 1229 | No | NA | 2 | 0 | 1 | Not-determined | Genome-fragment | NA | NA | 0 |
| S8_vs_k141_140236\|\|lt2gene | 1465 | No | NA | 1 | 1 | 0 | Low-quality | Genome-fragment | 3.78 | AAI-based (medium-confidence) | 0 |
| S1_extracted_k141_149033 | 1471 | No | NA | 1 | 0 | 1 | Not-determined | Genome-fragment | NA | NA | 0 |
| S11_extracted_k141_48520 | 1466 | No | NA | 2 | 0 | 2 | Low-quality | Genome-fragment | 1.91 | AAI-based (medium-confidence) | 0 |
| S1_extracted_k141_149037 | 1222 | No | NA | 2 | 0 | 2 | Not-determined | Genome-fragment | NA | NA | 0 |
| S1_extracted_k141_78265 | 1006 | No | NA | 1 | 0 | 1 | Not-determined | Genome-fragment | NA | NA | 0 |
| S1_extracted_k141_11757 | 1007 | No | NA | 2 | 0 | 1 | Not-determined | Genome-fragment | NA | NA | 0 |
| S11_vs_k141_16997\|\|full | 1466 | No | NA | 3 | 1 | 0 | Low-quality | Genome-fragment | 3.92 | AAI-based (medium-confidence) | 0 |
| S1_extracted_k141_111574 | 6376 | No | NA | 4 | 0 | 4 | Not-determined | Genome-fragment | NA | NA | 0 |
| S1_extracted_k141_156225 | 3096 | No | NA | 4 | 0 | 2 | Not-determined | Genome-fragment | NA | NA | 0 |
| S1_extracted_k141_3849 | 1124 | No | NA | 1 | 0 | 0 | Not-determined | Genome-fragment | NA | NA | 0 |
| S1_extracted_k141_58167 | 1194 | No | NA | 1 | 0 | 1 | Not-determined | Genome-fragment | NA | NA | 0 |
| S1_extracted_k141_111588 | 1588 | No | NA | 1 | 0 | 0 | Not-determined | Genome-fragment | NA | NA | 0 |
| S1_extracted_k141_131662 | 1318 | No | NA | 1 | 0 | 1 | Not-determined | Genome-fragment | NA | NA | 0 |
| S1_extracted_k141_44983 | 1040 | No | NA | 2 | 0 | 0 | Not-determined | Genome-fragment | NA | NA | 0 |
| S1_extracted_k141_138157 | 22249 | No | NA | 33 | 13 | 0 | Medium-quality | Genome-fragment | 50.23 | AAI-based (high-confidence) | 0 |
| S1_extracted_k141_18718 | 7642 | No | NA | 7 | 0 | 6 | Not-determined | Genome-fragment | NA | NA | 0 |
| S1_extracted_k141_37250 | 2337 | No | NA | 3 | 0 | 2 | Not-determined | Genome-fragment | NA | NA | 0 |
| S1_extracted_k141_118331 | 1078 | No | NA | 1 | 0 | 1 | Not-determined | Genome-fragment | NA | NA | 0 |
| S1_extracted_k141_144633 | 2427 | No | NA | 1 | 0 | 1 | Not-determined | Genome-fragment | NA | NA | 0 |
| S1_extracted_k141_91596 | 1016 | No | NA | 1 | 0 | 1 | Not-determined | Genome-fragment | NA | NA | 0 |
| S11_vs_k141_39884\|\|lt2gene | 1466 | No | NA | 1 | 1 | 0 | Low-quality | Genome-fragment | 4.56 | AAI-based (high-confidence) | 0 |
| S1_extracted_k141_24772 | 1951 | No | NA | 1 | 0 | 1 | Not-determined | Genome-fragment | NA | NA | 0 |
| S1_extracted_k141_3889 | 1217 | No | NA | 1 | 0 | 0 | Not-determined | Genome-fragment | NA | NA | 0 |
| S1_extracted_k141_78362 | 1265 | No | NA | 2 | 0 | 2 | Not-determined | Genome-fragment | NA | NA | 0 |
| S1_extracted_k141_138196 | 1681 | No | NA | 3 | 0 | 3 | Not-determined | Genome-fragment | NA | NA | 0 |
| S14_extracted_k141_15503 | 1466 | No | NA | 4 | 1 | 0 | Low-quality | Genome-fragment | 4.64 | AAI-based (high-confidence) | 0 |
| S1_extracted_k141_37313 | 1028 | No | NA | 1 | 0 | 0 | Not-determined | Genome-fragment | NA | NA | 0 |
| S1_extracted_k141_63682 | 1063 | No | NA | 2 | 0 | 2 | Not-determined | Genome-fragment | NA | NA | 0 |
| S1_extracted_k141_78386 | 1247 | No | NA | 3 | 0 | 1 | Not-determined | Genome-fragment | NA | NA | 0 |
| S1_extracted_k141_78388 | 1448 | No | NA | 1 | 0 | 1 | Not-determined | Genome-fragment | NA | NA | 0 |
| S1_extracted_k141_149159 | 1612 | No | NA | 3 | 0 | 0 | Not-determined | Genome-fragment | NA | NA | 0 |
| S1_extracted_k141_37327 | 1003 | No | NA | 1 | 0 | 0 | Not-determined | Genome-fragment | NA | NA | 0 |
| S3_extracted_k141_4270 | 1466 | No | NA | 2 | 2 | 0 | Low-quality | Genome-fragment | 3.21 | AAI-based (medium-confidence) | 0 |
| S1_extracted_k141_11934 | 1319 | No | NA | 2 | 0 | 0 | Not-determined | Genome-fragment | NA | NA | 0 |
| S1_extracted_k141_3972 | 1231 | No | NA | 1 | 0 | 1 | Not-determined | Genome-fragment | NA | NA | 0 |
| S5_vs_k141_211\|\|full | 1466 | No | NA | 3 | 0 | 1 | Low-quality | Genome-fragment | 0.76 | AAI-based (medium-confidence) | 0 |
| S5_vs_k141_2572\|\|full | 1466 | No | NA | 2 | 0 | 1 | Low-quality | Genome-fragment | 0.44 | AAI-based (medium-confidence) | 0 |
| S1_extracted_k141_31728 | 1294 | No | NA | 2 | 0 | 0 | Not-determined | Genome-fragment | NA | NA | 0 |
| S1_extracted_k141_85104 | 1270 | No | NA | 1 | 0 | 1 | Not-determined | Genome-fragment | NA | NA | 0 |
| S15_extracted_66367 | 1466 | No | NA | 1 | 0 | 0 | Low-quality | Genome-fragment | 2.38 | AAI-based (high-confidence) | 0 |
| S1_extracted_k141_118488 | 1121 | No | NA | 1 | 0 | 1 | Not-determined | Genome-fragment | NA | NA | 0 |
| S1_extracted_k141_144809 | 1124 | No | NA | 1 | 0 | 1 | Not-determined | Genome-fragment | NA | NA | 0 |
| S1_extracted_k141_63804 | 1369 | No | NA | 1 | 0 | 1 | Not-determined | Genome-fragment | NA | NA | 0 |
| S1_extracted_k141_24941 | 1063 | No | NA | 2 | 0 | 2 | Not-determined | Genome-fragment | NA | NA | 0 |
| S1_extracted_k141_52024 | 3853 | No | NA | 7 | 0 | 0 | Not-determined | Genome-fragment | NA | NA | 0 |
| S1_extracted_k141_78538 | 1126 | No | NA | 3 | 0 | 3 | Not-determined | Genome-fragment | NA | NA | 0 |
| S1_extracted_k141_18971 | 3369 | No | NA | 1 | 0 | 1 | Not-determined | Genome-fragment | NA | NA | 0 |
| S1_extracted_k141_52038 | 1189 | No | NA | 1 | 0 | 0 | Not-determined | Genome-fragment | NA | NA | 0 |
| S1_extracted_k141_52049 | 2445 | No | NA | 3 | 0 | 2 | Not-determined | Genome-fragment | NA | NA | 0 |
| S1_extracted_k141_58419 | 1028 | No | NA | 1 | 0 | 1 | Not-determined | Genome-fragment | NA | NA | 0 |
| S1_extracted_k141_149230 | 2043 | No | NA | 1 | 0 | 1 | Not-determined | Genome-fragment | NA | NA | 0 |
| S1_extracted_k141_24978 | 1473 | No | NA | 1 | 0 | 0 | Not-determined | Genome-fragment | NA | NA | 0 |
| S6_extracted_k141_51308 | 1466 | No | NA | 1 | 0 | 0 | Low-quality | Genome-fragment | 2.47 | AAI-based (high-confidence) | 0 |
| S1_extracted_k141_111848 | 2320 | No | NA | 1 | 0 | 1 | Not-determined | Genome-fragment | NA | NA | 0 |
| S1_extracted_k141_31822 | 1453 | No | NA | 2 | 0 | 2 | Not-determined | Genome-fragment | NA | NA | 0 |
| S1_extracted_k141_58440 | 3443 | No | NA | 3 | 0 | 0 | Not-determined | Genome-fragment | NA | NA | 0 |
| S1_extracted_k141_52080 | 1271 | No | NA | 1 | 0 | 1 | Not-determined | Genome-fragment | NA | NA | 0 |
| S1_extracted_k141_138338 | 1089 | No | NA | 2 | 0 | 1 | Not-determined | Genome-fragment | NA | NA | 0 |
| S1_extracted_k141_12072 | 1642 | No | NA | 1 | 0 | 1 | Not-determined | Genome-fragment | NA | NA | 0 |
| S8_vs_k141_138428\|\|lt2gene | 1466 | No | NA | 1 | 1 | 0 | Low-quality | Genome-fragment | 1.92 | HMM-based (lower-bound) | 0 |
| S17_vs_k141_127606\|\|full | 1467 | No | NA | 4 | 1 | 0 | Low-quality | Genome-fragment | 2.52 | AAI-based (high-confidence) | 0 |
| S1_extracted_k141_144915 | 1163 | No | NA | 1 | 0 | 0 | Not-determined | Genome-fragment | NA | NA | 0 |
| S1_extracted_k141_131996 | 1370 | No | NA | 3 | 0 | 0 | Not-determined | Genome-fragment | NA | NA | 0 |
| S1_extracted_k141_91890 | 1170 | No | NA | 1 | 0 | 1 | Not-determined | Genome-fragment | NA | NA | 0 |
| S14_extracted_k141_47220 | 1467 | No | NA | 4 | 3 | 0 | Low-quality | Genome-fragment | 4.84 | AAI-based (high-confidence) | 0 |
| S1_extracted_k141_4143 | 1762 | No | NA | 1 | 0 | 1 | Not-determined | Genome-fragment | NA | NA | 0 |
| S5_vs_k141_107933\|\|full | 1467 | No | NA | 3 | 0 | 1 | Low-quality | Genome-fragment | 0.68 | AAI-based (medium-confidence) | 0 |
| S7_vs_85869\|\|full | 1467 | No | NA | 2 | 1 | 0 | Low-quality | Genome-fragment | 2.19 | HMM-based (lower-bound) | 0 |
| S1_extracted_k141_25064 | 1077 | No | NA | 1 | 0 | 1 | Not-determined | Genome-fragment | NA | NA | 0 |
| S12_vs_k141_95655\|\|full | 1467 | No | NA | 3 | 3 | 0 | Low-quality | Genome-fragment | 1.96 | AAI-based (medium-confidence) | 0 |
| S1_extracted_k141_98744 | 2898 | No | NA | 2 | 0 | 2 | Not-determined | Genome-fragment | NA | NA | 0 |
| S12_vs_k141_1479\|\|full | 1467 | No | NA | 3 | 2 | 0 | Low-quality | Genome-fragment | 4.04 | AAI-based (high-confidence) | 0 |
| S1_extracted_k141_85261 | 2813 | No | NA | 1 | 0 | 1 | Not-determined | Genome-fragment | NA | NA | 0 |
| S1_extracted_k141_111915 | 3872 | No | NA | 4 | 0 | 3 | Not-determined | Genome-fragment | NA | NA | 0 |
| S1_extracted_k141_37502 | 1577 | No | NA | 1 | 0 | 0 | Not-determined | Genome-fragment | NA | NA | 0 |
| S12_vs_k141_69563\|\|full | 1467 | No | NA | 2 | 0 | 1 | Low-quality | Genome-fragment | 4.68 | AAI-based (medium-confidence) | 0 |
| S9_vs_k141_72464\|\|full | 1468 | No | NA | 3 | 2 | 0 | Low-quality | Genome-fragment | 1.97 | HMM-based (lower-bound) | 0 |
| S1_extracted_k141_118666 | 2738 | No | NA | 1 | 0 | 1 | Not-determined | Genome-fragment | NA | NA | 0 |
| S1_extracted_k141_105361 | 1851 | No | NA | 2 | 0 | 1 | Not-determined | Genome-fragment | NA | NA | 0 |
| S14_vs_k141_112128\|\|full | 1468 | No | NA | 4 | 0 | 0 | Low-quality | Genome-fragment | 3.18 | AAI-based (high-confidence) | 0 |
| S1_extracted_k141_95967 | 1468 | No | NA | 2 | 0 | 0 | Low-quality | Genome-fragment | 2.14 | AAI-based (medium-confidence) | 0 |
| S7_extracted_22000 | 1468 | No | NA | 3 | 0 | 0 | Low-quality | Genome-fragment | 2.81 | AAI-based (high-confidence) | 0 |
| S1_extracted_k141_25122 | 3073 | No | NA | 2 | 0 | 1 | Not-determined | Genome-fragment | NA | NA | 0 |
| S1_extracted_k141_138490 | 2185 | No | NA | 3 | 0 | 3 | Not-determined | Genome-fragment | NA | NA | 0 |
| S1_extracted_k141_31945 | 1295 | No | NA | 1 | 0 | 1 | Not-determined | Genome-fragment | NA | NA | 0 |
| S1_extracted_k141_105397 | 1481 | No | NA | 2 | 0 | 1 | Not-determined | Genome-fragment | NA | NA | 0 |
| S15_vs_10868\|\|full | 1468 | No | NA | 4 | 0 | 0 | Low-quality | Genome-fragment | 3.73 | AAI-based (high-confidence) | 0 |
| S1_extracted_k141_98804 | 1771 | No | NA | 1 | 0 | 0 | Not-determined | Genome-fragment | NA | NA | 0 |
| S17_vs_k141_151310\|\|full | 1469 | No | NA | 4 | 1 | 0 | Low-quality | Genome-fragment | 3.36 | AAI-based (high-confidence) | 0 |
| S17_vs_k141_139415\|\|full | 1469 | No | NA | 4 | 0 | 2 | Low-quality | Genome-fragment | 2.83 | AAI-based (medium-confidence) | 0 |
| S1_extracted_k141_12221 | 1327 | No | NA | 2 | 0 | 2 | Not-determined | Genome-fragment | NA | NA | 0 |
| S1_extracted_k141_31977 | 1114 | No | NA | 1 | 0 | 1 | Not-determined | Genome-fragment | NA | NA | 0 |
| S1_extracted_k141_118712 | 1090 | No | NA | 2 | 0 | 0 | Not-determined | Genome-fragment | NA | NA | 0 |
| S1_extracted_k141_71050 | 1719 | No | NA | 1 | 0 | 1 | Not-determined | Genome-fragment | NA | NA | 0 |
| S14_vs_k141_56044\|\|full | 1469 | No | NA | 3 | 0 | 1 | Low-quality | Genome-fragment | 2.77 | AAI-based (high-confidence) | 0 |
| S8_extracted_k141_139205 | 1469 | No | NA | 2 | 0 | 1 | Low-quality | Genome-fragment | 1.67 | AAI-based (medium-confidence) | 0 |
| S1_extracted_k141_145030 | 3532 | No | NA | 4 | 0 | 3 | Not-determined | Genome-fragment | NA | NA | 0 |
| S1_extracted_k141_138536 | 1189 | No | NA | 1 | 0 | 1 | Not-determined | Genome-fragment | NA | NA | 0 |
| S1_extracted_k141_45421 | 1921 | No | NA | 1 | 0 | 0 | Not-determined | Genome-fragment | NA | NA | 0 |
| S1_extracted_k141_32015 | 1011 | No | NA | 1 | 0 | 1 | Not-determined | Genome-fragment | NA | NA | 0 |
| S1_extracted_k141_12256 | 1717 | No | NA | 1 | 0 | 1 | Not-determined | Genome-fragment | NA | NA | 0 |
| S8_vs_k141_122457\|\|full | 1469 | No | NA | 2 | 0 | 0 | Low-quality | Genome-fragment | 3.08 | AAI-based (medium-confidence) | 0 |
| S1_extracted_k141_118757 | 1287 | No | NA | 1 | 0 | 1 | Not-determined | Genome-fragment | NA | NA | 0 |
| S8_vs_k141_112621\|\|full | 1469 | No | NA | 3 | 0 | 2 | Low-quality | Genome-fragment | 0.44 | AAI-based (medium-confidence) | 0 |
| S9_vs_k141_4865\|\|full | 1470 | No | NA | 3 | 2 | 0 | Low-quality | Genome-fragment | 3.19 | AAI-based (high-confidence) | 0 |
| S1_extracted_k141_138575 | 1233 | No | NA | 2 | 0 | 0 | Not-determined | Genome-fragment | NA | NA | 0 |
| S1_extracted_k141_32063 | 1184 | No | NA | 1 | 0 | 0 | Not-determined | Genome-fragment | NA | NA | 0 |
| S1_extracted_k141_118773 | 1145 | No | NA | 2 | 0 | 0 | Not-determined | Genome-fragment | NA | NA | 0 |
| S1_extracted_k141_92082 | 1053 | No | NA | 1 | 0 | 0 | Not-determined | Genome-fragment | NA | NA | 0 |
| S1_extracted_k141_58684 | 1077 | No | NA | 2 | 0 | 2 | Not-determined | Genome-fragment | NA | NA | 0 |
| S11_extracted_k141_68324 | 1470 | No | NA | 2 | 0 | 0 | Low-quality | Genome-fragment | 7.08 | AAI-based (high-confidence) | 0 |
| S1_extracted_k141_138591 | 3067 | No | NA | 5 | 0 | 5 | Not-determined | Genome-fragment | NA | NA | 0 |
| S13_vs_34708\|\|full | 1470 | No | NA | 5 | 1 | 0 | Low-quality | Genome-fragment | 2.51 | AAI-based (medium-confidence) | 0 |
| S1_extracted_k141_145085 | 1262 | No | NA | 1 | 0 | 1 | Not-determined | Genome-fragment | NA | NA | 0 |
| S1_extracted_k141_98878 | 1408 | No | NA | 1 | 0 | 0 | Not-determined | Genome-fragment | NA | NA | 0 |
| S13_vs_27970\|\|lt2gene | 1470 | No | NA | 2 | 0 | 1 | Low-quality | Genome-fragment | 3.34 | AAI-based (medium-confidence) | 0 |
| S1_extracted_k141_112099 | 1096 | No | NA | 2 | 0 | 0 | Not-determined | Genome-fragment | NA | NA | 0 |
| S3_extracted_k141_28089 | 1470 | No | NA | 3 | 0 | 1 | Low-quality | Genome-fragment | 0.74 | AAI-based (high-confidence) | 0 |
| S3_vs_k141_19681\|\|full | 1470 | No | NA | 6 | 0 | 0 | Low-quality | Genome-fragment | 1.87 | AAI-based (high-confidence) | 0 |
| S10_extracted_11536 | 1470 | No | NA | 2 | 1 | 0 | Low-quality | Genome-fragment | 1.35 | AAI-based (high-confidence) | 0 |
| S1_extracted_k141_32157 | 1096 | No | NA | 2 | 0 | 1 | Not-determined | Genome-fragment | NA | NA | 0 |
| S10_vs_28643\|\|lt2gene | 1470 | No | NA | 2 | 1 | 0 | Low-quality | Genome-fragment | 3.3 | AAI-based (high-confidence) | 0 |
| S1_extracted_k141_12378 | 1481 | No | NA | 1 | 0 | 1 | Not-determined | Genome-fragment | NA | NA | 0 |
| S12_extracted_k141_175821 | 1470 | No | NA | 3 | 3 | 0 | Low-quality | Genome-fragment | 2.84 | AAI-based (high-confidence) | 0 |
| S1_extracted_k141_12406 | 1172 | No | NA | 1 | 0 | 0 | Not-determined | Genome-fragment | NA | NA | 0 |
| S15_extracted_60755 | 1470 | No | NA | 1 | 0 | 1 | Low-quality | Genome-fragment | 4.15 | AAI-based (medium-confidence) | 0 |
| S1_extracted_k141_112147 | 5829 | No | NA | 2 | 0 | 2 | Not-determined | Genome-fragment | NA | NA | 0 |
| S1_extracted_k141_118907 | 3887 | No | NA | 2 | 0 | 2 | Not-determined | Genome-fragment | NA | NA | 0 |
| S1_extracted_k141_32211 | 1201 | No | NA | 1 | 0 | 1 | Not-determined | Genome-fragment | NA | NA | 0 |

| | | | | | | | | | | | |
|---|---|---|---|---|---|---|---|---|---|---|---|
| S1_extracted_k141_37651 | 1590 | No | NA | 2 | 0 | 1 | Not-determined | Genome-fragment | NA | NA | 0 |
| S6_vs_k141_27310\|\|full | 1470 | No | NA | 4 | 3 | 0 | Low-quality | Genome-fragment | 3.66 | AAI-based (high-confidence) | 0 |
| S8_vs_k141_81071\|\|full | 1470 | No | NA | 4 | 1 | 0 | Low-quality | Genome-fragment | 2.44 | AAI-based (high-confidence) | 0 |
| S1_extracted_k141_105605 | 2271 | No | NA | 2 | 0 | 1 | Not-determined | Genome-fragment | NA | NA | 0 |
| S1_extracted_k141_99022 | 1046 | No | NA | 2 | 0 | 0 | Not-determined | Genome-fragment | NA | NA | 0 |
| S17_vs_k141_68115\|\|full | 1471 | No | NA | 3 | 0 | 0 | Low-quality | Genome-fragment | 3.26 | AAI-based (medium-confidence) | 0 |
| S1_extracted_k141_156493 | 1540 | No | NA | 1 | 0 | 1 | Not-determined | Genome-fragment | NA | NA | 0 |
| S1_extracted_k141_125313 | 1401 | No | NA | 1 | 0 | 1 | Not-determined | Genome-fragment | NA | NA | 0 |
| S1_extracted_k141_145230 | 1626 | No | NA | 3 | 0 | 0 | Not-determined | Genome-fragment | NA | NA | 0 |
| S1_extracted_k141_25397 | 1599 | No | NA | 2 | 0 | 1 | Not-determined | Genome-fragment | NA | NA | 0 |
| S3_vs_k141_83743\|\|full | 1471 | No | NA | 3 | 0 | 2 | Low-quality | Genome-fragment | 2.37 | AAI-based (medium-confidence) | 0 |
| S1_extracted_k141_125321 | 1733 | No | NA | 1 | 0 | 1 | Not-determined | Genome-fragment | NA | NA | 0 |
| S1_extracted_k141_45665 | 4349 | No | NA | 2 | 0 | 2 | Not-determined | Genome-fragment | NA | NA | 0 |
| S1_extracted_k141_138728 | 1261 | No | NA | 2 | 0 | 0 | Not-determined | Genome-fragment | NA | NA | 0 |
| S12_vs_k141_186880\|\|full | 1471 | No | NA | 2 | 1 | 0 | Low-quality | Genome-fragment | 2.51 | AAI-based (high-confidence) | 0 |
| S1_extracted_k141_64357 | 1262 | No | NA | 1 | 0 | 1 | Not-determined | Genome-fragment | NA | NA | 0 |
| S1_extracted_k141_112259 | 1228 | No | NA | 2 | 0 | 2 | Not-determined | Genome-fragment | NA | NA | 0 |
| S1_extracted_k141_156547 | 1098 | No | NA | 1 | 0 | 1 | Not-determined | Genome-fragment | NA | NA | 0 |
| S1_extracted_k141_99107 | 1624 | No | NA | 1 | 0 | 1 | Not-determined | Genome-fragment | NA | NA | 0 |
| S1_extracted_k141_32343 | 1591 | No | NA | 3 | 0 | 2 | Not-determined | Genome-fragment | NA | NA | 0 |
| S1_extracted_k141_125396 | 1062 | No | NA | 1 | 0 | 1 | Not-determined | Genome-fragment | NA | NA | 0 |
| S1_extracted_k141_25490 | 2578 | No | NA | 1 | 0 | 1 | Not-determined | Genome-fragment | NA | NA | 0 |
| S1_extracted_k141_45764 | 3065 | No | NA | 1 | 0 | 1 | Not-determined | Genome-fragment | NA | NA | 0 |
| S1_extracted_k141_19520 | 1508 | No | NA | 1 | 0 | 1 | Not-determined | Genome-fragment | NA | NA | 0 |
| S1_extracted_k141_119050 | 3922 | No | NA | 3 | 0 | 2 | Not-determined | Genome-fragment | NA | NA | 0 |
| S1_extracted_k141_92360 | 1143 | No | NA | 1 | 0 | 1 | Not-determined | Genome-fragment | NA | NA | 0 |
| S6_extracted_k141_53314 | 1471 | No | NA | 2 | 2 | 0 | Low-quality | Genome-fragment | 3.52 | AAI-based (medium-confidence) | 0 |
| S8_vs_k141_185530\|\|full | 1471 | No | NA | 3 | 1 | 0 | Low-quality | Genome-fragment | 4.4 | HMM-based (lower-bound) | 0 |
| S1_extracted_k141_85754 | 1007 | No | NA | 1 | 0 | 0 | Not-determined | Genome-fragment | NA | NA | 0 |
| S1_extracted_k141_19545 | 1698 | No | NA | 2 | 0 | 1 | Not-determined | Genome-fragment | NA | NA | 0 |
| S17_vs_k141_122245\|\|full | 1472 | No | NA | 5 | 0 | 0 | Low-quality | Genome-fragment | 3.06 | AAI-based (high-confidence) | 0 |
| S1_extracted_k141_125466 | 1155 | No | NA | 2 | 0 | 0 | Not-determined | Genome-fragment | NA | NA | 0 |
| S9_vs_k141_73597\|\|full | 1472 | No | NA | 3 | 2 | 0 | Low-quality | Genome-fragment | 3.25 | AAI-based (high-confidence) | 0 |
| S9_vs_k141_7024\|\|full | 1472 | No | NA | 4 | 0 | 0 | Low-quality | Genome-fragment | 3.99 | AAI-based (high-confidence) | 0 |
| S1_extracted_k141_138858 | 1518 | No | NA | 1 | 0 | 1 | Not-determined | Genome-fragment | NA | NA | 0 |
| S3_vs_k141_52680\|\|full | 1472 | No | NA | 4 | 0 | 0 | Low-quality | Genome-fragment | 3.91 | AAI-based (high-confidence) | 0 |
| S1_extracted_k141_32448 | 1707 | No | NA | 1 | 0 | 1 | Not-determined | Genome-fragment | NA | NA | 0 |
| S1_extracted_k141_112384 | 3998 | No | NA | 4 | 0 | 3 | Not-determined | Genome-fragment | NA | NA | 0 |
| S5_extracted_k141_44642 | 1472 | No | NA | 4 | 0 | 0 | Low-quality | Genome-fragment | 2.32 | AAI-based (high-confidence) | 0 |
| S1_extracted_k141_132526 | 1232 | No | NA | 1 | 0 | 0 | Not-determined | Genome-fragment | NA | NA | 0 |
| S7_vs_3942\|\|full | 1472 | No | NA | 3 | 0 | 0 | Low-quality | Genome-fragment | 1.44 | AAI-based (high-confidence) | 0 |
| S7_vs_49690\|\|lt2gene | 1472 | No | NA | 2 | 2 | 0 | Low-quality | Genome-fragment | 2.52 | HMM-based (lower-bound) | 0 |
| S1_extracted_k141_119127 | 3274 | No | NA | 4 | 0 | 3 | Not-determined | Genome-fragment | NA | NA | 0 |
| S1_extracted_k141_145408 | 2024 | No | NA | 2 | 0 | 0 | Not-determined | Genome-fragment | NA | NA | 0 |
| S1_extracted_k141_25593 | 1004 | No | NA | 2 | 0 | 1 | Not-determined | Genome-fragment | NA | NA | 0 |
| S1_extracted_k141_19636 | 1605 | No | NA | 2 | 0 | 1 | Not-determined | Genome-fragment | NA | NA | 0 |
| S16_vs_121040\|\|full | 1472 | No | NA | 3 | 0 | 1 | Low-quality | Genome-fragment | 1.69 | AAI-based (high-confidence) | 0 |
| S16_vs_131978\|\|full | 1472 | No | NA | 3 | 0 | 0 | Low-quality | Genome-fragment | 2.4 | AAI-based (high-confidence) | 0 |
| S1_extracted_k141_99246 | 1987 | No | NA | 1 | 0 | 1 | Not-determined | Genome-fragment | NA | NA | 0 |
| S1_extracted_k141_25606 | 1329 | No | NA | 2 | 0 | 2 | Not-determined | Genome-fragment | NA | NA | 0 |
| S1_extracted_k141_25615 | 1755 | No | NA | 2 | 0 | 0 | Not-determined | Genome-fragment | NA | NA | 0 |
| S2_vs_k141_16349\|\|full | 1472 | No | NA | 3 | 0 | 0 | Low-quality | Genome-fragment | 1.53 | AAI-based (high-confidence) | 0 |
| S1_extracted_k141_59103 | 1101 | No | NA | 1 | 0 | 1 | Not-determined | Genome-fragment | NA | NA | 0 |
| S1_extracted_k141_19676 | 3625 | No | NA | 2 | 0 | 2 | Not-determined | Genome-fragment | NA | NA | 0 |
| S8_extracted_k141_53382 | 1472 | No | NA | 2 | 0 | 2 | Low-quality | Genome-fragment | 0.49 | AAI-based (medium-confidence) | 0 |
| S1_extracted_k141_19693 | 1090 | No | NA | 1 | 0 | 1 | Not-determined | Genome-fragment | NA | NA | 0 |
| S1_extracted_k141_92482 | 1472 | No | NA | 1 | 0 | 0 | Not-determined | Genome-fragment | NA | NA | 0 |
| S1_extracted_k141_138965 | 3393 | No | NA | 3 | 0 | 2 | Not-determined | Genome-fragment | NA | NA | 0 |
| S8_vs_k141_188400\|\|full | 1472 | No | NA | 4 | 2 | 0 | Low-quality | Genome-fragment | 2.48 | AAI-based (high-confidence) | 0 |
| S1_extracted_k141_37888 | 1312 | No | NA | 3 | 0 | 0 | Not-determined | Genome-fragment | NA | NA | 0 |
| S9_extracted_k141_83735 | 1473 | No | NA | 2 | 1 | 0 | Low-quality | Genome-fragment | 0.73 | AAI-based (high-confidence) | 0 |
| S9_extracted_k141_29961 | 1473 | No | NA | 2 | 1 | 0 | Low-quality | Genome-fragment | 3.39 | AAI-based (high-confidence) | 0 |
| S1_extracted_k141_32580 | 1148 | No | NA | 1 | 0 | 1 | Not-determined | Genome-fragment | NA | NA | 0 |
| S1_extracted_k141_59161 | 1040 | No | NA | 1 | 0 | 0 | Not-determined | Genome-fragment | NA | NA | 0 |
| S9_vs_k141_48179\|\|full | 1473 | No | NA | 5 | 0 | 0 | Low-quality | Genome-fragment | 2.93 | AAI-based (medium-confidence) | 0 |
| S11_vs_k141_95080\|\|full | 1473 | No | NA | 3 | 0 | 0 | Low-quality | Genome-fragment | 2.75 | AAI-based (medium-confidence) | 0 |
| S1_extracted_k141_37910 | 2215 | No | NA | 1 | 0 | 1 | Not-determined | Genome-fragment | NA | NA | 0 |
| S1_extracted_k141_156686 | 1812 | No | NA | 2 | 0 | 0 | Not-determined | Genome-fragment | NA | NA | 0 |
| S13_vs_22681\|\|full | 1473 | No | NA | 3 | 0 | 0 | Low-quality | Genome-fragment | 2.36 | AAI-based (medium-confidence) | 0 |
| S14_extracted_k141_8498 | 1473 | No | NA | 2 | 2 | 0 | Low-quality | Genome-fragment | 4.89 | AAI-based (high-confidence) | 0 |
| S3_vs_k141_96054\|\|full | 1473 | No | NA | 2 | 0 | 0 | Low-quality | Genome-fragment | 3.23 | AAI-based (medium-confidence) | 0 |
| S1_extracted_k141_59182 | 1870 | No | NA | 2 | 0 | 2 | Not-determined | Genome-fragment | NA | NA | 0 |
| S1_extracted_k141_45939 | 5340 | No | NA | 2 | 0 | 2 | Not-determined | Genome-fragment | NA | NA | 0 |
| S5_extracted_k141_16172 | 1473 | No | NA | 3 | 0 | 0 | Low-quality | Genome-fragment | 3.07 | AAI-based (medium-confidence) | 0 |
| S10_vs_50093\|\|full | 1473 | No | NA | 2 | 2 | 0 | Low-quality | Genome-fragment | 3.12 | AAI-based (high-confidence) | 0 |
| S1_extracted_k141_145531 | 1193 | No | NA | 1 | 0 | 0 | Not-determined | Genome-fragment | NA | NA | 0 |
| S12_vs_k141_15170\|\|full | 1473 | No | NA | 3 | 2 | 0 | Low-quality | Genome-fragment | 2.47 | HMM-based (lower-bound) | 0 |
| S1_extracted_k141_64620 | 4153 | No | NA | 2 | 0 | 2 | Not-determined | Genome-fragment | NA | NA | 0 |
| S1_extracted_k141_12788 | 1162 | No | NA | 1 | 0 | 1 | Not-determined | Genome-fragment | NA | NA | 0 |
| S1_extracted_k141_32637 | 1249 | No | NA | 1 | 0 | 0 | Not-determined | Genome-fragment | NA | NA | 0 |
| S6_vs_k141_232\|\|full | 1473 | No | NA | 3 | 1 | 0 | Low-quality | Genome-fragment | 1.2 | AAI-based (high-confidence) | 0 |
| S1_extracted_k141_125650 | 7776 | No | NA | 11 | 3 | 0 | Medium-quality | Genome-fragment | 56.76 | AAI-based (medium-confidence) | 0 |
| S1_extracted_k141_45982 | 4331 | No | NA | 6 | 0 | 1 | Not-determined | Genome-fragment | NA | NA | 0 |
| S8_vs_k141_138713\|\|full | 1473 | No | NA | 3 | 0 | 1 | Low-quality | Genome-fragment | 1.68 | AAI-based (high-confidence) | 0 |
| S1_extracted_k141_125673 | 1087 | No | NA | 1 | 0 | 1 | Not-determined | Genome-fragment | NA | NA | 0 |
| S1_extracted_k141_25789 | 1266 | No | NA | 1 | 0 | 1 | Not-determined | Genome-fragment | NA | NA | 0 |
| S1_extracted_k141_79459 | 1160 | No | NA | 1 | 0 | 1 | Not-determined | Genome-fragment | NA | NA | 0 |
| S13_extracted_10330 | 1474 | No | NA | 2 | 0 | 0 | Low-quality | Genome-fragment | 2.29 | AAI-based (high-confidence) | 0 |
| S1_extracted_k141_125709 | 1135 | No | NA | 1 | 0 | 1 | Not-determined | Genome-fragment | NA | NA | 0 |
| S1_extracted_k141_25824 | 1282 | No | NA | 1 | 0 | 0 | Not-determined | Genome-fragment | NA | NA | 0 |
| S14_extracted_k141_67335 | 1474 | No | NA | 6 | 5 | 0 | Low-quality | Genome-fragment | 4.86 | AAI-based (high-confidence) | 0 |
| S14_vs_k141_104719\|\|full | 1474 | No | NA | 3 | 1 | 0 | Low-quality | Genome-fragment | 3.16 | AAI-based (medium-confidence) | 0 |
| S1_extracted_k141_149707 | 3213 | No | NA | 1 | 0 | 1 | Not-determined | Genome-fragment | NA | NA | 0 |
| S1_extracted_k141_132812 | 1078 | No | NA | 2 | 0 | 0 | Not-determined | Genome-fragment | NA | NA | 0 |
| S1_extracted_k141_12899 | 4211 | No | NA | 4 | 0 | 4 | Not-determined | Genome-fragment | NA | NA | 0 |
| S1_extracted_k141_25839 | 5226 | No | NA | 10 | 0 | 2 | Not-determined | Genome-fragment | NA | NA | 0 |
| S3_extracted_k141_64850 | 1474 | No | NA | 5 | 3 | 0 | Low-quality | Genome-fragment | 6.73 | AAI-based (high-confidence) | 0 |
| S1_extracted_k141_106095 | 1024 | No | NA | 2 | 0 | 2 | Not-determined | Genome-fragment | NA | NA | 0 |
| S1_extracted_k141_112623 | 1511 | No | NA | 2 | 0 | 1 | Not-determined | Genome-fragment | NA | NA | 0 |
| S3_vs_k141_303\|\|lt2gene | 1474 | No | NA | 2 | 1 | 0 | Low-quality | Genome-fragment | 2.7 | AAI-based (high-confidence) | 0 |
| S1_extracted_k141_4747 | 3279 | No | NA | 1 | 0 | 1 | Not-determined | Genome-fragment | NA | NA | 0 |
| S1_extracted_k141_125782 | 1013 | No | NA | 1 | 0 | 1 | Not-determined | Genome-fragment | NA | NA | 0 |
| S1_extracted_k141_59346 | 13067 | No | NA | 12 | 0 | 3 | Not-determined | Genome-fragment | NA | NA | 0 |
| S1_extracted_k141_59355 | 1764 | No | NA | 1 | 0 | 1 | Not-determined | Genome-fragment | NA | NA | 0 |
| S1_extracted_k141_149767 | 1687 | No | NA | 2 | 0 | 1 | Not-determined | Genome-fragment | NA | NA | 0 |
| S5_extracted_k141_97240 | 1474 | No | NA | 3 | 3 | 0 | Low-quality | Genome-fragment | 3.64 | AAI-based (high-confidence) | 0 |
| S1_extracted_k141_99531 | 1155 | No | NA | 3 | 0 | 0 | Not-determined | Genome-fragment | NA | NA | 0 |
| S7_extracted_55836 | 1474 | No | NA | 2 | 0 | 0 | Low-quality | Genome-fragment | 0.88 | AAI-based (medium-confidence) | 0 |
| S1_extracted_k141_125817 | 2090 | No | NA | 1 | 0 | 0 | Not-determined | Genome-fragment | NA | NA | 0 |
| S1_extracted_k141_38111 | 3055 | No | NA | 2 | 0 | 2 | Not-determined | Genome-fragment | NA | NA | 0 |
| S1_extracted_k141_32810 | 1164 | No | NA | 1 | 0 | 1 | Not-determined | Genome-fragment | NA | NA | 0 |
| S7_vs_111300\|\|full | 1474 | No | NA | 3 | 1 | 0 | Low-quality | Genome-fragment | 3.65 | AAI-based (medium-confidence) | 0 |
| S7_vs_116651\|\|lt2gene | 1474 | No | NA | 1 | 1 | 0 | Low-quality | Genome-fragment | 3.11 | AAI-based (high-confidence) | 0 |
| S1_extracted_k141_149820 | 1006 | No | NA | 1 | 0 | 1 | Not-determined | Genome-fragment | NA | NA | 0 |
| S1_extracted_k141_64810 | 1048 | No | NA | 1 | 0 | 1 | Not-determined | Genome-fragment | NA | NA | 0 |
| S1_extracted_k141_145746 | 1306 | No | NA | 1 | 0 | 1 | Not-determined | Genome-fragment | NA | NA | 0 |
| S16_vs_88343\|\|full | 1474 | No | NA | 4 | 0 | 0 | Low-quality | Genome-fragment | 2.38 | AAI-based (high-confidence) | 0 |
| S12_vs_k141_185295\|\|full | 1474 | No | NA | 3 | 2 | 0 | Low-quality | Genome-fragment | 3.39 | AAI-based (high-confidence) | 0 |
| S1_extracted_k141_125901 | 1122 | No | NA | 1 | 0 | 0 | Not-determined | Genome-fragment | NA | NA | 0 |
| S12_vs_k141_174162\|\|lt2gene | 1474 | No | NA | 1 | 1 | 0 | Low-quality | Genome-fragment | 1.38 | HMM-based (lower-bound) | 0 |
| S6_vs_k141_1103\|\|lt2gene | 1474 | No | NA | 2 | 0 | 0 | Low-quality | Genome-fragment | 3.76 | AAI-based (high-confidence) | 0 |
| S1_extracted_k141_59434 | 5371 | No | NA | 6 | 0 | 5 | Not-determined | Genome-fragment | NA | NA | 0 |
| S8_extracted_k141_175408 | 1474 | No | NA | 1 | 0 | 0 | Low-quality | Genome-fragment | 1.45 | AAI-based (high-confidence) | 0 |
| S1_extracted_k141_149859 | 2142 | No | NA | 2 | 0 | 0 | Not-determined | Genome-fragment | NA | NA | 0 |
| S1_extracted_k141_59437 | 3184 | No | NA | 3 | 0 | 2 | Not-determined | Genome-fragment | NA | NA | 0 |
| S1_extracted_k141_4826 | 7257 | No | NA | 8 | 0 | 5 | Not-determined | Genome-fragment | NA | NA | 0 |
| S1_extracted_k141_112757 | 1198 | No | NA | 1 | 0 | 0 | Not-determined | Genome-fragment | NA | NA | 0 |
| S1_extracted_k141_79650 | 1228 | No | NA | 1 | 0 | 1 | Not-determined | Genome-fragment | NA | NA | 0 |
| S1_extracted_k141_106276 | 2004 | No | NA | 1 | 0 | 1 | Not-determined | Genome-fragment | NA | NA | 0 |
| S1_extracted_k141_106282 | 1636 | No | NA | 2 | 0 | 0 | Not-determined | Genome-fragment | NA | NA | 0 |
| S1_extracted_k141_112790 | 1240 | No | NA | 1 | 0 | 1 | Not-determined | Genome-fragment | NA | NA | 0 |
| S8_vs_k141_157306\|\|full | 1474 | No | NA | 2 | 0 | 0 | Low-quality | Genome-fragment | 0.88 | AAI-based (high-confidence) | 0 |
| S17_extracted_k141_43609 | 1475 | No | NA | 2 | 0 | 0 | Low-quality | Genome-fragment | 2.02 | AAI-based (high-confidence) | 0 |
| S14_vs_k141_33111\|\|full | 1475 | No | NA | 3 | 0 | 0 | Low-quality | Genome-fragment | 4.25 | AAI-based (high-confidence) | 0 |
| S1_extracted_k141_88445 | 1475 | No | NA | 3 | 0 | 0 | Low-quality | Genome-fragment | 3.67 | AAI-based (high-confidence) | 0 |
| S1_vs_k141_20466\|\|full | 1475 | No | NA | 3 | 0 | 0 | Low-quality | Genome-fragment | 1.99 | AAI-based (medium-confidence) | 0 |
| S1_extracted_k141_53184 | 1363 | No | NA | 2 | 0 | 1 | Not-determined | Genome-fragment | NA | NA | 0 |
| S1_extracted_k141_92898 | 1016 | No | NA | 1 | 0 | 1 | Not-determined | Genome-fragment | NA | NA | 0 |
| S1_extracted_k141_64935 | 3263 | No | NA | 1 | 0 | 1 | Not-determined | Genome-fragment | NA | NA | 0 |
| S1_extracted_k141_26069 | 1060 | No | NA | 1 | 0 | 0 | Not-determined | Genome-fragment | NA | NA | 0 |
| S1_extracted_k141_4907 | 1302 | No | NA | 2 | 0 | 1 | Not-determined | Genome-fragment | NA | NA | 0 |
| NC1_vf_k141_2039 | 1475 | No | NA | 3 | 0 | 0 | Low-quality | Genome-fragment | 3.13 | AAI-based (high-confidence) | 0 |
| S10_vs_51675\|\|full | 1475 | No | NA | 3 | 0 | 2 | Low-quality | Genome-fragment | 2.05 | AAI-based (medium-confidence) | 0 |
| S1_extracted_k141_38247 | 1019 | No | NA | 1 | 0 | 1 | Not-determined | Genome-fragment | NA | NA | 0 |
| S12_vs_k141_12437\|\|full | 1475 | No | NA | 3 | 1 | 0 | Low-quality | Genome-fragment | 2.63 | HMM-based (lower-bound) | 0 |
| S12_vs_k141_108751\|\|full | 1475 | No | NA | 2 | 0 | 0 | Low-quality | Genome-fragment | 1.14 | AAI-based (medium-confidence) | 0 |
| S13_vs_63037\|\|full | 1476 | No | NA | 2 | 0 | 0 | Low-quality | Genome-fragment | 2.31 | AAI-based (medium-confidence) | 0 |
| S1_vs_k141_155816\|\|full | 1476 | No | NA | 3 | 0 | 0 | Low-quality | Genome-fragment | 1.84 | AAI-based (medium-confidence) | 0 |
| S1_extracted_k141_13244 | 1058 | No | NA | 1 | 0 | 0 | Not-determined | Genome-fragment | NA | NA | 0 |
| S1_extracted_k141_106403 | 1036 | No | NA | 1 | 0 | 1 | Not-determined | Genome-fragment | NA | NA | 0 |
| S1_extracted_k141_53299 | 3656 | No | NA | 5 | 0 | 1 | Not-determined | Genome-fragment | NA | NA | 0 |
| S1_extracted_k141_119691 | 1602 | No | NA | 2 | 0 | 2 | Not-determined | Genome-fragment | NA | NA | 0 |
| S5_extracted_k141_4794 | 1476 | No | NA | 4 | 1 | 0 | Low-quality | Genome-fragment | 0.85 | HMM-based (lower-bound) | 0 |
| S1_extracted_k141_65027 | 1005 | No | NA | 1 | 0 | 0 | Not-determined | Genome-fragment | NA | NA | 0 |
| NC1_vf_k141_1196 | 1476 | No | NA | 4 | 0 | 0 | Low-quality | Genome-fragment | 3.25 | AAI-based (high-confidence) | 0 |
| S16_extracted_9466 | 1476 | No | NA | 2 | 1 | 0 | Low-quality | Genome-fragment | 3.46 | AAI-based (high-confidence) | 0 |
| S16_extracted_92050 | 1476 | No | NA | 2 | 0 | 0 | Low-quality | Genome-fragment | 1.02 | AAI-based (high-confidence) | 0 |
| S16_vs_17158\|\|full | 1476 | No | NA | 3 | 1 | 0 | Low-quality | Genome-fragment | 2.35 | AAI-based (high-confidence) | 0 |
| S16_vs_171000\|\|full | 1476 | No | NA | 3 | 1 | 0 | Low-quality | Genome-fragment | 3.03 | AAI-based (medium-confidence) | 0 |
| S1_extracted_k141_86477 | 2201 | No | NA | 2 | 0 | 0 | Not-determined | Genome-fragment | NA | NA | 0 |
| S1_extracted_k141_13349 | 1874 | No | NA | 2 | 0 | 2 | Not-determined | Genome-fragment | NA | NA | 0 |
| S1_extracted_k141_93050 | 1517 | No | NA | 2 | 0 | 2 | Not-determined | Genome-fragment | NA | NA | 0 |
| S15_extracted_23928 | 1476 | No | NA | 3 | 0 | 0 | Low-quality | Genome-fragment | 3.23 | AAI-based (medium-confidence) | 0 |
| S8_extracted_k141_75832 | 1476 | No | NA | 3 | 0 | 1 | Low-quality | Genome-fragment | 4.71 | AAI-based (medium-confidence) | 0 |
| S1_extracted_k141_26259 | 1434 | No | NA | 2 | 0 | 1 | Not-determined | Genome-fragment | NA | NA | 0 |
| S1_extracted_k141_145937 | 1593 | No | NA | 1 | 0 | 1 | Not-determined | Genome-fragment | NA | NA | 0 |
| S1_extracted_k141_126200 | 1056 | No | NA | 1 | 0 | 0 | Not-determined | Genome-fragment | NA | NA | 0 |
| S17_extracted_k141_12828 | 1477 | No | NA | 2 | 1 | 0 | Low-quality | Genome-fragment | 2.45 | AAI-based (high-confidence) | 0 |
| S11_extracted_k141_68278 | 1477 | No | NA | 2 | 0 | 0 | Low-quality | Genome-fragment | 3.1 | AAI-based (medium-confidence) | 0 |
| S11_vs_k141_34288\|\|full | 1477 | No | NA | 3 | 1 | 0 | Low-quality | Genome-fragment | 4.47 | AAI-based (medium-confidence) | 0 |
| S11_vs_k141_14637\|\|lt2gene | 1477 | No | NA | 1 | 1 | 0 | Low-quality | Genome-fragment | 1.38 | HMM-based (lower-bound) | 0 |
| S11_vs_k141_43555\|\|lt2gene | 1477 | No | NA | 2 | 2 | 0 | Low-quality | Genome-fragment | 3.83 | AAI-based (high-confidence) | 0 |
| S14_extracted_k141_32728 | 1477 | No | NA | 1 | 1 | 0 | Low-quality | Genome-fragment | 4.94 | AAI-based (high-confidence) | 0 |
| S1_extracted_k141_126210 | 5383 | No | NA | 2 | 0 | 2 | Not-determined | Genome-fragment | NA | NA | 0 |
| S1_extracted_k141_26305 | 1079 | No | NA | 3 | 0 | 0 | Not-determined | Genome-fragment | NA | NA | 0 |
| S1_extracted_k141_133332 | 1231 | No | NA | 4 | 0 | 1 | Not-determined | Genome-fragment | NA | NA | 0 |
| S1_extracted_k141_139654 | 1683 | No | NA | 1 | 0 | 1 | Not-determined | Genome-fragment | NA | NA | 0 |
| S1_extracted_k141_38425 | 1135 | No | NA | 1 | 0 | 1 | Not-determined | Genome-fragment | NA | NA | 0 |
| S1_extracted_k141_139663 | 1307 | No | NA | 1 | 0 | 0 | Not-determined | Genome-fragment | NA | NA | 0 |
| S1_extracted_k141_133352 | 5618 | No | NA | 4 | 0 | 3 | Not-determined | Genome-fragment | NA | NA | 0 |
| S1_extracted_k141_106579 | 1436 | No | NA | 1 | 0 | 1 | Not-determined | Genome-fragment | NA | NA | 0 |
| S1_vs_k141_138411\|\|full | 1477 | No | NA | 2 | 0 | 0 | Low-quality | Genome-fragment | 1.6 | AAI-based (high-confidence) | 0 |
| S16_vs_153993\|\|full | 1477 | No | NA | 3 | 1 | 0 | Low-quality | Genome-fragment | 3.37 | AAI-based (high-confidence) | 0 |
| S1_extracted_k141_139681 | 1530 | No | NA | 3 | 0 | 1 | Not-determined | Genome-fragment | NA | NA | 0 |
| S1_extracted_k141_33287 | 1549 | No | NA | 2 | 0 | 0 | Not-determined | Genome-fragment | NA | NA | 0 |
| S1_extracted_k141_93201 | 1013 | No | NA | 2 | 0 | 0 | Not-determined | Genome-fragment | NA | NA | 0 |
| S1_extracted_k141_133393 | 1320 | No | NA | 1 | 0 | 1 | Not-determined | Genome-fragment | NA | NA | 0 |
| S1_extracted_k141_65246 | 1248 | No | NA | 2 | 0 | 1 | Not-determined | Genome-fragment | NA | NA | 0 |
| S1_extracted_k141_26382 | 1283 | No | NA | 1 | 0 | 1 | Not-determined | Genome-fragment | NA | NA | 0 |
| S1_extracted_k141_139732 | 3395 | No | NA | 2 | 0 | 1 | Not-determined | Genome-fragment | NA | NA | 0 |
| S10_vs_96347\|\|full | 1477 | No | NA | 2 | 1 | 0 | Low-quality | Genome-fragment | 3.97 | AAI-based (high-confidence) | 0 |
| S12_vs_k141_134939\|\|full | 1477 | No | NA | 3 | 1 | 0 | Low-quality | Genome-fragment | 2.81 | AAI-based (high-confidence) | 0 |
| S1_extracted_k141_93270 | 1139 | No | NA | 2 | 0 | 0 | Not-determined | Genome-fragment | NA | NA | 0 |
| S6_vs_k141_1983\|\|full | 1477 | No | NA | 3 | 0 | 1 | Low-quality | Genome-fragment | 2.74 | AAI-based (medium-confidence) | 0 |
| S1_extracted_k141_119936 | 1064 | No | NA | 2 | 0 | 1 | Not-determined | Genome-fragment | NA | NA | 0 |
| S1_extracted_k141_65312 | 4007 | No | NA | 4 | 0 | 2 | Not-determined | Genome-fragment | NA | NA | 0 |
| S1_extracted_k141_146146 | 3672 | No | NA | 4 | 0 | 3 | Not-determined | Genome-fragment | NA | NA | 0 |
| S8_extracted_k141_160940 | 1477 | No | NA | 4 | 1 | 0 | Low-quality | Genome-fragment | 2.52 | AAI-based (high-confidence) | 0 |
| S1_extracted_k141_126373 | 2315 | No | NA | 2 | 0 | 2 | Not-determined | Genome-fragment | NA | NA | 0 |
| S1_extracted_k141_38561 | 1082 | No | NA | 1 | 0 | 1 | Not-determined | Genome-fragment | NA | NA | 0 |
| S1_extracted_k141_65352 | 1088 | No | NA | 2 | 0 | 0 | Not-determined | Genome-fragment | NA | NA | 0 |
| S1_extracted_k141_93375 | 1105 | No | NA | 2 | 0 | 1 | Not-determined | Genome-fragment | NA | NA | 0 |
| S1_extracted_k141_65356 | 1188 | No | NA | 1 | 0 | 0 | Not-determined | Genome-fragment | NA | NA | 0 |
| S8_extracted_k141_187194 | 1477 | No | NA | 3 | 0 | 1 | Low-quality | Genome-fragment | 2.33 | AAI-based (medium-confidence) | 0 |
| S1_extracted_k141_93383 | 2476 | No | NA | 1 | 0 | 1 | Not-determined | Genome-fragment | NA | NA | 0 |
| S1_extracted_k141_71832 | 1243 | No | NA | 1 | 0 | 1 | Not-determined | Genome-fragment | NA | NA | 0 |
| S1_extracted_k141_113310 | 1328 | No | NA | 1 | 0 | 1 | Not-determined | Genome-fragment | NA | NA | 0 |
| S8_vs_k141_113793\|\|full | 1477 | No | NA | 3 | 0 | 1 | Low-quality | Genome-fragment | 3.17 | AAI-based (high-confidence) | 0 |
| S1_extracted_k141_146170 | 1046 | No | NA | 2 | 0 | 1 | Not-determined | Genome-fragment | NA | NA | 0 |
| S1_extracted_k141_113321 | 1198 | No | NA | 1 | 0 | 0 | Not-determined | Genome-fragment | NA | NA | 0 |
| S11_vs_k141_60028\|\|full | 1478 | No | NA | 3 | 0 | 1 | Low-quality | Genome-fragment | 2.07 | AAI-based (medium-confidence) | 0 |
| S1_extracted_k141_126385 | 1104 | No | NA | 3 | 0 | 1 | Not-determined | Genome-fragment | NA | NA | 0 |
| S1_extracted_k141_120021 | 1503 | No | NA | 4 | 0 | 3 | Not-determined | Genome-fragment | NA | NA | 0 |
| S14_vs_k141_62586\|\|full | 1478 | No | NA | 3 | 1 | 0 | Low-quality | Genome-fragment | 3.87 | AAI-based (medium-confidence) | 0 |
| S1_extracted_k141_120035 | 1992 | No | NA | 1 | 0 | 1 | Not-determined | Genome-fragment | NA | NA | 0 |
| S14_vs_k141_12587\|\|full | 1478 | No | NA | 3 | 0 | 2 | Low-quality | Genome-fragment | 2.48 | AAI-based (medium-confidence) | 0 |
| S1_extracted_k141_150116 | 2244 | No | NA | 1 | 0 | 0 | Not-determined | Genome-fragment | NA | NA | 0 |
| S1_extracted_k141_26545 | 1144 | No | NA | 1 | 0 | 0 | Not-determined | Genome-fragment | NA | NA | 0 |
| S1_extracted_k141_150124 | 1002 | No | NA | 2 | 0 | 0 | Not-determined | Genome-fragment | NA | NA | 0 |
| S1_vs_k141_27448\|\|full | 1478 | No | NA | 4 | 0 | 0 | Low-quality | Genome-fragment | 3.16 | AAI-based (high-confidence) | 0 |
| S1_extracted_k141_65550 | 1781 | No | NA | 1 | 0 | 0 | Not-determined | Genome-fragment | NA | NA | 0 |
| S1_extracted_k141_146379 | 1542 | No | NA | 4 | 0 | 0 | Not-determined | Genome-fragment | NA | NA | 0 |
| S1_extracted_k141_38665 | 1926 | No | NA | 1 | 0 | 0 | Not-determined | Genome-fragment | NA | NA | 0 |
| S1_extracted_k141_38705 | 1340 | No | NA | 2 | 0 | 1 | Not-determined | Genome-fragment | NA | NA | 0 |
| S1_extracted_k141_5440 | 1536 | No | NA | 1 | 0 | 0 | Not-determined | Genome-fragment | NA | NA | 0 |
| S1_extracted_k141_65660 | 1104 | No | NA | 2 | 0 | 0 | Not-determined | Genome-fragment | NA | NA | 0 |
| S1_extracted_k141_150175 | 1897 | No | NA | 1 | 0 | 0 | Not-determined | Genome-fragment | NA | NA | 0 |
| S1_extracted_k141_38755 | 1735 | No | NA | 1 | 0 | 0 | Not-determined | Genome-fragment | NA | NA | 0 |
| S1_extracted_k141_140114 | 1055 | No | NA | 2 | 0 | 1 | Not-determined | Genome-fragment | NA | NA | 0 |
| S1_extracted_k141_65711 | 1183 | No | NA | 1 | 0 | 1 | Not-determined | Genome-fragment | NA | NA | 0 |
| S5_vs_k141_55372\|\|full | 1478 | No | NA | 3 | 0 | 0 | Low-quality | Genome-fragment | 2.87 | AAI-based (medium-confidence) | 0 |
| S1_extracted_k141_126677 | 1135 | No | NA | 2 | 0 | 1 | Not-determined | Genome-fragment | NA | NA | 0 |
| S1_extracted_k141_65767 | 1661 | No | NA | 2 | 0 | 1 | Not-determined | Genome-fragment | NA | NA | 0 |
| S5_vs_k141_1761\|\|full | 1478 | No | NA | 3 | 0 | 0 | Low-quality | Genome-fragment | 0.98 | AAI-based (medium-confidence) | 0 |
| S1_extracted_k141_5542 | 3754 | No | NA | 1 | 0 | 0 | Not-determined | Genome-fragment | NA | NA | 0 |
| S7_vs_6806\|\|full | 1478 | No | NA | 4 | 2 | 0 | Low-quality | Genome-fragment | 3.14 | AAI-based (medium-confidence) | 0 |
| S7_vs_39792\|\|full | 1478 | No | NA | 2 | 0 | 1 | Low-quality | Genome-fragment | 1.75 | AAI-based (medium-confidence) | 0 |
| S1_extracted_k141_126805 | 1920 | No | NA | 3 | 0 | 0 | Not-determined | Genome-fragment | NA | NA | 0 |
| S16_vs_8497\|\|full | 1478 | No | NA | 3 | 0 | 0 | Low-quality | Genome-fragment | 3 | AAI-based (medium-confidence) | 0 |
| S1_extracted_k141_65912 | 1834 | No | NA | 1 | 0 | 0 | Not-determined | Genome-fragment | NA | NA | 0 |
| S14_vs_k141_37210\|\|lt2gene | 1479 | No | NA | 2 | 2 | 0 | Low-quality | Genome-fragment | 2.74 | HMM-based (lower-bound) | 0 |
| S3_extracted_k141_81207 | 1479 | No | NA | 2 | 2 | 0 | Low-quality | Genome-fragment | 3.93 | AAI-based (high-confidence) | 0 |
| S1_extracted_k141_146819 | 1229 | No | NA | 2 | 0 | 0 | Not-determined | Genome-fragment | NA | NA | 0 |
| S1_extracted_k141_66029 | 1317 | No | NA | 2 | 0 | 1 | Not-determined | Genome-fragment | NA | NA | 0 |
| S1_extracted_k141_5759 | 3324 | No | NA | 1 | 0 | 1 | Not-determined | Genome-fragment | NA | NA | 0 |
| S1_extracted_k141_157421 | 1172 | No | NA | 1 | 0 | 0 | Not-determined | Genome-fragment | NA | NA | 0 |
| S1_extracted_k141_66193 | 1001 | No | NA | 3 | 0 | 0 | Not-determined | Genome-fragment | NA | NA | 0 |
| S1_extracted_k141_66254 | 1541 | No | NA | 2 | 0 | 2 | Not-determined | Genome-fragment | NA | NA | 0 |
| S3_vs_k141_107387\|\|full | 1479 | No | NA | 3 | 0 | 0 | Low-quality | Genome-fragment | 1.22 | AAI-based (high-confidence) | 0 |
| S1_extracted_k141_39177 | 1116 | No | NA | 2 | 0 | 1 | Not-determined | Genome-fragment | NA | NA | 0 |
| S1_extracted_k141_66296 | 3090 | No | NA | 3 | 0 | 1 | Not-determined | Genome-fragment | NA | NA | 0 |
| S1_extracted_k141_5947 | 1703 | No | NA | 1 | 0 | 0 | Not-determined | Genome-fragment | NA | NA | 0 |
| S3_vs_k141_49868\|\|lt2gene | 1479 | No | NA | 2 | 2 | 0 | Low-quality | Genome-fragment | 4.4 | AAI-based (high-confidence) | 0 |
| S1_extracted_k141_66370 | 1148 | No | NA | 1 | 0 | 0 | Not-determined | Genome-fragment | NA | NA | 0 |
| S1_extracted_k141_39238 | 1177 | No | NA | 2 | 0 | 0 | Not-determined | Genome-fragment | NA | NA | 0 |
| S16_vs_72346\|\|full | 1479 | No | NA | 4 | 1 | 0 | Low-quality | Genome-fragment | 3.75 | AAI-based (high-confidence) | 0 |
| S10_vs_49241\|\|full | 1479 | No | NA | 4 | 4 | 0 | Low-quality | Genome-fragment | 4.25 | AAI-based (high-confidence) | 0 |
| S10_vs_49267\|\|full | 1479 | No | NA | 3 | 0 | 0 | Low-quality | Genome-fragment | 3.59 | AAI-based (high-confidence) | 0 |
| S1_extracted_k141_6216 | 1304 | No | NA | 2 | 0 | 1 | Not-determined | Genome-fragment | NA | NA | 0 |
| S1_extracted_k141_66633 | 1193 | No | NA | 2 | 0 | 2 | Not-determined | Genome-fragment | NA | NA | 0 |
| S1_extracted_k141_6231 | 1352 | No | NA | 2 | 0 | 1 | Not-determined | Genome-fragment | NA | NA | 0 |
| S8_vs_k141_141770\|\|full | 1479 | No | NA | 3 | 0 | 0 | Low-quality | Genome-fragment | 2.51 | AAI-based (medium-confidence) | 0 |
| S9_vs_k141_45783\|\|full | 1480 | No | NA | 3 | 2 | 0 | Low-quality | Genome-fragment | 2.93 | AAI-based (medium-confidence) | 0 |
| S1_extracted_k141_6272 | 40918 | No | NA | 74 | 16 | 0 | Complete | High-quality | 100 | DTR (high-confidence) | 0 |
| S1_extracted_k141_6300 | 1029 | No | NA | 1 | 0 | 0 | Not-determined | Genome-fragment | NA | NA | 0 |
| S1_extracted_k141_157693 | 1033 | No | NA | 1 | 0 | 1 | Not-determined | Genome-fragment | NA | NA | 0 |
| S11_vs_k141_74333\|\|lt2gene | 1480 | No | NA | 2 | 1 | 0 | Low-quality | Genome-fragment | 3.98 | AAI-based (high-confidence) | 0 |
| S13_vs_120915\|\|full | 1480 | No | NA | 2 | 1 | 0 | Low-quality | Genome-fragment | 0.43 | HMM-based (lower-bound) | 0 |
| S1_extracted_k141_157715 | 1588 | No | NA | 1 | 0 | 1 | Not-determined | Genome-fragment | NA | NA | 0 |
| S14_extracted_k141_88403 | 1480 | No | NA | 4 | 3 | 0 | Low-quality | Genome-fragment | 4.87 | AAI-based (high-confidence) | 0 |
| S1_extracted_k141_157816 | 1418 | No | NA | 1 | 0 | 1 | Not-determined | Genome-fragment | NA | NA | 0 |
| S1_extracted_k141_6472 | 1116 | No | NA | 1 | 0 | 1 | Not-determined | Genome-fragment | NA | NA | 0 |
| S1_extracted_k141_39736 | 3754 | No | NA | 3 | 0 | 3 | Not-determined | Genome-fragment | NA | NA | 0 |
| S14_extracted_k141_9082 | 1480 | No | NA | 5 | 3 | 0 | Low-quality | Genome-fragment | 3.86 | AAI-based (high-confidence) | 0 |
| S1_extracted_k141_6578 | 1163 | No | NA | 1 | 0 | 0 | Not-determined | Genome-fragment | NA | NA | 0 |
| S1_extracted_k141_72902 | 1110 | No | NA | 1 | 0 | 1 | Not-determined | Genome-fragment | NA | NA | 0 |
| S14_vs_k141_44211\|\|full | 1480 | No | NA | 4 | 0 | 0 | Low-quality | Genome-fragment | 2.09 | AAI-based (high-confidence) | 0 |
| S1_extracted_k141_39854 | 1415 | No | NA | 2 | 0 | 0 | Not-determined | Genome-fragment | NA | NA | 0 |
| S14_vs_k141_67318\|\|full | 1480 | No | NA | 3 | 0 | 0 | Low-quality | Genome-fragment | 3.07 | AAI-based (high-confidence) | 0 |
| S1_extracted_k141_150750 | 1891 | No | NA | 1 | 0 | 1 | Not-determined | Genome-fragment | NA | NA | 0 |
| S1_extracted_k141_158015 | 1208 | No | NA | 1 | 0 | 1 | Not-determined | Genome-fragment | NA | NA | 0 |
| S1_extracted_k141_158033 | 1365 | No | NA | 1 | 0 | 1 | Not-determined | Genome-fragment | NA | NA | 0 |
| S1_extracted_k141_158087 | 1056 | No | NA | 1 | 0 | 1 | Not-determined | Genome-fragment | NA | NA | 0 |
| S1_vs_k141_87155\|\|full | 1480 | No | NA | 2 | 0 | 2 | Low-quality | Genome-fragment | 2.15 | AAI-based (high-confidence) | 0 |
| S5_extracted_k141_28089 | 1480 | No | NA | 5 | 1 | 0 | Low-quality | Genome-fragment | 2.53 | AAI-based (high-confidence) | 0 |
| S5_vs_k141_32176\|\|full | 1480 | No | NA | 5 | 0 | 0 | Low-quality | Genome-fragment | 3.72 | AAI-based (high-confidence) | 0 |
| S1_extracted_k141_73094 | 1035 | No | NA | 2 | 0 | 1 | Not-determined | Genome-fragment | NA | NA | 0 |
| S1_extracted_k141_73097 | 1006 | No | NA | 2 | 0 | 2 | Not-determined | Genome-fragment | NA | NA | 0 |
| S16_vs_86909\|\|full | 1480 | No | NA | 3 | 0 | 0 | Low-quality | Genome-fragment | 1.8 | AAI-based (high-confidence) | 0 |
| S12_vs_k141_152266\|\|full | 1480 | No | NA | 3 | 1 | 0 | Low-quality | Genome-fragment | 2.45 | HMM-based (lower-bound) | 0 |
| S6_vs_k141_22986\|\|full | 1480 | No | NA | 4 | 0 | 1 | Low-quality | Genome-fragment | 2.67 | AAI-based (medium-confidence) | 0 |
| S1_extracted_k141_73188 | 1126 | No | NA | 2 | 0 | 2 | Not-determined | Genome-fragment | NA | NA | 0 |
| S1_extracted_k141_151027 | 1028 | No | NA | 1 | 0 | 0 | Not-determined | Genome-fragment | NA | NA | 0 |
| S1_extracted_k141_158420 | 1486 | No | NA | 1 | 0 | 0 | Not-determined | Genome-fragment | NA | NA | 0 |
| S1_extracted_k141_158462 | 1035 | No | NA | 1 | 0 | 0 | Not-determined | Genome-fragment | NA | NA | 0 |
| S1_extracted_k141_151183 | 5309 | No | NA | 3 | 0 | 1 | Not-determined | Genome-fragment | NA | NA | 0 |
| S1_extracted_k141_151250 | 1557 | No | NA | 1 | 0 | 1 | Not-determined | Genome-fragment | NA | NA | 0 |
| S1_extracted_k141_158603 | 2164 | No | NA | 4 | 0 | 2 | Not-determined | Genome-fragment | NA | NA | 0 |
| S13_extracted_147516 | 1481 | No | NA | 3 | 1 | 0 | Low-quality | Genome-fragment | 3.67 | AAI-based (high-confidence) | 0 |
| S3_extracted_k141_41336 | 1481 | No | NA | 4 | 2 | 0 | Low-quality | Genome-fragment | 2.82 | AAI-based (high-confidence) | 0 |
| S1_extracted_k141_151391 | 1101 | No | NA | 1 | 0 | 1 | Not-determined | Genome-fragment | NA | NA | 0 |
| S1_extracted_k141_158693 | 2030 | No | NA | 2 | 0 | 1 | Not-determined | Genome-fragment | NA | NA | 0 |
| S1_extracted_k141_151485 | 1031 | No | NA | 1 | 0 | 1 | Not-determined | Genome-fragment | NA | NA | 0 |
| S3_vs_k141_96398\|\|full | 1481 | No | NA | 2 | 0 | 0 | Low-quality | Genome-fragment | 2.05 | AAI-based (high-confidence) | 0 |
| S1_extracted_k141_158890 | 1566 | No | NA | 1 | 0 | 1 | Not-determined | Genome-fragment | NA | NA | 0 |
| S1_extracted_k141_151678 | 1227 | No | NA | 1 | 0 | 0 | Not-determined | Genome-fragment | NA | NA | 0 |
| S5_extracted_k141_28971 | 1481 | No | NA | 1 | 1 | 0 | Low-quality | Genome-fragment | 0.92 | HMM-based (lower-bound) | 0 |
| S7_vs_63635\|\|full | 1481 | No | NA | 2 | 0 | 1 | Low-quality | Genome-fragment | 4.73 | AAI-based (high-confidence) | 0 |
| S1_extracted_k141_151780 | 2082 | No | NA | 3 | 0 | 2 | Not-determined | Genome-fragment | NA | NA | 0 |
| S1_extracted_k141_158961 | 43352 | No | NA | 66 | 9 | 1 | Medium-quality | Genome-fragment | 80.9 | AAI-based (high-confidence) | 0 |
| S1_extracted_k141_158985 | 2752 | No | NA | 2 | 0 | 1 | Not-determined | Genome-fragment | NA | NA | 0 |
| S1_extracted_k141_159144 | 1157 | No | NA | 3 | 0 | 2 | Not-determined | Genome-fragment | NA | NA | 0 |
| S16_extracted_64876 | 1481 | No | NA | 1 | 0 | 1 | Low-quality | Genome-fragment | 0.86 | AAI-based (medium-confidence) | 0 |
| S1_extracted_k141_152189 | 2109 | No | NA | 2 | 0 | 2 | Not-determined | Genome-fragment | NA | NA | 0 |
| S10_extracted_24939 | 1481 | No | NA | 3 | 0 | 1 | Low-quality | Genome-fragment | 3.98 | AAI-based (high-confidence) | 0 |
| S1_extracted_k141_159322 | 1771 | No | NA | 1 | 0 | 1 | Not-determined | Genome-fragment | NA | NA | 0 |
| S1_extracted_k141_159329 | 1177 | No | NA | 2 | 0 | 1 | Not-determined | Genome-fragment | NA | NA | 0 |
| S1_extracted_k141_159479 | 1036 | No | NA | 2 | 0 | 2 | Not-determined | Genome-fragment | NA | NA | 0 |
| S1_extracted_k141_159524 | 1279 | No | NA | 1 | 0 | 1 | Not-determined | Genome-fragment | NA | NA | 0 |
| S1_extracted_k141_159543 | 1948 | No | NA | 3 | 0 | 0 | Not-determined | Genome-fragment | NA | NA | 0 |
| S12_extracted_k141_40887 | 1481 | No | NA | 4 | 0 | 2 | Low-quality | Genome-fragment | 3.22 | AAI-based (medium-confidence) | 0 |
| S1_extracted_k141_152496 | 1277 | No | NA | 1 | 0 | 1 | Not-determined | Genome-fragment | NA | NA | 0 |
| S1_extracted_k141_159589 | 6712 | No | NA | 4 | 0 | 2 | Not-determined | Genome-fragment | NA | NA | 0 |
| S1_extracted_k141_159657 | 1097 | No | NA | 1 | 0 | 1 | Not-determined | Genome-fragment | NA | NA | 0 |
| S15_vs_28280\|\|full | 1481 | No | NA | 4 | 0 | 0 | Low-quality | Genome-fragment | 3.3 | AAI-based (high-confidence) | 0 |
| S8_extracted_k141_92549 | 1481 | No | NA | 3 | 0 | 0 | Low-quality | Genome-fragment | 1.64 | AAI-based (medium-confidence) | 0 |
| S1_extracted_k141_159785 | 1446 | No | NA | 1 | 0 | 1 | Not-determined | Genome-fragment | NA | NA | 0 |
| S1_extracted_k141_159839 | 1037 | No | NA | 1 | 0 | 0 | Not-determined | Genome-fragment | NA | NA | 0 |
| S8_vs_k141_8054\|\|full | 1481 | No | NA | 3 | 0 | 0 | Low-quality | Genome-fragment | 2.73 | AAI-based (medium-confidence) | 0 |
| S1_extracted_k141_153109 | 7731 | No | NA | 10 | 0 | 6 | Not-determined | Genome-fragment | NA | NA | 0 |
| S14_vs_k141_51106\|\|full | 1482 | No | NA | 3 | 0 | 1 | Low-quality | Genome-fragment | 0.79 | AAI-based (medium-confidence) | 0 |
| S1_extracted_k141_159951 | 101988 | No | NA | 124 | 17 | 3 | Complete | High-quality | 100 | DTR (high-confidence) | 0 |
| S1_extracted_k141_153189 | 1685 | No | NA | 4 | 0 | 1 | Not-determined | Genome-fragment | NA | NA | 0 |
| S1_extracted_k141_153230 | 1017 | No | NA | 1 | 0 | 1 | Not-determined | Genome-fragment | NA | NA | 0 |
| S1_extracted_k141_153237 | 1147 | No | NA | 1 | 0 | 1 | Not-determined | Genome-fragment | NA | NA | 0 |
| S1_extracted_k141_153397 | 1027 | No | NA | 2 | 0 | 2 | Not-determined | Genome-fragment | NA | NA | 0 |
| S1_extracted_k141_153500 | 2637 | No | NA | 1 | 0 | 1 | Not-determined | Genome-fragment | NA | NA | 0 |
| S1_vs_k141_143929\|\|full | 39654 | No | NA | 73 | 14 | 1 | High-quality | High-quality | 94.85 | AAI-based (high-confidence) | 0 |
| S1_vs_k141_66751\|\|full | 1760 | No | NA | 4 | 0 | 0 | Not-determined | Genome-fragment | NA | NA | 0 |
| S1_vs_k141_80114\|\|full | 1352 | No | NA | 2 | 0 | 2 | Not-determined | Genome-fragment | NA | NA | 0 |
| S1_extracted_k141_31875 | 1482 | No | NA | 3 | 1 | 0 | Low-quality | Genome-fragment | 1.85 | HMM-based (lower-bound) | 0 |
| S1_vs_k141_146858\|\|full | 3656 | No | NA | 3 | 0 | 3 | Not-determined | Genome-fragment | NA | NA | 0 |
| S3_extracted_k141_36917 | 1482 | No | NA | 1 | 1 | 0 | Low-quality | Genome-fragment | 2.55 | AAI-based (medium-confidence) | 0 |
| S1_vs_k141_33403\|\|full | 1012 | No | NA | 3 | 0 | 0 | Not-determined | Genome-fragment | NA | NA | 0 |
| S3_extracted_k141_28999 | 1482 | No | NA | 2 | 0 | 1 | Low-quality | Genome-fragment | 0.91 | AAI-based (medium-confidence) | 0 |
| S5_vs_k141_74854\|\|full | 1482 | No | NA | 3 | 1 | 2 | Low-quality | Genome-fragment | 0.24 | HMM-based (lower-bound) | 0 |
| S7_vs_26041\|\|full | 1482 | No | NA | 2 | 0 | 2 | Low-quality | Genome-fragment | 0.47 | AAI-based (medium-confidence) | 0 |
| S7_vs_133315\|\|full | 1482 | No | NA | 4 | 0 | 0 | Low-quality | Genome-fragment | 3.31 | AAI-based (medium-confidence) | 0 |
| S1_vs_k141_86808\|\|full | 1517 | No | NA | 5 | 0 | 0 | Not-determined | Genome-fragment | NA | NA | 0 |
| S1_vs_k141_26723\|\|full | 1304 | No | NA | 2 | 0 | 1 | Not-determined | Genome-fragment | NA | NA | 0 |
| S1_vs_k141_133544\|\|full | 1951 | No | NA | 2 | 0 | 2 | Not-determined | Genome-fragment | NA | NA | 0 |
| S1_vs_k141_73449\|\|full | 4289 | No | NA | 5 | 0 | 4 | Not-determined | Genome-fragment | NA | NA | 0 |
| S1_vs_k141_20064\|\|full | 6319 | No | NA | 7 | 0 | 5 | Not-determined | Genome-fragment | NA | NA | 0 |
| S16_vs_61133\|\|full | 1482 | No | NA | 4 | 0 | 0 | Low-quality | Genome-fragment | 2.46 | AAI-based (medium-confidence) | 0 |
| S1_vs_k141_113515\|\|full | 2691 | No | NA | 6 | 0 | 1 | Not-determined | Genome-fragment | NA | NA | 0 |
| S1_vs_k141_120200\|\|full | 2133 | No | NA | 3 | 0 | 1 | Not-determined | Genome-fragment | NA | NA | 0 |
| S16_vs_121885\|\|full | 1482 | No | NA | 4 | 1 | 0 | Low-quality | Genome-fragment | 3.23 | HMM-based (lower-bound) | 0 |
| S16_vs_146459\|\|full | 1482 | No | NA | 3 | 0 | 0 | Low-quality | Genome-fragment | 1.3 | AAI-based (medium-confidence) | 0 |
| S10_vs_58897\|\|full | 1482 | Yes | 999 | 3 | 1 | 1 | Low-quality | Genome-fragment | 0.69 | AAI-based (high-confidence) | 32.59 |
| S12_extracted_k141_183885 | 1482 | No | NA | 3 | 0 | 0 | Low-quality | Genome-fragment | 1.06 | AAI-based (high-confidence) | 0 |
| S12_extracted_k141_140828 | 1482 | No | NA | 2 | 0 | 1 | Low-quality | Genome-fragment | 0.27 | AAI-based (medium-confidence) | 0 |
| S12_extracted_k141_124648 | 1482 | No | NA | 3 | 1 | 0 | Low-quality | Genome-fragment | 1.7 | AAI-based (high-confidence) | 0 |
| S1_vs_k141_110\|\|full | 2157 | No | NA | 3 | 0 | 2 | Not-determined | Genome-fragment | NA | NA | 0 |
| S1_vs_k141_86859\|\|full | 1715 | No | NA | 3 | 0 | 3 | Not-determined | Genome-fragment | NA | NA | 0 |
| S1_vs_k141_66832\|\|full | 1189 | No | NA | 3 | 0 | 1 | Not-determined | Genome-fragment | NA | NA | 0 |
| S8_vs_k141_96715\|\|full | 1482 | No | NA | 3 | 1 | 0 | Low-quality | Genome-fragment | 1.37 | HMM-based (lower-bound) | 0 |
| S1_vs_k141_66837\|\|full | 4083 | No | NA | 7 | 0 | 0 | Not-determined | Genome-fragment | NA | NA | 0 |
| S17_vs_k141_44479\|\|full | 1483 | No | NA | 3 | 0 | 1 | Low-quality | Genome-fragment | 3.6 | AAI-based (medium-confidence) | 0 |
| S1_vs_k141_73513\|\|full | 2343 | No | NA | 3 | 0 | 2 | Not-determined | Genome-fragment | NA | NA | 0 |
| S1_extracted_k141_156374 | 1483 | Yes | 1048 | 4 | 1 | 1 | Low-quality | Genome-fragment | 2.29 | AAI-based (high-confidence) | 29.33 |
| S1_vs_k141_46837\|\|full | 2070 | No | NA | 3 | 0 | 2 | Not-determined | Genome-fragment | NA | NA | 0 |
| S1_vs_k141_140306\|\|full | 2429 | No | NA | 3 | 0 | 3 | Not-determined | Genome-fragment | NA | NA | 0 |
| S1_vs_k141_80245\|\|full | 2171 | No | NA | 3 | 0 | 2 | Not-determined | Genome-fragment | NA | NA | 0 |
| S1_vs_k141_133577\|\|full | 37683 | No | NA | 55 | 17 | 1 | High-quality | High-quality | 97.57 | AAI-based (high-confidence) | 0 |
| S1_vs_k141_73542\|\|full | 1200 | No | NA | 3 | 0 | 1 | Not-determined | Genome-fragment | NA | NA | 0 |
| S7_vs_36793\|\|lt2gene | 1483 | No | NA | 2 | 1 | 0 | Low-quality | Genome-fragment | 3.59 | AAI-based (medium-confidence) | 0 |
| S1_vs_k141_80273\|\|full | 1471 | No | NA | 3 | 0 | 2 | Not-determined | Genome-fragment | NA | NA | 0 |
| S1_vs_k141_113634\|\|full | 1099 | No | NA | 3 | 0 | 0 | Not-determined | Genome-fragment | NA | NA | 0 |
| S15_extracted_11555 | 1483 | No | NA | 2 | 0 | 2 | Low-quality | Genome-fragment | 0.67 | AAI-based (medium-confidence) | 0 |
| S4_vs_VIRSorter_31698-cat_3 | 1483 | No | NA | 4 | 1 | 0 | Low-quality | Genome-fragment | 3.83 | AAI-based (high-confidence) | 0 |
| S6_vs_k141_36798\|\|full | 1483 | No | NA | 3 | 0 | 0 | Low-quality | Genome-fragment | 4.69 | AAI-based (high-confidence) | 0 |
| S17_vs_k141_151326\|\|full | 1484 | No | NA | 5 | 1 | 0 | Low-quality | Genome-fragment | 2.8 | AAI-based (high-confidence) | 0 |
| S1_vs_k141_204\|\|full | 1044 | No | NA | 3 | 0 | 0 | Not-determined | Genome-fragment | NA | NA | 0 |
| S17_vs_k141_124312\|\|full | 1484 | No | NA | 2 | 0 | 2 | Low-quality | Genome-fragment | 0.81 | AAI-based (high-confidence) | 0 |
| S17_vs_k141_21395\|\|full | 1484 | No | NA | 3 | 2 | 0 | Low-quality | Genome-fragment | 3.46 | AAI-based (high-confidence) | 0 |
| S1_vs_k141_20257\|\|full | 3067 | No | NA | 3 | 0 | 0 | Not-determined | Genome-fragment | NA | NA | 0 |
| S14_vs_k141_20113\|\|full | 1484 | No | NA | 2 | 0 | 0 | Low-quality | Genome-fragment | 3.19 | AAI-based (medium-confidence) | 0 |
| S14_vs_k141_94513\|\|full | 1484 | No | NA | 3 | 0 | 1 | Low-quality | Genome-fragment | 1.97 | AAI-based (medium-confidence) | 0 |
| S1_vs_k141_87042\|\|full | 1981 | No | NA | 3 | 0 | 3 | Not-determined | Genome-fragment | NA | NA | 0 |
| S10_extracted_8869 | 1484 | No | NA | 3 | 0 | 2 | Low-quality | Genome-fragment | 0.45 | AAI-based (high-confidence) | 0 |
| S10_vs_79283\|\|full | 1484 | No | NA | 5 | 1 | 0 | Low-quality | Genome-fragment | 3.8 | AAI-based (high-confidence) | 0 |
| S8_vs_k141_175601\|\|full | 1484 | No | NA | 5 | 0 | 0 | Low-quality | Genome-fragment | 3 | AAI-based (medium-confidence) | 0 |
| S1_vs_k141_113719\|\|full | 2446 | No | NA | 3 | 0 | 1 | Not-determined | Genome-fragment | NA | NA | 0 |
| S1_vs_k141_127131\|\|full | 2151 | No | NA | 3 | 0 | 0 | Not-determined | Genome-fragment | NA | NA | 0 |
| S1_vs_k141_33617\|\|full | 1016 | No | NA | 4 | 0 | 0 | Not-determined | Genome-fragment | NA | NA | 0 |
| S1_vs_k141_66989\|\|full | 3099 | No | NA | 3 | 0 | 2 | Not-determined | Genome-fragment | NA | NA | 0 |
| S1_vs_k141_26890\|\|full | 1970 | No | NA | 3 | 0 | 1 | Not-determined | Genome-fragment | NA | NA | 0 |
| S8_vs_k141_48652\|\|full | 1484 | No | NA | 3 | 0 | 1 | Low-quality | Genome-fragment | 0.95 | AAI-based (medium-confidence) | 0 |
| S9_extracted_k141_29541 | 1485 | No | NA | 2 | 2 | 0 | Low-quality | Genome-fragment | 4.2 | AAI-based (high-confidence) | 0 |
| S1_vs_k141_66995\|\|full | 3974 | No | NA | 5 | 0 | 2 | Not-determined | Genome-fragment | NA | NA | 0 |
| S11_vs_k141_88004\|\|full | 1485 | No | NA | 2 | 0 | 0 | Low-quality | Genome-fragment | 3.61 | AAI-based (medium-confidence) | 0 |
| S1_vs_k141_113749\|\|full | 1680 | No | NA | 3 | 0 | 2 | Not-determined | Genome-fragment | NA | NA | 0 |
| S13_vs_74441\|\|full | 1485 | No | NA | 3 | 0 | 1 | Low-quality | Genome-fragment | 0.48 | AAI-based (medium-confidence) | 0 |
| S1_vs_k141_133731\|\|full | 1211 | No | NA | 2 | 0 | 0 | Not-determined | Genome-fragment | NA | NA | 0 |
| S1_vs_k141_127197\|\|full | 1173 | No | NA | 3 | 0 | 1 | Not-determined | Genome-fragment | NA | NA | 0 |
| S14_vs_k141_45208\|\|1_partial | 1485 | No | NA | 2 | 1 | 0 | Low-quality | Genome-fragment | 33.16 | AAI-based (high-confidence) | 0 |
| S5_extracted_k141_65432 | 1485 | No | NA | 1 | 1 | 0 | Low-quality | Genome-fragment | 2.53 | AAI-based (medium-confidence) | 0 |
| S1_vs_k141_120496\|\|full | 2292 | No | NA | 4 | 0 | 1 | Not-determined | Genome-fragment | NA | NA | 0 |
| S1_vs_k141_140495\|\|full | 2649 | No | NA | 3 | 0 | 2 | Not-determined | Genome-fragment | NA | NA | 0 |
| S1_vs_k141_350\|\|full | 1222 | No | NA | 2 | 0 | 1 | Not-determined | Genome-fragment | NA | NA | 0 |
| S1_vs_k141_73761\|\|full | 1303 | No | NA | 2 | 0 | 1 | Not-determined | Genome-fragment | NA | NA | 0 |
| S5_vs_k141_19719\|\|full | 1485 | No | NA | 2 | 0 | 0 | Low-quality | Genome-fragment | 1.26 | AAI-based (high-confidence) | 0 |
| S1_vs_k141_33692\|\|full | 1777 | No | NA | 3 | 0 | 1 | Not-determined | Genome-fragment | NA | NA | 0 |

| | | | | | | | | | | | |
|---|---|---|---|---|---|---|---|---|---|---|---|
| S5_vs_k141_16437\|\|full | 1485 | No | NA | 3 | 0 | 0 | Low-quality | Genome-fragment | 1.72 | AAI-based (medium-confidence) | 0 |
| S5_vs_k141_143980\|\|full | 1485 | No | NA | 4 | 0 | 0 | Low-quality | Genome-fragment | 3.12 | AAI-based (medium-confidence) | 0 |
| S10_vs_103735\|\|full | 1485 | No | NA | 3 | 1 | 0 | Low-quality | Genome-fragment | 1.3 | AAI-based (high-confidence) | 0 |
| S1_vs_k141_13759\|\|full | 1060 | No | NA | 2 | 0 | 1 | Not-determined | Genome-fragment | NA | NA | 0 |
| S12_vs_k141_136803\|\|full | 1485 | No | NA | 3 | 0 | 0 | Low-quality | Genome-fragment | 3.04 | AAI-based (medium-confidence) | 0 |
| S17_vs_k141_120010\|\|lt2gene | 1486 | No | NA | 2 | 1 | 0 | Low-quality | Genome-fragment | 2.51 | AAI-based (high-confidence) | 0 |
| S9_vs_k141_75120\|\|full | 1486 | No | NA | 3 | 0 | 1 | Low-quality | Genome-fragment | 1.31 | AAI-based (medium-confidence) | 0 |
| S1_vs_k141_53866\|\|full | 3531 | No | NA | 3 | 0 | 3 | Not-determined | Genome-fragment | NA | NA | 0 |
| S1_extracted_k141_17701 | 1486 | No | NA | 2 | 0 | 0 | Low-quality | Genome-fragment | 5.18 | AAI-based (high-confidence) | 0 |
| S1_vs_k141_31655\|\|full | 1486 | No | NA | 3 | 0 | 0 | Low-quality | Genome-fragment | 2.98 | AAI-based (medium-confidence) | 0 |
| S3_vs_k141_46246\|\|lt2gene | 1486 | No | NA | 1 | 1 | 0 | Low-quality | Genome-fragment | 1.03 | HMM-based (lower-bound) | 0 |
| S5_extracted_k141_80626 | 1486 | No | NA | 5 | 0 | 0 | Low-quality | Genome-fragment | 3.52 | AAI-based (high-confidence) | 0 |
| S1_vs_k141_47170\|\|full | 1260 | No | NA | 2 | 0 | 0 | Not-determined | Genome-fragment | NA | NA | 0 |
| S1_vs_k141_93967\|\|full | 2266 | No | NA | 3 | 0 | 1 | Not-determined | Genome-fragment | NA | NA | 0 |
| S1_vs_k141_113935\|\|full | 2251 | No | NA | 4 | 0 | 3 | Not-determined | Genome-fragment | NA | NA | 0 |
| S1_vs_k141_73917\|\|full | 1120 | No | NA | 2 | 0 | 2 | Not-determined | Genome-fragment | NA | NA | 0 |
| S1_vs_k141_120637\|\|full | 2543 | No | NA | 5 | 0 | 1 | Not-determined | Genome-fragment | NA | NA | 0 |
| S5_vs_k141_49268\|\|full | 1486 | No | NA | 3 | 1 | 0 | Low-quality | Genome-fragment | 4 | AAI-based (high-confidence) | 0 |
| S1_vs_k141_47221\|\|full | 1159 | No | NA | 2 | 0 | 1 | Not-determined | Genome-fragment | NA | NA | 0 |
| S5_vs_k141_59061\|\|full | 1486 | No | NA | 3 | 0 | 0 | Low-quality | Genome-fragment | 2.6 | AAI-based (medium-confidence) | 0 |
| S7_extracted_87035 | 1486 | No | NA | 4 | 2 | 0 | Low-quality | Genome-fragment | 3.84 | AAI-based (medium-confidence) | 0 |
| S16_extracted_101826 | 1486 | No | NA | 2 | 2 | 0 | Low-quality | Genome-fragment | 3.45 | AAI-based (high-confidence) | 0 |
| S1_vs_k141_140702\|\|full | 2976 | No | NA | 4 | 0 | 4 | Not-determined | Genome-fragment | NA | NA | 0 |
| S1_vs_k141_47236\|\|full | 2982 | No | NA | 3 | 0 | 2 | Not-determined | Genome-fragment | NA | NA | 0 |
| S16_vs_55677\|\|full | 1486 | No | NA | 7 | 0 | 0 | Low-quality | Genome-fragment | 0.86 | AAI-based (high-confidence) | 0 |
| S15_extracted_42392 | 1486 | No | NA | 2 | 0 | 2 | Low-quality | Genome-fragment | 1.05 | AAI-based (medium-confidence) | 0 |
| S1_vs_k141_27200\|\|full | 1000 | No | NA | 2 | 0 | 0 | Not-determined | Genome-fragment | NA | NA | 0 |
| S1_vs_k141_113975\|\|full | 2169 | No | NA | 4 | 0 | 0 | Not-determined | Genome-fragment | NA | NA | 0 |
| S15_vs_68973\|\|full | 1486 | No | NA | 4 | 1 | 0 | Low-quality | Genome-fragment | 1.14 | AAI-based (medium-confidence) | 0 |
| S1_vs_k141_20537\|\|full | 2182 | No | NA | 2 | 0 | 2 | Not-determined | Genome-fragment | NA | NA | 0 |
| S17_extracted_k141_175431 | 1487 | No | NA | 4 | 1 | 0 | Low-quality | Genome-fragment | 3.35 | AAI-based (high-confidence) | 0 |
| S1_vs_k141_87391\|\|full | 2513 | No | NA | 2 | 0 | 1 | Not-determined | Genome-fragment | NA | NA | 0 |
| S1_vs_k141_114040\|\|full | 1586 | No | NA | 3 | 0 | 1 | Not-determined | Genome-fragment | NA | NA | 0 |
| S14_vs_k141_4798\|\|full | 1487 | No | NA | 2 | 0 | 1 | Low-quality | Genome-fragment | 0.75 | AAI-based (medium-confidence) | 0 |
| S14_vs_k141_38233\|\|lt2gene | 1487 | No | NA | 1 | 1 | 0 | Low-quality | Genome-fragment | 3.03 | AAI-based (high-confidence) | 0 |
| S1_vs_k141_120731\|\|full | 1059 | No | NA | 2 | 0 | 1 | Not-determined | Genome-fragment | NA | NA | 0 |
| S7_vs_2747\|\|full | 1487 | No | NA | 4 | 0 | 0 | Low-quality | Genome-fragment | 1.73 | AAI-based (high-confidence) | 0 |
| S12_extracted_k141_71340 | 1487 | No | NA | 4 | 0 | 0 | Low-quality | Genome-fragment | 4.58 | AAI-based (high-confidence) | 0 |
| S8_vs_k141_19419\|\|lt2gene | 1487 | No | NA | 1 | 1 | 0 | Low-quality | Genome-fragment | 1.55 | HMM-based (lower-bound) | 0 |
| S14_extracted_k141_3424 | 1488 | No | NA | 2 | 0 | 2 | Low-quality | Genome-fragment | 3.58 | AAI-based (high-confidence) | 0 |
| S1_vs_k141_40755\|\|full | 1300 | No | NA | 3 | 0 | 0 | Not-determined | Genome-fragment | NA | NA | 0 |
| S3_vs_k141_68237\|\|full | 1488 | No | NA | 8 | 0 | 0 | Low-quality | Genome-fragment | 4.11 | AAI-based (high-confidence) | 0 |
| S1_vs_k141_14149\|\|full | 2304 | No | NA | 2 | 0 | 1 | Not-determined | Genome-fragment | NA | NA | 0 |
| S3_vs_k141_101414\|\|full | 1488 | No | NA | 3 | 1 | 0 | Low-quality | Genome-fragment | 3.26 | AAI-based (high-confidence) | 0 |
| S1_vs_k141_40790\|\|full | 1146 | No | NA | 3 | 0 | 0 | Not-determined | Genome-fragment | NA | NA | 0 |
| S1_vs_k141_94172\|\|full | 3815 | No | NA | 5 | 0 | 1 | Not-determined | Genome-fragment | NA | NA | 0 |
| S1_vs_k141_74138\|\|full | 1065 | No | NA | 3 | 0 | 0 | Not-determined | Genome-fragment | NA | NA | 0 |
| S3_vs_k141_46507\|\|full | 1488 | No | NA | 2 | 2 | 0 | Low-quality | Genome-fragment | 1.04 | AAI-based (high-confidence) | 0 |
| S5_vs_k141_98558\|\|full | 1488 | No | NA | 3 | 0 | 0 | Low-quality | Genome-fragment | 2.72 | AAI-based (high-confidence) | 0 |
| S1_vs_k141_67403\|\|full | 2032 | No | NA | 2 | 0 | 1 | Not-determined | Genome-fragment | NA | NA | 0 |
| S1_vs_k141_140932\|\|full | 2319 | No | NA | 2 | 0 | 2 | Not-determined | Genome-fragment | NA | NA | 0 |
| S7_extracted_48945 | 1488 | No | NA | 1 | 1 | 0 | Low-quality | Genome-fragment | 2.76 | AAI-based (medium-confidence) | 0 |
| S1_vs_k141_27422\|\|full | 2134 | No | NA | 2 | 0 | 2 | Not-determined | Genome-fragment | NA | NA | 0 |
| S16_extracted_152695 | 1488 | No | NA | 5 | 3 | 0 | Low-quality | Genome-fragment | 2.56 | AAI-based (high-confidence) | 0 |
| S1_vs_k141_107452\|\|full | 3933 | No | NA | 6 | 0 | 2 | Not-determined | Genome-fragment | NA | NA | 0 |
| S16_vs_124558\|\|lt2gene | 1488 | No | NA | 1 | 1 | 0 | Low-quality | Genome-fragment | 1.03 | HMM-based (lower-bound) | 0 |
| S12_vs_k141_72878\|\|full | 1488 | No | NA | 2 | 0 | 0 | Low-quality | Genome-fragment | 1.41 | AAI-based (high-confidence) | 0 |
| S15_extracted_29185 | 1488 | No | NA | 2 | 0 | 0 | Low-quality | Genome-fragment | 3.33 | AAI-based (high-confidence) | 0 |
| S1_vs_k141_94301\|\|full | 2758 | No | NA | 3 | 0 | 2 | Not-determined | Genome-fragment | NA | NA | 0 |
| S15_extracted_40218 | 1488 | No | NA | 2 | 0 | 1 | Low-quality | Genome-fragment | 0.99 | AAI-based (high-confidence) | 0 |
| S8_extracted_k141_54909 | 1488 | No | NA | 2 | 0 | 1 | Low-quality | Genome-fragment | 1.02 | AAI-based (medium-confidence) | 0 |
| S8_vs_k141_57783\|\|full | 1488 | No | NA | 2 | 0 | 2 | Low-quality | Genome-fragment | 3.85 | AAI-based (medium-confidence) | 0 |
| S1_vs_k141_140995\|\|full | 1007 | No | NA | 2 | 0 | 0 | Not-determined | Genome-fragment | NA | NA | 0 |
| S11_extracted_k141_103473 | 1489 | No | NA | 5 | 1 | 0 | Low-quality | Genome-fragment | 1.97 | AAI-based (medium-confidence) | 0 |
| S11_vs_k141_103627\|\|full | 1489 | No | NA | 2 | 0 | 0 | Low-quality | Genome-fragment | 1.4 | AAI-based (high-confidence) | 0 |
| S13_vs_92862\|\|full | 1489 | No | NA | 3 | 0 | 1 | Low-quality | Genome-fragment | 0.98 | AAI-based (medium-confidence) | 0 |
| S1_vs_k141_120880\|\|full | 1221 | No | NA | 2 | 0 | 1 | Not-determined | Genome-fragment | NA | NA | 0 |
| S1_vs_k141_40963\|\|full | 2059 | No | NA | 3 | 0 | 2 | Not-determined | Genome-fragment | NA | NA | 0 |
| S1_extracted_k141_155271 | 1489 | No | NA | 2 | 1 | 0 | Low-quality | Genome-fragment | 3.79 | AAI-based (high-confidence) | 0 |
| S1_vs_k141_67543\|\|full | 1661 | No | NA | 3 | 0 | 1 | Not-determined | Genome-fragment | NA | NA | 0 |
| S1_vs_k141_27524\|\|full | 3628 | No | NA | 2 | 0 | 2 | Not-determined | Genome-fragment | NA | NA | 0 |
| S16_extracted_119163 | 1489 | No | NA | 1 | 0 | 1 | Low-quality | Genome-fragment | 0.8 | AAI-based (medium-confidence) | 0 |
| S10_vs_96835\|\|full | 1489 | No | NA | 5 | 1 | 0 | Low-quality | Genome-fragment | 3.85 | AAI-based (high-confidence) | 0 |
| S12_vs_k141_151634\|\|full | 1489 | No | NA | 2 | 0 | 0 | Low-quality | Genome-fragment | 3.53 | AAI-based (high-confidence) | 0 |
| S15_extracted_69246 | 1489 | No | NA | 2 | 0 | 2 | Low-quality | Genome-fragment | 0.63 | AAI-based (medium-confidence) | 0 |
| S8_extracted_k141_6227 | 1489 | No | NA | 2 | 0 | 1 | Low-quality | Genome-fragment | 3.85 | AAI-based (medium-confidence) | 0 |
| S1_vs_k141_20910\|\|full | 2810 | No | NA | 4 | 0 | 2 | Not-determined | Genome-fragment | NA | NA | 0 |
| S17_extracted_k141_12846 | 1490 | No | NA | 3 | 1 | 0 | Low-quality | Genome-fragment | 2.35 | AAI-based (high-confidence) | 0 |
| S9_vs_k141_39685\|\|full | 1490 | No | NA | 3 | 1 | 0 | Low-quality | Genome-fragment | 3.61 | AAI-based (medium-confidence) | 0 |
| S13_vs_35590\|\|full | 1490 | No | NA | 3 | 1 | 0 | Low-quality | Genome-fragment | 3.16 | AAI-based (high-confidence) | 0 |
| S1_vs_k141_87711\|\|full | 1346 | No | NA | 3 | 0 | 1 | Not-determined | Genome-fragment | NA | NA | 0 |
| S13_vs_109253\|\|full | 1490 | No | NA | 3 | 0 | 0 | Low-quality | Genome-fragment | 2.72 | AAI-based (medium-confidence) | 0 |
| S3_vs_k141_39146\|\|full | 1490 | No | NA | 4 | 0 | 0 | Low-quality | Genome-fragment | 2.7 | AAI-based (medium-confidence) | 0 |
| S10_vs_52573\|\|full | 1490 | No | NA | 3 | 1 | 0 | Low-quality | Genome-fragment | 1.71 | HMM-based (lower-bound) | 0 |
| S12_vs_k141_127056\|\|full | 1490 | No | NA | 3 | 0 | 0 | Low-quality | Genome-fragment | 0.86 | AAI-based (medium-confidence) | 0 |
| S1_vs_k141_60613\|\|full | 3502 | No | NA | 6 | 0 | 1 | Not-determined | Genome-fragment | NA | NA | 0 |
| S1_vs_k141_114332\|\|full | 2050 | No | NA | 3 | 0 | 0 | Not-determined | Genome-fragment | NA | NA | 0 |
| S4_extracted_8279 | 1490 | No | NA | 3 | 0 | 0 | Low-quality | Genome-fragment | 1.07 | AAI-based (medium-confidence) | 0 |
| S11_vs_k141_77050\|\|full | 1491 | No | NA | 4 | 1 | 0 | Low-quality | Genome-fragment | 2.8 | AAI-based (medium-confidence) | 0 |
| S14_extracted_k141_31563 | 1491 | No | NA | 3 | 2 | 0 | Low-quality | Genome-fragment | 4.85 | AAI-based (high-confidence) | 0 |
| S1_vs_k141_94469\|\|full | 1830 | No | NA | 5 | 0 | 1 | Not-determined | Genome-fragment | NA | NA | 0 |
| S14_vs_k141_11773\|\|full | 1491 | No | NA | 4 | 1 | 0 | Low-quality | Genome-fragment | 3.43 | AAI-based (high-confidence) | 0 |
| S1_vs_k141_107613\|\|full | 2097 | No | NA | 3 | 0 | 0 | Not-determined | Genome-fragment | NA | NA | 0 |
| S1_vs_k141_47819\|\|full | 1491 | No | NA | 3 | 1 | 0 | Low-quality | Genome-fragment | 3.41 | AAI-based (high-confidence) | 0 |
| S3_extracted_k141_24182 | 1491 | No | NA | 2 | 0 | 1 | Low-quality | Genome-fragment | 1.01 | AAI-based (high-confidence) | 0 |
| S5_vs_k141_14094\|\|lt2gene | 1491 | No | NA | 1 | 1 | 0 | Low-quality | Genome-fragment | 3.02 | AAI-based (high-confidence) | 0 |
| S7_extracted_19384 | 1491 | No | NA | 5 | 0 | 0 | Low-quality | Genome-fragment | 2.95 | AAI-based (high-confidence) | 0 |
| S16_vs_65239\|\|full | 1491 | No | NA | 3 | 0 | 1 | Low-quality | Genome-fragment | 3.48 | AAI-based (medium-confidence) | 0 |
| S1_vs_k141_896\|\|full | 2093 | No | NA | 2 | 0 | 1 | Not-determined | Genome-fragment | NA | NA | 0 |
| S1_vs_k141_141202\|\|full | 1930 | No | NA | 3 | 0 | 2 | Not-determined | Genome-fragment | NA | NA | 0 |
| S16_vs_23137\|\|lt2gene | 1491 | No | NA | 2 | 2 | 0 | Low-quality | Genome-fragment | 3.27 | AAI-based (high-confidence) | 0 |
| S1_vs_k141_81207\|\|full | 6380 | No | NA | 7 | 0 | 7 | Not-determined | Genome-fragment | NA | NA | 0 |
| S6_vs_k141_7555\|\|full | 1491 | No | NA | 2 | 0 | 1 | Low-quality | Genome-fragment | 1.06 | AAI-based (medium-confidence) | 0 |
| S1_vs_k141_912\|\|full | 8990 | No | NA | 13 | 0 | 3 | Not-determined | Genome-fragment | NA | NA | 0 |
| S1_vs_k141_54491\|\|full | 1493 | No | NA | 2 | 0 | 1 | Not-determined | Genome-fragment | NA | NA | 0 |
| S6_vs_k141_45573\|\|full | 1491 | No | NA | 4 | 0 | 0 | Low-quality | Genome-fragment | 1.39 | AAI-based (medium-confidence) | 0 |
| S8_vs_k141_57429\|\|full | 1491 | No | NA | 2 | 0 | 1 | Low-quality | Genome-fragment | 0.97 | AAI-based (medium-confidence) | 0 |
| S1_vs_k141_21090\|\|full | 1042 | No | NA | 3 | 0 | 0 | Not-determined | Genome-fragment | NA | NA | 0 |
| S8_vs_k141_180910\|\|full | 1491 | No | NA | 2 | 0 | 0 | Low-quality | Genome-fragment | 1.62 | AAI-based (medium-confidence) | 0 |
| S17_vs_k141_28531\|\|full | 1492 | No | NA | 2 | 0 | 0 | Low-quality | Genome-fragment | 3.14 | AAI-based (medium-confidence) | 0 |
| S9_vs_k141_38768\|\|full | 1492 | No | NA | 3 | 1 | 0 | Low-quality | Genome-fragment | 3.34 | AAI-based (high-confidence) | 0 |
| S9_vs_k141_62719\|\|full | 1492 | No | NA | 3 | 0 | 0 | Low-quality | Genome-fragment | 3.14 | AAI-based (medium-confidence) | 0 |
| S14_vs_k141_8657\|\|full | 1492 | No | NA | 4 | 0 | 0 | Low-quality | Genome-fragment | 3.1 | AAI-based (medium-confidence) | 0 |
| S5_vs_k141_10183\|\|full | 1492 | No | NA | 4 | 1 | 0 | Low-quality | Genome-fragment | 2.24 | AAI-based (medium-confidence) | 0 |
| S1_vs_k141_94638\|\|full | 1940 | No | NA | 4 | 0 | 0 | Not-determined | Genome-fragment | NA | NA | 0 |
| S1_vs_k141_101269\|\|full | 2134 | No | NA | 4 | 0 | 0 | Not-determined | Genome-fragment | NA | NA | 0 |
| S7_vs_299\|\|full | 1492 | No | NA | 4 | 1 | 0 | Low-quality | Genome-fragment | 2.01 | HMM-based (lower-bound) | 0 |
| S1_vs_k141_47832\|\|full | 4015 | No | NA | 6 | 0 | 1 | Not-determined | Genome-fragment | NA | NA | 0 |
| S16_vs_59764\|\|full | 1492 | No | NA | 3 | 0 | 1 | Low-quality | Genome-fragment | 0.77 | AAI-based (medium-confidence) | 0 |
| S1_vs_k141_81314\|\|full | 1187 | No | NA | 2 | 0 | 1 | Not-determined | Genome-fragment | NA | NA | 0 |
| S15_vs_9780\|\|full | 1492 | No | NA | 3 | 2 | 0 | Low-quality | Genome-fragment | 3.3 | AAI-based (high-confidence) | 0 |
| S8_vs_k141_56168\|\|full | 1492 | No | NA | 3 | 1 | 0 | Low-quality | Genome-fragment | 0.86 | AAI-based (high-confidence) | 0 |
| S1_vs_k141_41290\|\|full | 2854 | No | NA | 4 | 0 | 0 | Not-determined | Genome-fragment | NA | NA | 0 |
| S1_vs_k141_141309\|\|full | 2304 | No | NA | 3 | 0 | 1 | Not-determined | Genome-fragment | NA | NA | 0 |
| S8_vs_k141_146953\|\|full | 1492 | No | NA | 3 | 1 | 0 | Low-quality | Genome-fragment | 3.33 | AAI-based (medium-confidence) | 0 |
| S17_vs_k141_32085\|\|full | 1493 | No | NA | 1 | 0 | 0 | Low-quality | Genome-fragment | 1.65 | AAI-based (high-confidence) | 0 |
| S1_vs_k141_27788\|\|full | 2289 | No | NA | 4 | 0 | 3 | Not-determined | Genome-fragment | NA | NA | 0 |
| S11_extracted_k141_37120 | 1493 | No | NA | 3 | 0 | 0 | Low-quality | Genome-fragment | 3.27 | AAI-based (high-confidence) | 0 |
| S11_vs_k141_92210\|\|full | 1493 | No | NA | 4 | 1 | 0 | Low-quality | Genome-fragment | 3.54 | AAI-based (high-confidence) | 0 |
| S1_vs_k141_41325\|\|full | 1244 | No | NA | 3 | 0 | 1 | Not-determined | Genome-fragment | NA | NA | 0 |
| S3_vs_k141_126263\|\|full | 1493 | No | NA | 5 | 0 | 0 | Low-quality | Genome-fragment | 3.7 | AAI-based (high-confidence) | 0 |
| S1_vs_k141_14853\|\|full | 1265 | No | NA | 3 | 0 | 1 | Not-determined | Genome-fragment | NA | NA | 0 |
| S1_vs_k141_1033\|\|full | 2538 | No | NA | 3 | 0 | 2 | Not-determined | Genome-fragment | NA | NA | 0 |
| S1_vs_k141_128046\|\|full | 7825 | No | NA | 3 | 0 | 3 | Not-determined | Genome-fragment | NA | NA | 0 |
| S1_vs_k141_27866\|\|full | 1126 | No | NA | 3 | 0 | 1 | Not-determined | Genome-fragment | NA | NA | 0 |
| S5_vs_k141_85760\|\|lt2gene | 1493 | No | NA | 2 | 2 | 0 | Low-quality | Genome-fragment | 2.5 | HMM-based (lower-bound) | 0 |
| S16_vs_134123\|\|full | 1493 | No | NA | 3 | 0 | 0 | Low-quality | Genome-fragment | 3.62 | AAI-based (medium-confidence) | 0 |
| S10_vs_3934\|\|full | 1493 | No | NA | 3 | 0 | 0 | Low-quality | Genome-fragment | 1.92 | AAI-based (high-confidence) | 0 |
| S6_extracted_k141_46715 | 1493 | No | NA | 3 | 0 | 1 | Low-quality | Genome-fragment | 3.32 | AAI-based (medium-confidence) | 0 |
| S1_vs_k141_21263\|\|full | 14418 | No | NA | 11 | 0 | 9 | Not-determined | Genome-fragment | NA | NA | 0 |
| S6_vs_k141_13215\|\|lt2gene | 1493 | No | NA | 1 | 1 | 0 | Low-quality | Genome-fragment | 2.51 | HMM-based (lower-bound) | 0 |
| S1_vs_k141_154248\|\|full | 1019 | No | NA | 2 | 0 | 2 | Not-determined | Genome-fragment | NA | NA | 0 |
| S8_extracted_k141_739 | 1493 | No | NA | 2 | 0 | 0 | Low-quality | Genome-fragment | 4.75 | AAI-based (high-confidence) | 0 |
| S1_vs_k141_48031\|\|full | 2285 | No | NA | 3 | 0 | 0 | Not-determined | Genome-fragment | NA | NA | 0 |
| S13_extracted_68522 | 1494 | No | NA | 2 | 0 | 0 | Low-quality | Genome-fragment | 2.01 | AAI-based (high-confidence) | 0 |
| S1_vs_k141_54705\|\|full | 1928 | No | NA | 3 | 0 | 1 | Not-determined | Genome-fragment | NA | NA | 0 |
| S7_vs_40601\|\|full | 1494 | No | NA | 4 | 1 | 0 | Low-quality | Genome-fragment | 2.02 | AAI-based (medium-confidence) | 0 |
| S1_vs_k141_54708\|\|full | 5273 | No | NA | 4 | 0 | 2 | Not-determined | Genome-fragment | NA | NA | 0 |
| S1_vs_k141_54717\|\|full | 1491 | No | NA | 4 | 0 | 0 | Not-determined | Genome-fragment | NA | NA | 0 |
| S7_vs_105989\|\|full | 1494 | No | NA | 3 | 1 | 0 | Low-quality | Genome-fragment | 0.89 | AAI-based (medium-confidence) | 0 |
| S1_vs_k141_141428\|\|full | 4318 | No | NA | 4 | 0 | 3 | Not-determined | Genome-fragment | NA | NA | 0 |
| S16_vs_11046\|\|full | 1494 | No | NA | 2 | 0 | 0 | Low-quality | Genome-fragment | 2.96 | AAI-based (high-confidence) | 0 |
| S1_vs_k141_41473\|\|full | 1684 | No | NA | 3 | 0 | 0 | Not-determined | Genome-fragment | NA | NA | 0 |
| S6_vs_k141_5624\|\|full | 1494 | No | NA | 2 | 0 | 0 | Low-quality | Genome-fragment | 2 | AAI-based (medium-confidence) | 0 |
| S8_extracted_k141_76891 | 1494 | No | NA | 1 | 0 | 1 | Low-quality | Genome-fragment | 0.8 | AAI-based (medium-confidence) | 0 |
| S8_vs_k141_38068\|\|full | 1494 | No | NA | 3 | 0 | 0 | Low-quality | Genome-fragment | 3.12 | AAI-based (medium-confidence) | 0 |
| S17_vs_k141_49140\|\|full | 1495 | No | NA | 3 | 0 | 0 | Low-quality | Genome-fragment | 1.95 | AAI-based (high-confidence) | 0 |
| S1_vs_k141_1139\|\|full | 2101 | No | NA | 3 | 0 | 2 | Not-determined | Genome-fragment | NA | NA | 0 |
| S1_vs_k141_74768\|\|full | 3032 | No | NA | 6 | 0 | 1 | Not-determined | Genome-fragment | NA | NA | 0 |
| S13_vs_25368\|\|lt2gene | 1495 | No | NA | 2 | 1 | 0 | Low-quality | Genome-fragment | 3.92 | AAI-based (high-confidence) | 0 |
| S1_vs_k141_81533\|\|full | 2360 | No | NA | 6 | 0 | 2 | Not-determined | Genome-fragment | NA | NA | 0 |
| S1_vs_k141_88156\|\|full | 2489 | No | NA | 4 | 0 | 0 | Not-determined | Genome-fragment | NA | NA | 0 |
| S1_vs_k141_101602\|\|full | 2085 | No | NA | 3 | 0 | 1 | Not-determined | Genome-fragment | NA | NA | 0 |
| S1_vs_k141_137457\|\|full | 1495 | No | NA | 4 | 2 | 0 | Low-quality | Genome-fragment | 3.95 | AAI-based (high-confidence) | 0 |
| S3_vs_k141_20255\|\|full | 1495 | No | NA | 4 | 0 | 0 | Low-quality | Genome-fragment | 3.02 | AAI-based (medium-confidence) | 0 |
| S5_extracted_k141_140153 | 1495 | No | NA | 1 | 1 | 0 | Low-quality | Genome-fragment | 23.95 | AAI-based (high-confidence) | 0 |
| S1_vs_k141_81575\|\|full | 3625 | No | NA | 2 | 0 | 2 | Not-determined | Genome-fragment | NA | NA | 0 |
| S5_extracted_k141_121069 | 1495 | No | NA | 4 | 0 | 0 | Low-quality | Genome-fragment | 3.18 | AAI-based (high-confidence) | 0 |
| S13_vs_111195\|\|full | 1496 | No | NA | 3 | 0 | 0 | Low-quality | Genome-fragment | 3.77 | AAI-based (medium-confidence) | 0 |
| S13_vs_110649\|\|lt2gene | 1496 | No | NA | 1 | 1 | 0 | Low-quality | Genome-fragment | 2.41 | AAI-based (high-confidence) | 0 |
| S1_vs_k141_68137\|\|full | 10139 | No | NA | 9 | 0 | 5 | Not-determined | Genome-fragment | NA | NA | 0 |
| S1_vs_k141_108132\|\|full | 2217 | No | NA | 3 | 0 | 0 | Not-determined | Genome-fragment | NA | NA | 0 |
| S1_vs_k141_154366\|\|full | 5645 | No | NA | 2 | 0 | 2 | Not-determined | Genome-fragment | NA | NA | 0 |
| S1_vs_k141_21439\|\|full | 32806 | No | NA | 56 | 15 | 0 | Medium-quality | Genome-fragment | 77.41 | AAI-based (high-confidence) | 0 |
| S3_vs_k141_58300\|\|full | 1496 | No | NA | 3 | 1 | 0 | Low-quality | Genome-fragment | 0.89 | HMM-based (lower-bound) | 0 |
| S12_vs_k141_168499\|\|full | 1496 | No | NA | 2 | 2 | 0 | Low-quality | Genome-fragment | 3.24 | AAI-based (high-confidence) | 0 |
| S1_vs_k141_54880\|\|full | 2549 | No | NA | 3 | 0 | 1 | Not-determined | Genome-fragment | NA | NA | 0 |
| S12_vs_k141_30225\|\|full | 1496 | No | NA | 2 | 0 | 0 | Low-quality | Genome-fragment | 0.74 | AAI-based (high-confidence) | 0 |
| S1_vs_k141_15193\|\|full | 1661 | No | NA | 3 | 0 | 2 | Not-determined | Genome-fragment | NA | NA | 0 |
| S15_extracted_57194 | 1496 | No | NA | 1 | 0 | 1 | Low-quality | Genome-fragment | 0.25 | AAI-based (high-confidence) | 0 |
| S2_extracted_k141_13597 | 1496 | No | NA | 2 | 2 | 0 | Low-quality | Genome-fragment | 4.94 | HMM-based (lower-bound) | 0 |
| S1_vs_k141_154403\|\|full | 2774 | No | NA | 5 | 0 | 0 | Not-determined | Genome-fragment | NA | NA | 0 |
| S1_vs_k141_134887\|\|full | 4360 | No | NA | 2 | 0 | 2 | Not-determined | Genome-fragment | NA | NA | 0 |
| S6_vs_k141_26066\|\|full | 1496 | No | NA | 5 | 0 | 2 | Low-quality | Genome-fragment | 1.23 | AAI-based (medium-confidence) | 0 |
| S1_vs_k141_95065\|\|full | 2223 | No | NA | 5 | 0 | 1 | Not-determined | Genome-fragment | NA | NA | 0 |
| S6_vs_k141_36924\|\|full | 1496 | No | NA | 1 | 0 | 0 | Low-quality | Genome-fragment | 2.61 | AAI-based (high-confidence) | 0 |
| S8_extracted_k141_180053 | 1496 | No | NA | 1 | 0 | 0 | Low-quality | Genome-fragment | 0.93 | AAI-based (medium-confidence) | 0 |
| S1_vs_k141_101747\|\|full | 1505 | No | NA | 2 | 0 | 0 | Not-determined | Genome-fragment | NA | NA | 0 |
| S9_extracted_k141_1832 | 1497 | No | NA | 4 | 0 | 0 | Low-quality | Genome-fragment | 2.49 | AAI-based (medium-confidence) | 0 |
| S9_vs_k141_65292\|\|full | 1497 | No | NA | 4 | 0 | 0 | Low-quality | Genome-fragment | 4.37 | AAI-based (medium-confidence) | 0 |
| S1_vs_k141_1144\|\|full | 1497 | No | NA | 4 | 3 | 0 | Low-quality | Genome-fragment | 4.31 | AAI-based (high-confidence) | 0 |
| S1_vs_k141_111313\|\|full | 1497 | No | NA | 2 | 1 | 0 | Low-quality | Genome-fragment | 4.01 | AAI-based (high-confidence) | 0 |
| S12_vs_k141_14922\|\|full | 1497 | No | NA | 4 | 0 | 1 | Low-quality | Genome-fragment | 1.22 | AAI-based (medium-confidence) | 0 |
| S1_vs_k141_134935\|\|full | 1340 | No | NA | 2 | 0 | 1 | Not-determined | Genome-fragment | NA | NA | 0 |
| S6_vs_k141_8702\|\|full | 1497 | No | NA | 4 | 2 | 0 | Low-quality | Genome-fragment | 2.67 | HMM-based (lower-bound) | 0 |
| S8_extracted_k141_174010 | 1497 | No | NA | 2 | 0 | 2 | Low-quality | Genome-fragment | 0.94 | AAI-based (high-confidence) | 0 |
| S1_vs_k141_55011\|\|full | 1348 | No | NA | 3 | 0 | 2 | Not-determined | Genome-fragment | NA | NA | 0 |
| S8_extracted_k141_192038 | 1497 | No | NA | 2 | 1 | 0 | Low-quality | Genome-fragment | 2.32 | AAI-based (high-confidence) | 0 |
| S9_vs_k141_49234\|\|full | 1498 | No | NA | 3 | 1 | 0 | Low-quality | Genome-fragment | 3.09 | AAI-based (medium-confidence) | 0 |
| S1_vs_k141_55032\|\|full | 3120 | No | NA | 3 | 0 | 1 | Not-determined | Genome-fragment | NA | NA | 0 |
| S1_vs_k141_108318\|\|full | 1653 | No | NA | 3 | 0 | 0 | Not-determined | Genome-fragment | NA | NA | 0 |
| S1_vs_k141_128440\|\|full | 1047 | No | NA | 2 | 0 | 1 | Not-determined | Genome-fragment | NA | NA | 0 |
| S1_vs_k141_41754\|\|full | 6172 | No | NA | 2 | 0 | 2 | Not-determined | Genome-fragment | NA | NA | 0 |
| S9_vs_k141_52995\|\|full | 1498 | No | NA | 3 | 2 | 0 | Low-quality | Genome-fragment | 3.2 | AAI-based (high-confidence) | 0 |
| S1_vs_k141_15337\|\|full | 6505 | No | NA | 2 | 0 | 2 | Not-determined | Genome-fragment | NA | NA | 0 |
| S1_vs_k141_34749\|\|full | 2061 | No | NA | 4 | 0 | 1 | Not-determined | Genome-fragment | NA | NA | 0 |
| S11_vs_k141_37716\|\|full | 1498 | No | NA | 3 | 0 | 1 | Low-quality | Genome-fragment | 1.87 | AAI-based (medium-confidence) | 0 |
| S14_vs_k141_109894\|\|full | 1498 | No | NA | 3 | 0 | 0 | Low-quality | Genome-fragment | 2.88 | AAI-based (high-confidence) | 0 |
| S1_vs_k141_121573\|\|full | 1491 | No | NA | 2 | 0 | 1 | Not-determined | Genome-fragment | NA | NA | 0 |
| S1_vs_k141_48428\|\|full | 2078 | No | NA | 3 | 0 | 2 | Not-determined | Genome-fragment | NA | NA | 0 |
| S14_vs_k141_28423\|\|full | 1498 | No | NA | 4 | 0 | 0 | Low-quality | Genome-fragment | 3.38 | AAI-based (medium-confidence) | 0 |
| S1_vs_k141_101835\|\|full | 1082 | No | NA | 3 | 0 | 1 | Not-determined | Genome-fragment | NA | NA | 0 |
| S1_extracted_k141_117652 | 1498 | No | NA | 2 | 0 | 1 | Low-quality | Genome-fragment | 1.96 | AAI-based (medium-confidence) | 0 |
| S1_vs_k141_37922\|\|full | 1498 | No | NA | 4 | 2 | 0 | Low-quality | Genome-fragment | 2.45 | AAI-based (high-confidence) | 0 |
| S5_extracted_k141_131615 | 1498 | No | NA | 2 | 0 | 1 | Low-quality | Genome-fragment | 1.77 | AAI-based (medium-confidence) | 0 |
| S1_vs_k141_8461\|\|full | 1740 | No | NA | 3 | 0 | 1 | Not-determined | Genome-fragment | NA | NA | 0 |
| S10_extracted_15889 | 1498 | No | NA | 2 | 0 | 1 | Low-quality | Genome-fragment | 3.82 | AAI-based (medium-confidence) | 0 |
| S17_extracted_k141_82683 | 1499 | No | NA | 2 | 0 | 2 | Low-quality | Genome-fragment | 0.71 | AAI-based (medium-confidence) | 0 |
| S9_vs_k141_45469\|\|full | 1499 | No | NA | 5 | 0 | 1 | Low-quality | Genome-fragment | 1.33 | AAI-based (medium-confidence) | 0 |
| S16_vs_109445\|\|full | 1499 | No | NA | 3 | 2 | 0 | Low-quality | Genome-fragment | 2.83 | AAI-based (medium-confidence) | 0 |
| S10_vs_38278\|\|full | 1499 | No | NA | 2 | 0 | 1 | Low-quality | Genome-fragment | 4.23 | AAI-based (medium-confidence) | 0 |
| S11_vs_k141_24819\|\|lt2gene | 1500 | No | NA | 2 | 1 | 0 | Low-quality | Genome-fragment | 3.79 | AAI-based (high-confidence) | 0 |
| S14_extracted_k141_62978 | 1500 | No | NA | 2 | 1 | 0 | Low-quality | Genome-fragment | 2.52 | AAI-based (high-confidence) | 0 |
| S1_vs_k141_90132\|\|lt2gene | 1500 | No | NA | 1 | 1 | 0 | Low-quality | Genome-fragment | 0.97 | AAI-based (high-confidence) | 0 |
| S1_vs_k141_15413\|\|full | 5143 | No | NA | 3 | 0 | 1 | Not-determined | Genome-fragment | NA | NA | 0 |
| S1_vs_k141_75149\|\|full | 8778 | No | NA | 8 | 0 | 6 | Not-determined | Genome-fragment | NA | NA | 0 |
| S3_vs_k141_80771\|\|full | 1500 | No | NA | 3 | 0 | 0 | Low-quality | Genome-fragment | 2.64 | AAI-based (medium-confidence) | 0 |
| S1_vs_k141_8517\|\|full | 1147 | No | NA | 3 | 0 | 0 | Not-determined | Genome-fragment | NA | NA | 0 |
| S3_vs_k141_103446\|\|lt2gene | 1500 | No | NA | 2 | 2 | 0 | Low-quality | Genome-fragment | 1.07 | AAI-based (high-confidence) | 0 |
| S1_vs_k141_15446\|\|full | 1306 | No | NA | 2 | 0 | 1 | Not-determined | Genome-fragment | NA | NA | 0 |
| S10_vs_4673\|\|full | 1500 | No | NA | 3 | 1 | 0 | Low-quality | Genome-fragment | 2.88 | AAI-based (high-confidence) | 0 |
| S1_vs_k141_75193\|\|full | 2576 | No | NA | 6 | 0 | 1 | Not-determined | Genome-fragment | NA | NA | 0 |
| S10_vs_24357\|\|full | 1500 | No | NA | 4 | 0 | 0 | Low-quality | Genome-fragment | 4.13 | AAI-based (medium-confidence) | 0 |
| S1_vs_k141_88467\|\|full | 1318 | No | NA | 2 | 0 | 1 | Not-determined | Genome-fragment | NA | NA | 0 |
| S1_vs_k141_154596\|\|full | 3147 | No | NA | 3 | 0 | 3 | Not-determined | Genome-fragment | NA | NA | 0 |
| S12_vs_k141_76139\|\|full | 1500 | No | NA | 4 | 0 | 0 | Low-quality | Genome-fragment | 2.89 | AAI-based (high-confidence) | 0 |
| S1_vs_k141_154608\|\|full | 5978 | No | NA | 6 | 0 | 6 | Not-determined | Genome-fragment | NA | NA | 0 |
| S17_vs_k141_129213\|\|lt2gene | 1501 | No | NA | 1 | 1 | 0 | Low-quality | Genome-fragment | 2.63 | HMM-based (lower-bound) | 0 |
| S1_vs_k141_88512\|\|full | 1020 | No | NA | 2 | 0 | 0 | Not-determined | Genome-fragment | NA | NA | 0 |
| S1_vs_k141_61059\|\|full | 1089 | No | NA | 2 | 0 | 0 | Not-determined | Genome-fragment | NA | NA | 0 |
| S11_vs_k141_79681\|\|full | 1501 | No | NA | 6 | 0 | 0 | Low-quality | Genome-fragment | 3.45 | AAI-based (medium-confidence) | 0 |
| S1_vs_k141_8591\|\|full | 5231 | No | NA | 5 | 0 | 3 | Not-determined | Genome-fragment | NA | NA | 0 |
| S1_vs_k141_68518\|\|full | 3310 | No | NA | 3 | 0 | 2 | Not-determined | Genome-fragment | NA | NA | 0 |
| S14_vs_k141_15631\|\|full | 1501 | No | NA | 3 | 0 | 0 | Low-quality | Genome-fragment | 3.71 | AAI-based (high-confidence) | 0 |
| S1_vs_k141_48597\|\|full | 1029 | No | NA | 2 | 0 | 0 | Not-determined | Genome-fragment | NA | NA | 0 |
| S1_vs_k141_15540\|\|full | 2061 | No | NA | 3 | 0 | 0 | Not-determined | Genome-fragment | NA | NA | 0 |
| S3_vs_k141_112710\|\|full | 1501 | No | NA | 3 | 1 | 0 | Low-quality | Genome-fragment | 3.6 | AAI-based (high-confidence) | 0 |
| S1_vs_k141_115256\|\|full | 2231 | No | NA | 5 | 0 | 0 | Not-determined | Genome-fragment | NA | NA | 0 |
| S5_vs_k141_156\|\|full | 1501 | No | NA | 3 | 2 | 0 | Low-quality | Genome-fragment | 2.41 | HMM-based (lower-bound) | 0 |
| S1_vs_k141_141964\|\|full | 2950 | No | NA | 3 | 0 | 3 | Not-determined | Genome-fragment | NA | NA | 0 |
| S7_vs_10561\|\|full | 1501 | No | NA | 2 | 0 | 1 | Low-quality | Genome-fragment | 1.26 | AAI-based (medium-confidence) | 0 |
| S1_vs_k141_21832\|\|full | 5391 | No | NA | 8 | 0 | 6 | Not-determined | Genome-fragment | NA | NA | 0 |
| S12_extracted_k141_14602 | 1501 | No | NA | 3 | 1 | 0 | Low-quality | Genome-fragment | 3.83 | AAI-based (high-confidence) | 0 |
| S12_vs_k141_91868\|\|full | 1501 | No | NA | 4 | 1 | 0 | Low-quality | Genome-fragment | 2.9 | AAI-based (high-confidence) | 0 |
| S12_vs_k141_147282\|\|full | 1501 | No | NA | 3 | 0 | 0 | Low-quality | Genome-fragment | 3.18 | AAI-based (high-confidence) | 0 |
| S12_vs_k141_148474\|\|full | 1501 | No | NA | 2 | 0 | 0 | Low-quality | Genome-fragment | 0.84 | AAI-based (high-confidence) | 0 |
| S1_vs_k141_61092\|\|full | 1014 | No | NA | 2 | 0 | 0 | Not-determined | Genome-fragment | NA | NA | 0 |
| S6_vs_k141_49906\|\|full | 1501 | No | NA | 4 | 1 | 0 | Low-quality | Genome-fragment | 2.62 | AAI-based (high-confidence) | 0 |
| S1_vs_k141_75354\|\|full | 1294 | No | NA | 2 | 0 | 1 | Not-determined | Genome-fragment | NA | NA | 0 |
| S1_vs_k141_28600\|\|full | 2250 | No | NA | 3 | 0 | 2 | Not-determined | Genome-fragment | NA | NA | 0 |
| S6_vs_k141_27445\|\|full | 1501 | No | NA | 3 | 0 | 0 | Low-quality | Genome-fragment | 4.26 | AAI-based (high-confidence) | 0 |
| S9_extracted_k141_70925 | 1502 | No | NA | 1 | 0 | 0 | Low-quality | Genome-fragment | 1.38 | AAI-based (high-confidence) | 0 |
| S9_vs_k141_18162\|\|full | 1502 | No | NA | 2 | 0 | 2 | Low-quality | Genome-fragment | 0.8 | AAI-based (medium-confidence) | 0 |
| S13_vs_34701\|\|full | 1502 | No | NA | 3 | 3 | 0 | Low-quality | Genome-fragment | 2.46 | AAI-based (high-confidence) | 0 |
| S13_vs_53690\|\|full | 1502 | No | NA | 2 | 1 | 0 | Low-quality | Genome-fragment | 3.63 | AAI-based (medium-confidence) | 0 |
| S1_vs_k141_68889\|\|full | 1502 | No | NA | 4 | 1 | 0 | Low-quality | Genome-fragment | 4.23 | AAI-based (high-confidence) | 0 |
| S3_vs_k141_126464\|\|full | 1502 | No | NA | 2 | 0 | 0 | Low-quality | Genome-fragment | 2.73 | AAI-based (medium-confidence) | 0 |
| S3_vs_k141_80041\|\|lt2gene | 1502 | No | NA | 2 | 2 | 0 | Low-quality | Genome-fragment | 3.21 | AAI-based (high-confidence) | 0 |
| S1_vs_k141_75403\|\|full | 8872 | No | NA | 8 | 0 | 5 | Not-determined | Genome-fragment | NA | NA | 0 |
| S5_extracted_k141_98794 | 1502 | No | NA | 1 | 1 | 0 | Low-quality | Genome-fragment | 4.17 | AAI-based (high-confidence) | 0 |
| S1_vs_k141_68686\|\|full | 1291 | No | NA | 3 | 0 | 3 | Not-determined | Genome-fragment | NA | NA | 0 |
| S1_vs_k141_8760\|\|full | 1073 | No | NA | 2 | 0 | 0 | Not-determined | Genome-fragment | NA | NA | 0 |
| S5_vs_k141_138995\|\|lt2gene | 1502 | No | NA | 1 | 0 | 0 | Low-quality | Genome-fragment | 4.61 | AAI-based (high-confidence) | 0 |
| S7_vs_70290\|\|full | 1502 | No | NA | 4 | 0 | 2 | Low-quality | Genome-fragment | 0.65 | AAI-based (medium-confidence) | 0 |
| S1_vs_k141_48783\|\|full | 1206 | No | NA | 2 | 0 | 1 | Not-determined | Genome-fragment | NA | NA | 0 |
| S1_vs_k141_21936\|\|full | 2382 | No | NA | 4 | 0 | 1 | Not-determined | Genome-fragment | NA | NA | 0 |
| S16_extracted_72283 | 1502 | No | NA | 3 | 0 | 0 | Low-quality | Genome-fragment | 2.36 | AAI-based (high-confidence) | 0 |
| S1_vs_k141_102196\|\|full | 3843 | No | NA | 6 | 0 | 5 | Not-determined | Genome-fragment | NA | NA | 0 |
| S1_vs_k141_48796\|\|full | 1422 | No | NA | 2 | 0 | 1 | Not-determined | Genome-fragment | NA | NA | 0 |
| S1_vs_k141_82190\|\|full | 2823 | No | NA | 3 | 0 | 2 | Not-determined | Genome-fragment | NA | NA | 0 |
| S10_extracted_85642 | 1502 | No | NA | 1 | 0 | 0 | Low-quality | Genome-fragment | 2.27 | AAI-based (high-confidence) | 0 |
| S1_vs_k141_1696\|\|full | 1403 | No | NA | 3 | 0 | 1 | Not-determined | Genome-fragment | NA | NA | 0 |
| S10_vs_40985\|\|full | 1502 | No | NA | 3 | 0 | 0 | Low-quality | Genome-fragment | 2.78 | AAI-based (medium-confidence) | 0 |
| S15_extracted_38566 | 1502 | No | NA | 1 | 0 | 1 | Low-quality | Genome-fragment | 0.56 | AAI-based (high-confidence) | 0 |
| S2_extracted_k141_18690 | 1502 | No | NA | 1 | 0 | 1 | Low-quality | Genome-fragment | 0.61 | AAI-based (medium-confidence) | 0 |
| S1_vs_k141_68723\|\|full | 2570 | No | NA | 3 | 0 | 2 | Not-determined | Genome-fragment | NA | NA | 0 |
| S1_vs_k141_88772\|\|full | 1381 | No | NA | 3 | 0 | 0 | Not-determined | Genome-fragment | NA | NA | 0 |
| S1_vs_k141_88777\|\|full | 4338 | No | NA | 3 | 0 | 2 | Not-determined | Genome-fragment | NA | NA | 0 |
| S6_extracted_k141_9626 | 1502 | No | NA | 6 | 2 | 0 | Low-quality | Genome-fragment | 2.78 | AAI-based (high-confidence) | 0 |
| S1_vs_k141_35057\|\|full | 1584 | No | NA | 3 | 0 | 0 | Not-determined | Genome-fragment | NA | NA | 0 |
| S6_vs_k141_48295\|\|full | 1502 | No | NA | 4 | 1 | 0 | Low-quality | Genome-fragment | 4.21 | AAI-based (high-confidence) | 0 |
| S1_vs_k141_95592\|\|full | 2377 | No | NA | 2 | 0 | 2 | Not-determined | Genome-fragment | NA | NA | 0 |
| S1_vs_k141_121990\|\|full | 1241 | No | NA | 3 | 0 | 0 | Not-determined | Genome-fragment | NA | NA | 0 |
| S8_vs_k141_176184\|\|full | 1502 | No | NA | 6 | 1 | 0 | Low-quality | Genome-fragment | 3.29 | AAI-based (high-confidence) | 0 |
| S1_vs_k141_75515\|\|full | 10470 | No | NA | 11 | 0 | 2 | Not-determined | Genome-fragment | NA | NA | 0 |
| S1_vs_k141_82263\|\|full | 1090 | No | NA | 2 | 0 | 1 | Not-determined | Genome-fragment | NA | NA | 0 |
| S9_vs_k141_82634\|\|full | 1503 | No | NA | 3 | 0 | 0 | Low-quality | Genome-fragment | 3.95 | AAI-based (medium-confidence) | 0 |
| S11_vs_k141_40002\|\|full | 1503 | No | NA | 5 | 1 | 0 | Low-quality | Genome-fragment | 3.22 | AAI-based (medium-confidence) | 0 |
| S1_vs_k141_154851\|\|full | 1186 | No | NA | 2 | 0 | 1 | Not-determined | Genome-fragment | NA | NA | 0 |
| S1_vs_k141_82307\|\|full | 5383 | No | NA | 4 | 0 | 3 | Not-determined | Genome-fragment | NA | NA | 0 |
| S1_vs_k141_28822\|\|full | 2160 | No | NA | 2 | 0 | 0 | Not-determined | Genome-fragment | NA | NA | 0 |
| S1_vs_k141_22105\|\|full | 3011 | No | NA | 4 | 0 | 0 | Not-determined | Genome-fragment | NA | NA | 0 |
| S1_vs_k141_35134\|\|full | 3044 | No | NA | 4 | 0 | 3 | Not-determined | Genome-fragment | NA | NA | 0 |
| S11_vs_k141_102210\|\|full | 1503 | No | NA | 3 | 1 | 0 | Low-quality | Genome-fragment | 3.41 | AAI-based (medium-confidence) | 0 |
| S12_extracted_k141_169020 | 1503 | No | NA | 2 | 0 | 1 | Low-quality | Genome-fragment | 0.99 | AAI-based (high-confidence) | 0 |
| S1_vs_k141_8955\|\|full | 3002 | No | NA | 2 | 0 | 1 | Not-determined | Genome-fragment | NA | NA | 0 |
| S12_vs_k141_238\|\|full | 1503 | No | NA | 4 | 3 | 0 | Low-quality | Genome-fragment | 1.88 | HMM-based (lower-bound) | 0 |
| S1_vs_k141_42301\|\|full | 1758 | No | NA | 2 | 0 | 1 | Not-determined | Genome-fragment | NA | NA | 0 |
| S1_vs_k141_35182\|\|full | 1350 | No | NA | 4 | 0 | 0 | Not-determined | Genome-fragment | NA | NA | 0 |
| S1_vs_k141_16041\|\|full | 5415 | No | NA | 2 | 0 | 2 | Not-determined | Genome-fragment | NA | NA | 0 |
| S12_vs_k141_14382\|\|full | 1503 | No | NA | 3 | 1 | 0 | Low-quality | Genome-fragment | 3.69 | AAI-based (high-confidence) | 0 |
| S1_vs_k141_75659\|\|full | 4347 | No | NA | 5 | 0 | 2 | Not-determined | Genome-fragment | NA | NA | 0 |
| S8_vs_k141_106204\|\|full | 1503 | No | NA | 5 | 0 | 0 | Low-quality | Genome-fragment | 3.57 | AAI-based (high-confidence) | 0 |
| S8_vs_k141_185701\|\|full | 1503 | No | NA | 2 | 0 | 1 | Low-quality | Genome-fragment | 1.76 | AAI-based (high-confidence) | 0 |
| S8_vs_k141_60058\|\|full | 1503 | No | NA | 2 | 0 | 1 | Low-quality | Genome-fragment | 0.36 | AAI-based (medium-confidence) | 0 |
| S9_extracted_k141_23833 | 1504 | No | NA | 2 | 0 | 1 | Low-quality | Genome-fragment | 3.11 | AAI-based (high-confidence) | 0 |
| S1_vs_k141_82405\|\|full | 3623 | No | NA | 3 | 0 | 2 | Not-determined | Genome-fragment | NA | NA | 0 |
| S1_vs_k141_122195\|\|full | 1387 | No | NA | 2 | 0 | 1 | Not-determined | Genome-fragment | NA | NA | 0 |
| S11_extracted_k141_22197 | 1504 | No | NA | 3 | 1 | 0 | Low-quality | Genome-fragment | 3.59 | AAI-based (high-confidence) | 0 |
| S1_vs_k141_122206\|\|full | 3004 | No | NA | 3 | 0 | 0 | Not-determined | Genome-fragment | NA | NA | 0 |
| S14_vs_k141_36876\|\|full | 1504 | No | NA | 4 | 2 | 0 | Low-quality | Genome-fragment | 2.64 | AAI-based (medium-confidence) | 0 |
| S14_vs_k141_99299\|\|full | 1504 | No | NA | 3 | 2 | 0 | Low-quality | Genome-fragment | 3.75 | AAI-based (medium-confidence) | 0 |
| S1_vs_k141_16117\|\|full | 1110 | No | NA | 3 | 0 | 0 | Not-determined | Genome-fragment | NA | NA | 0 |
| S14_vs_k141_62003\|\|lt2gene | 1504 | No | NA | 2 | 2 | 0 | Low-quality | Genome-fragment | 3.86 | AAI-based (high-confidence) | 0 |
| S16_vs_38912\|\|full | 1504 | No | NA | 4 | 0 | 0 | Low-quality | Genome-fragment | 4.18 | AAI-based (high-confidence) | 0 |
| S12_vs_k141_161809\|\|full | 1504 | No | NA | 3 | 0 | 0 | Low-quality | Genome-fragment | 2.21 | AAI-based (high-confidence) | 0 |
| S1_vs_k141_122257\|\|full | 1604 | No | NA | 3 | 0 | 0 | Not-determined | Genome-fragment | NA | NA | 0 |
| S9_extracted_k141_13348 | 1505 | No | NA | 3 | 2 | 0 | Low-quality | Genome-fragment | 0.86 | HMM-based (lower-bound) | 0 |
| S11_extracted_k141_30889 | 1505 | No | NA | 1 | 0 | 0 | Low-quality | Genome-fragment | 3.65 | AAI-based (medium-confidence) | 0 |
| S1_vs_k141_28994\|\|full | 10611 | No | NA | 9 | 0 | 9 | Not-determined | Genome-fragment | NA | NA | 0 |
| S1_vs_k141_102522\|\|full | 2342 | No | NA | 3 | 0 | 3 | Not-determined | Genome-fragment | NA | NA | 0 |
| S1_vs_k141_142408\|\|full | 3908 | No | NA | 3 | 0 | 2 | Not-determined | Genome-fragment | NA | NA | 0 |
| S1_vs_k141_22216\|\|full | 8041 | No | NA | 6 | 0 | 6 | Not-determined | Genome-fragment | NA | NA | 0 |
| S14_extracted_k141_2730 | 1505 | No | NA | 3 | 2 | 0 | Low-quality | Genome-fragment | 4.83 | AAI-based (high-confidence) | 0 |
| S3_extracted_k141_81982 | 1505 | No | NA | 3 | 1 | 0 | Low-quality | Genome-fragment | 0.9 | HMM-based (lower-bound) | 0 |
| S1_vs_k141_109039\|\|full | 1301 | No | NA | 2 | 0 | 1 | Not-determined | Genome-fragment | NA | NA | 0 |
| S1_vs_k141_16203\|\|full | 2647 | No | NA | 4 | 0 | 1 | Not-determined | Genome-fragment | NA | NA | 0 |
| S1_vs_k141_16216\|\|full | 1001 | No | NA | 3 | 0 | 0 | Not-determined | Genome-fragment | NA | NA | 0 |
| S1_vs_k141_75836\|\|full | 1568 | No | NA | 2 | 0 | 1 | Not-determined | Genome-fragment | NA | NA | 0 |
| S1_vs_k141_109088\|\|full | 3552 | No | NA | 7 | 0 | 0 | Not-determined | Genome-fragment | NA | NA | 0 |
| S10_extracted_15917 | 1505 | No | NA | 2 | 0 | 2 | Low-quality | Genome-fragment | 0.48 | AAI-based (medium-confidence) | 0 |
| S1_vs_k141_55858\|\|full | 1145 | No | NA | 2 | 0 | 0 | Not-determined | Genome-fragment | NA | NA | 0 |
| S6_extracted_k141_44267 | 1505 | No | NA | 4 | 4 | 0 | Low-quality | Genome-fragment | 3.78 | AAI-based (high-confidence) | 0 |
| S14_vs_k141_68515\|\|full | 1506 | No | NA | 4 | 0 | 0 | Low-quality | Genome-fragment | 3.54 | AAI-based (high-confidence) | 0 |
| S14_vs_k141_107992\|\|full | 1506 | No | NA | 3 | 3 | 0 | Low-quality | Genome-fragment | 3.07 | AAI-based (high-confidence) | 0 |
| S1_vs_k141_109128\|\|full | 1302 | No | NA | 2 | 0 | 0 | Not-determined | Genome-fragment | NA | NA | 0 |
| S1_extracted_k141_81250 | 1506 | No | NA | 3 | 1 | 0 | Low-quality | Genome-fragment | 1.91 | AAI-based (high-confidence) | 0 |
| S1_vs_k141_155039\|\|full | 2093 | No | NA | 4 | 0 | 2 | Not-determined | Genome-fragment | NA | NA | 0 |
| S7_vs_58691\|\|full | 1506 | No | NA | 5 | 0 | 0 | Low-quality | Genome-fragment | 2.29 | AAI-based (medium-confidence) | 0 |
| S7_vs_114260\|\|full | 1506 | No | NA | 3 | 2 | 0 | Low-quality | Genome-fragment | 3.31 | AAI-based (high-confidence) | 0 |
| S12_extracted_k141_24553 | 1506 | No | NA | 2 | 1 | 0 | Low-quality | Genome-fragment | 2.33 | AAI-based (high-confidence) | 0 |
| S1_vs_k141_82604\|\|full | 1872 | No | NA | 2 | 0 | 0 | Not-determined | Genome-fragment | NA | NA | 0 |
| S1_vs_k141_135847\|\|full | 4568 | No | NA | 3 | 0 | 2 | Not-determined | Genome-fragment | NA | NA | 0 |
| S1_vs_k141_16300\|\|full | 1514 | No | NA | 2 | 0 | 2 | Not-determined | Genome-fragment | NA | NA | 0 |
| S1_vs_k141_147943\|\|full | 1064 | No | NA | 2 | 0 | 1 | Not-determined | Genome-fragment | NA | NA | 0 |
| S1_vs_k141_129321\|\|full | 3811 | No | NA | 5 | 0 | 0 | Not-determined | Genome-fragment | NA | NA | 0 |
| S1_vs_k141_155058\|\|full | 1769 | No | NA | 4 | 0 | 0 | Not-determined | Genome-fragment | NA | NA | 0 |
| S12_vs_k141_187265\|\|full | 1506 | No | NA | 3 | 0 | 0 | Low-quality | Genome-fragment | 2.57 | AAI-based (medium-confidence) | 0 |
| S8_extracted_k141_73613 | 1506 | No | NA | 2 | 0 | 0 | Low-quality | Genome-fragment | 1.94 | AAI-based (high-confidence) | 0 |
| S1_vs_k141_82662\|\|full | 3142 | No | NA | 5 | 0 | 1 | Not-determined | Genome-fragment | NA | NA | 0 |
| S1_vs_k141_102687\|\|full | 1891 | No | NA | 3 | 0 | 2 | Not-determined | Genome-fragment | NA | NA | 0 |
| S1_vs_k141_22361\|\|full | 1904 | No | NA | 3 | 0 | 2 | Not-determined | Genome-fragment | NA | NA | 0 |
| S1_vs_k141_89189\|\|full | 1346 | No | NA | 2 | 0 | 0 | Not-determined | Genome-fragment | NA | NA | 0 |
| S1_vs_k141_22371\|\|full | 1896 | No | NA | 3 | 0 | 0 | Not-determined | Genome-fragment | NA | NA | 0 |
| S8_vs_k141_179623\|\|full | 1506 | No | NA | 3 | 0 | 0 | Low-quality | Genome-fragment | 0.5 | AAI-based (medium-confidence) | 0 |
| S1_vs_k141_82691\|\|full | 1412 | No | NA | 3 | 0 | 3 | Not-determined | Genome-fragment | NA | NA | 0 |
| S9_vs_k141_59994\|\|full | 1507 | No | NA | 3 | 0 | 1 | Low-quality | Genome-fragment | 2.67 | AAI-based (medium-confidence) | 0 |
| S14_vs_k141_113117\|\|full | 1507 | No | NA | 2 | 1 | 0 | Low-quality | Genome-fragment | 4.08 | AAI-based (high-confidence) | 0 |
| S14_vs_k141_67369\|\|lt2gene | 1507 | No | NA | 2 | 1 | 0 | Low-quality | Genome-fragment | 3.87 | AAI-based (high-confidence) | 0 |
| S1_vs_k141_9343\|\|full | 1024 | No | NA | 2 | 0 | 0 | Not-determined | Genome-fragment | NA | NA | 0 |
| S1_vs_k141_56020\|\|full | 1382 | No | NA | 3 | 0 | 0 | Not-determined | Genome-fragment | NA | NA | 0 |
| S1_extracted_k141_50205 | 1507 | No | NA | 1 | 0 | 1 | Low-quality | Genome-fragment | 0.26 | AAI-based (high-confidence) | 0 |
| S5_vs_k141_15042\|\|full | 1507 | No | NA | 3 | 0 | 0 | Low-quality | Genome-fragment | 3.38 | AAI-based (medium-confidence) | 0 |
| S1_vs_k141_69231\|\|full | 5411 | No | NA | 2 | 0 | 1 | Not-determined | Genome-fragment | NA | NA | 0 |
| S1_vs_k141_109289\|\|full | 1094 | No | NA | 2 | 0 | 1 | Not-determined | Genome-fragment | NA | NA | 0 |
| S5_vs_k141_99502\|\|full | 1507 | No | NA | 6 | 1 | 0 | Low-quality | Genome-fragment | 3.4 | AAI-based (medium-confidence) | 0 |
| S7_vs_20432\|\|full | 1507 | No | NA | 2 | 1 | 0 | Low-quality | Genome-fragment | 2.33 | HMM-based (lower-bound) | 0 |
| S10_extracted_83392 | 1507 | No | NA | 1 | 0 | 1 | Low-quality | Genome-fragment | 0.57 | AAI-based (high-confidence) | 0 |
| S15_vs_52164\|\|full | 1507 | No | NA | 3 | 0 | 0 | Low-quality | Genome-fragment | 5.22 | AAI-based (high-confidence) | 0 |
| S6_vs_k141_35734\|\|full | 1507 | No | NA | 4 | 0 | 2 | Low-quality | Genome-fragment | 1.13 | AAI-based (medium-confidence) | 0 |
| S6_vs_k141_24632\|\|full | 1507 | No | NA | 2 | 0 | 1 | Low-quality | Genome-fragment | 3.75 | AAI-based (medium-confidence) | 0 |
| S1_vs_k141_142716\|\|full | 1071 | No | NA | 3 | 0 | 1 | Not-determined | Genome-fragment | NA | NA | 0 |
| S1_vs_k141_9403\|\|full | 1876 | No | NA | 3 | 0 | 2 | Not-determined | Genome-fragment | NA | NA | 0 |
| S1_vs_k141_35492\|\|full | 1228 | No | NA | 3 | 0 | 0 | Not-determined | Genome-fragment | NA | NA | 0 |
| S17_vs_k141_17544\|\|full | 1508 | No | NA | 3 | 0 | 1 | Low-quality | Genome-fragment | 1.79 | AAI-based (high-confidence) | 0 |
| S9_extracted_k141_84567 | 1508 | No | NA | 2 | 2 | 0 | Low-quality | Genome-fragment | 3.36 | AAI-based (high-confidence) | 0 |
| S11_vs_k141_1011\|\|full | 1508 | No | NA | 3 | 3 | 0 | Low-quality | Genome-fragment | 3.41 | AAI-based (high-confidence) | 0 |
| S14_vs_k141_103462\|\|full | 1508 | No | NA | 3 | 0 | 1 | Low-quality | Genome-fragment | 1.45 | AAI-based (high-confidence) | 0 |
| S1_vs_k141_96192\|\|full | 3538 | No | NA | 2 | 0 | 1 | Not-determined | Genome-fragment | NA | NA | 0 |
| S3_vs_k141_48981\|\|full | 1508 | No | NA | 5 | 2 | 0 | Low-quality | Genome-fragment | 3.46 | AAI-based (high-confidence) | 0 |
| S1_vs_k141_76106\|\|full | 4018 | No | NA | 2 | 0 | 2 | Not-determined | Genome-fragment | NA | NA | 0 |
| S1_vs_k141_56110\|\|full | 1206 | No | NA | 3 | 0 | 0 | Not-determined | Genome-fragment | NA | NA | 0 |
| S3_vs_k141_30955\|\|full | 1508 | No | NA | 3 | 2 | 0 | Low-quality | Genome-fragment | 3.98 | AAI-based (high-confidence) | 0 |
| S1_vs_k141_96231\|\|full | 1314 | No | NA | 2 | 0 | 0 | Not-determined | Genome-fragment | NA | NA | 0 |
| S1_vs_k141_142793\|\|full | 3273 | No | NA | 4 | 0 | 3 | Not-determined | Genome-fragment | NA | NA | 0 |
| S1_vs_k141_49539\|\|full | 2576 | No | NA | 3 | 0 | 0 | Not-determined | Genome-fragment | NA | NA | 0 |
| S5_vs_k141_7756\|\|full | 1508 | No | NA | 4 | 0 | 0 | Low-quality | Genome-fragment | 2.59 | AAI-based (medium-confidence) | 0 |
| S16_vs_49456\|\|full | 1508 | No | NA | 3 | 0 | 1 | Low-quality | Genome-fragment | 1.04 | AAI-based (medium-confidence) | 0 |
| S1_vs_k141_2251\|\|full | 2079 | No | NA | 2 | 0 | 0 | Not-determined | Genome-fragment | NA | NA | 0 |
| S1_vs_k141_155183\|\|full | 30775 | No | NA | 51 | 12 | 0 | Medium-quality | Genome-fragment | 74.09 | AAI-based (high-confidence) | 0 |
| S10_extracted_95257 | 1508 | No | NA | 2 | 0 | 1 | Low-quality | Genome-fragment | 1.01 | AAI-based (high-confidence) | 0 |
| S8_vs_k141_127191\|\|full | 1508 | No | NA | 3 | 0 | 0 | Low-quality | Genome-fragment | 3.19 | AAI-based (medium-confidence) | 0 |
| S1_vs_k141_49565\|\|full | 1146 | No | NA | 3 | 0 | 0 | Not-determined | Genome-fragment | NA | NA | 0 |
| S8_vs_k141_207314\|\|full | 1508 | No | NA | 2 | 0 | 1 | Low-quality | Genome-fragment | 0.92 | AAI-based (high-confidence) | 0 |
| S1_vs_k141_102915\|\|full | 1119 | No | NA | 2 | 0 | 0 | Not-determined | Genome-fragment | NA | NA | 0 |
| S17_extracted_k141_33778 | 1509 | No | NA | 3 | 1 | 0 | Low-quality | Genome-fragment | 3.38 | AAI-based (medium-confidence) | 0 |
| S9_extracted_k141_76527 | 1509 | No | NA | 3 | 3 | 0 | Low-quality | Genome-fragment | 2.32 | AAI-based (high-confidence) | 0 |
| S11_vs_k141_53070\|\|lt2gene | 1509 | No | NA | 2 | 1 | 0 | Low-quality | Genome-fragment | 1.74 | HMM-based (lower-bound) | 0 |
| S14_extracted_k141_63150 | 1509 | No | NA | 2 | 1 | 0 | Low-quality | Genome-fragment | 1.18 | AAI-based (medium-confidence) | 0 |
| S1_vs_k141_42883\|\|full | 1064 | No | NA | 3 | 0 | 0 | Not-determined | Genome-fragment | NA | NA | 0 |
| S14_vs_k141_102751\|\|full | 1509 | No | NA | 3 | 1 | 0 | Low-quality | Genome-fragment | 4.42 | AAI-based (high-confidence) | 0 |
| S16_vs_71762\|\|full | 1509 | No | NA | 4 | 2 | 0 | Low-quality | Genome-fragment | 2.51 | AAI-based (high-confidence) | 0 |
| S10_extracted_15520 | 1509 | No | NA | 2 | 0 | 0 | Low-quality | Genome-fragment | 2.66 | AAI-based (high-confidence) | 0 |

| | | | | | | | | | | | |
|---|---|---|---|---|---|---|---|---|---|---|---|
| S1_vs_k141_49621\|\|full | 1270 | No | NA | 2 | 0 | 2 | Not-determined | Genome-fragment | NA | NA | 0 |
| S1_vs_k141_9574\|\|full | 2795 | No | NA | 3 | 0 | 3 | Not-determined | Genome-fragment | NA | NA | 0 |
| S1_vs_k141_9575\|\|full | 1167 | No | NA | 3 | 0 | 0 | Not-determined | Genome-fragment | NA | NA | 0 |
| S12_extracted_k141_119422 | 1509 | No | NA | 3 | 1 | 0 | Low-quality | Genome-fragment | 1.23 | AAI-based (medium-confidence) | 0 |
| S8_extracted_k141_14804 | 1509 | No | NA | 1 | 1 | 0 | Low-quality | Genome-fragment | 1.41 | HMM-based (lower-bound) | 0 |
| S1_vs_k141_9610\|\|full | 3585 | No | NA | 4 | 0 | 0 | Not-determined | Genome-fragment | NA | NA | 0 |
| S1_vs_k141_103022\|\|full | 1912 | No | NA | 2 | 0 | 1 | Not-determined | Genome-fragment | NA | NA | 0 |
| S8_vs_k141_151332\|\|lt2gene | 1509 | No | NA | 2 | 2 | 0 | Low-quality | Genome-fragment | 4.17 | AAI-based (medium-confidence) | 0 |
| S1_vs_k141_22687\|\|full | 2430 | No | NA | 2 | 0 | 0 | Not-determined | Genome-fragment | NA | NA | 0 |
| S1_vs_k141_83035\|\|full | 1463 | No | NA | 2 | 0 | 2 | Not-determined | Genome-fragment | NA | NA | 0 |
| S9_extracted_k141_86608 | 1510 | No | NA | 5 | 0 | 0 | Low-quality | Genome-fragment | 4.28 | AAI-based (high-confidence) | 0 |
| S9_vs_k141_58730\|\|full | 1510 | No | NA | 3 | 1 | 0 | Low-quality | Genome-fragment | 2.96 | AAI-based (high-confidence) | 0 |
| S14_extracted_k141_24935 | 1510 | No | NA | 2 | 1 | 0 | Low-quality | Genome-fragment | 3.38 | AAI-based (high-confidence) | 0 |
| S1_vs_k141_69490\|\|full | 1618 | No | NA | 3 | 0 | 0 | Not-determined | Genome-fragment | NA | NA | 0 |
| S3_extracted_k141_60749 | 1510 | No | NA | 5 | 0 | 0 | Low-quality | Genome-fragment | 1.45 | AAI-based (high-confidence) | 0 |
| S3_vs_k141_47443\|\|full | 1510 | No | NA | 3 | 0 | 0 | Low-quality | Genome-fragment | 2.68 | AAI-based (medium-confidence) | 0 |
| S1_vs_k141_22716\|\|full | 1264 | No | NA | 2 | 0 | 1 | Not-determined | Genome-fragment | NA | NA | 0 |
| S16_vs_40111\|\|full | 1510 | No | NA | 3 | 0 | 1 | Low-quality | Genome-fragment | 3.59 | AAI-based (medium-confidence) | 0 |
| S1_vs_k141_116400\|\|full | 2516 | No | NA | 2 | 0 | 2 | Not-determined | Genome-fragment | NA | NA | 0 |
| S1_vs_k141_49763\|\|full | 3479 | No | NA | 4 | 0 | 4 | Not-determined | Genome-fragment | NA | NA | 0 |
| S4_extracted_6674 | 1510 | No | NA | 2 | 0 | 2 | Low-quality | Genome-fragment | 0.8 | AAI-based (medium-confidence) | 0 |
| S6_vs_k141_33214\|\|full | 1510 | No | NA | 2 | 0 | 1 | Low-quality | Genome-fragment | 2.98 | AAI-based (medium-confidence) | 0 |
| S17_vs_k141_45968\|\|full | 1511 | No | NA | 3 | 1 | 0 | Low-quality | Genome-fragment | 3.94 | AAI-based (high-confidence) | 0 |
| S1_vs_k141_122805\|\|full | 2984 | No | NA | 3 | 0 | 0 | Not-determined | Genome-fragment | NA | NA | 0 |
| S11_extracted_k141_75426 | 1511 | No | NA | 2 | 0 | 2 | Low-quality | Genome-fragment | 2.54 | AAI-based (medium-confidence) | 0 |
| S13_extracted_87962 | 1511 | No | NA | 4 | 0 | 0 | Low-quality | Genome-fragment | 0.95 | AAI-based (high-confidence) | 0 |
| S14_extracted_k141_101845 | 1511 | No | NA | 1 | 0 | 0 | Low-quality | Genome-fragment | 28.43 | AAI-based (medium-confidence) | 0 |
| S7_vs_42464\|\|full | 1511 | No | NA | 4 | 1 | 0 | Low-quality | Genome-fragment | 2.4 | AAI-based (high-confidence) | 0 |
| S16_extracted_147795 | 1511 | No | NA | 1 | 0 | 0 | Low-quality | Genome-fragment | 1.78 | AAI-based (medium-confidence) | 0 |
| S1_vs_k141_148101\|\|full | 1629 | No | NA | 1 | 0 | 1 | Not-determined | Genome-fragment | NA | NA | 0 |
| S12_vs_k141_134462\|\|full | 1511 | No | NA | 2 | 0 | 2 | Low-quality | Genome-fragment | 2.98 | AAI-based (medium-confidence) | 0 |
| S1_vs_k141_89615\|\|full | 1596 | No | NA | 2 | 0 | 0 | Not-determined | Genome-fragment | NA | NA | 0 |
| S2_extracted_k141_16549 | 1511 | No | NA | 5 | 0 | 0 | Low-quality | Genome-fragment | 2.47 | AAI-based (high-confidence) | 0 |
| S11_extracted_k141_66322 | 1512 | No | NA | 1 | 0 | 1 | Low-quality | Genome-fragment | 0.44 | AAI-based (high-confidence) | 0 |
| S11_extracted_k141_34466 | 1512 | No | NA | 1 | 0 | 1 | Low-quality | Genome-fragment | 0.86 | AAI-based (medium-confidence) | 0 |
| S1_vs_k141_83124\|\|full | 1125 | No | NA | 2 | 0 | 0 | Not-determined | Genome-fragment | NA | NA | 0 |
| S11_vs_k141_94560\|\|full | 1512 | No | NA | 6 | 0 | 0 | Low-quality | Genome-fragment | 2.55 | AAI-based (high-confidence) | 0 |
| S1_vs_k141_61792\|\|full | 1716 | No | NA | 3 | 0 | 2 | Not-determined | Genome-fragment | NA | NA | 0 |
| S13_vs_477\|\|full | 1512 | No | NA | 3 | 1 | 0 | Low-quality | Genome-fragment | 3.01 | AAI-based (high-confidence) | 0 |
| S1_vs_k141_35758\|\|full | 1313 | No | NA | 3 | 0 | 2 | Not-determined | Genome-fragment | NA | NA | 0 |
| S14_extracted_k141_25133 | 1512 | No | NA | 4 | 4 | 0 | Low-quality | Genome-fragment | 4.99 | AAI-based (high-confidence) | 0 |
| S1_vs_k141_134022\|\|lt2gene | 1512 | No | NA | 1 | 1 | 0 | Low-quality | Genome-fragment | 3.5 | AAI-based (high-confidence) | 0 |
| S1_vs_k141_122871\|\|full | 6345 | No | NA | 5 | 0 | 2 | Not-determined | Genome-fragment | NA | NA | 0 |
| S1_vs_k141_136355\|\|full | 6839 | No | NA | 8 | 0 | 3 | Not-determined | Genome-fragment | NA | NA | 0 |
| S1_vs_k141_56370\|\|full | 2720 | No | NA | 3 | 0 | 1 | Not-determined | Genome-fragment | NA | NA | 0 |
| S1_vs_k141_16880\|\|full | 1065 | No | NA | 2 | 0 | 1 | Not-determined | Genome-fragment | NA | NA | 0 |
| S5_vs_k141_79812\|\|full | 1512 | No | NA | 3 | 1 | 0 | Low-quality | Genome-fragment | 2.8 | HMM-based (lower-bound) | 0 |
| S16_vs_80406\|\|full | 1512 | No | NA | 7 | 1 | 0 | Low-quality | Genome-fragment | 1.67 | HMM-based (lower-bound) | 0 |
| S1_vs_k141_76430\|\|full | 1136 | No | NA | 2 | 0 | 0 | Not-determined | Genome-fragment | NA | NA | 0 |
| S1_vs_k141_61824\|\|full | 3488 | No | NA | 3 | 0 | 1 | Not-determined | Genome-fragment | NA | NA | 0 |
| S16_vs_116577\|\|full | 1512 | No | NA | 2 | 0 | 1 | Low-quality | Genome-fragment | 0.99 | AAI-based (medium-confidence) | 0 |
| S1_vs_k141_35773\|\|full | 1465 | No | NA | 2 | 0 | 1 | Not-determined | Genome-fragment | NA | NA | 0 |
| S10_vs_55748\|\|full | 1512 | No | NA | 3 | 0 | 2 | Low-quality | Genome-fragment | 0.66 | AAI-based (high-confidence) | 0 |
| S8_vs_k141_85406\|\|full | 1512 | Yes | 1017 | 3 | 1 | 1 | Low-quality | Genome-fragment | 2.13 | AAI-based (medium-confidence) | 32.74 |
| S17_extracted_k141_106256 | 1513 | No | NA | 1 | 0 | 0 | Low-quality | Genome-fragment | 2.67 | AAI-based (high-confidence) | 0 |
| S1_vs_k141_2565\|\|full | 1699 | No | NA | 2 | 0 | 1 | Not-determined | Genome-fragment | NA | NA | 0 |
| S1_vs_k141_69668\|\|full | 1900 | No | NA | 3 | 0 | 3 | Not-determined | Genome-fragment | NA | NA | 0 |
| S17_vs_k141_138661\|\|full | 1513 | No | NA | 4 | 2 | 0 | Low-quality | Genome-fragment | 2.32 | AAI-based (high-confidence) | 0 |
| S1_vs_k141_89710\|\|full | 4505 | No | NA | 3 | 0 | 1 | Not-determined | Genome-fragment | NA | NA | 0 |
| S14_extracted_k141_39046 | 1513 | No | NA | 3 | 3 | 0 | Low-quality | Genome-fragment | 4.99 | AAI-based (high-confidence) | 0 |
| S14_extracted_k141_1919 | 1513 | No | NA | 1 | 0 | 1 | Low-quality | Genome-fragment | 0.28 | AAI-based (medium-confidence) | 0 |
| S1_vs_k141_61897\|\|full | 1454 | No | NA | 2 | 0 | 0 | Not-determined | Genome-fragment | NA | NA | 0 |
| S5_extracted_k141_135065 | 1513 | No | NA | 1 | 0 | 1 | Low-quality | Genome-fragment | 0.14 | AAI-based (medium-confidence) | 0 |
| S5_vs_k141_90054\|\|full | 1513 | No | NA | 1 | 0 | 0 | Low-quality | Genome-fragment | 11.18 | AAI-based (high-confidence) | 0 |
| S7_extracted_100045 | 1513 | No | NA | 3 | 2 | 0 | Low-quality | Genome-fragment | 1.09 | AAI-based (medium-confidence) | 0 |
| S1_vs_k141_103321\|\|full | 4678 | No | NA | 6 | 0 | 1 | Not-determined | Genome-fragment | NA | NA | 0 |
| S1_vs_k141_22905\|\|full | 2038 | No | NA | 3 | 0 | 3 | Not-determined | Genome-fragment | NA | NA | 0 |
| S7_vs_47417\|\|lt2gene | 1513 | No | NA | 2 | 2 | 0 | Low-quality | Genome-fragment | 3 | AAI-based (high-confidence) | 0 |
| S1_vs_k141_122999\|\|full | 1444 | No | NA | 2 | 0 | 1 | Not-determined | Genome-fragment | NA | NA | 0 |
| S10_vs_74095\|\|full | 1513 | No | NA | 5 | 0 | 1 | Low-quality | Genome-fragment | 3.27 | AAI-based (high-confidence) | 0 |
| S1_vs_k141_61947\|\|full | 5430 | No | NA | 3 | 0 | 2 | Not-determined | Genome-fragment | NA | NA | 0 |
| S11_extracted_k141_41664 | 1514 | No | NA | 4 | 0 | 0 | Low-quality | Genome-fragment | 2.44 | AAI-based (medium-confidence) | 0 |
| S1_vs_k141_29833\|\|full | 4086 | No | NA | 4 | 0 | 1 | Not-determined | Genome-fragment | NA | NA | 0 |
| S13_extracted_131317 | 1514 | No | NA | 3 | 0 | 3 | Low-quality | Genome-fragment | 0.26 | AAI-based (medium-confidence) | 0 |
| S1_vs_k141_129979\|\|full | 1565 | No | NA | 2 | 0 | 1 | Not-determined | Genome-fragment | NA | NA | 0 |
| S13_vs_131210\|\|full | 1514 | No | NA | 2 | 0 | 0 | Low-quality | Genome-fragment | 2.35 | AAI-based (high-confidence) | 0 |
| S14_vs_k141_25384\|\|full | 1514 | No | NA | 3 | 0 | 0 | Low-quality | Genome-fragment | 3.54 | AAI-based (medium-confidence) | 0 |
| S1_vs_k141_123034\|\|full | 1832 | No | NA | 2 | 0 | 2 | Not-determined | Genome-fragment | NA | NA | 0 |
| S1_vs_k141_116699\|\|full | 1267 | No | NA | 3 | 0 | 2 | Not-determined | Genome-fragment | NA | NA | 0 |
| S1_extracted_k141_102363 | 1514 | No | NA | 3 | 0 | 1 | Low-quality | Genome-fragment | 1.11 | AAI-based (medium-confidence) | 0 |
| S1_extracted_k141_147906 | 1514 | Yes | 1220 | 3 | 1 | 2 | Low-quality | Genome-fragment | 2.53 | AAI-based (medium-confidence) | 19.42 |
| S1_vs_k141_64779\|\|full | 1514 | No | NA | 4 | 1 | 0 | Low-quality | Genome-fragment | 3.64 | AAI-based (high-confidence) | 0 |
| S3_extracted_k141_89960 | 1514 | No | NA | 2 | 1 | 0 | Low-quality | Genome-fragment | 2.36 | AAI-based (high-confidence) | 0 |
| S1_vs_k141_62040\|\|full | 1807 | No | NA | 2 | 0 | 2 | Not-determined | Genome-fragment | NA | NA | 0 |
| S3_vs_k141_108852\|\|full | 1514 | Yes | 680 | 2 | 1 | 1 | Low-quality | Genome-fragment | 0.35 | HMM-based (lower-bound) | 55.09 |
| S1_vs_k141_123082\|\|full | 2195 | No | NA | 2 | 0 | 2 | Not-determined | Genome-fragment | NA | NA | 0 |
| S3_vs_k141_35714\|\|lt2gene | 1514 | No | NA | 2 | 1 | 0 | Low-quality | Genome-fragment | 3.2 | AAI-based (high-confidence) | 0 |
| S5_extracted_k141_108693 | 1514 | No | NA | 1 | 0 | 1 | Low-quality | Genome-fragment | 0.61 | AAI-based (medium-confidence) | 0 |
| S1_vs_k141_62045\|\|full | 9875 | No | NA | 9 | 0 | 4 | Not-determined | Genome-fragment | NA | NA | 0 |
| S5_extracted_k141_84705 | 1514 | No | NA | 1 | 0 | 1 | Low-quality | Genome-fragment | 1.91 | AAI-based (medium-confidence) | 0 |
| S5_vs_k141_69674\|\|lt2gene | 1514 | No | NA | 2 | 1 | 0 | Low-quality | Genome-fragment | 4.51 | AAI-based (high-confidence) | 0 |
| S1_vs_k141_123094\|\|full | 2269 | No | NA | 8 | 0 | 0 | Not-determined | Genome-fragment | NA | NA | 0 |
| S7_extracted_14624 | 1514 | No | NA | 1 | 0 | 1 | Low-quality | Genome-fragment | 0.4 | AAI-based (medium-confidence) | 0 |
| S1_vs_k141_83426\|\|full | 2031 | No | NA | 5 | 0 | 2 | Not-determined | Genome-fragment | NA | NA | 0 |
| S7_vs_4708\|\|full | 1514 | No | NA | 2 | 1 | 0 | Low-quality | Genome-fragment | 2.19 | HMM-based (lower-bound) | 0 |
| S16_vs_8589\|\|full | 1514 | No | NA | 3 | 0 | 0 | Low-quality | Genome-fragment | 3.42 | AAI-based (medium-confidence) | 0 |
| S1_vs_k141_143236\|\|full | 2718 | No | NA | 5 | 0 | 2 | Not-determined | Genome-fragment | NA | NA | 0 |
| S16_vs_27232\|\|full | 1514 | No | NA | 4 | 0 | 0 | Low-quality | Genome-fragment | 2.7 | AAI-based (medium-confidence) | 0 |
| S16_vs_169915\|\|full | 1514 | No | NA | 2 | 1 | 0 | Low-quality | Genome-fragment | 3.12 | AAI-based (high-confidence) | 0 |
| S1_vs_k141_2729\|\|full | 2271 | No | NA | 3 | 0 | 0 | Not-determined | Genome-fragment | NA | NA | 0 |
| S1_vs_k141_29992\|\|full | 3026 | No | NA | 3 | 0 | 1 | Not-determined | Genome-fragment | NA | NA | 0 |
| S10_vs_107526\|\|full | 1514 | No | NA | 2 | 0 | 0 | Low-quality | Genome-fragment | 2.87 | AAI-based (medium-confidence) | 0 |
| S6_vs_k141_15030\|\|full | 1514 | No | NA | 3 | 0 | 0 | Low-quality | Genome-fragment | 4.71 | AAI-based (high-confidence) | 0 |
| S1_vs_k141_56700\|\|full | 1210 | No | NA | 4 | 0 | 1 | Not-determined | Genome-fragment | NA | NA | 0 |
| S8_vs_k141_32265\|\|full | 1514 | No | NA | 1 | 0 | 0 | Low-quality | Genome-fragment | 2.33 | AAI-based (medium-confidence) | 0 |
| S17_vs_k141_59941\|\|lt2gene | 1515 | No | NA | 1 | 1 | 0 | Low-quality | Genome-fragment | 3.22 | AAI-based (medium-confidence) | 0 |
| S11_vs_k141_95401\|\|full | 1515 | No | NA | 3 | 1 | 0 | Low-quality | Genome-fragment | 3.73 | AAI-based (medium-confidence) | 0 |
| S14_extracted_k141_62996 | 1515 | No | NA | 2 | 1 | 0 | Low-quality | Genome-fragment | 2.54 | AAI-based (high-confidence) | 0 |
| S1_vs_k141_96904\|\|full | 4371 | No | NA | 8 | 0 | 0 | Not-determined | Genome-fragment | NA | NA | 0 |
| S1_vs_k141_56165\|\|full | 1515 | No | NA | 2 | 0 | 0 | Low-quality | Genome-fragment | 0.63 | AAI-based (medium-confidence) | 0 |
| S1_vs_k141_24066\|\|full | 1515 | No | NA | 3 | 0 | 0 | Low-quality | Genome-fragment | 3.14 | AAI-based (high-confidence) | 0 |
| S1_vs_k141_50243\|\|full | 1050 | No | NA | 2 | 0 | 0 | Not-determined | Genome-fragment | NA | NA | 0 |
| S5_extracted_k141_95237 | 1515 | No | NA | 2 | 2 | 0 | Low-quality | Genome-fragment | 2.59 | AAI-based (high-confidence) | 0 |
| S10_vs_84692\|\|full | 1515 | No | NA | 3 | 0 | 1 | Low-quality | Genome-fragment | 1.04 | AAI-based (medium-confidence) | 0 |
| S10_vs_121091\|\|full | 1515 | No | NA | 3 | 1 | 0 | Low-quality | Genome-fragment | 2 | AAI-based (medium-confidence) | 0 |
| S1_vs_k141_143302\|\|full | 5037 | No | NA | 5 | 0 | 3 | Not-determined | Genome-fragment | NA | NA | 0 |
| S1_vs_k141_123250\|\|full | 1030 | No | NA | 2 | 0 | 0 | Not-determined | Genome-fragment | NA | NA | 0 |
| S12_extracted_k141_91874 | 1515 | No | NA | 2 | 0 | 0 | Low-quality | Genome-fragment | 3 | AAI-based (high-confidence) | 0 |
| S6_vs_k141_36360\|\|full | 1515 | No | NA | 7 | 2 | 0 | Low-quality | Genome-fragment | 3.04 | AAI-based (high-confidence) | 0 |
| S1_vs_k141_83575\|\|full | 1568 | No | NA | 2 | 0 | 1 | Not-determined | Genome-fragment | NA | NA | 0 |
| S6_vs_k141_55606\|\|full | 1515 | No | NA | 4 | 0 | 0 | Low-quality | Genome-fragment | 2.57 | AAI-based (high-confidence) | 0 |
| S8_extracted_k141_65820 | 1515 | No | NA | 2 | 0 | 1 | Low-quality | Genome-fragment | 2.53 | AAI-based (high-confidence) | 0 |
| S1_vs_k141_96972\|\|full | 1080 | No | NA | 3 | 0 | 0 | Not-determined | Genome-fragment | NA | NA | 0 |
| S1_vs_k141_83594\|\|full | 1122 | No | NA | 2 | 0 | 0 | Not-determined | Genome-fragment | NA | NA | 0 |
| S1_vs_k141_136718\|\|full | 1743 | No | NA | 2 | 0 | 1 | Not-determined | Genome-fragment | NA | NA | 0 |
| S8_extracted_k141_137477 | 1515 | No | NA | 5 | 0 | 0 | Low-quality | Genome-fragment | 2.3 | AAI-based (high-confidence) | 0 |
| S1_vs_k141_30131\|\|full | 1692 | No | NA | 4 | 0 | 1 | Not-determined | Genome-fragment | NA | NA | 0 |
| S17_vs_k141_138439\|\|full | 1516 | No | NA | 2 | 1 | 0 | Low-quality | Genome-fragment | 3.18 | AAI-based (high-confidence) | 0 |
| S9_vs_k141_35276\|\|full | 1516 | No | NA | 2 | 2 | 0 | Low-quality | Genome-fragment | 4.48 | AAI-based (high-confidence) | 0 |
| S11_extracted_k141_59809 | 1516 | No | NA | 4 | 0 | 0 | Low-quality | Genome-fragment | 2.41 | AAI-based (medium-confidence) | 0 |
| S11_extracted_k141_79842 | 1516 | No | NA | 2 | 0 | 0 | Low-quality | Genome-fragment | 2.4 | AAI-based (high-confidence) | 0 |
| S13_vs_131640\|\|full | 1516 | No | NA | 2 | 0 | 0 | Low-quality | Genome-fragment | 2.92 | AAI-based (medium-confidence) | 0 |
| S1_vs_k141_83677\|\|full | 3369 | No | NA | 3 | 0 | 3 | Not-determined | Genome-fragment | NA | NA | 0 |
| S14_vs_k141_113430\|\|full | 1516 | No | NA | 5 | 2 | 0 | Low-quality | Genome-fragment | 4.02 | AAI-based (medium-confidence) | 0 |
| S1_vs_k141_136781\|\|full | 2173 | No | NA | 3 | 0 | 2 | Not-determined | Genome-fragment | NA | NA | 0 |
| S1_vs_k141_90107\|\|full | 1816 | No | NA | 3 | 0 | 3 | Not-determined | Genome-fragment | NA | NA | 0 |
| S1_vs_k141_9902\|\|full | 1516 | No | NA | 2 | 0 | 1 | Low-quality | Genome-fragment | 1 | AAI-based (high-confidence) | 0 |
| S1_vs_k141_83704\|\|full | 2296 | No | NA | 4 | 0 | 1 | Not-determined | Genome-fragment | NA | NA | 0 |
| S5_vs_k141_122627\|\|full | 1516 | No | NA | 5 | 0 | 0 | Low-quality | Genome-fragment | 3.87 | AAI-based (high-confidence) | 0 |
| S7_extracted_38846 | 1516 | No | NA | 2 | 0 | 1 | Low-quality | Genome-fragment | 1.19 | AAI-based (high-confidence) | 0 |
| S7_vs_37889\|\|full | 1516 | No | NA | 5 | 1 | 0 | Low-quality | Genome-fragment | 5.99 | AAI-based (medium-confidence) | 0 |
| S1_vs_k141_136821\|\|full | 1554 | No | NA | 2 | 0 | 2 | Not-determined | Genome-fragment | NA | NA | 0 |
| S16_extracted_82201 | 1516 | No | NA | 5 | 0 | 0 | Low-quality | Genome-fragment | 4.18 | AAI-based (high-confidence) | 0 |
| S12_extracted_k141_138605 | 1516 | No | NA | 1 | 1 | 0 | Low-quality | Genome-fragment | 2.22 | HMM-based (lower-bound) | 0 |
| S4_extracted_7485 | 1516 | No | NA | 5 | 5 | 0 | Low-quality | Genome-fragment | 5.01 | AAI-based (high-confidence) | 0 |
| S1_vs_k141_50446\|\|full | 2252 | No | NA | 4 | 0 | 2 | Not-determined | Genome-fragment | NA | NA | 0 |
| S4_extracted_18990 | 1516 | No | NA | 1 | 0 | 0 | Low-quality | Genome-fragment | 2.15 | AAI-based (high-confidence) | 0 |
| S1_vs_k141_136903\|\|full | 2033 | No | NA | 3 | 0 | 3 | Not-determined | Genome-fragment | NA | NA | 0 |
| S8_vs_k141_55708\|\|full | 1516 | No | NA | 2 | 0 | 1 | Low-quality | Genome-fragment | 1.78 | AAI-based (high-confidence) | 0 |
| S1_vs_k141_123489\|\|full | 1525 | No | NA | 4 | 0 | 1 | Not-determined | Genome-fragment | NA | NA | 0 |
| S1_vs_k141_103765\|\|full | 1406 | No | NA | 2 | 0 | 0 | Not-determined | Genome-fragment | NA | NA | 0 |
| S1_vs_k141_17466\|\|full | 6837 | No | NA | 4 | 0 | 1 | Not-determined | Genome-fragment | NA | NA | 0 |
| S17_vs_k141_104025\|\|full | 1517 | No | NA | 2 | 0 | 0 | Low-quality | Genome-fragment | 3.44 | AAI-based (medium-confidence) | 0 |
| S11_extracted_k141_86595 | 1517 | No | NA | 2 | 0 | 0 | Low-quality | Genome-fragment | 3.58 | AAI-based (medium-confidence) | 0 |
| S1_vs_k141_23408\|\|full | 1247 | No | NA | 2 | 0 | 2 | Not-determined | Genome-fragment | NA | NA | 0 |
| S1_vs_k141_62409\|\|full | 16910 | No | NA | 23 | 10 | 0 | Medium-quality | Genome-fragment | 50 | AAI-based (medium-confidence) | 0 |
| S1_vs_k141_90265\|\|full | 1958 | No | NA | 2 | 0 | 2 | Not-determined | Genome-fragment | NA | NA | 0 |
| S1_vs_k141_136968\|\|full | 2037 | No | NA | 4 | 0 | 1 | Not-determined | Genome-fragment | NA | NA | 0 |
| S11_vs_k141_42277\|\|full | 1517 | No | NA | 3 | 0 | 0 | Low-quality | Genome-fragment | 3.64 | AAI-based (medium-confidence) | 0 |
| S14_extracted_k141_56246 | 1517 | No | NA | 1 | 0 | 1 | Low-quality | Genome-fragment | 0.66 | AAI-based (medium-confidence) | 0 |
| S1_vs_k141_103839\|\|full | 2894 | No | NA | 2 | 0 | 1 | Not-determined | Genome-fragment | NA | NA | 0 |
| S1_vs_k141_76279\|\|full | 1517 | No | NA | 2 | 0 | 1 | Low-quality | Genome-fragment | 3.19 | AAI-based (high-confidence) | 0 |
| S1_vs_k141_136983\|\|full | 2299 | No | NA | 2 | 0 | 2 | Not-determined | Genome-fragment | NA | NA | 0 |
| S1_vs_k141_57082\|\|full | 2186 | No | NA | 3 | 0 | 2 | Not-determined | Genome-fragment | NA | NA | 0 |
| S1_vs_k141_90305\|\|full | 1158 | No | NA | 3 | 0 | 0 | Not-determined | Genome-fragment | NA | NA | 0 |
| S3_vs_k141_72060\|\|full | 1517 | No | NA | 2 | 0 | 0 | Low-quality | Genome-fragment | 3.83 | AAI-based (medium-confidence) | 0 |
| S3_vs_k141_90447\|\|full | 1517 | No | NA | 5 | 1 | 0 | Low-quality | Genome-fragment | 3.39 | AAI-based (high-confidence) | 0 |
| S1_vs_k141_23540\|\|full | 2698 | No | NA | 3 | 0 | 3 | Not-determined | Genome-fragment | NA | NA | 0 |
| S5_vs_k141_35102\|\|full | 1517 | No | NA | 2 | 0 | 0 | Low-quality | Genome-fragment | 2.53 | AAI-based (high-confidence) | 0 |
| S7_vs_93904\|\|lt2gene | 1517 | No | NA | 2 | 1 | 0 | Low-quality | Genome-fragment | 2.13 | HMM-based (lower-bound) | 0 |
| S1_vs_k141_97324\|\|full | 5349 | No | NA | 7 | 0 | 0 | Not-determined | Genome-fragment | NA | NA | 0 |
| S1_vs_k141_143608\|\|full | 1472 | No | NA | 3 | 0 | 1 | Not-determined | Genome-fragment | NA | NA | 0 |
| S1_vs_k141_117303\|\|full | 1673 | No | NA | 3 | 0 | 0 | Not-determined | Genome-fragment | NA | NA | 0 |
| S16_extracted_126825 | 1517 | No | NA | 2 | 0 | 0 | Low-quality | Genome-fragment | 0.99 | AAI-based (high-confidence) | 0 |
| S1_vs_k141_50650\|\|full | 1189 | No | NA | 4 | 0 | 0 | Not-determined | Genome-fragment | NA | NA | 0 |
| S16_extracted_159367 | 1517 | No | NA | 6 | 0 | 0 | Low-quality | Genome-fragment | 1 | AAI-based (medium-confidence) | 0 |
| S16_vs_105783\|\|full | 1517 | No | NA | 3 | 0 | 0 | Low-quality | Genome-fragment | 2.32 | AAI-based (medium-confidence) | 0 |
| S1_vs_k141_97370\|\|full | 1332 | No | NA | 3 | 0 | 1 | Not-determined | Genome-fragment | NA | NA | 0 |
| S10_vs_112725\|\|full | 1517 | No | NA | 2 | 0 | 0 | Low-quality | Genome-fragment | 2.54 | AAI-based (medium-confidence) | 0 |
| S1_vs_k141_70084\|\|full | 2742 | No | NA | 3 | 0 | 2 | Not-determined | Genome-fragment | NA | NA | 0 |
| S1_vs_k141_130612\|\|full | 1330 | No | NA | 3 | 0 | 0 | Not-determined | Genome-fragment | NA | NA | 0 |
| S1_vs_k141_117317\|\|full | 33680 | No | NA | 60 | 14 | 1 | Medium-quality | Genome-fragment | 81.81 | AAI-based (high-confidence) | 0 |
| S1_vs_k141_84004\|\|full | 1200 | No | NA | 2 | 0 | 1 | Not-determined | Genome-fragment | NA | NA | 0 |
| S12_extracted_k141_4720 | 1517 | No | NA | 2 | 0 | 2 | Low-quality | Genome-fragment | 1.19 | AAI-based (medium-confidence) | 0 |
| S15_vs_50157\|\|full | 1517 | No | NA | 5 | 0 | 0 | Low-quality | Genome-fragment | 3.3 | AAI-based (high-confidence) | 0 |
| S1_vs_k141_143727\|\|full | 1586 | No | NA | 2 | 0 | 2 | Not-determined | Genome-fragment | NA | NA | 0 |
| S1_vs_k141_137204\|\|full | 5350 | No | NA | 3 | 0 | 2 | Not-determined | Genome-fragment | NA | NA | 0 |
| S17_extracted_k141_134575 | 1518 | No | NA | 2 | 0 | 2 | Low-quality | Genome-fragment | 3.34 | AAI-based (medium-confidence) | 0 |
| S1_vs_k141_17717\|\|full | 1344 | No | NA | 2 | 0 | 1 | Not-determined | Genome-fragment | NA | NA | 0 |
| S9_vs_k141_66191\|\|lt2gene | 1518 | No | NA | 2 | 2 | 0 | Low-quality | Genome-fragment | 3.97 | AAI-based (high-confidence) | 0 |
| S13_extracted_122436 | 1518 | No | NA | 3 | 0 | 0 | Low-quality | Genome-fragment | 2.03 | AAI-based (high-confidence) | 0 |
| S1_vs_k141_62672\|\|full | 1864 | No | NA | 3 | 0 | 1 | Not-determined | Genome-fragment | NA | NA | 0 |
| S13_extracted_125844 | 1518 | No | NA | 1 | 1 | 0 | Low-quality | Genome-fragment | 0.86 | HMM-based (lower-bound) | 0 |
| S3_extracted_k141_29674 | 1518 | No | NA | 4 | 1 | 0 | Low-quality | Genome-fragment | 3.77 | AAI-based (high-confidence) | 0 |
| S16_vs_28928\|\|full | 1518 | No | NA | 3 | 0 | 0 | Low-quality | Genome-fragment | 3.15 | AAI-based (medium-confidence) | 0 |
| S10_vs_119265\|\|full | 1518 | No | NA | 3 | 0 | 0 | Low-quality | Genome-fragment | 0.9 | AAI-based (high-confidence) | 0 |
| S12_vs_k141_156235\|\|full | 1518 | No | NA | 3 | 0 | 0 | Low-quality | Genome-fragment | 3.22 | AAI-based (medium-confidence) | 0 |
| S6_vs_k141_26883\|\|full | 1518 | No | NA | 3 | 0 | 2 | Low-quality | Genome-fragment | 0.43 | AAI-based (medium-confidence) | 0 |
| S9_vs_k141_20884\|\|full | 1519 | No | NA | 2 | 0 | 2 | Low-quality | Genome-fragment | 1.36 | AAI-based (medium-confidence) | 0 |
| S13_extracted_141715 | 1519 | No | NA | 3 | 0 | 0 | Low-quality | Genome-fragment | 3.32 | AAI-based (high-confidence) | 0 |
| S1_vs_k141_70228\|\|full | 1086 | No | NA | 2 | 0 | 1 | Not-determined | Genome-fragment | NA | NA | 0 |
| S1_vs_k141_23682\|\|full | 44452 | No | NA | 76 | 18 | 0 | High-quality | High-quality | 100 | AAI-based (high-confidence) | 0 |
| S1_vs_k141_30728\|\|full | 2108 | No | NA | 2 | 0 | 2 | Not-determined | Genome-fragment | NA | NA | 0 |
| S1_vs_k141_99969\|\|full | 1519 | No | NA | 3 | 0 | 0 | Low-quality | Genome-fragment | 2.05 | AAI-based (medium-confidence) | 0 |
| S7_vs_34665\|\|full | 1519 | No | NA | 2 | 0 | 0 | Low-quality | Genome-fragment | 2.51 | AAI-based (high-confidence) | 0 |
| S1_vs_k141_148660\|\|full | 1240 | No | NA | 3 | 0 | 0 | Not-determined | Genome-fragment | NA | NA | 0 |
| S7_vs_109847\|\|full | 1519 | No | NA | 6 | 1 | 0 | Low-quality | Genome-fragment | 2.82 | AAI-based (medium-confidence) | 0 |
| S16_vs_144825\|\|full | 1519 | No | NA | 4 | 0 | 0 | Low-quality | Genome-fragment | 0.81 | AAI-based (high-confidence) | 0 |
| S8_vs_k141_195462\|\|lt2gene | 1519 | No | NA | 2 | 1 | 0 | Low-quality | Genome-fragment | 1.4 | HMM-based (lower-bound) | 0 |
| S1_vs_k141_3173\|\|full | 1220 | No | NA | 1 | 0 | 0 | Not-determined | Genome-fragment | NA | NA | 0 |
| S1_vs_k141_104138\|\|full | 1486 | No | NA | 3 | 0 | 2 | Not-determined | Genome-fragment | NA | NA | 0 |
| S1_vs_k141_10846\|\|full | 2422 | No | NA | 3 | 0 | 2 | Not-determined | Genome-fragment | NA | NA | 0 |
| S1_vs_k141_36520\|\|full | 2897 | No | NA | 2 | 0 | 2 | Not-determined | Genome-fragment | NA | NA | 0 |
| S1_vs_k141_143860\|\|full | 1510 | No | NA | 2 | 0 | 1 | Not-determined | Genome-fragment | NA | NA | 0 |
| S14_extracted_k141_114789 | 1520 | No | NA | 1 | 0 | 1 | Low-quality | Genome-fragment | 3.67 | AAI-based (medium-confidence) | 0 |
| S1_vs_k141_17870\|\|full | 4203 | No | NA | 4 | 0 | 3 | Not-determined | Genome-fragment | NA | NA | 0 |
| S14_vs_k141_116315\|\|full | 1520 | No | NA | 3 | 0 | 0 | Low-quality | Genome-fragment | 0.87 | AAI-based (medium-confidence) | 0 |
| S1_vs_k141_63844\|\|full | 1520 | No | NA | 3 | 0 | 2 | Low-quality | Genome-fragment | 3.39 | AAI-based (medium-confidence) | 0 |
| S3_extracted_k141_62122 | 1520 | No | NA | 3 | 0 | 0 | Low-quality | Genome-fragment | 1 | AAI-based (medium-confidence) | 0 |
| S3_vs_k141_113504\|\|full | 1520 | No | NA | 3 | 0 | 2 | Low-quality | Genome-fragment | 0.58 | AAI-based (medium-confidence) | 0 |
| S1_vs_k141_143887\|\|full | 1781 | No | NA | 2 | 0 | 2 | Not-determined | Genome-fragment | NA | NA | 0 |
| S5_vs_k141_9629\|\|full | 1520 | No | NA | 2 | 0 | 0 | Low-quality | Genome-fragment | 1.67 | AAI-based (high-confidence) | 0 |
| S7_vs_87071\|\|full | 1520 | No | NA | 4 | 2 | 0 | Low-quality | Genome-fragment | 3.31 | AAI-based (high-confidence) | 0 |
| S1_vs_k141_110783\|\|full | 3131 | No | NA | 4 | 0 | 4 | Not-determined | Genome-fragment | NA | NA | 0 |
| S7_vs_96117\|\|lt2gene | 1520 | No | NA | 2 | 2 | 0 | Low-quality | Genome-fragment | 3.19 | AAI-based (high-confidence) | 0 |
| S16_vs_17592\|\|full | 1520 | No | NA | 2 | 0 | 0 | Low-quality | Genome-fragment | 3.48 | AAI-based (medium-confidence) | 0 |
| S1_vs_k141_104200\|\|full | 9414 | No | NA | 9 | 0 | 7 | Not-determined | Genome-fragment | NA | NA | 0 |
| S1_vs_k141_97681\|\|full | 3520 | No | NA | 4 | 0 | 4 | Not-determined | Genome-fragment | NA | NA | 0 |
| S1_vs_k141_77509\|\|full | 2709 | No | NA | 2 | 0 | 1 | Not-determined | Genome-fragment | NA | NA | 0 |
| S1_vs_k141_36584\|\|full | 1304 | No | NA | 2 | 0 | 2 | Not-determined | Genome-fragment | NA | NA | 0 |
| S8_vs_k141_168086\|\|full | 1520 | No | NA | 2 | 0 | 1 | Low-quality | Genome-fragment | 0.93 | AAI-based (medium-confidence) | 0 |
| S1_vs_k141_10967\|\|full | 3492 | No | NA | 12 | 0 | 0 | Not-determined | Genome-fragment | NA | NA | 0 |
| S1_vs_k141_123896\|\|full | 2760 | No | NA | 4 | 0 | 2 | Not-determined | Genome-fragment | NA | NA | 0 |
| S8_vs_k141_172806\|\|full | 1520 | No | NA | 7 | 0 | 0 | Low-quality | Genome-fragment | 4.09 | AAI-based (high-confidence) | 0 |
| S14_vs_k141_55848\|\|full | 1521 | No | NA | 3 | 2 | 0 | Low-quality | Genome-fragment | 3.03 | AAI-based (medium-confidence) | 0 |
| S1_vs_k141_3267\|\|full | 4887 | No | NA | 9 | 6 | 0 | High-quality | High-quality | 100 | AAI-based (high-confidence) | 0 |
| S1_vs_k141_130882\|\|full | 4747 | No | NA | 4 | 0 | 1 | Not-determined | Genome-fragment | NA | NA | 0 |
| S14_vs_k141_15333\|\|lt2gene | 1521 | No | NA | 1 | 1 | 0 | Low-quality | Genome-fragment | 2.6 | AAI-based (high-confidence) | 0 |
| S1_vs_k141_10979\|\|full | 1150 | No | NA | 2 | 0 | 1 | Not-determined | Genome-fragment | NA | NA | 0 |
| S3_extracted_k141_3500 | 1521 | No | NA | 3 | 1 | 0 | Low-quality | Genome-fragment | 2.49 | HMM-based (lower-bound) | 0 |
| S3_extracted_k141_110388 | 1521 | No | NA | 2 | 0 | 0 | Low-quality | Genome-fragment | 4.32 | AAI-based (high-confidence) | 0 |
| S3_vs_k141_35215\|\|lt2gene | 1521 | No | NA | 2 | 2 | 0 | Low-quality | Genome-fragment | 2.64 | HMM-based (lower-bound) | 0 |
| S3_vs_k141_21523\|\|lt2gene | 1521 | No | NA | 1 | 1 | 0 | Low-quality | Genome-fragment | 1.05 | HMM-based (lower-bound) | 0 |
| S1_vs_k141_90786\|\|full | 2073 | No | NA | 5 | 0 | 0 | Not-determined | Genome-fragment | NA | NA | 0 |
| S1_vs_k141_123934\|\|full | 1227 | No | NA | 2 | 0 | 1 | Not-determined | Genome-fragment | NA | NA | 0 |
| S5_vs_k141_98521\|\|full | 1521 | No | NA | 3 | 1 | 0 | Low-quality | Genome-fragment | 4.84 | AAI-based (high-confidence) | 0 |
| S5_vs_k141_82157\|\|full | 1521 | No | NA | 3 | 0 | 2 | Low-quality | Genome-fragment | 0.87 | AAI-based (medium-confidence) | 0 |
| S1_vs_k141_84346\|\|full | 2157 | No | NA | 3 | 0 | 0 | Not-determined | Genome-fragment | NA | NA | 0 |
| S1_vs_k141_123953\|\|full | 5516 | No | NA | 6 | 0 | 2 | Not-determined | Genome-fragment | NA | NA | 0 |
| S7_extracted_30020 | 1521 | No | NA | 2 | 1 | 0 | Low-quality | Genome-fragment | 3.53 | AAI-based (high-confidence) | 0 |
| S7_vs_105470\|\|full | 1521 | No | NA | 5 | 0 | 0 | Low-quality | Genome-fragment | 3.42 | AAI-based (medium-confidence) | 0 |
| S1_vs_k141_23996\|\|full | 1090 | No | NA | 3 | 0 | 0 | Not-determined | Genome-fragment | NA | NA | 0 |
| S1_vs_k141_97796\|\|full | 3099 | No | NA | 4 | 0 | 1 | Not-determined | Genome-fragment | NA | NA | 0 |
| S16_extracted_61554 | 1521 | No | NA | 1 | 0 | 1 | Low-quality | Genome-fragment | 1.01 | AAI-based (medium-confidence) | 0 |
| S1_vs_k141_44358\|\|full | 3030 | No | NA | 3 | 0 | 1 | Not-determined | Genome-fragment | NA | NA | 0 |
| S16_vs_76370\|\|full | 1521 | No | NA | 3 | 0 | 0 | Low-quality | Genome-fragment | 3.57 | AAI-based (high-confidence) | 0 |
| S1_vs_k141_123963\|\|full | 1118 | No | NA | 2 | 0 | 1 | Not-determined | Genome-fragment | NA | NA | 0 |
| S15_extracted_33395 | 1521 | No | NA | 4 | 0 | 0 | Low-quality | Genome-fragment | 0.89 | AAI-based (high-confidence) | 0 |
| S8_vs_k141_183495\|\|full | 1521 | No | NA | 2 | 0 | 0 | Low-quality | Genome-fragment | 2.87 | AAI-based (medium-confidence) | 0 |
| S8_vs_k141_37134\|\|full | 1521 | No | NA | 5 | 0 | 0 | Low-quality | Genome-fragment | 3.55 | AAI-based (high-confidence) | 0 |
| S1_vs_k141_104399\|\|full | 2135 | No | NA | 3 | 0 | 1 | Not-determined | Genome-fragment | NA | NA | 0 |
| S1_vs_k141_131038\|\|full | 2444 | No | NA | 2 | 0 | 0 | Not-determined | Genome-fragment | NA | NA | 0 |
| S1_vs_k141_11152\|\|full | 1714 | No | NA | 2 | 0 | 2 | Not-determined | Genome-fragment | NA | NA | 0 |
| S8_vs_k141_2678\|\|full | 1521 | No | NA | 4 | 0 | 0 | Low-quality | Genome-fragment | 4.59 | AAI-based (high-confidence) | 0 |
| S1_vs_k141_44436\|\|full | 2012 | No | NA | 2 | 0 | 1 | Not-determined | Genome-fragment | NA | NA | 0 |
| S17_extracted_k141_129484 | 1522 | No | NA | 3 | 3 | 0 | Low-quality | Genome-fragment | 5.02 | AAI-based (high-confidence) | 0 |
| S1_vs_k141_137646\|\|full | 1135 | No | NA | 2 | 0 | 1 | Not-determined | Genome-fragment | NA | NA | 0 |
| S1_vs_k141_24058\|\|full | 4841 | No | NA | 3 | 0 | 3 | Not-determined | Genome-fragment | NA | NA | 0 |
| S17_vs_k141_82166\|\|full | 1522 | No | NA | 3 | 0 | 2 | Low-quality | Genome-fragment | 2.07 | AAI-based (medium-confidence) | 0 |
| S13_vs_3811\|\|lt2gene | 1522 | No | NA | 2 | 0 | 1 | Low-quality | Genome-fragment | 4.31 | AAI-based (medium-confidence) | 0 |
| S1_vs_k141_24074\|\|full | 1429 | No | NA | 3 | 0 | 1 | Not-determined | Genome-fragment | NA | NA | 0 |
| S1_vs_k141_131103\|\|full | 5200 | No | NA | 6 | 0 | 3 | Not-determined | Genome-fragment | NA | NA | 0 |
| S1_vs_k141_32423\|\|full | 1522 | No | NA | 2 | 0 | 0 | Low-quality | Genome-fragment | 2.49 | AAI-based (high-confidence) | 0 |
| S1_vs_k141_3435\|\|full | 3839 | No | NA | 4 | 2 | 0 | Medium-quality | Genome-fragment | 70.06 | AAI-based (high-confidence) | 0 |
| S1_vs_k141_137685\|\|full | 1724 | No | NA | 3 | 0 | 1 | Not-determined | Genome-fragment | NA | NA | 0 |
| S3_vs_k141_47056\|\|full | 1522 | No | NA | 5 | 1 | 0 | Low-quality | Genome-fragment | 3.22 | AAI-based (high-confidence) | 0 |
| S3_vs_k141_10435\|\|lt2gene | 1522 | No | NA | 1 | 1 | 0 | Low-quality | Genome-fragment | 4.41 | AAI-based (high-confidence) | 0 |
| S1_vs_k141_111093\|\|full | 2287 | No | NA | 3 | 0 | 3 | Not-determined | Genome-fragment | NA | NA | 0 |
| S16_extracted_118150 | 1522 | No | NA | 2 | 1 | 0 | Low-quality | Genome-fragment | 2.16 | AAI-based (high-confidence) | 0 |
| S1_vs_k141_91019\|\|full | 5820 | No | NA | 3 | 0 | 2 | Not-determined | Genome-fragment | NA | NA | 0 |
| S16_vs_101681\|\|full | 1522 | No | NA | 3 | 0 | 1 | Low-quality | Genome-fragment | 1.24 | AAI-based (medium-confidence) | 0 |
| S1_vs_k141_36848\|\|full | 2960 | No | NA | 3 | 0 | 1 | Not-determined | Genome-fragment | NA | NA | 0 |
| S1_vs_k141_77817\|\|full | 1058 | No | NA | 3 | 0 | 0 | Not-determined | Genome-fragment | NA | NA | 0 |
| S12_vs_k141_130225\|\|full | 1522 | No | NA | 2 | 0 | 0 | Low-quality | Genome-fragment | 4.26 | AAI-based (high-confidence) | 0 |
| S4_extracted_10388 | 1522 | No | NA | 2 | 2 | 0 | Low-quality | Genome-fragment | 4.13 | HMM-based (lower-bound) | 0 |
| S8_extracted_k141_188978 | 1522 | No | NA | 1 | 0 | 1 | Low-quality | Genome-fragment | 0.31 | AAI-based (medium-confidence) | 0 |
| S1_vs_k141_84524\|\|full | 1644 | No | NA | 3 | 0 | 2 | Not-determined | Genome-fragment | NA | NA | 0 |
| S17_extracted_k141_119911 | 1523 | No | NA | 2 | 2 | 0 | Low-quality | Genome-fragment | 4.91 | AAI-based (high-confidence) | 0 |
| S1_vs_k141_91074\|\|full | 3106 | No | NA | 2 | 0 | 1 | Not-determined | Genome-fragment | NA | NA | 0 |
| S1_vs_k141_36898\|\|full | 1448 | No | NA | 5 | 0 | 4 | Not-determined | Genome-fragment | NA | NA | 0 |
| S1_vs_k141_24203\|\|full | 1688 | No | NA | 3 | 0 | 1 | Not-determined | Genome-fragment | NA | NA | 0 |
| S1_vs_k141_148878\|\|full | 1857 | No | NA | 2 | 0 | 2 | Not-determined | Genome-fragment | NA | NA | 0 |
| S1_vs_k141_57750\|\|full | 1623 | No | NA | 2 | 0 | 2 | Not-determined | Genome-fragment | NA | NA | 0 |
| S17_vs_k141_160776\|\|full | 1523 | No | NA | 2 | 0 | 0 | Low-quality | Genome-fragment | 3.56 | AAI-based (medium-confidence) | 0 |
| S11_vs_k141_17483\|\|lt2gene | 1523 | No | NA | 1 | 0 | 0 | Low-quality | Genome-fragment | 3.45 | AAI-based (high-confidence) | 0 |
| S1_vs_k141_44585\|\|full | 1697 | No | NA | 2 | 0 | 2 | Not-determined | Genome-fragment | NA | NA | 0 |
| S1_vs_k141_111186\|\|full | 4673 | No | NA | 3 | 0 | 2 | Not-determined | Genome-fragment | NA | NA | 0 |
| S1_vs_k141_70560\|\|full | 2109 | No | NA | 2 | 0 | 1 | Not-determined | Genome-fragment | NA | NA | 0 |
| S11_vs_k141_69248\|\|lt2gene | 1523 | No | NA | 2 | 1 | 0 | Low-quality | Genome-fragment | 2.82 | HMM-based (lower-bound) | 0 |
| S1_vs_k141_18329\|\|full | 1737 | No | NA | 3 | 0 | 1 | Not-determined | Genome-fragment | NA | NA | 0 |
| S14_vs_k141_90846\|\|full | 1523 | No | NA | 3 | 1 | 0 | Low-quality | Genome-fragment | 3.24 | AAI-based (high-confidence) | 0 |
| S14_vs_k141_105967\|\|full | 1523 | No | NA | 5 | 1 | 0 | Low-quality | Genome-fragment | 3.3 | AAI-based (medium-confidence) | 0 |
| S3_vs_k141_101397\|\|full | 1523 | No | NA | 3 | 1 | 0 | Low-quality | Genome-fragment | 3.26 | AAI-based (medium-confidence) | 0 |
| S5_vs_k141_17888\|\|full | 1523 | No | NA | 3 | 0 | 2 | Low-quality | Genome-fragment | 1.03 | AAI-based (medium-confidence) | 0 |
| S7_extracted_28804 | 1523 | No | NA | 1 | 0 | 1 | Low-quality | Genome-fragment | 0.49 | AAI-based (medium-confidence) | 0 |
| S7_vs_50445\|\|full | 1523 | No | NA | 3 | 2 | 0 | Low-quality | Genome-fragment | 3.23 | AAI-based (high-confidence) | 0 |
| S16_vs_88394\|\|full | 1523 | No | NA | 3 | 0 | 2 | Low-quality | Genome-fragment | 1.67 | AAI-based (medium-confidence) | 0 |
| S10_extracted_108848 | 1523 | No | NA | 2 | 1 | 0 | Low-quality | Genome-fragment | 0.66 | AAI-based (high-confidence) | 0 |
| S12_extracted_k141_885 | 1523 | No | NA | 1 | 1 | 0 | Low-quality | Genome-fragment | 1.6 | AAI-based (high-confidence) | 0 |
| S12_vs_k141_103781\|\|full | 1523 | No | NA | 5 | 0 | 0 | Low-quality | Genome-fragment | 3.48 | AAI-based (high-confidence) | 0 |
| S1_vs_k141_18382\|\|full | 1430 | No | NA | 2 | 0 | 1 | Not-determined | Genome-fragment | NA | NA | 0 |
| S1_vs_k141_144309\|\|full | 1532 | No | NA | 2 | 0 | 0 | Not-determined | Genome-fragment | NA | NA | 0 |
| S12_vs_k141_86440\|\|lt2gene | 1523 | No | NA | 2 | 2 | 0 | Low-quality | Genome-fragment | 4.04 | AAI-based (high-confidence) | 0 |
| S1_vs_k141_144333\|\|full | 4378 | No | NA | 4 | 0 | 2 | Not-determined | Genome-fragment | NA | NA | 0 |
| S1_vs_k141_104662\|\|full | 2357 | No | NA | 4 | 0 | 0 | Not-determined | Genome-fragment | NA | NA | 0 |
| S17_extracted_k141_74802 | 1524 | No | NA | 2 | 1 | 0 | Low-quality | Genome-fragment | 6.96 | AAI-based (high-confidence) | 0 |
| S1_vs_k141_118032\|\|full | 3684 | No | NA | 4 | 0 | 2 | Not-determined | Genome-fragment | NA | NA | 0 |
| S1_vs_k141_137897\|\|full | 1250 | No | NA | 2 | 0 | 2 | Not-determined | Genome-fragment | NA | NA | 0 |
| S1_vs_k141_124297\|\|full | 2345 | No | NA | 2 | 0 | 2 | Not-determined | Genome-fragment | NA | NA | 0 |
| S17_vs_k141_22103\|\|full | 1524 | No | NA | 4 | 1 | 0 | Low-quality | Genome-fragment | 0.88 | AAI-based (high-confidence) | 0 |
| S11_vs_k141_80357\|\|full | 1524 | No | NA | 4 | 1 | 0 | Low-quality | Genome-fragment | 3.5 | AAI-based (medium-confidence) | 0 |
| S3_extracted_k141_404 | 1524 | No | NA | 2 | 1 | 0 | Low-quality | Genome-fragment | 2.19 | AAI-based (high-confidence) | 0 |
| S16_extracted_8505 | 1524 | No | NA | 4 | 0 | 0 | Low-quality | Genome-fragment | 2.73 | AAI-based (high-confidence) | 0 |
| S16_vs_116417\|\|full | 1524 | No | NA | 3 | 0 | 3 | Low-quality | Genome-fragment | 0.8 | AAI-based (high-confidence) | 0 |
| S1_vs_k141_24387\|\|full | 2405 | No | NA | 3 | 0 | 2 | Not-determined | Genome-fragment | NA | NA | 0 |
| S12_vs_k141_139479\|\|full | 1524 | No | NA | 3 | 1 | 0 | Low-quality | Genome-fragment | 3.1 | AAI-based (high-confidence) | 0 |
| S8_vs_k141_75379\|\|full | 1524 | No | NA | 3 | 0 | 0 | Low-quality | Genome-fragment | 1.55 | AAI-based (high-confidence) | 0 |
| S17_extracted_k141_59286 | 1525 | No | NA | 1 | 1 | 0 | Low-quality | Genome-fragment | 2.55 | AAI-based (medium-confidence) | 0 |
| S1_vs_k141_31365\|\|full | 1116 | No | NA | 2 | 0 | 1 | Not-determined | Genome-fragment | NA | NA | 0 |
| S9_vs_k141_2296\|\|full | 1525 | No | NA | 3 | 0 | 0 | Low-quality | Genome-fragment | 3.4 | AAI-based (high-confidence) | 0 |
| S1_vs_k141_91270\|\|full | 18290 | No | NA | 26 | 11 | 0 | Medium-quality | Genome-fragment | 53.58 | AAI-based (high-confidence) | 0 |
| S1_vs_k141_37065\|\|full | 2009 | No | NA | 4 | 0 | 2 | Not-determined | Genome-fragment | NA | NA | 0 |
| S1_vs_k141_78072\|\|full | 2707 | No | NA | 2 | 0 | 1 | Not-determined | Genome-fragment | NA | NA | 0 |
| S11_vs_k141_64993\|\|full | 1525 | No | NA | 2 | 0 | 0 | Low-quality | Genome-fragment | 3.39 | AAI-based (high-confidence) | 0 |
| S1_vs_k141_106515\|\|full | 1525 | No | NA | 5 | 0 | 0 | Low-quality | Genome-fragment | 2.99 | AAI-based (high-confidence) | 0 |
| S5_vs_k141_23192\|\|full | 1525 | No | NA | 5 | 3 | 0 | Low-quality | Genome-fragment | 4.28 | AAI-based (high-confidence) | 0 |
| S7_extracted_16739 | 1525 | No | NA | 3 | 0 | 0 | Low-quality | Genome-fragment | 3.29 | AAI-based (high-confidence) | 0 |
| S16_extracted_152213 | 1525 | No | NA | 1 | 0 | 1 | Low-quality | Genome-fragment | 0.15 | AAI-based (medium-confidence) | 0 |
| S16_vs_116233\|\|full | 1525 | No | NA | 3 | 0 | 0 | Low-quality | Genome-fragment | 3.73 | AAI-based (medium-confidence) | 0 |
| S12_vs_k141_154991\|\|full | 1525 | No | NA | 4 | 1 | 0 | Low-quality | Genome-fragment | 2.53 | AAI-based (high-confidence) | 0 |
| S1_vs_k141_98193\|\|full | 1791 | No | NA | 3 | 0 | 3 | Not-determined | Genome-fragment | NA | NA | 0 |
| S6_vs_k141_8754\|\|full | 1525 | No | NA | 4 | 0 | 0 | Low-quality | Genome-fragment | 4.08 | AAI-based (high-confidence) | 0 |
| S1_vs_k141_156131\|\|full | 1377 | No | NA | 4 | 0 | 1 | Not-determined | Genome-fragment | NA | NA | 0 |
| S1_vs_k141_98210\|\|full | 1599 | No | NA | 3 | 0 | 0 | Not-determined | Genome-fragment | NA | NA | 0 |
| S1_vs_k141_3742\|\|full | 1401 | No | NA | 3 | 0 | 0 | Not-determined | Genome-fragment | NA | NA | 0 |
| S1_vs_k141_111419\|\|full | 1431 | No | NA | 2 | 0 | 2 | Not-determined | Genome-fragment | NA | NA | 0 |
| S13_extracted_152195 | 1526 | No | NA | 2 | 0 | 0 | Low-quality | Genome-fragment | 1.62 | AAI-based (high-confidence) | 0 |
| S13_vs_56702\|\|full | 1526 | No | NA | 2 | 0 | 1 | Low-quality | Genome-fragment | 1.46 | AAI-based (high-confidence) | 0 |
| S1_vs_k141_98239\|\|full | 1160 | No | NA | 2 | 0 | 0 | Not-determined | Genome-fragment | NA | NA | 0 |
| S1_vs_k141_101707\|\|full | 1526 | No | NA | 3 | 0 | 1 | Low-quality | Genome-fragment | 3.35 | AAI-based (high-confidence) | 0 |
| S1_vs_k141_68634\|\|full | 1526 | No | NA | 2 | 0 | 1 | Low-quality | Genome-fragment | 0.8 | AAI-based (high-confidence) | 0 |
| S1_vs_k141_69082\|\|full | 1526 | No | NA | 5 | 2 | 0 | Low-quality | Genome-fragment | 2.51 | AAI-based (high-confidence) | 0 |
| S1_vs_k141_147045\|\|lt2gene | 1526 | No | NA | 2 | 1 | 0 | Low-quality | Genome-fragment | 3.38 | AAI-based (high-confidence) | 0 |
| S3_extracted_k141_9963 | 1526 | No | NA | 4 | 0 | 0 | Low-quality | Genome-fragment | 2.42 | AAI-based (high-confidence) | 0 |
| S1_vs_k141_37157\|\|full | 2008 | No | NA | 2 | 0 | 0 | Not-determined | Genome-fragment | NA | NA | 0 |
| S5_vs_k141_49100\|\|full | 1526 | No | NA | 2 | 0 | 0 | Low-quality | Genome-fragment | 4.29 | AAI-based (high-confidence) | 0 |
| S1_vs_k141_3779\|\|full | 1075 | No | NA | 3 | 0 | 0 | Not-determined | Genome-fragment | NA | NA | 0 |
| S16_extracted_137895 | 1526 | No | NA | 1 | 1 | 0 | Low-quality | Genome-fragment | 1.82 | HMM-based (lower-bound) | 0 |
| S16_vs_164904\|\|full | 1526 | No | NA | 2 | 0 | 1 | Low-quality | Genome-fragment | 0.91 | AAI-based (medium-confidence) | 0 |
| S11_vs_k141_79492\|\|full | 1527 | No | NA | 3 | 0 | 2 | Low-quality | Genome-fragment | 1.74 | AAI-based (medium-confidence) | 0 |
| S1_vs_k141_98292\|\|full | 3963 | No | NA | 4 | 0 | 1 | Not-determined | Genome-fragment | NA | NA | 0 |
| S1_vs_k141_144537\|\|full | 2849 | No | NA | 3 | 0 | 1 | Not-determined | Genome-fragment | NA | NA | 0 |
| S1_extracted_k141_44390 | 1527 | No | NA | 1 | 1 | 0 | Low-quality | Genome-fragment | 3.81 | AAI-based (high-confidence) | 0 |
| S1_vs_k141_18657\|\|full | 1865 | No | NA | 4 | 0 | 4 | Not-determined | Genome-fragment | NA | NA | 0 |
| S1_vs_k141_42994\|\|full | 1527 | No | NA | 2 | 0 | 1 | Low-quality | Genome-fragment | 1.49 | AAI-based (medium-confidence) | 0 |
| S1_vs_k141_44917\|\|full | 4719 | No | NA | 5 | 0 | 1 | Not-determined | Genome-fragment | NA | NA | 0 |
| S16_extracted_18151 | 1527 | No | NA | 1 | 0 | 1 | Low-quality | Genome-fragment | 0.73 | AAI-based (medium-confidence) | 0 |
| S12_vs_k141_157107\|\|full | 1527 | No | NA | 3 | 0 | 1 | Low-quality | Genome-fragment | 3.07 | AAI-based (medium-confidence) | 0 |
| S6_extracted_k141_32499 | 1527 | No | NA | 1 | 1 | 0 | Low-quality | Genome-fragment | 1.07 | AAI-based (high-confidence) | 0 |
| S6_vs_k141_10755\|\|full | 1527 | No | NA | 4 | 0 | 0 | Low-quality | Genome-fragment | 2.37 | AAI-based (high-confidence) | 0 |
| S11_vs_k141_28276\|\|full | 1528 | No | NA | 3 | 0 | 0 | Low-quality | Genome-fragment | 3.52 | AAI-based (medium-confidence) | 0 |
| S14_extracted_k141_115988 | 1528 | No | NA | 1 | 0 | 0 | Low-quality | Genome-fragment | 3.17 | AAI-based (medium-confidence) | 0 |
| S1_vs_k141_58139\|\|full | 3324 | No | NA | 2 | 0 | 1 | Not-determined | Genome-fragment | NA | NA | 0 |
| S1_vs_k141_104951\|\|full | 4773 | No | NA | 4 | 0 | 3 | Not-determined | Genome-fragment | NA | NA | 0 |
| S14_vs_k141_81076\|\|full | 1528 | No | NA | 3 | 0 | 0 | Low-quality | Genome-fragment | 3.21 | AAI-based (medium-confidence) | 0 |
| S1_vs_k141_118326\|\|full | 1390 | No | NA | 2 | 0 | 1 | Not-determined | Genome-fragment | NA | NA | 0 |
| S1_vs_k141_128880\|\|lt2gene | 1528 | No | NA | 1 | 1 | 0 | Low-quality | Genome-fragment | 1.43 | HMM-based (lower-bound) | 0 |
| S1_vs_k141_138167\|\|full | 2121 | No | NA | 3 | 0 | 1 | Not-determined | Genome-fragment | NA | NA | 0 |
| S3_extracted_k141_9243 | 1528 | No | NA | 2 | 0 | 0 | Low-quality | Genome-fragment | 0.85 | AAI-based (medium-confidence) | 0 |
| S1_vs_k141_131684\|\|full | 5340 | No | NA | 5 | 0 | 1 | Not-determined | Genome-fragment | NA | NA | 0 |
| S5_vs_k141_130924\|\|full | 1528 | No | NA | 4 | 0 | 0 | Low-quality | Genome-fragment | 3.03 | AAI-based (medium-confidence) | 0 |
| S7_vs_75579\|\|full | 1528 | No | NA | 2 | 0 | 0 | Low-quality | Genome-fragment | 4.14 | AAI-based (high-confidence) | 0 |
| S1_vs_k141_11821\|\|full | 3180 | No | NA | 7 | 0 | 3 | Not-determined | Genome-fragment | NA | NA | 0 |
| S10_vs_67258\|\|full | 1528 | No | NA | 3 | 0 | 0 | Low-quality | Genome-fragment | 3.34 | AAI-based (medium-confidence) | 0 |
| S10_vs_75993\|\|full | 1528 | No | NA | 3 | 0 | 0 | Low-quality | Genome-fragment | 2.86 | AAI-based (high-confidence) | 0 |
| S12_vs_k141_45635\|\|full | 1528 | No | NA | 4 | 0 | 3 | Low-quality | Genome-fragment | 0.39 | AAI-based (medium-confidence) | 0 |
| S1_vs_k141_45026\|\|full | 2038 | No | NA | 4 | 0 | 4 | Not-determined | Genome-fragment | NA | NA | 0 |
| S1_vs_k141_31597\|\|full | 3115 | No | NA | 3 | 0 | 3 | Not-determined | Genome-fragment | NA | NA | 0 |
| S12_vs_k141_45875\|\|full | 1528 | No | NA | 2 | 1 | 0 | Low-quality | Genome-fragment | 4.36 | AAI-based (high-confidence) | 0 |
| S8_vs_k141_59497\|\|full | 1528 | No | NA | 3 | 2 | 0 | Low-quality | Genome-fragment | 3.81 | AAI-based (high-confidence) | 0 |
| S17_vs_k141_16695\|\|full | 1529 | No | NA | 2 | 0 | 1 | Low-quality | Genome-fragment | 1.39 | AAI-based (medium-confidence) | 0 |
| S17_vs_k141_138410\|\|full | 1529 | No | NA | 2 | 0 | 1 | Low-quality | Genome-fragment | 2.08 | AAI-based (high-confidence) | 0 |
| S9_vs_k141_74839\|\|full | 1529 | No | NA | 2 | 0 | 2 | Low-quality | Genome-fragment | 0.82 | AAI-based (medium-confidence) | 0 |
| S11_vs_k141_84368\|\|full | 1529 | No | NA | 4 | 1 | 0 | Low-quality | Genome-fragment | 3.5 | AAI-based (medium-confidence) | 0 |
| S1_vs_k141_18824\|\|full | 1178 | No | NA | 3 | 0 | 0 | Not-determined | Genome-fragment | NA | NA | 0 |
| S3_extracted_k141_113602 | 1529 | No | NA | 3 | 0 | 0 | Low-quality | Genome-fragment | 1.88 | AAI-based (high-confidence) | 0 |
| S1_vs_k141_91663\|\|full | 1023 | No | NA | 3 | 0 | 0 | Not-determined | Genome-fragment | NA | NA | 0 |
| S3_vs_k141_119332\|\|full | 1529 | No | NA | 2 | 0 | 1 | Low-quality | Genome-fragment | 1.02 | AAI-based (high-confidence) | 0 |
| S7_extracted_41259 | 1529 | No | NA | 1 | 0 | 0 | Low-quality | Genome-fragment | 2.73 | AAI-based (medium-confidence) | 0 |
| S1_vs_k141_144725\|\|full | 4044 | No | NA | 5 | 0 | 0 | Not-determined | Genome-fragment | NA | NA | 0 |
| S16_extracted_137676 | 1529 | No | NA | 2 | 1 | 0 | Low-quality | Genome-fragment | 3.48 | AAI-based (medium-confidence) | 0 |
| S16_vs_25443\|\|full | 1529 | No | NA | 4 | 0 | 0 | Low-quality | Genome-fragment | 3.33 | AAI-based (high-confidence) | 0 |
| S16_vs_29840\|\|full | 1529 | No | NA | 3 | 0 | 2 | Low-quality | Genome-fragment | 0.49 | AAI-based (medium-confidence) | 0 |
| S12_vs_k141_30989\|\|full | 1529 | No | NA | 3 | 1 | 0 | Low-quality | Genome-fragment | 1.09 | HMM-based (lower-bound) | 0 |
| S9_vs_k141_65410\|\|full | 1530 | No | NA | 3 | 0 | 0 | Low-quality | Genome-fragment | 3.66 | AAI-based (high-confidence) | 0 |
| S11_vs_k141_58568\|\|full | 1530 | No | NA | 3 | 0 | 0 | Low-quality | Genome-fragment | 3.52 | AAI-based (medium-confidence) | 0 |
| S1_vs_k141_144791\|\|full | 1052 | No | NA | 2 | 0 | 1 | Not-determined | Genome-fragment | NA | NA | 0 |
| S14_vs_k141_89885\|\|full | 1530 | No | NA | 3 | 0 | 1 | Low-quality | Genome-fragment | 2.5 | AAI-based (high-confidence) | 0 |
| S14_vs_k141_41099\|\|full | 1530 | No | NA | 3 | 0 | 1 | Low-quality | Genome-fragment | 1.79 | AAI-based (high-confidence) | 0 |
| S1_vs_k141_31749\|\|full | 1735 | No | NA | 3 | 0 | 0 | Not-determined | Genome-fragment | NA | NA | 0 |
| S1_extracted_k141_113198 | 1530 | No | NA | 1 | 0 | 0 | Low-quality | Genome-fragment | 0.86 | AAI-based (high-confidence) | 0 |
| S1_vs_k141_4042\|\|full | 2285 | No | NA | 2 | 0 | 2 | Not-determined | Genome-fragment | NA | NA | 0 |
| S3_vs_k141_8859\|\|full | 1530 | No | NA | 3 | 1 | 0 | Low-quality | Genome-fragment | 1.77 | HMM-based (lower-bound) | 0 |

| | | | | | | | | | | | |
|---|---|---|---|---|---|---|---|---|---|---|---|
| S3_vs_k141_65206\|\|full | 1530 | No | NA | 2 | 0 | 1 | Low-quality | Genome-fragment | 2.08 | AAI-based (high-confidence) | 0 |
| S3_vs_k141_14503\|\|full | 1530 | No | NA | 5 | 0 | 0 | Low-quality | Genome-fragment | 3.31 | AAI-based (medium-confidence) | 0 |
| S7_extracted_28377 | 1530 | No | NA | 3 | 0 | 0 | Low-quality | Genome-fragment | 1.85 | AAI-based (medium-confidence) | 0 |
| S1_vs_k141_85180\|\|full | 2740 | No | NA | 3 | 0 | 1 | Not-determined | Genome-fragment | NA | NA | 0 |
| S7_extracted_52597 | 1530 | No | NA | 1 | 0 | 1 | Low-quality | Genome-fragment | 4.16 | AAI-based (medium-confidence) | 0 |
| S16_extracted_117186 | 1530 | No | NA | 1 | 0 | 0 | Low-quality | Genome-fragment | 2.63 | AAI-based (high-confidence) | 0 |
| S10_vs_9087\|\|full | 1530 | No | NA | 2 | 0 | 0 | Low-quality | Genome-fragment | 1.04 | AAI-based (high-confidence) | 0 |
| S12_extracted_k141_179806 | 1530 | No | NA | 5 | 0 | 0 | Low-quality | Genome-fragment | 3.48 | AAI-based (high-confidence) | 0 |
| S6_vs_k141_8488\|\|full | 1530 | No | NA | 4 | 1 | 0 | Low-quality | Genome-fragment | 0.84 | HMM-based (lower-bound) | 0 |
| S1_vs_k141_25000\|\|full | 1394 | No | NA | 2 | 0 | 2 | Not-determined | Genome-fragment | NA | NA | 0 |
| S1_vs_k141_25005\|\|full | 1579 | No | NA | 3 | 0 | 1 | Not-determined | Genome-fragment | NA | NA | 0 |
| S8_extracted_k141_107519 | 1530 | No | NA | 1 | 0 | 1 | Low-quality | Genome-fragment | 1.42 | AAI-based (medium-confidence) | 0 |
| S17_extracted_k141_83451 | 1531 | No | NA | 1 | 0 | 1 | Low-quality | Genome-fragment | 1.42 | AAI-based (medium-confidence) | 0 |
| S1_vs_k141_124942\|\|full | 3029 | No | NA | 4 | 0 | 3 | Not-determined | Genome-fragment | NA | NA | 0 |
| S1_vs_k141_111868\|\|full | 4981 | No | NA | 5 | 0 | 5 | Not-determined | Genome-fragment | NA | NA | 0 |
| S1_vs_k141_91864\|\|full | 1529 | No | NA | 3 | 0 | 1 | Not-determined | Genome-fragment | NA | NA | 0 |
| S9_vs_k141_22207\|\|lt2gene | 1531 | No | NA | 1 | 1 | 0 | Low-quality | Genome-fragment | 2.65 | AAI-based (medium-confidence) | 0 |
| S1_vs_k141_19045\|\|full | 1136 | No | NA | 2 | 0 | 0 | Not-determined | Genome-fragment | NA | NA | 0 |
| S11_extracted_k141_88911 | 1531 | No | NA | 2 | 0 | 1 | Low-quality | Genome-fragment | 1.6 | AAI-based (medium-confidence) | 0 |
| S11_extracted_k141_96926 | 1531 | No | NA | 4 | 2 | 0 | Low-quality | Genome-fragment | 24.12 | AAI-based (high-confidence) | 0 |
| S11_vs_k141_3294\|\|full | 1531 | No | NA | 2 | 0 | 0 | Low-quality | Genome-fragment | 3.43 | AAI-based (high-confidence) | 0 |
| S13_vs_11133\|\|full | 1531 | No | NA | 2 | 0 | 0 | Low-quality | Genome-fragment | 1.9 | AAI-based (medium-confidence) | 0 |
| S1_vs_k141_45325\|\|full | 1213 | No | NA | 3 | 0 | 0 | Not-determined | Genome-fragment | NA | NA | 0 |
| S14_vs_k141_45687\|\|full | 1531 | No | NA | 5 | 0 | 0 | Low-quality | Genome-fragment | 3.52 | AAI-based (high-confidence) | 0 |
| S1_extracted_k141_57597 | 1531 | No | NA | 1 | 0 | 0 | Low-quality | Genome-fragment | 0.66 | AAI-based (medium-confidence) | 0 |
| S3_extracted_k141_43696 | 1531 | No | NA | 3 | 3 | 0 | Low-quality | Genome-fragment | 7.01 | AAI-based (high-confidence) | 0 |
| S5_vs_k141_43546\|\|full | 1531 | No | NA | 4 | 2 | 0 | Low-quality | Genome-fragment | 4.15 | AAI-based (high-confidence) | 0 |
| S1_vs_k141_45396\|\|full | 6966 | No | NA | 6 | 0 | 4 | Not-determined | Genome-fragment | NA | NA | 0 |
| S1_vs_k141_125096\|\|full | 7099 | No | NA | 5 | 0 | 5 | Not-determined | Genome-fragment | NA | NA | 0 |
| S5_vs_k141_10113\|\|full | 1531 | No | NA | 5 | 1 | 0 | Low-quality | Genome-fragment | 4.43 | AAI-based (medium-confidence) | 0 |
| S16_vs_85405\|\|full | 1531 | No | NA | 4 | 0 | 1 | Low-quality | Genome-fragment | 2.44 | AAI-based (high-confidence) | 0 |
| S1_vs_k141_32013\|\|full | 1094 | No | NA | 1 | 0 | 1 | Not-determined | Genome-fragment | NA | NA | 0 |
| S1_vs_k141_145035\|\|full | 1721 | No | NA | 3 | 0 | 0 | Not-determined | Genome-fragment | NA | NA | 0 |
| S16_vs_139275\|\|lt2gene | 1531 | No | NA | 2 | 1 | 0 | Low-quality | Genome-fragment | 3.38 | AAI-based (high-confidence) | 0 |
| S1_vs_k141_45422\|\|full | 2844 | No | NA | 3 | 0 | 3 | Not-determined | Genome-fragment | NA | NA | 0 |
| S8_vs_k141_155582\|\|full | 1531 | No | NA | 2 | 0 | 0 | Low-quality | Genome-fragment | 1.09 | AAI-based (medium-confidence) | 0 |
| S1_vs_k141_105456\|\|full | 3382 | No | NA | 4 | 0 | 2 | Not-determined | Genome-fragment | NA | NA | 0 |
| S17_extracted_k141_28306 | 1532 | No | NA | 3 | 3 | 0 | Low-quality | Genome-fragment | 6.99 | AAI-based (high-confidence) | 0 |
| S14_extracted_k141_59826 | 1532 | No | NA | 1 | 1 | 0 | Low-quality | Genome-fragment | 3.29 | AAI-based (high-confidence) | 0 |
| S5_vs_k141_101592\|\|full | 1532 | No | NA | 3 | 1 | 0 | Low-quality | Genome-fragment | 4 | AAI-based (high-confidence) | 0 |
| S1_vs_k141_92090\|\|full | 1319 | No | NA | 2 | 0 | 0 | Not-determined | Genome-fragment | NA | NA | 0 |
| S1_vs_k141_37576\|\|full | 1945 | No | NA | 3 | 0 | 0 | Not-determined | Genome-fragment | NA | NA | 0 |
| S16_vs_32086\|\|full | 1532 | No | NA | 4 | 1 | 0 | Low-quality | Genome-fragment | 3.07 | AAI-based (medium-confidence) | 0 |
| S16_vs_157123\|\|full | 1532 | No | NA | 4 | 1 | 0 | Low-quality | Genome-fragment | 1.1 | HMM-based (lower-bound) | 0 |
| S12_extracted_k141_109633 | 1532 | No | NA | 3 | 0 | 1 | Low-quality | Genome-fragment | 3.32 | AAI-based (high-confidence) | 0 |
| S6_extracted_k141_34497 | 1532 | No | NA | 4 | 1 | 0 | Low-quality | Genome-fragment | 2.85 | AAI-based (medium-confidence) | 0 |
| S6_vs_k141_49295\|\|full | 1532 | No | NA | 2 | 0 | 1 | Low-quality | Genome-fragment | 0.91 | AAI-based (high-confidence) | 0 |
| S8_extracted_k141_173401 | 1532 | No | NA | 3 | 0 | 1 | Low-quality | Genome-fragment | 0.41 | AAI-based (medium-confidence) | 0 |
| S1_vs_k141_37617\|\|full | 3546 | No | NA | 8 | 0 | 0 | Not-determined | Genome-fragment | NA | NA | 0 |
| S1_vs_k141_138637\|\|full | 10188 | No | NA | 12 | 0 | 7 | Not-determined | Genome-fragment | NA | NA | 0 |
| S1_vs_k141_52363\|\|full | 1406 | No | NA | 2 | 0 | 1 | Not-determined | Genome-fragment | NA | NA | 0 |
| S9_extracted_k141_28072 | 1533 | No | NA | 4 | 1 | 0 | Low-quality | Genome-fragment | 2.59 | AAI-based (medium-confidence) | 0 |
| S11_vs_k141_94232\|\|full | 1533 | No | NA | 3 | 0 | 0 | Low-quality | Genome-fragment | 2.8 | AAI-based (medium-confidence) | 0 |
| S1_vs_k141_56323\|\|full | 1533 | No | NA | 5 | 1 | 0 | Low-quality | Genome-fragment | 3.52 | AAI-based (medium-confidence) | 0 |
| S12_extracted_k141_176886 | 1533 | No | NA | 4 | 1 | 0 | Low-quality | Genome-fragment | 2.67 | AAI-based (medium-confidence) | 0 |
| S12_vs_k141_72065\|\|full | 1533 | No | NA | 4 | 1 | 0 | Low-quality | Genome-fragment | 3.59 | AAI-based (medium-confidence) | 0 |
| S1_vs_k141_19378\|\|full | 4815 | No | NA | 3 | 0 | 2 | Not-determined | Genome-fragment | NA | NA | 0 |
| S12_vs_k141_67553\|\|full | 1533 | No | NA | 4 | 0 | 0 | Low-quality | Genome-fragment | 2.26 | AAI-based (medium-confidence) | 0 |
| S17_vs_k141_50520\|\|full | 1534 | No | NA | 5 | 3 | 0 | Low-quality | Genome-fragment | 2.61 | HMM-based (lower-bound) | 0 |
| S9_extracted_k141_57432 | 1534 | No | NA | 2 | 0 | 0 | Low-quality | Genome-fragment | 0.29 | AAI-based (medium-confidence) | 0 |
| S14_vs_k141_15072\|\|full | 1534 | No | NA | 5 | 1 | 0 | Low-quality | Genome-fragment | 4.43 | AAI-based (high-confidence) | 0 |
| S1_vs_k141_78976\|\|full | 1489 | No | NA | 3 | 0 | 0 | Not-determined | Genome-fragment | NA | NA | 0 |
| S1_vs_k141_94096\|\|full | 1534 | No | NA | 3 | 0 | 0 | Low-quality | Genome-fragment | 3.63 | AAI-based (medium-confidence) | 0 |
| S1_vs_k141_138837\|\|full | 1534 | No | NA | 4 | 1 | 0 | Low-quality | Genome-fragment | 3.93 | AAI-based (high-confidence) | 0 |
| S1_vs_k141_85583\|\|full | 1832 | No | NA | 5 | 0 | 0 | Not-determined | Genome-fragment | NA | NA | 0 |
| S1_vs_k141_46122\|\|full | 1534 | No | NA | 2 | 2 | 0 | Low-quality | Genome-fragment | 2.6 | AAI-based (high-confidence) | 0 |
| S3_extracted_k141_56840 | 1534 | No | NA | 2 | 1 | 0 | Low-quality | Genome-fragment | 1.71 | HMM-based (lower-bound) | 0 |
| S5_vs_k141_89116\|\|full | 1534 | No | NA | 3 | 0 | 0 | Low-quality | Genome-fragment | 46.34 | AAI-based (medium-confidence) | 0 |
| S7_extracted_9647 | 1534 | No | NA | 3 | 0 | 0 | Low-quality | Genome-fragment | 2.91 | AAI-based (high-confidence) | 0 |
| S7_vs_47010\|\|full | 1534 | No | NA | 2 | 0 | 1 | Low-quality | Genome-fragment | 1.62 | AAI-based (high-confidence) | 0 |
| S10_extracted_22518 | 1534 | No | NA | 1 | 0 | 0 | Low-quality | Genome-fragment | 1 | AAI-based (high-confidence) | 0 |
| S1_vs_k141_99061\|\|full | 1779 | No | NA | 2 | 0 | 2 | Not-determined | Genome-fragment | NA | NA | 0 |
| S1_vs_k141_79019\|\|full | 1478 | No | NA | 4 | 0 | 0 | Not-determined | Genome-fragment | NA | NA | 0 |
| S17_vs_k141_5749\|\|full | 1535 | No | NA | 6 | 0 | 0 | Low-quality | Genome-fragment | 3.25 | AAI-based (medium-confidence) | 0 |
| S14_extracted_k141_37988 | 1535 | No | NA | 1 | 0 | 0 | Low-quality | Genome-fragment | 2.78 | AAI-based (high-confidence) | 0 |
| S14_extracted_k141_86830 | 1535 | No | NA | 5 | 5 | 0 | Low-quality | Genome-fragment | 5.04 | AAI-based (high-confidence) | 0 |
| S1_vs_k141_124331\|\|full | 1535 | No | NA | 3 | 0 | 0 | Low-quality | Genome-fragment | 0.76 | AAI-based (medium-confidence) | 0 |
| S1_vs_k141_52546\|\|full | 4239 | No | NA | 3 | 0 | 1 | Not-determined | Genome-fragment | NA | NA | 0 |
| S3_vs_k141_71044\|\|full | 1535 | No | NA | 5 | 0 | 0 | Low-quality | Genome-fragment | 1.74 | AAI-based (medium-confidence) | 0 |
| S1_vs_k141_85684\|\|full | 1254 | No | NA | 2 | 0 | 1 | Not-determined | Genome-fragment | NA | NA | 0 |
| S1_vs_k141_138796\|\|full | 1105 | No | NA | 3 | 0 | 1 | Not-determined | Genome-fragment | NA | NA | 0 |
| S1_vs_k141_119028\|\|full | 1367 | No | NA | 2 | 0 | 2 | Not-determined | Genome-fragment | NA | NA | 0 |
| S5_extracted_k141_74729 | 1535 | No | NA | 4 | 0 | 0 | Low-quality | Genome-fragment | 4.35 | AAI-based (high-confidence) | 0 |
| S5_vs_k141_49890\|\|full | 1535 | No | NA | 2 | 0 | 0 | Low-quality | Genome-fragment | 3.73 | AAI-based (high-confidence) | 0 |
| S7_vs_28647\|\|full | 1535 | No | NA | 3 | 1 | 0 | Low-quality | Genome-fragment | 2.31 | HMM-based (lower-bound) | 0 |
| S16_vs_58938\|\|lt2gene | 1535 | No | NA | 2 | 1 | 0 | Low-quality | Genome-fragment | 3.41 | AAI-based (high-confidence) | 0 |
| S12_vs_k141_62509\|\|full | 1535 | No | NA | 4 | 1 | 0 | Low-quality | Genome-fragment | 3.85 | AAI-based (medium-confidence) | 0 |
| S8_extracted_k141_131124 | 1535 | No | NA | 2 | 0 | 0 | Low-quality | Genome-fragment | 0.81 | AAI-based (medium-confidence) | 0 |
| S17_extracted_k141_72980 | 1536 | No | NA | 2 | 0 | 0 | Low-quality | Genome-fragment | 4.88 | AAI-based (high-confidence) | 0 |
| S17_vs_k141_169490\|\|full | 1536 | No | NA | 2 | 0 | 1 | Low-quality | Genome-fragment | 3.86 | AAI-based (high-confidence) | 0 |
| S1_vs_k141_52600\|\|full | 1022 | No | NA | 2 | 0 | 0 | Not-determined | Genome-fragment | NA | NA | 0 |
| S1_vs_k141_99158\|\|full | 1563 | No | NA | 2 | 0 | 1 | Not-determined | Genome-fragment | NA | NA | 0 |
| S1_vs_k141_149979\|\|full | 1536 | No | NA | 4 | 1 | 0 | Low-quality | Genome-fragment | 3.55 | AAI-based (medium-confidence) | 0 |
| S3_extracted_k141_4659 | 1536 | No | NA | 5 | 0 | 0 | Low-quality | Genome-fragment | 0.75 | AAI-based (high-confidence) | 0 |
| S1_vs_k141_25524\|\|full | 3461 | No | NA | 3 | 0 | 2 | Not-determined | Genome-fragment | NA | NA | 0 |
| S3_vs_k141_120548\|\|full | 1536 | No | NA | 2 | 0 | 1 | Low-quality | Genome-fragment | 1.48 | AAI-based (medium-confidence) | 0 |
| S1_vs_k141_99175\|\|full | 2378 | No | NA | 2 | 0 | 2 | Not-determined | Genome-fragment | NA | NA | 0 |
| S5_extracted_k141_20966 | 1536 | No | NA | 3 | 0 | 0 | Low-quality | Genome-fragment | 3.37 | AAI-based (medium-confidence) | 0 |
| S5_vs_k141_106913\|\|lt2gene | 1536 | No | NA | 1 | 1 | 0 | Low-quality | Genome-fragment | 3.59 | AAI-based (medium-confidence) | 0 |
| S16_extracted_77914 | 1536 | No | NA | 3 | 0 | 0 | Low-quality | Genome-fragment | 1.25 | AAI-based (high-confidence) | 0 |
| S16_vs_9218\|\|full | 1536 | No | NA | 3 | 0 | 0 | Low-quality | Genome-fragment | 4.12 | AAI-based (high-confidence) | 0 |
| S10_extracted_1923 | 1536 | No | NA | 3 | 0 | 0 | Low-quality | Genome-fragment | 2.74 | AAI-based (high-confidence) | 0 |
| S15_vs_61073\|\|full | 1536 | No | NA | 3 | 0 | 0 | Low-quality | Genome-fragment | 2.57 | AAI-based (high-confidence) | 0 |
| S1_vs_k141_64468\|\|full | 3443 | No | NA | 4 | 0 | 1 | Not-determined | Genome-fragment | NA | NA | 0 |
| S1_vs_k141_52685\|\|full | 1683 | No | NA | 2 | 0 | 2 | Not-determined | Genome-fragment | NA | NA | 0 |
| S9_vs_k141_11713\|\|lt2gene | 1537 | No | NA | 1 | 1 | 0 | Low-quality | Genome-fragment | 3.5 | AAI-based (high-confidence) | 0 |
| S1_vs_k141_108320\|\|lt2gene | 1537 | No | NA | 2 | 2 | 0 | Low-quality | Genome-fragment | 3.93 | AAI-based (high-confidence) | 0 |
| S5_vs_k141_88736\|\|full | 1537 | No | NA | 3 | 1 | 0 | Low-quality | Genome-fragment | 3.43 | AAI-based (high-confidence) | 0 |
| S1_vs_k141_19670\|\|full | 1922 | No | NA | 2 | 0 | 0 | Not-determined | Genome-fragment | NA | NA | 0 |
| S1_vs_k141_92453\|\|full | 1197 | No | NA | 2 | 0 | 0 | Not-determined | Genome-fragment | NA | NA | 0 |
| S1_vs_k141_19675\|\|full | 1316 | No | NA | 3 | 0 | 0 | Not-determined | Genome-fragment | NA | NA | 0 |
| S1_vs_k141_92486\|\|full | 1323 | No | NA | 2 | 0 | 0 | Not-determined | Genome-fragment | NA | NA | 0 |
| S1_vs_k141_145456\|\|full | 1141 | No | NA | 3 | 0 | 1 | Not-determined | Genome-fragment | NA | NA | 0 |
| S8_extracted_k141_3736 | 1537 | No | NA | 3 | 2 | 0 | Low-quality | Genome-fragment | 4.99 | AAI-based (high-confidence) | 0 |
| S1_vs_k141_112467\|\|full | 1851 | No | NA | 4 | 0 | 2 | Not-determined | Genome-fragment | NA | NA | 0 |
| S7_vs_70016\|\|full | 1538 | No | NA | 3 | 2 | 0 | Low-quality | Genome-fragment | 3.82 | AAI-based (high-confidence) | 0 |
| S1_vs_k141_138973\|\|full | 1103 | No | NA | 2 | 0 | 1 | Not-determined | Genome-fragment | NA | NA | 0 |
| S16_vs_17468\|\|full | 1538 | No | NA | 3 | 1 | 0 | Low-quality | Genome-fragment | 3.52 | AAI-based (high-confidence) | 0 |
| S1_vs_k141_25680\|\|full | 1357 | No | NA | 2 | 0 | 0 | Not-determined | Genome-fragment | NA | NA | 0 |
| S1_vs_k141_105924\|\|full | 1341 | No | NA | 2 | 0 | 0 | Not-determined | Genome-fragment | NA | NA | 0 |
| S16_vs_53780\|\|lt2gene | 1538 | No | NA | 1 | 1 | 0 | Low-quality | Genome-fragment | 2.91 | HMM-based (lower-bound) | 0 |
| S10_extracted_18682 | 1538 | No | NA | 2 | 0 | 2 | Low-quality | Genome-fragment | 0.92 | AAI-based (medium-confidence) | 0 |
| S1_vs_k141_79353\|\|full | 1174 | No | NA | 3 | 0 | 2 | Not-determined | Genome-fragment | NA | NA | 0 |
| S1_vs_k141_149633\|\|full | 1300 | No | NA | 2 | 0 | 0 | Not-determined | Genome-fragment | NA | NA | 0 |
| S12_vs_k141_101417\|\|full | 1538 | No | NA | 3 | 0 | 0 | Low-quality | Genome-fragment | 1.65 | AAI-based (high-confidence) | 0 |
| S8_vs_k141_106344\|\|full | 1538 | No | NA | 3 | 0 | 0 | Low-quality | Genome-fragment | 3.38 | AAI-based (medium-confidence) | 0 |
| S8_vs_k141_40329\|\|lt2gene | 1538 | No | NA | 1 | 1 | 0 | Low-quality | Genome-fragment | 0.13 | HMM-based (lower-bound) | 0 |
| S1_vs_k141_112480\|\|full | 1024 | No | NA | 2 | 0 | 1 | Not-determined | Genome-fragment | NA | NA | 0 |
| S1_vs_k141_105944\|\|full | 1809 | No | NA | 2 | 0 | 1 | Not-determined | Genome-fragment | NA | NA | 0 |
| S1_vs_k141_145513\|\|full | 1550 | No | NA | 4 | 0 | 1 | Not-determined | Genome-fragment | NA | NA | 0 |
| S17_extracted_k141_24589 | 1539 | No | NA | 1 | 0 | 1 | Low-quality | Genome-fragment | 4.34 | AAI-based (medium-confidence) | 0 |
| S1_vs_k141_4629\|\|full | 1761 | No | NA | 3 | 0 | 1 | Not-determined | Genome-fragment | NA | NA | 0 |
| S11_vs_k141_26850\|\|full | 1539 | No | NA | 3 | 1 | 0 | Low-quality | Genome-fragment | 3.51 | AAI-based (high-confidence) | 0 |
| S1_vs_k141_45960\|\|full | 2318 | No | NA | 2 | 0 | 1 | Not-determined | Genome-fragment | NA | NA | 0 |
| S1_vs_k141_85990\|\|full | 2402 | No | NA | 2 | 0 | 2 | Not-determined | Genome-fragment | NA | NA | 0 |
| S14_extracted_k141_63198 | 1539 | No | NA | 3 | 2 | 0 | Low-quality | Genome-fragment | 2.68 | AAI-based (high-confidence) | 0 |
| S1_vs_k141_92575\|\|full | 2594 | No | NA | 3 | 0 | 2 | Not-determined | Genome-fragment | NA | NA | 0 |
| S1_vs_k141_25748\|\|full | 2035 | No | NA | 4 | 0 | 2 | Not-determined | Genome-fragment | NA | NA | 0 |
| S5_vs_k141_87211\|\|full | 1539 | No | NA | 2 | 1 | 0 | Low-quality | Genome-fragment | 4.23 | AAI-based (medium-confidence) | 0 |
| S5_vs_k141_42370\|\|full | 1539 | No | NA | 4 | 1 | 0 | Low-quality | Genome-fragment | 2.75 | AAI-based (high-confidence) | 0 |
| S1_vs_k141_32665\|\|full | 1817 | No | NA | 3 | 0 | 1 | Not-determined | Genome-fragment | NA | NA | 0 |
| S1_vs_k141_37979\|\|full | 2650 | No | NA | 4 | 0 | 0 | Not-determined | Genome-fragment | NA | NA | 0 |
| S1_vs_k141_86035\|\|full | 1142 | No | NA | 3 | 0 | 1 | Not-determined | Genome-fragment | NA | NA | 0 |
| S16_vs_56546\|\|full | 1539 | No | NA | 3 | 0 | 1 | Low-quality | Genome-fragment | 3.08 | AAI-based (medium-confidence) | 0 |
| S16_vs_1589\|\|lt2gene | 1539 | No | NA | 2 | 0 | 0 | Low-quality | Genome-fragment | 4.12 | AAI-based (medium-confidence) | 0 |
| S8_extracted_k141_33047 | 1539 | No | NA | 2 | 0 | 1 | Low-quality | Genome-fragment | 0.5 | AAI-based (medium-confidence) | 0 |
| S13_vs_13625\|\|lt2gene | 1540 | No | NA | 1 | 1 | 0 | Low-quality | Genome-fragment | 2.25 | HMM-based (lower-bound) | 0 |
| S1_extracted_k141_108678 | 1540 | No | NA | 1 | 0 | 1 | Low-quality | Genome-fragment | 1.15 | AAI-based (medium-confidence) | 0 |
| S1_vs_k141_120320\|\|full | 1540 | No | NA | 2 | 2 | 0 | Low-quality | Genome-fragment | 4.5 | AAI-based (high-confidence) | 0 |
| S3_extracted_k141_88809 | 1540 | No | NA | 3 | 1 | 0 | Low-quality | Genome-fragment | 3.04 | AAI-based (high-confidence) | 0 |
| S3_extracted_k141_61030 | 1540 | No | NA | 3 | 0 | 0 | Low-quality | Genome-fragment | 2.75 | AAI-based (high-confidence) | 0 |
| S3_vs_k141_91814\|\|full | 1540 | No | NA | 4 | 0 | 0 | Low-quality | Genome-fragment | 2.44 | AAI-based (high-confidence) | 0 |
| S5_extracted_k141_77617 | 1540 | No | NA | 1 | 0 | 1 | Low-quality | Genome-fragment | 0.75 | AAI-based (medium-confidence) | 0 |
| S7_vs_81552\|\|full | 1540 | No | NA | 3 | 1 | 0 | Low-quality | Genome-fragment | 3.52 | AAI-based (high-confidence) | 0 |
| S7_vs_43903\|\|lt2gene | 1540 | No | NA | 1 | 1 | 0 | Low-quality | Genome-fragment | 3.35 | AAI-based (high-confidence) | 0 |
| S1_vs_k141_71393\|\|full | 1116 | No | NA | 2 | 0 | 1 | Not-determined | Genome-fragment | NA | NA | 0 |
| S16_extracted_128427 | 1540 | No | NA | 4 | 2 | 0 | Low-quality | Genome-fragment | 3.48 | AAI-based (high-confidence) | 0 |
| S1_vs_k141_12922\|\|full | 1088 | No | NA | 2 | 0 | 1 | Not-determined | Genome-fragment | NA | NA | 0 |
| S16_extracted_160888 | 1540 | No | NA | 3 | 3 | 0 | Low-quality | Genome-fragment | 4.07 | AAI-based (high-confidence) | 0 |
| S10_vs_4519\|\|full | 1540 | No | NA | 3 | 1 | 0 | Low-quality | Genome-fragment | 0.55 | HMM-based (lower-bound) | 0 |
| S8_vs_k141_178128\|\|lt2gene | 1540 | No | NA | 2 | 1 | 0 | Low-quality | Genome-fragment | 3.37 | AAI-based (high-confidence) | 0 |
| S1_vs_k141_32764\|\|full | 6028 | No | NA | 8 | 0 | 1 | Not-determined | Genome-fragment | NA | NA | 0 |
| S11_vs_k141_35189\|\|full | 1541 | No | NA | 6 | 0 | 2 | Low-quality | Genome-fragment | 3.02 | AAI-based (high-confidence) | 0 |
| S13_vs_65024\|\|full | 1541 | No | NA | 3 | 0 | 0 | Low-quality | Genome-fragment | 2.69 | AAI-based (high-confidence) | 0 |
| S14_vs_k141_50639\|\|full | 1541 | No | NA | 4 | 1 | 0 | Low-quality | Genome-fragment | 3.84 | AAI-based (high-confidence) | 0 |
| S1_extracted_k141_30753 | 1541 | No | NA | 3 | 2 | 0 | Low-quality | Genome-fragment | 3.92 | AAI-based (high-confidence) | 0 |
| S1_vs_k141_71409\|\|full | 2070 | No | NA | 2 | 0 | 2 | Not-determined | Genome-fragment | NA | NA | 0 |
| S1_vs_k141_106123\|\|full | 1883 | No | NA | 3 | 0 | 0 | Not-determined | Genome-fragment | NA | NA | 0 |
| S1_extracted_k141_125607 | 1541 | No | NA | 1 | 0 | 1 | Low-quality | Genome-fragment | 2.63 | AAI-based (high-confidence) | 0 |
| S7_vs_67765\|\|full | 1541 | No | NA | 4 | [illegible] | 3 | Low-quality | Genome-fragment | 2.47 | AAI-based (high-confidence) | 0 |
| S1_vs_k141_53070\|\|full | 1164 | No | NA | 3 | 0 | 1 | Not-determined | Genome-fragment | NA | NA | 0 |
| S7_vs_88588\|\|lt2gene | 1541 | No | NA | 2 | 1 | 0 | Low-quality | Genome-fragment | 3.77 | AAI-based (high-confidence) | 0 |
| S1_vs_k141_132924\|\|full | 2446 | No | NA | 3 | 0 | 1 | Not-determined | Genome-fragment | NA | NA | 0 |
| S16_vs_172379\|\|full | 1541 | No | NA | 4 | 0 | 1 | Low-quality | Genome-fragment | 2.86 | AAI-based (high-confidence) | 0 |
| S10_extracted_74007 | 1541 | No | NA | 2 | 0 | 1 | Low-quality | Genome-fragment | 3.44 | AAI-based (medium-confidence) | 0 |
| S1_vs_k141_156821\|\|full | 2010 | No | NA | 2 | 0 | 1 | Not-determined | Genome-fragment | NA | NA | 0 |
| S10_vs_99459\|\|full | 1541 | No | NA | 3 | 2 | 0 | Low-quality | Genome-fragment | 3.54 | AAI-based (high-confidence) | 0 |
| S1_vs_k141_79577\|\|full | 22821 | No | NA | 22 | 0 | 13 | Not-determined | Genome-fragment | NA | NA | 0 |
| S12_vs_k141_179592\|\|full | 1541 | No | NA | 3 | 0 | 0 | Low-quality | Genome-fragment | 2.8 | AAI-based (medium-confidence) | 0 |
| S8_extracted_k141_139122 | 1541 | No | NA | 5 | 0 | 0 | Low-quality | Genome-fragment | 2.45 | AAI-based (high-confidence) | 0 |
| S8_vs_k141_110253\|\|full | 1541 | No | NA | 2 | 1 | 0 | Low-quality | Genome-fragment | 3.72 | AAI-based (medium-confidence) | 0 |
| S9_vs_k141_64095\|\|full | 1542 | No | NA | 3 | 0 | 1 | Low-quality | Genome-fragment | 2.38 | AAI-based (high-confidence) | 0 |
| S13_extracted_49964 | 1542 | No | NA | 1 | 0 | 0 | Low-quality | Genome-fragment | 0.26 | AAI-based (high-confidence) | 0 |
| S14_extracted_k141_48649 | 1542 | No | NA | 1 | 0 | 1 | Low-quality | Genome-fragment | 0.16 | AAI-based (medium-confidence) | 0 |
| S1_vs_k141_4830\|\|full | 1623 | No | NA | 3 | 0 | 0 | Not-determined | Genome-fragment | NA | NA | 0 |
| S14_vs_k141_116655\|\|full | 1542 | No | NA | 2 | 0 | 0 | Low-quality | Genome-fragment | 1.16 | AAI-based (high-confidence) | 0 |
| S1_vs_k141_4836\|\|full | 1662 | No | NA | 4 | 0 | 2 | Not-determined | Genome-fragment | NA | NA | 0 |
| S14_vs_k141_56680\|\|lt2gene | 1542 | No | NA | 2 | 1 | 0 | Low-quality | Genome-fragment | 3.47 | AAI-based (medium-confidence) | 0 |
| S1_vs_k141_79655\|\|full | 1190 | No | NA | 2 | 0 | 1 | Not-determined | Genome-fragment | NA | NA | 0 |
| S1_vs_k141_46993\|\|full | 1542 | No | NA | 3 | 0 | 1 | Low-quality | Genome-fragment | 0.7 | AAI-based (medium-confidence) | 0 |
| S3_vs_k141_76276\|\|full | 1542 | No | NA | 3 | 0 | 0 | Low-quality | Genome-fragment | 0.97 | AAI-based (high-confidence) | 0 |
| S7_vs_79139\|\|full | 1542 | No | NA | 3 | 1 | 0 | Low-quality | Genome-fragment | 4.11 | AAI-based (medium-confidence) | 0 |
| S16_extracted_148119 | 1542 | No | NA | 4 | 0 | 0 | Low-quality | Genome-fragment | 2.62 | AAI-based (high-confidence) | 0 |
| S1_vs_k141_112814\|\|full | 1506 | No | NA | 2 | 0 | 2 | Not-determined | Genome-fragment | NA | NA | 0 |
| S1_vs_k141_139386\|\|full | 1262 | No | NA | 3 | 0 | 2 | Not-determined | Genome-fragment | NA | NA | 0 |
| S16_vs_22254\|\|full | 1542 | No | NA | 4 | 0 | 0 | Low-quality | Genome-fragment | 2.62 | AAI-based (medium-confidence) | 0 |
| S16_vs_96268\|\|full | 1542 | No | NA | 3 | 0 | 0 | Low-quality | Genome-fragment | 2.91 | AAI-based (medium-confidence) | 0 |
| S10_vs_12626\|\|full | 1542 | No | NA | 3 | 0 | 1 | Low-quality | Genome-fragment | 2.13 | AAI-based (high-confidence) | 0 |
| S12_extracted_k141_172523 | 1542 | No | NA | 2 | 0 | 0 | Low-quality | Genome-fragment | 2.51 | AAI-based (high-confidence) | 0 |
| S8_vs_k141_117694\|\|full | 1542 | No | NA | 4 | 2 | 0 | Low-quality | Genome-fragment | 4.22 | AAI-based (high-confidence) | 0 |
| S1_vs_k141_53216\|\|full | 1821 | No | NA | 3 | 0 | 2 | Not-determined | Genome-fragment | NA | NA | 0 |
| S17_extracted_k141_37459 | 1543 | No | NA | 3 | 0 | 1 | Low-quality | Genome-fragment | 0.98 | AAI-based (medium-confidence) | 0 |
| S1_vs_k141_79734\|\|full | 1092 | No | NA | 2 | 0 | 0 | Not-determined | Genome-fragment | NA | NA | 0 |
| S17_vs_k141_78052\|\|full | 1543 | No | NA | 3 | 0 | 0 | Low-quality | Genome-fragment | 1.36 | AAI-based (medium-confidence) | 0 |
| S1_vs_k141_145792\|\|full | 23601 | No | NA | 22 | 1 | 1 | Medium-quality | Genome-fragment | 55.35 | AAI-based (high-confidence) | 0 |
| S1_vs_k141_46318\|\|full | 2042 | No | NA | 4 | 0 | 0 | Not-determined | Genome-fragment | NA | NA | 0 |
| S1_vs_k141_71554\|\|full | 1458 | No | NA | 2 | 0 | 0 | Not-determined | Genome-fragment | NA | NA | 0 |
| S11_vs_k141_42570\|\|full | 1543 | No | NA | 3 | 1 | 0 | Low-quality | Genome-fragment | 2.48 | HMM-based (lower-bound) | 0 |
| S1_vs_k141_46339\|\|full | 1461 | No | NA | 3 | 0 | 1 | Not-determined | Genome-fragment | NA | NA | 0 |
| S1_vs_k141_86382\|\|full | 3921 | No | NA | 5 | 0 | 0 | Not-determined | Genome-fragment | NA | NA | 0 |
| S1_vs_k141_71567\|\|full | 2388 | No | NA | 5 | 0 | 2 | Not-determined | Genome-fragment | NA | NA | 0 |
| S13_vs_52326\|\|full | 1543 | No | NA | 2 | 0 | 1 | Low-quality | Genome-fragment | 1.25 | AAI-based (high-confidence) | 0 |
| S1_vs_k141_13256\|\|full | 4005 | No | NA | 4 | 0 | 0 | Not-determined | Genome-fragment | NA | NA | 0 |
| S1_vs_k141_92969\|\|full | 11064 | No | NA | 13 | 0 | 11 | Not-determined | Genome-fragment | NA | NA | 0 |
| S14_vs_k141_65219\|\|full | 1543 | No | NA | 3 | 1 | 0 | Low-quality | Genome-fragment | 0.91 | AAI-based (high-confidence) | 0 |
| S1_vs_k141_106418\|\|full | 1791 | No | NA | 3 | 0 | 1 | Not-determined | Genome-fragment | NA | NA | 0 |
| S1_vs_k141_126102\|\|full | 2170 | No | NA | 5 | 0 | 0 | Not-determined | Genome-fragment | NA | NA | 0 |
| S3_extracted_k141_78393 | 1543 | No | NA | 4 | 1 | 0 | Low-quality | Genome-fragment | 1.98 | AAI-based (medium-confidence) | 0 |
| S1_vs_k141_79780\|\|full | 2413 | No | NA | 3 | 0 | 1 | Not-determined | Genome-fragment | NA | NA | 0 |
| S1_vs_k141_59633\|\|full | 2042 | No | NA | 3 | 0 | 3 | Not-determined | Genome-fragment | NA | NA | 0 |
| S1_vs_k141_13330\|\|full | 1717 | No | NA | 2 | 0 | 2 | Not-determined | Genome-fragment | NA | NA | 0 |
| S1_vs_k141_119722\|\|full | 2065 | No | NA | 4 | 0 | 0 | Not-determined | Genome-fragment | NA | NA | 0 |
| S10_vs_68211\|\|full | 1543 | No | NA | 6 | 1 | 0 | Low-quality | Genome-fragment | 3.74 | AAI-based (medium-confidence) | 0 |
| S12_extracted_k141_138560 | 1543 | No | NA | 1 | 0 | 0 | Low-quality | Genome-fragment | 1.58 | AAI-based (high-confidence) | 0 |
| S15_vs_48389\|\|full | 1543 | No | NA | 4 | 2 | 0 | Low-quality | Genome-fragment | 3 | AAI-based (medium-confidence) | 0 |
| S1_vs_k141_26229\|\|full | 1389 | No | NA | 3 | 0 | 0 | Not-determined | Genome-fragment | NA | NA | 0 |
| S1_vs_k141_38334\|\|full | 1773 | No | NA | 3 | 0 | 0 | Not-determined | Genome-fragment | NA | NA | 0 |
| S6_extracted_k141_47425 | 1543 | No | NA | 3 | 0 | 0 | Low-quality | Genome-fragment | 1.9 | AAI-based (high-confidence) | 0 |
| S14_vs_k141_110034\|\|full | 1544 | No | NA | 4 | 0 | 0 | Low-quality | Genome-fragment | 3.7 | AAI-based (high-confidence) | 0 |
| S3_vs_k141_3670\|\|full | 1544 | No | NA | 3 | 2 | 0 | Low-quality | Genome-fragment | 2.83 | AAI-based (high-confidence) | 0 |
| S1_vs_k141_126196\|\|full | 1494 | No | NA | 2 | 0 | 0 | Not-determined | Genome-fragment | NA | NA | 0 |
| S3_vs_k141_20573\|\|full | 1544 | No | NA | 4 | 1 | 0 | Low-quality | Genome-fragment | 2.84 | AAI-based (high-confidence) | 0 |
| S7_extracted_23903 | 1544 | No | NA | 3 | 0 | 0 | Low-quality | Genome-fragment | 1.01 | AAI-based (high-confidence) | 0 |
| S7_vs_37851\|\|full | 1544 | No | NA | 5 | 1 | 0 | Low-quality | Genome-fragment | 3.89 | AAI-based (medium-confidence) | 0 |
| S16_vs_123991\|\|full | 1544 | No | NA | 2 | 1 | 0 | Low-quality | Genome-fragment | 1.41 | AAI-based (high-confidence) | 0 |
| S16_vs_162490\|\|lt2gene | 1544 | No | NA | 2 | 2 | 0 | Low-quality | Genome-fragment | 3.04 | AAI-based (high-confidence) | 0 |
| S1_vs_k141_93112\|\|full | 2198 | No | NA | 3 | 0 | 1 | Not-determined | Genome-fragment | NA | NA | 0 |
| S10_extracted_93122 | 1544 | No | NA | 2 | 0 | 1 | Low-quality | Genome-fragment | 1.22 | AAI-based (medium-confidence) | 0 |
| S4_extracted_18145 | 1544 | No | NA | 4 | 4 | 0 | Low-quality | Genome-fragment | 5.09 | AAI-based (high-confidence) | 0 |
| S1_vs_k141_65158\|\|full | 1467 | No | NA | 2 | 0 | 1 | Not-determined | Genome-fragment | NA | NA | 0 |
| S1_vs_k141_26298\|\|full | 4596 | No | NA | 4 | 0 | 4 | Not-determined | Genome-fragment | NA | NA | 0 |
| S6_extracted_k141_51871 | 1544 | No | NA | 3 | 1 | 0 | Low-quality | Genome-fragment | 2.57 | AAI-based (high-confidence) | 0 |
| S1_vs_k141_119810\|\|full | 3511 | No | NA | 2 | 0 | 2 | Not-determined | Genome-fragment | NA | NA | 0 |
| S8_vs_k141_176074\|\|full | 1544 | No | NA | 3 | 1 | 0 | Low-quality | Genome-fragment | 2.56 | AAI-based (high-confidence) | 0 |
| S17_extracted_k141_58229 | 1545 | No | NA | 1 | 0 | 1 | Low-quality | Genome-fragment | 0.78 | AAI-based (medium-confidence) | 0 |
| S1_vs_k141_5102\|\|full | 4763 | No | NA | 5 | 0 | 2 | Not-determined | Genome-fragment | NA | NA | 0 |
| S1_vs_k141_38430\|\|full | 1953 | No | NA | 3 | 0 | 1 | Not-determined | Genome-fragment | NA | NA | 0 |
| S1_vs_k141_106573\|\|full | 1506 | No | NA | 3 | 0 | 2 | Not-determined | Genome-fragment | NA | NA | 0 |
| S17_vs_k141_59956\|\|full | 1545 | No | NA | 4 | 0 | 0 | Low-quality | Genome-fragment | 2.93 | AAI-based (high-confidence) | 0 |
| S9_vs_k141_6639\|\|full | 1545 | No | NA | 4 | 0 | 0 | Low-quality | Genome-fragment | 3.51 | AAI-based (medium-confidence) | 0 |
| S13_vs_130962\|\|full | 1545 | No | NA | 2 | 0 | 0 | Low-quality | Genome-fragment | 1.57 | AAI-based (medium-confidence) | 0 |
| S1_vs_k141_23244\|\|full | 1545 | No | NA | 4 | 0 | 0 | Low-quality | Genome-fragment | 0.88 | AAI-based (high-confidence) | 0 |
| S1_vs_k141_113156\|\|full | 1419 | No | NA | 2 | 0 | 2 | Not-determined | Genome-fragment | NA | NA | 0 |
| S3_vs_k141_53027\|\|full | 1545 | No | NA | 2 | 0 | 0 | Low-quality | Genome-fragment | 3.13 | AAI-based (medium-confidence) | 0 |
| S1_vs_k141_59804\|\|full | 1406 | No | NA | 3 | 0 | 0 | Not-determined | Genome-fragment | NA | NA | 0 |
| S1_vs_k141_100019\|\|full | 4858 | No | NA | 7 | 2 | 0 | Medium-quality | Genome-fragment | 87.87 | AAI-based (high-confidence) | 0 |
| S5_vs_k141_116379\|\|full | 1545 | No | NA | 1 | 0 | 1 | Low-quality | Genome-fragment | 2.24 | AAI-based (high-confidence) | 0 |
| S7_vs_25200\|\|full | 1545 | No | NA | 1 | 1 | 0 | Low-quality | Genome-fragment | 3.32 | AAI-based (medium-confidence) | 0 |
| S10_extracted_114349 | 1545 | No | NA | 2 | 2 | 0 | Low-quality | Genome-fragment | 0.92 | HMM-based (lower-bound) | 0 |
| S10_vs_93910\|\|full | 1545 | No | NA | 4 | 0 | 0 | Low-quality | Genome-fragment | 2.81 | AAI-based (high-confidence) | 0 |
| S1_vs_k141_38485\|\|full | 2010 | No | NA | 2 | 0 | 2 | Not-determined | Genome-fragment | NA | NA | 0 |
| S1_vs_k141_133422\|\|full | 3144 | No | NA | 4 | 0 | 2 | Not-determined | Genome-fragment | NA | NA | 0 |
| S12_vs_k141_68486\|\|full | 1545 | No | NA | 2 | 0 | 0 | Low-quality | Genome-fragment | 2.55 | AAI-based (medium-confidence) | 0 |
| S1_vs_k141_26418\|\|full | 2092 | No | NA | 3 | 0 | 2 | Not-determined | Genome-fragment | NA | NA | 0 |
| S1_vs_k141_26419\|\|full | 1746 | No | NA | 5 | 0 | 1 | Not-determined | Genome-fragment | NA | NA | 0 |
| S8_vs_k141_157573\|\|full | 1545 | No | NA | 2 | 0 | 0 | Low-quality | Genome-fragment | 5.55 | AAI-based (high-confidence) | 0 |
| S1_vs_k141_146096\|\|full | 5232 | No | NA | 3 | 0 | 3 | Not-determined | Genome-fragment | NA | NA | 0 |
| S9_vs_k141_55777\|\|full | 1546 | No | NA | 1 | 0 | 0 | Low-quality | Genome-fragment | 1.3 | AAI-based (medium-confidence) | 0 |
| S11_vs_k141_63940\|\|full | 1546 | No | NA | 6 | 1 | 0 | Low-quality | Genome-fragment | 3.96 | AAI-based (medium-confidence) | 0 |
| S14_extracted_k141_10246 | 1546 | No | NA | 2 | 2 | 0 | Low-quality | Genome-fragment | 4.99 | AAI-based (high-confidence) | 0 |
| S1_vs_k141_46676\|\|full | 3157 | No | NA | 4 | 0 | 3 | Not-determined | Genome-fragment | NA | NA | 0 |
| S14_vs_k141_75392\|\|full | 1546 | No | NA | 4 | 1 | 0 | Low-quality | Genome-fragment | 2.61 | AAI-based (medium-confidence) | 0 |
| S1_vs_k141_71787\|\|full | 1012 | No | NA | 3 | 0 | 1 | Not-determined | Genome-fragment | NA | NA | 0 |
| S1_vs_k141_86729\|\|full | 1699 | No | NA | 2 | 0 | 1 | Not-determined | Genome-fragment | NA | NA | 0 |
| S1_vs_k141_119949\|\|full | 1454 | No | NA | 3 | 0 | 0 | Not-determined | Genome-fragment | NA | NA | 0 |
| S1_vs_k141_124046\|\|full | 1546 | No | NA | 3 | 0 | 0 | Low-quality | Genome-fragment | 1.08 | AAI-based (medium-confidence) | 0 |
| S1_vs_k141_71799\|\|full | 1179 | No | NA | 3 | 0 | 2 | Not-determined | Genome-fragment | NA | NA | 0 |
| S7_vs_45494\|\|full | 1546 | No | NA | 2 | 0 | 0 | Low-quality | Genome-fragment | 2.93 | AAI-based (medium-confidence) | 0 |
| S16_extracted_14182 | 1546 | No | NA | 2 | 2 | 0 | Low-quality | Genome-fragment | 3.4 | AAI-based (high-confidence) | 0 |
| S16_vs_119381\|\|full | 1546 | No | NA | 3 | 1 | 0 | Low-quality | Genome-fragment | 3.55 | AAI-based (high-confidence) | 0 |
| S1_vs_k141_65361\|\|full | 1936 | No | NA | 3 | 0 | 2 | Not-determined | Genome-fragment | NA | NA | 0 |
| S1_vs_k141_46716\|\|full | 2368 | No | NA | 3 | 0 | 0 | Not-determined | Genome-fragment | NA | NA | 0 |
| S1_vs_k141_119974\|\|full | 4871 | No | NA | 5 | 0 | 4 | Not-determined | Genome-fragment | NA | NA | 0 |
| S10_vs_69242\|\|full | 1546 | No | NA | 3 | 2 | 0 | Low-quality | Genome-fragment | 8.61 | AAI-based (high-confidence) | 0 |
| S1_vs_k141_26455\|\|full | 1262 | No | NA | 2 | 0 | 1 | Not-determined | Genome-fragment | NA | NA | 0 |
| S12_vs_k141_104093\|\|full | 1546 | No | NA | 3 | 0 | 1 | Low-quality | Genome-fragment | 1.12 | AAI-based (medium-confidence) | 0 |
| S17_vs_k141_149240\|\|full | 1547 | No | NA | 6 | 1 | 0 | Low-quality | Genome-fragment | 3.45 | AAI-based (high-confidence) | 0 |
| S1_vs_k141_88069\|\|full | 1547 | Yes | 808 | 3 | 2 | 1 | Low-quality | Genome-fragment | 1.44 | AAI-based (medium-confidence) | 47.77 |
| S10_vs_13145\|\|full | 1547 | No | NA | 3 | 0 | 0 | Low-quality | Genome-fragment | 1.14 | AAI-based (medium-confidence) | 0 |
| S1_vs_k141_26586\|\|full | 2738 | No | NA | 5 | 0 | 2 | Not-determined | Genome-fragment | NA | NA | 0 |
| S1_vs_k141_146354\|\|full | 1824 | No | NA | 3 | 0 | 1 | Not-determined | Genome-fragment | NA | NA | 0 |
| S17_extracted_k141_49419 | 1548 | No | NA | 3 | 0 | 0 | Low-quality | Genome-fragment | 2.32 | AAI-based (high-confidence) | 0 |
| S1_vs_k141_26638\|\|full | 1688 | No | NA | 3 | 0 | 2 | Not-determined | Genome-fragment | NA | NA | 0 |
| S1_vs_k141_5432\|\|full | 3162 | No | NA | 4 | 0 | 2 | Not-determined | Genome-fragment | NA | NA | 0 |
| S1_vs_k141_146444\|\|full | 2065 | No | NA | 3 | 0 | 2 | Not-determined | Genome-fragment | NA | NA | 0 |
| S17_vs_k141_31423\|\|full | 1548 | No | NA | 4 | 2 | 0 | Low-quality | Genome-fragment | 3.54 | AAI-based (high-confidence) | 0 |
| S11_vs_k141_24880\|\|full | 1548 | No | NA | 3 | 1 | 0 | Low-quality | Genome-fragment | 2.61 | HMM-based (lower-bound) | 0 |
| S14_vs_k141_26346\|\|full | 1548 | No | NA | 2 | 0 | 1 | Low-quality | Genome-fragment | 0.96 | AAI-based (high-confidence) | 0 |
| S1_vs_k141_140167\|\|full | 1782 | No | NA | 2 | 0 | 0 | Not-determined | Genome-fragment | NA | NA | 0 |
| S1_vs_k141_5524\|\|full | 1537 | No | NA | 3 | 0 | 2 | Not-determined | Genome-fragment | NA | NA | 0 |
| S1_vs_k141_38795\|\|full | 2663 | No | NA | 5 | 0 | 0 | Not-determined | Genome-fragment | NA | NA | 0 |
| S1_vs_k141_47963\|\|lt2gene | 1548 | No | NA | 2 | 1 | 0 | Low-quality | Genome-fragment | 3.64 | AAI-based (high-confidence) | 0 |
| S5_vs_k141_115747\|\|full | 1548 | No | NA | 3 | 1 | 0 | Low-quality | Genome-fragment | 3.34 | AAI-based (high-confidence) | 0 |
| S7_extracted_25249 | 1548 | No | NA | 3 | 0 | 0 | Low-quality | Genome-fragment | 3.43 | AAI-based (medium-confidence) | 0 |
| S16_vs_12227\|\|lt2gene | 1548 | No | NA | 2 | 1 | 0 | Low-quality | Genome-fragment | 2.53 | HMM-based (lower-bound) | 0 |
| S1_vs_k141_126755\|\|full | 4255 | No | NA | 4 | 0 | 3 | Not-determined | Genome-fragment | NA | NA | 0 |
| S10_extracted_57712 | 1548 | No | NA | 2 | 0 | 1 | Low-quality | Genome-fragment | 1 | AAI-based (high-confidence) | 0 |
| S1_vs_k141_146644\|\|full | 2762 | No | NA | 6 | 0 | 0 | Not-determined | Genome-fragment | NA | NA | 0 |
| S12_vs_k141_156872\|\|full | 1548 | No | NA | 4 | 0 | 0 | Low-quality | Genome-fragment | 3.91 | AAI-based (high-confidence) | 0 |
| S17_extracted_k141_14811 | 1549 | No | NA | 4 | 4 | 0 | Low-quality | Genome-fragment | 4.04 | AAI-based (high-confidence) | 0 |
| S1_vs_k141_65899\|\|full | 2770 | No | NA | 3 | 0 | 0 | Not-determined | Genome-fragment | NA | NA | 0 |
| S1_vs_k141_5646\|\|full | 2684 | No | NA | 2 | 0 | 2 | Not-determined | Genome-fragment | NA | NA | 0 |
| S1_extracted_k141_125670 | 1549 | No | NA | 3 | 1 | 0 | Low-quality | Genome-fragment | 4.12 | AAI-based (medium-confidence) | 0 |
| S1_vs_k141_145528\|\|full | 1549 | No | NA | 4 | 0 | 0 | Low-quality | Genome-fragment | 3.65 | AAI-based (medium-confidence) | 0 |
| S3_vs_k141_91390\|\|full | 1549 | No | NA | 4 | 0 | 0 | Low-quality | Genome-fragment | 2.98 | AAI-based (high-confidence) | 0 |
| S1_vs_k141_146764\|\|full | 3654 | No | NA | 4 | 0 | 2 | Not-determined | Genome-fragment | NA | NA | 0 |
| S1_vs_k141_157283\|\|full | 1341 | No | NA | 3 | 0 | 0 | Not-determined | Genome-fragment | NA | NA | 0 |
| S1_vs_k141_65984\|\|full | 2765 | No | NA | 6 | 0 | 1 | Not-determined | Genome-fragment | NA | NA | 0 |
| S5_extracted_k141_86854 | 1549 | No | NA | 3 | 1 | 0 | Low-quality | Genome-fragment | 2.35 | AAI-based (high-confidence) | 0 |
| S1_vs_k141_66089\|\|full | 1532 | No | NA | 3 | 0 | 1 | Not-determined | Genome-fragment | NA | NA | 0 |
| S1_vs_k141_66182\|\|full | 1301 | No | NA | 2 | 0 | 2 | Not-determined | Genome-fragment | NA | NA | 0 |
| S1_vs_k141_39095\|\|full | 2231 | No | NA | 2 | 0 | 0 | Not-determined | Genome-fragment | NA | NA | 0 |
| S5_extracted_k141_124458 | 1549 | No | NA | 2 | 2 | 0 | Low-quality | Genome-fragment | 4.1 | AAI-based (high-confidence) | 0 |
| S5_vs_k141_123206\|\|full | 1549 | No | NA | 3 | 0 | 0 | Low-quality | Genome-fragment | 3.5 | AAI-based (medium-confidence) | 0 |
| S5_vs_k141_88318\|\|full | 1549 | No | NA | 5 | 1 | 0 | Low-quality | Genome-fragment | 3.82 | AAI-based (high-confidence) | 0 |
| S1_vs_k141_66340\|\|full | 1146 | No | NA | 3 | 0 | 1 | Not-determined | Genome-fragment | NA | NA | 0 |
| S1_vs_k141_39244\|\|full | 4216 | No | NA | 6 | 0 | 3 | Not-determined | Genome-fragment | NA | NA | 0 |
| S7_vs_94006\|\|full | 1549 | No | NA | 4 | 0 | 0 | Low-quality | Genome-fragment | 3.52 | AAI-based (medium-confidence) | 0 |
| S16_vs_19047\|\|full | 1549 | No | NA | 3 | 1 | 0 | Low-quality | Genome-fragment | 3.75 | HMM-based (lower-bound) | 0 |
| S1_vs_k141_6033\|\|full | 1672 | No | NA | 3 | 0 | 0 | Not-determined | Genome-fragment | NA | NA | 0 |
| S1_vs_k141_6037\|\|full | 2076 | No | NA | 3 | 0 | 1 | Not-determined | Genome-fragment | NA | NA | 0 |
| S1_vs_k141_157548\|\|full | 2228 | No | NA | 2 | 0 | 1 | Not-determined | Genome-fragment | NA | NA | 0 |
| S16_vs_97459\|\|full | 1549 | No | NA | 2 | 0 | 1 | Low-quality | Genome-fragment | 2.23 | AAI-based (medium-confidence) | 0 |
| S12_vs_k141_54209\|\|full | 1549 | No | NA | 2 | 1 | 0 | Low-quality | Genome-fragment | 3.63 | AAI-based (medium-confidence) | 0 |
| S15_vs_56252\|\|full | 1549 | No | NA | 2 | 0 | 1 | Low-quality | Genome-fragment | 2.46 | AAI-based (high-confidence) | 0 |
| S8_extracted_k141_4173 | 1549 | No | NA | 3 | 1 | 0 | Low-quality | Genome-fragment | 2.46 | AAI-based (high-confidence) | 0 |
| S1_vs_k141_6259\|\|full | 1530 | No | NA | 2 | 0 | 1 | Not-determined | Genome-fragment | NA | NA | 0 |
| S1_vs_k141_66685\|\|full | 1521 | No | NA | 4 | 0 | 0 | Not-determined | Genome-fragment | NA | NA | 0 |
| S1_vs_k141_72656\|\|full | 2848 | No | NA | 3 | 0 | 3 | Not-determined | Genome-fragment | NA | NA | 0 |
| S1_vs_k141_150530\|\|full | 1397 | No | NA | 2 | 0 | 2 | Not-determined | Genome-fragment | NA | NA | 0 |
| S8_vs_k141_74161\|\|full | 1549 | No | NA | 3 | 0 | 0 | Low-quality | Genome-fragment | 2.46 | AAI-based (high-confidence) | 0 |
| S1_vs_k141_6281\|\|full | 1211 | No | NA | 3 | 0 | 1 | Not-determined | Genome-fragment | NA | NA | 0 |
| S17_vs_k141_97588\|\|full | 1550 | No | NA | 3 | 0 | 0 | Low-quality | Genome-fragment | 2.59 | AAI-based (medium-confidence) | 0 |
| S1_vs_k141_72762\|\|full | 1998 | No | NA | 2 | 0 | 1 | Not-determined | Genome-fragment | NA | NA | 0 |
| S17_vs_k141_147618\|\|full | 1550 | No | NA | 3 | 0 | 0 | Low-quality | Genome-fragment | 0.91 | AAI-based (high-confidence) | 0 |
| S1_extracted_k141_103614 | 1550 | No | NA | 2 | 0 | 2 | Low-quality | Genome-fragment | 0.17 | AAI-based (medium-confidence) | 0 |
| S1_vs_k141_157764\|\|full | 5060 | No | NA | 5 | 0 | 2 | Not-determined | Genome-fragment | NA | NA | 0 |
| S16_vs_29977\|\|full | 1550 | No | NA | 2 | 0 | 0 | Low-quality | Genome-fragment | 1.81 | AAI-based (medium-confidence) | 0 |
| S16_vs_158572\|\|full | 1550 | No | NA | 3 | 1 | 0 | Low-quality | Genome-fragment | 3.74 | AAI-based (high-confidence) | 0 |
| S1_vs_k141_39730\|\|full | 1048 | No | NA | 2 | 0 | 2 | Not-determined | Genome-fragment | NA | NA | 0 |
| S2_vs_k141_17411\|\|full | 1550 | No | NA | 2 | 1 | 0 | Low-quality | Genome-fragment | 3.14 | AAI-based (high-confidence) | 0 |
| S1_vs_k141_39809\|\|full | 2467 | No | NA | 4 | 0 | 3 | Not-determined | Genome-fragment | NA | NA | 0 |
| S6_vs_k141_15257\|\|full | 1550 | No | NA | 3 | 0 | 0 | Low-quality | Genome-fragment | 0.91 | AAI-based (medium-confidence) | 0 |
| S1_vs_k141_115119\|\|full | 1551 | No | NA | 4 | 0 | 0 | Low-quality | Genome-fragment | 1.13 | AAI-based (high-confidence) | 0 |
| S1_vs_k141_39913\|\|full | 1406 | No | NA | 3 | 0 | 0 | Not-determined | Genome-fragment | NA | NA | 0 |
| S3_vs_k141_16184\|\|full | 1551 | No | NA | 4 | 0 | 0 | Low-quality | Genome-fragment | 2.72 | AAI-based (high-confidence) | 0 |
| S1_vs_k141_150865\|\|full | 2223 | No | NA | 4 | 0 | 2 | Not-determined | Genome-fragment | NA | NA | 0 |
| S1_vs_k141_158190\|\|full | 1002 | No | NA | 2 | 0 | 1 | Not-determined | Genome-fragment | NA | NA | 0 |
| S7_vs_7267\|\|full | 1551 | No | NA | 3 | 0 | 0 | Low-quality | Genome-fragment | 3.86 | AAI-based (high-confidence) | 0 |
| S1_vs_k141_150919\|\|full | 1655 | No | NA | 3 | 0 | 1 | Not-determined | Genome-fragment | NA | NA | 0 |
| S1_vs_k141_158362\|\|full | 1251 | No | NA | 3 | 0 | 0 | Not-determined | Genome-fragment | NA | NA | 0 |
| S7_vs_75084\|\|full | 1551 | No | NA | 4 | 0 | 0 | Low-quality | Genome-fragment | 3.62 | AAI-based (high-confidence) | 0 |
| S15_vs_45237\|\|lt2gene | 1551 | No | NA | 1 | 1 | 0 | Low-quality | Genome-fragment | 3.48 | AAI-based (high-confidence) | 0 |
| S1_vs_k141_158492\|\|full | 7107 | No | NA | 5 | 0 | 5 | Not-determined | Genome-fragment | NA | NA | 0 |
| S1_vs_k141_158493\|\|full | 1129 | No | NA | 2 | 0 | 1 | Not-determined | Genome-fragment | NA | NA | 0 |
| S4_extracted_4997 | 1551 | No | NA | 2 | 2 | 0 | Low-quality | Genome-fragment | 5.1 | AAI-based (high-confidence) | 0 |
| S4_extracted_13380 | 1551 | No | NA | 2 | 0 | 0 | Low-quality | Genome-fragment | 2.12 | AAI-based (high-confidence) | 0 |
| S6_vs_k141_7352\|\|full | 1551 | No | NA | 2 | 1 | 0 | Low-quality | Genome-fragment | 2.99 | HMM-based (lower-bound) | 0 |
| S1_vs_k141_151403\|\|full | 5241 | No | NA | 4 | 0 | 3 | Not-determined | Genome-fragment | NA | NA | 0 |
| S6_vs_k141_11485\|\|full | 1551 | No | NA | 4 | 0 | 2 | Low-quality | Genome-fragment | 3.28 | AAI-based (high-confidence) | 0 |
| S8_extracted_k141_152231 | 1551 | No | NA | 1 | 0 | 1 | Low-quality | Genome-fragment | 0.84 | AAI-based (medium-confidence) | 0 |
| S1_vs_k141_151692\|\|full | 1708 | No | NA | 3 | 0 | 0 | Not-determined | Genome-fragment | NA | NA | 0 |
| S8_vs_k141_8231\|\|full | 1551 | No | NA | 4 | 1 | 0 | Low-quality | Genome-fragment | 4.81 | AAI-based (high-confidence) | 0 |
| S1_vs_k141_159004\|\|full | 3978 | No | NA | 3 | 0 | 0 | Not-determined | Genome-fragment | NA | NA | 0 |
| S1_vs_k141_151840\|\|full | 3139 | No | NA | 7 | 0 | 2 | Not-determined | Genome-fragment | NA | NA | 0 |
| S1_vs_k141_151883\|\|full | 2246 | No | NA | 2 | 0 | 2 | Not-determined | Genome-fragment | NA | NA | 0 |
| S8_vs_k141_56314\|\|lt2gene | 1551 | No | NA | 2 | 2 | 0 | Low-quality | Genome-fragment | 1.18 | AAI-based (medium-confidence) | 0 |
| S17_extracted_k141_120249 | 1552 | No | NA | 3 | 1 | 0 | Low-quality | Genome-fragment | 2.51 | AAI-based (high-confidence) | 0 |
| S17_extracted_k141_71464 | 1552 | No | NA | 1 | 0 | 0 | Low-quality | Genome-fragment | 1.55 | AAI-based (medium-confidence) | 0 |
| S1_vs_k141_159244\|\|full | 2412 | No | NA | 2 | 0 | 2 | Not-determined | Genome-fragment | NA | NA | 0 |
| S9_vs_k141_27805\|\|full | 1552 | No | NA | 4 | 2 | 0 | Low-quality | Genome-fragment | 4.31 | AAI-based (high-confidence) | 0 |
| S1_vs_k141_159281\|\|full | 1530 | No | NA | 2 | 0 | 0 | Not-determined | Genome-fragment | NA | NA | 0 |
| S11_vs_k141_20453\|\|full | 1552 | No | NA | 3 | 2 | 0 | Low-quality | Genome-fragment | 1.91 | HMM-based (lower-bound) | 0 |
| S1_vs_k141_106624\|\|full | 1552 | No | NA | 4 | 1 | 0 | Low-quality | Genome-fragment | 3.96 | AAI-based (high-confidence) | 0 |
| S1_vs_k141_152288\|\|full | 1040 | No | NA | 2 | 0 | 0 | Not-determined | Genome-fragment | NA | NA | 0 |
| S1_vs_k141_152399\|\|full | 3204 | No | NA | 3 | 0 | 1 | Not-determined | Genome-fragment | NA | NA | 0 |
| S3_vs_k141_30745\|\|full | 1552 | No | NA | 2 | 0 | 1 | Low-quality | Genome-fragment | 2.27 | AAI-based (medium-confidence) | 0 |
| S3_vs_k141_7652\|\|full | 1552 | No | NA | 4 | 0 | 0 | Low-quality | Genome-fragment | 2.82 | AAI-based (high-confidence) | 0 |
| S7_extracted_44429 | 1552 | No | NA | 1 | 0 | 1 | Low-quality | Genome-fragment | 0.88 | AAI-based (medium-confidence) | 0 |
| S12_extracted_k141_170295 | 1552 | No | NA | 3 | 2 | 0 | Low-quality | Genome-fragment | 3.4 | AAI-based (medium-confidence) | 0 |
| S12_vs_k141_165477\|\|lt2gene | 1552 | No | NA | 2 | 1 | 0 | Low-quality | Genome-fragment | 3.77 | AAI-based (high-confidence) | 0 |
| S1_vs_k141_159736\|\|full | 1139 | No | NA | 2 | 0 | 2 | Not-determined | Genome-fragment | NA | NA | 0 |
| S1_vs_k141_152864\|\|full | 1201 | No | NA | 2 | 0 | 1 | Not-determined | Genome-fragment | NA | NA | 0 |
| S1_vs_k141_159927\|\|full | 3184 | No | NA | 3 | 0 | 0 | Not-determined | Genome-fragment | NA | NA | 0 |
| S8_vs_k141_162534\|\|full | 1552 | No | NA | 3 | 0 | 0 | Low-quality | Genome-fragment | 2.12 | AAI-based (high-confidence) | 0 |
| S17_vs_k141_133303\|\|full | 1553 | No | NA | 3 | 1 | 0 | Low-quality | Genome-fragment | 3 | AAI-based (high-confidence) | 0 |
| S1_vs_k141_153340\|\|full | 4479 | No | NA | 8 | 0 | 1 | Not-determined | Genome-fragment | NA | NA | 0 |
| S1_vs_k141_53468\|\|lt2gene | 1434 | No | NA | 2 | 0 | 0 | Not-determined | Genome-fragment | NA | NA | 0 |
| S17_vs_k141_15298\|\|full | 1553 | No | NA | 2 | 0 | 0 | Low-quality | Genome-fragment | 1.27 | AAI-based (high-confidence) | 0 |
| S17_vs_k141_150951\|\|full | 1553 | No | NA | 4 | 0 | 0 | Low-quality | Genome-fragment | 2.14 | AAI-based (medium-confidence) | 0 |
| S11_vs_k141_1973\|\|full | 1553 | No | NA | 4 | 0 | 0 | Low-quality | Genome-fragment | 3.35 | AAI-based (medium-confidence) | 0 |
| S14_vs_k141_22370\|\|full | 1553 | No | NA | 4 | 0 | 0 | Low-quality | Genome-fragment | 3.85 | AAI-based (high-confidence) | 0 |
| S1_vs_k141_73975\|\|lt2gene | 1142 | No | NA | 1 | 0 | 0 | Not-determined | Genome-fragment | NA | NA | 0 |
| S14_vs_k141_30059\|\|full | 1553 | No | NA | 3 | 0 | 2 | Low-quality | Genome-fragment | 1.12 | AAI-based (medium-confidence) | 0 |
| S1_vs_k141_153906\|\|lt2gene | 2950 | No | NA | 2 | 0 | 1 | Not-determined | Genome-fragment | NA | NA | 0 |
| S14_vs_k141_73802\|\|full | 1553 | No | NA | 2 | 0 | 0 | Low-quality | Genome-fragment | 3.6 | AAI-based (high-confidence) | 0 |
| S3_extracted_k141_114692 | 1553 | No | NA | 4 | 1 | 0 | Low-quality | Genome-fragment | 3.4 | AAI-based (high-confidence) | 0 |
| S3_vs_k141_22812\|\|full | 1553 | No | NA | 4 | 1 | 0 | Low-quality | Genome-fragment | 2.42 | AAI-based (high-confidence) | 0 |
| S7_extracted_29742 | 1553 | No | NA | 2 | 0 | 2 | Low-quality | Genome-fragment | 2.88 | AAI-based (medium-confidence) | 0 |
| S7_extracted_60692 | 1553 | No | NA | 5 | 1 | 0 | Low-quality | Genome-fragment | 2.93 | AAI-based (high-confidence) | 0 |
| S7_vs_125711\|\|full | 1553 | No | NA | 3 | 3 | 0 | Low-quality | Genome-fragment | 3.33 | AAI-based (high-confidence) | 0 |
| S16_vs_166471\|\|full | 1553 | No | NA | 3 | 0 | 2 | Low-quality | Genome-fragment | 0.76 | AAI-based (high-confidence) | 0 |
| S10_vs_67733\|\|lt2gene | 1553 | No | NA | 1 | 1 | 0 | Low-quality | Genome-fragment | 0.84 | HMM-based (lower-bound) | 0 |

| | | | | | | | | | | | |
|---|---|---|---|---|---|---|---|---|---|---|---|
| S12_extracted_k141_146725 | 1553 | No | NA | 5 | 0 | 0 | Low-quality | Genome-fragment | 4.13 | AAI-based (high-confidence) | 0 |
| S9_vs_k141_46896\|\|full | 1554 | No | NA | 4 | 2 | 0 | Low-quality | Genome-fragment | 4.63 | AAI-based (high-confidence) | 0 |
| S3_vs_k141_75706\|\|full | 1554 | No | NA | 3 | 1 | 0 | Low-quality | Genome-fragment | 2.73 | HMM-based (lower-bound) | 0 |
| S3_vs_k141_81639\|\|lt2gene | 1554 | No | NA | 1 | 1 | 0 | Low-quality | Genome-fragment | 1.49 | AAI-based (high-confidence) | 0 |
| S1_vs_k141_48439\|\|lt2gene | 1236 | No | NA | 1 | 0 | 0 | Not-determined | Genome-fragment | NA | NA | 0 |
| S16_vs_136042\|\|lt2gene | 1554 | No | NA | 2 | 1 | 0 | Low-quality | Genome-fragment | 4.09 | AAI-based (high-confidence) | 0 |
| S12_vs_k141_163346\|\|full | 1554 | No | NA | 3 | 2 | 0 | Low-quality | Genome-fragment | 3.96 | AAI-based (medium-confidence) | 0 |
| S13_vs_20158\|\|full | 1555 | No | NA | 2 | 0 | 0 | Low-quality | Genome-fragment | 2.25 | AAI-based (medium-confidence) | 0 |
| S7_vs_51535\|\|full | 1555 | No | NA | 2 | 0 | 1 | Low-quality | Genome-fragment | 1.97 | AAI-based (medium-confidence) | 0 |
| S16_vs_129975\|\|full | 1555 | No | NA | 4 | 2 | 0 | Low-quality | Genome-fragment | 2.38 | AAI-based (high-confidence) | 0 |
| S1_vs_k141_61144\|\|lt2gene | 1996 | No | NA | 2 | 0 | 1 | Not-determined | Genome-fragment | NA | NA | 0 |
| S17_vs_k141_146824\|\|full | 1556 | No | NA | 3 | 0 | 1 | Low-quality | Genome-fragment | 3.19 | AAI-based (medium-confidence) | 0 |
| S13_vs_24549\|\|full | 1556 | No | NA | 3 | 0 | 2 | Low-quality | Genome-fragment | 1.41 | AAI-based (medium-confidence) | 0 |
| S1_vs_k141_115502\|\|lt2gene | 1053 | No | NA | 1 | 0 | 0 | Not-determined | Genome-fragment | NA | NA | 0 |
| S13_vs_113751\|\|lt2gene | 1556 | No | NA | 1 | 1 | 0 | Low-quality | Genome-fragment | 2.95 | HMM-based (lower-bound) | 0 |
| S1_extracted_k141_109462 | 1556 | No | NA | 2 | 1 | 0 | Low-quality | Genome-fragment | 2.53 | AAI-based (high-confidence) | 0 |
| S1_extracted_k141_65477 | 1556 | No | NA | 1 | 1 | 0 | Low-quality | Genome-fragment | 1.57 | AAI-based (high-confidence) | 0 |
| S5_vs_k141_25057\|\|full | 1556 | No | NA | 3 | 0 | 1 | Low-quality | Genome-fragment | 0.49 | AAI-based (medium-confidence) | 0 |
| S5_vs_k141_25827\|\|full | 1556 | No | NA | 3 | 0 | 1 | Low-quality | Genome-fragment | 1 | AAI-based (medium-confidence) | 0 |
| S16_vs_25689\|\|full | 1556 | No | NA | 3 | 0 | 0 | Low-quality | Genome-fragment | 1.98 | AAI-based (medium-confidence) | 0 |
| S16_vs_136724\|\|full | 1556 | No | NA | 3 | 0 | 0 | Low-quality | Genome-fragment | 2.6 | AAI-based (high-confidence) | 0 |
| S12_extracted_k141_54184 | 1556 | No | NA | 4 | 2 | 0 | Low-quality | Genome-fragment | 3.49 | AAI-based (medium-confidence) | 0 |
| S1_vs_k141_55888\|\|lt2gene | 2453 | No | NA | 1 | 0 | 0 | Not-determined | Genome-fragment | NA | NA | 0 |
| S1_vs_k141_2162\|\|lt2gene | 2002 | No | NA | 2 | 0 | 0 | Not-determined | Genome-fragment | NA | NA | 0 |
| S12_vs_k141_118308\|\|full | 1556 | No | NA | 4 | 1 | 0 | Low-quality | Genome-fragment | 4.71 | AAI-based (high-confidence) | 0 |
| S8_vs_k141_199660\|\|lt2gene | 1556 | No | NA | 1 | 1 | 0 | Low-quality | Genome-fragment | 4.15 | AAI-based (medium-confidence) | 0 |
| S1_vs_k141_69370\|\|lt2gene | 2020 | No | NA | 2 | 0 | 0 | Not-determined | Genome-fragment | NA | NA | 0 |
| S1_vs_k141_96477\|\|lt2gene | 2064 | No | NA | 1 | 0 | 0 | Not-determined | Genome-fragment | NA | NA | 0 |
| S8_vs_k141_170500\|\|lt2gene | 1556 | No | NA | 2 | 1 | 0 | Low-quality | Genome-fragment | 3 | HMM-based (lower-bound) | 0 |
| S1_vs_k141_49970\|\|lt2gene | 1543 | No | NA | 2 | 0 | 1 | Not-determined | Genome-fragment | NA | NA | 0 |
| S17_vs_k141_91677\|\|full | 1557 | No | NA | 2 | 0 | 0 | Low-quality | Genome-fragment | 2.99 | AAI-based (medium-confidence) | 0 |
| S9_vs_k141_57782\|\|full | 1557 | No | NA | 3 | 2 | 0 | Low-quality | Genome-fragment | 3.83 | AAI-based (high-confidence) | 0 |
| S1_vs_k141_136636\|\|lt2gene | 1214 | No | NA | 1 | 0 | 1 | Not-determined | Genome-fragment | NA | NA | 0 |
| S9_vs_k141_9103\|\|full | 1557 | No | NA | 4 | 0 | 0 | Low-quality | Genome-fragment | 3.54 | AAI-based (high-confidence) | 0 |
| S14_extracted_k141_102728 | 1557 | No | NA | 4 | 3 | 0 | Low-quality | Genome-fragment | 5.04 | AAI-based (high-confidence) | 0 |
| S1_vs_k141_30448\|\|lt2gene | 1307 | No | NA | 2 | 0 | 0 | Not-determined | Genome-fragment | NA | NA | 0 |
| S1_vs_k141_97439\|\|lt2gene | 1771 | No | NA | 1 | 0 | 1 | Not-determined | Genome-fragment | NA | NA | 0 |
| S1_vs_k141_84240\|\|lt2gene | 1023 | No | NA | 2 | 0 | 1 | Not-determined | Genome-fragment | NA | NA | 0 |
| S1_vs_k141_155806\|\|full | 1557 | No | NA | 3 | 3 | 0 | Low-quality | Genome-fragment | 3.26 | AAI-based (high-confidence) | 0 |
| S1_vs_k141_90613\|\|full | 1557 | No | NA | 2 | 0 | 0 | Low-quality | Genome-fragment | 3.72 | AAI-based (high-confidence) | 0 |
| S3_vs_k141_96818\|\|full | 1557 | No | NA | 2 | 0 | 0 | Low-quality | Genome-fragment | 2.73 | AAI-based (high-confidence) | 0 |
| S1_vs_k141_51427\|\|lt2gene | 1344 | No | NA | 2 | 0 | 0 | Not-determined | Genome-fragment | NA | NA | 0 |
| S3_vs_k141_82821\|\|full | 1557 | No | NA | 3 | 0 | 0 | Low-quality | Genome-fragment | 1.16 | AAI-based (high-confidence) | 0 |
| S1_vs_k141_24465\|\|lt2gene | 1333 | No | NA | 1 | 0 | 0 | Not-determined | Genome-fragment | NA | NA | 0 |
| S5_vs_k141_130285\|\|full | 1557 | Yes | 802 | 4 | 1 | 1 | Low-quality | Genome-fragment | 1.6 | AAI-based (high-confidence) | 48.49 |
| S7_vs_117006\|\|full | 1557 | No | NA | 2 | 0 | 0 | Low-quality | Genome-fragment | 0.93 | AAI-based (medium-confidence) | 0 |
| S16_extracted_121629 | 1557 | No | NA | 1 | 0 | 0 | Low-quality | Genome-fragment | 0.97 | AAI-based (high-confidence) | 0 |
| S12_extracted_k141_75408 | 1557 | No | NA | 3 | 0 | 0 | Low-quality | Genome-fragment | 1.73 | AAI-based (high-confidence) | 0 |
| S8_vs_k141_173289\|\|full | 1557 | No | NA | 4 | 2 | 0 | Low-quality | Genome-fragment | 4.09 | AAI-based (medium-confidence) | 0 |
| S17_vs_k141_124083\|\|full | 1558 | No | NA | 2 | 0 | 2 | Low-quality | Genome-fragment | 1.75 | AAI-based (medium-confidence) | 0 |
| S1_vs_k141_37190\|\|lt2gene | 1915 | No | NA | 1 | 0 | 0 | Not-determined | Genome-fragment | NA | NA | 0 |
| S17_vs_k141_22026\|\|full | 1558 | No | NA | 6 | 0 | 0 | Low-quality | Genome-fragment | 3.46 | AAI-based (medium-confidence) | 0 |
| S1_vs_k141_144838\|\|lt2gene | 1157 | No | NA | 2 | 0 | 0 | Not-determined | Genome-fragment | NA | NA | 0 |
| S13_vs_123916\|\|full | 1558 | No | NA | 2 | 0 | 1 | Low-quality | Genome-fragment | 3.2 | AAI-based (high-confidence) | 0 |
| S1_vs_k141_145087\|\|lt2gene | 1162 | No | NA | 2 | 0 | 0 | Not-determined | Genome-fragment | NA | NA | 0 |
| S14_vs_k141_61176\|\|full | 1558 | No | NA | 5 | 0 | 0 | Low-quality | Genome-fragment | 3.6 | AAI-based (medium-confidence) | 0 |
| S3_extracted_k141_98639 | 1558 | No | NA | 3 | 0 | 0 | Low-quality | Genome-fragment | 2.45 | AAI-based (medium-confidence) | 0 |
| S3_extracted_k141_126731 | 1558 | No | NA | 4 | 0 | 0 | Low-quality | Genome-fragment | 2.37 | AAI-based (high-confidence) | 0 |
| S5_vs_k141_62141\|\|full | 1558 | No | NA | 4 | 0 | 4 | Low-quality | Genome-fragment | 0.44 | AAI-based (medium-confidence) | 0 |
| S16_vs_25681\|\|full | 1558 | No | NA | 3 | 0 | 1 | Low-quality | Genome-fragment | 0.57 | AAI-based (medium-confidence) | 0 |
| S16_vs_59861\|\|full | 1558 | No | NA | 2 | 0 | 2 | Low-quality | Genome-fragment | 2.88 | AAI-based (medium-confidence) | 0 |
| S10_vs_27922\|\|full | 1558 | No | NA | 3 | 1 | 0 | Low-quality | Genome-fragment | 2.95 | AAI-based (medium-confidence) | 0 |
| S10_vs_49307\|\|full | 1558 | No | NA | 3 | 0 | 1 | Low-quality | Genome-fragment | 2.1 | AAI-based (medium-confidence) | 0 |
| S8_extracted_k141_200820 | 1558 | No | NA | 1 | 0 | 1 | Low-quality | Genome-fragment | 3.71 | AAI-based (medium-confidence) | 0 |
| S8_extracted_k141_11231 | 1558 | No | NA | 3 | 0 | 2 | Low-quality | Genome-fragment | 1.97 | AAI-based (medium-confidence) | 0 |
| S9_vs_k141_43872\|\|full | 1559 | No | NA | 4 | 0 | 0 | Low-quality | Genome-fragment | 0.9 | AAI-based (high-confidence) | 0 |
| S9_vs_k141_47582\|\|full | 1559 | No | NA | 4 | 0 | 0 | Low-quality | Genome-fragment | 3.71 | AAI-based (high-confidence) | 0 |
| S1_vs_k141_149817\|\|lt2gene | 1085 | No | NA | 1 | 0 | 0 | Not-determined | Genome-fragment | NA | NA | 0 |
| S11_extracted_k141_96375 | 1559 | No | NA | 3 | 1 | 0 | Low-quality | Genome-fragment | 3.4 | AAI-based (high-confidence) | 0 |
| S14_vs_k141_81725\|\|full | 1559 | No | NA | 3 | 2 | 0 | Low-quality | Genome-fragment | 3.3 | AAI-based (medium-confidence) | 0 |
| S3_extracted_k141_98820 | 1559 | No | NA | 2 | 2 | 0 | Low-quality | Genome-fragment | 5.14 | AAI-based (high-confidence) | 0 |
| S3_vs_k141_44716\|\|full | 1559 | No | NA | 4 | 1 | 0 | Low-quality | Genome-fragment | 3.74 | AAI-based (high-confidence) | 0 |
| S1_vs_k141_146363\|\|lt2gene | 1120 | No | NA | 1 | 0 | 0 | Not-determined | Genome-fragment | NA | NA | 0 |
| S3_vs_k141_22171\|\|full | 1559 | No | NA | 2 | 0 | 1 | Low-quality | Genome-fragment | 1.84 | AAI-based (high-confidence) | 0 |
| S1_vs_k141_39058\|\|lt2gene | 1072 | No | NA | 1 | 0 | 0 | Not-determined | Genome-fragment | NA | NA | 0 |
| S1_vs_k141_150696\|\|lt2gene | 1661 | No | NA | 1 | 0 | 0 | Not-determined | Genome-fragment | NA | NA | 0 |
| S7_vs_87108\|\|full | 1559 | No | NA | 3 | 1 | 0 | Low-quality | Genome-fragment | 3.14 | AAI-based (high-confidence) | 0 |
| S16_extracted_57523 | 1559 | No | NA | 3 | 0 | 0 | Low-quality | Genome-fragment | 4.22 | AAI-based (high-confidence) | 0 |
| S1_vs_k141_158282\|\|lt2gene | 1084 | No | NA | 2 | 0 | 0 | Not-determined | Genome-fragment | NA | NA | 0 |
| S16_vs_27759\|\|full | 1559 | No | NA | 2 | 0 | 0 | Low-quality | Genome-fragment | 2.26 | AAI-based (high-confidence) | 0 |
| S16_vs_136280\|\|full | 1559 | No | NA | 2 | 0 | 0 | Low-quality | Genome-fragment | 3.18 | AAI-based (medium-confidence) | 0 |
| S3_extracted_k141_63761 | 6864 | No | NA | 7 | 0 | 1 | Not-determined | Genome-fragment | NA | NA | 0 |
| S3_extracted_k141_15957 | 1626 | No | NA | 2 | 0 | 0 | Not-determined | Genome-fragment | NA | NA | 0 |
| S3_extracted_k141_71750 | 1580 | No | NA | 1 | 0 | 0 | Not-determined | Genome-fragment | NA | NA | 0 |
| S16_vs_172320\|\|full | 1559 | No | NA | 3 | 2 | 0 | Low-quality | Genome-fragment | 3.31 | AAI-based (high-confidence) | 0 |
| S3_extracted_k141_63800 | 7289 | No | NA | 9 | 0 | 6 | Not-determined | Genome-fragment | NA | NA | 0 |
| S10_vs_49610\|\|full | 1559 | No | NA | 3 | 2 | 0 | Low-quality | Genome-fragment | 3.46 | AAI-based (high-confidence) | 0 |
| S15_extracted_64024 | 1559 | No | NA | 3 | 0 | 0 | Low-quality | Genome-fragment | 3.47 | AAI-based (high-confidence) | 0 |
| S3_extracted_k141_111632 | 3684 | No | NA | 3 | 0 | 3 | Not-determined | Genome-fragment | NA | NA | 0 |
| S3_extracted_k141_63814 | 2579 | No | NA | 3 | 0 | 1 | Not-determined | Genome-fragment | NA | NA | 0 |
| S3_extracted_k141_47927 | 2902 | No | NA | 2 | 0 | 2 | Not-determined | Genome-fragment | NA | NA | 0 |
| S6_vs_k141_51812\|\|full | 1559 | No | NA | 3 | 0 | 0 | Low-quality | Genome-fragment | 2.79 | AAI-based (medium-confidence) | 0 |
| S3_extracted_k141_24046 | 1520 | No | NA | 1 | 0 | 1 | Not-determined | Genome-fragment | NA | NA | 0 |
| S3_extracted_k141_63825 | 1100 | No | NA | 2 | 0 | 1 | Not-determined | Genome-fragment | NA | NA | 0 |
| S8_extracted_k141_106759 | 1559 | No | NA | 3 | 0 | 2 | Low-quality | Genome-fragment | 0.9 | AAI-based (high-confidence) | 0 |
| S3_extracted_k141_71819 | 1666 | No | NA | 2 | 0 | 2 | Not-determined | Genome-fragment | NA | NA | 0 |
| S8_vs_k141_155394\|\|full | 1559 | No | NA | 3 | 0 | 0 | Low-quality | Genome-fragment | 2.65 | AAI-based (high-confidence) | 0 |
| S3_extracted_k141_87692 | 15849 | No | NA | 16 | 0 | 7 | Not-determined | Genome-fragment | NA | NA | 0 |
| S3_extracted_k141_79771 | 1114 | No | NA | 1 | 0 | 1 | Not-determined | Genome-fragment | NA | NA | 0 |
| S3_extracted_k141_31936 | 1409 | No | NA | 1 | 0 | 1 | Not-determined | Genome-fragment | NA | NA | 0 |
| S1_extracted_k141_158613 | 1560 | No | NA | 3 | 0 | 0 | Low-quality | Genome-fragment | 2.23 | AAI-based (high-confidence) | 0 |
| S3_extracted_k141_71851 | 3053 | No | NA | 2 | 0 | 1 | Not-determined | Genome-fragment | NA | NA | 0 |
| S3_vs_k141_63581\|\|full | 1560 | No | NA | 4 | 2 | 0 | Low-quality | Genome-fragment | 4.03 | AAI-based (high-confidence) | 0 |
| S3_extracted_k141_8058 | 49145 | No | NA | 67 | 19 | 0 | High-quality | High-quality | 100 | AAI-based (high-confidence) | 0 |
| S16_vs_57802\|\|full | 1560 | No | NA | 3 | 0 | 0 | Low-quality | Genome-fragment | 3.49 | AAI-based (medium-confidence) | 0 |
| S3_extracted_k141_119629 | 1225 | No | NA | 2 | 0 | 1 | Not-determined | Genome-fragment | NA | NA | 0 |
| S3_extracted_k141_55923 | 1550 | No | NA | 2 | 0 | 1 | Not-determined | Genome-fragment | NA | NA | 0 |
| S16_vs_127579\|\|full | 1560 | No | NA | 3 | 1 | 0 | Low-quality | Genome-fragment | 3.18 | AAI-based (medium-confidence) | 0 |
| S12_extracted_k141_46816 | 1560 | No | NA | 5 | 3 | 0 | Low-quality | Genome-fragment | 1.02 | AAI-based (high-confidence) | 0 |
| S12_vs_k141_99141\|\|full | 1560 | No | NA | 2 | 0 | 0 | Low-quality | Genome-fragment | 0.6 | AAI-based (medium-confidence) | 0 |
| S3_extracted_k141_24136 | 1299 | No | NA | 1 | 0 | 0 | Not-determined | Genome-fragment | NA | NA | 0 |
| S3_extracted_k141_24139 | 1299 | No | NA | 1 | 0 | 0 | Not-determined | Genome-fragment | NA | NA | 0 |
| S8_vs_k141_89673\|\|full | 1560 | No | NA | 1 | 0 | 0 | Low-quality | Genome-fragment | 3.01 | AAI-based (medium-confidence) | 0 |
| S3_extracted_k141_87730 | 1617 | No | NA | 2 | 0 | 1 | Not-determined | Genome-fragment | NA | NA | 0 |
| S3_extracted_k141_8122 | 1035 | No | NA | 2 | 0 | 1 | Not-determined | Genome-fragment | NA | NA | 0 |
| S3_extracted_k141_8132 | 1020 | No | NA | 2 | 0 | 0 | Not-determined | Genome-fragment | NA | NA | 0 |
| S3_extracted_k141_79795 | 1490 | No | NA | 3 | 0 | 3 | Not-determined | Genome-fragment | NA | NA | 0 |
| S3_extracted_k141_55992 | 1464 | No | NA | 1 | 0 | 1 | Not-determined | Genome-fragment | NA | NA | 0 |
| S3_extracted_k141_154 | 1278 | No | NA | 2 | 0 | 0 | Not-determined | Genome-fragment | NA | NA | 0 |
| S3_extracted_k141_8157 | 2690 | No | NA | 7 | 0 | 0 | Not-determined | Genome-fragment | NA | NA | 0 |
| S3_extracted_k141_31988 | 5641 | No | NA | 3 | 0 | 3 | Not-determined | Genome-fragment | NA | NA | 0 |
| S8_vs_k141_152193\|\|full | 1560 | No | NA | 3 | 1 | 0 | Low-quality | Genome-fragment | 1.02 | AAI-based (high-confidence) | 0 |
| S17_extracted_k141_104599 | 1561 | No | NA | 2 | 0 | 0 | Low-quality | Genome-fragment | 1.68 | AAI-based (high-confidence) | 0 |
| S3_extracted_k141_56007 | 1473 | No | NA | 2 | 0 | 1 | Not-determined | Genome-fragment | NA | NA | 0 |
| S3_extracted_k141_111741 | 1041 | No | NA | 1 | 0 | 1 | Not-determined | Genome-fragment | NA | NA | 0 |
| S3_extracted_k141_8164 | 36393 | No | NA | 59 | 18 | 0 | High-quality | High-quality | 94.33 | AAI-based (high-confidence) | 0 |
| S3_extracted_k141_95782 | 4790 | No | NA | 3 | 0 | 3 | Not-determined | Genome-fragment | NA | NA | 0 |
| S14_extracted_k141_66092 | 1561 | No | NA | 3 | 2 | 0 | Low-quality | Genome-fragment | 5.15 | AAI-based (high-confidence) | 0 |
| S3_extracted_k141_79891 | 1205 | No | NA | 2 | 0 | 1 | Not-determined | Genome-fragment | NA | NA | 0 |
| S3_extracted_k141_39992 | 2000 | No | NA | 5 | 0 | 0 | Not-determined | Genome-fragment | NA | NA | 0 |
| S14_vs_k141_9817\|\|full | 1561 | No | NA | 3 | 1 | 0 | Low-quality | Genome-fragment | 3.91 | AAI-based (medium-confidence) | 0 |
| S3_extracted_k141_116727 | 1561 | No | NA | 2 | 0 | 0 | Low-quality | Genome-fragment | 1.55 | AAI-based (high-confidence) | 0 |
| S5_vs_k141_68051\|\|full | 1561 | No | NA | 4 | 1 | 0 | Low-quality | Genome-fragment | 3.04 | AAI-based (high-confidence) | 0 |
| S3_extracted_k141_48189 | 1190 | No | NA | 3 | 0 | 1 | Not-determined | Genome-fragment | NA | NA | 0 |
| S5_vs_k141_113538\|\|full | 1561 | No | NA | 2 | 0 | 0 | Low-quality | Genome-fragment | 3.58 | AAI-based (medium-confidence) | 0 |
| S3_extracted_k141_64013 | 2177 | No | NA | 2 | 0 | 1 | Not-determined | Genome-fragment | NA | NA | 0 |
| S3_extracted_k141_234 | 6907 | No | NA | 11 | 0 | 1 | Not-determined | Genome-fragment | NA | NA | 0 |
| S7_vs_112332\|\|full | 1561 | No | NA | 3 | 2 | 0 | Low-quality | Genome-fragment | 1.94 | AAI-based (high-confidence) | 0 |
| S3_extracted_k141_56047 | 3304 | No | NA | 5 | 0 | 2 | Not-determined | Genome-fragment | NA | NA | 0 |
| S3_extracted_k141_95866 | 5603 | No | NA | 6 | 0 | 2 | Not-determined | Genome-fragment | NA | NA | 0 |
| S3_extracted_k141_79945 | 1387 | No | NA | 2 | 0 | 1 | Not-determined | Genome-fragment | NA | NA | 0 |
| S16_vs_152420\|\|full | 1561 | No | NA | 3 | 0 | 1 | Low-quality | Genome-fragment | 1.18 | AAI-based (medium-confidence) | 0 |
| S10_vs_63017\|\|full | 1561 | No | NA | 3 | 0 | 1 | Low-quality | Genome-fragment | 1.4 | AAI-based (medium-confidence) | 0 |
| S17_vs_k141_104459\|\|full | 1562 | No | NA | 3 | 0 | 0 | Low-quality | Genome-fragment | 0.89 | AAI-based (high-confidence) | 0 |
| S3_extracted_k141_40064 | 1077 | No | NA | 2 | 0 | 0 | Not-determined | Genome-fragment | NA | NA | 0 |
| S3_extracted_k141_87845 | 1329 | No | NA | 1 | 0 | 1 | Not-determined | Genome-fragment | NA | NA | 0 |
| S3_extracted_k141_72023 | 9202 | No | NA | 5 | 0 | 3 | Not-determined | Genome-fragment | NA | NA | 0 |
| S3_extracted_k141_72024 | 1104 | No | NA | 1 | 0 | 1 | Not-determined | Genome-fragment | NA | NA | 0 |
| S3_extracted_k141_103806 | 1420 | No | NA | 1 | 0 | 1 | Not-determined | Genome-fragment | NA | NA | 0 |
| S9_vs_k141_26410\|\|full | 1562 | No | NA | 2 | 1 | 0 | Low-quality | Genome-fragment | 0.92 | AAI-based (high-confidence) | 0 |
| S3_extracted_k141_111776 | 16294 | No | NA | 18 | 8 | 0 | Medium-quality | Genome-fragment | 70.53 | AAI-based (medium-confidence) | 0 |
| S3_extracted_k141_119735 | 7142 | No | NA | 8 | 0 | 1 | Not-determined | Genome-fragment | NA | NA | 0 |
| S9_vs_k141_60417\|\|full | 1562 | No | NA | 3 | 0 | 0 | Low-quality | Genome-fragment | 3.59 | AAI-based (medium-confidence) | 0 |
| S3_extracted_k141_87878 | 2106 | No | NA | 4 | 0 | 0 | Not-determined | Genome-fragment | NA | NA | 0 |
| S3_extracted_k141_358 | 1009 | No | NA | 1 | 0 | 1 | Not-determined | Genome-fragment | NA | NA | 0 |
| S11_vs_k141_81191\|\|full | 1562 | No | NA | 3 | 1 | 0 | Low-quality | Genome-fragment | 3.92 | AAI-based (high-confidence) | 0 |
| S3_extracted_k141_56135 | 1026 | No | NA | 1 | 0 | 0 | Not-determined | Genome-fragment | NA | NA | 0 |
| S3_extracted_k141_72066 | 1352 | No | NA | 2 | 0 | 0 | Not-determined | Genome-fragment | NA | NA | 0 |
| S11_vs_k141_48927\|\|full | 1562 | No | NA | 2 | 1 | 0 | Low-quality | Genome-fragment | 3.35 | AAI-based (high-confidence) | 0 |
| S13_extracted_33519 | 1562 | No | NA | 3 | 0 | 0 | Low-quality | Genome-fragment | 2.95 | AAI-based (medium-confidence) | 0 |
| S3_extracted_k141_24303 | 1947 | No | NA | 1 | 0 | 1 | Not-determined | Genome-fragment | NA | NA | 0 |
| S3_extracted_k141_87896 | 14143 | No | NA | 21 | 1 | 1 | Medium-quality | Genome-fragment | 87.95 | AAI-based (medium-confidence) | 0 |
| S14_vs_k141_88862\|\|full | 1562 | No | NA | 3 | 1 | 0 | Low-quality | Genome-fragment | 3.2 | AAI-based (medium-confidence) | 0 |
| S14_vs_k141_35809\|\|full | 1562 | No | NA | 5 | 0 | 1 | Low-quality | Genome-fragment | 3.59 | AAI-based (medium-confidence) | 0 |
| S3_extracted_k141_56214 | 2303 | No | NA | 1 | 0 | 1 | Not-determined | Genome-fragment | NA | NA | 0 |
| S3_extracted_k141_8477 | 2063 | No | NA | 3 | 0 | 2 | Not-determined | Genome-fragment | NA | NA | 0 |
| S3_extracted_k141_103951 | 1216 | No | NA | 2 | 0 | 1 | Not-determined | Genome-fragment | NA | NA | 0 |
| S3_extracted_k141_96116 | 1030 | No | NA | 2 | 0 | 2 | Not-determined | Genome-fragment | NA | NA | 0 |
| S14_vs_k141_108940\|\|lt2gene | 1562 | No | NA | 2 | 1 | 0 | Low-quality | Genome-fragment | 3.89 | AAI-based (high-confidence) | 0 |
| S3_extracted_k141_80111 | 1396 | No | NA | 2 | 0 | 0 | Not-determined | Genome-fragment | NA | NA | 0 |
| S3_extracted_k141_445 | 3363 | No | NA | 2 | 0 | 1 | Not-determined | Genome-fragment | NA | NA | 0 |
| S3_extracted_k141_32316 | 1129 | No | NA | 2 | 0 | 0 | Not-determined | Genome-fragment | NA | NA | 0 |
| S3_extracted_k141_16146 | 3358 | No | NA | 7 | 0 | 0 | Not-determined | Genome-fragment | NA | NA | 0 |
| S3_extracted_k141_87970 | 1640 | No | NA | 3 | 0 | 2 | Not-determined | Genome-fragment | NA | NA | 0 |
| S3_extracted_k141_111893 | 34477 | No | NA | 51 | 24 | 0 | Complete | High-quality | 100 | DTR (high-confidence) | 0 |
| S3_extracted_k141_56240 | 1071 | No | NA | 1 | 0 | 1 | Not-determined | Genome-fragment | NA | NA | 0 |
| S3_vs_k141_78756\|\|full | 1562 | No | NA | 3 | 1 | 0 | Low-quality | Genome-fragment | 1.81 | AAI-based (medium-confidence) | 0 |
| S3_extracted_k141_40291 | 1436 | No | NA | 1 | 0 | 1 | Not-determined | Genome-fragment | NA | NA | 0 |
| S5_vs_k141_38295\|\|full | 1562 | Yes | 921 | 2 | 1 | 1 | Low-quality | Genome-fragment | 0.5 | HMM-based (lower-bound) | 41.04 |
| S3_extracted_k141_56251 | 1073 | No | NA | 1 | 0 | 1 | Not-determined | Genome-fragment | NA | NA | 0 |
| S3_extracted_k141_96141 | 1385 | No | NA | 2 | 0 | 0 | Not-determined | Genome-fragment | NA | NA | 0 |
| S3_extracted_k141_64175 | 12906 | No | NA | 7 | 0 | 5 | Not-determined | Genome-fragment | NA | NA | 0 |
| S3_extracted_k141_104028 | 2963 | No | NA | 1 | 0 | 0 | Not-determined | Genome-fragment | NA | NA | 0 |
| S7_vs_8853\|\|full | 1562 | No | NA | 4 | 0 | 0 | Low-quality | Genome-fragment | 2.75 | AAI-based (high-confidence) | 0 |
| S3_extracted_k141_119940 | 1020 | No | NA | 1 | 0 | 1 | Not-determined | Genome-fragment | NA | NA | 0 |
| S16_vs_152644\|\|full | 1562 | No | NA | 1 | 0 | 1 | Low-quality | Genome-fragment | 3.36 | AAI-based (medium-confidence) | 0 |
| S3_extracted_k141_96156 | 1671 | No | NA | 2 | 0 | 2 | Not-determined | Genome-fragment | NA | NA | 0 |
| S3_extracted_k141_40369 | 2906 | No | NA | 1 | 0 | 1 | Not-determined | Genome-fragment | NA | NA | 0 |
| S3_extracted_k141_119967 | 1356 | No | NA | 1 | 0 | 0 | Not-determined | Genome-fragment | NA | NA | 0 |
| S3_extracted_k141_56261 | 3569 | No | NA | 4 | 0 | 1 | Not-determined | Genome-fragment | NA | NA | 0 |
| S3_extracted_k141_119991 | 1492 | No | NA | 2 | 0 | 1 | Not-determined | Genome-fragment | NA | NA | 0 |
| S3_extracted_k141_104061 | 4266 | No | NA | 4 | 0 | 4 | Not-determined | Genome-fragment | NA | NA | 0 |
| S10_extracted_70720 | 1562 | No | NA | 4 | 0 | 1 | Low-quality | Genome-fragment | 4.39 | AAI-based (high-confidence) | 0 |
| S3_extracted_k141_16170 | 1077 | No | NA | 2 | 0 | 0 | Not-determined | Genome-fragment | NA | NA | 0 |
| S3_extracted_k141_64230 | 4332 | No | NA | 5 | 0 | 2 | Not-determined | Genome-fragment | NA | NA | 0 |
| S3_extracted_k141_72134 | 1588 | No | NA | 1 | 0 | 1 | Not-determined | Genome-fragment | NA | NA | 0 |
| S3_extracted_k141_64248 | 1301 | No | NA | 1 | 0 | 0 | Not-determined | Genome-fragment | NA | NA | 0 |
| S3_extracted_k141_8669 | 2134 | No | NA | 1 | 0 | 1 | Not-determined | Genome-fragment | NA | NA | 0 |
| S3_extracted_k141_72155 | 1429 | No | NA | 1 | 0 | 1 | Not-determined | Genome-fragment | NA | NA | 0 |
| S3_extracted_k141_40391 | 3241 | No | NA | 4 | 0 | 0 | Not-determined | Genome-fragment | NA | NA | 0 |
| S3_extracted_k141_569 | 1097 | No | NA | 1 | 0 | 1 | Not-determined | Genome-fragment | NA | NA | 0 |
| S3_extracted_k141_16193 | 1240 | No | NA | 2 | 0 | 0 | Not-determined | Genome-fragment | NA | NA | 0 |
| S3_extracted_k141_48433 | 2078 | No | NA | 1 | 0 | 1 | Not-determined | Genome-fragment | NA | NA | 0 |
| S3_extracted_k141_587 | 1073 | No | NA | 1 | 0 | 1 | Not-determined | Genome-fragment | NA | NA | 0 |
| S3_extracted_k141_88065 | 3070 | No | NA | 5 | 0 | 4 | Not-determined | Genome-fragment | NA | NA | 0 |
| S3_extracted_k141_32494 | 1302 | No | NA | 1 | 0 | 1 | Not-determined | Genome-fragment | NA | NA | 0 |
| S3_extracted_k141_598 | 1015 | No | NA | 1 | 0 | 1 | Not-determined | Genome-fragment | NA | NA | 0 |
| S17_vs_k141_96090\|\|full | 1563 | No | NA | 3 | 0 | 0 | Low-quality | Genome-fragment | 2.62 | AAI-based (high-confidence) | 0 |
| S3_extracted_k141_602 | 4744 | No | NA | 4 | 0 | 2 | Not-determined | Genome-fragment | NA | NA | 0 |
| S17_vs_k141_51117\|\|full | 1563 | No | NA | 3 | 3 | 0 | Low-quality | Genome-fragment | 4.32 | AAI-based (high-confidence) | 0 |
| S14_extracted_k141_88197 | 1563 | No | NA | 4 | 4 | 0 | Low-quality | Genome-fragment | 5.16 | AAI-based (high-confidence) | 0 |
| S3_extracted_k141_104128 | 1149 | No | NA | 2 | 0 | 1 | Not-determined | Genome-fragment | NA | NA | 0 |
| S3_extracted_k141_8789 | 1107 | No | NA | 1 | 0 | 0 | Not-determined | Genome-fragment | NA | NA | 0 |
| S3_extracted_k141_112004 | 1001 | No | NA | 1 | 0 | 0 | Not-determined | Genome-fragment | NA | NA | 0 |
| S14_extracted_k141_68516 | 1563 | No | NA | 1 | 0 | 0 | Low-quality | Genome-fragment | 0.59 | AAI-based (high-confidence) | 0 |
| S3_extracted_k141_112037 | 1016 | No | NA | 1 | 0 | 1 | Not-determined | Genome-fragment | NA | NA | 0 |
| S3_extracted_k141_8836 | 4175 | No | NA | 3 | 0 | 0 | Not-determined | Genome-fragment | NA | NA | 0 |
| S3_extracted_k141_96222 | 22007 | No | NA | 37 | 31 | 0 | Medium-quality | Genome-fragment | 72.48 | AAI-based (high-confidence) | 0 |
| S3_extracted_k141_96223 | 1084 | No | NA | 1 | 0 | 1 | Not-determined | Genome-fragment | NA | NA | 0 |
| S3_extracted_k141_16198 | 6117 | No | NA | 3 | 0 | 3 | Not-determined | Genome-fragment | NA | NA | 0 |
| S3_extracted_k141_32547 | 2933 | No | NA | 2 | 0 | 1 | Not-determined | Genome-fragment | NA | NA | 0 |
| S1_vs_k141_111010\|\|full | 1563 | No | NA | 2 | 0 | 1 | Low-quality | Genome-fragment | 0.88 | AAI-based (high-confidence) | 0 |
| S3_extracted_k141_88111 | 1435 | No | NA | 2 | 0 | 0 | Not-determined | Genome-fragment | NA | NA | 0 |
| S7_extracted_30034 | 1563 | No | NA | 2 | 0 | 0 | Low-quality | Genome-fragment | 2.14 | AAI-based (high-confidence) | 0 |
| S8_vs_k141_64366\|\|full | 1563 | No | NA | 3 | 0 | 0 | Low-quality | Genome-fragment | 3.62 | AAI-based (medium-confidence) | 0 |
| S8_vs_k141_109353\|\|full | 1563 | No | NA | 1 | 0 | 0 | Low-quality | Genome-fragment | 22.81 | AAI-based (medium-confidence) | 0 |
| S17_extracted_k141_49697 | 1564 | No | NA | 2 | 1 | 0 | Low-quality | Genome-fragment | 3.28 | AAI-based (medium-confidence) | 0 |
| S3_extracted_k141_32617 | 1719 | No | NA | 1 | 0 | 0 | Not-determined | Genome-fragment | NA | NA | 0 |
| S3_extracted_k141_728 | 1880 | No | NA | 1 | 0 | 1 | Not-determined | Genome-fragment | NA | NA | 0 |
| S3_extracted_k141_32655 | 1497 | No | NA | 6 | 0 | 0 | Not-determined | Genome-fragment | NA | NA | 0 |
| S3_extracted_k141_64287 | 1004 | No | NA | 1 | 0 | 1 | Not-determined | Genome-fragment | NA | NA | 0 |
| S11_vs_k141_13585\|\|full | 1564 | No | NA | 3 | 0 | 0 | Low-quality | Genome-fragment | 2.77 | AAI-based (high-confidence) | 0 |
| S3_extracted_k141_8875 | 1425 | No | NA | 1 | 0 | 1 | Not-determined | Genome-fragment | NA | NA | 0 |
| S3_extracted_k141_32677 | 3380 | No | NA | 4 | 0 | 1 | Not-determined | Genome-fragment | NA | NA | 0 |
| S3_extracted_k141_80287 | 44726 | No | NA | 61 | 31 | 0 | Complete | High-quality | 100 | DTR (high-confidence) | 0 |
| S3_extracted_k141_80292 | 1043 | No | NA | 1 | 0 | 0 | Not-determined | Genome-fragment | NA | NA | 0 |
| S3_extracted_k141_56363 | 34937 | No | NA | 48 | 20 | 1 | Medium-quality | Genome-fragment | 86.49 | AAI-based (high-confidence) | 0 |
| S3_extracted_k141_80303 | 1004 | No | NA | 1 | 0 | 0 | Not-determined | Genome-fragment | NA | NA | 0 |
| S3_extracted_k141_751 | 1233 | No | NA | 2 | 0 | 0 | Not-determined | Genome-fragment | NA | NA | 0 |
| S11_vs_k141_92310\|\|full | 1564 | No | NA | 3 | 0 | 1 | Low-quality | Genome-fragment | 3.61 | AAI-based (medium-confidence) | 0 |
| S3_extracted_k141_48670 | 4074 | No | NA | 5 | 0 | 1 | Not-determined | Genome-fragment | NA | NA | 0 |
| S3_extracted_k141_48684 | 2313 | No | NA | 1 | 0 | 1 | Not-determined | Genome-fragment | NA | NA | 0 |
| S11_vs_k141_88233\|\|full | 1564 | No | NA | 4 | 0 | 3 | Low-quality | Genome-fragment | 0.85 | AAI-based (medium-confidence) | 0 |
| S3_extracted_k141_56449 | 1385 | No | NA | 2 | 0 | 1 | Not-determined | Genome-fragment | NA | NA | 0 |
| S11_vs_k141_56715\|\|full | 1564 | No | NA | 3 | 0 | 1 | Low-quality | Genome-fragment | 3.39 | AAI-based (medium-confidence) | 0 |
| S14_extracted_k141_106255 | 1564 | No | NA | 3 | 2 | 0 | Low-quality | Genome-fragment | 5.16 | AAI-based (high-confidence) | 0 |
| S14_vs_k141_53833\|\|full | 1564 | No | NA | 3 | 2 | 0 | Low-quality | Genome-fragment | 3.51 | AAI-based (high-confidence) | 0 |
| S3_extracted_k141_96404 | 1081 | No | NA | 2 | 0 | 0 | Not-determined | Genome-fragment | NA | NA | 0 |
| S3_extracted_k141_104317 | 1684 | No | NA | 2 | 0 | 0 | Not-determined | Genome-fragment | NA | NA | 0 |
| S3_extracted_k141_9030 | 4413 | No | NA | 3 | 0 | 1 | Not-determined | Genome-fragment | NA | NA | 0 |
| S3_extracted_k141_56503 | 2356 | No | NA | 1 | 0 | 0 | Not-determined | Genome-fragment | NA | NA | 0 |
| S16_extracted_37937 | 1564 | No | NA | 1 | 0 | 0 | Low-quality | Genome-fragment | 1.38 | AAI-based (medium-confidence) | 0 |
| S3_extracted_k141_16269 | 4908 | No | NA | 5 | 0 | 3 | Not-determined | Genome-fragment | NA | NA | 0 |
| S3_extracted_k141_9046 | 6983 | No | NA | 6 | 0 | 2 | Not-determined | Genome-fragment | NA | NA | 0 |
| S3_extracted_k141_32819 | 1314 | No | NA | 2 | 0 | 2 | Not-determined | Genome-fragment | NA | NA | 0 |
| S3_extracted_k141_9074 | 2520 | No | NA | 4 | 0 | 1 | Not-determined | Genome-fragment | NA | NA | 0 |
| S16_extracted_172017 | 1564 | No | NA | 5 | 1 | 0 | Low-quality | Genome-fragment | 1.44 | AAI-based (high-confidence) | 0 |
| S3_extracted_k141_16328 | 7310 | No | NA | 6 | 0 | 6 | Not-determined | Genome-fragment | NA | NA | 0 |
| S16_vs_136361\|\|full | 1564 | No | NA | 3 | 1 | 0 | Low-quality | Genome-fragment | 3.78 | AAI-based (medium-confidence) | 0 |
| S10_extracted_50108 | 1564 | No | NA | 3 | 0 | 1 | Low-quality | Genome-fragment | 0.66 | AAI-based (medium-confidence) | 0 |
| S12_vs_k141_182183\|\|lt2gene | 1564 | No | NA | 2 | 1 | 0 | Low-quality | Genome-fragment | 2.85 | AAI-based (high-confidence) | 0 |
| S3_extracted_k141_112249 | 9942 | No | NA | 6 | 0 | 0 | Not-determined | Genome-fragment | NA | NA | 0 |
| S3_extracted_k141_56580 | 1199 | No | NA | 4 | 0 | 0 | Not-determined | Genome-fragment | NA | NA | 0 |
| S6_extracted_k141_27722 | 1564 | No | NA | 2 | 0 | 1 | Low-quality | Genome-fragment | 1.65 | AAI-based (high-confidence) | 0 |
| S17_vs_k141_39022\|\|full | 1565 | No | NA | 4 | 1 | 0 | Low-quality | Genome-fragment | 3.97 | AAI-based (high-confidence) | 0 |
| S3_extracted_k141_9147 | 5981 | No | NA | 9 | 0 | 4 | Not-determined | Genome-fragment | NA | NA | 0 |
| S3_extracted_k141_9154 | 3014 | No | NA | 3 | 0 | 3 | Not-determined | Genome-fragment | NA | NA | 0 |
| S3_extracted_k141_24676 | 1642 | No | NA | 2 | 0 | 1 | Not-determined | Genome-fragment | NA | NA | 0 |
| S11_extracted_k141_13170 | 1565 | No | NA | 1 | 1 | 0 | Low-quality | Genome-fragment | 4.08 | AAI-based (medium-confidence) | 0 |
| S3_extracted_k141_80446 | 1829 | No | NA | 3 | 0 | 2 | Not-determined | Genome-fragment | NA | NA | 0 |
| S3_extracted_k141_56613 | 1147 | No | NA | 2 | 0 | 1 | Not-determined | Genome-fragment | NA | NA | 0 |
| S3_extracted_k141_88306 | 1720 | No | NA | 2 | 0 | 0 | Not-determined | Genome-fragment | NA | NA | 0 |
| S3_extracted_k141_80463 | 2175 | No | NA | 3 | 0 | 0 | Not-determined | Genome-fragment | NA | NA | 0 |
| S3_extracted_k141_72769 | 1442 | No | NA | 2 | 0 | 0 | Not-determined | Genome-fragment | NA | NA | 0 |
| S14_extracted_k141_30535 | 1565 | No | NA | 1 | 1 | 0 | Low-quality | Genome-fragment | 5.02 | AAI-based (high-confidence) | 0 |
| S14_extracted_k141_54060 | 1565 | No | NA | 3 | 0 | 2 | Low-quality | Genome-fragment | 1.88 | AAI-based (medium-confidence) | 0 |
| S1_vs_k141_12945\|\|full | 1565 | No | NA | 4 | 2 | 0 | Low-quality | Genome-fragment | 3.47 | AAI-based (high-confidence) | 0 |
| S3_extracted_k141_96592 | 3391 | No | NA | 4 | 0 | 0 | Not-determined | Genome-fragment | NA | NA | 0 |
| S5_vs_k141_106468\|\|full | 1565 | No | NA | 3 | 0 | 1 | Low-quality | Genome-fragment | 0.58 | AAI-based (medium-confidence) | 0 |
| S10_vs_71985\|\|full | 1565 | No | NA | 3 | 2 | 0 | Low-quality | Genome-fragment | 3.95 | AAI-based (high-confidence) | 0 |
| S3_extracted_k141_56715 | 1188 | No | NA | 1 | 0 | 0 | Not-determined | Genome-fragment | NA | NA | 0 |
| S10_vs_88825\|\|lt2gene | 1565 | No | NA | 1 | 1 | 0 | Low-quality | Genome-fragment | 0.6 | HMM-based (lower-bound) | 0 |
| S3_extracted_k141_72853 | 6373 | No | NA | 4 | 0 | 0 | Not-determined | Genome-fragment | NA | NA | 0 |
| S12_extracted_k141_169647 | 1565 | No | NA | 2 | 0 | 0 | Low-quality | Genome-fragment | 2.12 | AAI-based (medium-confidence) | 0 |
| S3_extracted_k141_56726 | 14683 | No | NA | 19 | 0 | 0 | Not-determined | Genome-fragment | NA | NA | 0 |
| S12_vs_k141_9612\|\|full | 1565 | No | NA | 2 | 1 | 0 | Low-quality | Genome-fragment | 3.77 | AAI-based (medium-confidence) | 0 |
| S15_extracted_49118 | 1565 | No | NA | 1 | 0 | 1 | Low-quality | Genome-fragment | 1.35 | AAI-based (medium-confidence) | 0 |
| S4_extracted_684 | 1565 | No | NA | 3 | 3 | 0 | Low-quality | Genome-fragment | 5.16 | AAI-based (high-confidence) | 0 |
| S3_extracted_k141_96685 | 1169 | No | NA | 2 | 0 | 1 | Not-determined | Genome-fragment | NA | NA | 0 |
| S3_extracted_k141_72910 | 1069 | No | NA | 1 | 0 | 1 | Not-determined | Genome-fragment | NA | NA | 0 |
| S3_extracted_k141_9265 | 3041 | No | NA | 3 | 0 | 2 | Not-determined | Genome-fragment | NA | NA | 0 |
| S6_vs_k141_43058\|\|full | 1565 | No | NA | 3 | 0 | 0 | Low-quality | Genome-fragment | 5.26 | AAI-based (medium-confidence) | 0 |
| S3_extracted_k141_72923 | 4532 | No | NA | 4 | 0 | 0 | Not-determined | Genome-fragment | NA | NA | 0 |
| S8_vs_k141_36189\|\|full | 1565 | No | NA | 3 | 0 | 0 | Low-quality | Genome-fragment | 1.69 | AAI-based (medium-confidence) | 0 |
| S3_extracted_k141_72933 | 3320 | No | NA | 4 | 0 | 2 | Not-determined | Genome-fragment | NA | NA | 0 |
| S13_extracted_37066 | 1566 | No | NA | 1 | 0 | 1 | Low-quality | Genome-fragment | 0.67 | AAI-based (medium-confidence) | 0 |
| S3_extracted_k141_24780 | 5675 | No | NA | 4 | 0 | 4 | Not-determined | Genome-fragment | NA | NA | 0 |
| S14_vs_k141_2887\|\|lt2gene | 1566 | No | NA | 2 | 1 | 0 | Low-quality | Genome-fragment | 3.62 | AAI-based (high-confidence) | 0 |
| S3_vs_k141_76849\|\|full | 1566 | No | NA | 4 | 1 | 0 | Low-quality | Genome-fragment | 2.54 | AAI-based (high-confidence) | 0 |
| S3_extracted_k141_49022 | 1068 | No | NA | 2 | 0 | 0 | Not-determined | Genome-fragment | NA | NA | 0 |
| S7_vs_97206\|\|lt2gene | 1566 | No | NA | 2 | 1 | 0 | Low-quality | Genome-fragment | 3.63 | AAI-based (high-confidence) | 0 |
| S3_extracted_k141_120181 | 1350 | No | NA | 2 | 0 | 1 | Not-determined | Genome-fragment | NA | NA | 0 |
| S16_vs_150465\|\|full | 1566 | No | NA | 4 | 3 | 0 | Low-quality | Genome-fragment | 4.65 | AAI-based (high-confidence) | 0 |
| S3_extracted_k141_9291 | 1236 | No | NA | 1 | 0 | 1 | Not-determined | Genome-fragment | NA | NA | 0 |
| S16_vs_164342\|\|full | 1566 | No | NA | 3 | 0 | 2 | Low-quality | Genome-fragment | 1.23 | AAI-based (medium-confidence) | 0 |
| S12_vs_k141_13258\|\|full | 1566 | No | NA | 2 | 0 | 0 | Low-quality | Genome-fragment | 2.7 | AAI-based (high-confidence) | 0 |
| S12_vs_k141_45622\|\|lt2gene | 1566 | No | NA | 2 | 1 | 0 | Low-quality | Genome-fragment | 3 | AAI-based (high-confidence) | 0 |
| S3_extracted_k141_88484 | 1039 | No | NA | 5 | 0 | 3 | Not-determined | Genome-fragment | NA | NA | 0 |
| S3_extracted_k141_64507 | 1184 | No | NA | 1 | 0 | 0 | Not-determined | Genome-fragment | NA | NA | 0 |
| S3_extracted_k141_104465 | 1057 | No | NA | 1 | 0 | 0 | Not-determined | Genome-fragment | NA | NA | 0 |
| S17_vs_k141_45987\|\|full | 1567 | No | NA | 3 | 2 | 0 | Low-quality | Genome-fragment | 2.55 | AAI-based (high-confidence) | 0 |
| S3_extracted_k141_49045 | 1004 | No | NA | 1 | 0 | 0 | Not-determined | Genome-fragment | NA | NA | 0 |
| S17_vs_k141_33819\|\|full | 1567 | No | NA | 2 | 2 | 0 | Low-quality | Genome-fragment | 3.63 | AAI-based (high-confidence) | 0 |
| S3_extracted_k141_49050 | 2554 | No | NA | 1 | 0 | 1 | Not-determined | Genome-fragment | NA | NA | 0 |
| S11_vs_k141_45974\|\|full | 1567 | No | NA | 2 | 0 | 1 | Low-quality | Genome-fragment | 2.96 | AAI-based (medium-confidence) | 0 |
| S3_extracted_k141_56817 | 3562 | No | NA | 4 | 0 | 3 | Not-determined | Genome-fragment | NA | NA | 0 |
| S14_vs_k141_114806\|\|full | 1567 | No | NA | 4 | 1 | 0 | Low-quality | Genome-fragment | 5.12 | AAI-based (high-confidence) | 0 |
| S1_extracted_k141_114645 | 1567 | No | NA | 1 | 0 | 1 | Low-quality | Genome-fragment | 1.9 | AAI-based (medium-confidence) | 0 |
| S1_vs_k141_75646\|\|full | 1567 | No | NA | 4 | 1 | 0 | Low-quality | Genome-fragment | 1.81 | HMM-based (lower-bound) | 0 |
| S3_extracted_k141_56843 | 3455 | No | NA | 6 | 0 | 5 | Not-determined | Genome-fragment | NA | NA | 0 |
| S1_vs_k141_48705\|\|lt2gene | 1567 | No | NA | 2 | 1 | 0 | Low-quality | Genome-fragment | 3.74 | AAI-based (high-confidence) | 0 |
| S3_extracted_k141_56853 | 4420 | No | NA | 6 | 0 | 6 | Not-determined | Genome-fragment | NA | NA | 0 |
| S10_vs_46170\|\|full | 1567 | No | NA | 3 | 0 | 1 | Low-quality | Genome-fragment | 2.5 | AAI-based (medium-confidence) | 0 |
| S3_extracted_k141_24901 | 3995 | No | NA | 5 | 0 | 2 | Not-determined | Genome-fragment | NA | NA | 0 |
| S3_extracted_k141_24920 | 1019 | No | NA | 2 | 0 | 1 | Not-determined | Genome-fragment | NA | NA | 0 |
| S3_extracted_k141_73099 | 1897 | No | NA | 1 | 0 | 1 | Not-determined | Genome-fragment | NA | NA | 0 |
| S3_extracted_k141_9431 | 1239 | No | NA | 2 | 0 | 1 | Not-determined | Genome-fragment | NA | NA | 0 |
| S10_vs_62338\|\|full | 1567 | No | NA | 3 | 0 | 1 | Low-quality | Genome-fragment | 3.64 | AAI-based (high-confidence) | 0 |
| S12_vs_k141_181741\|\|full | 1567 | No | NA | 3 | 0 | 2 | Low-quality | Genome-fragment | 5.23 | AAI-based (medium-confidence) | 0 |
| S3_extracted_k141_104536 | 1168 | No | NA | 1 | 0 | 0 | Not-determined | Genome-fragment | NA | NA | 0 |
| S3_extracted_k141_104575 | 2186 | No | NA | 3 | 0 | 0 | Not-determined | Genome-fragment | NA | NA | 0 |
| S3_extracted_k141_9481 | 1071 | No | NA | 2 | 0 | 0 | Not-determined | Genome-fragment | NA | NA | 0 |
| S3_extracted_k141_49209 | 2214 | No | NA | 5 | 0 | 0 | Not-determined | Genome-fragment | NA | NA | 0 |
| S3_extracted_k141_16602 | 1023 | No | NA | 1 | 0 | 0 | Not-determined | Genome-fragment | NA | NA | 0 |
| S12_vs_k141_14678\|\|full | 1567 | No | NA | 6 | 0 | 0 | Low-quality | Genome-fragment | 3.12 | AAI-based (medium-confidence) | 0 |
| S3_extracted_k141_73208 | 10456 | No | NA | 16 | 0 | 0 | Not-determined | Genome-fragment | NA | NA | 0 |
| S9_vs_k141_62162\|\|full | 1568 | No | NA | 3 | 0 | 0 | Low-quality | Genome-fragment | 2.64 | AAI-based (high-confidence) | 0 |
| S3_extracted_k141_96883 | 1382 | No | NA | 1 | 0 | 0 | Not-determined | Genome-fragment | NA | NA | 0 |
| S3_extracted_k141_120266 | 2060 | No | NA | 4 | 0 | 3 | Not-determined | Genome-fragment | NA | NA | 0 |
| S13_vs_99394\|\|full | 1568 | No | NA | 3 | 0 | 0 | Low-quality | Genome-fragment | 0.95 | AAI-based (medium-confidence) | 0 |
| S5_extracted_k141_40840 | 1568 | No | NA | 3 | 1 | 0 | Low-quality | Genome-fragment | 4.53 | AAI-based (high-confidence) | 0 |
| S3_extracted_k141_64602 | 2167 | No | NA | 2 | 0 | 1 | Not-determined | Genome-fragment | NA | NA | 0 |
| S3_extracted_k141_1051 | 4374 | No | NA | 4 | 0 | 3 | Not-determined | Genome-fragment | NA | NA | 0 |
| S7_vs_3626\|\|full | 1568 | No | NA | 2 | 1 | 0 | Low-quality | Genome-fragment | 2.68 | AAI-based (medium-confidence) | 0 |
| S3_extracted_k141_73309 | 1529 | No | NA | 1 | 0 | 1 | Not-determined | Genome-fragment | NA | NA | 0 |
| S7_vs_62647\|\|full | 1568 | No | NA | 3 | 1 | 0 | Low-quality | Genome-fragment | 3.82 | AAI-based (high-confidence) | 0 |
| S16_extracted_139875 | 1568 | No | NA | 4 | 0 | 0 | Low-quality | Genome-fragment | 1.8 | AAI-based (medium-confidence) | 0 |
| S3_extracted_k141_80909 | 1595 | No | NA | 3 | 0 | 1 | Not-determined | Genome-fragment | NA | NA | 0 |
| S16_vs_45200\|\|full | 1568 | No | NA | 3 | 0 | 0 | Low-quality | Genome-fragment | 2.41 | AAI-based (medium-confidence) | 0 |
| S3_extracted_k141_97026 | 4003 | No | NA | 7 | 0 | 0 | Not-determined | Genome-fragment | NA | NA | 0 |
| S3_extracted_k141_40991 | 1141 | No | NA | 1 | 0 | 1 | Not-determined | Genome-fragment | NA | NA | 0 |
| S16_vs_82598\|\|full | 1568 | No | NA | 3 | 0 | 0 | Low-quality | Genome-fragment | 3.43 | AAI-based (medium-confidence) | 0 |
| S3_extracted_k141_64674 | 7822 | No | NA | 9 | 0 | 2 | Not-determined | Genome-fragment | NA | NA | 0 |
| S3_extracted_k141_88683 | 1372 | No | NA | 1 | 0 | 0 | Not-determined | Genome-fragment | NA | NA | 0 |
| S12_vs_k141_133877\|\|full | 1568 | No | NA | 2 | 0 | 0 | Low-quality | Genome-fragment | 2.41 | AAI-based (medium-confidence) | 0 |
| S3_extracted_k141_97053 | 6198 | No | NA | 5 | 0 | 3 | Not-determined | Genome-fragment | NA | NA | 0 |
| S15_vs_51281\|\|full | 1568 | No | NA | 4 | 4 | 0 | Low-quality | Genome-fragment | 1.95 | AAI-based (high-confidence) | 0 |
| S3_extracted_k141_112476 | 1473 | No | NA | 1 | 0 | 0 | Not-determined | Genome-fragment | NA | NA | 0 |
| S17_vs_k141_70768\|\|lt2gene | 1569 | Yes | 1107 | 2 | 1 | 1 | Low-quality | Genome-fragment | 2.94 | AAI-based (high-confidence) | 29.45 |
| S3_extracted_k141_104699 | 1216 | No | NA | 1 | 0 | 1 | Not-determined | Genome-fragment | NA | NA | 0 |
| S11_extracted_k141_21665 | 1569 | No | NA | 4 | 1 | 0 | Low-quality | Genome-fragment | 2.35 | AAI-based (high-confidence) | 0 |
| S13_vs_99716\|\|full | 1569 | No | NA | 3 | 1 | 0 | Low-quality | Genome-fragment | 1.18 | AAI-based (medium-confidence) | 0 |
| S3_extracted_k141_120401 | 1330 | No | NA | 2 | 0 | 1 | Not-determined | Genome-fragment | NA | NA | 0 |
| S5_vs_k141_24287\|\|full | 1569 | No | NA | 6 | 0 | 0 | Low-quality | Genome-fragment | 3.74 | AAI-based (high-confidence) | 0 |
| S16_vs_131552\|\|full | 1569 | No | NA | 2 | 0 | 0 | Low-quality | Genome-fragment | 3.56 | AAI-based (medium-confidence) | 0 |
| S12_extracted_k141_132728 | 1569 | No | NA | 2 | 2 | 0 | Low-quality | Genome-fragment | 3.44 | AAI-based (high-confidence) | 0 |
| S3_extracted_k141_120409 | 4421 | No | NA | 4 | 0 | 3 | Not-determined | Genome-fragment | NA | NA | 0 |
| S12_vs_k141_29766\|\|full | 1569 | No | NA | 1 | 0 | 1 | Low-quality | Genome-fragment | 2.55 | AAI-based (medium-confidence) | 0 |
| S15_vs_29613\|\|full | 1569 | No | NA | 4 | 1 | 0 | Low-quality | Genome-fragment | 3.22 | AAI-based (high-confidence) | 0 |
| S3_extracted_k141_64743 | 4930 | No | NA | 3 | 0 | 3 | Not-determined | Genome-fragment | NA | NA | 0 |
| S3_extracted_k141_81118 | 1028 | No | NA | 1 | 0 | 0 | Not-determined | Genome-fragment | NA | NA | 0 |
| S3_extracted_k141_104715 | 3951 | No | NA | 2 | 0 | 1 | Not-determined | Genome-fragment | NA | NA | 0 |
| S3_extracted_k141_33389 | 17765 | No | NA | 25 | 8 | 0 | High-quality | High-quality | 91.08 | AAI-based (medium-confidence) | 0 |
| S15_vs_73644\|\|full | 1569 | No | NA | 4 | 0 | 0 | Low-quality | Genome-fragment | 3.73 | AAI-based (medium-confidence) | 0 |
| S6_extracted_k141_39574 | 1569 | No | NA | 2 | 2 | 0 | Low-quality | Genome-fragment | 3.47 | AAI-based (high-confidence) | 0 |
| S8_extracted_k141_131057 | 1569 | No | NA | 1 | 1 | 0 | Low-quality | Genome-fragment | 2.94 | HMM-based (lower-bound) | 0 |
| S8_vs_k141_108852\|\|full | 1569 | No | NA | 3 | 0 | 0 | Low-quality | Genome-fragment | 3.32 | AAI-based (high-confidence) | 0 |
| S8_vs_k141_153546\|\|full | 1569 | No | NA | 5 | 0 | 0 | Low-quality | Genome-fragment | 3.3 | AAI-based (medium-confidence) | 0 |
| S14_extracted_k141_75158 | 1570 | No | NA | 2 | 1 | 0 | Low-quality | Genome-fragment | 3.58 | AAI-based (high-confidence) | 0 |
| S14_vs_k141_68035\|\|full | 1570 | No | NA | 3 | 0 | 0 | Low-quality | Genome-fragment | 3.26 | AAI-based (medium-confidence) | 0 |
| S3_extracted_k141_112585 | 2540 | No | NA | 3 | 0 | 2 | Not-determined | Genome-fragment | NA | NA | 0 |
| S14_vs_k141_15410\|\|lt2gene | 1570 | No | NA | 1 | 0 | 0 | Low-quality | Genome-fragment | 1.54 | AAI-based (medium-confidence) | 0 |
| S3_extracted_k141_81165 | 3387 | No | NA | 5 | 0 | 2 | Not-determined | Genome-fragment | NA | NA | 0 |
| S1_vs_k141_51445\|\|full | 1570 | No | NA | 3 | 0 | 1 | Low-quality | Genome-fragment | 0.99 | AAI-based (medium-confidence) | 0 |
| S3_vs_k141_95045\|\|full | 1570 | No | NA | 4 | 1 | 0 | Low-quality | Genome-fragment | 3.76 | AAI-based (high-confidence) | 0 |
| S3_extracted_k141_97131 | 1033 | No | NA | 1 | 0 | 0 | Not-determined | Genome-fragment | NA | NA | 0 |
| S5_vs_k141_44872\|\|full | 1570 | No | NA | 4 | 2 | 0 | Low-quality | Genome-fragment | 2.6 | AAI-based (medium-confidence) | 0 |
| S16_extracted_130686 | 1570 | No | NA | 4 | 0 | 0 | Low-quality | Genome-fragment | 1.99 | AAI-based (high-confidence) | 0 |
| S16_vs_27690\|\|full | 1570 | No | NA | 3 | 0 | 1 | Low-quality | Genome-fragment | 3.15 | AAI-based (medium-confidence) | 0 |
| S12_vs_k141_63288\|\|full | 1570 | No | NA | 5 | 1 | 0 | Low-quality | Genome-fragment | 4.15 | AAI-based (high-confidence) | 0 |
| S3_extracted_k141_81200 | 1170 | No | NA | 1 | 0 | 1 | Not-determined | Genome-fragment | NA | NA | 0 |
| S17_vs_k141_14875\|\|full | 1571 | No | NA | 3 | 0 | 0 | Low-quality | Genome-fragment | 3.24 | AAI-based (medium-confidence) | 0 |
| S3_extracted_k141_16969 | 3277 | No | NA | 4 | 0 | 2 | Not-determined | Genome-fragment | NA | NA | 0 |
| S3_extracted_k141_16983 | 1164 | No | NA | 1 | 0 | 1 | Not-determined | Genome-fragment | NA | NA | 0 |
| S3_extracted_k141_104799 | 1343 | No | NA | 2 | 0 | 0 | Not-determined | Genome-fragment | NA | NA | 0 |
| S17_vs_k141_30258\|\|full | 1571 | No | NA | 3 | 3 | 0 | Low-quality | Genome-fragment | 2.7 | AAI-based (high-confidence) | 0 |
| S3_extracted_k141_88868 | 1951 | No | NA | 1 | 0 | 0 | Not-determined | Genome-fragment | NA | NA | 0 |
| S17_vs_k141_165641\|\|full | 1571 | No | NA | 4 | 1 | 0 | Low-quality | Genome-fragment | 2.88 | HMM-based (lower-bound) | 0 |
| S3_extracted_k141_1335 | 4084 | No | NA | 5 | 0 | 2 | Not-determined | Genome-fragment | NA | NA | 0 |
| S3_extracted_k141_104842 | 3385 | No | NA | 2 | 0 | 2 | Not-determined | Genome-fragment | NA | NA | 0 |
| S3_extracted_k141_41207 | 3305 | No | NA | 3 | 0 | 1 | Not-determined | Genome-fragment | NA | NA | 0 |
| S3_extracted_k141_120562 | 4356 | No | NA | 2 | 0 | 2 | Not-determined | Genome-fragment | NA | NA | 0 |
| S9_vs_k141_52896\|\|full | 1571 | No | NA | 5 | 0 | 0 | Low-quality | Genome-fragment | 1.88 | AAI-based (high-confidence) | 0 |
| S3_extracted_k141_41221 | 3771 | No | NA | 5 | 0 | 1 | Not-determined | Genome-fragment | NA | NA | 0 |
| S14_vs_k141_68771\|\|full | 1571 | No | NA | 3 | 0 | 0 | Low-quality | Genome-fragment | 1.38 | AAI-based (high-confidence) | 0 |
| S3_extracted_k141_57197 | 6541 | No | NA | 5 | 0 | 3 | Not-determined | Genome-fragment | NA | NA | 0 |
| S14_vs_k141_54369\|\|lt2gene | 1571 | No | NA | 2 | 1 | 0 | Low-quality | Genome-fragment | 2.87 | HMM-based (lower-bound) | 0 |
| S3_extracted_k141_57208 | 5651 | No | NA | 6 | 0 | 2 | Not-determined | Genome-fragment | NA | NA | 0 |
| S3_extracted_k141_17011 | 1421 | No | NA | 1 | 0 | 0 | Not-determined | Genome-fragment | NA | NA | 0 |
| S16_vs_6415\|\|full | 1571 | No | NA | 5 | 0 | 0 | Low-quality | Genome-fragment | 2.6 | AAI-based (high-confidence) | 0 |
| S10_vs_40642\|\|full | 1571 | No | NA | 2 | 0 | 0 | Low-quality | Genome-fragment | 2.64 | AAI-based (medium-confidence) | 0 |
| S3_extracted_k141_64989 | 3887 | No | NA | 3 | 0 | 2 | Not-determined | Genome-fragment | NA | NA | 0 |
| S3_extracted_k141_81302 | 1078 | No | NA | 2 | 0 | 2 | Not-determined | Genome-fragment | NA | NA | 0 |

| | | | | | | | | | | | |
|---|---|---|---|---|---|---|---|---|---|---|---|
| S12_extracted_k141_64256 | 1571 | No | NA | 2 | 2 | 0 | Low-quality | Genome-fragment | 3.52 | AAI-based (high-confidence) | 0 |
| S15_vs_31634\|\|full | 1571 | No | NA | 2 | 0 | 1 | Low-quality | Genome-fragment | 2.77 | AAI-based (high-confidence) | 0 |
| S6_vs_k141_49511\|\|full | 1571 | No | NA | 3 | 1 | 0 | Low-quality | Genome-fragment | 3.1 | HMM-based (lower-bound) | 0 |
| S3_extracted_k141_73597 | 57020 | No | NA | 89 | 14 | 4 | Medium-quality | Genome-fragment | 82.14 | AAI-based (high-confidence) | 0 |
| S3_extracted_k141_112725 | 3898 | No | NA | 2 | 0 | 2 | Not-determined | Genome-fragment | NA | NA | 0 |
| S3_extracted_k141_17054 | 1107 | No | NA | 1 | 0 | 1 | Not-determined | Genome-fragment | NA | NA | 0 |
| S3_extracted_k141_57274 | 2988 | No | NA | 7 | 0 | 2 | Not-determined | Genome-fragment | NA | NA | 0 |
| S17_extracted_k141_8178 | 1572 | No | NA | 1 | 0 | 1 | Low-quality | Genome-fragment | 0.85 | AAI-based (medium-confidence) | 0 |
| S3_extracted_k141_25131 | 1164 | No | NA | 1 | 0 | 0 | Not-determined | Genome-fragment | NA | NA | 0 |
| S3_extracted_k141_41255 | 4606 | No | NA | 6 | 0 | 0 | Not-determined | Genome-fragment | NA | NA | 0 |
| S17_extracted_k141_36927 | 1572 | No | NA | 3 | 0 | 0 | Low-quality | Genome-fragment | 3.26 | AAI-based (high-confidence) | 0 |
| S3_extracted_k141_73718 | 1306 | No | NA | 4 | 0 | 0 | Not-determined | Genome-fragment | NA | NA | 0 |
| S3_extracted_k141_41300 | 1287 | No | NA | 1 | 0 | 1 | Not-determined | Genome-fragment | NA | NA | 0 |
| S3_extracted_k141_73732 | 1705 | No | NA | 3 | 0 | 1 | Not-determined | Genome-fragment | NA | NA | 0 |
| S9_extracted_k141_63775 | 1572 | No | NA | 6 | 1 | 0 | Low-quality | Genome-fragment | 2.56 | AAI-based (medium-confidence) | 0 |
| S3_extracted_k141_88934 | 2723 | No | NA | 2 | 0 | 0 | Not-determined | Genome-fragment | NA | NA | 0 |
| S3_extracted_k141_81426 | 3800 | No | NA | 4 | 0 | 2 | Not-determined | Genome-fragment | NA | NA | 0 |
| S3_extracted_k141_65092 | 1278 | No | NA | 0 | 0 | 0 | Not-determined | Genome-fragment | NA | NA | 0 |
| S3_extracted_k141_57359 | 2182 | No | NA | 1 | 0 | 0 | Not-determined | Genome-fragment | NA | NA | 0 |
| S14_extracted_k141_45070 | 1572 | No | NA | 1 | 1 | 0 | Low-quality | Genome-fragment | 1.93 | HMM-based (lower-bound) | 0 |
| S14_extracted_k141_35469 | 1572 | No | NA | 3 | 1 | 0 | Low-quality | Genome-fragment | 5.1 | AAI-based (high-confidence) | 0 |
| S3_extracted_k141_41373 | 1271 | No | NA | 2 | 0 | 1 | Not-determined | Genome-fragment | NA | NA | 0 |
| S3_extracted_k141_49680 | 1447 | No | NA | 2 | 0 | 1 | Not-determined | Genome-fragment | NA | NA | 0 |
| S7_vs_50853\|\|full | 1572 | No | NA | 3 | 1 | 0 | Low-quality | Genome-fragment | 3.55 | AAI-based (high-confidence) | 0 |
| S3_extracted_k141_57398 | 1082 | No | NA | 1 | 0 | 1 | Not-determined | Genome-fragment | NA | NA | 0 |
| S3_extracted_k141_120690 | 1337 | No | NA | 1 | 0 | 1 | Not-determined | Genome-fragment | NA | NA | 0 |
| S3_extracted_k141_25163 | 103478 | No | NA | 94 | 16 | 1 | Complete | High-quality | 100 | DTR (high-confidence) | 0 |
| S3_extracted_k141_25170 | 1434 | No | NA | 2 | 0 | 0 | Not-determined | Genome-fragment | NA | NA | 0 |
| S12_vs_k141_132413\|\|full | 1572 | No | NA | 2 | 0 | 0 | Low-quality | Genome-fragment | 3.17 | AAI-based (medium-confidence) | 0 |
| S3_extracted_k141_57422 | 15991 | No | NA | 21 | 8 | 0 | Medium-quality | Genome-fragment | 70.78 | AAI-based (medium-confidence) | 0 |
| S3_extracted_k141_73779 | 3335 | No | NA | 1 | 0 | 0 | Not-determined | Genome-fragment | NA | NA | 0 |
| S3_extracted_k141_49708 | 1243 | No | NA | 2 | 0 | 0 | Not-determined | Genome-fragment | NA | NA | 0 |
| S3_extracted_k141_73784 | 1008 | No | NA | 3 | 0 | 0 | Not-determined | Genome-fragment | NA | NA | 0 |
| S3_extracted_k141_1593 | 1003 | No | NA | 1 | 0 | 1 | Not-determined | Genome-fragment | NA | NA | 0 |
| S3_extracted_k141_97354 | 1248 | No | NA | 2 | 0 | 1 | Not-determined | Genome-fragment | NA | NA | 0 |
| S4_extracted_6598 | 1572 | No | NA | 4 | 4 | 0 | Low-quality | Genome-fragment | 5.19 | AAI-based (high-confidence) | 0 |
| S17_extracted_k141_66881 | 1573 | No | NA | 1 | 0 | 0 | Low-quality | Genome-fragment | 0.29 | AAI-based (medium-confidence) | 0 |
| S3_extracted_k141_73855 | 2670 | No | NA | 1 | 0 | 0 | Not-determined | Genome-fragment | NA | NA | 0 |
| S11_vs_k141_22322\|\|full | 1573 | No | NA | 3 | 1 | 0 | Low-quality | Genome-fragment | 3.59 | AAI-based (high-confidence) | 0 |
| S13_vs_9868\|\|full | 1573 | No | NA | 4 | 0 | 1 | Low-quality | Genome-fragment | 1.66 | AAI-based (medium-confidence) | 0 |
| S3_extracted_k141_47390 | 1573 | No | NA | 2 | 0 | 0 | Low-quality | Genome-fragment | 2.97 | AAI-based (medium-confidence) | 0 |
| S7_vs_11752\|\|full | 1573 | No | NA | 2 | 1 | 0 | Low-quality | Genome-fragment | 4.29 | AAI-based (high-confidence) | 0 |
| S3_extracted_k141_17230 | 7606 | No | NA | 6 | 0 | 3 | Not-determined | Genome-fragment | NA | NA | 0 |
| S3_extracted_k141_73893 | 1427 | No | NA | 1 | 0 | 1 | Not-determined | Genome-fragment | NA | NA | 0 |
| S7_vs_90253\|\|full | 1573 | No | NA | 5 | 2 | 0 | Low-quality | Genome-fragment | 1.93 | HMM-based (lower-bound) | 0 |
| S3_extracted_k141_112984 | 1158 | No | NA | 1 | 0 | 0 | Not-determined | Genome-fragment | NA | NA | 0 |
| S3_extracted_k141_57481 | 1168 | No | NA | 3 | 0 | 0 | Not-determined | Genome-fragment | NA | NA | 0 |
| S10_vs_18570\|\|full | 1573 | No | NA | 3 | 0 | 0 | Low-quality | Genome-fragment | 3.23 | AAI-based (medium-confidence) | 0 |
| S17_vs_k141_126823\|\|full | 1574 | No | NA | 2 | 0 | 0 | Low-quality | Genome-fragment | 3.03 | AAI-based (medium-confidence) | 0 |
| S3_extracted_k141_120799 | 1014 | No | NA | 1 | 0 | 0 | Not-determined | Genome-fragment | NA | NA | 0 |
| S3_extracted_k141_120817 | 1485 | No | NA | 3 | 0 | 2 | Not-determined | Genome-fragment | NA | NA | 0 |
| S3_extracted_k141_10258 | 1640 | No | NA | 3 | 0 | 0 | Not-determined | Genome-fragment | NA | NA | 0 |
| S3_extracted_k141_17280 | 1194 | No | NA | 1 | 0 | 1 | Not-determined | Genome-fragment | NA | NA | 0 |
| S3_extracted_k141_89209 | 4682 | No | NA | 3 | 0 | 1 | Not-determined | Genome-fragment | NA | NA | 0 |
| S9_vs_k141_74183\|\|lt2gene | 1574 | No | NA | 2 | 2 | 0 | Low-quality | Genome-fragment | 4.14 | AAI-based (high-confidence) | 0 |
| S7_extracted_118389 | 1574 | No | NA | 3 | 0 | 0 | Low-quality | Genome-fragment | 1.87 | AAI-based (high-confidence) | 0 |
| S16_vs_78756\|\|full | 1574 | No | NA | 5 | 2 | 0 | Low-quality | Genome-fragment | 3.74 | AAI-based (medium-confidence) | 0 |
| S3_extracted_k141_41508 | 3496 | No | NA | 9 | 0 | 9 | Not-determined | Genome-fragment | NA | NA | 0 |
| S3_extracted_k141_17284 | 2016 | No | NA | 2 | 0 | 0 | Not-determined | Genome-fragment | NA | NA | 0 |
| S3_extracted_k141_74010 | 1174 | No | NA | 2 | 0 | 2 | Not-determined | Genome-fragment | NA | NA | 0 |
| S12_vs_k141_142412\|\|full | 1574 | No | NA | 2 | 0 | 1 | Low-quality | Genome-fragment | 1.83 | AAI-based (high-confidence) | 0 |
| S1_vs_k141_126860\|\|full | 1575 | No | NA | 5 | 0 | 0 | Low-quality | Genome-fragment | 1.55 | AAI-based (medium-confidence) | 0 |
| S7_extracted_99362 | 1575 | No | NA | 3 | 0 | 3 | Low-quality | Genome-fragment | 4.44 | AAI-based (medium-confidence) | 0 |
| S16_extracted_140584 | 1575 | No | NA | 3 | 0 | 0 | Low-quality | Genome-fragment | 1.89 | AAI-based (high-confidence) | 0 |
| S10_extracted_31625 | 1575 | No | NA | 1 | 1 | 0 | Low-quality | Genome-fragment | 24.91 | AAI-based (high-confidence) | 0 |
| S10_vs_51880\|\|full | 1575 | No | NA | 3 | 0 | 1 | Low-quality | Genome-fragment | 0.84 | AAI-based (medium-confidence) | 0 |
| S8_vs_k141_154183\|\|lt2gene | 1575 | No | NA | 1 | 1 | 0 | Low-quality | Genome-fragment | 1.48 | AAI-based (high-confidence) | 0 |
| S3_extracted_k141_49904 | 1261 | No | NA | 1 | 0 | 0 | Not-determined | Genome-fragment | NA | NA | 0 |
| S3_extracted_k141_81538 | 5357 | No | NA | 3 | 0 | 2 | Not-determined | Genome-fragment | NA | NA | 0 |
| S3_extracted_k141_105014 | 2855 | No | NA | 5 | 0 | 0 | Not-determined | Genome-fragment | NA | NA | 0 |
| S3_extracted_k141_113154 | 1341 | No | NA | 2 | 0 | 1 | Not-determined | Genome-fragment | NA | NA | 0 |
| S11_extracted_k141_78694 | 1576 | No | NA | 4 | 0 | 0 | Low-quality | Genome-fragment | 2.35 | AAI-based (high-confidence) | 0 |
| S3_extracted_k141_1751 | 3119 | No | NA | 4 | 0 | 0 | Not-determined | Genome-fragment | NA | NA | 0 |
| S3_extracted_k141_41590 | 1124 | No | NA | 1 | 0 | 0 | Not-determined | Genome-fragment | NA | NA | 0 |
| S3_extracted_k141_81579 | 2219 | No | NA | 4 | 0 | 0 | Not-determined | Genome-fragment | NA | NA | 0 |
| S3_extracted_k141_57677 | 11276 | No | NA | 11 | 0 | 7 | Not-determined | Genome-fragment | NA | NA | 0 |
| S3_extracted_k141_17399 | 1120 | No | NA | 2 | 0 | 1 | Not-determined | Genome-fragment | NA | NA | 0 |
| S13_vs_26034\|\|full | 1576 | No | NA | 3 | 0 | 0 | Low-quality | Genome-fragment | 3.5 | AAI-based (medium-confidence) | 0 |
| S14_vs_k141_111784\|\|full | 1576 | No | NA | 4 | 1 | 0 | Low-quality | Genome-fragment | 3.12 | AAI-based (high-confidence) | 0 |
| S14_vs_k141_80302\|\|full | 1576 | No | NA | 3 | 1 | 0 | Low-quality | Genome-fragment | 3.83 | AAI-based (high-confidence) | 0 |
| S5_vs_k141_94805\|\|full | 1576 | No | NA | 3 | 0 | 1 | Low-quality | Genome-fragment | 0.84 | AAI-based (high-confidence) | 0 |
| S7_extracted_16045 | 1576 | No | NA | 3 | 0 | 1 | Low-quality | Genome-fragment | 1.83 | AAI-based (medium-confidence) | 0 |
| S3_extracted_k141_105111 | 1230 | No | NA | 1 | 0 | 0 | Not-determined | Genome-fragment | NA | NA | 0 |
| S10_vs_96780\|\|full | 1576 | No | NA | 5 | 0 | 0 | Low-quality | Genome-fragment | 3.75 | AAI-based (high-confidence) | 0 |
| S11_vs_k141_26421\|\|full | 1577 | No | NA | 2 | 0 | 0 | Low-quality | Genome-fragment | 3.94 | AAI-based (high-confidence) | 0 |
| S3_extracted_k141_105131 | 6126 | No | NA | 5 | 0 | 3 | Not-determined | Genome-fragment | NA | NA | 0 |
| S3_extracted_k141_50005 | 3826 | No | NA | 4 | 0 | 4 | Not-determined | Genome-fragment | NA | NA | 0 |
| S3_extracted_k141_25494 | 1018 | No | NA | 3 | 0 | 3 | Not-determined | Genome-fragment | NA | NA | 0 |
| S3_extracted_k141_113254 | 1232 | No | NA | 3 | 0 | 0 | Not-determined | Genome-fragment | NA | NA | 0 |
| S3_extracted_k141_1850 | 3856 | No | NA | 2 | 0 | 2 | Not-determined | Genome-fragment | NA | NA | 0 |
| S11_vs_k141_3750\|\|full | 1577 | No | NA | 4 | 0 | 0 | Low-quality | Genome-fragment | 2.72 | AAI-based (medium-confidence) | 0 |
| S3_extracted_k141_1868 | 1027 | No | NA | 2 | 0 | 1 | Not-determined | Genome-fragment | NA | NA | 0 |
| S3_extracted_k141_105168 | 1331 | No | NA | 2 | 0 | 1 | Not-determined | Genome-fragment | NA | NA | 0 |
| S3_extracted_k141_41703 | 1333 | No | NA | 2 | 0 | 1 | Not-determined | Genome-fragment | NA | NA | 0 |
| S14_extracted_k141_22950 | 1577 | No | NA | 6 | 2 | 0 | Low-quality | Genome-fragment | 4.55 | AAI-based (high-confidence) | 0 |
| S3_extracted_k141_120985 | 3233 | No | NA | 1 | 0 | 0 | Not-determined | Genome-fragment | NA | NA | 0 |
| S3_extracted_k141_105181 | 2174 | No | NA | 1 | 0 | 1 | Not-determined | Genome-fragment | NA | NA | 0 |
| S3_extracted_k141_113276 | 1032 | No | NA | 2 | 0 | 2 | Not-determined | Genome-fragment | NA | NA | 0 |
| S3_extracted_k141_25513 | 1786 | No | NA | 2 | 0 | 1 | Not-determined | Genome-fragment | NA | NA | 0 |
| S3_extracted_k141_33903 | 4718 | No | NA | 3 | 0 | 3 | Not-determined | Genome-fragment | NA | NA | 0 |
| S1_extracted_k141_100237 | 1577 | No | NA | 1 | 0 | 1 | Low-quality | Genome-fragment | 0.49 | AAI-based (medium-confidence) | 0 |
| S3_extracted_k141_105869 | 1577 | No | NA | 2 | 0 | 1 | Low-quality | Genome-fragment | 0.97 | AAI-based (high-confidence) | 0 |
| S3_extracted_k141_57780 | 2820 | No | NA | 1 | 0 | 0 | Not-determined | Genome-fragment | NA | NA | 0 |
| S3_extracted_k141_10539 | 8831 | No | NA | 4 | 0 | 2 | Not-determined | Genome-fragment | NA | NA | 0 |
| S3_vs_k141_77469\|\|full | 1577 | No | NA | 3 | 0 | 1 | Low-quality | Genome-fragment | 1.18 | AAI-based (medium-confidence) | 0 |
| S3_extracted_k141_41725 | 3297 | No | NA | 4 | 0 | 1 | Not-determined | Genome-fragment | NA | NA | 0 |
| S7_vs_45166\|\|full | 1577 | No | NA | 2 | 0 | 0 | Low-quality | Genome-fragment | 0.99 | AAI-based (medium-confidence) | 0 |
| S3_extracted_k141_25557 | 1688 | No | NA | 5 | 0 | 0 | Not-determined | Genome-fragment | NA | NA | 0 |
| S3_extracted_k141_57795 | 4432 | No | NA | 4 | 0 | 2 | Not-determined | Genome-fragment | NA | NA | 0 |
| S3_extracted_k141_57797 | 1234 | No | NA | 2 | 0 | 1 | Not-determined | Genome-fragment | NA | NA | 0 |
| S16_vs_64603\|\|full | 1577 | No | NA | 3 | 0 | 0 | Low-quality | Genome-fragment | 4.29 | AAI-based (high-confidence) | 0 |
| S15_extracted_73591 | 1577 | No | NA | 1 | 0 | 1 | Low-quality | Genome-fragment | 0.27 | AAI-based (medium-confidence) | 0 |
| S15_vs_76400\|\|full | 1577 | No | NA | 2 | 2 | 0 | Low-quality | Genome-fragment | 3.6 | AAI-based (medium-confidence) | 0 |
| S3_extracted_k141_74099 | 1058 | No | NA | 2 | 0 | 2 | Not-determined | Genome-fragment | NA | NA | 0 |
| S3_extracted_k141_25593 | 1593 | No | NA | 1 | 0 | 1 | Not-determined | Genome-fragment | NA | NA | 0 |
| S3_extracted_k141_81697 | 1092 | No | NA | 1 | 0 | 0 | Not-determined | Genome-fragment | NA | NA | 0 |
| S3_extracted_k141_34005 | 4998 | No | NA | 6 | 0 | 5 | Not-determined | Genome-fragment | NA | NA | 0 |
| S3_extracted_k141_34009 | 3154 | No | NA | 3 | 0 | 0 | Not-determined | Genome-fragment | NA | NA | 0 |
| S3_extracted_k141_105305 | 6712 | No | NA | 6 | 0 | 2 | Not-determined | Genome-fragment | NA | NA | 0 |
| S8_vs_k141_134720\|\|full | 1577 | No | NA | 6 | 0 | 0 | Low-quality | Genome-fragment | 3.68 | AAI-based (high-confidence) | 0 |
| S3_extracted_k141_25616 | 2531 | No | NA | 4 | 0 | 3 | Not-determined | Genome-fragment | NA | NA | 0 |
| S11_extracted_k141_78336 | 1578 | No | NA | 3 | 1 | 0 | Low-quality | Genome-fragment | 2.62 | AAI-based (high-confidence) | 0 |
| S3_extracted_k141_81720 | 1333 | No | NA | 2 | 0 | 1 | Not-determined | Genome-fragment | NA | NA | 0 |
| S3_extracted_k141_17487 | 2711 | No | NA | 6 | 0 | 4 | Not-determined | Genome-fragment | NA | NA | 0 |
| S3_extracted_k141_113441 | 8879 | No | NA | 4 | 0 | 2 | Not-determined | Genome-fragment | NA | NA | 0 |
| S13_extracted_133210 | 1578 | No | NA | 2 | 1 | 0 | Low-quality | Genome-fragment | 1.73 | AAI-based (high-confidence) | 0 |
| S1_extracted_k141_153122 | 1578 | No | NA | 2 | 1 | 0 | Low-quality | Genome-fragment | 5.35 | HMM-based (lower-bound) | 0 |
| S3_extracted_k141_121169 | 1405 | No | NA | 2 | 0 | 1 | Not-determined | Genome-fragment | NA | NA | 0 |
| S3_extracted_k141_65384 | 1035 | No | NA | 1 | 0 | 1 | Not-determined | Genome-fragment | NA | NA | 0 |
| S3_extracted_k141_34082 | 1540 | No | NA | 2 | 0 | 1 | Not-determined | Genome-fragment | NA | NA | 0 |
| S3_vs_k141_91974\|\|full | 1578 | No | NA | 4 | 0 | 0 | Low-quality | Genome-fragment | 2.78 | AAI-based (high-confidence) | 0 |
| S5_vs_k141_49727\|\|full | 1578 | No | NA | 5 | 2 | 0 | Low-quality | Genome-fragment | 2.42 | HMM-based (lower-bound) | 0 |
| S7_vs_75153\|\|full | 1578 | No | NA | 3 | 0 | 0 | Low-quality | Genome-fragment | 3.92 | AAI-based (high-confidence) | 0 |
| S3_extracted_k141_97659 | 5869 | No | NA | 3 | 0 | 3 | Not-determined | Genome-fragment | NA | NA | 0 |
| S10_vs_64349\|\|full | 1578 | No | NA | 4 | 1 | 0 | Low-quality | Genome-fragment | 2.77 | AAI-based (medium-confidence) | 0 |
| S12_extracted_k141_124205 | 1578 | No | NA | 5 | 2 | 0 | Low-quality | Genome-fragment | 2.49 | AAI-based (high-confidence) | 0 |
| S3_extracted_k141_89604 | 1996 | No | NA | 3 | 0 | 1 | Not-determined | Genome-fragment | NA | NA | 0 |
| S6_vs_k141_18315\|\|full | 1578 | No | NA | 2 | 0 | 1 | Low-quality | Genome-fragment | 0.72 | AAI-based (medium-confidence) | 0 |
| S8_vs_k141_58560\|\|full | 1578 | Yes | 632 | 3 | 1 | 1 | Low-quality | Genome-fragment | 1.11 | HMM-based (lower-bound) | 59.95 |
| S8_vs_k141_112563\|\|full | 1578 | No | NA | 5 | 0 | 0 | Low-quality | Genome-fragment | 3.07 | AAI-based (high-confidence) | 0 |
| S3_extracted_k141_57903 | 1428 | No | NA | 2 | 0 | 2 | Not-determined | Genome-fragment | NA | NA | 0 |
| S9_extracted_k141_38325 | 1579 | No | NA | 3 | 2 | 0 | Low-quality | Genome-fragment | 3.84 | AAI-based (high-confidence) | 0 |
| S3_extracted_k141_17559 | 1984 | No | NA | 1 | 0 | 0 | Not-determined | Genome-fragment | NA | NA | 0 |
| S13_extracted_5074 | 1579 | No | NA | 3 | 0 | 0 | Low-quality | Genome-fragment | 2.87 | AAI-based (high-confidence) | 0 |
| S13_vs_48542\|\|full | 1579 | No | NA | 5 | 0 | 0 | Low-quality | Genome-fragment | 1.77 | AAI-based (high-confidence) | 0 |
| S14_vs_k141_103571\|\|full | 1579 | No | NA | 4 | 2 | 0 | Low-quality | Genome-fragment | 3.68 | AAI-based (high-confidence) | 0 |
| S3_extracted_k141_34249 | 4827 | No | NA | 3 | 0 | 0 | Not-determined | Genome-fragment | NA | NA | 0 |
| S3_extracted_k141_121318 | 2013 | No | NA | 1 | 0 | 1 | Not-determined | Genome-fragment | NA | NA | 0 |
| S3_extracted_k141_34265 | 1885 | No | NA | 2 | 0 | 2 | Not-determined | Genome-fragment | NA | NA | 0 |
| S5_vs_k141_113514\|\|full | 1579 | No | NA | 2 | 1 | 0 | Low-quality | Genome-fragment | 3.97 | AAI-based (high-confidence) | 0 |
| S3_extracted_k141_17593 | 1000 | No | NA | 1 | 0 | 1 | Not-determined | Genome-fragment | NA | NA | 0 |
| S3_extracted_k141_17594 | 3747 | No | NA | 8 | 0 | 7 | Not-determined | Genome-fragment | NA | NA | 0 |
| S3_extracted_k141_57952 | 1417 | No | NA | 2 | 0 | 2 | Not-determined | Genome-fragment | NA | NA | 0 |
| S3_extracted_k141_17633 | 1359 | No | NA | 2 | 0 | 0 | Not-determined | Genome-fragment | NA | NA | 0 |
| S3_extracted_k141_65555 | 2342 | No | NA | 2 | 0 | 1 | Not-determined | Genome-fragment | NA | NA | 0 |
| S3_extracted_k141_10970 | 1094 | No | NA | 2 | 0 | 0 | Not-determined | Genome-fragment | NA | NA | 0 |
| S7_vs_75148\|\|full | 1579 | No | NA | 3 | 0 | 0 | Low-quality | Genome-fragment | 2.39 | AAI-based (high-confidence) | 0 |
| S3_extracted_k141_105472 | 14209 | No | NA | 18 | 0 | 3 | Not-determined | Genome-fragment | NA | NA | 0 |
| S12_extracted_k141_186906 | 1579 | No | NA | 3 | 0 | 2 | Low-quality | Genome-fragment | 0.69 | AAI-based (medium-confidence) | 0 |
| S3_extracted_k141_97723 | 3440 | No | NA | 5 | 0 | 2 | Not-determined | Genome-fragment | NA | NA | 0 |
| S12_vs_k141_112785\|\|full | 1579 | No | NA | 5 | 1 | 0 | Low-quality | Genome-fragment | 3.07 | AAI-based (medium-confidence) | 0 |
| S3_extracted_k141_121378 | 7330 | No | NA | 10 | 0 | 2 | Not-determined | Genome-fragment | NA | NA | 0 |
| S3_extracted_k141_34339 | 1090 | No | NA | 2 | 0 | 1 | Not-determined | Genome-fragment | NA | NA | 0 |
| S12_vs_k141_160231\|\|full | 1579 | No | NA | 2 | 0 | 1 | Low-quality | Genome-fragment | 1.1 | AAI-based (medium-confidence) | 0 |
| S3_extracted_k141_113607 | 1043 | No | NA | 2 | 0 | 0 | Not-determined | Genome-fragment | NA | NA | 0 |
| S3_extracted_k141_65592 | 1745 | No | NA | 2 | 0 | 0 | Not-determined | Genome-fragment | NA | NA | 0 |
| S11_extracted_k141_4768 | 1580 | No | NA | 3 | 2 | 0 | Low-quality | Genome-fragment | 3.73 | AAI-based (high-confidence) | 0 |
| S3_extracted_k141_89686 | 1507 | No | NA | 3 | 0 | 2 | Not-determined | Genome-fragment | NA | NA | 0 |
| S3_extracted_k141_121400 | 5972 | No | NA | 9 | 0 | 7 | Not-determined | Genome-fragment | NA | NA | 0 |
| S1_vs_k141_34995\|\|full | 1580 | No | NA | 4 | 1 | 0 | Low-quality | Genome-fragment | 2.73 | AAI-based (medium-confidence) | 0 |
| S5_extracted_k141_103482 | 1580 | No | NA | 2 | 2 | 0 | Low-quality | Genome-fragment | 4.08 | AAI-based (high-confidence) | 0 |
| S3_extracted_k141_105531 | 5403 | No | NA | 3 | 0 | 3 | Not-determined | Genome-fragment | NA | NA | 0 |
| S16_vs_79584\|\|lt2gene | 1580 | No | NA | 2 | 2 | 0 | Low-quality | Genome-fragment | 4.6 | AAI-based (high-confidence) | 0 |
| S16_vs_79860\|\|lt2gene | 1580 | No | NA | 2 | 2 | 0 | Low-quality | Genome-fragment | 4.6 | AAI-based (high-confidence) | 0 |
| S12_extracted_k141_134134 | 1580 | No | NA | 3 | 2 | 0 | Low-quality | Genome-fragment | 4.15 | AAI-based (medium-confidence) | 0 |
| S3_extracted_k141_11040 | 1100 | No | NA | 1 | 0 | 0 | Not-determined | Genome-fragment | NA | NA | 0 |
| S3_extracted_k141_50225 | 4181 | No | NA | 6 | 0 | 3 | Not-determined | Genome-fragment | NA | NA | 0 |
| S3_extracted_k141_50229 | 1805 | No | NA | 3 | 0 | 2 | Not-determined | Genome-fragment | NA | NA | 0 |
| S3_extracted_k141_11057 | 1165 | No | NA | 1 | 0 | 0 | Not-determined | Genome-fragment | NA | NA | 0 |
| S12_extracted_k141_166810 | 1580 | No | NA | 1 | 0 | 1 | Low-quality | Genome-fragment | 0.92 | AAI-based (medium-confidence) | 0 |
| S3_extracted_k141_113688 | 2346 | No | NA | 1 | 0 | 1 | Not-determined | Genome-fragment | NA | NA | 0 |
| S3_extracted_k141_65701 | 1078 | No | NA | 1 | 0 | 1 | Not-determined | Genome-fragment | NA | NA | 0 |
| S8_extracted_k141_200404 | 1580 | No | NA | 2 | 0 | 1 | Low-quality | Genome-fragment | 0.47 | AAI-based (medium-confidence) | 0 |
| S3_extracted_k141_58115 | 1144 | No | NA | 1 | 0 | 1 | Not-determined | Genome-fragment | NA | NA | 0 |
| S3_extracted_k141_42068 | 1028 | No | NA | 1 | 0 | 0 | Not-determined | Genome-fragment | NA | NA | 0 |
| S8_vs_k141_78583\|\|full | 1580 | No | NA | 6 | 1 | 0 | Low-quality | Genome-fragment | 3.92 | AAI-based (medium-confidence) | 0 |
| S17_vs_k141_158544\|\|full | 1581 | No | NA | 3 | 0 | 0 | Low-quality | Genome-fragment | 3.87 | AAI-based (high-confidence) | 0 |
| S3_extracted_k141_113743 | 1021 | No | NA | 1 | 0 | 0 | Not-determined | Genome-fragment | NA | NA | 0 |
| S11_vs_k141_67659\|\|full | 1581 | No | NA | 4 | 0 | 0 | Low-quality | Genome-fragment | 6.7 | AAI-based (high-confidence) | 0 |
| S13_vs_40355\|\|full | 1581 | No | NA | 3 | 0 | 0 | Low-quality | Genome-fragment | 1.88 | AAI-based (medium-confidence) | 0 |
| S14_extracted_k141_66403 | 1581 | No | NA | 4 | 3 | 0 | Low-quality | Genome-fragment | 5.21 | AAI-based (high-confidence) | 0 |
| S3_extracted_k141_81824 | 3111 | No | NA | 1 | 0 | 0 | Not-determined | Genome-fragment | NA | NA | 0 |
| S3_extracted_k141_80649 | 1581 | No | NA | 5 | 0 | 0 | Low-quality | Genome-fragment | 7.21 | AAI-based (high-confidence) | 0 |
| S3_extracted_k141_11156 | 1691 | No | NA | 3 | 0 | 1 | Not-determined | Genome-fragment | NA | NA | 0 |
| S7_vs_123556\|\|full | 1581 | No | NA | 5 | 0 | 0 | Low-quality | Genome-fragment | 3.15 | AAI-based (high-confidence) | 0 |
| S16_extracted_69261 | 1581 | No | NA | 2 | 0 | 0 | Low-quality | Genome-fragment | 3.68 | AAI-based (medium-confidence) | 0 |
| S3_extracted_k141_50256 | 1633 | No | NA | 2 | 0 | 0 | Not-determined | Genome-fragment | NA | NA | 0 |
| S3_extracted_k141_65744 | 2795 | No | NA | 2 | 0 | 0 | Not-determined | Genome-fragment | NA | NA | 0 |
| S14_extracted_k141_109960 | 1582 | No | NA | 3 | 2 | 0 | Low-quality | Genome-fragment | 5.22 | AAI-based (high-confidence) | 0 |
| S14_extracted_k141_15175 | 1582 | No | NA | 2 | 2 | 0 | Low-quality | Genome-fragment | 3.48 | AAI-based (high-confidence) | 0 |
| S3_extracted_k141_25934 | 1657 | No | NA | 2 | 0 | 2 | Not-determined | Genome-fragment | NA | NA | 0 |
| S3_extracted_k141_42108 | 5897 | No | NA | 4 | 0 | 1 | Not-determined | Genome-fragment | NA | NA | 0 |
| S3_extracted_k141_50268 | 7115 | No | NA | 7 | 0 | 4 | Not-determined | Genome-fragment | NA | NA | 0 |
| S3_extracted_k141_11191 | 1005 | No | NA | 2 | 0 | 0 | Not-determined | Genome-fragment | NA | NA | 0 |
| S3_vs_k141_95113\|\|full | 1582 | No | NA | 5 | 0 | 0 | Low-quality | Genome-fragment | 3.99 | AAI-based (medium-confidence) | 0 |
| S12_vs_k141_40043\|\|full | 1582 | No | NA | 2 | 0 | 2 | Low-quality | Genome-fragment | 3.29 | AAI-based (medium-confidence) | 0 |
| S3_extracted_k141_81868 | 1186 | No | NA | 1 | 0 | 1 | Not-determined | Genome-fragment | NA | NA | 0 |
| S3_extracted_k141_65799 | 3333 | No | NA | 3 | 0 | 2 | Not-determined | Genome-fragment | NA | NA | 0 |
| S3_extracted_k141_121460 | 1866 | No | NA | 3 | 0 | 0 | Not-determined | Genome-fragment | NA | NA | 0 |
| S12_vs_k141_19971\|\|full | 1582 | No | NA | 2 | 1 | 0 | Low-quality | Genome-fragment | 1.88 | HMM-based (lower-bound) | 0 |
| S3_extracted_k141_50328 | 1204 | No | NA | 2 | 0 | 2 | Not-determined | Genome-fragment | NA | NA | 0 |
| S3_extracted_k141_58218 | 4514 | No | NA | 3 | 0 | 3 | Not-determined | Genome-fragment | NA | NA | 0 |
| S6_extracted_k141_26003 | 1582 | No | NA | 2 | 0 | 0 | Low-quality | Genome-fragment | 2.86 | AAI-based (high-confidence) | 0 |
| S3_extracted_k141_65838 | 1730 | No | NA | 1 | 0 | 0 | Not-determined | Genome-fragment | NA | NA | 0 |
| S3_extracted_k141_11295 | 3831 | No | NA | 7 | 0 | 1 | Not-determined | Genome-fragment | NA | NA | 0 |
| S3_extracted_k141_34479 | 1287 | No | NA | 1 | 0 | 0 | Not-determined | Genome-fragment | NA | NA | 0 |
| S8_extracted_k141_56860 | 1582 | No | NA | 5 | 2 | 0 | Low-quality | Genome-fragment | 4.77 | AAI-based (high-confidence) | 0 |
| S9_vs_k141_74607\|\|full | 1583 | No | NA | 4 | 2 | 0 | Low-quality | Genome-fragment | 3.92 | AAI-based (high-confidence) | 0 |
| S13_vs_111027\|\|lt2gene | 1583 | No | NA | 2 | 1 | 0 | Low-quality | Genome-fragment | 4.13 | AAI-based (high-confidence) | 0 |
| S14_vs_k141_31088\|\|full | 1583 | No | NA | 4 | 1 | 0 | Low-quality | Genome-fragment | 3.47 | AAI-based (high-confidence) | 0 |
| S3_extracted_k141_17748 | 96667 | No | NA | 94 | 12 | 0 | Complete | High-quality | 100 | DTR (high-confidence) | 0 |
| S3_extracted_k141_113788 | 5231 | No | NA | 3 | 0 | 2 | Not-determined | Genome-fragment | NA | NA | 0 |
| S1_vs_k141_1837\|\|lt2gene | 1583 | No | NA | 2 | 1 | 0 | Low-quality | Genome-fragment | 3.27 | AAI-based (high-confidence) | 0 |
| S3_extracted_k141_17751 | 2472 | No | NA | 3 | 0 | 2 | Not-determined | Genome-fragment | NA | NA | 0 |
| S3_extracted_k141_2186 | 55789 | No | NA | 61 | 6 | 0 | Medium-quality | Genome-fragment | 77.64 | AAI-based (high-confidence) | 0 |
| S3_extracted_k141_89872 | 2686 | No | NA | 2 | 0 | 0 | Not-determined | Genome-fragment | NA | NA | 0 |
| S3_extracted_k141_50468 | 3198 | No | NA | 4 | 0 | 1 | Not-determined | Genome-fragment | NA | NA | 0 |
| S3_extracted_k141_121579 | 1395 | No | NA | 2 | 0 | 0 | Not-determined | Genome-fragment | NA | NA | 0 |
| S3_extracted_k141_74484 | 3196 | No | NA | 2 | 0 | 1 | Not-determined | Genome-fragment | NA | NA | 0 |
| S3_extracted_k141_113819 | 2649 | No | NA | 3 | 0 | 1 | Not-determined | Genome-fragment | NA | NA | 0 |
| S3_extracted_k141_65930 | 1385 | No | NA | 1 | 0 | 0 | Not-determined | Genome-fragment | NA | NA | 0 |
| S3_extracted_k141_121593 | 1035 | No | NA | 2 | 0 | 1 | Not-determined | Genome-fragment | NA | NA | 0 |
| S3_vs_k141_2878\|\|full | 1583 | No | NA | 3 | 1 | 0 | Low-quality | Genome-fragment | 2.35 | HMM-based (lower-bound) | 0 |
| S7_vs_74395\|\|full | 1583 | No | NA | 5 | 0 | 0 | Low-quality | Genome-fragment | 3.64 | AAI-based (high-confidence) | 0 |
| S3_extracted_k141_26015 | 5582 | No | NA | 8 | 0 | 3 | Not-determined | Genome-fragment | NA | NA | 0 |
| S3_extracted_k141_58272 | 1305 | No | NA | 3 | 0 | 3 | Not-determined | Genome-fragment | NA | NA | 0 |
| S16_extracted_120447 | 1583 | No | NA | 3 | 0 | 2 | Low-quality | Genome-fragment | 0.98 | AAI-based (high-confidence) | 0 |
| S16_vs_48478\|\|full | 1583 | No | NA | 2 | 1 | 0 | Low-quality | Genome-fragment | 3.59 | AAI-based (high-confidence) | 0 |
| S3_extracted_k141_11428 | 45436 | No | NA | 76 | 16 | 1 | High-quality | High-quality | 100 | AAI-based (high-confidence) | 0 |
| S16_vs_119362\|\|full | 1583 | No | NA | 2 | 0 | 2 | Low-quality | Genome-fragment | 2.65 | AAI-based (high-confidence) | 0 |
| S3_extracted_k141_81915 | 4764 | No | NA | 5 | 0 | 1 | Not-determined | Genome-fragment | NA | NA | 0 |
| S12_vs_k141_8187\|\|full | 1583 | No | NA | 2 | 0 | 1 | Low-quality | Genome-fragment | 0.42 | AAI-based (medium-confidence) | 0 |
| S3_extracted_k141_81935 | 1743 | No | NA | 2 | 0 | 1 | Not-determined | Genome-fragment | NA | NA | 0 |
| S3_extracted_k141_17786 | 1031 | No | NA | 1 | 0 | 1 | Not-determined | Genome-fragment | NA | NA | 0 |
| S3_extracted_k141_42234 | 1200 | No | NA | 1 | 0 | 0 | Not-determined | Genome-fragment | NA | NA | 0 |
| S3_extracted_k141_42245 | 2622 | No | NA | 3 | 0 | 0 | Not-determined | Genome-fragment | NA | NA | 0 |
| S17_extracted_k141_165546 | 1584 | No | NA | 4 | 0 | 0 | Low-quality | Genome-fragment | 2.01 | AAI-based (high-confidence) | 0 |
| S17_vs_k141_142256\|\|full | 1584 | No | NA | 4 | 2 | 0 | Low-quality | Genome-fragment | 2.44 | AAI-based (high-confidence) | 0 |
| S3_extracted_k141_97935 | 1661 | No | NA | 4 | 0 | 0 | Not-determined | Genome-fragment | NA | NA | 0 |
| S3_extracted_k141_74580 | 1039 | No | NA | 1 | 0 | 0 | Not-determined | Genome-fragment | NA | NA | 0 |
| S3_extracted_k141_26095 | 1629 | No | NA | 2 | 0 | 2 | Not-determined | Genome-fragment | NA | NA | 0 |
| S9_vs_k141_55122\|\|full | 1584 | No | NA | 3 | 2 | 0 | Low-quality | Genome-fragment | 3.28 | AAI-based (high-confidence) | 0 |
| S14_vs_k141_27716\|\|full | 1584 | No | NA | 3 | 0 | 0 | Low-quality | Genome-fragment | 2.37 | AAI-based (high-confidence) | 0 |
| S3_extracted_k141_105924 | 3616 | No | NA | 6 | 0 | 0 | Not-determined | Genome-fragment | NA | NA | 0 |
| S3_extracted_k141_42303 | 3515 | No | NA | 4 | 0 | 1 | Not-determined | Genome-fragment | NA | NA | 0 |
| S3_extracted_k141_42307 | 1856 | No | NA | 3 | 0 | 1 | Not-determined | Genome-fragment | NA | NA | 0 |
| S3_extracted_k141_105962 | 1712 | No | NA | 3 | 0 | 1 | Not-determined | Genome-fragment | NA | NA | 0 |
| S1_extracted_k141_22988 | 1584 | No | NA | 2 | 0 | 0 | Low-quality | Genome-fragment | 1.56 | AAI-based (high-confidence) | 0 |
| S1_vs_k141_131929\|\|full | 1584 | No | NA | 4 | 1 | 0 | Low-quality | Genome-fragment | 2.98 | AAI-based (high-confidence) | 0 |
| S3_extracted_k141_97973 | 3251 | No | NA | 6 | 0 | 5 | Not-determined | Genome-fragment | NA | NA | 0 |
| S3_extracted_k141_83202 | 1584 | No | NA | 4 | 1 | 0 | Low-quality | Genome-fragment | 3.26 | AAI-based (high-confidence) | 0 |
| S3_extracted_k141_2309 | 1539 | No | NA | 2 | 0 | 1 | Not-determined | Genome-fragment | NA | NA | 0 |
| S3_vs_k141_19104\|\|lt2gene | 1584 | No | NA | 2 | 2 | 0 | Low-quality | Genome-fragment | 1.07 | AAI-based (high-confidence) | 0 |
| S7_extracted_42607 | 1584 | No | NA | 2 | 0 | 2 | Low-quality | Genome-fragment | 0.91 | AAI-based (medium-confidence) | 0 |
| S3_extracted_k141_58381 | 1728 | No | NA | 1 | 0 | 0 | Not-determined | Genome-fragment | NA | NA | 0 |
| S7_extracted_118747 | 1584 | No | NA | 1 | 0 | 1 | Low-quality | Genome-fragment | 0.36 | AAI-based (medium-confidence) | 0 |
| S3_extracted_k141_74666 | 1070 | No | NA | 1 | 0 | 1 | Not-determined | Genome-fragment | NA | NA | 0 |
| S7_vs_22558\|\|full | 1584 | No | NA | 5 | 0 | 0 | Low-quality | Genome-fragment | 2.77 | AAI-based (high-confidence) | 0 |
| S3_extracted_k141_50608 | 5553 | No | NA | 5 | 0 | 5 | Not-determined | Genome-fragment | NA | NA | 0 |
| S3_extracted_k141_42430 | 1714 | No | NA | 2 | 0 | 0 | Not-determined | Genome-fragment | NA | NA | 0 |
| S3_extracted_k141_114029 | 2642 | No | NA | 1 | 0 | 0 | Not-determined | Genome-fragment | NA | NA | 0 |
| S3_extracted_k141_17848 | 1274 | No | NA | 1 | 0 | 0 | Not-determined | Genome-fragment | NA | NA | 0 |
| S16_vs_110371\|\|full | 1584 | No | NA | 2 | 0 | 1 | Low-quality | Genome-fragment | 2.01 | AAI-based (high-confidence) | 0 |
| S3_extracted_k141_11692 | 3129 | No | NA | 3 | 0 | 2 | Not-determined | Genome-fragment | NA | NA | 0 |
| S3_extracted_k141_74707 | 2689 | No | NA | 3 | 0 | 1 | Not-determined | Genome-fragment | NA | NA | 0 |
| S3_extracted_k141_17877 | 1720 | No | NA | 5 | 0 | 0 | Not-determined | Genome-fragment | NA | NA | 0 |
| S12_extracted_k141_54891 | 1584 | No | NA | 1 | 0 | 1 | Low-quality | Genome-fragment | 0.85 | AAI-based (medium-confidence) | 0 |
| S12_vs_k141_66576\|\|full | 1584 | No | NA | 5 | 1 | 0 | Low-quality | Genome-fragment | 2.53 | AAI-based (high-confidence) | 0 |
| S8_extracted_k141_154642 | 1584 | No | NA | 1 | 0 | 1 | Low-quality | Genome-fragment | 2.26 | AAI-based (medium-confidence) | 0 |
| S9_vs_k141_34769\|\|full | 1585 | No | NA | 3 | 2 | 0 | Low-quality | Genome-fragment | 1.94 | AAI-based (high-confidence) | 0 |
| S9_vs_k141_58390\|\|full | 1585 | No | NA | 4 | 1 | 0 | Low-quality | Genome-fragment | 1.02 | HMM-based (lower-bound) | 0 |
| S1_vs_k141_97950\|\|full | 1585 | No | NA | 4 | 1 | 0 | Low-quality | Genome-fragment | 2.76 | AAI-based (medium-confidence) | 0 |
| S3_extracted_k141_34587 | 1233 | No | NA | 1 | 0 | 0 | Not-determined | Genome-fragment | NA | NA | 0 |
| S3_extracted_k141_58438 | 3240 | No | NA | 5 | 0 | 2 | Not-determined | Genome-fragment | NA | NA | 0 |
| NC1_vf_k141_1922 | 1585 | No | NA | 2 | 0 | 0 | Low-quality | Genome-fragment | 2.26 | AAI-based (medium-confidence) | 0 |
| S3_extracted_k141_90146 | 5479 | No | NA | 3 | 0 | 3 | Not-determined | Genome-fragment | NA | NA | 0 |
| S16_vs_139186\|\|full | 1585 | No | NA | 1 | 0 | 0 | Low-quality | Genome-fragment | 3.57 | AAI-based (medium-confidence) | 0 |
| S3_extracted_k141_26326 | 2954 | No | NA | 2 | 0 | 2 | Not-determined | Genome-fragment | NA | NA | 0 |
| S3_extracted_k141_42501 | 1667 | No | NA | 2 | 0 | 0 | Not-determined | Genome-fragment | NA | NA | 0 |
| S3_extracted_k141_106100 | 1801 | No | NA | 3 | 0 | 2 | Not-determined | Genome-fragment | NA | NA | 0 |
| S8_vs_k141_41521\|\|lt2gene | 1585 | No | NA | 2 | 2 | 0 | Low-quality | Genome-fragment | 4.61 | AAI-based (high-confidence) | 0 |
| S3_extracted_k141_106112 | 1144 | No | NA | 1 | 0 | 1 | Not-determined | Genome-fragment | NA | NA | 0 |
| S9_vs_k141_77539\|\|full | 1586 | No | NA | 2 | 0 | 0 | Low-quality | Genome-fragment | 3.76 | AAI-based (high-confidence) | 0 |
| S3_extracted_k141_58530 | 1093 | No | NA | 3 | 0 | 3 | Not-determined | Genome-fragment | NA | NA | 0 |
| S14_vs_k141_11784\|\|full | 1586 | No | NA | 3 | 0 | 3 | Low-quality | Genome-fragment | 0.76 | AAI-based (medium-confidence) | 0 |
| S3_extracted_k141_90207 | 1241 | No | NA | 1 | 0 | 0 | Not-determined | Genome-fragment | NA | NA | 0 |
| S3_extracted_k141_50683 | 3400 | No | NA | 4 | 0 | 2 | Not-determined | Genome-fragment | NA | NA | 0 |
| S5_vs_k141_48051\|\|full | 1586 | No | NA | 4 | 1 | 0 | Low-quality | Genome-fragment | 3.5 | AAI-based (high-confidence) | 0 |
| S3_extracted_k141_2479 | 1645 | No | NA | 2 | 0 | 1 | Not-determined | Genome-fragment | NA | NA | 0 |
| S3_extracted_k141_34687 | 1168 | No | NA | 1 | 0 | 1 | Not-determined | Genome-fragment | NA | NA | 0 |
| S7_vs_75321\|\|full | 1586 | No | NA | 5 | 0 | 0 | Low-quality | Genome-fragment | 4.25 | AAI-based (high-confidence) | 0 |
| S3_extracted_k141_17976 | 1376 | No | NA | 3 | 0 | 0 | Not-determined | Genome-fragment | NA | NA | 0 |
| S3_extracted_k141_98191 | 1154 | No | NA | 1 | 0 | 0 | Not-determined | Genome-fragment | NA | NA | 0 |
| S7_vs_134529\|\|full | 1586 | No | NA | 4 | 0 | 0 | Low-quality | Genome-fragment | 4.79 | AAI-based (high-confidence) | 0 |
| S3_extracted_k141_106165 | 1084 | No | NA | 1 | 0 | 0 | Not-determined | Genome-fragment | NA | NA | 0 |
| S10_vs_112309\|\|full | 1586 | No | NA | 2 | 0 | 1 | Low-quality | Genome-fragment | 0.11 | AAI-based (medium-confidence) | 0 |
| S10_vs_119797\|\|full | 1586 | No | NA | 3 | 0 | 1 | Low-quality | Genome-fragment | 3.74 | AAI-based (high-confidence) | 0 |
| S3_extracted_k141_82322 | 1238 | No | NA | 3 | 0 | 3 | Not-determined | Genome-fragment | NA | NA | 0 |
| S3_extracted_k141_98231 | 7697 | No | NA | 7 | 0 | 4 | Not-determined | Genome-fragment | NA | NA | 0 |
| S3_extracted_k141_106197 | 1678 | No | NA | 2 | 0 | 2 | Not-determined | Genome-fragment | NA | NA | 0 |
| S8_vs_k141_48749\|\|full | 1586 | No | NA | 3 | 0 | 1 | Low-quality | Genome-fragment | 3.87 | AAI-based (high-confidence) | 0 |
| S17_extracted_k141_162053 | 1587 | No | NA | 2 | 0 | 0 | Low-quality | Genome-fragment | 2.93 | AAI-based (medium-confidence) | 0 |
| S3_extracted_k141_26454 | 1220 | No | NA | 1 | 0 | 1 | Not-determined | Genome-fragment | NA | NA | 0 |
| S3_extracted_k141_26461 | 1250 | No | NA | 2 | 0 | 0 | Not-determined | Genome-fragment | NA | NA | 0 |
| S3_extracted_k141_26462 | 1160 | No | NA | 3 | 0 | 1 | Not-determined | Genome-fragment | NA | NA | 0 |
| S3_extracted_k141_11779 | 1198 | No | NA | 1 | 0 | 1 | Not-determined | Genome-fragment | NA | NA | 0 |
| S3_extracted_k141_34731 | 7492 | No | NA | 7 | 0 | 6 | Not-determined | Genome-fragment | NA | NA | 0 |
| S11_vs_k141_46142\|\|full | 1587 | No | NA | 3 | 0 | 0 | Low-quality | Genome-fragment | 2.87 | AAI-based (high-confidence) | 0 |
| S11_vs_k141_24019\|\|full | 1587 | No | NA | 4 | 3 | 0 | Low-quality | Genome-fragment | 3.95 | AAI-based (medium-confidence) | 0 |
| S14_vs_k141_87850\|\|full | 1587 | No | NA | 5 | 0 | 0 | Low-quality | Genome-fragment | 2.98 | AAI-based (medium-confidence) | 0 |
| S1_vs_k141_58878\|\|full | 1587 | No | NA | 4 | 2 | 0 | Low-quality | Genome-fragment | 2.69 | AAI-based (high-confidence) | 0 |
| S3_extracted_k141_42636 | 3335 | No | NA | 1 | 0 | 1 | Not-determined | Genome-fragment | NA | NA | 0 |
| S3_extracted_k141_90550 | 1587 | No | NA | 1 | 0 | 0 | Low-quality | Genome-fragment | 0.5 | AAI-based (medium-confidence) | 0 |
| S3_vs_k141_114630\|\|full | 1587 | No | NA | 4 | 1 | 0 | Low-quality | Genome-fragment | 3.18 | AAI-based (high-confidence) | 0 |
| S3_extracted_k141_74915 | 1745 | No | NA | 2 | 0 | 2 | Not-determined | Genome-fragment | NA | NA | 0 |
| S7_vs_42026\|\|full | 1587 | No | NA | 3 | 3 | 0 | Low-quality | Genome-fragment | 2.98 | AAI-based (high-confidence) | 0 |
| S3_extracted_k141_11826 | 4132 | No | NA | 5 | 0 | 4 | Not-determined | Genome-fragment | NA | NA | 0 |
| S3_extracted_k141_121847 | 1262 | No | NA | 2 | 0 | 0 | Not-determined | Genome-fragment | NA | NA | 0 |
| S3_extracted_k141_106267 | 2927 | No | NA | 2 | 0 | 0 | Not-determined | Genome-fragment | NA | NA | 0 |
| S3_extracted_k141_26547 | 2641 | No | NA | 3 | 0 | 1 | Not-determined | Genome-fragment | NA | NA | 0 |
| S3_extracted_k141_74958 | 8252 | No | NA | 10 | 0 | 4 | Not-determined | Genome-fragment | NA | NA | 0 |
| S3_extracted_k141_74961 | 2448 | No | NA | 3 | 0 | 1 | Not-determined | Genome-fragment | NA | NA | 0 |
| S3_extracted_k141_34812 | 1128 | No | NA | 2 | 0 | 1 | Not-determined | Genome-fragment | NA | NA | 0 |
| S16_extracted_16281 | 1587 | No | NA | 2 | 0 | 0 | Low-quality | Genome-fragment | 2.87 | AAI-based (medium-confidence) | 0 |
| S3_extracted_k141_26589 | 3637 | No | NA | 1 | 0 | 1 | Not-determined | Genome-fragment | NA | NA | 0 |
| S12_vs_k141_119180\|\|full | 1587 | No | NA | 5 | 2 | 0 | Low-quality | Genome-fragment | 3.9 | AAI-based (high-confidence) | 0 |
| S12_vs_k141_3037\|\|full | 1587 | No | NA | 4 | 1 | 0 | Low-quality | Genome-fragment | 0.8 | HMM-based (lower-bound) | 0 |
| S3_extracted_k141_58845 | 1499 | No | NA | 1 | 0 | 1 | Not-determined | Genome-fragment | NA | NA | 0 |
| S3_extracted_k141_114260 | 1814 | No | NA | 4 | 0 | 3 | Not-determined | Genome-fragment | NA | NA | 0 |
| S3_extracted_k141_11876 | 1068 | No | NA | 1 | 0 | 1 | Not-determined | Genome-fragment | NA | NA | 0 |
| S3_extracted_k141_42759 | 1245 | No | NA | 2 | 0 | 2 | Not-determined | Genome-fragment | NA | NA | 0 |
| S3_extracted_k141_2770 | 1237 | No | NA | 2 | 0 | 1 | Not-determined | Genome-fragment | NA | NA | 0 |
| S3_extracted_k141_18080 | 46776 | No | NA | 64 | 11 | 1 | Medium-quality | Genome-fragment | 84.86 | AAI-based (high-confidence) | 0 |
| S3_extracted_k141_75046 | 1309 | No | NA | 1 | 0 | 0 | Not-determined | Genome-fragment | NA | NA | 0 |
| S3_extracted_k141_34847 | 1448 | No | NA | 2 | 0 | 1 | Not-determined | Genome-fragment | NA | NA | 0 |
| S3_extracted_k141_58913 | 1193 | No | NA | 2 | 0 | 0 | Not-determined | Genome-fragment | NA | NA | 0 |
| S6_vs_k141_36466\|\|full | 1587 | No | NA | 2 | 0 | 1 | Low-quality | Genome-fragment | 0.78 | AAI-based (high-confidence) | 0 |
| S3_extracted_k141_90380 | 2387 | No | NA | 4 | 0 | 1 | Not-determined | Genome-fragment | NA | NA | 0 |
| S3_extracted_k141_26656 | 4007 | No | NA | 4 | 0 | 2 | Not-determined | Genome-fragment | NA | NA | 0 |
| S3_extracted_k141_106401 | 39810 | Yes | 30429 | 52 | 19 | 6 | Medium-quality | Genome-fragment | 77.08 | AAI-based (high-confidence) | 23.56 |
| S8_extracted_k141_201475 | 1587 | No | NA | 3 | 0 | 0 | Low-quality | Genome-fragment | 1.33 | AAI-based (medium-confidence) | 0 |
| S11_extracted_k141_69475 | 1588 | No | NA | 2 | 0 | 2 | Low-quality | Genome-fragment | 0.79 | AAI-based (medium-confidence) | 0 |
| S3_extracted_k141_106405 | 3769 | No | NA | 3 | 0 | 3 | Not-determined | Genome-fragment | NA | NA | 0 |
| S3_extracted_k141_75082 | 1018 | No | NA | 1 | 0 | 1 | Not-determined | Genome-fragment | NA | NA | 0 |
| S1_vs_k141_69816\|\|full | 1588 | No | NA | 3 | 0 | 1 | Low-quality | Genome-fragment | 3.57 | AAI-based (medium-confidence) | 0 |
| S3_extracted_k141_121995 | 1062 | No | NA | 2 | 0 | 1 | Not-determined | Genome-fragment | NA | NA | 0 |
| S1_vs_k141_92278\|\|full | 1588 | No | NA | 4 | 0 | 0 | Low-quality | Genome-fragment | 3.51 | AAI-based (medium-confidence) | 0 |
| S3_extracted_k141_90416 | 1552 | No | NA | 2 | 0 | 2 | Not-determined | Genome-fragment | NA | NA | 0 |
| S3_extracted_k141_11997 | 6497 | No | NA | 5 | 0 | 2 | Not-determined | Genome-fragment | NA | NA | 0 |
| S3_extracted_k141_50987 | 1097 | No | NA | 1 | 0 | 1 | Not-determined | Genome-fragment | NA | NA | 0 |
| S7_vs_32990\|\|full | 1588 | No | NA | 3 | 0 | 1 | Low-quality | Genome-fragment | 3.02 | AAI-based (high-confidence) | 0 |
| S16_vs_126981\|\|full | 1588 | No | NA | 4 | 2 | 0 | Low-quality | Genome-fragment | 3.72 | AAI-based (medium-confidence) | 0 |
| S12_vs_k141_143470\|\|full | 1588 | No | NA | 3 | 0 | 0 | Low-quality | Genome-fragment | 3.15 | AAI-based (medium-confidence) | 0 |
| S3_extracted_k141_114355 | 1209 | No | NA | 1 | 0 | 1 | Not-determined | Genome-fragment | NA | NA | 0 |
| S3_extracted_k141_114368 | 3685 | No | NA | 3 | 0 | 3 | Not-determined | Genome-fragment | NA | NA | 0 |
| S3_extracted_k141_35008 | 1217 | No | NA | 1 | 0 | 0 | Not-determined | Genome-fragment | NA | NA | 0 |
| S3_extracted_k141_59078 | 5910 | No | NA | 7 | 2 | 0 | High-quality | High-quality | 95.34 | AAI-based (high-confidence) | 0 |
| S3_extracted_k141_82571 | 25954 | No | NA | 38 | 16 | 0 | Medium-quality | Genome-fragment | 80.6 | AAI-based (high-confidence) | 0 |
| S8_extracted_k141_168405 | 1588 | No | NA | 2 | 1 | 0 | Low-quality | Genome-fragment | 3.41 | AAI-based (high-confidence) | 0 |
| S17_vs_k141_86007\|\|full | 1589 | No | NA | 2 | 0 | 1 | Low-quality | Genome-fragment | 4.15 | AAI-based (medium-confidence) | 0 |
| S3_extracted_k141_106526 | 4408 | No | NA | 5 | 0 | 4 | Not-determined | Genome-fragment | NA | NA | 0 |
| S3_extracted_k141_82615 | 3940 | No | NA | 1 | 0 | 0 | Not-determined | Genome-fragment | NA | NA | 0 |
| S7_extracted_43862 | 1589 | No | NA | 2 | 0 | 1 | Low-quality | Genome-fragment | 2.76 | AAI-based (high-confidence) | 0 |
| S3_extracted_k141_98509 | 1174 | No | NA | 2 | 0 | 0 | Not-determined | Genome-fragment | NA | NA | 0 |
| S3_extracted_k141_26817 | 3136 | No | NA | 4 | 0 | 2 | Not-determined | Genome-fragment | NA | NA | 0 |
| S12_vs_k141_87745\|\|full | 1589 | No | NA | 5 | 0 | 0 | Low-quality | Genome-fragment | 3.13 | AAI-based (medium-confidence) | 0 |
| S3_extracted_k141_35073 | 59386 | No | NA | 71 | 33 | 0 | High-quality | High-quality | 97.39 | AAI-based (high-confidence) | 0 |
| S3_extracted_k141_90593 | 2469 | No | NA | 3 | 0 | 1 | Not-determined | Genome-fragment | NA | NA | 0 |
| S3_extracted_k141_12092 | 1051 | No | NA | 2 | 0 | 0 | Not-determined | Genome-fragment | NA | NA | 0 |
| S12_vs_k141_183161\|\|full | 1589 | No | NA | 4 | 0 | 0 | Low-quality | Genome-fragment | 5.59 | AAI-based (high-confidence) | 0 |
| S12_vs_k141_184491\|\|full | 1589 | No | NA | 2 | 0 | 1 | Low-quality | Genome-fragment | 1.66 | AAI-based (high-confidence) | 0 |
| S2_extracted_k141_6805 | 1589 | No | NA | 3 | 0 | 0 | Low-quality | Genome-fragment | 1.09 | AAI-based (high-confidence) | 0 |
| S3_extracted_k141_12116 | 15486 | No | NA | 12 | 0 | 3 | Not-determined | Genome-fragment | NA | NA | 0 |
| S3_extracted_k141_106604 | 5009 | No | NA | 4 | 0 | 2 | Not-determined | Genome-fragment | NA | NA | 0 |
| S3_extracted_k141_82741 | 1591 | No | NA | 2 | 0 | 2 | Not-determined | Genome-fragment | NA | NA | 0 |
| S3_extracted_k141_43010 | 1181 | No | NA | 1 | 0 | 1 | Not-determined | Genome-fragment | NA | NA | 0 |
| S3_extracted_k141_75346 | 1190 | No | NA | 1 | 0 | 0 | Not-determined | Genome-fragment | NA | NA | 0 |
| S3_extracted_k141_98588 | 3573 | No | NA | 3 | 0 | 1 | Not-determined | Genome-fragment | NA | NA | 0 |
| S6_vs_k141_9065\|\|full | 1589 | No | NA | 3 | 0 | 1 | Low-quality | Genome-fragment | 0.97 | AAI-based (medium-confidence) | 0 |
| S17_extracted_k141_24655 | 1590 | No | NA | 3 | 0 | 2 | Low-quality | Genome-fragment | 0.75 | AAI-based (medium-confidence) | 0 |
| S9_vs_k141_71857\|\|full | 1590 | Yes | 1362 | 3 | 1 | 1 | Low-quality | Genome-fragment | 0.83 | HMM-based (lower-bound) | 14.34 |
| S3_extracted_k141_2948 | 1404 | No | NA | 2 | 0 | 2 | Not-determined | Genome-fragment | NA | NA | 0 |
| S13_vs_24178\|\|full | 1590 | No | NA | 7 | 0 | 0 | Low-quality | Genome-fragment | 3.89 | AAI-based (high-confidence) | 0 |
| S3_extracted_k141_3012 | 1937 | No | NA | 2 | 0 | 2 | Not-determined | Genome-fragment | NA | NA | 0 |
| S3_extracted_k141_75400 | 1820 | No | NA | 3 | 0 | 2 | Not-determined | Genome-fragment | NA | NA | 0 |
| S14_vs_k141_105749\|\|full | 1590 | No | NA | 4 | 2 | 0 | Low-quality | Genome-fragment | 2.49 | AAI-based (medium-confidence) | 0 |
| S3_extracted_k141_3030 | 1145 | No | NA | 0 | 0 | 0 | Not-determined | Genome-fragment | NA | NA | 0 |
| S3_extracted_k141_98647 | 3590 | No | NA | 3 | 0 | 3 | Not-determined | Genome-fragment | NA | NA | 0 |
| S3_extracted_k141_12231 | 1908 | No | NA | 2 | 0 | 0 | Not-determined | Genome-fragment | NA | NA | 0 |
| S16_extracted_24727 | 1590 | No | NA | 3 | 0 | 0 | Low-quality | Genome-fragment | 3.17 | AAI-based (high-confidence) | 0 |
| S3_extracted_k141_90684 | 10747 | No | NA | 10 | 0 | 1 | Not-determined | Genome-fragment | NA | NA | 0 |
| S3_extracted_k141_3050 | 2362 | No | NA | 1 | 0 | 1 | Not-determined | Genome-fragment | NA | NA | 0 |
| S3_extracted_k141_106759 | 1416 | No | NA | 1 | 0 | 1 | Not-determined | Genome-fragment | NA | NA | 0 |
| S16_extracted_128965 | 1590 | No | NA | 1 | 0 | 0 | Low-quality | Genome-fragment | 18.68 | AAI-based (high-confidence) | 0 |
| S3_extracted_k141_75463 | 1629 | No | NA | 1 | 0 | 1 | Not-determined | Genome-fragment | NA | NA | 0 |
| S16_vs_89883\|\|lt2gene | 1590 | No | NA | 2 | 2 | 0 | Low-quality | Genome-fragment | 5.09 | AAI-based (high-confidence) | 0 |
| S3_extracted_k141_114537 | 1288 | No | NA | 2 | 0 | 2 | Not-determined | Genome-fragment | NA | NA | 0 |
| S3_extracted_k141_106789 | 5768 | No | NA | 5 | 0 | 2 | Not-determined | Genome-fragment | NA | NA | 0 |
| S3_extracted_k141_66373 | 1303 | No | NA | 2 | 0 | 1 | Not-determined | Genome-fragment | NA | NA | 0 |
| S3_extracted_k141_27065 | 1115 | No | NA | 2 | 0 | 2 | Not-determined | Genome-fragment | NA | NA | 0 |
| S3_extracted_k141_75476 | 1246 | No | NA | 3 | 0 | 1 | Not-determined | Genome-fragment | NA | NA | 0 |
| S3_extracted_k141_106805 | 1171 | No | NA | 1 | 0 | 0 | Not-determined | Genome-fragment | NA | NA | 0 |
| S3_extracted_k141_98677 | 1008 | No | NA | 1 | 0 | 0 | Not-determined | Genome-fragment | NA | NA | 0 |

| | | | | | | | | | | | |
|---|---|---|---|---|---|---|---|---|---|---|---|
| S3_extracted_k141_12294 | 1495 | No | NA | 2 | 0 | 1 | Not-determined | Genome-fragment | NA | NA | 0 |
| S3_extracted_k141_114576 | 4815 | No | NA | 4 | 0 | 0 | Not-determined | Genome-fragment | NA | NA | 0 |
| S12_extracted_k141_126405 | 1590 | No | NA | 2 | 0 | 0 | Low-quality | Genome-fragment | 2.93 | AAI-based (high-confidence) | 0 |
| S3_extracted_k141_98706 | 1071 | No | NA | 2 | 0 | 0 | Not-determined | Genome-fragment | NA | NA | 0 |
| S13_vs_153132\|\|full | 1591 | No | NA | 3 | 0 | 1 | Low-quality | Genome-fragment | 0.47 | AAI-based (high-confidence) | 0 |
| S14_extracted_k141_88830 | 1591 | No | NA | 2 | 2 | 0 | Low-quality | Genome-fragment | 5.23 | AAI-based (high-confidence) | 0 |
| S14_extracted_k141_56212 | 1591 | No | NA | 5 | 2 | 0 | Low-quality | Genome-fragment | 3.24 | AAI-based (high-confidence) | 0 |
| S14_vs_k141_20458\|\|full | 1591 | No | NA | 3 | 0 | 0 | Low-quality | Genome-fragment | 2.84 | AAI-based (medium-confidence) | 0 |
| S3_extracted_k141_98759 | 1079 | No | NA | 2 | 0 | 0 | Not-determined | Genome-fragment | NA | NA | 0 |
| S3_extracted_k141_66426 | 1242 | No | NA | 1 | 0 | 0 | Not-determined | Genome-fragment | NA | NA | 0 |
| S3_vs_k141_29033\|\|full | 1591 | No | NA | 4 | 4 | 0 | Low-quality | Genome-fragment | 2.98 | AAI-based (high-confidence) | 0 |
| S3_extracted_k141_3151 | 1055 | No | NA | 2 | 0 | 1 | Not-determined | Genome-fragment | NA | NA | 0 |
| S3_extracted_k141_98771 | 3031 | No | NA | 4 | 0 | 1 | Not-determined | Genome-fragment | NA | NA | 0 |
| S8_vs_k141_122503\|\|full | 1591 | No | NA | 3 | 0 | 1 | Low-quality | Genome-fragment | 0.72 | AAI-based (medium-confidence) | 0 |
| S3_extracted_k141_59351 | 1515 | No | NA | 1 | 0 | 1 | Not-determined | Genome-fragment | NA | NA | 0 |
| S3_extracted_k141_75625 | 1889 | No | NA | 2 | 0 | 1 | Not-determined | Genome-fragment | NA | NA | 0 |
| S11_vs_k141_54670\|\|full | 1592 | No | NA | 3 | 0 | 0 | Low-quality | Genome-fragment | 2.82 | AAI-based (medium-confidence) | 0 |
| S3_vs_k141_16642\|\|full | 1592 | No | NA | 3 | 0 | 0 | Low-quality | Genome-fragment | 1.02 | AAI-based (high-confidence) | 0 |
| S5_vs_k141_79537\|\|full | 1592 | No | NA | 4 | 0 | 0 | Low-quality | Genome-fragment | 1.44 | AAI-based (medium-confidence) | 0 |
| S3_extracted_k141_90820 | 1145 | No | NA | 3 | 0 | 0 | Not-determined | Genome-fragment | NA | NA | 0 |
| S3_extracted_k141_122228 | 1109 | No | NA | 1 | 0 | 0 | Not-determined | Genome-fragment | NA | NA | 0 |
| S3_extracted_k141_90833 | 1421 | No | NA | 3 | 0 | 0 | Not-determined | Genome-fragment | NA | NA | 0 |
| S8_extracted_k141_111477 | 1592 | No | NA | 2 | 0 | 1 | Low-quality | Genome-fragment | 1.43 | AAI-based (high-confidence) | 0 |
| S3_extracted_k141_114707 | 4627 | No | NA | 3 | 0 | 2 | Not-determined | Genome-fragment | NA | NA | 0 |
| S9_extracted_k141_39086 | 1593 | No | NA | 3 | 1 | 0 | Low-quality | Genome-fragment | 2.69 | AAI-based (medium-confidence) | 0 |
| S3_extracted_k141_106967 | 15363 | No | NA | 21 | 0 | 3 | Not-determined | Genome-fragment | NA | NA | 0 |
| S3_extracted_k141_90858 | 3983 | No | NA | 3 | 0 | 0 | Not-determined | Genome-fragment | NA | NA | 0 |
| S3_extracted_k141_12403 | 1228 | No | NA | 2 | 0 | 0 | Not-determined | Genome-fragment | NA | NA | 0 |
| S3_extracted_k141_98814 | 1066 | No | NA | 2 | 0 | 0 | Not-determined | Genome-fragment | NA | NA | 0 |
| S3_extracted_k141_12413 | 1160 | No | NA | 1 | 0 | 0 | Not-determined | Genome-fragment | NA | NA | 0 |
| S3_extracted_k141_90874 | 1085 | No | NA | 2 | 0 | 1 | Not-determined | Genome-fragment | NA | NA | 0 |
| S3_extracted_k141_35519 | 1172 | No | NA | 1 | 0 | 1 | Not-determined | Genome-fragment | NA | NA | 0 |
| S11_vs_k141_94561\|\|full | 1593 | No | NA | 3 | 0 | 0 | Low-quality | Genome-fragment | 2.57 | AAI-based (medium-confidence) | 0 |
| S3_extracted_k141_106993 | 1407 | No | NA | 3 | 0 | 2 | Not-determined | Genome-fragment | NA | NA | 0 |
| S1_vs_k141_63833\|\|full | 1593 | No | NA | 3 | 3 | 0 | Low-quality | Genome-fragment | 2.57 | AAI-based (high-confidence) | 0 |
| S3_extracted_k141_59402 | 1032 | No | NA | 2 | 0 | 0 | Not-determined | Genome-fragment | NA | NA | 0 |
| S3_extracted_k141_51543 | 3007 | No | NA | 4 | 0 | 2 | Not-determined | Genome-fragment | NA | NA | 0 |
| S3_extracted_k141_90937 | 2499 | No | NA | 4 | 0 | 0 | Not-determined | Genome-fragment | NA | NA | 0 |
| S3_extracted_k141_35583 | 1843 | No | NA | 2 | 0 | 2 | Not-determined | Genome-fragment | NA | NA | 0 |
| S7_extracted_12531 | 1593 | No | NA | 3 | 1 | 0 | Low-quality | Genome-fragment | 1.09 | AAI-based (medium-confidence) | 0 |
| S3_extracted_k141_51574 | 1436 | No | NA | 1 | 0 | 1 | Not-determined | Genome-fragment | NA | NA | 0 |
| S3_extracted_k141_122283 | 1494 | No | NA | 1 | 0 | 1 | Not-determined | Genome-fragment | NA | NA | 0 |
| S3_extracted_k141_82911 | 1770 | No | NA | 2 | 0 | 0 | Not-determined | Genome-fragment | NA | NA | 0 |
| S3_extracted_k141_66508 | 1220 | No | NA | 2 | 0 | 1 | Not-determined | Genome-fragment | NA | NA | 0 |
| S3_extracted_k141_27199 | 1023 | No | NA | 1 | 0 | 0 | Not-determined | Genome-fragment | NA | NA | 0 |
| S3_extracted_k141_51592 | 1981 | No | NA | 1 | 0 | 1 | Not-determined | Genome-fragment | NA | NA | 0 |
| S7_extracted_31035 | 1593 | No | NA | 4 | 2 | 0 | Low-quality | Genome-fragment | 11.57 | AAI-based (high-confidence) | 0 |
| S3_extracted_k141_82940 | 8311 | No | NA | 5 | 0 | 4 | Not-determined | Genome-fragment | NA | NA | 0 |
| S3_extracted_k141_18555 | 1083 | No | NA | 1 | 0 | 0 | Not-determined | Genome-fragment | NA | NA | 0 |
| S3_extracted_k141_12504 | 3792 | No | NA | 3 | 0 | 0 | Not-determined | Genome-fragment | NA | NA | 0 |
| S3_extracted_k141_51628 | 3598 | No | NA | 4 | 0 | 3 | Not-determined | Genome-fragment | NA | NA | 0 |
| S3_extracted_k141_43331 | 1926 | No | NA | 1 | 0 | 1 | Not-determined | Genome-fragment | NA | NA | 0 |
| S3_extracted_k141_35668 | 1083 | No | NA | 1 | 0 | 0 | Not-determined | Genome-fragment | NA | NA | 0 |
| S12_vs_k141_24489\|\|full | 1593 | No | NA | 4 | 0 | 0 | Low-quality | Genome-fragment | 2.11 | AAI-based (high-confidence) | 0 |
| S6_extracted_k141_160 | 1593 | No | NA | 2 | 0 | 1 | Low-quality | Genome-fragment | 1.29 | AAI-based (high-confidence) | 0 |
| S8_extracted_k141_120043 | 1593 | No | NA | 3 | 0 | 3 | Low-quality | Genome-fragment | 0.45 | AAI-based (medium-confidence) | 0 |
| S3_extracted_k141_27318 | 1383 | No | NA | 1 | 0 | 1 | Not-determined | Genome-fragment | NA | NA | 0 |
| S3_extracted_k141_99000 | 4445 | No | NA | 7 | 0 | 1 | Not-determined | Genome-fragment | NA | NA | 0 |
| S3_extracted_k141_91060 | 5014 | No | NA | 2 | 0 | 2 | Not-determined | Genome-fragment | NA | NA | 0 |
| S3_extracted_k141_99050 | 2754 | No | NA | 4 | 0 | 4 | Not-determined | Genome-fragment | NA | NA | 0 |
| S3_extracted_k141_12572 | 1611 | No | NA | 2 | 0 | 2 | Not-determined | Genome-fragment | NA | NA | 0 |
| S3_extracted_k141_18648 | 2100 | No | NA | 2 | 0 | 2 | Not-determined | Genome-fragment | NA | NA | 0 |
| S8_vs_k141_64113\|\|full | 1593 | No | NA | 4 | 0 | 0 | Low-quality | Genome-fragment | 3.53 | AAI-based (medium-confidence) | 0 |
| S17_extracted_k141_76601 | 1594 | No | NA | 2 | 1 | 0 | Low-quality | Genome-fragment | 6.85 | AAI-based (high-confidence) | 0 |
| S17_extracted_k141_88602 | 1594 | No | NA | 4 | 4 | 0 | Low-quality | Genome-fragment | 4.35 | AAI-based (high-confidence) | 0 |
| S3_extracted_k141_12598 | 7516 | No | NA | 7 | 0 | 1 | Not-determined | Genome-fragment | NA | NA | 0 |
| S3_extracted_k141_83068 | 1311 | No | NA | 1 | 0 | 1 | Not-determined | Genome-fragment | NA | NA | 0 |
| S3_extracted_k141_27350 | 1088 | No | NA | 2 | 0 | 1 | Not-determined | Genome-fragment | NA | NA | 0 |
| S3_extracted_k141_3288 | 1342 | No | NA | 1 | 0 | 0 | Not-determined | Genome-fragment | NA | NA | 0 |
| S3_extracted_k141_83077 | 1135 | No | NA | 2 | 0 | 0 | Not-determined | Genome-fragment | NA | NA | 0 |
| S3_extracted_k141_122516 | 1455 | No | NA | 1 | 0 | 1 | Not-determined | Genome-fragment | NA | NA | 0 |
| S17_vs_k141_84002\|\|full | 1594 | No | NA | 3 | 1 | 0 | Low-quality | Genome-fragment | 2.93 | HMM-based (lower-bound) | 0 |
| S3_extracted_k141_51711 | 1095 | No | NA | 1 | 0 | 1 | Not-determined | Genome-fragment | NA | NA | 0 |
| S9_vs_k141_80704\|\|full | 1594 | No | NA | 3 | 0 | 0 | Low-quality | Genome-fragment | 3.65 | AAI-based (medium-confidence) | 0 |
| S3_extracted_k141_107235 | 11821 | No | NA | 9 | 0 | 2 | Not-determined | Genome-fragment | NA | NA | 0 |
| S9_vs_k141_84831\|\|full | 1594 | No | NA | 4 | 0 | 0 | Low-quality | Genome-fragment | 3.51 | AAI-based (high-confidence) | 0 |
| S11_extracted_k141_34886 | 1594 | No | NA | 3 | 0 | 1 | Low-quality | Genome-fragment | 4.15 | AAI-based (medium-confidence) | 0 |
| S3_extracted_k141_99097 | 3039 | No | NA | 6 | 0 | 2 | Not-determined | Genome-fragment | NA | NA | 0 |
| S3_extracted_k141_107261 | 10076 | No | NA | 4 | 0 | 4 | Not-determined | Genome-fragment | NA | NA | 0 |
| S3_extracted_k141_7979 | 1594 | No | NA | 4 | 0 | 0 | Low-quality | Genome-fragment | 1.19 | AAI-based (high-confidence) | 0 |
| S3_extracted_k141_27433 | 2161 | No | NA | 3 | 0 | 2 | Not-determined | Genome-fragment | NA | NA | 0 |
| S3_extracted_k141_43442 | 1134 | No | NA | 2 | 0 | 2 | Not-determined | Genome-fragment | NA | NA | 0 |
| S3_extracted_k141_122572 | 1810 | No | NA | 2 | 0 | 2 | Not-determined | Genome-fragment | NA | NA | 0 |
| S3_extracted_k141_66696 | 1195 | No | NA | 1 | 0 | 0 | Not-determined | Genome-fragment | NA | NA | 0 |
| S3_extracted_k141_35764 | 1785 | No | NA | 2 | 0 | 2 | Not-determined | Genome-fragment | NA | NA | 0 |
| S5_vs_k141_89026\|\|full | 1594 | No | NA | 4 | 0 | 0 | Low-quality | Genome-fragment | 2.74 | AAI-based (medium-confidence) | 0 |
| S3_extracted_k141_107299 | 9491 | No | NA | 6 | 0 | 4 | Not-determined | Genome-fragment | NA | NA | 0 |
| S3_extracted_k141_91166 | 1005 | No | NA | 1 | 0 | 0 | Not-determined | Genome-fragment | NA | NA | 0 |
| S7_extracted_48754 | 1594 | No | NA | 2 | 1 | 0 | Low-quality | Genome-fragment | 1.18 | AAI-based (high-confidence) | 0 |
| S16_extracted_158733 | 1594 | No | NA | 1 | 0 | 1 | Low-quality | Genome-fragment | 0.43 | AAI-based (high-confidence) | 0 |
| S16_extracted_164579 | 1594 | No | NA | 5 | 0 | 0 | Low-quality | Genome-fragment | 1.46 | AAI-based (high-confidence) | 0 |
| S6_vs_k141_18056\|\|full | 1594 | No | NA | 3 | 1 | 0 | Low-quality | Genome-fragment | 2.59 | AAI-based (high-confidence) | 0 |
| S8_vs_k141_71575\|\|full | 1594 | No | NA | 4 | 1 | 0 | Low-quality | Genome-fragment | 4.47 | AAI-based (high-confidence) | 0 |
| S8_vs_k141_178789\|\|full | 1594 | No | NA | 4 | 0 | 0 | Low-quality | Genome-fragment | 2.61 | AAI-based (medium-confidence) | 0 |
| S3_extracted_k141_83256 | 1096 | No | NA | 2 | 0 | 0 | Not-determined | Genome-fragment | NA | NA | 0 |
| S3_extracted_k141_107327 | 1042 | No | NA | 2 | 0 | 2 | Not-determined | Genome-fragment | NA | NA | 0 |
| S3_extracted_k141_59691 | 5486 | No | NA | 4 | 0 | 2 | Not-determined | Genome-fragment | NA | NA | 0 |
| S3_extracted_k141_12744 | 4616 | No | NA | 3 | 0 | 1 | Not-determined | Genome-fragment | NA | NA | 0 |
| S3_extracted_k141_99164 | 2282 | No | NA | 4 | 0 | 1 | Not-determined | Genome-fragment | NA | NA | 0 |
| S3_extracted_k141_83296 | 3586 | No | NA | 5 | 0 | 0 | Not-determined | Genome-fragment | NA | NA | 0 |
| S3_extracted_k141_43531 | 1264 | No | NA | 1 | 0 | 1 | Not-determined | Genome-fragment | NA | NA | 0 |
| S3_extracted_k141_122622 | 2909 | No | NA | 4 | 0 | 1 | Not-determined | Genome-fragment | NA | NA | 0 |
| S3_extracted_k141_43538 | 1097 | No | NA | 1 | 0 | 1 | Not-determined | Genome-fragment | NA | NA | 0 |
| S8_vs_k141_66952\|\|full | 1594 | No | NA | 3 | 0 | 1 | Low-quality | Genome-fragment | 3.62 | AAI-based (medium-confidence) | 0 |
| S11_vs_k141_6190\|\|full | 1595 | No | NA | 4 | 1 | 0 | Low-quality | Genome-fragment | 3.75 | AAI-based (high-confidence) | 0 |
| S3_extracted_k141_122643 | 4665 | No | NA | 4 | 0 | 3 | Not-determined | Genome-fragment | NA | NA | 0 |
| S3_extracted_k141_12773 | 1667 | No | NA | 1 | 0 | 0 | Not-determined | Genome-fragment | NA | NA | 0 |
| S3_extracted_k141_43554 | 1136 | No | NA | 1 | 0 | 0 | Not-determined | Genome-fragment | NA | NA | 0 |
| S3_extracted_k141_115115 | 5094 | No | NA | 2 | 0 | 2 | Not-determined | Genome-fragment | NA | NA | 0 |
| S3_extracted_k141_12779 | 3131 | No | NA | 5 | 0 | 5 | Not-determined | Genome-fragment | NA | NA | 0 |
| S3_extracted_k141_91272 | 1331 | No | NA | 1 | 0 | 1 | Not-determined | Genome-fragment | NA | NA | 0 |
| S14_vs_k141_7910\|\|full | 1595 | No | NA | 3 | 0 | 0 | Low-quality | Genome-fragment | 3.06 | AAI-based (high-confidence) | 0 |
| S1_extracted_k141_43554 | 1595 | No | NA | 1 | 0 | 0 | Low-quality | Genome-fragment | 3.45 | AAI-based (high-confidence) | 0 |
| S1_extracted_k141_30484 | 1595 | No | NA | 5 | 1 | 0 | Low-quality | Genome-fragment | 2.69 | AAI-based (high-confidence) | 0 |
| S3_extracted_k141_122696 | 1093 | No | NA | 2 | 0 | 0 | Not-determined | Genome-fragment | NA | NA | 0 |
| S5_vs_k141_40455\|\|full | 1595 | No | NA | 3 | 0 | 2 | Low-quality | Genome-fragment | 1.83 | AAI-based (medium-confidence) | 0 |
| S3_extracted_k141_59818 | 3223 | No | NA | 3 | 0 | 1 | Not-determined | Genome-fragment | NA | NA | 0 |
| S3_extracted_k141_115164 | 4731 | No | NA | 4 | 0 | 3 | Not-determined | Genome-fragment | NA | NA | 0 |
| S3_extracted_k141_91357 | 6746 | No | NA | 5 | 0 | 4 | Not-determined | Genome-fragment | NA | NA | 0 |
| S7_vs_129901\|\|full | 1595 | No | NA | 5 | 2 | 0 | Low-quality | Genome-fragment | 3.09 | AAI-based (high-confidence) | 0 |
| S3_extracted_k141_3475 | 1209 | No | NA | 2 | 0 | 2 | Not-determined | Genome-fragment | NA | NA | 0 |
| S3_extracted_k141_51971 | 1558 | No | NA | 2 | 0 | 1 | Not-determined | Genome-fragment | NA | NA | 0 |
| S16_vs_133472\|\|full | 1595 | No | NA | 3 | 1 | 0 | Low-quality | Genome-fragment | 3.87 | HMM-based (lower-bound) | 0 |
| S16_vs_156625\|\|lt2gene | 1595 | No | NA | 2 | 2 | 0 | Low-quality | Genome-fragment | 4.18 | AAI-based (high-confidence) | 0 |
| S3_extracted_k141_66848 | 2915 | No | NA | 1 | 0 | 0 | Not-determined | Genome-fragment | NA | NA | 0 |
| S12_vs_k141_82831\|\|full | 1595 | No | NA | 3 | 1 | 0 | Low-quality | Genome-fragment | 2.74 | HMM-based (lower-bound) | 0 |
| S3_extracted_k141_18766 | 3638 | No | NA | 3 | 0 | 3 | Not-determined | Genome-fragment | NA | NA | 0 |
| S6_vs_k141_15196\|\|full | 1595 | No | NA | 6 | 0 | 0 | Low-quality | Genome-fragment | 4.18 | AAI-based (high-confidence) | 0 |
| S3_extracted_k141_3535 | 1033 | No | NA | 2 | 0 | 2 | Not-determined | Genome-fragment | NA | NA | 0 |
| S3_extracted_k141_122750 | 2311 | No | NA | 1 | 0 | 0 | Not-determined | Genome-fragment | NA | NA | 0 |
| S3_extracted_k141_52051 | 1069 | No | NA | 1 | 0 | 0 | Not-determined | Genome-fragment | NA | NA | 0 |
| S13_vs_5405\|\|lt2gene | 1596 | No | NA | 2 | 1 | 0 | Low-quality | Genome-fragment | 3.36 | AAI-based (medium-confidence) | 0 |
| S3_extracted_k141_52059 | 4653 | No | NA | 4 | 0 | 2 | Not-determined | Genome-fragment | NA | NA | 0 |
| S3_extracted_k141_52060 | 4530 | No | NA | 2 | 0 | 1 | Not-determined | Genome-fragment | NA | NA | 0 |
| S3_extracted_k141_83450 | 1234 | No | NA | 1 | 0 | 1 | Not-determined | Genome-fragment | NA | NA | 0 |
| S1_extracted_k141_106538 | 1596 | No | NA | 4 | 0 | 0 | Low-quality | Genome-fragment | 2.37 | AAI-based (high-confidence) | 0 |
| S5_extracted_k141_140268 | 1596 | No | NA | 4 | 1 | 0 | Low-quality | Genome-fragment | 4.36 | AAI-based (high-confidence) | 0 |
| S5_vs_k141_124891\|\|full | 1596 | No | NA | 3 | 1 | 0 | Low-quality | Genome-fragment | 3.15 | HMM-based (lower-bound) | 0 |
| S5_vs_k141_76580\|\|full | 1596 | No | NA | 4 | 0 | 1 | Low-quality | Genome-fragment | 4.78 | AAI-based (medium-confidence) | 0 |
| S16_extracted_39964 | 1596 | No | NA | 2 | 1 | 0 | Low-quality | Genome-fragment | 2.41 | HMM-based (lower-bound) | 0 |
| S3_extracted_k141_12893 | 1156 | No | NA | 1 | 0 | 0 | Not-determined | Genome-fragment | NA | NA | 0 |
| S16_vs_33610\|\|full | 1596 | No | NA | 3 | 0 | 0 | Low-quality | Genome-fragment | 4.44 | AAI-based (high-confidence) | 0 |
| S3_extracted_k141_27642 | 6009 | No | NA | 6 | 0 | 6 | Not-determined | Genome-fragment | NA | NA | 0 |
| S10_extracted_1019 | 1596 | No | NA | 4 | 0 | 0 | Low-quality | Genome-fragment | 3.09 | AAI-based (high-confidence) | 0 |
| S3_extracted_k141_83505 | 1238 | No | NA | 2 | 0 | 1 | Not-determined | Genome-fragment | NA | NA | 0 |
| S12_vs_k141_68273\|\|full | 1596 | No | NA | 3 | 1 | 0 | Low-quality | Genome-fragment | 2.58 | AAI-based (high-confidence) | 0 |
| S13_vs_45970\|\|full | 1597 | No | NA | 2 | 0 | 2 | Low-quality | Genome-fragment | 0.72 | AAI-based (medium-confidence) | 0 |
| S3_extracted_k141_36008 | 4645 | No | NA | 4 | 0 | 0 | Not-determined | Genome-fragment | NA | NA | 0 |
| S3_extracted_k141_83536 | 15430 | No | NA | 17 | 7 | 0 | Medium-quality | Genome-fragment | 66.36 | AAI-based (medium-confidence) | 0 |
| S3_extracted_k141_18829 | 86825 | No | NA | 111 | 15 | 2 | Complete | High-quality | 100 | DTR (high-confidence) | 0 |
| S3_extracted_k141_36063 | 3368 | No | NA | 3 | 0 | 1 | Not-determined | Genome-fragment | NA | NA | 0 |
| S13_vs_65500\|\|full | 1597 | No | NA | 2 | 0 | 1 | Low-quality | Genome-fragment | 1.47 | AAI-based (high-confidence) | 0 |
| S3_extracted_k141_36079 | 1978 | No | NA | 1 | 0 | 0 | Not-determined | Genome-fragment | NA | NA | 0 |
| S3_extracted_k141_83560 | 3157 | No | NA | 3 | 0 | 1 | Not-determined | Genome-fragment | NA | NA | 0 |
| S3_extracted_k141_91556 | 3824 | No | NA | 4 | 0 | 2 | Not-determined | Genome-fragment | NA | NA | 0 |
| S3_extracted_k141_83583 | 2090 | No | NA | 4 | 0 | 1 | Not-determined | Genome-fragment | NA | NA | 0 |
| S3_extracted_k141_91582 | 2241 | No | NA | 2 | 0 | 0 | Not-determined | Genome-fragment | NA | NA | 0 |
| S5_vs_k141_104893\|\|full | 1597 | No | NA | 3 | 1 | 0 | Low-quality | Genome-fragment | 2.21 | AAI-based (medium-confidence) | 0 |
| S3_extracted_k141_115304 | 100971 | No | NA | 106 | 16 | 1 | Complete | High-quality | 100 | DTR (high-confidence) | 0 |
| S3_extracted_k141_36093 | 14516 | No | NA | 16 | 0 | 3 | Not-determined | Genome-fragment | NA | NA | 0 |
| S3_extracted_k141_107482 | 7654 | No | NA | 5 | 0 | 3 | Not-determined | Genome-fragment | NA | NA | 0 |
| S3_extracted_k141_60053 | 2649 | No | NA | 3 | 0 | 1 | Not-determined | Genome-fragment | NA | NA | 0 |
| S5_vs_k141_48057\|\|full | 1597 | No | NA | 4 | 1 | 0 | Low-quality | Genome-fragment | 3.52 | AAI-based (high-confidence) | 0 |
| S3_extracted_k141_60075 | 3445 | No | NA | 2 | 0 | 2 | Not-determined | Genome-fragment | NA | NA | 0 |
| S3_extracted_k141_60085 | 1667 | No | NA | 1 | 0 | 1 | Not-determined | Genome-fragment | NA | NA | 0 |
| S7_vs_45085\|\|full | 1597 | No | NA | 3 | 1 | 0 | Low-quality | Genome-fragment | 3.89 | AAI-based (high-confidence) | 0 |
| S12_extracted_k141_72777 | 1597 | No | NA | 3 | 2 | 0 | Low-quality | Genome-fragment | 3.79 | HMM-based (lower-bound) | 0 |
| S3_extracted_k141_60095 | 3335 | No | NA | 2 | 0 | 2 | Not-determined | Genome-fragment | NA | NA | 0 |
| S3_extracted_k141_60112 | 1467 | No | NA | 2 | 0 | 2 | Not-determined | Genome-fragment | NA | NA | 0 |
| S3_extracted_k141_76243 | 1302 | No | NA | 2 | 0 | 1 | Not-determined | Genome-fragment | NA | NA | 0 |
| S12_vs_k141_18949\|\|full | 1597 | No | NA | 4 | 1 | 0 | Low-quality | Genome-fragment | 4.36 | AAI-based (high-confidence) | 0 |
| S3_extracted_k141_13075 | 4326 | No | NA | 7 | 0 | 3 | Not-determined | Genome-fragment | NA | NA | 0 |
| S3_extracted_k141_52386 | 1063 | No | NA | 1 | 0 | 1 | Not-determined | Genome-fragment | NA | NA | 0 |
| S3_extracted_k141_60147 | 5130 | No | NA | 3 | 0 | 2 | Not-determined | Genome-fragment | NA | NA | 0 |
| S2_extracted_k141_13392 | 1597 | No | NA | 1 | 0 | 1 | Low-quality | Genome-fragment | 2.43 | AAI-based (medium-confidence) | 0 |
| S3_extracted_k141_52418 | 1212 | No | NA | 1 | 0 | 0 | Not-determined | Genome-fragment | NA | NA | 0 |
| S3_extracted_k141_91677 | 5510 | No | NA | 1 | 0 | 1 | Not-determined | Genome-fragment | NA | NA | 0 |
| S6_extracted_k141_25454 | 1597 | No | NA | 5 | 1 | 0 | Low-quality | Genome-fragment | 3.77 | AAI-based (high-confidence) | 0 |
| S8_extracted_k141_71753 | 1597 | No | NA | 2 | 0 | 2 | Low-quality | Genome-fragment | 0.71 | AAI-based (medium-confidence) | 0 |
| S3_extracted_k141_122901 | 1028 | No | NA | 1 | 0 | 1 | Not-determined | Genome-fragment | NA | NA | 0 |
| S3_extracted_k141_107627 | 1858 | No | NA | 2 | 0 | 2 | Not-determined | Genome-fragment | NA | NA | 0 |
| S3_extracted_k141_13154 | 1014 | No | NA | 1 | 0 | 0 | Not-determined | Genome-fragment | NA | NA | 0 |
| S3_extracted_k141_3792 | 6349 | No | NA | 6 | 0 | 0 | Not-determined | Genome-fragment | NA | NA | 0 |
| S3_extracted_k141_52454 | 6126 | No | NA | 9 | 0 | 1 | Not-determined | Genome-fragment | NA | NA | 0 |
| S8_vs_k141_33497\|\|full | 1597 | No | NA | 2 | 1 | 0 | Low-quality | Genome-fragment | 4.35 | AAI-based (high-confidence) | 0 |
| S3_extracted_k141_60201 | 4682 | No | NA | 6 | 0 | 4 | Not-determined | Genome-fragment | NA | NA | 0 |
| S3_extracted_k141_107662 | 1266 | No | NA | 1 | 0 | 1 | Not-determined | Genome-fragment | NA | NA | 0 |
| S17_extracted_k141_102607 | 1598 | No | NA | 3 | 2 | 0 | Low-quality | Genome-fragment | 4.34 | AAI-based (high-confidence) | 0 |
| S3_extracted_k141_3841 | 1229 | No | NA | 2 | 0 | 0 | Not-determined | Genome-fragment | NA | NA | 0 |
| S17_vs_k141_19327\|\|full | 1598 | No | NA | 3 | 0 | 0 | Low-quality | Genome-fragment | 1.15 | AAI-based (medium-confidence) | 0 |
| S17_vs_k141_9849\|\|lt2gene | 1598 | No | NA | 1 | 1 | 0 | Low-quality | Genome-fragment | 4.44 | AAI-based (medium-confidence) | 0 |
| S14_extracted_k141_70559 | 1598 | No | NA | 3 | 0 | 0 | Low-quality | Genome-fragment | 1.02 | AAI-based (medium-confidence) | 0 |
| S3_vs_k141_91154\|\|full | 1598 | No | NA | 5 | 0 | 0 | Low-quality | Genome-fragment | 3.4 | AAI-based (high-confidence) | 0 |
| S3_vs_k141_76309\|\|full | 1598 | No | NA | 2 | 1 | 0 | Low-quality | Genome-fragment | 2.85 | AAI-based (high-confidence) | 0 |
| S3_extracted_k141_43921 | 2123 | No | NA | 1 | 0 | 1 | Not-determined | Genome-fragment | NA | NA | 0 |
| S3_extracted_k141_91782 | 1041 | No | NA | 4 | 0 | 0 | Not-determined | Genome-fragment | NA | NA | 0 |
| S5_vs_k141_63291\|\|lt2gene | 1598 | No | NA | 1 | 1 | 0 | Low-quality | Genome-fragment | 3.58 | AAI-based (medium-confidence) | 0 |
| S3_extracted_k141_67143 | 1018 | No | NA | 1 | 0 | 0 | Not-determined | Genome-fragment | NA | NA | 0 |
| S3_extracted_k141_3957 | 1705 | No | NA | 1 | 0 | 1 | Not-determined | Genome-fragment | NA | NA | 0 |
| S3_extracted_k141_91803 | 2250 | No | NA | 2 | 0 | 1 | Not-determined | Genome-fragment | NA | NA | 0 |
| S3_extracted_k141_36248 | 2671 | No | NA | 2 | 0 | 2 | Not-determined | Genome-fragment | NA | NA | 0 |
| S7_vs_65590\|\|full | 1598 | No | NA | 3 | 0 | 0 | Low-quality | Genome-fragment | 4.37 | AAI-based (high-confidence) | 0 |
| S15_vs_19881\|\|full | 1598 | No | NA | 3 | 1 | 0 | Low-quality | Genome-fragment | 3.54 | AAI-based (high-confidence) | 0 |
| S15_vs_74723\|\|full | 1598 | No | NA | 3 | 0 | 0 | Low-quality | Genome-fragment | 1.18 | AAI-based (medium-confidence) | 0 |
| S3_extracted_k141_123013 | 11838 | No | NA | 5 | 0 | 3 | Not-determined | Genome-fragment | NA | NA | 0 |
| S3_extracted_k141_107759 | 4800 | No | NA | 5 | 0 | 0 | Not-determined | Genome-fragment | NA | NA | 0 |
| S2_extracted_k141_19419 | 1598 | No | NA | 6 | 0 | 0 | Low-quality | Genome-fragment | 1.66 | AAI-based (high-confidence) | 0 |
| S8_extracted_k141_25157 | 1598 | No | NA | 2 | 0 | 0 | Low-quality | Genome-fragment | 3.76 | AAI-based (high-confidence) | 0 |
| S3_extracted_k141_91865 | 3274 | No | NA | 3 | 0 | 2 | Not-determined | Genome-fragment | NA | NA | 0 |
| S3_extracted_k141_107768 | 1180 | No | NA | 2 | 0 | 0 | Not-determined | Genome-fragment | NA | NA | 0 |
| S8_vs_k141_1943\|\|full | 1598 | No | NA | 3 | 0 | 0 | Low-quality | Genome-fragment | 18.16 | AAI-based (medium-confidence) | 0 |
| S11_vs_k141_32679\|\|full | 1599 | No | NA | 5 | 1 | 0 | Low-quality | Genome-fragment | 2.31 | AAI-based (medium-confidence) | 0 |
| S3_extracted_k141_107799 | 5072 | No | NA | 8 | 0 | 2 | Not-determined | Genome-fragment | NA | NA | 0 |
| S3_extracted_k141_13359 | 1431 | No | NA | 4 | 0 | 2 | Not-determined | Genome-fragment | NA | NA | 0 |
| S3_extracted_k141_19190 | 13533 | No | NA | 11 | 0 | 2 | Not-determined | Genome-fragment | NA | NA | 0 |
| S3_extracted_k141_91875 | 1129 | No | NA | 2 | 0 | 1 | Not-determined | Genome-fragment | NA | NA | 0 |
| S11_vs_k141_24669\|\|full | 1599 | No | NA | 3 | 2 | 0 | Low-quality | Genome-fragment | 2.4 | AAI-based (medium-confidence) | 0 |
| S1_extracted_k141_27994 | 1599 | No | NA | 1 | 0 | 1 | Low-quality | Genome-fragment | 0.6 | AAI-based (high-confidence) | 0 |
| S10_vs_57536\|\|full | 1599 | No | NA | 3 | 0 | 3 | Low-quality | Genome-fragment | 2.44 | AAI-based (high-confidence) | 0 |
| S3_extracted_k141_27984 | 5648 | No | NA | 4 | 0 | 1 | Not-determined | Genome-fragment | NA | NA | 0 |
| S3_extracted_k141_67270 | 5219 | No | NA | 3 | 0 | 0 | Not-determined | Genome-fragment | NA | NA | 0 |
| S3_extracted_k141_13438 | 1241 | No | NA | 2 | 0 | 0 | Not-determined | Genome-fragment | NA | NA | 0 |
| S10_vs_93700\|\|full | 1599 | No | NA | 3 | 1 | 0 | Low-quality | Genome-fragment | 3.4 | AAI-based (high-confidence) | 0 |
| S8_vs_k141_24812\|\|full | 1599 | No | NA | 4 | 1 | 0 | Low-quality | Genome-fragment | 3.78 | AAI-based (high-confidence) | 0 |
| S3_extracted_k141_52607 | 1152 | No | NA | 3 | 0 | 0 | Not-determined | Genome-fragment | NA | NA | 0 |
| S3_extracted_k141_36385 | 1158 | No | NA | 2 | 0 | 1 | Not-determined | Genome-fragment | NA | NA | 0 |
| S3_extracted_k141_4067 | 6741 | No | NA | 11 | 0 | 11 | Not-determined | Genome-fragment | NA | NA | 0 |
| S17_extracted_k141_155914 | 1600 | No | NA | 3 | 2 | 0 | Low-quality | Genome-fragment | 7.25 | AAI-based (high-confidence) | 0 |
| S9_extracted_k141_164 | 1600 | No | NA | 3 | 0 | 0 | Low-quality | Genome-fragment | 2.21 | AAI-based (high-confidence) | 0 |
| S13_vs_98755\|\|full | 1600 | No | NA | 3 | 0 | 0 | Low-quality | Genome-fragment | 3.98 | AAI-based (high-confidence) | 0 |
| S3_extracted_k141_4082 | 1751 | No | NA | 1 | 0 | 1 | Not-determined | Genome-fragment | NA | NA | 0 |
| S14_extracted_k141_79091 | 1600 | No | NA | 3 | 0 | 0 | Low-quality | Genome-fragment | 3.72 | AAI-based (high-confidence) | 0 |
| S14_vs_k141_13609\|\|full | 1600 | No | NA | 3 | 0 | 2 | Low-quality | Genome-fragment | 2.08 | AAI-based (medium-confidence) | 0 |
| S3_extracted_k141_13510 | 1749 | No | NA | 1 | 0 | 1 | Not-determined | Genome-fragment | NA | NA | 0 |
| S14_vs_k141_104305\|\|lt2gene | 1600 | No | NA | 1 | 1 | 0 | Low-quality | Genome-fragment | 3.01 | AAI-based (medium-confidence) | 0 |
| S3_extracted_k141_60543 | 4225 | No | NA | 10 | 0 | 10 | Not-determined | Genome-fragment | NA | NA | 0 |
| S1_extracted_k141_96105 | 1600 | No | NA | 1 | 0 | 0 | Low-quality | Genome-fragment | 1.72 | AAI-based (high-confidence) | 0 |
| S3_extracted_k141_123230 | 3562 | No | NA | 4 | 0 | 2 | Not-determined | Genome-fragment | NA | NA | 0 |
| S3_extracted_k141_67333 | 56683 | No | NA | 98 | 14 | 0 | Complete | High-quality | 100 | DTR (high-confidence) | 0 |
| S3_extracted_k141_60581 | 2060 | No | NA | 2 | 0 | 2 | Not-determined | Genome-fragment | NA | NA | 0 |
| S3_extracted_k141_99567 | 1116 | No | NA | 1 | 0 | 0 | Not-determined | Genome-fragment | NA | NA | 0 |
| S5_extracted_k141_12960 | 1600 | No | NA | 4 | 0 | 0 | Low-quality | Genome-fragment | 1.02 | AAI-based (medium-confidence) | 0 |
| S3_extracted_k141_52709 | 1027 | No | NA | 2 | 0 | 0 | Not-determined | Genome-fragment | NA | NA | 0 |
| S3_extracted_k141_4138 | 1231 | No | NA | 1 | 0 | 1 | Not-determined | Genome-fragment | NA | NA | 0 |
| S10_vs_35001\|\|full | 1600 | No | NA | 3 | 0 | 0 | Low-quality | Genome-fragment | 3.35 | AAI-based (medium-confidence) | 0 |
| S3_extracted_k141_44104 | 7909 | No | NA | 12 | 0 | 3 | Not-determined | Genome-fragment | NA | NA | 0 |
| S10_vs_72962\|\|full | 1600 | No | NA | 4 | 0 | 2 | Low-quality | Genome-fragment | 2.76 | AAI-based (high-confidence) | 0 |
| S10_vs_69130\|\|lt2gene | 1600 | No | NA | 2 | 2 | 0 | Low-quality | Genome-fragment | 3.5 | AAI-based (high-confidence) | 0 |
| S3_extracted_k141_36425 | 8709 | No | NA | 8 | 0 | 0 | Not-determined | Genome-fragment | NA | NA | 0 |
| S3_extracted_k141_13604 | 1120 | No | NA | 1 | 0 | 0 | Not-determined | Genome-fragment | NA | NA | 0 |
| S3_extracted_k141_123294 | 1211 | No | NA | 1 | 0 | 1 | Not-determined | Genome-fragment | NA | NA | 0 |
| S3_extracted_k141_60660 | 1566 | No | NA | 2 | 0 | 2 | Not-determined | Genome-fragment | NA | NA | 0 |
| S12_vs_k141_178076\|\|full | 1600 | No | NA | 4 | 0 | 0 | Low-quality | Genome-fragment | 3.89 | AAI-based (high-confidence) | 0 |
| S3_extracted_k141_52767 | 1305 | No | NA | 1 | 0 | 0 | Not-determined | Genome-fragment | NA | NA | 0 |
| S12_vs_k141_171571\|\|full | 1600 | No | NA | 2 | 0 | 0 | Low-quality | Genome-fragment | 2.08 | AAI-based (medium-confidence) | 0 |
| S3_extracted_k141_76729 | 6698 | No | NA | 7 | 0 | 3 | Not-determined | Genome-fragment | NA | NA | 0 |
| S3_extracted_k141_52772 | 3804 | No | NA | 3 | 0 | 3 | Not-determined | Genome-fragment | NA | NA | 0 |
| S3_extracted_k141_123321 | 2583 | No | NA | 2 | 0 | 2 | Not-determined | Genome-fragment | NA | NA | 0 |
| S8_extracted_k141_63596 | 1600 | No | NA | 1 | 0 | 0 | Low-quality | Genome-fragment | 1.14 | AAI-based (medium-confidence) | 0 |
| S3_extracted_k141_83839 | 1914 | No | NA | 6 | 0 | 6 | Not-determined | Genome-fragment | NA | NA | 0 |
| S3_extracted_k141_60682 | 1527 | No | NA | 1 | 0 | 1 | Not-determined | Genome-fragment | NA | NA | 0 |
| S9_vs_k141_21238\|\|full | 1601 | No | NA | 4 | 0 | 0 | Low-quality | Genome-fragment | 4.02 | AAI-based (high-confidence) | 0 |
| S3_extracted_k141_60692 | 4173 | No | NA | 2 | 0 | 1 | Not-determined | Genome-fragment | NA | NA | 0 |
| S3_extracted_k141_115815 | 1720 | No | NA | 4 | 0 | 1 | Not-determined | Genome-fragment | NA | NA | 0 |
| S3_extracted_k141_76762 | 7576 | No | NA | 5 | 0 | 3 | Not-determined | Genome-fragment | NA | NA | 0 |
| S13_vs_4991\|\|full | 1601 | No | NA | 3 | 0 | 0 | Low-quality | Genome-fragment | 4.43 | AAI-based (medium-confidence) | 0 |
| S3_extracted_k141_44145 | 1881 | No | NA | 3 | 0 | 0 | Not-determined | Genome-fragment | NA | NA | 0 |
| S14_extracted_k141_93760 | 1601 | No | NA | 4 | 3 | 0 | Low-quality | Genome-fragment | 3.55 | AAI-based (high-confidence) | 0 |
| S3_extracted_k141_115861 | 2082 | No | NA | 2 | 0 | 2 | Not-determined | Genome-fragment | NA | NA | 0 |
| S3_extracted_k141_99626 | 39745 | No | NA | 57 | 21 | 0 | Complete | High-quality | 100 | DTR (high-confidence) | 0 |
| S14_vs_k141_9012\|\|full | 1601 | No | NA | 3 | 0 | 0 | Low-quality | Genome-fragment | 3.76 | AAI-based (high-confidence) | 0 |
| S3_extracted_k141_67436 | 1697 | No | NA | 2 | 0 | 1 | Not-determined | Genome-fragment | NA | NA | 0 |
| S3_extracted_k141_13673 | 9090 | No | NA | 6 | 0 | 5 | Not-determined | Genome-fragment | NA | NA | 0 |
| S1_vs_k141_49044\|\|full | 1601 | No | NA | 4 | 1 | 0 | Low-quality | Genome-fragment | 3.47 | AAI-based (high-confidence) | 0 |
| S3_extracted_k141_123409 | 4145 | No | NA | 2 | 0 | 0 | Not-determined | Genome-fragment | NA | NA | 0 |
| S3_extracted_k141_64973 | 1601 | No | NA | 1 | 0 | 1 | Low-quality | Genome-fragment | 1 | AAI-based (medium-confidence) | 0 |
| S3_extracted_k141_115869 | 3204 | No | NA | 5 | 0 | 3 | Not-determined | Genome-fragment | NA | NA | 0 |
| S3_vs_k141_119193\|\|full | 1601 | No | NA | 3 | 1 | 0 | Low-quality | Genome-fragment | 3.84 | AAI-based (high-confidence) | 0 |
| S3_extracted_k141_28171 | 1540 | No | NA | 3 | 0 | 3 | Not-determined | Genome-fragment | NA | NA | 0 |
| S3_extracted_k141_115901 | 1650 | No | NA | 2 | 0 | 1 | Not-determined | Genome-fragment | NA | NA | 0 |
| S5_vs_k141_82186\|\|lt2gene | 1601 | No | NA | 1 | 1 | 0 | Low-quality | Genome-fragment | 2.34 | HMM-based (lower-bound) | 0 |
| S3_extracted_k141_36485 | 35347 | No | NA | 19 | 0 | 13 | Not-determined | Genome-fragment | NA | NA | 0 |
| S3_extracted_k141_83910 | 8049 | No | NA | 5 | 0 | 5 | Not-determined | Genome-fragment | NA | NA | 0 |
| S10_vs_92518\|\|full | 1601 | No | NA | 2 | 0 | 1 | Low-quality | Genome-fragment | 2.2 | AAI-based (medium-confidence) | 0 |
| S12_vs_k141_170641\|\|full | 1601 | No | NA | 3 | 0 | 0 | Low-quality | Genome-fragment | 2.88 | AAI-based (high-confidence) | 0 |
| S8_extracted_k141_132078 | 1601 | No | NA | 3 | 0 | 0 | Low-quality | Genome-fragment | 1 | AAI-based (high-confidence) | 0 |
| S9_extracted_k141_6490 | 1602 | No | NA | 4 | 1 | 0 | Low-quality | Genome-fragment | 1.86 | AAI-based (high-confidence) | 0 |
| S3_extracted_k141_108049 | 1098 | No | NA | 1 | 0 | 1 | Not-determined | Genome-fragment | NA | NA | 0 |
| S13_vs_96119\|\|full | 1602 | No | NA | 4 | 1 | 0 | Low-quality | Genome-fragment | 3.62 | AAI-based (medium-confidence) | 0 |
| S3_extracted_k141_67556 | 1334 | No | NA | 1 | 0 | 1 | Not-determined | Genome-fragment | NA | NA | 0 |
| S3_extracted_k141_36509 | 1626 | No | NA | 1 | 0 | 1 | Not-determined | Genome-fragment | NA | NA | 0 |
| S1_extracted_k141_47748 | 1602 | No | NA | 3 | 0 | 1 | Low-quality | Genome-fragment | 0.89 | AAI-based (medium-confidence) | 0 |
| S3_extracted_k141_13804 | 1495 | No | NA | 1 | 0 | 1 | Not-determined | Genome-fragment | NA | NA | 0 |
| S3_extracted_k141_92053 | 4291 | No | NA | 2 | 0 | 1 | Not-determined | Genome-fragment | NA | NA | 0 |
| S3_extracted_k141_52947 | 7764 | No | NA | 7 | 0 | 3 | Not-determined | Genome-fragment | NA | NA | 0 |
| S3_extracted_k141_76896 | 3253 | No | NA | 2 | 0 | 2 | Not-determined | Genome-fragment | NA | NA | 0 |
| S3_extracted_k141_36558 | 5958 | No | NA | 6 | 0 | 5 | Not-determined | Genome-fragment | NA | NA | 0 |
| S3_extracted_k141_13827 | 2098 | No | NA | 1 | 0 | 0 | Not-determined | Genome-fragment | NA | NA | 0 |
| S1_extracted_k141_104905 | 1602 | No | NA | 2 | 0 | 0 | Low-quality | Genome-fragment | 3.65 | AAI-based (high-confidence) | 0 |
| S1_vs_k141_42042\|\|full | 1602 | No | NA | 3 | 0 | 0 | Low-quality | Genome-fragment | 4 | AAI-based (high-confidence) | 0 |
| S3_extracted_k141_4272 | 1292 | No | NA | 3 | 0 | 3 | Not-determined | Genome-fragment | NA | NA | 0 |
| S3_vs_k141_111423\|\|full | 1602 | No | NA | 4 | 0 | 0 | Low-quality | Genome-fragment | 4.13 | AAI-based (high-confidence) | 0 |
| S16_vs_6261\|\|full | 1602 | No | NA | 3 | 0 | 0 | Low-quality | Genome-fragment | 3.69 | AAI-based (medium-confidence) | 0 |
| S3_extracted_k141_36577 | 3234 | No | NA | 2 | 0 | 2 | Not-determined | Genome-fragment | NA | NA | 0 |
| S3_extracted_k141_19536 | 1049 | No | NA | 1 | 0 | 0 | Not-determined | Genome-fragment | NA | NA | 0 |
| S3_extracted_k141_108122 | 4204 | No | NA | 4 | 0 | 3 | Not-determined | Genome-fragment | NA | NA | 0 |
| S16_vs_53753\|\|full | 1602 | No | NA | 3 | 0 | 0 | Low-quality | Genome-fragment | 2.93 | AAI-based (high-confidence) | 0 |
| S3_extracted_k141_108127 | 1297 | No | NA | 2 | 0 | 0 | Not-determined | Genome-fragment | NA | NA | 0 |
| S3_extracted_k141_19544 | 3193 | No | NA | 1 | 0 | 1 | Not-determined | Genome-fragment | NA | NA | 0 |
| S10_extracted_27357 | 1602 | No | NA | 3 | 2 | 0 | Low-quality | Genome-fragment | 2.08 | AAI-based (high-confidence) | 0 |
| S3_extracted_k141_44397 | 4338 | No | NA | 3 | 0 | 1 | Not-determined | Genome-fragment | NA | NA | 0 |
| S3_extracted_k141_28306 | 1000 | No | NA | 2 | 0 | 0 | Not-determined | Genome-fragment | NA | NA | 0 |
| S12_extracted_k141_86737 | 1602 | No | NA | 1 | 0 | 0 | Low-quality | Genome-fragment | 2.57 | AAI-based (high-confidence) | 0 |
| S8_vs_k141_86373\|\|full | 1602 | No | NA | 3 | 0 | 1 | Low-quality | Genome-fragment | 2.25 | AAI-based (medium-confidence) | 0 |
| S3_extracted_k141_28309 | 8674 | No | NA | 5 | 0 | 4 | Not-determined | Genome-fragment | NA | NA | 0 |
| S11_extracted_k141_98157 | 1603 | No | NA | 4 | 0 | 0 | Low-quality | Genome-fragment | 3.5 | AAI-based (high-confidence) | 0 |
| S3_extracted_k141_84066 | 1680 | No | NA | 1 | 0 | 0 | Not-determined | Genome-fragment | NA | NA | 0 |
| S3_extracted_k141_53053 | 3046 | No | NA | 4 | 0 | 4 | Not-determined | Genome-fragment | NA | NA | 0 |
| S11_vs_k141_74777\|\|full | 1603 | No | NA | 3 | 0 | 0 | Low-quality | Genome-fragment | 2.08 | AAI-based (medium-confidence) | 0 |
| S14_vs_k141_84166\|\|full | 1603 | No | NA | 4 | 0 | 0 | Low-quality | Genome-fragment | 1 | AAI-based (medium-confidence) | 0 |
| S3_extracted_k141_123575 | 4915 | No | NA | 3 | 0 | 3 | Not-determined | Genome-fragment | NA | NA | 0 |
| S3_extracted_k141_44405 | 38184 | No | NA | 52 | 26 | 0 | Medium-quality | Genome-fragment | 63.68 | AAI-based (medium-confidence) | 0 |
| S3_vs_k141_56834\|\|full | 1603 | No | NA | 4 | 1 | 0 | Low-quality | Genome-fragment | 0.71 | HMM-based (lower-bound) | 0 |
| S3_extracted_k141_19637 | 3081 | No | NA | 4 | 0 | 3 | Not-determined | Genome-fragment | NA | NA | 0 |
| S7_vs_84859\|\|full | 1603 | No | NA | 2 | 1 | 0 | Low-quality | Genome-fragment | 3.65 | AAI-based (high-confidence) | 0 |
| S3_extracted_k141_53108 | 3131 | No | NA | 7 | 0 | 1 | Not-determined | Genome-fragment | NA | NA | 0 |
| S7_vs_109895\|\|full | 1603 | No | NA | 6 | 0 | 0 | Low-quality | Genome-fragment | 2.99 | AAI-based (medium-confidence) | 0 |
| S3_extracted_k141_4359 | 2000 | No | NA | 1 | 0 | 0 | Not-determined | Genome-fragment | NA | NA | 0 |
| S16_vs_94972\|\|full | 1603 | No | NA | 2 | 0 | 0 | Low-quality | Genome-fragment | 3.16 | AAI-based (medium-confidence) | 0 |
| S10_vs_107588\|\|full | 1603 | No | NA | 3 | 0 | 0 | Low-quality | Genome-fragment | 0.65 | AAI-based (high-confidence) | 0 |
| S3_extracted_k141_99732 | 13105 | No | NA | 9 | 0 | 7 | Not-determined | Genome-fragment | NA | NA | 0 |
| S17_extracted_k141_44673 | 1604 | No | NA | 1 | 0 | 1 | Low-quality | Genome-fragment | 0.6 | AAI-based (high-confidence) | 0 |
| S3_extracted_k141_84179 | 1056 | No | NA | 2 | 0 | 2 | Not-determined | Genome-fragment | NA | NA | 0 |
| S3_extracted_k141_14060 | 1066 | No | NA | 1 | 0 | 1 | Not-determined | Genome-fragment | NA | NA | 0 |
| S9_vs_k141_2680\|\|full | 1604 | No | NA | 5 | 2 | 0 | Low-quality | Genome-fragment | 4.19 | AAI-based (high-confidence) | 0 |
| S3_extracted_k141_92218 | 3182 | No | NA | 5 | 0 | 0 | Not-determined | Genome-fragment | NA | NA | 0 |
| S3_extracted_k141_77117 | 5315 | No | NA | 5 | 0 | 3 | Not-determined | Genome-fragment | NA | NA | 0 |
| S3_extracted_k141_99734 | 46304 | No | NA | 69 | 18 | 0 | High-quality | High-quality | 98.26 | AAI-based (high-confidence) | 0 |
| S14_extracted_k141_39622 | 1604 | No | NA | 1 | 0 | 1 | Low-quality | Genome-fragment | 0.85 | AAI-based (medium-confidence) | 0 |
| S3_extracted_k141_4406 | 1185 | No | NA | 2 | 0 | 1 | Not-determined | Genome-fragment | NA | NA | 0 |
| S3_extracted_k141_19734 | 2449 | No | NA | 2 | 0 | 2 | Not-determined | Genome-fragment | NA | NA | 0 |
| S3_extracted_k141_99750 | 3513 | No | NA | 3 | 0 | 3 | Not-determined | Genome-fragment | NA | NA | 0 |
| S3_extracted_k141_19767 | 1070 | No | NA | 1 | 0 | 1 | Not-determined | Genome-fragment | NA | NA | 0 |
| S3_extracted_k141_36683 | 2340 | No | NA | 2 | 0 | 1 | Not-determined | Genome-fragment | NA | NA | 0 |
| S3_extracted_k141_28419 | 1460 | No | NA | 1 | 0 | 1 | Not-determined | Genome-fragment | NA | NA | 0 |
| S3_extracted_k141_67772 | 3582 | No | NA | 5 | 0 | 2 | Not-determined | Genome-fragment | NA | NA | 0 |
| S3_extracted_k141_92336 | 1550 | No | NA | 1 | 0 | 0 | Not-determined | Genome-fragment | NA | NA | 0 |
| S3_extracted_k141_67786 | 3212 | No | NA | 3 | 0 | 2 | Not-determined | Genome-fragment | NA | NA | 0 |
| S3_vs_k141_66207\|\|full | 1604 | No | NA | 3 | 0 | 0 | Low-quality | Genome-fragment | 1.2 | AAI-based (high-confidence) | 0 |
| S3_extracted_k141_44445 | 1009 | No | NA | 1 | 0 | 0 | Not-determined | Genome-fragment | NA | NA | 0 |
| S3_extracted_k141_77224 | 2318 | No | NA | 2 | 0 | 0 | Not-determined | Genome-fragment | NA | NA | 0 |
| S3_extracted_k141_77227 | 1923 | No | NA | 1 | 0 | 1 | Not-determined | Genome-fragment | NA | NA | 0 |
| S3_extracted_k141_28466 | 1620 | No | NA | 5 | 0 | 1 | Not-determined | Genome-fragment | NA | NA | 0 |
| S3_extracted_k141_77230 | 1600 | No | NA | 1 | 0 | 1 | Not-determined | Genome-fragment | NA | NA | 0 |
| S5_vs_k141_28743\|\|full | 1604 | No | NA | 2 | 0 | 0 | Low-quality | Genome-fragment | 2.08 | AAI-based (high-confidence) | 0 |
| S3_extracted_k141_4440 | 3503 | No | NA | 3 | 0 | 3 | Not-determined | Genome-fragment | NA | NA | 0 |
| S3_extracted_k141_61062 | 1969 | No | NA | 2 | 0 | 1 | Not-determined | Genome-fragment | NA | NA | 0 |
| S16_extracted_114946 | 1604 | No | NA | 6 | 1 | 0 | Low-quality | Genome-fragment | 2.47 | HMM-based (lower-bound) | 0 |
| S16_vs_112982\|\|full | 1604 | No | NA | 2 | 1 | 0 | Low-quality | Genome-fragment | 0.89 | HMM-based (lower-bound) | 0 |
| S3_extracted_k141_77242 | 1406 | No | NA | 1 | 0 | 1 | Not-determined | Genome-fragment | NA | NA | 0 |
| S16_vs_156641\|\|full | 1604 | No | NA | 2 | 0 | 0 | Low-quality | Genome-fragment | 2.75 | AAI-based (high-confidence) | 0 |
| S6_extracted_k141_55199 | 1604 | No | NA | 2 | 0 | 0 | Low-quality | Genome-fragment | 2.01 | AAI-based (high-confidence) | 0 |
| S3_extracted_k141_67828 | 6187 | No | NA | 10 | 2 | 0 | Complete | High-quality | 100 | DTR (high-confidence) | 0 |
| S6_vs_k141_22524\|\|full | 1604 | No | NA | 4 | 4 | 0 | Low-quality | Genome-fragment | 4.22 | AAI-based (high-confidence) | 0 |
| S8_extracted_k141_192470 | 1604 | No | NA | 3 | 0 | 0 | Low-quality | Genome-fragment | 0.92 | AAI-based (high-confidence) | 0 |
| S3_extracted_k141_84385 | 1327 | No | NA | 1 | 0 | 0 | Not-determined | Genome-fragment | NA | NA | 0 |
| S17_extracted_k141_61388 | 1605 | No | NA | 1 | 0 | 0 | Low-quality | Genome-fragment | 1.47 | AAI-based (medium-confidence) | 0 |
| S17_extracted_k141_100264 | 1605 | No | NA | 4 | 0 | 0 | Low-quality | Genome-fragment | 4.01 | AAI-based (high-confidence) | 0 |
| S3_extracted_k141_123731 | 2021 | No | NA | 5 | 0 | 1 | Not-determined | Genome-fragment | NA | NA | 0 |
| S13_vs_40441\|\|full | 1605 | No | NA | 5 | 0 | 0 | Low-quality | Genome-fragment | 1.61 | AAI-based (medium-confidence) | 0 |
| S14_extracted_k141_62820 | 1605 | No | NA | 3 | 0 | 1 | Low-quality | Genome-fragment | 1.43 | AAI-based (high-confidence) | 0 |
| S3_extracted_k141_92484 | 1001 | No | NA | 2 | 0 | 1 | Not-determined | Genome-fragment | NA | NA | 0 |
| S14_extracted_k141_64579 | 1605 | No | NA | 3 | 0 | 0 | Low-quality | Genome-fragment | 2.68 | AAI-based (high-confidence) | 0 |
| S14_vs_k141_98701\|\|full | 1605 | No | NA | 4 | 3 | 0 | Low-quality | Genome-fragment | 3.29 | AAI-based (medium-confidence) | 0 |
| S1_vs_k141_89447\|\|full | 1605 | No | NA | 4 | 0 | 0 | Low-quality | Genome-fragment | 3.8 | AAI-based (high-confidence) | 0 |
| S3_extracted_k141_4545 | 1765 | No | NA | 2 | 0 | 0 | Not-determined | Genome-fragment | NA | NA | 0 |
| S3_vs_k141_34524\|\|full | 1605 | No | NA | 2 | 0 | 1 | Low-quality | Genome-fragment | 1.54 | AAI-based (high-confidence) | 0 |
| S7_vs_38028\|\|full | 1605 | No | NA | 6 | 1 | 0 | Low-quality | Genome-fragment | 3.55 | AAI-based (high-confidence) | 0 |
| S3_extracted_k141_4574 | 2056 | No | NA | 2 | 0 | 1 | Not-determined | Genome-fragment | NA | NA | 0 |
| S3_extracted_k141_61332 | 1795 | No | NA | 2 | 0 | 1 | Not-determined | Genome-fragment | NA | NA | 0 |
| S3_extracted_k141_116362 | 3952 | No | NA | 2 | 0 | 2 | Not-determined | Genome-fragment | NA | NA | 0 |
| S16_extracted_48641 | 1605 | No | NA | 3 | 0 | 0 | Low-quality | Genome-fragment | 3.9 | AAI-based (high-confidence) | 0 |
| S3_extracted_k141_77346 | 1081 | No | NA | 2 | 0 | 2 | Not-determined | Genome-fragment | NA | NA | 0 |
| S3_extracted_k141_14361 | 3307 | No | NA | 3 | 0 | 0 | Not-determined | Genome-fragment | NA | NA | 0 |
| S3_extracted_k141_99990 | 2015 | No | NA | 2 | 0 | 1 | Not-determined | Genome-fragment | NA | NA | 0 |
| S3_extracted_k141_77365 | 1944 | No | NA | 1 | 0 | 1 | Not-determined | Genome-fragment | NA | NA | 0 |
| S16_vs_138721\|\|full | 1605 | No | NA | 3 | 0 | 0 | Low-quality | Genome-fragment | 3.42 | AAI-based (high-confidence) | 0 |
| S3_extracted_k141_44555 | 1662 | No | NA | 1 | 0 | 0 | Not-determined | Genome-fragment | NA | NA | 0 |
| S3_extracted_k141_123797 | 3312 | No | NA | 1 | 0 | 1 | Not-determined | Genome-fragment | NA | NA | 0 |
| S3_extracted_k141_67926 | 1368 | No | NA | 2 | 0 | 0 | Not-determined | Genome-fragment | NA | NA | 0 |
| S3_extracted_k141_123799 | 4080 | No | NA | 3 | 0 | 1 | Not-determined | Genome-fragment | NA | NA | 0 |
| S12_vs_k141_114960\|\|full | 1605 | No | NA | 3 | 3 | 0 | Low-quality | Genome-fragment | 4.68 | AAI-based (high-confidence) | 0 |
| S3_extracted_k141_116400 | 1278 | No | NA | 1 | 0 | 1 | Not-determined | Genome-fragment | NA | NA | 0 |
| S12_vs_k141_148410\|\|full | 1605 | No | NA | 3 | 1 | 0 | Low-quality | Genome-fragment | 2.22 | AAI-based (high-confidence) | 0 |
| S3_extracted_k141_108459 | 3113 | No | NA | 4 | 0 | 2 | Not-determined | Genome-fragment | NA | NA | 0 |
| S3_extracted_k141_123809 | 6021 | No | NA | 4 | 0 | 2 | Not-determined | Genome-fragment | NA | NA | 0 |
| S3_extracted_k141_84548 | 2310 | No | NA | 3 | 0 | 3 | Not-determined | Genome-fragment | NA | NA | 0 |
| S3_extracted_k141_4618 | 5458 | No | NA | 4 | 0 | 4 | Not-determined | Genome-fragment | NA | NA | 0 |
| S11_vs_k141_7972\|\|full | 1606 | No | NA | 5 | 3 | 0 | Low-quality | Genome-fragment | 3.52 | AAI-based (high-confidence) | 0 |
| S3_extracted_k141_116442 | 1005 | No | NA | 2 | 0 | 2 | Not-determined | Genome-fragment | NA | NA | 0 |
| S3_extracted_k141_116468 | 2687 | No | NA | 3 | 0 | 3 | Not-determined | Genome-fragment | NA | NA | 0 |
| S3_extracted_k141_67958 | 2369 | No | NA | 3 | 0 | 0 | Not-determined | Genome-fragment | NA | NA | 0 |
| S14_vs_k141_23148\|\|full | 1606 | No | NA | 4 | 0 | 0 | Low-quality | Genome-fragment | 2.75 | AAI-based (medium-confidence) | 0 |
| S3_extracted_k141_20071 | 1119 | No | NA | 1 | 0 | 0 | Not-determined | Genome-fragment | NA | NA | 0 |
| S3_extracted_k141_28704 | 1670 | No | NA | 4 | 0 | 2 | Not-determined | Genome-fragment | NA | NA | 0 |
| S5_vs_k141_81352\|\|full | 1606 | No | NA | 2 | 0 | 0 | Low-quality | Genome-fragment | 1.05 | AAI-based (high-confidence) | 0 |
| S5_vs_k141_123926\|\|lt2gene | 1606 | No | NA | 2 | 1 | 0 | Low-quality | Genome-fragment | 2.71 | AAI-based (high-confidence) | 0 |
| S7_vs_86435\|\|full | 1606 | No | NA | 3 | 2 | 0 | Low-quality | Genome-fragment | 4.7 | AAI-based (high-confidence) | 0 |
| S3_extracted_k141_14465 | 1010 | No | NA | 4 | 0 | 0 | Not-determined | Genome-fragment | NA | NA | 0 |
| S10_vs_115452\|\|full | 1606 | No | NA | 3 | 2 | 0 | Low-quality | Genome-fragment | 2.69 | AAI-based (medium-confidence) | 0 |
| S3_extracted_k141_123878 | 1845 | No | NA | 2 | 0 | 2 | Not-determined | Genome-fragment | NA | NA | 0 |
| S3_extracted_k141_123888 | 2133 | No | NA | 1 | 0 | 1 | Not-determined | Genome-fragment | NA | NA | 0 |
| S3_extracted_k141_36932 | 1085 | No | NA | 2 | 0 | 1 | Not-determined | Genome-fragment | NA | NA | 0 |

| | | | | | | | | | | | |
|---|---|---|---|---|---|---|---|---|---|---|---|
| S12_vs_k141_80270\|\|full | 1606 | No | NA | 3 | 0 | 1 | Low-quality | Genome-fragment | 0.76 | AAI-based (medium-confidence) | 0 |
| S3_extracted_k141_123890 | 1016 | No | NA | 1 | 0 | 0 | Not-determined | Genome-fragment | NA | NA | 0 |
| S12_vs_k141_148746\|\|full | 1606 | No | NA | 3 | 1 | 0 | Low-quality | Genome-fragment | 2.43 | HMM-based (lower-bound) | 0 |
| S6_vs_k141_17960\|\|full | 1606 | No | NA | 5 | 0 | 0 | Low-quality | Genome-fragment | 3.53 | AAI-based (high-confidence) | 0 |
| S8_vs_k141_78737\|\|full | 1606 | No | NA | 3 | 0 | 1 | Low-quality | Genome-fragment | 1.3 | AAI-based (high-confidence) | 0 |
| S3_extracted_k141_14489 | 3162 | No | NA | 4 | 0 | 1 | Not-determined | Genome-fragment | NA | NA | 0 |
| S3_extracted_k141_28722 | 1472 | No | NA | 4 | 0 | 0 | Not-determined | Genome-fragment | NA | NA | 0 |
| S8_vs_k141_165916\|\|full | 1606 | No | NA | 3 | 0 | 1 | Low-quality | Genome-fragment | 1.92 | AAI-based (high-confidence) | 0 |
| S17_extracted_k141_143228 | 1607 | No | NA | 4 | 1 | 0 | Low-quality | Genome-fragment | 2.6 | AAI-based (high-confidence) | 0 |
| S17_vs_k141_136591\|\|full | 1607 | No | NA | 4 | 1 | 0 | Low-quality | Genome-fragment | 2.4 | AAI-based (high-confidence) | 0 |
| S3_extracted_k141_61564 | 2115 | No | NA | 3 | 0 | 3 | Not-determined | Genome-fragment | NA | NA | 0 |
| S3_extracted_k141_108546 | 4668 | No | NA | 5 | 0 | 3 | Not-determined | Genome-fragment | NA | NA | 0 |
| S9_extracted_k141_12268 | 1607 | No | NA | 2 | 0 | 0 | Low-quality | Genome-fragment | 3.87 | AAI-based (medium-confidence) | 0 |
| S11_vs_k141_2150\|\|full | 1607 | No | NA | 3 | 0 | 1 | Low-quality | Genome-fragment | 1.35 | AAI-based (high-confidence) | 0 |
| S3_extracted_k141_100080 | 3058 | No | NA | 3 | 0 | 1 | Not-determined | Genome-fragment | NA | NA | 0 |
| S3_extracted_k141_61590 | 2355 | No | NA | 5 | 0 | 3 | Not-determined | Genome-fragment | NA | NA | 0 |
| S3_extracted_k141_53501 | 1065 | No | NA | 1 | 0 | 0 | Not-determined | Genome-fragment | NA | NA | 0 |
| S13_vs_88503\|\|full | 1607 | No | NA | 2 | 0 | 1 | Low-quality | Genome-fragment | 3.45 | AAI-based (high-confidence) | 0 |
| S14_extracted_k141_45652 | 1607 | No | NA | 3 | 2 | 0 | Low-quality | Genome-fragment | 5.31 | AAI-based (high-confidence) | 0 |
| S14_vs_k141_4310\|\|full | 1607 | No | NA | 3 | 0 | 0 | Low-quality | Genome-fragment | 3.79 | AAI-based (high-confidence) | 0 |
| S3_extracted_k141_84832 | 12719 | No | NA | 15 | 10 | 0 | Medium-quality | Genome-fragment | 54.33 | AAI-based (medium-confidence) | 0 |
| S3_extracted_k141_68038 | 1000 | No | NA | 2 | 0 | 1 | Not-determined | Genome-fragment | NA | NA | 0 |
| S3_vs_k141_27821\|\|full | 1607 | No | NA | 3 | 1 | 0 | Low-quality | Genome-fragment | 2.55 | AAI-based (high-confidence) | 0 |
| S3_extracted_k141_92779 | 1927 | No | NA | 5 | 0 | 0 | Not-determined | Genome-fragment | NA | NA | 0 |
| S3_extracted_k141_124049 | 1502 | No | NA | 3 | 0 | 0 | Not-determined | Genome-fragment | NA | NA | 0 |
| S3_extracted_k141_108597 | 2034 | No | NA | 2 | 0 | 2 | Not-determined | Genome-fragment | NA | NA | 0 |
| S3_extracted_k141_108599 | 1166 | No | NA | 3 | 0 | 0 | Not-determined | Genome-fragment | NA | NA | 0 |
| S3_extracted_k141_68059 | 1383 | No | NA | 2 | 0 | 1 | Not-determined | Genome-fragment | NA | NA | 0 |
| S5_vs_k141_3735\|\|full | 1607 | No | NA | 2 | 0 | 0 | Low-quality | Genome-fragment | 4.01 | AAI-based (high-confidence) | 0 |
| S16_extracted_29992 | 1607 | No | NA | 5 | 0 | 0 | Low-quality | Genome-fragment | 2.93 | AAI-based (high-confidence) | 0 |
| S3_extracted_k141_124077 | 1032 | No | NA | 1 | 0 | 1 | Not-determined | Genome-fragment | NA | NA | 0 |
| S12_vs_k141_71075\|\|full | 1607 | No | NA | 3 | 1 | 0 | Low-quality | Genome-fragment | 3.66 | AAI-based (high-confidence) | 0 |
| S6_vs_k141_8216\|\|full | 1607 | No | NA | 5 | 1 | 0 | Low-quality | Genome-fragment | 3.64 | AAI-based (high-confidence) | 0 |
| S3_extracted_k141_84866 | 1181 | No | NA | 1 | 0 | 1 | Not-determined | Genome-fragment | NA | NA | 0 |
| S3_extracted_k141_14672 | 1142 | No | NA | 2 | 0 | 1 | Not-determined | Genome-fragment | NA | NA | 0 |
| S3_extracted_k141_92798 | 1146 | No | NA | 2 | 0 | 1 | Not-determined | Genome-fragment | NA | NA | 0 |
| S3_extracted_k141_108641 | 1631 | No | NA | 1 | 0 | 1 | Not-determined | Genome-fragment | NA | NA | 0 |
| S8_vs_k141_200146\|\|full | 1607 | No | NA | 3 | 1 | 0 | Low-quality | Genome-fragment | 3.03 | AAI-based (medium-confidence) | 0 |
| S8_vs_k141_146133\|\|full | 1607 | No | NA | 2 | 0 | 1 | Low-quality | Genome-fragment | 1.55 | AAI-based (medium-confidence) | 0 |
| S3_extracted_k141_108662 | 1105 | No | NA | 3 | 0 | 1 | Not-determined | Genome-fragment | NA | NA | 0 |
| S11_vs_k141_22434\|\|full | 1608 | No | NA | 3 | 0 | 0 | Low-quality | Genome-fragment | 2.57 | AAI-based (medium-confidence) | 0 |
| S3_extracted_k141_92844 | 1635 | No | NA | 1 | 0 | 1 | Not-determined | Genome-fragment | NA | NA | 0 |
| S11_vs_k141_36954\|\|full | 1608 | No | NA | 2 | 0 | 0 | Low-quality | Genome-fragment | 4.62 | AAI-based (high-confidence) | 0 |
| S3_extracted_k141_68157 | 1295 | No | NA | 1 | 0 | 0 | Not-determined | Genome-fragment | NA | NA | 0 |
| S13_vs_141198\|\|full | 1608 | Yes | 1464 | 5 | 1 | 1 | Low-quality | Genome-fragment | 3.74 | AAI-based (medium-confidence) | 8.96 |
| S3_extracted_k141_4723 | 1050 | No | NA | 2 | 0 | 0 | Not-determined | Genome-fragment | NA | NA | 0 |
| S3_extracted_k141_68210 | 1243 | No | NA | 2 | 0 | 0 | Not-determined | Genome-fragment | NA | NA | 0 |
| S1_extracted_k141_49240 | 1608 | No | NA | 2 | 1 | 0 | Low-quality | Genome-fragment | 3.66 | AAI-based (high-confidence) | 0 |
| S3_extracted_k141_68220 | 1105 | No | NA | 2 | 0 | 2 | Not-determined | Genome-fragment | NA | NA | 0 |
| S1_extracted_k141_118586 | 1608 | No | NA | 2 | 1 | 0 | Low-quality | Genome-fragment | 0.92 | AAI-based (high-confidence) | 0 |
| S3_extracted_k141_92883 | 1160 | No | NA | 3 | 0 | 3 | Not-determined | Genome-fragment | NA | NA | 0 |
| S3_extracted_k141_14796 | 1344 | No | NA | 1 | 0 | 1 | Not-determined | Genome-fragment | NA | NA | 0 |
| S3_extracted_k141_4761 | 1088 | No | NA | 1 | 0 | 1 | Not-determined | Genome-fragment | NA | NA | 0 |
| S1_vs_k141_31294\|\|full | 1608 | No | NA | 5 | 0 | 0 | Low-quality | Genome-fragment | 1.13 | AAI-based (medium-confidence) | 0 |
| S3_extracted_k141_61795 | 1217 | No | NA | 2 | 0 | 0 | Not-determined | Genome-fragment | NA | NA | 0 |
| S7_vs_47569\|\|full | 1608 | No | NA | 2 | 0 | 1 | Low-quality | Genome-fragment | 1.06 | AAI-based (medium-confidence) | 0 |
| S3_extracted_k141_4783 | 1750 | No | NA | 3 | 0 | 0 | Not-determined | Genome-fragment | NA | NA | 0 |
| S16_vs_101328\|\|full | 1608 | No | NA | 4 | 0 | 0 | Low-quality | Genome-fragment | 0.94 | AAI-based (medium-confidence) | 0 |
| S3_extracted_k141_124184 | 1185 | No | NA | 1 | 0 | 0 | Not-determined | Genome-fragment | NA | NA | 0 |
| S3_extracted_k141_37218 | 7637 | No | NA | 6 | 0 | 6 | Not-determined | Genome-fragment | NA | NA | 0 |
| S3_extracted_k141_116704 | 1262 | No | NA | 1 | 0 | 0 | Not-determined | Genome-fragment | NA | NA | 0 |
| S10_vs_76332\|\|full | 1608 | No | NA | 4 | 0 | 1 | Low-quality | Genome-fragment | 1.09 | AAI-based (medium-confidence) | 0 |
| S3_extracted_k141_108841 | 2315 | No | NA | 2 | 0 | 2 | Not-determined | Genome-fragment | NA | NA | 0 |
| S6_vs_k141_23047\|\|full | 1608 | No | NA | 5 | 0 | 0 | Low-quality | Genome-fragment | 4.16 | AAI-based (high-confidence) | 0 |
| S8_vs_k141_177854\|\|full | 1608 | No | NA | 5 | 0 | 2 | Low-quality | Genome-fragment | 2.9 | AAI-based (medium-confidence) | 0 |
| S3_extracted_k141_14849 | 5705 | No | NA | 3 | 0 | 3 | Not-determined | Genome-fragment | NA | NA | 0 |
| S3_extracted_k141_85052 | 1561 | No | NA | 2 | 0 | 1 | Not-determined | Genome-fragment | NA | NA | 0 |
| S3_extracted_k141_37253 | 1298 | No | NA | 1 | 0 | 0 | Not-determined | Genome-fragment | NA | NA | 0 |
| S3_extracted_k141_77840 | 1489 | No | NA | 3 | 0 | 0 | Not-determined | Genome-fragment | NA | NA | 0 |
| S3_extracted_k141_108897 | 6123 | No | NA | 4 | 0 | 3 | Not-determined | Genome-fragment | NA | NA | 0 |
| S11_vs_k141_60283\|\|full | 1609 | No | NA | 3 | 0 | 0 | Low-quality | Genome-fragment | 3.76 | AAI-based (high-confidence) | 0 |
| S11_vs_k141_93141\|\|full | 1609 | No | NA | 3 | 0 | 1 | Low-quality | Genome-fragment | 2.51 | AAI-based (medium-confidence) | 0 |
| S3_extracted_k141_61860 | 67881 | No | NA | 92 | 27 | 0 | Complete | High-quality | 100 | DTR (high-confidence) | 0 |
| S3_extracted_k141_108936 | 1102 | No | NA | 2 | 0 | 0 | Not-determined | Genome-fragment | NA | NA | 0 |
| S3_extracted_k141_92980 | 3249 | No | NA | 4 | 0 | 0 | Not-determined | Genome-fragment | NA | NA | 0 |
| S3_extracted_k141_68317 | 1056 | No | NA | 2 | 0 | 0 | Not-determined | Genome-fragment | NA | NA | 0 |
| S11_vs_k141_79473\|\|lt2gene | 1609 | No | NA | 1 | 1 | 0 | Low-quality | Genome-fragment | 3.05 | HMM-based (lower-bound) | 0 |
| S3_extracted_k141_28981 | 2818 | No | NA | 4 | 0 | 3 | Not-determined | Genome-fragment | NA | NA | 0 |
| S3_extracted_k141_85104 | 1085 | No | NA | 1 | 0 | 1 | Not-determined | Genome-fragment | NA | NA | 0 |
| S1_extracted_k141_2973 | 1609 | No | NA | 1 | 0 | 0 | Low-quality | Genome-fragment | 2.77 | AAI-based (high-confidence) | 0 |
| S3_extracted_k141_29008 | 4575 | No | NA | 8 | 0 | 2 | Not-determined | Genome-fragment | NA | NA | 0 |
| S3_extracted_k141_68366 | 1239 | No | NA | 2 | 0 | 0 | Not-determined | Genome-fragment | NA | NA | 0 |
| S1_vs_k141_85839\|\|full | 1609 | No | NA | 4 | 0 | 0 | Low-quality | Genome-fragment | 4.44 | AAI-based (high-confidence) | 0 |
| S3_extracted_k141_100440 | 31270 | No | NA | 43 | 24 | 0 | Medium-quality | Genome-fragment | 68.24 | AAI-based (high-confidence) | 0 |
| S3_extracted_k141_37312 | 3304 | No | NA | 7 | 0 | 3 | Not-determined | Genome-fragment | NA | NA | 0 |
| S3_extracted_k141_5021 | 1514 | No | NA | 2 | 0 | 1 | Not-determined | Genome-fragment | NA | NA | 0 |
| S3_extracted_k141_108959 | 61482 | No | NA | 115 | 26 | 1 | Complete | High-quality | 100 | DTR (high-confidence) | 0 |
| S3_extracted_k141_61941 | 1720 | No | NA | 2 | 0 | 0 | Not-determined | Genome-fragment | NA | NA | 0 |
| S3_extracted_k141_68398 | 1429 | No | NA | 3 | 0 | 1 | Not-determined | Genome-fragment | NA | NA | 0 |
| S3_extracted_k141_61959 | 4813 | No | NA | 3 | 0 | 2 | Not-determined | Genome-fragment | NA | NA | 0 |
| S3_extracted_k141_5042 | 2385 | No | NA | 1 | 0 | 1 | Not-determined | Genome-fragment | NA | NA | 0 |
| S3_extracted_k141_68417 | 1476 | No | NA | 2 | 0 | 1 | Not-determined | Genome-fragment | NA | NA | 0 |
| S3_vs_k141_12119\|\|full | 1609 | No | NA | 2 | 0 | 1 | Low-quality | Genome-fragment | 1.54 | AAI-based (high-confidence) | 0 |
| S7_vs_113013\|\|full | 1609 | No | NA | 2 | 0 | 0 | Low-quality | Genome-fragment | 3.79 | AAI-based (medium-confidence) | 0 |
| S3_extracted_k141_68442 | 1051 | No | NA | 1 | 0 | 0 | Not-determined | Genome-fragment | NA | NA | 0 |
| S3_extracted_k141_5123 | 1000 | No | NA | 2 | 0 | 0 | Not-determined | Genome-fragment | NA | NA | 0 |
| S7_vs_126743\|\|full | 1609 | No | NA | 3 | 0 | 0 | Low-quality | Genome-fragment | 1.35 | AAI-based (high-confidence) | 0 |
| S7_vs_61046\|\|lt2gene | 1609 | No | NA | 1 | 1 | 0 | Low-quality | Genome-fragment | 4.02 | AAI-based (high-confidence) | 0 |
| S16_vs_112078\|\|full | 1609 | No | NA | 3 | 2 | 0 | Low-quality | Genome-fragment | 4.07 | AAI-based (high-confidence) | 0 |
| S3_extracted_k141_37410 | 1166 | No | NA | 2 | 0 | 0 | Not-determined | Genome-fragment | NA | NA | 0 |
| S16_vs_170711\|\|full | 1609 | Yes | 309 | 3 | 1 | 1 | Low-quality | Genome-fragment | 0.5 | HMM-based (lower-bound) | 80.8 |
| S3_extracted_k141_109090 | 1043 | No | NA | 2 | 0 | 2 | Not-determined | Genome-fragment | NA | NA | 0 |
| S10_extracted_42331 | 1609 | No | NA | 4 | 1 | 0 | Low-quality | Genome-fragment | 1.58 | AAI-based (high-confidence) | 0 |
| S3_extracted_k141_68523 | 1032 | No | NA | 2 | 0 | 1 | Not-determined | Genome-fragment | NA | NA | 0 |
| S3_extracted_k141_29099 | 4741 | No | NA | 3 | 0 | 1 | Not-determined | Genome-fragment | NA | NA | 0 |
| S3_extracted_k141_116927 | 1761 | No | NA | 2 | 0 | 0 | Not-determined | Genome-fragment | NA | NA | 0 |
| S3_extracted_k141_78056 | 3494 | No | NA | 6 | 0 | 1 | Not-determined | Genome-fragment | NA | NA | 0 |
| S3_extracted_k141_15003 | 5303 | No | NA | 3 | 0 | 1 | Not-determined | Genome-fragment | NA | NA | 0 |
| S3_extracted_k141_85324 | 1590 | No | NA | 1 | 0 | 0 | Not-determined | Genome-fragment | NA | NA | 0 |
| S3_extracted_k141_109151 | 3701 | No | NA | 6 | 0 | 4 | Not-determined | Genome-fragment | NA | NA | 0 |
| S10_vs_11420\|\|full | 1609 | No | NA | 3 | 3 | 0 | Low-quality | Genome-fragment | 1.52 | AAI-based (high-confidence) | 0 |
| S3_extracted_k141_15084 | 4963 | No | NA | 7 | 0 | 4 | Not-determined | Genome-fragment | NA | NA | 0 |
| S8_extracted_k141_195304 | 1609 | No | NA | 2 | 1 | 0 | Low-quality | Genome-fragment | 1.03 | AAI-based (high-confidence) | 0 |
| S3_extracted_k141_116976 | 1341 | No | NA | 1 | 0 | 1 | Not-determined | Genome-fragment | NA | NA | 0 |
| S17_vs_k141_123891\|\|full | 1610 | No | NA | 3 | 2 | 0 | Low-quality | Genome-fragment | 3.35 | HMM-based (lower-bound) | 0 |
| S14_vs_k141_41053\|\|full | 1610 | No | NA | 2 | 0 | 2 | Low-quality | Genome-fragment | 2.36 | AAI-based (medium-confidence) | 0 |
| S1_extracted_k141_120575 | 1610 | No | NA | 1 | 0 | 0 | Low-quality | Genome-fragment | 3.52 | AAI-based (high-confidence) | 0 |
| S3_extracted_k141_68675 | 1559 | No | NA | 2 | 0 | 1 | Not-determined | Genome-fragment | NA | NA | 0 |
| S1_vs_k141_21855\|\|full | 1610 | No | NA | 2 | 0 | 2 | Low-quality | Genome-fragment | 1.15 | AAI-based (high-confidence) | 0 |
| S3_extracted_k141_37563 | 1106 | No | NA | 1 | 0 | 0 | Not-determined | Genome-fragment | NA | NA | 0 |
| S3_extracted_k141_15218 | 1326 | No | NA | 4 | 0 | 0 | Not-determined | Genome-fragment | NA | NA | 0 |
| S3_extracted_k141_15221 | 3266 | No | NA | 3 | 0 | 3 | Not-determined | Genome-fragment | NA | NA | 0 |
| S8_extracted_k141_41297 | 1610 | No | NA | 3 | 1 | 0 | Low-quality | Genome-fragment | 2.57 | AAI-based (high-confidence) | 0 |
| S3_extracted_k141_124497 | 1093 | No | NA | 1 | 0 | 0 | Not-determined | Genome-fragment | NA | NA | 0 |
| S3_extracted_k141_109277 | 7138 | No | NA | 12 | 0 | 0 | Medium-quality | Genome-fragment | 60.06 | AAI-based (high-confidence) | 0 |
| S8_vs_k141_13980\|\|full | 1610 | No | NA | 2 | 0 | 1 | Low-quality | Genome-fragment | 1.68 | AAI-based (high-confidence) | 0 |
| S3_extracted_k141_78134 | 1801 | No | NA | 3 | 0 | 1 | Not-determined | Genome-fragment | NA | NA | 0 |
| S8_vs_k141_24603\|\|full | 1610 | No | NA | 3 | 0 | 1 | Low-quality | Genome-fragment | 1.42 | AAI-based (medium-confidence) | 0 |
| S7_vs_43884\|\|full | 1611 | No | NA | 3 | 1 | 0 | Low-quality | Genome-fragment | 3.9 | AAI-based (medium-confidence) | 0 |
| S16_extracted_124353 | 1611 | No | NA | 2 | 1 | 0 | Low-quality | Genome-fragment | 2.73 | AAI-based (high-confidence) | 0 |
| S10_vs_15728\|\|full | 1611 | No | NA | 3 | 0 | 1 | Low-quality | Genome-fragment | 3.82 | AAI-based (medium-confidence) | 0 |
| S3_extracted_k141_20739 | 1730 | No | NA | 2 | 0 | 2 | Not-determined | Genome-fragment | NA | NA | 0 |
| S12_extracted_k141_114703 | 1611 | No | NA | 2 | 0 | 0 | Low-quality | Genome-fragment | 2.38 | AAI-based (medium-confidence) | 0 |
| S11_vs_k141_54995\|\|full | 1612 | No | NA | 4 | 0 | 0 | Low-quality | Genome-fragment | 3.52 | AAI-based (high-confidence) | 0 |
| S14_vs_k141_112401\|\|full | 1612 | No | NA | 3 | 0 | 0 | Low-quality | Genome-fragment | 3.99 | AAI-based (medium-confidence) | 0 |
| S3_extracted_k141_85533 | 1182 | No | NA | 1 | 0 | 1 | Not-determined | Genome-fragment | NA | NA | 0 |
| S3_extracted_k141_20759 | 10910 | No | NA | 12 | 0 | 1 | Not-determined | Genome-fragment | NA | NA | 0 |
| S3_extracted_k141_37639 | 13259 | No | NA | 1 | 0 | 1 | Not-determined | Genome-fragment | NA | NA | 0 |
| S3_extracted_k141_37640 | 3671 | No | NA | 6 | 0 | 5 | Not-determined | Genome-fragment | NA | NA | 0 |
| S14_vs_k141_63729\|\|full | 1612 | No | NA | 2 | 1 | 0 | Low-quality | Genome-fragment | 3.81 | AAI-based (high-confidence) | 0 |
| S3_extracted_k141_37645 | 1358 | No | NA | 2 | 0 | 2 | Not-determined | Genome-fragment | NA | NA | 0 |
| S3_extracted_k141_109474 | 1158 | No | NA | 1 | 0 | 1 | Not-determined | Genome-fragment | NA | NA | 0 |
| S1_vs_k141_125111\|\|lt2gene | 1612 | No | NA | 1 | 1 | 0 | Low-quality | Genome-fragment | 3.73 | AAI-based (high-confidence) | 0 |
| S3_extracted_k141_15318 | 1102 | No | NA | 1 | 0 | 1 | Not-determined | Genome-fragment | NA | NA | 0 |
| S5_extracted_k141_20127 | 1612 | No | NA | 5 | 1 | 0 | Low-quality | Genome-fragment | 0.85 | AAI-based (medium-confidence) | 0 |
| S7_vs_112992\|\|full | 1612 | No | NA | 5 | 1 | 0 | Low-quality | Genome-fragment | 3.83 | AAI-based (medium-confidence) | 0 |
| S10_extracted_21376 | 1612 | No | NA | 3 | 0 | 0 | Low-quality | Genome-fragment | 2.8 | AAI-based (medium-confidence) | 0 |
| S4_extracted_15002 | 1612 | No | NA | 5 | 0 | 0 | Low-quality | Genome-fragment | 2.89 | AAI-based (medium-confidence) | 0 |
| S3_extracted_k141_29410 | 2097 | No | NA | 2 | 0 | 2 | Not-determined | Genome-fragment | NA | NA | 0 |
| S9_vs_k141_59739\|\|full | 1613 | No | NA | 4 | 1 | 0 | Low-quality | Genome-fragment | 0.98 | HMM-based (lower-bound) | 0 |
| S11_vs_k141_29327\|\|full | 1613 | No | NA | 3 | 1 | 0 | Low-quality | Genome-fragment | 2.96 | AAI-based (medium-confidence) | 0 |
| S3_extracted_k141_78265 | 14179 | No | NA | 19 | 0 | 2 | Not-determined | Genome-fragment | NA | NA | 0 |
| S3_extracted_k141_124596 | 1486 | No | NA | 2 | 0 | 1 | Not-determined | Genome-fragment | NA | NA | 0 |
| S3_extracted_k141_78281 | 1241 | No | NA | 2 | 0 | 2 | Not-determined | Genome-fragment | NA | NA | 0 |
| S1_vs_k141_146884\|\|full | 1613 | No | NA | 1 | 0 | 0 | Low-quality | Genome-fragment | 3.81 | AAI-based (medium-confidence) | 0 |
| S3_extracted_k141_78288 | 3002 | No | NA | 5 | 0 | 4 | Not-determined | Genome-fragment | NA | NA | 0 |
| S3_extracted_k141_117135 | 1519 | No | NA | 2 | 0 | 2 | Not-determined | Genome-fragment | NA | NA | 0 |
| S3_extracted_k141_29480 | 2188 | No | NA | 3 | 0 | 3 | Not-determined | Genome-fragment | NA | NA | 0 |
| S3_extracted_k141_45352 | 1835 | No | NA | 2 | 0 | 1 | Not-determined | Genome-fragment | NA | NA | 0 |
| S3_extracted_k141_15383 | 1333 | No | NA | 1 | 0 | 0 | Not-determined | Genome-fragment | NA | NA | 0 |
| S3_extracted_k141_93406 | 2767 | No | NA | 2 | 0 | 0 | Not-determined | Genome-fragment | NA | NA | 0 |
| S3_extracted_k141_32594 | 1613 | No | NA | 1 | 0 | 1 | Low-quality | Genome-fragment | 0.45 | AAI-based (medium-confidence) | 0 |
| S3_extracted_k141_109593 | 1159 | No | NA | 4 | 0 | 0 | Not-determined | Genome-fragment | NA | NA | 0 |
| S3_extracted_k141_38467 | 1613 | No | NA | 2 | 0 | 1 | Low-quality | Genome-fragment | 3.71 | AAI-based (high-confidence) | 0 |
| S5_extracted_k141_67368 | 1613 | No | NA | 3 | 0 | 0 | Low-quality | Genome-fragment | 2.88 | AAI-based (high-confidence) | 0 |
| S3_extracted_k141_109607 | 1062 | No | NA | 1 | 0 | 0 | Not-determined | Genome-fragment | NA | NA | 0 |
| S3_extracted_k141_29514 | 7426 | No | NA | 10 | 0 | 3 | Not-determined | Genome-fragment | NA | NA | 0 |
| S3_extracted_k141_29531 | 3323 | No | NA | 1 | 0 | 0 | Not-determined | Genome-fragment | NA | NA | 0 |
| S7_extracted_13765 | 1613 | No | NA | 2 | 1 | 0 | Low-quality | Genome-fragment | 6 | AAI-based (high-confidence) | 0 |
| S3_extracted_k141_5525 | 40068 | No | NA | 61 | 16 | 0 | High-quality | High-quality | 90.61 | AAI-based (high-confidence) | 0 |
| NC1_vf_k141_2670 | 1613 | No | NA | 3 | 0 | 1 | Low-quality | Genome-fragment | 2 | AAI-based (medium-confidence) | 0 |
| S12_vs_k141_24782\|\|full | 1613 | No | NA | 4 | 0 | 0 | Low-quality | Genome-fragment | 3.4 | AAI-based (high-confidence) | 0 |
| S3_extracted_k141_54185 | 2544 | No | NA | 1 | 0 | 0 | Not-determined | Genome-fragment | NA | NA | 0 |
| S3_extracted_k141_29581 | 1189 | No | NA | 2 | 0 | 1 | Not-determined | Genome-fragment | NA | NA | 0 |
| S3_extracted_k141_117190 | 1126 | No | NA | 2 | 0 | 0 | Not-determined | Genome-fragment | NA | NA | 0 |
| S3_extracted_k141_5569 | 1040 | No | NA | 2 | 0 | 1 | Not-determined | Genome-fragment | NA | NA | 0 |
| S9_extracted_k141_58524 | 1614 | No | NA | 2 | 1 | 0 | Low-quality | Genome-fragment | 4.07 | AAI-based (high-confidence) | 0 |
| S11_vs_k141_78333\|\|full | 1614 | No | NA | 3 | 2 | 0 | Low-quality | Genome-fragment | 4.22 | AAI-based (high-confidence) | 0 |
| S3_extracted_k141_45497 | 1392 | No | NA | 3 | 0 | 2 | Not-determined | Genome-fragment | NA | NA | 0 |
| S3_extracted_k141_45506 | 1008 | No | NA | 1 | 0 | 1 | Not-determined | Genome-fragment | NA | NA | 0 |
| S5_vs_k141_11307\|\|full | 1614 | Yes | 510 | 4 | 1 | 1 | Low-quality | Genome-fragment | 0.37 | HMM-based (lower-bound) | 68.4 |
| S3_extracted_k141_45540 | 1156 | No | NA | 1 | 0 | 0 | Not-determined | Genome-fragment | NA | NA | 0 |
| S12_vs_k141_37836\|\|lt2gene | 1614 | No | NA | 2 | 2 | 0 | Low-quality | Genome-fragment | 4.3 | AAI-based (high-confidence) | 0 |
| S3_extracted_k141_117272 | 1012 | No | NA | 1 | 0 | 0 | Not-determined | Genome-fragment | NA | NA | 0 |
| S3_extracted_k141_16045 | 1615 | No | NA | 3 | 0 | 0 | Low-quality | Genome-fragment | 2.7 | AAI-based (high-confidence) | 0 |
| S3_extracted_k141_93558 | 1367 | No | NA | 2 | 0 | 1 | Not-determined | Genome-fragment | NA | NA | 0 |
| S5_vs_k141_49783\|\|full | 1615 | No | NA | 2 | 0 | 0 | Low-quality | Genome-fragment | 3.7 | AAI-based (medium-confidence) | 0 |
| S3_extracted_k141_15477 | 1058 | No | NA | 2 | 0 | 2 | Not-determined | Genome-fragment | NA | NA | 0 |
| S3_extracted_k141_5641 | 1107 | No | NA | 2 | 0 | 2 | Not-determined | Genome-fragment | NA | NA | 0 |
| S3_extracted_k141_5642 | 1051 | No | NA | 1 | 0 | 1 | Not-determined | Genome-fragment | NA | NA | 0 |
| S3_extracted_k141_62507 | 1922 | No | NA | 3 | 0 | 2 | Not-determined | Genome-fragment | NA | NA | 0 |
| S3_extracted_k141_5659 | 1016 | No | NA | 2 | 0 | 0 | Not-determined | Genome-fragment | NA | NA | 0 |
| S7_extracted_111242 | 1615 | No | NA | 2 | 2 | 0 | Low-quality | Genome-fragment | 3.41 | AAI-based (high-confidence) | 0 |
| S7_vs_67596\|\|full | 1615 | No | NA | 4 | 0 | 2 | Low-quality | Genome-fragment | 2.74 | AAI-based (medium-confidence) | 0 |
| S7_vs_121348\|\|full | 1615 | No | NA | 4 | 0 | 0 | Low-quality | Genome-fragment | 3.88 | AAI-based (high-confidence) | 0 |
| S3_extracted_k141_15591 | 3038 | No | NA | 2 | 0 | 2 | Not-determined | Genome-fragment | NA | NA | 0 |
| S3_extracted_k141_85834 | 1909 | No | NA | 1 | 0 | 0 | Not-determined | Genome-fragment | NA | NA | 0 |
| S16_extracted_79515 | 1615 | No | NA | 4 | 1 | 0 | Low-quality | Genome-fragment | 2.8 | AAI-based (medium-confidence) | 0 |
| S3_extracted_k141_21052 | 1157 | No | NA | 2 | 0 | 2 | Not-determined | Genome-fragment | NA | NA | 0 |
| S3_extracted_k141_124835 | 1754 | No | NA | 2 | 0 | 0 | Not-determined | Genome-fragment | NA | NA | 0 |
| S3_extracted_k141_69003 | 3368 | No | NA | 4 | 0 | 1 | Not-determined | Genome-fragment | NA | NA | 0 |
| S3_extracted_k141_85852 | 10278 | No | NA | 7 | 0 | 4 | Not-determined | Genome-fragment | NA | NA | 0 |
| S3_extracted_k141_29796 | 1029 | No | NA | 1 | 0 | 1 | Not-determined | Genome-fragment | NA | NA | 0 |
| S3_extracted_k141_78594 | 4145 | No | NA | 3 | 0 | 3 | Not-determined | Genome-fragment | NA | NA | 0 |
| S3_extracted_k141_45761 | 3286 | No | NA | 2 | 0 | 2 | Not-determined | Genome-fragment | NA | NA | 0 |
| S3_extracted_k141_124956 | 1052 | No | NA | 3 | 0 | 0 | Not-determined | Genome-fragment | NA | NA | 0 |
| S16_extracted_126842 | 1615 | No | NA | 2 | 1 | 0 | Low-quality | Genome-fragment | 2.6 | AAI-based (high-confidence) | 0 |
| S3_extracted_k141_21140 | 1659 | No | NA | 2 | 0 | 2 | Not-determined | Genome-fragment | NA | NA | 0 |
| S3_extracted_k141_100917 | 3131 | No | NA | 1 | 0 | 1 | Not-determined | Genome-fragment | NA | NA | 0 |
| S10_extracted_76079 | 1615 | No | NA | 1 | 0 | 1 | Low-quality | Genome-fragment | 2.6 | AAI-based (medium-confidence) | 0 |
| S3_extracted_k141_54353 | 57601 | No | NA | 65 | 14 | 0 | High-quality | High-quality | 99.5 | AAI-based (medium-confidence) | 0 |
| S3_extracted_k141_45807 | 1920 | No | NA | 3 | 0 | 3 | Not-determined | Genome-fragment | NA | NA | 0 |
| S3_extracted_k141_15721 | 1625 | No | NA | 2 | 0 | 2 | Not-determined | Genome-fragment | NA | NA | 0 |
| S3_extracted_k141_117375 | 1854 | No | NA | 1 | 0 | 0 | Not-determined | Genome-fragment | NA | NA | 0 |
| S3_extracted_k141_109833 | 5846 | No | NA | 7 | 0 | 5 | Not-determined | Genome-fragment | NA | NA | 0 |
| S4_extracted_10417 | 1615 | No | NA | 6 | 4 | 0 | Low-quality | Genome-fragment | 5.33 | AAI-based (high-confidence) | 0 |
| S8_extracted_k141_170307 | 1615 | No | NA | 2 | 0 | 1 | Low-quality | Genome-fragment | 1.07 | AAI-based (high-confidence) | 0 |
| S8_vs_k141_186215\|\|full | 1615 | No | NA | 2 | 0 | 1 | Low-quality | Genome-fragment | 1.88 | AAI-based (high-confidence) | 0 |
| S3_extracted_k141_21221 | 1038 | No | NA | 3 | 0 | 1 | Not-determined | Genome-fragment | NA | NA | 0 |
| S3_extracted_k141_117389 | 3109 | No | NA | 2 | 0 | 1 | Not-determined | Genome-fragment | NA | NA | 0 |
| S3_extracted_k141_45909 | 2226 | No | NA | 1 | 0 | 0 | Not-determined | Genome-fragment | NA | NA | 0 |
| S3_extracted_k141_15791 | 1423 | No | NA | 2 | 0 | 2 | Not-determined | Genome-fragment | NA | NA | 0 |
| S3_extracted_k141_69051 | 2579 | No | NA | 2 | 0 | 1 | Not-determined | Genome-fragment | NA | NA | 0 |
| S3_extracted_k141_93912 | 1712 | No | NA | 3 | 0 | 2 | Not-determined | Genome-fragment | NA | NA | 0 |
| S3_extracted_k141_54498 | 5405 | No | NA | 7 | 2 | 0 | High-quality | High-quality | 93.06 | AAI-based (high-confidence) | 0 |
| S11_vs_k141_87940\|\|full | 1616 | No | NA | 4 | 0 | 1 | Low-quality | Genome-fragment | 2.74 | AAI-based (medium-confidence) | 0 |
| S7_vs_87048\|\|lt2gene | 1616 | No | NA | 1 | 1 | 0 | Low-quality | Genome-fragment | 1.12 | HMM-based (lower-bound) | 0 |
| S3_extracted_k141_29950 | 1015 | No | NA | 2 | 0 | 0 | Not-determined | Genome-fragment | NA | NA | 0 |
| S3_extracted_k141_54554 | 1135 | No | NA | 2 | 0 | 0 | Not-determined | Genome-fragment | NA | NA | 0 |
| S16_extracted_48384 | 1616 | No | NA | 2 | 0 | 1 | Low-quality | Genome-fragment | 3.61 | AAI-based (medium-confidence) | 0 |
| S16_vs_122887\|\|full | 1616 | No | NA | 3 | 0 | 0 | Low-quality | Genome-fragment | 1.02 | AAI-based (medium-confidence) | 0 |
| S4_extracted_1473 | 1616 | No | NA | 5 | 4 | 0 | Low-quality | Genome-fragment | 5.33 | AAI-based (high-confidence) | 0 |
| S3_extracted_k141_78779 | 2224 | No | NA | 3 | 0 | 1 | Not-determined | Genome-fragment | NA | NA | 0 |
| S6_vs_k141_3203\|\|full | 1616 | No | NA | 3 | 0 | 1 | Low-quality | Genome-fragment | 2.74 | AAI-based (medium-confidence) | 0 |
| S3_extracted_k141_125076 | 1801 | No | NA | 2 | 0 | 0 | Not-determined | Genome-fragment | NA | NA | 0 |
| S3_extracted_k141_69104 | 1731 | No | NA | 1 | 0 | 1 | Not-determined | Genome-fragment | NA | NA | 0 |
| S11_vs_k141_102076\|\|full | 1617 | No | NA | 6 | 1 | 0 | Low-quality | Genome-fragment | 4.02 | AAI-based (high-confidence) | 0 |
| S3_extracted_k141_78835 | 3533 | No | NA | 6 | 0 | 3 | Not-determined | Genome-fragment | NA | NA | 0 |
| S5_extracted_k141_16198 | 1617 | No | NA | 1 | 0 | 0 | Low-quality | Genome-fragment | 2.7 | AAI-based (high-confidence) | 0 |
| S3_extracted_k141_86319 | 1166 | No | NA | 1 | 0 | 0 | Not-determined | Genome-fragment | NA | NA | 0 |
| S3_extracted_k141_5872 | 3290 | No | NA | 3 | 0 | 1 | Not-determined | Genome-fragment | NA | NA | 0 |
| S5_vs_k141_115581\|\|full | 1617 | No | NA | 2 | 0 | 2 | Low-quality | Genome-fragment | 2.77 | AAI-based (medium-confidence) | 0 |
| S3_extracted_k141_78845 | 2339 | No | NA | 2 | 0 | 0 | Not-determined | Genome-fragment | NA | NA | 0 |
| S5_vs_k141_131726\|\|full | 1617 | No | NA | 2 | 0 | 2 | Low-quality | Genome-fragment | 1.86 | AAI-based (medium-confidence) | 0 |
| S3_extracted_k141_110042 | 9816 | No | NA | 7 | 0 | 6 | Not-determined | Genome-fragment | NA | NA | 0 |
| S3_extracted_k141_110077 | 1387 | No | NA | 2 | 0 | 1 | Not-determined | Genome-fragment | NA | NA | 0 |
| S3_extracted_k141_110094 | 1973 | No | NA | 1 | 0 | 1 | Not-determined | Genome-fragment | NA | NA | 0 |
| S3_extracted_k141_62907 | 1115 | No | NA | 1 | 0 | 0 | Not-determined | Genome-fragment | NA | NA | 0 |
| S7_vs_45206\|\|full | 1617 | No | NA | 2 | 0 | 1 | Low-quality | Genome-fragment | 0.57 | AAI-based (high-confidence) | 0 |
| S3_extracted_k141_46155 | 2718 | No | NA | 1 | 0 | 1 | Not-determined | Genome-fragment | NA | NA | 0 |
| S3_extracted_k141_78891 | 3217 | No | NA | 3 | 0 | 1 | Not-determined | Genome-fragment | NA | NA | 0 |
| S10_vs_71903\|\|lt2gene | 1617 | No | NA | 2 | 2 | 0 | Low-quality | Genome-fragment | 4.27 | AAI-based (high-confidence) | 0 |
| S3_extracted_k141_69162 | 4947 | No | NA | 4 | 0 | 3 | Not-determined | Genome-fragment | NA | NA | 0 |
| S6_vs_k141_7116\|\|full | 1617 | No | NA | 6 | 0 | 0 | Low-quality | Genome-fragment | 5.06 | AAI-based (high-confidence) | 0 |
| S13_vs_121968\|\|full | 1618 | No | NA | 3 | 0 | 0 | Low-quality | Genome-fragment | 0.71 | AAI-based (medium-confidence) | 0 |
| S3_extracted_k141_30206 | 1655 | No | NA | 1 | 0 | 1 | Not-determined | Genome-fragment | NA | NA | 0 |
| S3_extracted_k141_46218 | 3112 | No | NA | 4 | 0 | 2 | Not-determined | Genome-fragment | NA | NA | 0 |
| S3_extracted_k141_38234 | 1587 | No | NA | 2 | 0 | 1 | Not-determined | Genome-fragment | NA | NA | 0 |
| S14_extracted_k141_110058 | 1618 | No | NA | 3 | 0 | 0 | Low-quality | Genome-fragment | 3.98 | AAI-based (medium-confidence) | 0 |
| S3_extracted_k141_78953 | 1170 | No | NA | 2 | 0 | 0 | Not-determined | Genome-fragment | NA | NA | 0 |
| S3_extracted_k141_54726 | 8662 | No | NA | 5 | 0 | 4 | Not-determined | Genome-fragment | NA | NA | 0 |
| S14_extracted_k141_37862 | 1618 | No | NA | 6 | 6 | 0 | Low-quality | Genome-fragment | 5.34 | AAI-based (high-confidence) | 0 |
| S1_vs_k141_74319\|\|full | 1618 | No | NA | 3 | 2 | 0 | Low-quality | Genome-fragment | 4.09 | AAI-based (high-confidence) | 0 |
| S3_extracted_k141_46263 | 1374 | No | NA | 1 | 0 | 0 | Not-determined | Genome-fragment | NA | NA | 0 |
| S3_vs_k141_10496\|\|full | 1618 | No | NA | 5 | 3 | 0 | Low-quality | Genome-fragment | 4.27 | AAI-based (high-confidence) | 0 |
| S16_extracted_86998 | 1618 | No | NA | 1 | 0 | 1 | Low-quality | Genome-fragment | 3.14 | AAI-based (medium-confidence) | 0 |
| S6_vs_k141_16727\|\|full | 1618 | No | NA | 3 | 2 | 0 | Low-quality | Genome-fragment | 4.54 | AAI-based (high-confidence) | 0 |
| S13_extracted_121042 | 1619 | No | NA | 2 | 0 | 0 | Low-quality | Genome-fragment | 3.2 | AAI-based (high-confidence) | 0 |
| S3_extracted_k141_69216 | 1140 | No | NA | 2 | 0 | 0 | Not-determined | Genome-fragment | NA | NA | 0 |
| S14_vs_k141_101570\|\|full | 1619 | No | NA | 3 | 0 | 0 | Low-quality | Genome-fragment | 1.14 | AAI-based (high-confidence) | 0 |
| S3_extracted_k141_38348 | 2757 | No | NA | 4 | 0 | 2 | Not-determined | Genome-fragment | NA | NA | 0 |
| S1_vs_k141_2028\|\|full | 1619 | No | NA | 2 | 0 | 0 | Low-quality | Genome-fragment | 1.27 | AAI-based (high-confidence) | 0 |
| S3_extracted_k141_122731 | 1619 | No | NA | 2 | 0 | 2 | Low-quality | Genome-fragment | 0.6 | AAI-based (high-confidence) | 0 |
| S3_extracted_k141_63125 | 1157 | No | NA | 1 | 0 | 1 | Not-determined | Genome-fragment | NA | NA | 0 |
| S3_extracted_k141_94239 | 5601 | No | NA | 7 | 0 | 7 | Not-determined | Genome-fragment | NA | NA | 0 |
| S3_extracted_k141_54849 | 5339 | No | NA | 7 | 0 | 4 | Not-determined | Genome-fragment | NA | NA | 0 |
| S3_vs_k141_80974\|\|full | 1619 | No | NA | 4 | 1 | 0 | Low-quality | Genome-fragment | 1.74 | AAI-based (high-confidence) | 0 |
| S3_extracted_k141_125357 | 1101 | No | NA | 1 | 0 | 1 | Not-determined | Genome-fragment | NA | NA | 0 |
| S5_extracted_k141_7560 | 1619 | No | NA | 4 | 2 | 0 | Low-quality | Genome-fragment | 4.36 | AAI-based (high-confidence) | 0 |
| S3_extracted_k141_38389 | 1543 | No | NA | 2 | 0 | 2 | Not-determined | Genome-fragment | NA | NA | 0 |
| S3_extracted_k141_30312 | 2002 | No | NA | 2 | 0 | 2 | Not-determined | Genome-fragment | NA | NA | 0 |
| S3_extracted_k141_110312 | 9088 | No | NA | 2 | 0 | 1 | Not-determined | Genome-fragment | NA | NA | 0 |
| S5_vs_k141_49764\|\|full | 1619 | No | NA | 4 | 0 | 0 | Low-quality | Genome-fragment | 2.45 | AAI-based (high-confidence) | 0 |
| S10_extracted_25552 | 1619 | No | NA | 1 | 1 | 0 | Low-quality | Genome-fragment | 5.14 | AAI-based (high-confidence) | 0 |
| S3_extracted_k141_6137 | 2201 | No | NA | 3 | 0 | 2 | Not-determined | Genome-fragment | NA | NA | 0 |
| S3_extracted_k141_69304 | 1482 | No | NA | 1 | 0 | 1 | Not-determined | Genome-fragment | NA | NA | 0 |
| S3_extracted_k141_21549 | 1145 | No | NA | 1 | 0 | 1 | Not-determined | Genome-fragment | NA | NA | 0 |
| S10_extracted_107470 | 1619 | No | NA | 4 | 1 | 0 | Low-quality | Genome-fragment | 3.08 | HMM-based (lower-bound) | 0 |
| S15_vs_1056\|\|full | 1619 | No | NA | 2 | 1 | 0 | Low-quality | Genome-fragment | 0.92 | AAI-based (high-confidence) | 0 |
| S3_extracted_k141_6177 | 2203 | No | NA | 4 | 0 | 1 | Not-determined | Genome-fragment | NA | NA | 0 |
| S3_extracted_k141_110361 | 3902 | No | NA | 3 | 0 | 2 | Not-determined | Genome-fragment | NA | NA | 0 |
| S3_extracted_k141_110372 | 2072 | No | NA | 2 | 0 | 2 | Not-determined | Genome-fragment | NA | NA | 0 |
| S6_vs_k141_9256\|\|full | 1619 | No | NA | 6 | 1 | 0 | Low-quality | Genome-fragment | 3.43 | AAI-based (medium-confidence) | 0 |
| S3_extracted_k141_46419 | 4078 | No | NA | 3 | 0 | 3 | Not-determined | Genome-fragment | NA | NA | 0 |
| S9_vs_k141_36181\|\|full | 1620 | No | NA | 3 | 0 | 0 | Low-quality | Genome-fragment | 3.71 | AAI-based (high-confidence) | 0 |
| S3_extracted_k141_69389 | 1026 | No | NA | 2 | 0 | 0 | Not-determined | Genome-fragment | NA | NA | 0 |
| S1_vs_k141_74243\|\|full | 1620 | No | NA | 1 | 0 | 0 | Low-quality | Genome-fragment | 0.98 | AAI-based (medium-confidence) | 0 |
| S3_extracted_k141_86745 | 1402 | No | NA | 1 | 0 | 1 | Not-determined | Genome-fragment | NA | NA | 0 |
| S3_extracted_k141_117927 | 3007 | No | NA | 4 | 0 | 2 | Not-determined | Genome-fragment | NA | NA | 0 |
| S3_vs_k141_66384\|\|full | 1620 | No | NA | 3 | 1 | 0 | Low-quality | Genome-fragment | 3.6 | AAI-based (medium-confidence) | 0 |
| S3_extracted_k141_86753 | 12294 | No | NA | 10 | 0 | 6 | Not-determined | Genome-fragment | NA | NA | 0 |
| S7_vs_24874\|\|full | 1620 | No | NA | 5 | 0 | 0 | Low-quality | Genome-fragment | 3.36 | AAI-based (high-confidence) | 0 |
| S7_vs_107485\|\|full | 1620 | No | NA | 3 | 0 | 0 | Low-quality | Genome-fragment | 2.98 | AAI-based (high-confidence) | 0 |
| S3_extracted_k141_86766 | 1084 | No | NA | 2 | 0 | 1 | Not-determined | Genome-fragment | NA | NA | 0 |
| S16_extracted_11388 | 1620 | No | NA | 4 | 1 | 0 | Low-quality | Genome-fragment | 0.96 | AAI-based (high-confidence) | 0 |
| S3_extracted_k141_118004 | 1531 | No | NA | 3 | 0 | 0 | Not-determined | Genome-fragment | NA | NA | 0 |
| S3_extracted_k141_125466 | 1432 | No | NA | 1 | 0 | 0 | Not-determined | Genome-fragment | NA | NA | 0 |
| S16_extracted_167250 | 1620 | No | NA | 2 | 0 | 1 | Low-quality | Genome-fragment | 2.65 | AAI-based (high-confidence) | 0 |
| S16_vs_15312\|\|full | 1620 | No | NA | 3 | 0 | 0 | Low-quality | Genome-fragment | 2.44 | AAI-based (high-confidence) | 0 |
| S12_vs_k141_62044\|\|full | 1620 | No | NA | 3 | 0 | 0 | Low-quality | Genome-fragment | 3.59 | AAI-based (medium-confidence) | 0 |
| S3_extracted_k141_125480 | 12884 | No | NA | 11 | 0 | 9 | Not-determined | Genome-fragment | NA | NA | 0 |
| S2_vs_k141_3534\|\|full | 1620 | No | NA | 3 | 0 | 3 | Low-quality | Genome-fragment | 1.11 | AAI-based (medium-confidence) | 0 |
| S8_vs_k141_183232\|\|full | 1620 | No | NA | 3 | 0 | 0 | Low-quality | Genome-fragment | 3.04 | AAI-based (high-confidence) | 0 |
| S3_extracted_k141_63326 | 1051 | No | NA | 1 | 0 | 0 | Not-determined | Genome-fragment | NA | NA | 0 |
| S3_extracted_k141_69429 | 1301 | No | NA | 2 | 0 | 1 | Not-determined | Genome-fragment | NA | NA | 0 |
| S3_extracted_k141_21772 | 2228 | No | NA | 3 | 0 | 0 | Not-determined | Genome-fragment | NA | NA | 0 |
| S17_vs_k141_29696\|\|full | 1621 | No | NA | 3 | 0 | 0 | Low-quality | Genome-fragment | 1.47 | AAI-based (medium-confidence) | 0 |
| S3_extracted_k141_125574 | 1337 | No | NA | 2 | 0 | 2 | Not-determined | Genome-fragment | NA | NA | 0 |
| S17_vs_k141_82360\|\|full | 1621 | No | NA | 3 | 0 | 1 | Low-quality | Genome-fragment | 2.24 | AAI-based (high-confidence) | 0 |
| S3_extracted_k141_55082 | 1950 | No | NA | 4 | 0 | 0 | Not-determined | Genome-fragment | NA | NA | 0 |
| S17_vs_k141_97928\|\|full | 1621 | No | NA | 2 | 0 | 0 | Low-quality | Genome-fragment | 1.34 | AAI-based (medium-confidence) | 0 |
| S3_extracted_k141_69462 | 1077 | No | NA | 1 | 0 | 0 | Not-determined | Genome-fragment | NA | NA | 0 |
| S3_extracted_k141_125587 | 2002 | No | NA | 2 | 0 | 0 | Not-determined | Genome-fragment | NA | NA | 0 |
| S3_extracted_k141_86820 | 1040 | No | NA | 1 | 0 | 0 | Not-determined | Genome-fragment | NA | NA | 0 |
| S11_extracted_k141_58038 | 1621 | No | NA | 3 | 0 | 0 | Low-quality | Genome-fragment | 1.31 | AAI-based (high-confidence) | 0 |
| S3_extracted_k141_125609 | 3882 | No | NA | 7 | 0 | 2 | Not-determined | Genome-fragment | NA | NA | 0 |
| S3_extracted_k141_86846 | 1059 | No | NA | 2 | 0 | 0 | Not-determined | Genome-fragment | NA | NA | 0 |
| S3_extracted_k141_30516 | 2939 | No | NA | 2 | 0 | 2 | Not-determined | Genome-fragment | NA | NA | 0 |
| S3_extracted_k141_118140 | 1317 | No | NA | 3 | 0 | 0 | Not-determined | Genome-fragment | NA | NA | 0 |
| S11_vs_k141_33094\|\|lt2gene | 1621 | No | NA | 2 | 1 | 0 | Low-quality | Genome-fragment | 2.81 | AAI-based (medium-confidence) | 0 |
| S3_extracted_k141_94424 | 1371 | No | NA | 2 | 0 | 2 | Not-determined | Genome-fragment | NA | NA | 0 |
| S3_extracted_k141_86857 | 5642 | No | NA | 3 | 0 | 1 | Not-determined | Genome-fragment | NA | NA | 0 |
| S3_extracted_k141_79096 | 4328 | No | NA | 5 | 0 | 4 | Not-determined | Genome-fragment | NA | NA | 0 |
| S3_extracted_k141_79104 | 1042 | No | NA | 1 | 0 | 1 | Not-determined | Genome-fragment | NA | NA | 0 |
| S3_vs_k141_119029\|\|full | 1621 | No | NA | 3 | 1 | 0 | Low-quality | Genome-fragment | 2.96 | HMM-based (lower-bound) | 0 |
| S3_extracted_k141_21909 | 1328 | No | NA | 2 | 0 | 0 | Not-determined | Genome-fragment | NA | NA | 0 |
| S5_extracted_k141_137052 | 1621 | No | NA | 4 | 0 | 0 | Low-quality | Genome-fragment | 1.73 | AAI-based (medium-confidence) | 0 |
| S5_vs_k141_117378\|\|full | 1621 | No | NA | 6 | 0 | 0 | Low-quality | Genome-fragment | 3.58 | AAI-based (medium-confidence) | 0 |
| S3_extracted_k141_21931 | 1086 | No | NA | 2 | 0 | 0 | Not-determined | Genome-fragment | NA | NA | 0 |
| S5_vs_k141_111704\|\|full | 1621 | No | NA | 2 | 0 | 1 | Low-quality | Genome-fragment | 2.98 | AAI-based (medium-confidence) | 0 |
| S3_extracted_k141_21949 | 1570 | No | NA | 4 | 0 | 0 | Not-determined | Genome-fragment | NA | NA | 0 |
| S3_extracted_k141_30592 | 1353 | No | NA | 2 | 0 | 2 | Not-determined | Genome-fragment | NA | NA | 0 |
| S5_vs_k141_77080\|\|full | 1621 | No | NA | 4 | 1 | 0 | Low-quality | Genome-fragment | 3.03 | AAI-based (medium-confidence) | 0 |
| S3_extracted_k141_22018 | 1266 | No | NA | 3 | 0 | 0 | Not-determined | Genome-fragment | NA | NA | 0 |
| S3_extracted_k141_6510 | 1067 | No | NA | 2 | 0 | 0 | Not-determined | Genome-fragment | NA | NA | 0 |
| S7_vs_114917\|\|full | 1621 | No | NA | 3 | 2 | 0 | Low-quality | Genome-fragment | 4.12 | AAI-based (high-confidence) | 0 |
| S3_extracted_k141_94518 | 1009 | No | NA | 3 | 0 | 0 | Not-determined | Genome-fragment | NA | NA | 0 |
| S16_vs_146532\|\|full | 1621 | Yes | 489 | 4 | 1 | 1 | Low-quality | Genome-fragment | 2.97 | AAI-based (medium-confidence) | 69.83 |
| S1_extracted_k141_121438 | 1622 | No | NA | 1 | 1 | 0 | Low-quality | Genome-fragment | 3.13 | AAI-based (high-confidence) | 0 |
| S3_extracted_k141_6513 | 3571 | No | NA | 3 | 0 | 2 | Not-determined | Genome-fragment | NA | NA | 0 |
| S3_extracted_k141_101591 | 4782 | No | NA | 5 | 4 | 0 | Medium-quality | Genome-fragment | 84.94 | AAI-based (high-confidence) | 0 |
| S3_vs_k141_120316\|\|full | 1622 | No | NA | 2 | 0 | 0 | Low-quality | Genome-fragment | 3.96 | AAI-based (medium-confidence) | 0 |
| S3_extracted_k141_63459 | 3733 | No | NA | 1 | 0 | 1 | Not-determined | Genome-fragment | NA | NA | 0 |
| S3_vs_k141_1518\|\|full | 1622 | No | NA | 3 | 0 | 0 | Low-quality | Genome-fragment | 2.94 | AAI-based (high-confidence) | 0 |
| S3_extracted_k141_6536 | 3271 | No | NA | 4 | 0 | 3 | Not-determined | Genome-fragment | NA | NA | 0 |
| S3_extracted_k141_125734 | 1036 | No | NA | 2 | 0 | 2 | Not-determined | Genome-fragment | NA | NA | 0 |
| S3_vs_k141_80823\|\|full | 1623 | No | NA | 2 | 0 | 0 | Low-quality | Genome-fragment | 2.32 | AAI-based (high-confidence) | 0 |
| S3_extracted_k141_110664 | 4383 | No | NA | 4 | 0 | 2 | Not-determined | Genome-fragment | NA | NA | 0 |
| S3_extracted_k141_22087 | 1426 | No | NA | 1 | 0 | 1 | Not-determined | Genome-fragment | NA | NA | 0 |
| S3_vs_k141_107438\|\|full | 1623 | No | NA | 3 | 1 | 0 | Low-quality | Genome-fragment | 3.71 | AAI-based (high-confidence) | 0 |
| S3_extracted_k141_101597 | 1016 | No | NA | 2 | 0 | 0 | Not-determined | Genome-fragment | NA | NA | 0 |
| S16_vs_169934\|\|full | 1623 | No | NA | 5 | 2 | 0 | Low-quality | Genome-fragment | 2.8 | AAI-based (high-confidence) | 0 |
| S10_extracted_19900 | 1623 | No | NA | 4 | 1 | 0 | Low-quality | Genome-fragment | 2.46 | AAI-based (high-confidence) | 0 |
| S12_vs_k141_144247\|\|full | 1623 | Yes | 954 | 2 | 1 | 1 | Low-quality | Genome-fragment | 2.16 | AAI-based (medium-confidence) | 41.22 |
| S3_extracted_k141_94585 | 5039 | No | NA | 3 | 0 | 3 | Not-determined | Genome-fragment | NA | NA | 0 |
| S6_extracted_k141_31420 | 1623 | No | NA | 1 | 0 | 0 | Low-quality | Genome-fragment | 1.02 | AAI-based (medium-confidence) | 0 |
| S8_extracted_k141_9837 | 1623 | No | NA | 1 | 0 | 1 | Low-quality | Genome-fragment | 4.88 | AAI-based (medium-confidence) | 0 |
| S3_extracted_k141_118320 | 1386 | No | NA | 1 | 0 | 0 | Not-determined | Genome-fragment | NA | NA | 0 |
| S8_vs_k141_61681\|\|full | 1623 | No | NA | 2 | 0 | 1 | Low-quality | Genome-fragment | 1.6 | AAI-based (high-confidence) | 0 |
| S3_extracted_k141_118322 | 3128 | No | NA | 3 | 0 | 2 | Not-determined | Genome-fragment | NA | NA | 0 |
| S11_extracted_k141_69111 | 1624 | No | NA | 3 | 0 | 0 | Low-quality | Genome-fragment | 1.94 | AAI-based (high-confidence) | 0 |
| S3_extracted_k141_22158 | 1224 | No | NA | 1 | 0 | 1 | Not-determined | Genome-fragment | NA | NA | 0 |
| S3_extracted_k141_101611 | 40986 | No | NA | 50 | 24 | 3 | Medium-quality | Genome-fragment | 73.47 | HMM-based (lower-bound) | 0 |
| S3_extracted_k141_22166 | 1231 | No | NA | 2 | 0 | 0 | Not-determined | Genome-fragment | NA | NA | 0 |
| S3_extracted_k141_79418 | 1040 | No | NA | 2 | 0 | 0 | Not-determined | Genome-fragment | NA | NA | 0 |
| S3_extracted_k141_87113 | 1062 | No | NA | 2 | 0 | 1 | Not-determined | Genome-fragment | NA | NA | 0 |
| S3_extracted_k141_94650 | 1476 | No | NA | 2 | 0 | 2 | Not-determined | Genome-fragment | NA | NA | 0 |
| S14_vs_k141_73280\|\|full | 1624 | No | NA | 4 | 0 | 0 | Low-quality | Genome-fragment | 4.07 | AAI-based (medium-confidence) | 0 |
| S3_vs_k141_37332\|\|full | 1624 | No | NA | 4 | 0 | 0 | Low-quality | Genome-fragment | 3.02 | AAI-based (high-confidence) | 0 |
| S3_extracted_k141_94659 | 3925 | No | NA | 5 | 0 | 0 | Not-determined | Genome-fragment | NA | NA | 0 |
| S7_vs_96808\|\|full | 1624 | No | NA | 3 | 0 | 0 | Low-quality | Genome-fragment | 5.19 | AAI-based (high-confidence) | 0 |
| S3_extracted_k141_87133 | 4199 | No | NA | 2 | 0 | 2 | Not-determined | Genome-fragment | NA | NA | 0 |
| S3_extracted_k141_94665 | 1205 | No | NA | 2 | 0 | 2 | Not-determined | Genome-fragment | NA | NA | 0 |
| S3_extracted_k141_46787 | 1416 | No | NA | 3 | 0 | 0 | Not-determined | Genome-fragment | NA | NA | 0 |
| S3_extracted_k141_55262 | 5684 | No | NA | 3 | 0 | 1 | Not-determined | Genome-fragment | NA | NA | 0 |
| S16_extracted_153022 | 1624 | No | NA | 4 | 1 | 0 | Low-quality | Genome-fragment | 4.08 | AAI-based (high-confidence) | 0 |
| S3_extracted_k141_94721 | 1096 | No | NA | 2 | 0 | 1 | Not-determined | Genome-fragment | NA | NA | 0 |
| S3_extracted_k141_6609 | 1048 | No | NA | 2 | 0 | 0 | Not-determined | Genome-fragment | NA | NA | 0 |
| S12_vs_k141_90939\|\|full | 1624 | No | NA | 4 | 0 | 0 | Low-quality | Genome-fragment | 3.84 | AAI-based (high-confidence) | 0 |
| S8_extracted_k141_153417 | 1624 | No | NA | 2 | 0 | 0 | Low-quality | Genome-fragment | 3.35 | AAI-based (high-confidence) | 0 |
| S8_vs_k141_189191\|\|full | 1624 | No | NA | 1 | 0 | 1 | Low-quality | Genome-fragment | 2.26 | AAI-based (medium-confidence) | 0 |
| S17_vs_k141_135717\|\|full | 1625 | No | NA | 3 | 0 | 0 | Low-quality | Genome-fragment | 3.49 | AAI-based (medium-confidence) | 0 |
| S3_extracted_k141_46863 | 1621 | No | NA | 2 | 0 | 1 | Not-determined | Genome-fragment | NA | NA | 0 |
| S3_extracted_k141_94791 | 5466 | No | NA | 6 | 0 | 0 | Not-determined | Genome-fragment | NA | NA | 0 |
| S3_extracted_k141_38899 | 1036 | No | NA | 2 | 0 | 0 | Not-determined | Genome-fragment | NA | NA | 0 |
| S11_vs_k141_36386\|\|full | 1625 | No | NA | 4 | 2 | 0 | Low-quality | Genome-fragment | 3.77 | AAI-based (medium-confidence) | 0 |
| S3_extracted_k141_81238 | 1625 | No | NA | 2 | 0 | 0 | Low-quality | Genome-fragment | 3.24 | AAI-based (high-confidence) | 0 |
| S16_vs_71631\|\|full | 1625 | No | NA | 4 | 2 | 0 | Low-quality | Genome-fragment | 0.94 | AAI-based (high-confidence) | 0 |
| S3_extracted_k141_125868 | 3401 | No | NA | 1 | 0 | 1 | Not-determined | Genome-fragment | NA | NA | 0 |
| S3_extracted_k141_118417 | 1422 | No | NA | 2 | 0 | 1 | Not-determined | Genome-fragment | NA | NA | 0 |
| S8_extracted_k141_175243 | 1625 | No | NA | 3 | 0 | 0 | Low-quality | Genome-fragment | 2.18 | AAI-based (medium-confidence) | 0 |
| S17_extracted_k141_89237 | 1626 | No | NA | 2 | 0 | 1 | Low-quality | Genome-fragment | 4.15 | AAI-based (medium-confidence) | 0 |
| S3_extracted_k141_69921 | 3347 | No | NA | 5 | 0 | 2 | Not-determined | Genome-fragment | NA | NA | 0 |
| S17_extracted_k141_169250 | 1626 | No | NA | 4 | 1 | 0 | Low-quality | Genome-fragment | 2.42 | HMM-based (lower-bound) | 0 |
| S3_extracted_k141_118442 | 3637 | No | NA | 2 | 0 | 1 | Not-determined | Genome-fragment | NA | NA | 0 |
| S3_extracted_k141_94858 | 4201 | No | NA | 4 | 0 | 2 | Not-determined | Genome-fragment | NA | NA | 0 |
| S3_extracted_k141_94861 | 1694 | No | NA | 1 | 0 | 0 | Not-determined | Genome-fragment | NA | NA | 0 |
| S3_extracted_k141_101821 | 1759 | No | NA | 2 | 0 | 1 | Not-determined | Genome-fragment | NA | NA | 0 |
| S3_extracted_k141_125932 | 2579 | No | NA | 3 | 0 | 2 | Not-determined | Genome-fragment | NA | NA | 0 |
| S17_vs_k141_119048\|\|full | 1626 | No | NA | 3 | 1 | 0 | Low-quality | Genome-fragment | 2.91 | AAI-based (high-confidence) | 0 |
| S3_extracted_k141_63743 | 1445 | No | NA | 2 | 0 | 1 | Not-determined | Genome-fragment | NA | NA | 0 |
| S14_vs_k141_6363\|\|full | 1626 | No | NA | 3 | 0 | 2 | Low-quality | Genome-fragment | 1.41 | AAI-based (medium-confidence) | 0 |
| S3_extracted_k141_55394 | 1095 | No | NA | 3 | 0 | 0 | Not-determined | Genome-fragment | NA | NA | 0 |

| | | | | | | | | | | | |
|---|---|---|---|---|---|---|---|---|---|---|---|
| S3_extracted_k141_101853 | 1214 | No | NA | 2 | 0 | 0 | Not-determined | Genome-fragment | NA | NA | 0 |
| S3_extracted_k141_125956 | 2891 | No | NA | 2 | 0 | 2 | Not-determined | Genome-fragment | NA | NA | 0 |
| S3_extracted_k141_55410 | 1322 | No | NA | 2 | 0 | 1 | Not-determined | Genome-fragment | NA | NA | 0 |
| S3_extracted_k141_88471 | 1626 | No | NA | 3 | 0 | 0 | Low-quality | Genome-fragment | 1.24 | AAI-based (high-confidence) | 0 |
| S3_extracted_k141_6775 | 2236 | No | NA | 3 | 0 | 1 | Not-determined | Genome-fragment | NA | NA | 0 |
| S3_extracted_k141_6780 | 6559 | No | NA | 9 | 0 | 6 | Not-determined | Genome-fragment | NA | NA | 0 |
| S3_extracted_k141_70005 | 1096 | No | NA | 1 | 0 | 1 | Not-determined | Genome-fragment | NA | NA | 0 |
| S3_extracted_k141_125983 | 1444 | No | NA | 1 | 0 | 1 | Not-determined | Genome-fragment | NA | NA | 0 |
| S3_vs_k141_4719\|\|full | 1626 | No | NA | 2 | 0 | 0 | Low-quality | Genome-fragment | 1.79 | AAI-based (high-confidence) | 0 |
| S3_extracted_k141_55454 | 5054 | No | NA | 6 | 0 | 4 | Not-determined | Genome-fragment | NA | NA | 0 |
| S3_extracted_k141_94977 | 1760 | No | NA | 1 | 0 | 1 | Not-determined | Genome-fragment | NA | NA | 0 |
| S5_vs_k141_122530\|\|full | 1626 | No | NA | 3 | 1 | 0 | Low-quality | Genome-fragment | 3.61 | HMM-based (lower-bound) | 0 |
| S3_extracted_k141_47053 | 1636 | No | NA | 1 | 0 | 0 | Not-determined | Genome-fragment | NA | NA | 0 |
| S3_extracted_k141_94997 | 4342 | No | NA | 5 | 0 | 5 | Not-determined | Genome-fragment | NA | NA | 0 |
| S3_extracted_k141_70054 | 1289 | No | NA | 1 | 0 | 1 | Not-determined | Genome-fragment | NA | NA | 0 |
| S3_extracted_k141_126040 | 3295 | No | NA | 5 | 0 | 1 | Not-determined | Genome-fragment | NA | NA | 0 |
| S5_vs_k141_113385\|\|full | 1626 | No | NA | 4 | 0 | 0 | Low-quality | Genome-fragment | 3.28 | AAI-based (medium-confidence) | 0 |
| S3_extracted_k141_101896 | 3636 | No | NA | 4 | 0 | 4 | Not-determined | Genome-fragment | NA | NA | 0 |
| S7_extracted_86547 | 1626 | No | NA | 1 | 0 | 1 | Low-quality | Genome-fragment | 0.66 | AAI-based (high-confidence) | 0 |
| S3_extracted_k141_118599 | 1374 | No | NA | 2 | 0 | 1 | Not-determined | Genome-fragment | NA | NA | 0 |
| S3_extracted_k141_118603 | 1025 | No | NA | 2 | 0 | 0 | Not-determined | Genome-fragment | NA | NA | 0 |
| S3_extracted_k141_126087 | 1294 | No | NA | 2 | 0 | 1 | Not-determined | Genome-fragment | NA | NA | 0 |
| S3_extracted_k141_95036 | 3224 | No | NA | 4 | 0 | 2 | Not-determined | Genome-fragment | NA | NA | 0 |
| S10_vs_35219\|\|full | 1626 | No | NA | 2 | 0 | 1 | Low-quality | Genome-fragment | 1.31 | AAI-based (medium-confidence) | 0 |
| S3_extracted_k141_39144 | 1000 | No | NA | 2 | 0 | 2 | Not-determined | Genome-fragment | NA | NA | 0 |
| S3_extracted_k141_30972 | 1376 | No | NA | 1 | 0 | 1 | Not-determined | Genome-fragment | NA | NA | 0 |
| S8_vs_k141_43919\|\|full | 1626 | No | NA | 3 | 0 | 1 | Low-quality | Genome-fragment | 3.59 | AAI-based (medium-confidence) | 0 |
| S3_extracted_k141_39181 | 1019 | No | NA | 2 | 0 | 2 | Not-determined | Genome-fragment | NA | NA | 0 |
| S9_extracted_k141_23769 | 1627 | No | NA | 2 | 1 | 0 | Low-quality | Genome-fragment | 27.31 | AAI-based (high-confidence) | 0 |
| S3_extracted_k141_39187 | 2067 | No | NA | 1 | 0 | 0 | Not-determined | Genome-fragment | NA | NA | 0 |
| S3_extracted_k141_87351 | 3439 | No | NA | 1 | 0 | 1 | Not-determined | Genome-fragment | NA | NA | 0 |
| S3_extracted_k141_87357 | 1041 | No | NA | 1 | 0 | 0 | Not-determined | Genome-fragment | NA | NA | 0 |
| S3_extracted_k141_39193 | 1180 | No | NA | 1 | 0 | 0 | Not-determined | Genome-fragment | NA | NA | 0 |
| S3_extracted_k141_118695 | 1115 | No | NA | 1 | 0 | 1 | Not-determined | Genome-fragment | NA | NA | 0 |
| S3_extracted_k141_118710 | 3034 | No | NA | 2 | 0 | 2 | Not-determined | Genome-fragment | NA | NA | 0 |
| S3_extracted_k141_47186 | 2832 | No | NA | 1 | 0 | 0 | Not-determined | Genome-fragment | NA | NA | 0 |
| S14_vs_k141_54938\|\|full | 1627 | No | NA | 3 | 1 | 0 | Low-quality | Genome-fragment | 5.28 | AAI-based (medium-confidence) | 0 |
| S3_extracted_k141_111218 | 3530 | No | NA | 4 | 0 | 0 | Not-determined | Genome-fragment | NA | NA | 0 |
| S3_extracted_k141_126250 | 1626 | No | NA | 3 | 0 | 2 | Not-determined | Genome-fragment | NA | NA | 0 |
| S14_vs_k141_5315\|\|full | 1627 | No | NA | 6 | 5 | 0 | Low-quality | Genome-fragment | 5.37 | AAI-based (high-confidence) | 0 |
| S3_extracted_k141_70217 | 1019 | No | NA | 1 | 0 | 0 | Not-determined | Genome-fragment | NA | NA | 0 |
| S3_extracted_k141_89656 | 1627 | No | NA | 2 | 2 | 0 | Low-quality | Genome-fragment | 2.85 | AAI-based (high-confidence) | 0 |
| S7_vs_106350\|\|full | 1627 | No | NA | 3 | 0 | 2 | Low-quality | Genome-fragment | 3.26 | AAI-based (medium-confidence) | 0 |
| S3_extracted_k141_87433 | 1043 | No | NA | 1 | 0 | 0 | Not-determined | Genome-fragment | NA | NA | 0 |
| S16_extracted_63641 | 1627 | No | NA | 2 | 0 | 1 | Low-quality | Genome-fragment | 3.5 | AAI-based (high-confidence) | 0 |
| S3_extracted_k141_47258 | 1283 | No | NA | 2 | 0 | 1 | Not-determined | Genome-fragment | NA | NA | 0 |
| S3_extracted_k141_31108 | 1845 | No | NA | 2 | 0 | 0 | Not-determined | Genome-fragment | NA | NA | 0 |
| S3_extracted_k141_87484 | 1521 | No | NA | 3 | 0 | 0 | Not-determined | Genome-fragment | NA | NA | 0 |
| S16_vs_62132\|\|full | 1627 | No | NA | 3 | 1 | 0 | Low-quality | Genome-fragment | 0.96 | AAI-based (medium-confidence) | 0 |
| S3_extracted_k141_111330 | 2142 | No | NA | 3 | 0 | 0 | Not-determined | Genome-fragment | NA | NA | 0 |
| S3_extracted_k141_111334 | 1151 | No | NA | 2 | 0 | 1 | Not-determined | Genome-fragment | NA | NA | 0 |
| S3_extracted_k141_95195 | 1995 | No | NA | 2 | 0 | 1 | Not-determined | Genome-fragment | NA | NA | 0 |
| S16_vs_128797\|\|lt2gene | 1627 | No | NA | 1 | 1 | 0 | Low-quality | Genome-fragment | 1.25 | HMM-based (lower-bound) | 0 |
| S10_vs_34208\|\|full | 1627 | No | NA | 3 | 0 | 0 | Low-quality | Genome-fragment | 2.85 | AAI-based (medium-confidence) | 0 |
| S12_vs_k141_8287\|\|full | 1627 | No | NA | 4 | 2 | 0 | Low-quality | Genome-fragment | 4.04 | AAI-based (high-confidence) | 0 |
| S2_vs_k141_21411\|\|full | 1627 | No | NA | 1 | 0 | 0 | Low-quality | Genome-fragment | 2.41 | AAI-based (medium-confidence) | 0 |
| S3_extracted_k141_55665 | 5952 | No | NA | 10 | 0 | 4 | Not-determined | Genome-fragment | NA | NA | 0 |
| S3_extracted_k141_95214 | 7615 | No | NA | 9 | 0 | 3 | Not-determined | Genome-fragment | NA | NA | 0 |
| S3_extracted_k141_95217 | 1484 | No | NA | 1 | 0 | 1 | Not-determined | Genome-fragment | NA | NA | 0 |
| S6_vs_k141_5934\|\|full | 1627 | No | NA | 3 | 1 | 0 | Low-quality | Genome-fragment | 2.68 | AAI-based (high-confidence) | 0 |
| S3_extracted_k141_55673 | 3268 | No | NA | 4 | 0 | 4 | Not-determined | Genome-fragment | NA | NA | 0 |
| S8_extracted_k141_66007 | 1627 | No | NA | 1 | 1 | 0 | Low-quality | Genome-fragment | 0.85 | HMM-based (lower-bound) | 0 |
| S8_extracted_k141_110009 | 1627 | No | NA | 3 | 0 | 0 | Low-quality | Genome-fragment | 2.57 | AAI-based (high-confidence) | 0 |
| S3_extracted_k141_87578 | 8068 | No | NA | 3 | 0 | 2 | Not-determined | Genome-fragment | NA | NA | 0 |
| S8_extracted_k141_190567 | 1627 | No | NA | 2 | 0 | 1 | Low-quality | Genome-fragment | 3.08 | AAI-based (medium-confidence) | 0 |
| S11_extracted_k141_100215 | 1628 | No | NA | 4 | 0 | 0 | Low-quality | Genome-fragment | 1.46 | AAI-based (medium-confidence) | 0 |
| S3_extracted_k141_70305 | 1369 | No | NA | 2 | 0 | 0 | Not-determined | Genome-fragment | NA | NA | 0 |
| S3_extracted_k141_126375 | 3295 | No | NA | 2 | 0 | 1 | Not-determined | Genome-fragment | NA | NA | 0 |
| S3_extracted_k141_55719 | 2052 | No | NA | 1 | 0 | 1 | Not-determined | Genome-fragment | NA | NA | 0 |
| S3_extracted_k141_102061 | 10119 | No | NA | 8 | 0 | 7 | Not-determined | Genome-fragment | NA | NA | 0 |
| S11_vs_k141_35341\|\|full | 1628 | No | NA | 4 | 1 | 0 | Low-quality | Genome-fragment | 3.4 | AAI-based (high-confidence) | 0 |
| S3_extracted_k141_22683 | 4265 | No | NA | 11 | 0 | 0 | Not-determined | Genome-fragment | NA | NA | 0 |
| S3_extracted_k141_55759 | 2673 | No | NA | 3 | 0 | 1 | Not-determined | Genome-fragment | NA | NA | 0 |
| S3_extracted_k141_31282 | 1647 | No | NA | 2 | 0 | 2 | Not-determined | Genome-fragment | NA | NA | 0 |
| S3_extracted_k141_111426 | 3475 | No | NA | 5 | 0 | 1 | Not-determined | Genome-fragment | NA | NA | 0 |
| S3_extracted_k141_87623 | 5502 | No | NA | 5 | 0 | 4 | Not-determined | Genome-fragment | NA | NA | 0 |
| S3_extracted_k141_102115 | 4402 | No | NA | 6 | 0 | 4 | Not-determined | Genome-fragment | NA | NA | 0 |
| S13_extracted_139578 | 1628 | No | NA | 2 | 1 | 0 | Low-quality | Genome-fragment | 3.34 | HMM-based (lower-bound) | 0 |
| S3_extracted_k141_111441 | 3230 | No | NA | 2 | 0 | 2 | Not-determined | Genome-fragment | NA | NA | 0 |
| S3_extracted_k141_111442 | 1387 | No | NA | 1 | 0 | 1 | Not-determined | Genome-fragment | NA | NA | 0 |
| S3_extracted_k141_70368 | 2217 | No | NA | 2 | 0 | 1 | Not-determined | Genome-fragment | NA | NA | 0 |
| S3_extracted_k141_102130 | 6558 | No | NA | 4 | 0 | 3 | Not-determined | Genome-fragment | NA | NA | 0 |
| S3_extracted_k141_39433 | 1005 | No | NA | 2 | 0 | 1 | Not-determined | Genome-fragment | NA | NA | 0 |
| S13_vs_28456\|\|full | 1628 | No | NA | 2 | 0 | 0 | Low-quality | Genome-fragment | 1.99 | AAI-based (high-confidence) | 0 |
| S3_extracted_k141_47512 | 2140 | No | NA | 2 | 0 | 2 | Not-determined | Genome-fragment | NA | NA | 0 |
| S3_extracted_k141_31388 | 4643 | No | NA | 3 | 0 | 2 | Not-determined | Genome-fragment | NA | NA | 0 |
| S3_extracted_k141_95329 | 5302 | No | NA | 4 | 0 | 1 | Not-determined | Genome-fragment | NA | NA | 0 |
| S13_vs_117733\|\|full | 1628 | No | NA | 2 | 0 | 1 | Low-quality | Genome-fragment | 1.74 | AAI-based (medium-confidence) | 0 |
| S3_extracted_k141_118896 | 2108 | No | NA | 1 | 0 | 1 | Not-determined | Genome-fragment | NA | NA | 0 |
| S3_extracted_k141_7054 | 1535 | No | NA | 1 | 0 | 0 | Not-determined | Genome-fragment | NA | NA | 0 |
| S3_extracted_k141_7057 | 1176 | No | NA | 2 | 0 | 0 | Not-determined | Genome-fragment | NA | NA | 0 |
| S3_extracted_k141_47576 | 1107 | No | NA | 2 | 0 | 1 | Not-determined | Genome-fragment | NA | NA | 0 |
| S1_vs_k141_101712\|\|full | 1628 | No | NA | 3 | 1 | 0 | Low-quality | Genome-fragment | 2.86 | AAI-based (high-confidence) | 0 |
| S3_extracted_k141_126560 | 1320 | No | NA | 2 | 0 | 2 | Not-determined | Genome-fragment | NA | NA | 0 |
| S3_extracted_k141_47587 | 11361 | No | NA | 9 | 0 | 2 | Not-determined | Genome-fragment | NA | NA | 0 |
| S1_vs_k141_106246\|\|full | 1628 | No | NA | 1 | 0 | 0 | Low-quality | Genome-fragment | 2.73 | AAI-based (high-confidence) | 0 |
| S3_extracted_k141_22784 | 1270 | No | NA | 2 | 0 | 0 | Not-determined | Genome-fragment | NA | NA | 0 |
| S3_extracted_k141_102245 | 2070 | No | NA | 2 | 0 | 2 | Not-determined | Genome-fragment | NA | NA | 0 |
| S3_extracted_k141_111579 | 1576 | No | NA | 2 | 0 | 1 | Not-determined | Genome-fragment | NA | NA | 0 |
| S3_extracted_k141_47625 | 5116 | No | NA | 4 | 0 | 4 | Not-determined | Genome-fragment | NA | NA | 0 |
| S3_extracted_k141_39644 | 1313 | No | NA | 1 | 0 | 1 | Not-determined | Genome-fragment | NA | NA | 0 |
| S3_extracted_k141_7107 | 2983 | No | NA | 4 | 0 | 0 | Not-determined | Genome-fragment | NA | NA | 0 |
| S3_extracted_k141_126624 | 1042 | No | NA | 3 | 0 | 0 | Not-determined | Genome-fragment | NA | NA | 0 |
| S5_vs_k141_74381\|\|full | 1628 | No | NA | 3 | 0 | 0 | Low-quality | Genome-fragment | 2.22 | AAI-based (high-confidence) | 0 |
| S10_vs_107335\|\|full | 1628 | No | NA | 5 | 0 | 0 | Low-quality | Genome-fragment | 4.86 | AAI-based (high-confidence) | 0 |
| S3_extracted_k141_47709 | 1013 | No | NA | 2 | 0 | 0 | Not-determined | Genome-fragment | NA | NA | 0 |
| S3_extracted_k141_39713 | 1007 | No | NA | 2 | 0 | 2 | Not-determined | Genome-fragment | NA | NA | 0 |
| S3_extracted_k141_22874 | 1083 | No | NA | 1 | 0 | 0 | Not-determined | Genome-fragment | NA | NA | 0 |
| S3_extracted_k141_7137 | 15928 | No | NA | 18 | 5 | 0 | Medium-quality | Genome-fragment | 53.02 | HMM-based (lower-bound) | 0 |
| S3_extracted_k141_7145 | 1343 | No | NA | 2 | 0 | 1 | Not-determined | Genome-fragment | NA | NA | 0 |
| S3_extracted_k141_39750 | 1066 | No | NA | 3 | 0 | 2 | Not-determined | Genome-fragment | NA | NA | 0 |
| S3_extracted_k141_31551 | 2042 | No | NA | 2 | 0 | 0 | Not-determined | Genome-fragment | NA | NA | 0 |
| S3_extracted_k141_22968 | 3167 | No | NA | 5 | 2 | 0 | Medium-quality | Genome-fragment | 52.91 | AAI-based (high-confidence) | 0 |
| S3_extracted_k141_47796 | 3227 | No | NA | 5 | 0 | 3 | Not-determined | Genome-fragment | NA | NA | 0 |
| S12_vs_k141_163970\|\|full | 1628 | No | NA | 3 | 0 | 1 | Low-quality | Genome-fragment | 3.14 | AAI-based (medium-confidence) | 0 |
| S8_extracted_k141_8262 | 1628 | No | NA | 3 | 0 | 2 | Low-quality | Genome-fragment | 3.82 | AAI-based (medium-confidence) | 0 |
| S11_extracted_k141_27146 | 1629 | No | NA | 4 | 0 | 3 | Low-quality | Genome-fragment | 2.98 | AAI-based (medium-confidence) | 0 |
| S1_extracted_k141_33266 | 1629 | No | NA | 1 | 0 | 1 | Low-quality | Genome-fragment | 1.02 | AAI-based (medium-confidence) | 0 |
| S3_extracted_k141_23036 | 1005 | No | NA | 3 | 0 | 1 | Not-determined | Genome-fragment | NA | NA | 0 |
| S3_extracted_k141_23042 | 19014 | No | NA | 33 | 31 | 0 | Medium-quality | Genome-fragment | 62.67 | AAI-based (high-confidence) | 0 |
| S1_vs_k141_55861\|\|full | 1629 | No | NA | 4 | 0 | 4 | Low-quality | Genome-fragment | 1.2 | AAI-based (medium-confidence) | 0 |
| S3_extracted_k141_11696 | 1629 | No | NA | 2 | 0 | 1 | Low-quality | Genome-fragment | 2.46 | AAI-based (medium-confidence) | 0 |
| S3_vs_k141_98190\|\|full | 1629 | No | NA | 2 | 0 | 0 | Low-quality | Genome-fragment | 3.51 | AAI-based (medium-confidence) | 0 |
| S3_extracted_k141_23080 | 5718 | No | NA | 3 | 0 | 2 | Not-determined | Genome-fragment | NA | NA | 0 |
| S3_vs_k141_66103\|\|full | 1629 | No | NA | 3 | 1 | 0 | Low-quality | Genome-fragment | 4.83 | AAI-based (high-confidence) | 0 |
| S3_extracted_k141_7337 | 1120 | No | NA | 1 | 0 | 1 | Not-determined | Genome-fragment | NA | NA | 0 |
| S3_extracted_k141_31684 | 3127 | No | NA | 1 | 0 | 1 | Not-determined | Genome-fragment | NA | NA | 0 |
| S3_extracted_k141_23095 | 1380 | No | NA | 1 | 0 | 0 | Not-determined | Genome-fragment | NA | NA | 0 |
| S3_extracted_k141_7355 | 9233 | No | NA | 11 | 0 | 2 | Not-determined | Genome-fragment | NA | NA | 0 |
| S3_extracted_k141_95598 | 1112 | No | NA | 2 | 0 | 1 | Not-determined | Genome-fragment | NA | NA | 0 |
| S12_vs_k141_181245\|\|full | 1629 | No | NA | 3 | 0 | 2 | Low-quality | Genome-fragment | 2.89 | AAI-based (medium-confidence) | 0 |
| S3_extracted_k141_102514 | 3006 | No | NA | 4 | 0 | 2 | Not-determined | Genome-fragment | NA | NA | 0 |
| S3_extracted_k141_70746 | 30924 | No | NA | 52 | 19 | 0 | Medium-quality | Genome-fragment | 81.81 | AAI-based (high-confidence) | 0 |
| S3_extracted_k141_7466 | 1511 | No | NA | 2 | 0 | 1 | Not-determined | Genome-fragment | NA | NA | 0 |
| S3_extracted_k141_102589 | 3710 | No | NA | 3 | 0 | 2 | Not-determined | Genome-fragment | NA | NA | 0 |
| S8_extracted_k141_100834 | 1629 | No | NA | 2 | 0 | 1 | Low-quality | Genome-fragment | 3.64 | AAI-based (medium-confidence) | 0 |
| S17_vs_k141_140792\|\|full | 1630 | No | NA | 2 | 0 | 2 | Low-quality | Genome-fragment | 2.23 | AAI-based (medium-confidence) | 0 |
| S3_extracted_k141_7511 | 1630 | No | NA | 2 | 0 | 1 | Not-determined | Genome-fragment | NA | NA | 0 |
| S14_vs_k141_11900\|\|full | 1630 | No | NA | 3 | 1 | 0 | Low-quality | Genome-fragment | 2.2 | AAI-based (high-confidence) | 0 |
| S3_extracted_k141_126815 | 1131 | No | NA | 2 | 0 | 1 | Not-determined | Genome-fragment | NA | NA | 0 |
| S3_extracted_k141_70923 | 2214 | No | NA | 2 | 0 | 1 | Not-determined | Genome-fragment | NA | NA | 0 |
| S3_extracted_k141_126877 | 1000 | No | NA | 2 | 0 | 0 | Not-determined | Genome-fragment | NA | NA | 0 |
| S3_extracted_k141_126881 | 1032 | No | NA | 2 | 0 | 2 | Not-determined | Genome-fragment | NA | NA | 0 |
| S5_vs_k141_102902\|\|full | 1630 | No | NA | 2 | 0 | 0 | Low-quality | Genome-fragment | 2.14 | AAI-based (medium-confidence) | 0 |
| S3_extracted_k141_23288 | 1328 | No | NA | 2 | 0 | 1 | Not-determined | Genome-fragment | NA | NA | 0 |
| S7_vs_59065\|\|full | 1630 | No | NA | 3 | 1 | 0 | Low-quality | Genome-fragment | 4 | AAI-based (medium-confidence) | 0 |
| S10_vs_42214\|\|full | 1630 | No | NA | 4 | 0 | 0 | Low-quality | Genome-fragment | 2.27 | AAI-based (medium-confidence) | 0 |
| S3_extracted_k141_70983 | 1230 | No | NA | 2 | 0 | 0 | Not-determined | Genome-fragment | NA | NA | 0 |
| S3_extracted_k141_102783 | 1308 | No | NA | 2 | 0 | 2 | Not-determined | Genome-fragment | NA | NA | 0 |
| S3_extracted_k141_23351 | 3052 | No | NA | 3 | 0 | 0 | Not-determined | Genome-fragment | NA | NA | 0 |
| S17_vs_k141_78221\|\|full | 1631 | No | NA | 2 | 0 | 1 | Low-quality | Genome-fragment | 4.3 | AAI-based (high-confidence) | 0 |
| S3_extracted_k141_7721 | 1035 | No | NA | 2 | 0 | 2 | Not-determined | Genome-fragment | NA | NA | 0 |
| S3_extracted_k141_7728 | 1094 | No | NA | 4 | 0 | 4 | Not-determined | Genome-fragment | NA | NA | 0 |
| S11_extracted_k141_80551 | 1631 | No | NA | 4 | 0 | 0 | Low-quality | Genome-fragment | 3.79 | AAI-based (high-confidence) | 0 |
| S3_vs_k141_3433\|\|full | 1631 | No | NA | 5 | 1 | 0 | Low-quality | Genome-fragment | 2.71 | AAI-based (high-confidence) | 0 |
| S3_extracted_k141_7773 | 6855 | No | NA | 17 | 0 | 1 | Not-determined | Genome-fragment | NA | NA | 0 |
| S3_extracted_k141_119246 | 1405 | No | NA | 1 | 0 | 0 | Not-determined | Genome-fragment | NA | NA | 0 |
| S3_extracted_k141_127058 | 2838 | No | NA | 2 | 0 | 2 | Not-determined | Genome-fragment | NA | NA | 0 |
| S3_extracted_k141_71183 | 3802 | No | NA | 6 | 0 | 4 | Not-determined | Genome-fragment | NA | NA | 0 |
| S3_extracted_k141_23513 | 1225 | No | NA | 1 | 0 | 0 | Not-determined | Genome-fragment | NA | NA | 0 |
| S3_extracted_k141_127091 | 1568 | No | NA | 3 | 0 | 1 | Not-determined | Genome-fragment | NA | NA | 0 |
| S5_extracted_k141_18485 | 1631 | No | NA | 2 | 0 | 2 | Low-quality | Genome-fragment | 0.4 | AAI-based (medium-confidence) | 0 |
| S3_extracted_k141_127132 | 3128 | No | NA | 1 | 0 | 1 | Not-determined | Genome-fragment | NA | NA | 0 |
| S16_extracted_81717 | 1631 | No | NA | 2 | 0 | 0 | Low-quality | Genome-fragment | 1.87 | AAI-based (medium-confidence) | 0 |
| S8_vs_k141_206528\|\|full | 1631 | No | NA | 2 | 0 | 1 | Low-quality | Genome-fragment | 0.61 | AAI-based (medium-confidence) | 0 |
| S3_extracted_k141_127168 | 13910 | No | NA | 17 | 7 | 0 | Complete | High-quality | 100 | ITR (high-confidence) | 0 |
| S3_extracted_k141_119419 | 1929 | No | NA | 2 | 0 | 0 | Not-determined | Genome-fragment | NA | NA | 0 |
| S9_vs_k141_80189\|\|full | 1632 | No | NA | 5 | 2 | 0 | Low-quality | Genome-fragment | 4.19 | AAI-based (high-confidence) | 0 |
| S3_extracted_k141_119441 | 3095 | No | NA | 3 | 0 | 0 | Not-determined | Genome-fragment | NA | NA | 0 |
| S3_extracted_k141_103057 | 2799 | No | NA | 2 | 0 | 1 | Not-determined | Genome-fragment | NA | NA | 0 |
| S3_extracted_k141_71407 | 1350 | No | NA | 1 | 0 | 1 | Not-determined | Genome-fragment | NA | NA | 0 |
| S3_extracted_k141_71410 | 1365 | No | NA | 4 | 0 | 4 | Not-determined | Genome-fragment | NA | NA | 0 |
| S3_extracted_k141_127233 | 1401 | No | NA | 3 | 0 | 0 | Not-determined | Genome-fragment | NA | NA | 0 |
| S3_extracted_k141_23705 | 3889 | No | NA | 3 | 0 | 1 | Not-determined | Genome-fragment | NA | NA | 0 |
| S3_extracted_k141_127272 | 5804 | No | NA | 6 | 0 | 4 | Not-determined | Genome-fragment | NA | NA | 0 |
| S3_extracted_k141_71491 | 2150 | No | NA | 3 | 0 | 1 | Not-determined | Genome-fragment | NA | NA | 0 |
| S3_extracted_k141_127309 | 2855 | No | NA | 1 | 0 | 1 | Not-determined | Genome-fragment | NA | NA | 0 |
| S14_vs_k141_45002\|\|full | 1632 | No | NA | 2 | 0 | 1 | Low-quality | Genome-fragment | 1.08 | AAI-based (high-confidence) | 0 |
| S3_extracted_k141_103211 | 1011 | No | NA | 1 | 0 | 1 | Not-determined | Genome-fragment | NA | NA | 0 |
| S3_extracted_k141_23828 | 3997 | No | NA | 3 | 0 | 3 | Not-determined | Genome-fragment | NA | NA | 0 |
| S14_vs_k141_13664\|\|full | 1632 | No | NA | 2 | 1 | 0 | Low-quality | Genome-fragment | 0.65 | HMM-based (lower-bound) | 0 |
| S3_extracted_k141_127392 | 54915 | No | NA | 75 | 25 | 1 | Complete | High-quality | 100 | DTR (high-confidence) | 0 |
| S3_extracted_k141_103431 | 3141 | No | NA | 2 | 0 | 1 | Not-determined | Genome-fragment | NA | NA | 0 |
| S3_extracted_k141_64144 | 1632 | No | NA | 2 | 0 | 0 | Low-quality | Genome-fragment | 19.17 | AAI-based (high-confidence) | 0 |
| S3_extracted_k141_103451 | 1318 | No | NA | 2 | 0 | 2 | Not-determined | Genome-fragment | NA | NA | 0 |
| S3_extracted_k141_127399 | 97442 | No | NA | 117 | 15 | 3 | Complete | High-quality | 100 | DTR (high-confidence) | 0 |
| S3_extracted_k141_127400 | 42799 | No | NA | 74 | 19 | 0 | Complete | High-quality | 100 | DTR (high-confidence) | 0 |
| S7_extracted_57207 | 1632 | No | NA | 4 | 1 | 0 | Low-quality | Genome-fragment | 2.95 | AAI-based (medium-confidence) | 0 |
| S3_extracted_k141_103554 | 1818 | No | NA | 1 | 0 | 0 | Not-determined | Genome-fragment | NA | NA | 0 |
| S10_vs_104517\|\|full | 1632 | No | NA | 4 | 1 | 0 | Low-quality | Genome-fragment | 1.05 | HMM-based (lower-bound) | 0 |
| S3_extracted_k141_127405 | 39265 | No | NA | 57 | 19 | 1 | Complete | High-quality | 100 | DTR (high-confidence) | 0 |
| S11_vs_k141_31401\|\|lt2gene | 1633 | No | NA | 1 | 0 | 0 | Low-quality | Genome-fragment | 4.29 | AAI-based (high-confidence) | 0 |
| S3_extracted_k141_127406 | 78572 | No | NA | 106 | 14 | 1 | Complete | High-quality | 100 | DTR (high-confidence) | 0 |
| S3_extracted_k141_127408 | 29543 | No | NA | 33 | 10 | 0 | Complete | High-quality | 100 | DTR (high-confidence) | 0 |
| S3_extracted_k141_127409 | 44970 | No | NA | 61 | 32 | 1 | Complete | High-quality | 100 | DTR (high-confidence) | 0 |
| S3_vs_k141_56286\|\|full | 1633 | No | NA | 3 | 1 | 0 | Low-quality | Genome-fragment | 3.44 | HMM-based (lower-bound) | 0 |
| S3_extracted_k141_127423 | 48451 | No | NA | 56 | 14 | 1 | Complete | High-quality | 100 | DTR (high-confidence) | 0 |
| S3_extracted_k141_127426 | 3462 | No | NA | 7 | 4 | 0 | Medium-quality | Genome-fragment | 67.36 | AAI-based (high-confidence) | 0 |
| S3_extracted_k141_127427 | 2474 | No | NA | 1 | 0 | 0 | Not-determined | Genome-fragment | NA | NA | 0 |
| S3_extracted_k141_127431 | 5479 | No | NA | 11 | 0 | 0 | Complete | High-quality | 100 | DTR (high-confidence) | 0 |
| S3_extracted_k141_127447 | 101130 | No | NA | 117 | 23 | 0 | Complete | High-quality | 100 | DTR (high-confidence) | 0 |
| S3_extracted_k141_127450 | 31350 | No | NA | 38 | 12 | 0 | Complete | High-quality | 100 | DTR (high-confidence) | 0 |
| S3_extracted_k141_127464 | 4249 | No | NA | 4 | 0 | 0 | Not-determined | Genome-fragment | NA | NA | 0 |
| S3_vs_k141_119605\|\|full | 56346 | Yes | 45953 | 64 | 21 | 10 | High-quality | High-quality | 96.21 | AAI-based (high-confidence) | 18.44 |
| S3_vs_k141_80151\|\|full | 149101 | No | NA | 174 | 22 | 4 | High-quality | High-quality | 98.71 | AAI-based (high-confidence) | 0 |
| S3_vs_k141_16825\|\|full | 60023 | No | NA | 105 | 17 | 3 | High-quality | High-quality | 100 | AAI-based (high-confidence) | 0 |
| S3_vs_k141_49835\|\|full | 38726 | No | NA | 39 | 8 | 0 | High-quality | High-quality | 95.83 | AAI-based (high-confidence) | 0 |
| S3_vs_k141_114293\|\|full | 1633 | No | NA | 2 | 0 | 0 | Low-quality | Genome-fragment | 2.91 | AAI-based (high-confidence) | 0 |
| S3_vs_k141_89485\|\|full | 36153 | No | NA | 49 | 12 | 1 | High-quality | High-quality | 100 | AAI-based (high-confidence) | 0 |
| S3_vs_k141_81793\|\|full | 74426 | No | NA | 96 | 21 | 7 | High-quality | High-quality | 100 | AAI-based (high-confidence) | 0 |
| S3_vs_k141_90331\|\|full | 52928 | No | NA | 82 | 29 | 0 | High-quality | High-quality | 99.13 | AAI-based (high-confidence) | 0 |
| S3_vs_k141_90341\|\|full | 33590 | No | NA | 43 | 16 | 0 | High-quality | High-quality | 93.64 | AAI-based (high-confidence) | 0 |
| S3_vs_k141_50924\|\|full | 39215 | No | NA | 57 | 14 | 0 | High-quality | High-quality | 100 | AAI-based (high-confidence) | 0 |
| S3_vs_k141_98675\|\|full | 34112 | No | NA | 63 | 19 | 0 | High-quality | High-quality | 95.06 | AAI-based (high-confidence) | 0 |
| S3_vs_k141_82895\|\|full | 147228 | No | NA | 173 | 20 | 1 | High-quality | High-quality | 95.72 | AAI-based (high-confidence) | 0 |
| S3_vs_k141_122810\|\|full | 12399 | No | NA | 16 | 0 | 2 | Not-determined | Genome-fragment | NA | NA | 0 |
| S3_vs_k141_127371\|\|full | 1633 | No | NA | 5 | 0 | 0 | Low-quality | Genome-fragment | 1.07 | AAI-based (high-confidence) | 0 |
| S3_vs_k141_108076\|\|full | 39296 | No | NA | 54 | 33 | 1 | High-quality | High-quality | 90.63 | AAI-based (high-confidence) | 0 |
| S3_vs_k141_28402\|\|full | 34037 | No | NA | 55 | 19 | 0 | High-quality | High-quality | 98.13 | AAI-based (medium-confidence) | 0 |
| S5_vs_k141_69905\|\|lt2gene | 1633 | No | NA | 2 | 1 | 0 | Low-quality | Genome-fragment | 3.33 | AAI-based (high-confidence) | 0 |
| S3_vs_k141_127391\|\|full | 60441 | No | NA | 82 | 19 | 1 | High-quality | High-quality | 100 | AAI-based (high-confidence) | 0 |
| S3_vs_k141_127395\|\|full | 38336 | No | NA | 53 | 16 | 2 | High-quality | High-quality | 94.12 | AAI-based (high-confidence) | 0 |
| S3_vs_k141_127397\|\|full | 32948 | No | NA | 58 | 19 | 1 | High-quality | High-quality | 100 | AAI-based (high-confidence) | 0 |
| S3_vs_k141_127401\|\|full | 39557 | No | NA | 51 | 29 | 1 | High-quality | High-quality | 91.47 | AAI-based (high-confidence) | 0 |
| S16_vs_71872\|\|full | 1633 | No | NA | 3 | 1 | 0 | Low-quality | Genome-fragment | 1.47 | HMM-based (lower-bound) | 0 |
| S3_vs_k141_127404\|\|full | 45259 | No | NA | 60 | 5 | 0 | High-quality | High-quality | 96.12 | AAI-based (high-confidence) | 0 |
| S3_vs_k141_127410\|\|full | 34932 | No | NA | 58 | 21 | 3 | High-quality | High-quality | 100 | AAI-based (high-confidence) | 0 |
| S3_vs_k141_127411\|\|full | 15600 | No | NA | 26 | 2 | 0 | High-quality | High-quality | 100 | AAI-based (high-confidence) | 0 |
| S3_vs_k141_127412\|\|full | 7400 | No | NA | 13 | 0 | 1 | Not-determined | Genome-fragment | NA | NA | 0 |
| S3_vs_k141_127413\|\|full | 35912 | No | NA | 43 | 10 | 0 | Medium-quality | Genome-fragment | 79.48 | AAI-based (high-confidence) | 0 |
| S3_vs_k141_127421\|\|full | 41647 | No | NA | 78 | 19 | 1 | High-quality | High-quality | 98.86 | AAI-based (high-confidence) | 0 |
| S3_vs_k141_127442\|\|full | 5755 | No | NA | 8 | 3 | 0 | High-quality | High-quality | 99.17 | AAI-based (high-confidence) | 0 |
| S3_vs_k141_127445\|\|full | 76534 | No | NA | 101 | 9 | 1 | High-quality | High-quality | 98.6 | AAI-based (high-confidence) | 0 |
| S3_vs_k141_127448\|\|full | 3513 | No | NA | 4 | 0 | 0 | Not-determined | Genome-fragment | NA | NA | 0 |
| S3_vs_k141_63760\|\|full | 1522 | No | NA | 4 | 0 | 0 | Not-determined | Genome-fragment | NA | NA | 0 |
| S3_vs_k141_15941\|\|full | 2498 | No | NA | 2 | 0 | 2 | Not-determined | Genome-fragment | NA | NA | 0 |
| S3_vs_k141_103618\|\|full | 1103 | No | NA | 1 | 0 | 0 | Not-determined | Genome-fragment | NA | NA | 0 |
| S3_vs_k141_0\|\|full | 5769 | No | NA | 5 | 0 | 1 | Not-determined | Genome-fragment | NA | NA | 0 |
| S3_vs_k141_63768\|\|full | 1824 | No | NA | 3 | 0 | 1 | Not-determined | Genome-fragment | NA | NA | 0 |
| S16_vs_134968\|\|full | 1633 | No | NA | 3 | 0 | 1 | Low-quality | Genome-fragment | 1.04 | AAI-based (high-confidence) | 0 |
| S3_vs_k141_63782\|\|full | 2434 | No | NA | 3 | 0 | 2 | Not-determined | Genome-fragment | NA | NA | 0 |
| S3_vs_k141_111595\|\|full | 7388 | No | NA | 6 | 0 | 4 | Not-determined | Genome-fragment | NA | NA | 0 |
| S3_vs_k141_47858\|\|full | 3590 | No | NA | 4 | 0 | 1 | Not-determined | Genome-fragment | NA | NA | 0 |
| S16_vs_92874\|\|lt2gene | 1633 | No | NA | 1 | 0 | 0 | Low-quality | Genome-fragment | 2.69 | AAI-based (high-confidence) | 0 |
| S12_extracted_k141_56612 | 1633 | No | NA | 1 | 1 | 0 | Low-quality | Genome-fragment | 3.92 | AAI-based (medium-confidence) | 0 |
| S6_vs_k141_44092\|\|full | 1633 | No | NA | 3 | 0 | 0 | Low-quality | Genome-fragment | 3.78 | AAI-based (medium-confidence) | 0 |
| S8_vs_k141_64350\|\|full | 1633 | No | NA | 3 | 0 | 2 | Low-quality | Genome-fragment | 0.68 | AAI-based (medium-confidence) | 0 |
| S3_vs_k141_23996\|\|full | 3047 | No | NA | 4 | 0 | 1 | Not-determined | Genome-fragment | NA | NA | 0 |
| S11_vs_k141_65586\|\|full | 1634 | No | NA | 5 | 0 | 0 | Low-quality | Genome-fragment | 4.46 | AAI-based (medium-confidence) | 0 |
| S13_vs_9319\|\|lt2gene | 1634 | No | NA | 2 | 2 | 0 | Low-quality | Genome-fragment | 3.81 | AAI-based (high-confidence) | 0 |
| S14_extracted_k141_986 | 1634 | No | NA | 2 | 2 | 0 | Low-quality | Genome-fragment | 5.2 | AAI-based (high-confidence) | 0 |
| S3_vs_k141_79766\|\|full | 1024 | No | NA | 3 | 0 | 1 | Not-determined | Genome-fragment | NA | NA | 0 |
| S3_vs_k141_92\|\|full | 2016 | No | NA | 5 | 0 | 1 | Not-determined | Genome-fragment | NA | NA | 0 |
| S3_extracted_k141_106067 | 1634 | No | NA | 2 | 2 | 0 | Low-quality | Genome-fragment | 4.07 | AAI-based (high-confidence) | 0 |
| S3_vs_k141_63844\|\|full | 3442 | No | NA | 4 | 0 | 4 | Not-determined | Genome-fragment | NA | NA | 0 |
| S3_vs_k141_55912\|\|full | 2346 | No | NA | 3 | 0 | 0 | Not-determined | Genome-fragment | NA | NA | 0 |
| S3_vs_k141_31953\|\|full | 4528 | No | NA | 4 | 0 | 4 | Not-determined | Genome-fragment | NA | NA | 0 |
| S3_vs_k141_39930\|\|full | 6892 | No | NA | 4 | 0 | 4 | Not-determined | Genome-fragment | NA | NA | 0 |
| S5_extracted_k141_133037 | 1634 | No | NA | 1 | 0 | 0 | Low-quality | Genome-fragment | 3.3 | AAI-based (medium-confidence) | 0 |
| S16_vs_15001\|\|full | 1634 | No | NA | 2 | 0 | 1 | Low-quality | Genome-fragment | 4.3 | AAI-based (medium-confidence) | 0 |
| S3_vs_k141_71927\|\|full | 1093 | No | NA | 2 | 0 | 1 | Not-determined | Genome-fragment | NA | NA | 0 |
| S3_vs_k141_39940\|\|full | 20073 | No | NA | 39 | 7 | 0 | Medium-quality | Genome-fragment | 51.66 | AAI-based (high-confidence) | 0 |
| S3_vs_k141_87771\|\|full | 1432 | No | NA | 3 | 0 | 0 | Not-determined | Genome-fragment | NA | NA | 0 |
| S3_vs_k141_63915\|\|full | 2316 | No | NA | 4 | 0 | 0 | Not-determined | Genome-fragment | NA | NA | 0 |
| S16_vs_19781\|\|full | 1634 | No | NA | 3 | 0 | 0 | Low-quality | Genome-fragment | 3.46 | AAI-based (high-confidence) | 0 |
| S3_vs_k141_8161\|\|full | 2713 | No | NA | 4 | 0 | 0 | Not-determined | Genome-fragment | NA | NA | 0 |
| S3_vs_k141_119660\|\|full | 39852 | Yes | 33361 | 54 | 7 | 6 | Medium-quality | Genome-fragment | 77.89 | AAI-based (high-confidence) | 16.29 |
| S3_vs_k141_39945\|\|full | 2283 | No | NA | 4 | 0 | 0 | Not-determined | Genome-fragment | NA | NA | 0 |
| S3_vs_k141_103686\|\|full | 5220 | No | NA | 7 | 0 | 2 | Not-determined | Genome-fragment | NA | NA | 0 |
| S12_extracted_k141_110789 | 1634 | No | NA | 2 | 0 | 0 | Low-quality | Genome-fragment | 5.74 | AAI-based (high-confidence) | 0 |
| S4_extracted_8439 | 1634 | No | NA | 3 | 3 | 0 | Low-quality | Genome-fragment | 5.37 | AAI-based (high-confidence) | 0 |
| S3_vs_k141_63954\|\|full | 1797 | No | NA | 4 | 0 | 2 | Not-determined | Genome-fragment | NA | NA | 0 |
| S3_vs_k141_24158\|\|full | 1725 | No | NA | 3 | 0 | 0 | Not-determined | Genome-fragment | NA | NA | 0 |
| S11_vs_k141_49476\|\|full | 1635 | No | NA | 3 | 0 | 0 | Low-quality | Genome-fragment | 3.17 | AAI-based (high-confidence) | 0 |
| S3_vs_k141_79870\|\|full | 3755 | No | NA | 3 | 0 | 0 | Not-determined | Genome-fragment | NA | NA | 0 |
| S3_vs_k141_111755\|\|full | 1684 | No | NA | 3 | 0 | 3 | Not-determined | Genome-fragment | NA | NA | 0 |
| S3_vs_k141_48147\|\|full | 2391 | No | NA | 4 | 0 | 1 | Not-determined | Genome-fragment | NA | NA | 0 |
| S3_vs_k141_8172\|\|full | 10396 | No | NA | 10 | 0 | 0 | Not-determined | Genome-fragment | NA | NA | 0 |
| S11_vs_k141_57282\|\|full | 1635 | No | NA | 3 | 1 | 0 | Low-quality | Genome-fragment | 2.14 | HMM-based (lower-bound) | 0 |
| S3_vs_k141_24181\|\|full | 1835 | No | NA | 3 | 0 | 1 | Not-determined | Genome-fragment | NA | NA | 0 |
| S3_vs_k141_8176\|\|full | 1809 | No | NA | 2 | 0 | 1 | Not-determined | Genome-fragment | NA | NA | 0 |
| S1_extracted_k141_19918 | 1635 | No | NA | 2 | 2 | 0 | Low-quality | Genome-fragment | 2.53 | AAI-based (high-confidence) | 0 |
| S5_vs_k141_91473\|\|full | 1635 | No | NA | 4 | 0 | 0 | Low-quality | Genome-fragment | 2.92 | AAI-based (medium-confidence) | 0 |
| S5_vs_k141_81764\|\|full | 1635 | No | NA | 2 | 2 | 0 | Low-quality | Genome-fragment | 3.47 | AAI-based (medium-confidence) | 0 |
| S3_vs_k141_111773\|\|full | 1224 | No | NA | 2 | 0 | 0 | Not-determined | Genome-fragment | NA | NA | 0 |
| S7_vs_119162\|\|full | 1635 | No | NA | 3 | 0 | 0 | Low-quality | Genome-fragment | 3.28 | AAI-based (medium-confidence) | 0 |
| S3_vs_k141_8211\|\|full | 2344 | No | NA | 2 | 0 | 2 | Not-determined | Genome-fragment | NA | NA | 0 |
| S3_vs_k141_48198\|\|full | 1383 | No | NA | 3 | 0 | 0 | Not-determined | Genome-fragment | NA | NA | 0 |
| S3_vs_k141_95863\|\|full | 3297 | No | NA | 3 | 0 | 2 | Not-determined | Genome-fragment | NA | NA | 0 |
| S17_vs_k141_34808\|\|full | 1636 | No | NA | 3 | 2 | 0 | Low-quality | Genome-fragment | 3.79 | AAI-based (high-confidence) | 0 |
| S9_vs_k141_62755\|\|full | 1636 | No | NA | 3 | 0 | 3 | Low-quality | Genome-fragment | 2.42 | AAI-based (medium-confidence) | 0 |
| S11_extracted_k141_70736 | 1636 | No | NA | 3 | 2 | 0 | Low-quality | Genome-fragment | 3.43 | AAI-based (high-confidence) | 0 |
| S14_extracted_k141_112177 | 1636 | No | NA | 1 | 1 | 0 | Low-quality | Genome-fragment | 2.01 | HMM-based (lower-bound) | 0 |
| S14_vs_k141_62948\|\|full | 1636 | No | NA | 2 | 0 | 1 | Low-quality | Genome-fragment | 2.13 | AAI-based (medium-confidence) | 0 |
| S5_vs_k141_56864\|\|full | 1636 | No | NA | 4 | 1 | 0 | Low-quality | Genome-fragment | 4.08 | AAI-based (high-confidence) | 0 |
| S16_vs_78602\|\|full | 1636 | No | NA | 2 | 0 | 0 | Low-quality | Genome-fragment | 2.32 | AAI-based (medium-confidence) | 0 |
| S3_vs_k141_95945\|\|full | 1292 | No | NA | 3 | 0 | 1 | Not-determined | Genome-fragment | NA | NA | 0 |
| S16_vs_83831\|\|full | 1636 | No | NA | 4 | 1 | 0 | Low-quality | Genome-fragment | 3.07 | AAI-based (medium-confidence) | 0 |
| S12_vs_k141_24692\|\|full | 1636 | No | NA | 5 | 1 | 0 | Low-quality | Genome-fragment | 4.03 | AAI-based (high-confidence) | 0 |
| S15_extracted_15688 | 1636 | No | NA | 2 | 0 | 0 | Low-quality | Genome-fragment | 2.74 | AAI-based (high-confidence) | 0 |
| S8_vs_k141_204706\|\|full | 1636 | No | NA | 4 | 0 | 1 | Low-quality | Genome-fragment | 2.58 | AAI-based (medium-confidence) | 0 |
| S3_vs_k141_103805\|\|full | 2314 | No | NA | 4 | 0 | 4 | Not-determined | Genome-fragment | NA | NA | 0 |
| S3_vs_k141_8325\|\|full | 2653 | No | NA | 4 | 0 | 2 | Not-determined | Genome-fragment | NA | NA | 0 |
| S3_vs_k141_87855\|\|full | 6976 | No | NA | 7 | 0 | 4 | Not-determined | Genome-fragment | NA | NA | 0 |
| S3_vs_k141_111777\|\|full | 3676 | No | NA | 4 | 0 | 1 | Not-determined | Genome-fragment | NA | NA | 0 |
| S3_vs_k141_48282\|\|full | 5559 | No | NA | 7 | 0 | 4 | Not-determined | Genome-fragment | NA | NA | 0 |
| S17_extracted_k141_128181 | 1637 | No | NA | 1 | 0 | 0 | Low-quality | Genome-fragment | 2.56 | AAI-based (medium-confidence) | 0 |
| S5_vs_k141_106941\|\|lt2gene | 1637 | No | NA | 2 | 1 | 0 | Low-quality | Genome-fragment | 3.4 | AAI-based (medium-confidence) | 0 |
| S7_vs_27821\|\|full | 1637 | No | NA | 3 | 1 | 0 | Low-quality | Genome-fragment | 4.38 | AAI-based (high-confidence) | 0 |
| S7_vs_95330\|\|full | 1637 | No | NA | 3 | 1 | 0 | Low-quality | Genome-fragment | 3.37 | AAI-based (medium-confidence) | 0 |
| S16_vs_2647\|\|full | 1637 | No | NA | 4 | 2 | 0 | Low-quality | Genome-fragment | 4.83 | AAI-based (high-confidence) | 0 |
| S10_vs_61580\|\|full | 1637 | No | NA | 2 | 0 | 0 | Low-quality | Genome-fragment | 4.09 | AAI-based (high-confidence) | 0 |
| S3_vs_k141_96020\|\|full | 1172 | No | NA | 3 | 0 | 2 | Not-determined | Genome-fragment | NA | NA | 0 |
| S4_extracted_8048 | 1637 | No | NA | 3 | 2 | 0 | Low-quality | Genome-fragment | 5.38 | AAI-based (high-confidence) | 0 |
| S4_extracted_8705 | 1637 | No | NA | 3 | 3 | 0 | Low-quality | Genome-fragment | 5.39 | AAI-based (high-confidence) | 0 |
| S6_vs_k141_42950\|\|full | 1637 | No | NA | 2 | 1 | 0 | Low-quality | Genome-fragment | 3.8 | AAI-based (high-confidence) | 0 |
| S3_vs_k141_119771\|\|full | 29244 | No | NA | 39 | 12 | 0 | Medium-quality | Genome-fragment | 79 | AAI-based (high-confidence) | 0 |
| S17_vs_k141_125407\|\|full | 1638 | Yes | 1020 | 5 | 1 | 1 | Low-quality | Genome-fragment | 1.96 | AAI-based (medium-confidence) | 37.73 |
| S3_vs_k141_111827\|\|full | 3588 | No | NA | 6 | 0 | 3 | Not-determined | Genome-fragment | NA | NA | 0 |
| S11_vs_k141_62130\|\|full | 1638 | No | NA | 4 | 2 | 0 | Low-quality | Genome-fragment | 2.71 | HMM-based (lower-bound) | 0 |
| S1_extracted_k141_89058 | 1638 | No | NA | 2 | 1 | 0 | Low-quality | Genome-fragment | 0.93 | AAI-based (high-confidence) | 0 |
| S3_vs_k141_48365\|\|full | 1148 | No | NA | 3 | 0 | 0 | Not-determined | Genome-fragment | NA | NA | 0 |
| S3_vs_k141_40161\|\|full | 5268 | No | NA | 10 | 0 | 0 | Not-determined | Genome-fragment | NA | NA | 0 |
| S3_vs_k141_87897\|\|full | 7660 | No | NA | 5 | 0 | 4 | Not-determined | Genome-fragment | NA | NA | 0 |
| S1_extracted_k141_56866 | 1638 | No | NA | 2 | 0 | 0 | Low-quality | Genome-fragment | 1.24 | AAI-based (medium-confidence) | 0 |
| S3_vs_k141_103927\|\|full | 2104 | No | NA | 3 | 0 | 1 | Not-determined | Genome-fragment | NA | NA | 0 |
| S3_vs_k141_40187\|\|full | 9861 | No | NA | 8 | 0 | 0 | Not-determined | Genome-fragment | NA | NA | 0 |
| S3_vs_k141_96005\|\|full | 1638 | No | NA | 2 | 1 | 0 | Low-quality | Genome-fragment | 3.57 | AAI-based (high-confidence) | 0 |
| S3_vs_k141_101957\|\|full | 1638 | No | NA | 3 | 0 | 2 | Low-quality | Genome-fragment | 2.36 | AAI-based (medium-confidence) | 0 |
| S3_vs_k141_75792\|\|lt2gene | 1638 | No | NA | 2 | 0 | 0 | Low-quality | Genome-fragment | 3.55 | AAI-based (high-confidence) | 0 |
| S5_extracted_k141_59632 | 1638 | No | NA | 1 | 0 | 0 | Low-quality | Genome-fragment | 2.66 | AAI-based (high-confidence) | 0 |
| S7_vs_85001\|\|full | 1638 | No | NA | 3 | 1 | 0 | Low-quality | Genome-fragment | 3.43 | AAI-based (medium-confidence) | 0 |
| S16_extracted_4702 | 1638 | No | NA | 1 | 0 | 0 | Low-quality | Genome-fragment | 1.28 | AAI-based (medium-confidence) | 0 |
| S16_extracted_56025 | 1638 | No | NA | 3 | 0 | 1 | Low-quality | Genome-fragment | 0.93 | AAI-based (medium-confidence) | 0 |
| S3_vs_k141_48407\|\|full | 3063 | No | NA | 2 | 0 | 1 | Not-determined | Genome-fragment | NA | NA | 0 |
| S3_vs_k141_24401\|\|full | 3250 | No | NA | 4 | 0 | 0 | Not-determined | Genome-fragment | NA | NA | 0 |
| S16_vs_116854\|\|full | 1638 | No | NA | 2 | 0 | 0 | Low-quality | Genome-fragment | 3.05 | AAI-based (high-confidence) | 0 |
| S16_vs_116978\|\|full | 1638 | No | NA | 4 | 0 | 0 | Low-quality | Genome-fragment | 3.61 | AAI-based (high-confidence) | 0 |
| S3_vs_k141_40278\|\|full | 2127 | No | NA | 3 | 0 | 1 | Not-determined | Genome-fragment | NA | NA | 0 |
| S12_vs_k141_171481\|\|full | 1638 | No | NA | 3 | 0 | 0 | Low-quality | Genome-fragment | 3.26 | AAI-based (medium-confidence) | 0 |
| S4_extracted_24905 | 1638 | No | NA | 1 | 1 | 0 | Low-quality | Genome-fragment | 4.01 | AAI-based (medium-confidence) | 0 |
| S8_vs_k141_201666\|\|full | 1638 | No | NA | 3 | 2 | 0 | Low-quality | Genome-fragment | 1.07 | AAI-based (high-confidence) | 0 |
| S8_vs_k141_32766\|\|full | 1638 | No | NA | 1 | 0 | 1 | Low-quality | Genome-fragment | 2.51 | AAI-based (high-confidence) | 0 |
| S3_vs_k141_96148\|\|full | 15554 | No | NA | 14 | 0 | 12 | Not-determined | Genome-fragment | NA | NA | 0 |
| S3_vs_k141_32431\|\|full | 1991 | No | NA | 5 | 0 | 0 | Not-determined | Genome-fragment | NA | NA | 0 |
| S11_vs_k141_57136\|\|full | 1639 | No | NA | 4 | 3 | 0 | Low-quality | Genome-fragment | 3.46 | AAI-based (high-confidence) | 0 |
| S14_extracted_k141_50222 | 1639 | No | NA | 2 | 0 | 0 | Low-quality | Genome-fragment | 3.16 | AAI-based (medium-confidence) | 0 |
| S3_vs_k141_32438\|\|full | 5402 | No | NA | 3 | 0 | 2 | Not-determined | Genome-fragment | NA | NA | 0 |
| S3_vs_k141_32441\|\|full | 1008 | No | NA | 2 | 0 | 2 | Not-determined | Genome-fragment | NA | NA | 0 |
| S3_vs_k141_104054\|\|full | 2204 | No | NA | 4 | 0 | 0 | Not-determined | Genome-fragment | NA | NA | 0 |
| S3_vs_k141_72118\|\|full | 34453 | No | NA | 50 | 8 | 0 | Medium-quality | Genome-fragment | 87.8 | AAI-based (medium-confidence) | 0 |
| S3_vs_k141_16171\|\|full | 2213 | No | NA | 4 | 0 | 1 | Not-determined | Genome-fragment | NA | NA | 0 |
| S7_vs_30360\|\|full | 1639 | No | NA | 3 | 1 | 0 | Low-quality | Genome-fragment | 1.89 | HMM-based (lower-bound) | 0 |
| S3_vs_k141_32474\|\|full | 3674 | No | NA | 6 | 0 | 1 | Not-determined | Genome-fragment | NA | NA | 0 |
| S3_vs_k141_88048\|\|full | 4374 | No | NA | 3 | 0 | 3 | Not-determined | Genome-fragment | NA | NA | 0 |
| S16_vs_140237\|\|full | 1639 | No | NA | 2 | 0 | 1 | Low-quality | Genome-fragment | 1.31 | AAI-based (high-confidence) | 0 |
| S10_vs_30570\|\|full | 1639 | No | NA | 3 | 0 | 1 | Low-quality | Genome-fragment | 1.12 | AAI-based (high-confidence) | 0 |
| S3_vs_k141_40395\|\|full | 1574 | No | NA | 1 | 0 | 1 | Not-determined | Genome-fragment | NA | NA | 0 |
| S3_vs_k141_80179\|\|full | 2044 | No | NA | 3 | 0 | 1 | Not-determined | Genome-fragment | NA | NA | 0 |
| S8_extracted_k141_127516 | 1639 | No | NA | 4 | 0 | 0 | Low-quality | Genome-fragment | 3.34 | AAI-based (high-confidence) | 0 |
| S3_vs_k141_72213\|\|full | 1914 | No | NA | 3 | 0 | 2 | Not-determined | Genome-fragment | NA | NA | 0 |
| S8_vs_k141_85073\|\|full | 1639 | No | NA | 2 | 0 | 2 | Low-quality | Genome-fragment | 3.18 | AAI-based (medium-confidence) | 0 |
| S3_vs_k141_8745\|\|full | 1916 | No | NA | 2 | 0 | 0 | Not-determined | Genome-fragment | NA | NA | 0 |
| S17_vs_k141_22857\|\|full | 1640 | No | NA | 2 | 0 | 1 | Low-quality | Genome-fragment | 2.98 | AAI-based (medium-confidence) | 0 |
| S11_extracted_k141_8974 | 1640 | No | NA | 2 | 1 | 0 | Low-quality | Genome-fragment | 2.32 | AAI-based (high-confidence) | 0 |
| S3_vs_k141_111987\|\|full | 1526 | No | NA | 2 | 0 | 1 | Not-determined | Genome-fragment | NA | NA | 0 |
| S11_vs_k141_43665\|\|full | 1640 | No | NA | 2 | 0 | 0 | Low-quality | Genome-fragment | 1.47 | AAI-based (medium-confidence) | 0 |
| S3_vs_k141_48507\|\|full | 1855 | No | NA | 3 | 0 | 1 | Not-determined | Genome-fragment | NA | NA | 0 |
| S3_vs_k141_104138\|\|full | 1213 | No | NA | 5 | 0 | 0 | Not-determined | Genome-fragment | NA | NA | 0 |
| S3_vs_k141_689\|\|full | 1178 | No | NA | 3 | 0 | 0 | Not-determined | Genome-fragment | NA | NA | 0 |
| S3_vs_k141_8810\|\|full | 1764 | No | NA | 3 | 0 | 1 | Not-determined | Genome-fragment | NA | NA | 0 |
| S1_vs_k141_100517\|\|full | 1640 | No | NA | 2 | 1 | 0 | Low-quality | Genome-fragment | 3.58 | AAI-based (high-confidence) | 0 |
| S3_vs_k141_56292\|\|full | 52629 | No | NA | 57 | 13 | 2 | High-quality | High-quality | 100 | AAI-based (high-confidence) | 0 |
| S7_vs_81828\|\|full | 1640 | No | NA | 3 | 0 | 0 | Low-quality | Genome-fragment | 2.82 | AAI-based (high-confidence) | 0 |
| S12_vs_k141_178919\|\|full | 1640 | No | NA | 5 | 1 | 0 | Low-quality | Genome-fragment | 1 | HMM-based (lower-bound) | 0 |
| S3_vs_k141_718\|\|full | 6579 | No | NA | 6 | 0 | 3 | Not-determined | Genome-fragment | NA | NA | 0 |
| S3_vs_k141_96281\|\|full | 1103 | No | NA | 2 | 0 | 0 | Not-determined | Genome-fragment | NA | NA | 0 |
| S3_vs_k141_48591\|\|full | 4998 | No | NA | 4 | 0 | 3 | Not-determined | Genome-fragment | NA | NA | 0 |
| S17_vs_k141_3763\|\|lt2gene | 1641 | No | NA | 2 | 1 | 0 | Low-quality | Genome-fragment | 4.34 | AAI-based (high-confidence) | 0 |
| S9_extracted_k141_87446 | 1641 | No | NA | 2 | 2 | 0 | Low-quality | Genome-fragment | 2.42 | HMM-based (lower-bound) | 0 |
| S9_extracted_k141_47261 | 1641 | No | NA | 3 | 0 | 1 | Low-quality | Genome-fragment | 2.95 | AAI-based (high-confidence) | 0 |
| S11_vs_k141_82833\|\|full | 1641 | No | NA | 3 | 0 | 1 | Low-quality | Genome-fragment | 0.99 | AAI-based (high-confidence) | 0 |
| S5_vs_k141_100990\|\|full | 1641 | No | NA | 3 | 2 | 0 | Low-quality | Genome-fragment | 2.56 | AAI-based (high-confidence) | 0 |
| S3_vs_k141_32679\|\|full | 4566 | No | NA | 5 | 0 | 4 | Not-determined | Genome-fragment | NA | NA | 0 |
| S3_vs_k141_80301\|\|full | 1067 | No | NA | 2 | 0 | 1 | Not-determined | Genome-fragment | NA | NA | 0 |
| S7_vs_111011\|\|full | 1641 | No | NA | 3 | 1 | 0 | Low-quality | Genome-fragment | 3.22 | AAI-based (medium-confidence) | 0 |
| S16_extracted_59694 | 1641 | No | NA | 2 | 0 | 0 | Low-quality | Genome-fragment | 1.54 | AAI-based (high-confidence) | 0 |
| S16_extracted_97501 | 1641 | No | NA | 4 | 0 | 0 | Low-quality | Genome-fragment | 2.66 | AAI-based (high-confidence) | 0 |
| S10_vs_103423\|\|full | 1641 | No | NA | 3 | 0 | 2 | Low-quality | Genome-fragment | 0.73 | AAI-based (medium-confidence) | 0 |
| S3_vs_k141_48664\|\|full | 1105 | No | NA | 2 | 0 | 0 | Not-determined | Genome-fragment | NA | NA | 0 |
| S3_vs_k141_112078\|\|full | 3721 | No | NA | 4 | 0 | 1 | Not-determined | Genome-fragment | NA | NA | 0 |
| S12_vs_k141_73982\|\|full | 1641 | No | NA | 2 | 0 | 1 | Low-quality | Genome-fragment | 4.72 | AAI-based (medium-confidence) | 0 |
| S3_vs_k141_48675\|\|full | 2986 | No | NA | 4 | 0 | 2 | Not-determined | Genome-fragment | NA | NA | 0 |
| S9_vs_k141_83203\|\|full | 1642 | No | NA | 5 | 1 | 0 | Low-quality | Genome-fragment | 3.28 | AAI-based (high-confidence) | 0 |
| S11_vs_k141_3009\|\|full | 1642 | No | NA | 4 | 0 | 1 | Low-quality | Genome-fragment | 1.33 | AAI-based (medium-confidence) | 0 |
| S3_vs_k141_88202\|\|full | 1631 | No | NA | 3 | 0 | 1 | Not-determined | Genome-fragment | NA | NA | 0 |
| S3_vs_k141_796\|\|full | 1269 | No | NA | 2 | 0 | 0 | Not-determined | Genome-fragment | NA | NA | 0 |
| S3_vs_k141_120070\|\|full | 2881 | No | NA | 2 | 0 | 2 | Not-determined | Genome-fragment | NA | NA | 0 |
| S3_vs_k141_48702\|\|full | 3248 | No | NA | 4 | 0 | 2 | Not-determined | Genome-fragment | NA | NA | 0 |
| S14_vs_k141_41439\|\|full | 1642 | No | NA | 3 | 0 | 2 | Low-quality | Genome-fragment | 0.97 | AAI-based (medium-confidence) | 0 |
| S1_vs_k141_67547\|\|full | 1642 | No | NA | 3 | 0 | 0 | Low-quality | Genome-fragment | 3.01 | AAI-based (high-confidence) | 0 |
| S3_vs_k141_112160\|\|full | 3967 | No | NA | 5 | 0 | 0 | Not-determined | Genome-fragment | NA | NA | 0 |
| S3_vs_k141_48716\|\|full | 3009 | No | NA | 3 | 0 | 3 | Not-determined | Genome-fragment | NA | NA | 0 |
| S1_vs_k141_52551\|\|full | 1642 | No | NA | 3 | 2 | 0 | Low-quality | Genome-fragment | 4.81 | AAI-based (high-confidence) | 0 |
| S3_extracted_k141_6810 | 1642 | No | NA | 5 | 0 | 0 | Low-quality | Genome-fragment | 2.9 | AAI-based (high-confidence) | 0 |
| S5_vs_k141_108947\|\|full | 1642 | No | NA | 4 | 0 | 1 | Low-quality | Genome-fragment | 3.86 | AAI-based (medium-confidence) | 0 |
| S16_vs_92990\|\|full | 1642 | No | NA | 4 | 1 | 0 | Low-quality | Genome-fragment | 0.95 | AAI-based (high-confidence) | 0 |
| S12_vs_k141_21721\|\|full | 1642 | No | NA | 6 | 0 | 0 | Low-quality | Genome-fragment | 4.33 | AAI-based (medium-confidence) | 0 |
| S3_vs_k141_72538\|\|full | 1390 | No | NA | 2 | 0 | 2 | Not-determined | Genome-fragment | NA | NA | 0 |
| S12_vs_k141_3294\|\|lt2gene | 1642 | No | NA | 2 | 2 | 0 | Low-quality | Genome-fragment | 4.09 | AAI-based (high-confidence) | 0 |
| S3_vs_k141_32772\|\|full | 6542 | No | NA | 3 | 0 | 3 | Not-determined | Genome-fragment | NA | NA | 0 |
| S17_extracted_k141_111081 | 1643 | No | NA | 4 | 4 | 0 | Low-quality | Genome-fragment | 5.42 | AAI-based (high-confidence) | 0 |
| S3_vs_k141_48763\|\|full | 1262 | No | NA | 2 | 0 | 1 | Not-determined | Genome-fragment | NA | NA | 0 |

| | | | | | | | | | | | |
|---|---|---|---|---|---|---|---|---|---|---|---|
| S3_vs_k141_64335\|\|full | 5837 | No | NA | 5 | 0 | 4 | Not-determined | Genome-fragment | NA | NA | 0 |
| S17_vs_k141_170447\|\|full | 1643 | No | NA | 4 | 0 | 1 | Low-quality | Genome-fragment | 2.79 | AAI-based (high-confidence) | 0 |
| S9_vs_k141_25010\|\|full | 1643 | No | NA | 2 | 0 | 2 | Low-quality | Genome-fragment | 4.01 | AAI-based (medium-confidence) | 0 |
| S11_extracted_k141_30609 | 1643 | No | NA | 3 | 1 | 0 | Low-quality | Genome-fragment | 3.49 | AAI-based (high-confidence) | 0 |
| S3_vs_k141_32853\|\|full | 1320 | No | NA | 4 | 0 | 0 | Not-determined | Genome-fragment | NA | NA | 0 |
| S11_vs_k141_1394\|\|full | 1643 | No | NA | 2 | 2 | 0 | Low-quality | Genome-fragment | 2.54 | AAI-based (high-confidence) | 0 |
| S13_extracted_123204 | 1643 | No | NA | 1 | 0 | 0 | Low-quality | Genome-fragment | 2.89 | AAI-based (medium-confidence) | 0 |
| S13_vs_38485\|\|full | 1643 | No | NA | 4 | 0 | 0 | Low-quality | Genome-fragment | 4.12 | AAI-based (high-confidence) | 0 |
| S14_extracted_k141_100530 | 1643 | No | NA | 2 | 0 | 2 | Low-quality | Genome-fragment | 2.9 | AAI-based (high-confidence) | 0 |
| S14_vs_k141_33472\|\|full | 1643 | No | NA | 2 | 1 | 0 | Low-quality | Genome-fragment | 3.81 | AAI-based (medium-confidence) | 0 |
| S3_vs_k141_80424\|\|full | 2773 | No | NA | 3 | 0 | 0 | Not-determined | Genome-fragment | NA | NA | 0 |
| S1_extracted_k141_94964 | 1643 | No | NA | 2 | 2 | 0 | Low-quality | Genome-fragment | 1.66 | AAI-based (high-confidence) | 0 |
| S10_vs_51946\|\|full | 1643 | No | NA | 2 | 0 | 0 | Low-quality | Genome-fragment | 1.52 | AAI-based (medium-confidence) | 0 |
| S12_vs_k141_16942\|\|full | 1643 | No | NA | 3 | 0 | 0 | Low-quality | Genome-fragment | 1.08 | AAI-based (medium-confidence) | 0 |
| S3_vs_k141_64376\|\|full | 1872 | No | NA | 2 | 0 | 1 | Not-determined | Genome-fragment | NA | NA | 0 |
| S8_vs_k141_122447\|\|lt2gene | 1643 | No | NA | 2 | 2 | 0 | Low-quality | Genome-fragment | 4.36 | AAI-based (high-confidence) | 0 |
| S14_extracted_k141_50918 | 1644 | No | NA | 1 | 1 | 0 | Low-quality | Genome-fragment | 1.21 | AAI-based (medium-confidence) | 0 |
| S7_vs_27495\|\|full | 1644 | No | NA | 5 | 1 | 0 | Low-quality | Genome-fragment | 3.91 | AAI-based (medium-confidence) | 0 |
| S7_vs_55814\|\|full | 1644 | No | NA | 3 | 0 | 0 | Low-quality | Genome-fragment | 2.57 | AAI-based (high-confidence) | 0 |
| S3_vs_k141_104376\|\|full | 1374 | No | NA | 2 | 0 | 0 | Not-determined | Genome-fragment | NA | NA | 0 |
| S3_vs_k141_48813\|\|full | 2814 | No | NA | 2 | 0 | 2 | Not-determined | Genome-fragment | NA | NA | 0 |
| S3_vs_k141_88322\|\|full | 1682 | No | NA | 2 | 0 | 2 | Not-determined | Genome-fragment | NA | NA | 0 |
| S7_vs_40578\|\|lt2gene | 1644 | No | NA | 2 | 2 | 0 | Low-quality | Genome-fragment | 4.77 | AAI-based (high-confidence) | 0 |
| S16_vs_33663\|\|full | 1644 | No | NA | 4 | 1 | 0 | Low-quality | Genome-fragment | 1.47 | HMM-based (lower-bound) | 0 |
| S3_vs_k141_96557\|\|full | 2372 | No | NA | 2 | 0 | 1 | Not-determined | Genome-fragment | NA | NA | 0 |
| S3_vs_k141_32914\|\|full | 1468 | No | NA | 3 | 0 | 1 | Not-determined | Genome-fragment | NA | NA | 0 |
| S3_vs_k141_32915\|\|full | 1359 | No | NA | 3 | 0 | 1 | Not-determined | Genome-fragment | NA | NA | 0 |
| S3_vs_k141_96565\|\|full | 3380 | No | NA | 4 | 0 | 0 | Not-determined | Genome-fragment | NA | NA | 0 |
| S3_vs_k141_88328\|\|full | 3215 | No | NA | 3 | 0 | 1 | Not-determined | Genome-fragment | NA | NA | 0 |
| S3_vs_k141_104387\|\|full | 1655 | No | NA | 3 | 0 | 2 | Not-determined | Genome-fragment | NA | NA | 0 |
| S12_vs_k141_182122\|\|full | 1644 | No | NA | 5 | 0 | 0 | Low-quality | Genome-fragment | 3.13 | AAI-based (high-confidence) | 0 |
| S3_vs_k141_9158\|\|full | 61321 | Yes | 29689 | 62 | 9 | 24 | High-quality | High-quality | 100 | AAI-based (high-confidence) | 51.58 |
| S8_vs_k141_190996\|\|full | 1644 | No | NA | 5 | 3 | 0 | Low-quality | Genome-fragment | 3.82 | AAI-based (high-confidence) | 0 |
| S11_vs_k141_88646\|\|full | 1645 | No | NA | 3 | 2 | 0 | Low-quality | Genome-fragment | 1.93 | HMM-based (lower-bound) | 0 |
| S5_vs_k141_46462\|\|full | 1645 | No | NA | 3 | 0 | 0 | Low-quality | Genome-fragment | 0.59 | AAI-based (high-confidence) | 0 |
| S7_vs_36434\|\|full | 1645 | No | NA | 3 | 0 | 0 | Low-quality | Genome-fragment | 4.04 | AAI-based (high-confidence) | 0 |
| S3_vs_k141_88386\|\|full | 6549 | No | NA | 10 | 0 | 4 | Not-determined | Genome-fragment | NA | NA | 0 |
| S16_vs_5411\|\|full | 1645 | No | NA | 5 | 0 | 0 | Low-quality | Genome-fragment | 2.18 | AAI-based (medium-confidence) | 0 |
| S3_vs_k141_9194\|\|full | 4156 | No | NA | 3 | 0 | 2 | Not-determined | Genome-fragment | NA | NA | 0 |
| S3_vs_k141_64424\|\|full | 75252 | No | NA | 60 | 1 | 20 | High-quality | High-quality | 100 | AAI-based (high-confidence) | 0 |
| S10_vs_41831\|\|full | 1645 | No | NA | 2 | 0 | 0 | Low-quality | Genome-fragment | 4.2 | AAI-based (medium-confidence) | 0 |
| S12_extracted_k141_183839 | 1645 | No | NA | 2 | 0 | 1 | Low-quality | Genome-fragment | 0.73 | AAI-based (medium-confidence) | 0 |
| S12_vs_k141_58152\|\|full | 1645 | No | NA | 4 | 0 | 0 | Low-quality | Genome-fragment | 2.9 | AAI-based (medium-confidence) | 0 |
| S3_vs_k141_9260\|\|full | 1648 | No | NA | 3 | 0 | 1 | Not-determined | Genome-fragment | NA | NA | 0 |
| S3_vs_k141_48980\|\|full | 4622 | No | NA | 4 | 0 | 0 | Not-determined | Genome-fragment | NA | NA | 0 |
| S9_vs_k141_12913\|\|full | 1646 | No | NA | 2 | 0 | 1 | Low-quality | Genome-fragment | 2.74 | AAI-based (high-confidence) | 0 |
| S3_vs_k141_112370\|\|full | 4200 | No | NA | 8 | 0 | 1 | Not-determined | Genome-fragment | NA | NA | 0 |
| S3_vs_k141_80602\|\|full | 10591 | No | NA | 11 | 0 | 2 | Not-determined | Genome-fragment | NA | NA | 0 |
| S3_vs_k141_33050\|\|full | 2746 | No | NA | 3 | 0 | 3 | Not-determined | Genome-fragment | NA | NA | 0 |
| S3_vs_k141_9284\|\|full | 1747 | No | NA | 3 | 0 | 0 | Not-determined | Genome-fragment | NA | NA | 0 |
| S3_vs_k141_33069\|\|full | 2655 | No | NA | 3 | 0 | 2 | Not-determined | Genome-fragment | NA | NA | 0 |
| S3_vs_k141_9295\|\|full | 7376 | No | NA | 6 | 0 | 1 | Not-determined | Genome-fragment | NA | NA | 0 |
| S9_vs_k141_37600\|\|full | 1646 | No | NA | 3 | 0 | 2 | Low-quality | Genome-fragment | 0.51 | AAI-based (medium-confidence) | 0 |
| S11_vs_k141_86895\|\|full | 1646 | No | NA | 2 | 0 | 1 | Low-quality | Genome-fragment | 0.64 | AAI-based (high-confidence) | 0 |
| S13_extracted_149466 | 1646 | No | NA | 4 | 0 | 0 | Low-quality | Genome-fragment | 4.45 | AAI-based (medium-confidence) | 0 |
| S14_vs_k141_22761\|\|full | 1646 | No | NA | 3 | 0 | 0 | Low-quality | Genome-fragment | 3.13 | AAI-based (high-confidence) | 0 |
| S3_vs_k141_64533\|\|full | 1583 | No | NA | 3 | 0 | 0 | Not-determined | Genome-fragment | NA | NA | 0 |
| S1_vs_k141_143165\|\|full | 1646 | No | NA | 4 | 0 | 0 | Low-quality | Genome-fragment | 1.06 | AAI-based (high-confidence) | 0 |
| S3_vs_k141_49053\|\|full | 7143 | No | NA | 5 | 0 | 5 | Not-determined | Genome-fragment | NA | NA | 0 |
| S3_vs_k141_27872\|\|full | 1646 | No | NA | 3 | 0 | 0 | Low-quality | Genome-fragment | 2.69 | AAI-based (medium-confidence) | 0 |
| S7_vs_27285\|\|full | 1646 | No | NA | 6 | 0 | 0 | Low-quality | Genome-fragment | 4.77 | AAI-based (high-confidence) | 0 |
| S12_vs_k141_956\|\|full | 1646 | No | NA | 5 | 1 | 0 | Low-quality | Genome-fragment | 2.16 | HMM-based (lower-bound) | 0 |
| S17_extracted_k141_153151 | 1647 | No | NA | 2 | 0 | 1 | Low-quality | Genome-fragment | 0.97 | AAI-based (medium-confidence) | 0 |
| S3_vs_k141_80675\|\|full | 2357 | No | NA | 3 | 0 | 1 | Not-determined | Genome-fragment | NA | NA | 0 |
| S17_vs_k141_171446\|\|full | 1647 | No | NA | 5 | 1 | 0 | Low-quality | Genome-fragment | 2.8 | AAI-based (medium-confidence) | 0 |
| S3_vs_k141_49095\|\|full | 1374 | No | NA | 2 | 0 | 1 | Not-determined | Genome-fragment | NA | NA | 0 |
| S11_extracted_k141_18699 | 1647 | No | NA | 1 | 0 | 1 | Low-quality | Genome-fragment | 0.23 | AAI-based (medium-confidence) | 0 |
| S3_vs_k141_1031\|\|full | 14570 | No | NA | 23 | 10 | 0 | Medium-quality | Genome-fragment | 57.41 | AAI-based (medium-confidence) | 0 |
| S3_vs_k141_16537\|\|full | 7661 | No | NA | 7 | 0 | 3 | Not-determined | Genome-fragment | NA | NA | 0 |
| S3_vs_k141_56895\|\|full | 1993 | No | NA | 3 | 0 | 1 | Not-determined | Genome-fragment | NA | NA | 0 |
| S14_extracted_k141_62112 | 1647 | No | NA | 3 | 1 | 0 | Low-quality | Genome-fragment | 3.31 | AAI-based (high-confidence) | 0 |
| S1_vs_k141_145328\|\|full | 1647 | No | NA | 3 | 0 | 2 | Low-quality | Genome-fragment | 0.52 | AAI-based (medium-confidence) | 0 |
| S3_vs_k141_56906\|\|full | 1774 | No | NA | 3 | 0 | 1 | Not-determined | Genome-fragment | NA | NA | 0 |
| S3_vs_k141_49173\|\|full | 2108 | No | NA | 2 | 0 | 2 | Not-determined | Genome-fragment | NA | NA | 0 |
| S3_vs_k141_104542\|\|full | 2221 | No | NA | 2 | 0 | 1 | Not-determined | Genome-fragment | NA | NA | 0 |
| S7_vs_29074\|\|full | 1647 | No | NA | 6 | 0 | 0 | Low-quality | Genome-fragment | 4.75 | AAI-based (high-confidence) | 0 |
| S15_extracted_53854 | 1647 | No | NA | 3 | 1 | 0 | Low-quality | Genome-fragment | 0.99 | AAI-based (high-confidence) | 0 |
| S3_vs_k141_49200\|\|full | 14483 | No | NA | 9 | 0 | 8 | Not-determined | Genome-fragment | NA | NA | 0 |
| S17_extracted_k141_115356 | 1648 | No | NA | 2 | 0 | 0 | Low-quality | Genome-fragment | 1.05 | AAI-based (medium-confidence) | 0 |
| S17_vs_k141_30522\|\|full | 1648 | No | NA | 2 | 1 | 0 | Low-quality | Genome-fragment | 3.45 | AAI-based (high-confidence) | 0 |
| S14_vs_k141_10248\|\|full | 1648 | No | NA | 2 | 2 | 0 | Low-quality | Genome-fragment | 4.15 | AAI-based (high-confidence) | 0 |
| S14_vs_k141_101163\|\|full | 1648 | No | NA | 2 | 0 | 1 | Low-quality | Genome-fragment | 2.03 | AAI-based (high-confidence) | 0 |
| S3_vs_k141_119228\|\|full | 1648 | No | NA | 3 | 1 | 0 | Low-quality | Genome-fragment | 0.43 | HMM-based (lower-bound) | 0 |
| S5_vs_k141_44034\|\|full | 1648 | No | NA | 4 | 0 | 0 | Low-quality | Genome-fragment | 3.38 | AAI-based (medium-confidence) | 0 |
| S5_vs_k141_134600\|\|full | 1648 | No | NA | 3 | 0 | 0 | Low-quality | Genome-fragment | 3.95 | AAI-based (high-confidence) | 0 |
| S3_vs_k141_73225\|\|full | 1471 | No | NA | 3 | 0 | 0 | Not-determined | Genome-fragment | NA | NA | 0 |
| S16_vs_72248\|\|full | 1648 | No | NA | 3 | 2 | 0 | Low-quality | Genome-fragment | 4.38 | AAI-based (high-confidence) | 0 |
| S12_vs_k141_12877\|\|full | 1648 | No | NA | 3 | 1 | 0 | Low-quality | Genome-fragment | 3.76 | AAI-based (high-confidence) | 0 |
| S3_vs_k141_80832\|\|full | 2302 | No | NA | 2 | 0 | 1 | Not-determined | Genome-fragment | NA | NA | 0 |
| S12_vs_k141_132657\|\|full | 1648 | No | NA | 4 | 0 | 0 | Low-quality | Genome-fragment | 2.83 | AAI-based (high-confidence) | 0 |
| S3_vs_k141_9582\|\|full | 2344 | No | NA | 2 | 0 | 2 | Not-determined | Genome-fragment | NA | NA | 0 |
| S3_vs_k141_49309\|\|full | 2239 | No | NA | 2 | 0 | 2 | Not-determined | Genome-fragment | NA | NA | 0 |
| S15_vs_31088\|\|full | 1648 | No | NA | 3 | 0 | 0 | Low-quality | Genome-fragment | 4.19 | AAI-based (high-confidence) | 0 |
| S15_vs_45085\|\|full | 1648 | No | NA | 2 | 0 | 2 | Low-quality | Genome-fragment | 1.55 | AAI-based (medium-confidence) | 0 |
| S14_vs_k141_72769\|\|full | 1649 | No | NA | 6 | 1 | 0 | Low-quality | Genome-fragment | 2.09 | AAI-based (high-confidence) | 0 |
| S3_vs_k141_73307\|\|full | 2826 | No | NA | 3 | 0 | 0 | Not-determined | Genome-fragment | NA | NA | 0 |
| S5_extracted_k141_88494 | 1649 | No | NA | 5 | 4 | 0 | Low-quality | Genome-fragment | 4.07 | AAI-based (high-confidence) | 0 |
| S5_vs_k141_29243\|\|full | 1649 | No | NA | 3 | 1 | 0 | Low-quality | Genome-fragment | 2.63 | HMM-based (lower-bound) | 0 |
| S7_extracted_31051 | 1649 | No | NA | 2 | 1 | 0 | Low-quality | Genome-fragment | 3.56 | AAI-based (high-confidence) | 0 |
| S16_extracted_154645 | 1649 | Yes | 713 | 4 | 1 | 1 | Low-quality | Genome-fragment | 0.41 | AAI-based (high-confidence) | 56.76 |
| S3_vs_k141_73348\|\|full | 3037 | No | NA | 3 | 0 | 3 | Not-determined | Genome-fragment | NA | NA | 0 |
| S16_vs_121784\|\|full | 1649 | No | NA | 3 | 2 | 0 | Low-quality | Genome-fragment | 2.46 | HMM-based (lower-bound) | 0 |
| S12_extracted_k141_68699 | 1649 | No | NA | 2 | 0 | 2 | Low-quality | Genome-fragment | 0.93 | AAI-based (medium-confidence) | 0 |
| S3_vs_k141_57093\|\|full | 11866 | No | NA | 13 | 0 | 8 | Not-determined | Genome-fragment | NA | NA | 0 |
| S8_extracted_k141_149377 | 1649 | No | NA | 4 | 1 | 0 | Low-quality | Genome-fragment | 5.24 | AAI-based (high-confidence) | 0 |
| S8_extracted_k141_81437 | 1649 | No | NA | 3 | 0 | 0 | Low-quality | Genome-fragment | 3.74 | AAI-based (high-confidence) | 0 |
| S8_vs_k141_128987\|\|full | 1649 | No | NA | 4 | 0 | 0 | Low-quality | Genome-fragment | 4.07 | AAI-based (medium-confidence) | 0 |
| S9_vs_k141_7485\|\|full | 1650 | No | NA | 3 | 0 | 0 | Low-quality | Genome-fragment | 4.2 | AAI-based (medium-confidence) | 0 |
| S3_vs_k141_73361\|\|full | 33306 | No | NA | 62 | 12 | 0 | High-quality | High-quality | 93.08 | AAI-based (high-confidence) | 0 |
| S14_extracted_k141_29430 | 1650 | No | NA | 2 | 2 | 0 | Low-quality | Genome-fragment | 5.33 | AAI-based (high-confidence) | 0 |
| S3_vs_k141_9663\|\|full | 1657 | No | NA | 4 | 0 | 0 | Not-determined | Genome-fragment | NA | NA | 0 |
| S3_vs_k141_40745\|\|full | 1650 | No | NA | 2 | 2 | 0 | Low-quality | Genome-fragment | 3.64 | AAI-based (high-confidence) | 0 |
| S3_vs_k141_88676\|\|full | 1852 | No | NA | 3 | 0 | 0 | Not-determined | Genome-fragment | NA | NA | 0 |
| S3_vs_k141_1143\|\|full | 47753 | No | NA | 68 | 22 | 0 | High-quality | High-quality | 97.11 | AAI-based (high-confidence) | 0 |
| S7_vs_31711\|\|full | 1650 | No | NA | 2 | 0 | 0 | Low-quality | Genome-fragment | 3.8 | AAI-based (high-confidence) | 0 |
| S3_vs_k141_1148\|\|full | 1276 | No | NA | 3 | 0 | 1 | Not-determined | Genome-fragment | NA | NA | 0 |
| S16_vs_8596\|\|full | 1650 | No | NA | 3 | 2 | 0 | Low-quality | Genome-fragment | 2.96 | HMM-based (lower-bound) | 0 |
| S12_vs_k141_107906\|\|full | 1650 | No | NA | 3 | 0 | 1 | Low-quality | Genome-fragment | 1.23 | AAI-based (medium-confidence) | 0 |
| S8_vs_k141_108596\|\|full | 1650 | No | NA | 4 | 2 | 0 | Low-quality | Genome-fragment | 3.32 | AAI-based (medium-confidence) | 0 |
| S17_vs_k141_148482\|\|full | 1651 | No | NA | 2 | 2 | 0 | Low-quality | Genome-fragment | 4.21 | AAI-based (high-confidence) | 0 |
| S3_vs_k141_25003\|\|full | 1170 | No | NA | 3 | 0 | 0 | Not-determined | Genome-fragment | NA | NA | 0 |
| S3_vs_k141_112485\|\|full | 2100 | No | NA | 4 | 0 | 0 | Not-determined | Genome-fragment | NA | NA | 0 |
| S3_vs_k141_81027\|\|full | 12069 | No | NA | 12 | 0 | 3 | Not-determined | Genome-fragment | NA | NA | 0 |
| S1_vs_k141_9523\|\|full | 1651 | No | NA | 3 | 0 | 2 | Low-quality | Genome-fragment | 1.34 | AAI-based (medium-confidence) | 0 |
| S3_extracted_k141_42258 | 1651 | No | NA | 2 | 1 | 0 | Low-quality | Genome-fragment | 1.91 | AAI-based (high-confidence) | 0 |
| S3_vs_k141_104710\|\|full | 1075 | No | NA | 2 | 0 | 0 | Not-determined | Genome-fragment | NA | NA | 0 |
| S5_vs_k141_58398\|\|full | 1651 | No | NA | 2 | 0 | 0 | Low-quality | Genome-fragment | 1.02 | AAI-based (medium-confidence) | 0 |
| S16_extracted_92627 | 1651 | No | NA | 2 | 2 | 0 | Low-quality | Genome-fragment | 5.14 | AAI-based (high-confidence) | 0 |
| S2_extracted_k141_8144 | 1651 | No | NA | 4 | 1 | 0 | Low-quality | Genome-fragment | 4.2 | AAI-based (high-confidence) | 0 |
| S3_vs_k141_16808\|\|full | 4659 | No | NA | 5 | 0 | 3 | Not-determined | Genome-fragment | NA | NA | 0 |
| S8_extracted_k141_25276 | 1651 | No | NA | 1 | 0 | 1 | Low-quality | Genome-fragment | 2.08 | AAI-based (medium-confidence) | 0 |
| S3_vs_k141_25054\|\|full | 3286 | No | NA | 6 | 0 | 0 | Not-determined | Genome-fragment | NA | NA | 0 |
| S3_vs_k141_120417\|\|full | 1285 | No | NA | 2 | 0 | 0 | Not-determined | Genome-fragment | NA | NA | 0 |
| S3_vs_k141_9691\|\|full | 4469 | No | NA | 11 | 0 | 0 | Not-determined | Genome-fragment | NA | NA | 0 |
| S3_vs_k141_112528\|\|full | 5487 | No | NA | 6 | 0 | 3 | Not-determined | Genome-fragment | NA | NA | 0 |
| S3_vs_k141_81125\|\|full | 12048 | No | NA | 19 | 0 | 6 | Not-determined | Genome-fragment | NA | NA | 0 |
| S8_vs_k141_148611\|\|full | 1651 | No | NA | 5 | 0 | 0 | Low-quality | Genome-fragment | 4.26 | AAI-based (high-confidence) | 0 |
| S11_vs_k141_1334\|\|full | 1652 | No | NA | 2 | 0 | 1 | Low-quality | Genome-fragment | 1.88 | AAI-based (high-confidence) | 0 |
| S1_vs_k141_157534\|\|full | 1652 | No | NA | 2 | 0 | 0 | Low-quality | Genome-fragment | 1.04 | AAI-based (high-confidence) | 0 |
| S5_vs_k141_53361\|\|full | 1652 | No | NA | 3 | 2 | 0 | Low-quality | Genome-fragment | 3.61 | AAI-based (high-confidence) | 0 |
| S3_vs_k141_112563\|\|full | 2422 | No | NA | 3 | 0 | 2 | Not-determined | Genome-fragment | NA | NA | 0 |
| S3_vs_k141_33390\|\|full | 11860 | No | NA | 7 | 0 | 3 | Not-determined | Genome-fragment | NA | NA | 0 |
| S5_vs_k141_47264\|\|full | 1652 | No | NA | 3 | 0 | 0 | Low-quality | Genome-fragment | 2.95 | AAI-based (high-confidence) | 0 |
| S7_vs_37827\|\|full | 1652 | No | NA | 3 | 1 | 0 | Low-quality | Genome-fragment | 3.66 | AAI-based (high-confidence) | 0 |
| S3_vs_k141_16871\|\|full | 5026 | No | NA | 9 | 0 | 0 | Not-determined | Genome-fragment | NA | NA | 0 |
| S4_extracted_20052 | 1652 | No | NA | 2 | 1 | 0 | Low-quality | Genome-fragment | 1.77 | AAI-based (high-confidence) | 0 |
| S6_vs_k141_7608\|\|full | 1652 | No | NA | 3 | 0 | 0 | Low-quality | Genome-fragment | 2.56 | AAI-based (high-confidence) | 0 |
| S8_extracted_k141_120265 | 1652 | No | NA | 2 | 0 | 0 | Low-quality | Genome-fragment | 2.85 | AAI-based (medium-confidence) | 0 |
| S3_vs_k141_41108\|\|full | 1171 | No | NA | 2 | 0 | 1 | Not-determined | Genome-fragment | NA | NA | 0 |
| S11_vs_k141_13766\|\|full | 1653 | No | NA | 3 | 0 | 0 | Low-quality | Genome-fragment | 4.11 | AAI-based (high-confidence) | 0 |
| S3_vs_k141_73473\|\|full | 2701 | No | NA | 2 | 0 | 1 | Not-determined | Genome-fragment | NA | NA | 0 |
| S11_vs_k141_26567\|\|lt2gene | 1653 | No | NA | 2 | 2 | 0 | Low-quality | Genome-fragment | 4.37 | AAI-based (high-confidence) | 0 |
| S1_vs_k141_144589\|\|full | 1653 | No | NA | 3 | 3 | 0 | Low-quality | Genome-fragment | 3.58 | AAI-based (medium-confidence) | 0 |
| S3_vs_k141_111880\|\|full | 1653 | No | NA | 2 | 0 | 2 | Low-quality | Genome-fragment | 0.73 | AAI-based (medium-confidence) | 0 |
| S3_vs_k141_9757\|\|full | 1291 | No | NA | 2 | 0 | 2 | Not-determined | Genome-fragment | NA | NA | 0 |
| S3_vs_k141_73492\|\|full | 2370 | No | NA | 4 | 0 | 4 | Not-determined | Genome-fragment | NA | NA | 0 |
| S3_vs_k141_109507\|\|full | 1653 | No | NA | 2 | 0 | 1 | Low-quality | Genome-fragment | 1.03 | AAI-based (high-confidence) | 0 |
| S5_vs_k141_8067\|\|full | 1653 | No | NA | 5 | 1 | 0 | Low-quality | Genome-fragment | 3.96 | AAI-based (medium-confidence) | 0 |
| S8_extracted_k141_180480 | 1653 | No | NA | 2 | 0 | 0 | Low-quality | Genome-fragment | 3.82 | AAI-based (high-confidence) | 0 |
| S8_vs_k141_23290\|\|full | 1653 | No | NA | 3 | 0 | 0 | Low-quality | Genome-fragment | 3.86 | AAI-based (high-confidence) | 0 |
| S17_extracted_k141_75148 | 1654 | No | NA | 5 | 4 | 0 | Low-quality | Genome-fragment | 5.46 | AAI-based (high-confidence) | 0 |
| S9_extracted_k141_83456 | 1654 | No | NA | 1 | 0 | 1 | Low-quality | Genome-fragment | 1 | AAI-based (medium-confidence) | 0 |
| S9_vs_k141_9149\|\|full | 1654 | No | NA | 3 | 0 | 2 | Low-quality | Genome-fragment | 0.8 | AAI-based (high-confidence) | 0 |
| S11_extracted_k141_41720 | 1654 | No | NA | 3 | 0 | 0 | Low-quality | Genome-fragment | 3.69 | AAI-based (high-confidence) | 0 |
| S13_vs_70168\|\|full | 1654 | No | NA | 3 | 0 | 3 | Low-quality | Genome-fragment | 2.78 | AAI-based (medium-confidence) | 0 |
| S3_vs_k141_115637\|\|full | 1654 | No | NA | 4 | 0 | 0 | Low-quality | Genome-fragment | 3.46 | AAI-based (high-confidence) | 0 |
| S5_vs_k141_122106\|\|full | 1654 | No | NA | 5 | 4 | 0 | Low-quality | Genome-fragment | 2.74 | AAI-based (medium-confidence) | 0 |
| S16_extracted_27803 | 1654 | No | NA | 1 | 0 | 0 | Low-quality | Genome-fragment | 0.72 | AAI-based (medium-confidence) | 0 |
| S3_vs_k141_33554\|\|full | 1255 | No | NA | 2 | 0 | 1 | Not-determined | Genome-fragment | NA | NA | 0 |
| S3_vs_k141_73594\|\|full | 3397 | No | NA | 5 | 0 | 0 | Not-determined | Genome-fragment | NA | NA | 0 |
| S16_vs_34072\|\|full | 1654 | No | NA | 2 | 1 | 0 | Low-quality | Genome-fragment | 3.54 | HMM-based (lower-bound) | 0 |
| S10_extracted_56123 | 1654 | No | NA | 2 | 0 | 1 | Low-quality | Genome-fragment | 1.43 | AAI-based (medium-confidence) | 0 |
| S12_vs_k141_49187\|\|full | 1654 | No | NA | 3 | 0 | 2 | Low-quality | Genome-fragment | 1.05 | AAI-based (medium-confidence) | 0 |
| S3_vs_k141_104874\|\|full | 2057 | No | NA | 2 | 0 | 2 | Not-determined | Genome-fragment | NA | NA | 0 |
| S15_extracted_64759 | 1654 | No | NA | 6 | 0 | 0 | Low-quality | Genome-fragment | 3.67 | AAI-based (high-confidence) | 0 |
| S3_vs_k141_81263\|\|full | 1165 | No | NA | 2 | 0 | 1 | Not-determined | Genome-fragment | NA | NA | 0 |
| S15_vs_65456\|\|full | 1654 | No | NA | 3 | 0 | 0 | Low-quality | Genome-fragment | 3.16 | AAI-based (high-confidence) | 0 |
| S3_vs_k141_57243\|\|full | 7575 | No | NA | 5 | 0 | 4 | Not-determined | Genome-fragment | NA | NA | 0 |
| S15_vs_76804\|\|full | 1654 | Yes | 588 | 3 | 1 | 1 | Low-quality | Genome-fragment | 0.35 | HMM-based (lower-bound) | 64.45 |
| S8_vs_k141_45151\|\|full | 1654 | No | NA | 4 | 1 | 0 | Low-quality | Genome-fragment | 1.49 | HMM-based (lower-bound) | 0 |
| S8_vs_k141_110723\|\|full | 1654 | No | NA | 4 | 1 | 0 | Low-quality | Genome-fragment | 2.89 | AAI-based (medium-confidence) | 0 |
| S13_vs_50817\|\|full | 1655 | No | NA | 5 | 0 | 5 | Low-quality | Genome-fragment | 0.52 | AAI-based (medium-confidence) | 0 |
| S3_vs_k141_104900\|\|full | 2094 | No | NA | 2 | 0 | 0 | Not-determined | Genome-fragment | NA | NA | 0 |
| S1_extracted_k141_89180 | 1655 | No | NA | 2 | 1 | 0 | Low-quality | Genome-fragment | 7.4 | AAI-based (medium-confidence) | 0 |
| S3_vs_k141_112723\|\|full | 1850 | No | NA | 2 | 0 | 2 | Not-determined | Genome-fragment | NA | NA | 0 |
| S3_vs_k141_49634\|\|full | 1022 | No | NA | 2 | 0 | 0 | Not-determined | Genome-fragment | NA | NA | 0 |
| S1_vs_k141_86479\|\|full | 1655 | No | NA | 3 | 0 | 0 | Low-quality | Genome-fragment | 2.63 | AAI-based (high-confidence) | 0 |
| S5_extracted_k141_102465 | 1655 | No | NA | 5 | 2 | 0 | Low-quality | Genome-fragment | 3.41 | AAI-based (high-confidence) | 0 |
| S3_vs_k141_120634\|\|full | 2430 | No | NA | 3 | 0 | 1 | Not-determined | Genome-fragment | NA | NA | 0 |
| S3_vs_k141_17117\|\|full | 8580 | No | NA | 6 | 0 | 4 | Not-determined | Genome-fragment | NA | NA | 0 |
| S16_extracted_116183 | 1655 | No | NA | 2 | 1 | 0 | Low-quality | Genome-fragment | 0.92 | HMM-based (lower-bound) | 0 |
| S16_vs_40164\|\|full | 1655 | No | NA | 4 | 1 | 0 | Low-quality | Genome-fragment | 3.33 | AAI-based (high-confidence) | 0 |
| S16_vs_102792\|\|full | 1655 | No | NA | 3 | 1 | 0 | Low-quality | Genome-fragment | 0.94 | HMM-based (lower-bound) | 0 |
| S3_vs_k141_81383\|\|full | 2145 | No | NA | 2 | 0 | 0 | Not-determined | Genome-fragment | NA | NA | 0 |
| S10_vs_95731\|\|full | 1655 | Yes | 1019 | 4 | 1 | 1 | Low-quality | Genome-fragment | 2.44 | HMM-based (lower-bound) | 38.43 |
| S12_vs_k141_116020\|\|full | 1655 | No | NA | 5 | 1 | 0 | Low-quality | Genome-fragment | 3.49 | AAI-based (high-confidence) | 0 |
| S6_vs_k141_41726\|\|full | 1655 | No | NA | 4 | 1 | 0 | Low-quality | Genome-fragment | 4.59 | AAI-based (high-confidence) | 0 |
| S3_vs_k141_57348\|\|full | 3197 | No | NA | 3 | 0 | 0 | Not-determined | Genome-fragment | NA | NA | 0 |
| S9_extracted_k141_19176 | 1656 | No | NA | 3 | 0 | 0 | Low-quality | Genome-fragment | 3.71 | AAI-based (high-confidence) | 0 |
| S9_vs_k141_21769\|\|full | 1656 | No | NA | 2 | 0 | 1 | Low-quality | Genome-fragment | 3.62 | AAI-based (high-confidence) | 0 |
| S3_vs_k141_57358\|\|full | 3003 | No | NA | 3 | 0 | 2 | Not-determined | Genome-fragment | NA | NA | 0 |
| S11_extracted_k141_34218 | 1656 | No | NA | 2 | 2 | 0 | Low-quality | Genome-fragment | 4.06 | AAI-based (high-confidence) | 0 |
| S11_vs_k141_10540\|\|full | 1656 | No | NA | 3 | 0 | 1 | Low-quality | Genome-fragment | 2.58 | AAI-based (high-confidence) | 0 |
| S3_vs_k141_112812\|\|full | 3380 | No | NA | 4 | 0 | 2 | Not-determined | Genome-fragment | NA | NA | 0 |
| S11_vs_k141_49140\|\|lt2gene | 1656 | No | NA | 2 | 2 | 0 | Low-quality | Genome-fragment | 4.63 | AAI-based (medium-confidence) | 0 |
| S3_vs_k141_1563\|\|full | 3811 | No | NA | 3 | 0 | 2 | Not-determined | Genome-fragment | NA | NA | 0 |
| S7_extracted_8986 | 1656 | No | NA | 3 | 0 | 0 | Low-quality | Genome-fragment | 2.96 | AAI-based (high-confidence) | 0 |
| S3_vs_k141_57414\|\|full | 1816 | No | NA | 4 | 0 | 0 | Not-determined | Genome-fragment | NA | NA | 0 |
| S7_vs_71433\|\|full | 1656 | No | NA | 2 | 0 | 0 | Low-quality | Genome-fragment | 3.52 | AAI-based (medium-confidence) | 0 |
| S10_vs_119575\|\|full | 1656 | No | NA | 4 | 1 | 0 | Low-quality | Genome-fragment | 3.36 | AAI-based (high-confidence) | 0 |
| S3_vs_k141_17168\|\|full | 103365 | Yes | 31917 | 110 | 16 | 46 | Complete | High-quality | 100 | Provirus (medium-confidence) | 69.12 |
| S8_extracted_k141_194025 | 1656 | No | NA | 4 | 3 | 0 | Low-quality | Genome-fragment | 4.03 | AAI-based (medium-confidence) | 0 |
| S3_vs_k141_112879\|\|full | 2788 | No | NA | 3 | 0 | 2 | Not-determined | Genome-fragment | NA | NA | 0 |
| S8_vs_k141_69179\|\|full | 1656 | No | NA | 3 | 0 | 1 | Low-quality | Genome-fragment | 2.67 | AAI-based (medium-confidence) | 0 |
| S17_vs_k141_135031\|\|full | 1657 | No | NA | 5 | 1 | 0 | Low-quality | Genome-fragment | 3.67 | AAI-based (high-confidence) | 0 |
| S9_vs_k141_63142\|\|full | 1657 | No | NA | 3 | 1 | 0 | Low-quality | Genome-fragment | 4.34 | AAI-based (medium-confidence) | 0 |
| S1_vs_k141_98559\|\|full | 1657 | No | NA | 2 | 0 | 1 | Low-quality | Genome-fragment | 1.98 | AAI-based (high-confidence) | 0 |
| S3_vs_k141_81456\|\|full | 1217 | No | NA | 2 | 0 | 1 | Not-determined | Genome-fragment | NA | NA | 0 |
| S3_vs_k141_49754\|\|full | 1034 | No | NA | 2 | 0 | 2 | Not-determined | Genome-fragment | NA | NA | 0 |
| S3_vs_k141_112921\|\|full | 3701 | No | NA | 5 | 0 | 0 | Not-determined | Genome-fragment | NA | NA | 0 |
| S3_vs_k141_25242\|\|full | 1370 | No | NA | 3 | 0 | 2 | Not-determined | Genome-fragment | NA | NA | 0 |
| S5_extracted_k141_41455 | 1657 | No | NA | 2 | 1 | 0 | Low-quality | Genome-fragment | 0.47 | HMM-based (lower-bound) | 0 |
| S5_vs_k141_86174\|\|full | 1657 | No | NA | 3 | 1 | 0 | Low-quality | Genome-fragment | 3.92 | AAI-based (high-confidence) | 0 |
| S3_vs_k141_41448\|\|full | 3321 | No | NA | 3 | 0 | 1 | Not-determined | Genome-fragment | NA | NA | 0 |
| S3_vs_k141_89104\|\|full | 1444 | No | NA | 3 | 0 | 1 | Not-determined | Genome-fragment | NA | NA | 0 |
| S3_vs_k141_89108\|\|full | 1391 | No | NA | 2 | 0 | 2 | Not-determined | Genome-fragment | NA | NA | 0 |
| S3_vs_k141_17237\|\|full | 2676 | No | NA | 4 | 0 | 0 | Not-determined | Genome-fragment | NA | NA | 0 |
| S3_vs_k141_65167\|\|full | 2388 | No | NA | 2 | 0 | 1 | Not-determined | Genome-fragment | NA | NA | 0 |
| S3_vs_k141_120794\|\|full | 1729 | No | NA | 2 | 0 | 1 | Not-determined | Genome-fragment | NA | NA | 0 |
| S10_extracted_110728 | 1657 | No | NA | 3 | 0 | 1 | Low-quality | Genome-fragment | 0.65 | AAI-based (high-confidence) | 0 |
| S3_vs_k141_25323\|\|full | 2016 | No | NA | 3 | 0 | 1 | Not-determined | Genome-fragment | NA | NA | 0 |
| S3_vs_k141_89158\|\|full | 5695 | No | NA | 5 | 0 | 2 | Not-determined | Genome-fragment | NA | NA | 0 |
| S12_vs_k141_127493\|\|full | 1657 | No | NA | 3 | 2 | 0 | Low-quality | Genome-fragment | 4.38 | AAI-based (high-confidence) | 0 |
| S12_vs_k141_106180\|\|full | 1657 | No | NA | 3 | 0 | 2 | Low-quality | Genome-fragment | 2.36 | AAI-based (medium-confidence) | 0 |
| S2_vs_k141_13501\|\|lt2gene | 1657 | No | NA | 2 | 1 | 0 | Low-quality | Genome-fragment | 4.11 | AAI-based (high-confidence) | 0 |
| S9_vs_k141_12219\|\|full | 1658 | No | NA | 2 | 0 | 2 | Low-quality | Genome-fragment | 3.17 | AAI-based (medium-confidence) | 0 |
| S3_vs_k141_17275\|\|full | 1662 | No | NA | 3 | 0 | 0 | Not-determined | Genome-fragment | NA | NA | 0 |
| S3_vs_k141_10294\|\|full | 6670 | No | NA | 7 | 0 | 7 | Not-determined | Genome-fragment | NA | NA | 0 |
| S3_vs_k141_97435\|\|full | 1226 | No | NA | 3 | 0 | 0 | Not-determined | Genome-fragment | NA | NA | 0 |
| S13_vs_58993\|\|full | 1658 | No | NA | 3 | 0 | 0 | Low-quality | Genome-fragment | 1.16 | AAI-based (medium-confidence) | 0 |
| S3_vs_k141_89256\|\|full | 3910 | No | NA | 6 | 0 | 2 | Not-determined | Genome-fragment | NA | NA | 0 |
| S5_vs_k141_111975\|\|full | 1658 | No | NA | 2 | 0 | 1 | Low-quality | Genome-fragment | 3.09 | AAI-based (high-confidence) | 0 |
| S5_vs_k141_136229\|\|full | 1658 | No | NA | 5 | 1 | 0 | Low-quality | Genome-fragment | 3.36 | AAI-based (high-confidence) | 0 |
| S7_vs_88036\|\|lt2gene | 1658 | No | NA | 1 | 1 | 0 | Low-quality | Genome-fragment | 3.88 | AAI-based (high-confidence) | 0 |
| S16_extracted_67944 | 1658 | No | NA | 1 | 1 | 0 | Low-quality | Genome-fragment | 4.05 | AAI-based (high-confidence) | 0 |
| S16_extracted_75323 | 1658 | No | NA | 1 | 0 | 0 | Low-quality | Genome-fragment | 3.61 | AAI-based (high-confidence) | 0 |
| S10_extracted_22138 | 1658 | No | NA | 4 | 2 | 0 | Low-quality | Genome-fragment | 3.87 | AAI-based (high-confidence) | 0 |
| S12_vs_k141_13121\|\|full | 1658 | No | NA | 2 | 1 | 0 | Low-quality | Genome-fragment | 3.3 | AAI-based (medium-confidence) | 0 |
| S3_vs_k141_25421\|\|full | 2784 | No | NA | 3 | 0 | 1 | Not-determined | Genome-fragment | NA | NA | 0 |
| S3_vs_k141_113172\|\|full | 4067 | No | NA | 8 | 0 | 0 | Not-determined | Genome-fragment | NA | NA | 0 |
| S17_vs_k141_166400\|\|full | 1659 | No | NA | 5 | 0 | 0 | Low-quality | Genome-fragment | 2.81 | AAI-based (high-confidence) | 0 |
| S11_extracted_k141_32296 | 1659 | No | NA | 2 | 0 | 0 | Low-quality | Genome-fragment | 4.43 | AAI-based (medium-confidence) | 0 |
| S11_vs_k141_100396\|\|full | 1659 | No | NA | 4 | 1 | 0 | Low-quality | Genome-fragment | 2.43 | AAI-based (high-confidence) | 0 |
| S13_extracted_49286 | 1659 | No | NA | 2 | 0 | 1 | Low-quality | Genome-fragment | 1.4 | AAI-based (medium-confidence) | 0 |
| S3_vs_k141_96977\|\|full | 1659 | No | NA | 4 | 1 | 0 | Low-quality | Genome-fragment | 2.63 | AAI-based (high-confidence) | 0 |
| S3_vs_k141_81589\|\|full | 3874 | No | NA | 4 | 0 | 2 | Not-determined | Genome-fragment | NA | NA | 0 |
| S5_extracted_k141_64626 | 1659 | No | NA | 2 | 2 | 0 | Low-quality | Genome-fragment | 4.85 | AAI-based (high-confidence) | 0 |
| S5_vs_k141_10944\|\|full | 1659 | No | NA | 2 | 0 | 1 | Low-quality | Genome-fragment | 3.79 | AAI-based (medium-confidence) | 0 |
| S5_vs_k141_64645\|\|lt2gene | 1659 | No | NA | 2 | 2 | 0 | Low-quality | Genome-fragment | 4.85 | AAI-based (high-confidence) | 0 |
| S16_vs_10496\|\|full | 1659 | No | NA | 3 | 2 | 0 | Low-quality | Genome-fragment | 3.22 | AAI-based (high-confidence) | 0 |
| S3_vs_k141_10415\|\|full | 1679 | No | NA | 3 | 0 | 2 | Not-determined | Genome-fragment | NA | NA | 0 |
| S3_vs_k141_49990\|\|full | 2633 | No | NA | 4 | 0 | 0 | Not-determined | Genome-fragment | NA | NA | 0 |
| S12_vs_k141_145110\|\|full | 1659 | No | NA | 2 | 0 | 2 | Low-quality | Genome-fragment | 1.12 | AAI-based (medium-confidence) | 0 |
| S3_vs_k141_1830\|\|full | 4011 | No | NA | 3 | 0 | 1 | Not-determined | Genome-fragment | NA | NA | 0 |
| S8_vs_k141_165501\|\|full | 1659 | No | NA | 5 | 3 | 0 | Low-quality | Genome-fragment | 4.09 | AAI-based (medium-confidence) | 0 |
| S1_extracted_k141_129717 | 1660 | No | NA | 5 | 0 | 0 | Low-quality | Genome-fragment | 3.32 | AAI-based (high-confidence) | 0 |
| S3_vs_k141_50023\|\|full | 6264 | No | NA | 3 | 0 | 0 | Not-determined | Genome-fragment | NA | NA | 0 |
| S3_vs_k141_1853\|\|full | 4585 | No | NA | 5 | 0 | 0 | Not-determined | Genome-fragment | NA | NA | 0 |
| S1_vs_k141_50291\|\|full | 1660 | No | NA | 3 | 0 | 0 | Low-quality | Genome-fragment | 3.72 | AAI-based (high-confidence) | 0 |
| S1_vs_k141_57307\|\|full | 1660 | No | NA | 3 | 0 | 0 | Low-quality | Genome-fragment | 3.34 | AAI-based (high-confidence) | 0 |
| S5_extracted_k141_23336 | 1660 | No | NA | 4 | 0 | 2 | Low-quality | Genome-fragment | 2.1 | AAI-based (medium-confidence) | 0 |
| S3_vs_k141_10514\|\|full | 2786 | No | NA | 2 | 0 | 2 | Not-determined | Genome-fragment | NA | NA | 0 |
| S3_vs_k141_50070\|\|full | 1406 | No | NA | 2 | 0 | 2 | Not-determined | Genome-fragment | NA | NA | 0 |
| S5_vs_k141_43667\|\|full | 1660 | No | NA | 3 | 0 | 0 | Low-quality | Genome-fragment | 0.79 | AAI-based (medium-confidence) | 0 |
| S3_vs_k141_89411\|\|full | 2142 | No | NA | 3 | 0 | 1 | Not-determined | Genome-fragment | NA | NA | 0 |
| S5_vs_k141_83626\|\|full | 1660 | No | NA | 2 | 1 | 0 | Low-quality | Genome-fragment | 3.83 | AAI-based (high-confidence) | 0 |
| S7_vs_72467\|\|lt2gene | 1660 | No | NA | 1 | 1 | 0 | Low-quality | Genome-fragment | 2.44 | AAI-based (medium-confidence) | 0 |
| S7_vs_91935\|\|lt2gene | 1660 | No | NA | 2 | 0 | 0 | Low-quality | Genome-fragment | 3.96 | AAI-based (medium-confidence) | 0 |
| S3_vs_k141_41774\|\|full | 2165 | No | NA | 2 | 0 | 2 | Not-determined | Genome-fragment | NA | NA | 0 |
| S3_vs_k141_25612\|\|full | 21159 | No | NA | 18 | 0 | 18 | Not-determined | Genome-fragment | NA | NA | 0 |
| S3_vs_k141_81710\|\|full | 2931 | No | NA | 4 | 0 | 0 | Not-determined | Genome-fragment | NA | NA | 0 |
| S16_vs_28848\|\|full | 1660 | No | NA | 4 | 1 | 0 | Low-quality | Genome-fragment | 3.04 | HMM-based (lower-bound) | 0 |
| S3_vs_k141_25626\|\|full | 3436 | No | NA | 5 | 0 | 1 | Not-determined | Genome-fragment | NA | NA | 0 |
| S3_vs_k141_50099\|\|full | 3257 | No | NA | 6 | 0 | 4 | Not-determined | Genome-fragment | NA | NA | 0 |
| S16_vs_120363\|\|full | 1660 | No | NA | 3 | 0 | 2 | Low-quality | Genome-fragment | 0.99 | AAI-based (medium-confidence) | 0 |
| S17_extracted_k141_106024 | 1661 | No | NA | 2 | 0 | 0 | Low-quality | Genome-fragment | 4.18 | AAI-based (medium-confidence) | 0 |
| S3_vs_k141_105349\|\|full | 2824 | No | NA | 4 | 0 | 3 | Not-determined | Genome-fragment | NA | NA | 0 |
| S3_vs_k141_105354\|\|full | 1008 | No | NA | 2 | 0 | 0 | Not-determined | Genome-fragment | NA | NA | 0 |
| S3_vs_k141_89539\|\|full | 2938 | No | NA | 3 | 0 | 0 | Not-determined | Genome-fragment | NA | NA | 0 |
| S17_vs_k141_61694\|\|full | 1661 | No | NA | 3 | 0 | 0 | Low-quality | Genome-fragment | 1.99 | AAI-based (high-confidence) | 0 |
| S14_extracted_k141_83727 | 1661 | No | NA | 2 | 0 | 1 | Low-quality | Genome-fragment | 2.81 | AAI-based (high-confidence) | 0 |
| S14_vs_k141_98677\|\|full | 1661 | No | NA | 3 | 0 | 0 | Low-quality | Genome-fragment | 1.98 | AAI-based (high-confidence) | 0 |
| S5_vs_k141_48655\|\|full | 1661 | No | NA | 3 | 0 | 0 | Low-quality | Genome-fragment | 3.96 | AAI-based (medium-confidence) | 0 |
| S3_vs_k141_105410\|\|full | 1760 | No | NA | 2 | 0 | 1 | Not-determined | Genome-fragment | NA | NA | 0 |
| S7_extracted_68282 | 1661 | No | NA | 2 | 0 | 1 | Low-quality | Genome-fragment | 3.17 | AAI-based (medium-confidence) | 0 |
| S3_vs_k141_50120\|\|full | 7710 | No | NA | 12 | 0 | 4 | Not-determined | Genome-fragment | NA | NA | 0 |
| S16_vs_89033\|\|full | 1661 | No | NA | 3 | 0 | 0 | Low-quality | Genome-fragment | 3.91 | AAI-based (high-confidence) | 0 |
| S12_extracted_k141_94646 | 1661 | No | NA | 2 | 1 | 0 | Low-quality | Genome-fragment | 5.93 | HMM-based (lower-bound) | 0 |
| S2_vs_k141_7165\|\|full | 1661 | No | NA | 3 | 2 | 0 | Low-quality | Genome-fragment | 2.88 | AAI-based (high-confidence) | 0 |
| S3_vs_k141_89581\|\|full | 5302 | No | NA | 7 | 0 | 0 | Not-determined | Genome-fragment | NA | NA | 0 |
| S3_vs_k141_50135\|\|full | 5751 | No | NA | 4 | 0 | 3 | Not-determined | Genome-fragment | NA | NA | 0 |
| S3_vs_k141_10800\|\|full | 1313 | No | NA | 2 | 0 | 2 | Not-determined | Genome-fragment | NA | NA | 0 |
| S3_vs_k141_25725\|\|full | 2493 | No | NA | 3 | 0 | 0 | Not-determined | Genome-fragment | NA | NA | 0 |
| S8_extracted_k141_17038 | 1661 | No | NA | 3 | 0 | 0 | Low-quality | Genome-fragment | 3.84 | AAI-based (high-confidence) | 0 |
| S17_vs_k141_167712\|\|full | 1662 | No | NA | 2 | 0 | 0 | Low-quality | Genome-fragment | 2.06 | AAI-based (medium-confidence) | 0 |
| S9_vs_k141_61788\|\|full | 1662 | No | NA | 4 | 0 | 0 | Low-quality | Genome-fragment | 4.07 | AAI-based (high-confidence) | 0 |
| S3_vs_k141_25763\|\|full | 4676 | No | NA | 6 | 0 | 3 | Not-determined | Genome-fragment | NA | NA | 0 |
| S3_vs_k141_57932\|\|full | 2377 | No | NA | 4 | 0 | 1 | Not-determined | Genome-fragment | NA | NA | 0 |
| S3_vs_k141_10914\|\|full | 3417 | No | NA | 5 | 0 | 4 | Not-determined | Genome-fragment | NA | NA | 0 |
| S9_vs_k141_14562\|\|full | 1662 | No | NA | 3 | 0 | 2 | Low-quality | Genome-fragment | 2.96 | AAI-based (medium-confidence) | 0 |
| S3_extracted_k141_64493 | 1662 | No | NA | 4 | 0 | 0 | Low-quality | Genome-fragment | 3.26 | AAI-based (high-confidence) | 0 |
| S3_vs_k141_69844\|\|full | 1662 | No | NA | 3 | 0 | 0 | Low-quality | Genome-fragment | 3.33 | AAI-based (medium-confidence) | 0 |
| S3_vs_k141_34361\|\|full | 1225 | No | NA | 2 | 0 | 1 | Not-determined | Genome-fragment | NA | NA | 0 |
| S5_vs_k141_64011\|\|full | 1662 | No | NA | 3 | 2 | 0 | Low-quality | Genome-fragment | 1.47 | AAI-based (medium-confidence) | 0 |
| S7_vs_19748\|\|full | 1662 | No | NA | 3 | 0 | 0 | Low-quality | Genome-fragment | 3.05 | AAI-based (medium-confidence) | 0 |
| S3_vs_k141_25864\|\|full | 4853 | No | NA | 4 | 0 | 2 | Not-determined | Genome-fragment | NA | NA | 0 |
| S10_vs_88354\|\|full | 1662 | No | NA | 5 | 0 | 0 | Low-quality | Genome-fragment | 3.26 | AAI-based (high-confidence) | 0 |
| S3_vs_k141_97749\|\|full | 3834 | No | NA | 5 | 0 | 1 | Not-determined | Genome-fragment | NA | NA | 0 |
| S3_vs_k141_89706\|\|full | 2804 | No | NA | 2 | 0 | 1 | Not-determined | Genome-fragment | NA | NA | 0 |
| S11_vs_k141_44323\|\|full | 1663 | No | NA | 2 | 0 | 2 | Low-quality | Genome-fragment | 1.28 | AAI-based (medium-confidence) | 0 |
| S14_extracted_k141_15683 | 1663 | No | NA | 1 | 0 | 0 | Low-quality | Genome-fragment | 0.8 | AAI-based (medium-confidence) | 0 |
| S1_extracted_k141_36752 | 1663 | No | NA | 1 | 0 | 0 | Low-quality | Genome-fragment | 0.93 | AAI-based (high-confidence) | 0 |
| S3_vs_k141_17680\|\|full | 1206 | No | NA | 3 | 0 | 1 | Not-determined | Genome-fragment | NA | NA | 0 |
| S3_vs_k141_11093\|\|full | 1902 | No | NA | 3 | 0 | 0 | Not-determined | Genome-fragment | NA | NA | 0 |
| S1_vs_k141_135351\|\|full | 1663 | No | NA | 4 | 0 | 0 | Low-quality | Genome-fragment | 3.37 | AAI-based (high-confidence) | 0 |
| S5_extracted_k141_113433 | 1663 | No | NA | 2 | 0 | 0 | Low-quality | Genome-fragment | 2.82 | AAI-based (medium-confidence) | 0 |
| S3_vs_k141_2139\|\|full | 1118 | No | NA | 3 | 0 | 1 | Not-determined | Genome-fragment | NA | NA | 0 |
| S3_vs_k141_25896\|\|full | 45532 | No | NA | 63 | 6 | 2 | High-quality | High-quality | 100 | AAI-based (high-confidence) | 0 |
| S3_vs_k141_42074\|\|full | 7442 | No | NA | 13 | 0 | 1 | Not-determined | Genome-fragment | NA | NA | 0 |
| S7_vs_11769\|\|full | 1663 | No | NA | 4 | 0 | 0 | Low-quality | Genome-fragment | 1.56 | AAI-based (medium-confidence) | 0 |
| S7_vs_61389\|\|full | 1663 | Yes | 1144 | 3 | 1 | 1 | Low-quality | Genome-fragment | 2.19 | AAI-based (medium-confidence) | 31.21 |
| S12_extracted_k141_114026 | 1663 | No | NA | 3 | 0 | 3 | Low-quality | Genome-fragment | 2.84 | AAI-based (medium-confidence) | 0 |
| S15_vs_26968\|\|full | 1663 | No | NA | 4 | 0 | 1 | Low-quality | Genome-fragment | 2.79 | AAI-based (high-confidence) | 0 |
| S3_vs_k141_97766\|\|full | 5337 | No | NA | 4 | 0 | 2 | Not-determined | Genome-fragment | NA | NA | 0 |
| S6_vs_k141_27915\|\|full | 1663 | No | NA | 3 | 0 | 0 | Low-quality | Genome-fragment | 3.85 | AAI-based (medium-confidence) | 0 |
| S3_vs_k141_58154\|\|full | 5681 | No | NA | 6 | 0 | 4 | Not-determined | Genome-fragment | NA | NA | 0 |
| S8_vs_k141_165660\|\|full | 1663 | No | NA | 3 | 1 | 0 | Low-quality | Genome-fragment | 1.42 | AAI-based (high-confidence) | 0 |
| S8_vs_k141_7711\|\|full | 1663 | No | NA | 4 | 1 | 2 | Low-quality | Genome-fragment | 1.69 | AAI-based (medium-confidence) | 0 |
| S9_vs_k141_39181\|\|full | 1664 | No | NA | 3 | 0 | 3 | Low-quality | Genome-fragment | 1.95 | AAI-based (medium-confidence) | 0 |
| S3_vs_k141_105722\|\|full | 2730 | No | NA | 3 | 0 | 2 | Not-determined | Genome-fragment | NA | NA | 0 |
| S13_extracted_125787 | 1664 | No | NA | 3 | 0 | 0 | Low-quality | Genome-fragment | 2.5 | AAI-based (high-confidence) | 0 |
| S13_vs_46023\|\|full | 1664 | No | NA | 3 | 0 | 0 | Low-quality | Genome-fragment | 1.95 | AAI-based (high-confidence) | 0 |
| S5_vs_k141_5305\|\|full | 1664 | No | NA | 3 | 0 | 0 | Low-quality | Genome-fragment | 2.64 | AAI-based (high-confidence) | 0 |
| S10_extracted_87467 | 1664 | No | NA | 2 | 0 | 0 | Low-quality | Genome-fragment | 2.05 | AAI-based (high-confidence) | 0 |
| S12_extracted_k141_57920 | 1664 | No | NA | 2 | 1 | 0 | Low-quality | Genome-fragment | 2.34 | HMM-based (lower-bound) | 0 |
| S3_vs_k141_105768\|\|full | 1350 | No | NA | 2 | 0 | 0 | Not-determined | Genome-fragment | NA | NA | 0 |
| S3_vs_k141_74422\|\|full | 2473 | No | NA | 2 | 0 | 1 | Not-determined | Genome-fragment | NA | NA | 0 |
| S3_vs_k141_113764\|\|full | 4921 | No | NA | 7 | 0 | 1 | Not-determined | Genome-fragment | NA | NA | 0 |
| S3_vs_k141_74424\|\|full | 2225 | No | NA | 4 | 0 | 0 | Not-determined | Genome-fragment | NA | NA | 0 |
| S3_vs_k141_81881\|\|full | 2870 | No | NA | 5 | 0 | 1 | Not-determined | Genome-fragment | NA | NA | 0 |
| S3_vs_k141_2168\|\|full | 1037 | No | NA | 3 | 0 | 0 | Not-determined | Genome-fragment | NA | NA | 0 |
| S3_vs_k141_74428\|\|full | 10082 | No | NA | 10 | 0 | 0 | Not-determined | Genome-fragment | NA | NA | 0 |
| S3_vs_k141_113771\|\|full | 3188 | No | NA | 5 | 0 | 0 | Not-determined | Genome-fragment | NA | NA | 0 |
| S3_vs_k141_65886\|\|full | 2634 | No | NA | 2 | 0 | 1 | Not-determined | Genome-fragment | NA | NA | 0 |
| S3_vs_k141_74439\|\|full | 1710 | No | NA | 3 | 0 | 1 | Not-determined | Genome-fragment | NA | NA | 0 |
| S13_vs_98299\|\|full | 1665 | No | NA | 4 | 0 | 0 | Low-quality | Genome-fragment | 1.5 | AAI-based (high-confidence) | 0 |
| S3_vs_k141_97827\|\|full | 38749 | Yes | 25226 | 51 | 10 | 7 | Medium-quality | Genome-fragment | 63.89 | AAI-based (medium-confidence) | 34.9 |
| S14_vs_k141_105079\|\|full | 1665 | No | NA | 3 | 0 | 0 | Low-quality | Genome-fragment | 3.29 | AAI-based (medium-confidence) | 0 |
| S3_vs_k141_97831\|\|full | 2707 | No | NA | 4 | 0 | 0 | Not-determined | Genome-fragment | NA | NA | 0 |
| S3_vs_k141_50459\|\|full | 2417 | No | NA | 2 | 0 | 2 | Not-determined | Genome-fragment | NA | NA | 0 |
| S7_extracted_43168 | 1665 | No | NA | 2 | 0 | 0 | Low-quality | Genome-fragment | 2.76 | AAI-based (medium-confidence) | 0 |
| S16_vs_159828\|\|lt2gene | 1665 | No | NA | 1 | 1 | 0 | Low-quality | Genome-fragment | 4.74 | AAI-based (medium-confidence) | 0 |
| S15_vs_12437\|\|full | 1665 | No | NA | 2 | 0 | 1 | Low-quality | Genome-fragment | 1.19 | AAI-based (high-confidence) | 0 |
| S3_vs_k141_105863\|\|full | 1004 | No | NA | 3 | 0 | 0 | Not-determined | Genome-fragment | NA | NA | 0 |
| S15_vs_45631\|\|full | 1665 | No | NA | 5 | 0 | 0 | Low-quality | Genome-fragment | 2.71 | AAI-based (high-confidence) | 0 |
| S3_vs_k141_65941\|\|full | 1475 | No | NA | 2 | 0 | 1 | Not-determined | Genome-fragment | NA | NA | 0 |
| S6_vs_k141_38859\|\|full | 1665 | No | NA | 4 | 3 | 0 | Low-quality | Genome-fragment | 3.52 | AAI-based (high-confidence) | 0 |
| S3_vs_k141_74507\|\|full | 1024 | No | NA | 2 | 0 | 1 | Not-determined | Genome-fragment | NA | NA | 0 |
| S3_vs_k141_113837\|\|full | 2076 | No | NA | 2 | 0 | 2 | Not-determined | Genome-fragment | NA | NA | 0 |
| S8_vs_k141_183466\|\|full | 1665 | No | NA | 4 | 0 | 0 | Low-quality | Genome-fragment | 6.62 | AAI-based (high-confidence) | 0 |
| S8_vs_k141_164834\|\|full | 1665 | No | NA | 3 | 0 | 2 | Low-quality | Genome-fragment | 1.07 | AAI-based (medium-confidence) | 0 |
| S3_vs_k141_81928\|\|full | 2258 | No | NA | 3 | 0 | 1 | Not-determined | Genome-fragment | NA | NA | 0 |
| S3_vs_k141_121650\|\|full | 1701 | No | NA | 3 | 0 | 1 | Not-determined | Genome-fragment | NA | NA | 0 |
| S5_extracted_k141_44801 | 1666 | No | NA | 2 | 1 | 0 | Low-quality | Genome-fragment | 7.59 | AAI-based (high-confidence) | 0 |
| S7_vs_68516\|\|full | 1666 | No | NA | 3 | 0 | 0 | Low-quality | Genome-fragment | 3.88 | AAI-based (high-confidence) | 0 |
| S3_vs_k141_89898\|\|full | 47278 | No | NA | 56 | 7 | 1 | High-quality | High-quality | 100 | AAI-based (high-confidence) | 0 |
| S10_vs_101522\|\|full | 1666 | No | NA | 3 | 0 | 0 | Low-quality | Genome-fragment | 1.87 | AAI-based (medium-confidence) | 0 |
| S12_vs_k141_127298\|\|full | 1666 | No | NA | 2 | 0 | 0 | Low-quality | Genome-fragment | 3.74 | AAI-based (high-confidence) | 0 |
| S3_vs_k141_97913\|\|full | 10067 | No | NA | 6 | 0 | 6 | Not-determined | Genome-fragment | NA | NA | 0 |
| S3_vs_k141_50522\|\|full | 4094 | No | NA | 3 | 0 | 1 | Not-determined | Genome-fragment | NA | NA | 0 |
| S3_vs_k141_42283\|\|full | 2864 | No | NA | 3 | 0 | 1 | Not-determined | Genome-fragment | NA | NA | 0 |
| S12_vs_k141_128161\|\|lt2gene | 1666 | No | NA | 1 | 1 | 0 | Low-quality | Genome-fragment | 3.51 | AAI-based (medium-confidence) | 0 |
| S8_vs_k141_26498\|\|full | 1666 | No | NA | 4 | 1 | 0 | Low-quality | Genome-fragment | 4.03 | AAI-based (high-confidence) | 0 |
| S3_vs_k141_89977\|\|full | 1662 | No | NA | 3 | 0 | 0 | Not-determined | Genome-fragment | NA | NA | 0 |
| S17_extracted_k141_67876 | 1667 | No | NA | 3 | 3 | 0 | Low-quality | Genome-fragment | 7.87 | AAI-based (high-confidence) | 0 |
| S14_extracted_k141_53863 | 1667 | No | NA | 6 | 0 | 0 | Low-quality | Genome-fragment | 3.39 | AAI-based (medium-confidence) | 0 |
| S3_vs_k141_96742\|\|full | 1667 | No | NA | 1 | 0 | 1 | Low-quality | Genome-fragment | 3.55 | AAI-based (medium-confidence) | 0 |
| S3_vs_k141_98550\|\|full | 1667 | No | NA | 3 | 2 | 0 | Low-quality | Genome-fragment | 2.58 | AAI-based (high-confidence) | 0 |
| S3_vs_k141_105984\|\|full | 1357 | No | NA | 2 | 0 | 0 | Not-determined | Genome-fragment | NA | NA | 0 |
| S5_vs_k141_79094\|\|full | 1667 | No | NA | 2 | 0 | 1 | Low-quality | Genome-fragment | 0.98 | AAI-based (medium-confidence) | 0 |
| S3_vs_k141_50557\|\|full | 42085 | No | NA | 65 | 22 | 0 | High-quality | High-quality | 99.78 | AAI-based (high-confidence) | 0 |
| S3_vs_k141_42355\|\|full | 3412 | No | NA | 5 | 0 | 1 | Not-determined | Genome-fragment | NA | NA | 0 |
| S3_vs_k141_74657\|\|full | 8034 | No | NA | 11 | 0 | 2 | Not-determined | Genome-fragment | NA | NA | 0 |
| S3_vs_k141_50562\|\|full | 1682 | No | NA | 3 | 0 | 1 | Not-determined | Genome-fragment | NA | NA | 0 |
| S3_vs_k141_26232\|\|full | 4072 | No | NA | 6 | 0 | 1 | Not-determined | Genome-fragment | NA | NA | 0 |
| S5_vs_k141_114965\|\|full | 1667 | No | NA | 6 | 0 | 0 | Low-quality | Genome-fragment | 4.24 | AAI-based (high-confidence) | 0 |
| S3_vs_k141_114021\|\|full | 1283 | No | NA | 3 | 0 | 0 | Not-determined | Genome-fragment | NA | NA | 0 |
| S7_extracted_58665 | 1667 | No | NA | 6 | 6 | 0 | Low-quality | Genome-fragment | 5.5 | AAI-based (high-confidence) | 0 |
| S8_extracted_k141_103670 | 1667 | No | NA | 4 | 4 | 0 | Low-quality | Genome-fragment | 5.46 | AAI-based (high-confidence) | 0 |
| S3_vs_k141_50630\|\|full | 4907 | No | NA | 7 | 0 | 0 | Not-determined | Genome-fragment | NA | NA | 0 |
| S3_vs_k141_74684\|\|full | 1615 | No | NA | 2 | 0 | 2 | Not-determined | Genome-fragment | NA | NA | 0 |
| S9_extracted_k141_43167 | 1668 | No | NA | 3 | 3 | 0 | Low-quality | Genome-fragment | 2.78 | AAI-based (medium-confidence) | 0 |
| S3_vs_k141_17876\|\|full | 2767 | No | NA | 4 | 0 | 2 | Not-determined | Genome-fragment | NA | NA | 0 |
| S3_vs_k141_50650\|\|full | 3392 | No | NA | 3 | 0 | 3 | Not-determined | Genome-fragment | NA | NA | 0 |
| S9_vs_k141_62686\|\|full | 1668 | No | NA | 3 | 0 | 0 | Low-quality | Genome-fragment | 3.18 | AAI-based (high-confidence) | 0 |
| S3_vs_k141_50660\|\|full | 2292 | No | NA | 3 | 0 | 2 | Not-determined | Genome-fragment | NA | NA | 0 |
| S9_vs_k141_36357\|\|lt2gene | 1668 | No | NA | 2 | 2 | 0 | Low-quality | Genome-fragment | 4.32 | AAI-based (medium-confidence) | 0 |
| S14_extracted_k141_20955 | 1668 | No | NA | 2 | 0 | 1 | Low-quality | Genome-fragment | 3.62 | AAI-based (medium-confidence) | 0 |
| S3_vs_k141_82173\|\|full | 2150 | No | NA | 5 | 0 | 0 | Not-determined | Genome-fragment | NA | NA | 0 |
| S3_vs_k141_82190\|\|full | 2194 | No | NA | 6 | 0 | 2 | Not-determined | Genome-fragment | NA | NA | 0 |
| S3_vs_k141_94833\|\|full | 1668 | No | NA | 4 | 0 | 0 | Low-quality | Genome-fragment | 0.99 | AAI-based (high-confidence) | 0 |
| S16_extracted_100444 | 1668 | No | NA | 2 | 0 | 1 | Low-quality | Genome-fragment | 0.16 | AAI-based (medium-confidence) | 0 |
| S3_vs_k141_98090\|\|full | 1341 | No | NA | 3 | 0 | 1 | Not-determined | Genome-fragment | NA | NA | 0 |

| | | | | | | | | | | | |
|---|---|---|---|---|---|---|---|---|---|---|---|
| S12_extracted_k141_126490 | 1668 | No | NA | 2 | 0 | 0 | Low-quality | Genome-fragment | 3.28 | AAI-based (high-confidence) | 0 |
| S2_vs_k141_20681\|\|full | 1668 | No | NA | 1 | 0 | 0 | Low-quality | Genome-fragment | 3.77 | AAI-based (high-confidence) | 0 |
| S3_vs_k141_98102\|\|full | 1414 | No | NA | 3 | 0 | 0 | Not-determined | Genome-fragment | NA | NA | 0 |
| S6_vs_k141_21784\|\|full | 1668 | Yes | 968 | 4 | 1 | 1 | Low-quality | Genome-fragment | 0.42 | HMM-based (lower-bound) | 41.97 |
| S3_vs_k141_114081\|\|full | 1242 | No | NA | 2 | 0 | 0 | Not-determined | Genome-fragment | NA | NA | 0 |
| S8_vs_k141_38509\|\|full | 1668 | No | NA | 3 | 0 | 0 | Low-quality | Genome-fragment | 4.67 | AAI-based (medium-confidence) | 0 |
| S3_vs_k141_42535\|\|full | 1811 | No | NA | 3 | 0 | 0 | Not-determined | Genome-fragment | NA | NA | 0 |
| S3_vs_k141_58519\|\|full | 1229 | No | NA | 2 | 0 | 0 | Not-determined | Genome-fragment | NA | NA | 0 |
| S3_extracted_k141_87932 | 1669 | No | NA | 1 | 0 | 1 | Low-quality | Genome-fragment | 0.45 | AAI-based (high-confidence) | 0 |
| S7_extracted_114845 | 1669 | No | NA | 3 | 2 | 0 | Low-quality | Genome-fragment | 4.43 | AAI-based (high-confidence) | 0 |
| S7_vs_89818\|\|full | 1669 | No | NA | 4 | 0 | 0 | Low-quality | Genome-fragment | 3.43 | AAI-based (medium-confidence) | 0 |
| S16_extracted_31314 | 1669 | No | NA | 1 | 0 | 1 | Low-quality | Genome-fragment | 2.62 | AAI-based (medium-confidence) | 0 |
| S16_extracted_52638 | 1669 | No | NA | 6 | 1 | 0 | Low-quality | Genome-fragment | 2.78 | AAI-based (high-confidence) | 0 |
| S16_vs_38393\|\|full | 1669 | No | NA | 3 | 0 | 0 | Low-quality | Genome-fragment | 3.49 | AAI-based (medium-confidence) | 0 |
| S3_vs_k141_58610\|\|full | 1350 | No | NA | 2 | 0 | 2 | Not-determined | Genome-fragment | NA | NA | 0 |
| S3_vs_k141_98185\|\|full | 1460 | No | NA | 2 | 0 | 0 | Not-determined | Genome-fragment | NA | NA | 0 |
| S12_extracted_k141_34932 | 1669 | No | NA | 2 | 1 | 0 | Low-quality | Genome-fragment | 3.6 | AAI-based (high-confidence) | 0 |
| S12_vs_k141_159792\|\|full | 1669 | No | NA | 2 | 0 | 0 | Low-quality | Genome-fragment | 3.31 | AAI-based (high-confidence) | 0 |
| S3_vs_k141_58616\|\|full | 6014 | No | NA | 7 | 0 | 3 | Not-determined | Genome-fragment | NA | NA | 0 |
| S3_vs_k141_26427\|\|full | 4851 | No | NA | 5 | 0 | 2 | Not-determined | Genome-fragment | NA | NA | 0 |
| S12_vs_k141_50636\|\|full | 1669 | No | NA | 3 | 0 | 0 | Low-quality | Genome-fragment | 3.64 | AAI-based (high-confidence) | 0 |
| S3_vs_k141_58644\|\|full | 1734 | No | NA | 3 | 0 | 0 | Not-determined | Genome-fragment | NA | NA | 0 |
| S3_vs_k141_98202\|\|full | 13455 | No | NA | 11 | 0 | 2 | Not-determined | Genome-fragment | NA | NA | 0 |
| S12_vs_k141_46261\|\|full | 1669 | No | NA | 3 | 0 | 0 | Low-quality | Genome-fragment | 3.9 | AAI-based (high-confidence) | 0 |
| S2_extracted_k141_8859 | 1669 | No | NA | 2 | 1 | 0 | Low-quality | Genome-fragment | 5.39 | HMM-based (lower-bound) | 0 |
| S3_vs_k141_82324\|\|full | 5132 | No | NA | 7 | 0 | 1 | Not-determined | Genome-fragment | NA | NA | 0 |
| S3_vs_k141_106180\|\|full | 2129 | No | NA | 4 | 0 | 0 | Not-determined | Genome-fragment | NA | NA | 0 |
| S4_extracted_18099 | 1669 | No | NA | 3 | 0 | 0 | Low-quality | Genome-fragment | 4.18 | AAI-based (high-confidence) | 0 |
| S3_vs_k141_2565\|\|full | 1966 | No | NA | 3 | 0 | 1 | Not-determined | Genome-fragment | NA | NA | 0 |
| S3_vs_k141_74845\|\|full | 1699 | No | NA | 3 | 0 | 1 | Not-determined | Genome-fragment | NA | NA | 0 |
| S3_vs_k141_121821\|\|full | 1226 | No | NA | 2 | 0 | 0 | Not-determined | Genome-fragment | NA | NA | 0 |
| S3_vs_k141_42569\|\|full | 67795 | Yes | 55470 | 98 | 21 | 11 | High-quality | High-quality | 100 | AAI-based (high-confidence) | 18.18 |
| S3_vs_k141_42576\|\|full | 5532 | No | NA | 8 | 0 | 2 | Not-determined | Genome-fragment | NA | NA | 0 |
| S3_vs_k141_26472\|\|full | 4653 | No | NA | 5 | 0 | 3 | Not-determined | Genome-fragment | NA | NA | 0 |
| S8_vs_k141_75874\|\|full | 1669 | No | NA | 3 | 0 | 1 | Low-quality | Genome-fragment | 2.07 | AAI-based (medium-confidence) | 0 |
| S8_vs_k141_94294\|\|full | 1669 | No | NA | 3 | 0 | 3 | Low-quality | Genome-fragment | 1.9 | AAI-based (medium-confidence) | 0 |
| S14_vs_k141_14053\|\|full | 1670 | No | NA | 4 | 1 | 0 | Low-quality | Genome-fragment | 2.56 | AAI-based (medium-confidence) | 0 |
| S3_vs_k141_74869\|\|full | 1200 | No | NA | 3 | 0 | 1 | Not-determined | Genome-fragment | NA | NA | 0 |
| S3_vs_k141_98290\|\|full | 2861 | No | NA | 3 | 0 | 0 | Not-determined | Genome-fragment | NA | NA | 0 |
| S1_extracted_k141_159309 | 1670 | No | NA | 3 | 1 | 0 | Low-quality | Genome-fragment | 0.11 | HMM-based (lower-bound) | 0 |
| S3_vs_k141_82363\|\|full | 1220 | No | NA | 3 | 0 | 2 | Not-determined | Genome-fragment | NA | NA | 0 |
| S3_vs_k141_66062\|\|full | 1808 | No | NA | 4 | 0 | 1 | Not-determined | Genome-fragment | NA | NA | 0 |
| S3_vs_k141_114204\|\|full | 1597 | No | NA | 5 | 0 | 0 | Not-determined | Genome-fragment | NA | NA | 0 |
| S3_vs_k141_58796\|\|full | 1687 | No | NA | 2 | 0 | 1 | Not-determined | Genome-fragment | NA | NA | 0 |
| S1_vs_k141_84846\|\|full | 1670 | No | NA | 4 | 1 | 0 | Low-quality | Genome-fragment | 2.78 | AAI-based (high-confidence) | 0 |
| S3_vs_k141_72750\|\|full | 1670 | No | NA | 8 | 0 | 0 | Low-quality | Genome-fragment | 1.07 | AAI-based (high-confidence) | 0 |
| S3_vs_k141_42668\|\|full | 3592 | No | NA | 4 | 0 | 2 | Not-determined | Genome-fragment | NA | NA | 0 |
| S16_vs_40057\|\|full | 1670 | No | NA | 3 | 3 | 0 | Low-quality | Genome-fragment | 3.86 | AAI-based (high-confidence) | 0 |
| S16_vs_40944\|\|full | 1670 | No | NA | 2 | 2 | 0 | Low-quality | Genome-fragment | 2.97 | AAI-based (high-confidence) | 0 |
| S16_vs_46149\|\|full | 1670 | No | NA | 3 | 0 | 0 | Low-quality | Genome-fragment | 3.62 | AAI-based (high-confidence) | 0 |
| S14_vs_k141_41248\|\|full | 1671 | No | NA | 4 | 0 | 3 | Low-quality | Genome-fragment | 0.49 | AAI-based (medium-confidence) | 0 |
| S3_vs_k141_34799\|\|full | 2386 | No | NA | 4 | 0 | 1 | Not-determined | Genome-fragment | NA | NA | 0 |
| S1_vs_k141_156592\|\|lt2gene | 1671 | No | NA | 2 | 1 | 0 | Low-quality | Genome-fragment | 4.25 | AAI-based (high-confidence) | 0 |
| S3_vs_k141_98329\|\|full | 1043 | No | NA | 3 | 0 | 0 | Not-determined | Genome-fragment | NA | NA | 0 |
| S3_vs_k141_68445\|\|full | 1671 | No | NA | 3 | 1 | 0 | Low-quality | Genome-fragment | 0.9 | HMM-based (lower-bound) | 0 |
| S5_vs_k141_83106\|\|full | 1671 | No | NA | 5 | 0 | 0 | Low-quality | Genome-fragment | 1.59 | AAI-based (medium-confidence) | 0 |
| S7_vs_104825\|\|full | 1671 | No | NA | 5 | 0 | 0 | Low-quality | Genome-fragment | 3.29 | AAI-based (medium-confidence) | 0 |
| S16_vs_22643\|\|full | 1671 | No | NA | 4 | 1 | 0 | Low-quality | Genome-fragment | 4.02 | AAI-based (high-confidence) | 0 |
| S3_vs_k141_42767\|\|full | 3197 | No | NA | 3 | 0 | 3 | Not-determined | Genome-fragment | NA | NA | 0 |
| S3_vs_k141_106343\|\|full | 5244 | No | NA | 7 | 0 | 4 | Not-determined | Genome-fragment | NA | NA | 0 |
| S12_vs_k141_157689\|\|full | 1671 | No | NA | 3 | 0 | 0 | Low-quality | Genome-fragment | 2.87 | AAI-based (medium-confidence) | 0 |
| S6_vs_k141_9905\|\|full | 1671 | No | NA | 2 | 0 | 2 | Low-quality | Genome-fragment | 2.8 | AAI-based (medium-confidence) | 0 |
| S14_extracted_k141_29425 | 1672 | No | NA | 3 | 3 | 0 | Low-quality | Genome-fragment | 5.51 | AAI-based (high-confidence) | 0 |
| S3_vs_k141_114271\|\|full | 28459 | No | NA | 46 | 16 | 0 | Medium-quality | Genome-fragment | 88.52 | AAI-based (high-confidence) | 0 |
| NC3_vf_k141_1019 | 1672 | No | NA | 3 | 0 | 0 | Low-quality | Genome-fragment | 2.39 | AAI-based (high-confidence) | 0 |
| S16_extracted_104366 | 1672 | No | NA | 2 | 1 | 0 | Low-quality | Genome-fragment | 4.28 | AAI-based (medium-confidence) | 0 |
| S16_vs_99539\|\|lt2gene | 1672 | No | NA | 1 | 1 | 0 | Low-quality | Genome-fragment | 4.79 | AAI-based (high-confidence) | 0 |
| S10_extracted_44560 | 1672 | No | NA | 3 | 2 | 0 | Low-quality | Genome-fragment | 5.57 | AAI-based (high-confidence) | 0 |
| S3_vs_k141_50937\|\|full | 2784 | No | NA | 4 | 0 | 3 | Not-determined | Genome-fragment | NA | NA | 0 |
| S10_vs_113156\|\|lt2gene | 1672 | No | NA | 1 | 0 | 0 | Low-quality | Genome-fragment | 4.26 | AAI-based (high-confidence) | 0 |
| S15_extracted_69936 | 1672 | No | NA | 2 | 0 | 0 | Low-quality | Genome-fragment | 0.76 | AAI-based (high-confidence) | 0 |
| S17_vs_k141_72046\|\|full | 1673 | No | NA | 3 | 1 | 0 | Low-quality | Genome-fragment | 1.78 | AAI-based (high-confidence) | 0 |
| S14_vs_k141_34675\|\|full | 1673 | No | NA | 2 | 0 | 1 | Low-quality | Genome-fragment | 2.29 | AAI-based (high-confidence) | 0 |
| S3_extracted_k141_62823 | 1673 | No | NA | 6 | 2 | 0 | Low-quality | Genome-fragment | 4.07 | AAI-based (high-confidence) | 0 |
| S3_vs_k141_96479\|\|full | 1673 | No | NA | 5 | 0 | 1 | Low-quality | Genome-fragment | 2.87 | AAI-based (high-confidence) | 0 |
| S7_vs_3044\|\|full | 1673 | No | NA | 4 | 0 | 0 | Low-quality | Genome-fragment | 3.86 | AAI-based (medium-confidence) | 0 |
| S7_vs_19542\|\|full | 1673 | No | NA | 3 | 2 | 0 | Low-quality | Genome-fragment | 2.12 | HMM-based (lower-bound) | 0 |
| S7_vs_54954\|\|full | 1673 | No | NA | 3 | 0 | 1 | Low-quality | Genome-fragment | 1.23 | AAI-based (medium-confidence) | 0 |
| S3_vs_k141_11996\|\|full | 2802 | No | NA | 3 | 0 | 3 | Not-determined | Genome-fragment | NA | NA | 0 |
| S16_vs_14314\|\|full | 1673 | No | NA | 4 | 1 | 0 | Low-quality | Genome-fragment | 3.68 | AAI-based (high-confidence) | 0 |
| S8_extracted_k141_95131 | 1673 | No | NA | 1 | 0 | 0 | Low-quality | Genome-fragment | 1.76 | AAI-based (high-confidence) | 0 |
| S8_vs_k141_87021\|\|full | 1673 | No | NA | 3 | 0 | 0 | Low-quality | Genome-fragment | 3.22 | AAI-based (medium-confidence) | 0 |
| S8_vs_k141_140003\|\|full | 1673 | No | NA | 3 | 0 | 0 | Low-quality | Genome-fragment | 5.26 | AAI-based (high-confidence) | 0 |
| S8_vs_k141_180023\|\|full | 1673 | No | NA | 4 | 1 | 0 | Low-quality | Genome-fragment | 1.15 | HMM-based (lower-bound) | 0 |
| S14_vs_k141_86882\|\|full | 1674 | No | NA | 3 | 0 | 0 | Low-quality | Genome-fragment | 3.21 | AAI-based (high-confidence) | 0 |
| S3_vs_k141_50999\|\|full | 3196 | No | NA | 3 | 0 | 2 | Not-determined | Genome-fragment | NA | NA | 0 |
| S7_vs_5105\|\|full | 1674 | No | NA | 3 | 1 | 0 | Low-quality | Genome-fragment | 1.2 | HMM-based (lower-bound) | 0 |
| S7_vs_25811\|\|full | 1674 | No | NA | 3 | 0 | 0 | Low-quality | Genome-fragment | 3.49 | AAI-based (high-confidence) | 0 |
| S3_vs_k141_51005\|\|full | 2984 | No | NA | 7 | 3 | 0 | Medium-quality | Genome-fragment | 62.43 | AAI-based (high-confidence) | 0 |
| S7_vs_38833\|\|full | 1674 | No | NA | 3 | 1 | 0 | Low-quality | Genome-fragment | 2.51 | AAI-based (medium-confidence) | 0 |
| S3_vs_k141_98442\|\|full | 4258 | No | NA | 4 | 0 | 1 | Not-determined | Genome-fragment | NA | NA | 0 |
| S3_vs_k141_82550\|\|full | 4112 | No | NA | 4 | 0 | 3 | Not-determined | Genome-fragment | NA | NA | 0 |
| S7_vs_90003\|\|lt2gene | 1674 | No | NA | 2 | 1 | 0 | Low-quality | Genome-fragment | 4.38 | AAI-based (high-confidence) | 0 |
| S16_extracted_82420 | 1674 | No | NA | 4 | 0 | 0 | Low-quality | Genome-fragment | 2.79 | AAI-based (high-confidence) | 0 |
| S3_vs_k141_114351\|\|full | 2301 | No | NA | 3 | 0 | 0 | Not-determined | Genome-fragment | NA | NA | 0 |
| S3_vs_k141_75170\|\|full | 1788 | No | NA | 3 | 0 | 1 | Not-determined | Genome-fragment | NA | NA | 0 |
| S12_vs_k141_63747\|\|full | 1674 | No | NA | 4 | 0 | 1 | Low-quality | Genome-fragment | 3.44 | AAI-based (medium-confidence) | 0 |
| S3_vs_k141_12046\|\|full | 2279 | No | NA | 3 | 0 | 0 | Not-determined | Genome-fragment | NA | NA | 0 |
| S12_vs_k141_4653\|\|full | 1674 | No | NA | 4 | 2 | 0 | Low-quality | Genome-fragment | 4.02 | AAI-based (medium-confidence) | 0 |
| S3_vs_k141_42929\|\|full | 2594 | No | NA | 3 | 0 | 0 | Not-determined | Genome-fragment | NA | NA | 0 |
| S2_vs_k141_4229\|\|full | 1674 | No | NA | 4 | 0 | 0 | Low-quality | Genome-fragment | 3.91 | AAI-based (high-confidence) | 0 |
| S4_extracted_3757 | 1674 | No | NA | 2 | 0 | 1 | Low-quality | Genome-fragment | 1.81 | AAI-based (medium-confidence) | 0 |
| S3_vs_k141_75289\|\|full | 1425 | No | NA | 3 | 0 | 0 | Not-determined | Genome-fragment | NA | NA | 0 |
| S3_vs_k141_26836\|\|full | 1536 | No | NA | 2 | 0 | 2 | Not-determined | Genome-fragment | NA | NA | 0 |
| S8_extracted_k141_193142 | 1674 | No | NA | 5 | 3 | 0 | Low-quality | Genome-fragment | 5.32 | AAI-based (high-confidence) | 0 |
| S3_vs_k141_51163\|\|full | 8787 | No | NA | 8 | 0 | 6 | Not-determined | Genome-fragment | NA | NA | 0 |
| S9_vs_k141_34056\|\|full | 1675 | No | NA | 3 | 2 | 0 | Low-quality | Genome-fragment | 4.15 | AAI-based (high-confidence) | 0 |
| S10_extracted_13439 | 1675 | No | NA | 6 | 0 | 1 | Low-quality | Genome-fragment | 2.95 | AAI-based (high-confidence) | 0 |
| S12_extracted_k141_100464 | 1675 | No | NA | 2 | 2 | 0 | Low-quality | Genome-fragment | 4.13 | AAI-based (high-confidence) | 0 |
| S3_vs_k141_51243\|\|full | 2208 | No | NA | 3 | 0 | 0 | Not-determined | Genome-fragment | NA | NA | 0 |
| S3_vs_k141_82762\|\|full | 8501 | No | NA | 9 | 0 | 0 | Not-determined | Genome-fragment | NA | NA | 0 |
| S3_vs_k141_35142\|\|full | 1168 | No | NA | 2 | 0 | 0 | Not-determined | Genome-fragment | NA | NA | 0 |
| S3_vs_k141_18151\|\|full | 1588 | No | NA | 2 | 0 | 2 | Not-determined | Genome-fragment | NA | NA | 0 |
| S12_vs_k141_118579\|\|full | 1675 | No | NA | 4 | 0 | 0 | Low-quality | Genome-fragment | 3.33 | AAI-based (medium-confidence) | 0 |
| S1_extracted_k141_125601 | 1676 | No | NA | 4 | 0 | 0 | Low-quality | Genome-fragment | 1.69 | AAI-based (high-confidence) | 0 |
| S5_vs_k141_104716\|\|full | 1676 | No | NA | 3 | 3 | 0 | Low-quality | Genome-fragment | 2.17 | AAI-based (high-confidence) | 0 |
| S5_vs_k141_4644\|\|full | 1676 | No | NA | 4 | 0 | 1 | Low-quality | Genome-fragment | 0.89 | AAI-based (medium-confidence) | 0 |
| S10_vs_38179\|\|full | 1676 | No | NA | 4 | 0 | 1 | Low-quality | Genome-fragment | 3.58 | AAI-based (high-confidence) | 0 |
| S4_extracted_20406 | 1676 | No | NA | 3 | 0 | 0 | Low-quality | Genome-fragment | 3.73 | AAI-based (high-confidence) | 0 |
| S3_vs_k141_75390\|\|full | 1462 | No | NA | 2 | 0 | 2 | Not-determined | Genome-fragment | NA | NA | 0 |
| S8_vs_k141_179337\|\|full | 1676 | No | NA | 3 | 0 | 0 | Low-quality | Genome-fragment | 3.33 | AAI-based (high-confidence) | 0 |
| S13_vs_107676\|\|full | 1677 | No | NA | 3 | 0 | 1 | Low-quality | Genome-fragment | 1.44 | AAI-based (medium-confidence) | 0 |
| S3_vs_k141_98608\|\|full | 4163 | No | NA | 5 | 0 | 3 | Not-determined | Genome-fragment | NA | NA | 0 |
| S1_vs_k141_84724\|\|full | 1677 | No | NA | 3 | 0 | 1 | Low-quality | Genome-fragment | 1.3 | AAI-based (medium-confidence) | 0 |
| S3_vs_k141_114477\|\|full | 2554 | No | NA | 5 | 0 | 1 | Not-determined | Genome-fragment | NA | NA | 0 |
| S3_vs_k141_47879\|\|full | 1677 | No | NA | 2 | 0 | 1 | Low-quality | Genome-fragment | 3.06 | AAI-based (medium-confidence) | 0 |
| S3_vs_k141_114500\|\|full | 1077 | No | NA | 2 | 0 | 0 | Not-determined | Genome-fragment | NA | NA | 0 |
| S3_vs_k141_43080\|\|full | 2026 | No | NA | 3 | 0 | 1 | Not-determined | Genome-fragment | NA | NA | 0 |
| S3_vs_k141_91625\|\|lt2gene | 1677 | No | NA | 2 | 1 | 0 | Low-quality | Genome-fragment | 3.11 | AAI-based (medium-confidence) | 0 |
| S3_vs_k141_12219\|\|full | 8169 | No | NA | 11 | 0 | 3 | Not-determined | Genome-fragment | NA | NA | 0 |
| S5_vs_k141_107781\|\|full | 1677 | No | NA | 5 | 1 | 0 | Low-quality | Genome-fragment | 2.25 | HMM-based (lower-bound) | 0 |
| S7_vs_26569\|\|full | 1677 | No | NA | 3 | 0 | 2 | Low-quality | Genome-fragment | 0.86 | AAI-based (medium-confidence) | 0 |
| S3_vs_k141_35213\|\|full | 1608 | No | NA | 2 | 0 | 2 | Not-determined | Genome-fragment | NA | NA | 0 |
| S3_vs_k141_26941\|\|full | 8975 | No | NA | 11 | 0 | 8 | Not-determined | Genome-fragment | NA | NA | 0 |
| S10_extracted_22545 | 1677 | No | NA | 2 | 0 | 0 | Low-quality | Genome-fragment | 1.08 | AAI-based (high-confidence) | 0 |
| S6_extracted_k141_5356 | 1677 | No | NA | 4 | 2 | 0 | Low-quality | Genome-fragment | 3.16 | AAI-based (high-confidence) | 0 |
| S17_vs_k141_144280\|\|full | 1678 | No | NA | 2 | 0 | 0 | Low-quality | Genome-fragment | 4.79 | AAI-based (medium-confidence) | 0 |
| S3_vs_k141_59188\|\|full | 2751 | No | NA | 4 | 0 | 0 | Not-determined | Genome-fragment | NA | NA | 0 |
| S9_extracted_k141_23235 | 1678 | No | NA | 2 | 0 | 0 | Low-quality | Genome-fragment | 2.8 | AAI-based (high-confidence) | 0 |
| S14_vs_k141_37445\|\|full | 1678 | No | NA | 3 | 0 | 0 | Low-quality | Genome-fragment | 2.89 | AAI-based (medium-confidence) | 0 |
| S3_vs_k141_66362\|\|full | 5095 | No | NA | 4 | 0 | 2 | Not-determined | Genome-fragment | NA | NA | 0 |
| S3_vs_k141_66367\|\|full | 2450 | No | NA | 3 | 0 | 1 | Not-determined | Genome-fragment | NA | NA | 0 |
| S14_vs_k141_113670\|\|full | 1678 | No | NA | 3 | 2 | 0 | Low-quality | Genome-fragment | 4.06 | AAI-based (high-confidence) | 0 |
| S3_vs_k141_126991\|\|full | 1678 | No | NA | 4 | 0 | 1 | Low-quality | Genome-fragment | 0.84 | AAI-based (medium-confidence) | 0 |
| S3_vs_k141_66375\|\|full | 1449 | No | NA | 3 | 0 | 2 | Not-determined | Genome-fragment | NA | NA | 0 |
| S3_vs_k141_90742\|\|full | 3744 | No | NA | 4 | 0 | 2 | Not-determined | Genome-fragment | NA | NA | 0 |
| S5_vs_k141_55564\|\|full | 1678 | No | NA | 4 | 0 | 0 | Low-quality | Genome-fragment | 3.22 | AAI-based (high-confidence) | 0 |
| S5_vs_k141_20709\|\|full | 1678 | No | NA | 5 | 4 | 0 | Low-quality | Genome-fragment | 2.77 | AAI-based (medium-confidence) | 0 |
| S3_vs_k141_114623\|\|full | 1774 | No | NA | 3 | 0 | 3 | Not-determined | Genome-fragment | NA | NA | 0 |
| S7_vs_3761\|\|full | 1678 | No | NA | 3 | 1 | 0 | Low-quality | Genome-fragment | 1.13 | AAI-based (medium-confidence) | 0 |
| S3_vs_k141_18292\|\|full | 1448 | No | NA | 2 | 0 | 0 | Not-determined | Genome-fragment | NA | NA | 0 |
| S3_vs_k141_43185\|\|full | 1954 | No | NA | 3 | 0 | 0 | Not-determined | Genome-fragment | NA | NA | 0 |
| S7_vs_19316\|\|full | 1678 | No | NA | 7 | 0 | 0 | Low-quality | Genome-fragment | 4.32 | AAI-based (high-confidence) | 0 |
| S3_vs_k141_114652\|\|full | 1745 | No | NA | 3 | 0 | 1 | Not-determined | Genome-fragment | NA | NA | 0 |
| S3_vs_k141_59312\|\|full | 2109 | No | NA | 3 | 0 | 2 | Not-determined | Genome-fragment | NA | NA | 0 |
| S16_extracted_154219 | 1678 | No | NA | 2 | 0 | 0 | Low-quality | Genome-fragment | 3.34 | AAI-based (high-confidence) | 0 |
| S3_vs_k141_59362\|\|full | 5066 | No | NA | 3 | 0 | 0 | Not-determined | Genome-fragment | NA | NA | 0 |
| S12_vs_k141_98797\|\|full | 1678 | No | NA | 2 | 0 | 1 | Low-quality | Genome-fragment | 1.07 | AAI-based (medium-confidence) | 0 |
| S3_vs_k141_75655\|\|full | 1458 | No | NA | 3 | 0 | 0 | Not-determined | Genome-fragment | NA | NA | 0 |
| S3_vs_k141_18317\|\|full | 3829 | No | NA | 3 | 0 | 3 | Not-determined | Genome-fragment | NA | NA | 0 |
| S3_vs_k141_90819\|\|full | 5571 | No | NA | 9 | 0 | 0 | Not-determined | Genome-fragment | NA | NA | 0 |
| S6_vs_k141_14281\|\|full | 1678 | No | NA | 4 | 1 | 0 | Low-quality | Genome-fragment | 3.91 | AAI-based (high-confidence) | 0 |
| S3_vs_k141_122242\|\|full | 8434 | No | NA | 5 | 0 | 4 | Not-determined | Genome-fragment | NA | NA | 0 |
| S17_vs_k141_172195\|\|full | 1679 | No | NA | 2 | 0 | 0 | Low-quality | Genome-fragment | 0.85 | AAI-based (medium-confidence) | 0 |
| S11_vs_k141_68418\|\|full | 1679 | No | NA | 2 | 2 | 0 | Low-quality | Genome-fragment | 3.65 | AAI-based (high-confidence) | 0 |
| S3_vs_k141_75716\|\|full | 4613 | No | NA | 5 | 0 | 3 | Not-determined | Genome-fragment | NA | NA | 0 |
| S3_vs_k141_75786\|\|full | 1786 | No | NA | 3 | 0 | 1 | Not-determined | Genome-fragment | NA | NA | 0 |
| S14_vs_k141_72322\|\|lt2gene | 1679 | No | NA | 1 | 1 | 0 | Low-quality | Genome-fragment | 2.45 | HMM-based (lower-bound) | 0 |
| S3_vs_k141_98866\|\|full | 3373 | No | NA | 4 | 0 | 1 | Not-determined | Genome-fragment | NA | NA | 0 |
| S16_vs_69737\|\|full | 1679 | No | NA | 4 | 2 | 1 | Low-quality | Genome-fragment | 3.43 | AAI-based (high-confidence) | 0 |
| S3_vs_k141_59440\|\|full | 11277 | No | NA | 12 | 0 | 4 | Not-determined | Genome-fragment | NA | NA | 0 |
| S3_vs_k141_98876\|\|full | 11569 | No | NA | 11 | 0 | 5 | Not-determined | Genome-fragment | NA | NA | 0 |
| S3_vs_k141_114781\|\|full | 5344 | No | NA | 6 | 0 | 4 | Not-determined | Genome-fragment | NA | NA | 0 |
| S3_vs_k141_114789\|\|full | 2865 | No | NA | 5 | 0 | 0 | Not-determined | Genome-fragment | NA | NA | 0 |
| S3_vs_k141_107075\|\|full | 1600 | No | NA | 2 | 0 | 1 | Not-determined | Genome-fragment | NA | NA | 0 |
| S10_vs_93022\|\|full | 1679 | No | NA | 2 | 0 | 1 | Low-quality | Genome-fragment | 1.02 | AAI-based (high-confidence) | 0 |
| S9_extracted_k141_70029 | 1680 | No | NA | 6 | 1 | 0 | Low-quality | Genome-fragment | 1.01 | HMM-based (lower-bound) | 0 |
| S3_vs_k141_114803\|\|full | 3952 | No | NA | 5 | 0 | 0 | Not-determined | Genome-fragment | NA | NA | 0 |
| S9_vs_k141_42547\|\|full | 1680 | No | NA | 2 | 0 | 0 | Low-quality | Genome-fragment | 4.12 | AAI-based (high-confidence) | 0 |
| S11_vs_k141_68235\|\|full | 1680 | No | NA | 3 | 0 | 0 | Low-quality | Genome-fragment | 4.1 | AAI-based (medium-confidence) | 0 |
| S3_vs_k141_35620\|\|full | 1966 | No | NA | 3 | 0 | 1 | Not-determined | Genome-fragment | NA | NA | 0 |
| S3_vs_k141_18559\|\|full | 1057 | No | NA | 1 | 0 | 0 | Not-determined | Genome-fragment | NA | NA | 0 |
| S3_vs_k141_122343\|\|full | 6381 | No | NA | 9 | 0 | 1 | Not-determined | Genome-fragment | NA | NA | 0 |
| S1_vs_k141_21482\|\|full | 1680 | No | NA | 3 | 1 | 0 | Low-quality | Genome-fragment | 4 | HMM-based (lower-bound) | 0 |
| S3_vs_k141_59483\|\|full | 1480 | No | NA | 2 | 0 | 0 | Not-determined | Genome-fragment | NA | NA | 0 |
| S3_vs_k141_73451\|\|full | 1680 | No | NA | 4 | 1 | 0 | Low-quality | Genome-fragment | 0.93 | AAI-based (high-confidence) | 0 |
| S3_vs_k141_61599\|\|full | 1680 | No | NA | 3 | 1 | 0 | Low-quality | Genome-fragment | 4.62 | HMM-based (lower-bound) | 0 |
| S7_extracted_49353 | 1680 | No | NA | 2 | 1 | 0 | Low-quality | Genome-fragment | 3.57 | AAI-based (high-confidence) | 0 |
| S3_vs_k141_114891\|\|full | 1864 | No | NA | 5 | 0 | 1 | Not-determined | Genome-fragment | NA | NA | 0 |
| S7_vs_13842\|\|full | 1680 | No | NA | 4 | 1 | 0 | Low-quality | Genome-fragment | 4.16 | AAI-based (medium-confidence) | 0 |
| S3_vs_k141_114899\|\|full | 2557 | No | NA | 2 | 0 | 0 | Not-determined | Genome-fragment | NA | NA | 0 |
| S10_extracted_88202 | 1680 | No | NA | 5 | 1 | 0 | Low-quality | Genome-fragment | 0.94 | HMM-based (lower-bound) | 0 |
| S8_vs_k141_74480\|\|full | 1680 | No | NA | 4 | 1 | 0 | Low-quality | Genome-fragment | 1.03 | HMM-based (lower-bound) | 0 |
| S3_vs_k141_107162\|\|full | 2210 | No | NA | 3 | 0 | 2 | Not-determined | Genome-fragment | NA | NA | 0 |
| S8_vs_k141_77038\|\|full | 1680 | No | NA | 3 | 0 | 0 | Low-quality | Genome-fragment | 2.92 | AAI-based (medium-confidence) | 0 |
| S17_vs_k141_104678\|\|full | 1681 | No | NA | 6 | 1 | 0 | Low-quality | Genome-fragment | 1.58 | AAI-based (medium-confidence) | 0 |
| S17_vs_k141_95055\|\|full | 1681 | No | NA | 6 | 1 | 0 | Low-quality | Genome-fragment | 0.97 | HMM-based (lower-bound) | 0 |
| S9_vs_k141_54922\|\|lt2gene | 1681 | No | NA | 1 | 1 | 0 | Low-quality | Genome-fragment | 2.73 | AAI-based (high-confidence) | 0 |
| S3_vs_k141_75883\|\|full | 6607 | No | NA | 6 | 0 | 5 | Not-determined | Genome-fragment | NA | NA | 0 |
| S3_vs_k141_18669\|\|full | 5282 | No | NA | 7 | 0 | 3 | Not-determined | Genome-fragment | NA | NA | 0 |
| S9_vs_k141_54960\|\|lt2gene | 1681 | No | NA | 1 | 1 | 0 | Low-quality | Genome-fragment | 2.74 | AAI-based (high-confidence) | 0 |
| S16_vs_161446\|\|full | 1681 | No | NA | 3 | 0 | 0 | Low-quality | Genome-fragment | 4.67 | AAI-based (medium-confidence) | 0 |
| S3_vs_k141_51707\|\|full | 2093 | No | NA | 2 | 0 | 1 | Not-determined | Genome-fragment | NA | NA | 0 |
| S10_vs_120250\|\|full | 1681 | No | NA | 3 | 0 | 1 | Low-quality | Genome-fragment | 2.28 | AAI-based (medium-confidence) | 0 |
| S8_extracted_k141_70627 | 1681 | No | NA | 1 | 0 | 1 | Low-quality | Genome-fragment | 3.07 | AAI-based (high-confidence) | 0 |
| S17_vs_k141_171409\|\|full | 1682 | No | NA | 3 | 0 | 2 | Low-quality | Genome-fragment | 5.37 | AAI-based (high-confidence) | 0 |
| S17_vs_k141_150872\|\|full | 1682 | No | NA | 2 | 0 | 0 | Low-quality | Genome-fragment | 2.02 | AAI-based (medium-confidence) | 0 |
| S3_vs_k141_12674\|\|full | 2193 | No | NA | 3 | 0 | 2 | Not-determined | Genome-fragment | NA | NA | 0 |
| S3_vs_k141_66659\|\|full | 1452 | No | NA | 4 | 0 | 0 | Not-determined | Genome-fragment | NA | NA | 0 |
| S5_vs_k141_132945\|\|full | 1682 | No | NA | 3 | 3 | 0 | Low-quality | Genome-fragment | 3.59 | AAI-based (medium-confidence) | 0 |
| S16_vs_75901\|\|full | 1682 | No | NA | 3 | 1 | 0 | Low-quality | Genome-fragment | 4.21 | AAI-based (high-confidence) | 0 |
| S3_vs_k141_27403\|\|full | 2106 | No | NA | 4 | 0 | 4 | Not-determined | Genome-fragment | NA | NA | 0 |
| S6_vs_k141_48232\|\|full | 1682 | No | NA | 2 | 2 | 0 | Low-quality | Genome-fragment | 3.29 | AAI-based (high-confidence) | 0 |
| S8_vs_k141_123871\|\|full | 1682 | No | NA | 4 | 2 | 0 | Low-quality | Genome-fragment | 4.3 | AAI-based (high-confidence) | 0 |
| S3_vs_k141_83149\|\|full | 4065 | No | NA | 7 | 0 | 2 | Not-determined | Genome-fragment | NA | NA | 0 |
| S3_vs_k141_122575\|\|full | 1115 | No | NA | 2 | 0 | 0 | Not-determined | Genome-fragment | NA | NA | 0 |
| S17_extracted_k141_131021 | 1683 | No | NA | 2 | 0 | 2 | Low-quality | Genome-fragment | 1.11 | AAI-based (high-confidence) | 0 |
| S3_vs_k141_115056\|\|full | 1938 | No | NA | 2 | 0 | 0 | Not-determined | Genome-fragment | NA | NA | 0 |
| S17_vs_k141_96137\|\|full | 1683 | No | NA | 2 | 0 | 1 | Low-quality | Genome-fragment | 2.07 | AAI-based (medium-confidence) | 0 |
| S17_vs_k141_131698\|\|lt2gene | 1683 | No | NA | 1 | 1 | 0 | Low-quality | Genome-fragment | 3.9 | AAI-based (medium-confidence) | 0 |
| S11_vs_k141_92254\|\|full | 1683 | No | NA | 4 | 1 | 0 | Low-quality | Genome-fragment | 5.04 | HMM-based (lower-bound) | 0 |
| S11_vs_k141_44241\|\|full | 1683 | No | NA | 5 | 1 | 0 | Low-quality | Genome-fragment | 4.13 | AAI-based (medium-confidence) | 0 |
| S14_vs_k141_33717\|\|full | 1683 | No | NA | 3 | 0 | 1 | Low-quality | Genome-fragment | 2.01 | AAI-based (medium-confidence) | 0 |
| S3_vs_k141_107311\|\|full | 2481 | No | NA | 5 | 0 | 0 | Not-determined | Genome-fragment | NA | NA | 0 |
| S3_vs_k141_51808\|\|full | 2139 | No | NA | 5 | 0 | 0 | Not-determined | Genome-fragment | NA | NA | 0 |
| S14_vs_k141_100085\|\|full | 1683 | No | NA | 5 | 0 | 0 | Low-quality | Genome-fragment | 4.02 | AAI-based (high-confidence) | 0 |
| S5_extracted_k141_110479 | 1683 | No | NA | 2 | 2 | 0 | Low-quality | Genome-fragment | 5.54 | AAI-based (high-confidence) | 0 |
| S3_vs_k141_107380\|\|full | 1427 | No | NA | 3 | 0 | 1 | Not-determined | Genome-fragment | NA | NA | 0 |
| S3_vs_k141_122618\|\|full | 1819 | No | NA | 3 | 0 | 1 | Not-determined | Genome-fragment | NA | NA | 0 |
| S3_vs_k141_66732\|\|full | 1078 | No | NA | 4 | 0 | 0 | Not-determined | Genome-fragment | NA | NA | 0 |
| S7_extracted_26721 | 1683 | No | NA | 5 | 0 | 0 | Low-quality | Genome-fragment | 2.41 | AAI-based (medium-confidence) | 0 |
| S7_vs_84870\|\|full | 1683 | No | NA | 5 | 0 | 0 | Low-quality | Genome-fragment | 3.1 | AAI-based (high-confidence) | 0 |
| S3_vs_k141_43548\|\|full | 1485 | No | NA | 3 | 0 | 0 | Not-determined | Genome-fragment | NA | NA | 0 |
| S3_vs_k141_66752\|\|full | 10470 | Yes | 2661 | 7 | 1 | 4 | Not-determined | Genome-fragment | NA | NA | 74.58 |
| S3_vs_k141_75979\|\|full | 6260 | No | NA | 6 | 0 | 2 | Not-determined | Genome-fragment | NA | NA | 0 |
| S16_extracted_134485 | 1683 | No | NA | 2 | 0 | 1 | Low-quality | Genome-fragment | 2.95 | AAI-based (medium-confidence) | 0 |
| S16_vs_103570\|\|full | 1683 | No | NA | 3 | 0 | 1 | Low-quality | Genome-fragment | 1.56 | AAI-based (high-confidence) | 0 |
| S3_vs_k141_43562\|\|full | 6837 | No | NA | 8 | 0 | 5 | Not-determined | Genome-fragment | NA | NA | 0 |
| S10_vs_34309\|\|lt2gene | 1683 | No | NA | 2 | 1 | 0 | Low-quality | Genome-fragment | 4 | AAI-based (high-confidence) | 0 |
| S12_vs_k141_19603\|\|full | 1683 | No | NA | 3 | 1 | 0 | Low-quality | Genome-fragment | 3.92 | AAI-based (medium-confidence) | 0 |
| S15_extracted_9428 | 1683 | No | NA | 4 | 0 | 0 | Low-quality | Genome-fragment | 1.67 | AAI-based (high-confidence) | 0 |
| S11_vs_k141_4332\|\|full | 1684 | No | NA | 3 | 0 | 0 | Low-quality | Genome-fragment | 4.6 | AAI-based (medium-confidence) | 0 |
| S3_vs_k141_27577\|\|full | 1378 | No | NA | 3 | 0 | 1 | Not-determined | Genome-fragment | NA | NA | 0 |
| S3_vs_k141_122699\|\|full | 2217 | No | NA | 5 | 0 | 3 | Not-determined | Genome-fragment | NA | NA | 0 |
| S1_vs_k141_141347\|\|full | 1684 | No | NA | 4 | 1 | 0 | Low-quality | Genome-fragment | 0.51 | HMM-based (lower-bound) | 0 |
| S3_vs_k141_115139\|\|full | 1014 | No | NA | 2 | 0 | 0 | Not-determined | Genome-fragment | NA | NA | 0 |
| S3_vs_k141_12817\|\|full | 2738 | No | NA | 4 | 0 | 2 | Not-determined | Genome-fragment | NA | NA | 0 |
| S1_vs_k141_112740\|\|full | 1684 | No | NA | 6 | 1 | 0 | Low-quality | Genome-fragment | 3.88 | AAI-based (medium-confidence) | 0 |
| S3_vs_k141_115155\|\|full | 1745 | No | NA | 3 | 0 | 1 | Not-determined | Genome-fragment | NA | NA | 0 |
| S3_vs_k141_51931\|\|full | 3896 | No | NA | 7 | 6 | 0 | Medium-quality | Genome-fragment | 79.28 | AAI-based (high-confidence) | 0 |
| S3_vs_k141_122706\|\|full | 21591 | No | NA | 26 | 9 | 0 | Medium-quality | Genome-fragment | 62.56 | AAI-based (high-confidence) | 0 |
| S16_vs_60042\|\|full | 1684 | No | NA | 5 | 1 | 0 | Low-quality | Genome-fragment | 3.23 | HMM-based (lower-bound) | 0 |
| S3_vs_k141_3509\|\|full | 3307 | No | NA | 4 | 0 | 4 | Not-determined | Genome-fragment | NA | NA | 0 |
| S16_vs_124497\|\|full | 1684 | No | NA | 2 | 1 | 0 | Low-quality | Genome-fragment | 3.13 | HMM-based (lower-bound) | 0 |
| S3_vs_k141_51989\|\|full | 8285 | No | NA | 7 | 0 | 5 | Not-determined | Genome-fragment | NA | NA | 0 |
| S3_vs_k141_51994\|\|full | 1019 | No | NA | 3 | 0 | 0 | Not-determined | Genome-fragment | NA | NA | 0 |
| S10_extracted_22191 | 1684 | No | NA | 1 | 0 | 1 | Low-quality | Genome-fragment | 0.3 | AAI-based (medium-confidence) | 0 |
| S3_vs_k141_115217\|\|full | 1336 | No | NA | 3 | 0 | 2 | Not-determined | Genome-fragment | NA | NA | 0 |
| S10_vs_36790\|\|full | 1684 | No | NA | 4 | 0 | 1 | Low-quality | Genome-fragment | 2.64 | AAI-based (medium-confidence) | 0 |
| S3_vs_k141_115229\|\|full | 1226 | No | NA | 2 | 0 | 2 | Not-determined | Genome-fragment | NA | NA | 0 |
| S12_vs_k141_133821\|\|full | 1684 | No | NA | 3 | 0 | 0 | Low-quality | Genome-fragment | 3.16 | AAI-based (high-confidence) | 0 |
| S3_vs_k141_18772\|\|full | 1475 | No | NA | 3 | 0 | 1 | Not-determined | Genome-fragment | NA | NA | 0 |
| S3_vs_k141_27610\|\|full | 3175 | No | NA | 7 | 0 | 0 | Not-determined | Genome-fragment | NA | NA | 0 |
| S3_vs_k141_76101\|\|full | 1099 | No | NA | 2 | 0 | 2 | Not-determined | Genome-fragment | NA | NA | 0 |
| S9_extracted_k141_68149 | 1685 | No | NA | 3 | 2 | 0 | Low-quality | Genome-fragment | 3.87 | AAI-based (high-confidence) | 0 |
| S13_vs_76554\|\|full | 1685 | No | NA | 3 | 2 | 0 | Low-quality | Genome-fragment | 4.87 | AAI-based (high-confidence) | 0 |
| S3_vs_k141_66883\|\|full | 1631 | No | NA | 3 | 0 | 1 | Not-determined | Genome-fragment | NA | NA | 0 |
| S13_vs_96730\|\|full | 1685 | No | NA | 5 | 3 | 0 | Low-quality | Genome-fragment | 4.49 | AAI-based (high-confidence) | 0 |
| S3_vs_k141_15457\|\|full | 1685 | No | NA | 3 | 1 | 0 | Low-quality | Genome-fragment | 3.73 | AAI-based (medium-confidence) | 0 |
| S3_vs_k141_52112\|\|full | 2168 | No | NA | 4 | 0 | 0 | Not-determined | Genome-fragment | NA | NA | 0 |
| S3_vs_k141_59916\|\|full | 1386 | No | NA | 3 | 0 | 1 | Not-determined | Genome-fragment | NA | NA | 0 |
| S5_vs_k141_140343\|\|full | 1685 | No | NA | 3 | 1 | 0 | Low-quality | Genome-fragment | 4.18 | AAI-based (high-confidence) | 0 |
| S3_vs_k141_99223\|\|full | 1509 | No | NA | 2 | 0 | 1 | Not-determined | Genome-fragment | NA | NA | 0 |
| S10_vs_70275\|\|full | 1685 | No | NA | 3 | 2 | 0 | Low-quality | Genome-fragment | 4.06 | AAI-based (high-confidence) | 0 |
| S3_vs_k141_35975\|\|full | 1744 | No | NA | 2 | 0 | 1 | Not-determined | Genome-fragment | NA | NA | 0 |
| S11_vs_k141_98796\|\|full | 1686 | No | NA | 4 | 1 | 0 | Low-quality | Genome-fragment | 4.39 | AAI-based (high-confidence) | 0 |
| S3_vs_k141_35981\|\|full | 2369 | No | NA | 3 | 0 | 1 | Not-determined | Genome-fragment | NA | NA | 0 |
| S14_vs_k141_67781\|\|full | 1686 | No | NA | 3 | 0 | 0 | Low-quality | Genome-fragment | 4.04 | AAI-based (medium-confidence) | 0 |
| S3_vs_k141_12941\|\|full | 3091 | No | NA | 4 | 0 | 0 | Not-determined | Genome-fragment | NA | NA | 0 |
| S3_vs_k141_91500\|\|full | 5454 | No | NA | 5 | 0 | 0 | Not-determined | Genome-fragment | NA | NA | 0 |
| S3_vs_k141_27672\|\|full | 2813 | No | NA | 4 | 0 | 2 | Not-determined | Genome-fragment | NA | NA | 0 |
| S1_vs_k141_47305\|\|full | 1686 | Yes | 775 | 5 | 2 | 1 | Low-quality | Genome-fragment | 0.98 | AAI-based (high-confidence) | 54.03 |
| S3_vs_k141_18834\|\|full | 2073 | No | NA | 3 | 0 | 0 | Not-determined | Genome-fragment | NA | NA | 0 |
| S3_vs_k141_12973\|\|full | 20022 | No | NA | 35 | 4 | 1 | Medium-quality | Genome-fragment | 58.51 | AAI-based (high-confidence) | 0 |
| S5_extracted_k141_80757 | 1686 | No | NA | 3 | 1 | 0 | Low-quality | Genome-fragment | 7.69 | AAI-based (high-confidence) | 0 |
| S16_extracted_148659 | 1686 | No | NA | 4 | 2 | 0 | Low-quality | Genome-fragment | 2.9 | HMM-based (lower-bound) | 0 |
| S3_vs_k141_59977\|\|full | 1523 | No | NA | 3 | 0 | 2 | Not-determined | Genome-fragment | NA | NA | 0 |
| S10_vs_4027\|\|full | 1686 | No | NA | 3 | 0 | 0 | Low-quality | Genome-fragment | 3.66 | AAI-based (high-confidence) | 0 |
| S3_vs_k141_122823\|\|full | 5081 | No | NA | 3 | 0 | 0 | Not-determined | Genome-fragment | NA | NA | 0 |
| S10_vs_5745\|\|lt2gene | 1686 | No | NA | 1 | 1 | 0 | Low-quality | Genome-fragment | 2.93 | HMM-based (lower-bound) | 0 |
| S3_vs_k141_83571\|\|full | 1470 | No | NA | 3 | 0 | 0 | Not-determined | Genome-fragment | NA | NA | 0 |
| S3_vs_k141_43751\|\|full | 1720 | No | NA | 3 | 0 | 1 | Not-determined | Genome-fragment | NA | NA | 0 |
| S3_vs_k141_66962\|\|full | 2718 | No | NA | 3 | 0 | 2 | Not-determined | Genome-fragment | NA | NA | 0 |
| S12_vs_k141_91465\|\|full | 1686 | No | NA | 3 | 0 | 1 | Low-quality | Genome-fragment | 3.08 | AAI-based (medium-confidence) | 0 |
| S3_vs_k141_27740\|\|full | 1817 | No | NA | 3 | 0 | 0 | Not-determined | Genome-fragment | NA | NA | 0 |
| S3_vs_k141_107474\|\|full | 4291 | No | NA | 5 | 0 | 4 | Not-determined | Genome-fragment | NA | NA | 0 |
| S6_extracted_k141_28660 | 1686 | No | NA | 1 | 1 | 0 | Low-quality | Genome-fragment | 2.81 | AAI-based (high-confidence) | 0 |
| S8_vs_k141_17211\|\|full | 1686 | No | NA | 5 | 0 | 1 | Low-quality | Genome-fragment | 1.5 | AAI-based (medium-confidence) | 0 |
| S5_vs_k141_42930\|\|full | 1687 | No | NA | 2 | 1 | 0 | Low-quality | Genome-fragment | 3.43 | AAI-based (medium-confidence) | 0 |
| S3_vs_k141_115324\|\|full | 2754 | No | NA | 2 | 0 | 2 | Not-determined | Genome-fragment | NA | NA | 0 |
| S3_vs_k141_3688\|\|full | 4525 | No | NA | 6 | 0 | 0 | Not-determined | Genome-fragment | NA | NA | 0 |
| S7_extracted_59840 | 1687 | No | NA | 1 | 0 | 1 | Low-quality | Genome-fragment | 0.99 | AAI-based (medium-confidence) | 0 |
| S7_vs_88700\|\|full | 1687 | No | NA | 4 | 2 | 0 | Low-quality | Genome-fragment | 3.3 | AAI-based (high-confidence) | 0 |
| S3_vs_k141_122860\|\|full | 2393 | No | NA | 3 | 0 | 0 | Not-determined | Genome-fragment | NA | NA | 0 |
| S3_vs_k141_12985\|\|full | 19001 | No | NA | 22 | 13 | 1 | Medium-quality | Genome-fragment | 58.58 | AAI-based (high-confidence) | 0 |
| S3_vs_k141_52285\|\|full | 2812 | No | NA | 5 | 0 | 0 | Not-determined | Genome-fragment | NA | NA | 0 |
| S3_vs_k141_18858\|\|full | 40921 | Yes | 30395 | 47 | 9 | 6 | Medium-quality | Genome-fragment | 76.49 | AAI-based (high-confidence) | 25.72 |
| S16_extracted_4134 | 1687 | No | NA | 2 | 0 | 0 | Low-quality | Genome-fragment | 4.3 | AAI-based (high-confidence) | 0 |
| S10_vs_27805\|\|full | 1687 | No | NA | 5 | 1 | 0 | Low-quality | Genome-fragment | 2.27 | HMM-based (lower-bound) | 0 |
| S12_extracted_k141_6121 | 1687 | No | NA | 2 | 1 | 0 | Low-quality | Genome-fragment | 3.33 | HMM-based (lower-bound) | 0 |
| S3_vs_k141_36160\|\|full | 4792 | No | NA | 5 | 0 | 3 | Not-determined | Genome-fragment | NA | NA | 0 |
| S3_vs_k141_107548\|\|full | 1360 | No | NA | 2 | 0 | 1 | Not-determined | Genome-fragment | NA | NA | 0 |
| S3_vs_k141_115400\|\|full | 2232 | No | NA | 3 | 0 | 0 | Not-determined | Genome-fragment | NA | NA | 0 |
| S12_vs_k141_129795\|\|full | 1687 | No | NA | 2 | 0 | 1 | Low-quality | Genome-fragment | 4.11 | AAI-based (high-confidence) | 0 |
| S3_vs_k141_60146\|\|full | 1994 | No | NA | 2 | 0 | 1 | Not-determined | Genome-fragment | NA | NA | 0 |
| S3_vs_k141_115414\|\|full | 1286 | No | NA | 2 | 0 | 1 | Not-determined | Genome-fragment | NA | NA | 0 |
| S8_vs_k141_2597\|\|full | 1687 | No | NA | 4 | 0 | 0 | Low-quality | Genome-fragment | 3.1 | AAI-based (high-confidence) | 0 |
| S3_vs_k141_3727\|\|full | 12526 | No | NA | 15 | 0 | 4 | Not-determined | Genome-fragment | NA | NA | 0 |
| S17_vs_k141_163440\|\|full | 1688 | No | NA | 2 | 0 | 0 | Low-quality | Genome-fragment | 2.47 | AAI-based (high-confidence) | 0 |
| S11_extracted_k141_67322 | 1688 | No | NA | 6 | 0 | 0 | Low-quality | Genome-fragment | 3.09 | AAI-based (medium-confidence) | 0 |
| S11_extracted_k141_23201 | 1688 | No | NA | 2 | 0 | 0 | Low-quality | Genome-fragment | 1.81 | AAI-based (high-confidence) | 0 |
| S3_vs_k141_122869\|\|full | 1130 | No | NA | 3 | 0 | 1 | Not-determined | Genome-fragment | NA | NA | 0 |
| S14_vs_k141_16364\|\|full | 1688 | No | NA | 4 | 2 | 0 | Low-quality | Genome-fragment | 10.16 | AAI-based (high-confidence) | 0 |
| S3_vs_k141_91683\|\|full | 2278 | No | NA | 3 | 0 | 2 | Not-determined | Genome-fragment | NA | NA | 0 |
| S3_vs_k141_36975\|\|full | 1688 | No | NA | 2 | 2 | 0 | Low-quality | Genome-fragment | 4 | AAI-based (high-confidence) | 0 |
| S3_vs_k141_6334\|\|full | 1688 | No | NA | 3 | 1 | 0 | Low-quality | Genome-fragment | 1.78 | AAI-based (medium-confidence) | 0 |
| S3_vs_k141_13147\|\|full | 3197 | No | NA | 7 | 0 | 0 | Not-determined | Genome-fragment | NA | NA | 0 |
| S5_vs_k141_34890\|\|full | 1688 | No | NA | 5 | 2 | 0 | Low-quality | Genome-fragment | 3.41 | AAI-based (medium-confidence) | 0 |
| S10_vs_67667\|\|full | 1688 | No | NA | 3 | 0 | 0 | Low-quality | Genome-fragment | 1.67 | AAI-based (high-confidence) | 0 |
| S3_vs_k141_67016\|\|full | 3811 | No | NA | 4 | 0 | 4 | Not-determined | Genome-fragment | NA | NA | 0 |
| S3_vs_k141_115452\|\|full | 2299 | No | NA | 5 | 0 | 0 | Not-determined | Genome-fragment | NA | NA | 0 |
| S3_vs_k141_91690\|\|full | 1430 | No | NA | 3 | 0 | 1 | Not-determined | Genome-fragment | NA | NA | 0 |
| S12_vs_k141_160818\|\|lt2gene | 1688 | No | NA | 1 | 1 | 0 | Low-quality | Genome-fragment | 3.66 | AAI-based (high-confidence) | 0 |
| S6_vs_k141_39048\|\|full | 1688 | No | NA | 5 | 0 | 0 | Low-quality | Genome-fragment | 3.26 | AAI-based (high-confidence) | 0 |
| S3_vs_k141_76347\|\|full | 2959 | No | NA | 3 | 0 | 0 | Not-determined | Genome-fragment | NA | NA | 0 |
| S8_vs_k141_7975\|\|full | 1688 | Yes | 597 | 2 | 1 | 1 | Low-quality | Genome-fragment | 0.37 | HMM-based (lower-bound) | 64.63 |
| S3_vs_k141_107653\|\|full | 7962 | No | NA | 7 | 0 | 6 | Not-determined | Genome-fragment | NA | NA | 0 |
| S3_vs_k141_43861\|\|full | 1034 | No | NA | 3 | 0 | 0 | Not-determined | Genome-fragment | NA | NA | 0 |
| S3_vs_k141_91699\|\|full | 1658 | No | NA | 3 | 0 | 1 | Not-determined | Genome-fragment | NA | NA | 0 |
| S17_extracted_k141_15958 | 1689 | No | NA | 2 | 0 | 1 | Low-quality | Genome-fragment | 3.92 | AAI-based (medium-confidence) | 0 |
| S17_vs_k141_37880\|\|full | 1689 | No | NA | 3 | 0 | 2 | Low-quality | Genome-fragment | 0.78 | AAI-based (medium-confidence) | 0 |
| S3_vs_k141_115500\|\|full | 2288 | No | NA | 3 | 0 | 0 | Not-determined | Genome-fragment | NA | NA | 0 |
| S17_vs_k141_139260\|\|full | 1689 | No | NA | 2 | 0 | 0 | Low-quality | Genome-fragment | 1.31 | AAI-based (medium-confidence) | 0 |
| S3_vs_k141_107678\|\|full | 1860 | No | NA | 5 | 0 | 0 | Not-determined | Genome-fragment | NA | NA | 0 |
| S3_vs_k141_76398\|\|full | 1661 | No | NA | 5 | 0 | 0 | Not-determined | Genome-fragment | NA | NA | 0 |
| S1_vs_k141_129840\|\|full | 1689 | No | NA | 6 | 0 | 0 | Low-quality | Genome-fragment | 3.4 | AAI-based (high-confidence) | 0 |
| S7_vs_12063\|\|full | 1689 | No | NA | 2 | 2 | 0 | Low-quality | Genome-fragment | 3.08 | AAI-based (medium-confidence) | 0 |
| S16_extracted_89280 | 1689 | No | NA | 1 | 1 | 0 | Low-quality | Genome-fragment | 9.3 | AAI-based (high-confidence) | 0 |
| S16_extracted_103461 | 1689 | No | NA | 3 | 3 | 0 | Low-quality | Genome-fragment | 3.37 | AAI-based (medium-confidence) | 0 |
| S16_vs_78198\|\|full | 1689 | No | NA | 3 | 0 | 1 | Low-quality | Genome-fragment | 1.31 | AAI-based (high-confidence) | 0 |
| S10_vs_49445\|\|full | 1689 | No | NA | 4 | 0 | 0 | Low-quality | Genome-fragment | 2.56 | AAI-based (medium-confidence) | 0 |
| S12_vs_k141_150053\|\|full | 1689 | No | NA | 3 | 1 | 0 | Low-quality | Genome-fragment | 4.61 | AAI-based (medium-confidence) | 0 |
| S17_vs_k141_4085\|\|full | 1690 | No | NA | 5 | 0 | 0 | Low-quality | Genome-fragment | 2.66 | AAI-based (high-confidence) | 0 |
| S1_vs_k141_24653\|\|lt2gene | 1690 | No | NA | 2 | 2 | 0 | Low-quality | Genome-fragment | 4.02 | AAI-based (high-confidence) | 0 |
| S3_vs_k141_76455\|\|full | 1520 | No | NA | 4 | 0 | 0 | Not-determined | Genome-fragment | NA | NA | 0 |
| S3_vs_k141_3921\|\|full | 2625 | No | NA | 4 | 0 | 2 | Not-determined | Genome-fragment | NA | NA | 0 |
| S7_vs_3981\|\|full | 1690 | No | NA | 4 | 0 | 1 | Low-quality | Genome-fragment | 3.47 | AAI-based (medium-confidence) | 0 |
| S16_extracted_105213 | 1690 | No | NA | 3 | 2 | 0 | Low-quality | Genome-fragment | 3.49 | AAI-based (high-confidence) | 0 |
| S16_vs_100929\|\|full | 1690 | No | NA | 3 | 0 | 0 | Low-quality | Genome-fragment | 1.37 | AAI-based (medium-confidence) | 0 |
| S3_vs_k141_3966\|\|full | 4662 | No | NA | 6 | 0 | 5 | Not-determined | Genome-fragment | NA | NA | 0 |
| S16_vs_151531\|\|full | 1690 | No | NA | 3 | 1 | 0 | Low-quality | Genome-fragment | 4.17 | AAI-based (high-confidence) | 0 |
| S3_vs_k141_91811\|\|full | 1412 | No | NA | 2 | 0 | 0 | Not-determined | Genome-fragment | NA | NA | 0 |
| S10_vs_110074\|\|full | 1690 | No | NA | 7 | 0 | 0 | Low-quality | Genome-fragment | 3.96 | AAI-based (medium-confidence) | 0 |
| S3_vs_k141_36251\|\|full | 5855 | No | NA | 5 | 0 | 3 | Not-determined | Genome-fragment | NA | NA | 0 |
| S12_vs_k141_187099\|\|full | 1690 | No | NA | 3 | 0 | 0 | Low-quality | Genome-fragment | 2.92 | AAI-based (high-confidence) | 0 |
| S3_vs_k141_27954\|\|full | 23561 | No | NA | 19 | 0 | 2 | Not-determined | Genome-fragment | NA | NA | 0 |
| S8_extracted_k141_34032 | 1690 | No | NA | 5 | 1 | 0 | Low-quality | Genome-fragment | 0.31 | HMM-based (lower-bound) | 0 |
| S3_vs_k141_36272\|\|full | 3038 | No | NA | 3 | 0 | 2 | Not-determined | Genome-fragment | NA | NA | 0 |
| S3_vs_k141_91868\|\|full | 5661 | No | NA | 4 | 0 | 3 | Not-determined | Genome-fragment | NA | NA | 0 |
| S9_vs_k141_68182\|\|full | 1691 | No | NA | 5 | 3 | 0 | Low-quality | Genome-fragment | 4.46 | AAI-based (high-confidence) | 0 |
| S3_vs_k141_115585\|\|full | 1631 | No | NA | 2 | 0 | 0 | Not-determined | Genome-fragment | NA | NA | 0 |
| S14_extracted_k141_8531 | 1691 | No | NA | 4 | 4 | 0 | Low-quality | Genome-fragment | 2.5 | AAI-based (high-confidence) | 0 |
| S3_vs_k141_36300\|\|full | 1577 | No | NA | 2 | 0 | 0 | Not-determined | Genome-fragment | NA | NA | 0 |
| S3_vs_k141_49877\|\|full | 1691 | No | NA | 4 | 1 | 0 | Low-quality | Genome-fragment | 2.64 | HMM-based (lower-bound) | 0 |
| S3_vs_k141_76544\|\|full | 4323 | No | NA | 7 | 0 | 2 | Not-determined | Genome-fragment | NA | NA | 0 |
| S3_vs_k141_93495\|\|full | 1691 | No | NA | 3 | 0 | 0 | Low-quality | Genome-fragment | 1.19 | AAI-based (medium-confidence) | 0 |
| S3_vs_k141_4026\|\|full | 2730 | No | NA | 4 | 0 | 0 | Not-determined | Genome-fragment | NA | NA | 0 |
| S5_extracted_k141_82134 | 1691 | No | NA | 2 | 2 | 0 | Low-quality | Genome-fragment | 4.16 | AAI-based (high-confidence) | 0 |
| S5_extracted_k141_78315 | 1691 | No | NA | 3 | 0 | 2 | Low-quality | Genome-fragment | 1.36 | AAI-based (medium-confidence) | 0 |
| S3_vs_k141_123058\|\|full | 1260 | No | NA | 2 | 0 | 0 | Not-determined | Genome-fragment | NA | NA | 0 |
| S5_vs_k141_96028\|\|lt2gene | 1691 | No | NA | 2 | 2 | 0 | Low-quality | Genome-fragment | 4.16 | AAI-based (medium-confidence) | 0 |
| S7_extracted_57843 | 1691 | No | NA | 3 | 0 | 0 | Low-quality | Genome-fragment | 3.95 | AAI-based (high-confidence) | 0 |
| S3_vs_k141_91885\|\|full | 8231 | No | NA | 9 | 0 | 4 | Not-determined | Genome-fragment | NA | NA | 0 |
| S3_vs_k141_123078\|\|full | 1982 | No | NA | 3 | 0 | 0 | Not-determined | Genome-fragment | NA | NA | 0 |
| S6_vs_k141_39067\|\|full | 1691 | No | NA | 4 | 1 | 0 | Low-quality | Genome-fragment | 3.04 | AAI-based (medium-confidence) | 0 |
| S3_vs_k141_13419\|\|full | 1137 | No | NA | 3 | 0 | 1 | Not-determined | Genome-fragment | NA | NA | 0 |
| S17_extracted_k141_119017 | 1692 | No | NA | 1 | 0 | 1 | Low-quality | Genome-fragment | 0.62 | AAI-based (high-confidence) | 0 |
| S11_extracted_k141_79141 | 1692 | No | NA | 4 | 2 | 0 | Low-quality | Genome-fragment | 4.68 | AAI-based (high-confidence) | 0 |
| S3_vs_k141_76562\|\|full | 1954 | No | NA | 6 | 0 | 0 | Not-determined | Genome-fragment | NA | NA | 0 |
| S3_vs_k141_44031\|\|full | 4427 | No | NA | 3 | 0 | 3 | Not-determined | Genome-fragment | NA | NA | 0 |
| S3_vs_k141_67295\|\|full | 1380 | No | NA | 3 | 0 | 0 | Not-determined | Genome-fragment | NA | NA | 0 |
| S3_vs_k141_52602\|\|full | 6685 | No | NA | 12 | 0 | 4 | Not-determined | Genome-fragment | NA | NA | 0 |
| S13_vs_75788\|\|full | 1692 | No | NA | 4 | 2 | 0 | Low-quality | Genome-fragment | 2.81 | AAI-based (high-confidence) | 0 |
| S3_vs_k141_107293\|\|full | 1692 | No | NA | 3 | 0 | 2 | Low-quality | Genome-fragment | 0.81 | AAI-based (medium-confidence) | 0 |
| S5_vs_k141_112938\|\|full | 1692 | No | NA | 3 | 1 | 0 | Low-quality | Genome-fragment | 1.16 | HMM-based (lower-bound) | 0 |
| S3_vs_k141_107857\|\|full | 1977 | No | NA | 3 | 0 | 0 | Not-determined | Genome-fragment | NA | NA | 0 |
| S16_vs_123484\|\|full | 1692 | No | NA | 4 | 1 | 0 | Low-quality | Genome-fragment | 5.71 | AAI-based (high-confidence) | 0 |
| S3_vs_k141_52632\|\|full | 1511 | No | NA | 3 | 0 | 0 | Not-determined | Genome-fragment | NA | NA | 0 |
| S10_vs_24227\|\|full | 1692 | No | NA | 2 | 0 | 1 | Low-quality | Genome-fragment | 0.48 | AAI-based (medium-confidence) | 0 |
| S12_vs_k141_188774\|\|full | 1692 | No | NA | 5 | 0 | 0 | Low-quality | Genome-fragment | 2.98 | AAI-based (medium-confidence) | 0 |
| S2_extracted_k141_15983 | 1692 | No | NA | 8 | 0 | 0 | Low-quality | Genome-fragment | 1.76 | AAI-based (high-confidence) | 0 |
| S3_vs_k141_115708\|\|full | 6757 | No | NA | 7 | 0 | 3 | Not-determined | Genome-fragment | NA | NA | 0 |
| S3_vs_k141_91912\|\|full | 2976 | No | NA | 4 | 0 | 3 | Not-determined | Genome-fragment | NA | NA | 0 |
| S3_vs_k141_52674\|\|full | 2559 | No | NA | 4 | 0 | 0 | Not-determined | Genome-fragment | NA | NA | 0 |
| S6_vs_k141_29731\|\|lt2gene | 1692 | No | NA | 2 | 1 | 0 | Low-quality | Genome-fragment | 2.94 | HMM-based (lower-bound) | 0 |
| S8_vs_k141_143203\|\|lt2gene | 1692 | No | NA | 2 | 2 | 0 | Low-quality | Genome-fragment | 3.76 | AAI-based (high-confidence) | 0 |
| S17_vs_k141_14917\|\|lt2gene | 1693 | No | NA | 1 | 1 | 0 | Low-quality | Genome-fragment | 2.12 | AAI-based (high-confidence) | 0 |
| S3_vs_k141_99563\|\|full | 4396 | No | NA | 4 | 0 | 1 | Not-determined | Genome-fragment | NA | NA | 0 |
| S3_vs_k141_46739\|\|full | 1693 | No | NA | 3 | 0 | 0 | Low-quality | Genome-fragment | 3.45 | AAI-based (medium-confidence) | 0 |
| S3_vs_k141_91933\|\|full | 1742 | No | NA | 2 | 0 | 1 | Not-determined | Genome-fragment | NA | NA | 0 |
| S3_vs_k141_67367\|\|full | 1184 | No | NA | 3 | 0 | 1 | Not-determined | Genome-fragment | NA | NA | 0 |
| S5_vs_k141_142596\|\|full | 1693 | No | NA | 2 | 1 | 0 | Low-quality | Genome-fragment | 3.68 | AAI-based (high-confidence) | 0 |
| S3_vs_k141_123263\|\|full | 1896 | No | NA | 2 | 0 | 2 | Not-determined | Genome-fragment | NA | NA | 0 |

| | | | | | | | | | | | |
|---|---|---|---|---|---|---|---|---|---|---|---|
| S5_vs_k141_115304\|\|full | 1693 | No | NA | 4 | 2 | 0 | Low-quality | Genome-fragment | 4.39 | AAI-based (medium-confidence) | 0 |
| S3_vs_k141_123272\|\|full | 5652 | No | NA | 5 | 0 | 5 | Not-determined | Genome-fragment | NA | NA | 0 |
| S7_vs_13505\|\|full | 1693 | No | NA | 6 | 0 | 0 | Low-quality | Genome-fragment | 3.41 | AAI-based (high-confidence) | 0 |
| S16_extracted_112544 | 1693 | No | NA | 2 | 0 | 0 | Low-quality | Genome-fragment | 3.08 | AAI-based (high-confidence) | 0 |
| S16_vs_166190\|\|full | 1693 | No | NA | 4 | 0 | 0 | Low-quality | Genome-fragment | 3.93 | AAI-based (high-confidence) | 0 |
| S12_extracted_k141_83863 | 1693 | No | NA | 5 | 0 | 0 | Low-quality | Genome-fragment | 4.19 | AAI-based (medium-confidence) | 0 |
| S4_extracted_7732 | 1693 | No | NA | 4 | 0 | 0 | Low-quality | Genome-fragment | 3.07 | AAI-based (high-confidence) | 0 |
| S6_vs_k141_48689\|\|full | 1693 | No | NA | 5 | 1 | 0 | Low-quality | Genome-fragment | 3.09 | AAI-based (high-confidence) | 0 |
| S8_vs_k141_204192\|\|full | 1693 | No | NA | 2 | 0 | 1 | Low-quality | Genome-fragment | 1.33 | AAI-based (high-confidence) | 0 |
| S11_vs_k141_54260\|\|full | 1694 | No | NA | 6 | 1 | 0 | Low-quality | Genome-fragment | 2.99 | AAI-based (medium-confidence) | 0 |
| S3_vs_k141_44127\|\|full | 1266 | No | NA | 3 | 0 | 2 | Not-determined | Genome-fragment | NA | NA | 0 |
| S1_vs_k141_54513\|\|full | 1694 | No | NA | 4 | 1 | 0 | Low-quality | Genome-fragment | 3.79 | AAI-based (high-confidence) | 0 |
| S5_vs_k141_3151\|\|full | 1694 | No | NA | 3 | 0 | 0 | Low-quality | Genome-fragment | 0.41 | AAI-based (medium-confidence) | 0 |
| S3_vs_k141_115809\|\|full | 7091 | No | NA | 2 | 0 | 2 | Not-determined | Genome-fragment | NA | NA | 0 |
| S16_vs_14585\|\|full | 1694 | No | NA | 2 | 0 | 0 | Low-quality | Genome-fragment | 2.89 | AAI-based (high-confidence) | 0 |
| S3_vs_k141_13646\|\|full | 2266 | No | NA | 3 | 0 | 0 | Not-determined | Genome-fragment | NA | NA | 0 |
| S16_vs_88403\|\|full | 1694 | No | NA | 3 | 0 | 1 | Low-quality | Genome-fragment | 2.87 | AAI-based (medium-confidence) | 0 |
| S16_vs_91689\|\|full | 1694 | No | NA | 5 | 1 | 0 | Low-quality | Genome-fragment | 5.05 | AAI-based (high-confidence) | 0 |
| S12_extracted_k141_153598 | 1694 | No | NA | 1 | 0 | 1 | Low-quality | Genome-fragment | 1.64 | AAI-based (medium-confidence) | 0 |
| S3_vs_k141_115853\|\|full | 6481 | No | NA | 3 | 0 | 3 | Not-determined | Genome-fragment | NA | NA | 0 |
| S3_vs_k141_91983\|\|full | 2013 | No | NA | 3 | 0 | 2 | Not-determined | Genome-fragment | NA | NA | 0 |
| S12_extracted_k141_130222 | 1694 | No | NA | 4 | 1 | 0 | Low-quality | Genome-fragment | 1.21 | AAI-based (high-confidence) | 0 |
| S3_vs_k141_91986\|\|full | 1550 | No | NA | 3 | 0 | 0 | Not-determined | Genome-fragment | NA | NA | 0 |
| S3_vs_k141_76769\|\|full | 2166 | No | NA | 2 | 0 | 1 | Not-determined | Genome-fragment | NA | NA | 0 |
| S3_vs_k141_91987\|\|full | 1693 | No | NA | 3 | 0 | 1 | Not-determined | Genome-fragment | NA | NA | 0 |
| S3_vs_k141_52857\|\|full | 5689 | No | NA | 5 | 0 | 4 | Not-determined | Genome-fragment | NA | NA | 0 |
| S6_vs_k141_10591\|\|full | 1694 | No | NA | 5 | 0 | 0 | Low-quality | Genome-fragment | 4.1 | AAI-based (medium-confidence) | 0 |
| S3_vs_k141_52880\|\|full | 3658 | No | NA | 4 | 0 | 4 | Not-determined | Genome-fragment | NA | NA | 0 |
| S3_vs_k141_28165\|\|full | 7295 | No | NA | 5 | 0 | 0 | Not-determined | Genome-fragment | NA | NA | 0 |
| S3_vs_k141_44226\|\|full | 1039 | No | NA | 2 | 0 | 0 | Not-determined | Genome-fragment | NA | NA | 0 |
| S8_vs_k141_184380\|\|full | 1694 | No | NA | 3 | 0 | 0 | Low-quality | Genome-fragment | 3.28 | AAI-based (medium-confidence) | 0 |
| S3_vs_k141_92008\|\|full | 1328 | No | NA | 3 | 0 | 1 | Not-determined | Genome-fragment | NA | NA | 0 |
| S3_extracted_k141_101756 | 1695 | No | NA | 4 | 0 | 0 | Low-quality | Genome-fragment | 2.83 | AAI-based (high-confidence) | 0 |
| S3_vs_k141_9152\|\|full | 1695 | No | NA | 4 | 2 | 0 | Low-quality | Genome-fragment | 4.28 | AAI-based (high-confidence) | 0 |
| S3_vs_k141_67522\|\|full | 2038 | No | NA | 3 | 0 | 1 | Not-determined | Genome-fragment | NA | NA | 0 |
| S5_vs_k141_49297\|\|full | 1695 | No | NA | 5 | 0 | 0 | Low-quality | Genome-fragment | 3.5 | AAI-based (high-confidence) | 0 |
| S7_vs_47411\|\|full | 1695 | No | NA | 3 | 1 | 0 | Low-quality | Genome-fragment | 2.77 | HMM-based (lower-bound) | 0 |
| S3_vs_k141_123509\|\|full | 5112 | No | NA | 2 | 0 | 1 | Not-determined | Genome-fragment | NA | NA | 0 |
| S12_extracted_k141_16672 | 1695 | No | NA | 2 | 0 | 0 | Low-quality | Genome-fragment | 3.43 | AAI-based (high-confidence) | 0 |
| S3_vs_k141_19454\|\|full | 1205 | No | NA | 2 | 0 | 0 | Not-determined | Genome-fragment | NA | NA | 0 |
| S3_vs_k141_108062\|\|full | 1982 | No | NA | 3 | 0 | 0 | Not-determined | Genome-fragment | NA | NA | 0 |
| S3_vs_k141_4237\|\|full | 1413 | No | NA | 3 | 0 | 2 | Not-determined | Genome-fragment | NA | NA | 0 |
| S8_extracted_k141_10689 | 1695 | No | NA | 3 | 0 | 0 | Low-quality | Genome-fragment | 3.53 | AAI-based (medium-confidence) | 0 |
| S14_extracted_k141_79630 | 1696 | No | NA | 2 | 0 | 0 | Low-quality | Genome-fragment | 1.75 | AAI-based (medium-confidence) | 0 |
| S1_extracted_k141_143914 | 1696 | No | NA | 1 | 0 | 1 | Low-quality | Genome-fragment | 2.05 | AAI-based (high-confidence) | 0 |
| S8_extracted_k141_125943 | 1696 | No | NA | 2 | 0 | 0 | Low-quality | Genome-fragment | 4.07 | AAI-based (high-confidence) | 0 |
| S3_vs_k141_36538\|\|full | 5554 | No | NA | 4 | 0 | 3 | Not-determined | Genome-fragment | NA | NA | 0 |
| S17_vs_k141_14894\|\|lt2gene | 1697 | No | NA | 1 | 1 | 0 | Low-quality | Genome-fragment | 2.13 | AAI-based (high-confidence) | 0 |
| S11_vs_k141_58389\|\|lt2gene | 1697 | No | NA | 2 | 2 | 0 | Low-quality | Genome-fragment | 4.63 | AAI-based (high-confidence) | 0 |
| S14_vs_k141_30431\|\|full | 1697 | No | NA | 3 | 1 | 0 | Low-quality | Genome-fragment | 4.64 | AAI-based (high-confidence) | 0 |
| S3_vs_k141_108086\|\|full | 1622 | No | NA | 2 | 0 | 0 | Not-determined | Genome-fragment | NA | NA | 0 |
| S3_vs_k141_76950\|\|full | 1454 | No | NA | 3 | 0 | 0 | Not-determined | Genome-fragment | NA | NA | 0 |
| S1_vs_k141_13206\|\|full | 1697 | No | NA | 1 | 0 | 1 | Low-quality | Genome-fragment | 0.78 | AAI-based (high-confidence) | 0 |
| S3_vs_k141_44372\|\|full | 6323 | No | NA | 8 | 0 | 4 | Not-determined | Genome-fragment | NA | NA | 0 |
| S3_vs_k141_67582\|\|full | 3073 | No | NA | 4 | 0 | 3 | Not-determined | Genome-fragment | NA | NA | 0 |
| S3_extracted_k141_108412 | 1697 | No | NA | 1 | 1 | 0 | Low-quality | Genome-fragment | 0.93 | HMM-based (lower-bound) | 0 |
| S3_vs_k141_84035\|\|full | 1015 | No | NA | 3 | 0 | 0 | Not-determined | Genome-fragment | NA | NA | 0 |
| S3_vs_k141_51008\|\|full | 1697 | No | NA | 4 | 1 | 0 | Low-quality | Genome-fragment | 35.53 | AAI-based (high-confidence) | 0 |
| S3_vs_k141_36596\|\|full | 1629 | No | NA | 4 | 0 | 0 | Not-determined | Genome-fragment | NA | NA | 0 |
| S7_vs_110039\|\|full | 1697 | No | NA | 2 | 0 | 0 | Low-quality | Genome-fragment | 0.82 | AAI-based (medium-confidence) | 0 |
| S12_vs_k141_106785\|\|full | 1697 | No | NA | 5 | 0 | 0 | Low-quality | Genome-fragment | 3 | AAI-based (high-confidence) | 0 |
| S6_extracted_k141_681 | 1697 | No | NA | 2 | 0 | 0 | Low-quality | Genome-fragment | 1.6 | AAI-based (high-confidence) | 0 |
| S3_vs_k141_28333\|\|full | 2230 | No | NA | 3 | 0 | 0 | Not-determined | Genome-fragment | NA | NA | 0 |
| S3_vs_k141_4313\|\|full | 2729 | No | NA | 4 | 0 | 3 | Not-determined | Genome-fragment | NA | NA | 0 |
| S6_vs_k141_29353\|\|full | 1697 | No | NA | 4 | 0 | 4 | Low-quality | Genome-fragment | 2.66 | AAI-based (medium-confidence) | 0 |
| S8_vs_k141_56476\|\|lt2gene | 1697 | No | NA | 2 | 2 | 0 | Low-quality | Genome-fragment | 3.92 | AAI-based (medium-confidence) | 0 |
| S17_vs_k141_132658\|\|full | 1698 | No | NA | 4 | 0 | 0 | Low-quality | Genome-fragment | 3.61 | AAI-based (medium-confidence) | 0 |
| S9_vs_k141_39761\|\|full | 1698 | No | NA | 3 | 1 | 0 | Low-quality | Genome-fragment | 2.82 | AAI-based (high-confidence) | 0 |
| S11_vs_k141_63113\|\|full | 1698 | No | NA | 2 | 0 | 1 | Low-quality | Genome-fragment | 0.67 | AAI-based (medium-confidence) | 0 |
| S13_extracted_40401 | 1698 | Yes | 1188 | 3 | 1 | 1 | Low-quality | Genome-fragment | 0.68 | HMM-based (lower-bound) | 30.04 |
| S13_vs_114017\|\|full | 1698 | No | NA | 3 | 0 | 0 | Low-quality | Genome-fragment | 4.23 | AAI-based (high-confidence) | 0 |
| S14_vs_k141_74311\|\|lt2gene | 1698 | No | NA | 1 | 1 | 0 | Low-quality | Genome-fragment | 7.7 | AAI-based (medium-confidence) | 0 |
| S1_vs_k141_20806\|\|full | 1698 | No | NA | 3 | 0 | 0 | Low-quality | Genome-fragment | 1.22 | AAI-based (medium-confidence) | 0 |
| S3_vs_k141_77058\|\|full | 2250 | No | NA | 2 | 0 | 1 | Not-determined | Genome-fragment | NA | NA | 0 |
| S3_vs_k141_18770\|\|full | 1698 | No | NA | 4 | 1 | 0 | Low-quality | Genome-fragment | 0.96 | AAI-based (high-confidence) | 0 |
| S7_vs_52737\|\|full | 1698 | No | NA | 5 | 1 | 0 | Low-quality | Genome-fragment | 2.84 | AAI-based (medium-confidence) | 0 |
| S7_vs_81950\|\|full | 1698 | No | NA | 3 | 0 | 0 | Low-quality | Genome-fragment | 3.18 | AAI-based (high-confidence) | 0 |
| S3_vs_k141_116238\|\|full | 1198 | No | NA | 2 | 0 | 1 | Not-determined | Genome-fragment | NA | NA | 0 |
| S16_extracted_42264 | 1698 | No | NA | 4 | 1 | 0 | Low-quality | Genome-fragment | 4.69 | AAI-based (high-confidence) | 0 |
| S10_vs_56012\|\|full | 1698 | No | NA | 5 | 1 | 0 | Low-quality | Genome-fragment | 3.78 | AAI-based (medium-confidence) | 0 |
| S6_vs_k141_17521\|\|lt2gene | 1698 | No | NA | 2 | 2 | 0 | Low-quality | Genome-fragment | 4.45 | AAI-based (medium-confidence) | 0 |
| S8_extracted_k141_70868 | 1698 | No | NA | 4 | 2 | 0 | Low-quality | Genome-fragment | 2.1 | AAI-based (high-confidence) | 0 |
| S8_extracted_k141_167894 | 1698 | No | NA | 2 | 0 | 1 | Low-quality | Genome-fragment | 1.98 | AAI-based (medium-confidence) | 0 |
| S8_extracted_k141_161339 | 1698 | No | NA | 1 | 0 | 1 | Low-quality | Genome-fragment | 0.16 | AAI-based (medium-confidence) | 0 |
| S3_vs_k141_77156\|\|full | 5941 | No | NA | 9 | 3 | 0 | High-quality | High-quality | 100 | AAI-based (high-confidence) | 0 |
| S3_vs_k141_77163\|\|full | 1614 | No | NA | 4 | 0 | 0 | Not-determined | Genome-fragment | NA | NA | 0 |
| S3_vs_k141_14173\|\|full | 5863 | No | NA | 8 | 0 | 2 | Not-determined | Genome-fragment | NA | NA | 0 |
| S3_vs_k141_116288\|\|full | 2536 | No | NA | 4 | 0 | 0 | Not-determined | Genome-fragment | NA | NA | 0 |
| S8_vs_k141_65217\|\|full | 1698 | No | NA | 3 | 0 | 2 | Low-quality | Genome-fragment | 2.43 | AAI-based (medium-confidence) | 0 |
| S3_vs_k141_92317\|\|full | 1655 | No | NA | 3 | 0 | 0 | Not-determined | Genome-fragment | NA | NA | 0 |
| S3_vs_k141_99678\|\|full | 1699 | No | NA | 5 | 0 | 0 | Low-quality | Genome-fragment | 2.85 | AAI-based (high-confidence) | 0 |
| S5_vs_k141_99240\|\|full | 1699 | No | NA | 3 | 0 | 1 | Low-quality | Genome-fragment | 2.63 | AAI-based (medium-confidence) | 0 |
| S16_extracted_29646 | 1699 | No | NA | 3 | 0 | 2 | Low-quality | Genome-fragment | 1.04 | AAI-based (medium-confidence) | 0 |
| S16_vs_45973\|\|full | 1699 | No | NA | 4 | 0 | 0 | Low-quality | Genome-fragment | 2.85 | AAI-based (medium-confidence) | 0 |
| S3_vs_k141_123696\|\|full | 1641 | No | NA | 2 | 0 | 1 | Not-determined | Genome-fragment | NA | NA | 0 |
| S10_vs_58908\|\|full | 1699 | Yes | 1461 | 5 | 1 | 1 | Low-quality | Genome-fragment | 2.59 | AAI-based (high-confidence) | 14.01 |
| S3_vs_k141_84349\|\|full | 2900 | No | NA | 2 | 0 | 1 | Not-determined | Genome-fragment | NA | NA | 0 |
| S12_vs_k141_128863\|\|full | 1699 | No | NA | 5 | 1 | 0 | Low-quality | Genome-fragment | 3.76 | AAI-based (high-confidence) | 0 |
| S15_vs_70415\|\|full | 1699 | No | NA | 2 | 0 | 1 | Low-quality | Genome-fragment | 3.24 | AAI-based (medium-confidence) | 0 |
| S3_vs_k141_61189\|\|full | 1795 | No | NA | 3 | 0 | 2 | Not-determined | Genome-fragment | NA | NA | 0 |
| S2_vs_k141_7421\|\|full | 1699 | No | NA | 4 | 1 | 0 | Low-quality | Genome-fragment | 2.82 | AAI-based (high-confidence) | 0 |
| S3_vs_k141_92447\|\|full | 12356 | No | NA | 13 | 0 | 3 | Not-determined | Genome-fragment | NA | NA | 0 |
| S8_extracted_k141_54104 | 1699 | No | NA | 3 | 1 | 0 | Low-quality | Genome-fragment | 1.06 | AAI-based (high-confidence) | 0 |
| S8_vs_k141_61559\|\|full | 1699 | No | NA | 3 | 0 | 0 | Low-quality | Genome-fragment | 3.67 | AAI-based (medium-confidence) | 0 |
| S9_vs_k141_19803\|\|full | 1700 | No | NA | 4 | 3 | 0 | Low-quality | Genome-fragment | 2.78 | AAI-based (high-confidence) | 0 |
| S5_vs_k141_24045\|\|full | 1700 | No | NA | 6 | 1 | 0 | Low-quality | Genome-fragment | 4.94 | AAI-based (high-confidence) | 0 |
| S3_vs_k141_61269\|\|full | 2386 | No | NA | 4 | 0 | 2 | Not-determined | Genome-fragment | NA | NA | 0 |
| S3_vs_k141_77315\|\|full | 1006 | No | NA | 2 | 0 | 1 | Not-determined | Genome-fragment | NA | NA | 0 |
| S3_vs_k141_92488\|\|full | 1317 | No | NA | 4 | 0 | 0 | Not-determined | Genome-fragment | NA | NA | 0 |
| S3_vs_k141_99939\|\|full | 6144 | No | NA | 5 | 0 | 3 | Not-determined | Genome-fragment | NA | NA | 0 |
| S3_vs_k141_84481\|\|full | 1142 | No | NA | 2 | 0 | 1 | Not-determined | Genome-fragment | NA | NA | 0 |
| S3_vs_k141_99968\|\|full | 1906 | No | NA | 2 | 0 | 2 | Not-determined | Genome-fragment | NA | NA | 0 |
| S2_vs_k141_19995\|\|full | 1700 | No | NA | 2 | 0 | 2 | Low-quality | Genome-fragment | 1.5 | AAI-based (medium-confidence) | 0 |
| S3_vs_k141_61340\|\|full | 1148 | No | NA | 2 | 0 | 1 | Not-determined | Genome-fragment | NA | NA | 0 |
| S9_vs_k141_13590\|\|full | 1701 | No | NA | 3 | 0 | 0 | Low-quality | Genome-fragment | 0.67 | AAI-based (medium-confidence) | 0 |
| S3_vs_k141_123783\|\|full | 1575 | No | NA | 2 | 0 | 1 | Not-determined | Genome-fragment | NA | NA | 0 |
| S14_vs_k141_113479\|\|full | 1701 | No | NA | 2 | 1 | 0 | Low-quality | Genome-fragment | 4.24 | AAI-based (medium-confidence) | 0 |
| S1_vs_k141_7765\|\|lt2gene | 1701 | No | NA | 1 | 1 | 0 | Low-quality | Genome-fragment | 1.18 | HMM-based (lower-bound) | 0 |
| S7_extracted_21815 | 1701 | No | NA | 3 | 1 | 0 | Low-quality | Genome-fragment | 2.24 | HMM-based (lower-bound) | 0 |
| S3_vs_k141_116409\|\|full | 4535 | No | NA | 6 | 0 | 3 | Not-determined | Genome-fragment | NA | NA | 0 |
| S3_vs_k141_36826\|\|full | 3878 | No | NA | 3 | 0 | 2 | Not-determined | Genome-fragment | NA | NA | 0 |
| S3_vs_k141_84571\|\|full | 1944 | No | NA | 3 | 0 | 1 | Not-determined | Genome-fragment | NA | NA | 0 |
| S3_vs_k141_61493\|\|full | 3845 | No | NA | 5 | 0 | 4 | Not-determined | Genome-fragment | NA | NA | 0 |
| S7_extracted_56111 | 1701 | No | NA | 4 | 1 | 0 | Low-quality | Genome-fragment | 3.25 | AAI-based (medium-confidence) | 0 |
| S3_vs_k141_67962\|\|full | 6711 | No | NA | 4 | 0 | 2 | Not-determined | Genome-fragment | NA | NA | 0 |
| S16_vs_68118\|\|full | 1701 | No | NA | 3 | 0 | 0 | Low-quality | Genome-fragment | 3.01 | AAI-based (high-confidence) | 0 |
| S16_vs_98257\|\|full | 1701 | No | NA | 4 | 0 | 0 | Low-quality | Genome-fragment | 3.07 | AAI-based (high-confidence) | 0 |
| S3_vs_k141_123834\|\|full | 37186 | No | NA | 54 | 19 | 1 | High-quality | High-quality | 98.16 | AAI-based (high-confidence) | 0 |
| S10_vs_55138\|\|full | 1701 | No | NA | 2 | 0 | 1 | Low-quality | Genome-fragment | 1.03 | AAI-based (medium-confidence) | 0 |
| S15_vs_57434\|\|full | 1701 | No | NA | 3 | 0 | 0 | Low-quality | Genome-fragment | 2.93 | AAI-based (high-confidence) | 0 |
| S3_vs_k141_20099\|\|full | 3513 | No | NA | 2 | 0 | 1 | Not-determined | Genome-fragment | NA | NA | 0 |
| S3_vs_k141_61534\|\|full | 3530 | No | NA | 6 | 0 | 1 | Not-determined | Genome-fragment | NA | NA | 0 |
| S3_vs_k141_123889\|\|full | 3561 | No | NA | 4 | 0 | 3 | Not-determined | Genome-fragment | NA | NA | 0 |
| S2_extracted_k141_5217 | 1701 | No | NA | 4 | 4 | 0 | Low-quality | Genome-fragment | 5.6 | AAI-based (high-confidence) | 0 |
| S3_vs_k141_4654\|\|full | 1291 | No | NA | 3 | 0 | 1 | Not-determined | Genome-fragment | NA | NA | 0 |
| S2_vs_k141_1401\|\|full | 1701 | No | NA | 2 | 0 | 2 | Low-quality | Genome-fragment | 0.57 | AAI-based (medium-confidence) | 0 |
| S8_extracted_k141_129572 | 1701 | No | NA | 1 | 0 | 0 | Low-quality | Genome-fragment | 1.94 | AAI-based (medium-confidence) | 0 |
| S3_vs_k141_92587\|\|full | 1541 | No | NA | 3 | 0 | 0 | Not-determined | Genome-fragment | NA | NA | 0 |
| S11_vs_k141_85114\|\|full | 1702 | No | NA | 5 | 0 | 0 | Low-quality | Genome-fragment | 2.97 | AAI-based (medium-confidence) | 0 |
| S13_vs_65977\|\|full | 1702 | No | NA | 4 | 0 | 0 | Low-quality | Genome-fragment | 3.35 | AAI-based (high-confidence) | 0 |
| S14_vs_k141_6173\|\|full | 1702 | No | NA | 3 | 1 | 0 | Low-quality | Genome-fragment | 3.88 | AAI-based (medium-confidence) | 0 |
| S3_vs_k141_67976\|\|full | 2042 | No | NA | 2 | 0 | 1 | Not-determined | Genome-fragment | NA | NA | 0 |
| S1_extracted_k141_127729 | 1702 | No | NA | 2 | 0 | 0 | Low-quality | Genome-fragment | 1.29 | AAI-based (high-confidence) | 0 |
| S3_vs_k141_53425\|\|full | 2234 | No | NA | 3 | 0 | 1 | Not-determined | Genome-fragment | NA | NA | 0 |
| S3_vs_k141_123962\|\|full | 3388 | No | NA | 4 | 0 | 3 | Not-determined | Genome-fragment | NA | NA | 0 |
| S3_vs_k141_123965\|\|full | 1610 | No | NA | 2 | 0 | 2 | Not-determined | Genome-fragment | NA | NA | 0 |
| S3_vs_k141_49541\|\|full | 1702 | No | NA | 4 | 2 | 0 | Low-quality | Genome-fragment | 3.57 | AAI-based (high-confidence) | 0 |
| S3_vs_k141_93987\|\|full | 1702 | No | NA | 4 | 0 | 0 | Low-quality | Genome-fragment | 4.71 | AAI-based (medium-confidence) | 0 |
| S3_vs_k141_53444\|\|full | 3844 | No | NA | 2 | 0 | 0 | Not-determined | Genome-fragment | NA | NA | 0 |
| S3_vs_k141_123973\|\|full | 10858 | No | NA | 8 | 0 | 6 | Not-determined | Genome-fragment | NA | NA | 0 |
| S7_vs_24295\|\|full | 1702 | No | NA | 3 | 0 | 2 | Low-quality | Genome-fragment | 2.11 | AAI-based (high-confidence) | 0 |
| S3_vs_k141_84734\|\|full | 3213 | No | NA | 3 | 0 | 3 | Not-determined | Genome-fragment | NA | NA | 0 |
| S3_vs_k141_37033\|\|full | 6293 | No | NA | 6 | 0 | 3 | Not-determined | Genome-fragment | NA | NA | 0 |
| S3_vs_k141_100075\|\|full | 1300 | No | NA | 3 | 0 | 1 | Not-determined | Genome-fragment | NA | NA | 0 |
| S3_vs_k141_123981\|\|full | 1514 | No | NA | 3 | 0 | 0 | Not-determined | Genome-fragment | NA | NA | 0 |
| S16_vs_122928\|\|full | 1702 | No | NA | 2 | 0 | 0 | Low-quality | Genome-fragment | 4.84 | AAI-based (medium-confidence) | 0 |
| S4_extracted_20818 | 1702 | No | NA | 3 | 0 | 0 | Low-quality | Genome-fragment | 3.39 | AAI-based (high-confidence) | 0 |
| S6_vs_k141_5249\|\|full | 1702 | No | NA | 3 | 2 | 0 | Low-quality | Genome-fragment | 3.8 | AAI-based (high-confidence) | 0 |
| S8_extracted_k141_124903 | 1702 | No | NA | 4 | 0 | 0 | Low-quality | Genome-fragment | 1.42 | AAI-based (medium-confidence) | 0 |
| S11_vs_k141_69882\|\|full | 1703 | No | NA | 3 | 0 | 1 | Low-quality | Genome-fragment | 1.41 | AAI-based (medium-confidence) | 0 |
| S14_extracted_k141_36347 | 1703 | No | NA | 1 | 1 | 0 | Low-quality | Genome-fragment | 3.59 | AAI-based (high-confidence) | 0 |
| S3_vs_k141_14560\|\|full | 1155 | No | NA | 2 | 0 | 0 | Not-determined | Genome-fragment | NA | NA | 0 |
| S3_extracted_k141_125183 | 1703 | No | NA | 3 | 0 | 0 | Low-quality | Genome-fragment | 3.6 | AAI-based (medium-confidence) | 0 |
| S3_vs_k141_92728\|\|full | 1326 | No | NA | 5 | 0 | 0 | Not-determined | Genome-fragment | NA | NA | 0 |
| S3_vs_k141_100132\|\|full | 2550 | No | NA | 4 | 0 | 0 | Not-determined | Genome-fragment | NA | NA | 0 |
| S3_vs_k141_124019\|\|full | 5025 | No | NA | 6 | 0 | 3 | Not-determined | Genome-fragment | NA | NA | 0 |
| S3_vs_k141_100141\|\|full | 5544 | No | NA | 3 | 0 | 1 | Not-determined | Genome-fragment | NA | NA | 0 |
| S3_vs_k141_20315\|\|full | 36208 | No | NA | 46 | 15 | 1 | Medium-quality | Genome-fragment | 88.82 | AAI-based (high-confidence) | 0 |
| S3_vs_k141_124030\|\|full | 6703 | No | NA | 3 | 0 | 3 | Not-determined | Genome-fragment | NA | NA | 0 |
| S3_vs_k141_68031\|\|full | 80878 | Yes | 30744 | 109 | 15 | 37 | Medium-quality | Genome-fragment | 50.83 | AAI-based (high-confidence) | 61.99 |
| S3_vs_k141_100146\|\|full | 2312 | No | NA | 3 | 0 | 1 | Not-determined | Genome-fragment | NA | NA | 0 |
| S5_vs_k141_134286\|\|full | 1703 | No | NA | 3 | 1 | 0 | Low-quality | Genome-fragment | 4.9 | AAI-based (high-confidence) | 0 |
| S3_vs_k141_28793\|\|full | 3612 | No | NA | 3 | 0 | 2 | Not-determined | Genome-fragment | NA | NA | 0 |
| S3_vs_k141_37075\|\|full | 2094 | No | NA | 4 | 0 | 0 | Not-determined | Genome-fragment | NA | NA | 0 |
| S3_vs_k141_100157\|\|full | 1964 | No | NA | 2 | 0 | 0 | Not-determined | Genome-fragment | NA | NA | 0 |
| S7_vs_135483\|\|full | 1703 | No | NA | 3 | 0 | 0 | Low-quality | Genome-fragment | 4.6 | AAI-based (high-confidence) | 0 |
| S16_extracted_120732 | 1703 | Yes | 707 | 2 | 1 | 1 | Low-quality | Genome-fragment | 0.87 | AAI-based (medium-confidence) | 58.49 |
| S12_vs_k141_79047\|\|full | 1703 | No | NA | 2 | 0 | 1 | Low-quality | Genome-fragment | 1.01 | AAI-based (high-confidence) | 0 |
| S12_vs_k141_144629\|\|full | 1703 | No | NA | 8 | 1 | 0 | Low-quality | Genome-fragment | 1.64 | AAI-based (high-confidence) | 0 |
| S3_vs_k141_61673\|\|full | 1968 | No | NA | 3 | 0 | 2 | Not-determined | Genome-fragment | NA | NA | 0 |
| S3_vs_k141_84940\|\|full | 1968 | No | NA | 3 | 0 | 1 | Not-determined | Genome-fragment | NA | NA | 0 |
| S4_extracted_22291 | 1703 | No | NA | 3 | 1 | 0 | Low-quality | Genome-fragment | 4.27 | AAI-based (high-confidence) | 0 |
| S8_extracted_k141_157941 | 1703 | No | NA | 3 | 0 | 1 | Low-quality | Genome-fragment | 1.08 | AAI-based (high-confidence) | 0 |
| S8_vs_k141_96708\|\|full | 1703 | No | NA | 3 | 0 | 1 | Low-quality | Genome-fragment | 2.95 | AAI-based (medium-confidence) | 0 |
| S3_vs_k141_108658\|\|full | 2253 | No | NA | 3 | 0 | 0 | Not-determined | Genome-fragment | NA | NA | 0 |
| S3_vs_k141_108685\|\|full | 2410 | No | NA | 3 | 0 | 0 | Not-determined | Genome-fragment | NA | NA | 0 |
| S8_vs_k141_144090\|\|lt2gene | 1703 | No | NA | 2 | 2 | 0 | Low-quality | Genome-fragment | 5.07 | AAI-based (high-confidence) | 0 |
| S14_extracted_k141_101084 | 1704 | No | NA | 1 | 0 | 1 | Low-quality | Genome-fragment | 0.64 | AAI-based (high-confidence) | 0 |
| S3_extracted_k141_58058 | 1704 | No | NA | 3 | 1 | 0 | Low-quality | Genome-fragment | 7.77 | AAI-based (high-confidence) | 0 |
| S3_vs_k141_37159\|\|full | 4705 | No | NA | 6 | 0 | 0 | Not-determined | Genome-fragment | NA | NA | 0 |
| S5_vs_k141_141370\|\|full | 1704 | No | NA | 5 | 2 | 0 | Low-quality | Genome-fragment | 2.8 | AAI-based (high-confidence) | 0 |
| S3_vs_k141_53666\|\|full | 2642 | No | NA | 2 | 0 | 0 | Not-determined | Genome-fragment | NA | NA | 0 |
| S5_vs_k141_4649\|\|full | 1704 | No | NA | 4 | 0 | 0 | Low-quality | Genome-fragment | 2.16 | AAI-based (high-confidence) | 0 |
| S3_vs_k141_68204\|\|full | 2486 | No | NA | 3 | 0 | 1 | Not-determined | Genome-fragment | NA | NA | 0 |
| S16_extracted_101554 | 1704 | No | NA | 3 | 0 | 1 | Low-quality | Genome-fragment | 1.44 | AAI-based (high-confidence) | 0 |
| S3_vs_k141_20416\|\|full | 2294 | No | NA | 3 | 0 | 2 | Not-determined | Genome-fragment | NA | NA | 0 |
| S3_vs_k141_28857\|\|full | 2399 | No | NA | 2 | 0 | 2 | Not-determined | Genome-fragment | NA | NA | 0 |
| S3_vs_k141_84966\|\|full | 1779 | No | NA | 3 | 0 | 0 | Not-determined | Genome-fragment | NA | NA | 0 |
| S8_vs_k141_83859\|\|full | 1704 | No | NA | 5 | 0 | 0 | Low-quality | Genome-fragment | 3.07 | AAI-based (high-confidence) | 0 |
| S11_vs_k141_69001\|\|full | 1705 | No | NA | 3 | 1 | 0 | Low-quality | Genome-fragment | 2.25 | HMM-based (lower-bound) | 0 |
| S11_vs_k141_24722\|\|full | 1705 | No | NA | 3 | 2 | 0 | Low-quality | Genome-fragment | 4.32 | AAI-based (high-confidence) | 0 |
| S5_vs_k141_108342\|\|full | 1705 | No | NA | 3 | 0 | 2 | Low-quality | Genome-fragment | 0.98 | AAI-based (medium-confidence) | 0 |
| S16_extracted_36840 | 1705 | No | NA | 2 | 0 | 0 | Low-quality | Genome-fragment | 4.01 | AAI-based (medium-confidence) | 0 |
| S10_vs_55983\|\|full | 1705 | No | NA | 3 | 2 | 0 | Low-quality | Genome-fragment | 2.46 | HMM-based (lower-bound) | 0 |
| S10_vs_62248\|\|full | 1705 | No | NA | 4 | 1 | 0 | Low-quality | Genome-fragment | 3.01 | AAI-based (medium-confidence) | 0 |
| S12_vs_k141_43898\|\|full | 1705 | No | NA | 4 | 3 | 0 | Low-quality | Genome-fragment | 4.61 | AAI-based (high-confidence) | 0 |
| S6_vs_k141_38320\|\|full | 1705 | No | NA | 3 | 0 | 2 | Low-quality | Genome-fragment | 2.84 | AAI-based (medium-confidence) | 0 |
| S3_vs_k141_20447\|\|full | 2809 | No | NA | 4 | 0 | 1 | Not-determined | Genome-fragment | NA | NA | 0 |
| S3_vs_k141_100357\|\|full | 1785 | No | NA | 4 | 0 | 0 | Not-determined | Genome-fragment | NA | NA | 0 |
| S6_vs_k141_24187\|\|full | 1705 | No | NA | 5 | 0 | 0 | Low-quality | Genome-fragment | 3.8 | AAI-based (high-confidence) | 0 |
| S3_vs_k141_85019\|\|full | 1382 | No | NA | 3 | 0 | 0 | Not-determined | Genome-fragment | NA | NA | 0 |
| S3_vs_k141_116718\|\|full | 1324 | No | NA | 2 | 0 | 0 | Not-determined | Genome-fragment | NA | NA | 0 |
| S8_vs_k141_26712\|\|full | 1705 | No | NA | 4 | 1 | 0 | Low-quality | Genome-fragment | 3.6 | AAI-based (high-confidence) | 0 |
| S8_vs_k141_53664\|\|full | 1705 | No | NA | 2 | 0 | 1 | Low-quality | Genome-fragment | 2.43 | AAI-based (high-confidence) | 0 |
| S3_vs_k141_108863\|\|full | 6457 | No | NA | 12 | 0 | 1 | Not-determined | Genome-fragment | NA | NA | 0 |
| S3_vs_k141_21506\|\|full | 1706 | No | NA | 4 | 0 | 0 | Low-quality | Genome-fragment | 2.84 | AAI-based (high-confidence) | 0 |
| S5_vs_k141_89563\|\|full | 1706 | No | NA | 2 | 1 | 0 | Low-quality | Genome-fragment | 3.82 | AAI-based (high-confidence) | 0 |
| S12_vs_k141_140697\|\|full | 1706 | No | NA | 6 | 0 | 0 | Low-quality | Genome-fragment | 2.43 | AAI-based (medium-confidence) | 0 |
| S8_extracted_k141_175122 | 1706 | No | NA | 2 | 0 | 1 | Low-quality | Genome-fragment | 1.08 | AAI-based (medium-confidence) | 0 |
| S3_vs_k141_68319\|\|full | 1126 | No | NA | 3 | 0 | 1 | Not-determined | Genome-fragment | NA | NA | 0 |
| S8_extracted_k141_47742 | 1706 | No | NA | 3 | 1 | 0 | Low-quality | Genome-fragment | 2.49 | HMM-based (lower-bound) | 0 |
| S3_vs_k141_77921\|\|full | 1012 | No | NA | 3 | 0 | 2 | Not-determined | Genome-fragment | NA | NA | 0 |
| S3_vs_k141_4969\|\|full | 4241 | No | NA | 6 | 0 | 5 | Not-determined | Genome-fragment | NA | NA | 0 |
| S3_vs_k141_4972\|\|full | 1718 | No | NA | 3 | 0 | 1 | Not-determined | Genome-fragment | NA | NA | 0 |
| S8_extracted_k141_6298 | 1706 | No | NA | 5 | 0 | 0 | Low-quality | Genome-fragment | 5.27 | AAI-based (high-confidence) | 0 |
| S8_extracted_k141_128597 | 1706 | No | NA | 2 | 0 | 0 | Low-quality | Genome-fragment | 5.28 | AAI-based (high-confidence) | 0 |
| S3_vs_k141_68351\|\|full | 2256 | No | NA | 2 | 0 | 2 | Not-determined | Genome-fragment | NA | NA | 0 |
| S3_vs_k141_29006\|\|full | 1052 | No | NA | 3 | 0 | 1 | Not-determined | Genome-fragment | NA | NA | 0 |
| S3_vs_k141_68364\|\|full | 4906 | No | NA | 3 | 0 | 1 | Not-determined | Genome-fragment | NA | NA | 0 |
| S8_vs_k141_63840\|\|full | 1706 | No | NA | 3 | 0 | 2 | Low-quality | Genome-fragment | 0.65 | AAI-based (high-confidence) | 0 |
| S17_extracted_k141_37910 | 1707 | No | NA | 3 | 0 | 0 | Low-quality | Genome-fragment | 1.55 | AAI-based (high-confidence) | 0 |
| S11_extracted_k141_50940 | 1707 | No | NA | 4 | 1 | 0 | Low-quality | Genome-fragment | 3.03 | AAI-based (high-confidence) | 0 |
| S14_extracted_k141_47194 | 1707 | No | NA | 2 | 0 | 0 | Low-quality | Genome-fragment | 3.69 | AAI-based (high-confidence) | 0 |
| S3_vs_k141_116872\|\|full | 1255 | No | NA | 2 | 0 | 0 | Not-determined | Genome-fragment | NA | NA | 0 |
| S14_extracted_k141_13937 | 1707 | No | NA | 3 | 0 | 1 | Low-quality | Genome-fragment | 3.87 | AAI-based (high-confidence) | 0 |
| S3_vs_k141_119773\|\|lt2gene | 1707 | No | NA | 2 | 2 | 0 | Low-quality | Genome-fragment | 4.06 | AAI-based (high-confidence) | 0 |
| S7_vs_102087\|\|full | 1707 | No | NA | 4 | 0 | 0 | Low-quality | Genome-fragment | 2.83 | AAI-based (medium-confidence) | 0 |
| S3_vs_k141_20574\|\|full | 1627 | No | NA | 4 | 0 | 1 | Not-determined | Genome-fragment | NA | NA | 0 |
| S3_vs_k141_109009\|\|full | 2111 | No | NA | 5 | 0 | 1 | Not-determined | Genome-fragment | NA | NA | 0 |
| S16_vs_131717\|\|full | 1707 | No | NA | 4 | 1 | 0 | Low-quality | Genome-fragment | 3.35 | AAI-based (high-confidence) | 0 |
| S3_vs_k141_116887\|\|full | 3737 | No | NA | 4 | 0 | 4 | Not-determined | Genome-fragment | NA | NA | 0 |
| S3_vs_k141_20591\|\|full | 3291 | No | NA | 5 | 0 | 2 | Not-determined | Genome-fragment | NA | NA | 0 |
| S3_vs_k141_37376\|\|full | 2759 | No | NA | 3 | 0 | 1 | Not-determined | Genome-fragment | NA | NA | 0 |
| S3_vs_k141_68497\|\|full | 3026 | No | NA | 4 | 0 | 2 | Not-determined | Genome-fragment | NA | NA | 0 |
| S12_vs_k141_178148\|\|lt2gene | 1707 | No | NA | 2 | 2 | 0 | Low-quality | Genome-fragment | 4.46 | AAI-based (high-confidence) | 0 |
| S3_vs_k141_68507\|\|full | 3985 | No | NA | 8 | 0 | 0 | Not-determined | Genome-fragment | NA | NA | 0 |
| S3_vs_k141_85279\|\|full | 3067 | No | NA | 4 | 0 | 4 | Not-determined | Genome-fragment | NA | NA | 0 |
| S3_vs_k141_29077\|\|full | 28440 | No | NA | 44 | 19 | 0 | Medium-quality | Genome-fragment | 81.53 | AAI-based (high-confidence) | 0 |
| S3_vs_k141_85293\|\|full | 2611 | No | NA | 5 | 0 | 4 | Not-determined | Genome-fragment | NA | NA | 0 |
| S6_vs_k141_6405\|\|full | 1707 | Yes | 678 | 5 | 1 | 1 | Low-quality | Genome-fragment | 0.28 | HMM-based (lower-bound) | 60.28 |
| S8_extracted_k141_39726 | 1707 | No | NA | 3 | 0 | 0 | Low-quality | Genome-fragment | 3.75 | AAI-based (high-confidence) | 0 |
| S8_vs_k141_115794\|\|lt2gene | 1707 | No | NA | 2 | 1 | 0 | Low-quality | Genome-fragment | 4.5 | AAI-based (high-confidence) | 0 |
| S3_vs_k141_78067\|\|full | 1812 | No | NA | 4 | 0 | 1 | Not-determined | Genome-fragment | NA | NA | 0 |
| S3_vs_k141_53985\|\|full | 3519 | No | NA | 5 | 0 | 2 | Not-determined | Genome-fragment | NA | NA | 0 |
| S3_vs_k141_68556\|\|full | 2260 | No | NA | 2 | 0 | 1 | Not-determined | Genome-fragment | NA | NA | 0 |
| S3_vs_k141_5172\|\|full | 4439 | No | NA | 4 | 0 | 3 | Not-determined | Genome-fragment | NA | NA | 0 |
| S3_vs_k141_5211\|\|full | 1322 | No | NA | 3 | 0 | 0 | Not-determined | Genome-fragment | NA | NA | 0 |
| S3_vs_k141_5220\|\|full | 2162 | No | NA | 3 | 0 | 1 | Not-determined | Genome-fragment | NA | NA | 0 |
| S9_extracted_k141_48387 | 1708 | No | NA | 3 | 0 | 2 | Low-quality | Genome-fragment | 5.26 | AAI-based (high-confidence) | 0 |
| S9_vs_k141_64696\|\|full | 1708 | No | NA | 4 | 0 | 0 | Low-quality | Genome-fragment | 3.49 | AAI-based (medium-confidence) | 0 |
| S3_vs_k141_68591\|\|full | 2054 | No | NA | 3 | 0 | 0 | Not-determined | Genome-fragment | NA | NA | 0 |
| S3_vs_k141_116979\|\|full | 4934 | No | NA | 7 | 0 | 2 | Not-determined | Genome-fragment | NA | NA | 0 |
| S11_extracted_k141_90659 | 1708 | No | NA | 2 | 1 | 0 | Low-quality | Genome-fragment | 1.87 | HMM-based (lower-bound) | 0 |
| S3_vs_k141_15113\|\|full | 3345 | No | NA | 3 | 0 | 2 | Not-determined | Genome-fragment | NA | NA | 0 |
| S3_vs_k141_93084\|\|full | 3836 | No | NA | 4 | 0 | 1 | Not-determined | Genome-fragment | NA | NA | 0 |
| S13_vs_43911\|\|full | 1708 | No | NA | 2 | 0 | 1 | Low-quality | Genome-fragment | 2.2 | AAI-based (medium-confidence) | 0 |
| S14_extracted_k141_102820 | 1708 | No | NA | 4 | 4 | 0 | Low-quality | Genome-fragment | 5.64 | AAI-based (high-confidence) | 0 |
| S14_vs_k141_22667\|\|full | 1708 | No | NA | 3 | 0 | 1 | Low-quality | Genome-fragment | 4.02 | AAI-based (medium-confidence) | 0 |
| S3_vs_k141_62158\|\|full | 4733 | No | NA | 6 | 0 | 2 | Not-determined | Genome-fragment | NA | NA | 0 |
| S3_vs_k141_124412\|\|full | 3482 | No | NA | 5 | 0 | 4 | Not-determined | Genome-fragment | NA | NA | 0 |
| S3_vs_k141_93112\|\|full | 1502 | No | NA | 3 | 0 | 1 | Not-determined | Genome-fragment | NA | NA | 0 |
| S3_vs_k141_15176\|\|full | 1061 | No | NA | 2 | 0 | 1 | Not-determined | Genome-fragment | NA | NA | 0 |
| S16_extracted_23891 | 1708 | No | NA | 4 | 0 | 1 | Low-quality | Genome-fragment | 1.15 | AAI-based (high-confidence) | 0 |
| S16_vs_46579\|\|full | 1708 | No | NA | 3 | 0 | 0 | Low-quality | Genome-fragment | 0.97 | AAI-based (high-confidence) | 0 |
| S3_vs_k141_5287\|\|full | 1153 | No | NA | 3 | 0 | 0 | Not-determined | Genome-fragment | NA | NA | 0 |
| S16_vs_93161\|\|full | 1708 | No | NA | 4 | 0 | 0 | Low-quality | Genome-fragment | 1.25 | AAI-based (medium-confidence) | 0 |
| S16_vs_32070\|\|lt2gene | 1708 | No | NA | 2 | 2 | 0 | Low-quality | Genome-fragment | 5.77 | AAI-based (high-confidence) | 0 |
| S8_vs_k141_137048\|\|full | 1708 | No | NA | 3 | 1 | 0 | Low-quality | Genome-fragment | 34.28 | AAI-based (high-confidence) | 0 |
| S3_vs_k141_62246\|\|full | 1002 | No | NA | 3 | 0 | 2 | Not-determined | Genome-fragment | NA | NA | 0 |
| S3_vs_k141_85410\|\|full | 3217 | No | NA | 3 | 0 | 1 | Not-determined | Genome-fragment | NA | NA | 0 |
| S3_vs_k141_15223\|\|full | 1039 | No | NA | 2 | 0 | 0 | Not-determined | Genome-fragment | NA | NA | 0 |
| S11_vs_k141_87360\|\|full | 1709 | No | NA | 5 | 1 | 0 | Low-quality | Genome-fragment | 4.61 | AAI-based (medium-confidence) | 0 |
| S13_extracted_38612 | 1709 | No | NA | 2 | 0 | 0 | Low-quality | Genome-fragment | 1.14 | AAI-based (high-confidence) | 0 |
| S1_extracted_k141_32140 | 1709 | No | NA | 5 | 2 | 0 | Low-quality | Genome-fragment | 4.29 | AAI-based (high-confidence) | 0 |
| S3_vs_k141_124521\|\|full | 1556 | No | NA | 2 | 0 | 2 | Not-determined | Genome-fragment | NA | NA | 0 |
| S3_vs_k141_110714\|\|full | 1709 | No | NA | 2 | 0 | 0 | Low-quality | Genome-fragment | 1.06 | AAI-based (high-confidence) | 0 |
| S3_vs_k141_45147\|\|full | 90239 | No | NA | 141 | 16 | 1 | High-quality | High-quality | 100 | AAI-based (high-confidence) | 0 |
| S5_vs_k141_134558\|\|full | 1709 | No | NA | 3 | 0 | 0 | Low-quality | Genome-fragment | 1.69 | AAI-based (high-confidence) | 0 |
| S16_extracted_102596 | 1709 | No | NA | 3 | 1 | 0 | Low-quality | Genome-fragment | 3.81 | AAI-based (high-confidence) | 0 |
| S16_vs_52956\|\|full | 1709 | No | NA | 3 | 2 | 0 | Low-quality | Genome-fragment | 4.04 | AAI-based (high-confidence) | 0 |
| S3_vs_k141_20738\|\|full | 4046 | No | NA | 4 | 0 | 2 | Not-determined | Genome-fragment | NA | NA | 0 |
| S12_vs_k141_34084\|\|full | 1709 | No | NA | 3 | 1 | 0 | Low-quality | Genome-fragment | 4.12 | AAI-based (medium-confidence) | 0 |
| S15_extracted_45932 | 1709 | No | NA | 4 | 1 | 0 | Low-quality | Genome-fragment | 10.8 | AAI-based (medium-confidence) | 0 |
| S15_vs_21852\|\|full | 1709 | No | NA | 3 | 2 | 0 | Low-quality | Genome-fragment | 2.73 | HMM-based (lower-bound) | 0 |
| S3_vs_k141_124548\|\|full | 1062 | No | NA | 3 | 0 | 0 | Not-determined | Genome-fragment | NA | NA | 0 |
| S15_vs_51324\|\|full | 1709 | No | NA | 6 | 2 | 0 | Low-quality | Genome-fragment | 0.91 | AAI-based (high-confidence) | 0 |
| S3_vs_k141_29301\|\|full | 1665 | No | NA | 2 | 0 | 2 | Not-determined | Genome-fragment | NA | NA | 0 |
| S3_vs_k141_124568\|\|full | 1227 | No | NA | 2 | 0 | 1 | Not-determined | Genome-fragment | NA | NA | 0 |
| S3_vs_k141_68845\|\|full | 1071 | No | NA | 3 | 0 | 0 | Not-determined | Genome-fragment | NA | NA | 0 |
| S3_vs_k141_100582\|\|full | 1131 | No | NA | 3 | 0 | 1 | Not-determined | Genome-fragment | NA | NA | 0 |
| S2_vs_k141_4045\|\|full | 1709 | No | NA | 3 | 0 | 0 | Low-quality | Genome-fragment | 4.19 | AAI-based (high-confidence) | 0 |
| S3_vs_k141_29318\|\|full | 2808 | No | NA | 5 | 0 | 0 | Not-determined | Genome-fragment | NA | NA | 0 |
| S8_extracted_k141_124121 | 1709 | No | NA | 1 | 0 | 1 | Low-quality | Genome-fragment | 5.46 | AAI-based (high-confidence) | 0 |
| S3_vs_k141_124574\|\|full | 20388 | No | NA | 13 | 0 | 7 | Not-determined | Genome-fragment | NA | NA | 0 |
| S3_vs_k141_85586\|\|full | 2031 | No | NA | 2 | 0 | 0 | Not-determined | Genome-fragment | NA | NA | 0 |
| S17_extracted_k141_72831 | 1710 | No | NA | 2 | 2 | 0 | Low-quality | Genome-fragment | 7.8 | AAI-based (high-confidence) | 0 |
| S17_vs_k141_33216\|\|full | 1710 | No | NA | 3 | 0 | 2 | Low-quality | Genome-fragment | 0.48 | AAI-based (medium-confidence) | 0 |
| S3_vs_k141_97153\|\|full | 1710 | No | NA | 5 | 2 | 1 | Low-quality | Genome-fragment | 4.12 | AAI-based (high-confidence) | 0 |
| S5_vs_k141_130481\|\|full | 1710 | No | NA | 3 | 0 | 0 | Low-quality | Genome-fragment | 4.38 | AAI-based (high-confidence) | 0 |
| S3_vs_k141_29387\|\|full | 1482 | No | NA | 3 | 0 | 1 | Not-determined | Genome-fragment | NA | NA | 0 |
| S3_vs_k141_62410\|\|full | 33206 | No | NA | 62 | 20 | 1 | High-quality | High-quality | 91.88 | AAI-based (medium-confidence) | 0 |
| S7_extracted_687 | 1710 | No | NA | 5 | 3 | 0 | Low-quality | Genome-fragment | 3.9 | AAI-based (high-confidence) | 0 |
| S10_extracted_35430 | 1710 | No | NA | 4 | 2 | 0 | Low-quality | Genome-fragment | 2.95 | AAI-based (high-confidence) | 0 |
| S8_vs_k141_106724\|\|full | 1710 | No | NA | 3 | 0 | 2 | Low-quality | Genome-fragment | 2.32 | AAI-based (medium-confidence) | 0 |
| S3_vs_k141_100689\|\|full | 1430 | No | NA | 3 | 0 | 3 | Not-determined | Genome-fragment | NA | NA | 0 |
| S17_vs_k141_159558\|\|full | 1711 | No | NA | 4 | 3 | 0 | Low-quality | Genome-fragment | 2.81 | AAI-based (high-confidence) | 0 |
| S14_vs_k141_34258\|\|lt2gene | 1711 | No | NA | 2 | 1 | 0 | Low-quality | Genome-fragment | 3.68 | AAI-based (high-confidence) | 0 |
| S5_vs_k141_13823\|\|full | 1711 | No | NA | 2 | 0 | 0 | Low-quality | Genome-fragment | 1.07 | AAI-based (medium-confidence) | 0 |
| S7_vs_9127\|\|full | 1711 | No | NA | 2 | 2 | 0 | Low-quality | Genome-fragment | 2.77 | AAI-based (medium-confidence) | 0 |
| S16_extracted_150715 | 1711 | No | NA | 2 | 0 | 0 | Low-quality | Genome-fragment | 5.33 | AAI-based (high-confidence) | 0 |
| S3_vs_k141_109514\|\|full | 3848 | No | NA | 3 | 0 | 0 | Not-determined | Genome-fragment | NA | NA | 0 |
| S10_vs_36678\|\|full | 1711 | No | NA | 2 | 0 | 0 | Low-quality | Genome-fragment | 3.17 | AAI-based (medium-confidence) | 0 |
| S8_extracted_k141_164068 | 1711 | No | NA | 2 | 0 | 2 | Low-quality | Genome-fragment | 1.67 | AAI-based (medium-confidence) | 0 |
| S17_vs_k141_71183\|\|full | 1712 | No | NA | 4 | 0 | 1 | Low-quality | Genome-fragment | 1.34 | AAI-based (high-confidence) | 0 |
| S17_vs_k141_87026\|\|full | 1712 | No | NA | 2 | 0 | 0 | Low-quality | Genome-fragment | 2.49 | AAI-based (medium-confidence) | 0 |
| S9_vs_k141_77323\|\|full | 1712 | No | NA | 3 | 1 | 0 | Low-quality | Genome-fragment | 3.1 | AAI-based (medium-confidence) | 0 |
| S3_vs_k141_15382\|\|full | 2348 | No | NA | 3 | 0 | 0 | Not-determined | Genome-fragment | NA | NA | 0 |
| S3_vs_k141_54149\|\|full | 2622 | No | NA | 7 | 0 | 0 | Not-determined | Genome-fragment | NA | NA | 0 |
| S3_vs_k141_45367\|\|full | 2581 | No | NA | 3 | 0 | 3 | Not-determined | Genome-fragment | NA | NA | 0 |
| S11_extracted_k141_19638 | 1712 | No | NA | 4 | 0 | 0 | Low-quality | Genome-fragment | 3.82 | AAI-based (high-confidence) | 0 |
| S3_vs_k141_45396\|\|full | 1228 | No | NA | 2 | 0 | 1 | Not-determined | Genome-fragment | NA | NA | 0 |
| S3_vs_k141_109623\|\|full | 1143 | No | NA | 3 | 0 | 1 | Not-determined | Genome-fragment | NA | NA | 0 |
| S3_vs_k141_100779\|\|full | 1132 | No | NA | 2 | 0 | 0 | Not-determined | Genome-fragment | NA | NA | 0 |
| S11_vs_k141_46432\|\|full | 1712 | No | NA | 3 | 0 | 0 | Low-quality | Genome-fragment | 3.17 | AAI-based (high-confidence) | 0 |
| S1_vs_k141_59872\|\|lt2gene | 1712 | No | NA | 1 | 1 | 0 | Low-quality | Genome-fragment | 3.56 | AAI-based (high-confidence) | 0 |
| S3_vs_k141_124665\|\|full | 2586 | No | NA | 3 | 0 | 2 | Not-determined | Genome-fragment | NA | NA | 0 |
| S3_vs_k141_117154\|\|full | 62603 | No | NA | 63 | 0 | 18 | Not-determined | Genome-fragment | NA | NA | 0 |
| S3_vs_k141_36719\|\|lt2gene | 1712 | No | NA | 1 | 1 | 0 | Low-quality | Genome-fragment | 3.65 | AAI-based (high-confidence) | 0 |
| S16_vs_169594\|\|lt2gene | 1712 | No | NA | 1 | 1 | 0 | Low-quality | Genome-fragment | 2.89 | AAI-based (high-confidence) | 0 |
| S12_extracted_k141_126736 | 1712 | No | NA | 1 | 0 | 0 | Low-quality | Genome-fragment | 2.66 | AAI-based (high-confidence) | 0 |
| S3_vs_k141_78400\|\|full | 1091 | No | NA | 3 | 0 | 0 | Not-determined | Genome-fragment | NA | NA | 0 |
| S12_vs_k141_10980\|\|full | 1712 | No | NA | 4 | 3 | 0 | Low-quality | Genome-fragment | 4.32 | AAI-based (high-confidence) | 0 |
| S15_vs_61757\|\|full | 1712 | No | NA | 2 | 2 | 0 | Low-quality | Genome-fragment | 3.28 | AAI-based (high-confidence) | 0 |
| S3_vs_k141_20913\|\|full | 5706 | No | NA | 6 | 0 | 1 | Not-determined | Genome-fragment | NA | NA | 0 |
| S3_vs_k141_100785\|\|full | 38147 | No | NA | 56 | 15 | 0 | High-quality | High-quality | 98.45 | AAI-based (high-confidence) | 0 |
| S8_extracted_k141_117963 | 1712 | No | NA | 3 | 0 | 2 | Low-quality | Genome-fragment | 3.17 | AAI-based (medium-confidence) | 0 |
| S3_vs_k141_124716\|\|full | 1377 | No | NA | 3 | 0 | 0 | Not-determined | Genome-fragment | NA | NA | 0 |
| S3_vs_k141_85753\|\|full | 2991 | No | NA | 4 | 0 | 1 | Not-determined | Genome-fragment | NA | NA | 0 |
| S3_vs_k141_29654\|\|full | 1764 | No | NA | 2 | 0 | 1 | Not-determined | Genome-fragment | NA | NA | 0 |
| S8_vs_k141_194521\|\|full | 1712 | No | NA | 3 | 0 | 0 | Low-quality | Genome-fragment | 4.01 | AAI-based (high-confidence) | 0 |
| S8_vs_k141_92766\|\|full | 1712 | No | NA | 5 | 0 | 0 | Low-quality | Genome-fragment | 2.86 | AAI-based (medium-confidence) | 0 |
| S3_vs_k141_15450\|\|full | 44266 | No | NA | 63 | 18 | 0 | High-quality | High-quality | 99.79 | AAI-based (high-confidence) | 0 |
| S8_vs_k141_148509\|\|lt2gene | 1712 | No | NA | 2 | 1 | 0 | Low-quality | Genome-fragment | 4.11 | AAI-based (high-confidence) | 0 |
| S9_extracted_k141_80036 | 1713 | No | NA | 1 | 0 | 1 | Low-quality | Genome-fragment | 0.93 | AAI-based (medium-confidence) | 0 |
| S13_vs_83381\|\|full | 1713 | No | NA | 2 | 0 | 1 | Low-quality | Genome-fragment | 2.83 | AAI-based (medium-confidence) | 0 |
| S3_vs_k141_15487\|\|full | 2070 | No | NA | 3 | 0 | 2 | Not-determined | Genome-fragment | NA | NA | 0 |
| S3_vs_k141_12652\|\|full | 1713 | No | NA | 2 | 1 | 0 | Low-quality | Genome-fragment | 1.45 | AAI-based (high-confidence) | 0 |
| S5_vs_k141_140990\|\|full | 1713 | No | NA | 3 | 0 | 0 | Low-quality | Genome-fragment | 1.47 | AAI-based (medium-confidence) | 0 |
| S3_vs_k141_124783\|\|full | 2048 | No | NA | 2 | 0 | 2 | Not-determined | Genome-fragment | NA | NA | 0 |
| S8_extracted_k141_10500 | 1713 | No | NA | 3 | 1 | 0 | Low-quality | Genome-fragment | 3.72 | AAI-based (high-confidence) | 0 |
| S3_vs_k141_93634\|\|full | 1029 | No | NA | 2 | 0 | 0 | Not-determined | Genome-fragment | NA | NA | 0 |
| S8_extracted_k141_150162 | 1713 | No | NA | 1 | 0 | 0 | Low-quality | Genome-fragment | 4.62 | AAI-based (medium-confidence) | 0 |
| S17_vs_k141_29739\|\|full | 1714 | No | NA | 4 | 0 | 0 | Low-quality | Genome-fragment | 1.8 | AAI-based (high-confidence) | 0 |
| S9_extracted_k141_27580 | 1714 | No | NA | 1 | 0 | 0 | Low-quality | Genome-fragment | 0.99 | AAI-based (high-confidence) | 0 |
| S3_vs_k141_85838\|\|full | 2969 | No | NA | 2 | 0 | 1 | Not-determined | Genome-fragment | NA | NA | 0 |
| S9_vs_k141_1928\|\|lt2gene | 1714 | No | NA | 1 | 1 | 0 | Low-quality | Genome-fragment | 2.5 | HMM-based (lower-bound) | 0 |
| S3_vs_k141_54303\|\|full | 43708 | No | NA | 67 | 17 | 2 | High-quality | High-quality | 97.38 | AAI-based (high-confidence) | 0 |
| S3_vs_k141_54307\|\|full | 1605 | No | NA | 2 | 0 | 1 | Not-determined | Genome-fragment | NA | NA | 0 |
| S11_vs_k141_72764\|\|full | 1714 | No | NA | 3 | 1 | 0 | Low-quality | Genome-fragment | 3.23 | AAI-based (high-confidence) | 0 |
| S13_vs_151534\|\|full | 1714 | No | NA | 4 | 0 | 0 | Low-quality | Genome-fragment | 3.46 | AAI-based (high-confidence) | 0 |
| S3_vs_k141_111717\|\|full | 1714 | No | NA | 3 | 0 | 0 | Low-quality | Genome-fragment | 1.14 | AAI-based (medium-confidence) | 0 |
| S3_vs_k141_15646\|\|full | 1475 | No | NA | 3 | 0 | 2 | Not-determined | Genome-fragment | NA | NA | 0 |
| S5_vs_k141_29262\|\|full | 1714 | No | NA | 3 | 0 | 0 | Low-quality | Genome-fragment | 4.15 | AAI-based (medium-confidence) | 0 |
| S5_vs_k141_69334\|\|lt2gene | 1714 | No | NA | 1 | 1 | 0 | Low-quality | Genome-fragment | 4.23 | AAI-based (medium-confidence) | 0 |
| S16_vs_89326\|\|full | 1714 | No | NA | 5 | 0 | 0 | Low-quality | Genome-fragment | 3.54 | AAI-based (high-confidence) | 0 |
| S3_vs_k141_5687\|\|full | 1028 | No | NA | 2 | 0 | 0 | Not-determined | Genome-fragment | NA | NA | 0 |
| S3_vs_k141_21138\|\|full | 1635 | No | NA | 3 | 0 | 0 | Not-determined | Genome-fragment | NA | NA | 0 |
| S16_vs_145928\|\|full | 1714 | No | NA | 3 | 1 | 0 | Low-quality | Genome-fragment | 2.81 | AAI-based (high-confidence) | 0 |
| S12_vs_k141_157838\|\|full | 1714 | No | NA | 3 | 1 | 0 | Low-quality | Genome-fragment | 2.46 | HMM-based (lower-bound) | 0 |
| S12_vs_k141_26493\|\|full | 1714 | No | NA | 4 | 1 | 0 | Low-quality | Genome-fragment | 3.12 | AAI-based (medium-confidence) | 0 |
| S6_vs_k141_53162\|\|full | 1714 | No | NA | 3 | 2 | 0 | Low-quality | Genome-fragment | 2.85 | AAI-based (high-confidence) | 0 |
| S8_extracted_k141_44269 | 1714 | No | NA | 5 | 1 | 0 | Low-quality | Genome-fragment | 1.48 | AAI-based (medium-confidence) | 0 |
| S8_vs_k141_157007\|\|full | 1714 | No | NA | 3 | 0 | 0 | Low-quality | Genome-fragment | 2.8 | AAI-based (high-confidence) | 0 |
| S8_vs_k141_133540\|\|full | 1714 | No | NA | 5 | 0 | 0 | Low-quality | Genome-fragment | 3.45 | AAI-based (high-confidence) | 0 |
| S9_vs_k141_23043\|\|full | 1715 | No | NA | 2 | 0 | 1 | Low-quality | Genome-fragment | 0.78 | AAI-based (medium-confidence) | 0 |
| S3_vs_k141_15708\|\|full | 5857 | No | NA | 5 | 0 | 2 | Not-determined | Genome-fragment | NA | NA | 0 |
| S13_extracted_104172 | 1715 | No | NA | 1 | 0 | 0 | Low-quality | Genome-fragment | 3.54 | AAI-based (medium-confidence) | 0 |
| S3_vs_k141_62613\|\|full | 1697 | No | NA | 3 | 0 | 0 | Not-determined | Genome-fragment | NA | NA | 0 |
| S3_vs_k141_57757\|\|lt2gene | 1715 | No | NA | 1 | 1 | 0 | Low-quality | Genome-fragment | 4.05 | AAI-based (high-confidence) | 0 |
| S5_vs_k141_53\|\|full | 1715 | No | NA | 2 | 0 | 2 | Low-quality | Genome-fragment | 2.88 | AAI-based (medium-confidence) | 0 |
| S3_vs_k141_29835\|\|full | 1714 | No | NA | 2 | 0 | 1 | Not-determined | Genome-fragment | NA | NA | 0 |
| S12_vs_k141_1280\|\|full | 1715 | No | NA | 2 | 0 | 2 | Low-quality | Genome-fragment | 0.87 | AAI-based (medium-confidence) | 0 |
| S12_vs_k141_22854\|\|full | 1715 | No | NA | 3 | 0 | 1 | Low-quality | Genome-fragment | 2.52 | AAI-based (medium-confidence) | 0 |
| S2_vs_k141_4826\|\|full | 1715 | No | NA | 3 | 1 | 0 | Low-quality | Genome-fragment | 5.16 | AAI-based (high-confidence) | 0 |
| S3_vs_k141_93793\|\|full | 1460 | No | NA | 3 | 0 | 0 | Not-determined | Genome-fragment | NA | NA | 0 |
| S17_vs_k141_174433\|\|full | 1716 | No | NA | 3 | 0 | 0 | Low-quality | Genome-fragment | 3.59 | AAI-based (high-confidence) | 0 |
| S3_vs_k141_54410\|\|full | 1033 | No | NA | 2 | 0 | 0 | Not-determined | Genome-fragment | NA | NA | 0 |
| S3_vs_k141_93867\|\|full | 8246 | No | NA | 9 | 0 | 8 | Not-determined | Genome-fragment | NA | NA | 0 |
| S11_extracted_k141_40367 | 1716 | No | NA | 2 | 2 | 0 | Low-quality | Genome-fragment | 3.75 | AAI-based (high-confidence) | 0 |
| S3_vs_k141_29898\|\|full | 1889 | No | NA | 3 | 0 | 2 | Not-determined | Genome-fragment | NA | NA | 0 |
| S3_vs_k141_100970\|\|full | 2854 | No | NA | 5 | 0 | 0 | Not-determined | Genome-fragment | NA | NA | 0 |
| S13_vs_135157\|\|full | 1716 | No | NA | 2 | 0 | 0 | Low-quality | Genome-fragment | 3.79 | AAI-based (high-confidence) | 0 |
| S3_vs_k141_29907\|\|full | 2576 | No | NA | 4 | 0 | 0 | Not-determined | Genome-fragment | NA | NA | 0 |
| S3_vs_k141_29909\|\|full | 5501 | No | NA | 5 | 0 | 2 | Not-determined | Genome-fragment | NA | NA | 0 |
| S3_vs_k141_69059\|\|full | 2280 | No | NA | 3 | 0 | 2 | Not-determined | Genome-fragment | NA | NA | 0 |
| S3_vs_k141_86122\|\|full | 1227 | No | NA | 2 | 0 | 0 | Not-determined | Genome-fragment | NA | NA | 0 |
| S3_vs_k141_29921\|\|full | 1508 | No | NA | 2 | 0 | 0 | Not-determined | Genome-fragment | NA | NA | 0 |
| S14_vs_k141_8752\|\|full | 1716 | No | NA | 3 | 0 | 0 | Low-quality | Genome-fragment | 2.72 | AAI-based (high-confidence) | 0 |
| S3_vs_k141_29941\|\|full | 2812 | No | NA | 3 | 0 | 3 | Not-determined | Genome-fragment | NA | NA | 0 |
| S3_vs_k141_15828\|\|full | 1757 | No | NA | 3 | 0 | 0 | Not-determined | Genome-fragment | NA | NA | 0 |
| S3_vs_k141_86150\|\|full | 2534 | No | NA | 5 | 0 | 0 | Not-determined | Genome-fragment | NA | NA | 0 |
| S3_vs_k141_44962\|\|full | 1716 | No | NA | 2 | 0 | 0 | Low-quality | Genome-fragment | 1.06 | AAI-based (medium-confidence) | 0 |
| S7_vs_53044\|\|full | 1716 | No | NA | 4 | 1 | 0 | Low-quality | Genome-fragment | 4.1 | AAI-based (high-confidence) | 0 |

| | | | | | | | | | | | | | |
|---|---|---|---|---|---|---|---|---|---|---|---|---|---|
| S16_vs_132235\|\|full | 1716 | No | NA | | 2 | 0 | 0 | Low-quality | Genome-fragment | | 0.85 | AAI-based (medium-confidence) | 0 |
| S12_extracted_k141_188000 | 1716 | No | NA | | 3 | 0 | 0 | Low-quality | Genome-fragment | | 2.21 | AAI-based (medium-confidence) | 0 |
| S6_extracted_k141_2020 | 1716 | No | NA | | 1 | 0 | 0 | Low-quality | Genome-fragment | | 2.5 | AAI-based (medium-confidence) | 0 |
| S3_vs_k141_38105\|\|full | 1695 | No | NA | | 4 | 0 | 0 | Not-determined | Genome-fragment | NA | | NA | 0 |
| S3_vs_k141_109944\|\|full | 1552 | No | NA | | 4 | 0 | 0 | Not-determined | Genome-fragment | NA | | NA | 0 |
| S3_vs_k141_86250\|\|full | 3482 | No | NA | | 3 | 0 | 2 | Not-determined | Genome-fragment | NA | | NA | 0 |
| S11_vs_k141_87060\|\|full | 1717 | Yes | | 399 | 4 | 1 | 1 | Low-quality | Genome-fragment | | 0.82 | AAI-based (high-confidence) | 76.76 |
| S14_vs_k141_93323\|\|full | 1717 | No | NA | | 3 | 3 | 0 | Low-quality | Genome-fragment | | 4.25 | AAI-based (high-confidence) | 0 |
| S3_vs_k141_38129\|\|full | 1660 | No | NA | | 3 | 0 | 0 | Not-determined | Genome-fragment | NA | | NA | 0 |
| S1_vs_k141_156858\|\|full | 1717 | No | NA | | 3 | 0 | 0 | Low-quality | Genome-fragment | | 2.91 | AAI-based (high-confidence) | 0 |
| S3_vs_k141_5876\|\|full | 3578 | No | NA | | 5 | 0 | 4 | Not-determined | Genome-fragment | NA | | NA | 0 |
| S3_vs_k141_125134\|\|full | 1442 | No | NA | | 2 | 0 | 1 | Not-determined | Genome-fragment | NA | | NA | 0 |
| S5_vs_k141_61812\|\|full | 1717 | No | NA | | 4 | 4 | 0 | Low-quality | Genome-fragment | | 3.55 | AAI-based (medium-confidence) | 0 |
| S7_extracted_123324 | 1717 | No | NA | | 2 | 0 | 1 | Low-quality | Genome-fragment | | 2.69 | AAI-based (medium-confidence) | 0 |
| S16_vs_157485\|\|full | 1717 | No | NA | | 4 | 1 | 0 | Low-quality | Genome-fragment | | 1.33 | AAI-based (high-confidence) | 0 |
| S3_vs_k141_78876\|\|full | 2000 | No | NA | | 3 | 0 | 1 | Not-determined | Genome-fragment | NA | | NA | 0 |
| S17_extracted_k141_121113 | 1718 | Yes | | 776 | 4 | 1 | 1 | Low-quality | Genome-fragment | | 0.53 | AAI-based (medium-confidence) | 54.83 |
| S17_extracted_k141_20552 | 1718 | No | NA | | 2 | 0 | 0 | Low-quality | Genome-fragment | | 4.15 | AAI-based (high-confidence) | 0 |
| S9_extracted_k141_7671 | 1718 | No | NA | | 2 | 0 | 0 | Low-quality | Genome-fragment | | 2.04 | AAI-based (medium-confidence) | 0 |
| S3_vs_k141_62885\|\|full | 1074 | No | NA | | 4 | 0 | 3 | Not-determined | Genome-fragment | NA | | NA | 0 |
| S11_vs_k141_61885\|\|full | 1718 | No | NA | | 2 | 0 | 0 | Low-quality | Genome-fragment | | 3.61 | AAI-based (high-confidence) | 0 |
| S16_vs_78582\|\|full | 1718 | No | NA | | 3 | 0 | 0 | Low-quality | Genome-fragment | | 3.32 | AAI-based (medium-confidence) | 0 |
| S16_vs_89329\|\|full | 1718 | No | NA | | 2 | 0 | 0 | Low-quality | Genome-fragment | | 0.78 | AAI-based (high-confidence) | 0 |
| S3_vs_k141_117742\|\|full | 3782 | No | NA | | 3 | 0 | 3 | Not-determined | Genome-fragment | NA | | NA | 0 |
| S14_vs_k141_68490\|\|full | 1719 | No | NA | | 3 | 1 | 0 | Low-quality | Genome-fragment | | 2 | HMM-based (lower-bound) | 0 |
| S3_vs_k141_86443\|\|full | 3007 | No | NA | | 4 | 0 | 2 | Not-determined | Genome-fragment | NA | | NA | 0 |
| S1_extracted_k141_116117 | 1719 | No | NA | | 3 | 1 | 0 | Low-quality | Genome-fragment | | 5.82 | HMM-based (lower-bound) | 0 |
| S1_vs_k141_55698\|\|lt2gene | 1719 | No | NA | | 1 | 1 | 0 | Low-quality | Genome-fragment | | 3.28 | AAI-based (medium-confidence) | 0 |
| S7_vs_98023\|\|full | 1719 | No | NA | | 3 | 0 | 0 | Low-quality | Genome-fragment | | 3.93 | AAI-based (high-confidence) | 0 |
| S12_vs_k141_60676\|\|full | 1719 | No | NA | | 3 | 0 | 1 | Low-quality | Genome-fragment | | 1.31 | AAI-based (medium-confidence) | 0 |
| S3_vs_k141_5975\|\|full | 27273 | No | NA | | 25 | 0 | 6 | Not-determined | Genome-fragment | NA | | NA | 0 |
| S17_vs_k141_9943\|\|full | 1720 | No | NA | | 6 | 0 | 0 | Low-quality | Genome-fragment | | 2.73 | AAI-based (high-confidence) | 0 |
| S3_vs_k141_6017\|\|full | 2260 | No | NA | | 4 | 0 | 0 | Not-determined | Genome-fragment | NA | | NA | 0 |
| S13_vs_20205\|\|full | 1720 | No | NA | | 3 | 0 | 1 | Low-quality | Genome-fragment | | 1.27 | AAI-based (medium-confidence) | 0 |
| S13_vs_97379\|\|full | 1720 | No | NA | | 2 | 0 | 1 | Low-quality | Genome-fragment | | 2.08 | AAI-based (medium-confidence) | 0 |
| S3_vs_k141_117872\|\|full | 2037 | No | NA | | 3 | 0 | 0 | Not-determined | Genome-fragment | NA | | NA | 0 |
| S3_vs_k141_94171\|\|full | 1894 | No | NA | | 4 | 0 | 1 | Not-determined | Genome-fragment | NA | | NA | 0 |
| S3_vs_k141_94175\|\|full | 4180 | No | NA | | 3 | 0 | 3 | Not-determined | Genome-fragment | NA | | NA | 0 |
| S1_vs_k141_38864\|\|full | 1720 | No | NA | | 3 | 0 | 1 | Low-quality | Genome-fragment | | 3.84 | AAI-based (medium-confidence) | 0 |
| S3_vs_k141_72675\|\|full | 1720 | No | NA | | 4 | 0 | 1 | Low-quality | Genome-fragment | | 2.39 | AAI-based (medium-confidence) | 0 |
| S12_vs_k141_182150\|\|full | 1720 | No | NA | | 3 | 1 | 0 | Low-quality | Genome-fragment | | 5.55 | HMM-based (lower-bound) | 0 |
| S3_vs_k141_54801\|\|full | 1752 | No | NA | | 2 | 0 | 2 | Not-determined | Genome-fragment | NA | | NA | 0 |
| S3_vs_k141_63088\|\|full | 1261 | No | NA | | 3 | 0 | 1 | Not-determined | Genome-fragment | NA | | NA | 0 |
| S3_vs_k141_110254\|\|full | 1615 | No | NA | | 3 | 0 | 0 | Not-determined | Genome-fragment | NA | | NA | 0 |
| S8_vs_k141_77586\|\|full | 1720 | No | NA | | 2 | 0 | 2 | Low-quality | Genome-fragment | | 1.23 | AAI-based (medium-confidence) | 0 |
| S14_extracted_k141_61453 | 1721 | No | NA | | 2 | 2 | 0 | Low-quality | Genome-fragment | | 5.67 | AAI-based (high-confidence) | 0 |
| S3_vs_k141_102812\|\|full | 1721 | No | NA | | 4 | 1 | 0 | Low-quality | Genome-fragment | | 3.16 | AAI-based (high-confidence) | 0 |
| S3_vs_k141_24361\|\|lt2gene | 1721 | No | NA | | 2 | 2 | 0 | Low-quality | Genome-fragment | | 4.55 | AAI-based (high-confidence) | 0 |
| S5_vs_k141_39804\|\|lt2gene | 1721 | No | NA | | 1 | 1 | 0 | Low-quality | Genome-fragment | | 5.02 | AAI-based (high-confidence) | 0 |
| S7_vs_5291\|\|full | 1721 | No | NA | | 4 | 0 | 0 | Low-quality | Genome-fragment | | 4.05 | AAI-based (high-confidence) | 0 |
| S12_extracted_k141_144653 | 1721 | No | NA | | 3 | 0 | 1 | Low-quality | Genome-fragment | | 1.05 | AAI-based (medium-confidence) | 0 |
| S8_extracted_k141_51853 | 1721 | No | NA | | 1 | 0 | 1 | Low-quality | Genome-fragment | | 0.47 | AAI-based (medium-confidence) | 0 |
| S17_extracted_k141_104189 | 1722 | No | NA | | 4 | 4 | 0 | Low-quality | Genome-fragment | | 5.68 | AAI-based (high-confidence) | 0 |
| S9_vs_k141_18355\|\|full | 1722 | No | NA | | 3 | 2 | 0 | Low-quality | Genome-fragment | | 3.97 | AAI-based (high-confidence) | 0 |
| S3_vs_k141_110325\|\|full | 3167 | No | NA | | 2 | 0 | 1 | Not-determined | Genome-fragment | NA | | NA | 0 |
| S3_vs_k141_115341\|\|full | 1722 | No | NA | | 6 | 2 | 0 | Low-quality | Genome-fragment | | 4.47 | AAI-based (medium-confidence) | 0 |
| S3_vs_k141_54904\|\|full | 4046 | No | NA | | 6 | 0 | 5 | Not-determined | Genome-fragment | NA | | NA | 0 |
| S3_vs_k141_30318\|\|full | 37805 | No | NA | | 55 | 9 | 1 | High-quality | High-quality | | 92.84 | AAI-based (medium-confidence) | 0 |
| S3_vs_k141_110330\|\|full | 3315 | No | NA | | 4 | 0 | 3 | Not-determined | Genome-fragment | NA | | NA | 0 |
| S3_vs_k141_101159\|\|full | 1590 | No | NA | | 3 | 0 | 0 | Not-determined | Genome-fragment | NA | | NA | 0 |
| S3_vs_k141_38488\|\|full | 1531 | No | NA | | 3 | 0 | 0 | Not-determined | Genome-fragment | NA | | NA | 0 |
| S3_vs_k141_110345\|\|full | 1802 | No | NA | | 3 | 0 | 0 | Not-determined | Genome-fragment | NA | | NA | 0 |
| S3_vs_k141_38507\|\|full | 2294 | No | NA | | 3 | 0 | 2 | Not-determined | Genome-fragment | NA | | NA | 0 |
| S5_extracted_k141_106708 | 1722 | No | NA | | 6 | 1 | 0 | Low-quality | Genome-fragment | | 5.08 | AAI-based (high-confidence) | 0 |
| S5_vs_k141_113821\|\|full | 1722 | No | NA | | 3 | 1 | 0 | Low-quality | Genome-fragment | | 3.66 | AAI-based (high-confidence) | 0 |
| S5_vs_k141_57880\|\|full | 1722 | No | NA | | 4 | 0 | 0 | Low-quality | Genome-fragment | | 3.05 | AAI-based (high-confidence) | 0 |
| S15_vs_27464\|\|full | 1722 | No | NA | | 3 | 0 | 1 | Low-quality | Genome-fragment | | 2.82 | AAI-based (medium-confidence) | 0 |
| S17_vs_k141_77311\|\|full | 1723 | No | NA | | 3 | 0 | 2 | Low-quality | Genome-fragment | | 2.24 | AAI-based (medium-confidence) | 0 |
| S11_vs_k141_50017\|\|full | 1723 | No | NA | | 3 | 0 | 0 | Low-quality | Genome-fragment | | 2.79 | AAI-based (medium-confidence) | 0 |
| S3_vs_k141_117912\|\|full | 3068 | No | NA | | 2 | 0 | 2 | Not-determined | Genome-fragment | NA | | NA | 0 |
| S3_vs_k141_46414\|\|full | 2602 | No | NA | | 2 | 0 | 2 | Not-determined | Genome-fragment | NA | | NA | 0 |
| S14_extracted_k141_73669 | 1723 | No | NA | | 6 | 5 | 0 | Low-quality | Genome-fragment | | 5.68 | AAI-based (high-confidence) | 0 |
| S14_vs_k141_108062\|\|full | 1723 | No | NA | | 3 | 0 | 0 | Low-quality | Genome-fragment | | 3.04 | AAI-based (medium-confidence) | 0 |
| S5_vs_k141_114947\|\|full | 1723 | No | NA | | 3 | 0 | 0 | Low-quality | Genome-fragment | | 3.02 | AAI-based (medium-confidence) | 0 |
| S3_vs_k141_63225\|\|full | 8186 | No | NA | | 8 | 0 | 2 | Not-determined | Genome-fragment | NA | | NA | 0 |
| S12_vs_k141_127944\|\|full | 1723 | No | NA | | 3 | 1 | 0 | Low-quality | Genome-fragment | | 1.05 | AAI-based (high-confidence) | 0 |
| S8_vs_k141_48759\|\|full | 1723 | No | NA | | 5 | 1 | 0 | Low-quality | Genome-fragment | | 2.04 | AAI-based (medium-confidence) | 0 |
| S17_vs_k141_46553\|\|full | 1724 | No | NA | | 4 | 2 | 0 | Low-quality | Genome-fragment | | 4.25 | AAI-based (high-confidence) | 0 |
| S3_vs_k141_101277\|\|full | 1843 | No | NA | | 4 | 0 | 1 | Not-determined | Genome-fragment | NA | | NA | 0 |
| S14_extracted_k141_29414 | 1724 | No | NA | | 6 | 4 | 0 | Low-quality | Genome-fragment | | 5.69 | AAI-based (high-confidence) | 0 |
| S1_extracted_k141_95084 | 1724 | No | NA | | 3 | 0 | 0 | Low-quality | Genome-fragment | | 1.84 | AAI-based (high-confidence) | 0 |
| S3_vs_k141_110408\|\|full | 1171 | No | NA | | 2 | 0 | 1 | Not-determined | Genome-fragment | NA | | NA | 0 |
| S10_vs_1980\|\|full | 1724 | No | NA | | 2 | 0 | 0 | Low-quality | Genome-fragment | | 4.45 | AAI-based (high-confidence) | 0 |
| S3_vs_k141_118022\|\|full | 3220 | No | NA | | 4 | 0 | 1 | Not-determined | Genome-fragment | NA | | NA | 0 |
| S2_extracted_k141_10152 | 1724 | No | NA | | 4 | 0 | 1 | Low-quality | Genome-fragment | | 4.03 | AAI-based (high-confidence) | 0 |
| S17_extracted_k141_133527 | 1725 | No | NA | | 2 | 0 | 0 | Low-quality | Genome-fragment | | 1.07 | AAI-based (medium-confidence) | 0 |
| S9_vs_k141_28894\|\|full | 1725 | No | NA | | 2 | 1 | 0 | Low-quality | Genome-fragment | | 4.31 | AAI-based (high-confidence) | 0 |
| S3_vs_k141_86804\|\|full | 2144 | No | NA | | 2 | 0 | 0 | Not-determined | Genome-fragment | NA | | NA | 0 |
| S3_vs_k141_31324\|\|full | 1725 | No | NA | | 3 | 0 | 1 | Low-quality | Genome-fragment | | 1.12 | AAI-based (medium-confidence) | 0 |
| S3_vs_k141_46425\|\|full | 124817 | No | NA | | 132 | 0 | 38 | Not-determined | Genome-fragment | NA | | NA | 0 |
| S3_vs_k141_101396\|\|full | 1741 | No | NA | | 2 | 0 | 2 | Not-determined | Genome-fragment | NA | | NA | 0 |
| S7_vs_33363\|\|full | 1725 | No | NA | | 3 | 3 | 0 | Low-quality | Genome-fragment | | 5.68 | AAI-based (high-confidence) | 0 |
| S16_extracted_128144 | 1725 | No | NA | | 4 | 0 | 0 | Low-quality | Genome-fragment | | 4.68 | AAI-based (high-confidence) | 0 |
| S16_vs_165480\|\|full | 1725 | No | NA | | 2 | 0 | 0 | Low-quality | Genome-fragment | | 1 | AAI-based (high-confidence) | 0 |
| S3_vs_k141_55057\|\|full | 1395 | No | NA | | 3 | 0 | 0 | Not-determined | Genome-fragment | NA | | NA | 0 |
| S8_vs_k141_54152\|\|full | 1725 | No | NA | | 2 | 1 | 0 | Low-quality | Genome-fragment | | 2.44 | HMM-based (lower-bound) | 0 |
| S11_vs_k141_34231\|\|full | 1726 | No | NA | | 3 | 1 | 0 | Low-quality | Genome-fragment | | 0.7 | HMM-based (lower-bound) | 0 |
| S3_vs_k141_101442\|\|full | 1933 | No | NA | | 2 | 0 | 2 | Not-determined | Genome-fragment | NA | | NA | 0 |
| S3_vs_k141_86819\|\|full | 21762 | No | NA | | 22 | 0 | 4 | Not-determined | Genome-fragment | NA | | NA | 0 |
| S3_vs_k141_110535\|\|full | 2300 | No | NA | | 4 | 0 | 4 | Not-determined | Genome-fragment | NA | | NA | 0 |
| S3_vs_k141_21810\|\|full | 2488 | No | NA | | 3 | 0 | 2 | Not-determined | Genome-fragment | NA | | NA | 0 |
| S11_vs_k141_47659\|\|full | 1726 | No | NA | | 3 | 1 | 0 | Low-quality | Genome-fragment | | 1.07 | HMM-based (lower-bound) | 0 |
| S3_vs_k141_94407\|\|full | 1127 | No | NA | | 2 | 0 | 0 | Not-determined | Genome-fragment | NA | | NA | 0 |
| S11_vs_k141_9740\|\|full | 1726 | No | NA | | 4 | 0 | 0 | Low-quality | Genome-fragment | | 3.38 | AAI-based (high-confidence) | 0 |
| S11_vs_k141_43041\|\|lt2gene | 1726 | No | NA | | 2 | 1 | 0 | Low-quality | Genome-fragment | | 3.26 | HMM-based (lower-bound) | 0 |
| S14_vs_k141_31827\|\|full | 1726 | No | NA | | 3 | 2 | 0 | Low-quality | Genome-fragment | | 2.3 | AAI-based (high-confidence) | 0 |
| S3_extracted_k141_7489 | 1726 | No | NA | | 2 | 1 | 0 | Low-quality | Genome-fragment | | 3.33 | HMM-based (lower-bound) | 0 |
| S5_vs_k141_128008\|\|full | 1726 | No | NA | | 5 | 5 | 0 | Low-quality | Genome-fragment | | 3.27 | HMM-based (lower-bound) | 0 |
| S6_vs_k141_53628\|\|full | 1726 | No | NA | | 2 | 0 | 2 | Low-quality | Genome-fragment | | 2.26 | AAI-based (high-confidence) | 0 |
| S8_vs_k141_91841\|\|full | 1726 | No | NA | | 5 | 0 | 0 | Low-quality | Genome-fragment | | 4.11 | AAI-based (high-confidence) | 0 |
| S9_vs_k141_4617\|\|full | 1727 | No | NA | | 4 | 0 | 1 | Low-quality | Genome-fragment | | 3.04 | AAI-based (medium-confidence) | 0 |
| S3_vs_k141_101497\|\|full | 3934 | No | NA | | 3 | 0 | 3 | Not-determined | Genome-fragment | NA | | NA | 0 |
| S3_vs_k141_86961\|\|full | 3735 | No | NA | | 4 | 0 | 3 | Not-determined | Genome-fragment | NA | | NA | 0 |
| S3_vs_k141_94506\|\|full | 1196 | No | NA | | 3 | 0 | 1 | Not-determined | Genome-fragment | NA | | NA | 0 |
| S3_vs_k141_101533\|\|full | 1908 | No | NA | | 2 | 0 | 0 | Not-determined | Genome-fragment | NA | | NA | 0 |
| S3_vs_k141_55166\|\|full | 3716 | No | NA | | 4 | 0 | 3 | Not-determined | Genome-fragment | NA | | NA | 0 |
| S3_vs_k141_110617\|\|full | 4247 | No | NA | | 5 | 0 | 1 | Not-determined | Genome-fragment | NA | | NA | 0 |
| S3_vs_k141_125712\|\|full | 2023 | No | NA | | 4 | 0 | 1 | Not-determined | Genome-fragment | NA | | NA | 0 |
| S3_vs_k141_63433\|\|full | 2809 | No | NA | | 2 | 0 | 2 | Not-determined | Genome-fragment | NA | | NA | 0 |
| S3_vs_k141_110639\|\|full | 1164 | No | NA | | 2 | 0 | 0 | Not-determined | Genome-fragment | NA | | NA | 0 |
| S3_vs_k141_94542\|\|full | 1653 | No | NA | | 4 | 0 | 0 | Not-determined | Genome-fragment | NA | | NA | 0 |
| S3_vs_k141_63445\|\|full | 4112 | No | NA | | 3 | 0 | 2 | Not-determined | Genome-fragment | NA | | NA | 0 |
| S11_extracted_k141_75102 | 1727 | No | NA | | 4 | 1 | 0 | Low-quality | Genome-fragment | | 1.17 | HMM-based (lower-bound) | 0 |
| S1_extracted_k141_27903 | 1727 | No | NA | | 5 | 1 | 0 | Low-quality | Genome-fragment | | 3.32 | AAI-based (high-confidence) | 0 |
| S1_extracted_k141_135974 | 1727 | No | NA | | 2 | 0 | 1 | Low-quality | Genome-fragment | | 0.61 | AAI-based (medium-confidence) | 0 |
| S3_vs_k141_38781\|\|full | 7760 | No | NA | | 7 | 0 | 5 | Not-determined | Genome-fragment | NA | | NA | 0 |
| S3_vs_k141_22059\|\|full | 2118 | No | NA | | 2 | 0 | 2 | Not-determined | Genome-fragment | NA | | NA | 0 |
| S3_extracted_k141_67603 | 1727 | No | NA | | 2 | 0 | 2 | Low-quality | Genome-fragment | | 1.15 | AAI-based (high-confidence) | 0 |
| S3_vs_k141_79305\|\|full | 5508 | No | NA | | 4 | 0 | 4 | Not-determined | Genome-fragment | NA | | NA | 0 |
| S7_extracted_121157 | 1727 | No | NA | | 3 | 0 | 2 | Low-quality | Genome-fragment | | 1.28 | AAI-based (medium-confidence) | 0 |
| S3_vs_k141_118287\|\|full | 8310 | No | NA | | 8 | 0 | 0 | Not-determined | Genome-fragment | NA | | NA | 0 |
| S3_vs_k141_87055\|\|full | 9861 | No | NA | | 8 | 0 | 3 | Not-determined | Genome-fragment | NA | | NA | 0 |
| S3_vs_k141_94577\|\|full | 20384 | No | NA | | 33 | 18 | 0 | Medium-quality | Genome-fragment | | 61.58 | AAI-based (high-confidence) | 0 |
| S7_vs_36821\|\|full | 1727 | No | NA | | 2 | 1 | 0 | Low-quality | Genome-fragment | | 1.89 | HMM-based (lower-bound) | 0 |
| S7_vs_60348\|\|full | 1727 | No | NA | | 4 | 0 | 0 | Low-quality | Genome-fragment | | 3.89 | AAI-based (high-confidence) | 0 |
| S3_vs_k141_94590\|\|full | 1656 | No | NA | | 2 | 0 | 1 | Not-determined | Genome-fragment | NA | | NA | 0 |
| S16_vs_46359\|\|full | 1727 | No | NA | | 4 | 1 | 0 | Low-quality | Genome-fragment | | 1.27 | HMM-based (lower-bound) | 0 |
| S3_vs_k141_94612\|\|full | 2220 | No | NA | | 2 | 0 | 0 | Not-determined | Genome-fragment | NA | | NA | 0 |
| S12_vs_k141_56168\|\|full | 1727 | No | NA | | 2 | 0 | 2 | Low-quality | Genome-fragment | | 2.91 | AAI-based (medium-confidence) | 0 |
| S15_vs_36526\|\|full | 1727 | No | NA | | 3 | 0 | 0 | Low-quality | Genome-fragment | | 2 | AAI-based (high-confidence) | 0 |
| S4_extracted_7056 | 1727 | No | NA | | 4 | 3 | 0 | Low-quality | Genome-fragment | | 5.67 | AAI-based (high-confidence) | 0 |
| S3_vs_k141_110749\|\|full | 1712 | No | NA | | 3 | 0 | 1 | Not-determined | Genome-fragment | NA | | NA | 0 |
| S17_extracted_k141_84701 | 1728 | No | NA | | 3 | 3 | 0 | Low-quality | Genome-fragment | | 5.78 | AAI-based (high-confidence) | 0 |
| S3_vs_k141_63609\|\|full | 1558 | No | NA | | 3 | 0 | 0 | Not-determined | Genome-fragment | NA | | NA | 0 |
| S13_vs_24324\|\|full | 1728 | No | NA | | 2 | 0 | 1 | Low-quality | Genome-fragment | | 0.86 | AAI-based (medium-confidence) | 0 |
| S5_extracted_k141_42284 | 1728 | No | NA | | 4 | 0 | 1 | Low-quality | Genome-fragment | | 4 | AAI-based (high-confidence) | 0 |
| S12_extracted_k141_21852 | 1728 | No | NA | | 3 | 1 | 0 | Low-quality | Genome-fragment | | 1.22 | AAI-based (medium-confidence) | 0 |
| S3_vs_k141_38840\|\|full | 1817 | No | NA | | 2 | 0 | 2 | Not-determined | Genome-fragment | NA | | NA | 0 |
| S3_vs_k141_30658\|\|full | 4085 | No | NA | | 3 | 0 | 1 | Not-determined | Genome-fragment | NA | | NA | 0 |
| S3_vs_k141_6604\|\|full | 78643 | Yes | | 32409 | 81 | 3 | 26 | Medium-quality | Genome-fragment | | 74.74 | AAI-based (medium-confidence) | 58.79 |
| S15_vs_11738\|\|full | 1728 | No | NA | | 2 | 0 | 0 | Low-quality | Genome-fragment | | 4.12 | AAI-based (medium-confidence) | 0 |
| S8_vs_k141_39724\|\|full | 1728 | No | NA | | 2 | 0 | 1 | Low-quality | Genome-fragment | | 1.03 | AAI-based (high-confidence) | 0 |
| S8_vs_k141_33025\|\|full | 1728 | No | NA | | 2 | 0 | 1 | Low-quality | Genome-fragment | | 1.49 | AAI-based (high-confidence) | 0 |
| S3_vs_k141_6644\|\|full | 1113 | No | NA | | 2 | 0 | 0 | Not-determined | Genome-fragment | NA | | NA | 0 |
| S13_vs_128195\|\|full | 1729 | No | NA | | 5 | 1 | 0 | Low-quality | Genome-fragment | | 4.72 | AAI-based (medium-confidence) | 0 |
| S3_vs_k141_94776\|\|full | 1923 | No | NA | | 3 | 0 | 1 | Not-determined | Genome-fragment | NA | | NA | 0 |
| S3_vs_k141_69850\|\|full | 14836 | No | NA | | 21 | 0 | 4 | Not-determined | Genome-fragment | NA | | NA | 0 |
| S1_extracted_k141_110388 | 1729 | No | NA | | 3 | 2 | 0 | Low-quality | Genome-fragment | | 1.3 | AAI-based (high-confidence) | 0 |
| S3_vs_k141_94795\|\|full | 1904 | No | NA | | 3 | 0 | 0 | Not-determined | Genome-fragment | NA | | NA | 0 |
| S3_vs_k141_6671\|\|full | 1023 | No | NA | | 3 | 0 | 0 | Not-determined | Genome-fragment | NA | | NA | 0 |
| S3_vs_k141_101748\|\|full | 1917 | No | NA | | 3 | 0 | 1 | Not-determined | Genome-fragment | NA | | NA | 0 |
| S3_vs_k141_94806\|\|full | 6157 | No | NA | | 11 | 0 | 0 | Not-determined | Genome-fragment | NA | | NA | 0 |
| S5_vs_k141_38550\|\|full | 1729 | No | NA | | 2 | 1 | 0 | Low-quality | Genome-fragment | | 3 | HMM-based (lower-bound) | 0 |
| S3_vs_k141_94818\|\|full | 3884 | No | NA | | 4 | 0 | 0 | Not-determined | Genome-fragment | NA | | NA | 0 |
| S7_vs_104589\|\|lt2gene | 1729 | No | NA | | 2 | 0 | 0 | Low-quality | Genome-fragment | | 3.57 | AAI-based (high-confidence) | 0 |
| S16_vs_67646\|\|full | 1729 | No | NA | | 7 | 1 | 0 | Low-quality | Genome-fragment | | 2.89 | AAI-based (medium-confidence) | 0 |
| S3_vs_k141_101804\|\|full | 4441 | No | NA | | 5 | 0 | 5 | Not-determined | Genome-fragment | NA | | NA | 0 |
| S3_vs_k141_79471\|\|full | 1519 | No | NA | | 2 | 0 | 0 | Not-determined | Genome-fragment | NA | | NA | 0 |
| S3_vs_k141_63713\|\|full | 6892 | No | NA | | 8 | 0 | 3 | Not-determined | Genome-fragment | NA | | NA | 0 |
| S3_vs_k141_30789\|\|full | 1802 | No | NA | | 3 | 0 | 1 | Not-determined | Genome-fragment | NA | | NA | 0 |
| S3_vs_k141_118462\|\|full | 5711 | No | NA | | 5 | 0 | 3 | Not-determined | Genome-fragment | NA | | NA | 0 |
| S16_vs_155259\|\|full | 1729 | No | NA | | 3 | 0 | 1 | Low-quality | Genome-fragment | | 2.18 | AAI-based (high-confidence) | 0 |
| S10_extracted_3481 | 1729 | No | NA | | 5 | 1 | 0 | Low-quality | Genome-fragment | | 5.02 | AAI-based (high-confidence) | 0 |
| S3_vs_k141_55378\|\|full | 2792 | No | NA | | 3 | 0 | 1 | Not-determined | Genome-fragment | NA | | NA | 0 |
| S3_vs_k141_22397\|\|full | 3154 | No | NA | | 4 | 0 | 3 | Not-determined | Genome-fragment | NA | | NA | 0 |
| S10_extracted_83217 | 1729 | No | NA | | 3 | 1 | 0 | Low-quality | Genome-fragment | | 4.07 | AAI-based (high-confidence) | 0 |
| S3_vs_k141_46961\|\|full | 2558 | No | NA | | 5 | 0 | 1 | Not-determined | Genome-fragment | NA | | NA | 0 |
| S3_vs_k141_118501\|\|full | 1071 | No | NA | | 2 | 0 | 0 | Not-determined | Genome-fragment | NA | | NA | 0 |
| S10_vs_66542\|\|lt2gene | 1729 | No | NA | | 1 | 1 | 0 | Low-quality | Genome-fragment | | 4.6 | AAI-based (medium-confidence) | 0 |
| S3_vs_k141_46990\|\|full | 6364 | No | NA | | 5 | 0 | 3 | Not-determined | Genome-fragment | NA | | NA | 0 |
| S3_vs_k141_118517\|\|full | 1184 | No | NA | | 2 | 0 | 1 | Not-determined | Genome-fragment | NA | | NA | 0 |
| S3_vs_k141_94913\|\|full | 1461 | No | NA | | 4 | 0 | 2 | Not-determined | Genome-fragment | NA | | NA | 0 |
| S12_vs_k141_181802\|\|full | 1729 | No | NA | | 2 | 0 | 0 | Low-quality | Genome-fragment | | 3.64 | AAI-based (medium-confidence) | 0 |
| S8_vs_k141_107642\|\|full | 1729 | No | NA | | 3 | 0 | 0 | Low-quality | Genome-fragment | | 7.26 | AAI-based (medium-confidence) | 0 |
| S3_vs_k141_110910\|\|full | 1548 | No | NA | | 2 | 0 | 1 | Not-determined | Genome-fragment | NA | | NA | 0 |
| S3_vs_k141_110912\|\|full | 1254 | No | NA | | 2 | 0 | 0 | Not-determined | Genome-fragment | NA | | NA | 0 |
| S8_vs_k141_32388\|\|full | 1729 | No | NA | | 4 | 2 | 0 | Low-quality | Genome-fragment | | 5.01 | AAI-based (high-confidence) | 0 |
| S3_vs_k141_126004\|\|full | 1359 | No | NA | | 2 | 0 | 0 | Not-determined | Genome-fragment | NA | | NA | 0 |
| S3_vs_k141_22419\|\|full | 4900 | No | NA | | 5 | 0 | 2 | Not-determined | Genome-fragment | NA | | NA | 0 |
| S3_vs_k141_55453\|\|full | 5028 | No | NA | | 6 | 0 | 3 | Not-determined | Genome-fragment | NA | | NA | 0 |
| S3_vs_k141_79585\|\|full | 2042 | No | NA | | 3 | 0 | 1 | Not-determined | Genome-fragment | NA | | NA | 0 |
| S8_vs_k141_163994\|\|full | 1729 | No | NA | | 4 | 2 | 0 | Low-quality | Genome-fragment | | 6.29 | HMM-based (lower-bound) | 0 |
| S3_vs_k141_110971\|\|full | 1947 | No | NA | | 3 | 0 | 0 | Not-determined | Genome-fragment | NA | | NA | 0 |
| S14_extracted_k141_46798 | 1730 | No | NA | | 4 | 2 | 0 | Low-quality | Genome-fragment | | 3.85 | AAI-based (high-confidence) | 0 |
| S3_vs_k141_70049\|\|full | 2075 | No | NA | | 3 | 0 | 2 | Not-determined | Genome-fragment | NA | | NA | 0 |
| S3_vs_k141_70052\|\|full | 1355 | No | NA | | 2 | 0 | 2 | Not-determined | Genome-fragment | NA | | NA | 0 |
| S3_vs_k141_63384\|\|full | 1730 | No | NA | | 3 | 0 | 0 | Low-quality | Genome-fragment | | 1.11 | AAI-based (medium-confidence) | 0 |
| S3_vs_k141_79636\|\|full | 2138 | No | NA | | 2 | 0 | 0 | Not-determined | Genome-fragment | NA | | NA | 0 |
| S5_vs_k141_110552\|\|full | 1730 | No | NA | | 5 | 0 | 1 | Low-quality | Genome-fragment | | 3.64 | AAI-based (medium-confidence) | 0 |
| S16_vs_15204\|\|full | 1730 | No | NA | | 2 | 1 | 0 | Low-quality | Genome-fragment | | 2.97 | HMM-based (lower-bound) | 0 |
| S3_vs_k141_70093\|\|full | 20643 | No | NA | | 22 | 5 | 0 | Medium-quality | Genome-fragment | | 51.56 | AAI-based (high-confidence) | 0 |
| S3_vs_k141_95030\|\|full | 1576 | No | NA | | 4 | 0 | 0 | Not-determined | Genome-fragment | NA | | NA | 0 |
| S16_vs_28086\|\|full | 1730 | No | NA | | 3 | 0 | 0 | Low-quality | Genome-fragment | | 1.53 | AAI-based (high-confidence) | 0 |
| S16_vs_170187\|\|full | 1730 | No | NA | | 4 | 1 | 0 | Low-quality | Genome-fragment | | 4.31 | HMM-based (lower-bound) | 0 |
| S10_vs_97316\|\|full | 1730 | No | NA | | 3 | 0 | 0 | Low-quality | Genome-fragment | | 3.23 | AAI-based (high-confidence) | 0 |
| S12_vs_k141_960\|\|full | 1730 | No | NA | | 5 | 1 | 0 | Low-quality | Genome-fragment | | 2.27 | HMM-based (lower-bound) | 0 |
| S11_vs_k141_36568\|\|full | 1731 | No | NA | | 4 | 2 | 0 | Low-quality | Genome-fragment | | 4.05 | AAI-based (high-confidence) | 0 |
| S1_extracted_k141_87138 | 1731 | No | NA | | 1 | 0 | 1 | Low-quality | Genome-fragment | | 0.81 | AAI-based (medium-confidence) | 0 |
| S3_vs_k141_106172\|\|full | 1731 | No | NA | | 2 | 1 | 0 | Low-quality | Genome-fragment | | 4 | AAI-based (high-confidence) | 0 |
| S3_vs_k141_70007\|\|full | 1731 | No | NA | | 4 | 1 | 0 | Low-quality | Genome-fragment | | 3.75 | AAI-based (high-confidence) | 0 |
| S3_vs_k141_95051\|\|full | 4737 | No | NA | | 5 | 0 | 4 | Not-determined | Genome-fragment | NA | | NA | 0 |
| S3_vs_k141_118644\|\|full | 5368 | No | NA | | 6 | 0 | 4 | Not-determined | Genome-fragment | NA | | NA | 0 |
| S16_extracted_81551 | 1731 | No | NA | | 7 | 1 | 0 | Low-quality | Genome-fragment | | 1.11 | AAI-based (high-confidence) | 0 |
| S8_extracted_k141_131624 | 1731 | No | NA | | 3 | 0 | 2 | Low-quality | Genome-fragment | | 0.62 | AAI-based (medium-confidence) | 0 |
| S3_vs_k141_47153\|\|full | 1278 | No | NA | | 3 | 0 | 1 | Not-determined | Genome-fragment | NA | | NA | 0 |
| S3_vs_k141_118675\|\|full | 1618 | No | NA | | 2 | 0 | 0 | Not-determined | Genome-fragment | NA | | NA | 0 |
| S3_vs_k141_111188\|\|full | 5421 | No | NA | | 6 | 0 | 4 | Not-determined | Genome-fragment | NA | | NA | 0 |
| S8_vs_k141_19653\|\|full | 1731 | No | NA | | 1 | 0 | 0 | Low-quality | Genome-fragment | | 3.96 | AAI-based (medium-confidence) | 0 |
| S3_vs_k141_39220\|\|full | 1025 | No | NA | | 2 | 0 | 0 | Not-determined | Genome-fragment | NA | | NA | 0 |
| S17_vs_k141_70575\|\|full | 1732 | No | NA | | 3 | 0 | 0 | Low-quality | Genome-fragment | | 3.36 | AAI-based (medium-confidence) | 0 |
| S17_vs_k141_27722\|\|full | 1732 | No | NA | | 5 | 1 | 0 | Low-quality | Genome-fragment | | 3.79 | AAI-based (high-confidence) | 0 |
| S3_vs_k141_111241\|\|full | 2037 | No | NA | | 4 | 0 | 2 | Not-determined | Genome-fragment | NA | | NA | 0 |
| S14_vs_k141_31608\|\|full | 1732 | No | NA | | 4 | 0 | 1 | Low-quality | Genome-fragment | | 1.73 | AAI-based (medium-confidence) | 0 |
| S3_vs_k141_70191\|\|full | 3369 | No | NA | | 3 | 0 | 2 | Not-determined | Genome-fragment | NA | | NA | 0 |
| S5_vs_k141_33016\|\|full | 1732 | No | NA | | 2 | 0 | 0 | Low-quality | Genome-fragment | | 4.07 | AAI-based (medium-confidence) | 0 |
| S16_extracted_28997 | 1732 | No | NA | | 3 | 1 | 0 | Low-quality | Genome-fragment | | 4.3 | AAI-based (high-confidence) | 0 |
| S10_extracted_40558 | 1732 | No | NA | | 2 | 0 | 2 | Low-quality | Genome-fragment | | 0.65 | AAI-based (high-confidence) | 0 |
| S17_extracted_k141_111640 | 1733 | No | NA | | 4 | 1 | 0 | Low-quality | Genome-fragment | | 3.94 | AAI-based (high-confidence) | 0 |
| S3_vs_k141_111260\|\|full | 2594 | No | NA | | 3 | 0 | 1 | Not-determined | Genome-fragment | NA | | NA | 0 |
| S17_vs_k141_76379\|\|full | 1733 | No | NA | | 3 | 0 | 2 | Low-quality | Genome-fragment | | 0.78 | AAI-based (medium-confidence) | 0 |
| S3_vs_k141_111288\|\|full | 2748 | No | NA | | 3 | 0 | 0 | Not-determined | Genome-fragment | NA | | NA | 0 |
| S3_vs_k141_31047\|\|full | 2927 | No | NA | | 3 | 0 | 3 | Not-determined | Genome-fragment | NA | | NA | 0 |
| S9_extracted_k141_19394 | 1733 | No | NA | | 2 | 1 | 0 | Low-quality | Genome-fragment | | 2.53 | HMM-based (lower-bound) | 0 |
| S14_extracted_k141_111704 | 1733 | No | NA | | 3 | 2 | 0 | Low-quality | Genome-fragment | | 5.71 | AAI-based (high-confidence) | 0 |
| S14_vs_k141_7574\|\|full | 1733 | No | NA | | 5 | 1 | 0 | Low-quality | Genome-fragment | | 2.93 | AAI-based (high-confidence) | 0 |
| S14_vs_k141_35378\|\|full | 1733 | No | NA | | 3 | 1 | 0 | Low-quality | Genome-fragment | | 3.35 | AAI-based (medium-confidence) | 0 |
| S16_extracted_49214 | 1733 | No | NA | | 2 | 0 | 1 | Low-quality | Genome-fragment | | 2.75 | AAI-based (medium-confidence) | 0 |
| S10_vs_102891\|\|full | 1733 | No | NA | | 2 | 0 | 0 | Low-quality | Genome-fragment | | 2.76 | AAI-based (high-confidence) | 0 |
| S3_vs_k141_55664\|\|full | 2899 | No | NA | | 3 | 0 | 1 | Not-determined | Genome-fragment | NA | | NA | 0 |
| S3_vs_k141_95212\|\|full | 3725 | No | NA | | 4 | 0 | 0 | Not-determined | Genome-fragment | NA | | NA | 0 |
| S3_vs_k141_87579\|\|full | 1898 | No | NA | | 3 | 0 | 1 | Not-determined | Genome-fragment | NA | | NA | 0 |
| S12_extracted_k141_12534 | 1733 | No | NA | | 2 | 1 | 0 | Low-quality | Genome-fragment | | 2.72 | AAI-based (high-confidence) | 0 |
| S3_vs_k141_39328\|\|full | 1040 | No | NA | | 3 | 0 | 0 | Not-determined | Genome-fragment | NA | | NA | 0 |
| S3_vs_k141_39333\|\|full | 2222 | No | NA | | 4 | 0 | 2 | Not-determined | Genome-fragment | NA | | NA | 0 |
| S2_extracted_k141_4137 | 1733 | No | NA | | 4 | 0 | 0 | Low-quality | Genome-fragment | | 4.27 | AAI-based (high-confidence) | 0 |
| S17_extracted_k141_160634 | 1734 | No | NA | | 6 | 0 | 0 | Low-quality | Genome-fragment | | 0.95 | AAI-based (high-confidence) | 0 |
| S3_vs_k141_70299\|\|full | 2828 | No | NA | | 3 | 0 | 2 | Not-determined | Genome-fragment | NA | | NA | 0 |
| S3_vs_k141_31206\|\|full | 2522 | No | NA | | 4 | 0 | 2 | Not-determined | Genome-fragment | NA | | NA | 0 |
| S3_vs_k141_111411\|\|full | 3795 | No | NA | | 5 | 0 | 3 | Not-determined | Genome-fragment | NA | | NA | 0 |
| S11_extracted_k141_85140 | 1734 | No | NA | | 5 | 3 | 0 | Low-quality | Genome-fragment | | 4.63 | AAI-based (high-confidence) | 0 |
| S3_vs_k141_70311\|\|full | 1590 | No | NA | | 2 | 0 | 1 | Not-determined | Genome-fragment | NA | | NA | 0 |
| S3_vs_k141_22653\|\|full | 1192 | No | NA | | 4 | 0 | 0 | Not-determined | Genome-fragment | NA | | NA | 0 |
| S14_vs_k141_9626\|\|full | 1734 | No | NA | | 5 | 4 | 0 | Low-quality | Genome-fragment | | 3.34 | AAI-based (high-confidence) | 0 |
| S1_vs_k141_128007\|\|full | 1734 | No | NA | | 6 | 0 | 0 | Low-quality | Genome-fragment | | 4.16 | AAI-based (high-confidence) | 0 |
| S5_vs_k141_66966\|\|full | 1734 | No | NA | | 4 | 2 | 0 | Low-quality | Genome-fragment | | 3.8 | AAI-based (medium-confidence) | 0 |
| S7_vs_122483\|\|full | 1734 | No | NA | | 4 | 0 | 0 | Low-quality | Genome-fragment | | 3.64 | AAI-based (high-confidence) | 0 |
| S16_vs_138702\|\|full | 1734 | No | NA | | 6 | 0 | 0 | Low-quality | Genome-fragment | | 3.12 | AAI-based (medium-confidence) | 0 |
| S6_extracted_k141_18637 | 1734 | No | NA | | 2 | 0 | 1 | Low-quality | Genome-fragment | | 1.08 | AAI-based (high-confidence) | 0 |
| S3_vs_k141_55777\|\|full | 3936 | No | NA | | 3 | 0 | 0 | Not-determined | Genome-fragment | NA | | NA | 0 |
| S3_vs_k141_111767\|\|lt2gene | 1735 | No | NA | | 2 | 1 | 0 | Low-quality | Genome-fragment | | 4.5 | AAI-based (high-confidence) | 0 |
| S3_vs_k141_39383\|\|full | 1427 | No | NA | | 3 | 0 | 1 | Not-determined | Genome-fragment | NA | | NA | 0 |
| S16_vs_17119\|\|full | 1735 | No | NA | | 4 | 0 | 0 | Low-quality | Genome-fragment | | 2.77 | AAI-based (high-confidence) | 0 |
| S3_vs_k141_7012\|\|full | 6321 | No | NA | | 8 | 0 | 4 | Not-determined | Genome-fragment | NA | | NA | 0 |
| S10_vs_8087\|\|full | 1735 | No | NA | | 4 | 1 | 0 | Low-quality | Genome-fragment | | 4.09 | HMM-based (lower-bound) | 0 |
| S15_vs_48969\|\|full | 1735 | No | NA | | 4 | 0 | 0 | Low-quality | Genome-fragment | | 1.01 | AAI-based (high-confidence) | 0 |
| S8_vs_k141_190388\|\|full | 1735 | No | NA | | 3 | 1 | 0 | Low-quality | Genome-fragment | | 3.5 | AAI-based (high-confidence) | 0 |
| S17_vs_k141_167347\|\|full | 1736 | No | NA | | 4 | 0 | 0 | Low-quality | Genome-fragment | | 3.94 | AAI-based (high-confidence) | 0 |
| S3_vs_k141_39460\|\|full | 1556 | No | NA | | 3 | 0 | 0 | Not-determined | Genome-fragment | NA | | NA | 0 |
| S11_extracted_k141_80385 | 1736 | No | NA | | 5 | 2 | 0 | Low-quality | Genome-fragment | | 4.28 | AAI-based (high-confidence) | 0 |
| S11_vs_k141_87292\|\|lt2gene | 1736 | No | NA | | 2 | 1 | 0 | Low-quality | Genome-fragment | | 4.85 | AAI-based (high-confidence) | 0 |
| S2_extracted_k141_6337 | 1736 | No | NA | | 2 | 0 | 0 | Low-quality | Genome-fragment | | 2.63 | AAI-based (medium-confidence) | 0 |
| S14_vs_k141_28629\|\|full | 1737 | No | NA | | 6 | 3 | 0 | Low-quality | Genome-fragment | | 3.88 | AAI-based (high-confidence) | 0 |
| S3_vs_k141_111499\|\|full | 7778 | No | NA | | 11 | 0 | 4 | Not-determined | Genome-fragment | NA | | NA | 0 |
| S3_vs_k141_111511\|\|full | 2112 | No | NA | | 4 | 0 | 0 | Not-determined | Genome-fragment | NA | | NA | 0 |
| S3_vs_k141_102202\|\|full | 1368 | No | NA | | 2 | 0 | 2 | Not-determined | Genome-fragment | NA | | NA | 0 |
| S3_vs_k141_47566\|\|full | 1065 | No | NA | | 3 | 0 | 0 | Not-determined | Genome-fragment | NA | | NA | 0 |
| S1_vs_k141_58084\|\|full | 1737 | No | NA | | 2 | 1 | 0 | Low-quality | Genome-fragment | | 4.78 | AAI-based (high-confidence) | 0 |
| S1_vs_k141_152103\|\|full | 1737 | No | NA | | 2 | 0 | 0 | Low-quality | Genome-fragment | | 3.32 | AAI-based (medium-confidence) | 0 |
| S5_extracted_k141_85914 | 1737 | No | NA | | 1 | 1 | 0 | Low-quality | Genome-fragment | | 0.91 | HMM-based (lower-bound) | 0 |
| S3_vs_k141_22772\|\|full | 4441 | No | NA | | 6 | 0 | 2 | Not-determined | Genome-fragment | NA | | NA | 0 |
| S3_vs_k141_102219\|\|full | 11813 | No | NA | | 11 | 0 | 7 | Not-determined | Genome-fragment | NA | | NA | 0 |
| S16_vs_102985\|\|full | 1737 | No | NA | | 4 | 2 | 0 | Low-quality | Genome-fragment | | 2.9 | AAI-based (medium-confidence) | 0 |
| S3_vs_k141_102232\|\|full | 3266 | No | NA | | 3 | 0 | 1 | Not-determined | Genome-fragment | NA | | NA | 0 |
| S6_vs_k141_32817\|\|full | 1737 | No | NA | | 6 | 1 | 0 | Low-quality | Genome-fragment | | 3.8 | AAI-based (high-confidence) | 0 |
| S8_extracted_k141_96031 | 1737 | No | NA | | 3 | 0 | 1 | Low-quality | Genome-fragment | | 1.4 | AAI-based (medium-confidence) | 0 |
| S3_vs_k141_102238\|\|full | 2682 | No | NA | | 3 | 0 | 2 | Not-determined | Genome-fragment | NA | | NA | 0 |
| S8_extracted_k141_74196 | 1737 | No | NA | | 4 | 0 | 3 | Low-quality | Genome-fragment | | 0.73 | AAI-based (medium-confidence) | 0 |
| S8_vs_k141_90628\|\|full | 1737 | No | NA | | 3 | 2 | 0 | Low-quality | Genome-fragment | | 3.68 | AAI-based (medium-confidence) | 0 |
| S3_vs_k141_47629\|\|full | 5227 | No | NA | | 5 | 0 | 2 | Not-determined | Genome-fragment | NA | | NA | 0 |
| S9_vs_k141_90376\|\|full | 1738 | No | NA | | 2 | 0 | 0 | Low-quality | Genome-fragment | | 3.14 | AAI-based (medium-confidence) | 0 |
| S9_vs_k141_90638\|\|full | 1738 | No | NA | | 5 | 0 | 0 | Low-quality | Genome-fragment | | 4.02 | AAI-based (high-confidence) | 0 |
| S3_vs_k141_126608\|\|full | 2458 | No | NA | | 4 | 0 | 2 | Not-determined | Genome-fragment | NA | | NA | 0 |
| S3_vs_k141_39685\|\|full | 1926 | No | NA | | 5 | 0 | 1 | Not-determined | Genome-fragment | NA | | NA | 0 |
| S11_extracted_k141_31859 | 1738 | No | NA | | 4 | 1 | 0 | Low-quality | Genome-fragment | | 5.09 | AAI-based (high-confidence) | 0 |
| S5_vs_k141_38167\|\|full | 1738 | No | NA | | 3 | 1 | 0 | Low-quality | Genome-fragment | | 2.5 | HMM-based (lower-bound) | 0 |
| S3_vs_k141_95494\|\|full | 2442 | No | NA | | 3 | 0 | 2 | Not-determined | Genome-fragment | NA | | NA | 0 |
| S7_extracted_27410 | 1738 | No | NA | | 3 | 0 | 0 | Low-quality | Genome-fragment | | 4.23 | AAI-based (high-confidence) | 0 |
| S3_vs_k141_70508\|\|full | 57171 | No | NA | | 59 | 4 | 7 | High-quality | High-quality | | 100 | AAI-based (high-confidence) | 0 |
| S7_extracted_68279 | 1738 | No | NA | | 3 | 2 | 0 | Low-quality | Genome-fragment | | 7.91 | AAI-based (high-confidence) | 0 |
| S7_vs_40520\|\|full | 1738 | No | NA | | 3 | 1 | 0 | Low-quality | Genome-fragment | | 4.07 | AAI-based (high-confidence) | 0 |
| S3_vs_k141_47748\|\|full | 5247 | No | NA | | 7 | 0 | 1 | Not-determined | Genome-fragment | NA | | NA | 0 |
| S3_vs_k141_102349\|\|full | 7137 | No | NA | | 7 | 0 | 3 | Not-determined | Genome-fragment | NA | | NA | 0 |
| S16_vs_108467\|\|full | 1738 | No | NA | | 6 | 1 | 0 | Low-quality | Genome-fragment | | 4.84 | AAI-based (high-confidence) | 0 |
| S16_vs_144896\|\|full | 1738 | No | NA | | 3 | 0 | 2 | Low-quality | Genome-fragment | | 0.87 | AAI-based (medium-confidence) | 0 |
| S11_vs_k141_89985\|\|full | 1739 | No | NA | | 1 | 0 | 1 | Low-quality | Genome-fragment | | 2.34 | AAI-based (high-confidence) | 0 |
| S3_vs_k141_102391\|\|full | 2425 | No | NA | | 3 | 0 | 3 | Not-determined | Genome-fragment | NA | | NA | 0 |
| S3_vs_k141_126726\|\|full | 5577 | No | NA | | 3 | 0 | 2 | Not-determined | Genome-fragment | NA | | NA | 0 |
| S3_vs_k141_102410\|\|full | 2495 | No | NA | | 2 | 0 | 1 | Not-determined | Genome-fragment | NA | | NA | 0 |
| S14_extracted_k141_29387 | 1739 | No | NA | | 2 | 0 | 2 | Low-quality | Genome-fragment | | 0.71 | AAI-based (medium-confidence) | 0 |
| S3_vs_k141_39774\|\|full | 73767 | No | NA | | 99 | 6 | 5 | Medium-quality | Genome-fragment | | 66.49 | AAI-based (medium-confidence) | 0 |
| S3_vs_k141_95574\|\|full | 22530 | No | NA | | 21 | 6 | 1 | Medium-quality | Genome-fragment | | 53.28 | AAI-based (high-confidence) | 0 |
| S3_vs_k141_118967\|\|full | 4548 | No | NA | | 4 | 0 | 4 | Not-determined | Genome-fragment | NA | | NA | 0 |
| S3_vs_k141_23066\|\|full | 1754 | No | NA | | 3 | 0 | 2 | Not-determined | Genome-fragment | NA | | NA | 0 |
| S1_extracted_k141_78762 | 1739 | No | NA | | 1 | 0 | 1 | Low-quality | Genome-fragment | | 0.52 | AAI-based (medium-confidence) | 0 |
| S3_vs_k141_28909\|\|full | 1739 | No | NA | | 3 | 0 | 0 | Low-quality | Genome-fragment | | 4.09 | AAI-based (medium-confidence) | 0 |
| S3_vs_k141_118970\|\|full | 15654 | No | NA | | 12 | 0 | 6 | Not-determined | Genome-fragment | NA | | NA | 0 |
| S3_vs_k141_102451\|\|full | 7596 | No | NA | | 4 | 0 | 0 | Not-determined | Genome-fragment | NA | | NA | 0 |
| S3_vs_k141_102456\|\|full | 1693 | No | NA | | 3 | 0 | 2 | Not-determined | Genome-fragment | NA | | NA | 0 |
| S3_vs_k141_71453\|\|full | 1739 | No | NA | | 2 | 2 | 0 | Low-quality | Genome-fragment | | 4.43 | AAI-based (high-confidence) | 0 |
| S3_vs_k141_102465\|\|full | 1861 | No | NA | | 2 | 0 | 1 | Not-determined | Genome-fragment | NA | | NA | 0 |
| S7_vs_96042\|\|full | 1739 | No | NA | | 4 | 1 | 0 | Low-quality | Genome-fragment | | 3.72 | AAI-based (high-confidence) | 0 |
| S3_vs_k141_31700\|\|full | 3089 | No | NA | | 3 | 0 | 3 | Not-determined | Genome-fragment | NA | | NA | 0 |
| S12_vs_k141_186130\|\|full | 1739 | No | NA | | 7 | 0 | 0 | Low-quality | Genome-fragment | | 3.71 | AAI-based (high-confidence) | 0 |
| S4_extracted_4801 | 1739 | No | NA | | 1 | 0 | 0 | Low-quality | Genome-fragment | | 1.87 | AAI-based (high-confidence) | 0 |
| S3_vs_k141_31720\|\|full | 2352 | No | NA | | 3 | 0 | 0 | Not-determined | Genome-fragment | NA | | NA | 0 |
| S3_vs_k141_7437\|\|full | 2770 | No | NA | | 3 | 0 | 1 | Not-determined | Genome-fragment | NA | | NA | 0 |
| S8_extracted_k141_157561 | 1739 | No | NA | | 3 | 0 | 2 | Low-quality | Genome-fragment | | 0.8 | AAI-based (high-confidence) | 0 |
| S3_vs_k141_102608\|\|full | 5199 | No | NA | | 10 | 0 | 0 | Not-determined | Genome-fragment | NA | | NA | 0 |
| S8_vs_k141_72426\|\|full | 1739 | No | NA | | 2 | 0 | 0 | Low-quality | Genome-fragment | | 3.01 | AAI-based (medium-confidence) | 0 |
| S3_vs_k141_23194\|\|full | 37613 | No | NA | | 44 | 2 | 2 | Medium-quality | Genome-fragment | | 86.73 | AAI-based (medium-confidence) | 0 |
| S3_vs_k141_119082\|\|full | 1359 | No | NA | | 3 | 0 | 1 | Not-determined | Genome-fragment | NA | | NA | 0 |
| S5_vs_k141_73690\|\|full | 1740 | No | NA | | 3 | 3 | 0 | Low-quality | Genome-fragment | | 3.7 | AAI-based (medium-confidence) | 0 |
| S16_vs_33483\|\|full | 1740 | No | NA | | 3 | 2 | 0 | Low-quality | Genome-fragment | | 1.55 | AAI-based (high-confidence) | 0 |
| S3_vs_k141_7553\|\|full | 3628 | No | NA | | 3 | 0 | 2 | Not-determined | Genome-fragment | NA | | NA | 0 |
| S3_vs_k141_31813\|\|full | 1548 | No | NA | | 3 | 0 | 1 | Not-determined | Genome-fragment | NA | | NA | 0 |
| S10_extracted_24396 | 1740 | No | NA | | 2 | 0 | 0 | Low-quality | Genome-fragment | | 1.22 | AAI-based (high-confidence) | 0 |
| S8_vs_k141_125743\|\|full | 1740 | No | NA | | 4 | 1 | 0 | Low-quality | Genome-fragment | | 3.67 | HMM-based (lower-bound) | 0 |
| S8_vs_k141_93238\|\|full | 1740 | No | NA | | 5 | 0 | 0 | Low-quality | Genome-fragment | | 3.58 | AAI-based (medium-confidence) | 0 |
| S9_vs_k141_27970\|\|full | 1741 | No | NA | | 3 | 0 | 0 | Low-quality | Genome-fragment | | 4.11 | AAI-based (high-confidence) | 0 |
| S11_vs_k141_91665\|\|full | 1741 | No | NA | | 3 | 0 | 1 | Low-quality | Genome-fragment | | 1.1 | AAI-based (medium-confidence) | 0 |
| S5_vs_k141_15309\|\|lt2gene | 1741 | No | NA | | 2 | 2 | 0 | Low-quality | Genome-fragment | | 0.94 | HMM-based (lower-bound) | 0 |
| S3_vs_k141_71046\|\|full | 2903 | No | NA | | 4 | 0 | 3 | Not-determined | Genome-fragment | NA | | NA | 0 |
| S3_vs_k141_23364\|\|full | 6092 | No | NA | | 8 | 0 | 3 | Not-determined | Genome-fragment | NA | | NA | 0 |
| S7_extracted_16224 | 1741 | No | NA | | 1 | 0 | 0 | Low-quality | Genome-fragment | | 1.81 | AAI-based (high-confidence) | 0 |
| S12_vs_k141_139783\|\|full | 1741 | No | NA | | 2 | 0 | 0 | Low-quality | Genome-fragment | | 2.1 | AAI-based (high-confidence) | 0 |
| S5_extracted_k141_74748 | 1742 | No | NA | | 4 | 0 | 0 | Low-quality | Genome-fragment | | 4.95 | AAI-based (high-confidence) | 0 |
| S3_vs_k141_119216\|\|full | 4861 | No | NA | | 7 | 0 | 5 | Not-determined | Genome-fragment | NA | | NA | 0 |
| S7_vs_37434\|\|full | 1742 | No | NA | | 4 | 1 | 0 | Low-quality | Genome-fragment | | 5.21 | AAI-based (medium-confidence) | 0 |
| S7_vs_67644\|\|full | 1742 | No | NA | | 2 | 0 | 0 | Low-quality | Genome-fragment | | 1.16 | AAI-based (high-confidence) | 0 |
| S10_extracted_109253 | 1742 | No | NA | | 3 | 0 | 0 | Low-quality | Genome-fragment | | 2.27 | AAI-based (high-confidence) | 0 |
| S12_extracted_k141_128704 | 1742 | No | NA | | 2 | 0 | 2 | Low-quality | Genome-fragment | | 0.44 | AAI-based (high-confidence) | 0 |
| S4_extracted_28940 | 1742 | No | NA | | 1 | 0 | 1 | Low-quality | Genome-fragment | | 1.15 | AAI-based (high-confidence) | 0 |
| S6_vs_k141_25987\|\|full | 1742 | No | NA | | 4 | 0 | 1 | Low-quality | Genome-fragment | | 1.79 | AAI-based (medium-confidence) | 0 |
| S9_vs_k141_88332\|\|full | 1743 | No | NA | | 3 | 0 | 2 | Low-quality | Genome-fragment | | 0.59 | AAI-based (medium-confidence) | 0 |
| S11_vs_k141_32722\|\|full | 1743 | No | NA | | 5 | 0 | 0 | Low-quality | Genome-fragment | | 2.7 | AAI-based (high-confidence) | 0 |
| S14_extracted_k141_66199 | 1743 | No | NA | | 2 | 2 | 0 | Low-quality | Genome-fragment | | 5.76 | AAI-based (high-confidence) | 0 |
| S3_vs_k141_127088\|\|full | 4976 | No | NA | | 3 | 0 | 2 | Not-determined | Genome-fragment | NA | | NA | 0 |
| S3_vs_k141_127093\|\|full | 1224 | No | NA | | 2 | 0 | 1 | Not-determined | Genome-fragment | NA | | NA | 0 |
| S1_vs_k141_120115\|\|full | 1743 | No | NA | | 6 | 2 | 0 | Low-quality | Genome-fragment | | 4.5 | AAI-based (high-confidence) | 0 |
| S3_vs_k141_119306\|\|full | 1767 | No | NA | | 3 | 0 | 0 | Not-determined | Genome-fragment | NA | | NA | 0 |
| S3_vs_k141_99577\|\|full | 1743 | No | NA | | 3 | 0 | 0 | Low-quality | Genome-fragment | | 2.21 | AAI-based (medium-confidence) | 0 |
| S3_vs_k141_23544\|\|full | 5566 | No | NA | | 3 | 0 | 1 | Not-determined | Genome-fragment | NA | | NA | 0 |
| S5_vs_k141_29954\|\|full | 1743 | No | NA | | 4 | 0 | 0 | Low-quality | Genome-fragment | | 3.08 | AAI-based (high-confidence) | 0 |
| S16_extracted_160096 | 1743 | No | NA | | 2 | 0 | 0 | Low-quality | Genome-fragment | | 3.01 | AAI-based (medium-confidence) | 0 |
| S16_vs_36708\|\|full | 1743 | No | NA | | 1 | 0 | 1 | Low-quality | Genome-fragment | | 2.62 | AAI-based (medium-confidence) | 0 |
| S3_vs_k141_7948\|\|full | 5076 | No | NA | | 4 | 0 | 3 | Not-determined | Genome-fragment | NA | | NA | 0 |
| S3_vs_k141_23597\|\|full | 6554 | No | NA | | 5 | 0 | 4 | Not-determined | Genome-fragment | NA | | NA | 0 |
| S12_vs_k141_5790\|\|full | 1743 | No | NA | | 5 | 1 | 0 | Low-quality | Genome-fragment | | 2.94 | HMM-based (lower-bound) | 0 |
| S3_vs_k141_103061\|\|full | 1046 | No | NA | | 2 | 0 | 0 | Not-determined | Genome-fragment | NA | | NA | 0 |
| S8_extracted_k141_43481 | 1743 | No | NA | | 2 | 0 | 0 | Low-quality | Genome-fragment | | 3.87 | AAI-based (high-confidence) | 0 |
| S3_vs_k141_71439\|\|full | 2470 | No | NA | | 3 | 0 | 3 | Not-determined | Genome-fragment | NA | | NA | 0 |
| S8_extracted_k141_141002 | 1743 | No | NA | | 1 | 0 | 1 | Low-quality | Genome-fragment | | 2.79 | AAI-based (medium-confidence) | 0 |
| S8_vs_k141_91188\|\|full | 1743 | No | NA | | 3 | 0 | 0 | Low-quality | Genome-fragment | | 3.07 | AAI-based (medium-confidence) | 0 |
| S3_vs_k141_119479\|\|full | 2613 | No | NA | | 2 | 0 | 0 | Not-determined | Genome-fragment | NA | | NA | 0 |
| S3_vs_k141_127319\|\|full | 1975 | No | NA | | 4 | 0 | 0 | Not-determined | Genome-fragment | NA | | NA | 0 |
| S3_vs_k141_127360\|\|full | 1333 | No | NA | | 2 | 0 | 1 | Not-determined | Genome-fragment | NA | | NA | 0 |
| S17_vs_k141_63043\|\|full | 1744 | No | NA | | 4 | 0 | 0 | Low-quality | Genome-fragment | | 2.5 | AAI-based (high-confidence) | 0 |
| S3_vs_k141_127387\|\|full | 28569 | No | NA | | 40 | 12 | 0 | Medium-quality | Genome-fragment | | 74.37 | AAI-based (medium-confidence) | 0 |
| S3_vs_k141_127388\|\|full | 1797 | No | NA | | 2 | 0 | 1 | Not-determined | Genome-fragment | NA | | NA | 0 |
| S3_vs_k141_23831\|\|full | 5020 | No | NA | | 4 | 0 | 3 | Not-determined | Genome-fragment | NA | | NA | 0 |
| S3_vs_k141_103298\|\|full | 2917 | No | NA | | 5 | 0 | 2 | Not-determined | Genome-fragment | NA | | NA | 0 |
| S3_vs_k141_71620\|\|full | 2817 | No | NA | | 4 | 0 | 1 | Not-determined | Genome-fragment | NA | | NA | 0 |
| S11_vs_k141_53903\|\|full | 1744 | No | NA | | 5 | 1 | 0 | Low-quality | Genome-fragment | | 2.87 | AAI-based (high-confidence) | 0 |
| S3_vs_k141_103368\|\|full | 1683 | No | NA | | 2 | 0 | 1 | Not-determined | Genome-fragment | NA | | NA | 0 |
| S3_vs_k141_71690\|\|full | 1269 | No | NA | | 3 | 0 | 1 | Not-determined | Genome-fragment | NA | | NA | 0 |
| S3_vs_k141_103411\|\|full | 5302 | No | NA | | 7 | 0 | 4 | Not-determined | Genome-fragment | NA | | NA | 0 |
| S3_vs_k141_14191\|\|full | 1744 | No | NA | | 5 | 0 | 0 | Low-quality | Genome-fragment | | 1.66 | AAI-based (medium-confidence) | 0 |
| S5_vs_k141_87961\|\|full | 1744 | No | NA | | 3 | 1 | 0 | Low-quality | Genome-fragment | | 3.96 | AAI-based (medium-confidence) | 0 |
| S3_vs_k141_87728\|\|0_partial | 15881 | No | NA | | 20 | 3 | 1 | Medium-quality | Genome-fragment | | 55.35 | AAI-based (high-confidence) | 0 |
| S3_vs_k141_16671\|\|0_partial | 23004 | No | NA | | 27 | 0 | 17 | Not-determined | Genome-fragment | NA | | NA | 0 |
| S3_vs_k141_40939\|\|0_partial | 44878 | Yes | | 42098 | 47 | 3 | 7 | Medium-quality | Genome-fragment | | 68.69 | AAI-based (high-confidence) | 6.19 |
| S3_vs_k141_40939\|\|1_partial | 15072 | No | NA | | 8 | 0 | 6 | Not-determined | Genome-fragment | NA | | NA | 0 |
| S7_vs_46511\|\|lt2gene | 1744 | No | NA | | 1 | 1 | 0 | Low-quality | Genome-fragment | | 2.26 | AAI-based (medium-confidence) | 0 |
| S8_extracted_k141_13112 | 1744 | No | NA | | 3 | 0 | 0 | Low-quality | Genome-fragment | | 1.11 | AAI-based (medium-confidence) | 0 |
| S3_vs_k141_122047\|\|0_partial | 14833 | No | NA | | 10 | 0 | 6 | Not-determined | Genome-fragment | NA | | NA | 0 |
| S8_vs_k141_38072\|\|full | 1744 | No | NA | | 2 | 1 | 0 | Low-quality | Genome-fragment | | 1 | HMM-based (lower-bound) | 0 |
| S3_vs_k141_82962\|\|0_partial | 11132 | No | NA | | 14 | 0 | 7 | Not-determined | Genome-fragment | NA | | NA | 0 |
| S17_vs_k141_11203\|\|full | 1745 | No | NA | | 3 | 0 | 0 | Low-quality | Genome-fragment | | 2.15 | AAI-based (high-confidence) | 0 |
| S9_vs_k141_19428\|\|full | 1745 | No | NA | | 3 | 1 | 0 | Low-quality | Genome-fragment | | 3.16 | AAI-based (medium-confidence) | 0 |
| S3_vs_k141_3710\|\|0_partial | 14536 | No | NA | | 13 | 0 | 11 | Not-determined | Genome-fragment | NA | | NA | 0 |
| S3_vs_k141_43757\|\|0_partial | 17443 | No | NA | | 16 | 0 | 7 | Not-determined | Genome-fragment | NA | | NA | 0 |
| S3_vs_k141_19171\|\|0_partial | 13012 | No | NA | | 17 | 0 | 5 | Not-determined | Genome-fragment | NA | | NA | 0 |
| S11_vs_k141_74867\|\|full | 1745 | No | NA | | 5 | 1 | 0 | Low-quality | Genome-fragment | | 3.89 | AAI-based (high-confidence) | 0 |
| S3_vs_k141_36691\|\|0_partial | 12386 | No | NA | | 15 | 0 | 3 | Not-determined | Genome-fragment | NA | | NA | 0 |
| S7_vs_50406\|\|full | 1745 | No | NA | | 3 | 0 | 0 | Low-quality | Genome-fragment | | 3.51 | AAI-based (medium-confidence) | 0 |
| S3_vs_k141_68123\|\|0_partial | 23940 | No | NA | | 18 | 0 | 13 | Not-determined | Genome-fragment | NA | | NA | 0 |
| S3_vs_k141_85076\|\|0_partial | 11247 | No | NA | | 7 | 0 | 5 | Not-determined | Genome-fragment | NA | | NA | 0 |
| S3_vs_k141_124525\|\|0_partial | 21186 | No | NA | | 21 | 0 | 15 | Not-determined | Genome-fragment | NA | | NA | 0 |
| S3_vs_k141_117894\|\|0_partial | 56521 | No | NA | | 66 | 2 | 15 | Medium-quality | Genome-fragment | | 73.03 | HMM-based (lower-bound) | 0 |
| S16_vs_34608\|\|full | 1745 | No | NA | | 2 | 0 | 0 | Low-quality | Genome-fragment | | 1.44 | AAI-based (high-confidence) | 0 |
| S3_vs_k141_71320\|\|0_partial | 16672 | No | NA | | 16 | 0 | 2 | Not-determined | Genome-fragment | NA | | NA | 0 |
| S3_vs_k141_103464\|\|0_partial | 28172 | No | NA | | 30 | 0 | 10 | Not-determined | Genome-fragment | NA | | NA | 0 |
| S12_vs_k141_62362\|\|full | 1745 | No | NA | | 2 | 1 | 0 | Low-quality | Genome-fragment | | 32.82 | AAI-based (medium-confidence) | 0 |

| | | | | | | | | | | | |
|---|---|---|---|---|---|---|---|---|---|---|---|
| S8_extracted_k141_114192 | 1745 | No | NA | 2 | 0 | 0 | Low-quality | Genome-fragment | 4.11 | AAI-based (high-confidence) | 0 |
| S8_vs_k141_171477\|\|lt2gene | 1745 | No | NA | 1 | 0 | 0 | Low-quality | Genome-fragment | 4.11 | AAI-based (medium-confidence) | 0 |
| S11_extracted_k141_70135 | 1746 | No | NA | 2 | 0 | 2 | Low-quality | Genome-fragment | 1.16 | AAI-based (high-confidence) | 0 |
| S11_vs_k141_98506\|\|full | 1746 | No | NA | 5 | 1 | 0 | Low-quality | Genome-fragment | 3.04 | AAI-based (high-confidence) | 0 |
| S13_extracted_43087 | 1746 | No | NA | 5 | 0 | 0 | Low-quality | Genome-fragment | 1.81 | AAI-based (high-confidence) | 0 |
| S1_extracted_k141_6619 | 1746 | No | NA | 3 | 0 | 0 | Low-quality | Genome-fragment | 2.82 | AAI-based (high-confidence) | 0 |
| S3_vs_k141_96304\|\|lt2gene | 1157 | No | NA | 2 | 0 | 0 | Not-determined | Genome-fragment | NA | NA | 0 |
| S5_vs_k141_1146\|\|full | 1746 | No | NA | 3 | 2 | 0 | Low-quality | Genome-fragment | 4.19 | AAI-based (high-confidence) | 0 |
| S5_vs_k141_102354\|\|full | 1746 | No | NA | 4 | 2 | 0 | Low-quality | Genome-fragment | 3.79 | AAI-based (medium-confidence) | 0 |
| S16_vs_17901\|\|full | 1746 | No | NA | 4 | 0 | 0 | Low-quality | Genome-fragment | 3.3 | AAI-based (medium-confidence) | 0 |
| S12_vs_k141_82435\|\|full | 1746 | No | NA | 3 | 1 | 0 | Low-quality | Genome-fragment | 0.16 | HMM-based (lower-bound) | 0 |
| S4_extracted_17976 | 1746 | No | NA | 3 | 0 | 2 | Low-quality | Genome-fragment | 0.61 | AAI-based (medium-confidence) | 0 |
| S8_extracted_k141_23205 | 1746 | No | NA | 2 | 0 | 0 | Low-quality | Genome-fragment | 2.61 | AAI-based (high-confidence) | 0 |
| S9_extracted_k141_63070 | 1747 | No | NA | 5 | 2 | 0 | Low-quality | Genome-fragment | 4.31 | AAI-based (high-confidence) | 0 |
| S11_vs_k141_18152\|\|full | 1747 | No | NA | 3 | 0 | 0 | Low-quality | Genome-fragment | 1.19 | AAI-based (high-confidence) | 0 |
| S14_extracted_k141_80080 | 1747 | No | NA | 3 | 3 | 0 | Low-quality | Genome-fragment | 5.11 | AAI-based (high-confidence) | 0 |
| S1_vs_k141_144640\|\|full | 1747 | No | NA | 8 | 0 | 0 | Low-quality | Genome-fragment | 1.13 | AAI-based (high-confidence) | 0 |
| S1_vs_k141_59390\|\|full | 1747 | No | NA | 4 | 0 | 0 | Low-quality | Genome-fragment | 3.51 | AAI-based (high-confidence) | 0 |
| S3_vs_k141_41177\|\|lt2gene | 1284 | No | NA | 1 | 0 | 0 | Not-determined | Genome-fragment | NA | NA | 0 |
| S3_vs_k141_112709\|\|lt2gene | 1349 | No | NA | 1 | 0 | 0 | Not-determined | Genome-fragment | NA | NA | 0 |
| S7_vs_79327\|\|full | 1747 | No | NA | 3 | 0 | 0 | Low-quality | Genome-fragment | 1.02 | AAI-based (high-confidence) | 0 |
| S3_vs_k141_97482\|\|lt2gene | 1028 | No | NA | 1 | 0 | 0 | Not-determined | Genome-fragment | NA | NA | 0 |
| S16_extracted_79133 | 1747 | No | NA | 5 | 4 | 0 | Low-quality | Genome-fragment | 2.68 | AAI-based (high-confidence) | 0 |
| S12_vs_k141_186283\|\|full | 1747 | No | NA | 6 | 0 | 0 | Low-quality | Genome-fragment | 2.44 | AAI-based (high-confidence) | 0 |
| S13_vs_97403\|\|full | 1748 | No | NA | 3 | 0 | 2 | Low-quality | Genome-fragment | 0.9 | AAI-based (medium-confidence) | 0 |
| S14_vs_k141_59115\|\|full | 1748 | No | NA | 3 | 0 | 2 | Low-quality | Genome-fragment | 2.16 | AAI-based (medium-confidence) | 0 |
| S3_vs_k141_10538\|\|lt2gene | 1651 | No | NA | 2 | 0 | 0 | Not-determined | Genome-fragment | NA | NA | 0 |
| S1_extracted_k141_36324 | 1748 | No | NA | 3 | 0 | 0 | Low-quality | Genome-fragment | 1.21 | AAI-based (high-confidence) | 0 |
| S1_vs_k141_151566\|\|full | 1748 | No | NA | 6 | 1 | 0 | Low-quality | Genome-fragment | 3.62 | AAI-based (high-confidence) | 0 |
| S3_vs_k141_82061\|\|lt2gene | 1790 | No | NA | 1 | 0 | 0 | Not-determined | Genome-fragment | NA | NA | 0 |
| S5_vs_k141_135494\|\|full | 1748 | No | NA | 4 | 1 | 0 | Low-quality | Genome-fragment | 3.89 | AAI-based (high-confidence) | 0 |
| S3_vs_k141_50606\|\|lt2gene | 1214 | No | NA | 2 | 0 | 0 | Not-determined | Genome-fragment | NA | NA | 0 |
| S3_vs_k141_34668\|\|lt2gene | 1460 | No | NA | 2 | 0 | 0 | Not-determined | Genome-fragment | NA | NA | 0 |
| S3_vs_k141_74963\|\|lt2gene | 1225 | No | NA | 1 | 0 | 0 | Not-determined | Genome-fragment | NA | NA | 0 |
| S16_vs_119473\|\|full | 1748 | No | NA | 3 | 0 | 1 | Low-quality | Genome-fragment | 1.3 | AAI-based (high-confidence) | 0 |
| S8_extracted_k141_17388 | 1748 | No | NA | 2 | 0 | 1 | Low-quality | Genome-fragment | 0.19 | AAI-based (high-confidence) | 0 |
| S8_extracted_k141_191027 | 1748 | No | NA | 1 | 0 | 0 | Low-quality | Genome-fragment | 2.69 | AAI-based (medium-confidence) | 0 |
| S17_vs_k141_86876\|\|full | 1749 | No | NA | 5 | 0 | 0 | Low-quality | Genome-fragment | 3.32 | AAI-based (medium-confidence) | 0 |
| S14_vs_k141_28891\|\|full | 1749 | No | NA | 4 | 0 | 1 | Low-quality | Genome-fragment | 2.86 | AAI-based (medium-confidence) | 0 |
| S7_vs_54897\|\|full | 1749 | No | NA | 5 | 2 | 0 | Low-quality | Genome-fragment | 4.22 | AAI-based (high-confidence) | 0 |
| S16_vs_123691\|\|full | 1749 | No | NA | 3 | 0 | 0 | Low-quality | Genome-fragment | 4.26 | AAI-based (high-confidence) | 0 |
| S3_vs_k141_18582\|\|lt2gene | 1387 | No | NA | 2 | 0 | 0 | Not-determined | Genome-fragment | NA | NA | 0 |
| S15_extracted_77440 | 1749 | No | NA | 1 | 0 | 0 | Low-quality | Genome-fragment | 2.93 | AAI-based (medium-confidence) | 0 |
| S8_vs_k141_32925\|\|full | 1749 | No | NA | 2 | 0 | 0 | Low-quality | Genome-fragment | 5.59 | AAI-based (high-confidence) | 0 |
| S9_extracted_k141_59297 | 1750 | No | NA | 1 | 0 | 1 | Low-quality | Genome-fragment | 0.55 | AAI-based (medium-confidence) | 0 |
| S9_vs_k141_57504\|\|full | 1750 | No | NA | 2 | 0 | 1 | Low-quality | Genome-fragment | 3.33 | AAI-based (high-confidence) | 0 |
| S13_vs_16423\|\|full | 1750 | No | NA | 4 | 2 | 0 | Low-quality | Genome-fragment | 3.01 | AAI-based (high-confidence) | 0 |
| S3_extracted_k141_91764 | 1750 | No | NA | 7 | 0 | 0 | Low-quality | Genome-fragment | 0.97 | AAI-based (high-confidence) | 0 |
| S10_vs_37752\|\|full | 1750 | No | NA | 3 | 2 | 0 | Low-quality | Genome-fragment | 2.79 | AAI-based (high-confidence) | 0 |
| S10_vs_121252\|\|full | 1750 | No | NA | 3 | 1 | 0 | Low-quality | Genome-fragment | 1.52 | AAI-based (high-confidence) | 0 |
| S15_vs_38445\|\|full | 1750 | No | NA | 3 | 0 | 0 | Low-quality | Genome-fragment | 4.41 | AAI-based (high-confidence) | 0 |
| S6_extracted_k141_13931 | 1750 | No | NA | 4 | 3 | 0 | Low-quality | Genome-fragment | 11.77 | AAI-based (medium-confidence) | 0 |
| S11_vs_k141_79853\|\|full | 1751 | No | NA | 3 | 1 | 0 | Low-quality | Genome-fragment | 0.77 | HMM-based (lower-bound) | 0 |
| S14_vs_k141_101576\|\|full | 1751 | No | NA | 2 | 0 | 2 | Low-quality | Genome-fragment | 2.83 | AAI-based (medium-confidence) | 0 |
| S1_vs_k141_139452\|\|full | 1751 | No | NA | 2 | 1 | 0 | Low-quality | Genome-fragment | 6 | HMM-based (lower-bound) | 0 |
| S3_vs_k141_123228\|\|lt2gene | 1215 | No | NA | 1 | 0 | 0 | Not-determined | Genome-fragment | NA | NA | 0 |
| S3_vs_k141_28334\|\|lt2gene | 1753 | No | NA | 2 | 0 | 0 | Not-determined | Genome-fragment | NA | NA | 0 |
| S3_extracted_k141_27978 | 1751 | No | NA | 4 | 0 | 0 | Low-quality | Genome-fragment | 1.52 | AAI-based (medium-confidence) | 0 |
| S7_vs_38579\|\|full | 1751 | No | NA | 4 | 3 | 0 | Low-quality | Genome-fragment | 4.35 | AAI-based (high-confidence) | 0 |
| S16_vs_97608\|\|full | 1751 | No | NA | 3 | 2 | 0 | Low-quality | Genome-fragment | 3.57 | HMM-based (lower-bound) | 0 |
| S10_vs_82286\|\|full | 1751 | No | NA | 4 | 4 | 0 | Low-quality | Genome-fragment | 3.91 | AAI-based (high-confidence) | 0 |
| S8_extracted_k141_56621 | 1751 | No | NA | 2 | 0 | 2 | Low-quality | Genome-fragment | 0.5 | AAI-based (high-confidence) | 0 |
| S3_vs_k141_53356\|\|lt2gene | 1213 | No | NA | 2 | 0 | 0 | Not-determined | Genome-fragment | NA | NA | 0 |
| S8_extracted_k141_172207 | 1751 | No | NA | 2 | 0 | 1 | Low-quality | Genome-fragment | 0.48 | AAI-based (medium-confidence) | 0 |
| S3_vs_k141_20410\|\|lt2gene | 1432 | No | NA | 1 | 0 | 1 | Not-determined | Genome-fragment | NA | NA | 0 |
| S8_vs_k141_103756\|\|full | 1751 | No | NA | 3 | 0 | 0 | Low-quality | Genome-fragment | 3.31 | AAI-based (medium-confidence) | 0 |
| S9_vs_k141_84342\|\|full | 1752 | No | NA | 3 | 1 | 0 | Low-quality | Genome-fragment | 0.99 | AAI-based (medium-confidence) | 0 |
| S1_extracted_k141_116504 | 1752 | No | NA | 2 | 0 | 2 | Low-quality | Genome-fragment | 2.34 | AAI-based (medium-confidence) | 0 |
| S7_vs_15273\|\|full | 1752 | No | NA | 4 | 3 | 0 | Low-quality | Genome-fragment | 3.16 | AAI-based (medium-confidence) | 0 |
| S10_vs_73690\|\|lt2gene | 1752 | No | NA | 2 | 1 | 0 | Low-quality | Genome-fragment | 9.75 | AAI-based (high-confidence) | 0 |
| S12_extracted_k141_62499 | 1752 | No | NA | 3 | 0 | 0 | Low-quality | Genome-fragment | 4.4 | AAI-based (high-confidence) | 0 |
| S12_vs_k141_137557\|\|full | 1752 | No | NA | 3 | 0 | 1 | Low-quality | Genome-fragment | 1.79 | AAI-based (medium-confidence) | 0 |
| S6_vs_k141_4843\|\|full | 1752 | No | NA | 4 | 0 | 0 | Low-quality | Genome-fragment | 3.36 | AAI-based (medium-confidence) | 0 |
| S8_extracted_k141_156355 | 1752 | No | NA | 4 | 0 | 0 | Low-quality | Genome-fragment | 1.08 | AAI-based (high-confidence) | 0 |
| S9_extracted_k141_35002 | 1753 | No | NA | 1 | 1 | 0 | Low-quality | Genome-fragment | 4.96 | AAI-based (high-confidence) | 0 |
| S14_vs_k141_38206\|\|full | 1753 | No | NA | 3 | 0 | 0 | Low-quality | Genome-fragment | 3.95 | AAI-based (medium-confidence) | 0 |
| S3_vs_k141_33367\|\|full | 1753 | Yes | 1186 | 3 | 1 | 1 | Low-quality | Genome-fragment | 3.45 | AAI-based (high-confidence) | 32.34 |
| S16_extracted_19243 | 1753 | No | NA | 3 | 0 | 1 | Low-quality | Genome-fragment | 2.22 | AAI-based (medium-confidence) | 0 |
| S16_vs_33563\|\|full | 1753 | No | NA | 3 | 0 | 2 | Low-quality | Genome-fragment | 1.05 | AAI-based (high-confidence) | 0 |
| S10_vs_34954\|\|full | 1753 | No | NA | 5 | 0 | 0 | Low-quality | Genome-fragment | 2.95 | AAI-based (medium-confidence) | 0 |
| S10_vs_8607\|\|lt2gene | 1753 | No | NA | 2 | 1 | 0 | Low-quality | Genome-fragment | 0.63 | HMM-based (lower-bound) | 0 |
| S17_vs_k141_46783\|\|full | 1754 | No | NA | 4 | 1 | 0 | Low-quality | Genome-fragment | 3.87 | AAI-based (high-confidence) | 0 |
| S13_extracted_49943 | 1754 | No | NA | 3 | 1 | 0 | Low-quality | Genome-fragment | 4.18 | AAI-based (high-confidence) | 0 |
| S14_vs_k141_102681\|\|full | 1754 | No | NA | 2 | 0 | 2 | Low-quality | Genome-fragment | 2.49 | AAI-based (medium-confidence) | 0 |
| S3_extracted_k141_93349 | 1754 | No | NA | 5 | 0 | 0 | Low-quality | Genome-fragment | 4.52 | AAI-based (medium-confidence) | 0 |
| S7_extracted_53414 | 1754 | No | NA | 1 | 0 | 1 | Low-quality | Genome-fragment | 4.95 | AAI-based (medium-confidence) | 0 |
| S7_extracted_126276 | 1754 | No | NA | 2 | 0 | 0 | Low-quality | Genome-fragment | 2.71 | AAI-based (medium-confidence) | 0 |
| S10_vs_67195\|\|full | 1754 | No | NA | 4 | 1 | 0 | Low-quality | Genome-fragment | 4.48 | AAI-based (medium-confidence) | 0 |
| S12_vs_k141_126460\|\|full | 1754 | No | NA | 3 | 0 | 3 | Low-quality | Genome-fragment | 1.06 | AAI-based (medium-confidence) | 0 |
| S6_vs_k141_27191\|\|full | 1754 | No | NA | 5 | 0 | 0 | Low-quality | Genome-fragment | 3.72 | AAI-based (high-confidence) | 0 |
| S8_vs_k141_73300\|\|full | 1754 | No | NA | 5 | 0 | 3 | Low-quality | Genome-fragment | 0.51 | AAI-based (medium-confidence) | 0 |
| S3_vs_k141_7687\|\|lt2gene | 1178 | No | NA | 2 | 0 | 0 | Not-determined | Genome-fragment | NA | NA | 0 |
| S9_vs_k141_31440\|\|full | 1755 | No | NA | 2 | 0 | 2 | Low-quality | Genome-fragment | 0.88 | AAI-based (medium-confidence) | 0 |
| S5_vs_k141_130752\|\|full | 1755 | Yes | 1176 | 3 | 1 | 1 | Low-quality | Genome-fragment | 0.7 | AAI-based (medium-confidence) | 32.99 |
| S3_vs_k141_103605\|\|lt2gene | 3689 | No | NA | 1 | 0 | 1 | Not-determined | Genome-fragment | NA | NA | 0 |
| S5_extracted_k141_72860 | 2105 | No | NA | 1 | 0 | 0 | Not-determined | Genome-fragment | NA | NA | 0 |
| S7_vs_97060\|\|full | 1755 | No | NA | 4 | 1 | 0 | Low-quality | Genome-fragment | 3.66 | AAI-based (medium-confidence) | 0 |
| S5_extracted_k141_103226 | 2337 | No | NA | 3 | 0 | 2 | Not-determined | Genome-fragment | NA | NA | 0 |
| S5_extracted_k141_54650 | 5883 | No | NA | 5 | 0 | 5 | Not-determined | Genome-fragment | NA | NA | 0 |
| S10_vs_35875\|\|full | 1755 | No | NA | 3 | 2 | 0 | Low-quality | Genome-fragment | 2.62 | HMM-based (lower-bound) | 0 |
| S5_extracted_k141_42581 | 1799 | No | NA | 2 | 0 | 1 | Not-determined | Genome-fragment | NA | NA | 0 |
| S5_extracted_k141_85063 | 2874 | No | NA | 5 | 0 | 4 | Not-determined | Genome-fragment | NA | NA | 0 |
| S5_extracted_k141_54688 | 1174 | No | NA | 2 | 0 | 1 | Not-determined | Genome-fragment | NA | NA | 0 |
| S10_vs_83538\|\|full | 1755 | No | NA | 3 | 0 | 1 | Low-quality | Genome-fragment | 2.45 | AAI-based (medium-confidence) | 0 |
| S5_extracted_k141_24324 | 4805 | No | NA | 8 | 0 | 3 | Not-determined | Genome-fragment | NA | NA | 0 |
| S8_extracted_k141_128105 | 1755 | No | NA | 2 | 0 | 0 | Low-quality | Genome-fragment | 3.63 | AAI-based (medium-confidence) | 0 |
| S5_extracted_k141_12197 | 1640 | No | NA | 4 | 0 | 2 | Not-determined | Genome-fragment | NA | NA | 0 |
| S17_extracted_k141_61753 | 1756 | No | NA | 4 | 0 | 0 | Low-quality | Genome-fragment | 4.28 | AAI-based (high-confidence) | 0 |
| S17_extracted_k141_124501 | 1756 | No | NA | 5 | 1 | 0 | Low-quality | Genome-fragment | 2.63 | AAI-based (high-confidence) | 0 |
| S17_vs_k141_51869\|\|full | 1756 | Yes | 507 | 3 | 1 | 1 | Low-quality | Genome-fragment | 0.27 | HMM-based (lower-bound) | 71.13 |
| S5_extracted_k141_85071 | 1199 | No | NA | 2 | 0 | 1 | Not-determined | Genome-fragment | NA | NA | 0 |
| S9_vs_k141_38148\|\|full | 1756 | No | NA | 2 | 0 | 0 | Low-quality | Genome-fragment | 1.16 | AAI-based (medium-confidence) | 0 |
| S5_extracted_k141_139765 | 3163 | No | NA | 2 | 0 | 2 | Not-determined | Genome-fragment | NA | NA | 0 |
| S9_vs_k141_64223\|\|full | 1756 | No | NA | 4 | 0 | 1 | Low-quality | Genome-fragment | 4.21 | AAI-based (medium-confidence) | 0 |
| S5_extracted_k141_6233 | 3149 | No | NA | 4 | 0 | 1 | Not-determined | Genome-fragment | NA | NA | 0 |
| S1_vs_k141_127397\|\|full | 1756 | No | NA | 2 | 0 | 2 | Low-quality | Genome-fragment | 1.08 | AAI-based (medium-confidence) | 0 |
| S5_extracted_k141_60928 | 3426 | No | NA | 3 | 0 | 0 | Not-determined | Genome-fragment | NA | NA | 0 |
| S1_vs_k141_108858\|\|full | 1756 | No | NA | 4 | 0 | 1 | Low-quality | Genome-fragment | 3.71 | AAI-based (medium-confidence) | 0 |
| S5_extracted_k141_121571 | 1650 | No | NA | 2 | 0 | 1 | Not-determined | Genome-fragment | NA | NA | 0 |
| S5_extracted_k141_48695 | 1739 | No | NA | 1 | 0 | 1 | Not-determined | Genome-fragment | NA | NA | 0 |
| S5_extracted_k141_42740 | 1457 | No | NA | 2 | 0 | 1 | Not-determined | Genome-fragment | NA | NA | 0 |
| S5_extracted_k141_127640 | 3197 | No | NA | 4 | 0 | 0 | Not-determined | Genome-fragment | NA | NA | 0 |
| S1_vs_k141_4459\|\|full | 1756 | No | NA | 3 | 1 | 0 | Low-quality | Genome-fragment | 4.08 | AAI-based (medium-confidence) | 0 |
| S5_extracted_k141_133653 | 1140 | No | NA | 1 | 0 | 1 | Not-determined | Genome-fragment | NA | NA | 0 |
| S5_extracted_k141_133656 | 1311 | No | NA | 3 | 0 | 0 | Not-determined | Genome-fragment | NA | NA | 0 |
| S5_extracted_k141_97312 | 1216 | No | NA | 2 | 0 | 2 | Not-determined | Genome-fragment | NA | NA | 0 |
| S5_vs_k141_134549\|\|full | 1756 | No | NA | 2 | 1 | 0 | Low-quality | Genome-fragment | 2.66 | HMM-based (lower-bound) | 0 |
| S5_vs_k141_136564\|\|full | 1756 | No | NA | 4 | 1 | 0 | Low-quality | Genome-fragment | 2.33 | AAI-based (high-confidence) | 0 |
| S16_extracted_51657 | 1756 | No | NA | 1 | 0 | 1 | Low-quality | Genome-fragment | 3.43 | AAI-based (medium-confidence) | 0 |
| S5_extracted_k141_6338 | 3886 | No | NA | 3 | 0 | 1 | Not-determined | Genome-fragment | NA | NA | 0 |
| S16_vs_15233\|\|full | 1756 | Yes | 1082 | 3 | 1 | 1 | Low-quality | Genome-fragment | 2.28 | AAI-based (medium-confidence) | 38.38 |
| S5_extracted_k141_139892 | 1217 | No | NA | 1 | 0 | 0 | Not-determined | Genome-fragment | NA | NA | 0 |
| S5_extracted_k141_6349 | 6135 | No | NA | 4 | 0 | 4 | Not-determined | Genome-fragment | NA | NA | 0 |
| S16_vs_150061\|\|lt2gene | 1756 | No | NA | 1 | 1 | 0 | Low-quality | Genome-fragment | 3.32 | HMM-based (lower-bound) | 0 |
| S5_extracted_k141_18461 | 29821 | No | NA | 42 | 7 | 1 | Medium-quality | Genome-fragment | 56.09 | AAI-based (high-confidence) | 0 |
| S5_extracted_k141_48749 | 2179 | No | NA | 1 | 0 | 1 | Not-determined | Genome-fragment | NA | NA | 0 |
| S5_extracted_k141_127686 | 1074 | No | NA | 1 | 0 | 1 | Not-determined | Genome-fragment | NA | NA | 0 |
| S8_vs_k141_131525\|\|full | 1756 | No | NA | 4 | 0 | 3 | Low-quality | Genome-fragment | 2.82 | AAI-based (high-confidence) | 0 |
| S5_extracted_k141_18473 | 3679 | No | NA | 4 | 0 | 4 | Not-determined | Genome-fragment | NA | NA | 0 |
| S8_vs_k141_137612\|\|full | 1756 | No | NA | 5 | 1 | 0 | Low-quality | Genome-fragment | 2.63 | AAI-based (high-confidence) | 0 |
| S5_extracted_k141_73014 | 1020 | No | NA | 2 | 0 | 2 | Not-determined | Genome-fragment | NA | NA | 0 |
| S13_vs_98144\|\|full | 1757 | No | NA | 3 | 0 | 2 | Low-quality | Genome-fragment | 4.28 | AAI-based (medium-confidence) | 0 |
| S5_extracted_k141_115464 | 6837 | No | NA | 9 | 0 | 1 | Not-determined | Genome-fragment | NA | NA | 0 |
| S14_extracted_k141_33741 | 1757 | No | NA | 5 | 1 | 0 | Low-quality | Genome-fragment | 1.11 | HMM-based (lower-bound) | 0 |
| S5_extracted_k141_18487 | 1135 | No | NA | 2 | 0 | 1 | Not-determined | Genome-fragment | NA | NA | 0 |
| S16_extracted_4680 | 1757 | No | NA | 1 | 0 | 0 | Low-quality | Genome-fragment | 3.48 | AAI-based (medium-confidence) | 0 |
| S5_extracted_k141_61053 | 3447 | No | NA | 3 | 0 | 0 | Not-determined | Genome-fragment | NA | NA | 0 |
| S12_vs_k141_94923\|\|full | 1757 | No | NA | 2 | 0 | 1 | Low-quality | Genome-fragment | 2.93 | AAI-based (medium-confidence) | 0 |
| S5_extracted_k141_121816 | 1047 | No | NA | 2 | 0 | 0 | Not-determined | Genome-fragment | NA | NA | 0 |
| S5_extracted_k141_127742 | 3728 | No | NA | 1 | 0 | 1 | Not-determined | Genome-fragment | NA | NA | 0 |
| S5_extracted_k141_24462 | 1107 | No | NA | 2 | 0 | 2 | Not-determined | Genome-fragment | NA | NA | 0 |
| S5_extracted_k141_6500 | 4103 | No | NA | 6 | 0 | 1 | Not-determined | Genome-fragment | NA | NA | 0 |
| S5_extracted_k141_91311 | 2476 | No | NA | 2 | 0 | 0 | Not-determined | Genome-fragment | NA | NA | 0 |
| S5_extracted_k141_424 | 1025 | No | NA | 1 | 0 | 1 | Not-determined | Genome-fragment | NA | NA | 0 |
| S5_extracted_k141_24471 | 1889 | No | NA | 1 | 0 | 1 | Not-determined | Genome-fragment | NA | NA | 0 |
| S12_vs_k141_76072\|\|full | 1757 | No | NA | 6 | 0 | 0 | Low-quality | Genome-fragment | 5.05 | AAI-based (high-confidence) | 0 |
| S5_extracted_k141_6506 | 23722 | No | NA | 30 | 14 | 0 | Medium-quality | Genome-fragment | 50.94 | AAI-based (high-confidence) | 0 |
| S5_extracted_k141_451 | 1178 | No | NA | 1 | 0 | 1 | Not-determined | Genome-fragment | NA | NA | 0 |
| S3_extracted_k141_116361 | 1758 | No | NA | 2 | 0 | 0 | Low-quality | Genome-fragment | 1.26 | AAI-based (high-confidence) | 0 |
| S5_extracted_k141_43007 | 1982 | No | NA | 1 | 0 | 1 | Not-determined | Genome-fragment | NA | NA | 0 |
| S3_vs_k141_14766\|\|full | 1758 | No | NA | 5 | 1 | 0 | Low-quality | Genome-fragment | 2.53 | AAI-based (high-confidence) | 0 |
| S10_vs_86092\|\|full | 1758 | No | NA | 5 | 1 | 0 | Low-quality | Genome-fragment | 2.64 | AAI-based (high-confidence) | 0 |
| S5_extracted_k141_48879 | 1526 | No | NA | 3 | 0 | 3 | Not-determined | Genome-fragment | NA | NA | 0 |
| S8_extracted_k141_12103 | 1758 | No | NA | 2 | 2 | 0 | Low-quality | Genome-fragment | 6.18 | HMM-based (lower-bound) | 0 |
| S17_vs_k141_174964\|\|full | 1759 | No | NA | 3 | 0 | 0 | Low-quality | Genome-fragment | 1.65 | AAI-based (high-confidence) | 0 |
| S9_vs_k141_53854\|\|full | 1759 | No | NA | 5 | 4 | 0 | Low-quality | Genome-fragment | 4.64 | AAI-based (high-confidence) | 0 |
| S5_extracted_k141_6569 | 1078 | No | NA | 1 | 0 | 0 | Not-determined | Genome-fragment | NA | NA | 0 |
| S13_vs_5882\|\|full | 1759 | No | NA | 3 | 0 | 0 | Low-quality | Genome-fragment | 1.04 | AAI-based (high-confidence) | 0 |
| S5_extracted_k141_133744 | 2253 | No | NA | 2 | 0 | 0 | Not-determined | Genome-fragment | NA | NA | 0 |
| S5_extracted_k141_97443 | 9336 | No | NA | 9 | 0 | 4 | Not-determined | Genome-fragment | NA | NA | 0 |
| S13_vs_58884\|\|full | 1759 | No | NA | 3 | 0 | 0 | Low-quality | Genome-fragment | 3.81 | AAI-based (medium-confidence) | 0 |
| S14_vs_k141_66679\|\|full | 1759 | No | NA | 4 | 0 | 1 | Low-quality | Genome-fragment | 1.57 | AAI-based (medium-confidence) | 0 |
| S5_extracted_k141_115757 | 1437 | No | NA | 1 | 0 | 0 | Not-determined | Genome-fragment | NA | NA | 0 |
| S16_vs_2049\|\|full | 1759 | Yes | 843 | 2 | 1 | 1 | Low-quality | Genome-fragment | 1.44 | AAI-based (medium-confidence) | 52.08 |
| S8_vs_k141_122567\|\|full | 1759 | No | NA | 6 | 0 | 0 | Low-quality | Genome-fragment | 4.31 | AAI-based (medium-confidence) | 0 |
| S5_extracted_k141_43142 | 7712 | No | NA | 8 | 0 | 1 | Not-determined | Genome-fragment | NA | NA | 0 |
| S5_extracted_k141_659 | 3239 | No | NA | 1 | 0 | 0 | Not-determined | Genome-fragment | NA | NA | 0 |
| S5_extracted_k141_91509 | 3029 | No | NA | 5 | 0 | 0 | Not-determined | Genome-fragment | NA | NA | 0 |
| S3_vs_k141_59052\|\|full | 1760 | No | NA | 3 | 2 | 0 | Low-quality | Genome-fragment | 4.71 | AAI-based (high-confidence) | 0 |
| S5_extracted_k141_122091 | 1297 | No | NA | 2 | 0 | 1 | Not-determined | Genome-fragment | NA | NA | 0 |
| S5_extracted_k141_30830 | 1211 | No | NA | 2 | 0 | 0 | Not-determined | Genome-fragment | NA | NA | 0 |
| S5_extracted_k141_733 | 1141 | No | NA | 2 | 0 | 0 | Not-determined | Genome-fragment | NA | NA | 0 |
| S5_vs_k141_115631\|\|full | 1760 | No | NA | 2 | 0 | 0 | Low-quality | Genome-fragment | 3.86 | AAI-based (medium-confidence) | 0 |
| S7_vs_36876\|\|full | 1760 | No | NA | 6 | 0 | 0 | Low-quality | Genome-fragment | 3.82 | AAI-based (medium-confidence) | 0 |
| S12_extracted_k141_173141 | 1760 | No | NA | 4 | 1 | 0 | Low-quality | Genome-fragment | 5.1 | AAI-based (high-confidence) | 0 |
| S12_vs_k141_155966\|\|full | 1760 | No | NA | 3 | 0 | 0 | Low-quality | Genome-fragment | 3.28 | AAI-based (medium-confidence) | 0 |
| S8_extracted_k141_185214 | 1760 | No | NA | 1 | 0 | 1 | Low-quality | Genome-fragment | 0.46 | AAI-based (high-confidence) | 0 |
| S5_extracted_k141_779 | 1223 | No | NA | 2 | 0 | 0 | Not-determined | Genome-fragment | NA | NA | 0 |
| S5_extracted_k141_18665 | 1449 | No | NA | 1 | 0 | 1 | Not-determined | Genome-fragment | NA | NA | 0 |
| S5_extracted_k141_791 | 1002 | No | NA | 1 | 0 | 1 | Not-determined | Genome-fragment | NA | NA | 0 |
| S5_extracted_k141_30912 | 1231 | No | NA | 1 | 0 | 1 | Not-determined | Genome-fragment | NA | NA | 0 |
| S5_extracted_k141_36719 | 1828 | No | NA | 1 | 0 | 1 | Not-determined | Genome-fragment | NA | NA | 0 |
| S5_extracted_k141_61232 | 1188 | No | NA | 1 | 0 | 0 | Not-determined | Genome-fragment | NA | NA | 0 |
| S5_extracted_k141_18683 | 1080 | No | NA | 1 | 0 | 0 | Not-determined | Genome-fragment | NA | NA | 0 |
| S5_extracted_k141_122216 | 1130 | No | NA | 1 | 0 | 0 | Not-determined | Genome-fragment | NA | NA | 0 |
| S5_extracted_k141_115881 | 1571 | No | NA | 1 | 0 | 0 | Not-determined | Genome-fragment | NA | NA | 0 |
| S5_extracted_k141_49063 | 3124 | No | NA | 1 | 0 | 1 | Not-determined | Genome-fragment | NA | NA | 0 |
| S5_extracted_k141_122240 | 1279 | No | NA | 1 | 0 | 1 | Not-determined | Genome-fragment | NA | NA | 0 |
| S9_vs_k141_72295\|\|full | 1761 | No | NA | 3 | 0 | 0 | Low-quality | Genome-fragment | 4.04 | AAI-based (high-confidence) | 0 |
| S9_vs_k141_13599\|\|full | 1761 | No | NA | 4 | 0 | 0 | Low-quality | Genome-fragment | 3.46 | AAI-based (medium-confidence) | 0 |
| S5_extracted_k141_49080 | 1380 | No | NA | 1 | 0 | 1 | Not-determined | Genome-fragment | NA | NA | 0 |
| S5_vs_k141_21288\|\|full | 1761 | No | NA | 2 | 0 | 1 | Low-quality | Genome-fragment | 1.46 | AAI-based (high-confidence) | 0 |
| S7_extracted_103843 | 1761 | No | NA | 2 | 1 | 0 | Low-quality | Genome-fragment | 1.23 | HMM-based (lower-bound) | 0 |
| S7_vs_22679\|\|full | 1761 | No | NA | 2 | 0 | 0 | Low-quality | Genome-fragment | 3.91 | AAI-based (medium-confidence) | 0 |
| S5_extracted_k141_122289 | 4899 | No | NA | 7 | 0 | 1 | Not-determined | Genome-fragment | NA | NA | 0 |
| S7_vs_72921\|\|full | 1761 | No | NA | 3 | 2 | 0 | Low-quality | Genome-fragment | 2.81 | AAI-based (high-confidence) | 0 |
| S2_vs_k141_5173\|\|full | 1761 | No | NA | 4 | 0 | 1 | Low-quality | Genome-fragment | 1.36 | AAI-based (medium-confidence) | 0 |
| S5_extracted_k141_855 | 1037 | No | NA | 3 | 0 | 0 | Not-determined | Genome-fragment | NA | NA | 0 |
| S5_extracted_k141_85520 | 1083 | No | NA | 2 | 0 | 2 | Not-determined | Genome-fragment | NA | NA | 0 |
| S17_vs_k141_28233\|\|full | 1762 | No | NA | 2 | 0 | 0 | Low-quality | Genome-fragment | 2.13 | AAI-based (medium-confidence) | 0 |
| S9_extracted_k141_58333 | 1762 | No | NA | 3 | 3 | 0 | Low-quality | Genome-fragment | 5.86 | AAI-based (high-confidence) | 0 |
| S5_extracted_k141_85540 | 1876 | No | NA | 1 | 0 | 1 | Not-determined | Genome-fragment | NA | NA | 0 |
| S11_vs_k141_28477\|\|lt2gene | 1762 | No | NA | 2 | 1 | 0 | Low-quality | Genome-fragment | 4.35 | AAI-based (high-confidence) | 0 |
| S5_extracted_k141_67094 | 3435 | No | NA | 5 | 0 | 2 | Not-determined | Genome-fragment | NA | NA | 0 |
| S14_vs_k141_35746\|\|full | 1762 | No | NA | 4 | 0 | 0 | Low-quality | Genome-fragment | 3.48 | AAI-based (medium-confidence) | 0 |
| S1_vs_k141_132753\|\|full | 1762 | No | NA | 4 | 2 | 0 | Low-quality | Genome-fragment | 4.45 | AAI-based (high-confidence) | 0 |
| S5_extracted_k141_91615 | 3490 | No | NA | 4 | 0 | 0 | Not-determined | Genome-fragment | NA | NA | 0 |
| S5_extracted_k141_91616 | 1203 | No | NA | 2 | 0 | 2 | Not-determined | Genome-fragment | NA | NA | 0 |
| S1_vs_k141_39949\|\|lt2gene | 1762 | No | NA | 2 | 2 | 0 | Low-quality | Genome-fragment | 3.46 | AAI-based (high-confidence) | 0 |
| S5_extracted_k141_128133 | 1171 | No | NA | 1 | 0 | 1 | Not-determined | Genome-fragment | NA | NA | 0 |
| S5_extracted_k141_67186 | 3663 | No | NA | 2 | 0 | 1 | Not-determined | Genome-fragment | NA | NA | 0 |
| S3_vs_k141_42670\|\|full | 1762 | No | NA | 5 | 3 | 0 | Low-quality | Genome-fragment | 2.77 | HMM-based (lower-bound) | 0 |
| S10_vs_51\|\|full | 1762 | No | NA | 3 | 0 | 0 | Low-quality | Genome-fragment | 5.33 | AAI-based (high-confidence) | 0 |
| S5_extracted_k141_122441 | 1175 | No | NA | 2 | 0 | 2 | Not-determined | Genome-fragment | NA | NA | 0 |
| S5_extracted_k141_36940 | 1278 | No | NA | 1 | 0 | 1 | Not-determined | Genome-fragment | NA | NA | 0 |
| S10_vs_28742\|\|full | 1762 | No | NA | 5 | 1 | 0 | Low-quality | Genome-fragment | 3.7 | AAI-based (high-confidence) | 0 |
| S5_extracted_k141_122485 | 1203 | No | NA | 2 | 0 | 2 | Not-determined | Genome-fragment | NA | NA | 0 |
| S5_extracted_k141_6769 | 1527 | No | NA | 1 | 0 | 0 | Not-determined | Genome-fragment | NA | NA | 0 |
| S5_extracted_k141_122492 | 4152 | No | NA | 3 | 0 | 1 | Not-determined | Genome-fragment | NA | NA | 0 |
| S12_vs_k141_40358\|\|full | 1762 | No | NA | 4 | 0 | 1 | Low-quality | Genome-fragment | 1.25 | AAI-based (medium-confidence) | 0 |
| S5_extracted_k141_31187 | 3730 | No | NA | 7 | 0 | 0 | Not-determined | Genome-fragment | NA | NA | 0 |
| S5_extracted_k141_6794 | 10026 | No | NA | 13 | 0 | 0 | Not-determined | Genome-fragment | NA | NA | 0 |
| S5_extracted_k141_6815 | 1473 | No | NA | 2 | 0 | 0 | Not-determined | Genome-fragment | NA | NA | 0 |
| S5_extracted_k141_6836 | 2043 | No | NA | 1 | 0 | 1 | Not-determined | Genome-fragment | NA | NA | 0 |
| S12_vs_k141_58116\|\|full | 1762 | No | NA | 3 | 1 | 0 | Low-quality | Genome-fragment | 3.47 | AAI-based (high-confidence) | 0 |
| S17_vs_k141_160473\|\|full | 1763 | No | NA | 2 | 0 | 1 | Low-quality | Genome-fragment | 1.1 | AAI-based (medium-confidence) | 0 |
| S5_extracted_k141_37039 | 1373 | No | NA | 1 | 0 | 1 | Not-determined | Genome-fragment | NA | NA | 0 |
| S5_extracted_k141_43540 | 1813 | No | NA | 2 | 0 | 2 | Not-determined | Genome-fragment | NA | NA | 0 |
| S5_extracted_k141_43547 | 2570 | No | NA | 6 | 0 | 1 | Not-determined | Genome-fragment | NA | NA | 0 |
| S3_extracted_k141_28219 | 1763 | No | NA | 4 | 3 | 0 | Low-quality | Genome-fragment | 4.69 | AAI-based (high-confidence) | 0 |
| S5_extracted_k141_49387 | 1042 | No | NA | 1 | 0 | 1 | Not-determined | Genome-fragment | NA | NA | 0 |
| S7_extracted_15512 | 1763 | No | NA | 4 | 2 | 0 | Low-quality | Genome-fragment | 10.59 | AAI-based (medium-confidence) | 0 |
| S5_extracted_k141_79237 | 3229 | No | NA | 3 | 0 | 1 | Not-determined | Genome-fragment | NA | NA | 0 |
| S5_extracted_k141_122568 | 53264 | No | NA | 78 | 21 | 1 | Complete | High-quality | 100 | DTR (high-confidence) | 0 |
| S11_extracted_k141_42037 | 1764 | No | NA | 4 | 0 | 0 | Low-quality | Genome-fragment | 1.85 | AAI-based (medium-confidence) | 0 |
| S5_extracted_k141_6911 | 1393 | No | NA | 4 | 0 | 0 | Not-determined | Genome-fragment | NA | NA | 0 |
| S5_extracted_k141_140205 | 1172 | No | NA | 1 | 0 | 1 | Not-determined | Genome-fragment | NA | NA | 0 |
| S5_extracted_k141_79251 | 5802 | No | NA | 3 | 0 | 3 | Not-determined | Genome-fragment | NA | NA | 0 |
| S5_extracted_k141_49438 | 4737 | No | NA | 2 | 0 | 1 | Not-determined | Genome-fragment | NA | NA | 0 |
| S5_extracted_k141_116217 | 1240 | No | NA | 1 | 0 | 0 | Not-determined | Genome-fragment | NA | NA | 0 |
| S5_extracted_k141_134040 | 36207 | Yes | 24097 | 42 | 12 | 4 | Medium-quality | Genome-fragment | 67.33 | AAI-based (medium-confidence) | 33.45 |
| S11_extracted_k141_10052 | 1764 | No | NA | 2 | 1 | 0 | Low-quality | Genome-fragment | 1.19 | HMM-based (lower-bound) | 0 |
| S5_extracted_k141_12593 | 4331 | No | NA | 3 | 0 | 1 | Not-determined | Genome-fragment | NA | NA | 0 |
| S13_vs_74868\|\|full | 1764 | No | NA | 3 | 1 | 0 | Low-quality | Genome-fragment | 2.83 | HMM-based (lower-bound) | 0 |
| S5_extracted_k141_134076 | 2192 | No | NA | 3 | 0 | 3 | Not-determined | Genome-fragment | NA | NA | 0 |
| S14_vs_k141_419\|\|full | 1764 | Yes | 1362 | 4 | 1 | 1 | Low-quality | Genome-fragment | 3.39 | HMM-based (lower-bound) | 22.79 |
| S5_extracted_k141_122626 | 23241 | No | NA | 21 | 0 | 4 | Not-determined | Genome-fragment | NA | NA | 0 |
| S5_extracted_k141_67421 | 1194 | No | NA | 4 | 0 | 0 | Not-determined | Genome-fragment | NA | NA | 0 |
| S5_extracted_k141_121683 | 1764 | No | NA | 5 | 4 | 0 | Low-quality | Genome-fragment | 5.1 | AAI-based (high-confidence) | 0 |
| S5_extracted_k141_104075 | 1216 | No | NA | 1 | 0 | 0 | Not-determined | Genome-fragment | NA | NA | 0 |
| S5_extracted_k141_19016 | 1379 | No | NA | 3 | 0 | 2 | Not-determined | Genome-fragment | NA | NA | 0 |
| S5_vs_k141_47226\|\|full | 1764 | No | NA | 3 | 1 | 0 | Low-quality | Genome-fragment | 2.85 | HMM-based (lower-bound) | 0 |
| S7_extracted_45011 | 1764 | No | NA | 3 | 0 | 0 | Low-quality | Genome-fragment | 2.01 | AAI-based (high-confidence) | 0 |
| S5_extracted_k141_134130 | 1979 | No | NA | 4 | 0 | 0 | Not-determined | Genome-fragment | NA | NA | 0 |
| S5_extracted_k141_7051 | 6577 | No | NA | 6 | 0 | 1 | Not-determined | Genome-fragment | NA | NA | 0 |
| S5_extracted_k141_116300 | 20151 | Yes | 18965 | 39 | 5 | 3 | Medium-quality | Genome-fragment | 50.66 | AAI-based (high-confidence) | 5.89 |
| S16_extracted_2391 | 1764 | No | NA | 3 | 0 | 0 | Low-quality | Genome-fragment | 2.35 | AAI-based (high-confidence) | 0 |
| S5_extracted_k141_116320 | 1696 | No | NA | 1 | 0 | 0 | Not-determined | Genome-fragment | NA | NA | 0 |
| S16_extracted_71346 | 1764 | No | NA | 5 | 2 | 0 | Low-quality | Genome-fragment | 4.41 | AAI-based (high-confidence) | 0 |
| S5_extracted_k141_116322 | 1148 | No | NA | 4 | 0 | 0 | Not-determined | Genome-fragment | NA | NA | 0 |
| S12_vs_k141_53827\|\|full | 1764 | No | NA | 3 | 1 | 0 | Low-quality | Genome-fragment | 1.98 | HMM-based (lower-bound) | 0 |
| S5_extracted_k141_12747 | 1222 | No | NA | 2 | 0 | 0 | Not-determined | Genome-fragment | NA | NA | 0 |
| S5_extracted_k141_85761 | 4531 | No | NA | 2 | 0 | 0 | Not-determined | Genome-fragment | NA | NA | 0 |
| S5_extracted_k141_73299 | 1071 | No | NA | 1 | 0 | 1 | Not-determined | Genome-fragment | NA | NA | 0 |
| S5_extracted_k141_122856 | 4133 | No | NA | 6 | 0 | 2 | Not-determined | Genome-fragment | NA | NA | 0 |
| S5_extracted_k141_122858 | 1335 | No | NA | 3 | 0 | 0 | Not-determined | Genome-fragment | NA | NA | 0 |
| S8_vs_k141_138809\|\|full | 1764 | No | NA | 2 | 0 | 0 | Low-quality | Genome-fragment | 3.32 | AAI-based (medium-confidence) | 0 |
| S5_extracted_k141_67519 | 1409 | No | NA | 1 | 0 | 1 | Not-determined | Genome-fragment | NA | NA | 0 |
| S9_extracted_k141_81962 | 1765 | No | NA | 2 | 2 | 0 | Low-quality | Genome-fragment | 3.81 | AAI-based (high-confidence) | 0 |
| S11_vs_k141_47579\|\|full | 1765 | No | NA | 4 | 1 | 0 | Low-quality | Genome-fragment | 5.29 | AAI-based (high-confidence) | 0 |
| S11_vs_k141_49101\|\|full | 1765 | No | NA | 5 | 0 | 0 | Low-quality | Genome-fragment | 3.96 | AAI-based (high-confidence) | 0 |
| S14_extracted_k141_34257 | 1765 | No | NA | 1 | 0 | 1 | Low-quality | Genome-fragment | 0.61 | AAI-based (medium-confidence) | 0 |
| S1_vs_k141_29746\|\|full | 1765 | No | NA | 2 | 0 | 0 | Low-quality | Genome-fragment | 2.82 | AAI-based (medium-confidence) | 0 |
| S5_extracted_k141_122922 | 1375 | No | NA | 1 | 0 | 0 | Not-determined | Genome-fragment | NA | NA | 0 |
| S3_vs_k141_3292\|\|full | 1765 | No | NA | 4 | 0 | 0 | Low-quality | Genome-fragment | 2.52 | AAI-based (high-confidence) | 0 |
| S7_extracted_6047 | 1765 | No | NA | 2 | 1 | 0 | Low-quality | Genome-fragment | 7.59 | AAI-based (high-confidence) | 0 |
| S12_vs_k141_179844\|\|full | 1765 | No | NA | 2 | 0 | 0 | Low-quality | Genome-fragment | 4.37 | AAI-based (high-confidence) | 0 |
| S5_extracted_k141_31559 | 3726 | No | NA | 6 | 0 | 4 | Not-determined | Genome-fragment | NA | NA | 0 |
| S8_extracted_k141_33733 | 1765 | No | NA | 3 | 2 | 0 | Low-quality | Genome-fragment | 4.27 | AAI-based (high-confidence) | 0 |
| S8_vs_k141_182137\|\|full | 1765 | No | NA | 2 | 1 | 0 | Low-quality | Genome-fragment | 4.31 | AAI-based (high-confidence) | 0 |
| S17_vs_k141_358\|\|full | 1766 | No | NA | 3 | 0 | 0 | Low-quality | Genome-fragment | 1.25 | AAI-based (medium-confidence) | 0 |
| S5_extracted_k141_85955 | 3712 | No | NA | 5 | 0 | 0 | Not-determined | Genome-fragment | NA | NA | 0 |
| S9_vs_k141_36286\|\|full | 1766 | No | NA | 4 | 1 | 0 | Low-quality | Genome-fragment | 4.35 | AAI-based (high-confidence) | 0 |
| S9_vs_k141_70678\|\|full | 1766 | No | NA | 3 | 2 | 0 | Low-quality | Genome-fragment | 3.07 | AAI-based (high-confidence) | 0 |
| S11_vs_k141_53762\|\|full | 1766 | No | NA | 5 | 3 | 0 | Low-quality | Genome-fragment | 4.11 | HMM-based (lower-bound) | 0 |
| S7_vs_93734\|\|full | 1766 | No | NA | 4 | 1 | 0 | Low-quality | Genome-fragment | 4.92 | AAI-based (high-confidence) | 0 |
| S7_vs_94174\|\|full | 1766 | No | NA | 3 | 2 | 0 | Low-quality | Genome-fragment | 4.02 | AAI-based (medium-confidence) | 0 |
| S7_vs_114024\|\|full | 1766 | No | NA | 3 | 0 | 0 | Low-quality | Genome-fragment | 3.2 | AAI-based (medium-confidence) | 0 |
| S5_extracted_k141_67622 | 168282 | No | NA | 190 | 18 | 6 | High-quality | High-quality | 91.68 | AAI-based (high-confidence) | 0 |
| S16_vs_130588\|\|full | 1766 | No | NA | 2 | 0 | 2 | Low-quality | Genome-fragment | 1.44 | AAI-based (medium-confidence) | 0 |
| S5_extracted_k141_110171 | 47834 | No | NA | 70 | 11 | 0 | High-quality | High-quality | 100 | AAI-based (high-confidence) | 0 |
| S12_extracted_k141_176337 | 1766 | No | NA | 3 | 2 | 0 | Low-quality | Genome-fragment | 3.77 | AAI-based (high-confidence) | 0 |
| S5_extracted_k141_37460 | 1271 | No | NA | 1 | 0 | 0 | Not-determined | Genome-fragment | NA | NA | 0 |
| S2_extracted_k141_13995 | 1766 | No | NA | 6 | 5 | 0 | Low-quality | Genome-fragment | 5.83 | AAI-based (high-confidence) | 0 |
| S5_extracted_k141_67685 | 2259 | No | NA | 1 | 0 | 1 | Not-determined | Genome-fragment | NA | NA | 0 |
| S5_extracted_k141_7313 | 1155 | No | NA | 1 | 0 | 0 | Not-determined | Genome-fragment | NA | NA | 0 |
| S5_extracted_k141_37488 | 8022 | No | NA | 5 | 0 | 5 | Not-determined | Genome-fragment | NA | NA | 0 |
| S5_extracted_k141_97883 | 1336 | No | NA | 1 | 0 | 0 | Not-determined | Genome-fragment | NA | NA | 0 |
| S11_vs_k141_84433\|\|full | 1767 | No | NA | 3 | 0 | 0 | Low-quality | Genome-fragment | 2.06 | AAI-based (medium-confidence) | 0 |
| S5_extracted_k141_128315 | 2428 | No | NA | 4 | 0 | 0 | Not-determined | Genome-fragment | NA | NA | 0 |
| S3_vs_k141_62016\|\|lt2gene | 1767 | No | NA | 2 | 2 | 0 | Low-quality | Genome-fragment | 3.93 | AAI-based (medium-confidence) | 0 |
| S5_vs_k141_50610\|\|full | 1767 | No | NA | 3 | 0 | 0 | Low-quality | Genome-fragment | 3.15 | AAI-based (high-confidence) | 0 |
| S10_extracted_102941 | 1767 | No | NA | 1 | 0 | 0 | Low-quality | Genome-fragment | 2.3 | AAI-based (medium-confidence) | 0 |
| S5_extracted_k141_92502 | 1768 | No | NA | 2 | 2 | 0 | Low-quality | Genome-fragment | 4.42 | AAI-based (high-confidence) | 0 |
| S5_extracted_k141_61710 | 5673 | No | NA | 11 | 0 | 0 | Not-determined | Genome-fragment | NA | NA | 0 |
| S16_vs_82546\|\|full | 1768 | No | NA | 4 | 2 | 0 | Low-quality | Genome-fragment | 3.25 | AAI-based (medium-confidence) | 0 |
| S5_extracted_k141_13162 | 1149 | No | NA | 2 | 0 | 0 | Not-determined | Genome-fragment | NA | NA | 0 |
| S15_extracted_3377 | 1768 | No | NA | 5 | 1 | 0 | Low-quality | Genome-fragment | 0.95 | AAI-based (high-confidence) | 0 |
| S5_extracted_k141_79466 | 2442 | No | NA | 1 | 0 | 1 | Not-determined | Genome-fragment | NA | NA | 0 |
| S5_extracted_k141_73617 | 1160 | No | NA | 1 | 0 | 1 | Not-determined | Genome-fragment | NA | NA | 0 |
| S5_extracted_k141_123252 | 1041 | No | NA | 1 | 0 | 0 | Not-determined | Genome-fragment | NA | NA | 0 |
| S8_extracted_k141_108898 | 1768 | No | NA | 2 | 2 | 0 | Low-quality | Genome-fragment | 5.7 | AAI-based (high-confidence) | 0 |
| S8_extracted_k141_126939 | 1768 | No | NA | 1 | 0 | 0 | Low-quality | Genome-fragment | 1.86 | AAI-based (high-confidence) | 0 |
| S5_extracted_k141_92193 | 4438 | No | NA | 5 | 0 | 0 | Not-determined | Genome-fragment | NA | NA | 0 |
| S5_extracted_k141_67871 | 3823 | No | NA | 3 | 0 | 0 | Not-determined | Genome-fragment | NA | NA | 0 |
| S5_extracted_k141_116938 | 1786 | No | NA | 1 | 0 | 1 | Not-determined | Genome-fragment | NA | NA | 0 |
| S8_vs_k141_50223\|\|full | 1768 | No | NA | 4 | 0 | 0 | Low-quality | Genome-fragment | 2.44 | AAI-based (medium-confidence) | 0 |
| S5_extracted_k141_116965 | 4508 | No | NA | 8 | 0 | 1 | Not-determined | Genome-fragment | NA | NA | 0 |
| S17_extracted_k141_99310 | 1769 | No | NA | 4 | 4 | 0 | Low-quality | Genome-fragment | 5.84 | AAI-based (high-confidence) | 0 |
| S14_vs_k141_75742\|\|full | 1769 | No | NA | 5 | 3 | 0 | Low-quality | Genome-fragment | 3.89 | AAI-based (high-confidence) | 0 |
| S5_extracted_k141_140588 | 1154 | No | NA | 3 | 0 | 0 | Not-determined | Genome-fragment | NA | NA | 0 |
| S5_extracted_k141_49716 | 1748 | No | NA | 1 | 0 | 0 | Not-determined | Genome-fragment | NA | NA | 0 |
| S5_vs_k141_67617\|\|full | 1769 | No | NA | 6 | 0 | 0 | Low-quality | Genome-fragment | 4.92 | AAI-based (high-confidence) | 0 |
| S5_extracted_k141_73804 | 1506 | No | NA | 3 | 0 | 0 | Not-determined | Genome-fragment | NA | NA | 0 |
| S5_vs_k141_22587\|\|full | 1769 | No | NA | 2 | 0 | 0 | Low-quality | Genome-fragment | 2.14 | AAI-based (high-confidence) | 0 |
| S5_extracted_k141_128479 | 1160 | No | NA | 1 | 0 | 1 | Not-determined | Genome-fragment | NA | NA | 0 |
| S16_vs_100971\|\|full | 1769 | No | NA | 3 | 2 | 0 | Low-quality | Genome-fragment | 3.63 | AAI-based (high-confidence) | 0 |
| S12_extracted_k141_83895 | 1769 | No | NA | 4 | 0 | 0 | Low-quality | Genome-fragment | 2.48 | AAI-based (high-confidence) | 0 |
| S5_extracted_k141_104469 | 1138 | No | NA | 1 | 0 | 0 | Not-determined | Genome-fragment | NA | NA | 0 |
| S5_extracted_k141_67978 | 1108 | No | NA | 1 | 0 | 0 | Not-determined | Genome-fragment | NA | NA | 0 |
| S5_extracted_k141_37711 | 2843 | No | NA | 2 | 0 | 2 | Not-determined | Genome-fragment | NA | NA | 0 |
| S12_vs_k141_93173\|\|full | 1769 | No | NA | 5 | 3 | 0 | Low-quality | Genome-fragment | 5.13 | AAI-based (high-confidence) | 0 |
| S5_extracted_k141_110608 | 1000 | No | NA | 2 | 0 | 0 | Not-determined | Genome-fragment | NA | NA | 0 |
| S5_extracted_k141_32046 | 4362 | No | NA | 5 | 0 | 1 | Not-determined | Genome-fragment | NA | NA | 0 |
| S5_extracted_k141_92498 | 2192 | No | NA | 1 | 0 | 1 | Not-determined | Genome-fragment | NA | NA | 0 |
| S5_extracted_k141_55415 | 98442 | No | NA | 101 | 18 | 0 | High-quality | High-quality | 97.88 | AAI-based (high-confidence) | 0 |
| S8_vs_k141_144577\|\|full | 1769 | No | NA | 5 | 1 | 0 | Low-quality | Genome-fragment | 3.78 | AAI-based (medium-confidence) | 0 |
| S11_extracted_k141_33735 | 1770 | No | NA | 3 | 0 | 0 | Low-quality | Genome-fragment | 4.33 | AAI-based (high-confidence) | 0 |
| S5_extracted_k141_104518 | 3832 | No | NA | 4 | 0 | 0 | Not-determined | Genome-fragment | NA | NA | 0 |
| S14_vs_k141_49524\|\|full | 1770 | Yes | 990 | 2 | 1 | 1 | Low-quality | Genome-fragment | 1.12 | HMM-based (lower-bound) | 44.07 |
| S5_extracted_k141_68035 | 3099 | No | NA | 4 | 0 | 0 | Not-determined | Genome-fragment | NA | NA | 0 |
| S5_extracted_k141_32099 | 3188 | No | NA | 4 | 0 | 0 | Not-determined | Genome-fragment | NA | NA | 0 |
| S7_vs_31309\|\|full | 1770 | No | NA | 6 | 1 | 0 | Low-quality | Genome-fragment | 3.13 | AAI-based (medium-confidence) | 0 |
| S5_extracted_k141_140718 | 7808 | No | NA | 5 | 0 | 2 | Not-determined | Genome-fragment | NA | NA | 0 |
| S5_extracted_k141_128539 | 1625 | No | NA | 3 | 0 | 0 | Not-determined | Genome-fragment | NA | NA | 0 |
| S5_extracted_k141_110667 | 1403 | No | NA | 2 | 0 | 2 | Not-determined | Genome-fragment | NA | NA | 0 |
| S1_vs_k141_64047\|\|full | 1771 | No | NA | 3 | 0 | 2 | Low-quality | Genome-fragment | 2.6 | AAI-based (medium-confidence) | 0 |
| S15_extracted_47391 | 1771 | No | NA | 3 | 0 | 0 | Low-quality | Genome-fragment | 2.2 | AAI-based (high-confidence) | 0 |
| S5_extracted_k141_61932 | 18822 | No | NA | 12 | 0 | 9 | Not-determined | Genome-fragment | NA | NA | 0 |
| S5_extracted_k141_32170 | 1054 | No | NA | 1 | 0 | 1 | Not-determined | Genome-fragment | NA | NA | 0 |
| S5_extracted_k141_7552 | 3886 | No | NA | 4 | 0 | 0 | Not-determined | Genome-fragment | NA | NA | 0 |
| S8_extracted_k141_90033 | 1771 | No | NA | 3 | 0 | 2 | Low-quality | Genome-fragment | 1.91 | AAI-based (high-confidence) | 0 |
| S9_extracted_k141_82147 | 1772 | No | NA | 1 | 0 | 1 | Low-quality | Genome-fragment | 0.87 | AAI-based (medium-confidence) | 0 |
| S1_vs_k141_57065\|\|full | 1772 | No | NA | 4 | 0 | 2 | Low-quality | Genome-fragment | 3.37 | AAI-based (medium-confidence) | 0 |
| S5_extracted_k141_73891 | 5519 | No | NA | 5 | 0 | 4 | Not-determined | Genome-fragment | NA | NA | 0 |
| S5_extracted_k141_110733 | 1012 | No | NA | 2 | 0 | 0 | Not-determined | Genome-fragment | NA | NA | 0 |
| S3_vs_k141_120681\|\|full | 1772 | No | NA | 4 | 2 | 0 | Low-quality | Genome-fragment | 4.99 | AAI-based (high-confidence) | 0 |
| S5_extracted_k141_68174 | 3127 | No | NA | 3 | 0 | 0 | Not-determined | Genome-fragment | NA | NA | 0 |
| S5_extracted_k141_110791 | 1014 | No | NA | 1 | 0 | 0 | Not-determined | Genome-fragment | NA | NA | 0 |
| S5_extracted_k141_110793 | 3366 | No | NA | 5 | 0 | 2 | Not-determined | Genome-fragment | NA | NA | 0 |
| S5_extracted_k141_128693 | 2471 | No | NA | 2 | 0 | 0 | Not-determined | Genome-fragment | NA | NA | 0 |
| S5_extracted_k141_24958 | 8534 | No | NA | 4 | 0 | 2 | Not-determined | Genome-fragment | NA | NA | 0 |
| S5_extracted_k141_79581 | 1203 | No | NA | 2 | 0 | 1 | Not-determined | Genome-fragment | NA | NA | 0 |
| S5_extracted_k141_86344 | 1034 | No | NA | 1 | 0 | 1 | Not-determined | Genome-fragment | NA | NA | 0 |
| S5_extracted_k141_79602 | 1203 | No | NA | 2 | 0 | 1 | Not-determined | Genome-fragment | NA | NA | 0 |
| S5_extracted_k141_104747 | 1519 | No | NA | 1 | 0 | 1 | Not-determined | Genome-fragment | NA | NA | 0 |
| S5_extracted_k141_62010 | 1698 | No | NA | 1 | 0 | 0 | Not-determined | Genome-fragment | NA | NA | 0 |
| S5_extracted_k141_13500 | 1040 | No | NA | 1 | 0 | 0 | Not-determined | Genome-fragment | NA | NA | 0 |
| S6_vs_k141_55476\|\|lt2gene | 1772 | No | NA | 2 | 1 | 0 | Low-quality | Genome-fragment | 2.32 | HMM-based (lower-bound) | 0 |
| S5_extracted_k141_27921 | 5958 | No | NA | 7 | 0 | 4 | Not-determined | Genome-fragment | NA | NA | 0 |
| S10_extracted_9668 | 1773 | No | NA | 3 | 0 | 0 | Low-quality | Genome-fragment | 4.04 | AAI-based (medium-confidence) | 0 |
| S5_extracted_k141_86433 | 1307 | No | NA | 1 | 0 | 0 | Not-determined | Genome-fragment | NA | NA | 0 |
| S10_extracted_79062 | 1773 | No | NA | 4 | 2 | 0 | Low-quality | Genome-fragment | 1.26 | AAI-based (high-confidence) | 0 |
| S12_vs_k141_98145\|\|full | 1773 | No | NA | 3 | 2 | 0 | Low-quality | Genome-fragment | 3.97 | AAI-based (high-confidence) | 0 |
| S15_extracted_16245 | 1773 | No | NA | 3 | 0 | 0 | Low-quality | Genome-fragment | 2.14 | AAI-based (medium-confidence) | 0 |
| S9_vs_k141_33445\|\|lt2gene | 1774 | No | NA | 2 | 0 | 1 | Low-quality | Genome-fragment | 1.36 | AAI-based (medium-confidence) | 0 |
| S5_extracted_k141_38011 | 16427 | No | NA | 24 | 6 | 0 | Medium-quality | Genome-fragment | 68.39 | AAI-based (high-confidence) | 0 |
| S3_vs_k141_100050\|\|full | 1774 | No | NA | 3 | 0 | 0 | Low-quality | Genome-fragment | 3.32 | AAI-based (high-confidence) | 0 |
| S5_extracted_k141_19721 | 6983 | No | NA | 7 | 0 | 7 | Not-determined | Genome-fragment | NA | NA | 0 |
| S7_extracted_135777 | 1774 | No | NA | 3 | 3 | 0 | Low-quality | Genome-fragment | 3.1 | AAI-based (high-confidence) | 0 |
| S5_extracted_k141_32392 | 3851 | No | NA | 2 | 0 | 1 | Not-determined | Genome-fragment | NA | NA | 0 |
| S8_vs_k141_111855\|\|full | 1774 | No | NA | 3 | 1 | 0 | Low-quality | Genome-fragment | 3.57 | HMM-based (lower-bound) | 0 |
| S5_extracted_k141_50170 | 1368 | No | NA | 3 | 0 | 1 | Not-determined | Genome-fragment | NA | NA | 0 |
| S5_extracted_k141_7729 | 8903 | No | NA | 6 | 0 | 2 | Not-determined | Genome-fragment | NA | NA | 0 |
| S17_vs_k141_47387\|\|full | 1775 | No | NA | 3 | 0 | 0 | Low-quality | Genome-fragment | 1.22 | AAI-based (medium-confidence) | 0 |
| S5_extracted_k141_44437 | 1132 | No | NA | 1 | 0 | 1 | Not-determined | Genome-fragment | NA | NA | 0 |
| S11_vs_k141_43956\|\|full | 1775 | No | NA | 4 | 0 | 0 | Low-quality | Genome-fragment | 4.01 | AAI-based (high-confidence) | 0 |
| S5_extracted_k141_110899 | 1586 | No | NA | 3 | 0 | 0 | Not-determined | Genome-fragment | NA | NA | 0 |
| S1_vs_k141_125962\|\|full | 1775 | No | NA | 2 | 2 | 0 | Low-quality | Genome-fragment | 4.62 | AAI-based (high-confidence) | 0 |
| S5_extracted_k141_25147 | 3133 | No | NA | 1 | 0 | 0 | Not-determined | Genome-fragment | NA | NA | 0 |
| S12_vs_k141_75895\|\|full | 1775 | No | NA | 4 | 2 | 0 | Low-quality | Genome-fragment | 4.97 | AAI-based (high-confidence) | 0 |
| S6_vs_k141_19971\|\|full | 1775 | No | NA | 4 | 2 | 0 | Low-quality | Genome-fragment | 3.88 | AAI-based (high-confidence) | 0 |
| S5_extracted_k141_79671 | 2696 | No | NA | 2 | 0 | 0 | Not-determined | Genome-fragment | NA | NA | 0 |
| S5_extracted_k141_32475 | 1535 | No | NA | 2 | 0 | 0 | Not-determined | Genome-fragment | NA | NA | 0 |
| S5_extracted_k141_55687 | 14413 | No | NA | 16 | 12 | 0 | Medium-quality | Genome-fragment | 65.79 | AAI-based (high-confidence) | 0 |
| S5_extracted_k141_117489 | 1034 | No | NA | 2 | 0 | 0 | Not-determined | Genome-fragment | NA | NA | 0 |
| S5_extracted_k141_44480 | 15595 | No | NA | 13 | 0 | 10 | Not-determined | Genome-fragment | NA | NA | 0 |
| S13_vs_2690\|\|full | 1776 | No | NA | 4 | 0 | 0 | Low-quality | Genome-fragment | 4.04 | AAI-based (high-confidence) | 0 |
| S5_extracted_k141_134818 | 2205 | No | NA | 1 | 0 | 1 | Not-determined | Genome-fragment | NA | NA | 0 |
| S5_extracted_k141_2025 | 1115 | No | NA | 2 | 0 | 1 | Not-determined | Genome-fragment | NA | NA | 0 |
| S5_extracted_k141_68319 | 3642 | No | NA | 4 | 0 | 0 | Not-determined | Genome-fragment | NA | NA | 0 |
| S5_extracted_k141_44496 | 1009 | No | NA | 3 | 0 | 0 | Not-determined | Genome-fragment | NA | NA | 0 |
| S5_extracted_k141_111048 | 1131 | No | NA | 3 | 0 | 0 | Not-determined | Genome-fragment | NA | NA | 0 |

| | | | | | | | | | | | |
|---|---|---|---|---|---|---|---|---|---|---|---|
| S14_vs_k141_68255\|\|full | 1776 | No | NA | 4 | 0 | 0 | Low-quality | Genome-fragment | 2.4 | AAI-based (high-confidence) | 0 |
| S3_vs_k141_46557\|\|full | 1776 | No | NA | 3 | 0 | 0 | Low-quality | Genome-fragment | 3.98 | AAI-based (high-confidence) | 0 |
| S5_extracted_k141_139461 | 1776 | No | NA | 3 | 0 | 0 | Low-quality | Genome-fragment | 4.4 | AAI-based (medium-confidence) | 0 |
| S5_extracted_k141_32506 | 3402 | No | NA | 2 | 0 | 2 | Not-determined | Genome-fragment | NA | NA | 0 |
| S5_extracted_k141_55751 | 6090 | No | NA | 4 | 0 | 2 | Not-determined | Genome-fragment | NA | NA | 0 |
| S5_extracted_k141_86663 | 8069 | No | NA | 12 | 0 | 3 | Not-determined | Genome-fragment | NA | NA | 0 |
| S7_vs_119495\|\|full | 1776 | No | NA | 4 | 2 | 0 | Low-quality | Genome-fragment | 3.01 | AAI-based (high-confidence) | 0 |
| S16_vs_489\|\|full | 1776 | No | NA | 3 | 0 | 0 | Low-quality | Genome-fragment | 3.28 | AAI-based (high-confidence) | 0 |
| S5_extracted_k141_19874 | 1020 | No | NA | 1 | 0 | 0 | Not-determined | Genome-fragment | NA | NA | 0 |
| S5_extracted_k141_68420 | 3843 | No | NA | 5 | 0 | 3 | Not-determined | Genome-fragment | NA | NA | 0 |
| S10_extracted_42395 | 1776 | No | NA | 2 | 0 | 0 | Low-quality | Genome-fragment | 1.26 | AAI-based (high-confidence) | 0 |
| S12_vs_k141_143482\|\|full | 1776 | No | NA | 3 | 0 | 2 | Low-quality | Genome-fragment | 2 | AAI-based (medium-confidence) | 0 |
| S5_extracted_k141_13854 | 1699 | No | NA | 2 | 0 | 2 | Not-determined | Genome-fragment | NA | NA | 0 |
| S5_extracted_k141_8035 | 1646 | No | NA | 1 | 0 | 1 | Not-determined | Genome-fragment | NA | NA | 0 |
| S5_extracted_k141_134916 | 2313 | No | NA | 2 | 0 | 2 | Not-determined | Genome-fragment | NA | NA | 0 |
| S5_extracted_k141_140978 | 3684 | No | NA | 6 | 0 | 2 | Not-determined | Genome-fragment | NA | NA | 0 |
| S2_extracted_k141_20809 | 1776 | No | NA | 2 | 1 | 0 | Low-quality | Genome-fragment | 1.74 | AAI-based (high-confidence) | 0 |
| S4_extracted_13954 | 1776 | No | NA | 3 | 1 | 0 | Low-quality | Genome-fragment | 5.59 | AAI-based (high-confidence) | 0 |
| S5_extracted_k141_8061 | 3214 | No | NA | 2 | 0 | 0 | Not-determined | Genome-fragment | NA | NA | 0 |
| S6_vs_k141_29972\|\|full | 1776 | No | NA | 4 | 0 | 0 | Low-quality | Genome-fragment | 2.57 | AAI-based (high-confidence) | 0 |
| S5_extracted_k141_13945 | 1187 | No | NA | 1 | 0 | 1 | Not-determined | Genome-fragment | NA | NA | 0 |
| S5_extracted_k141_93025 | 1331 | No | NA | 1 | 0 | 0 | Not-determined | Genome-fragment | NA | NA | 0 |
| S8_extracted_k141_28711 | 1776 | No | NA | 3 | 1 | 0 | Low-quality | Genome-fragment | 0.5 | HMM-based (lower-bound) | 0 |
| S5_extracted_k141_2349 | 5500 | No | NA | 9 | 0 | 0 | Not-determined | Genome-fragment | NA | NA | 0 |
| S5_extracted_k141_38162 | 1066 | No | NA | 1 | 0 | 1 | Not-determined | Genome-fragment | NA | NA | 0 |
| S8_extracted_k141_17346 | 1776 | No | NA | 5 | 0 | 3 | Low-quality | Genome-fragment | 5.17 | AAI-based (medium-confidence) | 0 |
| S8_vs_k141_40889\|\|full | 1776 | No | NA | 3 | 3 | 0 | Low-quality | Genome-fragment | 4.04 | AAI-based (high-confidence) | 0 |
| S5_extracted_k141_129208 | 6543 | No | NA | 6 | 0 | 2 | Not-determined | Genome-fragment | NA | NA | 0 |
| S8_vs_k141_137451\|\|full | 1776 | No | NA | 3 | 3 | 0 | Low-quality | Genome-fragment | 4.05 | AAI-based (high-confidence) | 0 |
| S5_extracted_k141_93039 | 1973 | No | NA | 3 | 0 | 1 | Not-determined | Genome-fragment | NA | NA | 0 |
| S9_vs_k141_15794\|\|full | 1777 | No | NA | 2 | 2 | 0 | Low-quality | Genome-fragment | 4.73 | AAI-based (high-confidence) | 0 |
| S5_extracted_k141_50582 | 1079 | No | NA | 1 | 0 | 0 | Not-determined | Genome-fragment | NA | NA | 0 |
| S13_extracted_14967 | 1777 | No | NA | 2 | 0 | 1 | Low-quality | Genome-fragment | 1.89 | AAI-based (medium-confidence) | 0 |
| S5_extracted_k141_93102 | 1351 | No | NA | 2 | 0 | 0 | Not-determined | Genome-fragment | NA | NA | 0 |
| S5_extracted_k141_50600 | 1028 | No | NA | 1 | 0 | 1 | Not-determined | Genome-fragment | NA | NA | 0 |
| S14_extracted_k141_25724 | 1777 | No | NA | 7 | 7 | 0 | Low-quality | Genome-fragment | 5.84 | AAI-based (high-confidence) | 0 |
| S5_extracted_k141_20014 | 1283 | No | NA | 3 | 0 | 1 | Not-determined | Genome-fragment | NA | NA | 0 |
| S3_vs_k141_71128\|\|full | 1777 | No | NA | 3 | 0 | 0 | Low-quality | Genome-fragment | 4.58 | AAI-based (high-confidence) | 0 |
| S7_vs_81829\|\|lt2gene | 1777 | No | NA | 2 | 0 | 0 | Low-quality | Genome-fragment | 4.49 | AAI-based (high-confidence) | 0 |
| S5_extracted_k141_20057 | 1456 | No | NA | 2 | 0 | 1 | Not-determined | Genome-fragment | NA | NA | 0 |
| S5_extracted_k141_2490 | 1037 | No | NA | 2 | 0 | 0 | Not-determined | Genome-fragment | NA | NA | 0 |
| S5_extracted_k141_93158 | 4630 | No | NA | 4 | 0 | 0 | Not-determined | Genome-fragment | NA | NA | 0 |
| S5_extracted_k141_32700 | 6145 | No | NA | 5 | 0 | 4 | Not-determined | Genome-fragment | NA | NA | 0 |
| S12_extracted_k141_113076 | 1777 | No | NA | 2 | 0 | 0 | Low-quality | Genome-fragment | 3.69 | AAI-based (medium-confidence) | 0 |
| S6_extracted_k141_4733 | 1777 | No | NA | 3 | 1 | 0 | Low-quality | Genome-fragment | 5.73 | HMM-based (lower-bound) | 0 |
| S17_extracted_k141_53880 | 1778 | No | NA | 2 | 0 | 2 | Low-quality | Genome-fragment | 2.62 | AAI-based (high-confidence) | 0 |
| S9_vs_k141_34648\|\|full | 1778 | No | NA | 4 | 0 | 0 | Low-quality | Genome-fragment | 4.12 | AAI-based (high-confidence) | 0 |
| S5_extracted_k141_44773 | 1311 | No | NA | 2 | 0 | 2 | Not-determined | Genome-fragment | NA | NA | 0 |
| S5_extracted_k141_62379 | 1173 | No | NA | 3 | 0 | 2 | Not-determined | Genome-fragment | NA | NA | 0 |
| S5_extracted_k141_141199 | 1109 | No | NA | 1 | 0 | 0 | Not-determined | Genome-fragment | NA | NA | 0 |
| S11_vs_k141_44187\|\|full | 1778 | No | NA | 3 | 1 | 0 | Low-quality | Genome-fragment | 4.69 | AAI-based (high-confidence) | 0 |
| S3_vs_k141_7769\|\|full | 1778 | No | NA | 3 | 1 | 0 | Low-quality | Genome-fragment | 3.07 | AAI-based (high-confidence) | 0 |
| S5_extracted_k141_14195 | 4658 | No | NA | 9 | 0 | 9 | Not-determined | Genome-fragment | NA | NA | 0 |
| S7_vs_92557\|\|lt2gene | 1778 | No | NA | 1 | 1 | 0 | Low-quality | Genome-fragment | 4.14 | AAI-based (high-confidence) | 0 |
| S5_extracted_k141_93189 | 1133 | No | NA | 2 | 0 | 0 | Not-determined | Genome-fragment | NA | NA | 0 |
| S5_extracted_k141_105478 | 1616 | No | NA | 1 | 0 | 1 | Not-determined | Genome-fragment | NA | NA | 0 |
| S16_extracted_146968 | 1778 | No | NA | 2 | 0 | 1 | Low-quality | Genome-fragment | 2.86 | AAI-based (high-confidence) | 0 |
| S5_extracted_k141_141252 | 1009 | No | NA | 2 | 0 | 0 | Not-determined | Genome-fragment | NA | NA | 0 |
| S5_extracted_k141_93213 | 1182 | No | NA | 3 | 0 | 3 | Not-determined | Genome-fragment | NA | NA | 0 |
| S10_extracted_79115 | 1778 | No | NA | 1 | 0 | 1 | Low-quality | Genome-fragment | 0.48 | AAI-based (high-confidence) | 0 |
| S5_extracted_k141_25423 | 29056 | No | NA | 48 | 45 | 0 | High-quality | High-quality | 95.81 | AAI-based (high-confidence) | 0 |
| S10_extracted_120611 | 1778 | No | NA | 2 | 0 | 0 | Low-quality | Genome-fragment | 1 | AAI-based (high-confidence) | 0 |
| S10_vs_2435\|\|full | 1778 | No | NA | 3 | 1 | 0 | Low-quality | Genome-fragment | 1.6 | AAI-based (high-confidence) | 0 |
| S5_extracted_k141_86917 | 1398 | No | NA | 1 | 0 | 0 | Not-determined | Genome-fragment | NA | NA | 0 |
| S5_extracted_k141_123990 | 17185 | No | NA | 21 | 9 | 0 | Medium-quality | Genome-fragment | 75.2 | AAI-based (medium-confidence) | 0 |
| S5_extracted_k141_68631 | 1046 | No | NA | 2 | 0 | 1 | Not-determined | Genome-fragment | NA | NA | 0 |
| S8_vs_k141_11741\|\|full | 1778 | No | NA | 2 | 0 | 0 | Low-quality | Genome-fragment | 4.31 | AAI-based (high-confidence) | 0 |
| S8_vs_k141_46469\|\|full | 1778 | No | NA | 3 | 0 | 3 | Low-quality | Genome-fragment | 1.02 | AAI-based (medium-confidence) | 0 |
| S11_vs_k141_17857\|\|full | 1779 | No | NA | 4 | 0 | 1 | Low-quality | Genome-fragment | 2.66 | AAI-based (high-confidence) | 0 |
| S5_extracted_k141_68637 | 4845 | No | NA | 3 | 0 | 3 | Not-determined | Genome-fragment | NA | NA | 0 |
| S5_extracted_k141_50798 | 1705 | No | NA | 1 | 0 | 1 | Not-determined | Genome-fragment | NA | NA | 0 |
| S3_vs_k141_12560\|\|full | 1779 | No | NA | 5 | 1 | 0 | Low-quality | Genome-fragment | 3.91 | AAI-based (medium-confidence) | 0 |
| S5_vs_k141_57989\|\|full | 1779 | No | NA | 2 | 0 | 2 | Low-quality | Genome-fragment | 3.53 | AAI-based (medium-confidence) | 0 |
| S5_extracted_k141_20225 | 1444 | No | NA | 2 | 0 | 2 | Not-determined | Genome-fragment | NA | NA | 0 |
| S5_extracted_k141_62493 | 7225 | No | NA | 9 | 0 | 0 | Not-determined | Genome-fragment | NA | NA | 0 |
| S5_extracted_k141_129448 | 2707 | No | NA | 1 | 0 | 0 | Not-determined | Genome-fragment | NA | NA | 0 |
| S7_extracted_60031 | 1779 | No | NA | 3 | 0 | 1 | Low-quality | Genome-fragment | 1.62 | AAI-based (high-confidence) | 0 |
| S12_vs_k141_66192\|\|full | 1779 | No | NA | 3 | 0 | 0 | Low-quality | Genome-fragment | 3.71 | AAI-based (medium-confidence) | 0 |
| S5_extracted_k141_93318 | 1169 | No | NA | 1 | 0 | 0 | Not-determined | Genome-fragment | NA | NA | 0 |
| S5_extracted_k141_105728 | 36024 | No | NA | 72 | 16 | 0 | Medium-quality | Genome-fragment | 79.55 | AAI-based (high-confidence) | 0 |
| S6_vs_k141_54414\|\|full | 1779 | No | NA | 3 | 0 | 2 | Low-quality | Genome-fragment | 1.32 | AAI-based (high-confidence) | 0 |
| S8_vs_k141_11601\|\|full | 1779 | No | NA | 2 | 1 | 0 | Low-quality | Genome-fragment | 1.2 | HMM-based (lower-bound) | 0 |
| S5_extracted_k141_124133 | 3426 | No | NA | 6 | 0 | 1 | Not-determined | Genome-fragment | NA | NA | 0 |
| S5_extracted_k141_44857 | 1376 | No | NA | 1 | 0 | 1 | Not-determined | Genome-fragment | NA | NA | 0 |
| S5_extracted_k141_124178 | 3120 | No | NA | 7 | 0 | 1 | Not-determined | Genome-fragment | NA | NA | 0 |
| S5_extracted_k141_14416 | 12045 | No | NA | 10 | 0 | 3 | Not-determined | Genome-fragment | NA | NA | 0 |
| S3_vs_k141_91759\|\|full | 1780 | No | NA | 2 | 0 | 2 | Low-quality | Genome-fragment | 0.35 | AAI-based (medium-confidence) | 0 |
| S7_extracted_18847 | 1780 | No | NA | 3 | 1 | 0 | Low-quality | Genome-fragment | 3.53 | AAI-based (high-confidence) | 0 |
| S7_vs_94199\|\|full | 1780 | No | NA | 6 | 0 | 0 | Low-quality | Genome-fragment | 4.01 | AAI-based (high-confidence) | 0 |
| S12_vs_k141_51276\|\|full | 1780 | Yes | 828 | 2 | 1 | 1 | Low-quality | Genome-fragment | 0.23 | HMM-based (lower-bound) | 53.48 |
| S5_extracted_k141_98430 | 1186 | No | NA | 1 | 0 | 0 | Not-determined | Genome-fragment | NA | NA | 0 |
| S5_extracted_k141_38453 | 1012 | No | NA | 2 | 0 | 0 | Not-determined | Genome-fragment | NA | NA | 0 |
| S5_extracted_k141_105862 | 2761 | No | NA | 1 | 0 | 0 | Not-determined | Genome-fragment | NA | NA | 0 |
| S5_extracted_k141_68789 | 1394 | No | NA | 3 | 0 | 2 | Not-determined | Genome-fragment | NA | NA | 0 |
| S5_extracted_k141_51075 | 1035 | No | NA | 2 | 0 | 0 | Not-determined | Genome-fragment | NA | NA | 0 |
| S5_extracted_k141_2871 | 1686 | No | NA | 2 | 0 | 2 | Not-determined | Genome-fragment | NA | NA | 0 |
| S2_extracted_k141_13376 | 1780 | No | NA | 6 | 0 | 0 | Low-quality | Genome-fragment | 1.85 | AAI-based (high-confidence) | 0 |
| S5_extracted_k141_20393 | 1130 | No | NA | 1 | 0 | 1 | Not-determined | Genome-fragment | NA | NA | 0 |
| S2_vs_k141_8297\|\|full | 1780 | No | NA | 4 | 2 | 0 | Low-quality | Genome-fragment | 5 | AAI-based (high-confidence) | 0 |
| S17_vs_k141_52299\|\|full | 1781 | No | NA | 3 | 0 | 0 | Low-quality | Genome-fragment | 2.05 | AAI-based (medium-confidence) | 0 |
| S5_extracted_k141_124254 | 1358 | No | NA | 1 | 0 | 1 | Not-determined | Genome-fragment | NA | NA | 0 |
| S5_extracted_k141_68846 | 2573 | No | NA | 2 | 0 | 2 | Not-determined | Genome-fragment | NA | NA | 0 |
| S9_vs_k141_29282\|\|full | 1781 | No | NA | 3 | 0 | 1 | Low-quality | Genome-fragment | 3.71 | AAI-based (medium-confidence) | 0 |
| S5_extracted_k141_141459 | 3520 | No | NA | 2 | 0 | 2 | Not-determined | Genome-fragment | NA | NA | 0 |
| S13_vs_80826\|\|full | 1781 | No | NA | 3 | 1 | 0 | Low-quality | Genome-fragment | 4.03 | AAI-based (high-confidence) | 0 |
| S14_vs_k141_57197\|\|full | 1781 | No | NA | 4 | 0 | 0 | Low-quality | Genome-fragment | 2.02 | AAI-based (medium-confidence) | 0 |
| S5_extracted_k141_124299 | 1013 | No | NA | 1 | 0 | 0 | Not-determined | Genome-fragment | NA | NA | 0 |
| S7_vs_93545\|\|full | 1781 | No | NA | 3 | 1 | 0 | Low-quality | Genome-fragment | 3.94 | AAI-based (medium-confidence) | 0 |
| S5_extracted_k141_51129 | 1091 | No | NA | 1 | 0 | 0 | Not-determined | Genome-fragment | NA | NA | 0 |
| S17_extracted_k141_156128 | 1782 | No | NA | 4 | 1 | 0 | Low-quality | Genome-fragment | 1.91 | AAI-based (high-confidence) | 0 |
| S5_extracted_k141_14500 | 1846 | No | NA | 1 | 0 | 0 | Not-determined | Genome-fragment | NA | NA | 0 |
| S5_extracted_k141_80146 | 1533 | No | NA | 2 | 0 | 1 | Not-determined | Genome-fragment | NA | NA | 0 |
| S9_extracted_k141_36644 | 1782 | No | NA | 2 | 0 | 1 | Low-quality | Genome-fragment | 3.23 | AAI-based (high-confidence) | 0 |
| S5_extracted_k141_3013 | 2877 | No | NA | 6 | 0 | 5 | Not-determined | Genome-fragment | NA | NA | 0 |
| S5_extracted_k141_93615 | 1452 | No | NA | 2 | 0 | 0 | Not-determined | Genome-fragment | NA | NA | 0 |
| S9_vs_k141_42968\|\|full | 1782 | No | NA | 3 | 0 | 1 | Low-quality | Genome-fragment | 4.57 | AAI-based (high-confidence) | 0 |
| S11_vs_k141_16223\|\|full | 1782 | No | NA | 4 | 0 | 0 | Low-quality | Genome-fragment | 1.61 | AAI-based (high-confidence) | 0 |
| S5_extracted_k141_38547 | 1957 | No | NA | 3 | 0 | 1 | Not-determined | Genome-fragment | NA | NA | 0 |
| S14_extracted_k141_80627 | 1782 | No | NA | 3 | 2 | 0 | Low-quality | Genome-fragment | 6.49 | AAI-based (high-confidence) | 0 |
| S5_extracted_k141_129765 | 4796 | No | NA | 5 | 0 | 0 | Not-determined | Genome-fragment | NA | NA | 0 |
| S5_extracted_k141_135275 | 3094 | No | NA | 6 | 0 | 2 | Not-determined | Genome-fragment | NA | NA | 0 |
| S14_vs_k141_26782\|\|full | 1782 | No | NA | 3 | 2 | 0 | Low-quality | Genome-fragment | 4.53 | AAI-based (high-confidence) | 0 |
| S5_extracted_k141_8721 | 12692 | No | NA | 25 | 0 | 25 | Not-determined | Genome-fragment | NA | NA | 0 |
| S5_extracted_k141_106020 | 3452 | No | NA | 6 | 0 | 3 | Not-determined | Genome-fragment | NA | NA | 0 |
| S5_extracted_k141_118360 | 1757 | No | NA | 3 | 0 | 1 | Not-determined | Genome-fragment | NA | NA | 0 |
| S14_vs_k141_116993\|\|full | 1782 | No | NA | 3 | 0 | 0 | Low-quality | Genome-fragment | 2.83 | AAI-based (high-confidence) | 0 |
| S7_extracted_43909 | 1782 | No | NA | 2 | 0 | 0 | Low-quality | Genome-fragment | 1.36 | AAI-based (high-confidence) | 0 |
| S5_extracted_k141_124470 | 2458 | No | NA | 1 | 0 | 0 | Not-determined | Genome-fragment | NA | NA | 0 |
| S5_extracted_k141_62782 | 3653 | No | NA | 4 | 0 | 2 | Not-determined | Genome-fragment | NA | NA | 0 |
| S5_extracted_k141_62783 | 1303 | No | NA | 3 | 0 | 0 | Not-determined | Genome-fragment | NA | NA | 0 |
| S7_vs_32413\|\|full | 1782 | No | NA | 3 | 1 | 0 | Low-quality | Genome-fragment | 2.04 | AAI-based (high-confidence) | 0 |
| S5_extracted_k141_38606 | 1603 | No | NA | 2 | 0 | 1 | Not-determined | Genome-fragment | NA | NA | 0 |
| S5_extracted_k141_124490 | 6949 | No | NA | 9 | 0 | 2 | Not-determined | Genome-fragment | NA | NA | 0 |
| S7_vs_75771\|\|full | 1782 | No | NA | 3 | 1 | 0 | Low-quality | Genome-fragment | 3.28 | AAI-based (high-confidence) | 0 |
| S16_vs_152570\|\|full | 1782 | No | NA | 5 | 0 | 0 | Low-quality | Genome-fragment | 4.26 | AAI-based (high-confidence) | 0 |
| S15_vs_61269\|\|lt2gene | 1782 | No | NA | 1 | 1 | 0 | Low-quality | Genome-fragment | 1.67 | HMM-based (lower-bound) | 0 |
| S5_extracted_k141_62813 | 1470 | No | NA | 1 | 0 | 0 | Not-determined | Genome-fragment | NA | NA | 0 |
| S5_extracted_k141_3198 | 1115 | No | NA | 1 | 0 | 1 | Not-determined | Genome-fragment | NA | NA | 0 |
| S5_extracted_k141_51239 | 4978 | No | NA | 6 | 0 | 1 | Not-determined | Genome-fragment | NA | NA | 0 |
| S4_extracted_3612 | 1782 | No | NA | 6 | 5 | 0 | Low-quality | Genome-fragment | 5.85 | AAI-based (high-confidence) | 0 |
| S5_extracted_k141_68990 | 1254 | No | NA | 1 | 0 | 1 | Not-determined | Genome-fragment | NA | NA | 0 |
| S5_extracted_k141_106133 | 2726 | No | NA | 2 | 0 | 2 | Not-determined | Genome-fragment | NA | NA | 0 |
| S5_extracted_k141_51304 | 2896 | No | NA | 4 | 0 | 2 | Not-determined | Genome-fragment | NA | NA | 0 |
| S11_vs_k141_54108\|\|full | 1783 | No | NA | 5 | 1 | 0 | Low-quality | Genome-fragment | 3.26 | HMM-based (lower-bound) | 0 |
| S11_vs_k141_29364\|\|full | 1783 | No | NA | 4 | 1 | 0 | Low-quality | Genome-fragment | 3.58 | AAI-based (high-confidence) | 0 |
| S3_extracted_k141_106107 | 1783 | No | NA | 2 | 0 | 0 | Low-quality | Genome-fragment | 3.24 | AAI-based (high-confidence) | 0 |
| S5_extracted_k141_74412 | 1781 | No | NA | 4 | 0 | 0 | Not-determined | Genome-fragment | NA | NA | 0 |
| S5_extracted_k141_98551 | 1004 | No | NA | 2 | 0 | 0 | Not-determined | Genome-fragment | NA | NA | 0 |
| S3_vs_k141_93394\|\|full | 1783 | No | NA | 2 | 0 | 1 | Low-quality | Genome-fragment | 2.92 | AAI-based (medium-confidence) | 0 |
| S5_extracted_k141_69020 | 1011 | No | NA | 2 | 0 | 1 | Not-determined | Genome-fragment | NA | NA | 0 |
| S5_extracted_k141_124557 | 1777 | No | NA | 2 | 0 | 0 | Not-determined | Genome-fragment | NA | NA | 0 |
| S5_extracted_k141_98563 | 1509 | No | NA | 1 | 0 | 0 | Not-determined | Genome-fragment | NA | NA | 0 |
| S12_vs_k141_128307\|\|full | 1783 | No | NA | 3 | 1 | 0 | Low-quality | Genome-fragment | 1.4 | AAI-based (high-confidence) | 0 |
| S5_extracted_k141_20642 | 2570 | No | NA | 3 | 0 | 2 | Not-determined | Genome-fragment | NA | NA | 0 |
| S12_vs_k141_34310\|\|full | 1783 | No | NA | 2 | 0 | 0 | Low-quality | Genome-fragment | 4.95 | AAI-based (high-confidence) | 0 |
| S8_vs_k141_89569\|\|full | 1783 | No | NA | 3 | 0 | 0 | Low-quality | Genome-fragment | 2.71 | AAI-based (medium-confidence) | 0 |
| S1_vs_k141_109411\|\|full | 1784 | No | NA | 3 | 0 | 0 | Low-quality | Genome-fragment | 4.23 | AAI-based (high-confidence) | 0 |
| S5_extracted_k141_8808 | 22785 | No | NA | 31 | 15 | 0 | Medium-quality | Genome-fragment | 66.73 | AAI-based (high-confidence) | 0 |
| S5_vs_k141_106369\|\|full | 1784 | No | NA | 4 | 0 | 1 | Low-quality | Genome-fragment | 4.85 | AAI-based (high-confidence) | 0 |
| S5_vs_k141_75370\|\|full | 1784 | No | NA | 2 | 0 | 1 | Low-quality | Genome-fragment | 2.98 | AAI-based (medium-confidence) | 0 |
| S5_extracted_k141_14840 | 1066 | No | NA | 1 | 0 | 0 | Not-determined | Genome-fragment | NA | NA | 0 |
| S16_vs_126244\|\|full | 1784 | No | NA | 3 | 0 | 0 | Low-quality | Genome-fragment | 4.28 | AAI-based (high-confidence) | 0 |
| S5_extracted_k141_38759 | 1675 | No | NA | 3 | 0 | 2 | Not-determined | Genome-fragment | NA | NA | 0 |
| S5_extracted_k141_98631 | 1027 | No | NA | 1 | 0 | 0 | Not-determined | Genome-fragment | NA | NA | 0 |
| S16_vs_134570\|\|full | 1784 | No | NA | 6 | 1 | 0 | Low-quality | Genome-fragment | 1.65 | AAI-based (high-confidence) | 0 |
| S5_extracted_k141_38790 | 3059 | No | NA | 6 | 0 | 5 | Not-determined | Genome-fragment | NA | NA | 0 |
| S5_extracted_k141_93850 | 3345 | No | NA | 3 | 0 | 2 | Not-determined | Genome-fragment | NA | NA | 0 |
| S13_vs_64926\|\|full | 1785 | No | NA | 2 | 1 | 0 | Low-quality | Genome-fragment | 0.97 | HMM-based (lower-bound) | 0 |
| S3_vs_k141_126275\|\|full | 1785 | No | NA | 4 | 0 | 0 | Low-quality | Genome-fragment | 4.88 | AAI-based (high-confidence) | 0 |
| S5_vs_k141_29624\|\|full | 1785 | No | NA | 2 | 2 | 0 | Low-quality | Genome-fragment | 4.53 | AAI-based (medium-confidence) | 0 |
| S5_extracted_k141_93888 | 5936 | No | NA | 4 | 0 | 1 | Not-determined | Genome-fragment | NA | NA | 0 |
| S7_vs_88169\|\|full | 1785 | No | NA | 3 | 1 | 0 | Low-quality | Genome-fragment | 3.69 | AAI-based (high-confidence) | 0 |
| S7_vs_134374\|\|full | 1785 | No | NA | 3 | 0 | 0 | Low-quality | Genome-fragment | 4.58 | AAI-based (high-confidence) | 0 |
| S16_vs_29585\|\|full | 1785 | No | NA | 5 | 2 | 0 | Low-quality | Genome-fragment | 2.2 | HMM-based (lower-bound) | 0 |
| S16_vs_100932\|\|full | 1785 | No | NA | 3 | 0 | 3 | Low-quality | Genome-fragment | 0.75 | AAI-based (medium-confidence) | 0 |
| S5_extracted_k141_33201 | 1115 | No | NA | 2 | 0 | 2 | Not-determined | Genome-fragment | NA | NA | 0 |
| S5_extracted_k141_20772 | 3955 | No | NA | 11 | 0 | 0 | Not-determined | Genome-fragment | NA | NA | 0 |
| S8_extracted_k141_37850 | 1785 | No | NA | 4 | 0 | 0 | Low-quality | Genome-fragment | 1.34 | AAI-based (medium-confidence) | 0 |
| S17_extracted_k141_17706 | 1786 | No | NA | 1 | 1 | 0 | Low-quality | Genome-fragment | 2.74 | AAI-based (high-confidence) | 0 |
| S9_vs_k141_86066\|\|full | 1786 | No | NA | 4 | 2 | 0 | Low-quality | Genome-fragment | 2.28 | AAI-based (high-confidence) | 0 |
| S5_extracted_k141_51551 | 1717 | No | NA | 1 | 0 | 0 | Not-determined | Genome-fragment | NA | NA | 0 |
| S5_extracted_k141_111697 | 1419 | No | NA | 4 | 0 | 0 | Not-determined | Genome-fragment | NA | NA | 0 |
| S1_extracted_k141_134869 | 1786 | No | NA | 2 | 1 | 0 | Low-quality | Genome-fragment | 5.22 | AAI-based (high-confidence) | 0 |
| S5_vs_k141_67830\|\|full | 1786 | No | NA | 4 | 1 | 0 | Low-quality | Genome-fragment | 3.83 | AAI-based (medium-confidence) | 0 |
| S5_extracted_k141_8965 | 22925 | No | NA | 31 | 10 | 1 | Medium-quality | Genome-fragment | 70.63 | AAI-based (high-confidence) | 0 |
| S5_extracted_k141_8971 | 3382 | No | NA | 4 | 0 | 1 | Not-determined | Genome-fragment | NA | NA | 0 |
| S5_extracted_k141_98765 | 3384 | No | NA | 4 | 0 | 1 | Not-determined | Genome-fragment | NA | NA | 0 |
| S5_extracted_k141_51656 | 5823 | No | NA | 10 | 0 | 0 | Not-determined | Genome-fragment | NA | NA | 0 |
| S5_vs_k141_20238\|\|full | 1786 | No | NA | 3 | 0 | 0 | Low-quality | Genome-fragment | 4.34 | AAI-based (high-confidence) | 0 |
| S5_extracted_k141_69368 | 1177 | No | NA | 1 | 0 | 1 | Not-determined | Genome-fragment | NA | NA | 0 |
| S5_extracted_k141_38844 | 1141 | No | NA | 1 | 0 | 1 | Not-determined | Genome-fragment | NA | NA | 0 |
| S5_extracted_k141_20918 | 4200 | No | NA | 3 | 0 | 0 | Not-determined | Genome-fragment | NA | NA | 0 |
| S7_extracted_100494 | 1786 | No | NA | 2 | 0 | 0 | Low-quality | Genome-fragment | 4.76 | AAI-based (medium-confidence) | 0 |
| S5_extracted_k141_38859 | 1201 | No | NA | 1 | 0 | 0 | Not-determined | Genome-fragment | NA | NA | 0 |
| S7_extracted_122812 | 1786 | No | NA | 2 | 0 | 0 | Low-quality | Genome-fragment | 2.41 | AAI-based (medium-confidence) | 0 |
| S7_vs_39503\|\|full | 1786 | No | NA | 6 | 0 | 0 | Low-quality | Genome-fragment | 3.52 | AAI-based (medium-confidence) | 0 |
| S16_vs_69730\|\|full | 1786 | No | NA | 3 | 2 | 0 | Low-quality | Genome-fragment | 4.91 | AAI-based (high-confidence) | 0 |
| S16_vs_145532\|\|full | 1786 | No | NA | 2 | 0 | 1 | Low-quality | Genome-fragment | 1.36 | AAI-based (high-confidence) | 0 |
| S5_extracted_k141_118714 | 1970 | No | NA | 1 | 0 | 1 | Not-determined | Genome-fragment | NA | NA | 0 |
| S16_vs_156248\|\|full | 1786 | No | NA | 3 | 3 | 0 | Low-quality | Genome-fragment | 2.87 | HMM-based (lower-bound) | 0 |
| S13_extracted_88307 | 1787 | No | NA | 1 | 0 | 0 | Low-quality | Genome-fragment | 0.31 | AAI-based (medium-confidence) | 0 |
| S7_vs_88569\|\|full | 1787 | No | NA | 3 | 1 | 0 | Low-quality | Genome-fragment | 5.55 | AAI-based (high-confidence) | 0 |
| S5_extracted_k141_118734 | 1043 | No | NA | 1 | 0 | 1 | Not-determined | Genome-fragment | NA | NA | 0 |
| S5_extracted_k141_3755 | 1116 | No | NA | 2 | 0 | 0 | Not-determined | Genome-fragment | NA | NA | 0 |
| S12_vs_k141_23859\|\|full | 1787 | No | NA | 3 | 0 | 0 | Low-quality | Genome-fragment | 1.31 | AAI-based (medium-confidence) | 0 |
| S2_vs_k141_11865\|\|full | 1787 | No | NA | 1 | 0 | 1 | Low-quality | Genome-fragment | 2.62 | AAI-based (medium-confidence) | 0 |
| S5_extracted_k141_3768 | 1180 | No | NA | 1 | 0 | 0 | Not-determined | Genome-fragment | NA | NA | 0 |
| S6_vs_k141_40761\|\|full | 1787 | No | NA | 5 | 1 | 0 | Low-quality | Genome-fragment | 4.58 | AAI-based (high-confidence) | 0 |
| S5_extracted_k141_106472 | 1307 | No | NA | 1 | 0 | 0 | Not-determined | Genome-fragment | NA | NA | 0 |
| S8_vs_k141_193067\|\|full | 1787 | No | NA | 3 | 0 | 1 | Low-quality | Genome-fragment | 3.11 | AAI-based (medium-confidence) | 0 |
| S14_vs_k141_101338\|\|full | 1788 | No | NA | 4 | 4 | 0 | Low-quality | Genome-fragment | 3.51 | AAI-based (high-confidence) | 0 |
| S5_extracted_k141_69506 | 1017 | No | NA | 1 | 0 | 1 | Not-determined | Genome-fragment | NA | NA | 0 |
| S1_extracted_k141_803 | 1788 | No | NA | 1 | 0 | 0 | Low-quality | Genome-fragment | 1.47 | AAI-based (high-confidence) | 0 |
| S3_extracted_k141_48032 | 1788 | No | NA | 3 | 2 | 0 | Low-quality | Genome-fragment | 3.56 | AAI-based (high-confidence) | 0 |
| S3_vs_k141_95416\|\|full | 1788 | No | NA | 3 | 0 | 1 | Low-quality | Genome-fragment | 3.54 | AAI-based (medium-confidence) | 0 |
| S5_extracted_k141_25979 | 3817 | No | NA | 3 | 0 | 1 | Not-determined | Genome-fragment | NA | NA | 0 |
| S5_vs_k141_111026\|\|full | 1788 | No | NA | 7 | 1 | 0 | Low-quality | Genome-fragment | 1.22 | HMM-based (lower-bound) | 0 |
| S5_extracted_k141_33429 | 2300 | No | NA | 3 | 0 | 1 | Not-determined | Genome-fragment | NA | NA | 0 |
| S5_extracted_k141_130213 | 1645 | No | NA | 1 | 0 | 1 | Not-determined | Genome-fragment | NA | NA | 0 |
| S5_extracted_k141_118840 | 3075 | No | NA | 2 | 0 | 2 | Not-determined | Genome-fragment | NA | NA | 0 |
| S16_extracted_91505 | 1788 | No | NA | 5 | 0 | 0 | Low-quality | Genome-fragment | 1.51 | AAI-based (high-confidence) | 0 |
| S5_extracted_k141_26003 | 1649 | No | NA | 1 | 0 | 0 | Not-determined | Genome-fragment | NA | NA | 0 |
| S16_vs_66159\|\|full | 1788 | No | NA | 4 | 0 | 3 | Low-quality | Genome-fragment | 2.65 | AAI-based (medium-confidence) | 0 |
| S5_extracted_k141_118864 | 1868 | No | NA | 3 | 0 | 0 | Not-determined | Genome-fragment | NA | NA | 0 |
| S16_vs_79369\|\|full | 1788 | No | NA | 5 | 0 | 0 | Low-quality | Genome-fragment | 2.27 | AAI-based (high-confidence) | 0 |
| S5_extracted_k141_130262 | 1275 | No | NA | 3 | 0 | 3 | Not-determined | Genome-fragment | NA | NA | 0 |
| S6_vs_k141_34875\|\|lt2gene | 1788 | No | NA | 1 | 1 | 0 | Low-quality | Genome-fragment | 4.52 | AAI-based (high-confidence) | 0 |
| S5_extracted_k141_87314 | 2498 | No | NA | 1 | 0 | 0 | Not-determined | Genome-fragment | NA | NA | 0 |
| S5_extracted_k141_94087 | 2079 | No | NA | 6 | 0 | 0 | Not-determined | Genome-fragment | NA | NA | 0 |
| S5_extracted_k141_87335 | 1247 | No | NA | 2 | 0 | 0 | Not-determined | Genome-fragment | NA | NA | 0 |
| S5_extracted_k141_45427 | 1071 | No | NA | 2 | 0 | 2 | Not-determined | Genome-fragment | NA | NA | 0 |
| S5_extracted_k141_69568 | 1114 | No | NA | 1 | 0 | 0 | Not-determined | Genome-fragment | NA | NA | 0 |
| S5_extracted_k141_74590 | 2192 | No | NA | 3 | 0 | 0 | Not-determined | Genome-fragment | NA | NA | 0 |
| S5_extracted_k141_112127 | 1126 | No | NA | 2 | 0 | 1 | Not-determined | Genome-fragment | NA | NA | 0 |
| S8_vs_k141_207711\|\|full | 1788 | No | NA | 3 | 2 | 0 | Low-quality | Genome-fragment | 4.24 | AAI-based (high-confidence) | 0 |
| S17_vs_k141_138542\|\|full | 1789 | No | NA | 2 | 0 | 0 | Low-quality | Genome-fragment | 1.04 | AAI-based (high-confidence) | 0 |
| S5_extracted_k141_94131 | 1098 | No | NA | 3 | 0 | 2 | Not-determined | Genome-fragment | NA | NA | 0 |
| S5_extracted_k141_118971 | 1198 | No | NA | 1 | 0 | 1 | Not-determined | Genome-fragment | NA | NA | 0 |
| S5_extracted_k141_118973 | 1079 | No | NA | 2 | 0 | 0 | Not-determined | Genome-fragment | NA | NA | 0 |
| S5_extracted_k141_38978 | 1451 | No | NA | 1 | 0 | 1 | Not-determined | Genome-fragment | NA | NA | 0 |
| S5_extracted_k141_130325 | 1124 | No | NA | 1 | 0 | 0 | Not-determined | Genome-fragment | NA | NA | 0 |
| S5_extracted_k141_130332 | 3156 | No | NA | 3 | 0 | 0 | Not-determined | Genome-fragment | NA | NA | 0 |
| S5_extracted_k141_124960 | 3372 | No | NA | 1 | 0 | 1 | Not-determined | Genome-fragment | NA | NA | 0 |
| S11_vs_k141_99247\|\|full | 1789 | No | NA | 2 | 0 | 0 | Low-quality | Genome-fragment | 4.27 | AAI-based (high-confidence) | 0 |
| S5_extracted_k141_130343 | 8272 | No | NA | 11 | 0 | 2 | Not-determined | Genome-fragment | NA | NA | 0 |
| S11_vs_k141_67468\|\|full | 1789 | No | NA | 3 | 0 | 1 | Low-quality | Genome-fragment | 4.18 | AAI-based (medium-confidence) | 0 |
| S14_extracted_k141_62088 | 1789 | No | NA | 2 | 0 | 0 | Low-quality | Genome-fragment | 3.04 | AAI-based (high-confidence) | 0 |
| S5_extracted_k141_3988 | 29546 | No | NA | 24 | 0 | 13 | Not-determined | Genome-fragment | NA | NA | 0 |
| S3_vs_k141_35781\|\|full | 1789 | No | NA | 2 | 0 | 1 | Low-quality | Genome-fragment | 2.84 | AAI-based (medium-confidence) | 0 |
| S5_extracted_k141_135609 | 15998 | No | NA | 30 | 0 | 30 | Not-determined | Genome-fragment | NA | NA | 0 |
| S5_extracted_k141_106718 | 5682 | No | NA | 5 | 0 | 1 | Not-determined | Genome-fragment | NA | NA | 0 |
| S5_extracted_k141_124990 | 2787 | No | NA | 3 | 0 | 1 | Not-determined | Genome-fragment | NA | NA | 0 |
| S5_extracted_k141_106743 | 3498 | No | NA | 5 | 0 | 4 | Not-determined | Genome-fragment | NA | NA | 0 |
| S5_extracted_k141_106752 | 3985 | No | NA | 6 | 0 | 3 | Not-determined | Genome-fragment | NA | NA | 0 |
| S5_extracted_k141_70272 | 1789 | No | NA | 3 | 1 | 0 | Low-quality | Genome-fragment | 3.83 | AAI-based (high-confidence) | 0 |
| S7_vs_128266\|\|full | 1789 | No | NA | 3 | 1 | 0 | Low-quality | Genome-fragment | 2.5 | HMM-based (lower-bound) | 0 |
| S7_vs_95654\|\|lt2gene | 1789 | No | NA | 1 | 1 | 0 | Low-quality | Genome-fragment | 3.92 | AAI-based (high-confidence) | 0 |
| S5_extracted_k141_52074 | 1137 | No | NA | 1 | 0 | 0 | Not-determined | Genome-fragment | NA | NA | 0 |
| S5_extracted_k141_39086 | 2601 | No | NA | 4 | 0 | 0 | Not-determined | Genome-fragment | NA | NA | 0 |
| S16_extracted_43860 | 1789 | No | NA | 1 | 1 | 0 | Low-quality | Genome-fragment | 0.84 | AAI-based (high-confidence) | 0 |
| S5_extracted_k141_106838 | 1689 | No | NA | 4 | 0 | 4 | Not-determined | Genome-fragment | NA | NA | 0 |
| S10_vs_1884\|\|full | 1789 | No | NA | 3 | 0 | 0 | Low-quality | Genome-fragment | 0.43 | AAI-based (medium-confidence) | 0 |
| S5_extracted_k141_106872 | 1781 | No | NA | 4 | 0 | 0 | Not-determined | Genome-fragment | NA | NA | 0 |
| S4_extracted_1279 | 1789 | No | NA | 6 | 0 | 0 | Low-quality | Genome-fragment | 2.23 | AAI-based (high-confidence) | 0 |
| S5_extracted_k141_142279 | 1000 | No | NA | 2 | 0 | 0 | Not-determined | Genome-fragment | NA | NA | 0 |
| S6_vs_k141_9814\|\|full | 1789 | No | NA | 3 | 0 | 0 | Low-quality | Genome-fragment | 2.84 | AAI-based (medium-confidence) | 0 |
| S8_extracted_k141_39455 | 1789 | No | NA | 1 | 0 | 1 | Low-quality | Genome-fragment | 2.88 | AAI-based (high-confidence) | 0 |
| S11_extracted_k141_86115 | 1790 | No | NA | 2 | 2 | 0 | Low-quality | Genome-fragment | 4.25 | AAI-based (high-confidence) | 0 |
| S5_extracted_k141_125153 | 4511 | No | NA | 2 | 0 | 0 | Not-determined | Genome-fragment | NA | NA | 0 |
| S5_extracted_k141_142321 | 1324 | No | NA | 1 | 0 | 1 | Not-determined | Genome-fragment | NA | NA | 0 |
| S5_extracted_k141_87587 | 1244 | No | NA | 2 | 0 | 2 | Not-determined | Genome-fragment | NA | NA | 0 |
| S5_extracted_k141_45670 | 5870 | No | NA | 6 | 0 | 4 | Not-determined | Genome-fragment | NA | NA | 0 |
| S5_extracted_k141_99049 | 1198 | No | NA | 2 | 0 | 1 | Not-determined | Genome-fragment | NA | NA | 0 |
| S5_extracted_k141_142363 | 3135 | No | NA | 2 | 0 | 1 | Not-determined | Genome-fragment | NA | NA | 0 |
| S5_extracted_k141_74661 | 40404 | No | NA | 52 | 40 | 1 | High-quality | High-quality | 100 | AAI-based (high-confidence) | 0 |
| S5_extracted_k141_130570 | 1028 | No | NA | 1 | 0 | 0 | Not-determined | Genome-fragment | NA | NA | 0 |
| S3_vs_k141_30800\|\|full | 1790 | No | NA | 6 | 5 | 0 | Low-quality | Genome-fragment | 3.33 | AAI-based (high-confidence) | 0 |
| S5_vs_k141_128346\|\|full | 1790 | No | NA | 3 | 2 | 0 | Low-quality | Genome-fragment | 3.95 | AAI-based (high-confidence) | 0 |
| S16_extracted_36843 | 1790 | No | NA | 2 | 0 | 0 | Low-quality | Genome-fragment | 1.35 | AAI-based (high-confidence) | 0 |
| S5_extracted_k141_80629 | 6049 | No | NA | 7 | 0 | 3 | Not-determined | Genome-fragment | NA | NA | 0 |
| S5_extracted_k141_107039 | 1498 | No | NA | 1 | 0 | 0 | Not-determined | Genome-fragment | NA | NA | 0 |
| S5_extracted_k141_4305 | 1116 | No | NA | 2 | 0 | 1 | Not-determined | Genome-fragment | NA | NA | 0 |
| S5_extracted_k141_125300 | 1062 | No | NA | 2 | 0 | 0 | Not-determined | Genome-fragment | NA | NA | 0 |
| S5_extracted_k141_33893 | 1531 | No | NA | 1 | 0 | 0 | Not-determined | Genome-fragment | NA | NA | 0 |
| S12_vs_k141_125621\|\|full | 1790 | No | NA | 3 | 2 | 0 | Low-quality | Genome-fragment | 4.23 | AAI-based (high-confidence) | 0 |
| S17_extracted_k141_63625 | 1791 | No | NA | 4 | 0 | 0 | Low-quality | Genome-fragment | 2.74 | AAI-based (medium-confidence) | 0 |
| S5_extracted_k141_135765 | 1104 | No | NA | 1 | 0 | 0 | Not-determined | Genome-fragment | NA | NA | 0 |
| S5_extracted_k141_125333 | 1576 | No | NA | 2 | 0 | 0 | Not-determined | Genome-fragment | NA | NA | 0 |
| S5_extracted_k141_63697 | 7317 | No | NA | 6 | 0 | 6 | Not-determined | Genome-fragment | NA | NA | 0 |
| S5_extracted_k141_74695 | 1120 | No | NA | 2 | 0 | 2 | Not-determined | Genome-fragment | NA | NA | 0 |
| S11_extracted_k141_32723 | 1791 | No | NA | 3 | 1 | 0 | Low-quality | Genome-fragment | 4.51 | AAI-based (medium-confidence) | 0 |
| S11_vs_k141_27875\|\|full | 1791 | No | NA | 5 | 1 | 0 | Low-quality | Genome-fragment | 4.62 | AAI-based (high-confidence) | 0 |
| S5_extracted_k141_107098 | 1016 | No | NA | 1 | 0 | 0 | Not-determined | Genome-fragment | NA | NA | 0 |
| S5_extracted_k141_130648 | 2332 | No | NA | 1 | 0 | 1 | Not-determined | Genome-fragment | NA | NA | 0 |
| S5_extracted_k141_45780 | 4561 | No | NA | 3 | 0 | 3 | Not-determined | Genome-fragment | NA | NA | 0 |
| S3_extracted_k141_80608 | 1791 | No | NA | 2 | 0 | 1 | Low-quality | Genome-fragment | 2.54 | AAI-based (medium-confidence) | 0 |
| S3_vs_k141_107043\|\|full | 1791 | No | NA | 4 | 0 | 0 | Low-quality | Genome-fragment | 4.03 | AAI-based (high-confidence) | 0 |
| S5_extracted_k141_941 | 1791 | No | NA | 5 | 5 | 0 | Low-quality | Genome-fragment | 5.91 | AAI-based (high-confidence) | 0 |
| S5_vs_k141_86335\|\|full | 1791 | No | NA | 3 | 0 | 0 | Low-quality | Genome-fragment | 2.73 | AAI-based (medium-confidence) | 0 |
| S8_vs_k141_142022\|\|lt2gene | 1791 | No | NA | 2 | 0 | 1 | Low-quality | Genome-fragment | 1.44 | AAI-based (medium-confidence) | 0 |
| S5_extracted_k141_107112 | 1597 | No | NA | 2 | 0 | 1 | Not-determined | Genome-fragment | NA | NA | 0 |
| S11_vs_k141_6747\|\|full | 1792 | No | NA | 4 | 0 | 0 | Low-quality | Genome-fragment | 3.61 | AAI-based (high-confidence) | 0 |
| S5_extracted_k141_69636 | 1863 | No | NA | 2 | 0 | 2 | Not-determined | Genome-fragment | NA | NA | 0 |
| S16_vs_94793\|\|full | 1792 | No | NA | 3 | 2 | 0 | Low-quality | Genome-fragment | 4.55 | AAI-based (medium-confidence) | 0 |
| S5_extracted_k141_119137 | 4649 | No | NA | 4 | 0 | 3 | Not-determined | Genome-fragment | NA | NA | 0 |
| S12_vs_k141_61327\|\|full | 1792 | No | NA | 2 | 0 | 2 | Low-quality | Genome-fragment | 2.58 | AAI-based (medium-confidence) | 0 |
| S5_extracted_k141_125383 | 1975 | No | NA | 5 | 0 | 0 | Not-determined | Genome-fragment | NA | NA | 0 |
| S5_extracted_k141_130762 | 1527 | No | NA | 2 | 0 | 0 | Not-determined | Genome-fragment | NA | NA | 0 |
| S17_vs_k141_17594\|\|full | 1793 | No | NA | 3 | 1 | 0 | Low-quality | Genome-fragment | 4.54 | AAI-based (high-confidence) | 0 |
| S11_vs_k141_69219\|\|full | 1793 | No | NA | 3 | 0 | 0 | Low-quality | Genome-fragment | 4.61 | AAI-based (medium-confidence) | 0 |
| S5_extracted_k141_34065 | 1340 | No | NA | 1 | 0 | 0 | Not-determined | Genome-fragment | NA | NA | 0 |
| S14_extracted_k141_62496 | 1793 | No | NA | 1 | 0 | 0 | Low-quality | Genome-fragment | 4.44 | AAI-based (medium-confidence) | 0 |
| S5_extracted_k141_34092 | 1304 | No | NA | 1 | 0 | 0 | Not-determined | Genome-fragment | NA | NA | 0 |
| S5_extracted_k141_125473 | 4304 | No | NA | 3 | 0 | 0 | Not-determined | Genome-fragment | NA | NA | 0 |
| S5_extracted_k141_45927 | 1021 | No | NA | 1 | 0 | 0 | Not-determined | Genome-fragment | NA | NA | 0 |
| S14_vs_k141_22263\|\|full | 1793 | No | NA | 3 | 1 | 0 | Low-quality | Genome-fragment | 0.66 | HMM-based (lower-bound) | 0 |
| S1_extracted_k141_125558 | 1793 | No | NA | 3 | 0 | 1 | Low-quality | Genome-fragment | 3.7 | AAI-based (medium-confidence) | 0 |
| S5_extracted_k141_69705 | 5126 | No | NA | 9 | 0 | 1 | Not-determined | Genome-fragment | NA | NA | 0 |
| S3_extracted_k141_87084 | 1793 | No | NA | 5 | 0 | 0 | Low-quality | Genome-fragment | 1.17 | AAI-based (high-confidence) | 0 |
| S5_vs_k141_51790\|\|full | 1793 | No | NA | 2 | 0 | 0 | Low-quality | Genome-fragment | 1.02 | AAI-based (medium-confidence) | 0 |
| S10_extracted_55290 | 1793 | No | NA | 4 | 0 | 0 | Low-quality | Genome-fragment | 4.84 | AAI-based (high-confidence) | 0 |
| S5_extracted_k141_125563 | 1817 | No | NA | 1 | 0 | 1 | Not-determined | Genome-fragment | NA | NA | 0 |
| S5_extracted_k141_99306 | 1412 | No | NA | 1 | 0 | 1 | Not-determined | Genome-fragment | NA | NA | 0 |
| S10_vs_59767\|\|full | 1793 | No | NA | 3 | 0 | 1 | Low-quality | Genome-fragment | 0.6 | AAI-based (medium-confidence) | 0 |
| S5_extracted_k141_69757 | 3339 | No | NA | 2 | 0 | 1 | Not-determined | Genome-fragment | NA | NA | 0 |
| S5_extracted_k141_112516 | 1002 | No | NA | 1 | 0 | 1 | Not-determined | Genome-fragment | NA | NA | 0 |
| S5_extracted_k141_34185 | 1913 | No | NA | 1 | 0 | 1 | Not-determined | Genome-fragment | NA | NA | 0 |
| S8_vs_k141_133481\|\|full | 1793 | No | NA | 2 | 0 | 0 | Low-quality | Genome-fragment | 3.84 | AAI-based (medium-confidence) | 0 |
| S5_extracted_k141_125654 | 1041 | No | NA | 2 | 0 | 2 | Not-determined | Genome-fragment | NA | NA | 0 |
| S5_extracted_k141_74763 | 1559 | No | NA | 2 | 0 | 2 | Not-determined | Genome-fragment | NA | NA | 0 |
| S5_extracted_k141_119312 | 1354 | No | NA | 3 | 0 | 0 | Not-determined | Genome-fragment | NA | NA | 0 |
| S9_vs_k141_63602\|\|full | 1794 | No | NA | 4 | 0 | 0 | Low-quality | Genome-fragment | 2.84 | AAI-based (high-confidence) | 0 |
| S5_extracted_k141_69816 | 2439 | No | NA | 1 | 0 | 1 | Not-determined | Genome-fragment | NA | NA | 0 |
| S11_vs_k141_29955\|\|full | 1794 | No | NA | 3 | 0 | 0 | Low-quality | Genome-fragment | 1.09 | AAI-based (medium-confidence) | 0 |
| S7_extracted_91523 | 1794 | No | NA | 5 | 0 | 0 | Low-quality | Genome-fragment | 4.5 | AAI-based (medium-confidence) | 0 |
| S5_extracted_k141_125701 | 1631 | No | NA | 3 | 0 | 2 | Not-determined | Genome-fragment | NA | NA | 0 |
| S5_extracted_k141_63880 | 7335 | No | NA | 5 | 0 | 3 | Not-determined | Genome-fragment | NA | NA | 0 |
| S16_vs_100358\|\|full | 1794 | No | NA | 4 | 1 | 0 | Low-quality | Genome-fragment | 4.22 | AAI-based (high-confidence) | 0 |
| S10_vs_83265\|\|full | 1794 | No | NA | 2 | 0 | 0 | Low-quality | Genome-fragment | 2.28 | AAI-based (high-confidence) | 0 |
| S17_vs_k141_59211\|\|full | 1795 | No | NA | 6 | 1 | 0 | Low-quality | Genome-fragment | 4.25 | AAI-based (high-confidence) | 0 |
| S5_extracted_k141_63905 | 1561 | No | NA | 3 | 0 | 1 | Not-determined | Genome-fragment | NA | NA | 0 |
| S5_extracted_k141_130910 | 3058 | No | NA | 2 | 0 | 2 | Not-determined | Genome-fragment | NA | NA | 0 |
| S11_vs_k141_37351\|\|full | 1795 | No | NA | 4 | 0 | 0 | Low-quality | Genome-fragment | 1.91 | AAI-based (high-confidence) | 0 |
| S5_extracted_k141_130933 | 1091 | No | NA | 1 | 0 | 1 | Not-determined | Genome-fragment | NA | NA | 0 |
| S5_extracted_k141_125761 | 3413 | No | NA | 2 | 0 | 0 | Not-determined | Genome-fragment | NA | NA | 0 |
| S5_extracted_k141_125770 | 1053 | No | NA | 2 | 0 | 0 | Not-determined | Genome-fragment | NA | NA | 0 |
| S5_extracted_k141_26366 | 1177 | No | NA | 2 | 0 | 0 | Not-determined | Genome-fragment | NA | NA | 0 |
| S5_extracted_k141_4624 | 1163 | No | NA | 1 | 0 | 0 | Not-determined | Genome-fragment | NA | NA | 0 |
| S13_vs_39603\|\|full | 1795 | No | NA | 4 | 0 | 0 | Low-quality | Genome-fragment | 5.06 | AAI-based (medium-confidence) | 0 |
| S5_extracted_k141_4642 | 1010 | No | NA | 1 | 0 | 1 | Not-determined | Genome-fragment | NA | NA | 0 |
| S5_extracted_k141_125837 | 1137 | No | NA | 2 | 0 | 0 | Not-determined | Genome-fragment | NA | NA | 0 |
| S5_extracted_k141_136015 | 3327 | No | NA | 7 | 0 | 1 | Not-determined | Genome-fragment | NA | NA | 0 |
| S5_extracted_k141_52727 | 1893 | No | NA | 2 | 0 | 0 | Not-determined | Genome-fragment | NA | NA | 0 |
| S5_extracted_k141_63981 | 1013 | No | NA | 2 | 0 | 0 | Not-determined | Genome-fragment | NA | NA | 0 |
| S5_extracted_k141_99447 | 1135 | No | NA | 1 | 0 | 0 | Not-determined | Genome-fragment | NA | NA | 0 |
| S5_extracted_k141_39556 | 3584 | No | NA | 4 | 0 | 3 | Not-determined | Genome-fragment | NA | NA | 0 |
| S5_extracted_k141_94576 | 1863 | No | NA | 3 | 0 | 1 | Not-determined | Genome-fragment | NA | NA | 0 |
| S5_extracted_k141_94577 | 1106 | No | NA | 1 | 0 | 1 | Not-determined | Genome-fragment | NA | NA | 0 |
| S5_extracted_k141_136047 | 1670 | No | NA | 1 | 0 | 0 | Not-determined | Genome-fragment | NA | NA | 0 |
| S5_extracted_k141_99485 | 1287 | No | NA | 1 | 0 | 1 | Not-determined | Genome-fragment | NA | NA | 0 |
| S5_extracted_k141_56706 | 1067 | No | NA | 1 | 0 | 1 | Not-determined | Genome-fragment | NA | NA | 0 |
| S5_extracted_k141_99516 | 1785 | No | NA | 8 | 0 | 0 | Not-determined | Genome-fragment | NA | NA | 0 |
| S1_vs_k141_50028\|\|full | 1795 | No | NA | 2 | 2 | 0 | Low-quality | Genome-fragment | 4.41 | AAI-based (high-confidence) | 0 |
| S10_vs_24382\|\|full | 1795 | No | NA | 7 | 0 | 0 | Low-quality | Genome-fragment | 3.69 | AAI-based (high-confidence) | 0 |
| S11_vs_k141_31707\|\|full | 1796 | No | NA | 3 | 3 | 0 | Low-quality | Genome-fragment | 4.23 | AAI-based (high-confidence) | 0 |
| S5_extracted_k141_119460 | 9630 | No | NA | 8 | 0 | 8 | Not-determined | Genome-fragment | NA | NA | 0 |
| S5_extracted_k141_52795 | 2624 | No | NA | 1 | 0 | 0 | Not-determined | Genome-fragment | NA | NA | 0 |
| S5_extracted_k141_34383 | 1024 | No | NA | 2 | 0 | 1 | Not-determined | Genome-fragment | NA | NA | 0 |
| S13_vs_43981\|\|full | 1796 | No | NA | 2 | 0 | 0 | Low-quality | Genome-fragment | 3.36 | AAI-based (high-confidence) | 0 |
| S5_extracted_k141_15754 | 3672 | No | NA | 4 | 0 | 1 | Not-determined | Genome-fragment | NA | NA | 0 |
| S5_extracted_k141_131015 | 1005 | No | NA | 2 | 0 | 0 | Not-determined | Genome-fragment | NA | NA | 0 |
| S14_vs_k141_52574\|\|full | 1796 | No | NA | 4 | 1 | 0 | Low-quality | Genome-fragment | 4.57 | AAI-based (high-confidence) | 0 |
| S5_extracted_k141_26404 | 8679 | No | NA | 12 | 0 | 1 | Not-determined | Genome-fragment | NA | NA | 0 |
| S14_vs_k141_92775\|\|full | 1796 | No | NA | 5 | 4 | 0 | Low-quality | Genome-fragment | 3.86 | AAI-based (medium-confidence) | 0 |
| S5_extracted_k141_88231 | 1040 | No | NA | 2 | 0 | 0 | Not-determined | Genome-fragment | NA | NA | 0 |
| S5_extracted_k141_88233 | 1240 | No | NA | 1 | 0 | 0 | Not-determined | Genome-fragment | NA | NA | 0 |
| S5_extracted_k141_99567 | 1663 | No | NA | 7 | 0 | 0 | Not-determined | Genome-fragment | NA | NA | 0 |
| S1_vs_k141_153867\|\|full | 1796 | No | NA | 3 | 0 | 1 | Low-quality | Genome-fragment | 2.34 | AAI-based (medium-confidence) | 0 |
| S5_extracted_k141_64025 | 2631 | No | NA | 2 | 0 | 2 | Not-determined | Genome-fragment | NA | NA | 0 |
| S5_extracted_k141_136076 | 45466 | No | NA | 69 | 11 | 0 | High-quality | High-quality | 99 | AAI-based (high-confidence) | 0 |
| S5_extracted_k141_52856 | 1151 | No | NA | 2 | 0 | 0 | Not-determined | Genome-fragment | NA | NA | 0 |
| S5_extracted_k141_15780 | 1846 | No | NA | 1 | 0 | 1 | Not-determined | Genome-fragment | NA | NA | 0 |
| S5_vs_k141_96857\|\|full | 1796 | No | NA | 3 | 0 | 0 | Low-quality | Genome-fragment | 3.66 | AAI-based (medium-confidence) | 0 |
| S16_vs_18966\|\|full | 1796 | No | NA | 6 | 0 | 0 | Low-quality | Genome-fragment | 3 | AAI-based (high-confidence) | 0 |
| S16_vs_54734\|\|full | 1796 | No | NA | 5 | 1 | 0 | Low-quality | Genome-fragment | 2.68 | AAI-based (medium-confidence) | 0 |
| S16_vs_135475\|\|full | 1796 | No | NA | 3 | 0 | 1 | Low-quality | Genome-fragment | 0.97 | AAI-based (medium-confidence) | 0 |
| S5_extracted_k141_107678 | 2280 | No | NA | 1 | 0 | 0 | Not-determined | Genome-fragment | NA | NA | 0 |
| S5_extracted_k141_126059 | 1187 | No | NA | 2 | 0 | 1 | Not-determined | Genome-fragment | NA | NA | 0 |
| S5_extracted_k141_34586 | 1229 | No | NA | 2 | 0 | 2 | Not-determined | Genome-fragment | NA | NA | 0 |
| S5_extracted_k141_4979 | 2952 | No | NA | 1 | 0 | 0 | Not-determined | Genome-fragment | NA | NA | 0 |
| S5_extracted_k141_46055 | 1027 | No | NA | 2 | 0 | 2 | Not-determined | Genome-fragment | NA | NA | 0 |
| S12_extracted_k141_121377 | 1796 | No | NA | 5 | 2 | 0 | Low-quality | Genome-fragment | 4.26 | AAI-based (high-confidence) | 0 |
| S5_extracted_k141_52993 | 1220 | No | NA | 1 | 0 | 0 | Not-determined | Genome-fragment | NA | NA | 0 |
| S12_vs_k141_52766\|\|full | 1796 | No | NA | 3 | 0 | 0 | Low-quality | Genome-fragment | 1.7 | AAI-based (high-confidence) | 0 |
| S5_extracted_k141_56877 | 5708 | No | NA | 5 | 0 | 4 | Not-determined | Genome-fragment | NA | NA | 0 |
| S8_extracted_k141_99780 | 1796 | No | NA | 3 | 0 | 1 | Low-quality | Genome-fragment | 1.35 | AAI-based (high-confidence) | 0 |
| S8_vs_k141_89758\|\|full | 1796 | No | NA | 5 | 0 | 0 | Low-quality | Genome-fragment | 3.9 | AAI-based (high-confidence) | 0 |
| S5_extracted_k141_107744 | 1052 | No | NA | 1 | 0 | 0 | Not-determined | Genome-fragment | NA | NA | 0 |
| S5_extracted_k141_88363 | 1014 | No | NA | 1 | 0 | 0 | Not-determined | Genome-fragment | NA | NA | 0 |
| S5_extracted_k141_131154 | 1021 | No | NA | 3 | 0 | 0 | Not-determined | Genome-fragment | NA | NA | 0 |
| S5_extracted_k141_21748 | 1922 | No | NA | 1 | 0 | 0 | Not-determined | Genome-fragment | NA | NA | 0 |
| S5_extracted_k141_107776 | 1148 | No | NA | 1 | 0 | 1 | Not-determined | Genome-fragment | NA | NA | 0 |
| S5_extracted_k141_113030 | 1153 | No | NA | 1 | 0 | 1 | Not-determined | Genome-fragment | NA | NA | 0 |
| S8_vs_k141_50685\|\|full | 1796 | No | NA | 5 | 0 | 0 | Low-quality | Genome-fragment | 4.27 | AAI-based (high-confidence) | 0 |
| S5_extracted_k141_21767 | 5972 | No | NA | 8 | 0 | 6 | Not-determined | Genome-fragment | NA | NA | 0 |
| S5_extracted_k141_107802 | 1046 | No | NA | 1 | 0 | 1 | Not-determined | Genome-fragment | NA | NA | 0 |
| S11_extracted_k141_22345 | 1797 | No | NA | 3 | 1 | 0 | Low-quality | Genome-fragment | 4.15 | AAI-based (medium-confidence) | 0 |
| S5_extracted_k141_80989 | 1082 | No | NA | 2 | 0 | 1 | Not-determined | Genome-fragment | NA | NA | 0 |
| S13_vs_6499\|\|full | 1797 | No | NA | 3 | 0 | 0 | Low-quality | Genome-fragment | 1.32 | AAI-based (high-confidence) | 0 |
| S5_extracted_k141_56963 | 3478 | No | NA | 4 | 0 | 3 | Not-determined | Genome-fragment | NA | NA | 0 |
| S13_vs_24334\|\|full | 1797 | No | NA | 3 | 0 | 1 | Low-quality | Genome-fragment | 3.8 | AAI-based (high-confidence) | 0 |
| S5_extracted_k141_53140 | 1274 | No | NA | 1 | 0 | 0 | Not-determined | Genome-fragment | NA | NA | 0 |
| S13_vs_61324\|\|full | 1797 | No | NA | 5 | 1 | 0 | Low-quality | Genome-fragment | 32.52 | AAI-based (high-confidence) | 0 |

| | | | | | | | | | | | | |
|---|---|---|---|---|---|---|---|---|---|---|---|---|
| S5_extracted_k141_94937 | 1835 | No | NA | 1 | 0 | 0 | Not-determined | Genome-fragment | NA | | NA | 0 |
| S14_vs_k141_13958\|\|full | 1797 | No | NA | 2 | 0 | 1 | Low-quality | Genome-fragment | | 3.22 | AAI-based (high-confidence) | 0 |
| S5_extracted_k141_98455 | 1797 | No | NA | 3 | 3 | 0 | Low-quality | Genome-fragment | | 8.18 | AAI-based (high-confidence) | 0 |
| S7_vs_16789\|\|full | 1797 | No | NA | 7 | 0 | 0 | Low-quality | Genome-fragment | | 3.57 | AAI-based (medium-confidence) | 0 |
| S5_extracted_k141_16022 | 1043 | No | NA | 1 | 0 | 0 | Not-determined | Genome-fragment | NA | | NA | 0 |
| S16_extracted_115911 | 1797 | No | NA | 4 | 0 | 0 | Low-quality | Genome-fragment | | 1.64 | AAI-based (high-confidence) | 0 |
| S5_extracted_k141_99771 | 4957 | No | NA | 5 | 0 | 0 | Not-determined | Genome-fragment | NA | | NA | 0 |
| S16_vs_154049\|\|lt2gene | 1797 | No | NA | 2 | 1 | 0 | Low-quality | Genome-fragment | | 4.64 | AAI-based (high-confidence) | 0 |
| S12_extracted_k141_87355 | 1797 | No | NA | 1 | 0 | 0 | Low-quality | Genome-fragment | | 1.88 | AAI-based (high-confidence) | 0 |
| S5_extracted_k141_99817 | 1033 | No | NA | 2 | 0 | 1 | Not-determined | Genome-fragment | NA | | NA | 0 |
| S5_extracted_k141_136326 | 1018 | No | NA | 1 | 0 | 1 | Not-determined | Genome-fragment | NA | | NA | 0 |
| S8_extracted_k141_42902 | 1797 | No | NA | 2 | 1 | 0 | Low-quality | Genome-fragment | | 5.57 | AAI-based (high-confidence) | 0 |
| S5_extracted_k141_21801 | 1000 | No | NA | 1 | 0 | 1 | Not-determined | Genome-fragment | NA | | NA | 0 |
| S5_extracted_k141_34854 | 1343 | No | NA | 2 | 0 | 0 | Not-determined | Genome-fragment | NA | | NA | 0 |
| S8_vs_k141_120682\|\|full | 1797 | No | NA | 3 | 0 | 0 | Low-quality | Genome-fragment | | 4.68 | AAI-based (high-confidence) | 0 |
| S5_extracted_k141_5297 | 1270 | No | NA | 1 | 0 | 1 | Not-determined | Genome-fragment | NA | | NA | 0 |
| S5_extracted_k141_39748 | 1595 | No | NA | 1 | 0 | 1 | Not-determined | Genome-fragment | NA | | NA | 0 |
| S5_extracted_k141_39766 | 1081 | No | NA | 2 | 0 | 0 | Not-determined | Genome-fragment | NA | | NA | 0 |
| S1_extracted_k141_26460 | 1798 | No | NA | 1 | 0 | 1 | Low-quality | Genome-fragment | | 1.61 | AAI-based (medium-confidence) | 0 |
| S1_vs_k141_8332\|\|full | 1798 | No | NA | 5 | 2 | 0 | Low-quality | Genome-fragment | | 4.36 | AAI-based (medium-confidence) | 0 |
| S5_extracted_k141_95104 | 1358 | No | NA | 2 | 0 | 0 | Not-determined | Genome-fragment | NA | | NA | 0 |
| S7_vs_98916\|\|full | 1798 | No | NA | 5 | 1 | 0 | Low-quality | Genome-fragment | | 4.06 | AAI-based (medium-confidence) | 0 |
| S5_extracted_k141_142932 | 3032 | No | NA | 4 | 0 | 0 | Not-determined | Genome-fragment | NA | | NA | 0 |
| S16_vs_54277\|\|full | 1798 | No | NA | 5 | 1 | 0 | Low-quality | Genome-fragment | | 3.5 | AAI-based (high-confidence) | 0 |
| S5_extracted_k141_95119 | 1089 | No | NA | 1 | 0 | 0 | Not-determined | Genome-fragment | NA | | NA | 0 |
| S16_vs_99835\|\|full | 1798 | No | NA | 4 | 2 | 0 | Low-quality | Genome-fragment | | 3.75 | AAI-based (high-confidence) | 0 |
| S5_extracted_k141_108046 | 6977 | No | NA | 9 | 0 | 7 | Not-determined | Genome-fragment | NA | | NA | 0 |
| S16_vs_129870\|\|full | 1798 | No | NA | 3 | 1 | 0 | Low-quality | Genome-fragment | | 1.21 | HMM-based (lower-bound) | 0 |
| S16_vs_172000\|\|full | 1798 | No | NA | 3 | 0 | 0 | Low-quality | Genome-fragment | | 4.65 | AAI-based (medium-confidence) | 0 |
| S5_extracted_k141_21869 | 43235 | No | NA | 80 | 18 | 0 | Complete | High-quality | | 100 | DTR (high-confidence) | 0 |
| S1_vs_k141_147279\|\|full | 1799 | No | NA | 2 | 0 | 2 | Low-quality | Genome-fragment | | 0.44 | AAI-based (medium-confidence) | 0 |
| S5_extracted_k141_75075 | 17572 | No | NA | 22 | 0 | 21 | Not-determined | Genome-fragment | NA | | NA | 0 |
| S3_extracted_k141_97296 | 1799 | No | NA | 4 | 4 | 0 | Low-quality | Genome-fragment | | 5.93 | AAI-based (high-confidence) | 0 |
| S3_vs_k141_117325\|\|lt2gene | 1799 | No | NA | 2 | 2 | 0 | Low-quality | Genome-fragment | | 3.1 | AAI-based (high-confidence) | 0 |
| S5_extracted_k141_95186 | 3099 | No | NA | 2 | 0 | 1 | Not-determined | Genome-fragment | NA | | NA | 0 |
| S5_extracted_k141_136446 | 1117 | No | NA | 2 | 0 | 1 | Not-determined | Genome-fragment | NA | | NA | 0 |
| S5_extracted_k141_10296 | 1575 | No | NA | 2 | 0 | 0 | Not-determined | Genome-fragment | NA | | NA | 0 |
| S5_vs_k141_17271\|\|lt2gene | 1799 | No | NA | 2 | 2 | 0 | Low-quality | Genome-fragment | | 4.03 | AAI-based (medium-confidence) | 0 |
| S7_vs_51295\|\|full | 1799 | No | NA | 4 | 1 | 0 | Low-quality | Genome-fragment | | 4.14 | AAI-based (medium-confidence) | 0 |
| S5_extracted_k141_57108 | 5885 | No | NA | 5 | 0 | 5 | Not-determined | Genome-fragment | NA | | NA | 0 |
| S5_extracted_k141_21977 | 1058 | No | NA | 2 | 0 | 1 | Not-determined | Genome-fragment | NA | | NA | 0 |
| S5_extracted_k141_35049 | 1803 | No | NA | 1 | 0 | 1 | Not-determined | Genome-fragment | NA | | NA | 0 |
| S16_extracted_43581 | 1799 | No | NA | 4 | 1 | 0 | Low-quality | Genome-fragment | | 2.82 | HMM-based (lower-bound) | 0 |
| S5_extracted_k141_88853 | 1794 | No | NA | 2 | 0 | 0 | Not-determined | Genome-fragment | NA | | NA | 0 |
| S5_extracted_k141_53462 | 1430 | No | NA | 1 | 0 | 1 | Not-determined | Genome-fragment | NA | | NA | 0 |
| S5_extracted_k141_136504 | 1479 | No | NA | 2 | 0 | 1 | Not-determined | Genome-fragment | NA | | NA | 0 |
| S5_extracted_k141_53465 | 3435 | No | NA | 2 | 0 | 0 | Not-determined | Genome-fragment | NA | | NA | 0 |
| S10_vs_8076\|\|full | 1799 | No | NA | 3 | 0 | 1 | Low-quality | Genome-fragment | | 1.62 | AAI-based (medium-confidence) | 0 |
| S5_extracted_k141_75212 | 3186 | No | NA | 5 | 0 | 0 | Not-determined | Genome-fragment | NA | | NA | 0 |
| S5_extracted_k141_53506 | 1726 | No | NA | 3 | 0 | 1 | Not-determined | Genome-fragment | NA | | NA | 0 |
| S5_extracted_k141_10397 | 3802 | No | NA | 4 | 0 | 2 | Not-determined | Genome-fragment | NA | | NA | 0 |
| S12_vs_k141_75412\|\|full | 1799 | No | NA | 4 | 1 | 0 | Low-quality | Genome-fragment | | 3.2 | HMM-based (lower-bound) | 0 |
| S5_extracted_k141_16242 | 1028 | No | NA | 2 | 0 | 1 | Not-determined | Genome-fragment | NA | | NA | 0 |
| S5_extracted_k141_108227 | 1001 | No | NA | 2 | 0 | 2 | Not-determined | Genome-fragment | NA | | NA | 0 |
| S15_extracted_56969 | 1799 | No | NA | 2 | 0 | 0 | Low-quality | Genome-fragment | | 0.93 | AAI-based (medium-confidence) | 0 |
| S6_vs_k141_44033\|\|full | 1799 | No | NA | 2 | 0 | 2 | Low-quality | Genome-fragment | | 3.84 | AAI-based (high-confidence) | 0 |
| S5_extracted_k141_75285 | 1274 | No | NA | 1 | 0 | 1 | Not-determined | Genome-fragment | NA | | NA | 0 |
| S8_vs_k141_61148\|\|full | 1799 | No | NA | 4 | 0 | 0 | Low-quality | Genome-fragment | | 1.51 | AAI-based (medium-confidence) | 0 |
| S5_extracted_k141_16299 | 1135 | No | NA | 2 | 0 | 0 | Not-determined | Genome-fragment | NA | | NA | 0 |
| S5_extracted_k141_16302 | 1437 | No | NA | 1 | 0 | 0 | Not-determined | Genome-fragment | NA | | NA | 0 |
| S17_extracted_k141_88479 | 1800 | No | NA | 2 | 0 | 0 | Low-quality | Genome-fragment | | 4.57 | AAI-based (high-confidence) | 0 |
| S11_vs_k141_37311\|\|full | 1800 | No | NA | 5 | 0 | 0 | Low-quality | Genome-fragment | | 4.11 | AAI-based (medium-confidence) | 0 |
| S13_vs_63334\|\|full | 1800 | No | NA | 4 | 0 | 1 | Low-quality | Genome-fragment | | 2.16 | AAI-based (medium-confidence) | 0 |
| S5_extracted_k141_22030 | 1471 | No | NA | 2 | 0 | 0 | Not-determined | Genome-fragment | NA | | NA | 0 |
| S7_vs_128694\|\|full | 1800 | No | NA | 4 | 1 | 0 | Low-quality | Genome-fragment | | 2.88 | AAI-based (high-confidence) | 0 |
| S5_extracted_k141_88990 | 3973 | No | NA | 1 | 0 | 1 | Not-determined | Genome-fragment | NA | | NA | 0 |
| S5_extracted_k141_75333 | 1144 | No | NA | 2 | 0 | 2 | Not-determined | Genome-fragment | NA | | NA | 0 |
| S5_extracted_k141_126450 | 7901 | No | NA | 6 | 0 | 5 | Not-determined | Genome-fragment | NA | | NA | 0 |
| S7_vs_63409\|\|lt2gene | 1800 | No | NA | 2 | 1 | 0 | Low-quality | Genome-fragment | | 4.13 | AAI-based (medium-confidence) | 0 |
| S5_extracted_k141_113417 | 7325 | No | NA | 4 | 0 | 1 | Not-determined | Genome-fragment | NA | | NA | 0 |
| S8_extracted_k141_98324 | 1800 | No | NA | 5 | 0 | 0 | Low-quality | Genome-fragment | | 1.32 | AAI-based (medium-confidence) | 0 |
| S8_vs_k141_127858\|\|full | 1800 | No | NA | 4 | 0 | 0 | Low-quality | Genome-fragment | | 3.96 | AAI-based (high-confidence) | 0 |
| S1_vs_k141_32589\|\|full | 1801 | No | NA | 3 | 3 | 0 | Low-quality | Genome-fragment | | 4.12 | AAI-based (high-confidence) | 0 |
| S3_extracted_k141_122562 | 1801 | No | NA | 4 | 0 | 0 | Low-quality | Genome-fragment | | 2.14 | AAI-based (medium-confidence) | 0 |
| S5_extracted_k141_100031 | 1376 | No | NA | 1 | 0 | 1 | Not-determined | Genome-fragment | NA | | NA | 0 |
| S5_extracted_k141_143156 | 1123 | No | NA | 1 | 0 | 1 | Not-determined | Genome-fragment | NA | | NA | 0 |
| S5_extracted_k141_22864 | 1801 | No | NA | 1 | 0 | 1 | Low-quality | Genome-fragment | | 5.08 | AAI-based (medium-confidence) | 0 |
| S5_vs_k141_6555\|\|full | 1801 | No | NA | 4 | 1 | 0 | Low-quality | Genome-fragment | | 2.95 | HMM-based (lower-bound) | 0 |
| S5_vs_k141_44883\|\|full | 1801 | No | NA | 3 | 0 | 0 | Low-quality | Genome-fragment | | 5.27 | AAI-based (high-confidence) | 0 |
| S12_vs_k141_9311\|\|full | 1801 | No | NA | 2 | 0 | 0 | Low-quality | Genome-fragment | | 2.29 | AAI-based (medium-confidence) | 0 |
| S5_extracted_k141_95426 | 1334 | No | NA | 3 | 0 | 2 | Not-determined | Genome-fragment | NA | | NA | 0 |
| S8_vs_k141_139594\|\|full | 1801 | No | NA | 6 | 1 | 0 | Low-quality | Genome-fragment | | 5.34 | AAI-based (high-confidence) | 0 |
| S5_extracted_k141_95454 | 1753 | No | NA | 3 | 0 | 0 | Not-determined | Genome-fragment | NA | | NA | 0 |
| S11_extracted_k141_101211 | 1802 | No | NA | 3 | 0 | 0 | Low-quality | Genome-fragment | | 21.95 | AAI-based (medium-confidence) | 0 |
| S5_extracted_k141_64722 | 1872 | No | NA | 3 | 0 | 0 | Not-determined | Genome-fragment | NA | | NA | 0 |
| S5_extracted_k141_26901 | 1035 | No | NA | 1 | 0 | 0 | Not-determined | Genome-fragment | NA | | NA | 0 |
| S14_vs_k141_45423\|\|lt2gene | 1802 | No | NA | 2 | 2 | 0 | Low-quality | Genome-fragment | | 5.77 | AAI-based (high-confidence) | 0 |
| S5_extracted_k141_75479 | 1099 | No | NA | 1 | 0 | 1 | Not-determined | Genome-fragment | NA | | NA | 0 |
| S5_extracted_k141_70605 | 1949 | No | NA | 2 | 0 | 0 | Not-determined | Genome-fragment | NA | | NA | 0 |
| S5_extracted_k141_95477 | 1027 | No | NA | 1 | 0 | 1 | Not-determined | Genome-fragment | NA | | NA | 0 |
| S3_extracted_k141_4022 | 1802 | No | NA | 2 | 1 | 0 | Low-quality | Genome-fragment | | 1.1 | HMM-based (lower-bound) | 0 |
| S8_vs_k141_33907\|\|full | 1802 | No | NA | 5 | 1 | 0 | Low-quality | Genome-fragment | | 4.29 | AAI-based (high-confidence) | 0 |
| S5_extracted_k141_70615 | 11406 | No | NA | 7 | 0 | 4 | Not-determined | Genome-fragment | NA | | NA | 0 |
| S5_extracted_k141_53682 | 1020 | No | NA | 3 | 0 | 0 | Not-determined | Genome-fragment | NA | | NA | 0 |
| S5_extracted_k141_131630 | 7240 | No | NA | 7 | 0 | 6 | Not-determined | Genome-fragment | NA | | NA | 0 |
| S17_extracted_k141_126995 | 1803 | No | NA | 3 | 1 | 0 | Low-quality | Genome-fragment | | 2.55 | AAI-based (high-confidence) | 0 |
| S5_extracted_k141_89111 | 1546 | No | NA | 1 | 0 | 1 | Not-determined | Genome-fragment | NA | | NA | 0 |
| S5_extracted_k141_108522 | 1086 | No | NA | 1 | 0 | 0 | Not-determined | Genome-fragment | NA | | NA | 0 |
| S5_extracted_k141_70664 | 1145 | No | NA | 2 | 0 | 1 | Not-determined | Genome-fragment | NA | | NA | 0 |
| S5_extracted_k141_57251 | 2496 | No | NA | 6 | 0 | 1 | Not-determined | Genome-fragment | NA | | NA | 0 |
| S5_extracted_k141_100209 | 1739 | No | NA | 2 | 0 | 0 | Not-determined | Genome-fragment | NA | | NA | 0 |
| S5_extracted_k141_10458 | 5413 | No | NA | 4 | 0 | 4 | Not-determined | Genome-fragment | NA | | NA | 0 |
| S5_extracted_k141_75538 | 2414 | No | NA | 2 | 0 | 2 | Not-determined | Genome-fragment | NA | | NA | 0 |
| S5_extracted_k141_35371 | 1713 | No | NA | 1 | 0 | 1 | Not-determined | Genome-fragment | NA | | NA | 0 |
| S11_vs_k141_62267\|\|full | 1803 | No | NA | 4 | 0 | 0 | Low-quality | Genome-fragment | | 3.23 | AAI-based (medium-confidence) | 0 |
| S5_extracted_k141_95581 | 1093 | No | NA | 1 | 0 | 1 | Not-determined | Genome-fragment | NA | | NA | 0 |
| S14_vs_k141_112253\|\|full | 1803 | No | NA | 3 | 0 | 1 | Low-quality | Genome-fragment | | 1.56 | AAI-based (high-confidence) | 0 |
| S5_extracted_k141_64850 | 1312 | No | NA | 1 | 0 | 1 | Not-determined | Genome-fragment | NA | | NA | 0 |
| S1_vs_k141_142657\|\|full | 1803 | No | NA | 3 | 0 | 0 | Low-quality | Genome-fragment | | 3.84 | AAI-based (medium-confidence) | 0 |
| S5_extracted_k141_22219 | 4218 | No | NA | 6 | 0 | 4 | Not-determined | Genome-fragment | NA | | NA | 0 |
| S5_extracted_k141_126707 | 2496 | No | NA | 7 | 0 | 0 | Not-determined | Genome-fragment | NA | | NA | 0 |
| S5_extracted_k141_57308 | 3758 | No | NA | 4 | 0 | 3 | Not-determined | Genome-fragment | NA | | NA | 0 |
| S3_vs_k141_3992\|\|full | 1803 | No | NA | 4 | 0 | 0 | Low-quality | Genome-fragment | | 1.25 | AAI-based (high-confidence) | 0 |
| S5_extracted_k141_126744 | 2625 | No | NA | 5 | 0 | 1 | Not-determined | Genome-fragment | NA | | NA | 0 |
| S5_extracted_k141_16827 | 1204 | No | NA | 3 | 0 | 0 | Not-determined | Genome-fragment | NA | | NA | 0 |
| S10_vs_37169\|\|full | 1803 | No | NA | 6 | 1 | 0 | Low-quality | Genome-fragment | | 3.32 | AAI-based (high-confidence) | 0 |
| S5_extracted_k141_75688 | 1106 | No | NA | 3 | 0 | 0 | Not-determined | Genome-fragment | NA | | NA | 0 |
| S12_vs_k141_166045\|\|full | 1803 | No | NA | 5 | 1 | 0 | Low-quality | Genome-fragment | | 5.33 | AAI-based (high-confidence) | 0 |
| S5_extracted_k141_131804 | 6567 | No | NA | 9 | 0 | 7 | Not-determined | Genome-fragment | NA | | NA | 0 |
| S5_extracted_k141_131807 | 1021 | No | NA | 3 | 0 | 3 | Not-determined | Genome-fragment | NA | | NA | 0 |
| S6_vs_k141_14502\|\|full | 1803 | No | NA | 4 | 1 | 0 | Low-quality | Genome-fragment | | 1.17 | AAI-based (medium-confidence) | 0 |
| S8_vs_k141_92583\|\|full | 1803 | No | NA | 6 | 3 | 0 | Low-quality | Genome-fragment | | 4.66 | AAI-based (high-confidence) | 0 |
| S5_extracted_k141_75722 | 1168 | No | NA | 1 | 0 | 1 | Not-determined | Genome-fragment | NA | | NA | 0 |
| S5_extracted_k141_113718 | 1227 | No | NA | 3 | 0 | 0 | Not-determined | Genome-fragment | NA | | NA | 0 |
| S9_vs_k141_26888\|\|full | 1804 | No | NA | 5 | 1 | 0 | Low-quality | Genome-fragment | | 4.84 | AAI-based (high-confidence) | 0 |
| S9_vs_k141_54325\|\|full | 1804 | No | NA | 4 | 4 | 0 | Low-quality | Genome-fragment | | 3.06 | HMM-based (lower-bound) | 0 |
| S5_extracted_k141_16867 | 7779 | No | NA | 5 | 0 | 5 | Not-determined | Genome-fragment | NA | | NA | 0 |
| S5_extracted_k141_75743 | 4452 | No | NA | 3 | 0 | 3 | Not-determined | Genome-fragment | NA | | NA | 0 |
| S5_extracted_k141_5751 | 7951 | No | NA | 8 | 0 | 2 | Not-determined | Genome-fragment | NA | | NA | 0 |
| S14_extracted_k141_37778 | 1804 | No | NA | 2 | 1 | 0 | Low-quality | Genome-fragment | | 4.71 | AAI-based (medium-confidence) | 0 |
| S5_extracted_k141_113776 | 2227 | No | NA | 1 | 0 | 1 | Not-determined | Genome-fragment | NA | | NA | 0 |
| S3_extracted_k141_69697 | 1804 | No | NA | 1 | 0 | 1 | Low-quality | Genome-fragment | | 2.75 | AAI-based (medium-confidence) | 0 |
| S5_vs_k141_58070\|\|full | 1804 | No | NA | 3 | 0 | 0 | Low-quality | Genome-fragment | | 2.22 | AAI-based (medium-confidence) | 0 |
| S5_extracted_k141_108696 | 1043 | No | NA | 1 | 0 | 1 | Not-determined | Genome-fragment | NA | | NA | 0 |
| S7_vs_37009\|\|full | 1804 | No | NA | 3 | 0 | 0 | Low-quality | Genome-fragment | | 3.35 | AAI-based (medium-confidence) | 0 |
| S5_extracted_k141_113783 | 10797 | No | NA | 6 | 0 | 6 | Not-determined | Genome-fragment | NA | | NA | 0 |
| S7_vs_58453\|\|full | 1804 | No | NA | 3 | 1 | 0 | Low-quality | Genome-fragment | | 5.56 | AAI-based (high-confidence) | 0 |
| S5_extracted_k141_22281 | 1850 | No | NA | 2 | 0 | 0 | Not-determined | Genome-fragment | NA | | NA | 0 |
| S9_extracted_k141_50226 | 1805 | No | NA | 2 | 1 | 0 | Low-quality | Genome-fragment | | 4.24 | AAI-based (high-confidence) | 0 |
| S5_extracted_k141_108757 | 1599 | No | NA | 2 | 0 | 0 | Not-determined | Genome-fragment | NA | | NA | 0 |
| S11_extracted_k141_52646 | 1805 | No | NA | 2 | 1 | 0 | Low-quality | Genome-fragment | | 3.97 | AAI-based (high-confidence) | 0 |
| S11_vs_k141_1105\|\|lt2gene | 1805 | No | NA | 2 | 2 | 0 | Low-quality | Genome-fragment | | 2.79 | HMM-based (lower-bound) | 0 |
| S5_extracted_k141_89218 | 1216 | No | NA | 1 | 0 | 1 | Not-determined | Genome-fragment | NA | | NA | 0 |
| S5_extracted_k141_108770 | 1554 | No | NA | 2 | 0 | 1 | Not-determined | Genome-fragment | NA | | NA | 0 |
| S13_vs_77580\|\|lt2gene | 1805 | No | NA | 2 | 1 | 0 | Low-quality | Genome-fragment | | 4.42 | AAI-based (high-confidence) | 0 |
| S1_extracted_k141_154936 | 1805 | No | NA | 2 | 0 | 2 | Low-quality | Genome-fragment | | 0.93 | AAI-based (high-confidence) | 0 |
| S5_extracted_k141_10522 | 4365 | No | NA | 3 | 0 | 3 | Not-determined | Genome-fragment | NA | | NA | 0 |
| S3_extracted_k141_69196 | 1805 | No | NA | 5 | 1 | 0 | Low-quality | Genome-fragment | | 1.18 | AAI-based (high-confidence) | 0 |
| S5_extracted_k141_70994 | 1260 | No | NA | 2 | 0 | 0 | Not-determined | Genome-fragment | NA | | NA | 0 |
| S5_extracted_k141_126952 | 1169 | No | NA | 3 | 0 | 0 | Not-determined | Genome-fragment | NA | | NA | 0 |
| S5_extracted_k141_22384 | 1646 | No | NA | 4 | 0 | 0 | Not-determined | Genome-fragment | NA | | NA | 0 |
| S16_vs_94687\|\|full | 1805 | No | NA | 3 | 0 | 1 | Low-quality | Genome-fragment | | 1.74 | AAI-based (medium-confidence) | 0 |
| S5_extracted_k141_136869 | 1163 | No | NA | 2 | 0 | 0 | Not-determined | Genome-fragment | NA | | NA | 0 |
| S12_extracted_k141_90015 | 1805 | No | NA | 3 | 1 | 0 | Low-quality | Genome-fragment | | 1.63 | HMM-based (lower-bound) | 0 |
| S5_extracted_k141_46446 | 2689 | No | NA | 1 | 0 | 0 | Not-determined | Genome-fragment | NA | | NA | 0 |
| S8_vs_k141_9046\|\|full | 1805 | No | NA | 4 | 1 | 0 | Low-quality | Genome-fragment | | 3.91 | AAI-based (medium-confidence) | 0 |
| S5_extracted_k141_53888 | 11327 | No | NA | 7 | 0 | 5 | Not-determined | Genome-fragment | NA | | NA | 0 |
| S5_extracted_k141_17012 | 1094 | No | NA | 2 | 0 | 1 | Not-determined | Genome-fragment | NA | | NA | 0 |
| S5_extracted_k141_120547 | 1011 | No | NA | 3 | 0 | 0 | Not-determined | Genome-fragment | NA | | NA | 0 |
| S17_vs_k141_163244\|\|full | 1806 | No | NA | 3 | 0 | 0 | Low-quality | Genome-fragment | | 4.14 | AAI-based (medium-confidence) | 0 |
| S5_extracted_k141_71029 | 1039 | No | NA | 1 | 0 | 1 | Not-determined | Genome-fragment | NA | | NA | 0 |
| S5_extracted_k141_100460 | 1076 | No | NA | 2 | 0 | 1 | Not-determined | Genome-fragment | NA | | NA | 0 |
| S11_vs_k141_86527\|\|full | 1806 | No | NA | 4 | 2 | 0 | Low-quality | Genome-fragment | | 3.96 | AAI-based (high-confidence) | 0 |
| S11_vs_k141_18686\|\|full | 1806 | No | NA | 4 | 3 | 0 | Low-quality | Genome-fragment | | 2.94 | AAI-based (high-confidence) | 0 |
| S5_extracted_k141_136900 | 1331 | No | NA | 1 | 0 | 1 | Not-determined | Genome-fragment | NA | | NA | 0 |
| S5_extracted_k141_89372 | 12638 | No | NA | 14 | 0 | 10 | Not-determined | Genome-fragment | NA | | NA | 0 |
| S5_extracted_k141_46532 | 1071 | No | NA | 2 | 0 | 0 | Not-determined | Genome-fragment | NA | | NA | 0 |
| S14_vs_k141_79306\|\|full | 1806 | No | NA | 3 | 1 | 0 | Low-quality | Genome-fragment | | 1.22 | AAI-based (high-confidence) | 0 |
| S5_extracted_k141_53971 | 2303 | No | NA | 2 | 0 | 2 | Not-determined | Genome-fragment | NA | | NA | 0 |
| S5_extracted_k141_100504 | 1638 | No | NA | 2 | 0 | 2 | Not-determined | Genome-fragment | NA | | NA | 0 |
| S3_vs_k141_93696\|\|full | 1806 | No | NA | 7 | 0 | 0 | Low-quality | Genome-fragment | | 5.42 | AAI-based (high-confidence) | 0 |
| S5_extracted_k141_136940 | 1381 | No | NA | 2 | 0 | 1 | Not-determined | Genome-fragment | NA | | NA | 0 |
| S5_extracted_k141_40841 | 1806 | No | NA | 2 | 0 | 0 | Low-quality | Genome-fragment | | 5.67 | AAI-based (medium-confidence) | 0 |
| S5_extracted_k141_54025 | 2574 | No | NA | 1 | 0 | 1 | Not-determined | Genome-fragment | NA | | NA | 0 |
| S5_extracted_k141_17107 | 1460 | No | NA | 1 | 0 | 0 | Not-determined | Genome-fragment | NA | | NA | 0 |
| S5_extracted_k141_108911 | 1078 | No | NA | 1 | 0 | 1 | Not-determined | Genome-fragment | NA | | NA | 0 |
| S5_extracted_k141_46606 | 1250 | No | NA | 2 | 0 | 0 | Not-determined | Genome-fragment | NA | | NA | 0 |
| S5_extracted_k141_5965 | 1804 | No | NA | 1 | 0 | 1 | Not-determined | Genome-fragment | NA | | NA | 0 |
| S5_extracted_k141_120648 | 1042 | No | NA | 2 | 0 | 1 | Not-determined | Genome-fragment | NA | | NA | 0 |
| S5_vs_k141_92238\|\|lt2gene | 1806 | No | NA | 2 | 2 | 0 | Low-quality | Genome-fragment | | 4.31 | AAI-based (high-confidence) | 0 |
| S5_extracted_k141_137000 | 1000 | No | NA | 2 | 0 | 1 | Not-determined | Genome-fragment | NA | | NA | 0 |
| S16_extracted_4059 | 1806 | No | NA | 3 | 2 | 0 | Low-quality | Genome-fragment | | 1.54 | AAI-based (high-confidence) | 0 |
| S5_extracted_k141_127128 | 1054 | No | NA | 3 | 0 | 3 | Not-determined | Genome-fragment | NA | | NA | 0 |
| S10_vs_118923\|\|full | 1806 | No | NA | 4 | 0 | 0 | Low-quality | Genome-fragment | | 4.14 | AAI-based (high-confidence) | 0 |
| S12_vs_k141_96773\|\|full | 1806 | No | NA | 6 | 0 | 0 | Low-quality | Genome-fragment | | 3.55 | AAI-based (medium-confidence) | 0 |
| S5_extracted_k141_17228 | 1228 | No | NA | 1 | 0 | 0 | Not-determined | Genome-fragment | NA | | NA | 0 |
| S5_extracted_k141_96025 | 2075 | No | NA | 1 | 0 | 1 | Not-determined | Genome-fragment | NA | | NA | 0 |
| S12_vs_k141_65994\|\|lt2gene | 1806 | No | NA | 1 | 1 | 0 | Low-quality | Genome-fragment | | 2.64 | HMM-based (lower-bound) | 0 |
| S8_vs_k141_69664\|\|full | 1806 | No | NA | 3 | 0 | 0 | Low-quality | Genome-fragment | | 1.14 | AAI-based (medium-confidence) | 0 |
| S17_vs_k141_25528\|\|full | 1807 | No | NA | 4 | 1 | 0 | Low-quality | Genome-fragment | | 1.4 | AAI-based (high-confidence) | 0 |
| S5_extracted_k141_6032 | 31164 | No | NA | 43 | 15 | 0 | High-quality | High-quality | | 99.12 | AAI-based (high-confidence) | 0 |
| S5_extracted_k141_81732 | 2074 | No | NA | 3 | 0 | 0 | Not-determined | Genome-fragment | NA | | NA | 0 |
| S5_extracted_k141_57548 | 2124 | No | NA | 2 | 0 | 0 | Not-determined | Genome-fragment | NA | | NA | 0 |
| S13_vs_36630\|\|full | 1807 | No | NA | 3 | 0 | 1 | Low-quality | Genome-fragment | | 3 | AAI-based (medium-confidence) | 0 |
| S5_extracted_k141_40577 | 1556 | No | NA | 2 | 0 | 2 | Not-determined | Genome-fragment | NA | | NA | 0 |
| S5_extracted_k141_132196 | 1664 | No | NA | 5 | 0 | 3 | Not-determined | Genome-fragment | NA | | NA | 0 |
| S5_extracted_k141_10853 | 1031 | No | NA | 1 | 0 | 0 | Not-determined | Genome-fragment | NA | | NA | 0 |
| S5_extracted_k141_71163 | 1383 | No | NA | 1 | 0 | 1 | Not-determined | Genome-fragment | NA | | NA | 0 |
| S5_extracted_k141_89557 | 1592 | No | NA | 1 | 0 | 1 | Not-determined | Genome-fragment | NA | | NA | 0 |
| S14_vs_k141_53977\|\|full | 1807 | No | NA | 4 | 1 | 0 | Low-quality | Genome-fragment | | 2.99 | AAI-based (high-confidence) | 0 |
| S14_vs_k141_10390\|\|full | 1807 | No | NA | 4 | 0 | 1 | Low-quality | Genome-fragment | | 3.44 | AAI-based (medium-confidence) | 0 |
| S7_vs_101248\|\|full | 1807 | No | NA | 2 | 0 | 1 | Low-quality | Genome-fragment | | 3.38 | AAI-based (medium-confidence) | 0 |
| S5_extracted_k141_89612 | 1329 | No | NA | 2 | 0 | 0 | Not-determined | Genome-fragment | NA | | NA | 0 |
| S12_extracted_k141_4784 | 1807 | No | NA | 1 | 0 | 0 | Low-quality | Genome-fragment | | 4.45 | AAI-based (high-confidence) | 0 |
| S5_extracted_k141_114294 | 1011 | No | NA | 2 | 0 | 1 | Not-determined | Genome-fragment | NA | | NA | 0 |
| S5_extracted_k141_54358 | 1049 | No | NA | 1 | 0 | 1 | Not-determined | Genome-fragment | NA | | NA | 0 |
| S5_extracted_k141_17462 | 3126 | No | NA | 2 | 0 | 2 | Not-determined | Genome-fragment | NA | | NA | 0 |
| S12_vs_k141_35254\|\|full | 1807 | No | NA | 3 | 1 | 0 | Low-quality | Genome-fragment | | 4.21 | AAI-based (high-confidence) | 0 |
| S5_extracted_k141_57583 | 1898 | No | NA | 2 | 0 | 2 | Not-determined | Genome-fragment | NA | | NA | 0 |
| S5_extracted_k141_114336 | 1199 | No | NA | 1 | 0 | 0 | Not-determined | Genome-fragment | NA | | NA | 0 |
| S8_vs_k141_28173\|\|full | 1807 | No | NA | 2 | 0 | 1 | Low-quality | Genome-fragment | | 2.75 | AAI-based (medium-confidence) | 0 |
| S17_vs_k141_134127\|\|full | 1808 | No | NA | 6 | 0 | 0 | Low-quality | Genome-fragment | | 4.01 | AAI-based (high-confidence) | 0 |
| S5_extracted_k141_40750 | 10422 | No | NA | 10 | 0 | 10 | Not-determined | Genome-fragment | NA | | NA | 0 |
| S5_extracted_k141_65448 | 1223 | No | NA | 1 | 0 | 0 | Not-determined | Genome-fragment | NA | | NA | 0 |
| S5_extracted_k141_81853 | 1750 | No | NA | 2 | 0 | 1 | Not-determined | Genome-fragment | NA | | NA | 0 |
| S5_extracted_k141_109215 | 1388 | No | NA | 1 | 0 | 0 | Not-determined | Genome-fragment | NA | | NA | 0 |
| S5_extracted_k141_114389 | 1528 | No | NA | 1 | 0 | 0 | Not-determined | Genome-fragment | NA | | NA | 0 |
| S11_vs_k141_74517\|\|full | 1808 | No | NA | 3 | 0 | 0 | Low-quality | Genome-fragment | | 0.68 | AAI-based (medium-confidence) | 0 |
| S14_vs_k141_692\|\|full | 1808 | No | NA | 6 | 1 | 0 | Low-quality | Genome-fragment | | 5.47 | AAI-based (high-confidence) | 0 |
| S14_vs_k141_86210\|\|full | 1808 | No | NA | 4 | 0 | 0 | Low-quality | Genome-fragment | | 3.83 | AAI-based (medium-confidence) | 0 |
| S5_extracted_k141_114418 | 1155 | No | NA | 4 | 0 | 0 | Not-determined | Genome-fragment | NA | | NA | 0 |
| S5_vs_k141_50538\|\|full | 1808 | No | NA | 3 | 0 | 1 | Low-quality | Genome-fragment | | 0.39 | AAI-based (medium-confidence) | 0 |
| S16_vs_36394\|\|full | 1808 | No | NA | 3 | 0 | 1 | Low-quality | Genome-fragment | | 1.25 | AAI-based (medium-confidence) | 0 |
| S5_extracted_k141_47068 | 1057 | No | NA | 3 | 0 | 3 | Not-determined | Genome-fragment | NA | | NA | 0 |
| S5_extracted_k141_22925 | 1364 | No | NA | 2 | 0 | 1 | Not-determined | Genome-fragment | NA | | NA | 0 |
| S6_vs_k141_5687\|\|full | 1808 | No | NA | 4 | 0 | 0 | Low-quality | Genome-fragment | | 1.29 | AAI-based (medium-confidence) | 0 |
| S8_vs_k141_33879\|\|full | 1808 | Yes | 1500 | 5 | 1 | 1 | Low-quality | Genome-fragment | | 3.86 | AAI-based (medium-confidence) | 17.04 |
| S5_extracted_k141_10957 | 2094 | No | NA | 3 | 0 | 0 | Not-determined | Genome-fragment | NA | | NA | 0 |
| S9_extracted_k141_55276 | 1809 | No | NA | 4 | 3 | 0 | Low-quality | Genome-fragment | | 5.99 | AAI-based (high-confidence) | 0 |
| S5_extracted_k141_81895 | 1416 | No | NA | 4 | 0 | 0 | Not-determined | Genome-fragment | NA | | NA | 0 |
| S9_extracted_k141_55865 | 1809 | No | NA | 1 | 0 | 0 | Low-quality | Genome-fragment | | 4.06 | AAI-based (medium-confidence) | 0 |
| S9_vs_k141_88453\|\|full | 1809 | No | NA | 2 | 0 | 1 | Low-quality | Genome-fragment | | 1.08 | AAI-based (high-confidence) | 0 |
| S11_vs_k141_80092\|\|full | 1809 | No | NA | 4 | 3 | 0 | Low-quality | Genome-fragment | | 3.45 | AAI-based (high-confidence) | 0 |
| S5_extracted_k141_54528 | 1066 | No | NA | 1 | 0 | 1 | Not-determined | Genome-fragment | NA | | NA | 0 |
| S5_extracted_k141_17661 | 3924 | No | NA | 2 | 0 | 1 | Not-determined | Genome-fragment | NA | | NA | 0 |
| S5_extracted_k141_114535 | 1342 | No | NA | 2 | 0 | 2 | Not-determined | Genome-fragment | NA | | NA | 0 |
| S1_extracted_k141_94036 | 1809 | No | NA | 6 | 0 | 0 | Low-quality | Genome-fragment | | 3.42 | AAI-based (high-confidence) | 0 |
| S5_extracted_k141_89878 | 1040 | No | NA | 1 | 0 | 1 | Not-determined | Genome-fragment | NA | | NA | 0 |
| S1_vs_k141_45044\|\|full | 1809 | No | NA | 4 | 0 | 0 | Low-quality | Genome-fragment | | 1.68 | AAI-based (high-confidence) | 0 |
| S5_extracted_k141_47231 | 6791 | No | NA | 5 | 0 | 5 | Not-determined | Genome-fragment | NA | | NA | 0 |
| S5_extracted_k141_121079 | 1489 | No | NA | 3 | 0 | 0 | Not-determined | Genome-fragment | NA | | NA | 0 |
| S5_extracted_k141_137208 | 1345 | No | NA | 2 | 0 | 1 | Not-determined | Genome-fragment | NA | | NA | 0 |
| S3_extracted_k141_120872 | 1809 | No | NA | 4 | 1 | 0 | Low-quality | Genome-fragment | | 2.48 | AAI-based (high-confidence) | 0 |
| S5_extracted_k141_23054 | 1345 | No | NA | 2 | 0 | 0 | Not-determined | Genome-fragment | NA | | NA | 0 |
| S10_extracted_19819 | 1809 | No | NA | 2 | 0 | 0 | Low-quality | Genome-fragment | | 1.54 | AAI-based (high-confidence) | 0 |
| S2_vs_k141_9871\|\|full | 1809 | No | NA | 5 | 0 | 0 | Low-quality | Genome-fragment | | 4.41 | AAI-based (high-confidence) | 0 |
| S17_vs_k141_118428\|\|full | 1810 | No | NA | 3 | 1 | 0 | Low-quality | Genome-fragment | | 4.84 | AAI-based (high-confidence) | 0 |
| S5_extracted_k141_89950 | 2298 | No | NA | 2 | 0 | 2 | Not-determined | Genome-fragment | NA | | NA | 0 |
| S3_vs_k141_116546\|\|full | 1810 | No | NA | 2 | 0 | 0 | Low-quality | Genome-fragment | | 3.43 | AAI-based (high-confidence) | 0 |
| S3_vs_k141_93733\|\|full | 1810 | No | NA | 4 | 0 | 0 | Low-quality | Genome-fragment | | 3.14 | AAI-based (high-confidence) | 0 |
| S5_extracted_k141_137236 | 3442 | No | NA | 7 | 0 | 3 | Not-determined | Genome-fragment | NA | | NA | 0 |
| S12_vs_k141_163168\|\|full | 1810 | No | NA | 4 | 0 | 1 | Low-quality | Genome-fragment | | 1.4 | AAI-based (medium-confidence) | 0 |
| S5_extracted_k141_132395 | 1378 | No | NA | 2 | 0 | 0 | Not-determined | Genome-fragment | NA | | NA | 0 |
| S5_extracted_k141_89980 | 1648 | No | NA | 1 | 0 | 1 | Not-determined | Genome-fragment | NA | | NA | 0 |
| S9_vs_k141_69620\|\|full | 1811 | No | NA | 2 | 0 | 0 | Low-quality | Genome-fragment | | 4.06 | AAI-based (medium-confidence) | 0 |
| S11_vs_k141_69759\|\|full | 1811 | No | NA | 3 | 2 | 0 | Low-quality | Genome-fragment | | 4.42 | AAI-based (high-confidence) | 0 |
| S5_extracted_k141_143889 | 2421 | No | NA | 2 | 0 | 0 | Not-determined | Genome-fragment | NA | | NA | 0 |
| S5_extracted_k141_36386 | 3588 | No | NA | 4 | 0 | 2 | Not-determined | Genome-fragment | NA | | NA | 0 |
| S5_extracted_k141_137260 | 16081 | No | NA | 19 | 6 | 0 | Complete | High-quality | | 100 | ITR (medium-confidence) | 0 |
| S14_vs_k141_111651\|\|full | 1811 | No | NA | 4 | 1 | 0 | Low-quality | Genome-fragment | | 3.19 | AAI-based (high-confidence) | 0 |
| S5_extracted_k141_137289 | 1212 | No | NA | 1 | 0 | 0 | Not-determined | Genome-fragment | NA | | NA | 0 |
| S6_vs_k141_11180\|\|full | 1811 | No | NA | 4 | 0 | 2 | Low-quality | Genome-fragment | | 2.03 | AAI-based (medium-confidence) | 0 |
| S5_extracted_k141_100813 | 2702 | No | NA | 3 | 0 | 0 | Not-determined | Genome-fragment | NA | | NA | 0 |
| S5_extracted_k141_17901 | 2641 | No | NA | 1 | 0 | 0 | Not-determined | Genome-fragment | NA | | NA | 0 |
| S5_extracted_k141_132513 | 11950 | No | NA | 10 | 0 | 9 | Not-determined | Genome-fragment | NA | | NA | 0 |
| S17_extracted_k141_6965 | 1812 | No | NA | 2 | 1 | 0 | Low-quality | Genome-fragment | | 3.46 | AAI-based (high-confidence) | 0 |
| S1_vs_k141_53670\|\|full | 1812 | No | NA | 3 | 0 | 1 | Low-quality | Genome-fragment | | 1.79 | AAI-based (medium-confidence) | 0 |
| S5_extracted_k141_143946 | 3447 | No | NA | 3 | 0 | 2 | Not-determined | Genome-fragment | NA | | NA | 0 |
| S5_extracted_k141_114754 | 11172 | No | NA | 9 | 0 | 8 | Not-determined | Genome-fragment | NA | | NA | 0 |
| S16_extracted_154978 | 1812 | No | NA | 4 | 0 | 1 | Low-quality | Genome-fragment | | 4.03 | AAI-based (medium-confidence) | 0 |
| S5_extracted_k141_71465 | 2304 | No | NA | 3 | 0 | 0 | Not-determined | Genome-fragment | NA | | NA | 0 |
| S5_extracted_k141_71470 | 1517 | No | NA | 2 | 0 | 0 | Not-determined | Genome-fragment | NA | | NA | 0 |
| S16_vs_74986\|\|full | 1812 | No | NA | 3 | 1 | 0 | Low-quality | Genome-fragment | | 4.49 | AAI-based (high-confidence) | 0 |
| S5_extracted_k141_47482 | 1443 | No | NA | 5 | 0 | 5 | Not-determined | Genome-fragment | NA | | NA | 0 |
| S15_extracted_58254 | 1812 | No | NA | 4 | 2 | 0 | Low-quality | Genome-fragment | | 3.98 | AAI-based (high-confidence) | 0 |
| S17_vs_k141_159929\|\|full | 1813 | No | NA | 2 | 0 | 1 | Low-quality | Genome-fragment | | 0.55 | AAI-based (medium-confidence) | 0 |
| S5_extracted_k141_96464 | 2568 | No | NA | 1 | 0 | 1 | Not-determined | Genome-fragment | NA | | NA | 0 |
| S1_vs_k141_2233\|\|full | 1813 | No | NA | 2 | 0 | 0 | Low-quality | Genome-fragment | | 3.35 | AAI-based (medium-confidence) | 0 |
| S5_extracted_k141_27660 | 1212 | No | NA | 3 | 0 | 0 | Not-determined | Genome-fragment | NA | | NA | 0 |
| S5_extracted_k141_132620 | 1092 | No | NA | 1 | 0 | 0 | Not-determined | Genome-fragment | NA | | NA | 0 |
| S3_extracted_k141_61918 | 1813 | No | NA | 4 | 0 | 0 | Low-quality | Genome-fragment | | 2.29 | AAI-based (high-confidence) | 0 |
| S7_vs_40985\|\|full | 1813 | No | NA | 3 | 0 | 0 | Low-quality | Genome-fragment | | 4.06 | AAI-based (medium-confidence) | 0 |
| S16_vs_21455\|\|full | 1813 | No | NA | 2 | 0 | 2 | Low-quality | Genome-fragment | | 0.79 | AAI-based (medium-confidence) | 0 |
| S5_extracted_k141_27686 | 1106 | No | NA | 2 | 0 | 2 | Not-determined | Genome-fragment | NA | | NA | 0 |
| S5_extracted_k141_17987 | 1075 | No | NA | 2 | 0 | 1 | Not-determined | Genome-fragment | NA | | NA | 0 |
| S5_extracted_k141_47609 | 1816 | No | NA | 2 | 0 | 0 | Not-determined | Genome-fragment | NA | | NA | 0 |
| S5_extracted_k141_121402 | 1043 | No | NA | 1 | 0 | 0 | Not-determined | Genome-fragment | NA | | NA | 0 |
| S10_vs_25731\|\|full | 1813 | No | NA | 4 | 0 | 0 | Low-quality | Genome-fragment | | 0.99 | AAI-based (medium-confidence) | 0 |
| S5_extracted_k141_47661 | 1210 | No | NA | 1 | 0 | 0 | Not-determined | Genome-fragment | NA | | NA | 0 |
| S12_extracted_k141_58464 | 1813 | No | NA | 2 | 0 | 1 | Low-quality | Genome-fragment | | 1.21 | AAI-based (high-confidence) | 0 |
| S5_extracted_k141_100914 | 4276 | No | NA | 2 | 0 | 1 | Not-determined | Genome-fragment | NA | | NA | 0 |
| S8_extracted_k141_91783 | 1813 | No | NA | 2 | 2 | 0 | Low-quality | Genome-fragment | | 4.64 | AAI-based (high-confidence) | 0 |
| S8_extracted_k141_196106 | 1813 | No | NA | 2 | 0 | 1 | Low-quality | Genome-fragment | | 0.54 | AAI-based (medium-confidence) | 0 |
| S13_extracted_15577 | 1814 | No | NA | 1 | 0 | 1 | Low-quality | Genome-fragment | | 0.31 | AAI-based (medium-confidence) | 0 |
| S5_extracted_k141_47713 | 1152 | No | NA | 4 | 0 | 3 | Not-determined | Genome-fragment | NA | | NA | 0 |
| S5_extracted_k141_132780 | 1268 | No | NA | 1 | 0 | 1 | Not-determined | Genome-fragment | NA | | NA | 0 |
| S14_vs_k141_112054\|\|full | 1814 | No | NA | 2 | 0 | 0 | Low-quality | Genome-fragment | | 4.36 | AAI-based (medium-confidence) | 0 |
| S5_extracted_k141_71720 | 1146 | No | NA | 1 | 0 | 1 | Not-determined | Genome-fragment | NA | | NA | 0 |
| S7_vs_55314\|\|full | 1814 | No | NA | 4 | 2 | 0 | Low-quality | Genome-fragment | | 4.65 | AAI-based (high-confidence) | 0 |
| S5_extracted_k141_40939 | 4144 | No | NA | 4 | 0 | 1 | Not-determined | Genome-fragment | NA | | NA | 0 |
| S5_extracted_k141_90160 | 5201 | No | NA | 3 | 0 | 1 | Not-determined | Genome-fragment | NA | | NA | 0 |
| S5_extracted_k141_144119 | 1102 | No | NA | 2 | 0 | 0 | Not-determined | Genome-fragment | NA | | NA | 0 |
| S16_extracted_43239 | 1814 | No | NA | 2 | 0 | 2 | Low-quality | Genome-fragment | | 0.78 | AAI-based (medium-confidence) | 0 |
| S12_extracted_k141_17374 | 1814 | No | NA | 1 | 0 | 1 | Low-quality | Genome-fragment | | 0.27 | AAI-based (medium-confidence) | 0 |
| S5_extracted_k141_90181 | 2335 | No | NA | 1 | 0 | 1 | Not-determined | Genome-fragment | NA | | NA | 0 |
| S12_extracted_k141_168409 | 1814 | No | NA | 4 | 0 | 0 | Low-quality | Genome-fragment | | 5.02 | AAI-based (high-confidence) | 0 |
| S14_extracted_k141_74308 | 1815 | No | NA | 5 | 3 | 0 | Low-quality | Genome-fragment | | 4.09 | AAI-based (high-confidence) | 0 |
| S14_vs_k141_100897\|\|full | 1815 | No | NA | 4 | 2 | 0 | Low-quality | Genome-fragment | | 4.3 | AAI-based (high-confidence) | 0 |
| S3_vs_k141_16381\|\|full | 1815 | No | NA | 3 | 0 | 2 | Low-quality | Genome-fragment | | 3.78 | AAI-based (medium-confidence) | 0 |
| S5_extracted_k141_65681 | 3296 | No | NA | 6 | 0 | 1 | Not-determined | Genome-fragment | NA | | NA | 0 |
| S5_extracted_k141_967 | 1815 | No | NA | 3 | 2 | 0 | Low-quality | Genome-fragment | | 5.37 | AAI-based (high-confidence) | 0 |
| S5_extracted_k141_132961 | 1179 | No | NA | 1 | 0 | 1 | Not-determined | Genome-fragment | NA | | NA | 0 |
| S5_extracted_k141_132967 | 1093 | No | NA | 2 | 0 | 2 | Not-determined | Genome-fragment | NA | | NA | 0 |
| S5_extracted_k141_132969 | 1591 | No | NA | 3 | 0 | 0 | Not-determined | Genome-fragment | NA | | NA | 0 |
| S5_extracted_k141_96718 | 1029 | No | NA | 2 | 0 | 0 | Not-determined | Genome-fragment | NA | | NA | 0 |
| S5_extracted_k141_41005 | 2460 | No | NA | 3 | 0 | 0 | Not-determined | Genome-fragment | NA | | NA | 0 |
| S5_extracted_k141_98541 | 1815 | No | NA | 4 | 2 | 0 | Low-quality | Genome-fragment | | 5.24 | AAI-based (high-confidence) | 0 |
| S5_extracted_k141_90346 | 1331 | No | NA | 1 | 0 | 1 | Not-determined | Genome-fragment | NA | | NA | 0 |
| S5_vs_k141_2622\|\|lt2gene | 1815 | No | NA | 1 | 0 | 0 | Low-quality | Genome-fragment | | 4.35 | AAI-based (medium-confidence) | 0 |
| S5_extracted_k141_144159 | 1559 | No | NA | 2 | 0 | 0 | Not-determined | Genome-fragment | NA | | NA | 0 |
| S7_extracted_95958 | 1815 | No | NA | 2 | 0 | 1 | Low-quality | Genome-fragment | | 1.85 | AAI-based (medium-confidence) | 0 |
| S7_vs_59482\|\|full | 1815 | No | NA | 5 | 2 | 0 | Low-quality | Genome-fragment | | 3.52 | AAI-based (high-confidence) | 0 |
| S5_extracted_k141_18198 | 5711 | No | NA | 9 | 0 | 1 | Not-determined | Genome-fragment | NA | | NA | 0 |
| S5_extracted_k141_65815 | 1809 | No | NA | 4 | 0 | 0 | Not-determined | Genome-fragment | NA | | NA | 0 |
| S7_vs_83636\|\|full | 1815 | No | NA | 5 | 2 | 0 | Low-quality | Genome-fragment | | 3.67 | AAI-based (high-confidence) | 0 |
| S5_extracted_k141_27926 | 1289 | No | NA | 1 | 0 | 1 | Not-determined | Genome-fragment | NA | | NA | 0 |
| S10_vs_51581\|\|full | 1815 | No | NA | 5 | 0 | 0 | Low-quality | Genome-fragment | | 3.37 | AAI-based (high-confidence) | 0 |
| S5_extracted_k141_47946 | 1058 | No | NA | 1 | 0 | 1 | Not-determined | Genome-fragment | NA | | NA | 0 |
| S6_vs_k141_10631\|\|full | 1815 | No | NA | 5 | 3 | 0 | Low-quality | Genome-fragment | | 4.96 | AAI-based (high-confidence) | 0 |
| S8_extracted_k141_145355 | 1815 | No | NA | 4 | 0 | 2 | Low-quality | Genome-fragment | | 0.75 | AAI-based (medium-confidence) | 0 |
| S8_vs_k141_62392\|\|lt2gene | 1815 | No | NA | 1 | 0 | 0 | Low-quality | Genome-fragment | | 5.7 | AAI-based (high-confidence) | 0 |
| S11_vs_k141_96232\|\|full | 1816 | No | NA | 6 | 0 | 0 | Low-quality | Genome-fragment | | 4.33 | AAI-based (high-confidence) | 0 |
| S5_extracted_k141_41215 | 3064 | No | NA | 1 | 0 | 1 | Not-determined | Genome-fragment | NA | | NA | 0 |
| S14_extracted_k141_110917 | 1816 | No | NA | 4 | 1 | 0 | Low-quality | Genome-fragment | | 5.46 | AAI-based (high-confidence) | 0 |
| S5_extracted_k141_72029 | 1288 | No | NA | 2 | 0 | 0 | Not-determined | Genome-fragment | NA | | NA | 0 |
| S1_vs_k141_18625\|\|full | 1816 | No | NA | 3 | 3 | 0 | Low-quality | Genome-fragment | | 2.12 | HMM-based (lower-bound) | 0 |
| S5_extracted_k141_11367 | 1992 | No | NA | 1 | 0 | 1 | Not-determined | Genome-fragment | NA | | NA | 0 |
| S5_extracted_k141_23686 | 1176 | No | NA | 1 | 0 | 1 | Not-determined | Genome-fragment | NA | | NA | 0 |
| S5_extracted_k141_76832 | 1050 | No | NA | 1 | 0 | 1 | Not-determined | Genome-fragment | NA | | NA | 0 |
| S5_extracted_k141_115209 | 1441 | No | NA | 3 | 0 | 0 | Not-determined | Genome-fragment | NA | | NA | 0 |
| S17_vs_k141_76224\|\|full | 1817 | No | NA | 5 | 2 | 0 | Low-quality | Genome-fragment | | 3.74 | AAI-based (medium-confidence) | 0 |
| S5_extracted_k141_72098 | 1254 | No | NA | 2 | 0 | 1 | Not-determined | Genome-fragment | NA | | NA | 0 |
| S5_extracted_k141_115237 | 1839 | No | NA | 1 | 0 | 1 | Not-determined | Genome-fragment | NA | | NA | 0 |
| S5_extracted_k141_72117 | 6973 | No | NA | 8 | 0 | 6 | Not-determined | Genome-fragment | NA | | NA | 0 |
| S5_extracted_k141_97056 | 6000 | No | NA | 5 | 0 | 5 | Not-determined | Genome-fragment | NA | | NA | 0 |
| S5_extracted_k141_72140 | 1031 | No | NA | 2 | 0 | 0 | Not-determined | Genome-fragment | NA | | NA | 0 |
| S5_extracted_k141_133348 | 1597 | No | NA | 1 | 0 | 0 | Not-determined | Genome-fragment | NA | | NA | 0 |
| S5_extracted_k141_90691 | 5188 | No | NA | 4 | 0 | 4 | Not-determined | Genome-fragment | NA | | NA | 0 |
| S1_vs_k141_134223\|\|lt2gene | 1817 | No | NA | 2 | 1 | 0 | Low-quality | Genome-fragment | | 1.87 | AAI-based (high-confidence) | 0 |
| S5_extracted_k141_76900 | 2565 | No | NA | 2 | 0 | 1 | Not-determined | Genome-fragment | NA | | NA | 0 |
| S5_extracted_k141_115314 | 1289 | No | NA | 1 | 0 | 1 | Not-determined | Genome-fragment | NA | | NA | 0 |
| S5_extracted_k141_28022 | 1108 | No | NA | 2 | 0 | 0 | Not-determined | Genome-fragment | NA | | NA | 0 |
| S5_extracted_k141_101065 | 1671 | No | NA | 3 | 0 | 3 | Not-determined | Genome-fragment | NA | | NA | 0 |
| S3_vs_k141_93327\|\|full | 1817 | No | NA | 2 | 0 | 0 | Low-quality | Genome-fragment | | 3.18 | AAI-based (high-confidence) | 0 |
| S5_extracted_k141_133440 | 1302 | No | NA | 2 | 0 | 1 | Not-determined | Genome-fragment | NA | | NA | 0 |
| S5_extracted_k141_11450 | 32114 | No | NA | 48 | 18 | 1 | Complete | High-quality | | 100 | DTR (high-confidence) | 0 |
| S5_extracted_k141_11456 | 5272 | No | NA | 3 | 0 | 2 | Not-determined | Genome-fragment | NA | | NA | 0 |
| S5_extracted_k141_11460 | 1092 | No | NA | 1 | 0 | 1 | Not-determined | Genome-fragment | NA | | NA | 0 |
| S16_extracted_156788 | 1817 | No | NA | 3 | 0 | 2 | Low-quality | Genome-fragment | | 3.09 | AAI-based (medium-confidence) | 0 |
| S5_extracted_k141_11499 | 1652 | No | NA | 3 | 0 | 2 | Not-determined | Genome-fragment | NA | | NA | 0 |
| S16_vs_136451\|\|full | 1817 | No | NA | 4 | 0 | 0 | Low-quality | Genome-fragment | | 3.43 | AAI-based (medium-confidence) | 0 |
| S16_vs_148019\|\|full | 1817 | No | NA | 4 | 0 | 3 | Low-quality | Genome-fragment | | 0.51 | AAI-based (medium-confidence) | 0 |
| S5_extracted_k141_23846 | 1061 | No | NA | 3 | 0 | 0 | Not-determined | Genome-fragment | NA | | NA | 0 |
| S12_vs_k141_112061\|\|full | 1817 | No | NA | 6 | 0 | 0 | Low-quality | Genome-fragment | | 1.66 | AAI-based (high-confidence) | 0 |
| S12_vs_k141_82053\|\|lt2gene | 1817 | No | NA | 3 | 2 | 0 | Low-quality | Genome-fragment | | 3.97 | AAI-based (high-confidence) | 0 |
| S5_extracted_k141_28061 | 38334 | No | NA | 53 | 16 | 1 | Complete | High-quality | | 100 | DTR (medium-confidence) | 0 |
| S2_vs_k141_11089\|\|full | 1817 | No | NA | 3 | 1 | 0 | Low-quality | Genome-fragment | | 4.32 | HMM-based (lower-bound) | 0 |
| S8_extracted_k141_60430 | 1817 | No | NA | 1 | 0 | 0 | Low-quality | Genome-fragment | | 4.1 | AAI-based (medium-confidence) | 0 |
| S5_extracted_k141_77064 | 3297 | No | NA | 2 | 0 | 0 | Not-determined | Genome-fragment | NA | | NA | 0 |
| S5_extracted_k141_41416 | 1006 | No | NA | 3 | 0 | 2 | Not-determined | Genome-fragment | NA | | NA | 0 |
| S5_extracted_k141_101151 | 1133 | No | NA | 1 | 0 | 1 | Not-determined | Genome-fragment | NA | | NA | 0 |
| S14_vs_k141_14643\|\|full | 1818 | No | NA | 4 | 1 | 0 | Low-quality | Genome-fragment | | 4.06 | AAI-based (medium-confidence) | 0 |
| S1_vs_k141_36667\|\|full | 1818 | No | NA | 3 | 0 | 0 | Low-quality | Genome-fragment | | 3.09 | AAI-based (high-confidence) | 0 |
| S5_extracted_k141_137738 | 5517 | No | NA | 4 | 0 | 0 | Not-determined | Genome-fragment | NA | | NA | 0 |
| S5_extracted_k141_72383 | 3235 | No | NA | 1 | 0 | 0 | Not-determined | Genome-fragment | NA | | NA | 0 |
| S1_vs_k141_93347\|\|full | 1818 | No | NA | 4 | 2 | 0 | Low-quality | Genome-fragment | | 4.5 | AAI-based (high-confidence) | 0 |
| S5_extracted_k141_11620 | 1010 | No | NA | 2 | 0 | 1 | Not-determined | Genome-fragment | NA | | NA | 0 |
| S12_vs_k141_24114\|\|full | 1818 | No | NA | 3 | 0 | 1 | Low-quality | Genome-fragment | | 0.89 | AAI-based (medium-confidence) | 0 |
| S5_extracted_k141_11654 | 1023 | No | NA | 2 | 0 | 0 | Not-determined | Genome-fragment | NA | | NA | 0 |
| S5_extracted_k141_77224 | 1139 | No | NA | 2 | 0 | 1 | Not-determined | Genome-fragment | NA | | NA | 0 |
| S5_extracted_k141_90802 | 1032 | No | NA | 1 | 0 | 1 | Not-determined | Genome-fragment | NA | | NA | 0 |
| S5_extracted_k141_101275 | 6507 | No | NA | 4 | 0 | 4 | Not-determined | Genome-fragment | NA | | NA | 0 |
| S8_vs_k141_10227\|\|full | 1818 | No | NA | 3 | 0 | 0 | Low-quality | Genome-fragment | | 0.76 | AAI-based (high-confidence) | 0 |
| S5_extracted_k141_137805 | 3570 | No | NA | 3 | 0 | 2 | Not-determined | Genome-fragment | NA | | NA | 0 |
| S5_extracted_k141_137814 | 1363 | No | NA | 2 | 0 | 1 | Not-determined | Genome-fragment | NA | | NA | 0 |
| S8_vs_k141_141226\|\|full | 1818 | No | NA | 3 | 0 | 0 | Low-quality | Genome-fragment | | 3 | AAI-based (medium-confidence) | 0 |
| S5_extracted_k141_72483 | 7851 | No | NA | 5 | 0 | 3 | Not-determined | Genome-fragment | NA | | NA | 0 |
| S17_vs_k141_11842\|\|full | 1819 | No | NA | 3 | 0 | 0 | Low-quality | Genome-fragment | | 1.08 | AAI-based (high-confidence) | 0 |
| S11_vs_k141_65181\|\|full | 1819 | No | NA | 3 | 2 | 0 | Low-quality | Genome-fragment | | 4.44 | AAI-based (high-confidence) | 0 |
| S5_extracted_k141_137846 | 4070 | No | NA | 3 | 0 | 1 | Not-determined | Genome-fragment | NA | | NA | 0 |
| S5_extracted_k141_48466 | 1149 | No | NA | 1 | 0 | 0 | Not-determined | Genome-fragment | NA | | NA | 0 |
| S5_extracted_k141_137884 | 1188 | No | NA | 1 | 0 | 0 | Not-determined | Genome-fragment | NA | | NA | 0 |
| S5_extracted_k141_90893 | 1015 | No | NA | 1 | 0 | 1 | Not-determined | Genome-fragment | NA | | NA | 0 |
| S13_extracted_23690 | 1819 | No | NA | 2 | 0 | 0 | Low-quality | Genome-fragment | | 3.04 | AAI-based (medium-confidence) | 0 |
| S13_vs_41412\|\|full | 1819 | No | NA | 4 | 0 | 0 | Low-quality | Genome-fragment | | 6.27 | AAI-based (high-confidence) | 0 |
| S1_extracted_k141_34103 | 1819 | No | NA | 1 | 0 | 1 | Low-quality | Genome-fragment | | 0.32 | AAI-based (high-confidence) | 0 |
| S3_vs_k141_75949\|\|full | 1819 | No | NA | 3 | 0 | 2 | Low-quality | Genome-fragment | | 3.92 | AAI-based (high-confidence) | 0 |
| S5_extracted_k141_24229 | 1096 | No | NA | 2 | 0 | 0 | Not-determined | Genome-fragment | NA | | NA | 0 |
| S16_extracted_125591 | 1819 | No | NA | 2 | 0 | 2 | Low-quality | Genome-fragment | | 0.79 | AAI-based (medium-confidence) | 0 |
| S5_extracted_k141_72579 | 2045 | No | NA | 1 | 0 | 1 | Not-determined | Genome-fragment | NA | | NA | 0 |
| S5_extracted_k141_41732 | 1000 | No | NA | 1 | 0 | 0 | Not-determined | Genome-fragment | NA | | NA | 0 |
| S16_extracted_127074 | 1819 | No | NA | 2 | 2 | 0 | Low-quality | Genome-fragment | | 5.99 | AAI-based (high-confidence) | 0 |
| S5_extracted_k141_101380 | 1366 | No | NA | 1 | 0 | 1 | Not-determined | Genome-fragment | NA | | NA | 0 |
| S5_extracted_k141_66370 | 1673 | No | NA | 2 | 0 | 0 | Not-determined | Genome-fragment | NA | | NA | 0 |
| S5_extracted_k141_72643 | 1064 | No | NA | 1 | 0 | 0 | Not-determined | Genome-fragment | NA | | NA | 0 |
| S5_extracted_k141_58279 | 10670 | No | NA | 7 | 0 | 2 | Not-determined | Genome-fragment | NA | | NA | 0 |
| S5_extracted_k141_137935 | 1403 | No | NA | 2 | 0 | 2 | Not-determined | Genome-fragment | NA | | NA | 0 |
| S5_extracted_k141_28510 | 1061 | No | NA | 2 | 0 | 2 | Not-determined | Genome-fragment | NA | | NA | 0 |
| S5_extracted_k141_66406 | 2458 | No | NA | 2 | 0 | 1 | Not-determined | Genome-fragment | NA | | NA | 0 |
| S10_vs_45754\|\|full | 1819 | No | NA | 3 | 0 | 0 | Low-quality | Genome-fragment | | 3.56 | AAI-based (medium-confidence) | 0 |
| S5_extracted_k141_144833 | 1408 | No | NA | 1 | 0 | 1 | Not-determined | Genome-fragment | NA | | NA | 0 |
| S5_extracted_k141_137950 | 2762 | No | NA | 5 | 0 | 5 | Not-determined | Genome-fragment | NA | | NA | 0 |
| S5_extracted_k141_72743 | 1262 | No | NA | 1 | 0 | 1 | Not-determined | Genome-fragment | NA | | NA | 0 |

| | | | | | | | | | | | |
|---|---|---|---|---|---|---|---|---|---|---|---|
| S12_vs_k141_176706\|\|full | 1819 | No | NA | 3 | 0 | 2 | Low-quality | Genome-fragment | 0.87 | AAI-based (medium-confidence) | 0 |
| S5_extracted_k141_144910 | 6253 | No | NA | 6 | 0 | 4 | Not-determined | Genome-fragment | NA | NA | 0 |
| S5_extracted_k141_144922 | 1369 | No | NA | 1 | 0 | 1 | Not-determined | Genome-fragment | NA | NA | 0 |
| S5_extracted_k141_83018 | 3870 | No | NA | 1 | 0 | 1 | Not-determined | Genome-fragment | NA | NA | 0 |
| S5_extracted_k141_41968 | 5646 | No | NA | 8 | 0 | 4 | Not-determined | Genome-fragment | NA | NA | 0 |
| S5_extracted_k141_77412 | 1042 | No | NA | 2 | 0 | 2 | Not-determined | Genome-fragment | NA | NA | 0 |
| S5_extracted_k141_28629 | 3055 | No | NA | 4 | 0 | 3 | Not-determined | Genome-fragment | NA | NA | 0 |
| S3_vs_k141_98668\|\|full | 1820 | No | NA | 2 | 0 | 1 | Low-quality | Genome-fragment | 0.49 | AAI-based (medium-confidence) | 0 |
| S5_extracted_k141_83161 | 5422 | No | NA | 5 | 0 | 2 | Not-determined | Genome-fragment | NA | NA | 0 |
| S5_extracted_k141_83177 | 1106 | No | NA | 1 | 0 | 1 | Not-determined | Genome-fragment | NA | NA | 0 |
| S7_vs_48129\|\|full | 1820 | No | NA | 3 | 1 | 0 | Low-quality | Genome-fragment | 3.47 | AAI-based (high-confidence) | 0 |
| S5_extracted_k141_42163 | 1278 | No | NA | 2 | 0 | 1 | Not-determined | Genome-fragment | NA | NA | 0 |
| S5_extracted_k141_145033 | 1296 | No | NA | 1 | 0 | 0 | Not-determined | Genome-fragment | NA | NA | 0 |
| S5_extracted_k141_145044 | 1199 | No | NA | 3 | 0 | 2 | Not-determined | Genome-fragment | NA | NA | 0 |
| S8_extracted_k141_44550 | 1820 | No | NA | 1 | 1 | 0 | Low-quality | Genome-fragment | 0.96 | HMM-based (lower-bound) | 0 |
| S5_extracted_k141_66553 | 1833 | No | NA | 5 | 0 | 5 | Not-determined | Genome-fragment | NA | NA | 0 |
| S5_extracted_k141_138093 | 6607 | No | NA | 7 | 0 | 3 | Not-determined | Genome-fragment | NA | NA | 0 |
| S5_extracted_k141_138094 | 1218 | No | NA | 1 | 0 | 0 | Not-determined | Genome-fragment | NA | NA | 0 |
| S8_extracted_k141_31877 | 1820 | No | NA | 4 | 0 | 0 | Low-quality | Genome-fragment | 2.98 | AAI-based (high-confidence) | 0 |
| S8_vs_k141_52295\|\|full | 1820 | No | NA | 2 | 0 | 1 | Low-quality | Genome-fragment | 5.19 | AAI-based (medium-confidence) | 0 |
| S3_extracted_k141_100400 | 1821 | No | NA | 2 | 0 | 0 | Low-quality | Genome-fragment | 1.54 | AAI-based (high-confidence) | 0 |
| S5_extracted_k141_101494 | 16172 | No | NA | 14 | 0 | 12 | Not-determined | Genome-fragment | NA | NA | 0 |
| S5_extracted_k141_42406 | 2802 | No | NA | 2 | 0 | 0 | Not-determined | Genome-fragment | NA | NA | 0 |
| S5_extracted_k141_83453 | 1167 | No | NA | 3 | 0 | 0 | Not-determined | Genome-fragment | NA | NA | 0 |
| S5_vs_k141_66113\|\|full | 1821 | No | NA | 2 | 0 | 2 | Low-quality | Genome-fragment | 2.75 | AAI-based (medium-confidence) | 0 |
| S5_extracted_k141_77701 | 38591 | No | NA | 43 | 27 | 0 | Medium-quality | Genome-fragment | 64.37 | AAI-based (high-confidence) | 0 |
| S5_extracted_k141_83522 | 1971 | No | NA | 2 | 0 | 1 | Not-determined | Genome-fragment | NA | NA | 0 |
| S5_extracted_k141_145262 | 2328 | No | NA | 1 | 0 | 1 | Not-determined | Genome-fragment | NA | NA | 0 |
| S5_extracted_k141_101531 | 4695 | No | NA | 4 | 0 | 3 | Not-determined | Genome-fragment | NA | NA | 0 |
| S7_vs_31558\|\|full | 1821 | No | NA | 4 | 0 | 1 | Low-quality | Genome-fragment | 5.07 | AAI-based (medium-confidence) | 0 |
| S10_vs_60364\|\|full | 1821 | No | NA | 3 | 0 | 0 | Low-quality | Genome-fragment | 1.5 | AAI-based (medium-confidence) | 0 |
| S5_extracted_k141_145329 | 1238 | No | NA | 1 | 0 | 1 | Not-determined | Genome-fragment | NA | NA | 0 |
| S5_extracted_k141_138350 | 1805 | No | NA | 3 | 0 | 2 | Not-determined | Genome-fragment | NA | NA | 0 |
| S12_vs_k141_62957\|\|full | 1821 | No | NA | 4 | 0 | 0 | Low-quality | Genome-fragment | 4.58 | AAI-based (medium-confidence) | 0 |
| S8_extracted_k141_173746 | 1821 | No | NA | 2 | 0 | 1 | Low-quality | Genome-fragment | 4.06 | AAI-based (medium-confidence) | 0 |
| S17_vs_k141_176709\|\|full | 1822 | No | NA | 4 | 0 | 0 | Low-quality | Genome-fragment | 1.82 | AAI-based (high-confidence) | 0 |
| S5_extracted_k141_145417 | 1239 | No | NA | 2 | 0 | 2 | Not-determined | Genome-fragment | NA | NA | 0 |
| S11_vs_k141_17884\|\|full | 1822 | No | NA | 4 | 0 | 0 | Low-quality | Genome-fragment | 2.46 | AAI-based (high-confidence) | 0 |
| S5_extracted_k141_145445 | 3831 | No | NA | 3 | 0 | 3 | Not-determined | Genome-fragment | NA | NA | 0 |
| S11_vs_k141_84199\|\|lt2gene | 1822 | No | NA | 2 | 1 | 0 | Low-quality | Genome-fragment | 5.31 | AAI-based (high-confidence) | 0 |
| S5_extracted_k141_145481 | 3531 | No | NA | 2 | 0 | 2 | Not-determined | Genome-fragment | NA | NA | 0 |
| S14_vs_k141_40653\|\|full | 1822 | No | NA | 3 | 0 | 2 | Low-quality | Genome-fragment | 1.38 | AAI-based (medium-confidence) | 0 |
| S5_extracted_k141_101594 | 1128 | No | NA | 1 | 0 | 1 | Not-determined | Genome-fragment | NA | NA | 0 |
| S3_vs_k141_84792\|\|full | 1822 | No | NA | 3 | 1 | 0 | Low-quality | Genome-fragment | 3.06 | AAI-based (medium-confidence) | 0 |
| S7_vs_92547\|\|full | 1822 | No | NA | 3 | 0 | 0 | Low-quality | Genome-fragment | 1.49 | AAI-based (high-confidence) | 0 |
| S12_extracted_k141_37499 | 1822 | No | NA | 5 | 1 | 2 | Low-quality | Genome-fragment | 0.98 | HMM-based (lower-bound) | 0 |
| S12_vs_k141_126129\|\|full | 1822 | No | NA | 3 | 2 | 0 | Low-quality | Genome-fragment | 3.16 | AAI-based (medium-confidence) | 0 |
| S6_extracted_k141_1713 | 1822 | No | NA | 2 | 0 | 1 | Low-quality | Genome-fragment | 2.81 | AAI-based (high-confidence) | 0 |
| S5_extracted_k141_138571 | 1045 | No | NA | 2 | 0 | 0 | Not-determined | Genome-fragment | NA | NA | 0 |
| S5_extracted_k141_101699 | 4218 | No | NA | 12 | 0 | 0 | Not-determined | Genome-fragment | NA | NA | 0 |
| S9_extracted_k141_28820 | 1823 | No | NA | 4 | 4 | 0 | Low-quality | Genome-fragment | 6.02 | AAI-based (high-confidence) | 0 |
| S9_vs_k141_43849\|\|full | 1823 | No | NA | 6 | 0 | 0 | Low-quality | Genome-fragment | 4.01 | AAI-based (high-confidence) | 0 |
| S12_vs_k141_117835\|\|full | 1823 | No | NA | 3 | 0 | 0 | Low-quality | Genome-fragment | 2.67 | AAI-based (medium-confidence) | 0 |
| S5_extracted_k141_145579 | 186438 | No | NA | 207 | 28 | 2 | Complete | High-quality | 100 | DTR (high-confidence) | 0 |
| S5_extracted_k141_83968 | 6365 | No | NA | 3 | 0 | 2 | Not-determined | Genome-fragment | NA | NA | 0 |
| S5_extracted_k141_59249 | 1049 | No | NA | 1 | 0 | 1 | Not-determined | Genome-fragment | NA | NA | 0 |
| S6_extracted_k141_3405 | 1823 | No | NA | 4 | 2 | 0 | Low-quality | Genome-fragment | 2.82 | AAI-based (high-confidence) | 0 |
| S5_extracted_k141_101843 | 1008 | No | NA | 3 | 0 | 0 | Not-determined | Genome-fragment | NA | NA | 0 |
| S5_extracted_k141_77970 | 7713 | No | NA | 8 | 0 | 1 | Not-determined | Genome-fragment | NA | NA | 0 |
| S17_vs_k141_8433\|\|full | 1824 | No | NA | 1 | 0 | 1 | Low-quality | Genome-fragment | 3.87 | AAI-based (medium-confidence) | 0 |
| S5_extracted_k141_77984 | 1002 | No | NA | 1 | 0 | 1 | Not-determined | Genome-fragment | NA | NA | 0 |
| S5_extracted_k141_59387 | 3858 | No | NA | 3 | 0 | 2 | Not-determined | Genome-fragment | NA | NA | 0 |
| S11_vs_k141_84096\|\|full | 1824 | No | NA | 4 | 1 | 0 | Low-quality | Genome-fragment | 3.83 | HMM-based (lower-bound) | 0 |
| S3_extracted_k141_37600 | 1824 | No | NA | 2 | 2 | 0 | Low-quality | Genome-fragment | 4.28 | AAI-based (high-confidence) | 0 |
| S16_vs_97306\|\|full | 1824 | No | NA | 5 | 0 | 0 | Low-quality | Genome-fragment | 3.22 | AAI-based (high-confidence) | 0 |
| S5_extracted_k141_145583 | 38922 | No | NA | 66 | 22 | 1 | Complete | High-quality | 100 | DTR (high-confidence) | 0 |
| S5_extracted_k141_29332 | 37610 | No | NA | 56 | 20 | 0 | High-quality | High-quality | 99.7 | AAI-based (high-confidence) | 0 |
| S5_extracted_k141_78104 | 1128 | No | NA | 2 | 0 | 1 | Not-determined | Genome-fragment | NA | NA | 0 |
| S10_extracted_13049 | 1824 | No | NA | 3 | 0 | 0 | Low-quality | Genome-fragment | 1.63 | AAI-based (high-confidence) | 0 |
| S9_extracted_k141_4498 | 1825 | No | NA | 2 | 0 | 1 | Low-quality | Genome-fragment | 2.93 | AAI-based (high-confidence) | 0 |
| S11_vs_k141_75483\|\|full | 1825 | No | NA | 3 | 0 | 1 | Low-quality | Genome-fragment | 4.01 | AAI-based (medium-confidence) | 0 |
| S1_vs_k141_60810\|\|full | 1825 | No | NA | 3 | 3 | 0 | Low-quality | Genome-fragment | 4 | AAI-based (high-confidence) | 0 |
| S1_vs_k141_82570\|\|lt2gene | 1825 | No | NA | 2 | 1 | 0 | Low-quality | Genome-fragment | 3.1 | AAI-based (high-confidence) | 0 |
| S5_extracted_k141_145585 | 65185 | No | NA | 92 | 14 | 0 | Complete | High-quality | 100 | DTR (high-confidence) | 0 |
| S5_extracted_k141_84279 | 1402 | No | NA | 3 | 0 | 2 | Not-determined | Genome-fragment | NA | NA | 0 |
| S5_extracted_k141_78227 | 1417 | No | NA | 3 | 0 | 2 | Not-determined | Genome-fragment | NA | NA | 0 |
| S7_vs_3197\|\|full | 1825 | No | NA | 4 | 1 | 0 | Low-quality | Genome-fragment | 3.37 | AAI-based (medium-confidence) | 0 |
| S5_extracted_k141_145586 | 39392 | No | NA | 63 | 13 | 0 | Complete | High-quality | 100 | DTR (high-confidence) | 0 |
| S5_extracted_k141_102041 | 5817 | No | NA | 5 | 0 | 2 | Not-determined | Genome-fragment | NA | NA | 0 |
| S17_vs_k141_136343\|\|full | 1826 | No | NA | 3 | 0 | 2 | Low-quality | Genome-fragment | 0.73 | AAI-based (medium-confidence) | 0 |
| S11_vs_k141_85123\|\|full | 1826 | No | NA | 5 | 0 | 0 | Low-quality | Genome-fragment | 2.84 | AAI-based (high-confidence) | 0 |
| S5_extracted_k141_138835 | 1149 | No | NA | 1 | 0 | 0 | Not-determined | Genome-fragment | NA | NA | 0 |
| S5_extracted_k141_145588 | 46058 | No | NA | 81 | 19 | 1 | Complete | High-quality | 100 | DTR (high-confidence) | 0 |
| S5_extracted_k141_138857 | 1708 | No | NA | 3 | 0 | 3 | Not-determined | Genome-fragment | NA | NA | 0 |
| S5_extracted_k141_62622 | 1826 | No | NA | 5 | 1 | 0 | Low-quality | Genome-fragment | 4.62 | AAI-based (medium-confidence) | 0 |
| S5_vs_k141_96731\|\|full | 1826 | No | NA | 3 | 0 | 0 | Low-quality | Genome-fragment | 2.83 | AAI-based (medium-confidence) | 0 |
| S5_extracted_k141_78503 | 1058 | No | NA | 4 | 0 | 0 | Not-determined | Genome-fragment | NA | NA | 0 |
| S5_extracted_k141_138986 | 5380 | No | NA | 5 | 0 | 5 | Not-determined | Genome-fragment | NA | NA | 0 |
| S7_extracted_6037 | 1826 | No | NA | 4 | 1 | 0 | Low-quality | Genome-fragment | 3.8 | AAI-based (high-confidence) | 0 |
| S5_extracted_k141_102262 | 4495 | No | NA | 8 | 0 | 7 | Not-determined | Genome-fragment | NA | NA | 0 |
| S5_extracted_k141_59813 | 14769 | No | NA | 10 | 0 | 7 | Not-determined | Genome-fragment | NA | NA | 0 |
| S16_extracted_11470 | 1826 | No | NA | 3 | 0 | 0 | Low-quality | Genome-fragment | 2.94 | AAI-based (high-confidence) | 0 |
| S5_extracted_k141_78835 | 3544 | No | NA | 3 | 0 | 3 | Not-determined | Genome-fragment | NA | NA | 0 |
| S5_extracted_k141_102384 | 1229 | No | NA | 3 | 0 | 0 | Not-determined | Genome-fragment | NA | NA | 0 |
| S5_extracted_k141_145604 | 42930 | No | NA | 69 | 15 | 1 | Complete | High-quality | 100 | DTR (high-confidence) | 0 |
| S5_extracted_k141_139108 | 6809 | No | NA | 5 | 0 | 3 | Not-determined | Genome-fragment | NA | NA | 0 |
| S9_vs_k141_1097\|\|lt2gene | 1827 | No | NA | 2 | 1 | 0 | Low-quality | Genome-fragment | 3.46 | HMM-based (lower-bound) | 0 |
| S5_extracted_k141_102420 | 2404 | No | NA | 1 | 0 | 1 | Not-determined | Genome-fragment | NA | NA | 0 |
| S3_extracted_k141_122920 | 1827 | No | NA | 5 | 0 | 0 | Low-quality | Genome-fragment | 1.88 | AAI-based (high-confidence) | 0 |
| S3_extracted_k141_110260 | 1827 | Yes | 483 | 3 | 1 | 1 | Low-quality | Genome-fragment | 0.31 | HMM-based (lower-bound) | 73.56 |
| S5_extracted_k141_60005 | 5602 | No | NA | 5 | 0 | 5 | Not-determined | Genome-fragment | NA | NA | 0 |
| S3_vs_k141_123521\|\|full | 1827 | No | NA | 5 | 1 | 0 | Low-quality | Genome-fragment | 3.77 | AAI-based (high-confidence) | 0 |
| S5_extracted_k141_102491 | 11143 | No | NA | 14 | 0 | 6 | Not-determined | Genome-fragment | NA | NA | 0 |
| S5_extracted_k141_84793 | 1241 | No | NA | 1 | 0 | 0 | Not-determined | Genome-fragment | NA | NA | 0 |
| S5_extracted_k141_104549 | 1827 | No | NA | 4 | 4 | 0 | Low-quality | Genome-fragment | 6.01 | AAI-based (high-confidence) | 0 |
| S5_extracted_k141_84832 | 1967 | No | NA | 1 | 0 | 0 | Not-determined | Genome-fragment | NA | NA | 0 |
| S5_extracted_k141_102581 | 1047 | No | NA | 1 | 0 | 0 | Not-determined | Genome-fragment | NA | NA | 0 |
| S5_vs_k141_105210\|\|lt2gene | 1827 | No | NA | 1 | 1 | 0 | Low-quality | Genome-fragment | 4.77 | AAI-based (high-confidence) | 0 |
| S5_extracted_k141_29967 | 1529 | No | NA | 1 | 0 | 0 | Not-determined | Genome-fragment | NA | NA | 0 |
| S5_extracted_k141_102662 | 6048 | No | NA | 4 | 0 | 4 | Not-determined | Genome-fragment | NA | NA | 0 |
| S5_extracted_k141_30042 | 1056 | No | NA | 1 | 0 | 1 | Not-determined | Genome-fragment | NA | NA | 0 |
| S5_extracted_k141_60147 | 1190 | No | NA | 1 | 0 | 1 | Not-determined | Genome-fragment | NA | NA | 0 |
| S12_vs_k141_132503\|\|full | 1827 | No | NA | 8 | 1 | 0 | Low-quality | Genome-fragment | 4.98 | AAI-based (high-confidence) | 0 |
| S6_vs_k141_16470\|\|full | 1827 | No | NA | 3 | 2 | 0 | Low-quality | Genome-fragment | 3.58 | HMM-based (lower-bound) | 0 |
| S17_extracted_k141_94867 | 1828 | No | NA | 1 | 0 | 1 | Low-quality | Genome-fragment | 0.35 | AAI-based (medium-confidence) | 0 |
| S5_extracted_k141_102755 | 1644 | No | NA | 1 | 0 | 1 | Not-determined | Genome-fragment | NA | NA | 0 |
| S1_extracted_k141_34509 | 1828 | No | NA | 5 | 2 | 0 | Low-quality | Genome-fragment | 2.94 | AAI-based (high-confidence) | 0 |
| S3_vs_k141_81011\|\|full | 1828 | No | NA | 5 | 0 | 0 | Low-quality | Genome-fragment | 1 | AAI-based (high-confidence) | 0 |
| S5_extracted_k141_102858 | 1297 | No | NA | 2 | 0 | 0 | Not-determined | Genome-fragment | NA | NA | 0 |
| S7_vs_70638\|\|full | 1828 | No | NA | 4 | 0 | 2 | Low-quality | Genome-fragment | 2.56 | AAI-based (high-confidence) | 0 |
| S5_extracted_k141_139448 | 4957 | No | NA | 2 | 0 | 0 | Not-determined | Genome-fragment | NA | NA | 0 |
| S12_vs_k141_25861\|\|full | 1828 | No | NA | 3 | 0 | 0 | Low-quality | Genome-fragment | 2.52 | AAI-based (medium-confidence) | 0 |
| S12_vs_k141_151368\|\|full | 1828 | Yes | 1314 | 6 | 1 | 1 | Low-quality | Genome-fragment | 3.44 | AAI-based (high-confidence) | 28.12 |
| S5_extracted_k141_103119 | 1111 | No | NA | 1 | 0 | 1 | Not-determined | Genome-fragment | NA | NA | 0 |
| S8_extracted_k141_130769 | 1828 | No | NA | 4 | 1 | 0 | Low-quality | Genome-fragment | 5.97 | AAI-based (high-confidence) | 0 |
| S1_extracted_k141_143614 | 1829 | No | NA | 1 | 0 | 0 | Low-quality | Genome-fragment | 3.45 | AAI-based (high-confidence) | 0 |
| S5_extracted_k141_103152 | 1195 | No | NA | 3 | 0 | 1 | Not-determined | Genome-fragment | NA | NA | 0 |
| S5_vs_k141_6140\|\|full | 1829 | No | NA | 3 | 0 | 2 | Low-quality | Genome-fragment | 4.35 | AAI-based (medium-confidence) | 0 |
| S5_extracted_k141_139591 | 1161 | No | NA | 2 | 0 | 0 | Not-determined | Genome-fragment | NA | NA | 0 |
| S5_extracted_k141_103206 | 18577 | No | NA | 28 | 1 | 0 | High-quality | High-quality | 100 | AAI-based (medium-confidence) | 0 |
| S5_extracted_k141_60508 | 3586 | No | NA | 5 | 0 | 3 | Not-determined | Genome-fragment | NA | NA | 0 |
| S5_extracted_k141_60582 | 4539 | No | NA | 7 | 0 | 2 | Not-determined | Genome-fragment | NA | NA | 0 |
| S5_extracted_k141_60597 | 51608 | No | NA | 92 | 16 | 2 | Complete | High-quality | 100 | DTR (high-confidence) | 0 |
| S5_extracted_k141_60599 | 1829 | No | NA | 1 | 0 | 1 | Not-determined | Genome-fragment | NA | NA | 0 |
| S5_extracted_k141_60672 | 1355 | No | NA | 3 | 0 | 1 | Not-determined | Genome-fragment | NA | NA | 0 |
| S5_vs_k141_64164\|\|full | 1829 | No | NA | 4 | 0 | 1 | Low-quality | Genome-fragment | 0.63 | AAI-based (medium-confidence) | 0 |
| S5_vs_k141_60959\|\|full | 3547 | No | NA | 6 | 0 | 3 | Not-determined | Genome-fragment | NA | NA | 0 |
| S5_vs_k141_36605\|\|full | 39273 | No | NA | 63 | 17 | 1 | High-quality | High-quality | 97.19 | AAI-based (high-confidence) | 0 |
| S5_vs_k141_666\|\|full | 7517 | No | NA | 3 | 0 | 2 | Not-determined | Genome-fragment | NA | NA | 0 |
| S5_vs_k141_122344\|\|full | 36736 | No | NA | 57 | 21 | 0 | High-quality | High-quality | 100 | AAI-based (high-confidence) | 0 |
| S5_vs_k141_122618\|\|full | 40649 | No | NA | 66 | 20 | 0 | High-quality | High-quality | 100 | AAI-based (high-confidence) | 0 |
| S5_vs_k141_97574\|\|full | 177746 | No | NA | 210 | 42 | 4 | High-quality | High-quality | 100 | AAI-based (high-confidence) | 0 |
| S7_extracted_80725 | 1830 | No | NA | 3 | 1 | 0 | Low-quality | Genome-fragment | 3.95 | AAI-based (medium-confidence) | 0 |
| S5_vs_k141_37659\|\|full | 39786 | No | NA | 54 | 17 | 2 | Medium-quality | Genome-fragment | 58.01 | HMM-based (lower-bound) | 0 |
| S5_vs_k141_67958\|\|full | 38068 | No | NA | 58 | 11 | 0 | Medium-quality | Genome-fragment | 78.56 | AAI-based (high-confidence) | 0 |
| S5_vs_k141_68254\|\|full | 44015 | No | NA | 78 | 20 | 0 | High-quality | High-quality | 96.66 | AAI-based (high-confidence) | 0 |
| S5_vs_k141_123766\|\|full | 4785 | No | NA | 8 | 0 | 2 | Not-determined | Genome-fragment | NA | NA | 0 |
| S5_vs_k141_118434\|\|full | 3666 | No | NA | 2 | 0 | 0 | Not-determined | Genome-fragment | NA | NA | 0 |
| S5_vs_k141_38949\|\|full | 44115 | No | NA | 68 | 16 | 1 | High-quality | High-quality | 98.29 | AAI-based (high-confidence) | 0 |
| S14_vs_k141_7724\|\|full | 1831 | No | NA | 4 | 1 | 0 | Low-quality | Genome-fragment | 3.31 | AAI-based (medium-confidence) | 0 |
| S5_vs_k141_21164\|\|full | 33592 | No | NA | 37 | 0 | 13 | Not-determined | Genome-fragment | NA | NA | 0 |
| S5_vs_k141_120079\|\|full | 38193 | No | NA | 58 | 16 | 1 | Medium-quality | Genome-fragment | 71.74 | AAI-based (high-confidence) | 0 |
| S5_vs_k141_10428\|\|full | 49404 | No | NA | 77 | 21 | 0 | High-quality | High-quality | 90.03 | AAI-based (medium-confidence) | 0 |
| S5_vs_k141_136679\|\|full | 37872 | No | NA | 43 | 8 | 0 | High-quality | High-quality | 90.51 | AAI-based (high-confidence) | 0 |
| S5_vs_k141_35522\|\|full | 5132 | No | NA | 6 | 0 | 5 | Not-determined | Genome-fragment | NA | NA | 0 |
| S5_vs_k141_120584\|\|full | 62832 | No | NA | 103 | 29 | 1 | High-quality | High-quality | 100 | AAI-based (high-confidence) | 0 |
| S5_vs_k141_109085\|\|full | 36721 | No | NA | 51 | 22 | 1 | High-quality | High-quality | 97.59 | AAI-based (high-confidence) | 0 |
| S5_vs_k141_121411\|\|full | 38687 | No | NA | 55 | 16 | 0 | High-quality | High-quality | 97.45 | AAI-based (high-confidence) | 0 |
| S5_vs_k141_121419\|\|full | 37918 | No | NA | 51 | 7 | 0 | High-quality | High-quality | 98.27 | AAI-based (high-confidence) | 0 |
| S5_vs_k141_27918\|\|full | 43293 | No | NA | 61 | 13 | 1 | High-quality | High-quality | 100 | AAI-based (medium-confidence) | 0 |
| S5_vs_k141_82785\|\|full | 2289 | No | NA | 3 | 0 | 0 | Not-determined | Genome-fragment | NA | NA | 0 |
| S5_vs_k141_145578\|\|full | 39974 | No | NA | 57 | 23 | 1 | High-quality | High-quality | 100 | AAI-based (high-confidence) | 0 |
| S5_vs_k141_145581\|\|full | 38965 | No | NA | 67 | 19 | 1 | High-quality | High-quality | 94.18 | AAI-based (medium-confidence) | 0 |
| S5_vs_k141_145582\|\|full | 57891 | No | NA | 82 | 28 | 0 | High-quality | High-quality | 100 | AAI-based (high-confidence) | 0 |
| S5_vs_k141_145590\|\|full | 39101 | No | NA | 53 | 29 | 0 | High-quality | High-quality | 95.17 | AAI-based (high-confidence) | 0 |
| S5_vs_k141_145591\|\|full | 43141 | No | NA | 51 | 14 | 0 | High-quality | High-quality | 95.38 | AAI-based (high-confidence) | 0 |
| S5_vs_k141_145592\|\|full | 38088 | No | NA | 57 | 14 | 1 | Medium-quality | Genome-fragment | 81.4 | AAI-based (medium-confidence) | 0 |
| S5_vs_k141_145593\|\|full | 14496 | No | NA | 20 | 2 | 0 | High-quality | High-quality | 99.49 | AAI-based (high-confidence) | 0 |
| S1_extracted_k141_139383 | 1831 | No | NA | 3 | 0 | 0 | Low-quality | Genome-fragment | 2.32 | AAI-based (high-confidence) | 0 |
| S5_vs_k141_145597\|\|full | 32371 | No | NA | 52 | 20 | 2 | High-quality | High-quality | 94.87 | AAI-based (medium-confidence) | 0 |
| S5_vs_k141_145598\|\|full | 40137 | No | NA | 60 | 28 | 0 | Medium-quality | Genome-fragment | 87.35 | AAI-based (medium-confidence) | 0 |
| S5_vs_k141_145599\|\|full | 42481 | No | NA | 57 | 31 | 2 | High-quality | High-quality | 100 | AAI-based (high-confidence) | 0 |
| S5_vs_k141_145601\|\|full | 41896 | No | NA | 62 | 29 | 0 | High-quality | High-quality | 95.62 | AAI-based (high-confidence) | 0 |
| S5_vs_k141_145602\|\|full | 42207 | No | NA | 64 | 28 | 3 | High-quality | High-quality | 100 | AAI-based (high-confidence) | 0 |
| S5_vs_k141_145605\|\|full | 14670 | No | NA | 16 | 5 | 0 | Medium-quality | Genome-fragment | 64.18 | AAI-based (medium-confidence) | 0 |
| S5_vs_k141_145611\|\|full | 40032 | No | NA | 60 | 26 | 0 | High-quality | High-quality | 92.74 | AAI-based (high-confidence) | 0 |
| S5_vs_k141_145621\|\|full | 2783 | No | NA | 3 | 0 | 0 | Not-determined | Genome-fragment | NA | NA | 0 |
| S16_vs_132410\|\|full | 1831 | No | NA | 2 | 0 | 0 | Low-quality | Genome-fragment | 1.8 | AAI-based (medium-confidence) | 0 |
| S5_vs_k141_145633\|\|full | 9979 | No | NA | 15 | 1 | 0 | Medium-quality | Genome-fragment | 65.2 | AAI-based (medium-confidence) | 0 |
| S5_vs_k141_133569\|\|full | 1109 | No | NA | 2 | 0 | 1 | Not-determined | Genome-fragment | NA | NA | 0 |
| S5_vs_k141_36438\|\|full | 4957 | No | NA | 6 | 0 | 5 | Not-determined | Genome-fragment | NA | NA | 0 |
| S8_vs_k141_196075\|\|full | 1831 | No | NA | 3 | 1 | 0 | Low-quality | Genome-fragment | 4.09 | AAI-based (high-confidence) | 0 |
| S5_vs_k141_97162\|\|full | 2164 | No | NA | 2 | 0 | 1 | Not-determined | Genome-fragment | NA | NA | 0 |
| S9_vs_k141_2504\|\|full | 1832 | No | NA | 3 | 0 | 1 | Low-quality | Genome-fragment | 1.3 | AAI-based (high-confidence) | 0 |
| S3_vs_k141_66789\|\|full | 1832 | No | NA | 3 | 0 | 0 | Low-quality | Genome-fragment | 1.34 | AAI-based (medium-confidence) | 0 |
| S5_vs_k141_6084\|\|full | 1520 | No | NA | 3 | 0 | 1 | Not-determined | Genome-fragment | NA | NA | 0 |
| S7_extracted_44447 | 1832 | No | NA | 4 | 1 | 0 | Low-quality | Genome-fragment | 3.1 | AAI-based (high-confidence) | 0 |
| S7_vs_108304\|\|full | 1832 | No | NA | 3 | 0 | 0 | Low-quality | Genome-fragment | 4.28 | AAI-based (medium-confidence) | 0 |
| S5_vs_k141_18251\|\|full | 2337 | No | NA | 5 | 0 | 0 | Not-determined | Genome-fragment | NA | NA | 0 |
| S16_extracted_33624 | 1832 | No | NA | 2 | 0 | 0 | Low-quality | Genome-fragment | 1.64 | AAI-based (high-confidence) | 0 |
| S15_vs_74371\|\|full | 1832 | No | NA | 5 | 3 | 0 | Low-quality | Genome-fragment | 3.57 | AAI-based (high-confidence) | 0 |
| S5_vs_k141_69\|\|full | 2601 | No | NA | 2 | 0 | 2 | Not-determined | Genome-fragment | NA | NA | 0 |
| S5_vs_k141_91134\|\|full | 1421 | No | NA | 2 | 0 | 1 | Not-determined | Genome-fragment | NA | NA | 0 |
| S5_vs_k141_48611\|\|full | 1274 | No | NA | 3 | 0 | 1 | Not-determined | Genome-fragment | NA | NA | 0 |
| S5_vs_k141_6091\|\|full | 1102 | No | NA | 3 | 0 | 0 | Not-determined | Genome-fragment | NA | NA | 0 |
| S8_vs_k141_158146\|\|full | 1832 | No | NA | 4 | 0 | 0 | Low-quality | Genome-fragment | 3.61 | AAI-based (high-confidence) | 0 |
| S17_extracted_k141_83232 | 1833 | No | NA | 2 | 2 | 0 | Low-quality | Genome-fragment | 6.04 | AAI-based (high-confidence) | 0 |
| S5_vs_k141_109324\|\|full | 2004 | No | NA | 5 | 0 | 1 | Not-determined | Genome-fragment | NA | NA | 0 |
| S5_vs_k141_60798\|\|full | 6912 | No | NA | 7 | 0 | 3 | Not-determined | Genome-fragment | NA | NA | 0 |
| S5_vs_k141_12185\|\|full | 5351 | No | NA | 2 | 0 | 2 | Not-determined | Genome-fragment | NA | NA | 0 |
| S5_vs_k141_18299\|\|full | 1008 | No | NA | 2 | 0 | 1 | Not-determined | Genome-fragment | NA | NA | 0 |
| S5_vs_k141_127546\|\|full | 2947 | No | NA | 6 | 0 | 2 | Not-determined | Genome-fragment | NA | NA | 0 |
| S9_vs_k141_49612\|\|full | 1833 | No | NA | 2 | 0 | 2 | Low-quality | Genome-fragment | 1.72 | AAI-based (medium-confidence) | 0 |
| S5_vs_k141_121532\|\|full | 3464 | No | NA | 3 | 0 | 1 | Not-determined | Genome-fragment | NA | NA | 0 |
| S11_extracted_k141_36497 | 1833 | No | NA | 2 | 2 | 0 | Low-quality | Genome-fragment | 2.99 | HMM-based (lower-bound) | 0 |
| S7_extracted_21932 | 1833 | No | NA | 4 | 0 | 0 | Low-quality | Genome-fragment | 4.7 | AAI-based (high-confidence) | 0 |
| S5_vs_k141_127558\|\|full | 1009 | No | NA | 3 | 0 | 0 | Not-determined | Genome-fragment | NA | NA | 0 |
| S5_vs_k141_18322\|\|full | 4647 | No | NA | 8 | 0 | 0 | Not-determined | Genome-fragment | NA | NA | 0 |
| S5_vs_k141_103332\|\|full | 1045 | No | NA | 2 | 0 | 0 | Not-determined | Genome-fragment | NA | NA | 0 |
| S16_vs_139934\|\|full | 1833 | No | NA | 4 | 1 | 0 | Low-quality | Genome-fragment | 3.79 | AAI-based (medium-confidence) | 0 |
| S12_extracted_k141_107204 | 1833 | No | NA | 2 | 0 | 0 | Low-quality | Genome-fragment | 2.09 | AAI-based (medium-confidence) | 0 |
| S5_vs_k141_139743\|\|full | 1396 | No | NA | 3 | 0 | 1 | Not-determined | Genome-fragment | NA | NA | 0 |
| S5_vs_k141_54728\|\|full | 2431 | No | NA | 5 | 0 | 1 | Not-determined | Genome-fragment | NA | NA | 0 |
| S5_vs_k141_85065\|\|full | 57832 | No | NA | 93 | 34 | 0 | High-quality | High-quality | 99.94 | AAI-based (high-confidence) | 0 |
| S5_vs_k141_36514\|\|full | 44567 | No | NA | 33 | 0 | 20 | Not-determined | Genome-fragment | NA | NA | 0 |
| S5_vs_k141_54734\|\|full | 2038 | No | NA | 5 | 0 | 0 | Not-determined | Genome-fragment | NA | NA | 0 |
| S5_vs_k141_85067\|\|full | 5903 | No | NA | 6 | 0 | 0 | Not-determined | Genome-fragment | NA | NA | 0 |
| S12_vs_k141_134894\|\|full | 1833 | No | NA | 4 | 0 | 1 | Low-quality | Genome-fragment | 3.71 | AAI-based (medium-confidence) | 0 |
| S6_vs_k141_3545\|\|full | 1833 | No | NA | 5 | 1 | 0 | Low-quality | Genome-fragment | 3.01 | AAI-based (high-confidence) | 0 |
| S8_vs_k141_19011\|\|full | 1833 | No | NA | 3 | 2 | 0 | Low-quality | Genome-fragment | 4.33 | AAI-based (high-confidence) | 0 |
| S11_vs_k141_18428\|\|full | 1834 | No | NA | 4 | 0 | 0 | Low-quality | Genome-fragment | 4.11 | AAI-based (medium-confidence) | 0 |
| S5_vs_k141_60882\|\|full | 1483 | No | NA | 3 | 0 | 2 | Not-determined | Genome-fragment | NA | NA | 0 |
| S5_vs_k141_103404\|\|full | 2122 | No | NA | 5 | 0 | 4 | Not-determined | Genome-fragment | NA | NA | 0 |
| S5_vs_k141_6229\|\|full | 2287 | No | NA | 3 | 0 | 2 | Not-determined | Genome-fragment | NA | NA | 0 |
| S5_vs_k141_60905\|\|full | 2208 | No | NA | 2 | 0 | 2 | Not-determined | Genome-fragment | NA | NA | 0 |
| S5_vs_k141_60921\|\|full | 1200 | No | NA | 2 | 0 | 1 | Not-determined | Genome-fragment | NA | NA | 0 |
| S5_vs_k141_97253\|\|full | 1867 | No | NA | 3 | 0 | 1 | Not-determined | Genome-fragment | NA | NA | 0 |
| S5_vs_k141_127618\|\|full | 3371 | No | NA | 2 | 0 | 2 | Not-determined | Genome-fragment | NA | NA | 0 |
| S5_vs_k141_36563\|\|full | 3039 | No | NA | 6 | 0 | 0 | Not-determined | Genome-fragment | NA | NA | 0 |
| S5_vs_k141_30453\|\|full | 2053 | No | NA | 4 | 0 | 0 | Not-determined | Genome-fragment | NA | NA | 0 |
| S7_vs_44030\|\|full | 1834 | No | NA | 3 | 0 | 0 | Low-quality | Genome-fragment | 4.37 | AAI-based (high-confidence) | 0 |
| S5_vs_k141_139777\|\|full | 2041 | No | NA | 3 | 0 | 2 | Not-determined | Genome-fragment | NA | NA | 0 |
| S5_vs_k141_66839\|\|full | 4838 | No | NA | 6 | 0 | 2 | Not-determined | Genome-fragment | NA | NA | 0 |
| S16_vs_94519\|\|full | 1834 | No | NA | 3 | 0 | 0 | Low-quality | Genome-fragment | 3.9 | AAI-based (medium-confidence) | 0 |
| S5_vs_k141_48696\|\|full | 2472 | No | NA | 4 | 0 | 1 | Not-determined | Genome-fragment | NA | NA | 0 |
| S5_vs_k141_109453\|\|full | 4170 | No | NA | 5 | 0 | 0 | Not-determined | Genome-fragment | NA | NA | 0 |
| S5_vs_k141_121592\|\|full | 1130 | No | NA | 2 | 0 | 1 | Not-determined | Genome-fragment | NA | NA | 0 |
| S10_vs_80547\|\|full | 1834 | No | NA | 3 | 0 | 1 | Low-quality | Genome-fragment | 0.89 | AAI-based (medium-confidence) | 0 |
| S10_vs_108735\|\|full | 1834 | No | NA | 5 | 2 | 0 | Low-quality | Genome-fragment | 2.23 | AAI-based (medium-confidence) | 0 |
| S5_vs_k141_72979\|\|full | 2128 | No | NA | 4 | 0 | 1 | Not-determined | Genome-fragment | NA | NA | 0 |
| S12_extracted_k141_19818 | 1834 | No | NA | 3 | 2 | 0 | Low-quality | Genome-fragment | 4.92 | AAI-based (high-confidence) | 0 |
| S15_vs_43524\|\|full | 1834 | No | NA | 3 | 0 | 0 | Low-quality | Genome-fragment | 3.49 | AAI-based (high-confidence) | 0 |
| S5_vs_k141_60952\|\|full | 12369 | No | NA | 9 | 0 | 8 | Not-determined | Genome-fragment | NA | NA | 0 |
| S5_vs_k141_18376\|\|full | 5486 | No | NA | 5 | 0 | 4 | Not-determined | Genome-fragment | NA | NA | 0 |
| S4_extracted_11649 | 1834 | No | NA | 1 | 0 | 0 | Low-quality | Genome-fragment | 4.11 | AAI-based (high-confidence) | 0 |
| S4_extracted_26505 | 1834 | No | NA | 7 | 6 | 0 | Low-quality | Genome-fragment | 5.98 | AAI-based (high-confidence) | 0 |
| S1_vs_k141_139820\|\|full | 1835 | No | NA | 4 | 0 | 0 | Low-quality | Genome-fragment | 3.86 | AAI-based (high-confidence) | 0 |
| S5_vs_k141_42782\|\|full | 3242 | No | NA | 4 | 0 | 0 | Not-determined | Genome-fragment | NA | NA | 0 |
| S3_vs_k141_28216\|\|full | 1835 | No | NA | 3 | 3 | 0 | Low-quality | Genome-fragment | 4.32 | AAI-based (medium-confidence) | 0 |
| S5_extracted_k141_110184 | 1835 | No | NA | 5 | 0 | 0 | Low-quality | Genome-fragment | 2.48 | AAI-based (medium-confidence) | 0 |
| S5_extracted_k141_144861 | 1835 | No | NA | 1 | 0 | 1 | Low-quality | Genome-fragment | 2.94 | AAI-based (medium-confidence) | 0 |
| S7_vs_105809\|\|full | 1835 | No | NA | 6 | 3 | 0 | Low-quality | Genome-fragment | 4.8 | AAI-based (medium-confidence) | 0 |
| S7_vs_107280\|\|full | 1835 | No | NA | 2 | 1 | 0 | Low-quality | Genome-fragment | 1.15 | HMM-based (lower-bound) | 0 |
| S12_vs_k141_11131\|\|full | 1835 | No | NA | 2 | 0 | 2 | Low-quality | Genome-fragment | 4.79 | AAI-based (medium-confidence) | 0 |
| S12_vs_k141_161154\|\|full | 1835 | No | NA | 5 | 1 | 0 | Low-quality | Genome-fragment | 5.24 | AAI-based (high-confidence) | 0 |
| S5_vs_k141_30490\|\|full | 56501 | Yes | 33176 | 53 | 4 | 14 | Medium-quality | Genome-fragment | 86.22 | AAI-based (high-confidence) | 41.28 |
| S8_vs_k141_3418\|\|lt2gene | 1835 | No | NA | 2 | 1 | 0 | Low-quality | Genome-fragment | 4.54 | AAI-based (medium-confidence) | 0 |
| S14_extracted_k141_15592 | 1836 | No | NA | 2 | 2 | 0 | Low-quality | Genome-fragment | 6.14 | AAI-based (high-confidence) | 0 |
| S5_vs_k141_103509\|\|full | 2338 | No | NA | 5 | 0 | 0 | Not-determined | Genome-fragment | NA | NA | 0 |
| S14_vs_k141_1158\|\|full | 1836 | No | NA | 2 | 0 | 0 | Low-quality | Genome-fragment | 1 | AAI-based (medium-confidence) | 0 |
| S14_vs_k141_84071\|\|full | 1836 | No | NA | 6 | 0 | 0 | Low-quality | Genome-fragment | 4.04 | AAI-based (high-confidence) | 0 |
| S3_extracted_k141_20385 | 1836 | No | NA | 5 | 1 | 0 | Low-quality | Genome-fragment | 1.76 | AAI-based (high-confidence) | 0 |
| S5_vs_k141_30184\|\|lt2gene | 1836 | No | NA | 2 | 0 | 0 | Low-quality | Genome-fragment | 3.89 | AAI-based (medium-confidence) | 0 |
| S7_vs_3701\|\|full | 1836 | No | NA | 3 | 0 | 1 | Low-quality | Genome-fragment | 0.53 | AAI-based (medium-confidence) | 0 |
| S16_vs_131604\|\|full | 1836 | No | NA | 2 | 0 | 1 | Low-quality | Genome-fragment | 1.14 | AAI-based (medium-confidence) | 0 |
| S5_vs_k141_127662\|\|lt2gene | 1837 | No | NA | 2 | 2 | 0 | Low-quality | Genome-fragment | 4.63 | AAI-based (high-confidence) | 0 |
| S7_vs_92145\|\|lt2gene | 1837 | No | NA | 2 | 1 | 0 | Low-quality | Genome-fragment | 4.46 | AAI-based (high-confidence) | 0 |
| S5_vs_k141_61018\|\|full | 2450 | No | NA | 3 | 0 | 1 | Not-determined | Genome-fragment | NA | NA | 0 |
| S5_vs_k141_18510\|\|full | 1675 | No | NA | 3 | 0 | 0 | Not-determined | Genome-fragment | NA | NA | 0 |
| S16_vs_30293\|\|full | 1837 | No | NA | 3 | 0 | 1 | Low-quality | Genome-fragment | 4.12 | AAI-based (high-confidence) | 0 |
| S10_vs_41112\|\|full | 1837 | No | NA | 5 | 2 | 0 | Low-quality | Genome-fragment | 4.25 | AAI-based (medium-confidence) | 0 |
| S5_vs_k141_103594\|\|full | 1033 | No | NA | 2 | 0 | 1 | Not-determined | Genome-fragment | NA | NA | 0 |
| S11_vs_k141_71913\|\|full | 1838 | No | NA | 3 | 0 | 0 | Low-quality | Genome-fragment | 4.3 | AAI-based (high-confidence) | 0 |
| S1_vs_k141_19194\|\|full | 1838 | No | NA | 4 | 0 | 0 | Low-quality | Genome-fragment | 4.66 | AAI-based (medium-confidence) | 0 |
| S5_vs_k141_109484\|\|full | 1515 | No | NA | 3 | 0 | 0 | Not-determined | Genome-fragment | NA | NA | 0 |
| S5_vs_k141_115531\|\|full | 1045 | No | NA | 3 | 0 | 0 | Not-determined | Genome-fragment | NA | NA | 0 |
| S3_vs_k141_72462\|\|full | 1838 | No | NA | 5 | 0 | 0 | Low-quality | Genome-fragment | 5.53 | AAI-based (medium-confidence) | 0 |
| S3_vs_k141_74069\|\|full | 1838 | No | NA | 2 | 1 | 0 | Low-quality | Genome-fragment | 2.82 | AAI-based (high-confidence) | 0 |
| S3_vs_k141_124055\|\|full | 1838 | No | NA | 8 | 2 | 0 | Low-quality | Genome-fragment | 5.11 | AAI-based (high-confidence) | 0 |
| S5_vs_k141_103820\|\|full | 1838 | No | NA | 5 | 2 | 0 | Low-quality | Genome-fragment | 3.34 | AAI-based (high-confidence) | 0 |
| S5_vs_k141_143298\|\|full | 1838 | No | NA | 3 | 0 | 2 | Low-quality | Genome-fragment | 1.78 | AAI-based (high-confidence) | 0 |
| S5_vs_k141_138036\|\|full | 1838 | No | NA | 3 | 1 | 0 | Low-quality | Genome-fragment | 1.19 | HMM-based (lower-bound) | 0 |
| S10_vs_18009\|\|full | 1838 | No | NA | 2 | 0 | 1 | Low-quality | Genome-fragment | 3.71 | AAI-based (high-confidence) | 0 |
| S10_vs_60579\|\|full | 1838 | No | NA | 3 | 0 | 1 | Low-quality | Genome-fragment | 2.75 | AAI-based (medium-confidence) | 0 |
| S8_vs_k141_119346\|\|full | 1838 | No | NA | 5 | 0 | 0 | Low-quality | Genome-fragment | 4.3 | AAI-based (medium-confidence) | 0 |
| S13_vs_111363\|\|full | 1839 | No | NA | 3 | 0 | 2 | Low-quality | Genome-fragment | 0.84 | AAI-based (medium-confidence) | 0 |
| S3_extracted_k141_76777 | 1839 | No | NA | 3 | 0 | 0 | Low-quality | Genome-fragment | 1.26 | AAI-based (high-confidence) | 0 |
| S7_vs_91588\|\|full | 1839 | No | NA | 3 | 0 | 0 | Low-quality | Genome-fragment | 5.4 | AAI-based (high-confidence) | 0 |
| S17_vs_k141_119013\|\|full | 1840 | No | NA | 7 | 0 | 0 | Low-quality | Genome-fragment | 3.16 | AAI-based (high-confidence) | 0 |
| S17_vs_k141_167464\|\|full | 1840 | No | NA | 3 | 1 | 0 | Low-quality | Genome-fragment | 2.41 | HMM-based (lower-bound) | 0 |
| S5_vs_k141_127770\|\|full | 1521 | No | NA | 3 | 0 | 0 | Not-determined | Genome-fragment | NA | NA | 0 |
| S9_vs_k141_75502\|\|full | 1840 | No | NA | 3 | 0 | 2 | Low-quality | Genome-fragment | 3.22 | AAI-based (medium-confidence) | 0 |
| S5_vs_k141_6478\|\|full | 41423 | No | NA | 68 | 21 | 0 | High-quality | High-quality | 98 | AAI-based (high-confidence) | 0 |
| S5_vs_k141_61077\|\|full | 2461 | No | NA | 3 | 0 | 0 | Not-determined | Genome-fragment | NA | NA | 0 |
| S5_vs_k141_36630\|\|full | 5093 | No | NA | 5 | 0 | 4 | Not-determined | Genome-fragment | NA | NA | 0 |
| S11_vs_k141_80053\|\|full | 1840 | No | NA | 3 | 2 | 0 | Low-quality | Genome-fragment | 3.37 | AAI-based (medium-confidence) | 0 |
| S11_vs_k141_45063\|\|full | 1840 | No | NA | 2 | 0 | 0 | Low-quality | Genome-fragment | 1.79 | AAI-based (medium-confidence) | 0 |
| S13_extracted_43788 | 1840 | No | NA | 3 | 0 | 1 | Low-quality | Genome-fragment | 2.97 | AAI-based (medium-confidence) | 0 |
| S5_vs_k141_66996\|\|full | 1636 | No | NA | 3 | 0 | 2 | Not-determined | Genome-fragment | NA | NA | 0 |
| S5_vs_k141_399\|\|full | 3739 | No | NA | 7 | 0 | 0 | Not-determined | Genome-fragment | NA | NA | 0 |
| S14_vs_k141_6013\|\|full | 1840 | No | NA | 4 | 0 | 0 | Low-quality | Genome-fragment | 4.04 | AAI-based (medium-confidence) | 0 |
| S3_vs_k141_88526\|\|full | 1840 | No | NA | 2 | 0 | 1 | Low-quality | Genome-fragment | 2.86 | AAI-based (medium-confidence) | 0 |
| S3_vs_k141_41075\|\|full | 1840 | No | NA | 3 | 0 | 1 | Low-quality | Genome-fragment | 1.66 | AAI-based (medium-confidence) | 0 |
| S5_vs_k141_30610\|\|full | 1600 | No | NA | 2 | 0 | 0 | Not-determined | Genome-fragment | NA | NA | 0 |
| S5_vs_k141_454\|\|full | 1157 | No | NA | 2 | 0 | 1 | Not-determined | Genome-fragment | NA | NA | 0 |
| S5_vs_k141_63218\|\|full | 1840 | No | NA | 3 | 0 | 1 | Low-quality | Genome-fragment | 2.25 | AAI-based (medium-confidence) | 0 |
| S5_vs_k141_121927\|\|full | 1417 | No | NA | 2 | 0 | 0 | Not-determined | Genome-fragment | NA | NA | 0 |
| S7_extracted_16137 | 1840 | No | NA | 3 | 0 | 2 | Low-quality | Genome-fragment | 1.55 | AAI-based (high-confidence) | 0 |
| S5_vs_k141_115659\|\|full | 6211 | No | NA | 3 | 0 | 3 | Not-determined | Genome-fragment | NA | NA | 0 |
| S5_vs_k141_48876\|\|full | 1197 | No | NA | 3 | 0 | 0 | Not-determined | Genome-fragment | NA | NA | 0 |
| S16_extracted_16649 | 1840 | No | NA | 5 | 2 | 0 | Low-quality | Genome-fragment | 3.32 | AAI-based (high-confidence) | 0 |
| S5_vs_k141_73119\|\|full | 4649 | No | NA | 6 | 0 | 3 | Not-determined | Genome-fragment | NA | NA | 0 |
| S5_vs_k141_30666\|\|full | 7450 | No | NA | 5 | 0 | 2 | Not-determined | Genome-fragment | NA | NA | 0 |
| S17_vs_k141_24648\|\|full | 1841 | No | NA | 2 | 0 | 0 | Low-quality | Genome-fragment | 4.13 | AAI-based (high-confidence) | 0 |
| S5_vs_k141_85360\|\|full | 1038 | No | NA | 3 | 0 | 0 | Not-determined | Genome-fragment | NA | NA | 0 |
| S5_vs_k141_103764\|\|full | 16178 | No | NA | 18 | 0 | 9 | Not-determined | Genome-fragment | NA | NA | 0 |
| S9_extracted_k141_48938 | 1841 | No | NA | 1 | 1 | 0 | Low-quality | Genome-fragment | 2.69 | HMM-based (lower-bound) | 0 |
| S13_vs_114837\|\|lt2gene | 1841 | No | NA | 2 | 0 | 1 | Low-quality | Genome-fragment | 0.52 | AAI-based (medium-confidence) | 0 |
| S5_vs_k141_36671\|\|full | 2043 | No | NA | 4 | 0 | 0 | Not-determined | Genome-fragment | NA | NA | 0 |
| S1_extracted_k141_35843 | 1841 | No | NA | 1 | 0 | 0 | Low-quality | Genome-fragment | 1.87 | AAI-based (high-confidence) | 0 |
| S5_vs_k141_48919\|\|full | 1406 | No | NA | 2 | 0 | 0 | Not-determined | Genome-fragment | NA | NA | 0 |
| S7_extracted_107029 | 1841 | No | NA | 1 | 0 | 0 | Low-quality | Genome-fragment | 0.8 | AAI-based (medium-confidence) | 0 |
| S16_vs_115763\|\|full | 1841 | No | NA | 3 | 1 | 0 | Low-quality | Genome-fragment | 4.71 | AAI-based (high-confidence) | 0 |
| S5_vs_k141_48943\|\|full | 1142 | No | NA | 2 | 0 | 0 | Not-determined | Genome-fragment | NA | NA | 0 |
| S13_extracted_38635 | 1842 | No | NA | 2 | 0 | 1 | Low-quality | Genome-fragment | 0.93 | AAI-based (medium-confidence) | 0 |
| S5_vs_k141_79159\|\|full | 1587 | No | NA | 3 | 0 | 1 | Not-determined | Genome-fragment | NA | NA | 0 |
| S14_extracted_k141_67011 | 1842 | No | NA | 2 | 2 | 0 | Low-quality | Genome-fragment | 5.98 | AAI-based (high-confidence) | 0 |
| S5_vs_k141_109527\|\|full | 142384 | No | NA | 133 | 2 | 28 | Medium-quality | Genome-fragment | 76.5 | HMM-based (lower-bound) | 0 |
| S14_vs_k141_87103\|\|full | 1842 | No | NA | 6 | 1 | 0 | Low-quality | Genome-fragment | 1.1 | AAI-based (medium-confidence) | 0 |
| S7_vs_136399\|\|full | 1842 | No | NA | 6 | 0 | 0 | Low-quality | Genome-fragment | 4.96 | AAI-based (high-confidence) | 0 |
| S16_vs_134556\|\|full | 1842 | No | NA | 5 | 0 | 0 | Low-quality | Genome-fragment | 5.14 | AAI-based (high-confidence) | 0 |
| S16_vs_147705\|\|full | 1842 | No | NA | 3 | 1 | 0 | Low-quality | Genome-fragment | 2.09 | HMM-based (lower-bound) | 0 |
| S10_vs_91561\|\|full | 1842 | No | NA | 3 | 2 | 0 | Low-quality | Genome-fragment | 5.28 | AAI-based (medium-confidence) | 0 |
| S15_extracted_30515 | 1842 | No | NA | 5 | 1 | 0 | Low-quality | Genome-fragment | 2.35 | AAI-based (high-confidence) | 0 |
| S8_vs_k141_123360\|\|full | 1842 | No | NA | 3 | 0 | 0 | Low-quality | Genome-fragment | 4.35 | AAI-based (high-confidence) | 0 |
| S14_extracted_k141_37400 | 1843 | No | NA | 5 | 0 | 2 | Low-quality | Genome-fragment | 4.03 | AAI-based (medium-confidence) | 0 |
| S14_extracted_k141_27591 | 1843 | No | NA | 4 | 1 | 0 | Low-quality | Genome-fragment | 4.34 | AAI-based (high-confidence) | 0 |
| S14_vs_k141_108421\|\|full | 1843 | No | NA | 4 | 0 | 0 | Low-quality | Genome-fragment | 4.92 | AAI-based (medium-confidence) | 0 |
| S5_vs_k141_61174\|\|full | 2818 | No | NA | 3 | 0 | 1 | Not-determined | Genome-fragment | NA | NA | 0 |
| S5_vs_k141_97483\|\|full | 1924 | No | NA | 2 | 0 | 1 | Not-determined | Genome-fragment | NA | NA | 0 |
| S3_vs_k141_101358\|\|full | 1843 | No | NA | 3 | 0 | 1 | Low-quality | Genome-fragment | 0.81 | AAI-based (high-confidence) | 0 |
| S5_vs_k141_73137\|\|full | 1840 | No | NA | 4 | 0 | 2 | Not-determined | Genome-fragment | NA | NA | 0 |
| S10_extracted_109721 | 1843 | No | NA | 2 | 0 | 0 | Low-quality | Genome-fragment | 2.8 | AAI-based (medium-confidence) | 0 |
| S5_vs_k141_661\|\|full | 4580 | No | NA | 5 | 0 | 2 | Not-determined | Genome-fragment | NA | NA | 0 |
| S5_vs_k141_6582\|\|full | 1988 | No | NA | 2 | 0 | 1 | Not-determined | Genome-fragment | NA | NA | 0 |
| S10_vs_109134\|\|full | 1843 | No | NA | 3 | 0 | 1 | Low-quality | Genome-fragment | 2.41 | AAI-based (high-confidence) | 0 |
| S5_vs_k141_97522\|\|full | 2176 | No | NA | 3 | 0 | 0 | Not-determined | Genome-fragment | NA | NA | 0 |
| S5_vs_k141_49011\|\|full | 33303 | No | NA | 57 | 20 | 0 | High-quality | High-quality | 95.31 | AAI-based (high-confidence) | 0 |
| S5_vs_k141_18599\|\|full | 4665 | No | NA | 8 | 0 | 2 | Not-determined | Genome-fragment | NA | NA | 0 |
| S12_extracted_k141_116660 | 1843 | No | NA | 5 | 0 | 0 | Low-quality | Genome-fragment | 5.16 | AAI-based (high-confidence) | 0 |
| S15_vs_11949\|\|full | 1843 | No | NA | 4 | 0 | 0 | Low-quality | Genome-fragment | 1.11 | AAI-based (high-confidence) | 0 |
| S5_vs_k141_30842\|\|full | 2770 | No | NA | 2 | 0 | 2 | Not-determined | Genome-fragment | NA | NA | 0 |
| S8_extracted_k141_31115 | 1843 | No | NA | 5 | 1 | 0 | Low-quality | Genome-fragment | 11.85 | AAI-based (high-confidence) | 0 |
| S8_vs_k141_61154\|\|full | 1843 | No | NA | 5 | 1 | 0 | Low-quality | Genome-fragment | 4.43 | AAI-based (high-confidence) | 0 |
| S17_vs_k141_106862\|\|full | 1844 | No | NA | 6 | 0 | 0 | Low-quality | Genome-fragment | 5.15 | AAI-based (high-confidence) | 0 |
| S11_vs_k141_97641\|\|full | 1844 | No | NA | 4 | 0 | 0 | Low-quality | Genome-fragment | 4.55 | AAI-based (medium-confidence) | 0 |
| S1_extracted_k141_42622 | 1844 | No | NA | 1 | 0 | 0 | Low-quality | Genome-fragment | 1.03 | AAI-based (high-confidence) | 0 |
| S5_vs_k141_30886\|\|full | 1006 | No | NA | 3 | 0 | 0 | Not-determined | Genome-fragment | NA | NA | 0 |
| S1_extracted_k141_70256 | 1844 | No | NA | 6 | 0 | 0 | Low-quality | Genome-fragment | 4.16 | AAI-based (high-confidence) | 0 |
| S5_vs_k141_789\|\|full | 2982 | No | NA | 4 | 0 | 2 | Not-determined | Genome-fragment | NA | NA | 0 |
| S5_vs_k141_43207\|\|full | 37982 | No | NA | 49 | 15 | 0 | High-quality | High-quality | 92.44 | AAI-based (high-confidence) | 0 |
| S5_vs_k141_85432\|\|full | 12722 | No | NA | 9 | 0 | 0 | Not-determined | Genome-fragment | NA | NA | 0 |
| S5_vs_k141_36720\|\|full | 1064 | No | NA | 2 | 0 | 1 | Not-determined | Genome-fragment | NA | NA | 0 |
| S5_vs_k141_85442\|\|full | 1171 | No | NA | 3 | 0 | 0 | Not-determined | Genome-fragment | NA | NA | 0 |
| S5_vs_k141_30918\|\|full | 2323 | No | NA | 3 | 0 | 1 | Not-determined | Genome-fragment | NA | NA | 0 |
| S5_vs_k141_140040\|\|full | 2256 | No | NA | 2 | 0 | 2 | Not-determined | Genome-fragment | NA | NA | 0 |
| S1_vs_k141_5039\|\|full | 1844 | No | NA | 5 | 1 | 0 | Low-quality | Genome-fragment | 3.18 | HMM-based (lower-bound) | 0 |
| S3_extracted_k141_114111 | 1844 | No | NA | 1 | 0 | 0 | Low-quality | Genome-fragment | 4.63 | AAI-based (medium-confidence) | 0 |
| S5_extracted_k141_86473 | 1844 | No | NA | 6 | 1 | 1 | Low-quality | Genome-fragment | 2.9 | AAI-based (medium-confidence) | 0 |
| S5_vs_k141_127979\|\|full | 1309 | No | NA | 2 | 0 | 0 | Not-determined | Genome-fragment | NA | NA | 0 |
| S5_vs_k141_139684\|\|full | 1844 | No | NA | 6 | 1 | 0 | Low-quality | Genome-fragment | 3.03 | AAI-based (medium-confidence) | 0 |
| S16_vs_26169\|\|full | 1844 | No | NA | 4 | 0 | 0 | Low-quality | Genome-fragment | 3.59 | AAI-based (high-confidence) | 0 |
| S16_vs_167709\|\|full | 1844 | No | NA | 4 | 1 | 0 | Low-quality | Genome-fragment | 4.25 | AAI-based (high-confidence) | 0 |
| S5_vs_k141_49077\|\|full | 1832 | No | NA | 2 | 0 | 0 | Not-determined | Genome-fragment | NA | NA | 0 |
| S5_vs_k141_833\|\|full | 2111 | No | NA | 3 | 0 | 0 | Not-determined | Genome-fragment | NA | NA | 0 |
| S10_vs_101146\|\|full | 1844 | No | NA | 3 | 0 | 2 | Low-quality | Genome-fragment | 2.28 | AAI-based (medium-confidence) | 0 |
| S8_vs_k141_74798\|\|full | 1844 | No | NA | 2 | 0 | 0 | Low-quality | Genome-fragment | 3.84 | AAI-based (medium-confidence) | 0 |
| S9_extracted_k141_4275 | 1845 | No | NA | 1 | 0 | 1 | Low-quality | Genome-fragment | 1.01 | AAI-based (medium-confidence) | 0 |
| S3_vs_k141_83838\|\|full | 1845 | No | NA | 7 | 0 | 0 | Low-quality | Genome-fragment | 3.25 | AAI-based (high-confidence) | 0 |
| S7_vs_18159\|\|full | 1845 | No | NA | 5 | 1 | 0 | Low-quality | Genome-fragment | 3.08 | AAI-based (medium-confidence) | 0 |
| S5_vs_k141_12417\|\|full | 3176 | No | NA | 5 | 0 | 0 | Not-determined | Genome-fragment | NA | NA | 0 |
| S5_vs_k141_848\|\|full | 6135 | No | NA | 6 | 0 | 4 | Not-determined | Genome-fragment | NA | NA | 0 |
| S7_vs_96906\|\|full | 1845 | No | NA | 3 | 1 | 0 | Low-quality | Genome-fragment | 3.15 | AAI-based (medium-confidence) | 0 |
| S16_extracted_45418 | 1845 | No | NA | 3 | 1 | 0 | Low-quality | Genome-fragment | 3.08 | AAI-based (high-confidence) | 0 |
| S5_vs_k141_18703\|\|full | 5840 | No | NA | 6 | 0 | 0 | Not-determined | Genome-fragment | NA | NA | 0 |
| S16_vs_124965\|\|lt2gene | 1845 | No | NA | 2 | 1 | 0 | Low-quality | Genome-fragment | 4.47 | AAI-based (high-confidence) | 0 |
| S5_vs_k141_73179\|\|full | 1849 | No | NA | 2 | 0 | 0 | Not-determined | Genome-fragment | NA | NA | 0 |
| S5_vs_k141_122317\|\|full | 1927 | No | NA | 4 | 0 | 0 | Not-determined | Genome-fragment | NA | NA | 0 |
| S12_vs_k141_176295\|\|lt2gene | 1845 | No | NA | 2 | 1 | 0 | Low-quality | Genome-fragment | 3.19 | HMM-based (lower-bound) | 0 |
| S8_vs_k141_178377\|\|full | 1845 | No | NA | 5 | 1 | 0 | Low-quality | Genome-fragment | 5.34 | AAI-based (medium-confidence) | 0 |
| S8_vs_k141_134855\|\|full | 1845 | No | NA | 6 | 0 | 0 | Low-quality | Genome-fragment | 4.66 | AAI-based (high-confidence) | 0 |
| S5_vs_k141_850\|\|full | 44156 | No | NA | 57 | 20 | 1 | High-quality | High-quality | 96.5 | AAI-based (medium-confidence) | 0 |
| S5_vs_k141_107928\|\|full | 1846 | No | NA | 3 | 3 | 0 | Low-quality | Genome-fragment | 4.62 | AAI-based (high-confidence) | 0 |
| S5_vs_k141_36803\|\|full | 1175 | No | NA | 2 | 0 | 1 | Not-determined | Genome-fragment | NA | NA | 0 |
| S7_vs_89294\|\|lt2gene | 1846 | No | NA | 2 | 1 | 0 | Low-quality | Genome-fragment | 1.14 | HMM-based (lower-bound) | 0 |
| S5_vs_k141_6694\|\|full | 4142 | No | NA | 4 | 0 | 2 | Not-determined | Genome-fragment | NA | NA | 0 |
| S5_vs_k141_36852\|\|full | 3476 | No | NA | 3 | 0 | 3 | Not-determined | Genome-fragment | NA | NA | 0 |
| S5_vs_k141_115922\|\|full | 2268 | No | NA | 4 | 0 | 0 | Not-determined | Genome-fragment | NA | NA | 0 |
| S5_vs_k141_31038\|\|full | 1753 | No | NA | 3 | 0 | 2 | Not-determined | Genome-fragment | NA | NA | 0 |
| S5_vs_k141_36870\|\|full | 1467 | No | NA | 2 | 0 | 1 | Not-determined | Genome-fragment | NA | NA | 0 |
| S10_extracted_57472 | 1846 | No | NA | 2 | 0 | 1 | Low-quality | Genome-fragment | 1.64 | AAI-based (medium-confidence) | 0 |
| S5_vs_k141_67115\|\|full | 1261 | No | NA | 2 | 0 | 1 | Not-determined | Genome-fragment | NA | NA | 0 |
| S2_vs_k141_4893\|\|full | 1846 | No | NA | 2 | 0 | 2 | Low-quality | Genome-fragment | 1.77 | AAI-based (high-confidence) | 0 |
| S4_extracted_19265 | 1846 | No | NA | 2 | 1 | 0 | Low-quality | Genome-fragment | 4.1 | HMM-based (lower-bound) | 0 |
| S5_vs_k141_12497\|\|full | 1517 | No | NA | 3 | 0 | 0 | Not-determined | Genome-fragment | NA | NA | 0 |
| S6_vs_k141_11262\|\|full | 1846 | No | NA | 3 | 0 | 0 | Low-quality | Genome-fragment | 1.6 | AAI-based (medium-confidence) | 0 |
| S9_vs_k141_78881\|\|full | 1847 | No | NA | 3 | 0 | 0 | Low-quality | Genome-fragment | 4.63 | AAI-based (high-confidence) | 0 |
| S13_vs_107530\|\|full | 1847 | No | NA | 3 | 0 | 0 | Low-quality | Genome-fragment | 2.12 | AAI-based (medium-confidence) | 0 |
| S14_vs_k141_110525\|\|full | 1847 | No | NA | 5 | 1 | 0 | Low-quality | Genome-fragment | 4.82 | AAI-based (high-confidence) | 0 |
| S5_vs_k141_961\|\|full | 1753 | No | NA | 3 | 0 | 2 | Not-determined | Genome-fragment | NA | NA | 0 |
| S3_vs_k141_85854\|\|full | 1847 | No | NA | 3 | 0 | 0 | Low-quality | Genome-fragment | 13.74 | AAI-based (medium-confidence) | 0 |
| S12_vs_k141_178366\|\|full | 1847 | No | NA | 4 | 1 | 0 | Low-quality | Genome-fragment | 1.79 | HMM-based (lower-bound) | 0 |
| S17_vs_k141_134961\|\|full | 1848 | No | NA | 2 | 0 | 0 | Low-quality | Genome-fragment | 3.05 | AAI-based (medium-confidence) | 0 |

| | | | | | | | | | | | |
|---|---|---|---|---|---|---|---|---|---|---|---|
| S5_vs_k141_67203\|\|full | 2220 | No | NA | 5 | 0 | 0 | Not-determined | Genome-fragment | NA | NA | 0 |
| S5_vs_k141_140062\|\|full | 4276 | No | NA | 2 | 0 | 1 | Not-determined | Genome-fragment | NA | NA | 0 |
| S5_vs_k141_43410\|\|full | 1522 | No | NA | 2 | 0 | 1 | Not-determined | Genome-fragment | NA | NA | 0 |
| S5_vs_k141_36926\|\|full | 4015 | No | NA | 4 | 0 | 4 | Not-determined | Genome-fragment | NA | NA | 0 |
| S5_vs_k141_85626\|\|full | 1547 | No | NA | 2 | 0 | 0 | Not-determined | Genome-fragment | NA | NA | 0 |
| S11_extracted_k141_7693 | 1848 | No | NA | 3 | 1 | 0 | Low-quality | Genome-fragment | 2.58 | HMM-based (lower-bound) | 0 |
| S11_vs_k141_103924\|\|full | 1848 | No | NA | 3 | 0 | 0 | Low-quality | Genome-fragment | 3.4 | AAI-based (high-confidence) | 0 |
| S5_vs_k141_130823\|\|full | 1848 | No | NA | 4 | 1 | 0 | Low-quality | Genome-fragment | 3.1 | AAI-based (high-confidence) | 0 |
| S5_vs_k141_91689\|\|full | 2742 | No | NA | 2 | 0 | 2 | Not-determined | Genome-fragment | NA | NA | 0 |
| S5_vs_k141_11859\|\|full | 1848 | No | NA | 6 | 1 | 0 | Low-quality | Genome-fragment | 5.09 | AAI-based (medium-confidence) | 0 |
| S7_vs_106529\|\|full | 1848 | No | NA | 5 | 0 | 0 | Low-quality | Genome-fragment | 3.34 | AAI-based (high-confidence) | 0 |
| S12_extracted_k141_134484 | 1848 | No | NA | 2 | 0 | 0 | Low-quality | Genome-fragment | 3.33 | AAI-based (medium-confidence) | 0 |
| S5_vs_k141_31164\|\|full | 1565 | No | NA | 3 | 0 | 2 | Not-determined | Genome-fragment | NA | NA | 0 |
| S11_vs_k141_17601\|\|lt2gene | 1849 | No | NA | 2 | 2 | 0 | Low-quality | Genome-fragment | 4.66 | AAI-based (high-confidence) | 0 |
| S5_vs_k141_85659\|\|full | 1629 | No | NA | 3 | 0 | 2 | Not-determined | Genome-fragment | NA | NA | 0 |
| S13_vs_115836\|\|full | 1849 | No | NA | 4 | 0 | 0 | Low-quality | Genome-fragment | 4.35 | AAI-based (medium-confidence) | 0 |
| S5_vs_k141_1027\|\|full | 3034 | No | NA | 5 | 0 | 3 | Not-determined | Genome-fragment | NA | NA | 0 |
| S5_vs_k141_133971\|\|full | 1897 | No | NA | 3 | 0 | 0 | Not-determined | Genome-fragment | NA | NA | 0 |
| S5_vs_k141_43448\|\|full | 2384 | No | NA | 3 | 0 | 2 | Not-determined | Genome-fragment | NA | NA | 0 |
| S14_vs_k141_1996\|\|full | 1849 | No | NA | 3 | 0 | 1 | Low-quality | Genome-fragment | 4.29 | AAI-based (high-confidence) | 0 |
| S5_vs_k141_116119\|\|full | 2275 | No | NA | 4 | 0 | 3 | Not-determined | Genome-fragment | NA | NA | 0 |
| S12_extracted_k141_60501 | 1849 | No | NA | 2 | 1 | 0 | Low-quality | Genome-fragment | 5.45 | AAI-based (high-confidence) | 0 |
| S7_extracted_129708 | 1850 | No | NA | 3 | 0 | 1 | Low-quality | Genome-fragment | 0.82 | AAI-based (medium-confidence) | 0 |
| S16_vs_28088\|\|full | 1850 | No | NA | 3 | 0 | 2 | Low-quality | Genome-fragment | 0.48 | AAI-based (medium-confidence) | 0 |
| S10_vs_77848\|\|full | 1850 | No | NA | 2 | 0 | 1 | Low-quality | Genome-fragment | 2.35 | AAI-based (high-confidence) | 0 |
| S5_vs_k141_1118\|\|full | 1655 | No | NA | 3 | 0 | 3 | Not-determined | Genome-fragment | NA | NA | 0 |
| S17_vs_k141_53760\|\|full | 1851 | No | NA | 3 | 0 | 1 | Low-quality | Genome-fragment | 1.21 | AAI-based (medium-confidence) | 0 |
| S9_extracted_k141_80566 | 1851 | No | NA | 4 | 2 | 0 | Low-quality | Genome-fragment | 4.41 | AAI-based (high-confidence) | 0 |
| S14_extracted_k141_80879 | 1851 | No | NA | 2 | 0 | 0 | Low-quality | Genome-fragment | 3.98 | AAI-based (medium-confidence) | 0 |
| S5_vs_k141_116132\|\|full | 7564 | No | NA | 7 | 0 | 1 | Not-determined | Genome-fragment | NA | NA | 0 |
| S5_vs_k141_18893\|\|full | 1293 | No | NA | 2 | 0 | 1 | Not-determined | Genome-fragment | NA | NA | 0 |
| S14_vs_k141_94007\|\|full | 1851 | No | NA | 3 | 0 | 0 | Low-quality | Genome-fragment | 1.45 | AAI-based (high-confidence) | 0 |
| S3_extracted_k141_39224 | 1851 | No | NA | 2 | 0 | 0 | Low-quality | Genome-fragment | 2.78 | AAI-based (high-confidence) | 0 |
| S3_vs_k141_49438\|\|full | 1851 | No | NA | 4 | 0 | 2 | Low-quality | Genome-fragment | 1.64 | AAI-based (medium-confidence) | 0 |
| S5_vs_k141_140148\|\|full | 1477 | No | NA | 2 | 0 | 1 | Not-determined | Genome-fragment | NA | NA | 0 |
| S7_vs_67978\|\|full | 1851 | No | NA | 3 | 0 | 1 | Low-quality | Genome-fragment | 2.19 | AAI-based (medium-confidence) | 0 |
| S15_vs_70325\|\|full | 1851 | No | NA | 3 | 0 | 0 | Low-quality | Genome-fragment | 3.06 | AAI-based (medium-confidence) | 0 |
| S5_vs_k141_67322\|\|full | 1181 | No | NA | 2 | 0 | 1 | Not-determined | Genome-fragment | NA | NA | 0 |
| S5_vs_k141_140167\|\|full | 1839 | No | NA | 3 | 0 | 2 | Not-determined | Genome-fragment | NA | NA | 0 |
| S6_vs_k141_6731\|\|lt2gene | 1851 | No | NA | 2 | 2 | 0 | Low-quality | Genome-fragment | 4.79 | AAI-based (medium-confidence) | 0 |
| S5_vs_k141_1192\|\|full | 1114 | No | NA | 3 | 0 | 2 | Not-determined | Genome-fragment | NA | NA | 0 |
| S1_vs_k141_65303\|\|full | 1852 | No | NA | 6 | 0 | 0 | Low-quality | Genome-fragment | 4.7 | AAI-based (high-confidence) | 0 |
| S5_vs_k141_77926\|\|full | 1852 | No | NA | 3 | 0 | 0 | Low-quality | Genome-fragment | 4.51 | AAI-based (medium-confidence) | 0 |
| S5_vs_k141_1221\|\|full | 4249 | No | NA | 3 | 0 | 3 | Not-determined | Genome-fragment | NA | NA | 0 |
| S12_extracted_k141_87344 | 1852 | No | NA | 3 | 1 | 0 | Low-quality | Genome-fragment | 4.06 | AAI-based (high-confidence) | 0 |
| S5_vs_k141_122607\|\|full | 2211 | No | NA | 2 | 0 | 2 | Not-determined | Genome-fragment | NA | NA | 0 |
| S8_vs_k141_173818\|\|full | 1852 | No | NA | 3 | 1 | 0 | Low-quality | Genome-fragment | 3.37 | AAI-based (high-confidence) | 0 |
| S8_vs_k141_150338\|\|full | 1852 | No | NA | 8 | 2 | 0 | Low-quality | Genome-fragment | 3.75 | HMM-based (lower-bound) | 0 |
| S11_vs_k141_77390\|\|lt2gene | 1853 | No | NA | 2 | 2 | 0 | Low-quality | Genome-fragment | 4.31 | AAI-based (high-confidence) | 0 |
| S13_vs_86129\|\|full | 1853 | No | NA | 3 | 1 | 0 | Low-quality | Genome-fragment | 0.86 | HMM-based (lower-bound) | 0 |
| S7_vs_118033\|\|full | 1853 | No | NA | 4 | 1 | 0 | Low-quality | Genome-fragment | 3.92 | AAI-based (high-confidence) | 0 |
| S8_extracted_k141_193140 | 1853 | No | NA | 1 | 0 | 1 | Low-quality | Genome-fragment | 1 | AAI-based (medium-confidence) | 0 |
| S8_extracted_k141_81521 | 1853 | No | NA | 2 | 0 | 0 | Low-quality | Genome-fragment | 2.87 | AAI-based (high-confidence) | 0 |
| S5_vs_k141_79300\|\|full | 4391 | No | NA | 4 | 0 | 3 | Not-determined | Genome-fragment | NA | NA | 0 |
| S8_extracted_k141_57658 | 1853 | No | NA | 1 | 0 | 1 | Low-quality | Genome-fragment | 5.23 | AAI-based (medium-confidence) | 0 |
| S5_vs_k141_104048\|\|full | 1916 | No | NA | 3 | 0 | 3 | Not-determined | Genome-fragment | NA | NA | 0 |
| S17_vs_k141_111781\|\|full | 1854 | No | NA | 3 | 0 | 0 | Low-quality | Genome-fragment | 3.87 | AAI-based (medium-confidence) | 0 |
| S17_vs_k141_10985\|\|full | 1854 | No | NA | 4 | 2 | 0 | Low-quality | Genome-fragment | 4.72 | AAI-based (high-confidence) | 0 |
| S5_vs_k141_116235\|\|full | 37669 | No | NA | 52 | 19 | 1 | High-quality | High-quality | 98.06 | AAI-based (high-confidence) | 0 |
| S9_vs_k141_55023\|\|full | 1854 | No | NA | 3 | 1 | 0 | Low-quality | Genome-fragment | 1.03 | AAI-based (high-confidence) | 0 |
| S9_vs_k141_47885\|\|full | 1854 | No | NA | 3 | 0 | 0 | Low-quality | Genome-fragment | 2.29 | AAI-based (medium-confidence) | 0 |
| S5_vs_k141_31370\|\|full | 3021 | No | NA | 4 | 0 | 3 | Not-determined | Genome-fragment | NA | NA | 0 |
| S11_vs_k141_16986\|\|full | 1854 | No | NA | 4 | 1 | 0 | Low-quality | Genome-fragment | 4.82 | AAI-based (high-confidence) | 0 |
| S5_vs_k141_6999\|\|full | 4347 | No | NA | 6 | 0 | 1 | Not-determined | Genome-fragment | NA | NA | 0 |
| S1_extracted_k141_95241 | 1854 | No | NA | 1 | 0 | 1 | Low-quality | Genome-fragment | 1.28 | AAI-based (medium-confidence) | 0 |
| S5_vs_k141_19014\|\|full | 8474 | No | NA | 9 | 0 | 6 | Not-determined | Genome-fragment | NA | NA | 0 |
| S5_vs_k141_134108\|\|full | 3946 | No | NA | 5 | 0 | 2 | Not-determined | Genome-fragment | NA | NA | 0 |
| S5_vs_k141_91854\|\|full | 2806 | No | NA | 3 | 0 | 1 | Not-determined | Genome-fragment | NA | NA | 0 |
| S1_extracted_k141_130182 | 1854 | No | NA | 4 | 0 | 0 | Low-quality | Genome-fragment | 3.51 | AAI-based (high-confidence) | 0 |
| S5_vs_k141_1338\|\|full | 56857 | No | NA | 66 | 8 | 4 | Medium-quality | Genome-fragment | 51.1 | AAI-based (medium-confidence) | 0 |
| S1_vs_k141_52979\|\|full | 1854 | No | NA | 3 | 0 | 2 | Low-quality | Genome-fragment | 0.94 | AAI-based (medium-confidence) | 0 |
| S5_vs_k141_19069\|\|full | 1057 | No | NA | 2 | 0 | 1 | Not-determined | Genome-fragment | NA | NA | 0 |
| S1_vs_k141_86712\|\|full | 1854 | No | NA | 3 | 0 | 2 | Low-quality | Genome-fragment | 0.77 | AAI-based (medium-confidence) | 0 |
| S5_vs_k141_128281\|\|full | 1233 | No | NA | 2 | 0 | 0 | Not-determined | Genome-fragment | NA | NA | 0 |
| S5_vs_k141_73271\|\|full | 5580 | No | NA | 8 | 0 | 0 | Not-determined | Genome-fragment | NA | NA | 0 |
| S3_vs_k141_24518\|\|full | 1854 | No | NA | 4 | 2 | 0 | Low-quality | Genome-fragment | 3.85 | AAI-based (high-confidence) | 0 |
| S5_extracted_k141_8110 | 1854 | No | NA | 1 | 0 | 0 | Low-quality | Genome-fragment | 2.57 | AAI-based (medium-confidence) | 0 |
| S5_vs_k141_14775\|\|full | 1854 | No | NA | 4 | 0 | 1 | Low-quality | Genome-fragment | 3.89 | AAI-based (medium-confidence) | 0 |
| S5_vs_k141_97601\|\|full | 1901 | No | NA | 2 | 0 | 0 | Not-determined | Genome-fragment | NA | NA | 0 |
| S7_vs_81322\|\|full | 1854 | No | NA | 4 | 1 | 0 | Low-quality | Genome-fragment | 4.42 | AAI-based (high-confidence) | 0 |
| S5_vs_k141_122793\|\|full | 2893 | No | NA | 3 | 0 | 0 | Not-determined | Genome-fragment | NA | NA | 0 |
| S7_vs_101606\|\|full | 1854 | No | NA | 3 | 3 | 0 | Low-quality | Genome-fragment | 5.09 | AAI-based (high-confidence) | 0 |
| S5_vs_k141_7091\|\|full | 2807 | No | NA | 3 | 0 | 2 | Not-determined | Genome-fragment | NA | NA | 0 |
| S5_vs_k141_104159\|\|full | 1465 | No | NA | 3 | 0 | 0 | Not-determined | Genome-fragment | NA | NA | 0 |
| S5_vs_k141_116331\|\|full | 2798 | No | NA | 4 | 0 | 1 | Not-determined | Genome-fragment | NA | NA | 0 |
| S16_vs_25173\|\|full | 1854 | No | NA | 3 | 0 | 0 | Low-quality | Genome-fragment | 3.2 | AAI-based (high-confidence) | 0 |
| S6_vs_k141_52862\|\|full | 1854 | No | NA | 3 | 1 | 0 | Low-quality | Genome-fragment | 1.06 | HMM-based (lower-bound) | 0 |
| S5_vs_k141_104172\|\|full | 1754 | No | NA | 3 | 0 | 0 | Not-determined | Genome-fragment | NA | NA | 0 |
| S17_vs_k141_159358\|\|lt2gene | 1855 | No | NA | 2 | 2 | 0 | Low-quality | Genome-fragment | 4.52 | AAI-based (medium-confidence) | 0 |
| S7_extracted_80695 | 1855 | No | NA | 3 | 0 | 0 | Low-quality | Genome-fragment | 1.28 | AAI-based (high-confidence) | 0 |
| S7_vs_6077\|\|full | 1855 | No | NA | 4 | 2 | 0 | Low-quality | Genome-fragment | 3.62 | AAI-based (high-confidence) | 0 |
| S8_vs_k141_171024\|\|lt2gene | 1855 | No | NA | 1 | 1 | 0 | Low-quality | Genome-fragment | 4.61 | AAI-based (high-confidence) | 0 |
| S5_vs_k141_55253\|\|full | 2976 | No | NA | 5 | 0 | 0 | Not-determined | Genome-fragment | NA | NA | 0 |
| S5_vs_k141_134197\|\|full | 1050 | No | NA | 2 | 0 | 0 | Not-determined | Genome-fragment | NA | NA | 0 |
| S5_vs_k141_85817\|\|full | 3507 | No | NA | 5 | 0 | 2 | Not-determined | Genome-fragment | NA | NA | 0 |
| S5_vs_k141_97632\|\|full | 1169 | No | NA | 3 | 0 | 0 | Not-determined | Genome-fragment | NA | NA | 0 |
| S5_vs_k141_31498\|\|full | 1012 | No | NA | 2 | 0 | 1 | Not-determined | Genome-fragment | NA | NA | 0 |
| S5_vs_k141_61305\|\|full | 3360 | No | NA | 5 | 0 | 0 | Not-determined | Genome-fragment | NA | NA | 0 |
| S5_vs_k141_116411\|\|full | 2668 | No | NA | 3 | 0 | 1 | Not-determined | Genome-fragment | NA | NA | 0 |
| S17_vs_k141_89937\|\|full | 1856 | No | NA | 3 | 0 | 0 | Low-quality | Genome-fragment | 3.62 | AAI-based (medium-confidence) | 0 |
| S14_extracted_k141_62071 | 1856 | No | NA | 3 | 3 | 0 | Low-quality | Genome-fragment | 6.12 | AAI-based (high-confidence) | 0 |
| S7_vs_69728\|\|full | 1856 | No | NA | 4 | 1 | 0 | Low-quality | Genome-fragment | 4.5 | AAI-based (medium-confidence) | 0 |
| S7_vs_105288\|\|full | 1856 | No | NA | 2 | 0 | 1 | Low-quality | Genome-fragment | 2.96 | AAI-based (medium-confidence) | 0 |
| S5_vs_k141_109944\|\|full | 2643 | No | NA | 4 | 0 | 3 | Not-determined | Genome-fragment | NA | NA | 0 |
| S5_vs_k141_122898\|\|full | 2461 | No | NA | 3 | 0 | 1 | Not-determined | Genome-fragment | NA | NA | 0 |
| S5_vs_k141_67574\|\|full | 1017 | No | NA | 4 | 0 | 0 | Not-determined | Genome-fragment | NA | NA | 0 |
| S16_vs_153824\|\|full | 1856 | No | NA | 3 | 0 | 0 | Low-quality | Genome-fragment | 2.29 | AAI-based (high-confidence) | 0 |
| S5_vs_k141_109997\|\|full | 1085 | No | NA | 2 | 0 | 1 | Not-determined | Genome-fragment | NA | NA | 0 |
| S12_extracted_k141_180691 | 1856 | No | NA | 2 | 0 | 0 | Low-quality | Genome-fragment | 2.97 | AAI-based (high-confidence) | 0 |
| S4_extracted_19554 | 1856 | No | NA | 4 | 1 | 0 | Low-quality | Genome-fragment | 5.18 | AAI-based (high-confidence) | 0 |
| S5_vs_k141_134217\|\|full | 1282 | No | NA | 3 | 0 | 2 | Not-determined | Genome-fragment | NA | NA | 0 |
| S5_vs_k141_140330\|\|full | 6326 | No | NA | 6 | 0 | 0 | Not-determined | Genome-fragment | NA | NA | 0 |
| S3_extracted_k141_115045 | 1857 | No | NA | 3 | 3 | 0 | Low-quality | Genome-fragment | 6.12 | AAI-based (high-confidence) | 0 |
| S5_extracted_k141_106276 | 1857 | No | NA | 4 | 1 | 0 | Low-quality | Genome-fragment | 0.98 | HMM-based (lower-bound) | 0 |
| S16_vs_101029\|\|full | 1857 | No | NA | 4 | 0 | 1 | Low-quality | Genome-fragment | 1.75 | AAI-based (medium-confidence) | 0 |
| S10_vs_102982\|\|full | 1857 | No | NA | 4 | 0 | 0 | Low-quality | Genome-fragment | 3.76 | AAI-based (high-confidence) | 0 |
| S5_vs_k141_49480\|\|full | 30873 | No | NA | 56 | 21 | 1 | High-quality | High-quality | 92.04 | AAI-based (high-confidence) | 0 |
| S5_vs_k141_104195\|\|full | 2085 | No | NA | 3 | 0 | 1 | Not-determined | Genome-fragment | NA | NA | 0 |
| S5_vs_k141_85892\|\|full | 1781 | No | NA | 3 | 0 | 1 | Not-determined | Genome-fragment | NA | NA | 0 |
| S5_vs_k141_85893\|\|full | 1004 | No | NA | 2 | 0 | 2 | Not-determined | Genome-fragment | NA | NA | 0 |
| S5_vs_k141_7219\|\|full | 1918 | No | NA | 3 | 0 | 1 | Not-determined | Genome-fragment | NA | NA | 0 |
| S12_extracted_k141_9289 | 1857 | No | NA | 3 | 0 | 0 | Low-quality | Genome-fragment | 3.71 | AAI-based (high-confidence) | 0 |
| S6_vs_k141_37151\|\|full | 1857 | No | NA | 3 | 0 | 0 | Low-quality | Genome-fragment | 1.85 | AAI-based (high-confidence) | 0 |
| S5_vs_k141_140370\|\|full | 2308 | No | NA | 2 | 0 | 0 | Not-determined | Genome-fragment | NA | NA | 0 |
| S5_vs_k141_85934\|\|full | 2299 | No | NA | 3 | 0 | 0 | Not-determined | Genome-fragment | NA | NA | 0 |
| S8_vs_k141_86750\|\|full | 1857 | No | NA | 2 | 0 | 1 | Low-quality | Genome-fragment | 3.81 | AAI-based (high-confidence) | 0 |
| S5_vs_k141_31595\|\|full | 1948 | No | NA | 2 | 0 | 1 | Not-determined | Genome-fragment | NA | NA | 0 |
| S11_vs_k141_69875\|\|full | 1858 | No | NA | 3 | 0 | 1 | Low-quality | Genome-fragment | 3.99 | AAI-based (high-confidence) | 0 |
| S5_vs_k141_31598\|\|full | 1233 | No | NA | 2 | 0 | 0 | Not-determined | Genome-fragment | NA | NA | 0 |
| S5_vs_k141_12914\|\|full | 1171 | No | NA | 3 | 0 | 0 | Not-determined | Genome-fragment | NA | NA | 0 |
| S3_extracted_k141_26515 | 1858 | No | NA | 3 | 0 | 2 | Low-quality | Genome-fragment | 1.38 | AAI-based (medium-confidence) | 0 |
| S5_vs_k141_37309\|\|full | 1470 | No | NA | 2 | 0 | 1 | Not-determined | Genome-fragment | NA | NA | 0 |
| S5_vs_k141_85988\|\|full | 2064 | No | NA | 3 | 0 | 1 | Not-determined | Genome-fragment | NA | NA | 0 |
| S5_vs_k141_19309\|\|full | 4602 | No | NA | 9 | 0 | 6 | Not-determined | Genome-fragment | NA | NA | 0 |
| S5_vs_k141_42900\|\|lt2gene | 1858 | No | NA | 2 | 2 | 0 | Low-quality | Genome-fragment | 4.08 | AAI-based (high-confidence) | 0 |
| S7_extracted_35682 | 1858 | No | NA | 3 | 0 | 0 | Low-quality | Genome-fragment | 4.7 | AAI-based (high-confidence) | 0 |
| S7_vs_133051\|\|full | 1858 | No | NA | 6 | 1 | 0 | Low-quality | Genome-fragment | 3.1 | AAI-based (medium-confidence) | 0 |
| S16_extracted_159780 | 1858 | No | NA | 5 | 1 | 0 | Low-quality | Genome-fragment | 1.2 | HMM-based (lower-bound) | 0 |
| S12_vs_k141_159711\|\|full | 1858 | No | NA | 3 | 0 | 0 | Low-quality | Genome-fragment | 4.19 | AAI-based (medium-confidence) | 0 |
| S8_vs_k141_73629\|\|full | 1858 | No | NA | 2 | 0 | 0 | Low-quality | Genome-fragment | 2.01 | AAI-based (medium-confidence) | 0 |
| S17_vs_k141_45107\|\|full | 1859 | No | NA | 3 | 2 | 0 | Low-quality | Genome-fragment | 3.08 | AAI-based (high-confidence) | 0 |
| S5_vs_k141_19341\|\|full | 1147 | No | NA | 2 | 0 | 0 | Not-determined | Genome-fragment | NA | NA | 0 |
| S5_vs_k141_86014\|\|full | 3416 | No | NA | 4 | 0 | 4 | Not-determined | Genome-fragment | NA | NA | 0 |
| S5_vs_k141_31634\|\|full | 3780 | No | NA | 5 | 0 | 1 | Not-determined | Genome-fragment | NA | NA | 0 |
| S11_vs_k141_73005\|\|lt2gene | 1859 | No | NA | 1 | 1 | 0 | Low-quality | Genome-fragment | 4.58 | AAI-based (medium-confidence) | 0 |
| S5_vs_k141_24795\|\|full | 2001 | No | NA | 3 | 0 | 2 | Not-determined | Genome-fragment | NA | NA | 0 |
| S5_vs_k141_61564\|\|full | 6495 | No | NA | 7 | 0 | 3 | Not-determined | Genome-fragment | NA | NA | 0 |
| S13_vs_73394\|\|full | 1859 | No | NA | 3 | 0 | 0 | Low-quality | Genome-fragment | 4.21 | AAI-based (high-confidence) | 0 |
| S5_vs_k141_140519\|\|full | 1514 | No | NA | 2 | 0 | 1 | Not-determined | Genome-fragment | NA | NA | 0 |
| S10_vs_42011\|\|lt2gene | 1859 | No | NA | 1 | 1 | 0 | Low-quality | Genome-fragment | 5.91 | AAI-based (high-confidence) | 0 |
| S12_vs_k141_142707\|\|full | 1859 | No | NA | 4 | 0 | 0 | Low-quality | Genome-fragment | 2.07 | AAI-based (high-confidence) | 0 |
| S11_vs_k141_41393\|\|full | 1860 | No | NA | 2 | 0 | 1 | Low-quality | Genome-fragment | 0.87 | AAI-based (medium-confidence) | 0 |
| S13_extracted_150831 | 1860 | No | NA | 1 | 0 | 1 | Low-quality | Genome-fragment | 2.02 | AAI-based (medium-confidence) | 0 |
| S14_vs_k141_87049\|\|full | 1860 | No | NA | 4 | 2 | 0 | Low-quality | Genome-fragment | 4.12 | AAI-based (high-confidence) | 0 |
| S5_vs_k141_73498\|\|full | 1843 | No | NA | 3 | 0 | 2 | Not-determined | Genome-fragment | NA | NA | 0 |
| S3_vs_k141_92538\|\|full | 1860 | No | NA | 6 | 1 | 0 | Low-quality | Genome-fragment | 3.23 | AAI-based (high-confidence) | 0 |
| S3_vs_k141_117786\|\|lt2gene | 1860 | No | NA | 2 | 2 | 0 | Low-quality | Genome-fragment | 5.22 | AAI-based (high-confidence) | 0 |
| S10_vs_45314\|\|full | 1860 | No | NA | 5 | 1 | 0 | Low-quality | Genome-fragment | 3.38 | AAI-based (medium-confidence) | 0 |
| S5_vs_k141_37485\|\|full | 2016 | No | NA | 4 | 0 | 0 | Not-determined | Genome-fragment | NA | NA | 0 |
| S4_extracted_18279 | 1860 | No | NA | 5 | 1 | 0 | Low-quality | Genome-fragment | 3.43 | AAI-based (high-confidence) | 0 |
| S11_vs_k141_35443\|\|full | 1861 | No | NA | 4 | 2 | 0 | Low-quality | Genome-fragment | 4.16 | AAI-based (high-confidence) | 0 |
| S13_vs_144025\|\|full | 1861 | No | NA | 3 | 0 | 0 | Low-quality | Genome-fragment | 3.78 | AAI-based (high-confidence) | 0 |
| S5_vs_k141_31686\|\|full | 1132 | No | NA | 2 | 0 | 1 | Not-determined | Genome-fragment | NA | NA | 0 |
| S5_vs_k141_37490\|\|full | 2634 | No | NA | 4 | 0 | 0 | Not-determined | Genome-fragment | NA | NA | 0 |
| S14_extracted_k141_44767 | 1861 | No | NA | 6 | 5 | 0 | Low-quality | Genome-fragment | 6.13 | AAI-based (high-confidence) | 0 |
| S5_vs_k141_37498\|\|full | 2095 | No | NA | 2 | 0 | 1 | Not-determined | Genome-fragment | NA | NA | 0 |
| S14_extracted_k141_60198 | 1861 | Yes | 778 | 2 | 1 | 1 | Low-quality | Genome-fragment | 1.79 | AAI-based (high-confidence) | 58.19 |
| S5_vs_k141_97890\|\|full | 4995 | No | NA | 4 | 0 | 1 | Not-determined | Genome-fragment | NA | NA | 0 |
| S5_vs_k141_1446\|\|full | 1316 | No | NA | 3 | 0 | 3 | Not-determined | Genome-fragment | NA | NA | 0 |
| S5_vs_k141_43877\|\|full | 1769 | No | NA | 4 | 0 | 0 | Not-determined | Genome-fragment | NA | NA | 0 |
| S5_vs_k141_123137\|\|full | 5638 | No | NA | 6 | 0 | 4 | Not-determined | Genome-fragment | NA | NA | 0 |
| S5_vs_k141_13090\|\|full | 32057 | No | NA | 50 | 18 | 0 | High-quality | High-quality | 90.23 | AAI-based (high-confidence) | 0 |
| S1_extracted_k141_69514 | 1861 | No | NA | 2 | 2 | 0 | Low-quality | Genome-fragment | 5.18 | AAI-based (high-confidence) | 0 |
| S1_vs_k141_60999\|\|full | 1861 | No | NA | 2 | 0 | 2 | Low-quality | Genome-fragment | 0.74 | AAI-based (medium-confidence) | 0 |
| S5_vs_k141_67730\|\|full | 5125 | No | NA | 6 | 0 | 4 | Not-determined | Genome-fragment | NA | NA | 0 |
| S3_extracted_k141_85794 | 1861 | No | NA | 3 | 3 | 0 | Low-quality | Genome-fragment | 4.11 | AAI-based (high-confidence) | 0 |
| S3_vs_k141_37415\|\|full | 1861 | No | NA | 3 | 1 | 0 | Low-quality | Genome-fragment | 1.38 | AAI-based (medium-confidence) | 0 |
| S5_vs_k141_134375\|\|full | 1364 | No | NA | 3 | 0 | 1 | Not-determined | Genome-fragment | NA | NA | 0 |
| S12_vs_k141_119681\|\|full | 1861 | No | NA | 3 | 0 | 2 | Low-quality | Genome-fragment | 3.44 | AAI-based (medium-confidence) | 0 |
| S5_vs_k141_13150\|\|full | 2365 | No | NA | 3 | 0 | 3 | Not-determined | Genome-fragment | NA | NA | 0 |
| S8_extracted_k141_18773 | 1861 | No | NA | 6 | 0 | 0 | Low-quality | Genome-fragment | 1.45 | AAI-based (high-confidence) | 0 |
| S8_extracted_k141_146955 | 1861 | No | NA | 1 | 0 | 1 | Low-quality | Genome-fragment | 1.64 | AAI-based (medium-confidence) | 0 |
| S5_vs_k141_19522\|\|full | 1833 | No | NA | 3 | 0 | 1 | Not-determined | Genome-fragment | NA | NA | 0 |
| S5_vs_k141_1454\|\|full | 1952 | No | NA | 3 | 0 | 0 | Not-determined | Genome-fragment | NA | NA | 0 |
| S17_extracted_k141_88837 | 1862 | No | NA | 3 | 2 | 0 | Low-quality | Genome-fragment | 4.57 | AAI-based (high-confidence) | 0 |
| S9_extracted_k141_2748 | 1862 | No | NA | 6 | 1 | 0 | Low-quality | Genome-fragment | 1.2 | AAI-based (high-confidence) | 0 |
| S1_extracted_k141_88820 | 1862 | No | NA | 2 | 0 | 1 | Low-quality | Genome-fragment | 2.36 | AAI-based (medium-confidence) | 0 |
| S5_vs_k141_128347\|\|full | 2243 | No | NA | 2 | 0 | 2 | Not-determined | Genome-fragment | NA | NA | 0 |
| S5_vs_k141_134421\|\|full | 1451 | No | NA | 3 | 0 | 3 | Not-determined | Genome-fragment | NA | NA | 0 |
| S1_vs_k141_39723\|\|full | 1862 | No | NA | 2 | 0 | 1 | Low-quality | Genome-fragment | 2.89 | AAI-based (medium-confidence) | 0 |
| S5_vs_k141_61736\|\|full | 18902 | No | NA | 19 | 0 | 14 | Not-determined | Genome-fragment | NA | NA | 0 |
| S3_vs_k141_111575\|\|full | 1862 | No | NA | 5 | 1 | 0 | Low-quality | Genome-fragment | 4.42 | AAI-based (high-confidence) | 0 |
| S16_vs_98646\|\|full | 1862 | No | NA | 4 | 2 | 0 | Low-quality | Genome-fragment | 6.29 | AAI-based (high-confidence) | 0 |
| S5_vs_k141_92163\|\|full | 2652 | No | NA | 6 | 0 | 1 | Not-determined | Genome-fragment | NA | NA | 0 |
| S12_vs_k141_178236\|\|full | 1862 | No | NA | 3 | 1 | 0 | Low-quality | Genome-fragment | 3.15 | AAI-based (high-confidence) | 0 |
| S12_vs_k141_160749\|\|lt2gene | 1862 | No | NA | 2 | 2 | 0 | Low-quality | Genome-fragment | 4.15 | AAI-based (high-confidence) | 0 |
| S8_extracted_k141_151948 | 1862 | No | NA | 2 | 1 | 0 | Low-quality | Genome-fragment | 2.72 | HMM-based (lower-bound) | 0 |
| S17_vs_k141_148910\|\|full | 1863 | No | NA | 5 | 0 | 0 | Low-quality | Genome-fragment | 3.08 | AAI-based (high-confidence) | 0 |
| S5_vs_k141_73635\|\|full | 1077 | No | NA | 3 | 0 | 0 | Not-determined | Genome-fragment | NA | NA | 0 |
| S11_extracted_k141_25988 | 1863 | No | NA | 1 | 1 | 0 | Low-quality | Genome-fragment | 4.5 | AAI-based (medium-confidence) | 0 |
| S7_vs_68688\|\|full | 1863 | No | NA | 5 | 0 | 0 | Low-quality | Genome-fragment | 3.95 | AAI-based (medium-confidence) | 0 |
| S5_vs_k141_1480\|\|full | 2531 | No | NA | 4 | 0 | 4 | Not-determined | Genome-fragment | NA | NA | 0 |
| S5_vs_k141_128398\|\|full | 26290 | No | NA | 36 | 18 | 0 | Medium-quality | Genome-fragment | 55.6 | AAI-based (medium-confidence) | 0 |
| S5_vs_k141_110413\|\|full | 2221 | No | NA | 3 | 0 | 2 | Not-determined | Genome-fragment | NA | NA | 0 |
| S5_vs_k141_73676\|\|full | 1605 | No | NA | 3 | 0 | 0 | Not-determined | Genome-fragment | NA | NA | 0 |
| S10_vs_104198\|\|full | 1863 | No | NA | 4 | 2 | 0 | Low-quality | Genome-fragment | 4.78 | AAI-based (medium-confidence) | 0 |
| S5_vs_k141_134497\|\|full | 1320 | No | NA | 2 | 0 | 1 | Not-determined | Genome-fragment | NA | NA | 0 |
| S10_vs_119812\|\|full | 1863 | No | NA | 3 | 2 | 0 | Low-quality | Genome-fragment | 3.22 | AAI-based (medium-confidence) | 0 |
| S12_vs_k141_97559\|\|full | 1863 | No | NA | 4 | 0 | 0 | Low-quality | Genome-fragment | 3.76 | AAI-based (high-confidence) | 0 |
| S17_vs_k141_165181\|\|full | 1864 | No | NA | 3 | 1 | 0 | Low-quality | Genome-fragment | 3.21 | AAI-based (high-confidence) | 0 |
| S1_vs_k141_117929\|\|full | 1864 | No | NA | 3 | 0 | 0 | Low-quality | Genome-fragment | 2.43 | AAI-based (medium-confidence) | 0 |
| S5_vs_k141_49639\|\|full | 1580 | No | NA | 2 | 0 | 0 | Not-determined | Genome-fragment | NA | NA | 0 |
| S3_extracted_k141_57871 | 1864 | No | NA | 6 | 0 | 0 | Low-quality | Genome-fragment | 3.14 | AAI-based (high-confidence) | 0 |
| S5_vs_k141_123256\|\|full | 86610 | Yes | 31697 | 75 | 11 | 31 | Medium-quality | Genome-fragment | 51.46 | HMM-based (lower-bound) | 63.4 |
| S5_vs_k141_13254\|\|full | 31809 | No | NA | 45 | 13 | 1 | Medium-quality | Genome-fragment | 81.74 | AAI-based (medium-confidence) | 0 |
| S5_extracted_k141_58428 | 1864 | No | NA | 2 | 2 | 0 | Low-quality | Genome-fragment | 4.79 | AAI-based (medium-confidence) | 0 |
| S16_vs_158818\|\|full | 1864 | No | NA | 3 | 0 | 2 | Low-quality | Genome-fragment | 0.59 | AAI-based (medium-confidence) | 0 |
| S5_vs_k141_97999\|\|full | 2113 | No | NA | 3 | 0 | 3 | Not-determined | Genome-fragment | NA | NA | 0 |
| S5_vs_k141_134534\|\|full | 5765 | No | NA | 5 | 0 | 4 | Not-determined | Genome-fragment | NA | NA | 0 |
| S10_extracted_5501 | 1864 | No | NA | 2 | 0 | 0 | Low-quality | Genome-fragment | 2.22 | AAI-based (high-confidence) | 0 |
| S5_vs_k141_31905\|\|full | 1926 | No | NA | 6 | 0 | 0 | Not-determined | Genome-fragment | NA | NA | 0 |
| S5_vs_k141_1522\|\|full | 1264 | No | NA | 2 | 0 | 2 | Not-determined | Genome-fragment | NA | NA | 0 |
| S5_vs_k141_98006\|\|full | 1879 | No | NA | 4 | 0 | 1 | Not-determined | Genome-fragment | NA | NA | 0 |
| S5_vs_k141_117024\|\|full | 1108 | No | NA | 2 | 0 | 1 | Not-determined | Genome-fragment | NA | NA | 0 |
| S5_vs_k141_134540\|\|full | 1441 | No | NA | 3 | 0 | 0 | Not-determined | Genome-fragment | NA | NA | 0 |
| S12_vs_k141_111442\|\|full | 1864 | No | NA | 3 | 0 | 1 | Low-quality | Genome-fragment | 1.16 | AAI-based (high-confidence) | 0 |
| S12_vs_k141_83780\|\|full | 1864 | No | NA | 3 | 0 | 0 | Low-quality | Genome-fragment | 4.88 | AAI-based (high-confidence) | 0 |
| S5_vs_k141_1552\|\|full | 1130 | No | NA | 2 | 0 | 1 | Not-determined | Genome-fragment | NA | NA | 0 |
| S15_vs_6441\|\|full | 1864 | No | NA | 2 | 0 | 0 | Low-quality | Genome-fragment | 2.78 | AAI-based (medium-confidence) | 0 |
| S5_vs_k141_49689\|\|full | 5878 | No | NA | 10 | 0 | 4 | Not-determined | Genome-fragment | NA | NA | 0 |
| S5_vs_k141_20850\|\|full | 1865 | No | NA | 4 | 2 | 0 | Low-quality | Genome-fragment | 5.85 | AAI-based (high-confidence) | 0 |
| S7_vs_94157\|\|full | 1865 | No | NA | 6 | 0 | 0 | Low-quality | Genome-fragment | 3.67 | AAI-based (medium-confidence) | 0 |
| S5_vs_k141_104399\|\|full | 1539 | No | NA | 3 | 0 | 0 | Not-determined | Genome-fragment | NA | NA | 0 |
| S5_vs_k141_61857\|\|full | 1620 | No | NA | 3 | 0 | 0 | Not-determined | Genome-fragment | NA | NA | 0 |
| S16_extracted_53540 | 1865 | No | NA | 2 | 0 | 0 | Low-quality | Genome-fragment | 4.13 | AAI-based (high-confidence) | 0 |
| S5_vs_k141_140597\|\|full | 1676 | No | NA | 2 | 0 | 1 | Not-determined | Genome-fragment | NA | NA | 0 |
| S16_vs_128652\|\|lt2gene | 1865 | No | NA | 2 | 2 | 0 | Low-quality | Genome-fragment | 4.73 | AAI-based (high-confidence) | 0 |
| S10_vs_23468\|\|full | 1865 | Yes | 930 | 4 | 1 | 1 | Low-quality | Genome-fragment | 1.86 | HMM-based (lower-bound) | 50.13 |
| S12_vs_k141_141992\|\|full | 1865 | No | NA | 5 | 3 | 0 | Low-quality | Genome-fragment | 4.28 | AAI-based (medium-confidence) | 0 |
| S5_vs_k141_110537\|\|full | 1940 | No | NA | 3 | 0 | 1 | Not-determined | Genome-fragment | NA | NA | 0 |
| S5_vs_k141_13303\|\|full | 1443 | No | NA | 3 | 0 | 1 | Not-determined | Genome-fragment | NA | NA | 0 |
| S5_vs_k141_24897\|\|full | 3911 | No | NA | 4 | 0 | 2 | Not-determined | Genome-fragment | NA | NA | 0 |
| S12_vs_k141_111742\|\|full | 1865 | No | NA | 3 | 0 | 1 | Low-quality | Genome-fragment | 2.96 | AAI-based (high-confidence) | 0 |
| S5_vs_k141_92424\|\|full | 1046 | No | NA | 2 | 0 | 1 | Not-determined | Genome-fragment | NA | NA | 0 |
| S8_extracted_k141_47797 | 1865 | No | NA | 2 | 0 | 0 | Low-quality | Genome-fragment | 1.05 | AAI-based (medium-confidence) | 0 |
| S5_vs_k141_110559\|\|full | 6099 | No | NA | 11 | 0 | 0 | Not-determined | Genome-fragment | NA | NA | 0 |
| S17_extracted_k141_170745 | 1866 | No | NA | 5 | 5 | 0 | Low-quality | Genome-fragment | 6.15 | AAI-based (high-confidence) | 0 |
| S5_vs_k141_1572\|\|full | 37649 | No | NA | 48 | 7 | 1 | Medium-quality | Genome-fragment | 83.1 | AAI-based (high-confidence) | 0 |
| S17_extracted_k141_34085 | 1866 | No | NA | 6 | 6 | 0 | Low-quality | Genome-fragment | 6.15 | AAI-based (high-confidence) | 0 |
| S17_vs_k141_146571\|\|full | 1866 | No | NA | 3 | 1 | 0 | Low-quality | Genome-fragment | 32.62 | AAI-based (high-confidence) | 0 |
| S13_extracted_21591 | 1866 | No | NA | 3 | 0 | 0 | Low-quality | Genome-fragment | 2.94 | AAI-based (medium-confidence) | 0 |
| S1_vs_k141_117197\|\|full | 1866 | No | NA | 2 | 1 | 0 | Low-quality | Genome-fragment | 3.19 | AAI-based (high-confidence) | 0 |
| S1_vs_k141_105073\|\|full | 1866 | No | NA | 2 | 0 | 0 | Low-quality | Genome-fragment | 2.3 | AAI-based (high-confidence) | 0 |
| S5_vs_k141_67971\|\|full | 16082 | No | NA | 13 | 0 | 9 | Not-determined | Genome-fragment | NA | NA | 0 |
| S7_vs_42668\|\|full | 1866 | No | NA | 6 | 1 | 0 | Low-quality | Genome-fragment | 3.79 | AAI-based (medium-confidence) | 0 |
| S7_vs_96508\|\|full | 1866 | No | NA | 3 | 0 | 0 | Low-quality | Genome-fragment | 4.48 | AAI-based (high-confidence) | 0 |
| S12_vs_k141_11464\|\|full | 1866 | Yes | 729 | 5 | 1 | 1 | Low-quality | Genome-fragment | 0.45 | HMM-based (lower-bound) | 60.93 |
| S4_extracted_13336 | 1866 | No | NA | 4 | 3 | 0 | Low-quality | Genome-fragment | 6.15 | AAI-based (high-confidence) | 0 |
| S5_vs_k141_110613\|\|full | 1371 | No | NA | 1 | 0 | 0 | Not-determined | Genome-fragment | NA | NA | 0 |
| S5_vs_k141_49808\|\|full | 3458 | No | NA | 3 | 0 | 2 | Not-determined | Genome-fragment | NA | NA | 0 |
| S8_vs_k141_56158\|\|full | 1866 | No | NA | 5 | 1 | 0 | Low-quality | Genome-fragment | 4.47 | AAI-based (high-confidence) | 0 |
| S8_vs_k141_160863\|\|full | 1866 | No | NA | 3 | 0 | 1 | Low-quality | Genome-fragment | 1.29 | AAI-based (medium-confidence) | 0 |
| S14_vs_k141_56112\|\|full | 1867 | No | NA | 4 | 1 | 0 | Low-quality | Genome-fragment | 4.47 | AAI-based (high-confidence) | 0 |
| S5_vs_k141_115566\|\|full | 1867 | No | NA | 3 | 2 | 0 | Low-quality | Genome-fragment | 4.12 | AAI-based (high-confidence) | 0 |
| S16_extracted_53505 | 1867 | No | NA | 7 | 1 | 0 | Low-quality | Genome-fragment | 4.53 | AAI-based (high-confidence) | 0 |
| S5_vs_k141_49827\|\|full | 2779 | No | NA | 4 | 0 | 1 | Not-determined | Genome-fragment | NA | NA | 0 |
| S10_vs_114820\|\|full | 1867 | No | NA | 5 | 1 | 0 | Low-quality | Genome-fragment | 4.02 | AAI-based (high-confidence) | 0 |
| S5_vs_k141_104525\|\|full | 3135 | No | NA | 3 | 0 | 2 | Not-determined | Genome-fragment | NA | NA | 0 |
| S12_vs_k141_77044\|\|full | 1867 | No | NA | 4 | 0 | 0 | Low-quality | Genome-fragment | 4.05 | AAI-based (high-confidence) | 0 |
| S5_vs_k141_92533\|\|full | 2306 | No | NA | 3 | 0 | 2 | Not-determined | Genome-fragment | NA | NA | 0 |
| S5_vs_k141_55440\|\|full | 3098 | No | NA | 3 | 0 | 2 | Not-determined | Genome-fragment | NA | NA | 0 |
| S17_vs_k141_25729\|\|full | 1868 | No | NA | 4 | 1 | 0 | Low-quality | Genome-fragment | 4.47 | HMM-based (lower-bound) | 0 |
| S5_vs_k141_117173\|\|full | 2270 | No | NA | 7 | 0 | 0 | Not-determined | Genome-fragment | NA | NA | 0 |
| S14_extracted_k141_116845 | 1868 | No | NA | 4 | 0 | 0 | Low-quality | Genome-fragment | 3.19 | AAI-based (high-confidence) | 0 |
| S1_vs_k141_159646\|\|full | 1868 | No | NA | 4 | 0 | 3 | Low-quality | Genome-fragment | 0.83 | AAI-based (medium-confidence) | 0 |
| S12_vs_k141_12626\|\|full | 1868 | Yes | 987 | 2 | 1 | 1 | Low-quality | Genome-fragment | 33.09 | AAI-based (high-confidence) | 47.16 |
| S8_extracted_k141_71812 | 1868 | No | NA | 2 | 0 | 0 | Low-quality | Genome-fragment | 3.32 | AAI-based (medium-confidence) | 0 |
| S5_vs_k141_128499\|\|full | 3716 | No | NA | 5 | 0 | 2 | Not-determined | Genome-fragment | NA | NA | 0 |
| S5_vs_k141_98061\|\|full | 2441 | No | NA | 4 | 0 | 3 | Not-determined | Genome-fragment | NA | NA | 0 |
| S8_vs_k141_48938\|\|full | 1868 | No | NA | 3 | 0 | 0 | Low-quality | Genome-fragment | 0.79 | AAI-based (medium-confidence) | 0 |
| S5_vs_k141_49850\|\|full | 2182 | No | NA | 2 | 0 | 2 | Not-determined | Genome-fragment | NA | NA | 0 |
| S17_vs_k141_153979\|\|full | 1869 | No | NA | 3 | 0 | 2 | Low-quality | Genome-fragment | 2.34 | AAI-based (medium-confidence) | 0 |
| S1_vs_k141_27527\|\|full | 1869 | No | NA | 4 | 1 | 0 | Low-quality | Genome-fragment | 4.67 | AAI-based (high-confidence) | 0 |
| S5_vs_k141_128558\|\|full | 3050 | No | NA | 2 | 0 | 2 | Not-determined | Genome-fragment | NA | NA | 0 |
| S5_vs_k141_32157\|\|full | 1582 | No | NA | 2 | 0 | 2 | Not-determined | Genome-fragment | NA | NA | 0 |
| S5_vs_k141_92572\|\|full | 4132 | No | NA | 4 | 0 | 2 | Not-determined | Genome-fragment | NA | NA | 0 |
| S1_vs_k141_139406\|\|full | 1869 | No | NA | 5 | 1 | 0 | Low-quality | Genome-fragment | 4.14 | AAI-based (high-confidence) | 0 |
| S3_extracted_k141_69013 | 1869 | No | NA | 5 | 1 | 0 | Low-quality | Genome-fragment | 1.34 | HMM-based (lower-bound) | 0 |
| S7_extracted_112192 | 1869 | No | NA | 3 | 3 | 0 | Low-quality | Genome-fragment | 4.53 | AAI-based (high-confidence) | 0 |
| S5_vs_k141_117191\|\|full | 2207 | No | NA | 2 | 0 | 0 | Not-determined | Genome-fragment | NA | NA | 0 |
| S7_extracted_127030 | 1869 | No | NA | 3 | 0 | 0 | Low-quality | Genome-fragment | 4.13 | AAI-based (medium-confidence) | 0 |
| S5_vs_k141_92588\|\|full | 1725 | No | NA | 4 | 0 | 0 | Not-determined | Genome-fragment | NA | NA | 0 |
| S5_vs_k141_49905\|\|full | 6459 | No | NA | 6 | 0 | 6 | Not-determined | Genome-fragment | NA | NA | 0 |
| S5_vs_k141_24919\|\|full | 1934 | No | NA | 4 | 0 | 1 | Not-determined | Genome-fragment | NA | NA | 0 |
| S5_vs_k141_55512\|\|full | 1559 | No | NA | 2 | 0 | 2 | Not-determined | Genome-fragment | NA | NA | 0 |
| S5_vs_k141_128608\|\|full | 1908 | No | NA | 3 | 0 | 1 | Not-determined | Genome-fragment | NA | NA | 0 |
| S11_vs_k141_81732\|\|full | 1870 | No | NA | 5 | 2 | 0 | Low-quality | Genome-fragment | 3.89 | AAI-based (high-confidence) | 0 |
| S3_vs_k141_111829\|\|full | 1870 | Yes | 1101 | 4 | 1 | 1 | Low-quality | Genome-fragment | 0.75 | AAI-based (medium-confidence) | 41.12 |
| S5_vs_k141_44163\|\|full | 5065 | No | NA | 3 | 0 | 0 | Not-determined | Genome-fragment | NA | NA | 0 |
| S3_vs_k141_4364\|\|full | 1870 | No | NA | 3 | 0 | 0 | Low-quality | Genome-fragment | 1.22 | AAI-based (high-confidence) | 0 |
| S3_vs_k141_21511\|\|full | 1870 | No | NA | 6 | 1 | 0 | Low-quality | Genome-fragment | 4.13 | AAI-based (high-confidence) | 0 |
| S5_vs_k141_49951\|\|full | 2397 | No | NA | 4 | 0 | 0 | Not-determined | Genome-fragment | NA | NA | 0 |
| S5_vs_k141_134579\|\|full | 1610 | No | NA | 3 | 0 | 1 | Not-determined | Genome-fragment | NA | NA | 0 |
| S5_vs_k141_13434\|\|full | 1786 | No | NA | 3 | 0 | 2 | Not-determined | Genome-fragment | NA | NA | 0 |
| S16_vs_80046\|\|full | 1870 | No | NA | 3 | 1 | 0 | Low-quality | Genome-fragment | 3.48 | AAI-based (high-confidence) | 0 |
| S15_vs_59636\|\|full | 1870 | No | NA | 3 | 0 | 2 | Low-quality | Genome-fragment | 3.32 | AAI-based (medium-confidence) | 0 |
| S4_extracted_25051 | 1870 | No | NA | 2 | 2 | 0 | Low-quality | Genome-fragment | 3.18 | AAI-based (high-confidence) | 0 |
| S11_vs_k141_50328\|\|full | 1871 | No | NA | 4 | 2 | 0 | Low-quality | Genome-fragment | 3.03 | AAI-based (medium-confidence) | 0 |
| S5_vs_k141_1731\|\|full | 2222 | No | NA | 3 | 0 | 3 | Not-determined | Genome-fragment | NA | NA | 0 |
| S7_vs_49663\|\|full | 1871 | No | NA | 4 | 1 | 0 | Low-quality | Genome-fragment | 1.12 | AAI-based (medium-confidence) | 0 |
| S10_extracted_106184 | 1871 | No | NA | 1 | 1 | 0 | Low-quality | Genome-fragment | 1.43 | HMM-based (lower-bound) | 0 |
| S5_vs_k141_13482\|\|full | 8316 | No | NA | 8 | 0 | 4 | Not-determined | Genome-fragment | NA | NA | 0 |
| S10_vs_5588\|\|full | 1871 | No | NA | 4 | 0 | 0 | Low-quality | Genome-fragment | 1.26 | AAI-based (medium-confidence) | 0 |
| S5_vs_k141_7598\|\|full | 3521 | No | NA | 3 | 0 | 1 | Not-determined | Genome-fragment | NA | NA | 0 |
| S10_vs_108886\|\|full | 1871 | No | NA | 3 | 0 | 1 | Low-quality | Genome-fragment | 2.46 | AAI-based (medium-confidence) | 0 |
| S12_extracted_k141_142448 | 1871 | No | NA | 1 | 0 | 0 | Low-quality | Genome-fragment | 1.79 | AAI-based (high-confidence) | 0 |
| S5_vs_k141_140774\|\|full | 2221 | No | NA | 3 | 0 | 3 | Not-determined | Genome-fragment | NA | NA | 0 |
| S5_vs_k141_128710\|\|full | 2552 | No | NA | 3 | 0 | 1 | Not-determined | Genome-fragment | NA | NA | 0 |
| S5_vs_k141_140777\|\|full | 2067 | No | NA | 3 | 0 | 0 | Not-determined | Genome-fragment | NA | NA | 0 |
| S5_vs_k141_134618\|\|full | 3406 | No | NA | 3 | 0 | 1 | Not-determined | Genome-fragment | NA | NA | 0 |
| S11_vs_k141_46683\|\|lt2gene | 1872 | No | NA | 2 | 2 | 0 | Low-quality | Genome-fragment | 3.66 | HMM-based (lower-bound) | 0 |
| S13_vs_133522\|\|full | 1872 | No | NA | 4 | 0 | 1 | Low-quality | Genome-fragment | 3.19 | AAI-based (medium-confidence) | 0 |
| S12_vs_k141_165991\|\|full | 1872 | No | NA | 4 | 0 | 0 | Low-quality | Genome-fragment | 3.36 | AAI-based (medium-confidence) | 0 |
| S8_vs_k141_175590\|\|full | 1872 | No | NA | 3 | 0 | 3 | Low-quality | Genome-fragment | 1.53 | AAI-based (medium-confidence) | 0 |
| S8_vs_k141_90195\|\|full | 1872 | No | NA | 4 | 0 | 0 | Low-quality | Genome-fragment | 3.64 | AAI-based (medium-confidence) | 0 |
| S17_vs_k141_43733\|\|full | 1873 | No | NA | 2 | 2 | 0 | Low-quality | Genome-fragment | 3.99 | AAI-based (high-confidence) | 0 |
| S11_extracted_k141_67354 | 1873 | No | NA | 3 | 1 | 0 | Low-quality | Genome-fragment | 4.59 | AAI-based (high-confidence) | 0 |
| S5_vs_k141_50077\|\|full | 1843 | No | NA | 2 | 0 | 1 | Not-determined | Genome-fragment | NA | NA | 0 |
| S5_vs_k141_24973\|\|full | 1047 | No | NA | 2 | 0 | 1 | Not-determined | Genome-fragment | NA | NA | 0 |
| S11_vs_k141_6214\|\|full | 1873 | No | NA | 4 | 1 | 0 | Low-quality | Genome-fragment | 4.08 | AAI-based (high-confidence) | 0 |
| S5_vs_k141_86364\|\|full | 1247 | No | NA | 4 | 0 | 0 | Not-determined | Genome-fragment | NA | NA | 0 |
| S14_vs_k141_43328\|\|full | 1873 | No | NA | 3 | 1 | 0 | Low-quality | Genome-fragment | 5.58 | AAI-based (high-confidence) | 0 |
| S11_extracted_k141_77284 | 1874 | No | NA | 5 | 3 | 0 | Low-quality | Genome-fragment | 4.64 | AAI-based (high-confidence) | 0 |
| S5_vs_k141_123514\|\|full | 7382 | No | NA | 10 | 0 | 4 | Not-determined | Genome-fragment | NA | NA | 0 |
| S5_vs_k141_32255\|\|full | 1451 | No | NA | 3 | 0 | 1 | Not-determined | Genome-fragment | NA | NA | 0 |
| S5_vs_k141_13526\|\|full | 1969 | No | NA | 2 | 0 | 2 | Not-determined | Genome-fragment | NA | NA | 0 |
| S5_vs_k141_123532\|\|full | 1641 | No | NA | 3 | 0 | 0 | Not-determined | Genome-fragment | NA | NA | 0 |
| S5_vs_k141_7632\|\|full | 4726 | No | NA | 5 | 0 | 3 | Not-determined | Genome-fragment | NA | NA | 0 |
| S5_vs_k141_13534\|\|full | 3630 | No | NA | 6 | 0 | 2 | Not-determined | Genome-fragment | NA | NA | 0 |
| S5_vs_k141_37925\|\|full | 2000 | No | NA | 5 | 0 | 0 | Not-determined | Genome-fragment | NA | NA | 0 |
| S5_vs_k141_62040\|\|full | 3276 | No | NA | 3 | 0 | 2 | Not-determined | Genome-fragment | NA | NA | 0 |
| S5_vs_k141_44304\|\|full | 3434 | No | NA | 6 | 0 | 1 | Not-determined | Genome-fragment | NA | NA | 0 |
| S1_vs_k141_37703\|\|full | 1874 | No | NA | 3 | 0 | 0 | Low-quality | Genome-fragment | 3.18 | AAI-based (high-confidence) | 0 |
| S5_vs_k141_7657\|\|full | 1790 | No | NA | 2 | 0 | 2 | Not-determined | Genome-fragment | NA | NA | 0 |
| S7_vs_122197\|\|full | 1874 | No | NA | 3 | 2 | 0 | Low-quality | Genome-fragment | 4.25 | AAI-based (medium-confidence) | 0 |
| S5_vs_k141_134660\|\|full | 10154 | No | NA | 15 | 0 | 3 | Not-determined | Genome-fragment | NA | NA | 0 |
| S5_vs_k141_7675\|\|full | 1971 | No | NA | 2 | 0 | 1 | Not-determined | Genome-fragment | NA | NA | 0 |
| S5_vs_k141_32333\|\|full | 1990 | No | NA | 3 | 0 | 2 | Not-determined | Genome-fragment | NA | NA | 0 |
| S16_vs_72815\|\|full | 1874 | No | NA | 2 | 0 | 0 | Low-quality | Genome-fragment | 2.96 | AAI-based (medium-confidence) | 0 |
| S14_vs_k141_86284\|\|full | 1875 | No | NA | 2 | 0 | 0 | Low-quality | Genome-fragment | 3.22 | AAI-based (medium-confidence) | 0 |
| S16_extracted_12309 | 1875 | No | NA | 2 | 0 | 0 | Low-quality | Genome-fragment | 4.1 | AAI-based (medium-confidence) | 0 |
| S5_vs_k141_117319\|\|full | 108449 | Yes | 63947 | 138 | 32 | 31 | Complete | High-quality | 100 | Provirus (high-confidence) | 41.03 |
| S12_vs_k141_79861\|\|full | 1875 | No | NA | 4 | 2 | 0 | Low-quality | Genome-fragment | 5.37 | AAI-based (medium-confidence) | 0 |
| S5_vs_k141_13624\|\|full | 2692 | No | NA | 3 | 0 | 2 | Not-determined | Genome-fragment | NA | NA | 0 |
| S5_vs_k141_62096\|\|full | 8735 | No | NA | 11 | 0 | 1 | Not-determined | Genome-fragment | NA | NA | 0 |
| S5_vs_k141_13642\|\|full | 5815 | No | NA | 8 | 0 | 2 | Not-determined | Genome-fragment | NA | NA | 0 |
| S5_vs_k141_1897\|\|full | 4348 | No | NA | 3 | 0 | 1 | Not-determined | Genome-fragment | NA | NA | 0 |
| S5_vs_k141_19715\|\|full | 1159 | No | NA | 3 | 0 | 2 | Not-determined | Genome-fragment | NA | NA | 0 |
| S13_extracted_17989 | 1876 | No | NA | 1 | 0 | 1 | Low-quality | Genome-fragment | 1.74 | AAI-based (medium-confidence) | 0 |
| S7_vs_9934\|\|full | 1876 | No | NA | 3 | 1 | 0 | Low-quality | Genome-fragment | 4.42 | AAI-based (high-confidence) | 0 |
| S12_extracted_k141_111554 | 1876 | No | NA | 2 | 0 | 2 | Low-quality | Genome-fragment | 0.1 | AAI-based (medium-confidence) | 0 |
| S11_extracted_k141_90780 | 1877 | No | NA | 5 | 0 | 0 | Low-quality | Genome-fragment | 2.82 | AAI-based (high-confidence) | 0 |
| S11_vs_k141_95186\|\|full | 1877 | No | NA | 4 | 2 | 0 | Low-quality | Genome-fragment | 3.02 | AAI-based (high-confidence) | 0 |
| S13_vs_134140\|\|full | 1877 | No | NA | 3 | 0 | 2 | Low-quality | Genome-fragment | 2.44 | AAI-based (medium-confidence) | 0 |
| S14_vs_k141_41484\|\|full | 1877 | No | NA | 3 | 0 | 0 | Low-quality | Genome-fragment | 3.9 | AAI-based (high-confidence) | 0 |
| S14_vs_k141_25128\|\|lt2gene | 1877 | No | NA | 2 | 1 | 0 | Low-quality | Genome-fragment | 4.53 | AAI-based (high-confidence) | 0 |
| S5_vs_k141_62152\|\|full | 4449 | No | NA | 2 | 0 | 1 | Not-determined | Genome-fragment | NA | NA | 0 |
| S5_vs_k141_133813\|\|full | 1877 | No | NA | 3 | 0 | 0 | Low-quality | Genome-fragment | 3.84 | AAI-based (medium-confidence) | 0 |
| S5_vs_k141_55598\|\|full | 8259 | No | NA | 11 | 0 | 9 | Not-determined | Genome-fragment | NA | NA | 0 |
| S5_vs_k141_117417\|\|full | 19695 | No | NA | 32 | 11 | 0 | Medium-quality | Genome-fragment | 50.78 | AAI-based (medium-confidence) | 0 |
| S5_vs_k141_1973\|\|full | 1372 | No | NA | 3 | 0 | 1 | Not-determined | Genome-fragment | NA | NA | 0 |
| S7_vs_135111\|\|full | 1877 | No | NA | 4 | 0 | 0 | Low-quality | Genome-fragment | 3.28 | AAI-based (high-confidence) | 0 |
| S5_vs_k141_32419\|\|full | 2804 | No | NA | 3 | 0 | 3 | Not-determined | Genome-fragment | NA | NA | 0 |
| S5_vs_k141_140856\|\|full | 4985 | No | NA | 8 | 0 | 0 | Not-determined | Genome-fragment | NA | NA | 0 |
| S12_vs_k141_59622\|\|full | 1877 | No | NA | 6 | 5 | 0 | Low-quality | Genome-fragment | 5.37 | AAI-based (medium-confidence) | 0 |
| S3_vs_k141_124743\|\|full | 1878 | No | NA | 3 | 1 | 0 | Low-quality | Genome-fragment | 4.98 | AAI-based (high-confidence) | 0 |
| S5_vs_k141_62216\|\|full | 3101 | No | NA | 4 | 0 | 2 | Not-determined | Genome-fragment | NA | NA | 0 |
| S5_vs_k141_80165\|\|full | 1878 | No | NA | 5 | 1 | 0 | Low-quality | Genome-fragment | 3.47 | AAI-based (high-confidence) | 0 |
| S10_extracted_58364 | 1878 | No | NA | 3 | 0 | 0 | Low-quality | Genome-fragment | 3.83 | AAI-based (medium-confidence) | 0 |
| S10_extracted_79311 | 1878 | No | NA | 2 | 0 | 2 | Low-quality | Genome-fragment | 0.5 | AAI-based (medium-confidence) | 0 |
| S12_vs_k141_25761\|\|full | 1878 | No | NA | 4 | 0 | 0 | Low-quality | Genome-fragment | 4.57 | AAI-based (high-confidence) | 0 |
| S5_vs_k141_7785\|\|full | 2572 | No | NA | 2 | 0 | 2 | Not-determined | Genome-fragment | NA | NA | 0 |
| S5_vs_k141_128854\|\|full | 14458 | No | NA | 15 | 0 | 2 | Not-determined | Genome-fragment | NA | NA | 0 |
| S12_vs_k141_183312\|\|full | 1878 | No | NA | 3 | 0 | 2 | Low-quality | Genome-fragment | 2.94 | AAI-based (medium-confidence) | 0 |
| S5_vs_k141_128855\|\|full | 1205 | No | NA | 3 | 0 | 0 | Not-determined | Genome-fragment | NA | NA | 0 |
| S12_vs_k141_121816\|\|full | 1878 | No | NA | 5 | 1 | 0 | Low-quality | Genome-fragment | 3.76 | AAI-based (high-confidence) | 0 |
| S9_vs_k141_28964\|\|full | 1879 | No | NA | 5 | 2 | 0 | Low-quality | Genome-fragment | 2.7 | AAI-based (high-confidence) | 0 |
| S14_extracted_k141_30752 | 1879 | No | NA | 2 | 0 | 1 | Low-quality | Genome-fragment | 0.83 | AAI-based (medium-confidence) | 0 |
| S5_vs_k141_55666\|\|full | 1026 | No | NA | 1 | 0 | 1 | Not-determined | Genome-fragment | NA | NA | 0 |
| S1_vs_k141_103087\|\|full | 1879 | No | NA | 6 | 1 | 0 | Low-quality | Genome-fragment | 5.18 | AAI-based (high-confidence) | 0 |
| S1_vs_k141_157757\|\|full | 1879 | No | NA | 2 | 0 | 0 | Low-quality | Genome-fragment | 2.64 | AAI-based (high-confidence) | 0 |
| S7_vs_7065\|\|full | 1879 | No | NA | 4 | 1 | 0 | Low-quality | Genome-fragment | 3.64 | HMM-based (lower-bound) | 0 |
| S5_vs_k141_134795\|\|full | 1089 | No | NA | 3 | 0 | 0 | Not-determined | Genome-fragment | NA | NA | 0 |
| S5_vs_k141_68305\|\|full | 2459 | No | NA | 3 | 0 | 1 | Not-determined | Genome-fragment | NA | NA | 0 |
| S7_vs_49848\|\|full | 1879 | No | NA | 6 | 3 | 0 | Low-quality | Genome-fragment | 4.34 | AAI-based (medium-confidence) | 0 |
| S5_vs_k141_68316\|\|full | 2096 | No | NA | 3 | 0 | 2 | Not-determined | Genome-fragment | NA | NA | 0 |
| S5_vs_k141_25183\|\|full | 7822 | No | NA | 9 | 0 | 4 | Not-determined | Genome-fragment | NA | NA | 0 |
| S5_vs_k141_128881\|\|full | 26715 | No | NA | 45 | 6 | 0 | Medium-quality | Genome-fragment | 54.47 | AAI-based (high-confidence) | 0 |
| S5_vs_k141_92882\|\|full | 20826 | No | NA | 19 | 0 | 16 | Not-determined | Genome-fragment | NA | NA | 0 |
| S5_vs_k141_86562\|\|full | 37878 | No | NA | 51 | 7 | 1 | High-quality | High-quality | 100 | AAI-based (medium-confidence) | 0 |
| S7_vs_110362\|\|full | 1879 | No | NA | 5 | 1 | 0 | Low-quality | Genome-fragment | 3.43 | AAI-based (medium-confidence) | 0 |
| S12_vs_k141_88800\|\|full | 1879 | No | NA | 3 | 0 | 1 | Low-quality | Genome-fragment | 2.34 | AAI-based (high-confidence) | 0 |
| S8_extracted_k141_79223 | 1879 | No | NA | 5 | 2 | 0 | Low-quality | Genome-fragment | 5.79 | AAI-based (high-confidence) | 0 |
| S9_extracted_k141_59883 | 1880 | No | NA | 4 | 0 | 0 | Low-quality | Genome-fragment | 3.14 | AAI-based (medium-confidence) | 0 |
| S9_vs_k141_11866\|\|full | 1880 | No | NA | 4 | 0 | 1 | Low-quality | Genome-fragment | 4.74 | AAI-based (medium-confidence) | 0 |
| S3_vs_k141_63952\|\|full | 1880 | No | NA | 5 | 1 | 0 | Low-quality | Genome-fragment | 4.48 | AAI-based (medium-confidence) | 0 |
| S5_vs_k141_111011\|\|full | 3772 | No | NA | 5 | 0 | 2 | Not-determined | Genome-fragment | NA | NA | 0 |
| S5_vs_k141_2037\|\|full | 1423 | No | NA | 4 | 0 | 0 | Not-determined | Genome-fragment | NA | NA | 0 |
| S7_vs_31288\|\|full | 1880 | No | NA | 3 | 0 | 0 | Low-quality | Genome-fragment | 3.7 | AAI-based (high-confidence) | 0 |
| S10_extracted_43219 | 1880 | Yes | 587 | 3 | 1 | 1 | Low-quality | Genome-fragment | 0.34 | AAI-based (high-confidence) | 68.78 |
| S5_vs_k141_79720\|\|full | 4599 | No | NA | 5 | 0 | 3 | Not-determined | Genome-fragment | NA | NA | 0 |
| S8_vs_k141_154170\|\|full | 1880 | No | NA | 2 | 0 | 0 | Low-quality | Genome-fragment | 8.21 | AAI-based (high-confidence) | 0 |
| S5_vs_k141_140946\|\|full | 26678 | No | NA | 33 | 8 | 0 | Medium-quality | Genome-fragment | 68.08 | AAI-based (high-confidence) | 0 |

| | | | | | | | | | | | |
|---|---|---|---|---|---|---|---|---|---|---|---|
| S11_vs_k141_49437\|\|full | 1881 | No | NA | 3 | 1 | 0 | Low-quality | Genome-fragment | 1.92 | AAI-based (medium-confidence) | 0 |
| S5_vs_k141_128952\|\|full | 2358 | No | NA | 3 | 0 | 3 | Not-determined | Genome-fragment | NA | NA | 0 |
| S5_extracted_k141_126341 | 1881 | No | NA | 2 | 0 | 2 | Low-quality | Genome-fragment | 1.39 | AAI-based (medium-confidence) | 0 |
| S7_vs_21116\|\|full | 1881 | No | NA | 3 | 0 | 0 | Low-quality | Genome-fragment | 3.54 | AAI-based (high-confidence) | 0 |
| S16_vs_12331\|\|full | 1881 | No | NA | 4 | 1 | 0 | Low-quality | Genome-fragment | 2.79 | HMM-based (lower-bound) | 0 |
| S5_vs_k141_111038\|\|full | 1951 | No | NA | 2 | 0 | 0 | Not-determined | Genome-fragment | NA | NA | 0 |
| S5_vs_k141_68353\|\|full | 5182 | No | NA | 6 | 0 | 4 | Not-determined | Genome-fragment | NA | NA | 0 |
| S6_extracted_k141_15912 | 1881 | No | NA | 4 | 0 | 2 | Low-quality | Genome-fragment | 1.87 | AAI-based (medium-confidence) | 0 |
| S5_vs_k141_117553\|\|full | 1581 | No | NA | 3 | 0 | 1 | Not-determined | Genome-fragment | NA | NA | 0 |
| S5_vs_k141_92979\|\|full | 1913 | No | NA | 4 | 0 | 1 | Not-determined | Genome-fragment | NA | NA | 0 |
| S5_vs_k141_50368\|\|full | 2407 | No | NA | 2 | 0 | 0 | Not-determined | Genome-fragment | NA | NA | 0 |
| S8_extracted_k141_3859 | 1881 | No | NA | 2 | 0 | 1 | Low-quality | Genome-fragment | 0.49 | AAI-based (high-confidence) | 0 |
| S17_extracted_k141_99561 | 1882 | No | NA | 2 | 0 | 0 | Low-quality | Genome-fragment | 3.95 | AAI-based (medium-confidence) | 0 |
| S14_vs_k141_67214\|\|full | 1882 | No | NA | 2 | 1 | 0 | Low-quality | Genome-fragment | 4.81 | AAI-based (high-confidence) | 0 |
| S5_vs_k141_84356\|\|full | 1882 | No | NA | 3 | 0 | 0 | Low-quality | Genome-fragment | 3.34 | AAI-based (high-confidence) | 0 |
| S5_vs_k141_128994\|\|full | 1457 | No | NA | 3 | 0 | 1 | Not-determined | Genome-fragment | NA | NA | 0 |
| S5_vs_k141_3422\|\|lt2gene | 1882 | No | NA | 2 | 1 | 0 | Low-quality | Genome-fragment | 3.89 | AAI-based (high-confidence) | 0 |
| S5_vs_k141_25212\|\|full | 2353 | No | NA | 3 | 0 | 0 | Not-determined | Genome-fragment | NA | NA | 0 |
| S16_vs_119841\|\|full | 1882 | No | NA | 4 | 0 | 4 | Low-quality | Genome-fragment | 2.91 | AAI-based (medium-confidence) | 0 |
| S8_vs_k141_50368\|\|full | 1882 | No | NA | 4 | 0 | 0 | Low-quality | Genome-fragment | 5.97 | AAI-based (high-confidence) | 0 |
| S11_vs_k141_67327\|\|full | 1883 | No | NA | 2 | 0 | 1 | Low-quality | Genome-fragment | 3.36 | AAI-based (medium-confidence) | 0 |
| S14_extracted_k141_4344 | 1883 | No | NA | 3 | 0 | 1 | Low-quality | Genome-fragment | 1.82 | AAI-based (medium-confidence) | 0 |
| S14_vs_k141_4332\|\|full | 1883 | No | NA | 7 | 2 | 0 | Low-quality | Genome-fragment | 3.79 | AAI-based (medium-confidence) | 0 |
| S3_vs_k141_57297\|\|full | 1883 | No | NA | 4 | 0 | 0 | Low-quality | Genome-fragment | 3.58 | AAI-based (high-confidence) | 0 |
| S5_vs_k141_31315\|\|full | 1883 | No | NA | 2 | 0 | 0 | Low-quality | Genome-fragment | 1.24 | AAI-based (medium-confidence) | 0 |
| S5_vs_k141_81309\|\|full | 1883 | No | NA | 3 | 0 | 0 | Low-quality | Genome-fragment | 4.38 | AAI-based (high-confidence) | 0 |
| S7_extracted_74230 | 1883 | No | NA | 5 | 4 | 0 | Low-quality | Genome-fragment | 8.6 | AAI-based (high-confidence) | 0 |
| S5_vs_k141_134883\|\|full | 4598 | No | NA | 5 | 0 | 5 | Not-determined | Genome-fragment | NA | NA | 0 |
| S5_vs_k141_25279\|\|full | 2104 | No | NA | 3 | 0 | 1 | Not-determined | Genome-fragment | NA | NA | 0 |
| S7_extracted_117495 | 1883 | No | NA | 2 | 0 | 1 | Low-quality | Genome-fragment | 1.77 | AAI-based (medium-confidence) | 0 |
| S5_vs_k141_86714\|\|full | 2608 | No | NA | 5 | 0 | 1 | Not-determined | Genome-fragment | NA | NA | 0 |
| S5_vs_k141_2216\|\|full | 8417 | No | NA | 11 | 0 | 8 | Not-determined | Genome-fragment | NA | NA | 0 |
| S7_vs_6851\|\|full | 1883 | No | NA | 2 | 1 | 0 | Low-quality | Genome-fragment | 1.44 | HMM-based (lower-bound) | 0 |
| S5_vs_k141_129075\|\|full | 1561 | No | NA | 2 | 0 | 0 | Not-determined | Genome-fragment | NA | NA | 0 |
| S7_vs_36901\|\|full | 1883 | No | NA | 3 | 3 | 0 | Low-quality | Genome-fragment | 3.07 | HMM-based (lower-bound) | 0 |
| S7_vs_100992\|\|full | 1883 | No | NA | 3 | 0 | 1 | Low-quality | Genome-fragment | 2.81 | AAI-based (high-confidence) | 0 |
| S5_vs_k141_123696\|\|full | 11912 | No | NA | 19 | 0 | 2 | Not-determined | Genome-fragment | NA | NA | 0 |
| S5_vs_k141_32555\|\|full | 2276 | No | NA | 2 | 0 | 2 | Not-determined | Genome-fragment | NA | NA | 0 |
| S5_vs_k141_86738\|\|full | 7292 | No | NA | 7 | 0 | 0 | Not-determined | Genome-fragment | NA | NA | 0 |
| S5_vs_k141_62278\|\|full | 2428 | No | NA | 3 | 0 | 0 | Not-determined | Genome-fragment | NA | NA | 0 |
| S16_vs_53203\|\|full | 1883 | No | NA | 2 | 1 | 0 | Low-quality | Genome-fragment | 3.65 | AAI-based (medium-confidence) | 0 |
| S10_vs_43966\|\|full | 1883 | No | NA | 3 | 0 | 1 | Low-quality | Genome-fragment | 3.46 | AAI-based (medium-confidence) | 0 |
| S5_vs_k141_44573\|\|full | 2816 | No | NA | 3 | 0 | 2 | Not-determined | Genome-fragment | NA | NA | 0 |
| S12_extracted_k141_75583 | 1883 | No | NA | 4 | 2 | 0 | Low-quality | Genome-fragment | 3.03 | AAI-based (high-confidence) | 0 |
| S5_vs_k141_79810\|\|full | 5647 | No | NA | 4 | 0 | 3 | Not-determined | Genome-fragment | NA | NA | 0 |
| S12_vs_k141_116782\|\|full | 1883 | No | NA | 3 | 1 | 0 | Low-quality | Genome-fragment | 1.54 | AAI-based (high-confidence) | 0 |
| S5_vs_k141_13948\|\|full | 4347 | No | NA | 5 | 0 | 2 | Not-determined | Genome-fragment | NA | NA | 0 |
| S9_vs_k141_39096\|\|full | 1884 | No | NA | 3 | 3 | 0 | Low-quality | Genome-fragment | 4.66 | AAI-based (high-confidence) | 0 |
| S5_vs_k141_117726\|\|full | 3769 | No | NA | 8 | 0 | 1 | Not-determined | Genome-fragment | NA | NA | 0 |
| S11_extracted_k141_30023 | 1884 | No | NA | 2 | 1 | 0 | Low-quality | Genome-fragment | 4.61 | AAI-based (high-confidence) | 0 |
| S5_vs_k141_68464\|\|full | 3261 | No | NA | 2 | 0 | 2 | Not-determined | Genome-fragment | NA | NA | 0 |
| S3_vs_k141_97716\|\|full | 1884 | No | NA | 5 | 1 | 0 | Low-quality | Genome-fragment | 4.07 | AAI-based (medium-confidence) | 0 |
| S5_vs_k141_8125\|\|full | 40010 | No | NA | 54 | 14 | 1 | High-quality | High-quality | 94.63 | AAI-based (high-confidence) | 0 |
| S5_vs_k141_50535\|\|full | 3012 | No | NA | 3 | 0 | 2 | Not-determined | Genome-fragment | NA | NA | 0 |
| S5_vs_k141_10395\|\|full | 1884 | No | NA | 3 | 2 | 0 | Low-quality | Genome-fragment | 4.78 | AAI-based (high-confidence) | 0 |
| S5_vs_k141_79872\|\|full | 1136 | No | NA | 2 | 0 | 0 | Not-determined | Genome-fragment | NA | NA | 0 |
| S7_vs_113497\|\|full | 1884 | No | NA | 4 | 0 | 0 | Low-quality | Genome-fragment | 5.23 | AAI-based (medium-confidence) | 0 |
| S5_vs_k141_86758\|\|full | 1348 | No | NA | 3 | 0 | 1 | Not-determined | Genome-fragment | NA | NA | 0 |
| S7_vs_125064\|\|full | 1884 | No | NA | 4 | 0 | 0 | Low-quality | Genome-fragment | 4.73 | AAI-based (high-confidence) | 0 |
| S16_vs_23870\|\|full | 1884 | No | NA | 2 | 0 | 0 | Low-quality | Genome-fragment | 1.07 | AAI-based (medium-confidence) | 0 |
| S5_vs_k141_14008\|\|full | 1371 | No | NA | 3 | 0 | 2 | Not-determined | Genome-fragment | NA | NA | 0 |
| S5_vs_k141_19999\|\|full | 3813 | No | NA | 5 | 0 | 1 | Not-determined | Genome-fragment | NA | NA | 0 |
| S16_vs_61613\|\|full | 1884 | No | NA | 2 | 0 | 0 | Low-quality | Genome-fragment | 4.81 | AAI-based (high-confidence) | 0 |
| S12_vs_k141_7634\|\|full | 1884 | No | NA | 3 | 0 | 2 | Low-quality | Genome-fragment | 1.19 | AAI-based (medium-confidence) | 0 |
| S5_vs_k141_93073\|\|full | 2789 | No | NA | 3 | 0 | 2 | Not-determined | Genome-fragment | NA | NA | 0 |
| S8_extracted_k141_47023 | 1884 | No | NA | 2 | 1 | 0 | Low-quality | Genome-fragment | 3.82 | AAI-based (high-confidence) | 0 |
| S5_vs_k141_117777\|\|full | 1662 | No | NA | 3 | 0 | 2 | Not-determined | Genome-fragment | NA | NA | 0 |
| S5_vs_k141_93095\|\|full | 1235 | No | NA | 2 | 0 | 0 | Not-determined | Genome-fragment | NA | NA | 0 |
| S17_vs_k141_31982\|\|full | 1885 | No | NA | 2 | 0 | 0 | Low-quality | Genome-fragment | 2.26 | AAI-based (medium-confidence) | 0 |
| S5_vs_k141_2436\|\|full | 2408 | No | NA | 6 | 0 | 2 | Not-determined | Genome-fragment | NA | NA | 0 |
| S13_vs_88541\|\|full | 1885 | No | NA | 4 | 1 | 0 | Low-quality | Genome-fragment | 2.15 | AAI-based (high-confidence) | 0 |
| S5_vs_k141_62359\|\|full | 1042 | No | NA | 2 | 0 | 0 | Not-determined | Genome-fragment | NA | NA | 0 |
| S13_vs_124491\|\|lt2gene | 1885 | No | NA | 2 | 1 | 0 | Low-quality | Genome-fragment | 5.02 | AAI-based (high-confidence) | 0 |
| S5_vs_k141_68538\|\|full | 1653 | No | NA | 4 | 0 | 0 | Not-determined | Genome-fragment | NA | NA | 0 |
| S5_vs_k141_68544\|\|full | 1840 | No | NA | 3 | 0 | 0 | Not-determined | Genome-fragment | NA | NA | 0 |
| S5_vs_k141_32643\|\|full | 1665 | No | NA | 2 | 0 | 0 | Not-determined | Genome-fragment | NA | NA | 0 |
| S5_vs_k141_75123\|\|full | 1885 | No | NA | 5 | 1 | 0 | Low-quality | Genome-fragment | 4.68 | AAI-based (medium-confidence) | 0 |
| S5_vs_k141_137229\|\|lt2gene | 1885 | No | NA | 1 | 0 | 0 | Low-quality | Genome-fragment | 4.82 | AAI-based (medium-confidence) | 0 |
| S7_vs_46373\|\|full | 1885 | No | NA | 6 | 1 | 0 | Low-quality | Genome-fragment | 2.9 | AAI-based (medium-confidence) | 0 |
| S5_vs_k141_38184\|\|full | 4450 | No | NA | 5 | 0 | 0 | Not-determined | Genome-fragment | NA | NA | 0 |
| S16_extracted_92260 | 1885 | No | NA | 1 | 0 | 1 | Low-quality | Genome-fragment | 4.33 | AAI-based (high-confidence) | 0 |
| S6_vs_k141_55146\|\|full | 1885 | No | NA | 5 | 1 | 0 | Low-quality | Genome-fragment | 3.47 | AAI-based (high-confidence) | 0 |
| S8_extracted_k141_34980 | 1885 | No | NA | 3 | 0 | 1 | Low-quality | Genome-fragment | 5.49 | AAI-based (medium-confidence) | 0 |
| S17_vs_k141_112517\|\|full | 1886 | No | NA | 3 | 0 | 1 | Low-quality | Genome-fragment | 1.53 | AAI-based (high-confidence) | 0 |
| S5_vs_k141_105324\|\|full | 1698 | No | NA | 2 | 0 | 1 | Not-determined | Genome-fragment | NA | NA | 0 |
| S9_vs_k141_2408\|\|full | 1886 | No | NA | 6 | 3 | 0 | Low-quality | Genome-fragment | 4.98 | AAI-based (medium-confidence) | 0 |
| S3_extracted_k141_102936 | 1886 | No | NA | 2 | 1 | 0 | Low-quality | Genome-fragment | 3.76 | AAI-based (high-confidence) | 0 |
| S5_vs_k141_103938\|\|full | 1886 | No | NA | 3 | 0 | 1 | Low-quality | Genome-fragment | 1.28 | AAI-based (medium-confidence) | 0 |
| S5_vs_k141_93147\|\|full | 3262 | No | NA | 3 | 0 | 1 | Not-determined | Genome-fragment | NA | NA | 0 |
| S5_vs_k141_20051\|\|full | 2167 | No | NA | 3 | 0 | 3 | Not-determined | Genome-fragment | NA | NA | 0 |
| S7_vs_55176\|\|full | 1886 | No | NA | 3 | 0 | 0 | Low-quality | Genome-fragment | 3.72 | AAI-based (high-confidence) | 0 |
| S16_vs_11225\|\|full | 1886 | No | NA | 2 | 0 | 0 | Low-quality | Genome-fragment | 3.7 | AAI-based (high-confidence) | 0 |
| S10_extracted_69112 | 1886 | No | NA | 7 | 1 | 0 | Low-quality | Genome-fragment | 3.68 | AAI-based (high-confidence) | 0 |
| S5_vs_k141_38220\|\|full | 7253 | No | NA | 8 | 0 | 1 | Not-determined | Genome-fragment | NA | NA | 0 |
| S5_vs_k141_74121\|\|full | 1764 | No | NA | 4 | 0 | 0 | Not-determined | Genome-fragment | NA | NA | 0 |
| S5_vs_k141_2510\|\|full | 1393 | No | NA | 3 | 0 | 0 | Not-determined | Genome-fragment | NA | NA | 0 |
| S10_vs_30356\|\|full | 1886 | No | NA | 4 | 0 | 2 | Low-quality | Genome-fragment | 2.81 | AAI-based (medium-confidence) | 0 |
| S5_vs_k141_44751\|\|full | 2058 | No | NA | 3 | 0 | 0 | Not-determined | Genome-fragment | NA | NA | 0 |
| S12_vs_k141_32702\|\|full | 1886 | No | NA | 3 | 1 | 0 | Low-quality | Genome-fragment | 4.59 | AAI-based (high-confidence) | 0 |
| S8_extracted_k141_86205 | 1886 | No | NA | 2 | 0 | 1 | Low-quality | Genome-fragment | 2.27 | AAI-based (medium-confidence) | 0 |
| S5_vs_k141_38264\|\|full | 2577 | No | NA | 3 | 0 | 0 | Not-determined | Genome-fragment | NA | NA | 0 |
| S5_vs_k141_38265\|\|full | 1131 | No | NA | 2 | 0 | 1 | Not-determined | Genome-fragment | NA | NA | 0 |
| S9_vs_k141_31077\|\|full | 1887 | No | NA | 2 | 0 | 1 | Low-quality | Genome-fragment | 0.87 | AAI-based (high-confidence) | 0 |
| S5_vs_k141_38281\|\|full | 6876 | No | NA | 8 | 0 | 1 | Not-determined | Genome-fragment | NA | NA | 0 |
| S9_vs_k141_17768\|\|full | 1887 | No | NA | 3 | 0 | 0 | Low-quality | Genome-fragment | 1.93 | AAI-based (medium-confidence) | 0 |
| S13_extracted_22338 | 1887 | No | NA | 1 | 0 | 1 | Low-quality | Genome-fragment | 0.5 | AAI-based (high-confidence) | 0 |
| S13_extracted_132600 | 1887 | No | NA | 5 | 2 | 0 | Low-quality | Genome-fragment | 3.91 | AAI-based (high-confidence) | 0 |
| S1_vs_k141_105063\|\|full | 1887 | No | NA | 2 | 0 | 1 | Low-quality | Genome-fragment | 2.73 | AAI-based (medium-confidence) | 0 |
| S1_vs_k141_79887\|\|full | 1887 | No | NA | 5 | 0 | 0 | Low-quality | Genome-fragment | 3.95 | AAI-based (high-confidence) | 0 |
| S16_vs_34680\|\|full | 1887 | No | NA | 6 | 1 | 0 | Low-quality | Genome-fragment | 5.37 | AAI-based (high-confidence) | 0 |
| S5_vs_k141_62440\|\|full | 2048 | No | NA | 4 | 0 | 1 | Not-determined | Genome-fragment | NA | NA | 0 |
| S5_vs_k141_105481\|\|full | 2495 | No | NA | 5 | 0 | 2 | Not-determined | Genome-fragment | NA | NA | 0 |
| S5_vs_k141_2547\|\|full | 2159 | No | NA | 3 | 0 | 2 | Not-determined | Genome-fragment | NA | NA | 0 |
| S5_vs_k141_32790\|\|full | 1121 | No | NA | 2 | 0 | 1 | Not-determined | Genome-fragment | NA | NA | 0 |
| S10_extracted_30253 | 1887 | No | NA | 3 | 0 | 0 | Low-quality | Genome-fragment | 2.09 | AAI-based (high-confidence) | 0 |
| S5_vs_k141_117981\|\|full | 1293 | No | NA | 3 | 0 | 0 | Not-determined | Genome-fragment | NA | NA | 0 |
| S12_vs_k141_81884\|\|full | 1887 | No | NA | 3 | 0 | 0 | Low-quality | Genome-fragment | 3.31 | AAI-based (medium-confidence) | 0 |
| S5_vs_k141_32809\|\|full | 1338 | No | NA | 2 | 0 | 1 | Not-determined | Genome-fragment | NA | NA | 0 |
| S17_vs_k141_8356\|\|full | 1888 | No | NA | 3 | 0 | 0 | Low-quality | Genome-fragment | 2.14 | AAI-based (medium-confidence) | 0 |
| S5_vs_k141_93217\|\|full | 1697 | No | NA | 4 | 0 | 0 | Not-determined | Genome-fragment | NA | NA | 0 |
| S5_vs_k141_141282\|\|full | 1685 | No | NA | 2 | 0 | 0 | Not-determined | Genome-fragment | NA | NA | 0 |
| S14_vs_k141_10276\|\|full | 1888 | No | NA | 3 | 0 | 3 | Low-quality | Genome-fragment | 1.11 | AAI-based (medium-confidence) | 0 |
| S14_vs_k141_77769\|\|full | 1888 | No | NA | 4 | 3 | 0 | Low-quality | Genome-fragment | 4.64 | AAI-based (high-confidence) | 0 |
| S5_vs_k141_111230\|\|full | 1006 | No | NA | 2 | 0 | 0 | Not-determined | Genome-fragment | NA | NA | 0 |
| S5_vs_k141_32818\|\|full | 1415 | No | NA | 3 | 0 | 2 | Not-determined | Genome-fragment | NA | NA | 0 |
| S1_vs_k141_108439\|\|full | 1888 | No | NA | 2 | 0 | 0 | Low-quality | Genome-fragment | 4.6 | AAI-based (high-confidence) | 0 |
| S5_vs_k141_93228\|\|full | 1481 | No | NA | 2 | 0 | 2 | Not-determined | Genome-fragment | NA | NA | 0 |
| S3_extracted_k141_113065 | 1888 | No | NA | 2 | 0 | 0 | Low-quality | Genome-fragment | 0.61 | AAI-based (medium-confidence) | 0 |
| S5_vs_k141_105527\|\|full | 1823 | No | NA | 4 | 0 | 3 | Not-determined | Genome-fragment | NA | NA | 0 |
| S3_vs_k141_27572\|\|full | 1888 | No | NA | 5 | 2 | 0 | Low-quality | Genome-fragment | 5.29 | AAI-based (high-confidence) | 0 |
| S5_vs_k141_123981\|\|full | 2829 | No | NA | 5 | 0 | 0 | Not-determined | Genome-fragment | NA | NA | 0 |
| S3_vs_k141_36226\|\|full | 1888 | No | NA | 4 | 1 | 0 | Low-quality | Genome-fragment | 4.35 | AAI-based (high-confidence) | 0 |
| S5_extracted_k141_26919 | 1888 | No | NA | 4 | 3 | 0 | Low-quality | Genome-fragment | 6.23 | AAI-based (high-confidence) | 0 |
| S5_vs_k141_141325\|\|full | 1226 | No | NA | 2 | 0 | 1 | Not-determined | Genome-fragment | NA | NA | 0 |
| S5_vs_k141_118033\|\|full | 3204 | No | NA | 4 | 0 | 1 | Not-determined | Genome-fragment | NA | NA | 0 |
| S5_vs_k141_111251\|\|full | 6819 | No | NA | 8 | 0 | 1 | Not-determined | Genome-fragment | NA | NA | 0 |
| S5_vs_k141_86337\|\|full | 1888 | No | NA | 5 | 3 | 0 | Low-quality | Genome-fragment | 7.87 | AAI-based (high-confidence) | 0 |
| S5_vs_k141_8388\|\|full | 3840 | No | NA | 2 | 0 | 1 | Not-determined | Genome-fragment | NA | NA | 0 |
| S5_vs_k141_14309\|\|full | 2001 | No | NA | 2 | 0 | 1 | Not-determined | Genome-fragment | NA | NA | 0 |
| S5_vs_k141_8391\|\|full | 1126 | No | NA | 2 | 0 | 1 | Not-determined | Genome-fragment | NA | NA | 0 |
| S7_vs_19113\|\|full | 1888 | No | NA | 3 | 0 | 0 | Low-quality | Genome-fragment | 3.7 | AAI-based (high-confidence) | 0 |
| S12_extracted_k141_87731 | 1888 | No | NA | 3 | 1 | 0 | Low-quality | Genome-fragment | 5.72 | AAI-based (medium-confidence) | 0 |
| S5_vs_k141_105614\|\|full | 1114 | No | NA | 2 | 0 | 1 | Not-determined | Genome-fragment | NA | NA | 0 |
| S5_vs_k141_124019\|\|full | 2950 | No | NA | 3 | 0 | 3 | Not-determined | Genome-fragment | NA | NA | 0 |
| S6_vs_k141_20569\|\|full | 1888 | No | NA | 4 | 1 | 0 | Low-quality | Genome-fragment | 3.37 | AAI-based (medium-confidence) | 0 |
| S8_extracted_k141_128910 | 1888 | No | NA | 3 | 2 | 0 | Low-quality | Genome-fragment | 3.82 | AAI-based (high-confidence) | 0 |
| S5_vs_k141_124035\|\|full | 12251 | No | NA | 10 | 1 | 3 | High-quality | High-quality | 100 | HMM-based (lower-bound) | 0 |
| S5_vs_k141_50835\|\|full | 2670 | No | NA | 3 | 0 | 0 | Not-determined | Genome-fragment | NA | NA | 0 |
| S5_vs_k141_141328\|\|full | 1538 | No | NA | 3 | 0 | 0 | Not-determined | Genome-fragment | NA | NA | 0 |
| S9_vs_k141_80425\|\|full | 1889 | No | NA | 3 | 0 | 0 | Low-quality | Genome-fragment | 3.66 | AAI-based (high-confidence) | 0 |
| S7_vs_119685\|\|full | 1889 | No | NA | 3 | 2 | 0 | Low-quality | Genome-fragment | 6.01 | AAI-based (high-confidence) | 0 |
| S16_vs_53269\|\|full | 1889 | No | NA | 3 | 1 | 0 | Low-quality | Genome-fragment | 3.53 | HMM-based (lower-bound) | 0 |
| S12_vs_k141_172549\|\|full | 1889 | No | NA | 3 | 0 | 0 | Low-quality | Genome-fragment | 4.21 | AAI-based (medium-confidence) | 0 |
| S17_extracted_k141_45402 | 1890 | No | NA | 2 | 1 | 0 | Low-quality | Genome-fragment | 1.09 | HMM-based (lower-bound) | 0 |
| S5_vs_k141_25525\|\|full | 2715 | No | NA | 4 | 0 | 0 | Not-determined | Genome-fragment | NA | NA | 0 |
| S5_vs_k141_20244\|\|full | 5094 | No | NA | 4 | 0 | 4 | Not-determined | Genome-fragment | NA | NA | 0 |
| S17_extracted_k141_117786 | 1890 | No | NA | 4 | 0 | 0 | Low-quality | Genome-fragment | 2.83 | AAI-based (high-confidence) | 0 |
| S3_vs_k141_97050\|\|full | 1890 | No | NA | 6 | 0 | 0 | Low-quality | Genome-fragment | 1.28 | AAI-based (high-confidence) | 0 |
| S5_vs_k141_117501\|\|full | 1890 | No | NA | 3 | 2 | 0 | Low-quality | Genome-fragment | 5.2 | AAI-based (high-confidence) | 0 |
| S5_vs_k141_93292\|\|full | 2218 | No | NA | 2 | 0 | 1 | Not-determined | Genome-fragment | NA | NA | 0 |
| S5_vs_k141_38307\|\|full | 3885 | No | NA | 3 | 0 | 1 | Not-determined | Genome-fragment | NA | NA | 0 |
| S5_vs_k141_2656\|\|full | 3144 | No | NA | 4 | 0 | 4 | Not-determined | Genome-fragment | NA | NA | 0 |
| S16_extracted_5063 | 1890 | No | NA | 2 | 0 | 1 | Low-quality | Genome-fragment | 1.07 | AAI-based (medium-confidence) | 0 |
| S5_vs_k141_25547\|\|full | 1947 | No | NA | 2 | 0 | 2 | Not-determined | Genome-fragment | NA | NA | 0 |
| S5_vs_k141_20268\|\|full | 14771 | No | NA | 23 | 13 | 0 | Medium-quality | Genome-fragment | 60.47 | AAI-based (high-confidence) | 0 |
| S12_extracted_k141_114391 | 1890 | No | NA | 4 | 0 | 0 | Low-quality | Genome-fragment | 2.96 | AAI-based (high-confidence) | 0 |
| S12_vs_k141_6956\|\|full | 1890 | No | NA | 4 | 1 | 0 | Low-quality | Genome-fragment | 2.76 | AAI-based (medium-confidence) | 0 |
| S5_vs_k141_38347\|\|full | 5920 | No | NA | 8 | 0 | 1 | Not-determined | Genome-fragment | NA | NA | 0 |
| S5_vs_k141_79992\|\|full | 54138 | Yes | 36331 | 61 | 15 | 10 | High-quality | High-quality | 92.92 | AAI-based (medium-confidence) | 32.89 |
| S5_vs_k141_50984\|\|full | 2517 | No | NA | 3 | 0 | 0 | Not-determined | Genome-fragment | NA | NA | 0 |
| S8_extracted_k141_155608 | 1890 | No | NA | 4 | 2 | 0 | Low-quality | Genome-fragment | 3.03 | AAI-based (high-confidence) | 0 |
| S17_vs_k141_117127\|\|lt2gene | 1891 | No | NA | 1 | 1 | 0 | Low-quality | Genome-fragment | 6.04 | AAI-based (medium-confidence) | 0 |
| S5_vs_k141_141439\|\|full | 3606 | No | NA | 7 | 0 | 0 | Not-determined | Genome-fragment | NA | NA | 0 |
| S9_vs_k141_9725\|\|full | 1891 | No | NA | 2 | 1 | 0 | Low-quality | Genome-fragment | 4.06 | AAI-based (medium-confidence) | 0 |
| S5_vs_k141_20332\|\|full | 2107 | No | NA | 2 | 0 | 1 | Not-determined | Genome-fragment | NA | NA | 0 |
| S5_vs_k141_14415\|\|full | 6022 | No | NA | 9 | 0 | 0 | Not-determined | Genome-fragment | NA | NA | 0 |
| S5_vs_k141_62536\|\|full | 1935 | No | NA | 3 | 0 | 0 | Not-determined | Genome-fragment | NA | NA | 0 |
| S11_extracted_k141_96062 | 1891 | No | NA | 2 | 0 | 0 | Low-quality | Genome-fragment | 4.41 | AAI-based (medium-confidence) | 0 |
| S5_vs_k141_25704\|\|full | 2895 | No | NA | 2 | 0 | 1 | Not-determined | Genome-fragment | NA | NA | 0 |
| S5_vs_k141_80023\|\|full | 6036 | No | NA | 9 | 0 | 6 | Not-determined | Genome-fragment | NA | NA | 0 |
| S5_vs_k141_105791\|\|full | 1728 | No | NA | 2 | 0 | 1 | Not-determined | Genome-fragment | NA | NA | 0 |
| S13_vs_23669\|\|full | 1891 | No | NA | 5 | 0 | 3 | Low-quality | Genome-fragment | 4.22 | AAI-based (medium-confidence) | 0 |
| S13_vs_120202\|\|full | 1891 | No | NA | 5 | 2 | 0 | Low-quality | Genome-fragment | 3.22 | AAI-based (medium-confidence) | 0 |
| S14_vs_k141_58212\|\|full | 1891 | No | NA | 3 | 0 | 0 | Low-quality | Genome-fragment | 3.75 | AAI-based (medium-confidence) | 0 |
| S1_extracted_k141_25399 | 1891 | No | NA | 1 | 0 | 1 | Low-quality | Genome-fragment | 0.68 | AAI-based (medium-confidence) | 0 |
| S3_vs_k141_121237\|\|full | 1891 | No | NA | 3 | 1 | 0 | Low-quality | Genome-fragment | 1.22 | AAI-based (high-confidence) | 0 |
| S5_vs_k141_91149\|\|full | 1891 | No | NA | 3 | 0 | 2 | Low-quality | Genome-fragment | 1.98 | AAI-based (medium-confidence) | 0 |
| S5_vs_k141_56029\|\|full | 1309 | No | NA | 3 | 0 | 0 | Not-determined | Genome-fragment | NA | NA | 0 |
| S5_vs_k141_80047\|\|full | 1440 | No | NA | 2 | 0 | 2 | Not-determined | Genome-fragment | NA | NA | 0 |
| S5_vs_k141_2831\|\|full | 4934 | No | NA | 5 | 0 | 3 | Not-determined | Genome-fragment | NA | NA | 0 |
| S16_extracted_64427 | 1891 | No | NA | 2 | 0 | 2 | Low-quality | Genome-fragment | 2.01 | AAI-based (medium-confidence) | 0 |
| S5_vs_k141_20364\|\|full | 1134 | No | NA | 2 | 0 | 1 | Not-determined | Genome-fragment | NA | NA | 0 |
| S5_vs_k141_2844\|\|full | 1243 | No | NA | 3 | 0 | 2 | Not-determined | Genome-fragment | NA | NA | 0 |
| S5_vs_k141_105849\|\|full | 2134 | No | NA | 4 | 0 | 1 | Not-determined | Genome-fragment | NA | NA | 0 |
| S5_vs_k141_98439\|\|full | 2853 | No | NA | 3 | 0 | 2 | Not-determined | Genome-fragment | NA | NA | 0 |
| S5_vs_k141_51072\|\|full | 2810 | No | NA | 4 | 0 | 3 | Not-determined | Genome-fragment | NA | NA | 0 |
| S5_vs_k141_118168\|\|full | 34729 | No | NA | 61 | 17 | 0 | Medium-quality | Genome-fragment | 78.83 | AAI-based (high-confidence) | 0 |
| S5_vs_k141_20390\|\|full | 2832 | No | NA | 4 | 0 | 2 | Not-determined | Genome-fragment | NA | NA | 0 |
| S5_vs_k141_124252\|\|full | 5312 | No | NA | 8 | 0 | 4 | Not-determined | Genome-fragment | NA | NA | 0 |
| S5_vs_k141_56044\|\|full | 1193 | No | NA | 2 | 0 | 2 | Not-determined | Genome-fragment | NA | NA | 0 |
| S16_vs_122763\|\|lt2gene | 1891 | No | NA | 1 | 1 | 0 | Low-quality | Genome-fragment | 4.68 | AAI-based (medium-confidence) | 0 |
| S5_vs_k141_32885\|\|full | 39417 | No | NA | 58 | 13 | 1 | Medium-quality | Genome-fragment | 83.27 | AAI-based (medium-confidence) | 0 |
| S5_vs_k141_2876\|\|full | 22336 | No | NA | 26 | 18 | 2 | Medium-quality | Genome-fragment | 55.91 | AAI-based (high-confidence) | 0 |
| S8_extracted_k141_66158 | 1891 | No | NA | 3 | 0 | 0 | Low-quality | Genome-fragment | 3.01 | AAI-based (high-confidence) | 0 |
| S5_vs_k141_68818\|\|full | 3880 | No | NA | 4 | 0 | 3 | Not-determined | Genome-fragment | NA | NA | 0 |
| S17_extracted_k141_148601 | 1892 | No | NA | 1 | 0 | 1 | Low-quality | Genome-fragment | 2.33 | AAI-based (medium-confidence) | 0 |
| S3_vs_k141_47648\|\|lt2gene | 1892 | No | NA | 2 | 2 | 0 | Low-quality | Genome-fragment | 4.81 | AAI-based (high-confidence) | 0 |
| S5_vs_k141_68821\|\|full | 3159 | No | NA | 3 | 0 | 3 | Not-determined | Genome-fragment | NA | NA | 0 |
| S5_vs_k141_9570\|\|full | 1892 | No | NA | 6 | 1 | 0 | Low-quality | Genome-fragment | 4.56 | AAI-based (medium-confidence) | 0 |
| S7_extracted_123117 | 1892 | No | NA | 3 | 0 | 0 | Low-quality | Genome-fragment | 4.67 | AAI-based (high-confidence) | 0 |
| S7_vs_20516\|\|full | 1892 | No | NA | 3 | 1 | 0 | Low-quality | Genome-fragment | 0.69 | HMM-based (lower-bound) | 0 |
| S10_extracted_64080 | 1892 | No | NA | 2 | 0 | 2 | Low-quality | Genome-fragment | 0.69 | AAI-based (medium-confidence) | 0 |
| S10_vs_3137\|\|full | 1892 | No | NA | 3 | 0 | 1 | Low-quality | Genome-fragment | 0.55 | AAI-based (high-confidence) | 0 |
| S5_vs_k141_8569\|\|full | 1088 | No | NA | 2 | 0 | 0 | Not-determined | Genome-fragment | NA | NA | 0 |
| S9_vs_k141_3825\|\|lt2gene | 1893 | No | NA | 1 | 1 | 0 | Low-quality | Genome-fragment | 4.44 | AAI-based (high-confidence) | 0 |
| S5_vs_k141_135225\|\|full | 1043 | No | NA | 2 | 0 | 1 | Not-determined | Genome-fragment | NA | NA | 0 |
| S5_vs_k141_93521\|\|full | 1319 | No | NA | 2 | 0 | 1 | Not-determined | Genome-fragment | NA | NA | 0 |
| S16_vs_68257\|\|full | 1893 | No | NA | 3 | 3 | 0 | Low-quality | Genome-fragment | 4.05 | AAI-based (high-confidence) | 0 |
| S12_extracted_k141_81482 | 1893 | No | NA | 6 | 1 | 0 | Low-quality | Genome-fragment | 1.75 | AAI-based (high-confidence) | 0 |
| S12_vs_k141_179759\|\|full | 1893 | No | NA | 4 | 2 | 0 | Low-quality | Genome-fragment | 4.54 | AAI-based (medium-confidence) | 0 |
| S5_vs_k141_20464\|\|full | 6979 | No | NA | 6 | 0 | 6 | Not-determined | Genome-fragment | NA | NA | 0 |
| S5_vs_k141_2941\|\|full | 3737 | No | NA | 3 | 0 | 0 | Not-determined | Genome-fragment | NA | NA | 0 |
| S5_vs_k141_20469\|\|full | 1210 | No | NA | 2 | 0 | 2 | Not-determined | Genome-fragment | NA | NA | 0 |
| S5_vs_k141_2948\|\|full | 2519 | No | NA | 5 | 0 | 2 | Not-determined | Genome-fragment | NA | NA | 0 |
| S5_vs_k141_2952\|\|full | 3309 | No | NA | 3 | 0 | 1 | Not-determined | Genome-fragment | NA | NA | 0 |
| S14_vs_k141_10207\|\|full | 1894 | No | NA | 2 | 0 | 0 | Low-quality | Genome-fragment | 1.2 | AAI-based (medium-confidence) | 0 |
| S14_vs_k141_87380\|\|full | 1894 | No | NA | 3 | 1 | 0 | Low-quality | Genome-fragment | 4.63 | AAI-based (medium-confidence) | 0 |
| S3_vs_k141_104045\|\|full | 1894 | No | NA | 3 | 1 | 0 | Low-quality | Genome-fragment | 3.65 | HMM-based (lower-bound) | 0 |
| S5_vs_k141_2981\|\|full | 1533 | No | NA | 3 | 0 | 1 | Not-determined | Genome-fragment | NA | NA | 0 |
| S5_vs_k141_51430\|\|full | 1894 | No | NA | 4 | 2 | 0 | Low-quality | Genome-fragment | 4.38 | AAI-based (high-confidence) | 0 |
| S5_vs_k141_56083\|\|full | 32027 | No | NA | 55 | 16 | 0 | High-quality | High-quality | 99.67 | AAI-based (high-confidence) | 0 |
| S7_vs_114815\|\|full | 1894 | No | NA | 3 | 0 | 0 | Low-quality | Genome-fragment | 3.89 | AAI-based (medium-confidence) | 0 |
| S5_vs_k141_105904\|\|full | 3368 | No | NA | 3 | 0 | 0 | Not-determined | Genome-fragment | NA | NA | 0 |
| S12_vs_k141_32485\|\|full | 1894 | No | NA | 2 | 0 | 1 | Low-quality | Genome-fragment | 1.27 | AAI-based (medium-confidence) | 0 |
| S8_vs_k141_37098\|\|full | 1894 | No | NA | 4 | 0 | 0 | Low-quality | Genome-fragment | 5.09 | AAI-based (medium-confidence) | 0 |
| S5_vs_k141_67218\|\|full | 1895 | No | NA | 2 | 0 | 1 | Low-quality | Genome-fragment | 1.52 | AAI-based (medium-confidence) | 0 |
| S10_extracted_4592 | 1895 | No | NA | 3 | 2 | 0 | Low-quality | Genome-fragment | 4.02 | AAI-based (high-confidence) | 0 |
| S5_vs_k141_124369\|\|full | 2693 | No | NA | 5 | 0 | 0 | Not-determined | Genome-fragment | NA | NA | 0 |
| S10_vs_26768\|\|full | 1895 | No | NA | 3 | 2 | 0 | Low-quality | Genome-fragment | 4.36 | AAI-based (medium-confidence) | 0 |
| S12_vs_k141_99227\|\|full | 1895 | No | NA | 3 | 2 | 0 | Low-quality | Genome-fragment | 5.58 | AAI-based (medium-confidence) | 0 |
| S5_vs_k141_14558\|\|full | 1208 | No | NA | 3 | 0 | 0 | Not-determined | Genome-fragment | NA | NA | 0 |
| S8_extracted_k141_31492 | 1895 | No | NA | 4 | 0 | 0 | Low-quality | Genome-fragment | 5.95 | AAI-based (high-confidence) | 0 |
| S5_vs_k141_38540\|\|full | 2221 | No | NA | 4 | 0 | 0 | Not-determined | Genome-fragment | NA | NA | 0 |
| S17_vs_k141_147812\|\|full | 1896 | No | NA | 2 | 1 | 0 | Low-quality | Genome-fragment | 1.04 | AAI-based (medium-confidence) | 0 |
| S5_vs_k141_44877\|\|full | 1725 | No | NA | 2 | 0 | 2 | Not-determined | Genome-fragment | NA | NA | 0 |
| S5_vs_k141_8703\|\|full | 2756 | No | NA | 2 | 0 | 2 | Not-determined | Genome-fragment | NA | NA | 0 |
| S17_vs_k141_112032\|\|full | 1896 | No | NA | 4 | 0 | 0 | Low-quality | Genome-fragment | 3.58 | AAI-based (medium-confidence) | 0 |
| S11_extracted_k141_95911 | 1896 | No | NA | 1 | 0 | 0 | Low-quality | Genome-fragment | 1.39 | AAI-based (high-confidence) | 0 |
| S11_vs_k141_63523\|\|full | 1896 | No | NA | 7 | 2 | 0 | Low-quality | Genome-fragment | 3.36 | AAI-based (high-confidence) | 0 |
| S3_vs_k141_24628\|\|full | 1896 | No | NA | 2 | 0 | 1 | Low-quality | Genome-fragment | 2.99 | AAI-based (medium-confidence) | 0 |
| S5_vs_k141_62702\|\|full | 1108 | No | NA | 2 | 0 | 1 | Not-determined | Genome-fragment | NA | NA | 0 |
| S7_vs_103404\|\|full | 1896 | No | NA | 5 | 1 | 0 | Low-quality | Genome-fragment | 4.37 | AAI-based (high-confidence) | 0 |
| S5_vs_k141_20500\|\|full | 71704 | Yes | 33200 | 80 | 16 | 28 | Complete | High-quality | 100 | Provirus (high-confidence) | 53.7 |
| S5_vs_k141_106005\|\|full | 2030 | No | NA | 4 | 0 | 2 | Not-determined | Genome-fragment | NA | NA | 0 |
| S5_vs_k141_25844\|\|full | 2784 | No | NA | 2 | 0 | 2 | Not-determined | Genome-fragment | NA | NA | 0 |
| S5_vs_k141_80213\|\|full | 2276 | No | NA | 3 | 0 | 2 | Not-determined | Genome-fragment | NA | NA | 0 |
| S16_vs_151426\|\|full | 1896 | No | NA | 3 | 0 | 0 | Low-quality | Genome-fragment | 4.4 | AAI-based (high-confidence) | 0 |
| S5_vs_k141_56155\|\|full | 1824 | No | NA | 3 | 0 | 0 | Not-determined | Genome-fragment | NA | NA | 0 |
| S2_vs_k141_13425\|\|full | 1896 | No | NA | 3 | 0 | 0 | Low-quality | Genome-fragment | 3.13 | AAI-based (high-confidence) | 0 |
| S8_extracted_k141_19380 | 1896 | No | NA | 4 | 4 | 0 | Low-quality | Genome-fragment | 3.27 | AAI-based (high-confidence) | 0 |
| S17_extracted_k141_128979 | 1897 | No | NA | 4 | 4 | 0 | Low-quality | Genome-fragment | 6.26 | AAI-based (high-confidence) | 0 |
| S5_vs_k141_141543\|\|full | 3264 | No | NA | 5 | 0 | 4 | Not-determined | Genome-fragment | NA | NA | 0 |
| S13_vs_38651\|\|full | 1897 | No | NA | 5 | 3 | 0 | Low-quality | Genome-fragment | 3.16 | AAI-based (high-confidence) | 0 |
| S7_vs_81705\|\|full | 1897 | No | NA | 3 | 1 | 0 | Low-quality | Genome-fragment | 1.44 | AAI-based (medium-confidence) | 0 |
| S17_vs_k141_37082\|\|full | 1898 | No | NA | 4 | 0 | 0 | Low-quality | Genome-fragment | 2.43 | AAI-based (medium-confidence) | 0 |
| S11_vs_k141_86489\|\|full | 1898 | No | NA | 3 | 0 | 0 | Low-quality | Genome-fragment | 1.43 | AAI-based (high-confidence) | 0 |
| S11_vs_k141_69557\|\|full | 1898 | No | NA | 5 | 0 | 0 | Low-quality | Genome-fragment | 4.29 | AAI-based (high-confidence) | 0 |
| S5_vs_k141_100445\|\|full | 1898 | No | NA | 3 | 1 | 0 | Low-quality | Genome-fragment | 3.68 | AAI-based (high-confidence) | 0 |
| S5_vs_k141_106069\|\|full | 1259 | No | NA | 3 | 0 | 0 | Not-determined | Genome-fragment | NA | NA | 0 |
| S5_vs_k141_118408\|\|full | 2647 | No | NA | 3 | 0 | 2 | Not-determined | Genome-fragment | NA | NA | 0 |
| S7_vs_119980\|\|full | 1898 | No | NA | 3 | 0 | 1 | Low-quality | Genome-fragment | 2.46 | AAI-based (medium-confidence) | 0 |
| S5_vs_k141_25864\|\|full | 1571 | No | NA | 4 | 0 | 0 | Not-determined | Genome-fragment | NA | NA | 0 |
| S5_vs_k141_51217\|\|full | 4932 | No | NA | 3 | 0 | 3 | Not-determined | Genome-fragment | NA | NA | 0 |
| S5_vs_k141_98501\|\|full | 1942 | No | NA | 4 | 0 | 0 | Not-determined | Genome-fragment | NA | NA | 0 |
| S5_vs_k141_3183\|\|full | 18914 | No | NA | 27 | 12 | 1 | Medium-quality | Genome-fragment | 56.46 | AAI-based (high-confidence) | 0 |
| S12_vs_k141_33178\|\|full | 1898 | No | NA | 6 | 0 | 0 | Low-quality | Genome-fragment | 3.11 | AAI-based (high-confidence) | 0 |
| S5_vs_k141_124502\|\|full | 4286 | No | NA | 6 | 0 | 2 | Not-determined | Genome-fragment | NA | NA | 0 |
| S8_vs_k141_112678\|\|full | 1898 | No | NA | 4 | 0 | 0 | Low-quality | Genome-fragment | 1.22 | AAI-based (medium-confidence) | 0 |
| S17_extracted_k141_96034 | 1899 | No | NA | 3 | 0 | 2 | Low-quality | Genome-fragment | 3.36 | AAI-based (medium-confidence) | 0 |
| S5_vs_k141_48346\|\|full | 1899 | No | NA | 5 | 2 | 0 | Low-quality | Genome-fragment | 3.79 | AAI-based (medium-confidence) | 0 |
| S8_vs_k141_38507\|\|full | 1899 | No | NA | 6 | 2 | 0 | Low-quality | Genome-fragment | 2.45 | AAI-based (medium-confidence) | 0 |
| S5_vs_k141_124521\|\|full | 10601 | No | NA | 13 | 0 | 3 | Not-determined | Genome-fragment | NA | NA | 0 |
| S5_vs_k141_80272\|\|full | 4468 | No | NA | 8 | 0 | 1 | Not-determined | Genome-fragment | NA | NA | 0 |
| S5_vs_k141_80273\|\|full | 6764 | No | NA | 9 | 0 | 7 | Not-determined | Genome-fragment | NA | NA | 0 |
| S17_vs_k141_13813\|\|full | 1900 | No | NA | 2 | 0 | 0 | Low-quality | Genome-fragment | 2.57 | AAI-based (high-confidence) | 0 |
| S5_vs_k141_69452\|\|full | 1900 | No | NA | 2 | 0 | 2 | Low-quality | Genome-fragment | 1.55 | AAI-based (high-confidence) | 0 |
| S5_vs_k141_98915\|\|full | 1900 | No | NA | 3 | 1 | 0 | Low-quality | Genome-fragment | 4.87 | AAI-based (high-confidence) | 0 |
| S5_vs_k141_141621\|\|full | 18852 | No | NA | 27 | 16 | 1 | Medium-quality | Genome-fragment | 50.09 | AAI-based (high-confidence) | 0 |
| S5_vs_k141_62890\|\|full | 2448 | No | NA | 3 | 0 | 1 | Not-determined | Genome-fragment | NA | NA | 0 |
| S7_vs_122351\|\|full | 1900 | No | NA | 4 | 0 | 2 | Low-quality | Genome-fragment | 1.46 | AAI-based (medium-confidence) | 0 |
| S10_vs_91566\|\|full | 1900 | No | NA | 4 | 0 | 0 | Low-quality | Genome-fragment | 2.98 | AAI-based (high-confidence) | 0 |
| S5_vs_k141_106151\|\|full | 4292 | No | NA | 6 | 0 | 6 | Not-determined | Genome-fragment | NA | NA | 0 |
| S8_extracted_k141_194663 | 1900 | No | NA | 2 | 0 | 0 | Low-quality | Genome-fragment | 5.22 | AAI-based (medium-confidence) | 0 |
| S5_vs_k141_20600\|\|full | 6171 | No | NA | 4 | 0 | 2 | Not-determined | Genome-fragment | NA | NA | 0 |
| S14_vs_k141_25385\|\|full | 1901 | No | NA | 8 | 1 | 0 | Low-quality | Genome-fragment | 4.83 | AAI-based (high-confidence) | 0 |
| S5_vs_k141_3294\|\|full | 1434 | No | NA | 2 | 0 | 0 | Not-determined | Genome-fragment | NA | NA | 0 |
| S1_extracted_k141_147143 | 1901 | No | NA | 1 | 0 | 1 | Low-quality | Genome-fragment | 1.7 | AAI-based (medium-confidence) | 0 |
| S5_vs_k141_56224\|\|full | 1210 | No | NA | 3 | 0 | 1 | Not-determined | Genome-fragment | NA | NA | 0 |
| S7_extracted_40561 | 1901 | No | NA | 3 | 0 | 0 | Low-quality | Genome-fragment | 1.15 | AAI-based (high-confidence) | 0 |
| S16_extracted_140266 | 1901 | No | NA | 2 | 0 | 2 | Low-quality | Genome-fragment | 0.7 | AAI-based (high-confidence) | 0 |
| S5_vs_k141_69055\|\|full | 8578 | No | NA | 5 | 0 | 4 | Not-determined | Genome-fragment | NA | NA | 0 |
| S16_vs_72057\|\|full | 1901 | No | NA | 4 | 1 | 0 | Low-quality | Genome-fragment | 2.97 | HMM-based (lower-bound) | 0 |
| S10_vs_2595\|\|full | 1901 | No | NA | 2 | 0 | 1 | Low-quality | Genome-fragment | 0.53 | AAI-based (medium-confidence) | 0 |
| S5_vs_k141_111569\|\|full | 1180 | No | NA | 2 | 0 | 1 | Not-determined | Genome-fragment | NA | NA | 0 |
| S5_vs_k141_129899\|\|full | 1666 | No | NA | 3 | 0 | 2 | Not-determined | Genome-fragment | NA | NA | 0 |
| S8_vs_k141_73080\|\|full | 1901 | No | NA | 6 | 1 | 0 | Low-quality | Genome-fragment | 1.03 | AAI-based (high-confidence) | 0 |
| S5_vs_k141_38688\|\|full | 9822 | No | NA | 6 | 0 | 6 | Not-determined | Genome-fragment | NA | NA | 0 |
| S9_vs_k141_53066\|\|full | 1902 | No | NA | 3 | 0 | 0 | Low-quality | Genome-fragment | 5.21 | AAI-based (medium-confidence) | 0 |
| S5_vs_k141_3349\|\|full | 2243 | No | NA | 3 | 0 | 0 | Not-determined | Genome-fragment | NA | NA | 0 |
| S5_vs_k141_56302\|\|full | 1354 | No | NA | 2 | 0 | 0 | Not-determined | Genome-fragment | NA | NA | 0 |
| S16_vs_67030\|\|full | 1902 | No | NA | 3 | 2 | 0 | Low-quality | Genome-fragment | 2.96 | HMM-based (lower-bound) | 0 |
| S16_vs_113283\|\|full | 1902 | No | NA | 2 | 2 | 0 | Low-quality | Genome-fragment | 4.03 | AAI-based (high-confidence) | 0 |
| S5_vs_k141_3354\|\|full | 1431 | No | NA | 3 | 0 | 0 | Not-determined | Genome-fragment | NA | NA | 0 |
| S10_vs_76787\|\|full | 1902 | No | NA | 2 | 0 | 1 | Low-quality | Genome-fragment | 2.81 | AAI-based (medium-confidence) | 0 |
| S5_vs_k141_69083\|\|full | 32634 | No | NA | 44 | 12 | 0 | Medium-quality | Genome-fragment | 60.39 | HMM-based (lower-bound) | 0 |
| S5_vs_k141_111610\|\|full | 1299 | No | NA | 2 | 0 | 2 | Not-determined | Genome-fragment | NA | NA | 0 |
| S5_vs_k141_129920\|\|full | 6064 | No | NA | 10 | 0 | 2 | Not-determined | Genome-fragment | NA | NA | 0 |
| S5_vs_k141_69085\|\|full | 1878 | No | NA | 2 | 0 | 0 | Not-determined | Genome-fragment | NA | NA | 0 |
| S5_vs_k141_111616\|\|full | 1665 | No | NA | 2 | 0 | 1 | Not-determined | Genome-fragment | NA | NA | 0 |
| S12_vs_k141_87144\|\|full | 1902 | No | NA | 4 | 0 | 0 | Low-quality | Genome-fragment | 5.11 | AAI-based (medium-confidence) | 0 |
| S8_extracted_k141_114150 | 1902 | No | NA | 4 | 0 | 1 | Low-quality | Genome-fragment | 1.07 | AAI-based (medium-confidence) | 0 |
| S5_vs_k141_56326\|\|full | 8543 | No | NA | 14 | 0 | 2 | Not-determined | Genome-fragment | NA | NA | 0 |
| S9_vs_k141_54241\|\|full | 1903 | No | NA | 4 | 0 | 1 | Low-quality | Genome-fragment | 3.57 | AAI-based (medium-confidence) | 0 |
| S14_extracted_k141_113212 | 1903 | No | NA | 5 | 0 | 0 | Low-quality | Genome-fragment | 4.36 | AAI-based (high-confidence) | 0 |
| S5_vs_k141_8830\|\|full | 6023 | No | NA | 4 | 0 | 3 | Not-determined | Genome-fragment | NA | NA | 0 |
| S5_vs_k141_45126\|\|full | 4195 | No | NA | 5 | 0 | 0 | Not-determined | Genome-fragment | NA | NA | 0 |
| S14_vs_k141_20092\|\|full | 1903 | No | NA | 4 | 2 | 0 | Low-quality | Genome-fragment | 4.68 | AAI-based (medium-confidence) | 0 |
| S5_vs_k141_74476\|\|full | 1135 | No | NA | 3 | 0 | 0 | Not-determined | Genome-fragment | NA | NA | 0 |
| S5_vs_k141_74478\|\|full | 1616 | No | NA | 2 | 0 | 0 | Not-determined | Genome-fragment | NA | NA | 0 |
| S5_vs_k141_69150\|\|full | 1919 | No | NA | 2 | 0 | 0 | Not-determined | Genome-fragment | NA | NA | 0 |
| S5_vs_k141_51416\|\|full | 38157 | No | NA | 55 | 14 | 0 | High-quality | High-quality | 99.54 | AAI-based (high-confidence) | 0 |
| S5_vs_k141_38785\|\|full | 1636 | No | NA | 3 | 0 | 2 | Not-determined | Genome-fragment | NA | NA | 0 |
| S5_vs_k141_69158\|\|full | 12724 | No | NA | 12 | 0 | 6 | Not-determined | Genome-fragment | NA | NA | 0 |
| S7_vs_36177\|\|lt2gene | 1903 | No | NA | 2 | 2 | 0 | Low-quality | Genome-fragment | 3.05 | HMM-based (lower-bound) | 0 |
| S5_vs_k141_8847\|\|full | 31194 | No | NA | 37 | 14 | 3 | Medium-quality | Genome-fragment | 64.14 | AAI-based (high-confidence) | 0 |
| S5_vs_k141_56365\|\|full | 8770 | No | NA | 10 | 0 | 2 | Not-determined | Genome-fragment | NA | NA | 0 |
| S12_extracted_k141_183850 | 1903 | No | NA | 6 | 1 | 0 | Low-quality | Genome-fragment | 1.96 | AAI-based (medium-confidence) | 0 |
| S8_extracted_k141_204292 | 1903 | No | NA | 1 | 0 | 0 | Low-quality | Genome-fragment | 6.08 | AAI-based (medium-confidence) | 0 |
| S5_vs_k141_93839\|\|full | 30686 | No | NA | 41 | 7 | 0 | Medium-quality | Genome-fragment | 63.9 | HMM-based (lower-bound) | 0 |
| S11_vs_k141_103786\|\|full | 1904 | No | NA | 3 | 1 | 0 | Low-quality | Genome-fragment | 4.3 | AAI-based (high-confidence) | 0 |
| S5_vs_k141_93840\|\|full | 2380 | No | NA | 3 | 0 | 2 | Not-determined | Genome-fragment | NA | NA | 0 |
| S7_vs_36084\|\|full | 1904 | No | NA | 2 | 1 | 0 | Low-quality | Genome-fragment | 5.23 | AAI-based (high-confidence) | 0 |
| S12_vs_k141_167685\|\|full | 1904 | No | NA | 5 | 1 | 0 | Low-quality | Genome-fragment | 4.22 | AAI-based (medium-confidence) | 0 |
| S12_vs_k141_161265\|\|full | 1904 | No | NA | 2 | 0 | 2 | Low-quality | Genome-fragment | 3.62 | AAI-based (medium-confidence) | 0 |
| S6_vs_k141_20665\|\|full | 1904 | No | NA | 3 | 1 | 0 | Low-quality | Genome-fragment | 2.36 | AAI-based (high-confidence) | 0 |
| S5_vs_k141_3501\|\|full | 39219 | Yes | 23805 | 45 | 12 | 12 | Medium-quality | Genome-fragment | 57.93 | AAI-based (high-confidence) | 39.3 |
| S5_vs_k141_63007\|\|full | 2095 | No | NA | 3 | 0 | 1 | Not-determined | Genome-fragment | NA | NA | 0 |
| S5_vs_k141_141693\|\|full | 6908 | No | NA | 9 | 0 | 4 | Not-determined | Genome-fragment | NA | NA | 0 |
| S9_vs_k141_13080\|\|full | 1905 | No | NA | 4 | 0 | 1 | Low-quality | Genome-fragment | 5.42 | AAI-based (medium-confidence) | 0 |
| S5_vs_k141_33187\|\|full | 2827 | No | NA | 3 | 0 | 3 | Not-determined | Genome-fragment | NA | NA | 0 |
| S11_vs_k141_99246\|\|full | 1905 | No | NA | 2 | 0 | 1 | Low-quality | Genome-fragment | 2.93 | AAI-based (high-confidence) | 0 |
| S13_vs_92807\|\|full | 1905 | No | NA | 4 | 1 | 0 | Low-quality | Genome-fragment | 3.81 | AAI-based (high-confidence) | 0 |
| S14_extracted_k141_94580 | 1905 | No | NA | 2 | 2 | 0 | Low-quality | Genome-fragment | 6.25 | AAI-based (high-confidence) | 0 |
| S5_vs_k141_63025\|\|full | 1188 | No | NA | 3 | 0 | 1 | Not-determined | Genome-fragment | NA | NA | 0 |
| S5_vs_k141_8922\|\|full | 1653 | No | NA | 3 | 0 | 0 | Not-determined | Genome-fragment | NA | NA | 0 |
| S5_vs_k141_20793\|\|full | 2120 | No | NA | 2 | 0 | 1 | Not-determined | Genome-fragment | NA | NA | 0 |
| S5_vs_k141_129970\|\|full | 32525 | No | NA | 33 | 4 | 2 | Medium-quality | Genome-fragment | 70.81 | AAI-based (high-confidence) | 0 |
| S14_vs_k141_47199\|\|full | 1905 | No | NA | 3 | 0 | 0 | Low-quality | Genome-fragment | 5.9 | AAI-based (medium-confidence) | 0 |
| S14_vs_k141_3952\|\|full | 1905 | No | NA | 4 | 1 | 0 | Low-quality | Genome-fragment | 5.18 | AAI-based (high-confidence) | 0 |
| S5_vs_k141_45666\|\|full | 1905 | No | NA | 5 | 1 | 0 | Low-quality | Genome-fragment | 4.74 | AAI-based (medium-confidence) | 0 |
| S7_vs_44365\|\|full | 1905 | No | NA | 3 | 0 | 1 | Low-quality | Genome-fragment | 4.29 | AAI-based (high-confidence) | 0 |
| S12_vs_k141_103827\|\|full | 1905 | No | NA | 3 | 0 | 1 | Low-quality | Genome-fragment | 1.25 | AAI-based (high-confidence) | 0 |
| S12_vs_k141_105493\|\|full | 1905 | No | NA | 5 | 1 | 0 | Low-quality | Genome-fragment | 4.61 | AAI-based (medium-confidence) | 0 |
| S5_vs_k141_69273\|\|full | 1430 | No | NA | 2 | 0 | 2 | Not-determined | Genome-fragment | NA | NA | 0 |
| S5_vs_k141_124677\|\|full | 105125 | Yes | 42430 | 113 | 22 | 50 | Medium-quality | Genome-fragment | 81.67 | AAI-based (high-confidence) | 59.64 |
| S5_vs_k141_63060\|\|full | 1970 | No | NA | 2 | 0 | 0 | Not-determined | Genome-fragment | NA | NA | 0 |
| S17_extracted_k141_163607 | 1906 | No | NA | 2 | 1 | 0 | Low-quality | Genome-fragment | 4.09 | AAI-based (high-confidence) | 0 |
| S5_vs_k141_69292\|\|full | 2206 | No | NA | 4 | 0 | 1 | Not-determined | Genome-fragment | NA | NA | 0 |
| S11_extracted_k141_9720 | 1906 | No | NA | 4 | 1 | 0 | Low-quality | Genome-fragment | 4.54 | AAI-based (medium-confidence) | 0 |
| S5_vs_k141_118659\|\|full | 1902 | No | NA | 3 | 0 | 0 | Not-determined | Genome-fragment | NA | NA | 0 |
| S11_vs_k141_96657\|\|full | 1906 | No | NA | 3 | 0 | 0 | Low-quality | Genome-fragment | 3.84 | AAI-based (medium-confidence) | 0 |
| S5_vs_k141_129998\|\|full | 4268 | No | NA | 6 | 0 | 4 | Not-determined | Genome-fragment | NA | NA | 0 |
| S3_vs_k141_142\|\|full | 1906 | No | NA | 2 | 0 | 1 | Low-quality | Genome-fragment | 2.32 | AAI-based (medium-confidence) | 0 |
| S5_vs_k141_99605\|\|lt2gene | 1906 | No | NA | 2 | 2 | 0 | Low-quality | Genome-fragment | 5.05 | AAI-based (high-confidence) | 0 |
| S5_vs_k141_111745\|\|full | 2261 | No | NA | 4 | 0 | 1 | Not-determined | Genome-fragment | NA | NA | 0 |
| S5_vs_k141_111760\|\|full | 1062 | No | NA | 2 | 0 | 0 | Not-determined | Genome-fragment | NA | NA | 0 |
| S17_vs_k141_123414\|\|full | 1907 | No | NA | 5 | 1 | 0 | Low-quality | Genome-fragment | 4.9 | AAI-based (high-confidence) | 0 |
| S14_vs_k141_106490\|\|full | 1907 | No | NA | 3 | 0 | 0 | Low-quality | Genome-fragment | 2.71 | AAI-based (high-confidence) | 0 |
| S1_vs_k141_115682\|\|full | 1907 | No | NA | 3 | 0 | 0 | Low-quality | Genome-fragment | 3.7 | AAI-based (high-confidence) | 0 |
| S3_vs_k141_18617\|\|full | 1907 | No | NA | 3 | 0 | 0 | Low-quality | Genome-fragment | 3.53 | AAI-based (medium-confidence) | 0 |
| S7_vs_54072\|\|full | 1907 | No | NA | 3 | 0 | 0 | Low-quality | Genome-fragment | 2.95 | AAI-based (medium-confidence) | 0 |
| S10_vs_90661\|\|full | 1907 | No | NA | 4 | 0 | 0 | Low-quality | Genome-fragment | 5.07 | AAI-based (high-confidence) | 0 |
| S5_vs_k141_45370\|\|full | 1474 | No | NA | 3 | 0 | 0 | Not-determined | Genome-fragment | NA | NA | 0 |
| S8_extracted_k141_96922 | 1907 | No | NA | 1 | 0 | 0 | Low-quality | Genome-fragment | 1.36 | AAI-based (high-confidence) | 0 |
| S5_vs_k141_69361\|\|full | 4119 | No | NA | 12 | 0 | 0 | Not-determined | Genome-fragment | NA | NA | 0 |
| S17_extracted_k141_59314 | 1908 | No | NA | 2 | 0 | 2 | Low-quality | Genome-fragment | 1.25 | AAI-based (high-confidence) | 0 |
| S17_extracted_k141_135432 | 1908 | No | NA | 2 | 1 | 0 | Low-quality | Genome-fragment | 4.02 | AAI-based (high-confidence) | 0 |
| S5_vs_k141_130032\|\|full | 38770 | No | NA | 41 | 25 | 1 | Medium-quality | Genome-fragment | 83.11 | AAI-based (high-confidence) | 0 |
| S5_vs_k141_3665\|\|full | 2814 | No | NA | 4 | 0 | 0 | Not-determined | Genome-fragment | NA | NA | 0 |
| S17_vs_k141_44436\|\|full | 1908 | No | NA | 3 | 0 | 2 | Low-quality | Genome-fragment | 1.17 | AAI-based (medium-confidence) | 0 |
| S9_vs_k141_67315\|\|lt2gene | 1908 | No | NA | 1 | 1 | 0 | Low-quality | Genome-fragment | 3.26 | AAI-based (medium-confidence) | 0 |
| S5_vs_k141_141881\|\|full | 1248 | No | NA | 3 | 0 | 2 | Not-determined | Genome-fragment | NA | NA | 0 |
| S3_vs_k141_52327\|\|full | 1908 | No | NA | 7 | 1 | 0 | Low-quality | Genome-fragment | 3.63 | AAI-based (high-confidence) | 0 |
| S5_vs_k141_63211\|\|full | 1041 | No | NA | 3 | 0 | 0 | Not-determined | Genome-fragment | NA | NA | 0 |
| S5_vs_k141_141894\|\|full | 1331 | No | NA | 3 | 0 | 1 | Not-determined | Genome-fragment | NA | NA | 0 |
| S7_vs_41934\|\|full | 1908 | No | NA | 4 | 0 | 1 | Low-quality | Genome-fragment | 4.07 | AAI-based (medium-confidence) | 0 |
| S16_vs_79278\|\|full | 1908 | No | NA | 4 | 0 | 1 | Low-quality | Genome-fragment | 4.6 | AAI-based (high-confidence) | 0 |
| S5_vs_k141_33311\|\|full | 1809 | No | NA | 3 | 0 | 1 | Not-determined | Genome-fragment | NA | NA | 0 |
| S10_vs_14777\|\|full | 1908 | No | NA | 4 | 0 | 0 | Low-quality | Genome-fragment | 3.74 | AAI-based (high-confidence) | 0 |
| S5_vs_k141_51688\|\|full | 1343 | No | NA | 2 | 0 | 1 | Not-determined | Genome-fragment | NA | NA | 0 |
| S5_vs_k141_15176\|\|full | 1991 | No | NA | 4 | 0 | 2 | Not-determined | Genome-fragment | NA | NA | 0 |
| S5_vs_k141_3729\|\|full | 1794 | No | NA | 3 | 0 | 3 | Not-determined | Genome-fragment | NA | NA | 0 |
| S13_extracted_79874 | 1909 | No | NA | 3 | 0 | 1 | Low-quality | Genome-fragment | 2.99 | AAI-based (high-confidence) | 0 |
| S5_vs_k141_51708\|\|full | 3538 | No | NA | 3 | 0 | 1 | Not-determined | Genome-fragment | NA | NA | 0 |
| S3_vs_k141_13172\|\|full | 1909 | No | NA | 5 | 0 | 0 | Low-quality | Genome-fragment | 2.5 | AAI-based (high-confidence) | 0 |
| S4_extracted_24087 | 1909 | No | NA | 2 | 1 | 0 | Low-quality | Genome-fragment | 4.31 | AAI-based (high-confidence) | 0 |
| S8_extracted_k141_9996 | 1909 | No | NA | 4 | 0 | 1 | Low-quality | Genome-fragment | 4.72 | AAI-based (high-confidence) | 0 |
| S8_extracted_k141_180613 | 1909 | No | NA | 1 | 0 | 0 | Low-quality | Genome-fragment | 0.35 | AAI-based (medium-confidence) | 0 |
| S17_vs_k141_22852\|\|full | 1910 | No | NA | 4 | 2 | 0 | Low-quality | Genome-fragment | 3.41 | HMM-based (lower-bound) | 0 |
| S17_vs_k141_99972\|\|full | 1910 | No | NA | 3 | 0 | 1 | Low-quality | Genome-fragment | 1.03 | AAI-based (medium-confidence) | 0 |
| S5_vs_k141_38943\|\|full | 1060 | No | NA | 2 | 0 | 1 | Not-determined | Genome-fragment | NA | NA | 0 |
| S5_vs_k141_118755\|\|full | 2183 | No | NA | 3 | 0 | 0 | Not-determined | Genome-fragment | NA | NA | 0 |
| S5_vs_k141_106453\|\|full | 1538 | No | NA | 3 | 0 | 2 | Not-determined | Genome-fragment | NA | NA | 0 |

| | | | | | | | | | | | |
|---|---|---|---|---|---|---|---|---|---|---|---|
| S5_vs_k141_80466\|\|full | 2617 | No | NA | 3 | 0 | 1 | Not-determined | Genome-fragment | NA | NA | 0 |
| S17_vs_k141_177092\|\|full | 1910 | Yes | 1304 | 4 | 1 | 1 | Low-quality | Genome-fragment | 3.04 | AAI-based (medium-confidence) | 31.73 |
| S14_vs_k141_105880\|\|full | 1910 | No | NA | 5 | 0 | 2 | Low-quality | Genome-fragment | 2.79 | AAI-based (high-confidence) | 0 |
| S14_vs_k141_77800\|\|full | 1910 | No | NA | 7 | 1 | 0 | Low-quality | Genome-fragment | 4.91 | AAI-based (high-confidence) | 0 |
| S1_extracted_k141_64565 | 1910 | No | NA | 3 | 0 | 0 | Low-quality | Genome-fragment | 4.45 | AAI-based (high-confidence) | 0 |
| S1_vs_k141_29298\|\|full | 1910 | No | NA | 5 | 0 | 0 | Low-quality | Genome-fragment | 3.57 | AAI-based (medium-confidence) | 0 |
| S12_extracted_k141_8783 | 1910 | No | NA | 3 | 0 | 2 | Low-quality | Genome-fragment | 1.36 | AAI-based (medium-confidence) | 0 |
| S12_vs_k141_85855\|\|full | 1910 | No | NA | 2 | 1 | 0 | Low-quality | Genome-fragment | 2.68 | HMM-based (lower-bound) | 0 |
| S8_vs_k141_177123\|\|full | 1910 | No | NA | 3 | 0 | 2 | Low-quality | Genome-fragment | 1.36 | AAI-based (medium-confidence) | 0 |
| S7_vs_46828\|\|full | 1911 | No | NA | 5 | 1 | 0 | Low-quality | Genome-fragment | 3.43 | AAI-based (medium-confidence) | 0 |
| S5_vs_k141_25973\|\|full | 1249 | No | NA | 3 | 0 | 1 | Not-determined | Genome-fragment | NA | NA | 0 |
| S7_vs_108395\|\|full | 1911 | No | NA | 4 | 2 | 0 | Low-quality | Genome-fragment | 4.21 | AAI-based (high-confidence) | 0 |
| S5_vs_k141_3825\|\|full | 2572 | No | NA | 4 | 0 | 0 | Not-determined | Genome-fragment | NA | NA | 0 |
| S16_extracted_90401 | 1911 | No | NA | 1 | 0 | 1 | Low-quality | Genome-fragment | 0.19 | AAI-based (medium-confidence) | 0 |
| S16_vs_21034\|\|full | 1911 | No | NA | 2 | 0 | 1 | Low-quality | Genome-fragment | 2.07 | AAI-based (high-confidence) | 0 |
| S5_vs_k141_142039\|\|full | 3407 | No | NA | 4 | 0 | 3 | Not-determined | Genome-fragment | NA | NA | 0 |
| S10_extracted_74345 | 1911 | No | NA | 3 | 0 | 1 | Low-quality | Genome-fragment | 1.22 | AAI-based (high-confidence) | 0 |
| S10_vs_56737\|\|full | 1911 | No | NA | 3 | 0 | 0 | Low-quality | Genome-fragment | 4.09 | AAI-based (high-confidence) | 0 |
| S1_extracted_k141_30679 | 1912 | No | NA | 3 | 2 | 0 | Low-quality | Genome-fragment | 2.87 | AAI-based (high-confidence) | 0 |
| S1_vs_k141_5227\|\|full | 1912 | No | NA | 2 | 1 | 0 | Low-quality | Genome-fragment | 4.54 | AAI-based (medium-confidence) | 0 |
| S5_vs_k141_9046\|\|full | 4119 | No | NA | 3 | 0 | 0 | Not-determined | Genome-fragment | NA | NA | 0 |
| S5_vs_k141_51812\|\|full | 8348 | No | NA | 3 | 0 | 1 | Not-determined | Genome-fragment | NA | NA | 0 |
| S3_extracted_k141_84480 | 1912 | No | NA | 2 | 0 | 0 | Low-quality | Genome-fragment | 2.11 | AAI-based (high-confidence) | 0 |
| S5_vs_k141_26018\|\|full | 2053 | No | NA | 2 | 0 | 1 | Not-determined | Genome-fragment | NA | NA | 0 |
| S5_vs_k141_63372\|\|full | 2720 | No | NA | 3 | 0 | 1 | Not-determined | Genome-fragment | NA | NA | 0 |
| S5_vs_k141_112052\|\|full | 2772 | No | NA | 5 | 0 | 0 | Not-determined | Genome-fragment | NA | NA | 0 |
| S3_vs_k141_16988\|\|full | 1912 | No | NA | 5 | 1 | 0 | Low-quality | Genome-fragment | 5.44 | AAI-based (high-confidence) | 0 |
| S5_vs_k141_141628\|\|full | 1912 | No | NA | 4 | 0 | 0 | Low-quality | Genome-fragment | 3.87 | AAI-based (high-confidence) | 0 |
| S5_vs_k141_30088\|\|full | 1912 | No | NA | 2 | 1 | 0 | Low-quality | Genome-fragment | 5.4 | AAI-based (medium-confidence) | 0 |
| S16_extracted_100506 | 1912 | No | NA | 4 | 1 | 0 | Low-quality | Genome-fragment | 3.76 | AAI-based (high-confidence) | 0 |
| S5_vs_k141_135554\|\|full | 1736 | No | NA | 2 | 0 | 2 | Not-determined | Genome-fragment | NA | NA | 0 |
| S16_vs_39547\|\|full | 1912 | No | NA | 6 | 0 | 0 | Low-quality | Genome-fragment | 3.93 | AAI-based (high-confidence) | 0 |
| S16_vs_151199\|\|full | 1912 | No | NA | 3 | 2 | 0 | Low-quality | Genome-fragment | 3.19 | AAI-based (high-confidence) | 0 |
| S5_vs_k141_51859\|\|full | 6963 | No | NA | 9 | 0 | 1 | Not-determined | Genome-fragment | NA | NA | 0 |
| S5_vs_k141_45403\|\|full | 1030 | No | NA | 3 | 0 | 1 | Not-determined | Genome-fragment | NA | NA | 0 |
| S10_vs_70727\|\|full | 1912 | No | NA | 2 | 0 | 2 | Low-quality | Genome-fragment | 1.03 | AAI-based (medium-confidence) | 0 |
| S12_vs_k141_104199\|\|full | 1912 | No | NA | 2 | 0 | 0 | Low-quality | Genome-fragment | 10.25 | AAI-based (high-confidence) | 0 |
| S5_vs_k141_94085\|\|full | 2523 | No | NA | 4 | 0 | 1 | Not-determined | Genome-fragment | NA | NA | 0 |
| S5_vs_k141_74565\|\|full | 2532 | No | NA | 2 | 0 | 1 | Not-determined | Genome-fragment | NA | NA | 0 |
| S6_vs_k141_13378\|\|full | 1912 | No | NA | 3 | 0 | 1 | Low-quality | Genome-fragment | 3.65 | AAI-based (medium-confidence) | 0 |
| S5_vs_k141_3910\|\|full | 5660 | No | NA | 6 | 0 | 1 | Not-determined | Genome-fragment | NA | NA | 0 |
| S5_vs_k141_112104\|\|full | 2080 | No | NA | 3 | 0 | 0 | Not-determined | Genome-fragment | NA | NA | 0 |
| S6_vs_k141_43742\|\|full | 1912 | No | NA | 5 | 1 | 0 | Low-quality | Genome-fragment | 2.91 | HMM-based (lower-bound) | 0 |
| S5_vs_k141_33506\|\|full | 1328 | No | NA | 3 | 0 | 1 | Not-determined | Genome-fragment | NA | NA | 0 |
| S5_vs_k141_124889\|\|full | 1431 | No | NA | 4 | 0 | 0 | Not-determined | Genome-fragment | NA | NA | 0 |
| S9_vs_k141_36914\|\|lt2gene | 1913 | No | NA | 1 | 1 | 0 | Low-quality | Genome-fragment | 3.14 | AAI-based (high-confidence) | 0 |
| S11_vs_k141_102454\|\|full | 1913 | No | NA | 6 | 0 | 0 | Low-quality | Genome-fragment | 4.67 | AAI-based (high-confidence) | 0 |
| S5_vs_k141_45440\|\|full | 1566 | No | NA | 2 | 0 | 0 | Not-determined | Genome-fragment | NA | NA | 0 |
| S5_vs_k141_94117\|\|full | 2285 | No | NA | 7 | 0 | 2 | Not-determined | Genome-fragment | NA | NA | 0 |
| S5_vs_k141_124898\|\|full | 2172 | No | NA | 4 | 0 | 0 | Not-determined | Genome-fragment | NA | NA | 0 |
| S3_vs_k141_120013\|\|full | 1913 | No | NA | 3 | 0 | 0 | Low-quality | Genome-fragment | 4.2 | AAI-based (medium-confidence) | 0 |
| S5_vs_k141_69572\|\|full | 1040 | No | NA | 3 | 0 | 1 | Not-determined | Genome-fragment | NA | NA | 0 |
| S5_vs_k141_69574\|\|full | 2411 | No | NA | 3 | 0 | 0 | Not-determined | Genome-fragment | NA | NA | 0 |
| S3_vs_k141_86189\|\|full | 1913 | No | NA | 3 | 0 | 0 | Low-quality | Genome-fragment | 1.14 | AAI-based (high-confidence) | 0 |
| S5_extracted_k141_73174 | 1913 | No | NA | 3 | 3 | 0 | Low-quality | Genome-fragment | 6.31 | AAI-based (high-confidence) | 0 |
| S5_extracted_k141_73193 | 1913 | No | NA | 3 | 3 | 0 | Low-quality | Genome-fragment | 6.31 | AAI-based (high-confidence) | 0 |
| S5_vs_k141_124913\|\|full | 2049 | No | NA | 3 | 0 | 0 | Not-determined | Genome-fragment | NA | NA | 0 |
| S16_vs_13491\|\|full | 1913 | No | NA | 3 | 0 | 1 | Low-quality | Genome-fragment | 2.36 | AAI-based (high-confidence) | 0 |
| S5_vs_k141_130294\|\|full | 43034 | No | NA | 40 | 3 | 16 | High-quality | High-quality | 100 | AAI-based (high-confidence) | 0 |
| S5_vs_k141_98931\|\|full | 2111 | No | NA | 4 | 0 | 3 | Not-determined | Genome-fragment | NA | NA | 0 |
| S5_vs_k141_56587\|\|full | 2368 | No | NA | 3 | 0 | 1 | Not-determined | Genome-fragment | NA | NA | 0 |
| S16_vs_83077\|\|full | 1913 | No | NA | 3 | 1 | 0 | Low-quality | Genome-fragment | 4.47 | AAI-based (high-confidence) | 0 |
| S8_vs_k141_177745\|\|full | 1913 | No | NA | 3 | 0 | 0 | Low-quality | Genome-fragment | 4.61 | AAI-based (medium-confidence) | 0 |
| S5_vs_k141_56590\|\|full | 2199 | No | NA | 3 | 0 | 0 | Not-determined | Genome-fragment | NA | NA | 0 |
| S5_vs_k141_142130\|\|full | 2815 | No | NA | 2 | 0 | 2 | Not-determined | Genome-fragment | NA | NA | 0 |
| S5_vs_k141_63483\|\|full | 1558 | No | NA | 3 | 0 | 1 | Not-determined | Genome-fragment | NA | NA | 0 |
| S9_extracted_k141_35619 | 1914 | No | NA | 1 | 0 | 1 | Low-quality | Genome-fragment | 1.06 | AAI-based (high-confidence) | 0 |
| S1_extracted_k141_123806 | 1914 | No | NA | 4 | 1 | 0 | Low-quality | Genome-fragment | 2.25 | AAI-based (medium-confidence) | 0 |
| S5_vs_k141_124957\|\|full | 3054 | No | NA | 4 | 0 | 2 | Not-determined | Genome-fragment | NA | NA | 0 |
| S5_vs_k141_39027\|\|full | 6598 | No | NA | 8 | 0 | 5 | Not-determined | Genome-fragment | NA | NA | 0 |
| S7_vs_78774\|\|full | 1914 | No | NA | 5 | 0 | 0 | Low-quality | Genome-fragment | 4.39 | AAI-based (high-confidence) | 0 |
| S16_vs_146080\|\|full | 1914 | No | NA | 5 | 1 | 0 | Low-quality | Genome-fragment | 2.93 | AAI-based (high-confidence) | 0 |
| S5_vs_k141_94159\|\|full | 1017 | No | NA | 2 | 0 | 0 | Not-determined | Genome-fragment | NA | NA | 0 |
| S12_vs_k141_84139\|\|full | 1914 | No | NA | 3 | 1 | 0 | Low-quality | Genome-fragment | 5.84 | AAI-based (medium-confidence) | 0 |
| S5_vs_k141_124991\|\|full | 4070 | No | NA | 5 | 0 | 4 | Not-determined | Genome-fragment | NA | NA | 0 |
| S12_vs_k141_133524\|\|full | 1914 | No | NA | 5 | 0 | 0 | Low-quality | Genome-fragment | 3.44 | AAI-based (high-confidence) | 0 |
| S5_vs_k141_9252\|\|full | 4000 | No | NA | 4 | 0 | 1 | Not-determined | Genome-fragment | NA | NA | 0 |
| S5_vs_k141_33625\|\|full | 1386 | No | NA | 3 | 0 | 0 | Not-determined | Genome-fragment | NA | NA | 0 |
| S12_vs_k141_161198\|\|lt2gene | 1914 | No | NA | 2 | 2 | 0 | Low-quality | Genome-fragment | 3.11 | HMM-based (lower-bound) | 0 |
| S5_vs_k141_33641\|\|full | 2107 | No | NA | 7 | 0 | 0 | Not-determined | Genome-fragment | NA | NA | 0 |
| S5_vs_k141_80496\|\|full | 150464 | Yes | 44475 | 151 | 14 | 41 | Medium-quality | Genome-fragment | 68.78 | AAI-based (high-confidence) | 70.44 |
| S5_vs_k141_87496\|\|full | 8279 | No | NA | 6 | 0 | 3 | Not-determined | Genome-fragment | NA | NA | 0 |
| S8_vs_k141_142076\|\|full | 1914 | No | NA | 4 | 0 | 1 | Low-quality | Genome-fragment | 1.66 | AAI-based (medium-confidence) | 0 |
| S5_vs_k141_106807\|\|full | 2666 | No | NA | 3 | 0 | 2 | Not-determined | Genome-fragment | NA | NA | 0 |
| S5_vs_k141_9308\|\|full | 3054 | No | NA | 7 | 0 | 1 | Not-determined | Genome-fragment | NA | NA | 0 |
| S17_extracted_k141_119838 | 1915 | No | NA | 2 | 0 | 2 | Low-quality | Genome-fragment | 0.71 | AAI-based (medium-confidence) | 0 |
| S5_vs_k141_45552\|\|full | 1472 | No | NA | 3 | 0 | 2 | Not-determined | Genome-fragment | NA | NA | 0 |
| S14_extracted_k141_81461 | 1915 | No | NA | 5 | 4 | 0 | Low-quality | Genome-fragment | 6.31 | AAI-based (high-confidence) | 0 |
| S5_vs_k141_52075\|\|full | 1215 | No | NA | 3 | 0 | 0 | Not-determined | Genome-fragment | NA | NA | 0 |
| S14_vs_k141_111659\|\|lt2gene | 1915 | No | NA | 1 | 1 | 0 | Low-quality | Genome-fragment | 6.03 | AAI-based (high-confidence) | 0 |
| S7_vs_120164\|\|full | 1915 | No | NA | 2 | 1 | 0 | Low-quality | Genome-fragment | 4.65 | AAI-based (medium-confidence) | 0 |
| S5_vs_k141_130471\|\|full | 15081 | No | NA | 14 | 0 | 7 | Not-determined | Genome-fragment | NA | NA | 0 |
| S5_vs_k141_142195\|\|full | 84815 | Yes | 43797 | 63 | 5 | 22 | Medium-quality | Genome-fragment | 72.35 | AAI-based (medium-confidence) | 48.36 |
| S16_vs_15245\|\|full | 1915 | No | NA | 3 | 1 | 0 | Low-quality | Genome-fragment | 4.43 | AAI-based (medium-confidence) | 0 |
| S16_vs_22322\|\|full | 1915 | No | NA | 6 | 0 | 0 | Low-quality | Genome-fragment | 2.68 | AAI-based (high-confidence) | 0 |
| S5_vs_k141_80498\|\|full | 24008 | No | NA | 29 | 20 | 0 | Medium-quality | Genome-fragment | 59.39 | AAI-based (medium-confidence) | 0 |
| S5_vs_k141_125060\|\|full | 1009 | No | NA | 3 | 0 | 0 | Not-determined | Genome-fragment | NA | NA | 0 |
| S8_vs_k141_146601\|\|full | 1915 | No | NA | 6 | 2 | 0 | Low-quality | Genome-fragment | 3.1 | AAI-based (high-confidence) | 0 |
| S17_extracted_k141_85729 | 1916 | No | NA | 3 | 0 | 0 | Low-quality | Genome-fragment | 3.59 | AAI-based (high-confidence) | 0 |
| S5_vs_k141_142213\|\|full | 3149 | No | NA | 5 | 0 | 4 | Not-determined | Genome-fragment | NA | NA | 0 |
| S5_vs_k141_39118\|\|full | 2836 | No | NA | 4 | 0 | 3 | Not-determined | Genome-fragment | NA | NA | 0 |
| S5_vs_k141_130476\|\|full | 20990 | No | NA | 38 | 4 | 0 | Medium-quality | Genome-fragment | 54.82 | AAI-based (medium-confidence) | 0 |
| S3_vs_k141_91746\|\|full | 1916 | No | NA | 5 | 0 | 0 | Low-quality | Genome-fragment | 3.14 | AAI-based (high-confidence) | 0 |
| S16_vs_124876\|\|full | 1916 | No | NA | 6 | 3 | 0 | Low-quality | Genome-fragment | 5.83 | AAI-based (high-confidence) | 0 |
| S16_vs_125462\|\|full | 1916 | No | NA | 4 | 0 | 0 | Low-quality | Genome-fragment | 4.11 | AAI-based (high-confidence) | 0 |
| S5_vs_k141_125099\|\|full | 1191 | No | NA | 3 | 0 | 0 | Not-determined | Genome-fragment | NA | NA | 0 |
| S10_extracted_23928 | 1916 | No | NA | 5 | 1 | 0 | Low-quality | Genome-fragment | 5.37 | AAI-based (high-confidence) | 0 |
| S5_vs_k141_45618\|\|full | 1716 | No | NA | 3 | 0 | 2 | Not-determined | Genome-fragment | NA | NA | 0 |
| S5_vs_k141_52180\|\|full | 1992 | No | NA | 4 | 0 | 1 | Not-determined | Genome-fragment | NA | NA | 0 |
| S5_vs_k141_56627\|\|full | 2097 | No | NA | 3 | 0 | 0 | Not-determined | Genome-fragment | NA | NA | 0 |
| S5_vs_k141_106923\|\|full | 3262 | No | NA | 4 | 0 | 0 | Not-determined | Genome-fragment | NA | NA | 0 |
| S5_vs_k141_9412\|\|full | 1713 | No | NA | 2 | 0 | 2 | Not-determined | Genome-fragment | NA | NA | 0 |
| S17_vs_k141_32390\|\|full | 1917 | No | NA | 4 | 0 | 0 | Low-quality | Genome-fragment | 4.39 | AAI-based (high-confidence) | 0 |
| S9_vs_k141_36411\|\|full | 1917 | No | NA | 5 | 1 | 0 | Low-quality | Genome-fragment | 3.96 | AAI-based (high-confidence) | 0 |
| S5_vs_k141_106943\|\|full | 3611 | No | NA | 5 | 0 | 0 | Not-determined | Genome-fragment | NA | NA | 0 |
| S5_vs_k141_39181\|\|full | 1411 | No | NA | 2 | 0 | 1 | Not-determined | Genome-fragment | NA | NA | 0 |
| S11_vs_k141_66930\|\|full | 1917 | No | NA | 3 | 0 | 0 | Low-quality | Genome-fragment | 2.19 | AAI-based (medium-confidence) | 0 |
| S5_vs_k141_99026\|\|full | 1839 | No | NA | 3 | 0 | 1 | Not-determined | Genome-fragment | NA | NA | 0 |
| S5_vs_k141_4161\|\|full | 1580 | No | NA | 2 | 0 | 2 | Not-determined | Genome-fragment | NA | NA | 0 |
| S14_extracted_k141_71630 | 1917 | No | NA | 3 | 0 | 0 | Low-quality | Genome-fragment | 5.38 | AAI-based (high-confidence) | 0 |
| S5_vs_k141_142295\|\|full | 1013 | No | NA | 3 | 0 | 0 | Not-determined | Genome-fragment | NA | NA | 0 |
| S5_vs_k141_4171\|\|full | 2992 | No | NA | 2 | 0 | 2 | Not-determined | Genome-fragment | NA | NA | 0 |
| S3_extracted_k141_31863 | 1917 | No | NA | 2 | 2 | 0 | Low-quality | Genome-fragment | 3.64 | HMM-based (lower-bound) | 0 |
| S5_vs_k141_45657\|\|full | 1116 | No | NA | 2 | 0 | 1 | Not-determined | Genome-fragment | NA | NA | 0 |
| S5_vs_k141_26196\|\|full | 3619 | No | NA | 5 | 0 | 1 | Not-determined | Genome-fragment | NA | NA | 0 |
| S3_vs_k141_115626\|\|full | 1917 | No | NA | 5 | 3 | 0 | Low-quality | Genome-fragment | 5.02 | AAI-based (high-confidence) | 0 |
| S5_vs_k141_142326\|\|full | 3653 | No | NA | 4 | 0 | 2 | Not-determined | Genome-fragment | NA | NA | 0 |
| S5_vs_k141_4212\|\|full | 2030 | No | NA | 3 | 0 | 1 | Not-determined | Genome-fragment | NA | NA | 0 |
| S5_vs_k141_87590\|\|full | 4848 | No | NA | 5 | 0 | 3 | Not-determined | Genome-fragment | NA | NA | 0 |
| S16_vs_87868\|\|full | 1917 | No | NA | 4 | 0 | 1 | Low-quality | Genome-fragment | 5.77 | AAI-based (high-confidence) | 0 |
| S6_vs_k141_38340\|\|full | 1917 | No | NA | 5 | 0 | 2 | Low-quality | Genome-fragment | 0.56 | AAI-based (medium-confidence) | 0 |
| S5_vs_k141_142361\|\|full | 1692 | No | NA | 3 | 0 | 0 | Not-determined | Genome-fragment | NA | NA | 0 |
| S5_vs_k141_33846\|\|full | 2277 | No | NA | 3 | 0 | 2 | Not-determined | Genome-fragment | NA | NA | 0 |
| S5_vs_k141_130541\|\|full | 2829 | No | NA | 4 | 0 | 3 | Not-determined | Genome-fragment | NA | NA | 0 |
| S5_vs_k141_107005\|\|full | 1174 | No | NA | 3 | 0 | 1 | Not-determined | Genome-fragment | NA | NA | 0 |
| S8_vs_k141_203154\|\|full | 1917 | Yes | 1473 | 3 | 1 | 1 | Low-quality | Genome-fragment | 0.86 | AAI-based (medium-confidence) | 23.16 |
| S1_vs_k141_8460\|\|full | 1918 | No | NA | 5 | 5 | 0 | Low-quality | Genome-fragment | 2.88 | AAI-based (high-confidence) | 0 |
| S5_vs_k141_112395\|\|full | 3809 | No | NA | 4 | 0 | 3 | Not-determined | Genome-fragment | NA | NA | 0 |
| S5_vs_k141_52285\|\|full | 2201 | No | NA | 3 | 0 | 2 | Not-determined | Genome-fragment | NA | NA | 0 |
| S5_extracted_k141_64666 | 1918 | No | NA | 4 | 2 | 0 | Low-quality | Genome-fragment | 4.48 | AAI-based (high-confidence) | 0 |
| S5_vs_k141_4298\|\|full | 1669 | No | NA | 3 | 0 | 2 | Not-determined | Genome-fragment | NA | NA | 0 |
| S7_vs_68805\|\|full | 1918 | No | NA | 2 | 0 | 0 | Low-quality | Genome-fragment | 4.39 | AAI-based (high-confidence) | 0 |
| S10_vs_7686\|\|full | 1918 | No | NA | 2 | 2 | 0 | Low-quality | Genome-fragment | 5.49 | AAI-based (high-confidence) | 0 |
| S10_vs_88082\|\|full | 1918 | No | NA | 3 | 0 | 1 | Low-quality | Genome-fragment | 1.25 | AAI-based (medium-confidence) | 0 |
| S8_vs_k141_51109\|\|full | 1918 | No | NA | 4 | 1 | 0 | Low-quality | Genome-fragment | 3.72 | AAI-based (high-confidence) | 0 |
| S1_extracted_k141_27004 | 1919 | No | NA | 4 | 0 | 0 | Low-quality | Genome-fragment | 2.38 | AAI-based (high-confidence) | 0 |
| S5_vs_k141_142430\|\|full | 3131 | No | NA | 4 | 0 | 3 | Not-determined | Genome-fragment | NA | NA | 0 |
| S7_vs_120954\|\|full | 1919 | No | NA | 2 | 1 | 0 | Low-quality | Genome-fragment | 4.14 | AAI-based (high-confidence) | 0 |
| S16_vs_150853\|\|full | 1919 | No | NA | 2 | 0 | 0 | Low-quality | Genome-fragment | 4.53 | AAI-based (high-confidence) | 0 |
| S5_vs_k141_4368\|\|full | 2981 | No | NA | 3 | 0 | 0 | Not-determined | Genome-fragment | NA | NA | 0 |
| S5_vs_k141_74726\|\|full | 1206 | No | NA | 2 | 0 | 1 | Not-determined | Genome-fragment | NA | NA | 0 |
| S10_extracted_25361 | 1919 | No | NA | 2 | 0 | 0 | Low-quality | Genome-fragment | 1.74 | AAI-based (high-confidence) | 0 |
| S5_vs_k141_107105\|\|full | 26809 | No | NA | 38 | 16 | 1 | Medium-quality | Genome-fragment | 89.33 | AAI-based (medium-confidence) | 0 |
| S5_vs_k141_142450\|\|full | 13654 | No | NA | 18 | 6 | 0 | Medium-quality | Genome-fragment | 68.89 | AAI-based (medium-confidence) | 0 |
| S5_vs_k141_74738\|\|full | 1515 | No | NA | 3 | 0 | 2 | Not-determined | Genome-fragment | NA | NA | 0 |
| S5_vs_k141_135795\|\|full | 1604 | No | NA | 2 | 0 | 1 | Not-determined | Genome-fragment | NA | NA | 0 |
| S6_vs_k141_46404\|\|lt2gene | 1919 | No | NA | 1 | 1 | 0 | Low-quality | Genome-fragment | 3.58 | AAI-based (medium-confidence) | 0 |
| S17_vs_k141_31819\|\|full | 1920 | No | NA | 3 | 1 | 0 | Low-quality | Genome-fragment | 3.47 | AAI-based (high-confidence) | 0 |
| S11_vs_k141_56370\|\|full | 1920 | No | NA | 5 | 1 | 0 | Low-quality | Genome-fragment | 2.56 | AAI-based (high-confidence) | 0 |
| S5_vs_k141_99116\|\|full | 2026 | No | NA | 2 | 0 | 0 | Not-determined | Genome-fragment | NA | NA | 0 |
| S11_vs_k141_43610\|\|full | 1920 | No | NA | 4 | 3 | 0 | Low-quality | Genome-fragment | 4.34 | AAI-based (medium-confidence) | 0 |
| S5_vs_k141_107129\|\|full | 2073 | No | NA | 3 | 0 | 2 | Not-determined | Genome-fragment | NA | NA | 0 |
| S14_vs_k141_20374\|\|full | 1920 | No | NA | 3 | 1 | 0 | Low-quality | Genome-fragment | 4.51 | AAI-based (high-confidence) | 0 |
| S5_vs_k141_99122\|\|full | 1920 | No | NA | 4 | 0 | 0 | Low-quality | Genome-fragment | 4.88 | AAI-based (high-confidence) | 0 |
| S5_vs_k141_15505\|\|full | 3164 | No | NA | 3 | 0 | 1 | Not-determined | Genome-fragment | NA | NA | 0 |
| S5_vs_k141_15508\|\|full | 1715 | No | NA | 2 | 0 | 1 | Not-determined | Genome-fragment | NA | NA | 0 |
| S16_extracted_138838 | 1920 | No | NA | 6 | 0 | 0 | Low-quality | Genome-fragment | 2.06 | AAI-based (high-confidence) | 0 |
| S16_vs_126813\|\|full | 1920 | No | NA | 3 | 0 | 0 | Low-quality | Genome-fragment | 2.63 | AAI-based (medium-confidence) | 0 |
| S5_vs_k141_87766\|\|full | 6397 | No | NA | 6 | 0 | 4 | Not-determined | Genome-fragment | NA | NA | 0 |
| S16_vs_165674\|\|full | 1920 | Yes | 768 | 2 | 1 | 1 | Low-quality | Genome-fragment | 0.97 | AAI-based (medium-confidence) | 60 |
| S5_vs_k141_130765\|\|full | 1062 | No | NA | 2 | 0 | 1 | Not-determined | Genome-fragment | NA | NA | 0 |
| S11_vs_k141_76807\|\|full | 1921 | No | NA | 4 | 3 | 0 | Low-quality | Genome-fragment | 4.38 | AAI-based (high-confidence) | 0 |
| S5_vs_k141_46106\|\|full | 1921 | No | NA | 6 | 0 | 0 | Low-quality | Genome-fragment | 3.57 | AAI-based (medium-confidence) | 0 |
| S5_vs_k141_52488\|\|full | 1264 | No | NA | 2 | 0 | 0 | Not-determined | Genome-fragment | NA | NA | 0 |
| S5_vs_k141_80731\|\|full | 1936 | No | NA | 7 | 0 | 0 | Not-determined | Genome-fragment | NA | NA | 0 |
| S5_vs_k141_112450\|\|full | 1493 | No | NA | 2 | 0 | 2 | Not-determined | Genome-fragment | NA | NA | 0 |
| S7_vs_9759\|\|full | 1921 | No | NA | 5 | 1 | 0 | Low-quality | Genome-fragment | 4.45 | AAI-based (medium-confidence) | 0 |
| S5_vs_k141_99224\|\|full | 1900 | No | NA | 3 | 0 | 1 | Not-determined | Genome-fragment | NA | NA | 0 |
| S7_vs_89896\|\|full | 1921 | No | NA | 6 | 1 | 0 | Low-quality | Genome-fragment | 4.33 | AAI-based (medium-confidence) | 0 |
| S5_vs_k141_87830\|\|full | 19755 | No | NA | 17 | 0 | 13 | Not-determined | Genome-fragment | NA | NA | 0 |
| S8_vs_k141_108206\|\|full | 1921 | No | NA | 4 | 2 | 1 | Low-quality | Genome-fragment | 5.4 | AAI-based (medium-confidence) | 0 |
| S17_extracted_k141_39793 | 1922 | No | NA | 2 | 0 | 2 | Low-quality | Genome-fragment | 0.64 | AAI-based (high-confidence) | 0 |
| S9_extracted_k141_48488 | 1922 | No | NA | 1 | 1 | 0 | Low-quality | Genome-fragment | 2.96 | HMM-based (lower-bound) | 0 |
| S9_vs_k141_3459\|\|full | 1922 | No | NA | 5 | 2 | 0 | Low-quality | Genome-fragment | 5.35 | AAI-based (medium-confidence) | 0 |
| S1_vs_k141_86613\|\|lt2gene | 1922 | No | NA | 1 | 1 | 0 | Low-quality | Genome-fragment | 5.15 | AAI-based (medium-confidence) | 0 |
| S7_vs_64669\|\|full | 1922 | No | NA | 2 | 0 | 0 | Low-quality | Genome-fragment | 4.51 | AAI-based (high-confidence) | 0 |
| S5_vs_k141_39339\|\|full | 2501 | No | NA | 3 | 0 | 2 | Not-determined | Genome-fragment | NA | NA | 0 |
| S5_vs_k141_112452\|\|full | 23378 | No | NA | 33 | 22 | 1 | Medium-quality | Genome-fragment | 63.62 | AAI-based (medium-confidence) | 0 |
| S5_vs_k141_80744\|\|full | 3002 | No | NA | 3 | 0 | 1 | Not-determined | Genome-fragment | NA | NA | 0 |
| S7_vs_95713\|\|full | 1922 | No | NA | 3 | 1 | 0 | Low-quality | Genome-fragment | 4.71 | AAI-based (high-confidence) | 0 |
| S10_vs_102488\|\|full | 1922 | No | NA | 3 | 0 | 3 | Low-quality | Genome-fragment | 0.76 | AAI-based (medium-confidence) | 0 |
| S12_vs_k141_65297\|\|full | 1922 | No | NA | 3 | 1 | 0 | Low-quality | Genome-fragment | 3.37 | AAI-based (medium-confidence) | 0 |
| S5_vs_k141_112470\|\|full | 2332 | No | NA | 5 | 0 | 1 | Not-determined | Genome-fragment | NA | NA | 0 |
| S4_extracted_5586 | 1922 | No | NA | 3 | 0 | 0 | Low-quality | Genome-fragment | 3.16 | AAI-based (high-confidence) | 0 |
| S6_vs_k141_9601\|\|full | 1922 | No | NA | 5 | 2 | 0 | Low-quality | Genome-fragment | 4.21 | AAI-based (high-confidence) | 0 |
| S5_vs_k141_125594\|\|full | 1118 | No | NA | 2 | 0 | 0 | Not-determined | Genome-fragment | NA | NA | 0 |
| S5_vs_k141_112496\|\|full | 1997 | No | NA | 3 | 0 | 2 | Not-determined | Genome-fragment | NA | NA | 0 |
| S5_vs_k141_69751\|\|full | 3407 | No | NA | 3 | 0 | 0 | Not-determined | Genome-fragment | NA | NA | 0 |
| S17_vs_k141_134773\|\|full | 1923 | No | NA | 4 | 0 | 0 | Low-quality | Genome-fragment | 4.22 | AAI-based (high-confidence) | 0 |
| S9_vs_k141_26879\|\|full | 1923 | No | NA | 6 | 0 | 0 | Low-quality | Genome-fragment | 4.41 | AAI-based (high-confidence) | 0 |
| S11_vs_k141_69815\|\|full | 1923 | Yes | 153 | 3 | 1 | 1 | Low-quality | Genome-fragment | 0.09 | HMM-based (lower-bound) | 92.04 |
| S13_vs_118808\|\|full | 1923 | Yes | 411 | 3 | 1 | 1 | Low-quality | Genome-fragment | 0.69 | AAI-based (medium-confidence) | 78.63 |
| S5_vs_k141_3306\|\|full | 1923 | No | NA | 4 | 0 | 0 | Low-quality | Genome-fragment | 2.24 | AAI-based (high-confidence) | 0 |
| S5_vs_k141_87860\|\|full | 2172 | No | NA | 3 | 0 | 2 | Not-determined | Genome-fragment | NA | NA | 0 |
| S5_vs_k141_99342\|\|full | 9228 | No | NA | 9 | 0 | 3 | Not-determined | Genome-fragment | NA | NA | 0 |
| S10_vs_10188\|\|full | 1923 | No | NA | 3 | 0 | 0 | Low-quality | Genome-fragment | 3.23 | AAI-based (medium-confidence) | 0 |
| S5_vs_k141_87878\|\|full | 5716 | No | NA | 5 | 0 | 4 | Not-determined | Genome-fragment | NA | NA | 0 |
| S12_vs_k141_161827\|\|full | 1923 | No | NA | 3 | 0 | 0 | Low-quality | Genome-fragment | 3.72 | AAI-based (medium-confidence) | 0 |
| S12_vs_k141_85512\|\|full | 1923 | No | NA | 2 | 0 | 0 | Low-quality | Genome-fragment | 3.44 | AAI-based (medium-confidence) | 0 |
| S5_vs_k141_112556\|\|full | 1096 | No | NA | 2 | 0 | 1 | Not-determined | Genome-fragment | NA | NA | 0 |
| S12_vs_k141_157288\|\|full | 1923 | No | NA | 3 | 1 | 0 | Low-quality | Genome-fragment | 3.31 | AAI-based (medium-confidence) | 0 |
| S8_vs_k141_141299\|\|full | 1923 | No | NA | 3 | 0 | 1 | Low-quality | Genome-fragment | 3.21 | AAI-based (medium-confidence) | 0 |
| S8_vs_k141_141949\|\|full | 1923 | No | NA | 3 | 0 | 1 | Low-quality | Genome-fragment | 4.93 | AAI-based (medium-confidence) | 0 |
| S14_extracted_k141_55197 | 1924 | No | NA | 4 | 1 | 0 | Low-quality | Genome-fragment | 6.42 | AAI-based (high-confidence) | 0 |
| S5_vs_k141_107409\|\|full | 2472 | No | NA | 3 | 0 | 0 | Not-determined | Genome-fragment | NA | NA | 0 |
| S5_vs_k141_99362\|\|full | 1163 | No | NA | 3 | 0 | 0 | Not-determined | Genome-fragment | NA | NA | 0 |
| S5_vs_k141_9649\|\|full | 2053 | No | NA | 3 | 0 | 2 | Not-determined | Genome-fragment | NA | NA | 0 |
| S5_vs_k141_69830\|\|full | 5750 | No | NA | 7 | 0 | 5 | Not-determined | Genome-fragment | NA | NA | 0 |
| S5_vs_k141_135919\|\|full | 1207 | No | NA | 1 | 0 | 1 | Not-determined | Genome-fragment | NA | NA | 0 |
| S3_vs_k141_23101\|\|full | 1924 | No | NA | 6 | 1 | 0 | Low-quality | Genome-fragment | 1.24 | AAI-based (high-confidence) | 0 |
| S5_vs_k141_87930\|\|full | 13152 | No | NA | 10 | 0 | 10 | Not-determined | Genome-fragment | NA | NA | 0 |
| S5_vs_k141_112610\|\|full | 1922 | No | NA | 3 | 0 | 0 | Not-determined | Genome-fragment | NA | NA | 0 |
| S5_vs_k141_9660\|\|full | 16823 | No | NA | 15 | 0 | 0 | Not-determined | Genome-fragment | NA | NA | 0 |
| S5_vs_k141_119387\|\|full | 1007 | No | NA | 3 | 0 | 1 | Not-determined | Genome-fragment | NA | NA | 0 |
| S5_vs_k141_19092\|\|lt2gene | 1924 | No | NA | 2 | 1 | 0 | Low-quality | Genome-fragment | 1.16 | HMM-based (lower-bound) | 0 |
| S5_vs_k141_135938\|\|full | 2629 | No | NA | 3 | 0 | 2 | Not-determined | Genome-fragment | NA | NA | 0 |
| S5_vs_k141_107451\|\|full | 2012 | No | NA | 3 | 0 | 1 | Not-determined | Genome-fragment | NA | NA | 0 |
| S5_vs_k141_63886\|\|full | 1269 | No | NA | 3 | 0 | 0 | Not-determined | Genome-fragment | NA | NA | 0 |
| S10_vs_86082\|\|full | 1924 | No | NA | 4 | 0 | 0 | Low-quality | Genome-fragment | 2.79 | AAI-based (medium-confidence) | 0 |
| S5_vs_k141_52617\|\|full | 3171 | No | NA | 3 | 0 | 1 | Not-determined | Genome-fragment | NA | NA | 0 |
| S5_vs_k141_9679\|\|full | 1516 | No | NA | 2 | 0 | 1 | Not-determined | Genome-fragment | NA | NA | 0 |
| S12_vs_k141_154626\|\|full | 1924 | No | NA | 5 | 2 | 0 | Low-quality | Genome-fragment | 2.73 | HMM-based (lower-bound) | 0 |
| S15_vs_19647\|\|full | 1924 | No | NA | 4 | 0 | 0 | Low-quality | Genome-fragment | 1.17 | AAI-based (high-confidence) | 0 |
| S5_vs_k141_15596\|\|full | 2929 | No | NA | 3 | 0 | 2 | Not-determined | Genome-fragment | NA | NA | 0 |
| S8_vs_k141_95567\|\|full | 1924 | No | NA | 3 | 0 | 2 | Low-quality | Genome-fragment | 1.07 | AAI-based (medium-confidence) | 0 |
| S17_vs_k141_16177\|\|full | 1925 | No | NA | 3 | 0 | 2 | Low-quality | Genome-fragment | 0.73 | AAI-based (medium-confidence) | 0 |
| S11_vs_k141_55670\|\|full | 1925 | No | NA | 3 | 0 | 0 | Low-quality | Genome-fragment | 4.87 | AAI-based (high-confidence) | 0 |
| S5_vs_k141_94497\|\|full | 1298 | No | NA | 1 | 0 | 0 | Not-determined | Genome-fragment | NA | NA | 0 |
| S5_vs_k141_52696\|\|full | 3987 | No | NA | 5 | 0 | 1 | Not-determined | Genome-fragment | NA | NA | 0 |
| S5_vs_k141_135986\|\|full | 1030 | No | NA | 3 | 0 | 0 | Not-determined | Genome-fragment | NA | NA | 0 |
| S1_vs_k141_16820\|\|full | 1925 | No | NA | 5 | 3 | 0 | Low-quality | Genome-fragment | 4.39 | AAI-based (medium-confidence) | 0 |
| S7_vs_118650\|\|full | 1925 | No | NA | 5 | 3 | 0 | Low-quality | Genome-fragment | 3.16 | AAI-based (medium-confidence) | 0 |
| S5_vs_k141_34260\|\|full | 2302 | No | NA | 3 | 0 | 3 | Not-determined | Genome-fragment | NA | NA | 0 |
| S5_vs_k141_63924\|\|full | 35637 | No | NA | 58 | 24 | 0 | High-quality | High-quality | 100 | AAI-based (high-confidence) | 0 |
| S7_vs_125548\|\|full | 1925 | No | NA | 4 | 3 | 0 | Low-quality | Genome-fragment | 4.23 | AAI-based (high-confidence) | 0 |
| S16_vs_115441\|\|full | 1925 | No | NA | 2 | 0 | 2 | Low-quality | Genome-fragment | 0.52 | AAI-based (medium-confidence) | 0 |
| S5_vs_k141_34263\|\|full | 3199 | No | NA | 3 | 0 | 2 | Not-determined | Genome-fragment | NA | NA | 0 |
| S5_vs_k141_9745\|\|full | 1767 | No | NA | 2 | 0 | 2 | Not-determined | Genome-fragment | NA | NA | 0 |
| S8_extracted_k141_40516 | 1925 | No | NA | 2 | 0 | 0 | Low-quality | Genome-fragment | 2.86 | AAI-based (high-confidence) | 0 |
| S5_vs_k141_125828\|\|full | 1065 | No | NA | 2 | 0 | 1 | Not-determined | Genome-fragment | NA | NA | 0 |
| S14_vs_k141_74195\|\|full | 1926 | No | NA | 4 | 0 | 0 | Low-quality | Genome-fragment | 2.75 | AAI-based (high-confidence) | 0 |
| S5_vs_k141_69869\|\|full | 3063 | No | NA | 3 | 0 | 1 | Not-determined | Genome-fragment | NA | NA | 0 |
| S14_vs_k141_74875\|\|full | 1926 | No | NA | 2 | 0 | 1 | Low-quality | Genome-fragment | 3.22 | AAI-based (medium-confidence) | 0 |
| S7_vs_13716\|\|full | 1926 | No | NA | 4 | 1 | 0 | Low-quality | Genome-fragment | 3.87 | AAI-based (high-confidence) | 0 |
| S16_vs_47661\|\|full | 1926 | No | NA | 2 | 0 | 1 | Low-quality | Genome-fragment | 0.91 | AAI-based (medium-confidence) | 0 |
| S5_vs_k141_107532\|\|full | 1796 | No | NA | 3 | 0 | 3 | Not-determined | Genome-fragment | NA | NA | 0 |
| S5_vs_k141_107534\|\|full | 1143 | No | NA | 2 | 0 | 2 | Not-determined | Genome-fragment | NA | NA | 0 |
| S5_vs_k141_94536\|\|full | 1313 | No | NA | 3 | 0 | 2 | Not-determined | Genome-fragment | NA | NA | 0 |
| S8_extracted_k141_91962 | 1926 | No | NA | 3 | 1 | 0 | Low-quality | Genome-fragment | 5.44 | AAI-based (medium-confidence) | 0 |
| S9_vs_k141_55915\|\|full | 1927 | No | NA | 3 | 1 | 0 | Low-quality | Genome-fragment | 2.16 | HMM-based (lower-bound) | 0 |
| S5_vs_k141_99449\|\|full | 2210 | No | NA | 2 | 0 | 1 | Not-determined | Genome-fragment | NA | NA | 0 |
| S5_vs_k141_94544\|\|full | 1095 | No | NA | 3 | 0 | 0 | Not-determined | Genome-fragment | NA | NA | 0 |
| S5_vs_k141_130958\|\|full | 36390 | No | NA | 49 | 15 | 1 | Medium-quality | Genome-fragment | 65.93 | AAI-based (high-confidence) | 0 |
| S3_vs_k141_68350\|\|full | 1927 | No | NA | 5 | 0 | 0 | Low-quality | Genome-fragment | 3.92 | AAI-based (high-confidence) | 0 |
| S5_vs_k141_56677\|\|full | 2582 | No | NA | 2 | 0 | 1 | Not-determined | Genome-fragment | NA | NA | 0 |
| S5_vs_k141_34322\|\|full | 2879 | No | NA | 6 | 0 | 0 | Not-determined | Genome-fragment | NA | NA | 0 |
| S16_extracted_151822 | 1927 | No | NA | 3 | 0 | 0 | Low-quality | Genome-fragment | 1.51 | AAI-based (high-confidence) | 0 |
| S5_vs_k141_9747\|\|full | 64437 | Yes | 46640 | 66 | 13 | 5 | High-quality | High-quality | 100 | AAI-based (high-confidence) | 27.62 |
| S5_vs_k141_39566\|\|full | 1117 | No | NA | 3 | 0 | 2 | Not-determined | Genome-fragment | NA | NA | 0 |
| S5_vs_k141_39570\|\|full | 3208 | No | NA | 6 | 0 | 3 | Not-determined | Genome-fragment | NA | NA | 0 |
| S16_vs_97578\|\|full | 1927 | No | NA | 3 | 0 | 0 | Low-quality | Genome-fragment | 4.43 | AAI-based (high-confidence) | 0 |
| S12_vs_k141_102478\|\|full | 1927 | No | NA | 4 | 0 | 0 | Low-quality | Genome-fragment | 5.21 | AAI-based (medium-confidence) | 0 |
| S12_vs_k141_143566\|\|full | 1927 | No | NA | 4 | 2 | 0 | Low-quality | Genome-fragment | 3.47 | AAI-based (high-confidence) | 0 |
| S12_vs_k141_177496\|\|full | 1927 | No | NA | 4 | 4 | 0 | Low-quality | Genome-fragment | 4.78 | AAI-based (high-confidence) | 0 |
| S12_vs_k141_47172\|\|full | 1927 | No | NA | 3 | 0 | 0 | Low-quality | Genome-fragment | 3.99 | AAI-based (high-confidence) | 0 |
| S5_vs_k141_99510\|\|full | 1154 | No | NA | 3 | 0 | 0 | Not-determined | Genome-fragment | NA | NA | 0 |
| S9_vs_k141_43567\|\|full | 1928 | No | NA | 5 | 0 | 0 | Low-quality | Genome-fragment | 4.38 | AAI-based (medium-confidence) | 0 |
| S5_vs_k141_34375\|\|full | 2450 | No | NA | 3 | 0 | 2 | Not-determined | Genome-fragment | NA | NA | 0 |
| S5_vs_k141_39630\|\|full | 1223 | No | NA | 3 | 0 | 1 | Not-determined | Genome-fragment | NA | NA | 0 |
| S5_vs_k141_80841\|\|full | 4444 | No | NA | 3 | 0 | 2 | Not-determined | Genome-fragment | NA | NA | 0 |
| S5_vs_k141_4737\|\|full | 37808 | No | NA | 56 | 15 | 1 | Medium-quality | Genome-fragment | 63.15 | HMM-based (lower-bound) | 0 |
| S5_vs_k141_130971\|\|full | 4580 | No | NA | 5 | 0 | 2 | Not-determined | Genome-fragment | NA | NA | 0 |
| S6_vs_k141_24001\|\|full | 1928 | No | NA | 3 | 2 | 0 | Low-quality | Genome-fragment | 3.16 | AAI-based (medium-confidence) | 0 |
| S5_vs_k141_52796\|\|full | 13838 | No | NA | 16 | 0 | 11 | Not-determined | Genome-fragment | NA | NA | 0 |
| S17_vs_k141_74691\|\|full | 1929 | No | NA | 3 | 0 | 2 | Low-quality | Genome-fragment | 0.63 | AAI-based (medium-confidence) | 0 |
| S9_vs_k141_62323\|\|full | 1929 | No | NA | 4 | 0 | 0 | Low-quality | Genome-fragment | 3.63 | AAI-based (high-confidence) | 0 |
| S11_vs_k141_43626\|\|full | 1929 | No | NA | 5 | 1 | 0 | Low-quality | Genome-fragment | 2.22 | AAI-based (high-confidence) | 0 |
| S5_vs_k141_125976\|\|full | 3063 | No | NA | 3 | 0 | 1 | Not-determined | Genome-fragment | NA | NA | 0 |
| S5_vs_k141_9822\|\|full | 9698 | No | NA | 11 | 0 | 1 | Not-determined | Genome-fragment | NA | NA | 0 |
| S14_vs_k141_97841\|\|full | 1929 | No | NA | 5 | 1 | 0 | Low-quality | Genome-fragment | 2.59 | AAI-based (high-confidence) | 0 |
| S5_vs_k141_88207\|\|full | 12966 | No | NA | 18 | 0 | 7 | Not-determined | Genome-fragment | NA | NA | 0 |
| S5_extracted_k141_137161 | 1929 | No | NA | 1 | 0 | 1 | Low-quality | Genome-fragment | 1.43 | AAI-based (medium-confidence) | 0 |
| S5_vs_k141_2825\|\|lt2gene | 1929 | No | NA | 2 | 1 | 0 | Low-quality | Genome-fragment | 3.04 | HMM-based (lower-bound) | 0 |
| S7_vs_116771\|\|full | 1929 | No | NA | 3 | 0 | 0 | Low-quality | Genome-fragment | 4.92 | AAI-based (medium-confidence) | 0 |
| S5_vs_k141_34429\|\|full | 3500 | No | NA | 4 | 0 | 2 | Not-determined | Genome-fragment | NA | NA | 0 |
| S17_vs_k141_164861\|\|full | 1930 | No | NA | 3 | 0 | 0 | Low-quality | Genome-fragment | 3.39 | AAI-based (high-confidence) | 0 |
| S9_extracted_k141_64838 | 1930 | No | NA | 3 | 0 | 0 | Low-quality | Genome-fragment | 1.11 | AAI-based (high-confidence) | 0 |
| S9_extracted_k141_4979 | 1930 | No | NA | 4 | 0 | 0 | Low-quality | Genome-fragment | 1.74 | AAI-based (high-confidence) | 0 |
| S5_vs_k141_74824\|\|full | 1691 | No | NA | 3 | 0 | 1 | Not-determined | Genome-fragment | NA | NA | 0 |
| S5_vs_k141_9871\|\|full | 1535 | No | NA | 4 | 0 | 0 | Not-determined | Genome-fragment | NA | NA | 0 |
| S5_vs_k141_94631\|\|full | 1615 | No | NA | 3 | 0 | 2 | Not-determined | Genome-fragment | NA | NA | 0 |
| S7_extracted_31341 | 1930 | No | NA | 3 | 1 | 0 | Low-quality | Genome-fragment | 3.26 | HMM-based (lower-bound) | 0 |
| S5_vs_k141_64026\|\|full | 2516 | No | NA | 3 | 0 | 1 | Not-determined | Genome-fragment | NA | NA | 0 |
| S5_vs_k141_34472\|\|full | 1100 | No | NA | 2 | 0 | 0 | Not-determined | Genome-fragment | NA | NA | 0 |
| S5_vs_k141_64034\|\|full | 1759 | No | NA | 4 | 0 | 3 | Not-determined | Genome-fragment | NA | NA | 0 |
| S16_vs_117328\|\|full | 1930 | No | NA | 4 | 0 | 0 | Low-quality | Genome-fragment | 2.33 | AAI-based (medium-confidence) | 0 |
| S5_vs_k141_107642\|\|full | 1620 | No | NA | 4 | 0 | 0 | Not-determined | Genome-fragment | NA | NA | 0 |
| S5_vs_k141_64041\|\|full | 2383 | No | NA | 3 | 0 | 1 | Not-determined | Genome-fragment | NA | NA | 0 |
| S16_vs_150474\|\|full | 1930 | No | NA | 3 | 2 | 0 | Low-quality | Genome-fragment | 4.75 | AAI-based (high-confidence) | 0 |
| S8_vs_k141_44277\|\|full | 1930 | No | NA | 3 | 2 | 0 | Low-quality | Genome-fragment | 4.6 | AAI-based (medium-confidence) | 0 |
| S5_vs_k141_80911\|\|full | 36909 | No | NA | 64 | 19 | 0 | High-quality | High-quality | 93.74 | AAI-based (medium-confidence) | 0 |
| S5_vs_k141_112929\|\|full | 4817 | No | NA | 6 | 0 | 5 | Not-determined | Genome-fragment | NA | NA | 0 |
| S5_vs_k141_80916\|\|full | 1336 | No | NA | 3 | 0 | 0 | Not-determined | Genome-fragment | NA | NA | 0 |
| S13_vs_32966\|\|full | 1931 | No | NA | 3 | 2 | 0 | Low-quality | Genome-fragment | 4.97 | AAI-based (high-confidence) | 0 |
| S14_vs_k141_5091\|\|full | 1931 | No | NA | 3 | 0 | 0 | Low-quality | Genome-fragment | 2.33 | AAI-based (medium-confidence) | 0 |
| S5_vs_k141_107658\|\|full | 1257 | No | NA | 3 | 0 | 2 | Not-determined | Genome-fragment | NA | NA | 0 |
| S7_vs_91997\|\|full | 1931 | No | NA | 3 | 0 | 0 | Low-quality | Genome-fragment | 1.25 | AAI-based (medium-confidence) | 0 |
| S3_vs_k141_124978\|\|full | 1932 | No | NA | 3 | 0 | 0 | Low-quality | Genome-fragment | 1.37 | AAI-based (high-confidence) | 0 |
| S7_extracted_25376 | 1932 | No | NA | 2 | 0 | 0 | Low-quality | Genome-fragment | 0.23 | AAI-based (medium-confidence) | 0 |
| S7_vs_67540\|\|full | 1932 | No | NA | 5 | 0 | 0 | Low-quality | Genome-fragment | 5.18 | AAI-based (high-confidence) | 0 |
| S5_vs_k141_99616\|\|full | 1462 | No | NA | 3 | 0 | 1 | Not-determined | Genome-fragment | NA | NA | 0 |
| S5_vs_k141_64074\|\|full | 2118 | No | NA | 3 | 0 | 1 | Not-determined | Genome-fragment | NA | NA | 0 |
| S5_vs_k141_34563\|\|full | 2593 | No | NA | 2 | 0 | 0 | Not-determined | Genome-fragment | NA | NA | 0 |
| S16_vs_172373\|\|full | 1932 | No | NA | 3 | 0 | 0 | Low-quality | Genome-fragment | 2.14 | AAI-based (medium-confidence) | 0 |
| S5_vs_k141_21681\|\|full | 2030 | No | NA | 2 | 0 | 0 | Not-determined | Genome-fragment | NA | NA | 0 |
| S10_vs_61009\|\|full | 1932 | No | NA | 3 | 0 | 1 | Low-quality | Genome-fragment | 1.35 | AAI-based (medium-confidence) | 0 |
| S12_extracted_k141_178033 | 1932 | No | NA | 1 | 0 | 0 | Low-quality | Genome-fragment | 1.41 | AAI-based (high-confidence) | 0 |
| S2_vs_k141_13777\|\|full | 1932 | No | NA | 5 | 1 | 0 | Low-quality | Genome-fragment | 3.15 | AAI-based (high-confidence) | 0 |
| S14_vs_k141_36785\|\|full | 1933 | No | NA | 2 | 0 | 1 | Low-quality | Genome-fragment | 3.08 | AAI-based (high-confidence) | 0 |
| S5_vs_k141_56849\|\|full | 2883 | No | NA | 3 | 0 | 2 | Not-determined | Genome-fragment | NA | NA | 0 |
| S1_extracted_k141_140879 | 1933 | No | NA | 1 | 1 | 0 | Low-quality | Genome-fragment | 1.95 | AAI-based (high-confidence) | 0 |
| S5_vs_k141_46060\|\|full | 5459 | Yes | 1671 | 3 | 1 | 2 | Not-determined | Genome-fragment | NA | NA | 69.39 |
| S3_vs_k141_101187\|\|full | 1933 | No | NA | 4 | 1 | 0 | Low-quality | Genome-fragment | 4.21 | AAI-based (high-confidence) | 0 |
| S5_vs_k141_17769\|\|full | 1933 | No | NA | 2 | 0 | 0 | Low-quality | Genome-fragment | 2.92 | AAI-based (high-confidence) | 0 |
| S7_extracted_24485 | 1933 | No | NA | 2 | 0 | 0 | Low-quality | Genome-fragment | 3.11 | AAI-based (medium-confidence) | 0 |
| S16_extracted_53974 | 1933 | No | NA | 3 | 3 | 0 | Low-quality | Genome-fragment | 4.12 | AAI-based (high-confidence) | 0 |
| S5_vs_k141_112984\|\|full | 1163 | No | NA | 2 | 0 | 1 | Not-determined | Genome-fragment | NA | NA | 0 |
| S5_vs_k141_69980\|\|full | 22516 | No | NA | 30 | 12 | 0 | Medium-quality | Genome-fragment | 57.76 | AAI-based (high-confidence) | 0 |
| S16_vs_3677\|\|full | 1933 | No | NA | 2 | 0 | 0 | Low-quality | Genome-fragment | 4.1 | AAI-based (high-confidence) | 0 |
| S10_vs_35468\|\|full | 1933 | No | NA | 3 | 3 | 0 | Low-quality | Genome-fragment | 3.17 | AAI-based (high-confidence) | 0 |
| S5_vs_k141_53048\|\|full | 1138 | No | NA | 3 | 0 | 0 | Not-determined | Genome-fragment | NA | NA | 0 |
| S8_extracted_k141_114950 | 1933 | No | NA | 5 | 0 | 0 | Low-quality | Genome-fragment | 2.36 | AAI-based (high-confidence) | 0 |
| S17_vs_k141_60602\|\|full | 1934 | No | NA | 3 | 2 | 0 | Low-quality | Genome-fragment | 3.69 | AAI-based (medium-confidence) | 0 |
| S9_vs_k141_27717\|\|lt2gene | 1934 | No | NA | 2 | 2 | 0 | Low-quality | Genome-fragment | 5.38 | AAI-based (high-confidence) | 0 |
| S3_extracted_k141_77528 | 1934 | No | NA | 2 | 0 | 0 | Low-quality | Genome-fragment | 4.01 | AAI-based (high-confidence) | 0 |
| S5_vs_k141_126121\|\|full | 1745 | No | NA | 3 | 0 | 2 | Not-determined | Genome-fragment | NA | NA | 0 |
| S5_vs_k141_136129\|\|full | 2000 | No | NA | 3 | 0 | 1 | Not-determined | Genome-fragment | NA | NA | 0 |
| S5_vs_k141_126131\|\|full | 7826 | No | NA | 5 | 0 | 3 | Not-determined | Genome-fragment | NA | NA | 0 |
| S5_vs_k141_113028\|\|full | 1506 | No | NA | 3 | 0 | 2 | Not-determined | Genome-fragment | NA | NA | 0 |
| S3_vs_k141_66314\|\|full | 1934 | No | NA | 3 | 0 | 0 | Low-quality | Genome-fragment | 3.55 | AAI-based (high-confidence) | 0 |
| S3_vs_k141_123109\|\|lt2gene | 1934 | No | NA | 2 | 2 | 0 | Low-quality | Genome-fragment | 4.42 | AAI-based (high-confidence) | 0 |
| S5_vs_k141_99725\|\|full | 3236 | No | NA | 7 | 0 | 0 | Not-determined | Genome-fragment | NA | NA | 0 |
| S5_vs_k141_131184\|\|full | 6900 | No | NA | 8 | 0 | 6 | Not-determined | Genome-fragment | NA | NA | 0 |
| S10_vs_106453\|\|full | 1934 | No | NA | 4 | 0 | 1 | Low-quality | Genome-fragment | 4.64 | AAI-based (medium-confidence) | 0 |
| S5_vs_k141_5077\|\|full | 4986 | No | NA | 4 | 0 | 2 | Not-determined | Genome-fragment | NA | NA | 0 |
| S12_extracted_k141_179012 | 1934 | No | NA | 2 | 0 | 0 | Low-quality | Genome-fragment | 3.2 | AAI-based (high-confidence) | 0 |
| S8_vs_k141_80668\|\|full | 1934 | No | NA | 3 | 2 | 0 | Low-quality | Genome-fragment | 3.75 | AAI-based (high-confidence) | 0 |
| S5_vs_k141_64168\|\|full | 2098 | No | NA | 3 | 0 | 0 | Not-determined | Genome-fragment | NA | NA | 0 |
| S8_vs_k141_20037\|\|full | 1934 | No | NA | 3 | 0 | 2 | Low-quality | Genome-fragment | 6.18 | AAI-based (high-confidence) | 0 |
| S17_vs_k141_105944\|\|full | 1935 | No | NA | 4 | 0 | 0 | Low-quality | Genome-fragment | 3.23 | AAI-based (medium-confidence) | 0 |
| S5_vs_k141_34722\|\|full | 1553 | No | NA | 2 | 0 | 0 | Not-determined | Genome-fragment | NA | NA | 0 |
| S3_extracted_k141_59766 | 1935 | No | NA | 5 | 0 | 0 | Low-quality | Genome-fragment | 3.87 | AAI-based (medium-confidence) | 0 |
| S5_vs_k141_29753\|\|lt2gene | 1935 | No | NA | 2 | 2 | 0 | Low-quality | Genome-fragment | 4.03 | AAI-based (high-confidence) | 0 |
| S7_vs_126702\|\|full | 1935 | No | NA | 3 | 2 | 0 | Low-quality | Genome-fragment | 3.03 | AAI-based (high-confidence) | 0 |
| S5_vs_k141_64195\|\|full | 1405 | No | NA | 2 | 0 | 2 | Not-determined | Genome-fragment | NA | NA | 0 |
| S5_vs_k141_136185\|\|full | 2423 | No | NA | 6 | 0 | 0 | Not-determined | Genome-fragment | NA | NA | 0 |
| S7_vs_136749\|\|full | 1935 | No | NA | 6 | 0 | 0 | Low-quality | Genome-fragment | 3.64 | AAI-based (medium-confidence) | 0 |
| S5_vs_k141_53113\|\|full | 9934 | No | NA | 8 | 0 | 4 | Not-determined | Genome-fragment | NA | NA | 0 |
| S5_vs_k141_46142\|\|full | 2358 | No | NA | 4 | 0 | 3 | Not-determined | Genome-fragment | NA | NA | 0 |
| S16_extracted_85804 | 1935 | No | NA | 3 | 0 | 2 | Low-quality | Genome-fragment | 0.67 | AAI-based (high-confidence) | 0 |
| S16_vs_64541\|\|full | 1935 | No | NA | 3 | 0 | 0 | Low-quality | Genome-fragment | 3.94 | AAI-based (high-confidence) | 0 |
| S8_extracted_k141_64792 | 1935 | No | NA | 3 | 0 | 2 | Low-quality | Genome-fragment | 2.62 | AAI-based (medium-confidence) | 0 |
| S5_vs_k141_46155\|\|full | 1813 | No | NA | 3 | 0 | 1 | Not-determined | Genome-fragment | NA | NA | 0 |
| S5_vs_k141_56971\|\|full | 5388 | No | NA | 7 | 0 | 5 | Not-determined | Genome-fragment | NA | NA | 0 |
| S17_vs_k141_20254\|\|full | 1936 | No | NA | 3 | 0 | 1 | Low-quality | Genome-fragment | 3.32 | AAI-based (high-confidence) | 0 |
| S5_vs_k141_70096\|\|full | 1610 | No | NA | 2 | 0 | 0 | Not-determined | Genome-fragment | NA | NA | 0 |
| S5_vs_k141_5153\|\|full | 1316 | No | NA | 2 | 0 | 1 | Not-determined | Genome-fragment | NA | NA | 0 |
| S5_vs_k141_94934\|\|full | 1472 | No | NA | 3 | 0 | 1 | Not-determined | Genome-fragment | NA | NA | 0 |
| S5_vs_k141_46183\|\|full | 1771 | No | NA | 4 | 0 | 1 | Not-determined | Genome-fragment | NA | NA | 0 |
| S11_vs_k141_80047\|\|full | 1936 | No | NA | 2 | 0 | 0 | Low-quality | Genome-fragment | 2.29 | AAI-based (high-confidence) | 0 |
| S3_vs_k141_64968\|\|full | 1936 | No | NA | 7 | 0 | 0 | Low-quality | Genome-fragment | 5.08 | AAI-based (high-confidence) | 0 |
| S3_vs_k141_12460\|\|full | 1936 | No | NA | 6 | 1 | 0 | Low-quality | Genome-fragment | 4.71 | AAI-based (medium-confidence) | 0 |
| S7_vs_5717\|\|full | 1936 | No | NA | 4 | 0 | 3 | Low-quality | Genome-fragment | 2.14 | AAI-based (medium-confidence) | 0 |
| S5_vs_k141_10073\|\|full | 1067 | No | NA | 2 | 0 | 2 | Not-determined | Genome-fragment | NA | NA | 0 |
| S12_extracted_k141_41433 | 1936 | No | NA | 4 | 0 | 2 | Low-quality | Genome-fragment | 2.22 | AAI-based (medium-confidence) | 0 |
| S6_vs_k141_21003\|\|full | 1936 | No | NA | 4 | 0 | 0 | Low-quality | Genome-fragment | 3.46 | AAI-based (high-confidence) | 0 |
| S5_vs_k141_26551\|\|full | 4005 | No | NA | 7 | 0 | 1 | Not-determined | Genome-fragment | NA | NA | 0 |
| S6_vs_k141_11015\|\|lt2gene | 1936 | No | NA | 1 | 1 | 0 | Low-quality | Genome-fragment | 6.37 | AAI-based (high-confidence) | 0 |
| S9_vs_k141_31471\|\|full | 1937 | No | NA | 2 | 0 | 1 | Low-quality | Genome-fragment | 2.35 | AAI-based (high-confidence) | 0 |
| S14_extracted_k141_116445 | 1937 | No | NA | 7 | 2 | 0 | Low-quality | Genome-fragment | 7.21 | AAI-based (high-confidence) | 0 |
| S14_vs_k141_76573\|\|full | 1937 | No | NA | 2 | 0 | 0 | Low-quality | Genome-fragment | 0.54 | AAI-based (medium-confidence) | 0 |
| S5_vs_k141_70162\|\|full | 12920 | No | NA | 15 | 0 | 8 | Not-determined | Genome-fragment | NA | NA | 0 |
| S4_extracted_23325 | 1937 | No | NA | 4 | 0 | 0 | Low-quality | Genome-fragment | 2.01 | AAI-based (high-confidence) | 0 |
| S8_extracted_k141_55364 | 1937 | No | NA | 1 | 0 | 0 | Low-quality | Genome-fragment | 1.96 | AAI-based (high-confidence) | 0 |
| S5_vs_k141_53224\|\|full | 1112 | No | NA | 2 | 0 | 0 | Not-determined | Genome-fragment | NA | NA | 0 |
| S8_vs_k141_69612\|\|lt2gene | 1937 | No | NA | 2 | 1 | 0 | Low-quality | Genome-fragment | 3.66 | HMM-based (lower-bound) | 0 |
| S17_vs_k141_166833\|\|full | 1938 | No | NA | 3 | 0 | 1 | Low-quality | Genome-fragment | 2.63 | AAI-based (high-confidence) | 0 |
| S5_vs_k141_88543\|\|full | 7671 | No | NA | 7 | 0 | 2 | Not-determined | Genome-fragment | NA | NA | 0 |
| S5_vs_k141_99794\|\|full | 3045 | No | NA | 3 | 0 | 1 | Not-determined | Genome-fragment | NA | NA | 0 |
| S9_extracted_k141_88633 | 1938 | No | NA | 3 | 1 | 0 | Low-quality | Genome-fragment | 1.1 | AAI-based (medium-confidence) | 0 |
| S5_vs_k141_136316\|\|full | 2296 | No | NA | 3 | 0 | 2 | Not-determined | Genome-fragment | NA | NA | 0 |
| S5_vs_k141_21778\|\|full | 157098 | No | NA | 151 | 4 | 27 | High-quality | High-quality | 92.73 | HMM-based (lower-bound) | 0 |
| S5_vs_k141_10109\|\|full | 2327 | No | NA | 4 | 0 | 3 | Not-determined | Genome-fragment | NA | NA | 0 |
| S14_extracted_k141_5409 | 1938 | No | NA | 3 | 3 | 0 | Low-quality | Genome-fragment | 6.57 | AAI-based (high-confidence) | 0 |
| S3_vs_k141_90470\|\|full | 1938 | No | NA | 7 | 1 | 0 | Low-quality | Genome-fragment | 3.39 | AAI-based (high-confidence) | 0 |
| S5_vs_k141_39721\|\|full | 1996 | No | NA | 3 | 0 | 2 | Not-determined | Genome-fragment | NA | NA | 0 |
| S5_vs_k141_107968\|\|full | 3545 | No | NA | 6 | 0 | 1 | Not-determined | Genome-fragment | NA | NA | 0 |
| S5_vs_k141_39741\|\|full | 1223 | No | NA | 2 | 0 | 1 | Not-determined | Genome-fragment | NA | NA | 0 |
| S7_vs_18225\|\|full | 1938 | No | NA | 4 | 1 | 0 | Low-quality | Genome-fragment | 2.4 | HMM-based (lower-bound) | 0 |
| S12_vs_k141_32851\|\|full | 1938 | No | NA | 4 | 0 | 0 | Low-quality | Genome-fragment | 4.09 | AAI-based (high-confidence) | 0 |
| S5_vs_k141_136342\|\|full | 8562 | No | NA | 8 | 0 | 3 | Not-determined | Genome-fragment | NA | NA | 0 |
| S5_extracted_k141_80781 | 1939 | No | NA | 3 | 1 | 0 | Low-quality | Genome-fragment | 8.84 | AAI-based (high-confidence) | 0 |
| S5_vs_k141_113187\|\|full | 2155 | No | NA | 2 | 0 | 1 | Not-determined | Genome-fragment | NA | NA | 0 |

| | | | | | | | | | | | |
|---|---|---|---|---|---|---|---|---|---|---|---|
| S5_vs_k141_99862\|\|full | 1539 | No | NA | 2 | 0 | 2 | Not-determined | Genome-fragment | NA | NA | 0 |
| S16_vs_153946\|\|full | 1939 | No | NA | 3 | 3 | 0 | Low-quality | Genome-fragment | 4.22 | AAI-based (medium-confidence) | 0 |
| S8_vs_k141_104140\|\|full | 1939 | No | NA | 3 | 3 | 0 | Low-quality | Genome-fragment | 3.51 | AAI-based (high-confidence) | 0 |
| S8_vs_k141_22551\|\|full | 1939 | No | NA | 2 | 0 | 0 | Low-quality | Genome-fragment | 3.03 | AAI-based (high-confidence) | 0 |
| S8_vs_k141_51593\|\|full | 1939 | No | NA | 6 | 0 | 0 | Low-quality | Genome-fragment | 4.83 | AAI-based (medium-confidence) | 0 |
| S9_extracted_k141_30984 | 1940 | Yes | 646 | 5 | 1 | 2 | Low-quality | Genome-fragment | 0.93 | HMM-based (lower-bound) | 66.7 |
| S5_vs_k141_39790\|\|full | 1332 | No | NA | 2 | 0 | 2 | Not-determined | Genome-fragment | NA | NA | 0 |
| S9_vs_k141_69148\|\|full | 1940 | No | NA | 3 | 0 | 1 | Low-quality | Genome-fragment | 0.81 | AAI-based (medium-confidence) | 0 |
| S5_vs_k141_21855\|\|full | 1477 | No | NA | 2 | 0 | 2 | Not-determined | Genome-fragment | NA | NA | 0 |
| S1_extracted_k141_132007 | 1940 | No | NA | 1 | 1 | 0 | Low-quality | Genome-fragment | 3.38 | AAI-based (high-confidence) | 0 |
| S3_extracted_k141_122396 | 1940 | No | NA | 4 | 1 | 0 | Low-quality | Genome-fragment | 1.09 | AAI-based (high-confidence) | 0 |
| S5_vs_k141_119866\|\|full | 1227 | No | NA | 2 | 0 | 1 | Not-determined | Genome-fragment | NA | NA | 0 |
| S5_vs_k141_16142\|\|full | 3345 | No | NA | 2 | 0 | 1 | Not-determined | Genome-fragment | NA | NA | 0 |
| S3_vs_k141_79679\|\|full | 1940 | No | NA | 3 | 1 | 0 | Low-quality | Genome-fragment | 0.16 | HMM-based (lower-bound) | 0 |
| S5_vs_k141_108040\|\|full | 1016 | No | NA | 2 | 0 | 0 | Not-determined | Genome-fragment | NA | NA | 0 |
| S5_extracted_k141_45599 | 1940 | No | NA | 3 | 3 | 0 | Low-quality | Genome-fragment | 5.17 | AAI-based (high-confidence) | 0 |
| S7_extracted_9058 | 1940 | No | NA | 3 | 0 | 0 | Low-quality | Genome-fragment | 1.33 | AAI-based (high-confidence) | 0 |
| S12_vs_k141_185328\|\|full | 1940 | No | NA | 3 | 0 | 0 | Low-quality | Genome-fragment | 3 | AAI-based (high-confidence) | 0 |
| S5_vs_k141_75067\|\|full | 2396 | No | NA | 3 | 0 | 3 | Not-determined | Genome-fragment | NA | NA | 0 |
| S5_vs_k141_142954\|\|full | 2609 | No | NA | 3 | 0 | 3 | Not-determined | Genome-fragment | NA | NA | 0 |
| S5_vs_k141_95155\|\|full | 3078 | No | NA | 3 | 0 | 0 | Not-determined | Genome-fragment | NA | NA | 0 |
| S15_vs_11255\|\|full | 1940 | No | NA | 6 | 0 | 0 | Low-quality | Genome-fragment | 4.84 | AAI-based (high-confidence) | 0 |
| S8_extracted_k141_86777 | 1940 | No | NA | 3 | 0 | 2 | Low-quality | Genome-fragment | 0.85 | AAI-based (medium-confidence) | 0 |
| S17_extracted_k141_85009 | 1941 | No | NA | 5 | 5 | 0 | Low-quality | Genome-fragment | 6.4 | AAI-based (high-confidence) | 0 |
| S9_vs_k141_25455\|\|full | 1941 | No | NA | 4 | 0 | 1 | Low-quality | Genome-fragment | 4.29 | AAI-based (high-confidence) | 0 |
| S5_vs_k141_34914\|\|full | 1370 | No | NA | 2 | 0 | 1 | Not-determined | Genome-fragment | NA | NA | 0 |
| S14_vs_k141_42414\|\|full | 1941 | No | NA | 3 | 0 | 0 | Low-quality | Genome-fragment | 4.9 | AAI-based (medium-confidence) | 0 |
| S3_extracted_k141_39271 | 1941 | No | NA | 2 | 2 | 0 | Low-quality | Genome-fragment | 4.12 | AAI-based (high-confidence) | 0 |
| S5_vs_k141_10244\|\|full | 1490 | No | NA | 4 | 0 | 0 | Not-determined | Genome-fragment | NA | NA | 0 |
| S7_vs_93634\|\|full | 1941 | No | NA | 5 | 1 | 0 | Low-quality | Genome-fragment | 4.07 | AAI-based (high-confidence) | 0 |
| S16_extracted_163570 | 1941 | No | NA | 1 | 0 | 1 | Low-quality | Genome-fragment | 0.71 | AAI-based (high-confidence) | 0 |
| S12_vs_k141_50559\|\|full | 1941 | No | NA | 3 | 0 | 0 | Low-quality | Genome-fragment | 2.95 | AAI-based (medium-confidence) | 0 |
| S5_vs_k141_26651\|\|full | 5427 | No | NA | 6 | 0 | 4 | Not-determined | Genome-fragment | NA | NA | 0 |
| S5_vs_k141_81208\|\|full | 3356 | No | NA | 3 | 0 | 2 | Not-determined | Genome-fragment | NA | NA | 0 |
| S5_vs_k141_81215\|\|full | 2268 | No | NA | 3 | 0 | 0 | Not-determined | Genome-fragment | NA | NA | 0 |
| S5_vs_k141_5450\|\|full | 1728 | No | NA | 2 | 0 | 2 | Not-determined | Genome-fragment | NA | NA | 0 |
| S5_vs_k141_75111\|\|full | 1719 | No | NA | 4 | 0 | 0 | Not-determined | Genome-fragment | NA | NA | 0 |
| S12_vs_k141_13628\|\|full | 1941 | No | NA | 3 | 0 | 0 | Low-quality | Genome-fragment | 4.7 | AAI-based (medium-confidence) | 0 |
| S17_vs_k141_136714\|\|full | 1942 | No | NA | 3 | 0 | 0 | Low-quality | Genome-fragment | 3.38 | AAI-based (high-confidence) | 0 |
| S17_vs_k141_85840\|\|full | 1942 | No | NA | 2 | 0 | 1 | Low-quality | Genome-fragment | 7 | AAI-based (medium-confidence) | 0 |
| S5_vs_k141_81231\|\|full | 2698 | No | NA | 2 | 0 | 1 | Not-determined | Genome-fragment | NA | NA | 0 |
| S5_vs_k141_39889\|\|full | 1726 | No | NA | 3 | 0 | 0 | Not-determined | Genome-fragment | NA | NA | 0 |
| S9_vs_k141_72605\|\|full | 1942 | No | NA | 4 | 2 | 0 | Low-quality | Genome-fragment | 3.52 | AAI-based (high-confidence) | 0 |
| S5_vs_k141_131412\|\|full | 2812 | No | NA | 3 | 0 | 3 | Not-determined | Genome-fragment | NA | NA | 0 |
| S5_vs_k141_88794\|\|full | 1081 | No | NA | 1 | 0 | 0 | Not-determined | Genome-fragment | NA | NA | 0 |
| S12_extracted_k141_24201 | 1942 | No | NA | 2 | 0 | 0 | Low-quality | Genome-fragment | 1.27 | AAI-based (high-confidence) | 0 |
| S5_vs_k141_64380\|\|full | 5051 | No | NA | 4 | 0 | 3 | Not-determined | Genome-fragment | NA | NA | 0 |
| S5_vs_k141_10314\|\|full | 2405 | No | NA | 2 | 0 | 1 | Not-determined | Genome-fragment | NA | NA | 0 |
| S12_vs_k141_54433\|\|full | 1942 | Yes | 1762 | 5 | 1 | 1 | Low-quality | Genome-fragment | 2.96 | AAI-based (high-confidence) | 9.27 |
| S6_vs_k141_20165\|\|full | 1942 | No | NA | 5 | 0 | 0 | Low-quality | Genome-fragment | 3.31 | AAI-based (medium-confidence) | 0 |
| S8_vs_k141_66352\|\|full | 1942 | No | NA | 3 | 1 | 0 | Low-quality | Genome-fragment | 2.79 | HMM-based (lower-bound) | 0 |
| S11_vs_k141_101783\|\|full | 1943 | No | NA | 4 | 3 | 0 | Low-quality | Genome-fragment | 3.98 | AAI-based (high-confidence) | 0 |
| S5_vs_k141_21989\|\|full | 1923 | No | NA | 3 | 0 | 0 | Not-determined | Genome-fragment | NA | NA | 0 |
| S5_vs_k141_95264\|\|full | 1750 | No | NA | 3 | 0 | 0 | Not-determined | Genome-fragment | NA | NA | 0 |
| S5_vs_k141_5500\|\|full | 2820 | No | NA | 3 | 0 | 2 | Not-determined | Genome-fragment | NA | NA | 0 |
| S5_vs_k141_35072\|\|full | 1038 | No | NA | 3 | 0 | 1 | Not-determined | Genome-fragment | NA | NA | 0 |
| S5_vs_k141_64415\|\|full | 4449 | No | NA | 4 | 0 | 1 | Not-determined | Genome-fragment | NA | NA | 0 |
| S11_vs_k141_83524\|\|full | 1943 | No | NA | 3 | 2 | 0 | Low-quality | Genome-fragment | 3.94 | HMM-based (lower-bound) | 0 |
| S5_vs_k141_5521\|\|full | 1152 | No | NA | 3 | 0 | 0 | Not-determined | Genome-fragment | NA | NA | 0 |
| S13_extracted_14901 | 1943 | No | NA | 1 | 0 | 0 | Low-quality | Genome-fragment | 1.44 | AAI-based (medium-confidence) | 0 |
| S7_vs_21638\|\|full | 1943 | No | NA | 2 | 0 | 0 | Low-quality | Genome-fragment | 4.95 | AAI-based (medium-confidence) | 0 |
| S5_vs_k141_99970\|\|full | 3809 | No | NA | 7 | 0 | 1 | Not-determined | Genome-fragment | NA | NA | 0 |
| S5_vs_k141_120062\|\|full | 2185 | No | NA | 4 | 0 | 0 | Not-determined | Genome-fragment | NA | NA | 0 |
| S5_vs_k141_39945\|\|full | 1193 | No | NA | 2 | 0 | 2 | Not-determined | Genome-fragment | NA | NA | 0 |
| S16_extracted_42603 | 1943 | No | NA | 5 | 0 | 0 | Low-quality | Genome-fragment | 3.2 | AAI-based (high-confidence) | 0 |
| S16_vs_68619\|\|full | 1943 | No | NA | 5 | 1 | 0 | Low-quality | Genome-fragment | 5.69 | AAI-based (high-confidence) | 0 |
| S5_vs_k141_108179\|\|full | 2809 | No | NA | 3 | 0 | 1 | Not-determined | Genome-fragment | NA | NA | 0 |
| S10_vs_108044\|\|full | 1943 | No | NA | 4 | 1 | 0 | Low-quality | Genome-fragment | 1.26 | HMM-based (lower-bound) | 0 |
| S5_vs_k141_53480\|\|full | 1196 | No | NA | 3 | 0 | 0 | Not-determined | Genome-fragment | NA | NA | 0 |
| S5_vs_k141_57128\|\|full | 2304 | No | NA | 3 | 0 | 0 | Not-determined | Genome-fragment | NA | NA | 0 |
| S5_vs_k141_113386\|\|full | 20152 | No | NA | 42 | 13 | 0 | Medium-quality | Genome-fragment | 57.35 | AAI-based (medium-confidence) | 0 |
| S5_vs_k141_113389\|\|full | 2213 | No | NA | 3 | 0 | 3 | Not-determined | Genome-fragment | NA | NA | 0 |
| S13_vs_16211\|\|full | 1944 | No | NA | 5 | 1 | 0 | Low-quality | Genome-fragment | 6.66 | HMM-based (lower-bound) | 0 |
| S3_vs_k141_16406\|\|full | 1944 | No | NA | 3 | 0 | 0 | Low-quality | Genome-fragment | 3.92 | AAI-based (high-confidence) | 0 |
| S3_vs_k141_26033\|\|full | 1944 | No | NA | 4 | 1 | 0 | Low-quality | Genome-fragment | 4 | AAI-based (high-confidence) | 0 |
| S5_vs_k141_95307\|\|full | 33203 | No | NA | 53 | 21 | 0 | Medium-quality | Genome-fragment | 83.67 | AAI-based (high-confidence) | 0 |
| S3_vs_k141_15702\|\|full | 1944 | No | NA | 5 | 3 | 0 | Low-quality | Genome-fragment | 5.48 | AAI-based (medium-confidence) | 0 |
| S5_vs_k141_16258\|\|full | 1148 | No | NA | 2 | 0 | 0 | Not-determined | Genome-fragment | NA | NA | 0 |
| S5_vs_k141_136552\|\|full | 2268 | No | NA | 2 | 0 | 0 | Not-determined | Genome-fragment | NA | NA | 0 |
| S7_extracted_53035 | 1944 | No | NA | 2 | 0 | 2 | Low-quality | Genome-fragment | 0.52 | AAI-based (medium-confidence) | 0 |
| S12_vs_k141_141421\|\|lt2gene | 1944 | No | NA | 2 | 0 | 0 | Low-quality | Genome-fragment | 4.13 | AAI-based (medium-confidence) | 0 |
| S5_vs_k141_120108\|\|full | 1662 | No | NA | 4 | 0 | 0 | Not-determined | Genome-fragment | NA | NA | 0 |
| S5_vs_k141_75284\|\|full | 3193 | No | NA | 4 | 0 | 2 | Not-determined | Genome-fragment | NA | NA | 0 |
| S8_extracted_k141_95381 | 1944 | No | NA | 2 | 0 | 0 | Low-quality | Genome-fragment | 1.46 | AAI-based (medium-confidence) | 0 |
| S5_vs_k141_26723\|\|full | 2254 | No | NA | 4 | 0 | 0 | Not-determined | Genome-fragment | NA | NA | 0 |
| S8_vs_k141_22660\|\|full | 1944 | No | NA | 4 | 0 | 2 | Low-quality | Genome-fragment | 2.8 | AAI-based (medium-confidence) | 0 |
| S16_extracted_24198 | 1945 | No | NA | 4 | 0 | 0 | Low-quality | Genome-fragment | 6.04 | AAI-based (high-confidence) | 0 |
| S12_extracted_k141_109556 | 1945 | No | NA | 3 | 0 | 0 | Low-quality | Genome-fragment | 3.05 | AAI-based (high-confidence) | 0 |
| S5_vs_k141_120136\|\|full | 3585 | No | NA | 4 | 0 | 3 | Not-determined | Genome-fragment | NA | NA | 0 |
| S5_vs_k141_57203\|\|full | 3951 | No | NA | 4 | 0 | 4 | Not-determined | Genome-fragment | NA | NA | 0 |
| S8_vs_k141_170421\|\|lt2gene | 1945 | No | NA | 2 | 1 | 0 | Low-quality | Genome-fragment | 4.93 | AAI-based (medium-confidence) | 0 |
| S11_vs_k141_2628\|\|full | 1946 | No | NA | 3 | 1 | 0 | Low-quality | Genome-fragment | 4.36 | AAI-based (medium-confidence) | 0 |
| S5_vs_k141_26777\|\|full | 2448 | No | NA | 4 | 0 | 4 | Not-determined | Genome-fragment | NA | NA | 0 |
| S5_vs_k141_22019\|\|full | 3558 | No | NA | 4 | 0 | 2 | Not-determined | Genome-fragment | NA | NA | 0 |
| S14_vs_k141_63637\|\|full | 1946 | No | NA | 2 | 0 | 2 | Low-quality | Genome-fragment | 0.86 | AAI-based (medium-confidence) | 0 |
| S16_vs_34862\|\|full | 1946 | No | NA | 4 | 2 | 0 | Low-quality | Genome-fragment | 6.57 | AAI-based (high-confidence) | 0 |
| S12_vs_k141_130723\|\|full | 1946 | No | NA | 4 | 0 | 1 | Low-quality | Genome-fragment | 2.27 | AAI-based (high-confidence) | 0 |
| S5_vs_k141_126449\|\|full | 1768 | No | NA | 2 | 0 | 1 | Not-determined | Genome-fragment | NA | NA | 0 |
| S15_extracted_72739 | 1946 | No | NA | 3 | 1 | 0 | Low-quality | Genome-fragment | 4.29 | AAI-based (high-confidence) | 0 |
| S5_vs_k141_22040\|\|full | 1616 | No | NA | 2 | 0 | 2 | Not-determined | Genome-fragment | NA | NA | 0 |
| S5_vs_k141_26787\|\|full | 12807 | No | NA | 15 | 0 | 4 | Not-determined | Genome-fragment | NA | NA | 0 |
| S11_extracted_k141_67519 | 1947 | No | NA | 7 | 1 | 0 | Low-quality | Genome-fragment | 4.36 | AAI-based (high-confidence) | 0 |
| S5_vs_k141_75344\|\|full | 1222 | No | NA | 3 | 0 | 1 | Not-determined | Genome-fragment | NA | NA | 0 |
| S11_extracted_k141_96973 | 1947 | No | NA | 5 | 0 | 0 | Low-quality | Genome-fragment | 3.27 | AAI-based (medium-confidence) | 0 |
| S14_vs_k141_89896\|\|full | 1947 | No | NA | 3 | 1 | 0 | Low-quality | Genome-fragment | 4.6 | AAI-based (high-confidence) | 0 |
| S3_extracted_k141_125755 | 1947 | Yes | 730 | 3 | 1 | 1 | Low-quality | Genome-fragment | 1.56 | AAI-based (medium-confidence) | 62.51 |
| S3_vs_k141_17926\|\|full | 1947 | No | NA | 6 | 0 | 0 | Low-quality | Genome-fragment | 4.29 | AAI-based (high-confidence) | 0 |
| S5_vs_k141_40114\|\|full | 1185 | No | NA | 2 | 0 | 1 | Not-determined | Genome-fragment | NA | NA | 0 |
| S16_vs_106816\|\|full | 1947 | No | NA | 3 | 0 | 1 | Low-quality | Genome-fragment | 3.96 | AAI-based (medium-confidence) | 0 |
| S5_vs_k141_126492\|\|full | 3002 | No | NA | 4 | 0 | 2 | Not-determined | Genome-fragment | NA | NA | 0 |
| S5_vs_k141_143148\|\|full | 3562 | No | NA | 4 | 0 | 1 | Not-determined | Genome-fragment | NA | NA | 0 |
| S10_extracted_32795 | 1947 | No | NA | 1 | 0 | 1 | Low-quality | Genome-fragment | 1.04 | AAI-based (medium-confidence) | 0 |
| S5_vs_k141_5634\|\|full | 1034 | No | NA | 3 | 0 | 0 | Not-determined | Genome-fragment | NA | NA | 0 |
| S5_vs_k141_131510\|\|full | 169132 | Yes | 38937 | 134 | 7 | 71 | Complete | High-quality | 100 | Provirus (high-confidence) | 76.98 |
| S5_vs_k141_35301\|\|full | 1050 | No | NA | 3 | 0 | 1 | Not-determined | Genome-fragment | NA | NA | 0 |
| S8_vs_k141_197401\|\|full | 1947 | No | NA | 3 | 1 | 0 | Low-quality | Genome-fragment | 0.75 | HMM-based (lower-bound) | 0 |
| S17_vs_k141_159501\|\|lt2gene | 1948 | No | NA | 2 | 2 | 0 | Low-quality | Genome-fragment | 3.19 | AAI-based (high-confidence) | 0 |
| S9_vs_k141_72212\|\|full | 1948 | No | NA | 2 | 0 | 2 | Low-quality | Genome-fragment | 0.86 | AAI-based (medium-confidence) | 0 |
| S5_vs_k141_126528\|\|full | 1318 | No | NA | 3 | 0 | 1 | Not-determined | Genome-fragment | NA | NA | 0 |
| S5_vs_k141_53587\|\|full | 2225 | No | NA | 3 | 0 | 1 | Not-determined | Genome-fragment | NA | NA | 0 |
| S13_vs_85351\|\|full | 1948 | No | NA | 3 | 1 | 0 | Low-quality | Genome-fragment | 2.19 | HMM-based (lower-bound) | 0 |
| S5_vs_k141_143179\|\|full | 6465 | No | NA | 6 | 0 | 4 | Not-determined | Genome-fragment | NA | NA | 0 |
| S5_vs_k141_126543\|\|full | 8177 | No | NA | 12 | 0 | 1 | Not-determined | Genome-fragment | NA | NA | 0 |
| S5_vs_k141_53599\|\|full | 2475 | No | NA | 3 | 0 | 0 | Not-determined | Genome-fragment | NA | NA | 0 |
| S5_vs_k141_53600\|\|full | 1652 | No | NA | 3 | 0 | 3 | Not-determined | Genome-fragment | NA | NA | 0 |
| S14_vs_k141_92766\|\|lt2gene | 1948 | No | NA | 2 | 2 | 0 | Low-quality | Genome-fragment | 4.54 | AAI-based (high-confidence) | 0 |
| S5_vs_k141_95449\|\|full | 1813 | No | NA | 2 | 0 | 0 | Not-determined | Genome-fragment | NA | NA | 0 |
| S12_extracted_k141_96274 | 1948 | No | NA | 2 | 0 | 0 | Low-quality | Genome-fragment | 4.21 | AAI-based (medium-confidence) | 0 |
| S5_vs_k141_53649\|\|full | 2701 | No | NA | 4 | 0 | 3 | Not-determined | Genome-fragment | NA | NA | 0 |
| S5_vs_k141_95486\|\|full | 1898 | No | NA | 3 | 0 | 1 | Not-determined | Genome-fragment | NA | NA | 0 |
| S5_vs_k141_126600\|\|full | 2023 | No | NA | 3 | 0 | 0 | Not-determined | Genome-fragment | NA | NA | 0 |
| S5_vs_k141_108490\|\|full | 2197 | No | NA | 3 | 0 | 0 | Not-determined | Genome-fragment | NA | NA | 0 |
| S17_vs_k141_94860\|\|full | 1949 | No | NA | 3 | 3 | 0 | Low-quality | Genome-fragment | 4.11 | AAI-based (medium-confidence) | 0 |
| S9_vs_k141_53736\|\|full | 1949 | No | NA | 4 | 3 | 0 | Low-quality | Genome-fragment | 4.28 | AAI-based (high-confidence) | 0 |
| S5_vs_k141_131642\|\|full | 2974 | No | NA | 2 | 0 | 2 | Not-determined | Genome-fragment | NA | NA | 0 |
| S11_vs_k141_62288\|\|full | 1949 | No | NA | 3 | 3 | 0 | Low-quality | Genome-fragment | 4.63 | AAI-based (high-confidence) | 0 |
| S5_vs_k141_16558\|\|full | 2256 | No | NA | 3 | 0 | 3 | Not-determined | Genome-fragment | NA | NA | 0 |
| S5_vs_k141_126635\|\|full | 3881 | No | NA | 2 | 0 | 1 | Not-determined | Genome-fragment | NA | NA | 0 |
| S13_extracted_31285 | 1949 | No | NA | 2 | 0 | 0 | Low-quality | Genome-fragment | 2.87 | AAI-based (high-confidence) | 0 |
| S3_extracted_k141_82857 | 1949 | No | NA | 4 | 0 | 0 | Low-quality | Genome-fragment | 7.02 | AAI-based (high-confidence) | 0 |
| S5_vs_k141_113504\|\|full | 1369 | No | NA | 2 | 0 | 2 | Not-determined | Genome-fragment | NA | NA | 0 |
| S9_vs_k141_40430\|\|full | 1950 | No | NA | 2 | 0 | 0 | Low-quality | Genome-fragment | 3.56 | AAI-based (medium-confidence) | 0 |
| S9_vs_k141_32772\|\|full | 1950 | No | NA | 2 | 2 | 0 | Low-quality | Genome-fragment | 5.08 | AAI-based (high-confidence) | 0 |
| S10_vs_56413\|\|full | 1950 | No | NA | 4 | 1 | 0 | Low-quality | Genome-fragment | 1.79 | AAI-based (high-confidence) | 0 |
| S10_vs_93879\|\|full | 1950 | No | NA | 2 | 0 | 0 | Low-quality | Genome-fragment | 3.49 | AAI-based (high-confidence) | 0 |
| S5_vs_k141_143268\|\|full | 1375 | No | NA | 2 | 0 | 1 | Not-determined | Genome-fragment | NA | NA | 0 |
| S15_extracted_16191 | 1950 | No | NA | 3 | 1 | 0 | Low-quality | Genome-fragment | 2.43 | AAI-based (high-confidence) | 0 |
| S3_extracted_k141_51054 | 1951 | No | NA | 4 | 0 | 0 | Low-quality | Genome-fragment | 3.91 | AAI-based (medium-confidence) | 0 |
| S7_extracted_11011 | 1951 | No | NA | 6 | 0 | 0 | Low-quality | Genome-fragment | 4.59 | AAI-based (high-confidence) | 0 |
| S5_vs_k141_143273\|\|full | 1723 | No | NA | 3 | 0 | 1 | Not-determined | Genome-fragment | NA | NA | 0 |
| S5_vs_k141_89162\|\|full | 2253 | No | NA | 4 | 0 | 0 | Not-determined | Genome-fragment | NA | NA | 0 |
| S5_vs_k141_108607\|\|full | 1210 | No | NA | 3 | 0 | 0 | Not-determined | Genome-fragment | NA | NA | 0 |
| S10_vs_46195\|\|full | 1951 | No | NA | 3 | 0 | 0 | Low-quality | Genome-fragment | 2.27 | AAI-based (medium-confidence) | 0 |
| S11_vs_k141_40328\|\|full | 1952 | No | NA | 4 | 1 | 0 | Low-quality | Genome-fragment | 1.23 | AAI-based (medium-confidence) | 0 |
| S13_vs_24106\|\|full | 1952 | No | NA | 4 | 0 | 0 | Low-quality | Genome-fragment | 4.08 | AAI-based (medium-confidence) | 0 |
| S14_vs_k141_11400\|\|full | 1952 | No | NA | 1 | 1 | 0 | Low-quality | Genome-fragment | 4.44 | AAI-based (high-confidence) | 0 |
| S5_vs_k141_70707\|\|full | 6377 | No | NA | 4 | 0 | 1 | Not-determined | Genome-fragment | NA | NA | 0 |
| S1_vs_k141_120494\|\|full | 1952 | No | NA | 3 | 0 | 1 | Low-quality | Genome-fragment | 3.53 | AAI-based (medium-confidence) | 0 |
| S7_vs_65205\|\|full | 1952 | No | NA | 4 | 2 | 0 | Low-quality | Genome-fragment | 4.09 | AAI-based (high-confidence) | 0 |
| S5_vs_k141_143324\|\|full | 1019 | No | NA | 3 | 0 | 0 | Not-determined | Genome-fragment | NA | NA | 0 |
| S5_vs_k141_95588\|\|full | 1307 | No | NA | 3 | 0 | 0 | Not-determined | Genome-fragment | NA | NA | 0 |
| S10_vs_120079\|\|full | 1952 | No | NA | 5 | 1 | 0 | Low-quality | Genome-fragment | 3.58 | AAI-based (high-confidence) | 0 |
| S5_vs_k141_113611\|\|full | 1354 | No | NA | 2 | 0 | 0 | Not-determined | Genome-fragment | NA | NA | 0 |
| S5_vs_k141_57272\|\|full | 2706 | No | NA | 2 | 0 | 2 | Not-determined | Genome-fragment | NA | NA | 0 |
| S8_vs_k141_164087\|\|full | 1952 | No | NA | 6 | 2 | 0 | Low-quality | Genome-fragment | 3.76 | AAI-based (medium-confidence) | 0 |
| S5_vs_k141_26997\|\|full | 1736 | No | NA | 2 | 0 | 2 | Not-determined | Genome-fragment | NA | NA | 0 |
| S9_extracted_k141_37339 | 1953 | No | NA | 1 | 1 | 0 | Low-quality | Genome-fragment | 1.12 | AAI-based (high-confidence) | 0 |
| S5_vs_k141_35431\|\|full | 2160 | No | NA | 4 | 0 | 2 | Not-determined | Genome-fragment | NA | NA | 0 |
| S9_vs_k141_17555\|\|full | 1953 | No | NA | 3 | 2 | 0 | Low-quality | Genome-fragment | 5.35 | AAI-based (medium-confidence) | 0 |
| S5_vs_k141_136704\|\|full | 1252 | No | NA | 2 | 0 | 0 | Not-determined | Genome-fragment | NA | NA | 0 |
| S14_vs_k141_95859\|\|full | 1953 | No | NA | 3 | 0 | 0 | Low-quality | Genome-fragment | 1.4 | AAI-based (medium-confidence) | 0 |
| S5_vs_k141_40269\|\|full | 1517 | No | NA | 3 | 0 | 0 | Not-determined | Genome-fragment | NA | NA | 0 |
| S5_vs_k141_95637\|\|full | 1451 | No | NA | 2 | 0 | 1 | Not-determined | Genome-fragment | NA | NA | 0 |
| S5_vs_k141_40281\|\|full | 1376 | No | NA | 3 | 0 | 3 | Not-determined | Genome-fragment | NA | NA | 0 |
| S3_extracted_k141_46199 | 1953 | No | NA | 4 | 1 | 0 | Low-quality | Genome-fragment | 3.18 | AAI-based (high-confidence) | 0 |
| S5_vs_k141_22210\|\|full | 12964 | No | NA | 21 | 0 | 4 | Not-determined | Genome-fragment | NA | NA | 0 |
| S3_vs_k141_40451\|\|full | 1953 | No | NA | 6 | 2 | 0 | Low-quality | Genome-fragment | 3.26 | AAI-based (high-confidence) | 0 |
| S3_vs_k141_101091\|\|full | 1953 | No | NA | 5 | 3 | 0 | Low-quality | Genome-fragment | 3.69 | AAI-based (medium-confidence) | 0 |
| S16_vs_65731\|\|full | 1953 | No | NA | 4 | 2 | 0 | Low-quality | Genome-fragment | 4.15 | HMM-based (lower-bound) | 0 |
| S5_vs_k141_70804\|\|full | 6067 | No | NA | 7 | 0 | 5 | Not-determined | Genome-fragment | NA | NA | 0 |
| S10_vs_35919\|\|full | 1953 | No | NA | 3 | 1 | 0 | Low-quality | Genome-fragment | 3.76 | AAI-based (high-confidence) | 0 |
| S5_vs_k141_126726\|\|full | 1747 | No | NA | 4 | 0 | 0 | Not-determined | Genome-fragment | NA | NA | 0 |
| S5_vs_k141_75644\|\|full | 13262 | No | NA | 12 | 0 | 9 | Not-determined | Genome-fragment | NA | NA | 0 |
| S5_vs_k141_131759\|\|full | 1332 | No | NA | 2 | 0 | 0 | Not-determined | Genome-fragment | NA | NA | 0 |
| S12_vs_k141_83505\|\|full | 1953 | No | NA | 2 | 0 | 2 | Low-quality | Genome-fragment | 1.25 | AAI-based (medium-confidence) | 0 |
| S5_vs_k141_131792\|\|full | 1078 | No | NA | 2 | 0 | 1 | Not-determined | Genome-fragment | NA | NA | 0 |
| S8_extracted_k141_87607 | 1953 | No | NA | 4 | 0 | 1 | Low-quality | Genome-fragment | 0.58 | AAI-based (medium-confidence) | 0 |
| S5_vs_k141_57351\|\|full | 1548 | No | NA | 3 | 0 | 0 | Not-determined | Genome-fragment | NA | NA | 0 |
| S5_vs_k141_46306\|\|full | 4267 | No | NA | 11 | 0 | 2 | Not-determined | Genome-fragment | NA | NA | 0 |
| S17_vs_k141_60574\|\|full | 1954 | No | NA | 3 | 2 | 0 | Low-quality | Genome-fragment | 3.97 | AAI-based (high-confidence) | 0 |
| S5_vs_k141_95697\|\|full | 10300 | No | NA | 9 | 0 | 9 | Not-determined | Genome-fragment | NA | NA | 0 |
| S14_vs_k141_111272\|\|full | 1954 | No | NA | 7 | 0 | 0 | Low-quality | Genome-fragment | 1.12 | AAI-based (high-confidence) | 0 |
| S5_vs_k141_126813\|\|full | 1565 | No | NA | 3 | 0 | 1 | Not-determined | Genome-fragment | NA | NA | 0 |
| S5_vs_k141_120297\|\|full | 1630 | No | NA | 2 | 0 | 1 | Not-determined | Genome-fragment | NA | NA | 0 |
| S9_vs_k141_8000\|\|full | 1955 | No | NA | 3 | 0 | 0 | Low-quality | Genome-fragment | 4.64 | AAI-based (high-confidence) | 0 |
| S5_vs_k141_113780\|\|full | 1253 | No | NA | 3 | 0 | 0 | Not-determined | Genome-fragment | NA | NA | 0 |
| S5_vs_k141_143405\|\|full | 7063 | No | NA | 6 | 0 | 4 | Not-determined | Genome-fragment | NA | NA | 0 |
| S5_vs_k141_131887\|\|full | 3018 | No | NA | 3 | 0 | 2 | Not-determined | Genome-fragment | NA | NA | 0 |
| S5_vs_k141_35626\|\|full | 1447 | No | NA | 3 | 0 | 0 | Not-determined | Genome-fragment | NA | NA | 0 |
| S5_vs_k141_5772\|\|full | 4797 | No | NA | 3 | 0 | 1 | Not-determined | Genome-fragment | NA | NA | 0 |
| S7_vs_64081\|\|full | 1955 | No | NA | 5 | 0 | 0 | Low-quality | Genome-fragment | 4.92 | AAI-based (medium-confidence) | 0 |
| S16_vs_74062\|\|full | 1955 | No | NA | 3 | 0 | 2 | Low-quality | Genome-fragment | 3.58 | AAI-based (medium-confidence) | 0 |
| S8_extracted_k141_188486 | 1955 | No | NA | 7 | 0 | 0 | Low-quality | Genome-fragment | 6.22 | AAI-based (high-confidence) | 0 |
| S5_vs_k141_100310\|\|full | 1453 | No | NA | 2 | 0 | 0 | Not-determined | Genome-fragment | NA | NA | 0 |
| S5_vs_k141_143471\|\|full | 5595 | No | NA | 6 | 0 | 6 | Not-determined | Genome-fragment | NA | NA | 0 |
| S5_vs_k141_76758\|\|full | 1956 | No | NA | 4 | 0 | 0 | Low-quality | Genome-fragment | 5.06 | AAI-based (medium-confidence) | 0 |
| S5_vs_k141_46394\|\|full | 60490 | No | NA | 70 | 6 | 32 | High-quality | High-quality | 100 | AAI-based (medium-confidence) | 0 |
| S7_vs_125365\|\|lt2gene | 1956 | No | NA | 1 | 1 | 0 | Low-quality | Genome-fragment | 2.86 | HMM-based (lower-bound) | 0 |
| S12_vs_k141_142437\|\|full | 1956 | No | NA | 4 | 3 | 0 | Low-quality | Genome-fragment | 1.25 | HMM-based (lower-bound) | 0 |
| S6_vs_k141_39938\|\|full | 1956 | No | NA | 2 | 0 | 2 | Low-quality | Genome-fragment | 6.65 | AAI-based (medium-confidence) | 0 |
| S5_vs_k141_136818\|\|full | 3538 | No | NA | 4 | 0 | 2 | Not-determined | Genome-fragment | NA | NA | 0 |
| S5_vs_k141_143512\|\|full | 1070 | No | NA | 2 | 0 | 1 | Not-determined | Genome-fragment | NA | NA | 0 |
| S5_vs_k141_75843\|\|full | 7638 | No | NA | 4 | 0 | 3 | Not-determined | Genome-fragment | NA | NA | 0 |
| S5_vs_k141_143521\|\|full | 1906 | No | NA | 4 | 0 | 0 | Not-determined | Genome-fragment | NA | NA | 0 |
| S5_vs_k141_27182\|\|full | 1035 | No | NA | 2 | 0 | 0 | Not-determined | Genome-fragment | NA | NA | 0 |
| S9_extracted_k141_44323 | 1957 | No | NA | 1 | 1 | 0 | Low-quality | Genome-fragment | 3.59 | AAI-based (high-confidence) | 0 |
| S14_extracted_k141_76062 | 1957 | No | NA | 5 | 1 | 0 | Low-quality | Genome-fragment | 7.29 | AAI-based (high-confidence) | 0 |
| S14_extracted_k141_49812 | 1957 | No | NA | 2 | 1 | 0 | Low-quality | Genome-fragment | 1.21 | HMM-based (lower-bound) | 0 |
| S5_vs_k141_5841\|\|full | 1212 | No | NA | 3 | 0 | 1 | Not-determined | Genome-fragment | NA | NA | 0 |
| S5_vs_k141_27197\|\|full | 4357 | No | NA | 7 | 0 | 0 | Not-determined | Genome-fragment | NA | NA | 0 |
| S14_vs_k141_89015\|\|full | 1957 | No | NA | 3 | 0 | 2 | Low-quality | Genome-fragment | 0.57 | AAI-based (high-confidence) | 0 |
| S14_vs_k141_45664\|\|full | 1957 | No | NA | 5 | 0 | 1 | Low-quality | Genome-fragment | 1.99 | AAI-based (medium-confidence) | 0 |
| S5_vs_k141_75866\|\|full | 2327 | No | NA | 3 | 0 | 0 | Not-determined | Genome-fragment | NA | NA | 0 |
| S1_extracted_k141_22752 | 1957 | No | NA | 1 | 0 | 0 | Low-quality | Genome-fragment | 4.46 | AAI-based (high-confidence) | 0 |
| S7_vs_102401\|\|full | 1957 | No | NA | 2 | 0 | 1 | Low-quality | Genome-fragment | 2.26 | AAI-based (medium-confidence) | 0 |
| S7_vs_112490\|\|full | 1957 | No | NA | 4 | 0 | 0 | Low-quality | Genome-fragment | 2.83 | AAI-based (medium-confidence) | 0 |
| S5_vs_k141_22380\|\|full | 1099 | No | NA | 3 | 0 | 2 | Not-determined | Genome-fragment | NA | NA | 0 |
| S5_vs_k141_46436\|\|full | 1017 | No | NA | 2 | 0 | 0 | Not-determined | Genome-fragment | NA | NA | 0 |
| S5_vs_k141_100392\|\|full | 3724 | No | NA | 4 | 0 | 3 | Not-determined | Genome-fragment | NA | NA | 0 |
| S5_vs_k141_143572\|\|full | 2481 | No | NA | 3 | 0 | 2 | Not-determined | Genome-fragment | NA | NA | 0 |
| S5_vs_k141_89317\|\|full | 1598 | No | NA | 2 | 0 | 1 | Not-determined | Genome-fragment | NA | NA | 0 |
| S5_vs_k141_27251\|\|full | 3250 | No | NA | 3 | 0 | 2 | Not-determined | Genome-fragment | NA | NA | 0 |
| S5_vs_k141_65125\|\|full | 1305 | No | NA | 3 | 0 | 2 | Not-determined | Genome-fragment | NA | NA | 0 |
| S5_vs_k141_40362\|\|full | 1182 | No | NA | 3 | 0 | 3 | Not-determined | Genome-fragment | NA | NA | 0 |
| S16_vs_108588\|\|full | 1957 | No | NA | 6 | 1 | 0 | Low-quality | Genome-fragment | 4.52 | AAI-based (high-confidence) | 0 |
| S5_vs_k141_81598\|\|full | 3428 | No | NA | 4 | 0 | 2 | Not-determined | Genome-fragment | NA | NA | 0 |
| S10_vs_69189\|\|full | 1957 | No | NA | 4 | 0 | 0 | Low-quality | Genome-fragment | 2.63 | AAI-based (medium-confidence) | 0 |
| S12_vs_k141_50526\|\|full | 1957 | No | NA | 3 | 0 | 1 | Low-quality | Genome-fragment | 3.95 | AAI-based (medium-confidence) | 0 |
| S12_vs_k141_50658\|\|full | 1957 | No | NA | 4 | 1 | 0 | Low-quality | Genome-fragment | 3.67 | AAI-based (high-confidence) | 0 |
| S15_extracted_13416 | 1957 | Yes | 415 | 3 | 1 | 1 | Low-quality | Genome-fragment | 0.34 | AAI-based (high-confidence) | 78.79 |
| S4_extracted_22392 | 1957 | No | NA | 6 | 6 | 0 | Low-quality | Genome-fragment | 6.46 | AAI-based (high-confidence) | 0 |
| S8_vs_k141_182354\|\|full | 1957 | No | NA | 3 | 0 | 1 | Low-quality | Genome-fragment | 1.13 | AAI-based (medium-confidence) | 0 |
| S5_vs_k141_35789\|\|full | 4782 | No | NA | 5 | 0 | 3 | Not-determined | Genome-fragment | NA | NA | 0 |
| S5_vs_k141_100459\|\|full | 2639 | No | NA | 3 | 0 | 0 | Not-determined | Genome-fragment | NA | NA | 0 |
| S5_vs_k141_120566\|\|full | 2906 | No | NA | 6 | 0 | 0 | Not-determined | Genome-fragment | NA | NA | 0 |
| S8_vs_k141_134601\|\|full | 1957 | No | NA | 3 | 0 | 2 | Low-quality | Genome-fragment | 0.9 | AAI-based (high-confidence) | 0 |
| S17_vs_k141_111126\|\|full | 1958 | No | NA | 2 | 0 | 1 | Low-quality | Genome-fragment | 1.35 | AAI-based (medium-confidence) | 0 |
| S5_vs_k141_114002\|\|full | 4389 | No | NA | 5 | 0 | 1 | Not-determined | Genome-fragment | NA | NA | 0 |
| S5_vs_k141_95918\|\|full | 1440 | No | NA | 3 | 0 | 2 | Not-determined | Genome-fragment | NA | NA | 0 |
| S5_vs_k141_35796\|\|full | 2861 | No | NA | 3 | 0 | 2 | Not-determined | Genome-fragment | NA | NA | 0 |
| S11_vs_k141_15194\|\|full | 1958 | No | NA | 2 | 0 | 1 | Low-quality | Genome-fragment | 1.07 | AAI-based (medium-confidence) | 0 |
| S13_vs_29250\|\|full | 1958 | No | NA | 3 | 0 | 1 | Low-quality | Genome-fragment | 3.88 | AAI-based (medium-confidence) | 0 |
| S14_extracted_k141_17592 | 1958 | No | NA | 3 | 2 | 0 | Low-quality | Genome-fragment | 6.45 | AAI-based (high-confidence) | 0 |
| S5_vs_k141_10760\|\|full | 2918 | No | NA | 4 | 0 | 1 | Not-determined | Genome-fragment | NA | NA | 0 |
| S14_vs_k141_63588\|\|full | 1958 | No | NA | 6 | 1 | 0 | Low-quality | Genome-fragment | 2.23 | AAI-based (medium-confidence) | 0 |
| S5_vs_k141_10776\|\|full | 3647 | No | NA | 5 | 0 | 3 | Not-determined | Genome-fragment | NA | NA | 0 |
| S14_vs_k141_21874\|\|full | 1958 | No | NA | 6 | 1 | 0 | Low-quality | Genome-fragment | 4.6 | AAI-based (high-confidence) | 0 |
| S5_vs_k141_22537\|\|full | 1107 | No | NA | 3 | 0 | 0 | Not-determined | Genome-fragment | NA | NA | 0 |
| S5_vs_k141_76043\|\|full | 3910 | No | NA | 6 | 0 | 0 | Not-determined | Genome-fragment | NA | NA | 0 |
| S5_vs_k141_46620\|\|full | 1409 | No | NA | 4 | 0 | 1 | Not-determined | Genome-fragment | NA | NA | 0 |
| S5_vs_k141_95973\|\|full | 2837 | No | NA | 2 | 0 | 1 | Not-determined | Genome-fragment | NA | NA | 0 |
| S5_vs_k141_35856\|\|full | 5357 | No | NA | 4 | 0 | 4 | Not-determined | Genome-fragment | NA | NA | 0 |
| S5_vs_k141_120612\|\|full | 1946 | No | NA | 3 | 0 | 1 | Not-determined | Genome-fragment | NA | NA | 0 |
| S5_vs_k141_35859\|\|full | 1461 | No | NA | 3 | 0 | 0 | Not-determined | Genome-fragment | NA | NA | 0 |
| S5_vs_k141_132089\|\|full | 5490 | No | NA | 7 | 0 | 3 | Not-determined | Genome-fragment | NA | NA | 0 |
| S5_vs_k141_22202\|\|full | 1958 | No | NA | 4 | 0 | 0 | Low-quality | Genome-fragment | 5.48 | AAI-based (medium-confidence) | 0 |
| S7_extracted_67116 | 1958 | No | NA | 3 | 0 | 0 | Low-quality | Genome-fragment | 6.85 | AAI-based (high-confidence) | 0 |
| S5_vs_k141_40431\|\|full | 1851 | No | NA | 3 | 0 | 0 | Not-determined | Genome-fragment | NA | NA | 0 |
| S5_vs_k141_76063\|\|full | 2671 | No | NA | 6 | 0 | 1 | Not-determined | Genome-fragment | NA | NA | 0 |
| S4_extracted_15892 | 1958 | No | NA | 5 | 3 | 0 | Low-quality | Genome-fragment | 6.46 | AAI-based (high-confidence) | 0 |
| S5_vs_k141_114082\|\|full | 2549 | No | NA | 3 | 0 | 2 | Not-determined | Genome-fragment | NA | NA | 0 |
| S1_vs_k141_87087\|\|full | 1959 | No | NA | 4 | 0 | 0 | Low-quality | Genome-fragment | 4.72 | AAI-based (high-confidence) | 0 |
| S5_vs_k141_136973\|\|full | 1679 | No | NA | 3 | 0 | 2 | Not-determined | Genome-fragment | NA | NA | 0 |
| S1_vs_k141_101872\|\|full | 1959 | No | NA | 7 | 1 | 0 | Low-quality | Genome-fragment | 4.39 | AAI-based (medium-confidence) | 0 |
| S7_vs_73106\|\|full | 1959 | No | NA | 4 | 1 | 0 | Low-quality | Genome-fragment | 4.23 | AAI-based (high-confidence) | 0 |
| S7_vs_85175\|\|full | 1959 | No | NA | 2 | 1 | 0 | Low-quality | Genome-fragment | 1.72 | HMM-based (lower-bound) | 0 |
| S10_vs_102460\|\|full | 1959 | No | NA | 3 | 0 | 0 | Low-quality | Genome-fragment | 1.26 | AAI-based (medium-confidence) | 0 |
| S15_vs_63143\|\|full | 1959 | No | NA | 3 | 0 | 0 | Low-quality | Genome-fragment | 4.39 | AAI-based (high-confidence) | 0 |
| S3_extracted_k141_100293 | 1960 | No | NA | 5 | 0 | 2 | Low-quality | Genome-fragment | 4.33 | AAI-based (medium-confidence) | 0 |
| S5_vs_k141_22652\|\|full | 1114 | No | NA | 2 | 0 | 0 | Not-determined | Genome-fragment | NA | NA | 0 |
| S5_vs_k141_46669\|\|full | 2005 | No | NA | 4 | 0 | 0 | Not-determined | Genome-fragment | NA | NA | 0 |
| S3_extracted_k141_111402 | 1960 | No | NA | 3 | 0 | 0 | Low-quality | Genome-fragment | 3.88 | AAI-based (high-confidence) | 0 |
| S3_vs_k141_29102\|\|full | 1960 | No | NA | 3 | 0 | 1 | Low-quality | Genome-fragment | 2.95 | AAI-based (high-confidence) | 0 |
| S5_extracted_k141_88786 | 1960 | No | NA | 5 | 1 | 0 | Low-quality | Genome-fragment | 3.45 | AAI-based (medium-confidence) | 0 |
| S5_vs_k141_96021\|\|full | 1583 | No | NA | 3 | 0 | 0 | Not-determined | Genome-fragment | NA | NA | 0 |
| S5_vs_k141_81687\|\|full | 3182 | No | NA | 3 | 0 | 2 | Not-determined | Genome-fragment | NA | NA | 0 |
| S5_vs_k141_6021\|\|full | 6938 | No | NA | 8 | 0 | 2 | Not-determined | Genome-fragment | NA | NA | 0 |
| S5_vs_k141_22685\|\|full | 2339 | No | NA | 4 | 0 | 1 | Not-determined | Genome-fragment | NA | NA | 0 |
| S17_vs_k141_82106\|\|full | 1961 | No | NA | 3 | 0 | 1 | Low-quality | Genome-fragment | 1.72 | AAI-based (high-confidence) | 0 |
| S5_vs_k141_54170\|\|full | 1153 | No | NA | 3 | 0 | 1 | Not-determined | Genome-fragment | NA | NA | 0 |
| S1_extracted_k141_28694 | 1961 | No | NA | 2 | 0 | 1 | Low-quality | Genome-fragment | 1.48 | AAI-based (high-confidence) | 0 |
| S5_vs_k141_114161\|\|full | 1050 | No | NA | 2 | 0 | 1 | Not-determined | Genome-fragment | NA | NA | 0 |
| S5_vs_k141_114168\|\|full | 2111 | No | NA | 2 | 0 | 1 | Not-determined | Genome-fragment | NA | NA | 0 |
| S7_vs_70112\|\|full | 1961 | No | NA | 6 | 0 | 0 | Low-quality | Genome-fragment | 2.19 | AAI-based (high-confidence) | 0 |
| S5_vs_k141_109078\|\|full | 5367 | No | NA | 8 | 0 | 0 | Not-determined | Genome-fragment | NA | NA | 0 |
| S16_extracted_151398 | 1961 | No | NA | 4 | 2 | 0 | Low-quality | Genome-fragment | 5.33 | AAI-based (high-confidence) | 0 |
| S6_vs_k141_12066\|\|full | 1961 | No | NA | 4 | 0 | 0 | Low-quality | Genome-fragment | 3.24 | AAI-based (high-confidence) | 0 |
| S17_extracted_k141_132421 | 1962 | No | NA | 2 | 0 | 0 | Low-quality | Genome-fragment | 0.9 | AAI-based (medium-confidence) | 0 |
| S17_vs_k141_99780\|\|full | 1962 | No | NA | 3 | 0 | 0 | Low-quality | Genome-fragment | 4.29 | AAI-based (medium-confidence) | 0 |
| S11_vs_k141_70083\|\|full | 1962 | No | NA | 4 | 3 | 0 | Low-quality | Genome-fragment | 4.49 | AAI-based (high-confidence) | 0 |
| S14_extracted_k141_31439 | 1962 | No | NA | 1 | 0 | 1 | Low-quality | Genome-fragment | 0.51 | AAI-based (high-confidence) | 0 |
| S5_vs_k141_114241\|\|full | 2213 | No | NA | 3 | 0 | 0 | Not-determined | Genome-fragment | NA | NA | 0 |
| S5_vs_k141_77460\|\|full | 1962 | No | NA | 5 | 1 | 0 | Low-quality | Genome-fragment | 3.66 | AAI-based (medium-confidence) | 0 |
| S5_vs_k141_89540\|\|full | 22259 | No | NA | 30 | 20 | 0 | Medium-quality | Genome-fragment | 59.98 | AAI-based (medium-confidence) | 0 |
| S12_vs_k141_55572\|\|full | 1962 | No | NA | 3 | 3 | 0 | Low-quality | Genome-fragment | 3.2 | HMM-based (lower-bound) | 0 |
| S5_vs_k141_40617\|\|full | 1709 | No | NA | 2 | 0 | 0 | Not-determined | Genome-fragment | NA | NA | 0 |
| S5_vs_k141_89547\|\|full | 1826 | No | NA | 4 | 0 | 3 | Not-determined | Genome-fragment | NA | NA | 0 |
| S8_vs_k141_178054\|\|full | 1962 | No | NA | 3 | 0 | 0 | Low-quality | Genome-fragment | 1.3 | AAI-based (medium-confidence) | 0 |
| S11_vs_k141_14475\|\|lt2gene | 1963 | No | NA | 2 | 2 | 0 | Low-quality | Genome-fragment | 5.08 | AAI-based (medium-confidence) | 0 |
| S5_vs_k141_89551\|\|full | 1289 | No | NA | 3 | 0 | 0 | Not-determined | Genome-fragment | NA | NA | 0 |
| S5_vs_k141_40635\|\|full | 1427 | No | NA | 2 | 0 | 1 | Not-determined | Genome-fragment | NA | NA | 0 |
| S5_vs_k141_89554\|\|full | 1721 | No | NA | 2 | 0 | 2 | Not-determined | Genome-fragment | NA | NA | 0 |
| S5_vs_k141_65378\|\|full | 1396 | No | NA | 3 | 0 | 0 | Not-determined | Genome-fragment | NA | NA | 0 |
| S13_vs_53462\|\|full | 1963 | No | NA | 3 | 2 | 0 | Low-quality | Genome-fragment | 4.5 | AAI-based (medium-confidence) | 0 |
| S3_extracted_k141_17709 | 1963 | No | NA | 2 | 0 | 0 | Low-quality | Genome-fragment | 4.24 | AAI-based (medium-confidence) | 0 |
| S3_vs_k141_22108\|\|full | 1963 | No | NA | 2 | 0 | 1 | Low-quality | Genome-fragment | 1.06 | AAI-based (high-confidence) | 0 |
| S7_vs_17170\|\|full | 1963 | No | NA | 5 | 2 | 0 | Low-quality | Genome-fragment | 4.51 | AAI-based (medium-confidence) | 0 |
| S5_vs_k141_40663\|\|full | 2905 | No | NA | 3 | 0 | 1 | Not-determined | Genome-fragment | NA | NA | 0 |
| S16_extracted_98641 | 1963 | No | NA | 3 | 2 | 0 | Low-quality | Genome-fragment | 10.79 | AAI-based (high-confidence) | 0 |
| S5_vs_k141_76165\|\|full | 1327 | No | NA | 2 | 0 | 0 | Not-determined | Genome-fragment | NA | NA | 0 |
| S5_vs_k141_132227\|\|full | 2266 | No | NA | 3 | 0 | 3 | Not-determined | Genome-fragment | NA | NA | 0 |
| S6_vs_k141_28253\|\|full | 1963 | No | NA | 6 | 1 | 0 | Low-quality | Genome-fragment | 3.79 | AAI-based (high-confidence) | 0 |
| S5_vs_k141_137059\|\|full | 1361 | No | NA | 4 | 0 | 0 | Not-determined | Genome-fragment | NA | NA | 0 |
| S8_vs_k141_22945\|\|full | 1963 | No | NA | 5 | 1 | 0 | Low-quality | Genome-fragment | 4.84 | AAI-based (medium-confidence) | 0 |
| S5_vs_k141_109148\|\|full | 2738 | No | NA | 4 | 0 | 2 | Not-determined | Genome-fragment | NA | NA | 0 |
| S17_vs_k141_28929\|\|full | 1964 | No | NA | 4 | 1 | 0 | Low-quality | Genome-fragment | 5.26 | AAI-based (high-confidence) | 0 |
| S5_vs_k141_143658\|\|full | 3671 | No | NA | 4 | 0 | 2 | Not-determined | Genome-fragment | NA | NA | 0 |
| S9_vs_k141_29099\|\|lt2gene | 1964 | No | NA | 1 | 1 | 0 | Low-quality | Genome-fragment | 3.46 | AAI-based (medium-confidence) | 0 |
| S5_vs_k141_76194\|\|full | 3378 | No | NA | 6 | 0 | 0 | Not-determined | Genome-fragment | NA | NA | 0 |
| S5_vs_k141_114283\|\|full | 1502 | No | NA | 7 | 0 | 0 | Not-determined | Genome-fragment | NA | NA | 0 |
| S9_vs_k141_49208\|\|lt2gene | 1964 | No | NA | 2 | 1 | 0 | Low-quality | Genome-fragment | 4.54 | AAI-based (medium-confidence) | 0 |
| S5_vs_k141_46904\|\|full | 1069 | No | NA | 3 | 0 | 1 | Not-determined | Genome-fragment | NA | NA | 0 |
| S11_vs_k141_90954\|\|full | 1964 | No | NA | 5 | 3 | 0 | Low-quality | Genome-fragment | 0.89 | HMM-based (lower-bound) | 0 |
| S5_vs_k141_22801\|\|full | 1221 | No | NA | 3 | 0 | 0 | Not-determined | Genome-fragment | NA | NA | 0 |
| S13_extracted_92382 | 1964 | No | NA | 2 | 0 | 1 | Low-quality | Genome-fragment | 0.9 | AAI-based (medium-confidence) | 0 |
| S5_vs_k141_81841\|\|full | 3333 | No | NA | 4 | 0 | 3 | Not-determined | Genome-fragment | NA | NA | 0 |
| S5_vs_k141_65464\|\|full | 1580 | No | NA | 3 | 0 | 1 | Not-determined | Genome-fragment | NA | NA | 0 |
| S5_vs_k141_76285\|\|full | 3924 | No | NA | 5 | 0 | 2 | Not-determined | Genome-fragment | NA | NA | 0 |
| S13_extracted_134548 | 1964 | No | NA | 1 | 0 | 1 | Low-quality | Genome-fragment | 2.18 | AAI-based (medium-confidence) | 0 |
| S5_vs_k141_89690\|\|full | 1922 | No | NA | 2 | 0 | 0 | Not-determined | Genome-fragment | NA | NA | 0 |
| S13_vs_98656\|\|full | 1964 | No | NA | 3 | 0 | 1 | Low-quality | Genome-fragment | 4.11 | AAI-based (medium-confidence) | 0 |
| S3_vs_k141_15993\|\|full | 1964 | No | NA | 3 | 0 | 1 | Low-quality | Genome-fragment | 5.33 | AAI-based (medium-confidence) | 0 |
| S5_vs_k141_125630\|\|full | 1964 | No | NA | 3 | 0 | 0 | Low-quality | Genome-fragment | 2.39 | AAI-based (medium-confidence) | 0 |
| S7_vs_89853\|\|full | 1964 | No | NA | 3 | 0 | 0 | Low-quality | Genome-fragment | 6.05 | AAI-based (high-confidence) | 0 |
| S5_vs_k141_17520\|\|full | 27566 | No | NA | 40 | 17 | 0 | Medium-quality | Genome-fragment | 73.4 | AAI-based (high-confidence) | 0 |
| S5_vs_k141_114465\|\|full | 2088 | No | NA | 3 | 0 | 1 | Not-determined | Genome-fragment | NA | NA | 0 |
| S5_vs_k141_22914\|\|full | 6452 | No | NA | 11 | 0 | 2 | Not-determined | Genome-fragment | NA | NA | 0 |
| S5_vs_k141_36112\|\|full | 1489 | No | NA | 3 | 0 | 2 | Not-determined | Genome-fragment | NA | NA | 0 |
| S16_vs_169560\|\|full | 1964 | No | NA | 2 | 0 | 1 | Low-quality | Genome-fragment | 0.77 | AAI-based (medium-confidence) | 0 |
| S5_vs_k141_132291\|\|full | 1354 | No | NA | 2 | 0 | 1 | Not-determined | Genome-fragment | NA | NA | 0 |
| S10_vs_78603\|\|full | 1964 | No | NA | 3 | 3 | 0 | Low-quality | Genome-fragment | 3.22 | AAI-based (medium-confidence) | 0 |
| S2_vs_k141_15781\|\|full | 1964 | No | NA | 3 | 1 | 0 | Low-quality | Genome-fragment | 3.61 | AAI-based (high-confidence) | 0 |
| S5_vs_k141_17582\|\|full | 2649 | No | NA | 4 | 0 | 4 | Not-determined | Genome-fragment | NA | NA | 0 |
| S3_extracted_k141_55263 | 1965 | No | NA | 4 | 1 | 0 | Low-quality | Genome-fragment | 3.18 | AAI-based (high-confidence) | 0 |
| S5_vs_k141_19875\|\|full | 1965 | No | NA | 2 | 0 | 2 | Low-quality | Genome-fragment | 2.38 | AAI-based (high-confidence) | 0 |
| S5_vs_k141_71275\|\|full | 2486 | No | NA | 4 | 0 | 1 | Not-determined | Genome-fragment | NA | NA | 0 |
| S5_vs_k141_54481\|\|full | 1468 | No | NA | 3 | 0 | 1 | Not-determined | Genome-fragment | NA | NA | 0 |
| S10_extracted_42544 | 1965 | No | NA | 1 | 0 | 1 | Low-quality | Genome-fragment | 0.34 | AAI-based (medium-confidence) | 0 |
| S5_vs_k141_114502\|\|full | 3140 | No | NA | 4 | 0 | 1 | Not-determined | Genome-fragment | NA | NA | 0 |
| S8_extracted_k141_103604 | 1965 | No | NA | 3 | 1 | 0 | Low-quality | Genome-fragment | 9.95 | AAI-based (high-confidence) | 0 |
| S5_vs_k141_36150\|\|full | 1365 | No | NA | 3 | 0 | 0 | Not-determined | Genome-fragment | NA | NA | 0 |
| S3_extracted_k141_119402 | 1966 | No | NA | 4 | 1 | 0 | Low-quality | Genome-fragment | 4.04 | AAI-based (high-confidence) | 0 |
| S5_vs_k141_60055\|\|lt2gene | 1966 | No | NA | 2 | 2 | 0 | Low-quality | Genome-fragment | 5.08 | AAI-based (high-confidence) | 0 |
| S5_vs_k141_22951\|\|full | 1079 | No | NA | 2 | 0 | 0 | Not-determined | Genome-fragment | NA | NA | 0 |
| S12_vs_k141_156464\|\|full | 1966 | No | NA | 3 | 2 | 0 | Low-quality | Genome-fragment | 5.56 | AAI-based (high-confidence) | 0 |
| S5_vs_k141_137154\|\|full | 18227 | No | NA | 23 | 14 | 1 | Medium-quality | Genome-fragment | 52.9 | AAI-based (high-confidence) | 0 |
| S5_vs_k141_71311\|\|full | 1224 | No | NA | 3 | 0 | 2 | Not-determined | Genome-fragment | NA | NA | 0 |
| S5_vs_k141_137166\|\|full | 1307 | No | NA | 3 | 0 | 0 | Not-determined | Genome-fragment | NA | NA | 0 |
| S8_vs_k141_139817\|\|full | 1966 | No | NA | 3 | 0 | 0 | Low-quality | Genome-fragment | 1.04 | AAI-based (medium-confidence) | 0 |
| S14_vs_k141_51852\|\|full | 1967 | No | NA | 3 | 1 | 0 | Low-quality | Genome-fragment | 4.13 | AAI-based (high-confidence) | 0 |
| S3_vs_k141_19017\|\|full | 1967 | No | NA | 2 | 1 | 0 | Low-quality | Genome-fragment | 3.41 | HMM-based (lower-bound) | 0 |
| S5_vs_k141_121052\|\|full | 1312 | No | NA | 3 | 0 | 0 | Not-determined | Genome-fragment | NA | NA | 0 |
| S5_vs_k141_17679\|\|full | 1292 | No | NA | 2 | 0 | 1 | Not-determined | Genome-fragment | NA | NA | 0 |
| S5_vs_k141_54554\|\|full | 1595 | No | NA | 3 | 0 | 0 | Not-determined | Genome-fragment | NA | NA | 0 |
| S5_vs_k141_23021\|\|full | 8282 | No | NA | 9 | 0 | 2 | Not-determined | Genome-fragment | NA | NA | 0 |
| S10_vs_81618\|\|full | 1967 | No | NA | 3 | 1 | 0 | Low-quality | Genome-fragment | 4.15 | AAI-based (high-confidence) | 0 |
| S8_extracted_k141_150759 | 1967 | No | NA | 1 | 0 | 1 | Low-quality | Genome-fragment | 0.52 | AAI-based (high-confidence) | 0 |
| S5_vs_k141_127241\|\|full | 1322 | No | NA | 2 | 0 | 0 | Not-determined | Genome-fragment | NA | NA | 0 |
| S5_vs_k141_17722\|\|full | 1419 | No | NA | 3 | 0 | 0 | Not-determined | Genome-fragment | NA | NA | 0 |
| S5_vs_k141_121074\|\|full | 18227 | No | NA | 12 | 0 | 10 | Not-determined | Genome-fragment | NA | NA | 0 |
| S5_vs_k141_27519\|\|full | 4171 | No | NA | 5 | 0 | 3 | Not-determined | Genome-fragment | NA | NA | 0 |
| S16_vs_40508\|\|full | 1968 | No | NA | 4 | 2 | 0 | Low-quality | Genome-fragment | 4.43 | AAI-based (medium-confidence) | 0 |
| S5_vs_k141_114558\|\|full | 2041 | No | NA | 3 | 0 | 1 | Not-determined | Genome-fragment | NA | NA | 0 |
| S10_extracted_65546 | 1968 | No | NA | 4 | 0 | 0 | Low-quality | Genome-fragment | 3.75 | AAI-based (medium-confidence) | 0 |
| S10_vs_119874\|\|full | 1968 | No | NA | 4 | 0 | 3 | Low-quality | Genome-fragment | 2.65 | AAI-based (medium-confidence) | 0 |
| S11_vs_k141_98673\|\|full | 1969 | No | NA | 3 | 0 | 2 | Low-quality | Genome-fragment | 2.56 | AAI-based (medium-confidence) | 0 |
| S5_vs_k141_47267\|\|full | 2144 | No | NA | 2 | 0 | 1 | Not-determined | Genome-fragment | NA | NA | 0 |
| S14_vs_k141_96545\|\|full | 1969 | No | NA | 2 | 1 | 0 | Low-quality | Genome-fragment | 6.43 | AAI-based (high-confidence) | 0 |
| S5_vs_k141_17774\|\|full | 2223 | No | NA | 3 | 0 | 0 | Not-determined | Genome-fragment | NA | NA | 0 |
| S5_vs_k141_89955\|\|full | 1153 | No | NA | 2 | 0 | 1 | Not-determined | Genome-fragment | NA | NA | 0 |
| S5_vs_k141_71313\|\|full | 2059 | No | NA | 3 | 0 | 2 | Not-determined | Genome-fragment | NA | NA | 0 |
| S16_vs_62255\|\|full | 1969 | No | NA | 2 | 1 | 0 | Low-quality | Genome-fragment | 3.77 | AAI-based (medium-confidence) | 0 |
| S5_vs_k141_17796\|\|full | 1933 | No | NA | 3 | 0 | 1 | Not-determined | Genome-fragment | NA | NA | 0 |
| S5_vs_k141_82010\|\|full | 1560 | No | NA | 3 | 0 | 3 | Not-determined | Genome-fragment | NA | NA | 0 |
| S14_extracted_k141_73543 | 1970 | No | NA | 5 | 5 | 0 | Low-quality | Genome-fragment | 6.5 | AAI-based (high-confidence) | 0 |
| S1_extracted_k141_46748 | 1970 | No | NA | 2 | 0 | 2 | Low-quality | Genome-fragment | 1.27 | AAI-based (high-confidence) | 0 |
| S16_vs_156187\|\|full | 1970 | No | NA | 3 | 0 | 2 | Low-quality | Genome-fragment | 2.68 | AAI-based (medium-confidence) | 0 |
| S15_vs_62764\|\|full | 1970 | No | NA | 4 | 0 | 0 | Low-quality | Genome-fragment | 36.45 | AAI-based (medium-confidence) | 0 |
| S5_vs_k141_114626\|\|full | 1330 | No | NA | 3 | 0 | 0 | Not-determined | Genome-fragment | NA | NA | 0 |
| S5_vs_k141_17834\|\|full | 1081 | No | NA | 2 | 0 | 0 | Not-determined | Genome-fragment | NA | NA | 0 |
| S5_vs_k141_132447\|\|full | 2398 | No | NA | 3 | 0 | 1 | Not-determined | Genome-fragment | NA | NA | 0 |
| S6_vs_k141_43818\|\|full | 1970 | No | NA | 6 | 0 | 0 | Low-quality | Genome-fragment | 5.61 | AAI-based (high-confidence) | 0 |
| S5_vs_k141_96327\|\|full | 2240 | No | NA | 6 | 0 | 0 | Not-determined | Genome-fragment | NA | NA | 0 |
| S8_vs_k141_119112\|\|full | 1970 | No | NA | 5 | 0 | 2 | Low-quality | Genome-fragment | 0.61 | AAI-based (high-confidence) | 0 |
| S5_vs_k141_17844\|\|full | 3930 | No | NA | 7 | 0 | 1 | Not-determined | Genome-fragment | NA | NA | 0 |
| S11_vs_k141_61233\|\|full | 1971 | No | NA | 4 | 1 | 0 | Low-quality | Genome-fragment | 4.31 | AAI-based (high-confidence) | 0 |
| S13_extracted_6612 | 1971 | No | NA | 2 | 0 | 1 | Low-quality | Genome-fragment | 0.98 | AAI-based (medium-confidence) | 0 |
| S13_vs_33706\|\|full | 1971 | No | NA | 3 | 0 | 1 | Low-quality | Genome-fragment | 4.97 | AAI-based (medium-confidence) | 0 |
| S14_vs_k141_71332\|\|full | 1971 | No | NA | 3 | 0 | 0 | Low-quality | Genome-fragment | 3.14 | AAI-based (high-confidence) | 0 |
| S14_vs_k141_21478\|\|full | 1971 | No | NA | 5 | 1 | 0 | Low-quality | Genome-fragment | 4.48 | AAI-based (medium-confidence) | 0 |
| S1_vs_k141_136423\|\|lt2gene | 1971 | No | NA | 1 | 1 | 0 | Low-quality | Genome-fragment | 2.45 | AAI-based (high-confidence) | 0 |
| S5_vs_k141_100777\|\|full | 4534 | No | NA | 6 | 0 | 1 | Not-determined | Genome-fragment | NA | NA | 0 |
| S5_vs_k141_139902\|\|full | 1971 | No | NA | 3 | 1 | 0 | Low-quality | Genome-fragment | 3.08 | HMM-based (lower-bound) | 0 |

| | | | | | | | | | | | |
|---|---|---|---|---|---|---|---|---|---|---|---|
| S5_vs_k141_27582\|\|full | 3120 | No | NA | 5 | 0 | 0 | Not-determined | Genome-fragment | NA | NA | 0 |
| S7_vs_112228\|\|full | 1971 | No | NA | 3 | 1 | 0 | Low-quality | Genome-fragment | 5.87 | AAI-based (high-confidence) | 0 |
| S16_extracted_58368 | 1971 | No | NA | 5 | 1 | 0 | Low-quality | Genome-fragment | 3.11 | AAI-based (high-confidence) | 0 |
| S5_vs_k141_127419\|\|full | 2620 | No | NA | 4 | 0 | 4 | Not-determined | Genome-fragment | NA | NA | 0 |
| S10_vs_64951\|\|full | 1971 | No | NA | 3 | 0 | 1 | Low-quality | Genome-fragment | 3.32 | AAI-based (medium-confidence) | 0 |
| S5_vs_k141_71409\|\|full | 1405 | No | NA | 2 | 0 | 0 | Not-determined | Genome-fragment | NA | NA | 0 |
| S10_vs_76491\|\|full | 1971 | No | NA | 3 | 2 | 0 | Low-quality | Genome-fragment | 3.36 | AAI-based (high-confidence) | 0 |
| S5_vs_k141_127455\|\|full | 2247 | No | NA | 2 | 0 | 1 | Not-determined | Genome-fragment | NA | NA | 0 |
| S5_vs_k141_127460\|\|full | 1615 | No | NA | 4 | 0 | 0 | Not-determined | Genome-fragment | NA | NA | 0 |
| S1_extracted_k141_26056 | 1972 | No | NA | 2 | 0 | 1 | Low-quality | Genome-fragment | 1.2 | AAI-based (medium-confidence) | 0 |
| S5_vs_k141_23197\|\|full | 2108 | No | NA | 4 | 0 | 1 | Not-determined | Genome-fragment | NA | NA | 0 |
| S3_extracted_k141_88259 | 1972 | No | NA | 1 | 1 | 0 | Low-quality | Genome-fragment | 3.65 | AAI-based (high-confidence) | 0 |
| S15_extracted_60845 | 1972 | No | NA | 3 | 0 | 0 | Low-quality | Genome-fragment | 3.69 | AAI-based (medium-confidence) | 0 |
| S5_vs_k141_47480\|\|full | 1140 | No | NA | 3 | 0 | 1 | Not-determined | Genome-fragment | NA | NA | 0 |
| S8_extracted_k141_155184 | 1972 | No | NA | 1 | 0 | 1 | Low-quality | Genome-fragment | 3.22 | AAI-based (medium-confidence) | 0 |
| S17_extracted_k141_150765 | 1973 | No | NA | 4 | 0 | 1 | Low-quality | Genome-fragment | 3.69 | AAI-based (high-confidence) | 0 |
| S5_vs_k141_64400\|\|full | 1973 | No | NA | 3 | 1 | 0 | Low-quality | Genome-fragment | 2.78 | HMM-based (lower-bound) | 0 |
| S5_vs_k141_40848\|\|full | 233641 | Yes | 62183 | 234 | 25 | 111 | High-quality | High-quality | 100 | AAI-based (high-confidence) | 73.39 |
| S16_extracted_47419 | 1973 | No | NA | 4 | 0 | 0 | Low-quality | Genome-fragment | 2.11 | AAI-based (high-confidence) | 0 |
| S16_vs_141786\|\|full | 1973 | No | NA | 4 | 0 | 4 | Low-quality | Genome-fragment | 1.77 | AAI-based (medium-confidence) | 0 |
| S4_extracted_19596 | 1973 | No | NA | 3 | 2 | 0 | Low-quality | Genome-fragment | 5.45 | AAI-based (high-confidence) | 0 |
| S6_vs_k141_52172\|\|full | 1973 | No | NA | 5 | 0 | 0 | Low-quality | Genome-fragment | 5.14 | AAI-based (medium-confidence) | 0 |
| S5_vs_k141_17979\|\|full | 2992 | No | NA | 3 | 0 | 0 | Not-determined | Genome-fragment | NA | NA | 0 |
| S17_vs_k141_50914\|\|full | 1974 | No | NA | 3 | 1 | 0 | Low-quality | Genome-fragment | 1.24 | HMM-based (lower-bound) | 0 |
| S9_vs_k141_77467\|\|full | 1974 | No | NA | 4 | 1 | 0 | Low-quality | Genome-fragment | 4.73 | AAI-based (high-confidence) | 0 |
| S5_vs_k141_137339\|\|full | 1421 | No | NA | 2 | 0 | 0 | Not-determined | Genome-fragment | NA | NA | 0 |
| S3_vs_k141_88796\|\|full | 1974 | No | NA | 4 | 0 | 0 | Low-quality | Genome-fragment | 1.29 | AAI-based (high-confidence) | 0 |
| S5_vs_k141_132690\|\|full | 2009 | No | NA | 2 | 0 | 1 | Not-determined | Genome-fragment | NA | NA | 0 |
| S5_vs_k141_47647\|\|full | 1358 | No | NA | 7 | 0 | 0 | Not-determined | Genome-fragment | NA | NA | 0 |
| S5_vs_k141_7352\|\|full | 1974 | No | NA | 4 | 0 | 0 | Low-quality | Genome-fragment | 2.8 | AAI-based (medium-confidence) | 0 |
| S12_vs_k141_173547\|\|full | 1974 | No | NA | 3 | 0 | 1 | Low-quality | Genome-fragment | 2.22 | AAI-based (medium-confidence) | 0 |
| S12_vs_k141_102764\|\|full | 1974 | No | NA | 1 | 0 | 1 | Low-quality | Genome-fragment | 2.53 | AAI-based (medium-confidence) | 0 |
| S17_extracted_k141_105127 | 1975 | Yes | 1145 | 2 | 1 | 1 | Low-quality | Genome-fragment | 2.19 | AAI-based (medium-confidence) | 42.03 |
| S13_vs_27342\|\|full | 1975 | No | NA | 2 | 0 | 1 | Low-quality | Genome-fragment | 2.39 | AAI-based (medium-confidence) | 0 |
| S7_vs_59881\|\|full | 1975 | No | NA | 3 | 0 | 0 | Low-quality | Genome-fragment | 4.39 | AAI-based (medium-confidence) | 0 |
| S5_vs_k141_47708\|\|full | 2494 | No | NA | 3 | 0 | 3 | Not-determined | Genome-fragment | NA | NA | 0 |
| S8_extracted_k141_29508 | 1975 | No | NA | 3 | 0 | 1 | Low-quality | Genome-fragment | 6.31 | AAI-based (high-confidence) | 0 |
| S11_vs_k141_636\|\|full | 1976 | No | NA | 3 | 1 | 0 | Low-quality | Genome-fragment | 1.34 | HMM-based (lower-bound) | 0 |
| S5_vs_k141_141367\|\|full | 1976 | No | NA | 4 | 0 | 0 | Low-quality | Genome-fragment | 3.71 | AAI-based (medium-confidence) | 0 |
| S5_vs_k141_89697\|\|full | 1976 | No | NA | 5 | 1 | 0 | Low-quality | Genome-fragment | 2.96 | HMM-based (lower-bound) | 0 |
| S5_vs_k141_18106\|\|full | 1623 | No | NA | 3 | 0 | 1 | Not-determined | Genome-fragment | NA | NA | 0 |
| S7_extracted_79967 | 1976 | No | NA | 2 | 2 | 0 | Low-quality | Genome-fragment | 3.82 | HMM-based (lower-bound) | 0 |
| S7_vs_98721\|\|full | 1976 | No | NA | 2 | 0 | 2 | Low-quality | Genome-fragment | 3.08 | AAI-based (medium-confidence) | 0 |
| S5_vs_k141_144103\|\|full | 2500 | No | NA | 3 | 0 | 2 | Not-determined | Genome-fragment | NA | NA | 0 |
| S10_extracted_23779 | 1976 | No | NA | 1 | 0 | 1 | Low-quality | Genome-fragment | 0.34 | AAI-based (medium-confidence) | 0 |
| S10_extracted_116949 | 1976 | No | NA | 3 | 1 | 0 | Low-quality | Genome-fragment | 3.92 | AAI-based (high-confidence) | 0 |
| S10_vs_101747\|\|full | 1976 | No | NA | 3 | 0 | 2 | Low-quality | Genome-fragment | 0.78 | AAI-based (medium-confidence) | 0 |
| S12_extracted_k141_175062 | 1976 | No | NA | 3 | 0 | 0 | Low-quality | Genome-fragment | 3.15 | AAI-based (high-confidence) | 0 |
| S12_vs_k141_64650\|\|full | 1976 | No | NA | 3 | 1 | 0 | Low-quality | Genome-fragment | 5.48 | AAI-based (medium-confidence) | 0 |
| S6_vs_k141_10726\|\|full | 1976 | No | NA | 4 | 0 | 0 | Low-quality | Genome-fragment | 3.25 | AAI-based (high-confidence) | 0 |
| S8_vs_k141_152625\|\|full | 1976 | No | NA | 4 | 0 | 0 | Low-quality | Genome-fragment | 3.83 | AAI-based (high-confidence) | 0 |
| S17_vs_k141_29551\|\|full | 1977 | No | NA | 3 | 1 | 0 | Low-quality | Genome-fragment | 5.57 | AAI-based (high-confidence) | 0 |
| S11_vs_k141_5242\|\|full | 1977 | No | NA | 5 | 2 | 0 | Low-quality | Genome-fragment | 5.52 | AAI-based (medium-confidence) | 0 |
| S14_vs_k141_110049\|\|full | 1977 | No | NA | 5 | 1 | 0 | Low-quality | Genome-fragment | 3.49 | AAI-based (high-confidence) | 0 |
| S14_vs_k141_63623\|\|lt2gene | 1977 | No | NA | 2 | 2 | 0 | Low-quality | Genome-fragment | 5.3 | AAI-based (high-confidence) | 0 |
| S1_vs_k141_112942\|\|full | 1977 | No | NA | 5 | 0 | 0 | Low-quality | Genome-fragment | 2.87 | AAI-based (high-confidence) | 0 |
| S5_vs_k141_76627\|\|full | 1695 | No | NA | 2 | 0 | 1 | Not-determined | Genome-fragment | NA | NA | 0 |
| S5_vs_k141_137416\|\|full | 1479 | No | NA | 2 | 0 | 0 | Not-determined | Genome-fragment | NA | NA | 0 |
| S3_extracted_k141_87340 | 1977 | No | NA | 4 | 2 | 0 | Low-quality | Genome-fragment | 4.1 | AAI-based (high-confidence) | 0 |
| S5_vs_k141_137418\|\|full | 2488 | No | NA | 4 | 0 | 3 | Not-determined | Genome-fragment | NA | NA | 0 |
| S16_vs_109645\|\|lt2gene | 1977 | No | NA | 2 | 1 | 0 | Low-quality | Genome-fragment | 3.21 | HMM-based (lower-bound) | 0 |
| S5_vs_k141_40956\|\|full | 1766 | No | NA | 5 | 0 | 0 | Not-determined | Genome-fragment | NA | NA | 0 |
| S5_vs_k141_23408\|\|full | 1151 | No | NA | 3 | 0 | 1 | Not-determined | Genome-fragment | NA | NA | 0 |
| S5_vs_k141_23414\|\|full | 8547 | No | NA | 5 | 0 | 1 | Not-determined | Genome-fragment | NA | NA | 0 |
| S6_vs_k141_11203\|\|full | 1977 | No | NA | 3 | 0 | 0 | Low-quality | Genome-fragment | 3.16 | AAI-based (medium-confidence) | 0 |
| S13_vs_6293\|\|full | 1978 | No | NA | 3 | 0 | 0 | Low-quality | Genome-fragment | 5.23 | AAI-based (high-confidence) | 0 |
| S14_extracted_k141_66649 | 1978 | No | NA | 2 | 2 | 0 | Low-quality | Genome-fragment | 3.06 | HMM-based (lower-bound) | 0 |
| S5_vs_k141_47883\|\|full | 1127 | No | NA | 4 | 0 | 0 | Not-determined | Genome-fragment | NA | NA | 0 |
| S14_vs_k141_8466\|\|full | 1978 | No | NA | 3 | 3 | 0 | Low-quality | Genome-fragment | 6.24 | AAI-based (high-confidence) | 0 |
| S1_vs_k141_152282\|\|full | 1978 | No | NA | 3 | 2 | 0 | Low-quality | Genome-fragment | 4.68 | AAI-based (high-confidence) | 0 |
| S3_vs_k141_62216\|\|full | 1978 | No | NA | 3 | 0 | 0 | Low-quality | Genome-fragment | 1.12 | AAI-based (high-confidence) | 0 |
| S7_vs_82969\|\|lt2gene | 1978 | No | NA | 1 | 1 | 0 | Low-quality | Genome-fragment | 4.29 | AAI-based (medium-confidence) | 0 |
| S16_extracted_38017 | 1978 | No | NA | 6 | 3 | 0 | Low-quality | Genome-fragment | 3.6 | HMM-based (lower-bound) | 0 |
| S5_vs_k141_96733\|\|full | 1737 | No | NA | 5 | 0 | 4 | Not-determined | Genome-fragment | NA | NA | 0 |
| S12_vs_k141_124474\|\|full | 1978 | No | NA | 2 | 1 | 0 | Low-quality | Genome-fragment | 3.31 | AAI-based (medium-confidence) | 0 |
| S5_vs_k141_71858\|\|full | 1184 | No | NA | 3 | 0 | 0 | Not-determined | Genome-fragment | NA | NA | 0 |
| S5_vs_k141_11225\|\|full | 1646 | No | NA | 4 | 0 | 3 | Not-determined | Genome-fragment | NA | NA | 0 |
| S15_extracted_65883 | 1978 | No | NA | 3 | 0 | 0 | Low-quality | Genome-fragment | 4.02 | AAI-based (high-confidence) | 0 |
| S5_vs_k141_114951\|\|full | 1876 | No | NA | 3 | 0 | 3 | Not-determined | Genome-fragment | NA | NA | 0 |
| S15_vs_70116\|\|full | 1978 | No | NA | 4 | 0 | 0 | Low-quality | Genome-fragment | 3.2 | AAI-based (medium-confidence) | 0 |
| S8_extracted_k141_61697 | 1978 | No | NA | 2 | 0 | 2 | Low-quality | Genome-fragment | 6.32 | AAI-based (high-confidence) | 0 |
| S5_vs_k141_65778\|\|full | 3671 | No | NA | 3 | 0 | 1 | Not-determined | Genome-fragment | NA | NA | 0 |
| S8_extracted_k141_28519 | 1978 | No | NA | 4 | 0 | 1 | Low-quality | Genome-fragment | 2.46 | AAI-based (high-confidence) | 0 |
| S14_extracted_k141_90574 | 1979 | No | NA | 3 | 1 | 0 | Low-quality | Genome-fragment | 3.14 | AAI-based (medium-confidence) | 0 |
| S5_vs_k141_11227\|\|full | 2638 | No | NA | 2 | 0 | 1 | Not-determined | Genome-fragment | NA | NA | 0 |
| S5_vs_k141_41067\|\|full | 9031 | No | NA | 11 | 0 | 9 | Not-determined | Genome-fragment | NA | NA | 0 |
| S5_vs_k141_23526\|\|full | 1549 | No | NA | 2 | 0 | 1 | Not-determined | Genome-fragment | NA | NA | 0 |
| S5_vs_k141_133055\|\|full | 1729 | No | NA | 2 | 0 | 2 | Not-determined | Genome-fragment | NA | NA | 0 |
| S12_vs_k141_36962\|\|full | 1979 | No | NA | 8 | 1 | 0 | Low-quality | Genome-fragment | 2.66 | AAI-based (medium-confidence) | 0 |
| S5_vs_k141_41089\|\|full | 1467 | No | NA | 3 | 0 | 1 | Not-determined | Genome-fragment | NA | NA | 0 |
| S8_extracted_k141_57693 | 1979 | No | NA | 2 | 0 | 0 | Low-quality | Genome-fragment | 1.35 | AAI-based (high-confidence) | 0 |
| S17_extracted_k141_69762 | 1980 | No | NA | 4 | 4 | 0 | Low-quality | Genome-fragment | 6.53 | AAI-based (high-confidence) | 0 |
| S5_vs_k141_144183\|\|full | 13930 | No | NA | 17 | 0 | 0 | Not-determined | Genome-fragment | NA | NA | 0 |
| S11_extracted_k141_100752 | 1980 | No | NA | 4 | 0 | 0 | Low-quality | Genome-fragment | 3.41 | AAI-based (high-confidence) | 0 |
| S5_vs_k141_144193\|\|full | 1456 | No | NA | 3 | 0 | 2 | Not-determined | Genome-fragment | NA | NA | 0 |
| S5_vs_k141_65842\|\|full | 2026 | No | NA | 3 | 0 | 0 | Not-determined | Genome-fragment | NA | NA | 0 |
| S5_vs_k141_82321\|\|full | 4925 | No | NA | 3 | 0 | 2 | Not-determined | Genome-fragment | NA | NA | 0 |
| S11_vs_k141_77451\|\|full | 1980 | No | NA | 3 | 0 | 1 | Low-quality | Genome-fragment | 1.97 | AAI-based (high-confidence) | 0 |
| S1_extracted_k141_74136 | 1980 | No | NA | 3 | 1 | 0 | Low-quality | Genome-fragment | 4.45 | AAI-based (high-confidence) | 0 |
| S5_extracted_k141_75474 | 1980 | No | NA | 2 | 2 | 0 | Low-quality | Genome-fragment | 3.35 | AAI-based (high-confidence) | 0 |
| S5_vs_k141_60977\|\|full | 1980 | No | NA | 3 | 0 | 0 | Low-quality | Genome-fragment | 3.27 | AAI-based (high-confidence) | 0 |
| S7_vs_36487\|\|lt2gene | 1980 | No | NA | 1 | 1 | 0 | Low-quality | Genome-fragment | 5.02 | AAI-based (high-confidence) | 0 |
| S5_vs_k141_41161\|\|full | 2076 | No | NA | 2 | 0 | 1 | Not-determined | Genome-fragment | NA | NA | 0 |
| S10_vs_1759\|\|full | 1980 | No | NA | 5 | 0 | 0 | Low-quality | Genome-fragment | 3.18 | AAI-based (medium-confidence) | 0 |
| S5_vs_k141_115150\|\|full | 5989 | No | NA | 5 | 0 | 3 | Not-determined | Genome-fragment | NA | NA | 0 |
| S12_vs_k141_74013\|\|lt2gene | 1980 | No | NA | 2 | 2 | 0 | Low-quality | Genome-fragment | 4.99 | AAI-based (high-confidence) | 0 |
| S6_extracted_k141_49018 | 1980 | No | NA | 2 | 1 | 0 | Low-quality | Genome-fragment | 1.27 | AAI-based (high-confidence) | 0 |
| S5_vs_k141_76805\|\|full | 1319 | No | NA | 3 | 0 | 1 | Not-determined | Genome-fragment | NA | NA | 0 |
| S11_vs_k141_7744\|\|full | 1981 | No | NA | 6 | 1 | 0 | Low-quality | Genome-fragment | 4.67 | AAI-based (high-confidence) | 0 |
| S11_vs_k141_61817\|\|full | 1981 | No | NA | 3 | 1 | 0 | Low-quality | Genome-fragment | 0.81 | AAI-based (medium-confidence) | 0 |
| S5_vs_k141_23642\|\|full | 3150 | No | NA | 4 | 0 | 1 | Not-determined | Genome-fragment | NA | NA | 0 |
| S5_vs_k141_72020\|\|full | 2154 | No | NA | 3 | 0 | 0 | Not-determined | Genome-fragment | NA | NA | 0 |
| S14_vs_k141_93806\|\|full | 1981 | No | NA | 5 | 1 | 0 | Low-quality | Genome-fragment | 2.94 | HMM-based (lower-bound) | 0 |
| S5_vs_k141_96988\|\|full | 1712 | No | NA | 3 | 0 | 2 | Not-determined | Genome-fragment | NA | NA | 0 |
| S5_vs_k141_121860\|\|full | 1981 | No | NA | 4 | 2 | 0 | Low-quality | Genome-fragment | 4.58 | AAI-based (medium-confidence) | 0 |
| S5_vs_k141_82456\|\|full | 1512 | No | NA | 3 | 0 | 3 | Not-determined | Genome-fragment | NA | NA | 0 |
| S5_vs_k141_27475\|\|full | 1981 | No | NA | 4 | 1 | 0 | Low-quality | Genome-fragment | 3.91 | HMM-based (lower-bound) | 0 |
| S5_vs_k141_82238\|\|full | 1981 | No | NA | 6 | 0 | 1 | Low-quality | Genome-fragment | 4.64 | AAI-based (medium-confidence) | 0 |
| S16_extracted_18382 | 1981 | No | NA | 3 | 0 | 0 | Low-quality | Genome-fragment | 2.55 | AAI-based (high-confidence) | 0 |
| S5_vs_k141_90659\|\|full | 1148 | No | NA | 2 | 0 | 2 | Not-determined | Genome-fragment | NA | NA | 0 |
| S5_vs_k141_90670\|\|full | 2040 | No | NA | 2 | 0 | 0 | Not-determined | Genome-fragment | NA | NA | 0 |
| S16_vs_171236\|\|full | 1981 | No | NA | 6 | 0 | 0 | Low-quality | Genome-fragment | 4.4 | AAI-based (high-confidence) | 0 |
| S5_vs_k141_76881\|\|full | 1735 | No | NA | 2 | 0 | 1 | Not-determined | Genome-fragment | NA | NA | 0 |
| S12_vs_k141_130380\|\|full | 1981 | No | NA | 4 | 0 | 0 | Low-quality | Genome-fragment | 3.68 | AAI-based (medium-confidence) | 0 |
| S8_vs_k141_45587\|\|full | 1981 | No | NA | 2 | 0 | 2 | Low-quality | Genome-fragment | 1.64 | AAI-based (medium-confidence) | 0 |
| S11_vs_k141_23311\|\|full | 1982 | No | NA | 3 | 2 | 0 | Low-quality | Genome-fragment | 6.82 | AAI-based (high-confidence) | 0 |
| S5_vs_k141_9682\|\|full | 1982 | No | NA | 2 | 2 | 0 | Low-quality | Genome-fragment | 4.84 | AAI-based (high-confidence) | 0 |
| S5_vs_k141_133418\|\|full | 2387 | No | NA | 3 | 0 | 0 | Not-determined | Genome-fragment | NA | NA | 0 |
| S5_vs_k141_41305\|\|full | 3001 | No | NA | 3 | 0 | 2 | Not-determined | Genome-fragment | NA | NA | 0 |
| S7_vs_1906\|\|full | 1982 | No | NA | 3 | 0 | 0 | Low-quality | Genome-fragment | 1.49 | AAI-based (high-confidence) | 0 |
| S12_vs_k141_164347\|\|full | 1982 | No | NA | 3 | 0 | 0 | Low-quality | Genome-fragment | 2.2 | AAI-based (medium-confidence) | 0 |
| S17_vs_k141_74165\|\|full | 1983 | No | NA | 4 | 0 | 0 | Low-quality | Genome-fragment | 4.5 | AAI-based (medium-confidence) | 0 |
| S11_vs_k141_79444\|\|lt2gene | 1983 | No | NA | 2 | 2 | 0 | Low-quality | Genome-fragment | 4.87 | AAI-based (medium-confidence) | 0 |
| S14_extracted_k141_96234 | 1983 | No | NA | 7 | 7 | 0 | Low-quality | Genome-fragment | 6.55 | AAI-based (high-confidence) | 0 |
| S14_vs_k141_35551\|\|full | 1983 | No | NA | 4 | 0 | 1 | Low-quality | Genome-fragment | 2.22 | AAI-based (medium-confidence) | 0 |
| S1_extracted_k141_42747 | 1983 | No | NA | 3 | 2 | 0 | Low-quality | Genome-fragment | 2.47 | AAI-based (high-confidence) | 0 |
| S5_vs_k141_65999\|\|full | 2958 | No | NA | 6 | 0 | 1 | Not-determined | Genome-fragment | NA | NA | 0 |
| S5_vs_k141_66015\|\|full | 1065 | No | NA | 2 | 0 | 0 | Not-determined | Genome-fragment | NA | NA | 0 |
| S1_vs_k141_137892\|\|full | 1983 | No | NA | 3 | 1 | 0 | Low-quality | Genome-fragment | 2.66 | HMM-based (lower-bound) | 0 |
| S5_vs_k141_23806\|\|full | 3497 | No | NA | 4 | 0 | 3 | Not-determined | Genome-fragment | NA | NA | 0 |
| S3_extracted_k141_100656 | 1983 | No | NA | 6 | 2 | 0 | Low-quality | Genome-fragment | 1.84 | AAI-based (high-confidence) | 0 |
| S5_vs_k141_48137\|\|full | 1191 | No | NA | 2 | 0 | 1 | Not-determined | Genome-fragment | NA | NA | 0 |
| S7_extracted_11971 | 1983 | No | NA | 2 | 2 | 0 | Low-quality | Genome-fragment | 4.83 | AAI-based (high-confidence) | 0 |
| S5_vs_k141_144419\|\|full | 3635 | No | NA | 2 | 0 | 2 | Not-determined | Genome-fragment | NA | NA | 0 |
| S7_extracted_27207 | 1983 | No | NA | 3 | 2 | 0 | Low-quality | Genome-fragment | 4.21 | AAI-based (high-confidence) | 0 |
| S5_vs_k141_144424\|\|full | 4946 | No | NA | 4 | 0 | 2 | Not-determined | Genome-fragment | NA | NA | 0 |
| S7_extracted_113775 | 1983 | No | NA | 3 | 0 | 0 | Low-quality | Genome-fragment | 5.02 | AAI-based (medium-confidence) | 0 |
| S5_vs_k141_23910\|\|full | 4137 | No | NA | 6 | 0 | 4 | Not-determined | Genome-fragment | NA | NA | 0 |
| S5_vs_k141_144446\|\|full | 1882 | No | NA | 3 | 0 | 3 | Not-determined | Genome-fragment | NA | NA | 0 |
| S7_vs_98129\|\|full | 1983 | No | NA | 5 | 0 | 0 | Low-quality | Genome-fragment | 4.16 | AAI-based (high-confidence) | 0 |
| S8_vs_k141_174989\|\|full | 1983 | No | NA | 4 | 0 | 0 | Low-quality | Genome-fragment | 1.16 | AAI-based (high-confidence) | 0 |
| S3_extracted_k141_67621 | 1984 | No | NA | 4 | 2 | 0 | Low-quality | Genome-fragment | 3.99 | AAI-based (high-confidence) | 0 |
| S5_extracted_k141_40515 | 1984 | No | NA | 6 | 1 | 0 | Low-quality | Genome-fragment | 0.96 | AAI-based (high-confidence) | 0 |
| S5_vs_k141_137719\|\|full | 7485 | No | NA | 16 | 0 | 0 | Not-determined | Genome-fragment | NA | NA | 0 |
| S5_vs_k141_23949\|\|full | 1381 | No | NA | 2 | 0 | 2 | Not-determined | Genome-fragment | NA | NA | 0 |
| S8_vs_k141_6824\|\|full | 1984 | No | NA | 3 | 0 | 0 | Low-quality | Genome-fragment | 4.56 | AAI-based (medium-confidence) | 0 |
| S5_vs_k141_137730\|\|full | 2329 | No | NA | 2 | 0 | 1 | Not-determined | Genome-fragment | NA | NA | 0 |
| S5_vs_k141_66132\|\|full | 1074 | No | NA | 2 | 0 | 2 | Not-determined | Genome-fragment | NA | NA | 0 |
| S5_vs_k141_23954\|\|full | 1835 | No | NA | 3 | 0 | 3 | Not-determined | Genome-fragment | NA | NA | 0 |
| S9_vs_k141_27455\|\|full | 1985 | No | NA | 4 | 1 | 0 | Low-quality | Genome-fragment | 5.55 | AAI-based (high-confidence) | 0 |
| S13_vs_32133\|\|full | 1985 | No | NA | 2 | 0 | 0 | Low-quality | Genome-fragment | 3.12 | AAI-based (high-confidence) | 0 |
| S5_vs_k141_31538\|\|full | 1985 | No | NA | 3 | 0 | 2 | Low-quality | Genome-fragment | 6.34 | AAI-based (high-confidence) | 0 |
| S5_vs_k141_93619\|\|lt2gene | 1985 | No | NA | 2 | 1 | 0 | Low-quality | Genome-fragment | 5.62 | AAI-based (high-confidence) | 0 |
| S5_vs_k141_11634\|\|full | 1309 | No | NA | 2 | 0 | 0 | Not-determined | Genome-fragment | NA | NA | 0 |
| S5_vs_k141_66183\|\|full | 3342 | No | NA | 3 | 0 | 1 | Not-determined | Genome-fragment | NA | NA | 0 |
| S7_vs_33462\|\|full | 1985 | No | NA | 4 | 2 | 0 | Low-quality | Genome-fragment | 4.39 | AAI-based (high-confidence) | 0 |
| S17_vs_k141_75215\|\|full | 1986 | No | NA | 2 | 0 | 0 | Low-quality | Genome-fragment | 3.18 | AAI-based (medium-confidence) | 0 |
| S5_vs_k141_137788\|\|full | 1186 | No | NA | 2 | 0 | 0 | Not-determined | Genome-fragment | NA | NA | 0 |
| S7_extracted_11390 | 1986 | No | NA | 4 | 4 | 0 | Low-quality | Genome-fragment | 6.51 | AAI-based (high-confidence) | 0 |
| S10_extracted_16567 | 1986 | No | NA | 2 | 1 | 0 | Low-quality | Genome-fragment | 3.21 | AAI-based (high-confidence) | 0 |
| S10_vs_7651\|\|full | 1986 | No | NA | 5 | 1 | 0 | Low-quality | Genome-fragment | 1.19 | AAI-based (medium-confidence) | 0 |
| S5_vs_k141_77192\|\|full | 2044 | No | NA | 3 | 0 | 1 | Not-determined | Genome-fragment | NA | NA | 0 |
| S5_vs_k141_41554\|\|full | 10880 | No | NA | 11 | 0 | 4 | Not-determined | Genome-fragment | NA | NA | 0 |
| S8_extracted_k141_58490 | 1986 | No | NA | 3 | 0 | 0 | Low-quality | Genome-fragment | 3.13 | AAI-based (high-confidence) | 0 |
| S5_vs_k141_77222\|\|full | 1148 | No | NA | 3 | 0 | 1 | Not-determined | Genome-fragment | NA | NA | 0 |
| S5_vs_k141_41562\|\|full | 2533 | No | NA | 2 | 0 | 2 | Not-determined | Genome-fragment | NA | NA | 0 |
| S5_vs_k141_72451\|\|full | 1680 | No | NA | 3 | 0 | 2 | Not-determined | Genome-fragment | NA | NA | 0 |
| S9_vs_k141_32419\|\|full | 1987 | No | NA | 3 | 3 | 0 | Low-quality | Genome-fragment | 4.93 | AAI-based (high-confidence) | 0 |
| S5_vs_k141_77242\|\|full | 1271 | No | NA | 5 | 0 | 0 | Not-determined | Genome-fragment | NA | NA | 0 |
| S3_vs_k141_21745\|\|full | 1987 | No | NA | 7 | 1 | 0 | Low-quality | Genome-fragment | 4.76 | AAI-based (high-confidence) | 0 |
| S7_extracted_33700 | 1987 | No | NA | 4 | 0 | 0 | Low-quality | Genome-fragment | 6.01 | AAI-based (medium-confidence) | 0 |
| S12_extracted_k141_154005 | 1987 | No | NA | 2 | 0 | 1 | Low-quality | Genome-fragment | 5.23 | AAI-based (medium-confidence) | 0 |
| S11_vs_k141_76373\|\|full | 1988 | No | NA | 5 | 1 | 0 | Low-quality | Genome-fragment | 2.95 | HMM-based (lower-bound) | 0 |
| S5_vs_k141_48458\|\|full | 4869 | No | NA | 5 | 0 | 3 | Not-determined | Genome-fragment | NA | NA | 0 |
| S5_vs_k141_66234\|\|full | 3152 | No | NA | 2 | 0 | 1 | Not-determined | Genome-fragment | NA | NA | 0 |
| S13_vs_114202\|\|full | 1988 | No | NA | 5 | 0 | 0 | Low-quality | Genome-fragment | 4.54 | AAI-based (medium-confidence) | 0 |
| S5_vs_k141_66237\|\|full | 2366 | No | NA | 4 | 0 | 2 | Not-determined | Genome-fragment | NA | NA | 0 |
| S5_vs_k141_28263\|\|full | 29578 | No | NA | 42 | 15 | 0 | Medium-quality | Genome-fragment | 85.3 | AAI-based (high-confidence) | 0 |
| S5_vs_k141_66244\|\|full | 1559 | No | NA | 2 | 0 | 1 | Not-determined | Genome-fragment | NA | NA | 0 |
| S5_vs_k141_24111\|\|full | 2668 | No | NA | 4 | 0 | 0 | Not-determined | Genome-fragment | NA | NA | 0 |
| S3_vs_k141_72116\|\|full | 1988 | No | NA | 3 | 1 | 0 | Low-quality | Genome-fragment | 3.14 | AAI-based (medium-confidence) | 0 |
| S3_vs_k141_126935\|\|full | 1988 | No | NA | 3 | 0 | 0 | Low-quality | Genome-fragment | 4.25 | AAI-based (high-confidence) | 0 |
| S5_vs_k141_28288\|\|full | 2029 | No | NA | 3 | 0 | 1 | Not-determined | Genome-fragment | NA | NA | 0 |
| S5_vs_k141_82717\|\|full | 1682 | No | NA | 3 | 0 | 1 | Not-determined | Genome-fragment | NA | NA | 0 |
| S5_vs_k141_82721\|\|full | 1984 | No | NA | 3 | 0 | 1 | Not-determined | Genome-fragment | NA | NA | 0 |
| S16_vs_103582\|\|lt2gene | 1988 | No | NA | 2 | 1 | 0 | Low-quality | Genome-fragment | 1.23 | HMM-based (lower-bound) | 0 |
| S5_vs_k141_58153\|\|full | 1198 | No | NA | 3 | 0 | 0 | Not-determined | Genome-fragment | NA | NA | 0 |
| S5_vs_k141_72532\|\|full | 53583 | Yes | 36298 | 68 | 8 | 11 | Medium-quality | Genome-fragment | 70.51 | AAI-based (high-confidence) | 32.26 |
| S5_vs_k141_41663\|\|full | 60673 | No | NA | 91 | 23 | 4 | High-quality | High-quality | 95.52 | AAI-based (medium-confidence) | 0 |
| S5_vs_k141_90932\|\|full | 2550 | No | NA | 4 | 0 | 1 | Not-determined | Genome-fragment | NA | NA | 0 |
| S8_vs_k141_93667\|\|full | 1988 | No | NA | 3 | 0 | 1 | Low-quality | Genome-fragment | 1.21 | AAI-based (medium-confidence) | 0 |
| S13_vs_96099\|\|full | 1989 | No | NA | 3 | 0 | 0 | Low-quality | Genome-fragment | 2.33 | AAI-based (high-confidence) | 0 |
| S1_vs_k141_57990\|\|full | 1989 | No | NA | 3 | 2 | 0 | Low-quality | Genome-fragment | 1.36 | HMM-based (lower-bound) | 0 |
| S5_extracted_k141_47796 | 1989 | No | NA | 5 | 1 | 0 | Low-quality | Genome-fragment | 1.19 | HMM-based (lower-bound) | 0 |
| S5_vs_k141_74525\|\|full | 1989 | No | NA | 5 | 0 | 0 | Low-quality | Genome-fragment | 4.64 | AAI-based (medium-confidence) | 0 |
| S10_extracted_97263 | 1989 | No | NA | 1 | 0 | 1 | Low-quality | Genome-fragment | 0.52 | AAI-based (high-confidence) | 0 |
| S10_vs_113330\|\|full | 1989 | No | NA | 4 | 1 | 0 | Low-quality | Genome-fragment | 4.22 | AAI-based (medium-confidence) | 0 |
| S5_vs_k141_24269\|\|full | 1558 | No | NA | 3 | 0 | 2 | Not-determined | Genome-fragment | NA | NA | 0 |
| S6_vs_k141_44165\|\|full | 1989 | No | NA | 3 | 1 | 0 | Low-quality | Genome-fragment | 3.27 | AAI-based (high-confidence) | 0 |
| S5_vs_k141_144749\|\|full | 2877 | No | NA | 4 | 0 | 1 | Not-determined | Genome-fragment | NA | NA | 0 |
| S11_vs_k141_60807\|\|full | 1990 | No | NA | 4 | 1 | 0 | Low-quality | Genome-fragment | 3.34 | HMM-based (lower-bound) | 0 |
| S5_vs_k141_82818\|\|full | 2519 | No | NA | 4 | 0 | 0 | Not-determined | Genome-fragment | NA | NA | 0 |
| S13_vs_105920\|\|full | 1990 | No | NA | 3 | 0 | 3 | Low-quality | Genome-fragment | 1.92 | AAI-based (high-confidence) | 0 |
| S5_vs_k141_72631\|\|full | 8882 | No | NA | 9 | 0 | 5 | Not-determined | Genome-fragment | NA | NA | 0 |
| S5_vs_k141_28380\|\|full | 1086 | No | NA | 4 | 0 | 0 | Not-determined | Genome-fragment | NA | NA | 0 |
| S14_extracted_k141_77853 | 1990 | No | NA | 2 | 0 | 1 | Low-quality | Genome-fragment | 0.96 | AAI-based (medium-confidence) | 0 |
| S5_vs_k141_41776\|\|full | 1429 | No | NA | 3 | 0 | 3 | Not-determined | Genome-fragment | NA | NA | 0 |
| S3_vs_k141_62603\|\|full | 1990 | No | NA | 5 | 0 | 0 | Low-quality | Genome-fragment | 5.27 | AAI-based (high-confidence) | 0 |
| S5_vs_k141_82909\|\|full | 2027 | No | NA | 3 | 0 | 0 | Not-determined | Genome-fragment | NA | NA | 0 |
| S5_vs_k141_28491\|\|full | 1084 | No | NA | 2 | 0 | 0 | Not-determined | Genome-fragment | NA | NA | 0 |
| S5_vs_k141_41898\|\|full | 2814 | No | NA | 4 | 0 | 2 | Not-determined | Genome-fragment | NA | NA | 0 |
| S15_vs_18399\|\|full | 1990 | No | NA | 3 | 0 | 0 | Low-quality | Genome-fragment | 5.23 | AAI-based (high-confidence) | 0 |
| S8_extracted_k141_78298 | 1990 | No | NA | 3 | 0 | 0 | Low-quality | Genome-fragment | 4.77 | AAI-based (medium-confidence) | 0 |
| S11_extracted_k141_74348 | 1991 | No | NA | 6 | 0 | 0 | Low-quality | Genome-fragment | 3.08 | AAI-based (high-confidence) | 0 |
| S5_vs_k141_144846\|\|full | 1767 | No | NA | 2 | 0 | 2 | Not-determined | Genome-fragment | NA | NA | 0 |
| S14_vs_k141_89278\|\|full | 1991 | No | NA | 2 | 0 | 0 | Low-quality | Genome-fragment | 1.08 | AAI-based (medium-confidence) | 0 |
| S5_vs_k141_144855\|\|full | 2444 | No | NA | 4 | 0 | 2 | Not-determined | Genome-fragment | NA | NA | 0 |
| S1_extracted_k141_53048 | 1991 | No | NA | 1 | 0 | 0 | Low-quality | Genome-fragment | 3.28 | AAI-based (high-confidence) | 0 |
| S5_vs_k141_66468\|\|full | 2567 | No | NA | 3 | 0 | 2 | Not-determined | Genome-fragment | NA | NA | 0 |
| S5_vs_k141_137979\|\|full | 2229 | No | NA | 4 | 0 | 2 | Not-determined | Genome-fragment | NA | NA | 0 |
| S7_extracted_54593 | 1991 | No | NA | 1 | 1 | 0 | Low-quality | Genome-fragment | 1.2 | AAI-based (high-confidence) | 0 |
| S5_vs_k141_66517\|\|full | 1228 | No | NA | 2 | 0 | 2 | Not-determined | Genome-fragment | NA | NA | 0 |
| S5_vs_k141_72774\|\|full | 29785 | No | NA | 19 | 0 | 16 | Not-determined | Genome-fragment | NA | NA | 0 |
| S6_extracted_k141_10958 | 1991 | No | NA | 3 | 0 | 0 | Low-quality | Genome-fragment | 1.98 | AAI-based (medium-confidence) | 0 |
| S5_vs_k141_83048\|\|full | 4301 | No | NA | 5 | 0 | 4 | Not-determined | Genome-fragment | NA | NA | 0 |
| S17_extracted_k141_117369 | 1992 | No | NA | 3 | 0 | 0 | Low-quality | Genome-fragment | 3.36 | AAI-based (high-confidence) | 0 |
| S5_vs_k141_83071\|\|full | 1824 | No | NA | 2 | 0 | 2 | Not-determined | Genome-fragment | NA | NA | 0 |
| S5_vs_k141_83075\|\|full | 1311 | No | NA | 3 | 0 | 2 | Not-determined | Genome-fragment | NA | NA | 0 |
| S5_vs_k141_58407\|\|full | 1956 | No | NA | 4 | 0 | 1 | Not-determined | Genome-fragment | NA | NA | 0 |
| S9_vs_k141_27863\|\|full | 1992 | No | NA | 2 | 0 | 0 | Low-quality | Genome-fragment | 1.15 | AAI-based (high-confidence) | 0 |
| S11_vs_k141_37764\|\|full | 1992 | No | NA | 5 | 1 | 0 | Low-quality | Genome-fragment | 4.11 | AAI-based (high-confidence) | 0 |
| S3_vs_k141_24865\|\|lt2gene | 1992 | No | NA | 2 | 2 | 0 | Low-quality | Genome-fragment | 5.02 | AAI-based (medium-confidence) | 0 |
| S5_extracted_k141_84954 | 1992 | No | NA | 5 | 2 | 0 | Low-quality | Genome-fragment | 4.35 | AAI-based (high-confidence) | 0 |
| S5_vs_k141_83167\|\|full | 1721 | No | NA | 3 | 0 | 3 | Not-determined | Genome-fragment | NA | NA | 0 |
| S5_vs_k141_77497\|\|full | 2694 | No | NA | 2 | 0 | 2 | Not-determined | Genome-fragment | NA | NA | 0 |
| S5_vs_k141_42109\|\|full | 3669 | No | NA | 4 | 0 | 1 | Not-determined | Genome-fragment | NA | NA | 0 |
| S7_vs_70894\|\|full | 1992 | No | NA | 6 | 0 | 1 | Low-quality | Genome-fragment | 4.06 | AAI-based (medium-confidence) | 0 |
| S17_vs_k141_161757\|\|full | 1993 | No | NA | 3 | 0 | 1 | Low-quality | Genome-fragment | 4.43 | AAI-based (medium-confidence) | 0 |
| S5_vs_k141_58512\|\|full | 1240 | No | NA | 3 | 0 | 1 | Not-determined | Genome-fragment | NA | NA | 0 |
| S14_extracted_k141_39830 | 1993 | No | NA | 4 | 1 | 0 | Low-quality | Genome-fragment | 2.92 | AAI-based (medium-confidence) | 0 |
| S14_vs_k141_26617\|\|full | 1993 | No | NA | 4 | 1 | 0 | Low-quality | Genome-fragment | 3.93 | AAI-based (medium-confidence) | 0 |
| S5_vs_k141_145026\|\|full | 41474 | No | NA | 60 | 12 | 1 | High-quality | High-quality | 96.6 | AAI-based (high-confidence) | 0 |
| S5_vs_k141_138004\|\|full | 2684 | No | NA | 3 | 0 | 0 | Not-determined | Genome-fragment | NA | NA | 0 |
| S7_extracted_7448 | 1993 | No | NA | 1 | 0 | 0 | Low-quality | Genome-fragment | 2.44 | AAI-based (high-confidence) | 0 |
| S5_vs_k141_28717\|\|full | 7463 | No | NA | 6 | 0 | 5 | Not-determined | Genome-fragment | NA | NA | 0 |
| S7_vs_68889\|\|full | 1993 | No | NA | 5 | 0 | 0 | Low-quality | Genome-fragment | 4.42 | AAI-based (medium-confidence) | 0 |
| S7_vs_37235\|\|lt2gene | 1993 | No | NA | 2 | 2 | 0 | Low-quality | Genome-fragment | 2.23 | HMM-based (lower-bound) | 0 |
| S16_extracted_109253 | 1993 | No | NA | 5 | 1 | 0 | Low-quality | Genome-fragment | 3.21 | AAI-based (high-confidence) | 0 |
| S16_vs_38272\|\|full | 1993 | No | NA | 4 | 0 | 0 | Low-quality | Genome-fragment | 1.15 | AAI-based (high-confidence) | 0 |
| S5_vs_k141_77593\|\|full | 135431 | Yes | 45565 | 166 | 30 | 83 | Medium-quality | Genome-fragment | 69.58 | HMM-based (lower-bound) | 66.36 |
| S5_vs_k141_66581\|\|full | 2491 | No | NA | 4 | 0 | 2 | Not-determined | Genome-fragment | NA | NA | 0 |
| S5_vs_k141_138123\|\|full | 1134 | No | NA | 3 | 0 | 1 | Not-determined | Genome-fragment | NA | NA | 0 |
| S5_vs_k141_77626\|\|full | 1018 | No | NA | 2 | 0 | 0 | Not-determined | Genome-fragment | NA | NA | 0 |
| S5_vs_k141_138130\|\|full | 4475 | No | NA | 5 | 0 | 2 | Not-determined | Genome-fragment | NA | NA | 0 |
| S15_vs_41539\|\|full | 1993 | No | NA | 5 | 0 | 0 | Low-quality | Genome-fragment | 5.34 | AAI-based (medium-confidence) | 0 |
| S5_vs_k141_77646\|\|full | 2706 | No | NA | 3 | 0 | 3 | Not-determined | Genome-fragment | NA | NA | 0 |
| S8_vs_k141_54350\|\|full | 1993 | No | NA | 2 | 0 | 0 | Low-quality | Genome-fragment | 1.39 | AAI-based (high-confidence) | 0 |
| S8_vs_k141_81650\|\|full | 1993 | No | NA | 5 | 0 | 0 | Low-quality | Genome-fragment | 2.42 | AAI-based (medium-confidence) | 0 |
| S8_vs_k141_195909\|\|full | 1993 | Yes | 946 | 3 | 1 | 1 | Low-quality | Genome-fragment | 0.51 | HMM-based (lower-bound) | 52.53 |
| S17_vs_k141_157711\|\|full | 1994 | No | NA | 2 | 0 | 0 | Low-quality | Genome-fragment | 5.31 | AAI-based (medium-confidence) | 0 |
| S13_vs_60776\|\|full | 1994 | Yes | 1575 | 8 | 1 | 1 | Low-quality | Genome-fragment | 4.12 | AAI-based (high-confidence) | 21.01 |
| S14_vs_k141_2065\|\|full | 1994 | No | NA | 3 | 1 | 0 | Low-quality | Genome-fragment | 3.25 | HMM-based (lower-bound) | 0 |
| S5_vs_k141_83434\|\|full | 1279 | No | NA | 4 | 0 | 2 | Not-determined | Genome-fragment | NA | NA | 0 |
| S5_vs_k141_58744\|\|full | 13312 | No | NA | 13 | 0 | 9 | Not-determined | Genome-fragment | NA | NA | 0 |
| S5_vs_k141_42418\|\|full | 1550 | No | NA | 3 | 0 | 1 | Not-determined | Genome-fragment | NA | NA | 0 |
| S5_vs_k141_77686\|\|full | 1663 | No | NA | 3 | 0 | 1 | Not-determined | Genome-fragment | NA | NA | 0 |
| S1_vs_k141_118219\|\|lt2gene | 1994 | No | NA | 2 | 1 | 0 | Low-quality | Genome-fragment | 37.73 | AAI-based (high-confidence) | 0 |
| S8_vs_k141_17803\|\|full | 1994 | No | NA | 5 | 0 | 0 | Low-quality | Genome-fragment | 5.12 | AAI-based (high-confidence) | 0 |
| S3_vs_k141_54465\|\|full | 1995 | No | NA | 2 | 1 | 0 | Low-quality | Genome-fragment | 3.46 | AAI-based (high-confidence) | 0 |
| S7_vs_91920\|\|full | 1995 | No | NA | 3 | 2 | 0 | Low-quality | Genome-fragment | 5.82 | AAI-based (high-confidence) | 0 |
| S5_vs_k141_138231\|\|full | 2580 | No | NA | 4 | 0 | 1 | Not-determined | Genome-fragment | NA | NA | 0 |
| S5_vs_k141_42455\|\|full | 4310 | No | NA | 4 | 0 | 1 | Not-determined | Genome-fragment | NA | NA | 0 |
| S12_extracted_k141_172626 | 1995 | No | NA | 2 | 2 | 0 | Low-quality | Genome-fragment | 3.82 | AAI-based (high-confidence) | 0 |
| S5_vs_k141_77705\|\|full | 2769 | No | NA | 3 | 0 | 2 | Not-determined | Genome-fragment | NA | NA | 0 |
| S5_vs_k141_58766\|\|full | 20940 | No | NA | 25 | 12 | 0 | Medium-quality | Genome-fragment | 51.31 | AAI-based (high-confidence) | 0 |
| S5_vs_k141_58775\|\|full | 1511 | No | NA | 3 | 0 | 1 | Not-determined | Genome-fragment | NA | NA | 0 |
| S5_vs_k141_145267\|\|full | 2129 | No | NA | 4 | 0 | 0 | Not-determined | Genome-fragment | NA | NA | 0 |
| S8_vs_k141_32430\|\|full | 1995 | No | NA | 6 | 0 | 0 | Low-quality | Genome-fragment | 3.64 | AAI-based (high-confidence) | 0 |
| S5_vs_k141_138276\|\|full | 3795 | No | NA | 4 | 0 | 4 | Not-determined | Genome-fragment | NA | NA | 0 |
| S5_vs_k141_28901\|\|full | 6608 | No | NA | 5 | 0 | 4 | Not-determined | Genome-fragment | NA | NA | 0 |
| S5_vs_k141_42501\|\|full | 3020 | No | NA | 4 | 0 | 3 | Not-determined | Genome-fragment | NA | NA | 0 |
| S11_vs_k141_48729\|\|full | 1996 | No | NA | 2 | 2 | 0 | Low-quality | Genome-fragment | 5.6 | AAI-based (high-confidence) | 0 |
| S13_vs_76157\|\|lt2gene | 1996 | No | NA | 2 | 1 | 0 | Low-quality | Genome-fragment | 3.88 | AAI-based (high-confidence) | 0 |
| S3_extracted_k141_10785 | 1996 | No | NA | 4 | 2 | 0 | Low-quality | Genome-fragment | 2.83 | AAI-based (high-confidence) | 0 |
| S5_extracted_k141_99667 | 1996 | No | NA | 2 | 2 | 0 | Low-quality | Genome-fragment | 5.89 | AAI-based (medium-confidence) | 0 |
| S5_vs_k141_58859\|\|full | 21538 | No | NA | 32 | 18 | 0 | Medium-quality | Genome-fragment | 62.86 | AAI-based (high-confidence) | 0 |
| S6_vs_k141_20329\|\|full | 1996 | No | NA | 3 | 1 | 0 | Low-quality | Genome-fragment | 4.29 | AAI-based (high-confidence) | 0 |
| S5_vs_k141_145398\|\|full | 3260 | No | NA | 6 | 0 | 0 | Not-determined | Genome-fragment | NA | NA | 0 |
| S5_vs_k141_77820\|\|full | 3174 | No | NA | 3 | 0 | 3 | Not-determined | Genome-fragment | NA | NA | 0 |
| S14_vs_k141_37619\|\|full | 1997 | No | NA | 4 | 1 | 0 | Low-quality | Genome-fragment | 1.41 | HMM-based (lower-bound) | 0 |
| S5_vs_k141_77828\|\|full | 1481 | No | NA | 2 | 0 | 1 | Not-determined | Genome-fragment | NA | NA | 0 |
| S5_vs_k141_28993\|\|full | 10059 | No | NA | 9 | 0 | 6 | Not-determined | Genome-fragment | NA | NA | 0 |
| S1_vs_k141_105669\|\|full | 1997 | No | NA | 2 | 0 | 0 | Low-quality | Genome-fragment | 2.62 | AAI-based (medium-confidence) | 0 |
| S5_vs_k141_134837\|\|full | 1997 | No | NA | 5 | 0 | 1 | Low-quality | Genome-fragment | 2.6 | AAI-based (high-confidence) | 0 |
| S5_vs_k141_29027\|\|full | 1126 | No | NA | 2 | 0 | 1 | Not-determined | Genome-fragment | NA | NA | 0 |
| S5_vs_k141_138450\|\|full | 4521 | No | NA | 6 | 0 | 5 | Not-determined | Genome-fragment | NA | NA | 0 |
| S7_vs_60391\|\|full | 1997 | No | NA | 6 | 1 | 0 | Low-quality | Genome-fragment | 2.24 | HMM-based (lower-bound) | 0 |
| S16_vs_75517\|\|full | 1997 | No | NA | 3 | 0 | 0 | Low-quality | Genome-fragment | 3.2 | AAI-based (medium-confidence) | 0 |
| S10_vs_51294\|\|lt2gene | 1997 | No | NA | 2 | 2 | 0 | Low-quality | Genome-fragment | 3.89 | AAI-based (medium-confidence) | 0 |
| S5_vs_k141_58989\|\|full | 1284 | No | NA | 2 | 0 | 1 | Not-determined | Genome-fragment | NA | NA | 0 |
| S5_vs_k141_145569\|\|full | 3658 | No | NA | 5 | 0 | 2 | Not-determined | Genome-fragment | NA | NA | 0 |
| S5_vs_k141_138511\|\|full | 2245 | No | NA | 3 | 0 | 1 | Not-determined | Genome-fragment | NA | NA | 0 |
| S12_extracted_k141_70916 | 1997 | No | NA | 2 | 1 | 0 | Low-quality | Genome-fragment | 4.1 | AAI-based (high-confidence) | 0 |
| S5_vs_k141_29113\|\|full | 1060 | No | NA | 2 | 0 | 0 | Not-determined | Genome-fragment | NA | NA | 0 |
| S3_vs_k141_117280\|\|full | 1998 | No | NA | 3 | 3 | 0 | Low-quality | Genome-fragment | 3.11 | HMM-based (lower-bound) | 0 |
| S5_vs_k141_83886\|\|full | 24786 | No | NA | 29 | 6 | 0 | Medium-quality | Genome-fragment | 60.38 | AAI-based (high-confidence) | 0 |
| S5_vs_k141_101736\|\|full | 1474 | No | NA | 2 | 0 | 2 | Not-determined | Genome-fragment | NA | NA | 0 |
| S7_vs_97091\|\|full | 1998 | No | NA | 7 | 1 | 0 | Low-quality | Genome-fragment | 3.33 | AAI-based (medium-confidence) | 0 |
| S7_vs_125533\|\|full | 1998 | No | NA | 6 | 0 | 0 | Low-quality | Genome-fragment | 1.07 | AAI-based (medium-confidence) | 0 |
| S5_vs_k141_101829\|\|full | 1137 | No | NA | 2 | 0 | 0 | Not-determined | Genome-fragment | NA | NA | 0 |
| S10_vs_85920\|\|full | 1998 | No | NA | 3 | 1 | 0 | Low-quality | Genome-fragment | 5.79 | AAI-based (high-confidence) | 0 |
| S2_vs_k141_7460\|\|full | 1998 | No | NA | 5 | 1 | 0 | Low-quality | Genome-fragment | 4.56 | AAI-based (high-confidence) | 0 |
| S5_vs_k141_77887\|\|full | 1811 | No | NA | 3 | 0 | 1 | Not-determined | Genome-fragment | NA | NA | 0 |
| S6_extracted_k141_21422 | 1998 | No | NA | 5 | 5 | 0 | Low-quality | Genome-fragment | 6.59 | AAI-based (high-confidence) | 0 |
| S5_vs_k141_77920\|\|full | 1884 | No | NA | 4 | 0 | 3 | Not-determined | Genome-fragment | NA | NA | 0 |
| S13_vs_85087\|\|full | 1999 | No | NA | 2 | 0 | 0 | Low-quality | Genome-fragment | 3.46 | AAI-based (medium-confidence) | 0 |
| S5_vs_k141_11890\|\|full | 2681 | No | NA | 3 | 0 | 1 | Not-determined | Genome-fragment | NA | NA | 0 |
| S14_extracted_k141_67156 | 1999 | No | NA | 1 | 1 | 0 | Low-quality | Genome-fragment | 6.58 | AAI-based (high-confidence) | 0 |
| S5_vs_k141_84187\|\|full | 1154 | No | NA | 3 | 0 | 1 | Not-determined | Genome-fragment | NA | NA | 0 |
| S1_extracted_k141_81888 | 1999 | No | NA | 1 | 1 | 0 | Low-quality | Genome-fragment | 3.42 | AAI-based (medium-confidence) | 0 |
| S1_vs_k141_29610\|\|full | 1999 | No | NA | 9 | 1 | 0 | Low-quality | Genome-fragment | 5.73 | AAI-based (high-confidence) | 0 |
| S5_vs_k141_59417\|\|full | 2074 | No | NA | 3 | 0 | 2 | Not-determined | Genome-fragment | NA | NA | 0 |
| S1_vs_k141_121070\|\|lt2gene | 1999 | No | NA | 1 | 1 | 0 | Low-quality | Genome-fragment | 2.11 | AAI-based (medium-confidence) | 0 |
| S3_extracted_k141_116484 | 1999 | No | NA | 2 | 0 | 0 | Low-quality | Genome-fragment | 2.3 | AAI-based (high-confidence) | 0 |
| S7_vs_72189\|\|full | 1999 | No | NA | 4 | 0 | 0 | Low-quality | Genome-fragment | 3.72 | AAI-based (medium-confidence) | 0 |
| S11_extracted_k141_259 | 2000 | No | NA | 6 | 0 | 0 | Low-quality | Genome-fragment | 2.81 | AAI-based (high-confidence) | 0 |
| S13_vs_154095\|\|full | 2000 | No | NA | 5 | 0 | 0 | Low-quality | Genome-fragment | 3.81 | AAI-based (high-confidence) | 0 |
| S14_extracted_k141_99476 | 2000 | No | NA | 5 | 3 | 0 | Low-quality | Genome-fragment | 4.31 | AAI-based (high-confidence) | 0 |
| S5_extracted_k141_144416 | 2000 | No | NA | 2 | 1 | 0 | Low-quality | Genome-fragment | 2.81 | HMM-based (lower-bound) | 0 |
| S5_vs_k141_78173\|\|full | 1794 | No | NA | 3 | 0 | 3 | Not-determined | Genome-fragment | NA | NA | 0 |
| S5_vs_k141_29368\|\|full | 2845 | No | NA | 3 | 0 | 0 | Not-determined | Genome-fragment | NA | NA | 0 |
| S5_vs_k141_59598\|\|full | 11598 | No | NA | 16 | 0 | 0 | Not-determined | Genome-fragment | NA | NA | 0 |
| S16_vs_114076\|\|full | 2000 | No | NA | 4 | 0 | 0 | Low-quality | Genome-fragment | 4.14 | AAI-based (medium-confidence) | 0 |
| S5_vs_k141_138819\|\|full | 2416 | No | NA | 4 | 0 | 3 | Not-determined | Genome-fragment | NA | NA | 0 |
| S10_vs_33293\|\|full | 2000 | No | NA | 4 | 3 | 0 | Low-quality | Genome-fragment | 4.16 | AAI-based (high-confidence) | 0 |
| S5_vs_k141_84333\|\|full | 1120 | No | NA | 3 | 0 | 0 | Not-determined | Genome-fragment | NA | NA | 0 |
| S10_vs_105862\|\|full | 2000 | No | NA | 3 | 0 | 1 | Low-quality | Genome-fragment | 4.48 | AAI-based (medium-confidence) | 0 |
| S9_vs_k141_32984\|\|full | 2001 | No | NA | 4 | 0 | 2 | Low-quality | Genome-fragment | 1.03 | AAI-based (medium-confidence) | 0 |
| S5_vs_k141_78445\|\|full | 1932 | No | NA | 2 | 0 | 1 | Not-determined | Genome-fragment | NA | NA | 0 |
| S14_vs_k141_45791\|\|full | 2001 | No | NA | 5 | 4 | 0 | Low-quality | Genome-fragment | 6.49 | AAI-based (high-confidence) | 0 |
| S5_vs_k141_138919\|\|full | 1909 | No | NA | 5 | 0 | 0 | Not-determined | Genome-fragment | NA | NA | 0 |
| S7_extracted_122133 | 2001 | No | NA | 5 | 2 | 0 | Low-quality | Genome-fragment | 6.17 | AAI-based (high-confidence) | 0 |
| S5_vs_k141_29405\|\|full | 2597 | No | NA | 3 | 0 | 0 | Not-determined | Genome-fragment | NA | NA | 0 |
| S7_vs_89473\|\|full | 2001 | No | NA | 5 | 1 | 0 | Low-quality | Genome-fragment | 4.2 | AAI-based (high-confidence) | 0 |
| S5_vs_k141_138980\|\|full | 54544 | No | NA | 84 | 19 | 7 | High-quality | High-quality | 100 | AAI-based (high-confidence) | 0 |
| S7_vs_99003\|\|full | 2001 | No | NA | 3 | 2 | 0 | Low-quality | Genome-fragment | 4.67 | AAI-based (medium-confidence) | 0 |
| S5_vs_k141_29444\|\|full | 1970 | No | NA | 4 | 0 | 1 | Not-determined | Genome-fragment | NA | NA | 0 |
| S12_extracted_k141_72899 | 2001 | No | NA | 2 | 0 | 0 | Low-quality | Genome-fragment | 0.37 | AAI-based (medium-confidence) | 0 |
| S5_vs_k141_78660\|\|full | 1547 | No | NA | 2 | 0 | 1 | Not-determined | Genome-fragment | NA | NA | 0 |
| S8_vs_k141_49480\|\|full | 2001 | No | NA | 5 | 4 | 0 | Low-quality | Genome-fragment | 4.6 | AAI-based (high-confidence) | 0 |
| S17_vs_k141_13650\|\|full | 2002 | No | NA | 4 | 0 | 2 | Low-quality | Genome-fragment | 0.94 | AAI-based (medium-confidence) | 0 |
| S14_vs_k141_68986\|\|lt2gene | 2002 | No | NA | 2 | 1 | 0 | Low-quality | Genome-fragment | 4.17 | AAI-based (high-confidence) | 0 |
| S5_vs_k141_29506\|\|full | 2626 | No | NA | 2 | 0 | 2 | Not-determined | Genome-fragment | NA | NA | 0 |
| S1_vs_k141_16608\|\|full | 2002 | No | NA | 3 | 0 | 0 | Low-quality | Genome-fragment | 1.5 | AAI-based (high-confidence) | 0 |
| S5_extracted_k141_761 | 2002 | No | NA | 3 | 2 | 0 | Low-quality | Genome-fragment | 5.8 | AAI-based (high-confidence) | 0 |
| S5_vs_k141_59798\|\|full | 40187 | No | NA | 58 | 26 | 0 | Medium-quality | Genome-fragment | 87.96 | AAI-based (high-confidence) | 0 |
| S7_extracted_109880 | 2002 | No | NA | 4 | 1 | 0 | Low-quality | Genome-fragment | 4.66 | AAI-based (medium-confidence) | 0 |
| S7_vs_23759\|\|full | 2002 | No | NA | 2 | 0 | 1 | Low-quality | Genome-fragment | 3.38 | AAI-based (medium-confidence) | 0 |
| S5_vs_k141_59826\|\|full | 1160 | No | NA | 2 | 0 | 1 | Not-determined | Genome-fragment | NA | NA | 0 |
| S5_vs_k141_102337\|\|full | 1939 | No | NA | 4 | 0 | 1 | Not-determined | Genome-fragment | NA | NA | 0 |
| S5_vs_k141_29622\|\|full | 1847 | No | NA | 3 | 0 | 1 | Not-determined | Genome-fragment | NA | NA | 0 |
| S7_vs_24695\|\|full | 2002 | No | NA | 4 | 0 | 0 | Low-quality | Genome-fragment | 3.42 | AAI-based (high-confidence) | 0 |
| S7_vs_57139\|\|full | 2002 | No | NA | 5 | 0 | 0 | Low-quality | Genome-fragment | 4.75 | AAI-based (medium-confidence) | 0 |
| S7_vs_58623\|\|full | 2002 | No | NA | 6 | 1 | 0 | Low-quality | Genome-fragment | 5.08 | AAI-based (high-confidence) | 0 |
| S7_vs_59575\|\|lt2gene | 2002 | No | NA | 1 | 1 | 0 | Low-quality | Genome-fragment | 3.87 | AAI-based (high-confidence) | 0 |
| S6_vs_k141_16604\|\|lt2gene | 2002 | No | NA | 2 | 2 | 0 | Low-quality | Genome-fragment | 4.57 | AAI-based (high-confidence) | 0 |
| S8_vs_k141_44610\|\|full | 2002 | No | NA | 3 | 0 | 2 | Low-quality | Genome-fragment | 1.27 | AAI-based (medium-confidence) | 0 |
| S5_vs_k141_59890\|\|full | 2530 | No | NA | 3 | 0 | 2 | Not-determined | Genome-fragment | NA | NA | 0 |
| S5_vs_k141_84574\|\|full | 36270 | No | NA | 41 | 6 | 2 | High-quality | High-quality | 96 | AAI-based (medium-confidence) | 0 |
| S5_vs_k141_78892\|\|full | 5521 | No | NA | 12 | 0 | 1 | Not-determined | Genome-fragment | NA | NA | 0 |
| S5_vs_k141_84580\|\|full | 1073 | No | NA | 2 | 0 | 0 | Not-determined | Genome-fragment | NA | NA | 0 |
| S8_vs_k141_94644\|\|full | 2002 | No | NA | 3 | 0 | 1 | Low-quality | Genome-fragment | 3.23 | AAI-based (medium-confidence) | 0 |
| S11_extracted_k141_52076 | 2003 | No | NA | 1 | 1 | 0 | Low-quality | Genome-fragment | 0.69 | HMM-based (lower-bound) | 0 |
| S1_extracted_k141_141441 | 2003 | No | NA | 2 | 0 | 0 | Low-quality | Genome-fragment | 4.89 | AAI-based (high-confidence) | 0 |
| S5_vs_k141_139130\|\|full | 4487 | No | NA | 5 | 0 | 0 | Not-determined | Genome-fragment | NA | NA | 0 |
| S1_vs_k141_85294\|\|full | 2003 | No | NA | 3 | 0 | 2 | Low-quality | Genome-fragment | 0.97 | AAI-based (medium-confidence) | 0 |
| S1_vs_k141_33138\|\|full | 2003 | No | NA | 7 | 1 | 0 | Low-quality | Genome-fragment | 4.03 | AAI-based (high-confidence) | 0 |
| S4_extracted_12575 | 2003 | No | NA | 5 | 0 | 0 | Low-quality | Genome-fragment | 4.38 | AAI-based (high-confidence) | 0 |
| S5_vs_k141_60028\|\|full | 1697 | No | NA | 2 | 0 | 2 | Not-determined | Genome-fragment | NA | NA | 0 |
| S6_vs_k141_28484\|\|full | 2003 | No | NA | 5 | 2 | 0 | Low-quality | Genome-fragment | 4.63 | AAI-based (medium-confidence) | 0 |
| S5_vs_k141_102558\|\|full | 1619 | No | NA | 2 | 0 | 0 | Not-determined | Genome-fragment | NA | NA | 0 |
| S11_vs_k141_69309\|\|full | 2004 | No | NA | 4 | 1 | 0 | Low-quality | Genome-fragment | 4.76 | AAI-based (medium-confidence) | 0 |
| S5_vs_k141_29911\|\|full | 1071 | No | NA | 3 | 0 | 0 | Not-determined | Genome-fragment | NA | NA | 0 |
| S1_extracted_k141_129436 | 2004 | No | NA | 5 | 1 | 0 | Low-quality | Genome-fragment | 4.17 | AAI-based (high-confidence) | 0 |
| S1_vs_k141_122821\|\|full | 2004 | No | NA | 3 | 0 | 1 | Low-quality | Genome-fragment | 3.86 | AAI-based (medium-confidence) | 0 |
| S5_vs_k141_132308\|\|full | 2004 | No | NA | 4 | 1 | 0 | Low-quality | Genome-fragment | 3.06 | AAI-based (medium-confidence) | 0 |
| S8_extracted_k141_62923 | 2004 | No | NA | 1 | 0 | 0 | Low-quality | Genome-fragment | 3.14 | AAI-based (medium-confidence) | 0 |
| S11_vs_k141_87095\|\|full | 2005 | No | NA | 5 | 1 | 0 | Low-quality | Genome-fragment | 4.82 | AAI-based (high-confidence) | 0 |
| S1_vs_k141_77264\|\|full | 2005 | No | NA | 2 | 0 | 1 | Low-quality | Genome-fragment | 2.76 | AAI-based (high-confidence) | 0 |
| S5_vs_k141_30074\|\|full | 1053 | No | NA | 3 | 0 | 0 | Not-determined | Genome-fragment | NA | NA | 0 |
| S16_vs_59858\|\|full | 2005 | No | NA | 3 | 0 | 1 | Low-quality | Genome-fragment | 1.55 | AAI-based (medium-confidence) | 0 |
| S12_vs_k141_85420\|\|full | 2005 | No | NA | 4 | 0 | 0 | Low-quality | Genome-fragment | 3.13 | AAI-based (high-confidence) | 0 |
| S5_vs_k141_60163\|\|full | 3720 | No | NA | 4 | 0 | 2 | Not-determined | Genome-fragment | NA | NA | 0 |
| S17_extracted_k141_116040 | 2006 | No | NA | 2 | 2 | 0 | Low-quality | Genome-fragment | 6.61 | AAI-based (high-confidence) | 0 |
| S5_vs_k141_84946\|\|full | 35711 | No | NA | 62 | 19 | 2 | High-quality | High-quality | 100 | AAI-based (high-confidence) | 0 |
| S5_vs_k141_30094\|\|full | 2323 | No | NA | 3 | 0 | 1 | Not-determined | Genome-fragment | NA | NA | 0 |
| S9_vs_k141_60754\|\|full | 2006 | No | NA | 3 | 0 | 0 | Low-quality | Genome-fragment | 4.78 | AAI-based (medium-confidence) | 0 |
| S5_vs_k141_84982\|\|full | 8391 | No | NA | 8 | 0 | 5 | Not-determined | Genome-fragment | NA | NA | 0 |
| S5_vs_k141_102781\|\|full | 1342 | No | NA | 3 | 0 | 1 | Not-determined | Genome-fragment | NA | NA | 0 |
| S5_vs_k141_60230\|\|full | 1423 | No | NA | 2 | 0 | 1 | Not-determined | Genome-fragment | NA | NA | 0 |
| S5_vs_k141_84996\|\|full | 1290 | No | NA | 2 | 0 | 0 | Not-determined | Genome-fragment | NA | NA | 0 |
| S11_extracted_k141_93996 | 2006 | No | NA | 5 | 0 | 1 | Low-quality | Genome-fragment | 2.2 | AAI-based (medium-confidence) | 0 |
| S13_extracted_108426 | 2006 | No | NA | 3 | 0 | 1 | Low-quality | Genome-fragment | 3.67 | AAI-based (medium-confidence) | 0 |

| | | | | | | | | | | | |
|---|---|---|---|---|---|---|---|---|---|---|---|
| S14_extracted_k141_26258 | 2006 | No | NA | 5 | 5 | 0 | Low-quality | Genome-fragment | 6.59 | AAI-based (high-confidence) | 0 |
| S14_extracted_k141_4769 | 2006 | No | NA | 4 | 1 | 0 | Low-quality | Genome-fragment | 5.2 | AAI-based (high-confidence) | 0 |
| S3_vs_k141_31299\|\|full | 2006 | No | NA | 2 | 0 | 0 | Low-quality | Genome-fragment | 1.08 | AAI-based (high-confidence) | 0 |
| S10_extracted_34322 | 2006 | No | NA | 2 | 0 | 1 | Low-quality | Genome-fragment | 2.49 | AAI-based (high-confidence) | 0 |
| S5_vs_k141_60280\|\|full | 10987 | No | NA | 10 | 0 | 5 | Not-determined | Genome-fragment | NA | NA | 0 |
| S4_extracted_9130 | 2006 | No | NA | 4 | 3 | 0 | Low-quality | Genome-fragment | 5 | AAI-based (high-confidence) | 0 |
| S5_vs_k141_139401\|\|full | 28466 | No | NA | 45 | 19 | 2 | Medium-quality | Genome-fragment | 80.85 | AAI-based (medium-confidence) | 0 |
| S5_vs_k141_60343\|\|full | 1421 | No | NA | 3 | 0 | 1 | Not-determined | Genome-fragment | NA | NA | 0 |
| S9_vs_k141_55710\|\|lt2gene | 2007 | No | NA | 2 | 2 | 0 | Low-quality | Genome-fragment | 5.7 | AAI-based (high-confidence) | 0 |
| S5_vs_k141_103055\|\|full | 5602 | No | NA | 6 | 0 | 1 | Not-determined | Genome-fragment | NA | NA | 0 |
| S5_vs_k141_139454\|\|full | 1642 | No | NA | 3 | 0 | 0 | Not-determined | Genome-fragment | NA | NA | 0 |
| S1_vs_k141_130216\|\|full | 2007 | No | NA | 3 | 2 | 0 | Low-quality | Genome-fragment | 5.59 | AAI-based (high-confidence) | 0 |
| S5_vs_k141_139530\|\|full | 6777 | No | NA | 7 | 0 | 4 | Not-determined | Genome-fragment | NA | NA | 0 |
| S5_extracted_k141_87837 | 2007 | No | NA | 4 | 2 | 0 | Low-quality | Genome-fragment | 2.76 | HMM-based (lower-bound) | 0 |
| S5_vs_k141_139554\|\|full | 3078 | No | NA | 4 | 0 | 1 | Not-determined | Genome-fragment | NA | NA | 0 |
| S5_vs_k141_139555\|\|full | 1193 | No | NA | 2 | 0 | 1 | Not-determined | Genome-fragment | NA | NA | 0 |
| S10_vs_82334\|\|full | 2007 | No | NA | 5 | 0 | 1 | Low-quality | Genome-fragment | 4.33 | AAI-based (medium-confidence) | 0 |
| S5_vs_k141_139636\|\|full | 4444 | No | NA | 7 | 0 | 1 | Not-determined | Genome-fragment | NA | NA | 0 |
| S8_extracted_k141_40746 | 2007 | No | NA | 3 | 0 | 0 | Low-quality | Genome-fragment | 4.26 | AAI-based (high-confidence) | 0 |
| S5_vs_k141_60507\|\|full | 27241 | No | NA | 38 | 28 | 0 | Medium-quality | Genome-fragment | 76.93 | AAI-based (high-confidence) | 0 |
| S5_vs_k141_60647\|\|full | 1876 | No | NA | 5 | 0 | 2 | Not-determined | Genome-fragment | NA | NA | 0 |
| S5_vs_k141_60667\|\|full | 2795 | No | NA | 4 | 0 | 1 | Not-determined | Genome-fragment | NA | NA | 0 |
| S5_vs_k141_24363\|\|0_partial | 10172 | No | NA | 11 | 0 | 6 | Not-determined | Genome-fragment | NA | NA | 0 |
| S8_vs_k141_135251\|\|full | 2007 | No | NA | 3 | 0 | 0 | Low-quality | Genome-fragment | 5.45 | AAI-based (medium-confidence) | 0 |
| S9_vs_k141_14100\|\|full | 2008 | No | NA | 3 | 0 | 0 | Low-quality | Genome-fragment | 3.49 | AAI-based (medium-confidence) | 0 |
| S5_vs_k141_55140\|\|0_partial | 11257 | No | NA | 8 | 0 | 3 | Not-determined | Genome-fragment | NA | NA | 0 |
| S5_vs_k141_61284\|\|0_partial | 11592 | No | NA | 16 | 0 | 2 | Not-determined | Genome-fragment | NA | NA | 0 |
| S5_extracted_k141_41356 | 2008 | No | NA | 1 | 0 | 1 | Low-quality | Genome-fragment | 0.56 | AAI-based (high-confidence) | 0 |
| S5_vs_k141_89744\|\|full | 2008 | No | NA | 6 | 3 | 0 | Low-quality | Genome-fragment | 6.04 | AAI-based (high-confidence) | 0 |
| S5_vs_k141_7444\|\|0_partial | 5735 | Yes | 3063 | 6 | 1 | 1 | Medium-quality | Genome-fragment | 73.61 | HMM-based (lower-bound) | 46.59 |
| S5_vs_k141_24905\|\|0_partial | 13230 | No | NA | 16 | 0 | 6 | Not-determined | Genome-fragment | NA | NA | 0 |
| S16_extracted_88075 | 2008 | No | NA | 3 | 2 | 0 | Low-quality | Genome-fragment | 3.94 | AAI-based (high-confidence) | 0 |
| S5_vs_k141_25023\|\|0_partial | 15558 | No | NA | 12 | 0 | 3 | Not-determined | Genome-fragment | NA | NA | 0 |
| S5_vs_k141_117564\|\|0_partial | 15713 | No | NA | 22 | 0 | 4 | Not-determined | Genome-fragment | NA | NA | 0 |
| S5_vs_k141_117625\|\|0_partial | 18877 | No | NA | 16 | 0 | 7 | Not-determined | Genome-fragment | NA | NA | 0 |
| S16_vs_170805\|\|full | 2008 | No | NA | 5 | 0 | 0 | Low-quality | Genome-fragment | 9.17 | AAI-based (medium-confidence) | 0 |
| S5_vs_k141_86751\|\|0_partial | 12590 | No | NA | 12 | 0 | 7 | Not-determined | Genome-fragment | NA | NA | 0 |
| S10_vs_19451\|\|full | 2008 | No | NA | 3 | 0 | 3 | Low-quality | Genome-fragment | 2.32 | AAI-based (medium-confidence) | 0 |
| S12_vs_k141_148103\|\|full | 2008 | No | NA | 4 | 1 | 0 | Low-quality | Genome-fragment | 4.79 | AAI-based (high-confidence) | 0 |
| S5_vs_k141_32820\|\|0_partial | 19334 | No | NA | 12 | 0 | 6 | Not-determined | Genome-fragment | NA | NA | 0 |
| S5_vs_k141_38380\|\|0_partial | 15548 | No | NA | 11 | 0 | 5 | Not-determined | Genome-fragment | NA | NA | 0 |
| S5_vs_k141_74199\|\|0_partial | 30526 | Yes | 25854 | 27 | 3 | 4 | Medium-quality | Genome-fragment | 62.06 | AAI-based (medium-confidence) | 15.3 |
| S5_vs_k141_93704\|\|0_partial | 14816 | No | NA | 15 | 0 | 6 | Not-determined | Genome-fragment | NA | NA | 0 |
| S5_vs_k141_51394\|\|0_partial | 33314 | No | NA | 25 | 0 | 21 | Not-determined | Genome-fragment | NA | NA | 0 |
| S6_vs_k141_47675\|\|full | 2008 | No | NA | 3 | 0 | 1 | Low-quality | Genome-fragment | 1.88 | AAI-based (high-confidence) | 0 |
| S5_vs_k141_141661\|\|0_partial | 13979 | No | NA | 11 | 0 | 2 | Not-determined | Genome-fragment | NA | NA | 0 |
| S17_vs_k141_31144\|\|full | 2009 | No | NA | 4 | 1 | 0 | Low-quality | Genome-fragment | 3.58 | AAI-based (medium-confidence) | 0 |
| S17_vs_k141_54326\|\|full | 2009 | No | NA | 5 | 1 | 0 | Low-quality | Genome-fragment | 4.8 | AAI-based (high-confidence) | 0 |
| S14_extracted_k141_78432 | 2009 | No | NA | 3 | 2 | 0 | Low-quality | Genome-fragment | 4.86 | AAI-based (high-confidence) | 0 |
| S1_extracted_k141_130411 | 2009 | No | NA | 1 | 0 | 1 | Low-quality | Genome-fragment | 1.01 | AAI-based (medium-confidence) | 0 |
| S3_extracted_k141_89613 | 2009 | No | NA | 5 | 2 | 0 | Low-quality | Genome-fragment | 5.11 | AAI-based (high-confidence) | 0 |
| S5_vs_k141_108624\|\|0_partial | 13968 | No | NA | 11 | 0 | 8 | Not-determined | Genome-fragment | NA | NA | 0 |
| S5_vs_k141_27258\|\|0_partial | 20858 | No | NA | 21 | 0 | 6 | Not-determined | Genome-fragment | NA | NA | 0 |
| S7_extracted_108648 | 2009 | No | NA | 2 | 0 | 0 | Low-quality | Genome-fragment | 3.55 | AAI-based (high-confidence) | 0 |
| S7_vs_69414\|\|full | 2009 | No | NA | 3 | 0 | 1 | Low-quality | Genome-fragment | 4.5 | AAI-based (medium-confidence) | 0 |
| S5_vs_k141_65490\|\|0_partial | 35560 | Yes | 19595 | 32 | 2 | 8 | High-quality | High-quality | 100 | AAI-based (high-confidence) | 44.9 |
| S10_vs_50362\|\|full | 2009 | No | NA | 4 | 1 | 0 | Low-quality | Genome-fragment | 3.16 | AAI-based (high-confidence) | 0 |
| S5_vs_k141_76500\|\|0_partial | 64407 | No | NA | 56 | 7 | 7 | Medium-quality | Genome-fragment | 85.82 | HMM-based (lower-bound) | 0 |
| S8_vs_k141_138749\|\|full | 2009 | No | NA | 5 | 1 | 0 | Low-quality | Genome-fragment | 4.79 | AAI-based (high-confidence) | 0 |
| S17_extracted_k141_126331 | 2010 | No | NA | 3 | 1 | 0 | Low-quality | Genome-fragment | 2.48 | AAI-based (high-confidence) | 0 |
| S17_vs_k141_77210\|\|full | 2010 | No | NA | 3 | 0 | 0 | Low-quality | Genome-fragment | 4.22 | AAI-based (medium-confidence) | 0 |
| S5_vs_k141_30407\|\|lt2gene | 1838 | No | NA | 1 | 0 | 1 | Not-determined | Genome-fragment | NA | NA | 0 |
| S3_extracted_k141_81117 | 2010 | No | NA | 4 | 0 | 0 | Low-quality | Genome-fragment | 3.46 | AAI-based (high-confidence) | 0 |
| S5_vs_k141_97302\|\|full | 2010 | No | NA | 4 | 0 | 0 | Low-quality | Genome-fragment | 4.54 | AAI-based (medium-confidence) | 0 |
| S6_vs_k141_45927\|\|full | 2010 | No | NA | 3 | 0 | 1 | Low-quality | Genome-fragment | 1.93 | AAI-based (high-confidence) | 0 |
| S8_vs_k141_160296\|\|full | 2010 | No | NA | 3 | 3 | 0 | Low-quality | Genome-fragment | 4.46 | AAI-based (high-confidence) | 0 |
| S17_vs_k141_148653\|\|full | 2011 | No | NA | 5 | 0 | 0 | Low-quality | Genome-fragment | 2.15 | AAI-based (high-confidence) | 0 |
| S9_vs_k141_76102\|\|full | 2011 | No | NA | 4 | 4 | 0 | Low-quality | Genome-fragment | 4.35 | AAI-based (high-confidence) | 0 |
| S11_vs_k141_77599\|\|full | 2011 | No | NA | 2 | 0 | 1 | Low-quality | Genome-fragment | 2.28 | AAI-based (medium-confidence) | 0 |
| S11_vs_k141_101689\|\|lt2gene | 2011 | No | NA | 1 | 1 | 0 | Low-quality | Genome-fragment | 1.39 | HMM-based (lower-bound) | 0 |
| S14_vs_k141_26901\|\|full | 2011 | No | NA | 5 | 2 | 0 | Low-quality | Genome-fragment | 5.48 | AAI-based (medium-confidence) | 0 |
| S3_extracted_k141_120291 | 2011 | No | NA | 5 | 0 | 3 | Low-quality | Genome-fragment | 4.39 | AAI-based (medium-confidence) | 0 |
| S7_vs_15526\|\|full | 2011 | No | NA | 3 | 0 | 1 | Low-quality | Genome-fragment | 0.45 | AAI-based (medium-confidence) | 0 |
| S16_vs_27411\|\|full | 2011 | No | NA | 3 | 0 | 1 | Low-quality | Genome-fragment | 3.29 | AAI-based (medium-confidence) | 0 |
| S10_vs_44037\|\|full | 2011 | No | NA | 4 | 0 | 3 | Low-quality | Genome-fragment | 2.3 | AAI-based (medium-confidence) | 0 |
| S12_extracted_k141_155477 | 2011 | No | NA | 1 | 0 | 1 | Low-quality | Genome-fragment | 1.28 | AAI-based (medium-confidence) | 0 |
| S8_vs_k141_104446\|\|full | 2011 | No | NA | 2 | 0 | 1 | Low-quality | Genome-fragment | 1.46 | AAI-based (high-confidence) | 0 |
| S17_vs_k141_99213\|\|full | 2012 | No | NA | 2 | 1 | 0 | Low-quality | Genome-fragment | 1.19 | AAI-based (high-confidence) | 0 |
| S11_vs_k141_48382\|\|full | 2012 | No | NA | 3 | 2 | 0 | Low-quality | Genome-fragment | 3.58 | AAI-based (high-confidence) | 0 |
| S13_extracted_54766 | 2012 | No | NA | 4 | 0 | 0 | Low-quality | Genome-fragment | 3.55 | AAI-based (medium-confidence) | 0 |
| S13_vs_20454\|\|full | 2012 | No | NA | 4 | 0 | 1 | Low-quality | Genome-fragment | 1.98 | AAI-based (medium-confidence) | 0 |
| S14_vs_k141_102673\|\|full | 2012 | No | NA | 3 | 0 | 1 | Low-quality | Genome-fragment | 4.06 | AAI-based (medium-confidence) | 0 |
| S5_vs_k141_107814\|\|full | 2012 | No | NA | 3 | 3 | 0 | Low-quality | Genome-fragment | 5.86 | AAI-based (medium-confidence) | 0 |
| S7_vs_102895\|\|full | 2012 | No | NA | 5 | 1 | 0 | Low-quality | Genome-fragment | 3.84 | AAI-based (high-confidence) | 0 |
| S11_vs_k141_52747\|\|full | 2013 | No | NA | 5 | 0 | 0 | Low-quality | Genome-fragment | 4.72 | AAI-based (medium-confidence) | 0 |
| S11_vs_k141_11540\|\|full | 2013 | No | NA | 3 | 0 | 0 | Low-quality | Genome-fragment | 4.49 | AAI-based (high-confidence) | 0 |
| S14_extracted_k141_25409 | 2013 | No | NA | 3 | 3 | 0 | Low-quality | Genome-fragment | 6.64 | AAI-based (high-confidence) | 0 |
| S4_extracted_7889 | 2013 | No | NA | 5 | 1 | 0 | Low-quality | Genome-fragment | 5.39 | AAI-based (high-confidence) | 0 |
| S17_vs_k141_171548\|\|full | 2014 | No | NA | 3 | 0 | 2 | Low-quality | Genome-fragment | 0.9 | AAI-based (medium-confidence) | 0 |
| S9_vs_k141_90864\|\|full | 2014 | No | NA | 2 | 0 | 0 | Low-quality | Genome-fragment | 4.11 | AAI-based (high-confidence) | 0 |
| S11_vs_k141_67844\|\|full | 2014 | No | NA | 4 | 0 | 0 | Low-quality | Genome-fragment | 4.69 | AAI-based (high-confidence) | 0 |
| S11_vs_k141_57048\|\|full | 2014 | No | NA | 4 | 1 | 0 | Low-quality | Genome-fragment | 5.03 | AAI-based (medium-confidence) | 0 |
| S14_vs_k141_45217\|\|full | 2014 | No | NA | 5 | 0 | 0 | Low-quality | Genome-fragment | 1.19 | AAI-based (medium-confidence) | 0 |
| S5_vs_k141_134276\|\|lt2gene | 3084 | No | NA | 1 | 0 | 0 | Not-determined | Genome-fragment | NA | NA | 0 |
| S3_vs_k141_113871\|\|full | 2014 | No | NA | 3 | 0 | 0 | Low-quality | Genome-fragment | 1.03 | AAI-based (high-confidence) | 0 |
| S5_vs_k141_134311\|\|lt2gene | 1661 | No | NA | 1 | 0 | 0 | Not-determined | Genome-fragment | NA | NA | 0 |
| S5_vs_k141_61546\|\|lt2gene | 1296 | No | NA | 2 | 0 | 0 | Not-determined | Genome-fragment | NA | NA | 0 |
| S7_vs_97151\|\|lt2gene | 2014 | No | NA | 2 | 2 | 0 | Low-quality | Genome-fragment | 0.77 | HMM-based (lower-bound) | 0 |
| S10_vs_57385\|\|full | 2014 | No | NA | 3 | 0 | 0 | Low-quality | Genome-fragment | 3.9 | AAI-based (medium-confidence) | 0 |
| S12_extracted_k141_67267 | 2014 | No | NA | 2 | 0 | 1 | Low-quality | Genome-fragment | 4.37 | AAI-based (medium-confidence) | 0 |
| S12_vs_k141_52171\|\|full | 2014 | No | NA | 3 | 0 | 2 | Low-quality | Genome-fragment | 6.43 | AAI-based (medium-confidence) | 0 |
| S12_vs_k141_176366\|\|lt2gene | 2014 | No | NA | 2 | 1 | 0 | Low-quality | Genome-fragment | 3.54 | HMM-based (lower-bound) | 0 |
| S8_extracted_k141_119552 | 2014 | No | NA | 1 | 1 | 0 | Low-quality | Genome-fragment | 1.39 | AAI-based (medium-confidence) | 0 |
| S9_extracted_k141_78432 | 2015 | No | NA | 5 | 1 | 0 | Low-quality | Genome-fragment | 4.96 | AAI-based (high-confidence) | 0 |
| S1_vs_k141_143264\|\|full | 2015 | No | NA | 3 | 2 | 0 | Low-quality | Genome-fragment | 3.22 | HMM-based (lower-bound) | 0 |
| S12_extracted_k141_153426 | 2015 | No | NA | 3 | 0 | 1 | Low-quality | Genome-fragment | 4.03 | AAI-based (medium-confidence) | 0 |
| S12_vs_k141_180598\|\|full | 2015 | No | NA | 4 | 1 | 0 | Low-quality | Genome-fragment | 5.18 | AAI-based (high-confidence) | 0 |
| S6_extracted_k141_30507 | 2015 | No | NA | 1 | 1 | 0 | Low-quality | Genome-fragment | 1.38 | HMM-based (lower-bound) | 0 |
| S8_vs_k141_102517\|\|full | 2015 | No | NA | 5 | 3 | 0 | Low-quality | Genome-fragment | 5.46 | AAI-based (medium-confidence) | 0 |
| S17_vs_k141_93321\|\|full | 2016 | Yes | 1451 | 4 | 1 | 1 | Low-quality | Genome-fragment | 2.09 | AAI-based (medium-confidence) | 28.03 |
| S5_vs_k141_2215\|\|lt2gene | 1940 | No | NA | 1 | 0 | 0 | Not-determined | Genome-fragment | NA | NA | 0 |
| S14_extracted_k141_100342 | 2016 | No | NA | 7 | 7 | 0 | Low-quality | Genome-fragment | 6.66 | AAI-based (high-confidence) | 0 |
| S14_vs_k141_85272\|\|full | 2016 | No | NA | 5 | 1 | 0 | Low-quality | Genome-fragment | 4.18 | AAI-based (high-confidence) | 0 |
| S5_vs_k141_17092\|\|full | 2016 | No | NA | 3 | 0 | 0 | Low-quality | Genome-fragment | 3.85 | AAI-based (high-confidence) | 0 |
| S7_vs_89006\|\|full | 2016 | No | NA | 4 | 0 | 0 | Low-quality | Genome-fragment | 2.97 | AAI-based (high-confidence) | 0 |
| S10_vs_57054\|\|full | 2016 | No | NA | 5 | 0 | 0 | Low-quality | Genome-fragment | 2.48 | AAI-based (high-confidence) | 0 |
| S5_vs_k141_62439\|\|lt2gene | 1034 | No | NA | 1 | 0 | 0 | Not-determined | Genome-fragment | NA | NA | 0 |
| S2_extracted_k141_16277 | 2016 | No | NA | 5 | 0 | 0 | Low-quality | Genome-fragment | 4.71 | AAI-based (high-confidence) | 0 |
| S6_vs_k141_24999\|\|full | 2016 | No | NA | 4 | 0 | 0 | Low-quality | Genome-fragment | 3.82 | AAI-based (high-confidence) | 0 |
| S17_extracted_k141_105439 | 2017 | No | NA | 1 | 0 | 0 | Low-quality | Genome-fragment | 0.88 | AAI-based (medium-confidence) | 0 |
| S17_vs_k141_10946\|\|full | 2017 | No | NA | 2 | 0 | 2 | Low-quality | Genome-fragment | 3.42 | AAI-based (medium-confidence) | 0 |
| S13_vs_17497\|\|full | 2017 | No | NA | 5 | 1 | 0 | Low-quality | Genome-fragment | 3.19 | AAI-based (high-confidence) | 0 |
| S1_vs_k141_63791\|\|lt2gene | 2017 | No | NA | 1 | 0 | 0 | Low-quality | Genome-fragment | 4.77 | AAI-based (medium-confidence) | 0 |
| S5_vs_k141_56056\|\|lt2gene | 1235 | No | NA | 2 | 0 | 0 | Not-determined | Genome-fragment | NA | NA | 0 |
| S5_vs_k141_124261\|\|lt2gene | 1678 | No | NA | 1 | 0 | 0 | Not-determined | Genome-fragment | NA | NA | 0 |
| S5_vs_k141_13363\|\|full | 2017 | No | NA | 4 | 4 | 0 | Low-quality | Genome-fragment | 3.02 | HMM-based (lower-bound) | 0 |
| S5_vs_k141_105977\|\|lt2gene | 1004 | No | NA | 1 | 0 | 0 | Not-determined | Genome-fragment | NA | NA | 0 |
| S5_vs_k141_118353\|\|lt2gene | 1177 | No | NA | 1 | 0 | 0 | Not-determined | Genome-fragment | NA | NA | 0 |
| S16_extracted_162565 | 2017 | No | NA | 5 | 0 | 0 | Low-quality | Genome-fragment | 3.54 | AAI-based (high-confidence) | 0 |
| S10_extracted_58834 | 2017 | No | NA | 2 | 0 | 2 | Low-quality | Genome-fragment | 1.23 | AAI-based (medium-confidence) | 0 |
| S5_vs_k141_20643\|\|lt2gene | 1464 | No | NA | 1 | 0 | 0 | Not-determined | Genome-fragment | NA | NA | 0 |
| S12_vs_k141_72744\|\|full | 2017 | No | NA | 2 | 0 | 0 | Low-quality | Genome-fragment | 3.99 | AAI-based (medium-confidence) | 0 |
| S8_extracted_k141_68617 | 2017 | No | NA | 3 | 0 | 1 | Low-quality | Genome-fragment | 3.78 | AAI-based (high-confidence) | 0 |
| S9_vs_k141_2747\|\|full | 2018 | No | NA | 2 | 1 | 0 | Low-quality | Genome-fragment | 4.56 | AAI-based (high-confidence) | 0 |
| S9_vs_k141_83671\|\|lt2gene | 2018 | No | NA | 2 | 1 | 0 | Low-quality | Genome-fragment | 4.52 | AAI-based (medium-confidence) | 0 |
| S5_vs_k141_136355\|\|full | 2018 | No | NA | 2 | 0 | 1 | Low-quality | Genome-fragment | 2.44 | AAI-based (high-confidence) | 0 |
| S7_vs_63719\|\|full | 2018 | No | NA | 3 | 0 | 1 | Low-quality | Genome-fragment | 1.06 | AAI-based (medium-confidence) | 0 |
| S12_vs_k141_114198\|\|full | 2018 | No | NA | 4 | 1 | 0 | Low-quality | Genome-fragment | 0.18 | HMM-based (lower-bound) | 0 |
| S11_vs_k141_72737\|\|full | 2019 | No | NA | 3 | 0 | 1 | Low-quality | Genome-fragment | 3.07 | AAI-based (medium-confidence) | 0 |
| S5_extracted_k141_33313 | 2019 | No | NA | 4 | 1 | 0 | Low-quality | Genome-fragment | 5.14 | AAI-based (high-confidence) | 0 |
| S5_vs_k141_51816\|\|lt2gene | 1056 | No | NA | 1 | 0 | 0 | Not-determined | Genome-fragment | NA | NA | 0 |
| S5_vs_k141_94053\|\|lt2gene | 1068 | No | NA | 1 | 0 | 0 | Not-determined | Genome-fragment | NA | NA | 0 |
| S7_vs_68702\|\|full | 2019 | No | NA | 2 | 0 | 1 | Low-quality | Genome-fragment | 0.51 | AAI-based (medium-confidence) | 0 |
| S10_vs_67549\|\|full | 2019 | No | NA | 3 | 1 | 0 | Low-quality | Genome-fragment | 0.86 | AAI-based (high-confidence) | 0 |
| S8_extracted_k141_74649 | 2019 | No | NA | 2 | 0 | 0 | Low-quality | Genome-fragment | 6.71 | AAI-based (high-confidence) | 0 |
| S1_vs_k141_98032\|\|full | 2020 | No | NA | 5 | 2 | 0 | Low-quality | Genome-fragment | 4.65 | AAI-based (high-confidence) | 0 |
| S3_vs_k141_33954\|\|full | 2020 | No | NA | 4 | 0 | 0 | Low-quality | Genome-fragment | 2.87 | AAI-based (high-confidence) | 0 |
| S5_vs_k141_393\|\|full | 2020 | No | NA | 2 | 0 | 0 | Low-quality | Genome-fragment | 2.06 | AAI-based (medium-confidence) | 0 |
| S7_vs_497\|\|full | 2020 | No | NA | 5 | 0 | 0 | Low-quality | Genome-fragment | 2.97 | AAI-based (high-confidence) | 0 |
| S7_vs_102671\|\|full | 2020 | No | NA | 3 | 0 | 2 | Low-quality | Genome-fragment | 4.15 | AAI-based (medium-confidence) | 0 |
| S16_vs_170213\|\|full | 2020 | No | NA | 6 | 2 | 0 | Low-quality | Genome-fragment | 3.91 | AAI-based (high-confidence) | 0 |
| S12_vs_k141_117972\|\|full | 2020 | No | NA | 3 | 1 | 0 | Low-quality | Genome-fragment | 3.3 | HMM-based (lower-bound) | 0 |
| S2_extracted_k141_14988 | 2020 | No | NA | 9 | 9 | 0 | Low-quality | Genome-fragment | 6.67 | AAI-based (high-confidence) | 0 |
| S9_vs_k141_45624\|\|full | 2021 | No | NA | 4 | 0 | 2 | Low-quality | Genome-fragment | 2.75 | AAI-based (high-confidence) | 0 |
| S13_vs_52521\|\|full | 2021 | No | NA | 4 | 0 | 1 | Low-quality | Genome-fragment | 3.56 | AAI-based (high-confidence) | 0 |
| S14_vs_k141_59267\|\|full | 2021 | No | NA | 4 | 1 | 0 | Low-quality | Genome-fragment | 1.25 | HMM-based (lower-bound) | 0 |
| S5_vs_k141_52337\|\|lt2gene | 1474 | No | NA | 2 | 0 | 1 | Not-determined | Genome-fragment | NA | NA | 0 |
| S3_vs_k141_109761\|\|full | 2021 | No | NA | 3 | 0 | 0 | Low-quality | Genome-fragment | 1.08 | AAI-based (medium-confidence) | 0 |
| S5_vs_k141_34126\|\|lt2gene | 1312 | No | NA | 1 | 0 | 0 | Not-determined | Genome-fragment | NA | NA | 0 |
| S7_vs_124950\|\|full | 2021 | No | NA | 8 | 0 | 0 | Low-quality | Genome-fragment | 5.8 | AAI-based (high-confidence) | 0 |
| S10_vs_105091\|\|full | 2021 | No | NA | 5 | 4 | 0 | Low-quality | Genome-fragment | 3.06 | AAI-based (high-confidence) | 0 |
| S14_vs_k141_78961\|\|full | 2022 | No | NA | 4 | 0 | 0 | Low-quality | Genome-fragment | 15.92 | AAI-based (high-confidence) | 0 |
| S3_vs_k141_103971\|\|full | 2022 | No | NA | 7 | 0 | 0 | Low-quality | Genome-fragment | 6.05 | AAI-based (high-confidence) | 0 |
| S7_vs_88901\|\|full | 2022 | No | NA | 4 | 1 | 0 | Low-quality | Genome-fragment | 4.61 | AAI-based (high-confidence) | 0 |
| S7_vs_108795\|\|full | 2022 | No | NA | 6 | 1 | 0 | Low-quality | Genome-fragment | 4.94 | AAI-based (medium-confidence) | 0 |
| S5_vs_k141_63896\|\|lt2gene | 2307 | No | NA | 1 | 0 | 0 | Not-determined | Genome-fragment | NA | NA | 0 |
| S7_vs_122405\|\|full | 2022 | No | NA | 3 | 1 | 0 | Low-quality | Genome-fragment | 3.99 | HMM-based (lower-bound) | 0 |
| S16_vs_41334\|\|full | 2022 | No | NA | 3 | 0 | 1 | Low-quality | Genome-fragment | 2.44 | AAI-based (medium-confidence) | 0 |
| S10_extracted_23347 | 2022 | No | NA | 2 | 0 | 1 | Low-quality | Genome-fragment | 3.36 | AAI-based (high-confidence) | 0 |
| S5_vs_k141_88254\|\|lt2gene | 1447 | No | NA | 1 | 0 | 0 | Not-determined | Genome-fragment | NA | NA | 0 |
| S12_vs_k141_131880\|\|full | 2022 | No | NA | 3 | 2 | 0 | Low-quality | Genome-fragment | 3.13 | HMM-based (lower-bound) | 0 |
| S8_extracted_k141_118417 | 2022 | No | NA | 2 | 0 | 2 | Low-quality | Genome-fragment | 0.74 | AAI-based (medium-confidence) | 0 |
| S5_vs_k141_74897\|\|lt2gene | 1154 | No | NA | 1 | 0 | 0 | Not-determined | Genome-fragment | NA | NA | 0 |
| S9_vs_k141_87334\|\|full | 2023 | No | NA | 2 | 0 | 0 | Low-quality | Genome-fragment | 2.5 | AAI-based (high-confidence) | 0 |
| S17_extracted_k141_5427 | 2024 | No | NA | 4 | 0 | 3 | Low-quality | Genome-fragment | 0.61 | AAI-based (medium-confidence) | 0 |
| S5_vs_k141_107853\|\|lt2gene | 1413 | No | NA | 1 | 0 | 0 | Not-determined | Genome-fragment | NA | NA | 0 |
| S1_vs_k141_83913\|\|full | 2024 | No | NA | 3 | 0 | 0 | Low-quality | Genome-fragment | 5.65 | AAI-based (high-confidence) | 0 |
| S5_vs_k141_10091\|\|lt2gene | 1531 | No | NA | 1 | 0 | 0 | Not-determined | Genome-fragment | NA | NA | 0 |
| S3_extracted_k141_121671 | 2024 | No | NA | 3 | 0 | 1 | Low-quality | Genome-fragment | 2.25 | AAI-based (high-confidence) | 0 |
| S3_extracted_k141_100649 | 2024 | No | NA | 3 | 0 | 1 | Low-quality | Genome-fragment | 2.56 | AAI-based (high-confidence) | 0 |
| S3_vs_k141_87656\|\|full | 2024 | No | NA | 5 | 1 | 0 | Low-quality | Genome-fragment | 3.24 | HMM-based (lower-bound) | 0 |
| S5_vs_k141_114094\|\|full | 2024 | No | NA | 5 | 1 | 0 | Low-quality | Genome-fragment | 5.39 | AAI-based (high-confidence) | 0 |
| S16_extracted_24269 | 2024 | No | NA | 3 | 0 | 0 | Low-quality | Genome-fragment | 3.67 | AAI-based (medium-confidence) | 0 |
| S2_extracted_k141_12705 | 2024 | No | NA | 2 | 1 | 0 | Low-quality | Genome-fragment | 4.82 | HMM-based (lower-bound) | 0 |
| S5_vs_k141_108224\|\|lt2gene | 1395 | No | NA | 1 | 0 | 0 | Not-determined | Genome-fragment | NA | NA | 0 |
| S4_vs_VIRSorter_13276-cat_2 | 2024 | No | NA | 5 | 5 | 0 | Low-quality | Genome-fragment | 6.63 | AAI-based (high-confidence) | 0 |
| S5_vs_k141_99997\|\|lt2gene | 1232 | No | NA | 2 | 0 | 1 | Not-determined | Genome-fragment | NA | NA | 0 |
| S8_vs_k141_91136\|\|full | 2024 | No | NA | 4 | 0 | 2 | Low-quality | Genome-fragment | 2.17 | AAI-based (medium-confidence) | 0 |
| S1_vs_k141_81008\|\|full | 2025 | No | NA | 3 | 1 | 0 | Low-quality | Genome-fragment | 6.86 | HMM-based (lower-bound) | 0 |
| S5_extracted_k141_1865 | 2025 | No | NA | 7 | 5 | 0 | Low-quality | Genome-fragment | 5.02 | AAI-based (high-confidence) | 0 |
| S5_vs_k141_117774\|\|full | 2025 | No | NA | 4 | 0 | 0 | Low-quality | Genome-fragment | 5.36 | AAI-based (high-confidence) | 0 |
| S7_vs_23336\|\|full | 2025 | No | NA | 3 | 0 | 2 | Low-quality | Genome-fragment | 2.16 | AAI-based (medium-confidence) | 0 |
| S12_vs_k141_156522\|\|full | 2025 | No | NA | 4 | 0 | 0 | Low-quality | Genome-fragment | 4.2 | AAI-based (medium-confidence) | 0 |
| S9_vs_k141_9285\|\|full | 2026 | No | NA | 5 | 3 | 0 | Low-quality | Genome-fragment | 3.4 | AAI-based (medium-confidence) | 0 |
| S1_vs_k141_68419\|\|lt2gene | 2026 | No | NA | 1 | 0 | 0 | Low-quality | Genome-fragment | 4.43 | AAI-based (high-confidence) | 0 |
| S5_extracted_k141_29959 | 2026 | No | NA | 5 | 0 | 0 | Low-quality | Genome-fragment | 4.1 | AAI-based (medium-confidence) | 0 |
| S7_vs_65491\|\|lt2gene | 2026 | No | NA | 2 | 1 | 0 | Low-quality | Genome-fragment | 6.56 | AAI-based (high-confidence) | 0 |
| S12_vs_k141_121004\|\|full | 2026 | No | NA | 3 | 1 | 0 | Low-quality | Genome-fragment | 1.43 | AAI-based (medium-confidence) | 0 |
| S6_extracted_k141_56470 | 2026 | No | NA | 2 | 0 | 1 | Low-quality | Genome-fragment | 1.14 | AAI-based (high-confidence) | 0 |
| S6_vs_k141_5597\|\|full | 2026 | No | NA | 4 | 0 | 0 | Low-quality | Genome-fragment | 0.64 | AAI-based (medium-confidence) | 0 |
| S17_extracted_k141_160657 | 2027 | No | NA | 2 | 0 | 2 | Low-quality | Genome-fragment | 1.07 | AAI-based (medium-confidence) | 0 |
| S14_extracted_k141_69318 | 2027 | No | NA | 2 | 0 | 2 | Low-quality | Genome-fragment | 0.46 | AAI-based (medium-confidence) | 0 |
| S7_vs_2699\|\|full | 2027 | No | NA | 7 | 1 | 0 | Low-quality | Genome-fragment | 4.71 | AAI-based (medium-confidence) | 0 |
| S5_vs_k141_100942\|\|lt2gene | 1752 | No | NA | 1 | 1 | 0 | Not-determined | Genome-fragment | NA | NA | 0 |
| S7_vs_137427\|\|full | 2027 | No | NA | 3 | 0 | 0 | Low-quality | Genome-fragment | 3.8 | AAI-based (medium-confidence) | 0 |
| S16_vs_7832\|\|full | 2027 | No | NA | 3 | 0 | 2 | Low-quality | Genome-fragment | 0.96 | AAI-based (medium-confidence) | 0 |
| S10_vs_62408\|\|full | 2027 | No | NA | 3 | 0 | 0 | Low-quality | Genome-fragment | 5.42 | AAI-based (high-confidence) | 0 |
| S5_vs_k141_90353\|\|lt2gene | 1107 | No | NA | 1 | 0 | 0 | Not-determined | Genome-fragment | NA | NA | 0 |
| S11_extracted_k141_66647 | 2028 | No | NA | 1 | 0 | 0 | Low-quality | Genome-fragment | 1.24 | AAI-based (medium-confidence) | 0 |
| S11_vs_k141_9090\|\|full | 2028 | No | NA | 5 | 2 | 0 | Low-quality | Genome-fragment | 4.98 | AAI-based (medium-confidence) | 0 |
| S11_vs_k141_10578\|\|full | 2028 | No | NA | 5 | 0 | 0 | Low-quality | Genome-fragment | 5.92 | AAI-based (high-confidence) | 0 |
| S5_vs_k141_137601\|\|lt2gene | 1468 | No | NA | 2 | 0 | 1 | Not-determined | Genome-fragment | NA | NA | 0 |
| S5_vs_k141_137619\|\|lt2gene | 1162 | No | NA | 1 | 0 | 1 | Not-determined | Genome-fragment | NA | NA | 0 |
| S3_vs_k141_108645\|\|full | 2028 | No | NA | 6 | 2 | 0 | Low-quality | Genome-fragment | 5.03 | AAI-based (high-confidence) | 0 |
| S5_vs_k141_48240\|\|lt2gene | 1041 | No | NA | 1 | 0 | 0 | Not-determined | Genome-fragment | NA | NA | 0 |
| S5_vs_k141_72359\|\|lt2gene | 1319 | No | NA | 2 | 0 | 0 | Not-determined | Genome-fragment | NA | NA | 0 |
| S5_vs_k141_137058\|\|full | 2028 | No | NA | 3 | 0 | 0 | Low-quality | Genome-fragment | 1.09 | AAI-based (medium-confidence) | 0 |
| S16_extracted_123353 | 2028 | No | NA | 2 | 0 | 2 | Low-quality | Genome-fragment | 1.65 | AAI-based (high-confidence) | 0 |
| S14_extracted_k141_4818 | 2029 | No | NA | 7 | 0 | 1 | Low-quality | Genome-fragment | 3.55 | AAI-based (high-confidence) | 0 |
| S14_vs_k141_83995\|\|full | 2029 | No | NA | 7 | 0 | 0 | Low-quality | Genome-fragment | 5.28 | AAI-based (high-confidence) | 0 |
| S16_extracted_159243 | 2029 | No | NA | 2 | 0 | 0 | Low-quality | Genome-fragment | 2.51 | AAI-based (high-confidence) | 0 |
| S10_vs_43284\|\|full | 2029 | No | NA | 4 | 0 | 1 | Low-quality | Genome-fragment | 0.61 | AAI-based (medium-confidence) | 0 |
| S9_vs_k141_19864\|\|full | 2030 | No | NA | 3 | 0 | 0 | Low-quality | Genome-fragment | 5.55 | AAI-based (high-confidence) | 0 |
| S1_vs_k141_24019\|\|full | 2030 | No | NA | 6 | 1 | 0 | Low-quality | Genome-fragment | 5.15 | AAI-based (high-confidence) | 0 |
| S5_vs_k141_29333\|\|lt2gene | 2366 | No | NA | 2 | 0 | 1 | Not-determined | Genome-fragment | NA | NA | 0 |
| S5_vs_k141_55715\|\|full | 2030 | No | NA | 5 | 4 | 0 | Low-quality | Genome-fragment | 3.32 | AAI-based (high-confidence) | 0 |
| S5_vs_k141_59626\|\|lt2gene | 1655 | No | NA | 1 | 0 | 0 | Not-determined | Genome-fragment | NA | NA | 0 |
| S7_extracted_751 | 2030 | No | NA | 4 | 0 | 2 | Low-quality | Genome-fragment | 1.37 | AAI-based (medium-confidence) | 0 |
| S12_vs_k141_169828\|\|full | 2030 | No | NA | 4 | 0 | 0 | Low-quality | Genome-fragment | 3.99 | AAI-based (high-confidence) | 0 |
| S12_vs_k141_15561\|\|full | 2030 | No | NA | 3 | 0 | 2 | Low-quality | Genome-fragment | 1.61 | AAI-based (medium-confidence) | 0 |
| S12_vs_k141_168927\|\|lt2gene | 2030 | No | NA | 2 | 1 | 0 | Low-quality | Genome-fragment | 4.88 | AAI-based (medium-confidence) | 0 |
| S11_extracted_k141_3084 | 2031 | No | NA | 3 | 0 | 0 | Low-quality | Genome-fragment | 1.28 | AAI-based (medium-confidence) | 0 |
| S16_extracted_121594 | 2031 | No | NA | 5 | 1 | 0 | Low-quality | Genome-fragment | 3.93 | AAI-based (high-confidence) | 0 |
| S7_extracted_220 | 1498 | No | NA | 1 | 0 | 1 | Not-determined | Genome-fragment | NA | NA | 0 |
| S16_vs_78512\|\|full | 2031 | No | NA | 3 | 0 | 2 | Low-quality | Genome-fragment | 1.45 | AAI-based (high-confidence) | 0 |
| S7_extracted_364 | 1697 | No | NA | 1 | 0 | 1 | Not-determined | Genome-fragment | NA | NA | 0 |
| S7_extracted_370 | 4615 | No | NA | 6 | 0 | 3 | Not-determined | Genome-fragment | NA | NA | 0 |
| S7_extracted_388 | 2375 | No | NA | 2 | 0 | 1 | Not-determined | Genome-fragment | NA | NA | 0 |
| S7_extracted_430 | 1102 | No | NA | 1 | 0 | 0 | Not-determined | Genome-fragment | NA | NA | 0 |
| S10_extracted_90660 | 2031 | No | NA | 5 | 0 | 0 | Low-quality | Genome-fragment | 2.52 | AAI-based (high-confidence) | 0 |
| S1_vs_k141_133765\|\|full | 2032 | No | NA | 4 | 1 | 0 | Low-quality | Genome-fragment | 4.18 | AAI-based (high-confidence) | 0 |
| S1_vs_k141_156096\|\|full | 2032 | No | NA | 3 | 0 | 1 | Low-quality | Genome-fragment | 4.6 | AAI-based (medium-confidence) | 0 |
| S7_vs_99993\|\|full | 2032 | No | NA | 4 | 0 | 0 | Low-quality | Genome-fragment | 6.26 | AAI-based (high-confidence) | 0 |
| S10_extracted_57053 | 2032 | No | NA | 2 | 0 | 0 | Low-quality | Genome-fragment | 1.53 | AAI-based (high-confidence) | 0 |
| S7_extracted_700 | 1396 | No | NA | 2 | 0 | 1 | Not-determined | Genome-fragment | NA | NA | 0 |
| S7_extracted_748 | 2238 | No | NA | 3 | 0 | 0 | Not-determined | Genome-fragment | NA | NA | 0 |
| S2_extracted_k141_12999 | 2032 | No | NA | 6 | 1 | 0 | Low-quality | Genome-fragment | 5.22 | AAI-based (medium-confidence) | 0 |
| S8_extracted_k141_191556 | 2032 | No | NA | 7 | 7 | 0 | Low-quality | Genome-fragment | 6.68 | AAI-based (high-confidence) | 0 |
| S9_vs_k141_32243\|\|full | 2033 | No | NA | 2 | 0 | 2 | Low-quality | Genome-fragment | 1.44 | AAI-based (high-confidence) | 0 |
| S7_extracted_1108 | 4484 | No | NA | 8 | 0 | 1 | Not-determined | Genome-fragment | NA | NA | 0 |
| S7_extracted_1124 | 1115 | No | NA | 1 | 0 | 0 | Not-determined | Genome-fragment | NA | NA | 0 |
| S7_extracted_1194 | 4740 | No | NA | 5 | 0 | 1 | Not-determined | Genome-fragment | NA | NA | 0 |
| S9_vs_k141_79493\|\|full | 2033 | No | NA | 3 | 0 | 0 | Low-quality | Genome-fragment | 4.6 | AAI-based (medium-confidence) | 0 |
| S7_extracted_1483 | 1187 | No | NA | 2 | 0 | 1 | Not-determined | Genome-fragment | NA | NA | 0 |
| S7_extracted_1646 | 1479 | No | NA | 3 | 0 | 0 | Not-determined | Genome-fragment | NA | NA | 0 |
| S7_extracted_1652 | 3675 | No | NA | 5 | 0 | 2 | Not-determined | Genome-fragment | NA | NA | 0 |
| S11_extracted_k141_11701 | 2033 | No | NA | 5 | 0 | 0 | Low-quality | Genome-fragment | 4.54 | AAI-based (medium-confidence) | 0 |
| S7_extracted_1792 | 5243 | No | NA | 6 | 0 | 0 | Not-determined | Genome-fragment | NA | NA | 0 |
| S7_extracted_1811 | 2588 | No | NA | 2 | 0 | 0 | Not-determined | Genome-fragment | NA | NA | 0 |
| S11_vs_k141_75480\|\|full | 2033 | No | NA | 4 | 1 | 0 | Low-quality | Genome-fragment | 6.09 | HMM-based (lower-bound) | 0 |
| S16_vs_11647\|\|full | 2033 | No | NA | 3 | 3 | 0 | Low-quality | Genome-fragment | 4.68 | AAI-based (high-confidence) | 0 |
| S7_extracted_2096 | 3428 | No | NA | 3 | 0 | 3 | Not-determined | Genome-fragment | NA | NA | 0 |
| S7_extracted_2234 | 2351 | No | NA | 4 | 0 | 3 | Not-determined | Genome-fragment | NA | NA | 0 |
| S10_vs_51415\|\|full | 2033 | No | NA | 5 | 0 | 0 | Low-quality | Genome-fragment | 3.2 | AAI-based (medium-confidence) | 0 |
| S7_extracted_2390 | 5170 | No | NA | 5 | 0 | 5 | Not-determined | Genome-fragment | NA | NA | 0 |
| S7_extracted_2703 | 1571 | No | NA | 3 | 0 | 2 | Not-determined | Genome-fragment | NA | NA | 0 |
| S8_extracted_k141_162884 | 2033 | No | NA | 2 | 1 | 0 | Low-quality | Genome-fragment | 4.67 | AAI-based (high-confidence) | 0 |
| S7_extracted_2846 | 1053 | No | NA | 1 | 0 | 1 | Not-determined | Genome-fragment | NA | NA | 0 |
| S7_extracted_2873 | 1285 | No | NA | 3 | 0 | 1 | Not-determined | Genome-fragment | NA | NA | 0 |
| S7_extracted_2876 | 12800 | No | NA | 12 | 0 | 9 | Not-determined | Genome-fragment | NA | NA | 0 |
| S7_extracted_2903 | 1062 | No | NA | 2 | 0 | 2 | Not-determined | Genome-fragment | NA | NA | 0 |
| S7_extracted_3040 | 4378 | No | NA | 9 | 0 | 0 | Not-determined | Genome-fragment | NA | NA | 0 |
| S3_vs_k141_21012\|\|full | 2034 | No | NA | 3 | 1 | 0 | Low-quality | Genome-fragment | 3.21 | AAI-based (high-confidence) | 0 |
| S7_extracted_3118 | 1260 | No | NA | 1 | 0 | 0 | Not-determined | Genome-fragment | NA | NA | 0 |
| S7_vs_64849\|\|full | 2034 | No | NA | 4 | 1 | 0 | Low-quality | Genome-fragment | 4.52 | AAI-based (medium-confidence) | 0 |
| S7_extracted_3255 | 1235 | No | NA | 1 | 0 | 0 | Not-determined | Genome-fragment | NA | NA | 0 |
| S7_extracted_3298 | 1233 | No | NA | 1 | 0 | 1 | Not-determined | Genome-fragment | NA | NA | 0 |
| S7_extracted_3482 | 1174 | No | NA | 1 | 0 | 0 | Not-determined | Genome-fragment | NA | NA | 0 |
| S7_extracted_3593 | 2177 | No | NA | 2 | 0 | 2 | Not-determined | Genome-fragment | NA | NA | 0 |
| S7_extracted_3719 | 1403 | No | NA | 2 | 0 | 1 | Not-determined | Genome-fragment | NA | NA | 0 |
| S16_extracted_60770 | 2034 | No | NA | 2 | 0 | 0 | Low-quality | Genome-fragment | 3.24 | AAI-based (high-confidence) | 0 |
| S7_extracted_3785 | 1032 | No | NA | 1 | 0 | 1 | Not-determined | Genome-fragment | NA | NA | 0 |
| S7_extracted_3815 | 8161 | No | NA | 8 | 0 | 7 | Not-determined | Genome-fragment | NA | NA | 0 |
| S16_vs_53620\|\|full | 2034 | No | NA | 4 | 1 | 0 | Low-quality | Genome-fragment | 3.26 | HMM-based (lower-bound) | 0 |
| S7_extracted_4093 | 1158 | No | NA | 2 | 0 | 0 | Not-determined | Genome-fragment | NA | NA | 0 |
| S12_vs_k141_126316\|\|full | 2034 | No | NA | 4 | 1 | 0 | Low-quality | Genome-fragment | 5.59 | HMM-based (lower-bound) | 0 |
| S7_extracted_4294 | 2938 | No | NA | 2 | 0 | 2 | Not-determined | Genome-fragment | NA | NA | 0 |
| S7_extracted_4407 | 1285 | No | NA | 3 | 0 | 0 | Not-determined | Genome-fragment | NA | NA | 0 |
| S12_vs_k141_35171\|\|full | 2034 | No | NA | 2 | 0 | 2 | Low-quality | Genome-fragment | 1.02 | AAI-based (medium-confidence) | 0 |
| S7_extracted_4720 | 5635 | No | NA | 3 | 0 | 0 | Not-determined | Genome-fragment | NA | NA | 0 |
| S7_extracted_4803 | 3305 | No | NA | 4 | 0 | 1 | Not-determined | Genome-fragment | NA | NA | 0 |
| S7_extracted_4815 | 2142 | No | NA | 3 | 0 | 0 | Not-determined | Genome-fragment | NA | NA | 0 |
| S7_extracted_4819 | 3199 | No | NA | 2 | 0 | 2 | Not-determined | Genome-fragment | NA | NA | 0 |
| S7_extracted_4887 | 2031 | No | NA | 2 | 0 | 2 | Not-determined | Genome-fragment | NA | NA | 0 |
| S7_extracted_4999 | 7560 | No | NA | 5 | 0 | 4 | Not-determined | Genome-fragment | NA | NA | 0 |
| S8_extracted_k141_199617 | 2034 | No | NA | 4 | 0 | 1 | Low-quality | Genome-fragment | 2.61 | AAI-based (medium-confidence) | 0 |
| S9_extracted_k141_1798 | 2035 | No | NA | 2 | 2 | 0 | Low-quality | Genome-fragment | 3.36 | AAI-based (medium-confidence) | 0 |
| S7_extracted_5375 | 1420 | No | NA | 2 | 0 | 2 | Not-determined | Genome-fragment | NA | NA | 0 |
| S9_extracted_k141_45010 | 2035 | No | NA | 6 | 5 | 0 | Low-quality | Genome-fragment | 5.89 | AAI-based (high-confidence) | 0 |
| S7_extracted_5439 | 50770 | No | NA | 72 | 22 | 0 | Complete | High-quality | 100 | DTR (high-confidence) | 0 |
| S7_extracted_5462 | 1701 | No | NA | 2 | 0 | 0 | Not-determined | Genome-fragment | NA | NA | 0 |
| S9_vs_k141_76517\|\|full | 2035 | No | NA | 3 | 0 | 0 | Low-quality | Genome-fragment | 3.44 | AAI-based (medium-confidence) | 0 |
| S7_extracted_5533 | 1481 | No | NA | 3 | 0 | 1 | Not-determined | Genome-fragment | NA | NA | 0 |
| S7_extracted_5638 | 2417 | No | NA | 1 | 0 | 1 | Not-determined | Genome-fragment | NA | NA | 0 |
| S7_extracted_5645 | 5615 | No | NA | 11 | 0 | 1 | Not-determined | Genome-fragment | NA | NA | 0 |
| S14_extracted_k141_110930 | 2035 | No | NA | 3 | 0 | 1 | Low-quality | Genome-fragment | 2.12 | AAI-based (medium-confidence) | 0 |
| S7_extracted_5801 | 63941 | No | NA | 92 | 19 | 4 | Complete | High-quality | 100 | DTR (high-confidence) | 0 |
| S7_vs_109649\|\|full | 2035 | No | NA | 3 | 0 | 0 | Low-quality | Genome-fragment | 0.85 | AAI-based (medium-confidence) | 0 |
| S7_extracted_5827 | 1352 | No | NA | 1 | 0 | 1 | Not-determined | Genome-fragment | NA | NA | 0 |
| S7_extracted_5843 | 14473 | No | NA | 23 | 7 | 0 | Medium-quality | Genome-fragment | 84.07 | AAI-based (high-confidence) | 0 |
| S16_extracted_23469 | 2035 | No | NA | 2 | 0 | 0 | Low-quality | Genome-fragment | 3.87 | AAI-based (high-confidence) | 0 |
| S8_extracted_k141_88684 | 2035 | No | NA | 2 | 2 | 0 | Low-quality | Genome-fragment | 6.71 | HMM-based (lower-bound) | 0 |
| S8_vs_k141_89805\|\|full | 2035 | No | NA | 3 | 0 | 2 | Low-quality | Genome-fragment | 3.18 | AAI-based (medium-confidence) | 0 |
| S7_extracted_6461 | 1009 | No | NA | 2 | 0 | 0 | Not-determined | Genome-fragment | NA | NA | 0 |
| S7_extracted_6559 | 4153 | No | NA | 2 | 0 | 1 | Not-determined | Genome-fragment | NA | NA | 0 |
| S17_vs_k141_120473\|\|full | 2036 | No | NA | 3 | 0 | 2 | Low-quality | Genome-fragment | 1.32 | AAI-based (medium-confidence) | 0 |
| S7_extracted_6673 | 1112 | No | NA | 1 | 0 | 0 | Not-determined | Genome-fragment | NA | NA | 0 |
| S7_extracted_6699 | 1353 | No | NA | 2 | 0 | 0 | Not-determined | Genome-fragment | NA | NA | 0 |
| S7_extracted_6808 | 2257 | No | NA | 4 | 0 | 2 | Not-determined | Genome-fragment | NA | NA | 0 |
| S7_extracted_7003 | 4803 | No | NA | 4 | 0 | 3 | Not-determined | Genome-fragment | NA | NA | 0 |
| S13_extracted_58631 | 2036 | No | NA | 3 | 0 | 1 | Low-quality | Genome-fragment | 2.18 | AAI-based (medium-confidence) | 0 |
| S16_vs_66051\|\|full | 2036 | No | NA | 4 | 0 | 1 | Low-quality | Genome-fragment | 4.64 | AAI-based (medium-confidence) | 0 |
| S7_extracted_7175 | 4064 | No | NA | 4 | 0 | 1 | Not-determined | Genome-fragment | NA | NA | 0 |
| S3_extracted_k141_48561 | 2037 | No | NA | 4 | 0 | 0 | Low-quality | Genome-fragment | 3.97 | AAI-based (medium-confidence) | 0 |
| S7_extracted_7323 | 3861 | No | NA | 4 | 0 | 1 | Not-determined | Genome-fragment | NA | NA | 0 |
| S7_vs_65518\|\|full | 2037 | No | NA | 3 | 1 | 0 | Low-quality | Genome-fragment | 4.78 | AAI-based (high-confidence) | 0 |
| S8_vs_k141_97417\|\|full | 2037 | No | NA | 3 | 1 | 0 | Low-quality | Genome-fragment | 4.38 | AAI-based (high-confidence) | 0 |
| S7_extracted_7583 | 1022 | No | NA | 1 | 0 | 0 | Not-determined | Genome-fragment | NA | NA | 0 |
| S7_extracted_7599 | 1007 | No | NA | 2 | 0 | 0 | Not-determined | Genome-fragment | NA | NA | 0 |
| S7_extracted_7647 | 1042 | No | NA | 1 | 0 | 1 | Not-determined | Genome-fragment | NA | NA | 0 |
| S7_extracted_7716 | 1237 | No | NA | 2 | 0 | 2 | Not-determined | Genome-fragment | NA | NA | 0 |
| S11_vs_k141_37795\|\|full | 2038 | No | NA | 3 | 1 | 0 | Low-quality | Genome-fragment | 5.43 | AAI-based (high-confidence) | 0 |
| S7_extracted_7760 | 1241 | No | NA | 1 | 0 | 1 | Not-determined | Genome-fragment | NA | NA | 0 |
| S7_extracted_7795 | 2945 | No | NA | 2 | 0 | 1 | Not-determined | Genome-fragment | NA | NA | 0 |
| S7_extracted_7839 | 1683 | No | NA | 2 | 0 | 1 | Not-determined | Genome-fragment | NA | NA | 0 |
| S7_extracted_7891 | 1378 | No | NA | 2 | 0 | 0 | Not-determined | Genome-fragment | NA | NA | 0 |
| S1_vs_k141_70597\|\|full | 2038 | No | NA | 4 | 0 | 0 | Low-quality | Genome-fragment | 3.43 | AAI-based (medium-confidence) | 0 |
| S7_extracted_7969 | 4856 | No | NA | 3 | 0 | 2 | Not-determined | Genome-fragment | NA | NA | 0 |
| S7_extracted_7988 | 1529 | No | NA | 2 | 0 | 2 | Not-determined | Genome-fragment | NA | NA | 0 |
| S7_extracted_8069 | 4867 | No | NA | 8 | 0 | 2 | Not-determined | Genome-fragment | NA | NA | 0 |
| S7_extracted_8104 | 1759 | No | NA | 3 | 0 | 0 | Not-determined | Genome-fragment | NA | NA | 0 |
| S5_vs_k141_47555\|\|full | 2038 | No | NA | 4 | 1 | 0 | Low-quality | Genome-fragment | 4.08 | HMM-based (lower-bound) | 0 |
| S7_extracted_8155 | 1018 | No | NA | 1 | 0 | 0 | Not-determined | Genome-fragment | NA | NA | 0 |
| S7_extracted_8332 | 31505 | No | NA | 50 | 17 | 0 | Medium-quality | Genome-fragment | 81.6 | AAI-based (high-confidence) | 0 |
| S7_extracted_8518 | 1155 | No | NA | 2 | 0 | 2 | Not-determined | Genome-fragment | NA | NA | 0 |
| S7_extracted_8522 | 1261 | No | NA | 1 | 0 | 0 | Not-determined | Genome-fragment | NA | NA | 0 |
| S7_extracted_8557 | 1014 | No | NA | 3 | 0 | 0 | Not-determined | Genome-fragment | NA | NA | 0 |
| S7_extracted_8578 | 3675 | No | NA | 4 | 0 | 0 | Not-determined | Genome-fragment | NA | NA | 0 |
| S16_vs_55071\|\|full | 2038 | Yes | 1018 | 4 | 1 | 1 | Low-quality | Genome-fragment | 1.81 | AAI-based (high-confidence) | 50.05 |
| S7_extracted_8648 | 4176 | No | NA | 5 | 0 | 0 | Not-determined | Genome-fragment | NA | NA | 0 |
| S7_extracted_8922 | 1340 | No | NA | 1 | 0 | 0 | Not-determined | Genome-fragment | NA | NA | 0 |
| S7_extracted_8959 | 1562 | No | NA | 3 | 0 | 1 | Not-determined | Genome-fragment | NA | NA | 0 |
| S12_vs_k141_102720\|\|full | 2038 | No | NA | 2 | 0 | 1 | Low-quality | Genome-fragment | 1.64 | AAI-based (medium-confidence) | 0 |
| S11_extracted_k141_90235 | 2039 | No | NA | 1 | 0 | 0 | Low-quality | Genome-fragment | 3.3 | AAI-based (high-confidence) | 0 |
| S13_vs_92621\|\|full | 2039 | No | NA | 4 | 0 | 0 | Low-quality | Genome-fragment | 4.94 | AAI-based (high-confidence) | 0 |
| S10_vs_12882\|\|full | 2039 | No | NA | 2 | 0 | 1 | Low-quality | Genome-fragment | 1.42 | AAI-based (high-confidence) | 0 |
| S7_extracted_9334 | 40566 | No | NA | 48 | 18 | 0 | Complete | High-quality | 100 | DTR (high-confidence) | 0 |
| S12_vs_k141_149941\|\|full | 2039 | No | NA | 3 | 1 | 0 | Low-quality | Genome-fragment | 1.57 | AAI-based (high-confidence) | 0 |
| S7_extracted_9393 | 1701 | No | NA | 1 | 0 | 1 | Not-determined | Genome-fragment | NA | NA | 0 |
| S2_extracted_k141_3627 | 2039 | No | NA | 2 | 0 | 0 | Low-quality | Genome-fragment | 4.59 | AAI-based (high-confidence) | 0 |
| S7_extracted_9544 | 5052 | No | NA | 5 | 0 | 0 | Not-determined | Genome-fragment | NA | NA | 0 |
| S7_extracted_9608 | 1012 | No | NA | 1 | 0 | 0 | Not-determined | Genome-fragment | NA | NA | 0 |
| S11_vs_k141_43757\|\|full | 2040 | No | NA | 4 | 2 | 0 | Low-quality | Genome-fragment | 3.22 | AAI-based (high-confidence) | 0 |
| S7_extracted_9695 | 3633 | No | NA | 3 | 0 | 0 | Not-determined | Genome-fragment | NA | NA | 0 |
| S7_extracted_9703 | 1039 | No | NA | 1 | 0 | 1 | Not-determined | Genome-fragment | NA | NA | 0 |
| S7_extracted_9929 | 1715 | No | NA | 1 | 0 | 1 | Not-determined | Genome-fragment | NA | NA | 0 |
| S7_vs_445\|\|full | 2040 | No | NA | 5 | 0 | 0 | Low-quality | Genome-fragment | 3.94 | AAI-based (high-confidence) | 0 |
| S7_extracted_10088 | 1028 | No | NA | 2 | 0 | 0 | Not-determined | Genome-fragment | NA | NA | 0 |
| S7_extracted_10127 | 1049 | No | NA | 1 | 0 | 1 | Not-determined | Genome-fragment | NA | NA | 0 |
| S7_extracted_10174 | 1479 | No | NA | 1 | 0 | 1 | Not-determined | Genome-fragment | NA | NA | 0 |
| S7_extracted_10182 | 1137 | No | NA | 2 | 0 | 1 | Not-determined | Genome-fragment | NA | NA | 0 |
| S7_extracted_10230 | 1017 | No | NA | 1 | 0 | 0 | Not-determined | Genome-fragment | NA | NA | 0 |
| S7_extracted_10258 | 1284 | No | NA | 3 | 0 | 1 | Not-determined | Genome-fragment | NA | NA | 0 |
| S7_extracted_10285 | 1037 | No | NA | 1 | 0 | 1 | Not-determined | Genome-fragment | NA | NA | 0 |
| S7_extracted_10304 | 1291 | No | NA | 2 | 0 | 1 | Not-determined | Genome-fragment | NA | NA | 0 |
| S7_extracted_10317 | 1045 | No | NA | 1 | 0 | 0 | Not-determined | Genome-fragment | NA | NA | 0 |
| S7_extracted_10582 | 3332 | No | NA | 4 | 0 | 2 | Not-determined | Genome-fragment | NA | NA | 0 |
| S7_extracted_10823 | 2080 | No | NA | 2 | 0 | 2 | Not-determined | Genome-fragment | NA | NA | 0 |
| S7_extracted_10854 | 1021 | No | NA | 1 | 0 | 0 | Not-determined | Genome-fragment | NA | NA | 0 |
| S2_vs_k141_8371\|\|full | 2040 | No | NA | 3 | 1 | 0 | Low-quality | Genome-fragment | 6.91 | HMM-based (lower-bound) | 0 |
| S8_extracted_k141_41446 | 2040 | No | NA | 2 | 0 | 1 | Low-quality | Genome-fragment | 2.46 | AAI-based (high-confidence) | 0 |
| S7_extracted_11162 | 1805 | No | NA | 1 | 0 | 1 | Not-determined | Genome-fragment | NA | NA | 0 |
| S7_extracted_11203 | 2098 | No | NA | 2 | 0 | 0 | Not-determined | Genome-fragment | NA | NA | 0 |
| S8_vs_k141_131469\|\|full | 2040 | No | NA | 3 | 0 | 0 | Low-quality | Genome-fragment | 3.86 | AAI-based (medium-confidence) | 0 |
| S9_extracted_k141_31551 | 2041 | No | NA | 6 | 0 | 1 | Low-quality | Genome-fragment | 3.41 | AAI-based (medium-confidence) | 0 |
| S3_vs_k141_53407\|\|full | 2041 | No | NA | 3 | 0 | 1 | Low-quality | Genome-fragment | 2.33 | AAI-based (medium-confidence) | 0 |
| S7_extracted_11417 | 1416 | No | NA | 1 | 0 | 0 | Not-determined | Genome-fragment | NA | NA | 0 |
| S7_extracted_11458 | 2064 | No | NA | 1 | 0 | 0 | Not-determined | Genome-fragment | NA | NA | 0 |
| S7_extracted_11460 | 4591 | No | NA | 5 | 0 | 5 | Not-determined | Genome-fragment | NA | NA | 0 |
| S7_extracted_103924 | 2041 | No | NA | 2 | 2 | 0 | Low-quality | Genome-fragment | 3.59 | AAI-based (high-confidence) | 0 |
| S15_vs_76010\|\|full | 2041 | No | NA | 4 | 0 | 2 | Low-quality | Genome-fragment | 2.86 | AAI-based (high-confidence) | 0 |
| S7_extracted_11726 | 1916 | No | NA | 1 | 0 | 1 | Not-determined | Genome-fragment | NA | NA | 0 |
| S7_extracted_11744 | 19870 | No | NA | 26 | 7 | 0 | Medium-quality | Genome-fragment | 61.7 | AAI-based (medium-confidence) | 0 |
| S7_extracted_11902 | 1388 | No | NA | 2 | 0 | 0 | Not-determined | Genome-fragment | NA | NA | 0 |
| S7_extracted_11936 | 4497 | No | NA | 2 | 0 | 2 | Not-determined | Genome-fragment | NA | NA | 0 |
| S9_vs_k141_3263\|\|full | 2042 | No | NA | 3 | 3 | 0 | Low-quality | Genome-fragment | 5.46 | AAI-based (medium-confidence) | 0 |
| S7_extracted_12173 | 1383 | No | NA | 2 | 0 | 2 | Not-determined | Genome-fragment | NA | NA | 0 |
| S7_extracted_12324 | 4172 | No | NA | 5 | 0 | 1 | Not-determined | Genome-fragment | NA | NA | 0 |
| S7_extracted_12378 | 3942 | No | NA | 3 | 0 | 2 | Not-determined | Genome-fragment | NA | NA | 0 |
| S9_vs_k141_64122\|\|full | 2042 | No | NA | 4 | 1 | 0 | Low-quality | Genome-fragment | 4.66 | AAI-based (high-confidence) | 0 |
| S7_extracted_12485 | 3548 | No | NA | 2 | 0 | 1 | Not-determined | Genome-fragment | NA | NA | 0 |
| S11_extracted_k141_566 | 2042 | No | NA | 3 | 3 | 0 | Low-quality | Genome-fragment | 5.04 | AAI-based (high-confidence) | 0 |
| S7_extracted_12621 | 1348 | No | NA | 1 | 0 | 1 | Not-determined | Genome-fragment | NA | NA | 0 |
| S11_vs_k141_95537\|\|full | 2042 | No | NA | 4 | 2 | 0 | Low-quality | Genome-fragment | 15.48 | AAI-based (high-confidence) | 0 |
| S7_extracted_12738 | 1041 | No | NA | 2 | 0 | 1 | Not-determined | Genome-fragment | NA | NA | 0 |
| S7_extracted_12855 | 1201 | No | NA | 2 | 0 | 1 | Not-determined | Genome-fragment | NA | NA | 0 |
| S7_extracted_12919 | 5106 | No | NA | 2 | 0 | 1 | Not-determined | Genome-fragment | NA | NA | 0 |
| S14_vs_k141_84002\|\|full | 2042 | No | NA | 4 | 0 | 0 | Low-quality | Genome-fragment | 5.47 | AAI-based (medium-confidence) | 0 |
| S7_extracted_12932 | 1342 | No | NA | 1 | 0 | 1 | Not-determined | Genome-fragment | NA | NA | 0 |
| S7_extracted_13044 | 1662 | No | NA | 1 | 0 | 1 | Not-determined | Genome-fragment | NA | NA | 0 |
| S7_extracted_13074 | 1472 | No | NA | 2 | 0 | 1 | Not-determined | Genome-fragment | NA | NA | 0 |
| S7_extracted_13192 | 1178 | No | NA | 2 | 0 | 0 | Not-determined | Genome-fragment | NA | NA | 0 |
| S7_extracted_13269 | 4252 | No | NA | 5 | 0 | 3 | Not-determined | Genome-fragment | NA | NA | 0 |

| | | | | | | | | | | | |
|---|---|---|---|---|---|---|---|---|---|---|---|
| S7_extracted_13368 | 1032 | No | NA | 1 | 0 | 0 | Not-determined | Genome-fragment | NA | NA | 0 |
| S7_extracted_13376 | 1398 | No | NA | 2 | 0 | 2 | Not-determined | Genome-fragment | NA | NA | 0 |
| S7_extracted_13425 | 3398 | No | NA | 3 | 0 | 2 | Not-determined | Genome-fragment | NA | NA | 0 |
| S1_extracted_k141_26863 | 2042 | No | NA | 3 | 3 | 0 | Low-quality | Genome-fragment | 3.39 | AAI-based (high-confidence) | 0 |
| S7_extracted_13559 | 2129 | No | NA | 1 | 0 | 1 | Not-determined | Genome-fragment | NA | NA | 0 |
| S7_extracted_13579 | 2457 | No | NA | 2 | 0 | 1 | Not-determined | Genome-fragment | NA | NA | 0 |
| S7_extracted_13614 | 34819 | No | NA | 50 | 22 | 1 | Complete | High-quality | 100 | DTR (high-confidence) | 0 |
| S7_extracted_13694 | 5930 | No | NA | 7 | 0 | 5 | Not-determined | Genome-fragment | NA | NA | 0 |
| S7_extracted_13715 | 1544 | No | NA | 3 | 0 | 2 | Not-determined | Genome-fragment | NA | NA | 0 |
| S7_extracted_13740 | 4383 | No | NA | 7 | 0 | 2 | Not-determined | Genome-fragment | NA | NA | 0 |
| S3_vs_k141_93276\|\|lt2gene | 2042 | No | NA | 2 | 2 | 0 | Low-quality | Genome-fragment | 3.08 | HMM-based (lower-bound) | 0 |
| S7_vs_48843\|\|full | 2042 | No | NA | 8 | 2 | 0 | Low-quality | Genome-fragment | 5.13 | AAI-based (medium-confidence) | 0 |
| S7_extracted_14013 | 3528 | No | NA | 4 | 0 | 3 | Not-determined | Genome-fragment | NA | NA | 0 |
| S10_extracted_6344 | 2042 | No | NA | 3 | 1 | 0 | Low-quality | Genome-fragment | 3.07 | AAI-based (high-confidence) | 0 |
| S7_extracted_14297 | 2230 | No | NA | 3 | 0 | 0 | Not-determined | Genome-fragment | NA | NA | 0 |
| S7_extracted_14301 | 1175 | No | NA | 2 | 0 | 0 | Not-determined | Genome-fragment | NA | NA | 0 |
| S7_extracted_14558 | 1088 | No | NA | 2 | 0 | 2 | Not-determined | Genome-fragment | NA | NA | 0 |
| S10_extracted_63815 | 2042 | No | NA | 1 | 0 | 0 | Low-quality | Genome-fragment | 2.78 | AAI-based (high-confidence) | 0 |
| S7_extracted_14694 | 1743 | No | NA | 2 | 0 | 2 | Not-determined | Genome-fragment | NA | NA | 0 |
| S7_extracted_14716 | 1175 | No | NA | 2 | 0 | 0 | Not-determined | Genome-fragment | NA | NA | 0 |
| S7_extracted_14936 | 2511 | No | NA | 1 | 0 | 1 | Not-determined | Genome-fragment | NA | NA | 0 |
| S7_extracted_15000 | 1580 | No | NA | 2 | 0 | 2 | Not-determined | Genome-fragment | NA | NA | 0 |
| S7_extracted_15028 | 2014 | No | NA | 5 | 0 | 0 | Not-determined | Genome-fragment | NA | NA | 0 |
| S10_vs_102292\|\|full | 2042 | No | NA | 4 | 0 | 0 | Low-quality | Genome-fragment | 4.45 | AAI-based (medium-confidence) | 0 |
| S7_extracted_15117 | 1479 | No | NA | 1 | 0 | 1 | Not-determined | Genome-fragment | NA | NA | 0 |
| S7_extracted_15283 | 1147 | No | NA | 1 | 0 | 0 | Not-determined | Genome-fragment | NA | NA | 0 |
| S7_extracted_15316 | 3224 | No | NA | 2 | 0 | 1 | Not-determined | Genome-fragment | NA | NA | 0 |
| S7_extracted_15367 | 1221 | No | NA | 1 | 0 | 0 | Not-determined | Genome-fragment | NA | NA | 0 |
| S7_extracted_15447 | 1587 | No | NA | 4 | 0 | 3 | Not-determined | Genome-fragment | NA | NA | 0 |
| S8_vs_k141_26037\|\|full | 2042 | No | NA | 2 | 0 | 2 | Low-quality | Genome-fragment | 1.49 | AAI-based (high-confidence) | 0 |
| S17_vs_k141_134039\|\|full | 2043 | No | NA | 3 | 0 | 1 | Low-quality | Genome-fragment | 2.5 | AAI-based (medium-confidence) | 0 |
| S17_vs_k141_93369\|\|full | 2043 | No | NA | 3 | 0 | 2 | Low-quality | Genome-fragment | 1.55 | AAI-based (medium-confidence) | 0 |
| S7_extracted_15597 | 1155 | No | NA | 1 | 0 | 1 | Not-determined | Genome-fragment | NA | NA | 0 |
| S14_vs_k141_13027\|\|full | 2043 | No | NA | 4 | 0 | 1 | Low-quality | Genome-fragment | 3.5 | AAI-based (medium-confidence) | 0 |
| S7_extracted_15669 | 5405 | No | NA | 9 | 0 | 1 | Not-determined | Genome-fragment | NA | NA | 0 |
| S1_vs_k141_70371\|\|full | 2043 | No | NA | 3 | 0 | 1 | Low-quality | Genome-fragment | 3.28 | AAI-based (medium-confidence) | 0 |
| S12_vs_k141_58894\|\|full | 2043 | No | NA | 3 | 0 | 2 | Low-quality | Genome-fragment | 4.66 | AAI-based (medium-confidence) | 0 |
| S7_extracted_15829 | 1379 | No | NA | 1 | 0 | 1 | Not-determined | Genome-fragment | NA | NA | 0 |
| S8_extracted_k141_64409 | 2043 | No | NA | 2 | 2 | 0 | Low-quality | Genome-fragment | 6.74 | HMM-based (lower-bound) | 0 |
| S7_extracted_15980 | 1011 | No | NA | 1 | 0 | 0 | Not-determined | Genome-fragment | NA | NA | 0 |
| S8_vs_k141_60814\|\|full | 2043 | No | NA | 6 | 0 | 0 | Low-quality | Genome-fragment | 1.83 | AAI-based (medium-confidence) | 0 |
| S7_extracted_16061 | 8934 | No | NA | 10 | 0 | 3 | Not-determined | Genome-fragment | NA | NA | 0 |
| S9_vs_k141_10435\|\|lt2gene | 2044 | No | NA | 2 | 2 | 0 | Low-quality | Genome-fragment | 5.2 | AAI-based (high-confidence) | 0 |
| S11_extracted_k141_66572 | 2044 | No | NA | 5 | 1 | 0 | Low-quality | Genome-fragment | 3.16 | AAI-based (high-confidence) | 0 |
| S7_extracted_16138 | 3556 | No | NA | 2 | 0 | 1 | Not-determined | Genome-fragment | NA | NA | 0 |
| S7_extracted_16141 | 1027 | No | NA | 2 | 0 | 2 | Not-determined | Genome-fragment | NA | NA | 0 |
| S11_extracted_k141_74444 | 2044 | No | NA | 5 | 0 | 0 | Low-quality | Genome-fragment | 2.12 | AAI-based (medium-confidence) | 0 |
| S7_extracted_16204 | 5350 | No | NA | 4 | 0 | 1 | Not-determined | Genome-fragment | NA | NA | 0 |
| S7_extracted_16212 | 3856 | No | NA | 3 | 0 | 3 | Not-determined | Genome-fragment | NA | NA | 0 |
| S14_vs_k141_16570\|\|full | 2044 | No | NA | 4 | 0 | 0 | Low-quality | Genome-fragment | 4.82 | AAI-based (medium-confidence) | 0 |
| S7_extracted_16246 | 1948 | No | NA | 3 | 0 | 0 | Not-determined | Genome-fragment | NA | NA | 0 |
| S1_vs_k141_40934\|\|full | 2044 | No | NA | 3 | 1 | 0 | Low-quality | Genome-fragment | 3.41 | AAI-based (high-confidence) | 0 |
| S7_vs_105498\|\|full | 2044 | No | NA | 9 | 0 | 0 | Low-quality | Genome-fragment | 4.71 | AAI-based (medium-confidence) | 0 |
| S7_extracted_16409 | 15221 | No | NA | 14 | 0 | 2 | Not-determined | Genome-fragment | NA | NA | 0 |
| S15_vs_9532\|\|full | 2044 | No | NA | 5 | 0 | 0 | Low-quality | Genome-fragment | 4.18 | AAI-based (medium-confidence) | 0 |
| S15_vs_76633\|\|full | 2044 | Yes | 1468 | 3 | 1 | 1 | Low-quality | Genome-fragment | 0.98 | AAI-based (high-confidence) | 28.18 |
| S7_extracted_16533 | 5700 | No | NA | 8 | 0 | 2 | Not-determined | Genome-fragment | NA | NA | 0 |
| S8_vs_k141_67469\|\|full | 2044 | No | NA | 4 | 0 | 0 | Low-quality | Genome-fragment | 4.78 | AAI-based (medium-confidence) | 0 |
| S8_extracted_k141_169751 | 2045 | No | NA | 2 | 0 | 1 | Low-quality | Genome-fragment | 3.42 | AAI-based (medium-confidence) | 0 |
| S17_extracted_k141_174629 | 2046 | No | NA | 2 | 0 | 0 | Low-quality | Genome-fragment | 3.42 | AAI-based (high-confidence) | 0 |
| S7_extracted_16865 | 1171 | No | NA | 1 | 0 | 1 | Not-determined | Genome-fragment | NA | NA | 0 |
| S7_extracted_17127 | 2269 | No | NA | 1 | 0 | 1 | Not-determined | Genome-fragment | NA | NA | 0 |
| S17_extracted_k141_28924 | 2046 | No | NA | 4 | 1 | 0 | Low-quality | Genome-fragment | 2.04 | AAI-based (high-confidence) | 0 |
| S17_vs_k141_10749\|\|full | 2046 | No | NA | 3 | 0 | 0 | Low-quality | Genome-fragment | 3.32 | AAI-based (high-confidence) | 0 |
| S14_vs_k141_108002\|\|full | 2046 | No | NA | 8 | 1 | 0 | Low-quality | Genome-fragment | 5.27 | AAI-based (high-confidence) | 0 |
| S3_vs_k141_39079\|\|full | 2046 | No | NA | 3 | 0 | 0 | Low-quality | Genome-fragment | 2.27 | AAI-based (high-confidence) | 0 |
| S7_extracted_17470 | 3538 | No | NA | 3 | 0 | 2 | Not-determined | Genome-fragment | NA | NA | 0 |
| S7_extracted_17628 | 1196 | No | NA | 2 | 0 | 2 | Not-determined | Genome-fragment | NA | NA | 0 |
| S7_extracted_17652 | 1663 | No | NA | 2 | 0 | 0 | Not-determined | Genome-fragment | NA | NA | 0 |
| S7_extracted_17677 | 1141 | No | NA | 2 | 0 | 0 | Not-determined | Genome-fragment | NA | NA | 0 |
| S7_extracted_17808 | 6070 | No | NA | 9 | 0 | 3 | Not-determined | Genome-fragment | NA | NA | 0 |
| S5_vs_k141_14135\|\|full | 2046 | No | NA | 3 | 2 | 0 | Low-quality | Genome-fragment | 5.51 | AAI-based (medium-confidence) | 0 |
| S7_extracted_18002 | 1665 | No | NA | 2 | 0 | 1 | Not-determined | Genome-fragment | NA | NA | 0 |
| S7_extracted_18021 | 1085 | No | NA | 3 | 0 | 0 | Not-determined | Genome-fragment | NA | NA | 0 |
| S7_extracted_18063 | 6885 | No | NA | 8 | 0 | 5 | Not-determined | Genome-fragment | NA | NA | 0 |
| S7_extracted_18066 | 1602 | No | NA | 1 | 0 | 1 | Not-determined | Genome-fragment | NA | NA | 0 |
| S7_extracted_18100 | 3990 | No | NA | 3 | 0 | 3 | Not-determined | Genome-fragment | NA | NA | 0 |
| S10_vs_2672\|\|full | 2046 | No | NA | 3 | 0 | 0 | Low-quality | Genome-fragment | 3.08 | AAI-based (high-confidence) | 0 |
| S7_extracted_18183 | 4291 | No | NA | 4 | 0 | 1 | Not-determined | Genome-fragment | NA | NA | 0 |
| S7_extracted_18260 | 3539 | No | NA | 5 | 0 | 3 | Not-determined | Genome-fragment | NA | NA | 0 |
| S10_vs_55810\|\|full | 2046 | No | NA | 5 | 0 | 0 | Low-quality | Genome-fragment | 4.95 | AAI-based (high-confidence) | 0 |
| S7_extracted_18620 | 3055 | No | NA | 2 | 0 | 0 | Not-determined | Genome-fragment | NA | NA | 0 |
| S8_extracted_k141_185559 | 2046 | No | NA | 3 | 0 | 2 | Low-quality | Genome-fragment | 1.27 | AAI-based (medium-confidence) | 0 |
| S17_extracted_k141_154126 | 2047 | No | NA | 4 | 0 | 0 | Low-quality | Genome-fragment | 3.07 | AAI-based (high-confidence) | 0 |
| S17_vs_k141_116691\|\|full | 2047 | No | NA | 3 | 0 | 1 | Low-quality | Genome-fragment | 4.7 | AAI-based (medium-confidence) | 0 |
| S7_extracted_18970 | 2178 | No | NA | 2 | 0 | 2 | Not-determined | Genome-fragment | NA | NA | 0 |
| S7_extracted_19040 | 1359 | No | NA | 2 | 0 | 0 | Not-determined | Genome-fragment | NA | NA | 0 |
| S7_extracted_19254 | 1445 | No | NA | 1 | 0 | 1 | Not-determined | Genome-fragment | NA | NA | 0 |
| S7_extracted_19376 | 1912 | No | NA | 2 | 0 | 1 | Not-determined | Genome-fragment | NA | NA | 0 |
| S1_vs_k141_21661\|\|full | 2047 | No | NA | 3 | 2 | 0 | Low-quality | Genome-fragment | 4.87 | AAI-based (high-confidence) | 0 |
| S7_extracted_19415 | 1026 | No | NA | 1 | 0 | 1 | Not-determined | Genome-fragment | NA | NA | 0 |
| S7_extracted_19508 | 1346 | No | NA | 1 | 0 | 1 | Not-determined | Genome-fragment | NA | NA | 0 |
| S7_extracted_19762 | 1527 | No | NA | 2 | 0 | 1 | Not-determined | Genome-fragment | NA | NA | 0 |
| S3_extracted_k141_81097 | 2047 | No | NA | 4 | 1 | 0 | Low-quality | Genome-fragment | 6.12 | AAI-based (high-confidence) | 0 |
| S3_vs_k141_94924\|\|full | 2047 | No | NA | 2 | 0 | 2 | Low-quality | Genome-fragment | 1.15 | AAI-based (medium-confidence) | 0 |
| S7_extracted_19866 | 16490 | No | NA | 27 | 9 | 0 | Medium-quality | Genome-fragment | 74.58 | AAI-based (medium-confidence) | 0 |
| S7_extracted_20005 | 3148 | No | NA | 4 | 0 | 3 | Not-determined | Genome-fragment | NA | NA | 0 |
| S7_extracted_20090 | 5567 | No | NA | 3 | 0 | 1 | Not-determined | Genome-fragment | NA | NA | 0 |
| S7_extracted_20129 | 16300 | No | NA | 19 | 8 | 0 | Medium-quality | Genome-fragment | 53.36 | HMM-based (lower-bound) | 0 |
| S7_extracted_20156 | 1078 | No | NA | 1 | 0 | 1 | Not-determined | Genome-fragment | NA | NA | 0 |
| S16_vs_18969\|\|full | 2047 | No | NA | 8 | 2 | 0 | Low-quality | Genome-fragment | 1.46 | HMM-based (lower-bound) | 0 |
| S7_extracted_20304 | 2757 | No | NA | 3 | 0 | 2 | Not-determined | Genome-fragment | NA | NA | 0 |
| S7_extracted_20328 | 1438 | No | NA | 1 | 0 | 1 | Not-determined | Genome-fragment | NA | NA | 0 |
| S7_extracted_20368 | 1361 | No | NA | 2 | 0 | 1 | Not-determined | Genome-fragment | NA | NA | 0 |
| S7_extracted_20645 | 1853 | No | NA | 1 | 0 | 0 | Not-determined | Genome-fragment | NA | NA | 0 |
| S7_extracted_20655 | 2442 | No | NA | 3 | 0 | 2 | Not-determined | Genome-fragment | NA | NA | 0 |
| S7_extracted_20679 | 1975 | No | NA | 2 | 0 | 1 | Not-determined | Genome-fragment | NA | NA | 0 |
| S7_extracted_20702 | 1337 | No | NA | 1 | 0 | 1 | Not-determined | Genome-fragment | NA | NA | 0 |
| S7_extracted_20840 | 1455 | No | NA | 2 | 0 | 2 | Not-determined | Genome-fragment | NA | NA | 0 |
| S7_extracted_20998 | 3176 | No | NA | 4 | 0 | 1 | Not-determined | Genome-fragment | NA | NA | 0 |
| S7_extracted_21028 | 1344 | No | NA | 1 | 0 | 1 | Not-determined | Genome-fragment | NA | NA | 0 |
| S7_extracted_21061 | 1101 | No | NA | 2 | 0 | 1 | Not-determined | Genome-fragment | NA | NA | 0 |
| S7_extracted_21066 | 1196 | No | NA | 2 | 0 | 1 | Not-determined | Genome-fragment | NA | NA | 0 |
| S6_vs_k141_35143\|\|full | 2047 | No | NA | 3 | 0 | 0 | Low-quality | Genome-fragment | 2.96 | AAI-based (medium-confidence) | 0 |
| S6_vs_k141_44678\|\|full | 2047 | No | NA | 6 | 2 | 0 | Low-quality | Genome-fragment | 4.95 | AAI-based (high-confidence) | 0 |
| S7_extracted_21164 | 2294 | No | NA | 1 | 0 | 1 | Not-determined | Genome-fragment | NA | NA | 0 |
| S17_extracted_k141_21354 | 2048 | No | NA | 2 | 0 | 1 | Low-quality | Genome-fragment | 2.16 | AAI-based (high-confidence) | 0 |
| S7_extracted_21319 | 1428 | No | NA | 1 | 0 | 0 | Not-determined | Genome-fragment | NA | NA | 0 |
| S7_extracted_21387 | 1118 | No | NA | 1 | 0 | 1 | Not-determined | Genome-fragment | NA | NA | 0 |
| S11_vs_k141_97937\|\|full | 2048 | No | NA | 4 | 1 | 0 | Low-quality | Genome-fragment | 2.42 | AAI-based (medium-confidence) | 0 |
| S3_vs_k141_47455\|\|full | 2048 | No | NA | 2 | 1 | 0 | Low-quality | Genome-fragment | 1.19 | AAI-based (medium-confidence) | 0 |
| S7_vs_137149\|\|full | 2048 | No | NA | 3 | 1 | 0 | Low-quality | Genome-fragment | 4.93 | AAI-based (high-confidence) | 0 |
| S16_extracted_70891 | 2048 | No | NA | 5 | 0 | 1 | Low-quality | Genome-fragment | 0.87 | AAI-based (medium-confidence) | 0 |
| S10_extracted_6838 | 2048 | No | NA | 4 | 1 | 0 | Low-quality | Genome-fragment | 2.11 | AAI-based (high-confidence) | 0 |
| S12_extracted_k141_181789 | 2048 | No | NA | 1 | 0 | 1 | Low-quality | Genome-fragment | 3.1 | AAI-based (medium-confidence) | 0 |
| S7_extracted_21954 | 1186 | No | NA | 2 | 0 | 2 | Not-determined | Genome-fragment | NA | NA | 0 |
| S7_extracted_21966 | 1912 | No | NA | 1 | 0 | 1 | Not-determined | Genome-fragment | NA | NA | 0 |
| S7_extracted_21996 | 3510 | No | NA | 3 | 0 | 2 | Not-determined | Genome-fragment | NA | NA | 0 |
| S12_vs_k141_79725\|\|full | 2048 | No | NA | 6 | 2 | 0 | Low-quality | Genome-fragment | 2.01 | AAI-based (medium-confidence) | 0 |
| S7_extracted_22111 | 5289 | No | NA | 4 | 0 | 2 | Not-determined | Genome-fragment | NA | NA | 0 |
| S6_extracted_k141_41431 | 2048 | No | NA | 2 | 0 | 0 | Low-quality | Genome-fragment | 2.28 | AAI-based (medium-confidence) | 0 |
| S7_extracted_22151 | 6888 | No | NA | 6 | 0 | 5 | Not-determined | Genome-fragment | NA | NA | 0 |
| S6_extracted_k141_6776 | 2048 | No | NA | 2 | 0 | 2 | Low-quality | Genome-fragment | 5.23 | AAI-based (medium-confidence) | 0 |
| S7_extracted_22509 | 1022 | No | NA | 2 | 0 | 1 | Not-determined | Genome-fragment | NA | NA | 0 |
| S7_extracted_22758 | 1237 | No | NA | 2 | 0 | 0 | Not-determined | Genome-fragment | NA | NA | 0 |
| S17_vs_k141_170748\|\|full | 2049 | No | NA | 2 | 0 | 1 | Low-quality | Genome-fragment | 1.24 | AAI-based (high-confidence) | 0 |
| S7_extracted_22962 | 2057 | No | NA | 2 | 0 | 1 | Not-determined | Genome-fragment | NA | NA | 0 |
| S7_extracted_22984 | 1294 | No | NA | 1 | 0 | 0 | Not-determined | Genome-fragment | NA | NA | 0 |
| S7_extracted_23061 | 1616 | No | NA | 2 | 0 | 1 | Not-determined | Genome-fragment | NA | NA | 0 |
| S7_extracted_23306 | 1667 | No | NA | 2 | 0 | 2 | Not-determined | Genome-fragment | NA | NA | 0 |
| S7_extracted_23372 | 1020 | No | NA | 2 | 0 | 1 | Not-determined | Genome-fragment | NA | NA | 0 |
| S7_extracted_23471 | 1254 | No | NA | 1 | 0 | 1 | Not-determined | Genome-fragment | NA | NA | 0 |
| S7_extracted_23497 | 2028 | No | NA | 1 | 0 | 1 | Not-determined | Genome-fragment | NA | NA | 0 |
| S7_extracted_23504 | 1618 | No | NA | 1 | 0 | 1 | Not-determined | Genome-fragment | NA | NA | 0 |
| S7_extracted_23695 | 3309 | No | NA | 3 | 0 | 3 | Not-determined | Genome-fragment | NA | NA | 0 |
| S7_extracted_23801 | 4885 | No | NA | 6 | 0 | 6 | Not-determined | Genome-fragment | NA | NA | 0 |
| S7_vs_48278\|\|full | 2049 | No | NA | 3 | 0 | 1 | Low-quality | Genome-fragment | 2.62 | AAI-based (high-confidence) | 0 |
| S7_vs_123671\|\|full | 2049 | No | NA | 4 | 0 | 0 | Low-quality | Genome-fragment | 4.91 | AAI-based (medium-confidence) | 0 |
| S10_vs_81791\|\|full | 2049 | No | NA | 4 | 0 | 1 | Low-quality | Genome-fragment | 5.87 | AAI-based (medium-confidence) | 0 |
| S7_extracted_23955 | 4319 | No | NA | 10 | 0 | 0 | Not-determined | Genome-fragment | NA | NA | 0 |
| S7_extracted_23987 | 1151 | No | NA | 2 | 0 | 1 | Not-determined | Genome-fragment | NA | NA | 0 |
| S7_extracted_24276 | 3919 | No | NA | 2 | 0 | 2 | Not-determined | Genome-fragment | NA | NA | 0 |
| S7_extracted_24334 | 1866 | No | NA | 2 | 0 | 0 | Not-determined | Genome-fragment | NA | NA | 0 |
| S15_vs_21947\|\|full | 2049 | No | NA | 4 | 0 | 0 | Low-quality | Genome-fragment | 4.09 | AAI-based (high-confidence) | 0 |
| S11_vs_k141_5463\|\|full | 2050 | No | NA | 6 | 1 | 0 | Low-quality | Genome-fragment | 4.09 | AAI-based (medium-confidence) | 0 |
| S1_extracted_k141_36045 | 2050 | No | NA | 1 | 1 | 0 | Low-quality | Genome-fragment | 5.34 | AAI-based (high-confidence) | 0 |
| S7_extracted_24509 | 9463 | No | NA | 12 | 0 | 9 | Not-determined | Genome-fragment | NA | NA | 0 |
| S7_extracted_24724 | 5005 | No | NA | 4 | 0 | 3 | Not-determined | Genome-fragment | NA | NA | 0 |
| S7_extracted_24820 | 10024 | No | NA | 12 | 0 | 6 | Not-determined | Genome-fragment | NA | NA | 0 |
| S7_extracted_24868 | 1526 | No | NA | 3 | 0 | 1 | Not-determined | Genome-fragment | NA | NA | 0 |
| S7_extracted_24891 | 12818 | No | NA | 26 | 0 | 26 | Not-determined | Genome-fragment | NA | NA | 0 |
| S7_extracted_24934 | 1682 | No | NA | 1 | 0 | 0 | Not-determined | Genome-fragment | NA | NA | 0 |
| S3_vs_k141_44585\|\|full | 2050 | No | NA | 5 | 1 | 0 | Low-quality | Genome-fragment | 1.56 | AAI-based (high-confidence) | 0 |
| S5_vs_k141_112847\|\|full | 2050 | No | NA | 3 | 3 | 0 | Low-quality | Genome-fragment | 6.06 | AAI-based (high-confidence) | 0 |
| S5_vs_k141_9630\|\|lt2gene | 2050 | No | NA | 2 | 2 | 0 | Low-quality | Genome-fragment | 3.19 | HMM-based (lower-bound) | 0 |
| S7_extracted_25570 | 1269 | No | NA | 1 | 0 | 0 | Not-determined | Genome-fragment | NA | NA | 0 |
| S7_extracted_25688 | 1429 | No | NA | 2 | 0 | 1 | Not-determined | Genome-fragment | NA | NA | 0 |
| S7_extracted_25723 | 1069 | No | NA | 2 | 0 | 0 | Not-determined | Genome-fragment | NA | NA | 0 |
| S16_vs_1801\|\|full | 2050 | No | NA | 4 | 0 | 1 | Low-quality | Genome-fragment | 1.57 | AAI-based (medium-confidence) | 0 |
| S7_extracted_25810 | 1725 | No | NA | 2 | 0 | 1 | Not-determined | Genome-fragment | NA | NA | 0 |
| S7_extracted_25820 | 1191 | No | NA | 1 | 0 | 1 | Not-determined | Genome-fragment | NA | NA | 0 |
| S7_extracted_25836 | 2675 | No | NA | 2 | 0 | 2 | Not-determined | Genome-fragment | NA | NA | 0 |
| S16_vs_62763\|\|full | 2050 | No | NA | 6 | 0 | 0 | Low-quality | Genome-fragment | 1.39 | AAI-based (high-confidence) | 0 |
| S7_extracted_25997 | 7783 | No | NA | 6 | 0 | 4 | Not-determined | Genome-fragment | NA | NA | 0 |
| S17_extracted_k141_54755 | 2051 | No | NA | 4 | 1 | 0 | Low-quality | Genome-fragment | 3.07 | AAI-based (high-confidence) | 0 |
| S7_extracted_26225 | 1719 | No | NA | 1 | 0 | 0 | Not-determined | Genome-fragment | NA | NA | 0 |
| S7_extracted_26339 | 1728 | No | NA | 2 | 0 | 2 | Not-determined | Genome-fragment | NA | NA | 0 |
| S7_extracted_26442 | 3195 | No | NA | 2 | 0 | 2 | Not-determined | Genome-fragment | NA | NA | 0 |
| S7_extracted_26445 | 1427 | No | NA | 2 | 0 | 1 | Not-determined | Genome-fragment | NA | NA | 0 |
| S7_extracted_26587 | 1462 | No | NA | 1 | 0 | 0 | Not-determined | Genome-fragment | NA | NA | 0 |
| S1_vs_k141_114323\|\|full | 2051 | No | NA | 3 | 2 | 0 | Low-quality | Genome-fragment | 4.87 | HMM-based (lower-bound) | 0 |
| S7_extracted_26683 | 1197 | No | NA | 1 | 0 | 1 | Not-determined | Genome-fragment | NA | NA | 0 |
| S3_vs_k141_63927\|\|full | 2051 | No | NA | 5 | 3 | 0 | Low-quality | Genome-fragment | 3.35 | AAI-based (high-confidence) | 0 |
| S7_extracted_26725 | 1662 | No | NA | 3 | 0 | 2 | Not-determined | Genome-fragment | NA | NA | 0 |
| S7_extracted_26933 | 4672 | No | NA | 7 | 0 | 2 | Not-determined | Genome-fragment | NA | NA | 0 |
| S3_vs_k141_57475\|\|lt2gene | 2051 | No | NA | 1 | 0 | 0 | Low-quality | Genome-fragment | 5.57 | AAI-based (medium-confidence) | 0 |
| S7_extracted_27194 | 1605 | No | NA | 2 | 0 | 2 | Not-determined | Genome-fragment | NA | NA | 0 |
| S16_extracted_44898 | 2051 | No | NA | 2 | 0 | 2 | Low-quality | Genome-fragment | 0.25 | AAI-based (medium-confidence) | 0 |
| S7_extracted_27309 | 34140 | No | NA | 49 | 13 | 0 | Complete | High-quality | 100 | DTR (high-confidence) | 0 |
| S7_extracted_27321 | 1828 | No | NA | 3 | 0 | 0 | Not-determined | Genome-fragment | NA | NA | 0 |
| S7_extracted_27326 | 1005 | No | NA | 2 | 0 | 1 | Not-determined | Genome-fragment | NA | NA | 0 |
| S7_extracted_27336 | 1161 | No | NA | 1 | 0 | 1 | Not-determined | Genome-fragment | NA | NA | 0 |
| S7_extracted_27360 | 1942 | No | NA | 3 | 0 | 0 | Not-determined | Genome-fragment | NA | NA | 0 |
| S7_extracted_27388 | 1187 | No | NA | 1 | 0 | 1 | Not-determined | Genome-fragment | NA | NA | 0 |
| S7_extracted_27401 | 1364 | No | NA | 2 | 0 | 1 | Not-determined | Genome-fragment | NA | NA | 0 |
| S16_vs_111606\|\|full | 2051 | No | NA | 3 | 0 | 0 | Low-quality | Genome-fragment | 5.23 | AAI-based (medium-confidence) | 0 |
| S7_extracted_27437 | 13570 | No | NA | 21 | 6 | 0 | Complete | High-quality | 100 | ITR (high-confidence) | 0 |
| S7_extracted_27469 | 2557 | No | NA | 3 | 0 | 2 | Not-determined | Genome-fragment | NA | NA | 0 |
| S7_extracted_27577 | 4213 | No | NA | 8 | 0 | 2 | Not-determined | Genome-fragment | NA | NA | 0 |
| S7_extracted_27611 | 12590 | No | NA | 7 | 0 | 2 | Not-determined | Genome-fragment | NA | NA | 0 |
| S7_extracted_27721 | 2490 | No | NA | 1 | 0 | 0 | Not-determined | Genome-fragment | NA | NA | 0 |
| S7_extracted_27750 | 1058 | No | NA | 2 | 0 | 1 | Not-determined | Genome-fragment | NA | NA | 0 |
| S7_extracted_27767 | 1711 | No | NA | 3 | 0 | 0 | Not-determined | Genome-fragment | NA | NA | 0 |
| S4_extracted_5349 | 2051 | No | NA | 5 | 4 | 0 | Low-quality | Genome-fragment | 6.66 | AAI-based (high-confidence) | 0 |
| S17_extracted_k141_61083 | 2052 | No | NA | 6 | 5 | 0 | Low-quality | Genome-fragment | 6.77 | AAI-based (high-confidence) | 0 |
| S7_extracted_28038 | 2184 | No | NA | 2 | 0 | 2 | Not-determined | Genome-fragment | NA | NA | 0 |
| S7_extracted_28077 | 1031 | No | NA | 1 | 0 | 1 | Not-determined | Genome-fragment | NA | NA | 0 |
| S7_extracted_28282 | 2104 | No | NA | 4 | 0 | 1 | Not-determined | Genome-fragment | NA | NA | 0 |
| S7_extracted_28313 | 1029 | No | NA | 1 | 0 | 1 | Not-determined | Genome-fragment | NA | NA | 0 |
| S17_extracted_k141_2471 | 2052 | No | NA | 1 | 0 | 1 | Low-quality | Genome-fragment | 1.25 | AAI-based (medium-confidence) | 0 |
| S17_extracted_k141_153803 | 2052 | No | NA | 3 | 0 | 0 | Low-quality | Genome-fragment | 44.01 | AAI-based (medium-confidence) | 0 |
| S7_extracted_28380 | 2011 | No | NA | 3 | 0 | 0 | Not-determined | Genome-fragment | NA | NA | 0 |
| S7_extracted_28423 | 3515 | No | NA | 2 | 0 | 2 | Not-determined | Genome-fragment | NA | NA | 0 |
| S7_extracted_28429 | 156691 | No | NA | 185 | 17 | 3 | Complete | High-quality | 100 | DTR (high-confidence) | 0 |
| S7_extracted_28431 | 1191 | No | NA | 3 | 0 | 1 | Not-determined | Genome-fragment | NA | NA | 0 |
| S17_vs_k141_90029\|\|full | 2052 | No | NA | 1 | 0 | 1 | Low-quality | Genome-fragment | 3.31 | AAI-based (medium-confidence) | 0 |
| S7_extracted_28693 | 1099 | No | NA | 2 | 0 | 2 | Not-determined | Genome-fragment | NA | NA | 0 |
| S9_vs_k141_51862\|\|full | 2052 | No | NA | 3 | 0 | 0 | Low-quality | Genome-fragment | 5.49 | AAI-based (medium-confidence) | 0 |
| S11_extracted_k141_80344 | 2052 | No | NA | 3 | 0 | 0 | Low-quality | Genome-fragment | 2.48 | AAI-based (high-confidence) | 0 |
| S16_extracted_143578 | 2052 | No | NA | 2 | 0 | 2 | Low-quality | Genome-fragment | 0.56 | AAI-based (medium-confidence) | 0 |
| S7_extracted_29019 | 3125 | No | NA | 3 | 0 | 3 | Not-determined | Genome-fragment | NA | NA | 0 |
| S7_extracted_29026 | 1679 | No | NA | 1 | 0 | 0 | Not-determined | Genome-fragment | NA | NA | 0 |
| S7_extracted_29196 | 28259 | No | NA | 36 | 11 | 1 | Medium-quality | Genome-fragment | 50.97 | AAI-based (high-confidence) | 0 |
| S7_extracted_29282 | 3097 | No | NA | 3 | 0 | 1 | Not-determined | Genome-fragment | NA | NA | 0 |
| S7_extracted_29407 | 2231 | No | NA | 4 | 0 | 2 | Not-determined | Genome-fragment | NA | NA | 0 |
| S7_extracted_29408 | 2112 | No | NA | 2 | 0 | 1 | Not-determined | Genome-fragment | NA | NA | 0 |
| S7_extracted_29439 | 1162 | No | NA | 1 | 0 | 1 | Not-determined | Genome-fragment | NA | NA | 0 |
| S7_extracted_29645 | 1319 | No | NA | 2 | 0 | 1 | Not-determined | Genome-fragment | NA | NA | 0 |
| S12_extracted_k141_17310 | 2052 | No | NA | 2 | 1 | 0 | Low-quality | Genome-fragment | 1.27 | HMM-based (lower-bound) | 0 |
| S7_extracted_29785 | 1681 | No | NA | 1 | 0 | 1 | Not-determined | Genome-fragment | NA | NA | 0 |
| S7_extracted_29808 | 1025 | No | NA | 2 | 0 | 1 | Not-determined | Genome-fragment | NA | NA | 0 |
| S12_vs_k141_35260\|\|full | 2052 | No | NA | 3 | 0 | 1 | Low-quality | Genome-fragment | 2.34 | AAI-based (medium-confidence) | 0 |
| S8_vs_k141_23043\|\|full | 2052 | No | NA | 3 | 2 | 0 | Low-quality | Genome-fragment | 4.55 | AAI-based (high-confidence) | 0 |
| S9_extracted_k141_88100 | 2053 | No | NA | 4 | 2 | 1 | Low-quality | Genome-fragment | 4.82 | AAI-based (high-confidence) | 0 |
| S9_vs_k141_58834\|\|full | 2053 | Yes | 1478 | 4 | 1 | 1 | Low-quality | Genome-fragment | 4.9 | AAI-based (medium-confidence) | 28.01 |
| S7_extracted_29941 | 19166 | No | NA | 37 | 36 | 0 | Medium-quality | Genome-fragment | 63.15 | AAI-based (high-confidence) | 0 |
| S7_extracted_29997 | 30505 | No | NA | 28 | 0 | 8 | Not-determined | Genome-fragment | NA | NA | 0 |
| S1_extracted_k141_137865 | 2053 | Yes | 822 | 6 | 2 | 1 | Low-quality | Genome-fragment | 1.42 | AAI-based (high-confidence) | 59.96 |
| S7_extracted_30025 | 2730 | No | NA | 1 | 0 | 0 | Not-determined | Genome-fragment | NA | NA | 0 |
| S1_extracted_k141_73026 | 2053 | No | NA | 5 | 0 | 0 | Low-quality | Genome-fragment | 2.1 | AAI-based (high-confidence) | 0 |
| S3_extracted_k141_95490 | 2053 | No | NA | 4 | 0 | 1 | Low-quality | Genome-fragment | 4.05 | AAI-based (medium-confidence) | 0 |
| S7_extracted_30083 | 1827 | No | NA | 4 | 0 | 0 | Not-determined | Genome-fragment | NA | NA | 0 |
| S5_vs_k141_13893\|\|full | 2053 | No | NA | 4 | 0 | 2 | Low-quality | Genome-fragment | 2.85 | AAI-based (medium-confidence) | 0 |
| S12_vs_k141_63568\|\|full | 2053 | No | NA | 2 | 0 | 0 | Low-quality | Genome-fragment | 2.32 | AAI-based (medium-confidence) | 0 |
| S12_vs_k141_103144\|\|full | 2053 | No | NA | 3 | 0 | 0 | Low-quality | Genome-fragment | 1.22 | AAI-based (high-confidence) | 0 |
| S7_extracted_30546 | 1774 | No | NA | 4 | 0 | 0 | Not-determined | Genome-fragment | NA | NA | 0 |
| S7_extracted_30575 | 1750 | No | NA | 3 | 0 | 3 | Not-determined | Genome-fragment | NA | NA | 0 |
| S12_vs_k141_185321\|\|full | 2053 | No | NA | 7 | 0 | 0 | Low-quality | Genome-fragment | 4.21 | AAI-based (high-confidence) | 0 |
| S7_extracted_30762 | 1107 | No | NA | 1 | 0 | 1 | Not-determined | Genome-fragment | NA | NA | 0 |
| S7_extracted_30903 | 1748 | No | NA | 3 | 0 | 1 | Not-determined | Genome-fragment | NA | NA | 0 |
| S9_vs_k141_12323\|\|full | 2054 | No | NA | 6 | 0 | 0 | Low-quality | Genome-fragment | 5.67 | AAI-based (high-confidence) | 0 |
| S9_vs_k141_70490\|\|full | 2054 | No | NA | 4 | 0 | 0 | Low-quality | Genome-fragment | 2.45 | AAI-based (high-confidence) | 0 |
| S7_extracted_31126 | 1058 | No | NA | 2 | 0 | 1 | Not-determined | Genome-fragment | NA | NA | 0 |
| S7_extracted_31276 | 7476 | No | NA | 5 | 0 | 2 | Not-determined | Genome-fragment | NA | NA | 0 |
| S13_vs_18213\|\|full | 2054 | No | NA | 3 | 2 | 0 | Low-quality | Genome-fragment | 3.62 | HMM-based (lower-bound) | 0 |
| S7_extracted_31499 | 1382 | No | NA | 1 | 0 | 1 | Not-determined | Genome-fragment | NA | NA | 0 |
| S7_extracted_31546 | 5439 | No | NA | 3 | 0 | 3 | Not-determined | Genome-fragment | NA | NA | 0 |
| S7_extracted_31851 | 1312 | No | NA | 2 | 0 | 1 | Not-determined | Genome-fragment | NA | NA | 0 |
| S7_extracted_32040 | 1394 | No | NA | 1 | 0 | 1 | Not-determined | Genome-fragment | NA | NA | 0 |
| S13_vs_99623\|\|full | 2054 | No | NA | 3 | 0 | 2 | Low-quality | Genome-fragment | 0.98 | AAI-based (medium-confidence) | 0 |
| S7_extracted_32237 | 1402 | No | NA | 2 | 0 | 2 | Not-determined | Genome-fragment | NA | NA | 0 |
| S5_vs_k141_17680\|\|lt2gene | 2054 | No | NA | 1 | 1 | 0 | Low-quality | Genome-fragment | 2.98 | AAI-based (high-confidence) | 0 |
| S7_extracted_32396 | 16486 | No | NA | 23 | 7 | 0 | Medium-quality | Genome-fragment | 72.1 | AAI-based (medium-confidence) | 0 |
| S7_extracted_32475 | 1217 | No | NA | 2 | 0 | 1 | Not-determined | Genome-fragment | NA | NA | 0 |
| S7_extracted_32559 | 14766 | No | NA | 20 | 7 | 0 | Medium-quality | Genome-fragment | 68.66 | AAI-based (medium-confidence) | 0 |
| S7_extracted_32764 | 1772 | No | NA | 4 | 0 | 1 | Not-determined | Genome-fragment | NA | NA | 0 |
| S16_vs_14701\|\|full | 2054 | No | NA | 5 | 0 | 0 | Low-quality | Genome-fragment | 4.17 | AAI-based (medium-confidence) | 0 |
| S7_extracted_32874 | 1435 | No | NA | 2 | 0 | 1 | Not-determined | Genome-fragment | NA | NA | 0 |
| S7_extracted_33221 | 1447 | No | NA | 1 | 0 | 1 | Not-determined | Genome-fragment | NA | NA | 0 |
| S7_extracted_33332 | 2068 | No | NA | 3 | 0 | 0 | Not-determined | Genome-fragment | NA | NA | 0 |
| S7_extracted_33501 | 2042 | No | NA | 2 | 0 | 0 | Not-determined | Genome-fragment | NA | NA | 0 |
| S7_extracted_33627 | 3816 | No | NA | 1 | 0 | 1 | Not-determined | Genome-fragment | NA | NA | 0 |
| S16_vs_105854\|\|lt2gene | 2054 | No | NA | 1 | 1 | 0 | Low-quality | Genome-fragment | 1.42 | HMM-based (lower-bound) | 0 |
| S12_extracted_k141_154364 | 2054 | No | NA | 2 | 0 | 2 | Low-quality | Genome-fragment | 1.34 | AAI-based (medium-confidence) | 0 |
| S7_extracted_33716 | 5728 | No | NA | 3 | 0 | 2 | Not-determined | Genome-fragment | NA | NA | 0 |
| S7_extracted_33756 | 2659 | No | NA | 3 | 0 | 2 | Not-determined | Genome-fragment | NA | NA | 0 |
| S15_extracted_41340 | 2054 | No | NA | 4 | 0 | 0 | Low-quality | Genome-fragment | 6.53 | AAI-based (medium-confidence) | 0 |
| S6_vs_k141_41721\|\|full | 2054 | No | NA | 2 | 0 | 1 | Low-quality | Genome-fragment | 0.85 | AAI-based (medium-confidence) | 0 |
| S7_extracted_34008 | 1630 | No | NA | 2 | 0 | 0 | Not-determined | Genome-fragment | NA | NA | 0 |
| S7_extracted_34013 | 3417 | No | NA | 4 | 0 | 3 | Not-determined | Genome-fragment | NA | NA | 0 |
| S7_extracted_34024 | 1441 | No | NA | 1 | 0 | 0 | Not-determined | Genome-fragment | NA | NA | 0 |
| S7_extracted_34060 | 1033 | No | NA | 1 | 0 | 1 | Not-determined | Genome-fragment | NA | NA | 0 |
| S7_extracted_34064 | 3110 | No | NA | 2 | 0 | 2 | Not-determined | Genome-fragment | NA | NA | 0 |
| S7_extracted_34084 | 1136 | No | NA | 1 | 0 | 1 | Not-determined | Genome-fragment | NA | NA | 0 |
| S8_vs_k141_206416\|\|full | 2054 | No | NA | 3 | 0 | 1 | Low-quality | Genome-fragment | 1.33 | AAI-based (high-confidence) | 0 |
| S7_extracted_34262 | 1767 | No | NA | 2 | 0 | 1 | Not-determined | Genome-fragment | NA | NA | 0 |
| S14_vs_k141_112635\|\|full | 2055 | No | NA | 3 | 0 | 2 | Low-quality | Genome-fragment | 2.55 | AAI-based (medium-confidence) | 0 |
| S3_vs_k141_19642\|\|full | 2055 | No | NA | 2 | 0 | 0 | Low-quality | Genome-fragment | 4.77 | AAI-based (medium-confidence) | 0 |
| S5_vs_k141_30476\|\|full | 2055 | No | NA | 6 | 1 | 0 | Low-quality | Genome-fragment | 4.58 | AAI-based (high-confidence) | 0 |
| S7_extracted_34821 | 2768 | No | NA | 1 | 0 | 0 | Not-determined | Genome-fragment | NA | NA | 0 |
| S7_extracted_35102 | 1019 | No | NA | 1 | 0 | 1 | Not-determined | Genome-fragment | NA | NA | 0 |
| S16_vs_78657\|\|full | 2055 | No | NA | 4 | 1 | 0 | Low-quality | Genome-fragment | 4.16 | AAI-based (high-confidence) | 0 |
| S7_extracted_35257 | 9748 | No | NA | 7 | 0 | 2 | Not-determined | Genome-fragment | NA | NA | 0 |
| S7_extracted_35294 | 1643 | No | NA | 1 | 0 | 0 | Not-determined | Genome-fragment | NA | NA | 0 |
| S7_extracted_35303 | 7508 | No | NA | 11 | 0 | 2 | Not-determined | Genome-fragment | NA | NA | 0 |
| S7_extracted_35324 | 1070 | No | NA | 3 | 0 | 0 | Not-determined | Genome-fragment | NA | NA | 0 |
| S7_extracted_35453 | 5988 | No | NA | 13 | 0 | 8 | Not-determined | Genome-fragment | NA | NA | 0 |
| S7_extracted_35464 | 2583 | No | NA | 2 | 0 | 2 | Not-determined | Genome-fragment | NA | NA | 0 |
| S7_extracted_35528 | 1024 | No | NA | 2 | 0 | 1 | Not-determined | Genome-fragment | NA | NA | 0 |
| S8_vs_k141_117583\|\|full | 2055 | No | NA | 2 | 0 | 2 | Low-quality | Genome-fragment | 2.45 | AAI-based (high-confidence) | 0 |
| S11_vs_k141_48912\|\|full | 2056 | No | NA | 4 | 0 | 0 | Low-quality | Genome-fragment | 5.2 | AAI-based (high-confidence) | 0 |
| S10_vs_91730\|\|full | 2056 | No | NA | 3 | 0 | 1 | Low-quality | Genome-fragment | 1.14 | AAI-based (high-confidence) | 0 |
| S7_extracted_36290 | 1225 | No | NA | 1 | 0 | 1 | Not-determined | Genome-fragment | NA | NA | 0 |
| S10_vs_105239\|\|full | 2056 | No | NA | 5 | 1 | 0 | Low-quality | Genome-fragment | 4.09 | AAI-based (high-confidence) | 0 |
| S7_extracted_36333 | 1508 | No | NA | 1 | 0 | 1 | Not-determined | Genome-fragment | NA | NA | 0 |
| S8_vs_k141_133932\|\|full | 2056 | No | NA | 3 | 2 | 0 | Low-quality | Genome-fragment | 4.14 | AAI-based (high-confidence) | 0 |
| S7_extracted_36469 | 4389 | No | NA | 6 | 0 | 3 | Not-determined | Genome-fragment | NA | NA | 0 |
| S7_extracted_36627 | 14887 | No | NA | 21 | 7 | 1 | Medium-quality | Genome-fragment | 51.36 | HMM-based (lower-bound) | 0 |
| S7_extracted_36716 | 9277 | No | NA | 5 | 0 | 4 | Not-determined | Genome-fragment | NA | NA | 0 |
| S7_extracted_36775 | 1708 | No | NA | 1 | 0 | 1 | Not-determined | Genome-fragment | NA | NA | 0 |
| S8_vs_k141_42990\|\|full | 2056 | No | NA | 4 | 1 | 0 | Low-quality | Genome-fragment | 1.3 | HMM-based (lower-bound) | 0 |
| S14_vs_k141_97316\|\|full | 2057 | No | NA | 4 | 4 | 0 | Low-quality | Genome-fragment | 5.69 | AAI-based (high-confidence) | 0 |
| S7_extracted_36933 | 4409 | No | NA | 2 | 0 | 2 | Not-determined | Genome-fragment | NA | NA | 0 |
| S7_extracted_37066 | 1298 | No | NA | 1 | 0 | 0 | Not-determined | Genome-fragment | NA | NA | 0 |
| S7_extracted_37091 | 3323 | No | NA | 3 | 0 | 0 | Not-determined | Genome-fragment | NA | NA | 0 |
| S7_extracted_37132 | 1886 | No | NA | 2 | 0 | 1 | Not-determined | Genome-fragment | NA | NA | 0 |
| S7_extracted_37279 | 80069 | No | NA | 127 | 16 | 1 | Complete | High-quality | 100 | DTR (high-confidence) | 0 |
| S7_extracted_37293 | 53167 | No | NA | 74 | 27 | 0 | High-quality | High-quality | 94.3 | AAI-based (high-confidence) | 0 |
| S7_extracted_37727 | 3081 | No | NA | 1 | 0 | 1 | Not-determined | Genome-fragment | NA | NA | 0 |
| S7_extracted_37741 | 1030 | No | NA | 2 | 0 | 2 | Not-determined | Genome-fragment | NA | NA | 0 |
| S1_vs_k141_769\|\|full | 2057 | No | NA | 3 | 1 | 0 | Low-quality | Genome-fragment | 4.65 | AAI-based (high-confidence) | 0 |
| S7_extracted_37962 | 2429 | No | NA | 2 | 0 | 1 | Not-determined | Genome-fragment | NA | NA | 0 |
| S7_extracted_38012 | 1614 | No | NA | 3 | 0 | 3 | Not-determined | Genome-fragment | NA | NA | 0 |
| S1_vs_k141_78458\|\|full | 2057 | No | NA | 3 | 0 | 0 | Low-quality | Genome-fragment | 4.76 | AAI-based (high-confidence) | 0 |
| S7_extracted_38253 | 1068 | No | NA | 1 | 0 | 0 | Not-determined | Genome-fragment | NA | NA | 0 |
| S3_extracted_k141_87790 | 2057 | No | NA | 5 | 0 | 0 | Low-quality | Genome-fragment | 2.19 | AAI-based (high-confidence) | 0 |
| S5_vs_k141_4635\|\|full | 2057 | No | NA | 2 | 0 | 2 | Low-quality | Genome-fragment | 1.45 | AAI-based (high-confidence) | 0 |
| S7_extracted_38494 | 1135 | No | NA | 1 | 0 | 1 | Not-determined | Genome-fragment | NA | NA | 0 |
| S7_extracted_38546 | 4044 | No | NA | 5 | 0 | 1 | Not-determined | Genome-fragment | NA | NA | 0 |
| S7_extracted_38554 | 1073 | No | NA | 1 | 0 | 1 | Not-determined | Genome-fragment | NA | NA | 0 |
| S7_extracted_38697 | 60640 | No | NA | 84 | 33 | 1 | Complete | High-quality | 100 | DTR (high-confidence) | 0 |
| S16_vs_50059\|\|full | 2057 | No | NA | 3 | 0 | 0 | Low-quality | Genome-fragment | 2.49 | AAI-based (medium-confidence) | 0 |
| S10_extracted_109594 | 2057 | No | NA | 3 | 2 | 0 | Low-quality | Genome-fragment | 1.47 | HMM-based (lower-bound) | 0 |
| S7_extracted_39222 | 13069 | No | NA | 20 | 0 | 5 | Not-determined | Genome-fragment | NA | NA | 0 |
| S7_extracted_39255 | 1356 | No | NA | 3 | 0 | 0 | Not-determined | Genome-fragment | NA | NA | 0 |
| S7_extracted_39305 | 1025 | No | NA | 1 | 0 | 0 | Not-determined | Genome-fragment | NA | NA | 0 |
| S7_extracted_39311 | 1089 | No | NA | 2 | 0 | 1 | Not-determined | Genome-fragment | NA | NA | 0 |
| S7_extracted_39334 | 1233 | No | NA | 2 | 0 | 1 | Not-determined | Genome-fragment | NA | NA | 0 |
| S7_extracted_39379 | 1343 | No | NA | 1 | 0 | 1 | Not-determined | Genome-fragment | NA | NA | 0 |
| S7_extracted_39664 | 1131 | No | NA | 2 | 0 | 1 | Not-determined | Genome-fragment | NA | NA | 0 |
| S7_extracted_39724 | 37389 | No | NA | 52 | 14 | 0 | High-quality | High-quality | 100 | AAI-based (medium-confidence) | 0 |
| S7_extracted_39854 | 1473 | No | NA | 1 | 0 | 1 | Not-determined | Genome-fragment | NA | NA | 0 |
| S7_extracted_39972 | 1095 | No | NA | 4 | 0 | 1 | Not-determined | Genome-fragment | NA | NA | 0 |
| S17_vs_k141_92496\|\|full | 2058 | No | NA | 3 | 2 | 0 | Low-quality | Genome-fragment | 3.2 | HMM-based (lower-bound) | 0 |
| S7_extracted_40018 | 1611 | No | NA | 4 | 0 | 0 | Not-determined | Genome-fragment | NA | NA | 0 |
| S7_extracted_40046 | 1287 | No | NA | 2 | 0 | 2 | Not-determined | Genome-fragment | NA | NA | 0 |
| S7_extracted_40143 | 1086 | No | NA | 1 | 0 | 0 | Not-determined | Genome-fragment | NA | NA | 0 |
| S7_extracted_40241 | 1776 | No | NA | 1 | 0 | 0 | Not-determined | Genome-fragment | NA | NA | 0 |
| S1_vs_k141_146273\|\|full | 2058 | No | NA | 3 | 0 | 0 | Low-quality | Genome-fragment | 4.45 | AAI-based (high-confidence) | 0 |
| S7_vs_90681\|\|full | 2058 | No | NA | 4 | 0 | 0 | Low-quality | Genome-fragment | 5.36 | AAI-based (high-confidence) | 0 |
| S7_extracted_40432 | 5088 | No | NA | 5 | 0 | 2 | Not-determined | Genome-fragment | NA | NA | 0 |
| S7_extracted_40433 | 5710 | No | NA | 4 | 0 | 3 | Not-determined | Genome-fragment | NA | NA | 0 |
| S7_extracted_40441 | 1376 | No | NA | 1 | 0 | 1 | Not-determined | Genome-fragment | NA | NA | 0 |
| S7_extracted_40480 | 1156 | No | NA | 1 | 0 | 1 | Not-determined | Genome-fragment | NA | NA | 0 |
| S15_vs_58209\|\|full | 2058 | No | NA | 7 | 0 | 0 | Low-quality | Genome-fragment | 3.61 | AAI-based (high-confidence) | 0 |
| S7_extracted_40563 | 5704 | No | NA | 6 | 0 | 0 | Not-determined | Genome-fragment | NA | NA | 0 |
| S7_extracted_40648 | 4048 | No | NA | 4 | 0 | 0 | Not-determined | Genome-fragment | NA | NA | 0 |
| S7_extracted_40683 | 1255 | No | NA | 2 | 0 | 1 | Not-determined | Genome-fragment | NA | NA | 0 |
| S5_extracted_k141_128988 | 2059 | No | NA | 2 | 2 | 0 | Low-quality | Genome-fragment | 5.6 | AAI-based (high-confidence) | 0 |
| S7_extracted_40832 | 1346 | No | NA | 1 | 0 | 1 | Not-determined | Genome-fragment | NA | NA | 0 |
| S7_extracted_40865 | 1136 | No | NA | 1 | 0 | 0 | Not-determined | Genome-fragment | NA | NA | 0 |
| S12_vs_k141_131720\|\|full | 2059 | No | NA | 6 | 1 | 0 | Low-quality | Genome-fragment | 4.42 | AAI-based (high-confidence) | 0 |
| S7_extracted_40912 | 1261 | No | NA | 2 | 0 | 1 | Not-determined | Genome-fragment | NA | NA | 0 |
| S6_vs_k141_11589\|\|full | 2059 | No | NA | 7 | 1 | 0 | Low-quality | Genome-fragment | 4.95 | AAI-based (high-confidence) | 0 |
| S8_vs_k141_16140\|\|full | 2059 | No | NA | 3 | 0 | 2 | Low-quality | Genome-fragment | 2.76 | AAI-based (medium-confidence) | 0 |
| S7_vs_63786\|\|full | 2060 | No | NA | 3 | 0 | 1 | Low-quality | Genome-fragment | 3.79 | AAI-based (medium-confidence) | 0 |
| S7_extracted_41272 | 1452 | No | NA | 3 | 0 | 0 | Not-determined | Genome-fragment | NA | NA | 0 |
| S7_extracted_41413 | 1304 | No | NA | 1 | 0 | 0 | Not-determined | Genome-fragment | NA | NA | 0 |
| S7_extracted_41540 | 3460 | No | NA | 7 | 0 | 1 | Not-determined | Genome-fragment | NA | NA | 0 |
| S7_extracted_41771 | 8400 | No | NA | 6 | 0 | 4 | Not-determined | Genome-fragment | NA | NA | 0 |
| S7_extracted_41780 | 1156 | No | NA | 1 | 0 | 1 | Not-determined | Genome-fragment | NA | NA | 0 |
| S7_extracted_41988 | 1017 | No | NA | 1 | 0 | 1 | Not-determined | Genome-fragment | NA | NA | 0 |
| S7_extracted_42121 | 3201 | No | NA | 3 | 0 | 0 | Not-determined | Genome-fragment | NA | NA | 0 |
| S7_vs_72743\|\|full | 2060 | No | NA | 3 | 0 | 0 | Low-quality | Genome-fragment | 4.65 | AAI-based (high-confidence) | 0 |
| S16_vs_52772\|\|full | 2060 | No | NA | 4 | 0 | 0 | Low-quality | Genome-fragment | 4.17 | AAI-based (high-confidence) | 0 |
| S7_extracted_42189 | 1035 | No | NA | 1 | 0 | 0 | Not-determined | Genome-fragment | NA | NA | 0 |
| S7_extracted_42215 | 5199 | No | NA | 3 | 0 | 3 | Not-determined | Genome-fragment | NA | NA | 0 |
| S7_extracted_42269 | 4139 | No | NA | 6 | 0 | 1 | Not-determined | Genome-fragment | NA | NA | 0 |
| S7_extracted_42294 | 1532 | No | NA | 2 | 0 | 2 | Not-determined | Genome-fragment | NA | NA | 0 |
| S7_extracted_42318 | 2175 | No | NA | 1 | 0 | 1 | Not-determined | Genome-fragment | NA | NA | 0 |
| S7_extracted_42363 | 1785 | No | NA | 1 | 0 | 0 | Not-determined | Genome-fragment | NA | NA | 0 |
| S7_extracted_42534 | 1309 | No | NA | 2 | 0 | 0 | Not-determined | Genome-fragment | NA | NA | 0 |
| S12_vs_k141_54391\|\|full | 2060 | No | NA | 3 | 0 | 1 | Low-quality | Genome-fragment | 1.83 | AAI-based (medium-confidence) | 0 |
| S7_extracted_42691 | 3622 | No | NA | 3 | 0 | 3 | Not-determined | Genome-fragment | NA | NA | 0 |
| S7_extracted_42704 | 3717 | No | NA | 4 | 0 | 1 | Not-determined | Genome-fragment | NA | NA | 0 |
| S7_extracted_42739 | 2481 | No | NA | 3 | 0 | 1 | Not-determined | Genome-fragment | NA | NA | 0 |
| S7_extracted_42742 | 1097 | No | NA | 1 | 0 | 0 | Not-determined | Genome-fragment | NA | NA | 0 |
| S7_extracted_42800 | 23850 | No | NA | 19 | 0 | 1 | Not-determined | Genome-fragment | NA | NA | 0 |
| S6_vs_k141_20863\|\|full | 2060 | No | NA | 5 | 2 | 0 | Low-quality | Genome-fragment | 2.86 | HMM-based (lower-bound) | 0 |
| S14_vs_k141_53569\|\|full | 2061 | No | NA | 4 | 3 | 0 | Low-quality | Genome-fragment | 4.54 | AAI-based (high-confidence) | 0 |
| S7_extracted_124986 | 2061 | No | NA | 3 | 0 | 0 | Low-quality | Genome-fragment | 3.03 | AAI-based (medium-confidence) | 0 |
| S7_vs_132643\|\|full | 2061 | No | NA | 3 | 0 | 2 | Low-quality | Genome-fragment | 1.28 | AAI-based (medium-confidence) | 0 |
| S7_extracted_43240 | 1311 | No | NA | 2 | 0 | 0 | Not-determined | Genome-fragment | NA | NA | 0 |
| S7_extracted_43375 | 4207 | No | NA | 4 | 0 | 0 | Not-determined | Genome-fragment | NA | NA | 0 |
| S7_extracted_43425 | 3267 | No | NA | 8 | 0 | 1 | Not-determined | Genome-fragment | NA | NA | 0 |
| S7_extracted_43471 | 1375 | No | NA | 1 | 0 | 1 | Not-determined | Genome-fragment | NA | NA | 0 |
| S7_extracted_43581 | 3525 | No | NA | 6 | 0 | 2 | Not-determined | Genome-fragment | NA | NA | 0 |
| S7_extracted_43615 | 1354 | No | NA | 1 | 0 | 1 | Not-determined | Genome-fragment | NA | NA | 0 |
| S7_extracted_43663 | 1108 | No | NA | 1 | 0 | 1 | Not-determined | Genome-fragment | NA | NA | 0 |
| S7_extracted_43716 | 1374 | No | NA | 1 | 0 | 0 | Not-determined | Genome-fragment | NA | NA | 0 |
| S7_extracted_43798 | 7581 | No | NA | 11 | 0 | 2 | Not-determined | Genome-fragment | NA | NA | 0 |
| S16_extracted_111936 | 2061 | No | NA | 5 | 3 | 0 | Low-quality | Genome-fragment | 4.63 | AAI-based (high-confidence) | 0 |
| S16_vs_133986\|\|full | 2061 | No | NA | 3 | 2 | 0 | Low-quality | Genome-fragment | 3.6 | AAI-based (high-confidence) | 0 |
| S7_extracted_43835 | 1322 | No | NA | 1 | 0 | 1 | Not-determined | Genome-fragment | NA | NA | 0 |
| S16_vs_106825\|\|lt2gene | 2061 | No | NA | 2 | 2 | 0 | Low-quality | Genome-fragment | 3.82 | HMM-based (lower-bound) | 0 |
| S11_vs_k141_86209\|\|full | 2062 | No | NA | 4 | 1 | 0 | Low-quality | Genome-fragment | 3.56 | AAI-based (high-confidence) | 0 |
| S7_extracted_43942 | 1737 | No | NA | 2 | 0 | 0 | Not-determined | Genome-fragment | NA | NA | 0 |
| S7_extracted_43983 | 1723 | No | NA | 3 | 0 | 3 | Not-determined | Genome-fragment | NA | NA | 0 |
| S7_extracted_43984 | 3742 | No | NA | 3 | 0 | 2 | Not-determined | Genome-fragment | NA | NA | 0 |
| S7_extracted_44131 | 2670 | No | NA | 1 | 0 | 1 | Not-determined | Genome-fragment | NA | NA | 0 |
| S7_extracted_44344 | 2168 | No | NA | 2 | 0 | 1 | Not-determined | Genome-fragment | NA | NA | 0 |
| S7_extracted_44391 | 1272 | No | NA | 1 | 0 | 1 | Not-determined | Genome-fragment | NA | NA | 0 |
| S3_vs_k141_69893\|\|full | 2062 | No | NA | 5 | 2 | 0 | Low-quality | Genome-fragment | 4.63 | AAI-based (high-confidence) | 0 |

| | | | | | | | | | | | | |
|---|---|---|---|---|---|---|---|---|---|---|---|---|
| S6_vs_k141_25395\|\|full | 2062 | No | NA | 3 | 0 | 1 | Low-quality | Genome-fragment | | 1.94 | AAI-based (high-confidence) | 0 |
| S9_vs_k141_46328\|\|full | 2063 | No | NA | 3 | 2 | 0 | Low-quality | Genome-fragment | | 5.05 | AAI-based (high-confidence) | 0 |
| S3_vs_k141_64919\|\|full | 2063 | No | NA | 3 | 0 | 0 | Low-quality | Genome-fragment | | 2.52 | AAI-based (high-confidence) | 0 |
| S7_extracted_44625 | 1312 | No | NA | 3 | 0 | 1 | Not-determined | Genome-fragment | NA | | NA | 0 |
| S7_extracted_44634 | 1032 | No | NA | 1 | 0 | 1 | Not-determined | Genome-fragment | NA | | NA | 0 |
| S7_extracted_44968 | 2001 | No | NA | 5 | 0 | 4 | Not-determined | Genome-fragment | NA | | NA | 0 |
| S16_vs_41722\|\|full | 2063 | No | NA | 6 | 4 | 0 | Low-quality | Genome-fragment | | 3.17 | AAI-based (high-confidence) | 0 |
| S7_extracted_45314 | 1553 | No | NA | 2 | 0 | 1 | Not-determined | Genome-fragment | NA | | NA | 0 |
| S7_extracted_45565 | 3312 | No | NA | 3 | 0 | 2 | Not-determined | Genome-fragment | NA | | NA | 0 |
| S12_extracted_k141_91070 | 2063 | No | NA | 3 | 0 | 0 | Low-quality | Genome-fragment | | 4 | AAI-based (high-confidence) | 0 |
| S12_vs_k141_29739\|\|full | 2063 | No | NA | 4 | 0 | 0 | Low-quality | Genome-fragment | | 5.11 | AAI-based (medium-confidence) | 0 |
| S7_extracted_45744 | 9902 | No | NA | 9 | 0 | 8 | Not-determined | Genome-fragment | NA | | NA | 0 |
| S7_extracted_46093 | 1101 | No | NA | 1 | 0 | 0 | Not-determined | Genome-fragment | NA | | NA | 0 |
| S7_extracted_46509 | 1460 | No | NA | 1 | 0 | 1 | Not-determined | Genome-fragment | NA | | NA | 0 |
| S6_vs_k141_33811\|\|full | 2063 | No | NA | 4 | 0 | 2 | Low-quality | Genome-fragment | | 0.72 | AAI-based (medium-confidence) | 0 |
| S8_vs_k141_374\|\|full | 2063 | No | NA | 3 | 1 | 0 | Low-quality | Genome-fragment | | 1.28 | AAI-based (high-confidence) | 0 |
| S7_extracted_46765 | 2042 | No | NA | 2 | 0 | 1 | Not-determined | Genome-fragment | NA | | NA | 0 |
| S7_extracted_46818 | 1021 | No | NA | 1 | 0 | 1 | Not-determined | Genome-fragment | NA | | NA | 0 |
| S7_extracted_46889 | 1442 | No | NA | 2 | 0 | 0 | Not-determined | Genome-fragment | NA | | NA | 0 |
| S7_extracted_47171 | 1589 | No | NA | 2 | 0 | 2 | Not-determined | Genome-fragment | NA | | NA | 0 |
| S7_extracted_47179 | 1097 | No | NA | 1 | 0 | 1 | Not-determined | Genome-fragment | NA | | NA | 0 |
| S17_vs_k141_24259\|\|full | 2064 | No | NA | 3 | 0 | 1 | Low-quality | Genome-fragment | | 3.36 | AAI-based (medium-confidence) | 0 |
| S14_vs_k141_78885\|\|full | 2064 | No | NA | 3 | 1 | 0 | Low-quality | Genome-fragment | | 46.7 | AAI-based (high-confidence) | 0 |
| S7_extracted_47391 | 1524 | No | NA | 1 | 0 | 1 | Not-determined | Genome-fragment | NA | | NA | 0 |
| S7_extracted_47523 | 1369 | No | NA | 2 | 0 | 2 | Not-determined | Genome-fragment | NA | | NA | 0 |
| S7_extracted_47777 | 3020 | No | NA | 4 | 0 | 1 | Not-determined | Genome-fragment | NA | | NA | 0 |
| S16_extracted_74821 | 2064 | No | NA | 3 | 2 | 0 | Low-quality | Genome-fragment | | 5.62 | AAI-based (high-confidence) | 0 |
| S7_extracted_47908 | 3279 | No | NA | 4 | 0 | 0 | Not-determined | Genome-fragment | NA | | NA | 0 |
| S6_vs_k141_42475\|\|full | 2064 | No | NA | 3 | 0 | 0 | Low-quality | Genome-fragment | | 2.97 | AAI-based (medium-confidence) | 0 |
| S7_extracted_48099 | 2214 | No | NA | 2 | 0 | 2 | Not-determined | Genome-fragment | NA | | NA | 0 |
| S7_extracted_48117 | 1385 | No | NA | 3 | 0 | 1 | Not-determined | Genome-fragment | NA | | NA | 0 |
| S7_extracted_48304 | 2205 | No | NA | 2 | 0 | 1 | Not-determined | Genome-fragment | NA | | NA | 0 |
| S7_extracted_48547 | 3002 | No | NA | 3 | 0 | 0 | Not-determined | Genome-fragment | NA | | NA | 0 |
| S7_extracted_48574 | 1119 | No | NA | 2 | 0 | 0 | Not-determined | Genome-fragment | NA | | NA | 0 |
| S7_extracted_48593 | 1362 | No | NA | 1 | 0 | 1 | Not-determined | Genome-fragment | NA | | NA | 0 |
| S17_extracted_k141_72209 | 2065 | No | NA | 2 | 0 | 1 | Low-quality | Genome-fragment | | 1.44 | AAI-based (medium-confidence) | 0 |
| S9_vs_k141_41136\|\|full | 2065 | No | NA | 4 | 1 | 0 | Low-quality | Genome-fragment | | 4.47 | AAI-based (medium-confidence) | 0 |
| S7_extracted_48908 | 5122 | No | NA | 5 | 0 | 2 | Not-determined | Genome-fragment | NA | | NA | 0 |
| S7_extracted_48933 | 3255 | No | NA | 2 | 0 | 1 | Not-determined | Genome-fragment | NA | | NA | 0 |
| S11_extracted_k141_48484 | 2065 | No | NA | 2 | 0 | 0 | Low-quality | Genome-fragment | | 3.21 | AAI-based (high-confidence) | 0 |
| S7_extracted_49007 | 46929 | No | NA | 62 | 22 | 0 | Medium-quality | Genome-fragment | | 80.75 | AAI-based (high-confidence) | 0 |
| S7_extracted_49107 | 1757 | No | NA | 1 | 0 | 0 | Not-determined | Genome-fragment | NA | | NA | 0 |
| S7_extracted_49164 | 1230 | No | NA | 1 | 0 | 1 | Not-determined | Genome-fragment | NA | | NA | 0 |
| S7_extracted_49239 | 1266 | No | NA | 2 | 0 | 0 | Not-determined | Genome-fragment | NA | | NA | 0 |
| S11_vs_k141_102134\|\|full | 2065 | No | NA | 4 | 0 | 0 | Low-quality | Genome-fragment | | 4.29 | AAI-based (high-confidence) | 0 |
| S7_extracted_49287 | 2577 | No | NA | 1 | 0 | 0 | Not-determined | Genome-fragment | NA | | NA | 0 |
| S7_vs_103612\|\|full | 2065 | No | NA | 2 | 2 | 0 | Low-quality | Genome-fragment | | 4.49 | AAI-based (high-confidence) | 0 |
| S7_vs_108368\|\|full | 2065 | No | NA | 4 | 0 | 0 | Low-quality | Genome-fragment | | 5.54 | AAI-based (high-confidence) | 0 |
| S7_extracted_49454 | 1672 | No | NA | 1 | 0 | 1 | Not-determined | Genome-fragment | NA | | NA | 0 |
| S7_extracted_49485 | 38429 | No | NA | 54 | 13 | 0 | Medium-quality | Genome-fragment | | 86.12 | AAI-based (high-confidence) | 0 |
| S7_extracted_49627 | 3408 | No | NA | 3 | 0 | 2 | Not-determined | Genome-fragment | NA | | NA | 0 |
| S7_extracted_49634 | 1240 | No | NA | 3 | 0 | 1 | Not-determined | Genome-fragment | NA | | NA | 0 |
| S7_extracted_49835 | 2054 | No | NA | 1 | 0 | 1 | Not-determined | Genome-fragment | NA | | NA | 0 |
| S7_extracted_49954 | 1346 | No | NA | 3 | 0 | 1 | Not-determined | Genome-fragment | NA | | NA | 0 |
| S7_extracted_49964 | 1329 | No | NA | 1 | 0 | 0 | Not-determined | Genome-fragment | NA | | NA | 0 |
| S7_extracted_50025 | 4240 | No | NA | 5 | 0 | 4 | Not-determined | Genome-fragment | NA | | NA | 0 |
| S7_extracted_50091 | 1302 | No | NA | 1 | 0 | 0 | Not-determined | Genome-fragment | NA | | NA | 0 |
| S7_extracted_50115 | 1699 | No | NA | 1 | 0 | 1 | Not-determined | Genome-fragment | NA | | NA | 0 |
| S7_extracted_50175 | 1505 | No | NA | 3 | 0 | 0 | Not-determined | Genome-fragment | NA | | NA | 0 |
| S7_extracted_50360 | 1186 | No | NA | 1 | 0 | 0 | Not-determined | Genome-fragment | NA | | NA | 0 |
| S7_extracted_50591 | 1206 | No | NA | 1 | 0 | 0 | Not-determined | Genome-fragment | NA | | NA | 0 |
| S7_extracted_50594 | 1108 | No | NA | 2 | 0 | 2 | Not-determined | Genome-fragment | NA | | NA | 0 |
| S10_vs_5214\|\|full | 2065 | No | NA | 3 | 2 | 0 | Low-quality | Genome-fragment | | 3.79 | HMM-based (lower-bound) | 0 |
| S7_extracted_50747 | 1643 | No | NA | 1 | 0 | 1 | Not-determined | Genome-fragment | NA | | NA | 0 |
| S7_extracted_50812 | 1324 | No | NA | 1 | 0 | 0 | Not-determined | Genome-fragment | NA | | NA | 0 |
| S7_extracted_50867 | 4750 | No | NA | 10 | 0 | 9 | Not-determined | Genome-fragment | NA | | NA | 0 |
| S7_extracted_50996 | 1466 | No | NA | 1 | 0 | 1 | Not-determined | Genome-fragment | NA | | NA | 0 |
| S7_extracted_51139 | 2599 | No | NA | 3 | 0 | 3 | Not-determined | Genome-fragment | NA | | NA | 0 |
| S10_vs_30001\|\|full | 2065 | No | NA | 3 | 0 | 1 | Low-quality | Genome-fragment | | 4.99 | AAI-based (medium-confidence) | 0 |
| S7_extracted_51408 | 1228 | No | NA | 2 | 0 | 2 | Not-determined | Genome-fragment | NA | | NA | 0 |
| S7_extracted_51495 | 1543 | No | NA | 2 | 0 | 0 | Not-determined | Genome-fragment | NA | | NA | 0 |
| S7_extracted_51542 | 1017 | No | NA | 1 | 0 | 0 | Not-determined | Genome-fragment | NA | | NA | 0 |
| S17_vs_k141_112654\|\|full | 2066 | No | NA | 5 | 1 | 0 | Low-quality | Genome-fragment | | 3.33 | AAI-based (high-confidence) | 0 |
| S7_extracted_51632 | 2060 | No | NA | 1 | 0 | 1 | Not-determined | Genome-fragment | NA | | NA | 0 |
| S7_extracted_51653 | 1105 | No | NA | 2 | 0 | 0 | Not-determined | Genome-fragment | NA | | NA | 0 |
| S7_extracted_51785 | 1550 | No | NA | 2 | 0 | 1 | Not-determined | Genome-fragment | NA | | NA | 0 |
| S9_vs_k141_86242\|\|full | 2066 | No | NA | 3 | 0 | 2 | Low-quality | Genome-fragment | | 4.77 | AAI-based (medium-confidence) | 0 |
| S7_extracted_51918 | 1051 | No | NA | 1 | 0 | 1 | Not-determined | Genome-fragment | NA | | NA | 0 |
| S7_extracted_51928 | 6539 | No | NA | 8 | 0 | 6 | Not-determined | Genome-fragment | NA | | NA | 0 |
| S7_extracted_51947 | 1106 | No | NA | 3 | 0 | 3 | Not-determined | Genome-fragment | NA | | NA | 0 |
| S1_extracted_k141_34031 | 2066 | No | NA | 2 | 0 | 1 | Low-quality | Genome-fragment | | 0.77 | AAI-based (medium-confidence) | 0 |
| S14_vs_k141_113963\|\|full | 2067 | No | NA | 4 | 0 | 0 | Low-quality | Genome-fragment | | 5.54 | AAI-based (high-confidence) | 0 |
| S7_vs_100535\|\|full | 2067 | No | NA | 6 | 1 | 0 | Low-quality | Genome-fragment | | 2.52 | AAI-based (medium-confidence) | 0 |
| S7_extracted_52225 | 1560 | No | NA | 3 | 0 | 1 | Not-determined | Genome-fragment | NA | | NA | 0 |
| S7_extracted_52231 | 1626 | No | NA | 4 | 0 | 1 | Not-determined | Genome-fragment | NA | | NA | 0 |
| S7_extracted_52451 | 1197 | No | NA | 2 | 0 | 0 | Not-determined | Genome-fragment | NA | | NA | 0 |
| S16_vs_91439\|\|full | 2067 | No | NA | 4 | 0 | 0 | Low-quality | Genome-fragment | | 3.7 | AAI-based (medium-confidence) | 0 |
| S7_extracted_52628 | 1293 | No | NA | 1 | 0 | 1 | Not-determined | Genome-fragment | NA | | NA | 0 |
| S7_extracted_52631 | 1840 | No | NA | 1 | 0 | 1 | Not-determined | Genome-fragment | NA | | NA | 0 |
| S7_extracted_52640 | 2281 | No | NA | 3 | 0 | 0 | Not-determined | Genome-fragment | NA | | NA | 0 |
| S7_extracted_52856 | 1626 | No | NA | 3 | 0 | 1 | Not-determined | Genome-fragment | NA | | NA | 0 |
| S7_extracted_52893 | 1104 | No | NA | 2 | 0 | 2 | Not-determined | Genome-fragment | NA | | NA | 0 |
| S7_extracted_52916 | 3725 | No | NA | 6 | 0 | 5 | Not-determined | Genome-fragment | NA | | NA | 0 |
| S7_extracted_53025 | 1812 | No | NA | 1 | 0 | 0 | Not-determined | Genome-fragment | NA | | NA | 0 |
| S6_extracted_k141_391 | 2067 | No | NA | 3 | 3 | 0 | Low-quality | Genome-fragment | | 6.79 | AAI-based (high-confidence) | 0 |
| S7_vs_17410\|\|full | 2068 | No | NA | 4 | 3 | 0 | Low-quality | Genome-fragment | | 4.22 | AAI-based (medium-confidence) | 0 |
| S7_extracted_53050 | 1019 | No | NA | 1 | 0 | 1 | Not-determined | Genome-fragment | NA | | NA | 0 |
| S16_extracted_74014 | 2068 | No | NA | 3 | 0 | 0 | Low-quality | Genome-fragment | | 2.28 | AAI-based (high-confidence) | 0 |
| S7_extracted_53284 | 1332 | No | NA | 6 | 0 | 1 | Not-determined | Genome-fragment | NA | | NA | 0 |
| S7_extracted_53390 | 3391 | No | NA | 2 | 0 | 2 | Not-determined | Genome-fragment | NA | | NA | 0 |
| S16_vs_48805\|\|full | 2068 | No | NA | 4 | 0 | 0 | Low-quality | Genome-fragment | | 2.97 | AAI-based (high-confidence) | 0 |
| S12_vs_k141_174375\|\|full | 2068 | No | NA | 6 | 2 | 0 | Low-quality | Genome-fragment | | 2.6 | HMM-based (lower-bound) | 0 |
| S7_extracted_53558 | 1261 | No | NA | 5 | 0 | 2 | Not-determined | Genome-fragment | NA | | NA | 0 |
| S7_extracted_53694 | 1296 | No | NA | 2 | 0 | 2 | Not-determined | Genome-fragment | NA | | NA | 0 |
| S7_extracted_53841 | 2118 | No | NA | 2 | 0 | 1 | Not-determined | Genome-fragment | NA | | NA | 0 |
| S7_extracted_54011 | 1094 | No | NA | 2 | 0 | 2 | Not-determined | Genome-fragment | NA | | NA | 0 |
| S7_extracted_54053 | 2043 | No | NA | 3 | 0 | 0 | Not-determined | Genome-fragment | NA | | NA | 0 |
| S2_extracted_k141_13483 | 2068 | No | NA | 4 | 1 | 0 | Low-quality | Genome-fragment | | 4.92 | HMM-based (lower-bound) | 0 |
| S8_extracted_k141_79859 | 2068 | No | NA | 2 | 0 | 1 | Low-quality | Genome-fragment | | 0.88 | AAI-based (high-confidence) | 0 |
| S7_extracted_54572 | 2409 | No | NA | 1 | 0 | 1 | Not-determined | Genome-fragment | NA | | NA | 0 |
| S11_vs_k141_10249\|\|full | 2069 | No | NA | 4 | 0 | 0 | Low-quality | Genome-fragment | | 3.26 | AAI-based (high-confidence) | 0 |
| S7_extracted_54636 | 1079 | No | NA | 1 | 0 | 0 | Not-determined | Genome-fragment | NA | | NA | 0 |
| S7_extracted_54718 | 2520 | No | NA | 3 | 0 | 0 | Not-determined | Genome-fragment | NA | | NA | 0 |
| S7_extracted_54823 | 1221 | No | NA | 1 | 0 | 1 | Not-determined | Genome-fragment | NA | | NA | 0 |
| S14_vs_k141_8877\|\|full | 2069 | No | NA | 5 | 2 | 0 | Low-quality | Genome-fragment | | 4.15 | AAI-based (high-confidence) | 0 |
| S7_extracted_54911 | 1099 | No | NA | 3 | 0 | 0 | Not-determined | Genome-fragment | NA | | NA | 0 |
| S7_extracted_54950 | 2979 | No | NA | 5 | 0 | 1 | Not-determined | Genome-fragment | NA | | NA | 0 |
| S7_extracted_54989 | 1175 | No | NA | 2 | 0 | 2 | Not-determined | Genome-fragment | NA | | NA | 0 |
| S16_vs_93811\|\|full | 2069 | No | NA | 2 | 0 | 0 | Low-quality | Genome-fragment | | 4.29 | AAI-based (medium-confidence) | 0 |
| S7_extracted_55112 | 3831 | No | NA | 2 | 0 | 2 | Not-determined | Genome-fragment | NA | | NA | 0 |
| S12_vs_k141_45781\|\|full | 2069 | No | NA | 4 | 0 | 0 | Low-quality | Genome-fragment | | 4.93 | AAI-based (medium-confidence) | 0 |
| S7_extracted_55375 | 3869 | No | NA | 2 | 0 | 1 | Not-determined | Genome-fragment | NA | | NA | 0 |
| S7_extracted_55643 | 2546 | No | NA | 2 | 0 | 1 | Not-determined | Genome-fragment | NA | | NA | 0 |
| S9_vs_k141_12385\|\|full | 2070 | No | NA | 4 | 0 | 0 | Low-quality | Genome-fragment | | 3.93 | AAI-based (high-confidence) | 0 |
| S14_vs_k141_24370\|\|full | 2070 | No | NA | 4 | 2 | 0 | Low-quality | Genome-fragment | | 4.78 | AAI-based (high-confidence) | 0 |
| S1_vs_k141_21761\|\|full | 2070 | No | NA | 3 | 2 | 0 | Low-quality | Genome-fragment | | 4.86 | AAI-based (high-confidence) | 0 |
| S5_vs_k141_108021\|\|full | 2070 | No | NA | 4 | 0 | 3 | Low-quality | Genome-fragment | | 1.64 | AAI-based (medium-confidence) | 0 |
| S7_vs_125907\|\|full | 2070 | No | NA | 3 | 1 | 0 | Low-quality | Genome-fragment | | 5.18 | AAI-based (high-confidence) | 0 |
| S7_extracted_56489 | 1074 | No | NA | 2 | 0 | 0 | Not-determined | Genome-fragment | NA | | NA | 0 |
| S7_extracted_56544 | 1869 | No | NA | 2 | 0 | 1 | Not-determined | Genome-fragment | NA | | NA | 0 |
| S7_extracted_56664 | 3030 | No | NA | 2 | 0 | 1 | Not-determined | Genome-fragment | NA | | NA | 0 |
| S7_extracted_56689 | 3090 | No | NA | 1 | 0 | 1 | Not-determined | Genome-fragment | NA | | NA | 0 |
| S7_vs_34094\|\|lt2gene | 2070 | No | NA | 2 | 2 | 0 | Low-quality | Genome-fragment | | 5.19 | AAI-based (high-confidence) | 0 |
| S11_vs_k141_78057\|\|full | 2071 | No | NA | 3 | 2 | 0 | Low-quality | Genome-fragment | | 3.32 | AAI-based (high-confidence) | 0 |
| S7_extracted_56955 | 1521 | No | NA | 2 | 0 | 1 | Not-determined | Genome-fragment | NA | | NA | 0 |
| S7_extracted_57009 | 3958 | No | NA | 3 | 0 | 3 | Not-determined | Genome-fragment | NA | | NA | 0 |
| S7_extracted_57097 | 2476 | No | NA | 3 | 0 | 2 | Not-determined | Genome-fragment | NA | | NA | 0 |
| S7_extracted_57115 | 1882 | No | NA | 1 | 0 | 1 | Not-determined | Genome-fragment | NA | | NA | 0 |
| S1_vs_k141_75076\|\|full | 2071 | No | NA | 4 | 2 | 0 | Low-quality | Genome-fragment | | 5.58 | AAI-based (high-confidence) | 0 |
| S7_extracted_57147 | 1643 | No | NA | 2 | 0 | 1 | Not-determined | Genome-fragment | NA | | NA | 0 |
| S7_extracted_57149 | 1945 | No | NA | 4 | 0 | 0 | Not-determined | Genome-fragment | NA | | NA | 0 |
| S3_vs_k141_73336\|\|full | 2071 | No | NA | 4 | 2 | 0 | Low-quality | Genome-fragment | | 3.33 | HMM-based (lower-bound) | 0 |
| S7_extracted_57237 | 1129 | No | NA | 1 | 0 | 0 | Not-determined | Genome-fragment | NA | | NA | 0 |
| S7_extracted_57619 | 1180 | No | NA | 1 | 0 | 1 | Not-determined | Genome-fragment | NA | | NA | 0 |
| S7_vs_24529\|\|full | 2071 | No | NA | 3 | 1 | 0 | Low-quality | Genome-fragment | | 3.49 | AAI-based (high-confidence) | 0 |
| S7_extracted_57764 | 1339 | No | NA | 1 | 0 | 0 | Not-determined | Genome-fragment | NA | | NA | 0 |
| S7_extracted_57787 | 2692 | No | NA | 2 | 0 | 2 | Not-determined | Genome-fragment | NA | | NA | 0 |
| S7_extracted_57831 | 7131 | No | NA | 3 | 0 | 2 | Not-determined | Genome-fragment | NA | | NA | 0 |
| S8_extracted_k141_62705 | 2071 | No | NA | 6 | 2 | 0 | Low-quality | Genome-fragment | | 4.08 | AAI-based (high-confidence) | 0 |
| S8_extracted_k141_41327 | 2071 | No | NA | 3 | 0 | 2 | Low-quality | Genome-fragment | | 0.99 | AAI-based (medium-confidence) | 0 |
| S9_extracted_k141_65843 | 2072 | No | NA | 1 | 1 | 0 | Low-quality | Genome-fragment | | 1.44 | AAI-based (high-confidence) | 0 |
| S7_extracted_57952 | 3105 | No | NA | 6 | 0 | 0 | Not-determined | Genome-fragment | NA | | NA | 0 |
| S7_extracted_58081 | 1680 | No | NA | 2 | 0 | 0 | Not-determined | Genome-fragment | NA | | NA | 0 |
| S7_extracted_58097 | 1999 | No | NA | 2 | 0 | 1 | Not-determined | Genome-fragment | NA | | NA | 0 |
| S7_extracted_58141 | 2096 | No | NA | 2 | 0 | 1 | Not-determined | Genome-fragment | NA | | NA | 0 |
| S7_extracted_58321 | 5560 | No | NA | 1 | 0 | 0 | Not-determined | Genome-fragment | NA | | NA | 0 |
| S1_vs_k141_84944\|\|full | 2072 | No | NA | 4 | 0 | 0 | Low-quality | Genome-fragment | | 0.99 | AAI-based (medium-confidence) | 0 |
| S7_extracted_58930 | 1231 | No | NA | 2 | 0 | 2 | Not-determined | Genome-fragment | NA | | NA | 0 |
| S7_extracted_58997 | 3578 | No | NA | 5 | 0 | 1 | Not-determined | Genome-fragment | NA | | NA | 0 |
| S7_extracted_59074 | 1198 | No | NA | 2 | 0 | 2 | Not-determined | Genome-fragment | NA | | NA | 0 |
| S7_extracted_59097 | 3660 | No | NA | 1 | 0 | 1 | Not-determined | Genome-fragment | NA | | NA | 0 |
| S7_extracted_59109 | 1326 | No | NA | 3 | 0 | 1 | Not-determined | Genome-fragment | NA | | NA | 0 |
| S7_extracted_59131 | 2590 | No | NA | 2 | 0 | 0 | Not-determined | Genome-fragment | NA | | NA | 0 |
| S7_extracted_59141 | 1005 | No | NA | 2 | 0 | 0 | Not-determined | Genome-fragment | NA | | NA | 0 |
| S7_vs_5578\|\|full | 2072 | No | NA | 3 | 3 | 0 | Low-quality | Genome-fragment | | 4.48 | AAI-based (high-confidence) | 0 |
| S10_vs_44498\|\|full | 2072 | No | NA | 3 | 0 | 2 | Low-quality | Genome-fragment | | 0.86 | AAI-based (medium-confidence) | 0 |
| S7_extracted_59549 | 3458 | No | NA | 3 | 0 | 1 | Not-determined | Genome-fragment | NA | | NA | 0 |
| S7_extracted_59640 | 1360 | No | NA | 2 | 0 | 2 | Not-determined | Genome-fragment | NA | | NA | 0 |
| S8_extracted_k141_120657 | 2072 | No | NA | 2 | 0 | 1 | Low-quality | Genome-fragment | | 2.2 | AAI-based (medium-confidence) | 0 |
| S9_vs_k141_25200\|\|full | 2073 | No | NA | 4 | 0 | 2 | Low-quality | Genome-fragment | | 1.35 | AAI-based (medium-confidence) | 0 |
| S7_extracted_60158 | 1251 | No | NA | 2 | 0 | 0 | Not-determined | Genome-fragment | NA | | NA | 0 |
| S11_vs_k141_61179\|\|full | 2073 | No | NA | 3 | 1 | 0 | Low-quality | Genome-fragment | | 2.29 | AAI-based (medium-confidence) | 0 |
| S7_extracted_60412 | 1069 | No | NA | 2 | 0 | 1 | Not-determined | Genome-fragment | NA | | NA | 0 |
| S7_extracted_60492 | 1363 | No | NA | 1 | 0 | 0 | Not-determined | Genome-fragment | NA | | NA | 0 |
| S16_vs_170208\|\|full | 2073 | No | NA | 8 | 1 | 0 | Low-quality | Genome-fragment | | 2.32 | HMM-based (lower-bound) | 0 |
| S7_extracted_60701 | 1070 | No | NA | 1 | 0 | 1 | Not-determined | Genome-fragment | NA | | NA | 0 |
| S7_extracted_60968 | 41855 | No | NA | 64 | 22 | 1 | Complete | High-quality | | 100 | DTR (high-confidence) | 0 |
| S7_extracted_60997 | 4845 | No | NA | 4 | 0 | 4 | Not-determined | Genome-fragment | NA | | NA | 0 |
| S7_extracted_61029 | 1307 | No | NA | 3 | 0 | 0 | Not-determined | Genome-fragment | NA | | NA | 0 |
| S7_extracted_61057 | 4115 | No | NA | 9 | 0 | 3 | Not-determined | Genome-fragment | NA | | NA | 0 |
| S7_extracted_61095 | 1131 | No | NA | 3 | 0 | 0 | Not-determined | Genome-fragment | NA | | NA | 0 |
| S12_vs_k141_154508\|\|full | 2073 | No | NA | 4 | 1 | 0 | Low-quality | Genome-fragment | | 3.71 | AAI-based (high-confidence) | 0 |
| S7_extracted_61311 | 1208 | No | NA | 3 | 0 | 2 | Not-determined | Genome-fragment | NA | | NA | 0 |
| S7_extracted_61416 | 2253 | No | NA | 1 | 0 | 1 | Not-determined | Genome-fragment | NA | | NA | 0 |
| S12_vs_k141_7409\|\|full | 2073 | No | NA | 4 | 0 | 4 | Low-quality | Genome-fragment | | 3.03 | AAI-based (medium-confidence) | 0 |
| S7_extracted_61559 | 1501 | No | NA | 2 | 0 | 2 | Not-determined | Genome-fragment | NA | | NA | 0 |
| S7_extracted_61596 | 2009 | No | NA | 3 | 0 | 0 | Not-determined | Genome-fragment | NA | | NA | 0 |
| S7_extracted_61685 | 2235 | No | NA | 2 | 0 | 2 | Not-determined | Genome-fragment | NA | | NA | 0 |
| S7_extracted_61722 | 1283 | No | NA | 3 | 0 | 0 | Not-determined | Genome-fragment | NA | | NA | 0 |
| S7_extracted_61744 | 1224 | No | NA | 2 | 0 | 1 | Not-determined | Genome-fragment | NA | | NA | 0 |
| S7_extracted_61803 | 3557 | No | NA | 7 | 0 | 0 | Not-determined | Genome-fragment | NA | | NA | 0 |
| S7_extracted_61848 | 1444 | No | NA | 2 | 0 | 2 | Not-determined | Genome-fragment | NA | | NA | 0 |
| S7_extracted_61922 | 1187 | No | NA | 1 | 0 | 1 | Not-determined | Genome-fragment | NA | | NA | 0 |
| S7_extracted_61977 | 1358 | No | NA | 2 | 0 | 1 | Not-determined | Genome-fragment | NA | | NA | 0 |
| S15_extracted_2598 | 2073 | No | NA | 4 | 0 | 0 | Low-quality | Genome-fragment | | 2.94 | AAI-based (medium-confidence) | 0 |
| S9_extracted_k141_11699 | 2074 | No | NA | 2 | 2 | 0 | Low-quality | Genome-fragment | | 4.57 | AAI-based (high-confidence) | 0 |
| S9_vs_k141_84909\|\|full | 2074 | No | NA | 4 | 0 | 0 | Low-quality | Genome-fragment | | 2.41 | AAI-based (high-confidence) | 0 |
| S7_extracted_62265 | 1477 | No | NA | 1 | 0 | 1 | Not-determined | Genome-fragment | NA | | NA | 0 |
| S7_extracted_62393 | 1089 | No | NA | 1 | 0 | 0 | Not-determined | Genome-fragment | NA | | NA | 0 |
| S7_extracted_62663 | 3059 | No | NA | 3 | 0 | 1 | Not-determined | Genome-fragment | NA | | NA | 0 |
| S7_extracted_62800 | 3057 | No | NA | 2 | 0 | 2 | Not-determined | Genome-fragment | NA | | NA | 0 |
| S7_extracted_62830 | 1734 | No | NA | 1 | 0 | 1 | Not-determined | Genome-fragment | NA | | NA | 0 |
| S7_extracted_62847 | 1029 | No | NA | 1 | 0 | 1 | Not-determined | Genome-fragment | NA | | NA | 0 |
| S7_extracted_62916 | 1440 | No | NA | 2 | 0 | 2 | Not-determined | Genome-fragment | NA | | NA | 0 |
| S13_extracted_41848 | 2074 | No | NA | 3 | 0 | 2 | Low-quality | Genome-fragment | | 1.34 | AAI-based (high-confidence) | 0 |
| S3_vs_k141_112274\|\|full | 2074 | No | NA | 5 | 2 | 0 | Low-quality | Genome-fragment | | 3.59 | HMM-based (lower-bound) | 0 |
| S7_extracted_63109 | 11275 | No | NA | 8 | 0 | 7 | Not-determined | Genome-fragment | NA | | NA | 0 |
| S7_vs_64391\|\|full | 2074 | No | NA | 4 | 0 | 0 | Low-quality | Genome-fragment | | 4.92 | AAI-based (high-confidence) | 0 |
| S6_vs_k141_44882\|\|full | 2074 | No | NA | 3 | 0 | 0 | Low-quality | Genome-fragment | | 1.64 | AAI-based (high-confidence) | 0 |
| S1_vs_k141_140782\|\|full | 2075 | No | NA | 5 | 1 | 0 | Low-quality | Genome-fragment | | 3.55 | AAI-based (medium-confidence) | 0 |
| S7_extracted_63513 | 1763 | No | NA | 1 | 0 | 1 | Not-determined | Genome-fragment | NA | | NA | 0 |
| S7_extracted_63648 | 1051 | No | NA | 1 | 0 | 0 | Not-determined | Genome-fragment | NA | | NA | 0 |
| S7_extracted_63690 | 1367 | No | NA | 2 | 0 | 0 | Not-determined | Genome-fragment | NA | | NA | 0 |
| S7_extracted_63731 | 2114 | No | NA | 3 | 0 | 0 | Not-determined | Genome-fragment | NA | | NA | 0 |
| S7_extracted_63805 | 1155 | No | NA | 2 | 0 | 1 | Not-determined | Genome-fragment | NA | | NA | 0 |
| S7_extracted_63859 | 1422 | No | NA | 2 | 0 | 2 | Not-determined | Genome-fragment | NA | | NA | 0 |
| S7_extracted_63883 | 2330 | No | NA | 3 | 0 | 3 | Not-determined | Genome-fragment | NA | | NA | 0 |
| S7_extracted_63894 | 27117 | No | NA | 41 | 18 | 0 | High-quality | High-quality | | 94.05 | AAI-based (high-confidence) | 0 |
| S7_extracted_63953 | 1053 | No | NA | 2 | 0 | 0 | Not-determined | Genome-fragment | NA | | NA | 0 |
| S7_extracted_64202 | 2107 | No | NA | 6 | 0 | 0 | Not-determined | Genome-fragment | NA | | NA | 0 |
| S1_vs_k141_126290\|\|full | 2075 | No | NA | 2 | 0 | 0 | Low-quality | Genome-fragment | | 3.59 | AAI-based (high-confidence) | 0 |
| S7_extracted_64320 | 1617 | No | NA | 2 | 0 | 1 | Not-determined | Genome-fragment | NA | | NA | 0 |
| S7_extracted_64375 | 5613 | No | NA | 4 | 0 | 3 | Not-determined | Genome-fragment | NA | | NA | 0 |
| S7_extracted_64486 | 1680 | No | NA | 2 | 0 | 1 | Not-determined | Genome-fragment | NA | | NA | 0 |
| S7_extracted_64571 | 6023 | No | NA | 7 | 0 | 3 | Not-determined | Genome-fragment | NA | | NA | 0 |
| S7_extracted_64609 | 3206 | No | NA | 2 | 0 | 2 | Not-determined | Genome-fragment | NA | | NA | 0 |
| S16_vs_17474\|\|full | 2075 | No | NA | 3 | 0 | 0 | Low-quality | Genome-fragment | | 5.03 | AAI-based (medium-confidence) | 0 |
| S7_extracted_65058 | 1067 | No | NA | 1 | 0 | 1 | Not-determined | Genome-fragment | NA | | NA | 0 |
| S15_vs_68169\|\|full | 2075 | No | NA | 6 | 0 | 0 | Low-quality | Genome-fragment | | 4.54 | AAI-based (high-confidence) | 0 |
| S7_extracted_65549 | 1325 | No | NA | 2 | 0 | 0 | Not-determined | Genome-fragment | NA | | NA | 0 |
| S7_extracted_65569 | 1150 | No | NA | 2 | 0 | 1 | Not-determined | Genome-fragment | NA | | NA | 0 |
| S7_extracted_65585 | 1097 | No | NA | 1 | 0 | 1 | Not-determined | Genome-fragment | NA | | NA | 0 |
| S7_extracted_65630 | 1614 | No | NA | 2 | 0 | 2 | Not-determined | Genome-fragment | NA | | NA | 0 |
| S7_extracted_65662 | 1314 | No | NA | 2 | 0 | 2 | Not-determined | Genome-fragment | NA | | NA | 0 |
| S17_vs_k141_101638\|\|full | 2076 | No | NA | 2 | 0 | 1 | Low-quality | Genome-fragment | | 0.63 | AAI-based (medium-confidence) | 0 |
| S7_extracted_65832 | 1705 | No | NA | 1 | 0 | 0 | Not-determined | Genome-fragment | NA | | NA | 0 |
| S9_vs_k141_40559\|\|full | 2076 | No | NA | 6 | 0 | 0 | Low-quality | Genome-fragment | | 5.49 | AAI-based (medium-confidence) | 0 |
| S11_vs_k141_9044\|\|full | 2076 | No | NA | 5 | 1 | 0 | Low-quality | Genome-fragment | | 5.56 | AAI-based (medium-confidence) | 0 |
| S7_extracted_66052 | 1285 | No | NA | 3 | 0 | 0 | Not-determined | Genome-fragment | NA | | NA | 0 |
| S13_vs_92270\|\|full | 2076 | No | NA | 3 | 2 | 0 | Low-quality | Genome-fragment | | 5.61 | AAI-based (high-confidence) | 0 |
| S7_extracted_66230 | 2120 | No | NA | 3 | 0 | 3 | Not-determined | Genome-fragment | NA | | NA | 0 |
| S7_extracted_66262 | 1012 | No | NA | 1 | 0 | 1 | Not-determined | Genome-fragment | NA | | NA | 0 |
| S3_extracted_k141_68622 | 2076 | No | NA | 3 | 0 | 0 | Low-quality | Genome-fragment | | 4 | AAI-based (medium-confidence) | 0 |
| S7_extracted_66401 | 1297 | No | NA | 1 | 0 | 0 | Not-determined | Genome-fragment | NA | | NA | 0 |
| S7_extracted_66604 | 3261 | No | NA | 3 | 0 | 1 | Not-determined | Genome-fragment | NA | | NA | 0 |
| S7_extracted_66629 | 1324 | No | NA | 2 | 0 | 1 | Not-determined | Genome-fragment | NA | | NA | 0 |
| S7_extracted_66643 | 2211 | No | NA | 1 | 0 | 0 | Not-determined | Genome-fragment | NA | | NA | 0 |
| S3_vs_k141_77076\|\|full | 2076 | No | NA | 2 | 0 | 1 | Low-quality | Genome-fragment | | 0.97 | AAI-based (high-confidence) | 0 |
| S16_vs_144303\|\|full | 2076 | No | NA | 3 | 0 | 2 | Low-quality | Genome-fragment | | 1.2 | AAI-based (medium-confidence) | 0 |
| S17_vs_k141_11585\|\|full | 2077 | No | NA | 3 | 0 | 0 | Low-quality | Genome-fragment | | 3.47 | AAI-based (medium-confidence) | 0 |
| S7_extracted_66920 | 2577 | No | NA | 2 | 0 | 0 | Not-determined | Genome-fragment | NA | | NA | 0 |
| S7_extracted_67013 | 4876 | No | NA | 6 | 0 | 6 | Not-determined | Genome-fragment | NA | | NA | 0 |
| S17_vs_k141_63520\|\|full | 2077 | No | NA | 4 | 0 | 0 | Low-quality | Genome-fragment | | 4.19 | AAI-based (medium-confidence) | 0 |
| S7_extracted_67246 | 1199 | No | NA | 2 | 0 | 1 | Not-determined | Genome-fragment | NA | | NA | 0 |
| S14_extracted_k141_36863 | 2077 | No | NA | 8 | 6 | 0 | Low-quality | Genome-fragment | | 6.84 | AAI-based (high-confidence) | 0 |
| S14_vs_k141_17713\|\|full | 2077 | No | NA | 5 | 3 | 0 | Low-quality | Genome-fragment | | 4.57 | AAI-based (medium-confidence) | 0 |
| S7_extracted_67538 | 5151 | No | NA | 2 | 0 | 1 | Not-determined | Genome-fragment | NA | | NA | 0 |
| S7_extracted_67552 | 1184 | No | NA | 1 | 0 | 1 | Not-determined | Genome-fragment | NA | | NA | 0 |
| S14_vs_k141_33606\|\|full | 2077 | No | NA | 6 | 0 | 0 | Low-quality | Genome-fragment | | 5.76 | AAI-based (high-confidence) | 0 |
| S7_extracted_67576 | 1093 | No | NA | 3 | 0 | 0 | Not-determined | Genome-fragment | NA | | NA | 0 |
| S7_extracted_67677 | 1165 | No | NA | 1 | 0 | 1 | Not-determined | Genome-fragment | NA | | NA | 0 |
| S7_extracted_67702 | 1495 | No | NA | 2 | 0 | 0 | Not-determined | Genome-fragment | NA | | NA | 0 |
| S7_vs_101259\|\|full | 2077 | No | NA | 5 | 1 | 0 | Low-quality | Genome-fragment | | 4.6 | AAI-based (high-confidence) | 0 |
| S7_extracted_67863 | 1131 | No | NA | 1 | 0 | 1 | Not-determined | Genome-fragment | NA | | NA | 0 |
| S7_extracted_67910 | 33641 | No | NA | 51 | 16 | 0 | High-quality | High-quality | | 100 | AAI-based (medium-confidence) | 0 |
| S7_extracted_67964 | 3127 | No | NA | 4 | 0 | 4 | Not-determined | Genome-fragment | NA | | NA | 0 |
| S7_extracted_67968 | 1104 | No | NA | 1 | 0 | 1 | Not-determined | Genome-fragment | NA | | NA | 0 |
| S7_extracted_68018 | 3626 | No | NA | 2 | 0 | 0 | Not-determined | Genome-fragment | NA | | NA | 0 |
| S7_extracted_68238 | 1489 | No | NA | 3 | 0 | 1 | Not-determined | Genome-fragment | NA | | NA | 0 |
| S7_extracted_68239 | 35810 | No | NA | 75 | 41 | 0 | High-quality | High-quality | | 100 | AAI-based (high-confidence) | 0 |
| S7_extracted_68256 | 1094 | No | NA | 2 | 0 | 1 | Not-determined | Genome-fragment | NA | | NA | 0 |
| S8_vs_k141_20920\|\|full | 2077 | No | NA | 3 | 0 | 1 | Low-quality | Genome-fragment | | 3.12 | AAI-based (medium-confidence) | 0 |
| S17_vs_k141_90414\|\|full | 2078 | No | NA | 6 | 0 | 0 | Low-quality | Genome-fragment | | 3.87 | AAI-based (medium-confidence) | 0 |
| S5_vs_k141_127726\|\|full | 2078 | No | NA | 3 | 0 | 0 | Low-quality | Genome-fragment | | 4.02 | AAI-based (medium-confidence) | 0 |
| S10_extracted_97568 | 2078 | No | NA | 2 | 0 | 0 | Low-quality | Genome-fragment | | 0.86 | AAI-based (high-confidence) | 0 |
| S7_extracted_68507 | 2279 | No | NA | 2 | 0 | 0 | Not-determined | Genome-fragment | NA | | NA | 0 |
| S7_extracted_68546 | 32399 | No | NA | 53 | 21 | 0 | Medium-quality | Genome-fragment | | 65.12 | AAI-based (high-confidence) | 0 |
| S7_extracted_68670 | 1307 | No | NA | 1 | 0 | 1 | Not-determined | Genome-fragment | NA | | NA | 0 |
| S13_vs_39455\|\|full | 2079 | No | NA | 3 | 0 | 0 | Low-quality | Genome-fragment | | 3.19 | AAI-based (high-confidence) | 0 |
| S7_extracted_68806 | 6704 | No | NA | 6 | 0 | 2 | Not-determined | Genome-fragment | NA | | NA | 0 |
| S7_extracted_68970 | 1028 | No | NA | 1 | 0 | 1 | Not-determined | Genome-fragment | NA | | NA | 0 |
| S13_vs_132460\|\|full | 2079 | No | NA | 4 | 0 | 2 | Low-quality | Genome-fragment | | 1.31 | AAI-based (medium-confidence) | 0 |
| S7_extracted_69348 | 15428 | No | NA | 20 | 8 | 0 | Medium-quality | Genome-fragment | | 62.87 | AAI-based (medium-confidence) | 0 |
| S7_extracted_69404 | 1010 | No | NA | 2 | 0 | 2 | Not-determined | Genome-fragment | NA | | NA | 0 |
| S7_extracted_69514 | 5611 | No | NA | 7 | 0 | 6 | Not-determined | Genome-fragment | NA | | NA | 0 |
| S14_extracted_k141_8616 | 2079 | No | NA | 5 | 5 | 0 | Low-quality | Genome-fragment | | 6.85 | AAI-based (high-confidence) | 0 |
| S1_vs_k141_135076\|\|full | 2079 | No | NA | 3 | 1 | 0 | Low-quality | Genome-fragment | | 4.11 | AAI-based (high-confidence) | 0 |
| S3_vs_k141_120452\|\|full | 2079 | No | NA | 7 | 2 | 0 | Low-quality | Genome-fragment | | 3.62 | AAI-based (high-confidence) | 0 |
| S7_extracted_69965 | 1351 | No | NA | 2 | 0 | 1 | Not-determined | Genome-fragment | NA | | NA | 0 |
| S7_extracted_70123 | 1963 | No | NA | 2 | 0 | 2 | Not-determined | Genome-fragment | NA | | NA | 0 |
| S7_extracted_70583 | 1617 | No | NA | 1 | 0 | 1 | Not-determined | Genome-fragment | NA | | NA | 0 |
| S5_extracted_k141_139502 | 2079 | No | NA | 7 | 0 | 0 | Low-quality | Genome-fragment | | 4.34 | AAI-based (medium-confidence) | 0 |
| S7_extracted_70869 | 4149 | No | NA | 5 | 0 | 4 | Not-determined | Genome-fragment | NA | | NA | 0 |
| S7_extracted_70893 | 1727 | No | NA | 2 | 0 | 1 | Not-determined | Genome-fragment | NA | | NA | 0 |
| S7_extracted_70987 | 11883 | No | NA | 7 | 0 | 4 | Not-determined | Genome-fragment | NA | | NA | 0 |
| S5_vs_k141_75017\|\|lt2gene | 2079 | No | NA | 2 | 0 | 1 | Low-quality | Genome-fragment | | 3.58 | AAI-based (medium-confidence) | 0 |
| S7_extracted_71195 | 1117 | No | NA | 1 | 0 | 1 | Not-determined | Genome-fragment | NA | | NA | 0 |
| S7_extracted_71217 | 1129 | No | NA | 4 | 0 | 0 | Not-determined | Genome-fragment | NA | | NA | 0 |
| S7_extracted_71268 | 1162 | No | NA | 1 | 0 | 0 | Not-determined | Genome-fragment | NA | | NA | 0 |
| S7_extracted_71321 | 1196 | No | NA | 1 | 0 | 1 | Not-determined | Genome-fragment | NA | | NA | 0 |
| S7_vs_17574\|\|full | 2079 | No | NA | 3 | 3 | 0 | Low-quality | Genome-fragment | | 4.57 | AAI-based (high-confidence) | 0 |
| S7_extracted_71527 | 12797 | No | NA | 16 | 1 | 2 | High-quality | High-quality | | 100 | AAI-based (high-confidence) | 0 |
| S7_extracted_71705 | 1408 | No | NA | 1 | 0 | 1 | Not-determined | Genome-fragment | NA | | NA | 0 |
| S7_extracted_71766 | 1113 | No | NA | 2 | 0 | 1 | Not-determined | Genome-fragment | NA | | NA | 0 |
| S12_vs_k141_142252\|\|full | 2079 | No | NA | 6 | 1 | 0 | Low-quality | Genome-fragment | | 5.42 | AAI-based (high-confidence) | 0 |
| S6_extracted_k141_17103 | 2079 | No | NA | 4 | 2 | 0 | Low-quality | Genome-fragment | | 0.99 | AAI-based (medium-confidence) | 0 |
| S7_extracted_72048 | 1437 | No | NA | 3 | 0 | 2 | Not-determined | Genome-fragment | NA | | NA | 0 |
| S7_extracted_72166 | 1665 | No | NA | 2 | 0 | 1 | Not-determined | Genome-fragment | NA | | NA | 0 |
| S7_extracted_72204 | 1034 | No | NA | 2 | 0 | 1 | Not-determined | Genome-fragment | NA | | NA | 0 |
| S11_extracted_k141_27848 | 2080 | No | NA | 2 | 0 | 0 | Low-quality | Genome-fragment | | 3.02 | AAI-based (medium-confidence) | 0 |
| S7_extracted_72320 | 3284 | No | NA | 4 | 0 | 2 | Not-determined | Genome-fragment | NA | | NA | 0 |
| S7_extracted_72446 | 3091 | No | NA | 2 | 0 | 0 | Not-determined | Genome-fragment | NA | | NA | 0 |
| S7_extracted_72493 | 22981 | No | NA | 20 | 5 | 0 | Medium-quality | Genome-fragment | | 54.26 | AAI-based (high-confidence) | 0 |
| S7_extracted_72682 | 1392 | No | NA | 1 | 0 | 1 | Not-determined | Genome-fragment | NA | | NA | 0 |
| S1_extracted_k141_61606 | 2080 | No | NA | 6 | 0 | 0 | Low-quality | Genome-fragment | | 2.05 | AAI-based (high-confidence) | 0 |
| S1_vs_k141_84410\|\|full | 2080 | No | NA | 3 | 0 | 3 | Low-quality | Genome-fragment | | 1.02 | AAI-based (medium-confidence) | 0 |
| S7_extracted_72777 | 1151 | No | NA | 1 | 0 | 1 | Not-determined | Genome-fragment | NA | | NA | 0 |
| S7_extracted_72922 | 2963 | No | NA | 5 | 0 | 5 | Not-determined | Genome-fragment | NA | | NA | 0 |
| S7_extracted_73108 | 1685 | No | NA | 2 | 0 | 0 | Not-determined | Genome-fragment | NA | | NA | 0 |
| S7_extracted_73156 | 4433 | No | NA | 1 | 0 | 1 | Not-determined | Genome-fragment | NA | | NA | 0 |
| S5_extracted_k141_13338 | 2080 | No | NA | 1 | 0 | 0 | Low-quality | Genome-fragment | | 4.13 | AAI-based (medium-confidence) | 0 |
| S5_vs_k141_101398\|\|full | 2080 | No | NA | 4 | 0 | 0 | Low-quality | Genome-fragment | | 4.01 | AAI-based (medium-confidence) | 0 |
| S7_extracted_73327 | 1007 | No | NA | 2 | 0 | 2 | Not-determined | Genome-fragment | NA | | NA | 0 |
| S7_extracted_73559 | 9703 | No | NA | 6 | 0 | 1 | Not-determined | Genome-fragment | NA | | NA | 0 |
| S7_extracted_73591 | 1017 | No | NA | 1 | 0 | 1 | Not-determined | Genome-fragment | NA | | NA | 0 |
| S7_extracted_73782 | 1068 | No | NA | 1 | 0 | 1 | Not-determined | Genome-fragment | NA | | NA | 0 |
| S16_vs_12254\|\|full | 2080 | No | NA | 2 | 2 | 0 | Low-quality | Genome-fragment | | 5.16 | AAI-based (high-confidence) | 0 |
| S7_extracted_73893 | 3359 | No | NA | 1 | 0 | 0 | Not-determined | Genome-fragment | NA | | NA | 0 |
| S16_vs_143722\|\|full | 2080 | No | NA | 8 | 1 | 0 | Low-quality | Genome-fragment | | 3.26 | HMM-based (lower-bound) | 0 |
| S13_vs_147838\|\|full | 2081 | No | NA | 4 | 0 | 0 | Low-quality | Genome-fragment | | 1.45 | AAI-based (medium-confidence) | 0 |
| S7_extracted_74440 | 1141 | No | NA | 2 | 0 | 0 | Not-determined | Genome-fragment | NA | | NA | 0 |
| S14_extracted_k141_46698 | 2081 | No | NA | 1 | 0 | 1 | Low-quality | Genome-fragment | | 2.2 | AAI-based (medium-confidence) | 0 |
| S7_extracted_74507 | 1122 | No | NA | 1 | 0 | 0 | Not-determined | Genome-fragment | NA | | NA | 0 |
| S7_extracted_74605 | 1030 | No | NA | 2 | 0 | 1 | Not-determined | Genome-fragment | NA | | NA | 0 |
| S7_extracted_74669 | 1086 | No | NA | 1 | 0 | 1 | Not-determined | Genome-fragment | NA | | NA | 0 |
| S3_extracted_k141_53839 | 2081 | No | NA | 2 | 1 | 0 | Low-quality | Genome-fragment | | 1.44 | HMM-based (lower-bound) | 0 |
| S7_extracted_74800 | 1435 | No | NA | 2 | 0 | 1 | Not-determined | Genome-fragment | NA | | NA | 0 |
| S5_vs_k141_79398\|\|full | 2081 | No | NA | 3 | 0 | 2 | Low-quality | Genome-fragment | | 2.65 | AAI-based (medium-confidence) | 0 |
| S7_extracted_75040 | 7462 | No | NA | 10 | 0 | 5 | Not-determined | Genome-fragment | NA | | NA | 0 |
| S7_extracted_75111 | 1735 | No | NA | 3 | 0 | 1 | Not-determined | Genome-fragment | NA | | NA | 0 |
| S7_extracted_75137 | 3326 | No | NA | 5 | 0 | 2 | Not-determined | Genome-fragment | NA | | NA | 0 |
| S7_extracted_75298 | 1018 | No | NA | 1 | 0 | 1 | Not-determined | Genome-fragment | NA | | NA | 0 |
| S8_vs_k141_1528\|\|full | 2081 | No | NA | 3 | 0 | 1 | Low-quality | Genome-fragment | | 2.64 | AAI-based (high-confidence) | 0 |
| S7_extracted_75538 | 1682 | No | NA | 1 | 0 | 0 | Not-determined | Genome-fragment | NA | | NA | 0 |
| S5_extracted_k141_53781 | 2082 | No | NA | 2 | 0 | 0 | Low-quality | Genome-fragment | | 3.43 | AAI-based (high-confidence) | 0 |
| S7_extracted_75660 | 1741 | No | NA | 2 | 0 | 1 | Not-determined | Genome-fragment | NA | | NA | 0 |
| S7_extracted_75701 | 1081 | No | NA | 1 | 0 | 1 | Not-determined | Genome-fragment | NA | | NA | 0 |
| S7_extracted_75744 | 1380 | No | NA | 1 | 0 | 1 | Not-determined | Genome-fragment | NA | | NA | 0 |
| S16_vs_127718\|\|full | 2082 | No | NA | 5 | 2 | 0 | Low-quality | Genome-fragment | | 5.73 | AAI-based (high-confidence) | 0 |
| S7_extracted_75899 | 2420 | No | NA | 2 | 0 | 2 | Not-determined | Genome-fragment | NA | | NA | 0 |
| S12_vs_k141_67732\|\|full | 2082 | No | NA | 3 | 0 | 0 | Low-quality | Genome-fragment | | 2.26 | AAI-based (high-confidence) | 0 |
| S7_extracted_75974 | 8041 | No | NA | 9 | 0 | 6 | Not-determined | Genome-fragment | NA | | NA | 0 |
| S12_vs_k141_163429\|\|full | 2082 | No | NA | 4 | 0 | 0 | Low-quality | Genome-fragment | | 4.71 | AAI-based (medium-confidence) | 0 |
| S9_extracted_k141_11908 | 2083 | No | NA | 2 | 1 | 0 | Low-quality | Genome-fragment | | 1.2 | AAI-based (high-confidence) | 0 |
| S7_extracted_76101 | 3163 | No | NA | 3 | 0 | 3 | Not-determined | Genome-fragment | NA | | NA | 0 |
| S9_extracted_k141_59693 | 2083 | No | NA | 5 | 1 | 0 | Low-quality | Genome-fragment | | 4.88 | AAI-based (high-confidence) | 0 |
| S7_extracted_76279 | 2573 | No | NA | 1 | 0 | 1 | Not-determined | Genome-fragment | NA | | NA | 0 |
| S7_extracted_76327 | 3465 | No | NA | 4 | 0 | 0 | Not-determined | Genome-fragment | NA | | NA | 0 |
| S7_extracted_76365 | 1464 | No | NA | 1 | 0 | 0 | Not-determined | Genome-fragment | NA | | NA | 0 |
| S7_extracted_76382 | 17054 | No | NA | 22 | 8 | 0 | Medium-quality | Genome-fragment | | 78.25 | AAI-based (medium-confidence) | 0 |
| S7_extracted_76463 | 46228 | No | NA | 72 | 14 | 0 | High-quality | High-quality | | 100 | AAI-based (high-confidence) | 0 |
| S1_vs_k141_2107\|\|full | 2083 | No | NA | 4 | 2 | 0 | Low-quality | Genome-fragment | | 5.3 | AAI-based (high-confidence) | 0 |
| S7_extracted_76517 | 1918 | No | NA | 2 | 0 | 1 | Not-determined | Genome-fragment | NA | | NA | 0 |
| S7_extracted_76586 | 3686 | No | NA | 6 | 0 | 0 | Not-determined | Genome-fragment | NA | | NA | 0 |
| S10_extracted_94395 | 2083 | No | NA | 3 | 1 | 0 | Low-quality | Genome-fragment | | 1.45 | AAI-based (medium-confidence) | 0 |
| S10_vs_118957\|\|full | 2083 | No | NA | 3 | 0 | 3 | Low-quality | Genome-fragment | | 1.2 | AAI-based (medium-confidence) | 0 |
| S7_extracted_76881 | 4586 | No | NA | 6 | 0 | 3 | Not-determined | Genome-fragment | NA | | NA | 0 |
| S7_extracted_77061 | 1083 | No | NA | 2 | 0 | 1 | Not-determined | Genome-fragment | NA | | NA | 0 |
| S8_vs_k141_174069\|\|full | 2083 | No | NA | 2 | 2 | 0 | Low-quality | Genome-fragment | | 4.61 | AAI-based (medium-confidence) | 0 |
| S7_extracted_77243 | 1138 | No | NA | 1 | 0 | 1 | Not-determined | Genome-fragment | NA | | NA | 0 |
| S7_extracted_77363 | 4229 | No | NA | 4 | 0 | 2 | Not-determined | Genome-fragment | NA | | NA | 0 |
| S11_vs_k141_42973\|\|full | 2084 | No | NA | 5 | 0 | 0 | Low-quality | Genome-fragment | | 5 | AAI-based (high-confidence) | 0 |
| S7_extracted_77436 | 1188 | No | NA | 1 | 0 | 0 | Not-determined | Genome-fragment | NA | | NA | 0 |
| S7_extracted_77704 | 2439 | No | NA | 2 | 0 | 2 | Not-determined | Genome-fragment | NA | | NA | 0 |
| S7_extracted_77719 | 1336 | No | NA | 1 | 0 | 1 | Not-determined | Genome-fragment | NA | | NA | 0 |
| S14_extracted_k141_90086 | 2084 | No | NA | 5 | 5 | 0 | Low-quality | Genome-fragment | | 6.87 | AAI-based (high-confidence) | 0 |
| S7_extracted_77864 | 1600 | No | NA | 1 | 0 | 1 | Not-determined | Genome-fragment | NA | | NA | 0 |
| S7_extracted_78089 | 2366 | No | NA | 2 | 0 | 2 | Not-determined | Genome-fragment | NA | | NA | 0 |
| S7_extracted_78149 | 4235 | No | NA | 3 | 0 | 3 | Not-determined | Genome-fragment | NA | | NA | 0 |
| S7_extracted_78327 | 3815 | No | NA | 4 | 0 | 2 | Not-determined | Genome-fragment | NA | | NA | 0 |
| S7_extracted_78501 | 2841 | No | NA | 2 | 0 | 1 | Not-determined | Genome-fragment | NA | | NA | 0 |
| S3_vs_k141_9802\|\|full | 2084 | No | NA | 3 | 1 | 0 | Low-quality | Genome-fragment | | 4.22 | AAI-based (high-confidence) | 0 |
| S7_extracted_78649 | 1524 | No | NA | 1 | 0 | 1 | Not-determined | Genome-fragment | NA | | NA | 0 |
| S7_extracted_78656 | 3098 | No | NA | 3 | 0 | 2 | Not-determined | Genome-fragment | NA | | NA | 0 |
| S7_extracted_78963 | 1119 | No | NA | 1 | 0 | 1 | Not-determined | Genome-fragment | NA | | NA | 0 |
| S7_extracted_79100 | 1726 | No | NA | 1 | 0 | 1 | Not-determined | Genome-fragment | NA | | NA | 0 |
| S7_extracted_79165 | 1331 | No | NA | 1 | 0 | 0 | Not-determined | Genome-fragment | NA | | NA | 0 |
| S3_vs_k141_92941\|\|full | 2084 | No | NA | 4 | 0 | 0 | Low-quality | Genome-fragment | | 5.71 | AAI-based (high-confidence) | 0 |
| S7_extracted_79285 | 1643 | No | NA | 4 | 0 | 1 | Not-determined | Genome-fragment | NA | | NA | 0 |
| S5_vs_k141_35827\|\|full | 2084 | No | NA | 3 | 0 | 0 | Low-quality | Genome-fragment | | 4.26 | AAI-based (high-confidence) | 0 |
| S12_extracted_k141_70517 | 2084 | No | NA | 1 | 0 | 1 | Low-quality | Genome-fragment | | 2.09 | AAI-based (medium-confidence) | 0 |
| S7_extracted_79753 | 1132 | No | NA | 3 | 0 | 1 | Not-determined | Genome-fragment | NA | | NA | 0 |
| S7_extracted_79813 | 2053 | No | NA | 1 | 0 | 1 | Not-determined | Genome-fragment | NA | | NA | 0 |
| S7_extracted_79830 | 9902 | No | NA | 3 | 0 | 1 | Not-determined | Genome-fragment | NA | | NA | 0 |
| S6_vs_k141_3395\|\|full | 2084 | No | NA | 4 | 0 | 1 | Low-quality | Genome-fragment | | 4.08 | AAI-based (high-confidence) | 0 |
| S7_extracted_80149 | 1215 | No | NA | 2 | 0 | 1 | Not-determined | Genome-fragment | NA | | NA | 0 |
| S7_extracted_80186 | 1086 | No | NA | 1 | 0 | 0 | Not-determined | Genome-fragment | NA | | NA | 0 |
| S8_vs_k141_175395\|\|full | 2084 | No | NA | 3 | 2 | 0 | Low-quality | Genome-fragment | | 3.57 | AAI-based (high-confidence) | 0 |
| S7_extracted_80326 | 1813 | No | NA | 2 | 0 | 0 | Not-determined | Genome-fragment | NA | | NA | 0 |
| S7_extracted_80365 | 1253 | No | NA | 1 | 0 | 0 | Not-determined | Genome-fragment | NA | | NA | 0 |
| S7_extracted_80391 | 1938 | No | NA | 2 | 0 | 1 | Not-determined | Genome-fragment | NA | | NA | 0 |
| S17_vs_k141_158851\|\|full | 2085 | No | NA | 3 | 0 | 0 | Low-quality | Genome-fragment | | 2.81 | AAI-based (high-confidence) | 0 |
| S7_extracted_80598 | 1294 | No | NA | 2 | 0 | 0 | Not-determined | Genome-fragment | NA | | NA | 0 |
| S11_vs_k141_101529\|\|full | 2085 | No | NA | 4 | 1 | 0 | Low-quality | Genome-fragment | | 4.98 | AAI-based (high-confidence) | 0 |
| S3_extracted_k141_79342 | 2085 | No | NA | 1 | 0 | 0 | Low-quality | Genome-fragment | | 1.36 | AAI-based (high-confidence) | 0 |
| S7_extracted_80768 | 1264 | No | NA | 1 | 0 | 1 | Not-determined | Genome-fragment | NA | | NA | 0 |
| S5_vs_k141_119054\|\|full | 2085 | No | NA | 4 | 1 | 0 | Low-quality | Genome-fragment | | 1.27 | HMM-based (lower-bound) | 0 |
| S7_extracted_81043 | 6735 | No | NA | 5 | 0 | 4 | Not-determined | Genome-fragment | NA | | NA | 0 |
| S7_extracted_81137 | 3308 | No | NA | 2 | 0 | 2 | Not-determined | Genome-fragment | NA | | NA | 0 |
| S7_extracted_81254 | 2387 | No | NA | 2 | 0 | 1 | Not-determined | Genome-fragment | NA | | NA | 0 |
| S7_extracted_81265 | 1140 | No | NA | 2 | 0 | 0 | Not-determined | Genome-fragment | NA | | NA | 0 |
| S7_extracted_81306 | 1697 | No | NA | 3 | 0 | 3 | Not-determined | Genome-fragment | NA | | NA | 0 |
| S7_extracted_81310 | 1352 | No | NA | 1 | 0 | 1 | Not-determined | Genome-fragment | NA | | NA | 0 |

| | | | | | | | | | | | |
|---|---|---|---|---|---|---|---|---|---|---|---|
| S16_vs_51439\|\|full | 2085 | No | NA | 4 | 2 | 0 | Low-quality | Genome-fragment | 5.17 | AAI-based (high-confidence) | 0 |
| S12_extracted_k141_165420 | 2085 | No | NA | 2 | 0 | 1 | Low-quality | Genome-fragment | 4.26 | AAI-based (medium-confidence) | 0 |
| S7_extracted_81579 | 1244 | No | NA | 1 | 0 | 1 | Not-determined | Genome-fragment | NA | NA | 0 |
| S8_extracted_k141_49105 | 2085 | No | NA | 3 | 1 | 0 | Low-quality | Genome-fragment | 2.2 | AAI-based (medium-confidence) | 0 |
| S7_extracted_81733 | 1424 | No | NA | 2 | 0 | 1 | Not-determined | Genome-fragment | NA | NA | 0 |
| S14_extracted_k141_577 | 2086 | No | NA | 3 | 2 | 0 | Low-quality | Genome-fragment | 5.05 | AAI-based (medium-confidence) | 0 |
| S7_extracted_81836 | 1232 | No | NA | 1 | 0 | 0 | Not-determined | Genome-fragment | NA | NA | 0 |
| S14_vs_k141_15505\|\|full | 2086 | No | NA | 5 | 0 | 0 | Low-quality | Genome-fragment | 5.09 | AAI-based (medium-confidence) | 0 |
| S7_extracted_81853 | 1229 | No | NA | 3 | 0 | 0 | Not-determined | Genome-fragment | NA | NA | 0 |
| S7_extracted_81959 | 1271 | No | NA | 2 | 0 | 1 | Not-determined | Genome-fragment | NA | NA | 0 |
| S7_extracted_82016 | 2018 | No | NA | 1 | 0 | 1 | Not-determined | Genome-fragment | NA | NA | 0 |
| S7_extracted_82100 | 2495 | No | NA | 3 | 0 | 0 | Not-determined | Genome-fragment | NA | NA | 0 |
| S14_vs_k141_100079\|\|full | 2086 | No | NA | 3 | 1 | 0 | Low-quality | Genome-fragment | 3.51 | HMM-based (lower-bound) | 0 |
| S7_extracted_82375 | 1012 | No | NA | 1 | 0 | 1 | Not-determined | Genome-fragment | NA | NA | 0 |
| S7_extracted_82377 | 2323 | No | NA | 2 | 0 | 2 | Not-determined | Genome-fragment | NA | NA | 0 |
| S1_extracted_k141_117310 | 2086 | No | NA | 1 | 0 | 1 | Low-quality | Genome-fragment | 0.18 | AAI-based (medium-confidence) | 0 |
| S7_extracted_82810 | 3206 | No | NA | 4 | 0 | 3 | Not-determined | Genome-fragment | NA | NA | 0 |
| S7_extracted_82899 | 4306 | No | NA | 4 | 0 | 2 | Not-determined | Genome-fragment | NA | NA | 0 |
| S7_extracted_83046 | 3106 | No | NA | 5 | 0 | 4 | Not-determined | Genome-fragment | NA | NA | 0 |
| S12_vs_k141_114142\|\|full | 2086 | No | NA | 6 | 2 | 0 | Low-quality | Genome-fragment | 4.82 | AAI-based (high-confidence) | 0 |
| S7_extracted_83329 | 1432 | No | NA | 1 | 0 | 0 | Not-determined | Genome-fragment | NA | NA | 0 |
| S7_extracted_83380 | 1503 | No | NA | 2 | 0 | 2 | Not-determined | Genome-fragment | NA | NA | 0 |
| S7_extracted_83478 | 3367 | No | NA | 3 | 0 | 2 | Not-determined | Genome-fragment | NA | NA | 0 |
| S7_extracted_83490 | 17786 | No | NA | 17 | 0 | 3 | Not-determined | Genome-fragment | NA | NA | 0 |
| S7_extracted_83624 | 16244 | No | NA | 24 | 7 | 0 | Medium-quality | Genome-fragment | 77.28 | AAI-based (medium-confidence) | 0 |
| S11_vs_k141_10401\|\|full | 2087 | No | NA | 6 | 3 | 0 | Low-quality | Genome-fragment | 3.39 | HMM-based (lower-bound) | 0 |
| S7_extracted_83654 | 1066 | No | NA | 2 | 0 | 1 | Not-determined | Genome-fragment | NA | NA | 0 |
| S7_extracted_83739 | 1287 | No | NA | 2 | 0 | 1 | Not-determined | Genome-fragment | NA | NA | 0 |
| S3_vs_k141_68587\|\|full | 2087 | No | NA | 3 | 1 | 0 | Low-quality | Genome-fragment | 4.95 | AAI-based (medium-confidence) | 0 |
| S7_extracted_83852 | 3398 | No | NA | 4 | 0 | 2 | Not-determined | Genome-fragment | NA | NA | 0 |
| S7_extracted_83858 | 1012 | No | NA | 2 | 0 | 2 | Not-determined | Genome-fragment | NA | NA | 0 |
| S7_extracted_83875 | 3696 | No | NA | 3 | 0 | 3 | Not-determined | Genome-fragment | NA | NA | 0 |
| S7_extracted_83927 | 2329 | No | NA | 3 | 0 | 2 | Not-determined | Genome-fragment | NA | NA | 0 |
| S7_extracted_83990 | 6118 | No | NA | 6 | 0 | 1 | Not-determined | Genome-fragment | NA | NA | 0 |
| S8_vs_k141_71936\|\|full | 2087 | No | NA | 3 | 2 | 0 | Low-quality | Genome-fragment | 4.63 | AAI-based (high-confidence) | 0 |
| S7_extracted_84314 | 2193 | No | NA | 1 | 0 | 1 | Not-determined | Genome-fragment | NA | NA | 0 |
| S17_extracted_k141_177157 | 2088 | No | NA | 6 | 0 | 0 | Low-quality | Genome-fragment | 2.47 | AAI-based (high-confidence) | 0 |
| S7_extracted_84360 | 1051 | No | NA | 1 | 0 | 0 | Not-determined | Genome-fragment | NA | NA | 0 |
| S7_extracted_84432 | 1014 | No | NA | 1 | 0 | 0 | Not-determined | Genome-fragment | NA | NA | 0 |
| S7_extracted_84557 | 1138 | No | NA | 2 | 0 | 0 | Not-determined | Genome-fragment | NA | NA | 0 |
| S7_extracted_84689 | 1019 | No | NA | 1 | 0 | 0 | Not-determined | Genome-fragment | NA | NA | 0 |
| S7_extracted_84792 | 2036 | No | NA | 1 | 0 | 1 | Not-determined | Genome-fragment | NA | NA | 0 |
| S7_extracted_84801 | 3646 | No | NA | 8 | 0 | 7 | Not-determined | Genome-fragment | NA | NA | 0 |
| S7_extracted_84953 | 7931 | No | NA | 9 | 0 | 6 | Not-determined | Genome-fragment | NA | NA | 0 |
| S7_extracted_84974 | 80171 | No | NA | 97 | 22 | 1 | Medium-quality | Genome-fragment | 55.23 | AAI-based (high-confidence) | 0 |
| S7_extracted_85031 | 4719 | No | NA | 6 | 0 | 4 | Not-determined | Genome-fragment | NA | NA | 0 |
| S17_vs_k141_142349\|\|full | 2088 | No | NA | 3 | 0 | 0 | Low-quality | Genome-fragment | 5.72 | AAI-based (medium-confidence) | 0 |
| S7_extracted_85118 | 1341 | No | NA | 2 | 0 | 0 | Not-determined | Genome-fragment | NA | NA | 0 |
| S7_extracted_85199 | 1842 | No | NA | 2 | 0 | 0 | Not-determined | Genome-fragment | NA | NA | 0 |
| S7_extracted_85229 | 8111 | No | NA | 11 | 0 | 2 | Not-determined | Genome-fragment | NA | NA | 0 |
| S7_extracted_85472 | 1053 | No | NA | 1 | 0 | 0 | Not-determined | Genome-fragment | NA | NA | 0 |
| S7_extracted_85678 | 10562 | No | NA | 9 | 0 | 4 | Not-determined | Genome-fragment | NA | NA | 0 |
| S7_extracted_85860 | 1680 | No | NA | 2 | 0 | 0 | Not-determined | Genome-fragment | NA | NA | 0 |
| S7_extracted_86229 | 1410 | No | NA | 2 | 0 | 0 | Not-determined | Genome-fragment | NA | NA | 0 |
| S7_extracted_86339 | 1140 | No | NA | 3 | 0 | 0 | Not-determined | Genome-fragment | NA | NA | 0 |
| S9_vs_k141_32213\|\|full | 2088 | No | NA | 3 | 0 | 1 | Low-quality | Genome-fragment | 2.49 | AAI-based (high-confidence) | 0 |
| S7_extracted_86433 | 1172 | No | NA | 1 | 0 | 1 | Not-determined | Genome-fragment | NA | NA | 0 |
| S7_extracted_86448 | 3693 | No | NA | 3 | 0 | 3 | Not-determined | Genome-fragment | NA | NA | 0 |
| S5_vs_k141_139521\|\|full | 2088 | No | NA | 2 | 0 | 0 | Low-quality | Genome-fragment | 3.87 | AAI-based (medium-confidence) | 0 |
| S7_vs_44806\|\|full | 2088 | Yes | 927 | 2 | 1 | 1 | Low-quality | Genome-fragment | 1.86 | HMM-based (lower-bound) | 55.6 |
| S7_extracted_86605 | 1954 | No | NA | 2 | 0 | 1 | Not-determined | Genome-fragment | NA | NA | 0 |
| S7_vs_94856\|\|full | 2088 | No | NA | 4 | 1 | 0 | Low-quality | Genome-fragment | 2.03 | HMM-based (lower-bound) | 0 |
| S7_extracted_86762 | 1661 | No | NA | 1 | 0 | 0 | Not-determined | Genome-fragment | NA | NA | 0 |
| S7_extracted_86803 | 1126 | No | NA | 2 | 0 | 0 | Not-determined | Genome-fragment | NA | NA | 0 |
| S7_extracted_86977 | 2476 | No | NA | 7 | 0 | 1 | Not-determined | Genome-fragment | NA | NA | 0 |
| S9_vs_k141_48044\|\|full | 2089 | No | NA | 6 | 2 | 0 | Low-quality | Genome-fragment | 3.78 | AAI-based (medium-confidence) | 0 |
| S7_extracted_87085 | 1381 | No | NA | 1 | 0 | 1 | Not-determined | Genome-fragment | NA | NA | 0 |
| S7_extracted_87250 | 3540 | No | NA | 2 | 0 | 2 | Not-determined | Genome-fragment | NA | NA | 0 |
| S11_vs_k141_93100\|\|full | 2089 | No | NA | 3 | 1 | 0 | Low-quality | Genome-fragment | 3.95 | AAI-based (high-confidence) | 0 |
| S7_extracted_87276 | 4856 | No | NA | 5 | 0 | 0 | Not-determined | Genome-fragment | NA | NA | 0 |
| S7_extracted_87338 | 1450 | No | NA | 1 | 0 | 1 | Not-determined | Genome-fragment | NA | NA | 0 |
| S13_vs_99104\|\|full | 2089 | No | NA | 5 | 1 | 0 | Low-quality | Genome-fragment | 4.19 | AAI-based (medium-confidence) | 0 |
| S7_extracted_87493 | 3845 | No | NA | 2 | 0 | 2 | Not-determined | Genome-fragment | NA | NA | 0 |
| S7_extracted_87637 | 3037 | No | NA | 2 | 0 | 2 | Not-determined | Genome-fragment | NA | NA | 0 |
| S7_extracted_87821 | 1183 | No | NA | 1 | 0 | 1 | Not-determined | Genome-fragment | NA | NA | 0 |
| S7_vs_29598\|\|full | 2089 | No | NA | 3 | 0 | 0 | Low-quality | Genome-fragment | 4.89 | AAI-based (medium-confidence) | 0 |
| S7_extracted_88256 | 1248 | No | NA | 1 | 0 | 1 | Not-determined | Genome-fragment | NA | NA | 0 |
| S12_vs_k141_82809\|\|full | 2089 | No | NA | 3 | 0 | 1 | Low-quality | Genome-fragment | 6.52 | AAI-based (high-confidence) | 0 |
| S7_extracted_88530 | 2137 | No | NA | 4 | 0 | 1 | Not-determined | Genome-fragment | NA | NA | 0 |
| S7_extracted_88664 | 1133 | No | NA | 2 | 0 | 2 | Not-determined | Genome-fragment | NA | NA | 0 |
| S7_extracted_88691 | 1089 | No | NA | 2 | 0 | 2 | Not-determined | Genome-fragment | NA | NA | 0 |
| S7_extracted_88795 | 1082 | No | NA | 1 | 0 | 0 | Not-determined | Genome-fragment | NA | NA | 0 |
| S7_extracted_88846 | 24184 | No | NA | 18 | 0 | 13 | Not-determined | Genome-fragment | NA | NA | 0 |
| S6_vs_k141_48631\|\|full | 2089 | No | NA | 5 | 1 | 0 | Low-quality | Genome-fragment | 2.52 | AAI-based (medium-confidence) | 0 |
| S7_extracted_89143 | 2968 | No | NA | 3 | 0 | 2 | Not-determined | Genome-fragment | NA | NA | 0 |
| S7_extracted_89197 | 16972 | No | NA | 33 | 19 | 0 | Medium-quality | Genome-fragment | 77.43 | AAI-based (high-confidence) | 0 |
| S11_vs_k141_28250\|\|full | 2090 | No | NA | 2 | 0 | 0 | Low-quality | Genome-fragment | 3.18 | AAI-based (medium-confidence) | 0 |
| S7_extracted_89344 | 1426 | No | NA | 1 | 0 | 1 | Not-determined | Genome-fragment | NA | NA | 0 |
| S7_extracted_89387 | 3232 | No | NA | 5 | 0 | 2 | Not-determined | Genome-fragment | NA | NA | 0 |
| S7_extracted_89413 | 3363 | No | NA | 3 | 0 | 3 | Not-determined | Genome-fragment | NA | NA | 0 |
| S7_extracted_89420 | 2101 | No | NA | 2 | 0 | 0 | Not-determined | Genome-fragment | NA | NA | 0 |
| S7_extracted_89431 | 1398 | No | NA | 2 | 0 | 1 | Not-determined | Genome-fragment | NA | NA | 0 |
| S5_extracted_k141_129981 | 2090 | No | NA | 3 | 2 | 0 | Low-quality | Genome-fragment | 6.11 | AAI-based (high-confidence) | 0 |
| S7_extracted_89546 | 3983 | No | NA | 4 | 0 | 2 | Not-determined | Genome-fragment | NA | NA | 0 |
| S7_extracted_89603 | 1559 | No | NA | 2 | 0 | 2 | Not-determined | Genome-fragment | NA | NA | 0 |
| S7_extracted_89633 | 3263 | No | NA | 3 | 0 | 3 | Not-determined | Genome-fragment | NA | NA | 0 |
| S7_extracted_89668 | 16930 | No | NA | 27 | 8 | 0 | Medium-quality | Genome-fragment | 72.64 | AAI-based (medium-confidence) | 0 |
| S7_extracted_89680 | 5114 | No | NA | 5 | 0 | 1 | Not-determined | Genome-fragment | NA | NA | 0 |
| S7_extracted_89750 | 1711 | No | NA | 3 | 0 | 2 | Not-determined | Genome-fragment | NA | NA | 0 |
| S7_extracted_89835 | 1107 | No | NA | 2 | 0 | 1 | Not-determined | Genome-fragment | NA | NA | 0 |
| S7_extracted_90206 | 3936 | No | NA | 7 | 0 | 1 | Not-determined | Genome-fragment | NA | NA | 0 |
| S7_extracted_90252 | 1094 | No | NA | 2 | 0 | 2 | Not-determined | Genome-fragment | NA | NA | 0 |
| S5_vs_k141_2397\|\|full | 2090 | No | NA | 3 | 0 | 0 | Low-quality | Genome-fragment | 4.3 | AAI-based (medium-confidence) | 0 |
| S7_extracted_90294 | 4498 | No | NA | 3 | 0 | 1 | Not-determined | Genome-fragment | NA | NA | 0 |
| S7_extracted_90308 | 2307 | No | NA | 2 | 0 | 1 | Not-determined | Genome-fragment | NA | NA | 0 |
| S7_extracted_90341 | 8216 | No | NA | 6 | 0 | 0 | Not-determined | Genome-fragment | NA | NA | 0 |
| S7_extracted_90356 | 1719 | No | NA | 3 | 0 | 0 | Not-determined | Genome-fragment | NA | NA | 0 |
| S7_extracted_90612 | 1113 | No | NA | 1 | 0 | 1 | Not-determined | Genome-fragment | NA | NA | 0 |
| S7_extracted_90748 | 1191 | No | NA | 1 | 0 | 0 | Not-determined | Genome-fragment | NA | NA | 0 |
| S7_extracted_90970 | 2282 | No | NA | 3 | 0 | 0 | Not-determined | Genome-fragment | NA | NA | 0 |
| S7_extracted_90989 | 1192 | No | NA | 1 | 0 | 1 | Not-determined | Genome-fragment | NA | NA | 0 |
| S7_vs_37811\|\|full | 2090 | No | NA | 5 | 0 | 0 | Low-quality | Genome-fragment | 5.2 | AAI-based (medium-confidence) | 0 |
| S16_vs_76849\|\|full | 2090 | No | NA | 5 | 1 | 0 | Low-quality | Genome-fragment | 4.04 | AAI-based (high-confidence) | 0 |
| S7_extracted_91408 | 1192 | No | NA | 2 | 0 | 1 | Not-determined | Genome-fragment | NA | NA | 0 |
| S7_extracted_91521 | 1317 | No | NA | 2 | 0 | 2 | Not-determined | Genome-fragment | NA | NA | 0 |
| S10_vs_91839\|\|full | 2090 | No | NA | 4 | 1 | 0 | Low-quality | Genome-fragment | 7.08 | HMM-based (lower-bound) | 0 |
| S7_extracted_91546 | 3898 | No | NA | 6 | 0 | 2 | Not-determined | Genome-fragment | NA | NA | 0 |
| S4_extracted_14369 | 2090 | No | NA | 3 | 2 | 0 | Low-quality | Genome-fragment | 4.77 | AAI-based (high-confidence) | 0 |
| S7_extracted_91815 | 3644 | No | NA | 4 | 0 | 1 | Not-determined | Genome-fragment | NA | NA | 0 |
| S6_vs_k141_12162\|\|full | 2090 | No | NA | 4 | 1 | 0 | Low-quality | Genome-fragment | 2.35 | HMM-based (lower-bound) | 0 |
| S17_vs_k141_33828\|\|full | 2091 | No | NA | 4 | 1 | 0 | Low-quality | Genome-fragment | 1.28 | AAI-based (medium-confidence) | 0 |
| S1_vs_k141_116503\|\|full | 2091 | No | NA | 4 | 0 | 1 | Low-quality | Genome-fragment | 1.31 | AAI-based (medium-confidence) | 0 |
| S7_extracted_92156 | 1076 | No | NA | 2 | 0 | 1 | Not-determined | Genome-fragment | NA | NA | 0 |
| S7_vs_2952\|\|full | 2091 | No | NA | 3 | 0 | 0 | Low-quality | Genome-fragment | 4.23 | AAI-based (high-confidence) | 0 |
| S7_extracted_92275 | 1288 | No | NA | 1 | 0 | 1 | Not-determined | Genome-fragment | NA | NA | 0 |
| S7_extracted_92310 | 1192 | No | NA | 2 | 0 | 0 | Not-determined | Genome-fragment | NA | NA | 0 |
| S7_extracted_92327 | 1408 | No | NA | 1 | 0 | 0 | Not-determined | Genome-fragment | NA | NA | 0 |
| S7_extracted_92455 | 2320 | No | NA | 2 | 0 | 0 | Not-determined | Genome-fragment | NA | NA | 0 |
| S7_extracted_92456 | 1243 | No | NA | 3 | 0 | 1 | Not-determined | Genome-fragment | NA | NA | 0 |
| S7_vs_127796\|\|full | 2091 | No | NA | 2 | 0 | 2 | Low-quality | Genome-fragment | 2.15 | AAI-based (medium-confidence) | 0 |
| S7_extracted_92518 | 1130 | No | NA | 1 | 0 | 0 | Not-determined | Genome-fragment | NA | NA | 0 |
| S7_extracted_92621 | 1355 | No | NA | 2 | 0 | 0 | Not-determined | Genome-fragment | NA | NA | 0 |
| S7_extracted_92642 | 3613 | No | NA | 4 | 0 | 4 | Not-determined | Genome-fragment | NA | NA | 0 |
| S16_vs_82002\|\|full | 2091 | No | NA | 5 | 2 | 0 | Low-quality | Genome-fragment | 5.1 | AAI-based (high-confidence) | 0 |
| S7_extracted_92759 | 3658 | No | NA | 3 | 0 | 3 | Not-determined | Genome-fragment | NA | NA | 0 |
| S7_extracted_92843 | 1407 | No | NA | 1 | 0 | 0 | Not-determined | Genome-fragment | NA | NA | 0 |
| S7_extracted_92875 | 1528 | No | NA | 2 | 0 | 2 | Not-determined | Genome-fragment | NA | NA | 0 |
| S7_extracted_92934 | 1777 | No | NA | 2 | 0 | 2 | Not-determined | Genome-fragment | NA | NA | 0 |
| S7_extracted_92941 | 1129 | No | NA | 2 | 0 | 1 | Not-determined | Genome-fragment | NA | NA | 0 |
| S12_vs_k141_113421\|\|full | 2091 | No | NA | 2 | 0 | 2 | Low-quality | Genome-fragment | 2.91 | AAI-based (high-confidence) | 0 |
| S7_extracted_93165 | 9786 | No | NA | 9 | 0 | 4 | Not-determined | Genome-fragment | NA | NA | 0 |
| S7_extracted_93423 | 2827 | No | NA | 3 | 0 | 3 | Not-determined | Genome-fragment | NA | NA | 0 |
| S7_extracted_93532 | 1109 | No | NA | 1 | 0 | 0 | Not-determined | Genome-fragment | NA | NA | 0 |
| S8_vs_k141_21745\|\|full | 2091 | No | NA | 5 | 1 | 0 | Low-quality | Genome-fragment | 1.84 | AAI-based (medium-confidence) | 0 |
| S9_vs_k141_24095\|\|full | 2092 | No | NA | 2 | 0 | 0 | Low-quality | Genome-fragment | 2.81 | AAI-based (medium-confidence) | 0 |
| S11_vs_k141_12406\|\|full | 2092 | No | NA | 4 | 2 | 0 | Low-quality | Genome-fragment | 6.24 | AAI-based (medium-confidence) | 0 |
| S7_extracted_93871 | 1500 | No | NA | 2 | 0 | 0 | Not-determined | Genome-fragment | NA | NA | 0 |
| S7_extracted_93876 | 2694 | No | NA | 1 | 0 | 0 | Not-determined | Genome-fragment | NA | NA | 0 |
| S7_extracted_93927 | 1035 | No | NA | 1 | 0 | 1 | Not-determined | Genome-fragment | NA | NA | 0 |
| S7_extracted_93994 | 1116 | No | NA | 1 | 0 | 1 | Not-determined | Genome-fragment | NA | NA | 0 |
| S7_extracted_94176 | 1011 | No | NA | 2 | 0 | 1 | Not-determined | Genome-fragment | NA | NA | 0 |
| S7_extracted_94337 | 1915 | No | NA | 1 | 0 | 1 | Not-determined | Genome-fragment | NA | NA | 0 |
| S7_extracted_94349 | 1952 | No | NA | 1 | 0 | 1 | Not-determined | Genome-fragment | NA | NA | 0 |
| S7_extracted_94474 | 3197 | No | NA | 1 | 0 | 1 | Not-determined | Genome-fragment | NA | NA | 0 |
| S11_vs_k141_98978\|\|lt2gene | 2092 | No | NA | 2 | 1 | 0 | Low-quality | Genome-fragment | 6.42 | AAI-based (medium-confidence) | 0 |
| S7_extracted_94518 | 1578 | No | NA | 2 | 0 | 0 | Not-determined | Genome-fragment | NA | NA | 0 |
| S7_extracted_94535 | 2137 | No | NA | 1 | 0 | 1 | Not-determined | Genome-fragment | NA | NA | 0 |
| S7_extracted_94547 | 2278 | No | NA | 2 | 0 | 1 | Not-determined | Genome-fragment | NA | NA | 0 |
| S7_extracted_94570 | 1036 | No | NA | 1 | 0 | 1 | Not-determined | Genome-fragment | NA | NA | 0 |
| S7_extracted_94662 | 1053 | No | NA | 2 | 0 | 0 | Not-determined | Genome-fragment | NA | NA | 0 |
| S7_extracted_94935 | 1130 | No | NA | 1 | 0 | 0 | Not-determined | Genome-fragment | NA | NA | 0 |
| S13_vs_90245\|\|full | 2092 | No | NA | 2 | 0 | 1 | Low-quality | Genome-fragment | 0.93 | AAI-based (medium-confidence) | 0 |
| S7_extracted_94954 | 3214 | No | NA | 4 | 0 | 3 | Not-determined | Genome-fragment | NA | NA | 0 |
| S7_extracted_94964 | 1217 | No | NA | 2 | 0 | 1 | Not-determined | Genome-fragment | NA | NA | 0 |
| S14_vs_k141_83896\|\|full | 2092 | No | NA | 3 | 0 | 2 | Low-quality | Genome-fragment | 7.02 | AAI-based (high-confidence) | 0 |
| S7_vs_66545\|\|full | 2092 | No | NA | 4 | 0 | 0 | Low-quality | Genome-fragment | 3.89 | AAI-based (medium-confidence) | 0 |
| S7_extracted_95220 | 3611 | No | NA | 5 | 0 | 0 | Not-determined | Genome-fragment | NA | NA | 0 |
| S7_vs_68252\|\|full | 2092 | No | NA | 3 | 0 | 0 | Low-quality | Genome-fragment | 4.65 | AAI-based (high-confidence) | 0 |
| S7_extracted_95386 | 3656 | No | NA | 4 | 0 | 3 | Not-determined | Genome-fragment | NA | NA | 0 |
| S7_vs_124003\|\|full | 2092 | No | NA | 7 | 0 | 1 | Low-quality | Genome-fragment | 4.81 | AAI-based (high-confidence) | 0 |
| S7_extracted_95636 | 1399 | No | NA | 2 | 0 | 0 | Not-determined | Genome-fragment | NA | NA | 0 |
| S7_extracted_95645 | 4648 | No | NA | 3 | 0 | 2 | Not-determined | Genome-fragment | NA | NA | 0 |
| S7_extracted_95714 | 1935 | No | NA | 2 | 0 | 2 | Not-determined | Genome-fragment | NA | NA | 0 |
| S16_vs_44679\|\|full | 2092 | No | NA | 4 | 1 | 0 | Low-quality | Genome-fragment | 5.36 | AAI-based (high-confidence) | 0 |
| S6_extracted_k141_36855 | 2092 | No | NA | 3 | 0 | 1 | Low-quality | Genome-fragment | 4.67 | AAI-based (medium-confidence) | 0 |
| S7_extracted_96067 | 3182 | No | NA | 4 | 0 | 2 | Not-determined | Genome-fragment | NA | NA | 0 |
| S7_extracted_96194 | 4537 | No | NA | 3 | 0 | 3 | Not-determined | Genome-fragment | NA | NA | 0 |
| S7_extracted_96207 | 3127 | No | NA | 3 | 0 | 2 | Not-determined | Genome-fragment | NA | NA | 0 |
| S7_extracted_96411 | 1216 | No | NA | 1 | 0 | 1 | Not-determined | Genome-fragment | NA | NA | 0 |
| S7_extracted_96451 | 6038 | No | NA | 5 | 0 | 2 | Not-determined | Genome-fragment | NA | NA | 0 |
| S9_vs_k141_76434\|\|full | 2093 | No | NA | 4 | 3 | 0 | Low-quality | Genome-fragment | 5.15 | AAI-based (high-confidence) | 0 |
| S7_extracted_96748 | 1010 | No | NA | 1 | 0 | 0 | Not-determined | Genome-fragment | NA | NA | 0 |
| S3_vs_k141_29477\|\|full | 2093 | No | NA | 3 | 1 | 0 | Low-quality | Genome-fragment | 3.59 | AAI-based (high-confidence) | 0 |
| S16_vs_70549\|\|full | 2093 | No | NA | 3 | 1 | 0 | Low-quality | Genome-fragment | 0.85 | HMM-based (lower-bound) | 0 |
| S2_vs_k141_16159\|\|full | 2093 | No | NA | 5 | 1 | 0 | Low-quality | Genome-fragment | 4.87 | AAI-based (high-confidence) | 0 |
| S7_extracted_97117 | 1083 | No | NA | 2 | 0 | 1 | Not-determined | Genome-fragment | NA | NA | 0 |
| S6_extracted_k141_4397 | 2093 | No | NA | 2 | 0 | 0 | Low-quality | Genome-fragment | 2.25 | AAI-based (high-confidence) | 0 |
| S7_extracted_97186 | 27695 | No | NA | 38 | 0 | 33 | Not-determined | Genome-fragment | NA | NA | 0 |
| S7_extracted_97240 | 1143 | No | NA | 2 | 0 | 1 | Not-determined | Genome-fragment | NA | NA | 0 |
| S7_extracted_97244 | 3228 | No | NA | 6 | 0 | 0 | Not-determined | Genome-fragment | NA | NA | 0 |
| S8_vs_k141_148553\|\|full | 2093 | No | NA | 4 | 0 | 0 | Low-quality | Genome-fragment | 3.78 | AAI-based (medium-confidence) | 0 |
| S14_vs_k141_40365\|\|full | 2094 | No | NA | 4 | 2 | 0 | Low-quality | Genome-fragment | 5.02 | AAI-based (high-confidence) | 0 |
| S7_extracted_97911 | 3773 | No | NA | 4 | 0 | 3 | Not-determined | Genome-fragment | NA | NA | 0 |
| S7_extracted_97961 | 6849 | No | NA | 8 | 0 | 2 | Not-determined | Genome-fragment | NA | NA | 0 |
| S7_extracted_98052 | 3766 | No | NA | 3 | 0 | 2 | Not-determined | Genome-fragment | NA | NA | 0 |
| S1_extracted_k141_3327 | 2094 | No | NA | 4 | 1 | 0 | Low-quality | Genome-fragment | 5.84 | AAI-based (high-confidence) | 0 |
| S7_extracted_15074 | 2094 | No | NA | 5 | 0 | 0 | Low-quality | Genome-fragment | 1.48 | AAI-based (high-confidence) | 0 |
| S7_vs_40576\|\|full | 2094 | No | NA | 3 | 0 | 3 | Low-quality | Genome-fragment | 1.26 | AAI-based (medium-confidence) | 0 |
| S7_vs_123563\|\|full | 2094 | No | NA | 4 | 3 | 0 | Low-quality | Genome-fragment | 4.62 | AAI-based (high-confidence) | 0 |
| S12_vs_k141_183420\|\|full | 2094 | No | NA | 4 | 0 | 0 | Low-quality | Genome-fragment | 3.76 | AAI-based (high-confidence) | 0 |
| S7_extracted_98613 | 1999 | No | NA | 3 | 0 | 0 | Not-determined | Genome-fragment | NA | NA | 0 |
| S7_extracted_98634 | 1136 | No | NA | 2 | 0 | 1 | Not-determined | Genome-fragment | NA | NA | 0 |
| S7_extracted_98711 | 1281 | No | NA | 1 | 0 | 1 | Not-determined | Genome-fragment | NA | NA | 0 |
| S7_extracted_98759 | 1257 | No | NA | 3 | 0 | 1 | Not-determined | Genome-fragment | NA | NA | 0 |
| S7_extracted_98838 | 1264 | No | NA | 2 | 0 | 2 | Not-determined | Genome-fragment | NA | NA | 0 |
| S7_extracted_98881 | 1248 | No | NA | 1 | 0 | 0 | Not-determined | Genome-fragment | NA | NA | 0 |
| S7_extracted_98885 | 4700 | No | NA | 3 | 0 | 1 | Not-determined | Genome-fragment | NA | NA | 0 |
| S7_extracted_98978 | 1203 | No | NA | 2 | 0 | 0 | Not-determined | Genome-fragment | NA | NA | 0 |
| S17_extracted_k141_47059 | 2095 | No | NA | 4 | 1 | 0 | Low-quality | Genome-fragment | 1.8 | HMM-based (lower-bound) | 0 |
| S7_extracted_99062 | 7177 | No | NA | 6 | 0 | 5 | Not-determined | Genome-fragment | NA | NA | 0 |
| S7_extracted_99070 | 1729 | No | NA | 3 | 0 | 3 | Not-determined | Genome-fragment | NA | NA | 0 |
| S17_vs_k141_92137\|\|full | 2095 | No | NA | 3 | 0 | 1 | Low-quality | Genome-fragment | 2.9 | AAI-based (medium-confidence) | 0 |
| S14_vs_k141_30891\|\|full | 2095 | No | NA | 5 | 1 | 0 | Low-quality | Genome-fragment | 3.37 | AAI-based (high-confidence) | 0 |
| S7_extracted_99204 | 1203 | No | NA | 1 | 0 | 1 | Not-determined | Genome-fragment | NA | NA | 0 |
| S7_extracted_99223 | 2174 | No | NA | 1 | 0 | 0 | Not-determined | Genome-fragment | NA | NA | 0 |
| S16_vs_107563\|\|full | 2095 | No | NA | 3 | 0 | 2 | Low-quality | Genome-fragment | 1.21 | AAI-based (medium-confidence) | 0 |
| S7_extracted_99413 | 2042 | No | NA | 3 | 0 | 0 | Not-determined | Genome-fragment | NA | NA | 0 |
| S7_extracted_99627 | 1070 | No | NA | 1 | 0 | 1 | Not-determined | Genome-fragment | NA | NA | 0 |
| S7_extracted_99806 | 1604 | No | NA | 2 | 0 | 2 | Not-determined | Genome-fragment | NA | NA | 0 |
| S8_extracted_k141_197342 | 2095 | No | NA | 2 | 2 | 0 | Low-quality | Genome-fragment | 4.19 | AAI-based (high-confidence) | 0 |
| S7_extracted_100090 | 1311 | No | NA | 1 | 0 | 1 | Not-determined | Genome-fragment | NA | NA | 0 |
| S7_extracted_100289 | 5241 | No | NA | 6 | 0 | 2 | Not-determined | Genome-fragment | NA | NA | 0 |
| S7_extracted_100422 | 4651 | No | NA | 6 | 0 | 6 | Not-determined | Genome-fragment | NA | NA | 0 |
| S8_vs_k141_53377\|\|full | 2095 | No | NA | 3 | 1 | 0 | Low-quality | Genome-fragment | 2.37 | AAI-based (medium-confidence) | 0 |
| S7_extracted_100604 | 1833 | No | NA | 2 | 0 | 0 | Not-determined | Genome-fragment | NA | NA | 0 |
| S17_extracted_k141_13321 | 2096 | No | NA | 3 | 0 | 1 | Low-quality | Genome-fragment | 4.35 | AAI-based (high-confidence) | 0 |
| S13_extracted_11911 | 2096 | No | NA | 2 | 0 | 1 | Low-quality | Genome-fragment | 1.08 | AAI-based (medium-confidence) | 0 |
| S7_extracted_100690 | 1389 | No | NA | 2 | 0 | 1 | Not-determined | Genome-fragment | NA | NA | 0 |
| S7_extracted_100750 | 1510 | No | NA | 2 | 0 | 2 | Not-determined | Genome-fragment | NA | NA | 0 |
| S7_extracted_100932 | 1628 | No | NA | 1 | 0 | 1 | Not-determined | Genome-fragment | NA | NA | 0 |
| S5_extracted_k141_14964 | 2096 | No | NA | 2 | 0 | 0 | Low-quality | Genome-fragment | 4.8 | AAI-based (medium-confidence) | 0 |
| S7_extracted_101000 | 3297 | No | NA | 4 | 0 | 1 | Not-determined | Genome-fragment | NA | NA | 0 |
| S7_extracted_101179 | 1025 | No | NA | 1 | 0 | 0 | Not-determined | Genome-fragment | NA | NA | 0 |
| S7_extracted_101245 | 1382 | No | NA | 2 | 0 | 0 | Not-determined | Genome-fragment | NA | NA | 0 |
| S7_extracted_101391 | 1284 | No | NA | 1 | 0 | 0 | Not-determined | Genome-fragment | NA | NA | 0 |
| S16_extracted_139264 | 2096 | No | NA | 2 | 2 | 0 | Low-quality | Genome-fragment | 11.53 | AAI-based (high-confidence) | 0 |
| S7_extracted_101497 | 7515 | No | NA | 10 | 0 | 9 | Not-determined | Genome-fragment | NA | NA | 0 |
| S7_extracted_101629 | 1245 | No | NA | 1 | 0 | 0 | Not-determined | Genome-fragment | NA | NA | 0 |
| S16_vs_142922\|\|full | 2096 | No | NA | 4 | 2 | 0 | Low-quality | Genome-fragment | 5.89 | AAI-based (high-confidence) | 0 |
| S12_vs_k141_62023\|\|full | 2096 | No | NA | 8 | 1 | 0 | Low-quality | Genome-fragment | 3.24 | HMM-based (lower-bound) | 0 |
| S7_extracted_102151 | 2356 | No | NA | 4 | 0 | 0 | Not-determined | Genome-fragment | NA | NA | 0 |
| S17_vs_k141_25382\|\|full | 2097 | No | NA | 3 | 0 | 0 | Low-quality | Genome-fragment | 4.73 | AAI-based (medium-confidence) | 0 |
| S9_vs_k141_79520\|\|full | 2097 | No | NA | 6 | 2 | 0 | Low-quality | Genome-fragment | 5.03 | AAI-based (medium-confidence) | 0 |
| S7_extracted_102394 | 2306 | No | NA | 4 | 0 | 1 | Not-determined | Genome-fragment | NA | NA | 0 |
| S7_extracted_102478 | 1823 | No | NA | 1 | 0 | 1 | Not-determined | Genome-fragment | NA | NA | 0 |
| S7_extracted_102646 | 1142 | No | NA | 2 | 0 | 1 | Not-determined | Genome-fragment | NA | NA | 0 |
| S7_extracted_102977 | 1414 | No | NA | 1 | 0 | 1 | Not-determined | Genome-fragment | NA | NA | 0 |
| S7_extracted_103054 | 2634 | No | NA | 2 | 0 | 1 | Not-determined | Genome-fragment | NA | NA | 0 |
| S7_extracted_103064 | 1179 | No | NA | 1 | 0 | 1 | Not-determined | Genome-fragment | NA | NA | 0 |
| S7_extracted_103326 | 1059 | No | NA | 2 | 0 | 1 | Not-determined | Genome-fragment | NA | NA | 0 |
| S7_extracted_103344 | 1194 | No | NA | 2 | 0 | 0 | Not-determined | Genome-fragment | NA | NA | 0 |
| S11_vs_k141_69329\|\|full | 2097 | No | NA | 3 | 2 | 0 | Low-quality | Genome-fragment | 4 | HMM-based (lower-bound) | 0 |
| S7_extracted_103409 | 5416 | No | NA | 12 | 0 | 0 | Not-determined | Genome-fragment | NA | NA | 0 |
| S7_vs_116510\|\|full | 2097 | No | NA | 3 | 0 | 0 | Low-quality | Genome-fragment | 3.17 | AAI-based (medium-confidence) | 0 |
| S7_extracted_103529 | 1079 | No | NA | 2 | 0 | 0 | Not-determined | Genome-fragment | NA | NA | 0 |
| S7_extracted_103636 | 1013 | No | NA | 1 | 0 | 1 | Not-determined | Genome-fragment | NA | NA | 0 |
| S7_extracted_103766 | 1027 | No | NA | 1 | 0 | 0 | Not-determined | Genome-fragment | NA | NA | 0 |
| S7_extracted_103767 | 1359 | No | NA | 1 | 0 | 1 | Not-determined | Genome-fragment | NA | NA | 0 |
| S7_extracted_103801 | 1298 | No | NA | 1 | 0 | 1 | Not-determined | Genome-fragment | NA | NA | 0 |
| S6_vs_k141_11570\|\|full | 2097 | No | NA | 2 | 0 | 0 | Low-quality | Genome-fragment | 4.81 | AAI-based (medium-confidence) | 0 |
| S7_extracted_103911 | 1193 | No | NA | 1 | 0 | 1 | Not-determined | Genome-fragment | NA | NA | 0 |
| S7_extracted_103920 | 1149 | No | NA | 2 | 0 | 2 | Not-determined | Genome-fragment | NA | NA | 0 |
| S9_vs_k141_71018\|\|full | 2098 | No | NA | 4 | 1 | 0 | Low-quality | Genome-fragment | 0.9 | HMM-based (lower-bound) | 0 |
| S7_extracted_103964 | 1584 | No | NA | 1 | 0 | 1 | Not-determined | Genome-fragment | NA | NA | 0 |
| S7_extracted_104021 | 1006 | No | NA | 1 | 0 | 0 | Not-determined | Genome-fragment | NA | NA | 0 |
| S7_extracted_104038 | 1142 | No | NA | 4 | 0 | 0 | Not-determined | Genome-fragment | NA | NA | 0 |
| S7_extracted_104181 | 1737 | No | NA | 1 | 0 | 1 | Not-determined | Genome-fragment | NA | NA | 0 |
| S7_extracted_104328 | 1069 | No | NA | 3 | 0 | 0 | Not-determined | Genome-fragment | NA | NA | 0 |
| S7_extracted_104698 | 3134 | No | NA | 2 | 0 | 1 | Not-determined | Genome-fragment | NA | NA | 0 |
| S7_extracted_104774 | 1089 | No | NA | 1 | 0 | 0 | Not-determined | Genome-fragment | NA | NA | 0 |
| S7_extracted_104833 | 4951 | No | NA | 4 | 0 | 3 | Not-determined | Genome-fragment | NA | NA | 0 |
| S7_extracted_104890 | 6313 | No | NA | 5 | 0 | 3 | Not-determined | Genome-fragment | NA | NA | 0 |
| S7_extracted_104899 | 9124 | No | NA | 6 | 0 | 3 | Not-determined | Genome-fragment | NA | NA | 0 |
| S7_extracted_104942 | 2643 | No | NA | 2 | 0 | 1 | Not-determined | Genome-fragment | NA | NA | 0 |
| S1_vs_k141_118317\|\|full | 2098 | No | NA | 3 | 0 | 1 | Low-quality | Genome-fragment | 3.96 | AAI-based (medium-confidence) | 0 |
| S7_vs_6963\|\|full | 2098 | No | NA | 3 | 0 | 2 | Low-quality | Genome-fragment | 2.6 | AAI-based (medium-confidence) | 0 |
| S7_extracted_105121 | 1223 | No | NA | 1 | 0 | 0 | Not-determined | Genome-fragment | NA | NA | 0 |
| S7_extracted_105291 | 1430 | No | NA | 1 | 0 | 0 | Not-determined | Genome-fragment | NA | NA | 0 |
| S7_extracted_105312 | 1500 | No | NA | 6 | 0 | 0 | Not-determined | Genome-fragment | NA | NA | 0 |
| S7_extracted_105559 | 1444 | No | NA | 2 | 0 | 2 | Not-determined | Genome-fragment | NA | NA | 0 |
| S16_vs_109874\|\|full | 2098 | No | NA | 3 | 0 | 0 | Low-quality | Genome-fragment | 3.34 | AAI-based (medium-confidence) | 0 |
| S7_extracted_105984 | 1059 | No | NA | 1 | 0 | 1 | Not-determined | Genome-fragment | NA | NA | 0 |
| S7_extracted_105986 | 6481 | No | NA | 9 | 0 | 7 | Not-determined | Genome-fragment | NA | NA | 0 |
| S7_extracted_106192 | 1869 | No | NA | 2 | 0 | 1 | Not-determined | Genome-fragment | NA | NA | 0 |
| S7_extracted_106381 | 2566 | No | NA | 1 | 0 | 1 | Not-determined | Genome-fragment | NA | NA | 0 |
| S7_extracted_106497 | 5086 | No | NA | 8 | 0 | 1 | Not-determined | Genome-fragment | NA | NA | 0 |
| S7_extracted_106718 | 1191 | No | NA | 1 | 0 | 1 | Not-determined | Genome-fragment | NA | NA | 0 |
| S11_extracted_k141_53747 | 2099 | No | NA | 1 | 0 | 1 | Low-quality | Genome-fragment | 0.19 | AAI-based (medium-confidence) | 0 |
| S16_extracted_120865 | 2099 | No | NA | 3 | 0 | 1 | Low-quality | Genome-fragment | 2.99 | AAI-based (high-confidence) | 0 |
| S7_extracted_107158 | 4361 | No | NA | 3 | 0 | 2 | Not-determined | Genome-fragment | NA | NA | 0 |
| S7_extracted_107195 | 4220 | No | NA | 3 | 0 | 2 | Not-determined | Genome-fragment | NA | NA | 0 |
| S7_extracted_107291 | 1376 | No | NA | 2 | 0 | 0 | Not-determined | Genome-fragment | NA | NA | 0 |
| S7_extracted_107366 | 1175 | No | NA | 2 | 0 | 1 | Not-determined | Genome-fragment | NA | NA | 0 |
| S7_extracted_107654 | 4239 | No | NA | 3 | 0 | 2 | Not-determined | Genome-fragment | NA | NA | 0 |
| S7_extracted_107792 | 1947 | No | NA | 3 | 0 | 0 | Not-determined | Genome-fragment | NA | NA | 0 |
| S7_extracted_107801 | 3924 | No | NA | 4 | 0 | 4 | Not-determined | Genome-fragment | NA | NA | 0 |
| S7_extracted_107882 | 3760 | No | NA | 4 | 0 | 4 | Not-determined | Genome-fragment | NA | NA | 0 |
| S8_vs_k141_163506\|\|full | 2099 | No | NA | 4 | 1 | 0 | Low-quality | Genome-fragment | 4.2 | HMM-based (lower-bound) | 0 |
| S7_extracted_108155 | 8819 | No | NA | 12 | 0 | 3 | Not-determined | Genome-fragment | NA | NA | 0 |
| S9_vs_k141_59895\|\|full | 2100 | No | NA | 3 | 0 | 1 | Low-quality | Genome-fragment | 0.84 | AAI-based (medium-confidence) | 0 |
| S10_vs_117351\|\|full | 2100 | No | NA | 5 | 0 | 3 | Low-quality | Genome-fragment | 0.61 | AAI-based (medium-confidence) | 0 |
| S7_extracted_108324 | 5416 | No | NA | 6 | 0 | 5 | Not-determined | Genome-fragment | NA | NA | 0 |
| S12_vs_k141_74070\|\|full | 2100 | No | NA | 4 | 1 | 0 | Low-quality | Genome-fragment | 5.72 | AAI-based (high-confidence) | 0 |
| S8_extracted_k141_24428 | 2100 | No | NA | 2 | 1 | 0 | Low-quality | Genome-fragment | 1.97 | HMM-based (lower-bound) | 0 |
| S14_extracted_k141_67892 | 2101 | No | NA | 3 | 0 | 0 | Low-quality | Genome-fragment | 1.63 | AAI-based (medium-confidence) | 0 |
| S7_vs_75087\|\|full | 2101 | Yes | 1627 | 3 | 1 | 1 | Low-quality | Genome-fragment | 1.99 | AAI-based (high-confidence) | 22.56 |
| S16_vs_95892\|\|full | 2101 | No | NA | 3 | 0 | 1 | Low-quality | Genome-fragment | 1.19 | AAI-based (high-confidence) | 0 |
| S7_extracted_108663 | 1979 | No | NA | 3 | 0 | 2 | Not-determined | Genome-fragment | NA | NA | 0 |
| S16_vs_154881\|\|full | 2101 | No | NA | 4 | 1 | 0 | Low-quality | Genome-fragment | 3.66 | AAI-based (medium-confidence) | 0 |
| S7_extracted_108883 | 22864 | No | NA | 17 | 0 | 15 | Not-determined | Genome-fragment | NA | NA | 0 |
| S7_extracted_109011 | 6675 | No | NA | 8 | 0 | 1 | Not-determined | Genome-fragment | NA | NA | 0 |
| S7_extracted_109054 | 1103 | No | NA | 1 | 0 | 1 | Not-determined | Genome-fragment | NA | NA | 0 |
| S8_extracted_k141_380 | 2101 | No | NA | 3 | 0 | 0 | Low-quality | Genome-fragment | 4.35 | AAI-based (high-confidence) | 0 |
| S7_extracted_109266 | 1247 | No | NA | 1 | 0 | 1 | Not-determined | Genome-fragment | NA | NA | 0 |
| S9_extracted_k141_49113 | 2102 | No | NA | 1 | 0 | 0 | Low-quality | Genome-fragment | 4.83 | AAI-based (high-confidence) | 0 |
| S11_vs_k141_58558\|\|full | 2102 | No | NA | 5 | 0 | 0 | Low-quality | Genome-fragment | 5.68 | AAI-based (medium-confidence) | 0 |
| S17_vs_k141_153627\|\|full | 2103 | No | NA | 3 | 0 | 0 | Low-quality | Genome-fragment | 4.68 | AAI-based (high-confidence) | 0 |
| S7_extracted_109814 | 1432 | No | NA | 2 | 0 | 1 | Not-determined | Genome-fragment | NA | NA | 0 |
| S12_vs_k141_35342\|\|full | 2103 | No | NA | 4 | 0 | 0 | Low-quality | Genome-fragment | 3.48 | AAI-based (medium-confidence) | 0 |
| S7_extracted_109893 | 3649 | No | NA | 7 | 0 | 2 | Not-determined | Genome-fragment | NA | NA | 0 |
| S7_extracted_109906 | 1483 | No | NA | 2 | 0 | 2 | Not-determined | Genome-fragment | NA | NA | 0 |
| S7_extracted_109926 | 1264 | No | NA | 1 | 0 | 1 | Not-determined | Genome-fragment | NA | NA | 0 |
| S7_extracted_110011 | 1671 | No | NA | 1 | 0 | 1 | Not-determined | Genome-fragment | NA | NA | 0 |
| S8_extracted_k141_137562 | 2103 | No | NA | 4 | 0 | 0 | Low-quality | Genome-fragment | 1.95 | AAI-based (high-confidence) | 0 |
| S7_extracted_110148 | 2633 | No | NA | 3 | 0 | 1 | Not-determined | Genome-fragment | NA | NA | 0 |
| S8_extracted_k141_190895 | 2103 | No | NA | 2 | 0 | 0 | Low-quality | Genome-fragment | 0.2 | AAI-based (medium-confidence) | 0 |
| S7_extracted_110665 | 2322 | No | NA | 1 | 0 | 0 | Not-determined | Genome-fragment | NA | NA | 0 |
| S11_vs_k141_10807\|\|full | 2104 | No | NA | 3 | 0 | 0 | Low-quality | Genome-fragment | 1.61 | AAI-based (medium-confidence) | 0 |
| S7_extracted_110841 | 1194 | No | NA | 1 | 0 | 1 | Not-determined | Genome-fragment | NA | NA | 0 |
| S7_extracted_110867 | 1093 | No | NA | 1 | 0 | 1 | Not-determined | Genome-fragment | NA | NA | 0 |
| S13_vs_132543\|\|full | 2104 | No | NA | 2 | 0 | 2 | Low-quality | Genome-fragment | 1.58 | AAI-based (medium-confidence) | 0 |
| S7_extracted_111367 | 1265 | No | NA | 1 | 0 | 1 | Not-determined | Genome-fragment | NA | NA | 0 |
| S7_extracted_111375 | 4346 | No | NA | 2 | 0 | 0 | Not-determined | Genome-fragment | NA | NA | 0 |
| S7_extracted_111396 | 5907 | No | NA | 6 | 0 | 1 | Not-determined | Genome-fragment | NA | NA | 0 |
| S7_extracted_111417 | 2350 | No | NA | 2 | 0 | 2 | Not-determined | Genome-fragment | NA | NA | 0 |
| S7_extracted_111655 | 1351 | No | NA | 3 | 0 | 1 | Not-determined | Genome-fragment | NA | NA | 0 |
| S7_extracted_111668 | 3322 | No | NA | 4 | 0 | 0 | Not-determined | Genome-fragment | NA | NA | 0 |
| S3_vs_k141_67876\|\|full | 2104 | No | NA | 2 | 0 | 0 | Low-quality | Genome-fragment | 5.48 | AAI-based (high-confidence) | 0 |
| S7_extracted_112068 | 7592 | No | NA | 9 | 0 | 5 | Not-determined | Genome-fragment | NA | NA | 0 |
| S5_vs_k141_34980\|\|full | 2104 | No | NA | 4 | 1 | 0 | Low-quality | Genome-fragment | 3.36 | AAI-based (high-confidence) | 0 |
| S7_vs_97542\|\|full | 2104 | No | NA | 4 | 2 | 0 | Low-quality | Genome-fragment | 3.85 | AAI-based (medium-confidence) | 0 |
| S7_extracted_112229 | 1036 | No | NA | 2 | 0 | 0 | Not-determined | Genome-fragment | NA | NA | 0 |
| S13_extracted_89176 | 2105 | No | NA | 2 | 0 | 2 | Low-quality | Genome-fragment | 0.59 | AAI-based (high-confidence) | 0 |
| S7_extracted_112465 | 1162 | No | NA | 3 | 0 | 1 | Not-determined | Genome-fragment | NA | NA | 0 |
| S14_vs_k141_51201\|\|full | 2105 | No | NA | 6 | 1 | 0 | Low-quality | Genome-fragment | 3.55 | HMM-based (lower-bound) | 0 |
| S10_vs_111067\|\|full | 2105 | No | NA | 4 | 0 | 0 | Low-quality | Genome-fragment | 0.61 | AAI-based (medium-confidence) | 0 |
| S7_extracted_112683 | 1369 | No | NA | 1 | 0 | 1 | Not-determined | Genome-fragment | NA | NA | 0 |
| S7_extracted_113199 | 1334 | No | NA | 2 | 0 | 2 | Not-determined | Genome-fragment | NA | NA | 0 |
| S12_extracted_k141_122530 | 2105 | No | NA | 3 | 1 | 0 | Low-quality | Genome-fragment | 6.16 | AAI-based (medium-confidence) | 0 |
| S12_vs_k141_10419\|\|full | 2105 | No | NA | 5 | 0 | 0 | Low-quality | Genome-fragment | 4.68 | AAI-based (medium-confidence) | 0 |
| S7_extracted_113702 | 2518 | No | NA | 3 | 0 | 2 | Not-determined | Genome-fragment | NA | NA | 0 |
| S7_extracted_113756 | 1028 | No | NA | 1 | 0 | 0 | Not-determined | Genome-fragment | NA | NA | 0 |
| S9_vs_k141_55942\|\|full | 2106 | No | NA | 5 | 1 | 0 | Low-quality | Genome-fragment | 1.73 | HMM-based (lower-bound) | 0 |
| S11_extracted_k141_27602 | 2106 | No | NA | 6 | 0 | 0 | Low-quality | Genome-fragment | 4.64 | AAI-based (high-confidence) | 0 |
| S11_vs_k141_66204\|\|full | 2106 | No | NA | 4 | 0 | 0 | Low-quality | Genome-fragment | 3.54 | AAI-based (high-confidence) | 0 |
| S14_extracted_k141_22313 | 2106 | No | NA | 8 | 8 | 0 | Low-quality | Genome-fragment | 6.97 | AAI-based (high-confidence) | 0 |
| S7_extracted_114068 | 1279 | No | NA | 1 | 0 | 1 | Not-determined | Genome-fragment | NA | NA | 0 |
| S7_extracted_114128 | 3618 | No | NA | 3 | 0 | 2 | Not-determined | Genome-fragment | NA | NA | 0 |
| S7_extracted_114164 | 6751 | No | NA | 4 | 0 | 0 | Not-determined | Genome-fragment | NA | NA | 0 |
| S7_extracted_114206 | 1347 | No | NA | 2 | 0 | 1 | Not-determined | Genome-fragment | NA | NA | 0 |
| S3_vs_k141_117944\|\|full | 2106 | No | NA | 3 | 1 | 0 | Low-quality | Genome-fragment | 3.29 | AAI-based (high-confidence) | 0 |
| S7_extracted_114476 | 24797 | No | NA | 27 | 12 | 0 | Medium-quality | Genome-fragment | 59.98 | AAI-based (high-confidence) | 0 |
| S7_extracted_114479 | 3432 | No | NA | 4 | 0 | 3 | Not-determined | Genome-fragment | NA | NA | 0 |
| S5_vs_k141_41761\|\|full | 2106 | No | NA | 5 | 3 | 0 | Low-quality | Genome-fragment | 3.64 | AAI-based (high-confidence) | 0 |
| S12_vs_k141_22274\|\|full | 2106 | No | NA | 4 | 0 | 3 | Low-quality | Genome-fragment | 0.73 | AAI-based (medium-confidence) | 0 |
| S4_extracted_11304 | 2106 | No | NA | 6 | 0 | 0 | Low-quality | Genome-fragment | 5.34 | AAI-based (high-confidence) | 0 |
| S7_extracted_115096 | 1946 | No | NA | 2 | 0 | 2 | Not-determined | Genome-fragment | NA | NA | 0 |
| S7_extracted_115195 | 1405 | No | NA | 2 | 0 | 0 | Not-determined | Genome-fragment | NA | NA | 0 |
| S7_extracted_115433 | 3035 | No | NA | 2 | 0 | 2 | Not-determined | Genome-fragment | NA | NA | 0 |
| S7_extracted_115536 | 1031 | No | NA | 2 | 0 | 2 | Not-determined | Genome-fragment | NA | NA | 0 |
| S3_vs_k141_47238\|\|full | 2107 | No | NA | 6 | 1 | 0 | Low-quality | Genome-fragment | 5.03 | AAI-based (medium-confidence) | 0 |
| S7_extracted_115745 | 5470 | No | NA | 4 | 0 | 1 | Not-determined | Genome-fragment | NA | NA | 0 |
| S7_vs_90842\|\|full | 2107 | No | NA | 5 | 0 | 0 | Low-quality | Genome-fragment | 3.52 | AAI-based (medium-confidence) | 0 |
| S16_extracted_156214 | 2107 | No | NA | 2 | 2 | 0 | Low-quality | Genome-fragment | 2.54 | HMM-based (lower-bound) | 0 |
| S7_extracted_115989 | 2243 | No | NA | 4 | 0 | 0 | Not-determined | Genome-fragment | NA | NA | 0 |
| S7_extracted_116110 | 1824 | No | NA | 2 | 0 | 0 | Not-determined | Genome-fragment | NA | NA | 0 |
| S7_extracted_116117 | 1760 | No | NA | 2 | 0 | 0 | Not-determined | Genome-fragment | NA | NA | 0 |
| S7_extracted_116229 | 1259 | No | NA | 2 | 0 | 2 | Not-determined | Genome-fragment | NA | NA | 0 |
| S7_extracted_116266 | 1044 | No | NA | 1 | 0 | 1 | Not-determined | Genome-fragment | NA | NA | 0 |
| S7_extracted_116454 | 3275 | No | NA | 3 | 0 | 0 | Not-determined | Genome-fragment | NA | NA | 0 |
| S7_extracted_116475 | 3918 | No | NA | 4 | 0 | 2 | Not-determined | Genome-fragment | NA | NA | 0 |
| S7_extracted_116585 | 1016 | No | NA | 2 | 0 | 0 | Not-determined | Genome-fragment | NA | NA | 0 |
| S7_extracted_116668 | 2365 | No | NA | 1 | 0 | 1 | Not-determined | Genome-fragment | NA | NA | 0 |
| S7_extracted_116793 | 1848 | No | NA | 3 | 0 | 0 | Not-determined | Genome-fragment | NA | NA | 0 |
| S7_extracted_116913 | 1566 | No | NA | 2 | 0 | 2 | Not-determined | Genome-fragment | NA | NA | 0 |
| S7_extracted_116994 | 11810 | No | NA | 7 | 0 | 3 | Not-determined | Genome-fragment | NA | NA | 0 |
| S7_extracted_117215 | 1319 | No | NA | 1 | 0 | 1 | Not-determined | Genome-fragment | NA | NA | 0 |
| S16_vs_144786\|\|full | 2107 | No | NA | 4 | 1 | 0 | Low-quality | Genome-fragment | 0.59 | HMM-based (lower-bound) | 0 |
| S8_extracted_k141_192151 | 2107 | No | NA | 3 | 0 | 0 | Low-quality | Genome-fragment | 0.2 | AAI-based (medium-confidence) | 0 |
| S17_vs_k141_147571\|\|full | 2108 | No | NA | 2 | 0 | 0 | Low-quality | Genome-fragment | 6.6 | AAI-based (high-confidence) | 0 |
| S7_extracted_117697 | 40769 | No | NA | 54 | 9 | 1 | High-quality | High-quality | 100 | AAI-based (high-confidence) | 0 |
| S9_vs_k141_28152\|\|full | 2108 | No | NA | 3 | 1 | 0 | Low-quality | Genome-fragment | 2.81 | AAI-based (medium-confidence) | 0 |
| S7_extracted_118106 | 1976 | No | NA | 2 | 0 | 2 | Not-determined | Genome-fragment | NA | NA | 0 |
| S7_extracted_118171 | 1443 | No | NA | 3 | 0 | 0 | Not-determined | Genome-fragment | NA | NA | 0 |
| S7_extracted_118312 | 1146 | No | NA | 2 | 0 | 1 | Not-determined | Genome-fragment | NA | NA | 0 |
| S7_extracted_118346 | 3477 | No | NA | 4 | 0 | 1 | Not-determined | Genome-fragment | NA | NA | 0 |
| S3_extracted_k141_71608 | 2108 | No | NA | 1 | 0 | 0 | Low-quality | Genome-fragment | 1.57 | AAI-based (high-confidence) | 0 |
| S8_extracted_k141_27237 | 2108 | No | NA | 5 | 1 | 0 | Low-quality | Genome-fragment | 4.59 | AAI-based (high-confidence) | 0 |
| S7_extracted_118411 | 1953 | No | NA | 3 | 0 | 0 | Not-determined | Genome-fragment | NA | NA | 0 |
| S7_extracted_118482 | 1533 | No | NA | 2 | 0 | 1 | Not-determined | Genome-fragment | NA | NA | 0 |
| S7_extracted_118483 | 1164 | No | NA | 1 | 0 | 0 | Not-determined | Genome-fragment | NA | NA | 0 |
| S7_extracted_118667 | 1137 | No | NA | 2 | 0 | 1 | Not-determined | Genome-fragment | NA | NA | 0 |
| S7_extracted_118727 | 1483 | No | NA | 2 | 0 | 2 | Not-determined | Genome-fragment | NA | NA | 0 |
| S8_extracted_k141_171521 | 2108 | No | NA | 4 | 0 | 0 | Low-quality | Genome-fragment | 4.79 | AAI-based (medium-confidence) | 0 |
| S8_vs_k141_43142\|\|lt2gene | 2108 | No | NA | 2 | 2 | 0 | Low-quality | Genome-fragment | 5.65 | AAI-based (high-confidence) | 0 |
| S9_vs_k141_2442\|\|full | 2109 | No | NA | 3 | 0 | 3 | Low-quality | Genome-fragment | 1.12 | AAI-based (medium-confidence) | 0 |
| S14_vs_k141_83398\|\|full | 2109 | No | NA | 6 | 0 | 0 | Low-quality | Genome-fragment | 4.5 | AAI-based (high-confidence) | 0 |
| S14_vs_k141_35653\|\|full | 2109 | No | NA | 5 | 2 | 0 | Low-quality | Genome-fragment | 4.53 | AAI-based (medium-confidence) | 0 |
| S7_extracted_119367 | 1174 | No | NA | 1 | 0 | 1 | Not-determined | Genome-fragment | NA | NA | 0 |
| S7_extracted_119410 | 1122 | No | NA | 1 | 0 | 1 | Not-determined | Genome-fragment | NA | NA | 0 |
| S7_vs_50633\|\|lt2gene | 2109 | No | NA | 2 | 2 | 0 | Low-quality | Genome-fragment | 6.16 | AAI-based (medium-confidence) | 0 |
| S7_extracted_119573 | 1011 | No | NA | 2 | 0 | 2 | Not-determined | Genome-fragment | NA | NA | 0 |
| S7_extracted_119622 | 7182 | No | NA | 6 | 0 | 2 | Not-determined | Genome-fragment | NA | NA | 0 |

| | | | | | | | | | | | | |
|---|---|---|---|---|---|---|---|---|---|---|---|---|
| S9_vs_k141_66204\|\|full | 2110 | No | NA | 6 | 0 | 0 | Low-quality | Genome-fragment | | 2.99 | AAI-based (high-confidence) | 0 |
| S7_extracted_119805 | 1033 | No | NA | 1 | 0 | 1 | Not-determined | Genome-fragment | NA | | NA | 0 |
| S5_extracted_k141_133163 | 2110 | No | NA | 4 | 2 | 0 | Low-quality | Genome-fragment | | 5.13 | AAI-based (high-confidence) | 0 |
| S7_extracted_119897 | 1974 | No | NA | 1 | 0 | 1 | Not-determined | Genome-fragment | NA | | NA | 0 |
| S7_extracted_119974 | 1045 | No | NA | 1 | 0 | 0 | Not-determined | Genome-fragment | NA | | NA | 0 |
| S16_vs_13777\|\|full | 2110 | No | NA | 3 | 0 | 3 | Low-quality | Genome-fragment | | 5.42 | AAI-based (medium-confidence) | 0 |
| S16_vs_27554\|\|full | 2110 | No | NA | 3 | 0 | 0 | Low-quality | Genome-fragment | | 2.92 | AAI-based (high-confidence) | 0 |
| S7_extracted_120452 | 1353 | No | NA | 3 | 0 | 0 | Not-determined | Genome-fragment | NA | | NA | 0 |
| S7_extracted_120534 | 1601 | No | NA | 3 | 0 | 1 | Not-determined | Genome-fragment | NA | | NA | 0 |
| S7_extracted_120617 | 1351 | No | NA | 1 | 0 | 0 | Not-determined | Genome-fragment | NA | | NA | 0 |
| S7_extracted_120791 | 5487 | No | NA | 4 | 0 | 1 | Not-determined | Genome-fragment | NA | | NA | 0 |
| S7_extracted_120896 | 4948 | No | NA | 7 | 0 | 1 | Not-determined | Genome-fragment | NA | | NA | 0 |
| S7_extracted_120936 | 3036 | No | NA | 3 | 0 | 0 | Not-determined | Genome-fragment | NA | | NA | 0 |
| S7_extracted_120953 | 9388 | No | NA | 6 | 0 | 2 | Not-determined | Genome-fragment | NA | | NA | 0 |
| S7_extracted_120999 | 1029 | No | NA | 1 | 0 | 1 | Not-determined | Genome-fragment | NA | | NA | 0 |
| S8_vs_k141_208148\|\|full | 2110 | No | NA | 2 | 1 | 0 | Low-quality | Genome-fragment | | 0.69 | HMM-based (lower-bound) | 0 |
| S7_extracted_121158 | 1072 | No | NA | 2 | 0 | 2 | Not-determined | Genome-fragment | NA | | NA | 0 |
| S14_extracted_k141_95505 | 2111 | No | NA | 2 | 1 | 0 | Low-quality | Genome-fragment | | 1.3 | HMM-based (lower-bound) | 0 |
| S7_extracted_121168 | 1495 | No | NA | 1 | 0 | 0 | Not-determined | Genome-fragment | NA | | NA | 0 |
| S14_extracted_k141_99933 | 2111 | No | NA | 2 | 0 | 0 | Low-quality | Genome-fragment | | 2.88 | AAI-based (high-confidence) | 0 |
| S1_vs_k141_11508\|\|full | 2111 | No | NA | 2 | 0 | 0 | Low-quality | Genome-fragment | | 3.61 | AAI-based (high-confidence) | 0 |
| S7_extracted_121301 | 1177 | No | NA | 2 | 0 | 0 | Not-determined | Genome-fragment | NA | | NA | 0 |
| S7_extracted_121350 | 1828 | No | NA | 2 | 0 | 0 | Not-determined | Genome-fragment | NA | | NA | 0 |
| S7_extracted_121442 | 2475 | No | NA | 3 | 0 | 2 | Not-determined | Genome-fragment | NA | | NA | 0 |
| S7_extracted_121495 | 3715 | No | NA | 4 | 0 | 1 | Not-determined | Genome-fragment | NA | | NA | 0 |
| S7_extracted_121511 | 1079 | No | NA | 1 | 0 | 1 | Not-determined | Genome-fragment | NA | | NA | 0 |
| S7_extracted_121522 | 1216 | No | NA | 1 | 0 | 1 | Not-determined | Genome-fragment | NA | | NA | 0 |
| S7_extracted_121530 | 1188 | No | NA | 3 | 0 | 0 | Not-determined | Genome-fragment | NA | | NA | 0 |
| S3_extracted_k141_1198 | 2111 | No | NA | 4 | 1 | 0 | Low-quality | Genome-fragment | | 1.38 | AAI-based (high-confidence) | 0 |
| S7_extracted_121815 | 1044 | No | NA | 2 | 0 | 2 | Not-determined | Genome-fragment | NA | | NA | 0 |
| S7_extracted_121848 | 1478 | No | NA | 1 | 0 | 0 | Not-determined | Genome-fragment | NA | | NA | 0 |
| S5_extracted_k141_140737 | 2111 | No | NA | 1 | 1 | 0 | Low-quality | Genome-fragment | | 1.47 | AAI-based (high-confidence) | 0 |
| S7_extracted_121981 | 1121 | No | NA | 2 | 0 | 1 | Not-determined | Genome-fragment | NA | | NA | 0 |
| S7_extracted_122010 | 1373 | No | NA | 3 | 0 | 0 | Not-determined | Genome-fragment | NA | | NA | 0 |
| S7_extracted_122063 | 1017 | No | NA | 2 | 0 | 1 | Not-determined | Genome-fragment | NA | | NA | 0 |
| S7_extracted_122078 | 1781 | No | NA | 2 | 0 | 2 | Not-determined | Genome-fragment | NA | | NA | 0 |
| S5_extracted_k141_51838 | 2111 | No | NA | 5 | 1 | 0 | Low-quality | Genome-fragment | | 6.03 | AAI-based (medium-confidence) | 0 |
| S7_extracted_122208 | 1057 | No | NA | 1 | 0 | 0 | Not-determined | Genome-fragment | NA | | NA | 0 |
| S7_extracted_122277 | 1618 | No | NA | 2 | 0 | 0 | Not-determined | Genome-fragment | NA | | NA | 0 |
| S7_extracted_122376 | 1789 | No | NA | 1 | 0 | 0 | Not-determined | Genome-fragment | NA | | NA | 0 |
| S5_vs_k141_139790\|\|full | 2111 | No | NA | 3 | 0 | 1 | Low-quality | Genome-fragment | | 1.47 | AAI-based (medium-confidence) | 0 |
| S7_extracted_122484 | 1398 | No | NA | 3 | 0 | 3 | Not-determined | Genome-fragment | NA | | NA | 0 |
| S7_extracted_122576 | 1035 | No | NA | 1 | 0 | 1 | Not-determined | Genome-fragment | NA | | NA | 0 |
| S7_extracted_122718 | 5413 | No | NA | 4 | 0 | 2 | Not-determined | Genome-fragment | NA | | NA | 0 |
| S7_extracted_122753 | 9910 | No | NA | 5 | 0 | 3 | Not-determined | Genome-fragment | NA | | NA | 0 |
| S7_extracted_122760 | 1134 | No | NA | 1 | 0 | 0 | Not-determined | Genome-fragment | NA | | NA | 0 |
| S5_vs_k141_90192\|\|full | 2111 | No | NA | 2 | 0 | 2 | Low-quality | Genome-fragment | | 1.37 | AAI-based (medium-confidence) | 0 |
| S7_extracted_829 | 2111 | No | NA | 4 | 0 | 1 | Low-quality | Genome-fragment | | 4.06 | AAI-based (high-confidence) | 0 |
| S7_extracted_123090 | 1066 | No | NA | 1 | 0 | 0 | Not-determined | Genome-fragment | NA | | NA | 0 |
| S7_vs_7542\|\|lt2gene | 2111 | No | NA | 2 | 2 | 0 | Low-quality | Genome-fragment | | 5.47 | AAI-based (high-confidence) | 0 |
| S10_vs_94600\|\|full | 2111 | No | NA | 4 | 0 | 0 | Low-quality | Genome-fragment | | 4.69 | AAI-based (high-confidence) | 0 |
| S7_extracted_123212 | 1311 | No | NA | 2 | 0 | 1 | Not-determined | Genome-fragment | NA | | NA | 0 |
| S15_extracted_70071 | 2111 | No | NA | 2 | 0 | 1 | Low-quality | Genome-fragment | | 1.18 | AAI-based (high-confidence) | 0 |
| S8_vs_k141_157532\|\|full | 2111 | No | NA | 4 | 0 | 0 | Low-quality | Genome-fragment | | 5.23 | AAI-based (medium-confidence) | 0 |
| S7_extracted_123407 | 1413 | No | NA | 1 | 0 | 0 | Not-determined | Genome-fragment | NA | | NA | 0 |
| S7_extracted_123462 | 1021 | No | NA | 1 | 0 | 1 | Not-determined | Genome-fragment | NA | | NA | 0 |
| S7_extracted_123498 | 1106 | No | NA | 1 | 0 | 0 | Not-determined | Genome-fragment | NA | | NA | 0 |
| S1_extracted_k141_126162 | 2112 | No | NA | 2 | 0 | 0 | Low-quality | Genome-fragment | | 2.13 | AAI-based (high-confidence) | 0 |
| S7_extracted_123541 | 1965 | No | NA | 2 | 0 | 1 | Not-determined | Genome-fragment | NA | | NA | 0 |
| S7_extracted_123606 | 3219 | No | NA | 4 | 0 | 1 | Not-determined | Genome-fragment | NA | | NA | 0 |
| S3_vs_k141_25099\|\|full | 2112 | No | NA | 3 | 0 | 1 | Low-quality | Genome-fragment | | 4.18 | AAI-based (medium-confidence) | 0 |
| S7_extracted_123815 | 4213 | No | NA | 3 | 0 | 2 | Not-determined | Genome-fragment | NA | | NA | 0 |
| S7_extracted_123822 | 1151 | No | NA | 1 | 0 | 1 | Not-determined | Genome-fragment | NA | | NA | 0 |
| S7_extracted_124254 | 3940 | No | NA | 3 | 0 | 0 | Not-determined | Genome-fragment | NA | | NA | 0 |
| S7_extracted_124412 | 4698 | No | NA | 5 | 0 | 3 | Not-determined | Genome-fragment | NA | | NA | 0 |
| S3_vs_k141_30155\|\|full | 2112 | No | NA | 4 | 1 | 0 | Low-quality | Genome-fragment | | 5.29 | AAI-based (high-confidence) | 0 |
| S7_extracted_124521 | 6070 | No | NA | 8 | 0 | 6 | Not-determined | Genome-fragment | NA | | NA | 0 |
| S7_extracted_124547 | 1015 | No | NA | 1 | 0 | 0 | Not-determined | Genome-fragment | NA | | NA | 0 |
| S7_extracted_124602 | 1599 | No | NA | 2 | 0 | 2 | Not-determined | Genome-fragment | NA | | NA | 0 |
| S7_extracted_124604 | 1008 | No | NA | 1 | 0 | 1 | Not-determined | Genome-fragment | NA | | NA | 0 |
| S7_extracted_124775 | 1444 | No | NA | 5 | 0 | 5 | Not-determined | Genome-fragment | NA | | NA | 0 |
| S7_extracted_124807 | 2650 | No | NA | 4 | 0 | 1 | Not-determined | Genome-fragment | NA | | NA | 0 |
| S7_extracted_124866 | 3700 | No | NA | 5 | 0 | 2 | Not-determined | Genome-fragment | NA | | NA | 0 |
| S7_extracted_124903 | 2240 | No | NA | 2 | 0 | 1 | Not-determined | Genome-fragment | NA | | NA | 0 |
| S7_extracted_21427 | 2112 | No | NA | 1 | 0 | 1 | Low-quality | Genome-fragment | | 0.19 | AAI-based (medium-confidence) | 0 |
| S7_extracted_124990 | 4036 | No | NA | 2 | 0 | 2 | Not-determined | Genome-fragment | NA | | NA | 0 |
| S10_vs_34260\|\|full | 2112 | No | NA | 4 | 2 | 0 | Low-quality | Genome-fragment | | 5.4 | AAI-based (high-confidence) | 0 |
| S7_extracted_125160 | 1504 | No | NA | 1 | 0 | 1 | Not-determined | Genome-fragment | NA | | NA | 0 |
| S7_extracted_125166 | 3300 | No | NA | 5 | 0 | 0 | Not-determined | Genome-fragment | NA | | NA | 0 |
| S7_vs_87443\|\|full | 2113 | No | NA | 2 | 0 | 0 | Low-quality | Genome-fragment | | 3.86 | AAI-based (medium-confidence) | 0 |
| S7_extracted_125352 | 1034 | No | NA | 1 | 0 | 1 | Not-determined | Genome-fragment | NA | | NA | 0 |
| S7_vs_93647\|\|full | 2113 | No | NA | 5 | 0 | 0 | Low-quality | Genome-fragment | | 3.16 | AAI-based (high-confidence) | 0 |
| S7_extracted_125601 | 1150 | No | NA | 2 | 0 | 1 | Not-determined | Genome-fragment | NA | | NA | 0 |
| S7_extracted_125677 | 1322 | No | NA | 1 | 0 | 0 | Not-determined | Genome-fragment | NA | | NA | 0 |
| S7_extracted_125793 | 2167 | No | NA | 2 | 0 | 1 | Not-determined | Genome-fragment | NA | | NA | 0 |
| S16_vs_155240\|\|full | 2113 | No | NA | 4 | 2 | 0 | Low-quality | Genome-fragment | | 2.57 | AAI-based (high-confidence) | 0 |
| S16_vs_167553\|\|full | 2113 | No | NA | 5 | 0 | 0 | Low-quality | Genome-fragment | | 6.13 | AAI-based (high-confidence) | 0 |
| S17_vs_k141_8941\|\|full | 2114 | No | NA | 3 | 0 | 2 | Low-quality | Genome-fragment | | 3.34 | AAI-based (high-confidence) | 0 |
| S9_vs_k141_64591\|\|full | 2114 | No | NA | 6 | 1 | 0 | Low-quality | Genome-fragment | | 6.1 | AAI-based (high-confidence) | 0 |
| S7_extracted_126326 | 4102 | No | NA | 3 | 0 | 2 | Not-determined | Genome-fragment | NA | | NA | 0 |
| S7_extracted_126369 | 1155 | No | NA | 3 | 0 | 0 | Not-determined | Genome-fragment | NA | | NA | 0 |
| S5_vs_k141_21692\|\|full | 2114 | No | NA | 2 | 1 | 0 | Low-quality | Genome-fragment | | 6.03 | AAI-based (high-confidence) | 0 |
| S7_extracted_126884 | 1591 | No | NA | 1 | 0 | 1 | Not-determined | Genome-fragment | NA | | NA | 0 |
| S7_extracted_108442 | 2114 | No | NA | 9 | 2 | 0 | Low-quality | Genome-fragment | | 6.14 | AAI-based (high-confidence) | 0 |
| S7_vs_36582\|\|full | 2114 | No | NA | 3 | 1 | 0 | Low-quality | Genome-fragment | | 1.62 | AAI-based (medium-confidence) | 0 |
| S7_extracted_127056 | 5036 | No | NA | 3 | 0 | 1 | Not-determined | Genome-fragment | NA | | NA | 0 |
| S7_extracted_127082 | 1250 | No | NA | 1 | 0 | 1 | Not-determined | Genome-fragment | NA | | NA | 0 |
| S7_extracted_127092 | 1159 | No | NA | 3 | 0 | 1 | Not-determined | Genome-fragment | NA | | NA | 0 |
| S7_extracted_127194 | 2670 | No | NA | 3 | 0 | 2 | Not-determined | Genome-fragment | NA | | NA | 0 |
| S7_extracted_127529 | 3346 | No | NA | 2 | 0 | 2 | Not-determined | Genome-fragment | NA | | NA | 0 |
| S7_extracted_127556 | 1212 | No | NA | 2 | 0 | 1 | Not-determined | Genome-fragment | NA | | NA | 0 |
| S7_extracted_127599 | 3409 | No | NA | 1 | 0 | 1 | Not-determined | Genome-fragment | NA | | NA | 0 |
| S7_extracted_127602 | 2732 | No | NA | 8 | 0 | 1 | Not-determined | Genome-fragment | NA | | NA | 0 |
| S7_extracted_127608 | 1322 | No | NA | 1 | 0 | 0 | Not-determined | Genome-fragment | NA | | NA | 0 |
| S7_extracted_127623 | 3193 | No | NA | 3 | 0 | 0 | Not-determined | Genome-fragment | NA | | NA | 0 |
| S7_vs_118478\|\|full | 2114 | No | NA | 5 | 0 | 0 | Low-quality | Genome-fragment | | 4.94 | AAI-based (high-confidence) | 0 |
| S7_extracted_127717 | 1424 | No | NA | 1 | 0 | 1 | Not-determined | Genome-fragment | NA | | NA | 0 |
| S16_vs_2273\|\|full | 2114 | No | NA | 5 | 0 | 0 | Low-quality | Genome-fragment | | 4.74 | AAI-based (medium-confidence) | 0 |
| S7_extracted_127985 | 1213 | No | NA | 1 | 0 | 1 | Not-determined | Genome-fragment | NA | | NA | 0 |
| S8_vs_k141_37191\|\|full | 2114 | No | NA | 2 | 0 | 1 | Low-quality | Genome-fragment | | 4.99 | AAI-based (medium-confidence) | 0 |
| S7_extracted_128053 | 3758 | No | NA | 2 | 0 | 2 | Not-determined | Genome-fragment | NA | | NA | 0 |
| S7_extracted_128320 | 1372 | No | NA | 2 | 0 | 1 | Not-determined | Genome-fragment | NA | | NA | 0 |
| S8_vs_k141_38438\|\|full | 2114 | No | NA | 5 | 2 | 0 | Low-quality | Genome-fragment | | 13.38 | AAI-based (high-confidence) | 0 |
| S11_extracted_k141_32290 | 2115 | No | NA | 4 | 1 | 0 | Low-quality | Genome-fragment | | 3.16 | AAI-based (high-confidence) | 0 |
| S5_vs_k141_91196\|\|full | 2115 | No | NA | 6 | 3 | 0 | Low-quality | Genome-fragment | | 3.17 | AAI-based (high-confidence) | 0 |
| S7_extracted_128779 | 1090 | No | NA | 2 | 0 | 2 | Not-determined | Genome-fragment | NA | | NA | 0 |
| S7_extracted_129094 | 6206 | No | NA | 3 | 0 | 3 | Not-determined | Genome-fragment | NA | | NA | 0 |
| S7_extracted_129119 | 1144 | No | NA | 1 | 0 | 1 | Not-determined | Genome-fragment | NA | | NA | 0 |
| S7_extracted_129232 | 1229 | No | NA | 3 | 0 | 2 | Not-determined | Genome-fragment | NA | | NA | 0 |
| S7_extracted_129249 | 1050 | No | NA | 2 | 0 | 2 | Not-determined | Genome-fragment | NA | | NA | 0 |
| S7_extracted_129274 | 1779 | No | NA | 1 | 0 | 0 | Not-determined | Genome-fragment | NA | | NA | 0 |
| S16_vs_170041\|\|full | 2115 | No | NA | 5 | 0 | 0 | Low-quality | Genome-fragment | | 5.49 | AAI-based (high-confidence) | 0 |
| S7_extracted_129303 | 1336 | No | NA | 3 | 0 | 0 | Not-determined | Genome-fragment | NA | | NA | 0 |
| S7_extracted_129390 | 1544 | No | NA | 1 | 0 | 0 | Not-determined | Genome-fragment | NA | | NA | 0 |
| S12_vs_k141_24236\|\|full | 2115 | No | NA | 3 | 0 | 0 | Low-quality | Genome-fragment | | 1.43 | AAI-based (high-confidence) | 0 |
| S7_extracted_129535 | 1169 | No | NA | 1 | 0 | 0 | Not-determined | Genome-fragment | NA | | NA | 0 |
| S12_vs_k141_112223\|\|full | 2115 | No | NA | 4 | 0 | 1 | Low-quality | Genome-fragment | | 1.05 | AAI-based (high-confidence) | 0 |
| S7_extracted_129604 | 1044 | No | NA | 2 | 0 | 0 | Not-determined | Genome-fragment | NA | | NA | 0 |
| S7_extracted_129617 | 3214 | No | NA | 2 | 0 | 0 | Not-determined | Genome-fragment | NA | | NA | 0 |
| S12_vs_k141_130823\|\|full | 2115 | No | NA | 3 | 0 | 2 | Low-quality | Genome-fragment | | 1.04 | AAI-based (medium-confidence) | 0 |
| S6_vs_k141_42395\|\|full | 2115 | No | NA | 4 | 0 | 1 | Low-quality | Genome-fragment | | 2.11 | AAI-based (medium-confidence) | 0 |
| S11_vs_k141_65772\|\|full | 2116 | No | NA | 4 | 2 | 0 | Low-quality | Genome-fragment | | 4.39 | HMM-based (lower-bound) | 0 |
| S1_vs_k141_90886\|\|lt2gene | 2116 | No | NA | 1 | 1 | 0 | Low-quality | Genome-fragment | | 3.85 | AAI-based (medium-confidence) | 0 |
| S7_extracted_129806 | 1031 | No | NA | 2 | 0 | 0 | Not-determined | Genome-fragment | NA | | NA | 0 |
| S3_vs_k141_68607\|\|full | 2116 | No | NA | 3 | 0 | 0 | Low-quality | Genome-fragment | | 2.77 | AAI-based (high-confidence) | 0 |
| S7_extracted_129831 | 1018 | No | NA | 1 | 0 | 1 | Not-determined | Genome-fragment | NA | | NA | 0 |
| S7_extracted_129887 | 7025 | No | NA | 7 | 0 | 6 | Not-determined | Genome-fragment | NA | | NA | 0 |
| S7_extracted_130044 | 1376 | No | NA | 1 | 0 | 1 | Not-determined | Genome-fragment | NA | | NA | 0 |
| S7_extracted_130077 | 1307 | No | NA | 2 | 0 | 1 | Not-determined | Genome-fragment | NA | | NA | 0 |
| S7_extracted_130181 | 3695 | No | NA | 3 | 0 | 3 | Not-determined | Genome-fragment | NA | | NA | 0 |
| S5_vs_k141_15121\|\|full | 2116 | No | NA | 3 | 1 | 0 | Low-quality | Genome-fragment | | 3.49 | AAI-based (high-confidence) | 0 |
| S7_extracted_130588 | 1130 | No | NA | 1 | 0 | 1 | Not-determined | Genome-fragment | NA | | NA | 0 |
| S7_extracted_130616 | 20482 | No | NA | 17 | 0 | 14 | Not-determined | Genome-fragment | NA | | NA | 0 |
| S7_extracted_130664 | 1728 | No | NA | 1 | 0 | 0 | Not-determined | Genome-fragment | NA | | NA | 0 |
| S12_extracted_k141_76582 | 2116 | No | NA | 5 | 1 | 0 | Low-quality | Genome-fragment | | 3.52 | AAI-based (high-confidence) | 0 |
| S7_extracted_130723 | 1018 | No | NA | 2 | 0 | 0 | Not-determined | Genome-fragment | NA | | NA | 0 |
| S15_extracted_56213 | 2116 | No | NA | 3 | 2 | 0 | Low-quality | Genome-fragment | | 4.89 | AAI-based (high-confidence) | 0 |
| S7_extracted_130941 | 1106 | No | NA | 1 | 0 | 0 | Not-determined | Genome-fragment | NA | | NA | 0 |
| S7_extracted_130945 | 1805 | No | NA | 2 | 0 | 2 | Not-determined | Genome-fragment | NA | | NA | 0 |
| S7_extracted_131004 | 1578 | No | NA | 4 | 0 | 0 | Not-determined | Genome-fragment | NA | | NA | 0 |
| S7_extracted_131213 | 1279 | No | NA | 2 | 0 | 2 | Not-determined | Genome-fragment | NA | | NA | 0 |
| S7_extracted_131247 | 10231 | No | NA | 24 | 0 | 1 | Not-determined | Genome-fragment | NA | | NA | 0 |
| S7_extracted_131353 | 3801 | No | NA | 3 | 0 | 2 | Not-determined | Genome-fragment | NA | | NA | 0 |
| S7_extracted_131370 | 1076 | No | NA | 1 | 0 | 1 | Not-determined | Genome-fragment | NA | | NA | 0 |
| S7_extracted_131664 | 1048 | No | NA | 1 | 0 | 1 | Not-determined | Genome-fragment | NA | | NA | 0 |
| S7_extracted_131665 | 1265 | No | NA | 1 | 0 | 1 | Not-determined | Genome-fragment | NA | | NA | 0 |
| S7_extracted_131712 | 3467 | No | NA | 7 | 0 | 2 | Not-determined | Genome-fragment | NA | | NA | 0 |
| S7_extracted_131962 | 3410 | No | NA | 4 | 0 | 4 | Not-determined | Genome-fragment | NA | | NA | 0 |
| S7_extracted_132043 | 1322 | No | NA | 2 | 0 | 1 | Not-determined | Genome-fragment | NA | | NA | 0 |
| S11_vs_k141_2704\|\|full | 2117 | No | NA | 4 | 0 | 0 | Low-quality | Genome-fragment | | 4.37 | AAI-based (high-confidence) | 0 |
| S7_extracted_132317 | 1164 | No | NA | 2 | 0 | 1 | Not-determined | Genome-fragment | NA | | NA | 0 |
| S7_extracted_132459 | 1393 | No | NA | 1 | 0 | 0 | Not-determined | Genome-fragment | NA | | NA | 0 |
| S7_extracted_132475 | 3967 | No | NA | 4 | 0 | 0 | Not-determined | Genome-fragment | NA | | NA | 0 |
| S10_vs_114417\|\|full | 2117 | No | NA | 3 | 0 | 0 | Low-quality | Genome-fragment | | 5.65 | AAI-based (high-confidence) | 0 |
| S7_extracted_132548 | 1657 | No | NA | 1 | 0 | 0 | Not-determined | Genome-fragment | NA | | NA | 0 |
| S7_extracted_132562 | 7865 | No | NA | 8 | 0 | 0 | Not-determined | Genome-fragment | NA | | NA | 0 |
| S7_extracted_132615 | 1888 | No | NA | 1 | 0 | 1 | Not-determined | Genome-fragment | NA | | NA | 0 |
| S15_extracted_51548 | 2117 | No | NA | 9 | 0 | 0 | Low-quality | Genome-fragment | | 6.12 | AAI-based (high-confidence) | 0 |
| S6_extracted_k141_48593 | 2117 | No | NA | 3 | 0 | 1 | Low-quality | Genome-fragment | | 1.04 | AAI-based (medium-confidence) | 0 |
| S6_vs_k141_26110\|\|full | 2117 | No | NA | 4 | 1 | 0 | Low-quality | Genome-fragment | | 3.49 | AAI-based (high-confidence) | 0 |
| S8_vs_k141_142542\|\|full | 2117 | No | NA | 3 | 0 | 2 | Low-quality | Genome-fragment | | 0.61 | AAI-based (medium-confidence) | 0 |
| S7_extracted_132968 | 2483 | No | NA | 6 | 0 | 1 | Not-determined | Genome-fragment | NA | | NA | 0 |
| S7_extracted_133089 | 1002 | No | NA | 1 | 0 | 1 | Not-determined | Genome-fragment | NA | | NA | 0 |
| S1_extracted_k141_50555 | 2118 | No | NA | 3 | 2 | 0 | Low-quality | Genome-fragment | | 6.1 | AAI-based (high-confidence) | 0 |
| S3_vs_k141_33118\|\|full | 2118 | No | NA | 6 | 3 | 0 | Low-quality | Genome-fragment | | 5.19 | AAI-based (high-confidence) | 0 |
| S7_extracted_133156 | 1266 | No | NA | 2 | 0 | 2 | Not-determined | Genome-fragment | NA | | NA | 0 |
| S7_extracted_133205 | 5594 | No | NA | 4 | 0 | 4 | Not-determined | Genome-fragment | NA | | NA | 0 |
| S7_extracted_133347 | 4876 | No | NA | 5 | 0 | 3 | Not-determined | Genome-fragment | NA | | NA | 0 |
| S7_extracted_133362 | 1443 | No | NA | 2 | 0 | 2 | Not-determined | Genome-fragment | NA | | NA | 0 |
| S7_extracted_133463 | 3650 | No | NA | 4 | 0 | 3 | Not-determined | Genome-fragment | NA | | NA | 0 |
| S7_extracted_133516 | 1487 | No | NA | 2 | 0 | 1 | Not-determined | Genome-fragment | NA | | NA | 0 |
| S7_extracted_133554 | 1398 | No | NA | 1 | 0 | 1 | Not-determined | Genome-fragment | NA | | NA | 0 |
| S7_vs_125655\|\|full | 2118 | No | NA | 4 | 0 | 0 | Low-quality | Genome-fragment | | 4.5 | AAI-based (medium-confidence) | 0 |
| S7_extracted_133577 | 1682 | No | NA | 1 | 0 | 1 | Not-determined | Genome-fragment | NA | | NA | 0 |
| S7_extracted_133722 | 1063 | No | NA | 1 | 0 | 1 | Not-determined | Genome-fragment | NA | | NA | 0 |
| S7_extracted_133746 | 3573 | No | NA | 3 | 0 | 1 | Not-determined | Genome-fragment | NA | | NA | 0 |
| S7_extracted_133761 | 1752 | No | NA | 3 | 0 | 0 | Not-determined | Genome-fragment | NA | | NA | 0 |
| S7_extracted_133849 | 3044 | No | NA | 4 | 0 | 1 | Not-determined | Genome-fragment | NA | | NA | 0 |
| S7_extracted_133861 | 3491 | No | NA | 6 | 0 | 0 | Not-determined | Genome-fragment | NA | | NA | 0 |
| S7_extracted_134014 | 1115 | No | NA | 1 | 0 | 0 | Not-determined | Genome-fragment | NA | | NA | 0 |
| S7_extracted_134023 | 41645 | No | NA | 73 | 16 | 1 | Complete | High-quality | | 100 | DTR (high-confidence) | 0 |
| S7_extracted_134061 | 4591 | No | NA | 2 | 0 | 0 | Not-determined | Genome-fragment | NA | | NA | 0 |
| S7_extracted_134201 | 1010 | No | NA | 1 | 0 | 0 | Not-determined | Genome-fragment | NA | | NA | 0 |
| S3_extracted_k141_102443 | 2119 | No | NA | 2 | 1 | 0 | Low-quality | Genome-fragment | | 3.38 | AAI-based (high-confidence) | 0 |
| S10_vs_40791\|\|full | 2119 | No | NA | 4 | 0 | 0 | Low-quality | Genome-fragment | | 4.56 | AAI-based (high-confidence) | 0 |
| S7_extracted_134434 | 1633 | No | NA | 2 | 0 | 0 | Not-determined | Genome-fragment | NA | | NA | 0 |
| S10_vs_90917\|\|full | 2119 | No | NA | 4 | 2 | 0 | Low-quality | Genome-fragment | | 3.87 | AAI-based (high-confidence) | 0 |
| S7_extracted_134488 | 1193 | No | NA | 1 | 0 | 1 | Not-determined | Genome-fragment | NA | | NA | 0 |
| S7_extracted_134528 | 1342 | No | NA | 2 | 0 | 1 | Not-determined | Genome-fragment | NA | | NA | 0 |
| S10_vs_104380\|\|full | 2119 | No | NA | 3 | 1 | 0 | Low-quality | Genome-fragment | | 38.84 | AAI-based (high-confidence) | 0 |
| S7_extracted_134718 | 1366 | No | NA | 1 | 0 | 1 | Not-determined | Genome-fragment | NA | | NA | 0 |
| S7_extracted_134793 | 3493 | No | NA | 3 | 0 | 3 | Not-determined | Genome-fragment | NA | | NA | 0 |
| S7_extracted_134850 | 1157 | No | NA | 2 | 0 | 0 | Not-determined | Genome-fragment | NA | | NA | 0 |
| S7_extracted_134876 | 1011 | No | NA | 1 | 0 | 0 | Not-determined | Genome-fragment | NA | | NA | 0 |
| S8_vs_k141_168147\|\|full | 2119 | No | NA | 5 | 0 | 0 | Low-quality | Genome-fragment | | 4.91 | AAI-based (high-confidence) | 0 |
| S7_extracted_135056 | 2431 | No | NA | 2 | 0 | 1 | Not-determined | Genome-fragment | NA | | NA | 0 |
| S7_extracted_135081 | 1978 | No | NA | 3 | 0 | 0 | Not-determined | Genome-fragment | NA | | NA | 0 |
| S7_extracted_135084 | 1455 | No | NA | 2 | 0 | 0 | Not-determined | Genome-fragment | NA | | NA | 0 |
| S7_extracted_135183 | 1386 | No | NA | 2 | 0 | 1 | Not-determined | Genome-fragment | NA | | NA | 0 |
| S7_extracted_135197 | 2087 | No | NA | 2 | 0 | 1 | Not-determined | Genome-fragment | NA | | NA | 0 |
| S7_extracted_135462 | 2305 | No | NA | 2 | 0 | 0 | Not-determined | Genome-fragment | NA | | NA | 0 |
| S11_extracted_k141_21016 | 2120 | No | NA | 2 | 1 | 0 | Low-quality | Genome-fragment | | 1.32 | HMM-based (lower-bound) | 0 |
| S7_extracted_135520 | 4503 | No | NA | 8 | 0 | 2 | Not-determined | Genome-fragment | NA | | NA | 0 |
| S7_extracted_135572 | 1132 | No | NA | 2 | 0 | 0 | Not-determined | Genome-fragment | NA | | NA | 0 |
| S7_extracted_135580 | 1115 | No | NA | 1 | 0 | 0 | Not-determined | Genome-fragment | NA | | NA | 0 |
| S14_vs_k141_31036\|\|full | 2120 | No | NA | 4 | 3 | 0 | Low-quality | Genome-fragment | | 4.25 | AAI-based (medium-confidence) | 0 |
| S7_extracted_135778 | 14133 | No | NA | 19 | 6 | 1 | Complete | High-quality | | 100 | DTR (high-confidence) | 0 |
| S1_vs_k141_88745\|\|full | 2120 | No | NA | 4 | 1 | 0 | Low-quality | Genome-fragment | | 5.04 | HMM-based (lower-bound) | 0 |
| S7_extracted_135980 | 2692 | No | NA | 3 | 0 | 1 | Not-determined | Genome-fragment | NA | | NA | 0 |
| S7_vs_13059\|\|full | 2120 | No | NA | 4 | 2 | 1 | Low-quality | Genome-fragment | | 3.69 | HMM-based (lower-bound) | 0 |
| S7_extracted_136152 | 3889 | No | NA | 3 | 0 | 1 | Not-determined | Genome-fragment | NA | | NA | 0 |
| S7_extracted_136199 | 3115 | No | NA | 3 | 0 | 1 | Not-determined | Genome-fragment | NA | | NA | 0 |
| S2_vs_k141_12419\|\|full | 2120 | No | NA | 5 | 0 | 0 | Low-quality | Genome-fragment | | 3.55 | AAI-based (high-confidence) | 0 |
| S7_extracted_136348 | 1136 | No | NA | 3 | 0 | 0 | Not-determined | Genome-fragment | NA | | NA | 0 |
| S7_extracted_136362 | 3110 | No | NA | 2 | 0 | 1 | Not-determined | Genome-fragment | NA | | NA | 0 |
| S7_extracted_136427 | 1097 | No | NA | 2 | 0 | 1 | Not-determined | Genome-fragment | NA | | NA | 0 |
| S7_extracted_136544 | 1213 | No | NA | 2 | 0 | 1 | Not-determined | Genome-fragment | NA | | NA | 0 |
| S7_extracted_136632 | 2268 | No | NA | 3 | 0 | 0 | Not-determined | Genome-fragment | NA | | NA | 0 |
| S7_extracted_136837 | 1183 | No | NA | 2 | 0 | 0 | Not-determined | Genome-fragment | NA | | NA | 0 |
| S7_extracted_136858 | 1295 | No | NA | 1 | 0 | 0 | Not-determined | Genome-fragment | NA | | NA | 0 |
| S7_extracted_137206 | 3419 | No | NA | 3 | 0 | 2 | Not-determined | Genome-fragment | NA | | NA | 0 |
| S8_vs_k141_129979\|\|full | 2120 | No | NA | 4 | 0 | 0 | Low-quality | Genome-fragment | | 2.89 | AAI-based (medium-confidence) | 0 |
| S5_extracted_k141_135558 | 2121 | No | NA | 2 | 2 | 0 | Low-quality | Genome-fragment | | 1.82 | HMM-based (lower-bound) | 0 |
| S7_extracted_137599 | 2187 | No | NA | 1 | 0 | 1 | Not-determined | Genome-fragment | NA | | NA | 0 |
| S7_vs_96214\|\|full | 2121 | No | NA | 2 | 2 | 0 | Low-quality | Genome-fragment | | 5.32 | AAI-based (high-confidence) | 0 |
| S7_extracted_137726 | 1097 | No | NA | 2 | 0 | 0 | Not-determined | Genome-fragment | NA | | NA | 0 |
| S7_extracted_137744 | 1294 | No | NA | 1 | 0 | 1 | Not-determined | Genome-fragment | NA | | NA | 0 |
| S7_extracted_137797 | 1170 | No | NA | 2 | 0 | 0 | Not-determined | Genome-fragment | NA | | NA | 0 |
| S10_vs_17292\|\|full | 2121 | No | NA | 3 | 0 | 0 | Low-quality | Genome-fragment | | 1.61 | AAI-based (high-confidence) | 0 |
| S12_vs_k141_33431\|\|full | 2121 | No | NA | 3 | 0 | 1 | Low-quality | Genome-fragment | | 1.88 | AAI-based (medium-confidence) | 0 |
| S7_extracted_137977 | 1113 | No | NA | 2 | 0 | 1 | Not-determined | Genome-fragment | NA | | NA | 0 |
| S7_extracted_138019 | 10319 | No | NA | 22 | 2 | 0 | Medium-quality | Genome-fragment | | 76.65 | AAI-based (medium-confidence) | 0 |
| S7_extracted_138045 | 3411 | No | NA | 4 | 0 | 2 | Not-determined | Genome-fragment | NA | | NA | 0 |
| S7_extracted_138069 | 59463 | No | NA | 83 | 17 | 0 | Complete | High-quality | | 100 | DTR (high-confidence) | 0 |
| S7_extracted_138106 | 1408 | No | NA | 1 | 0 | 1 | Not-determined | Genome-fragment | NA | | NA | 0 |
| S7_extracted_138114 | 1013 | No | NA | 2 | 0 | 1 | Not-determined | Genome-fragment | NA | | NA | 0 |
| S7_extracted_138143 | 1276 | No | NA | 1 | 0 | 0 | Not-determined | Genome-fragment | NA | | NA | 0 |
| S7_extracted_138149 | 1131 | No | NA | 1 | 0 | 1 | Not-determined | Genome-fragment | NA | | NA | 0 |
| S7_extracted_138201 | 6278 | No | NA | 12 | 0 | 2 | Not-determined | Genome-fragment | NA | | NA | 0 |
| S17_vs_k141_100573\|\|full | 2122 | No | NA | 2 | 0 | 1 | Low-quality | Genome-fragment | | 2.31 | AAI-based (medium-confidence) | 0 |
| S7_extracted_138225 | 40099 | No | NA | 46 | 19 | 0 | Complete | High-quality | | 100 | DTR (high-confidence) | 0 |
| S7_extracted_138237 | 5056 | No | NA | 7 | 0 | 0 | Not-determined | Genome-fragment | NA | | NA | 0 |
| S7_extracted_138246 | 1902 | No | NA | 2 | 0 | 0 | Not-determined | Genome-fragment | NA | | NA | 0 |
| S7_vs_557\|\|full | 37167 | No | NA | 55 | 10 | 1 | High-quality | High-quality | | 93.88 | AAI-based (medium-confidence) | 0 |
| S7_vs_2599\|\|full | 46176 | No | NA | 53 | 6 | 2 | High-quality | High-quality | | 100 | AAI-based (high-confidence) | 0 |
| S7_vs_2692\|\|full | 5934 | No | NA | 10 | 0 | 2 | Not-determined | Genome-fragment | NA | | NA | 0 |
| S7_vs_4956\|\|full | 44351 | No | NA | 66 | 17 | 1 | High-quality | High-quality | | 98.81 | AAI-based (high-confidence) | 0 |
| S7_vs_5236\|\|full | 32721 | No | NA | 49 | 17 | 1 | High-quality | High-quality | | 100 | AAI-based (high-confidence) | 0 |
| S13_vs_45909\|\|full | 2122 | No | NA | 4 | 0 | 1 | Low-quality | Genome-fragment | | 0.98 | AAI-based (medium-confidence) | 0 |
| S7_vs_27530\|\|full | 41445 | No | NA | 66 | 15 | 2 | High-quality | High-quality | | 98.98 | AAI-based (medium-confidence) | 0 |
| S7_vs_36656\|\|full | 41765 | No | NA | 68 | 32 | 2 | Medium-quality | Genome-fragment | | 85.59 | AAI-based (medium-confidence) | 0 |
| S7_vs_42563\|\|full | 37834 | No | NA | 41 | 4 | 1 | High-quality | High-quality | | 98.7 | AAI-based (high-confidence) | 0 |
| S7_vs_54322\|\|full | 56805 | No | NA | 81 | 13 | 2 | High-quality | High-quality | | 100 | AAI-based (high-confidence) | 0 |
| S7_vs_60775\|\|full | 36609 | No | NA | 51 | 23 | 0 | High-quality | High-quality | | 96.77 | AAI-based (high-confidence) | 0 |
| S7_vs_65719\|\|full | 40735 | No | NA | 78 | 17 | 1 | Medium-quality | Genome-fragment | | 76.86 | AAI-based (high-confidence) | 0 |
| S7_vs_75613\|\|full | 7686 | No | NA | 10 | 0 | 0 | Not-determined | Genome-fragment | NA | | NA | 0 |
| S13_vs_66243\|\|full | 2122 | No | NA | 4 | 1 | 0 | Low-quality | Genome-fragment | | 3.62 | AAI-based (high-confidence) | 0 |
| S7_vs_94738\|\|full | 39859 | No | NA | 77 | 20 | 0 | High-quality | High-quality | | 97.69 | AAI-based (high-confidence) | 0 |
| S7_vs_95276\|\|full | 41741 | No | NA | 54 | 13 | 2 | High-quality | High-quality | | 100 | AAI-based (high-confidence) | 0 |
| S7_vs_103530\|\|full | 53326 | No | NA | 88 | 18 | 2 | High-quality | High-quality | | 94.18 | AAI-based (high-confidence) | 0 |
| S7_vs_129271\|\|full | 35566 | Yes | 32266 | 54 | 15 | 2 | Medium-quality | Genome-fragment | | 54.18 | HMM-based (lower-bound) | 9.28 |
| S7_vs_134391\|\|full | 36975 | No | NA | 47 | 21 | 0 | Medium-quality | Genome-fragment | | 80.3 | AAI-based (high-confidence) | 0 |
| S7_vs_134784\|\|full | 35044 | No | NA | 42 | 16 | 0 | High-quality | High-quality | | 100 | AAI-based (high-confidence) | 0 |
| S7_vs_135322\|\|full | 55799 | No | NA | 89 | 23 | 0 | High-quality | High-quality | | 100 | AAI-based (high-confidence) | 0 |
| S7_vs_135679\|\|full | 41488 | No | NA | 59 | 15 | 1 | High-quality | High-quality | | 100 | AAI-based (high-confidence) | 0 |
| S7_vs_135828\|\|full | 6008 | No | NA | 6 | 3 | 0 | High-quality | High-quality | | 98.9 | AAI-based (high-confidence) | 0 |
| S7_vs_136164\|\|full | 34652 | No | NA | 49 | 12 | 1 | High-quality | High-quality | | 90.63 | AAI-based (medium-confidence) | 0 |
| S7_vs_137611\|\|full | 142455 | No | NA | 177 | 31 | 6 | Medium-quality | Genome-fragment | | 81.41 | AAI-based (high-confidence) | 0 |
| S7_vs_137915\|\|full | 101806 | No | NA | 120 | 19 | 4 | High-quality | High-quality | | 99.08 | AAI-based (high-confidence) | 0 |
| S7_vs_137998\|\|full | 87575 | No | NA | 143 | 27 | 3 | High-quality | High-quality | | 100 | AAI-based (high-confidence) | 0 |
| S7_vs_138096\|\|full | 34972 | No | NA | 44 | 7 | 0 | High-quality | High-quality | | 90.47 | AAI-based (medium-confidence) | 0 |
| S7_vs_138109\|\|full | 42405 | No | NA | 55 | 22 | 0 | High-quality | High-quality | | 100 | AAI-based (high-confidence) | 0 |
| S7_vs_138116\|\|full | 10338 | No | NA | 16 | 3 | 1 | Medium-quality | Genome-fragment | | 80.72 | AAI-based (high-confidence) | 0 |
| S7_vs_138140\|\|full | 39653 | No | NA | 50 | 10 | 1 | High-quality | High-quality | | 100 | AAI-based (high-confidence) | 0 |
| S7_vs_138173\|\|full | 48528 | Yes | 45350 | 65 | 10 | 2 | High-quality | High-quality | | 91.19 | AAI-based (high-confidence) | 6.55 |
| S7_vs_138193\|\|full | 4972 | No | NA | 9 | 0 | 1 | Not-determined | Genome-fragment | NA | | NA | 0 |
| S7_vs_138260\|\|full | 1354 | No | NA | 3 | 0 | 0 | Not-determined | Genome-fragment | NA | | NA | 0 |
| S13_vs_75349\|\|full | 2122 | No | NA | 4 | 0 | 0 | Low-quality | Genome-fragment | | 3.97 | AAI-based (medium-confidence) | 0 |
| S7_vs_119\|\|full | 2460 | No | NA | 4 | 0 | 1 | Not-determined | Genome-fragment | NA | | NA | 0 |
| S7_vs_149\|\|full | 3523 | No | NA | 3 | 0 | 3 | Not-determined | Genome-fragment | NA | | NA | 0 |
| S7_vs_151\|\|full | 6439 | No | NA | 6 | 0 | 3 | Not-determined | Genome-fragment | NA | | NA | 0 |
| S7_vs_152\|\|full | 1208 | No | NA | 2 | 0 | 0 | Not-determined | Genome-fragment | NA | | NA | 0 |
| S7_vs_277\|\|full | 1013 | No | NA | 2 | 0 | 1 | Not-determined | Genome-fragment | NA | | NA | 0 |
| S7_vs_282\|\|full | 1192 | No | NA | 4 | 0 | 0 | Not-determined | Genome-fragment | NA | | NA | 0 |
| S3_vs_k141_18341\|\|full | 2122 | No | NA | 4 | 0 | 0 | Low-quality | Genome-fragment | | 4.3 | AAI-based (high-confidence) | 0 |
| S7_vs_48876\|\|full | 2122 | No | NA | 7 | 2 | 0 | Low-quality | Genome-fragment | | 6.23 | AAI-based (high-confidence) | 0 |
| S7_vs_366\|\|full | 1731 | No | NA | 3 | 0 | 2 | Not-determined | Genome-fragment | NA | | NA | 0 |
| S16_vs_26758\|\|full | 2122 | No | NA | 7 | 2 | 0 | Low-quality | Genome-fragment | | 4.6 | AAI-based (high-confidence) | 0 |
| S10_extracted_22083 | 2122 | No | NA | 4 | 0 | 3 | Low-quality | Genome-fragment | | 1.65 | AAI-based (medium-confidence) | 0 |
| S10_vs_30560\|\|full | 2122 | No | NA | 3 | 0 | 1 | Low-quality | Genome-fragment | | 1.73 | AAI-based (medium-confidence) | 0 |
| S8_extracted_k141_48051 | 2122 | No | NA | 2 | 0 | 0 | Low-quality | Genome-fragment | | 0.39 | AAI-based (medium-confidence) | 0 |
| S9_vs_k141_51223\|\|full | 2123 | No | NA | 2 | 0 | 0 | Low-quality | Genome-fragment | | 5.39 | AAI-based (high-confidence) | 0 |
| S13_extracted_119923 | 2123 | No | NA | 3 | 0 | 2 | Low-quality | Genome-fragment | | 4.24 | AAI-based (medium-confidence) | 0 |
| S16_vs_56700\|\|full | 2123 | No | NA | 5 | 1 | 0 | Low-quality | Genome-fragment | | 2.15 | AAI-based (high-confidence) | 0 |
| S7_vs_944\|\|full | 23257 | No | NA | 27 | 13 | 0 | Medium-quality | Genome-fragment | | 57.89 | AAI-based (high-confidence) | 0 |
| S10_vs_83731\|\|full | 2123 | No | NA | 2 | 0 | 1 | Low-quality | Genome-fragment | | 7.15 | AAI-based (high-confidence) | 0 |
| S12_vs_k141_2649\|\|full | 2123 | No | NA | 4 | 1 | 0 | Low-quality | Genome-fragment | | 4.89 | AAI-based (medium-confidence) | 0 |
| S17_extracted_k141_102519 | 2124 | No | NA | 3 | 0 | 0 | Low-quality | Genome-fragment | | 1.44 | AAI-based (medium-confidence) | 0 |
| S14_vs_k141_63952\|\|full | 2124 | No | NA | 5 | 0 | 1 | Low-quality | Genome-fragment | | 4.96 | AAI-based (medium-confidence) | 0 |
| S5_vs_k141_130256\|\|full | 2124 | No | NA | 2 | 0 | 1 | Low-quality | Genome-fragment | | 1.25 | AAI-based (medium-confidence) | 0 |
| S7_extracted_22423 | 2124 | No | NA | 6 | 0 | 2 | Low-quality | Genome-fragment | | 4.1 | AAI-based (medium-confidence) | 0 |
| S16_vs_146544\|\|full | 2124 | No | NA | 3 | 1 | 0 | Low-quality | Genome-fragment | | 3.35 | AAI-based (high-confidence) | 0 |
| S8_extracted_k141_116220 | 2124 | No | NA | 6 | 1 | 0 | Low-quality | Genome-fragment | | 6.75 | AAI-based (high-confidence) | 0 |
| S7_vs_1727\|\|full | 1728 | No | NA | 3 | 0 | 3 | Not-determined | Genome-fragment | NA | | NA | 0 |
| S11_vs_k141_67259\|\|full | 2125 | No | NA | 4 | 0 | 0 | Low-quality | Genome-fragment | | 4.14 | AAI-based (medium-confidence) | 0 |
| S5_extracted_k141_26258 | 2125 | No | NA | 5 | 4 | 0 | Low-quality | Genome-fragment | | 6.16 | AAI-based (high-confidence) | 0 |
| S7_vs_1907\|\|full | 7939 | No | NA | 10 | 0 | 6 | Not-determined | Genome-fragment | NA | | NA | 0 |
| S7_vs_103838\|\|full | 2125 | No | NA | 4 | 1 | 0 | Low-quality | Genome-fragment | | 4.28 | AAI-based (medium-confidence) | 0 |
| S8_extracted_k141_135243 | 2125 | No | NA | 5 | 0 | 2 | Low-quality | Genome-fragment | | 4.26 | AAI-based (high-confidence) | 0 |
| S2_extracted_k141_13197 | 2126 | No | NA | 3 | 2 | 0 | Low-quality | Genome-fragment | | 2.07 | AAI-based (high-confidence) | 0 |
| S7_vs_2406\|\|full | 7399 | No | NA | 7 | 0 | 6 | Not-determined | Genome-fragment | NA | | NA | 0 |
| S7_vs_2456\|\|full | 1044 | No | NA | 2 | 0 | 0 | Not-determined | Genome-fragment | NA | | NA | 0 |
| S7_vs_2490\|\|full | 1399 | No | NA | 1 | 0 | 1 | Not-determined | Genome-fragment | NA | | NA | 0 |
| S7_vs_2589\|\|full | 2780 | No | NA | 3 | 0 | 3 | Not-determined | Genome-fragment | NA | | NA | 0 |
| S7_vs_2632\|\|full | 1470 | No | NA | 3 | 0 | 1 | Not-determined | Genome-fragment | NA | | NA | 0 |
| S8_extracted_k141_16282 | 2126 | No | NA | 5 | 1 | 0 | Low-quality | Genome-fragment | | 5.42 | AAI-based (high-confidence) | 0 |
| S8_vs_k141_41036\|\|full | 2126 | No | NA | 4 | 0 | 0 | Low-quality | Genome-fragment | | 3.51 | AAI-based (high-confidence) | 0 |
| S9_vs_k141_5350\|\|full | 2127 | No | NA | 6 | 3 | 0 | Low-quality | Genome-fragment | | 5.63 | AAI-based (medium-confidence) | 0 |
| S14_vs_k141_92323\|\|full | 2127 | No | NA | 3 | 1 | 0 | Low-quality | Genome-fragment | | 5 | AAI-based (high-confidence) | 0 |
| S3_extracted_k141_16878 | 2127 | No | NA | 4 | 1 | 0 | Low-quality | Genome-fragment | | 7.29 | HMM-based (lower-bound) | 0 |
| S3_extracted_k141_81809 | 2127 | No | NA | 4 | 1 | 0 | Low-quality | Genome-fragment | | 7.29 | HMM-based (lower-bound) | 0 |
| S7_vs_78828\|\|full | 2127 | No | NA | 5 | 0 | 0 | Low-quality | Genome-fragment | | 4.06 | AAI-based (high-confidence) | 0 |
| S7_vs_2830\|\|full | 1461 | No | NA | 3 | 0 | 1 | Not-determined | Genome-fragment | NA | | NA | 0 |
| S7_vs_2928\|\|full | 1844 | No | NA | 2 | 0 | 1 | Not-determined | Genome-fragment | NA | | NA | 0 |
| S12_extracted_k141_57887 | 2127 | No | NA | 2 | 0 | 0 | Low-quality | Genome-fragment | | 3.69 | AAI-based (medium-confidence) | 0 |
| S12_vs_k141_128099\|\|full | 2127 | No | NA | 2 | 0 | 2 | Low-quality | Genome-fragment | | 1.28 | AAI-based (medium-confidence) | 0 |
| S12_vs_k141_97489\|\|full | 2127 | No | NA | 3 | 1 | 0 | Low-quality | Genome-fragment | | 5.86 | AAI-based (high-confidence) | 0 |
| S2_extracted_k141_15227 | 2127 | No | NA | 2 | 0 | 1 | Low-quality | Genome-fragment | | 1.59 | AAI-based (medium-confidence) | 0 |
| S6_vs_k141_78\|\|full | 2127 | No | NA | 3 | 0 | 2 | Low-quality | Genome-fragment | | 2.26 | AAI-based (medium-confidence) | 0 |
| S7_vs_3209\|\|full | 1682 | No | NA | 4 | 0 | 2 | Not-determined | Genome-fragment | NA | | NA | 0 |
| S8_extracted_k141_87935 | 2127 | No | NA | 1 | 0 | 1 | Low-quality | Genome-fragment | | 6.79 | AAI-based (medium-confidence) | 0 |
| S7_vs_3287\|\|full | 3041 | No | NA | 4 | 0 | 4 | Not-determined | Genome-fragment | NA | | NA | 0 |
| S7_vs_3391\|\|full | 5284 | No | NA | 2 | 0 | 2 | Not-determined | Genome-fragment | NA | | NA | 0 |
| S7_vs_3425\|\|full | 1269 | No | NA | 2 | 0 | 0 | Not-determined | Genome-fragment | NA | | NA | 0 |
| S8_vs_k141_41303\|\|full | 2127 | No | NA | 3 | 0 | 0 | Low-quality | Genome-fragment | | 4.63 | AAI-based (high-confidence) | 0 |
| S7_vs_3685\|\|full | 2913 | No | NA | 2 | 0 | 2 | Not-determined | Genome-fragment | NA | | NA | 0 |
| S11_vs_k141_23162\|\|full | 2128 | No | NA | 4 | 1 | 0 | Low-quality | Genome-fragment | | 5.91 | AAI-based (medium-confidence) | 0 |
| S7_vs_3729\|\|full | 1398 | No | NA | 1 | 0 | 0 | Not-determined | Genome-fragment | NA | | NA | 0 |
| S5_vs_k141_10485\|\|full | 2128 | No | NA | 4 | 0 | 0 | Low-quality | Genome-fragment | | 5.13 | AAI-based (medium-confidence) | 0 |
| S7_vs_3797\|\|full | 4509 | No | NA | 6 | 0 | 2 | Not-determined | Genome-fragment | NA | | NA | 0 |
| S7_vs_97785\|\|full | 2128 | No | NA | 2 | 0 | 2 | Low-quality | Genome-fragment | | 0.93 | AAI-based (medium-confidence) | 0 |
| S7_vs_3952\|\|full | 5985 | No | NA | 9 | 0 | 1 | Not-determined | Genome-fragment | NA | | NA | 0 |
| S10_vs_103550\|\|full | 2128 | No | NA | 5 | 0 | 0 | Low-quality | Genome-fragment | | 3.51 | AAI-based (high-confidence) | 0 |
| S7_vs_4114\|\|full | 36060 | No | NA | 68 | 11 | 1 | Medium-quality | Genome-fragment | | 87.94 | AAI-based (high-confidence) | 0 |
| S7_vs_4129\|\|full | 4192 | No | NA | 8 | 0 | 0 | Not-determined | Genome-fragment | NA | | NA | 0 |
| S7_vs_4158\|\|full | 3241 | No | NA | 3 | 0 | 1 | Not-determined | Genome-fragment | NA | | NA | 0 |
| S7_vs_4164\|\|full | 4276 | No | NA | 5 | 0 | 4 | Not-determined | Genome-fragment | NA | | NA | 0 |
| S7_vs_4165\|\|full | 2644 | No | NA | 4 | 0 | 1 | Not-determined | Genome-fragment | NA | | NA | 0 |
| S7_vs_4189\|\|full | 4577 | No | NA | 3 | 0 | 2 | Not-determined | Genome-fragment | NA | | NA | 0 |
| S7_vs_4200\|\|full | 1194 | No | NA | 3 | 0 | 0 | Not-determined | Genome-fragment | NA | | NA | 0 |
| S17_vs_k141_163922\|\|full | 2129 | No | NA | 3 | 0 | 0 | Low-quality | Genome-fragment | | 5.32 | AAI-based (high-confidence) | 0 |
| S17_vs_k141_167283\|\|full | 2129 | No | NA | 6 | 1 | 0 | Low-quality | Genome-fragment | | 2.39 | AAI-based (high-confidence) | 0 |
| S1_extracted_k141_47536 | 2129 | No | NA | 3 | 0 | 0 | Low-quality | Genome-fragment | | 1.21 | AAI-based (high-confidence) | 0 |
| S7_vs_4412\|\|full | 1251 | No | NA | 2 | 0 | 2 | Not-determined | Genome-fragment | NA | | NA | 0 |
| S3_vs_k141_122639\|\|full | 2129 | No | NA | 4 | 0 | 0 | Low-quality | Genome-fragment | | 5.96 | AAI-based (high-confidence) | 0 |
| S17_vs_k141_96433\|\|full | 2130 | No | NA | 3 | 0 | 1 | Low-quality | Genome-fragment | | 1.23 | AAI-based (medium-confidence) | 0 |
| S14_vs_k141_6985\|\|lt2gene | 2130 | No | NA | 1 | 0 | 0 | Low-quality | Genome-fragment | | 5.42 | AAI-based (medium-confidence) | 0 |
| S7_vs_4556\|\|full | 3163 | No | NA | 4 | 0 | 3 | Not-determined | Genome-fragment | NA | | NA | 0 |
| S7_vs_4564\|\|full | 2940 | No | NA | 3 | 0 | 3 | Not-determined | Genome-fragment | NA | | NA | 0 |
| S7_vs_37190\|\|full | 2130 | No | NA | 5 | 0 | 1 | Low-quality | Genome-fragment | | 4.63 | AAI-based (high-confidence) | 0 |
| S7_vs_4709\|\|full | 1562 | No | NA | 3 | 0 | 0 | Not-determined | Genome-fragment | NA | | NA | 0 |
| S7_vs_56616\|\|full | 2130 | No | NA | 2 | 0 | 2 | Low-quality | Genome-fragment | | 3.94 | AAI-based (medium-confidence) | 0 |
| S7_vs_4745\|\|full | 2220 | No | NA | 2 | 0 | 0 | Not-determined | Genome-fragment | NA | | NA | 0 |
| S7_vs_70498\|\|full | 2130 | No | NA | 4 | 0 | 1 | Low-quality | Genome-fragment | | 2.36 | AAI-based (medium-confidence) | 0 |
| S16_vs_43159\|\|full | 2130 | No | NA | 5 | 1 | 0 | Low-quality | Genome-fragment | | 1.91 | HMM-based (lower-bound) | 0 |
| S7_vs_4941\|\|full | 2695 | No | NA | 4 | 0 | 0 | Not-determined | Genome-fragment | NA | | NA | 0 |
| S16_vs_81219\|\|full | 2130 | No | NA | 6 | 2 | 0 | Low-quality | Genome-fragment | | 5.1 | AAI-based (medium-confidence) | 0 |
| S7_vs_4982\|\|full | 4288 | No | NA | 9 | 0 | 9 | Not-determined | Genome-fragment | NA | | NA | 0 |
| S4_extracted_11584 | 2130 | No | NA | 4 | 0 | 0 | Low-quality | Genome-fragment | | 3.21 | AAI-based (high-confidence) | 0 |
| S14_vs_k141_62268\|\|full | 2131 | No | NA | 5 | 1 | 0 | Low-quality | Genome-fragment | | 3.24 | HMM-based (lower-bound) | 0 |
| S5_vs_k141_39809\|\|lt2gene | 2131 | No | NA | 2 | 1 | 0 | Low-quality | Genome-fragment | | 5.33 | AAI-based (medium-confidence) | 0 |
| S17_vs_k141_158961\|\|full | 2132 | No | NA | 6 | 0 | 0 | Low-quality | Genome-fragment | | 3.52 | AAI-based (high-confidence) | 0 |
| S5_vs_k141_101018\|\|lt2gene | 2132 | No | NA | 2 | 2 | 0 | Low-quality | Genome-fragment | | 4.88 | AAI-based (high-confidence) | 0 |
| S7_vs_75368\|\|full | 2132 | No | NA | 3 | 0 | 0 | Low-quality | Genome-fragment | | 1.51 | AAI-based (high-confidence) | 0 |
| S7_vs_5283\|\|full | 4401 | No | NA | 5 | 0 | 4 | Not-determined | Genome-fragment | NA | | NA | 0 |
| S4_vs_VIRSorter_9973-cat_2 | 2132 | No | NA | 5 | 0 | 0 | Low-quality | Genome-fragment | | 4.47 | AAI-based (high-confidence) | 0 |
| S11_extracted_k141_6326 | 2133 | No | NA | 2 | 2 | 0 | Low-quality | Genome-fragment | | 3.68 | AAI-based (medium-confidence) | 0 |
| S13_vs_80984\|\|full | 2133 | No | NA | 3 | 0 | 1 | Low-quality | Genome-fragment | | 1.08 | AAI-based (high-confidence) | 0 |
| S3_extracted_k141_3348 | 2133 | No | NA | 5 | 1 | 0 | Low-quality | Genome-fragment | | 5.27 | AAI-based (high-confidence) | 0 |
| S3_vs_k141_92367\|\|full | 2133 | No | NA | 5 | 0 | 0 | Low-quality | Genome-fragment | | 3.57 | AAI-based (high-confidence) | 0 |
| S7_vs_43023\|\|full | 2133 | No | NA | 4 | 2 | 0 | Low-quality | Genome-fragment | | 4.81 | AAI-based (high-confidence) | 0 |
| S12_vs_k141_142295\|\|full | 2133 | No | NA | 3 | 1 | 0 | Low-quality | Genome-fragment | | 0.74 | HMM-based (lower-bound) | 0 |
| S6_vs_k141_1586\|\|full | 2133 | No | NA | 2 | 0 | 2 | Low-quality | Genome-fragment | | 1.26 | AAI-based (medium-confidence) | 0 |
| S7_vs_5708\|\|full | 1871 | No | NA | 5 | 0 | 0 | Not-determined | Genome-fragment | NA | | NA | 0 |
| S11_vs_k141_100964\|\|full | 2134 | No | NA | 3 | 0 | 2 | Low-quality | Genome-fragment | | 1.29 | AAI-based (medium-confidence) | 0 |
| S14_vs_k141_68303\|\|lt2gene | 2134 | No | NA | 2 | 2 | 0 | Low-quality | Genome-fragment | | 5.49 | AAI-based (high-confidence) | 0 |
| S5_vs_k141_76444\|\|lt2gene | 2134 | No | NA | 2 | 0 | 0 | Low-quality | Genome-fragment | | 4.58 | AAI-based (medium-confidence) | 0 |
| S7_vs_5850\|\|full | 1941 | No | NA | 3 | 0 | 1 | Not-determined | Genome-fragment | NA | | NA | 0 |
| S16_extracted_94422 | 2134 | No | NA | 4 | 1 | 0 | Low-quality | Genome-fragment | | 3.43 | HMM-based (lower-bound) | 0 |
| S7_vs_5930\|\|full | 1806 | No | NA | 3 | 0 | 0 | Not-determined | Genome-fragment | NA | | NA | 0 |
| S10_vs_56043\|\|full | 2134 | No | NA | 4 | 2 | 0 | Low-quality | Genome-fragment | | 1.21 | AAI-based (medium-confidence) | 0 |
| S7_vs_6298\|\|full | 4798 | No | NA | 5 | 0 | 3 | Not-determined | Genome-fragment | NA | | NA | 0 |
| S7_vs_6334\|\|full | 3241 | No | NA | 5 | 0 | 3 | Not-determined | Genome-fragment | NA | | NA | 0 |
| S7_vs_6428\|\|full | 1067 | No | NA | 2 | 0 | 0 | Not-determined | Genome-fragment | NA | | NA | 0 |
| S12_vs_k141_125663\|\|full | 2134 | No | NA | 4 | 0 | 0 | Low-quality | Genome-fragment | | 5.82 | AAI-based (high-confidence) | 0 |
| S7_vs_6626\|\|full | 61993 | No | NA | 109 | 21 | 1 | High-quality | High-quality | | 100 | AAI-based (high-confidence) | 0 |
| S7_vs_6638\|\|full | 1336 | No | NA | 3 | 0 | 0 | Not-determined | Genome-fragment | NA | | NA | 0 |
| S1_vs_k141_149048\|\|full | 2135 | No | NA | 4 | 0 | 0 | Low-quality | Genome-fragment | | 2.9 | AAI-based (medium-confidence) | 0 |
| S7_vs_131329\|\|lt2gene | 2135 | No | NA | 2 | 1 | 0 | Low-quality | Genome-fragment | | 1.41 | AAI-based (high-confidence) | 0 |
| S16_vs_130379\|\|full | 2135 | No | NA | 3 | 0 | 0 | Low-quality | Genome-fragment | | 1.62 | AAI-based (medium-confidence) | 0 |
| S4_extracted_4120 | 2135 | No | NA | 4 | 1 | 0 | Low-quality | Genome-fragment | | 5.27 | AAI-based (high-confidence) | 0 |
| S8_extracted_k141_108521 | 2135 | No | NA | 2 | 0 | 1 | Low-quality | Genome-fragment | | 2.09 | AAI-based (medium-confidence) | 0 |
| S8_vs_k141_95801\|\|full | 2135 | No | NA | 2 | 0 | 0 | Low-quality | Genome-fragment | | 1.3 | AAI-based (high-confidence) | 0 |

| | | | | | | | | | | | |
|---|---|---|---|---|---|---|---|---|---|---|---|
| S11_extracted_k141_98456 | 2136 | No | NA | 2 | 1 | 0 | Low-quality | Genome-fragment | 4.66 | AAI-based (high-confidence) | 0 |
| S7_vs_7038\|\|full | 2210 | No | NA | 2 | 0 | 1 | Not-determined | Genome-fragment | NA | NA | 0 |
| S11_extracted_k141_100933 | 2136 | No | NA | 5 | 2 | 0 | Low-quality | Genome-fragment | 1.72 | AAI-based (high-confidence) | 0 |
| S7_vs_7135\|\|full | 2095 | No | NA | 3 | 0 | 2 | Not-determined | Genome-fragment | NA | NA | 0 |
| S7_vs_7202\|\|full | 1513 | No | NA | 3 | 0 | 1 | Not-determined | Genome-fragment | NA | NA | 0 |
| S1_vs_k141_136772\|\|full | 2136 | No | NA | 5 | 1 | 0 | Low-quality | Genome-fragment | 4.67 | AAI-based (high-confidence) | 0 |
| S3_vs_k141_35670\|\|full | 2136 | No | NA | 5 | 1 | 0 | Low-quality | Genome-fragment | 4.99 | AAI-based (high-confidence) | 0 |
| S7_vs_7331\|\|full | 1868 | No | NA | 3 | 0 | 3 | Not-determined | Genome-fragment | NA | NA | 0 |
| S7_vs_7378\|\|full | 1844 | No | NA | 4 | 0 | 0 | Not-determined | Genome-fragment | NA | NA | 0 |
| S7_vs_7761\|\|full | 20219 | No | NA | 28 | 0 | 4 | Not-determined | Genome-fragment | NA | NA | 0 |
| S7_vs_7944\|\|full | 3335 | No | NA | 4 | 0 | 2 | Not-determined | Genome-fragment | NA | NA | 0 |
| S5_extracted_k141_133075 | 2136 | Yes | 730 | 4 | 1 | 1 | Low-quality | Genome-fragment | 2.05 | AAI-based (high-confidence) | 65.82 |
| S12_extracted_k141_151475 | 2136 | No | NA | 2 | 1 | 0 | Low-quality | Genome-fragment | 3.77 | AAI-based (high-confidence) | 0 |
| S7_vs_8084\|\|full | 1284 | No | NA | 2 | 0 | 1 | Not-determined | Genome-fragment | NA | NA | 0 |
| S9_vs_k141_69592\|\|full | 2137 | No | NA | 4 | 2 | 0 | Low-quality | Genome-fragment | 4.69 | AAI-based (medium-confidence) | 0 |
| S7_vs_8121\|\|full | 1043 | No | NA | 2 | 0 | 1 | Not-determined | Genome-fragment | NA | NA | 0 |
| S7_vs_8130\|\|full | 1521 | No | NA | 3 | 0 | 0 | Not-determined | Genome-fragment | NA | NA | 0 |
| S14_extracted_k141_30300 | 2137 | No | NA | 6 | 6 | 0 | Low-quality | Genome-fragment | 7.05 | AAI-based (high-confidence) | 0 |
| S5_vs_k141_138876\|\|full | 2137 | No | NA | 2 | 2 | 0 | Low-quality | Genome-fragment | 5.93 | AAI-based (high-confidence) | 0 |
| S5_vs_k141_19741\|\|full | 2138 | No | NA | 4 | 2 | 0 | Low-quality | Genome-fragment | 5.28 | AAI-based (high-confidence) | 0 |
| S7_vs_8292\|\|full | 1660 | No | NA | 3 | 0 | 2 | Not-determined | Genome-fragment | NA | NA | 0 |
| S5_vs_k141_75624\|\|full | 2138 | No | NA | 4 | 3 | 0 | Low-quality | Genome-fragment | 5.46 | AAI-based (medium-confidence) | 0 |
| S7_vs_8413\|\|full | 1756 | No | NA | 3 | 0 | 1 | Not-determined | Genome-fragment | NA | NA | 0 |
| S7_vs_8453\|\|full | 1455 | No | NA | 3 | 0 | 0 | Not-determined | Genome-fragment | NA | NA | 0 |
| S10_extracted_6273 | 2138 | No | NA | 4 | 1 | 0 | Low-quality | Genome-fragment | 4.72 | AAI-based (high-confidence) | 0 |
| S7_vs_8507\|\|full | 3475 | No | NA | 9 | 0 | 0 | Not-determined | Genome-fragment | NA | NA | 0 |
| S8_vs_k141_1238\|\|full | 2138 | No | NA | 4 | 0 | 0 | Low-quality | Genome-fragment | 6.41 | AAI-based (medium-confidence) | 0 |
| S14_vs_k141_108359\|\|lt2gene | 2139 | No | NA | 1 | 1 | 0 | Low-quality | Genome-fragment | 3.97 | AAI-based (medium-confidence) | 0 |
| S7_vs_8632\|\|full | 2800 | No | NA | 4 | 0 | 0 | Not-determined | Genome-fragment | NA | NA | 0 |
| S7_vs_8697\|\|full | 1203 | No | NA | 4 | 0 | 0 | Not-determined | Genome-fragment | NA | NA | 0 |
| S5_vs_k141_105444\|\|full | 2139 | No | NA | 3 | 2 | 0 | Low-quality | Genome-fragment | 3.33 | HMM-based (lower-bound) | 0 |
| S7_vs_8892\|\|full | 3502 | No | NA | 3 | 0 | 1 | Not-determined | Genome-fragment | NA | NA | 0 |
| S7_vs_59006\|\|full | 2139 | No | NA | 4 | 1 | 0 | Low-quality | Genome-fragment | 4.31 | AAI-based (medium-confidence) | 0 |
| S7_vs_9020\|\|full | 2719 | No | NA | 2 | 0 | 0 | Not-determined | Genome-fragment | NA | NA | 0 |
| S11_vs_k141_55425\|\|full | 2140 | No | NA | 2 | 0 | 1 | Low-quality | Genome-fragment | 7.21 | AAI-based (high-confidence) | 0 |
| S1_extracted_k141_100960 | 2140 | No | NA | 1 | 0 | 1 | Low-quality | Genome-fragment | 1.22 | AAI-based (medium-confidence) | 0 |
| S1_vs_k141_34430\|\|full | 2140 | No | NA | 3 | 0 | 0 | Low-quality | Genome-fragment | 4.04 | AAI-based (high-confidence) | 0 |
| S5_vs_k141_8209\|\|full | 2140 | No | NA | 4 | 0 | 1 | Low-quality | Genome-fragment | 5.03 | AAI-based (medium-confidence) | 0 |
| S16_vs_90382\|\|full | 2140 | No | NA | 3 | 3 | 0 | Low-quality | Genome-fragment | 6.21 | AAI-based (high-confidence) | 0 |
| S10_vs_77726\|\|full | 2140 | No | NA | 3 | 0 | 0 | Low-quality | Genome-fragment | 3.72 | AAI-based (high-confidence) | 0 |
| S12_vs_k141_133222\|\|full | 2140 | No | NA | 4 | 0 | 0 | Low-quality | Genome-fragment | 4.94 | AAI-based (high-confidence) | 0 |
| S7_vs_9364\|\|full | 4510 | No | NA | 3 | 0 | 3 | Not-determined | Genome-fragment | NA | NA | 0 |
| S8_vs_k141_184543\|\|full | 2140 | No | NA | 6 | 2 | 0 | Low-quality | Genome-fragment | 3.45 | AAI-based (high-confidence) | 0 |
| S7_vs_9659\|\|full | 1170 | No | NA | 3 | 0 | 0 | Not-determined | Genome-fragment | NA | NA | 0 |
| S7_vs_9687\|\|full | 4509 | No | NA | 3 | 0 | 1 | Not-determined | Genome-fragment | NA | NA | 0 |
| S17_extracted_k141_16787 | 2141 | No | NA | 3 | 0 | 3 | Low-quality | Genome-fragment | 1.09 | AAI-based (medium-confidence) | 0 |
| S5_vs_k141_77153\|\|lt2gene | 2141 | No | NA | 2 | 2 | 0 | Low-quality | Genome-fragment | 4.89 | AAI-based (medium-confidence) | 0 |
| S7_vs_10000\|\|full | 13588 | No | NA | 7 | 0 | 5 | Not-determined | Genome-fragment | NA | NA | 0 |
| S10_vs_78666\|\|full | 2141 | No | NA | 2 | 0 | 0 | Low-quality | Genome-fragment | 1.45 | AAI-based (high-confidence) | 0 |
| S8_vs_k141_185063\|\|full | 2141 | No | NA | 3 | 0 | 1 | Low-quality | Genome-fragment | 3.28 | AAI-based (medium-confidence) | 0 |
| S7_vs_10199\|\|full | 3355 | No | NA | 3 | 0 | 2 | Not-determined | Genome-fragment | NA | NA | 0 |
| S7_vs_10294\|\|full | 2249 | No | NA | 3 | 0 | 2 | Not-determined | Genome-fragment | NA | NA | 0 |
| S7_vs_10396\|\|full | 14874 | No | NA | 10 | 0 | 5 | Not-determined | Genome-fragment | NA | NA | 0 |
| S7_vs_10477\|\|full | 4760 | No | NA | 3 | 0 | 2 | Not-determined | Genome-fragment | NA | NA | 0 |
| S7_vs_10480\|\|full | 4101 | No | NA | 5 | 0 | 3 | Not-determined | Genome-fragment | NA | NA | 0 |
| S14_vs_k141_9671\|\|full | 2142 | No | NA | 5 | 0 | 0 | Low-quality | Genome-fragment | 2.29 | AAI-based (medium-confidence) | 0 |
| S5_vs_k141_17081\|\|full | 2142 | No | NA | 7 | 2 | 0 | Low-quality | Genome-fragment | 3.53 | HMM-based (lower-bound) | 0 |
| S7_vs_10657\|\|full | 1727 | No | NA | 2 | 0 | 1 | Not-determined | Genome-fragment | NA | NA | 0 |
| S16_vs_109716\|\|full | 2142 | No | NA | 4 | 0 | 1 | Low-quality | Genome-fragment | 1.13 | AAI-based (high-confidence) | 0 |
| S7_vs_10832\|\|full | 2430 | No | NA | 7 | 0 | 0 | Not-determined | Genome-fragment | NA | NA | 0 |
| S8_vs_k141_201841\|\|full | 2142 | No | NA | 3 | 0 | 0 | Low-quality | Genome-fragment | 5.53 | AAI-based (high-confidence) | 0 |
| S5_vs_k141_42811\|\|full | 2143 | No | NA | 3 | 2 | 0 | Low-quality | Genome-fragment | 5.2 | AAI-based (high-confidence) | 0 |
| S7_vs_10986\|\|full | 2427 | No | NA | 2 | 0 | 0 | Not-determined | Genome-fragment | NA | NA | 0 |
| S7_vs_11014\|\|full | 1260 | No | NA | 1 | 0 | 0 | Not-determined | Genome-fragment | NA | NA | 0 |
| S12_extracted_k141_33221 | 2143 | No | NA | 2 | 1 | 0 | Low-quality | Genome-fragment | 4.94 | AAI-based (medium-confidence) | 0 |
| S7_vs_11081\|\|full | 57891 | No | NA | 62 | 8 | 3 | High-quality | High-quality | 100 | AAI-based (high-confidence) | 0 |
| S7_vs_11138\|\|full | 5152 | No | NA | 3 | 0 | 2 | Not-determined | Genome-fragment | NA | NA | 0 |
| S15_extracted_17606 | 2143 | No | NA | 4 | 0 | 0 | Low-quality | Genome-fragment | 2.47 | AAI-based (high-confidence) | 0 |
| S7_vs_11314\|\|full | 1988 | No | NA | 6 | 0 | 0 | Not-determined | Genome-fragment | NA | NA | 0 |
| S11_vs_k141_18011\|\|full | 2144 | No | NA | 4 | 0 | 1 | Low-quality | Genome-fragment | 3.28 | AAI-based (medium-confidence) | 0 |
| S14_vs_k141_44175\|\|full | 2144 | No | NA | 4 | 0 | 0 | Low-quality | Genome-fragment | 4 | AAI-based (high-confidence) | 0 |
| S7_vs_11593\|\|full | 2453 | No | NA | 4 | 0 | 2 | Not-determined | Genome-fragment | NA | NA | 0 |
| S5_vs_k141_82956\|\|full | 2144 | No | NA | 3 | 1 | 0 | Low-quality | Genome-fragment | 3.95 | HMM-based (lower-bound) | 0 |
| S7_vs_11668\|\|full | 1440 | No | NA | 3 | 0 | 0 | Not-determined | Genome-fragment | NA | NA | 0 |
| S7_vs_2666\|\|full | 2144 | No | NA | 4 | 1 | 0 | Low-quality | Genome-fragment | 4.25 | AAI-based (high-confidence) | 0 |
| S7_vs_136759\|\|full | 2144 | No | NA | 8 | 1 | 0 | Low-quality | Genome-fragment | 4.78 | AAI-based (medium-confidence) | 0 |
| S16_vs_123619\|\|full | 2144 | No | NA | 3 | 2 | 0 | Low-quality | Genome-fragment | 4.47 | AAI-based (high-confidence) | 0 |
| S7_vs_11834\|\|full | 1050 | No | NA | 2 | 0 | 0 | Not-determined | Genome-fragment | NA | NA | 0 |
| S8_extracted_k141_83846 | 2144 | No | NA | 2 | 0 | 2 | Low-quality | Genome-fragment | 0.66 | AAI-based (medium-confidence) | 0 |
| S7_vs_11901\|\|full | 9402 | No | NA | 5 | 0 | 5 | Not-determined | Genome-fragment | NA | NA | 0 |
| S7_vs_11918\|\|full | 1393 | No | NA | 2 | 0 | 0 | Not-determined | Genome-fragment | NA | NA | 0 |
| S8_vs_k141_174800\|\|full | 2144 | No | NA | 4 | 2 | 0 | Low-quality | Genome-fragment | 5.28 | AAI-based (medium-confidence) | 0 |
| S3_extracted_k141_118010 | 2145 | No | NA | 2 | 0 | 1 | Low-quality | Genome-fragment | 3.68 | AAI-based (medium-confidence) | 0 |
| S5_extracted_k141_4899 | 2145 | No | NA | 3 | 0 | 0 | Low-quality | Genome-fragment | 2.17 | AAI-based (high-confidence) | 0 |
| S7_vs_12132\|\|full | 3727 | No | NA | 3 | 0 | 2 | Not-determined | Genome-fragment | NA | NA | 0 |
| S6_extracted_k141_16619 | 2145 | No | NA | 1 | 0 | 1 | Low-quality | Genome-fragment | 1.15 | AAI-based (medium-confidence) | 0 |
| S13_extracted_121857 | 2146 | No | NA | 5 | 3 | 0 | Low-quality | Genome-fragment | 3.66 | AAI-based (high-confidence) | 0 |
| S7_vs_12298\|\|full | 1166 | No | NA | 2 | 0 | 0 | Not-determined | Genome-fragment | NA | NA | 0 |
| S7_vs_12384\|\|full | 1440 | No | NA | 4 | 0 | 0 | Not-determined | Genome-fragment | NA | NA | 0 |
| S14_vs_k141_82380\|\|full | 2146 | No | NA | 3 | 0 | 0 | Low-quality | Genome-fragment | 18.03 | AAI-based (medium-confidence) | 0 |
| S7_vs_12405\|\|full | 2456 | No | NA | 3 | 0 | 2 | Not-determined | Genome-fragment | NA | NA | 0 |
| S7_vs_12428\|\|full | 2885 | No | NA | 4 | 0 | 1 | Not-determined | Genome-fragment | NA | NA | 0 |
| S7_vs_12482\|\|full | 2669 | No | NA | 3 | 0 | 0 | Not-determined | Genome-fragment | NA | NA | 0 |
| S14_vs_k141_10657\|\|full | 2146 | No | NA | 2 | 0 | 1 | Low-quality | Genome-fragment | 4.15 | AAI-based (medium-confidence) | 0 |
| S5_vs_k141_80344\|\|full | 2146 | No | NA | 4 | 0 | 0 | Low-quality | Genome-fragment | 4.63 | AAI-based (medium-confidence) | 0 |
| S16_vs_159618\|\|full | 2146 | No | NA | 4 | 0 | 3 | Low-quality | Genome-fragment | 0.69 | AAI-based (medium-confidence) | 0 |
| S12_vs_k141_172020\|\|full | 2146 | No | NA | 3 | 0 | 1 | Low-quality | Genome-fragment | 1.72 | AAI-based (high-confidence) | 0 |
| S6_extracted_k141_55599 | 2146 | No | NA | 3 | 1 | 0 | Low-quality | Genome-fragment | 0.18 | HMM-based (lower-bound) | 0 |
| S17_vs_k141_154218\|\|full | 2147 | No | NA | 3 | 0 | 0 | Low-quality | Genome-fragment | 3.62 | AAI-based (medium-confidence) | 0 |
| S7_vs_13041\|\|full | 31516 | No | NA | 57 | 16 | 0 | High-quality | High-quality | 97.9 | AAI-based (high-confidence) | 0 |
| S9_vs_k141_19675\|\|full | 2147 | No | NA | 5 | 0 | 1 | Low-quality | Genome-fragment | 3.25 | AAI-based (high-confidence) | 0 |
| S7_vs_13154\|\|full | 2704 | No | NA | 5 | 0 | 3 | Not-determined | Genome-fragment | NA | NA | 0 |
| S7_vs_13217\|\|full | 2610 | No | NA | 3 | 0 | 0 | Not-determined | Genome-fragment | NA | NA | 0 |
| S7_vs_13370\|\|full | 31005 | No | NA | 33 | 1 | 19 | Medium-quality | Genome-fragment | 53.41 | HMM-based (lower-bound) | 0 |
| S1_vs_k141_42416\|\|full | 2147 | No | NA | 5 | 1 | 0 | Low-quality | Genome-fragment | 5.34 | AAI-based (medium-confidence) | 0 |
| S7_vs_13388\|\|full | 1011 | No | NA | 2 | 0 | 1 | Not-determined | Genome-fragment | NA | NA | 0 |
| S7_vs_13397\|\|full | 1881 | No | NA | 4 | 0 | 0 | Not-determined | Genome-fragment | NA | NA | 0 |
| S7_vs_13454\|\|full | 2348 | No | NA | 2 | 0 | 1 | Not-determined | Genome-fragment | NA | NA | 0 |
| S7_vs_13501\|\|full | 3330 | No | NA | 3 | 0 | 0 | Not-determined | Genome-fragment | NA | NA | 0 |
| S1_vs_k141_104779\|\|full | 2147 | No | NA | 3 | 1 | 0 | Low-quality | Genome-fragment | 1.91 | HMM-based (lower-bound) | 0 |
| S7_vs_13578\|\|full | 2259 | No | NA | 3 | 0 | 3 | Not-determined | Genome-fragment | NA | NA | 0 |
| S7_vs_13636\|\|full | 2735 | No | NA | 3 | 0 | 0 | Not-determined | Genome-fragment | NA | NA | 0 |
| S7_vs_13640\|\|full | 4835 | No | NA | 4 | 0 | 4 | Not-determined | Genome-fragment | NA | NA | 0 |
| S7_vs_13653\|\|full | 2312 | No | NA | 2 | 0 | 0 | Not-determined | Genome-fragment | NA | NA | 0 |
| S6_vs_k141_25769\|\|full | 2147 | No | NA | 3 | 0 | 0 | Low-quality | Genome-fragment | 4.78 | AAI-based (medium-confidence) | 0 |
| S13_vs_151723\|\|full | 2148 | No | NA | 3 | 1 | 0 | Low-quality | Genome-fragment | 5.22 | AAI-based (medium-confidence) | 0 |
| S1_extracted_k141_108154 | 2148 | No | NA | 2 | 0 | 2 | Low-quality | Genome-fragment | 0.6 | AAI-based (high-confidence) | 0 |
| S7_vs_13773\|\|full | 7965 | No | NA | 4 | 0 | 4 | Not-determined | Genome-fragment | NA | NA | 0 |
| S7_vs_13799\|\|full | 39151 | No | NA | 60 | 26 | 0 | High-quality | High-quality | 97.95 | AAI-based (high-confidence) | 0 |
| S3_vs_k141_8277\|\|full | 2148 | No | NA | 6 | 1 | 0 | Low-quality | Genome-fragment | 4.57 | AAI-based (medium-confidence) | 0 |
| S12_vs_k141_34627\|\|full | 2148 | No | NA | 7 | 0 | 0 | Low-quality | Genome-fragment | 4.43 | AAI-based (high-confidence) | 0 |
| S7_vs_13903\|\|full | 15940 | No | NA | 19 | 7 | 0 | High-quality | High-quality | 100 | AAI-based (high-confidence) | 0 |
| S8_vs_k141_198966\|\|full | 2148 | No | NA | 3 | 0 | 2 | Low-quality | Genome-fragment | 1.86 | AAI-based (medium-confidence) | 0 |
| S7_vs_14037\|\|full | 1694 | No | NA | 2 | 0 | 0 | Not-determined | Genome-fragment | NA | NA | 0 |
| S7_vs_14040\|\|full | 3226 | No | NA | 4 | 0 | 4 | Not-determined | Genome-fragment | NA | NA | 0 |
| S7_vs_14140\|\|full | 2173 | No | NA | 3 | 0 | 3 | Not-determined | Genome-fragment | NA | NA | 0 |
| S14_vs_k141_90513\|\|full | 2149 | No | NA | 4 | 1 | 0 | Low-quality | Genome-fragment | 4.78 | AAI-based (high-confidence) | 0 |
| S7_vs_14199\|\|full | 1263 | No | NA | 3 | 0 | 0 | Not-determined | Genome-fragment | NA | NA | 0 |
| S7_vs_14232\|\|full | 1003 | No | NA | 3 | 0 | 1 | Not-determined | Genome-fragment | NA | NA | 0 |
| S7_vs_14242\|\|full | 1155 | No | NA | 3 | 0 | 1 | Not-determined | Genome-fragment | NA | NA | 0 |
| S7_vs_14275\|\|full | 37694 | No | NA | 58 | 11 | 2 | High-quality | High-quality | 100 | AAI-based (high-confidence) | 0 |
| S7_vs_14469\|\|full | 1518 | No | NA | 3 | 0 | 1 | Not-determined | Genome-fragment | NA | NA | 0 |
| S1_extracted_k141_25573 | 2149 | No | NA | 10 | 1 | 0 | Low-quality | Genome-fragment | 5.4 | AAI-based (high-confidence) | 0 |
| S7_vs_23316\|\|full | 2149 | No | NA | 6 | 6 | 0 | Low-quality | Genome-fragment | 4.73 | AAI-based (medium-confidence) | 0 |
| S7_vs_14618\|\|full | 10011 | No | NA | 8 | 0 | 7 | Not-determined | Genome-fragment | NA | NA | 0 |
| S7_vs_14645\|\|full | 3456 | No | NA | 4 | 0 | 2 | Not-determined | Genome-fragment | NA | NA | 0 |
| S7_vs_14669\|\|full | 1617 | No | NA | 3 | 0 | 2 | Not-determined | Genome-fragment | NA | NA | 0 |
| S10_vs_58615\|\|full | 2149 | No | NA | 2 | 0 | 2 | Low-quality | Genome-fragment | 1.28 | AAI-based (medium-confidence) | 0 |
| S12_vs_k141_57575\|\|full | 2149 | No | NA | 3 | 0 | 1 | Low-quality | Genome-fragment | 5.14 | AAI-based (medium-confidence) | 0 |
| S7_vs_14804\|\|full | 1341 | No | NA | 2 | 0 | 0 | Not-determined | Genome-fragment | NA | NA | 0 |
| S12_vs_k141_101813\|\|full | 2149 | No | NA | 2 | 0 | 2 | Low-quality | Genome-fragment | 4.97 | AAI-based (medium-confidence) | 0 |
| S4_extracted_13834 | 2149 | No | NA | 3 | 3 | 0 | Low-quality | Genome-fragment | 5.58 | AAI-based (medium-confidence) | 0 |
| S17_extracted_k141_112383 | 2150 | No | NA | 3 | 1 | 0 | Low-quality | Genome-fragment | 4.41 | HMM-based (lower-bound) | 0 |
| S11_extracted_k141_102572 | 2150 | No | NA | 2 | 0 | 0 | Low-quality | Genome-fragment | 8.97 | AAI-based (high-confidence) | 0 |
| S11_vs_k141_59977\|\|full | 2150 | No | NA | 6 | 6 | 0 | Low-quality | Genome-fragment | 5.26 | AAI-based (high-confidence) | 0 |
| S7_vs_14969\|\|full | 1952 | No | NA | 3 | 0 | 0 | Not-determined | Genome-fragment | NA | NA | 0 |
| S7_vs_15045\|\|full | 1197 | No | NA | 2 | 0 | 1 | Not-determined | Genome-fragment | NA | NA | 0 |
| S16_extracted_34198 | 2150 | No | NA | 5 | 1 | 0 | Low-quality | Genome-fragment | 6.58 | AAI-based (medium-confidence) | 0 |
| S7_vs_15101\|\|full | 1545 | No | NA | 2 | 0 | 0 | Not-determined | Genome-fragment | NA | NA | 0 |
| S7_vs_15236\|\|full | 1666 | No | NA | 3 | 0 | 1 | Not-determined | Genome-fragment | NA | NA | 0 |
| S16_vs_45193\|\|full | 2150 | Yes | 1250 | 3 | 2 | 1 | Low-quality | Genome-fragment | 3.06 | AAI-based (high-confidence) | 41.86 |
| S7_vs_15285\|\|full | 2953 | No | NA | 5 | 0 | 3 | Not-determined | Genome-fragment | NA | NA | 0 |
| S7_vs_15288\|\|full | 1140 | No | NA | 3 | 0 | 0 | Not-determined | Genome-fragment | NA | NA | 0 |
| S7_vs_15332\|\|full | 4349 | No | NA | 4 | 0 | 3 | Not-determined | Genome-fragment | NA | NA | 0 |
| S12_vs_k141_33923\|\|full | 2150 | No | NA | 4 | 2 | 0 | Low-quality | Genome-fragment | 4.75 | AAI-based (medium-confidence) | 0 |
| S17_extracted_k141_56036 | 2151 | No | NA | 3 | 0 | 0 | Low-quality | Genome-fragment | 1.5 | AAI-based (high-confidence) | 0 |
| S9_vs_k141_56262\|\|full | 2151 | No | NA | 3 | 2 | 0 | Low-quality | Genome-fragment | 4.36 | AAI-based (high-confidence) | 0 |
| S11_extracted_k141_16102 | 2151 | No | NA | 2 | 0 | 0 | Low-quality | Genome-fragment | 1.58 | AAI-based (high-confidence) | 0 |
| S7_vs_15675\|\|full | 5489 | No | NA | 6 | 0 | 4 | Not-determined | Genome-fragment | NA | NA | 0 |
| S7_vs_15694\|\|full | 2322 | No | NA | 3 | 0 | 2 | Not-determined | Genome-fragment | NA | NA | 0 |
| S11_vs_k141_83559\|\|full | 2151 | No | NA | 4 | 0 | 0 | Low-quality | Genome-fragment | 5.67 | AAI-based (medium-confidence) | 0 |
| S7_vs_16038\|\|full | 2559 | No | NA | 4 | 0 | 0 | Not-determined | Genome-fragment | NA | NA | 0 |
| S14_extracted_k141_56427 | 2151 | No | NA | 3 | 0 | 0 | Low-quality | Genome-fragment | 1.47 | AAI-based (high-confidence) | 0 |
| S3_vs_k141_15509\|\|full | 2151 | No | NA | 5 | 1 | 0 | Low-quality | Genome-fragment | 4.34 | AAI-based (medium-confidence) | 0 |
| S12_extracted_k141_114556 | 2151 | No | NA | 2 | 2 | 0 | Low-quality | Genome-fragment | 4.79 | AAI-based (high-confidence) | 0 |
| S7_vs_16334\|\|full | 1080 | No | NA | 3 | 0 | 2 | Not-determined | Genome-fragment | NA | NA | 0 |
| S7_vs_16344\|\|full | 171746 | Yes | 27695 | 164 | 17 | 95 | Medium-quality | Genome-fragment | 51.53 | HMM-based (lower-bound) | 83.87 |
| S2_vs_k141_11878\|\|full | 2151 | No | NA | 6 | 0 | 0 | Low-quality | Genome-fragment | 5.53 | AAI-based (medium-confidence) | 0 |
| S7_vs_16381\|\|full | 1721 | No | NA | 4 | 0 | 2 | Not-determined | Genome-fragment | NA | NA | 0 |
| S7_vs_16531\|\|full | 1425 | No | NA | 2 | 0 | 0 | Not-determined | Genome-fragment | NA | NA | 0 |
| S7_vs_16635\|\|full | 1978 | No | NA | 6 | 0 | 0 | Not-determined | Genome-fragment | NA | NA | 0 |
| S17_vs_k141_152700\|\|full | 2152 | No | NA | 2 | 2 | 0 | Low-quality | Genome-fragment | 4.45 | AAI-based (high-confidence) | 0 |
| S11_vs_k141_37778\|\|full | 2152 | No | NA | 3 | 0 | 3 | Low-quality | Genome-fragment | 1.14 | AAI-based (medium-confidence) | 0 |
| S7_vs_16823\|\|full | 1460 | No | NA | 2 | 0 | 1 | Not-determined | Genome-fragment | NA | NA | 0 |
| S7_vs_16855\|\|full | 1391 | No | NA | 3 | 0 | 0 | Not-determined | Genome-fragment | NA | NA | 0 |
| S1_vs_k141_125506\|\|full | 2152 | No | NA | 5 | 0 | 1 | Low-quality | Genome-fragment | 3.97 | AAI-based (medium-confidence) | 0 |
| S7_vs_17076\|\|full | 3170 | No | NA | 6 | 0 | 1 | Not-determined | Genome-fragment | NA | NA | 0 |
| S7_vs_17118\|\|full | 5947 | No | NA | 5 | 0 | 3 | Not-determined | Genome-fragment | NA | NA | 0 |
| S5_extracted_k141_105667 | 2152 | No | NA | 2 | 2 | 0 | Low-quality | Genome-fragment | 3.32 | HMM-based (lower-bound) | 0 |
| S7_vs_17181\|\|full | 1580 | No | NA | 2 | 0 | 1 | Not-determined | Genome-fragment | NA | NA | 0 |
| S7_vs_17370\|\|full | 2152 | Yes | 519 | 3 | 1 | 2 | Low-quality | Genome-fragment | 0.32 | HMM-based (lower-bound) | 75.88 |
| S16_vs_65579\|\|full | 2152 | No | NA | 4 | 0 | 0 | Low-quality | Genome-fragment | 4.24 | AAI-based (high-confidence) | 0 |
| S16_vs_86822\|\|full | 2152 | No | NA | 3 | 0 | 0 | Low-quality | Genome-fragment | 5.16 | AAI-based (medium-confidence) | 0 |
| S7_vs_17371\|\|full | 5918 | No | NA | 4 | 0 | 2 | Not-determined | Genome-fragment | NA | NA | 0 |
| S10_vs_4714\|\|full | 2152 | No | NA | 3 | 2 | 0 | Low-quality | Genome-fragment | 4.63 | AAI-based (medium-confidence) | 0 |
| S12_extracted_k141_45804 | 2152 | No | NA | 3 | 0 | 0 | Low-quality | Genome-fragment | 3.65 | AAI-based (medium-confidence) | 0 |
| S7_vs_17655\|\|full | 1323 | No | NA | 2 | 0 | 0 | Not-determined | Genome-fragment | NA | NA | 0 |
| S7_vs_17695\|\|full | 5076 | No | NA | 8 | 0 | 0 | Not-determined | Genome-fragment | NA | NA | 0 |
| S7_vs_17696\|\|full | 2279 | No | NA | 2 | 0 | 2 | Not-determined | Genome-fragment | NA | NA | 0 |
| S7_vs_17743\|\|full | 4900 | No | NA | 5 | 0 | 2 | Not-determined | Genome-fragment | NA | NA | 0 |
| S12_vs_k141_75960\|\|full | 2152 | No | NA | 2 | 0 | 0 | Low-quality | Genome-fragment | 1.15 | AAI-based (medium-confidence) | 0 |
| S7_vs_18014\|\|full | 1956 | No | NA | 2 | 0 | 0 | Not-determined | Genome-fragment | NA | NA | 0 |
| S15_extracted_71987 | 2152 | No | NA | 4 | 0 | 0 | Low-quality | Genome-fragment | 1.63 | AAI-based (high-confidence) | 0 |
| S8_vs_k141_122158\|\|full | 2152 | No | NA | 3 | 3 | 0 | Low-quality | Genome-fragment | 6 | AAI-based (high-confidence) | 0 |
| S7_vs_18193\|\|full | 1592 | No | NA | 2 | 0 | 0 | Not-determined | Genome-fragment | NA | NA | 0 |
| S17_vs_k141_88622\|\|full | 2153 | No | NA | 4 | 0 | 0 | Low-quality | Genome-fragment | 5.61 | AAI-based (medium-confidence) | 0 |
| S7_vs_18234\|\|full | 2929 | No | NA | 4 | 0 | 2 | Not-determined | Genome-fragment | NA | NA | 0 |
| S7_vs_18367\|\|full | 2714 | No | NA | 4 | 0 | 0 | Not-determined | Genome-fragment | NA | NA | 0 |
| S9_vs_k141_83436\|\|full | 2153 | Yes | 1405 | 6 | 2 | 1 | Low-quality | Genome-fragment | 3.21 | AAI-based (high-confidence) | 34.74 |
| S11_extracted_k141_26480 | 2153 | No | NA | 4 | 0 | 0 | Low-quality | Genome-fragment | 1.64 | AAI-based (medium-confidence) | 0 |
| S16_vs_62671\|\|full | 2153 | Yes | 948 | 2 | 1 | 1 | Low-quality | Genome-fragment | 0.96 | AAI-based (high-confidence) | 55.97 |
| S7_vs_18709\|\|full | 4365 | No | NA | 4 | 0 | 4 | Not-determined | Genome-fragment | NA | NA | 0 |
| S7_vs_18802\|\|full | 3499 | No | NA | 5 | 0 | 1 | Not-determined | Genome-fragment | NA | NA | 0 |
| S7_vs_18837\|\|full | 2457 | No | NA | 3 | 0 | 1 | Not-determined | Genome-fragment | NA | NA | 0 |
| S10_extracted_115474 | 2153 | No | NA | 3 | 0 | 1 | Low-quality | Genome-fragment | 4.76 | AAI-based (medium-confidence) | 0 |
| S7_vs_18925\|\|full | 3056 | No | NA | 4 | 0 | 0 | Not-determined | Genome-fragment | NA | NA | 0 |
| S7_vs_19001\|\|full | 1424 | No | NA | 5 | 0 | 0 | Not-determined | Genome-fragment | NA | NA | 0 |
| S6_extracted_k141_4279 | 2153 | No | NA | 3 | 0 | 3 | Low-quality | Genome-fragment | 1.71 | AAI-based (medium-confidence) | 0 |
| S7_vs_19016\|\|full | 12271 | No | NA | 9 | 0 | 2 | Not-determined | Genome-fragment | NA | NA | 0 |
| S8_vs_k141_27690\|\|full | 2153 | No | NA | 2 | 1 | 0 | Low-quality | Genome-fragment | 4.88 | AAI-based (medium-confidence) | 0 |
| S8_vs_k141_123424\|\|full | 2153 | No | NA | 3 | 0 | 2 | Low-quality | Genome-fragment | 0.62 | AAI-based (high-confidence) | 0 |
| S1_extracted_k141_85456 | 2154 | No | NA | 5 | 0 | 0 | Low-quality | Genome-fragment | 3.73 | AAI-based (high-confidence) | 0 |
| S7_vs_19302\|\|full | 4760 | No | NA | 3 | 0 | 3 | Not-determined | Genome-fragment | NA | NA | 0 |
| S7_vs_19303\|\|full | 2156 | No | NA | 3 | 0 | 2 | Not-determined | Genome-fragment | NA | NA | 0 |
| S7_vs_108436\|\|full | 2154 | No | NA | 6 | 1 | 0 | Low-quality | Genome-fragment | 3.49 | AAI-based (high-confidence) | 0 |
| S12_vs_k141_114573\|\|full | 2154 | No | NA | 4 | 0 | 1 | Low-quality | Genome-fragment | 2.86 | AAI-based (high-confidence) | 0 |
| S7_vs_19466\|\|full | 1485 | No | NA | 3 | 0 | 0 | Not-determined | Genome-fragment | NA | NA | 0 |
| S7_vs_19481\|\|full | 2166 | No | NA | 2 | 0 | 2 | Not-determined | Genome-fragment | NA | NA | 0 |
| S7_vs_19511\|\|full | 1979 | No | NA | 2 | 0 | 2 | Not-determined | Genome-fragment | NA | NA | 0 |
| S17_vs_k141_75057\|\|full | 2155 | No | NA | 4 | 0 | 0 | Low-quality | Genome-fragment | 3.33 | AAI-based (high-confidence) | 0 |
| S7_vs_19569\|\|full | 12241 | No | NA | 10 | 0 | 6 | Not-determined | Genome-fragment | NA | NA | 0 |
| S7_vs_19742\|\|full | 2798 | No | NA | 3 | 0 | 3 | Not-determined | Genome-fragment | NA | NA | 0 |
| S1_vs_k141_122526\|\|full | 2155 | No | NA | 5 | 0 | 0 | Low-quality | Genome-fragment | 3.66 | AAI-based (high-confidence) | 0 |
| S7_extracted_91077 | 2155 | No | NA | 3 | 0 | 1 | Low-quality | Genome-fragment | 2.78 | AAI-based (medium-confidence) | 0 |
| S7_vs_17231\|\|full | 2155 | No | NA | 3 | 2 | 0 | Low-quality | Genome-fragment | 4.98 | AAI-based (high-confidence) | 0 |
| S7_vs_19805\|\|full | 3208 | No | NA | 7 | 0 | 1 | Not-determined | Genome-fragment | NA | NA | 0 |
| S10_vs_117048\|\|full | 2155 | No | NA | 2 | 0 | 1 | Low-quality | Genome-fragment | 1.03 | AAI-based (medium-confidence) | 0 |
| S7_vs_20046\|\|full | 1854 | No | NA | 3 | 0 | 2 | Not-determined | Genome-fragment | NA | NA | 0 |
| S7_vs_20067\|\|full | 2981 | No | NA | 5 | 0 | 3 | Not-determined | Genome-fragment | NA | NA | 0 |
| S7_vs_20200\|\|full | 39503 | No | NA | 49 | 9 | 0 | High-quality | High-quality | 93.87 | AAI-based (medium-confidence) | 0 |
| S7_vs_20217\|\|full | 9819 | No | NA | 9 | 0 | 8 | Not-determined | Genome-fragment | NA | NA | 0 |
| S7_vs_20240\|\|full | 2968 | No | NA | 3 | 0 | 1 | Not-determined | Genome-fragment | NA | NA | 0 |
| S7_vs_20253\|\|full | 1842 | No | NA | 3 | 0 | 1 | Not-determined | Genome-fragment | NA | NA | 0 |
| S7_vs_20286\|\|full | 11126 | No | NA | 13 | 0 | 3 | Not-determined | Genome-fragment | NA | NA | 0 |
| S17_extracted_k141_142126 | 2156 | Yes | 1238 | 4 | 1 | 1 | Low-quality | Genome-fragment | 0.72 | AAI-based (high-confidence) | 42.58 |
| S14_vs_k141_1089\|\|full | 2156 | No | NA | 4 | 0 | 0 | Low-quality | Genome-fragment | 5.96 | AAI-based (high-confidence) | 0 |
| S7_vs_20511\|\|full | 1565 | No | NA | 4 | 0 | 0 | Not-determined | Genome-fragment | NA | NA | 0 |
| S1_extracted_k141_17015 | 2156 | No | NA | 5 | 0 | 0 | Low-quality | Genome-fragment | 3.74 | AAI-based (high-confidence) | 0 |
| S3_vs_k141_70605\|\|full | 2156 | No | NA | 4 | 1 | 0 | Low-quality | Genome-fragment | 4.88 | AAI-based (high-confidence) | 0 |
| S7_vs_20737\|\|full | 1384 | No | NA | 3 | 0 | 0 | Not-determined | Genome-fragment | NA | NA | 0 |
| S7_vs_20781\|\|full | 1425 | No | NA | 2 | 0 | 1 | Not-determined | Genome-fragment | NA | NA | 0 |
| S7_vs_20847\|\|full | 2181 | No | NA | 4 | 0 | 0 | Not-determined | Genome-fragment | NA | NA | 0 |
| S16_extracted_33653 | 2156 | No | NA | 2 | 0 | 1 | Low-quality | Genome-fragment | 1.65 | AAI-based (high-confidence) | 0 |
| S16_vs_86080\|\|full | 2156 | No | NA | 2 | 0 | 2 | Low-quality | Genome-fragment | 4.56 | AAI-based (medium-confidence) | 0 |
| S7_vs_21056\|\|full | 1577 | No | NA | 3 | 0 | 0 | Not-determined | Genome-fragment | NA | NA | 0 |
| S16_vs_136928\|\|full | 2156 | No | NA | 4 | 1 | 0 | Low-quality | Genome-fragment | 7.69 | HMM-based (lower-bound) | 0 |
| S6_extracted_k141_53761 | 2156 | No | NA | 3 | 0 | 2 | Low-quality | Genome-fragment | 1.33 | AAI-based (high-confidence) | 0 |
| S6_vs_k141_47634\|\|full | 2156 | No | NA | 3 | 0 | 0 | Low-quality | Genome-fragment | 3.8 | AAI-based (medium-confidence) | 0 |
| S17_vs_k141_106271\|\|full | 2157 | No | NA | 3 | 0 | 0 | Low-quality | Genome-fragment | 5.49 | AAI-based (high-confidence) | 0 |
| S12_vs_k141_115153\|\|full | 2157 | No | NA | 2 | 0 | 2 | Low-quality | Genome-fragment | 1.19 | AAI-based (medium-confidence) | 0 |
| S2_vs_k141_8230\|\|full | 2157 | No | NA | 3 | 0 | 0 | Low-quality | Genome-fragment | 5.01 | AAI-based (high-confidence) | 0 |
| S8_extracted_k141_174695 | 2157 | No | NA | 3 | 0 | 2 | Low-quality | Genome-fragment | 3.09 | AAI-based (medium-confidence) | 0 |
| S7_vs_21450\|\|full | 1582 | No | NA | 3 | 0 | 3 | Not-determined | Genome-fragment | NA | NA | 0 |
| S7_vs_21521\|\|full | 55699 | No | NA | 54 | 1 | 27 | Medium-quality | Genome-fragment | 63.58 | HMM-based (lower-bound) | 0 |
| S7_vs_21555\|\|full | 3179 | No | NA | 3 | 0 | 2 | Not-determined | Genome-fragment | NA | NA | 0 |
| S3_vs_k141_51114\|\|full | 2158 | No | NA | 4 | 1 | 0 | Low-quality | Genome-fragment | 4.55 | AAI-based (high-confidence) | 0 |
| S7_vs_21613\|\|full | 3653 | No | NA | 5 | 0 | 0 | Not-determined | Genome-fragment | NA | NA | 0 |
| S7_vs_21618\|\|full | 1840 | No | NA | 4 | 0 | 0 | Not-determined | Genome-fragment | NA | NA | 0 |
| S16_vs_1094\|\|full | 2158 | No | NA | 3 | 0 | 2 | Low-quality | Genome-fragment | 1.03 | AAI-based (medium-confidence) | 0 |
| S9_vs_k141_83625\|\|full | 2159 | No | NA | 4 | 3 | 0 | Low-quality | Genome-fragment | 6.28 | AAI-based (medium-confidence) | 0 |
| S11_vs_k141_96737\|\|full | 2159 | Yes | 1712 | 2 | 1 | 1 | Low-quality | Genome-fragment | 3.46 | AAI-based (medium-confidence) | 20.7 |
| S5_vs_k141_41463\|\|full | 2159 | No | NA | 3 | 0 | 1 | Low-quality | Genome-fragment | 4.42 | AAI-based (medium-confidence) | 0 |
| S7_vs_21985\|\|full | 1007 | No | NA | 2 | 0 | 0 | Not-determined | Genome-fragment | NA | NA | 0 |
| S7_vs_22037\|\|full | 1241 | No | NA | 4 | 0 | 2 | Not-determined | Genome-fragment | NA | NA | 0 |
| S7_vs_22172\|\|full | 7492 | No | NA | 6 | 0 | 3 | Not-determined | Genome-fragment | NA | NA | 0 |
| S7_extracted_40011 | 2159 | No | NA | 3 | 0 | 1 | Low-quality | Genome-fragment | 3.44 | AAI-based (medium-confidence) | 0 |
| S7_vs_22243\|\|full | 8913 | No | NA | 11 | 0 | 1 | Not-determined | Genome-fragment | NA | NA | 0 |
| S7_vs_22277\|\|full | 1573 | No | NA | 2 | 0 | 2 | Not-determined | Genome-fragment | NA | NA | 0 |
| S7_vs_22315\|\|full | 2894 | No | NA | 6 | 0 | 1 | Not-determined | Genome-fragment | NA | NA | 0 |
| S7_vs_22325\|\|full | 2948 | No | NA | 4 | 0 | 1 | Not-determined | Genome-fragment | NA | NA | 0 |
| S7_vs_22337\|\|full | 2212 | No | NA | 3 | 0 | 1 | Not-determined | Genome-fragment | NA | NA | 0 |
| S7_vs_22383\|\|full | 1451 | No | NA | 2 | 0 | 0 | Not-determined | Genome-fragment | NA | NA | 0 |
| S10_vs_17465\|\|full | 2159 | No | NA | 3 | 0 | 0 | Low-quality | Genome-fragment | 1.43 | AAI-based (medium-confidence) | 0 |
| S7_vs_22443\|\|full | 12647 | No | NA | 5 | 0 | 1 | Not-determined | Genome-fragment | NA | NA | 0 |
| S11_vs_k141_4135\|\|full | 2160 | No | NA | 3 | 1 | 0 | Low-quality | Genome-fragment | 4.35 | AAI-based (medium-confidence) | 0 |
| S7_vs_22503\|\|full | 7887 | No | NA | 8 | 0 | 5 | Not-determined | Genome-fragment | NA | NA | 0 |
| S11_vs_k141_17014\|\|full | 2160 | No | NA | 5 | 1 | 0 | Low-quality | Genome-fragment | 5.42 | AAI-based (high-confidence) | 0 |
| S14_extracted_k141_12734 | 2160 | No | NA | 5 | 2 | 0 | Low-quality | Genome-fragment | 3.74 | AAI-based (medium-confidence) | 0 |
| S7_vs_22759\|\|full | 1332 | No | NA | 2 | 0 | 0 | Not-determined | Genome-fragment | NA | NA | 0 |
| S7_vs_22791\|\|full | 8324 | No | NA | 7 | 0 | 2 | Not-determined | Genome-fragment | NA | NA | 0 |
| S7_vs_22954\|\|full | 5486 | No | NA | 3 | 0 | 1 | Not-determined | Genome-fragment | NA | NA | 0 |
| S7_vs_23011\|\|full | 1947 | No | NA | 3 | 0 | 2 | Not-determined | Genome-fragment | NA | NA | 0 |
| S7_vs_104695\|\|full | 2160 | No | NA | 6 | 1 | 0 | Low-quality | Genome-fragment | 6.13 | AAI-based (medium-confidence) | 0 |
| S7_vs_23033\|\|full | 2116 | No | NA | 2 | 0 | 2 | Not-determined | Genome-fragment | NA | NA | 0 |
| S7_vs_23066\|\|full | 6415 | No | NA | 3 | 0 | 3 | Not-determined | Genome-fragment | NA | NA | 0 |
| S7_vs_131252\|\|full | 2160 | No | NA | 4 | 2 | 1 | Low-quality | Genome-fragment | 4.15 | AAI-based (high-confidence) | 0 |
| S16_vs_105931\|\|full | 2160 | No | NA | 3 | 0 | 1 | Low-quality | Genome-fragment | 4.38 | AAI-based (medium-confidence) | 0 |
| S9_vs_k141_3515\|\|full | 2161 | No | NA | 3 | 2 | 0 | Low-quality | Genome-fragment | 3.61 | HMM-based (lower-bound) | 0 |
| S7_vs_23291\|\|full | 1246 | No | NA | 3 | 0 | 0 | Not-determined | Genome-fragment | NA | NA | 0 |
| S13_vs_62806\|\|full | 2161 | No | NA | 6 | 0 | 0 | Low-quality | Genome-fragment | 4.19 | AAI-based (medium-confidence) | 0 |
| S10_vs_116576\|\|full | 2161 | No | NA | 4 | 0 | 0 | Low-quality | Genome-fragment | 3.32 | AAI-based (medium-confidence) | 0 |
| S7_vs_23346\|\|full | 1187 | No | NA | 3 | 0 | 0 | Not-determined | Genome-fragment | NA | NA | 0 |
| S12_extracted_k141_90521 | 2161 | No | NA | 6 | 3 | 0 | Low-quality | Genome-fragment | 4.35 | HMM-based (lower-bound) | 0 |
| S6_extracted_k141_10905 | 2161 | No | NA | 2 | 0 | 1 | Low-quality | Genome-fragment | 1.12 | AAI-based (high-confidence) | 0 |
| S11_extracted_k141_39753 | 2162 | No | NA | 4 | 0 | 0 | Low-quality | Genome-fragment | 3.33 | AAI-based (high-confidence) | 0 |
| S11_vs_k141_86847\|\|full | 2162 | No | NA | 4 | 1 | 0 | Low-quality | Genome-fragment | 1.33 | AAI-based (high-confidence) | 0 |
| S16_vs_34870\|\|full | 2162 | No | NA | 3 | 0 | 3 | Low-quality | Genome-fragment | 0.72 | AAI-based (high-confidence) | 0 |
| S6_vs_k141_2894\|\|full | 2162 | No | NA | 5 | 1 | 0 | Low-quality | Genome-fragment | 4.28 | AAI-based (high-confidence) | 0 |
| S16_vs_100294\|\|full | 2163 | No | NA | 5 | 0 | 0 | Low-quality | Genome-fragment | 4.05 | AAI-based (medium-confidence) | 0 |
| S7_vs_23770\|\|full | 23399 | No | NA | 31 | 17 | 0 | Medium-quality | Genome-fragment | 65.59 | AAI-based (high-confidence) | 0 |
| S17_vs_k141_19620\|\|full | 2164 | No | NA | 5 | 2 | 0 | Low-quality | Genome-fragment | 5.44 | AAI-based (high-confidence) | 0 |
| S7_vs_23799\|\|full | 4399 | No | NA | 4 | 0 | 0 | Not-determined | Genome-fragment | NA | NA | 0 |
| S7_vs_23922\|\|full | 1844 | No | NA | 2 | 0 | 1 | Not-determined | Genome-fragment | NA | NA | 0 |
| S3_vs_k141_113000\|\|full | 2164 | No | NA | 3 | 1 | 0 | Low-quality | Genome-fragment | 1.54 | AAI-based (high-confidence) | 0 |
| S7_vs_23981\|\|full | 13009 | No | NA | 16 | 0 | 0 | Not-determined | Genome-fragment | NA | NA | 0 |
| S7_vs_24070\|\|full | 3819 | No | NA | 4 | 0 | 2 | Not-determined | Genome-fragment | NA | NA | 0 |
| S16_extracted_58592 | 2164 | No | NA | 3 | 1 | 0 | Low-quality | Genome-fragment | 2.56 | AAI-based (high-confidence) | 0 |
| S16_extracted_142717 | 2164 | No | NA | 7 | 3 | 0 | Low-quality | Genome-fragment | 3.77 | AAI-based (high-confidence) | 0 |
| S7_vs_24199\|\|full | 2279 | No | NA | 4 | 0 | 2 | Not-determined | Genome-fragment | NA | NA | 0 |
| S10_extracted_98469 | 2164 | Yes | 1573 | 3 | 1 | 1 | Low-quality | Genome-fragment | 3.74 | HMM-based (lower-bound) | 27.31 |
| S7_vs_24352\|\|full | 1089 | No | NA | 2 | 0 | 0 | Not-determined | Genome-fragment | NA | NA | 0 |
| S6_vs_k141_23613\|\|full | 2164 | No | NA | 4 | 0 | 0 | Low-quality | Genome-fragment | 3.37 | AAI-based (medium-confidence) | 0 |
| S6_vs_k141_41135\|\|full | 2164 | No | NA | 2 | 0 | 0 | Low-quality | Genome-fragment | 5.62 | AAI-based (high-confidence) | 0 |
| S8_vs_k141_10832\|\|full | 2164 | No | NA | 3 | 0 | 1 | Low-quality | Genome-fragment | 3.46 | AAI-based (medium-confidence) | 0 |
| S7_vs_24550\|\|full | 1147 | No | NA | 2 | 0 | 0 | Not-determined | Genome-fragment | NA | NA | 0 |
| S13_vs_93547\|\|full | 2165 | No | NA | 4 | 0 | 1 | Low-quality | Genome-fragment | 2.88 | AAI-based (medium-confidence) | 0 |
| S1_extracted_k141_110316 | 2165 | No | NA | 3 | 1 | 0 | Low-quality | Genome-fragment | 6.1 | AAI-based (high-confidence) | 0 |
| S7_vs_24714\|\|full | 1334 | No | NA | 2 | 0 | 0 | Not-determined | Genome-fragment | NA | NA | 0 |
| S7_vs_24805\|\|full | 2812 | No | NA | 4 | 0 | 1 | Not-determined | Genome-fragment | NA | NA | 0 |
| S1_vs_k141_65833\|\|full | 2165 | No | NA | 4 | 3 | 0 | Low-quality | Genome-fragment | 4.76 | AAI-based (high-confidence) | 0 |
| S7_vs_24843\|\|full | 1646 | No | NA | 2 | 0 | 1 | Not-determined | Genome-fragment | NA | NA | 0 |
| S7_extracted_29894 | 2165 | No | NA | 1 | 0 | 0 | Low-quality | Genome-fragment | 4.08 | AAI-based (high-confidence) | 0 |
| S7_vs_46382\|\|full | 2165 | No | NA | 4 | 3 | 0 | Low-quality | Genome-fragment | 3.57 | HMM-based (lower-bound) | 0 |
| S16_vs_129540\|\|full | 2165 | No | NA | 4 | 1 | 0 | Low-quality | Genome-fragment | 3.18 | HMM-based (lower-bound) | 0 |
| S7_vs_25035\|\|full | 2909 | No | NA | 3 | 0 | 1 | Not-determined | Genome-fragment | NA | NA | 0 |
| S14_vs_k141_93711\|\|full | 2166 | No | NA | 8 | 1 | 0 | Low-quality | Genome-fragment | 5.99 | AAI-based (medium-confidence) | 0 |
| S16_vs_798\|\|full | 2166 | No | NA | 3 | 1 | 0 | Low-quality | Genome-fragment | 1.29 | AAI-based (high-confidence) | 0 |
| S7_vs_25215\|\|full | 1028 | No | NA | 3 | 0 | 0 | Not-determined | Genome-fragment | NA | NA | 0 |
| S12_vs_k141_107436\|\|full | 2166 | No | NA | 3 | 1 | 0 | Low-quality | Genome-fragment | 0.18 | HMM-based (lower-bound) | 0 |
| S7_vs_25281\|\|full | 1834 | No | NA | 3 | 0 | 2 | Not-determined | Genome-fragment | NA | NA | 0 |
| S8_vs_k141_105412\|\|full | 2166 | No | NA | 4 | 2 | 0 | Low-quality | Genome-fragment | 5.33 | AAI-based (medium-confidence) | 0 |
| S17_vs_k141_143433\|\|full | 2167 | No | NA | 2 | 0 | 1 | Low-quality | Genome-fragment | 1.42 | AAI-based (high-confidence) | 0 |
| S14_vs_k141_61836\|\|full | 2167 | No | NA | 3 | 0 | 0 | Low-quality | Genome-fragment | 3.74 | AAI-based (medium-confidence) | 0 |
| S7_vs_25466\|\|full | 1533 | No | NA | 3 | 0 | 0 | Not-determined | Genome-fragment | NA | NA | 0 |
| S7_vs_25565\|\|full | 1541 | No | NA | 2 | 0 | 1 | Not-determined | Genome-fragment | NA | NA | 0 |
| S7_vs_25618\|\|full | 6740 | No | NA | 3 | 0 | 1 | Not-determined | Genome-fragment | NA | NA | 0 |
| S3_extracted_k141_19547 | 2167 | No | NA | 1 | 0 | 1 | Low-quality | Genome-fragment | 0.58 | AAI-based (high-confidence) | 0 |
| S7_vs_104650\|\|full | 2167 | No | NA | 4 | 0 | 0 | Low-quality | Genome-fragment | 5.49 | AAI-based (medium-confidence) | 0 |
| S7_vs_25746\|\|full | 3381 | No | NA | 3 | 0 | 0 | Not-determined | Genome-fragment | NA | NA | 0 |
| S7_vs_25759\|\|full | 1509 | No | NA | 3 | 0 | 1 | Not-determined | Genome-fragment | NA | NA | 0 |
| S12_vs_k141_9990\|\|full | 2167 | No | NA | 2 | 0 | 0 | Low-quality | Genome-fragment | 3.52 | AAI-based (medium-confidence) | 0 |
| S7_vs_25933\|\|full | 3002 | No | NA | 4 | 0 | 3 | Not-determined | Genome-fragment | NA | NA | 0 |
| S7_vs_26007\|\|full | 3891 | No | NA | 8 | 0 | 0 | Not-determined | Genome-fragment | NA | NA | 0 |
| S6_vs_k141_19921\|\|full | 2167 | No | NA | 3 | 0 | 0 | Low-quality | Genome-fragment | 7.23 | AAI-based (high-confidence) | 0 |
| S13_vs_94225\|\|full | 2168 | No | NA | 3 | 0 | 1 | Low-quality | Genome-fragment | 0.57 | AAI-based (medium-confidence) | 0 |
| S14_extracted_k141_56768 | 2168 | No | NA | 3 | 1 | 0 | Low-quality | Genome-fragment | 1.2 | HMM-based (lower-bound) | 0 |
| S7_vs_26235\|\|full | 30810 | No | NA | 51 | 16 | 2 | Medium-quality | Genome-fragment | 82.23 | AAI-based (high-confidence) | 0 |
| S3_vs_k141_4645\|\|full | 2168 | No | NA | 4 | 3 | 0 | Low-quality | Genome-fragment | 5.36 | AAI-based (high-confidence) | 0 |
| S10_vs_71823\|\|full | 2168 | No | NA | 6 | 3 | 0 | Low-quality | Genome-fragment | 5.27 | AAI-based (high-confidence) | 0 |
| S7_vs_26330\|\|full | 8257 | No | NA | 9 | 0 | 8 | Not-determined | Genome-fragment | NA | NA | 0 |
| S7_vs_26487\|\|full | 3584 | No | NA | 4 | 0 | 1 | Not-determined | Genome-fragment | NA | NA | 0 |
| S17_extracted_k141_892 | 2169 | No | NA | 2 | 2 | 0 | Low-quality | Genome-fragment | 7.15 | AAI-based (high-confidence) | 0 |
| S7_vs_26571\|\|full | 7227 | No | NA | 6 | 0 | 5 | Not-determined | Genome-fragment | NA | NA | 0 |
| S7_vs_26608\|\|full | 2626 | No | NA | 3 | 0 | 2 | Not-determined | Genome-fragment | NA | NA | 0 |
| S7_vs_26714\|\|full | 1403 | No | NA | 4 | 0 | 0 | Not-determined | Genome-fragment | NA | NA | 0 |
| S14_vs_k141_61124\|\|full | 2169 | No | NA | 4 | 0 | 0 | Low-quality | Genome-fragment | 4.13 | AAI-based (high-confidence) | 0 |
| S7_vs_26757\|\|full | 5493 | No | NA | 6 | 0 | 1 | Medium-quality | Genome-fragment | 62.42 | AAI-based (medium-confidence) | 0 |
| S5_vs_k141_96849\|\|full | 2169 | No | NA | 3 | 2 | 0 | Low-quality | Genome-fragment | 3.15 | HMM-based (lower-bound) | 0 |
| S7_vs_26842\|\|full | 1717 | No | NA | 3 | 0 | 1 | Not-determined | Genome-fragment | NA | NA | 0 |
| S10_vs_68967\|\|full | 2169 | No | NA | 2 | 0 | 2 | Low-quality | Genome-fragment | 0.73 | AAI-based (medium-confidence) | 0 |
| S9_extracted_k141_78873 | 2170 | No | NA | 4 | 0 | 0 | Low-quality | Genome-fragment | 2.87 | AAI-based (high-confidence) | 0 |
| S9_vs_k141_44581\|\|full | 2170 | No | NA | 3 | 0 | 0 | Low-quality | Genome-fragment | 3.56 | AAI-based (high-confidence) | 0 |
| S14_extracted_k141_25323 | 2170 | No | NA | 1 | 1 | 0 | Low-quality | Genome-fragment | 1.61 | HMM-based (lower-bound) | 0 |
| S7_vs_27458\|\|full | 1471 | No | NA | 3 | 0 | 0 | Not-determined | Genome-fragment | NA | NA | 0 |
| S7_vs_27471\|\|full | 3939 | No | NA | 5 | 0 | 2 | Not-determined | Genome-fragment | NA | NA | 0 |
| S5_vs_k141_92535\|\|full | 2170 | No | NA | 3 | 0 | 2 | Low-quality | Genome-fragment | 0.89 | AAI-based (medium-confidence) | 0 |
| S5_vs_k141_101329\|\|full | 2170 | No | NA | 2 | 0 | 0 | Low-quality | Genome-fragment | 4.94 | AAI-based (high-confidence) | 0 |
| S7_vs_27590\|\|full | 2790 | No | NA | 3 | 0 | 3 | Not-determined | Genome-fragment | NA | NA | 0 |
| S7_vs_27675\|\|full | 1905 | No | NA | 3 | 0 | 2 | Not-determined | Genome-fragment | NA | NA | 0 |
| S7_vs_27677\|\|full | 1442 | No | NA | 2 | 0 | 0 | Not-determined | Genome-fragment | NA | NA | 0 |
| S8_vs_k141_190521\|\|full | 2170 | No | NA | 5 | 0 | 0 | Low-quality | Genome-fragment | 5.05 | AAI-based (medium-confidence) | 0 |
| S7_extracted_113982 | 2171 | No | NA | 1 | 0 | 0 | Low-quality | Genome-fragment | 5.43 | AAI-based (high-confidence) | 0 |
| S16_extracted_125810 | 2171 | No | NA | 6 | 0 | 0 | Low-quality | Genome-fragment | 4.41 | AAI-based (medium-confidence) | 0 |
| S7_vs_27882\|\|full | 22230 | No | NA | 8 | 0 | 6 | Not-determined | Genome-fragment | NA | NA | 0 |
| S10_vs_117254\|\|full | 2171 | No | NA | 3 | 0 | 1 | Low-quality | Genome-fragment | 4.49 | AAI-based (medium-confidence) | 0 |
| S7_vs_27927\|\|full | 3071 | No | NA | 4 | 0 | 2 | Not-determined | Genome-fragment | NA | NA | 0 |
| S7_vs_27965\|\|full | 1755 | No | NA | 4 | 0 | 0 | Not-determined | Genome-fragment | NA | NA | 0 |
| S17_vs_k141_120892\|\|full | 2172 | No | NA | 3 | 2 | 0 | Low-quality | Genome-fragment | 6.07 | AAI-based (high-confidence) | 0 |
| S7_vs_28009\|\|full | 3109 | No | NA | 3 | 0 | 2 | Not-determined | Genome-fragment | NA | NA | 0 |
| S7_vs_28072\|\|full | 1093 | No | NA | 2 | 0 | 0 | Not-determined | Genome-fragment | NA | NA | 0 |
| S12_vs_k141_21225\|\|full | 2172 | No | NA | 3 | 0 | 2 | Low-quality | Genome-fragment | 1.38 | AAI-based (high-confidence) | 0 |
| S7_vs_28150\|\|full | 33434 | No | NA | 52 | 13 | 1 | High-quality | High-quality | 90.86 | AAI-based (high-confidence) | 0 |
| S12_vs_k141_77863\|\|full | 2172 | No | NA | 3 | 0 | 1 | Low-quality | Genome-fragment | 1.75 | AAI-based (high-confidence) | 0 |
| S7_vs_28285\|\|full | 2296 | No | NA | 3 | 0 | 0 | Not-determined | Genome-fragment | NA | NA | 0 |
| S17_extracted_k141_164731 | 2173 | No | NA | 4 | 0 | 0 | Low-quality | Genome-fragment | 5.68 | AAI-based (high-confidence) | 0 |
| S16_extracted_90565 | 2173 | No | NA | 2 | 0 | 0 | Low-quality | Genome-fragment | 5.15 | AAI-based (high-confidence) | 0 |
| S7_vs_28372\|\|full | 1246 | No | NA | 3 | 0 | 0 | Not-determined | Genome-fragment | NA | NA | 0 |
| S7_vs_28458\|\|full | 2579 | No | NA | 4 | 0 | 1 | Not-determined | Genome-fragment | NA | NA | 0 |
| S7_vs_28463\|\|full | 1472 | No | NA | 3 | 0 | 3 | Not-determined | Genome-fragment | NA | NA | 0 |
| S10_vs_80355\|\|full | 2173 | No | NA | 5 | 1 | 0 | Low-quality | Genome-fragment | 3.24 | AAI-based (high-confidence) | 0 |
| S7_vs_28536\|\|full | 1413 | No | NA | 4 | 0 | 3 | Not-determined | Genome-fragment | NA | NA | 0 |
| S8_vs_k141_131492\|\|full | 2173 | No | NA | 3 | 1 | 0 | Low-quality | Genome-fragment | 3.86 | AAI-based (medium-confidence) | 0 |
| S11_vs_k141_31578\|\|full | 2174 | No | NA | 3 | 0 | 0 | Low-quality | Genome-fragment | 5.08 | AAI-based (medium-confidence) | 0 |
| S7_vs_28648\|\|full | 1530 | No | NA | 2 | 0 | 1 | Not-determined | Genome-fragment | NA | NA | 0 |
| S7_vs_28657\|\|full | 1770 | No | NA | 3 | 0 | 1 | Not-determined | Genome-fragment | NA | NA | 0 |
| S7_vs_28733\|\|full | 28764 | No | NA | 45 | 16 | 0 | High-quality | High-quality | 94.61 | AAI-based (high-confidence) | 0 |
| S14_vs_k141_47560\|\|full | 2174 | No | NA | 2 | 0 | 1 | Low-quality | Genome-fragment | 0.97 | AAI-based (medium-confidence) | 0 |
| S5_vs_k141_2133\|\|full | 2174 | No | NA | 6 | 0 | 0 | Low-quality | Genome-fragment | 5.76 | AAI-based (medium-confidence) | 0 |
| S7_vs_12051\|\|full | 2174 | No | NA | 3 | 0 | 0 | Low-quality | Genome-fragment | 4.14 | AAI-based (medium-confidence) | 0 |
| S7_vs_79181\|\|full | 2174 | No | NA | 2 | 2 | 0 | Low-quality | Genome-fragment | 5.81 | AAI-based (medium-confidence) | 0 |
| S7_vs_28973\|\|full | 1799 | No | NA | 2 | 0 | 0 | Not-determined | Genome-fragment | NA | NA | 0 |
| S7_vs_28996\|\|full | 2289 | No | NA | 2 | 0 | 2 | Not-determined | Genome-fragment | NA | NA | 0 |
| S7_vs_29001\|\|full | 1307 | No | NA | 2 | 0 | 1 | Not-determined | Genome-fragment | NA | NA | 0 |
| S16_vs_70156\|\|full | 2174 | No | NA | 3 | 0 | 1 | Low-quality | Genome-fragment | 5.1 | AAI-based (medium-confidence) | 0 |
| S10_extracted_29507 | 2174 | No | NA | 1 | 0 | 1 | Low-quality | Genome-fragment | 2.37 | AAI-based (medium-confidence) | 0 |
| S8_vs_k141_3597\|\|full | 2174 | No | NA | 2 | 0 | 0 | Low-quality | Genome-fragment | 1.16 | AAI-based (high-confidence) | 0 |
| S9_extracted_k141_63063 | 2175 | No | NA | 2 | 2 | 0 | Low-quality | Genome-fragment | 6.15 | AAI-based (high-confidence) | 0 |
| S9_vs_k141_40322\|\|full | 2175 | No | NA | 3 | 0 | 1 | Low-quality | Genome-fragment | 5.93 | AAI-based (high-confidence) | 0 |
| S13_extracted_61108 | 2175 | No | NA | 3 | 0 | 2 | Low-quality | Genome-fragment | 1.42 | AAI-based (medium-confidence) | 0 |
| S6_vs_k141_48086\|\|full | 2175 | No | NA | 5 | 2 | 0 | Low-quality | Genome-fragment | 4.95 | AAI-based (high-confidence) | 0 |
| S7_vs_29595\|\|full | 71684 | No | NA | 87 | 14 | 0 | Medium-quality | Genome-fragment | 82.12 | AAI-based (high-confidence) | 0 |
| S8_vs_k141_88752\|\|full | 2175 | No | NA | 4 | 0 | 1 | Low-quality | Genome-fragment | 1.51 | AAI-based (high-confidence) | 0 |
| S7_vs_29707\|\|full | 1514 | No | NA | 5 | 0 | 0 | Not-determined | Genome-fragment | NA | NA | 0 |

| | | | | | | | | | | | |
|---|---|---|---|---|---|---|---|---|---|---|---|
| S7_vs_29718\|\|full | 2649 | No | NA | 2 | 0 | 0 | Not-determined | Genome-fragment | NA | NA | 0 |
| S8_vs_k141_197959\|\|full | 2175 | No | NA | 3 | 0 | 1 | Low-quality | Genome-fragment | 1.29 | AAI-based (medium-confidence) | 0 |
| S7_vs_29844\|\|full | 1523 | No | NA | 3 | 0 | 0 | Not-determined | Genome-fragment | NA | NA | 0 |
| S7_vs_29912\|\|full | 3565 | No | NA | 5 | 0 | 0 | Not-determined | Genome-fragment | NA | NA | 0 |
| S7_vs_30074\|\|full | 1043 | No | NA | 2 | 0 | 0 | Not-determined | Genome-fragment | NA | NA | 0 |
| S9_extracted_k141_16711 | 2176 | No | NA | 2 | 2 | 0 | Low-quality | Genome-fragment | 3.63 | AAI-based (high-confidence) | 0 |
| S1_vs_k141_36607\|\|full | 2176 | No | NA | 3 | 1 | 0 | Low-quality | Genome-fragment | 2.72 | HMM-based (lower-bound) | 0 |
| S7_vs_38565\|\|full | 2176 | Yes | 1645 | 4 | 1 | 1 | Low-quality | Genome-fragment | 3.17 | AAI-based (medium-confidence) | 24.4 |
| S16_vs_114402\|\|lt2gene | 2176 | No | NA | 2 | 1 | 0 | Low-quality | Genome-fragment | 5.04 | AAI-based (high-confidence) | 0 |
| S8_vs_k141_103539\|\|full | 2176 | No | NA | 3 | 1 | 0 | Low-quality | Genome-fragment | 5.57 | AAI-based (high-confidence) | 0 |
| S16_vs_95931\|\|full | 2177 | No | NA | 4 | 1 | 0 | Low-quality | Genome-fragment | 5.49 | AAI-based (high-confidence) | 0 |
| S6_extracted_k141_40453 | 2177 | No | NA | 5 | 0 | 1 | Low-quality | Genome-fragment | 2.22 | AAI-based (medium-confidence) | 0 |
| S7_vs_30505\|\|full | 2545 | No | NA | 3 | 0 | 0 | Not-determined | Genome-fragment | NA | NA | 0 |
| S7_vs_30591\|\|full | 8932 | No | NA | 8 | 0 | 1 | Not-determined | Genome-fragment | NA | NA | 0 |
| S8_vs_k141_10221\|\|full | 2177 | No | NA | 4 | 0 | 1 | Low-quality | Genome-fragment | 2.19 | AAI-based (medium-confidence) | 0 |
| S7_vs_30640\|\|full | 30658 | No | NA | 43 | 16 | 0 | Medium-quality | Genome-fragment | 81.83 | AAI-based (high-confidence) | 0 |
| S7_vs_30665\|\|full | 9811 | No | NA | 16 | 6 | 1 | Medium-quality | Genome-fragment | 70.37 | AAI-based (high-confidence) | 0 |
| S7_vs_125016\|\|full | 2178 | No | NA | 5 | 3 | 0 | Low-quality | Genome-fragment | 5.98 | AAI-based (medium-confidence) | 0 |
| S8_vs_k141_182178\|\|full | 2178 | No | NA | 3 | 0 | 2 | Low-quality | Genome-fragment | 2.39 | AAI-based (medium-confidence) | 0 |
| S7_vs_30771\|\|full | 1668 | No | NA | 3 | 0 | 2 | Not-determined | Genome-fragment | NA | NA | 0 |
| S17_vs_k141_54095\|\|full | 2179 | No | NA | 4 | 1 | 0 | Low-quality | Genome-fragment | 3.48 | AAI-based (high-confidence) | 0 |
| S7_vs_31151\|\|full | 1174 | No | NA | 3 | 0 | 1 | Not-determined | Genome-fragment | NA | NA | 0 |
| S9_extracted_k141_77469 | 2179 | No | NA | 5 | 0 | 0 | Low-quality | Genome-fragment | 1.26 | AAI-based (high-confidence) | 0 |
| S7_vs_31271\|\|full | 1286 | No | NA | 2 | 0 | 0 | Not-determined | Genome-fragment | NA | NA | 0 |
| S12_vs_k141_125488\|\|full | 2179 | No | NA | 4 | 0 | 1 | Low-quality | Genome-fragment | 4.45 | AAI-based (medium-confidence) | 0 |
| S11_extracted_k141_92590 | 2180 | No | NA | 4 | 0 | 0 | Low-quality | Genome-fragment | 5.47 | AAI-based (high-confidence) | 0 |
| S5_extracted_k141_124627 | 2180 | No | NA | 3 | 3 | 0 | Low-quality | Genome-fragment | 5.75 | AAI-based (high-confidence) | 0 |
| S5_vs_k141_107028\|\|full | 2180 | No | NA | 5 | 1 | 0 | Low-quality | Genome-fragment | 1.41 | HMM-based (lower-bound) | 0 |
| S7_vs_31431\|\|full | 3914 | No | NA | 5 | 0 | 2 | Not-determined | Genome-fragment | NA | NA | 0 |
| S7_vs_31473\|\|full | 5648 | No | NA | 6 | 0 | 2 | Not-determined | Genome-fragment | NA | NA | 0 |
| S7_vs_31535\|\|full | 13243 | No | NA | 17 | 0 | 0 | Not-determined | Genome-fragment | NA | NA | 0 |
| S5_vs_k141_21440\|\|full | 2180 | No | NA | 8 | 1 | 0 | Low-quality | Genome-fragment | 4.75 | AAI-based (medium-confidence) | 0 |
| S5_vs_k141_22372\|\|full | 2180 | No | NA | 4 | 2 | 0 | Low-quality | Genome-fragment | 4.45 | AAI-based (medium-confidence) | 0 |
| S5_vs_k141_66646\|\|full | 2180 | No | NA | 7 | 0 | 0 | Low-quality | Genome-fragment | 4.61 | AAI-based (medium-confidence) | 0 |
| S16_vs_42948\|\|full | 2180 | No | NA | 3 | 0 | 0 | Low-quality | Genome-fragment | 4.68 | AAI-based (high-confidence) | 0 |
| S16_vs_151788\|\|full | 2180 | No | NA | 3 | 1 | 0 | Low-quality | Genome-fragment | 5.62 | AAI-based (high-confidence) | 0 |
| S7_vs_31714\|\|full | 3320 | No | NA | 4 | 0 | 2 | Not-determined | Genome-fragment | NA | NA | 0 |
| S7_vs_31721\|\|full | 2136 | No | NA | 2 | 0 | 1 | Not-determined | Genome-fragment | NA | NA | 0 |
| S10_extracted_22224 | 2180 | No | NA | 2 | 0 | 0 | Low-quality | Genome-fragment | 3.39 | AAI-based (medium-confidence) | 0 |
| S10_vs_75336\|\|full | 2180 | No | NA | 3 | 3 | 0 | Low-quality | Genome-fragment | 5.13 | AAI-based (high-confidence) | 0 |
| S7_vs_31843\|\|full | 2812 | No | NA | 4 | 0 | 0 | Not-determined | Genome-fragment | NA | NA | 0 |
| S7_vs_31848\|\|full | 6904 | No | NA | 5 | 0 | 3 | Not-determined | Genome-fragment | NA | NA | 0 |
| S7_vs_31890\|\|full | 1515 | No | NA | 2 | 0 | 1 | Not-determined | Genome-fragment | NA | NA | 0 |
| S12_extracted_k141_105160 | 2180 | No | NA | 3 | 0 | 2 | Low-quality | Genome-fragment | 0.36 | AAI-based (high-confidence) | 0 |
| S7_vs_32043\|\|full | 3177 | No | NA | 4 | 0 | 0 | Not-determined | Genome-fragment | NA | NA | 0 |
| S7_vs_32096\|\|full | 1040 | No | NA | 2 | 0 | 1 | Not-determined | Genome-fragment | NA | NA | 0 |
| S7_vs_32184\|\|full | 2022 | No | NA | 3 | 0 | 2 | Not-determined | Genome-fragment | NA | NA | 0 |
| S12_vs_k141_32849\|\|full | 2180 | No | NA | 5 | 3 | 0 | Low-quality | Genome-fragment | 3.62 | HMM-based (lower-bound) | 0 |
| S11_extracted_k141_28778 | 2181 | No | NA | 5 | 2 | 0 | Low-quality | Genome-fragment | 5.6 | AAI-based (medium-confidence) | 0 |
| S7_vs_32256\|\|full | 1933 | No | NA | 3 | 0 | 2 | Not-determined | Genome-fragment | NA | NA | 0 |
| S7_vs_32308\|\|full | 6450 | No | NA | 7 | 0 | 3 | Not-determined | Genome-fragment | NA | NA | 0 |
| S1_extracted_k141_79145 | 2181 | No | NA | 2 | 0 | 2 | Low-quality | Genome-fragment | 0.58 | AAI-based (medium-confidence) | 0 |
| S3_vs_k141_104152\|\|full | 2181 | No | NA | 3 | 0 | 0 | Low-quality | Genome-fragment | 5.38 | AAI-based (high-confidence) | 0 |
| S9_vs_k141_83584\|\|full | 2182 | No | NA | 7 | 1 | 0 | Low-quality | Genome-fragment | 16.92 | AAI-based (high-confidence) | 0 |
| S14_vs_k141_55728\|\|full | 2182 | No | NA | 6 | 1 | 0 | Low-quality | Genome-fragment | 1.6 | AAI-based (medium-confidence) | 0 |
| S1_vs_k141_59837\|\|full | 2182 | No | NA | 2 | 0 | 1 | Low-quality | Genome-fragment | 2.67 | AAI-based (high-confidence) | 0 |
| S8_vs_k141_74774\|\|full | 2182 | No | NA | 3 | 3 | 0 | Low-quality | Genome-fragment | 6.05 | AAI-based (medium-confidence) | 0 |
| S13_vs_146543\|\|full | 2183 | No | NA | 2 | 0 | 1 | Low-quality | Genome-fragment | 2.74 | AAI-based (medium-confidence) | 0 |
| S7_vs_32822\|\|full | 1508 | No | NA | 3 | 0 | 0 | Not-determined | Genome-fragment | NA | NA | 0 |
| S7_vs_32840\|\|full | 5605 | No | NA | 5 | 0 | 1 | Not-determined | Genome-fragment | NA | NA | 0 |
| S7_vs_32964\|\|full | 5795 | No | NA | 7 | 0 | 3 | Not-determined | Genome-fragment | NA | NA | 0 |
| S7_vs_32973\|\|full | 1347 | No | NA | 6 | 0 | 0 | Not-determined | Genome-fragment | NA | NA | 0 |
| S16_vs_144518\|\|full | 2183 | No | NA | 4 | 0 | 0 | Low-quality | Genome-fragment | 2.05 | AAI-based (medium-confidence) | 0 |
| S12_vs_k141_112743\|\|full | 2183 | No | NA | 3 | 0 | 0 | Low-quality | Genome-fragment | 4.85 | AAI-based (high-confidence) | 0 |
| S7_vs_33067\|\|full | 3073 | No | NA | 4 | 0 | 2 | Not-determined | Genome-fragment | NA | NA | 0 |
| S7_vs_33076\|\|full | 5878 | No | NA | 7 | 0 | 4 | Not-determined | Genome-fragment | NA | NA | 0 |
| S7_vs_33085\|\|full | 2880 | No | NA | 3 | 0 | 3 | Not-determined | Genome-fragment | NA | NA | 0 |
| S7_vs_33180\|\|full | 2390 | No | NA | 5 | 0 | 3 | Not-determined | Genome-fragment | NA | NA | 0 |
| S4_extracted_17347 | 2183 | No | NA | 4 | 2 | 0 | Low-quality | Genome-fragment | 4.3 | AAI-based (high-confidence) | 0 |
| S8_vs_k141_20803\|\|full | 2183 | No | NA | 6 | 2 | 0 | Low-quality | Genome-fragment | 5.02 | AAI-based (high-confidence) | 0 |
| S9_extracted_k141_60460 | 2184 | No | NA | 3 | 3 | 0 | Low-quality | Genome-fragment | 5.04 | AAI-based (high-confidence) | 0 |
| S9_vs_k141_16114\|\|lt2gene | 2184 | No | NA | 2 | 2 | 0 | Low-quality | Genome-fragment | 4.89 | AAI-based (high-confidence) | 0 |
| S14_vs_k141_18735\|\|full | 2184 | No | NA | 3 | 0 | 0 | Low-quality | Genome-fragment | 3.7 | AAI-based (medium-confidence) | 0 |
| S16_vs_146301\|\|full | 2184 | No | NA | 3 | 0 | 0 | Low-quality | Genome-fragment | 5.01 | AAI-based (medium-confidence) | 0 |
| S8_extracted_k141_29190 | 2184 | No | NA | 2 | 0 | 2 | Low-quality | Genome-fragment | 1.04 | AAI-based (medium-confidence) | 0 |
| S7_vs_32452\|\|full | 2185 | No | NA | 6 | 0 | 0 | Low-quality | Genome-fragment | 5.46 | AAI-based (medium-confidence) | 0 |
| S7_vs_34124\|\|full | 1905 | No | NA | 2 | 0 | 0 | Not-determined | Genome-fragment | NA | NA | 0 |
| S7_vs_34130\|\|full | 4192 | No | NA | 6 | 0 | 0 | Not-determined | Genome-fragment | NA | NA | 0 |
| S12_vs_k141_80046\|\|full | 2185 | No | NA | 3 | 1 | 0 | Low-quality | Genome-fragment | 3.31 | HMM-based (lower-bound) | 0 |
| S7_vs_34226\|\|full | 6675 | No | NA | 3 | 0 | 2 | Not-determined | Genome-fragment | NA | NA | 0 |
| S12_vs_k141_78140\|\|full | 2185 | No | NA | 3 | 0 | 1 | Low-quality | Genome-fragment | 2.9 | AAI-based (medium-confidence) | 0 |
| S7_vs_34495\|\|full | 9623 | No | NA | 16 | 0 | 1 | Not-determined | Genome-fragment | NA | NA | 0 |
| S4_extracted_10895 | 2185 | No | NA | 2 | 2 | 0 | Low-quality | Genome-fragment | 7.21 | HMM-based (lower-bound) | 0 |
| S8_extracted_k141_30558 | 2185 | No | NA | 4 | 2 | 0 | Low-quality | Genome-fragment | 6.78 | AAI-based (high-confidence) | 0 |
| S11_vs_k141_78211\|\|full | 2186 | No | NA | 8 | 0 | 0 | Low-quality | Genome-fragment | 2.73 | AAI-based (medium-confidence) | 0 |
| S7_vs_521\|\|full | 2186 | No | NA | 5 | 2 | 0 | Low-quality | Genome-fragment | 6.21 | AAI-based (medium-confidence) | 0 |
| S7_vs_34787\|\|full | 6138 | No | NA | 6 | 0 | 3 | Not-determined | Genome-fragment | NA | NA | 0 |
| S7_vs_20420\|\|full | 2186 | No | NA | 1 | 0 | 0 | Low-quality | Genome-fragment | 2.4 | AAI-based (medium-confidence) | 0 |
| S16_extracted_111518 | 2186 | No | NA | 2 | 0 | 1 | Low-quality | Genome-fragment | 1.95 | AAI-based (medium-confidence) | 0 |
| S10_vs_19955\|\|full | 2186 | No | NA | 4 | 0 | 2 | Low-quality | Genome-fragment | 3.5 | AAI-based (high-confidence) | 0 |
| S7_vs_34966\|\|full | 1197 | No | NA | 3 | 0 | 2 | Not-determined | Genome-fragment | NA | NA | 0 |
| S7_vs_34997\|\|full | 2222 | No | NA | 3 | 0 | 1 | Not-determined | Genome-fragment | NA | NA | 0 |
| S10_vs_55742\|\|full | 2186 | No | NA | 4 | 0 | 1 | Low-quality | Genome-fragment | 5.17 | AAI-based (medium-confidence) | 0 |
| S15_vs_71673\|\|full | 2186 | No | NA | 3 | 0 | 0 | Low-quality | Genome-fragment | 0.62 | AAI-based (medium-confidence) | 0 |
| S7_vs_35091\|\|full | 6084 | No | NA | 4 | 0 | 3 | Not-determined | Genome-fragment | NA | NA | 0 |
| S17_vs_k141_122569\|\|full | 2187 | No | NA | 3 | 1 | 0 | Low-quality | Genome-fragment | 4.7 | AAI-based (high-confidence) | 0 |
| S7_vs_35121\|\|full | 1879 | No | NA | 5 | 0 | 1 | Not-determined | Genome-fragment | NA | NA | 0 |
| S3_vs_k141_98087\|\|full | 2187 | No | NA | 5 | 0 | 0 | Low-quality | Genome-fragment | 4.33 | AAI-based (medium-confidence) | 0 |
| S7_vs_35295\|\|full | 1115 | No | NA | 2 | 0 | 1 | Not-determined | Genome-fragment | NA | NA | 0 |
| S7_vs_35351\|\|full | 2417 | No | NA | 5 | 0 | 0 | Not-determined | Genome-fragment | NA | NA | 0 |
| S7_vs_35393\|\|full | 3105 | No | NA | 3 | 0 | 3 | Not-determined | Genome-fragment | NA | NA | 0 |
| S7_vs_35420\|\|full | 5484 | No | NA | 2 | 0 | 1 | Not-determined | Genome-fragment | NA | NA | 0 |
| S7_extracted_54859 | 2187 | No | NA | 3 | 1 | 0 | Low-quality | Genome-fragment | 33.7 | AAI-based (high-confidence) | 0 |
| S7_vs_35513\|\|full | 3171 | No | NA | 4 | 0 | 0 | Not-determined | Genome-fragment | NA | NA | 0 |
| S7_vs_35521\|\|full | 3409 | No | NA | 4 | 0 | 2 | Not-determined | Genome-fragment | NA | NA | 0 |
| S16_extracted_31444 | 2187 | No | NA | 1 | 0 | 1 | Low-quality | Genome-fragment | 1.1 | AAI-based (medium-confidence) | 0 |
| S16_vs_167097\|\|full | 2187 | No | NA | 4 | 0 | 3 | Low-quality | Genome-fragment | 1.34 | AAI-based (medium-confidence) | 0 |
| S7_vs_35740\|\|full | 7866 | No | NA | 7 | 0 | 4 | Not-determined | Genome-fragment | NA | NA | 0 |
| S7_vs_35777\|\|full | 4401 | No | NA | 6 | 0 | 5 | Not-determined | Genome-fragment | NA | NA | 0 |
| S10_vs_116968\|\|full | 2187 | No | NA | 4 | 0 | 0 | Low-quality | Genome-fragment | 3.55 | AAI-based (medium-confidence) | 0 |
| S12_vs_k141_17527\|\|full | 2187 | No | NA | 3 | 1 | 0 | Low-quality | Genome-fragment | 2.76 | HMM-based (lower-bound) | 0 |
| S11_vs_k141_11129\|\|full | 2188 | No | NA | 4 | 0 | 0 | Low-quality | Genome-fragment | 3.72 | AAI-based (high-confidence) | 0 |
| S1_vs_k141_137804\|\|full | 2188 | No | NA | 3 | 0 | 0 | Low-quality | Genome-fragment | 5.02 | AAI-based (medium-confidence) | 0 |
| S7_vs_36164\|\|full | 1034 | No | NA | 2 | 0 | 1 | Not-determined | Genome-fragment | NA | NA | 0 |
| S7_vs_36358\|\|full | 8450 | No | NA | 9 | 0 | 4 | Not-determined | Genome-fragment | NA | NA | 0 |
| S12_vs_k141_127842\|\|full | 2188 | No | NA | 4 | 1 | 0 | Low-quality | Genome-fragment | 3.3 | HMM-based (lower-bound) | 0 |
| S8_vs_k141_134084\|\|full | 2188 | No | NA | 2 | 0 | 2 | Low-quality | Genome-fragment | 1.52 | AAI-based (medium-confidence) | 0 |
| S9_vs_k141_6769\|\|full | 2189 | No | NA | 3 | 0 | 1 | Low-quality | Genome-fragment | 1.61 | AAI-based (medium-confidence) | 0 |
| S7_extracted_5273 | 2189 | No | NA | 4 | 3 | 0 | Low-quality | Genome-fragment | 7.72 | HMM-based (lower-bound) | 0 |
| S7_vs_36751\|\|full | 4447 | No | NA | 3 | 0 | 0 | Not-determined | Genome-fragment | NA | NA | 0 |
| S7_vs_36820\|\|full | 4948 | No | NA | 2 | 0 | 2 | Not-determined | Genome-fragment | NA | NA | 0 |
| S7_extracted_92011 | 2189 | No | NA | 4 | 4 | 0 | Low-quality | Genome-fragment | 9.98 | AAI-based (high-confidence) | 0 |
| S7_vs_12921\|\|full | 2189 | No | NA | 4 | 2 | 0 | Low-quality | Genome-fragment | 5.03 | AAI-based (medium-confidence) | 0 |
| S12_extracted_k141_28826 | 2189 | No | NA | 4 | 1 | 0 | Low-quality | Genome-fragment | 1.84 | AAI-based (high-confidence) | 0 |
| S5_vs_k141_57107\|\|full | 2190 | No | NA | 3 | 0 | 0 | Low-quality | Genome-fragment | 3.91 | AAI-based (high-confidence) | 0 |
| S16_vs_63008\|\|full | 2190 | No | NA | 3 | 0 | 1 | Low-quality | Genome-fragment | 4 | AAI-based (medium-confidence) | 0 |
| S12_vs_k141_150255\|\|full | 2190 | No | NA | 3 | 1 | 0 | Low-quality | Genome-fragment | 4 | AAI-based (high-confidence) | 0 |
| S17_extracted_k141_35974 | 2191 | No | NA | 3 | 1 | 0 | Low-quality | Genome-fragment | 3.28 | AAI-based (high-confidence) | 0 |
| S17_vs_k141_100928\|\|full | 2191 | No | NA | 6 | 0 | 0 | Low-quality | Genome-fragment | 5.22 | AAI-based (medium-confidence) | 0 |
| S7_vs_37202\|\|full | 3498 | No | NA | 4 | 0 | 1 | Not-determined | Genome-fragment | NA | NA | 0 |
| S14_vs_k141_36442\|\|full | 2191 | No | NA | 4 | 2 | 0 | Low-quality | Genome-fragment | 4.08 | AAI-based (high-confidence) | 0 |
| S7_vs_37233\|\|full | 1674 | No | NA | 2 | 0 | 1 | Not-determined | Genome-fragment | NA | NA | 0 |
| S7_vs_37317\|\|full | 4542 | No | NA | 3 | 0 | 0 | Not-determined | Genome-fragment | NA | NA | 0 |
| S5_vs_k141_63239\|\|full | 2191 | No | NA | 3 | 1 | 0 | Low-quality | Genome-fragment | 6.29 | AAI-based (high-confidence) | 0 |
| S5_vs_k141_130727\|\|full | 2191 | No | NA | 4 | 4 | 0 | Low-quality | Genome-fragment | 6.53 | AAI-based (high-confidence) | 0 |
| S7_vs_37492\|\|full | 1001 | No | NA | 2 | 0 | 1 | Not-determined | Genome-fragment | NA | NA | 0 |
| S16_extracted_138167 | 2191 | No | NA | 2 | 0 | 2 | Low-quality | Genome-fragment | 1.8 | AAI-based (high-confidence) | 0 |
| S7_vs_37570\|\|full | 6406 | No | NA | 6 | 0 | 1 | Not-determined | Genome-fragment | NA | NA | 0 |
| S7_vs_37616\|\|full | 16300 | No | NA | 17 | 0 | 17 | Not-determined | Genome-fragment | NA | NA | 0 |
| S7_vs_37728\|\|full | 1091 | No | NA | 2 | 0 | 1 | Not-determined | Genome-fragment | NA | NA | 0 |
| S10_vs_90284\|\|full | 2191 | No | NA | 4 | 2 | 0 | Low-quality | Genome-fragment | 2.72 | AAI-based (high-confidence) | 0 |
| S8_vs_k141_67588\|\|full | 2191 | No | NA | 3 | 0 | 0 | Low-quality | Genome-fragment | 5.31 | AAI-based (high-confidence) | 0 |
| S17_extracted_k141_102365 | 2192 | No | NA | 1 | 0 | 0 | Low-quality | Genome-fragment | 2.24 | AAI-based (high-confidence) | 0 |
| S9_vs_k141_13219\|\|full | 2192 | No | NA | 6 | 3 | 0 | Low-quality | Genome-fragment | 5.51 | AAI-based (high-confidence) | 0 |
| S16_vs_67867\|\|full | 2192 | No | NA | 5 | 0 | 1 | Low-quality | Genome-fragment | 2.53 | AAI-based (medium-confidence) | 0 |
| S12_vs_k141_141900\|\|full | 2192 | No | NA | 6 | 1 | 0 | Low-quality | Genome-fragment | 4.93 | AAI-based (medium-confidence) | 0 |
| S14_vs_k141_116187\|\|full | 2193 | No | NA | 3 | 0 | 2 | Low-quality | Genome-fragment | 1.16 | AAI-based (medium-confidence) | 0 |
| S1_vs_k141_142023\|\|full | 2193 | No | NA | 4 | 1 | 0 | Low-quality | Genome-fragment | 5.13 | AAI-based (high-confidence) | 0 |
| S7_vs_38113\|\|full | 17045 | No | NA | 21 | 15 | 0 | Medium-quality | Genome-fragment | 50.08 | AAI-based (high-confidence) | 0 |
| S7_vs_38123\|\|full | 1235 | No | NA | 3 | 0 | 0 | Not-determined | Genome-fragment | NA | NA | 0 |
| S1_vs_k141_9386\|\|full | 2193 | No | NA | 6 | 2 | 0 | Low-quality | Genome-fragment | 2.68 | AAI-based (medium-confidence) | 0 |
| S7_vs_38254\|\|full | 2053 | No | NA | 3 | 0 | 0 | Not-determined | Genome-fragment | NA | NA | 0 |
| S7_vs_38308\|\|full | 2353 | No | NA | 3 | 0 | 1 | Not-determined | Genome-fragment | NA | NA | 0 |
| S7_vs_38322\|\|full | 1005 | No | NA | 2 | 0 | 1 | Not-determined | Genome-fragment | NA | NA | 0 |
| S7_vs_38408\|\|full | 1244 | No | NA | 2 | 0 | 0 | Not-determined | Genome-fragment | NA | NA | 0 |
| S5_vs_k141_66070\|\|full | 2193 | No | NA | 5 | 5 | 0 | Low-quality | Genome-fragment | 3.58 | AAI-based (high-confidence) | 0 |
| S7_vs_38437\|\|full | 1589 | No | NA | 3 | 0 | 3 | Not-determined | Genome-fragment | NA | NA | 0 |
| S12_vs_k141_77993\|\|full | 2193 | No | NA | 3 | 0 | 1 | Low-quality | Genome-fragment | 1.24 | AAI-based (medium-confidence) | 0 |
| S7_vs_38500\|\|full | 1241 | No | NA | 3 | 0 | 1 | Not-determined | Genome-fragment | NA | NA | 0 |
| S6_vs_k141_42642\|\|full | 2193 | No | NA | 3 | 0 | 0 | Low-quality | Genome-fragment | 3.88 | AAI-based (high-confidence) | 0 |
| S8_vs_k141_95076\|\|full | 2193 | No | NA | 4 | 3 | 0 | Low-quality | Genome-fragment | 5.38 | AAI-based (high-confidence) | 0 |
| S7_vs_38675\|\|full | 15100 | No | NA | 16 | 0 | 3 | Not-determined | Genome-fragment | NA | NA | 0 |
| S8_vs_k141_52025\|\|full | 2193 | No | NA | 3 | 2 | 0 | Low-quality | Genome-fragment | 4.59 | AAI-based (high-confidence) | 0 |
| S14_extracted_k141_85977 | 2194 | No | NA | 5 | 3 | 0 | Low-quality | Genome-fragment | 3.18 | AAI-based (medium-confidence) | 0 |
| S7_vs_38720\|\|full | 1316 | No | NA | 2 | 0 | 1 | Not-determined | Genome-fragment | NA | NA | 0 |
| S15_vs_63661\|\|full | 2194 | No | NA | 4 | 1 | 0 | Low-quality | Genome-fragment | 1.25 | AAI-based (high-confidence) | 0 |
| S8_vs_k141_38195\|\|full | 2194 | No | NA | 3 | 3 | 0 | Low-quality | Genome-fragment | 4.8 | AAI-based (high-confidence) | 0 |
| S7_vs_38810\|\|full | 1598 | No | NA | 3 | 0 | 1 | Not-determined | Genome-fragment | NA | NA | 0 |
| S9_vs_k141_49504\|\|full | 2195 | No | NA | 5 | 0 | 2 | Low-quality | Genome-fragment | 4.42 | AAI-based (medium-confidence) | 0 |
| S7_vs_38834\|\|full | 20668 | No | NA | 23 | 13 | 0 | Medium-quality | Genome-fragment | 58.53 | AAI-based (medium-confidence) | 0 |
| S7_vs_38838\|\|full | 1594 | No | NA | 2 | 0 | 0 | Not-determined | Genome-fragment | NA | NA | 0 |
| S7_vs_38880\|\|full | 2045 | No | NA | 4 | 0 | 1 | Not-determined | Genome-fragment | NA | NA | 0 |
| S7_vs_24481\|\|full | 2195 | No | NA | 7 | 5 | 0 | Low-quality | Genome-fragment | 5.92 | AAI-based (medium-confidence) | 0 |
| S7_vs_132972\|\|full | 2195 | No | NA | 3 | 0 | 0 | Low-quality | Genome-fragment | 2.79 | AAI-based (medium-confidence) | 0 |
| S16_vs_61402\|\|full | 2195 | No | NA | 9 | 2 | 0 | Low-quality | Genome-fragment | 3.17 | HMM-based (lower-bound) | 0 |
| S4_extracted_2612 | 2195 | No | NA | 3 | 0 | 3 | Low-quality | Genome-fragment | 0.94 | AAI-based (medium-confidence) | 0 |
| S7_vs_39120\|\|full | 2446 | No | NA | 4 | 0 | 3 | Not-determined | Genome-fragment | NA | NA | 0 |
| S10_vs_73129\|\|full | 2196 | No | NA | 3 | 0 | 1 | Low-quality | Genome-fragment | 4.22 | AAI-based (medium-confidence) | 0 |
| S12_vs_k141_93516\|\|full | 2196 | No | NA | 3 | 0 | 0 | Low-quality | Genome-fragment | 5.65 | AAI-based (high-confidence) | 0 |
| S1_vs_k141_152148\|\|full | 2197 | No | NA | 4 | 1 | 0 | Low-quality | Genome-fragment | 4.4 | HMM-based (lower-bound) | 0 |
| S16_vs_109402\|\|lt2gene | 2197 | No | NA | 2 | 1 | 0 | Low-quality | Genome-fragment | 4.03 | HMM-based (lower-bound) | 0 |
| S7_vs_39356\|\|full | 2238 | No | NA | 3 | 0 | 2 | Not-determined | Genome-fragment | NA | NA | 0 |
| S7_vs_39373\|\|full | 1802 | No | NA | 4 | 0 | 1 | Not-determined | Genome-fragment | NA | NA | 0 |
| S7_vs_39382\|\|full | 1944 | No | NA | 3 | 0 | 1 | Not-determined | Genome-fragment | NA | NA | 0 |
| S7_vs_39393\|\|full | 1104 | No | NA | 3 | 0 | 0 | Not-determined | Genome-fragment | NA | NA | 0 |
| S10_vs_7647\|\|full | 2197 | No | NA | 5 | 0 | 1 | Low-quality | Genome-fragment | 5.37 | AAI-based (high-confidence) | 0 |
| S7_vs_39456\|\|full | 2128 | No | NA | 2 | 0 | 0 | Not-determined | Genome-fragment | NA | NA | 0 |
| S12_vs_k141_37691\|\|full | 2197 | No | NA | 4 | 0 | 0 | Low-quality | Genome-fragment | 3.88 | AAI-based (high-confidence) | 0 |
| S7_vs_39631\|\|full | 3105 | No | NA | 5 | 0 | 3 | Not-determined | Genome-fragment | NA | NA | 0 |
| S6_vs_k141_52928\|\|full | 2197 | No | NA | 8 | 2 | 0 | Low-quality | Genome-fragment | 3.65 | AAI-based (high-confidence) | 0 |
| S17_vs_k141_106962\|\|full | 2198 | No | NA | 5 | 0 | 0 | Low-quality | Genome-fragment | 3.33 | AAI-based (high-confidence) | 0 |
| S3_vs_k141_113526\|\|full | 2198 | No | NA | 2 | 1 | 0 | Low-quality | Genome-fragment | 2.91 | HMM-based (lower-bound) | 0 |
| S7_vs_39740\|\|full | 3068 | No | NA | 4 | 0 | 0 | Not-determined | Genome-fragment | NA | NA | 0 |
| S3_vs_k141_60068\|\|full | 2198 | No | NA | 3 | 1 | 0 | Low-quality | Genome-fragment | 4.8 | AAI-based (high-confidence) | 0 |
| S16_vs_93295\|\|full | 2198 | No | NA | 2 | 1 | 0 | Low-quality | Genome-fragment | 5.62 | AAI-based (high-confidence) | 0 |
| S7_vs_40050\|\|full | 2069 | No | NA | 3 | 0 | 1 | Not-determined | Genome-fragment | NA | NA | 0 |
| S10_vs_67322\|\|full | 2198 | No | NA | 5 | 0 | 3 | Low-quality | Genome-fragment | 1.75 | AAI-based (medium-confidence) | 0 |
| S7_vs_40197\|\|full | 2857 | No | NA | 2 | 0 | 0 | Not-determined | Genome-fragment | NA | NA | 0 |
| S11_vs_k141_12764\|\|full | 2199 | No | NA | 6 | 5 | 0 | Low-quality | Genome-fragment | 5.72 | AAI-based (medium-confidence) | 0 |
| S7_vs_40259\|\|full | 41109 | No | NA | 57 | 24 | 0 | Medium-quality | Genome-fragment | 88.55 | AAI-based (medium-confidence) | 0 |
| S7_vs_40328\|\|full | 10906 | No | NA | 11 | 0 | 5 | Not-determined | Genome-fragment | NA | NA | 0 |
| S7_vs_40464\|\|full | 2307 | No | NA | 4 | 0 | 1 | Not-determined | Genome-fragment | NA | NA | 0 |
| S7_vs_28543\|\|full | 2199 | No | NA | 3 | 1 | 0 | Low-quality | Genome-fragment | 2.36 | AAI-based (medium-confidence) | 0 |
| S7_vs_40552\|\|full | 1724 | No | NA | 2 | 0 | 1 | Not-determined | Genome-fragment | NA | NA | 0 |
| S7_vs_40557\|\|full | 2130 | No | NA | 2 | 0 | 2 | Not-determined | Genome-fragment | NA | NA | 0 |
| S7_vs_40574\|\|full | 2241 | No | NA | 5 | 0 | 0 | Not-determined | Genome-fragment | NA | NA | 0 |
| S12_vs_k141_83867\|\|full | 2199 | No | NA | 5 | 1 | 0 | Low-quality | Genome-fragment | 5.37 | AAI-based (medium-confidence) | 0 |
| S7_vs_40591\|\|full | 1736 | No | NA | 5 | 0 | 0 | Not-determined | Genome-fragment | NA | NA | 0 |
| S8_vs_k141_21857\|\|full | 2199 | No | NA | 3 | 0 | 2 | Low-quality | Genome-fragment | 5.08 | AAI-based (medium-confidence) | 0 |
| S7_vs_40626\|\|full | 3565 | No | NA | 2 | 0 | 1 | Not-determined | Genome-fragment | NA | NA | 0 |
| S11_extracted_k141_102315 | 2200 | No | NA | 2 | 0 | 1 | Low-quality | Genome-fragment | 2.77 | AAI-based (medium-confidence) | 0 |
| S7_vs_40769\|\|full | 1187 | No | NA | 3 | 0 | 2 | Not-determined | Genome-fragment | NA | NA | 0 |
| S7_vs_40892\|\|full | 1686 | No | NA | 3 | 0 | 1 | Not-determined | Genome-fragment | NA | NA | 0 |
| S7_vs_40930\|\|full | 1032 | No | NA | 3 | 0 | 0 | Not-determined | Genome-fragment | NA | NA | 0 |
| S14_vs_k141_48149\|\|full | 2200 | No | NA | 4 | 0 | 0 | Low-quality | Genome-fragment | 5.23 | AAI-based (high-confidence) | 0 |
| S1_extracted_k141_22138 | 2200 | No | NA | 3 | 1 | 0 | Low-quality | Genome-fragment | 5.94 | AAI-based (medium-confidence) | 0 |
| S7_vs_41098\|\|full | 60067 | No | NA | 89 | 32 | 0 | High-quality | High-quality | 99.27 | AAI-based (high-confidence) | 0 |
| S7_vs_41275\|\|full | 1259 | No | NA | 4 | 0 | 0 | Not-determined | Genome-fragment | NA | NA | 0 |
| S7_vs_41397\|\|full | 1677 | No | NA | 2 | 0 | 2 | Not-determined | Genome-fragment | NA | NA | 0 |
| S1_extracted_k141_143203 | 2200 | No | NA | 7 | 0 | 0 | Low-quality | Genome-fragment | 4.16 | AAI-based (medium-confidence) | 0 |
| S7_vs_41534\|\|full | 15804 | No | NA | 23 | 7 | 0 | Medium-quality | Genome-fragment | 72.22 | AAI-based (medium-confidence) | 0 |
| S16_vs_83223\|\|full | 2200 | No | NA | 3 | 0 | 2 | Low-quality | Genome-fragment | 0.61 | AAI-based (medium-confidence) | 0 |
| S8_vs_k141_29611\|\|full | 2200 | No | NA | 3 | 2 | 0 | Low-quality | Genome-fragment | 1.18 | AAI-based (high-confidence) | 0 |
| S7_vs_41816\|\|full | 15799 | No | NA | 17 | 0 | 10 | Not-determined | Genome-fragment | NA | NA | 0 |
| S8_vs_k141_57275\|\|full | 2200 | No | NA | 7 | 0 | 0 | Low-quality | Genome-fragment | 3.73 | AAI-based (medium-confidence) | 0 |
| S3_vs_k141_26929\|\|full | 2201 | No | NA | 5 | 2 | 0 | Low-quality | Genome-fragment | 4.69 | AAI-based (high-confidence) | 0 |
| S7_vs_67042\|\|full | 2201 | No | NA | 2 | 0 | 0 | Low-quality | Genome-fragment | 5.24 | AAI-based (high-confidence) | 0 |
| S7_vs_42051\|\|full | 2582 | No | NA | 3 | 0 | 1 | Not-determined | Genome-fragment | NA | NA | 0 |
| S6_vs_k141_12787\|\|full | 2201 | No | NA | 3 | 1 | 0 | Low-quality | Genome-fragment | 0.94 | HMM-based (lower-bound) | 0 |
| S11_extracted_k141_99114 | 2202 | No | NA | 5 | 0 | 0 | Low-quality | Genome-fragment | 5.3 | AAI-based (high-confidence) | 0 |
| S14_vs_k141_83502\|\|full | 2202 | No | NA | 5 | 1 | 0 | Low-quality | Genome-fragment | 3.18 | AAI-based (medium-confidence) | 0 |
| S7_vs_42299\|\|full | 3665 | No | NA | 3 | 0 | 2 | Not-determined | Genome-fragment | NA | NA | 0 |
| S7_vs_42372\|\|full | 4123 | No | NA | 5 | 0 | 1 | Not-determined | Genome-fragment | NA | NA | 0 |
| S7_vs_42419\|\|full | 2996 | No | NA | 2 | 0 | 2 | Not-determined | Genome-fragment | NA | NA | 0 |
| S7_vs_42453\|\|full | 1133 | No | NA | 2 | 0 | 0 | Not-determined | Genome-fragment | NA | NA | 0 |
| S1_vs_k141_120522\|\|full | 2202 | No | NA | 2 | 0 | 0 | Low-quality | Genome-fragment | 4.57 | AAI-based (medium-confidence) | 0 |
| S1_vs_k141_95466\|\|full | 2202 | No | NA | 3 | 1 | 0 | Low-quality | Genome-fragment | 4.63 | AAI-based (high-confidence) | 0 |
| S3_extracted_k141_71895 | 2202 | No | NA | 2 | 0 | 0 | Low-quality | Genome-fragment | 5.09 | AAI-based (high-confidence) | 0 |
| S8_vs_k141_99267\|\|full | 2202 | Yes | 969 | 2 | 1 | 1 | Low-quality | Genome-fragment | 1.33 | AAI-based (medium-confidence) | 55.99 |
| S7_vs_42824\|\|full | 1341 | No | NA | 2 | 0 | 2 | Not-determined | Genome-fragment | NA | NA | 0 |
| S7_vs_42840\|\|full | 1127 | No | NA | 3 | 0 | 0 | Not-determined | Genome-fragment | NA | NA | 0 |
| S15_vs_36432\|\|full | 2203 | No | NA | 3 | 0 | 0 | Low-quality | Genome-fragment | 1.78 | AAI-based (high-confidence) | 0 |
| S7_vs_43330\|\|full | 1259 | No | NA | 2 | 0 | 1 | Not-determined | Genome-fragment | NA | NA | 0 |
| S7_vs_43385\|\|full | 1569 | No | NA | 2 | 0 | 1 | Not-determined | Genome-fragment | NA | NA | 0 |
| S7_vs_43515\|\|full | 1611 | No | NA | 2 | 0 | 1 | Not-determined | Genome-fragment | NA | NA | 0 |
| S7_vs_43529\|\|full | 34062 | No | NA | 54 | 17 | 0 | Medium-quality | Genome-fragment | 69.5 | AAI-based (high-confidence) | 0 |
| S7_vs_43538\|\|full | 1106 | No | NA | 3 | 0 | 0 | Not-determined | Genome-fragment | NA | NA | 0 |
| S7_vs_43545\|\|full | 14591 | No | NA | 15 | 0 | 6 | Not-determined | Genome-fragment | NA | NA | 0 |
| S8_vs_k141_27236\|\|full | 2203 | No | NA | 3 | 0 | 3 | Low-quality | Genome-fragment | 6.24 | AAI-based (medium-confidence) | 0 |
| S7_vs_43601\|\|full | 2194 | No | NA | 3 | 0 | 2 | Not-determined | Genome-fragment | NA | NA | 0 |
| S7_vs_43832\|\|full | 7929 | No | NA | 8 | 0 | 7 | Not-determined | Genome-fragment | NA | NA | 0 |
| S17_vs_k141_679\|\|full | 2204 | Yes | 1766 | 4 | 1 | 1 | Low-quality | Genome-fragment | 1.06 | HMM-based (lower-bound) | 19.87 |
| S7_vs_43876\|\|full | 4950 | No | NA | 5 | 0 | 0 | Not-determined | Genome-fragment | NA | NA | 0 |
| S9_extracted_k141_89204 | 2204 | No | NA | 3 | 0 | 3 | Low-quality | Genome-fragment | 1.45 | AAI-based (high-confidence) | 0 |
| S1_vs_k141_118193\|\|full | 2204 | No | NA | 10 | 0 | 0 | Low-quality | Genome-fragment | 5.61 | AAI-based (high-confidence) | 0 |
| S7_vs_44012\|\|full | 3099 | No | NA | 2 | 0 | 2 | Not-determined | Genome-fragment | NA | NA | 0 |
| S7_extracted_53474 | 2204 | No | NA | 3 | 0 | 0 | Low-quality | Genome-fragment | 2.9 | AAI-based (medium-confidence) | 0 |
| S7_vs_44079\|\|full | 1557 | No | NA | 5 | 0 | 0 | Not-determined | Genome-fragment | NA | NA | 0 |
| S7_vs_44106\|\|full | 3742 | No | NA | 3 | 0 | 1 | Not-determined | Genome-fragment | NA | NA | 0 |
| S7_vs_44191\|\|full | 2522 | No | NA | 3 | 0 | 2 | Not-determined | Genome-fragment | NA | NA | 0 |
| S7_vs_44265\|\|full | 2586 | No | NA | 3 | 0 | 1 | Not-determined | Genome-fragment | NA | NA | 0 |
| S7_vs_44301\|\|full | 1260 | No | NA | 3 | 0 | 1 | Not-determined | Genome-fragment | NA | NA | 0 |
| S8_extracted_k141_153132 | 2204 | No | NA | 6 | 0 | 0 | Low-quality | Genome-fragment | 2.24 | AAI-based (high-confidence) | 0 |
| S17_vs_k141_149970\|\|full | 2205 | No | NA | 4 | 0 | 0 | Low-quality | Genome-fragment | 5.44 | AAI-based (medium-confidence) | 0 |
| S7_vs_44423\|\|full | 3343 | No | NA | 3 | 0 | 3 | Not-determined | Genome-fragment | NA | NA | 0 |
| S11_vs_k141_52403\|\|full | 2205 | No | NA | 8 | 0 | 0 | Low-quality | Genome-fragment | 6.27 | AAI-based (medium-confidence) | 0 |
| S7_vs_44612\|\|full | 3263 | No | NA | 4 | 0 | 3 | Not-determined | Genome-fragment | NA | NA | 0 |
| S7_vs_44629\|\|full | 3886 | No | NA | 3 | 0 | 1 | Not-determined | Genome-fragment | NA | NA | 0 |
| S1_extracted_k141_113884 | 2205 | No | NA | 5 | 1 | 0 | Low-quality | Genome-fragment | 3.74 | AAI-based (high-confidence) | 0 |
| S16_extracted_83975 | 2205 | No | NA | 2 | 0 | 2 | Low-quality | Genome-fragment | 2.33 | AAI-based (medium-confidence) | 0 |
| S16_vs_29235\|\|full | 2205 | No | NA | 3 | 0 | 0 | Low-quality | Genome-fragment | 2.99 | AAI-based (high-confidence) | 0 |
| S12_extracted_k141_32360 | 2205 | No | NA | 2 | 0 | 0 | Low-quality | Genome-fragment | 3.44 | AAI-based (medium-confidence) | 0 |
| S8_vs_k141_33002\|\|full | 2205 | No | NA | 3 | 0 | 2 | Low-quality | Genome-fragment | 0.74 | AAI-based (medium-confidence) | 0 |
| S17_extracted_k141_29547 | 2206 | No | NA | 4 | 4 | 0 | Low-quality | Genome-fragment | 7.27 | AAI-based (high-confidence) | 0 |
| S5_vs_k141_109426\|\|lt2gene | 2206 | No | NA | 1 | 1 | 0 | Low-quality | Genome-fragment | 4.59 | AAI-based (medium-confidence) | 0 |
| S7_vs_45505\|\|full | 2340 | No | NA | 4 | 0 | 0 | Not-determined | Genome-fragment | NA | NA | 0 |
| S16_vs_89295\|\|full | 2206 | No | NA | 4 | 0 | 1 | Low-quality | Genome-fragment | 5.53 | AAI-based (high-confidence) | 0 |
| S16_vs_154803\|\|full | 2206 | No | NA | 3 | 0 | 0 | Low-quality | Genome-fragment | 3.94 | AAI-based (high-confidence) | 0 |
| S10_vs_15894\|\|full | 2206 | No | NA | 4 | 0 | 3 | Low-quality | Genome-fragment | 0.68 | AAI-based (medium-confidence) | 0 |
| S7_vs_45605\|\|full | 1287 | No | NA | 2 | 0 | 1 | Not-determined | Genome-fragment | NA | NA | 0 |
| S7_vs_45635\|\|full | 20669 | No | NA | 26 | 0 | 8 | Not-determined | Genome-fragment | NA | NA | 0 |
| S7_vs_45866\|\|full | 2079 | No | NA | 3 | 0 | 1 | Not-determined | Genome-fragment | NA | NA | 0 |
| S7_vs_45904\|\|full | 1976 | No | NA | 3 | 0 | 2 | Not-determined | Genome-fragment | NA | NA | 0 |
| S7_vs_45965\|\|full | 2431 | No | NA | 4 | 0 | 1 | Not-determined | Genome-fragment | NA | NA | 0 |
| S17_vs_k141_135810\|\|full | 2207 | No | NA | 2 | 0 | 0 | Low-quality | Genome-fragment | 2.7 | AAI-based (medium-confidence) | 0 |
| S13_vs_129760\|\|full | 2207 | No | NA | 9 | 0 | 0 | Low-quality | Genome-fragment | 4.2 | AAI-based (medium-confidence) | 0 |
| S7_vs_46051\|\|full | 2110 | No | NA | 2 | 0 | 2 | Not-determined | Genome-fragment | NA | NA | 0 |
| S7_vs_46064\|\|full | 2184 | No | NA | 3 | 0 | 0 | Not-determined | Genome-fragment | NA | NA | 0 |
| S5_extracted_k141_59557 | 2207 | No | NA | 3 | 2 | 0 | Low-quality | Genome-fragment | 5.43 | AAI-based (high-confidence) | 0 |
| S7_vs_46145\|\|full | 1664 | No | NA | 2 | 0 | 0 | Not-determined | Genome-fragment | NA | NA | 0 |
| S7_vs_46182\|\|full | 1907 | No | NA | 3 | 0 | 0 | Not-determined | Genome-fragment | NA | NA | 0 |
| S5_vs_k141_62605\|\|full | 2207 | No | NA | 2 | 1 | 0 | Low-quality | Genome-fragment | 4.96 | AAI-based (high-confidence) | 0 |
| S7_vs_14145\|\|full | 2207 | No | NA | 6 | 1 | 0 | Low-quality | Genome-fragment | 6.01 | AAI-based (high-confidence) | 0 |
| S7_vs_46270\|\|full | 1835 | No | NA | 4 | 0 | 0 | Not-determined | Genome-fragment | NA | NA | 0 |
| S12_extracted_k141_1695 | 2207 | No | NA | 4 | 0 | 0 | Low-quality | Genome-fragment | 3.16 | AAI-based (high-confidence) | 0 |
| S8_extracted_k141_3360 | 2207 | No | NA | 2 | 0 | 1 | Low-quality | Genome-fragment | 4.21 | AAI-based (medium-confidence) | 0 |
| S8_extracted_k141_39078 | 2207 | No | NA | 3 | 0 | 2 | Low-quality | Genome-fragment | 3.12 | AAI-based (medium-confidence) | 0 |
| S7_vs_46406\|\|full | 2626 | No | NA | 2 | 0 | 2 | Not-determined | Genome-fragment | NA | NA | 0 |
| S7_vs_46436\|\|full | 5205 | No | NA | 3 | 0 | 2 | Not-determined | Genome-fragment | NA | NA | 0 |
| S17_vs_k141_147350\|\|full | 2208 | Yes | 1665 | 6 | 1 | 1 | Low-quality | Genome-fragment | 2.06 | AAI-based (high-confidence) | 24.59 |
| S7_vs_46680\|\|full | 2680 | No | NA | 2 | 0 | 2 | Not-determined | Genome-fragment | NA | NA | 0 |
| S7_vs_46733\|\|full | 2928 | No | NA | 3 | 0 | 2 | Not-determined | Genome-fragment | NA | NA | 0 |
| S7_vs_46759\|\|full | 4871 | No | NA | 3 | 0 | 0 | Not-determined | Genome-fragment | NA | NA | 0 |
| S7_vs_46769\|\|full | 2750 | No | NA | 3 | 0 | 2 | Not-determined | Genome-fragment | NA | NA | 0 |
| S7_vs_46787\|\|full | 3051 | No | NA | 3 | 0 | 1 | Not-determined | Genome-fragment | NA | NA | 0 |
| S5_extracted_k141_107397 | 2208 | No | NA | 4 | 0 | 0 | Low-quality | Genome-fragment | 2.68 | AAI-based (medium-confidence) | 0 |
| S7_vs_46950\|\|full | 1460 | No | NA | 2 | 0 | 1 | Not-determined | Genome-fragment | NA | NA | 0 |
| S7_vs_46987\|\|full | 3457 | No | NA | 4 | 0 | 2 | Not-determined | Genome-fragment | NA | NA | 0 |
| S8_vs_k141_183075\|\|full | 2208 | No | NA | 3 | 0 | 1 | Low-quality | Genome-fragment | 5.21 | AAI-based (medium-confidence) | 0 |
| S7_vs_47042\|\|full | 2380 | No | NA | 3 | 0 | 0 | Not-determined | Genome-fragment | NA | NA | 0 |
| S7_vs_47050\|\|full | 39088 | Yes | 34528 | 64 | 17 | 4 | High-quality | High-quality | 99.51 | AAI-based (high-confidence) | 11.67 |
| S10_extracted_30295 | 2209 | No | NA | 4 | 0 | 1 | Low-quality | Genome-fragment | 3.78 | AAI-based (medium-confidence) | 0 |
| S7_vs_47183\|\|full | 2320 | No | NA | 3 | 0 | 2 | Not-determined | Genome-fragment | NA | NA | 0 |
| S10_extracted_91084 | 2209 | No | NA | 6 | 2 | 0 | Low-quality | Genome-fragment | 2.12 | AAI-based (medium-confidence) | 0 |
| S7_vs_47288\|\|full | 4753 | No | NA | 5 | 0 | 0 | Not-determined | Genome-fragment | NA | NA | 0 |
| S10_vs_72239\|\|full | 2209 | No | NA | 3 | 0 | 3 | Low-quality | Genome-fragment | 5.32 | AAI-based (medium-confidence) | 0 |
| S7_vs_87865\|\|full | 2210 | No | NA | 4 | 0 | 0 | Low-quality | Genome-fragment | 4.1 | AAI-based (medium-confidence) | 0 |
| S7_vs_47504\|\|full | 1493 | No | NA | 3 | 0 | 1 | Not-determined | Genome-fragment | NA | NA | 0 |
| S7_vs_119825\|\|full | 2210 | No | NA | 3 | 0 | 0 | Low-quality | Genome-fragment | 5.98 | AAI-based (medium-confidence) | 0 |
| S16_extracted_82180 | 2210 | No | NA | 2 | 0 | 2 | Low-quality | Genome-fragment | 1.44 | AAI-based (medium-confidence) | 0 |
| S16_vs_63717\|\|full | 2210 | No | NA | 5 | 0 | 0 | Low-quality | Genome-fragment | 5.33 | AAI-based (medium-confidence) | 0 |
| S7_vs_47656\|\|full | 6238 | No | NA | 7 | 0 | 5 | Not-determined | Genome-fragment | NA | NA | 0 |
| S7_vs_47673\|\|full | 2838 | No | NA | 3 | 0 | 2 | Not-determined | Genome-fragment | NA | NA | 0 |
| S12_extracted_k141_85881 | 2210 | No | NA | 6 | 3 | 0 | Low-quality | Genome-fragment | 3.68 | AAI-based (high-confidence) | 0 |
| S15_vs_44947\|\|full | 2210 | No | NA | 3 | 0 | 1 | Low-quality | Genome-fragment | 3.27 | AAI-based (medium-confidence) | 0 |
| S8_vs_k141_78906\|\|full | 2210 | No | NA | 4 | 1 | 0 | Low-quality | Genome-fragment | 3.66 | AAI-based (high-confidence) | 0 |
| S7_vs_47909\|\|full | 1717 | No | NA | 3 | 0 | 1 | Not-determined | Genome-fragment | NA | NA | 0 |
| S7_vs_47922\|\|full | 1964 | No | NA | 3 | 0 | 0 | Not-determined | Genome-fragment | NA | NA | 0 |
| S9_vs_k141_27194\|\|full | 2211 | No | NA | 3 | 0 | 0 | Low-quality | Genome-fragment | 4.44 | AAI-based (high-confidence) | 0 |
| S3_extracted_k141_12363 | 2211 | No | NA | 5 | 0 | 0 | Low-quality | Genome-fragment | 1.27 | AAI-based (high-confidence) | 0 |
| S7_vs_48010\|\|full | 1196 | No | NA | 2 | 0 | 1 | Not-determined | Genome-fragment | NA | NA | 0 |
| S3_vs_k141_64527\|\|full | 2211 | No | NA | 4 | 1 | 0 | Low-quality | Genome-fragment | 3.8 | AAI-based (high-confidence) | 0 |
| S5_extracted_k141_51368 | 2211 | No | NA | 5 | 1 | 0 | Low-quality | Genome-fragment | 5.06 | AAI-based (high-confidence) | 0 |
| S10_extracted_90685 | 2211 | No | NA | 4 | 0 | 1 | Low-quality | Genome-fragment | 3.75 | AAI-based (medium-confidence) | 0 |
| S12_vs_k141_108065\|\|full | 2211 | No | NA | 4 | 2 | 0 | Low-quality | Genome-fragment | 4.53 | AAI-based (high-confidence) | 0 |
| S7_vs_48174\|\|full | 2424 | No | NA | 2 | 0 | 2 | Not-determined | Genome-fragment | NA | NA | 0 |
| S4_extracted_7858 | 2211 | No | NA | 6 | 3 | 0 | Low-quality | Genome-fragment | 5.64 | AAI-based (high-confidence) | 0 |
| S7_vs_48328\|\|full | 4341 | No | NA | 5 | 0 | 3 | Not-determined | Genome-fragment | NA | NA | 0 |
| S7_vs_48334\|\|full | 1495 | No | NA | 3 | 0 | 0 | Not-determined | Genome-fragment | NA | NA | 0 |
| S7_vs_48456\|\|full | 1758 | No | NA | 3 | 0 | 0 | Not-determined | Genome-fragment | NA | NA | 0 |
| S7_vs_48493\|\|full | 2712 | No | NA | 4 | 0 | 3 | Not-determined | Genome-fragment | NA | NA | 0 |
| S7_vs_48523\|\|full | 44450 | No | NA | 81 | 24 | 0 | High-quality | High-quality | 100 | AAI-based (high-confidence) | 0 |
| S7_vs_48671\|\|full | 1189 | No | NA | 2 | 0 | 0 | Not-determined | Genome-fragment | NA | NA | 0 |
| S7_vs_48678\|\|full | 2359 | No | NA | 3 | 0 | 0 | Not-determined | Genome-fragment | NA | NA | 0 |
| S17_vs_k141_9575\|\|full | 2212 | No | NA | 4 | 2 | 0 | Low-quality | Genome-fragment | 6.95 | AAI-based (high-confidence) | 0 |
| S7_vs_48708\|\|full | 68692 | No | NA | 98 | 25 | 4 | High-quality | High-quality | 100 | AAI-based (high-confidence) | 0 |
| S13_extracted_133535 | 2212 | No | NA | 3 | 1 | 0 | Low-quality | Genome-fragment | 3.64 | AAI-based (medium-confidence) | 0 |
| S7_vs_48832\|\|full | 37951 | No | NA | 50 | 28 | 0 | High-quality | High-quality | 91.91 | AAI-based (high-confidence) | 0 |
| S5_vs_k141_67527\|\|full | 2212 | No | NA | 4 | 0 | 0 | Low-quality | Genome-fragment | 4.46 | AAI-based (high-confidence) | 0 |
| S5_vs_k141_19336\|\|full | 2212 | No | NA | 3 | 0 | 0 | Low-quality | Genome-fragment | 1.98 | AAI-based (medium-confidence) | 0 |
| S7_vs_48963\|\|full | 1356 | No | NA | 3 | 0 | 0 | Not-determined | Genome-fragment | NA | NA | 0 |
| S5_vs_k141_98907\|\|lt2gene | 2212 | No | NA | 2 | 2 | 0 | Low-quality | Genome-fragment | 6.43 | AAI-based (high-confidence) | 0 |
| S7_vs_49033\|\|full | 1370 | No | NA | 2 | 0 | 0 | Not-determined | Genome-fragment | NA | NA | 0 |
| S10_extracted_101942 | 2212 | No | NA | 2 | 1 | 0 | Low-quality | Genome-fragment | 5.26 | HMM-based (lower-bound) | 0 |
| S7_vs_49195\|\|full | 1432 | No | NA | 3 | 0 | 2 | Not-determined | Genome-fragment | NA | NA | 0 |
| S7_vs_49278\|\|full | 5581 | No | NA | 5 | 0 | 3 | Not-determined | Genome-fragment | NA | NA | 0 |
| S7_vs_49426\|\|full | 57556 | Yes | 34483 | 75 | 13 | 22 | High-quality | High-quality | 98.72 | AAI-based (high-confidence) | 40.09 |
| S7_vs_49440\|\|full | 1773 | No | NA | 2 | 0 | 1 | Not-determined | Genome-fragment | NA | NA | 0 |
| S12_vs_k141_181512\|\|full | 2212 | No | NA | 4 | 0 | 0 | Low-quality | Genome-fragment | 4.91 | AAI-based (high-confidence) | 0 |
| S11_vs_k141_11294\|\|full | 2213 | No | NA | 3 | 0 | 1 | Low-quality | Genome-fragment | 4.77 | AAI-based (high-confidence) | 0 |
| S3_vs_k141_124828\|\|full | 2213 | No | NA | 4 | 2 | 0 | Low-quality | Genome-fragment | 3.93 | AAI-based (high-confidence) | 0 |
| S7_vs_61844\|\|lt2gene | 2213 | No | NA | 1 | 1 | 0 | Low-quality | Genome-fragment | 4.56 | AAI-based (medium-confidence) | 0 |
| S7_vs_49694\|\|full | 1995 | No | NA | 3 | 0 | 0 | Not-determined | Genome-fragment | NA | NA | 0 |
| S16_extracted_49221 | 2213 | No | NA | 4 | 2 | 0 | Low-quality | Genome-fragment | 5.98 | AAI-based (high-confidence) | 0 |
| S7_vs_50036\|\|full | 1740 | No | NA | 3 | 0 | 0 | Not-determined | Genome-fragment | NA | NA | 0 |
| S11_vs_k141_33623\|\|full | 2214 | No | NA | 2 | 1 | 0 | Low-quality | Genome-fragment | 5.78 | AAI-based (high-confidence) | 0 |
| S7_vs_50194\|\|full | 38824 | No | NA | 52 | 7 | 2 | High-quality | High-quality | 98.16 | AAI-based (high-confidence) | 0 |
| S5_vs_k141_144916\|\|full | 2214 | No | NA | 5 | 1 | 0 | Low-quality | Genome-fragment | 5.65 | AAI-based (high-confidence) | 0 |
| S7_vs_50438\|\|full | 2467 | No | NA | 5 | 0 | 0 | Not-determined | Genome-fragment | NA | NA | 0 |
| S7_vs_55723\|\|full | 2214 | No | NA | 7 | 0 | 1 | Low-quality | Genome-fragment | 4.8 | AAI-based (medium-confidence) | 0 |
| S7_vs_132305\|\|full | 2214 | No | NA | 3 | 1 | 0 | Low-quality | Genome-fragment | 6.03 | AAI-based (medium-confidence) | 0 |
| S7_vs_50489\|\|full | 1482 | No | NA | 3 | 0 | 0 | Not-determined | Genome-fragment | NA | NA | 0 |
| S7_vs_50588\|\|full | 1276 | No | NA | 2 | 0 | 0 | Not-determined | Genome-fragment | NA | NA | 0 |
| S7_vs_50836\|\|full | 1505 | No | NA | 3 | 0 | 0 | Not-determined | Genome-fragment | NA | NA | 0 |
| S8_vs_k141_94622\|\|full | 2214 | No | NA | 3 | 0 | 0 | Low-quality | Genome-fragment | 1.95 | AAI-based (medium-confidence) | 0 |
| S7_vs_50875\|\|full | 3761 | No | NA | 2 | 0 | 0 | Not-determined | Genome-fragment | NA | NA | 0 |
| S13_extracted_140926 | 2215 | No | NA | 3 | 0 | 2 | Low-quality | Genome-fragment | 1.47 | AAI-based (high-confidence) | 0 |
| S7_vs_50917\|\|full | 2271 | No | NA | 3 | 0 | 0 | Not-determined | Genome-fragment | NA | NA | 0 |
| S14_extracted_k141_116714 | 2215 | No | NA | 3 | 0 | 0 | Low-quality | Genome-fragment | 3.62 | AAI-based (high-confidence) | 0 |
| S12_extracted_k141_28921 | 2215 | No | NA | 1 | 0 | 0 | Low-quality | Genome-fragment | 2.41 | AAI-based (medium-confidence) | 0 |
| S7_vs_51000\|\|full | 4884 | No | NA | 6 | 0 | 6 | Not-determined | Genome-fragment | NA | NA | 0 |
| S2_extracted_k141_13731 | 2215 | No | NA | 1 | 0 | 0 | Low-quality | Genome-fragment | 2.31 | AAI-based (high-confidence) | 0 |
| S7_vs_51190\|\|full | 1024 | No | NA | 2 | 0 | 1 | Not-determined | Genome-fragment | NA | NA | 0 |
| S9_vs_k141_16881\|\|lt2gene | 2216 | No | NA | 1 | 1 | 0 | Low-quality | Genome-fragment | 3.73 | HMM-based (lower-bound) | 0 |
| S7_vs_51332\|\|full | 47629 | No | NA | 45 | 2 | 19 | Medium-quality | Genome-fragment | 51.81 | HMM-based (lower-bound) | 0 |
| S7_vs_51365\|\|full | 2666 | No | NA | 4 | 0 | 0 | Not-determined | Genome-fragment | NA | NA | 0 |
| S7_vs_51393\|\|full | 1451 | No | NA | 2 | 0 | 2 | Not-determined | Genome-fragment | NA | NA | 0 |
| S11_extracted_k141_65798 | 2216 | No | NA | 1 | 1 | 0 | Low-quality | Genome-fragment | 4.19 | HMM-based (lower-bound) | 0 |
| S11_vs_k141_70253\|\|full | 2216 | No | NA | 4 | 0 | 1 | Low-quality | Genome-fragment | 1.19 | AAI-based (medium-confidence) | 0 |
| S7_vs_51603\|\|full | 1492 | No | NA | 3 | 0 | 0 | Not-determined | Genome-fragment | NA | NA | 0 |
| S7_vs_51761\|\|full | 1096 | No | NA | 2 | 0 | 1 | Not-determined | Genome-fragment | NA | NA | 0 |
| S13_vs_17708\|\|full | 2216 | No | NA | 3 | 0 | 2 | Low-quality | Genome-fragment | 1.83 | AAI-based (medium-confidence) | 0 |
| S7_vs_23348\|\|full | 2216 | No | NA | 3 | 2 | 0 | Low-quality | Genome-fragment | 3.64 | AAI-based (high-confidence) | 0 |
| S8_extracted_k141_145300 | 2216 | No | NA | 1 | 0 | 0 | Low-quality | Genome-fragment | 3.46 | AAI-based (high-confidence) | 0 |
| S1_extracted_k141_159546 | 2217 | No | NA | 2 | 0 | 1 | Low-quality | Genome-fragment | 4.11 | AAI-based (medium-confidence) | 0 |
| S11_extracted_k141_29077 | 2218 | No | NA | 3 | 1 | 0 | Low-quality | Genome-fragment | 4.6 | AAI-based (high-confidence) | 0 |
| S14_extracted_k141_96715 | 2218 | No | NA | 5 | 4 | 0 | Low-quality | Genome-fragment | 7.19 | AAI-based (high-confidence) | 0 |
| S1_extracted_k141_9745 | 2218 | No | NA | 7 | 2 | 0 | Low-quality | Genome-fragment | 4.62 | AAI-based (high-confidence) | 0 |
| S3_vs_k141_100531\|\|full | 2218 | No | NA | 4 | 3 | 0 | Low-quality | Genome-fragment | 3.8 | AAI-based (high-confidence) | 0 |
| S3_vs_k141_31295\|\|full | 2218 | No | NA | 2 | 0 | 2 | Low-quality | Genome-fragment | 2.64 | AAI-based (medium-confidence) | 0 |
| S7_vs_52634\|\|full | 3908 | No | NA | 4 | 0 | 3 | Not-determined | Genome-fragment | NA | NA | 0 |
| S7_vs_71392\|\|full | 2218 | No | NA | 3 | 0 | 1 | Low-quality | Genome-fragment | 1.56 | AAI-based (medium-confidence) | 0 |
| S4_extracted_30455 | 2218 | No | NA | 5 | 3 | 0 | Low-quality | Genome-fragment | 7.22 | AAI-based (high-confidence) | 0 |
| S7_vs_52995\|\|full | 4144 | No | NA | 4 | 0 | 4 | Not-determined | Genome-fragment | NA | NA | 0 |
| S14_vs_k141_29434\|\|full | 2219 | No | NA | 7 | 6 | 0 | Low-quality | Genome-fragment | 7.32 | AAI-based (high-confidence) | 0 |
| S7_vs_53065\|\|full | 1295 | No | NA | 2 | 0 | 2 | Not-determined | Genome-fragment | NA | NA | 0 |
| S7_vs_53134\|\|full | 1331 | No | NA | 2 | 0 | 0 | Not-determined | Genome-fragment | NA | NA | 0 |
| S1_extracted_k141_47543 | 2219 | No | NA | 1 | 0 | 1 | Low-quality | Genome-fragment | 1.12 | AAI-based (medium-confidence) | 0 |
| S5_vs_k141_89280\|\|full | 2219 | No | NA | 2 | 0 | 1 | Low-quality | Genome-fragment | 3.49 | AAI-based (high-confidence) | 0 |
| S16_vs_14430\|\|full | 2219 | No | NA | 4 | 0 | 1 | Low-quality | Genome-fragment | 3.12 | AAI-based (high-confidence) | 0 |
| S7_vs_53456\|\|full | 1662 | No | NA | 5 | 0 | 1 | Not-determined | Genome-fragment | NA | NA | 0 |
| S7_vs_53591\|\|full | 2175 | No | NA | 9 | 0 | 0 | Not-determined | Genome-fragment | NA | NA | 0 |
| S7_vs_53604\|\|full | 1719 | No | NA | 3 | 0 | 0 | Not-determined | Genome-fragment | NA | NA | 0 |

| | | | | | | | | | | | |
|---|---|---|---|---|---|---|---|---|---|---|---|
| S7_vs_53609\|\|full | 1286 | No | NA | 2 | 0 | 0 | Not-determined | Genome-fragment | NA | NA | 0 |
| S7_vs_53921\|\|full | 2501 | No | NA | 2 | 0 | 1 | Not-determined | Genome-fragment | NA | NA | 0 |
| S7_vs_53926\|\|full | 1718 | No | NA | 3 | 0 | 1 | Not-determined | Genome-fragment | NA | NA | 0 |
| S8_vs_k141_207749\|\|full | 2219 | No | NA | 3 | 0 | 2 | Low-quality | Genome-fragment | 0.81 | AAI-based (medium-confidence) | 0 |
| S5_vs_k141_117897\|\|full | 2220 | No | NA | 3 | 1 | 0 | Low-quality | Genome-fragment | 4.05 | AAI-based (medium-confidence) | 0 |
| S8_vs_k141_126596\|\|lt2gene | 2220 | No | NA | 1 | 1 | 0 | Low-quality | Genome-fragment | 6.07 | AAI-based (high-confidence) | 0 |
| S3_vs_k141_84416\|\|full | 2221 | No | NA | 2 | 2 | 0 | Low-quality | Genome-fragment | 2.74 | AAI-based (medium-confidence) | 0 |
| S7_vs_54146\|\|full | 5333 | No | NA | 7 | 0 | 4 | Not-determined | Genome-fragment | NA | NA | 0 |
| S7_vs_54151\|\|full | 2625 | No | NA | 6 | 0 | 1 | Not-determined | Genome-fragment | NA | NA | 0 |
| S7_vs_54249\|\|full | 4027 | No | NA | 4 | 0 | 3 | Not-determined | Genome-fragment | NA | NA | 0 |
| S5_vs_k141_7931\|\|full | 2221 | No | NA | 4 | 1 | 0 | Low-quality | Genome-fragment | 5.32 | AAI-based (high-confidence) | 0 |
| S7_vs_54507\|\|full | 1604 | No | NA | 2 | 0 | 0 | Not-determined | Genome-fragment | NA | NA | 0 |
| S16_vs_28039\|\|full | 2221 | No | NA | 3 | 0 | 1 | Low-quality | Genome-fragment | 2.09 | AAI-based (high-confidence) | 0 |
| S16_vs_138032\|\|full | 2221 | No | NA | 4 | 0 | 0 | Low-quality | Genome-fragment | 4.27 | AAI-based (medium-confidence) | 0 |
| S7_vs_54738\|\|full | 3099 | No | NA | 7 | 0 | 2 | Not-determined | Genome-fragment | NA | NA | 0 |
| S10_vs_67070\|\|full | 2221 | No | NA | 3 | 0 | 2 | Low-quality | Genome-fragment | 3.42 | AAI-based (medium-confidence) | 0 |
| S5_extracted_k141_131538 | 2222 | No | NA | 3 | 0 | 3 | Low-quality | Genome-fragment | 1.44 | AAI-based (high-confidence) | 0 |
| S16_vs_72982\|\|full | 2222 | No | NA | 4 | 3 | 0 | Low-quality | Genome-fragment | 5.28 | AAI-based (high-confidence) | 0 |
| S10_vs_58406\|\|full | 2222 | No | NA | 4 | 0 | 0 | Low-quality | Genome-fragment | 5.93 | AAI-based (high-confidence) | 0 |
| S7_vs_54974\|\|full | 1558 | No | NA | 3 | 0 | 1 | Not-determined | Genome-fragment | NA | NA | 0 |
| S7_vs_55031\|\|full | 36725 | No | NA | 54 | 11 | 0 | Medium-quality | Genome-fragment | 79.12 | AAI-based (high-confidence) | 0 |
| S7_vs_55097\|\|full | 1789 | No | NA | 3 | 0 | 2 | Not-determined | Genome-fragment | NA | NA | 0 |
| S17_vs_k141_24010\|\|full | 2223 | No | NA | 4 | 0 | 2 | Low-quality | Genome-fragment | 0.69 | AAI-based (medium-confidence) | 0 |
| S9_vs_k141_63215\|\|full | 2223 | No | NA | 6 | 1 | 0 | Low-quality | Genome-fragment | 1.13 | HMM-based (lower-bound) | 0 |
| S3_vs_k141_81536\|\|full | 2223 | No | NA | 3 | 1 | 0 | Low-quality | Genome-fragment | 1.09 | AAI-based (high-confidence) | 0 |
| S7_vs_66486\|\|full | 2223 | No | NA | 3 | 0 | 2 | Low-quality | Genome-fragment | 1.81 | AAI-based (medium-confidence) | 0 |
| S7_vs_55409\|\|full | 31512 | No | NA | 46 | 12 | 1 | Medium-quality | Genome-fragment | 88.89 | AAI-based (medium-confidence) | 0 |
| S7_vs_114775\|\|full | 2223 | No | NA | 3 | 0 | 0 | Low-quality | Genome-fragment | 5.29 | AAI-based (high-confidence) | 0 |
| S7_vs_55513\|\|full | 1159 | No | NA | 3 | 0 | 0 | Not-determined | Genome-fragment | NA | NA | 0 |
| S7_vs_55526\|\|full | 28778 | No | NA | 38 | 13 | 0 | Medium-quality | Genome-fragment | 74.98 | AAI-based (high-confidence) | 0 |
| S7_vs_55530\|\|full | 1934 | No | NA | 3 | 0 | 3 | Not-determined | Genome-fragment | NA | NA | 0 |
| S7_vs_55565\|\|full | 3994 | No | NA | 3 | 0 | 0 | Not-determined | Genome-fragment | NA | NA | 0 |
| S7_vs_55624\|\|full | 2510 | No | NA | 3 | 0 | 3 | Not-determined | Genome-fragment | NA | NA | 0 |
| S7_vs_55647\|\|full | 1008 | No | NA | 2 | 0 | 1 | Not-determined | Genome-fragment | NA | NA | 0 |
| S7_vs_55685\|\|full | 5303 | No | NA | 9 | 0 | 2 | Not-determined | Genome-fragment | NA | NA | 0 |
| S12_vs_k141_98669\|\|full | 2223 | No | NA | 3 | 0 | 2 | Low-quality | Genome-fragment | 2.66 | AAI-based (medium-confidence) | 0 |
| S12_vs_k141_38294\|\|full | 2223 | No | NA | 5 | 5 | 0 | Low-quality | Genome-fragment | 6.14 | AAI-based (high-confidence) | 0 |
| S8_vs_k141_175680\|\|full | 2223 | No | NA | 3 | 1 | 0 | Low-quality | Genome-fragment | 5.16 | AAI-based (medium-confidence) | 0 |
| S9_vs_k141_74858\|\|full | 2224 | No | NA | 4 | 0 | 0 | Low-quality | Genome-fragment | 2.87 | AAI-based (high-confidence) | 0 |
| S7_vs_55926\|\|full | 11566 | No | NA | 12 | 0 | 1 | Not-determined | Genome-fragment | NA | NA | 0 |
| S7_vs_56176\|\|full | 4908 | No | NA | 3 | 0 | 3 | Not-determined | Genome-fragment | NA | NA | 0 |
| S7_vs_56260\|\|full | 2125 | No | NA | 3 | 0 | 3 | Not-determined | Genome-fragment | NA | NA | 0 |
| S13_vs_76716\|\|full | 2224 | No | NA | 3 | 0 | 1 | Low-quality | Genome-fragment | 3.35 | AAI-based (medium-confidence) | 0 |
| S3_extracted_k141_93180 | 2224 | No | NA | 5 | 1 | 0 | Low-quality | Genome-fragment | 4.4 | AAI-based (high-confidence) | 0 |
| S7_vs_56322\|\|full | 2286 | No | NA | 3 | 0 | 2 | Not-determined | Genome-fragment | NA | NA | 0 |
| S7_vs_56335\|\|full | 2848 | No | NA | 4 | 0 | 0 | Not-determined | Genome-fragment | NA | NA | 0 |
| S7_vs_56529\|\|full | 32265 | No | NA | 45 | 12 | 1 | High-quality | High-quality | 100 | AAI-based (high-confidence) | 0 |
| S14_vs_k141_114781\|\|full | 2225 | No | NA | 4 | 0 | 0 | Low-quality | Genome-fragment | 14.98 | AAI-based (high-confidence) | 0 |
| S5_vs_k141_142708\|\|full | 2225 | No | NA | 3 | 2 | 0 | Low-quality | Genome-fragment | 6.36 | AAI-based (high-confidence) | 0 |
| S7_vs_56668\|\|full | 2455 | No | NA | 5 | 0 | 0 | Not-determined | Genome-fragment | NA | NA | 0 |
| S16_extracted_119190 | 2225 | No | NA | 1 | 0 | 1 | Low-quality | Genome-fragment | 5.35 | AAI-based (high-confidence) | 0 |
| S16_vs_98148\|\|full | 2225 | No | NA | 4 | 0 | 0 | Low-quality | Genome-fragment | 1.9 | AAI-based (high-confidence) | 0 |
| S12_vs_k141_168161\|\|full | 2225 | No | NA | 5 | 3 | 0 | Low-quality | Genome-fragment | 5.36 | AAI-based (medium-confidence) | 0 |
| S11_vs_k141_87692\|\|full | 2226 | No | NA | 3 | 0 | 0 | Low-quality | Genome-fragment | 3.05 | AAI-based (medium-confidence) | 0 |
| S7_vs_57152\|\|full | 1555 | No | NA | 3 | 0 | 1 | Not-determined | Genome-fragment | NA | NA | 0 |
| S14_extracted_k141_9242 | 2226 | No | NA | 6 | 6 | 0 | Low-quality | Genome-fragment | 7.32 | AAI-based (high-confidence) | 0 |
| S14_vs_k141_40734\|\|full | 2226 | No | NA | 4 | 0 | 0 | Low-quality | Genome-fragment | 4.03 | AAI-based (high-confidence) | 0 |
| S14_vs_k141_67440\|\|lt2gene | 2226 | No | NA | 2 | 2 | 0 | Low-quality | Genome-fragment | 3.44 | HMM-based (lower-bound) | 0 |
| S7_vs_57428\|\|full | 1105 | No | NA | 2 | 0 | 0 | Not-determined | Genome-fragment | NA | NA | 0 |
| S16_vs_58887\|\|full | 2226 | No | NA | 3 | 0 | 0 | Low-quality | Genome-fragment | 2.44 | AAI-based (high-confidence) | 0 |
| S10_vs_108991\|\|full | 2226 | No | NA | 3 | 3 | 0 | Low-quality | Genome-fragment | 5.21 | AAI-based (medium-confidence) | 0 |
| S7_vs_57702\|\|full | 1475 | No | NA | 3 | 0 | 1 | Not-determined | Genome-fragment | NA | NA | 0 |
| S7_vs_57715\|\|full | 4306 | No | NA | 3 | 0 | 2 | Not-determined | Genome-fragment | NA | NA | 0 |
| S7_vs_57731\|\|full | 4218 | No | NA | 5 | 0 | 3 | Not-determined | Genome-fragment | NA | NA | 0 |
| S11_vs_k141_71801\|\|full | 2227 | No | NA | 6 | 0 | 0 | Low-quality | Genome-fragment | 4.92 | AAI-based (high-confidence) | 0 |
| S7_vs_57866\|\|full | 1260 | No | NA | 3 | 0 | 1 | Not-determined | Genome-fragment | NA | NA | 0 |
| S14_vs_k141_81967\|\|full | 2227 | No | NA | 3 | 0 | 1 | Low-quality | Genome-fragment | 0.63 | AAI-based (high-confidence) | 0 |
| S5_vs_k141_136792\|\|full | 2227 | No | NA | 2 | 0 | 1 | Low-quality | Genome-fragment | 4.09 | AAI-based (medium-confidence) | 0 |
| S7_vs_11324\|\|full | 2227 | No | NA | 6 | 1 | 0 | Low-quality | Genome-fragment | 4.96 | AAI-based (high-confidence) | 0 |
| S16_extracted_164974 | 2227 | Yes | 1559 | 5 | 1 | 1 | Low-quality | Genome-fragment | 4.04 | AAI-based (high-confidence) | 30 |
| S7_vs_58335\|\|full | 1462 | No | NA | 2 | 0 | 1 | Not-determined | Genome-fragment | NA | NA | 0 |
| S4_extracted_14197 | 2227 | No | NA | 2 | 0 | 2 | Low-quality | Genome-fragment | 0.97 | AAI-based (high-confidence) | 0 |
| S7_vs_58462\|\|full | 1145 | No | NA | 3 | 0 | 0 | Not-determined | Genome-fragment | NA | NA | 0 |
| S7_vs_58487\|\|full | 2906 | No | NA | 4 | 0 | 4 | Not-determined | Genome-fragment | NA | NA | 0 |
| S6_vs_k141_7739\|\|full | 2227 | No | NA | 4 | 2 | 0 | Low-quality | Genome-fragment | 3.5 | AAI-based (high-confidence) | 0 |
| S6_vs_k141_38816\|\|full | 2227 | No | NA | 5 | 0 | 0 | Low-quality | Genome-fragment | 2.97 | AAI-based (medium-confidence) | 0 |
| S7_vs_58771\|\|full | 1534 | No | NA | 3 | 0 | 1 | Not-determined | Genome-fragment | NA | NA | 0 |
| S9_vs_k141_6852\|\|full | 2228 | No | NA | 7 | 2 | 0 | Low-quality | Genome-fragment | 1.92 | HMM-based (lower-bound) | 0 |
| S7_vs_58881\|\|full | 1563 | No | NA | 3 | 0 | 2 | Not-determined | Genome-fragment | NA | NA | 0 |
| S13_vs_3598\|\|full | 2228 | No | NA | 3 | 2 | 0 | Low-quality | Genome-fragment | 2.03 | HMM-based (lower-bound) | 0 |
| S3_vs_k141_99237\|\|full | 2228 | No | NA | 3 | 0 | 1 | Low-quality | Genome-fragment | 1.49 | AAI-based (high-confidence) | 0 |
| S7_vs_75220\|\|full | 2228 | No | NA | 4 | 0 | 0 | Low-quality | Genome-fragment | 6.14 | AAI-based (medium-confidence) | 0 |
| S7_vs_59372\|\|full | 3418 | No | NA | 2 | 0 | 2 | Not-determined | Genome-fragment | NA | NA | 0 |
| S7_vs_116044\|\|full | 2228 | No | NA | 3 | 1 | 0 | Low-quality | Genome-fragment | 4.38 | AAI-based (high-confidence) | 0 |
| S10_vs_107213\|\|full | 2228 | No | NA | 3 | 0 | 0 | Low-quality | Genome-fragment | 3.46 | AAI-based (high-confidence) | 0 |
| S7_vs_59465\|\|full | 37931 | No | NA | 58 | 25 | 2 | Medium-quality | Genome-fragment | 82.37 | AAI-based (medium-confidence) | 0 |
| S12_vs_k141_45938\|\|full | 2228 | No | NA | 3 | 0 | 0 | Low-quality | Genome-fragment | 3.92 | AAI-based (medium-confidence) | 0 |
| S7_vs_59708\|\|full | 1377 | No | NA | 3 | 0 | 2 | Not-determined | Genome-fragment | NA | NA | 0 |
| S17_vs_k141_76435\|\|full | 2229 | No | NA | 3 | 0 | 1 | Low-quality | Genome-fragment | 4.01 | AAI-based (medium-confidence) | 0 |
| S9_vs_k141_18337\|\|full | 2229 | No | NA | 6 | 0 | 1 | Low-quality | Genome-fragment | 5.35 | AAI-based (medium-confidence) | 0 |
| S5_vs_k141_124358\|\|full | 2229 | No | NA | 6 | 1 | 0 | Low-quality | Genome-fragment | 4.73 | AAI-based (medium-confidence) | 0 |
| S7_vs_59960\|\|full | 1176 | No | NA | 2 | 0 | 2 | Not-determined | Genome-fragment | NA | NA | 0 |
| S7_vs_60023\|\|full | 9387 | No | NA | 9 | 0 | 4 | Not-determined | Genome-fragment | NA | NA | 0 |
| S10_vs_114073\|\|full | 2229 | No | NA | 2 | 0 | 0 | Low-quality | Genome-fragment | 4.75 | AAI-based (high-confidence) | 0 |
| S7_vs_60196\|\|full | 1719 | No | NA | 3 | 0 | 0 | Not-determined | Genome-fragment | NA | NA | 0 |
| S7_vs_60258\|\|full | 1429 | No | NA | 3 | 0 | 0 | Not-determined | Genome-fragment | NA | NA | 0 |
| S7_vs_60320\|\|full | 1843 | No | NA | 4 | 0 | 3 | Not-determined | Genome-fragment | NA | NA | 0 |
| S8_vs_k141_118244\|\|full | 2229 | No | NA | 3 | 0 | 0 | Low-quality | Genome-fragment | 5.7 | AAI-based (high-confidence) | 0 |
| S7_vs_60358\|\|full | 2944 | No | NA | 3 | 0 | 3 | Not-determined | Genome-fragment | NA | NA | 0 |
| S7_vs_60386\|\|full | 1636 | No | NA | 3 | 0 | 1 | Not-determined | Genome-fragment | NA | NA | 0 |
| S9_vs_k141_30929\|\|full | 2230 | No | NA | 4 | 1 | 0 | Low-quality | Genome-fragment | 5.36 | AAI-based (medium-confidence) | 0 |
| S11_vs_k141_48112\|\|full | 2230 | No | NA | 3 | 0 | 0 | Low-quality | Genome-fragment | 4.67 | AAI-based (medium-confidence) | 0 |
| S7_vs_60764\|\|full | 1064 | No | NA | 3 | 0 | 0 | Not-determined | Genome-fragment | NA | NA | 0 |
| S7_vs_60779\|\|full | 1247 | No | NA | 2 | 0 | 1 | Not-determined | Genome-fragment | NA | NA | 0 |
| S5_vs_k141_36168\|\|full | 2230 | No | NA | 2 | 0 | 0 | Low-quality | Genome-fragment | 4.28 | AAI-based (high-confidence) | 0 |
| S7_vs_60802\|\|full | 3039 | No | NA | 3 | 0 | 2 | Not-determined | Genome-fragment | NA | NA | 0 |
| S7_vs_39422\|\|full | 2230 | No | NA | 6 | 0 | 0 | Low-quality | Genome-fragment | 2.94 | AAI-based (medium-confidence) | 0 |
| S7_vs_58906\|\|full | 2230 | No | NA | 5 | 0 | 0 | Low-quality | Genome-fragment | 4.85 | AAI-based (high-confidence) | 0 |
| S12_vs_k141_144541\|\|full | 2230 | No | NA | 3 | 3 | 0 | Low-quality | Genome-fragment | 5.75 | AAI-based (high-confidence) | 0 |
| S7_vs_60904\|\|full | 4615 | No | NA | 4 | 0 | 2 | Not-determined | Genome-fragment | NA | NA | 0 |
| S7_vs_60947\|\|full | 3104 | No | NA | 5 | 0 | 1 | Not-determined | Genome-fragment | NA | NA | 0 |
| S14_vs_k141_93104\|\|full | 2231 | No | NA | 2 | 0 | 0 | Low-quality | Genome-fragment | 1.01 | AAI-based (medium-confidence) | 0 |
| S7_vs_4451\|\|full | 2231 | No | NA | 8 | 0 | 0 | Low-quality | Genome-fragment | 4.89 | AAI-based (high-confidence) | 0 |
| S7_vs_61147\|\|full | 2915 | No | NA | 5 | 0 | 0 | Not-determined | Genome-fragment | NA | NA | 0 |
| S7_vs_61162\|\|full | 4344 | No | NA | 3 | 0 | 0 | Not-determined | Genome-fragment | NA | NA | 0 |
| S12_extracted_k141_173600 | 2231 | No | NA | 3 | 0 | 0 | Low-quality | Genome-fragment | 1.75 | AAI-based (high-confidence) | 0 |
| S7_vs_61262\|\|full | 1381 | No | NA | 2 | 0 | 0 | Not-determined | Genome-fragment | NA | NA | 0 |
| S12_vs_k141_8422\|\|full | 2231 | No | NA | 2 | 0 | 1 | Low-quality | Genome-fragment | 1.41 | AAI-based (medium-confidence) | 0 |
| S7_vs_61296\|\|full | 7491 | No | NA | 8 | 0 | 4 | Not-determined | Genome-fragment | NA | NA | 0 |
| S8_vs_k141_167876\|\|full | 2231 | No | NA | 3 | 1 | 0 | Low-quality | Genome-fragment | 3.93 | AAI-based (medium-confidence) | 0 |
| S9_vs_k141_86612\|\|lt2gene | 2232 | No | NA | 2 | 1 | 0 | Low-quality | Genome-fragment | 5.37 | AAI-based (high-confidence) | 0 |
| S7_vs_40736\|\|full | 2232 | No | NA | 4 | 0 | 0 | Low-quality | Genome-fragment | 5.25 | AAI-based (high-confidence) | 0 |
| S12_vs_k141_161882\|\|full | 2232 | No | NA | 5 | 2 | 1 | Low-quality | Genome-fragment | 3.63 | HMM-based (lower-bound) | 0 |
| S11_vs_k141_4009\|\|full | 2233 | No | NA | 4 | 0 | 0 | Low-quality | Genome-fragment | 1.82 | AAI-based (high-confidence) | 0 |
| S7_vs_61522\|\|full | 3993 | No | NA | 5 | 0 | 0 | Not-determined | Genome-fragment | NA | NA | 0 |
| S3_vs_k141_112713\|\|full | 2233 | No | NA | 4 | 0 | 0 | Low-quality | Genome-fragment | 2.56 | AAI-based (medium-confidence) | 0 |
| S7_vs_61548\|\|full | 1198 | No | NA | 2 | 0 | 1 | Not-determined | Genome-fragment | NA | NA | 0 |
| S7_vs_61550\|\|full | 2204 | No | NA | 2 | 0 | 2 | Not-determined | Genome-fragment | NA | NA | 0 |
| S7_vs_61601\|\|full | 1745 | No | NA | 3 | 0 | 2 | Not-determined | Genome-fragment | NA | NA | 0 |
| S7_vs_61610\|\|full | 3111 | Yes | 1870 | 3 | 1 | 1 | Not-determined | Genome-fragment | NA | NA | 39.89 |
| S5_vs_k141_137943\|\|full | 2233 | No | NA | 2 | 1 | 0 | Low-quality | Genome-fragment | 4.78 | AAI-based (high-confidence) | 0 |
| S7_vs_61866\|\|full | 1729 | No | NA | 5 | 0 | 0 | Not-determined | Genome-fragment | NA | NA | 0 |
| S7_vs_62025\|\|full | 1207 | No | NA | 2 | 0 | 1 | Not-determined | Genome-fragment | NA | NA | 0 |
| S7_vs_62056\|\|full | 2342 | No | NA | 5 | 0 | 1 | Not-determined | Genome-fragment | NA | NA | 0 |
| S7_vs_62060\|\|full | 1973 | No | NA | 4 | 0 | 1 | Not-determined | Genome-fragment | NA | NA | 0 |
| S12_vs_k141_25328\|\|full | 2233 | No | NA | 6 | 1 | 0 | Low-quality | Genome-fragment | 2.59 | HMM-based (lower-bound) | 0 |
| S7_vs_62068\|\|full | 1600 | No | NA | 4 | 0 | 0 | Not-determined | Genome-fragment | NA | NA | 0 |
| S7_vs_62125\|\|full | 1717 | No | NA | 3 | 0 | 0 | Not-determined | Genome-fragment | NA | NA | 0 |
| S7_vs_62158\|\|full | 1267 | No | NA | 2 | 0 | 1 | Not-determined | Genome-fragment | NA | NA | 0 |
| S7_vs_62182\|\|full | 1292 | No | NA | 3 | 0 | 0 | Not-determined | Genome-fragment | NA | NA | 0 |
| S7_vs_62304\|\|full | 1022 | No | NA | 2 | 0 | 1 | Not-determined | Genome-fragment | NA | NA | 0 |
| S8_vs_k141_105613\|\|full | 2233 | No | NA | 4 | 4 | 0 | Low-quality | Genome-fragment | 3.58 | AAI-based (high-confidence) | 0 |
| S7_vs_62458\|\|full | 2245 | No | NA | 2 | 0 | 0 | Not-determined | Genome-fragment | NA | NA | 0 |
| S1_vs_k141_54376\|\|full | 2234 | No | NA | 3 | 1 | 0 | Low-quality | Genome-fragment | 2.16 | HMM-based (lower-bound) | 0 |
| S3_vs_k141_117776\|\|full | 2234 | No | NA | 4 | 0 | 0 | Low-quality | Genome-fragment | 1.4 | AAI-based (high-confidence) | 0 |
| S16_vs_127690\|\|full | 2234 | No | NA | 3 | 0 | 0 | Low-quality | Genome-fragment | 2.04 | AAI-based (high-confidence) | 0 |
| S8_extracted_k141_8338 | 2234 | No | NA | 5 | 0 | 0 | Low-quality | Genome-fragment | 3.61 | AAI-based (high-confidence) | 0 |
| S7_vs_101689\|\|full | 2235 | No | NA | 3 | 0 | 0 | Low-quality | Genome-fragment | 4.24 | AAI-based (medium-confidence) | 0 |
| S7_vs_62677\|\|full | 1966 | No | NA | 2 | 0 | 0 | Not-determined | Genome-fragment | NA | NA | 0 |
| S7_vs_62692\|\|full | 3010 | No | NA | 3 | 0 | 0 | Not-determined | Genome-fragment | NA | NA | 0 |
| S7_vs_62733\|\|full | 1246 | No | NA | 2 | 0 | 0 | Not-determined | Genome-fragment | NA | NA | 0 |
| S16_extracted_7798 | 2235 | No | NA | 2 | 2 | 0 | Low-quality | Genome-fragment | 3.48 | AAI-based (high-confidence) | 0 |
| S7_vs_62775\|\|full | 7082 | No | NA | 6 | 0 | 2 | Not-determined | Genome-fragment | NA | NA | 0 |
| S17_extracted_k141_63065 | 2236 | No | NA | 4 | 3 | 0 | Low-quality | Genome-fragment | 1.3 | AAI-based (high-confidence) | 0 |
| S17_vs_k141_47300\|\|lt2gene | 2236 | No | NA | 2 | 1 | 0 | Low-quality | Genome-fragment | 3.73 | AAI-based (high-confidence) | 0 |
| S11_extracted_k141_60637 | 2236 | No | NA | 2 | 1 | 0 | Low-quality | Genome-fragment | 5.4 | AAI-based (high-confidence) | 0 |
| S11_vs_k141_87339\|\|full | 2236 | No | NA | 3 | 0 | 0 | Low-quality | Genome-fragment | 3.92 | AAI-based (high-confidence) | 0 |
| S7_vs_8105\|\|full | 2236 | No | NA | 4 | 1 | 0 | Low-quality | Genome-fragment | 3.63 | AAI-based (medium-confidence) | 0 |
| S7_vs_71310\|\|full | 2236 | No | NA | 3 | 1 | 0 | Low-quality | Genome-fragment | 2.96 | HMM-based (lower-bound) | 0 |
| S17_extracted_k141_6122 | 2237 | Yes | 1108 | 3 | 1 | 1 | Low-quality | Genome-fragment | 0.4 | AAI-based (medium-confidence) | 50.47 |
| S7_vs_63203\|\|full | 1493 | No | NA | 3 | 0 | 2 | Not-determined | Genome-fragment | NA | NA | 0 |
| S7_vs_63263\|\|full | 3429 | No | NA | 3 | 0 | 2 | Not-determined | Genome-fragment | NA | NA | 0 |
| S7_vs_63284\|\|full | 1732 | No | NA | 3 | 0 | 3 | Not-determined | Genome-fragment | NA | NA | 0 |
| S7_vs_63339\|\|full | 1011 | No | NA | 3 | 0 | 0 | Not-determined | Genome-fragment | NA | NA | 0 |
| S7_vs_63341\|\|full | 1268 | No | NA | 3 | 0 | 1 | Not-determined | Genome-fragment | NA | NA | 0 |
| S7_vs_63373\|\|full | 5064 | No | NA | 10 | 0 | 0 | Not-determined | Genome-fragment | NA | NA | 0 |
| S7_vs_63393\|\|full | 4124 | No | NA | 5 | 0 | 0 | Not-determined | Genome-fragment | NA | NA | 0 |
| S7_vs_63408\|\|full | 2903 | No | NA | 5 | 0 | 2 | Not-determined | Genome-fragment | NA | NA | 0 |
| S7_vs_63532\|\|full | 1999 | No | NA | 2 | 0 | 0 | Not-determined | Genome-fragment | NA | NA | 0 |
| S7_vs_63565\|\|full | 2212 | No | NA | 2 | 0 | 2 | Not-determined | Genome-fragment | NA | NA | 0 |
| S13_vs_63851\|\|full | 2237 | No | NA | 3 | 0 | 0 | Low-quality | Genome-fragment | 3.18 | AAI-based (high-confidence) | 0 |
| S7_vs_63664\|\|full | 1579 | No | NA | 3 | 0 | 1 | Not-determined | Genome-fragment | NA | NA | 0 |
| S5_vs_k141_92068\|\|full | 2237 | No | NA | 4 | 0 | 0 | Low-quality | Genome-fragment | 1.1 | AAI-based (medium-confidence) | 0 |
| S10_vs_110361\|\|full | 2237 | No | NA | 2 | 0 | 0 | Low-quality | Genome-fragment | 18.81 | AAI-based (high-confidence) | 0 |
| S14_vs_k141_43229\|\|full | 2238 | No | NA | 4 | 1 | 0 | Low-quality | Genome-fragment | 5.11 | AAI-based (medium-confidence) | 0 |
| S7_vs_30183\|\|full | 2238 | No | NA | 5 | 2 | 0 | Low-quality | Genome-fragment | 5.63 | AAI-based (high-confidence) | 0 |
| S7_vs_63961\|\|full | 2175 | No | NA | 3 | 0 | 0 | Not-determined | Genome-fragment | NA | NA | 0 |
| S7_vs_64017\|\|full | 1269 | No | NA | 3 | 0 | 0 | Not-determined | Genome-fragment | NA | NA | 0 |
| S12_extracted_k141_40848 | 2238 | No | NA | 2 | 0 | 1 | Low-quality | Genome-fragment | 4.39 | AAI-based (medium-confidence) | 0 |
| S7_extracted_112681 | 2239 | No | NA | 1 | 0 | 1 | Low-quality | Genome-fragment | 0.9 | AAI-based (medium-confidence) | 0 |
| S7_vs_64165\|\|full | 1750 | No | NA | 2 | 0 | 1 | Not-determined | Genome-fragment | NA | NA | 0 |
| S7_vs_64280\|\|full | 2302 | No | NA | 3 | 0 | 1 | Not-determined | Genome-fragment | NA | NA | 0 |
| S8_vs_k141_39704\|\|full | 2239 | No | NA | 3 | 1 | 0 | Low-quality | Genome-fragment | 5.38 | AAI-based (high-confidence) | 0 |
| S1_vs_k141_22506\|\|full | 2240 | No | NA | 3 | 0 | 1 | Low-quality | Genome-fragment | 1.54 | AAI-based (high-confidence) | 0 |
| S5_extracted_k141_122272 | 2240 | No | NA | 3 | 2 | 0 | Low-quality | Genome-fragment | 6.59 | AAI-based (high-confidence) | 0 |
| S16_vs_52016\|\|full | 2240 | No | NA | 4 | 2 | 0 | Low-quality | Genome-fragment | 4.84 | AAI-based (medium-confidence) | 0 |
| S12_vs_k141_105202\|\|full | 2240 | No | NA | 4 | 0 | 0 | Low-quality | Genome-fragment | 6.05 | AAI-based (high-confidence) | 0 |
| S7_vs_64616\|\|full | 1004 | No | NA | 4 | 0 | 0 | Not-determined | Genome-fragment | NA | NA | 0 |
| S9_vs_k141_76664\|\|full | 2241 | No | NA | 3 | 0 | 0 | Low-quality | Genome-fragment | 4.9 | AAI-based (high-confidence) | 0 |
| S7_vs_64688\|\|full | 1201 | No | NA | 2 | 0 | 2 | Not-determined | Genome-fragment | NA | NA | 0 |
| S7_vs_64796\|\|full | 5514 | No | NA | 5 | 0 | 1 | Not-determined | Genome-fragment | NA | NA | 0 |
| S11_vs_k141_43230\|\|full | 2241 | No | NA | 5 | 2 | 0 | Low-quality | Genome-fragment | 5.52 | AAI-based (medium-confidence) | 0 |
| S11_vs_k141_82238\|\|full | 2241 | No | NA | 2 | 0 | 1 | Low-quality | Genome-fragment | 4.52 | AAI-based (medium-confidence) | 0 |
| S7_vs_64985\|\|full | 7197 | No | NA | 7 | 0 | 4 | Not-determined | Genome-fragment | NA | NA | 0 |
| S7_vs_65174\|\|full | 2659 | No | NA | 3 | 0 | 0 | Not-determined | Genome-fragment | NA | NA | 0 |
| S3_vs_k141_29825\|\|full | 2241 | Yes | 1968 | 3 | 1 | 1 | Low-quality | Genome-fragment | 1.6 | HMM-based (lower-bound) | 12.18 |
| S16_vs_121485\|\|full | 2241 | No | NA | 4 | 1 | 0 | Low-quality | Genome-fragment | 4.44 | AAI-based (high-confidence) | 0 |
| S2_extracted_k141_9896 | 2241 | No | NA | 2 | 0 | 0 | Low-quality | Genome-fragment | 2.33 | AAI-based (high-confidence) | 0 |
| S7_vs_65310\|\|full | 21775 | No | NA | 26 | 0 | 10 | Not-determined | Genome-fragment | NA | NA | 0 |
| S7_vs_65362\|\|full | 1842 | No | NA | 3 | 0 | 1 | Not-determined | Genome-fragment | NA | NA | 0 |
| S7_vs_65438\|\|full | 6902 | No | NA | 7 | 0 | 0 | Not-determined | Genome-fragment | NA | NA | 0 |
| S7_vs_65465\|\|full | 3350 | No | NA | 4 | 0 | 3 | Not-determined | Genome-fragment | NA | NA | 0 |
| S7_vs_65471\|\|full | 1909 | No | NA | 3 | 0 | 0 | Not-determined | Genome-fragment | NA | NA | 0 |
| S8_vs_k141_202839\|\|full | 2241 | No | NA | 3 | 2 | 0 | Low-quality | Genome-fragment | 5.15 | AAI-based (high-confidence) | 0 |
| S7_vs_65557\|\|full | 11627 | No | NA | 9 | 0 | 2 | Not-determined | Genome-fragment | NA | NA | 0 |
| S13_vs_40625\|\|full | 2242 | No | NA | 5 | 1 | 0 | Low-quality | Genome-fragment | 3.16 | AAI-based (medium-confidence) | 0 |
| S13_vs_121186\|\|full | 2242 | No | NA | 4 | 2 | 0 | Low-quality | Genome-fragment | 6.49 | AAI-based (high-confidence) | 0 |
| S3_vs_k141_44557\|\|full | 2242 | No | NA | 2 | 0 | 2 | Low-quality | Genome-fragment | 1.05 | AAI-based (medium-confidence) | 0 |
| S7_vs_48013\|\|full | 2242 | No | NA | 4 | 0 | 2 | Low-quality | Genome-fragment | 4.1 | AAI-based (medium-confidence) | 0 |
| NC3_vf_k141_505 | 2242 | No | NA | 4 | 0 | 0 | Low-quality | Genome-fragment | 4.31 | AAI-based (medium-confidence) | 0 |
| S7_vs_65753\|\|full | 2540 | No | NA | 2 | 0 | 0 | Not-determined | Genome-fragment | NA | NA | 0 |
| S10_vs_8236\|\|full | 2242 | No | NA | 6 | 0 | 0 | Low-quality | Genome-fragment | 4.71 | AAI-based (high-confidence) | 0 |
| S17_vs_k141_115430\|\|full | 2243 | No | NA | 3 | 0 | 1 | Low-quality | Genome-fragment | 1.95 | AAI-based (high-confidence) | 0 |
| S7_vs_66086\|\|full | 3281 | No | NA | 5 | 0 | 0 | Not-determined | Genome-fragment | NA | NA | 0 |
| S7_vs_66211\|\|full | 1695 | No | NA | 2 | 0 | 1 | Not-determined | Genome-fragment | NA | NA | 0 |
| S7_vs_66229\|\|full | 1638 | No | NA | 3 | 0 | 0 | Not-determined | Genome-fragment | NA | NA | 0 |
| S14_vs_k141_71998\|\|full | 2243 | No | NA | 6 | 5 | 0 | Low-quality | Genome-fragment | 6.53 | AAI-based (high-confidence) | 0 |
| S7_vs_66319\|\|full | 1062 | No | NA | 3 | 0 | 1 | Not-determined | Genome-fragment | NA | NA | 0 |
| S7_vs_66404\|\|full | 1426 | No | NA | 2 | 0 | 2 | Not-determined | Genome-fragment | NA | NA | 0 |
| S3_vs_k141_56459\|\|full | 2243 | No | NA | 3 | 0 | 1 | Low-quality | Genome-fragment | 2.51 | AAI-based (medium-confidence) | 0 |
| S5_vs_k141_126614\|\|full | 2243 | No | NA | 5 | 0 | 0 | Low-quality | Genome-fragment | 4.49 | AAI-based (medium-confidence) | 0 |
| S8_extracted_k141_108511 | 2243 | No | NA | 3 | 0 | 1 | Low-quality | Genome-fragment | 3.52 | AAI-based (high-confidence) | 0 |
| S7_vs_66573\|\|full | 1050 | No | NA | 3 | 0 | 2 | Not-determined | Genome-fragment | NA | NA | 0 |
| S7_vs_66574\|\|full | 2829 | No | NA | 5 | 0 | 0 | Not-determined | Genome-fragment | NA | NA | 0 |
| S7_vs_66593\|\|full | 1562 | No | NA | 3 | 0 | 2 | Not-determined | Genome-fragment | NA | NA | 0 |
| S7_vs_66598\|\|full | 1028 | No | NA | 2 | 0 | 0 | Not-determined | Genome-fragment | NA | NA | 0 |
| S12_vs_k141_134829\|\|full | 2244 | No | NA | 4 | 0 | 2 | Low-quality | Genome-fragment | 1.36 | AAI-based (medium-confidence) | 0 |
| S7_vs_66618\|\|full | 2190 | No | NA | 3 | 0 | 3 | Not-determined | Genome-fragment | NA | NA | 0 |
| S7_vs_66631\|\|full | 2901 | No | NA | 3 | 0 | 3 | Not-determined | Genome-fragment | NA | NA | 0 |
| S17_extracted_k141_67269 | 2245 | No | NA | 7 | 0 | 0 | Low-quality | Genome-fragment | 2.8 | AAI-based (high-confidence) | 0 |
| S7_vs_66651\|\|full | 1221 | No | NA | 2 | 0 | 0 | Not-determined | Genome-fragment | NA | NA | 0 |
| S7_vs_66655\|\|full | 3567 | No | NA | 4 | 0 | 3 | Not-determined | Genome-fragment | NA | NA | 0 |
| S1_vs_k141_42888\|\|full | 2245 | No | NA | 3 | 0 | 1 | Low-quality | Genome-fragment | 2.09 | AAI-based (medium-confidence) | 0 |
| S7_vs_66750\|\|full | 1579 | No | NA | 3 | 0 | 1 | Not-determined | Genome-fragment | NA | NA | 0 |
| S3_vs_k141_75042\|\|full | 2245 | No | NA | 3 | 0 | 0 | Low-quality | Genome-fragment | 4.45 | AAI-based (high-confidence) | 0 |
| S7_vs_66865\|\|full | 1959 | No | NA | 4 | 0 | 0 | Not-determined | Genome-fragment | NA | NA | 0 |
| S7_vs_66979\|\|full | 7696 | No | NA | 6 | 0 | 3 | Not-determined | Genome-fragment | NA | NA | 0 |
| S7_vs_32186\|\|full | 2245 | No | NA | 3 | 1 | 0 | Low-quality | Genome-fragment | 4.84 | AAI-based (medium-confidence) | 0 |
| S7_vs_67065\|\|full | 5651 | No | NA | 4 | 0 | 2 | Not-determined | Genome-fragment | NA | NA | 0 |
| S3_extracted_k141_79922 | 2246 | No | NA | 4 | 0 | 1 | Low-quality | Genome-fragment | 4.93 | AAI-based (medium-confidence) | 0 |
| S7_vs_67128\|\|full | 2899 | No | NA | 5 | 0 | 3 | Not-determined | Genome-fragment | NA | NA | 0 |
| S7_vs_67152\|\|full | 1190 | No | NA | 2 | 0 | 0 | Not-determined | Genome-fragment | NA | NA | 0 |
| S7_vs_67209\|\|full | 2853 | No | NA | 3 | 0 | 2 | Not-determined | Genome-fragment | NA | NA | 0 |
| S3_vs_k141_85718\|\|full | 2246 | No | NA | 6 | 0 | 0 | Low-quality | Genome-fragment | 6.61 | AAI-based (high-confidence) | 0 |
| S10_extracted_80812 | 2246 | No | NA | 2 | 0 | 1 | Low-quality | Genome-fragment | 3.55 | AAI-based (high-confidence) | 0 |
| S12_extracted_k141_96300 | 2246 | No | NA | 2 | 1 | 0 | Low-quality | Genome-fragment | 4.6 | HMM-based (lower-bound) | 0 |
| S12_extracted_k141_36312 | 2246 | No | NA | 2 | 1 | 0 | Low-quality | Genome-fragment | 0.73 | HMM-based (lower-bound) | 0 |
| S12_vs_k141_130992\|\|full | 2246 | No | NA | 3 | 0 | 0 | Low-quality | Genome-fragment | 3.06 | AAI-based (high-confidence) | 0 |
| S7_vs_67534\|\|full | 1929 | No | NA | 3 | 0 | 3 | Not-determined | Genome-fragment | NA | NA | 0 |
| S11_vs_k141_79602\|\|full | 2247 | No | NA | 6 | 0 | 2 | Low-quality | Genome-fragment | 4.54 | AAI-based (medium-confidence) | 0 |
| S8_extracted_k141_96431 | 2247 | No | NA | 2 | 0 | 1 | Low-quality | Genome-fragment | 0.7 | AAI-based (medium-confidence) | 0 |
| S7_vs_67642\|\|full | 2120 | No | NA | 3 | 0 | 0 | Not-determined | Genome-fragment | NA | NA | 0 |
| S1_vs_k141_45677\|\|full | 2248 | No | NA | 3 | 2 | 0 | Low-quality | Genome-fragment | 5.52 | AAI-based (high-confidence) | 0 |
| S7_vs_67652\|\|full | 1607 | No | NA | 2 | 0 | 1 | Not-determined | Genome-fragment | NA | NA | 0 |
| S7_vs_67661\|\|full | 2656 | No | NA | 1 | 0 | 1 | Not-determined | Genome-fragment | NA | NA | 0 |
| S12_vs_k141_181931\|\|full | 2248 | No | NA | 5 | 1 | 0 | Low-quality | Genome-fragment | 2.94 | HMM-based (lower-bound) | 0 |
| S8_extracted_k141_7838 | 2248 | No | NA | 3 | 0 | 1 | Low-quality | Genome-fragment | 3.43 | AAI-based (medium-confidence) | 0 |
| S7_vs_67846\|\|full | 2940 | No | NA | 3 | 0 | 3 | Not-determined | Genome-fragment | NA | NA | 0 |
| S17_vs_k141_1769\|\|full | 2249 | No | NA | 3 | 0 | 1 | Low-quality | Genome-fragment | 1.58 | AAI-based (medium-confidence) | 0 |
| S7_vs_68175\|\|full | 3176 | No | NA | 5 | 0 | 0 | Not-determined | Genome-fragment | NA | NA | 0 |
| S11_vs_k141_21672\|\|full | 2249 | No | NA | 4 | 0 | 0 | Low-quality | Genome-fragment | 5.5 | AAI-based (high-confidence) | 0 |
| S1_vs_k141_10517\|\|full | 2249 | No | NA | 5 | 1 | 0 | Low-quality | Genome-fragment | 4.73 | AAI-based (high-confidence) | 0 |
| S7_vs_68278\|\|full | 1347 | No | NA | 3 | 0 | 0 | Not-determined | Genome-fragment | NA | NA | 0 |
| S7_vs_68294\|\|full | 2614 | No | NA | 3 | 0 | 2 | Not-determined | Genome-fragment | NA | NA | 0 |
| S7_vs_30605\|\|full | 2249 | No | NA | 5 | 1 | 0 | Low-quality | Genome-fragment | 6.38 | AAI-based (high-confidence) | 0 |
| S16_extracted_45896 | 2249 | No | NA | 6 | 0 | 0 | Low-quality | Genome-fragment | 4.85 | AAI-based (medium-confidence) | 0 |
| S12_vs_k141_177362\|\|lt2gene | 2249 | No | NA | 1 | 0 | 0 | Low-quality | Genome-fragment | 4.02 | AAI-based (high-confidence) | 0 |
| S7_vs_68453\|\|full | 3312 | No | NA | 3 | 0 | 3 | Not-determined | Genome-fragment | NA | NA | 0 |
| S8_vs_k141_201721\|\|full | 2249 | No | NA | 4 | 0 | 1 | Low-quality | Genome-fragment | 6.42 | AAI-based (medium-confidence) | 0 |
| S7_vs_68572\|\|full | 1093 | No | NA | 3 | 0 | 1 | Not-determined | Genome-fragment | NA | NA | 0 |
| S3_vs_k141_16372\|\|full | 2250 | No | NA | 4 | 3 | 0 | Low-quality | Genome-fragment | 3.88 | HMM-based (lower-bound) | 0 |
| S3_vs_k141_117681\|\|full | 2250 | No | NA | 4 | 2 | 0 | Low-quality | Genome-fragment | 1.38 | AAI-based (high-confidence) | 0 |
| S3_vs_k141_103537\|\|full | 2250 | No | NA | 4 | 0 | 2 | Low-quality | Genome-fragment | 4.94 | AAI-based (medium-confidence) | 0 |
| S7_vs_68808\|\|full | 2203 | No | NA | 3 | 0 | 1 | Not-determined | Genome-fragment | NA | NA | 0 |
| S3_vs_k141_98964\|\|lt2gene | 2250 | Yes | 1021 | 2 | 1 | 1 | Low-quality | Genome-fragment | 0.62 | HMM-based (lower-bound) | 54.62 |
| S7_vs_69072\|\|full | 1288 | No | NA | 1 | 0 | 1 | Not-determined | Genome-fragment | NA | NA | 0 |
| S5_vs_k141_130798\|\|full | 2250 | No | NA | 5 | 1 | 0 | Low-quality | Genome-fragment | 3.85 | AAI-based (high-confidence) | 0 |
| S7_vs_69238\|\|full | 5508 | No | NA | 3 | 0 | 3 | Not-determined | Genome-fragment | NA | NA | 0 |
| S11_vs_k141_74571\|\|full | 2251 | No | NA | 4 | 0 | 0 | Low-quality | Genome-fragment | 3.06 | AAI-based (high-confidence) | 0 |
| S5_vs_k141_45991\|\|full | 2251 | No | NA | 4 | 2 | 0 | Low-quality | Genome-fragment | 4.95 | AAI-based (medium-confidence) | 0 |
| S7_vs_69415\|\|full | 1614 | No | NA | 3 | 0 | 0 | Not-determined | Genome-fragment | NA | NA | 0 |
| S7_vs_69435\|\|full | 2123 | No | NA | 2 | 0 | 2 | Not-determined | Genome-fragment | NA | NA | 0 |
| S7_vs_83337\|\|full | 2251 | No | NA | 3 | 0 | 1 | Low-quality | Genome-fragment | 1.03 | AAI-based (medium-confidence) | 0 |
| S11_vs_k141_38812\|\|full | 2252 | No | NA | 4 | 3 | 0 | Low-quality | Genome-fragment | 5.68 | AAI-based (high-confidence) | 0 |
| S10_vs_88576\|\|full | 2252 | No | NA | 3 | 0 | 0 | Low-quality | Genome-fragment | 4.87 | AAI-based (high-confidence) | 0 |
| S7_vs_69630\|\|full | 39417 | No | NA | 60 | 26 | 0 | High-quality | High-quality | 100 | AAI-based (high-confidence) | 0 |
| S7_vs_69644\|\|full | 2598 | No | NA | 3 | 0 | 1 | Not-determined | Genome-fragment | NA | NA | 0 |
| S11_extracted_k141_79418 | 2253 | No | NA | 4 | 0 | 0 | Low-quality | Genome-fragment | 1.12 | AAI-based (medium-confidence) | 0 |
| S7_vs_69717\|\|full | 2746 | No | NA | 3 | 0 | 2 | Not-determined | Genome-fragment | NA | NA | 0 |
| S11_vs_k141_17397\|\|full | 2253 | No | NA | 3 | 0 | 0 | Low-quality | Genome-fragment | 5.71 | AAI-based (high-confidence) | 0 |
| S7_vs_69901\|\|full | 3000 | No | NA | 2 | 0 | 2 | Not-determined | Genome-fragment | NA | NA | 0 |
| S7_vs_69947\|\|full | 30996 | No | NA | 31 | 0 | 3 | Not-determined | Genome-fragment | NA | NA | 0 |
| S7_vs_69951\|\|full | 2082 | No | NA | 2 | 0 | 2 | Not-determined | Genome-fragment | NA | NA | 0 |
| S7_vs_69970\|\|full | 5363 | No | NA | 5 | 0 | 3 | Not-determined | Genome-fragment | NA | NA | 0 |
| S3_extracted_k141_113629 | 2253 | No | NA | 4 | 2 | 0 | Low-quality | Genome-fragment | 3.48 | AAI-based (high-confidence) | 0 |
| S3_vs_k141_85419\|\|full | 2253 | No | NA | 3 | 0 | 1 | Low-quality | Genome-fragment | 1.78 | AAI-based (medium-confidence) | 0 |
| S7_vs_14917\|\|full | 2253 | No | NA | 2 | 0 | 2 | Low-quality | Genome-fragment | 1.58 | AAI-based (medium-confidence) | 0 |
| S16_extracted_76158 | 2253 | No | NA | 5 | 2 | 0 | Low-quality | Genome-fragment | 2.39 | AAI-based (high-confidence) | 0 |
| S12_extracted_k141_187023 | 2253 | No | NA | 2 | 0 | 0 | Low-quality | Genome-fragment | 0.41 | AAI-based (medium-confidence) | 0 |
| S15_vs_51984\|\|full | 2253 | Yes | 534 | 3 | 1 | 2 | Low-quality | Genome-fragment | 0.61 | AAI-based (medium-confidence) | 76.3 |
| S6_extracted_k141_12567 | 2253 | No | NA | 3 | 0 | 3 | Low-quality | Genome-fragment | 3.68 | AAI-based (medium-confidence) | 0 |
| S7_vs_70265\|\|full | 17875 | No | NA | 25 | 9 | 0 | Medium-quality | Genome-fragment | 52.56 | HMM-based (lower-bound) | 0 |
| S17_vs_k141_169065\|\|full | 2254 | Yes | 1623 | 5 | 1 | 1 | Low-quality | Genome-fragment | 3.45 | AAI-based (high-confidence) | 27.99 |
| S14_extracted_k141_9243 | 2254 | No | NA | 7 | 7 | 0 | Low-quality | Genome-fragment | 7.43 | AAI-based (high-confidence) | 0 |
| S7_vs_70374\|\|full | 5575 | No | NA | 7 | 0 | 6 | Not-determined | Genome-fragment | NA | NA | 0 |
| S14_vs_k141_58189\|\|full | 2254 | No | NA | 4 | 0 | 1 | Low-quality | Genome-fragment | 0.88 | AAI-based (medium-confidence) | 0 |
| S16_vs_167159\|\|full | 2254 | No | NA | 6 | 1 | 0 | Low-quality | Genome-fragment | 3.94 | AAI-based (high-confidence) | 0 |
| S10_vs_107312\|\|full | 2254 | No | NA | 7 | 2 | 0 | Low-quality | Genome-fragment | 5.29 | AAI-based (high-confidence) | 0 |
| S7_vs_70537\|\|full | 8912 | No | NA | 14 | 0 | 5 | Not-determined | Genome-fragment | NA | NA | 0 |
| S17_extracted_k141_116830 | 2255 | No | NA | 3 | 0 | 1 | Low-quality | Genome-fragment | 3.26 | AAI-based (medium-confidence) | 0 |
| S7_vs_70659\|\|full | 1808 | No | NA | 3 | 0 | 1 | Not-determined | Genome-fragment | NA | NA | 0 |
| S7_vs_70665\|\|full | 12370 | No | NA | 12 | 0 | 8 | Not-determined | Genome-fragment | NA | NA | 0 |
| S7_vs_70716\|\|full | 1634 | No | NA | 2 | 0 | 0 | Not-determined | Genome-fragment | NA | NA | 0 |
| S7_vs_70725\|\|full | 2275 | No | NA | 2 | 0 | 0 | Not-determined | Genome-fragment | NA | NA | 0 |
| S7_vs_70803\|\|full | 1157 | No | NA | 3 | 0 | 0 | Not-determined | Genome-fragment | NA | NA | 0 |
| S3_vs_k141_40121\|\|full | 2255 | No | NA | 4 | 0 | 2 | Low-quality | Genome-fragment | 1.14 | AAI-based (medium-confidence) | 0 |
| S5_vs_k141_46133\|\|full | 2255 | No | NA | 5 | 1 | 0 | Low-quality | Genome-fragment | 2.81 | AAI-based (medium-confidence) | 0 |
| S16_vs_20922\|\|full | 2255 | No | NA | 3 | 0 | 2 | Low-quality | Genome-fragment | 1 | AAI-based (medium-confidence) | 0 |
| S12_vs_k141_178379\|\|full | 2255 | No | NA | 3 | 0 | 0 | Low-quality | Genome-fragment | 5.81 | AAI-based (medium-confidence) | 0 |
| S7_vs_71170\|\|full | 2666 | No | NA | 4 | 0 | 0 | Not-determined | Genome-fragment | NA | NA | 0 |
| S8_vs_k141_90694\|\|full | 2255 | No | NA | 3 | 1 | 2 | Low-quality | Genome-fragment | 3.59 | HMM-based (lower-bound) | 0 |
| S14_extracted_k141_47085 | 2256 | No | NA | 5 | 3 | 0 | Low-quality | Genome-fragment | 7.26 | AAI-based (high-confidence) | 0 |
| S11_extracted_k141_94644 | 2257 | No | NA | 4 | 0 | 0 | Low-quality | Genome-fragment | 4.14 | AAI-based (medium-confidence) | 0 |
| S5_extracted_k141_10824 | 2257 | No | NA | 1 | 1 | 0 | Low-quality | Genome-fragment | 4.81 | AAI-based (medium-confidence) | 0 |
| S16_extracted_60090 | 2257 | No | NA | 1 | 0 | 1 | Low-quality | Genome-fragment | 3.9 | AAI-based (medium-confidence) | 0 |
| S16_extracted_107765 | 2257 | No | NA | 5 | 0 | 0 | Low-quality | Genome-fragment | 12.4 | AAI-based (high-confidence) | 0 |
| S7_vs_71521\|\|full | 2072 | No | NA | 3 | 0 | 2 | Not-determined | Genome-fragment | NA | NA | 0 |
| S7_vs_71526\|\|full | 1603 | No | NA | 2 | 0 | 1 | Not-determined | Genome-fragment | NA | NA | 0 |
| S7_vs_71614\|\|full | 5941 | No | NA | 10 | 0 | 0 | Not-determined | Genome-fragment | NA | NA | 0 |
| S8_vs_k141_26775\|\|full | 2257 | No | NA | 2 | 0 | 1 | Low-quality | Genome-fragment | 2.97 | AAI-based (medium-confidence) | 0 |
| S7_vs_71836\|\|full | 1121 | No | NA | 3 | 0 | 0 | Not-determined | Genome-fragment | NA | NA | 0 |
| S7_vs_71854\|\|full | 1287 | No | NA | 3 | 0 | 0 | Not-determined | Genome-fragment | NA | NA | 0 |
| S13_vs_26641\|\|full | 2258 | No | NA | 3 | 1 | 0 | Low-quality | Genome-fragment | 1.21 | HMM-based (lower-bound) | 0 |
| S14_vs_k141_110390\|\|full | 2258 | No | NA | 3 | 0 | 0 | Low-quality | Genome-fragment | 5.07 | AAI-based (medium-confidence) | 0 |
| S7_vs_72007\|\|full | 1377 | No | NA | 3 | 0 | 0 | Not-determined | Genome-fragment | NA | NA | 0 |
| S7_vs_72012\|\|full | 1388 | No | NA | 4 | 0 | 0 | Not-determined | Genome-fragment | NA | NA | 0 |
| S14_vs_k141_114748\|\|full | 2258 | No | NA | 3 | 0 | 2 | Low-quality | Genome-fragment | 3.9 | AAI-based (medium-confidence) | 0 |
| S12_vs_k141_143613\|\|full | 2258 | No | NA | 4 | 0 | 1 | Low-quality | Genome-fragment | 2.32 | AAI-based (medium-confidence) | 0 |
| S12_vs_k141_139368\|\|full | 2258 | No | NA | 6 | 1 | 0 | Low-quality | Genome-fragment | 4.96 | AAI-based (medium-confidence) | 0 |
| S7_vs_72203\|\|full | 1628 | No | NA | 2 | 0 | 2 | Not-determined | Genome-fragment | NA | NA | 0 |
| S7_vs_72213\|\|full | 2149 | No | NA | 4 | 0 | 0 | Not-determined | Genome-fragment | NA | NA | 0 |
| S7_vs_72336\|\|full | 4062 | No | NA | 7 | 0 | 0 | Not-determined | Genome-fragment | NA | NA | 0 |
| S7_vs_72415\|\|full | 1180 | No | NA | 1 | 0 | 1 | Not-determined | Genome-fragment | NA | NA | 0 |
| S12_vs_k141_171493\|\|full | 2258 | No | NA | 4 | 0 | 1 | Low-quality | Genome-fragment | 1.09 | AAI-based (medium-confidence) | 0 |
| S13_vs_28449\|\|full | 2259 | No | NA | 3 | 0 | 2 | Low-quality | Genome-fragment | 1.96 | AAI-based (medium-confidence) | 0 |
| S7_vs_72628\|\|full | 2137 | No | NA | 3 | 0 | 0 | Not-determined | Genome-fragment | NA | NA | 0 |
| S14_vs_k141_98601\|\|full | 2259 | No | NA | 3 | 3 | 0 | Low-quality | Genome-fragment | 5.75 | AAI-based (medium-confidence) | 0 |
| S12_vs_k141_152958\|\|full | 2259 | No | NA | 3 | 1 | 0 | Low-quality | Genome-fragment | 1.42 | HMM-based (lower-bound) | 0 |
| S7_vs_72858\|\|full | 3995 | No | NA | 4 | 0 | 2 | Not-determined | Genome-fragment | NA | NA | 0 |
| S8_vs_k141_97317\|\|full | 2259 | No | NA | 8 | 2 | 0 | Low-quality | Genome-fragment | 4.08 | AAI-based (high-confidence) | 0 |
| S7_vs_72927\|\|full | 1218 | No | NA | 2 | 0 | 1 | Not-determined | Genome-fragment | NA | NA | 0 |
| S7_vs_72930\|\|full | 4653 | No | NA | 4 | 0 | 2 | Not-determined | Genome-fragment | NA | NA | 0 |
| S3_extracted_k141_45426 | 2260 | No | NA | 3 | 0 | 3 | Low-quality | Genome-fragment | 1.36 | AAI-based (medium-confidence) | 0 |
| S7_vs_124320\|\|full | 2260 | No | NA | 4 | 0 | 0 | Low-quality | Genome-fragment | 1.91 | AAI-based (high-confidence) | 0 |
| S7_vs_73028\|\|full | 4453 | No | NA | 6 | 0 | 1 | Not-determined | Genome-fragment | NA | NA | 0 |
| S10_extracted_58574 | 2260 | No | NA | 7 | 0 | 0 | Low-quality | Genome-fragment | 4.17 | AAI-based (high-confidence) | 0 |
| S12_vs_k141_34775\|\|full | 2260 | No | NA | 6 | 3 | 0 | Low-quality | Genome-fragment | 1.48 | AAI-based (high-confidence) | 0 |
| S4_extracted_6023 | 2260 | No | NA | 5 | 3 | 0 | Low-quality | Genome-fragment | 7.25 | AAI-based (high-confidence) | 0 |
| S7_vs_73280\|\|full | 39043 | No | NA | 62 | 17 | 0 | High-quality | High-quality | 97.58 | AAI-based (high-confidence) | 0 |
| S7_vs_73287\|\|full | 5585 | No | NA | 5 | 0 | 4 | Not-determined | Genome-fragment | NA | NA | 0 |
| S7_vs_73305\|\|full | 1601 | No | NA | 3 | 0 | 0 | Not-determined | Genome-fragment | NA | NA | 0 |
| S14_extracted_k141_55996 | 2261 | No | NA | 6 | 2 | 0 | Low-quality | Genome-fragment | 5.59 | AAI-based (high-confidence) | 0 |
| S7_vs_73437\|\|full | 4110 | No | NA | 7 | 0 | 0 | Not-determined | Genome-fragment | NA | NA | 0 |
| S7_vs_57153\|\|full | 2261 | No | NA | 4 | 1 | 0 | Low-quality | Genome-fragment | 2.38 | HMM-based (lower-bound) | 0 |
| S7_vs_73475\|\|full | 1591 | No | NA | 3 | 0 | 0 | Not-determined | Genome-fragment | NA | NA | 0 |
| S16_vs_173136\|\|full | 2261 | No | NA | 4 | 0 | 0 | Low-quality | Genome-fragment | 41.84 | AAI-based (medium-confidence) | 0 |
| S7_vs_73628\|\|full | 1350 | No | NA | 2 | 0 | 1 | Not-determined | Genome-fragment | NA | NA | 0 |
| S7_vs_73647\|\|full | 1883 | No | NA | 4 | 0 | 0 | Not-determined | Genome-fragment | NA | NA | 0 |
| S4_extracted_22116 | 2261 | No | NA | 5 | 0 | 1 | Low-quality | Genome-fragment | 1.42 | AAI-based (high-confidence) | 0 |
| S7_vs_73746\|\|full | 2946 | No | NA | 2 | 0 | 0 | Not-determined | Genome-fragment | NA | NA | 0 |
| S7_vs_73758\|\|full | 2800 | No | NA | 4 | 0 | 1 | Not-determined | Genome-fragment | NA | NA | 0 |
| S7_vs_73804\|\|full | 1435 | No | NA | 3 | 0 | 0 | Not-determined | Genome-fragment | NA | NA | 0 |
| S7_vs_73827\|\|full | 6890 | No | NA | 8 | 0 | 0 | Not-determined | Genome-fragment | NA | NA | 0 |
| S7_vs_73867\|\|full | 2056 | No | NA | 2 | 0 | 0 | Not-determined | Genome-fragment | NA | NA | 0 |
| S7_vs_73892\|\|full | 2713 | No | NA | 2 | 0 | 0 | Not-determined | Genome-fragment | NA | NA | 0 |
| S7_vs_73898\|\|full | 3478 | No | NA | 5 | 0 | 2 | Not-determined | Genome-fragment | NA | NA | 0 |
| S7_vs_73974\|\|full | 2310 | No | NA | 3 | 0 | 0 | Not-determined | Genome-fragment | NA | NA | 0 |
| S14_extracted_k141_61940 | 2262 | No | NA | 4 | 0 | 0 | Low-quality | Genome-fragment | 1.52 | AAI-based (medium-confidence) | 0 |
| S7_vs_74229\|\|full | 1383 | No | NA | 3 | 0 | 1 | Not-determined | Genome-fragment | NA | NA | 0 |
| S1_extracted_k141_110542 | 2262 | No | NA | 4 | 1 | 0 | Low-quality | Genome-fragment | 3.95 | AAI-based (high-confidence) | 0 |
| S1_extracted_k141_11471 | 2262 | No | NA | 3 | 0 | 2 | Low-quality | Genome-fragment | 0.62 | AAI-based (medium-confidence) | 0 |
| S1_vs_k141_69477\|\|full | 2262 | No | NA | 3 | 0 | 2 | Low-quality | Genome-fragment | 2.7 | AAI-based (high-confidence) | 0 |
| S7_vs_74388\|\|full | 1778 | No | NA | 2 | 0 | 0 | Not-determined | Genome-fragment | NA | NA | 0 |
| S3_vs_k141_37562\|\|full | 2262 | No | NA | 4 | 1 | 0 | Low-quality | Genome-fragment | 1.52 | HMM-based (lower-bound) | 0 |
| S7_vs_74511\|\|full | 2981 | No | NA | 3 | 0 | 2 | Not-determined | Genome-fragment | NA | NA | 0 |
| S7_vs_74545\|\|full | 1381 | No | NA | 3 | 0 | 1 | Not-determined | Genome-fragment | NA | NA | 0 |
| S7_vs_74583\|\|full | 4256 | No | NA | 5 | 0 | 1 | Not-determined | Genome-fragment | NA | NA | 0 |
| S7_vs_39678\|\|full | 2262 | No | NA | 3 | 1 | 0 | Low-quality | Genome-fragment | 3.31 | HMM-based (lower-bound) | 0 |
| S7_vs_74874\|\|full | 4062 | No | NA | 3 | 0 | 3 | Not-determined | Genome-fragment | NA | NA | 0 |
| S7_vs_74985\|\|full | 3074 | No | NA | 3 | 0 | 1 | Not-determined | Genome-fragment | NA | NA | 0 |
| S7_vs_74998\|\|full | 1204 | No | NA | 3 | 0 | 1 | Not-determined | Genome-fragment | NA | NA | 0 |
| S7_vs_75057\|\|full | 1385 | No | NA | 3 | 0 | 0 | Not-determined | Genome-fragment | NA | NA | 0 |
| S10_vs_54237\|\|full | 2262 | No | NA | 4 | 2 | 0 | Low-quality | Genome-fragment | 3.68 | AAI-based (high-confidence) | 0 |
| S9_vs_k141_77121\|\|full | 2263 | No | NA | 5 | 1 | 0 | Low-quality | Genome-fragment | 5.43 | AAI-based (high-confidence) | 0 |
| S7_vs_63031\|\|full | 2263 | No | NA | 3 | 2 | 0 | Low-quality | Genome-fragment | 5.42 | AAI-based (high-confidence) | 0 |
| S7_vs_75106\|\|full | 1352 | No | NA | 2 | 0 | 1 | Not-determined | Genome-fragment | NA | NA | 0 |
| S16_vs_59654\|\|full | 2263 | No | NA | 2 | 0 | 1 | Low-quality | Genome-fragment | 1.06 | AAI-based (medium-confidence) | 0 |
| S8_vs_k141_19940\|\|full | 2263 | No | NA | 5 | 3 | 0 | Low-quality | Genome-fragment | 6.03 | AAI-based (high-confidence) | 0 |
| S8_vs_k141_153648\|\|full | 2263 | No | NA | 5 | 0 | 0 | Low-quality | Genome-fragment | 4.13 | AAI-based (high-confidence) | 0 |
| S14_vs_k141_68050\|\|full | 2264 | No | NA | 5 | 5 | 0 | Low-quality | Genome-fragment | 4.86 | AAI-based (high-confidence) | 0 |
| S5_extracted_k141_108763 | 2264 | No | NA | 2 | 1 | 0 | Low-quality | Genome-fragment | 1.47 | HMM-based (lower-bound) | 0 |
| S16_vs_87243\|\|full | 2264 | No | NA | 5 | 1 | 0 | Low-quality | Genome-fragment | 5.22 | AAI-based (medium-confidence) | 0 |
| S3_vs_k141_53363\|\|full | 2266 | No | NA | 3 | 2 | 0 | Low-quality | Genome-fragment | 5.93 | AAI-based (high-confidence) | 0 |
| S5_vs_k141_140405\|\|full | 2266 | No | NA | 2 | 0 | 1 | Low-quality | Genome-fragment | 5.58 | AAI-based (medium-confidence) | 0 |
| S16_extracted_136635 | 2266 | No | NA | 1 | 1 | 0 | Low-quality | Genome-fragment | 3.74 | AAI-based (high-confidence) | 0 |
| S15_vs_76232\|\|full | 2266 | No | NA | 4 | 0 | 2 | Low-quality | Genome-fragment | 1 | AAI-based (medium-confidence) | 0 |
| S7_vs_75813\|\|full | 8158 | No | NA | 6 | 0 | 5 | Not-determined | Genome-fragment | NA | NA | 0 |
| S7_vs_76052\|\|full | 5882 | No | NA | 8 | 0 | 1 | Not-determined | Genome-fragment | NA | NA | 0 |
| S17_vs_k141_61435\|\|full | 2267 | No | NA | 3 | 0 | 3 | Low-quality | Genome-fragment | 0.65 | AAI-based (high-confidence) | 0 |
| S7_vs_76150\|\|full | 7023 | No | NA | 7 | 0 | 4 | Not-determined | Genome-fragment | NA | NA | 0 |
| S11_extracted_k141_83868 | 2267 | No | NA | 4 | 0 | 0 | Low-quality | Genome-fragment | 2.56 | AAI-based (medium-confidence) | 0 |
| S14_vs_k141_112830\|\|full | 2267 | No | NA | 6 | 2 | 0 | Low-quality | Genome-fragment | 6.27 | AAI-based (high-confidence) | 0 |
| S16_extracted_20237 | 2267 | No | NA | 4 | 0 | 3 | Low-quality | Genome-fragment | 3.78 | AAI-based (medium-confidence) | 0 |

| | | | | | | | | | | | |
|---|---|---|---|---|---|---|---|---|---|---|---|
| S12_extracted_k141_15667 | 2267 | No | NA | 4 | 1 | 0 | Low-quality | Genome-fragment | 3.55 | AAI-based (high-confidence) | 0 |
| S12_vs_k141_84746\|\|full | 2267 | No | NA | 7 | 1 | 0 | Low-quality | Genome-fragment | 5.27 | HMM-based (lower-bound) | 0 |
| S7_vs_76464\|\|full | 1531 | No | NA | 3 | 0 | 1 | Not-determined | Genome-fragment | NA | NA | 0 |
| S9_vs_k141_88751\|\|full | 2268 | No | NA | 6 | 2 | 0 | Low-quality | Genome-fragment | 5.88 | AAI-based (high-confidence) | 0 |
| S16_vs_6953\|\|full | 2268 | No | NA | 3 | 0 | 1 | Low-quality | Genome-fragment | 1.1 | AAI-based (medium-confidence) | 0 |
| S7_vs_76757\|\|full | 3716 | No | NA | 4 | 0 | 3 | Not-determined | Genome-fragment | NA | NA | 0 |
| S7_vs_76811\|\|full | 1315 | No | NA | 3 | 0 | 1 | Not-determined | Genome-fragment | NA | NA | 0 |
| S7_vs_76886\|\|full | 3154 | No | NA | 4 | 0 | 4 | Not-determined | Genome-fragment | NA | NA | 0 |
| S9_vs_k141_13038\|\|full | 2269 | No | NA | 5 | 2 | 0 | Low-quality | Genome-fragment | 5.72 | AAI-based (high-confidence) | 0 |
| S14_extracted_k141_106906 | 2269 | No | NA | 4 | 3 | 0 | Low-quality | Genome-fragment | 6.03 | AAI-based (high-confidence) | 0 |
| S7_vs_18838\|\|full | 2269 | No | NA | 3 | 0 | 0 | Low-quality | Genome-fragment | 4.42 | AAI-based (high-confidence) | 0 |
| S17_extracted_k141_141936 | 2270 | No | NA | 2 | 0 | 0 | Low-quality | Genome-fragment | 2.21 | AAI-based (high-confidence) | 0 |
| S7_vs_77494\|\|full | 3907 | No | NA | 3 | 0 | 3 | Not-determined | Genome-fragment | NA | NA | 0 |
| S7_extracted_57140 | 2270 | No | NA | 3 | 0 | 2 | Low-quality | Genome-fragment | 2.99 | AAI-based (medium-confidence) | 0 |
| S8_vs_k141_32554\|\|full | 2270 | No | NA | 3 | 1 | 0 | Low-quality | Genome-fragment | 1.23 | HMM-based (lower-bound) | 0 |
| S7_vs_77842\|\|full | 6203 | No | NA | 5 | 0 | 0 | Not-determined | Genome-fragment | NA | NA | 0 |
| S11_extracted_k141_100385 | 2271 | No | NA | 2 | 1 | 0 | Low-quality | Genome-fragment | 0.78 | HMM-based (lower-bound) | 0 |
| S14_vs_k141_47242\|\|full | 2271 | No | NA | 2 | 0 | 1 | Low-quality | Genome-fragment | 5.82 | AAI-based (medium-confidence) | 0 |
| S7_vs_77941\|\|full | 1505 | No | NA | 3 | 0 | 1 | Not-determined | Genome-fragment | NA | NA | 0 |
| S3_vs_k141_73476\|\|full | 2271 | No | NA | 3 | 0 | 2 | Low-quality | Genome-fragment | 0.52 | AAI-based (medium-confidence) | 0 |
| S7_vs_78034\|\|full | 2261 | No | NA | 2 | 0 | 1 | Not-determined | Genome-fragment | NA | NA | 0 |
| S7_vs_23282\|\|full | 2271 | No | NA | 3 | 0 | 2 | Low-quality | Genome-fragment | 1.64 | AAI-based (medium-confidence) | 0 |
| S15_vs_46242\|\|full | 2271 | No | NA | 3 | 0 | 0 | Low-quality | Genome-fragment | 2.34 | AAI-based (medium-confidence) | 0 |
| S17_extracted_k141_74884 | 2272 | No | NA | 2 | 0 | 1 | Low-quality | Genome-fragment | 1.08 | AAI-based (medium-confidence) | 0 |
| S14_extracted_k141_744 | 2272 | No | NA | 8 | 7 | 0 | Low-quality | Genome-fragment | 7.45 | AAI-based (high-confidence) | 0 |
| S7_vs_78284\|\|full | 1732 | No | NA | 5 | 0 | 0 | Not-determined | Genome-fragment | NA | NA | 0 |
| S7_vs_78453\|\|full | 1522 | No | NA | 3 | 0 | 0 | Not-determined | Genome-fragment | NA | NA | 0 |
| S16_vs_118349\|\|full | 2272 | No | NA | 3 | 3 | 0 | Low-quality | Genome-fragment | 6.24 | AAI-based (high-confidence) | 0 |
| S7_vs_78507\|\|full | 40891 | No | NA | 67 | 15 | 0 | Medium-quality | Genome-fragment | 88.01 | AAI-based (high-confidence) | 0 |
| S7_vs_78536\|\|full | 7532 | No | NA | 9 | 0 | 2 | Not-determined | Genome-fragment | NA | NA | 0 |
| S10_vs_36169\|\|full | 2272 | No | NA | 4 | 0 | 1 | Low-quality | Genome-fragment | 6.29 | AAI-based (medium-confidence) | 0 |
| S10_vs_75238\|\|full | 2272 | Yes | 1893 | 6 | 2 | 1 | Low-quality | Genome-fragment | 5.17 | AAI-based (high-confidence) | 16.68 |
| S11_extracted_k141_38567 | 2273 | No | NA | 2 | 0 | 2 | Low-quality | Genome-fragment | 3.31 | AAI-based (medium-confidence) | 0 |
| S7_vs_78693\|\|full | 9573 | No | NA | 9 | 0 | 4 | Not-determined | Genome-fragment | NA | NA | 0 |
| S7_vs_78704\|\|full | 1876 | No | NA | 4 | 0 | 2 | Not-determined | Genome-fragment | NA | NA | 0 |
| S11_vs_k141_1282\|\|full | 2273 | No | NA | 3 | 1 | 0 | Low-quality | Genome-fragment | 5.97 | AAI-based (high-confidence) | 0 |
| S13_vs_37534\|\|full | 2273 | Yes | 598 | 4 | 1 | 1 | Low-quality | Genome-fragment | 0.84 | AAI-based (high-confidence) | 73.69 |
| S10_vs_94482\|\|full | 2273 | No | NA | 5 | 2 | 0 | Low-quality | Genome-fragment | 5.31 | AAI-based (medium-confidence) | 0 |
| S7_vs_78888\|\|full | 4027 | No | NA | 6 | 0 | 0 | Not-determined | Genome-fragment | NA | NA | 0 |
| S12_vs_k141_167085\|\|full | 2273 | No | NA | 4 | 3 | 0 | Low-quality | Genome-fragment | 5.59 | AAI-based (high-confidence) | 0 |
| S7_vs_78950\|\|full | 1328 | No | NA | 2 | 0 | 2 | Not-determined | Genome-fragment | NA | NA | 0 |
| S15_extracted_3153 | 2273 | No | NA | 3 | 0 | 0 | Low-quality | Genome-fragment | 3.67 | AAI-based (high-confidence) | 0 |
| S4_extracted_10878 | 2273 | No | NA | 4 | 2 | 0 | Low-quality | Genome-fragment | 1.92 | HMM-based (lower-bound) | 0 |
| S1_vs_k141_61752\|\|full | 2274 | No | NA | 4 | 1 | 0 | Low-quality | Genome-fragment | 3.77 | AAI-based (high-confidence) | 0 |
| S3_vs_k141_102367\|\|full | 2274 | No | NA | 2 | 0 | 0 | Low-quality | Genome-fragment | 4.28 | AAI-based (medium-confidence) | 0 |
| S5_vs_k141_33328\|\|full | 2274 | No | NA | 2 | 0 | 1 | Low-quality | Genome-fragment | 4.22 | AAI-based (high-confidence) | 0 |
| S16_vs_85336\|\|lt2gene | 2274 | No | NA | 2 | 2 | 0 | Low-quality | Genome-fragment | 5.68 | AAI-based (high-confidence) | 0 |
| S10_vs_32599\|\|lt2gene | 2274 | No | NA | 1 | 1 | 0 | Low-quality | Genome-fragment | 4.14 | AAI-based (high-confidence) | 0 |
| S12_vs_k141_28124\|\|full | 2274 | No | NA | 5 | 2 | 0 | Low-quality | Genome-fragment | 4.29 | AAI-based (medium-confidence) | 0 |
| S8_vs_k141_70644\|\|full | 2274 | No | NA | 5 | 0 | 0 | Low-quality | Genome-fragment | 5.62 | AAI-based (high-confidence) | 0 |
| S17_vs_k141_47511\|\|full | 2275 | No | NA | 2 | 0 | 1 | Low-quality | Genome-fragment | 1.05 | AAI-based (medium-confidence) | 0 |
| S7_vs_79526\|\|full | 2248 | No | NA | 3 | 0 | 0 | Not-determined | Genome-fragment | NA | NA | 0 |
| S14_vs_k141_31778\|\|full | 2275 | No | NA | 3 | 0 | 0 | Low-quality | Genome-fragment | 3.81 | AAI-based (high-confidence) | 0 |
| S7_vs_79636\|\|full | 2749 | No | NA | 3 | 0 | 1 | Not-determined | Genome-fragment | NA | NA | 0 |
| S7_vs_79795\|\|full | 1196 | No | NA | 3 | 0 | 0 | Not-determined | Genome-fragment | NA | NA | 0 |
| S7_vs_79815\|\|full | 1685 | No | NA | 4 | 0 | 0 | Not-determined | Genome-fragment | NA | NA | 0 |
| S7_vs_79906\|\|full | 2839 | No | NA | 2 | 0 | 1 | Not-determined | Genome-fragment | NA | NA | 0 |
| S11_vs_k141_31521\|\|lt2gene | 2276 | No | NA | 1 | 1 | 0 | Low-quality | Genome-fragment | 6.23 | AAI-based (high-confidence) | 0 |
| S7_vs_80006\|\|full | 1389 | No | NA | 3 | 0 | 3 | Not-determined | Genome-fragment | NA | NA | 0 |
| S7_vs_80009\|\|full | 5932 | No | NA | 5 | 0 | 1 | Not-determined | Genome-fragment | NA | NA | 0 |
| S7_vs_80010\|\|full | 2213 | No | NA | 6 | 0 | 1 | Not-determined | Genome-fragment | NA | NA | 0 |
| S7_vs_96307\|\|full | 2276 | No | NA | 6 | 1 | 0 | Low-quality | Genome-fragment | 5.41 | AAI-based (high-confidence) | 0 |
| S7_vs_80095\|\|full | 1260 | No | NA | 4 | 0 | 0 | Not-determined | Genome-fragment | NA | NA | 0 |
| S6_extracted_k141_55416 | 2276 | No | NA | 7 | 6 | 0 | Low-quality | Genome-fragment | 7.5 | AAI-based (high-confidence) | 0 |
| S6_vs_k141_38230\|\|full | 2276 | No | NA | 3 | 3 | 0 | Low-quality | Genome-fragment | 6.04 | AAI-based (high-confidence) | 0 |
| S14_extracted_k141_43415 | 2277 | No | NA | 4 | 3 | 0 | Low-quality | Genome-fragment | 3.5 | HMM-based (lower-bound) | 0 |
| S7_vs_80214\|\|full | 3569 | No | NA | 3 | 0 | 1 | Not-determined | Genome-fragment | NA | NA | 0 |
| S7_vs_30742\|\|lt2gene | 2277 | No | NA | 1 | 1 | 0 | Low-quality | Genome-fragment | 5.08 | AAI-based (high-confidence) | 0 |
| S7_vs_80315\|\|full | 2117 | No | NA | 4 | 0 | 0 | Not-determined | Genome-fragment | NA | NA | 0 |
| S11_vs_k141_47406\|\|full | 2278 | No | NA | 4 | 0 | 0 | Low-quality | Genome-fragment | 3.58 | AAI-based (high-confidence) | 0 |
| S7_vs_80432\|\|full | 14357 | No | NA | 7 | 0 | 5 | Not-determined | Genome-fragment | NA | NA | 0 |
| S14_vs_k141_35460\|\|full | 2278 | No | NA | 3 | 0 | 2 | Low-quality | Genome-fragment | 3.01 | AAI-based (medium-confidence) | 0 |
| S7_vs_80652\|\|full | 2313 | No | NA | 6 | 0 | 1 | Not-determined | Genome-fragment | NA | NA | 0 |
| S14_vs_k141_87214\|\|full | 2279 | No | NA | 4 | 0 | 0 | Low-quality | Genome-fragment | 1.8 | AAI-based (medium-confidence) | 0 |
| S5_vs_k141_18659\|\|full | 2279 | No | NA | 6 | 1 | 0 | Low-quality | Genome-fragment | 4.06 | AAI-based (medium-confidence) | 0 |
| S7_vs_80826\|\|full | 7118 | No | NA | 18 | 0 | 0 | Not-determined | Genome-fragment | NA | NA | 0 |
| S7_vs_80835\|\|full | 1260 | No | NA | 3 | 0 | 3 | Not-determined | Genome-fragment | NA | NA | 0 |
| S5_vs_k141_116373\|\|full | 2279 | No | NA | 3 | 0 | 0 | Low-quality | Genome-fragment | 2.77 | AAI-based (high-confidence) | 0 |
| S7_vs_80892\|\|full | 4353 | No | NA | 7 | 0 | 0 | Not-determined | Genome-fragment | NA | NA | 0 |
| S5_vs_k141_63140\|\|full | 2279 | No | NA | 3 | 0 | 2 | Low-quality | Genome-fragment | 3.8 | AAI-based (medium-confidence) | 0 |
| S7_vs_81123\|\|full | 2362 | No | NA | 3 | 0 | 1 | Not-determined | Genome-fragment | NA | NA | 0 |
| S7_vs_81141\|\|full | 2343 | No | NA | 2 | 0 | 2 | Not-determined | Genome-fragment | NA | NA | 0 |
| S7_vs_30827\|\|full | 2279 | No | NA | 3 | 3 | 0 | Low-quality | Genome-fragment | 5.59 | AAI-based (high-confidence) | 0 |
| S7_vs_59914\|\|full | 2279 | No | NA | 7 | 0 | 1 | Low-quality | Genome-fragment | 6.62 | AAI-based (high-confidence) | 0 |
| S7_vs_81498\|\|full | 1458 | No | NA | 3 | 0 | 0 | Not-determined | Genome-fragment | NA | NA | 0 |
| S7_vs_81551\|\|full | 1303 | No | NA | 3 | 0 | 0 | Not-determined | Genome-fragment | NA | NA | 0 |
| S16_vs_16450\|\|full | 2279 | No | NA | 4 | 0 | 0 | Low-quality | Genome-fragment | 4.43 | AAI-based (high-confidence) | 0 |
| S7_vs_81553\|\|full | 3706 | No | NA | 4 | 0 | 0 | Not-determined | Genome-fragment | NA | NA | 0 |
| S7_vs_81677\|\|full | 1855 | No | NA | 3 | 0 | 2 | Not-determined | Genome-fragment | NA | NA | 0 |
| S12_vs_k141_175672\|\|full | 2279 | No | NA | 5 | 0 | 0 | Low-quality | Genome-fragment | 3.98 | AAI-based (medium-confidence) | 0 |
| S7_vs_81712\|\|full | 2080 | No | NA | 3 | 0 | 2 | Not-determined | Genome-fragment | NA | NA | 0 |
| S7_vs_81823\|\|full | 2386 | No | NA | 3 | 0 | 0 | Not-determined | Genome-fragment | NA | NA | 0 |
| S12_vs_k141_60869\|\|full | 2279 | No | NA | 5 | 0 | 0 | Low-quality | Genome-fragment | 5.64 | AAI-based (high-confidence) | 0 |
| S7_vs_81871\|\|full | 5815 | No | NA | 7 | 0 | 2 | Not-determined | Genome-fragment | NA | NA | 0 |
| S12_vs_k141_37622\|\|full | 2279 | No | NA | 3 | 0 | 1 | Low-quality | Genome-fragment | 1.49 | AAI-based (high-confidence) | 0 |
| S8_extracted_k141_77302 | 2279 | No | NA | 3 | 0 | 0 | Low-quality | Genome-fragment | 5.9 | AAI-based (medium-confidence) | 0 |
| S7_vs_81990\|\|full | 22604 | No | NA | 26 | 0 | 6 | Not-determined | Genome-fragment | NA | NA | 0 |
| S8_vs_k141_185418\|\|full | 2279 | No | NA | 7 | 2 | 0 | Low-quality | Genome-fragment | 3.94 | AAI-based (high-confidence) | 0 |
| S7_vs_82092\|\|full | 1382 | No | NA | 2 | 0 | 0 | Not-determined | Genome-fragment | NA | NA | 0 |
| S7_vs_82140\|\|full | 22206 | Yes | 19525 | 28 | 12 | 3 | Medium-quality | Genome-fragment | 51.21 | AAI-based (high-confidence) | 12.07 |
| S1_vs_k141_72278\|\|full | 2280 | No | NA | 3 | 0 | 1 | Low-quality | Genome-fragment | 3.9 | AAI-based (medium-confidence) | 0 |
| S7_vs_82383\|\|full | 2249 | No | NA | 3 | 0 | 2 | Not-determined | Genome-fragment | NA | NA | 0 |
| S3_vs_k141_107409\|\|full | 2280 | No | NA | 4 | 0 | 4 | Low-quality | Genome-fragment | 2.31 | AAI-based (medium-confidence) | 0 |
| S5_vs_k141_14513\|\|full | 2280 | No | NA | 5 | 1 | 0 | Low-quality | Genome-fragment | 6.32 | AAI-based (high-confidence) | 0 |
| S8_vs_k141_177021\|\|full | 2280 | No | NA | 3 | 0 | 2 | Low-quality | Genome-fragment | 6.25 | AAI-based (medium-confidence) | 0 |
| S7_vs_82471\|\|full | 2430 | No | NA | 2 | 0 | 2 | Not-determined | Genome-fragment | NA | NA | 0 |
| S7_vs_82534\|\|full | 2198 | No | NA | 2 | 0 | 2 | Not-determined | Genome-fragment | NA | NA | 0 |
| S13_vs_85462\|\|full | 2281 | No | NA | 3 | 0 | 0 | Low-quality | Genome-fragment | 1.19 | AAI-based (medium-confidence) | 0 |
| S7_vs_82610\|\|full | 1865 | No | NA | 2 | 0 | 2 | Not-determined | Genome-fragment | NA | NA | 0 |
| S7_vs_82658\|\|full | 1147 | No | NA | 2 | 0 | 1 | Not-determined | Genome-fragment | NA | NA | 0 |
| S7_vs_82663\|\|full | 2365 | No | NA | 3 | 0 | 1 | Not-determined | Genome-fragment | NA | NA | 0 |
| S3_vs_k141_91277\|\|full | 2281 | No | NA | 2 | 0 | 1 | Low-quality | Genome-fragment | 1.18 | AAI-based (medium-confidence) | 0 |
| S12_vs_k141_73442\|\|full | 2281 | No | NA | 5 | 2 | 0 | Low-quality | Genome-fragment | 4.42 | AAI-based (high-confidence) | 0 |
| S7_vs_82753\|\|full | 1874 | No | NA | 3 | 0 | 1 | Not-determined | Genome-fragment | NA | NA | 0 |
| S8_vs_k141_38334\|\|full | 2281 | No | NA | 3 | 3 | 0 | Low-quality | Genome-fragment | 7.42 | AAI-based (high-confidence) | 0 |
| S17_vs_k141_16213\|\|full | 2282 | No | NA | 7 | 0 | 0 | Low-quality | Genome-fragment | 2.43 | AAI-based (high-confidence) | 0 |
| S7_vs_83100\|\|full | 43538 | No | NA | 67 | 30 | 0 | High-quality | High-quality | 100 | AAI-based (medium-confidence) | 0 |
| S7_vs_83166\|\|full | 1592 | No | NA | 4 | 0 | 0 | Not-determined | Genome-fragment | NA | NA | 0 |
| S6_vs_k141_45672\|\|full | 2282 | No | NA | 6 | 1 | 0 | Low-quality | Genome-fragment | 6.57 | AAI-based (medium-confidence) | 0 |
| S7_vs_83322\|\|full | 1461 | No | NA | 3 | 0 | 1 | Not-determined | Genome-fragment | NA | NA | 0 |
| S17_vs_k141_165857\|\|full | 2283 | No | NA | 2 | 0 | 1 | Low-quality | Genome-fragment | 0.51 | AAI-based (medium-confidence) | 0 |
| S7_vs_83339\|\|full | 2514 | No | NA | 3 | 0 | 2 | Not-determined | Genome-fragment | NA | NA | 0 |
| S9_vs_k141_7784\|\|full | 2283 | No | NA | 5 | 2 | 0 | Low-quality | Genome-fragment | 5.01 | AAI-based (medium-confidence) | 0 |
| S9_vs_k141_47124\|\|full | 2283 | No | NA | 5 | 0 | 0 | Low-quality | Genome-fragment | 4.78 | AAI-based (medium-confidence) | 0 |
| S7_vs_83474\|\|full | 21040 | No | NA | 36 | 6 | 1 | Medium-quality | Genome-fragment | 51.99 | AAI-based (high-confidence) | 0 |
| S11_vs_k141_88564\|\|full | 2283 | No | NA | 5 | 1 | 0 | Low-quality | Genome-fragment | 5.07 | AAI-based (high-confidence) | 0 |
| S7_vs_83598\|\|full | 2136 | No | NA | 2 | 0 | 1 | Not-determined | Genome-fragment | NA | NA | 0 |
| S1_extracted_k141_158144 | 2283 | No | NA | 7 | 1 | 0 | Low-quality | Genome-fragment | 1.3 | AAI-based (high-confidence) | 0 |
| S7_vs_85461\|\|full | 2283 | No | NA | 5 | 2 | 0 | Low-quality | Genome-fragment | 6.17 | AAI-based (high-confidence) | 0 |
| S7_vs_83730\|\|full | 1920 | No | NA | 4 | 0 | 2 | Not-determined | Genome-fragment | NA | NA | 0 |
| S7_vs_83778\|\|full | 2806 | No | NA | 3 | 0 | 2 | Not-determined | Genome-fragment | NA | NA | 0 |
| S12_vs_k141_114630\|\|full | 2283 | No | NA | 3 | 1 | 0 | Low-quality | Genome-fragment | 4.24 | HMM-based (lower-bound) | 0 |
| S7_vs_83817\|\|full | 3445 | No | NA | 4 | 0 | 2 | Not-determined | Genome-fragment | NA | NA | 0 |
| S8_vs_k141_132572\|\|full | 2283 | No | NA | 6 | 0 | 0 | Low-quality | Genome-fragment | 4.68 | AAI-based (medium-confidence) | 0 |
| S8_vs_k141_57195\|\|full | 2283 | No | NA | 4 | 1 | 0 | Low-quality | Genome-fragment | 2.83 | HMM-based (lower-bound) | 0 |
| S7_vs_83910\|\|full | 1529 | No | NA | 2 | 0 | 1 | Not-determined | Genome-fragment | NA | NA | 0 |
| S7_vs_83957\|\|full | 1910 | No | NA | 4 | 0 | 0 | Not-determined | Genome-fragment | NA | NA | 0 |
| S7_vs_83983\|\|full | 1821 | No | NA | 3 | 0 | 0 | Not-determined | Genome-fragment | NA | NA | 0 |
| S9_vs_k141_40891\|\|full | 2284 | No | NA | 3 | 2 | 0 | Low-quality | Genome-fragment | 5.74 | AAI-based (high-confidence) | 0 |
| S7_vs_84289\|\|full | 2191 | No | NA | 3 | 0 | 3 | Not-determined | Genome-fragment | NA | NA | 0 |
| S7_vs_84290\|\|full | 1694 | No | NA | 3 | 0 | 1 | Not-determined | Genome-fragment | NA | NA | 0 |
| S7_vs_84346\|\|full | 1304 | No | NA | 2 | 0 | 1 | Not-determined | Genome-fragment | NA | NA | 0 |
| S7_vs_84456\|\|full | 2357 | No | NA | 4 | 0 | 2 | Not-determined | Genome-fragment | NA | NA | 0 |
| S11_vs_k141_4638\|\|full | 2284 | No | NA | 7 | 2 | 0 | Low-quality | Genome-fragment | 4.23 | AAI-based (medium-confidence) | 0 |
| S3_vs_k141_121250\|\|full | 2284 | No | NA | 3 | 0 | 3 | Low-quality | Genome-fragment | 1 | AAI-based (medium-confidence) | 0 |
| S7_vs_84583\|\|full | 6138 | No | NA | 4 | 0 | 4 | Not-determined | Genome-fragment | NA | NA | 0 |
| S16_vs_154796\|\|full | 2284 | No | NA | 3 | 0 | 2 | Low-quality | Genome-fragment | 0.71 | AAI-based (medium-confidence) | 0 |
| S7_vs_84612\|\|full | 1647 | No | NA | 3 | 0 | 0 | Not-determined | Genome-fragment | NA | NA | 0 |
| S7_vs_84618\|\|full | 1156 | No | NA | 2 | 0 | 0 | Not-determined | Genome-fragment | NA | NA | 0 |
| S4_vs_VIRSorter_18816-cat_2 | 2284 | No | NA | 5 | 3 | 0 | Low-quality | Genome-fragment | 6.55 | AAI-based (high-confidence) | 0 |
| S8_vs_k141_165832\|\|full | 2284 | No | NA | 5 | 2 | 0 | Low-quality | Genome-fragment | 7.78 | AAI-based (high-confidence) | 0 |
| S17_vs_k141_150829\|\|full | 2285 | No | NA | 4 | 1 | 0 | Low-quality | Genome-fragment | 3.43 | AAI-based (high-confidence) | 0 |
| S7_vs_84752\|\|full | 2313 | No | NA | 3 | 0 | 2 | Not-determined | Genome-fragment | NA | NA | 0 |
| S7_vs_84779\|\|full | 2516 | No | NA | 4 | 0 | 1 | Not-determined | Genome-fragment | NA | NA | 0 |
| S13_vs_52244\|\|full | 2285 | No | NA | 3 | 0 | 0 | Low-quality | Genome-fragment | 5.36 | AAI-based (high-confidence) | 0 |
| S3_vs_k141_43898\|\|full | 2285 | No | NA | 3 | 0 | 0 | Low-quality | Genome-fragment | 5.21 | AAI-based (high-confidence) | 0 |
| S5_vs_k141_77027\|\|full | 2285 | No | NA | 2 | 0 | 0 | Low-quality | Genome-fragment | 5.65 | AAI-based (medium-confidence) | 0 |
| S12_extracted_k141_23191 | 2285 | No | NA | 4 | 0 | 1 | Low-quality | Genome-fragment | 1.29 | AAI-based (medium-confidence) | 0 |
| S7_vs_85011\|\|full | 2734 | No | NA | 3 | 0 | 0 | Not-determined | Genome-fragment | NA | NA | 0 |
| S7_vs_85068\|\|full | 1641 | No | NA | 3 | 0 | 1 | Not-determined | Genome-fragment | NA | NA | 0 |
| S7_vs_85078\|\|full | 1462 | No | NA | 2 | 0 | 1 | Not-determined | Genome-fragment | NA | NA | 0 |
| S9_vs_k141_90868\|\|full | 2286 | No | NA | 2 | 0 | 1 | Low-quality | Genome-fragment | 1.47 | AAI-based (medium-confidence) | 0 |
| S3_vs_k141_81229\|\|full | 2286 | No | NA | 5 | 0 | 0 | Low-quality | Genome-fragment | 4.33 | AAI-based (high-confidence) | 0 |
| S5_vs_k141_9450\|\|full | 2286 | No | NA | 4 | 1 | 0 | Low-quality | Genome-fragment | 2.57 | HMM-based (lower-bound) | 0 |
| S9_vs_k141_83593\|\|full | 2287 | No | NA | 3 | 2 | 0 | Low-quality | Genome-fragment | 4.78 | AAI-based (medium-confidence) | 0 |
| S14_extracted_k141_111483 | 2287 | No | NA | 3 | 2 | 0 | Low-quality | Genome-fragment | 3.08 | HMM-based (lower-bound) | 0 |
| S7_vs_85488\|\|full | 1875 | No | NA | 3 | 0 | 0 | Not-determined | Genome-fragment | NA | NA | 0 |
| S7_vs_111555\|\|full | 2287 | No | NA | 5 | 0 | 0 | Low-quality | Genome-fragment | 1.88 | AAI-based (medium-confidence) | 0 |
| S7_vs_85688\|\|full | 6208 | No | NA | 7 | 0 | 5 | Not-determined | Genome-fragment | NA | NA | 0 |
| S6_vs_k141_47010\|\|lt2gene | 2287 | No | NA | 1 | 1 | 0 | Low-quality | Genome-fragment | 6 | AAI-based (high-confidence) | 0 |
| S8_vs_k141_202274\|\|full | 2287 | No | NA | 5 | 1 | 0 | Low-quality | Genome-fragment | 2.9 | AAI-based (high-confidence) | 0 |
| S7_vs_85764\|\|full | 4467 | No | NA | 3 | 0 | 1 | Not-determined | Genome-fragment | NA | NA | 0 |
| S3_vs_k141_80651\|\|full | 2288 | No | NA | 2 | 2 | 0 | Low-quality | Genome-fragment | 3.97 | AAI-based (high-confidence) | 0 |
| S7_extracted_23953 | 2288 | No | NA | 3 | 0 | 0 | Low-quality | Genome-fragment | 7.47 | AAI-based (high-confidence) | 0 |
| S7_vs_85922\|\|full | 3091 | No | NA | 2 | 0 | 2 | Not-determined | Genome-fragment | NA | NA | 0 |
| S13_vs_118292\|\|full | 2289 | No | NA | 7 | 0 | 1 | Low-quality | Genome-fragment | 2.42 | AAI-based (medium-confidence) | 0 |
| S1_extracted_k141_27863 | 2289 | No | NA | 2 | 0 | 1 | Low-quality | Genome-fragment | 0.4 | AAI-based (high-confidence) | 0 |
| S7_vs_86377\|\|full | 33765 | No | NA | 59 | 21 | 0 | High-quality | High-quality | 96.73 | AAI-based (high-confidence) | 0 |
| S3_vs_k141_65285\|\|full | 2289 | No | NA | 3 | 0 | 0 | Low-quality | Genome-fragment | 1.37 | AAI-based (high-confidence) | 0 |
| S5_vs_k141_86705\|\|full | 2289 | No | NA | 2 | 1 | 0 | Low-quality | Genome-fragment | 1.53 | AAI-based (high-confidence) | 0 |
| S10_vs_35524\|\|lt2gene | 2289 | No | NA | 2 | 1 | 0 | Low-quality | Genome-fragment | 5.49 | AAI-based (high-confidence) | 0 |
| S8_vs_k141_141787\|\|full | 2289 | No | NA | 4 | 0 | 0 | Low-quality | Genome-fragment | 1.39 | AAI-based (medium-confidence) | 0 |
| S3_vs_k141_20077\|\|full | 2290 | No | NA | 3 | 0 | 1 | Low-quality | Genome-fragment | 1.02 | AAI-based (high-confidence) | 0 |
| S16_vs_88511\|\|full | 2290 | No | NA | 3 | 0 | 0 | Low-quality | Genome-fragment | 1.91 | AAI-based (medium-confidence) | 0 |
| S7_vs_86818\|\|full | 1600 | No | NA | 3 | 0 | 2 | Not-determined | Genome-fragment | NA | NA | 0 |
| S11_extracted_k141_32880 | 2291 | No | NA | 7 | 1 | 0 | Low-quality | Genome-fragment | 3.66 | AAI-based (high-confidence) | 0 |
| S13_extracted_149123 | 2291 | No | NA | 5 | 0 | 0 | Low-quality | Genome-fragment | 4.07 | AAI-based (high-confidence) | 0 |
| S7_vs_86976\|\|full | 2641 | No | NA | 3 | 0 | 0 | Not-determined | Genome-fragment | NA | NA | 0 |
| S14_vs_k141_94118\|\|full | 2291 | No | NA | 6 | 0 | 0 | Low-quality | Genome-fragment | 1.31 | AAI-based (high-confidence) | 0 |
| S1_vs_k141_125235\|\|full | 2291 | No | NA | 2 | 0 | 1 | Low-quality | Genome-fragment | 1.02 | AAI-based (medium-confidence) | 0 |
| S7_vs_87116\|\|full | 2232 | No | NA | 2 | 0 | 1 | Not-determined | Genome-fragment | NA | NA | 0 |
| S5_vs_k141_103140\|\|full | 2291 | No | NA | 3 | 1 | 0 | Low-quality | Genome-fragment | 1.57 | HMM-based (lower-bound) | 0 |
| S7_vs_87269\|\|full | 1678 | No | NA | 3 | 0 | 1 | Not-determined | Genome-fragment | NA | NA | 0 |
| S7_vs_87279\|\|full | 54131 | No | NA | 87 | 27 | 0 | Medium-quality | Genome-fragment | 89.5 | AAI-based (high-confidence) | 0 |
| S10_vs_42592\|\|full | 2291 | No | NA | 3 | 0 | 2 | Low-quality | Genome-fragment | 1.76 | AAI-based (medium-confidence) | 0 |
| S8_vs_k141_166077\|\|full | 2291 | No | NA | 3 | 0 | 1 | Low-quality | Genome-fragment | 1.44 | AAI-based (high-confidence) | 0 |
| S7_vs_87439\|\|full | 23882 | No | NA | 44 | 7 | 0 | Medium-quality | Genome-fragment | 59.67 | AAI-based (high-confidence) | 0 |
| S7_vs_54271\|\|full | 2292 | No | NA | 5 | 0 | 1 | Low-quality | Genome-fragment | 2.44 | AAI-based (medium-confidence) | 0 |
| S12_vs_k141_133251\|\|full | 2292 | No | NA | 3 | 1 | 0 | Low-quality | Genome-fragment | 4.45 | AAI-based (high-confidence) | 0 |
| S15_vs_22040\|\|full | 2292 | No | NA | 4 | 0 | 0 | Low-quality | Genome-fragment | 4.47 | AAI-based (high-confidence) | 0 |
| S8_vs_k141_119978\|\|full | 2292 | No | NA | 4 | 0 | 0 | Low-quality | Genome-fragment | 5.45 | AAI-based (medium-confidence) | 0 |
| S14_extracted_k141_77175 | 2293 | No | NA | 1 | 1 | 0 | Low-quality | Genome-fragment | 5.85 | AAI-based (high-confidence) | 0 |
| S3_extracted_k141_61838 | 2293 | No | NA | 4 | 0 | 0 | Low-quality | Genome-fragment | 6.15 | AAI-based (high-confidence) | 0 |
| S15_vs_10181\|\|full | 2293 | No | NA | 2 | 0 | 0 | Low-quality | Genome-fragment | 4.38 | AAI-based (medium-confidence) | 0 |
| S7_vs_87660\|\|full | 1985 | No | NA | 3 | 0 | 2 | Not-determined | Genome-fragment | NA | NA | 0 |
| S9_extracted_k141_60584 | 2294 | No | NA | 5 | 2 | 0 | Low-quality | Genome-fragment | 6.55 | AAI-based (high-confidence) | 0 |
| S7_vs_87824\|\|full | 3503 | No | NA | 2 | 0 | 2 | Not-determined | Genome-fragment | NA | NA | 0 |
| S16_vs_48622\|\|full | 2294 | No | NA | 3 | 0 | 2 | Low-quality | Genome-fragment | 5.99 | AAI-based (medium-confidence) | 0 |
| S7_vs_87866\|\|full | 1837 | No | NA | 3 | 0 | 1 | Not-determined | Genome-fragment | NA | NA | 0 |
| S17_vs_k141_74468\|\|full | 2295 | No | NA | 6 | 1 | 0 | Low-quality | Genome-fragment | 4.57 | AAI-based (high-confidence) | 0 |
| S9_extracted_k141_30827 | 2295 | No | NA | 5 | 5 | 0 | Low-quality | Genome-fragment | 7.53 | AAI-based (high-confidence) | 0 |
| S11_extracted_k141_49097 | 2295 | No | NA | 3 | 1 | 0 | Low-quality | Genome-fragment | 0.89 | AAI-based (medium-confidence) | 0 |
| S14_extracted_k141_64411 | 2295 | No | NA | 2 | 0 | 2 | Low-quality | Genome-fragment | 5.46 | AAI-based (high-confidence) | 0 |
| S5_vs_k141_65679\|\|full | 2295 | No | NA | 3 | 2 | 0 | Low-quality | Genome-fragment | 5.5 | AAI-based (high-confidence) | 0 |
| S12_vs_k141_152693\|\|full | 2295 | No | NA | 5 | 2 | 0 | Low-quality | Genome-fragment | 6.1 | AAI-based (high-confidence) | 0 |
| S7_vs_88206\|\|full | 1887 | No | NA | 2 | 0 | 1 | Not-determined | Genome-fragment | NA | NA | 0 |
| S7_vs_88266\|\|full | 2480 | No | NA | 4 | 0 | 1 | Not-determined | Genome-fragment | NA | NA | 0 |
| S15_extracted_74557 | 2295 | No | NA | 2 | 2 | 0 | Low-quality | Genome-fragment | 5.18 | AAI-based (high-confidence) | 0 |
| S7_vs_88344\|\|full | 15282 | No | NA | 11 | 0 | 5 | Not-determined | Genome-fragment | NA | NA | 0 |
| S7_vs_88373\|\|full | 1402 | No | NA | 2 | 0 | 1 | Not-determined | Genome-fragment | NA | NA | 0 |
| S14_vs_k141_73809\|\|lt2gene | 2296 | No | NA | 2 | 1 | 0 | Low-quality | Genome-fragment | 2.1 | HMM-based (lower-bound) | 0 |
| S7_vs_88598\|\|full | 5135 | No | NA | 6 | 0 | 4 | Not-determined | Genome-fragment | NA | NA | 0 |
| S7_vs_8076\|\|full | 2296 | No | NA | 5 | 0 | 0 | Low-quality | Genome-fragment | 6.64 | AAI-based (high-confidence) | 0 |
| S7_vs_51783\|\|full | 2296 | No | NA | 7 | 3 | 0 | Low-quality | Genome-fragment | 4.2 | HMM-based (lower-bound) | 0 |
| S7_vs_88779\|\|full | 3659 | No | NA | 6 | 0 | 1 | Not-determined | Genome-fragment | NA | NA | 0 |
| S7_vs_88782\|\|full | 1800 | No | NA | 2 | 0 | 0 | Not-determined | Genome-fragment | NA | NA | 0 |
| S7_vs_88873\|\|full | 1520 | No | NA | 3 | 0 | 0 | Not-determined | Genome-fragment | NA | NA | 0 |
| S7_vs_126864\|\|lt2gene | 2296 | No | NA | 2 | 1 | 0 | Low-quality | Genome-fragment | 5.53 | AAI-based (high-confidence) | 0 |
| S7_vs_88912\|\|full | 2444 | No | NA | 4 | 0 | 2 | Not-determined | Genome-fragment | NA | NA | 0 |
| S7_vs_88984\|\|full | 1347 | No | NA | 2 | 0 | 1 | Not-determined | Genome-fragment | NA | NA | 0 |
| S16_vs_98178\|\|full | 2296 | No | NA | 3 | 0 | 2 | Low-quality | Genome-fragment | 1.38 | AAI-based (high-confidence) | 0 |
| S7_vs_89090\|\|full | 4914 | No | NA | 6 | 0 | 4 | Not-determined | Genome-fragment | NA | NA | 0 |
| S7_vs_89176\|\|full | 5075 | No | NA | 3 | 0 | 2 | Not-determined | Genome-fragment | NA | NA | 0 |
| S8_extracted_k141_94235 | 2296 | No | NA | 4 | 0 | 3 | Low-quality | Genome-fragment | 5.82 | AAI-based (medium-confidence) | 0 |
| S8_vs_k141_205508\|\|full | 2296 | No | NA | 3 | 0 | 0 | Low-quality | Genome-fragment | 1.07 | AAI-based (medium-confidence) | 0 |
| S14_vs_k141_32345\|\|full | 2297 | No | NA | 2 | 0 | 0 | Low-quality | Genome-fragment | 1.25 | AAI-based (medium-confidence) | 0 |
| S14_vs_k141_70109\|\|full | 2297 | No | NA | 4 | 0 | 1 | Low-quality | Genome-fragment | 4.19 | AAI-based (medium-confidence) | 0 |
| S3_extracted_k141_80653 | 2297 | No | NA | 2 | 1 | 0 | Low-quality | Genome-fragment | 1.63 | AAI-based (high-confidence) | 0 |
| S3_vs_k141_14263\|\|full | 2297 | No | NA | 7 | 2 | 0 | Low-quality | Genome-fragment | 6.63 | AAI-based (high-confidence) | 0 |
| S7_vs_89601\|\|full | 2688 | No | NA | 3 | 0 | 1 | Not-determined | Genome-fragment | NA | NA | 0 |
| S7_vs_89618\|\|full | 1704 | No | NA | 2 | 0 | 1 | Not-determined | Genome-fragment | NA | NA | 0 |
| S16_vs_161186\|\|full | 2297 | No | NA | 3 | 0 | 0 | Low-quality | Genome-fragment | 6.43 | AAI-based (high-confidence) | 0 |
| S7_vs_89746\|\|full | 3429 | No | NA | 3 | 0 | 3 | Not-determined | Genome-fragment | NA | NA | 0 |
| S10_vs_89371\|\|full | 2297 | No | NA | 3 | 0 | 0 | Low-quality | Genome-fragment | 6.16 | AAI-based (medium-confidence) | 0 |
| S15_extracted_9664 | 2297 | No | NA | 2 | 0 | 1 | Low-quality | Genome-fragment | 1.29 | AAI-based (high-confidence) | 0 |
| S8_vs_k141_150652\|\|full | 2297 | No | NA | 2 | 1 | 0 | Low-quality | Genome-fragment | 1.38 | HMM-based (lower-bound) | 0 |
| S1_vs_k141_148662\|\|full | 2298 | No | NA | 4 | 0 | 0 | Low-quality | Genome-fragment | 4.52 | AAI-based (high-confidence) | 0 |
| S14_vs_k141_63521\|\|full | 2299 | No | NA | 3 | 1 | 0 | Low-quality | Genome-fragment | 2.21 | AAI-based (medium-confidence) | 0 |
| S7_vs_90128\|\|full | 1483 | No | NA | 3 | 0 | 1 | Not-determined | Genome-fragment | NA | NA | 0 |
| S7_vs_86675\|\|full | 2299 | No | NA | 6 | 0 | 0 | Low-quality | Genome-fragment | 4.68 | AAI-based (medium-confidence) | 0 |
| S12_vs_k141_155639\|\|full | 2299 | No | NA | 4 | 0 | 0 | Low-quality | Genome-fragment | 3.43 | AAI-based (high-confidence) | 0 |
| S5_vs_k141_18610\|\|full | 2300 | No | NA | 5 | 1 | 0 | Low-quality | Genome-fragment | 4.45 | HMM-based (lower-bound) | 0 |
| S7_vs_90391\|\|full | 1398 | No | NA | 2 | 0 | 2 | Not-determined | Genome-fragment | NA | NA | 0 |
| S7_vs_90468\|\|full | 3160 | No | NA | 5 | 0 | 2 | Not-determined | Genome-fragment | NA | NA | 0 |
| S7_vs_90565\|\|full | 4821 | No | NA | 4 | 0 | 3 | Not-determined | Genome-fragment | NA | NA | 0 |
| S6_vs_k141_39628\|\|full | 2300 | No | NA | 2 | 1 | 0 | Low-quality | Genome-fragment | 0.93 | HMM-based (lower-bound) | 0 |
| S7_vs_90627\|\|full | 2158 | No | NA | 5 | 0 | 0 | Not-determined | Genome-fragment | NA | NA | 0 |
| S8_vs_k141_45352\|\|full | 2300 | No | NA | 8 | 0 | 0 | Low-quality | Genome-fragment | 6.42 | AAI-based (medium-confidence) | 0 |
| S7_vs_90763\|\|full | 1123 | No | NA | 2 | 0 | 0 | Not-determined | Genome-fragment | NA | NA | 0 |
| S16_vs_165769\|\|full | 2301 | No | NA | 4 | 1 | 0 | Low-quality | Genome-fragment | 5.11 | HMM-based (lower-bound) | 0 |
| S10_vs_59025\|\|full | 2301 | No | NA | 4 | 1 | 0 | Low-quality | Genome-fragment | 3.79 | AAI-based (high-confidence) | 0 |
| S7_vs_90911\|\|full | 1651 | No | NA | 2 | 0 | 2 | Not-determined | Genome-fragment | NA | NA | 0 |
| S7_vs_90915\|\|full | 45811 | No | NA | 80 | 30 | 0 | High-quality | High-quality | 100 | AAI-based (high-confidence) | 0 |
| S8_extracted_k141_11654 | 2301 | No | NA | 3 | 3 | 0 | Low-quality | Genome-fragment | 8.12 | HMM-based (lower-bound) | 0 |
| S8_extracted_k141_205835 | 2301 | No | NA | 4 | 3 | 0 | Low-quality | Genome-fragment | 5.1 | AAI-based (high-confidence) | 0 |
| S7_vs_91036\|\|full | 1491 | No | NA | 4 | 0 | 0 | Not-determined | Genome-fragment | NA | NA | 0 |
| S7_vs_91134\|\|full | 2325 | No | NA | 2 | 0 | 2 | Not-determined | Genome-fragment | NA | NA | 0 |
| S1_vs_k141_105277\|\|full | 2302 | No | NA | 2 | 0 | 2 | Low-quality | Genome-fragment | 1.25 | AAI-based (medium-confidence) | 0 |
| S3_vs_k141_104664\|\|full | 2302 | No | NA | 3 | 1 | 0 | Low-quality | Genome-fragment | 4.46 | AAI-based (medium-confidence) | 0 |
| S7_vs_91332\|\|full | 1237 | No | NA | 2 | 0 | 2 | Not-determined | Genome-fragment | NA | NA | 0 |
| S5_vs_k141_123815\|\|full | 2302 | No | NA | 2 | 0 | 1 | Low-quality | Genome-fragment | 0.74 | AAI-based (medium-confidence) | 0 |
| S7_vs_91445\|\|full | 5810 | No | NA | 4 | 0 | 3 | Not-determined | Genome-fragment | NA | NA | 0 |
| S7_vs_42567\|\|full | 2302 | No | NA | 6 | 2 | 0 | Low-quality | Genome-fragment | 6.14 | AAI-based (medium-confidence) | 0 |
| S7_vs_91464\|\|full | 1370 | No | NA | 3 | 0 | 1 | Not-determined | Genome-fragment | NA | NA | 0 |
| S7_vs_91496\|\|full | 4091 | No | NA | 5 | 0 | 1 | Not-determined | Genome-fragment | NA | NA | 0 |
| S12_vs_k141_41936\|\|full | 2302 | No | NA | 6 | 0 | 1 | Low-quality | Genome-fragment | 5.49 | AAI-based (medium-confidence) | 0 |
| S3_vs_k141_124515\|\|full | 2303 | No | NA | 3 | 0 | 1 | Low-quality | Genome-fragment | 2.87 | AAI-based (high-confidence) | 0 |
| S7_vs_91928\|\|full | 1896 | No | NA | 4 | 0 | 0 | Not-determined | Genome-fragment | NA | NA | 0 |
| S2_vs_k141_20306\|\|full | 2303 | No | NA | 6 | 1 | 0 | Low-quality | Genome-fragment | 3.87 | AAI-based (high-confidence) | 0 |
| S1_vs_k141_1928\|\|full | 2304 | No | NA | 5 | 0 | 0 | Low-quality | Genome-fragment | 1.21 | AAI-based (medium-confidence) | 0 |
| S7_vs_92014\|\|full | 1466 | No | NA | 4 | 0 | 0 | Not-determined | Genome-fragment | NA | NA | 0 |
| S10_vs_78312\|\|full | 2304 | No | NA | 5 | 2 | 0 | Low-quality | Genome-fragment | 6.35 | AAI-based (medium-confidence) | 0 |
| S10_vs_49029\|\|lt2gene | 2304 | No | NA | 2 | 2 | 0 | Low-quality | Genome-fragment | 3.86 | AAI-based (high-confidence) | 0 |
| S7_vs_92109\|\|full | 2599 | No | NA | 3 | 0 | 0 | Not-determined | Genome-fragment | NA | NA | 0 |
| S17_vs_k141_140639\|\|full | 2305 | No | NA | 4 | 0 | 2 | Low-quality | Genome-fragment | 3.17 | AAI-based (medium-confidence) | 0 |
| S17_vs_k141_173157\|\|full | 2305 | No | NA | 5 | 2 | 0 | Low-quality | Genome-fragment | 3.09 | AAI-based (high-confidence) | 0 |
| S17_vs_k141_79768\|\|full | 2305 | No | NA | 6 | 0 | 0 | Low-quality | Genome-fragment | 3.32 | AAI-based (high-confidence) | 0 |
| S7_vs_92303\|\|full | 1109 | No | NA | 3 | 0 | 1 | Not-determined | Genome-fragment | NA | NA | 0 |
| S7_vs_92500\|\|full | 1777 | No | NA | 2 | 0 | 1 | Not-determined | Genome-fragment | NA | NA | 0 |
| S7_vs_122160\|\|full | 2305 | No | NA | 6 | 2 | 0 | Low-quality | Genome-fragment | 3.86 | AAI-based (medium-confidence) | 0 |
| S7_vs_92560\|\|full | 2960 | No | NA | 3 | 0 | 2 | Not-determined | Genome-fragment | NA | NA | 0 |
| S10_extracted_54842 | 2305 | No | NA | 2 | 0 | 0 | Low-quality | Genome-fragment | 6.19 | AAI-based (high-confidence) | 0 |
| S12_vs_k141_77962\|\|full | 2305 | No | NA | 3 | 0 | 0 | Low-quality | Genome-fragment | 4.21 | AAI-based (high-confidence) | 0 |
| S15_vs_45391\|\|full | 2305 | No | NA | 3 | 0 | 0 | Low-quality | Genome-fragment | 5.05 | AAI-based (high-confidence) | 0 |
| S7_vs_92826\|\|full | 2063 | No | NA | 2 | 0 | 1 | Not-determined | Genome-fragment | NA | NA | 0 |
| S7_vs_92923\|\|full | 7287 | No | NA | 5 | 0 | 4 | Not-determined | Genome-fragment | NA | NA | 0 |
| S8_vs_k141_142396\|\|full | 2305 | No | NA | 3 | 0 | 1 | Low-quality | Genome-fragment | 0.67 | AAI-based (high-confidence) | 0 |
| S10_vs_83963\|\|full | 2306 | No | NA | 3 | 0 | 1 | Low-quality | Genome-fragment | 1.25 | AAI-based (medium-confidence) | 0 |
| S7_vs_93037\|\|full | 1753 | No | NA | 3 | 0 | 0 | Not-determined | Genome-fragment | NA | NA | 0 |
| S12_vs_k141_114590\|\|full | 2306 | No | NA | 3 | 0 | 1 | Low-quality | Genome-fragment | 2.03 | AAI-based (high-confidence) | 0 |
| S15_vs_55584\|\|full | 2306 | No | NA | 4 | 0 | 0 | Low-quality | Genome-fragment | 1.38 | AAI-based (high-confidence) | 0 |
| S7_vs_93098\|\|full | 8627 | No | NA | 9 | 0 | 9 | Not-determined | Genome-fragment | NA | NA | 0 |
| S7_vs_93103\|\|full | 1999 | No | NA | 6 | 0 | 0 | Not-determined | Genome-fragment | NA | NA | 0 |
| S7_vs_93513\|\|full | 2765 | No | NA | 3 | 0 | 3 | Not-determined | Genome-fragment | NA | NA | 0 |
| S8_extracted_k141_138998 | 2306 | No | NA | 5 | 0 | 0 | Low-quality | Genome-fragment | 5.41 | AAI-based (high-confidence) | 0 |
| S9_vs_k141_32769\|\|full | 2307 | No | NA | 5 | 0 | 0 | Low-quality | Genome-fragment | 3.96 | AAI-based (high-confidence) | 0 |
| S11_vs_k141_22782\|\|full | 2307 | No | NA | 5 | 2 | 0 | Low-quality | Genome-fragment | 3.4 | HMM-based (lower-bound) | 0 |
| S12_extracted_k141_67601 | 2307 | No | NA | 4 | 3 | 0 | Low-quality | Genome-fragment | 5.28 | AAI-based (high-confidence) | 0 |
| S7_vs_93650\|\|full | 1948 | No | NA | 4 | 0 | 0 | Not-determined | Genome-fragment | NA | NA | 0 |
| S8_vs_k141_192967\|\|full | 2307 | No | NA | 5 | 0 | 2 | Low-quality | Genome-fragment | 1.26 | AAI-based (medium-confidence) | 0 |
| S8_vs_k141_207007\|\|full | 2307 | No | NA | 2 | 0 | 2 | Low-quality | Genome-fragment | 1.26 | AAI-based (medium-confidence) | 0 |
| S3_extracted_k141_32176 | 2308 | No | NA | 2 | 0 | 1 | Low-quality | Genome-fragment | 1.25 | AAI-based (medium-confidence) | 0 |
| S7_vs_93948\|\|full | 5382 | No | NA | 6 | 0 | 4 | Not-determined | Genome-fragment | NA | NA | 0 |
| S16_extracted_118713 | 2308 | No | NA | 3 | 0 | 0 | Low-quality | Genome-fragment | 5.27 | AAI-based (high-confidence) | 0 |
| S16_vs_135033\|\|full | 2308 | No | NA | 6 | 0 | 0 | Low-quality | Genome-fragment | 3.98 | AAI-based (high-confidence) | 0 |
| S12_extracted_k141_99986 | 2308 | No | NA | 5 | 0 | 0 | Low-quality | Genome-fragment | 6.51 | AAI-based (medium-confidence) | 0 |
| S7_vs_94107\|\|full | 7429 | No | NA | 9 | 0 | 6 | Not-determined | Genome-fragment | NA | NA | 0 |
| S12_vs_k141_76024\|\|full | 2308 | No | NA | 4 | 0 | 1 | Low-quality | Genome-fragment | 4.39 | AAI-based (medium-confidence) | 0 |
| S8_vs_k141_46982\|\|full | 2308 | No | NA | 8 | 1 | 0 | Low-quality | Genome-fragment | 5.42 | AAI-based (high-confidence) | 0 |
| S17_vs_k141_132303\|\|full | 2309 | No | NA | 2 | 0 | 0 | Low-quality | Genome-fragment | 2.08 | AAI-based (high-confidence) | 0 |
| S9_vs_k141_24073\|\|full | 2309 | No | NA | 3 | 0 | 1 | Low-quality | Genome-fragment | 0.21 | AAI-based (medium-confidence) | 0 |
| S7_vs_94222\|\|full | 3071 | No | NA | 3 | 0 | 0 | Not-determined | Genome-fragment | NA | NA | 0 |
| S9_vs_k141_32331\|\|full | 2309 | No | NA | 4 | 0 | 0 | Low-quality | Genome-fragment | 3.23 | AAI-based (medium-confidence) | 0 |
| S12_vs_k141_24623\|\|full | 2309 | No | NA | 3 | 2 | 0 | Low-quality | Genome-fragment | 1.28 | HMM-based (lower-bound) | 0 |
| S14_vs_k141_77384\|\|full | 2310 | No | NA | 4 | 0 | 2 | Low-quality | Genome-fragment | 2.42 | AAI-based (high-confidence) | 0 |
| S12_vs_k141_136568\|\|lt2gene | 2310 | No | NA | 1 | 1 | 0 | Low-quality | Genome-fragment | 4.45 | AAI-based (medium-confidence) | 0 |
| S8_extracted_k141_121482 | 2310 | No | NA | 1 | 0 | 1 | Low-quality | Genome-fragment | 4.91 | AAI-based (high-confidence) | 0 |
| S7_vs_94543\|\|full | 1628 | No | NA | 2 | 0 | 1 | Not-determined | Genome-fragment | NA | NA | 0 |
| S7_vs_94649\|\|full | 3048 | No | NA | 6 | 0 | 0 | Not-determined | Genome-fragment | NA | NA | 0 |
| S17_vs_k141_2845\|\|full | 2311 | No | NA | 3 | 0 | 1 | Low-quality | Genome-fragment | 1.45 | AAI-based (medium-confidence) | 0 |
| S1_vs_k141_28093\|\|full | 2311 | No | NA | 3 | 0 | 1 | Low-quality | Genome-fragment | 1.93 | AAI-based (medium-confidence) | 0 |
| S3_extracted_k141_73490 | 2311 | No | NA | 2 | 1 | 0 | Low-quality | Genome-fragment | 3.98 | AAI-based (high-confidence) | 0 |
| S16_vs_87965\|\|full | 2311 | No | NA | 4 | 4 | 0 | Low-quality | Genome-fragment | 4.22 | AAI-based (medium-confidence) | 0 |
| S7_vs_94938\|\|full | 3798 | No | NA | 7 | 0 | 0 | Not-determined | Genome-fragment | NA | NA | 0 |
| S15_extracted_46223 | 2311 | No | NA | 3 | 0 | 0 | Low-quality | Genome-fragment | 2.36 | AAI-based (medium-confidence) | 0 |
| S7_vs_95072\|\|full | 2886 | No | NA | 3 | 0 | 2 | Not-determined | Genome-fragment | NA | NA | 0 |
| S6_vs_k141_37781\|\|full | 2311 | No | NA | 8 | 1 | 0 | Low-quality | Genome-fragment | 3.63 | AAI-based (high-confidence) | 0 |
| S7_vs_95186\|\|full | 1273 | No | NA | 3 | 0 | 1 | Not-determined | Genome-fragment | NA | NA | 0 |
| S7_vs_95274\|\|full | 3700 | No | NA | 5 | 0 | 3 | Not-determined | Genome-fragment | NA | NA | 0 |
| S6_vs_k141_52864\|\|full | 2311 | No | NA | 3 | 0 | 1 | Low-quality | Genome-fragment | 5.11 | AAI-based (medium-confidence) | 0 |
| S7_vs_95413\|\|full | 3854 | No | NA | 4 | 0 | 4 | Not-determined | Genome-fragment | NA | NA | 0 |
| S7_vs_95438\|\|full | 1235 | No | NA | 4 | 0 | 0 | Not-determined | Genome-fragment | NA | NA | 0 |
| S17_vs_k141_75709\|\|full | 2312 | No | NA | 4 | 0 | 1 | Low-quality | Genome-fragment | 0.6 | AAI-based (medium-confidence) | 0 |
| S7_vs_95761\|\|full | 2023 | No | NA | 3 | 0 | 0 | Not-determined | Genome-fragment | NA | NA | 0 |
| S7_vs_95794\|\|full | 6879 | No | NA | 9 | 0 | 5 | Not-determined | Genome-fragment | NA | NA | 0 |
| S7_vs_96036\|\|full | 1172 | No | NA | 3 | 0 | 2 | Not-determined | Genome-fragment | NA | NA | 0 |
| S7_vs_96038\|\|full | 3228 | No | NA | 4 | 0 | 2 | Not-determined | Genome-fragment | NA | NA | 0 |
| S9_vs_k141_17830\|\|full | 2312 | No | NA | 5 | 2 | 0 | Low-quality | Genome-fragment | 5.96 | AAI-based (high-confidence) | 0 |
| S14_extracted_k141_108048 | 2312 | No | NA | 3 | 3 | 0 | Low-quality | Genome-fragment | 7.62 | AAI-based (high-confidence) | 0 |
| S1_vs_k141_159698\|\|full | 2312 | No | NA | 3 | 1 | 0 | Low-quality | Genome-fragment | 1.59 | AAI-based (high-confidence) | 0 |
| S7_vs_50886\|\|full | 2312 | No | NA | 5 | 1 | 0 | Low-quality | Genome-fragment | 1.46 | HMM-based (lower-bound) | 0 |
| S7_vs_72036\|\|full | 2312 | No | NA | 3 | 3 | 0 | Low-quality | Genome-fragment | 5.06 | AAI-based (high-confidence) | 0 |
| S7_vs_96549\|\|full | 1315 | No | NA | 2 | 0 | 0 | Not-determined | Genome-fragment | NA | NA | 0 |
| S7_vs_96582\|\|full | 1524 | No | NA | 2 | 0 | 1 | Not-determined | Genome-fragment | NA | NA | 0 |
| S10_extracted_52379 | 2312 | No | NA | 2 | 0 | 0 | Low-quality | Genome-fragment | 4.91 | AAI-based (high-confidence) | 0 |
| S12_vs_k141_2117\|\|full | 2312 | No | NA | 3 | 0 | 1 | Low-quality | Genome-fragment | 3.89 | AAI-based (medium-confidence) | 0 |
| S7_vs_96846\|\|full | 1002 | No | NA | 3 | 0 | 1 | Not-determined | Genome-fragment | NA | NA | 0 |
| S1_vs_k141_116811\|\|full | 2313 | No | NA | 3 | 0 | 1 | Low-quality | Genome-fragment | 6.48 | AAI-based (medium-confidence) | 0 |
| S7_vs_97017\|\|full | 2484 | No | NA | 3 | 0 | 3 | Not-determined | Genome-fragment | NA | NA | 0 |
| S3_vs_k141_73484\|\|full | 2313 | No | NA | 6 | 1 | 0 | Low-quality | Genome-fragment | 4.2 | AAI-based (high-confidence) | 0 |
| S8_extracted_k141_79807 | 2313 | No | NA | 2 | 0 | 0 | Low-quality | Genome-fragment | 8.98 | AAI-based (high-confidence) | 0 |
| S7_vs_97312\|\|full | 2271 | No | NA | 3 | 0 | 1 | Not-determined | Genome-fragment | NA | NA | 0 |
| S8_vs_k141_39804\|\|full | 2313 | No | NA | 4 | 0 | 1 | Low-quality | Genome-fragment | 1.32 | AAI-based (medium-confidence) | 0 |
| S8_vs_k141_129488\|\|full | 2313 | No | NA | 3 | 0 | 2 | Low-quality | Genome-fragment | 1.4 | AAI-based (medium-confidence) | 0 |
| S7_vs_97447\|\|full | 3363 | No | NA | 4 | 0 | 2 | Not-determined | Genome-fragment | NA | NA | 0 |
| S7_vs_97507\|\|full | 3351 | No | NA | 3 | 0 | 3 | Not-determined | Genome-fragment | NA | NA | 0 |
| S13_vs_103896\|\|full | 2314 | No | NA | 3 | 0 | 1 | Low-quality | Genome-fragment | 0.56 | AAI-based (medium-confidence) | 0 |
| S1_vs_k141_60210\|\|full | 2314 | No | NA | 3 | 2 | 0 | Low-quality | Genome-fragment | 4.98 | AAI-based (high-confidence) | 0 |
| S7_vs_97705\|\|full | 2499 | No | NA | 3 | 0 | 2 | Not-determined | Genome-fragment | NA | NA | 0 |
| S7_vs_97737\|\|full | 74627 | No | NA | 66 | 3 | 36 | High-quality | High-quality | 100 | AAI-based (medium-confidence) | 0 |
| S7_vs_90586\|\|full | 2314 | No | NA | 3 | 1 | 0 | Low-quality | Genome-fragment | 5.58 | AAI-based (high-confidence) | 0 |
| S10_vs_1341\|\|full | 2314 | No | NA | 4 | 4 | 0 | Low-quality | Genome-fragment | 6.55 | AAI-based (medium-confidence) | 0 |
| S12_vs_k141_62134\|\|full | 2314 | No | NA | 3 | 3 | 0 | Low-quality | Genome-fragment | 3.87 | AAI-based (high-confidence) | 0 |
| S7_vs_97884\|\|full | 9709 | No | NA | 5 | 0 | 4 | Not-determined | Genome-fragment | NA | NA | 0 |
| S7_vs_97938\|\|full | 4303 | No | NA | 7 | 0 | 0 | Not-determined | Genome-fragment | NA | NA | 0 |
| S17_extracted_k141_116745 | 2315 | No | NA | 3 | 0 | 1 | Low-quality | Genome-fragment | 14.38 | AAI-based (medium-confidence) | 0 |
| S11_vs_k141_99074\|\|full | 2315 | No | NA | 2 | 0 | 1 | Low-quality | Genome-fragment | 5.18 | AAI-based (high-confidence) | 0 |
| S1_extracted_k141_145209 | 2315 | No | NA | 2 | 2 | 0 | Low-quality | Genome-fragment | 6.72 | AAI-based (high-confidence) | 0 |
| S7_vs_98142\|\|full | 4209 | No | NA | 7 | 0 | 3 | Not-determined | Genome-fragment | NA | NA | 0 |
| S3_extracted_k141_82528 | 2315 | No | NA | 6 | 2 | 0 | Low-quality | Genome-fragment | 5.97 | AAI-based (medium-confidence) | 0 |
| S7_vs_98179\|\|full | 2828 | No | NA | 3 | 0 | 1 | Not-determined | Genome-fragment | NA | NA | 0 |
| S10_extracted_75228 | 2315 | No | NA | 5 | 0 | 1 | Low-quality | Genome-fragment | 4.25 | AAI-based (high-confidence) | 0 |
| S12_vs_k141_127051\|\|full | 2316 | No | NA | 2 | 0 | 0 | Low-quality | Genome-fragment | 1.59 | AAI-based (medium-confidence) | 0 |
| S9_vs_k141_2520\|\|full | 2317 | No | NA | 3 | 0 | 1 | Low-quality | Genome-fragment | 3.63 | AAI-based (medium-confidence) | 0 |
| S7_vs_98638\|\|full | 1184 | No | NA | 3 | 0 | 1 | Not-determined | Genome-fragment | NA | NA | 0 |
| S7_vs_98642\|\|full | 1451 | No | NA | 2 | 0 | 2 | Not-determined | Genome-fragment | NA | NA | 0 |
| S1_vs_k141_128316\|\|full | 2317 | No | NA | 4 | 2 | 0 | Low-quality | Genome-fragment | 5.53 | AAI-based (medium-confidence) | 0 |
| S5_extracted_k141_30614 | 2317 | No | NA | 1 | 1 | 0 | Low-quality | Genome-fragment | 4.39 | HMM-based (lower-bound) | 0 |
| S5_vs_k141_104917\|\|full | 2317 | No | NA | 3 | 0 | 1 | Low-quality | Genome-fragment | 4.6 | AAI-based (high-confidence) | 0 |
| S7_vs_136381\|\|full | 2317 | No | NA | 5 | 1 | 0 | Low-quality | Genome-fragment | 1.44 | HMM-based (lower-bound) | 0 |
| S12_extracted_k141_184009 | 2317 | No | NA | 1 | 0 | 1 | Low-quality | Genome-fragment | 1.39 | AAI-based (medium-confidence) | 0 |
| S8_extracted_k141_107189 | 2317 | No | NA | 2 | 0 | 0 | Low-quality | Genome-fragment | 0.91 | AAI-based (medium-confidence) | 0 |
| S9_extracted_k141_73776 | 2318 | No | NA | 5 | 3 | 0 | Low-quality | Genome-fragment | 7.64 | AAI-based (high-confidence) | 0 |
| S7_vs_99014\|\|full | 1382 | No | NA | 2 | 0 | 0 | Not-determined | Genome-fragment | NA | NA | 0 |
| S14_extracted_k141_85206 | 2318 | No | NA | 1 | 1 | 0 | Low-quality | Genome-fragment | 5.43 | HMM-based (lower-bound) | 0 |
| S8_vs_k141_117262\|\|full | 2318 | No | NA | 5 | 2 | 0 | Low-quality | Genome-fragment | 4.66 | AAI-based (medium-confidence) | 0 |
| S7_vs_99189\|\|full | 1171 | No | NA | 2 | 0 | 1 | Not-determined | Genome-fragment | NA | NA | 0 |
| S7_vs_99275\|\|full | 3804 | No | NA | 3 | 0 | 2 | Not-determined | Genome-fragment | NA | NA | 0 |
| S7_vs_99310\|\|full | 1098 | No | NA | 4 | 0 | 0 | Not-determined | Genome-fragment | NA | NA | 0 |
| S7_vs_99361\|\|full | 1003 | No | NA | 3 | 0 | 0 | Not-determined | Genome-fragment | NA | NA | 0 |
| S7_vs_84142\|\|full | 2319 | No | NA | 6 | 1 | 0 | Low-quality | Genome-fragment | 11.85 | AAI-based (medium-confidence) | 0 |
| S7_vs_90891\|\|full | 2319 | Yes | 602 | 3 | 1 | 1 | Low-quality | Genome-fragment | 0.69 | HMM-based (lower-bound) | 74.04 |
| S7_vs_99598\|\|full | 1803 | No | NA | 2 | 0 | 1 | Not-determined | Genome-fragment | NA | NA | 0 |
| S7_vs_99674\|\|full | 3456 | No | NA | 5 | 0 | 2 | Not-determined | Genome-fragment | NA | NA | 0 |
| S8_vs_k141_18454\|\|full | 2319 | No | NA | 2 | 0 | 2 | Low-quality | Genome-fragment | 3.89 | AAI-based (medium-confidence) | 0 |
| S11_vs_k141_77759\|\|full | 2320 | No | NA | 6 | 3 | 0 | Low-quality | Genome-fragment | 5.45 | AAI-based (medium-confidence) | 0 |
| S13_vs_94564\|\|full | 2320 | No | NA | 3 | 1 | 0 | Low-quality | Genome-fragment | 4.92 | AAI-based (high-confidence) | 0 |
| S14_extracted_k141_33851 | 2320 | No | NA | 3 | 3 | 0 | Low-quality | Genome-fragment | 7.66 | AAI-based (high-confidence) | 0 |
| S14_vs_k141_29966\|\|full | 2320 | No | NA | 4 | 1 | 0 | Low-quality | Genome-fragment | 5.23 | AAI-based (high-confidence) | 0 |
| S14_vs_k141_19784\|\|full | 2320 | No | NA | 4 | 2 | 0 | Low-quality | Genome-fragment | 5.33 | AAI-based (high-confidence) | 0 |
| S1_vs_k141_16823\|\|full | 2320 | No | NA | 2 | 0 | 0 | Low-quality | Genome-fragment | 5.92 | AAI-based (high-confidence) | 0 |
| S7_vs_100285\|\|full | 1023 | No | NA | 2 | 0 | 0 | Not-determined | Genome-fragment | NA | NA | 0 |
| S8_extracted_k141_85037 | 2320 | No | NA | 2 | 0 | 2 | Low-quality | Genome-fragment | 0.12 | AAI-based (medium-confidence) | 0 |

| | | | | | | | | | | | |
|---|---|---|---|---|---|---|---|---|---|---|---|
| S8_vs_k141_52945||full | 2320 | No | NA | 3 | 0 | 2 | Low-quality | Genome-fragment | 3.42 | AAI-based (medium-confidence) | 0 |
| S17_vs_k141_105909||full | 2321 | No | NA | 6 | 1 | 0 | Low-quality | Genome-fragment | 4.67 | AAI-based (high-confidence) | 0 |
| S7_vs_100640||full | 19185 | No | NA | 28 | 15 | 0 | Medium-quality | Genome-fragment | 54.87 | AAI-based (high-confidence) | 0 |
| S13_extracted_45492 | 2321 | No | NA | 1 | 0 | 0 | Low-quality | Genome-fragment | 2.49 | AAI-based (high-confidence) | 0 |
| S13_extracted_84005 | 2321 | No | NA | 2 | 0 | 1 | Low-quality | Genome-fragment | 5.03 | AAI-based (medium-confidence) | 0 |
| S7_vs_100677||full | 2378 | No | NA | 3 | 0 | 3 | Not-determined | Genome-fragment | NA | NA | 0 |
| S13_vs_123076||full | 2321 | No | NA | 7 | 0 | 0 | Low-quality | Genome-fragment | 3.83 | AAI-based (high-confidence) | 0 |
| NC1_vf_k141_3092 | 2321 | No | NA | 4 | 0 | 0 | Low-quality | Genome-fragment | 4.33 | AAI-based (medium-confidence) | 0 |
| S16_vs_132294||full | 2321 | No | NA | 3 | 0 | 2 | Low-quality | Genome-fragment | 1.01 | AAI-based (medium-confidence) | 0 |
| S7_vs_100830||full | 37520 | No | NA | 57 | 0 | 10 | Not-determined | Genome-fragment | NA | NA | 0 |
| S9_vs_k141_79890||full | 2322 | No | NA | 8 | 0 | 0 | Low-quality | Genome-fragment | 4.9 | AAI-based (medium-confidence) | 0 |
| S7_vs_101006||full | 1145 | No | NA | 3 | 0 | 0 | Not-determined | Genome-fragment | NA | NA | 0 |
| S13_vs_19376||full | 2322 | No | NA | 2 | 0 | 1 | Low-quality | Genome-fragment | 1.75 | AAI-based (high-confidence) | 0 |
| S7_vs_101078||full | 2755 | No | NA | 3 | 0 | 2 | Not-determined | Genome-fragment | NA | NA | 0 |
| S14_extracted_k141_73587 | 2322 | No | NA | 2 | 2 | 0 | Low-quality | Genome-fragment | 5.48 | AAI-based (high-confidence) | 0 |
| S7_vs_101104||full | 2366 | No | NA | 4 | 0 | 2 | Not-determined | Genome-fragment | NA | NA | 0 |
| S7_vs_101221||full | 1949 | No | NA | 2 | 0 | 1 | Not-determined | Genome-fragment | NA | NA | 0 |
| S7_vs_101231||full | 1469 | No | NA | 4 | 0 | 1 | Not-determined | Genome-fragment | NA | NA | 0 |
| S16_vs_147941||full | 2322 | No | NA | 3 | 3 | 0 | Low-quality | Genome-fragment | 5.16 | AAI-based (high-confidence) | 0 |
| S10_vs_83898||full | 2322 | No | NA | 3 | 0 | 0 | Low-quality | Genome-fragment | 3.12 | AAI-based (medium-confidence) | 0 |
| S7_vs_101267||full | 2515 | No | NA | 2 | 0 | 1 | Not-determined | Genome-fragment | NA | NA | 0 |
| S10_vs_96174||full | 2322 | No | NA | 7 | 1 | 0 | Low-quality | Genome-fragment | 3.15 | AAI-based (high-confidence) | 0 |
| S6_vs_k141_48163||full | 2322 | No | NA | 3 | 2 | 0 | Low-quality | Genome-fragment | 4.42 | AAI-based (medium-confidence) | 0 |
| S7_vs_101456||full | 1668 | No | NA | 3 | 0 | 0 | Not-determined | Genome-fragment | NA | NA | 0 |
| S7_vs_101474||full | 2268 | No | NA | 3 | 0 | 2 | Not-determined | Genome-fragment | NA | NA | 0 |
| S8_vs_k141_40861||full | 2322 | No | NA | 5 | 2 | 0 | Low-quality | Genome-fragment | 2.84 | HMM-based (lower-bound) | 0 |
| S7_vs_101564||full | 1109 | No | NA | 3 | 0 | 0 | Not-determined | Genome-fragment | NA | NA | 0 |
| S14_vs_k141_64465||full | 2323 | No | NA | 5 | 0 | 0 | Low-quality | Genome-fragment | 4.25 | AAI-based (high-confidence) | 0 |
| S3_vs_k141_116184||full | 2323 | No | NA | 3 | 0 | 2 | Low-quality | Genome-fragment | 1.84 | AAI-based (medium-confidence) | 0 |
| S5_extracted_k141_27652 | 2323 | No | NA | 4 | 4 | 0 | Low-quality | Genome-fragment | 5.25 | AAI-based (high-confidence) | 0 |
| S17_extracted_k141_1457 | 2324 | No | NA | 7 | 6 | 0 | Low-quality | Genome-fragment | 7.64 | AAI-based (high-confidence) | 0 |
| S7_vs_101714||full | 2082 | No | NA | 2 | 0 | 1 | Not-determined | Genome-fragment | NA | NA | 0 |
| S7_vs_101728||full | 6067 | No | NA | 5 | 0 | 3 | Not-determined | Genome-fragment | NA | NA | 0 |
| S7_vs_101751||full | 91169 | No | NA | 154 | 31 | 2 | High-quality | High-quality | 100 | AAI-based (high-confidence) | 0 |
| S11_vs_k141_9904||full | 2324 | No | NA | 4 | 1 | 0 | Low-quality | Genome-fragment | 1.56 | HMM-based (lower-bound) | 0 |
| S7_vs_101815||full | 1130 | No | NA | 2 | 0 | 1 | Not-determined | Genome-fragment | NA | NA | 0 |
| S7_vs_101919||full | 9678 | No | NA | 6 | 0 | 4 | Not-determined | Genome-fragment | NA | NA | 0 |
| S14_vs_k141_49018||full | 2324 | No | NA | 2 | 0 | 1 | Low-quality | Genome-fragment | 1.95 | AAI-based (medium-confidence) | 0 |
| S3_extracted_k141_65253 | 2324 | No | NA | 4 | 2 | 0 | Low-quality | Genome-fragment | 36.07 | AAI-based (high-confidence) | 0 |
| S7_vs_102072||full | 1941 | No | NA | 3 | 0 | 2 | Not-determined | Genome-fragment | NA | NA | 0 |
| S8_vs_k141_3564||full | 2324 | No | NA | 5 | 0 | 0 | Low-quality | Genome-fragment | 2.71 | AAI-based (medium-confidence) | 0 |
| S7_vs_102125||full | 19310 | No | NA | 21 | 0 | 3 | Not-determined | Genome-fragment | NA | NA | 0 |
| S7_vs_102283||full | 2514 | No | NA | 4 | 0 | 2 | Not-determined | Genome-fragment | NA | NA | 0 |
| S17_vs_k141_143010||lt2gene | 2325 | No | NA | 1 | 1 | 0 | Low-quality | Genome-fragment | 3.08 | HMM-based (lower-bound) | 0 |
| S14_vs_k141_95659||full | 2325 | No | NA | 5 | 5 | 0 | Low-quality | Genome-fragment | 3.91 | AAI-based (high-confidence) | 0 |
| S3_vs_k141_14434||full | 2325 | No | NA | 1 | 1 | 0 | Low-quality | Genome-fragment | 1.38 | AAI-based (medium-confidence) | 0 |
| S7_vs_102388||full | 2153 | No | NA | 3 | 0 | 1 | Not-determined | Genome-fragment | NA | NA | 0 |
| S5_extracted_k141_108741 | 2325 | No | NA | 2 | 0 | 2 | Low-quality | Genome-fragment | 0.86 | AAI-based (high-confidence) | 0 |
| S7_vs_102499||full | 6501 | No | NA | 9 | 0 | 0 | Not-determined | Genome-fragment | NA | NA | 0 |
| S5_vs_k141_33938||full | 2325 | No | NA | 3 | 0 | 2 | Low-quality | Genome-fragment | 3.36 | AAI-based (medium-confidence) | 0 |
| S8_extracted_k141_123728 | 2325 | No | NA | 3 | 3 | 0 | Low-quality | Genome-fragment | 8.2 | HMM-based (lower-bound) | 0 |
| S13_vs_75626||full | 2326 | No | NA | 2 | 0 | 0 | Low-quality | Genome-fragment | 4.59 | AAI-based (medium-confidence) | 0 |
| S1_extracted_k141_3375 | 2326 | No | NA | 1 | 0 | 0 | Low-quality | Genome-fragment | 1.82 | AAI-based (medium-confidence) | 0 |
| S3_vs_k141_84199||full | 2326 | No | NA | 2 | 0 | 1 | Low-quality | Genome-fragment | 1.02 | AAI-based (medium-confidence) | 0 |
| S7_vs_102807||full | 1573 | No | NA | 3 | 0 | 1 | Not-determined | Genome-fragment | NA | NA | 0 |
| S7_vs_102844||full | 1423 | No | NA | 4 | 0 | 0 | Not-determined | Genome-fragment | NA | NA | 0 |
| S5_vs_k141_64211||full | 2326 | No | NA | 5 | 3 | 0 | Low-quality | Genome-fragment | 4.01 | HMM-based (lower-bound) | 0 |
| S5_vs_k141_108213||full | 2326 | No | NA | 3 | 0 | 2 | Low-quality | Genome-fragment | 3.58 | AAI-based (medium-confidence) | 0 |
| S12_extracted_k141_146946 | 2326 | No | NA | 4 | 0 | 0 | Low-quality | Genome-fragment | 3.91 | AAI-based (high-confidence) | 0 |
| S7_vs_103071||full | 4431 | No | NA | 7 | 0 | 4 | Not-determined | Genome-fragment | NA | NA | 0 |
| S7_vs_103093||full | 1130 | No | NA | 2 | 0 | 0 | Not-determined | Genome-fragment | NA | NA | 0 |
| S7_vs_103205||full | 3598 | No | NA | 11 | 0 | 0 | Not-determined | Genome-fragment | NA | NA | 0 |
| S7_vs_103208||full | 1427 | No | NA | 4 | 0 | 0 | Not-determined | Genome-fragment | NA | NA | 0 |
| S8_vs_k141_11347||full | 2326 | No | NA | 3 | 0 | 2 | Low-quality | Genome-fragment | 1.46 | AAI-based (medium-confidence) | 0 |
| S17_vs_k141_168555||full | 2327 | No | NA | 2 | 0 | 2 | Low-quality | Genome-fragment | 5.67 | AAI-based (medium-confidence) | 0 |
| S7_vs_103428||full | 4090 | No | NA | 10 | 0 | 0 | Not-determined | Genome-fragment | NA | NA | 0 |
| S9_vs_k141_76913||full | 2327 | No | NA | 5 | 1 | 0 | Low-quality | Genome-fragment | 1.51 | AAI-based (medium-confidence) | 0 |
| S16_extracted_138590 | 2327 | No | NA | 2 | 2 | 0 | Low-quality | Genome-fragment | 7.11 | AAI-based (high-confidence) | 0 |
| S16_vs_144438||full | 2327 | No | NA | 2 | 0 | 0 | Low-quality | Genome-fragment | 5.86 | AAI-based (high-confidence) | 0 |
| S7_vs_103812||full | 2194 | No | NA | 3 | 0 | 3 | Not-determined | Genome-fragment | NA | NA | 0 |
| S6_vs_k141_50769||full | 2327 | No | NA | 3 | 0 | 0 | Low-quality | Genome-fragment | 1.55 | AAI-based (high-confidence) | 0 |
| S7_vs_103880||full | 1632 | No | NA | 2 | 0 | 0 | Not-determined | Genome-fragment | NA | NA | 0 |
| S17_vs_k141_141866||full | 2328 | No | NA | 4 | 1 | 0 | Low-quality | Genome-fragment | 10.61 | AAI-based (high-confidence) | 0 |
| S9_extracted_k141_44891 | 2328 | No | NA | 4 | 1 | 0 | Low-quality | Genome-fragment | 3.34 | AAI-based (high-confidence) | 0 |
| S7_vs_104193||full | 9686 | No | NA | 11 | 0 | 0 | Not-determined | Genome-fragment | NA | NA | 0 |
| S7_vs_104195||full | 2262 | No | NA | 3 | 0 | 0 | Not-determined | Genome-fragment | NA | NA | 0 |
| S7_vs_104204||full | 19616 | No | NA | 22 | 0 | 6 | Not-determined | Genome-fragment | NA | NA | 0 |
| S1_vs_k141_157181||full | 2328 | No | NA | 4 | 1 | 0 | Low-quality | Genome-fragment | 3.69 | AAI-based (high-confidence) | 0 |
| S11_vs_k141_13896||full | 2329 | No | NA | 3 | 0 | 0 | Low-quality | Genome-fragment | 3.65 | AAI-based (medium-confidence) | 0 |
| S7_vs_104392||full | 1452 | No | NA | 3 | 0 | 0 | Not-determined | Genome-fragment | NA | NA | 0 |
| S7_vs_104493||full | 1009 | No | NA | 2 | 0 | 1 | Not-determined | Genome-fragment | NA | NA | 0 |
| S7_vs_104533||full | 5503 | No | NA | 4 | 0 | 3 | Not-determined | Genome-fragment | NA | NA | 0 |
| S7_vs_104555||full | 1635 | No | NA | 3 | 0 | 1 | Not-determined | Genome-fragment | NA | NA | 0 |
| S3_vs_k141_36707||full | 2329 | No | NA | 5 | 1 | 0 | Low-quality | Genome-fragment | 5.72 | AAI-based (medium-confidence) | 0 |
| S5_vs_k141_34577||full | 2329 | No | NA | 3 | 0 | 0 | Low-quality | Genome-fragment | 1.49 | AAI-based (medium-confidence) | 0 |
| S7_vs_34485||full | 2329 | No | NA | 3 | 0 | 2 | Low-quality | Genome-fragment | 1.09 | AAI-based (high-confidence) | 0 |
| S8_vs_k141_203581||full | 2329 | No | NA | 4 | 0 | 0 | Low-quality | Genome-fragment | 5.5 | AAI-based (medium-confidence) | 0 |
| S7_vs_104779||full | 1326 | No | NA | 2 | 0 | 0 | Not-determined | Genome-fragment | NA | NA | 0 |
| S13_vs_120103||full | 2330 | Yes | 1326 | 6 | 2 | 1 | Low-quality | Genome-fragment | 3.35 | AAI-based (high-confidence) | 43.09 |
| S7_vs_104986||full | 1285 | No | NA | 2 | 0 | 2 | Not-determined | Genome-fragment | NA | NA | 0 |
| S7_vs_105009||full | 3321 | No | NA | 4 | 0 | 1 | Not-determined | Genome-fragment | NA | NA | 0 |
| S5_vs_k141_43665||full | 2330 | No | NA | 4 | 0 | 0 | Low-quality | Genome-fragment | 1.49 | AAI-based (medium-confidence) | 0 |
| S7_vs_105147||full | 1322 | No | NA | 3 | 0 | 2 | Not-determined | Genome-fragment | NA | NA | 0 |
| S7_vs_105210||full | 2646 | No | NA | 3 | 0 | 1 | Not-determined | Genome-fragment | NA | NA | 0 |
| S16_vs_133019||full | 2330 | Yes | 892 | 5 | 1 | 1 | Low-quality | Genome-fragment | 1.86 | AAI-based (medium-confidence) | 61.72 |
| S7_vs_105245||full | 1571 | No | NA | 1 | 0 | 1 | Not-determined | Genome-fragment | NA | NA | 0 |
| S12_vs_k141_8539||full | 2330 | No | NA | 3 | 2 | 0 | Low-quality | Genome-fragment | 5.37 | AAI-based (high-confidence) | 0 |
| S12_vs_k141_17157||full | 2330 | No | NA | 3 | 0 | 0 | Low-quality | Genome-fragment | 2.64 | AAI-based (medium-confidence) | 0 |
| S6_extracted_k141_7145 | 2330 | No | NA | 2 | 0 | 2 | Low-quality | Genome-fragment | 2.37 | AAI-based (medium-confidence) | 0 |
| S8_extracted_k141_186205 | 2330 | No | NA | 1 | 0 | 1 | Low-quality | Genome-fragment | 4.35 | AAI-based (medium-confidence) | 0 |
| S3_vs_k141_111242||full | 2331 | No | NA | 3 | 0 | 0 | Low-quality | Genome-fragment | 7.47 | AAI-based (medium-confidence) | 0 |
| S7_vs_105546||full | 2575 | No | NA | 3 | 0 | 1 | Not-determined | Genome-fragment | NA | NA | 0 |
| S7_vs_105752||full | 1813 | No | NA | 5 | 0 | 0 | Not-determined | Genome-fragment | NA | NA | 0 |
| S5_vs_k141_92112||full | 2331 | No | NA | 5 | 1 | 0 | Low-quality | Genome-fragment | 5.97 | AAI-based (medium-confidence) | 0 |
| S7_vs_105855||full | 2609 | No | NA | 3 | 0 | 3 | Not-determined | Genome-fragment | NA | NA | 0 |
| S5_vs_k141_21844||full | 2331 | No | NA | 5 | 0 | 0 | Low-quality | Genome-fragment | 5.87 | AAI-based (high-confidence) | 0 |
| S7_vs_105921||full | 5417 | No | NA | 6 | 0 | 5 | Not-determined | Genome-fragment | NA | NA | 0 |
| S8_extracted_k141_73201 | 2331 | No | NA | 4 | 4 | 0 | Low-quality | Genome-fragment | 5.37 | AAI-based (high-confidence) | 0 |
| S7_vs_105965||full | 2475 | No | NA | 3 | 0 | 2 | Not-determined | Genome-fragment | NA | NA | 0 |
| S8_extracted_k141_120257 | 2331 | No | NA | 3 | 0 | 2 | Low-quality | Genome-fragment | 1.36 | AAI-based (medium-confidence) | 0 |
| S11_extracted_k141_45503 | 2332 | No | NA | 1 | 0 | 0 | Low-quality | Genome-fragment | 3.46 | AAI-based (high-confidence) | 0 |
| S3_vs_k141_55436||full | 2332 | No | NA | 5 | 0 | 0 | Low-quality | Genome-fragment | 1.44 | AAI-based (medium-confidence) | 0 |
| S7_vs_106108||full | 5145 | No | NA | 6 | 0 | 1 | Not-determined | Genome-fragment | NA | NA | 0 |
| S7_vs_106248||full | 5263 | No | NA | 4 | 0 | 4 | Not-determined | Genome-fragment | NA | NA | 0 |
| S17_extracted_k141_8672 | 2333 | No | NA | 6 | 3 | 0 | Low-quality | Genome-fragment | 7.57 | AAI-based (high-confidence) | 0 |
| S14_extracted_k141_82496 | 2333 | No | NA | 5 | 5 | 0 | Low-quality | Genome-fragment | 7.67 | AAI-based (high-confidence) | 0 |
| S7_extracted_95277 | 2333 | No | NA | 1 | 0 | 1 | Low-quality | Genome-fragment | 0.64 | AAI-based (medium-confidence) | 0 |
| S7_vs_106377||full | 1147 | No | NA | 2 | 0 | 0 | Not-determined | Genome-fragment | NA | NA | 0 |
| S7_vs_106454||full | 2485 | No | NA | 3 | 0 | 2 | Not-determined | Genome-fragment | NA | NA | 0 |
| S7_vs_106464||full | 3041 | No | NA | 3 | 0 | 2 | Not-determined | Genome-fragment | NA | NA | 0 |
| S16_extracted_40905 | 2333 | No | NA | 5 | 0 | 1 | Low-quality | Genome-fragment | 3.16 | AAI-based (medium-confidence) | 0 |
| S16_vs_23810||full | 2333 | No | NA | 3 | 0 | 2 | Low-quality | Genome-fragment | 3.29 | AAI-based (medium-confidence) | 0 |
| S7_vs_106651||full | 4113 | No | NA | 5 | 0 | 4 | Not-determined | Genome-fragment | NA | NA | 0 |
| S7_vs_106704||full | 5116 | No | NA | 4 | 0 | 4 | Not-determined | Genome-fragment | NA | NA | 0 |
| S7_vs_106740||full | 2229 | No | NA | 3 | 0 | 1 | Not-determined | Genome-fragment | NA | NA | 0 |
| S12_extracted_k141_162123 | 2333 | No | NA | 3 | 0 | 0 | Low-quality | Genome-fragment | 3.66 | AAI-based (high-confidence) | 0 |
| S7_vs_106924||full | 1047 | No | NA | 2 | 0 | 0 | Not-determined | Genome-fragment | NA | NA | 0 |
| S7_vs_106963||full | 3008 | No | NA | 5 | 0 | 1 | Not-determined | Genome-fragment | NA | NA | 0 |
| S7_vs_107104||full | 1899 | No | NA | 4 | 0 | 1 | Not-determined | Genome-fragment | NA | NA | 0 |
| S7_vs_107107||full | 4641 | No | NA | 4 | 0 | 0 | Not-determined | Genome-fragment | NA | NA | 0 |
| S7_vs_107164||full | 1559 | No | NA | 3 | 0 | 1 | Not-determined | Genome-fragment | NA | NA | 0 |
| S8_vs_k141_74503||full | 2333 | No | NA | 3 | 2 | 0 | Low-quality | Genome-fragment | 6.9 | HMM-based (lower-bound) | 0 |
| S7_vs_107250||full | 1401 | No | NA | 3 | 0 | 0 | Not-determined | Genome-fragment | NA | NA | 0 |
| S10_extracted_57552 | 2334 | Yes | 918 | 6 | 1 | 1 | Low-quality | Genome-fragment | 0.96 | AAI-based (high-confidence) | 60.67 |
| S7_vs_107369||full | 1423 | No | NA | 3 | 0 | 0 | Not-determined | Genome-fragment | NA | NA | 0 |
| S4_extracted_7905 | 2334 | No | NA | 3 | 0 | 3 | Low-quality | Genome-fragment | 0.81 | AAI-based (high-confidence) | 0 |
| S7_vs_107497||full | 4462 | No | NA | 5 | 0 | 0 | Not-determined | Genome-fragment | NA | NA | 0 |
| S13_vs_135042||full | 2335 | No | NA | 5 | 0 | 0 | Low-quality | Genome-fragment | 5.09 | AAI-based (high-confidence) | 0 |
| S7_vs_107715||full | 3346 | No | NA | 2 | 0 | 1 | Not-determined | Genome-fragment | NA | NA | 0 |
| S7_vs_107820||full | 1825 | No | NA | 2 | 0 | 1 | Not-determined | Genome-fragment | NA | NA | 0 |
| S14_vs_k141_110394||full | 2335 | No | NA | 3 | 0 | 1 | Low-quality | Genome-fragment | 3.91 | AAI-based (high-confidence) | 0 |
| S7_vs_107876||full | 16348 | No | NA | 22 | 7 | 0 | High-quality | High-quality | 95.98 | AAI-based (medium-confidence) | 0 |
| S7_vs_108000||full | 1262 | No | NA | 3 | 0 | 1 | Not-determined | Genome-fragment | NA | NA | 0 |
| S12_extracted_k141_73314 | 2335 | No | NA | 2 | 0 | 2 | Low-quality | Genome-fragment | 1.29 | AAI-based (medium-confidence) | 0 |
| S7_vs_108075||full | 1637 | No | NA | 2 | 0 | 1 | Not-determined | Genome-fragment | NA | NA | 0 |
| S2_extracted_k141_20194 | 2335 | No | NA | 2 | 0 | 0 | Low-quality | Genome-fragment | 5.36 | AAI-based (high-confidence) | 0 |
| S7_vs_108345||full | 2548 | No | NA | 6 | 0 | 0 | Not-determined | Genome-fragment | NA | NA | 0 |
| S9_vs_k141_28470||full | 2336 | No | NA | 5 | 0 | 0 | Low-quality | Genome-fragment | 4.5 | AAI-based (high-confidence) | 0 |
| S7_vs_108378||full | 9838 | No | NA | 19 | 0 | 0 | Not-determined | Genome-fragment | NA | NA | 0 |
| S8_extracted_k141_168031 | 2336 | No | NA | 2 | 0 | 1 | Low-quality | Genome-fragment | 0.64 | AAI-based (high-confidence) | 0 |
| S17_vs_k141_145620||full | 2337 | No | NA | 4 | 2 | 0 | Low-quality | Genome-fragment | 5.49 | AAI-based (medium-confidence) | 0 |
| S1_vs_k141_122895||full | 2337 | No | NA | 4 | 0 | 0 | Low-quality | Genome-fragment | 4.84 | AAI-based (high-confidence) | 0 |
| S1_vs_k141_125188||full | 2337 | No | NA | 3 | 0 | 0 | Low-quality | Genome-fragment | 2.25 | AAI-based (medium-confidence) | 0 |
| S7_vs_125063||full | 2337 | No | NA | 5 | 0 | 1 | Low-quality | Genome-fragment | 4.84 | AAI-based (medium-confidence) | 0 |
| S12_vs_k141_88763||full | 2337 | No | NA | 8 | 1 | 0 | Low-quality | Genome-fragment | 5.34 | AAI-based (medium-confidence) | 0 |
| S7_vs_108612||full | 2694 | No | NA | 3 | 0 | 2 | Not-determined | Genome-fragment | NA | NA | 0 |
| S9_vs_k141_66562||full | 2338 | No | NA | 3 | 0 | 1 | Low-quality | Genome-fragment | 1.39 | AAI-based (high-confidence) | 0 |
| S17_vs_k141_147428||full | 2339 | No | NA | 6 | 1 | 0 | Low-quality | Genome-fragment | 5.83 | AAI-based (high-confidence) | 0 |
| S11_vs_k141_33272||full | 2339 | No | NA | 6 | 1 | 0 | Low-quality | Genome-fragment | 4.09 | AAI-based (high-confidence) | 0 |
| S7_vs_108833||full | 3236 | No | NA | 6 | 0 | 0 | Not-determined | Genome-fragment | NA | NA | 0 |
| S7_vs_108887||full | 6639 | No | NA | 5 | 0 | 1 | Not-determined | Genome-fragment | NA | NA | 0 |
| S14_extracted_k141_112738 | 2339 | No | NA | 3 | 2 | 0 | Low-quality | Genome-fragment | 3.9 | AAI-based (high-confidence) | 0 |
| S7_vs_108958||full | 2193 | No | NA | 3 | 0 | 0 | Not-determined | Genome-fragment | NA | NA | 0 |
| S14_vs_k141_44973||lt2gene | 2339 | No | NA | 2 | 2 | 0 | Low-quality | Genome-fragment | 5.3 | HMM-based (lower-bound) | 0 |
| S1_vs_k141_120598||full | 2339 | No | NA | 4 | 1 | 0 | Low-quality | Genome-fragment | 3.98 | AAI-based (high-confidence) | 0 |
| S5_vs_k141_46125||full | 2339 | No | NA | 3 | 0 | 3 | Low-quality | Genome-fragment | 0.84 | AAI-based (medium-confidence) | 0 |
| S8_extracted_k141_177554 | 2339 | No | NA | 1 | 0 | 1 | Low-quality | Genome-fragment | 2.26 | AAI-based (medium-confidence) | 0 |
| S3_vs_k141_63113||full | 2340 | No | NA | 4 | 0 | 1 | Low-quality | Genome-fragment | 4.57 | AAI-based (high-confidence) | 0 |
| S7_vs_109354||full | 2624 | No | NA | 3 | 0 | 2 | Not-determined | Genome-fragment | NA | NA | 0 |
| S7_vs_109489||full | 3488 | No | NA | 6 | 0 | 2 | Not-determined | Genome-fragment | NA | NA | 0 |
| S7_vs_109582||full | 1308 | No | NA | 3 | 0 | 0 | Not-determined | Genome-fragment | NA | NA | 0 |
| S5_vs_k141_22410||full | 2340 | No | NA | 6 | 2 | 0 | Low-quality | Genome-fragment | 4.37 | AAI-based (medium-confidence) | 0 |
| S7_vs_101069||full | 2340 | No | NA | 3 | 0 | 0 | Low-quality | Genome-fragment | 5.58 | AAI-based (medium-confidence) | 0 |
| S16_vs_119128||full | 2340 | No | NA | 3 | 0 | 2 | Low-quality | Genome-fragment | 6.24 | AAI-based (high-confidence) | 0 |
| S10_vs_90069||full | 2340 | No | NA | 8 | 2 | 0 | Low-quality | Genome-fragment | 1.67 | HMM-based (lower-bound) | 0 |
| S7_vs_109887||full | 1217 | No | NA | 4 | 0 | 1 | Not-determined | Genome-fragment | NA | NA | 0 |
| S8_vs_k141_131207||full | 2340 | No | NA | 2 | 2 | 0 | Low-quality | Genome-fragment | 1.46 | AAI-based (high-confidence) | 0 |
| S17_vs_k141_42418||full | 2341 | No | NA | 3 | 0 | 2 | Low-quality | Genome-fragment | 1.66 | AAI-based (medium-confidence) | 0 |
| S7_vs_109919||full | 11032 | No | NA | 8 | 0 | 2 | Not-determined | Genome-fragment | NA | NA | 0 |
| S7_vs_110024||full | 2650 | No | NA | 4 | 0 | 1 | Not-determined | Genome-fragment | NA | NA | 0 |
| S13_vs_70940||full | 2341 | No | NA | 6 | 0 | 5 | Low-quality | Genome-fragment | 0.87 | AAI-based (medium-confidence) | 0 |
| S7_vs_110141||full | 1668 | No | NA | 4 | 0 | 0 | Not-determined | Genome-fragment | NA | NA | 0 |
| S5_vs_k141_31520||full | 2341 | No | NA | 4 | 0 | 0 | Low-quality | Genome-fragment | 3.7 | AAI-based (high-confidence) | 0 |
| S13_vs_19427||full | 2342 | No | NA | 3 | 0 | 1 | Low-quality | Genome-fragment | 1.14 | AAI-based (medium-confidence) | 0 |
| S7_vs_110291||full | 4189 | No | NA | 11 | 0 | 0 | Not-determined | Genome-fragment | NA | NA | 0 |
| S7_vs_110321||full | 1462 | No | NA | 3 | 0 | 0 | Not-determined | Genome-fragment | NA | NA | 0 |
| S12_vs_k141_24929||full | 2342 | No | NA | 3 | 0 | 2 | Low-quality | Genome-fragment | 2.79 | AAI-based (medium-confidence) | 0 |
| S15_vs_40326||full | 2342 | No | NA | 3 | 0 | 2 | Low-quality | Genome-fragment | 1.07 | AAI-based (medium-confidence) | 0 |
| S6_vs_k141_31634||full | 2342 | No | NA | 5 | 0 | 1 | Low-quality | Genome-fragment | 1.49 | AAI-based (medium-confidence) | 0 |
| S3_vs_k141_95043||full | 2343 | No | NA | 7 | 1 | 0 | Low-quality | Genome-fragment | 4.66 | AAI-based (medium-confidence) | 0 |
| S7_vs_110778||full | 3718 | No | NA | 4 | 0 | 3 | Not-determined | Genome-fragment | NA | NA | 0 |
| S7_vs_110826||full | 2710 | No | NA | 2 | 0 | 1 | Not-determined | Genome-fragment | NA | NA | 0 |
| S9_extracted_k141_73062 | 2344 | No | NA | 5 | 4 | 0 | Low-quality | Genome-fragment | 7.72 | AAI-based (high-confidence) | 0 |
| S7_vs_110932||full | 3231 | No | NA | 3 | 0 | 3 | Not-determined | Genome-fragment | NA | NA | 0 |
| S1_vs_k141_58289||full | 2344 | No | NA | 4 | 0 | 0 | Low-quality | Genome-fragment | 5.25 | AAI-based (high-confidence) | 0 |
| S5_vs_k141_22806||full | 2344 | No | NA | 9 | 1 | 0 | Low-quality | Genome-fragment | 5.55 | AAI-based (medium-confidence) | 0 |
| S10_vs_76493||full | 2344 | No | NA | 5 | 0 | 0 | Low-quality | Genome-fragment | 4.23 | AAI-based (high-confidence) | 0 |
| S8_vs_k141_152503||full | 2344 | No | NA | 3 | 3 | 0 | Low-quality | Genome-fragment | 6.21 | AAI-based (high-confidence) | 0 |
| S7_vs_111205||full | 1555 | No | NA | 3 | 0 | 2 | Not-determined | Genome-fragment | NA | NA | 0 |
| S14_extracted_k141_80575 | 2345 | No | NA | 3 | 3 | 0 | Low-quality | Genome-fragment | 7.7 | AAI-based (high-confidence) | 0 |
| S7_vs_111545||full | 1278 | No | NA | 2 | 0 | 1 | Not-determined | Genome-fragment | NA | NA | 0 |
| S5_extracted_k141_66337 | 2345 | No | NA | 3 | 2 | 0 | Low-quality | Genome-fragment | 1.25 | HMM-based (lower-bound) | 0 |
| S6_vs_k141_26835||full | 2345 | No | NA | 3 | 2 | 0 | Low-quality | Genome-fragment | 6.46 | AAI-based (high-confidence) | 0 |
| S7_vs_111617||full | 1435 | No | NA | 3 | 0 | 0 | Not-determined | Genome-fragment | NA | NA | 0 |
| S11_extracted_k141_10104 | 2346 | No | NA | 4 | 3 | 0 | Low-quality | Genome-fragment | 4.66 | AAI-based (high-confidence) | 0 |
| S7_vs_111831||full | 34066 | No | NA | 51 | 21 | 1 | High-quality | High-quality | 97.46 | AAI-based (high-confidence) | 0 |
| S7_vs_111863||full | 2005 | No | NA | 4 | 0 | 0 | Not-determined | Genome-fragment | NA | NA | 0 |
| S7_vs_111879||full | 2595 | No | NA | 4 | 0 | 1 | Not-determined | Genome-fragment | NA | NA | 0 |
| S13_vs_14338||full | 2346 | No | NA | 3 | 0 | 0 | Low-quality | Genome-fragment | 5.49 | AAI-based (high-confidence) | 0 |
| S7_vs_112015||full | 1970 | No | NA | 3 | 0 | 1 | Not-determined | Genome-fragment | NA | NA | 0 |
| S5_vs_k141_42797||full | 2346 | No | NA | 7 | 4 | 0 | Low-quality | Genome-fragment | 4.64 | AAI-based (medium-confidence) | 0 |
| S16_vs_6459||full | 2346 | No | NA | 4 | 1 | 0 | Low-quality | Genome-fragment | 5.83 | AAI-based (high-confidence) | 0 |
| S7_vs_112328||full | 2781 | No | NA | 4 | 0 | 0 | Not-determined | Genome-fragment | NA | NA | 0 |
| S11_vs_k141_102860||full | 2347 | No | NA | 7 | 2 | 0 | Low-quality | Genome-fragment | 3.63 | AAI-based (high-confidence) | 0 |
| S7_vs_112419||full | 2172 | No | NA | 2 | 0 | 1 | Not-determined | Genome-fragment | NA | NA | 0 |
| S14_vs_k141_84319||full | 2347 | No | NA | 4 | 0 | 3 | Low-quality | Genome-fragment | 5.68 | AAI-based (medium-confidence) | 0 |
| S16_vs_21843||full | 2347 | No | NA | 7 | 1 | 0 | Low-quality | Genome-fragment | 5.99 | AAI-based (high-confidence) | 0 |
| S7_vs_112689||full | 2046 | No | NA | 4 | 0 | 0 | Not-determined | Genome-fragment | NA | NA | 0 |
| S7_vs_112762||full | 1405 | No | NA | 2 | 0 | 0 | Not-determined | Genome-fragment | NA | NA | 0 |
| S16_vs_129478||full | 2347 | No | NA | 6 | 0 | 0 | Low-quality | Genome-fragment | 4.55 | AAI-based (high-confidence) | 0 |
| S10_vs_103958||full | 2347 | No | NA | 3 | 0 | 3 | Low-quality | Genome-fragment | 1.78 | AAI-based (medium-confidence) | 0 |
| S12_extracted_k141_141604 | 2347 | No | NA | 4 | 0 | 1 | Low-quality | Genome-fragment | 3.61 | AAI-based (medium-confidence) | 0 |
| S7_vs_112960||full | 1254 | No | NA | 3 | 0 | 0 | Not-determined | Genome-fragment | NA | NA | 0 |
| S9_vs_k141_6474||full | 2348 | No | NA | 5 | 1 | 0 | Low-quality | Genome-fragment | 4.77 | AAI-based (medium-confidence) | 0 |
| S11_vs_k141_65657||full | 2348 | No | NA | 3 | 0 | 3 | Low-quality | Genome-fragment | 1.08 | AAI-based (medium-confidence) | 0 |
| S7_vs_54716||full | 2348 | No | NA | 3 | 0 | 0 | Low-quality | Genome-fragment | 0.86 | AAI-based (medium-confidence) | 0 |
| S12_vs_k141_164578||lt2gene | 2348 | No | NA | 1 | 1 | 0 | Low-quality | Genome-fragment | 5.36 | AAI-based (high-confidence) | 0 |
| S7_vs_113142||full | 3488 | No | NA | 6 | 0 | 0 | Not-determined | Genome-fragment | NA | NA | 0 |
| S7_vs_25169||full | 2349 | No | NA | 3 | 1 | 0 | Low-quality | Genome-fragment | 6.79 | AAI-based (high-confidence) | 0 |
| S7_vs_113221||full | 1355 | No | NA | 3 | 0 | 0 | Not-determined | Genome-fragment | NA | NA | 0 |
| S16_extracted_126545 | 2349 | No | NA | 5 | 1 | 0 | Low-quality | Genome-fragment | 3.86 | AAI-based (medium-confidence) | 0 |
| S16_vs_74175||full | 2349 | No | NA | 3 | 0 | 2 | Low-quality | Genome-fragment | 1.12 | AAI-based (medium-confidence) | 0 |
| S7_vs_113482||full | 9921 | No | NA | 7 | 0 | 4 | Not-determined | Genome-fragment | NA | NA | 0 |
| S10_vs_62141||full | 2349 | No | NA | 2 | 0 | 0 | Low-quality | Genome-fragment | 2.78 | AAI-based (medium-confidence) | 0 |
| S7_vs_113547||full | 3180 | No | NA | 3 | 0 | 1 | Not-determined | Genome-fragment | NA | NA | 0 |
| S12_extracted_k141_118891 | 2349 | No | NA | 8 | 2 | 0 | Low-quality | Genome-fragment | 3.93 | AAI-based (high-confidence) | 0 |
| S7_vs_113750||full | 1688 | No | NA | 2 | 0 | 0 | Not-determined | Genome-fragment | NA | NA | 0 |
| S12_extracted_k141_92898 | 2349 | No | NA | 3 | 2 | 0 | Low-quality | Genome-fragment | 7.74 | AAI-based (high-confidence) | 0 |
| S8_vs_k141_202064||lt2gene | 2349 | No | NA | 2 | 1 | 0 | Low-quality | Genome-fragment | 5.44 | AAI-based (high-confidence) | 0 |
| S7_vs_113889||full | 1383 | No | NA | 3 | 0 | 1 | Not-determined | Genome-fragment | NA | NA | 0 |
| S1_extracted_k141_25101 | 2350 | No | NA | 4 | 0 | 0 | Low-quality | Genome-fragment | 1.28 | AAI-based (high-confidence) | 0 |
| S7_vs_118089||full | 2350 | No | NA | 3 | 0 | 0 | Low-quality | Genome-fragment | 1 | AAI-based (medium-confidence) | 0 |
| S7_vs_114238||full | 2071 | No | NA | 6 | 0 | 1 | Not-determined | Genome-fragment | NA | NA | 0 |
| S16_vs_77554||full | 2350 | No | NA | 4 | 0 | 0 | Low-quality | Genome-fragment | 4.47 | AAI-based (high-confidence) | 0 |
| S12_vs_k141_43222||full | 2350 | No | NA | 4 | 1 | 0 | Low-quality | Genome-fragment | 5.88 | AAI-based (medium-confidence) | 0 |
| S7_extracted_125045 | 2351 | No | NA | 5 | 2 | 0 | Low-quality | Genome-fragment | 5.88 | AAI-based (high-confidence) | 0 |
| S7_vs_114492||full | 2515 | No | NA | 2 | 0 | 1 | Not-determined | Genome-fragment | NA | NA | 0 |
| S7_vs_114588||full | 1025 | No | NA | 2 | 0 | 0 | Not-determined | Genome-fragment | NA | NA | 0 |
| S7_vs_114590||full | 1133 | No | NA | 3 | 0 | 0 | Not-determined | Genome-fragment | NA | NA | 0 |
| S7_vs_114633||full | 1542 | No | NA | 4 | 0 | 3 | Not-determined | Genome-fragment | NA | NA | 0 |
| S7_vs_114700||full | 1350 | No | NA | 3 | 0 | 0 | Not-determined | Genome-fragment | NA | NA | 0 |
| S16_vs_101203||full | 2351 | No | NA | 3 | 1 | 0 | Low-quality | Genome-fragment | 39.98 | AAI-based (high-confidence) | 0 |
| S17_vs_k141_55825||full | 2352 | No | NA | 2 | 0 | 1 | Low-quality | Genome-fragment | 1.07 | AAI-based (medium-confidence) | 0 |
| S7_vs_114809||full | 3233 | No | NA | 3 | 0 | 3 | Not-determined | Genome-fragment | NA | NA | 0 |
| S1_extracted_k141_157133 | 2352 | No | NA | 1 | 0 | 0 | Low-quality | Genome-fragment | 0.62 | AAI-based (high-confidence) | 0 |
| S16_extracted_55150 | 2352 | No | NA | 3 | 0 | 2 | Low-quality | Genome-fragment | 1.3 | AAI-based (medium-confidence) | 0 |
| S10_vs_14839||full | 2352 | No | NA | 4 | 0 | 0 | Low-quality | Genome-fragment | 4.29 | AAI-based (medium-confidence) | 0 |
| S7_vs_114943||full | 5441 | No | NA | 2 | 0 | 1 | Not-determined | Genome-fragment | NA | NA | 0 |
| S7_vs_115095||full | 4527 | No | NA | 6 | 0 | 1 | Not-determined | Genome-fragment | NA | NA | 0 |
| S7_vs_115098||full | 5095 | No | NA | 5 | 0 | 5 | Not-determined | Genome-fragment | NA | NA | 0 |
| S7_vs_115102||full | 4453 | No | NA | 5 | 0 | 4 | Not-determined | Genome-fragment | NA | NA | 0 |
| S7_vs_115139||full | 1205 | No | NA | 2 | 0 | 1 | Not-determined | Genome-fragment | NA | NA | 0 |
| S10_vs_104037||full | 2352 | No | NA | 6 | 0 | 0 | Low-quality | Genome-fragment | 2.41 | AAI-based (high-confidence) | 0 |
| S8_vs_k141_58570||full | 2352 | No | NA | 2 | 0 | 2 | Low-quality | Genome-fragment | 2.84 | AAI-based (high-confidence) | 0 |
| S17_vs_k141_55692||full | 2353 | No | NA | 3 | 0 | 0 | Low-quality | Genome-fragment | 4.04 | AAI-based (high-confidence) | 0 |
| S7_vs_115421||full | 1867 | No | NA | 3 | 0 | 0 | Not-determined | Genome-fragment | NA | NA | 0 |
| S7_vs_115641||full | 3458 | No | NA | 4 | 0 | 1 | Not-determined | Genome-fragment | NA | NA | 0 |
| S7_vs_115653||full | 2644 | No | NA | 3 | 0 | 0 | Not-determined | Genome-fragment | NA | NA | 0 |
| S16_extracted_64680 | 2353 | No | NA | 3 | 2 | 0 | Low-quality | Genome-fragment | 4.1 | HMM-based (lower-bound) | 0 |
| S7_vs_115900||full | 1063 | No | NA | 2 | 0 | 0 | Not-determined | Genome-fragment | NA | NA | 0 |
| S16_vs_35255||full | 2353 | No | NA | 3 | 1 | 0 | Low-quality | Genome-fragment | 6.14 | AAI-based (high-confidence) | 0 |
| S7_vs_116018||full | 1097 | No | NA | 2 | 0 | 1 | Not-determined | Genome-fragment | NA | NA | 0 |
| S16_vs_42319||full | 2353 | No | NA | 4 | 4 | 0 | Low-quality | Genome-fragment | 4.77 | AAI-based (high-confidence) | 0 |
| S4_extracted_11978 | 2353 | No | NA | 3 | 2 | 0 | Low-quality | Genome-fragment | 7.75 | AAI-based (high-confidence) | 0 |
| S7_vs_116353||full | 5675 | No | NA | 3 | 0 | 3 | Not-determined | Genome-fragment | NA | NA | 0 |
| S7_vs_116446||full | 1378 | No | NA | 3 | 0 | 0 | Not-determined | Genome-fragment | NA | NA | 0 |
| S9_vs_k141_18505||full | 2354 | No | NA | 4 | 0 | 1 | Low-quality | Genome-fragment | 4.14 | AAI-based (medium-confidence) | 0 |
| S11_extracted_k141_65795 | 2354 | No | NA | 2 | 0 | 0 | Low-quality | Genome-fragment | 0.91 | AAI-based (medium-confidence) | 0 |
| S7_vs_116553||full | 2293 | No | NA | 4 | 0 | 0 | Not-determined | Genome-fragment | NA | NA | 0 |
| S7_vs_47228||full | 2354 | No | NA | 8 | 2 | 0 | Low-quality | Genome-fragment | 6.54 | AAI-based (high-confidence) | 0 |
| S7_vs_116704||full | 3839 | No | NA | 3 | 0 | 2 | Not-determined | Genome-fragment | NA | NA | 0 |
| S7_vs_67286||full | 2354 | No | NA | 7 | 1 | 0 | Low-quality | Genome-fragment | 5.8 | AAI-based (high-confidence) | 0 |
| S7_vs_116801||full | 1314 | No | NA | 2 | 0 | 2 | Not-determined | Genome-fragment | NA | NA | 0 |
| S7_vs_116816||full | 2866 | No | NA | 6 | 0 | 1 | Not-determined | Genome-fragment | NA | NA | 0 |
| S7_vs_116965||full | 3143 | No | NA | 3 | 0 | 1 | Not-determined | Genome-fragment | NA | NA | 0 |
| S16_vs_79760||full | 2354 | No | NA | 6 | 2 | 0 | Low-quality | Genome-fragment | 6.58 | AAI-based (medium-confidence) | 0 |
| S7_vs_117104||full | 6591 | No | NA | 7 | 0 | 4 | Not-determined | Genome-fragment | NA | NA | 0 |
| S7_vs_117140||full | 6040 | No | NA | 4 | 0 | 2 | Not-determined | Genome-fragment | NA | NA | 0 |
| S7_vs_117304||full | 1078 | No | NA | 2 | 0 | 1 | Not-determined | Genome-fragment | NA | NA | 0 |
| S7_vs_117378||full | 1511 | No | NA | 2 | 0 | 1 | Not-determined | Genome-fragment | NA | NA | 0 |
| S13_vs_97066||full | 2355 | No | NA | 3 | 1 | 0 | Low-quality | Genome-fragment | 1.8 | HMM-based (lower-bound) | 0 |
| S7_vs_117474||full | 1862 | No | NA | 3 | 0 | 1 | Not-determined | Genome-fragment | NA | NA | 0 |
| S7_vs_117574||full | 2024 | No | NA | 2 | 0 | 2 | Not-determined | Genome-fragment | NA | NA | 0 |
| S1_extracted_k141_22669 | 2355 | No | NA | 1 | 0 | 1 | Low-quality | Genome-fragment | 0.65 | AAI-based (medium-confidence) | 0 |
| S3_extracted_k141_4161 | 2355 | No | NA | 5 | 5 | 0 | Low-quality | Genome-fragment | 7.77 | AAI-based (high-confidence) | 0 |
| S8_extracted_k141_147052 | 2355 | No | NA | 3 | 3 | 0 | Low-quality | Genome-fragment | 10.72 | AAI-based (high-confidence) | 0 |
| S8_vs_k141_147289||full | 2355 | No | NA | 3 | 0 | 1 | Low-quality | Genome-fragment | 1.48 | AAI-based (medium-confidence) | 0 |
| S7_vs_117804||full | 2256 | No | NA | 2 | 0 | 1 | Not-determined | Genome-fragment | NA | NA | 0 |
| S7_vs_117875||full | 5396 | No | NA | 5 | 0 | 3 | Not-determined | Genome-fragment | NA | NA | 0 |
| S3_vs_k141_26476||full | 2356 | No | NA | 4 | 2 | 0 | Low-quality | Genome-fragment | 5 | AAI-based (medium-confidence) | 0 |
| S3_vs_k141_15002||full | 2356 | No | NA | 5 | 1 | 0 | Low-quality | Genome-fragment | 5.19 | AAI-based (high-confidence) | 0 |
| S11_vs_k141_33102||full | 2357 | No | NA | 3 | 0 | 1 | Low-quality | Genome-fragment | 4.83 | AAI-based (high-confidence) | 0 |
| S1_extracted_k141_76618 | 2357 | No | NA | 3 | 1 | 0 | Low-quality | Genome-fragment | 3.81 | AAI-based (high-confidence) | 0 |
| S8_vs_k141_116519||full | 2357 | No | NA | 7 | 2 | 0 | Low-quality | Genome-fragment | 4.04 | AAI-based (medium-confidence) | 0 |
| S7_vs_118229||full | 1565 | No | NA | 4 | 0 | 0 | Not-determined | Genome-fragment | NA | NA | 0 |
| S7_vs_118295||full | 26388 | No | NA | 42 | 14 | 1 | Medium-quality | Genome-fragment | 61.89 | AAI-based (high-confidence) | 0 |
| S17_vs_k141_89302||full | 2358 | No | NA | 3 | 0 | 0 | Low-quality | Genome-fragment | 3.66 | AAI-based (high-confidence) | 0 |
| S14_vs_k141_56865||full | 2358 | No | NA | 2 | 0 | 1 | Low-quality | Genome-fragment | 4.73 | AAI-based (medium-confidence) | 0 |
| S5_extracted_k141_103912 | 2358 | No | NA | 5 | 4 | 0 | Low-quality | Genome-fragment | 3.72 | AAI-based (high-confidence) | 0 |
| S7_vs_118538||full | 2632 | No | NA | 3 | 0 | 1 | Not-determined | Genome-fragment | NA | NA | 0 |
| S7_vs_118648||full | 5824 | No | NA | 4 | 0 | 4 | Not-determined | Genome-fragment | NA | NA | 0 |
| S7_extracted_83836 | 2358 | No | NA | 5 | 4 | 0 | Low-quality | Genome-fragment | 10.75 | AAI-based (high-confidence) | 0 |
| S16_extracted_146390 | 2358 | No | NA | 5 | 0 | 0 | Low-quality | Genome-fragment | 6.59 | AAI-based (high-confidence) | 0 |
| S7_vs_118705||full | 1247 | No | NA | 2 | 0 | 1 | Not-determined | Genome-fragment | NA | NA | 0 |
| S8_extracted_k141_72486 | 2358 | No | NA | 4 | 2 | 0 | Low-quality | Genome-fragment | 6.21 | HMM-based (lower-bound) | 0 |
| S11_vs_k141_55283||full | 2359 | No | NA | 8 | 0 | 0 | Low-quality | Genome-fragment | 3.97 | AAI-based (medium-confidence) | 0 |
| S1_vs_k141_110672||full | 2359 | No | NA | 3 | 0 | 0 | Low-quality | Genome-fragment | 6.19 | AAI-based (high-confidence) | 0 |
| S7_extracted_51207 | 2359 | No | NA | 1 | 0 | 1 | Low-quality | Genome-fragment | 0.58 | AAI-based (medium-confidence) | 0 |
| S12_extracted_k141_81307 | 2359 | No | NA | 4 | 1 | 0 | Low-quality | Genome-fragment | 5.68 | AAI-based (high-confidence) | 0 |
| S7_vs_118959||full | 1733 | No | NA | 2 | 0 | 1 | Not-determined | Genome-fragment | NA | NA | 0 |
| S7_vs_119097||full | 2994 | No | NA | 5 | 0 | 0 | Not-determined | Genome-fragment | NA | NA | 0 |
| S7_vs_119103||full | 2550 | No | NA | 3 | 0 | 2 | Not-determined | Genome-fragment | NA | NA | 0 |
| S17_vs_k141_92876||full | 2360 | No | NA | 5 | 0 | 0 | Low-quality | Genome-fragment | 3.16 | AAI-based (medium-confidence) | 0 |
| S7_vs_119191||full | 2774 | No | NA | 3 | 0 | 1 | Not-determined | Genome-fragment | NA | NA | 0 |
| S1_vs_k141_34904||full | 2360 | No | NA | 3 | 0 | 1 | Low-quality | Genome-fragment | 1.46 | AAI-based (medium-confidence) | 0 |
| S5_vs_k141_84708||full | 2360 | No | NA | 3 | 1 | 0 | Low-quality | Genome-fragment | 6.15 | AAI-based (high-confidence) | 0 |
| S7_vs_119469||full | 1133 | No | NA | 2 | 0 | 1 | Not-determined | Genome-fragment | NA | NA | 0 |
| S7_vs_114424||full | 2360 | No | NA | 3 | 3 | 0 | Low-quality | Genome-fragment | 5.31 | AAI-based (high-confidence) | 0 |
| S7_vs_119492||full | 13185 | No | NA | 18 | 6 | 0 | Medium-quality | Genome-fragment | 66.91 | AAI-based (medium-confidence) | 0 |
| S16_vs_123820||full | 2360 | No | NA | 6 | 0 | 0 | Low-quality | Genome-fragment | 3.57 | AAI-based (high-confidence) | 0 |
| S7_vs_119507||full | 2529 | No | NA | 3 | 0 | 3 | Not-determined | Genome-fragment | NA | NA | 0 |
| S7_vs_119679||full | 51206 | Yes | 40433 | 60 | 14 | 8 | High-quality | High-quality | 100 | AAI-based (medium-confidence) | 21.04 |
| S8_vs_k141_75484||full | 2360 | No | NA | 3 | 0 | 0 | Low-quality | Genome-fragment | 3.08 | AAI-based (medium-confidence) | 0 |
| S7_vs_119759||full | 1733 | No | NA | 3 | 0 | 0 | Not-determined | Genome-fragment | NA | NA | 0 |
| S7_vs_119779||full | 1491 | No | NA | 2 | 0 | 2 | Not-determined | Genome-fragment | NA | NA | 0 |
| S9_vs_k141_19046||full | 2361 | No | NA | 4 | 1 | 0 | Low-quality | Genome-fragment | 1.24 | HMM-based (lower-bound) | 0 |
| S16_extracted_7282 | 2361 | No | NA | 3 | 0 | 0 | Low-quality | Genome-fragment | 2.17 | AAI-based (medium-confidence) | 0 |
| S7_vs_119916||full | 5627 | No | NA | 6 | 0 | 3 | Not-determined | Genome-fragment | NA | NA | 0 |
| S7_vs_119930||full | 5967 | No | NA | 6 | 0 | 3 | Not-determined | Genome-fragment | NA | NA | 0 |
| S11_vs_k141_79955||full | 2362 | No | NA | 2 | 0 | 2 | Low-quality | Genome-fragment | 4.46 | AAI-based (medium-confidence) | 0 |
| S11_vs_k141_22668||full | 2362 | No | NA | 3 | 0 | 1 | Low-quality | Genome-fragment | 1.64 | AAI-based (medium-confidence) | 0 |
| S7_vs_33566||full | 2362 | No | NA | 3 | 0 | 0 | Low-quality | Genome-fragment | 5.26 | AAI-based (medium-confidence) | 0 |
| S16_extracted_39280 | 2362 | No | NA | 1 | 0 | 1 | Low-quality | Genome-fragment | 5.07 | AAI-based (high-confidence) | 0 |
| S12_vs_k141_27754||full | 2362 | No | NA | 4 | 0 | 0 | Low-quality | Genome-fragment | 4.28 | AAI-based (high-confidence) | 0 |
| S7_vs_120207||full | 2006 | No | NA | 2 | 0 | 0 | Not-determined | Genome-fragment | NA | NA | 0 |
| S8_vs_k141_137896||full | 2362 | No | NA | 5 | 0 | 0 | Low-quality | Genome-fragment | 4.92 | AAI-based (high-confidence) | 0 |
| S7_vs_120402||full | 1287 | No | NA | 3 | 0 | 3 | Not-determined | Genome-fragment | NA | NA | 0 |
| S17_vs_k141_31100||full | 2363 | No | NA | 3 | 1 | 0 | Low-quality | Genome-fragment | 3.52 | AAI-based (high-confidence) | 0 |
| S7_vs_120520||full | 1426 | No | NA | 4 | 0 | 0 | Not-determined | Genome-fragment | NA | NA | 0 |
| S7_vs_120600||full | 1562 | No | NA | 2 | 0 | 1 | Not-determined | Genome-fragment | NA | NA | 0 |
| S7_vs_120951||full | 1083 | No | NA | 4 | 0 | 0 | Not-determined | Genome-fragment | NA | NA | 0 |
| S3_vs_k141_71992||full | 2363 | No | NA | 5 | 0 | 0 | Low-quality | Genome-fragment | 1.5 | AAI-based (high-confidence) | 0 |
| S7_vs_120988||full | 4189 | No | NA | 2 | 0 | 0 | Not-determined | Genome-fragment | NA | NA | 0 |
| S7_vs_121153||full | 1213 | No | NA | 2 | 0 | 1 | Not-determined | Genome-fragment | NA | NA | 0 |
| S8_extracted_k141_71153 | 2363 | Yes | 662 | 2 | 1 | 1 | Low-quality | Genome-fragment | 0.36 | HMM-based (lower-bound) | 71.98 |
| S5_vs_k141_56699||full | 2364 | No | NA | 7 | 2 | 0 | Low-quality | Genome-fragment | 7.5 | AAI-based (high-confidence) | 0 |
| S8_vs_k141_43985||full | 2364 | No | NA | 3 | 0 | 3 | Low-quality | Genome-fragment | 1.11 | AAI-based (medium-confidence) | 0 |
| S8_vs_k141_183160||full | 2364 | No | NA | 2 | 0 | 0 | Low-quality | Genome-fragment | 1.76 | AAI-based (medium-confidence) | 0 |
| S7_vs_121568||full | 1557 | No | NA | 3 | 0 | 0 | Not-determined | Genome-fragment | NA | NA | 0 |
| S7_vs_121569||full | 1896 | No | NA | 3 | 0 | 0 | Not-determined | Genome-fragment | NA | NA | 0 |
| S7_vs_121611||full | 1253 | No | NA | 2 | 0 | 1 | Not-determined | Genome-fragment | NA | NA | 0 |
| S8_vs_k141_24203||full | 2364 | No | NA | 5 | 1 | 0 | Low-quality | Genome-fragment | 7.29 | AAI-based (high-confidence) | 0 |
| S7_vs_121718||full | 2898 | No | NA | 3 | 0 | 2 | Not-determined | Genome-fragment | NA | NA | 0 |
| S7_vs_121764||full | 3820 | No | NA | 3 | 0 | 1 | Not-determined | Genome-fragment | NA | NA | 0 |
| S7_vs_121865||full | 1764 | No | NA | 4 | 0 | 0 | Not-determined | Genome-fragment | NA | NA | 0 |
| S11_extracted_k141_20760 | 2365 | No | NA | 2 | 1 | 0 | Low-quality | Genome-fragment | 3.37 | AAI-based (high-confidence) | 0 |
| S7_vs_121964||full | 2770 | No | NA | 2 | 0 | 2 | Not-determined | Genome-fragment | NA | NA | 0 |
| S7_vs_122151||full | 33897 | No | NA | 59 | 19 | 0 | High-quality | High-quality | 99.99 | AAI-based (medium-confidence) | 0 |
| S7_vs_113077||full | 2365 | No | NA | 3 | 0 | 0 | Low-quality | Genome-fragment | 6.37 | AAI-based (medium-confidence) | 0 |
| S7_vs_122165||full | 2989 | No | NA | 6 | 0 | 1 | Not-determined | Genome-fragment | NA | NA | 0 |
| S7_vs_122169||full | 4750 | No | NA | 5 | 0 | 2 | Not-determined | Genome-fragment | NA | NA | 0 |
| S15_vs_67510||full | 2365 | No | NA | 5 | 1 | 0 | Low-quality | Genome-fragment | 5.26 | AAI-based (high-confidence) | 0 |
| S8_vs_k141_92948||full | 2365 | No | NA | 5 | 0 | 0 | Low-quality | Genome-fragment | 4.93 | AAI-based (medium-confidence) | 0 |
| S7_vs_122334||full | 2056 | No | NA | 3 | 0 | 0 | Not-determined | Genome-fragment | NA | NA | 0 |
| S12_vs_k141_185480||full | 2366 | No | NA | 4 | 3 | 0 | Low-quality | Genome-fragment | 6.07 | AAI-based (high-confidence) | 0 |
| S8_vs_k141_149786||full | 2366 | No | NA | 4 | 1 | 0 | Low-quality | Genome-fragment | 5.69 | AAI-based (medium-confidence) | 0 |
| S7_vs_122424||full | 1232 | No | NA | 2 | 0 | 1 | Not-determined | Genome-fragment | NA | NA | 0 |
| S14_vs_k141_321||lt2gene | 2367 | No | NA | 1 | 1 | 0 | Low-quality | Genome-fragment | 1.5 | AAI-based (medium-confidence) | 0 |
| S1_vs_k141_140356||full | 2367 | No | NA | 3 | 2 | 0 | Low-quality | Genome-fragment | 7.81 | HMM-based (lower-bound) | 0 |
| S3_vs_k141_56082||full | 2367 | No | NA | 4 | 1 | 0 | Low-quality | Genome-fragment | 3.79 | AAI-based (high-confidence) | 0 |
| S10_extracted_51451 | 2367 | No | NA | 2 | 0 | 1 | Low-quality | Genome-fragment | 0.4 | AAI-based (high-confidence) | 0 |
| S12_extracted_k141_6511 | 2367 | No | NA | 3 | 1 | 0 | Low-quality | Genome-fragment | 2.54 | AAI-based (high-confidence) | 0 |
| S7_vs_122603||full | 1862 | No | NA | 4 | 0 | 3 | Not-determined | Genome-fragment | NA | NA | 0 |
| S7_vs_122627||full | 1506 | No | NA | 3 | 0 | 0 | Not-determined | Genome-fragment | NA | NA | 0 |
| S7_vs_122653||full | 1417 | No | NA | 2 | 0 | 0 | Not-determined | Genome-fragment | NA | NA | 0 |
| S7_vs_122761||full | 1771 | No | NA | 6 | 0 | 0 | Not-determined | Genome-fragment | NA | NA | 0 |
| S12_vs_k141_34234||full | 2367 | No | NA | 6 | 0 | 0 | Low-quality | Genome-fragment | 5.32 | AAI-based (medium-confidence) | 0 |
| S8_extracted_k141_26575 | 2367 | No | NA | 5 | 0 | 0 | Low-quality | Genome-fragment | 2.37 | AAI-based (high-confidence) | 0 |
| S17_vs_k141_140215||full | 2368 | No | NA | 3 | 0 | 0 | Low-quality | Genome-fragment | 4.25 | AAI-based (high-confidence) | 0 |
| S7_vs_123070||full | 2360 | No | NA | 2 | 0 | 0 | Not-determined | Genome-fragment | NA | NA | 0 |
| S6_vs_k141_30917||full | 2368 | Yes | 1004 | 3 | 1 | 1 | Low-quality | Genome-fragment | 0.68 | AAI-based (high-confidence) | 57.6 |
| S7_vs_123202||full | 5844 | No | NA | 4 | 0 | 3 | Not-determined | Genome-fragment | NA | NA | 0 |
| S7_vs_123216||full | 1166 | No | NA | 2 | 0 | 0 | Not-determined | Genome-fragment | NA | NA | 0 |
| S7_vs_123265||full | 2615 | No | NA | 3 | 0 | 0 | Not-determined | Genome-fragment | NA | NA | 0 |
| S7_vs_123266||full | 8560 | No | NA | 14 | 0 | 3 | Not-determined | Genome-fragment | NA | NA | 0 |
| S10_vs_116925||full | 2369 | No | NA | 3 | 2 | 0 | Low-quality | Genome-fragment | 3.83 | AAI-based (medium-confidence) | 0 |
| S7_vs_123348||full | 4330 | No | NA | 5 | 0 | 1 | Not-determined | Genome-fragment | NA | NA | 0 |
| S15_vs_61638||full | 2369 | No | NA | 3 | 0 | 3 | Low-quality | Genome-fragment | 1.07 | AAI-based (medium-confidence) | 0 |
| S8_vs_k141_104668||full | 2369 | No | NA | 2 | 0 | 1 | Low-quality | Genome-fragment | 1.15 | AAI-based (medium-confidence) | 0 |
| S17_extracted_k141_163901 | 2370 | No | NA | 2 | 0 | 1 | Low-quality | Genome-fragment | 0.27 | AAI-based (medium-confidence) | 0 |
| S11_vs_k141_37179||full | 2370 | No | NA | 8 | 0 | 0 | Low-quality | Genome-fragment | 5.51 | AAI-based (medium-confidence) | 0 |
| S3_vs_k141_109671||full | 2370 | No | NA | 5 | 4 | 0 | Low-quality | Genome-fragment | 6.59 | AAI-based (high-confidence) | 0 |
| S7_vs_123761||full | 2728 | No | NA | 3 | 0 | 0 | Not-determined | Genome-fragment | NA | NA | 0 |
| S7_vs_123802||full | 2077 | No | NA | 3 | 0 | 0 | Not-determined | Genome-fragment | NA | NA | 0 |
| S7_vs_123807||full | 2031 | No | NA | 3 | 0 | 2 | Not-determined | Genome-fragment | NA | NA | 0 |
| S7_vs_123940||full | 4257 | No | NA | 3 | 0 | 2 | Not-determined | Genome-fragment | NA | NA | 0 |
| S7_vs_123960||full | 3223 | No | NA | 4 | 0 | 0 | Not-determined | Genome-fragment | NA | NA | 0 |
| S16_vs_76599||full | 2370 | Yes | 1000 | 4 | 2 | 1 | Low-quality | Genome-fragment | 2.07 | AAI-based (medium-confidence) | 57.81 |
| S7_vs_124016||full | 1373 | No | NA | 2 | 0 | 2 | Not-determined | Genome-fragment | NA | NA | 0 |
| S7_vs_124230||full | 1170 | No | NA | 2 | 0 | 2 | Not-determined | Genome-fragment | NA | NA | 0 |
| S3_vs_k141_41413||full | 2371 | No | NA | 4 | 0 | 0 | Low-quality | Genome-fragment | 4.38 | AAI-based (high-confidence) | 0 |
| S7_vs_124460||full | 3384 | No | NA | 2 | 0 | 1 | Not-determined | Genome-fragment | NA | NA | 0 |
| S7_vs_124461||full | 3179 | No | NA | 3 | 0 | 1 | Not-determined | Genome-fragment | NA | NA | 0 |

| | | | | | | | | | | | |
|---|---|---|---|---|---|---|---|---|---|---|---|
| S7_vs_124501\|\|full | 1817 | No | NA | 4 | 0 | 0 | Not-determined | Genome-fragment | NA | NA | 0 |
| S7_vs_124631\|\|full | 1632 | No | NA | 3 | 0 | 1 | Not-determined | Genome-fragment | NA | NA | 0 |
| S7_vs_124650\|\|full | 1660 | No | NA | 3 | 0 | 1 | Not-determined | Genome-fragment | NA | NA | 0 |
| S7_vs_124687\|\|full | 4588 | No | NA | 7 | 0 | 1 | Not-determined | Genome-fragment | NA | NA | 0 |
| S7_vs_124737\|\|full | 1185 | No | NA | 3 | 0 | 0 | Not-determined | Genome-fragment | NA | NA | 0 |
| S5_vs_k141_132436\|\|full | 2371 | Yes | 2125 | 4 | 1 | 1 | Low-quality | Genome-fragment | 1.37 | AAI-based (medium-confidence) | 10.38 |
| S7_vs_124822\|\|full | 1734 | No | NA | 2 | 0 | 0 | Not-determined | Genome-fragment | NA | NA | 0 |
| S7_vs_124936\|\|full | 2474 | No | NA | 2 | 0 | 0 | Not-determined | Genome-fragment | NA | NA | 0 |
| S17_vs_k141_39780\|\|full | 2372 | No | NA | 5 | 1 | 0 | Low-quality | Genome-fragment | 3.5 | AAI-based (high-confidence) | 0 |
| S7_vs_124967\|\|full | 2198 | No | NA | 3 | 0 | 3 | Not-determined | Genome-fragment | NA | NA | 0 |
| S9_vs_k141_33736\|\|full | 2372 | Yes | 1290 | 6 | 1 | 1 | Low-quality | Genome-fragment | 1.6 | HMM-based (lower-bound) | 45.62 |
| S11_vs_k141_18049\|\|full | 2372 | No | NA | 5 | 5 | 0 | Low-quality | Genome-fragment | 5.96 | AAI-based (medium-confidence) | 0 |
| S13_vs_116541\|\|full | 2372 | No | NA | 4 | 0 | 1 | Low-quality | Genome-fragment | 2.22 | AAI-based (medium-confidence) | 0 |
| S1_vs_k141_122638\|\|full | 2372 | No | NA | 5 | 4 | 0 | Low-quality | Genome-fragment | 5.42 | AAI-based (high-confidence) | 0 |
| S7_vs_125101\|\|full | 17593 | No | NA | 12 | 0 | 12 | Not-determined | Genome-fragment | NA | NA | 0 |
| S8_extracted_k141_135532 | 2372 | No | NA | 6 | 3 | 1 | Low-quality | Genome-fragment | 5.52 | AAI-based (high-confidence) | 0 |
| S9_extracted_k141_3740 | 2373 | No | NA | 6 | 5 | 0 | Low-quality | Genome-fragment | 5.77 | AAI-based (high-confidence) | 0 |
| S7_vs_125296\|\|full | 1396 | No | NA | 3 | 0 | 0 | Not-determined | Genome-fragment | NA | NA | 0 |
| S7_vs_125478\|\|full | 2802 | No | NA | 5 | 0 | 0 | Not-determined | Genome-fragment | NA | NA | 0 |
| S9_vs_k141_5657\|\|full | 2373 | No | NA | 7 | 2 | 0 | Low-quality | Genome-fragment | 5.46 | AAI-based (high-confidence) | 0 |
| S13_vs_97545\|\|full | 2373 | No | NA | 3 | 1 | 0 | Low-quality | Genome-fragment | 4.09 | AAI-based (medium-confidence) | 0 |
| S7_vs_125546\|\|full | 7342 | No | NA | 6 | 0 | 4 | Not-determined | Genome-fragment | NA | NA | 0 |
| S1_vs_k141_15081\|\|full | 2373 | No | NA | 5 | 3 | 0 | Low-quality | Genome-fragment | 5.18 | AAI-based (high-confidence) | 0 |
| S3_vs_k141_99584\|\|full | 2373 | No | NA | 2 | 0 | 1 | Low-quality | Genome-fragment | 1.05 | AAI-based (medium-confidence) | 0 |
| S7_vs_125653\|\|full | 1430 | No | NA | 2 | 0 | 1 | Not-determined | Genome-fragment | NA | NA | 0 |
| S3_extracted_k141_27681 | 2374 | No | NA | 2 | 0 | 0 | Low-quality | Genome-fragment | 3.67 | AAI-based (high-confidence) | 0 |
| S3_extracted_k141_126153 | 2374 | No | NA | 2 | 0 | 1 | Low-quality | Genome-fragment | 1.14 | AAI-based (high-confidence) | 0 |
| S7_vs_125808\|\|full | 1374 | No | NA | 3 | 0 | 0 | Not-determined | Genome-fragment | NA | NA | 0 |
| S3_vs_k141_16609\|\|full | 2374 | No | NA | 3 | 0 | 1 | Low-quality | Genome-fragment | 1.53 | AAI-based (medium-confidence) | 0 |
| S7_extracted_96506 | 2374 | No | NA | 3 | 0 | 2 | Low-quality | Genome-fragment | 1.02 | AAI-based (medium-confidence) | 0 |
| S8_vs_k141_130693\|\|full | 2374 | No | NA | 5 | 1 | 0 | Low-quality | Genome-fragment | 3.52 | HMM-based (lower-bound) | 0 |
| S8_vs_k141_160512\|\|full | 2374 | No | NA | 6 | 1 | 0 | Low-quality | Genome-fragment | 7.46 | AAI-based (high-confidence) | 0 |
| S7_vs_126075\|\|full | 6899 | No | NA | 6 | 0 | 4 | Not-determined | Genome-fragment | NA | NA | 0 |
| S7_vs_126138\|\|full | 1103 | No | NA | 2 | 0 | 1 | Not-determined | Genome-fragment | NA | NA | 0 |
| S11_extracted_k141_12658 | 2375 | No | NA | 6 | 2 | 0 | Low-quality | Genome-fragment | 5.4 | AAI-based (medium-confidence) | 0 |
| S13_vs_105479\|\|full | 2375 | No | NA | 5 | 0 | 0 | Low-quality | Genome-fragment | 5.75 | AAI-based (high-confidence) | 0 |
| S16_vs_32606\|\|full | 2375 | Yes | 818 | 4 | 1 | 1 | Low-quality | Genome-fragment | 2.05 | AAI-based (medium-confidence) | 65.56 |
| S4_extracted_120 | 2375 | No | NA | 8 | 3 | 0 | Low-quality | Genome-fragment | 5.11 | AAI-based (medium-confidence) | 0 |
| S3_vs_k141_10908\|\|full | 2376 | No | NA | 5 | 0 | 0 | Low-quality | Genome-fragment | 4.64 | AAI-based (medium-confidence) | 0 |
| S6_vs_k141_4467\|\|full | 2376 | No | NA | 5 | 1 | 0 | Low-quality | Genome-fragment | 5.46 | AAI-based (high-confidence) | 0 |
| S7_vs_126786\|\|full | 1836 | No | NA | 2 | 0 | 2 | Not-determined | Genome-fragment | NA | NA | 0 |
| S7_vs_126808\|\|full | 14720 | No | NA | 14 | 0 | 9 | Not-determined | Genome-fragment | NA | NA | 0 |
| S7_vs_126883\|\|full | 2445 | No | NA | 3 | 0 | 0 | Not-determined | Genome-fragment | NA | NA | 0 |
| S7_vs_127089\|\|full | 6290 | No | NA | 6 | 0 | 6 | Not-determined | Genome-fragment | NA | NA | 0 |
| S7_vs_127192\|\|full | 2773 | No | NA | 3 | 0 | 2 | Not-determined | Genome-fragment | NA | NA | 0 |
| S7_vs_127257\|\|full | 1511 | No | NA | 3 | 0 | 1 | Not-determined | Genome-fragment | NA | NA | 0 |
| S17_vs_k141_143540\|\|full | 2377 | No | NA | 3 | 1 | 0 | Low-quality | Genome-fragment | 5.23 | AAI-based (high-confidence) | 0 |
| S7_vs_127366\|\|full | 1856 | No | NA | 2 | 0 | 2 | Not-determined | Genome-fragment | NA | NA | 0 |
| S7_vs_127737\|\|full | 2312 | No | NA | 4 | 0 | 2 | Not-determined | Genome-fragment | NA | NA | 0 |
| S7_vs_127769\|\|full | 1021 | No | NA | 2 | 0 | 1 | Not-determined | Genome-fragment | NA | NA | 0 |
| S11_vs_k141_58503\|\|full | 2377 | No | NA | 3 | 0 | 1 | Low-quality | Genome-fragment | 1.8 | AAI-based (medium-confidence) | 0 |
| S14_extracted_k141_7323 | 2377 | No | NA | 5 | 4 | 0 | Low-quality | Genome-fragment | 7.85 | AAI-based (high-confidence) | 0 |
| S10_vs_79912\|\|full | 2377 | No | NA | 5 | 0 | 0 | Low-quality | Genome-fragment | 2.92 | AAI-based (high-confidence) | 0 |
| S12_extracted_k141_100672 | 2377 | No | NA | 4 | 0 | 0 | Low-quality | Genome-fragment | 1.99 | AAI-based (high-confidence) | 0 |
| S15_extracted_5501 | 2377 | No | NA | 1 | 0 | 1 | Low-quality | Genome-fragment | 2.47 | AAI-based (high-confidence) | 0 |
| S7_vs_128375\|\|full | 4304 | No | NA | 4 | 0 | 4 | Not-determined | Genome-fragment | NA | NA | 0 |
| S7_vs_128387\|\|full | 1527 | No | NA | 3 | 0 | 0 | Not-determined | Genome-fragment | NA | NA | 0 |
| S7_vs_128459\|\|full | 4267 | No | NA | 3 | 0 | 3 | Not-determined | Genome-fragment | NA | NA | 0 |
| S7_vs_128504\|\|full | 23155 | No | NA | 18 | 0 | 16 | Not-determined | Genome-fragment | NA | NA | 0 |
| S7_vs_128509\|\|full | 1441 | No | NA | 3 | 0 | 2 | Not-determined | Genome-fragment | NA | NA | 0 |
| S7_vs_128643\|\|full | 2386 | No | NA | 5 | 0 | 0 | Not-determined | Genome-fragment | NA | NA | 0 |
| S14_extracted_k141_80701 | 2378 | No | NA | 5 | 5 | 0 | Low-quality | Genome-fragment | 5.12 | AAI-based (medium-confidence) | 0 |
| S7_vs_128752\|\|full | 1745 | No | NA | 3 | 0 | 1 | Not-determined | Genome-fragment | NA | NA | 0 |
| S1_vs_k141_100678\|\|full | 2378 | No | NA | 4 | 0 | 0 | Low-quality | Genome-fragment | 4.54 | AAI-based (medium-confidence) | 0 |
| S16_vs_103020\|\|lt2gene | 2378 | No | NA | 2 | 1 | 0 | Low-quality | Genome-fragment | 7.17 | AAI-based (high-confidence) | 0 |
| S8_vs_k141_38817\|\|full | 2378 | No | NA | 3 | 1 | 0 | Low-quality | Genome-fragment | 6.26 | AAI-based (medium-confidence) | 0 |
| S7_vs_129297\|\|full | 2742 | No | NA | 3 | 0 | 3 | Not-determined | Genome-fragment | NA | NA | 0 |
| S7_extracted_114808 | 2379 | No | NA | 3 | 2 | 0 | Low-quality | Genome-fragment | 4.41 | AAI-based (high-confidence) | 0 |
| S7_vs_129344\|\|full | 3425 | No | NA | 5 | 0 | 0 | Not-determined | Genome-fragment | NA | NA | 0 |
| S16_vs_101193\|\|full | 2379 | No | NA | 8 | 1 | 0 | Low-quality | Genome-fragment | 1.41 | AAI-based (high-confidence) | 0 |
| S10_vs_7049\|\|full | 2379 | No | NA | 8 | 0 | 0 | Low-quality | Genome-fragment | 2.17 | AAI-based (high-confidence) | 0 |
| S7_vs_129858\|\|full | 23771 | No | NA | 33 | 11 | 0 | Medium-quality | Genome-fragment | 66.82 | AAI-based (high-confidence) | 0 |
| S17_vs_k141_66781\|\|full | 2380 | Yes | 2182 | 5 | 1 | 1 | Low-quality | Genome-fragment | 1.23 | HMM-based (lower-bound) | 8.32 |
| S17_vs_k141_91917\|\|full | 2380 | No | NA | 2 | 1 | 0 | Low-quality | Genome-fragment | 3.76 | HMM-based (lower-bound) | 0 |
| S11_extracted_k141_54216 | 2380 | Yes | 2059 | 3 | 1 | 1 | Low-quality | Genome-fragment | 0.91 | AAI-based (medium-confidence) | 13.49 |
| S14_vs_k141_100095\|\|full | 2380 | No | NA | 3 | 1 | 0 | Low-quality | Genome-fragment | 5.14 | AAI-based (medium-confidence) | 0 |
| S1_vs_k141_93492\|\|full | 2380 | No | NA | 6 | 2 | 0 | Low-quality | Genome-fragment | 5.56 | AAI-based (high-confidence) | 0 |
| S1_vs_k141_48592\|\|full | 2380 | No | NA | 2 | 0 | 1 | Low-quality | Genome-fragment | 0.65 | AAI-based (medium-confidence) | 0 |
| S7_vs_130335\|\|full | 2101 | No | NA | 2 | 0 | 2 | Not-determined | Genome-fragment | NA | NA | 0 |
| S7_vs_130463\|\|full | 1281 | No | NA | 3 | 0 | 1 | Not-determined | Genome-fragment | NA | NA | 0 |
| S7_vs_130567\|\|full | 3495 | No | NA | 3 | 0 | 2 | Not-determined | Genome-fragment | NA | NA | 0 |
| S7_vs_130586\|\|full | 6841 | No | NA | 8 | 0 | 2 | Not-determined | Genome-fragment | NA | NA | 0 |
| S7_vs_54901\|\|full | 2380 | No | NA | 1 | 0 | 0 | Low-quality | Genome-fragment | 1.07 | AAI-based (medium-confidence) | 0 |
| S7_vs_130678\|\|full | 1586 | No | NA | 2 | 0 | 1 | Not-determined | Genome-fragment | NA | NA | 0 |
| S7_vs_130716\|\|full | 2091 | No | NA | 4 | 0 | 3 | Not-determined | Genome-fragment | NA | NA | 0 |
| S7_vs_130773\|\|full | 5029 | No | NA | 5 | 0 | 5 | Not-determined | Genome-fragment | NA | NA | 0 |
| S7_vs_130784\|\|full | 2183 | No | NA | 2 | 0 | 1 | Not-determined | Genome-fragment | NA | NA | 0 |
| S7_vs_130802\|\|full | 1426 | No | NA | 1 | 0 | 0 | Not-determined | Genome-fragment | NA | NA | 0 |
| S7_vs_130810\|\|full | 9150 | No | NA | 10 | 0 | 4 | Not-determined | Genome-fragment | NA | NA | 0 |
| S8_vs_k141_21220\|\|full | 2380 | No | NA | 5 | 0 | 0 | Low-quality | Genome-fragment | 2.28 | AAI-based (medium-confidence) | 0 |
| S7_vs_130901\|\|full | 2840 | No | NA | 3 | 0 | 3 | Not-determined | Genome-fragment | NA | NA | 0 |
| S8_vs_k141_181424\|\|full | 2380 | No | NA | 4 | 0 | 0 | Low-quality | Genome-fragment | 4.48 | AAI-based (medium-confidence) | 0 |
| S9_vs_k141_74415\|\|full | 2381 | No | NA | 4 | 0 | 2 | Low-quality | Genome-fragment | 2.81 | AAI-based (high-confidence) | 0 |
| S7_vs_131040\|\|full | 6629 | No | NA | 10 | 0 | 3 | Not-determined | Genome-fragment | NA | NA | 0 |
| S7_vs_131053\|\|full | 2958 | No | NA | 4 | 0 | 0 | Not-determined | Genome-fragment | NA | NA | 0 |
| S7_vs_131217\|\|full | 3179 | No | NA | 3 | 0 | 3 | Not-determined | Genome-fragment | NA | NA | 0 |
| S11_vs_k141_5503\|\|full | 2381 | No | NA | 6 | 0 | 0 | Low-quality | Genome-fragment | 2.67 | AAI-based (high-confidence) | 0 |
| S12_extracted_k141_62127 | 2381 | No | NA | 5 | 0 | 2 | Low-quality | Genome-fragment | 1.65 | AAI-based (medium-confidence) | 0 |
| S2_vs_k141_479\|\|full | 2381 | No | NA | 5 | 1 | 0 | Low-quality | Genome-fragment | 5.54 | AAI-based (high-confidence) | 0 |
| S7_vs_131416\|\|full | 1884 | No | NA | 2 | 0 | 0 | Not-determined | Genome-fragment | NA | NA | 0 |
| S7_vs_131443\|\|full | 1875 | No | NA | 3 | 0 | 2 | Not-determined | Genome-fragment | NA | NA | 0 |
| S7_vs_131489\|\|full | 3864 | No | NA | 4 | 0 | 3 | Not-determined | Genome-fragment | NA | NA | 0 |
| S7_vs_131553\|\|full | 5717 | No | NA | 9 | 0 | 3 | Not-determined | Genome-fragment | NA | NA | 0 |
| S7_vs_131561\|\|full | 1548 | No | NA | 3 | 0 | 3 | Not-determined | Genome-fragment | NA | NA | 0 |
| S7_vs_131573\|\|full | 6160 | No | NA | 5 | 0 | 3 | Not-determined | Genome-fragment | NA | NA | 0 |
| S7_vs_131698\|\|full | 3164 | No | NA | 3 | 0 | 2 | Not-determined | Genome-fragment | NA | NA | 0 |
| S11_vs_k141_88150\|\|full | 2382 | No | NA | 4 | 2 | 0 | Low-quality | Genome-fragment | 6.6 | AAI-based (high-confidence) | 0 |
| S14_extracted_k141_34491 | 2382 | No | NA | 2 | 1 | 0 | Low-quality | Genome-fragment | 8.79 | AAI-based (high-confidence) | 0 |
| S7_vs_131859\|\|full | 1190 | No | NA | 2 | 0 | 2 | Not-determined | Genome-fragment | NA | NA | 0 |
| S7_vs_131910\|\|full | 1082 | No | NA | 3 | 0 | 0 | Not-determined | Genome-fragment | NA | NA | 0 |
| S1_vs_k141_82130\|\|full | 2382 | No | NA | 3 | 0 | 2 | Low-quality | Genome-fragment | 1.63 | AAI-based (medium-confidence) | 0 |
| S16_vs_138537\|\|full | 2382 | No | NA | 7 | 1 | 0 | Low-quality | Genome-fragment | 2.35 | AAI-based (medium-confidence) | 0 |
| S7_vs_132026\|\|full | 1220 | No | NA | 4 | 0 | 0 | Not-determined | Genome-fragment | NA | NA | 0 |
| S8_extracted_k141_197679 | 2382 | No | NA | 3 | 0 | 2 | Low-quality | Genome-fragment | 4.2 | AAI-based (medium-confidence) | 0 |
| S13_extracted_62471 | 2383 | No | NA | 2 | 0 | 0 | Low-quality | Genome-fragment | 1.09 | AAI-based (high-confidence) | 0 |
| S7_vs_132463\|\|full | 4478 | No | NA | 3 | 0 | 3 | Not-determined | Genome-fragment | NA | NA | 0 |
| S14_vs_k141_89014\|\|full | 2383 | No | NA | 7 | 2 | 0 | Low-quality | Genome-fragment | 5.81 | AAI-based (high-confidence) | 0 |
| S16_vs_110499\|\|full | 2383 | No | NA | 3 | 0 | 2 | Low-quality | Genome-fragment | 5.51 | AAI-based (medium-confidence) | 0 |
| S10_vs_31670\|\|full | 2383 | No | NA | 7 | 0 | 0 | Low-quality | Genome-fragment | 5.6 | AAI-based (high-confidence) | 0 |
| S7_vs_132680\|\|full | 2012 | No | NA | 7 | 0 | 0 | Not-determined | Genome-fragment | NA | NA | 0 |
| S17_vs_k141_165302\|\|full | 2384 | No | NA | 4 | 1 | 0 | Low-quality | Genome-fragment | 1.22 | HMM-based (lower-bound) | 0 |
| S13_extracted_126990 | 2384 | No | NA | 8 | 0 | 0 | Low-quality | Genome-fragment | 3.88 | AAI-based (high-confidence) | 0 |
| S5_vs_k141_60141\|\|full | 2384 | No | NA | 3 | 0 | 2 | Low-quality | Genome-fragment | 1.38 | AAI-based (medium-confidence) | 0 |
| S7_vs_41849\|\|full | 2384 | No | NA | 3 | 2 | 0 | Low-quality | Genome-fragment | 6.24 | AAI-based (high-confidence) | 0 |
| S7_vs_109191\|\|full | 2384 | No | NA | 3 | 0 | 0 | Low-quality | Genome-fragment | 1.63 | AAI-based (high-confidence) | 0 |
| S12_vs_k141_78012\|\|full | 2384 | No | NA | 3 | 0 | 1 | Low-quality | Genome-fragment | 2.07 | AAI-based (high-confidence) | 0 |
| S7_vs_133216\|\|full | 1444 | No | NA | 3 | 0 | 2 | Not-determined | Genome-fragment | NA | NA | 0 |
| S8_vs_k141_13979\|\|full | 2384 | No | NA | 4 | 1 | 0 | Low-quality | Genome-fragment | 4.14 | AAI-based (medium-confidence) | 0 |
| S17_vs_k141_18812\|\|full | 2385 | No | NA | 4 | 1 | 0 | Low-quality | Genome-fragment | 1.37 | AAI-based (medium-confidence) | 0 |
| S11_extracted_k141_55533 | 2385 | No | NA | 4 | 0 | 0 | Low-quality | Genome-fragment | 4.83 | AAI-based (high-confidence) | 0 |
| S3_vs_k141_22474\|\|full | 2385 | No | NA | 2 | 0 | 2 | Low-quality | Genome-fragment | 1.15 | AAI-based (medium-confidence) | 0 |
| S7_vs_133838\|\|full | 2217 | No | NA | 4 | 0 | 1 | Not-determined | Genome-fragment | NA | NA | 0 |
| S7_vs_133935\|\|full | 12138 | No | NA | 6 | 0 | 5 | Not-determined | Genome-fragment | NA | NA | 0 |
| S7_vs_133943\|\|full | 7387 | No | NA | 10 | 0 | 2 | Not-determined | Genome-fragment | NA | NA | 0 |
| S7_vs_67334\|\|full | 2385 | No | NA | 6 | 0 | 0 | Low-quality | Genome-fragment | 6.85 | AAI-based (high-confidence) | 0 |
| S2_vs_k141_8593\|\|full | 2385 | No | NA | 6 | 2 | 0 | Low-quality | Genome-fragment | 3.96 | AAI-based (high-confidence) | 0 |
| S8_extracted_k141_134535 | 2386 | No | NA | 5 | 1 | 0 | Low-quality | Genome-fragment | 5.68 | HMM-based (lower-bound) | 0 |
| S16_vs_17500\|\|full | 2387 | No | NA | 7 | 1 | 0 | Low-quality | Genome-fragment | 3.41 | AAI-based (medium-confidence) | 0 |
| S7_vs_134070\|\|full | 4725 | No | NA | 6 | 0 | 5 | Not-determined | Genome-fragment | NA | NA | 0 |
| S7_vs_134144\|\|full | 2289 | No | NA | 3 | 0 | 1 | Not-determined | Genome-fragment | NA | NA | 0 |
| S8_extracted_k141_179018 | 2387 | No | NA | 6 | 0 | 0 | Low-quality | Genome-fragment | 2.39 | AAI-based (high-confidence) | 0 |
| S7_vs_134299\|\|full | 8172 | No | NA | 8 | 0 | 3 | Not-determined | Genome-fragment | NA | NA | 0 |
| S8_vs_k141_9232\|\|full | 2387 | No | NA | 3 | 0 | 0 | Low-quality | Genome-fragment | 6.18 | AAI-based (high-confidence) | 0 |
| S11_extracted_k141_103567 | 2388 | No | NA | 6 | 3 | 0 | Low-quality | Genome-fragment | 4.55 | AAI-based (medium-confidence) | 0 |
| S14_extracted_k141_25651 | 2388 | No | NA | 8 | 1 | 0 | Low-quality | Genome-fragment | 3.09 | AAI-based (high-confidence) | 0 |
| S7_vs_134450\|\|full | 1140 | No | NA | 2 | 0 | 0 | Not-determined | Genome-fragment | NA | NA | 0 |
| S7_vs_134493\|\|full | 1467 | No | NA | 2 | 0 | 1 | Not-determined | Genome-fragment | NA | NA | 0 |
| S14_vs_k141_17266\|\|full | 2388 | No | NA | 5 | 4 | 0 | Low-quality | Genome-fragment | 6.37 | AAI-based (medium-confidence) | 0 |
| S7_vs_134583\|\|full | 48947 | No | NA | 71 | 21 | 0 | High-quality | High-quality | 97.57 | AAI-based (high-confidence) | 0 |
| S7_vs_134636\|\|full | 1234 | No | NA | 3 | 0 | 2 | Not-determined | Genome-fragment | NA | NA | 0 |
| S3_vs_k141_105089\|\|full | 2388 | No | NA | 3 | 0 | 1 | Low-quality | Genome-fragment | 0.69 | AAI-based (high-confidence) | 0 |
| S12_vs_k141_6038\|\|full | 2388 | No | NA | 3 | 0 | 2 | Low-quality | Genome-fragment | 7.63 | AAI-based (high-confidence) | 0 |
| S7_vs_134722\|\|full | 1800 | No | NA | 3 | 0 | 1 | Not-determined | Genome-fragment | NA | NA | 0 |
| S7_vs_134771\|\|full | 1832 | No | NA | 3 | 0 | 0 | Not-determined | Genome-fragment | NA | NA | 0 |
| S8_vs_k141_169708\|\|full | 2388 | No | NA | 3 | 0 | 0 | Low-quality | Genome-fragment | 5.2 | AAI-based (high-confidence) | 0 |
| S13_vs_59388\|\|full | 2389 | No | NA | 9 | 1 | 0 | Low-quality | Genome-fragment | 2.94 | HMM-based (lower-bound) | 0 |
| S7_extracted_110016 | 2389 | No | NA | 3 | 1 | 0 | Low-quality | Genome-fragment | 1.51 | HMM-based (lower-bound) | 0 |
| S12_extracted_k141_162055 | 2389 | No | NA | 3 | 0 | 0 | Low-quality | Genome-fragment | 1.12 | AAI-based (high-confidence) | 0 |
| S14_vs_k141_76294\|\|full | 2390 | No | NA | 4 | 0 | 0 | Low-quality | Genome-fragment | 6.33 | AAI-based (high-confidence) | 0 |
| S14_vs_k141_11662\|\|full | 2390 | No | NA | 3 | 1 | 0 | Low-quality | Genome-fragment | 4.13 | AAI-based (high-confidence) | 0 |
| S16_vs_111413\|\|full | 2390 | No | NA | 3 | 2 | 0 | Low-quality | Genome-fragment | 6.14 | AAI-based (high-confidence) | 0 |
| S7_vs_135416\|\|full | 2140 | No | NA | 7 | 0 | 0 | Not-determined | Genome-fragment | NA | NA | 0 |
| S10_vs_54478\|\|full | 2390 | No | NA | 7 | 1 | 0 | Low-quality | Genome-fragment | 7.51 | AAI-based (high-confidence) | 0 |
| S7_vs_135928\|\|full | 2039 | No | NA | 2 | 0 | 1 | Not-determined | Genome-fragment | NA | NA | 0 |
| S7_vs_136090\|\|full | 2004 | No | NA | 2 | 0 | 2 | Not-determined | Genome-fragment | NA | NA | 0 |
| S7_vs_136139\|\|full | 2515 | No | NA | 6 | 0 | 0 | Not-determined | Genome-fragment | NA | NA | 0 |
| S7_vs_136181\|\|full | 1110 | No | NA | 4 | 0 | 0 | Not-determined | Genome-fragment | NA | NA | 0 |
| S7_vs_136207\|\|full | 4963 | No | NA | 4 | 0 | 1 | Not-determined | Genome-fragment | NA | NA | 0 |
| S7_vs_57625\|\|full | 2391 | No | NA | 5 | 1 | 0 | Low-quality | Genome-fragment | 2.67 | AAI-based (high-confidence) | 0 |
| S7_vs_136258\|\|full | 1614 | No | NA | 3 | 0 | 0 | Not-determined | Genome-fragment | NA | NA | 0 |
| S12_vs_k141_111721\|\|full | 2391 | No | NA | 3 | 0 | 0 | Low-quality | Genome-fragment | 4.74 | AAI-based (high-confidence) | 0 |
| S7_vs_136295\|\|full | 3247 | No | NA | 3 | 0 | 2 | Not-determined | Genome-fragment | NA | NA | 0 |
| S7_vs_136311\|\|full | 3926 | No | NA | 6 | 0 | 2 | Not-determined | Genome-fragment | NA | NA | 0 |
| S7_vs_136316\|\|full | 1685 | No | NA | 3 | 0 | 1 | Not-determined | Genome-fragment | NA | NA | 0 |
| S7_vs_136358\|\|full | 1006 | No | NA | 3 | 0 | 0 | Not-determined | Genome-fragment | NA | NA | 0 |
| S15_extracted_71816 | 2391 | No | NA | 7 | 0 | 6 | Low-quality | Genome-fragment | 0.99 | AAI-based (medium-confidence) | 0 |
| S6_vs_k141_36164\|\|full | 2391 | No | NA | 2 | 0 | 1 | Low-quality | Genome-fragment | 4.51 | AAI-based (medium-confidence) | 0 |
| S8_vs_k141_134178\|\|full | 2391 | No | NA | 4 | 0 | 0 | Low-quality | Genome-fragment | 2.66 | AAI-based (medium-confidence) | 0 |
| S7_vs_136447\|\|full | 4476 | No | NA | 5 | 0 | 4 | Not-determined | Genome-fragment | NA | NA | 0 |
| S8_vs_k141_164524\|\|full | 2391 | No | NA | 7 | 1 | 0 | Low-quality | Genome-fragment | 6.9 | AAI-based (medium-confidence) | 0 |
| S7_vs_136540\|\|full | 3626 | No | NA | 3 | 0 | 2 | Not-determined | Genome-fragment | NA | NA | 0 |
| S9_extracted_k141_43946 | 2392 | No | NA | 4 | 1 | 0 | Low-quality | Genome-fragment | 5.84 | AAI-based (high-confidence) | 0 |
| S9_vs_k141_79328\|\|full | 2392 | No | NA | 4 | 0 | 0 | Low-quality | Genome-fragment | 3.39 | AAI-based (high-confidence) | 0 |
| S11_extracted_k141_88582 | 2392 | No | NA | 4 | 0 | 0 | Low-quality | Genome-fragment | 5.26 | AAI-based (high-confidence) | 0 |
| S3_vs_k141_59594\|\|full | 2392 | No | NA | 2 | 0 | 1 | Low-quality | Genome-fragment | 3.9 | AAI-based (medium-confidence) | 0 |
| S7_vs_136816\|\|full | 2018 | No | NA | 3 | 0 | 3 | Not-determined | Genome-fragment | NA | NA | 0 |
| S7_vs_136945\|\|full | 1671 | No | NA | 5 | 0 | 1 | Not-determined | Genome-fragment | NA | NA | 0 |
| S7_vs_136965\|\|full | 2636 | No | NA | 5 | 0 | 0 | Not-determined | Genome-fragment | NA | NA | 0 |
| S7_vs_136979\|\|full | 6584 | No | NA | 6 | 0 | 3 | Not-determined | Genome-fragment | NA | NA | 0 |
| S5_vs_k141_2358\|\|full | 2392 | No | NA | 6 | 2 | 0 | Low-quality | Genome-fragment | 6.97 | AAI-based (medium-confidence) | 0 |
| S7_vs_137062\|\|full | 5051 | No | NA | 5 | 0 | 5 | Not-determined | Genome-fragment | NA | NA | 0 |
| S7_vs_6501\|\|full | 2392 | No | NA | 7 | 2 | 0 | Low-quality | Genome-fragment | 5.57 | AAI-based (high-confidence) | 0 |
| S7_vs_137152\|\|full | 6008 | No | NA | 7 | 0 | 1 | Not-determined | Genome-fragment | NA | NA | 0 |
| S7_vs_137197\|\|full | 1699 | No | NA | 4 | 0 | 0 | Not-determined | Genome-fragment | NA | NA | 0 |
| S7_vs_137240\|\|full | 2747 | No | NA | 4 | 0 | 2 | Not-determined | Genome-fragment | NA | NA | 0 |
| S16_extracted_168906 | 2392 | No | NA | 3 | 0 | 1 | Low-quality | Genome-fragment | 5.93 | AAI-based (medium-confidence) | 0 |
| S7_vs_137282\|\|full | 1025 | No | NA | 2 | 0 | 1 | Not-determined | Genome-fragment | NA | NA | 0 |
| S7_vs_137283\|\|full | 1171 | No | NA | 2 | 0 | 0 | Not-determined | Genome-fragment | NA | NA | 0 |
| S7_vs_137338\|\|full | 4444 | No | NA | 2 | 0 | 1 | Not-determined | Genome-fragment | NA | NA | 0 |
| S15_vs_69229\|\|full | 2392 | No | NA | 3 | 0 | 2 | Low-quality | Genome-fragment | 1.42 | AAI-based (high-confidence) | 0 |
| S17_vs_k141_105979\|\|full | 2393 | No | NA | 3 | 1 | 0 | Low-quality | Genome-fragment | 5.94 | AAI-based (high-confidence) | 0 |
| S7_vs_137439\|\|full | 2255 | No | NA | 3 | 0 | 1 | Not-determined | Genome-fragment | NA | NA | 0 |
| S7_vs_137466\|\|full | 2564 | No | NA | 4 | 0 | 1 | Not-determined | Genome-fragment | NA | NA | 0 |
| S7_vs_137484\|\|full | 2711 | No | NA | 10 | 0 | 0 | Not-determined | Genome-fragment | NA | NA | 0 |
| S7_vs_137486\|\|full | 1993 | No | NA | 3 | 0 | 0 | Not-determined | Genome-fragment | NA | NA | 0 |
| S7_vs_137553\|\|full | 1609 | No | NA | 3 | 0 | 1 | Not-determined | Genome-fragment | NA | NA | 0 |
| S7_vs_137574\|\|full | 3677 | No | NA | 4 | 0 | 1 | Not-determined | Genome-fragment | NA | NA | 0 |
| S7_vs_137612\|\|full | 5581 | No | NA | 8 | 0 | 4 | Not-determined | Genome-fragment | NA | NA | 0 |
| S7_vs_137613\|\|full | 1982 | No | NA | 2 | 0 | 2 | Not-determined | Genome-fragment | NA | NA | 0 |
| S7_vs_137639\|\|full | 1516 | No | NA | 3 | 0 | 1 | Not-determined | Genome-fragment | NA | NA | 0 |
| S7_vs_137739\|\|full | 1246 | No | NA | 2 | 0 | 1 | Not-determined | Genome-fragment | NA | NA | 0 |
| S7_vs_137793\|\|full | 5875 | No | NA | 4 | 0 | 0 | Not-determined | Genome-fragment | NA | NA | 0 |
| S7_vs_137863\|\|full | 9946 | No | NA | 9 | 0 | 8 | Not-determined | Genome-fragment | NA | NA | 0 |
| S7_vs_137866\|\|full | 4523 | No | NA | 9 | 0 | 1 | Not-determined | Genome-fragment | NA | NA | 0 |
| S7_vs_137913\|\|full | 2718 | No | NA | 4 | 0 | 0 | Not-determined | Genome-fragment | NA | NA | 0 |
| S7_vs_137993\|\|full | 2930 | No | NA | 3 | 0 | 0 | Not-determined | Genome-fragment | NA | NA | 0 |
| S3_vs_k141_65915\|\|full | 2393 | No | NA | 2 | 1 | 0 | Low-quality | Genome-fragment | 3.62 | HMM-based (lower-bound) | 0 |
| S7_vs_12150\|\|0_partial | 22205 | No | NA | 22 | 0 | 11 | Not-determined | Genome-fragment | NA | NA | 0 |
| S7_vs_15170\|\|0_partial | 12925 | No | NA | 5 | 0 | 4 | Not-determined | Genome-fragment | NA | NA | 0 |
| S7_vs_83867\|\|full | 2393 | No | NA | 4 | 1 | 0 | Low-quality | Genome-fragment | 1.47 | HMM-based (lower-bound) | 0 |
| S7_vs_47488\|\|0_partial | 12026 | No | NA | 14 | 0 | 8 | Not-determined | Genome-fragment | NA | NA | 0 |
| S7_vs_47978\|\|0_partial | 16561 | No | NA | 12 | 0 | 3 | Not-determined | Genome-fragment | NA | NA | 0 |
| S7_vs_72737\|\|0_partial | 33496 | No | NA | 33 | 1 | 14 | High-quality | High-quality | 100 | HMM-based (lower-bound) | 0 |
| S16_vs_82514\|\|full | 2393 | No | NA | 4 | 0 | 0 | Low-quality | Genome-fragment | 5.75 | AAI-based (medium-confidence) | 0 |
| S7_vs_118374\|\|0_partial | 13751 | No | NA | 10 | 0 | 2 | Not-determined | Genome-fragment | NA | NA | 0 |
| S6_vs_k141_7426\|\|full | 2393 | No | NA | 2 | 0 | 2 | Low-quality | Genome-fragment | 5.35 | AAI-based (medium-confidence) | 0 |
| S9_vs_k141_88443\|\|full | 2394 | No | NA | 3 | 0 | 0 | Low-quality | Genome-fragment | 5.61 | AAI-based (medium-confidence) | 0 |
| S7_vs_1369\|\|lt2gene | 1156 | No | NA | 2 | 0 | 0 | Not-determined | Genome-fragment | NA | NA | 0 |
| S1_extracted_k141_152792 | 2394 | No | NA | 5 | 1 | 0 | Low-quality | Genome-fragment | 12.08 | AAI-based (medium-confidence) | 0 |
| S7_vs_6139\|\|lt2gene | 2028 | No | NA | 2 | 0 | 0 | Not-determined | Genome-fragment | NA | NA | 0 |
| S7_vs_6310\|\|lt2gene | 2100 | No | NA | 2 | 0 | 0 | Not-determined | Genome-fragment | NA | NA | 0 |
| S16_vs_70687\|\|full | 2394 | No | NA | 6 | 0 | 1 | Low-quality | Genome-fragment | 5.66 | AAI-based (medium-confidence) | 0 |
| S7_vs_7054\|\|lt2gene | 1175 | No | NA | 1 | 0 | 1 | Not-determined | Genome-fragment | NA | NA | 0 |
| S10_vs_20419\|\|full | 2394 | No | NA | 5 | 0 | 1 | Low-quality | Genome-fragment | 5.51 | AAI-based (medium-confidence) | 0 |
| S1_extracted_k141_89072 | 2395 | No | NA | 5 | 1 | 0 | Low-quality | Genome-fragment | 6.93 | AAI-based (high-confidence) | 0 |
| S3_vs_k141_106427\|\|full | 2395 | No | NA | 5 | 0 | 0 | Low-quality | Genome-fragment | 5.29 | AAI-based (high-confidence) | 0 |
| S7_vs_8883\|\|lt2gene | 3101 | No | NA | 1 | 0 | 0 | Not-determined | Genome-fragment | NA | NA | 0 |
| S7_vs_9599\|\|lt2gene | 1029 | No | NA | 1 | 0 | 0 | Not-determined | Genome-fragment | NA | NA | 0 |
| S5_vs_k141_2883\|\|full | 2395 | No | NA | 4 | 1 | 0 | Low-quality | Genome-fragment | 2.14 | HMM-based (lower-bound) | 0 |
| S7_vs_13707\|\|lt2gene | 1233 | No | NA | 1 | 0 | 0 | Not-determined | Genome-fragment | NA | NA | 0 |
| S12_vs_k141_53318\|\|lt2gene | 2395 | No | NA | 1 | 1 | 0 | Low-quality | Genome-fragment | 5.29 | AAI-based (high-confidence) | 0 |
| S14_vs_k141_114204\|\|full | 2396 | No | NA | 2 | 0 | 0 | Low-quality | Genome-fragment | 1.62 | AAI-based (high-confidence) | 0 |
| S7_vs_18434\|\|lt2gene | 2078 | No | NA | 1 | 0 | 0 | Not-determined | Genome-fragment | NA | NA | 0 |
| S5_vs_k141_56829\|\|full | 2396 | No | NA | 4 | 2 | 0 | Low-quality | Genome-fragment | 1.48 | AAI-based (high-confidence) | 0 |
| S7_vs_20346\|\|lt2gene | 1200 | No | NA | 1 | 0 | 1 | Not-determined | Genome-fragment | NA | NA | 0 |
| S7_vs_20373\|\|lt2gene | 1838 | No | NA | 1 | 0 | 0 | Not-determined | Genome-fragment | NA | NA | 0 |
| S16_vs_52010\|\|full | 2396 | No | NA | 4 | 0 | 4 | Low-quality | Genome-fragment | 3.26 | AAI-based (high-confidence) | 0 |
| S12_extracted_k141_119265 | 2396 | No | NA | 6 | 3 | 0 | Low-quality | Genome-fragment | 5.97 | AAI-based (high-confidence) | 0 |
| S7_vs_29251\|\|lt2gene | 2169 | No | NA | 2 | 0 | 1 | Not-determined | Genome-fragment | NA | NA | 0 |
| S12_vs_k141_162142\|\|full | 2396 | No | NA | 3 | 0 | 1 | Low-quality | Genome-fragment | 7.65 | AAI-based (medium-confidence) | 0 |
| S17_vs_k141_135206\|\|full | 2397 | No | NA | 3 | 1 | 0 | Low-quality | Genome-fragment | 6.89 | AAI-based (high-confidence) | 0 |
| S17_vs_k141_12377\|\|full | 2397 | No | NA | 5 | 0 | 0 | Low-quality | Genome-fragment | 4.08 | AAI-based (high-confidence) | 0 |
| S13_vs_45566\|\|full | 2397 | No | NA | 2 | 0 | 1 | Low-quality | Genome-fragment | 3.02 | AAI-based (medium-confidence) | 0 |
| S1_extracted_k141_97881 | 2397 | No | NA | 4 | 2 | 0 | Low-quality | Genome-fragment | 8.43 | HMM-based (lower-bound) | 0 |
| S7_vs_32986\|\|lt2gene | 1141 | No | NA | 2 | 0 | 0 | Not-determined | Genome-fragment | NA | NA | 0 |
| S10_vs_100937\|\|full | 2397 | No | NA | 4 | 0 | 1 | Low-quality | Genome-fragment | 2.55 | AAI-based (medium-confidence) | 0 |
| S9_vs_k141_38455\|\|full | 2398 | No | NA | 5 | 0 | 2 | Low-quality | Genome-fragment | 4.37 | AAI-based (medium-confidence) | 0 |
| S1_vs_k141_59312\|\|full | 2398 | No | NA | 3 | 0 | 1 | Low-quality | Genome-fragment | 4.48 | AAI-based (medium-confidence) | 0 |
| S1_vs_k141_139158\|\|full | 2398 | No | NA | 3 | 0 | 3 | Low-quality | Genome-fragment | 0.69 | AAI-based (medium-confidence) | 0 |
| S5_vs_k141_125151\|\|full | 2398 | No | NA | 7 | 6 | 0 | Low-quality | Genome-fragment | 7.02 | AAI-based (high-confidence) | 0 |
| S16_extracted_102593 | 2398 | No | NA | 4 | 3 | 0 | Low-quality | Genome-fragment | 4.75 | AAI-based (medium-confidence) | 0 |
| S16_vs_123804\|\|full | 2398 | No | NA | 3 | 0 | 3 | Low-quality | Genome-fragment | 3.23 | AAI-based (medium-confidence) | 0 |
| S7_vs_56270\|\|full | 2399 | No | NA | 4 | 0 | 1 | Low-quality | Genome-fragment | 3.18 | AAI-based (medium-confidence) | 0 |
| S12_vs_k141_172224\|\|full | 2399 | No | NA | 5 | 1 | 0 | Low-quality | Genome-fragment | 1.47 | AAI-based (high-confidence) | 0 |
| S13_vs_5788\|\|full | 2400 | Yes | 1474 | 5 | 2 | 1 | Low-quality | Genome-fragment | 3.06 | AAI-based (medium-confidence) | 38.58 |
| S14_vs_k141_69536\|\|full | 2400 | No | NA | 3 | 0 | 0 | Low-quality | Genome-fragment | 4.8 | AAI-based (high-confidence) | 0 |
| S14_vs_k141_14169\|\|full | 2400 | No | NA | 4 | 2 | 0 | Low-quality | Genome-fragment | 5.54 | AAI-based (high-confidence) | 0 |
| S7_vs_115787\|\|full | 2400 | No | NA | 4 | 0 | 0 | Low-quality | Genome-fragment | 5.99 | AAI-based (medium-confidence) | 0 |
| S16_extracted_825 | 2400 | No | NA | 2 | 1 | 0 | Low-quality | Genome-fragment | 1.37 | AAI-based (high-confidence) | 0 |
| S16_vs_52187\|\|full | 2400 | No | NA | 4 | 0 | 0 | Low-quality | Genome-fragment | 3.51 | AAI-based (medium-confidence) | 0 |
| S12_vs_k141_130229\|\|full | 2400 | No | NA | 3 | 1 | 0 | Low-quality | Genome-fragment | 5.95 | AAI-based (high-confidence) | 0 |
| S8_extracted_k141_17539 | 2400 | No | NA | 7 | 5 | 0 | Low-quality | Genome-fragment | 7.92 | AAI-based (high-confidence) | 0 |
| S8_vs_k141_154512\|\|full | 2400 | No | NA | 4 | 3 | 0 | Low-quality | Genome-fragment | 2.27 | AAI-based (high-confidence) | 0 |
| S7_vs_46936\|\|lt2gene | 1397 | No | NA | 1 | 0 | 1 | Not-determined | Genome-fragment | NA | NA | 0 |
| S11_vs_k141_15876\|\|full | 2401 | No | NA | 3 | 0 | 0 | Low-quality | Genome-fragment | 2.31 | AAI-based (medium-confidence) | 0 |
| S16_vs_112756\|\|full | 2401 | No | NA | 7 | 1 | 0 | Low-quality | Genome-fragment | 1.91 | HMM-based (lower-bound) | 0 |
| S10_vs_86716\|\|full | 2401 | No | NA | 9 | 1 | 0 | Low-quality | Genome-fragment | 5.24 | AAI-based (medium-confidence) | 0 |
| S7_vs_48662\|\|lt2gene | 1671 | No | NA | 2 | 0 | 0 | Not-determined | Genome-fragment | NA | NA | 0 |
| S17_vs_k141_37417\|\|lt2gene | 2402 | No | NA | 2 | 1 | 0 | Low-quality | Genome-fragment | 4.22 | AAI-based (high-confidence) | 0 |
| S5_vs_k141_107694\|\|full | 2402 | Yes | 2015 | 3 | 1 | 1 | Low-quality | Genome-fragment | 3.4 | AAI-based (medium-confidence) | 16.11 |
| S10_vs_32445\|\|full | 2402 | No | NA | 4 | 0 | 0 | Low-quality | Genome-fragment | 4.41 | AAI-based (medium-confidence) | 0 |
| S10_vs_104554\|\|full | 2402 | No | NA | 4 | 1 | 0 | Low-quality | Genome-fragment | 0.98 | HMM-based (lower-bound) | 0 |
| S8_vs_k141_206587\|\|full | 2402 | No | NA | 2 | 0 | 1 | Low-quality | Genome-fragment | 1.08 | AAI-based (medium-confidence) | 0 |
| S13_vs_145888\|\|full | 2403 | No | NA | 3 | 0 | 0 | Low-quality | Genome-fragment | 4.26 | AAI-based (high-confidence) | 0 |
| S1_extracted_k141_113822 | 2403 | No | NA | 5 | 5 | 0 | Low-quality | Genome-fragment | 5.26 | AAI-based (high-confidence) | 0 |
| S8_extracted_k141_157523 | 2403 | No | NA | 2 | 0 | 1 | Low-quality | Genome-fragment | 5.17 | AAI-based (high-confidence) | 0 |
| S8_vs_k141_38410\|\|full | 2403 | No | NA | 6 | 1 | 0 | Low-quality | Genome-fragment | 5.95 | AAI-based (medium-confidence) | 0 |
| S11_vs_k141_39191\|\|full | 2404 | No | NA | 4 | 3 | 0 | Low-quality | Genome-fragment | 6.52 | AAI-based (high-confidence) | 0 |
| S11_vs_k141_2758\|\|full | 2404 | No | NA | 6 | 0 | 0 | Low-quality | Genome-fragment | 2.83 | AAI-based (medium-confidence) | 0 |
| S7_vs_62988\|\|lt2gene | 1186 | No | NA | 2 | 0 | 1 | Not-determined | Genome-fragment | NA | NA | 0 |
| S14_extracted_k141_7754 | 2404 | No | NA | 2 | 2 | 0 | Low-quality | Genome-fragment | 7.9 | AAI-based (high-confidence) | 0 |
| S1_extracted_k141_50693 | 2404 | No | NA | 5 | 3 | 0 | Low-quality | Genome-fragment | 5.3 | AAI-based (medium-confidence) | 0 |
| S3_vs_k141_83175\|\|full | 2404 | No | NA | 4 | 0 | 1 | Low-quality | Genome-fragment | 1.45 | AAI-based (high-confidence) | 0 |
| S8_vs_k141_207030\|\|full | 2404 | No | NA | 3 | 0 | 0 | Low-quality | Genome-fragment | 4.49 | AAI-based (medium-confidence) | 0 |
| S11_vs_k141_102705\|\|full | 2405 | No | NA | 2 | 0 | 1 | Low-quality | Genome-fragment | 0.72 | AAI-based (high-confidence) | 0 |
| S5_extracted_k141_129270 | 2405 | No | NA | 9 | 4 | 0 | Low-quality | Genome-fragment | 10.98 | AAI-based (high-confidence) | 0 |
| S16_extracted_106541 | 2405 | No | NA | 5 | 0 | 0 | Low-quality | Genome-fragment | 6.22 | AAI-based (high-confidence) | 0 |
| S12_vs_k141_180712\|\|full | 2405 | No | NA | 3 | 0 | 1 | Low-quality | Genome-fragment | 1.26 | AAI-based (medium-confidence) | 0 |
| S6_vs_k141_34813\|\|full | 2405 | No | NA | 2 | 0 | 1 | Low-quality | Genome-fragment | 1.86 | AAI-based (high-confidence) | 0 |
| S9_extracted_k141_75201 | 2406 | No | NA | 3 | 0 | 1 | Low-quality | Genome-fragment | 5.66 | AAI-based (high-confidence) | 0 |
| S1_extracted_k141_143539 | 2406 | No | NA | 3 | 0 | 1 | Low-quality | Genome-fragment | 2.45 | AAI-based (medium-confidence) | 0 |
| S5_vs_k141_6652\|\|full | 2407 | No | NA | 4 | 0 | 0 | Low-quality | Genome-fragment | 4.47 | AAI-based (medium-confidence) | 0 |
| S5_vs_k141_45950\|\|full | 2407 | No | NA | 6 | 2 | 0 | Low-quality | Genome-fragment | 5.58 | AAI-based (medium-confidence) | 0 |
| S10_vs_80411\|\|full | 2407 | No | NA | 4 | 0 | 2 | Low-quality | Genome-fragment | 2.31 | AAI-based (high-confidence) | 0 |
| S4_extracted_5438 | 2407 | No | NA | 4 | 1 | 0 | Low-quality | Genome-fragment | 6.93 | AAI-based (high-confidence) | 0 |
| S8_vs_k141_188476\|\|full | 2407 | No | NA | 4 | 0 | 0 | Low-quality | Genome-fragment | 5.85 | AAI-based (high-confidence) | 0 |
| S3_extracted_k141_25458 | 2408 | No | NA | 3 | 3 | 0 | Low-quality | Genome-fragment | 10.98 | AAI-based (high-confidence) | 0 |
| S3_vs_k141_80470\|\|full | 2408 | No | NA | 6 | 1 | 0 | Low-quality | Genome-fragment | 3.53 | AAI-based (high-confidence) | 0 |
| S13_vs_55805\|\|full | 2409 | No | NA | 6 | 1 | 0 | Low-quality | Genome-fragment | 4.41 | HMM-based (lower-bound) | 0 |
| S1_vs_k141_86086\|\|full | 2409 | No | NA | 3 | 1 | 0 | Low-quality | Genome-fragment | 3.65 | HMM-based (lower-bound) | 0 |
| S7_vs_77401\|\|lt2gene | 1023 | No | NA | 1 | 0 | 0 | Not-determined | Genome-fragment | NA | NA | 0 |
| S16_vs_118170\|\|full | 2409 | No | NA | 3 | 3 | 0 | Low-quality | Genome-fragment | 7.01 | AAI-based (high-confidence) | 0 |
| S10_extracted_60063 | 2409 | No | NA | 3 | 1 | 0 | Low-quality | Genome-fragment | 4.85 | AAI-based (high-confidence) | 0 |
| S7_vs_77921\|\|lt2gene | 1016 | No | NA | 1 | 0 | 0 | Not-determined | Genome-fragment | NA | NA | 0 |
| S8_extracted_k141_179106 | 2409 | No | NA | 2 | 0 | 2 | Low-quality | Genome-fragment | 0.53 | AAI-based (medium-confidence) | 0 |
| S17_extracted_k141_118658 | 2410 | No | NA | 5 | 4 | 0 | Low-quality | Genome-fragment | 7.95 | AAI-based (high-confidence) | 0 |
| S9_extracted_k141_27059 | 2410 | No | NA | 2 | 0 | 0 | Low-quality | Genome-fragment | 5.25 | AAI-based (high-confidence) | 0 |
| S11_vs_k141_100848\|\|full | 2410 | No | NA | 5 | 0 | 2 | Low-quality | Genome-fragment | 1.42 | AAI-based (medium-confidence) | 0 |
| S11_vs_k141_89317\|\|full | 2410 | No | NA | 6 | 0 | 0 | Low-quality | Genome-fragment | 6.44 | AAI-based (high-confidence) | 0 |
| S3_extracted_k141_28143 | 2410 | No | NA | 7 | 1 | 0 | Low-quality | Genome-fragment | 3.65 | AAI-based (high-confidence) | 0 |
| S5_extracted_k141_97172 | 2410 | No | NA | 2 | 0 | 2 | Low-quality | Genome-fragment | 0.89 | AAI-based (high-confidence) | 0 |
| S16_vs_59857\|\|full | 2411 | No | NA | 5 | 0 | 0 | Low-quality | Genome-fragment | 3.74 | AAI-based (high-confidence) | 0 |
| S13_vs_121022\|\|full | 2412 | No | NA | 6 | 1 | 0 | Low-quality | Genome-fragment | 3.24 | HMM-based (lower-bound) | 0 |
| S3_extracted_k141_33597 | 2412 | No | NA | 4 | 0 | 0 | Low-quality | Genome-fragment | 5.89 | AAI-based (high-confidence) | 0 |
| S5_vs_k141_104937\|\|full | 2412 | No | NA | 5 | 1 | 0 | Low-quality | Genome-fragment | 4.99 | HMM-based (lower-bound) | 0 |
| S12_extracted_k141_118399 | 2412 | No | NA | 4 | 1 | 0 | Low-quality | Genome-fragment | 1.79 | HMM-based (lower-bound) | 0 |
| S14_vs_k141_50862\|\|full | 2413 | No | NA | 5 | 1 | 0 | Low-quality | Genome-fragment | 5.71 | AAI-based (high-confidence) | 0 |
| S7_extracted_17200 | 2413 | No | NA | 4 | 4 | 0 | Low-quality | Genome-fragment | 7.96 | AAI-based (high-confidence) | 0 |
| S16_vs_32803\|\|lt2gene | 2413 | No | NA | 2 | 1 | 0 | Low-quality | Genome-fragment | 3.16 | HMM-based (lower-bound) | 0 |
| S6_vs_k141_7596\|\|full | 2413 | No | NA | 4 | 0 | 1 | Low-quality | Genome-fragment | 4.37 | AAI-based (high-confidence) | 0 |
| S8_vs_k141_12200\|\|full | 2413 | No | NA | 7 | 1 | 0 | Low-quality | Genome-fragment | 5.17 | AAI-based (medium-confidence) | 0 |
| S9_extracted_k141_34578 | 2414 | No | NA | 4 | 0 | 0 | Low-quality | Genome-fragment | 1.39 | AAI-based (high-confidence) | 0 |
| S1_vs_k141_50029\|\|full | 2414 | No | NA | 4 | 1 | 0 | Low-quality | Genome-fragment | 4.1 | AAI-based (high-confidence) | 0 |
| S12_vs_k141_114557\|\|full | 2414 | No | NA | 6 | 4 | 0 | Low-quality | Genome-fragment | 9.6 | AAI-based (high-confidence) | 0 |
| S7_vs_92341\|\|lt2gene | 1203 | No | NA | 1 | 0 | 1 | Not-determined | Genome-fragment | NA | NA | 0 |
| S8_extracted_k141_145059 | 2414 | No | NA | 3 | 0 | 3 | Low-quality | Genome-fragment | 0.98 | AAI-based (medium-confidence) | 0 |
| S9_vs_k141_21662\|\|full | 2415 | No | NA | 4 | 1 | 0 | Low-quality | Genome-fragment | 3.53 | HMM-based (lower-bound) | 0 |
| S5_vs_k141_70714\|\|full | 2415 | No | NA | 9 | 2 | 0 | Low-quality | Genome-fragment | 4.41 | AAI-based (high-confidence) | 0 |
| S15_vs_74374\|\|full | 2415 | No | NA | 3 | 0 | 3 | Low-quality | Genome-fragment | 1.83 | AAI-based (medium-confidence) | 0 |
| S14_vs_k141_66572\|\|full | 2416 | No | NA | 3 | 0 | 0 | Low-quality | Genome-fragment | 4.69 | AAI-based (medium-confidence) | 0 |
| S1_vs_k141_18438\|\|full | 2416 | No | NA | 6 | 4 | 0 | Low-quality | Genome-fragment | 6.19 | AAI-based (high-confidence) | 0 |
| S5_vs_k141_52746\|\|full | 2416 | No | NA | 6 | 2 | 0 | Low-quality | Genome-fragment | 3.34 | AAI-based (medium-confidence) | 0 |
| S13_vs_56539\|\|full | 2417 | No | NA | 3 | 0 | 1 | Low-quality | Genome-fragment | 1.23 | AAI-based (medium-confidence) | 0 |
| S1_vs_k141_119481\|\|full | 2417 | No | NA | 7 | 1 | 0 | Low-quality | Genome-fragment | 5.07 | AAI-based (medium-confidence) | 0 |
| S5_vs_k141_78501\|\|full | 2417 | No | NA | 2 | 0 | 1 | Low-quality | Genome-fragment | 0.68 | AAI-based (high-confidence) | 0 |
| S7_vs_22454\|\|full | 2417 | No | NA | 4 | 0 | 0 | Low-quality | Genome-fragment | 5.67 | AAI-based (medium-confidence) | 0 |
| S10_vs_77300\|\|lt2gene | 2417 | No | NA | 2 | 1 | 0 | Low-quality | Genome-fragment | 4.15 | AAI-based (high-confidence) | 0 |
| S12_vs_k141_89108\|\|full | 2417 | No | NA | 3 | 0 | 2 | Low-quality | Genome-fragment | 1.86 | AAI-based (medium-confidence) | 0 |
| S8_extracted_k141_113233 | 2417 | No | NA | 2 | 0 | 0 | Low-quality | Genome-fragment | 1.38 | AAI-based (high-confidence) | 0 |
| S14_vs_k141_12708\|\|full | 2418 | No | NA | 3 | 0 | 0 | Low-quality | Genome-fragment | 5.8 | AAI-based (medium-confidence) | 0 |
| S1_vs_k141_7725\|\|full | 2418 | No | NA | 2 | 0 | 1 | Low-quality | Genome-fragment | 1.2 | AAI-based (medium-confidence) | 0 |
| S5_extracted_k141_106680 | 2418 | No | NA | 6 | 2 | 0 | Low-quality | Genome-fragment | 7.25 | AAI-based (high-confidence) | 0 |
| S16_vs_149706\|\|lt2gene | 2418 | No | NA | 2 | 0 | 0 | Low-quality | Genome-fragment | 6.23 | AAI-based (high-confidence) | 0 |
| S10_vs_73745\|\|full | 2418 | No | NA | 4 | 1 | 0 | Low-quality | Genome-fragment | 3.62 | AAI-based (high-confidence) | 0 |
| S7_vs_111527\|\|lt2gene | 1173 | No | NA | 1 | 0 | 0 | Not-determined | Genome-fragment | NA | NA | 0 |
| S7_vs_112095\|\|lt2gene | 1102 | No | NA | 1 | 0 | 0 | Not-determined | Genome-fragment | NA | NA | 0 |
| S3_vs_k141_95185\|\|full | 2419 | No | NA | 3 | 2 | 0 | Low-quality | Genome-fragment | 5.07 | AAI-based (medium-confidence) | 0 |
| S16_extracted_167887 | 2419 | No | NA | 2 | 0 | 2 | Low-quality | Genome-fragment | 7.73 | AAI-based (medium-confidence) | 0 |
| S16_vs_138438\|\|full | 2419 | No | NA | 4 | 1 | 0 | Low-quality | Genome-fragment | 2.35 | AAI-based (medium-confidence) | 0 |
| S12_extracted_k141_134290 | 2419 | No | NA | 2 | 0 | 0 | Low-quality | Genome-fragment | 1.84 | AAI-based (high-confidence) | 0 |
| S5_extracted_k141_107788 | 2420 | No | NA | 3 | 0 | 2 | Low-quality | Genome-fragment | 1.35 | AAI-based (high-confidence) | 0 |
| S8_vs_k141_188326\|\|full | 2420 | No | NA | 5 | 1 | 0 | Low-quality | Genome-fragment | 4.03 | AAI-based (medium-confidence) | 0 |
| S17_extracted_k141_15302 | 2421 | No | NA | 3 | 0 | 0 | Low-quality | Genome-fragment | 4.04 | AAI-based (medium-confidence) | 0 |
| S17_vs_k141_151316\|\|full | 2421 | No | NA | 1 | 0 | 1 | Low-quality | Genome-fragment | 1.07 | AAI-based (medium-confidence) | 0 |
| S17_vs_k141_79720\|\|full | 2421 | No | NA | 3 | 1 | 0 | Low-quality | Genome-fragment | 4.03 | HMM-based (lower-bound) | 0 |
| S7_vs_30426\|\|full | 2421 | No | NA | 7 | 2 | 0 | Low-quality | Genome-fragment | 6.72 | AAI-based (high-confidence) | 0 |
| S7_vs_125217\|\|lt2gene | 1498 | No | NA | 1 | 0 | 0 | Not-determined | Genome-fragment | NA | NA | 0 |
| S6_extracted_k141_52122 | 2421 | No | NA | 5 | 2 | 0 | Low-quality | Genome-fragment | 5.72 | AAI-based (high-confidence) | 0 |
| S14_extracted_k141_24930 | 2422 | No | NA | 5 | 4 | 0 | Low-quality | Genome-fragment | 7.9 | AAI-based (high-confidence) | 0 |
| S1_extracted_k141_33559 | 2422 | No | NA | 2 | 2 | 0 | Low-quality | Genome-fragment | 3.96 | AAI-based (high-confidence) | 0 |
| S7_extracted_51575 | 2422 | Yes | 1398 | 6 | 1 | 1 | Low-quality | Genome-fragment | 4.29 | AAI-based (high-confidence) | 42.28 |
| S7_vs_127657\|\|lt2gene | 1031 | No | NA | 1 | 0 | 0 | Not-determined | Genome-fragment | NA | NA | 0 |
| S10_extracted_69030 | 2422 | No | NA | 4 | 0 | 0 | Low-quality | Genome-fragment | 1.08 | AAI-based (medium-confidence) | 0 |
| S15_vs_61456\|\|full | 2422 | No | NA | 4 | 4 | 0 | Low-quality | Genome-fragment | 3.9 | AAI-based (high-confidence) | 0 |
| S8_extracted_k141_159984 | 2423 | No | NA | 2 | 0 | 0 | Low-quality | Genome-fragment | 5.45 | AAI-based (medium-confidence) | 0 |
| S11_vs_k141_81426\|\|lt2gene | 2424 | No | NA | 2 | 2 | 0 | Low-quality | Genome-fragment | 6.06 | AAI-based (medium-confidence) | 0 |
| S10_extracted_51357 | 2424 | No | NA | 4 | 1 | 0 | Low-quality | Genome-fragment | 2.98 | AAI-based (high-confidence) | 0 |
| S14_vs_k141_47282\|\|full | 2425 | No | NA | 6 | 4 | 0 | Low-quality | Genome-fragment | 7.89 | AAI-based (high-confidence) | 0 |
| S14_vs_k141_35775\|\|full | 2425 | No | NA | 6 | 0 | 0 | Low-quality | Genome-fragment | 6.19 | AAI-based (high-confidence) | 0 |
| S3_vs_k141_15680\|\|full | 2425 | No | NA | 3 | 0 | 2 | Low-quality | Genome-fragment | 4.19 | AAI-based (medium-confidence) | 0 |
| S7_vs_6762\|\|full | 2425 | No | NA | 3 | 0 | 3 | Low-quality | Genome-fragment | 1.69 | AAI-based (medium-confidence) | 0 |
| S7_vs_61286\|\|full | 2425 | No | NA | 3 | 2 | 0 | Low-quality | Genome-fragment | 4.85 | AAI-based (high-confidence) | 0 |
| S12_extracted_k141_181146 | 2425 | No | NA | 2 | 0 | 2 | Low-quality | Genome-fragment | 1.45 | AAI-based (medium-confidence) | 0 |
| S12_vs_k141_57182\|\|full | 2425 | No | NA | 4 | 0 | 1 | Low-quality | Genome-fragment | 1.27 | AAI-based (medium-confidence) | 0 |
| S8_extracted_k141_66256 | 2425 | Yes | 1311 | 5 | 1 | 2 | Low-quality | Genome-fragment | 1.68 | AAI-based (medium-confidence) | 45.94 |
| S14_vs_k141_78630\|\|lt2gene | 2426 | No | NA | 2 | 0 | 0 | Low-quality | Genome-fragment | 7.34 | AAI-based (high-confidence) | 0 |
| S3_vs_k141_121994\|\|full | 2426 | No | NA | 8 | 1 | 0 | Low-quality | Genome-fragment | 3.9 | AAI-based (high-confidence) | 0 |
| S5_vs_k141_70390\|\|full | 2426 | No | NA | 5 | 0 | 0 | Low-quality | Genome-fragment | 5.43 | AAI-based (medium-confidence) | 0 |
| S11_vs_k141_673\|\|full | 2427 | No | NA | 3 | 1 | 0 | Low-quality | Genome-fragment | 0.97 | AAI-based (medium-confidence) | 0 |
| NC1_vf_k141_3157 | 1097 | No | NA | 3 | 0 | 1 | Not-determined | Genome-fragment | NA | NA | 0 |
| S1_vs_k141_79633\|\|full | 2427 | No | NA | 3 | 0 | 2 | Low-quality | Genome-fragment | 0.79 | AAI-based (high-confidence) | 0 |
| NC1_vf_k141_1594 | 1970 | No | NA | 3 | 0 | 1 | Not-determined | Genome-fragment | NA | NA | 0 |
| NC1_vf_k141_3178 | 1433 | No | NA | 1 | 0 | 1 | Not-determined | Genome-fragment | NA | NA | 0 |
| NC1_vf_k141_2719 | 1279 | No | NA | 1 | 0 | 0 | Not-determined | Genome-fragment | NA | NA | 0 |
| NC1_vf_k141_3404 | 1387 | No | NA | 2 | 0 | 0 | Not-determined | Genome-fragment | NA | NA | 0 |
| NC1_vf_k141_2276 | 1094 | No | NA | 1 | 0 | 1 | Not-determined | Genome-fragment | NA | NA | 0 |
| NC1_vf_k141_263 | 1863 | No | NA | 4 | 0 | 0 | Not-determined | Genome-fragment | NA | NA | 0 |
| NC1_vf_k141_487 | 1604 | No | NA | 1 | 0 | 0 | Not-determined | Genome-fragment | NA | NA | 0 |
| NC1_vf_k141_2069 | 1020 | No | NA | 3 | 0 | 0 | Not-determined | Genome-fragment | NA | NA | 0 |
| NC1_vf_k141_3200 | 1560 | No | NA | 3 | 0 | 2 | Not-determined | Genome-fragment | NA | NA | 0 |
| NC1_vf_k141_740 | 1263 | No | NA | 1 | 0 | 0 | Not-determined | Genome-fragment | NA | NA | 0 |
| NC1_vf_k141_743 | 2853 | No | NA | 4 | 0 | 2 | Not-determined | Genome-fragment | NA | NA | 0 |
| NC1_vf_k141_3444 | 1720 | No | NA | 2 | 0 | 0 | Not-determined | Genome-fragment | NA | NA | 0 |
| NC1_vf_k141_2993 | 1240 | No | NA | 2 | 0 | 0 | Not-determined | Genome-fragment | NA | NA | 0 |
| S3_extracted_k141_37934 | 2427 | No | NA | 7 | 1 | 0 | Low-quality | Genome-fragment | 4.27 | AAI-based (high-confidence) | 0 |
| NC1_vf_k141_103 | 1400 | No | NA | 2 | 0 | 1 | Not-determined | Genome-fragment | NA | NA | 0 |
| NC1_vf_k141_105 | 1133 | No | NA | 2 | 0 | 2 | Not-determined | Genome-fragment | NA | NA | 0 |
| NC1_vf_k141_2126 | 1004 | No | NA | 2 | 0 | 0 | Not-determined | Genome-fragment | NA | NA | 0 |
| NC1_vf_k141_112 | 2316 | No | NA | 3 | 0 | 3 | Not-determined | Genome-fragment | NA | NA | 0 |
| NC1_vf_k141_2791 | 1063 | No | NA | 1 | 0 | 1 | Not-determined | Genome-fragment | NA | NA | 0 |
| NC1_vf_k141_1229 | 2937 | No | NA | 4 | 0 | 0 | Not-determined | Genome-fragment | NA | NA | 0 |
| [illegible] | [illegible] | [illegible] | [illegible] | [illegible] | [illegible] | [illegible] | [illegible] | [illegible] | [illegible] | [illegible] | [illegible] |

| | | | | | | | | | | | |
|---|---|---|---|---|---|---|---|---|---|---|---|
| S3_vs_k141_64203\|\|full | 2427 | No | NA | 3 | 3 | 0 | Low-quality | Genome-fragment | 5.59 | AAI-based (high-confidence) | 0 |
| NC1_vf_k141_2373 | 1567 | No | NA | 3 | 0 | 2 | Not-determined | Genome-fragment | NA | NA | 0 |
| NC1_vf_k141_3050 | 1186 | No | NA | 2 | 0 | 1 | Not-determined | Genome-fragment | NA | NA | 0 |
| NC1_vf_k141_181 | 1382 | No | NA | 1 | 0 | 0 | Not-determined | Genome-fragment | NA | NA | 0 |
| NC1_vf_k141_3067 | 2356 | No | NA | 5 | 0 | 3 | Not-determined | Genome-fragment | NA | NA | 0 |
| NC1_vf_k141_827 | 1894 | No | NA | 2 | 0 | 0 | Not-determined | Genome-fragment | NA | NA | 0 |
| NC1_vf_k141_3524 | 2057 | No | NA | 4 | 0 | 0 | Not-determined | Genome-fragment | NA | NA | 0 |
| NC1_vf_k141_2395 | 1145 | No | NA | 1 | 0 | 1 | Not-determined | Genome-fragment | NA | NA | 0 |
| NC1_vf_k141_1766 | 1588 | No | NA | 2 | 0 | 1 | Not-determined | Genome-fragment | NA | NA | 0 |
| NC1_vf_k141_3534 | 1170 | No | NA | 2 | 0 | 0 | Not-determined | Genome-fragment | NA | NA | 0 |
| NC1_vf_k141_2854 | 1159 | No | NA | 1 | 0 | 1 | Not-determined | Genome-fragment | NA | NA | 0 |
| NC1_vf_k141_1524 | 1168 | No | NA | 2 | 0 | 1 | Not-determined | Genome-fragment | NA | NA | 0 |
| NC1_vf_k141_212 | 2250 | No | NA | 3 | 0 | 2 | Not-determined | Genome-fragment | NA | NA | 0 |
| NC1_vf_k141_850 | 2817 | No | NA | 3 | 0 | 2 | Not-determined | Genome-fragment | NA | NA | 0 |
| S6_extracted_k141_15879 | 2427 | No | NA | 3 | 0 | 0 | Low-quality | Genome-fragment | 1.6 | AAI-based (medium-confidence) | 0 |
| NC1_vf_k141_422 | 1197 | No | NA | 2 | 0 | 2 | Not-determined | Genome-fragment | NA | NA | 0 |
| S8_vs_k141_63851\|\|full | 2427 | No | NA | 5 | 1 | 0 | Low-quality | Genome-fragment | 5.31 | AAI-based (high-confidence) | 0 |
| NC1_vf_k141_1325 | 1116 | No | NA | 1 | 0 | 0 | Not-determined | Genome-fragment | NA | NA | 0 |
| NC1_vf_k141_1560 | 1050 | No | NA | 3 | 0 | 3 | Not-determined | Genome-fragment | NA | NA | 0 |
| NC1_vf_k141_2681 | 1186 | No | NA | 1 | 0 | 0 | Not-determined | Genome-fragment | NA | NA | 0 |
| NC1_vf_k141_2247 | 3416 | No | NA | 3 | 0 | 3 | Not-determined | Genome-fragment | NA | NA | 0 |
| NC3_vf_k141_798 | 1175 | No | NA | 3 | 0 | 0 | Not-determined | Genome-fragment | NA | NA | 0 |
| NC3_vf_k141_143 | 1455 | No | NA | 1 | 0 | 0 | Not-determined | Genome-fragment | NA | NA | 0 |
| NC3_vf_k141_546 | 1142 | No | NA | 2 | 0 | 1 | Not-determined | Genome-fragment | NA | NA | 0 |
| NC3_vf_k141_2001 | 3765 | No | NA | 5 | 0 | 4 | Not-determined | Genome-fragment | NA | NA | 0 |
| NC3_vf_k141_149 | 1431 | No | NA | 2 | 0 | 2 | Not-determined | Genome-fragment | NA | NA | 0 |
| NC3_vf_k141_2012 | 1081 | No | NA | 2 | 0 | 1 | Not-determined | Genome-fragment | NA | NA | 0 |
| NC3_vf_k141_566 | 1064 | No | NA | 1 | 0 | 1 | Not-determined | Genome-fragment | NA | NA | 0 |
| NC3_vf_k141_42 | 1022 | No | NA | 3 | 0 | 0 | Not-determined | Genome-fragment | NA | NA | 0 |
| NC3_vf_k141_1212 | 1354 | No | NA | 3 | 0 | 2 | Not-determined | Genome-fragment | NA | NA | 0 |
| NC3_vf_k141_299 | 1148 | No | NA | 2 | 0 | 1 | Not-determined | Genome-fragment | NA | NA | 0 |
| NC3_vf_k141_1904 | 3220 | No | NA | 1 | 0 | 0 | Not-determined | Genome-fragment | NA | NA | 0 |
| NC3_vf_k141_2033 | 1050 | No | NA | 2 | 0 | 1 | Not-determined | Genome-fragment | NA | NA | 0 |
| S14_vs_k141_76788\|\|full | 2428 | No | NA | 6 | 2 | 0 | Low-quality | Genome-fragment | 5.1 | AAI-based (high-confidence) | 0 |
| NC3_vf_k141_308 | 1857 | No | NA | 3 | 0 | 2 | Not-determined | Genome-fragment | NA | NA | 0 |
| NC3_vf_k141_2039 | 2248 | No | NA | 3 | 0 | 2 | Not-determined | Genome-fragment | NA | NA | 0 |
| NC3_vf_k141_187 | 2510 | No | NA | 3 | 0 | 2 | Not-determined | Genome-fragment | NA | NA | 0 |
| NC3_vf_k141_1647 | 2447 | No | NA | 3 | 0 | 3 | Not-determined | Genome-fragment | NA | NA | 0 |
| NC3_vf_k141_1513 | 5056 | No | NA | 6 | 0 | 6 | Not-determined | Genome-fragment | NA | NA | 0 |
| NC3_vf_k141_1916 | 2136 | No | NA | 3 | 0 | 1 | Not-determined | Genome-fragment | NA | NA | 0 |
| NC3_vf_k141_587 | 2113 | No | NA | 3 | 0 | 3 | Not-determined | Genome-fragment | NA | NA | 0 |
| NC3_vf_k141_2046 | 2152 | No | NA | 3 | 0 | 3 | Not-determined | Genome-fragment | NA | NA | 0 |
| NC3_vf_k141_327 | 1382 | No | NA | 2 | 0 | 0 | Not-determined | Genome-fragment | NA | NA | 0 |
| NC3_vf_k141_2047 | 1339 | No | NA | 2 | 0 | 1 | Not-determined | Genome-fragment | NA | NA | 0 |
| NC3_vf_k141_853 | 1123 | No | NA | 1 | 0 | 1 | Not-determined | Genome-fragment | NA | NA | 0 |
| NC3_vf_k141_600 | 1108 | No | NA | 2 | 0 | 0 | Not-determined | Genome-fragment | NA | NA | 0 |
| NC3_vf_k141_1390 | 1270 | No | NA | 3 | 0 | 2 | Not-determined | Genome-fragment | NA | NA | 0 |
| NC3_vf_k141_1243 | 1175 | No | NA | 2 | 0 | 1 | Not-determined | Genome-fragment | NA | NA | 0 |
| NC3_vf_k141_605 | 1069 | No | NA | 2 | 0 | 2 | Not-determined | Genome-fragment | NA | NA | 0 |
| NC3_vf_k141_1821 | 1587 | No | NA | 4 | 0 | 2 | Not-determined | Genome-fragment | NA | NA | 0 |
| NC3_vf_k141_460 | 1334 | No | NA | 3 | 0 | 2 | Not-determined | Genome-fragment | NA | NA | 0 |
| NC3_vf_k141_1951 | 1128 | No | NA | 2 | 0 | 0 | Not-determined | Genome-fragment | NA | NA | 0 |
| NC3_vf_k141_734 | 2943 | No | NA | 4 | 0 | 3 | Not-determined | Genome-fragment | NA | NA | 0 |
| S5_vs_k141_27806\|\|full | 2428 | No | NA | 4 | 0 | 1 | Low-quality | Genome-fragment | 1.17 | AAI-based (medium-confidence) | 0 |
| NC3_vf_k141_228 | 1613 | No | NA | 4 | 0 | 0 | Not-determined | Genome-fragment | NA | NA | 0 |
| NC3_vf_k141_735 | 1626 | No | NA | 2 | 0 | 1 | Not-determined | Genome-fragment | NA | NA | 0 |
| NC3_vf_k141_1406 | 1031 | No | NA | 3 | 0 | 0 | Not-determined | Genome-fragment | NA | NA | 0 |
| NC3_vf_k141_1557 | 1029 | No | NA | 1 | 0 | 1 | Not-determined | Genome-fragment | NA | NA | 0 |
| NC3_vf_k141_989 | 1412 | No | NA | 2 | 0 | 1 | Not-determined | Genome-fragment | NA | NA | 0 |
| NC3_vf_k141_473 | 1090 | No | NA | 1 | 0 | 1 | Not-determined | Genome-fragment | NA | NA | 0 |
| NC3_vf_k141_626 | 1138 | No | NA | 2 | 0 | 1 | Not-determined | Genome-fragment | NA | NA | 0 |
| NC3_vf_k141_115 | 2969 | No | NA | 5 | 0 | 2 | Not-determined | Genome-fragment | NA | NA | 0 |
| NC3_vf_k141_738 | 2777 | No | NA | 5 | 0 | 2 | Not-determined | Genome-fragment | NA | NA | 0 |
| NC3_vf_k141_1417 | 1066 | No | NA | 2 | 0 | 1 | Not-determined | Genome-fragment | NA | NA | 0 |
| NC3_vf_k141_2105 | 1058 | No | NA | 1 | 0 | 1 | Not-determined | Genome-fragment | NA | NA | 0 |
| NC3_vf_k141_1190 | 5560 | No | NA | 6 | 0 | 5 | Not-determined | Genome-fragment | NA | NA | 0 |
| NC3_vf_k141_494 | 1815 | No | NA | 1 | 0 | 0 | Not-determined | Genome-fragment | NA | NA | 0 |
| NC3_vf_k141_1193 | 1122 | No | NA | 3 | 0 | 1 | Not-determined | Genome-fragment | NA | NA | 0 |
| NC3_vf_k141_1011 | 1027 | No | NA | 2 | 0 | 0 | Not-determined | Genome-fragment | NA | NA | 0 |
| NC3_vf_k141_1196 | 3020 | No | NA | 4 | 0 | 4 | Not-determined | Genome-fragment | NA | NA | 0 |
| NC3_vf_k141_2109 | 8080 | No | NA | 4 | 0 | 3 | Not-determined | Genome-fragment | NA | NA | 0 |
| NC3_vf_k141_768 | 1804 | No | NA | 2 | 0 | 0 | Not-determined | Genome-fragment | NA | NA | 0 |
| NC3_vf_k141_906 | 1583 | No | NA | 2 | 0 | 2 | Not-determined | Genome-fragment | NA | NA | 0 |
| S8_vs_k141_109406\|\|full | 2428 | No | NA | 7 | 0 | 0 | Low-quality | Genome-fragment | 5.64 | AAI-based (high-confidence) | 0 |
| S11_vs_k141_24066\|\|full | 2429 | No | NA | 5 | 1 | 0 | Low-quality | Genome-fragment | 6.24 | AAI-based (high-confidence) | 0 |
| NC3_vf_k141_1296 | 1643 | No | NA | 3 | 0 | 0 | Not-determined | Genome-fragment | NA | NA | 0 |
| NC3_vf_k141_1039 | 1655 | No | NA | 1 | 0 | 0 | Not-determined | Genome-fragment | NA | NA | 0 |
| NC3_vf_k141_1054 | 1155 | No | NA | 2 | 0 | 2 | Not-determined | Genome-fragment | NA | NA | 0 |
| S16_extracted_50 | 2143 | No | NA | 2 | 0 | 0 | Not-determined | Genome-fragment | NA | NA | 0 |
| S16_extracted_121 | 1183 | No | NA | 1 | 0 | 0 | Not-determined | Genome-fragment | NA | NA | 0 |
| S7_vs_965\|\|full | 2429 | No | NA | 3 | 0 | 0 | Low-quality | Genome-fragment | 5.63 | AAI-based (medium-confidence) | 0 |
| S16_extracted_326 | 2452 | No | NA | 1 | 0 | 0 | Not-determined | Genome-fragment | NA | NA | 0 |
| S7_vs_69240\|\|lt2gene | 2429 | No | NA | 2 | 1 | 0 | Low-quality | Genome-fragment | 4.09 | HMM-based (lower-bound) | 0 |
| S16_extracted_416 | 6561 | No | NA | 3 | 0 | 1 | Not-determined | Genome-fragment | NA | NA | 0 |
| S16_extracted_465 | 5263 | No | NA | 2 | 0 | 1 | Not-determined | Genome-fragment | NA | NA | 0 |
| S16_extracted_548 | 1234 | No | NA | 2 | 0 | 1 | Not-determined | Genome-fragment | NA | NA | 0 |
| S16_extracted_560 | 1235 | No | NA | 2 | 0 | 0 | Not-determined | Genome-fragment | NA | NA | 0 |
| S16_extracted_575 | 1067 | No | NA | 2 | 0 | 1 | Not-determined | Genome-fragment | NA | NA | 0 |
| S16_extracted_8567 | 2429 | No | NA | 1 | 0 | 1 | Low-quality | Genome-fragment | 1.31 | AAI-based (medium-confidence) | 0 |
| S16_extracted_914 | 1441 | No | NA | 1 | 0 | 1 | Not-determined | Genome-fragment | NA | NA | 0 |
| S3_vs_k141_104371\|\|full | 2430 | No | NA | 4 | 0 | 0 | Low-quality | Genome-fragment | 5.76 | AAI-based (medium-confidence) | 0 |
| S16_extracted_1037 | 58665 | No | NA | 101 | 29 | 2 | Complete | High-quality | 100 | DTR (high-confidence) | 0 |
| S10_vs_81025\|\|full | 2430 | No | NA | 3 | 0 | 0 | Low-quality | Genome-fragment | 4.59 | AAI-based (medium-confidence) | 0 |
| S16_extracted_1090 | 1174 | No | NA | 1 | 0 | 0 | Not-determined | Genome-fragment | NA | NA | 0 |
| S16_extracted_1101 | 1013 | No | NA | 2 | 0 | 2 | Not-determined | Genome-fragment | NA | NA | 0 |
| S16_extracted_1289 | 1791 | No | NA | 1 | 0 | 1 | Not-determined | Genome-fragment | NA | NA | 0 |
| S16_extracted_1572 | 1103 | No | NA | 2 | 0 | 0 | Not-determined | Genome-fragment | NA | NA | 0 |
| S16_extracted_1591 | 1495 | No | NA | 2 | 0 | 2 | Not-determined | Genome-fragment | NA | NA | 0 |
| S12_vs_k141_139329\|\|lt2gene | 2430 | No | NA | 2 | 2 | 0 | Low-quality | Genome-fragment | 7.08 | AAI-based (medium-confidence) | 0 |
| S16_extracted_1763 | 1016 | No | NA | 3 | 0 | 0 | Not-determined | Genome-fragment | NA | NA | 0 |
| S16_extracted_1798 | 2199 | No | NA | 2 | 0 | 2 | Not-determined | Genome-fragment | NA | NA | 0 |
| S1_vs_k141_136354\|\|full | 2431 | No | NA | 4 | 0 | 0 | Low-quality | Genome-fragment | 1.46 | AAI-based (medium-confidence) | 0 |
| S16_extracted_1850 | 1045 | No | NA | 1 | 0 | 0 | Not-determined | Genome-fragment | NA | NA | 0 |
| S16_extracted_1959 | 1980 | No | NA | 2 | 0 | 0 | Not-determined | Genome-fragment | NA | NA | 0 |
| S16_extracted_2039 | 1382 | No | NA | 2 | 0 | 1 | Not-determined | Genome-fragment | NA | NA | 0 |
| S3_vs_k141_91373\|\|full | 2431 | No | NA | 4 | 0 | 0 | Low-quality | Genome-fragment | 4.96 | AAI-based (medium-confidence) | 0 |
| S3_vs_k141_23167\|\|full | 2431 | No | NA | 4 | 2 | 0 | Low-quality | Genome-fragment | 6.75 | AAI-based (high-confidence) | 0 |
| S5_vs_k141_76831\|\|full | 2431 | No | NA | 7 | 1 | 0 | Low-quality | Genome-fragment | 5.99 | AAI-based (medium-confidence) | 0 |
| S16_extracted_2463 | 3454 | No | NA | 1 | 0 | 1 | Not-determined | Genome-fragment | NA | NA | 0 |
| S16_extracted_2477 | 2847 | No | NA | 7 | 0 | 6 | Not-determined | Genome-fragment | NA | NA | 0 |
| S16_extracted_2496 | 1002 | No | NA | 1 | 0 | 0 | Not-determined | Genome-fragment | NA | NA | 0 |
| S9_vs_k141_85790\|\|full | 2432 | No | NA | 3 | 0 | 3 | Low-quality | Genome-fragment | 0.97 | AAI-based (medium-confidence) | 0 |
| S16_extracted_2750 | 1135 | No | NA | 1 | 0 | 1 | Not-determined | Genome-fragment | NA | NA | 0 |
| S16_extracted_2862 | 1573 | No | NA | 2 | 0 | 1 | Not-determined | Genome-fragment | NA | NA | 0 |
| S16_extracted_2976 | 1537 | No | NA | 1 | 0 | 1 | Not-determined | Genome-fragment | NA | NA | 0 |
| S16_extracted_3085 | 1853 | No | NA | 1 | 0 | 1 | Not-determined | Genome-fragment | NA | NA | 0 |
| S16_extracted_3151 | 5358 | No | NA | 5 | 0 | 3 | Not-determined | Genome-fragment | NA | NA | 0 |
| S16_extracted_3162 | 22704 | No | NA | 2 | 0 | 1 | Not-determined | Genome-fragment | NA | NA | 0 |
| S10_vs_29127\|\|lt2gene | 2432 | No | NA | 2 | 2 | 0 | Low-quality | Genome-fragment | 4.8 | AAI-based (medium-confidence) | 0 |
| S16_extracted_3336 | 1432 | No | NA | 1 | 0 | 1 | Not-determined | Genome-fragment | NA | NA | 0 |
| S17_vs_k141_128025\|\|full | 2433 | No | NA | 3 | 0 | 2 | Low-quality | Genome-fragment | 4.29 | AAI-based (medium-confidence) | 0 |
| S16_extracted_3452 | 1080 | No | NA | 2 | 0 | 2 | Not-determined | Genome-fragment | NA | NA | 0 |
| S1_extracted_k141_142458 | 2433 | No | NA | 3 | 1 | 0 | Low-quality | Genome-fragment | 2.46 | AAI-based (high-confidence) | 0 |
| S16_extracted_3543 | 3378 | No | NA | 2 | 0 | 1 | Not-determined | Genome-fragment | NA | NA | 0 |
| S16_extracted_3590 | 1610 | No | NA | 2 | 0 | 0 | Not-determined | Genome-fragment | NA | NA | 0 |
| S16_extracted_3637 | 3159 | No | NA | 2 | 0 | 2 | Not-determined | Genome-fragment | NA | NA | 0 |
| S16_extracted_3639 | 1063 | No | NA | 3 | 0 | 0 | Not-determined | Genome-fragment | NA | NA | 0 |
| S16_extracted_3680 | 2974 | No | NA | 3 | 0 | 2 | Not-determined | Genome-fragment | NA | NA | 0 |
| S16_extracted_3852 | 1562 | No | NA | 2 | 0 | 0 | Not-determined | Genome-fragment | NA | NA | 0 |
| S1_vs_k141_144075\|\|full | 2433 | No | NA | 4 | 3 | 0 | Low-quality | Genome-fragment | 5 | AAI-based (high-confidence) | 0 |
| S16_extracted_3940 | 4781 | No | NA | 4 | 0 | 2 | Not-determined | Genome-fragment | NA | NA | 0 |
| S1_vs_k141_150798\|\|full | 2433 | Yes | 898 | 3 | 2 | 1 | Low-quality | Genome-fragment | 1.27 | HMM-based (lower-bound) | 63.09 |
| S16_extracted_4061 | 4032 | No | NA | 7 | 0 | 0 | Not-determined | Genome-fragment | NA | NA | 0 |
| S3_extracted_k141_43660 | 2433 | No | NA | 2 | 1 | 0 | Low-quality | Genome-fragment | 4.21 | HMM-based (lower-bound) | 0 |
| S10_extracted_99672 | 2433 | No | NA | 3 | 2 | 0 | Low-quality | Genome-fragment | 4.61 | AAI-based (high-confidence) | 0 |
| S16_extracted_4205 | 4728 | No | NA | 5 | 0 | 4 | Not-determined | Genome-fragment | NA | NA | 0 |
| S16_extracted_4263 | 3334 | No | NA | 4 | 0 | 4 | Not-determined | Genome-fragment | NA | NA | 0 |
| S16_extracted_4305 | 1658 | No | NA | 1 | 0 | 1 | Not-determined | Genome-fragment | NA | NA | 0 |
| S16_extracted_4323 | 5750 | No | NA | 5 | 0 | 2 | Not-determined | Genome-fragment | NA | NA | 0 |
| S16_extracted_4418 | 1203 | No | NA | 1 | 0 | 1 | Not-determined | Genome-fragment | NA | NA | 0 |
| S16_extracted_4421 | 1149 | No | NA | 1 | 0 | 0 | Not-determined | Genome-fragment | NA | NA | 0 |
| S16_extracted_4513 | 3859 | No | NA | 4 | 0 | 2 | Not-determined | Genome-fragment | NA | NA | 0 |
| S16_extracted_4571 | 1351 | No | NA | 1 | 0 | 0 | Not-determined | Genome-fragment | NA | NA | 0 |
| S16_extracted_4643 | 1931 | No | NA | 2 | 0 | 1 | Not-determined | Genome-fragment | NA | NA | 0 |
| S14_extracted_k141_52676 | 2434 | No | NA | 4 | 4 | 0 | Low-quality | Genome-fragment | 8.02 | AAI-based (high-confidence) | 0 |
| S14_vs_k141_50360\|\|full | 2434 | No | NA | 3 | 0 | 2 | Low-quality | Genome-fragment | 1.29 | AAI-based (medium-confidence) | 0 |
| S3_vs_k141_112824\|\|full | 2434 | No | NA | 5 | 0 | 3 | Low-quality | Genome-fragment | 3.51 | AAI-based (medium-confidence) | 0 |
| S16_extracted_4698 | 2645 | No | NA | 4 | 0 | 4 | Not-determined | Genome-fragment | NA | NA | 0 |
| S5_vs_k141_114345\|\|full | 2434 | No | NA | 3 | 0 | 0 | Low-quality | Genome-fragment | 1.25 | AAI-based (medium-confidence) | 0 |
| S16_extracted_4731 | 25679 | No | NA | 12 | 0 | 8 | Not-determined | Genome-fragment | NA | NA | 0 |
| S16_extracted_4762 | 1093 | No | NA | 2 | 0 | 2 | Not-determined | Genome-fragment | NA | NA | 0 |
| S16_extracted_4763 | 2864 | No | NA | 3 | 0 | 3 | Not-determined | Genome-fragment | NA | NA | 0 |
| S16_extracted_4773 | 1235 | No | NA | 2 | 0 | 1 | Not-determined | Genome-fragment | NA | NA | 0 |
| S16_extracted_4779 | 1294 | No | NA | 3 | 0 | 0 | Not-determined | Genome-fragment | NA | NA | 0 |
| S16_extracted_4819 | 1350 | No | NA | 2 | 0 | 0 | Not-determined | Genome-fragment | NA | NA | 0 |
| S16_extracted_4999 | 1224 | No | NA | 2 | 0 | 2 | Not-determined | Genome-fragment | NA | NA | 0 |
| S16_vs_21384\|\|full | 2434 | No | NA | 3 | 0 | 2 | Low-quality | Genome-fragment | 2.9 | AAI-based (medium-confidence) | 0 |
| S16_extracted_5099 | 1019 | No | NA | 1 | 0 | 1 | Not-determined | Genome-fragment | NA | NA | 0 |
| S16_extracted_5121 | 1458 | No | NA | 2 | 0 | 2 | Not-determined | Genome-fragment | NA | NA | 0 |
| S16_extracted_5153 | 1375 | No | NA | 3 | 0 | 2 | Not-determined | Genome-fragment | NA | NA | 0 |
| S16_extracted_5240 | 6193 | No | NA | 6 | 0 | 4 | Not-determined | Genome-fragment | NA | NA | 0 |
| S16_vs_69182\|\|full | 2434 | No | NA | 7 | 1 | 0 | Low-quality | Genome-fragment | 4.95 | AAI-based (medium-confidence) | 0 |
| S16_extracted_5263 | 1267 | No | NA | 1 | 0 | 1 | Not-determined | Genome-fragment | NA | NA | 0 |
| S16_extracted_5270 | 56086 | No | NA | 86 | 23 | 0 | Medium-quality | Genome-fragment | 89.07 | AAI-based (medium-confidence) | 0 |
| S16_extracted_5273 | 4109 | No | NA | 4 | 0 | 3 | Not-determined | Genome-fragment | NA | NA | 0 |
| S16_extracted_5289 | 1302 | No | NA | 1 | 0 | 1 | Not-determined | Genome-fragment | NA | NA | 0 |
| S12_extracted_k141_171710 | 2434 | No | NA | 2 | 0 | 2 | Low-quality | Genome-fragment | 1.28 | AAI-based (medium-confidence) | 0 |
| S16_extracted_5373 | 3153 | No | NA | 2 | 0 | 1 | Not-determined | Genome-fragment | NA | NA | 0 |
| S16_vs_101407\|\|full | 2435 | No | NA | 2 | 0 | 1 | Low-quality | Genome-fragment | 1.27 | AAI-based (medium-confidence) | 0 |
| S16_extracted_5447 | 2707 | No | NA | 1 | 0 | 1 | Not-determined | Genome-fragment | NA | NA | 0 |
| S16_extracted_5630 | 1400 | No | NA | 2 | 0 | 0 | Not-determined | Genome-fragment | NA | NA | 0 |
| S16_extracted_5809 | 36823 | No | NA | 56 | 16 | 0 | Medium-quality | Genome-fragment | 62.36 | AAI-based (high-confidence) | 0 |
| S16_extracted_5896 | 17910 | No | NA | 18 | 0 | 6 | Not-determined | Genome-fragment | NA | NA | 0 |
| S10_extracted_81570 | 2435 | No | NA | 7 | 3 | 0 | Low-quality | Genome-fragment | 8.44 | AAI-based (high-confidence) | 0 |
| S16_extracted_5964 | 4334 | No | NA | 6 | 0 | 1 | Not-determined | Genome-fragment | NA | NA | 0 |
| S16_extracted_6024 | 2007 | No | NA | 1 | 0 | 1 | Not-determined | Genome-fragment | NA | NA | 0 |
| S16_extracted_6080 | 1079 | No | NA | 1 | 0 | 0 | Not-determined | Genome-fragment | NA | NA | 0 |
| S16_extracted_6102 | 51464 | No | NA | 72 | 19 | 0 | High-quality | High-quality | 97.58 | AAI-based (high-confidence) | 0 |
| S16_extracted_6115 | 1035 | No | NA | 2 | 0 | 0 | Not-determined | Genome-fragment | NA | NA | 0 |
| S6_vs_k141_4713\|\|full | 2436 | No | NA | 8 | 0 | 0 | Low-quality | Genome-fragment | 7.27 | AAI-based (high-confidence) | 0 |
| S17_vs_k141_21462\|\|full | 2437 | No | NA | 3 | 0 | 3 | Low-quality | Genome-fragment | 0.86 | AAI-based (medium-confidence) | 0 |
| S16_extracted_6521 | 1266 | No | NA | 2 | 0 | 0 | Not-determined | Genome-fragment | NA | NA | 0 |
| S16_extracted_6579 | 1062 | No | NA | 2 | 0 | 0 | Not-determined | Genome-fragment | NA | NA | 0 |
| S16_extracted_6649 | 3410 | No | NA | 5 | 0 | 2 | Not-determined | Genome-fragment | NA | NA | 0 |
| S16_extracted_6771 | 1242 | No | NA | 1 | 0 | 0 | Not-determined | Genome-fragment | NA | NA | 0 |
| S16_extracted_6814 | 1107 | No | NA | 5 | 0 | 0 | Not-determined | Genome-fragment | NA | NA | 0 |
| S16_extracted_6822 | 1661 | No | NA | 2 | 0 | 2 | Not-determined | Genome-fragment | NA | NA | 0 |
| S16_extracted_6920 | 4340 | No | NA | 1 | 0 | 1 | Not-determined | Genome-fragment | NA | NA | 0 |
| S16_extracted_6926 | 1776 | No | NA | 1 | 0 | 0 | Not-determined | Genome-fragment | NA | NA | 0 |
| S16_extracted_6977 | 1599 | No | NA | 3 | 0 | 1 | Not-determined | Genome-fragment | NA | NA | 0 |
| S3_extracted_k141_92974 | 2437 | No | NA | 3 | 0 | 0 | Low-quality | Genome-fragment | 5.6 | AAI-based (medium-confidence) | 0 |
| S16_extracted_7021 | 1455 | No | NA | 1 | 0 | 1 | Not-determined | Genome-fragment | NA | NA | 0 |
| S16_extracted_7025 | 1236 | No | NA | 2 | 0 | 1 | Not-determined | Genome-fragment | NA | NA | 0 |
| S16_vs_61112\|\|full | 2437 | No | NA | 2 | 0 | 2 | Low-quality | Genome-fragment | 3.9 | AAI-based (medium-confidence) | 0 |
| S16_extracted_7071 | 1039 | No | NA | 1 | 0 | 1 | Not-determined | Genome-fragment | NA | NA | 0 |
| S12_extracted_k141_129066 | 2438 | No | NA | 2 | 1 | 0 | Low-quality | Genome-fragment | 4.22 | AAI-based (high-confidence) | 0 |
| S8_vs_k141_5592\|\|full | 2438 | No | NA | 2 | 0 | 2 | Low-quality | Genome-fragment | 4.93 | AAI-based (medium-confidence) | 0 |
| S16_extracted_7164 | 1221 | No | NA | 1 | 0 | 1 | Not-determined | Genome-fragment | NA | NA | 0 |
| S14_vs_k141_32105\|\|full | 2439 | No | NA | 6 | 2 | 0 | Low-quality | Genome-fragment | 3.03 | AAI-based (high-confidence) | 0 |
| S16_extracted_7314 | 11100 | No | NA | 14 | 0 | 5 | Not-determined | Genome-fragment | NA | NA | 0 |
| S16_extracted_7319 | 1128 | No | NA | 2 | 0 | 1 | Not-determined | Genome-fragment | NA | NA | 0 |
| S16_extracted_7335 | 9297 | No | NA | 10 | 0 | 8 | Not-determined | Genome-fragment | NA | NA | 0 |
| S16_extracted_7372 | 1640 | No | NA | 3 | 0 | 2 | Not-determined | Genome-fragment | NA | NA | 0 |
| S16_extracted_7374 | 23085 | No | NA | 23 | 0 | 6 | Not-determined | Genome-fragment | NA | NA | 0 |
| S16_extracted_7387 | 1909 | No | NA | 2 | 0 | 1 | Not-determined | Genome-fragment | NA | NA | 0 |
| S16_extracted_7429 | 1045 | No | NA | 1 | 0 | 1 | Not-determined | Genome-fragment | NA | NA | 0 |
| S16_extracted_7448 | 3624 | No | NA | 1 | 0 | 1 | Not-determined | Genome-fragment | NA | NA | 0 |
| S7_extracted_32050 | 2439 | No | NA | 6 | 2 | 0 | Low-quality | Genome-fragment | 7.04 | AAI-based (high-confidence) | 0 |
| S16_extracted_7548 | 4061 | No | NA | 4 | 0 | 2 | Not-determined | Genome-fragment | NA | NA | 0 |
| S16_vs_157089\|\|full | 2439 | No | NA | 8 | 1 | 0 | Low-quality | Genome-fragment | 5.87 | AAI-based (high-confidence) | 0 |
| S16_extracted_7653 | 4242 | No | NA | 7 | 0 | 2 | Not-determined | Genome-fragment | NA | NA | 0 |
| S8_vs_k141_169514\|\|full | 2439 | No | NA | 3 | 0 | 1 | Low-quality | Genome-fragment | 1.93 | AAI-based (medium-confidence) | 0 |
| S14_vs_k141_49179\|\|full | 2440 | Yes | 844 | 3 | 1 | 1 | Low-quality | Genome-fragment | 1.48 | AAI-based (medium-confidence) | 65.41 |
| S16_extracted_7882 | 2957 | No | NA | 2 | 0 | 2 | Not-determined | Genome-fragment | NA | NA | 0 |
| S16_extracted_8157 | 1422 | No | NA | 1 | 0 | 1 | Not-determined | Genome-fragment | NA | NA | 0 |
| S16_extracted_8260 | 1049 | No | NA | 1 | 0 | 0 | Not-determined | Genome-fragment | NA | NA | 0 |
| S16_extracted_8289 | 1831 | No | NA | 2 | 0 | 1 | Not-determined | Genome-fragment | NA | NA | 0 |
| S5_vs_k141_72838\|\|full | 2440 | No | NA | 4 | 1 | 0 | Low-quality | Genome-fragment | 5.05 | AAI-based (medium-confidence) | 0 |
| S16_extracted_8360 | 4012 | No | NA | 4 | 0 | 3 | Not-determined | Genome-fragment | NA | NA | 0 |
| S16_extracted_8446 | 2426 | No | NA | 2 | 0 | 2 | Not-determined | Genome-fragment | NA | NA | 0 |
| S10_vs_34717\|\|full | 2440 | No | NA | 3 | 0 | 1 | Low-quality | Genome-fragment | 2.13 | AAI-based (high-confidence) | 0 |
| S17_vs_k141_107891\|\|full | 2441 | No | NA | 3 | 0 | 2 | Low-quality | Genome-fragment | 2.18 | AAI-based (medium-confidence) | 0 |
| S16_extracted_8582 | 4708 | No | NA | 5 | 0 | 4 | Not-determined | Genome-fragment | NA | NA | 0 |
| S16_extracted_8660 | 3097 | No | NA | 3 | 0 | 2 | Not-determined | Genome-fragment | NA | NA | 0 |
| S14_vs_k141_54607\|\|full | 2441 | No | NA | 4 | 1 | 0 | Low-quality | Genome-fragment | 5.71 | AAI-based (medium-confidence) | 0 |
| S16_extracted_8789 | 1666 | No | NA | 3 | 0 | 1 | Not-determined | Genome-fragment | NA | NA | 0 |
| S16_extracted_8807 | 1150 | No | NA | 1 | 0 | 1 | Not-determined | Genome-fragment | NA | NA | 0 |
| S16_extracted_8825 | 1599 | No | NA | 2 | 0 | 0 | Not-determined | Genome-fragment | NA | NA | 0 |
| S3_extracted_k141_74234 | 2441 | No | NA | 4 | 2 | 0 | Low-quality | Genome-fragment | 1.86 | AAI-based (high-confidence) | 0 |
| S3_vs_k141_12252\|\|full | 2441 | No | NA | 6 | 1 | 0 | Low-quality | Genome-fragment | 6.99 | AAI-based (high-confidence) | 0 |
| S16_extracted_8902 | 6950 | No | NA | 12 | 0 | 1 | Not-determined | Genome-fragment | NA | NA | 0 |
| S16_extracted_8909 | 1305 | No | NA | 1 | 0 | 0 | Not-determined | Genome-fragment | NA | NA | 0 |
| S16_vs_35842\|\|full | 2441 | No | NA | 2 | 0 | 1 | Low-quality | Genome-fragment | 4.27 | AAI-based (medium-confidence) | 0 |
| S16_extracted_9088 | 1185 | No | NA | 1 | 0 | 0 | Not-determined | Genome-fragment | NA | NA | 0 |
| S16_extracted_9128 | 3952 | No | NA | 3 | 0 | 1 | Not-determined | Genome-fragment | NA | NA | 0 |
| S16_extracted_9241 | 8712 | No | NA | 6 | 0 | 1 | Not-determined | Genome-fragment | NA | NA | 0 |
| S16_extracted_9260 | 1683 | No | NA | 3 | 0 | 1 | Not-determined | Genome-fragment | NA | NA | 0 |
| S16_extracted_9270 | 64521 | No | NA | 90 | 9 | 2 | Complete | High-quality | 100 | DTR (high-confidence) | 0 |
| S16_extracted_9378 | 3459 | No | NA | 2 | 0 | 2 | Not-determined | Genome-fragment | NA | NA | 0 |
| S10_extracted_13081 | 2441 | No | NA | 4 | 2 | 0 | Low-quality | Genome-fragment | 4.04 | AAI-based (high-confidence) | 0 |
| S16_extracted_9398 | 1085 | No | NA | 3 | 0 | 0 | Not-determined | Genome-fragment | NA | NA | 0 |
| S16_extracted_9411 | 2283 | No | NA | 3 | 0 | 1 | Not-determined | Genome-fragment | NA | NA | 0 |
| S10_vs_1890\|\|full | 2441 | No | NA | 3 | 0 | 0 | Low-quality | Genome-fragment | 0.61 | AAI-based (medium-confidence) | 0 |
| S15_vs_73891\|\|full | 2441 | No | NA | 4 | 0 | 3 | Low-quality | Genome-fragment | 1.62 | AAI-based (high-confidence) | 0 |
| S9_vs_k141_29255\|\|full | 2442 | No | NA | 4 | 3 | 0 | Low-quality | Genome-fragment | 5.07 | AAI-based (high-confidence) | 0 |
| S16_extracted_9617 | 1461 | No | NA | 2 | 0 | 0 | Not-determined | Genome-fragment | NA | NA | 0 |
| S16_extracted_9762 | 4627 | No | NA | 1 | 0 | 1 | Not-determined | Genome-fragment | NA | NA | 0 |
| S17_extracted_k141_127344 | 2443 | No | NA | 4 | 0 | 0 | Low-quality | Genome-fragment | 1.87 | AAI-based (high-confidence) | 0 |
| S16_extracted_9896 | 9239 | No | NA | 5 | 0 | 1 | Not-determined | Genome-fragment | NA | NA | 0 |
| S16_extracted_9954 | 1330 | No | NA | 1 | 0 | 0 | Not-determined | Genome-fragment | NA | NA | 0 |
| S13_vs_137372\|\|full | 2443 | No | NA | 2 | 1 | 0 | Low-quality | Genome-fragment | 1.49 | AAI-based (high-confidence) | 0 |
| S14_vs_k141_98571\|\|full | 2443 | No | NA | 6 | 1 | 0 | Low-quality | Genome-fragment | 1.64 | HMM-based (lower-bound) | 0 |
| S1_extracted_k141_66853 | 2443 | No | NA | 7 | 2 | 0 | Low-quality | Genome-fragment | 4.26 | AAI-based (high-confidence) | 0 |
| S16_extracted_10185 | 1481 | No | NA | 1 | 0 | 0 | Not-determined | Genome-fragment | NA | NA | 0 |
| S16_extracted_10388 | 1459 | No | NA | 1 | 0 | 1 | Not-determined | Genome-fragment | NA | NA | 0 |
| S16_extracted_10428 | 1455 | No | NA | 2 | 0 | 2 | Not-determined | Genome-fragment | NA | NA | 0 |
| S1_vs_k141_46049\|\|full | 2443 | No | NA | 4 | 0 | 0 | Low-quality | Genome-fragment | 7.4 | AAI-based (high-confidence) | 0 |
| S16_extracted_10467 | 1011 | No | NA | 1 | 0 | 0 | Not-determined | Genome-fragment | NA | NA | 0 |
| S8_vs_k141_207268\|\|full | 2443 | No | NA | 5 | 0 | 0 | Low-quality | Genome-fragment | 2.72 | AAI-based (medium-confidence) | 0 |
| S16_extracted_10567 | 3415 | No | NA | 2 | 0 | 1 | Not-determined | Genome-fragment | NA | NA | 0 |
| S9_vs_k141_88372\|\|full | 2444 | No | NA | 3 | 1 | 0 | Low-quality | Genome-fragment | 3.46 | HMM-based (lower-bound) | 0 |
| S16_extracted_10665 | 1808 | No | NA | 1 | 0 | 1 | Not-determined | Genome-fragment | NA | NA | 0 |
| S16_extracted_10778 | 2030 | No | NA | 2 | 0 | 2 | Not-determined | Genome-fragment | NA | NA | 0 |
| S16_extracted_10812 | 5464 | No | NA | 9 | 0 | 6 | Not-determined | Genome-fragment | NA | NA | 0 |
| S14_vs_k141_25606\|\|full | 2444 | No | NA | 3 | 1 | 0 | Low-quality | Genome-fragment | 4.56 | AAI-based (high-confidence) | 0 |
| S16_vs_95806\|\|full | 2444 | No | NA | 5 | 2 | 0 | Low-quality | Genome-fragment | 5.73 | AAI-based (high-confidence) | 0 |
| S17_extracted_k141_34485 | 2445 | No | NA | 10 | 1 | 0 | Low-quality | Genome-fragment | 5.47 | AAI-based (high-confidence) | 0 |
| S16_extracted_10943 | 1186 | No | NA | 1 | 0 | 1 | Not-determined | Genome-fragment | NA | NA | 0 |
| S16_extracted_11041 | 1894 | No | NA | 2 | 0 | 2 | Not-determined | Genome-fragment | NA | NA | 0 |
| S16_vs_156438\|\|full | 2445 | No | NA | 3 | 0 | 1 | Low-quality | Genome-fragment | 1.25 | AAI-based (medium-confidence) | 0 |
| S16_extracted_11083 | 3429 | No | NA | 2 | 0 | 1 | Not-determined | Genome-fragment | NA | NA | 0 |
| S12_vs_k141_29457\|\|full | 2445 | No | NA | 3 | 0 | 0 | Low-quality | Genome-fragment | 0.57 | AAI-based (medium-confidence) | 0 |
| S16_extracted_11114 | 3308 | No | NA | 5 | 0 | 2 | Not-determined | Genome-fragment | NA | NA | 0 |
| S16_extracted_11148 | 1712 | No | NA | 2 | 0 | 0 | Not-determined | Genome-fragment | NA | NA | 0 |
| S16_extracted_11204 | 1961 | No | NA | 1 | 0 | 0 | Not-determined | Genome-fragment | NA | NA | 0 |
| S16_extracted_11324 | 1205 | No | NA | 1 | 0 | 1 | Not-determined | Genome-fragment | NA | NA | 0 |
| S16_extracted_11350 | 2080 | No | NA | 2 | 0 | 0 | Not-determined | Genome-fragment | NA | NA | 0 |
| S15_extracted_59874 | 2445 | No | NA | 3 | 0 | 1 | Low-quality | Genome-fragment | 2.24 | AAI-based (medium-confidence) | 0 |
| S16_extracted_11446 | 1004 | No | NA | 1 | 0 | 1 | Not-determined | Genome-fragment | NA | NA | 0 |
| S8_extracted_k141_194525 | 2445 | No | NA | 1 | 1 | 0 | Low-quality | Genome-fragment | 3.24 | HMM-based (lower-bound) | 0 |
| S16_extracted_11524 | 1367 | No | NA | 3 | 0 | 1 | Not-determined | Genome-fragment | NA | NA | 0 |
| S16_extracted_11611 | 6296 | No | NA | 7 | 0 | 1 | Not-determined | Genome-fragment | NA | NA | 0 |
| S14_vs_k141_85054\|\|full | 2446 | No | NA | 3 | 0 | 1 | Low-quality | Genome-fragment | 5.21 | AAI-based (medium-confidence) | 0 |
| S16_extracted_11674 | 1529 | No | NA | 1 | 0 | 0 | Not-determined | Genome-fragment | NA | NA | 0 |
| S3_vs_k141_69503\|\|full | 2446 | No | NA | 6 | 1 | 0 | Low-quality | Genome-fragment | 4.83 | AAI-based (high-confidence) | 0 |
| S16_extracted_11812 | 1592 | No | NA | 1 | 0 | 1 | Not-determined | Genome-fragment | NA | NA | 0 |
| S16_extracted_11815 | 1422 | No | NA | 1 | 0 | 1 | Not-determined | Genome-fragment | NA | NA | 0 |
| S16_extracted_11828 | 1593 | No | NA | 1 | 0 | 1 | Not-determined | Genome-fragment | NA | NA | 0 |
| S10_extracted_11304 | 2446 | No | NA | 4 | 2 | 0 | Low-quality | Genome-fragment | 3.39 | HMM-based (lower-bound) | 0 |
| S16_extracted_11896 | 1833 | No | NA | 2 | 0 | 2 | Not-determined | Genome-fragment | NA | NA | 0 |
| S16_extracted_11974 | 1108 | No | NA | 2 | 0 | 1 | Not-determined | Genome-fragment | NA | NA | 0 |
| S9_vs_k141_54372\|\|full | 2447 | No | NA | 4 | 0 | 1 | Low-quality | Genome-fragment | 3.02 | AAI-based (high-confidence) | 0 |
| S16_extracted_12277 | 2035 | No | NA | 4 | 0 | 0 | Not-determined | Genome-fragment | NA | NA | 0 |
| S9_vs_k141_20326\|\|full | 2447 | No | NA | 5 | 3 | 0 | Low-quality | Genome-fragment | 5.29 | AAI-based (high-confidence) | 0 |
| S14_vs_k141_106533\|\|full | 2447 | No | NA | 4 | 0 | 0 | Low-quality | Genome-fragment | 7.17 | AAI-based (high-confidence) | 0 |
| S1_vs_k141_56364\|\|full | 2447 | No | NA | 3 | 0 | 2 | Low-quality | Genome-fragment | 1.92 | AAI-based (high-confidence) | 0 |
| S16_extracted_12551 | 37871 | No | NA | 71 | 14 | 0 | High-quality | High-quality | 91.98 | AAI-based (high-confidence) | 0 |
| S16_extracted_12592 | 1084 | No | NA | 2 | 0 | 0 | Not-determined | Genome-fragment | NA | NA | 0 |
| S16_extracted_12630 | 4364 | No | NA | 4 | 0 | 3 | Not-determined | Genome-fragment | NA | NA | 0 |
| S12_vs_k141_162721\|\|full | 2447 | No | NA | 5 | 0 | 0 | Low-quality | Genome-fragment | 4.17 | AAI-based (high-confidence) | 0 |
| S16_extracted_12820 | 1467 | No | NA | 1 | 0 | 1 | Not-determined | Genome-fragment | NA | NA | 0 |
| S16_extracted_12854 | 6174 | No | NA | 5 | 0 | 4 | Not-determined | Genome-fragment | NA | NA | 0 |
| S16_extracted_12856 | 1089 | No | NA | 1 | 0 | 0 | Not-determined | Genome-fragment | NA | NA | 0 |
| S16_extracted_12881 | 1665 | No | NA | 2 | 0 | 0 | Not-determined | Genome-fragment | NA | NA | 0 |
| S16_extracted_12903 | 1839 | No | NA | 3 | 0 | 2 | Not-determined | Genome-fragment | NA | NA | 0 |
| S16_extracted_12944 | 1278 | No | NA | 2 | 0 | 1 | Not-determined | Genome-fragment | NA | NA | 0 |
| S16_extracted_12945 | 1351 | No | NA | 3 | 0 | 1 | Not-determined | Genome-fragment | NA | NA | 0 |
| S16_extracted_12976 | 1724 | No | NA | 2 | 0 | 1 | Not-determined | Genome-fragment | NA | NA | 0 |
| S9_vs_k141_3590\|\|full | 2448 | No | NA | 4 | 0 | 0 | Low-quality | Genome-fragment | 6.44 | AAI-based (medium-confidence) | 0 |
| S16_extracted_13333 | 5184 | No | NA | 1 | 0 | 1 | Not-determined | Genome-fragment | NA | NA | 0 |
| S16_extracted_13354 | 1689 | No | NA | 1 | 0 | 0 | Not-determined | Genome-fragment | NA | NA | 0 |
| S16_extracted_13401 | 1290 | No | NA | 3 | 0 | 0 | Not-determined | Genome-fragment | NA | NA | 0 |
| S16_extracted_13417 | 5332 | No | NA | 4 | 0 | 1 | Not-determined | Genome-fragment | NA | NA | 0 |
| S1_vs_k141_19104\|\|full | 2448 | No | NA | 3 | 3 | 0 | Low-quality | Genome-fragment | 5.08 | AAI-based (high-confidence) | 0 |
| S7_vs_84526\|\|full | 2448 | No | NA | 6 | 1 | 0 | Low-quality | Genome-fragment | 4.8 | AAI-based (high-confidence) | 0 |
| S8_vs_k141_49407\|\|full | 2448 | No | NA | 7 | 0 | 0 | Low-quality | Genome-fragment | 42.81 | AAI-based (medium-confidence) | 0 |
| S16_extracted_13755 | 6198 | No | NA | 8 | 0 | 3 | Not-determined | Genome-fragment | NA | NA | 0 |
| S9_vs_k141_58564\|\|full | 2449 | No | NA | 4 | 0 | 2 | Low-quality | Genome-fragment | 4.58 | AAI-based (high-confidence) | 0 |
| S16_extracted_14017 | 3105 | No | NA | 5 | 0 | 1 | Not-determined | Genome-fragment | NA | NA | 0 |
| S16_extracted_14030 | 7116 | No | NA | 10 | 0 | 6 | Not-determined | Genome-fragment | NA | NA | 0 |
| S16_extracted_14106 | 4167 | No | NA | 4 | 0 | 4 | Not-determined | Genome-fragment | NA | NA | 0 |
| S16_extracted_14123 | 1198 | No | NA | 2 | 0 | 1 | Not-determined | Genome-fragment | NA | NA | 0 |
| S5_vs_k141_140723\|\|full | 2449 | No | NA | 2 | 0 | 1 | Low-quality | Genome-fragment | 1.35 | AAI-based (medium-confidence) | 0 |
| S12_vs_k141_44667\|\|lt2gene | 2449 | No | NA | 2 | 1 | 0 | Low-quality | Genome-fragment | 0.99 | HMM-based (lower-bound) | 0 |
| S11_vs_k141_59592\|\|full | 2450 | No | NA | 3 | 0 | 1 | Low-quality | Genome-fragment | 4.07 | AAI-based (medium-confidence) | 0 |
| S16_extracted_14263 | 5172 | No | NA | 5 | 0 | 1 | Not-determined | Genome-fragment | NA | NA | 0 |
| S16_extracted_14358 | 1395 | No | NA | 1 | 0 | 1 | Not-determined | Genome-fragment | NA | NA | 0 |
| S16_extracted_14387 | 1637 | No | NA | 4 | 0 | 1 | Not-determined | Genome-fragment | NA | NA | 0 |
| S16_extracted_14468 | 1335 | No | NA | 1 | 0 | 1 | Not-determined | Genome-fragment | NA | NA | 0 |
| S1_extracted_k141_67361 | 2450 | No | NA | 5 | 3 | 0 | Low-quality | Genome-fragment | 4.08 | AAI-based (high-confidence) | 0 |
| S16_extracted_14670 | 1050 | No | NA | 3 | 0 | 0 | Not-determined | Genome-fragment | NA | NA | 0 |
| S16_extracted_14705 | 1812 | No | NA | 1 | 0 | 0 | Not-determined | Genome-fragment | NA | NA | 0 |
| S3_vs_k141_18237\|\|full | 2450 | No | NA | 3 | 2 | 0 | Low-quality | Genome-fragment | 5.27 | AAI-based (high-confidence) | 0 |
| S16_extracted_14745 | 4566 | No | NA | 5 | 0 | 0 | Not-determined | Genome-fragment | NA | NA | 0 |
| S16_extracted_14796 | 1199 | No | NA | 1 | 0 | 0 | Not-determined | Genome-fragment | NA | NA | 0 |
| S16_extracted_14888 | 4755 | No | NA | 4 | 0 | 0 | Not-determined | Genome-fragment | NA | NA | 0 |
| S16_extracted_14923 | 1151 | No | NA | 1 | 0 | 0 | Not-determined | Genome-fragment | NA | NA | 0 |
| S16_extracted_15078 | 4069 | No | NA | 4 | 0 | 2 | Not-determined | Genome-fragment | NA | NA | 0 |
| S16_extracted_15278 | 1026 | No | NA | 1 | 0 | 0 | Not-determined | Genome-fragment | NA | NA | 0 |
| S16_extracted_15293 | 1201 | No | NA | 1 | 0 | 0 | Not-determined | Genome-fragment | NA | NA | 0 |
| S16_extracted_15359 | 3414 | No | NA | 4 | 0 | 2 | Not-determined | Genome-fragment | NA | NA | 0 |
| S10_vs_15103\|\|full | 2450 | No | NA | 3 | 1 | 0 | Low-quality | Genome-fragment | 0.94 | HMM-based (lower-bound) | 0 |
| S16_extracted_15391 | 1147 | No | NA | 1 | 0 | 0 | Not-determined | Genome-fragment | NA | NA | 0 |
| S16_extracted_15399 | 7043 | No | NA | 7 | 0 | 6 | Not-determined | Genome-fragment | NA | NA | 0 |
| S16_extracted_15441 | 1266 | No | NA | 1 | 0 | 0 | Not-determined | Genome-fragment | NA | NA | 0 |
| S16_extracted_15497 | 3363 | No | NA | 5 | 0 | 5 | Not-determined | Genome-fragment | NA | NA | 0 |
| S8_vs_k141_180928\|\|full | 2450 | No | NA | 3 | 0 | 1 | Low-quality | Genome-fragment | 5.49 | AAI-based (high-confidence) | 0 |
| S11_vs_k141_10725\|\|full | 2451 | No | NA | 3 | 1 | 0 | Low-quality | Genome-fragment | 3.74 | AAI-based (high-confidence) | 0 |
| S16_extracted_15600 | 1129 | No | NA | 1 | 0 | 1 | Not-determined | Genome-fragment | NA | NA | 0 |
| S16_extracted_15805 | 1169 | No | NA | 2 | 0 | 1 | Not-determined | Genome-fragment | NA | NA | 0 |
| S11_vs_k141_38353\|\|full | 2451 | No | NA | 4 | 2 | 0 | Low-quality | Genome-fragment | 5.18 | AAI-based (high-confidence) | 0 |
| S16_extracted_15881 | 1041 | No | NA | 1 | 0 | 1 | Not-determined | Genome-fragment | NA | NA | 0 |
| S16_extracted_15951 | 1155 | No | NA | 1 | 0 | 1 | Not-determined | Genome-fragment | NA | NA | 0 |
| S16_extracted_15980 | 8810 | No | NA | 7 | 0 | 4 | Not-determined | Genome-fragment | NA | NA | 0 |
| S1_vs_k141_131168\|\|full | 2451 | No | NA | 3 | 1 | 0 | Low-quality | Genome-fragment | 5.63 | AAI-based (high-confidence) | 0 |
| S3_extracted_k141_76755 | 2451 | No | NA | 5 | 1 | 0 | Low-quality | Genome-fragment | 5.41 | AAI-based (high-confidence) | 0 |
| S3_vs_k141_51280\|\|full | 2451 | No | NA | 7 | 1 | 0 | Low-quality | Genome-fragment | 4.26 | AAI-based (high-confidence) | 0 |
| S16_extracted_16182 | 33206 | No | NA | 44 | 22 | 0 | Medium-quality | Genome-fragment | 55.4 | AAI-based (medium-confidence) | 0 |
| S7_vs_111049\|\|full | 2451 | No | NA | 4 | 0 | 0 | Low-quality | Genome-fragment | 3.52 | AAI-based (medium-confidence) | 0 |
| S16_extracted_16430 | 4739 | No | NA | 5 | 0 | 0 | Not-determined | Genome-fragment | NA | NA | 0 |
| S14_extracted_k141_67066 | 2452 | No | NA | 5 | 0 | 0 | Low-quality | Genome-fragment | 3.36 | AAI-based (medium-confidence) | 0 |
| S16_extracted_16588 | 1393 | No | NA | 1 | 0 | 1 | Not-determined | Genome-fragment | NA | NA | 0 |
| S16_extracted_16620 | 2524 | No | NA | 3 | 0 | 3 | Not-determined | Genome-fragment | NA | NA | 0 |
| S16_extracted_16635 | 3058 | No | NA | 5 | 0 | 2 | Not-determined | Genome-fragment | NA | NA | 0 |
| S3_vs_k141_87825\|\|full | 2452 | No | NA | 4 | 1 | 0 | Low-quality | Genome-fragment | 1.86 | AAI-based (high-confidence) | 0 |
| S16_extracted_16725 | 1206 | No | NA | 1 | 0 | 1 | Not-determined | Genome-fragment | NA | NA | 0 |
| S5_vs_k141_97502\|\|full | 2452 | No | NA | 4 | 4 | 0 | Low-quality | Genome-fragment | 5.5 | AAI-based (medium-confidence) | 0 |
| S16_extracted_16856 | 1370 | No | NA | 1 | 0 | 1 | Not-determined | Genome-fragment | NA | NA | 0 |
| S16_extracted_16861 | 1635 | No | NA | 1 | 0 | 0 | Not-determined | Genome-fragment | NA | NA | 0 |
| S16_extracted_16910 | 1108 | No | NA | 1 | 0 | 1 | Not-determined | Genome-fragment | NA | NA | 0 |
| S16_extracted_17190 | 1062 | No | NA | 2 | 0 | 1 | Not-determined | Genome-fragment | NA | NA | 0 |
| S16_extracted_17245 | 1647 | No | NA | 1 | 0 | 1 | Not-determined | Genome-fragment | NA | NA | 0 |
| S16_extracted_17286 | 1714 | No | NA | 2 | 0 | 2 | Not-determined | Genome-fragment | NA | NA | 0 |
| S16_extracted_17366 | 16047 | No | NA | 19 | 0 | 10 | Not-determined | Genome-fragment | NA | NA | 0 |
| S16_extracted_17404 | 1295 | No | NA | 1 | 0 | 1 | Not-determined | Genome-fragment | NA | NA | 0 |
| S16_extracted_17641 | 2691 | No | NA | 3 | 0 | 0 | Not-determined | Genome-fragment | NA | NA | 0 |
| S16_extracted_17801 | 1835 | No | NA | 3 | 0 | 3 | Not-determined | Genome-fragment | NA | NA | 0 |
| S16_extracted_17814 | 4319 | No | NA | 2 | 0 | 2 | Not-determined | Genome-fragment | NA | NA | 0 |
| S16_extracted_17888 | 2027 | No | NA | 4 | 0 | 0 | Not-determined | Genome-fragment | NA | NA | 0 |
| S7_vs_15473\|\|full | 2452 | No | NA | 4 | 0 | 2 | Low-quality | Genome-fragment | 5.49 | AAI-based (medium-confidence) | 0 |
| S16_extracted_17938 | 1237 | No | NA | 3 | 0 | 2 | Not-determined | Genome-fragment | NA | NA | 0 |
| S16_extracted_17986 | 1472 | No | NA | 3 | 0 | 1 | Not-determined | Genome-fragment | NA | NA | 0 |
| S16_extracted_18041 | 1571 | No | NA | 3 | 0 | 2 | Not-determined | Genome-fragment | NA | NA | 0 |
| S16_extracted_18093 | 1410 | No | NA | 3 | 0 | 1 | Not-determined | Genome-fragment | NA | NA | 0 |
| S16_vs_51492\|\|full | 2452 | No | NA | 3 | 1 | 0 | Low-quality | Genome-fragment | 3.51 | AAI-based (high-confidence) | 0 |
| S16_extracted_18185 | 2020 | No | NA | 4 | 0 | 4 | Not-determined | Genome-fragment | NA | NA | 0 |
| S16_extracted_18194 | 2022 | No | NA | 3 | 0 | 1 | Not-determined | Genome-fragment | NA | NA | 0 |
| S16_extracted_18218 | 1290 | No | NA | 1 | 0 | 0 | Not-determined | Genome-fragment | NA | NA | 0 |
| S10_vs_58482\|\|full | 2452 | No | NA | 8 | 3 | 0 | Low-quality | Genome-fragment | 5.63 | AAI-based (high-confidence) | 0 |
| S16_extracted_18393 | 1165 | No | NA | 2 | 0 | 0 | Not-determined | Genome-fragment | NA | NA | 0 |
| S16_extracted_18487 | 1006 | No | NA | 2 | 0 | 0 | Not-determined | Genome-fragment | NA | NA | 0 |
| S16_extracted_18513 | 1915 | No | NA | 1 | 0 | 1 | Not-determined | Genome-fragment | NA | NA | 0 |
| S8_vs_k141_136782\|\|full | 2452 | No | NA | 3 | 2 | 0 | Low-quality | Genome-fragment | 5.6 | AAI-based (high-confidence) | 0 |
| S16_extracted_18573 | 3837 | No | NA | 4 | 0 | 3 | Not-determined | Genome-fragment | NA | NA | 0 |
| S8_vs_k141_181497\|\|full | 2452 | No | NA | 5 | 1 | 0 | Low-quality | Genome-fragment | 3.07 | AAI-based (high-confidence) | 0 |
| S16_extracted_18860 | 2927 | No | NA | 2 | 0 | 2 | Not-determined | Genome-fragment | NA | NA | 0 |
| S16_extracted_18864 | 1513 | No | NA | 1 | 0 | 0 | Not-determined | Genome-fragment | NA | NA | 0 |
| S7_vs_26748\|\|full | 2453 | No | NA | 3 | 1 | 0 | Low-quality | Genome-fragment | 5.57 | AAI-based (medium-confidence) | 0 |
| S16_extracted_51452 | 2453 | No | NA | 7 | 1 | 0 | Low-quality | Genome-fragment | 2.23 | AAI-based (high-confidence) | 0 |
| S8_vs_k141_90709\|\|full | 2453 | No | NA | 6 | 0 | 0 | Low-quality | Genome-fragment | 3.97 | AAI-based (high-confidence) | 0 |
| S16_extracted_18967 | 1017 | No | NA | 2 | 0 | 0 | Not-determined | Genome-fragment | NA | NA | 0 |
| S8_vs_k141_12732\|\|full | 2453 | No | NA | 5 | 0 | 1 | Low-quality | Genome-fragment | 4.61 | AAI-based (medium-confidence) | 0 |
| S16_extracted_19135 | 2350 | No | NA | 3 | 0 | 3 | Not-determined | Genome-fragment | NA | NA | 0 |
| S16_extracted_19162 | 2563 | No | NA | 2 | 0 | 1 | Not-determined | Genome-fragment | NA | NA | 0 |
| S16_extracted_19180 | 1040 | No | NA | 1 | 0 | 0 | Not-determined | Genome-fragment | NA | NA | 0 |
| S16_extracted_19191 | 2875 | No | NA | 3 | 0 | 1 | Not-determined | Genome-fragment | NA | NA | 0 |
| S11_vs_k141_88062\|\|lt2gene | 2454 | No | NA | 2 | 1 | 0 | Low-quality | Genome-fragment | 5.16 | HMM-based (lower-bound) | 0 |
| S1_vs_k141_35740\|\|full | 2454 | No | NA | 5 | 0 | 0 | Low-quality | Genome-fragment | 4.31 | AAI-based (high-confidence) | 0 |
| S17_vs_k141_28009\|\|full | 2455 | No | NA | 3 | 0 | 0 | Low-quality | Genome-fragment | 6.92 | AAI-based (medium-confidence) | 0 |
| S9_vs_k141_69538\|\|full | 2455 | No | NA | 3 | 0 | 0 | Low-quality | Genome-fragment | 4.33 | AAI-based (medium-confidence) | 0 |
| S16_extracted_19516 | 1637 | No | NA | 2 | 0 | 2 | Not-determined | Genome-fragment | NA | NA | 0 |
| S16_extracted_19789 | 1832 | No | NA | 2 | 0 | 1 | Not-determined | Genome-fragment | NA | NA | 0 |
| S16_extracted_19812 | 10877 | No | NA | 11 | 0 | 7 | Not-determined | Genome-fragment | NA | NA | 0 |

| | | | | | | | | | | | |
|---|---|---|---|---|---|---|---|---|---|---|---|
| S16_extracted_19839 | 1006 | No | NA | 2 | 0 | 2 | Not-determined | Genome-fragment | NA | NA | 0 |
| S11_vs_k141_33184\|\|full | 2455 | No | NA | 3 | 1 | 0 | Low-quality | Genome-fragment | 5.61 | AAI-based (medium-confidence) | 0 |
| S11_vs_k141_15994\|\|full | 2455 | No | NA | 4 | 2 | 0 | Low-quality | Genome-fragment | 4.21 | AAI-based (high-confidence) | 0 |
| S16_extracted_19915 | 9180 | No | NA | 9 | 0 | 3 | Not-determined | Genome-fragment | NA | NA | 0 |
| S16_extracted_20013 | 5699 | No | NA | 8 | 0 | 4 | Not-determined | Genome-fragment | NA | NA | 0 |
| S13_extracted_108832 | 2455 | No | NA | 3 | 1 | 0 | Low-quality | Genome-fragment | 2.56 | AAI-based (high-confidence) | 0 |
| S1_vs_k141_139644\|\|full | 2455 | No | NA | 5 | 0 | 0 | Low-quality | Genome-fragment | 4.07 | AAI-based (high-confidence) | 0 |
| S16_extracted_20082 | 3285 | No | NA | 3 | 0 | 2 | Not-determined | Genome-fragment | NA | NA | 0 |
| S5_vs_k141_100985\|\|full | 2455 | No | NA | 4 | 0 | 0 | Low-quality | Genome-fragment | 4.67 | AAI-based (high-confidence) | 0 |
| S5_vs_k141_85842\|\|lt2gene | 2455 | No | NA | 1 | 1 | 0 | Low-quality | Genome-fragment | 6.14 | AAI-based (medium-confidence) | 0 |
| S16_extracted_20277 | 1366 | No | NA | 2 | 0 | 0 | Not-determined | Genome-fragment | NA | NA | 0 |
| S16_extracted_20534 | 1042 | No | NA | 1 | 0 | 1 | Not-determined | Genome-fragment | NA | NA | 0 |
| S16_extracted_20633 | 4195 | No | NA | 5 | 0 | 4 | Not-determined | Genome-fragment | NA | NA | 0 |
| S16_vs_122877\|\|full | 2455 | No | NA | 3 | 2 | 0 | Low-quality | Genome-fragment | 5.74 | AAI-based (high-confidence) | 0 |
| S10_vs_45624\|\|full | 2455 | No | NA | 4 | 0 | 0 | Low-quality | Genome-fragment | 3.55 | AAI-based (high-confidence) | 0 |
| S16_extracted_20822 | 5994 | No | NA | 5 | 0 | 0 | Not-determined | Genome-fragment | NA | NA | 0 |
| S16_extracted_20901 | 7412 | No | NA | 6 | 0 | 2 | Not-determined | Genome-fragment | NA | NA | 0 |
| S16_extracted_20918 | 1443 | No | NA | 3 | 0 | 2 | Not-determined | Genome-fragment | NA | NA | 0 |
| S16_extracted_20920 | 1072 | No | NA | 1 | 0 | 0 | Not-determined | Genome-fragment | NA | NA | 0 |
| S16_extracted_20936 | 3657 | No | NA | 5 | 0 | 0 | Not-determined | Genome-fragment | NA | NA | 0 |
| S16_extracted_20966 | 3282 | No | NA | 4 | 0 | 3 | Not-determined | Genome-fragment | NA | NA | 0 |
| S16_extracted_21002 | 1026 | No | NA | 2 | 0 | 0 | Not-determined | Genome-fragment | NA | NA | 0 |
| S8_vs_k141_157866\|\|full | 2455 | No | NA | 3 | 0 | 1 | Low-quality | Genome-fragment | 0.87 | AAI-based (medium-confidence) | 0 |
| S16_extracted_21137 | 1236 | No | NA | 1 | 0 | 0 | Not-determined | Genome-fragment | NA | NA | 0 |
| S16_extracted_21176 | 4223 | No | NA | 4 | 0 | 0 | Not-determined | Genome-fragment | NA | NA | 0 |
| S16_extracted_21295 | 1564 | No | NA | 2 | 0 | 2 | Not-determined | Genome-fragment | NA | NA | 0 |
| S17_vs_k141_6639\|\|full | 2456 | No | NA | 4 | 0 | 2 | Low-quality | Genome-fragment | 1.53 | AAI-based (medium-confidence) | 0 |
| S16_extracted_21441 | 1395 | No | NA | 1 | 0 | 1 | Not-determined | Genome-fragment | NA | NA | 0 |
| S16_extracted_21473 | 1472 | No | NA | 2 | 0 | 0 | Not-determined | Genome-fragment | NA | NA | 0 |
| S16_extracted_21508 | 6199 | No | NA | 9 | 0 | 2 | Not-determined | Genome-fragment | NA | NA | 0 |
| S16_extracted_21521 | 1054 | No | NA | 1 | 0 | 1 | Not-determined | Genome-fragment | NA | NA | 0 |
| S16_extracted_21594 | 1059 | No | NA | 2 | 0 | 0 | Not-determined | Genome-fragment | NA | NA | 0 |
| S16_extracted_21601 | 4626 | No | NA | 1 | 0 | 1 | Not-determined | Genome-fragment | NA | NA | 0 |
| S16_extracted_21705 | 5086 | No | NA | 4 | 0 | 3 | Not-determined | Genome-fragment | NA | NA | 0 |
| S16_extracted_21734 | 1241 | No | NA | 1 | 0 | 1 | Not-determined | Genome-fragment | NA | NA | 0 |
| S16_extracted_21737 | 5504 | No | NA | 4 | 0 | 4 | Not-determined | Genome-fragment | NA | NA | 0 |
| S16_extracted_21754 | 3068 | No | NA | 1 | 0 | 1 | Not-determined | Genome-fragment | NA | NA | 0 |
| S16_extracted_21886 | 1052 | No | NA | 2 | 0 | 1 | Not-determined | Genome-fragment | NA | NA | 0 |
| S16_extracted_22163 | 3475 | No | NA | 3 | 0 | 3 | Not-determined | Genome-fragment | NA | NA | 0 |
| S14_extracted_k141_39786 | 2456 | No | NA | 7 | 5 | 0 | Low-quality | Genome-fragment | 8.01 | AAI-based (high-confidence) | 0 |
| S14_vs_k141_106153\|\|full | 2456 | No | NA | 5 | 0 | 0 | Low-quality | Genome-fragment | 11.55 | AAI-based (medium-confidence) | 0 |
| S16_extracted_22262 | 4313 | No | NA | 3 | 0 | 1 | Not-determined | Genome-fragment | NA | NA | 0 |
| S16_extracted_22287 | 1064 | No | NA | 2 | 0 | 1 | Not-determined | Genome-fragment | NA | NA | 0 |
| S16_extracted_22418 | 3772 | No | NA | 2 | 0 | 2 | Not-determined | Genome-fragment | NA | NA | 0 |
| S16_extracted_22482 | 1701 | No | NA | 3 | 0 | 3 | Not-determined | Genome-fragment | NA | NA | 0 |
| S12_extracted_k141_8451 | 2457 | No | NA | 3 | 1 | 2 | Low-quality | Genome-fragment | 1.42 | HMM-based (lower-bound) | 0 |
| S1_extracted_k141_159877 | 2458 | No | NA | 3 | 0 | 0 | Low-quality | Genome-fragment | 2.49 | AAI-based (high-confidence) | 0 |
| S16_extracted_22519 | 8852 | No | NA | 12 | 0 | 3 | Not-determined | Genome-fragment | NA | NA | 0 |
| S16_extracted_22712 | 1351 | No | NA | 2 | 0 | 1 | Not-determined | Genome-fragment | NA | NA | 0 |
| S16_extracted_22771 | 1478 | No | NA | 2 | 0 | 0 | Not-determined | Genome-fragment | NA | NA | 0 |
| S16_extracted_22849 | 1932 | No | NA | 2 | 0 | 2 | Not-determined | Genome-fragment | NA | NA | 0 |
| S16_extracted_22903 | 1265 | No | NA | 2 | 0 | 2 | Not-determined | Genome-fragment | NA | NA | 0 |
| S5_vs_k141_78161\|\|full | 2458 | No | NA | 3 | 2 | 0 | Low-quality | Genome-fragment | 6.51 | AAI-based (high-confidence) | 0 |
| S16_extracted_22933 | 3444 | No | NA | 3 | 0 | 2 | Not-determined | Genome-fragment | NA | NA | 0 |
| S16_extracted_22975 | 1010 | No | NA | 1 | 0 | 0 | Not-determined | Genome-fragment | NA | NA | 0 |
| S16_extracted_23011 | 5047 | No | NA | 6 | 0 | 3 | Not-determined | Genome-fragment | NA | NA | 0 |
| S16_extracted_23015 | 3226 | No | NA | 3 | 0 | 2 | Not-determined | Genome-fragment | NA | NA | 0 |
| S16_extracted_23086 | 1053 | No | NA | 2 | 0 | 0 | Not-determined | Genome-fragment | NA | NA | 0 |
| S16_extracted_23195 | 1390 | No | NA | 1 | 0 | 1 | Not-determined | Genome-fragment | NA | NA | 0 |
| S16_extracted_23240 | 1421 | No | NA | 2 | 0 | 2 | Not-determined | Genome-fragment | NA | NA | 0 |
| S16_extracted_23282 | 1340 | No | NA | 1 | 0 | 1 | Not-determined | Genome-fragment | NA | NA | 0 |
| S16_extracted_23346 | 2279 | No | NA | 1 | 0 | 1 | Not-determined | Genome-fragment | NA | NA | 0 |
| S16_extracted_23351 | 1194 | No | NA | 2 | 0 | 1 | Not-determined | Genome-fragment | NA | NA | 0 |
| S16_extracted_23363 | 1499 | No | NA | 2 | 0 | 1 | Not-determined | Genome-fragment | NA | NA | 0 |
| S2_vs_k141_11654\|\|full | 2458 | No | NA | 4 | 2 | 0 | Low-quality | Genome-fragment | 3.45 | AAI-based (high-confidence) | 0 |
| S16_extracted_23513 | 1088 | No | NA | 1 | 0 | 0 | Not-determined | Genome-fragment | NA | NA | 0 |
| S16_extracted_23519 | 5143 | No | NA | 4 | 0 | 4 | Not-determined | Genome-fragment | NA | NA | 0 |
| S16_extracted_23618 | 2569 | No | NA | 1 | 0 | 0 | Not-determined | Genome-fragment | NA | NA | 0 |
| S16_extracted_23653 | 3088 | No | NA | 1 | 0 | 0 | Not-determined | Genome-fragment | NA | NA | 0 |
| S1_extracted_k141_121087 | 2459 | No | NA | 2 | 2 | 0 | Low-quality | Genome-fragment | 2.49 | AAI-based (high-confidence) | 0 |
| S12_vs_k141_24377\|\|full | 2459 | No | NA | 2 | 0 | 1 | Low-quality | Genome-fragment | 1.36 | AAI-based (medium-confidence) | 0 |
| S16_extracted_23785 | 1075 | No | NA | 1 | 0 | 1 | Not-determined | Genome-fragment | NA | NA | 0 |
| S16_extracted_23787 | 1555 | No | NA | 1 | 0 | 0 | Not-determined | Genome-fragment | NA | NA | 0 |
| S3_vs_k141_49439\|\|lt2gene | 2460 | No | NA | 1 | 1 | 0 | Low-quality | Genome-fragment | 4.31 | AAI-based (high-confidence) | 0 |
| S5_vs_k141_83993\|\|full | 2460 | No | NA | 3 | 1 | 0 | Low-quality | Genome-fragment | 5.74 | AAI-based (medium-confidence) | 0 |
| S16_extracted_24047 | 3207 | No | NA | 5 | 0 | 0 | Not-determined | Genome-fragment | NA | NA | 0 |
| S16_extracted_24126 | 2845 | No | NA | 1 | 0 | 0 | Not-determined | Genome-fragment | NA | NA | 0 |
| S17_extracted_k141_40470 | 2461 | No | NA | 4 | 0 | 0 | Low-quality | Genome-fragment | 2.98 | AAI-based (high-confidence) | 0 |
| S16_extracted_24237 | 1604 | No | NA | 1 | 0 | 1 | Not-determined | Genome-fragment | NA | NA | 0 |
| S12_extracted_k141_36445 | 2461 | No | NA | 5 | 0 | 0 | Low-quality | Genome-fragment | 5.4 | AAI-based (medium-confidence) | 0 |
| S16_extracted_24274 | 1094 | No | NA | 1 | 0 | 1 | Not-determined | Genome-fragment | NA | NA | 0 |
| S8_vs_k141_110513\|\|full | 2461 | No | NA | 4 | 0 | 4 | Low-quality | Genome-fragment | 0.7 | AAI-based (medium-confidence) | 0 |
| S16_extracted_24315 | 2893 | No | NA | 3 | 0 | 3 | Not-determined | Genome-fragment | NA | NA | 0 |
| S16_extracted_24327 | 4739 | No | NA | 4 | 0 | 1 | Not-determined | Genome-fragment | NA | NA | 0 |
| S16_extracted_24347 | 1067 | No | NA | 2 | 0 | 1 | Not-determined | Genome-fragment | NA | NA | 0 |
| S16_extracted_24410 | 10972 | No | NA | 4 | 0 | 3 | Not-determined | Genome-fragment | NA | NA | 0 |
| S16_extracted_24503 | 2835 | No | NA | 2 | 0 | 0 | Not-determined | Genome-fragment | NA | NA | 0 |
| S16_extracted_24508 | 3398 | No | NA | 4 | 0 | 3 | Not-determined | Genome-fragment | NA | NA | 0 |
| S11_vs_k141_49433\|\|full | 2462 | No | NA | 3 | 0 | 0 | Low-quality | Genome-fragment | 4.47 | AAI-based (medium-confidence) | 0 |
| S5_vs_k141_70025\|\|full | 2462 | No | NA | 6 | 4 | 0 | Low-quality | Genome-fragment | 8.28 | AAI-based (high-confidence) | 0 |
| S16_extracted_24663 | 4125 | No | NA | 5 | 0 | 3 | Not-determined | Genome-fragment | NA | NA | 0 |
| S16_extracted_24721 | 1266 | No | NA | 2 | 0 | 1 | Not-determined | Genome-fragment | NA | NA | 0 |
| S12_vs_k141_79749\|\|full | 2462 | No | NA | 4 | 0 | 1 | Low-quality | Genome-fragment | 1.76 | AAI-based (high-confidence) | 0 |
| S9_vs_k141_7676\|\|full | 2463 | No | NA | 4 | 0 | 0 | Low-quality | Genome-fragment | 5.4 | AAI-based (high-confidence) | 0 |
| S9_vs_k141_28360\|\|full | 2463 | No | NA | 2 | 0 | 1 | Low-quality | Genome-fragment | 2.28 | AAI-based (medium-confidence) | 0 |
| S16_extracted_24908 | 1045 | No | NA | 1 | 0 | 1 | Not-determined | Genome-fragment | NA | NA | 0 |
| S11_vs_k141_101966\|\|full | 2463 | No | NA | 4 | 0 | 0 | Low-quality | Genome-fragment | 1.6 | AAI-based (high-confidence) | 0 |
| S16_extracted_25019 | 2199 | No | NA | 3 | 0 | 0 | Not-determined | Genome-fragment | NA | NA | 0 |
| S16_extracted_25026 | 9427 | No | NA | 5 | 0 | 3 | Not-determined | Genome-fragment | NA | NA | 0 |
| S16_extracted_25140 | 1084 | No | NA | 2 | 0 | 0 | Not-determined | Genome-fragment | NA | NA | 0 |
| S16_extracted_25243 | 4202 | No | NA | 6 | 0 | 0 | Not-determined | Genome-fragment | NA | NA | 0 |
| S16_extracted_25261 | 1140 | No | NA | 2 | 0 | 1 | Not-determined | Genome-fragment | NA | NA | 0 |
| S14_vs_k141_5902\|\|full | 2463 | No | NA | 3 | 0 | 0 | Low-quality | Genome-fragment | 1.23 | AAI-based (medium-confidence) | 0 |
| S10_vs_101293\|\|full | 2463 | No | NA | 4 | 0 | 3 | Low-quality | Genome-fragment | 5.87 | AAI-based (medium-confidence) | 0 |
| S17_vs_k141_158228\|\|full | 2464 | No | NA | 4 | 2 | 0 | Low-quality | Genome-fragment | 3.7 | AAI-based (high-confidence) | 0 |
| S14_vs_k141_11616\|\|full | 2464 | No | NA | 4 | 0 | 1 | Low-quality | Genome-fragment | 3.06 | AAI-based (high-confidence) | 0 |
| S16_extracted_25609 | 1387 | No | NA | 4 | 0 | 0 | Not-determined | Genome-fragment | NA | NA | 0 |
| S16_extracted_25647 | 3090 | No | NA | 3 | 0 | 2 | Not-determined | Genome-fragment | NA | NA | 0 |
| S16_extracted_25706 | 1322 | No | NA | 1 | 0 | 1 | Not-determined | Genome-fragment | NA | NA | 0 |
| S16_extracted_25826 | 5575 | No | NA | 9 | 0 | 0 | Not-determined | Genome-fragment | NA | NA | 0 |
| S16_extracted_25869 | 1060 | No | NA | 1 | 0 | 1 | Not-determined | Genome-fragment | NA | NA | 0 |
| S16_extracted_25891 | 1134 | No | NA | 1 | 0 | 1 | Not-determined | Genome-fragment | NA | NA | 0 |
| S16_extracted_35813 | 2464 | No | NA | 8 | 2 | 0 | Low-quality | Genome-fragment | 6.28 | AAI-based (high-confidence) | 0 |
| S16_extracted_26021 | 1815 | No | NA | 3 | 0 | 1 | Not-determined | Genome-fragment | NA | NA | 0 |
| S16_extracted_26068 | 5949 | No | NA | 5 | 0 | 2 | Not-determined | Genome-fragment | NA | NA | 0 |
| S16_vs_133725\|\|full | 2464 | No | NA | 5 | 0 | 0 | Low-quality | Genome-fragment | 2.7 | AAI-based (high-confidence) | 0 |
| S16_extracted_26310 | 4653 | No | NA | 6 | 0 | 1 | Not-determined | Genome-fragment | NA | NA | 0 |
| S16_extracted_26321 | 1475 | No | NA | 1 | 0 | 0 | Not-determined | Genome-fragment | NA | NA | 0 |
| S16_extracted_26447 | 1609 | No | NA | 3 | 0 | 0 | Not-determined | Genome-fragment | NA | NA | 0 |
| S16_extracted_26535 | 1095 | No | NA | 2 | 0 | 0 | Not-determined | Genome-fragment | NA | NA | 0 |
| S16_extracted_26640 | 1172 | No | NA | 1 | 0 | 1 | Not-determined | Genome-fragment | NA | NA | 0 |
| S16_extracted_26707 | 3388 | No | NA | 5 | 0 | 4 | Not-determined | Genome-fragment | NA | NA | 0 |
| S16_extracted_26712 | 1190 | No | NA | 2 | 0 | 0 | Not-determined | Genome-fragment | NA | NA | 0 |
| S16_extracted_26723 | 1259 | No | NA | 1 | 0 | 0 | Not-determined | Genome-fragment | NA | NA | 0 |
| S16_extracted_26725 | 6900 | No | NA | 5 | 0 | 0 | Not-determined | Genome-fragment | NA | NA | 0 |
| S16_extracted_26781 | 2428 | No | NA | 1 | 0 | 1 | Not-determined | Genome-fragment | NA | NA | 0 |
| S16_extracted_26816 | 2797 | No | NA | 2 | 0 | 1 | Not-determined | Genome-fragment | NA | NA | 0 |
| S16_extracted_26817 | 3395 | No | NA | 6 | 0 | 0 | Not-determined | Genome-fragment | NA | NA | 0 |
| S16_extracted_26930 | 3822 | No | NA | 3 | 0 | 3 | Not-determined | Genome-fragment | NA | NA | 0 |
| S8_vs_k141_40173\|\|full | 2464 | No | NA | 3 | 0 | 1 | Low-quality | Genome-fragment | 1.93 | AAI-based (medium-confidence) | 0 |
| S5_vs_k141_88621\|\|full | 2465 | No | NA | 8 | 2 | 0 | Low-quality | Genome-fragment | 4.57 | AAI-based (medium-confidence) | 0 |
| S16_extracted_27020 | 1104 | No | NA | 1 | 0 | 1 | Not-determined | Genome-fragment | NA | NA | 0 |
| S16_extracted_27069 | 3532 | No | NA | 6 | 0 | 5 | Not-determined | Genome-fragment | NA | NA | 0 |
| S16_extracted_27194 | 1285 | No | NA | 2 | 0 | 2 | Not-determined | Genome-fragment | NA | NA | 0 |
| S8_vs_k141_123481\|\|full | 2465 | No | NA | 5 | 0 | 0 | Low-quality | Genome-fragment | 4.69 | AAI-based (medium-confidence) | 0 |
| S16_extracted_27485 | 8808 | No | NA | 12 | 0 | 12 | Not-determined | Genome-fragment | NA | NA | 0 |
| S16_extracted_27708 | 1271 | No | NA | 2 | 0 | 1 | Not-determined | Genome-fragment | NA | NA | 0 |
| S16_extracted_27717 | 1063 | No | NA | 1 | 0 | 0 | Not-determined | Genome-fragment | NA | NA | 0 |
| S16_extracted_27754 | 1283 | No | NA | 1 | 0 | 0 | Not-determined | Genome-fragment | NA | NA | 0 |
| S11_vs_k141_41422\|\|full | 2466 | No | NA | 3 | 0 | 3 | Low-quality | Genome-fragment | 1.09 | AAI-based (medium-confidence) | 0 |
| S14_vs_k141_36960\|\|lt2gene | 2466 | No | NA | 2 | 1 | 0 | Low-quality | Genome-fragment | 6.44 | AAI-based (high-confidence) | 0 |
| S16_extracted_27844 | 2859 | No | NA | 2 | 0 | 1 | Not-determined | Genome-fragment | NA | NA | 0 |
| S16_extracted_27848 | 1440 | No | NA | 1 | 0 | 0 | Not-determined | Genome-fragment | NA | NA | 0 |
| S5_extracted_k141_96277 | 2466 | No | NA | 5 | 3 | 0 | Low-quality | Genome-fragment | 7.29 | AAI-based (high-confidence) | 0 |
| S16_extracted_27862 | 1431 | No | NA | 1 | 0 | 1 | Not-determined | Genome-fragment | NA | NA | 0 |
| S7_vs_54094\|\|full | 2466 | No | NA | 4 | 3 | 0 | Low-quality | Genome-fragment | 5.78 | AAI-based (medium-confidence) | 0 |
| S16_extracted_27979 | 1650 | No | NA | 2 | 0 | 1 | Not-determined | Genome-fragment | NA | NA | 0 |
| S16_extracted_28118 | 1164 | No | NA | 2 | 0 | 1 | Not-determined | Genome-fragment | NA | NA | 0 |
| S16_vs_149467\|\|lt2gene | 2466 | No | NA | 2 | 1 | 0 | Low-quality | Genome-fragment | 4.15 | HMM-based (lower-bound) | 0 |
| S16_extracted_28203 | 4958 | No | NA | 1 | 0 | 1 | Not-determined | Genome-fragment | NA | NA | 0 |
| S14_vs_k141_88848\|\|full | 2467 | No | NA | 3 | 1 | 0 | Low-quality | Genome-fragment | 4.94 | HMM-based (lower-bound) | 0 |
| S16_extracted_28253 | 1278 | No | NA | 2 | 0 | 2 | Not-determined | Genome-fragment | NA | NA | 0 |
| S16_extracted_28375 | 2151 | No | NA | 1 | 0 | 0 | Not-determined | Genome-fragment | NA | NA | 0 |
| S16_extracted_28426 | 2093 | No | NA | 1 | 0 | 1 | Not-determined | Genome-fragment | NA | NA | 0 |
| S14_vs_k141_25251\|\|full | 2467 | No | NA | 5 | 1 | 0 | Low-quality | Genome-fragment | 5.4 | AAI-based (high-confidence) | 0 |
| S16_extracted_28580 | 5406 | No | NA | 10 | 0 | 1 | Not-determined | Genome-fragment | NA | NA | 0 |
| S16_extracted_28643 | 3495 | No | NA | 5 | 0 | 2 | Not-determined | Genome-fragment | NA | NA | 0 |
| S16_extracted_28801 | 1123 | No | NA | 1 | 0 | 1 | Not-determined | Genome-fragment | NA | NA | 0 |
| S4_extracted_2523 | 2467 | No | NA | 2 | 2 | 0 | Low-quality | Genome-fragment | 6.01 | AAI-based (high-confidence) | 0 |
| S16_extracted_28971 | 1800 | No | NA | 2 | 0 | 1 | Not-determined | Genome-fragment | NA | NA | 0 |
| S17_vs_k141_67959\|\|full | 2468 | No | NA | 3 | 0 | 2 | Low-quality | Genome-fragment | 0.56 | AAI-based (medium-confidence) | 0 |
| S16_extracted_29042 | 3241 | No | NA | 5 | 0 | 3 | Not-determined | Genome-fragment | NA | NA | 0 |
| S16_extracted_29159 | 1924 | No | NA | 1 | 0 | 0 | Not-determined | Genome-fragment | NA | NA | 0 |
| S16_extracted_29169 | 3056 | No | NA | 3 | 0 | 0 | Not-determined | Genome-fragment | NA | NA | 0 |
| S16_extracted_29296 | 2364 | No | NA | 4 | 0 | 2 | Not-determined | Genome-fragment | NA | NA | 0 |
| S16_extracted_29344 | 1645 | No | NA | 2 | 0 | 1 | Not-determined | Genome-fragment | NA | NA | 0 |
| S16_extracted_29392 | 6116 | No | NA | 7 | 0 | 2 | Not-determined | Genome-fragment | NA | NA | 0 |
| S16_extracted_29393 | 3677 | No | NA | 2 | 0 | 2 | Not-determined | Genome-fragment | NA | NA | 0 |
| S16_extracted_29597 | 4273 | No | NA | 2 | 0 | 2 | Not-determined | Genome-fragment | NA | NA | 0 |
| S16_extracted_29610 | 1444 | No | NA | 2 | 0 | 1 | Not-determined | Genome-fragment | NA | NA | 0 |
| S16_extracted_13715 | 2468 | No | NA | 5 | 0 | 0 | Low-quality | Genome-fragment | 4.1 | AAI-based (medium-confidence) | 0 |
| S16_vs_116073\|\|full | 2468 | No | NA | 4 | 0 | 0 | Low-quality | Genome-fragment | 5.52 | AAI-based (high-confidence) | 0 |
| S16_extracted_29736 | 1059 | No | NA | 2 | 0 | 2 | Not-determined | Genome-fragment | NA | NA | 0 |
| S16_extracted_29804 | 6443 | No | NA | 7 | 0 | 5 | Not-determined | Genome-fragment | NA | NA | 0 |
| S12_extracted_k141_175209 | 2468 | No | NA | 3 | 0 | 0 | Low-quality | Genome-fragment | 2.99 | AAI-based (high-confidence) | 0 |
| S16_extracted_29879 | 5452 | No | NA | 7 | 0 | 4 | Not-determined | Genome-fragment | NA | NA | 0 |
| S16_extracted_29974 | 3853 | No | NA | 2 | 0 | 0 | Not-determined | Genome-fragment | NA | NA | 0 |
| S11_vs_k141_63707\|\|full | 2469 | No | NA | 5 | 3 | 0 | Low-quality | Genome-fragment | 4.89 | AAI-based (high-confidence) | 0 |
| S16_extracted_30062 | 1131 | No | NA | 2 | 0 | 1 | Not-determined | Genome-fragment | NA | NA | 0 |
| S7_vs_62785\|\|full | 2469 | No | NA | 5 | 0 | 0 | Low-quality | Genome-fragment | 4.43 | AAI-based (high-confidence) | 0 |
| S16_extracted_30285 | 3249 | No | NA | 1 | 0 | 1 | Not-determined | Genome-fragment | NA | NA | 0 |
| S16_extracted_30299 | 1036 | No | NA | 1 | 0 | 1 | Not-determined | Genome-fragment | NA | NA | 0 |
| S10_vs_87667\|\|full | 2469 | No | NA | 8 | 0 | 0 | Low-quality | Genome-fragment | 6.31 | AAI-based (high-confidence) | 0 |
| S16_extracted_30322 | 1370 | No | NA | 2 | 0 | 0 | Not-determined | Genome-fragment | NA | NA | 0 |
| S16_extracted_30329 | 2302 | No | NA | 4 | 0 | 0 | Not-determined | Genome-fragment | NA | NA | 0 |
| S16_extracted_30349 | 3892 | No | NA | 4 | 0 | 4 | Not-determined | Genome-fragment | NA | NA | 0 |
| S16_extracted_30356 | 1256 | No | NA | 3 | 0 | 1 | Not-determined | Genome-fragment | NA | NA | 0 |
| S16_extracted_30432 | 1008 | No | NA | 1 | 0 | 1 | Not-determined | Genome-fragment | NA | NA | 0 |
| S16_extracted_30435 | 1427 | No | NA | 3 | 0 | 0 | Not-determined | Genome-fragment | NA | NA | 0 |
| S16_extracted_30470 | 1722 | No | NA | 4 | 0 | 2 | Not-determined | Genome-fragment | NA | NA | 0 |
| S16_extracted_30566 | 2464 | No | NA | 2 | 0 | 0 | Not-determined | Genome-fragment | NA | NA | 0 |
| S16_extracted_30691 | 3001 | No | NA | 5 | 0 | 1 | Not-determined | Genome-fragment | NA | NA | 0 |
| S16_extracted_30739 | 1671 | No | NA | 1 | 0 | 0 | Not-determined | Genome-fragment | NA | NA | 0 |
| S16_extracted_30749 | 1013 | No | NA | 2 | 0 | 2 | Not-determined | Genome-fragment | NA | NA | 0 |
| S16_extracted_30750 | 1226 | No | NA | 2 | 0 | 1 | Not-determined | Genome-fragment | NA | NA | 0 |
| S16_extracted_30919 | 1204 | No | NA | 2 | 0 | 0 | Not-determined | Genome-fragment | NA | NA | 0 |
| S16_extracted_30994 | 1903 | No | NA | 1 | 0 | 1 | Not-determined | Genome-fragment | NA | NA | 0 |
| S16_extracted_31060 | 1257 | No | NA | 1 | 0 | 1 | Not-determined | Genome-fragment | NA | NA | 0 |
| S16_extracted_31173 | 1232 | No | NA | 2 | 0 | 1 | Not-determined | Genome-fragment | NA | NA | 0 |
| S16_extracted_31219 | 8577 | No | NA | 11 | 0 | 2 | Not-determined | Genome-fragment | NA | NA | 0 |
| S16_extracted_31240 | 1000 | No | NA | 1 | 0 | 1 | Not-determined | Genome-fragment | NA | NA | 0 |
| S16_extracted_31272 | 1144 | No | NA | 2 | 0 | 1 | Not-determined | Genome-fragment | NA | NA | 0 |
| S16_extracted_31296 | 3831 | No | NA | 2 | 0 | 0 | Not-determined | Genome-fragment | NA | NA | 0 |
| S16_extracted_31299 | 4406 | No | NA | 6 | 0 | 3 | Not-determined | Genome-fragment | NA | NA | 0 |
| S8_vs_k141_152932\|\|full | 2469 | No | NA | 6 | 0 | 0 | Low-quality | Genome-fragment | 5.14 | AAI-based (medium-confidence) | 0 |
| S9_vs_k141_90504\|\|full | 2470 | No | NA | 3 | 0 | 1 | Low-quality | Genome-fragment | 3.04 | AAI-based (medium-confidence) | 0 |
| S16_extracted_31366 | 4805 | No | NA | 8 | 0 | 1 | Not-determined | Genome-fragment | NA | NA | 0 |
| S16_extracted_31392 | 1108 | No | NA | 1 | 0 | 0 | Not-determined | Genome-fragment | NA | NA | 0 |
| S14_vs_k141_106363\|\|full | 2470 | No | NA | 6 | 0 | 0 | Low-quality | Genome-fragment | 4.96 | AAI-based (medium-confidence) | 0 |
| S16_extracted_31409 | 1023 | No | NA | 1 | 0 | 0 | Not-determined | Genome-fragment | NA | NA | 0 |
| S16_extracted_31417 | 1053 | No | NA | 1 | 0 | 1 | Not-determined | Genome-fragment | NA | NA | 0 |
| S16_extracted_31435 | 1132 | No | NA | 1 | 0 | 1 | Not-determined | Genome-fragment | NA | NA | 0 |
| S16_extracted_31439 | 1543 | No | NA | 3 | 0 | 2 | Not-determined | Genome-fragment | NA | NA | 0 |
| S12_vs_k141_91309\|\|full | 2470 | No | NA | 2 | 0 | 1 | Low-quality | Genome-fragment | 1.64 | AAI-based (medium-confidence) | 0 |
| S16_extracted_31535 | 1345 | No | NA | 1 | 0 | 1 | Not-determined | Genome-fragment | NA | NA | 0 |
| S16_extracted_31680 | 1048 | No | NA | 1 | 0 | 1 | Not-determined | Genome-fragment | NA | NA | 0 |
| S16_extracted_31791 | 1145 | No | NA | 2 | 0 | 1 | Not-determined | Genome-fragment | NA | NA | 0 |
| S16_extracted_31797 | 7939 | No | NA | 6 | 0 | 5 | Not-determined | Genome-fragment | NA | NA | 0 |
| S16_extracted_31848 | 1255 | No | NA | 1 | 0 | 1 | Not-determined | Genome-fragment | NA | NA | 0 |
| S16_extracted_31863 | 1050 | No | NA | 1 | 0 | 0 | Not-determined | Genome-fragment | NA | NA | 0 |
| S16_extracted_31913 | 1826 | No | NA | 3 | 0 | 1 | Not-determined | Genome-fragment | NA | NA | 0 |
| S16_extracted_31956 | 1624 | No | NA | 2 | 0 | 2 | Not-determined | Genome-fragment | NA | NA | 0 |
| S16_extracted_32010 | 1001 | No | NA | 1 | 0 | 0 | Not-determined | Genome-fragment | NA | NA | 0 |
| S16_extracted_32051 | 1581 | No | NA | 2 | 0 | 2 | Not-determined | Genome-fragment | NA | NA | 0 |
| S16_extracted_32101 | 4760 | No | NA | 11 | 0 | 0 | Not-determined | Genome-fragment | NA | NA | 0 |
| S16_extracted_32140 | 1389 | No | NA | 2 | 0 | 2 | Not-determined | Genome-fragment | NA | NA | 0 |
| S16_extracted_32235 | 2083 | No | NA | 3 | 0 | 2 | Not-determined | Genome-fragment | NA | NA | 0 |
| S8_vs_k141_76884\|\|full | 2470 | No | NA | 4 | 0 | 0 | Low-quality | Genome-fragment | 3.2 | AAI-based (high-confidence) | 0 |
| S16_extracted_32359 | 28101 | No | NA | 53 | 10 | 0 | Medium-quality | Genome-fragment | 59.6 | AAI-based (medium-confidence) | 0 |
| S11_vs_k141_75257\|\|full | 2471 | No | NA | 3 | 0 | 0 | Low-quality | Genome-fragment | 5.44 | AAI-based (medium-confidence) | 0 |
| S16_extracted_32558 | 1778 | No | NA | 1 | 0 | 0 | Not-determined | Genome-fragment | NA | NA | 0 |
| S16_extracted_32686 | 1197 | No | NA | 1 | 0 | 0 | Not-determined | Genome-fragment | NA | NA | 0 |
| S1_vs_k141_26062\|\|full | 2471 | No | NA | 2 | 1 | 0 | Low-quality | Genome-fragment | 5.44 | AAI-based (medium-confidence) | 0 |
| S17_vs_k141_37011\|\|full | 2472 | No | NA | 6 | 1 | 0 | Low-quality | Genome-fragment | 5.07 | AAI-based (high-confidence) | 0 |
| S16_extracted_32858 | 1551 | No | NA | 3 | 0 | 1 | Not-determined | Genome-fragment | NA | NA | 0 |
| S16_extracted_32886 | 3235 | No | NA | 5 | 0 | 0 | Not-determined | Genome-fragment | NA | NA | 0 |
| S16_extracted_32921 | 1432 | No | NA | 1 | 0 | 0 | Not-determined | Genome-fragment | NA | NA | 0 |
| S16_extracted_32952 | 1725 | No | NA | 1 | 0 | 1 | Not-determined | Genome-fragment | NA | NA | 0 |
| S16_extracted_32957 | 1028 | No | NA | 1 | 0 | 0 | Not-determined | Genome-fragment | NA | NA | 0 |
| S16_extracted_32994 | 2142 | No | NA | 1 | 0 | 0 | Not-determined | Genome-fragment | NA | NA | 0 |
| S16_extracted_33118 | 1870 | No | NA | 2 | 0 | 0 | Not-determined | Genome-fragment | NA | NA | 0 |
| S16_extracted_33218 | 17281 | No | NA | 33 | 29 | 0 | Medium-quality | Genome-fragment | 57.01 | AAI-based (high-confidence) | 0 |
| S7_vs_49693\|\|full | 2472 | No | NA | 4 | 0 | 3 | Low-quality | Genome-fragment | 3.25 | AAI-based (medium-confidence) | 0 |
| S16_extracted_33534 | 4345 | No | NA | 4 | 0 | 2 | Not-determined | Genome-fragment | NA | NA | 0 |
| S16_extracted_33559 | 3320 | No | NA | 3 | 0 | 1 | Not-determined | Genome-fragment | NA | NA | 0 |
| S7_vs_80208\|\|full | 2472 | No | NA | 4 | 1 | 0 | Low-quality | Genome-fragment | 5.98 | AAI-based (medium-confidence) | 0 |
| S7_vs_90935\|\|full | 2472 | No | NA | 3 | 0 | 1 | Low-quality | Genome-fragment | 4.01 | AAI-based (medium-confidence) | 0 |
| S16_vs_57366\|\|full | 2472 | No | NA | 2 | 0 | 1 | Low-quality | Genome-fragment | 1.09 | AAI-based (medium-confidence) | 0 |
| S16_extracted_33698 | 1672 | No | NA | 2 | 0 | 1 | Not-determined | Genome-fragment | NA | NA | 0 |
| S16_extracted_33705 | 1038 | No | NA | 1 | 0 | 1 | Not-determined | Genome-fragment | NA | NA | 0 |
| S16_vs_66279\|\|full | 2472 | No | NA | 5 | 2 | 0 | Low-quality | Genome-fragment | 3.11 | HMM-based (lower-bound) | 0 |
| S16_extracted_33808 | 7170 | No | NA | 5 | 0 | 2 | Not-determined | Genome-fragment | NA | NA | 0 |
| S10_vs_63318\|\|full | 2472 | No | NA | 3 | 0 | 2 | Low-quality | Genome-fragment | 1.48 | AAI-based (medium-confidence) | 0 |
| S16_extracted_34062 | 1791 | No | NA | 1 | 0 | 1 | Not-determined | Genome-fragment | NA | NA | 0 |
| S16_extracted_34115 | 3822 | No | NA | 6 | 0 | 2 | Not-determined | Genome-fragment | NA | NA | 0 |
| S16_extracted_34118 | 1116 | No | NA | 1 | 0 | 0 | Not-determined | Genome-fragment | NA | NA | 0 |
| S9_vs_k141_48075\|\|full | 2473 | No | NA | 4 | 2 | 0 | Low-quality | Genome-fragment | 5.99 | AAI-based (high-confidence) | 0 |
| S16_extracted_34323 | 7062 | No | NA | 4 | 0 | 3 | Not-determined | Genome-fragment | NA | NA | 0 |
| S16_extracted_34344 | 1177 | No | NA | 1 | 0 | 1 | Not-determined | Genome-fragment | NA | NA | 0 |
| S16_extracted_34410 | 5898 | No | NA | 7 | 0 | 6 | Not-determined | Genome-fragment | NA | NA | 0 |
| S5_vs_k141_114150\|\|full | 2473 | No | NA | 5 | 1 | 1 | Low-quality | Genome-fragment | 1.34 | HMM-based (lower-bound) | 0 |
| S16_extracted_34475 | 1415 | No | NA | 1 | 0 | 0 | Not-determined | Genome-fragment | NA | NA | 0 |
| S16_extracted_34624 | 1171 | No | NA | 1 | 0 | 0 | Not-determined | Genome-fragment | NA | NA | 0 |
| S16_extracted_34627 | 1153 | No | NA | 1 | 0 | 0 | Not-determined | Genome-fragment | NA | NA | 0 |
| S7_vs_130273\|\|lt2gene | 2473 | No | NA | 2 | 0 | 0 | Low-quality | Genome-fragment | 6.85 | AAI-based (medium-confidence) | 0 |
| S8_vs_k141_114783\|\|full | 2473 | No | NA | 3 | 0 | 0 | Low-quality | Genome-fragment | 3.06 | AAI-based (high-confidence) | 0 |
| S8_vs_k141_179635\|\|full | 2473 | No | NA | 3 | 0 | 0 | Low-quality | Genome-fragment | 5.59 | AAI-based (high-confidence) | 0 |
| S16_extracted_34883 | 4839 | No | NA | 9 | 0 | 1 | Not-determined | Genome-fragment | NA | NA | 0 |
| S16_extracted_34900 | 1189 | No | NA | 2 | 0 | 2 | Not-determined | Genome-fragment | NA | NA | 0 |
| S16_extracted_34904 | 1146 | No | NA | 1 | 0 | 1 | Not-determined | Genome-fragment | NA | NA | 0 |
| S16_extracted_34966 | 5863 | No | NA | 10 | 0 | 1 | Not-determined | Genome-fragment | NA | NA | 0 |
| S16_extracted_35055 | 1441 | No | NA | 2 | 0 | 2 | Not-determined | Genome-fragment | NA | NA | 0 |
| S16_extracted_35103 | 1960 | No | NA | 3 | 0 | 1 | Not-determined | Genome-fragment | NA | NA | 0 |
| S14_vs_k141_89250\|\|full | 2474 | No | NA | 6 | 1 | 0 | Low-quality | Genome-fragment | 4.9 | HMM-based (lower-bound) | 0 |
| S16_extracted_35298 | 2813 | No | NA | 2 | 0 | 2 | Not-determined | Genome-fragment | NA | NA | 0 |
| S16_extracted_35417 | 1076 | No | NA | 2 | 0 | 1 | Not-determined | Genome-fragment | NA | NA | 0 |
| S14_vs_k141_42941\|\|full | 2474 | No | NA | 6 | 6 | 0 | Low-quality | Genome-fragment | 7.09 | AAI-based (high-confidence) | 0 |
| S16_extracted_35493 | 1188 | No | NA | 2 | 0 | 1 | Not-determined | Genome-fragment | NA | NA | 0 |
| S16_extracted_35551 | 1207 | No | NA | 2 | 0 | 2 | Not-determined | Genome-fragment | NA | NA | 0 |
| S16_extracted_35566 | 3729 | No | NA | 3 | 0 | 3 | Not-determined | Genome-fragment | NA | NA | 0 |
| S16_extracted_35582 | 1666 | No | NA | 4 | 0 | 0 | Not-determined | Genome-fragment | NA | NA | 0 |
| S1_vs_k141_4688\|\|full | 2474 | No | NA | 5 | 1 | 0 | Low-quality | Genome-fragment | 5.59 | AAI-based (high-confidence) | 0 |
| S16_extracted_35684 | 1043 | No | NA | 1 | 0 | 1 | Not-determined | Genome-fragment | NA | NA | 0 |
| S16_extracted_35769 | 2298 | No | NA | 3 | 0 | 2 | Not-determined | Genome-fragment | NA | NA | 0 |
| S7_vs_39127\|\|full | 2474 | No | NA | 2 | 2 | 0 | Low-quality | Genome-fragment | 6.2 | AAI-based (high-confidence) | 0 |
| S6_vs_k141_55910\|\|full | 2474 | No | NA | 4 | 0 | 2 | Low-quality | Genome-fragment | 1.61 | AAI-based (medium-confidence) | 0 |
| S16_extracted_35907 | 2334 | No | NA | 3 | 0 | 1 | Not-determined | Genome-fragment | NA | NA | 0 |
| S16_extracted_35962 | 3495 | No | NA | 4 | 0 | 3 | Not-determined | Genome-fragment | NA | NA | 0 |
| S3_vs_k141_50885\|\|full | 2475 | No | NA | 5 | 1 | 0 | Low-quality | Genome-fragment | 38.69 | AAI-based (high-confidence) | 0 |
| S16_vs_2121\|\|full | 2475 | No | NA | 5 | 0 | 0 | Low-quality | Genome-fragment | 6.49 | AAI-based (high-confidence) | 0 |
| S16_extracted_36095 | 1546 | No | NA | 1 | 0 | 1 | Not-determined | Genome-fragment | NA | NA | 0 |
| S16_vs_113841\|\|full | 2475 | No | NA | 5 | 0 | 5 | Low-quality | Genome-fragment | 0.84 | AAI-based (medium-confidence) | 0 |
| S16_extracted_36130 | 1905 | No | NA | 4 | 0 | 2 | Not-determined | Genome-fragment | NA | NA | 0 |
| S10_vs_33354\|\|full | 2475 | No | NA | 9 | 8 | 0 | Low-quality | Genome-fragment | 6.28 | AAI-based (medium-confidence) | 0 |
| S16_extracted_36142 | 1094 | No | NA | 2 | 0 | 1 | Not-determined | Genome-fragment | NA | NA | 0 |
| S16_extracted_36146 | 28348 | No | NA | 36 | 24 | 0 | Medium-quality | Genome-fragment | 61.87 | AAI-based (high-confidence) | 0 |
| S16_extracted_36224 | 1270 | No | NA | 1 | 0 | 0 | Not-determined | Genome-fragment | NA | NA | 0 |
| S16_extracted_36303 | 2663 | No | NA | 1 | 0 | 1 | Not-determined | Genome-fragment | NA | NA | 0 |
| S16_extracted_36365 | 2122 | No | NA | 4 | 0 | 0 | Not-determined | Genome-fragment | NA | NA | 0 |
| S13_vs_72030\|\|full | 2476 | No | NA | 8 | 1 | 0 | Low-quality | Genome-fragment | 5.62 | AAI-based (medium-confidence) | 0 |
| S1_vs_k141_62187\|\|full | 2476 | No | NA | 6 | 0 | 0 | Low-quality | Genome-fragment | 6.18 | AAI-based (high-confidence) | 0 |
| S16_extracted_36548 | 4893 | No | NA | 5 | 0 | 4 | Not-determined | Genome-fragment | NA | NA | 0 |
| S16_extracted_36629 | 1497 | No | NA | 1 | 0 | 1 | Not-determined | Genome-fragment | NA | NA | 0 |
| S5_extracted_k141_90929 | 2476 | No | NA | 3 | 0 | 2 | Low-quality | Genome-fragment | 1.24 | AAI-based (medium-confidence) | 0 |
| S8_vs_k141_167418\|\|full | 2476 | No | NA | 3 | 1 | 0 | Low-quality | Genome-fragment | 4.45 | AAI-based (high-confidence) | 0 |
| S16_extracted_36736 | 1182 | No | NA | 3 | 0 | 0 | Not-determined | Genome-fragment | NA | NA | 0 |
| S5_vs_k141_42534\|\|full | 2477 | No | NA | 5 | 0 | 4 | Low-quality | Genome-fragment | 5.18 | AAI-based (medium-confidence) | 0 |
| S9_extracted_k141_20809 | 2478 | No | NA | 2 | 0 | 0 | Low-quality | Genome-fragment | 1.39 | AAI-based (high-confidence) | 0 |
| S11_vs_k141_68220\|\|full | 2478 | No | NA | 8 | 0 | 0 | Low-quality | Genome-fragment | 5.15 | AAI-based (high-confidence) | 0 |
| S16_extracted_36860 | 1308 | No | NA | 2 | 0 | 2 | Not-determined | Genome-fragment | NA | NA | 0 |
| S3_vs_k141_120910\|\|full | 2478 | No | NA | 2 | 0 | 2 | Low-quality | Genome-fragment | 1.98 | AAI-based (high-confidence) | 0 |
| S16_extracted_36958 | 1081 | No | NA | 2 | 0 | 1 | Not-determined | Genome-fragment | NA | NA | 0 |
| S16_extracted_37018 | 1386 | No | NA | 1 | 0 | 1 | Not-determined | Genome-fragment | NA | NA | 0 |
| S16_extracted_37073 | 5557 | No | NA | 3 | 0 | 3 | Not-determined | Genome-fragment | NA | NA | 0 |
| S16_extracted_37105 | 1223 | No | NA | 2 | 0 | 1 | Not-determined | Genome-fragment | NA | NA | 0 |
| S16_extracted_37126 | 4893 | No | NA | 3 | 0 | 0 | Not-determined | Genome-fragment | NA | NA | 0 |
| S16_extracted_37141 | 1757 | No | NA | 2 | 0 | 1 | Not-determined | Genome-fragment | NA | NA | 0 |
| S16_extracted_37218 | 2198 | No | NA | 3 | 0 | 0 | Not-determined | Genome-fragment | NA | NA | 0 |
| S16_extracted_37275 | 1983 | No | NA | 2 | 0 | 1 | Not-determined | Genome-fragment | NA | NA | 0 |
| S16_extracted_37392 | 1025 | No | NA | 1 | 0 | 0 | Not-determined | Genome-fragment | NA | NA | 0 |
| S16_extracted_37408 | 1682 | No | NA | 1 | 0 | 1 | Not-determined | Genome-fragment | NA | NA | 0 |
| S16_extracted_37529 | 1005 | No | NA | 1 | 0 | 1 | Not-determined | Genome-fragment | NA | NA | 0 |
| S16_extracted_37754 | 1038 | No | NA | 1 | 0 | 0 | Not-determined | Genome-fragment | NA | NA | 0 |
| S16_extracted_37796 | 1261 | No | NA | 2 | 0 | 0 | Not-determined | Genome-fragment | NA | NA | 0 |
| S16_extracted_37802 | 1151 | No | NA | 2 | 0 | 1 | Not-determined | Genome-fragment | NA | NA | 0 |
| S16_extracted_37912 | 4327 | No | NA | 5 | 0 | 0 | Not-determined | Genome-fragment | NA | NA | 0 |
| S7_vs_28000\|\|full | 2478 | No | NA | 6 | 1 | 0 | Low-quality | Genome-fragment | 5.53 | AAI-based (medium-confidence) | 0 |
| S16_vs_47759\|\|full | 2478 | No | NA | 3 | 0 | 0 | Low-quality | Genome-fragment | 1.96 | AAI-based (high-confidence) | 0 |
| S16_extracted_38069 | 1967 | No | NA | 3 | 0 | 1 | Not-determined | Genome-fragment | NA | NA | 0 |
| S16_extracted_38208 | 6251 | No | NA | 5 | 0 | 3 | Not-determined | Genome-fragment | NA | NA | 0 |
| S16_extracted_38257 | 1141 | No | NA | 3 | 0 | 2 | Not-determined | Genome-fragment | NA | NA | 0 |
| S16_extracted_38297 | 1174 | No | NA | 1 | 0 | 1 | Not-determined | Genome-fragment | NA | NA | 0 |
| S16_extracted_38315 | 12937 | No | NA | 11 | 0 | 11 | Not-determined | Genome-fragment | NA | NA | 0 |
| S16_extracted_38392 | 5887 | No | NA | 2 | 0 | 2 | Not-determined | Genome-fragment | NA | NA | 0 |
| S16_extracted_38471 | 2071 | No | NA | 3 | 0 | 2 | Not-determined | Genome-fragment | NA | NA | 0 |
| S10_vs_63479\|\|full | 2478 | No | NA | 3 | 2 | 0 | Low-quality | Genome-fragment | 5.53 | AAI-based (medium-confidence) | 0 |
| S8_vs_k141_43558\|\|full | 2478 | No | NA | 2 | 0 | 0 | Low-quality | Genome-fragment | 6.25 | AAI-based (medium-confidence) | 0 |
| S16_extracted_38649 | 1528 | No | NA | 4 | 0 | 0 | Not-determined | Genome-fragment | NA | NA | 0 |
| S16_extracted_38667 | 1493 | No | NA | 1 | 0 | 1 | Not-determined | Genome-fragment | NA | NA | 0 |
| S16_extracted_38747 | 1022 | No | NA | 2 | 0 | 0 | Not-determined | Genome-fragment | NA | NA | 0 |
| S16_extracted_38750 | 12616 | No | NA | 17 | 0 | 10 | Not-determined | Genome-fragment | NA | NA | 0 |
| S16_extracted_38762 | 2015 | No | NA | 2 | 0 | 1 | Not-determined | Genome-fragment | NA | NA | 0 |
| S16_extracted_38821 | 4632 | No | NA | 2 | 0 | 2 | Not-determined | Genome-fragment | NA | NA | 0 |
| S16_extracted_38902 | 3375 | No | NA | 6 | 0 | 1 | Not-determined | Genome-fragment | NA | NA | 0 |
| S16_extracted_38932 | 1773 | No | NA | 1 | 0 | 1 | Not-determined | Genome-fragment | NA | NA | 0 |
| S16_extracted_38949 | 4418 | No | NA | 7 | 0 | 5 | Not-determined | Genome-fragment | NA | NA | 0 |
| S10_vs_49336\|\|full | 2479 | No | NA | 4 | 1 | 0 | Low-quality | Genome-fragment | 6.8 | AAI-based (high-confidence) | 0 |
| S8_vs_k141_134776\|\|full | 2479 | No | NA | 5 | 2 | 0 | Low-quality | Genome-fragment | 5.94 | AAI-based (medium-confidence) | 0 |
| S16_extracted_39062 | 1959 | No | NA | 4 | 0 | 0 | Not-determined | Genome-fragment | NA | NA | 0 |
| S16_extracted_39135 | 1389 | No | NA | 1 | 0 | 1 | Not-determined | Genome-fragment | NA | NA | 0 |
| S16_extracted_39145 | 1895 | No | NA | 2 | 0 | 1 | Not-determined | Genome-fragment | NA | NA | 0 |
| S16_extracted_39211 | 1400 | No | NA | 1 | 0 | 0 | Not-determined | Genome-fragment | NA | NA | 0 |
| S16_extracted_39265 | 1559 | No | NA | 1 | 0 | 1 | Not-determined | Genome-fragment | NA | NA | 0 |
| S3_vs_k141_64715\|\|full | 2480 | No | NA | 3 | 0 | 1 | Low-quality | Genome-fragment | 5.24 | AAI-based (medium-confidence) | 0 |
| S16_extracted_39387 | 15623 | No | NA | 15 | 0 | 8 | Not-determined | Genome-fragment | NA | NA | 0 |
| S16_extracted_39610 | 5046 | No | NA | 5 | 0 | 0 | Not-determined | Genome-fragment | NA | NA | 0 |
| S16_extracted_39644 | 1049 | No | NA | 1 | 0 | 1 | Not-determined | Genome-fragment | NA | NA | 0 |
| S16_extracted_39734 | 2261 | No | NA | 2 | 0 | 1 | Not-determined | Genome-fragment | NA | NA | 0 |
| S16_extracted_39741 | 1104 | No | NA | 1 | 0 | 1 | Not-determined | Genome-fragment | NA | NA | 0 |
| S16_extracted_39759 | 3184 | No | NA | 2 | 0 | 1 | Not-determined | Genome-fragment | NA | NA | 0 |
| S5_vs_k141_44599\|\|full | 2480 | No | NA | 3 | 0 | 2 | Low-quality | Genome-fragment | 1.14 | AAI-based (medium-confidence) | 0 |
| S16_extracted_39817 | 1022 | No | NA | 2 | 0 | 0 | Not-determined | Genome-fragment | NA | NA | 0 |
| S5_vs_k141_16023\|\|full | 2480 | No | NA | 9 | 0 | 0 | Low-quality | Genome-fragment | 6.28 | AAI-based (medium-confidence) | 0 |
| S16_extracted_39947 | 1337 | No | NA | 1 | 0 | 0 | Not-determined | Genome-fragment | NA | NA | 0 |
| S7_vs_28818\|\|full | 2480 | No | NA | 5 | 0 | 0 | Low-quality | Genome-fragment | 3.37 | AAI-based (high-confidence) | 0 |
| S16_extracted_39965 | 4041 | No | NA | 7 | 0 | 2 | Not-determined | Genome-fragment | NA | NA | 0 |
| S16_extracted_40043 | 1359 | No | NA | 1 | 0 | 1 | Not-determined | Genome-fragment | NA | NA | 0 |
| S16_extracted_40247 | 1623 | No | NA | 2 | 0 | 1 | Not-determined | Genome-fragment | NA | NA | 0 |
| S16_extracted_40301 | 1103 | No | NA | 2 | 0 | 1 | Not-determined | Genome-fragment | NA | NA | 0 |
| S16_extracted_40319 | 1083 | No | NA | 1 | 0 | 1 | Not-determined | Genome-fragment | NA | NA | 0 |
| S16_extracted_40428 | 1210 | No | NA | 3 | 0 | 0 | Not-determined | Genome-fragment | NA | NA | 0 |
| S16_extracted_40479 | 1018 | No | NA | 2 | 0 | 1 | Not-determined | Genome-fragment | NA | NA | 0 |
| S16_extracted_40499 | 1339 | No | NA | 2 | 0 | 1 | Not-determined | Genome-fragment | NA | NA | 0 |
| S16_extracted_40520 | 1102 | No | NA | 1 | 0 | 0 | Not-determined | Genome-fragment | NA | NA | 0 |
| S10_vs_37121\|\|full | 2480 | No | NA | 4 | 0 | 0 | Low-quality | Genome-fragment | 3.13 | AAI-based (medium-confidence) | 0 |
| S16_extracted_40743 | 3205 | No | NA | 2 | 0 | 1 | Not-determined | Genome-fragment | NA | NA | 0 |
| S16_extracted_40754 | 1732 | No | NA | 1 | 0 | 0 | Not-determined | Genome-fragment | NA | NA | 0 |
| S16_extracted_40802 | 1879 | No | NA | 2 | 0 | 0 | Not-determined | Genome-fragment | NA | NA | 0 |
| S16_extracted_40890 | 1224 | No | NA | 2 | 0 | 0 | Not-determined | Genome-fragment | NA | NA | 0 |
| S8_vs_k141_1327\|\|full | 2480 | No | NA | 6 | 0 | 0 | Low-quality | Genome-fragment | 5.43 | AAI-based (high-confidence) | 0 |
| S3_extracted_k141_116909 | 2481 | No | NA | 3 | 3 | 0 | Low-quality | Genome-fragment | 1.47 | AAI-based (high-confidence) | 0 |
| S16_extracted_41118 | 1102 | No | NA | 2 | 0 | 1 | Not-determined | Genome-fragment | NA | NA | 0 |
| S16_extracted_41128 | 1184 | No | NA | 4 | 0 | 1 | Not-determined | Genome-fragment | NA | NA | 0 |
| S8_extracted_k141_118917 | 2481 | No | NA | 3 | 0 | 2 | Low-quality | Genome-fragment | 0.56 | AAI-based (medium-confidence) | 0 |
| S16_extracted_41250 | 3460 | No | NA | 1 | 0 | 1 | Not-determined | Genome-fragment | NA | NA | 0 |
| S16_extracted_41251 | 1258 | No | NA | 1 | 0 | 0 | Not-determined | Genome-fragment | NA | NA | 0 |
| S16_extracted_41316 | 2025 | No | NA | 2 | 0 | 1 | Not-determined | Genome-fragment | NA | NA | 0 |
| S16_extracted_41321 | 1306 | No | NA | 1 | 0 | 1 | Not-determined | Genome-fragment | NA | NA | 0 |
| S16_extracted_41434 | 1103 | No | NA | 1 | 0 | 0 | Not-determined | Genome-fragment | NA | NA | 0 |
| S1_extracted_k141_102211 | 2482 | No | NA | 2 | 0 | 0 | Low-quality | Genome-fragment | 4.35 | AAI-based (high-confidence) | 0 |
| S1_vs_k141_17838\|\|full | 2482 | No | NA | 6 | 0 | 0 | Low-quality | Genome-fragment | 5.09 | AAI-based (medium-confidence) | 0 |
| S1_vs_k141_106197\|\|full | 2482 | No | NA | 6 | 2 | 0 | Low-quality | Genome-fragment | 4.75 | AAI-based (medium-confidence) | 0 |
| S16_extracted_41648 | 1480 | No | NA | 1 | 0 | 1 | Not-determined | Genome-fragment | NA | NA | 0 |
| S5_vs_k141_9121\|\|full | 2482 | No | NA | 6 | 1 | 0 | Low-quality | Genome-fragment | 7.16 | AAI-based (high-confidence) | 0 |
| S16_extracted_41712 | 1279 | No | NA | 1 | 0 | 1 | Not-determined | Genome-fragment | NA | NA | 0 |
| S16_extracted_41762 | 1411 | No | NA | 2 | 0 | 2 | Not-determined | Genome-fragment | NA | NA | 0 |
| S16_extracted_41783 | 1386 | No | NA | 1 | 0 | 1 | Not-determined | Genome-fragment | NA | NA | 0 |
| S7_vs_134647\|\|full | 2482 | No | NA | 5 | 1 | 0 | Low-quality | Genome-fragment | 5.95 | AAI-based (high-confidence) | 0 |
| S17_extracted_k141_38239 | 2483 | No | NA | 6 | 3 | 0 | Low-quality | Genome-fragment | 5.51 | AAI-based (high-confidence) | 0 |
| S16_extracted_42101 | 1224 | No | NA | 2 | 0 | 0 | Not-determined | Genome-fragment | NA | NA | 0 |
| S16_extracted_42126 | 1824 | No | NA | 2 | 0 | 2 | Not-determined | Genome-fragment | NA | NA | 0 |
| S16_extracted_42137 | 1194 | No | NA | 2 | 0 | 1 | Not-determined | Genome-fragment | NA | NA | 0 |
| S1_vs_k141_144853\|\|full | 2483 | No | NA | 5 | 1 | 0 | Low-quality | Genome-fragment | 6.34 | AAI-based (high-confidence) | 0 |
| S16_extracted_42280 | 1071 | No | NA | 1 | 0 | 1 | Not-determined | Genome-fragment | NA | NA | 0 |
| S16_extracted_42322 | 1294 | No | NA | 1 | 0 | 1 | Not-determined | Genome-fragment | NA | NA | 0 |
| S16_extracted_42431 | 2107 | No | NA | 3 | 0 | 2 | Not-determined | Genome-fragment | NA | NA | 0 |
| S16_extracted_42455 | 1446 | No | NA | 1 | 0 | 1 | Not-determined | Genome-fragment | NA | NA | 0 |
| S3_extracted_k141_88004 | 2483 | No | NA | 5 | 2 | 0 | Low-quality | Genome-fragment | 5.72 | AAI-based (high-confidence) | 0 |
| S16_extracted_42708 | 1398 | No | NA | 1 | 0 | 1 | Not-determined | Genome-fragment | NA | NA | 0 |
| S16_extracted_42758 | 3439 | No | NA | 3 | 0 | 3 | Not-determined | Genome-fragment | NA | NA | 0 |
| S16_extracted_42827 | 1297 | No | NA | 2 | 0 | 1 | Not-determined | Genome-fragment | NA | NA | 0 |
| S16_extracted_42839 | 1995 | No | NA | 2 | 0 | 1 | Not-determined | Genome-fragment | NA | NA | 0 |
| S16_extracted_42892 | 5453 | No | NA | 7 | 0 | 1 | Not-determined | Genome-fragment | NA | NA | 0 |

| | | | | | | | | | | | |
|---|---|---|---|---|---|---|---|---|---|---|---|
| S3_vs_k141_80678\|\|full | 2483 | No | NA | 2 | 0 | 0 | Low-quality | Genome-fragment | 6.55 | AAI-based (medium-confidence) | 0 |
| S16_extracted_42930 | 1657 | No | NA | 2 | 0 | 1 | Not-determined | Genome-fragment | NA | NA | 0 |
| S16_extracted_42980 | 1308 | No | NA | 3 | 0 | 0 | Not-determined | Genome-fragment | NA | NA | 0 |
| S16_extracted_43044 | 4474 | No | NA | 4 | 0 | 2 | Not-determined | Genome-fragment | NA | NA | 0 |
| S3_vs_k141_3413\|\|full | 2483 | No | NA | 3 | 0 | 2 | Low-quality | Genome-fragment | 1.45 | AAI-based (medium-confidence) | 0 |
| S16_extracted_43145 | 1146 | No | NA | 1 | 0 | 1 | Not-determined | Genome-fragment | NA | NA | 0 |
| S16_extracted_43200 | 1262 | No | NA | 1 | 0 | 1 | Not-determined | Genome-fragment | NA | NA | 0 |
| S16_extracted_43201 | 3566 | No | NA | 4 | 0 | 4 | Not-determined | Genome-fragment | NA | NA | 0 |
| S5_extracted_k141_142011 | 2483 | No | NA | 2 | 2 | 0 | Low-quality | Genome-fragment | 1.78 | AAI-based (medium-confidence) | 0 |
| S5_vs_k141_84843\|\|full | 2483 | No | NA | 5 | 0 | 0 | Low-quality | Genome-fragment | 7.4 | AAI-based (high-confidence) | 0 |
| S16_extracted_43288 | 1042 | No | NA | 1 | 0 | 1 | Not-determined | Genome-fragment | NA | NA | 0 |
| S16_extracted_43293 | 3704 | No | NA | 7 | 0 | 2 | Not-determined | Genome-fragment | NA | NA | 0 |
| S16_extracted_20056 | 2483 | No | NA | 3 | 0 | 1 | Low-quality | Genome-fragment | 3.71 | AAI-based (medium-confidence) | 0 |
| S16_extracted_43409 | 1794 | No | NA | 1 | 0 | 1 | Not-determined | Genome-fragment | NA | NA | 0 |
| S16_extracted_43547 | 3174 | No | NA | 2 | 0 | 1 | Not-determined | Genome-fragment | NA | NA | 0 |
| S16_extracted_43576 | 3674 | No | NA | 2 | 0 | 1 | Not-determined | Genome-fragment | NA | NA | 0 |
| S16_vs_2763\|\|full | 2483 | No | NA | 3 | 0 | 0 | Low-quality | Genome-fragment | 1.94 | AAI-based (high-confidence) | 0 |
| S16_vs_155864\|\|full | 2483 | No | NA | 9 | 0 | 0 | Low-quality | Genome-fragment | 3.08 | AAI-based (high-confidence) | 0 |
| S10_vs_115935\|\|full | 2483 | No | NA | 6 | 4 | 0 | Low-quality | Genome-fragment | 6.1 | AAI-based (high-confidence) | 0 |
| S16_extracted_43779 | 9747 | No | NA | 16 | 0 | 2 | Not-determined | Genome-fragment | NA | NA | 0 |
| S16_extracted_43780 | 4653 | No | NA | 6 | 0 | 1 | Not-determined | Genome-fragment | NA | NA | 0 |
| S16_extracted_43823 | 6242 | No | NA | 7 | 0 | 3 | Not-determined | Genome-fragment | NA | NA | 0 |
| S11_vs_k141_79993\|\|full | 2484 | No | NA | 3 | 3 | 0 | Low-quality | Genome-fragment | 6.1 | AAI-based (high-confidence) | 0 |
| S14_extracted_k141_52280 | 2484 | No | NA | 6 | 6 | 0 | Low-quality | Genome-fragment | 8.17 | AAI-based (high-confidence) | 0 |
| S16_extracted_43997 | 1198 | No | NA | 2 | 0 | 1 | Not-determined | Genome-fragment | NA | NA | 0 |
| S16_extracted_44071 | 2151 | No | NA | 1 | 0 | 0 | Not-determined | Genome-fragment | NA | NA | 0 |
| S16_extracted_44222 | 2062 | No | NA | 3 | 0 | 3 | Not-determined | Genome-fragment | NA | NA | 0 |
| S16_extracted_44259 | 3071 | No | NA | 4 | 0 | 2 | Not-determined | Genome-fragment | NA | NA | 0 |
| S3_extracted_k141_25733 | 2484 | No | NA | 4 | 3 | 0 | Low-quality | Genome-fragment | 11.33 | AAI-based (high-confidence) | 0 |
| S16_extracted_44302 | 3905 | No | NA | 2 | 0 | 1 | Not-determined | Genome-fragment | NA | NA | 0 |
| S16_extracted_44372 | 5128 | No | NA | 6 | 0 | 3 | Not-determined | Genome-fragment | NA | NA | 0 |
| S16_extracted_44485 | 1071 | No | NA | 1 | 0 | 0 | Not-determined | Genome-fragment | NA | NA | 0 |
| S12_vs_k141_19011\|\|full | 2484 | Yes | 1998 | 4 | 1 | 2 | Low-quality | Genome-fragment | 0.96 | AAI-based (medium-confidence) | 19.57 |
| S14_vs_k141_45526\|\|full | 2485 | No | NA | 5 | 0 | 0 | Low-quality | Genome-fragment | 6.82 | AAI-based (medium-confidence) | 0 |
| S16_extracted_44695 | 1110 | No | NA | 1 | 0 | 0 | Not-determined | Genome-fragment | NA | NA | 0 |
| S16_extracted_44815 | 8912 | No | NA | 9 | 0 | 5 | Not-determined | Genome-fragment | NA | NA | 0 |
| S16_extracted_44849 | 1212 | No | NA | 2 | 0 | 0 | Not-determined | Genome-fragment | NA | NA | 0 |
| S1_vs_k141_107970\|\|full | 2485 | No | NA | 3 | 0 | 1 | Low-quality | Genome-fragment | 5.34 | AAI-based (medium-confidence) | 0 |
| S16_extracted_45070 | 1222 | No | NA | 1 | 0 | 0 | Not-determined | Genome-fragment | NA | NA | 0 |
| S3_vs_k141_120529\|\|lt2gene | 2485 | No | NA | 2 | 0 | 1 | Low-quality | Genome-fragment | 2.49 | AAI-based (medium-confidence) | 0 |
| S16_extracted_45142 | 1889 | No | NA | 2 | 0 | 2 | Not-determined | Genome-fragment | NA | NA | 0 |
| S16_vs_26903\|\|full | 2486 | No | NA | 2 | 0 | 2 | Low-quality | Genome-fragment | 0.6 | AAI-based (medium-confidence) | 0 |
| S16_extracted_45185 | 1309 | No | NA | 1 | 0 | 1 | Not-determined | Genome-fragment | NA | NA | 0 |
| S16_extracted_45305 | 1057 | No | NA | 2 | 0 | 1 | Not-determined | Genome-fragment | NA | NA | 0 |
| S16_extracted_45342 | 1559 | No | NA | 1 | 0 | 0 | Not-determined | Genome-fragment | NA | NA | 0 |
| S16_extracted_45371 | 24589 | No | NA | 12 | 0 | 6 | Not-determined | Genome-fragment | NA | NA | 0 |
| S9_vs_k141_61431\|\|full | 2487 | No | NA | 3 | 0 | 1 | Low-quality | Genome-fragment | 5.8 | AAI-based (high-confidence) | 0 |
| S13_extracted_109815 | 2487 | No | NA | 7 | 1 | 0 | Low-quality | Genome-fragment | 9.15 | HMM-based (lower-bound) | 0 |
| S14_vs_k141_107416\|\|full | 2487 | No | NA | 7 | 4 | 0 | Low-quality | Genome-fragment | 4.68 | AAI-based (high-confidence) | 0 |
| S16_extracted_45616 | 1509 | No | NA | 2 | 0 | 2 | Not-determined | Genome-fragment | NA | NA | 0 |
| S3_vs_k141_100083\|\|full | 2487 | No | NA | 5 | 1 | 0 | Low-quality | Genome-fragment | 2.58 | AAI-based (medium-confidence) | 0 |
| S16_extracted_45676 | 2419 | No | NA | 3 | 0 | 2 | Not-determined | Genome-fragment | NA | NA | 0 |
| S16_extracted_45736 | 1015 | No | NA | 2 | 0 | 0 | Not-determined | Genome-fragment | NA | NA | 0 |
| S9_extracted_k141_66428 | 2488 | No | NA | 5 | 1 | 0 | Low-quality | Genome-fragment | 5.24 | AAI-based (high-confidence) | 0 |
| S16_extracted_45776 | 1401 | No | NA | 4 | 0 | 0 | Not-determined | Genome-fragment | NA | NA | 0 |
| S16_extracted_45797 | 1456 | No | NA | 1 | 0 | 1 | Not-determined | Genome-fragment | NA | NA | 0 |
| S9_vs_k141_73896\|\|full | 2489 | No | NA | 2 | 0 | 2 | Low-quality | Genome-fragment | 2.32 | AAI-based (medium-confidence) | 0 |
| S16_extracted_45955 | 1181 | No | NA | 2 | 0 | 2 | Not-determined | Genome-fragment | NA | NA | 0 |
| S11_vs_k141_38289\|\|full | 2489 | No | NA | 2 | 0 | 0 | Low-quality | Genome-fragment | 3.88 | AAI-based (high-confidence) | 0 |
| S16_extracted_45979 | 1201 | No | NA | 1 | 0 | 1 | Not-determined | Genome-fragment | NA | NA | 0 |
| S16_vs_21967\|\|full | 2489 | No | NA | 3 | 0 | 1 | Low-quality | Genome-fragment | 1.28 | AAI-based (medium-confidence) | 0 |
| S16_extracted_46192 | 1009 | No | NA | 2 | 0 | 1 | Not-determined | Genome-fragment | NA | NA | 0 |
| S16_extracted_46213 | 3882 | No | NA | 2 | 0 | 1 | Not-determined | Genome-fragment | NA | NA | 0 |
| S16_extracted_46650 | 1031 | No | NA | 1 | 0 | 0 | Not-determined | Genome-fragment | NA | NA | 0 |
| S16_extracted_46653 | 3973 | No | NA | 7 | 0 | 3 | Not-determined | Genome-fragment | NA | NA | 0 |
| S16_extracted_46659 | 5300 | No | NA | 3 | 0 | 2 | Not-determined | Genome-fragment | NA | NA | 0 |
| S16_extracted_46761 | 3069 | No | NA | 1 | 0 | 1 | Not-determined | Genome-fragment | NA | NA | 0 |
| S16_extracted_46800 | 1126 | No | NA | 2 | 0 | 1 | Not-determined | Genome-fragment | NA | NA | 0 |
| S16_extracted_46839 | 4233 | No | NA | 5 | 0 | 3 | Not-determined | Genome-fragment | NA | NA | 0 |
| S10_vs_3328\|\|full | 2489 | No | NA | 4 | 0 | 1 | Low-quality | Genome-fragment | 1.34 | AAI-based (medium-confidence) | 0 |
| S16_extracted_46953 | 1545 | No | NA | 2 | 0 | 0 | Not-determined | Genome-fragment | NA | NA | 0 |
| S16_extracted_46987 | 1198 | No | NA | 1 | 0 | 0 | Not-determined | Genome-fragment | NA | NA | 0 |
| S16_extracted_47104 | 1732 | No | NA | 3 | 0 | 2 | Not-determined | Genome-fragment | NA | NA | 0 |
| S16_extracted_47179 | 2158 | No | NA | 3 | 0 | 1 | Not-determined | Genome-fragment | NA | NA | 0 |
| S16_extracted_47270 | 2487 | No | NA | 3 | 0 | 0 | Not-determined | Genome-fragment | NA | NA | 0 |
| S16_extracted_47277 | 1869 | No | NA | 3 | 0 | 1 | Not-determined | Genome-fragment | NA | NA | 0 |
| S16_extracted_47323 | 2974 | No | NA | 3 | 0 | 0 | Not-determined | Genome-fragment | NA | NA | 0 |
| S17_vs_k141_174324\|\|full | 2490 | No | NA | 3 | 0 | 2 | Low-quality | Genome-fragment | 1.99 | AAI-based (high-confidence) | 0 |
| S16_extracted_47400 | 2344 | No | NA | 2 | 0 | 1 | Not-determined | Genome-fragment | NA | NA | 0 |
| S16_extracted_47412 | 1222 | No | NA | 2 | 0 | 1 | Not-determined | Genome-fragment | NA | NA | 0 |
| S3_extracted_k141_84166 | 2490 | No | NA | 2 | 0 | 0 | Low-quality | Genome-fragment | 4.57 | AAI-based (high-confidence) | 0 |
| S16_extracted_47757 | 1023 | No | NA | 1 | 0 | 1 | Not-determined | Genome-fragment | NA | NA | 0 |
| S5_vs_k141_54877\|\|full | 2490 | No | NA | 7 | 1 | 0 | Low-quality | Genome-fragment | 1.7 | HMM-based (lower-bound) | 0 |
| S16_extracted_47819 | 1165 | No | NA | 3 | 0 | 0 | Not-determined | Genome-fragment | NA | NA | 0 |
| S16_extracted_47851 | 1122 | No | NA | 3 | 0 | 0 | Not-determined | Genome-fragment | NA | NA | 0 |
| S16_extracted_47949 | 1161 | No | NA | 2 | 0 | 2 | Not-determined | Genome-fragment | NA | NA | 0 |
| S16_extracted_47983 | 1615 | No | NA | 2 | 0 | 0 | Not-determined | Genome-fragment | NA | NA | 0 |
| S5_vs_k141_123865\|\|full | 2490 | No | NA | 5 | 0 | 3 | Low-quality | Genome-fragment | 2.01 | AAI-based (medium-confidence) | 0 |
| S16_extracted_48016 | 4926 | No | NA | 7 | 0 | 3 | Not-determined | Genome-fragment | NA | NA | 0 |
| S10_extracted_24763 | 2490 | No | NA | 4 | 0 | 0 | Low-quality | Genome-fragment | 5.44 | AAI-based (high-confidence) | 0 |
| S16_extracted_48247 | 6170 | No | NA | 8 | 0 | 5 | Not-determined | Genome-fragment | NA | NA | 0 |
| S10_vs_112570\|\|full | 2491 | No | NA | 3 | 2 | 0 | Low-quality | Genome-fragment | 5.27 | AAI-based (high-confidence) | 0 |
| S16_extracted_48319 | 3449 | No | NA | 2 | 0 | 2 | Not-determined | Genome-fragment | NA | NA | 0 |
| S16_extracted_48379 | 2191 | No | NA | 2 | 0 | 1 | Not-determined | Genome-fragment | NA | NA | 0 |
| S12_vs_k141_24396\|\|full | 2491 | No | NA | 4 | 1 | 0 | Low-quality | Genome-fragment | 4.56 | HMM-based (lower-bound) | 0 |
| S12_vs_k141_27934\|\|full | 2491 | No | NA | 4 | 0 | 0 | Low-quality | Genome-fragment | 2.82 | AAI-based (medium-confidence) | 0 |
| S15_vs_71538\|\|full | 2491 | No | NA | 4 | 0 | 0 | Low-quality | Genome-fragment | 1.44 | AAI-based (high-confidence) | 0 |
| S5_vs_k141_86735\|\|full | 2492 | No | NA | 5 | 1 | 0 | Low-quality | Genome-fragment | 4.32 | AAI-based (high-confidence) | 0 |
| S5_vs_k141_134972\|\|full | 2492 | No | NA | 6 | 5 | 0 | Low-quality | Genome-fragment | 5.89 | AAI-based (high-confidence) | 0 |
| S16_extracted_48697 | 7574 | No | NA | 7 | 0 | 4 | Not-determined | Genome-fragment | NA | NA | 0 |
| S16_extracted_48809 | 1156 | No | NA | 1 | 0 | 0 | Not-determined | Genome-fragment | NA | NA | 0 |
| S16_extracted_48923 | 1419 | No | NA | 1 | 0 | 1 | Not-determined | Genome-fragment | NA | NA | 0 |
| S16_extracted_49006 | 1782 | No | NA | 4 | 0 | 1 | Not-determined | Genome-fragment | NA | NA | 0 |
| S16_extracted_49090 | 9129 | No | NA | 8 | 0 | 6 | Not-determined | Genome-fragment | NA | NA | 0 |
| S7_vs_77963\|\|full | 2492 | No | NA | 3 | 2 | 0 | Low-quality | Genome-fragment | 5.23 | AAI-based (high-confidence) | 0 |
| S7_vs_81106\|\|full | 2492 | No | NA | 6 | 1 | 0 | Low-quality | Genome-fragment | 7.67 | AAI-based (high-confidence) | 0 |
| S16_extracted_49360 | 1083 | No | NA | 2 | 0 | 2 | Not-determined | Genome-fragment | NA | NA | 0 |
| S16_extracted_49376 | 1289 | No | NA | 1 | 0 | 1 | Not-determined | Genome-fragment | NA | NA | 0 |
| S16_extracted_49409 | 6689 | No | NA | 4 | 0 | 3 | Not-determined | Genome-fragment | NA | NA | 0 |
| S7_vs_97779\|\|full | 2492 | No | NA | 5 | 0 | 0 | Low-quality | Genome-fragment | 1.53 | AAI-based (high-confidence) | 0 |
| S16_extracted_49534 | 1363 | No | NA | 3 | 0 | 0 | Not-determined | Genome-fragment | NA | NA | 0 |
| S8_vs_k141_3894\|\|full | 2492 | No | NA | 2 | 0 | 2 | Low-quality | Genome-fragment | 3.02 | AAI-based (high-confidence) | 0 |
| S16_extracted_49608 | 1091 | No | NA | 1 | 0 | 1 | Not-determined | Genome-fragment | NA | NA | 0 |
| S16_extracted_49720 | 1475 | No | NA | 1 | 0 | 0 | Not-determined | Genome-fragment | NA | NA | 0 |
| S16_extracted_49789 | 1700 | No | NA | 3 | 0 | 3 | Not-determined | Genome-fragment | NA | NA | 0 |
| S5_extracted_k141_73344 | 2493 | No | NA | 4 | 1 | 0 | Low-quality | Genome-fragment | 6.16 | AAI-based (medium-confidence) | 0 |
| S16_extracted_50080 | 1003 | No | NA | 1 | 0 | 1 | Not-determined | Genome-fragment | NA | NA | 0 |
| S16_extracted_50125 | 4081 | No | NA | 4 | 0 | 1 | Not-determined | Genome-fragment | NA | NA | 0 |
| S16_extracted_50166 | 1582 | No | NA | 1 | 0 | 1 | Not-determined | Genome-fragment | NA | NA | 0 |
| S16_extracted_50205 | 1559 | No | NA | 2 | 0 | 0 | Not-determined | Genome-fragment | NA | NA | 0 |
| S16_extracted_50295 | 2705 | No | NA | 3 | 0 | 1 | Not-determined | Genome-fragment | NA | NA | 0 |
| S16_extracted_50309 | 4116 | No | NA | 4 | 0 | 4 | Not-determined | Genome-fragment | NA | NA | 0 |
| S16_extracted_50354 | 4146 | No | NA | 4 | 0 | 2 | Not-determined | Genome-fragment | NA | NA | 0 |
| S16_extracted_50360 | 1018 | No | NA | 1 | 0 | 1 | Not-determined | Genome-fragment | NA | NA | 0 |
| S16_extracted_50503 | 1744 | No | NA | 2 | 0 | 1 | Not-determined | Genome-fragment | NA | NA | 0 |
| S5_vs_k141_140279\|\|full | 2493 | No | NA | 5 | 1 | 0 | Low-quality | Genome-fragment | 2.69 | AAI-based (medium-confidence) | 0 |
| S16_extracted_50671 | 1002 | No | NA | 2 | 0 | 0 | Not-determined | Genome-fragment | NA | NA | 0 |
| S10_vs_100440\|\|full | 2493 | No | NA | 3 | 1 | 0 | Low-quality | Genome-fragment | 1.93 | HMM-based (lower-bound) | 0 |
| S16_extracted_50780 | 1661 | No | NA | 3 | 0 | 0 | Not-determined | Genome-fragment | NA | NA | 0 |
| S16_extracted_50945 | 3102 | No | NA | 3 | 0 | 0 | Not-determined | Genome-fragment | NA | NA | 0 |
| S14_vs_k141_7943\|\|full | 2494 | Yes | 1030 | 5 | 1 | 1 | Low-quality | Genome-fragment | 0.63 | HMM-based (lower-bound) | 58.7 |
| S16_extracted_51054 | 1178 | No | NA | 1 | 0 | 1 | Not-determined | Genome-fragment | NA | NA | 0 |
| S16_extracted_51057 | 1438 | No | NA | 1 | 0 | 0 | Not-determined | Genome-fragment | NA | NA | 0 |
| S16_extracted_51101 | 3587 | No | NA | 4 | 0 | 1 | Not-determined | Genome-fragment | NA | NA | 0 |
| S16_extracted_51242 | 4409 | No | NA | 5 | 0 | 2 | Not-determined | Genome-fragment | NA | NA | 0 |
| S16_extracted_51307 | 1153 | No | NA | 1 | 0 | 1 | Not-determined | Genome-fragment | NA | NA | 0 |
| S16_extracted_51363 | 1101 | No | NA | 2 | 0 | 0 | Not-determined | Genome-fragment | NA | NA | 0 |
| S16_extracted_51367 | 1314 | No | NA | 1 | 0 | 1 | Not-determined | Genome-fragment | NA | NA | 0 |
| S1_vs_k141_99146\|\|full | 2494 | No | NA | 5 | 1 | 0 | Low-quality | Genome-fragment | 6.4 | AAI-based (medium-confidence) | 0 |
| S16_extracted_51543 | 2190 | No | NA | 2 | 0 | 0 | Not-determined | Genome-fragment | NA | NA | 0 |
| S16_extracted_51554 | 5126 | No | NA | 4 | 0 | 3 | Not-determined | Genome-fragment | NA | NA | 0 |
| S5_extracted_k141_101556 | 2494 | No | NA | 3 | 3 | 0 | Low-quality | Genome-fragment | 6.22 | AAI-based (high-confidence) | 0 |
| S16_extracted_51755 | 1669 | No | NA | 3 | 0 | 3 | Not-determined | Genome-fragment | NA | NA | 0 |
| S16_extracted_51774 | 1718 | No | NA | 1 | 0 | 1 | Not-determined | Genome-fragment | NA | NA | 0 |
| S16_vs_119956\|\|full | 2494 | No | NA | 3 | 0 | 0 | Low-quality | Genome-fragment | 4.21 | AAI-based (medium-confidence) | 0 |
| S16_extracted_51988 | 1814 | No | NA | 1 | 0 | 0 | Not-determined | Genome-fragment | NA | NA | 0 |
| S16_extracted_52104 | 2484 | No | NA | 2 | 0 | 2 | Not-determined | Genome-fragment | NA | NA | 0 |
| S16_extracted_52141 | 5518 | No | NA | 3 | 0 | 1 | Not-determined | Genome-fragment | NA | NA | 0 |
| S16_extracted_52186 | 1248 | No | NA | 2 | 0 | 1 | Not-determined | Genome-fragment | NA | NA | 0 |
| S16_extracted_52197 | 6483 | No | NA | 7 | 0 | 1 | Not-determined | Genome-fragment | NA | NA | 0 |
| S16_extracted_52242 | 1055 | No | NA | 1 | 0 | 1 | Not-determined | Genome-fragment | NA | NA | 0 |
| S16_extracted_52322 | 1873 | No | NA | 2 | 0 | 2 | Not-determined | Genome-fragment | NA | NA | 0 |
| S12_vs_k141_75054\|\|full | 2494 | No | NA | 6 | 2 | 0 | Low-quality | Genome-fragment | 5.9 | AAI-based (high-confidence) | 0 |
| S16_extracted_52417 | 4621 | No | NA | 8 | 0 | 0 | Not-determined | Genome-fragment | NA | NA | 0 |
| S16_extracted_52442 | 1796 | No | NA | 1 | 0 | 0 | Not-determined | Genome-fragment | NA | NA | 0 |
| S16_extracted_52513 | 40335 | No | NA | 75 | 14 | 2 | High-quality | High-quality | 90.74 | AAI-based (high-confidence) | 0 |
| S16_extracted_52550 | 1841 | No | NA | 2 | 0 | 1 | Not-determined | Genome-fragment | NA | NA | 0 |
| S16_extracted_52564 | 1571 | No | NA | 2 | 0 | 1 | Not-determined | Genome-fragment | NA | NA | 0 |
| S13_vs_37633\|\|full | 2495 | No | NA | 3 | 0 | 2 | Low-quality | Genome-fragment | 1.98 | AAI-based (medium-confidence) | 0 |
| S1_extracted_k141_36402 | 2495 | No | NA | 4 | 0 | 0 | Low-quality | Genome-fragment | 3.1 | AAI-based (high-confidence) | 0 |
| S5_vs_k141_42921\|\|full | 2495 | No | NA | 3 | 0 | 0 | Low-quality | Genome-fragment | 2.19 | AAI-based (high-confidence) | 0 |
| S16_extracted_52727 | 1407 | No | NA | 1 | 0 | 1 | Not-determined | Genome-fragment | NA | NA | 0 |
| S16_extracted_52779 | 1079 | No | NA | 2 | 0 | 2 | Not-determined | Genome-fragment | NA | NA | 0 |
| S16_extracted_52814 | 4230 | No | NA | 5 | 0 | 3 | Not-determined | Genome-fragment | NA | NA | 0 |
| S16_extracted_52821 | 1088 | No | NA | 4 | 0 | 0 | Not-determined | Genome-fragment | NA | NA | 0 |
| S16_extracted_52882 | 1714 | No | NA | 3 | 0 | 1 | Not-determined | Genome-fragment | NA | NA | 0 |
| S16_extracted_52908 | 2730 | No | NA | 4 | 0 | 2 | Not-determined | Genome-fragment | NA | NA | 0 |
| S16_extracted_53243 | 8151 | No | NA | 8 | 0 | 3 | Not-determined | Genome-fragment | NA | NA | 0 |
| S16_extracted_53274 | 1331 | No | NA | 1 | 0 | 1 | Not-determined | Genome-fragment | NA | NA | 0 |
| S16_extracted_53301 | 1993 | No | NA | 3 | 0 | 0 | Not-determined | Genome-fragment | NA | NA | 0 |
| S16_extracted_53334 | 1035 | No | NA | 1 | 0 | 1 | Not-determined | Genome-fragment | NA | NA | 0 |
| S16_extracted_53366 | 1320 | No | NA | 1 | 0 | 1 | Not-determined | Genome-fragment | NA | NA | 0 |
| S16_extracted_53417 | 1176 | No | NA | 2 | 0 | 1 | Not-determined | Genome-fragment | NA | NA | 0 |
| S6_vs_k141_7675\|\|full | 2495 | No | NA | 6 | 0 | 0 | Low-quality | Genome-fragment | 6.55 | AAI-based (high-confidence) | 0 |
| S16_extracted_53507 | 3234 | No | NA | 4 | 0 | 1 | Not-determined | Genome-fragment | NA | NA | 0 |
| S16_extracted_53525 | 1350 | No | NA | 1 | 0 | 1 | Not-determined | Genome-fragment | NA | NA | 0 |
| S16_extracted_53537 | 1496 | No | NA | 2 | 0 | 1 | Not-determined | Genome-fragment | NA | NA | 0 |
| S13_vs_121819\|\|full | 2496 | No | NA | 7 | 0 | 0 | Low-quality | Genome-fragment | 5.83 | AAI-based (high-confidence) | 0 |
| S16_extracted_53541 | 1401 | No | NA | 3 | 0 | 2 | Not-determined | Genome-fragment | NA | NA | 0 |
| S16_extracted_53555 | 1016 | No | NA | 1 | 0 | 0 | Not-determined | Genome-fragment | NA | NA | 0 |
| S16_extracted_53642 | 2433 | No | NA | 2 | 0 | 1 | Not-determined | Genome-fragment | NA | NA | 0 |
| S16_extracted_53673 | 1633 | No | NA | 3 | 0 | 2 | Not-determined | Genome-fragment | NA | NA | 0 |
| S16_extracted_53758 | 2090 | No | NA | 1 | 0 | 0 | Not-determined | Genome-fragment | NA | NA | 0 |
| S16_extracted_53766 | 1366 | No | NA | 1 | 0 | 1 | Not-determined | Genome-fragment | NA | NA | 0 |
| S16_extracted_53961 | 1531 | No | NA | 1 | 0 | 1 | Not-determined | Genome-fragment | NA | NA | 0 |
| S5_vs_k141_113313\|\|full | 2496 | No | NA | 7 | 2 | 0 | Low-quality | Genome-fragment | 5.22 | AAI-based (medium-confidence) | 0 |
| S16_extracted_54052 | 1049 | No | NA | 1 | 0 | 0 | Not-determined | Genome-fragment | NA | NA | 0 |
| S16_extracted_54089 | 1629 | No | NA | 2 | 0 | 2 | Not-determined | Genome-fragment | NA | NA | 0 |
| S10_vs_22903\|\|full | 2496 | No | NA | 4 | 0 | 0 | Low-quality | Genome-fragment | 3.57 | AAI-based (high-confidence) | 0 |
| S16_extracted_54314 | 2272 | No | NA | 2 | 0 | 2 | Not-determined | Genome-fragment | NA | NA | 0 |
| S16_extracted_54315 | 1310 | No | NA | 1 | 0 | 1 | Not-determined | Genome-fragment | NA | NA | 0 |
| S16_extracted_54329 | 1318 | No | NA | 2 | 0 | 1 | Not-determined | Genome-fragment | NA | NA | 0 |
| S16_extracted_54335 | 1070 | No | NA | 2 | 0 | 1 | Not-determined | Genome-fragment | NA | NA | 0 |
| S16_extracted_54519 | 4242 | No | NA | 5 | 0 | 0 | Not-determined | Genome-fragment | NA | NA | 0 |
| S16_extracted_54711 | 3000 | No | NA | 2 | 0 | 1 | Not-determined | Genome-fragment | NA | NA | 0 |
| S8_vs_k141_167463\|\|full | 2496 | No | NA | 4 | 1 | 0 | Low-quality | Genome-fragment | 4.82 | AAI-based (medium-confidence) | 0 |
| S16_extracted_54864 | 1145 | No | NA | 1 | 0 | 1 | Not-determined | Genome-fragment | NA | NA | 0 |
| S16_extracted_54980 | 1664 | No | NA | 2 | 0 | 1 | Not-determined | Genome-fragment | NA | NA | 0 |
| S16_extracted_55120 | 1220 | No | NA | 1 | 0 | 1 | Not-determined | Genome-fragment | NA | NA | 0 |
| S9_vs_k141_43191\|\|full | 2497 | No | NA | 4 | 3 | 0 | Low-quality | Genome-fragment | 4.1 | HMM-based (lower-bound) | 0 |
| S16_extracted_55167 | 1299 | No | NA | 2 | 0 | 0 | Not-determined | Genome-fragment | NA | NA | 0 |
| S16_extracted_55263 | 1036 | No | NA | 2 | 0 | 0 | Not-determined | Genome-fragment | NA | NA | 0 |
| S16_extracted_55459 | 2088 | No | NA | 4 | 0 | 3 | Not-determined | Genome-fragment | NA | NA | 0 |
| S16_extracted_55657 | 8344 | No | NA | 7 | 0 | 4 | Not-determined | Genome-fragment | NA | NA | 0 |
| S16_extracted_55696 | 5619 | No | NA | 4 | 0 | 3 | Not-determined | Genome-fragment | NA | NA | 0 |
| S16_extracted_55701 | 1076 | No | NA | 2 | 0 | 1 | Not-determined | Genome-fragment | NA | NA | 0 |
| S1_extracted_k141_98807 | 2497 | No | NA | 1 | 1 | 0 | Low-quality | Genome-fragment | 6.96 | AAI-based (high-confidence) | 0 |
| S16_extracted_55752 | 1002 | No | NA | 2 | 0 | 0 | Not-determined | Genome-fragment | NA | NA | 0 |
| S16_vs_132040\|\|full | 2497 | No | NA | 4 | 0 | 1 | Low-quality | Genome-fragment | 1.37 | AAI-based (medium-confidence) | 0 |
| S16_extracted_55988 | 1005 | No | NA | 1 | 0 | 0 | Not-determined | Genome-fragment | NA | NA | 0 |
| S16_extracted_55998 | 2967 | No | NA | 3 | 0 | 1 | Not-determined | Genome-fragment | NA | NA | 0 |
| S8_vs_k141_64170\|\|full | 2497 | No | NA | 4 | 0 | 1 | Low-quality | Genome-fragment | 2.43 | AAI-based (high-confidence) | 0 |
| S16_extracted_56028 | 1121 | No | NA | 2 | 0 | 0 | Not-determined | Genome-fragment | NA | NA | 0 |
| S8_vs_k141_30514\|\|full | 2497 | No | NA | 2 | 0 | 2 | Low-quality | Genome-fragment | 2.19 | AAI-based (medium-confidence) | 0 |
| S16_extracted_56311 | 1358 | No | NA | 1 | 0 | 0 | Not-determined | Genome-fragment | NA | NA | 0 |
| S16_extracted_56354 | 5385 | No | NA | 6 | 0 | 2 | Not-determined | Genome-fragment | NA | NA | 0 |
| S16_extracted_56384 | 1802 | No | NA | 2 | 0 | 0 | Not-determined | Genome-fragment | NA | NA | 0 |
| S16_extracted_56550 | 1624 | No | NA | 2 | 0 | 0 | Not-determined | Genome-fragment | NA | NA | 0 |
| S16_extracted_56732 | 1265 | No | NA | 2 | 0 | 1 | Not-determined | Genome-fragment | NA | NA | 0 |
| S16_extracted_56787 | 3567 | No | NA | 6 | 0 | 5 | Not-determined | Genome-fragment | NA | NA | 0 |
| S16_extracted_56788 | 1079 | No | NA | 1 | 0 | 0 | Not-determined | Genome-fragment | NA | NA | 0 |
| S16_extracted_56856 | 3330 | No | NA | 6 | 0 | 6 | Not-determined | Genome-fragment | NA | NA | 0 |
| S16_extracted_56913 | 3006 | No | NA | 4 | 0 | 2 | Not-determined | Genome-fragment | NA | NA | 0 |
| S16_extracted_56931 | 1110 | No | NA | 1 | 0 | 0 | Not-determined | Genome-fragment | NA | NA | 0 |
| S16_extracted_56984 | 1247 | No | NA | 2 | 0 | 2 | Not-determined | Genome-fragment | NA | NA | 0 |
| S16_extracted_57191 | 2397 | No | NA | 2 | 0 | 1 | Not-determined | Genome-fragment | NA | NA | 0 |
| S16_extracted_57232 | 5983 | No | NA | 6 | 0 | 1 | Not-determined | Genome-fragment | NA | NA | 0 |
| S17_vs_k141_102941\|\|full | 2498 | No | NA | 3 | 0 | 2 | Low-quality | Genome-fragment | 5.36 | AAI-based (high-confidence) | 0 |
| S5_vs_k141_122002\|\|full | 2498 | No | NA | 3 | 1 | 0 | Low-quality | Genome-fragment | 6 | AAI-based (medium-confidence) | 0 |
| S16_extracted_57450 | 1008 | No | NA | 1 | 0 | 0 | Not-determined | Genome-fragment | NA | NA | 0 |
| S10_extracted_109174 | 2498 | No | NA | 1 | 0 | 0 | Low-quality | Genome-fragment | 1.02 | AAI-based (high-confidence) | 0 |
| S12_vs_k141_158866\|\|full | 2498 | No | NA | 6 | 2 | 0 | Low-quality | Genome-fragment | 6.63 | AAI-based (medium-confidence) | 0 |
| S16_extracted_57497 | 3116 | No | NA | 4 | 0 | 2 | Not-determined | Genome-fragment | NA | NA | 0 |
| S8_vs_k141_55525\|\|full | 2498 | No | NA | 2 | 1 | 0 | Low-quality | Genome-fragment | 1.57 | AAI-based (high-confidence) | 0 |
| S16_extracted_57539 | 1536 | No | NA | 2 | 0 | 1 | Not-determined | Genome-fragment | NA | NA | 0 |
| S8_extracted_k141_20132 | 2499 | No | NA | 4 | 0 | 0 | Low-quality | Genome-fragment | 1.37 | AAI-based (high-confidence) | 0 |
| S16_extracted_57723 | 1194 | No | NA | 1 | 0 | 0 | Not-determined | Genome-fragment | NA | NA | 0 |
| S17_vs_k141_117611\|\|full | 2500 | No | NA | 4 | 0 | 1 | Low-quality | Genome-fragment | 1.41 | AAI-based (medium-confidence) | 0 |
| S16_extracted_57814 | 3175 | No | NA | 1 | 0 | 0 | Not-determined | Genome-fragment | NA | NA | 0 |
| S16_extracted_58019 | 5304 | No | NA | 6 | 0 | 5 | Not-determined | Genome-fragment | NA | NA | 0 |
| S16_extracted_58237 | 3695 | No | NA | 2 | 0 | 2 | Not-determined | Genome-fragment | NA | NA | 0 |
| S16_extracted_58282 | 1704 | No | NA | 1 | 0 | 1 | Not-determined | Genome-fragment | NA | NA | 0 |
| S16_extracted_58338 | 11177 | No | NA | 11 | 0 | 7 | Not-determined | Genome-fragment | NA | NA | 0 |
| S5_vs_k141_110789\|\|full | 2500 | No | NA | 5 | 1 | 0 | Low-quality | Genome-fragment | 6.69 | AAI-based (medium-confidence) | 0 |
| S16_extracted_58551 | 3419 | No | NA | 5 | 0 | 1 | Not-determined | Genome-fragment | NA | NA | 0 |
| S5_vs_k141_80289\|\|full | 2500 | No | NA | 5 | 0 | 1 | Low-quality | Genome-fragment | 2.66 | AAI-based (medium-confidence) | 0 |
| S17_extracted_k141_110066 | 2501 | No | NA | 3 | 0 | 1 | Low-quality | Genome-fragment | 2.19 | AAI-based (medium-confidence) | 0 |
| S13_vs_99806\|\|full | 2501 | No | NA | 3 | 0 | 2 | Low-quality | Genome-fragment | 1.77 | AAI-based (high-confidence) | 0 |
| S16_extracted_58850 | 2046 | No | NA | 2 | 0 | 0 | Not-determined | Genome-fragment | NA | NA | 0 |
| S16_extracted_59493 | 1173 | No | NA | 1 | 0 | 1 | Not-determined | Genome-fragment | NA | NA | 0 |
| S16_extracted_59550 | 1136 | No | NA | 2 | 0 | 1 | Not-determined | Genome-fragment | NA | NA | 0 |
| S16_extracted_59586 | 12076 | No | NA | 9 | 0 | 7 | Not-determined | Genome-fragment | NA | NA | 0 |
| S16_extracted_59680 | 1604 | No | NA | 3 | 0 | 0 | Not-determined | Genome-fragment | NA | NA | 0 |
| S11_vs_k141_92574\|\|full | 2502 | No | NA | 6 | 0 | 0 | Low-quality | Genome-fragment | 3.93 | AAI-based (high-confidence) | 0 |
| S16_extracted_59919 | 1067 | No | NA | 1 | 0 | 1 | Not-determined | Genome-fragment | NA | NA | 0 |
| S16_extracted_60058 | 3388 | No | NA | 1 | 0 | 1 | Not-determined | Genome-fragment | NA | NA | 0 |
| S16_extracted_60070 | 1010 | No | NA | 2 | 0 | 1 | Not-determined | Genome-fragment | NA | NA | 0 |
| S3_vs_k141_36521\|\|full | 2502 | No | NA | 3 | 0 | 3 | Low-quality | Genome-fragment | 0.69 | AAI-based (medium-confidence) | 0 |
| S16_extracted_60129 | 1131 | No | NA | 1 | 0 | 0 | Not-determined | Genome-fragment | NA | NA | 0 |
| S7_vs_5673\|\|full | 2502 | No | NA | 5 | 1 | 0 | Low-quality | Genome-fragment | 5.14 | AAI-based (high-confidence) | 0 |
| S7_vs_120037\|\|full | 2502 | No | NA | 5 | 0 | 0 | Low-quality | Genome-fragment | 4.33 | AAI-based (medium-confidence) | 0 |
| S16_extracted_60320 | 1070 | No | NA | 2 | 0 | 0 | Not-determined | Genome-fragment | NA | NA | 0 |
| S16_extracted_60481 | 3411 | No | NA | 3 | 0 | 2 | Not-determined | Genome-fragment | NA | NA | 0 |
| S16_extracted_60661 | 3024 | No | NA | 4 | 0 | 1 | Not-determined | Genome-fragment | NA | NA | 0 |
| S13_extracted_129200 | 2503 | No | NA | 2 | 0 | 1 | Low-quality | Genome-fragment | 8.72 | AAI-based (medium-confidence) | 0 |
| S16_extracted_131878 | 2504 | No | NA | 1 | 0 | 1 | Low-quality | Genome-fragment | 2.26 | AAI-based (medium-confidence) | 0 |
| S16_extracted_60953 | 3077 | No | NA | 4 | 0 | 0 | Not-determined | Genome-fragment | NA | NA | 0 |
| S16_extracted_60966 | 1229 | No | NA | 1 | 0 | 1 | Not-determined | Genome-fragment | NA | NA | 0 |
| S16_extracted_60973 | 5637 | No | NA | 9 | 0 | 7 | Not-determined | Genome-fragment | NA | NA | 0 |
| S12_vs_k141_19081\|\|full | 2504 | No | NA | 4 | 0 | 2 | Low-quality | Genome-fragment | 1.52 | AAI-based (medium-confidence) | 0 |
| S16_extracted_61125 | 1097 | No | NA | 2 | 0 | 1 | Not-determined | Genome-fragment | NA | NA | 0 |
| S16_extracted_61229 | 3331 | No | NA | 7 | 0 | 0 | Not-determined | Genome-fragment | NA | NA | 0 |
| S16_extracted_61309 | 1114 | No | NA | 1 | 0 | 0 | Not-determined | Genome-fragment | NA | NA | 0 |
| S16_extracted_61322 | 4494 | No | NA | 4 | 0 | 4 | Not-determined | Genome-fragment | NA | NA | 0 |
| S16_extracted_61361 | 6305 | No | NA | 12 | 0 | 1 | Not-determined | Genome-fragment | NA | NA | 0 |
| S16_extracted_61478 | 2197 | No | NA | 3 | 0 | 1 | Not-determined | Genome-fragment | NA | NA | 0 |
| S16_extracted_61489 | 1284 | No | NA | 1 | 0 | 1 | Not-determined | Genome-fragment | NA | NA | 0 |
| S16_extracted_61496 | 8783 | No | NA | 7 | 0 | 6 | Not-determined | Genome-fragment | NA | NA | 0 |
| S6_extracted_k141_6194 | 2504 | No | NA | 3 | 2 | 0 | Low-quality | Genome-fragment | 41.03 | AAI-based (high-confidence) | 0 |
| S16_extracted_61603 | 1143 | No | NA | 2 | 0 | 0 | Not-determined | Genome-fragment | NA | NA | 0 |
| S9_vs_k141_58202\|\|full | 2505 | No | NA | 3 | 3 | 0 | Low-quality | Genome-fragment | 6.11 | AAI-based (high-confidence) | 0 |
| S16_extracted_61830 | 1044 | No | NA | 2 | 0 | 1 | Not-determined | Genome-fragment | NA | NA | 0 |
| S16_extracted_61947 | 1437 | No | NA | 2 | 0 | 0 | Not-determined | Genome-fragment | NA | NA | 0 |
| S16_extracted_62000 | 1004 | No | NA | 1 | 0 | 1 | Not-determined | Genome-fragment | NA | NA | 0 |
| S16_extracted_62109 | 2660 | No | NA | 2 | 0 | 2 | Not-determined | Genome-fragment | NA | NA | 0 |
| S16_extracted_62203 | 2671 | No | NA | 7 | 0 | 6 | Not-determined | Genome-fragment | NA | NA | 0 |
| S11_vs_k141_50935\|\|full | 2505 | No | NA | 7 | 2 | 0 | Low-quality | Genome-fragment | 6.84 | AAI-based (high-confidence) | 0 |
| S16_extracted_62595 | 1409 | No | NA | 2 | 0 | 0 | Not-determined | Genome-fragment | NA | NA | 0 |
| S16_extracted_62618 | 3580 | No | NA | 4 | 0 | 2 | Not-determined | Genome-fragment | NA | NA | 0 |
| S16_extracted_62672 | 1102 | No | NA | 1 | 0 | 1 | Not-determined | Genome-fragment | NA | NA | 0 |
| S3_extracted_k141_19688 | 2505 | No | NA | 4 | 0 | 0 | Low-quality | Genome-fragment | 3.55 | AAI-based (high-confidence) | 0 |
| S16_extracted_62708 | 1071 | No | NA | 1 | 0 | 1 | Not-determined | Genome-fragment | NA | NA | 0 |
| S16_vs_85209\|\|full | 2505 | No | NA | 3 | 1 | 0 | Low-quality | Genome-fragment | 4.32 | AAI-based (high-confidence) | 0 |
| S16_extracted_62835 | 1570 | No | NA | 1 | 0 | 1 | Not-determined | Genome-fragment | NA | NA | 0 |
| S6_vs_k141_44325\|\|full | 2505 | No | NA | 3 | 3 | 0 | Low-quality | Genome-fragment | 5.12 | AAI-based (high-confidence) | 0 |
| S16_extracted_63195 | 2172 | No | NA | 1 | 0 | 0 | Not-determined | Genome-fragment | NA | NA | 0 |
| S16_extracted_63269 | 1198 | No | NA | 1 | 0 | 1 | Not-determined | Genome-fragment | NA | NA | 0 |
| S16_extracted_63336 | 1542 | No | NA | 1 | 0 | 0 | Not-determined | Genome-fragment | NA | NA | 0 |
| S16_extracted_63376 | 1088 | No | NA | 2 | 0 | 0 | Not-determined | Genome-fragment | NA | NA | 0 |
| S17_vs_k141_26333\|\|full | 2506 | No | NA | 3 | 0 | 1 | Low-quality | Genome-fragment | 8 | AAI-based (high-confidence) | 0 |
| S1_vs_k141_28299\|\|full | 2506 | No | NA | 5 | 0 | 2 | Low-quality | Genome-fragment | 1.12 | AAI-based (medium-confidence) | 0 |
| S16_extracted_63460 | 1075 | No | NA | 1 | 0 | 1 | Not-determined | Genome-fragment | NA | NA | 0 |
| S16_vs_12730\|\|full | 2506 | No | NA | 3 | 0 | 0 | Low-quality | Genome-fragment | 4.05 | AAI-based (high-confidence) | 0 |
| S16_extracted_63557 | 1030 | No | NA | 2 | 0 | 1 | Not-determined | Genome-fragment | NA | NA | 0 |
| S16_extracted_63599 | 1158 | No | NA | 1 | 0 | 1 | Not-determined | Genome-fragment | NA | NA | 0 |
| S14_vs_k141_22051\|\|full | 2507 | No | NA | 3 | 0 | 2 | Low-quality | Genome-fragment | 2.41 | AAI-based (medium-confidence) | 0 |
| S16_extracted_63798 | 1796 | No | NA | 1 | 0 | 0 | Not-determined | Genome-fragment | NA | NA | 0 |
| S16_extracted_63880 | 3024 | No | NA | 1 | 0 | 0 | Not-determined | Genome-fragment | NA | NA | 0 |
| S16_extracted_63968 | 2341 | No | NA | 7 | 0 | 6 | Not-determined | Genome-fragment | NA | NA | 0 |
| S16_extracted_64041 | 1526 | No | NA | 2 | 0 | 1 | Not-determined | Genome-fragment | NA | NA | 0 |
| S16_extracted_64200 | 1034 | No | NA | 1 | 0 | 1 | Not-determined | Genome-fragment | NA | NA | 0 |
| S16_extracted_64341 | 8805 | No | NA | 14 | 0 | 2 | Not-determined | Genome-fragment | NA | NA | 0 |
| S16_extracted_64393 | 3079 | No | NA | 3 | 0 | 0 | Not-determined | Genome-fragment | NA | NA | 0 |
| S16_extracted_64399 | 1669 | No | NA | 1 | 0 | 0 | Not-determined | Genome-fragment | NA | NA | 0 |
| S16_vs_65080\|\|full | 2507 | No | NA | 3 | 0 | 0 | Low-quality | Genome-fragment | 4.93 | AAI-based (medium-confidence) | 0 |
| S16_extracted_64566 | 1166 | No | NA | 1 | 0 | 0 | Not-determined | Genome-fragment | NA | NA | 0 |
| S11_vs_k141_51701\|\|full | 2508 | No | NA | 5 | 0 | 0 | Low-quality | Genome-fragment | 4.24 | AAI-based (high-confidence) | 0 |
| S3_vs_k141_7941\|\|full | 2508 | No | NA | 5 | 0 | 0 | Low-quality | Genome-fragment | 3.16 | AAI-based (high-confidence) | 0 |
| S16_extracted_24661 | 2508 | No | NA | 2 | 0 | 2 | Low-quality | Genome-fragment | 0.61 | AAI-based (high-confidence) | 0 |
| S16_extracted_64700 | 2204 | No | NA | 3 | 0 | 2 | Not-determined | Genome-fragment | NA | NA | 0 |
| S16_extracted_64725 | 3805 | No | NA | 5 | 0 | 2 | Not-determined | Genome-fragment | NA | NA | 0 |
| S16_extracted_64729 | 1045 | No | NA | 2 | 0 | 1 | Not-determined | Genome-fragment | NA | NA | 0 |
| S16_extracted_64781 | 8516 | No | NA | 6 | 0 | 5 | Not-determined | Genome-fragment | NA | NA | 0 |
| S16_extracted_64815 | 3758 | No | NA | 2 | 0 | 2 | Not-determined | Genome-fragment | NA | NA | 0 |
| S16_vs_56301\|\|full | 2508 | No | NA | 5 | 0 | 1 | Low-quality | Genome-fragment | 5.35 | AAI-based (high-confidence) | 0 |
| S16_extracted_64927 | 2520 | No | NA | 5 | 0 | 2 | Not-determined | Genome-fragment | NA | NA | 0 |
| S10_extracted_90618 | 2508 | No | NA | 3 | 0 | 3 | Low-quality | Genome-fragment | 0.27 | AAI-based (high-confidence) | 0 |
| S16_extracted_65059 | 1726 | No | NA | 2 | 0 | 2 | Not-determined | Genome-fragment | NA | NA | 0 |
| S16_extracted_65070 | 7807 | No | NA | 8 | 0 | 1 | Not-determined | Genome-fragment | NA | NA | 0 |
| S16_extracted_65334 | 1244 | No | NA | 2 | 0 | 1 | Not-determined | Genome-fragment | NA | NA | 0 |
| S16_extracted_65364 | 1095 | No | NA | 1 | 0 | 1 | Not-determined | Genome-fragment | NA | NA | 0 |
| S16_extracted_65374 | 2483 | No | NA | 2 | 0 | 2 | Not-determined | Genome-fragment | NA | NA | 0 |
| S16_extracted_65410 | 1536 | No | NA | 2 | 0 | 0 | Not-determined | Genome-fragment | NA | NA | 0 |
| S16_extracted_65523 | 1246 | No | NA | 4 | 0 | 0 | Not-determined | Genome-fragment | NA | NA | 0 |
| S16_extracted_65542 | 1713 | No | NA | 2 | 0 | 2 | Not-determined | Genome-fragment | NA | NA | 0 |
| S16_extracted_65592 | 4483 | No | NA | 4 | 0 | 2 | Not-determined | Genome-fragment | NA | NA | 0 |
| S16_extracted_65596 | 1478 | No | NA | 2 | 0 | 1 | Not-determined | Genome-fragment | NA | NA | 0 |
| S16_extracted_65599 | 3144 | No | NA | 2 | 0 | 2 | Not-determined | Genome-fragment | NA | NA | 0 |
| S16_extracted_65607 | 1900 | No | NA | 1 | 0 | 1 | Not-determined | Genome-fragment | NA | NA | 0 |
| S16_extracted_65805 | 1617 | No | NA | 2 | 0 | 1 | Not-determined | Genome-fragment | NA | NA | 0 |
| S16_extracted_65862 | 2550 | No | NA | 2 | 0 | 0 | Not-determined | Genome-fragment | NA | NA | 0 |
| S16_extracted_65957 | 1660 | No | NA | 2 | 0 | 2 | Not-determined | Genome-fragment | NA | NA | 0 |
| S5_vs_k141_95340\|\|full | 2509 | No | NA | 5 | 0 | 0 | Low-quality | Genome-fragment | 6.51 | AAI-based (high-confidence) | 0 |
| S8_extracted_k141_182078 | 2510 | No | NA | 4 | 0 | 4 | Low-quality | Genome-fragment | 0.68 | AAI-based (medium-confidence) | 0 |
| S16_extracted_66358 | 1021 | No | NA | 1 | 0 | 0 | Not-determined | Genome-fragment | NA | NA | 0 |
| S11_extracted_k141_99953 | 2511 | No | NA | 4 | 3 | 0 | Low-quality | Genome-fragment | 6.07 | AAI-based (high-confidence) | 0 |
| S16_extracted_66516 | 1201 | No | NA | 1 | 0 | 0 | Not-determined | Genome-fragment | NA | NA | 0 |
| S7_extracted_73839 | 2511 | No | NA | 3 | 2 | 0 | Low-quality | Genome-fragment | 5.42 | AAI-based (high-confidence) | 0 |
| S16_extracted_66594 | 2829 | No | NA | 1 | 0 | 0 | Not-determined | Genome-fragment | NA | NA | 0 |
| S7_extracted_120015 | 2511 | No | NA | 8 | 1 | 0 | Low-quality | Genome-fragment | 4.81 | AAI-based (high-confidence) | 0 |
| S16_extracted_66794 | 1466 | No | NA | 1 | 0 | 1 | Not-determined | Genome-fragment | NA | NA | 0 |
| S16_extracted_66816 | 2802 | No | NA | 1 | 0 | 0 | Not-determined | Genome-fragment | NA | NA | 0 |
| S16_extracted_66829 | 1498 | No | NA | 2 | 0 | 0 | Not-determined | Genome-fragment | NA | NA | 0 |
| S16_vs_152680\|\|full | 2511 | No | NA | 4 | 0 | 0 | Low-quality | Genome-fragment | 5.92 | AAI-based (medium-confidence) | 0 |
| S16_extracted_66911 | 1845 | No | NA | 1 | 0 | 0 | Not-determined | Genome-fragment | NA | NA | 0 |
| S16_extracted_66930 | 1079 | No | NA | 1 | 0 | 0 | Not-determined | Genome-fragment | NA | NA | 0 |
| S16_extracted_66944 | 1093 | No | NA | 2 | 0 | 0 | Not-determined | Genome-fragment | NA | NA | 0 |
| S16_extracted_66974 | 2734 | No | NA | 1 | 0 | 0 | Not-determined | Genome-fragment | NA | NA | 0 |
| S16_extracted_67148 | 1023 | No | NA | 2 | 0 | 0 | Not-determined | Genome-fragment | NA | NA | 0 |
| S16_extracted_67214 | 1343 | No | NA | 1 | 0 | 1 | Not-determined | Genome-fragment | NA | NA | 0 |
| S16_extracted_67230 | 2323 | No | NA | 3 | 0 | 2 | Not-determined | Genome-fragment | NA | NA | 0 |
| S16_extracted_67262 | 1060 | No | NA | 2 | 0 | 0 | Not-determined | Genome-fragment | NA | NA | 0 |
| S6_vs_k141_26769\|\|full | 2511 | No | NA | 2 | 0 | 1 | Low-quality | Genome-fragment | 1.19 | AAI-based (medium-confidence) | 0 |
| S16_extracted_67373 | 11023 | No | NA | 4 | 0 | 0 | Not-determined | Genome-fragment | NA | NA | 0 |
| S16_extracted_67416 | 1994 | No | NA | 1 | 0 | 1 | Not-determined | Genome-fragment | NA | NA | 0 |
| S16_extracted_67461 | 6025 | No | NA | 9 | 0 | 3 | Not-determined | Genome-fragment | NA | NA | 0 |
| S13_vs_61110\|\|full | 2513 | No | NA | 7 | 0 | 0 | Low-quality | Genome-fragment | 2.31 | AAI-based (medium-confidence) | 0 |
| S16_extracted_67706 | 2841 | No | NA | 2 | 0 | 1 | Not-determined | Genome-fragment | NA | NA | 0 |
| S16_extracted_67713 | 9494 | No | NA | 14 | 0 | 0 | Not-determined | Genome-fragment | NA | NA | 0 |
| S16_extracted_67718 | 1116 | No | NA | 1 | 0 | 0 | Not-determined | Genome-fragment | NA | NA | 0 |
| S16_extracted_67840 | 1604 | No | NA | 3 | 0 | 0 | Not-determined | Genome-fragment | NA | NA | 0 |
| S16_extracted_67845 | 1352 | No | NA | 3 | 0 | 2 | Not-determined | Genome-fragment | NA | NA | 0 |
| S5_vs_k141_46221\|\|full | 2513 | No | NA | 6 | 2 | 0 | Low-quality | Genome-fragment | 2.61 | HMM-based (lower-bound) | 0 |
| S16_extracted_67895 | 6149 | No | NA | 5 | 0 | 2 | Not-determined | Genome-fragment | NA | NA | 0 |
| S16_extracted_67909 | 1184 | No | NA | 2 | 0 | 1 | Not-determined | Genome-fragment | NA | NA | 0 |
| S15_extracted_6564 | 2513 | No | NA | 5 | 2 | 0 | Low-quality | Genome-fragment | 1.72 | AAI-based (high-confidence) | 0 |
| S16_extracted_67950 | 3109 | No | NA | 4 | 0 | 3 | Not-determined | Genome-fragment | NA | NA | 0 |
| S16_extracted_68139 | 1729 | No | NA | 1 | 0 | 0 | Not-determined | Genome-fragment | NA | NA | 0 |
| S16_extracted_68167 | 5019 | No | NA | 2 | 0 | 0 | Not-determined | Genome-fragment | NA | NA | 0 |
| S8_extracted_k141_21847 | 2513 | No | NA | 2 | 0 | 0 | Low-quality | Genome-fragment | 8.72 | AAI-based (medium-confidence) | 0 |
| S16_extracted_68301 | 8093 | No | NA | 7 | 0 | 4 | Not-determined | Genome-fragment | NA | NA | 0 |
| S16_extracted_68364 | 1109 | No | NA | 2 | 0 | 0 | Not-determined | Genome-fragment | NA | NA | 0 |
| S16_extracted_68425 | 2409 | No | NA | 3 | 0 | 2 | Not-determined | Genome-fragment | NA | NA | 0 |
| S16_extracted_68460 | 3668 | No | NA | 3 | 0 | 3 | Not-determined | Genome-fragment | NA | NA | 0 |
| S16_extracted_68570 | 1123 | No | NA | 2 | 0 | 0 | Not-determined | Genome-fragment | NA | NA | 0 |
| S7_vs_110215\|\|full | 2514 | No | NA | 3 | 0 | 1 | Low-quality | Genome-fragment | 1.49 | AAI-based (medium-confidence) | 0 |
| S16_extracted_68727 | 3057 | No | NA | 3 | 0 | 0 | Not-determined | Genome-fragment | NA | NA | 0 |
| S16_extracted_68977 | 1337 | No | NA | 2 | 0 | 2 | Not-determined | Genome-fragment | NA | NA | 0 |
| S16_extracted_126755 | 2514 | No | NA | 7 | 1 | 0 | Low-quality | Genome-fragment | 5.96 | AAI-based (high-confidence) | 0 |
| S16_extracted_69163 | 4461 | No | NA | 5 | 0 | 4 | Not-determined | Genome-fragment | NA | NA | 0 |
| S8_extracted_k141_194481 | 2514 | No | NA | 1 | 0 | 1 | Low-quality | Genome-fragment | 1.01 | AAI-based (medium-confidence) | 0 |
| S9_extracted_k141_44053 | 2515 | No | NA | 4 | 1 | 0 | Low-quality | Genome-fragment | 5.92 | AAI-based (medium-confidence) | 0 |
| S3_vs_k141_114108\|\|full | 2515 | No | NA | 4 | 0 | 0 | Low-quality | Genome-fragment | 1.77 | AAI-based (medium-confidence) | 0 |
| S16_extracted_69364 | 1361 | No | NA | 2 | 0 | 0 | Not-determined | Genome-fragment | NA | NA | 0 |
| S16_extracted_69580 | 1451 | No | NA | 2 | 0 | 0 | Not-determined | Genome-fragment | NA | NA | 0 |
| S16_extracted_69703 | 1264 | No | NA | 1 | 0 | 1 | Not-determined | Genome-fragment | NA | NA | 0 |
| S16_extracted_69714 | 2234 | No | NA | 1 | 0 | 0 | Not-determined | Genome-fragment | NA | NA | 0 |
| S16_extracted_69804 | 3280 | No | NA | 3 | 0 | 1 | Not-determined | Genome-fragment | NA | NA | 0 |
| S16_extracted_70080 | 1776 | No | NA | 3 | 0 | 0 | Not-determined | Genome-fragment | NA | NA | 0 |
| S16_vs_20308\|\|full | 2515 | No | NA | 3 | 2 | 0 | Low-quality | Genome-fragment | 6.14 | AAI-based (high-confidence) | 0 |
| S16_extracted_70212 | 1030 | No | NA | 1 | 0 | 1 | Not-determined | Genome-fragment | NA | NA | 0 |
| S16_extracted_70266 | 1136 | No | NA | 2 | 0 | 1 | Not-determined | Genome-fragment | NA | NA | 0 |
| S16_extracted_70289 | 3999 | No | NA | 2 | 0 | 0 | Not-determined | Genome-fragment | NA | NA | 0 |
| S16_extracted_70295 | 1651 | No | NA | 3 | 0 | 0 | Not-determined | Genome-fragment | NA | NA | 0 |
| S16_extracted_70338 | 1049 | No | NA | 2 | 0 | 1 | Not-determined | Genome-fragment | NA | NA | 0 |
| S3_extracted_k141_112584 | 2516 | No | NA | 2 | 0 | 2 | Low-quality | Genome-fragment | 1.89 | AAI-based (medium-confidence) | 0 |
| S16_extracted_70435 | 3683 | No | NA | 2 | 0 | 2 | Not-determined | Genome-fragment | NA | NA | 0 |
| S16_extracted_70458 | 3019 | No | NA | 4 | 0 | 2 | Not-determined | Genome-fragment | NA | NA | 0 |
| S16_extracted_70579 | 3016 | No | NA | 5 | 0 | 0 | Not-determined | Genome-fragment | NA | NA | 0 |
| S16_extracted_70612 | 1081 | No | NA | 1 | 0 | 1 | Not-determined | Genome-fragment | NA | NA | 0 |
| S16_extracted_70686 | 1720 | No | NA | 2 | 0 | 1 | Not-determined | Genome-fragment | NA | NA | 0 |
| S16_extracted_70692 | 9872 | No | NA | 2 | 0 | 2 | Not-determined | Genome-fragment | NA | NA | 0 |
| S16_extracted_70784 | 2905 | No | NA | 2 | 0 | 2 | Not-determined | Genome-fragment | NA | NA | 0 |

| | | | | | | | | | | | |
|---|---|---|---|---|---|---|---|---|---|---|---|
| S16_extracted_70817 | 1045 | No | NA | 1 | 0 | 1 | Not-determined | Genome-fragment | NA | NA | 0 |
| S16_extracted_70825 | 1003 | No | NA | 1 | 0 | 0 | Not-determined | Genome-fragment | NA | NA | 0 |
| S7_vs_88168\|\|full | 2516 | No | NA | 5 | 3 | 0 | Low-quality | Genome-fragment | 5.73 | AAI-based (high-confidence) | 0 |
| S16_extracted_70921 | 1424 | No | NA | 3 | 0 | 1 | Not-determined | Genome-fragment | NA | NA | 0 |
| S16_extracted_70969 | 4013 | No | NA | 2 | 0 | 1 | Not-determined | Genome-fragment | NA | NA | 0 |
| S16_extracted_71043 | 1383 | No | NA | 1 | 0 | 0 | Not-determined | Genome-fragment | NA | NA | 0 |
| S16_vs_76605\|\|full | 2516 | No | NA | 3 | 0 | 0 | Low-quality | Genome-fragment | 3.33 | AAI-based (medium-confidence) | 0 |
| S8_extracted_k141_43599 | 2516 | No | NA | 3 | 2 | 0 | Low-quality | Genome-fragment | 4.31 | AAI-based (high-confidence) | 0 |
| S17_vs_k141_95143\|\|full | 2517 | No | NA | 2 | 2 | 0 | Low-quality | Genome-fragment | 6.34 | AAI-based (medium-confidence) | 0 |
| S16_extracted_71378 | 2524 | No | NA | 1 | 0 | 1 | Not-determined | Genome-fragment | NA | NA | 0 |
| S16_extracted_71398 | 2936 | No | NA | 4 | 0 | 3 | Not-determined | Genome-fragment | NA | NA | 0 |
| S14_vs_k141_23213\|\|full | 2517 | No | NA | 5 | 3 | 0 | Low-quality | Genome-fragment | 5.97 | AAI-based (high-confidence) | 0 |
| S1_extracted_k141_87569 | 2517 | No | NA | 3 | 0 | 0 | Low-quality | Genome-fragment | 4.75 | AAI-based (high-confidence) | 0 |
| S16_extracted_71665 | 3546 | No | NA | 1 | 0 | 1 | Not-determined | Genome-fragment | NA | NA | 0 |
| S16_extracted_71766 | 1591 | No | NA | 2 | 0 | 2 | Not-determined | Genome-fragment | NA | NA | 0 |
| S16_extracted_71861 | 1698 | No | NA | 2 | 0 | 0 | Not-determined | Genome-fragment | NA | NA | 0 |
| S16_extracted_71893 | 2480 | No | NA | 2 | 0 | 2 | Not-determined | Genome-fragment | NA | NA | 0 |
| S16_extracted_71925 | 1358 | No | NA | 2 | 0 | 0 | Not-determined | Genome-fragment | NA | NA | 0 |
| S16_extracted_71937 | 4073 | No | NA | 4 | 0 | 1 | Not-determined | Genome-fragment | NA | NA | 0 |
| S16_extracted_71943 | 5402 | No | NA | 3 | 0 | 3 | Not-determined | Genome-fragment | NA | NA | 0 |
| S5_vs_k141_20980\|\|full | 2517 | No | NA | 4 | 0 | 2 | Low-quality | Genome-fragment | 3.26 | AAI-based (high-confidence) | 0 |
| S16_extracted_72121 | 5391 | No | NA | 5 | 0 | 4 | Not-determined | Genome-fragment | NA | NA | 0 |
| S16_extracted_72238 | 5190 | No | NA | 5 | 0 | 2 | Not-determined | Genome-fragment | NA | NA | 0 |
| S5_vs_k141_90792\|\|full | 2517 | No | NA | 3 | 0 | 1 | Low-quality | Genome-fragment | 2.9 | AAI-based (high-confidence) | 0 |
| S16_extracted_72461 | 1235 | No | NA | 1 | 0 | 0 | Not-determined | Genome-fragment | NA | NA | 0 |
| S16_extracted_72744 | 1076 | No | NA | 1 | 0 | 0 | Not-determined | Genome-fragment | NA | NA | 0 |
| S16_extracted_72771 | 1298 | No | NA | 2 | 0 | 0 | Not-determined | Genome-fragment | NA | NA | 0 |
| S7_extracted_130812 | 2517 | No | NA | 3 | 2 | 0 | Low-quality | Genome-fragment | 4.82 | AAI-based (medium-confidence) | 0 |
| S16_extracted_73046 | 3002 | No | NA | 2 | 0 | 1 | Not-determined | Genome-fragment | NA | NA | 0 |
| S16_extracted_73209 | 1495 | No | NA | 2 | 0 | 1 | Not-determined | Genome-fragment | NA | NA | 0 |
| S16_extracted_73232 | 3657 | No | NA | 5 | 0 | 0 | Not-determined | Genome-fragment | NA | NA | 0 |
| S16_extracted_73251 | 1038 | No | NA | 2 | 0 | 0 | Not-determined | Genome-fragment | NA | NA | 0 |
| S16_extracted_73264 | 1886 | No | NA | 1 | 0 | 0 | Not-determined | Genome-fragment | NA | NA | 0 |
| S10_vs_48389\|\|full | 2517 | No | NA | 6 | 3 | 0 | Low-quality | Genome-fragment | 6.05 | AAI-based (high-confidence) | 0 |
| S16_extracted_73307 | 1026 | No | NA | 2 | 0 | 0 | Not-determined | Genome-fragment | NA | NA | 0 |
| S16_extracted_73333 | 1431 | No | NA | 2 | 0 | 0 | Not-determined | Genome-fragment | NA | NA | 0 |
| S16_extracted_73395 | 1392 | No | NA | 1 | 0 | 1 | Not-determined | Genome-fragment | NA | NA | 0 |
| S16_extracted_73526 | 1319 | No | NA | 1 | 0 | 0 | Not-determined | Genome-fragment | NA | NA | 0 |
| S16_extracted_73546 | 1076 | No | NA | 3 | 0 | 1 | Not-determined | Genome-fragment | NA | NA | 0 |
| S16_extracted_73656 | 1901 | No | NA | 3 | 0 | 2 | Not-determined | Genome-fragment | NA | NA | 0 |
| S16_extracted_73743 | 1022 | No | NA | 1 | 0 | 1 | Not-determined | Genome-fragment | NA | NA | 0 |
| S6_vs_k141_38338\|\|full | 2517 | No | NA | 3 | 0 | 0 | Low-quality | Genome-fragment | 5.98 | AAI-based (high-confidence) | 0 |
| S9_extracted_k141_34931 | 2518 | No | NA | 3 | 0 | 3 | Low-quality | Genome-fragment | 1.64 | AAI-based (medium-confidence) | 0 |
| S11_vs_k141_7828\|\|full | 2518 | No | NA | 2 | 0 | 0 | Low-quality | Genome-fragment | 4.12 | AAI-based (high-confidence) | 0 |
| S16_extracted_74045 | 3195 | No | NA | 3 | 0 | 1 | Not-determined | Genome-fragment | NA | NA | 0 |
| S16_extracted_74112 | 1113 | No | NA | 1 | 0 | 0 | Not-determined | Genome-fragment | NA | NA | 0 |
| S16_extracted_74276 | 1285 | No | NA | 1 | 0 | 1 | Not-determined | Genome-fragment | NA | NA | 0 |
| S16_extracted_74343 | 1083 | No | NA | 1 | 0 | 1 | Not-determined | Genome-fragment | NA | NA | 0 |
| S16_extracted_74376 | 1831 | No | NA | 3 | 0 | 1 | Not-determined | Genome-fragment | NA | NA | 0 |
| S16_extracted_74556 | 5758 | No | NA | 5 | 0 | 1 | Not-determined | Genome-fragment | NA | NA | 0 |
| S16_extracted_74579 | 1433 | No | NA | 3 | 0 | 0 | Not-determined | Genome-fragment | NA | NA | 0 |
| S16_extracted_74728 | 1085 | No | NA | 1 | 0 | 0 | Not-determined | Genome-fragment | NA | NA | 0 |
| S1_vs_k141_52486\|\|full | 2518 | No | NA | 4 | 1 | 0 | Low-quality | Genome-fragment | 5.54 | AAI-based (high-confidence) | 0 |
| S16_extracted_74852 | 1756 | No | NA | 1 | 0 | 1 | Not-determined | Genome-fragment | NA | NA | 0 |
| S16_extracted_74866 | 5554 | No | NA | 2 | 0 | 2 | Not-determined | Genome-fragment | NA | NA | 0 |
| S16_extracted_74904 | 2360 | No | NA | 1 | 0 | 0 | Not-determined | Genome-fragment | NA | NA | 0 |
| S16_extracted_74954 | 1068 | No | NA | 1 | 0 | 0 | Not-determined | Genome-fragment | NA | NA | 0 |
| S3_vs_k141_88338\|\|full | 2518 | No | NA | 3 | 3 | 0 | Low-quality | Genome-fragment | 5.66 | AAI-based (high-confidence) | 0 |
| S7_vs_70269\|\|full | 2518 | No | NA | 5 | 2 | 0 | Low-quality | Genome-fragment | 6.88 | AAI-based (high-confidence) | 0 |
| S17_vs_k141_71422\|\|full | 2519 | No | NA | 3 | 0 | 0 | Low-quality | Genome-fragment | 5.92 | AAI-based (medium-confidence) | 0 |
| S13_vs_127672\|\|full | 2519 | No | NA | 8 | 0 | 0 | Low-quality | Genome-fragment | 6.37 | AAI-based (medium-confidence) | 0 |
| S16_extracted_75355 | 1271 | No | NA | 2 | 0 | 1 | Not-determined | Genome-fragment | NA | NA | 0 |
| S10_vs_110502\|\|full | 2519 | No | NA | 6 | 1 | 0 | Low-quality | Genome-fragment | 3.13 | AAI-based (high-confidence) | 0 |
| S16_extracted_75390 | 5604 | No | NA | 4 | 0 | 4 | Not-determined | Genome-fragment | NA | NA | 0 |
| S16_extracted_75398 | 1037 | No | NA | 1 | 0 | 1 | Not-determined | Genome-fragment | NA | NA | 0 |
| S16_extracted_75438 | 1174 | No | NA | 1 | 0 | 0 | Not-determined | Genome-fragment | NA | NA | 0 |
| S16_extracted_75482 | 1117 | No | NA | 1 | 0 | 1 | Not-determined | Genome-fragment | NA | NA | 0 |
| S16_extracted_75531 | 5029 | No | NA | 10 | 0 | 10 | Not-determined | Genome-fragment | NA | NA | 0 |
| S16_extracted_75591 | 1558 | No | NA | 2 | 0 | 0 | Not-determined | Genome-fragment | NA | NA | 0 |
| S8_extracted_k141_179121 | 2519 | No | NA | 1 | 0 | 0 | Low-quality | Genome-fragment | 2.52 | AAI-based (medium-confidence) | 0 |
| S16_extracted_75797 | 3229 | No | NA | 7 | 0 | 0 | Not-determined | Genome-fragment | NA | NA | 0 |
| S16_extracted_75806 | 4722 | No | NA | 3 | 0 | 3 | Not-determined | Genome-fragment | NA | NA | 0 |
| S16_extracted_75863 | 3420 | No | NA | 5 | 0 | 0 | Not-determined | Genome-fragment | NA | NA | 0 |
| S16_extracted_75900 | 1375 | No | NA | 1 | 0 | 0 | Not-determined | Genome-fragment | NA | NA | 0 |
| S16_extracted_75956 | 2610 | No | NA | 4 | 0 | 3 | Not-determined | Genome-fragment | NA | NA | 0 |
| S16_extracted_76005 | 2678 | No | NA | 3 | 0 | 1 | Not-determined | Genome-fragment | NA | NA | 0 |
| S16_extracted_76006 | 3256 | No | NA | 1 | 0 | 1 | Not-determined | Genome-fragment | NA | NA | 0 |
| S16_extracted_76089 | 5042 | No | NA | 3 | 0 | 3 | Not-determined | Genome-fragment | NA | NA | 0 |
| S16_extracted_76092 | 5031 | No | NA | 6 | 0 | 1 | Not-determined | Genome-fragment | NA | NA | 0 |
| S16_extracted_76102 | 8192 | No | NA | 6 | 0 | 5 | Not-determined | Genome-fragment | NA | NA | 0 |
| S16_extracted_76112 | 2611 | No | NA | 3 | 0 | 2 | Not-determined | Genome-fragment | NA | NA | 0 |
| S16_extracted_76115 | 4641 | No | NA | 3 | 0 | 3 | Not-determined | Genome-fragment | NA | NA | 0 |
| S16_extracted_76147 | 1437 | No | NA | 1 | 0 | 0 | Not-determined | Genome-fragment | NA | NA | 0 |
| S17_extracted_k141_89425 | 2520 | No | NA | 4 | 0 | 0 | Low-quality | Genome-fragment | 1.46 | AAI-based (high-confidence) | 0 |
| S16_extracted_76226 | 8575 | No | NA | 2 | 0 | 0 | Not-determined | Genome-fragment | NA | NA | 0 |
| S16_extracted_76309 | 1312 | No | NA | 1 | 0 | 0 | Not-determined | Genome-fragment | NA | NA | 0 |
| S16_extracted_76824 | 1284 | No | NA | 1 | 0 | 1 | Not-determined | Genome-fragment | NA | NA | 0 |
| S16_extracted_76956 | 1210 | No | NA | 1 | 0 | 1 | Not-determined | Genome-fragment | NA | NA | 0 |
| S16_extracted_77042 | 5598 | No | NA | 5 | 0 | 2 | Not-determined | Genome-fragment | NA | NA | 0 |
| S11_extracted_k141_52276 | 2520 | No | NA | 4 | 0 | 0 | Low-quality | Genome-fragment | 5.99 | AAI-based (high-confidence) | 0 |
| S16_extracted_77208 | 1146 | No | NA | 2 | 0 | 1 | Not-determined | Genome-fragment | NA | NA | 0 |
| S16_extracted_77212 | 1221 | No | NA | 2 | 0 | 2 | Not-determined | Genome-fragment | NA | NA | 0 |
| S16_extracted_77267 | 1156 | No | NA | 1 | 0 | 1 | Not-determined | Genome-fragment | NA | NA | 0 |
| S16_extracted_77289 | 4482 | No | NA | 2 | 0 | 2 | Not-determined | Genome-fragment | NA | NA | 0 |
| S16_extracted_77337 | 3764 | No | NA | 5 | 0 | 1 | Not-determined | Genome-fragment | NA | NA | 0 |
| S16_extracted_77362 | 1069 | No | NA | 2 | 0 | 0 | Not-determined | Genome-fragment | NA | NA | 0 |
| S16_extracted_77647 | 5507 | No | NA | 2 | 0 | 2 | Not-determined | Genome-fragment | NA | NA | 0 |
| S16_extracted_77658 | 1396 | No | NA | 1 | 0 | 1 | Not-determined | Genome-fragment | NA | NA | 0 |
| S16_extracted_77661 | 1005 | No | NA | 1 | 0 | 1 | Not-determined | Genome-fragment | NA | NA | 0 |
| S3_vs_k141_86201\|\|full | 2520 | No | NA | 8 | 6 | 0 | Low-quality | Genome-fragment | 7.22 | AAI-based (medium-confidence) | 0 |
| S16_extracted_77778 | 6466 | No | NA | 4 | 0 | 4 | Not-determined | Genome-fragment | NA | NA | 0 |
| S16_extracted_77803 | 1489 | No | NA | 3 | 0 | 0 | Not-determined | Genome-fragment | NA | NA | 0 |
| S16_extracted_77810 | 1084 | No | NA | 2 | 0 | 1 | Not-determined | Genome-fragment | NA | NA | 0 |
| S16_extracted_77840 | 1221 | No | NA | 1 | 0 | 1 | Not-determined | Genome-fragment | NA | NA | 0 |
| S7_vs_11647\|\|full | 2520 | No | NA | 3 | 0 | 1 | Low-quality | Genome-fragment | 4.61 | AAI-based (high-confidence) | 0 |
| S16_extracted_77867 | 2252 | No | NA | 2 | 0 | 2 | Not-determined | Genome-fragment | NA | NA | 0 |
| S16_extracted_77900 | 1512 | No | NA | 2 | 0 | 0 | Not-determined | Genome-fragment | NA | NA | 0 |
| S1_vs_k141_17301\|\|full | 2521 | No | NA | 2 | 0 | 1 | Low-quality | Genome-fragment | 3.39 | AAI-based (high-confidence) | 0 |
| S16_extracted_77980 | 1277 | No | NA | 1 | 0 | 1 | Not-determined | Genome-fragment | NA | NA | 0 |
| S5_vs_k141_49514\|\|full | 2521 | No | NA | 6 | 2 | 0 | Low-quality | Genome-fragment | 5.81 | AAI-based (medium-confidence) | 0 |
| S16_extracted_78037 | 1694 | No | NA | 2 | 0 | 0 | Not-determined | Genome-fragment | NA | NA | 0 |
| S16_extracted_78240 | 1835 | No | NA | 2 | 0 | 1 | Not-determined | Genome-fragment | NA | NA | 0 |
| S16_extracted_78250 | 4969 | No | NA | 4 | 0 | 2 | Not-determined | Genome-fragment | NA | NA | 0 |
| S16_extracted_78333 | 1104 | No | NA | 2 | 0 | 1 | Not-determined | Genome-fragment | NA | NA | 0 |
| S5_vs_k141_21331\|\|full | 2521 | No | NA | 6 | 0 | 0 | Low-quality | Genome-fragment | 7.97 | AAI-based (high-confidence) | 0 |
| S16_extracted_78381 | 1324 | No | NA | 4 | 0 | 2 | Not-determined | Genome-fragment | NA | NA | 0 |
| S16_extracted_78402 | 1026 | No | NA | 1 | 0 | 1 | Not-determined | Genome-fragment | NA | NA | 0 |
| S16_extracted_78520 | 3398 | No | NA | 3 | 0 | 1 | Not-determined | Genome-fragment | NA | NA | 0 |
| S16_extracted_78600 | 1229 | No | NA | 2 | 0 | 0 | Not-determined | Genome-fragment | NA | NA | 0 |
| S16_extracted_78724 | 1578 | No | NA | 1 | 0 | 0 | Not-determined | Genome-fragment | NA | NA | 0 |
| S16_extracted_78848 | 1147 | No | NA | 2 | 0 | 1 | Not-determined | Genome-fragment | NA | NA | 0 |
| S16_extracted_78886 | 2898 | No | NA | 3 | 0 | 1 | Not-determined | Genome-fragment | NA | NA | 0 |
| S16_extracted_79086 | 3884 | No | NA | 4 | 0 | 2 | Not-determined | Genome-fragment | NA | NA | 0 |
| S10_vs_21058\|\|full | 2521 | No | NA | 4 | 1 | 0 | Low-quality | Genome-fragment | 4.56 | AAI-based (medium-confidence) | 0 |
| S10_vs_32972\|\|full | 2521 | No | NA | 6 | 0 | 1 | Low-quality | Genome-fragment | 4.71 | AAI-based (medium-confidence) | 0 |
| S16_extracted_79265 | 1002 | No | NA | 2 | 0 | 1 | Not-determined | Genome-fragment | NA | NA | 0 |
| S11_vs_k141_53380\|\|full | 2522 | No | NA | 5 | 2 | 0 | Low-quality | Genome-fragment | 5.85 | AAI-based (medium-confidence) | 0 |
| S3_vs_k141_97694\|\|full | 2522 | No | NA | 5 | 1 | 0 | Low-quality | Genome-fragment | 1.49 | AAI-based (high-confidence) | 0 |
| S16_extracted_79426 | 3374 | No | NA | 3 | 0 | 1 | Not-determined | Genome-fragment | NA | NA | 0 |
| S16_extracted_79498 | 1535 | No | NA | 1 | 0 | 0 | Not-determined | Genome-fragment | NA | NA | 0 |
| S16_vs_7730\|\|lt2gene | 2522 | No | NA | 2 | 0 | 0 | Low-quality | Genome-fragment | 5.61 | AAI-based (high-confidence) | 0 |
| S12_extracted_k141_33652 | 2522 | No | NA | 4 | 0 | 0 | Low-quality | Genome-fragment | 3.99 | AAI-based (high-confidence) | 0 |
| S16_extracted_79618 | 6437 | No | NA | 3 | 0 | 3 | Not-determined | Genome-fragment | NA | NA | 0 |
| S16_extracted_79724 | 10132 | No | NA | 8 | 0 | 4 | Not-determined | Genome-fragment | NA | NA | 0 |
| S8_vs_k141_182797\|\|full | 2522 | No | NA | 6 | 1 | 0 | Low-quality | Genome-fragment | 2.36 | HMM-based (lower-bound) | 0 |
| S17_extracted_k141_145248 | 2523 | No | NA | 3 | 0 | 0 | Low-quality | Genome-fragment | 4.05 | AAI-based (medium-confidence) | 0 |
| S9_vs_k141_60353\|\|full | 2523 | No | NA | 7 | 0 | 0 | Low-quality | Genome-fragment | 6.06 | AAI-based (medium-confidence) | 0 |
| S5_vs_k141_79164\|\|full | 2523 | No | NA | 4 | 0 | 2 | Low-quality | Genome-fragment | 1.68 | AAI-based (medium-confidence) | 0 |
| S5_vs_k141_110456\|\|full | 2523 | No | NA | 4 | 2 | 0 | Low-quality | Genome-fragment | 5.62 | AAI-based (high-confidence) | 0 |
| S16_extracted_80353 | 20856 | No | NA | 13 | 0 | 6 | Not-determined | Genome-fragment | NA | NA | 0 |
| S16_extracted_80385 | 5421 | No | NA | 3 | 0 | 2 | Not-determined | Genome-fragment | NA | NA | 0 |
| S16_extracted_80394 | 1326 | No | NA | 1 | 0 | 1 | Not-determined | Genome-fragment | NA | NA | 0 |
| S16_extracted_80466 | 5800 | No | NA | 6 | 0 | 3 | Not-determined | Genome-fragment | NA | NA | 0 |
| S16_extracted_80484 | 3443 | No | NA | 3 | 0 | 1 | Not-determined | Genome-fragment | NA | NA | 0 |
| S8_extracted_k141_50703 | 2523 | No | NA | 1 | 1 | 0 | Low-quality | Genome-fragment | 2.51 | AAI-based (high-confidence) | 0 |
| S9_vs_k141_50124\|\|full | 2524 | No | NA | 4 | 3 | 0 | Low-quality | Genome-fragment | 4.2 | AAI-based (high-confidence) | 0 |
| S16_extracted_80595 | 2052 | No | NA | 4 | 0 | 0 | Not-determined | Genome-fragment | NA | NA | 0 |
| S16_extracted_80747 | 1337 | No | NA | 2 | 0 | 0 | Not-determined | Genome-fragment | NA | NA | 0 |
| S16_extracted_80963 | 7325 | No | NA | 6 | 0 | 6 | Not-determined | Genome-fragment | NA | NA | 0 |
| S16_extracted_80977 | 1095 | No | NA | 2 | 0 | 2 | Not-determined | Genome-fragment | NA | NA | 0 |
| S16_extracted_81020 | 1129 | No | NA | 1 | 0 | 0 | Not-determined | Genome-fragment | NA | NA | 0 |
| S13_vs_85777\|\|full | 2524 | No | NA | 5 | 0 | 3 | Low-quality | Genome-fragment | 1.86 | AAI-based (high-confidence) | 0 |
| S16_extracted_81210 | 21170 | No | NA | 13 | 0 | 8 | Not-determined | Genome-fragment | NA | NA | 0 |
| S16_extracted_81217 | 2271 | No | NA | 3 | 0 | 2 | Not-determined | Genome-fragment | NA | NA | 0 |
| S16_extracted_81270 | 3492 | No | NA | 3 | 0 | 0 | Not-determined | Genome-fragment | NA | NA | 0 |
| S16_extracted_81357 | 1456 | No | NA | 4 | 0 | 0 | Not-determined | Genome-fragment | NA | NA | 0 |
| S16_extracted_81523 | 2412 | No | NA | 4 | 0 | 3 | Not-determined | Genome-fragment | NA | NA | 0 |
| S16_extracted_81547 | 3031 | No | NA | 4 | 0 | 2 | Not-determined | Genome-fragment | NA | NA | 0 |
| S13_vs_133554\|\|full | 2524 | No | NA | 4 | 4 | 0 | Low-quality | Genome-fragment | 4.14 | AAI-based (high-confidence) | 0 |
| S16_extracted_81696 | 1210 | No | NA | 3 | 0 | 0 | Not-determined | Genome-fragment | NA | NA | 0 |
| S5_vs_k141_87372\|\|full | 2524 | No | NA | 5 | 0 | 0 | Low-quality | Genome-fragment | 7.11 | AAI-based (high-confidence) | 0 |
| S16_extracted_81758 | 1035 | No | NA | 1 | 0 | 1 | Not-determined | Genome-fragment | NA | NA | 0 |
| S16_extracted_81823 | 1948 | No | NA | 3 | 0 | 3 | Not-determined | Genome-fragment | NA | NA | 0 |
| S16_extracted_81989 | 4318 | No | NA | 4 | 0 | 1 | Not-determined | Genome-fragment | NA | NA | 0 |
| S16_vs_73135\|\|full | 2524 | No | NA | 5 | 0 | 0 | Low-quality | Genome-fragment | 4.87 | AAI-based (high-confidence) | 0 |
| S16_extracted_82047 | 5877 | No | NA | 5 | 0 | 0 | Not-determined | Genome-fragment | NA | NA | 0 |
| S16_extracted_82110 | 7421 | No | NA | 5 | 0 | 4 | Not-determined | Genome-fragment | NA | NA | 0 |
| S1_vs_k141_126944\|\|full | 2525 | No | NA | 3 | 0 | 1 | Low-quality | Genome-fragment | 3.92 | AAI-based (medium-confidence) | 0 |
| S16_extracted_82181 | 4341 | No | NA | 6 | 0 | 2 | Not-determined | Genome-fragment | NA | NA | 0 |
| S16_extracted_82199 | 1627 | No | NA | 2 | 0 | 0 | Not-determined | Genome-fragment | NA | NA | 0 |
| S5_extracted_k141_94026 | 2525 | No | NA | 5 | 1 | 0 | Low-quality | Genome-fragment | 2.2 | AAI-based (medium-confidence) | 0 |
| S16_extracted_82219 | 2043 | No | NA | 3 | 0 | 0 | Not-determined | Genome-fragment | NA | NA | 0 |
| S16_extracted_82245 | 2616 | No | NA | 3 | 0 | 0 | Not-determined | Genome-fragment | NA | NA | 0 |
| S16_extracted_82246 | 1365 | No | NA | 2 | 0 | 0 | Not-determined | Genome-fragment | NA | NA | 0 |
| S16_extracted_82308 | 3767 | No | NA | 3 | 0 | 3 | Not-determined | Genome-fragment | NA | NA | 0 |
| S16_extracted_82379 | 1038 | No | NA | 1 | 0 | 1 | Not-determined | Genome-fragment | NA | NA | 0 |
| S16_vs_5956\|\|full | 2525 | No | NA | 7 | 3 | 0 | Low-quality | Genome-fragment | 7.04 | AAI-based (high-confidence) | 0 |
| S16_extracted_82473 | 1096 | No | NA | 2 | 0 | 0 | Not-determined | Genome-fragment | NA | NA | 0 |
| S16_extracted_82493 | 2514 | No | NA | 1 | 0 | 0 | Not-determined | Genome-fragment | NA | NA | 0 |
| S16_extracted_82496 | 1131 | No | NA | 2 | 0 | 1 | Not-determined | Genome-fragment | NA | NA | 0 |
| S16_extracted_82629 | 1177 | No | NA | 2 | 0 | 0 | Not-determined | Genome-fragment | NA | NA | 0 |
| S16_vs_141994\|\|full | 2525 | No | NA | 7 | 3 | 0 | Low-quality | Genome-fragment | 7.11 | AAI-based (high-confidence) | 0 |
| S16_extracted_82657 | 4844 | No | NA | 4 | 0 | 2 | Not-determined | Genome-fragment | NA | NA | 0 |
| S16_extracted_82700 | 1087 | No | NA | 1 | 0 | 1 | Not-determined | Genome-fragment | NA | NA | 0 |
| S16_extracted_82833 | 5708 | No | NA | 4 | 0 | 2 | Not-determined | Genome-fragment | NA | NA | 0 |
| S16_extracted_82879 | 4069 | No | NA | 3 | 0 | 2 | Not-determined | Genome-fragment | NA | NA | 0 |
| S16_extracted_82933 | 1244 | No | NA | 1 | 0 | 1 | Not-determined | Genome-fragment | NA | NA | 0 |
| S16_extracted_82990 | 4602 | No | NA | 2 | 0 | 0 | Not-determined | Genome-fragment | NA | NA | 0 |
| S16_extracted_83006 | 1002 | No | NA | 2 | 0 | 1 | Not-determined | Genome-fragment | NA | NA | 0 |
| S16_extracted_83118 | 2176 | No | NA | 3 | 0 | 0 | Not-determined | Genome-fragment | NA | NA | 0 |
| S16_extracted_83222 | 1807 | No | NA | 3 | 0 | 1 | Not-determined | Genome-fragment | NA | NA | 0 |
| S16_extracted_83432 | 1032 | No | NA | 1 | 0 | 1 | Not-determined | Genome-fragment | NA | NA | 0 |
| S16_extracted_83489 | 2285 | No | NA | 3 | 0 | 3 | Not-determined | Genome-fragment | NA | NA | 0 |
| S16_extracted_83521 | 2198 | No | NA | 1 | 0 | 1 | Not-determined | Genome-fragment | NA | NA | 0 |
| S16_extracted_83525 | 1219 | No | NA | 1 | 0 | 0 | Not-determined | Genome-fragment | NA | NA | 0 |
| S16_extracted_83535 | 1114 | No | NA | 1 | 0 | 0 | Not-determined | Genome-fragment | NA | NA | 0 |
| S16_extracted_83669 | 5265 | No | NA | 2 | 0 | 1 | Not-determined | Genome-fragment | NA | NA | 0 |
| S16_extracted_83761 | 1019 | No | NA | 1 | 0 | 1 | Not-determined | Genome-fragment | NA | NA | 0 |
| S16_extracted_83769 | 1291 | No | NA | 2 | 0 | 1 | Not-determined | Genome-fragment | NA | NA | 0 |
| S4_extracted_14118 | 2525 | No | NA | 3 | 2 | 0 | Low-quality | Genome-fragment | 8.32 | AAI-based (high-confidence) | 0 |
| S16_extracted_83918 | 1974 | No | NA | 2 | 0 | 0 | Not-determined | Genome-fragment | NA | NA | 0 |
| S16_extracted_83949 | 20838 | No | NA | 20 | 0 | 5 | Not-determined | Genome-fragment | NA | NA | 0 |
| S8_extracted_k141_109251 | 2525 | No | NA | 1 | 1 | 0 | Low-quality | Genome-fragment | 4.73 | HMM-based (lower-bound) | 0 |
| S11_vs_k141_6998\|\|full | 2526 | No | NA | 7 | 1 | 0 | Low-quality | Genome-fragment | 5.83 | AAI-based (medium-confidence) | 0 |
| S16_extracted_84001 | 7608 | No | NA | 7 | 0 | 1 | Not-determined | Genome-fragment | NA | NA | 0 |
| S16_extracted_84175 | 1049 | No | NA | 2 | 0 | 0 | Not-determined | Genome-fragment | NA | NA | 0 |
| S16_extracted_84283 | 3851 | No | NA | 6 | 0 | 4 | Not-determined | Genome-fragment | NA | NA | 0 |
| S16_extracted_84375 | 1260 | No | NA | 4 | 0 | 0 | Not-determined | Genome-fragment | NA | NA | 0 |
| S16_extracted_84432 | 1005 | No | NA | 1 | 0 | 1 | Not-determined | Genome-fragment | NA | NA | 0 |
| S16_extracted_84457 | 9275 | No | NA | 5 | 0 | 3 | Not-determined | Genome-fragment | NA | NA | 0 |
| S16_extracted_84462 | 54255 | No | NA | 96 | 21 | 1 | Complete | High-quality | 100 | DTR (high-confidence) | 0 |
| S11_vs_k141_79630\|\|full | 2527 | No | NA | 6 | 6 | 0 | Low-quality | Genome-fragment | 6.52 | AAI-based (high-confidence) | 0 |
| S16_extracted_84499 | 2116 | No | NA | 4 | 0 | 0 | Not-determined | Genome-fragment | NA | NA | 0 |
| S16_extracted_84507 | 1149 | No | NA | 2 | 0 | 2 | Not-determined | Genome-fragment | NA | NA | 0 |
| S16_extracted_84522 | 1312 | No | NA | 2 | 0 | 1 | Not-determined | Genome-fragment | NA | NA | 0 |
| S13_vs_5559\|\|full | 2527 | No | NA | 3 | 0 | 3 | Low-quality | Genome-fragment | 3.07 | AAI-based (medium-confidence) | 0 |
| S16_extracted_84662 | 4166 | No | NA | 3 | 0 | 3 | Not-determined | Genome-fragment | NA | NA | 0 |
| S16_extracted_84708 | 1461 | No | NA | 1 | 0 | 1 | Not-determined | Genome-fragment | NA | NA | 0 |
| S16_extracted_84734 | 1004 | No | NA | 1 | 0 | 0 | Not-determined | Genome-fragment | NA | NA | 0 |
| S16_extracted_84896 | 3217 | No | NA | 2 | 0 | 2 | Not-determined | Genome-fragment | NA | NA | 0 |
| S5_vs_k141_47799\|\|full | 2527 | No | NA | 4 | 1 | 0 | Low-quality | Genome-fragment | 7.03 | AAI-based (high-confidence) | 0 |
| S16_extracted_84916 | 3271 | No | NA | 3 | 0 | 2 | Not-determined | Genome-fragment | NA | NA | 0 |
| S16_extracted_84994 | 8283 | No | NA | 6 | 0 | 3 | Not-determined | Genome-fragment | NA | NA | 0 |
| S16_extracted_85061 | 2178 | No | NA | 1 | 0 | 0 | Not-determined | Genome-fragment | NA | NA | 0 |
| S16_vs_161146\|\|full | 2527 | No | NA | 4 | 0 | 0 | Low-quality | Genome-fragment | 6.17 | AAI-based (medium-confidence) | 0 |
| S16_extracted_85244 | 1598 | No | NA | 1 | 0 | 0 | Not-determined | Genome-fragment | NA | NA | 0 |
| S16_vs_41164\|\|full | 2528 | No | NA | 3 | 0 | 3 | Low-quality | Genome-fragment | 1.13 | AAI-based (medium-confidence) | 0 |
| S16_extracted_85428 | 1990 | No | NA | 1 | 0 | 1 | Not-determined | Genome-fragment | NA | NA | 0 |
| S16_extracted_85573 | 2079 | No | NA | 3 | 0 | 1 | Not-determined | Genome-fragment | NA | NA | 0 |
| S16_extracted_85579 | 4014 | No | NA | 5 | 0 | 2 | Not-determined | Genome-fragment | NA | NA | 0 |
| S16_extracted_85631 | 3147 | No | NA | 3 | 0 | 2 | Not-determined | Genome-fragment | NA | NA | 0 |
| S16_extracted_85738 | 2019 | No | NA | 2 | 0 | 2 | Not-determined | Genome-fragment | NA | NA | 0 |
| S16_extracted_85748 | 3503 | No | NA | 3 | 0 | 1 | Not-determined | Genome-fragment | NA | NA | 0 |
| S16_extracted_85771 | 1004 | No | NA | 1 | 0 | 1 | Not-determined | Genome-fragment | NA | NA | 0 |
| S12_vs_k141_125739\|\|full | 2528 | No | NA | 3 | 0 | 1 | Low-quality | Genome-fragment | 4.16 | AAI-based (medium-confidence) | 0 |
| S16_extracted_85854 | 5631 | No | NA | 6 | 0 | 1 | Not-determined | Genome-fragment | NA | NA | 0 |
| S16_extracted_85857 | 9130 | No | NA | 16 | 0 | 1 | Not-determined | Genome-fragment | NA | NA | 0 |
| S16_extracted_85892 | 1056 | No | NA | 1 | 0 | 1 | Not-determined | Genome-fragment | NA | NA | 0 |
| S16_extracted_85935 | 2641 | No | NA | 4 | 0 | 0 | Not-determined | Genome-fragment | NA | NA | 0 |
| S13_vs_95629\|\|full | 2529 | No | NA | 4 | 1 | 0 | Low-quality | Genome-fragment | 3.88 | AAI-based (medium-confidence) | 0 |
| S1_vs_k141_117848\|\|full | 2529 | No | NA | 3 | 0 | 0 | Low-quality | Genome-fragment | 6.42 | AAI-based (medium-confidence) | 0 |
| S16_vs_157694\|\|full | 2529 | No | NA | 4 | 3 | 0 | Low-quality | Genome-fragment | 5.4 | AAI-based (medium-confidence) | 0 |
| S16_extracted_86083 | 6769 | No | NA | 7 | 0 | 6 | Not-determined | Genome-fragment | NA | NA | 0 |
| S16_extracted_86128 | 1014 | No | NA | 2 | 0 | 1 | Not-determined | Genome-fragment | NA | NA | 0 |
| S16_extracted_86129 | 1310 | No | NA | 2 | 0 | 2 | Not-determined | Genome-fragment | NA | NA | 0 |
| S16_extracted_86227 | 4325 | No | NA | 5 | 0 | 5 | Not-determined | Genome-fragment | NA | NA | 0 |
| S2_vs_k141_2979\|\|full | 2530 | No | NA | 3 | 0 | 1 | Low-quality | Genome-fragment | 0.95 | AAI-based (high-confidence) | 0 |
| S16_extracted_86367 | 1018 | No | NA | 1 | 0 | 1 | Not-determined | Genome-fragment | NA | NA | 0 |
| S16_extracted_86474 | 3254 | No | NA | 3 | 0 | 2 | Not-determined | Genome-fragment | NA | NA | 0 |
| S16_extracted_86510 | 1065 | No | NA | 2 | 0 | 0 | Not-determined | Genome-fragment | NA | NA | 0 |
| S16_extracted_86515 | 4735 | No | NA | 6 | 0 | 2 | Not-determined | Genome-fragment | NA | NA | 0 |
| S16_extracted_86592 | 3465 | No | NA | 2 | 0 | 2 | Not-determined | Genome-fragment | NA | NA | 0 |
| S16_extracted_86702 | 1567 | No | NA | 3 | 0 | 1 | Not-determined | Genome-fragment | NA | NA | 0 |
| S16_extracted_86730 | 2052 | No | NA | 2 | 0 | 2 | Not-determined | Genome-fragment | NA | NA | 0 |
| S16_extracted_86797 | 1315 | No | NA | 2 | 0 | 1 | Not-determined | Genome-fragment | NA | NA | 0 |
| S4_extracted_19526 | 2530 | No | NA | 6 | 3 | 0 | Low-quality | Genome-fragment | 2.46 | AAI-based (high-confidence) | 0 |
| S17_extracted_k141_141346 | 2531 | No | NA | 1 | 0 | 0 | Low-quality | Genome-fragment | 3.81 | AAI-based (high-confidence) | 0 |
| S3_extracted_k141_113479 | 2531 | No | NA | 4 | 4 | 0 | Low-quality | Genome-fragment | 11.55 | AAI-based (high-confidence) | 0 |
| S16_extracted_87141 | 1250 | No | NA | 1 | 0 | 1 | Not-determined | Genome-fragment | NA | NA | 0 |
| S16_extracted_87190 | 67710 | No | NA | 85 | 10 | 1 | Medium-quality | Genome-fragment | 63.7 | AAI-based (high-confidence) | 0 |
| S16_extracted_87220 | 9470 | No | NA | 7 | 0 | 5 | Not-determined | Genome-fragment | NA | NA | 0 |
| S16_extracted_87321 | 1014 | No | NA | 2 | 0 | 0 | Not-determined | Genome-fragment | NA | NA | 0 |
| S16_extracted_87325 | 6330 | No | NA | 9 | 0 | 7 | Not-determined | Genome-fragment | NA | NA | 0 |
| S16_extracted_87477 | 1291 | No | NA | 2 | 0 | 0 | Not-determined | Genome-fragment | NA | NA | 0 |
| S16_extracted_87534 | 8840 | No | NA | 10 | 0 | 3 | Not-determined | Genome-fragment | NA | NA | 0 |
| S16_extracted_87580 | 1138 | No | NA | 1 | 0 | 1 | Not-determined | Genome-fragment | NA | NA | 0 |
| S7_vs_47899\|\|full | 2531 | No | NA | 5 | 2 | 0 | Low-quality | Genome-fragment | 6.16 | AAI-based (high-confidence) | 0 |
| S16_extracted_87754 | 3682 | No | NA | 4 | 0 | 1 | Not-determined | Genome-fragment | NA | NA | 0 |
| S8_extracted_k141_31565 | 2531 | No | NA | 2 | 0 | 2 | Low-quality | Genome-fragment | 2.26 | AAI-based (medium-confidence) | 0 |
| S16_extracted_88090 | 1880 | No | NA | 3 | 0 | 0 | Not-determined | Genome-fragment | NA | NA | 0 |
| S16_extracted_88214 | 2263 | No | NA | 2 | 0 | 1 | Not-determined | Genome-fragment | NA | NA | 0 |
| S16_extracted_88262 | 2174 | No | NA | 1 | 0 | 1 | Not-determined | Genome-fragment | NA | NA | 0 |
| S16_extracted_88263 | 2402 | No | NA | 3 | 0 | 0 | Not-determined | Genome-fragment | NA | NA | 0 |
| S16_extracted_88303 | 1048 | No | NA | 1 | 0 | 1 | Not-determined | Genome-fragment | NA | NA | 0 |
| S16_extracted_88337 | 14147 | No | NA | 17 | 0 | 8 | Not-determined | Genome-fragment | NA | NA | 0 |
| S17_extracted_k141_90022 | 2532 | No | NA | 5 | 1 | 0 | Low-quality | Genome-fragment | 5.74 | AAI-based (high-confidence) | 0 |
| S14_vs_k141_99812\|\|full | 2532 | No | NA | 4 | 0 | 1 | Low-quality | Genome-fragment | 1.22 | AAI-based (medium-confidence) | 0 |
| S16_extracted_88494 | 1080 | No | NA | 1 | 0 | 1 | Not-determined | Genome-fragment | NA | NA | 0 |
| S16_extracted_88528 | 3152 | No | NA | 3 | 0 | 3 | Not-determined | Genome-fragment | NA | NA | 0 |
| S16_extracted_88616 | 8974 | No | NA | 7 | 0 | 4 | Not-determined | Genome-fragment | NA | NA | 0 |
| S16_extracted_88678 | 5128 | No | NA | 3 | 0 | 1 | Not-determined | Genome-fragment | NA | NA | 0 |
| S16_extracted_88721 | 1273 | No | NA | 2 | 0 | 1 | Not-determined | Genome-fragment | NA | NA | 0 |
| S16_extracted_88751 | 6020 | No | NA | 3 | 0 | 3 | Not-determined | Genome-fragment | NA | NA | 0 |
| S16_extracted_88853 | 1142 | No | NA | 4 | 0 | 3 | Not-determined | Genome-fragment | NA | NA | 0 |
| S16_extracted_89014 | 1093 | No | NA | 1 | 0 | 1 | Not-determined | Genome-fragment | NA | NA | 0 |
| S1_vs_k141_125413\|\|full | 2532 | No | NA | 4 | 3 | 0 | Low-quality | Genome-fragment | 4.64 | AAI-based (high-confidence) | 0 |
| S7_extracted_77425 | 2532 | No | NA | 6 | 2 | 0 | Low-quality | Genome-fragment | 9.41 | AAI-based (high-confidence) | 0 |
| S16_vs_155956\|\|full | 2532 | No | NA | 7 | 1 | 0 | Low-quality | Genome-fragment | 1.47 | AAI-based (high-confidence) | 0 |
| S16_extracted_89396 | 1364 | No | NA | 3 | 0 | 1 | Not-determined | Genome-fragment | NA | NA | 0 |
| S16_extracted_89405 | 5252 | No | NA | 3 | 0 | 3 | Not-determined | Genome-fragment | NA | NA | 0 |
| S16_extracted_89450 | 6211 | No | NA | 6 | 0 | 5 | Not-determined | Genome-fragment | NA | NA | 0 |
| S4_extracted_17514 | 2532 | No | NA | 7 | 5 | 0 | Low-quality | Genome-fragment | 8.35 | AAI-based (high-confidence) | 0 |
| S11_vs_k141_89517\|\|full | 2533 | No | NA | 6 | 2 | 0 | Low-quality | Genome-fragment | 5.98 | AAI-based (high-confidence) | 0 |
| S16_extracted_89674 | 1114 | No | NA | 1 | 0 | 0 | Not-determined | Genome-fragment | NA | NA | 0 |
| S16_extracted_89702 | 1288 | No | NA | 2 | 0 | 2 | Not-determined | Genome-fragment | NA | NA | 0 |
| S14_extracted_k141_104113 | 2534 | No | NA | 5 | 0 | 1 | Low-quality | Genome-fragment | 6.01 | AAI-based (medium-confidence) | 0 |
| S16_extracted_89902 | 2407 | No | NA | 4 | 0 | 2 | Not-determined | Genome-fragment | NA | NA | 0 |
| S16_extracted_90014 | 1099 | No | NA | 1 | 0 | 0 | Not-determined | Genome-fragment | NA | NA | 0 |
| S16_extracted_90043 | 2959 | No | NA | 2 | 0 | 2 | Not-determined | Genome-fragment | NA | NA | 0 |
| S16_extracted_90137 | 1684 | No | NA | 2 | 0 | 2 | Not-determined | Genome-fragment | NA | NA | 0 |
| S16_extracted_90183 | 2447 | No | NA | 2 | 0 | 0 | Not-determined | Genome-fragment | NA | NA | 0 |
| S1_vs_k141_19645\|\|full | 2534 | No | NA | 7 | 0 | 0 | Low-quality | Genome-fragment | 5.34 | AAI-based (high-confidence) | 0 |
| S16_extracted_90202 | 1029 | No | NA | 2 | 0 | 2 | Not-determined | Genome-fragment | NA | NA | 0 |
| S16_extracted_90208 | 2529 | No | NA | 3 | 0 | 2 | Not-determined | Genome-fragment | NA | NA | 0 |
| S16_extracted_90328 | 26593 | No | NA | 42 | 17 | 0 | Medium-quality | Genome-fragment | 71.71 | AAI-based (high-confidence) | 0 |
| S16_extracted_90352 | 1453 | No | NA | 2 | 0 | 2 | Not-determined | Genome-fragment | NA | NA | 0 |
| S16_extracted_90380 | 4560 | No | NA | 3 | 0 | 2 | Not-determined | Genome-fragment | NA | NA | 0 |
| S16_vs_78286\|\|lt2gene | 2534 | No | NA | 1 | 1 | 0 | Low-quality | Genome-fragment | 4.14 | AAI-based (high-confidence) | 0 |
| S16_extracted_90407 | 1005 | No | NA | 2 | 0 | 0 | Not-determined | Genome-fragment | NA | NA | 0 |
| S16_extracted_90470 | 3715 | No | NA | 1 | 0 | 0 | Not-determined | Genome-fragment | NA | NA | 0 |
| S16_extracted_90493 | 3590 | No | NA | 4 | 0 | 4 | Not-determined | Genome-fragment | NA | NA | 0 |
| S15_vs_68802\|\|full | 2534 | No | NA | 7 | 0 | 1 | Low-quality | Genome-fragment | 2.04 | AAI-based (high-confidence) | 0 |
| S3_vs_k141_69498\|\|full | 2535 | No | NA | 4 | 0 | 0 | Low-quality | Genome-fragment | 5.07 | AAI-based (high-confidence) | 0 |
| S8_vs_k141_118399\|\|full | 2535 | No | NA | 5 | 0 | 0 | Low-quality | Genome-fragment | 1.46 | AAI-based (high-confidence) | 0 |
| S9_vs_k141_36903\|\|full | 2536 | No | NA | 5 | 0 | 0 | Low-quality | Genome-fragment | 1.47 | AAI-based (medium-confidence) | 0 |
| S11_vs_k141_13340\|\|full | 2536 | No | NA | 3 | 3 | 0 | Low-quality | Genome-fragment | 6.41 | AAI-based (high-confidence) | 0 |
| S16_extracted_90781 | 2000 | No | NA | 2 | 0 | 2 | Not-determined | Genome-fragment | NA | NA | 0 |
| S16_extracted_90850 | 4154 | No | NA | 5 | 0 | 3 | Not-determined | Genome-fragment | NA | NA | 0 |
| S16_extracted_90859 | 1057 | No | NA | 2 | 0 | 1 | Not-determined | Genome-fragment | NA | NA | 0 |
| S16_extracted_91307 | 3267 | No | NA | 3 | 0 | 3 | Not-determined | Genome-fragment | NA | NA | 0 |
| S16_extracted_91310 | 1431 | No | NA | 1 | 0 | 0 | Not-determined | Genome-fragment | NA | NA | 0 |
| S3_vs_k141_26007\|\|full | 2536 | No | NA | 3 | 0 | 0 | Low-quality | Genome-fragment | 1.82 | AAI-based (high-confidence) | 0 |
| S16_extracted_91355 | 1007 | No | NA | 1 | 0 | 0 | Not-determined | Genome-fragment | NA | NA | 0 |
| S10_vs_14940\|\|full | 2536 | No | NA | 2 | 2 | 0 | Low-quality | Genome-fragment | 5.07 | AAI-based (high-confidence) | 0 |
| S12_vs_k141_170676\|\|full | 2536 | No | NA | 4 | 2 | 0 | Low-quality | Genome-fragment | 4.11 | AAI-based (medium-confidence) | 0 |
| S16_extracted_91457 | 2050 | No | NA | 3 | 0 | 0 | Not-determined | Genome-fragment | NA | NA | 0 |
| S16_extracted_91500 | 1410 | No | NA | 1 | 0 | 0 | Not-determined | Genome-fragment | NA | NA | 0 |
| S16_extracted_91501 | 2019 | No | NA | 4 | 0 | 2 | Not-determined | Genome-fragment | NA | NA | 0 |
| S4_extracted_3420 | 2536 | No | NA | 2 | 2 | 0 | Low-quality | Genome-fragment | 8.08 | AAI-based (high-confidence) | 0 |
| S16_extracted_91559 | 1484 | No | NA | 1 | 0 | 1 | Not-determined | Genome-fragment | NA | NA | 0 |
| S11_vs_k141_35085\|\|full | 2537 | No | NA | 3 | 0 | 3 | Low-quality | Genome-fragment | 1.3 | AAI-based (high-confidence) | 0 |
| S16_extracted_91754 | 1067 | No | NA | 2 | 0 | 0 | Not-determined | Genome-fragment | NA | NA | 0 |
| S16_extracted_91809 | 1431 | No | NA | 3 | 0 | 2 | Not-determined | Genome-fragment | NA | NA | 0 |
| S16_extracted_91831 | 1218 | No | NA | 1 | 0 | 0 | Not-determined | Genome-fragment | NA | NA | 0 |
| S16_vs_5854\|\|full | 2537 | No | NA | 5 | 0 | 0 | Low-quality | Genome-fragment | 3.92 | AAI-based (high-confidence) | 0 |
| S16_extracted_91938 | 1203 | No | NA | 1 | 0 | 0 | Not-determined | Genome-fragment | NA | NA | 0 |
| S16_extracted_91975 | 1873 | No | NA | 2 | 0 | 1 | Not-determined | Genome-fragment | NA | NA | 0 |
| S8_extracted_k141_168542 | 2537 | No | NA | 4 | 1 | 0 | Low-quality | Genome-fragment | 6.4 | AAI-based (high-confidence) | 0 |
| S16_extracted_92115 | 1677 | No | NA | 2 | 0 | 0 | Not-determined | Genome-fragment | NA | NA | 0 |
| S8_vs_k141_188848\|\|full | 2538 | No | NA | 2 | 0 | 2 | Low-quality | Genome-fragment | 2.43 | AAI-based (medium-confidence) | 0 |
| S1_vs_k141_136005\|\|full | 2539 | No | NA | 5 | 1 | 0 | Low-quality | Genome-fragment | 1.99 | AAI-based (high-confidence) | 0 |
| S16_extracted_92263 | 1276 | No | NA | 3 | 0 | 1 | Not-determined | Genome-fragment | NA | NA | 0 |
| S16_extracted_92364 | 5231 | No | NA | 3 | 0 | 2 | Not-determined | Genome-fragment | NA | NA | 0 |
| S16_extracted_92368 | 1044 | No | NA | 2 | 0 | 0 | Not-determined | Genome-fragment | NA | NA | 0 |
| S16_extracted_92414 | 1528 | No | NA | 2 | 0 | 1 | Not-determined | Genome-fragment | NA | NA | 0 |
| S16_extracted_92537 | 1076 | No | NA | 1 | 0 | 0 | Not-determined | Genome-fragment | NA | NA | 0 |
| S16_extracted_92560 | 1105 | No | NA | 1 | 0 | 1 | Not-determined | Genome-fragment | NA | NA | 0 |
| S7_extracted_119425 | 2539 | No | NA | 5 | 0 | 3 | Low-quality | Genome-fragment | 1.67 | AAI-based (high-confidence) | 0 |
| S16_extracted_92827 | 3472 | No | NA | 1 | 0 | 1 | Not-determined | Genome-fragment | NA | NA | 0 |
| S16_extracted_92853 | 1478 | No | NA | 1 | 0 | 0 | Not-determined | Genome-fragment | NA | NA | 0 |
| S16_extracted_92952 | 1103 | No | NA | 1 | 0 | 1 | Not-determined | Genome-fragment | NA | NA | 0 |
| S16_extracted_92975 | 5738 | No | NA | 6 | 0 | 3 | Not-determined | Genome-fragment | NA | NA | 0 |
| S7_vs_108010\|\|full | 2539 | No | NA | 4 | 0 | 0 | Low-quality | Genome-fragment | 5.02 | AAI-based (high-confidence) | 0 |
| S1_vs_k141_156922\|\|full | 2540 | No | NA | 3 | 1 | 0 | Low-quality | Genome-fragment | 9.34 | HMM-based (lower-bound) | 0 |
| S16_extracted_93073 | 1057 | No | NA | 1 | 0 | 0 | Not-determined | Genome-fragment | NA | NA | 0 |
| S16_extracted_93103 | 5541 | No | NA | 8 | 0 | 2 | Not-determined | Genome-fragment | NA | NA | 0 |
| S16_extracted_93143 | 1112 | No | NA | 1 | 0 | 0 | Not-determined | Genome-fragment | NA | NA | 0 |
| S16_extracted_93266 | 1221 | No | NA | 4 | 0 | 4 | Not-determined | Genome-fragment | NA | NA | 0 |
| S5_extracted_k141_75706 | 2540 | No | NA | 3 | 2 | 0 | Low-quality | Genome-fragment | 8.43 | AAI-based (high-confidence) | 0 |
| S16_extracted_93414 | 1008 | No | NA | 1 | 0 | 0 | Not-determined | Genome-fragment | NA | NA | 0 |
| S16_extracted_93513 | 1396 | No | NA | 2 | 0 | 1 | Not-determined | Genome-fragment | NA | NA | 0 |
| S16_extracted_93678 | 1950 | No | NA | 2 | 0 | 0 | Not-determined | Genome-fragment | NA | NA | 0 |
| S5_vs_k141_89129\|\|full | 2540 | No | NA | 3 | 0 | 1 | Low-quality | Genome-fragment | 4.77 | AAI-based (medium-confidence) | 0 |
| S16_extracted_93803 | 4534 | No | NA | 3 | 0 | 1 | Not-determined | Genome-fragment | NA | NA | 0 |
| S16_extracted_93877 | 2395 | No | NA | 4 | 0 | 2 | Not-determined | Genome-fragment | NA | NA | 0 |
| S16_extracted_93954 | 5158 | No | NA | 4 | 0 | 2 | Not-determined | Genome-fragment | NA | NA | 0 |
| S16_extracted_94034 | 4830 | No | NA | 6 | 0 | 4 | Not-determined | Genome-fragment | NA | NA | 0 |
| S16_vs_145295\|\|full | 2540 | No | NA | 4 | 0 | 0 | Low-quality | Genome-fragment | 5.04 | AAI-based (medium-confidence) | 0 |
| S16_vs_159413\|\|full | 2540 | No | NA | 3 | 0 | 2 | Low-quality | Genome-fragment | 1.33 | AAI-based (high-confidence) | 0 |
| S16_extracted_94212 | 4994 | No | NA | 8 | 0 | 2 | Not-determined | Genome-fragment | NA | NA | 0 |
| S16_extracted_94267 | 1106 | No | NA | 5 | 0 | 0 | Not-determined | Genome-fragment | NA | NA | 0 |
| S12_vs_k141_105548\|\|full | 2540 | No | NA | 3 | 0 | 1 | Low-quality | Genome-fragment | 2.06 | AAI-based (high-confidence) | 0 |
| S16_extracted_94291 | 1049 | No | NA | 1 | 0 | 0 | Not-determined | Genome-fragment | NA | NA | 0 |
| S16_extracted_94352 | 2174 | No | NA | 2 | 0 | 1 | Not-determined | Genome-fragment | NA | NA | 0 |
| S17_extracted_k141_113104 | 2541 | No | NA | 5 | 1 | 0 | Low-quality | Genome-fragment | 4.52 | HMM-based (lower-bound) | 0 |
| S16_extracted_94539 | 1330 | No | NA | 1 | 0 | 1 | Not-determined | Genome-fragment | NA | NA | 0 |
| S16_extracted_94615 | 1047 | No | NA | 1 | 0 | 1 | Not-determined | Genome-fragment | NA | NA | 0 |
| S16_extracted_94684 | 1079 | No | NA | 1 | 0 | 1 | Not-determined | Genome-fragment | NA | NA | 0 |
| S16_extracted_94695 | 3014 | No | NA | 2 | 1 | 0 | Not-determined | Genome-fragment | NA | NA | 0 |
| S16_extracted_94710 | 1045 | No | NA | 1 | 0 | 1 | Not-determined | Genome-fragment | NA | NA | 0 |
| S17_vs_k141_122758\|\|full | 2541 | No | NA | 4 | 0 | 2 | Low-quality | Genome-fragment | 2.84 | AAI-based (medium-confidence) | 0 |
| S16_extracted_94866 | 16805 | No | NA | 27 | 7 | 0 | Medium-quality | Genome-fragment | 76.85 | AAI-based (medium-confidence) | 0 |
| S16_extracted_94883 | 1071 | No | NA | 1 | 0 | 0 | Not-determined | Genome-fragment | NA | NA | 0 |
| S11_vs_k141_26714\|\|full | 2541 | No | NA | 3 | 2 | 0 | Low-quality | Genome-fragment | 4.55 | AAI-based (medium-confidence) | 0 |
| S16_extracted_94989 | 1340 | No | NA | 1 | 0 | 1 | Not-determined | Genome-fragment | NA | NA | 0 |
| S5_vs_k141_1990\|\|full | 2541 | No | NA | 5 | 0 | 0 | Low-quality | Genome-fragment | 4.79 | AAI-based (medium-confidence) | 0 |
| S8_vs_k141_103010\|\|full | 2541 | No | NA | 4 | 0 | 0 | Low-quality | Genome-fragment | 4.5 | AAI-based (medium-confidence) | 0 |
| S16_extracted_95344 | 1314 | No | NA | 2 | 0 | 0 | Not-determined | Genome-fragment | NA | NA | 0 |
| S13_extracted_62492 | 2542 | No | NA | 7 | 0 | 0 | Low-quality | Genome-fragment | 1.56 | AAI-based (high-confidence) | 0 |
| S1_vs_k141_131690\|\|full | 2542 | No | NA | 2 | 0 | 2 | Low-quality | Genome-fragment | 2.09 | AAI-based (high-confidence) | 0 |
| S16_extracted_95628 | 1524 | No | NA | 3 | 0 | 1 | Not-determined | Genome-fragment | NA | NA | 0 |
| S16_extracted_95661 | 3131 | No | NA | 3 | 0 | 1 | Not-determined | Genome-fragment | NA | NA | 0 |
| S16_extracted_95677 | 1326 | No | NA | 3 | 0 | 0 | Not-determined | Genome-fragment | NA | NA | 0 |
| S16_extracted_95679 | 3014 | No | NA | 1 | 0 | 1 | Not-determined | Genome-fragment | NA | NA | 0 |
| S16_extracted_95848 | 1180 | No | NA | 1 | 0 | 1 | Not-determined | Genome-fragment | NA | NA | 0 |
| S16_extracted_95913 | 1040 | No | NA | 2 | 0 | 0 | Not-determined | Genome-fragment | NA | NA | 0 |
| S16_extracted_95942 | 1109 | No | NA | 2 | 0 | 0 | Not-determined | Genome-fragment | NA | NA | 0 |
| S16_extracted_96049 | 1114 | No | NA | 1 | 0 | 1 | Not-determined | Genome-fragment | NA | NA | 0 |
| S16_extracted_96056 | 2649 | No | NA | 2 | 0 | 0 | Not-determined | Genome-fragment | NA | NA | 0 |
| S16_extracted_96073 | 1043 | No | NA | 1 | 0 | 0 | Not-determined | Genome-fragment | NA | NA | 0 |
| S16_extracted_96102 | 1120 | No | NA | 1 | 0 | 1 | Not-determined | Genome-fragment | NA | NA | 0 |
| S3_vs_k141_44006\|\|full | 2542 | No | NA | 5 | 0 | 0 | Low-quality | Genome-fragment | 4.93 | AAI-based (medium-confidence) | 0 |
| S12_vs_k141_82275\|\|full | 2542 | No | NA | 3 | 0 | 1 | Low-quality | Genome-fragment | 3.69 | AAI-based (medium-confidence) | 0 |
| S14_vs_k141_102337\|\|full | 2543 | No | NA | 5 | 1 | 0 | Low-quality | Genome-fragment | 17.12 | AAI-based (high-confidence) | 0 |
| S16_extracted_96344 | 1115 | No | NA | 1 | 0 | 0 | Not-determined | Genome-fragment | NA | NA | 0 |
| S16_extracted_96370 | 2768 | No | NA | 2 | 0 | 1 | Not-determined | Genome-fragment | NA | NA | 0 |
| S16_vs_27812\|\|full | 2543 | No | NA | 3 | 2 | 0 | Low-quality | Genome-fragment | 7.9 | AAI-based (high-confidence) | 0 |
| S16_extracted_96483 | 52402 | No | NA | 86 | 12 | 3 | Medium-quality | Genome-fragment | 66.24 | AAI-based (high-confidence) | 0 |
| S8_vs_k141_13988\|\|full | 2543 | No | NA | 3 | 0 | 0 | Low-quality | Genome-fragment | 4.47 | AAI-based (high-confidence) | 0 |
| S16_extracted_96564 | 1378 | No | NA | 3 | 0 | 2 | Not-determined | Genome-fragment | NA | NA | 0 |
| S16_extracted_96620 | 1201 | No | NA | 3 | 0 | 0 | Not-determined | Genome-fragment | NA | NA | 0 |
| S16_extracted_96763 | 1154 | No | NA | 1 | 0 | 0 | Not-determined | Genome-fragment | NA | NA | 0 |
| S16_extracted_96838 | 1488 | Yes | 820 | 2 | 1 | 1 | Not-determined | Genome-fragment | NA | NA | 44.89 |

| | | | | | | | | | | | |
|---|---|---|---|---|---|---|---|---|---|---|---|
| S16_extracted_96910 | 4007 | No | NA | 4 | 0 | 2 | Not-determined | Genome-fragment | NA | NA | 0 |
| S16_extracted_97094 | 3490 | No | NA | 4 | 0 | 0 | Not-determined | Genome-fragment | NA | NA | 0 |
| S16_extracted_97207 | 5060 | No | NA | 5 | 0 | 2 | Not-determined | Genome-fragment | NA | NA | 0 |
| S16_extracted_97242 | 1414 | No | NA | 1 | 0 | 0 | Not-determined | Genome-fragment | NA | NA | 0 |
| S16_extracted_97249 | 1216 | No | NA | 1 | 0 | 1 | Not-determined | Genome-fragment | NA | NA | 0 |
| S16_extracted_97279 | 48249 | No | NA | 70 | 19 | 0 | High-quality | High-quality | 99.38 | AAI-based (high-confidence) | 0 |
| S16_extracted_97308 | 1761 | No | NA | 3 | 0 | 0 | Not-determined | Genome-fragment | NA | NA | 0 |
| S5_vs_k141_45695\|\|full | 2544 | No | NA | 6 | 0 | 0 | Low-quality | Genome-fragment | 6.02 | AAI-based (medium-confidence) | 0 |
| S16_extracted_97346 | 1473 | No | NA | 3 | 0 | 2 | Not-determined | Genome-fragment | NA | NA | 0 |
| S16_extracted_97389 | 1584 | No | NA | 2 | 0 | 1 | Not-determined | Genome-fragment | NA | NA | 0 |
| S16_extracted_97390 | 1035 | No | NA | 1 | 0 | 0 | Not-determined | Genome-fragment | NA | NA | 0 |
| S5_vs_k141_137496\|\|full | 2544 | No | NA | 1 | 1 | 0 | Low-quality | Genome-fragment | 3.81 | HMM-based (lower-bound) | 0 |
| S16_extracted_97514 | 2499 | No | NA | 3 | 0 | 0 | Not-determined | Genome-fragment | NA | NA | 0 |
| S14_vs_k141_52357\|\|full | 2545 | No | NA | 4 | 1 | 0 | Low-quality | Genome-fragment | 4.29 | AAI-based (high-confidence) | 0 |
| S16_extracted_97639 | 6909 | No | NA | 8 | 0 | 0 | Not-determined | Genome-fragment | NA | NA | 0 |
| S16_extracted_97641 | 1204 | No | NA | 1 | 0 | 0 | Not-determined | Genome-fragment | NA | NA | 0 |
| S6_vs_k141_34271\|\|full | 2546 | No | NA | 4 | 0 | 0 | Low-quality | Genome-fragment | 3.57 | AAI-based (high-confidence) | 0 |
| S16_extracted_97707 | 1189 | No | NA | 1 | 0 | 0 | Not-determined | Genome-fragment | NA | NA | 0 |
| S16_extracted_97731 | 10717 | No | NA | 16 | 0 | 11 | Not-determined | Genome-fragment | NA | NA | 0 |
| S16_extracted_97749 | 1626 | No | NA | 2 | 0 | 1 | Not-determined | Genome-fragment | NA | NA | 0 |
| S16_extracted_97846 | 1233 | No | NA | 2 | 0 | 2 | Not-determined | Genome-fragment | NA | NA | 0 |
| S16_extracted_97847 | 3373 | No | NA | 1 | 0 | 1 | Not-determined | Genome-fragment | NA | NA | 0 |
| S16_extracted_97882 | 1718 | No | NA | 2 | 0 | 1 | Not-determined | Genome-fragment | NA | NA | 0 |
| S16_extracted_97885 | 3619 | No | NA | 5 | 0 | 4 | Not-determined | Genome-fragment | NA | NA | 0 |
| S16_extracted_97929 | 1230 | No | NA | 2 | 0 | 1 | Not-determined | Genome-fragment | NA | NA | 0 |
| S16_extracted_98021 | 1276 | No | NA | 1 | 0 | 1 | Not-determined | Genome-fragment | NA | NA | 0 |
| S16_extracted_98052 | 2186 | No | NA | 1 | 0 | 1 | Not-determined | Genome-fragment | NA | NA | 0 |
| S16_extracted_98153 | 1058 | No | NA | 1 | 0 | 1 | Not-determined | Genome-fragment | NA | NA | 0 |
| S16_extracted_98251 | 1193 | No | NA | 2 | 0 | 0 | Not-determined | Genome-fragment | NA | NA | 0 |
| S16_extracted_98341 | 3420 | No | NA | 2 | 0 | 2 | Not-determined | Genome-fragment | NA | NA | 0 |
| S16_extracted_98614 | 3101 | No | NA | 2 | 0 | 1 | Not-determined | Genome-fragment | NA | NA | 0 |
| S8_vs_k141_166568\|\|full | 2546 | No | NA | 3 | 1 | 0 | Low-quality | Genome-fragment | 6.06 | HMM-based (lower-bound) | 0 |
| S16_extracted_98736 | 1131 | No | NA | 1 | 0 | 0 | Not-determined | Genome-fragment | NA | NA | 0 |
| S16_extracted_98760 | 4071 | No | NA | 5 | 0 | 3 | Not-determined | Genome-fragment | NA | NA | 0 |
| S16_extracted_98788 | 1113 | No | NA | 2 | 0 | 2 | Not-determined | Genome-fragment | NA | NA | 0 |
| S16_extracted_98869 | 1633 | No | NA | 1 | 0 | 1 | Not-determined | Genome-fragment | NA | NA | 0 |
| S16_extracted_98933 | 1403 | No | NA | 2 | 0 | 2 | Not-determined | Genome-fragment | NA | NA | 0 |
| S16_extracted_99017 | 3630 | No | NA | 8 | 0 | 7 | Not-determined | Genome-fragment | NA | NA | 0 |
| S16_extracted_99025 | 1391 | No | NA | 2 | 0 | 1 | Not-determined | Genome-fragment | NA | NA | 0 |
| S16_extracted_99056 | 4593 | No | NA | 3 | 0 | 3 | Not-determined | Genome-fragment | NA | NA | 0 |
| S16_extracted_99094 | 1691 | No | NA | 2 | 0 | 0 | Not-determined | Genome-fragment | NA | NA | 0 |
| S16_extracted_99169 | 1336 | No | NA | 2 | 0 | 1 | Not-determined | Genome-fragment | NA | NA | 0 |
| S17_vs_k141_7232\|\|full | 2547 | No | NA | 3 | 0 | 0 | Low-quality | Genome-fragment | 2.67 | AAI-based (high-confidence) | 0 |
| S16_extracted_99332 | 1380 | No | NA | 1 | 0 | 1 | Not-determined | Genome-fragment | NA | NA | 0 |
| S16_extracted_99374 | 6072 | No | NA | 6 | 0 | 3 | Not-determined | Genome-fragment | NA | NA | 0 |
| S16_extracted_99442 | 3555 | No | NA | 4 | 0 | 2 | Not-determined | Genome-fragment | NA | NA | 0 |
| S16_extracted_99582 | 1391 | No | NA | 1 | 0 | 1 | Not-determined | Genome-fragment | NA | NA | 0 |
| S16_extracted_99731 | 1388 | No | NA | 1 | 0 | 1 | Not-determined | Genome-fragment | NA | NA | 0 |
| S16_extracted_99814 | 3556 | No | NA | 3 | 0 | 1 | Not-determined | Genome-fragment | NA | NA | 0 |
| S16_extracted_100001 | 1406 | No | NA | 2 | 0 | 0 | Not-determined | Genome-fragment | NA | NA | 0 |
| S16_extracted_100088 | 4508 | No | NA | 4 | 0 | 2 | Not-determined | Genome-fragment | NA | NA | 0 |
| S16_extracted_100235 | 18057 | No | NA | 23 | 9 | 0 | Medium-quality | Genome-fragment | 84.79 | AAI-based (medium-confidence) | 0 |
| S16_extracted_100237 | 1690 | No | NA | 1 | 0 | 0 | Not-determined | Genome-fragment | NA | NA | 0 |
| S16_extracted_100378 | 31039 | No | NA | 40 | 23 | 0 | Medium-quality | Genome-fragment | 51.79 | AAI-based (medium-confidence) | 0 |
| S16_extracted_100392 | 5073 | No | NA | 2 | 0 | 2 | Not-determined | Genome-fragment | NA | NA | 0 |
| S7_vs_90087\|\|full | 2547 | No | NA | 8 | 0 | 1 | Low-quality | Genome-fragment | 6.15 | AAI-based (high-confidence) | 0 |
| S16_extracted_100473 | 16067 | No | NA | 21 | 6 | 0 | Medium-quality | Genome-fragment | 72.45 | AAI-based (medium-confidence) | 0 |
| S8_extracted_k141_168753 | 2547 | No | NA | 6 | 5 | 0 | Low-quality | Genome-fragment | 4.16 | HMM-based (lower-bound) | 0 |
| S16_extracted_100637 | 1478 | No | NA | 1 | 0 | 1 | Not-determined | Genome-fragment | NA | NA | 0 |
| S9_vs_k141_58395\|\|full | 2548 | No | NA | 7 | 0 | 5 | Low-quality | Genome-fragment | 1 | AAI-based (medium-confidence) | 0 |
| S16_extracted_100934 | 1190 | No | NA | 1 | 0 | 1 | Not-determined | Genome-fragment | NA | NA | 0 |
| S16_extracted_100944 | 1001 | No | NA | 2 | 0 | 1 | Not-determined | Genome-fragment | NA | NA | 0 |
| S16_extracted_100958 | 1422 | No | NA | 2 | 0 | 0 | Not-determined | Genome-fragment | NA | NA | 0 |
| S11_vs_k141_95696\|\|full | 2549 | No | NA | 4 | 0 | 0 | Low-quality | Genome-fragment | 4.43 | AAI-based (high-confidence) | 0 |
| S8_extracted_k141_103583 | 2549 | No | NA | 8 | 0 | 0 | Low-quality | Genome-fragment | 1.49 | AAI-based (high-confidence) | 0 |
| S3_vs_k141_119100\|\|full | 2550 | No | NA | 3 | 0 | 1 | Low-quality | Genome-fragment | 4.48 | AAI-based (medium-confidence) | 0 |
| S12_vs_k141_150512\|\|full | 2550 | No | NA | 6 | 0 | 0 | Low-quality | Genome-fragment | 6.11 | AAI-based (medium-confidence) | 0 |
| S3_extracted_k141_67904 | 2551 | No | NA | 3 | 1 | 0 | Low-quality | Genome-fragment | 1.52 | AAI-based (high-confidence) | 0 |
| S7_vs_73965\|\|lt2gene | 2551 | No | NA | 1 | 1 | 0 | Low-quality | Genome-fragment | 18.3 | AAI-based (high-confidence) | 0 |
| S16_extracted_101434 | 1478 | No | NA | 3 | 0 | 0 | Not-determined | Genome-fragment | NA | NA | 0 |
| S11_vs_k141_99117\|\|full | 2552 | No | NA | 6 | 2 | 0 | Low-quality | Genome-fragment | 2.3 | HMM-based (lower-bound) | 0 |
| S16_extracted_101453 | 2191 | No | NA | 1 | 0 | 1 | Not-determined | Genome-fragment | NA | NA | 0 |
| S16_extracted_101540 | 1674 | No | NA | 1 | 0 | 1 | Not-determined | Genome-fragment | NA | NA | 0 |
| S16_extracted_101544 | 1263 | No | NA | 1 | 0 | 1 | Not-determined | Genome-fragment | NA | NA | 0 |
| S1_vs_k141_117076\|\|full | 2552 | No | NA | 4 | 2 | 0 | Low-quality | Genome-fragment | 5.86 | AAI-based (high-confidence) | 0 |
| S16_extracted_101586 | 1627 | No | NA | 4 | 0 | 2 | Not-determined | Genome-fragment | NA | NA | 0 |
| S16_extracted_101590 | 1166 | No | NA | 1 | 0 | 1 | Not-determined | Genome-fragment | NA | NA | 0 |
| S16_extracted_101792 | 1140 | No | NA | 2 | 0 | 0 | Not-determined | Genome-fragment | NA | NA | 0 |
| S16_extracted_101795 | 5110 | No | NA | 4 | 0 | 3 | Not-determined | Genome-fragment | NA | NA | 0 |
| S16_vs_155195\|\|full | 2553 | No | NA | 2 | 0 | 2 | Low-quality | Genome-fragment | 3.08 | AAI-based (medium-confidence) | 0 |
| S16_extracted_101898 | 5145 | No | NA | 6 | 0 | 4 | Not-determined | Genome-fragment | NA | NA | 0 |
| S16_extracted_101905 | 3949 | No | NA | 9 | 0 | 0 | Not-determined | Genome-fragment | NA | NA | 0 |
| S16_extracted_102162 | 8865 | No | NA | 9 | 0 | 4 | Not-determined | Genome-fragment | NA | NA | 0 |
| S16_extracted_102178 | 8321 | No | NA | 5 | 0 | 1 | Not-determined | Genome-fragment | NA | NA | 0 |
| S16_extracted_102289 | 3173 | No | NA | 3 | 0 | 3 | Not-determined | Genome-fragment | NA | NA | 0 |
| S16_extracted_102421 | 1400 | No | NA | 1 | 0 | 1 | Not-determined | Genome-fragment | NA | NA | 0 |
| S16_extracted_102439 | 1513 | No | NA | 1 | 0 | 0 | Not-determined | Genome-fragment | NA | NA | 0 |
| S16_extracted_102463 | 5472 | No | NA | 5 | 0 | 4 | Not-determined | Genome-fragment | NA | NA | 0 |
| S16_extracted_102466 | 1503 | No | NA | 2 | 0 | 2 | Not-determined | Genome-fragment | NA | NA | 0 |
| S9_vs_k141_4649\|\|full | 2554 | No | NA | 7 | 3 | 0 | Low-quality | Genome-fragment | 4.93 | HMM-based (lower-bound) | 0 |
| S1_vs_k141_115882\|\|full | 2554 | No | NA | 5 | 5 | 0 | Low-quality | Genome-fragment | 3.18 | AAI-based (high-confidence) | 0 |
| S7_extracted_132505 | 2554 | No | NA | 3 | 2 | 0 | Low-quality | Genome-fragment | 4.49 | AAI-based (high-confidence) | 0 |
| S16_extracted_102599 | 1392 | No | NA | 1 | 0 | 1 | Not-determined | Genome-fragment | NA | NA | 0 |
| S16_extracted_102642 | 2424 | No | NA | 3 | 0 | 3 | Not-determined | Genome-fragment | NA | NA | 0 |
| S16_extracted_102737 | 1415 | No | NA | 1 | 0 | 0 | Not-determined | Genome-fragment | NA | NA | 0 |
| S16_extracted_102751 | 1038 | No | NA | 1 | 0 | 0 | Not-determined | Genome-fragment | NA | NA | 0 |
| S16_extracted_102833 | 3221 | No | NA | 3 | 0 | 0 | Not-determined | Genome-fragment | NA | NA | 0 |
| S10_vs_27794\|\|lt2gene | 2554 | No | NA | 1 | 1 | 0 | Low-quality | Genome-fragment | 6.1 | AAI-based (medium-confidence) | 0 |
| S16_extracted_103213 | 2428 | No | NA | 3 | 0 | 0 | Not-determined | Genome-fragment | NA | NA | 0 |
| S16_extracted_103309 | 1093 | No | NA | 4 | 0 | 0 | Not-determined | Genome-fragment | NA | NA | 0 |
| S16_extracted_103311 | 1393 | No | NA | 5 | 0 | 0 | Not-determined | Genome-fragment | NA | NA | 0 |
| S16_extracted_103418 | 2553 | No | NA | 3 | 0 | 2 | Not-determined | Genome-fragment | NA | NA | 0 |
| S17_vs_k141_65219\|\|full | 2555 | No | NA | 5 | 3 | 0 | Low-quality | Genome-fragment | 5.86 | AAI-based (high-confidence) | 0 |
| S16_extracted_103668 | 1845 | No | NA | 1 | 0 | 0 | Not-determined | Genome-fragment | NA | NA | 0 |
| S16_extracted_103694 | 3134 | No | NA | 1 | 0 | 1 | Not-determined | Genome-fragment | NA | NA | 0 |
| S7_vs_129609\|\|full | 2555 | No | NA | 8 | 1 | 0 | Low-quality | Genome-fragment | 4.85 | AAI-based (medium-confidence) | 0 |
| S9_vs_k141_52922\|\|full | 2556 | No | NA | 3 | 0 | 0 | Low-quality | Genome-fragment | 1.3 | AAI-based (high-confidence) | 0 |
| S16_extracted_103859 | 1471 | No | NA | 1 | 0 | 1 | Not-determined | Genome-fragment | NA | NA | 0 |
| S16_extracted_104094 | 3762 | No | NA | 4 | 0 | 4 | Not-determined | Genome-fragment | NA | NA | 0 |
| S3_vs_k141_75992\|\|full | 2557 | No | NA | 5 | 4 | 0 | Low-quality | Genome-fragment | 6.34 | AAI-based (medium-confidence) | 0 |
| S16_extracted_104115 | 1504 | No | NA | 1 | 0 | 1 | Not-determined | Genome-fragment | NA | NA | 0 |
| S7_vs_27343\|\|full | 2557 | No | NA | 4 | 0 | 0 | Low-quality | Genome-fragment | 5.99 | AAI-based (medium-confidence) | 0 |
| S16_extracted_104157 | 2057 | No | NA | 4 | 0 | 1 | Not-determined | Genome-fragment | NA | NA | 0 |
| S16_extracted_104223 | 4378 | No | NA | 4 | 0 | 0 | Not-determined | Genome-fragment | NA | NA | 0 |
| S16_extracted_104341 | 1594 | No | NA | 1 | 0 | 0 | Not-determined | Genome-fragment | NA | NA | 0 |
| S7_vs_71762\|\|full | 2557 | No | NA | 3 | 0 | 2 | Low-quality | Genome-fragment | 1.23 | AAI-based (medium-confidence) | 0 |
| S16_extracted_104415 | 1315 | No | NA | 1 | 0 | 0 | Not-determined | Genome-fragment | NA | NA | 0 |
| S7_vs_87526\|\|full | 2557 | No | NA | 8 | 2 | 0 | Low-quality | Genome-fragment | 5.42 | AAI-based (high-confidence) | 0 |
| S11_vs_k141_7871\|\|full | 2558 | No | NA | 2 | 0 | 2 | Low-quality | Genome-fragment | 2.99 | AAI-based (medium-confidence) | 0 |
| S14_vs_k141_41509\|\|full | 2558 | No | NA | 4 | 1 | 0 | Low-quality | Genome-fragment | 1.91 | AAI-based (medium-confidence) | 0 |
| S5_vs_k141_141366\|\|full | 2558 | No | NA | 2 | 0 | 2 | Low-quality | Genome-fragment | 0.63 | AAI-based (medium-confidence) | 0 |
| S16_extracted_104555 | 1652 | No | NA | 1 | 0 | 1 | Not-determined | Genome-fragment | NA | NA | 0 |
| S16_extracted_104618 | 3055 | No | NA | 5 | 0 | 1 | Not-determined | Genome-fragment | NA | NA | 0 |
| S7_vs_922\|\|full | 2559 | No | NA | 5 | 1 | 0 | Low-quality | Genome-fragment | 9.41 | HMM-based (lower-bound) | 0 |
| S16_extracted_104871 | 1638 | No | NA | 2 | 0 | 0 | Not-determined | Genome-fragment | NA | NA | 0 |
| S16_extracted_105042 | 4668 | No | NA | 4 | 0 | 4 | Not-determined | Genome-fragment | NA | NA | 0 |
| S16_extracted_105144 | 1025 | No | NA | 2 | 0 | 0 | Not-determined | Genome-fragment | NA | NA | 0 |
| S16_vs_8470\|\|full | 2559 | No | NA | 3 | 3 | 0 | Low-quality | Genome-fragment | 1.9 | HMM-based (lower-bound) | 0 |
| S8_vs_k141_175725\|\|full | 2559 | No | NA | 3 | 0 | 0 | Low-quality | Genome-fragment | 4.47 | AAI-based (medium-confidence) | 0 |
| S3_vs_k141_54843\|\|full | 2560 | No | NA | 3 | 3 | 0 | Low-quality | Genome-fragment | 5.64 | AAI-based (high-confidence) | 0 |
| S16_extracted_105346 | 1378 | No | NA | 3 | 0 | 0 | Not-determined | Genome-fragment | NA | NA | 0 |
| S16_extracted_105442 | 1211 | No | NA | 1 | 0 | 0 | Not-determined | Genome-fragment | NA | NA | 0 |
| S7_vs_32335\|\|full | 2560 | No | NA | 6 | 0 | 0 | Low-quality | Genome-fragment | 5.25 | AAI-based (medium-confidence) | 0 |
| S16_extracted_105482 | 1294 | No | NA | 2 | 0 | 0 | Not-determined | Genome-fragment | NA | NA | 0 |
| S16_extracted_105520 | 1271 | No | NA | 3 | 0 | 0 | Not-determined | Genome-fragment | NA | NA | 0 |
| S16_extracted_105558 | 1096 | No | NA | 3 | 0 | 2 | Not-determined | Genome-fragment | NA | NA | 0 |
| S16_extracted_105574 | 1235 | No | NA | 1 | 0 | 1 | Not-determined | Genome-fragment | NA | NA | 0 |
| S16_extracted_105751 | 1853 | No | NA | 3 | 0 | 2 | Not-determined | Genome-fragment | NA | NA | 0 |
| S16_vs_119113\|\|full | 2560 | No | NA | 2 | 1 | 0 | Low-quality | Genome-fragment | 1.97 | HMM-based (lower-bound) | 0 |
| S12_vs_k141_169202\|\|lt2gene | 2560 | No | NA | 1 | 1 | 0 | Low-quality | Genome-fragment | 48.93 | AAI-based (high-confidence) | 0 |
| S16_extracted_106076 | 3215 | No | NA | 3 | 0 | 1 | Not-determined | Genome-fragment | NA | NA | 0 |
| S16_extracted_106370 | 1321 | No | NA | 1 | 0 | 1 | Not-determined | Genome-fragment | NA | NA | 0 |
| S16_extracted_106436 | 1742 | No | NA | 1 | 0 | 1 | Not-determined | Genome-fragment | NA | NA | 0 |
| S16_extracted_106445 | 2117 | No | NA | 2 | 0 | 1 | Not-determined | Genome-fragment | NA | NA | 0 |
| S8_vs_k141_186925\|\|full | 2560 | No | NA | 4 | 2 | 0 | Low-quality | Genome-fragment | 6.62 | AAI-based (high-confidence) | 0 |
| S16_extracted_106559 | 4338 | No | NA | 4 | 0 | 4 | Not-determined | Genome-fragment | NA | NA | 0 |
| S16_extracted_106583 | 1591 | No | NA | 1 | 0 | 1 | Not-determined | Genome-fragment | NA | NA | 0 |
| S16_extracted_106696 | 2098 | No | NA | 3 | 0 | 3 | Not-determined | Genome-fragment | NA | NA | 0 |
| S16_extracted_106708 | 3413 | No | NA | 1 | 0 | 1 | Not-determined | Genome-fragment | NA | NA | 0 |
| S16_extracted_106750 | 6953 | No | NA | 2 | 0 | 2 | Not-determined | Genome-fragment | NA | NA | 0 |
| S16_extracted_106763 | 1584 | No | NA | 1 | 0 | 1 | Not-determined | Genome-fragment | NA | NA | 0 |
| S16_extracted_106802 | 1741 | No | NA | 2 | 0 | 1 | Not-determined | Genome-fragment | NA | NA | 0 |
| S16_extracted_106871 | 3515 | No | NA | 4 | 0 | 0 | Not-determined | Genome-fragment | NA | NA | 0 |
| S16_extracted_106897 | 4134 | No | NA | 10 | 0 | 0 | Not-determined | Genome-fragment | NA | NA | 0 |
| S16_extracted_106919 | 1050 | No | NA | 2 | 0 | 1 | Not-determined | Genome-fragment | NA | NA | 0 |
| S9_vs_k141_49478\|\|full | 2561 | No | NA | 3 | 0 | 1 | Low-quality | Genome-fragment | 3.52 | AAI-based (high-confidence) | 0 |
| S16_extracted_106949 | 3383 | No | NA | 4 | 0 | 0 | Not-determined | Genome-fragment | NA | NA | 0 |
| S16_extracted_106951 | 2231 | No | NA | 3 | 0 | 1 | Not-determined | Genome-fragment | NA | NA | 0 |
| S16_extracted_106985 | 3899 | No | NA | 2 | 0 | 2 | Not-determined | Genome-fragment | NA | NA | 0 |
| S16_extracted_107013 | 2482 | No | NA | 2 | 0 | 2 | Not-determined | Genome-fragment | NA | NA | 0 |
| S5_vs_k141_55578\|\|full | 2562 | No | NA | 3 | 0 | 0 | Low-quality | Genome-fragment | 1.77 | AAI-based (medium-confidence) | 0 |
| S16_extracted_107065 | 1015 | No | NA | 2 | 0 | 0 | Not-determined | Genome-fragment | NA | NA | 0 |
| S16_extracted_107080 | 2408 | No | NA | 2 | 0 | 1 | Not-determined | Genome-fragment | NA | NA | 0 |
| S16_extracted_107270 | 1504 | No | NA | 1 | 0 | 1 | Not-determined | Genome-fragment | NA | NA | 0 |
| S16_extracted_107366 | 1836 | No | NA | 2 | 0 | 0 | Not-determined | Genome-fragment | NA | NA | 0 |
| S16_extracted_107428 | 4273 | No | NA | 4 | 0 | 0 | Not-determined | Genome-fragment | NA | NA | 0 |
| S16_extracted_107559 | 1234 | No | NA | 2 | 0 | 1 | Not-determined | Genome-fragment | NA | NA | 0 |
| S16_extracted_107561 | 8680 | No | NA | 7 | 0 | 3 | Not-determined | Genome-fragment | NA | NA | 0 |
| S10_extracted_34002 | 2562 | No | NA | 5 | 2 | 0 | Low-quality | Genome-fragment | 3.43 | AAI-based (high-confidence) | 0 |
| S16_extracted_107594 | 3141 | No | NA | 1 | 0 | 1 | Not-determined | Genome-fragment | NA | NA | 0 |
| S11_vs_k141_21255\|\|full | 2563 | No | NA | 6 | 1 | 0 | Low-quality | Genome-fragment | 4.31 | AAI-based (high-confidence) | 0 |
| S14_vs_k141_39828\|\|full | 2563 | No | NA | 6 | 2 | 0 | Low-quality | Genome-fragment | 9.54 | AAI-based (high-confidence) | 0 |
| S16_extracted_107697 | 1339 | No | NA | 2 | 0 | 2 | Not-determined | Genome-fragment | NA | NA | 0 |
| S16_extracted_107759 | 1030 | No | NA | 1 | 0 | 0 | Not-determined | Genome-fragment | NA | NA | 0 |
| S14_vs_k141_62043\|\|full | 2563 | No | NA | 5 | 0 | 1 | Low-quality | Genome-fragment | 1.8 | AAI-based (medium-confidence) | 0 |
| S9_vs_k141_19862\|\|full | 2564 | No | NA | 3 | 3 | 0 | Low-quality | Genome-fragment | 6.55 | AAI-based (high-confidence) | 0 |
| S11_vs_k141_33198\|\|full | 2564 | No | NA | 3 | 3 | 0 | Low-quality | Genome-fragment | 7.88 | AAI-based (medium-confidence) | 0 |
| S16_extracted_107955 | 1070 | No | NA | 1 | 0 | 1 | Not-determined | Genome-fragment | NA | NA | 0 |
| S3_vs_k141_85661\|\|full | 2564 | No | NA | 4 | 0 | 3 | Low-quality | Genome-fragment | 3.15 | AAI-based (medium-confidence) | 0 |
| S16_extracted_107977 | 5162 | No | NA | 5 | 0 | 3 | Not-determined | Genome-fragment | NA | NA | 0 |
| S16_extracted_107993 | 1598 | No | NA | 2 | 0 | 1 | Not-determined | Genome-fragment | NA | NA | 0 |
| S16_extracted_108030 | 1981 | No | NA | 3 | 0 | 0 | Not-determined | Genome-fragment | NA | NA | 0 |
| S16_extracted_108071 | 1167 | No | NA | 1 | 0 | 1 | Not-determined | Genome-fragment | NA | NA | 0 |
| S16_extracted_108169 | 1149 | No | NA | 2 | 0 | 2 | Not-determined | Genome-fragment | NA | NA | 0 |
| S16_extracted_108177 | 3622 | No | NA | 3 | 0 | 1 | Not-determined | Genome-fragment | NA | NA | 0 |
| S12_extracted_k141_4810 | 2564 | No | NA | 2 | 1 | 0 | Low-quality | Genome-fragment | 5.86 | AAI-based (medium-confidence) | 0 |
| S16_extracted_108260 | 1381 | No | NA | 1 | 0 | 1 | Not-determined | Genome-fragment | NA | NA | 0 |
| S16_extracted_108262 | 2943 | No | NA | 5 | 0 | 2 | Not-determined | Genome-fragment | NA | NA | 0 |
| S16_extracted_108342 | 8223 | No | NA | 9 | 0 | 2 | Not-determined | Genome-fragment | NA | NA | 0 |
| S16_extracted_108528 | 4105 | No | NA | 2 | 0 | 0 | Not-determined | Genome-fragment | NA | NA | 0 |
| S16_extracted_108554 | 57027 | No | NA | 104 | 19 | 2 | High-quality | High-quality | 98.93 | AAI-based (high-confidence) | 0 |
| S16_extracted_108582 | 1124 | No | NA | 1 | 0 | 1 | Not-determined | Genome-fragment | NA | NA | 0 |
| S16_extracted_108749 | 1107 | No | NA | 1 | 0 | 0 | Not-determined | Genome-fragment | NA | NA | 0 |
| S12_vs_k141_81227\|\|full | 2564 | No | NA | 6 | 1 | 0 | Low-quality | Genome-fragment | 4.33 | HMM-based (lower-bound) | 0 |
| S16_extracted_108922 | 2111 | No | NA | 3 | 0 | 1 | Not-determined | Genome-fragment | NA | NA | 0 |
| S16_extracted_109062 | 1143 | No | NA | 1 | 0 | 0 | Not-determined | Genome-fragment | NA | NA | 0 |
| S13_extracted_139681 | 2565 | No | NA | 2 | 0 | 2 | Low-quality | Genome-fragment | 0.7 | AAI-based (medium-confidence) | 0 |
| S1_vs_k141_8486\|\|full | 2565 | No | NA | 2 | 0 | 1 | Low-quality | Genome-fragment | 1.19 | AAI-based (high-confidence) | 0 |
| S3_vs_k141_25090\|\|full | 2565 | No | NA | 4 | 0 | 1 | Low-quality | Genome-fragment | 9.01 | AAI-based (medium-confidence) | 0 |
| S16_extracted_109415 | 1403 | No | NA | 2 | 0 | 1 | Not-determined | Genome-fragment | NA | NA | 0 |
| S3_vs_k141_63056\|\|lt2gene | 2565 | No | NA | 2 | 1 | 0 | Low-quality | Genome-fragment | 2.76 | HMM-based (lower-bound) | 0 |
| S12_vs_k141_35806\|\|full | 2565 | No | NA | 6 | 5 | 0 | Low-quality | Genome-fragment | 6.16 | AAI-based (high-confidence) | 0 |
| S17_vs_k141_53302\|\|full | 2566 | No | NA | 3 | 0 | 2 | Low-quality | Genome-fragment | 0.59 | AAI-based (high-confidence) | 0 |
| S16_extracted_109477 | 1298 | No | NA | 1 | 0 | 0 | Not-determined | Genome-fragment | NA | NA | 0 |
| S16_extracted_109639 | 1065 | No | NA | 2 | 0 | 0 | Not-determined | Genome-fragment | NA | NA | 0 |
| S16_extracted_109667 | 22177 | No | NA | 23 | 0 | 8 | Not-determined | Genome-fragment | NA | NA | 0 |
| S16_extracted_109699 | 1402 | No | NA | 1 | 0 | 0 | Not-determined | Genome-fragment | NA | NA | 0 |
| S16_extracted_109731 | 5075 | No | NA | 5 | 0 | 3 | Not-determined | Genome-fragment | NA | NA | 0 |
| S16_extracted_109825 | 1569 | No | NA | 1 | 0 | 0 | Not-determined | Genome-fragment | NA | NA | 0 |
| S16_extracted_109881 | 1065 | No | NA | 2 | 0 | 2 | Not-determined | Genome-fragment | NA | NA | 0 |
| S1_extracted_k141_72186 | 2566 | No | NA | 4 | 0 | 0 | Low-quality | Genome-fragment | 4.4 | AAI-based (high-confidence) | 0 |
| S16_extracted_110168 | 1530 | No | NA | 2 | 0 | 1 | Not-determined | Genome-fragment | NA | NA | 0 |
| S16_extracted_110187 | 1577 | No | NA | 2 | 0 | 0 | Not-determined | Genome-fragment | NA | NA | 0 |
| S16_extracted_110230 | 1097 | No | NA | 2 | 0 | 2 | Not-determined | Genome-fragment | NA | NA | 0 |
| S1_vs_k141_18356\|\|full | 2566 | No | NA | 6 | 1 | 0 | Low-quality | Genome-fragment | 3.54 | AAI-based (high-confidence) | 0 |
| S3_vs_k141_113180\|\|full | 2566 | No | NA | 5 | 1 | 0 | Low-quality | Genome-fragment | 3.25 | AAI-based (high-confidence) | 0 |
| S16_extracted_110485 | 4600 | No | NA | 4 | 0 | 4 | Not-determined | Genome-fragment | NA | NA | 0 |
| S16_extracted_110711 | 6739 | No | NA | 8 | 0 | 6 | Not-determined | Genome-fragment | NA | NA | 0 |
| S16_extracted_110713 | 1168 | No | NA | 1 | 0 | 1 | Not-determined | Genome-fragment | NA | NA | 0 |
| S16_extracted_110737 | 4660 | No | NA | 5 | 0 | 3 | Not-determined | Genome-fragment | NA | NA | 0 |
| S16_extracted_110757 | 1012 | No | NA | 2 | 0 | 1 | Not-determined | Genome-fragment | NA | NA | 0 |
| S16_extracted_110863 | 1274 | No | NA | 1 | 0 | 1 | Not-determined | Genome-fragment | NA | NA | 0 |
| S16_extracted_110886 | 1072 | No | NA | 1 | 0 | 1 | Not-determined | Genome-fragment | NA | NA | 0 |
| S16_extracted_110905 | 1331 | No | NA | 3 | 0 | 1 | Not-determined | Genome-fragment | NA | NA | 0 |
| S10_extracted_107012 | 2566 | No | NA | 4 | 0 | 3 | Low-quality | Genome-fragment | 0.82 | AAI-based (medium-confidence) | 0 |
| S16_extracted_110993 | 1006 | No | NA | 2 | 0 | 2 | Not-determined | Genome-fragment | NA | NA | 0 |
| S16_extracted_111036 | 1007 | No | NA | 2 | 0 | 1 | Not-determined | Genome-fragment | NA | NA | 0 |
| S16_extracted_111062 | 2062 | No | NA | 2 | 0 | 1 | Not-determined | Genome-fragment | NA | NA | 0 |
| S16_extracted_111077 | 1635 | No | NA | 2 | 0 | 1 | Not-determined | Genome-fragment | NA | NA | 0 |
| S16_extracted_111127 | 3009 | No | NA | 1 | 0 | 0 | Not-determined | Genome-fragment | NA | NA | 0 |
| S16_extracted_111290 | 1420 | No | NA | 1 | 0 | 1 | Not-determined | Genome-fragment | NA | NA | 0 |
| S16_extracted_111355 | 2241 | No | NA | 1 | 0 | 1 | Not-determined | Genome-fragment | NA | NA | 0 |
| S16_extracted_111383 | 3301 | No | NA | 3 | 0 | 2 | Not-determined | Genome-fragment | NA | NA | 0 |
| S12_vs_k141_170315\|\|full | 2566 | No | NA | 6 | 1 | 0 | Low-quality | Genome-fragment | 7.19 | AAI-based (high-confidence) | 0 |
| S16_extracted_111656 | 1525 | No | NA | 1 | 0 | 1 | Not-determined | Genome-fragment | NA | NA | 0 |
| S9_vs_k141_24118\|\|full | 2567 | No | NA | 3 | 2 | 0 | Low-quality | Genome-fragment | 5.82 | AAI-based (high-confidence) | 0 |
| S14_extracted_k141_23575 | 2567 | No | NA | 8 | 7 | 0 | Low-quality | Genome-fragment | 8.46 | AAI-based (high-confidence) | 0 |
| S16_vs_23375\|\|full | 2567 | No | NA | 9 | 0 | 0 | Low-quality | Genome-fragment | 5.38 | AAI-based (high-confidence) | 0 |
| S15_extracted_22508 | 2567 | No | NA | 6 | 1 | 0 | Low-quality | Genome-fragment | 5.74 | AAI-based (medium-confidence) | 0 |
| S17_vs_k141_88905\|\|full | 2568 | No | NA | 9 | 0 | 0 | Low-quality | Genome-fragment | 4.63 | AAI-based (high-confidence) | 0 |
| S16_extracted_112051 | 3154 | No | NA | 3 | 0 | 3 | Not-determined | Genome-fragment | NA | NA | 0 |
| S16_extracted_112202 | 4018 | No | NA | 4 | 0 | 3 | Not-determined | Genome-fragment | NA | NA | 0 |
| S11_vs_k141_3462\|\|full | 2568 | No | NA | 3 | 2 | 0 | Low-quality | Genome-fragment | 7.64 | AAI-based (medium-confidence) | 0 |
| S16_extracted_112377 | 1029 | No | NA | 1 | 0 | 1 | Not-determined | Genome-fragment | NA | NA | 0 |
| S5_vs_k141_4754\|\|full | 2568 | No | NA | 4 | 3 | 0 | Low-quality | Genome-fragment | 1.18 | AAI-based (high-confidence) | 0 |
| S16_extracted_112643 | 1250 | No | NA | 2 | 0 | 1 | Not-determined | Genome-fragment | NA | NA | 0 |
| S16_extracted_112681 | 1346 | No | NA | 2 | 0 | 1 | Not-determined | Genome-fragment | NA | NA | 0 |
| S16_extracted_112737 | 1340 | No | NA | 4 | 0 | 2 | Not-determined | Genome-fragment | NA | NA | 0 |
| S7_vs_105946\|\|full | 2568 | No | NA | 5 | 2 | 0 | Low-quality | Genome-fragment | 6 | AAI-based (high-confidence) | 0 |
| S7_vs_114083\|\|full | 2568 | No | NA | 4 | 0 | 0 | Low-quality | Genome-fragment | 5.09 | AAI-based (high-confidence) | 0 |
| S16_extracted_113074 | 2524 | No | NA | 2 | 0 | 1 | Not-determined | Genome-fragment | NA | NA | 0 |
| S16_extracted_113089 | 1337 | No | NA | 3 | 0 | 0 | Not-determined | Genome-fragment | NA | NA | 0 |
| S16_extracted_113207 | 1558 | No | NA | 3 | 0 | 2 | Not-determined | Genome-fragment | NA | NA | 0 |
| S16_extracted_113434 | 1192 | No | NA | 1 | 0 | 0 | Not-determined | Genome-fragment | NA | NA | 0 |
| S10_extracted_70606 | 2568 | No | NA | 5 | 1 | 0 | Low-quality | Genome-fragment | 1.3 | AAI-based (medium-confidence) | 0 |
| S16_extracted_113453 | 1327 | No | NA | 2 | 0 | 1 | Not-determined | Genome-fragment | NA | NA | 0 |
| S16_extracted_113528 | 1726 | No | NA | 3 | 0 | 2 | Not-determined | Genome-fragment | NA | NA | 0 |
| S16_extracted_113561 | 1001 | No | NA | 1 | 0 | 0 | Not-determined | Genome-fragment | NA | NA | 0 |
| S16_extracted_113585 | 5861 | No | NA | 6 | 0 | 0 | Not-determined | Genome-fragment | NA | NA | 0 |
| S16_extracted_113682 | 5332 | No | NA | 5 | 0 | 3 | Not-determined | Genome-fragment | NA | NA | 0 |
| S16_extracted_113705 | 6128 | No | NA | 7 | 0 | 5 | Not-determined | Genome-fragment | NA | NA | 0 |
| S1_extracted_k141_72781 | 2569 | No | NA | 3 | 2 | 0 | Low-quality | Genome-fragment | 5.85 | AAI-based (high-confidence) | 0 |
| S7_vs_25293\|\|full | 2569 | No | NA | 4 | 0 | 1 | Low-quality | Genome-fragment | 3.55 | AAI-based (medium-confidence) | 0 |
| S16_vs_36866\|\|full | 2569 | No | NA | 3 | 0 | 1 | Low-quality | Genome-fragment | 0.63 | AAI-based (medium-confidence) | 0 |
| S15_vs_16407\|\|full | 2569 | No | NA | 4 | 0 | 0 | Low-quality | Genome-fragment | 5.4 | AAI-based (high-confidence) | 0 |
| S16_extracted_114042 | 3713 | No | NA | 8 | 0 | 3 | Not-determined | Genome-fragment | NA | NA | 0 |
| S17_extracted_k141_38040 | 2570 | No | NA | 6 | 5 | 0 | Low-quality | Genome-fragment | 8.48 | AAI-based (high-confidence) | 0 |
| S17_vs_k141_165686\|\|full | 2570 | Yes | 1153 | 4 | 1 | 1 | Low-quality | Genome-fragment | 2.33 | AAI-based (medium-confidence) | 55.14 |
| S16_vs_112484\|\|full | 2570 | No | NA | 2 | 1 | 0 | Low-quality | Genome-fragment | 6.49 | AAI-based (medium-confidence) | 0 |
| S16_extracted_114236 | 4741 | No | NA | 5 | 0 | 2 | Not-determined | Genome-fragment | NA | NA | 0 |
| S16_extracted_114265 | 1673 | No | NA | 2 | 0 | 2 | Not-determined | Genome-fragment | NA | NA | 0 |
| S8_vs_k141_12482\|\|full | 2570 | No | NA | 3 | 0 | 1 | Low-quality | Genome-fragment | 1.32 | AAI-based (medium-confidence) | 0 |
| S16_extracted_114286 | 3239 | No | NA | 3 | 0 | 0 | Not-determined | Genome-fragment | NA | NA | 0 |
| S16_extracted_114367 | 1174 | No | NA | 2 | 0 | 0 | Not-determined | Genome-fragment | NA | NA | 0 |
| S11_extracted_k141_76146 | 2571 | No | NA | 3 | 0 | 0 | Low-quality | Genome-fragment | 1.61 | AAI-based (medium-confidence) | 0 |
| S16_extracted_114502 | 1470 | No | NA | 2 | 0 | 0 | Not-determined | Genome-fragment | NA | NA | 0 |
| S3_vs_k141_40071\|\|full | 2571 | No | NA | 5 | 1 | 0 | Low-quality | Genome-fragment | 1.62 | HMM-based (lower-bound) | 0 |
| S16_extracted_114526 | 4927 | No | NA | 1 | 0 | 1 | Not-determined | Genome-fragment | NA | NA | 0 |
| S16_extracted_114531 | 1082 | No | NA | 2 | 0 | 2 | Not-determined | Genome-fragment | NA | NA | 0 |
| S16_extracted_114555 | 3199 | No | NA | 3 | 0 | 2 | Not-determined | Genome-fragment | NA | NA | 0 |
| S16_extracted_114653 | 3315 | No | NA | 3 | 0 | 2 | Not-determined | Genome-fragment | NA | NA | 0 |
| S16_extracted_114787 | 1258 | No | NA | 2 | 0 | 2 | Not-determined | Genome-fragment | NA | NA | 0 |
| S16_extracted_114814 | 3892 | No | NA | 7 | 0 | 0 | Not-determined | Genome-fragment | NA | NA | 0 |
| S16_extracted_114845 | 1226 | No | NA | 2 | 0 | 1 | Not-determined | Genome-fragment | NA | NA | 0 |
| S7_vs_74514\|\|lt2gene | 2571 | No | NA | 2 | 1 | 0 | Low-quality | Genome-fragment | 7.47 | AAI-based (high-confidence) | 0 |
| S16_vs_86919\|\|full | 2571 | No | NA | 7 | 0 | 0 | Low-quality | Genome-fragment | 5.99 | AAI-based (medium-confidence) | 0 |
| S16_extracted_115015 | 1195 | No | NA | 3 | 0 | 1 | Not-determined | Genome-fragment | NA | NA | 0 |
| S16_extracted_115021 | 1769 | No | NA | 2 | 0 | 0 | Not-determined | Genome-fragment | NA | NA | 0 |
| S12_vs_k141_172942\|\|full | 2571 | No | NA | 3 | 0 | 0 | Low-quality | Genome-fragment | 2.92 | AAI-based (medium-confidence) | 0 |
| S16_vs_88112\|\|full | 2572 | No | NA | 3 | 1 | 0 | Low-quality | Genome-fragment | 4.72 | AAI-based (high-confidence) | 0 |
| S16_extracted_115204 | 1558 | No | NA | 1 | 0 | 1 | Not-determined | Genome-fragment | NA | NA | 0 |
| S16_extracted_115345 | 1020 | No | NA | 2 | 0 | 0 | Not-determined | Genome-fragment | NA | NA | 0 |
| S16_extracted_115375 | 14345 | No | NA | 11 | 0 | 10 | Not-determined | Genome-fragment | NA | NA | 0 |
| S16_extracted_115483 | 1187 | No | NA | 2 | 0 | 0 | Not-determined | Genome-fragment | NA | NA | 0 |
| S16_extracted_115565 | 1192 | No | NA | 3 | 0 | 0 | Not-determined | Genome-fragment | NA | NA | 0 |
| S16_extracted_115589 | 6273 | No | NA | 3 | 0 | 3 | Not-determined | Genome-fragment | NA | NA | 0 |
| S16_extracted_115679 | 20057 | No | NA | 19 | 0 | 8 | Not-determined | Genome-fragment | NA | NA | 0 |
| S16_extracted_115692 | 1251 | No | NA | 2 | 0 | 1 | Not-determined | Genome-fragment | NA | NA | 0 |
| S16_extracted_115793 | 1204 | No | NA | 2 | 0 | 0 | Not-determined | Genome-fragment | NA | NA | 0 |
| S16_extracted_115799 | 3680 | No | NA | 4 | 0 | 3 | Not-determined | Genome-fragment | NA | NA | 0 |
| S16_extracted_115879 | 5335 | No | NA | 10 | 0 | 0 | Not-determined | Genome-fragment | NA | NA | 0 |
| S16_extracted_115896 | 1585 | No | NA | 3 | 0 | 1 | Not-determined | Genome-fragment | NA | NA | 0 |
| S16_vs_102597\|\|full | 2572 | No | NA | 4 | 0 | 0 | Low-quality | Genome-fragment | 3.22 | AAI-based (medium-confidence) | 0 |
| S16_extracted_115969 | 4178 | No | NA | 8 | 0 | 1 | Not-determined | Genome-fragment | NA | NA | 0 |
| S16_extracted_116042 | 1004 | No | NA | 1 | 0 | 0 | Not-determined | Genome-fragment | NA | NA | 0 |
| S16_extracted_116072 | 4429 | No | NA | 3 | 0 | 1 | Not-determined | Genome-fragment | NA | NA | 0 |
| S16_extracted_116102 | 1297 | No | NA | 3 | 0 | 1 | Not-determined | Genome-fragment | NA | NA | 0 |
| S16_extracted_116135 | 3280 | No | NA | 2 | 0 | 2 | Not-determined | Genome-fragment | NA | NA | 0 |
| S10_vs_70402\|\|full | 2572 | No | NA | 4 | 3 | 0 | Low-quality | Genome-fragment | 1.88 | HMM-based (lower-bound) | 0 |
| S16_extracted_116198 | 4654 | No | NA | 4 | 0 | 2 | Not-determined | Genome-fragment | NA | NA | 0 |
| S16_extracted_116443 | 12819 | No | NA | 13 | 0 | 10 | Not-determined | Genome-fragment | NA | NA | 0 |
| S16_extracted_116454 | 1251 | No | NA | 1 | 0 | 1 | Not-determined | Genome-fragment | NA | NA | 0 |
| S16_extracted_116511 | 1130 | No | NA | 2 | 0 | 0 | Not-determined | Genome-fragment | NA | NA | 0 |
| S12_vs_k141_67275\|\|full | 2572 | No | NA | 4 | 0 | 0 | Low-quality | Genome-fragment | 6.21 | AAI-based (medium-confidence) | 0 |
| S16_extracted_116551 | 2774 | No | NA | 2 | 0 | 2 | Not-determined | Genome-fragment | NA | NA | 0 |
| S11_vs_k141_39411\|\|full | 2573 | No | NA | 6 | 1 | 0 | Low-quality | Genome-fragment | 3.79 | HMM-based (lower-bound) | 0 |
| S16_extracted_116599 | 4744 | No | NA | 6 | 0 | 5 | Not-determined | Genome-fragment | NA | NA | 0 |
| S16_extracted_116668 | 5615 | No | NA | 3 | 0 | 1 | Not-determined | Genome-fragment | NA | NA | 0 |
| S8_vs_k141_132669\|\|full | 2573 | No | NA | 4 | 0 | 0 | Low-quality | Genome-fragment | 6.02 | AAI-based (high-confidence) | 0 |
| S16_extracted_116803 | 1576 | No | NA | 2 | 0 | 0 | Not-determined | Genome-fragment | NA | NA | 0 |
| S16_extracted_116860 | 3335 | No | NA | 3 | 0 | 2 | Not-determined | Genome-fragment | NA | NA | 0 |
| S16_extracted_116965 | 1150 | No | NA | 1 | 0 | 1 | Not-determined | Genome-fragment | NA | NA | 0 |
| S16_extracted_117020 | 3272 | No | NA | 3 | 0 | 2 | Not-determined | Genome-fragment | NA | NA | 0 |
| S7_vs_68431\|\|full | 2574 | No | NA | 5 | 3 | 0 | Low-quality | Genome-fragment | 5.32 | AAI-based (medium-confidence) | 0 |
| S12_vs_k141_164821\|\|full | 2574 | No | NA | 3 | 2 | 0 | Low-quality | Genome-fragment | 7.6 | AAI-based (high-confidence) | 0 |
| S16_extracted_117240 | 1096 | No | NA | 2 | 0 | 0 | Not-determined | Genome-fragment | NA | NA | 0 |
| S16_extracted_117372 | 1213 | No | NA | 1 | 0 | 1 | Not-determined | Genome-fragment | NA | NA | 0 |
| S1_extracted_k141_101599 | 2575 | No | NA | 5 | 0 | 0 | Low-quality | Genome-fragment | 3.46 | AAI-based (high-confidence) | 0 |
| S16_extracted_117695 | 1888 | No | NA | 3 | 0 | 1 | Not-determined | Genome-fragment | NA | NA | 0 |
| S5_vs_k141_106405\|\|full | 2575 | No | NA | 6 | 6 | 0 | Low-quality | Genome-fragment | 4.47 | HMM-based (lower-bound) | 0 |
| S16_extracted_117830 | 1038 | No | NA | 4 | 0 | 0 | Not-determined | Genome-fragment | NA | NA | 0 |
| S16_extracted_118099 | 1001 | No | NA | 1 | 0 | 1 | Not-determined | Genome-fragment | NA | NA | 0 |
| S16_extracted_118130 | 8627 | No | NA | 1 | 0 | 1 | Not-determined | Genome-fragment | NA | NA | 0 |
| S9_vs_k141_43638\|\|full | 2576 | No | NA | 4 | 1 | 0 | Low-quality | Genome-fragment | 3.37 | HMM-based (lower-bound) | 0 |
| S16_extracted_118211 | 1919 | No | NA | 3 | 0 | 2 | Not-determined | Genome-fragment | NA | NA | 0 |
| S9_vs_k141_55292\|\|full | 2576 | No | NA | 6 | 3 | 0 | Low-quality | Genome-fragment | 8.1 | AAI-based (high-confidence) | 0 |
| S14_vs_k141_13208\|\|full | 2576 | No | NA | 6 | 3 | 0 | Low-quality | Genome-fragment | 5.37 | AAI-based (high-confidence) | 0 |
| S16_extracted_118347 | 1750 | No | NA | 1 | 0 | 0 | Not-determined | Genome-fragment | NA | NA | 0 |
| S5_extracted_k141_144717 | 2576 | No | NA | 2 | 2 | 0 | Low-quality | Genome-fragment | 5.57 | AAI-based (high-confidence) | 0 |
| S16_extracted_118418 | 3189 | No | NA | 4 | 0 | 2 | Not-determined | Genome-fragment | NA | NA | 0 |
| S16_extracted_118427 | 1105 | No | NA | 1 | 0 | 1 | Not-determined | Genome-fragment | NA | NA | 0 |
| S16_extracted_118496 | 1038 | No | NA | 1 | 0 | 0 | Not-determined | Genome-fragment | NA | NA | 0 |
| S5_vs_k141_9147\|\|full | 2576 | No | NA | 5 | 3 | 0 | Low-quality | Genome-fragment | 4.72 | AAI-based (medium-confidence) | 0 |
| S16_vs_45517\|\|full | 2576 | No | NA | 3 | 0 | 1 | Low-quality | Genome-fragment | 3.6 | AAI-based (medium-confidence) | 0 |
| S11_extracted_k141_90409 | 2577 | No | NA | 6 | 0 | 1 | Low-quality | Genome-fragment | 8.41 | AAI-based (medium-confidence) | 0 |
| S13_extracted_107871 | 2577 | No | NA | 3 | 0 | 1 | Low-quality | Genome-fragment | 2.54 | AAI-based (high-confidence) | 0 |
| S16_extracted_118708 | 1320 | No | NA | 1 | 0 | 1 | Not-determined | Genome-fragment | NA | NA | 0 |
| S5_extracted_k141_10410 | 2577 | No | NA | 4 | 3 | 0 | Low-quality | Genome-fragment | 6.29 | AAI-based (high-confidence) | 0 |
| S16_extracted_118751 | 4696 | No | NA | 5 | 0 | 5 | Not-determined | Genome-fragment | NA | NA | 0 |
| S16_extracted_118777 | 1012 | No | NA | 1 | 0 | 0 | Not-determined | Genome-fragment | NA | NA | 0 |
| S16_extracted_118925 | 1014 | No | NA | 2 | 0 | 2 | Not-determined | Genome-fragment | NA | NA | 0 |
| S16_extracted_119019 | 3428 | No | NA | 3 | 0 | 3 | Not-determined | Genome-fragment | NA | NA | 0 |
| S5_vs_k141_10297\|\|full | 2577 | No | NA | 3 | 0 | 3 | Low-quality | Genome-fragment | 1.66 | AAI-based (medium-confidence) | 0 |
| S7_vs_21784\|\|full | 2577 | No | NA | 4 | 0 | 2 | Low-quality | Genome-fragment | 3.36 | AAI-based (high-confidence) | 0 |
| S6_vs_k141_32951\|\|full | 2577 | No | NA | 8 | 2 | 0 | Low-quality | Genome-fragment | 5.29 | AAI-based (medium-confidence) | 0 |
| S9_vs_k141_70109\|\|full | 2578 | No | NA | 3 | 2 | 0 | Low-quality | Genome-fragment | 5.57 | AAI-based (high-confidence) | 0 |
| S16_extracted_119276 | 1201 | No | NA | 2 | 0 | 2 | Not-determined | Genome-fragment | NA | NA | 0 |
| S16_extracted_119364 | 7268 | No | NA | 7 | 0 | 1 | Not-determined | Genome-fragment | NA | NA | 0 |
| S16_extracted_119428 | 3535 | No | NA | 3 | 0 | 3 | Not-determined | Genome-fragment | NA | NA | 0 |
| S16_extracted_119489 | 1082 | No | NA | 1 | 0 | 1 | Not-determined | Genome-fragment | NA | NA | 0 |
| S16_extracted_119589 | 1155 | No | NA | 2 | 0 | 0 | Not-determined | Genome-fragment | NA | NA | 0 |
| S16_extracted_119675 | 1131 | No | NA | 1 | 0 | 1 | Not-determined | Genome-fragment | NA | NA | 0 |
| S5_vs_k141_15892\|\|full | 2578 | No | NA | 5 | 0 | 0 | Low-quality | Genome-fragment | 5.87 | AAI-based (medium-confidence) | 0 |
| S4_extracted_8476 | 2578 | No | NA | 3 | 3 | 0 | Low-quality | Genome-fragment | 8.5 | AAI-based (high-confidence) | 0 |
| S16_extracted_119738 | 1125 | No | NA | 1 | 0 | 1 | Not-determined | Genome-fragment | NA | NA | 0 |
| S16_extracted_119783 | 1255 | No | NA | 2 | 0 | 1 | Not-determined | Genome-fragment | NA | NA | 0 |
| S16_extracted_119823 | 1002 | No | NA | 1 | 0 | 1 | Not-determined | Genome-fragment | NA | NA | 0 |
| S16_extracted_119834 | 15826 | No | NA | 18 | 7 | 0 | Medium-quality | Genome-fragment | 78.38 | AAI-based (medium-confidence) | 0 |
| S16_extracted_119873 | 1289 | No | NA | 2 | 0 | 1 | Not-determined | Genome-fragment | NA | NA | 0 |
| S6_vs_k141_41743\|\|full | 2578 | No | NA | 5 | 1 | 0 | Low-quality | Genome-fragment | 6.51 | AAI-based (high-confidence) | 0 |
| S16_extracted_119928 | 2643 | No | NA | 2 | 0 | 1 | Not-determined | Genome-fragment | NA | NA | 0 |
| S17_vs_k141_104899\|\|full | 2579 | No | NA | 3 | 0 | 2 | Low-quality | Genome-fragment | 5.26 | AAI-based (medium-confidence) | 0 |
| S16_extracted_120095 | 1013 | No | NA | 1 | 0 | 0 | Not-determined | Genome-fragment | NA | NA | 0 |
| S16_extracted_120184 | 4470 | No | NA | 3 | 0 | 1 | Not-determined | Genome-fragment | NA | NA | 0 |
| S16_extracted_120206 | 6138 | No | NA | 7 | 0 | 3 | Not-determined | Genome-fragment | NA | NA | 0 |
| S16_extracted_120319 | 1626 | No | NA | 2 | 0 | 1 | Not-determined | Genome-fragment | NA | NA | 0 |
| S16_extracted_120356 | 1632 | No | NA | 1 | 0 | 0 | Not-determined | Genome-fragment | NA | NA | 0 |
| S1_extracted_k141_86758 | 2579 | No | NA | 5 | 0 | 0 | Low-quality | Genome-fragment | 1.47 | AAI-based (high-confidence) | 0 |
| S16_extracted_120474 | 1165 | No | NA | 3 | 0 | 0 | Not-determined | Genome-fragment | NA | NA | 0 |
| S16_extracted_120544 | 1153 | No | NA | 1 | 0 | 0 | Not-determined | Genome-fragment | NA | NA | 0 |
| S16_extracted_120581 | 1127 | No | NA | 1 | 0 | 1 | Not-determined | Genome-fragment | NA | NA | 0 |
| S16_extracted_120658 | 1094 | No | NA | 1 | 0 | 1 | Not-determined | Genome-fragment | NA | NA | 0 |
| S16_extracted_120674 | 1084 | No | NA | 1 | 0 | 1 | Not-determined | Genome-fragment | NA | NA | 0 |
| S16_extracted_120677 | 1438 | No | NA | 1 | 0 | 1 | Not-determined | Genome-fragment | NA | NA | 0 |
| S16_extracted_120687 | 5612 | No | NA | 10 | 0 | 1 | Not-determined | Genome-fragment | NA | NA | 0 |
| S16_extracted_120696 | 5595 | No | NA | 9 | 0 | 1 | Not-determined | Genome-fragment | NA | NA | 0 |
| S5_vs_k141_102885\|\|full | 2579 | No | NA | 4 | 0 | 0 | Low-quality | Genome-fragment | 4.57 | AAI-based (high-confidence) | 0 |
| S16_extracted_120783 | 1009 | No | NA | 1 | 0 | 1 | Not-determined | Genome-fragment | NA | NA | 0 |
| S16_extracted_120804 | 1700 | No | NA | 2 | 0 | 0 | Not-determined | Genome-fragment | NA | NA | 0 |
| S7_extracted_129816 | 2579 | No | NA | 7 | 4 | 0 | Low-quality | Genome-fragment | 4.53 | AAI-based (high-confidence) | 0 |
| S9_extracted_k141_12811 | 2580 | No | NA | 2 | 0 | 2 | Low-quality | Genome-fragment | 0.64 | AAI-based (medium-confidence) | 0 |
| S16_extracted_120944 | 1195 | No | NA | 1 | 0 | 1 | Not-determined | Genome-fragment | NA | NA | 0 |
| S16_extracted_121013 | 3181 | No | NA | 5 | 0 | 0 | Not-determined | Genome-fragment | NA | NA | 0 |
| S14_vs_k141_72612\|\|full | 2580 | No | NA | 8 | 0 | 0 | Low-quality | Genome-fragment | 5.11 | AAI-based (high-confidence) | 0 |
| S1_vs_k141_106513\|\|full | 2580 | No | NA | 2 | 0 | 0 | Low-quality | Genome-fragment | 4.13 | AAI-based (medium-confidence) | 0 |
| S16_extracted_121222 | 1606 | No | NA | 3 | 0 | 0 | Not-determined | Genome-fragment | NA | NA | 0 |
| S3_vs_k141_95345\|\|full | 2580 | No | NA | 5 | 2 | 0 | Low-quality | Genome-fragment | 1.85 | HMM-based (lower-bound) | 0 |
| S5_vs_k141_3516\|\|full | 2580 | No | NA | 7 | 0 | 0 | Low-quality | Genome-fragment | 6 | AAI-based (medium-confidence) | 0 |
| S16_extracted_121275 | 1085 | No | NA | 1 | 0 | 1 | Not-determined | Genome-fragment | NA | NA | 0 |
| S16_extracted_121278 | 1397 | No | NA | 2 | 0 | 0 | Not-determined | Genome-fragment | NA | NA | 0 |
| S16_extracted_121409 | 1003 | No | NA | 2 | 0 | 2 | Not-determined | Genome-fragment | NA | NA | 0 |
| S16_extracted_121434 | 1100 | No | NA | 1 | 0 | 0 | Not-determined | Genome-fragment | NA | NA | 0 |
| S16_extracted_121540 | 2300 | No | NA | 1 | 0 | 0 | Not-determined | Genome-fragment | NA | NA | 0 |
| S7_vs_134906\|\|full | 2580 | No | NA | 4 | 0 | 0 | Low-quality | Genome-fragment | 4.03 | AAI-based (medium-confidence) | 0 |
| S8_vs_k141_115812\|\|full | 2580 | No | NA | 5 | 0 | 1 | Low-quality | Genome-fragment | 6.75 | AAI-based (medium-confidence) | 0 |
| S16_extracted_121887 | 1226 | No | NA | 1 | 0 | 0 | Not-determined | Genome-fragment | NA | NA | 0 |
| S16_extracted_121908 | 1420 | No | NA | 4 | 0 | 0 | Not-determined | Genome-fragment | NA | NA | 0 |
| S16_extracted_121924 | 1200 | No | NA | 2 | 0 | 2 | Not-determined | Genome-fragment | NA | NA | 0 |
| S16_extracted_121956 | 1372 | No | NA | 3 | 0 | 0 | Not-determined | Genome-fragment | NA | NA | 0 |
| S14_vs_k141_97928\|\|full | 2581 | No | NA | 4 | 0 | 0 | Low-quality | Genome-fragment | 5.29 | AAI-based (high-confidence) | 0 |
| S16_extracted_122052 | 1146 | No | NA | 2 | 0 | 1 | Not-determined | Genome-fragment | NA | NA | 0 |
| S16_extracted_122137 | 4142 | No | NA | 4 | 0 | 2 | Not-determined | Genome-fragment | NA | NA | 0 |
| S16_extracted_122195 | 1108 | No | NA | 2 | 0 | 0 | Not-determined | Genome-fragment | NA | NA | 0 |

| | | | | | | | | | | | |
|---|---|---|---|---|---|---|---|---|---|---|---|
| S16_extracted_122276 | 1221 | No | NA | 2 | 0 | 2 | Not-determined | Genome-fragment | NA | NA | 0 |
| S16_extracted_122306 | 5914 | No | NA | 4 | 0 | 2 | Not-determined | Genome-fragment | NA | NA | 0 |
| S16_extracted_122327 | 1781 | No | NA | 1 | 0 | 0 | Not-determined | Genome-fragment | NA | NA | 0 |
| S16_extracted_122480 | 1284 | No | NA | 2 | 0 | 1 | Not-determined | Genome-fragment | NA | NA | 0 |
| S16_extracted_122502 | 1586 | No | NA | 3 | 0 | 1 | Not-determined | Genome-fragment | NA | NA | 0 |
| S16_extracted_122606 | 6258 | No | NA | 8 | 0 | 1 | Not-determined | Genome-fragment | NA | NA | 0 |
| S7_vs_84519\|\|full | 2581 | No | NA | 3 | 0 | 0 | Low-quality | Genome-fragment | 5.99 | AAI-based (medium-confidence) | 0 |
| S16_extracted_122646 | 3689 | No | NA | 3 | 0 | 2 | Not-determined | Genome-fragment | NA | NA | 0 |
| S7_vs_94108\|\|full | 2581 | No | NA | 7 | 3 | 0 | Low-quality | Genome-fragment | 5.51 | AAI-based (medium-confidence) | 0 |
| S16_vs_41220\|\|full | 2581 | No | NA | 4 | 0 | 0 | Low-quality | Genome-fragment | 6.87 | AAI-based (medium-confidence) | 0 |
| S16_extracted_122808 | 1146 | No | NA | 1 | 0 | 0 | Not-determined | Genome-fragment | NA | NA | 0 |
| S6_vs_k141_16838\|\|full | 2581 | Yes | 1340 | 5 | 1 | 1 | Low-quality | Genome-fragment | 0.81 | HMM-based (lower-bound) | 48.08 |
| S16_extracted_122988 | 1742 | No | NA | 1 | 0 | 1 | Not-determined | Genome-fragment | NA | NA | 0 |
| S16_vs_154070\|\|full | 2582 | No | NA | 2 | 0 | 1 | Low-quality | Genome-fragment | 2.53 | AAI-based (medium-confidence) | 0 |
| S16_extracted_123152 | 4105 | No | NA | 2 | 0 | 2 | Not-determined | Genome-fragment | NA | NA | 0 |
| S16_extracted_123181 | 1006 | No | NA | 1 | 0 | 1 | Not-determined | Genome-fragment | NA | NA | 0 |
| S17_vs_k141_59474\|\|full | 2583 | No | NA | 6 | 1 | 0 | Low-quality | Genome-fragment | 5.63 | AAI-based (high-confidence) | 0 |
| S16_extracted_123307 | 1012 | No | NA | 1 | 0 | 1 | Not-determined | Genome-fragment | NA | NA | 0 |
| S13_vs_41325\|\|full | 2583 | No | NA | 3 | 0 | 1 | Low-quality | Genome-fragment | 5.02 | AAI-based (high-confidence) | 0 |
| S16_extracted_123423 | 3735 | No | NA | 1 | 0 | 0 | Not-determined | Genome-fragment | NA | NA | 0 |
| S16_extracted_123501 | 1086 | No | NA | 1 | 0 | 0 | Not-determined | Genome-fragment | NA | NA | 0 |
| S13_vs_52088\|\|full | 2583 | No | NA | 4 | 0 | 0 | Low-quality | Genome-fragment | 5.84 | AAI-based (medium-confidence) | 0 |
| S16_extracted_123565 | 5847 | No | NA | 6 | 0 | 3 | Not-determined | Genome-fragment | NA | NA | 0 |
| S16_extracted_123609 | 1036 | No | NA | 1 | 0 | 0 | Not-determined | Genome-fragment | NA | NA | 0 |
| S16_extracted_123676 | 1286 | No | NA | 2 | 0 | 1 | Not-determined | Genome-fragment | NA | NA | 0 |
| S16_extracted_123720 | 1359 | No | NA | 2 | 0 | 1 | Not-determined | Genome-fragment | NA | NA | 0 |
| S16_extracted_123799 | 3477 | No | NA | 2 | 0 | 2 | Not-determined | Genome-fragment | NA | NA | 0 |
| S16_extracted_123828 | 1550 | No | NA | 1 | 0 | 1 | Not-determined | Genome-fragment | NA | NA | 0 |
| S16_extracted_123843 | 3175 | No | NA | 4 | 0 | 2 | Not-determined | Genome-fragment | NA | NA | 0 |
| S14_vs_k141_72063\|\|full | 2583 | No | NA | 4 | 1 | 0 | Low-quality | Genome-fragment | 4.56 | AAI-based (high-confidence) | 0 |
| S16_vs_152268\|\|full | 2583 | No | NA | 7 | 0 | 0 | Low-quality | Genome-fragment | 3.18 | AAI-based (high-confidence) | 0 |
| S16_extracted_124029 | 1182 | No | NA | 1 | 0 | 1 | Not-determined | Genome-fragment | NA | NA | 0 |
| S9_extracted_k141_70017 | 2584 | No | NA | 2 | 2 | 0 | Low-quality | Genome-fragment | 8.51 | AAI-based (high-confidence) | 0 |
| S16_extracted_124099 | 1247 | No | NA | 2 | 0 | 0 | Not-determined | Genome-fragment | NA | NA | 0 |
| S14_vs_k141_14937\|\|full | 2584 | No | NA | 5 | 2 | 0 | Low-quality | Genome-fragment | 6.69 | AAI-based (high-confidence) | 0 |
| S16_extracted_124277 | 1654 | No | NA | 2 | 0 | 1 | Not-determined | Genome-fragment | NA | NA | 0 |
| S16_extracted_124300 | 3674 | No | NA | 3 | 0 | 3 | Not-determined | Genome-fragment | NA | NA | 0 |
| S16_extracted_124310 | 1179 | No | NA | 1 | 0 | 0 | Not-determined | Genome-fragment | NA | NA | 0 |
| S1_vs_k141_105184\|\|full | 2584 | No | NA | 3 | 3 | 0 | Low-quality | Genome-fragment | 5.49 | AAI-based (high-confidence) | 0 |
| S17_vs_k141_27913\|\|full | 2585 | No | NA | 5 | 0 | 0 | Low-quality | Genome-fragment | 3.58 | AAI-based (high-confidence) | 0 |
| S16_extracted_124565 | 1787 | No | NA | 3 | 0 | 1 | Not-determined | Genome-fragment | NA | NA | 0 |
| S16_extracted_124573 | 1028 | No | NA | 1 | 0 | 0 | Not-determined | Genome-fragment | NA | NA | 0 |
| S16_extracted_124577 | 4102 | No | NA | 3 | 0 | 3 | Not-determined | Genome-fragment | NA | NA | 0 |
| S16_extracted_124744 | 2178 | No | NA | 2 | 0 | 2 | Not-determined | Genome-fragment | NA | NA | 0 |
| S16_extracted_124860 | 1287 | No | NA | 1 | 0 | 1 | Not-determined | Genome-fragment | NA | NA | 0 |
| S3_extracted_k141_96599 | 2585 | No | NA | 5 | 0 | 0 | Low-quality | Genome-fragment | 1.93 | AAI-based (high-confidence) | 0 |
| S3_extracted_k141_65492 | 2585 | No | NA | 4 | 2 | 0 | Low-quality | Genome-fragment | 11.8 | AAI-based (high-confidence) | 0 |
| S16_extracted_125183 | 1332 | No | NA | 2 | 0 | 0 | Not-determined | Genome-fragment | NA | NA | 0 |
| S8_vs_k141_1271\|\|full | 2585 | No | NA | 7 | 4 | 0 | Low-quality | Genome-fragment | 3.71 | HMM-based (lower-bound) | 0 |
| S16_extracted_125303 | 5915 | No | NA | 3 | 0 | 2 | Not-determined | Genome-fragment | NA | NA | 0 |
| S8_vs_k141_25504\|\|full | 2585 | No | NA | 3 | 0 | 1 | Low-quality | Genome-fragment | 1.26 | AAI-based (high-confidence) | 0 |
| S13_vs_74834\|\|full | 2586 | No | NA | 5 | 0 | 0 | Low-quality | Genome-fragment | 7.31 | AAI-based (high-confidence) | 0 |
| S16_extracted_125475 | 1111 | No | NA | 1 | 0 | 0 | Not-determined | Genome-fragment | NA | NA | 0 |
| S3_vs_k141_78103\|\|full | 2586 | No | NA | 3 | 1 | 0 | Low-quality | Genome-fragment | 5.19 | AAI-based (high-confidence) | 0 |
| S6_vs_k141_26165\|\|full | 2586 | No | NA | 3 | 0 | 2 | Low-quality | Genome-fragment | 2.07 | AAI-based (high-confidence) | 0 |
| S8_vs_k141_48074\|\|lt2gene | 2586 | No | NA | 1 | 1 | 0 | Low-quality | Genome-fragment | 2.33 | AAI-based (high-confidence) | 0 |
| S9_vs_k141_64649\|\|full | 2587 | No | NA | 3 | 0 | 1 | Low-quality | Genome-fragment | 5.12 | AAI-based (high-confidence) | 0 |
| S16_extracted_125868 | 2627 | No | NA | 2 | 0 | 1 | Not-determined | Genome-fragment | NA | NA | 0 |
| S16_extracted_125897 | 1000 | No | NA | 2 | 0 | 0 | Not-determined | Genome-fragment | NA | NA | 0 |
| S16_extracted_126030 | 1636 | No | NA | 1 | 0 | 1 | Not-determined | Genome-fragment | NA | NA | 0 |
| S16_extracted_126050 | 3310 | No | NA | 4 | 0 | 1 | Not-determined | Genome-fragment | NA | NA | 0 |
| S16_extracted_126221 | 1302 | No | NA | 1 | 0 | 1 | Not-determined | Genome-fragment | NA | NA | 0 |
| S9_vs_k141_30647\|\|full | 2587 | No | NA | 4 | 0 | 0 | Low-quality | Genome-fragment | 4.25 | AAI-based (medium-confidence) | 0 |
| S16_extracted_126407 | 1051 | No | NA | 1 | 0 | 1 | Not-determined | Genome-fragment | NA | NA | 0 |
| S16_extracted_126431 | 4602 | No | NA | 4 | 0 | 3 | Not-determined | Genome-fragment | NA | NA | 0 |
| S16_extracted_48607 | 2587 | No | NA | 3 | 0 | 0 | Low-quality | Genome-fragment | 4.48 | AAI-based (medium-confidence) | 0 |
| S16_extracted_126748 | 7178 | No | NA | 3 | 0 | 0 | Not-determined | Genome-fragment | NA | NA | 0 |
| S13_extracted_111086 | 2588 | No | NA | 3 | 1 | 0 | Low-quality | Genome-fragment | 3.89 | AAI-based (high-confidence) | 0 |
| S16_extracted_126814 | 1299 | No | NA | 1 | 0 | 1 | Not-determined | Genome-fragment | NA | NA | 0 |
| S14_vs_k141_50390\|\|full | 2588 | No | NA | 2 | 0 | 0 | Low-quality | Genome-fragment | 4.84 | AAI-based (medium-confidence) | 0 |
| S5_vs_k141_32833\|\|full | 2588 | No | NA | 7 | 6 | 0 | Low-quality | Genome-fragment | 6.16 | AAI-based (high-confidence) | 0 |
| S16_extracted_126997 | 4842 | No | NA | 7 | 0 | 1 | Not-determined | Genome-fragment | NA | NA | 0 |
| S12_vs_k141_131017\|\|full | 2588 | No | NA | 6 | 4 | 0 | Low-quality | Genome-fragment | 6.75 | AAI-based (medium-confidence) | 0 |
| S16_extracted_127098 | 1506 | No | NA | 3 | 0 | 0 | Not-determined | Genome-fragment | NA | NA | 0 |
| S16_extracted_127169 | 2385 | No | NA | 2 | 0 | 2 | Not-determined | Genome-fragment | NA | NA | 0 |
| S16_extracted_127315 | 1373 | No | NA | 1 | 0 | 0 | Not-determined | Genome-fragment | NA | NA | 0 |
| S16_extracted_127320 | 1486 | No | NA | 2 | 0 | 2 | Not-determined | Genome-fragment | NA | NA | 0 |
| S16_extracted_127352 | 3751 | No | NA | 3 | 0 | 2 | Not-determined | Genome-fragment | NA | NA | 0 |
| S16_extracted_127386 | 1214 | No | NA | 2 | 0 | 2 | Not-determined | Genome-fragment | NA | NA | 0 |
| S16_extracted_127612 | 1629 | No | NA | 1 | 0 | 1 | Not-determined | Genome-fragment | NA | NA | 0 |
| S16_extracted_127632 | 7650 | No | NA | 5 | 0 | 5 | Not-determined | Genome-fragment | NA | NA | 0 |
| S16_extracted_127673 | 1625 | No | NA | 1 | 0 | 1 | Not-determined | Genome-fragment | NA | NA | 0 |
| S16_extracted_127762 | 3142 | No | NA | 4 | 0 | 4 | Not-determined | Genome-fragment | NA | NA | 0 |
| S14_extracted_k141_61173 | 2589 | No | NA | 3 | 3 | 0 | Low-quality | Genome-fragment | 8.55 | AAI-based (high-confidence) | 0 |
| S16_extracted_127844 | 1146 | No | NA | 2 | 0 | 1 | Not-determined | Genome-fragment | NA | NA | 0 |
| S16_extracted_127886 | 1960 | No | NA | 4 | 0 | 3 | Not-determined | Genome-fragment | NA | NA | 0 |
| S16_extracted_127963 | 1396 | No | NA | 3 | 0 | 0 | Not-determined | Genome-fragment | NA | NA | 0 |
| S16_extracted_128082 | 1052 | No | NA | 2 | 0 | 0 | Not-determined | Genome-fragment | NA | NA | 0 |
| S3_vs_k141_109485\|\|full | 2589 | No | NA | 4 | 0 | 3 | Low-quality | Genome-fragment | 0.82 | AAI-based (medium-confidence) | 0 |
| S5_vs_k141_45348\|\|full | 2589 | No | NA | 7 | 4 | 0 | Low-quality | Genome-fragment | 1.91 | HMM-based (lower-bound) | 0 |
| S16_extracted_128270 | 1907 | No | NA | 3 | 0 | 0 | Not-determined | Genome-fragment | NA | NA | 0 |
| S16_extracted_128323 | 1429 | No | NA | 2 | 0 | 0 | Not-determined | Genome-fragment | NA | NA | 0 |
| S16_extracted_128335 | 2490 | No | NA | 4 | 0 | 4 | Not-determined | Genome-fragment | NA | NA | 0 |
| S16_extracted_128336 | 1291 | No | NA | 3 | 0 | 0 | Not-determined | Genome-fragment | NA | NA | 0 |
| S16_vs_79589\|\|full | 2589 | No | NA | 3 | 0 | 0 | Low-quality | Genome-fragment | 4.48 | AAI-based (high-confidence) | 0 |
| S2_vs_k141_10834\|\|lt2gene | 2589 | No | NA | 2 | 2 | 0 | Low-quality | Genome-fragment | 2.86 | AAI-based (high-confidence) | 0 |
| S16_extracted_128479 | 9274 | No | NA | 10 | 0 | 5 | Not-determined | Genome-fragment | NA | NA | 0 |
| S16_extracted_128524 | 1768 | No | NA | 2 | 0 | 2 | Not-determined | Genome-fragment | NA | NA | 0 |
| S16_extracted_128544 | 5268 | No | NA | 3 | 0 | 2 | Not-determined | Genome-fragment | NA | NA | 0 |
| S16_extracted_128598 | 1488 | No | NA | 2 | 0 | 2 | Not-determined | Genome-fragment | NA | NA | 0 |
| S16_extracted_128768 | 7011 | No | NA | 8 | 0 | 6 | Not-determined | Genome-fragment | NA | NA | 0 |
| S16_extracted_128814 | 1209 | No | NA | 2 | 0 | 1 | Not-determined | Genome-fragment | NA | NA | 0 |
| S16_extracted_128829 | 15514 | No | NA | 27 | 0 | 26 | Not-determined | Genome-fragment | NA | NA | 0 |
| S16_extracted_128834 | 1865 | No | NA | 2 | 0 | 1 | Not-determined | Genome-fragment | NA | NA | 0 |
| S16_extracted_128894 | 1176 | No | NA | 1 | 0 | 1 | Not-determined | Genome-fragment | NA | NA | 0 |
| S16_extracted_128916 | 1063 | No | NA | 2 | 0 | 1 | Not-determined | Genome-fragment | NA | NA | 0 |
| S16_extracted_128920 | 1760 | No | NA | 3 | 0 | 1 | Not-determined | Genome-fragment | NA | NA | 0 |
| S9_vs_k141_76216\|\|full | 2590 | No | NA | 6 | 2 | 0 | Low-quality | Genome-fragment | 3.96 | AAI-based (high-confidence) | 0 |
| S14_vs_k141_69206\|\|full | 2590 | No | NA | 2 | 0 | 0 | Low-quality | Genome-fragment | 4.03 | AAI-based (medium-confidence) | 0 |
| S16_extracted_129175 | 2549 | No | NA | 2 | 0 | 1 | Not-determined | Genome-fragment | NA | NA | 0 |
| S16_extracted_129206 | 9833 | No | NA | 10 | 0 | 3 | Not-determined | Genome-fragment | NA | NA | 0 |
| S16_extracted_129284 | 1030 | No | NA | 1 | 0 | 0 | Not-determined | Genome-fragment | NA | NA | 0 |
| S14_vs_k141_86476\|\|full | 2591 | No | NA | 3 | 0 | 0 | Low-quality | Genome-fragment | 4.07 | AAI-based (medium-confidence) | 0 |
| S16_extracted_129404 | 4518 | No | NA | 6 | 0 | 4 | Not-determined | Genome-fragment | NA | NA | 0 |
| S16_extracted_129528 | 1700 | No | NA | 2 | 0 | 1 | Not-determined | Genome-fragment | NA | NA | 0 |
| S16_extracted_129629 | 3502 | No | NA | 3 | 0 | 3 | Not-determined | Genome-fragment | NA | NA | 0 |
| S16_extracted_129641 | 11864 | No | NA | 11 | 0 | 8 | Not-determined | Genome-fragment | NA | NA | 0 |
| S3_extracted_k141_69038 | 2591 | No | NA | 1 | 0 | 1 | Low-quality | Genome-fragment | 0.7 | AAI-based (high-confidence) | 0 |
| S16_extracted_129715 | 6356 | No | NA | 4 | 0 | 3 | Not-determined | Genome-fragment | NA | NA | 0 |
| S8_extracted_k141_199875 | 2592 | No | NA | 2 | 0 | 1 | Low-quality | Genome-fragment | 1.57 | AAI-based (medium-confidence) | 0 |
| S16_extracted_129769 | 3047 | No | NA | 3 | 0 | 1 | Not-determined | Genome-fragment | NA | NA | 0 |
| S16_extracted_129777 | 1297 | No | NA | 1 | 0 | 0 | Not-determined | Genome-fragment | NA | NA | 0 |
| S16_extracted_129839 | 2120 | No | NA | 4 | 0 | 1 | Not-determined | Genome-fragment | NA | NA | 0 |
| S16_extracted_130052 | 4689 | No | NA | 8 | 0 | 6 | Not-determined | Genome-fragment | NA | NA | 0 |
| S16_extracted_130126 | 3695 | No | NA | 1 | 0 | 1 | Not-determined | Genome-fragment | NA | NA | 0 |
| S16_extracted_130280 | 1211 | No | NA | 2 | 0 | 2 | Not-determined | Genome-fragment | NA | NA | 0 |
| S16_extracted_130389 | 1039 | No | NA | 1 | 0 | 0 | Not-determined | Genome-fragment | NA | NA | 0 |
| S16_extracted_130428 | 4705 | No | NA | 2 | 0 | 0 | Not-determined | Genome-fragment | NA | NA | 0 |
| S17_vs_k141_48178\|\|full | 2593 | No | NA | 3 | 2 | 0 | Low-quality | Genome-fragment | 1.5 | AAI-based (high-confidence) | 0 |
| S16_extracted_130602 | 6409 | No | NA | 9 | 0 | 3 | Not-determined | Genome-fragment | NA | NA | 0 |
| S9_vs_k141_4637\|\|full | 2593 | No | NA | 6 | 1 | 0 | Low-quality | Genome-fragment | 5.75 | AAI-based (high-confidence) | 0 |
| S14_vs_k141_5116\|\|full | 2593 | No | NA | 5 | 0 | 0 | Low-quality | Genome-fragment | 5.96 | AAI-based (medium-confidence) | 0 |
| S16_extracted_130732 | 1193 | No | NA | 1 | 0 | 0 | Not-determined | Genome-fragment | NA | NA | 0 |
| S1_vs_k141_5014\|\|full | 2593 | No | NA | 4 | 1 | 0 | Low-quality | Genome-fragment | 1.63 | HMM-based (lower-bound) | 0 |
| S16_extracted_130906 | 1489 | No | NA | 4 | 0 | 2 | Not-determined | Genome-fragment | NA | NA | 0 |
| S16_extracted_130923 | 9787 | No | NA | 11 | 0 | 5 | Not-determined | Genome-fragment | NA | NA | 0 |
| S16_extracted_130978 | 1091 | No | NA | 1 | 0 | 1 | Not-determined | Genome-fragment | NA | NA | 0 |
| S7_vs_112445\|\|lt2gene | 2593 | No | NA | 2 | 2 | 0 | Low-quality | Genome-fragment | 5.59 | AAI-based (high-confidence) | 0 |
| S16_extracted_131200 | 1367 | No | NA | 4 | 0 | 0 | Not-determined | Genome-fragment | NA | NA | 0 |
| S8_extracted_k141_202318 | 2593 | No | NA | 2 | 0 | 1 | Low-quality | Genome-fragment | 1.14 | AAI-based (medium-confidence) | 0 |
| S16_extracted_131338 | 1171 | No | NA | 1 | 0 | 0 | Not-determined | Genome-fragment | NA | NA | 0 |
| S16_extracted_131367 | 1204 | No | NA | 3 | 0 | 1 | Not-determined | Genome-fragment | NA | NA | 0 |
| S16_extracted_131377 | 1232 | No | NA | 3 | 0 | 2 | Not-determined | Genome-fragment | NA | NA | 0 |
| S16_extracted_131422 | 1948 | No | NA | 3 | 0 | 1 | Not-determined | Genome-fragment | NA | NA | 0 |
| S5_vs_k141_30749\|\|full | 2594 | No | NA | 3 | 0 | 0 | Low-quality | Genome-fragment | 0.47 | AAI-based (medium-confidence) | 0 |
| S16_extracted_131476 | 1001 | No | NA | 2 | 0 | 1 | Not-determined | Genome-fragment | NA | NA | 0 |
| S16_extracted_131589 | 3180 | No | NA | 2 | 0 | 2 | Not-determined | Genome-fragment | NA | NA | 0 |
| S16_extracted_131714 | 1783 | No | NA | 1 | 0 | 1 | Not-determined | Genome-fragment | NA | NA | 0 |
| S16_extracted_131749 | 2671 | No | NA | 4 | 0 | 1 | Not-determined | Genome-fragment | NA | NA | 0 |
| S16_extracted_131828 | 1796 | No | NA | 2 | 0 | 0 | Not-determined | Genome-fragment | NA | NA | 0 |
| S16_extracted_131856 | 1093 | No | NA | 2 | 0 | 2 | Not-determined | Genome-fragment | NA | NA | 0 |
| S5_vs_k141_135864\|\|full | 2594 | No | NA | 6 | 2 | 0 | Low-quality | Genome-fragment | 6.54 | AAI-based (high-confidence) | 0 |
| S7_vs_125946\|\|full | 2594 | No | NA | 3 | 0 | 0 | Low-quality | Genome-fragment | 4.72 | AAI-based (high-confidence) | 0 |
| S11_vs_k141_14493\|\|full | 2595 | No | NA | 3 | 0 | 2 | Low-quality | Genome-fragment | 3.17 | AAI-based (medium-confidence) | 0 |
| S14_vs_k141_95115\|\|full | 2595 | No | NA | 4 | 0 | 1 | Low-quality | Genome-fragment | 4.84 | AAI-based (high-confidence) | 0 |
| S16_extracted_131969 | 1606 | No | NA | 1 | 0 | 1 | Not-determined | Genome-fragment | NA | NA | 0 |
| S16_extracted_132258 | 1250 | No | NA | 3 | 0 | 0 | Not-determined | Genome-fragment | NA | NA | 0 |
| S5_vs_k141_129312\|\|full | 2595 | No | NA | 3 | 0 | 2 | Low-quality | Genome-fragment | 1.13 | AAI-based (medium-confidence) | 0 |
| S16_extracted_132892 | 1023 | No | NA | 1 | 0 | 1 | Not-determined | Genome-fragment | NA | NA | 0 |
| S16_extracted_132898 | 6702 | No | NA | 9 | 0 | 0 | Not-determined | Genome-fragment | NA | NA | 0 |
| S15_vs_18620\|\|full | 2595 | No | NA | 6 | 0 | 3 | Low-quality | Genome-fragment | 2.66 | AAI-based (high-confidence) | 0 |
| S16_extracted_132990 | 2949 | No | NA | 2 | 0 | 2 | Not-determined | Genome-fragment | NA | NA | 0 |
| S16_extracted_133017 | 2292 | No | NA | 3 | 0 | 3 | Not-determined | Genome-fragment | NA | NA | 0 |
| S16_extracted_133066 | 6619 | No | NA | 4 | 0 | 4 | Not-determined | Genome-fragment | NA | NA | 0 |
| S16_extracted_133155 | 3744 | No | NA | 3 | 0 | 2 | Not-determined | Genome-fragment | NA | NA | 0 |
| S9_vs_k141_46491\|\|full | 2596 | No | NA | 6 | 3 | 0 | Low-quality | Genome-fragment | 6.4 | AAI-based (medium-confidence) | 0 |
| S16_extracted_133342 | 1892 | No | NA | 3 | 0 | 0 | Not-determined | Genome-fragment | NA | NA | 0 |
| S16_extracted_133347 | 1201 | No | NA | 2 | 0 | 0 | Not-determined | Genome-fragment | NA | NA | 0 |
| S16_extracted_133448 | 1196 | No | NA | 3 | 0 | 2 | Not-determined | Genome-fragment | NA | NA | 0 |
| S16_extracted_133499 | 1010 | No | NA | 1 | 0 | 1 | Not-determined | Genome-fragment | NA | NA | 0 |
| S16_extracted_133588 | 2115 | No | NA | 2 | 0 | 2 | Not-determined | Genome-fragment | NA | NA | 0 |
| S16_extracted_133688 | 4714 | No | NA | 5 | 0 | 1 | Not-determined | Genome-fragment | NA | NA | 0 |
| S10_vs_66700\|\|full | 2596 | No | NA | 4 | 0 | 0 | Low-quality | Genome-fragment | 1.06 | AAI-based (high-confidence) | 0 |
| S16_extracted_133801 | 1231 | No | NA | 1 | 0 | 1 | Not-determined | Genome-fragment | NA | NA | 0 |
| S16_extracted_133862 | 1148 | No | NA | 2 | 0 | 1 | Not-determined | Genome-fragment | NA | NA | 0 |
| S16_extracted_133863 | 2086 | No | NA | 3 | 0 | 0 | Not-determined | Genome-fragment | NA | NA | 0 |
| S16_extracted_133880 | 1092 | No | NA | 1 | 0 | 0 | Not-determined | Genome-fragment | NA | NA | 0 |
| S16_extracted_134243 | 1587 | No | NA | 1 | 0 | 0 | Not-determined | Genome-fragment | NA | NA | 0 |
| S16_extracted_134271 | 2259 | No | NA | 4 | 0 | 3 | Not-determined | Genome-fragment | NA | NA | 0 |
| S10_vs_109569\|\|full | 2596 | No | NA | 4 | 0 | 0 | Low-quality | Genome-fragment | 0.86 | AAI-based (medium-confidence) | 0 |
| S16_extracted_134622 | 2601 | No | NA | 1 | 0 | 0 | Not-determined | Genome-fragment | NA | NA | 0 |
| S6_vs_k141_352\|\|full | 2596 | No | NA | 2 | 0 | 1 | Low-quality | Genome-fragment | 4.51 | AAI-based (medium-confidence) | 0 |
| S16_extracted_134975 | 1167 | No | NA | 2 | 0 | 0 | Not-determined | Genome-fragment | NA | NA | 0 |
| S14_vs_k141_68668\|\|full | 2597 | No | NA | 6 | 2 | 0 | Low-quality | Genome-fragment | 6.83 | AAI-based (medium-confidence) | 0 |
| S16_extracted_135657 | 1045 | No | NA | 1 | 0 | 1 | Not-determined | Genome-fragment | NA | NA | 0 |
| S16_vs_83928\|\|full | 2597 | No | NA | 6 | 1 | 0 | Low-quality | Genome-fragment | 4.02 | AAI-based (high-confidence) | 0 |
| S16_extracted_135791 | 1399 | No | NA | 2 | 0 | 0 | Not-determined | Genome-fragment | NA | NA | 0 |
| S16_extracted_135796 | 1332 | No | NA | 1 | 0 | 0 | Not-determined | Genome-fragment | NA | NA | 0 |
| S16_extracted_135928 | 1868 | No | NA | 1 | 0 | 1 | Not-determined | Genome-fragment | NA | NA | 0 |
| S16_extracted_135964 | 6027 | No | NA | 2 | 0 | 1 | Not-determined | Genome-fragment | NA | NA | 0 |
| S16_extracted_135969 | 1933 | No | NA | 1 | 0 | 1 | Not-determined | Genome-fragment | NA | NA | 0 |
| S16_extracted_136125 | 1019 | No | NA | 1 | 0 | 1 | Not-determined | Genome-fragment | NA | NA | 0 |
| S12_vs_k141_179505\|\|full | 2597 | No | NA | 3 | 1 | 0 | Low-quality | Genome-fragment | 0.93 | HMM-based (lower-bound) | 0 |
| S8_vs_k141_66314\|\|full | 2597 | No | NA | 3 | 0 | 3 | Low-quality | Genome-fragment | 1.24 | AAI-based (high-confidence) | 0 |
| S1_vs_k141_114509\|\|full | 2598 | No | NA | 3 | 1 | 0 | Low-quality | Genome-fragment | 1.67 | AAI-based (medium-confidence) | 0 |
| S5_vs_k141_92996\|\|full | 2598 | No | NA | 6 | 1 | 0 | Low-quality | Genome-fragment | 4.59 | AAI-based (high-confidence) | 0 |
| S5_vs_k141_28196\|\|full | 2598 | No | NA | 6 | 0 | 3 | Low-quality | Genome-fragment | 0.82 | AAI-based (medium-confidence) | 0 |
| S7_vs_58357\|\|lt2gene | 2598 | No | NA | 1 | 1 | 0 | Low-quality | Genome-fragment | 7.07 | AAI-based (high-confidence) | 0 |
| S16_extracted_136680 | 3381 | No | NA | 5 | 0 | 2 | Not-determined | Genome-fragment | NA | NA | 0 |
| S16_extracted_136756 | 1343 | No | NA | 3 | 0 | 0 | Not-determined | Genome-fragment | NA | NA | 0 |
| S16_extracted_136791 | 1753 | No | NA | 2 | 0 | 0 | Not-determined | Genome-fragment | NA | NA | 0 |
| S16_extracted_137260 | 1434 | No | NA | 1 | 0 | 1 | Not-determined | Genome-fragment | NA | NA | 0 |
| S16_extracted_137341 | 1014 | No | NA | 1 | 0 | 1 | Not-determined | Genome-fragment | NA | NA | 0 |
| S16_extracted_137417 | 1131 | No | NA | 1 | 0 | 0 | Not-determined | Genome-fragment | NA | NA | 0 |
| S16_extracted_137489 | 1243 | No | NA | 2 | 0 | 2 | Not-determined | Genome-fragment | NA | NA | 0 |
| S16_extracted_7147 | 2598 | No | NA | 3 | 0 | 0 | Low-quality | Genome-fragment | 4.03 | AAI-based (medium-confidence) | 0 |
| S16_extracted_137577 | 2039 | No | NA | 1 | 0 | 1 | Not-determined | Genome-fragment | NA | NA | 0 |
| S17_vs_k141_89851\|\|full | 2599 | No | NA | 3 | 0 | 2 | Low-quality | Genome-fragment | 2.01 | AAI-based (high-confidence) | 0 |
| S16_extracted_137785 | 4094 | No | NA | 5 | 0 | 1 | Not-determined | Genome-fragment | NA | NA | 0 |
| S16_extracted_137787 | 2268 | No | NA | 6 | 0 | 0 | Not-determined | Genome-fragment | NA | NA | 0 |
| S16_extracted_137823 | 1092 | No | NA | 2 | 0 | 0 | Not-determined | Genome-fragment | NA | NA | 0 |
| S9_vs_k141_81544\|\|full | 2599 | No | NA | 7 | 0 | 0 | Low-quality | Genome-fragment | 6.2 | AAI-based (medium-confidence) | 0 |
| S16_extracted_137927 | 1321 | No | NA | 2 | 0 | 2 | Not-determined | Genome-fragment | NA | NA | 0 |
| S16_extracted_138065 | 1268 | No | NA | 2 | 0 | 1 | Not-determined | Genome-fragment | NA | NA | 0 |
| S16_extracted_138140 | 1141 | No | NA | 2 | 0 | 1 | Not-determined | Genome-fragment | NA | NA | 0 |
| S14_vs_k141_22893\|\|full | 2599 | No | NA | 4 | 0 | 1 | Low-quality | Genome-fragment | 4.89 | AAI-based (medium-confidence) | 0 |
| S16_extracted_138477 | 1009 | No | NA | 3 | 0 | 2 | Not-determined | Genome-fragment | NA | NA | 0 |
| S16_extracted_138493 | 10670 | No | NA | 13 | 0 | 3 | Not-determined | Genome-fragment | NA | NA | 0 |
| S8_vs_k141_19777\|\|full | 2599 | No | NA | 2 | 1 | 0 | Low-quality | Genome-fragment | 5.76 | AAI-based (medium-confidence) | 0 |
| S16_extracted_138642 | 2085 | No | NA | 3 | 0 | 1 | Not-determined | Genome-fragment | NA | NA | 0 |
| S16_extracted_138652 | 3548 | No | NA | 5 | 0 | 4 | Not-determined | Genome-fragment | NA | NA | 0 |
| S16_extracted_138717 | 1144 | No | NA | 2 | 0 | 1 | Not-determined | Genome-fragment | NA | NA | 0 |
| S8_extracted_k141_113649 | 2600 | No | NA | 3 | 1 | 0 | Low-quality | Genome-fragment | 8.08 | AAI-based (high-confidence) | 0 |
| S16_extracted_138812 | 1011 | No | NA | 1 | 0 | 1 | Not-determined | Genome-fragment | NA | NA | 0 |
| S17_extracted_k141_151034 | 2601 | No | NA | 2 | 0 | 0 | Low-quality | Genome-fragment | 1.43 | AAI-based (high-confidence) | 0 |
| S9_vs_k141_33780\|\|full | 2601 | No | NA | 6 | 1 | 0 | Low-quality | Genome-fragment | 3.69 | AAI-based (high-confidence) | 0 |
| S16_extracted_138954 | 1041 | No | NA | 1 | 0 | 0 | Not-determined | Genome-fragment | NA | NA | 0 |
| S3_vs_k141_1676\|\|full | 2601 | No | NA | 6 | 0 | 0 | Low-quality | Genome-fragment | 4.48 | AAI-based (high-confidence) | 0 |
| S16_extracted_139188 | 5254 | No | NA | 4 | 0 | 3 | Not-determined | Genome-fragment | NA | NA | 0 |
| S16_extracted_139189 | 3000 | No | NA | 3 | 0 | 2 | Not-determined | Genome-fragment | NA | NA | 0 |
| S16_extracted_139196 | 1355 | No | NA | 2 | 0 | 1 | Not-determined | Genome-fragment | NA | NA | 0 |
| S7_vs_98857\|\|full | 2601 | No | NA | 4 | 1 | 0 | Low-quality | Genome-fragment | 6.23 | AAI-based (medium-confidence) | 0 |
| S9_extracted_k141_3671 | 2602 | No | NA | 3 | 2 | 0 | Low-quality | Genome-fragment | 43.39 | AAI-based (high-confidence) | 0 |
| S16_extracted_139353 | 1602 | No | NA | 2 | 0 | 1 | Not-determined | Genome-fragment | NA | NA | 0 |
| S16_extracted_139441 | 1613 | No | NA | 4 | 0 | 2 | Not-determined | Genome-fragment | NA | NA | 0 |
| S16_extracted_139560 | 1155 | No | NA | 1 | 0 | 0 | Not-determined | Genome-fragment | NA | NA | 0 |
| S16_extracted_139613 | 1328 | No | NA | 1 | 0 | 1 | Not-determined | Genome-fragment | NA | NA | 0 |
| S16_extracted_139720 | 1885 | No | NA | 2 | 0 | 0 | Not-determined | Genome-fragment | NA | NA | 0 |
| S16_extracted_139763 | 1340 | No | NA | 4 | 0 | 0 | Not-determined | Genome-fragment | NA | NA | 0 |
| S16_extracted_139798 | 1320 | No | NA | 1 | 0 | 1 | Not-determined | Genome-fragment | NA | NA | 0 |
| S16_extracted_139809 | 1131 | No | NA | 1 | 0 | 1 | Not-determined | Genome-fragment | NA | NA | 0 |
| S16_extracted_139859 | 1511 | No | NA | 1 | 0 | 1 | Not-determined | Genome-fragment | NA | NA | 0 |
| S14_vs_k141_55275\|\|full | 2602 | No | NA | 4 | 0 | 0 | Low-quality | Genome-fragment | 6.02 | AAI-based (high-confidence) | 0 |
| S16_extracted_139885 | 1742 | No | NA | 3 | 0 | 1 | Not-determined | Genome-fragment | NA | NA | 0 |
| S16_extracted_139944 | 5405 | No | NA | 5 | 0 | 3 | Not-determined | Genome-fragment | NA | NA | 0 |
| S8_vs_k141_30441\|\|full | 2602 | No | NA | 6 | 1 | 0 | Low-quality | Genome-fragment | 4.53 | AAI-based (high-confidence) | 0 |
| S16_extracted_140029 | 2002 | No | NA | 1 | 0 | 1 | Not-determined | Genome-fragment | NA | NA | 0 |
| S16_extracted_140164 | 2174 | No | NA | 2 | 0 | 1 | Not-determined | Genome-fragment | NA | NA | 0 |
| S16_extracted_140224 | 2486 | No | NA | 2 | 0 | 2 | Not-determined | Genome-fragment | NA | NA | 0 |
| S8_vs_k141_196345\|\|full | 2602 | No | NA | 4 | 0 | 1 | Low-quality | Genome-fragment | 3.93 | AAI-based (medium-confidence) | 0 |
| S14_extracted_k141_10660 | 2603 | No | NA | 5 | 1 | 0 | Low-quality | Genome-fragment | 4.51 | AAI-based (high-confidence) | 0 |
| S16_extracted_140398 | 14341 | No | NA | 7 | 0 | 5 | Not-determined | Genome-fragment | NA | NA | 0 |
| S16_extracted_140440 | 1019 | No | NA | 1 | 0 | 1 | Not-determined | Genome-fragment | NA | NA | 0 |
| S7_vs_39262\|\|full | 2603 | No | NA | 3 | 0 | 1 | Low-quality | Genome-fragment | 6.8 | AAI-based (medium-confidence) | 0 |
| S16_extracted_140455 | 1055 | No | NA | 1 | 0 | 0 | Not-determined | Genome-fragment | NA | NA | 0 |
| S16_extracted_140471 | 1487 | No | NA | 3 | 0 | 2 | Not-determined | Genome-fragment | NA | NA | 0 |
| S16_extracted_140502 | 1092 | No | NA | 1 | 0 | 0 | Not-determined | Genome-fragment | NA | NA | 0 |
| S16_extracted_140540 | 1318 | No | NA | 3 | 0 | 0 | Not-determined | Genome-fragment | NA | NA | 0 |
| S16_extracted_140583 | 9674 | No | NA | 8 | 0 | 6 | Not-determined | Genome-fragment | NA | NA | 0 |
| S16_extracted_85355 | 2603 | No | NA | 5 | 4 | 0 | Low-quality | Genome-fragment | 8.51 | AAI-based (high-confidence) | 0 |
| S16_extracted_140685 | 1227 | No | NA | 1 | 0 | 0 | Not-determined | Genome-fragment | NA | NA | 0 |
| S16_extracted_140708 | 1155 | No | NA | 2 | 0 | 2 | Not-determined | Genome-fragment | NA | NA | 0 |
| S16_extracted_140727 | 1392 | No | NA | 1 | 0 | 0 | Not-determined | Genome-fragment | NA | NA | 0 |
| S16_extracted_140760 | 2139 | No | NA | 2 | 0 | 1 | Not-determined | Genome-fragment | NA | NA | 0 |
| S16_extracted_140778 | 1075 | No | NA | 1 | 0 | 0 | Not-determined | Genome-fragment | NA | NA | 0 |
| S16_extracted_140910 | 3792 | No | NA | 4 | 0 | 3 | Not-determined | Genome-fragment | NA | NA | 0 |
| S16_extracted_141105 | 1785 | No | NA | 2 | 0 | 2 | Not-determined | Genome-fragment | NA | NA | 0 |
| S16_extracted_141147 | 3225 | No | NA | 3 | 0 | 3 | Not-determined | Genome-fragment | NA | NA | 0 |
| S16_extracted_141241 | 7847 | No | NA | 15 | 0 | 14 | Not-determined | Genome-fragment | NA | NA | 0 |
| S16_extracted_141265 | 2781 | No | NA | 2 | 0 | 1 | Not-determined | Genome-fragment | NA | NA | 0 |
| S16_extracted_141531 | 1252 | No | NA | 1 | 0 | 1 | Not-determined | Genome-fragment | NA | NA | 0 |
| S16_extracted_141535 | 3562 | No | NA | 6 | 0 | 1 | Not-determined | Genome-fragment | NA | NA | 0 |
| S10_vs_68674\|\|full | 2603 | No | NA | 6 | 1 | 0 | Low-quality | Genome-fragment | 4.54 | AAI-based (high-confidence) | 0 |
| S16_extracted_141659 | 5292 | No | NA | 5 | 0 | 5 | Not-determined | Genome-fragment | NA | NA | 0 |
| S12_vs_k141_150413\|\|full | 2603 | Yes | 1629 | 5 | 1 | 1 | Low-quality | Genome-fragment | 1.94 | HMM-based (lower-bound) | 27.42 |
| S16_extracted_141896 | 1197 | No | NA | 1 | 0 | 1 | Not-determined | Genome-fragment | NA | NA | 0 |
| S8_vs_k141_118211\|\|full | 2603 | No | NA | 2 | 0 | 2 | Low-quality | Genome-fragment | 2.5 | AAI-based (medium-confidence) | 0 |
| S8_vs_k141_7374\|\|full | 2603 | No | NA | 6 | 1 | 0 | Low-quality | Genome-fragment | 5.32 | AAI-based (medium-confidence) | 0 |
| S16_extracted_142140 | 1674 | No | NA | 4 | 0 | 0 | Not-determined | Genome-fragment | NA | NA | 0 |
| S16_extracted_89593 | 2604 | No | NA | 3 | 2 | 0 | Low-quality | Genome-fragment | 4.19 | AAI-based (high-confidence) | 0 |
| S16_extracted_142217 | 2196 | No | NA | 2 | 0 | 0 | Not-determined | Genome-fragment | NA | NA | 0 |
| S16_extracted_142280 | 1000 | No | NA | 2 | 0 | 2 | Not-determined | Genome-fragment | NA | NA | 0 |
| S16_extracted_142299 | 4688 | No | NA | 4 | 0 | 2 | Not-determined | Genome-fragment | NA | NA | 0 |
| S16_extracted_142342 | 2395 | No | NA | 3 | 0 | 0 | Not-determined | Genome-fragment | NA | NA | 0 |
| S16_extracted_142376 | 2056 | No | NA | 3 | 0 | 3 | Not-determined | Genome-fragment | NA | NA | 0 |
| S12_vs_k141_52623\|\|full | 2604 | No | NA | 5 | 1 | 0 | Low-quality | Genome-fragment | 3.5 | HMM-based (lower-bound) | 0 |
| S15_vs_64027\|\|full | 2605 | No | NA | 3 | 0 | 0 | Low-quality | Genome-fragment | 1.32 | AAI-based (medium-confidence) | 0 |
| S16_extracted_142435 | 1100 | No | NA | 1 | 0 | 0 | Not-determined | Genome-fragment | NA | NA | 0 |
| S16_extracted_142483 | 1605 | No | NA | 4 | 0 | 0 | Not-determined | Genome-fragment | NA | NA | 0 |
| S16_extracted_142485 | 1269 | No | NA | 2 | 0 | 2 | Not-determined | Genome-fragment | NA | NA | 0 |
| S10_vs_45978\|\|full | 2606 | No | NA | 6 | 0 | 0 | Low-quality | Genome-fragment | 2.39 | AAI-based (high-confidence) | 0 |
| S9_vs_k141_60928\|\|full | 2607 | No | NA | 2 | 0 | 0 | Low-quality | Genome-fragment | 1.63 | AAI-based (medium-confidence) | 0 |
| S16_extracted_142640 | 1011 | No | NA | 1 | 0 | 0 | Not-determined | Genome-fragment | NA | NA | 0 |
| S14_vs_k141_9229\|\|full | 2607 | No | NA | 8 | 7 | 0 | Low-quality | Genome-fragment | 8.58 | AAI-based (high-confidence) | 0 |
| S16_extracted_142850 | 3747 | No | NA | 2 | 0 | 0 | Not-determined | Genome-fragment | NA | NA | 0 |
| S1_extracted_k141_140234 | 2607 | No | NA | 1 | 0 | 0 | Low-quality | Genome-fragment | 1.4 | AAI-based (high-confidence) | 0 |
| S16_extracted_142961 | 1572 | No | NA | 2 | 0 | 0 | Not-determined | Genome-fragment | NA | NA | 0 |
| S16_extracted_143017 | 1085 | No | NA | 2 | 0 | 1 | Not-determined | Genome-fragment | NA | NA | 0 |
| S16_extracted_143181 | 1539 | No | NA | 3 | 0 | 0 | Not-determined | Genome-fragment | NA | NA | 0 |
| S16_extracted_143259 | 1241 | No | NA | 1 | 0 | 0 | Not-determined | Genome-fragment | NA | NA | 0 |
| S16_extracted_143260 | 9656 | No | NA | 8 | 0 | 7 | Not-determined | Genome-fragment | NA | NA | 0 |
| S16_extracted_143275 | 5156 | No | NA | 4 | 0 | 1 | Not-determined | Genome-fragment | NA | NA | 0 |
| S16_extracted_143286 | 1520 | No | NA | 2 | 0 | 1 | Not-determined | Genome-fragment | NA | NA | 0 |
| S16_extracted_143346 | 1568 | No | NA | 2 | 0 | 1 | Not-determined | Genome-fragment | NA | NA | 0 |
| S16_extracted_143408 | 5306 | No | NA | 3 | 0 | 3 | Not-determined | Genome-fragment | NA | NA | 0 |
| S16_extracted_143410 | 3489 | No | NA | 4 | 0 | 3 | Not-determined | Genome-fragment | NA | NA | 0 |
| S7_vs_88195\|\|full | 2607 | No | NA | 3 | 3 | 0 | Low-quality | Genome-fragment | 7.61 | AAI-based (high-confidence) | 0 |
| S16_extracted_143448 | 1470 | No | NA | 2 | 0 | 0 | Not-determined | Genome-fragment | NA | NA | 0 |
| S12_extracted_k141_85714 | 2607 | No | NA | 3 | 0 | 0 | Low-quality | Genome-fragment | 14.27 | AAI-based (medium-confidence) | 0 |
| S8_vs_k141_87683\|\|full | 2607 | No | NA | 3 | 2 | 0 | Low-quality | Genome-fragment | 6.54 | AAI-based (high-confidence) | 0 |
| S9_vs_k141_82185\|\|full | 2608 | No | NA | 6 | 3 | 0 | Low-quality | Genome-fragment | 6.22 | AAI-based (medium-confidence) | 0 |
| S16_extracted_143767 | 1108 | No | NA | 2 | 0 | 1 | Not-determined | Genome-fragment | NA | NA | 0 |
| S16_extracted_143784 | 1101 | No | NA | 2 | 0 | 2 | Not-determined | Genome-fragment | NA | NA | 0 |
| S16_extracted_143797 | 1395 | No | NA | 2 | 0 | 2 | Not-determined | Genome-fragment | NA | NA | 0 |
| S7_vs_86551\|\|full | 2608 | No | NA | 6 | 1 | 0 | Low-quality | Genome-fragment | 6.02 | AAI-based (medium-confidence) | 0 |
| S16_extracted_143984 | 3158 | No | NA | 2 | 0 | 1 | Not-determined | Genome-fragment | NA | NA | 0 |
| S1_extracted_k141_132859 | 2609 | No | NA | 2 | 0 | 1 | Low-quality | Genome-fragment | 2.32 | AAI-based (medium-confidence) | 0 |
| S16_extracted_144195 | 1090 | No | NA | 1 | 0 | 1 | Not-determined | Genome-fragment | NA | NA | 0 |
| S16_extracted_144324 | 1319 | No | NA | 3 | 0 | 0 | Not-determined | Genome-fragment | NA | NA | 0 |
| S7_vs_100555\|\|full | 2609 | No | NA | 7 | 0 | 0 | Low-quality | Genome-fragment | 6.7 | AAI-based (high-confidence) | 0 |
| S16_extracted_144352 | 15759 | No | NA | 8 | 0 | 3 | Not-determined | Genome-fragment | NA | NA | 0 |
| S16_extracted_144395 | 1209 | No | NA | 2 | 0 | 0 | Not-determined | Genome-fragment | NA | NA | 0 |
| S16_extracted_144406 | 3692 | No | NA | 2 | 0 | 1 | Not-determined | Genome-fragment | NA | NA | 0 |
| S16_extracted_144562 | 5638 | No | NA | 8 | 0 | 3 | Not-determined | Genome-fragment | NA | NA | 0 |
| S16_extracted_144565 | 1432 | No | NA | 2 | 0 | 1 | Not-determined | Genome-fragment | NA | NA | 0 |
| S6_extracted_k141_46156 | 2609 | No | NA | 7 | 1 | 0 | Low-quality | Genome-fragment | 3.81 | AAI-based (high-confidence) | 0 |
| S9_vs_k141_81996\|\|full | 2610 | No | NA | 5 | 4 | 0 | Low-quality | Genome-fragment | 3.65 | HMM-based (lower-bound) | 0 |
| S16_extracted_144748 | 1271 | No | NA | 2 | 0 | 2 | Not-determined | Genome-fragment | NA | NA | 0 |
| S1_vs_k141_152110\|\|full | 2610 | No | NA | 4 | 1 | 0 | Low-quality | Genome-fragment | 1.4 | HMM-based (lower-bound) | 0 |
| S16_extracted_144920 | 1409 | No | NA | 2 | 0 | 1 | Not-determined | Genome-fragment | NA | NA | 0 |
| S16_extracted_145119 | 1723 | No | NA | 2 | 0 | 1 | Not-determined | Genome-fragment | NA | NA | 0 |
| S7_extracted_132883 | 2610 | No | NA | 2 | 0 | 2 | Low-quality | Genome-fragment | 0.7 | AAI-based (high-confidence) | 0 |
| S16_extracted_145173 | 1156 | No | NA | 1 | 0 | 1 | Not-determined | Genome-fragment | NA | NA | 0 |
| S16_extracted_145273 | 1152 | No | NA | 2 | 0 | 1 | Not-determined | Genome-fragment | NA | NA | 0 |
| S16_extracted_145339 | 4983 | No | NA | 5 | 0 | 3 | Not-determined | Genome-fragment | NA | NA | 0 |
| S8_extracted_k141_68343 | 2610 | No | NA | 2 | 0 | 0 | Low-quality | Genome-fragment | 2.12 | AAI-based (high-confidence) | 0 |
| S16_extracted_145421 | 11778 | No | NA | 12 | 0 | 7 | Not-determined | Genome-fragment | NA | NA | 0 |
| S16_extracted_145467 | 1634 | No | NA | 2 | 0 | 1 | Not-determined | Genome-fragment | NA | NA | 0 |
| S16_extracted_145478 | 9106 | No | NA | 7 | 0 | 4 | Not-determined | Genome-fragment | NA | NA | 0 |
| S16_extracted_145514 | 1599 | No | NA | 1 | 0 | 1 | Not-determined | Genome-fragment | NA | NA | 0 |
| S16_extracted_145526 | 3123 | No | NA | 3 | 0 | 1 | Not-determined | Genome-fragment | NA | NA | 0 |
| S8_vs_k141_11705\|\|full | 2610 | No | NA | 3 | 1 | 0 | Low-quality | Genome-fragment | 4.29 | AAI-based (medium-confidence) | 0 |
| S17_vs_k141_109926\|\|full | 2611 | No | NA | 2 | 0 | 0 | Low-quality | Genome-fragment | 2.29 | AAI-based (high-confidence) | 0 |
| S16_extracted_145695 | 1000 | No | NA | 1 | 0 | 0 | Not-determined | Genome-fragment | NA | NA | 0 |
| S16_extracted_145820 | 1540 | No | NA | 2 | 0 | 1 | Not-determined | Genome-fragment | NA | NA | 0 |
| S16_extracted_145925 | 1397 | No | NA | 2 | 0 | 2 | Not-determined | Genome-fragment | NA | NA | 0 |
| S16_extracted_145989 | 1181 | No | NA | 2 | 0 | 1 | Not-determined | Genome-fragment | NA | NA | 0 |
| S5_vs_k141_16360\|\|full | 2611 | No | NA | 5 | 3 | 0 | Low-quality | Genome-fragment | 7.1 | AAI-based (high-confidence) | 0 |
| S16_extracted_146024 | 3000 | No | NA | 2 | 0 | 1 | Not-determined | Genome-fragment | NA | NA | 0 |
| S3_vs_k141_96040\|\|full | 2612 | Yes | 592 | 5 | 1 | 2 | Low-quality | Genome-fragment | 1.02 | AAI-based (high-confidence) | 77.34 |
| S16_extracted_146181 | 1457 | No | NA | 2 | 0 | 0 | Not-determined | Genome-fragment | NA | NA | 0 |
| S16_extracted_146214 | 1245 | No | NA | 2 | 0 | 0 | Not-determined | Genome-fragment | NA | NA | 0 |
| S16_extracted_146244 | 5117 | No | NA | 6 | 0 | 5 | Not-determined | Genome-fragment | NA | NA | 0 |
| S16_extracted_146357 | 1003 | No | NA | 2 | 0 | 0 | Not-determined | Genome-fragment | NA | NA | 0 |
| S2_vs_k141_7028\|\|full | 2612 | No | NA | 7 | 1 | 0 | Low-quality | Genome-fragment | 0.69 | HMM-based (lower-bound) | 0 |
| S11_extracted_k141_94000 | 2613 | No | NA | 2 | 0 | 0 | Low-quality | Genome-fragment | 5.92 | AAI-based (medium-confidence) | 0 |
| S16_extracted_146500 | 2079 | No | NA | 1 | 0 | 0 | Not-determined | Genome-fragment | NA | NA | 0 |
| S16_extracted_146538 | 3938 | No | NA | 2 | 0 | 1 | Not-determined | Genome-fragment | NA | NA | 0 |
| S16_extracted_146600 | 1075 | No | NA | 1 | 0 | 0 | Not-determined | Genome-fragment | NA | NA | 0 |
| S16_extracted_146635 | 3678 | No | NA | 3 | 0 | 2 | Not-determined | Genome-fragment | NA | NA | 0 |
| S16_extracted_146720 | 4550 | No | NA | 7 | 0 | 2 | Not-determined | Genome-fragment | NA | NA | 0 |
| S16_extracted_146922 | 13192 | No | NA | 12 | 0 | 4 | Not-determined | Genome-fragment | NA | NA | 0 |
| S11_vs_k141_58713\|\|full | 2613 | No | NA | 4 | 0 | 0 | Low-quality | Genome-fragment | 6.19 | AAI-based (high-confidence) | 0 |
| S16_extracted_146993 | 1129 | No | NA | 3 | 0 | 0 | Not-determined | Genome-fragment | NA | NA | 0 |
| S1_vs_k141_79959\|\|full | 2613 | No | NA | 6 | 0 | 0 | Low-quality | Genome-fragment | 5.69 | AAI-based (medium-confidence) | 0 |
| S16_extracted_147104 | 1118 | No | NA | 1 | 0 | 0 | Not-determined | Genome-fragment | NA | NA | 0 |
| S16_extracted_147326 | 15321 | No | NA | 19 | 8 | 0 | Medium-quality | Genome-fragment | 77.66 | AAI-based (medium-confidence) | 0 |
| S16_extracted_147349 | 8137 | No | NA | 7 | 0 | 6 | Not-determined | Genome-fragment | NA | NA | 0 |
| S16_extracted_147369 | 1003 | No | NA | 2 | 0 | 0 | Not-determined | Genome-fragment | NA | NA | 0 |
| S16_extracted_147635 | 1892 | No | NA | 3 | 0 | 1 | Not-determined | Genome-fragment | NA | NA | 0 |
| S16_extracted_147679 | 1054 | No | NA | 1 | 0 | 1 | Not-determined | Genome-fragment | NA | NA | 0 |
| S16_extracted_147713 | 1007 | No | NA | 1 | 0 | 1 | Not-determined | Genome-fragment | NA | NA | 0 |
| S16_extracted_147714 | 1272 | No | NA | 1 | 0 | 1 | Not-determined | Genome-fragment | NA | NA | 0 |
| S16_extracted_147715 | 1085 | No | NA | 1 | 0 | 1 | Not-determined | Genome-fragment | NA | NA | 0 |
| S5_vs_k141_97313\|\|full | 2613 | No | NA | 8 | 0 | 0 | Low-quality | Genome-fragment | 5.15 | AAI-based (medium-confidence) | 0 |
| S12_vs_k141_26309\|\|full | 2613 | No | NA | 3 | 0 | 2 | Low-quality | Genome-fragment | 1.22 | AAI-based (medium-confidence) | 0 |
| S16_extracted_147994 | 1379 | No | NA | 3 | 0 | 0 | Not-determined | Genome-fragment | NA | NA | 0 |
| S8_extracted_k141_7139 | 2613 | No | NA | 2 | 1 | 0 | Low-quality | Genome-fragment | 1.43 | HMM-based (lower-bound) | 0 |
| S16_extracted_148277 | 3002 | No | NA | 3 | 0 | 2 | Not-determined | Genome-fragment | NA | NA | 0 |
| S8_vs_k141_197867\|\|full | 2613 | No | NA | 6 | 5 | 0 | Low-quality | Genome-fragment | 7.67 | AAI-based (high-confidence) | 0 |
| S16_extracted_148417 | 3266 | No | NA | 2 | 0 | 2 | Not-determined | Genome-fragment | NA | NA | 0 |
| S16_extracted_148536 | 2013 | No | NA | 2 | 0 | 2 | Not-determined | Genome-fragment | NA | NA | 0 |
| S16_extracted_148585 | 1013 | No | NA | 1 | 0 | 1 | Not-determined | Genome-fragment | NA | NA | 0 |
| S16_extracted_148640 | 2722 | No | NA | 2 | 0 | 2 | Not-determined | Genome-fragment | NA | NA | 0 |
| S14_extracted_k141_23333 | 2614 | No | NA | 8 | 5 | 0 | Low-quality | Genome-fragment | 8.55 | AAI-based (high-confidence) | 0 |
| S16_extracted_148700 | 1291 | No | NA | 3 | 0 | 2 | Not-determined | Genome-fragment | NA | NA | 0 |
| S16_extracted_148732 | 6348 | No | NA | 3 | 0 | 3 | Not-determined | Genome-fragment | NA | NA | 0 |
| S16_extracted_148833 | 1379 | No | NA | 2 | 0 | 0 | Not-determined | Genome-fragment | NA | NA | 0 |
| S16_extracted_148835 | 1032 | No | NA | 2 | 0 | 2 | Not-determined | Genome-fragment | NA | NA | 0 |
| S16_extracted_148906 | 5711 | No | NA | 6 | 0 | 5 | Not-determined | Genome-fragment | NA | NA | 0 |
| S16_extracted_148946 | 1503 | No | NA | 2 | 0 | 0 | Not-determined | Genome-fragment | NA | NA | 0 |
| S7_vs_38728\|\|full | 2614 | No | NA | 5 | 0 | 0 | Low-quality | Genome-fragment | 6.48 | AAI-based (high-confidence) | 0 |
| S16_extracted_149025 | 1116 | No | NA | 2 | 0 | 1 | Not-determined | Genome-fragment | NA | NA | 0 |
| S16_extracted_147843 | 2614 | No | NA | 5 | 3 | 0 | Low-quality | Genome-fragment | 4.51 | AAI-based (high-confidence) | 0 |
| S16_extracted_149126 | 1311 | No | NA | 2 | 0 | 1 | Not-determined | Genome-fragment | NA | NA | 0 |
| S16_extracted_149171 | 1125 | No | NA | 2 | 0 | 1 | Not-determined | Genome-fragment | NA | NA | 0 |
| S6_vs_k141_47596\|\|full | 2614 | No | NA | 5 | 3 | 0 | Low-quality | Genome-fragment | 4.74 | AAI-based (high-confidence) | 0 |
| S16_extracted_149287 | 3137 | No | NA | 6 | 0 | 1 | Not-determined | Genome-fragment | NA | NA | 0 |
| S16_extracted_149307 | 1336 | No | NA | 2 | 0 | 1 | Not-determined | Genome-fragment | NA | NA | 0 |
| S8_extracted_k141_96988 | 2614 | Yes | 620 | 3 | 1 | 2 | Low-quality | Genome-fragment | 0.29 | HMM-based (lower-bound) | 76.28 |
| S16_extracted_149527 | 4886 | No | NA | 5 | 0 | 1 | Not-determined | Genome-fragment | NA | NA | 0 |
| S5_vs_k141_25516\|\|full | 2615 | No | NA | 3 | 1 | 0 | Low-quality | Genome-fragment | 3.13 | HMM-based (lower-bound) | 0 |
| S16_extracted_149558 | 5489 | No | NA | 5 | 0 | 4 | Not-determined | Genome-fragment | NA | NA | 0 |
| S16_vs_14544\|\|full | 2615 | No | NA | 4 | 1 | 0 | Low-quality | Genome-fragment | 6.39 | AAI-based (high-confidence) | 0 |
| S16_extracted_149694 | 1247 | No | NA | 1 | 0 | 1 | Not-determined | Genome-fragment | NA | NA | 0 |
| S16_vs_60397\|\|full | 2615 | No | NA | 3 | 0 | 1 | Low-quality | Genome-fragment | 4.44 | AAI-based (medium-confidence) | 0 |
| S16_extracted_149940 | 1317 | No | NA | 2 | 0 | 1 | Not-determined | Genome-fragment | NA | NA | 0 |
| S16_extracted_149965 | 1271 | No | NA | 1 | 0 | 1 | Not-determined | Genome-fragment | NA | NA | 0 |
| S15_vs_70096\|\|full | 2615 | No | NA | 7 | 0 | 0 | Low-quality | Genome-fragment | 5.97 | AAI-based (high-confidence) | 0 |
| S16_extracted_150077 | 3000 | No | NA | 2 | 0 | 2 | Not-determined | Genome-fragment | NA | NA | 0 |
| S16_extracted_150239 | 3235 | No | NA | 3 | 0 | 2 | Not-determined | Genome-fragment | NA | NA | 0 |
| S16_extracted_150298 | 1048 | No | NA | 2 | 0 | 0 | Not-determined | Genome-fragment | NA | NA | 0 |
| S16_extracted_150422 | 1750 | No | NA | 1 | 0 | 0 | Not-determined | Genome-fragment | NA | NA | 0 |

| | | | | | | | | | | | |
|---|---|---|---|---|---|---|---|---|---|---|---|
| S16_extracted_150482 | 1227 | No | NA | 1 | 0 | 0 | Not-determined | Genome-fragment | NA | NA | 0 |
| S14_extracted_k141_24314 | 2616 | No | NA | 6 | 5 | 0 | Low-quality | Genome-fragment | 8.53 | AAI-based (high-confidence) | 0 |
| S16_extracted_150814 | 1401 | No | NA | 1 | 0 | 0 | Not-determined | Genome-fragment | NA | NA | 0 |
| S16_extracted_150849 | 1075 | No | NA | 1 | 0 | 1 | Not-determined | Genome-fragment | NA | NA | 0 |
| S16_extracted_150936 | 1254 | No | NA | 1 | 0 | 1 | Not-determined | Genome-fragment | NA | NA | 0 |
| S16_extracted_150968 | 1305 | No | NA | 2 | 0 | 1 | Not-determined | Genome-fragment | NA | NA | 0 |
| S16_extracted_151099 | 2683 | No | NA | 3 | 0 | 1 | Not-determined | Genome-fragment | NA | NA | 0 |
| S16_extracted_151122 | 1640 | No | NA | 2 | 0 | 1 | Not-determined | Genome-fragment | NA | NA | 0 |
| S16_extracted_151136 | 1575 | No | NA | 2 | 0 | 1 | Not-determined | Genome-fragment | NA | NA | 0 |
| S16_extracted_151171 | 1152 | No | NA | 2 | 0 | 2 | Not-determined | Genome-fragment | NA | NA | 0 |
| S5_vs_k141_136351\|\|full | 2616 | No | NA | 4 | 0 | 0 | Low-quality | Genome-fragment | 6.35 | AAI-based (medium-confidence) | 0 |
| S7_vs_78650\|\|full | 2616 | No | NA | 5 | 3 | 0 | Low-quality | Genome-fragment | 6.79 | AAI-based (medium-confidence) | 0 |
| S16_extracted_151397 | 1521 | No | NA | 2 | 0 | 2 | Not-determined | Genome-fragment | NA | NA | 0 |
| S5_extracted_k141_102318 | 2617 | No | NA | 3 | 2 | 0 | Low-quality | Genome-fragment | 7.68 | AAI-based (high-confidence) | 0 |
| S16_extracted_151450 | 95263 | No | NA | 92 | 11 | 6 | Complete | High-quality | 100 | DTR (high-confidence) | 0 |
| S16_extracted_151484 | 1366 | No | NA | 1 | 0 | 1 | Not-determined | Genome-fragment | NA | NA | 0 |
| S16_extracted_151488 | 1418 | No | NA | 2 | 0 | 0 | Not-determined | Genome-fragment | NA | NA | 0 |
| S16_extracted_151505 | 4103 | No | NA | 6 | 0 | 2 | Not-determined | Genome-fragment | NA | NA | 0 |
| S16_vs_113875\|\|full | 2617 | No | NA | 5 | 3 | 0 | Low-quality | Genome-fragment | 4.21 | AAI-based (medium-confidence) | 0 |
| S16_extracted_151607 | 1000 | No | NA | 2 | 0 | 1 | Not-determined | Genome-fragment | NA | NA | 0 |
| S16_extracted_151663 | 1368 | No | NA | 1 | 0 | 1 | Not-determined | Genome-fragment | NA | NA | 0 |
| S14_vs_k141_108072\|\|full | 2618 | No | NA | 4 | 4 | 0 | Low-quality | Genome-fragment | 5.97 | AAI-based (high-confidence) | 0 |
| S1_vs_k141_42979\|\|full | 2618 | No | NA | 5 | 0 | 0 | Low-quality | Genome-fragment | 6.21 | AAI-based (medium-confidence) | 0 |
| S16_extracted_151753 | 1224 | No | NA | 1 | 0 | 1 | Not-determined | Genome-fragment | NA | NA | 0 |
| S16_extracted_151796 | 1304 | No | NA | 3 | 0 | 2 | Not-determined | Genome-fragment | NA | NA | 0 |
| S7_vs_29541\|\|full | 2618 | No | NA | 4 | 0 | 2 | Low-quality | Genome-fragment | 2.98 | AAI-based (medium-confidence) | 0 |
| S16_extracted_151888 | 2946 | No | NA | 3 | 0 | 2 | Not-determined | Genome-fragment | NA | NA | 0 |
| S16_extracted_151982 | 2589 | No | NA | 3 | 0 | 2 | Not-determined | Genome-fragment | NA | NA | 0 |
| S16_extracted_152018 | 1137 | No | NA | 2 | 0 | 0 | Not-determined | Genome-fragment | NA | NA | 0 |
| S16_extracted_152052 | 5506 | No | NA | 9 | 0 | 2 | Not-determined | Genome-fragment | NA | NA | 0 |
| S6_vs_k141_37686\|\|full | 2618 | No | NA | 3 | 0 | 2 | Low-quality | Genome-fragment | 1.83 | AAI-based (medium-confidence) | 0 |
| S17_vs_k141_81488\|\|full | 2619 | No | NA | 4 | 0 | 1 | Low-quality | Genome-fragment | 2.35 | AAI-based (high-confidence) | 0 |
| S3_extracted_k141_102045 | 2619 | No | NA | 2 | 0 | 2 | Low-quality | Genome-fragment | 1.45 | AAI-based (medium-confidence) | 0 |
| S16_extracted_152332 | 1398 | No | NA | 2 | 0 | 1 | Not-determined | Genome-fragment | NA | NA | 0 |
| S7_vs_26116\|\|full | 2619 | No | NA | 3 | 0 | 2 | Low-quality | Genome-fragment | 1.79 | AAI-based (high-confidence) | 0 |
| S16_extracted_152469 | 3206 | No | NA | 5 | 0 | 3 | Not-determined | Genome-fragment | NA | NA | 0 |
| S16_extracted_152677 | 3294 | No | NA | 3 | 0 | 0 | Not-determined | Genome-fragment | NA | NA | 0 |
| S16_extracted_152692 | 4200 | No | NA | 2 | 0 | 0 | Not-determined | Genome-fragment | NA | NA | 0 |
| S1_vs_k141_46300\|\|full | 2620 | No | NA | 7 | 1 | 0 | Low-quality | Genome-fragment | 2.25 | HMM-based (lower-bound) | 0 |
| S12_vs_k141_160390\|\|full | 2620 | No | NA | 3 | 0 | 1 | Low-quality | Genome-fragment | 1.19 | AAI-based (medium-confidence) | 0 |
| S16_extracted_152817 | 1036 | No | NA | 1 | 0 | 0 | Not-determined | Genome-fragment | NA | NA | 0 |
| S16_extracted_152824 | 1313 | No | NA | 1 | 0 | 1 | Not-determined | Genome-fragment | NA | NA | 0 |
| S16_extracted_152843 | 1460 | No | NA | 1 | 0 | 0 | Not-determined | Genome-fragment | NA | NA | 0 |
| S16_extracted_152852 | 3021 | No | NA | 4 | 0 | 3 | Not-determined | Genome-fragment | NA | NA | 0 |
| S16_extracted_153017 | 1783 | No | NA | 2 | 0 | 2 | Not-determined | Genome-fragment | NA | NA | 0 |
| S7_vs_30726\|\|full | 2621 | No | NA | 2 | 0 | 2 | Low-quality | Genome-fragment | 2.38 | AAI-based (medium-confidence) | 0 |
| S16_extracted_153087 | 3201 | No | NA | 5 | 0 | 1 | Not-determined | Genome-fragment | NA | NA | 0 |
| S16_extracted_153132 | 2354 | No | NA | 1 | 0 | 1 | Not-determined | Genome-fragment | NA | NA | 0 |
| S16_extracted_153230 | 2817 | No | NA | 3 | 0 | 1 | Not-determined | Genome-fragment | NA | NA | 0 |
| S16_extracted_153378 | 5819 | No | NA | 6 | 0 | 0 | Not-determined | Genome-fragment | NA | NA | 0 |
| S16_vs_74805\|\|full | 2621 | No | NA | 3 | 1 | 0 | Low-quality | Genome-fragment | 7.67 | AAI-based (medium-confidence) | 0 |
| S16_extracted_153562 | 4371 | No | NA | 8 | 0 | 5 | Not-determined | Genome-fragment | NA | NA | 0 |
| S8_vs_k141_132016\|\|full | 2621 | No | NA | 4 | 0 | 0 | Low-quality | Genome-fragment | 5.85 | AAI-based (medium-confidence) | 0 |
| S16_extracted_153695 | 1353 | No | NA | 3 | 0 | 1 | Not-determined | Genome-fragment | NA | NA | 0 |
| S16_extracted_153730 | 1912 | No | NA | 1 | 0 | 1 | Not-determined | Genome-fragment | NA | NA | 0 |
| S16_extracted_153964 | 1688 | No | NA | 5 | 0 | 1 | Not-determined | Genome-fragment | NA | NA | 0 |
| S17_vs_k141_150547\|\|full | 2623 | No | NA | 4 | 1 | 0 | Low-quality | Genome-fragment | 5.61 | HMM-based (lower-bound) | 0 |
| S16_extracted_154393 | 1425 | No | NA | 2 | 0 | 1 | Not-determined | Genome-fragment | NA | NA | 0 |
| S16_extracted_154558 | 2336 | No | NA | 3 | 0 | 2 | Not-determined | Genome-fragment | NA | NA | 0 |
| S16_extracted_154560 | 1109 | No | NA | 1 | 0 | 0 | Not-determined | Genome-fragment | NA | NA | 0 |
| S16_extracted_154566 | 1156 | No | NA | 1 | 0 | 0 | Not-determined | Genome-fragment | NA | NA | 0 |
| S16_extracted_154641 | 1491 | No | NA | 2 | 0 | 1 | Not-determined | Genome-fragment | NA | NA | 0 |
| S16_extracted_154644 | 2515 | No | NA | 1 | 0 | 0 | Not-determined | Genome-fragment | NA | NA | 0 |
| S11_extracted_k141_71142 | 2623 | No | NA | 3 | 0 | 2 | Low-quality | Genome-fragment | 2.03 | AAI-based (medium-confidence) | 0 |
| S16_vs_8018\|\|full | 2623 | No | NA | 3 | 0 | 1 | Low-quality | Genome-fragment | 4.67 | AAI-based (high-confidence) | 0 |
| S16_extracted_154800 | 1077 | No | NA | 2 | 0 | 0 | Not-determined | Genome-fragment | NA | NA | 0 |
| S16_extracted_154805 | 3247 | No | NA | 3 | 0 | 2 | Not-determined | Genome-fragment | NA | NA | 0 |
| S9_vs_k141_61546\|\|full | 2624 | No | NA | 9 | 1 | 0 | Low-quality | Genome-fragment | 5.07 | AAI-based (high-confidence) | 0 |
| S16_extracted_154931 | 1179 | No | NA | 2 | 0 | 1 | Not-determined | Genome-fragment | NA | NA | 0 |
| S16_extracted_154970 | 13214 | No | NA | 16 | 0 | 5 | Not-determined | Genome-fragment | NA | NA | 0 |
| S11_vs_k141_86652\|\|full | 2624 | No | NA | 6 | 0 | 0 | Low-quality | Genome-fragment | 3.71 | AAI-based (high-confidence) | 0 |
| S16_extracted_155149 | 1367 | No | NA | 3 | 0 | 0 | Not-determined | Genome-fragment | NA | NA | 0 |
| S16_extracted_155287 | 2602 | No | NA | 4 | 0 | 0 | Not-determined | Genome-fragment | NA | NA | 0 |
| S16_extracted_155303 | 1078 | No | NA | 2 | 0 | 1 | Not-determined | Genome-fragment | NA | NA | 0 |
| S16_extracted_155474 | 6551 | No | NA | 7 | 0 | 2 | Not-determined | Genome-fragment | NA | NA | 0 |
| S16_extracted_155551 | 2973 | No | NA | 2 | 0 | 1 | Not-determined | Genome-fragment | NA | NA | 0 |
| S12_vs_k141_90526\|\|full | 2624 | No | NA | 6 | 2 | 0 | Low-quality | Genome-fragment | 4.72 | AAI-based (medium-confidence) | 0 |
| S16_extracted_155739 | 6563 | No | NA | 6 | 0 | 5 | Not-determined | Genome-fragment | NA | NA | 0 |
| S8_extracted_k141_191762 | 2624 | No | NA | 5 | 4 | 0 | Low-quality | Genome-fragment | 4.05 | HMM-based (lower-bound) | 0 |
| S16_extracted_155783 | 1550 | No | NA | 2 | 0 | 2 | Not-determined | Genome-fragment | NA | NA | 0 |
| S8_vs_k141_98420\|\|full | 2624 | No | NA | 4 | 0 | 2 | Low-quality | Genome-fragment | 1.4 | AAI-based (medium-confidence) | 0 |
| S16_extracted_155940 | 3531 | No | NA | 3 | 0 | 1 | Not-determined | Genome-fragment | NA | NA | 0 |
| S16_extracted_156105 | 1047 | No | NA | 1 | 0 | 0 | Not-determined | Genome-fragment | NA | NA | 0 |
| S16_extracted_156193 | 2373 | No | NA | 2 | 0 | 1 | Not-determined | Genome-fragment | NA | NA | 0 |
| S17_extracted_k141_87173 | 2625 | No | NA | 6 | 0 | 0 | Low-quality | Genome-fragment | 3.31 | AAI-based (medium-confidence) | 0 |
| S16_extracted_156250 | 1721 | No | NA | 4 | 0 | 3 | Not-determined | Genome-fragment | NA | NA | 0 |
| S16_extracted_156314 | 1672 | No | NA | 1 | 0 | 1 | Not-determined | Genome-fragment | NA | NA | 0 |
| S16_extracted_156381 | 3220 | No | NA | 1 | 0 | 0 | Not-determined | Genome-fragment | NA | NA | 0 |
| S16_extracted_156391 | 5364 | No | NA | 5 | 0 | 3 | Not-determined | Genome-fragment | NA | NA | 0 |
| S16_extracted_156465 | 1434 | No | NA | 1 | 0 | 0 | Not-determined | Genome-fragment | NA | NA | 0 |
| S11_vs_k141_8584\|\|full | 2625 | No | NA | 3 | 1 | 0 | Low-quality | Genome-fragment | 5.27 | AAI-based (high-confidence) | 0 |
| S16_extracted_156503 | 1089 | No | NA | 1 | 0 | 1 | Not-determined | Genome-fragment | NA | NA | 0 |
| S14_vs_k141_71855\|\|full | 2625 | No | NA | 8 | 1 | 0 | Low-quality | Genome-fragment | 7.54 | AAI-based (high-confidence) | 0 |
| S16_extracted_156551 | 10631 | No | NA | 2 | 0 | 2 | Not-determined | Genome-fragment | NA | NA | 0 |
| S16_extracted_156658 | 8722 | No | NA | 9 | 0 | 3 | Not-determined | Genome-fragment | NA | NA | 0 |
| S16_extracted_156690 | 2203 | No | NA | 3 | 0 | 1 | Not-determined | Genome-fragment | NA | NA | 0 |
| S16_extracted_156729 | 1475 | No | NA | 1 | 0 | 1 | Not-determined | Genome-fragment | NA | NA | 0 |
| S3_vs_k141_11909\|\|full | 2625 | No | NA | 8 | 4 | 0 | Low-quality | Genome-fragment | 6.59 | AAI-based (high-confidence) | 0 |
| S16_extracted_156951 | 1275 | No | NA | 1 | 0 | 1 | Not-determined | Genome-fragment | NA | NA | 0 |
| S16_extracted_157103 | 1408 | No | NA | 2 | 0 | 1 | Not-determined | Genome-fragment | NA | NA | 0 |
| S16_extracted_157191 | 7079 | No | NA | 9 | 0 | 0 | Not-determined | Genome-fragment | NA | NA | 0 |
| S16_extracted_157304 | 1113 | No | NA | 1 | 0 | 0 | Not-determined | Genome-fragment | NA | NA | 0 |
| S16_extracted_157353 | 1118 | No | NA | 1 | 0 | 1 | Not-determined | Genome-fragment | NA | NA | 0 |
| S16_extracted_157355 | 1018 | No | NA | 2 | 0 | 1 | Not-determined | Genome-fragment | NA | NA | 0 |
| S16_extracted_157378 | 1008 | No | NA | 2 | 0 | 0 | Not-determined | Genome-fragment | NA | NA | 0 |
| S16_extracted_157392 | 16838 | No | NA | 9 | 0 | 6 | Not-determined | Genome-fragment | NA | NA | 0 |
| S16_extracted_157396 | 1152 | No | NA | 1 | 0 | 1 | Not-determined | Genome-fragment | NA | NA | 0 |
| S16_extracted_157434 | 1601 | No | NA | 3 | 0 | 0 | Not-determined | Genome-fragment | NA | NA | 0 |
| S16_extracted_157457 | 1092 | No | NA | 2 | 0 | 1 | Not-determined | Genome-fragment | NA | NA | 0 |
| S16_extracted_157476 | 1088 | No | NA | 2 | 0 | 0 | Not-determined | Genome-fragment | NA | NA | 0 |
| S16_extracted_157506 | 1081 | No | NA | 1 | 0 | 0 | Not-determined | Genome-fragment | NA | NA | 0 |
| S16_extracted_157671 | 3576 | No | NA | 5 | 0 | 4 | Not-determined | Genome-fragment | NA | NA | 0 |
| S16_extracted_157731 | 1309 | No | NA | 2 | 0 | 1 | Not-determined | Genome-fragment | NA | NA | 0 |
| S7_vs_137036\|\|full | 2625 | No | NA | 2 | 0 | 1 | Low-quality | Genome-fragment | 3.53 | AAI-based (high-confidence) | 0 |
| S16_extracted_157795 | 1473 | No | NA | 2 | 0 | 0 | Not-determined | Genome-fragment | NA | NA | 0 |
| S16_extracted_157872 | 1337 | No | NA | 2 | 0 | 2 | Not-determined | Genome-fragment | NA | NA | 0 |
| S16_extracted_157978 | 1653 | No | NA | 3 | 0 | 2 | Not-determined | Genome-fragment | NA | NA | 0 |
| S16_extracted_158090 | 1268 | No | NA | 1 | 0 | 0 | Not-determined | Genome-fragment | NA | NA | 0 |
| S16_extracted_158219 | 1191 | No | NA | 3 | 0 | 0 | Not-determined | Genome-fragment | NA | NA | 0 |
| S16_extracted_158254 | 7650 | No | NA | 7 | 0 | 4 | Not-determined | Genome-fragment | NA | NA | 0 |
| S16_vs_95790\|\|full | 2625 | No | NA | 4 | 4 | 0 | Low-quality | Genome-fragment | 6.66 | AAI-based (medium-confidence) | 0 |
| S16_extracted_158406 | 1219 | No | NA | 2 | 0 | 0 | Not-determined | Genome-fragment | NA | NA | 0 |
| S12_vs_k141_32589\|\|full | 2625 | No | NA | 3 | 2 | 0 | Low-quality | Genome-fragment | 7.07 | AAI-based (high-confidence) | 0 |
| S16_extracted_158596 | 1310 | No | NA | 2 | 0 | 0 | Not-determined | Genome-fragment | NA | NA | 0 |
| S16_extracted_158673 | 1691 | No | NA | 2 | 0 | 1 | Not-determined | Genome-fragment | NA | NA | 0 |
| S11_vs_k141_42778\|\|full | 2626 | No | NA | 6 | 1 | 0 | Low-quality | Genome-fragment | 6.29 | AAI-based (medium-confidence) | 0 |
| S11_vs_k141_55901\|\|full | 2626 | No | NA | 2 | 0 | 0 | Low-quality | Genome-fragment | 5.86 | AAI-based (medium-confidence) | 0 |
| S16_extracted_158815 | 1902 | No | NA | 3 | 0 | 2 | Not-determined | Genome-fragment | NA | NA | 0 |
| S2_vs_k141_12993\|\|full | 2626 | No | NA | 4 | 2 | 0 | Low-quality | Genome-fragment | 6.39 | AAI-based (high-confidence) | 0 |
| S16_extracted_158995 | 2031 | No | NA | 3 | 0 | 3 | Not-determined | Genome-fragment | NA | NA | 0 |
| S16_extracted_159003 | 5689 | No | NA | 5 | 0 | 1 | Not-determined | Genome-fragment | NA | NA | 0 |
| S8_vs_k141_75230\|\|full | 2626 | No | NA | 3 | 1 | 0 | Low-quality | Genome-fragment | 3.42 | HMM-based (lower-bound) | 0 |
| S14_vs_k141_115203\|\|full | 2627 | No | NA | 4 | 0 | 1 | Low-quality | Genome-fragment | 4.37 | AAI-based (high-confidence) | 0 |
| S16_extracted_159301 | 1413 | No | NA | 1 | 0 | 1 | Not-determined | Genome-fragment | NA | NA | 0 |
| S16_extracted_159314 | 1517 | No | NA | 1 | 0 | 0 | Not-determined | Genome-fragment | NA | NA | 0 |
| S1_vs_k141_97713\|\|full | 2627 | No | NA | 5 | 0 | 0 | Low-quality | Genome-fragment | 1.71 | AAI-based (high-confidence) | 0 |
| S16_extracted_159592 | 1432 | No | NA | 2 | 0 | 1 | Not-determined | Genome-fragment | NA | NA | 0 |
| S16_extracted_159595 | 1166 | No | NA | 1 | 0 | 1 | Not-determined | Genome-fragment | NA | NA | 0 |
| S16_extracted_159600 | 1674 | No | NA | 3 | 0 | 2 | Not-determined | Genome-fragment | NA | NA | 0 |
| S16_extracted_159613 | 1081 | No | NA | 2 | 0 | 0 | Not-determined | Genome-fragment | NA | NA | 0 |
| S3_vs_k141_95884\|\|full | 2627 | No | NA | 3 | 1 | 0 | Low-quality | Genome-fragment | 6.28 | AAI-based (high-confidence) | 0 |
| S5_vs_k141_72554\|\|full | 2627 | No | NA | 4 | 1 | 0 | Low-quality | Genome-fragment | 6.08 | AAI-based (medium-confidence) | 0 |
| S16_extracted_160016 | 5611 | No | NA | 4 | 0 | 0 | Not-determined | Genome-fragment | NA | NA | 0 |
| S9_vs_k141_76622\|\|full | 2628 | No | NA | 4 | 0 | 0 | Low-quality | Genome-fragment | 4.87 | AAI-based (high-confidence) | 0 |
| S16_extracted_160147 | 1303 | No | NA | 1 | 0 | 1 | Not-determined | Genome-fragment | NA | NA | 0 |
| S16_extracted_160207 | 4753 | No | NA | 4 | 0 | 0 | Not-determined | Genome-fragment | NA | NA | 0 |
| S16_extracted_160365 | 1576 | No | NA | 4 | 0 | 1 | Not-determined | Genome-fragment | NA | NA | 0 |
| S11_vs_k141_45954\|\|full | 2629 | No | NA | 3 | 0 | 1 | Low-quality | Genome-fragment | 1.25 | AAI-based (high-confidence) | 0 |
| S16_extracted_70166 | 2629 | No | NA | 6 | 3 | 0 | Low-quality | Genome-fragment | 4.33 | AAI-based (high-confidence) | 0 |
| S12_extracted_k141_21226 | 2629 | No | NA | 4 | 2 | 0 | Low-quality | Genome-fragment | 6.37 | AAI-based (medium-confidence) | 0 |
| S17_extracted_k141_168812 | 2630 | No | NA | 3 | 0 | 0 | Low-quality | Genome-fragment | 9.03 | AAI-based (medium-confidence) | 0 |
| S16_extracted_160798 | 3367 | No | NA | 1 | 0 | 1 | Not-determined | Genome-fragment | NA | NA | 0 |
| S16_extracted_160863 | 6930 | No | NA | 4 | 0 | 2 | Not-determined | Genome-fragment | NA | NA | 0 |
| S1_vs_k141_104171\|\|full | 2630 | No | NA | 7 | 1 | 0 | Low-quality | Genome-fragment | 6.81 | AAI-based (high-confidence) | 0 |
| S5_vs_k141_125903\|\|full | 2630 | No | NA | 5 | 1 | 0 | Low-quality | Genome-fragment | 4.95 | AAI-based (medium-confidence) | 0 |
| S16_extracted_160974 | 3700 | No | NA | 3 | 0 | 0 | Not-determined | Genome-fragment | NA | NA | 0 |
| S16_extracted_161010 | 2690 | No | NA | 3 | 0 | 0 | Not-determined | Genome-fragment | NA | NA | 0 |
| S16_extracted_161090 | 3343 | No | NA | 7 | 0 | 1 | Not-determined | Genome-fragment | NA | NA | 0 |
| S16_extracted_161285 | 5112 | No | NA | 4 | 0 | 1 | Not-determined | Genome-fragment | NA | NA | 0 |
| S16_extracted_161321 | 1156 | No | NA | 1 | 0 | 1 | Not-determined | Genome-fragment | NA | NA | 0 |
| S16_extracted_161358 | 2335 | No | NA | 2 | 0 | 1 | Not-determined | Genome-fragment | NA | NA | 0 |
| S16_extracted_161489 | 1642 | No | NA | 3 | 0 | 1 | Not-determined | Genome-fragment | NA | NA | 0 |
| S16_extracted_161593 | 1035 | No | NA | 2 | 0 | 2 | Not-determined | Genome-fragment | NA | NA | 0 |
| S16_extracted_161656 | 3548 | No | NA | 6 | 0 | 1 | Not-determined | Genome-fragment | NA | NA | 0 |
| S16_extracted_161679 | 1966 | No | NA | 2 | 0 | 2 | Not-determined | Genome-fragment | NA | NA | 0 |
| S16_extracted_161709 | 1750 | No | NA | 2 | 0 | 0 | Not-determined | Genome-fragment | NA | NA | 0 |
| S5_extracted_k141_108143 | 2631 | No | NA | 7 | 2 | 0 | Low-quality | Genome-fragment | 8.16 | AAI-based (high-confidence) | 0 |
| S16_extracted_161730 | 1254 | No | NA | 2 | 0 | 1 | Not-determined | Genome-fragment | NA | NA | 0 |
| S16_extracted_161733 | 1118 | No | NA | 2 | 0 | 0 | Not-determined | Genome-fragment | NA | NA | 0 |
| S16_extracted_161734 | 1109 | No | NA | 2 | 0 | 0 | Not-determined | Genome-fragment | NA | NA | 0 |
| S16_extracted_161741 | 2137 | No | NA | 1 | 0 | 1 | Not-determined | Genome-fragment | NA | NA | 0 |
| S16_extracted_161759 | 1054 | No | NA | 2 | 0 | 1 | Not-determined | Genome-fragment | NA | NA | 0 |
| S16_extracted_161820 | 1545 | No | NA | 3 | 0 | 1 | Not-determined | Genome-fragment | NA | NA | 0 |
| S16_extracted_161878 | 9158 | No | NA | 12 | 0 | 2 | Not-determined | Genome-fragment | NA | NA | 0 |
| S16_extracted_161951 | 4092 | No | NA | 2 | 0 | 1 | Not-determined | Genome-fragment | NA | NA | 0 |
| S16_extracted_162063 | 2718 | No | NA | 1 | 0 | 0 | Not-determined | Genome-fragment | NA | NA | 0 |
| S16_extracted_162086 | 7472 | No | NA | 9 | 0 | 7 | Not-determined | Genome-fragment | NA | NA | 0 |
| S16_extracted_162191 | 2113 | No | NA | 2 | 0 | 0 | Not-determined | Genome-fragment | NA | NA | 0 |
| S16_extracted_162261 | 1147 | No | NA | 2 | 0 | 1 | Not-determined | Genome-fragment | NA | NA | 0 |
| S16_extracted_162378 | 3098 | No | NA | 2 | 0 | 1 | Not-determined | Genome-fragment | NA | NA | 0 |
| S5_vs_k141_90149\|\|full | 2631 | No | NA | 6 | 3 | 0 | Low-quality | Genome-fragment | 6.13 | AAI-based (high-confidence) | 0 |
| S11_vs_k141_85274\|\|full | 2632 | No | NA | 3 | 0 | 1 | Low-quality | Genome-fragment | 3.72 | AAI-based (medium-confidence) | 0 |
| S3_vs_k141_54858\|\|full | 2632 | No | NA | 3 | 3 | 0 | Low-quality | Genome-fragment | 6.45 | AAI-based (high-confidence) | 0 |
| S16_extracted_162634 | 1234 | No | NA | 1 | 0 | 1 | Not-determined | Genome-fragment | NA | NA | 0 |
| S5_vs_k141_23741\|\|full | 2632 | No | NA | 9 | 0 | 0 | Low-quality | Genome-fragment | 4.05 | AAI-based (medium-confidence) | 0 |
| S16_extracted_162661 | 1619 | No | NA | 3 | 0 | 2 | Not-determined | Genome-fragment | NA | NA | 0 |
| S16_extracted_162665 | 1096 | No | NA | 3 | 0 | 1 | Not-determined | Genome-fragment | NA | NA | 0 |
| S12_vs_k141_8463\|\|full | 2632 | No | NA | 3 | 0 | 0 | Low-quality | Genome-fragment | 5.68 | AAI-based (medium-confidence) | 0 |
| S12_vs_k141_114666\|\|full | 2632 | No | NA | 4 | 1 | 0 | Low-quality | Genome-fragment | 5.71 | AAI-based (high-confidence) | 0 |
| S16_extracted_162877 | 1160 | No | NA | 2 | 0 | 0 | Not-determined | Genome-fragment | NA | NA | 0 |
| S8_extracted_k141_93905 | 2632 | No | NA | 3 | 0 | 3 | Low-quality | Genome-fragment | 0.99 | AAI-based (high-confidence) | 0 |
| S16_extracted_163111 | 3081 | No | NA | 3 | 0 | 3 | Not-determined | Genome-fragment | NA | NA | 0 |
| S8_extracted_k141_146894 | 2632 | No | NA | 3 | 0 | 2 | Low-quality | Genome-fragment | 6.02 | AAI-based (high-confidence) | 0 |
| S16_extracted_163118 | 3191 | No | NA | 4 | 0 | 1 | Not-determined | Genome-fragment | NA | NA | 0 |
| S16_extracted_163221 | 1167 | No | NA | 1 | 0 | 0 | Not-determined | Genome-fragment | NA | NA | 0 |
| S14_extracted_k141_104685 | 2633 | No | NA | 3 | 1 | 0 | Low-quality | Genome-fragment | 1.47 | HMM-based (lower-bound) | 0 |
| S3_extracted_k141_67581 | 2633 | No | NA | 2 | 0 | 0 | Low-quality | Genome-fragment | 2.9 | AAI-based (high-confidence) | 0 |
| S16_extracted_163493 | 3438 | No | NA | 1 | 0 | 0 | Not-determined | Genome-fragment | NA | NA | 0 |
| S16_extracted_163545 | 4913 | No | NA | 3 | 0 | 1 | Not-determined | Genome-fragment | NA | NA | 0 |
| S16_extracted_163558 | 1761 | No | NA | 2 | 0 | 2 | Not-determined | Genome-fragment | NA | NA | 0 |
| S5_vs_k141_20234\|\|full | 2633 | No | NA | 3 | 1 | 0 | Low-quality | Genome-fragment | 3.8 | AAI-based (high-confidence) | 0 |
| S16_extracted_163592 | 3281 | No | NA | 3 | 0 | 2 | Not-determined | Genome-fragment | NA | NA | 0 |
| S16_extracted_163674 | 3356 | No | NA | 5 | 0 | 1 | Not-determined | Genome-fragment | NA | NA | 0 |
| S16_extracted_163702 | 1314 | No | NA | 1 | 0 | 0 | Not-determined | Genome-fragment | NA | NA | 0 |
| S16_extracted_163765 | 1036 | No | NA | 2 | 0 | 2 | Not-determined | Genome-fragment | NA | NA | 0 |
| S16_extracted_163847 | 2184 | No | NA | 1 | 0 | 0 | Not-determined | Genome-fragment | NA | NA | 0 |
| S16_vs_167035\|\|full | 2633 | No | NA | 5 | 3 | 0 | Low-quality | Genome-fragment | 7.97 | AAI-based (high-confidence) | 0 |
| S16_extracted_163921 | 3177 | No | NA | 4 | 0 | 3 | Not-determined | Genome-fragment | NA | NA | 0 |
| S16_extracted_164037 | 2277 | No | NA | 3 | 0 | 0 | Not-determined | Genome-fragment | NA | NA | 0 |
| S16_extracted_164087 | 1023 | No | NA | 1 | 0 | 1 | Not-determined | Genome-fragment | NA | NA | 0 |
| S16_extracted_164146 | 1296 | No | NA | 1 | 0 | 1 | Not-determined | Genome-fragment | NA | NA | 0 |
| S16_extracted_164162 | 1149 | No | NA | 2 | 0 | 0 | Not-determined | Genome-fragment | NA | NA | 0 |
| S16_extracted_164236 | 2215 | No | NA | 2 | 0 | 1 | Not-determined | Genome-fragment | NA | NA | 0 |
| S16_extracted_164353 | 1130 | No | NA | 1 | 0 | 1 | Not-determined | Genome-fragment | NA | NA | 0 |
| S9_vs_k141_15495\|\|full | 2634 | No | NA | 6 | 5 | 0 | Low-quality | Genome-fragment | 6.27 | AAI-based (medium-confidence) | 0 |
| S11_extracted_k141_70180 | 2634 | No | NA | 1 | 0 | 0 | Low-quality | Genome-fragment | 0.95 | AAI-based (medium-confidence) | 0 |
| S16_extracted_164647 | 1010 | No | NA | 2 | 0 | 1 | Not-determined | Genome-fragment | NA | NA | 0 |
| S16_extracted_164667 | 1255 | No | NA | 2 | 0 | 2 | Not-determined | Genome-fragment | NA | NA | 0 |
| S16_extracted_164704 | 1830 | No | NA | 1 | 0 | 1 | Not-determined | Genome-fragment | NA | NA | 0 |
| S16_extracted_164732 | 5160 | No | NA | 5 | 0 | 3 | Not-determined | Genome-fragment | NA | NA | 0 |
| S16_extracted_164823 | 1104 | No | NA | 4 | 0 | 0 | Not-determined | Genome-fragment | NA | NA | 0 |
| S16_extracted_164913 | 17025 | Yes | 15002 | 21 | 6 | 2 | High-quality | High-quality | 100 | AAI-based (high-confidence) | 11.88 |
| S3_vs_k141_67709\|\|full | 2634 | No | NA | 3 | 2 | 0 | Low-quality | Genome-fragment | 1.45 | HMM-based (lower-bound) | 0 |
| S16_extracted_164994 | 1194 | No | NA | 2 | 0 | 1 | Not-determined | Genome-fragment | NA | NA | 0 |
| S16_extracted_165020 | 3566 | No | NA | 11 | 0 | 0 | Not-determined | Genome-fragment | NA | NA | 0 |
| S7_extracted_43807 | 2634 | No | NA | 4 | 1 | 0 | Low-quality | Genome-fragment | 2.74 | AAI-based (high-confidence) | 0 |
| S16_extracted_165087 | 3193 | No | NA | 1 | 0 | 1 | Not-determined | Genome-fragment | NA | NA | 0 |
| S16_extracted_165101 | 7282 | No | NA | 5 | 0 | 2 | Not-determined | Genome-fragment | NA | NA | 0 |
| S16_extracted_165129 | 1245 | No | NA | 1 | 0 | 1 | Not-determined | Genome-fragment | NA | NA | 0 |
| S16_extracted_165238 | 4100 | No | NA | 5 | 0 | 4 | Not-determined | Genome-fragment | NA | NA | 0 |
| S16_extracted_165411 | 3953 | No | NA | 5 | 0 | 4 | Not-determined | Genome-fragment | NA | NA | 0 |
| S16_extracted_165504 | 1226 | No | NA | 1 | 0 | 1 | Not-determined | Genome-fragment | NA | NA | 0 |
| S10_vs_1280\|\|full | 2634 | No | NA | 6 | 0 | 0 | Low-quality | Genome-fragment | 3.46 | AAI-based (high-confidence) | 0 |
| S16_extracted_165604 | 1141 | No | NA | 1 | 0 | 1 | Not-determined | Genome-fragment | NA | NA | 0 |
| S16_extracted_165627 | 3355 | No | NA | 2 | 0 | 2 | Not-determined | Genome-fragment | NA | NA | 0 |
| S8_extracted_k141_203223 | 2634 | No | NA | 3 | 1 | 0 | Low-quality | Genome-fragment | 5.24 | AAI-based (medium-confidence) | 0 |
| S16_extracted_165820 | 5528 | No | NA | 4 | 0 | 3 | Not-determined | Genome-fragment | NA | NA | 0 |
| S17_extracted_k141_81594 | 2635 | No | NA | 3 | 3 | 0 | Low-quality | Genome-fragment | 8.51 | AAI-based (high-confidence) | 0 |
| S11_vs_k141_32245\|\|full | 2635 | Yes | 2430 | 4 | 3 | 1 | Low-quality | Genome-fragment | 3.63 | HMM-based (lower-bound) | 7.78 |
| S16_extracted_165993 | 4521 | No | NA | 5 | 0 | 2 | Not-determined | Genome-fragment | NA | NA | 0 |
| S16_extracted_166046 | 3466 | No | NA | 4 | 0 | 4 | Not-determined | Genome-fragment | NA | NA | 0 |
| S16_extracted_166145 | 1586 | No | NA | 3 | 0 | 2 | Not-determined | Genome-fragment | NA | NA | 0 |
| S4_extracted_18567 | 2635 | No | NA | 7 | 7 | 0 | Low-quality | Genome-fragment | 8.67 | AAI-based (high-confidence) | 0 |
| S16_extracted_166258 | 1034 | No | NA | 2 | 0 | 2 | Not-determined | Genome-fragment | NA | NA | 0 |
| S8_vs_k141_59002\|\|full | 2635 | No | NA | 6 | 1 | 0 | Low-quality | Genome-fragment | 5.57 | AAI-based (medium-confidence) | 0 |
| S16_extracted_166293 | 2585 | No | NA | 3 | 0 | 2 | Not-determined | Genome-fragment | NA | NA | 0 |
| S17_vs_k141_126467\|\|full | 2637 | No | NA | 5 | 1 | 0 | Low-quality | Genome-fragment | 4.74 | AAI-based (high-confidence) | 0 |
| S16_extracted_166511 | 1834 | No | NA | 1 | 0 | 1 | Not-determined | Genome-fragment | NA | NA | 0 |
| S5_vs_k141_60859\|\|full | 2637 | No | NA | 3 | 1 | 0 | Low-quality | Genome-fragment | 6.25 | AAI-based (high-confidence) | 0 |
| S16_extracted_166667 | 1709 | No | NA | 1 | 0 | 1 | Not-determined | Genome-fragment | NA | NA | 0 |
| S16_extracted_166677 | 5073 | No | NA | 6 | 0 | 2 | Not-determined | Genome-fragment | NA | NA | 0 |
| S16_extracted_166802 | 1032 | No | NA | 2 | 0 | 2 | Not-determined | Genome-fragment | NA | NA | 0 |
| S16_extracted_166845 | 1162 | No | NA | 1 | 0 | 0 | Not-determined | Genome-fragment | NA | NA | 0 |
| S16_extracted_166923 | 1000 | No | NA | 1 | 0 | 0 | Not-determined | Genome-fragment | NA | NA | 0 |
| S16_extracted_166974 | 1495 | No | NA | 1 | 0 | 0 | Not-determined | Genome-fragment | NA | NA | 0 |
| S16_extracted_167027 | 1573 | No | NA | 3 | 0 | 1 | Not-determined | Genome-fragment | NA | NA | 0 |
| S16_extracted_167132 | 1308 | No | NA | 1 | 0 | 1 | Not-determined | Genome-fragment | NA | NA | 0 |
| S16_extracted_167144 | 1018 | No | NA | 1 | 0 | 0 | Not-determined | Genome-fragment | NA | NA | 0 |
| S5_vs_k141_84612\|\|full | 2637 | No | NA | 4 | 0 | 2 | Low-quality | Genome-fragment | 1.21 | AAI-based (high-confidence) | 0 |
| S12_vs_k141_154406\|\|full | 2637 | No | NA | 5 | 2 | 0 | Low-quality | Genome-fragment | 6.3 | AAI-based (medium-confidence) | 0 |
| S16_extracted_167312 | 1074 | No | NA | 2 | 0 | 0 | Not-determined | Genome-fragment | NA | NA | 0 |
| S16_extracted_167321 | 3108 | No | NA | 3 | 0 | 0 | Not-determined | Genome-fragment | NA | NA | 0 |
| S16_extracted_167441 | 1688 | No | NA | 1 | 0 | 0 | Not-determined | Genome-fragment | NA | NA | 0 |
| S5_vs_k141_59789\|\|full | 2638 | No | NA | 3 | 0 | 1 | Low-quality | Genome-fragment | 4.46 | AAI-based (medium-confidence) | 0 |
| S16_extracted_167580 | 1846 | No | NA | 1 | 0 | 1 | Not-determined | Genome-fragment | NA | NA | 0 |
| S16_extracted_167624 | 1330 | No | NA | 2 | 0 | 2 | Not-determined | Genome-fragment | NA | NA | 0 |
| S16_extracted_167674 | 1037 | No | NA | 1 | 0 | 1 | Not-determined | Genome-fragment | NA | NA | 0 |
| S16_extracted_167848 | 1382 | No | NA | 2 | 0 | 1 | Not-determined | Genome-fragment | NA | NA | 0 |
| S7_extracted_69574 | 2638 | No | NA | 3 | 0 | 3 | Low-quality | Genome-fragment | 1.74 | AAI-based (high-confidence) | 0 |
| S16_vs_3440\|\|full | 2638 | No | NA | 4 | 1 | 0 | Low-quality | Genome-fragment | 1.63 | HMM-based (lower-bound) | 0 |
| S16_extracted_167983 | 3577 | No | NA | 1 | 0 | 1 | Not-determined | Genome-fragment | NA | NA | 0 |
| S16_extracted_167986 | 2483 | No | NA | 3 | 0 | 0 | Not-determined | Genome-fragment | NA | NA | 0 |
| S16_extracted_167988 | 1153 | No | NA | 3 | 0 | 2 | Not-determined | Genome-fragment | NA | NA | 0 |
| S16_extracted_168092 | 4092 | No | NA | 4 | 0 | 2 | Not-determined | Genome-fragment | NA | NA | 0 |
| S14_vs_k141_86955\|\|full | 2639 | No | NA | 6 | 1 | 0 | Low-quality | Genome-fragment | 2.27 | HMM-based (lower-bound) | 0 |
| S10_extracted_21093 | 2639 | No | NA | 3 | 0 | 1 | Low-quality | Genome-fragment | 5.68 | AAI-based (medium-confidence) | 0 |
| S16_extracted_168243 | 3507 | No | NA | 3 | 0 | 1 | Not-determined | Genome-fragment | NA | NA | 0 |
| S16_extracted_168418 | 1360 | No | NA | 2 | 0 | 1 | Not-determined | Genome-fragment | NA | NA | 0 |
| S6_extracted_k141_3133 | 2639 | No | NA | 5 | 4 | 0 | Low-quality | Genome-fragment | 4.33 | AAI-based (high-confidence) | 0 |
| S16_extracted_168527 | 3015 | No | NA | 1 | 0 | 1 | Not-determined | Genome-fragment | NA | NA | 0 |
| S16_extracted_168629 | 1215 | No | NA | 2 | 0 | 1 | Not-determined | Genome-fragment | NA | NA | 0 |
| S17_vs_k141_108622\|\|full | 2640 | No | NA | 4 | 0 | 3 | Low-quality | Genome-fragment | 0.91 | AAI-based (medium-confidence) | 0 |
| S16_extracted_168837 | 5225 | No | NA | 5 | 0 | 3 | Not-determined | Genome-fragment | NA | NA | 0 |
| S13_vs_55811\|\|full | 2640 | No | NA | 3 | 0 | 0 | Low-quality | Genome-fragment | 1.62 | AAI-based (high-confidence) | 0 |
| S14_vs_k141_94997\|\|lt2gene | 2640 | No | NA | 2 | 1 | 0 | Low-quality | Genome-fragment | 8.1 | HMM-based (lower-bound) | 0 |
| S16_extracted_169022 | 1644 | No | NA | 3 | 0 | 1 | Not-determined | Genome-fragment | NA | NA | 0 |
| S16_extracted_169054 | 1953 | No | NA | 1 | 0 | 1 | Not-determined | Genome-fragment | NA | NA | 0 |
| S16_extracted_169077 | 3162 | No | NA | 2 | 0 | 2 | Not-determined | Genome-fragment | NA | NA | 0 |
| S16_extracted_169119 | 3140 | No | NA | 3 | 0 | 2 | Not-determined | Genome-fragment | NA | NA | 0 |
| S16_extracted_169151 | 3008 | No | NA | 4 | 0 | 1 | Not-determined | Genome-fragment | NA | NA | 0 |
| S16_extracted_169245 | 1324 | No | NA | 2 | 0 | 1 | Not-determined | Genome-fragment | NA | NA | 0 |
| S1_extracted_k141_117142 | 2640 | No | NA | 1 | 0 | 1 | Low-quality | Genome-fragment | 5.68 | AAI-based (high-confidence) | 0 |
| S16_extracted_169365 | 1342 | No | NA | 2 | 0 | 0 | Not-determined | Genome-fragment | NA | NA | 0 |
| S16_extracted_169377 | 4271 | No | NA | 6 | 0 | 1 | Not-determined | Genome-fragment | NA | NA | 0 |
| S16_extracted_169433 | 5238 | No | NA | 5 | 0 | 1 | Not-determined | Genome-fragment | NA | NA | 0 |
| S16_extracted_169545 | 3510 | No | NA | 4 | 0 | 1 | Not-determined | Genome-fragment | NA | NA | 0 |
| S16_extracted_169555 | 1090 | No | NA | 2 | 0 | 1 | Not-determined | Genome-fragment | NA | NA | 0 |
| S16_extracted_169694 | 1263 | No | NA | 2 | 0 | 2 | Not-determined | Genome-fragment | NA | NA | 0 |
| S16_extracted_169776 | 1392 | No | NA | 2 | 0 | 0 | Not-determined | Genome-fragment | NA | NA | 0 |
| S16_extracted_169788 | 1314 | No | NA | 1 | 0 | 1 | Not-determined | Genome-fragment | NA | NA | 0 |
| S16_extracted_169789 | 1063 | No | NA | 3 | 0 | 3 | Not-determined | Genome-fragment | NA | NA | 0 |
| S16_extracted_169797 | 1021 | No | NA | 1 | 0 | 0 | Not-determined | Genome-fragment | NA | NA | 0 |
| S16_extracted_169864 | 2023 | No | NA | 1 | 0 | 1 | Not-determined | Genome-fragment | NA | NA | 0 |
| S16_extracted_169895 | 17342 | No | NA | 26 | 8 | 0 | Medium-quality | Genome-fragment | 86.69 | AAI-based (medium-confidence) | 0 |
| S16_extracted_169901 | 1031 | No | NA | 2 | 0 | 0 | Not-determined | Genome-fragment | NA | NA | 0 |
| S16_extracted_169914 | 3701 | No | NA | 2 | 0 | 2 | Not-determined | Genome-fragment | NA | NA | 0 |
| S16_extracted_169948 | 1238 | No | NA | 2 | 0 | 2 | Not-determined | Genome-fragment | NA | NA | 0 |
| S16_extracted_169985 | 85365 | No | NA | 109 | 13 | 2 | Complete | High-quality | 100 | DTR (high-confidence) | 0 |
| S16_extracted_170032 | 5878 | No | NA | 3 | 0 | 1 | Not-determined | Genome-fragment | NA | NA | 0 |
| S16_extracted_152082 | 2640 | No | NA | 5 | 0 | 3 | Low-quality | Genome-fragment | 2.83 | AAI-based (medium-confidence) | 0 |
| S16_extracted_170185 | 5183 | No | NA | 5 | 0 | 2 | Not-determined | Genome-fragment | NA | NA | 0 |
| S8_extracted_k141_61757 | 2640 | No | NA | 3 | 3 | 0 | Low-quality | Genome-fragment | 8.71 | AAI-based (high-confidence) | 0 |
| S16_extracted_170264 | 1605 | No | NA | 2 | 0 | 2 | Not-determined | Genome-fragment | NA | NA | 0 |
| S14_extracted_k141_59396 | 2641 | No | NA | 5 | 3 | 0 | Low-quality | Genome-fragment | 8.71 | AAI-based (high-confidence) | 0 |
| S16_extracted_170305 | 1729 | No | NA | 1 | 0 | 1 | Not-determined | Genome-fragment | NA | NA | 0 |
| S16_extracted_170443 | 2153 | No | NA | 1 | 0 | 0 | Not-determined | Genome-fragment | NA | NA | 0 |
| S16_extracted_170508 | 1530 | No | NA | 1 | 0 | 0 | Not-determined | Genome-fragment | NA | NA | 0 |
| S16_extracted_170543 | 3976 | No | NA | 5 | 0 | 5 | Not-determined | Genome-fragment | NA | NA | 0 |
| S16_extracted_170548 | 1686 | No | NA | 1 | 0 | 0 | Not-determined | Genome-fragment | NA | NA | 0 |
| S16_extracted_170592 | 1218 | No | NA | 2 | 0 | 0 | Not-determined | Genome-fragment | NA | NA | 0 |
| S16_extracted_170799 | 1085 | No | NA | 1 | 0 | 0 | Not-determined | Genome-fragment | NA | NA | 0 |
| S16_extracted_170834 | 4711 | No | NA | 4 | 0 | 3 | Not-determined | Genome-fragment | NA | NA | 0 |
| S14_extracted_k141_67476 | 2641 | No | NA | 6 | 6 | 0 | Low-quality | Genome-fragment | 8.69 | AAI-based (high-confidence) | 0 |
| S16_extracted_170930 | 2238 | No | NA | 3 | 0 | 1 | Not-determined | Genome-fragment | NA | NA | 0 |
| S16_extracted_170995 | 3905 | No | NA | 4 | 0 | 3 | Not-determined | Genome-fragment | NA | NA | 0 |
| S16_extracted_171001 | 2199 | No | NA | 3 | 0 | 1 | Not-determined | Genome-fragment | NA | NA | 0 |
| S16_extracted_171048 | 1202 | No | NA | 2 | 0 | 1 | Not-determined | Genome-fragment | NA | NA | 0 |
| S1_vs_k141_118035\|\|full | 2641 | No | NA | 3 | 1 | 0 | Low-quality | Genome-fragment | 5.43 | AAI-based (medium-confidence) | 0 |
| S16_extracted_171127 | 4776 | No | NA | 1 | 0 | 1 | Not-determined | Genome-fragment | NA | NA | 0 |
| S16_extracted_171205 | 1345 | No | NA | 1 | 0 | 0 | Not-determined | Genome-fragment | NA | NA | 0 |
| S7_vs_60187\|\|full | 2641 | No | NA | 3 | 0 | 2 | Low-quality | Genome-fragment | 2.07 | AAI-based (high-confidence) | 0 |
| S16_extracted_171586 | 3361 | No | NA | 3 | 0 | 3 | Not-determined | Genome-fragment | NA | NA | 0 |
| S16_extracted_171588 | 1214 | No | NA | 2 | 0 | 2 | Not-determined | Genome-fragment | NA | NA | 0 |
| S16_extracted_171649 | 2528 | No | NA | 2 | 0 | 1 | Not-determined | Genome-fragment | NA | NA | 0 |
| S16_extracted_171654 | 4331 | No | NA | 4 | 0 | 2 | Not-determined | Genome-fragment | NA | NA | 0 |
| S16_extracted_171748 | 1000 | No | NA | 1 | 0 | 1 | Not-determined | Genome-fragment | NA | NA | 0 |
| S16_extracted_171755 | 17782 | No | NA | 10 | 0 | 8 | Not-determined | Genome-fragment | NA | NA | 0 |
| S16_extracted_171787 | 2178 | No | NA | 3 | 0 | 3 | Not-determined | Genome-fragment | NA | NA | 0 |
| S16_extracted_171926 | 3722 | No | NA | 3 | 0 | 2 | Not-determined | Genome-fragment | NA | NA | 0 |
| S17_vs_k141_135473\|\|full | 2642 | No | NA | 10 | 0 | 0 | Low-quality | Genome-fragment | 5.92 | AAI-based (high-confidence) | 0 |
| S16_extracted_172081 | 1590 | No | NA | 1 | 0 | 0 | Not-determined | Genome-fragment | NA | NA | 0 |
| S11_vs_k141_61520\|\|full | 2642 | No | NA | 3 | 2 | 0 | Low-quality | Genome-fragment | 7.73 | AAI-based (high-confidence) | 0 |
| S16_extracted_172223 | 1169 | No | NA | 1 | 0 | 0 | Not-determined | Genome-fragment | NA | NA | 0 |
| S3_vs_k141_47123\|\|full | 2642 | No | NA | 4 | 2 | 0 | Low-quality | Genome-fragment | 2.8 | HMM-based (lower-bound) | 0 |
| S16_extracted_172419 | 1088 | No | NA | 2 | 0 | 2 | Not-determined | Genome-fragment | NA | NA | 0 |
| S16_extracted_172544 | 1885 | No | NA | 3 | 0 | 1 | Not-determined | Genome-fragment | NA | NA | 0 |
| S16_extracted_172550 | 1025 | No | NA | 1 | 0 | 0 | Not-determined | Genome-fragment | NA | NA | 0 |
| S16_extracted_172552 | 1559 | No | NA | 3 | 0 | 1 | Not-determined | Genome-fragment | NA | NA | 0 |
| S16_extracted_172588 | 2463 | No | NA | 3 | 0 | 0 | Not-determined | Genome-fragment | NA | NA | 0 |
| S16_extracted_172748 | 1002 | No | NA | 1 | 0 | 0 | Not-determined | Genome-fragment | NA | NA | 0 |
| S16_extracted_172782 | 1090 | No | NA | 2 | 0 | 1 | Not-determined | Genome-fragment | NA | NA | 0 |
| S16_extracted_172785 | 3258 | No | NA | 2 | 0 | 2 | Not-determined | Genome-fragment | NA | NA | 0 |
| S16_extracted_172791 | 1455 | No | NA | 1 | 0 | 1 | Not-determined | Genome-fragment | NA | NA | 0 |
| S16_extracted_172830 | 1346 | No | NA | 2 | 0 | 1 | Not-determined | Genome-fragment | NA | NA | 0 |
| S16_extracted_172846 | 1463 | No | NA | 1 | 0 | 1 | Not-determined | Genome-fragment | NA | NA | 0 |
| S16_extracted_172877 | 1837 | No | NA | 1 | 0 | 0 | Not-determined | Genome-fragment | NA | NA | 0 |
| S16_extracted_172897 | 36447 | No | NA | 53 | 22 | 1 | Complete | High-quality | 100 | DTR (high-confidence) | 0 |
| S16_vs_81611\|\|full | 2642 | No | NA | 3 | 1 | 0 | Low-quality | Genome-fragment | 7 | AAI-based (high-confidence) | 0 |
| S16_extracted_173000 | 6383 | No | NA | 5 | 0 | 5 | Not-determined | Genome-fragment | NA | NA | 0 |
| S16_extracted_173095 | 96033 | No | NA | 80 | 10 | 0 | Complete | High-quality | 100 | DTR (high-confidence) | 0 |
| S16_extracted_173099 | 77224 | No | NA | 104 | 22 | 4 | Complete | High-quality | 100 | DTR (high-confidence) | 0 |
| S16_extracted_173103 | 3994 | No | NA | 4 | 0 | 0 | Not-determined | Genome-fragment | NA | NA | 0 |
| S10_vs_101681\|\|full | 2642 | No | NA | 5 | 0 | 2 | Low-quality | Genome-fragment | 1.47 | AAI-based (medium-confidence) | 0 |
| S16_extracted_173109 | 28358 | No | NA | 47 | 12 | 1 | Complete | High-quality | 100 | DTR (high-confidence) | 0 |
| S16_extracted_173114 | 3343 | No | NA | 4 | 0 | 0 | Not-determined | Genome-fragment | NA | NA | 0 |
| S16_extracted_173133 | 12950 | Yes | 12101 | 23 | 3 | 2 | Complete | High-quality | 100 | DTR (medium-confidence) | 6.56 |
| S16_vs_8916\|\|full | 8185 | No | NA | 13 | 0 | 0 | Not-determined | Genome-fragment | NA | NA | 0 |
| S16_vs_15093\|\|full | 52784 | No | NA | 94 | 17 | 1 | High-quality | High-quality | 100 | AAI-based (high-confidence) | 0 |
| S16_vs_20060\|\|full | 41359 | No | NA | 70 | 14 | 0 | High-quality | High-quality | 98.23 | AAI-based (high-confidence) | 0 |
| S12_vs_k141_157771\|\|full | 2642 | No | NA | 7 | 1 | 0 | Low-quality | Genome-fragment | 6.1 | AAI-based (medium-confidence) | 0 |
| S16_vs_59697\|\|full | 43120 | No | NA | 51 | 16 | 1 | High-quality | High-quality | 97.47 | AAI-based (high-confidence) | 0 |
| S16_vs_75108\|\|full | 41188 | No | NA | 48 | 7 | 0 | High-quality | High-quality | 100 | AAI-based (high-confidence) | 0 |
| S16_vs_76823\|\|full | 54530 | No | NA | 89 | 18 | 1 | High-quality | High-quality | 100 | AAI-based (high-confidence) | 0 |
| S16_vs_78361\|\|full | 37771 | No | NA | 55 | 20 | 2 | High-quality | High-quality | 99.76 | AAI-based (high-confidence) | 0 |
| S16_vs_78429\|\|full | 39776 | No | NA | 76 | 15 | 1 | High-quality | High-quality | 100 | AAI-based (high-confidence) | 0 |
| S16_vs_127905\|\|full | 63611 | No | NA | 103 | 20 | 0 | High-quality | High-quality | 100 | AAI-based (high-confidence) | 0 |
| S16_vs_128557\|\|full | 3427 | No | NA | 3 | 0 | 1 | Not-determined | Genome-fragment | NA | NA | 0 |
| S16_vs_139034\|\|full | 5752 | No | NA | 6 | 0 | 0 | Not-determined | Genome-fragment | NA | NA | 0 |
| S16_vs_172749\|\|full | 37714 | No | NA | 49 | 22 | 0 | High-quality | High-quality | 96.52 | AAI-based (high-confidence) | 0 |
| S16_vs_173079\|\|full | 61575 | No | NA | 88 | 22 | 2 | High-quality | High-quality | 99.07 | AAI-based (high-confidence) | 0 |
| S16_vs_173096\|\|full | 34364 | No | NA | 48 | 9 | 0 | High-quality | High-quality | 100 | AAI-based (high-confidence) | 0 |
| S16_vs_173102\|\|full | 43714 | No | NA | 55 | 10 | 0 | High-quality | High-quality | 96.76 | AAI-based (high-confidence) | 0 |
| S16_vs_173113\|\|full | 2615 | No | NA | 4 | 0 | 0 | Not-determined | Genome-fragment | NA | NA | 0 |
| S16_vs_173117\|\|full | 3494 | No | NA | 5 | 0 | 0 | Not-determined | Genome-fragment | NA | NA | 0 |
| S8_vs_k141_80237\|\|full | 2642 | No | NA | 5 | 0 | 0 | Low-quality | Genome-fragment | 7.22 | AAI-based (medium-confidence) | 0 |
| S16_vs_173128\|\|full | 2446 | No | NA | 3 | 0 | 0 | Not-determined | Genome-fragment | NA | NA | 0 |
| S1_extracted_k141_64894 | 2643 | No | NA | 4 | 0 | 0 | Low-quality | Genome-fragment | 3.37 | AAI-based (high-confidence) | 0 |
| S16_vs_173141\|\|full | 3774 | No | NA | 3 | 0 | 0 | Medium-quality | Genome-fragment | 57.26 | AAI-based (medium-confidence) | 0 |
| S16_vs_173146\|\|full | 1852 | No | NA | 3 | 0 | 0 | Not-determined | Genome-fragment | NA | NA | 0 |
| S13_extracted_78368 | 2644 | No | NA | 3 | 0 | 0 | Low-quality | Genome-fragment | 3.37 | AAI-based (medium-confidence) | 0 |
| S16_vs_97\|\|full | 6469 | No | NA | 5 | 0 | 4 | Not-determined | Genome-fragment | NA | NA | 0 |
| S3_vs_k141_80648\|\|full | 2644 | Yes | 1103 | 5 | 1 | 1 | Low-quality | Genome-fragment | 2.51 | AAI-based (medium-confidence) | 58.28 |
| S16_vs_241\|\|full | 1357 | No | NA | 2 | 0 | 1 | Not-determined | Genome-fragment | NA | NA | 0 |
| S16_vs_438\|\|full | 1790 | No | NA | 3 | 0 | 1 | Not-determined | Genome-fragment | NA | NA | 0 |
| S8_vs_k141_120582\|\|full | 2644 | No | NA | 5 | 2 | 0 | Low-quality | Genome-fragment | 8.12 | AAI-based (high-confidence) | 0 |
| S3_vs_k141_2833\|\|full | 2645 | No | NA | 4 | 1 | 0 | Low-quality | Genome-fragment | 2.37 | HMM-based (lower-bound) | 0 |
| S3_vs_k141_77795\|\|full | 2645 | No | NA | 4 | 2 | 0 | Low-quality | Genome-fragment | 6.19 | AAI-based (high-confidence) | 0 |
| S14_extracted_k141_8044 | 2646 | No | NA | 9 | 7 | 0 | Low-quality | Genome-fragment | 8.72 | AAI-based (high-confidence) | 0 |
| S14_extracted_k141_54880 | 2646 | No | NA | 6 | 4 | 0 | Low-quality | Genome-fragment | 6.28 | AAI-based (high-confidence) | 0 |
| S14_vs_k141_74550\|\|full | 2646 | No | NA | 3 | 3 | 0 | Low-quality | Genome-fragment | 7.2 | AAI-based (high-confidence) | 0 |
| S16_vs_748\|\|full | 1712 | No | NA | 3 | 0 | 1 | Not-determined | Genome-fragment | NA | NA | 0 |
| S16_vs_754\|\|full | 4692 | No | NA | 5 | 0 | 2 | Not-determined | Genome-fragment | NA | NA | 0 |
| S16_vs_780\|\|full | 1465 | No | NA | 2 | 0 | 1 | Not-determined | Genome-fragment | NA | NA | 0 |
| S14_vs_k141_94423\|\|full | 2646 | No | NA | 3 | 0 | 1 | Low-quality | Genome-fragment | 3.2 | AAI-based (high-confidence) | 0 |
| S16_vs_838\|\|full | 3966 | No | NA | 3 | 0 | 1 | Not-determined | Genome-fragment | NA | NA | 0 |
| S16_vs_847\|\|full | 2140 | No | NA | 2 | 0 | 1 | Not-determined | Genome-fragment | NA | NA | 0 |
| S12_vs_k141_174850\|\|full | 2646 | No | NA | 3 | 2 | 0 | Low-quality | Genome-fragment | 6 | AAI-based (high-confidence) | 0 |
| S16_vs_990\|\|full | 4004 | No | NA | 5 | 0 | 4 | Not-determined | Genome-fragment | NA | NA | 0 |
| S16_vs_1025\|\|full | 4174 | No | NA | 4 | 0 | 2 | Not-determined | Genome-fragment | NA | NA | 0 |
| S1_vs_k141_64384\|\|full | 2647 | No | NA | 3 | 0 | 0 | Low-quality | Genome-fragment | 1.47 | AAI-based (high-confidence) | 0 |
| S3_vs_k141_2789\|\|full | 2647 | No | NA | 8 | 2 | 0 | Low-quality | Genome-fragment | 6.18 | AAI-based (medium-confidence) | 0 |
| S16_vs_1203\|\|full | 4784 | No | NA | 6 | 0 | 1 | Not-determined | Genome-fragment | NA | NA | 0 |
| S16_vs_1321\|\|full | 1735 | No | NA | 2 | 0 | 0 | Not-determined | Genome-fragment | NA | NA | 0 |

| | | | | | | | | | | | |
|---|---|---|---|---|---|---|---|---|---|---|---|
| S16_vs_1355\|\|full | 3780 | No | NA | 3 | 0 | 3 | Not-determined | Genome-fragment | NA | NA | 0 |
| S16_vs_1368\|\|full | 3910 | No | NA | 7 | 0 | 3 | Not-determined | Genome-fragment | NA | NA | 0 |
| S16_vs_1386\|\|full | 2251 | No | NA | 2 | 0 | 2 | Not-determined | Genome-fragment | NA | NA | 0 |
| S16_vs_1402\|\|full | 1189 | No | NA | 3 | 0 | 1 | Not-determined | Genome-fragment | NA | NA | 0 |
| S16_vs_1407\|\|full | 2616 | No | NA | 3 | 0 | 3 | Not-determined | Genome-fragment | NA | NA | 0 |
| S16_vs_1434\|\|full | 1420 | No | NA | 3 | 0 | 2 | Not-determined | Genome-fragment | NA | NA | 0 |
| S16_vs_1473\|\|full | 1730 | No | NA | 2 | 0 | 0 | Not-determined | Genome-fragment | NA | NA | 0 |
| S1_vs_k141_120953\|\|full | 2648 | No | NA | 2 | 0 | 2 | Low-quality | Genome-fragment | 5.61 | AAI-based (medium-confidence) | 0 |
| S16_vs_1554\|\|full | 1615 | No | NA | 3 | 0 | 1 | Not-determined | Genome-fragment | NA | NA | 0 |
| S16_vs_1626\|\|full | 2561 | No | NA | 4 | 0 | 1 | Not-determined | Genome-fragment | NA | NA | 0 |
| S16_vs_1634\|\|full | 1150 | No | NA | 2 | 0 | 2 | Not-determined | Genome-fragment | NA | NA | 0 |
| S5_vs_k141_78650\|\|full | 2648 | No | NA | 6 | 4 | 0 | Low-quality | Genome-fragment | 7.12 | AAI-based (high-confidence) | 0 |
| S7_vs_2697\|\|full | 2648 | No | NA | 5 | 0 | 0 | Low-quality | Genome-fragment | 5.97 | AAI-based (medium-confidence) | 0 |
| S7_vs_35053\|\|full | 2648 | No | NA | 5 | 0 | 0 | Low-quality | Genome-fragment | 5.77 | AAI-based (medium-confidence) | 0 |
| S4_extracted_3735 | 2648 | No | NA | 8 | 6 | 0 | Low-quality | Genome-fragment | 8.74 | AAI-based (high-confidence) | 0 |
| S16_vs_2031\|\|full | 4215 | No | NA | 5 | 0 | 4 | Not-determined | Genome-fragment | NA | NA | 0 |
| S5_vs_k141_30944\|\|full | 2649 | No | NA | 6 | 1 | 0 | Low-quality | Genome-fragment | 4.48 | AAI-based (high-confidence) | 0 |
| S5_vs_k141_61346\|\|full | 2649 | No | NA | 4 | 1 | 0 | Low-quality | Genome-fragment | 0.95 | HMM-based (lower-bound) | 0 |
| S12_vs_k141_22502\|\|full | 2649 | No | NA | 6 | 3 | 0 | Low-quality | Genome-fragment | 6.08 | AAI-based (high-confidence) | 0 |
| S16_vs_2220\|\|full | 5239 | No | NA | 6 | 0 | 3 | Not-determined | Genome-fragment | NA | NA | 0 |
| S6_vs_k141_39021\|\|full | 2649 | No | NA | 9 | 0 | 0 | Low-quality | Genome-fragment | 4.36 | AAI-based (high-confidence) | 0 |
| S17_vs_k141_114473\|\|full | 2650 | No | NA | 3 | 1 | 0 | Low-quality | Genome-fragment | 4.08 | AAI-based (high-confidence) | 0 |
| S1_extracted_k141_133107 | 2650 | No | NA | 4 | 2 | 0 | Low-quality | Genome-fragment | 4.58 | AAI-based (high-confidence) | 0 |
| S16_vs_2488\|\|full | 2436 | No | NA | 3 | 0 | 2 | Not-determined | Genome-fragment | NA | NA | 0 |
| S5_extracted_k141_145230 | 2650 | No | NA | 3 | 0 | 2 | Low-quality | Genome-fragment | 4.3 | AAI-based (medium-confidence) | 0 |
| S16_vs_2521\|\|full | 3392 | No | NA | 3 | 0 | 0 | Not-determined | Genome-fragment | NA | NA | 0 |
| S16_vs_2564\|\|full | 1854 | No | NA | 2 | 0 | 0 | Not-determined | Genome-fragment | NA | NA | 0 |
| S16_vs_2578\|\|full | 3246 | No | NA | 3 | 0 | 3 | Not-determined | Genome-fragment | NA | NA | 0 |
| S7_vs_86942\|\|full | 2650 | No | NA | 3 | 1 | 0 | Low-quality | Genome-fragment | 6.34 | AAI-based (medium-confidence) | 0 |
| S16_vs_93040\|\|full | 2650 | No | NA | 5 | 1 | 0 | Low-quality | Genome-fragment | 6.07 | AAI-based (medium-confidence) | 0 |
| S12_extracted_k141_185164 | 2650 | No | NA | 8 | 0 | 0 | Low-quality | Genome-fragment | 6.41 | AAI-based (high-confidence) | 0 |
| S16_vs_2800\|\|full | 2815 | No | NA | 4 | 0 | 4 | Not-determined | Genome-fragment | NA | NA | 0 |
| S6_vs_k141_2768\|\|full | 2650 | No | NA | 3 | 1 | 0 | Low-quality | Genome-fragment | 2.13 | HMM-based (lower-bound) | 0 |
| S16_vs_2929\|\|full | 1103 | No | NA | 2 | 0 | 1 | Not-determined | Genome-fragment | NA | NA | 0 |
| S9_vs_k141_89547\|\|full | 2651 | No | NA | 5 | 2 | 0 | Low-quality | Genome-fragment | 6.08 | AAI-based (high-confidence) | 0 |
| S16_vs_3046\|\|full | 3699 | No | NA | 4 | 0 | 1 | Not-determined | Genome-fragment | NA | NA | 0 |
| S16_vs_3058\|\|full | 3651 | No | NA | 4 | 0 | 3 | Not-determined | Genome-fragment | NA | NA | 0 |
| S16_vs_3138\|\|full | 3620 | No | NA | 3 | 0 | 2 | Not-determined | Genome-fragment | NA | NA | 0 |
| S16_vs_3330\|\|full | 4842 | No | NA | 9 | 0 | 0 | Not-determined | Genome-fragment | NA | NA | 0 |
| S1_vs_k141_19935\|\|full | 2651 | No | NA | 6 | 2 | 0 | Low-quality | Genome-fragment | 6.24 | AAI-based (high-confidence) | 0 |
| S16_vs_116091\|\|full | 2651 | No | NA | 4 | 1 | 0 | Low-quality | Genome-fragment | 1.06 | AAI-based (medium-confidence) | 0 |
| S6_vs_k141_16077\|\|full | 2651 | No | NA | 5 | 0 | 1 | Low-quality | Genome-fragment | 5.06 | AAI-based (medium-confidence) | 0 |
| S16_vs_3856\|\|full | 2388 | No | NA | 3 | 0 | 1 | Not-determined | Genome-fragment | NA | NA | 0 |
| S16_vs_3872\|\|full | 2499 | No | NA | 2 | 0 | 1 | Not-determined | Genome-fragment | NA | NA | 0 |
| S16_vs_3965\|\|full | 1073 | No | NA | 2 | 0 | 0 | Not-determined | Genome-fragment | NA | NA | 0 |
| S16_vs_4004\|\|full | 1644 | No | NA | 3 | 0 | 1 | Not-determined | Genome-fragment | NA | NA | 0 |
| S8_vs_k141_164422\|\|full | 2651 | No | NA | 7 | 1 | 0 | Low-quality | Genome-fragment | 4.58 | AAI-based (medium-confidence) | 0 |
| S16_vs_4147\|\|full | 8671 | No | NA | 7 | 0 | 1 | Not-determined | Genome-fragment | NA | NA | 0 |
| S9_vs_k141_26724\|\|full | 2652 | Yes | 449 | 3 | 1 | 2 | Low-quality | Genome-fragment | 1.43 | AAI-based (high-confidence) | 83.07 |
| S3_extracted_k141_17702 | 2652 | No | NA | 4 | 1 | 0 | Low-quality | Genome-fragment | 2.7 | AAI-based (high-confidence) | 0 |
| S16_vs_4229\|\|full | 3353 | No | NA | 3 | 0 | 1 | Not-determined | Genome-fragment | NA | NA | 0 |
| S16_vs_4261\|\|full | 1248 | No | NA | 3 | 0 | 2 | Not-determined | Genome-fragment | NA | NA | 0 |
| S6_vs_k141_53640\|\|full | 2652 | No | NA | 4 | 3 | 0 | Low-quality | Genome-fragment | 4.41 | AAI-based (high-confidence) | 0 |
| S9_vs_k141_68738\|\|full | 2653 | No | NA | 5 | 1 | 0 | Low-quality | Genome-fragment | 5.82 | AAI-based (high-confidence) | 0 |
| S16_vs_104052\|\|full | 2653 | No | NA | 5 | 0 | 0 | Low-quality | Genome-fragment | 3.22 | AAI-based (high-confidence) | 0 |
| S16_vs_4548\|\|full | 2334 | No | NA | 4 | 0 | 1 | Not-determined | Genome-fragment | NA | NA | 0 |
| S16_vs_4558\|\|full | 1795 | No | NA | 2 | 0 | 2 | Not-determined | Genome-fragment | NA | NA | 0 |
| S16_vs_4588\|\|full | 1181 | No | NA | 2 | 0 | 0 | Not-determined | Genome-fragment | NA | NA | 0 |
| S16_vs_4604\|\|full | 1914 | No | NA | 3 | 0 | 1 | Not-determined | Genome-fragment | NA | NA | 0 |
| S16_vs_4662\|\|full | 28941 | No | NA | 47 | 11 | 2 | Medium-quality | Genome-fragment | 76.8 | AAI-based (high-confidence) | 0 |
| S16_vs_4733\|\|full | 2371 | No | NA | 2 | 0 | 2 | Not-determined | Genome-fragment | NA | NA | 0 |
| S16_vs_4801\|\|full | 2230 | No | NA | 4 | 0 | 2 | Not-determined | Genome-fragment | NA | NA | 0 |
| S16_vs_4860\|\|full | 1221 | No | NA | 2 | 0 | 1 | Not-determined | Genome-fragment | NA | NA | 0 |
| S16_vs_5087\|\|full | 10547 | No | NA | 10 | 0 | 5 | Not-determined | Genome-fragment | NA | NA | 0 |
| S16_vs_5088\|\|full | 10379 | No | NA | 6 | 0 | 4 | Not-determined | Genome-fragment | NA | NA | 0 |
| S16_vs_5258\|\|full | 4299 | No | NA | 6 | 0 | 2 | Not-determined | Genome-fragment | NA | NA | 0 |
| S16_vs_5261\|\|full | 2527 | No | NA | 3 | 0 | 2 | Not-determined | Genome-fragment | NA | NA | 0 |
| S6_vs_k141_22525\|\|full | 2654 | No | NA | 3 | 0 | 1 | Low-quality | Genome-fragment | 5.6 | AAI-based (medium-confidence) | 0 |
| S14_vs_k141_15071\|\|full | 2656 | No | NA | 3 | 0 | 2 | Low-quality | Genome-fragment | 1.86 | AAI-based (medium-confidence) | 0 |
| S8_vs_k141_88052\|\|full | 2656 | No | NA | 4 | 3 | 0 | Low-quality | Genome-fragment | 5.77 | AAI-based (medium-confidence) | 0 |
| S17_vs_k141_141981\|\|full | 2657 | No | NA | 5 | 0 | 4 | Low-quality | Genome-fragment | 3.41 | AAI-based (high-confidence) | 0 |
| S9_vs_k141_19331\|\|full | 2657 | No | NA | 3 | 1 | 0 | Low-quality | Genome-fragment | 4.87 | HMM-based (lower-bound) | 0 |
| S16_vs_5808\|\|full | 3487 | No | NA | 6 | 0 | 4 | Not-determined | Genome-fragment | NA | NA | 0 |
| S1_extracted_k141_81828 | 2657 | No | NA | 5 | 4 | 0 | Low-quality | Genome-fragment | 7.23 | AAI-based (high-confidence) | 0 |
| S16_vs_5925\|\|full | 1253 | No | NA | 3 | 0 | 2 | Not-determined | Genome-fragment | NA | NA | 0 |
| S7_vs_14964\|\|full | 2657 | No | NA | 3 | 1 | 0 | Low-quality | Genome-fragment | 1.72 | HMM-based (lower-bound) | 0 |
| S16_vs_6050\|\|full | 1614 | No | NA | 2 | 0 | 2 | Not-determined | Genome-fragment | NA | NA | 0 |
| S4_extracted_16388 | 2657 | No | NA | 2 | 1 | 0 | Low-quality | Genome-fragment | 2.81 | AAI-based (high-confidence) | 0 |
| S16_vs_6158\|\|full | 2965 | No | NA | 7 | 0 | 0 | Not-determined | Genome-fragment | NA | NA | 0 |
| S8_vs_k141_196400\|\|full | 2657 | No | NA | 4 | 0 | 0 | Low-quality | Genome-fragment | 4.8 | AAI-based (high-confidence) | 0 |
| S16_vs_6222\|\|full | 1304 | No | NA | 3 | 0 | 0 | Not-determined | Genome-fragment | NA | NA | 0 |
| S3_vs_k141_20509\|\|full | 2658 | No | NA | 4 | 0 | 0 | Low-quality | Genome-fragment | 3.95 | AAI-based (high-confidence) | 0 |
| S16_vs_6393\|\|full | 1407 | No | NA | 2 | 0 | 2 | Not-determined | Genome-fragment | NA | NA | 0 |
| S8_vs_k141_172596\|\|full | 2658 | No | NA | 5 | 0 | 0 | Low-quality | Genome-fragment | 3.79 | AAI-based (medium-confidence) | 0 |
| S17_vs_k141_126012\|\|full | 2659 | No | NA | 5 | 1 | 0 | Low-quality | Genome-fragment | 5.39 | AAI-based (medium-confidence) | 0 |
| S3_extracted_k141_74911 | 2659 | No | NA | 4 | 1 | 0 | Low-quality | Genome-fragment | 3.37 | AAI-based (high-confidence) | 0 |
| S16_vs_6526\|\|full | 1505 | No | NA | 2 | 0 | 1 | Not-determined | Genome-fragment | NA | NA | 0 |
| S7_vs_132679\|\|full | 2659 | No | NA | 7 | 1 | 0 | Low-quality | Genome-fragment | 7.28 | AAI-based (medium-confidence) | 0 |
| S16_vs_163138\|\|full | 2659 | No | NA | 3 | 1 | 0 | Low-quality | Genome-fragment | 1.91 | AAI-based (medium-confidence) | 0 |
| S12_vs_k141_17109\|\|full | 2659 | No | NA | 7 | 1 | 0 | Low-quality | Genome-fragment | 6.53 | AAI-based (high-confidence) | 0 |
| S16_vs_6585\|\|full | 1445 | No | NA | 3 | 0 | 0 | Not-determined | Genome-fragment | NA | NA | 0 |
| S16_vs_6601\|\|full | 7268 | No | NA | 9 | 0 | 5 | Not-determined | Genome-fragment | NA | NA | 0 |
| S16_vs_6605\|\|full | 1453 | No | NA | 3 | 0 | 0 | Not-determined | Genome-fragment | NA | NA | 0 |
| S6_vs_k141_17497\|\|full | 2659 | No | NA | 4 | 0 | 1 | Low-quality | Genome-fragment | 3.06 | AAI-based (high-confidence) | 0 |
| S16_vs_6882\|\|full | 1790 | No | NA | 3 | 0 | 0 | Not-determined | Genome-fragment | NA | NA | 0 |
| S16_vs_6889\|\|full | 2130 | No | NA | 3 | 0 | 2 | Not-determined | Genome-fragment | NA | NA | 0 |
| S16_vs_6905\|\|full | 1795 | No | NA | 3 | 0 | 0 | Not-determined | Genome-fragment | NA | NA | 0 |
| S3_vs_k141_89702\|\|full | 2660 | No | NA | 3 | 1 | 0 | Low-quality | Genome-fragment | 1.55 | AAI-based (medium-confidence) | 0 |
| S16_vs_7050\|\|full | 4075 | No | NA | 7 | 0 | 0 | Not-determined | Genome-fragment | NA | NA | 0 |
| S16_vs_7094\|\|full | 1748 | No | NA | 2 | 0 | 2 | Not-determined | Genome-fragment | NA | NA | 0 |
| S12_extracted_k141_183552 | 2660 | No | NA | 5 | 0 | 0 | Low-quality | Genome-fragment | 2.45 | AAI-based (high-confidence) | 0 |
| S16_vs_7379\|\|full | 2721 | No | NA | 3 | 0 | 2 | Not-determined | Genome-fragment | NA | NA | 0 |
| S16_vs_7501\|\|full | 4307 | No | NA | 3 | 0 | 2 | Not-determined | Genome-fragment | NA | NA | 0 |
| S16_vs_7566\|\|full | 3064 | No | NA | 2 | 0 | 1 | Not-determined | Genome-fragment | NA | NA | 0 |
| S16_vs_7583\|\|full | 7780 | No | NA | 7 | 0 | 4 | Not-determined | Genome-fragment | NA | NA | 0 |
| S16_vs_7756\|\|full | 4375 | No | NA | 6 | 0 | 1 | Not-determined | Genome-fragment | NA | NA | 0 |
| S14_vs_k141_18163\|\|lt2gene | 2661 | No | NA | 2 | 2 | 0 | Low-quality | Genome-fragment | 5.83 | AAI-based (medium-confidence) | 0 |
| S12_extracted_k141_25707 | 2661 | No | NA | 5 | 0 | 0 | Low-quality | Genome-fragment | 2.51 | AAI-based (high-confidence) | 0 |
| S12_vs_k141_169783\|\|full | 2661 | No | NA | 8 | 1 | 0 | Low-quality | Genome-fragment | 0.92 | HMM-based (lower-bound) | 0 |
| S9_vs_k141_53457\|\|full | 2662 | No | NA | 3 | 0 | 0 | Low-quality | Genome-fragment | 4.33 | AAI-based (medium-confidence) | 0 |
| S9_vs_k141_49551\|\|full | 2663 | No | NA | 6 | 3 | 0 | Low-quality | Genome-fragment | 7.15 | AAI-based (medium-confidence) | 0 |
| S5_vs_k141_108291\|\|full | 2663 | No | NA | 6 | 2 | 0 | Low-quality | Genome-fragment | 1.66 | HMM-based (lower-bound) | 0 |
| S16_extracted_25482 | 2663 | No | NA | 2 | 2 | 0 | Low-quality | Genome-fragment | 3.93 | HMM-based (lower-bound) | 0 |
| S17_extracted_k141_172087 | 2664 | No | NA | 3 | 0 | 2 | Low-quality | Genome-fragment | 0.99 | AAI-based (medium-confidence) | 0 |
| S16_vs_8412\|\|full | 2064 | No | NA | 3 | 0 | 0 | Not-determined | Genome-fragment | NA | NA | 0 |
| S16_vs_8413\|\|full | 4596 | No | NA | 5 | 0 | 1 | Not-determined | Genome-fragment | NA | NA | 0 |
| S9_vs_k141_961\|\|full | 2664 | No | NA | 2 | 0 | 0 | Low-quality | Genome-fragment | 6.11 | AAI-based (high-confidence) | 0 |
| S11_extracted_k141_3018 | 2664 | No | NA | 1 | 0 | 0 | Low-quality | Genome-fragment | 2.41 | AAI-based (medium-confidence) | 0 |
| S1_vs_k141_86950\|\|full | 2664 | No | NA | 3 | 1 | 0 | Low-quality | Genome-fragment | 6.38 | HMM-based (lower-bound) | 0 |
| S5_vs_k141_69736\|\|full | 2664 | No | NA | 5 | 1 | 0 | Low-quality | Genome-fragment | 7.33 | HMM-based (lower-bound) | 0 |
| S7_extracted_21872 | 2664 | No | NA | 3 | 3 | 0 | Low-quality | Genome-fragment | 9.4 | HMM-based (lower-bound) | 0 |
| S16_vs_5726\|\|full | 2664 | Yes | 2355 | 4 | 1 | 1 | Low-quality | Genome-fragment | 3.53 | AAI-based (medium-confidence) | 11.6 |
| S16_vs_8685\|\|full | 1365 | No | NA | 2 | 0 | 1 | Not-determined | Genome-fragment | NA | NA | 0 |
| S17_extracted_k141_75280 | 2665 | No | NA | 3 | 0 | 3 | Low-quality | Genome-fragment | 0.98 | AAI-based (high-confidence) | 0 |
| S17_extracted_k141_82586 | 2666 | No | NA | 3 | 0 | 0 | Low-quality | Genome-fragment | 6.69 | AAI-based (medium-confidence) | 0 |
| S16_vs_9052\|\|full | 1089 | No | NA | 3 | 0 | 1 | Not-determined | Genome-fragment | NA | NA | 0 |
| S3_vs_k141_43272\|\|full | 2666 | No | NA | 9 | 0 | 0 | Low-quality | Genome-fragment | 1.47 | AAI-based (high-confidence) | 0 |
| S16_vs_9193\|\|full | 5512 | No | NA | 6 | 0 | 3 | Not-determined | Genome-fragment | NA | NA | 0 |
| S5_vs_k141_76680\|\|full | 2666 | No | NA | 6 | 0 | 0 | Low-quality | Genome-fragment | 7.12 | AAI-based (high-confidence) | 0 |
| S16_vs_115135\|\|full | 2666 | No | NA | 3 | 3 | 0 | Low-quality | Genome-fragment | 7.72 | AAI-based (high-confidence) | 0 |
| S12_vs_k141_1686\|\|full | 2666 | No | NA | 7 | 2 | 0 | Low-quality | Genome-fragment | 4.5 | AAI-based (high-confidence) | 0 |
| S7_vs_1896\|\|full | 2667 | No | NA | 9 | 1 | 0 | Low-quality | Genome-fragment | 7.55 | AAI-based (high-confidence) | 0 |
| S16_vs_9269\|\|full | 5349 | No | NA | 4 | 0 | 2 | Not-determined | Genome-fragment | NA | NA | 0 |
| S3_vs_k141_108481\|\|full | 2668 | No | NA | 3 | 0 | 1 | Low-quality | Genome-fragment | 1.64 | AAI-based (medium-confidence) | 0 |
| S16_vs_9429\|\|full | 2910 | No | NA | 3 | 0 | 3 | Not-determined | Genome-fragment | NA | NA | 0 |
| S16_vs_9530\|\|full | 1496 | No | NA | 2 | 0 | 1 | Not-determined | Genome-fragment | NA | NA | 0 |
| S16_vs_39354\|\|full | 2668 | No | NA | 4 | 0 | 1 | Low-quality | Genome-fragment | 1.35 | AAI-based (high-confidence) | 0 |
| S16_vs_9722\|\|full | 6645 | No | NA | 7 | 0 | 5 | Not-determined | Genome-fragment | NA | NA | 0 |
| S6_vs_k141_8642\|\|lt2gene | 2668 | No | NA | 2 | 2 | 0 | Low-quality | Genome-fragment | 7.78 | AAI-based (high-confidence) | 0 |
| S16_vs_9869\|\|full | 9320 | No | NA | 7 | 0 | 2 | Not-determined | Genome-fragment | NA | NA | 0 |
| S16_vs_9906\|\|full | 1568 | No | NA | 5 | 0 | 0 | Not-determined | Genome-fragment | NA | NA | 0 |
| S16_vs_9918\|\|full | 5335 | No | NA | 3 | 0 | 0 | Not-determined | Genome-fragment | NA | NA | 0 |
| S16_vs_9981\|\|full | 2641 | No | NA | 3 | 0 | 0 | Not-determined | Genome-fragment | NA | NA | 0 |
| S17_extracted_k141_70564 | 2669 | No | NA | 2 | 0 | 0 | Low-quality | Genome-fragment | 6.31 | AAI-based (high-confidence) | 0 |
| S16_vs_10146\|\|full | 1091 | No | NA | 2 | 0 | 1 | Not-determined | Genome-fragment | NA | NA | 0 |
| S14_vs_k141_37495\|\|full | 2669 | No | NA | 5 | 0 | 0 | Low-quality | Genome-fragment | 7.16 | AAI-based (medium-confidence) | 0 |
| S16_vs_10252\|\|full | 9795 | No | NA | 9 | 0 | 6 | Not-determined | Genome-fragment | NA | NA | 0 |
| S16_vs_10277\|\|full | 1693 | No | NA | 2 | 0 | 1 | Not-determined | Genome-fragment | NA | NA | 0 |
| S14_vs_k141_27412\|\|full | 2669 | No | NA | 7 | 2 | 0 | Low-quality | Genome-fragment | 3.99 | AAI-based (high-confidence) | 0 |
| S16_vs_10333\|\|full | 1601 | No | NA | 2 | 0 | 2 | Not-determined | Genome-fragment | NA | NA | 0 |
| S16_vs_10432\|\|full | 1270 | No | NA | 3 | 0 | 0 | Not-determined | Genome-fragment | NA | NA | 0 |
| S7_vs_52295\|\|full | 2669 | No | NA | 7 | 1 | 0 | Low-quality | Genome-fragment | 6.07 | AAI-based (high-confidence) | 0 |
| S16_vs_10506\|\|full | 5269 | No | NA | 7 | 0 | 0 | Not-determined | Genome-fragment | NA | NA | 0 |
| S16_vs_10569\|\|full | 1966 | No | NA | 3 | 0 | 0 | Not-determined | Genome-fragment | NA | NA | 0 |
| S16_vs_10622\|\|full | 35184 | No | NA | 39 | 9 | 1 | Medium-quality | Genome-fragment | 65.41 | HMM-based (lower-bound) | 0 |
| S16_vs_10830\|\|full | 2377 | No | NA | 3 | 0 | 1 | Not-determined | Genome-fragment | NA | NA | 0 |
| S7_vs_89479\|\|full | 2669 | No | NA | 4 | 0 | 0 | Low-quality | Genome-fragment | 4.32 | AAI-based (medium-confidence) | 0 |
| S12_vs_k141_80959\|\|full | 2669 | No | NA | 4 | 1 | 0 | Low-quality | Genome-fragment | 6.48 | AAI-based (high-confidence) | 0 |
| S1_extracted_k141_92361 | 2670 | No | NA | 3 | 2 | 0 | Low-quality | Genome-fragment | 4.42 | AAI-based (high-confidence) | 0 |
| S1_vs_k141_111865\|\|full | 2670 | Yes | 1144 | 4 | 2 | 1 | Low-quality | Genome-fragment | 3.03 | AAI-based (high-confidence) | 57.15 |
| S5_vs_k141_79560\|\|lt2gene | 2670 | No | NA | 2 | 2 | 0 | Low-quality | Genome-fragment | 7.27 | AAI-based (high-confidence) | 0 |
| S16_vs_11165\|\|full | 3013 | No | NA | 2 | 0 | 2 | Not-determined | Genome-fragment | NA | NA | 0 |
| S8_extracted_k141_117428 | 2670 | No | NA | 1 | 1 | 0 | Low-quality | Genome-fragment | 3.9 | HMM-based (lower-bound) | 0 |
| S17_extracted_k141_51466 | 2671 | No | NA | 2 | 0 | 2 | Low-quality | Genome-fragment | 0.6 | AAI-based (medium-confidence) | 0 |
| S16_vs_11244\|\|full | 4985 | No | NA | 8 | 0 | 1 | Not-determined | Genome-fragment | NA | NA | 0 |
| S16_vs_11557\|\|full | 3373 | No | NA | 3 | 0 | 2 | Not-determined | Genome-fragment | NA | NA | 0 |
| S3_vs_k141_124735\|\|full | 2671 | No | NA | 4 | 3 | 0 | Low-quality | Genome-fragment | 6.92 | AAI-based (high-confidence) | 0 |
| S16_vs_11649\|\|full | 46564 | No | NA | 57 | 12 | 1 | High-quality | High-quality | 91.75 | AAI-based (high-confidence) | 0 |
| S5_extracted_k141_23503 | 2671 | No | NA | 2 | 1 | 0 | Low-quality | Genome-fragment | 6.64 | AAI-based (high-confidence) | 0 |
| S16_vs_11747\|\|full | 1212 | No | NA | 2 | 0 | 1 | Not-determined | Genome-fragment | NA | NA | 0 |
| S5_vs_k141_71125\|\|full | 2671 | No | NA | 3 | 3 | 0 | Low-quality | Genome-fragment | 6.77 | AAI-based (high-confidence) | 0 |
| S16_vs_11885\|\|full | 8894 | No | NA | 10 | 0 | 8 | Not-determined | Genome-fragment | NA | NA | 0 |
| S16_vs_12141\|\|full | 6920 | No | NA | 3 | 0 | 3 | Not-determined | Genome-fragment | NA | NA | 0 |
| S16_vs_12175\|\|full | 1153 | No | NA | 3 | 0 | 2 | Not-determined | Genome-fragment | NA | NA | 0 |
| S16_vs_12228\|\|full | 1733 | No | NA | 3 | 0 | 0 | Not-determined | Genome-fragment | NA | NA | 0 |
| S16_vs_12232\|\|full | 5971 | No | NA | 5 | 0 | 3 | Not-determined | Genome-fragment | NA | NA | 0 |
| S7_extracted_137616 | 2671 | No | NA | 6 | 1 | 0 | Low-quality | Genome-fragment | 2.78 | AAI-based (high-confidence) | 0 |
| S1_vs_k141_137264\|\|full | 2672 | No | NA | 5 | 0 | 0 | Low-quality | Genome-fragment | 5.43 | AAI-based (medium-confidence) | 0 |
| S7_vs_63001\|\|full | 2672 | Yes | 1715 | 4 | 1 | 1 | Low-quality | Genome-fragment | 1.33 | AAI-based (medium-confidence) | 35.82 |
| S16_vs_12477\|\|full | 2877 | No | NA | 4 | 0 | 0 | Not-determined | Genome-fragment | NA | NA | 0 |
| S16_vs_101370\|\|full | 2672 | No | NA | 2 | 0 | 1 | Low-quality | Genome-fragment | 1.49 | AAI-based (medium-confidence) | 0 |
| S16_vs_12620\|\|full | 3604 | No | NA | 3 | 0 | 2 | Not-determined | Genome-fragment | NA | NA | 0 |
| S9_extracted_k141_2053 | 2673 | No | NA | 2 | 0 | 1 | Low-quality | Genome-fragment | 0.95 | AAI-based (medium-confidence) | 0 |
| S16_vs_12736\|\|full | 1097 | No | NA | 2 | 0 | 1 | Not-determined | Genome-fragment | NA | NA | 0 |
| S16_vs_12741\|\|full | 11429 | No | NA | 5 | 0 | 4 | Not-determined | Genome-fragment | NA | NA | 0 |
| S10_vs_65050\|\|full | 2673 | No | NA | 2 | 1 | 0 | Low-quality | Genome-fragment | 1.12 | AAI-based (high-confidence) | 0 |
| S16_vs_12805\|\|full | 2144 | No | NA | 2 | 0 | 2 | Not-determined | Genome-fragment | NA | NA | 0 |
| S10_vs_80589\|\|full | 2673 | No | NA | 3 | 0 | 0 | Low-quality | Genome-fragment | 2.93 | AAI-based (high-confidence) | 0 |
| S16_vs_12863\|\|full | 5959 | No | NA | 10 | 0 | 0 | Not-determined | Genome-fragment | NA | NA | 0 |
| S16_vs_12884\|\|full | 3164 | No | NA | 3 | 0 | 1 | Not-determined | Genome-fragment | NA | NA | 0 |
| S10_vs_117499\|\|full | 2673 | No | NA | 4 | 2 | 0 | Low-quality | Genome-fragment | 1.67 | AAI-based (high-confidence) | 0 |
| S16_vs_12949\|\|full | 6946 | No | NA | 3 | 0 | 3 | Not-determined | Genome-fragment | NA | NA | 0 |
| S16_vs_12953\|\|full | 15423 | No | NA | 16 | 0 | 10 | Not-determined | Genome-fragment | NA | NA | 0 |
| S16_vs_12990\|\|full | 1940 | No | NA | 5 | 0 | 0 | Not-determined | Genome-fragment | NA | NA | 0 |
| S16_vs_13039\|\|full | 1601 | No | NA | 2 | 0 | 0 | Not-determined | Genome-fragment | NA | NA | 0 |
| S16_vs_13146\|\|full | 3599 | No | NA | 4 | 0 | 0 | Not-determined | Genome-fragment | NA | NA | 0 |
| S17_vs_k141_83583\|\|full | 2674 | No | NA | 3 | 0 | 1 | Low-quality | Genome-fragment | 5.81 | AAI-based (medium-confidence) | 0 |
| S16_vs_13242\|\|full | 4749 | No | NA | 3 | 0 | 2 | Not-determined | Genome-fragment | NA | NA | 0 |
| S16_vs_13438\|\|full | 2304 | No | NA | 3 | 0 | 1 | Not-determined | Genome-fragment | NA | NA | 0 |
| S13_vs_134737\|\|full | 2675 | No | NA | 5 | 2 | 0 | Low-quality | Genome-fragment | 5.92 | AAI-based (high-confidence) | 0 |
| S14_vs_k141_67965\|\|full | 2675 | No | NA | 6 | 3 | 0 | Low-quality | Genome-fragment | 6.84 | AAI-based (medium-confidence) | 0 |
| S16_vs_13489\|\|full | 1221 | No | NA | 2 | 0 | 0 | Not-determined | Genome-fragment | NA | NA | 0 |
| S3_extracted_k141_23043 | 2675 | No | NA | 7 | 6 | 0 | Low-quality | Genome-fragment | 8.77 | AAI-based (high-confidence) | 0 |
| S7_vs_50981\|\|full | 2675 | No | NA | 4 | 1 | 0 | Low-quality | Genome-fragment | 6.51 | AAI-based (high-confidence) | 0 |
| S16_vs_13677\|\|full | 10188 | No | NA | 17 | 0 | 0 | Not-determined | Genome-fragment | NA | NA | 0 |
| S8_vs_k141_26501\|\|full | 2675 | No | NA | 4 | 0 | 2 | Low-quality | Genome-fragment | 0.77 | AAI-based (high-confidence) | 0 |
| S16_vs_13708\|\|full | 1800 | No | NA | 4 | 0 | 1 | Not-determined | Genome-fragment | NA | NA | 0 |
| S11_vs_k141_102230\|\|full | 2676 | No | NA | 8 | 0 | 0 | Low-quality | Genome-fragment | 3.97 | AAI-based (high-confidence) | 0 |
| S16_vs_13782\|\|full | 1745 | No | NA | 2 | 0 | 0 | Not-determined | Genome-fragment | NA | NA | 0 |
| S16_vs_13815\|\|full | 4568 | No | NA | 4 | 0 | 3 | Not-determined | Genome-fragment | NA | NA | 0 |
| S12_extracted_k141_13941 | 2676 | No | NA | 6 | 2 | 0 | Low-quality | Genome-fragment | 5.05 | AAI-based (high-confidence) | 0 |
| S17_extracted_k141_35809 | 2677 | No | NA | 3 | 0 | 3 | Low-quality | Genome-fragment | 2.07 | AAI-based (medium-confidence) | 0 |
| S16_vs_14032\|\|full | 6386 | No | NA | 8 | 0 | 1 | Not-determined | Genome-fragment | NA | NA | 0 |
| S16_vs_14057\|\|full | 8073 | No | NA | 6 | 0 | 2 | Not-determined | Genome-fragment | NA | NA | 0 |
| S5_vs_k141_63960\|\|full | 2677 | No | NA | 4 | 0 | 0 | Low-quality | Genome-fragment | 9.28 | AAI-based (medium-confidence) | 0 |
| S16_vs_14178\|\|full | 2062 | No | NA | 3 | 0 | 0 | Not-determined | Genome-fragment | NA | NA | 0 |
| S16_extracted_43612 | 2677 | No | NA | 2 | 0 | 0 | Low-quality | Genome-fragment | 4.47 | AAI-based (high-confidence) | 0 |
| S16_vs_145080\|\|full | 2677 | No | NA | 4 | 1 | 0 | Low-quality | Genome-fragment | 5.63 | HMM-based (lower-bound) | 0 |
| S17_extracted_k141_24654 | 2678 | No | NA | 3 | 0 | 3 | Low-quality | Genome-fragment | 6.29 | AAI-based (high-confidence) | 0 |
| S17_vs_k141_126040\|\|full | 2678 | No | NA | 3 | 1 | 0 | Low-quality | Genome-fragment | 7.36 | AAI-based (high-confidence) | 0 |
| S15_extracted_35860 | 2678 | No | NA | 3 | 1 | 0 | Low-quality | Genome-fragment | 6.9 | AAI-based (high-confidence) | 0 |
| S16_vs_14662\|\|full | 3021 | No | NA | 4 | 0 | 1 | Not-determined | Genome-fragment | NA | NA | 0 |
| S5_vs_k141_45551\|\|full | 2679 | No | NA | 3 | 0 | 1 | Low-quality | Genome-fragment | 5.74 | AAI-based (medium-confidence) | 0 |
| S7_extracted_69527 | 2679 | No | NA | 5 | 4 | 0 | Low-quality | Genome-fragment | 4.78 | AAI-based (high-confidence) | 0 |
| S7_vs_2939\|\|full | 2679 | No | NA | 4 | 0 | 0 | Low-quality | Genome-fragment | 4.41 | AAI-based (high-confidence) | 0 |
| S10_vs_45670\|\|full | 2679 | No | NA | 4 | 0 | 0 | Low-quality | Genome-fragment | 5.31 | AAI-based (high-confidence) | 0 |
| S11_vs_k141_48979\|\|full | 2680 | No | NA | 4 | 1 | 0 | Low-quality | Genome-fragment | 3.77 | AAI-based (medium-confidence) | 0 |
| S1_extracted_k141_102733 | 2680 | No | NA | 5 | 0 | 0 | Low-quality | Genome-fragment | 5.03 | AAI-based (medium-confidence) | 0 |
| S16_vs_15193\|\|full | 3500 | No | NA | 2 | 0 | 2 | Not-determined | Genome-fragment | NA | NA | 0 |
| S14_vs_k141_92918\|\|full | 2681 | No | NA | 6 | 3 | 0 | Low-quality | Genome-fragment | 6.33 | AAI-based (medium-confidence) | 0 |
| S16_vs_15229\|\|full | 10594 | No | NA | 11 | 0 | 10 | Not-determined | Genome-fragment | NA | NA | 0 |
| S12_vs_k141_187336\|\|full | 2681 | No | NA | 5 | 0 | 0 | Low-quality | Genome-fragment | 10.6 | AAI-based (high-confidence) | 0 |
| S13_vs_55937\|\|full | 2682 | No | NA | 4 | 0 | 2 | Low-quality | Genome-fragment | 1.89 | AAI-based (high-confidence) | 0 |
| S16_vs_15257\|\|full | 1530 | No | NA | 3 | 0 | 0 | Not-determined | Genome-fragment | NA | NA | 0 |
| S16_vs_15272\|\|full | 1996 | No | NA | 3 | 0 | 0 | Not-determined | Genome-fragment | NA | NA | 0 |
| S16_vs_15280\|\|full | 1886 | No | NA | 3 | 0 | 3 | Not-determined | Genome-fragment | NA | NA | 0 |
| S16_vs_15309\|\|full | 1813 | No | NA | 2 | 0 | 2 | Not-determined | Genome-fragment | NA | NA | 0 |
| S5_vs_k141_34475\|\|full | 2682 | Yes | 1019 | 4 | 1 | 1 | Low-quality | Genome-fragment | 1.76 | AAI-based (medium-confidence) | 62.01 |
| S16_vs_23791\|\|full | 2682 | No | NA | 4 | 0 | 0 | Low-quality | Genome-fragment | 4.48 | AAI-based (high-confidence) | 0 |
| S8_vs_k141_183412\|\|full | 2682 | No | NA | 4 | 0 | 1 | Low-quality | Genome-fragment | 3.16 | AAI-based (medium-confidence) | 0 |
| S8_vs_k141_147173\|\|full | 2682 | No | NA | 3 | 0 | 2 | Low-quality | Genome-fragment | 0.95 | AAI-based (medium-confidence) | 0 |
| S16_vs_15542\|\|full | 1844 | No | NA | 3 | 0 | 1 | Not-determined | Genome-fragment | NA | NA | 0 |
| S16_vs_15751\|\|full | 2810 | No | NA | 3 | 0 | 2 | Not-determined | Genome-fragment | NA | NA | 0 |
| S17_vs_k141_65031\|\|full | 2683 | No | NA | 4 | 0 | 1 | Low-quality | Genome-fragment | 5.6 | AAI-based (medium-confidence) | 0 |
| S16_vs_15790\|\|full | 4431 | No | NA | 5 | 0 | 4 | Not-determined | Genome-fragment | NA | NA | 0 |
| S5_vs_k141_23483\|\|full | 2683 | No | NA | 6 | 3 | 0 | Low-quality | Genome-fragment | 5.92 | AAI-based (high-confidence) | 0 |
| S16_vs_15931\|\|full | 1216 | No | NA | 2 | 0 | 0 | Not-determined | Genome-fragment | NA | NA | 0 |
| S16_vs_15948\|\|full | 1562 | No | NA | 3 | 0 | 1 | Not-determined | Genome-fragment | NA | NA | 0 |
| S16_extracted_28174 | 2683 | No | NA | 3 | 2 | 0 | Low-quality | Genome-fragment | 8.35 | AAI-based (high-confidence) | 0 |
| S16_vs_16008\|\|full | 1159 | No | NA | 3 | 0 | 3 | Not-determined | Genome-fragment | NA | NA | 0 |
| S8_vs_k141_107162\|\|full | 2683 | Yes | 1834 | 5 | 1 | 1 | Low-quality | Genome-fragment | 2.95 | AAI-based (medium-confidence) | 31.64 |
| S16_vs_16113\|\|full | 1633 | No | NA | 3 | 0 | 1 | Not-determined | Genome-fragment | NA | NA | 0 |
| S13_extracted_135436 | 2684 | No | NA | 3 | 1 | 0 | Low-quality | Genome-fragment | 1.79 | AAI-based (high-confidence) | 0 |
| S3_extracted_k141_121611 | 2684 | No | NA | 2 | 1 | 0 | Low-quality | Genome-fragment | 2.67 | AAI-based (medium-confidence) | 0 |
| S3_vs_k141_9164\|\|full | 2684 | No | NA | 4 | 1 | 0 | Low-quality | Genome-fragment | 7.17 | AAI-based (high-confidence) | 0 |
| S16_vs_16210\|\|full | 6940 | No | NA | 10 | 0 | 4 | Not-determined | Genome-fragment | NA | NA | 0 |
| S3_vs_k141_76871\|\|full | 2684 | No | NA | 9 | 0 | 0 | Low-quality | Genome-fragment | 1.51 | AAI-based (high-confidence) | 0 |
| S13_extracted_20436 | 2685 | No | NA | 1 | 0 | 1 | Low-quality | Genome-fragment | 2.8 | AAI-based (medium-confidence) | 0 |
| S16_vs_16492\|\|full | 1694 | No | NA | 4 | 0 | 1 | Not-determined | Genome-fragment | NA | NA | 0 |
| S16_vs_16510\|\|full | 9147 | No | NA | 9 | 0 | 4 | Not-determined | Genome-fragment | NA | NA | 0 |
| S3_vs_k141_117884\|\|full | 2685 | No | NA | 7 | 0 | 0 | Low-quality | Genome-fragment | 5.45 | AAI-based (high-confidence) | 0 |
| S16_vs_16628\|\|full | 1405 | No | NA | 4 | 0 | 0 | Not-determined | Genome-fragment | NA | NA | 0 |
| S4_vs_VIRSorter_22024-cat_2 | 2685 | No | NA | 8 | 8 | 0 | Low-quality | Genome-fragment | 8.83 | AAI-based (high-confidence) | 0 |
| S16_vs_16771\|\|full | 2093 | No | NA | 2 | 0 | 1 | Not-determined | Genome-fragment | NA | NA | 0 |
| S16_vs_16785\|\|full | 8443 | No | NA | 5 | 0 | 3 | Not-determined | Genome-fragment | NA | NA | 0 |
| S16_vs_16847\|\|full | 6777 | No | NA | 3 | 0 | 0 | Not-determined | Genome-fragment | NA | NA | 0 |
| S16_vs_17010\|\|full | 1102 | No | NA | 2 | 0 | 1 | Not-determined | Genome-fragment | NA | NA | 0 |
| S16_vs_17072\|\|full | 2852 | No | NA | 3 | 0 | 1 | Not-determined | Genome-fragment | NA | NA | 0 |
| S11_vs_k141_83516\|\|full | 2686 | No | NA | 7 | 1 | 0 | Low-quality | Genome-fragment | 4.47 | AAI-based (high-confidence) | 0 |
| S14_extracted_k141_90297 | 2686 | No | NA | 7 | 5 | 0 | Low-quality | Genome-fragment | 8.79 | AAI-based (high-confidence) | 0 |
| S11_vs_k141_38322\|\|full | 2687 | No | NA | 6 | 1 | 0 | Low-quality | Genome-fragment | 6.19 | HMM-based (lower-bound) | 0 |
| S16_vs_17318\|\|full | 1328 | No | NA | 2 | 0 | 1 | Not-determined | Genome-fragment | NA | NA | 0 |
| S14_vs_k141_94036\|\|full | 2687 | No | NA | 4 | 0 | 0 | Low-quality | Genome-fragment | 2.55 | AAI-based (medium-confidence) | 0 |
| S16_vs_17473\|\|full | 1260 | No | NA | 3 | 0 | 1 | Not-determined | Genome-fragment | NA | NA | 0 |
| S14_vs_k141_101524\|\|full | 2687 | No | NA | 3 | 0 | 0 | Low-quality | Genome-fragment | 2.47 | AAI-based (medium-confidence) | 0 |
| S16_vs_8431\|\|full | 2687 | No | NA | 2 | 0 | 2 | Low-quality | Genome-fragment | 9.01 | AAI-based (medium-confidence) | 0 |
| S16_vs_17493\|\|full | 30692 | No | NA | 32 | 6 | 0 | Medium-quality | Genome-fragment | 74.31 | AAI-based (high-confidence) | 0 |
| S10_vs_27785\|\|full | 2687 | No | NA | 6 | 0 | 0 | Low-quality | Genome-fragment | 4.42 | AAI-based (medium-confidence) | 0 |
| S16_vs_17502\|\|full | 1884 | No | NA | 3 | 0 | 2 | Not-determined | Genome-fragment | NA | NA | 0 |
| S12_vs_k141_43091\|\|full | 2687 | No | NA | 3 | 0 | 2 | Low-quality | Genome-fragment | 2.18 | AAI-based (high-confidence) | 0 |
| S16_vs_17634\|\|full | 1054 | No | NA | 2 | 0 | 0 | Not-determined | Genome-fragment | NA | NA | 0 |
| S16_vs_17722\|\|full | 1131 | No | NA | 3 | 0 | 0 | Not-determined | Genome-fragment | NA | NA | 0 |
| S16_vs_17817\|\|full | 14787 | No | NA | 12 | 0 | 5 | Not-determined | Genome-fragment | NA | NA | 0 |
| S16_vs_17898\|\|full | 2180 | No | NA | 2 | 0 | 2 | Not-determined | Genome-fragment | NA | NA | 0 |
| S13_extracted_38433 | 2688 | No | NA | 3 | 0 | 1 | Low-quality | Genome-fragment | 5.08 | AAI-based (high-confidence) | 0 |
| S16_vs_17920\|\|full | 7831 | No | NA | 9 | 0 | 7 | Not-determined | Genome-fragment | NA | NA | 0 |
| S8_extracted_k141_115790 | 2688 | No | NA | 7 | 2 | 0 | Low-quality | Genome-fragment | 6.95 | AAI-based (high-confidence) | 0 |
| S16_vs_18189\|\|full | 2251 | No | NA | 4 | 0 | 2 | Not-determined | Genome-fragment | NA | NA | 0 |
| S16_vs_18207\|\|full | 1200 | No | NA | 2 | 0 | 0 | Not-determined | Genome-fragment | NA | NA | 0 |
| S9_extracted_k141_17742 | 2689 | No | NA | 8 | 1 | 0 | Low-quality | Genome-fragment | 5.62 | AAI-based (medium-confidence) | 0 |
| S16_vs_18293\|\|full | 31073 | No | NA | 31 | 15 | 1 | Medium-quality | Genome-fragment | 65.09 | AAI-based (high-confidence) | 0 |
| S7_extracted_17364 | 2689 | No | NA | 8 | 1 | 0 | Low-quality | Genome-fragment | 4.7 | AAI-based (high-confidence) | 0 |
| S14_extracted_k141_8328 | 2690 | No | NA | 2 | 0 | 2 | Low-quality | Genome-fragment | 1.45 | AAI-based (medium-confidence) | 0 |
| S16_vs_18440\|\|full | 50518 | No | NA | 75 | 20 | 1 | High-quality | High-quality | 96.99 | AAI-based (high-confidence) | 0 |
| S3_vs_k141_41522\|\|full | 2690 | No | NA | 5 | 2 | 0 | Low-quality | Genome-fragment | 5.47 | AAI-based (medium-confidence) | 0 |
| S16_vs_18563\|\|full | 5183 | No | NA | 7 | 0 | 4 | Not-determined | Genome-fragment | NA | NA | 0 |
| S5_vs_k141_16073\|\|full | 2690 | No | NA | 4 | 0 | 1 | Low-quality | Genome-fragment | 9.43 | AAI-based (medium-confidence) | 0 |
| S7_vs_134950\|\|full | 2690 | No | NA | 5 | 2 | 0 | Low-quality | Genome-fragment | 6.68 | AAI-based (high-confidence) | 0 |
| S16_vs_18744\|\|full | 1392 | No | NA | 2 | 0 | 2 | Not-determined | Genome-fragment | NA | NA | 0 |
| S12_vs_k141_121181\|\|full | 2690 | No | NA | 3 | 0 | 3 | Low-quality | Genome-fragment | 1.3 | AAI-based (high-confidence) | 0 |
| S15_vs_32113\|\|full | 2690 | No | NA | 5 | 0 | 1 | Low-quality | Genome-fragment | 5.19 | AAI-based (medium-confidence) | 0 |
| S9_vs_k141_73328\|\|lt2gene | 2691 | No | NA | 2 | 2 | 0 | Low-quality | Genome-fragment | 6.35 | AAI-based (high-confidence) | 0 |
| S16_vs_18885\|\|full | 1263 | No | NA | 3 | 0 | 0 | Not-determined | Genome-fragment | NA | NA | 0 |
| S9_vs_k141_55636\|\|full | 2692 | No | NA | 6 | 0 | 0 | Low-quality | Genome-fragment | 6.45 | AAI-based (high-confidence) | 0 |
| S9_vs_k141_71737\|\|full | 2692 | No | NA | 3 | 2 | 0 | Low-quality | Genome-fragment | 6.61 | AAI-based (high-confidence) | 0 |
| S11_vs_k141_13824\|\|lt2gene | 2692 | No | NA | 2 | 0 | 0 | Low-quality | Genome-fragment | 1.76 | AAI-based (high-confidence) | 0 |
| S16_vs_19049\|\|full | 13123 | No | NA | 15 | 0 | 8 | Not-determined | Genome-fragment | NA | NA | 0 |
| S5_vs_k141_125051\|\|full | 2692 | No | NA | 7 | 1 | 0 | Low-quality | Genome-fragment | 2.04 | HMM-based (lower-bound) | 0 |
| S16_vs_19159\|\|full | 9212 | No | NA | 7 | 0 | 5 | Not-determined | Genome-fragment | NA | NA | 0 |
| S16_vs_19199\|\|full | 10679 | No | NA | 16 | 0 | 3 | Not-determined | Genome-fragment | NA | NA | 0 |
| S16_vs_19320\|\|full | 29199 | No | NA | 39 | 10 | 0 | Medium-quality | Genome-fragment | 69.62 | AAI-based (medium-confidence) | 0 |
| S9_vs_k141_88083\|\|full | 2693 | No | NA | 4 | 0 | 3 | Low-quality | Genome-fragment | 3.94 | AAI-based (high-confidence) | 0 |
| S3_vs_k141_76793\|\|full | 2693 | No | NA | 4 | 2 | 0 | Low-quality | Genome-fragment | 6.32 | AAI-based (high-confidence) | 0 |
| S12_vs_k141_73967\|\|full | 2693 | No | NA | 3 | 0 | 2 | Low-quality | Genome-fragment | 0.89 | AAI-based (medium-confidence) | 0 |
| S16_vs_19502\|\|full | 3230 | No | NA | 2 | 0 | 2 | Not-determined | Genome-fragment | NA | NA | 0 |
| S16_vs_19607\|\|full | 6181 | No | NA | 7 | 0 | 5 | Not-determined | Genome-fragment | NA | NA | 0 |
| S17_vs_k141_162938\|\|full | 2694 | No | NA | 4 | 0 | 0 | Low-quality | Genome-fragment | 6.15 | AAI-based (medium-confidence) | 0 |
| S1_extracted_k141_128172 | 2694 | No | NA | 2 | 0 | 2 | Low-quality | Genome-fragment | 1 | AAI-based (high-confidence) | 0 |
| S16_vs_19879\|\|full | 3241 | No | NA | 8 | 0 | 2 | Not-determined | Genome-fragment | NA | NA | 0 |
| S16_vs_19910\|\|full | 6688 | No | NA | 5 | 0 | 3 | Not-determined | Genome-fragment | NA | NA | 0 |
| S16_vs_19968\|\|full | 54417 | Yes | 34564 | 61 | 10 | 14 | Medium-quality | Genome-fragment | 83 | AAI-based (high-confidence) | 36.48 |
| S10_vs_61582\|\|full | 2694 | No | NA | 4 | 0 | 0 | Low-quality | Genome-fragment | 4.87 | AAI-based (medium-confidence) | 0 |
| S17_extracted_k141_177090 | 2695 | No | NA | 3 | 0 | 0 | Low-quality | Genome-fragment | 5.13 | AAI-based (medium-confidence) | 0 |
| S17_vs_k141_128771\|\|lt2gene | 2695 | No | NA | 2 | 1 | 0 | Low-quality | Genome-fragment | 6.2 | AAI-based (high-confidence) | 0 |
| S16_vs_20046\|\|full | 1350 | No | NA | 1 | 0 | 0 | Not-determined | Genome-fragment | NA | NA | 0 |
| S9_vs_k141_22470\|\|full | 2695 | No | NA | 3 | 0 | 1 | Low-quality | Genome-fragment | 5.96 | AAI-based (high-confidence) | 0 |
| S16_vs_20164\|\|full | 3349 | No | NA | 4 | 0 | 2 | Not-determined | Genome-fragment | NA | NA | 0 |
| S8_vs_k141_175248\|\|full | 2695 | No | NA | 3 | 3 | 0 | Low-quality | Genome-fragment | 6.14 | AAI-based (high-confidence) | 0 |
| S16_vs_20356\|\|full | 1944 | No | NA | 3 | 0 | 2 | Not-determined | Genome-fragment | NA | NA | 0 |
| S16_vs_20394\|\|full | 2520 | No | NA | 5 | 0 | 2 | Not-determined | Genome-fragment | NA | NA | 0 |
| S16_vs_20420\|\|full | 2647 | No | NA | 5 | 0 | 0 | Not-determined | Genome-fragment | NA | NA | 0 |
| S1_vs_k141_130164\|\|full | 2696 | No | NA | 5 | 5 | 0 | Low-quality | Genome-fragment | 6.31 | AAI-based (medium-confidence) | 0 |
| S16_vs_20453\|\|full | 1163 | No | NA | 2 | 0 | 1 | Not-determined | Genome-fragment | NA | NA | 0 |
| S16_vs_20470\|\|full | 1481 | No | NA | 3 | 0 | 0 | Not-determined | Genome-fragment | NA | NA | 0 |
| S16_vs_20715\|\|full | 3312 | No | NA | 2 | 0 | 2 | Not-determined | Genome-fragment | NA | NA | 0 |
| S16_vs_20721\|\|full | 3305 | No | NA | 4 | 0 | 3 | Not-determined | Genome-fragment | NA | NA | 0 |
| S7_vs_21984\|\|lt2gene | 2696 | No | NA | 2 | 2 | 0 | Low-quality | Genome-fragment | 5.8 | AAI-based (medium-confidence) | 0 |
| S16_vs_20781\|\|full | 1911 | No | NA | 3 | 0 | 0 | Not-determined | Genome-fragment | NA | NA | 0 |
| S16_vs_20814\|\|full | 1667 | No | NA | 3 | 0 | 1 | Not-determined | Genome-fragment | NA | NA | 0 |
| S11_vs_k141_5387\|\|full | 2697 | No | NA | 6 | 0 | 0 | Low-quality | Genome-fragment | 3.7 | AAI-based (medium-confidence) | 0 |
| S13_vs_125508\|\|full | 2697 | No | NA | 4 | 0 | 1 | Low-quality | Genome-fragment | 1.83 | AAI-based (high-confidence) | 0 |
| S16_vs_20957\|\|full | 3155 | No | NA | 2 | 0 | 2 | Not-determined | Genome-fragment | NA | NA | 0 |
| S16_vs_84575\|\|full | 2697 | No | NA | 4 | 0 | 0 | Low-quality | Genome-fragment | 2.1 | AAI-based (high-confidence) | 0 |
| S16_vs_21055\|\|full | 3031 | No | NA | 7 | 0 | 0 | Not-determined | Genome-fragment | NA | NA | 0 |
| S16_vs_21098\|\|full | 3751 | No | NA | 3 | 0 | 1 | Not-determined | Genome-fragment | NA | NA | 0 |
| S8_extracted_k141_13392 | 2697 | No | NA | 6 | 0 | 0 | Low-quality | Genome-fragment | 5.73 | AAI-based (high-confidence) | 0 |
| S16_vs_21328\|\|full | 1136 | No | NA | 2 | 0 | 1 | Not-determined | Genome-fragment | NA | NA | 0 |
| S16_vs_21352\|\|full | 6078 | No | NA | 7 | 0 | 3 | Not-determined | Genome-fragment | NA | NA | 0 |
| S16_vs_21361\|\|full | 3496 | No | NA | 5 | 0 | 0 | Not-determined | Genome-fragment | NA | NA | 0 |
| S8_vs_k141_180392\|\|full | 2697 | No | NA | 4 | 1 | 0 | Low-quality | Genome-fragment | 6.9 | AAI-based (medium-confidence) | 0 |
| S16_vs_21408\|\|full | 2924 | No | NA | 5 | 0 | 1 | Not-determined | Genome-fragment | NA | NA | 0 |
| S16_vs_21414\|\|full | 4318 | No | NA | 5 | 0 | 1 | Not-determined | Genome-fragment | NA | NA | 0 |
| S16_vs_21438\|\|full | 1551 | No | NA | 2 | 0 | 0 | Not-determined | Genome-fragment | NA | NA | 0 |
| S17_vs_k141_147818\|\|full | 2698 | No | NA | 6 | 0 | 2 | Low-quality | Genome-fragment | 1.78 | AAI-based (high-confidence) | 0 |
| S16_vs_21627\|\|full | 2221 | No | NA | 2 | 0 | 1 | Not-determined | Genome-fragment | NA | NA | 0 |
| S16_vs_21639\|\|full | 1780 | No | NA | 2 | 0 | 1 | Not-determined | Genome-fragment | NA | NA | 0 |
| S16_vs_21674\|\|full | 1719 | No | NA | 3 | 0 | 3 | Not-determined | Genome-fragment | NA | NA | 0 |
| S16_vs_21750\|\|full | 1472 | No | NA | 3 | 0 | 0 | Not-determined | Genome-fragment | NA | NA | 0 |
| S16_vs_21775\|\|full | 6902 | No | NA | 7 | 0 | 4 | Not-determined | Genome-fragment | NA | NA | 0 |
| S11_vs_k141_86979\|\|full | 2698 | No | NA | 4 | 0 | 0 | Low-quality | Genome-fragment | 4.86 | AAI-based (high-confidence) | 0 |
| S3_extracted_k141_26019 | 2698 | No | NA | 6 | 6 | 0 | Low-quality | Genome-fragment | 8.89 | AAI-based (high-confidence) | 0 |
| S16_vs_21979\|\|full | 1898 | No | NA | 3 | 0 | 1 | Not-determined | Genome-fragment | NA | NA | 0 |
| S16_vs_21994\|\|full | 2373 | No | NA | 3 | 0 | 0 | Not-determined | Genome-fragment | NA | NA | 0 |
| S16_vs_166642\|\|full | 2698 | No | NA | 3 | 2 | 0 | Low-quality | Genome-fragment | 6.68 | AAI-based (high-confidence) | 0 |
| S16_vs_22321\|\|full | 18630 | No | NA | 19 | 0 | 8 | Not-determined | Genome-fragment | NA | NA | 0 |
| S12_vs_k141_33247\|\|full | 2698 | No | NA | 5 | 5 | 0 | Low-quality | Genome-fragment | 5.91 | AAI-based (high-confidence) | 0 |
| S16_vs_22404\|\|full | 2106 | No | NA | 3 | 0 | 0 | Not-determined | Genome-fragment | NA | NA | 0 |
| S6_extracted_k141_53780 | 2698 | No | NA | 5 | 5 | 0 | Low-quality | Genome-fragment | 8.9 | AAI-based (high-confidence) | 0 |
| S16_vs_22447\|\|full | 1377 | No | NA | 2 | 0 | 2 | Not-determined | Genome-fragment | NA | NA | 0 |
| S16_vs_22637\|\|full | 3716 | No | NA | 1 | 0 | 0 | Not-determined | Genome-fragment | NA | NA | 0 |
| S16_extracted_23758 | 2699 | No | NA | 6 | 0 | 0 | Low-quality | Genome-fragment | 4.54 | AAI-based (high-confidence) | 0 |
| S16_vs_22702\|\|full | 1404 | No | NA | 2 | 0 | 0 | Not-determined | Genome-fragment | NA | NA | 0 |
| S16_vs_22808\|\|full | 1244 | No | NA | 1 | 0 | 0 | Not-determined | Genome-fragment | NA | NA | 0 |
| S16_vs_22869\|\|full | 2676 | No | NA | 4 | 0 | 0 | Not-determined | Genome-fragment | NA | NA | 0 |
| S12_vs_k141_26645\|\|full | 2699 | No | NA | 4 | 0 | 0 | Low-quality | Genome-fragment | 4.49 | AAI-based (high-confidence) | 0 |
| S16_vs_23058\|\|full | 1255 | No | NA | 2 | 0 | 2 | Not-determined | Genome-fragment | NA | NA | 0 |
| S16_vs_23115\|\|full | 1141 | No | NA | 3 | 0 | 0 | Not-determined | Genome-fragment | NA | NA | 0 |
| S8_vs_k141_153251\|\|full | 2700 | No | NA | 6 | 2 | 0 | Low-quality | Genome-fragment | 6.74 | AAI-based (high-confidence) | 0 |
| S16_vs_23269\|\|full | 1629 | No | NA | 2 | 0 | 0 | Not-determined | Genome-fragment | NA | NA | 0 |
| S8_vs_k141_173276\|\|full | 2700 | No | NA | 5 | 1 | 0 | Low-quality | Genome-fragment | 2.53 | AAI-based (high-confidence) | 0 |
| S16_vs_23374\|\|full | 2900 | No | NA | 5 | 0 | 0 | Not-determined | Genome-fragment | NA | NA | 0 |
| S12_vs_k141_14062\|\|full | 2701 | No | NA | 4 | 0 | 0 | Low-quality | Genome-fragment | 4.41 | AAI-based (medium-confidence) | 0 |
| S16_vs_23405\|\|full | 1518 | No | NA | 3 | 0 | 0 | Not-determined | Genome-fragment | NA | NA | 0 |
| S16_vs_23511\|\|full | 2238 | No | NA | 2 | 0 | 1 | Not-determined | Genome-fragment | NA | NA | 0 |
| S16_vs_23550\|\|full | 3183 | No | NA | 3 | 0 | 3 | Not-determined | Genome-fragment | NA | NA | 0 |
| S16_vs_23565\|\|full | 3611 | No | NA | 3 | 0 | 2 | Not-determined | Genome-fragment | NA | NA | 0 |
| S16_vs_23649\|\|full | 2745 | No | NA | 4 | 0 | 3 | Not-determined | Genome-fragment | NA | NA | 0 |
| S8_vs_k141_3336\|\|full | 2701 | No | NA | 3 | 0 | 0 | Low-quality | Genome-fragment | 1.91 | AAI-based (medium-confidence) | 0 |
| S16_vs_23808\|\|full | 1547 | No | NA | 2 | 0 | 0 | Not-determined | Genome-fragment | NA | NA | 0 |
| S5_vs_k141_49357\|\|full | 2702 | No | NA | 5 | 1 | 0 | Low-quality | Genome-fragment | 6.97 | AAI-based (high-confidence) | 0 |
| S7_vs_50090\|\|full | 2702 | No | NA | 7 | 2 | 0 | Low-quality | Genome-fragment | 5.64 | AAI-based (high-confidence) | 0 |
| S16_vs_23853\|\|full | 6546 | No | NA | 6 | 0 | 0 | Not-determined | Genome-fragment | NA | NA | 0 |
| S7_vs_133787\|\|full | 2702 | No | NA | 3 | 0 | 1 | Low-quality | Genome-fragment | 1.61 | AAI-based (medium-confidence) | 0 |
| S16_vs_23981\|\|full | 1231 | No | NA | 3 | 0 | 1 | Not-determined | Genome-fragment | NA | NA | 0 |
| S16_vs_24017\|\|full | 2478 | No | NA | 3 | 0 | 0 | Not-determined | Genome-fragment | NA | NA | 0 |
| S16_vs_117299\|\|full | 2702 | No | NA | 4 | 1 | 0 | Low-quality | Genome-fragment | 4.33 | HMM-based (lower-bound) | 0 |
| S8_vs_k141_151863\|\|full | 2702 | No | NA | 6 | 0 | 1 | Low-quality | Genome-fragment | 6.71 | AAI-based (high-confidence) | 0 |
| S14_extracted_k141_82227 | 2703 | No | NA | 2 | 2 | 0 | Low-quality | Genome-fragment | 8.89 | AAI-based (high-confidence) | 0 |
| S5_vs_k141_82932\|\|full | 2703 | No | NA | 2 | 1 | 0 | Low-quality | Genome-fragment | 1.93 | AAI-based (medium-confidence) | 0 |
| S7_extracted_56922 | 2703 | No | NA | 4 | 0 | 0 | Low-quality | Genome-fragment | 3.97 | AAI-based (high-confidence) | 0 |
| S16_extracted_395 | 2703 | No | NA | 1 | 1 | 0 | Low-quality | Genome-fragment | 5.06 | HMM-based (lower-bound) | 0 |
| S16_vs_24486\|\|full | 1303 | No | NA | 2 | 0 | 2 | Not-determined | Genome-fragment | NA | NA | 0 |
| S12_extracted_k141_145998 | 2703 | No | NA | 4 | 2 | 0 | Low-quality | Genome-fragment | 1.92 | AAI-based (high-confidence) | 0 |
| S7_vs_80843\|\|full | 2704 | No | NA | 4 | 3 | 0 | Low-quality | Genome-fragment | 5.86 | AAI-based (high-confidence) | 0 |
| S16_vs_24785\|\|full | 2449 | No | NA | 2 | 0 | 2 | Not-determined | Genome-fragment | NA | NA | 0 |
| S16_vs_24891\|\|full | 4096 | No | NA | 2 | 0 | 0 | Not-determined | Genome-fragment | NA | NA | 0 |
| S16_vs_24999\|\|full | 5231 | No | NA | 8 | 0 | 5 | Not-determined | Genome-fragment | NA | NA | 0 |
| S16_vs_25001\|\|full | 61588 | No | NA | 77 | 0 | 12 | Not-determined | Genome-fragment | NA | NA | 0 |
| S16_vs_25059\|\|full | 1734 | No | NA | 3 | 0 | 0 | Not-determined | Genome-fragment | NA | NA | 0 |
| S7_vs_132756\|\|lt2gene | 2704 | No | NA | 2 | 2 | 0 | Low-quality | Genome-fragment | 6.18 | AAI-based (medium-confidence) | 0 |
| S4_extracted_23191 | 2704 | No | NA | 4 | 4 | 0 | Low-quality | Genome-fragment | 6.9 | AAI-based (high-confidence) | 0 |
| S16_vs_25225\|\|full | 10580 | No | NA | 18 | 0 | 1 | Not-determined | Genome-fragment | NA | NA | 0 |
| S3_extracted_k141_70403 | 2705 | No | NA | 3 | 2 | 0 | Low-quality | Genome-fragment | 6.72 | AAI-based (medium-confidence) | 0 |

| | | | | | | | | | | | |
|---|---|---|---|---|---|---|---|---|---|---|---|
| S5_vs_k141_35672\|\|full | 2705 | No | NA | 4 | 1 | 0 | Low-quality | Genome-fragment | 5.38 | AAI-based (high-confidence) | 0 |
| S16_vs_25499\|\|full | 1711 | No | NA | 3 | 0 | 1 | Not-determined | Genome-fragment | NA | NA | 0 |
| S16_vs_25541\|\|full | 1497 | No | NA | 2 | 0 | 1 | Not-determined | Genome-fragment | NA | NA | 0 |
| S7_vs_101586\|\|full | 2705 | No | NA | 5 | 1 | 0 | Low-quality | Genome-fragment | 6.8 | AAI-based (high-confidence) | 0 |
| S10_vs_54875\|\|full | 2705 | No | NA | 4 | 1 | 0 | Low-quality | Genome-fragment | 1.13 | AAI-based (high-confidence) | 0 |
| S16_vs_25582\|\|full | 4898 | No | NA | 4 | 0 | 0 | Not-determined | Genome-fragment | NA | NA | 0 |
| S7_vs_104013\|\|full | 2706 | No | NA | 4 | 0 | 1 | Low-quality | Genome-fragment | 2.9 | AAI-based (medium-confidence) | 0 |
| S7_vs_136668\|\|full | 2706 | No | NA | 3 | 2 | 0 | Low-quality | Genome-fragment | 4.95 | AAI-based (high-confidence) | 0 |
| S16_vs_25808\|\|full | 2004 | No | NA | 3 | 0 | 0 | Not-determined | Genome-fragment | NA | NA | 0 |
| S9_vs_k141_19804\|\|full | 2707 | No | NA | 3 | 0 | 2 | Low-quality | Genome-fragment | 1.2 | AAI-based (medium-confidence) | 0 |
| S16_vs_25982\|\|full | 1529 | No | NA | 3 | 0 | 0 | Not-determined | Genome-fragment | NA | NA | 0 |
| S13_vs_87321\|\|full | 2707 | No | NA | 2 | 0 | 1 | Low-quality | Genome-fragment | 3.43 | AAI-based (medium-confidence) | 0 |
| S16_vs_26062\|\|full | 5685 | No | NA | 7 | 0 | 7 | Not-determined | Genome-fragment | NA | NA | 0 |
| S16_vs_26078\|\|full | 21179 | No | NA | 25 | 12 | 0 | Medium-quality | Genome-fragment | 52.75 | AAI-based (high-confidence) | 0 |
| S7_vs_120264\|\|full | 2707 | No | NA | 5 | 1 | 0 | Low-quality | Genome-fragment | 5.08 | AAI-based (medium-confidence) | 0 |
| S16_extracted_79774 | 2707 | No | NA | 2 | 0 | 2 | Low-quality | Genome-fragment | 1.51 | AAI-based (medium-confidence) | 0 |
| S16_vs_26176\|\|full | 2862 | No | NA | 3 | 0 | 0 | Not-determined | Genome-fragment | NA | NA | 0 |
| S16_vs_26201\|\|full | 6409 | No | NA | 2 | 0 | 2 | Not-determined | Genome-fragment | NA | NA | 0 |
| S16_vs_26319\|\|full | 4813 | No | NA | 6 | 0 | 6 | Not-determined | Genome-fragment | NA | NA | 0 |
| S16_vs_26372\|\|full | 6033 | No | NA | 7 | 0 | 4 | Not-determined | Genome-fragment | NA | NA | 0 |
| S16_vs_26459\|\|full | 3146 | No | NA | 4 | 0 | 3 | Not-determined | Genome-fragment | NA | NA | 0 |
| S16_vs_26499\|\|full | 2450 | No | NA | 4 | 0 | 0 | Not-determined | Genome-fragment | NA | NA | 0 |
| S16_vs_26634\|\|full | 1015 | No | NA | 3 | 0 | 0 | Not-determined | Genome-fragment | NA | NA | 0 |
| S16_vs_26744\|\|full | 1731 | No | NA | 3 | 0 | 1 | Not-determined | Genome-fragment | NA | NA | 0 |
| S1_extracted_k141_4973 | 2708 | No | NA | 3 | 0 | 3 | Low-quality | Genome-fragment | 0.99 | AAI-based (medium-confidence) | 0 |
| S16_vs_26837\|\|full | 4768 | No | NA | 5 | 0 | 5 | Not-determined | Genome-fragment | NA | NA | 0 |
| S16_vs_15405\|\|full | 2708 | No | NA | 6 | 1 | 0 | Low-quality | Genome-fragment | 5.62 | AAI-based (high-confidence) | 0 |
| S16_vs_27006\|\|full | 1808 | No | NA | 2 | 0 | 1 | Not-determined | Genome-fragment | NA | NA | 0 |
| S16_vs_27124\|\|full | 1255 | No | NA | 3 | 0 | 0 | Not-determined | Genome-fragment | NA | NA | 0 |
| S16_vs_148580\|\|full | 2708 | No | NA | 5 | 1 | 0 | Low-quality | Genome-fragment | 7.82 | AAI-based (high-confidence) | 0 |
| S16_vs_27209\|\|full | 3411 | No | NA | 5 | 0 | 4 | Not-determined | Genome-fragment | NA | NA | 0 |
| S16_vs_27210\|\|full | 1632 | No | NA | 3 | 0 | 1 | Not-determined | Genome-fragment | NA | NA | 0 |
| S16_vs_150252\|\|full | 2708 | No | NA | 4 | 2 | 0 | Low-quality | Genome-fragment | 9.52 | HMM-based (lower-bound) | 0 |
| S16_vs_27250\|\|full | 5700 | No | NA | 6 | 0 | 0 | Not-determined | Genome-fragment | NA | NA | 0 |
| S16_vs_27361\|\|full | 2913 | No | NA | 3 | 0 | 3 | Not-determined | Genome-fragment | NA | NA | 0 |
| S16_vs_27387\|\|full | 1985 | No | NA | 2 | 0 | 0 | Not-determined | Genome-fragment | NA | NA | 0 |
| S16_vs_164221\|\|full | 2708 | No | NA | 2 | 0 | 1 | Low-quality | Genome-fragment | 1.56 | AAI-based (medium-confidence) | 0 |
| S16_vs_27420\|\|full | 6273 | No | NA | 6 | 0 | 4 | Not-determined | Genome-fragment | NA | NA | 0 |
| S12_vs_k141_38912\|\|full | 2708 | No | NA | 3 | 1 | 0 | Low-quality | Genome-fragment | 6.97 | AAI-based (medium-confidence) | 0 |
| S16_vs_27598\|\|full | 3095 | No | NA | 2 | 0 | 1 | Not-determined | Genome-fragment | NA | NA | 0 |
| S15_vs_17228\|\|full | 2708 | No | NA | 3 | 1 | 0 | Low-quality | Genome-fragment | 5.88 | AAI-based (medium-confidence) | 0 |
| S3_vs_k141_80725\|\|full | 2709 | No | NA | 7 | 1 | 0 | Low-quality | Genome-fragment | 1.77 | AAI-based (high-confidence) | 0 |
| S3_vs_k141_6079\|\|lt2gene | 2709 | No | NA | 2 | 1 | 0 | Low-quality | Genome-fragment | 8 | AAI-based (high-confidence) | 0 |
| S7_vs_60881\|\|full | 2709 | No | NA | 2 | 1 | 0 | Low-quality | Genome-fragment | 6.99 | AAI-based (high-confidence) | 0 |
| S16_vs_27777\|\|full | 1847 | No | NA | 2 | 0 | 0 | Not-determined | Genome-fragment | NA | NA | 0 |
| S11_vs_k141_69560\|\|lt2gene | 2711 | No | NA | 2 | 0 | 0 | Low-quality | Genome-fragment | 4.43 | AAI-based (high-confidence) | 0 |
| S16_vs_27829\|\|full | 2387 | No | NA | 3 | 0 | 1 | Not-determined | Genome-fragment | NA | NA | 0 |
| S13_vs_114670\|\|full | 2711 | No | NA | 6 | 1 | 0 | Low-quality | Genome-fragment | 1.82 | HMM-based (lower-bound) | 0 |
| S7_vs_43575\|\|full | 2711 | No | NA | 3 | 0 | 2 | Low-quality | Genome-fragment | 4.09 | AAI-based (medium-confidence) | 0 |
| S11_extracted_k141_43211 | 2712 | No | NA | 2 | 0 | 0 | Low-quality | Genome-fragment | 1.05 | AAI-based (medium-confidence) | 0 |
| S3_vs_k141_97029\|\|full | 2712 | No | NA | 7 | 2 | 0 | Low-quality | Genome-fragment | 3.08 | HMM-based (lower-bound) | 0 |
| S16_vs_159724\|\|full | 2712 | No | NA | 7 | 0 | 1 | Low-quality | Genome-fragment | 1.57 | AAI-based (high-confidence) | 0 |
| S16_vs_28106\|\|full | 1413 | No | NA | 2 | 0 | 1 | Not-determined | Genome-fragment | NA | NA | 0 |
| S16_vs_28175\|\|full | 2777 | No | NA | 3 | 0 | 0 | Not-determined | Genome-fragment | NA | NA | 0 |
| S16_vs_28237\|\|full | 2588 | No | NA | 2 | 0 | 2 | Not-determined | Genome-fragment | NA | NA | 0 |
| S10_vs_46593\|\|full | 2712 | No | NA | 4 | 3 | 0 | Low-quality | Genome-fragment | 4.57 | AAI-based (high-confidence) | 0 |
| S8_extracted_k141_2037 | 2712 | No | NA | 3 | 1 | 0 | Low-quality | Genome-fragment | 3.82 | HMM-based (lower-bound) | 0 |
| S16_vs_28249\|\|full | 6056 | No | NA | 6 | 0 | 4 | Not-determined | Genome-fragment | NA | NA | 0 |
| S7_vs_21264\|\|full | 2713 | No | NA | 4 | 0 | 0 | Low-quality | Genome-fragment | 2.05 | AAI-based (high-confidence) | 0 |
| S16_vs_28257\|\|full | 3467 | No | NA | 5 | 0 | 3 | Not-determined | Genome-fragment | NA | NA | 0 |
| S16_vs_32314\|\|full | 2713 | No | NA | 2 | 0 | 0 | Low-quality | Genome-fragment | 18.46 | AAI-based (high-confidence) | 0 |
| S16_vs_28368\|\|full | 4529 | No | NA | 5 | 0 | 1 | Not-determined | Genome-fragment | NA | NA | 0 |
| S16_vs_28416\|\|full | 1185 | No | NA | 3 | 0 | 0 | Not-determined | Genome-fragment | NA | NA | 0 |
| S16_vs_28540\|\|full | 4598 | No | NA | 7 | 0 | 1 | Not-determined | Genome-fragment | NA | NA | 0 |
| S16_vs_28723\|\|full | 14671 | No | NA | 19 | 0 | 2 | Not-determined | Genome-fragment | NA | NA | 0 |
| S3_extracted_k141_123846 | 2714 | No | NA | 7 | 4 | 0 | Low-quality | Genome-fragment | 4.49 | AAI-based (high-confidence) | 0 |
| S8_vs_k141_65976\|\|full | 2714 | No | NA | 3 | 0 | 0 | Low-quality | Genome-fragment | 7.81 | AAI-based (high-confidence) | 0 |
| S16_vs_28882\|\|full | 2560 | No | NA | 2 | 0 | 1 | Not-determined | Genome-fragment | NA | NA | 0 |
| S8_vs_k141_190617\|\|full | 2714 | No | NA | 6 | 0 | 1 | Low-quality | Genome-fragment | 4.28 | AAI-based (medium-confidence) | 0 |
| S16_vs_28941\|\|full | 3584 | No | NA | 3 | 0 | 1 | Not-determined | Genome-fragment | NA | NA | 0 |
| S17_vs_k141_54577\|\|full | 2715 | No | NA | 3 | 0 | 2 | Low-quality | Genome-fragment | 2.65 | AAI-based (medium-confidence) | 0 |
| S16_vs_29023\|\|full | 2318 | No | NA | 3 | 0 | 0 | Not-determined | Genome-fragment | NA | NA | 0 |
| S16_vs_29137\|\|full | 6502 | No | NA | 6 | 0 | 0 | Not-determined | Genome-fragment | NA | NA | 0 |
| S16_vs_29172\|\|full | 5185 | No | NA | 3 | 0 | 2 | Not-determined | Genome-fragment | NA | NA | 0 |
| S16_vs_29184\|\|full | 4344 | No | NA | 3 | 0 | 3 | Not-determined | Genome-fragment | NA | NA | 0 |
| S17_vs_k141_4657\|\|full | 2715 | No | NA | 4 | 0 | 0 | Low-quality | Genome-fragment | 7.59 | AAI-based (high-confidence) | 0 |
| S13_extracted_153205 | 2715 | No | NA | 2 | 0 | 2 | Low-quality | Genome-fragment | 0.85 | AAI-based (medium-confidence) | 0 |
| S16_vs_29521\|\|full | 1811 | No | NA | 3 | 0 | 1 | Not-determined | Genome-fragment | NA | NA | 0 |
| S16_vs_29552\|\|full | 1914 | No | NA | 3 | 0 | 2 | Not-determined | Genome-fragment | NA | NA | 0 |
| S14_extracted_k141_109827 | 2715 | No | NA | 4 | 2 | 0 | Low-quality | Genome-fragment | 1.47 | HMM-based (lower-bound) | 0 |
| S16_vs_29587\|\|full | 10213 | No | NA | 7 | 0 | 5 | Not-determined | Genome-fragment | NA | NA | 0 |
| S16_vs_29606\|\|full | 2863 | No | NA | 3 | 0 | 0 | Not-determined | Genome-fragment | NA | NA | 0 |
| S14_vs_k141_65390\|\|full | 2715 | No | NA | 4 | 1 | 0 | Low-quality | Genome-fragment | 6.74 | AAI-based (medium-confidence) | 0 |
| S16_vs_29720\|\|full | 5112 | No | NA | 5 | 0 | 0 | Not-determined | Genome-fragment | NA | NA | 0 |
| S16_vs_29737\|\|full | 3651 | No | NA | 4 | 0 | 0 | Not-determined | Genome-fragment | NA | NA | 0 |
| S16_vs_29743\|\|full | 4031 | No | NA | 6 | 0 | 3 | Not-determined | Genome-fragment | NA | NA | 0 |
| S5_vs_k141_99157\|\|full | 2715 | No | NA | 5 | 1 | 0 | Low-quality | Genome-fragment | 4.26 | AAI-based (high-confidence) | 0 |
| S12_vs_k141_909\|\|full | 2715 | No | NA | 2 | 0 | 1 | Low-quality | Genome-fragment | 1.3 | AAI-based (medium-confidence) | 0 |
| S9_vs_k141_62288\|\|full | 2716 | No | NA | 7 | 0 | 0 | Low-quality | Genome-fragment | 7.66 | AAI-based (high-confidence) | 0 |
| S16_vs_30029\|\|full | 1197 | No | NA | 3 | 0 | 0 | Not-determined | Genome-fragment | NA | NA | 0 |
| S16_vs_30034\|\|full | 13765 | No | NA | 14 | 0 | 10 | Not-determined | Genome-fragment | NA | NA | 0 |
| S16_vs_30110\|\|full | 1238 | No | NA | 2 | 0 | 2 | Not-determined | Genome-fragment | NA | NA | 0 |
| S12_vs_k141_95725\|\|full | 2716 | No | NA | 7 | 2 | 0 | Low-quality | Genome-fragment | 6.51 | AAI-based (high-confidence) | 0 |
| S14_vs_k141_17694\|\|full | 2717 | No | NA | 2 | 0 | 1 | Low-quality | Genome-fragment | 4.33 | AAI-based (medium-confidence) | 0 |
| S16_vs_122866\|\|full | 2717 | No | NA | 4 | 0 | 0 | Low-quality | Genome-fragment | 2.18 | AAI-based (high-confidence) | 0 |
| S8_vs_k141_158727\|\|full | 2717 | No | NA | 3 | 0 | 2 | Low-quality | Genome-fragment | 1.69 | AAI-based (medium-confidence) | 0 |
| S17_vs_k141_155779\|\|full | 2718 | No | NA | 3 | 0 | 2 | Low-quality | Genome-fragment | 2.57 | AAI-based (medium-confidence) | 0 |
| S3_vs_k141_91973\|\|full | 2718 | No | NA | 7 | 1 | 0 | Low-quality | Genome-fragment | 4.28 | HMM-based (lower-bound) | 0 |
| S16_vs_30597\|\|full | 4601 | No | NA | 6 | 0 | 4 | Not-determined | Genome-fragment | NA | NA | 0 |
| S16_vs_30700\|\|full | 1836 | No | NA | 3 | 0 | 0 | Not-determined | Genome-fragment | NA | NA | 0 |
| S16_vs_30809\|\|full | 11274 | No | NA | 9 | 0 | 6 | Not-determined | Genome-fragment | NA | NA | 0 |
| S16_vs_30929\|\|full | 2530 | No | NA | 3 | 0 | 2 | Not-determined | Genome-fragment | NA | NA | 0 |
| S16_vs_31017\|\|full | 1177 | No | NA | 3 | 0 | 3 | Not-determined | Genome-fragment | NA | NA | 0 |
| S16_vs_31155\|\|full | 3982 | No | NA | 8 | 0 | 0 | Not-determined | Genome-fragment | NA | NA | 0 |
| S7_vs_64585\|\|full | 2718 | No | NA | 5 | 1 | 0 | Low-quality | Genome-fragment | 7.91 | AAI-based (high-confidence) | 0 |
| S5_extracted_k141_68776 | 2719 | No | NA | 3 | 3 | 0 | Low-quality | Genome-fragment | 4.2 | HMM-based (lower-bound) | 0 |
| S16_vs_31347\|\|full | 1847 | No | NA | 3 | 0 | 2 | Not-determined | Genome-fragment | NA | NA | 0 |
| S16_vs_31456\|\|full | 1161 | No | NA | 2 | 0 | 1 | Not-determined | Genome-fragment | NA | NA | 0 |
| S8_vs_k141_119558\|\|full | 2719 | No | NA | 3 | 0 | 1 | Low-quality | Genome-fragment | 1.56 | AAI-based (high-confidence) | 0 |
| S7_vs_31280\|\|full | 2720 | No | NA | 7 | 2 | 0 | Low-quality | Genome-fragment | 6.13 | AAI-based (high-confidence) | 0 |
| S16_vs_31832\|\|full | 1406 | No | NA | 3 | 0 | 1 | Not-determined | Genome-fragment | NA | NA | 0 |
| S16_vs_31948\|\|full | 2158 | No | NA | 3 | 0 | 0 | Not-determined | Genome-fragment | NA | NA | 0 |
| S8_vs_k141_37805\|\|full | 2720 | No | NA | 8 | 3 | 0 | Low-quality | Genome-fragment | 6.61 | AAI-based (high-confidence) | 0 |
| S17_vs_k141_106833\|\|full | 2721 | No | NA | 5 | 2 | 0 | Low-quality | Genome-fragment | 2.35 | HMM-based (lower-bound) | 0 |
| S7_vs_13726\|\|full | 2721 | No | NA | 7 | 0 | 0 | Low-quality | Genome-fragment | 5.71 | AAI-based (medium-confidence) | 0 |
| S11_extracted_k141_42022 | 2723 | No | NA | 5 | 2 | 0 | Low-quality | Genome-fragment | 2.91 | AAI-based (medium-confidence) | 0 |
| S16_vs_32151\|\|full | 1770 | No | NA | 3 | 0 | 0 | Not-determined | Genome-fragment | NA | NA | 0 |
| S16_vs_32196\|\|full | 1733 | No | NA | 3 | 0 | 0 | Not-determined | Genome-fragment | NA | NA | 0 |
| S14_vs_k141_22112\|\|full | 2723 | No | NA | 6 | 5 | 0 | Low-quality | Genome-fragment | 6.46 | AAI-based (high-confidence) | 0 |
| S17_extracted_k141_80211 | 2724 | No | NA | 1 | 1 | 0 | Low-quality | Genome-fragment | 5.22 | AAI-based (medium-confidence) | 0 |
| S16_vs_32575\|\|full | 2747 | No | NA | 3 | 0 | 1 | Not-determined | Genome-fragment | NA | NA | 0 |
| S7_vs_57825\|\|full | 2724 | No | NA | 3 | 0 | 0 | Low-quality | Genome-fragment | 6.25 | AAI-based (medium-confidence) | 0 |
| S16_vs_32613\|\|full | 1368 | No | NA | 2 | 0 | 2 | Not-determined | Genome-fragment | NA | NA | 0 |
| S10_extracted_105331 | 2724 | No | NA | 6 | 4 | 0 | Low-quality | Genome-fragment | 4.71 | HMM-based (lower-bound) | 0 |
| S16_vs_32761\|\|full | 2334 | No | NA | 4 | 0 | 1 | Not-determined | Genome-fragment | NA | NA | 0 |
| S16_vs_32764\|\|full | 55386 | No | NA | 45 | 1 | 19 | Medium-quality | Genome-fragment | 82.86 | HMM-based (lower-bound) | 0 |
| S16_vs_32861\|\|full | 1574 | No | NA | 2 | 0 | 1 | Not-determined | Genome-fragment | NA | NA | 0 |
| S16_vs_32871\|\|full | 1214 | No | NA | 3 | 0 | 1 | Not-determined | Genome-fragment | NA | NA | 0 |
| S16_vs_32889\|\|full | 1579 | No | NA | 3 | 0 | 0 | Not-determined | Genome-fragment | NA | NA | 0 |
| S12_vs_k141_167576\|\|full | 2724 | No | NA | 3 | 0 | 3 | Low-quality | Genome-fragment | 2.55 | AAI-based (medium-confidence) | 0 |
| S16_vs_33009\|\|full | 1786 | No | NA | 2 | 0 | 1 | Not-determined | Genome-fragment | NA | NA | 0 |
| S12_extracted_k141_60439 | 2725 | No | NA | 3 | 1 | 0 | Low-quality | Genome-fragment | 8.35 | AAI-based (medium-confidence) | 0 |
| S16_vs_33109\|\|full | 1727 | No | NA | 3 | 0 | 0 | Not-determined | Genome-fragment | NA | NA | 0 |
| S16_vs_33149\|\|full | 10646 | No | NA | 9 | 0 | 3 | Not-determined | Genome-fragment | NA | NA | 0 |
| S16_vs_33180\|\|full | 1384 | No | NA | 3 | 0 | 0 | Not-determined | Genome-fragment | NA | NA | 0 |
| S16_vs_33198\|\|full | 1605 | No | NA | 2 | 0 | 2 | Not-determined | Genome-fragment | NA | NA | 0 |
| S16_vs_33270\|\|full | 8311 | No | NA | 7 | 0 | 3 | Not-determined | Genome-fragment | NA | NA | 0 |
| S16_vs_33281\|\|full | 36592 | No | NA | 35 | 1 | 22 | Medium-quality | Genome-fragment | 55.64 | HMM-based (lower-bound) | 0 |
| S16_vs_33282\|\|full | 1515 | No | NA | 2 | 0 | 0 | Not-determined | Genome-fragment | NA | NA | 0 |
| S8_extracted_k141_200611 | 2725 | No | NA | 9 | 0 | 0 | Low-quality | Genome-fragment | 2.92 | AAI-based (high-confidence) | 0 |
| S16_vs_33393\|\|full | 1362 | No | NA | 2 | 0 | 1 | Not-determined | Genome-fragment | NA | NA | 0 |
| S16_vs_33476\|\|full | 2391 | No | NA | 2 | 0 | 0 | Not-determined | Genome-fragment | NA | NA | 0 |
| S8_vs_k141_167709\|\|full | 2725 | No | NA | 5 | 2 | 0 | Low-quality | Genome-fragment | 4.75 | AAI-based (high-confidence) | 0 |
| S16_vs_33524\|\|full | 2803 | No | NA | 3 | 0 | 2 | Not-determined | Genome-fragment | NA | NA | 0 |
| S3_vs_k141_104315\|\|full | 2726 | No | NA | 5 | 0 | 0 | Low-quality | Genome-fragment | 1.78 | AAI-based (high-confidence) | 0 |
| S16_vs_33582\|\|full | 7247 | No | NA | 7 | 0 | 5 | Not-determined | Genome-fragment | NA | NA | 0 |
| S16_vs_33607\|\|full | 3514 | No | NA | 5 | 0 | 2 | Not-determined | Genome-fragment | NA | NA | 0 |
| S13_vs_37759\|\|full | 2728 | No | NA | 5 | 0 | 1 | Low-quality | Genome-fragment | 2.82 | AAI-based (medium-confidence) | 0 |
| S14_extracted_k141_93593 | 2728 | No | NA | 3 | 1 | 0 | Low-quality | Genome-fragment | 6.6 | AAI-based (high-confidence) | 0 |
| S11_vs_k141_28933\|\|full | 2729 | No | NA | 3 | 0 | 3 | Low-quality | Genome-fragment | 1.6 | AAI-based (medium-confidence) | 0 |
| S1_extracted_k141_58502 | 2729 | No | NA | 1 | 0 | 1 | Low-quality | Genome-fragment | 0.24 | AAI-based (medium-confidence) | 0 |
| S16_vs_33793\|\|full | 3208 | No | NA | 3 | 0 | 1 | Not-determined | Genome-fragment | NA | NA | 0 |
| S9_vs_k141_23174\|\|full | 2730 | No | NA | 8 | 1 | 0 | Low-quality | Genome-fragment | 7.93 | AAI-based (high-confidence) | 0 |
| S16_vs_34019\|\|full | 4521 | No | NA | 9 | 0 | 1 | Not-determined | Genome-fragment | NA | NA | 0 |
| S16_vs_34021\|\|full | 81863 | Yes | 31484 | 95 | 14 | 34 | Medium-quality | Genome-fragment | 58.14 | HMM-based (lower-bound) | 61.54 |
| S8_vs_k141_74927\|\|full | 2730 | No | NA | 4 | 0 | 0 | Low-quality | Genome-fragment | 4.63 | AAI-based (high-confidence) | 0 |
| S16_vs_34131\|\|full | 2416 | No | NA | 3 | 0 | 1 | Not-determined | Genome-fragment | NA | NA | 0 |
| S16_vs_34236\|\|full | 3427 | No | NA | 5 | 0 | 0 | Not-determined | Genome-fragment | NA | NA | 0 |
| S16_vs_34241\|\|full | 1893 | No | NA | 3 | 0 | 1 | Not-determined | Genome-fragment | NA | NA | 0 |
| S9_vs_k141_16452\|\|full | 2732 | No | NA | 4 | 1 | 0 | Low-quality | Genome-fragment | 4.44 | HMM-based (lower-bound) | 0 |
| S5_vs_k141_69187\|\|full | 2732 | No | NA | 5 | 0 | 0 | Low-quality | Genome-fragment | 6.08 | AAI-based (medium-confidence) | 0 |
| S16_vs_34621\|\|full | 2976 | No | NA | 4 | 0 | 2 | Not-determined | Genome-fragment | NA | NA | 0 |
| S7_vs_44412\|\|full | 2732 | No | NA | 6 | 0 | 1 | Low-quality | Genome-fragment | 6.53 | AAI-based (medium-confidence) | 0 |
| S16_vs_34751\|\|full | 19964 | No | NA | 33 | 5 | 0 | Medium-quality | Genome-fragment | 51.9 | AAI-based (medium-confidence) | 0 |
| S6_extracted_k141_20542 | 2732 | No | NA | 5 | 3 | 0 | Low-quality | Genome-fragment | 7.56 | AAI-based (high-confidence) | 0 |
| S8_extracted_k141_135867 | 2732 | No | NA | 4 | 0 | 1 | Low-quality | Genome-fragment | 2.07 | AAI-based (medium-confidence) | 0 |
| S11_vs_k141_59547\|\|full | 2733 | No | NA | 3 | 0 | 0 | Low-quality | Genome-fragment | 2.51 | AAI-based (medium-confidence) | 0 |
| S7_extracted_595 | 2733 | No | NA | 1 | 0 | 0 | Low-quality | Genome-fragment | 1.38 | AAI-based (high-confidence) | 0 |
| S16_vs_34885\|\|full | 1238 | No | NA | 3 | 0 | 0 | Not-determined | Genome-fragment | NA | NA | 0 |
| S16_extracted_131857 | 2733 | No | NA | 5 | 0 | 0 | Low-quality | Genome-fragment | 1.59 | AAI-based (high-confidence) | 0 |
| S16_vs_35031\|\|full | 31021 | No | NA | 51 | 14 | 1 | Medium-quality | Genome-fragment | 71.38 | AAI-based (high-confidence) | 0 |
| S16_vs_35145\|\|full | 3302 | No | NA | 5 | 0 | 0 | Not-determined | Genome-fragment | NA | NA | 0 |
| S16_vs_35211\|\|full | 8387 | No | NA | 5 | 0 | 1 | Not-determined | Genome-fragment | NA | NA | 0 |
| S16_vs_97552\|\|full | 2733 | No | NA | 4 | 0 | 1 | Low-quality | Genome-fragment | 3 | AAI-based (medium-confidence) | 0 |
| S16_vs_35320\|\|full | 4418 | No | NA | 7 | 0 | 3 | Not-determined | Genome-fragment | NA | NA | 0 |
| S16_vs_35340\|\|full | 4974 | No | NA | 2 | 0 | 2 | Not-determined | Genome-fragment | NA | NA | 0 |
| S16_vs_35379\|\|full | 1537 | No | NA | 2 | 0 | 1 | Not-determined | Genome-fragment | NA | NA | 0 |
| S14_vs_k141_52288\|\|full | 2734 | No | NA | 4 | 0 | 0 | Low-quality | Genome-fragment | 5.59 | AAI-based (high-confidence) | 0 |
| S16_vs_35709\|\|full | 1147 | No | NA | 3 | 0 | 1 | Not-determined | Genome-fragment | NA | NA | 0 |
| S16_vs_35732\|\|full | 3701 | No | NA | 5 | 0 | 0 | Not-determined | Genome-fragment | NA | NA | 0 |
| S16_vs_35740\|\|full | 2521 | No | NA | 4 | 0 | 2 | Not-determined | Genome-fragment | NA | NA | 0 |
| S3_vs_k141_100535\|\|full | 2734 | No | NA | 2 | 0 | 1 | Low-quality | Genome-fragment | 0.91 | AAI-based (high-confidence) | 0 |
| S16_vs_35889\|\|full | 2236 | No | NA | 2 | 0 | 2 | Not-determined | Genome-fragment | NA | NA | 0 |
| S16_vs_36028\|\|full | 3333 | No | NA | 3 | 0 | 3 | Not-determined | Genome-fragment | NA | NA | 0 |
| S16_vs_36064\|\|full | 2032 | No | NA | 3 | 0 | 0 | Not-determined | Genome-fragment | NA | NA | 0 |
| S16_vs_36092\|\|full | 4777 | No | NA | 5 | 0 | 4 | Not-determined | Genome-fragment | NA | NA | 0 |
| S16_vs_36133\|\|full | 1271 | No | NA | 3 | 0 | 0 | Not-determined | Genome-fragment | NA | NA | 0 |
| S16_vs_36197\|\|full | 2130 | No | NA | 3 | 0 | 3 | Not-determined | Genome-fragment | NA | NA | 0 |
| S5_vs_k141_142559\|\|full | 2734 | No | NA | 4 | 0 | 1 | Low-quality | Genome-fragment | 1.58 | AAI-based (high-confidence) | 0 |
| S16_vs_36287\|\|full | 1850 | No | NA | 4 | 0 | 0 | Not-determined | Genome-fragment | NA | NA | 0 |
| S8_extracted_k141_87089 | 2734 | No | NA | 4 | 0 | 2 | Low-quality | Genome-fragment | 1.68 | AAI-based (medium-confidence) | 0 |
| S16_vs_36294\|\|full | 4761 | No | NA | 5 | 0 | 1 | Not-determined | Genome-fragment | NA | NA | 0 |
| S17_vs_k141_13310\|\|full | 2735 | No | NA | 5 | 0 | 0 | Low-quality | Genome-fragment | 3.38 | AAI-based (high-confidence) | 0 |
| S16_vs_36339\|\|full | 1425 | No | NA | 3 | 0 | 0 | Not-determined | Genome-fragment | NA | NA | 0 |
| S9_extracted_k141_90721 | 2735 | No | NA | 5 | 3 | 0 | Low-quality | Genome-fragment | 6.71 | AAI-based (high-confidence) | 0 |
| S16_vs_36481\|\|full | 1927 | No | NA | 3 | 0 | 1 | Not-determined | Genome-fragment | NA | NA | 0 |
| S16_vs_36500\|\|full | 3849 | No | NA | 3 | 0 | 2 | Not-determined | Genome-fragment | NA | NA | 0 |
| S7_vs_86840\|\|full | 2735 | No | NA | 3 | 3 | 0 | Low-quality | Genome-fragment | 6.09 | AAI-based (high-confidence) | 0 |
| S12_vs_k141_68061\|\|full | 2735 | No | NA | 4 | 3 | 0 | Low-quality | Genome-fragment | 7.22 | AAI-based (high-confidence) | 0 |
| S16_vs_36664\|\|full | 4483 | No | NA | 5 | 0 | 2 | Not-determined | Genome-fragment | NA | NA | 0 |
| S2_vs_k141_293\|\|full | 2735 | No | NA | 4 | 0 | 0 | Low-quality | Genome-fragment | 4.98 | AAI-based (high-confidence) | 0 |
| S9_vs_k141_63863\|\|full | 2736 | No | NA | 5 | 0 | 1 | Low-quality | Genome-fragment | 3.98 | AAI-based (high-confidence) | 0 |
| S16_vs_36799\|\|full | 2881 | No | NA | 3 | 0 | 0 | Not-determined | Genome-fragment | NA | NA | 0 |
| S7_vs_10943\|\|full | 2736 | No | NA | 5 | 1 | 0 | Low-quality | Genome-fragment | 6.51 | AAI-based (high-confidence) | 0 |
| S16_vs_16142\|\|full | 2736 | No | NA | 5 | 0 | 0 | Low-quality | Genome-fragment | 7.63 | AAI-based (high-confidence) | 0 |
| S17_vs_k141_48870\|\|full | 2737 | No | NA | 7 | 0 | 0 | Low-quality | Genome-fragment | 5.21 | AAI-based (high-confidence) | 0 |
| S16_vs_36914\|\|full | 10177 | No | NA | 10 | 0 | 8 | Not-determined | Genome-fragment | NA | NA | 0 |
| S16_vs_36942\|\|full | 3004 | No | NA | 4 | 0 | 1 | Not-determined | Genome-fragment | NA | NA | 0 |
| S16_vs_37159\|\|full | 1114 | No | NA | 3 | 0 | 0 | Not-determined | Genome-fragment | NA | NA | 0 |
| S16_vs_37207\|\|full | 9097 | No | NA | 6 | 0 | 2 | Not-determined | Genome-fragment | NA | NA | 0 |
| S16_vs_37233\|\|full | 3535 | No | NA | 4 | 0 | 3 | Not-determined | Genome-fragment | NA | NA | 0 |
| S14_vs_k141_31048\|\|full | 2737 | No | NA | 4 | 0 | 3 | Low-quality | Genome-fragment | 4.12 | AAI-based (medium-confidence) | 0 |
| S16_vs_37300\|\|full | 16153 | No | NA | 5 | 0 | 2 | Not-determined | Genome-fragment | NA | NA | 0 |
| S16_vs_37307\|\|full | 1902 | No | NA | 5 | 0 | 0 | Not-determined | Genome-fragment | NA | NA | 0 |
| S16_vs_37361\|\|full | 6381 | No | NA | 10 | 0 | 1 | Not-determined | Genome-fragment | NA | NA | 0 |
| S16_vs_37448\|\|full | 6379 | No | NA | 7 | 0 | 0 | Not-determined | Genome-fragment | NA | NA | 0 |
| S1_vs_k141_91841\|\|full | 2737 | No | NA | 5 | 0 | 0 | Low-quality | Genome-fragment | 1.76 | AAI-based (medium-confidence) | 0 |
| S16_vs_37599\|\|full | 1109 | No | NA | 2 | 0 | 1 | Not-determined | Genome-fragment | NA | NA | 0 |
| S3_vs_k141_11708\|\|full | 2737 | No | NA | 4 | 1 | 0 | Low-quality | Genome-fragment | 6 | AAI-based (high-confidence) | 0 |
| S16_vs_37685\|\|full | 1521 | No | NA | 2 | 0 | 1 | Not-determined | Genome-fragment | NA | NA | 0 |
| S10_vs_48809\|\|full | 2737 | No | NA | 4 | 0 | 1 | Low-quality | Genome-fragment | 1.88 | AAI-based (medium-confidence) | 0 |
| S16_vs_37697\|\|full | 1557 | No | NA | 3 | 0 | 2 | Not-determined | Genome-fragment | NA | NA | 0 |
| S16_vs_37766\|\|full | 2696 | No | NA | 2 | 0 | 2 | Not-determined | Genome-fragment | NA | NA | 0 |
| S16_vs_37780\|\|full | 1078 | No | NA | 3 | 0 | 1 | Not-determined | Genome-fragment | NA | NA | 0 |
| S6_vs_k141_30048\|\|full | 2738 | No | NA | 4 | 0 | 0 | Low-quality | Genome-fragment | 5.43 | AAI-based (medium-confidence) | 0 |
| S16_vs_37919\|\|full | 1049 | No | NA | 2 | 0 | 1 | Not-determined | Genome-fragment | NA | NA | 0 |
| S16_vs_38105\|\|full | 1005 | No | NA | 2 | 0 | 0 | Not-determined | Genome-fragment | NA | NA | 0 |
| S8_vs_k141_75504\|\|full | 2738 | No | NA | 4 | 0 | 0 | Low-quality | Genome-fragment | 5.76 | AAI-based (medium-confidence) | 0 |
| S11_vs_k141_87531\|\|full | 2739 | No | NA | 10 | 1 | 0 | Low-quality | Genome-fragment | 5.28 | AAI-based (medium-confidence) | 0 |
| S11_vs_k141_68442\|\|full | 2740 | No | NA | 5 | 3 | 0 | Low-quality | Genome-fragment | 3.72 | AAI-based (high-confidence) | 0 |
| S14_vs_k141_82611\|\|full | 2740 | No | NA | 8 | 1 | 0 | Low-quality | Genome-fragment | 5.89 | AAI-based (high-confidence) | 0 |
| S16_vs_38466\|\|full | 1018 | No | NA | 3 | 0 | 0 | Not-determined | Genome-fragment | NA | NA | 0 |
| S16_vs_38503\|\|full | 1871 | No | NA | 2 | 0 | 1 | Not-determined | Genome-fragment | NA | NA | 0 |
| S11_vs_k141_69962\|\|full | 2741 | No | NA | 2 | 0 | 2 | Low-quality | Genome-fragment | 4.7 | AAI-based (medium-confidence) | 0 |
| S16_vs_38533\|\|full | 3050 | No | NA | 3 | 0 | 1 | Not-determined | Genome-fragment | NA | NA | 0 |
| S16_vs_38536\|\|full | 1393 | No | NA | 2 | 0 | 2 | Not-determined | Genome-fragment | NA | NA | 0 |
| S16_vs_38577\|\|full | 2870 | No | NA | 2 | 0 | 1 | Not-determined | Genome-fragment | NA | NA | 0 |
| S3_vs_k141_51685\|\|full | 2741 | No | NA | 5 | 0 | 0 | Low-quality | Genome-fragment | 6 | AAI-based (medium-confidence) | 0 |
| S5_vs_k141_37623\|\|full | 2741 | No | NA | 3 | 0 | 0 | Low-quality | Genome-fragment | 7.04 | AAI-based (high-confidence) | 0 |
| S16_vs_38711\|\|full | 2211 | No | NA | 4 | 0 | 2 | Not-determined | Genome-fragment | NA | NA | 0 |
| S16_vs_38734\|\|full | 6023 | No | NA | 6 | 0 | 4 | Not-determined | Genome-fragment | NA | NA | 0 |
| S16_vs_39518\|\|full | 2741 | No | NA | 5 | 0 | 0 | Low-quality | Genome-fragment | 3.82 | AAI-based (medium-confidence) | 0 |
| S11_vs_k141_84750\|\|full | 2743 | No | NA | 5 | 0 | 0 | Low-quality | Genome-fragment | 6.38 | AAI-based (medium-confidence) | 0 |
| S16_vs_38863\|\|full | 34794 | No | NA | 50 | 26 | 0 | Medium-quality | Genome-fragment | 68.17 | AAI-based (high-confidence) | 0 |
| S12_extracted_k141_165995 | 2743 | No | NA | 6 | 1 | 0 | Low-quality | Genome-fragment | 5.48 | AAI-based (medium-confidence) | 0 |
| S16_vs_39028\|\|full | 3740 | No | NA | 7 | 0 | 0 | Not-determined | Genome-fragment | NA | NA | 0 |
| S6_vs_k141_47358\|\|full | 2743 | No | NA | 5 | 1 | 0 | Low-quality | Genome-fragment | 7.68 | AAI-based (high-confidence) | 0 |
| S16_vs_39244\|\|full | 4489 | No | NA | 4 | 0 | 0 | Not-determined | Genome-fragment | NA | NA | 0 |
| S16_vs_39300\|\|full | 1365 | No | NA | 2 | 0 | 1 | Not-determined | Genome-fragment | NA | NA | 0 |
| S17_vs_k141_128460\|\|full | 2744 | No | NA | 8 | 0 | 0 | Low-quality | Genome-fragment | 3.99 | AAI-based (high-confidence) | 0 |
| S14_vs_k141_1313\|\|full | 2744 | No | NA | 3 | 0 | 0 | Low-quality | Genome-fragment | 6.07 | AAI-based (medium-confidence) | 0 |
| S16_vs_39419\|\|full | 6370 | No | NA | 5 | 0 | 2 | Not-determined | Genome-fragment | NA | NA | 0 |
| S8_vs_k141_206229\|\|full | 2744 | No | NA | 5 | 0 | 0 | Low-quality | Genome-fragment | 5.47 | AAI-based (high-confidence) | 0 |
| S11_extracted_k141_1942 | 2745 | No | NA | 4 | 1 | 0 | Low-quality | Genome-fragment | 1.73 | HMM-based (lower-bound) | 0 |
| S11_vs_k141_90379\|\|full | 2745 | No | NA | 5 | 4 | 0 | Low-quality | Genome-fragment | 3.82 | AAI-based (high-confidence) | 0 |
| S8_vs_k141_120041\|\|full | 2745 | Yes | 1430 | 3 | 1 | 1 | Low-quality | Genome-fragment | 0.6 | HMM-based (lower-bound) | 47.91 |
| S16_vs_39550\|\|full | 5303 | No | NA | 8 | 0 | 0 | Not-determined | Genome-fragment | NA | NA | 0 |
| S16_vs_39552\|\|full | 1363 | No | NA | 3 | 0 | 0 | Not-determined | Genome-fragment | NA | NA | 0 |
| S14_vs_k141_89537\|\|full | 2747 | No | NA | 3 | 1 | 0 | Low-quality | Genome-fragment | 4.16 | HMM-based (lower-bound) | 0 |
| S10_vs_96631\|\|full | 2747 | No | NA | 4 | 0 | 0 | Low-quality | Genome-fragment | 5.81 | AAI-based (medium-confidence) | 0 |
| S16_vs_39858\|\|full | 1611 | No | NA | 3 | 0 | 1 | Not-determined | Genome-fragment | NA | NA | 0 |
| S16_vs_39865\|\|full | 1172 | No | NA | 3 | 0 | 0 | Not-determined | Genome-fragment | NA | NA | 0 |
| S17_vs_k141_48480\|\|full | 2748 | No | NA | 4 | 0 | 0 | Low-quality | Genome-fragment | 5.16 | AAI-based (medium-confidence) | 0 |
| S17_vs_k141_42848\|\|full | 2748 | No | NA | 4 | 0 | 1 | Low-quality | Genome-fragment | 5.12 | AAI-based (high-confidence) | 0 |
| S16_vs_40137\|\|full | 2730 | No | NA | 3 | 0 | 1 | Not-determined | Genome-fragment | NA | NA | 0 |
| S16_vs_40144\|\|full | 1678 | No | NA | 2 | 0 | 2 | Not-determined | Genome-fragment | NA | NA | 0 |
| S14_vs_k141_96192\|\|full | 2748 | No | NA | 5 | 0 | 0 | Low-quality | Genome-fragment | 6.77 | AAI-based (high-confidence) | 0 |
| S7_vs_36109\|\|full | 2748 | No | NA | 3 | 0 | 1 | Low-quality | Genome-fragment | 3.22 | AAI-based (high-confidence) | 0 |
| S16_vs_40398\|\|full | 1779 | No | NA | 3 | 0 | 2 | Not-determined | Genome-fragment | NA | NA | 0 |
| S16_vs_40469\|\|full | 2450 | No | NA | 2 | 0 | 2 | Not-determined | Genome-fragment | NA | NA | 0 |
| S11_vs_k141_72719\|\|full | 2749 | No | NA | 4 | 0 | 1 | Low-quality | Genome-fragment | 3.85 | AAI-based (high-confidence) | 0 |
| S11_vs_k141_15112\|\|full | 2749 | No | NA | 2 | 0 | 2 | Low-quality | Genome-fragment | 4.18 | AAI-based (medium-confidence) | 0 |
| S16_vs_40791\|\|full | 1184 | No | NA | 3 | 0 | 0 | Not-determined | Genome-fragment | NA | NA | 0 |
| S16_vs_47440\|\|full | 2750 | No | NA | 4 | 0 | 0 | Low-quality | Genome-fragment | 5.76 | AAI-based (medium-confidence) | 0 |
| S8_extracted_k141_66488 | 2750 | No | NA | 6 | 1 | 0 | Low-quality | Genome-fragment | 5.04 | AAI-based (medium-confidence) | 0 |
| S16_vs_41158\|\|full | 1745 | No | NA | 4 | 0 | 1 | Not-determined | Genome-fragment | NA | NA | 0 |
| S8_vs_k141_121492\|\|full | 2750 | No | NA | 6 | 0 | 0 | Low-quality | Genome-fragment | 4.34 | AAI-based (high-confidence) | 0 |
| S16_vs_41166\|\|full | 1025 | No | NA | 2 | 0 | 0 | Not-determined | Genome-fragment | NA | NA | 0 |
| S16_vs_41175\|\|full | 1796 | No | NA | 3 | 0 | 2 | Not-determined | Genome-fragment | NA | NA | 0 |
| S16_vs_41181\|\|full | 1874 | No | NA | 3 | 0 | 2 | Not-determined | Genome-fragment | NA | NA | 0 |
| S16_vs_41195\|\|full | 1688 | No | NA | 2 | 0 | 2 | Not-determined | Genome-fragment | NA | NA | 0 |
| S14_vs_k141_82079\|\|full | 2751 | No | NA | 4 | 2 | 0 | Low-quality | Genome-fragment | 7.37 | AAI-based (high-confidence) | 0 |
| S14_vs_k141_62034\|\|full | 2751 | No | NA | 4 | 0 | 3 | Low-quality | Genome-fragment | 1.16 | AAI-based (medium-confidence) | 0 |
| S16_vs_41428\|\|full | 2871 | No | NA | 3 | 0 | 1 | Not-determined | Genome-fragment | NA | NA | 0 |
| S16_vs_41437\|\|full | 7865 | No | NA | 9 | 0 | 2 | Not-determined | Genome-fragment | NA | NA | 0 |
| S7_vs_124818\|\|full | 2751 | No | NA | 4 | 3 | 0 | Low-quality | Genome-fragment | 7.67 | AAI-based (high-confidence) | 0 |
| S10_vs_17661\|\|full | 2751 | No | NA | 6 | 1 | 0 | Low-quality | Genome-fragment | 6.41 | AAI-based (medium-confidence) | 0 |
| S15_extracted_18695 | 2751 | No | NA | 6 | 0 | 0 | Low-quality | Genome-fragment | 4.92 | AAI-based (medium-confidence) | 0 |
| S16_vs_41810\|\|full | 2549 | No | NA | 5 | 0 | 0 | Not-determined | Genome-fragment | NA | NA | 0 |
| S16_vs_42081\|\|full | 3245 | No | NA | 3 | 0 | 2 | Not-determined | Genome-fragment | NA | NA | 0 |
| S16_vs_42102\|\|full | 1514 | No | NA | 3 | 0 | 0 | Not-determined | Genome-fragment | NA | NA | 0 |
| S9_vs_k141_52332\|\|full | 2752 | No | NA | 4 | 0 | 2 | Low-quality | Genome-fragment | 0.91 | AAI-based (medium-confidence) | 0 |
| S16_vs_42206\|\|full | 1158 | No | NA | 3 | 0 | 0 | Not-determined | Genome-fragment | NA | NA | 0 |
| S7_vs_64529\|\|full | 2752 | No | NA | 5 | 0 | 0 | Low-quality | Genome-fragment | 6.32 | AAI-based (high-confidence) | 0 |
| S16_vs_42228\|\|full | 1783 | No | NA | 2 | 0 | 1 | Not-determined | Genome-fragment | NA | NA | 0 |
| S16_vs_42250\|\|full | 2961 | No | NA | 5 | 0 | 1 | Not-determined | Genome-fragment | NA | NA | 0 |
| S7_vs_103674\|\|full | 2752 | No | NA | 5 | 0 | 0 | Low-quality | Genome-fragment | 6.53 | AAI-based (medium-confidence) | 0 |
| S13_vs_87851\|\|full | 2753 | No | NA | 5 | 1 | 0 | Low-quality | Genome-fragment | 7 | AAI-based (high-confidence) | 0 |
| S8_vs_k141_101183\|\|full | 2753 | No | NA | 8 | 0 | 0 | Low-quality | Genome-fragment | 4.89 | AAI-based (high-confidence) | 0 |
| S5_vs_k141_131374\|\|full | 2754 | No | NA | 11 | 0 | 0 | Low-quality | Genome-fragment | 6.63 | AAI-based (high-confidence) | 0 |
| S16_vs_42677\|\|full | 5925 | No | NA | 3 | 0 | 3 | Not-determined | Genome-fragment | NA | NA | 0 |
| S16_vs_42695\|\|full | 1274 | No | NA | 2 | 0 | 1 | Not-determined | Genome-fragment | NA | NA | 0 |
| S16_vs_42769\|\|full | 3819 | No | NA | 3 | 0 | 3 | Not-determined | Genome-fragment | NA | NA | 0 |
| S10_vs_3778\|\|full | 2754 | No | NA | 3 | 0 | 0 | Low-quality | Genome-fragment | 3.43 | AAI-based (high-confidence) | 0 |
| S16_vs_42990\|\|full | 1500 | No | NA | 2 | 0 | 2 | Not-determined | Genome-fragment | NA | NA | 0 |
| S11_vs_k141_78975\|\|full | 2756 | No | NA | 8 | 2 | 0 | Low-quality | Genome-fragment | 5.91 | AAI-based (high-confidence) | 0 |
| S16_vs_43055\|\|full | 2144 | No | NA | 2 | 0 | 1 | Not-determined | Genome-fragment | NA | NA | 0 |
| S7_vs_64603\|\|full | 2757 | No | NA | 4 | 0 | 0 | Low-quality | Genome-fragment | 2.01 | AAI-based (high-confidence) | 0 |
| S16_vs_43241\|\|full | 1729 | No | NA | 3 | 0 | 1 | Not-determined | Genome-fragment | NA | NA | 0 |
| S10_extracted_15314 | 2757 | No | NA | 6 | 0 | 0 | Low-quality | Genome-fragment | 9.56 | AAI-based (high-confidence) | 0 |
| S16_vs_43305\|\|full | 3765 | No | NA | 4 | 0 | 0 | Not-determined | Genome-fragment | NA | NA | 0 |
| S9_vs_k141_24381\|\|lt2gene | 2758 | No | NA | 2 | 2 | 0 | Low-quality | Genome-fragment | 9.05 | AAI-based (high-confidence) | 0 |
| S16_vs_43472\|\|full | 2765 | No | NA | 3 | 0 | 3 | Not-determined | Genome-fragment | NA | NA | 0 |
| S16_vs_43599\|\|full | 1526 | No | NA | 2 | 0 | 2 | Not-determined | Genome-fragment | NA | NA | 0 |
| S16_vs_43638\|\|full | 1758 | No | NA | 2 | 0 | 0 | Not-determined | Genome-fragment | NA | NA | 0 |
| S16_vs_43651\|\|full | 4623 | No | NA | 3 | 0 | 2 | Not-determined | Genome-fragment | NA | NA | 0 |
| S16_vs_43673\|\|full | 2746 | No | NA | 7 | 0 | 1 | Not-determined | Genome-fragment | NA | NA | 0 |
| S16_vs_43689\|\|full | 1006 | No | NA | 3 | 0 | 0 | Not-determined | Genome-fragment | NA | NA | 0 |
| S16_vs_43728\|\|full | 2847 | No | NA | 4 | 0 | 0 | Not-determined | Genome-fragment | NA | NA | 0 |
| S6_vs_k141_35972\|\|full | 2758 | No | NA | 3 | 0 | 2 | Low-quality | Genome-fragment | 4.2 | AAI-based (medium-confidence) | 0 |
| S3_vs_k141_101834\|\|full | 2759 | No | NA | 6 | 2 | 0 | Low-quality | Genome-fragment | 6.31 | AAI-based (medium-confidence) | 0 |
| S16_vs_43925\|\|full | 7419 | No | NA | 6 | 0 | 2 | Not-determined | Genome-fragment | NA | NA | 0 |
| S10_vs_3974\|\|full | 2759 | No | NA | 7 | 0 | 0 | Low-quality | Genome-fragment | 3.15 | AAI-based (medium-confidence) | 0 |
| S16_vs_44064\|\|full | 2013 | No | NA | 3 | 0 | 2 | Not-determined | Genome-fragment | NA | NA | 0 |
| S17_vs_k141_7730\|\|full | 2760 | No | NA | 5 | 0 | 2 | Low-quality | Genome-fragment | 2.94 | AAI-based (medium-confidence) | 0 |
| S11_vs_k141_85799\|\|full | 2760 | No | NA | 6 | 2 | 0 | Low-quality | Genome-fragment | 5.49 | AAI-based (high-confidence) | 0 |
| S14_vs_k141_41522\|\|full | 2760 | No | NA | 8 | 1 | 0 | Low-quality | Genome-fragment | 3.28 | AAI-based (high-confidence) | 0 |
| S10_vs_60135\|\|full | 2760 | Yes | 1757 | 6 | 1 | 1 | Low-quality | Genome-fragment | 2.76 | AAI-based (high-confidence) | 36.34 |
| S16_vs_44330\|\|full | 3637 | No | NA | 2 | 0 | 2 | Not-determined | Genome-fragment | NA | NA | 0 |
| S12_vs_k141_23392\|\|full | 2760 | No | NA | 4 | 1 | 0 | Low-quality | Genome-fragment | 6.86 | AAI-based (high-confidence) | 0 |
| S16_vs_44483\|\|full | 2151 | No | NA | 4 | 0 | 1 | Not-determined | Genome-fragment | NA | NA | 0 |
| S16_vs_44514\|\|full | 1684 | No | NA | 2 | 0 | 0 | Not-determined | Genome-fragment | NA | NA | 0 |
| S5_extracted_k141_109918 | 2761 | No | NA | 9 | 1 | 0 | Low-quality | Genome-fragment | 1.29 | HMM-based (lower-bound) | 0 |
| S16_vs_44687\|\|full | 3385 | No | NA | 3 | 0 | 0 | Not-determined | Genome-fragment | NA | NA | 0 |
| S5_vs_k141_67742\|\|full | 2761 | No | NA | 4 | 1 | 0 | Low-quality | Genome-fragment | 5.76 | AAI-based (high-confidence) | 0 |
| S4_extracted_2544 | 2761 | No | NA | 6 | 5 | 0 | Low-quality | Genome-fragment | 9.11 | AAI-based (high-confidence) | 0 |
| S16_vs_44868\|\|full | 1141 | No | NA | 3 | 0 | 1 | Not-determined | Genome-fragment | NA | NA | 0 |
| S8_vs_k141_19242\|\|full | 2761 | No | NA | 2 | 0 | 1 | Low-quality | Genome-fragment | 1.31 | AAI-based (medium-confidence) | 0 |
| S8_vs_k141_103423\|\|full | 2761 | No | NA | 4 | 0 | 2 | Low-quality | Genome-fragment | 3.75 | AAI-based (medium-confidence) | 0 |
| S5_vs_k141_117184\|\|full | 2762 | No | NA | 2 | 0 | 1 | Low-quality | Genome-fragment | 0.86 | AAI-based (medium-confidence) | 0 |
| S8_extracted_k141_24265 | 2762 | No | NA | 5 | 1 | 0 | Low-quality | Genome-fragment | 1.58 | AAI-based (high-confidence) | 0 |
| S16_vs_45098\|\|full | 3147 | No | NA | 4 | 0 | 0 | Not-determined | Genome-fragment | NA | NA | 0 |
| S9_vs_k141_84739\|\|full | 2763 | No | NA | 6 | 0 | 0 | Low-quality | Genome-fragment | 6.86 | AAI-based (high-confidence) | 0 |
| S13_extracted_61392 | 2763 | No | NA | 9 | 0 | 0 | Low-quality | Genome-fragment | 1.76 | AAI-based (high-confidence) | 0 |
| S14_vs_k141_99430\|\|full | 2765 | No | NA | 6 | 0 | 1 | Low-quality | Genome-fragment | 7.25 | AAI-based (medium-confidence) | 0 |
| S16_vs_45403\|\|full | 3012 | No | NA | 7 | 0 | 0 | Not-determined | Genome-fragment | NA | NA | 0 |
| S16_vs_45443\|\|full | 4293 | No | NA | 6 | 0 | 0 | Not-determined | Genome-fragment | NA | NA | 0 |
| S16_vs_45493\|\|full | 1593 | No | NA | 2 | 0 | 0 | Not-determined | Genome-fragment | NA | NA | 0 |
| S16_vs_45499\|\|full | 3944 | No | NA | 3 | 0 | 3 | Not-determined | Genome-fragment | NA | NA | 0 |
| S3_vs_k141_53813\|\|full | 2765 | No | NA | 5 | 2 | 0 | Low-quality | Genome-fragment | 3.54 | HMM-based (lower-bound) | 0 |
| S16_vs_45684\|\|full | 1237 | No | NA | 2 | 0 | 1 | Not-determined | Genome-fragment | NA | NA | 0 |
| S16_vs_45685\|\|full | 1689 | No | NA | 3 | 0 | 3 | Not-determined | Genome-fragment | NA | NA | 0 |
| S4_extracted_9428 | 2765 | No | NA | 3 | 3 | 0 | Low-quality | Genome-fragment | 9.12 | AAI-based (high-confidence) | 0 |
| S1_vs_k141_66658\|\|full | 2766 | No | NA | 4 | 0 | 3 | Low-quality | Genome-fragment | 6.63 | AAI-based (medium-confidence) | 0 |
| S9_vs_k141_73775\|\|full | 2767 | No | NA | 2 | 2 | 0 | Low-quality | Genome-fragment | 6.52 | AAI-based (high-confidence) | 0 |
| S11_vs_k141_100885\|\|full | 2767 | No | NA | 4 | 1 | 0 | Low-quality | Genome-fragment | 4.6 | AAI-based (high-confidence) | 0 |
| S9_vs_k141_75287\|\|full | 2768 | No | NA | 4 | 0 | 2 | Low-quality | Genome-fragment | 4.99 | AAI-based (medium-confidence) | 0 |
| S16_vs_46202\|\|full | 2170 | No | NA | 3 | 0 | 0 | Not-determined | Genome-fragment | NA | NA | 0 |
| S16_vs_46303\|\|full | 4430 | No | NA | 4 | 0 | 3 | Not-determined | Genome-fragment | NA | NA | 0 |
| S10_extracted_8611 | 2768 | No | NA | 7 | 3 | 0 | Low-quality | Genome-fragment | 4.5 | AAI-based (high-confidence) | 0 |
| S7_vs_108433\|\|full | 2769 | No | NA | 5 | 0 | 1 | Low-quality | Genome-fragment | 5.53 | AAI-based (medium-confidence) | 0 |
| S16_vs_46409\|\|full | 2086 | No | NA | 6 | 0 | 0 | Not-determined | Genome-fragment | NA | NA | 0 |
| S3_vs_k141_91903\|\|full | 2770 | No | NA | 3 | 3 | 0 | Low-quality | Genome-fragment | 4.56 | HMM-based (lower-bound) | 0 |
| S16_vs_12536\|\|full | 2770 | No | NA | 3 | 0 | 0 | Low-quality | Genome-fragment | 1.44 | AAI-based (high-confidence) | 0 |
| S16_vs_46704\|\|full | 1976 | No | NA | 2 | 0 | 2 | Not-determined | Genome-fragment | NA | NA | 0 |
| S16_vs_46825\|\|full | 3047 | No | NA | 4 | 0 | 0 | Not-determined | Genome-fragment | NA | NA | 0 |
| S16_vs_46870\|\|full | 2531 | No | NA | 3 | 0 | 1 | Not-determined | Genome-fragment | NA | NA | 0 |
| S16_vs_47053\|\|full | 3447 | No | NA | 6 | 0 | 0 | Not-determined | Genome-fragment | NA | NA | 0 |
| S16_vs_47117\|\|full | 1784 | No | NA | 2 | 0 | 0 | Not-determined | Genome-fragment | NA | NA | 0 |
| S16_vs_47250\|\|full | 1851 | No | NA | 3 | 0 | 1 | Not-determined | Genome-fragment | NA | NA | 0 |
| S16_vs_47287\|\|full | 1392 | No | NA | 3 | 0 | 2 | Not-determined | Genome-fragment | NA | NA | 0 |
| S16_vs_47332\|\|full | 3458 | No | NA | 3 | 0 | 3 | Not-determined | Genome-fragment | NA | NA | 0 |
| S15_extracted_22393 | 2770 | No | NA | 5 | 2 | 0 | Low-quality | Genome-fragment | 4.51 | AAI-based (high-confidence) | 0 |
| S16_vs_47549\|\|full | 6855 | No | NA | 7 | 0 | 5 | Not-determined | Genome-fragment | NA | NA | 0 |
| S16_vs_47622\|\|full | 2701 | No | NA | 3 | 0 | 1 | Not-determined | Genome-fragment | NA | NA | 0 |
| S17_vs_k141_155465\|\|full | 2771 | No | NA | 4 | 0 | 0 | Low-quality | Genome-fragment | 6.1 | AAI-based (high-confidence) | 0 |
| S16_extracted_167537 | 2771 | No | NA | 5 | 2 | 0 | Low-quality | Genome-fragment | 5.06 | HMM-based (lower-bound) | 0 |
| S16_vs_47712\|\|full | 1586 | No | NA | 3 | 0 | 2 | Not-determined | Genome-fragment | NA | NA | 0 |
| S1_extracted_k141_85965 | 2772 | No | NA | 6 | 5 | 0 | Low-quality | Genome-fragment | 6.41 | AAI-based (high-confidence) | 0 |
| S1_vs_k141_16965\|\|full | 2772 | No | NA | 9 | 0 | 0 | Low-quality | Genome-fragment | 5.66 | AAI-based (high-confidence) | 0 |
| S16_vs_47795\|\|full | 2963 | No | NA | 4 | 0 | 2 | Not-determined | Genome-fragment | NA | NA | 0 |
| S6_vs_k141_13938\|\|lt2gene | 2772 | No | NA | 2 | 0 | 0 | Low-quality | Genome-fragment | 3.42 | AAI-based (high-confidence) | 0 |
| S16_vs_47926\|\|full | 1125 | No | NA | 2 | 0 | 0 | Not-determined | Genome-fragment | NA | NA | 0 |
| S11_vs_k141_65858\|\|full | 2773 | No | NA | 7 | 3 | 0 | Low-quality | Genome-fragment | 3.93 | AAI-based (high-confidence) | 0 |
| S16_vs_48070\|\|full | 3503 | No | NA | 6 | 0 | 4 | Not-determined | Genome-fragment | NA | NA | 0 |
| S16_vs_48075\|\|full | 4490 | No | NA | 5 | 0 | 3 | Not-determined | Genome-fragment | NA | NA | 0 |
| S7_vs_16863\|\|full | 2773 | No | NA | 5 | 1 | 0 | Low-quality | Genome-fragment | 7.35 | AAI-based (high-confidence) | 0 |
| S16_vs_48169\|\|full | 5202 | No | NA | 4 | 0 | 0 | Not-determined | Genome-fragment | NA | NA | 0 |
| S8_vs_k141_70063\|\|full | 2773 | No | NA | 3 | 0 | 2 | Low-quality | Genome-fragment | 0.75 | AAI-based (medium-confidence) | 0 |
| S13_extracted_90346 | 2774 | No | NA | 6 | 4 | 0 | Low-quality | Genome-fragment | 4.56 | AAI-based (high-confidence) | 0 |
| S16_vs_48425\|\|full | 7147 | No | NA | 2 | 0 | 2 | Not-determined | Genome-fragment | NA | NA | 0 |
| S16_vs_94259\|\|full | 2774 | No | NA | 5 | 0 | 0 | Low-quality | Genome-fragment | 5.91 | AAI-based (medium-confidence) | 0 |
| S16_vs_48530\|\|full | 2673 | No | NA | 4 | 0 | 1 | Not-determined | Genome-fragment | NA | NA | 0 |
| S16_vs_48559\|\|full | 2958 | No | NA | 3 | 0 | 1 | Not-determined | Genome-fragment | NA | NA | 0 |
| S16_vs_112562\|\|full | 2774 | No | NA | 3 | 0 | 0 | Low-quality | Genome-fragment | 1.84 | AAI-based (medium-confidence) | 0 |
| S12_vs_k141_71090\|\|full | 2774 | No | NA | 4 | 0 | 3 | Low-quality | Genome-fragment | 1.27 | AAI-based (medium-confidence) | 0 |
| S17_vs_k141_140415\|\|full | 2775 | No | NA | 4 | 0 | 1 | Low-quality | Genome-fragment | 7.61 | AAI-based (medium-confidence) | 0 |
| S16_vs_48637\|\|full | 1349 | No | NA | 3 | 0 | 2 | Not-determined | Genome-fragment | NA | NA | 0 |
| S16_extracted_31326 | 2775 | No | NA | 7 | 0 | 0 | Low-quality | Genome-fragment | 2.88 | AAI-based (medium-confidence) | 0 |
| S16_vs_48714\|\|full | 23424 | No | NA | 29 | 0 | 5 | Medium-quality | Genome-fragment | 62.31 | AAI-based (high-confidence) | 0 |
| S2_vs_k141_5858\|\|full | 2775 | No | NA | 6 | 4 | 0 | Low-quality | Genome-fragment | 4.6 | AAI-based (high-confidence) | 0 |
| S8_extracted_k141_148861 | 2775 | No | NA | 9 | 1 | 0 | Low-quality | Genome-fragment | 8.6 | AAI-based (high-confidence) | 0 |
| S16_vs_49139\|\|full | 1314 | No | NA | 3 | 0 | 1 | Not-determined | Genome-fragment | NA | NA | 0 |
| S16_vs_49142\|\|full | 11726 | No | NA | 11 | 0 | 9 | Not-determined | Genome-fragment | NA | NA | 0 |
| S1_vs_k141_120654\|\|full | 2776 | No | NA | 2 | 1 | 0 | Low-quality | Genome-fragment | 3.93 | HMM-based (lower-bound) | 0 |
| S1_vs_k141_143634\|\|full | 2776 | No | NA | 6 | 0 | 0 | Low-quality | Genome-fragment | 6.48 | AAI-based (high-confidence) | 0 |
| S16_vs_49349\|\|full | 2404 | No | NA | 3 | 0 | 2 | Not-determined | Genome-fragment | NA | NA | 0 |
| S1_vs_k141_151567\|\|full | 2776 | No | NA | 3 | 2 | 0 | Low-quality | Genome-fragment | 7.93 | AAI-based (high-confidence) | 0 |
| S16_vs_49395\|\|full | 2530 | No | NA | 4 | 0 | 2 | Not-determined | Genome-fragment | NA | NA | 0 |
| S8_vs_k141_58181\|\|full | 2776 | No | NA | 5 | 0 | 1 | Low-quality | Genome-fragment | 5.61 | AAI-based (medium-confidence) | 0 |
| S12_vs_k141_95488\|\|full | 2778 | No | NA | 3 | 0 | 1 | Low-quality | Genome-fragment | 2.41 | AAI-based (high-confidence) | 0 |
| S11_extracted_k141_71769 | 2779 | No | NA | 6 | 0 | 0 | Low-quality | Genome-fragment | 6.36 | AAI-based (high-confidence) | 0 |
| S16_vs_49848\|\|full | 9531 | No | NA | 11 | 0 | 2 | Not-determined | Genome-fragment | NA | NA | 0 |

| | | | | | | | | | | | |
|---|---|---|---|---|---|---|---|---|---|---|---|
| S16_vs_49862\|\|full | 3208 | No | NA | 3 | 0 | 1 | Not-determined | Genome-fragment | NA | NA | 0 |
| S7_vs_130060\|\|full | 2779 | No | NA | 3 | 0 | 2 | Low-quality | Genome-fragment | 2.27 | AAI-based (high-confidence) | 0 |
| S17_vs_k141_148712\|\|full | 2780 | No | NA | 5 | 0 | 2 | Low-quality | Genome-fragment | 1.43 | AAI-based (medium-confidence) | 0 |
| S11_vs_k141_47609\|\|full | 2780 | No | NA | 3 | 2 | 0 | Low-quality | Genome-fragment | 2.78 | AAI-based (high-confidence) | 0 |
| S16_vs_50063\|\|full | 1815 | No | NA | 3 | 0 | 2 | Not-determined | Genome-fragment | NA | NA | 0 |
| S13_vs_113324\|\|full | 2780 | No | NA | 4 | 0 | 1 | Low-quality | Genome-fragment | 1.91 | AAI-based (high-confidence) | 0 |
| S7_vs_84898\|\|full | 2780 | No | NA | 3 | 3 | 0 | Low-quality | Genome-fragment | 6.81 | AAI-based (high-confidence) | 0 |
| S8_vs_k141_140518\|\|full | 2780 | No | NA | 2 | 0 | 2 | Low-quality | Genome-fragment | 0.86 | AAI-based (medium-confidence) | 0 |
| S17_vs_k141_92785\|\|full | 2781 | No | NA | 3 | 0 | 3 | Low-quality | Genome-fragment | 1.64 | AAI-based (medium-confidence) | 0 |
| S16_vs_50358\|\|full | 5068 | No | NA | 6 | 0 | 2 | Not-determined | Genome-fragment | NA | NA | 0 |
| S16_vs_50368\|\|full | 3453 | No | NA | 5 | 0 | 2 | Not-determined | Genome-fragment | NA | NA | 0 |
| S16_vs_50372\|\|full | 1607 | No | NA | 2 | 0 | 2 | Not-determined | Genome-fragment | NA | NA | 0 |
| S11_vs_k141_38554\|\|full | 2781 | No | NA | 5 | 0 | 1 | Low-quality | Genome-fragment | 4.64 | AAI-based (medium-confidence) | 0 |
| S16_vs_50434\|\|full | 3192 | No | NA | 2 | 0 | 2 | Not-determined | Genome-fragment | NA | NA | 0 |
| S16_vs_50569\|\|full | 3208 | No | NA | 3 | 0 | 3 | Not-determined | Genome-fragment | NA | NA | 0 |
| S14_extracted_k141_67830 | 2781 | No | NA | 7 | 6 | 0 | Low-quality | Genome-fragment | 9.11 | AAI-based (high-confidence) | 0 |
| S16_vs_50594\|\|full | 4685 | No | NA | 4 | 0 | 3 | Not-determined | Genome-fragment | NA | NA | 0 |
| S16_vs_50666\|\|full | 8928 | No | NA | 8 | 0 | 6 | Not-determined | Genome-fragment | NA | NA | 0 |
| S16_vs_50739\|\|full | 6575 | No | NA | 6 | 0 | 0 | Not-determined | Genome-fragment | NA | NA | 0 |
| S16_vs_50748\|\|full | 1818 | No | NA | 2 | 0 | 0 | Not-determined | Genome-fragment | NA | NA | 0 |
| S14_vs_k141_1651\|\|full | 2781 | No | NA | 3 | 2 | 0 | Low-quality | Genome-fragment | 6.66 | AAI-based (medium-confidence) | 0 |
| S16_vs_51005\|\|full | 1736 | No | NA | 3 | 0 | 0 | Not-determined | Genome-fragment | NA | NA | 0 |
| S16_vs_51025\|\|full | 7053 | No | NA | 5 | 0 | 2 | Not-determined | Genome-fragment | NA | NA | 0 |
| S16_vs_51264\|\|full | 4959 | No | NA | 5 | 0 | 3 | Not-determined | Genome-fragment | NA | NA | 0 |
| S3_vs_k141_126546\|\|full | 2781 | No | NA | 7 | 1 | 0 | Low-quality | Genome-fragment | 4.86 | AAI-based (high-confidence) | 0 |
| S15_vs_61677\|\|full | 2781 | No | NA | 6 | 0 | 0 | Low-quality | Genome-fragment | 6.56 | AAI-based (medium-confidence) | 0 |
| S16_vs_51474\|\|full | 3598 | No | NA | 5 | 0 | 3 | Not-determined | Genome-fragment | NA | NA | 0 |
| S6_vs_k141_11813\|\|full | 2781 | No | NA | 4 | 1 | 0 | Low-quality | Genome-fragment | 44.84 | AAI-based (high-confidence) | 0 |
| S16_vs_51512\|\|full | 4306 | No | NA | 3 | 0 | 2 | Not-determined | Genome-fragment | NA | NA | 0 |
| S8_extracted_k141_135685 | 2781 | No | NA | 2 | 1 | 0 | Low-quality | Genome-fragment | 4.65 | AAI-based (high-confidence) | 0 |
| S12_vs_k141_130138\|\|full | 2782 | No | NA | 4 | 1 | 0 | Low-quality | Genome-fragment | 6.33 | AAI-based (medium-confidence) | 0 |
| S16_vs_51713\|\|full | 1057 | No | NA | 2 | 0 | 2 | Not-determined | Genome-fragment | NA | NA | 0 |
| S16_vs_51727\|\|full | 2107 | No | NA | 5 | 0 | 4 | Not-determined | Genome-fragment | NA | NA | 0 |
| S16_vs_51771\|\|full | 2031 | No | NA | 2 | 0 | 1 | Not-determined | Genome-fragment | NA | NA | 0 |
| S5_vs_k141_2412\|\|full | 2783 | No | NA | 4 | 0 | 2 | Low-quality | Genome-fragment | 0.91 | AAI-based (medium-confidence) | 0 |
| S16_vs_51901\|\|full | 37473 | No | NA | 64 | 7 | 0 | High-quality | High-quality | 100 | AAI-based (high-confidence) | 0 |
| S5_vs_k141_88018\|\|full | 2783 | No | NA | 6 | 0 | 0 | Low-quality | Genome-fragment | 5.38 | AAI-based (medium-confidence) | 0 |
| S16_vs_52012\|\|full | 2001 | No | NA | 3 | 0 | 1 | Not-determined | Genome-fragment | NA | NA | 0 |
| S9_extracted_k141_81798 | 2784 | No | NA | 5 | 4 | 0 | Low-quality | Genome-fragment | 6.32 | AAI-based (high-confidence) | 0 |
| S16_vs_52045\|\|full | 2774 | No | NA | 4 | 0 | 0 | Not-determined | Genome-fragment | NA | NA | 0 |
| S16_vs_52090\|\|full | 1905 | No | NA | 3 | 0 | 0 | Not-determined | Genome-fragment | NA | NA | 0 |
| S16_vs_52177\|\|full | 1461 | No | NA | 2 | 0 | 0 | Not-determined | Genome-fragment | NA | NA | 0 |
| S11_extracted_k141_36302 | 2784 | No | NA | 2 | 2 | 0 | Low-quality | Genome-fragment | 5.54 | AAI-based (high-confidence) | 0 |
| S16_vs_52209\|\|full | 7250 | No | NA | 10 | 0 | 6 | Not-determined | Genome-fragment | NA | NA | 0 |
| S16_vs_52214\|\|full | 2170 | No | NA | 3 | 0 | 2 | Not-determined | Genome-fragment | NA | NA | 0 |
| S16_vs_52237\|\|full | 2452 | No | NA | 3 | 0 | 3 | Not-determined | Genome-fragment | NA | NA | 0 |
| S16_vs_52278\|\|full | 1581 | No | NA | 2 | 0 | 2 | Not-determined | Genome-fragment | NA | NA | 0 |
| S11_extracted_k141_51864 | 2784 | No | NA | 2 | 2 | 0 | Low-quality | Genome-fragment | 1.06 | HMM-based (lower-bound) | 0 |
| S16_vs_52457\|\|full | 2362 | No | NA | 3 | 0 | 1 | Not-determined | Genome-fragment | NA | NA | 0 |
| S16_vs_52479\|\|full | 2333 | No | NA | 6 | 0 | 0 | Not-determined | Genome-fragment | NA | NA | 0 |
| S16_vs_52567\|\|full | 3270 | No | NA | 4 | 0 | 1 | Not-determined | Genome-fragment | NA | NA | 0 |
| S16_vs_52604\|\|full | 1521 | No | NA | 2 | 0 | 0 | Not-determined | Genome-fragment | NA | NA | 0 |
| S14_vs_k141_103026\|\|full | 2784 | No | NA | 2 | 1 | 0 | Low-quality | Genome-fragment | 6.8 | AAI-based (medium-confidence) | 0 |
| S14_vs_k141_78370\|\|full | 2785 | No | NA | 2 | 0 | 1 | Low-quality | Genome-fragment | 0.88 | AAI-based (high-confidence) | 0 |
| S14_vs_k141_21588\|\|full | 2785 | No | NA | 3 | 0 | 0 | Low-quality | Genome-fragment | 2.2 | AAI-based (high-confidence) | 0 |
| S7_vs_65649\|\|full | 2785 | No | NA | 5 | 3 | 0 | Low-quality | Genome-fragment | 6.9 | AAI-based (medium-confidence) | 0 |
| S16_vs_52865\|\|full | 2358 | No | NA | 3 | 0 | 2 | Not-determined | Genome-fragment | NA | NA | 0 |
| S16_vs_52914\|\|full | 1563 | No | NA | 4 | 0 | 0 | Not-determined | Genome-fragment | NA | NA | 0 |
| S16_vs_52917\|\|full | 2902 | No | NA | 6 | 0 | 0 | Not-determined | Genome-fragment | NA | NA | 0 |
| S10_vs_43707\|\|full | 2785 | No | NA | 3 | 0 | 3 | Low-quality | Genome-fragment | 4.61 | AAI-based (medium-confidence) | 0 |
| S16_vs_52980\|\|full | 1855 | No | NA | 4 | 0 | 0 | Not-determined | Genome-fragment | NA | NA | 0 |
| S11_vs_k141_81870\|\|full | 2786 | No | NA | 5 | 0 | 0 | Low-quality | Genome-fragment | 4.9 | AAI-based (high-confidence) | 0 |
| S16_vs_53061\|\|full | 1309 | No | NA | 2 | 0 | 1 | Not-determined | Genome-fragment | NA | NA | 0 |
| S16_vs_53109\|\|full | 3245 | No | NA | 4 | 0 | 0 | Not-determined | Genome-fragment | NA | NA | 0 |
| S16_vs_53174\|\|full | 1035 | No | NA | 3 | 0 | 0 | Not-determined | Genome-fragment | NA | NA | 0 |
| S7_extracted_123515 | 2786 | No | NA | 6 | 5 | 0 | Low-quality | Genome-fragment | 7.66 | AAI-based (high-confidence) | 0 |
| S7_vs_98637\|\|full | 2786 | No | NA | 3 | 0 | 3 | Low-quality | Genome-fragment | 1.27 | AAI-based (medium-confidence) | 0 |
| S16_vs_53210\|\|full | 3644 | No | NA | 4 | 0 | 1 | Not-determined | Genome-fragment | NA | NA | 0 |
| S16_vs_140885\|\|full | 2786 | Yes | 531 | 3 | 1 | 1 | Low-quality | Genome-fragment | 1.24 | HMM-based (lower-bound) | 80.94 |
| S12_vs_k141_110978\|\|full | 2786 | No | NA | 3 | 0 | 2 | Low-quality | Genome-fragment | 1.35 | AAI-based (medium-confidence) | 0 |
| S16_vs_53300\|\|full | 1781 | No | NA | 2 | 0 | 1 | Not-determined | Genome-fragment | NA | NA | 0 |
| S1_extracted_k141_124692 | 2787 | No | NA | 5 | 3 | 0 | Low-quality | Genome-fragment | 7.12 | AAI-based (high-confidence) | 0 |
| S16_vs_53340\|\|full | 1274 | No | NA | 2 | 0 | 0 | Not-determined | Genome-fragment | NA | NA | 0 |
| S16_vs_53452\|\|full | 3229 | No | NA | 5 | 0 | 0 | Not-determined | Genome-fragment | NA | NA | 0 |
| S16_vs_53483\|\|full | 5552 | No | NA | 4 | 0 | 4 | Not-determined | Genome-fragment | NA | NA | 0 |
| S8_vs_k141_31098\|\|full | 2787 | No | NA | 3 | 0 | 0 | Low-quality | Genome-fragment | 6.25 | AAI-based (high-confidence) | 0 |
| S16_vs_53623\|\|full | 4706 | No | NA | 4 | 0 | 2 | Not-determined | Genome-fragment | NA | NA | 0 |
| S8_vs_k141_13523\|\|full | 2787 | No | NA | 3 | 0 | 2 | Low-quality | Genome-fragment | 2.36 | AAI-based (medium-confidence) | 0 |
| S16_vs_53927\|\|full | 4965 | No | NA | 10 | 0 | 4 | Not-determined | Genome-fragment | NA | NA | 0 |
| S16_vs_53938\|\|full | 1139 | No | NA | 2 | 0 | 0 | Not-determined | Genome-fragment | NA | NA | 0 |
| S11_vs_k141_26400\|\|full | 2788 | No | NA | 6 | 0 | 0 | Low-quality | Genome-fragment | 3 | AAI-based (high-confidence) | 0 |
| S10_extracted_2820 | 2788 | No | NA | 6 | 0 | 2 | Low-quality | Genome-fragment | 9.08 | AAI-based (medium-confidence) | 0 |
| S17_vs_k141_19837\|\|full | 2789 | No | NA | 2 | 0 | 1 | Low-quality | Genome-fragment | 7.47 | AAI-based (medium-confidence) | 0 |
| S9_extracted_k141_4655 | 2790 | No | NA | 7 | 6 | 0 | Low-quality | Genome-fragment | 9.11 | AAI-based (high-confidence) | 0 |
| S16_vs_54250\|\|full | 23594 | No | NA | 47 | 17 | 0 | Medium-quality | Genome-fragment | 62.46 | AAI-based (high-confidence) | 0 |
| S9_vs_k141_76314\|\|full | 2790 | No | NA | 6 | 0 | 0 | Low-quality | Genome-fragment | 7.37 | AAI-based (high-confidence) | 0 |
| S16_vs_54378\|\|full | 1628 | No | NA | 2 | 0 | 1 | Not-determined | Genome-fragment | NA | NA | 0 |
| S13_vs_31514\|\|full | 2790 | No | NA | 3 | 2 | 0 | Low-quality | Genome-fragment | 4.92 | HMM-based (lower-bound) | 0 |
| S8_vs_k141_189410\|\|full | 2790 | No | NA | 5 | 0 | 0 | Low-quality | Genome-fragment | 5.12 | AAI-based (high-confidence) | 0 |
| S3_vs_k141_58143\|\|full | 2791 | No | NA | 5 | 0 | 0 | Low-quality | Genome-fragment | 3.36 | AAI-based (high-confidence) | 0 |
| S16_vs_54482\|\|full | 4098 | No | NA | 3 | 0 | 1 | Not-determined | Genome-fragment | NA | NA | 0 |
| S7_vs_42287\|\|full | 2791 | No | NA | 7 | 1 | 0 | Low-quality | Genome-fragment | 7.45 | AAI-based (high-confidence) | 0 |
| S16_vs_54616\|\|full | 17792 | No | NA | 20 | 17 | 0 | Medium-quality | Genome-fragment | 52.97 | AAI-based (medium-confidence) | 0 |
| S16_vs_54669\|\|full | 2749 | No | NA | 5 | 0 | 4 | Not-determined | Genome-fragment | NA | NA | 0 |
| S16_vs_64530\|\|full | 2791 | No | NA | 2 | 0 | 0 | Low-quality | Genome-fragment | 1.75 | AAI-based (high-confidence) | 0 |
| S12_vs_k141_47251\|\|full | 2791 | No | NA | 4 | 0 | 1 | Low-quality | Genome-fragment | 1.48 | AAI-based (medium-confidence) | 0 |
| S12_extracted_k141_58675 | 2792 | No | NA | 11 | 1 | 0 | Low-quality | Genome-fragment | 4.66 | AAI-based (high-confidence) | 0 |
| S3_vs_k141_123770\|\|full | 2794 | No | NA | 5 | 0 | 1 | Low-quality | Genome-fragment | 1.73 | AAI-based (high-confidence) | 0 |
| S16_vs_55104\|\|full | 4087 | No | NA | 5 | 0 | 4 | Not-determined | Genome-fragment | NA | NA | 0 |
| S16_vs_55131\|\|full | 8040 | No | NA | 7 | 0 | 4 | Not-determined | Genome-fragment | NA | NA | 0 |
| S16_vs_55133\|\|full | 1601 | No | NA | 3 | 0 | 2 | Not-determined | Genome-fragment | NA | NA | 0 |
| S16_vs_55164\|\|full | 12845 | No | NA | 15 | 0 | 3 | Not-determined | Genome-fragment | NA | NA | 0 |
| S16_vs_55223\|\|full | 2965 | No | NA | 3 | 0 | 2 | Not-determined | Genome-fragment | NA | NA | 0 |
| S16_vs_55275\|\|full | 3267 | No | NA | 3 | 0 | 1 | Not-determined | Genome-fragment | NA | NA | 0 |
| S16_vs_55306\|\|full | 4264 | No | NA | 5 | 0 | 1 | Not-determined | Genome-fragment | NA | NA | 0 |
| S16_vs_55360\|\|full | 1229 | No | NA | 3 | 0 | 1 | Not-determined | Genome-fragment | NA | NA | 0 |
| S12_vs_k141_121671\|\|full | 2794 | No | NA | 3 | 0 | 2 | Low-quality | Genome-fragment | 2.24 | AAI-based (medium-confidence) | 0 |
| S8_vs_k141_72346\|\|full | 2794 | No | NA | 4 | 0 | 2 | Low-quality | Genome-fragment | 3.54 | AAI-based (high-confidence) | 0 |
| S16_vs_55500\|\|full | 3602 | No | NA | 3 | 0 | 2 | Not-determined | Genome-fragment | NA | NA | 0 |
| S16_vs_55527\|\|full | 1335 | No | NA | 2 | 0 | 2 | Not-determined | Genome-fragment | NA | NA | 0 |
| S16_vs_55600\|\|full | 2364 | No | NA | 3 | 0 | 3 | Not-determined | Genome-fragment | NA | NA | 0 |
| S16_vs_55604\|\|full | 1170 | No | NA | 2 | 0 | 2 | Not-determined | Genome-fragment | NA | NA | 0 |
| S16_vs_55610\|\|full | 1805 | No | NA | 2 | 0 | 2 | Not-determined | Genome-fragment | NA | NA | 0 |
| S16_vs_55615\|\|full | 2701 | No | NA | 2 | 0 | 2 | Not-determined | Genome-fragment | NA | NA | 0 |
| S17_vs_k141_107041\|\|full | 2795 | No | NA | 3 | 0 | 2 | Low-quality | Genome-fragment | 7.3 | AAI-based (medium-confidence) | 0 |
| S13_vs_108376\|\|full | 2795 | No | NA | 3 | 0 | 3 | Low-quality | Genome-fragment | 4.92 | AAI-based (medium-confidence) | 0 |
| S3_extracted_k141_58121 | 2795 | No | NA | 9 | 0 | 0 | Low-quality | Genome-fragment | 1.83 | AAI-based (high-confidence) | 0 |
| S5_vs_k141_18489\|\|full | 2795 | No | NA | 3 | 3 | 0 | Low-quality | Genome-fragment | 6.36 | AAI-based (high-confidence) | 0 |
| S16_vs_55892\|\|full | 7637 | No | NA | 7 | 0 | 5 | Not-determined | Genome-fragment | NA | NA | 0 |
| S16_vs_49875\|\|full | 2796 | No | NA | 3 | 0 | 1 | Low-quality | Genome-fragment | 1 | AAI-based (medium-confidence) | 0 |
| S16_vs_56004\|\|full | 5778 | No | NA | 2 | 0 | 2 | Not-determined | Genome-fragment | NA | NA | 0 |
| S16_vs_56042\|\|full | 2156 | No | NA | 5 | 0 | 3 | Not-determined | Genome-fragment | NA | NA | 0 |
| S16_vs_56089\|\|full | 3362 | No | NA | 4 | 0 | 0 | Not-determined | Genome-fragment | NA | NA | 0 |
| S16_vs_56095\|\|full | 2646 | No | NA | 3 | 0 | 2 | Not-determined | Genome-fragment | NA | NA | 0 |
| S16_vs_56239\|\|full | 2684 | No | NA | 2 | 0 | 0 | Not-determined | Genome-fragment | NA | NA | 0 |
| S16_vs_56244\|\|full | 7114 | No | NA | 10 | 0 | 1 | Not-determined | Genome-fragment | NA | NA | 0 |
| S14_vs_k141_94281\|\|full | 2797 | No | NA | 5 | 0 | 0 | Low-quality | Genome-fragment | 2 | AAI-based (medium-confidence) | 0 |
| S15_vs_37645\|\|full | 2797 | No | NA | 3 | 0 | 2 | Low-quality | Genome-fragment | 1.63 | AAI-based (medium-confidence) | 0 |
| S8_vs_k141_74021\|\|full | 2797 | No | NA | 3 | 1 | 0 | Low-quality | Genome-fragment | 1.61 | AAI-based (high-confidence) | 0 |
| S14_vs_k141_40194\|\|full | 2798 | No | NA | 3 | 2 | 0 | Low-quality | Genome-fragment | 6.47 | AAI-based (high-confidence) | 0 |
| S16_vs_56719\|\|full | 8478 | No | NA | 3 | 0 | 2 | Not-determined | Genome-fragment | NA | NA | 0 |
| S16_vs_56738\|\|full | 2025 | No | NA | 3 | 0 | 3 | Not-determined | Genome-fragment | NA | NA | 0 |
| S16_vs_56808\|\|full | 1693 | No | NA | 2 | 0 | 2 | Not-determined | Genome-fragment | NA | NA | 0 |
| S6_vs_k141_11037\|\|full | 2798 | No | NA | 7 | 0 | 0 | Low-quality | Genome-fragment | 5.93 | AAI-based (medium-confidence) | 0 |
| S16_vs_56858\|\|full | 2489 | No | NA | 2 | 0 | 0 | Not-determined | Genome-fragment | NA | NA | 0 |
| S17_vs_k141_100836\|\|full | 2799 | No | NA | 2 | 0 | 1 | Low-quality | Genome-fragment | 4.69 | AAI-based (high-confidence) | 0 |
| S16_vs_56899\|\|full | 2116 | No | NA | 2 | 0 | 2 | Not-determined | Genome-fragment | NA | NA | 0 |
| S14_vs_k141_79686\|\|full | 2799 | No | NA | 3 | 3 | 0 | Low-quality | Genome-fragment | 6.83 | AAI-based (high-confidence) | 0 |
| S16_vs_56940\|\|full | 2091 | No | NA | 2 | 0 | 0 | Not-determined | Genome-fragment | NA | NA | 0 |
| S16_vs_56969\|\|full | 2763 | No | NA | 3 | 0 | 3 | Not-determined | Genome-fragment | NA | NA | 0 |
| S5_vs_k141_120052\|\|full | 2799 | No | NA | 5 | 1 | 0 | Low-quality | Genome-fragment | 1.81 | HMM-based (lower-bound) | 0 |
| S10_vs_8532\|\|full | 2799 | No | NA | 4 | 0 | 0 | Low-quality | Genome-fragment | 4.6 | AAI-based (medium-confidence) | 0 |
| S2_vs_k141_3426\|\|full | 2799 | No | NA | 6 | 3 | 0 | Low-quality | Genome-fragment | 9.21 | AAI-based (high-confidence) | 0 |
| S16_vs_57609\|\|full | 3862 | No | NA | 4 | 0 | 3 | Not-determined | Genome-fragment | NA | NA | 0 |
| S16_vs_57672\|\|full | 1572 | No | NA | 5 | 0 | 0 | Not-determined | Genome-fragment | NA | NA | 0 |
| S16_vs_57735\|\|full | 1290 | No | NA | 2 | 0 | 2 | Not-determined | Genome-fragment | NA | NA | 0 |
| S8_vs_k141_128244\|\|full | 2799 | No | NA | 3 | 3 | 0 | Low-quality | Genome-fragment | 8.12 | AAI-based (high-confidence) | 0 |
| S16_vs_57838\|\|full | 2439 | No | NA | 3 | 0 | 1 | Not-determined | Genome-fragment | NA | NA | 0 |
| S16_vs_57874\|\|full | 3820 | No | NA | 6 | 0 | 3 | Not-determined | Genome-fragment | NA | NA | 0 |
| S16_vs_57923\|\|full | 2476 | No | NA | 5 | 0 | 1 | Not-determined | Genome-fragment | NA | NA | 0 |
| S16_vs_57984\|\|full | 2920 | No | NA | 4 | 0 | 0 | Not-determined | Genome-fragment | NA | NA | 0 |
| S17_vs_k141_90625\|\|full | 2800 | No | NA | 3 | 0 | 0 | Low-quality | Genome-fragment | 0.48 | AAI-based (medium-confidence) | 0 |
| S16_vs_58144\|\|full | 2117 | No | NA | 2 | 0 | 2 | Not-determined | Genome-fragment | NA | NA | 0 |
| S16_vs_58186\|\|full | 1245 | No | NA | 3 | 0 | 0 | Not-determined | Genome-fragment | NA | NA | 0 |
| S16_vs_58327\|\|full | 1907 | No | NA | 5 | 0 | 3 | Not-determined | Genome-fragment | NA | NA | 0 |
| S17_vs_k141_110041\|\|full | 2800 | No | NA | 5 | 0 | 1 | Low-quality | Genome-fragment | 3.27 | AAI-based (high-confidence) | 0 |
| S16_vs_58359\|\|full | 1136 | No | NA | 3 | 0 | 0 | Not-determined | Genome-fragment | NA | NA | 0 |
| S16_vs_58402\|\|full | 1468 | No | NA | 2 | 0 | 2 | Not-determined | Genome-fragment | NA | NA | 0 |
| S16_vs_58403\|\|full | 7123 | No | NA | 9 | 0 | 7 | Not-determined | Genome-fragment | NA | NA | 0 |
| S14_vs_k141_15590\|\|full | 2800 | No | NA | 5 | 1 | 0 | Low-quality | Genome-fragment | 4.17 | AAI-based (high-confidence) | 0 |
| S16_vs_58507\|\|full | 1271 | No | NA | 3 | 0 | 0 | Not-determined | Genome-fragment | NA | NA | 0 |
| S14_vs_k141_84784\|\|full | 2800 | No | NA | 4 | 1 | 0 | Low-quality | Genome-fragment | 1.22 | HMM-based (lower-bound) | 0 |
| S16_vs_58549\|\|full | 1860 | No | NA | 4 | 0 | 2 | Not-determined | Genome-fragment | NA | NA | 0 |
| S16_vs_58623\|\|full | 2654 | No | NA | 2 | 0 | 1 | Not-determined | Genome-fragment | NA | NA | 0 |
| S3_extracted_k141_69683 | 2800 | No | NA | 6 | 0 | 0 | Low-quality | Genome-fragment | 1.59 | AAI-based (high-confidence) | 0 |
| S7_vs_94746\|\|full | 2800 | No | NA | 6 | 1 | 0 | Low-quality | Genome-fragment | 7.27 | AAI-based (medium-confidence) | 0 |
| S16_vs_58876\|\|full | 7831 | No | NA | 6 | 0 | 6 | Not-determined | Genome-fragment | NA | NA | 0 |
| S16_vs_93131\|\|full | 2800 | No | NA | 4 | 0 | 0 | Low-quality | Genome-fragment | 3.38 | AAI-based (high-confidence) | 0 |
| S10_extracted_121009 | 2800 | No | NA | 3 | 0 | 0 | Low-quality | Genome-fragment | 2.72 | AAI-based (high-confidence) | 0 |
| S10_vs_1559\|\|lt2gene | 2800 | No | NA | 2 | 2 | 0 | Low-quality | Genome-fragment | 6.41 | AAI-based (medium-confidence) | 0 |
| S16_vs_59076\|\|full | 1336 | No | NA | 3 | 0 | 1 | Not-determined | Genome-fragment | NA | NA | 0 |
| S16_vs_59151\|\|full | 1069 | No | NA | 3 | 0 | 0 | Not-determined | Genome-fragment | NA | NA | 0 |
| S16_vs_59235\|\|full | 2111 | No | NA | 3 | 0 | 2 | Not-determined | Genome-fragment | NA | NA | 0 |
| S16_vs_59259\|\|full | 1411 | No | NA | 3 | 0 | 0 | Not-determined | Genome-fragment | NA | NA | 0 |
| S16_vs_59306\|\|full | 1845 | No | NA | 3 | 0 | 1 | Not-determined | Genome-fragment | NA | NA | 0 |
| S16_vs_59340\|\|full | 1261 | No | NA | 2 | 0 | 1 | Not-determined | Genome-fragment | NA | NA | 0 |
| S16_vs_59360\|\|full | 2283 | No | NA | 2 | 0 | 1 | Not-determined | Genome-fragment | NA | NA | 0 |
| S8_vs_k141_122387\|\|full | 2800 | No | NA | 5 | 0 | 0 | Low-quality | Genome-fragment | 6.11 | AAI-based (high-confidence) | 0 |
| S17_vs_k141_41708\|\|full | 2801 | No | NA | 4 | 0 | 2 | Low-quality | Genome-fragment | 7.31 | AAI-based (high-confidence) | 0 |
| S16_vs_59593\|\|full | 2096 | No | NA | 3 | 0 | 1 | Not-determined | Genome-fragment | NA | NA | 0 |
| S11_extracted_k141_55934 | 2801 | No | NA | 7 | 1 | 0 | Low-quality | Genome-fragment | 3.98 | AAI-based (high-confidence) | 0 |
| S16_vs_59687\|\|full | 1125 | No | NA | 1 | 0 | 0 | Not-determined | Genome-fragment | NA | NA | 0 |
| S14_vs_k141_100556\|\|full | 2801 | No | NA | 3 | 0 | 0 | Low-quality | Genome-fragment | 3.93 | AAI-based (high-confidence) | 0 |
| S16_vs_59787\|\|full | 13249 | No | NA | 18 | 0 | 0 | Not-determined | Genome-fragment | NA | NA | 0 |
| S16_vs_59856\|\|full | 6749 | No | NA | 5 | 0 | 5 | Not-determined | Genome-fragment | NA | NA | 0 |
| S1_vs_k141_61460\|\|full | 2801 | No | NA | 3 | 0 | 0 | Low-quality | Genome-fragment | 8.42 | AAI-based (high-confidence) | 0 |
| S3_vs_k141_52572\|\|full | 2801 | No | NA | 4 | 3 | 0 | Low-quality | Genome-fragment | 7.17 | AAI-based (high-confidence) | 0 |
| S10_vs_67611\|\|full | 2801 | No | NA | 7 | 2 | 0 | Low-quality | Genome-fragment | 4.44 | AAI-based (high-confidence) | 0 |
| S16_vs_59946\|\|full | 3681 | No | NA | 10 | 0 | 0 | Not-determined | Genome-fragment | NA | NA | 0 |
| S16_vs_60019\|\|full | 4123 | No | NA | 3 | 0 | 0 | Not-determined | Genome-fragment | NA | NA | 0 |
| S16_vs_60039\|\|full | 5252 | No | NA | 5 | 0 | 3 | Not-determined | Genome-fragment | NA | NA | 0 |
| S10_vs_111340\|\|full | 2801 | No | NA | 3 | 0 | 1 | Low-quality | Genome-fragment | 9.16 | AAI-based (medium-confidence) | 0 |
| S16_vs_60186\|\|full | 2504 | No | NA | 4 | 0 | 0 | Not-determined | Genome-fragment | NA | NA | 0 |
| S17_extracted_k141_52301 | 2802 | No | NA | 2 | 0 | 2 | Low-quality | Genome-fragment | 6.67 | AAI-based (high-confidence) | 0 |
| S16_vs_60401\|\|full | 12116 | No | NA | 11 | 0 | 7 | Not-determined | Genome-fragment | NA | NA | 0 |
| S16_vs_60475\|\|full | 1092 | No | NA | 3 | 0 | 1 | Not-determined | Genome-fragment | NA | NA | 0 |
| S5_extracted_k141_6377 | 2802 | Yes | 1380 | 4 | 1 | 1 | Low-quality | Genome-fragment | 2.1 | HMM-based (lower-bound) | 50.75 |
| S16_vs_60544\|\|full | 1985 | No | NA | 3 | 0 | 2 | Not-determined | Genome-fragment | NA | NA | 0 |
| S7_vs_39143\|\|full | 2802 | No | NA | 6 | 1 | 0 | Low-quality | Genome-fragment | 6.08 | AAI-based (high-confidence) | 0 |
| S16_vs_60588\|\|full | 1956 | No | NA | 3 | 0 | 2 | Not-determined | Genome-fragment | NA | NA | 0 |
| S16_extracted_108202 | 2802 | No | NA | 3 | 0 | 2 | Low-quality | Genome-fragment | 2.39 | AAI-based (medium-confidence) | 0 |
| S10_extracted_76831 | 2802 | No | NA | 6 | 1 | 0 | Low-quality | Genome-fragment | 7.06 | AAI-based (high-confidence) | 0 |
| S16_vs_60768\|\|full | 1896 | No | NA | 3 | 0 | 2 | Not-determined | Genome-fragment | NA | NA | 0 |
| S16_vs_60875\|\|full | 1049 | No | NA | 3 | 0 | 0 | Not-determined | Genome-fragment | NA | NA | 0 |
| S16_vs_60877\|\|full | 5835 | No | NA | 4 | 0 | 3 | Not-determined | Genome-fragment | NA | NA | 0 |
| S16_vs_60977\|\|full | 1734 | No | NA | 2 | 0 | 1 | Not-determined | Genome-fragment | NA | NA | 0 |
| S16_vs_61042\|\|full | 2459 | No | NA | 3 | 0 | 2 | Not-determined | Genome-fragment | NA | NA | 0 |
| S12_extracted_k141_78208 | 2802 | No | NA | 6 | 0 | 0 | Low-quality | Genome-fragment | 6.77 | AAI-based (high-confidence) | 0 |
| S11_vs_k141_7257\|\|full | 2804 | No | NA | 5 | 1 | 0 | Low-quality | Genome-fragment | 4.86 | AAI-based (high-confidence) | 0 |
| S13_vs_77349\|\|full | 2804 | No | NA | 3 | 1 | 0 | Low-quality | Genome-fragment | 5.08 | HMM-based (lower-bound) | 0 |
| S1_vs_k141_134038\|\|full | 2804 | No | NA | 6 | 1 | 0 | Low-quality | Genome-fragment | 6.13 | AAI-based (high-confidence) | 0 |
| S16_vs_61219\|\|full | 2666 | No | NA | 3 | 0 | 2 | Not-determined | Genome-fragment | NA | NA | 0 |
| S16_vs_61250\|\|full | 3535 | No | NA | 2 | 0 | 0 | Not-determined | Genome-fragment | NA | NA | 0 |
| S16_vs_61311\|\|full | 1556 | No | NA | 3 | 0 | 1 | Not-determined | Genome-fragment | NA | NA | 0 |
| S5_vs_k141_109104\|\|full | 2804 | No | NA | 9 | 2 | 0 | Low-quality | Genome-fragment | 6.4 | AAI-based (medium-confidence) | 0 |
| S16_vs_61425\|\|full | 8275 | No | NA | 16 | 0 | 3 | Not-determined | Genome-fragment | NA | NA | 0 |
| S16_vs_61511\|\|full | 6963 | No | NA | 5 | 0 | 4 | Not-determined | Genome-fragment | NA | NA | 0 |
| S7_extracted_38476 | 2804 | No | NA | 9 | 2 | 0 | Low-quality | Genome-fragment | 1.46 | AAI-based (high-confidence) | 0 |
| S16_vs_61648\|\|full | 1053 | No | NA | 3 | 0 | 1 | Not-determined | Genome-fragment | NA | NA | 0 |
| S16_vs_61753\|\|full | 2422 | No | NA | 3 | 0 | 0 | Not-determined | Genome-fragment | NA | NA | 0 |
| S16_vs_61794\|\|full | 1888 | No | NA | 2 | 0 | 2 | Not-determined | Genome-fragment | NA | NA | 0 |
| S8_vs_k141_199698\|\|full | 2804 | No | NA | 6 | 0 | 0 | Low-quality | Genome-fragment | 4.67 | AAI-based (medium-confidence) | 0 |
| S16_vs_61834\|\|full | 1002 | No | NA | 2 | 0 | 1 | Not-determined | Genome-fragment | NA | NA | 0 |
| S16_vs_61951\|\|full | 2254 | No | NA | 3 | 0 | 3 | Not-determined | Genome-fragment | NA | NA | 0 |
| S16_vs_62064\|\|full | 1389 | No | NA | 5 | 0 | 0 | Not-determined | Genome-fragment | NA | NA | 0 |
| S8_vs_k141_12715\|\|full | 2805 | No | NA | 2 | 0 | 1 | Low-quality | Genome-fragment | 1.52 | AAI-based (medium-confidence) | 0 |
| S9_vs_k141_14806\|\|full | 2807 | No | NA | 7 | 3 | 0 | Low-quality | Genome-fragment | 6.11 | AAI-based (high-confidence) | 0 |
| S16_vs_62089\|\|full | 2207 | No | NA | 3 | 0 | 2 | Not-determined | Genome-fragment | NA | NA | 0 |
| S16_vs_62112\|\|full | 4938 | No | NA | 5 | 0 | 3 | Not-determined | Genome-fragment | NA | NA | 0 |
| S3_extracted_k141_17921 | 2808 | No | NA | 10 | 0 | 0 | Low-quality | Genome-fragment | 6.07 | AAI-based (high-confidence) | 0 |
| S16_vs_62229\|\|full | 2586 | No | NA | 2 | 0 | 2 | Not-determined | Genome-fragment | NA | NA | 0 |
| S16_vs_62239\|\|full | 5311 | No | NA | 5 | 0 | 2 | Not-determined | Genome-fragment | NA | NA | 0 |
| S17_extracted_k141_126238 | 2809 | No | NA | 5 | 3 | 0 | Low-quality | Genome-fragment | 4.58 | AAI-based (high-confidence) | 0 |
| S16_vs_62273\|\|full | 2421 | No | NA | 4 | 0 | 1 | Not-determined | Genome-fragment | NA | NA | 0 |
| S16_vs_62316\|\|full | 1190 | No | NA | 6 | 0 | 0 | Not-determined | Genome-fragment | NA | NA | 0 |
| S16_vs_62446\|\|full | 1312 | No | NA | 2 | 0 | 0 | Not-determined | Genome-fragment | NA | NA | 0 |
| S16_vs_62450\|\|full | 1181 | No | NA | 2 | 0 | 0 | Not-determined | Genome-fragment | NA | NA | 0 |
| S16_vs_62495\|\|full | 3382 | No | NA | 2 | 0 | 1 | Not-determined | Genome-fragment | NA | NA | 0 |
| S3_vs_k141_27753\|\|full | 2809 | No | NA | 9 | 1 | 0 | Low-quality | Genome-fragment | 4.93 | AAI-based (high-confidence) | 0 |
| S16_vs_62580\|\|full | 3864 | No | NA | 4 | 0 | 1 | Not-determined | Genome-fragment | NA | NA | 0 |
| S16_vs_62597\|\|full | 3029 | No | NA | 3 | 0 | 2 | Not-determined | Genome-fragment | NA | NA | 0 |
| S16_vs_62599\|\|full | 3262 | No | NA | 3 | 0 | 1 | Not-determined | Genome-fragment | NA | NA | 0 |
| S3_vs_k141_12684\|\|full | 2810 | No | NA | 3 | 0 | 3 | Low-quality | Genome-fragment | 1.68 | AAI-based (medium-confidence) | 0 |
| S8_vs_k141_589\|\|full | 2810 | No | NA | 8 | 1 | 0 | Low-quality | Genome-fragment | 7.85 | AAI-based (high-confidence) | 0 |
| S16_vs_62797\|\|full | 8501 | No | NA | 5 | 0 | 2 | Not-determined | Genome-fragment | NA | NA | 0 |
| S16_vs_62892\|\|full | 5497 | No | NA | 4 | 0 | 1 | Not-determined | Genome-fragment | NA | NA | 0 |
| S8_vs_k141_37162\|\|full | 2810 | No | NA | 5 | 1 | 0 | Low-quality | Genome-fragment | 4.17 | HMM-based (lower-bound) | 0 |
| S16_vs_62934\|\|full | 3202 | No | NA | 5 | 0 | 3 | Not-determined | Genome-fragment | NA | NA | 0 |
| S16_vs_62989\|\|full | 9140 | No | NA | 9 | 0 | 6 | Not-determined | Genome-fragment | NA | NA | 0 |
| S14_vs_k141_47349\|\|full | 2811 | No | NA | 2 | 0 | 2 | Low-quality | Genome-fragment | 0.95 | AAI-based (high-confidence) | 0 |
| S12_vs_k141_164580\|\|full | 2811 | No | NA | 9 | 1 | 0 | Low-quality | Genome-fragment | 4.73 | AAI-based (high-confidence) | 0 |
| S16_vs_63345\|\|full | 1527 | No | NA | 3 | 0 | 0 | Not-determined | Genome-fragment | NA | NA | 0 |
| S16_vs_63426\|\|full | 3006 | No | NA | 3 | 0 | 0 | Not-determined | Genome-fragment | NA | NA | 0 |
| S16_vs_63674\|\|full | 1258 | No | NA | 2 | 0 | 0 | Not-determined | Genome-fragment | NA | NA | 0 |
| S16_vs_63690\|\|full | 1239 | No | NA | 2 | 0 | 1 | Not-determined | Genome-fragment | NA | NA | 0 |
| S5_vs_k141_144077\|\|full | 2812 | No | NA | 5 | 0 | 5 | Low-quality | Genome-fragment | 1.81 | AAI-based (medium-confidence) | 0 |
| S11_vs_k141_68276\|\|full | 2813 | No | NA | 6 | 0 | 0 | Low-quality | Genome-fragment | 6.42 | AAI-based (high-confidence) | 0 |
| S5_vs_k141_32980\|\|full | 2814 | No | NA | 7 | 4 | 0 | Low-quality | Genome-fragment | 6.91 | AAI-based (high-confidence) | 0 |
| S17_vs_k141_50185\|\|full | 2815 | No | NA | 4 | 1 | 0 | Low-quality | Genome-fragment | 1.74 | HMM-based (lower-bound) | 0 |
| S16_vs_63816\|\|full | 10407 | No | NA | 10 | 0 | 7 | Not-determined | Genome-fragment | NA | NA | 0 |
| S1_vs_k141_79215\|\|full | 2815 | No | NA | 7 | 4 | 0 | Low-quality | Genome-fragment | 4.78 | AAI-based (high-confidence) | 0 |
| S17_vs_k141_149647\|\|full | 2816 | No | NA | 3 | 0 | 3 | Low-quality | Genome-fragment | 9.36 | AAI-based (high-confidence) | 0 |
| S16_vs_64062\|\|full | 2841 | No | NA | 5 | 0 | 0 | Not-determined | Genome-fragment | NA | NA | 0 |
| S11_vs_k141_31819\|\|full | 2816 | No | NA | 7 | 0 | 0 | Low-quality | Genome-fragment | 4.55 | AAI-based (high-confidence) | 0 |
| S3_extracted_k141_111362 | 2816 | No | NA | 4 | 0 | 0 | Low-quality | Genome-fragment | 1.76 | AAI-based (high-confidence) | 0 |
| S16_vs_64183\|\|full | 6385 | No | NA | 9 | 0 | 3 | Not-determined | Genome-fragment | NA | NA | 0 |
| S16_vs_64184\|\|full | 6376 | No | NA | 6 | 0 | 4 | Not-determined | Genome-fragment | NA | NA | 0 |
| S16_vs_64273\|\|full | 6253 | No | NA | 2 | 0 | 2 | Not-determined | Genome-fragment | NA | NA | 0 |
| S16_vs_64289\|\|full | 1267 | No | NA | 4 | 0 | 0 | Not-determined | Genome-fragment | NA | NA | 0 |
| S16_vs_64308\|\|full | 8618 | No | NA | 11 | 0 | 6 | Not-determined | Genome-fragment | NA | NA | 0 |
| S16_vs_64478\|\|full | 5874 | No | NA | 3 | 0 | 3 | Not-determined | Genome-fragment | NA | NA | 0 |
| S12_extracted_k141_139653 | 2817 | Yes | 1956 | 3 | 1 | 1 | Low-quality | Genome-fragment | 4.73 | AAI-based (high-confidence) | 30.56 |
| S2_extracted_k141_14150 | 2817 | No | NA | 5 | 2 | 0 | Low-quality | Genome-fragment | 2.76 | AAI-based (high-confidence) | 0 |
| S16_vs_64560\|\|full | 1403 | No | NA | 2 | 0 | 2 | Not-determined | Genome-fragment | NA | NA | 0 |
| S16_vs_64584\|\|full | 26658 | No | NA | 38 | 12 | 0 | Medium-quality | Genome-fragment | 78 | AAI-based (high-confidence) | 0 |
| S10_vs_82090\|\|full | 2818 | No | NA | 6 | 0 | 0 | Low-quality | Genome-fragment | 4.91 | AAI-based (high-confidence) | 0 |
| S17_vs_k141_152199\|\|full | 2819 | No | NA | 4 | 3 | 0 | Low-quality | Genome-fragment | 8.12 | AAI-based (high-confidence) | 0 |
| S17_vs_k141_143193\|\|lt2gene | 2819 | No | NA | 2 | 2 | 0 | Low-quality | Genome-fragment | 4.65 | AAI-based (high-confidence) | 0 |
| S16_vs_65403\|\|full | 3551 | No | NA | 5 | 0 | 2 | Not-determined | Genome-fragment | NA | NA | 0 |
| S16_vs_65530\|\|full | 5618 | No | NA | 7 | 0 | 2 | Not-determined | Genome-fragment | NA | NA | 0 |
| S11_vs_k141_65758\|\|full | 2819 | No | NA | 4 | 2 | 0 | Low-quality | Genome-fragment | 7.15 | AAI-based (high-confidence) | 0 |
| S14_vs_k141_115329\|\|full | 2819 | No | NA | 9 | 1 | 0 | Low-quality | Genome-fragment | 5.1 | AAI-based (high-confidence) | 0 |
| S6_vs_k141_46990\|\|full | 2819 | No | NA | 2 | 0 | 0 | Low-quality | Genome-fragment | 1.75 | AAI-based (high-confidence) | 0 |
| S14_vs_k141_31905\|\|full | 2820 | No | NA | 5 | 1 | 0 | Low-quality | Genome-fragment | 4.88 | AAI-based (medium-confidence) | 0 |
| S16_vs_65924\|\|full | 5665 | No | NA | 4 | 0 | 2 | Not-determined | Genome-fragment | NA | NA | 0 |
| S16_vs_154460\|\|full | 2820 | No | NA | 7 | 3 | 0 | Low-quality | Genome-fragment | 3.38 | AAI-based (high-confidence) | 0 |
| S16_vs_65968\|\|full | 1421 | No | NA | 2 | 0 | 1 | Not-determined | Genome-fragment | NA | NA | 0 |
| S1_vs_k141_4265\|\|full | 2821 | No | NA | 2 | 0 | 0 | Low-quality | Genome-fragment | 3.07 | AAI-based (high-confidence) | 0 |
| S16_vs_66083\|\|full | 2565 | No | NA | 4 | 0 | 1 | Not-determined | Genome-fragment | NA | NA | 0 |
| S7_extracted_66380 | 2821 | No | NA | 4 | 3 | 0 | Low-quality | Genome-fragment | 4.79 | AAI-based (high-confidence) | 0 |
| S12_vs_k141_37231\|\|lt2gene | 2821 | No | NA | 1 | 1 | 0 | Low-quality | Genome-fragment | 8.04 | AAI-based (high-confidence) | 0 |
| S13_extracted_111491 | 2822 | No | NA | 3 | 0 | 0 | Low-quality | Genome-fragment | 2.65 | AAI-based (high-confidence) | 0 |
| S3_vs_k141_35908\|\|full | 2822 | No | NA | 8 | 1 | 0 | Low-quality | Genome-fragment | 7.8 | AAI-based (high-confidence) | 0 |
| S5_vs_k141_98585\|\|full | 2822 | No | NA | 2 | 0 | 1 | Low-quality | Genome-fragment | 3.61 | AAI-based (medium-confidence) | 0 |
| S16_vs_66543\|\|full | 2980 | No | NA | 2 | 0 | 1 | Not-determined | Genome-fragment | NA | NA | 0 |
| S16_vs_66577\|\|full | 1438 | No | NA | 4 | 0 | 0 | Not-determined | Genome-fragment | NA | NA | 0 |
| S16_vs_66592\|\|full | 1890 | No | NA | 2 | 0 | 1 | Not-determined | Genome-fragment | NA | NA | 0 |
| S16_vs_66634\|\|full | 1162 | No | NA | 3 | 0 | 0 | Not-determined | Genome-fragment | NA | NA | 0 |
| S12_vs_k141_166097\|\|full | 2822 | No | NA | 3 | 0 | 2 | Low-quality | Genome-fragment | 0.68 | AAI-based (high-confidence) | 0 |
| S14_vs_k141_88130\|\|full | 2823 | No | NA | 4 | 2 | 0 | Low-quality | Genome-fragment | 5.04 | HMM-based (lower-bound) | 0 |
| S16_vs_66700\|\|full | 2864 | No | NA | 4 | 0 | 2 | Not-determined | Genome-fragment | NA | NA | 0 |
| S1_extracted_k141_109394 | 2823 | No | NA | 6 | 1 | 0 | Low-quality | Genome-fragment | 5.33 | AAI-based (high-confidence) | 0 |
| S16_vs_66766\|\|full | 2905 | No | NA | 3 | 0 | 2 | Not-determined | Genome-fragment | NA | NA | 0 |
| S16_vs_66949\|\|full | 2976 | No | NA | 4 | 0 | 1 | Not-determined | Genome-fragment | NA | NA | 0 |
| S16_vs_66963\|\|full | 3101 | No | NA | 2 | 0 | 1 | Not-determined | Genome-fragment | NA | NA | 0 |
| S7_vs_94481\|\|full | 2823 | No | NA | 3 | 2 | 0 | Low-quality | Genome-fragment | 5.74 | AAI-based (medium-confidence) | 0 |
| S16_vs_67073\|\|full | 6428 | No | NA | 5 | 0 | 2 | Not-determined | Genome-fragment | NA | NA | 0 |
| S16_vs_67232\|\|full | 3230 | No | NA | 3 | 0 | 2 | Not-determined | Genome-fragment | NA | NA | 0 |
| S16_vs_67294\|\|full | 4198 | No | NA | 8 | 0 | 3 | Not-determined | Genome-fragment | NA | NA | 0 |
| S5_vs_k141_106815\|\|full | 2824 | No | NA | 5 | 3 | 0 | Low-quality | Genome-fragment | 6.99 | AAI-based (high-confidence) | 0 |
| S7_vs_4335\|\|full | 2824 | Yes | 2462 | 6 | 1 | 1 | Low-quality | Genome-fragment | 3.45 | HMM-based (lower-bound) | 12.82 |
| S16_vs_110107\|\|full | 2824 | No | NA | 5 | 0 | 1 | Low-quality | Genome-fragment | 3.43 | AAI-based (medium-confidence) | 0 |
| S16_vs_67638\|\|full | 1499 | No | NA | 2 | 0 | 0 | Not-determined | Genome-fragment | NA | NA | 0 |
| S3_vs_k141_125024\|\|full | 2825 | No | NA | 5 | 1 | 0 | Low-quality | Genome-fragment | 1.84 | AAI-based (medium-confidence) | 0 |
| S5_vs_k141_54660\|\|full | 2825 | No | NA | 3 | 2 | 0 | Low-quality | Genome-fragment | 8.19 | AAI-based (high-confidence) | 0 |
| S16_vs_127937\|\|full | 2825 | No | NA | 4 | 0 | 0 | Low-quality | Genome-fragment | 4.84 | AAI-based (medium-confidence) | 0 |
| S16_vs_67740\|\|full | 2029 | No | NA | 3 | 0 | 0 | Not-determined | Genome-fragment | NA | NA | 0 |
| S8_extracted_k141_163323 | 2825 | No | NA | 3 | 0 | 1 | Low-quality | Genome-fragment | 1.08 | AAI-based (medium-confidence) | 0 |
| S9_vs_k141_22454\|\|full | 2826 | No | NA | 2 | 2 | 0 | Low-quality | Genome-fragment | 4.69 | AAI-based (high-confidence) | 0 |
| S16_vs_67896\|\|full | 2085 | No | NA | 3 | 0 | 2 | Not-determined | Genome-fragment | NA | NA | 0 |
| S14_vs_k141_115030\|\|full | 2826 | No | NA | 5 | 0 | 0 | Low-quality | Genome-fragment | 7.21 | AAI-based (high-confidence) | 0 |
| S16_vs_67990\|\|full | 1899 | No | NA | 2 | 0 | 2 | Not-determined | Genome-fragment | NA | NA | 0 |
| S16_vs_68036\|\|full | 4472 | No | NA | 4 | 0 | 0 | Not-determined | Genome-fragment | NA | NA | 0 |
| S16_vs_68117\|\|full | 6891 | No | NA | 5 | 0 | 4 | Not-determined | Genome-fragment | NA | NA | 0 |
| S1_vs_k141_82421\|\|full | 2826 | No | NA | 3 | 2 | 0 | Low-quality | Genome-fragment | 7.33 | AAI-based (high-confidence) | 0 |
| S16_vs_68160\|\|full | 4980 | No | NA | 6 | 0 | 3 | Not-determined | Genome-fragment | NA | NA | 0 |
| S1_vs_k141_126342\|\|full | 2826 | No | NA | 3 | 1 | 0 | Low-quality | Genome-fragment | 4.75 | AAI-based (high-confidence) | 0 |
| S16_vs_68226\|\|full | 9624 | No | NA | 13 | 0 | 6 | Not-determined | Genome-fragment | NA | NA | 0 |
| S16_vs_68254\|\|full | 1222 | No | NA | 3 | 0 | 0 | Not-determined | Genome-fragment | NA | NA | 0 |
| S3_vs_k141_17922\|\|full | 2826 | No | NA | 3 | 2 | 0 | Low-quality | Genome-fragment | 4.91 | AAI-based (high-confidence) | 0 |
| S16_vs_68327\|\|full | 3356 | No | NA | 4 | 0 | 2 | Not-determined | Genome-fragment | NA | NA | 0 |
| S16_vs_68331\|\|full | 1319 | No | NA | 3 | 0 | 1 | Not-determined | Genome-fragment | NA | NA | 0 |
| S16_vs_68350\|\|full | 2807 | No | NA | 3 | 0 | 0 | Not-determined | Genome-fragment | NA | NA | 0 |
| S16_vs_68511\|\|full | 1095 | No | NA | 2 | 0 | 0 | Not-determined | Genome-fragment | NA | NA | 0 |
| S5_vs_k141_59875\|\|full | 2826 | No | NA | 2 | 0 | 1 | Low-quality | Genome-fragment | 1.52 | AAI-based (medium-confidence) | 0 |
| S7_vs_959\|\|full | 2826 | No | NA | 4 | 3 | 0 | Low-quality | Genome-fragment | 7.02 | AAI-based (high-confidence) | 0 |
| S16_vs_68696\|\|full | 9914 | No | NA | 7 | 0 | 5 | Not-determined | Genome-fragment | NA | NA | 0 |
| S16_vs_68715\|\|full | 2268 | No | NA | 5 | 0 | 2 | Not-determined | Genome-fragment | NA | NA | 0 |
| S11_vs_k141_67575\|\|full | 2827 | No | NA | 7 | 1 | 0 | Low-quality | Genome-fragment | 6.02 | AAI-based (medium-confidence) | 0 |
| S16_vs_68774\|\|full | 8932 | No | NA | 5 | 0 | 3 | Not-determined | Genome-fragment | NA | NA | 0 |
| S1_extracted_k141_46607 | 2827 | No | NA | 4 | 2 | 0 | Low-quality | Genome-fragment | 7.06 | AAI-based (high-confidence) | 0 |
| S16_vs_68899\|\|full | 2108 | No | NA | 3 | 0 | 2 | Not-determined | Genome-fragment | NA | NA | 0 |
| S16_vs_69013\|\|full | 2033 | No | NA | 2 | 0 | 0 | Not-determined | Genome-fragment | NA | NA | 0 |
| S10_vs_38434\|\|full | 2827 | No | NA | 4 | 0 | 2 | Low-quality | Genome-fragment | 0.81 | AAI-based (medium-confidence) | 0 |
| S16_vs_69230\|\|full | 6232 | No | NA | 11 | 0 | 2 | Not-determined | Genome-fragment | NA | NA | 0 |
| S16_vs_69336\|\|full | 1057 | No | NA | 2 | 0 | 0 | Not-determined | Genome-fragment | NA | NA | 0 |
| S16_vs_69402\|\|full | 3315 | No | NA | 4 | 0 | 1 | Not-determined | Genome-fragment | NA | NA | 0 |
| S16_vs_69520\|\|full | 2912 | No | NA | 2 | 0 | 2 | Not-determined | Genome-fragment | NA | NA | 0 |
| S16_vs_69660\|\|full | 10217 | No | NA | 8 | 0 | 7 | Not-determined | Genome-fragment | NA | NA | 0 |
| S15_vs_64025\|\|full | 2827 | No | NA | 6 | 0 | 0 | Low-quality | Genome-fragment | 1.5 | AAI-based (high-confidence) | 0 |
| S8_vs_k141_190779\|\|full | 2827 | No | NA | 2 | 0 | 2 | Low-quality | Genome-fragment | 1.3 | AAI-based (medium-confidence) | 0 |
| S16_vs_69929\|\|full | 1109 | No | NA | 2 | 0 | 0 | Not-determined | Genome-fragment | NA | NA | 0 |
| S13_vs_52002\|\|full | 2828 | No | NA | 3 | 0 | 0 | Low-quality | Genome-fragment | 2 | AAI-based (medium-confidence) | 0 |
| S3_extracted_k141_64725 | 2828 | No | NA | 4 | 3 | 0 | Low-quality | Genome-fragment | 9.32 | AAI-based (high-confidence) | 0 |
| S16_vs_70196\|\|full | 1907 | No | NA | 2 | 0 | 1 | Not-determined | Genome-fragment | NA | NA | 0 |
| S16_vs_70245\|\|full | 1398 | No | NA | 4 | 0 | 0 | Not-determined | Genome-fragment | NA | NA | 0 |
| S16_vs_70251\|\|full | 1283 | No | NA | 2 | 0 | 0 | Not-determined | Genome-fragment | NA | NA | 0 |
| S16_vs_70256\|\|full | 1708 | No | NA | 2 | 0 | 0 | Not-determined | Genome-fragment | NA | NA | 0 |
| S3_extracted_k141_106445 | 2828 | No | NA | 7 | 2 | 0 | Low-quality | Genome-fragment | 1.8 | AAI-based (high-confidence) | 0 |
| S16_vs_70377\|\|full | 1823 | No | NA | 2 | 0 | 1 | Not-determined | Genome-fragment | NA | NA | 0 |
| S16_vs_70381\|\|full | 1114 | No | NA | 3 | 0 | 0 | Not-determined | Genome-fragment | NA | NA | 0 |
| S16_vs_60570\|\|full | 2828 | No | NA | 4 | 1 | 0 | Low-quality | Genome-fragment | 3.26 | HMM-based (lower-bound) | 0 |
| S16_vs_70436\|\|full | 2808 | No | NA | 3 | 0 | 1 | Not-determined | Genome-fragment | NA | NA | 0 |
| S10_extracted_83083 | 2828 | No | NA | 4 | 0 | 0 | Low-quality | Genome-fragment | 1.25 | AAI-based (high-confidence) | 0 |
| S16_vs_70504\|\|full | 1569 | No | NA | 2 | 0 | 2 | Not-determined | Genome-fragment | NA | NA | 0 |
| S17_vs_k141_23778\|\|full | 2830 | No | NA | 3 | 0 | 2 | Low-quality | Genome-fragment | 4.45 | AAI-based (medium-confidence) | 0 |
| S16_vs_70599\|\|full | 1918 | No | NA | 3 | 0 | 2 | Not-determined | Genome-fragment | NA | NA | 0 |
| S3_vs_k141_27721\|\|full | 2830 | No | NA | 3 | 0 | 2 | Low-quality | Genome-fragment | 0.67 | AAI-based (medium-confidence) | 0 |
| S7_vs_2686\|\|full | 2831 | No | NA | 3 | 0 | 0 | Low-quality | Genome-fragment | 6.68 | AAI-based (medium-confidence) | 0 |
| S16_vs_70709\|\|full | 2163 | No | NA | 2 | 0 | 1 | Not-determined | Genome-fragment | NA | NA | 0 |
| S12_vs_k141_167929\|\|full | 2831 | No | NA | 8 | 1 | 0 | Low-quality | Genome-fragment | 2.94 | HMM-based (lower-bound) | 0 |
| S16_vs_70933\|\|full | 3614 | No | NA | 4 | 0 | 2 | Not-determined | Genome-fragment | NA | NA | 0 |
| S16_vs_71000\|\|full | 1518 | No | NA | 3 | 0 | 1 | Not-determined | Genome-fragment | NA | NA | 0 |
| S16_vs_71046\|\|full | 1594 | No | NA | 2 | 0 | 1 | Not-determined | Genome-fragment | NA | NA | 0 |
| S16_vs_71136\|\|full | 1709 | No | NA | 4 | 0 | 3 | Not-determined | Genome-fragment | NA | NA | 0 |
| S16_vs_71154\|\|full | 7432 | No | NA | 4 | 0 | 2 | Not-determined | Genome-fragment | NA | NA | 0 |
| S5_vs_k141_100540\|\|full | 2832 | No | NA | 4 | 0 | 4 | Low-quality | Genome-fragment | 4.4 | AAI-based (medium-confidence) | 0 |
| S16_vs_71187\|\|full | 3200 | No | NA | 3 | 0 | 2 | Not-determined | Genome-fragment | NA | NA | 0 |
| S16_vs_49579\|\|full | 2832 | No | NA | 8 | 1 | 0 | Low-quality | Genome-fragment | 5.64 | AAI-based (high-confidence) | 0 |
| S16_vs_71451\|\|full | 5431 | No | NA | 5 | 0 | 4 | Not-determined | Genome-fragment | NA | NA | 0 |
| S12_vs_k141_137474\|\|full | 2832 | No | NA | 5 | 2 | 0 | Low-quality | Genome-fragment | 2.71 | AAI-based (high-confidence) | 0 |
| S16_vs_71473\|\|full | 1172 | No | NA | 3 | 0 | 1 | Not-determined | Genome-fragment | NA | NA | 0 |
| S16_vs_71598\|\|full | 1990 | No | NA | 3 | 0 | 3 | Not-determined | Genome-fragment | NA | NA | 0 |
| S11_vs_k141_111\|\|full | 2833 | No | NA | 5 | 5 | 0 | Low-quality | Genome-fragment | 7.51 | AAI-based (high-confidence) | 0 |
| S8_vs_k141_42907\|\|full | 2833 | No | NA | 9 | 2 | 0 | Low-quality | Genome-fragment | 9.01 | AAI-based (high-confidence) | 0 |
| S16_vs_36316\|\|full | 2834 | No | NA | 10 | 1 | 0 | Low-quality | Genome-fragment | 1.62 | HMM-based (lower-bound) | 0 |
| S16_vs_71835\|\|full | 4414 | No | NA | 8 | 0 | 2 | Not-determined | Genome-fragment | NA | NA | 0 |
| S16_vs_71856\|\|full | 2692 | No | NA | 6 | 0 | 1 | Not-determined | Genome-fragment | NA | NA | 0 |
| S7_vs_110430\|\|full | 2835 | No | NA | 8 | 0 | 0 | Low-quality | Genome-fragment | 5.97 | AAI-based (medium-confidence) | 0 |
| S16_vs_71973\|\|full | 16784 | No | NA | 23 | 0 | 7 | Not-determined | Genome-fragment | NA | NA | 0 |
| S9_vs_k141_48489\|\|full | 2836 | No | NA | 4 | 3 | 0 | Low-quality | Genome-fragment | 4.07 | HMM-based (lower-bound) | 0 |
| S16_vs_72073\|\|full | 1252 | No | NA | 4 | 0 | 0 | Not-determined | Genome-fragment | NA | NA | 0 |
| S16_vs_72111\|\|full | 3060 | No | NA | 2 | 0 | 0 | Not-determined | Genome-fragment | NA | NA | 0 |
| S7_vs_113708\|\|full | 2836 | No | NA | 3 | 0 | 2 | Low-quality | Genome-fragment | 1.97 | AAI-based (medium-confidence) | 0 |
| S16_vs_72285\|\|full | 3209 | No | NA | 3 | 0 | 2 | Not-determined | Genome-fragment | NA | NA | 0 |
| S10_vs_38112\|\|full | 2836 | No | NA | 3 | 0 | 3 | Low-quality | Genome-fragment | 2.29 | AAI-based (high-confidence) | 0 |
| S16_vs_72395\|\|full | 1279 | No | NA | 3 | 0 | 1 | Not-determined | Genome-fragment | NA | NA | 0 |
| S16_vs_72405\|\|full | 8181 | No | NA | 9 | 0 | 6 | Not-determined | Genome-fragment | NA | NA | 0 |
| S16_vs_72517\|\|full | 1360 | No | NA | 3 | 0 | 0 | Not-determined | Genome-fragment | NA | NA | 0 |
| S16_vs_72631\|\|full | 3142 | No | NA | 2 | 0 | 2 | Not-determined | Genome-fragment | NA | NA | 0 |
| S16_vs_72766\|\|full | 1384 | No | NA | 2 | 0 | 0 | Not-determined | Genome-fragment | NA | NA | 0 |
| S10_vs_9376\|\|full | 2837 | No | NA | 4 | 1 | 0 | Low-quality | Genome-fragment | 5.31 | AAI-based (high-confidence) | 0 |
| S16_vs_72827\|\|full | 3015 | No | NA | 2 | 0 | 1 | Not-determined | Genome-fragment | NA | NA | 0 |
| S16_vs_72853\|\|full | 1539 | No | NA | 3 | 0 | 1 | Not-determined | Genome-fragment | NA | NA | 0 |
| S16_vs_72916\|\|full | 1494 | No | NA | 2 | 0 | 0 | Not-determined | Genome-fragment | NA | NA | 0 |
| S10_vs_10488\|\|full | 2837 | No | NA | 3 | 0 | 0 | Low-quality | Genome-fragment | 1.74 | AAI-based (high-confidence) | 0 |
| S16_vs_72974\|\|full | 1588 | No | NA | 3 | 0 | 1 | Not-determined | Genome-fragment | NA | NA | 0 |
| S8_extracted_k141_187457 | 2837 | No | NA | 3 | 3 | 0 | Low-quality | Genome-fragment | 4.63 | HMM-based (lower-bound) | 0 |
| S16_vs_154246\|\|full | 2838 | No | NA | 3 | 3 | 0 | Low-quality | Genome-fragment | 6.09 | AAI-based (medium-confidence) | 0 |
| S10_vs_48098\|\|full | 2838 | No | NA | 5 | 0 | 0 | Low-quality | Genome-fragment | 4.15 | AAI-based (high-confidence) | 0 |
| S16_vs_73083\|\|full | 1904 | No | NA | 3 | 0 | 2 | Not-determined | Genome-fragment | NA | NA | 0 |
| S9_vs_k141_84681\|\|full | 2839 | No | NA | 4 | 1 | 0 | Low-quality | Genome-fragment | 6.97 | AAI-based (high-confidence) | 0 |
| S16_vs_73349\|\|full | 6714 | No | NA | 3 | 0 | 1 | Not-determined | Genome-fragment | NA | NA | 0 |

| | | | | | | | | | | | |
|---|---|---|---|---|---|---|---|---|---|---|---|
| S16_vs_73360\|\|full | 4358 | No | NA | 3 | 0 | 3 | Not-determined | Genome-fragment | NA | NA | 0 |
| S16_vs_73394\|\|full | 2382 | No | NA | 2 | 0 | 1 | Not-determined | Genome-fragment | NA | NA | 0 |
| S14_extracted_k141_7830 | 2839 | No | NA | 7 | 6 | 0 | Low-quality | Genome-fragment | 9.36 | AAI-based (high-confidence) | 0 |
| S5_vs_k141_43200\|\|full | 2839 | No | NA | 3 | 3 | 0 | Low-quality | Genome-fragment | 7.69 | AAI-based (high-confidence) | 0 |
| S16_vs_73851\|\|full | 3183 | No | NA | 4 | 0 | 2 | Not-determined | Genome-fragment | NA | NA | 0 |
| S16_vs_73889\|\|full | 3127 | No | NA | 3 | 0 | 2 | Not-determined | Genome-fragment | NA | NA | 0 |
| S16_vs_64143\|\|full | 2839 | No | NA | 5 | 0 | 1 | Low-quality | Genome-fragment | 1.95 | AAI-based (high-confidence) | 0 |
| S16_vs_45780\|\|lt2gene | 2839 | No | NA | 2 | 2 | 0 | Low-quality | Genome-fragment | 8.05 | AAI-based (high-confidence) | 0 |
| S8_extracted_k141_54374 | 2839 | No | NA | 3 | 0 | 1 | Low-quality | Genome-fragment | 4.32 | AAI-based (high-confidence) | 0 |
| S14_vs_k141_10308\|\|full | 2840 | No | NA | 6 | 0 | 0 | Low-quality | Genome-fragment | 8.04 | AAI-based (high-confidence) | 0 |
| S3_vs_k141_99229\|\|full | 2840 | No | NA | 8 | 0 | 0 | Low-quality | Genome-fragment | 6.88 | AAI-based (medium-confidence) | 0 |
| S16_vs_74781\|\|full | 4961 | No | NA | 5 | 0 | 0 | Not-determined | Genome-fragment | NA | NA | 0 |
| S7_vs_57210\|\|full | 2840 | No | NA | 3 | 0 | 1 | Low-quality | Genome-fragment | 0.94 | AAI-based (medium-confidence) | 0 |
| S6_vs_k141_31915\|\|full | 2840 | No | NA | 6 | 0 | 0 | Low-quality | Genome-fragment | 6.79 | AAI-based (high-confidence) | 0 |
| S16_vs_74859\|\|full | 2847 | No | NA | 5 | 0 | 5 | Not-determined | Genome-fragment | NA | NA | 0 |
| S8_extracted_k141_9979 | 2840 | No | NA | 5 | 1 | 0 | Low-quality | Genome-fragment | 6.53 | AAI-based (high-confidence) | 0 |
| S11_vs_k141_635\|\|full | 2841 | No | NA | 8 | 0 | 0 | Low-quality | Genome-fragment | 7.24 | AAI-based (medium-confidence) | 0 |
| S16_vs_75079\|\|full | 1810 | No | NA | 4 | 0 | 0 | Not-determined | Genome-fragment | NA | NA | 0 |
| S16_vs_75084\|\|full | 3106 | No | NA | 3 | 0 | 3 | Not-determined | Genome-fragment | NA | NA | 0 |
| S16_vs_75220\|\|full | 1090 | No | NA | 2 | 0 | 1 | Not-determined | Genome-fragment | NA | NA | 0 |
| S16_vs_75224\|\|full | 10762 | No | NA | 6 | 0 | 6 | Not-determined | Genome-fragment | NA | NA | 0 |
| S12_vs_k141_6731\|\|full | 2841 | Yes | 1014 | 2 | 1 | 1 | Low-quality | Genome-fragment | 0.68 | HMM-based (lower-bound) | 64.31 |
| S17_vs_k141_141223\|\|full | 2842 | No | NA | 3 | 0 | 2 | Low-quality | Genome-fragment | 6.19 | AAI-based (medium-confidence) | 0 |
| S16_vs_75430\|\|full | 2641 | No | NA | 4 | 0 | 1 | Not-determined | Genome-fragment | NA | NA | 0 |
| S8_vs_k141_51954\|\|full | 2843 | No | NA | 4 | 0 | 0 | Low-quality | Genome-fragment | 6.02 | AAI-based (medium-confidence) | 0 |
| S12_vs_k141_155715\|\|full | 2844 | No | NA | 4 | 0 | 0 | Low-quality | Genome-fragment | 5.94 | AAI-based (medium-confidence) | 0 |
| S3_vs_k141_19435\|\|full | 2845 | No | NA | 5 | 1 | 0 | Low-quality | Genome-fragment | 4.96 | AAI-based (medium-confidence) | 0 |
| S8_vs_k141_132476\|\|full | 2845 | No | NA | 6 | 1 | 0 | Low-quality | Genome-fragment | 1.52 | HMM-based (lower-bound) | 0 |
| S16_vs_75540\|\|full | 10043 | No | NA | 7 | 0 | 5 | Not-determined | Genome-fragment | NA | NA | 0 |
| S16_vs_75747\|\|full | 1338 | No | NA | 4 | 0 | 1 | Not-determined | Genome-fragment | NA | NA | 0 |
| S9_vs_k141_40253\|\|full | 2846 | No | NA | 8 | 0 | 0 | Low-quality | Genome-fragment | 7.03 | AAI-based (high-confidence) | 0 |
| S16_vs_75849\|\|full | 1860 | No | NA | 3 | 0 | 1 | Not-determined | Genome-fragment | NA | NA | 0 |
| S16_vs_75870\|\|full | 1635 | No | NA | 3 | 0 | 0 | Not-determined | Genome-fragment | NA | NA | 0 |
| S9_vs_k141_3385\|\|full | 2846 | No | NA | 2 | 0 | 1 | Low-quality | Genome-fragment | 2.08 | AAI-based (medium-confidence) | 0 |
| S16_vs_75950\|\|full | 2153 | No | NA | 4 | 0 | 0 | Not-determined | Genome-fragment | NA | NA | 0 |
| S16_vs_6442\|\|full | 2846 | No | NA | 4 | 1 | 0 | Low-quality | Genome-fragment | 3.53 | AAI-based (high-confidence) | 0 |
| S12_vs_k141_35894\|\|full | 2846 | No | NA | 4 | 0 | 1 | Low-quality | Genome-fragment | 1.4 | AAI-based (medium-confidence) | 0 |
| S8_vs_k141_8278\|\|full | 2846 | No | NA | 4 | 0 | 2 | Low-quality | Genome-fragment | 1.48 | AAI-based (medium-confidence) | 0 |
| S16_vs_76327\|\|full | 1554 | No | NA | 3 | 0 | 0 | Not-determined | Genome-fragment | NA | NA | 0 |
| S16_vs_76333\|\|full | 3340 | No | NA | 4 | 0 | 3 | Not-determined | Genome-fragment | NA | NA | 0 |
| S16_vs_76362\|\|full | 7726 | No | NA | 8 | 0 | 1 | Not-determined | Genome-fragment | NA | NA | 0 |
| S17_extracted_k141_104476 | 2847 | No | NA | 4 | 0 | 0 | Low-quality | Genome-fragment | 4.76 | AAI-based (high-confidence) | 0 |
| S16_vs_76422\|\|full | 2690 | No | NA | 4 | 0 | 2 | Not-determined | Genome-fragment | NA | NA | 0 |
| S16_vs_76587\|\|full | 3066 | No | NA | 3 | 0 | 3 | Not-determined | Genome-fragment | NA | NA | 0 |
| S16_vs_76591\|\|full | 1017 | No | NA | 2 | 0 | 0 | Not-determined | Genome-fragment | NA | NA | 0 |
| S3_vs_k141_2841\|\|full | 2847 | No | NA | 4 | 0 | 0 | Low-quality | Genome-fragment | 9.68 | AAI-based (medium-confidence) | 0 |
| S12_vs_k141_141067\|\|full | 2847 | No | NA | 3 | 0 | 2 | Low-quality | Genome-fragment | 0.61 | AAI-based (medium-confidence) | 0 |
| S9_vs_k141_30862\|\|full | 2848 | No | NA | 3 | 0 | 2 | Low-quality | Genome-fragment | 0.44 | AAI-based (medium-confidence) | 0 |
| S13_vs_71094\|\|full | 2848 | No | NA | 7 | 0 | 0 | Low-quality | Genome-fragment | 3.23 | AAI-based (medium-confidence) | 0 |
| S16_vs_76778\|\|full | 1207 | No | NA | 3 | 0 | 1 | Not-determined | Genome-fragment | NA | NA | 0 |
| S8_vs_k141_91058\|\|full | 2848 | No | NA | 8 | 0 | 0 | Low-quality | Genome-fragment | 8.12 | AAI-based (high-confidence) | 0 |
| S14_vs_k141_54197\|\|full | 2849 | No | NA | 3 | 2 | 0 | Low-quality | Genome-fragment | 4.93 | AAI-based (high-confidence) | 0 |
| S16_vs_77020\|\|full | 10191 | No | NA | 8 | 0 | 3 | Not-determined | Genome-fragment | NA | NA | 0 |
| S16_vs_77044\|\|full | 1165 | No | NA | 2 | 0 | 0 | Not-determined | Genome-fragment | NA | NA | 0 |
| S16_vs_77061\|\|full | 10720 | No | NA | 14 | 0 | 1 | Not-determined | Genome-fragment | NA | NA | 0 |
| S16_vs_77073\|\|full | 6920 | No | NA | 4 | 0 | 2 | Not-determined | Genome-fragment | NA | NA | 0 |
| S14_vs_k141_14271\|\|full | 2849 | No | NA | 3 | 0 | 1 | Low-quality | Genome-fragment | 4.88 | AAI-based (high-confidence) | 0 |
| S3_vs_k141_52743\|\|full | 2849 | No | NA | 4 | 0 | 1 | Low-quality | Genome-fragment | 4.41 | AAI-based (high-confidence) | 0 |
| S16_vs_77436\|\|full | 2717 | No | NA | 4 | 0 | 3 | Not-determined | Genome-fragment | NA | NA | 0 |
| S16_vs_77471\|\|full | 1379 | No | NA | 3 | 0 | 0 | Not-determined | Genome-fragment | NA | NA | 0 |
| S9_extracted_k141_36708 | 2850 | No | NA | 3 | 3 | 0 | Low-quality | Genome-fragment | 8.14 | AAI-based (medium-confidence) | 0 |
| S1_extracted_k141_100844 | 2850 | No | NA | 5 | 3 | 0 | Low-quality | Genome-fragment | 6.6 | AAI-based (high-confidence) | 0 |
| S12_vs_k141_4435\|\|full | 2850 | No | NA | 3 | 0 | 2 | Low-quality | Genome-fragment | 1.85 | AAI-based (high-confidence) | 0 |
| S16_vs_77656\|\|full | 4925 | No | NA | 6 | 0 | 0 | Not-determined | Genome-fragment | NA | NA | 0 |
| S16_vs_77704\|\|full | 2233 | No | NA | 4 | 0 | 0 | Not-determined | Genome-fragment | NA | NA | 0 |
| S16_vs_77808\|\|full | 2116 | No | NA | 2 | 0 | 0 | Not-determined | Genome-fragment | NA | NA | 0 |
| S16_vs_77812\|\|full | 1031 | No | NA | 3 | 0 | 1 | Not-determined | Genome-fragment | NA | NA | 0 |
| S8_extracted_k141_1844 | 2850 | No | NA | 4 | 1 | 0 | Low-quality | Genome-fragment | 6.98 | AAI-based (high-confidence) | 0 |
| S16_vs_77931\|\|full | 3399 | No | NA | 4 | 0 | 4 | Not-determined | Genome-fragment | NA | NA | 0 |
| S16_vs_78011\|\|full | 4196 | No | NA | 7 | 0 | 0 | Not-determined | Genome-fragment | NA | NA | 0 |
| S11_vs_k141_71854\|\|full | 2851 | No | NA | 3 | 3 | 0 | Low-quality | Genome-fragment | 8.24 | AAI-based (high-confidence) | 0 |
| S16_vs_78089\|\|full | 2256 | No | NA | 4 | 0 | 1 | Not-determined | Genome-fragment | NA | NA | 0 |
| S16_vs_78101\|\|full | 3158 | No | NA | 5 | 0 | 2 | Not-determined | Genome-fragment | NA | NA | 0 |
| S1_vs_k141_130675\|\|full | 2851 | No | NA | 4 | 0 | 1 | Low-quality | Genome-fragment | 1.34 | AAI-based (medium-confidence) | 0 |
| S16_vs_78218\|\|full | 1022 | No | NA | 3 | 0 | 0 | Not-determined | Genome-fragment | NA | NA | 0 |
| S16_vs_78241\|\|full | 5766 | No | NA | 6 | 0 | 3 | Not-determined | Genome-fragment | NA | NA | 0 |
| S16_vs_78350\|\|full | 1726 | No | NA | 3 | 0 | 1 | Not-determined | Genome-fragment | NA | NA | 0 |
| S16_vs_78500\|\|full | 1873 | No | NA | 2 | 0 | 2 | Not-determined | Genome-fragment | NA | NA | 0 |
| S3_vs_k141_118601\|\|full | 2851 | No | NA | 3 | 2 | 0 | Low-quality | Genome-fragment | 1.74 | AAI-based (high-confidence) | 0 |
| S16_vs_42505\|\|full | 2851 | No | NA | 5 | 3 | 0 | Low-quality | Genome-fragment | 5.33 | AAI-based (high-confidence) | 0 |
| S16_vs_117133\|\|full | 2851 | No | NA | 3 | 1 | 0 | Low-quality | Genome-fragment | 3.9 | HMM-based (lower-bound) | 0 |
| S10_vs_86052\|\|full | 2851 | No | NA | 3 | 0 | 1 | Low-quality | Genome-fragment | 5.14 | AAI-based (medium-confidence) | 0 |
| S14_vs_k141_48346\|\|full | 2852 | No | NA | 6 | 0 | 0 | Low-quality | Genome-fragment | 5.3 | AAI-based (medium-confidence) | 0 |
| S16_vs_78757\|\|full | 1236 | No | NA | 2 | 0 | 0 | Not-determined | Genome-fragment | NA | NA | 0 |
| S5_extracted_k141_54945 | 2852 | No | NA | 5 | 0 | 1 | Low-quality | Genome-fragment | 4.76 | AAI-based (high-confidence) | 0 |
| S16_vs_79123\|\|full | 1875 | No | NA | 2 | 0 | 0 | Not-determined | Genome-fragment | NA | NA | 0 |
| S16_vs_79127\|\|full | 1354 | No | NA | 2 | 0 | 2 | Not-determined | Genome-fragment | NA | NA | 0 |
| S12_extracted_k141_140766 | 2852 | No | NA | 7 | 1 | 0 | Low-quality | Genome-fragment | 6.47 | AAI-based (high-confidence) | 0 |
| S5_extracted_k141_143079 | 2853 | Yes | 2081 | 4 | 1 | 1 | Low-quality | Genome-fragment | 0.74 | HMM-based (lower-bound) | 27.06 |
| S5_vs_k141_36759\|\|full | 2853 | No | NA | 4 | 2 | 0 | Low-quality | Genome-fragment | 5.82 | AAI-based (high-confidence) | 0 |
| S16_vs_50242\|\|full | 2853 | No | NA | 4 | 0 | 1 | Low-quality | Genome-fragment | 3.71 | AAI-based (medium-confidence) | 0 |
| S16_vs_79396\|\|full | 1060 | No | NA | 2 | 0 | 0 | Not-determined | Genome-fragment | NA | NA | 0 |
| S1_vs_k141_45260\|\|full | 2854 | No | NA | 4 | 3 | 0 | Low-quality | Genome-fragment | 6.27 | AAI-based (high-confidence) | 0 |
| S3_extracted_k141_17198 | 2854 | No | NA | 7 | 2 | 0 | Low-quality | Genome-fragment | 4.47 | AAI-based (high-confidence) | 0 |
| S3_extracted_k141_97667 | 2855 | No | NA | 6 | 1 | 0 | Low-quality | Genome-fragment | 4.75 | AAI-based (high-confidence) | 0 |
| S3_vs_k141_42231\|\|full | 2855 | No | NA | 9 | 0 | 0 | Low-quality | Genome-fragment | 1.67 | AAI-based (medium-confidence) | 0 |
| S16_vs_79658\|\|full | 1031 | No | NA | 3 | 0 | 0 | Not-determined | Genome-fragment | NA | NA | 0 |
| S3_vs_k141_93258\|\|full | 2855 | No | NA | 3 | 0 | 0 | Low-quality | Genome-fragment | 6.25 | AAI-based (medium-confidence) | 0 |
| S16_vs_80038\|\|full | 4773 | No | NA | 6 | 0 | 4 | Not-determined | Genome-fragment | NA | NA | 0 |
| S10_vs_48131\|\|full | 2855 | No | NA | 7 | 2 | 0 | Low-quality | Genome-fragment | 7.25 | AAI-based (medium-confidence) | 0 |
| S4_vs_VIRSorter_9937-cat_2 | 2855 | No | NA | 7 | 6 | 0 | Low-quality | Genome-fragment | 9.41 | AAI-based (high-confidence) | 0 |
| S16_vs_80190\|\|full | 2555 | No | NA | 3 | 0 | 1 | Not-determined | Genome-fragment | NA | NA | 0 |
| S16_vs_80244\|\|full | 54066 | No | NA | 66 | 15 | 4 | High-quality | High-quality | 93.8 | AAI-based (high-confidence) | 0 |
| S13_vs_128033\|\|full | 2856 | No | NA | 4 | 0 | 0 | Low-quality | Genome-fragment | 7.13 | AAI-based (medium-confidence) | 0 |
| S16_vs_80672\|\|full | 3908 | No | NA | 6 | 0 | 0 | Not-determined | Genome-fragment | NA | NA | 0 |
| S7_vs_50465\|\|full | 2856 | No | NA | 3 | 3 | 0 | Low-quality | Genome-fragment | 6.43 | AAI-based (high-confidence) | 0 |
| S8_extracted_k141_175998 | 2856 | No | NA | 5 | 5 | 0 | Low-quality | Genome-fragment | 6.14 | AAI-based (high-confidence) | 0 |
| S16_vs_80993\|\|full | 1960 | No | NA | 4 | 0 | 2 | Not-determined | Genome-fragment | NA | NA | 0 |
| S16_vs_80999\|\|full | 1297 | No | NA | 3 | 0 | 1 | Not-determined | Genome-fragment | NA | NA | 0 |
| S16_vs_81032\|\|full | 2342 | No | NA | 4 | 0 | 1 | Not-determined | Genome-fragment | NA | NA | 0 |
| S2_vs_k141_472\|\|full | 2857 | No | NA | 8 | 0 | 0 | Low-quality | Genome-fragment | 6.22 | AAI-based (high-confidence) | 0 |
| S16_vs_81071\|\|full | 2195 | No | NA | 2 | 0 | 2 | Not-determined | Genome-fragment | NA | NA | 0 |
| S16_vs_81077\|\|full | 5136 | No | NA | 9 | 0 | 0 | Not-determined | Genome-fragment | NA | NA | 0 |
| S16_vs_81188\|\|full | 4304 | No | NA | 5 | 0 | 3 | Not-determined | Genome-fragment | NA | NA | 0 |
| S16_vs_81199\|\|full | 7000 | No | NA | 5 | 0 | 4 | Not-determined | Genome-fragment | NA | NA | 0 |
| S5_vs_k141_124308\|\|full | 2858 | No | NA | 4 | 3 | 0 | Low-quality | Genome-fragment | 6.21 | AAI-based (high-confidence) | 0 |
| S9_vs_k141_78069\|\|full | 2859 | No | NA | 5 | 4 | 0 | Low-quality | Genome-fragment | 5.94 | AAI-based (medium-confidence) | 0 |
| S13_extracted_1145 | 2859 | No | NA | 3 | 1 | 0 | Low-quality | Genome-fragment | 7.01 | AAI-based (medium-confidence) | 0 |
| S16_vs_81457\|\|full | 1919 | No | NA | 3 | 0 | 1 | Not-determined | Genome-fragment | NA | NA | 0 |
| S16_vs_81492\|\|full | 2618 | No | NA | 7 | 0 | 0 | Not-determined | Genome-fragment | NA | NA | 0 |
| S16_vs_81549\|\|full | 1996 | No | NA | 2 | 0 | 2 | Not-determined | Genome-fragment | NA | NA | 0 |
| S12_vs_k141_34612\|\|full | 2859 | No | NA | 8 | 2 | 0 | Low-quality | Genome-fragment | 7.94 | AAI-based (medium-confidence) | 0 |
| S3_vs_k141_35724\|\|lt2gene | 2861 | No | NA | 2 | 1 | 0 | Low-quality | Genome-fragment | 4.17 | HMM-based (lower-bound) | 0 |
| S16_extracted_166272 | 2861 | No | NA | 6 | 1 | 0 | Low-quality | Genome-fragment | 7.87 | AAI-based (high-confidence) | 0 |
| S16_vs_81805\|\|full | 32483 | No | NA | 44 | 17 | 0 | Medium-quality | Genome-fragment | 88.09 | AAI-based (high-confidence) | 0 |
| S10_vs_52918\|\|full | 2861 | No | NA | 8 | 1 | 0 | Low-quality | Genome-fragment | 5.71 | AAI-based (medium-confidence) | 0 |
| S16_vs_82004\|\|full | 6450 | No | NA | 7 | 0 | 3 | Not-determined | Genome-fragment | NA | NA | 0 |
| S17_extracted_k141_44122 | 2862 | No | NA | 4 | 2 | 0 | Low-quality | Genome-fragment | 4.92 | AAI-based (medium-confidence) | 0 |
| S16_vs_82087\|\|full | 1042 | No | NA | 1 | 0 | 0 | Not-determined | Genome-fragment | NA | NA | 0 |
| S16_vs_82094\|\|full | 1245 | No | NA | 2 | 0 | 0 | Not-determined | Genome-fragment | NA | NA | 0 |
| S16_vs_82116\|\|full | 113606 | Yes | 40122 | 131 | 19 | 56 | High-quality | High-quality | 100 | AAI-based (high-confidence) | 64.68 |
| S11_vs_k141_68202\|\|full | 2862 | No | NA | 3 | 2 | 0 | Low-quality | Genome-fragment | 7.42 | AAI-based (high-confidence) | 0 |
| S16_vs_82424\|\|full | 6889 | No | NA | 9 | 0 | 2 | Not-determined | Genome-fragment | NA | NA | 0 |
| S11_vs_k141_58397\|\|full | 2863 | No | NA | 5 | 1 | 0 | Low-quality | Genome-fragment | 8.37 | AAI-based (high-confidence) | 0 |
| S12_vs_k141_35497\|\|full | 2863 | No | NA | 4 | 2 | 0 | Low-quality | Genome-fragment | 5.72 | AAI-based (high-confidence) | 0 |
| S2_vs_k141_4182\|\|full | 2863 | No | NA | 3 | 2 | 0 | Low-quality | Genome-fragment | 1.78 | AAI-based (medium-confidence) | 0 |
| S16_vs_82633\|\|full | 2097 | No | NA | 3 | 0 | 0 | Not-determined | Genome-fragment | NA | NA | 0 |
| S16_vs_82642\|\|full | 3369 | No | NA | 4 | 0 | 3 | Not-determined | Genome-fragment | NA | NA | 0 |
| S16_vs_82663\|\|full | 10582 | No | NA | 10 | 0 | 7 | Not-determined | Genome-fragment | NA | NA | 0 |
| S16_vs_82716\|\|full | 6461 | No | NA | 6 | 0 | 3 | Not-determined | Genome-fragment | NA | NA | 0 |
| S5_extracted_k141_13111 | 2864 | No | NA | 6 | 4 | 0 | Low-quality | Genome-fragment | 6.88 | AAI-based (high-confidence) | 0 |
| S7_vs_122837\|\|full | 2864 | No | NA | 10 | 2 | 0 | Low-quality | Genome-fragment | 7.3 | AAI-based (medium-confidence) | 0 |
| S16_vs_82858\|\|full | 2122 | No | NA | 3 | 0 | 1 | Not-determined | Genome-fragment | NA | NA | 0 |
| S16_vs_82953\|\|full | 1458 | No | NA | 2 | 0 | 1 | Not-determined | Genome-fragment | NA | NA | 0 |
| S16_vs_82959\|\|full | 2088 | No | NA | 3 | 0 | 1 | Not-determined | Genome-fragment | NA | NA | 0 |
| S4_extracted_9902 | 2864 | No | NA | 7 | 1 | 0 | Low-quality | Genome-fragment | 1.55 | HMM-based (lower-bound) | 0 |
| S16_vs_83120\|\|full | 2461 | No | NA | 5 | 0 | 1 | Not-determined | Genome-fragment | NA | NA | 0 |
| S17_vs_k141_167536\|\|full | 2865 | No | NA | 5 | 1 | 0 | Low-quality | Genome-fragment | 5.51 | AAI-based (medium-confidence) | 0 |
| S13_vs_16848\|\|full | 2865 | No | NA | 4 | 0 | 1 | Low-quality | Genome-fragment | 1.89 | AAI-based (medium-confidence) | 0 |
| S16_vs_83234\|\|full | 4473 | No | NA | 5 | 0 | 3 | Not-determined | Genome-fragment | NA | NA | 0 |
| S4_extracted_1970 | 2865 | No | NA | 9 | 9 | 0 | Low-quality | Genome-fragment | 9.45 | AAI-based (high-confidence) | 0 |
| S16_vs_83375\|\|full | 2180 | No | NA | 4 | 0 | 1 | Not-determined | Genome-fragment | NA | NA | 0 |
| S16_vs_83442\|\|full | 1141 | No | NA | 2 | 0 | 1 | Not-determined | Genome-fragment | NA | NA | 0 |
| S16_vs_83508\|\|full | 1446 | No | NA | 3 | 0 | 3 | Not-determined | Genome-fragment | NA | NA | 0 |
| S16_vs_83516\|\|full | 2442 | No | NA | 4 | 0 | 1 | Not-determined | Genome-fragment | NA | NA | 0 |
| S16_vs_83518\|\|full | 5740 | No | NA | 4 | 0 | 3 | Not-determined | Genome-fragment | NA | NA | 0 |
| S16_vs_83668\|\|full | 1244 | No | NA | 3 | 0 | 1 | Not-determined | Genome-fragment | NA | NA | 0 |
| S17_vs_k141_30859\|\|full | 2866 | No | NA | 5 | 0 | 0 | Low-quality | Genome-fragment | 3.86 | AAI-based (high-confidence) | 0 |
| S3_vs_k141_33713\|\|full | 2866 | No | NA | 7 | 1 | 0 | Low-quality | Genome-fragment | 7 | AAI-based (medium-confidence) | 0 |
| S16_vs_83900\|\|full | 1883 | No | NA | 2 | 0 | 0 | Not-determined | Genome-fragment | NA | NA | 0 |
| S16_vs_83905\|\|full | 2367 | No | NA | 4 | 0 | 1 | Not-determined | Genome-fragment | NA | NA | 0 |
| S3_vs_k141_100276\|\|full | 2866 | No | NA | 4 | 1 | 0 | Low-quality | Genome-fragment | 1.77 | HMM-based (lower-bound) | 0 |
| S16_vs_83934\|\|full | 1720 | No | NA | 3 | 0 | 0 | Not-determined | Genome-fragment | NA | NA | 0 |
| S16_vs_83945\|\|full | 1421 | No | NA | 2 | 0 | 0 | Not-determined | Genome-fragment | NA | NA | 0 |
| S16_vs_84125\|\|full | 6281 | No | NA | 10 | 0 | 2 | Not-determined | Genome-fragment | NA | NA | 0 |
| S16_vs_158000\|\|full | 2866 | No | NA | 3 | 0 | 2 | Low-quality | Genome-fragment | 1.18 | AAI-based (medium-confidence) | 0 |
| S12_extracted_k141_125655 | 2866 | No | NA | 7 | 2 | 0 | Low-quality | Genome-fragment | 7.41 | AAI-based (high-confidence) | 0 |
| S16_vs_84376\|\|full | 1737 | No | NA | 3 | 0 | 2 | Not-determined | Genome-fragment | NA | NA | 0 |
| S16_vs_84398\|\|full | 4879 | No | NA | 6 | 0 | 0 | Not-determined | Genome-fragment | NA | NA | 0 |
| S16_vs_84489\|\|full | 2352 | No | NA | 4 | 0 | 0 | Not-determined | Genome-fragment | NA | NA | 0 |
| S12_vs_k141_50565\|\|full | 2866 | No | NA | 3 | 1 | 0 | Low-quality | Genome-fragment | 6.91 | AAI-based (high-confidence) | 0 |
| S17_vs_k141_99797\|\|full | 2867 | No | NA | 4 | 1 | 0 | Low-quality | Genome-fragment | 4.13 | AAI-based (high-confidence) | 0 |
| S16_vs_84788\|\|full | 1670 | No | NA | 2 | 0 | 1 | Not-determined | Genome-fragment | NA | NA | 0 |
| S9_vs_k141_20932\|\|full | 2867 | No | NA | 3 | 0 | 0 | Low-quality | Genome-fragment | 2.87 | AAI-based (medium-confidence) | 0 |
| S16_vs_84821\|\|full | 4250 | No | NA | 8 | 0 | 0 | Not-determined | Genome-fragment | NA | NA | 0 |
| S16_vs_85128\|\|full | 3201 | No | NA | 6 | 0 | 5 | Not-determined | Genome-fragment | NA | NA | 0 |
| S14_vs_k141_81436\|\|full | 2867 | No | NA | 7 | 0 | 0 | Low-quality | Genome-fragment | 5.26 | AAI-based (high-confidence) | 0 |
| S7_vs_134050\|\|full | 2867 | No | NA | 8 | 4 | 0 | Low-quality | Genome-fragment | 6.06 | AAI-based (high-confidence) | 0 |
| S8_extracted_k141_90555 | 2867 | No | NA | 3 | 0 | 2 | Low-quality | Genome-fragment | 0.79 | AAI-based (medium-confidence) | 0 |
| S11_vs_k141_79235\|\|full | 2869 | No | NA | 6 | 2 | 0 | Low-quality | Genome-fragment | 5.13 | AAI-based (medium-confidence) | 0 |
| S16_vs_85529\|\|full | 11675 | No | NA | 7 | 0 | 4 | Not-determined | Genome-fragment | NA | NA | 0 |
| S16_vs_85652\|\|full | 5687 | No | NA | 6 | 0 | 4 | Not-determined | Genome-fragment | NA | NA | 0 |
| S16_vs_85844\|\|full | 2717 | No | NA | 3 | 0 | 3 | Not-determined | Genome-fragment | NA | NA | 0 |
| S5_vs_k141_11189\|\|full | 2869 | No | NA | 6 | 3 | 0 | Low-quality | Genome-fragment | 7.2 | AAI-based (medium-confidence) | 0 |
| S16_vs_85882\|\|full | 1895 | No | NA | 3 | 0 | 2 | Not-determined | Genome-fragment | NA | NA | 0 |
| S16_vs_85966\|\|full | 4851 | No | NA | 6 | 0 | 1 | Not-determined | Genome-fragment | NA | NA | 0 |
| S16_vs_86034\|\|full | 1866 | No | NA | 3 | 0 | 1 | Not-determined | Genome-fragment | NA | NA | 0 |
| S16_vs_86039\|\|full | 2120 | No | NA | 4 | 0 | 1 | Not-determined | Genome-fragment | NA | NA | 0 |
| S7_vs_53966\|\|full | 2869 | No | NA | 3 | 2 | 0 | Low-quality | Genome-fragment | 4.66 | HMM-based (lower-bound) | 0 |
| S16_vs_86116\|\|full | 47323 | No | NA | 64 | 19 | 0 | High-quality | High-quality | 98.78 | AAI-based (high-confidence) | 0 |
| S16_vs_86136\|\|full | 1974 | No | NA | 5 | 0 | 0 | Not-determined | Genome-fragment | NA | NA | 0 |
| S16_vs_86207\|\|full | 10277 | No | NA | 8 | 0 | 8 | Not-determined | Genome-fragment | NA | NA | 0 |
| S16_vs_86259\|\|full | 3551 | No | NA | 4 | 0 | 3 | Not-determined | Genome-fragment | NA | NA | 0 |
| S16_vs_86400\|\|full | 2915 | No | NA | 5 | 0 | 5 | Not-determined | Genome-fragment | NA | NA | 0 |
| S10_extracted_58741 | 2869 | No | NA | 4 | 1 | 0 | Low-quality | Genome-fragment | 1.21 | AAI-based (high-confidence) | 0 |
| S15_vs_3575\|\|full | 2869 | No | NA | 5 | 0 | 3 | Low-quality | Genome-fragment | 2.83 | AAI-based (medium-confidence) | 0 |
| S16_vs_86634\|\|full | 1928 | No | NA | 4 | 0 | 1 | Not-determined | Genome-fragment | NA | NA | 0 |
| S16_vs_86715\|\|full | 1277 | No | NA | 3 | 0 | 1 | Not-determined | Genome-fragment | NA | NA | 0 |
| S11_vs_k141_26868\|\|full | 2870 | No | NA | 5 | 0 | 0 | Low-quality | Genome-fragment | 5.79 | AAI-based (high-confidence) | 0 |
| S17_vs_k141_98155\|\|full | 2871 | No | NA | 2 | 0 | 2 | Low-quality | Genome-fragment | 0.96 | AAI-based (medium-confidence) | 0 |
| S3_vs_k141_88052\|\|full | 2871 | No | NA | 9 | 1 | 0 | Low-quality | Genome-fragment | 4.68 | AAI-based (high-confidence) | 0 |
| S10_vs_35128\|\|full | 2871 | No | NA | 3 | 3 | 0 | Low-quality | Genome-fragment | 6.73 | AAI-based (high-confidence) | 0 |
| S16_vs_86955\|\|full | 4892 | No | NA | 6 | 0 | 0 | Not-determined | Genome-fragment | NA | NA | 0 |
| S16_vs_87120\|\|full | 1249 | No | NA | 3 | 0 | 2 | Not-determined | Genome-fragment | NA | NA | 0 |
| S16_vs_87173\|\|full | 1795 | No | NA | 2 | 0 | 0 | Not-determined | Genome-fragment | NA | NA | 0 |
| S1_extracted_k141_102301 | 2872 | No | NA | 3 | 0 | 1 | Low-quality | Genome-fragment | 1.92 | AAI-based (high-confidence) | 0 |
| S3_extracted_k141_51124 | 2872 | No | NA | 6 | 0 | 0 | Low-quality | Genome-fragment | 2.15 | AAI-based (high-confidence) | 0 |
| S8_vs_k141_187841\|\|full | 2872 | No | NA | 2 | 0 | 1 | Low-quality | Genome-fragment | 0.88 | AAI-based (medium-confidence) | 0 |
| S11_vs_k141_17774\|\|full | 2873 | No | NA | 4 | 2 | 0 | Low-quality | Genome-fragment | 7.72 | AAI-based (high-confidence) | 0 |
| S16_vs_87622\|\|full | 1147 | No | NA | 4 | 0 | 0 | Not-determined | Genome-fragment | NA | NA | 0 |
| S16_vs_87645\|\|full | 5121 | No | NA | 5 | 0 | 5 | Not-determined | Genome-fragment | NA | NA | 0 |
| S16_vs_87677\|\|full | 2193 | No | NA | 2 | 0 | 1 | Not-determined | Genome-fragment | NA | NA | 0 |
| S16_vs_87753\|\|full | 1595 | No | NA | 3 | 0 | 1 | Not-determined | Genome-fragment | NA | NA | 0 |
| S16_vs_87783\|\|full | 4550 | No | NA | 6 | 0 | 2 | Not-determined | Genome-fragment | NA | NA | 0 |
| S3_vs_k141_124404\|\|full | 2873 | No | NA | 6 | 2 | 0 | Low-quality | Genome-fragment | 20.06 | AAI-based (high-confidence) | 0 |
| S16_vs_87898\|\|full | 1396 | No | NA | 3 | 0 | 1 | Not-determined | Genome-fragment | NA | NA | 0 |
| S16_vs_87902\|\|full | 1349 | No | NA | 3 | 0 | 2 | Not-determined | Genome-fragment | NA | NA | 0 |
| S16_vs_87948\|\|full | 1947 | No | NA | 3 | 0 | 2 | Not-determined | Genome-fragment | NA | NA | 0 |
| S4_extracted_525 | 2873 | No | NA | 7 | 6 | 0 | Low-quality | Genome-fragment | 9.47 | AAI-based (high-confidence) | 0 |
| S17_extracted_k141_162965 | 2874 | No | NA | 4 | 0 | 0 | Low-quality | Genome-fragment | 2.74 | AAI-based (high-confidence) | 0 |
| S16_vs_88101\|\|full | 4736 | No | NA | 3 | 0 | 2 | Not-determined | Genome-fragment | NA | NA | 0 |
| S14_vs_k141_69644\|\|full | 2874 | No | NA | 4 | 4 | 0 | Low-quality | Genome-fragment | 9 | AAI-based (high-confidence) | 0 |
| S16_vs_88140\|\|full | 1225 | No | NA | 2 | 0 | 1 | Not-determined | Genome-fragment | NA | NA | 0 |
| S7_vs_133263\|\|full | 2874 | No | NA | 3 | 0 | 0 | Low-quality | Genome-fragment | 7.33 | AAI-based (medium-confidence) | 0 |
| S5_vs_k141_133698\|\|full | 2875 | No | NA | 3 | 0 | 2 | Low-quality | Genome-fragment | 1.58 | AAI-based (medium-confidence) | 0 |
| S16_vs_88373\|\|full | 11952 | No | NA | 11 | 0 | 4 | Not-determined | Genome-fragment | NA | NA | 0 |
| S5_vs_k141_123825\|\|full | 2875 | No | NA | 3 | 1 | 0 | Low-quality | Genome-fragment | 4.26 | HMM-based (lower-bound) | 0 |
| S5_vs_k141_132710\|\|full | 2875 | No | NA | 8 | 1 | 0 | Low-quality | Genome-fragment | 7.16 | AAI-based (medium-confidence) | 0 |
| S7_vs_8229\|\|full | 2876 | No | NA | 3 | 0 | 0 | Low-quality | Genome-fragment | 5.75 | AAI-based (medium-confidence) | 0 |
| S16_vs_88558\|\|full | 2171 | No | NA | 3 | 0 | 1 | Not-determined | Genome-fragment | NA | NA | 0 |
| S16_vs_88691\|\|full | 7521 | No | NA | 6 | 0 | 5 | Not-determined | Genome-fragment | NA | NA | 0 |
| S16_vs_88923\|\|full | 1134 | No | NA | 2 | 0 | 1 | Not-determined | Genome-fragment | NA | NA | 0 |
| S2_vs_k141_13473\|\|full | 2876 | No | NA | 5 | 0 | 0 | Low-quality | Genome-fragment | 6.78 | AAI-based (medium-confidence) | 0 |
| S8_extracted_k141_160149 | 2876 | No | NA | 5 | 2 | 0 | Low-quality | Genome-fragment | 8.5 | HMM-based (lower-bound) | 0 |
| S16_vs_89291\|\|full | 1079 | No | NA | 2 | 0 | 2 | Not-determined | Genome-fragment | NA | NA | 0 |
| S11_vs_k141_30073\|\|full | 2877 | No | NA | 4 | 2 | 0 | Low-quality | Genome-fragment | 7.32 | AAI-based (high-confidence) | 0 |
| S3_extracted_k141_89407 | 2877 | No | NA | 4 | 3 | 0 | Low-quality | Genome-fragment | 9.48 | AAI-based (high-confidence) | 0 |
| S8_extracted_k141_148248 | 2877 | No | NA | 3 | 0 | 1 | Low-quality | Genome-fragment | 1.8 | AAI-based (medium-confidence) | 0 |
| S16_vs_89332\|\|full | 1580 | No | NA | 2 | 0 | 0 | Not-determined | Genome-fragment | NA | NA | 0 |
| S16_vs_89346\|\|full | 3637 | No | NA | 4 | 0 | 3 | Not-determined | Genome-fragment | NA | NA | 0 |
| S16_vs_89360\|\|full | 4185 | No | NA | 3 | 0 | 2 | Not-determined | Genome-fragment | NA | NA | 0 |
| S8_extracted_k141_31256 | 2877 | No | NA | 3 | 2 | 0 | Low-quality | Genome-fragment | 9.15 | AAI-based (high-confidence) | 0 |
| S11_vs_k141_29929\|\|full | 2878 | No | NA | 3 | 0 | 2 | Low-quality | Genome-fragment | 3.39 | AAI-based (medium-confidence) | 0 |
| S16_vs_89511\|\|full | 84541 | Yes | 63605 | 114 | 21 | 15 | High-quality | High-quality | 92.55 | HMM-based (lower-bound) | 24.76 |
| S16_vs_90229\|\|full | 4711 | No | NA | 5 | 0 | 2 | Not-determined | Genome-fragment | NA | NA | 0 |
| S16_vs_90314\|\|full | 1063 | No | NA | 2 | 0 | 2 | Not-determined | Genome-fragment | NA | NA | 0 |
| S14_extracted_k141_26393 | 2878 | No | NA | 4 | 0 | 0 | Low-quality | Genome-fragment | 3.9 | AAI-based (medium-confidence) | 0 |
| S5_vs_k141_77696\|\|full | 2878 | No | NA | 3 | 0 | 3 | Low-quality | Genome-fragment | 1.26 | AAI-based (medium-confidence) | 0 |
| S5_vs_k141_145025\|\|full | 2879 | No | NA | 7 | 1 | 0 | Low-quality | Genome-fragment | 5.43 | AAI-based (medium-confidence) | 0 |
| S7_vs_35676\|\|full | 2880 | No | NA | 9 | 0 | 0 | Low-quality | Genome-fragment | 6.37 | AAI-based (medium-confidence) | 0 |
| S12_vs_k141_122154\|\|full | 2880 | No | NA | 3 | 2 | 1 | Low-quality | Genome-fragment | 4.47 | HMM-based (lower-bound) | 0 |
| S16_vs_90938\|\|full | 7331 | No | NA | 6 | 0 | 6 | Not-determined | Genome-fragment | NA | NA | 0 |
| S12_vs_k141_78437\|\|full | 2880 | No | NA | 7 | 0 | 0 | Low-quality | Genome-fragment | 5.25 | AAI-based (medium-confidence) | 0 |
| S16_vs_90962\|\|full | 2090 | No | NA | 3 | 0 | 0 | Not-determined | Genome-fragment | NA | NA | 0 |
| S2_vs_k141_7449\|\|full | 2880 | No | NA | 7 | 1 | 0 | Low-quality | Genome-fragment | 6.37 | AAI-based (medium-confidence) | 0 |
| S16_vs_91008\|\|full | 1429 | No | NA | 2 | 0 | 1 | Not-determined | Genome-fragment | NA | NA | 0 |
| S17_vs_k141_143453\|\|full | 2881 | No | NA | 2 | 0 | 2 | Low-quality | Genome-fragment | 1.58 | AAI-based (medium-confidence) | 0 |
| S16_vs_91096\|\|full | 1853 | No | NA | 2 | 0 | 1 | Not-determined | Genome-fragment | NA | NA | 0 |
| S8_extracted_k141_16186 | 2881 | No | NA | 3 | 2 | 0 | Low-quality | Genome-fragment | 8.88 | AAI-based (high-confidence) | 0 |
| S16_vs_91324\|\|full | 1417 | No | NA | 3 | 0 | 0 | Not-determined | Genome-fragment | NA | NA | 0 |
| S17_vs_k141_17043\|\|full | 2882 | No | NA | 9 | 0 | 0 | Low-quality | Genome-fragment | 7.2 | AAI-based (high-confidence) | 0 |
| S13_vs_140998\|\|full | 2882 | No | NA | 4 | 0 | 0 | Low-quality | Genome-fragment | 6.38 | AAI-based (high-confidence) | 0 |
| S16_vs_91448\|\|full | 1689 | No | NA | 4 | 0 | 1 | Not-determined | Genome-fragment | NA | NA | 0 |
| S16_vs_91496\|\|full | 3485 | No | NA | 2 | 0 | 1 | Not-determined | Genome-fragment | NA | NA | 0 |
| S16_vs_91534\|\|full | 6814 | No | NA | 6 | 0 | 2 | Not-determined | Genome-fragment | NA | NA | 0 |
| S1_vs_k141_50229\|\|full | 2882 | No | NA | 9 | 1 | 0 | Low-quality | Genome-fragment | 1.86 | HMM-based (lower-bound) | 0 |
| S3_vs_k141_29327\|\|full | 2882 | No | NA | 4 | 0 | 0 | Low-quality | Genome-fragment | 6.4 | AAI-based (high-confidence) | 0 |
| S16_vs_91795\|\|full | 2648 | No | NA | 3 | 0 | 1 | Not-determined | Genome-fragment | NA | NA | 0 |
| S16_vs_91796\|\|full | 1230 | No | NA | 2 | 0 | 0 | Not-determined | Genome-fragment | NA | NA | 0 |
| S16_vs_91861\|\|full | 1755 | No | NA | 4 | 0 | 1 | Not-determined | Genome-fragment | NA | NA | 0 |
| S16_vs_91932\|\|full | 2327 | No | NA | 2 | 0 | 1 | Not-determined | Genome-fragment | NA | NA | 0 |
| S16_vs_92043\|\|full | 4217 | No | NA | 4 | 0 | 0 | Not-determined | Genome-fragment | NA | NA | 0 |
| S7_vs_106506\|\|full | 2882 | No | NA | 2 | 1 | 0 | Low-quality | Genome-fragment | 6.93 | AAI-based (medium-confidence) | 0 |
| S16_vs_92150\|\|full | 29970 | No | NA | 42 | 16 | 0 | Medium-quality | Genome-fragment | 87.3 | AAI-based (medium-confidence) | 0 |
| S16_vs_92323\|\|full | 1403 | No | NA | 2 | 0 | 1 | Not-determined | Genome-fragment | NA | NA | 0 |
| S16_vs_92353\|\|full | 5576 | No | NA | 3 | 0 | 2 | Not-determined | Genome-fragment | NA | NA | 0 |
| S6_vs_k141_49257\|\|full | 2882 | No | NA | 6 | 1 | 0 | Low-quality | Genome-fragment | 5.95 | AAI-based (medium-confidence) | 0 |
| S1_vs_k141_84082\|\|full | 2883 | No | NA | 7 | 1 | 0 | Low-quality | Genome-fragment | 6.39 | AAI-based (high-confidence) | 0 |
| S16_vs_92650\|\|full | 1865 | No | NA | 4 | 0 | 0 | Not-determined | Genome-fragment | NA | NA | 0 |
| S16_vs_92870\|\|full | 1170 | No | NA | 4 | 0 | 1 | Not-determined | Genome-fragment | NA | NA | 0 |
| S7_vs_80386\|\|full | 2883 | Yes | 1575 | 6 | 1 | 1 | Low-quality | Genome-fragment | 4.5 | AAI-based (high-confidence) | 45.37 |
| S8_vs_k141_29550\|\|full | 2883 | Yes | 405 | 4 | 1 | 1 | Low-quality | Genome-fragment | 0.26 | HMM-based (lower-bound) | 85.95 |
| S1_vs_k141_54992\|\|full | 2884 | No | NA | 6 | 2 | 0 | Low-quality | Genome-fragment | 8.28 | AAI-based (high-confidence) | 0 |
| S1_extracted_k141_38306 | 2885 | No | NA | 3 | 2 | 0 | Low-quality | Genome-fragment | 2.92 | AAI-based (high-confidence) | 0 |
| S16_vs_93167\|\|full | 2764 | No | NA | 4 | 0 | 0 | Not-determined | Genome-fragment | NA | NA | 0 |
| S16_vs_93206\|\|full | 2213 | No | NA | 3 | 0 | 3 | Not-determined | Genome-fragment | NA | NA | 0 |
| S16_vs_93216\|\|full | 1233 | No | NA | 2 | 0 | 1 | Not-determined | Genome-fragment | NA | NA | 0 |
| S1_vs_k141_19209\|\|full | 2885 | No | NA | 4 | 0 | 3 | Low-quality | Genome-fragment | 5.47 | AAI-based (medium-confidence) | 0 |
| S5_extracted_k141_103128 | 2885 | No | NA | 3 | 1 | 0 | Low-quality | Genome-fragment | 13.16 | AAI-based (high-confidence) | 0 |
| S4_vs_VIRSorter_11382-cat_2 | 2885 | No | NA | 9 | 9 | 0 | Low-quality | Genome-fragment | 9.49 | AAI-based (high-confidence) | 0 |
| S8_extracted_k141_139224 | 2885 | No | NA | 3 | 0 | 2 | Low-quality | Genome-fragment | 0.9 | AAI-based (medium-confidence) | 0 |
| S16_vs_93396\|\|full | 1867 | No | NA | 3 | 0 | 1 | Not-determined | Genome-fragment | NA | NA | 0 |
| S7_vs_58850\|\|full | 2886 | No | NA | 4 | 3 | 0 | Low-quality | Genome-fragment | 1.39 | AAI-based (high-confidence) | 0 |
| S16_vs_93484\|\|full | 1946 | No | NA | 6 | 0 | 0 | Not-determined | Genome-fragment | NA | NA | 0 |
| S14_vs_k141_20114\|\|full | 2887 | No | NA | 6 | 0 | 0 | Low-quality | Genome-fragment | 6.55 | AAI-based (high-confidence) | 0 |
| S16_vs_93544\|\|full | 1479 | No | NA | 2 | 0 | 0 | Not-determined | Genome-fragment | NA | NA | 0 |
| S16_vs_93776\|\|full | 1285 | No | NA | 3 | 0 | 1 | Not-determined | Genome-fragment | NA | NA | 0 |
| S8_vs_k141_173702\|\|full | 2887 | No | NA | 3 | 0 | 3 | Low-quality | Genome-fragment | 2.05 | AAI-based (medium-confidence) | 0 |
| S11_extracted_k141_75466 | 2888 | No | NA | 5 | 3 | 0 | Low-quality | Genome-fragment | 5.24 | AAI-based (high-confidence) | 0 |
| S16_vs_93949\|\|full | 1184 | No | NA | 3 | 0 | 2 | Not-determined | Genome-fragment | NA | NA | 0 |
| S11_vs_k141_96790\|\|full | 2888 | Yes | 914 | 6 | 2 | 2 | Low-quality | Genome-fragment | 1.82 | AAI-based (high-confidence) | 68.35 |
| S16_vs_93995\|\|full | 1117 | No | NA | 3 | 0 | 0 | Not-determined | Genome-fragment | NA | NA | 0 |
| S16_vs_94037\|\|full | 1357 | No | NA | 2 | 0 | 0 | Not-determined | Genome-fragment | NA | NA | 0 |
| S16_vs_94075\|\|full | 4033 | No | NA | 4 | 0 | 3 | Not-determined | Genome-fragment | NA | NA | 0 |
| S1_vs_k141_53416\|\|full | 2888 | No | NA | 4 | 0 | 0 | Low-quality | Genome-fragment | 6.89 | AAI-based (high-confidence) | 0 |
| S16_vs_94176\|\|full | 1429 | No | NA | 2 | 0 | 2 | Not-determined | Genome-fragment | NA | NA | 0 |
| S1_vs_k141_156744\|\|lt2gene | 2888 | No | NA | 1 | 1 | 0 | Low-quality | Genome-fragment | 8.32 | AAI-based (high-confidence) | 0 |
| S5_vs_k141_5609\|\|full | 2888 | No | NA | 5 | 2 | 0 | Low-quality | Genome-fragment | 4.56 | AAI-based (medium-confidence) | 0 |
| S16_vs_94340\|\|full | 8311 | No | NA | 8 | 0 | 3 | Not-determined | Genome-fragment | NA | NA | 0 |
| S16_vs_94357\|\|full | 3751 | No | NA | 3 | 0 | 1 | Not-determined | Genome-fragment | NA | NA | 0 |
| S15_vs_4664\|\|full | 2888 | No | NA | 5 | 0 | 2 | Low-quality | Genome-fragment | 1.01 | AAI-based (medium-confidence) | 0 |
| S16_vs_94400\|\|full | 7615 | No | NA | 7 | 0 | 5 | Not-determined | Genome-fragment | NA | NA | 0 |
| S3_vs_k141_95995\|\|full | 2889 | No | NA | 3 | 1 | 0 | Low-quality | Genome-fragment | 5.95 | AAI-based (high-confidence) | 0 |
| S16_vs_94531\|\|full | 1598 | No | NA | 2 | 0 | 2 | Not-determined | Genome-fragment | NA | NA | 0 |
| S3_vs_k141_25941\|\|full | 2889 | No | NA | 7 | 1 | 0 | Low-quality | Genome-fragment | 3.68 | AAI-based (medium-confidence) | 0 |
| S5_vs_k141_3805\|\|full | 2889 | No | NA | 5 | 2 | 0 | Low-quality | Genome-fragment | 6.6 | AAI-based (medium-confidence) | 0 |
| S2_vs_k141_15307\|\|full | 2889 | No | NA | 3 | 0 | 0 | Low-quality | Genome-fragment | 8.67 | AAI-based (high-confidence) | 0 |
| S16_vs_94841\|\|full | 3021 | No | NA | 3 | 0 | 1 | Not-determined | Genome-fragment | NA | NA | 0 |
| S13_vs_102116\|\|full | 2890 | No | NA | 3 | 0 | 0 | Low-quality | Genome-fragment | 7.02 | AAI-based (medium-confidence) | 0 |
| S16_vs_94948\|\|full | 1766 | No | NA | 3 | 0 | 0 | Not-determined | Genome-fragment | NA | NA | 0 |
| S16_vs_162336\|\|full | 2890 | No | NA | 4 | 0 | 1 | Low-quality | Genome-fragment | 3.97 | AAI-based (medium-confidence) | 0 |
| S12_vs_k141_39800\|\|full | 2890 | No | NA | 7 | 0 | 0 | Low-quality | Genome-fragment | 6.6 | AAI-based (high-confidence) | 0 |
| S12_vs_k141_81569\|\|full | 2890 | Yes | 1882 | 3 | 1 | 1 | Low-quality | Genome-fragment | 0.67 | HMM-based (lower-bound) | 34.88 |
| S5_vs_k141_135027\|\|full | 2891 | No | NA | 5 | 5 | 0 | Low-quality | Genome-fragment | 6.74 | AAI-based (medium-confidence) | 0 |
| S16_vs_115588\|\|full | 2891 | No | NA | 5 | 0 | 0 | Low-quality | Genome-fragment | 6.96 | AAI-based (medium-confidence) | 0 |
| S10_vs_68786\|\|lt2gene | 2891 | No | NA | 2 | 0 | 0 | Low-quality | Genome-fragment | 7.5 | AAI-based (medium-confidence) | 0 |
| S16_vs_95272\|\|full | 1330 | No | NA | 2 | 0 | 1 | Not-determined | Genome-fragment | NA | NA | 0 |
| S16_vs_95309\|\|full | 2198 | No | NA | 4 | 0 | 2 | Not-determined | Genome-fragment | NA | NA | 0 |
| S8_vs_k141_167080\|\|full | 2891 | No | NA | 4 | 0 | 0 | Low-quality | Genome-fragment | 7.25 | AAI-based (medium-confidence) | 0 |
| S16_vs_95582\|\|full | 1985 | No | NA | 2 | 0 | 1 | Not-determined | Genome-fragment | NA | NA | 0 |
| S14_extracted_k141_75015 | 2892 | No | NA | 7 | 7 | 0 | Low-quality | Genome-fragment | 9.5 | AAI-based (high-confidence) | 0 |
| S16_vs_95742\|\|full | 3142 | No | NA | 4 | 0 | 3 | Not-determined | Genome-fragment | NA | NA | 0 |
| S1_vs_k141_69299\|\|full | 2892 | Yes | 1517 | 6 | 1 | 1 | Low-quality | Genome-fragment | 0.4 | HMM-based (lower-bound) | 47.54 |
| S7_vs_125219\|\|full | 2892 | No | NA | 5 | 0 | 0 | Low-quality | Genome-fragment | 5.14 | AAI-based (medium-confidence) | 0 |
| S10_vs_107823\|\|full | 2892 | No | NA | 9 | 0 | 0 | Low-quality | Genome-fragment | 8.46 | AAI-based (high-confidence) | 0 |
| S16_vs_95854\|\|full | 3325 | No | NA | 5 | 0 | 1 | Not-determined | Genome-fragment | NA | NA | 0 |
| S12_vs_k141_166321\|\|full | 2892 | No | NA | 4 | 3 | 0 | Low-quality | Genome-fragment | 1.74 | AAI-based (medium-confidence) | 0 |
| S17_extracted_k141_81389 | 2893 | No | NA | 6 | 5 | 0 | Low-quality | Genome-fragment | 9.54 | AAI-based (high-confidence) | 0 |
| S16_vs_95909\|\|full | 4278 | No | NA | 4 | 0 | 2 | Not-determined | Genome-fragment | NA | NA | 0 |
| S16_vs_95910\|\|full | 5472 | No | NA | 6 | 0 | 1 | Not-determined | Genome-fragment | NA | NA | 0 |
| S16_vs_95915\|\|full | 1509 | No | NA | 4 | 0 | 1 | Not-determined | Genome-fragment | NA | NA | 0 |
| S13_vs_35055\|\|lt2gene | 2893 | No | NA | 1 | 1 | 0 | Low-quality | Genome-fragment | 5.47 | AAI-based (medium-confidence) | 0 |
| S16_vs_95937\|\|full | 2038 | No | NA | 4 | 0 | 1 | Not-determined | Genome-fragment | NA | NA | 0 |
| S16_vs_96050\|\|full | 2361 | No | NA | 3 | 0 | 0 | Not-determined | Genome-fragment | NA | NA | 0 |
| S16_vs_96051\|\|full | 1080 | No | NA | 3 | 0 | 0 | Not-determined | Genome-fragment | NA | NA | 0 |
| S16_vs_96072\|\|full | 4381 | No | NA | 4 | 0 | 3 | Not-determined | Genome-fragment | NA | NA | 0 |
| S16_vs_96124\|\|full | 1442 | No | NA | 2 | 0 | 1 | Not-determined | Genome-fragment | NA | NA | 0 |
| S16_vs_96263\|\|full | 8824 | No | NA | 7 | 0 | 5 | Not-determined | Genome-fragment | NA | NA | 0 |
| S3_extracted_k141_50859 | 2893 | No | NA | 4 | 1 | 0 | Low-quality | Genome-fragment | 1.96 | AAI-based (high-confidence) | 0 |
| S16_vs_96278\|\|full | 2921 | No | NA | 7 | 0 | 0 | Not-determined | Genome-fragment | NA | NA | 0 |
| S16_vs_96340\|\|full | 3317 | No | NA | 5 | 0 | 3 | Not-determined | Genome-fragment | NA | NA | 0 |
| S16_vs_96400\|\|full | 2682 | No | NA | 3 | 0 | 2 | Not-determined | Genome-fragment | NA | NA | 0 |
| S4_extracted_4023 | 2893 | No | NA | 6 | 4 | 0 | Low-quality | Genome-fragment | 9.53 | AAI-based (high-confidence) | 0 |
| S16_vs_96584\|\|full | 3707 | No | NA | 5 | 0 | 2 | Not-determined | Genome-fragment | NA | NA | 0 |
| S16_vs_96607\|\|full | 1265 | No | NA | 3 | 0 | 0 | Not-determined | Genome-fragment | NA | NA | 0 |
| S16_vs_96671\|\|full | 4359 | No | NA | 7 | 0 | 6 | Not-determined | Genome-fragment | NA | NA | 0 |
| S16_vs_96822\|\|full | 8380 | No | NA | 6 | 0 | 4 | Not-determined | Genome-fragment | NA | NA | 0 |
| S7_vs_67326\|\|full | 2894 | No | NA | 4 | 0 | 2 | Low-quality | Genome-fragment | 2.01 | AAI-based (medium-confidence) | 0 |
| S16_vs_96975\|\|full | 19342 | No | NA | 21 | 0 | 7 | Not-determined | Genome-fragment | NA | NA | 0 |
| S16_vs_97121\|\|full | 1045 | No | NA | 2 | 0 | 2 | Not-determined | Genome-fragment | NA | NA | 0 |
| S3_vs_k141_50817\|\|full | 2895 | No | NA | 3 | 2 | 0 | Low-quality | Genome-fragment | 2.17 | AAI-based (high-confidence) | 0 |
| S14_extracted_k141_1955 | 2896 | No | NA | 6 | 0 | 0 | Low-quality | Genome-fragment | 4.96 | AAI-based (high-confidence) | 0 |
| S16_vs_97312\|\|full | 1054 | No | NA | 2 | 0 | 0 | Not-determined | Genome-fragment | NA | NA | 0 |
| S16_vs_97324\|\|full | 2637 | No | NA | 4 | 0 | 4 | Not-determined | Genome-fragment | NA | NA | 0 |
| S16_vs_97409\|\|full | 3141 | No | NA | 3 | 0 | 1 | Not-determined | Genome-fragment | NA | NA | 0 |
| S1_extracted_k141_16062 | 2896 | No | NA | 7 | 2 | 0 | Low-quality | Genome-fragment | 6.5 | AAI-based (high-confidence) | 0 |
| S1_vs_k141_59036\|\|full | 2896 | No | NA | 5 | 1 | 0 | Low-quality | Genome-fragment | 2.91 | AAI-based (high-confidence) | 0 |
| S5_vs_k141_2427\|\|full | 2896 | No | NA | 6 | 3 | 0 | Low-quality | Genome-fragment | 6.16 | AAI-based (high-confidence) | 0 |
| S12_vs_k141_149394\|\|full | 2896 | No | NA | 6 | 1 | 0 | Low-quality | Genome-fragment | 6.43 | HMM-based (lower-bound) | 0 |
| S8_vs_k141_193476\|\|full | 2896 | No | NA | 8 | 2 | 0 | Low-quality | Genome-fragment | 7.02 | AAI-based (medium-confidence) | 0 |
| S9_vs_k141_61145\|\|full | 2897 | No | NA | 3 | 0 | 0 | Low-quality | Genome-fragment | 7.29 | AAI-based (high-confidence) | 0 |
| S11_vs_k141_34720\|\|full | 2898 | No | NA | 3 | 1 | 0 | Low-quality | Genome-fragment | 6.08 | AAI-based (medium-confidence) | 0 |
| S14_vs_k141_78588\|\|full | 2898 | No | NA | 9 | 1 | 0 | Low-quality | Genome-fragment | 22.82 | AAI-based (high-confidence) | 0 |
| S16_vs_94379\|\|full | 2898 | No | NA | 7 | 0 | 0 | Low-quality | Genome-fragment | 5.8 | AAI-based (high-confidence) | 0 |
| S16_vs_116900\|\|full | 2899 | No | NA | 3 | 0 | 0 | Low-quality | Genome-fragment | 5.76 | AAI-based (high-confidence) | 0 |
| S16_vs_161955\|\|full | 2899 | No | NA | 4 | 0 | 1 | Low-quality | Genome-fragment | 3.38 | AAI-based (medium-confidence) | 0 |
| S16_vs_98174\|\|full | 2923 | No | NA | 4 | 0 | 3 | Not-determined | Genome-fragment | NA | NA | 0 |
| S8_vs_k141_43584\|\|full | 2899 | No | NA | 4 | 0 | 0 | Low-quality | Genome-fragment | 4.56 | AAI-based (medium-confidence) | 0 |
| S16_vs_98184\|\|full | 1601 | No | NA | 2 | 0 | 2 | Not-determined | Genome-fragment | NA | NA | 0 |
| S14_vs_k141_72600\|\|full | 2900 | No | NA | 7 | 1 | 0 | Low-quality | Genome-fragment | 5.7 | AAI-based (high-confidence) | 0 |
| S5_vs_k141_42932\|\|lt2gene | 2900 | No | NA | 1 | 1 | 0 | Low-quality | Genome-fragment | 6.88 | AAI-based (high-confidence) | 0 |
| S10_vs_105117\|\|full | 2900 | No | NA | 4 | 0 | 1 | Low-quality | Genome-fragment | 6.61 | AAI-based (high-confidence) | 0 |
| S13_vs_36291\|\|full | 2901 | No | NA | 2 | 0 | 2 | Low-quality | Genome-fragment | 2.35 | AAI-based (medium-confidence) | 0 |
| S14_extracted_k141_113525 | 2901 | No | NA | 10 | 1 | 0 | Low-quality | Genome-fragment | 8.16 | AAI-based (medium-confidence) | 0 |
| S3_vs_k141_7085\|\|full | 2901 | No | NA | 4 | 1 | 0 | Low-quality | Genome-fragment | 3.26 | HMM-based (lower-bound) | 0 |
| S16_vs_98750\|\|full | 1101 | No | NA | 4 | 0 | 1 | Not-determined | Genome-fragment | NA | NA | 0 |
| S16_vs_99124\|\|full | 2226 | No | NA | 3 | 0 | 1 | Not-determined | Genome-fragment | NA | NA | 0 |
| S16_vs_99172\|\|full | 1807 | No | NA | 3 | 0 | 3 | Not-determined | Genome-fragment | NA | NA | 0 |
| S16_vs_99248\|\|full | 2380 | No | NA | 3 | 0 | 0 | Not-determined | Genome-fragment | NA | NA | 0 |
| S16_vs_99438\|\|full | 2110 | No | NA | 4 | 0 | 0 | Not-determined | Genome-fragment | NA | NA | 0 |
| S16_vs_13839\|\|full | 2901 | No | NA | 5 | 0 | 0 | Low-quality | Genome-fragment | 6 | AAI-based (medium-confidence) | 0 |
| S16_vs_99460\|\|full | 1939 | No | NA | 3 | 0 | 3 | Not-determined | Genome-fragment | NA | NA | 0 |
| S16_vs_99701\|\|full | 1697 | No | NA | 2 | 0 | 1 | Not-determined | Genome-fragment | NA | NA | 0 |
| S16_vs_99718\|\|full | 1607 | No | NA | 3 | 0 | 0 | Not-determined | Genome-fragment | NA | NA | 0 |
| S16_vs_99773\|\|full | 2724 | No | NA | 2 | 0 | 0 | Not-determined | Genome-fragment | NA | NA | 0 |
| S16_vs_99797\|\|full | 1862 | No | NA | 3 | 0 | 0 | Not-determined | Genome-fragment | NA | NA | 0 |
| S16_vs_59552\|\|full | 2901 | No | NA | 3 | 0 | 0 | Low-quality | Genome-fragment | 7.39 | AAI-based (high-confidence) | 0 |
| S16_vs_100064\|\|full | 1185 | No | NA | 2 | 0 | 0 | Not-determined | Genome-fragment | NA | NA | 0 |
| S16_vs_100214\|\|full | 1759 | No | NA | 3 | 0 | 2 | Not-determined | Genome-fragment | NA | NA | 0 |
| S3_extracted_k141_46593 | 2902 | No | NA | 6 | 6 | 0 | Low-quality | Genome-fragment | 9.56 | AAI-based (high-confidence) | 0 |
| S3_vs_k141_85471\|\|full | 2902 | No | NA | 7 | 2 | 0 | Low-quality | Genome-fragment | 6.78 | AAI-based (high-confidence) | 0 |
| S16_vs_100337\|\|full | 1270 | No | NA | 2 | 0 | 2 | Not-determined | Genome-fragment | NA | NA | 0 |
| S3_vs_k141_126603\|\|full | 2902 | No | NA | 6 | 0 | 0 | Low-quality | Genome-fragment | 5.33 | AAI-based (high-confidence) | 0 |
| S16_vs_100381\|\|full | 8344 | No | NA | 8 | 0 | 5 | Not-determined | Genome-fragment | NA | NA | 0 |
| S16_vs_100405\|\|full | 3023 | No | NA | 5 | 0 | 2 | Not-determined | Genome-fragment | NA | NA | 0 |
| S16_vs_100525\|\|full | 1605 | No | NA | 3 | 0 | 2 | Not-determined | Genome-fragment | NA | NA | 0 |
| S16_vs_100596\|\|full | 2264 | No | NA | 3 | 0 | 2 | Not-determined | Genome-fragment | NA | NA | 0 |
| S16_vs_100684\|\|full | 4972 | No | NA | 7 | 0 | 1 | Not-determined | Genome-fragment | NA | NA | 0 |
| S16_vs_100703\|\|full | 2396 | No | NA | 2 | 0 | 2 | Not-determined | Genome-fragment | NA | NA | 0 |
| S16_vs_100825\|\|full | 2845 | No | NA | 3 | 0 | 2 | Not-determined | Genome-fragment | NA | NA | 0 |
| S7_vs_85878\|\|full | 2902 | No | NA | 11 | 2 | 0 | Low-quality | Genome-fragment | 6.57 | AAI-based (medium-confidence) | 0 |
| S16_extracted_142498 | 2902 | No | NA | 3 | 0 | 1 | Low-quality | Genome-fragment | 3.61 | AAI-based (medium-confidence) | 0 |
| S16_vs_100935\|\|full | 6194 | No | NA | 5 | 0 | 3 | Not-determined | Genome-fragment | NA | NA | 0 |

| | | | | | | | | | | | | |
|---|---|---|---|---|---|---|---|---|---|---|---|---|
| S16_vs_100947\|\|full | 1307 | No | NA | | 3 | 0 | 0 | Not-determined | Genome-fragment | NA | NA | 0 |
| S1_vs_k141_104839\|\|full | 2903 | No | NA | | 5 | 4 | 0 | Low-quality | Genome-fragment | 5.94 | AAI-based (high-confidence) | 0 |
| S16_vs_101028\|\|full | 5017 | No | NA | | 2 | 0 | 1 | Not-determined | Genome-fragment | NA | NA | 0 |
| S8_extracted_k141_161544 | 2903 | No | NA | | 10 | 1 | 0 | Low-quality | Genome-fragment | 2.5 | AAI-based (high-confidence) | 0 |
| S8_vs_k141_12316\|\|full | 2903 | No | NA | | 6 | 0 | 0 | Low-quality | Genome-fragment | 2.83 | AAI-based (high-confidence) | 0 |
| S16_vs_101037\|\|full | 5036 | No | NA | | 5 | 0 | 3 | Not-determined | Genome-fragment | NA | NA | 0 |
| S16_vs_101072\|\|full | 31451 | Yes | | 20004 | 42 | 14 | 5 | Medium-quality | Genome-fragment | 50.63 | AAI-based (high-confidence) | 36.4 |
| S16_vs_101188\|\|full | 2578 | No | NA | | 3 | 0 | 0 | Not-determined | Genome-fragment | NA | NA | 0 |
| S9_vs_k141_41792\|\|full | 2904 | No | NA | | 5 | 3 | 0 | Low-quality | Genome-fragment | 5.32 | AAI-based (medium-confidence) | 0 |
| S12_vs_k141_130770\|\|full | 2904 | No | NA | | 2 | 0 | 2 | Low-quality | Genome-fragment | 1.46 | AAI-based (medium-confidence) | 0 |
| S16_vs_101300\|\|full | 2283 | No | NA | | 4 | 0 | 2 | Not-determined | Genome-fragment | NA | NA | 0 |
| S6_vs_k141_22957\|\|full | 2904 | No | NA | | 7 | 0 | 0 | Low-quality | Genome-fragment | 4.78 | AAI-based (high-confidence) | 0 |
| S17_extracted_k141_76718 | 2905 | No | NA | | 1 | 1 | 0 | Low-quality | Genome-fragment | 6.8 | HMM-based (lower-bound) | 0 |
| S11_vs_k141_49220\|\|full | 2905 | No | NA | | 3 | 0 | 0 | Low-quality | Genome-fragment | 4.5 | AAI-based (high-confidence) | 0 |
| S16_vs_101494\|\|full | 2476 | No | NA | | 3 | 0 | 2 | Not-determined | Genome-fragment | NA | NA | 0 |
| S16_vs_101542\|\|full | 2060 | No | NA | | 3 | 0 | 0 | Not-determined | Genome-fragment | NA | NA | 0 |
| S13_vs_126944\|\|full | 2905 | No | NA | | 4 | 0 | 2 | Low-quality | Genome-fragment | 1.43 | AAI-based (medium-confidence) | 0 |
| S16_vs_101614\|\|full | 1494 | No | NA | | 2 | 0 | 0 | Not-determined | Genome-fragment | NA | NA | 0 |
| S16_vs_101671\|\|full | 3287 | No | NA | | 8 | 4 | 1 | Medium-quality | Genome-fragment | 68.8 | AAI-based (high-confidence) | 0 |
| S10_vs_104660\|\|full | 2905 | No | NA | | 7 | 1 | 0 | Low-quality | Genome-fragment | 1.65 | HMM-based (lower-bound) | 0 |
| S16_vs_101799\|\|full | 1481 | No | NA | | 3 | 0 | 2 | Not-determined | Genome-fragment | NA | NA | 0 |
| S17_vs_k141_168359\|\|full | 2906 | No | NA | | 5 | 0 | 1 | Low-quality | Genome-fragment | 7.98 | AAI-based (medium-confidence) | 0 |
| S16_vs_102052\|\|full | 2889 | No | NA | | 4 | 0 | 1 | Not-determined | Genome-fragment | NA | NA | 0 |
| S7_vs_16814\|\|full | 2906 | No | NA | | 9 | 2 | 0 | Low-quality | Genome-fragment | 5.51 | AAI-based (medium-confidence) | 0 |
| S8_vs_k141_142480\|\|full | 2906 | No | NA | | 6 | 1 | 0 | Low-quality | Genome-fragment | 5.5 | AAI-based (medium-confidence) | 0 |
| S16_vs_102474\|\|full | 5148 | No | NA | | 6 | 0 | 1 | Not-determined | Genome-fragment | NA | NA | 0 |
| S16_vs_102506\|\|full | 1243 | No | NA | | 2 | 0 | 2 | Not-determined | Genome-fragment | NA | NA | 0 |
| S12_vs_k141_159680\|\|full | 2907 | No | NA | | 7 | 2 | 1 | Low-quality | Genome-fragment | 6.13 | AAI-based (high-confidence) | 0 |
| S7_vs_130845\|\|full | 2908 | No | NA | | 3 | 0 | 3 | Low-quality | Genome-fragment | 2.27 | AAI-based (medium-confidence) | 0 |
| S14_extracted_k141_97764 | 2909 | No | NA | | 8 | 5 | 0 | Low-quality | Genome-fragment | 9.45 | AAI-based (high-confidence) | 0 |
| S3_extracted_k141_54035 | 2909 | No | NA | | 4 | 2 | 0 | Low-quality | Genome-fragment | 7.62 | AAI-based (high-confidence) | 0 |
| S3_vs_k141_124074\|\|full | 2909 | No | NA | | 2 | 0 | 0 | Low-quality | Genome-fragment | 1.05 | AAI-based (medium-confidence) | 0 |
| S7_extracted_92513 | 2909 | No | NA | | 2 | 0 | 1 | Low-quality | Genome-fragment | 5.3 | AAI-based (medium-confidence) | 0 |
| S16_vs_103288\|\|full | 2661 | No | NA | | 3 | 0 | 3 | Not-determined | Genome-fragment | NA | NA | 0 |
| S16_vs_103295\|\|full | 4758 | No | NA | | 5 | 0 | 3 | Not-determined | Genome-fragment | NA | NA | 0 |
| S16_vs_103315\|\|full | 2681 | No | NA | | 2 | 0 | 0 | Not-determined | Genome-fragment | NA | NA | 0 |
| S16_vs_103404\|\|full | 1604 | No | NA | | 6 | 0 | 0 | Not-determined | Genome-fragment | NA | NA | 0 |
| S16_vs_103477\|\|full | 1009 | No | NA | | 2 | 0 | 2 | Not-determined | Genome-fragment | NA | NA | 0 |
| S15_vs_44386\|\|full | 2909 | No | NA | | 6 | 1 | 0 | Low-quality | Genome-fragment | 2.1 | HMM-based (lower-bound) | 0 |
| S16_vs_103663\|\|full | 2021 | No | NA | | 2 | 0 | 1 | Not-determined | Genome-fragment | NA | NA | 0 |
| S16_vs_103726\|\|full | 33933 | No | NA | | 57 | 20 | 0 | High-quality | High-quality | 99.89 | AAI-based (high-confidence) | 0 |
| S6_vs_k141_13287\|\|full | 2911 | No | NA | | 3 | 1 | 0 | Low-quality | Genome-fragment | 8.4 | AAI-based (high-confidence) | 0 |
| S9_vs_k141_73001\|\|full | 2912 | No | NA | | 7 | 0 | 1 | Low-quality | Genome-fragment | 4.86 | AAI-based (high-confidence) | 0 |
| S16_vs_103900\|\|full | 3306 | No | NA | | 4 | 0 | 4 | Not-determined | Genome-fragment | NA | NA | 0 |
| S14_vs_k141_73974\|\|full | 2912 | No | NA | | 4 | 1 | 0 | Low-quality | Genome-fragment | 9.96 | AAI-based (high-confidence) | 0 |
| S12_vs_k141_178580\|\|full | 2912 | No | NA | | 3 | 0 | 1 | Low-quality | Genome-fragment | 5.54 | AAI-based (medium-confidence) | 0 |
| S1_vs_k141_87839\|\|full | 2913 | No | NA | | 2 | 0 | 1 | Low-quality | Genome-fragment | 1.91 | AAI-based (medium-confidence) | 0 |
| S16_vs_114187\|\|full | 2913 | No | NA | | 4 | 0 | 3 | Low-quality | Genome-fragment | 0.89 | AAI-based (medium-confidence) | 0 |
| S16_vs_104693\|\|full | 1712 | No | NA | | 4 | 0 | 0 | Not-determined | Genome-fragment | NA | NA | 0 |
| S16_vs_104811\|\|full | 6765 | No | NA | | 7 | 0 | 0 | Not-determined | Genome-fragment | NA | NA | 0 |
| S11_vs_k141_48262\|\|full | 2914 | No | NA | | 11 | 1 | 0 | Low-quality | Genome-fragment | 6.73 | AAI-based (medium-confidence) | 0 |
| S11_vs_k141_29746\|\|full | 2914 | No | NA | | 10 | 0 | 0 | Low-quality | Genome-fragment | 7.08 | AAI-based (high-confidence) | 0 |
| S16_vs_105064\|\|full | 5094 | No | NA | | 9 | 0 | 0 | Not-determined | Genome-fragment | NA | NA | 0 |
| S16_vs_105065\|\|full | 1177 | No | NA | | 2 | 0 | 0 | Not-determined | Genome-fragment | NA | NA | 0 |
| S16_vs_105082\|\|full | 1318 | No | NA | | 2 | 0 | 0 | Not-determined | Genome-fragment | NA | NA | 0 |
| S16_vs_105227\|\|full | 1158 | No | NA | | 3 | 0 | 1 | Not-determined | Genome-fragment | NA | NA | 0 |
| S16_vs_105256\|\|full | 1748 | No | NA | | 2 | 0 | 0 | Not-determined | Genome-fragment | NA | NA | 0 |
| S16_vs_105314\|\|full | 1190 | No | NA | | 3 | 0 | 1 | Not-determined | Genome-fragment | NA | NA | 0 |
| S14_vs_k141_38464\|\|full | 2914 | No | NA | | 2 | 2 | 0 | Low-quality | Genome-fragment | 6.88 | AAI-based (medium-confidence) | 0 |
| S16_vs_105588\|\|full | 2373 | No | NA | | 3 | 0 | 1 | Not-determined | Genome-fragment | NA | NA | 0 |
| S16_vs_105639\|\|full | 1469 | No | NA | | 2 | 0 | 0 | Not-determined | Genome-fragment | NA | NA | 0 |
| S16_vs_105672\|\|full | 1510 | No | NA | | 5 | 0 | 1 | Not-determined | Genome-fragment | NA | NA | 0 |
| S16_vs_105705\|\|full | 1335 | No | NA | | 3 | 0 | 1 | Not-determined | Genome-fragment | NA | NA | 0 |
| S8_vs_k141_101276\|\|full | 2914 | No | NA | | 3 | 0 | 0 | Low-quality | Genome-fragment | 8.86 | AAI-based (high-confidence) | 0 |
| S3_vs_k141_5480\|\|full | 2915 | No | NA | | 3 | 0 | 2 | Low-quality | Genome-fragment | 1.27 | AAI-based (medium-confidence) | 0 |
| S16_vs_134105\|\|full | 2915 | No | NA | | 3 | 0 | 3 | Low-quality | Genome-fragment | 2.25 | AAI-based (medium-confidence) | 0 |
| S12_vs_k141_160546\|\|full | 2915 | No | NA | | 4 | 0 | 0 | Low-quality | Genome-fragment | 5.27 | AAI-based (high-confidence) | 0 |
| S16_vs_106028\|\|full | 2229 | No | NA | | 4 | 0 | 1 | Not-determined | Genome-fragment | NA | NA | 0 |
| S14_vs_k141_47931\|\|full | 2916 | No | NA | | 7 | 0 | 0 | Low-quality | Genome-fragment | 3.11 | AAI-based (medium-confidence) | 0 |
| S16_vs_106188\|\|full | 1003 | No | NA | | 3 | 0 | 0 | Not-determined | Genome-fragment | NA | NA | 0 |
| S16_vs_106366\|\|full | 1546 | No | NA | | 3 | 0 | 1 | Not-determined | Genome-fragment | NA | NA | 0 |
| S1_vs_k141_85054\|\|full | 2916 | No | NA | | 5 | 0 | 0 | Low-quality | Genome-fragment | 5.42 | AAI-based (high-confidence) | 0 |
| S16_vs_106388\|\|full | 1474 | No | NA | | 3 | 0 | 2 | Not-determined | Genome-fragment | NA | NA | 0 |
| S16_vs_106394\|\|full | 3631 | No | NA | | 5 | 0 | 1 | Not-determined | Genome-fragment | NA | NA | 0 |
| S16_vs_106427\|\|full | 1470 | No | NA | | 3 | 0 | 1 | Not-determined | Genome-fragment | NA | NA | 0 |
| S16_vs_106508\|\|full | 1463 | No | NA | | 2 | 0 | 1 | Not-determined | Genome-fragment | NA | NA | 0 |
| S16_vs_106588\|\|full | 3486 | No | NA | | 6 | 0 | 0 | Not-determined | Genome-fragment | NA | NA | 0 |
| S16_vs_106652\|\|full | 9633 | No | NA | | 10 | 0 | 1 | Not-determined | Genome-fragment | NA | NA | 0 |
| S12_extracted_k141_118420 | 2916 | No | NA | | 1 | 0 | 1 | Low-quality | Genome-fragment | 0.25 | AAI-based (medium-confidence) | 0 |
| S16_vs_106836\|\|full | 1174 | No | NA | | 3 | 0 | 0 | Not-determined | Genome-fragment | NA | NA | 0 |
| S16_vs_106843\|\|full | 2971 | No | NA | | 4 | 0 | 1 | Not-determined | Genome-fragment | NA | NA | 0 |
| S12_vs_k141_87177\|\|full | 2916 | No | NA | | 3 | 1 | 0 | Low-quality | Genome-fragment | 3.65 | HMM-based (lower-bound) | 0 |
| S16_vs_106942\|\|full | 3944 | No | NA | | 3 | 0 | 0 | Not-determined | Genome-fragment | NA | NA | 0 |
| S16_vs_107022\|\|full | 1274 | No | NA | | 2 | 0 | 1 | Not-determined | Genome-fragment | NA | NA | 0 |
| S16_vs_107035\|\|full | 1232 | No | NA | | 3 | 0 | 0 | Not-determined | Genome-fragment | NA | NA | 0 |
| S16_vs_107060\|\|full | 1524 | No | NA | | 3 | 0 | 2 | Not-determined | Genome-fragment | NA | NA | 0 |
| S16_vs_107235\|\|full | 56373 | Yes | | 28357 | 64 | 18 | 17 | Medium-quality | Genome-fragment | 80.07 | AAI-based (high-confidence) | 49.7 |
| S16_vs_107274\|\|full | 1060 | No | NA | | 2 | 0 | 1 | Not-determined | Genome-fragment | NA | NA | 0 |
| S16_vs_107347\|\|full | 1199 | No | NA | | 2 | 0 | 1 | Not-determined | Genome-fragment | NA | NA | 0 |
| S16_vs_107363\|\|full | 4967 | No | NA | | 5 | 0 | 1 | Not-determined | Genome-fragment | NA | NA | 0 |
| S6_vs_k141_7198\|\|full | 2916 | No | NA | | 4 | 0 | 2 | Low-quality | Genome-fragment | 1.74 | AAI-based (medium-confidence) | 0 |
| S8_vs_k141_9251\|\|full | 2916 | No | NA | | 4 | 3 | 0 | Low-quality | Genome-fragment | 6.32 | AAI-based (medium-confidence) | 0 |
| S16_vs_107628\|\|full | 5555 | No | NA | | 3 | 0 | 3 | Not-determined | Genome-fragment | NA | NA | 0 |
| S16_vs_107684\|\|full | 1211 | No | NA | | 2 | 0 | 2 | Not-determined | Genome-fragment | NA | NA | 0 |
| S16_vs_107771\|\|full | 2157 | No | NA | | 4 | 0 | 0 | Not-determined | Genome-fragment | NA | NA | 0 |
| S16_vs_107790\|\|full | 8318 | No | NA | | 7 | 0 | 4 | Not-determined | Genome-fragment | NA | NA | 0 |
| S8_vs_k141_187803\|\|full | 2916 | No | NA | | 3 | 0 | 2 | Low-quality | Genome-fragment | 1.4 | AAI-based (medium-confidence) | 0 |
| S10_vs_75810\|\|full | 2917 | No | NA | | 4 | 2 | 0 | Low-quality | Genome-fragment | 6.94 | AAI-based (medium-confidence) | 0 |
| S17_vs_k141_139644\|\|full | 2918 | No | NA | | 2 | 0 | 1 | Low-quality | Genome-fragment | 2.39 | AAI-based (medium-confidence) | 0 |
| S16_vs_107979\|\|full | 1527 | No | NA | | 3 | 0 | 1 | Not-determined | Genome-fragment | NA | NA | 0 |
| S16_vs_108009\|\|full | 2094 | No | NA | | 4 | 0 | 2 | Not-determined | Genome-fragment | NA | NA | 0 |
| S11_vs_k141_89100\|\|full | 2918 | No | NA | | 8 | 1 | 0 | Low-quality | Genome-fragment | 4.16 | AAI-based (medium-confidence) | 0 |
| S2_extracted_k141_16915 | 2918 | No | NA | | 4 | 0 | 0 | Low-quality | Genome-fragment | 7.62 | AAI-based (high-confidence) | 0 |
| S17_vs_k141_2837\|\|full | 2919 | No | NA | | 7 | 1 | 0 | Low-quality | Genome-fragment | 4.92 | AAI-based (high-confidence) | 0 |
| S14_vs_k141_4143\|\|full | 2919 | No | NA | | 6 | 1 | 0 | Low-quality | Genome-fragment | 7.65 | AAI-based (medium-confidence) | 0 |
| S16_vs_108743\|\|full | 1812 | No | NA | | 2 | 0 | 2 | Not-determined | Genome-fragment | NA | NA | 0 |
| S16_vs_108767\|\|full | 1531 | No | NA | | 2 | 0 | 0 | Not-determined | Genome-fragment | NA | NA | 0 |
| S16_vs_108778\|\|full | 1524 | No | NA | | 3 | 0 | 0 | Not-determined | Genome-fragment | NA | NA | 0 |
| S1_vs_k141_142963\|\|full | 2919 | No | NA | | 5 | 0 | 0 | Low-quality | Genome-fragment | 4.68 | AAI-based (high-confidence) | 0 |
| S16_vs_108828\|\|full | 5020 | No | NA | | 4 | 0 | 1 | Not-determined | Genome-fragment | NA | NA | 0 |
| S16_vs_108949\|\|full | 2048 | No | NA | | 2 | 0 | 0 | Not-determined | Genome-fragment | NA | NA | 0 |
| S16_vs_108983\|\|full | 4233 | No | NA | | 2 | 0 | 1 | Not-determined | Genome-fragment | NA | NA | 0 |
| S16_vs_109093\|\|full | 1988 | No | NA | | 4 | 0 | 0 | Not-determined | Genome-fragment | NA | NA | 0 |
| S16_vs_109161\|\|full | 3810 | No | NA | | 7 | 0 | 0 | Not-determined | Genome-fragment | NA | NA | 0 |
| S16_vs_90347\|\|full | 2919 | No | NA | | 5 | 1 | 0 | Low-quality | Genome-fragment | 2.09 | HMM-based (lower-bound) | 0 |
| S16_vs_109271\|\|full | 2128 | No | NA | | 4 | 0 | 1 | Not-determined | Genome-fragment | NA | NA | 0 |
| S5_vs_k141_111216\|\|full | 2920 | No | NA | | 9 | 1 | 0 | Low-quality | Genome-fragment | 6.32 | AAI-based (high-confidence) | 0 |
| S7_vs_113812\|\|full | 2920 | No | NA | | 8 | 2 | 0 | Low-quality | Genome-fragment | 4.8 | AAI-based (high-confidence) | 0 |
| S16_vs_109465\|\|full | 5853 | No | NA | | 5 | 0 | 5 | Not-determined | Genome-fragment | NA | NA | 0 |
| S12_vs_k141_38295\|\|full | 2920 | No | NA | | 5 | 2 | 0 | Low-quality | Genome-fragment | 5.68 | AAI-based (high-confidence) | 0 |
| S16_vs_109576\|\|full | 3088 | No | NA | | 5 | 0 | 2 | Not-determined | Genome-fragment | NA | NA | 0 |
| S16_vs_109617\|\|full | 1291 | No | NA | | 3 | 0 | 1 | Not-determined | Genome-fragment | NA | NA | 0 |
| S16_vs_109630\|\|full | 2148 | No | NA | | 4 | 0 | 1 | Not-determined | Genome-fragment | NA | NA | 0 |
| S14_vs_k141_8844\|\|full | 2921 | No | NA | | 7 | 6 | 0 | Low-quality | Genome-fragment | 7.28 | AAI-based (high-confidence) | 0 |
| S16_vs_109815\|\|full | 2882 | No | NA | | 4 | 0 | 4 | Not-determined | Genome-fragment | NA | NA | 0 |
| S16_vs_109842\|\|full | 1268 | No | NA | | 3 | 0 | 1 | Not-determined | Genome-fragment | NA | NA | 0 |
| S5_vs_k141_48064\|\|full | 2921 | No | NA | | 6 | 1 | 0 | Low-quality | Genome-fragment | 1.77 | HMM-based (lower-bound) | 0 |
| S7_extracted_5988 | 2921 | No | NA | | 3 | 0 | 2 | Low-quality | Genome-fragment | 5.13 | AAI-based (medium-confidence) | 0 |
| S16_vs_109980\|\|full | 3435 | No | NA | | 4 | 0 | 0 | Not-determined | Genome-fragment | NA | NA | 0 |
| S16_vs_110035\|\|full | 3469 | No | NA | | 4 | 0 | 3 | Not-determined | Genome-fragment | NA | NA | 0 |
| S16_vs_110082\|\|full | 3250 | No | NA | | 3 | 0 | 2 | Not-determined | Genome-fragment | NA | NA | 0 |
| S7_vs_135284\|\|full | 2921 | No | NA | | 5 | 0 | 1 | Low-quality | Genome-fragment | 5.47 | AAI-based (medium-confidence) | 0 |
| S16_vs_110136\|\|full | 7117 | No | NA | | 4 | 0 | 4 | Not-determined | Genome-fragment | NA | NA | 0 |
| S9_extracted_k141_26069 | 2922 | No | NA | | 6 | 0 | 0 | Low-quality | Genome-fragment | 48.85 | AAI-based (medium-confidence) | 0 |
| S8_vs_k141_135743\|\|full | 2922 | No | NA | | 4 | 0 | 0 | Low-quality | Genome-fragment | 7.2 | AAI-based (medium-confidence) | 0 |
| S13_extracted_149344 | 2923 | No | NA | | 2 | 0 | 1 | Low-quality | Genome-fragment | 4.94 | AAI-based (medium-confidence) | 0 |
| S16_vs_110268\|\|full | 2221 | No | NA | | 3 | 0 | 3 | Not-determined | Genome-fragment | NA | NA | 0 |
| S3_vs_k141_98434\|\|full | 2923 | No | NA | | 3 | 0 | 0 | Low-quality | Genome-fragment | 2.19 | AAI-based (high-confidence) | 0 |
| S15_vs_76553\|\|full | 2923 | No | NA | | 6 | 1 | 1 | Low-quality | Genome-fragment | 6.82 | AAI-based (high-confidence) | 0 |
| S14_extracted_k141_9149 | 2924 | No | NA | | 1 | 1 | 0 | Low-quality | Genome-fragment | 1.71 | HMM-based (lower-bound) | 0 |
| S14_extracted_k141_21281 | 2924 | No | NA | | 4 | 0 | 0 | Low-quality | Genome-fragment | 3.22 | AAI-based (high-confidence) | 0 |
| S16_vs_110573\|\|full | 1322 | No | NA | | 2 | 0 | 2 | Not-determined | Genome-fragment | NA | NA | 0 |
| S16_vs_110682\|\|full | 3274 | No | NA | | 2 | 0 | 1 | Not-determined | Genome-fragment | NA | NA | 0 |
| S16_vs_110746\|\|full | 4722 | No | NA | | 9 | 0 | 0 | Not-determined | Genome-fragment | NA | NA | 0 |
| S16_vs_111136\|\|full | 2702 | No | NA | | 5 | 0 | 1 | Not-determined | Genome-fragment | NA | NA | 0 |
| S7_vs_133950\|\|full | 2924 | No | NA | | 5 | 5 | 0 | Low-quality | Genome-fragment | 6.42 | AAI-based (high-confidence) | 0 |
| S8_extracted_k141_123570 | 2924 | No | NA | | 6 | 1 | 0 | Low-quality | Genome-fragment | 6.04 | AAI-based (high-confidence) | 0 |
| S16_vs_111351\|\|full | 3411 | No | NA | | 6 | 0 | 2 | Not-determined | Genome-fragment | NA | NA | 0 |
| S16_vs_111365\|\|full | 1760 | No | NA | | 4 | 0 | 0 | Not-determined | Genome-fragment | NA | NA | 0 |
| S13_extracted_88136 | 2925 | No | NA | | 4 | 0 | 3 | Low-quality | Genome-fragment | 1.4 | AAI-based (medium-confidence) | 0 |
| S14_extracted_k141_74053 | 2925 | No | NA | | 3 | 3 | 0 | Low-quality | Genome-fragment | 9.72 | AAI-based (high-confidence) | 0 |
| S16_vs_111597\|\|full | 2334 | No | NA | | 2 | 0 | 1 | Not-determined | Genome-fragment | NA | NA | 0 |
| S7_vs_112917\|\|full | 2925 | No | NA | | 4 | 3 | 0 | Low-quality | Genome-fragment | 7.23 | AAI-based (high-confidence) | 0 |
| S5_vs_k141_48809\|\|full | 2926 | No | NA | | 5 | 0 | 2 | Low-quality | Genome-fragment | 4.81 | AAI-based (high-confidence) | 0 |
| S10_vs_86036\|\|full | 2926 | No | NA | | 3 | 3 | 0 | Low-quality | Genome-fragment | 7.37 | AAI-based (high-confidence) | 0 |
| S15_extracted_56231 | 2926 | No | NA | | 3 | 3 | 0 | Low-quality | Genome-fragment | 1.57 | AAI-based (high-confidence) | 0 |
| S9_vs_k141_85831\|\|full | 2927 | No | NA | | 9 | 8 | 0 | Low-quality | Genome-fragment | 9.62 | AAI-based (high-confidence) | 0 |
| S3_vs_k141_107961\|\|full | 2927 | No | NA | | 2 | 0 | 1 | Low-quality | Genome-fragment | 3.93 | AAI-based (medium-confidence) | 0 |
| S3_vs_k141_77187\|\|full | 2927 | No | NA | | 5 | 0 | 0 | Low-quality | Genome-fragment | 1.81 | AAI-based (high-confidence) | 0 |
| S16_vs_112513\|\|full | 2376 | No | NA | | 4 | 0 | 1 | Not-determined | Genome-fragment | NA | NA | 0 |
| S12_vs_k141_142726\|\|full | 2927 | No | NA | | 8 | 1 | 0 | Low-quality | Genome-fragment | 7.57 | HMM-based (lower-bound) | 0 |
| S16_vs_112646\|\|full | 2403 | No | NA | | 3 | 0 | 2 | Not-determined | Genome-fragment | NA | NA | 0 |
| S16_vs_112672\|\|full | 5249 | No | NA | | 5 | 0 | 4 | Not-determined | Genome-fragment | NA | NA | 0 |
| S16_vs_112711\|\|full | 6388 | No | NA | | 3 | 0 | 1 | Not-determined | Genome-fragment | NA | NA | 0 |
| S12_vs_k141_133349\|\|full | 2927 | No | NA | | 6 | 1 | 0 | Low-quality | Genome-fragment | 2.25 | HMM-based (lower-bound) | 0 |
| S16_vs_112790\|\|full | 2076 | No | NA | | 3 | 0 | 2 | Not-determined | Genome-fragment | NA | NA | 0 |
| S16_vs_112830\|\|full | 1932 | No | NA | | 4 | 0 | 0 | Not-determined | Genome-fragment | NA | NA | 0 |
| S8_vs_k141_47550\|\|full | 2927 | No | NA | | 4 | 3 | 0 | Low-quality | Genome-fragment | 6.04 | AAI-based (high-confidence) | 0 |
| S13_vs_116220\|\|full | 2928 | No | NA | | 3 | 2 | 0 | Low-quality | Genome-fragment | 8.16 | AAI-based (high-confidence) | 0 |
| S12_extracted_k141_24349 | 2928 | No | NA | | 5 | 4 | 0 | Low-quality | Genome-fragment | 4.92 | AAI-based (medium-confidence) | 0 |
| S16_vs_113002\|\|full | 1329 | No | NA | | 2 | 0 | 1 | Not-determined | Genome-fragment | NA | NA | 0 |
| S16_vs_113019\|\|full | 1223 | No | NA | | 3 | 0 | 0 | Not-determined | Genome-fragment | NA | NA | 0 |
| S16_vs_113101\|\|full | 1798 | No | NA | | 4 | 0 | 2 | Not-determined | Genome-fragment | NA | NA | 0 |
| S11_vs_k141_20084\|\|full | 2929 | No | NA | | 4 | 4 | 0 | Low-quality | Genome-fragment | 6.55 | AAI-based (medium-confidence) | 0 |
| S16_vs_113216\|\|full | 1954 | No | NA | | 2 | 0 | 1 | Not-determined | Genome-fragment | NA | NA | 0 |
| S16_vs_113249\|\|full | 1079 | No | NA | | 3 | 0 | 0 | Not-determined | Genome-fragment | NA | NA | 0 |
| S13_vs_33785\|\|full | 2929 | No | NA | | 6 | 5 | 0 | Low-quality | Genome-fragment | 7 | AAI-based (high-confidence) | 0 |
| S16_vs_113442\|\|full | 1328 | No | NA | | 2 | 0 | 0 | Not-determined | Genome-fragment | NA | NA | 0 |
| S16_vs_113462\|\|full | 1567 | No | NA | | 3 | 0 | 0 | Not-determined | Genome-fragment | NA | NA | 0 |
| S16_vs_113493\|\|full | 1753 | No | NA | | 3 | 0 | 0 | Not-determined | Genome-fragment | NA | NA | 0 |
| S16_vs_113535\|\|full | 5353 | No | NA | | 6 | 0 | 1 | Not-determined | Genome-fragment | NA | NA | 0 |
| S14_vs_k141_13072\|\|lt2gene | 2929 | No | NA | | 2 | 2 | 0 | Low-quality | Genome-fragment | 4.52 | AAI-based (high-confidence) | 0 |
| S7_vs_76122\|\|full | 2930 | No | NA | | 4 | 0 | 3 | Low-quality | Genome-fragment | 3.01 | AAI-based (medium-confidence) | 0 |
| S16_vs_113809\|\|full | 1595 | No | NA | | 3 | 0 | 2 | Not-determined | Genome-fragment | NA | NA | 0 |
| S4_extracted_641 | 2930 | No | NA | | 4 | 0 | 3 | Low-quality | Genome-fragment | 1.91 | AAI-based (medium-confidence) | 0 |
| S14_vs_k141_21728\|\|full | 2931 | No | NA | | 5 | 3 | 0 | Low-quality | Genome-fragment | 4.26 | AAI-based (high-confidence) | 0 |
| S16_vs_114001\|\|full | 2090 | No | NA | | 3 | 0 | 0 | Not-determined | Genome-fragment | NA | NA | 0 |
| S1_vs_k141_159643\|\|full | 2931 | No | NA | | 4 | 1 | 0 | Low-quality | Genome-fragment | 6.35 | AAI-based (high-confidence) | 0 |
| S16_vs_114044\|\|full | 7164 | No | NA | | 8 | 0 | 1 | Not-determined | Genome-fragment | NA | NA | 0 |
| S7_vs_39314\|\|full | 2932 | No | NA | | 5 | 2 | 0 | Low-quality | Genome-fragment | 5.69 | AAI-based (medium-confidence) | 0 |
| S10_vs_58865\|\|full | 2932 | No | NA | | 5 | 3 | 0 | Low-quality | Genome-fragment | 7.09 | AAI-based (high-confidence) | 0 |
| S16_vs_114192\|\|full | 2092 | No | NA | | 4 | 0 | 0 | Not-determined | Genome-fragment | NA | NA | 0 |
| S16_vs_114206\|\|full | 2774 | No | NA | | 5 | 0 | 0 | Not-determined | Genome-fragment | NA | NA | 0 |
| S12_vs_k141_163478\|\|full | 2932 | No | NA | | 3 | 0 | 2 | Low-quality | Genome-fragment | 2.63 | AAI-based (medium-confidence) | 0 |
| S16_vs_114333\|\|full | 3449 | No | NA | | 3 | 0 | 2 | Not-determined | Genome-fragment | NA | NA | 0 |
| S8_vs_k141_47215\|\|full | 2932 | No | NA | | 7 | 0 | 1 | Low-quality | Genome-fragment | 7.31 | AAI-based (medium-confidence) | 0 |
| S16_vs_114444\|\|full | 1076 | No | NA | | 2 | 0 | 1 | Not-determined | Genome-fragment | NA | NA | 0 |
| S16_vs_114568\|\|full | 2934 | No | NA | | 2 | 0 | 2 | Not-determined | Genome-fragment | NA | NA | 0 |
| S3_extracted_k141_53877 | 2933 | No | NA | | 4 | 4 | 0 | Low-quality | Genome-fragment | 7.82 | AAI-based (high-confidence) | 0 |
| S16_vs_114778\|\|full | 1200 | No | NA | | 3 | 0 | 0 | Not-determined | Genome-fragment | NA | NA | 0 |
| S16_vs_114785\|\|full | 2232 | No | NA | | 3 | 0 | 0 | Not-determined | Genome-fragment | NA | NA | 0 |
| S16_vs_13702\|\|full | 2933 | No | NA | | 3 | 0 | 2 | Low-quality | Genome-fragment | 0.83 | AAI-based (high-confidence) | 0 |
| S16_vs_114943\|\|full | 2761 | No | NA | | 3 | 0 | 1 | Not-determined | Genome-fragment | NA | NA | 0 |
| S16_vs_114983\|\|full | 1516 | No | NA | | 2 | 0 | 1 | Not-determined | Genome-fragment | NA | NA | 0 |
| S3_vs_k141_116000\|\|full | 2934 | No | NA | | 8 | 0 | 0 | Low-quality | Genome-fragment | 5.14 | AAI-based (high-confidence) | 0 |
| S16_vs_115068\|\|full | 3693 | No | NA | | 3 | 0 | 2 | Not-determined | Genome-fragment | NA | NA | 0 |
| S16_vs_115104\|\|full | 1429 | No | NA | | 3 | 0 | 2 | Not-determined | Genome-fragment | NA | NA | 0 |
| S6_vs_k141_1232\|\|full | 2934 | No | NA | | 3 | 0 | 1 | Low-quality | Genome-fragment | 0.87 | AAI-based (medium-confidence) | 0 |
| S3_extracted_k141_90295 | 2935 | No | NA | | 6 | 5 | 0 | Low-quality | Genome-fragment | 9.68 | AAI-based (high-confidence) | 0 |
| S7_vs_123308\|\|full | 2935 | No | NA | | 5 | 0 | 0 | Low-quality | Genome-fragment | 5.46 | AAI-based (high-confidence) | 0 |
| S16_vs_115374\|\|full | 2038 | No | NA | | 2 | 0 | 2 | Not-determined | Genome-fragment | NA | NA | 0 |
| S14_vs_k141_103660\|\|full | 2936 | No | NA | | 3 | 0 | 0 | Low-quality | Genome-fragment | 6.42 | AAI-based (high-confidence) | 0 |
| S1_vs_k141_93765\|\|full | 2936 | No | NA | | 5 | 0 | 0 | Low-quality | Genome-fragment | 6.56 | AAI-based (medium-confidence) | 0 |
| S16_vs_115478\|\|full | 2120 | No | NA | | 3 | 0 | 2 | Not-determined | Genome-fragment | NA | NA | 0 |
| S6_extracted_k141_14693 | 2937 | No | NA | | 4 | 0 | 4 | Low-quality | Genome-fragment | 0.93 | AAI-based (medium-confidence) | 0 |
| S8_extracted_k141_138276 | 2938 | No | NA | | 2 | 0 | 2 | Low-quality | Genome-fragment | 1.34 | AAI-based (medium-confidence) | 0 |
| S16_vs_115609\|\|full | 1162 | No | NA | | 2 | 0 | 1 | Not-determined | Genome-fragment | NA | NA | 0 |
| S16_vs_115712\|\|full | 1619 | No | NA | | 2 | 0 | 0 | Not-determined | Genome-fragment | NA | NA | 0 |
| S16_vs_115731\|\|full | 2776 | No | NA | | 2 | 0 | 2 | Not-determined | Genome-fragment | NA | NA | 0 |
| S16_vs_115743\|\|full | 5271 | No | NA | | 3 | 0 | 2 | Not-determined | Genome-fragment | NA | NA | 0 |
| S1_extracted_k141_158926 | 2939 | No | NA | | 4 | 2 | 0 | Low-quality | Genome-fragment | 8.62 | HMM-based (lower-bound) | 0 |
| S16_vs_115791\|\|full | 1127 | No | NA | | 3 | 0 | 3 | Not-determined | Genome-fragment | NA | NA | 0 |
| S16_vs_115795\|\|full | 4154 | No | NA | | 6 | 0 | 0 | Not-determined | Genome-fragment | NA | NA | 0 |
| S16_vs_115848\|\|full | 1570 | No | NA | | 3 | 0 | 0 | Not-determined | Genome-fragment | NA | NA | 0 |
| S16_vs_115856\|\|full | 3110 | No | NA | | 4 | 0 | 1 | Not-determined | Genome-fragment | NA | NA | 0 |
| S8_vs_k141_122001\|\|full | 2939 | No | NA | | 5 | 0 | 0 | Low-quality | Genome-fragment | 4.99 | AAI-based (medium-confidence) | 0 |
| S16_vs_116023\|\|full | 1595 | No | NA | | 2 | 0 | 0 | Not-determined | Genome-fragment | NA | NA | 0 |
| S6_vs_k141_41669\|\|full | 2940 | No | NA | | 2 | 0 | 1 | Low-quality | Genome-fragment | 1.41 | AAI-based (medium-confidence) | 0 |
| S1_extracted_k141_22541 | 2941 | No | NA | | 3 | 0 | 1 | Low-quality | Genome-fragment | 2.68 | AAI-based (medium-confidence) | 0 |
| S14_extracted_k141_74168 | 2942 | No | NA | | 6 | 5 | 0 | Low-quality | Genome-fragment | 9.71 | AAI-based (high-confidence) | 0 |
| S16_vs_116128\|\|full | 1304 | No | NA | | 3 | 0 | 0 | Not-determined | Genome-fragment | NA | NA | 0 |
| S7_vs_101084\|\|full | 2942 | No | NA | | 3 | 0 | 1 | Low-quality | Genome-fragment | 1.36 | AAI-based (medium-confidence) | 0 |
| S16_vs_116248\|\|full | 1251 | No | NA | | 3 | 0 | 2 | Not-determined | Genome-fragment | NA | NA | 0 |
| S16_vs_116364\|\|full | 1348 | No | NA | | 3 | 0 | 1 | Not-determined | Genome-fragment | NA | NA | 0 |
| S16_vs_116393\|\|full | 4330 | No | NA | | 3 | 0 | 2 | Not-determined | Genome-fragment | NA | NA | 0 |
| S10_vs_89521\|\|full | 2942 | No | NA | | 4 | 1 | 0 | Low-quality | Genome-fragment | 6.68 | AAI-based (high-confidence) | 0 |
| S16_vs_116468\|\|full | 1454 | No | NA | | 3 | 0 | 1 | Not-determined | Genome-fragment | NA | NA | 0 |
| S12_vs_k141_182423\|\|full | 2942 | No | NA | | 2 | 1 | 0 | Low-quality | Genome-fragment | 1.17 | HMM-based (lower-bound) | 0 |
| S17_vs_k141_143133\|\|full | 2943 | No | NA | | 4 | 0 | 4 | Low-quality | Genome-fragment | 1.12 | AAI-based (medium-confidence) | 0 |
| S16_vs_116604\|\|full | 2241 | No | NA | | 4 | 0 | 2 | Not-determined | Genome-fragment | NA | NA | 0 |
| S14_vs_k141_40368\|\|full | 2943 | No | NA | | 3 | 0 | 3 | Low-quality | Genome-fragment | 3.55 | AAI-based (medium-confidence) | 0 |
| S8_vs_k141_187632\|\|full | 2943 | No | NA | | 3 | 0 | 2 | Low-quality | Genome-fragment | 1.29 | AAI-based (medium-confidence) | 0 |
| S16_vs_116789\|\|full | 3267 | No | NA | | 4 | 0 | 3 | Not-determined | Genome-fragment | NA | NA | 0 |
| S16_vs_116834\|\|full | 2565 | No | NA | | 3 | 0 | 1 | Not-determined | Genome-fragment | NA | NA | 0 |
| S5_vs_k141_4153\|\|full | 2945 | No | NA | | 3 | 3 | 0 | Low-quality | Genome-fragment | 7.24 | AAI-based (high-confidence) | 0 |
| S8_extracted_k141_33535 | 2945 | No | NA | | 3 | 0 | 0 | Low-quality | Genome-fragment | 3.59 | AAI-based (high-confidence) | 0 |
| S14_extracted_k141_98159 | 2946 | No | NA | | 5 | 1 | 0 | Low-quality | Genome-fragment | 4.54 | HMM-based (lower-bound) | 0 |
| S7_vs_108949\|\|full | 2946 | No | NA | | 3 | 0 | 0 | Low-quality | Genome-fragment | 9.12 | AAI-based (high-confidence) | 0 |
| S16_vs_73012\|\|full | 2946 | No | NA | | 5 | 0 | 2 | Low-quality | Genome-fragment | 4.55 | AAI-based (medium-confidence) | 0 |
| S16_vs_116932\|\|full | 5135 | No | NA | | 5 | 0 | 3 | Not-determined | Genome-fragment | NA | NA | 0 |
| S16_vs_116936\|\|full | 2994 | No | NA | | 3 | 0 | 3 | Not-determined | Genome-fragment | NA | NA | 0 |
| S16_vs_116956\|\|full | 33562 | No | NA | | 50 | 2 | 4 | Medium-quality | Genome-fragment | 53.31 | AAI-based (high-confidence) | 0 |
| S12_extracted_k141_182128 | 2946 | No | NA | | 2 | 0 | 1 | Low-quality | Genome-fragment | 4.88 | AAI-based (medium-confidence) | 0 |
| S16_vs_117035\|\|full | 1595 | No | NA | | 3 | 0 | 2 | Not-determined | Genome-fragment | NA | NA | 0 |
| S16_vs_117074\|\|full | 2083 | No | NA | | 4 | 0 | 2 | Not-determined | Genome-fragment | NA | NA | 0 |
| S16_vs_117093\|\|full | 3764 | No | NA | | 3 | 0 | 2 | Not-determined | Genome-fragment | NA | NA | 0 |
| S11_extracted_k141_34576 | 2948 | No | NA | | 3 | 3 | 0 | Low-quality | Genome-fragment | 6.27 | AAI-based (high-confidence) | 0 |
| S5_vs_k141_29438\|\|full | 2948 | No | NA | | 6 | 0 | 0 | Low-quality | Genome-fragment | 5.63 | AAI-based (medium-confidence) | 0 |
| S9_vs_k141_70132\|\|full | 2949 | No | NA | | 5 | 1 | 0 | Low-quality | Genome-fragment | 7.15 | AAI-based (high-confidence) | 0 |
| S16_vs_67908\|\|full | 2949 | No | NA | | 5 | 1 | 0 | Low-quality | Genome-fragment | 5.73 | AAI-based (high-confidence) | 0 |
| S12_vs_k141_128068\|\|lt2gene | 2949 | No | NA | | 1 | 1 | 0 | Low-quality | Genome-fragment | 5.83 | AAI-based (medium-confidence) | 0 |
| S16_vs_117379\|\|full | 4452 | No | NA | | 5 | 0 | 2 | Not-determined | Genome-fragment | NA | NA | 0 |
| S16_vs_117434\|\|full | 4231 | No | NA | | 5 | 0 | 2 | Not-determined | Genome-fragment | NA | NA | 0 |
| S16_vs_117463\|\|full | 6796 | No | NA | | 10 | 0 | 1 | Not-determined | Genome-fragment | NA | NA | 0 |
| S16_vs_117568\|\|full | 2443 | No | NA | | 4 | 0 | 0 | Not-determined | Genome-fragment | NA | NA | 0 |
| S16_vs_117592\|\|full | 1470 | No | NA | | 3 | 0 | 1 | Not-determined | Genome-fragment | NA | NA | 0 |
| S16_vs_117649\|\|full | 2167 | No | NA | | 4 | 0 | 0 | Not-determined | Genome-fragment | NA | NA | 0 |
| S16_vs_125437\|\|full | 2950 | No | NA | | 4 | 0 | 0 | Low-quality | Genome-fragment | 5.58 | AAI-based (medium-confidence) | 0 |
| S16_vs_142909\|\|lt2gene | 2950 | No | NA | | 2 | 2 | 0 | Low-quality | Genome-fragment | 4.84 | HMM-based (lower-bound) | 0 |
| S16_vs_117914\|\|full | 1989 | No | NA | | 3 | 0 | 3 | Not-determined | Genome-fragment | NA | NA | 0 |
| S16_vs_117939\|\|full | 1863 | No | NA | | 3 | 0 | 2 | Not-determined | Genome-fragment | NA | NA | 0 |
| S16_vs_117993\|\|full | 2687 | No | NA | | 4 | 0 | 2 | Not-determined | Genome-fragment | NA | NA | 0 |
| S10_extracted_1359 | 2950 | Yes | | 2621 | 6 | 2 | 1 | Low-quality | Genome-fragment | 3.6 | AAI-based (high-confidence) | 11.15 |
| S16_vs_118029\|\|full | 37809 | No | NA | | 66 | 25 | 0 | High-quality | High-quality | 100 | AAI-based (high-confidence) | 0 |
| S16_vs_118070\|\|full | 3333 | No | NA | | 6 | 0 | 1 | Not-determined | Genome-fragment | NA | NA | 0 |
| S10_extracted_32390 | 2950 | No | NA | | 5 | 1 | 0 | Low-quality | Genome-fragment | 7.02 | HMM-based (lower-bound) | 0 |
| S5_extracted_k141_118482 | 2952 | No | NA | | 8 | 8 | 0 | Low-quality | Genome-fragment | 9.72 | AAI-based (high-confidence) | 0 |
| S16_vs_118424\|\|full | 4074 | No | NA | | 3 | 0 | 2 | Not-determined | Genome-fragment | NA | NA | 0 |
| S16_extracted_151604 | 2952 | No | NA | | 4 | 3 | 0 | Low-quality | Genome-fragment | 7.24 | AAI-based (high-confidence) | 0 |
| S16_vs_118550\|\|full | 2276 | No | NA | | 3 | 0 | 0 | Not-determined | Genome-fragment | NA | NA | 0 |
| S16_vs_118618\|\|full | 2161 | No | NA | | 3 | 0 | 1 | Not-determined | Genome-fragment | NA | NA | 0 |
| S16_vs_118663\|\|full | 3208 | No | NA | | 3 | 0 | 2 | Not-determined | Genome-fragment | NA | NA | 0 |
| S16_vs_118669\|\|full | 1446 | No | NA | | 3 | 0 | 1 | Not-determined | Genome-fragment | NA | NA | 0 |
| S16_vs_118677\|\|full | 2449 | No | NA | | 6 | 0 | 0 | Not-determined | Genome-fragment | NA | NA | 0 |
| S16_vs_118684\|\|full | 14636 | No | NA | | 14 | 0 | 8 | Not-determined | Genome-fragment | NA | NA | 0 |
| S16_vs_118811\|\|full | 1787 | No | NA | | 3 | 0 | 1 | Not-determined | Genome-fragment | NA | NA | 0 |
| S16_vs_118883\|\|full | 2233 | No | NA | | 3 | 0 | 3 | Not-determined | Genome-fragment | NA | NA | 0 |
| S16_vs_119037\|\|full | 1559 | No | NA | | 4 | 0 | 2 | Not-determined | Genome-fragment | NA | NA | 0 |
| S12_extracted_k141_157895 | 2952 | No | NA | | 3 | 1 | 0 | Low-quality | Genome-fragment | 4.38 | AAI-based (high-confidence) | 0 |
| S5_vs_k141_132148\|\|full | 2953 | No | NA | | 4 | 1 | 0 | Low-quality | Genome-fragment | 7.73 | AAI-based (high-confidence) | 0 |
| S10_vs_36718\|\|full | 2954 | No | NA | | 2 | 0 | 1 | Low-quality | Genome-fragment | 2.33 | AAI-based (medium-confidence) | 0 |
| S16_vs_119130\|\|full | 1177 | No | NA | | 2 | 0 | 0 | Not-determined | Genome-fragment | NA | NA | 0 |
| S16_vs_119152\|\|full | 2795 | No | NA | | 3 | 0 | 1 | Not-determined | Genome-fragment | NA | NA | 0 |
| S16_vs_119250\|\|full | 1296 | No | NA | | 3 | 0 | 0 | Not-determined | Genome-fragment | NA | NA | 0 |
| S10_vs_76928\|\|full | 2954 | No | NA | | 4 | 0 | 0 | Low-quality | Genome-fragment | 4.46 | AAI-based (medium-confidence) | 0 |
| S16_vs_119323\|\|full | 2171 | No | NA | | 3 | 0 | 0 | Not-determined | Genome-fragment | NA | NA | 0 |
| S8_extracted_k141_23529 | 2954 | No | NA | | 2 | 2 | 0 | Low-quality | Genome-fragment | 5.12 | AAI-based (medium-confidence) | 0 |
| S3_vs_k141_110097\|\|full | 2955 | No | NA | | 3 | 0 | 2 | Low-quality | Genome-fragment | 4.59 | AAI-based (medium-confidence) | 0 |
| S7_vs_78201\|\|full | 2955 | No | NA | | 9 | 0 | 0 | Low-quality | Genome-fragment | 8.73 | AAI-based (high-confidence) | 0 |
| S16_vs_114227\|\|full | 2955 | No | NA | | 6 | 1 | 0 | Low-quality | Genome-fragment | 6.09 | AAI-based (medium-confidence) | 0 |
| S9_vs_k141_15145\|\|lt2gene | 2956 | No | NA | | 2 | 2 | 0 | Low-quality | Genome-fragment | 3.82 | AAI-based (high-confidence) | 0 |
| S16_vs_119747\|\|full | 5663 | No | NA | | 6 | 0 | 3 | Not-determined | Genome-fragment | NA | NA | 0 |
| S11_vs_k141_79435\|\|full | 2956 | Yes | | 1738 | 6 | 1 | 1 | Low-quality | Genome-fragment | 2.73 | HMM-based (lower-bound) | 41.2 |
| S16_vs_119842\|\|full | 7258 | No | NA | | 4 | 0 | 4 | Not-determined | Genome-fragment | NA | NA | 0 |
| S16_vs_119859\|\|full | 4032 | No | NA | | 4 | 0 | 3 | Not-determined | Genome-fragment | NA | NA | 0 |
| S3_vs_k141_101573\|\|full | 2956 | No | NA | | 5 | 0 | 0 | Low-quality | Genome-fragment | 1.59 | AAI-based (high-confidence) | 0 |
| S16_vs_169944\|\|full | 2956 | No | NA | | 4 | 0 | 0 | Low-quality | Genome-fragment | 4.57 | AAI-based (high-confidence) | 0 |
| S16_vs_120116\|\|full | 1656 | No | NA | | 3 | 0 | 1 | Not-determined | Genome-fragment | NA | NA | 0 |
| S16_vs_120149\|\|full | 20723 | No | NA | | 31 | 8 | 1 | Medium-quality | Genome-fragment | 58.55 | AAI-based (medium-confidence) | 0 |
| S1_extracted_k141_72542 | 2957 | No | NA | | 1 | 1 | 0 | Low-quality | Genome-fragment | 1.18 | AAI-based (high-confidence) | 0 |
| S16_vs_120409\|\|full | 1024 | No | NA | | 2 | 0 | 1 | Not-determined | Genome-fragment | NA | NA | 0 |
| S16_vs_120572\|\|full | 4133 | No | NA | | 6 | 0 | 3 | Not-determined | Genome-fragment | NA | NA | 0 |
| S16_vs_120574\|\|full | 3857 | No | NA | | 4 | 0 | 0 | Not-determined | Genome-fragment | NA | NA | 0 |
| S16_vs_120605\|\|full | 3674 | No | NA | | 4 | 2 | 0 | Medium-quality | Genome-fragment | 60.7 | AAI-based (high-confidence) | 0 |
| S16_vs_120622\|\|full | 54160 | No | NA | | 78 | 28 | 0 | High-quality | High-quality | 100 | AAI-based (high-confidence) | 0 |
| S16_vs_120635\|\|full | 3738 | No | NA | | 3 | 0 | 1 | Not-determined | Genome-fragment | NA | NA | 0 |
| S16_vs_120788\|\|full | 1998 | No | NA | | 3 | 0 | 1 | Not-determined | Genome-fragment | NA | NA | 0 |
| S9_vs_k141_25610\|\|full | 2958 | No | NA | | 5 | 1 | 0 | Low-quality | Genome-fragment | 7.25 | AAI-based (high-confidence) | 0 |
| S16_vs_120895\|\|full | 2995 | No | NA | | 4 | 0 | 1 | Not-determined | Genome-fragment | NA | NA | 0 |
| S13_vs_99845\|\|full | 2958 | No | NA | | 5 | 2 | 0 | Low-quality | Genome-fragment | 4.89 | AAI-based (high-confidence) | 0 |
| S16_vs_120953\|\|full | 2073 | No | NA | | 2 | 0 | 0 | Not-determined | Genome-fragment | NA | NA | 0 |
| S7_vs_34069\|\|full | 2958 | No | NA | | 5 | 0 | 0 | Low-quality | Genome-fragment | 7.6 | AAI-based (high-confidence) | 0 |
| S12_vs_k141_89744\|\|full | 2958 | No | NA | | 8 | 3 | 0 | Low-quality | Genome-fragment | 7.12 | AAI-based (high-confidence) | 0 |
| S16_vs_121046\|\|full | 5142 | No | NA | | 4 | 0 | 3 | Not-determined | Genome-fragment | NA | NA | 0 |
| S5_vs_k141_55335\|\|full | 2959 | Yes | | 2395 | 6 | 1 | 1 | Low-quality | Genome-fragment | 6.25 | AAI-based (high-confidence) | 19.06 |
| S7_vs_38047\|\|full | 2959 | No | NA | | 5 | 2 | 0 | Low-quality | Genome-fragment | 6.74 | AAI-based (medium-confidence) | 0 |
| S16_vs_121290\|\|full | 2565 | No | NA | | 3 | 0 | 2 | Not-determined | Genome-fragment | NA | NA | 0 |
| S16_vs_121439\|\|full | 5126 | No | NA | | 9 | 0 | 5 | Not-determined | Genome-fragment | NA | NA | 0 |
| S12_vs_k141_142844\|\|full | 2959 | Yes | | 1327 | 3 | 1 | 1 | Low-quality | Genome-fragment | 0.58 | HMM-based (lower-bound) | 55.15 |
| S16_vs_121697\|\|full | 3789 | No | NA | | 3 | 0 | 2 | Not-determined | Genome-fragment | NA | NA | 0 |
| S14_extracted_k141_105050 | 2960 | No | NA | | 6 | 3 | 0 | Low-quality | Genome-fragment | 5.12 | AAI-based (high-confidence) | 0 |
| S16_vs_121730\|\|full | 12240 | No | NA | | 13 | 0 | 3 | Not-determined | Genome-fragment | NA | NA | 0 |
| S7_vs_77834\|\|full | 2960 | No | NA | | 3 | 0 | 2 | Low-quality | Genome-fragment | 3.57 | AAI-based (medium-confidence) | 0 |
| S16_vs_121843\|\|full | 3913 | No | NA | | 10 | 0 | 0 | Not-determined | Genome-fragment | NA | NA | 0 |
| S16_vs_73565\|\|full | 2960 | No | NA | | 3 | 0 | 0 | Low-quality | Genome-fragment | 5.34 | AAI-based (high-confidence) | 0 |
| S10_extracted_29609 | 2960 | No | NA | | 9 | 0 | 0 | Low-quality | Genome-fragment | 16.48 | AAI-based (high-confidence) | 0 |
| S16_vs_121967\|\|full | 1885 | No | NA | | 3 | 0 | 0 | Not-determined | Genome-fragment | NA | NA | 0 |
| S17_extracted_k141_1874 | 2961 | No | NA | | 8 | 7 | 0 | Low-quality | Genome-fragment | 9.74 | AAI-based (high-confidence) | 0 |
| S16_vs_122157\|\|full | 2872 | No | NA | | 3 | 0 | 1 | Not-determined | Genome-fragment | NA | NA | 0 |
| S16_vs_122173\|\|full | 3723 | No | NA | | 4 | 0 | 2 | Not-determined | Genome-fragment | NA | NA | 0 |
| S12_vs_k141_104088\|\|full | 2961 | No | NA | | 2 | 0 | 1 | Low-quality | Genome-fragment | 4.93 | AAI-based (medium-confidence) | 0 |
| S16_vs_122311\|\|full | 3823 | No | NA | | 4 | 0 | 4 | Not-determined | Genome-fragment | NA | NA | 0 |
| S16_vs_122329\|\|full | 3353 | No | NA | | 5 | 0 | 3 | Not-determined | Genome-fragment | NA | NA | 0 |
| S4_extracted_14750 | 2961 | No | NA | | 2 | 2 | 0 | Low-quality | Genome-fragment | 9.48 | AAI-based (high-confidence) | 0 |
| S11_vs_k141_27068\|\|full | 2962 | No | NA | | 6 | 1 | 0 | Low-quality | Genome-fragment | 2.18 | AAI-based (high-confidence) | 0 |
| S3_vs_k141_105696\|\|full | 2962 | No | NA | | 8 | 1 | 0 | Low-quality | Genome-fragment | 6.9 | AAI-based (high-confidence) | 0 |
| S16_vs_122458\|\|full | 4831 | No | NA | | 6 | 0 | 5 | Not-determined | Genome-fragment | NA | NA | 0 |
| S5_vs_k141_2895\|\|full | 2962 | No | NA | | 3 | 0 | 2 | Low-quality | Genome-fragment | 1.07 | AAI-based (medium-confidence) | 0 |
| S16_vs_122604\|\|full | 5227 | No | NA | | 4 | 0 | 4 | Not-determined | Genome-fragment | NA | NA | 0 |
| S16_vs_122639\|\|full | 3206 | No | NA | | 3 | 0 | 1 | Not-determined | Genome-fragment | NA | NA | 0 |
| S16_vs_122762\|\|full | 1759 | No | NA | | 2 | 0 | 0 | Not-determined | Genome-fragment | NA | NA | 0 |
| S7_vs_69534\|\|full | 2965 | No | NA | | 3 | 3 | 0 | Low-quality | Genome-fragment | 7.74 | AAI-based (high-confidence) | 0 |
| S10_vs_20208\|\|full | 2965 | No | NA | | 4 | 2 | 0 | Low-quality | Genome-fragment | 7.13 | AAI-based (medium-confidence) | 0 |
| S8_vs_k141_143322\|\|full | 2965 | No | NA | | 4 | 0 | 2 | Low-quality | Genome-fragment | 2.05 | AAI-based (medium-confidence) | 0 |
| S13_vs_90752\|\|full | 2966 | No | NA | | 4 | 0 | 0 | Low-quality | Genome-fragment | 6.17 | AAI-based (medium-confidence) | 0 |
| S16_vs_123067\|\|full | 2376 | No | NA | | 3 | 0 | 1 | Not-determined | Genome-fragment | NA | NA | 0 |
| S14_vs_k141_5679\|\|full | 2966 | No | NA | | 11 | 1 | 0 | Low-quality | Genome-fragment | 7.14 | AAI-based (high-confidence) | 0 |
| S16_vs_123114\|\|full | 9092 | No | NA | | 8 | 0 | 5 | Not-determined | Genome-fragment | NA | NA | 0 |
| S16_vs_123117\|\|full | 2850 | No | NA | | 4 | 0 | 4 | Not-determined | Genome-fragment | NA | NA | 0 |
| S16_vs_123128\|\|full | 1030 | No | NA | | 2 | 0 | 0 | Not-determined | Genome-fragment | NA | NA | 0 |
| S16_vs_123133\|\|full | 1494 | No | NA | | 2 | 0 | 1 | Not-determined | Genome-fragment | NA | NA | 0 |
| S16_vs_123183\|\|full | 1668 | No | NA | | 2 | 0 | 0 | Not-determined | Genome-fragment | NA | NA | 0 |
| S16_vs_123223\|\|full | 3205 | No | NA | | 3 | 0 | 2 | Not-determined | Genome-fragment | NA | NA | 0 |
| S16_vs_123229\|\|full | 1701 | No | NA | | 4 | 0 | 0 | Not-determined | Genome-fragment | NA | NA | 0 |
| S16_vs_123231\|\|full | 1060 | No | NA | | 3 | 0 | 0 | Not-determined | Genome-fragment | NA | NA | 0 |
| S5_vs_k141_26382\|\|full | 2966 | No | NA | | 5 | 2 | 0 | Low-quality | Genome-fragment | 6.35 | AAI-based (high-confidence) | 0 |
| S16_vs_123342\|\|full | 1127 | No | NA | | 2 | 0 | 2 | Not-determined | Genome-fragment | NA | NA | 0 |
| S16_vs_123435\|\|full | 1523 | No | NA | | 2 | 0 | 0 | Not-determined | Genome-fragment | NA | NA | 0 |
| S16_vs_123439\|\|full | 2634 | No | NA | | 3 | 0 | 3 | Not-determined | Genome-fragment | NA | NA | 0 |
| S16_vs_123450\|\|full | 3975 | No | NA | | 3 | 0 | 3 | Not-determined | Genome-fragment | NA | NA | 0 |
| S5_vs_k141_84649\|\|full | 2966 | No | NA | | 5 | 0 | 0 | Low-quality | Genome-fragment | 3.88 | AAI-based (medium-confidence) | 0 |
| S12_extracted_k141_113009 | 2966 | No | NA | | 4 | 0 | 1 | Low-quality | Genome-fragment | 3.04 | AAI-based (medium-confidence) | 0 |
| S12_vs_k141_140048\|\|full | 2966 | No | NA | | 10 | 1 | 0 | Low-quality | Genome-fragment | 5.12 | AAI-based (high-confidence) | 0 |
| S8_vs_k141_201164\|\|full | 2966 | No | NA | | 9 | 0 | 0 | Low-quality | Genome-fragment | 8.52 | AAI-based (medium-confidence) | 0 |
| S5_vs_k141_73484\|\|full | 2967 | No | NA | | 6 | 0 | 0 | Low-quality | Genome-fragment | 8.34 | AAI-based (high-confidence) | 0 |
| S16_extracted_119061 | 2967 | No | NA | | 3 | 0 | 0 | Low-quality | Genome-fragment | 2.79 | AAI-based (medium-confidence) | 0 |
| S16_vs_123825\|\|full | 1251 | No | NA | | 3 | 0 | 1 | Not-determined | Genome-fragment | NA | NA | 0 |
| S16_vs_123983\|\|full | 2466 | No | NA | | 3 | 0 | 2 | Not-determined | Genome-fragment | NA | NA | 0 |
| S6_vs_k141_8104\|\|full | 2967 | No | NA | | 2 | 0 | 2 | Low-quality | Genome-fragment | 2.62 | AAI-based (medium-confidence) | 0 |
| S16_vs_124120\|\|full | 2273 | No | NA | | 3 | 0 | 2 | Not-determined | Genome-fragment | NA | NA | 0 |
| S3_vs_k141_45963\|\|full | 2968 | No | NA | | 3 | 0 | 0 | Low-quality | Genome-fragment | 4.44 | AAI-based (medium-confidence) | 0 |
| S16_vs_124146\|\|full | 3217 | No | NA | | 3 | 0 | 3 | Not-determined | Genome-fragment | NA | NA | 0 |
| S2_vs_k141_15140\|\|full | 2968 | No | NA | | 5 | 0 | 0 | Low-quality | Genome-fragment | 6.93 | AAI-based (high-confidence) | 0 |
| S16_vs_124355\|\|full | 3029 | No | NA | | 4 | 0 | 3 | Not-determined | Genome-fragment | NA | NA | 0 |
| S16_vs_124373\|\|full | 2366 | No | NA | | 3 | 0 | 1 | Not-determined | Genome-fragment | NA | NA | 0 |
| S16_vs_124401\|\|full | 4409 | No | NA | | 5 | 0 | 1 | Not-determined | Genome-fragment | NA | NA | 0 |
| S16_extracted_62928 | 2969 | No | NA | | 5 | 0 | 0 | Low-quality | Genome-fragment | 3.78 | AAI-based (high-confidence) | 0 |
| S16_vs_124511\|\|full | 1447 | No | NA | | 2 | 0 | 2 | Not-determined | Genome-fragment | NA | NA | 0 |
| S16_vs_124513\|\|full | 3377 | No | NA | | 4 | 0 | 3 | Not-determined | Genome-fragment | NA | NA | 0 |
| S16_vs_124634\|\|full | 1238 | No | NA | | 2 | 0 | 0 | Not-determined | Genome-fragment | NA | NA | 0 |
| S16_vs_124681\|\|full | 14102 | No | NA | | 7 | 0 | 6 | Not-determined | Genome-fragment | NA | NA | 0 |
| S16_vs_124686\|\|full | 1223 | No | NA | | 3 | 0 | 2 | Not-determined | Genome-fragment | NA | NA | 0 |
| S16_vs_124703\|\|full | 33417 | No | NA | | 50 | 15 | 0 | Medium-quality | Genome-fragment | 89.63 | AAI-based (high-confidence) | 0 |
| S16_vs_124760\|\|full | 4592 | No | NA | | 7 | 0 | 7 | Not-determined | Genome-fragment | NA | NA | 0 |
| S16_vs_114031\|\|full | 2969 | No | NA | | 4 | 0 | 1 | Low-quality | Genome-fragment | 4.59 | AAI-based (medium-confidence) | 0 |
| S16_vs_124936\|\|full | 2848 | No | NA | | 3 | 0 | 3 | Not-determined | Genome-fragment | NA | NA | 0 |
| S16_vs_124951\|\|full | 2238 | No | NA | | 3 | 0 | 1 | Not-determined | Genome-fragment | NA | NA | 0 |
| S1_extracted_k141_27105 | 2970 | No | NA | | 4 | 1 | 0 | Low-quality | Genome-fragment | 7.07 | HMM-based (lower-bound) | 0 |
| S8_vs_k141_29636\|\|lt2gene | 2970 | No | NA | | 1 | 1 | 0 | Low-quality | Genome-fragment | 7.03 | AAI-based (medium-confidence) | 0 |
| S9_vs_k141_6440\|\|full | 2971 | No | NA | | 4 | 3 | 0 | Low-quality | Genome-fragment | 7.76 | AAI-based (high-confidence) | 0 |
| S16_vs_125272\|\|full | 1682 | No | NA | | 2 | 0 | 0 | Not-determined | Genome-fragment | NA | NA | 0 |
| S11_vs_k141_15418\|\|full | 2971 | No | NA | | 3 | 3 | 0 | Low-quality | Genome-fragment | 7.1 | AAI-based (medium-confidence) | 0 |
| S11_vs_k141_49831\|\|full | 2971 | No | NA | | 5 | 1 | 0 | Low-quality | Genome-fragment | 7.83 | AAI-based (high-confidence) | 0 |
| S3_extracted_k141_111028 | 2971 | No | NA | | 6 | 0 | 0 | Low-quality | Genome-fragment | 1.91 | AAI-based (high-confidence) | 0 |
| S3_vs_k141_47506\|\|full | 2971 | No | NA | | 7 | 0 | 0 | Low-quality | Genome-fragment | 7.4 | AAI-based (high-confidence) | 0 |
| S16_vs_125472\|\|full | 6513 | No | NA | | 4 | 0 | 3 | Not-determined | Genome-fragment | NA | NA | 0 |
| S10_vs_31086\|\|full | 2972 | No | NA | | 3 | 0 | 2 | Low-quality | Genome-fragment | 1.67 | AAI-based (medium-confidence) | 0 |
| S16_vs_125546\|\|full | 1467 | No | NA | | 4 | 0 | 1 | Not-determined | Genome-fragment | NA | NA | 0 |
| S16_vs_125620\|\|full | 2816 | No | NA | | 4 | 0 | 1 | Not-determined | Genome-fragment | NA | NA | 0 |
| S2_vs_k141_1572\|\|full | 2972 | No | NA | | 7 | 1 | 0 | Low-quality | Genome-fragment | 7.38 | AAI-based (high-confidence) | 0 |
| S11_vs_k141_81174\|\|full | 2973 | No | NA | | 3 | 0 | 0 | Low-quality | Genome-fragment | 2.84 | AAI-based (medium-confidence) | 0 |
| S16_vs_125911\|\|full | 1251 | No | NA | | 6 | 0 | 0 | Not-determined | Genome-fragment | NA | NA | 0 |
| S16_vs_125912\|\|full | 1282 | No | NA | | 4 | 0 | 3 | Not-determined | Genome-fragment | NA | NA | 0 |

| | | | | | | | | | | | | | |
|---|---|---|---|---|---|---|---|---|---|---|---|---|---|
| S14_extracted_k141_9979 | 2973 | No | NA | | 5 | 0 | 0 | Low-quality | Genome-fragment | | 1.92 | AAI-based (high-confidence) | 0 |
| S16_vs_126149\|\|full | 1366 | No | NA | | 3 | 0 | 0 | Not-determined | Genome-fragment | NA | | NA | 0 |
| S17_vs_k141_69317\|\|full | 2974 | No | NA | | 3 | 0 | 2 | Low-quality | Genome-fragment | | 0.76 | AAI-based (medium-confidence) | 0 |
| S16_vs_126243\|\|full | 1984 | No | NA | | 3 | 0 | 3 | Not-determined | Genome-fragment | NA | | NA | 0 |
| S3_vs_k141_123320\|\|full | 2974 | No | NA | | 4 | 0 | 3 | Low-quality | Genome-fragment | | 1.14 | AAI-based (medium-confidence) | 0 |
| S16_vs_126247\|\|full | 1369 | No | NA | | 3 | 0 | 0 | Not-determined | Genome-fragment | NA | | NA | 0 |
| S5_extracted_k141_83912 | 2974 | No | NA | | 10 | 2 | 0 | Low-quality | Genome-fragment | | 5.44 | AAI-based (high-confidence) | 0 |
| S16_vs_126368\|\|full | 1891 | No | NA | | 4 | 0 | 0 | Not-determined | Genome-fragment | NA | | NA | 0 |
| S16_vs_126613\|\|full | 3123 | No | NA | | 3 | 0 | 1 | Not-determined | Genome-fragment | NA | | NA | 0 |
| S16_vs_126704\|\|full | 1135 | No | NA | | 2 | 0 | 0 | Not-determined | Genome-fragment | NA | | NA | 0 |
| S7_vs_8996\|\|full | 2974 | No | NA | | 3 | 0 | 3 | Low-quality | Genome-fragment | | 1.46 | AAI-based (medium-confidence) | 0 |
| S10_vs_105202\|\|full | 2974 | No | NA | | 3 | 3 | 0 | Low-quality | Genome-fragment | | 8.67 | AAI-based (high-confidence) | 0 |
| S16_vs_126965\|\|full | 1118 | No | NA | | 3 | 0 | 0 | Not-determined | Genome-fragment | NA | | NA | 0 |
| S9_vs_k141_33063\|\|full | 2975 | No | NA | | 5 | 0 | 1 | Low-quality | Genome-fragment | | 8.06 | AAI-based (high-confidence) | 0 |
| S16_vs_136312\|\|full | 2975 | No | NA | | 3 | 0 | 2 | Low-quality | Genome-fragment | | 1.07 | AAI-based (medium-confidence) | 0 |
| S16_vs_127056\|\|full | 15042 | No | NA | | 10 | 0 | 1 | Not-determined | Genome-fragment | NA | | NA | 0 |
| S16_vs_127174\|\|full | 3108 | No | NA | | 5 | 0 | 1 | Not-determined | Genome-fragment | NA | | NA | 0 |
| S16_vs_127357\|\|full | 1665 | No | NA | | 2 | 0 | 1 | Not-determined | Genome-fragment | NA | | NA | 0 |
| S16_vs_127406\|\|full | 1298 | No | NA | | 3 | 0 | 1 | Not-determined | Genome-fragment | NA | | NA | 0 |
| S16_vs_127505\|\|full | 13847 | No | NA | | 7 | 0 | 3 | Not-determined | Genome-fragment | NA | | NA | 0 |
| S10_extracted_30493 | 2975 | No | NA | | 4 | 0 | 0 | Low-quality | Genome-fragment | | 4.09 | AAI-based (medium-confidence) | 0 |
| S16_vs_127594\|\|full | 2653 | No | NA | | 2 | 0 | 2 | Not-determined | Genome-fragment | NA | | NA | 0 |
| S16_vs_127637\|\|full | 4137 | No | NA | | 4 | 0 | 4 | Not-determined | Genome-fragment | NA | | NA | 0 |
| S16_vs_127669\|\|full | 81984 | Yes | | 76460 | 116 | 15 | 6 | High-quality | High-quality | | 100 | AAI-based (high-confidence) | 6.74 |
| S16_vs_127674\|\|full | 4612 | No | NA | | 5 | 0 | 4 | Not-determined | Genome-fragment | NA | | NA | 0 |
| S2_extracted_k141_8521 | 2975 | No | NA | | 3 | 1 | 0 | Low-quality | Genome-fragment | | 10.94 | HMM-based (lower-bound) | 0 |
| S16_vs_94619\|\|full | 2976 | No | NA | | 4 | 0 | 2 | Low-quality | Genome-fragment | | 5.49 | AAI-based (medium-confidence) | 0 |
| S16_vs_173044\|\|full | 2977 | No | NA | | 6 | 0 | 0 | Low-quality | Genome-fragment | | 5.09 | AAI-based (medium-confidence) | 0 |
| S13_extracted_30419 | 2978 | No | NA | | 3 | 0 | 1 | Low-quality | Genome-fragment | | 6.88 | AAI-based (medium-confidence) | 0 |
| S16_vs_127971\|\|full | 1966 | No | NA | | 3 | 0 | 1 | Not-determined | Genome-fragment | NA | | NA | 0 |
| S16_vs_128031\|\|full | 5371 | No | NA | | 6 | 0 | 3 | Not-determined | Genome-fragment | NA | | NA | 0 |
| S16_vs_128068\|\|full | 7666 | No | NA | | 7 | 0 | 4 | Not-determined | Genome-fragment | NA | | NA | 0 |
| S5_vs_k141_37704\|\|full | 2978 | No | NA | | 8 | 2 | 0 | Low-quality | Genome-fragment | | 7.39 | AAI-based (medium-confidence) | 0 |
| S16_vs_128080\|\|full | 1875 | No | NA | | 5 | 0 | 0 | Not-determined | Genome-fragment | NA | | NA | 0 |
| S16_vs_128129\|\|full | 1162 | No | NA | | 3 | 0 | 0 | Not-determined | Genome-fragment | NA | | NA | 0 |
| S6_vs_k141_40353\|\|full | 2979 | No | NA | | 5 | 1 | 0 | Low-quality | Genome-fragment | | 3.47 | AAI-based (medium-confidence) | 0 |
| S11_vs_k141_56928\|\|full | 2981 | No | NA | | 5 | 1 | 0 | Low-quality | Genome-fragment | | 4.91 | AAI-based (medium-confidence) | 0 |
| S16_vs_128490\|\|full | 1186 | No | NA | | 3 | 0 | 2 | Not-determined | Genome-fragment | NA | | NA | 0 |
| S16_vs_128517\|\|full | 1084 | No | NA | | 2 | 0 | 1 | Not-determined | Genome-fragment | NA | | NA | 0 |
| S16_vs_128563\|\|full | 1034 | No | NA | | 4 | 0 | 0 | Not-determined | Genome-fragment | NA | | NA | 0 |
| S16_vs_128572\|\|full | 1171 | No | NA | | 3 | 0 | 0 | Not-determined | Genome-fragment | NA | | NA | 0 |
| S13_vs_124403\|\|full | 2981 | No | NA | | 5 | 0 | 3 | Low-quality | Genome-fragment | | 5.84 | AAI-based (medium-confidence) | 0 |
| S14_vs_k141_47079\|\|full | 2981 | No | NA | | 4 | 1 | 0 | Low-quality | Genome-fragment | | 5.59 | AAI-based (high-confidence) | 0 |
| S16_vs_128706\|\|full | 6527 | No | NA | | 7 | 0 | 3 | Not-determined | Genome-fragment | NA | | NA | 0 |
| S16_vs_128813\|\|full | 8787 | No | NA | | 10 | 0 | 2 | Not-determined | Genome-fragment | NA | | NA | 0 |
| S16_vs_128849\|\|full | 1456 | No | NA | | 4 | 0 | 0 | Not-determined | Genome-fragment | NA | | NA | 0 |
| S10_vs_51319\|\|full | 2981 | No | NA | | 2 | 0 | 1 | Low-quality | Genome-fragment | | 4.95 | AAI-based (high-confidence) | 0 |
| S16_vs_129049\|\|full | 8243 | No | NA | | 7 | 0 | 4 | Not-determined | Genome-fragment | NA | | NA | 0 |
| S1_vs_k141_68966\|\|full | 2982 | No | NA | | 6 | 2 | 0 | Low-quality | Genome-fragment | | 3.71 | AAI-based (high-confidence) | 0 |
| S16_vs_129249\|\|full | 1164 | No | NA | | 3 | 0 | 0 | Not-determined | Genome-fragment | NA | | NA | 0 |
| S1_vs_k141_112417\|\|full | 2982 | No | NA | | 6 | 3 | 0 | Low-quality | Genome-fragment | | 8.03 | AAI-based (high-confidence) | 0 |
| S2_vs_k141_3508\|\|full | 2982 | No | NA | | 3 | 2 | 0 | Low-quality | Genome-fragment | | 7.82 | AAI-based (high-confidence) | 0 |
| S5_extracted_k141_40050 | 2983 | No | NA | | 7 | 3 | 0 | Low-quality | Genome-fragment | | 6.57 | AAI-based (high-confidence) | 0 |
| S16_vs_129412\|\|full | 1053 | No | NA | | 2 | 0 | 1 | Not-determined | Genome-fragment | NA | | NA | 0 |
| S16_vs_129452\|\|full | 2023 | No | NA | | 3 | 0 | 2 | Not-determined | Genome-fragment | NA | | NA | 0 |
| S5_vs_k141_31009\|\|full | 2983 | No | NA | | 4 | 0 | 2 | Low-quality | Genome-fragment | | 2.28 | AAI-based (medium-confidence) | 0 |
| S16_vs_129485\|\|full | 7912 | No | NA | | 11 | 0 | 7 | Not-determined | Genome-fragment | NA | | NA | 0 |
| S9_vs_k141_8401\|\|full | 2984 | No | NA | | 2 | 0 | 0 | Low-quality | Genome-fragment | | 4.65 | AAI-based (high-confidence) | 0 |
| S16_vs_129551\|\|full | 2447 | No | NA | | 2 | 0 | 1 | Not-determined | Genome-fragment | NA | | NA | 0 |
| S3_vs_k141_68077\|\|full | 2984 | No | NA | | 3 | 0 | 1 | Low-quality | Genome-fragment | | 1.85 | AAI-based (medium-confidence) | 0 |
| S16_vs_129779\|\|full | 4677 | No | NA | | 12 | 0 | 1 | Not-determined | Genome-fragment | NA | | NA | 0 |
| S10_vs_89468\|\|full | 2984 | No | NA | | 5 | 0 | 0 | Low-quality | Genome-fragment | | 4.13 | AAI-based (medium-confidence) | 0 |
| S16_vs_129890\|\|full | 1228 | No | NA | | 4 | 0 | 0 | Not-determined | Genome-fragment | NA | | NA | 0 |
| S16_vs_129925\|\|full | 1112 | No | NA | | 4 | 0 | 0 | Not-determined | Genome-fragment | NA | | NA | 0 |
| S15_extracted_13861 | 2984 | No | NA | | 5 | 0 | 0 | Low-quality | Genome-fragment | | 2.97 | AAI-based (high-confidence) | 0 |
| S16_vs_130037\|\|full | 3219 | No | NA | | 4 | 0 | 2 | Not-determined | Genome-fragment | NA | | NA | 0 |
| S17_vs_k141_19700\|\|full | 2985 | No | NA | | 6 | 1 | 0 | Low-quality | Genome-fragment | | 5.87 | AAI-based (high-confidence) | 0 |
| S16_vs_130135\|\|full | 1643 | No | NA | | 3 | 0 | 0 | Not-determined | Genome-fragment | NA | | NA | 0 |
| S16_vs_130141\|\|full | 8912 | No | NA | | 8 | 0 | 3 | Not-determined | Genome-fragment | NA | | NA | 0 |
| S11_vs_k141_53204\|\|full | 2985 | No | NA | | 3 | 1 | 0 | Low-quality | Genome-fragment | | 3.26 | HMM-based (lower-bound) | 0 |
| S16_vs_130332\|\|full | 2939 | No | NA | | 4 | 0 | 0 | Not-determined | Genome-fragment | NA | | NA | 0 |
| S17_extracted_k141_82695 | 2986 | No | NA | | 2 | 0 | 1 | Low-quality | Genome-fragment | | 4.74 | AAI-based (medium-confidence) | 0 |
| S16_vs_130408\|\|full | 2159 | No | NA | | 4 | 0 | 2 | Not-determined | Genome-fragment | NA | | NA | 0 |
| S5_vs_k141_50838\|\|full | 2986 | No | NA | | 4 | 3 | 0 | Low-quality | Genome-fragment | | 5 | AAI-based (high-confidence) | 0 |
| S14_vs_k141_70763\|\|full | 2987 | No | NA | | 5 | 3 | 0 | Low-quality | Genome-fragment | | 7.27 | AAI-based (medium-confidence) | 0 |
| S16_extracted_79858 | 2987 | No | NA | | 3 | 0 | 3 | Low-quality | Genome-fragment | | 2.13 | AAI-based (medium-confidence) | 0 |
| S14_vs_k141_53966\|\|full | 2988 | No | NA | | 4 | 0 | 3 | Low-quality | Genome-fragment | | 1.23 | AAI-based (medium-confidence) | 0 |
| S16_vs_130843\|\|full | 3076 | No | NA | | 4 | 0 | 2 | Not-determined | Genome-fragment | NA | | NA | 0 |
| S16_vs_130860\|\|full | 1675 | No | NA | | 4 | 0 | 0 | Not-determined | Genome-fragment | NA | | NA | 0 |
| S16_vs_130887\|\|full | 6266 | No | NA | | 5 | 0 | 3 | Not-determined | Genome-fragment | NA | | NA | 0 |
| S16_vs_130896\|\|full | 2724 | No | NA | | 2 | 0 | 0 | Not-determined | Genome-fragment | NA | | NA | 0 |
| S16_vs_130969\|\|full | 1153 | No | NA | | 2 | 0 | 1 | Not-determined | Genome-fragment | NA | | NA | 0 |
| S16_vs_130987\|\|full | 1666 | No | NA | | 3 | 0 | 2 | Not-determined | Genome-fragment | NA | | NA | 0 |
| S16_vs_131036\|\|full | 2812 | No | NA | | 3 | 0 | 3 | Not-determined | Genome-fragment | NA | | NA | 0 |
| S3_vs_k141_1086\|\|full | 2990 | No | NA | | 5 | 0 | 0 | Low-quality | Genome-fragment | | 5.01 | AAI-based (high-confidence) | 0 |
| S16_vs_131118\|\|full | 1979 | No | NA | | 3 | 0 | 0 | Not-determined | Genome-fragment | NA | | NA | 0 |
| S16_vs_131144\|\|full | 1304 | No | NA | | 2 | 0 | 0 | Not-determined | Genome-fragment | NA | | NA | 0 |
| S16_vs_131275\|\|full | 4650 | No | NA | | 7 | 0 | 1 | Not-determined | Genome-fragment | NA | | NA | 0 |
| S11_vs_k141_91104\|\|full | 2991 | No | NA | | 4 | 3 | 0 | Low-quality | Genome-fragment | | 2.42 | HMM-based (lower-bound) | 0 |
| S16_vs_111940\|\|full | 2991 | No | NA | | 5 | 0 | 0 | Low-quality | Genome-fragment | | 5.4 | AAI-based (medium-confidence) | 0 |
| S16_vs_131502\|\|full | 14542 | No | NA | | 9 | 0 | 7 | Not-determined | Genome-fragment | NA | | NA | 0 |
| S16_vs_131524\|\|full | 1188 | No | NA | | 2 | 0 | 1 | Not-determined | Genome-fragment | NA | | NA | 0 |
| S16_vs_131542\|\|full | 8146 | No | NA | | 5 | 0 | 3 | Not-determined | Genome-fragment | NA | | NA | 0 |
| S7_vs_109197\|\|full | 2992 | No | NA | | 6 | 2 | 0 | Low-quality | Genome-fragment | | 4.98 | HMM-based (lower-bound) | 0 |
| S11_vs_k141_78365\|\|full | 2995 | No | NA | | 4 | 2 | 0 | Low-quality | Genome-fragment | | 7.2 | AAI-based (high-confidence) | 0 |
| S16_vs_131621\|\|full | 1062 | No | NA | | 2 | 0 | 1 | Not-determined | Genome-fragment | NA | | NA | 0 |
| S1_vs_k141_2771\|\|full | 2995 | No | NA | | 3 | 2 | 0 | Low-quality | Genome-fragment | | 5.24 | AAI-based (high-confidence) | 0 |
| S16_vs_131666\|\|full | 7468 | No | NA | | 8 | 0 | 6 | Not-determined | Genome-fragment | NA | | NA | 0 |
| S16_vs_131713\|\|full | 2171 | No | NA | | 3 | 0 | 0 | Not-determined | Genome-fragment | NA | | NA | 0 |
| S7_vs_11888\|\|full | 2995 | Yes | | 2267 | 6 | 1 | 1 | Low-quality | Genome-fragment | | 3.02 | AAI-based (medium-confidence) | 24.31 |
| S16_vs_131720\|\|full | 1557 | No | NA | | 3 | 0 | 1 | Not-determined | Genome-fragment | NA | | NA | 0 |
| S16_vs_131797\|\|full | 2176 | No | NA | | 4 | 0 | 0 | Not-determined | Genome-fragment | NA | | NA | 0 |
| S16_vs_131810\|\|full | 3849 | No | NA | | 3 | 0 | 1 | Not-determined | Genome-fragment | NA | | NA | 0 |
| S14_extracted_k141_83313 | 2996 | No | NA | | 2 | 0 | 1 | Low-quality | Genome-fragment | | 1.97 | AAI-based (medium-confidence) | 0 |
| S16_vs_131986\|\|full | 2267 | No | NA | | 2 | 0 | 2 | Not-determined | Genome-fragment | NA | | NA | 0 |
| S1_extracted_k141_80503 | 2996 | No | NA | | 4 | 0 | 0 | Low-quality | Genome-fragment | | 5.19 | AAI-based (high-confidence) | 0 |
| S7_vs_102700\|\|full | 2996 | No | NA | | 4 | 1 | 0 | Low-quality | Genome-fragment | | 7.43 | AAI-based (high-confidence) | 0 |
| S16_vs_132069\|\|full | 2837 | No | NA | | 2 | 0 | 2 | Not-determined | Genome-fragment | NA | | NA | 0 |
| S12_vs_k141_163199\|\|full | 2996 | No | NA | | 4 | 1 | 0 | Low-quality | Genome-fragment | | 6.38 | AAI-based (high-confidence) | 0 |
| S1_vs_k141_140147\|\|full | 2997 | No | NA | | 5 | 3 | 0 | Low-quality | Genome-fragment | | 4.95 | AAI-based (high-confidence) | 0 |
| S16_vs_132273\|\|full | 1010 | No | NA | | 2 | 0 | 1 | Not-determined | Genome-fragment | NA | | NA | 0 |
| S5_vs_k141_67781\|\|full | 2997 | No | NA | | 7 | 1 | 1 | Low-quality | Genome-fragment | | 1.22 | HMM-based (lower-bound) | 0 |
| S16_vs_132345\|\|full | 5129 | No | NA | | 8 | 0 | 2 | Not-determined | Genome-fragment | NA | | NA | 0 |
| S7_extracted_133131 | 2997 | No | NA | | 6 | 4 | 0 | Low-quality | Genome-fragment | | 6.83 | AAI-based (high-confidence) | 0 |
| S16_vs_132412\|\|full | 1722 | No | NA | | 3 | 0 | 1 | Not-determined | Genome-fragment | NA | | NA | 0 |
| S17_extracted_k141_63132 | 2998 | No | NA | | 3 | 3 | 0 | Low-quality | Genome-fragment | | 9.54 | AAI-based (high-confidence) | 0 |
| S16_vs_132449\|\|full | 3883 | No | NA | | 5 | 0 | 2 | Not-determined | Genome-fragment | NA | | NA | 0 |
| S16_vs_132705\|\|full | 2919 | No | NA | | 4 | 0 | 1 | Not-determined | Genome-fragment | NA | | NA | 0 |
| S16_vs_132716\|\|full | 1804 | No | NA | | 4 | 0 | 1 | Not-determined | Genome-fragment | NA | | NA | 0 |
| S16_vs_132793\|\|full | 2382 | No | NA | | 3 | 0 | 0 | Not-determined | Genome-fragment | NA | | NA | 0 |
| S16_vs_132904\|\|full | 4609 | No | NA | | 4 | 0 | 3 | Not-determined | Genome-fragment | NA | | NA | 0 |
| S16_vs_132979\|\|full | 31106 | Yes | | 17760 | 38 | 9 | 5 | Medium-quality | Genome-fragment | | 51.15 | AAI-based (high-confidence) | 42.9 |
| S3_vs_k141_41711\|\|full | 2999 | Yes | | 1465 | 6 | 1 | 1 | Low-quality | Genome-fragment | | 2.47 | HMM-based (lower-bound) | 51.15 |
| S4_extracted_10330 | 2999 | No | NA | | 6 | 0 | 5 | Low-quality | Genome-fragment | | 2.04 | AAI-based (medium-confidence) | 0 |
| S16_vs_133208\|\|full | 1956 | No | NA | | 4 | 0 | 0 | Not-determined | Genome-fragment | NA | | NA | 0 |
| S16_vs_133392\|\|full | 1968 | No | NA | | 3 | 0 | 1 | Not-determined | Genome-fragment | NA | | NA | 0 |
| S9_vs_k141_66481\|\|full | 3000 | No | NA | | 5 | 0 | 4 | Low-quality | Genome-fragment | | 4.63 | AAI-based (medium-confidence) | 0 |
| S14_vs_k141_13399\|\|full | 3000 | No | NA | | 5 | 0 | 1 | Low-quality | Genome-fragment | | 3.09 | AAI-based (medium-confidence) | 0 |
| S16_vs_133519\|\|full | 6276 | No | NA | | 9 | 0 | 1 | Not-determined | Genome-fragment | NA | | NA | 0 |
| S16_vs_133579\|\|full | 3724 | No | NA | | 5 | 0 | 3 | Not-determined | Genome-fragment | NA | | NA | 0 |
| S16_vs_133598\|\|full | 4084 | No | NA | | 5 | 0 | 2 | Not-determined | Genome-fragment | NA | | NA | 0 |
| S1_extracted_k141_42674 | 3000 | No | NA | | 2 | 0 | 2 | Low-quality | Genome-fragment | | 0.8 | AAI-based (high-confidence) | 0 |
| S16_vs_133794\|\|full | 7786 | No | NA | | 13 | 0 | 3 | Not-determined | Genome-fragment | NA | | NA | 0 |
| S5_extracted_k141_54757 | 3001 | No | NA | | 6 | 1 | 0 | Low-quality | Genome-fragment | | 6.18 | AAI-based (medium-confidence) | 0 |
| S16_vs_133929\|\|full | 1957 | No | NA | | 4 | 0 | 0 | Not-determined | Genome-fragment | NA | | NA | 0 |
| S16_vs_133934\|\|full | 3832 | No | NA | | 5 | 0 | 1 | Not-determined | Genome-fragment | NA | | NA | 0 |
| S7_vs_107862\|\|full | 3001 | No | NA | | 9 | 5 | 0 | Low-quality | Genome-fragment | | 4.95 | AAI-based (high-confidence) | 0 |
| S10_vs_12619\|\|full | 3001 | No | NA | | 4 | 1 | 0 | Low-quality | Genome-fragment | | 8.62 | AAI-based (high-confidence) | 0 |
| S14_vs_k141_11809\|\|full | 3002 | No | NA | | 5 | 1 | 0 | Low-quality | Genome-fragment | | 4.77 | AAI-based (high-confidence) | 0 |
| S12_vs_k141_16067\|\|full | 3002 | No | NA | | 4 | 0 | 0 | Low-quality | Genome-fragment | | 1.88 | AAI-based (medium-confidence) | 0 |
| S9_vs_k141_27396\|\|full | 3003 | No | NA | | 3 | 0 | 0 | Low-quality | Genome-fragment | | 3.58 | AAI-based (medium-confidence) | 0 |
| S16_vs_134144\|\|full | 2445 | No | NA | | 2 | 0 | 2 | Not-determined | Genome-fragment | NA | | NA | 0 |
| S7_vs_17784\|\|full | 3003 | No | NA | | 7 | 0 | 0 | Low-quality | Genome-fragment | | 7.71 | AAI-based (high-confidence) | 0 |
| S16_vs_134374\|\|full | 2763 | No | NA | | 4 | 0 | 1 | Not-determined | Genome-fragment | NA | | NA | 0 |
| S16_vs_134421\|\|full | 2268 | No | NA | | 3 | 0 | 2 | Not-determined | Genome-fragment | NA | | NA | 0 |
| S10_extracted_8554 | 3003 | No | NA | | 5 | 1 | 0 | Low-quality | Genome-fragment | | 3.81 | AAI-based (high-confidence) | 0 |
| S14_vs_k141_33023\|\|full | 3004 | No | NA | | 3 | 0 | 3 | Low-quality | Genome-fragment | | 1.78 | AAI-based (medium-confidence) | 0 |
| S16_vs_134550\|\|full | 5407 | No | NA | | 4 | 0 | 3 | Not-determined | Genome-fragment | NA | | NA | 0 |
| S3_vs_k141_73969\|\|full | 3004 | No | NA | | 4 | 3 | 0 | Low-quality | Genome-fragment | | 6.41 | AAI-based (high-confidence) | 0 |
| S16_extracted_16583 | 3004 | No | NA | | 5 | 0 | 3 | Low-quality | Genome-fragment | | 1.35 | AAI-based (medium-confidence) | 0 |
| S16_vs_134649\|\|full | 1639 | No | NA | | 2 | 0 | 1 | Not-determined | Genome-fragment | NA | | NA | 0 |
| S5_vs_k141_69372\|\|full | 3005 | No | NA | | 3 | 1 | 0 | Low-quality | Genome-fragment | | 1.86 | HMM-based (lower-bound) | 0 |
| S6_extracted_k141_31880 | 3005 | No | NA | | 3 | 3 | 0 | Low-quality | Genome-fragment | | 9.9 | AAI-based (high-confidence) | 0 |
| S8_extracted_k141_143015 | 3005 | No | NA | | 11 | 2 | 0 | Low-quality | Genome-fragment | | 2.4 | AAI-based (high-confidence) | 0 |
| S16_vs_134840\|\|full | 1897 | No | NA | | 2 | 0 | 1 | Not-determined | Genome-fragment | NA | | NA | 0 |
| S16_vs_134923\|\|full | 3255 | No | NA | | 2 | 0 | 0 | Not-determined | Genome-fragment | NA | | NA | 0 |
| S16_vs_134942\|\|full | 1133 | No | NA | | 2 | 0 | 1 | Not-determined | Genome-fragment | NA | | NA | 0 |
| S13_extracted_14388 | 3007 | No | NA | | 3 | 0 | 1 | Low-quality | Genome-fragment | | 9.04 | AAI-based (medium-confidence) | 0 |
| S14_vs_k141_98431\|\|full | 3007 | No | NA | | 3 | 1 | 0 | Low-quality | Genome-fragment | | 6.02 | HMM-based (lower-bound) | 0 |
| S16_vs_135193\|\|full | 1521 | No | NA | | 3 | 0 | 1 | Not-determined | Genome-fragment | NA | | NA | 0 |
| S1_extracted_k141_59430 | 3007 | No | NA | | 7 | 0 | 0 | Low-quality | Genome-fragment | | 10.49 | AAI-based (medium-confidence) | 0 |
| S14_extracted_k141_16085 | 3008 | No | NA | | 4 | 0 | 1 | Low-quality | Genome-fragment | | 3.8 | AAI-based (high-confidence) | 0 |
| S16_vs_135501\|\|full | 4442 | No | NA | | 6 | 0 | 1 | Not-determined | Genome-fragment | NA | | NA | 0 |
| S16_vs_135782\|\|full | 5746 | No | NA | | 7 | 0 | 0 | Not-determined | Genome-fragment | NA | | NA | 0 |
| S16_vs_135874\|\|full | 1720 | No | NA | | 2 | 0 | 2 | Not-determined | Genome-fragment | NA | | NA | 0 |
| S14_vs_k141_71453\|\|full | 3008 | No | NA | | 7 | 1 | 0 | Low-quality | Genome-fragment | | 6.6 | AAI-based (high-confidence) | 0 |
| S16_vs_55487\|\|full | 3008 | No | NA | | 4 | 0 | 3 | Low-quality | Genome-fragment | | 5.11 | AAI-based (medium-confidence) | 0 |
| S16_vs_136062\|\|full | 4392 | No | NA | | 4 | 0 | 2 | Not-determined | Genome-fragment | NA | | NA | 0 |
| S16_vs_136117\|\|full | 2321 | No | NA | | 2 | 0 | 2 | Not-determined | Genome-fragment | NA | | NA | 0 |
| S1_extracted_k141_108493 | 3009 | No | NA | | 4 | 0 | 0 | Low-quality | Genome-fragment | | 1.73 | AAI-based (high-confidence) | 0 |
| S16_vs_136203\|\|full | 5201 | No | NA | | 8 | 0 | 3 | Not-determined | Genome-fragment | NA | | NA | 0 |
| S16_vs_47892\|\|full | 3009 | No | NA | | 4 | 1 | 0 | Low-quality | Genome-fragment | | 2.06 | HMM-based (lower-bound) | 0 |
| S8_extracted_k141_154577 | 3009 | No | NA | | 3 | 0 | 2 | Low-quality | Genome-fragment | | 4.98 | AAI-based (medium-confidence) | 0 |
| S16_vs_136333\|\|full | 1280 | No | NA | | 2 | 0 | 1 | Not-determined | Genome-fragment | NA | | NA | 0 |
| S8_extracted_k141_68319 | 3009 | No | NA | | 7 | 4 | 0 | Low-quality | Genome-fragment | | 13.73 | AAI-based (high-confidence) | 0 |
| S3_vs_k141_97255\|\|full | 3010 | No | NA | | 6 | 3 | 0 | Low-quality | Genome-fragment | | 7.2 | AAI-based (high-confidence) | 0 |
| S16_vs_136465\|\|full | 1798 | No | NA | | 2 | 0 | 1 | Not-determined | Genome-fragment | NA | | NA | 0 |
| S16_vs_136559\|\|full | 1004 | No | NA | | 2 | 0 | 1 | Not-determined | Genome-fragment | NA | | NA | 0 |
| S6_extracted_k141_43110 | 3010 | No | NA | | 3 | 0 | 0 | Low-quality | Genome-fragment | | 6.88 | AAI-based (medium-confidence) | 0 |
| S5_vs_k141_115701\|\|full | 3011 | No | NA | | 4 | 0 | 0 | Low-quality | Genome-fragment | | 7.58 | AAI-based (medium-confidence) | 0 |
| S11_vs_k141_15479\|\|full | 3012 | No | NA | | 9 | 5 | 0 | Low-quality | Genome-fragment | | 6.14 | HMM-based (lower-bound) | 0 |
| S16_vs_136935\|\|full | 40309 | No | NA | | 50 | 1 | 2 | Medium-quality | Genome-fragment | | 66.07 | AAI-based (high-confidence) | 0 |
| S16_vs_136980\|\|full | 4045 | No | NA | | 5 | 0 | 2 | Not-determined | Genome-fragment | NA | | NA | 0 |
| S1_extracted_k141_16415 | 3012 | No | NA | | 2 | 0 | 0 | Low-quality | Genome-fragment | | 6.66 | AAI-based (high-confidence) | 0 |
| S8_vs_k141_111021\|\|full | 3013 | No | NA | | 5 | 1 | 0 | Low-quality | Genome-fragment | | 4.56 | HMM-based (lower-bound) | 0 |
| S16_vs_137222\|\|full | 44270 | No | NA | | 77 | 20 | 0 | High-quality | High-quality | | 94.11 | AAI-based (medium-confidence) | 0 |
| S1_extracted_k141_20304 | 3014 | No | NA | | 5 | 1 | 0 | Low-quality | Genome-fragment | | 6.59 | AAI-based (high-confidence) | 0 |
| S16_vs_137321\|\|full | 2058 | No | NA | | 3 | 0 | 2 | Not-determined | Genome-fragment | NA | | NA | 0 |
| S3_extracted_k141_77284 | 3014 | No | NA | | 11 | 0 | 0 | Low-quality | Genome-fragment | | 3.21 | AAI-based (high-confidence) | 0 |
| S16_vs_137450\|\|full | 5110 | No | NA | | 4 | 0 | 1 | Not-determined | Genome-fragment | NA | | NA | 0 |
| S7_vs_31790\|\|full | 3014 | No | NA | | 10 | 1 | 0 | Low-quality | Genome-fragment | | 1.56 | HMM-based (lower-bound) | 0 |
| S16_vs_137828\|\|full | 2269 | No | NA | | 3 | 0 | 0 | Not-determined | Genome-fragment | NA | | NA | 0 |
| S16_vs_137880\|\|full | 2019 | No | NA | | 3 | 0 | 0 | Not-determined | Genome-fragment | NA | | NA | 0 |
| S16_vs_137889\|\|full | 2073 | No | NA | | 3 | 0 | 2 | Not-determined | Genome-fragment | NA | | NA | 0 |
| S16_vs_137960\|\|full | 21076 | No | NA | | 17 | 0 | 15 | Not-determined | Genome-fragment | NA | | NA | 0 |
| S12_vs_k141_17821\|\|full | 3014 | No | NA | | 6 | 1 | 0 | Low-quality | Genome-fragment | | 4.03 | AAI-based (high-confidence) | 0 |
| S16_vs_138062\|\|full | 2814 | No | NA | | 4 | 0 | 2 | Not-determined | Genome-fragment | NA | | NA | 0 |
| S16_vs_138077\|\|full | 2520 | No | NA | | 3 | 0 | 3 | Not-determined | Genome-fragment | NA | | NA | 0 |
| S16_vs_138323\|\|full | 2884 | No | NA | | 3 | 0 | 2 | Not-determined | Genome-fragment | NA | | NA | 0 |
| S16_vs_138346\|\|full | 1750 | No | NA | | 2 | 0 | 1 | Not-determined | Genome-fragment | NA | | NA | 0 |
| S16_vs_138373\|\|full | 5695 | No | NA | | 5 | 0 | 3 | Not-determined | Genome-fragment | NA | | NA | 0 |
| S16_vs_138410\|\|full | 1550 | No | NA | | 2 | 0 | 1 | Not-determined | Genome-fragment | NA | | NA | 0 |
| S16_vs_138419\|\|full | 1187 | No | NA | | 3 | 0 | 2 | Not-determined | Genome-fragment | NA | | NA | 0 |
| S9_vs_k141_90674\|\|full | 3015 | No | NA | | 4 | 3 | 0 | Low-quality | Genome-fragment | | 8.71 | AAI-based (high-confidence) | 0 |
| S11_extracted_k141_36360 | 3015 | No | NA | | 7 | 2 | 0 | Low-quality | Genome-fragment | | 4.78 | AAI-based (high-confidence) | 0 |
| S16_vs_138561\|\|full | 1554 | No | NA | | 2 | 0 | 0 | Not-determined | Genome-fragment | NA | | NA | 0 |
| S16_vs_18528\|\|full | 3015 | No | NA | | 4 | 2 | 0 | Low-quality | Genome-fragment | | 4.74 | AAI-based (high-confidence) | 0 |
| S12_vs_k141_8262\|\|full | 3015 | No | NA | | 4 | 1 | 0 | Low-quality | Genome-fragment | | 6.29 | AAI-based (high-confidence) | 0 |
| S16_vs_138801\|\|full | 2487 | No | NA | | 4 | 0 | 2 | Not-determined | Genome-fragment | NA | | NA | 0 |
| S16_vs_138861\|\|full | 3750 | No | NA | | 4 | 0 | 2 | Not-determined | Genome-fragment | NA | | NA | 0 |
| S16_vs_139176\|\|full | 4908 | No | NA | | 5 | 0 | 2 | Not-determined | Genome-fragment | NA | | NA | 0 |
| S8_extracted_k141_73289 | 3015 | No | NA | | 3 | 0 | 2 | Low-quality | Genome-fragment | | 1.65 | AAI-based (medium-confidence) | 0 |
| S16_vs_139242\|\|full | 2329 | No | NA | | 3 | 0 | 3 | Not-determined | Genome-fragment | NA | | NA | 0 |
| S16_vs_139321\|\|full | 3931 | No | NA | | 4 | 0 | 2 | Not-determined | Genome-fragment | NA | | NA | 0 |
| S16_vs_139375\|\|full | 1180 | No | NA | | 2 | 0 | 2 | Not-determined | Genome-fragment | NA | | NA | 0 |
| S17_extracted_k141_147639 | 3016 | No | NA | | 5 | 5 | 0 | Low-quality | Genome-fragment | | 4.88 | AAI-based (high-confidence) | 0 |
| S16_vs_139475\|\|full | 1358 | No | NA | | 4 | 0 | 0 | Not-determined | Genome-fragment | NA | | NA | 0 |
| S16_vs_139476\|\|full | 3607 | No | NA | | 3 | 0 | 3 | Not-determined | Genome-fragment | NA | | NA | 0 |
| S16_vs_139510\|\|full | 1714 | No | NA | | 4 | 0 | 0 | Not-determined | Genome-fragment | NA | | NA | 0 |
| S7_extracted_128006 | 3016 | No | NA | | 6 | 0 | 1 | Low-quality | Genome-fragment | | 5.68 | AAI-based (medium-confidence) | 0 |
| S7_vs_80196\|\|full | 3016 | No | NA | | 4 | 3 | 0 | Low-quality | Genome-fragment | | 7.98 | AAI-based (high-confidence) | 0 |
| S10_vs_56319\|\|full | 3016 | No | NA | | 4 | 0 | 0 | Low-quality | Genome-fragment | | 6.95 | AAI-based (medium-confidence) | 0 |
| S14_extracted_k141_72017 | 3017 | No | NA | | 4 | 3 | 0 | Low-quality | Genome-fragment | | 7.42 | AAI-based (high-confidence) | 0 |
| S1_vs_k141_146659\|\|full | 3017 | No | NA | | 11 | 2 | 0 | Low-quality | Genome-fragment | | 7.81 | AAI-based (high-confidence) | 0 |
| S16_vs_139922\|\|full | 4975 | No | NA | | 6 | 0 | 4 | Not-determined | Genome-fragment | NA | | NA | 0 |
| S16_vs_34942\|\|full | 3017 | No | NA | | 2 | 0 | 1 | Low-quality | Genome-fragment | | 1.12 | AAI-based (medium-confidence) | 0 |
| S8_vs_k141_3713\|\|full | 3017 | No | NA | | 3 | 3 | 0 | Low-quality | Genome-fragment | | 8.78 | AAI-based (high-confidence) | 0 |
| S16_vs_140027\|\|full | 1493 | No | NA | | 4 | 0 | 2 | Not-determined | Genome-fragment | NA | | NA | 0 |
| S16_vs_140116\|\|full | 1201 | No | NA | | 3 | 0 | 0 | Not-determined | Genome-fragment | NA | | NA | 0 |
| S16_vs_140119\|\|full | 1582 | No | NA | | 2 | 0 | 1 | Not-determined | Genome-fragment | NA | | NA | 0 |
| S16_vs_140128\|\|full | 6392 | No | NA | | 6 | 0 | 5 | Not-determined | Genome-fragment | NA | | NA | 0 |
| S7_extracted_126288 | 3018 | No | NA | | 4 | 3 | 0 | Low-quality | Genome-fragment | | 9.32 | AAI-based (high-confidence) | 0 |
| S16_vs_83232\|\|full | 3018 | No | NA | | 2 | 0 | 1 | Low-quality | Genome-fragment | | 1.52 | AAI-based (medium-confidence) | 0 |
| S16_vs_140343\|\|full | 4473 | No | NA | | 5 | 0 | 3 | Not-determined | Genome-fragment | NA | | NA | 0 |
| S10_extracted_46766 | 3018 | No | NA | | 3 | 0 | 1 | Low-quality | Genome-fragment | | 1.25 | AAI-based (medium-confidence) | 0 |
| S16_vs_140372\|\|full | 2461 | No | NA | | 3 | 0 | 0 | Not-determined | Genome-fragment | NA | | NA | 0 |
| S8_extracted_k141_163460 | 3018 | No | NA | | 4 | 1 | 0 | Low-quality | Genome-fragment | | 16.2 | AAI-based (medium-confidence) | 0 |
| S16_vs_140422\|\|full | 6012 | No | NA | | 7 | 0 | 3 | Not-determined | Genome-fragment | NA | | NA | 0 |
| S16_vs_140468\|\|full | 20506 | No | NA | | 33 | 3 | 1 | Medium-quality | Genome-fragment | | 52.97 | AAI-based (high-confidence) | 0 |
| S17_extracted_k141_148097 | 3019 | No | NA | | 4 | 0 | 2 | Low-quality | Genome-fragment | | 2.55 | AAI-based (high-confidence) | 0 |
| S16_vs_140581\|\|full | 3732 | No | NA | | 5 | 0 | 4 | Not-determined | Genome-fragment | NA | | NA | 0 |
| S16_vs_140653\|\|full | 5811 | No | NA | | 5 | 0 | 3 | Not-determined | Genome-fragment | NA | | NA | 0 |
| S16_vs_140670\|\|full | 1394 | No | NA | | 2 | 0 | 1 | Not-determined | Genome-fragment | NA | | NA | 0 |
| S16_vs_140709\|\|full | 40751 | No | NA | | 71 | 24 | 2 | High-quality | High-quality | | 100 | AAI-based (high-confidence) | 0 |
| S16_vs_140738\|\|full | 3046 | No | NA | | 3 | 0 | 1 | Not-determined | Genome-fragment | NA | | NA | 0 |
| S16_vs_140822\|\|full | 7094 | No | NA | | 4 | 0 | 3 | Not-determined | Genome-fragment | NA | | NA | 0 |
| S1_extracted_k141_35763 | 3019 | No | NA | | 4 | 0 | 0 | Low-quality | Genome-fragment | | 1.96 | AAI-based (high-confidence) | 0 |
| S16_vs_140934\|\|full | 1206 | No | NA | | 3 | 0 | 0 | Not-determined | Genome-fragment | NA | | NA | 0 |
| S16_vs_140952\|\|full | 1481 | No | NA | | 2 | 0 | 1 | Not-determined | Genome-fragment | NA | | NA | 0 |
| S16_vs_140970\|\|full | 2969 | No | NA | | 3 | 0 | 1 | Not-determined | Genome-fragment | NA | | NA | 0 |
| S16_vs_141099\|\|full | 4333 | No | NA | | 4 | 0 | 4 | Not-determined | Genome-fragment | NA | | NA | 0 |
| S7_extracted_74457 | 3019 | No | NA | | 9 | 0 | 0 | Low-quality | Genome-fragment | | 7.12 | AAI-based (medium-confidence) | 0 |
| S16_vs_141218\|\|full | 49177 | Yes | | 35609 | 71 | 12 | 9 | Medium-quality | Genome-fragment | | 59.04 | HMM-based (lower-bound) | 27.59 |
| S16_vs_141291\|\|full | 1054 | No | NA | | 2 | 0 | 0 | Not-determined | Genome-fragment | NA | | NA | 0 |
| S16_vs_141368\|\|full | 3525 | No | NA | | 3 | 0 | 2 | Not-determined | Genome-fragment | NA | | NA | 0 |
| S16_vs_141664\|\|full | 7667 | No | NA | | 4 | 0 | 3 | Not-determined | Genome-fragment | NA | | NA | 0 |
| S10_extracted_63724 | 3019 | No | NA | | 5 | 4 | 0 | Low-quality | Genome-fragment | | 5.43 | AAI-based (high-confidence) | 0 |
| S4_extracted_26150 | 3019 | No | NA | | 2 | 2 | 0 | Low-quality | Genome-fragment | | 7.84 | AAI-based (high-confidence) | 0 |
| S16_vs_142012\|\|full | 1417 | No | NA | | 3 | 0 | 1 | Not-determined | Genome-fragment | NA | | NA | 0 |
| S16_vs_142090\|\|full | 2800 | No | NA | | 2 | 0 | 1 | Not-determined | Genome-fragment | NA | | NA | 0 |
| S17_extracted_k141_28324 | 3020 | No | NA | | 4 | 0 | 0 | Low-quality | Genome-fragment | | 5.78 | AAI-based (medium-confidence) | 0 |
| S16_vs_142271\|\|full | 8039 | No | NA | | 5 | 0 | 0 | Not-determined | Genome-fragment | NA | | NA | 0 |
| S9_vs_k141_44943\|\|full | 3021 | No | NA | | 3 | 0 | 0 | Low-quality | Genome-fragment | | 1.16 | AAI-based (medium-confidence) | 0 |
| S16_vs_85856\|\|full | 3021 | No | NA | | 3 | 0 | 0 | Low-quality | Genome-fragment | | 4.5 | AAI-based (high-confidence) | 0 |
| S16_vs_142414\|\|full | 1879 | No | NA | | 2 | 0 | 2 | Not-determined | Genome-fragment | NA | | NA | 0 |
| S10_vs_67436\|\|full | 3021 | No | NA | | 3 | 0 | 1 | Low-quality | Genome-fragment | | 1.95 | AAI-based (medium-confidence) | 0 |
| S16_vs_142545\|\|full | 3791 | No | NA | | 2 | 0 | 2 | Not-determined | Genome-fragment | NA | | NA | 0 |
| S16_vs_142561\|\|full | 82728 | Yes | | 35882 | 90 | 16 | 30 | High-quality | High-quality | | 97.4 | AAI-based (high-confidence) | 56.63 |
| S17_extracted_k141_33513 | 3022 | No | NA | | 2 | 0 | 2 | Low-quality | Genome-fragment | | 3.63 | AAI-based (medium-confidence) | 0 |
| S16_vs_142809\|\|full | 1328 | No | NA | | 3 | 0 | 1 | Not-determined | Genome-fragment | NA | | NA | 0 |
| S16_vs_142917\|\|full | 1063 | No | NA | | 2 | 0 | 0 | Not-determined | Genome-fragment | NA | | NA | 0 |
| S5_extracted_k141_60262 | 3022 | No | NA | | 3 | 0 | 2 | Low-quality | Genome-fragment | | 1.14 | AAI-based (high-confidence) | 0 |
| S16_vs_142980\|\|full | 1222 | No | NA | | 2 | 0 | 1 | Not-determined | Genome-fragment | NA | | NA | 0 |
| S7_vs_119345\|\|full | 3022 | No | NA | | 3 | 2 | 0 | Low-quality | Genome-fragment | | 7.24 | AAI-based (high-confidence) | 0 |
| S17_extracted_k141_117414 | 3023 | No | NA | | 2 | 0 | 2 | Low-quality | Genome-fragment | | 0.8 | AAI-based (medium-confidence) | 0 |
| S16_vs_143106\|\|full | 7500 | No | NA | | 11 | 0 | 4 | Not-determined | Genome-fragment | NA | | NA | 0 |
| S16_vs_143123\|\|full | 1153 | No | NA | | 2 | 0 | 0 | Not-determined | Genome-fragment | NA | | NA | 0 |
| S16_vs_143147\|\|full | 2346 | No | NA | | 2 | 0 | 1 | Not-determined | Genome-fragment | NA | | NA | 0 |
| S16_vs_143173\|\|full | 1770 | No | NA | | 2 | 0 | 1 | Not-determined | Genome-fragment | NA | | NA | 0 |
| S17_vs_k141_140711\|\|full | 3023 | Yes | | 1105 | 3 | 1 | 1 | Low-quality | Genome-fragment | | 0.69 | HMM-based (lower-bound) | 63.45 |
| S1_extracted_k141_70458 | 3023 | No | NA | | 5 | 1 | 0 | Low-quality | Genome-fragment | | 6.35 | AAI-based (high-confidence) | 0 |
| S7_extracted_71490 | 3023 | No | NA | | 6 | 2 | 0 | Low-quality | Genome-fragment | | 4.93 | AAI-based (medium-confidence) | 0 |
| S16_vs_143361\|\|full | 1024 | No | NA | | 3 | 0 | 1 | Not-determined | Genome-fragment | NA | | NA | 0 |
| S16_vs_143446\|\|full | 7069 | No | NA | | 6 | 0 | 5 | Not-determined | Genome-fragment | NA | | NA | 0 |
| S16_vs_143482\|\|full | 1719 | No | NA | | 3 | 0 | 0 | Not-determined | Genome-fragment | NA | | NA | 0 |
| S16_vs_143518\|\|full | 2572 | No | NA | | 2 | 0 | 1 | Not-determined | Genome-fragment | NA | | NA | 0 |
| S16_vs_143585\|\|full | 3028 | No | NA | | 7 | 0 | 0 | Not-determined | Genome-fragment | NA | | NA | 0 |
| S16_vs_143641\|\|full | 4288 | No | NA | | 3 | 0 | 2 | Not-determined | Genome-fragment | NA | | NA | 0 |
| S10_vs_39493\|\|full | 3023 | No | NA | | 7 | 3 | 0 | Low-quality | Genome-fragment | | 7.2 | AAI-based (high-confidence) | 0 |
| S7_vs_9361\|\|full | 3024 | No | NA | | 8 | 0 | 0 | Low-quality | Genome-fragment | | 6.74 | AAI-based (high-confidence) | 0 |
| S16_vs_143856\|\|full | 3740 | No | NA | | 3 | 0 | 0 | Not-determined | Genome-fragment | NA | | NA | 0 |
| S16_vs_143871\|\|full | 1526 | No | NA | | 3 | 0 | 1 | Not-determined | Genome-fragment | NA | | NA | 0 |
| S16_vs_143942\|\|full | 1010 | No | NA | | 2 | 0 | 1 | Not-determined | Genome-fragment | NA | | NA | 0 |
| S16_vs_143955\|\|full | 2489 | No | NA | | 4 | 0 | 0 | Not-determined | Genome-fragment | NA | | NA | 0 |
| S16_vs_143986\|\|full | 2766 | No | NA | | 3 | 0 | 3 | Not-determined | Genome-fragment | NA | | NA | 0 |
| S10_extracted_117571 | 3024 | No | NA | | 5 | 2 | 0 | Low-quality | Genome-fragment | | 8.35 | AAI-based (high-confidence) | 0 |
| S6_extracted_k141_42993 | 3025 | Yes | | 2273 | 7 | 1 | 1 | Low-quality | Genome-fragment | | 5.32 | HMM-based (lower-bound) | 24.86 |
| S13_vs_137108\|\|full | 3026 | Yes | | 1218 | 3 | 1 | 1 | Low-quality | Genome-fragment | | 3.59 | AAI-based (high-confidence) | 59.75 |
| S8_extracted_k141_135629 | 3027 | No | NA | | 3 | 0 | 2 | Low-quality | Genome-fragment | | 1.86 | AAI-based (medium-confidence) | 0 |
| S7_vs_51015\|\|full | 3028 | No | NA | | 5 | 2 | 0 | Low-quality | Genome-fragment | | 6.35 | AAI-based (high-confidence) | 0 |
| S16_extracted_52403 | 3028 | No | NA | | 4 | 0 | 0 | Low-quality | Genome-fragment | | 5.6 | AAI-based (medium-confidence) | 0 |
| S16_vs_144378\|\|full | 2203 | No | NA | | 4 | 0 | 0 | Not-determined | Genome-fragment | NA | | NA | 0 |
| S17_extracted_k141_7606 | 3029 | No | NA | | 2 | 0 | 0 | Low-quality | Genome-fragment | | 4.73 | AAI-based (medium-confidence) | 0 |
| S16_vs_144392\|\|full | 1541 | No | NA | | 4 | 0 | 0 | Not-determined | Genome-fragment | NA | | NA | 0 |
| S17_vs_k141_29541\|\|full | 3029 | No | NA | | 3 | 1 | 0 | Low-quality | Genome-fragment | | 5.69 | AAI-based (medium-confidence) | 0 |
| S16_vs_144477\|\|full | 4558 | No | NA | | 8 | 0 | 2 | Not-determined | Genome-fragment | NA | | NA | 0 |
| S16_vs_144492\|\|full | 5128 | No | NA | | 6 | 0 | 4 | Not-determined | Genome-fragment | NA | | NA | 0 |
| S11_vs_k141_68390\|\|full | 3029 | No | NA | | 6 | 0 | 0 | Low-quality | Genome-fragment | | 4.99 | AAI-based (medium-confidence) | 0 |
| S16_vs_144539\|\|full | 5596 | No | NA | | 3 | 0 | 2 | Not-determined | Genome-fragment | NA | | NA | 0 |
| S12_extracted_k141_95805 | 3029 | No | NA | | 3 | 0 | 0 | Low-quality | Genome-fragment | | 8.87 | AAI-based (medium-confidence) | 0 |
| S16_vs_144749\|\|full | 2894 | No | NA | | 4 | 0 | 3 | Not-determined | Genome-fragment | NA | | NA | 0 |
| S6_vs_k141_15635\|\|full | 3029 | No | NA | | 4 | 0 | 4 | Low-quality | Genome-fragment | | 2.16 | AAI-based (medium-confidence) | 0 |
| S7_vs_65258\|\|full | 3030 | No | NA | | 3 | 0 | 1 | Low-quality | Genome-fragment | | 7.33 | AAI-based (medium-confidence) | 0 |
| S13_extracted_44422 | 3032 | No | NA | | 2 | 0 | 0 | Low-quality | Genome-fragment | | 0.28 | AAI-based (medium-confidence) | 0 |
| S16_vs_144847\|\|full | 2105 | No | NA | | 4 | 0 | 2 | Not-determined | Genome-fragment | NA | | NA | 0 |
| S1_extracted_k141_76047 | 3032 | No | NA | | 6 | 0 | 0 | Low-quality | Genome-fragment | | 5.26 | AAI-based (high-confidence) | 0 |
| S16_vs_144985\|\|full | 4969 | No | NA | | 7 | 0 | 0 | Not-determined | Genome-fragment | NA | | NA | 0 |
| S16_vs_144989\|\|full | 2486 | No | NA | | 3 | 0 | 2 | Not-determined | Genome-fragment | NA | | NA | 0 |
| S1_vs_k141_25603\|\|full | 3032 | No | NA | | 4 | 2 | 0 | Low-quality | Genome-fragment | | 7.49 | AAI-based (high-confidence) | 0 |
| S16_extracted_25435 | 3032 | No | NA | | 3 | 0 | 3 | Low-quality | Genome-fragment | | 0.52 | AAI-based (high-confidence) | 0 |
| S16_extracted_29634 | 3032 | No | NA | | 5 | 0 | 4 | Low-quality | Genome-fragment | | 1.99 | AAI-based (high-confidence) | 0 |
| S10_extracted_23735 | 3032 | No | NA | | 5 | 1 | 0 | Low-quality | Genome-fragment | | 1.16 | HMM-based (lower-bound) | 0 |
| S11_vs_k141_79129\|\|full | 3033 | No | NA | | 6 | 2 | 0 | Low-quality | Genome-fragment | | 7.54 | AAI-based (high-confidence) | 0 |
| S16_vs_145383\|\|full | 1143 | No | NA | | 3 | 0 | 1 | Not-determined | Genome-fragment | NA | | NA | 0 |
| S16_vs_145461\|\|full | 2516 | No | NA | | 3 | 0 | 1 | Not-determined | Genome-fragment | NA | | NA | 0 |
| S3_extracted_k141_99580 | 3034 | No | NA | | 3 | 3 | 0 | Low-quality | Genome-fragment | | 4.98 | AAI-based (high-confidence) | 0 |
| S7_vs_78830\|\|full | 3034 | No | NA | | 10 | 1 | 0 | Low-quality | Genome-fragment | | 3.52 | HMM-based (lower-bound) | 0 |
| S16_vs_110403\|\|full | 3034 | No | NA | | 4 | 0 | 3 | Low-quality | Genome-fragment | | 4.37 | AAI-based (medium-confidence) | 0 |
| S16_vs_145564\|\|full | 3408 | No | NA | | 5 | 0 | 4 | Not-determined | Genome-fragment | NA | | NA | 0 |
| S16_vs_145733\|\|full | 3512 | No | NA | | 5 | 0 | 2 | Not-determined | Genome-fragment | NA | | NA | 0 |
| S10_extracted_113856 | 3034 | Yes | | 772 | 5 | 1 | 2 | Low-quality | Genome-fragment | | 0.44 | HMM-based (lower-bound) | 74.56 |
| S12_extracted_k141_151539 | 3034 | No | NA | | 1 | 0 | 0 | Low-quality | Genome-fragment | | 2.83 | AAI-based (high-confidence) | 0 |
| S16_vs_145793\|\|full | 1609 | No | NA | | 3 | 0 | 2 | Not-determined | Genome-fragment | NA | | NA | 0 |
| S16_vs_145796\|\|full | 1930 | No | NA | | 3 | 0 | 3 | Not-determined | Genome-fragment | NA | | NA | 0 |
| S16_vs_145819\|\|full | 12179 | No | NA | | 12 | 0 | 8 | Not-determined | Genome-fragment | NA | | NA | 0 |
| S16_vs_145907\|\|full | 2093 | No | NA | | 5 | 0 | 0 | Not-determined | Genome-fragment | NA | | NA | 0 |
| S2_vs_k141_10380\|\|full | 3034 | No | NA | | 5 | 1 | 0 | Low-quality | Genome-fragment | | 9.22 | AAI-based (high-confidence) | 0 |
| S8_vs_k141_52971\|\|full | 3034 | No | NA | | 4 | 0 | 0 | Low-quality | Genome-fragment | | 6.06 | AAI-based (medium-confidence) | 0 |
| S16_vs_146221\|\|full | 2844 | No | NA | | 5 | 0 | 0 | Not-determined | Genome-fragment | NA | | NA | 0 |
| S9_vs_k141_79380\|\|full | 3035 | No | NA | | 11 | 0 | 0 | Low-quality | Genome-fragment | | 8.02 | AAI-based (high-confidence) | 0 |
| S16_vs_146267\|\|full | 10222 | No | NA | | 16 | 0 | 2 | Not-determined | Genome-fragment | NA | | NA | 0 |
| S1_extracted_k141_60719 | 3035 | No | NA | | 4 | 0 | 0 | Low-quality | Genome-fragment | | 7.29 | AAI-based (high-confidence) | 0 |
| S3_vs_k141_88406\|\|full | 3035 | No | NA | | 3 | 1 | 0 | Low-quality | Genome-fragment | | 1.65 | HMM-based (lower-bound) | 0 |
| S16_vs_146454\|\|full | 2550 | No | NA | | 5 | 0 | 1 | Not-determined | Genome-fragment | NA | | NA | 0 |
| S8_vs_k141_137090\|\|lt2gene | 3035 | No | NA | | 2 | 2 | 0 | Low-quality | Genome-fragment | | 7.95 | AAI-based (high-confidence) | 0 |
| S17_vs_k141_97840\|\|full | 3036 | No | NA | | 2 | 0 | 0 | Low-quality | Genome-fragment | | 3.43 | AAI-based (medium-confidence) | 0 |
| S14_extracted_k141_96330 | 3036 | No | NA | | 3 | 3 | 0 | Low-quality | Genome-fragment | | 10.01 | AAI-based (high-confidence) | 0 |
| S16_vs_146611\|\|full | 4729 | No | NA | | 7 | 0 | 2 | Not-determined | Genome-fragment | NA | | NA | 0 |
| S16_vs_146612\|\|full | 1701 | No | NA | | 2 | 0 | 0 | Not-determined | Genome-fragment | NA | | NA | 0 |
| S16_vs_146657\|\|full | 1803 | No | NA | | 2 | 0 | 1 | Not-determined | Genome-fragment | NA | | NA | 0 |
| S3_vs_k141_121489\|\|full | 3036 | No | NA | | 3 | 0 | 0 | Low-quality | Genome-fragment | | 5.68 | AAI-based (high-confidence) | 0 |
| S16_vs_146873\|\|full | 6622 | No | NA | | 6 | 0 | 5 | Not-determined | Genome-fragment | NA | | NA | 0 |
| S16_vs_147023\|\|full | 1109 | No | NA | | 2 | 0 | 0 | Not-determined | Genome-fragment | NA | | NA | 0 |
| S16_vs_147213\|\|full | 2616 | No | NA | | 3 | 0 | 1 | Not-determined | Genome-fragment | NA | | NA | 0 |
| S16_vs_147366\|\|full | 1062 | No | NA | | 2 | 0 | 0 | Not-determined | Genome-fragment | NA | | NA | 0 |
| S16_vs_147388\|\|full | 1939 | No | NA | | 3 | 0 | 2 | Not-determined | Genome-fragment | NA | | NA | 0 |
| S16_vs_147492\|\|full | 7294 | No | NA | | 8 | 0 | 2 | Not-determined | Genome-fragment | NA | | NA | 0 |
| S16_vs_147503\|\|full | 1606 | No | NA | | 4 | 0 | 0 | Not-determined | Genome-fragment | NA | | NA | 0 |
| S16_vs_147603\|\|full | 4076 | No | NA | | 3 | 0 | 2 | Not-determined | Genome-fragment | NA | | NA | 0 |
| S7_extracted_55919 | 3036 | No | NA | | 2 | 0 | 2 | Low-quality | Genome-fragment | | 2.79 | AAI-based (medium-confidence) | 0 |
| S16_vs_147868\|\|full | 1386 | No | NA | | 3 | 0 | 0 | Not-determined | Genome-fragment | NA | | NA | 0 |
| S16_vs_147882\|\|full | 1798 | No | NA | | 3 | 0 | 1 | Not-determined | Genome-fragment | NA | | NA | 0 |
| S16_vs_147891\|\|full | 2666 | No | NA | | 3 | 0 | 3 | Not-determined | Genome-fragment | NA | | NA | 0 |
| S17_extracted_k141_98119 | 3037 | No | NA | | 7 | 5 | 0 | Low-quality | Genome-fragment | | 6.81 | AAI-based (high-confidence) | 0 |
| S15_vs_8517\|\|full | 3037 | No | NA | | 3 | 0 | 0 | Low-quality | Genome-fragment | | 2.01 | AAI-based (high-confidence) | 0 |
| S10_vs_120542\|\|full | 3038 | No | NA | | 5 | 0 | 2 | Low-quality | Genome-fragment | | 8.16 | AAI-based (medium-confidence) | 0 |
| S16_vs_148537\|\|full | 2964 | No | NA | | 7 | 0 | 6 | Not-determined | Genome-fragment | NA | | NA | 0 |
| S12_vs_k141_109432\|\|full | 3038 | No | NA | | 3 | 3 | 0 | Low-quality | Genome-fragment | | 5.53 | AAI-based (high-confidence) | 0 |
| S8_vs_k141_207669\|\|full | 3038 | No | NA | | 6 | 1 | 0 | Low-quality | Genome-fragment | | 7.31 | AAI-based (high-confidence) | 0 |
| S9_vs_k141_6544\|\|full | 3039 | No | NA | | 6 | 1 | 0 | Low-quality | Genome-fragment | | 5.64 | AAI-based (high-confidence) | 0 |
| S16_vs_148792\|\|full | 3600 | No | NA | | 4 | 0 | 0 | Not-determined | Genome-fragment | NA | | NA | 0 |
| S16_vs_148886\|\|full | 1293 | No | NA | | 2 | 0 | 2 | Not-determined | Genome-fragment | NA | | NA | 0 |
| S16_vs_149059\|\|full | 5660 | No | NA | | 5 | 0 | 3 | Not-determined | Genome-fragment | NA | | NA | 0 |
| S16_vs_149070\|\|full | 11576 | No | NA | | 8 | 0 | 1 | Not-determined | Genome-fragment | NA | | NA | 0 |
| S8_extracted_k141_20493 | 3039 | No | NA | | 3 | 1 | 0 | Low-quality | Genome-fragment | | 10.84 | HMM-based (lower-bound) | 0 |
| S16_vs_149152\|\|full | 2206 | No | NA | | 4 | 0 | 0 | Not-determined | Genome-fragment | NA | | NA | 0 |
| S16_vs_149234\|\|full | 6006 | No | NA | | 9 | 0 | 4 | Not-determined | Genome-fragment | NA | | NA | 0 |
| S16_vs_149381\|\|full | 1121 | No | NA | | 2 | 0 | 1 | Not-determined | Genome-fragment | NA | | NA | 0 |
| S16_vs_149454\|\|full | 1405 | No | NA | | 2 | 0 | 2 | Not-determined | Genome-fragment | NA | | NA | 0 |
| S16_vs_149487\|\|full | 1557 | No | NA | | 3 | 0 | 1 | Not-determined | Genome-fragment | NA | | NA | 0 |
| S16_vs_149750\|\|full | 2711 | No | NA | | 5 | 0 | 1 | Not-determined | Genome-fragment | NA | | NA | 0 |
| S16_vs_149812\|\|full | 2855 | No | NA | | 5 | 0 | 2 | Not-determined | Genome-fragment | NA | | NA | 0 |
| S16_vs_150155\|\|full | 8064 | No | NA | | 10 | 0 | 2 | Not-determined | Genome-fragment | NA | | NA | 0 |
| S16_vs_150229\|\|full | 2115 | No | NA | | 3 | 0 | 1 | Not-determined | Genome-fragment | NA | | NA | 0 |
| S16_vs_150247\|\|full | 1781 | No | NA | | 2 | 0 | 1 | Not-determined | Genome-fragment | NA | | NA | 0 |
| S8_extracted_k141_22524 | 3039 | No | NA | | 6 | 0 | 0 | Low-quality | Genome-fragment | | 9.37 | AAI-based (high-confidence) | 0 |
| S16_vs_150295\|\|full | 12958 | No | NA | | 15 | 0 | 1 | Not-determined | Genome-fragment | NA | | NA | 0 |
| S11_extracted_k141_79488 | 3040 | No | NA | | 2 | 0 | 1 | Low-quality | Genome-fragment | | 2.8 | AAI-based (medium-confidence) | 0 |
| S5_extracted_k141_37991 | 3040 | No | NA | | 3 | 2 | 0 | Low-quality | Genome-fragment | | 8.56 | AAI-based (high-confidence) | 0 |
| S10_extracted_45771 | 3040 | No | NA | | 3 | 0 | 1 | Low-quality | Genome-fragment | | 1.53 | AAI-based (medium-confidence) | 0 |
| S7_extracted_119093 | 3041 | No | NA | | 8 | 2 | 0 | Low-quality | Genome-fragment | | 5.62 | AAI-based (medium-confidence) | 0 |
| S10_extracted_40069 | 3041 | No | NA | | 5 | 1 | 0 | Low-quality | Genome-fragment | | 3.02 | AAI-based (medium-confidence) | 0 |
| S16_vs_150709\|\|full | 2178 | No | NA | | 4 | 0 | 0 | Not-determined | Genome-fragment | NA | | NA | 0 |
| S16_vs_150763\|\|full | 1374 | No | NA | | 2 | 0 | 1 | Not-determined | Genome-fragment | NA | | NA | 0 |
| S10_vs_85896\|\|full | 3041 | No | NA | | 2 | 0 | 1 | Low-quality | Genome-fragment | | 5.3 | AAI-based (medium-confidence) | 0 |
| S12_extracted_k141_167379 | 3041 | No | NA | | 5 | 0 | 1 | Low-quality | Genome-fragment | | 10.41 | AAI-based (medium-confidence) | 0 |
| S3_extracted_k141_33478 | 3042 | No | NA | | 5 | 0 | 1 | Low-quality | Genome-fragment | | 5.03 | AAI-based (high-confidence) | 0 |
| S16_vs_150919\|\|full | 1304 | No | NA | | 3 | 0 | 0 | Not-determined | Genome-fragment | NA | | NA | 0 |
| S8_vs_k141_202089\|\|full | 3042 | No | NA | | 3 | 0 | 2 | Low-quality | Genome-fragment | | 0.69 | AAI-based (medium-confidence) | 0 |
| S17_vs_k141_151398\|\|full | 3043 | No | NA | | 9 | 4 | 0 | Low-quality | Genome-fragment | | 4.57 | AAI-based (high-confidence) | 0 |
| S16_vs_151028\|\|full | 2173 | No | NA | | 3 | 0 | 0 | Not-determined | Genome-fragment | NA | | NA | 0 |
| S1_vs_k141_156258\|\|full | 3043 | No | NA | | 3 | 0 | 3 | Low-quality | Genome-fragment | | 1.4 | AAI-based (medium-confidence) | 0 |
| S16_vs_151135\|\|full | 1498 | No | NA | | 3 | 0 | 0 | Not-determined | Genome-fragment | NA | | NA | 0 |
| S10_extracted_91179 | 3043 | No | NA | | 4 | 0 | 1 | Low-quality | Genome-fragment | | 7.16 | AAI-based (high-confidence) | 0 |
| S17_vs_k141_27147\|\|full | 3045 | No | NA | | 3 | 0 | 2 | Low-quality | Genome-fragment | | 5.14 | AAI-based (medium-confidence) | 0 |
| S16_vs_151291\|\|full | 6304 | No | NA | | 8 | 0 | 5 | Not-determined | Genome-fragment | NA | | NA | 0 |
| S16_vs_178\|\|full | 3045 | No | NA | | 6 | 0 | 1 | Low-quality | Genome-fragment | | 7.12 | AAI-based (medium-confidence) | 0 |
| S12_extracted_k141_38282 | 3045 | No | NA | | 5 | 0 | 4 | Low-quality | Genome-fragment | | 2.45 | AAI-based (medium-confidence) | 0 |
| S16_vs_67366\|\|full | 3046 | No | NA | | 6 | 1 | 0 | Low-quality | Genome-fragment | | 0.99 | HMM-based (lower-bound) | 0 |
| S16_vs_151538\|\|full | 8234 | No | NA | | 5 | 0 | 5 | Not-determined | Genome-fragment | NA | | NA | 0 |
| S16_vs_151588\|\|full | 1437 | No | NA | | 2 | 0 | 0 | Not-determined | Genome-fragment | NA | | NA | 0 |
| S12_vs_k141_82718\|\|full | 3046 | No | NA | | 6 | 0 | 0 | Low-quality | Genome-fragment | | 3.29 | AAI-based (medium-confidence) | 0 |
| S16_vs_151638\|\|full | 1938 | No | NA | | 3 | 0 | 1 | Not-determined | Genome-fragment | NA | | NA | 0 |

| | | | | | | | | | | | |
|---|---|---|---|---|---|---|---|---|---|---|---|
| S4_extracted_24926 | 3046 | No | NA | 5 | 1 | 0 | Low-quality | Genome-fragment | 6.77 | AAI-based (high-confidence) | 0 |
| S16_vs_151857\|\|full | 3063 | No | NA | 3 | 0 | 3 | Not-determined | Genome-fragment | NA | NA | 0 |
| S16_vs_152034\|\|full | 1514 | No | NA | 3 | 0 | 1 | Not-determined | Genome-fragment | NA | NA | 0 |
| S11_extracted_k141_31999 | 3048 | No | NA | 6 | 3 | 0 | Low-quality | Genome-fragment | 3.28 | HMM-based (lower-bound) | 0 |
| S7_extracted_5770 | 3048 | No | NA | 4 | 2 | 0 | Low-quality | Genome-fragment | 9.01 | HMM-based (lower-bound) | 0 |
| S16_vs_152323\|\|full | 2517 | No | NA | 3 | 0 | 2 | Not-determined | Genome-fragment | NA | NA | 0 |
| S9_extracted_k141_81271 | 3049 | No | NA | 3 | 0 | 1 | Low-quality | Genome-fragment | 10.57 | AAI-based (medium-confidence) | 0 |
| S16_vs_152414\|\|full | 1828 | No | NA | 2 | 0 | 0 | Not-determined | Genome-fragment | NA | NA | 0 |
| S9_extracted_k141_10826 | 3049 | No | NA | 7 | 6 | 0 | Low-quality | Genome-fragment | 8.69 | AAI-based (high-confidence) | 0 |
| S16_vs_152487\|\|full | 1581 | No | NA | 3 | 0 | 0 | Not-determined | Genome-fragment | NA | NA | 0 |
| S3_extracted_k141_75088 | 3049 | No | NA | 3 | 1 | 0 | Low-quality | Genome-fragment | 6.76 | AAI-based (high-confidence) | 0 |
| S3_extracted_k141_46003 | 3049 | No | NA | 6 | 2 | 0 | Low-quality | Genome-fragment | 7.16 | AAI-based (high-confidence) | 0 |
| S16_extracted_67848 | 3049 | No | NA | 5 | 2 | 0 | Low-quality | Genome-fragment | 7.09 | AAI-based (high-confidence) | 0 |
| S10_extracted_30328 | 3049 | No | NA | 4 | 2 | 0 | Low-quality | Genome-fragment | 5.98 | AAI-based (high-confidence) | 0 |
| S16_vs_152728\|\|full | 3860 | No | NA | 4 | 0 | 2 | Not-determined | Genome-fragment | NA | NA | 0 |
| S16_vs_152833\|\|full | 3883 | No | NA | 5 | 0 | 4 | Not-determined | Genome-fragment | NA | NA | 0 |
| S8_vs_k141_45424\|\|full | 3049 | No | NA | 7 | 1 | 0 | Low-quality | Genome-fragment | 6.6 | AAI-based (medium-confidence) | 0 |
| S17_extracted_k141_60794 | 3050 | No | NA | 5 | 0 | 0 | Low-quality | Genome-fragment | 4.47 | AAI-based (medium-confidence) | 0 |
| S16_vs_153074\|\|full | 6567 | No | NA | 5 | 0 | 4 | Not-determined | Genome-fragment | NA | NA | 0 |
| S14_vs_k141_1849\|\|full | 3050 | No | NA | 5 | 2 | 0 | Low-quality | Genome-fragment | 6.43 | AAI-based (medium-confidence) | 0 |
| S16_vs_153373\|\|full | 1485 | No | NA | 3 | 0 | 0 | Not-determined | Genome-fragment | NA | NA | 0 |
| S3_extracted_k141_17564 | 3050 | No | NA | 3 | 1 | 0 | Low-quality | Genome-fragment | 3.71 | AAI-based (medium-confidence) | 0 |
| S16_vs_153494\|\|full | 5132 | No | NA | 7 | 0 | 1 | Not-determined | Genome-fragment | NA | NA | 0 |
| S5_vs_k141_15305\|\|full | 3050 | No | NA | 3 | 0 | 3 | Low-quality | Genome-fragment | 2.32 | AAI-based (medium-confidence) | 0 |
| S16_vs_153545\|\|full | 1216 | No | NA | 2 | 0 | 0 | Not-determined | Genome-fragment | NA | NA | 0 |
| S16_vs_153564\|\|full | 1748 | No | NA | 3 | 0 | 0 | Not-determined | Genome-fragment | NA | NA | 0 |
| S16_vs_153622\|\|full | 4601 | No | NA | 6 | 0 | 3 | Not-determined | Genome-fragment | NA | NA | 0 |
| S16_vs_153636\|\|full | 2470 | No | NA | 2 | 0 | 1 | Not-determined | Genome-fragment | NA | NA | 0 |
| S7_vs_77451\|\|full | 3050 | No | NA | 4 | 3 | 0 | Low-quality | Genome-fragment | 7.83 | AAI-based (high-confidence) | 0 |
| S2_vs_k141_11817\|\|full | 3050 | No | NA | 6 | 2 | 0 | Low-quality | Genome-fragment | 8.04 | HMM-based (lower-bound) | 0 |
| S17_vs_k141_56044\|\|full | 3051 | No | NA | 5 | 0 | 4 | Low-quality | Genome-fragment | 1.33 | AAI-based (medium-confidence) | 0 |
| S16_vs_154045\|\|full | 3171 | No | NA | 3 | 0 | 2 | Not-determined | Genome-fragment | NA | NA | 0 |
| S11_vs_k141_45807\|\|full | 3051 | No | NA | 5 | 0 | 0 | Low-quality | Genome-fragment | 6.09 | AAI-based (medium-confidence) | 0 |
| S16_vs_154179\|\|full | 3452 | No | NA | 4 | 0 | 3 | Not-determined | Genome-fragment | NA | NA | 0 |
| S14_vs_k141_99954\|\|full | 3051 | No | NA | 4 | 3 | 0 | Low-quality | Genome-fragment | 7.27 | AAI-based (high-confidence) | 0 |
| S12_extracted_k141_76197 | 3051 | No | NA | 5 | 0 | 1 | Low-quality | Genome-fragment | 5.1 | AAI-based (high-confidence) | 0 |
| S16_vs_154450\|\|full | 5014 | No | NA | 8 | 0 | 2 | Not-determined | Genome-fragment | NA | NA | 0 |
| S11_extracted_k141_63538 | 3052 | No | NA | 5 | 0 | 0 | Low-quality | Genome-fragment | 5.55 | AAI-based (high-confidence) | 0 |
| S16_vs_154544\|\|full | 1948 | No | NA | 2 | 0 | 2 | Not-determined | Genome-fragment | NA | NA | 0 |
| S16_vs_154556\|\|full | 1131 | No | NA | 2 | 0 | 0 | Not-determined | Genome-fragment | NA | NA | 0 |
| S16_vs_154693\|\|full | 1534 | No | NA | 4 | 0 | 0 | Not-determined | Genome-fragment | NA | NA | 0 |
| S3_extracted_k141_112987 | 3052 | No | NA | 5 | 0 | 0 | Low-quality | Genome-fragment | 11.72 | AAI-based (high-confidence) | 0 |
| S5_extracted_k141_133496 | 3052 | No | NA | 3 | 0 | 3 | Low-quality | Genome-fragment | 2.34 | AAI-based (medium-confidence) | 0 |
| S7_extracted_79358 | 3052 | No | NA | 2 | 0 | 1 | Low-quality | Genome-fragment | 7.11 | AAI-based (medium-confidence) | 0 |
| S7_vs_29425\|\|full | 3052 | No | NA | 4 | 0 | 0 | Low-quality | Genome-fragment | 6.28 | AAI-based (medium-confidence) | 0 |
| S16_vs_155042\|\|full | 1380 | No | NA | 2 | 0 | 1 | Not-determined | Genome-fragment | NA | NA | 0 |
| S12_extracted_k141_136656 | 3052 | No | NA | 4 | 0 | 3 | Low-quality | Genome-fragment | 1.89 | AAI-based (medium-confidence) | 0 |
| S16_vs_155161\|\|full | 1096 | No | NA | 2 | 0 | 2 | Not-determined | Genome-fragment | NA | NA | 0 |
| S16_vs_155171\|\|full | 5579 | No | NA | 7 | 0 | 1 | Not-determined | Genome-fragment | NA | NA | 0 |
| S16_vs_155178\|\|full | 1255 | No | NA | 2 | 0 | 2 | Not-determined | Genome-fragment | NA | NA | 0 |
| S14_extracted_k141_27936 | 3053 | No | NA | 9 | 1 | 0 | Low-quality | Genome-fragment | 3.85 | AAI-based (medium-confidence) | 0 |
| S3_extracted_k141_97488 | 3053 | No | NA | 4 | 0 | 3 | Low-quality | Genome-fragment | 0.95 | AAI-based (medium-confidence) | 0 |
| S16_extracted_24629 | 3053 | No | NA | 2 | 0 | 0 | Low-quality | Genome-fragment | 6.25 | AAI-based (medium-confidence) | 0 |
| S16_vs_155316\|\|full | 1334 | No | NA | 2 | 0 | 0 | Not-determined | Genome-fragment | NA | NA | 0 |
| S16_vs_155317\|\|full | 2314 | No | NA | 4 | 0 | 1 | Not-determined | Genome-fragment | NA | NA | 0 |
| S3_vs_k141_40239\|\|full | 3054 | No | NA | 6 | 0 | 1 | Low-quality | Genome-fragment | 1.88 | AAI-based (high-confidence) | 0 |
| S16_vs_155426\|\|full | 2357 | No | NA | 5 | 0 | 0 | Not-determined | Genome-fragment | NA | NA | 0 |
| S16_vs_155485\|\|full | 1452 | No | NA | 3 | 0 | 0 | Not-determined | Genome-fragment | NA | NA | 0 |
| S16_vs_155505\|\|full | 9656 | No | NA | 8 | 0 | 1 | Not-determined | Genome-fragment | NA | NA | 0 |
| S16_vs_155547\|\|full | 1331 | No | NA | 3 | 0 | 0 | Not-determined | Genome-fragment | NA | NA | 0 |
| S16_vs_155810\|\|full | 2879 | No | NA | 7 | 0 | 1 | Not-determined | Genome-fragment | NA | NA | 0 |
| S16_vs_92104\|\|full | 3054 | No | NA | 3 | 0 | 0 | Low-quality | Genome-fragment | 1.93 | AAI-based (high-confidence) | 0 |
| S7_vs_34529\|\|full | 3055 | No | NA | 5 | 0 | 2 | Low-quality | Genome-fragment | 5.47 | AAI-based (high-confidence) | 0 |
| S16_vs_155965\|\|full | 2935 | No | NA | 3 | 0 | 3 | Not-determined | Genome-fragment | NA | NA | 0 |
| S16_extracted_92978 | 3055 | No | NA | 9 | 0 | 1 | Low-quality | Genome-fragment | 2.46 | AAI-based (high-confidence) | 0 |
| S10_extracted_18055 | 3057 | No | NA | 3 | 0 | 2 | Low-quality | Genome-fragment | 3.68 | AAI-based (medium-confidence) | 0 |
| S16_vs_156096\|\|full | 1402 | No | NA | 2 | 0 | 1 | Not-determined | Genome-fragment | NA | NA | 0 |
| S16_vs_156134\|\|full | 10702 | No | NA | 10 | 0 | 7 | Not-determined | Genome-fragment | NA | NA | 0 |
| S17_vs_k141_110669\|\|full | 3058 | No | NA | 3 | 0 | 2 | Low-quality | Genome-fragment | 0.97 | AAI-based (medium-confidence) | 0 |
| S16_vs_156199\|\|full | 3063 | No | NA | 3 | 0 | 0 | Not-determined | Genome-fragment | NA | NA | 0 |
| S5_extracted_k141_30858 | 3058 | No | NA | 6 | 1 | 0 | Low-quality | Genome-fragment | 6.37 | AAI-based (high-confidence) | 0 |
| S16_vs_156266\|\|full | 4025 | No | NA | 5 | 0 | 3 | Not-determined | Genome-fragment | NA | NA | 0 |
| S7_vs_89593\|\|full | 3058 | No | NA | 7 | 0 | 1 | Low-quality | Genome-fragment | 7.46 | AAI-based (high-confidence) | 0 |
| S12_extracted_k141_41087 | 3058 | No | NA | 3 | 0 | 2 | Low-quality | Genome-fragment | 3.28 | AAI-based (medium-confidence) | 0 |
| S16_vs_156442\|\|full | 2024 | No | NA | 5 | 0 | 0 | Not-determined | Genome-fragment | NA | NA | 0 |
| S16_vs_156461\|\|full | 4602 | No | NA | 5 | 0 | 4 | Not-determined | Genome-fragment | NA | NA | 0 |
| S6_vs_k141_52783\|\|full | 3058 | No | NA | 5 | 1 | 0 | Low-quality | Genome-fragment | 5.49 | AAI-based (high-confidence) | 0 |
| S16_vs_156549\|\|full | 3619 | No | NA | 4 | 0 | 3 | Not-determined | Genome-fragment | NA | NA | 0 |
| S16_vs_156573\|\|full | 5050 | No | NA | 6 | 0 | 1 | Not-determined | Genome-fragment | NA | NA | 0 |
| S16_vs_156584\|\|full | 1554 | No | NA | 2 | 0 | 0 | Not-determined | Genome-fragment | NA | NA | 0 |
| S16_vs_156592\|\|full | 12545 | No | NA | 12 | 0 | 6 | Not-determined | Genome-fragment | NA | NA | 0 |
| S17_extracted_k141_4843 | 3059 | No | NA | 5 | 0 | 0 | Low-quality | Genome-fragment | 4.67 | AAI-based (high-confidence) | 0 |
| S17_vs_k141_29489\|\|full | 3059 | No | NA | 3 | 0 | 1 | Low-quality | Genome-fragment | 0.76 | AAI-based (medium-confidence) | 0 |
| S16_vs_156660\|\|full | 1076 | No | NA | 3 | 0 | 2 | Not-determined | Genome-fragment | NA | NA | 0 |
| S16_vs_156750\|\|full | 1010 | No | NA | 2 | 0 | 0 | Not-determined | Genome-fragment | NA | NA | 0 |
| S16_vs_156845\|\|full | 2668 | No | NA | 3 | 0 | 1 | Not-determined | Genome-fragment | NA | NA | 0 |
| S16_vs_156916\|\|full | 5086 | No | NA | 4 | 0 | 1 | Not-determined | Genome-fragment | NA | NA | 0 |
| S16_vs_156921\|\|full | 2273 | No | NA | 3 | 0 | 0 | Not-determined | Genome-fragment | NA | NA | 0 |
| S9_vs_k141_82552\|\|full | 3059 | No | NA | 5 | 0 | 0 | Low-quality | Genome-fragment | 6.08 | AAI-based (medium-confidence) | 0 |
| S16_vs_157107\|\|full | 2891 | No | NA | 5 | 0 | 2 | Not-determined | Genome-fragment | NA | NA | 0 |
| S1_extracted_k141_140255 | 3059 | No | NA | 7 | 4 | 0 | Low-quality | Genome-fragment | 7.73 | AAI-based (high-confidence) | 0 |
| S16_vs_157193\|\|full | 3064 | No | NA | 5 | 0 | 1 | Not-determined | Genome-fragment | NA | NA | 0 |
| S16_vs_157204\|\|full | 4847 | No | NA | 5 | 0 | 4 | Not-determined | Genome-fragment | NA | NA | 0 |
| S16_vs_157324\|\|full | 3838 | No | NA | 4 | 0 | 0 | Not-determined | Genome-fragment | NA | NA | 0 |
| S8_extracted_k141_50999 | 3059 | No | NA | 3 | 0 | 1 | Low-quality | Genome-fragment | 1.07 | AAI-based (medium-confidence) | 0 |
| S16_vs_157380\|\|full | 2522 | No | NA | 3 | 0 | 2 | Not-determined | Genome-fragment | NA | NA | 0 |
| S13_extracted_147568 | 3060 | No | NA | 5 | 1 | 0 | Low-quality | Genome-fragment | 4.78 | HMM-based (lower-bound) | 0 |
| S16_vs_124331\|\|full | 3060 | No | NA | 4 | 1 | 0 | Low-quality | Genome-fragment | 1.92 | HMM-based (lower-bound) | 0 |
| S12_extracted_k141_173426 | 3060 | No | NA | 3 | 0 | 2 | Low-quality | Genome-fragment | 9.77 | AAI-based (medium-confidence) | 0 |
| S6_vs_k141_30474\|\|full | 3060 | No | NA | 3 | 2 | 0 | Low-quality | Genome-fragment | 8.23 | AAI-based (high-confidence) | 0 |
| S16_vs_157747\|\|full | 1323 | No | NA | 3 | 0 | 1 | Not-determined | Genome-fragment | NA | NA | 0 |
| S16_vs_157764\|\|full | 1414 | No | NA | 2 | 0 | 1 | Not-determined | Genome-fragment | NA | NA | 0 |
| S16_vs_157797\|\|full | 6898 | No | NA | 6 | 0 | 3 | Not-determined | Genome-fragment | NA | NA | 0 |
| S1_vs_k141_115378\|\|full | 3061 | No | NA | 3 | 0 | 1 | Low-quality | Genome-fragment | 2.04 | AAI-based (high-confidence) | 0 |
| S12_extracted_k141_184294 | 3061 | No | NA | 3 | 0 | 2 | Low-quality | Genome-fragment | 5.86 | AAI-based (medium-confidence) | 0 |
| S16_vs_158098\|\|full | 5473 | No | NA | 6 | 0 | 1 | Not-determined | Genome-fragment | NA | NA | 0 |
| S16_vs_158220\|\|full | 3715 | No | NA | 4 | 0 | 1 | Not-determined | Genome-fragment | NA | NA | 0 |
| S12_extracted_k141_131599 | 3061 | No | NA | 3 | 0 | 2 | Low-quality | Genome-fragment | 0.44 | AAI-based (medium-confidence) | 0 |
| S16_vs_158446\|\|full | 2403 | No | NA | 3 | 0 | 1 | Not-determined | Genome-fragment | NA | NA | 0 |
| S12_vs_k141_18523\|\|full | 3061 | No | NA | 4 | 3 | 0 | Low-quality | Genome-fragment | 7.21 | AAI-based (high-confidence) | 0 |
| S8_vs_k141_127625\|\|full | 3061 | No | NA | 8 | 2 | 0 | Low-quality | Genome-fragment | 7.86 | AAI-based (high-confidence) | 0 |
| S14_extracted_k141_40014 | 3062 | No | NA | 6 | 1 | 0 | Low-quality | Genome-fragment | 2.06 | HMM-based (lower-bound) | 0 |
| S16_vs_158718\|\|full | 1020 | No | NA | 3 | 0 | 3 | Not-determined | Genome-fragment | NA | NA | 0 |
| S5_extracted_k141_49241 | 3062 | No | NA | 8 | 1 | 0 | Low-quality | Genome-fragment | 4.84 | AAI-based (high-confidence) | 0 |
| S8_extracted_k141_88757 | 3062 | No | NA | 3 | 2 | 0 | Low-quality | Genome-fragment | 5.11 | AAI-based (high-confidence) | 0 |
| S14_extracted_k141_16360 | 3063 | Yes | 693 | 6 | 1 | 1 | Low-quality | Genome-fragment | 1.68 | AAI-based (high-confidence) | 77.38 |
| S16_vs_158850\|\|full | 2167 | No | NA | 3 | 0 | 0 | Not-determined | Genome-fragment | NA | NA | 0 |
| S12_vs_k141_137839\|\|full | 3063 | No | NA | 6 | 1 | 0 | Low-quality | Genome-fragment | 8.49 | AAI-based (high-confidence) | 0 |
| S16_vs_158978\|\|full | 1124 | No | NA | 2 | 0 | 1 | Not-determined | Genome-fragment | NA | NA | 0 |
| S16_vs_159019\|\|full | 3207 | No | NA | 3 | 0 | 1 | Not-determined | Genome-fragment | NA | NA | 0 |
| S15_extracted_68494 | 3063 | No | NA | 3 | 2 | 0 | Low-quality | Genome-fragment | 4.97 | AAI-based (high-confidence) | 0 |
| S16_vs_159089\|\|full | 1841 | No | NA | 2 | 0 | 0 | Not-determined | Genome-fragment | NA | NA | 0 |
| S16_vs_159111\|\|full | 2321 | No | NA | 4 | 0 | 2 | Not-determined | Genome-fragment | NA | NA | 0 |
| S16_vs_159133\|\|full | 1956 | No | NA | 3 | 0 | 2 | Not-determined | Genome-fragment | NA | NA | 0 |
| S16_vs_159193\|\|full | 1025 | No | NA | 2 | 0 | 1 | Not-determined | Genome-fragment | NA | NA | 0 |
| S16_vs_159355\|\|full | 1578 | No | NA | 3 | 0 | 1 | Not-determined | Genome-fragment | NA | NA | 0 |
| S6_extracted_k141_44608 | 3063 | No | NA | 3 | 3 | 0 | Low-quality | Genome-fragment | 10.68 | HMM-based (lower-bound) | 0 |
| S16_vs_159439\|\|full | 1740 | No | NA | 4 | 0 | 3 | Not-determined | Genome-fragment | NA | NA | 0 |
| S9_extracted_k141_15656 | 3064 | No | NA | 6 | 2 | 0 | Low-quality | Genome-fragment | 4.55 | AAI-based (medium-confidence) | 0 |
| S8_extracted_k141_141498 | 3065 | No | NA | 4 | 0 | 2 | Low-quality | Genome-fragment | 2.99 | AAI-based (medium-confidence) | 0 |
| S14_vs_k141_105897\|\|full | 3066 | No | NA | 4 | 2 | 0 | Low-quality | Genome-fragment | 7.1 | AAI-based (high-confidence) | 0 |
| S16_vs_159772\|\|full | 2647 | No | NA | 3 | 0 | 3 | Not-determined | Genome-fragment | NA | NA | 0 |
| S16_vs_159832\|\|full | 1842 | No | NA | 3 | 0 | 0 | Not-determined | Genome-fragment | NA | NA | 0 |
| S8_vs_k141_13659\|\|full | 3066 | No | NA | 3 | 0 | 3 | Low-quality | Genome-fragment | 1.03 | AAI-based (medium-confidence) | 0 |
| S5_vs_k141_30249\|\|full | 3067 | No | NA | 4 | 0 | 0 | Low-quality | Genome-fragment | 1.17 | AAI-based (medium-confidence) | 0 |
| S7_vs_56531\|\|full | 3067 | No | NA | 6 | 0 | 0 | Low-quality | Genome-fragment | 4.27 | AAI-based (high-confidence) | 0 |
| S16_vs_160109\|\|full | 3363 | No | NA | 5 | 0 | 2 | Not-determined | Genome-fragment | NA | NA | 0 |
| S16_vs_160243\|\|full | 2470 | No | NA | 3 | 0 | 0 | Not-determined | Genome-fragment | NA | NA | 0 |
| S16_vs_160577\|\|full | 1774 | No | NA | 2 | 0 | 2 | Not-determined | Genome-fragment | NA | NA | 0 |
| S16_vs_160650\|\|full | 1418 | No | NA | 2 | 0 | 2 | Not-determined | Genome-fragment | NA | NA | 0 |
| S10_vs_56118\|\|full | 3067 | No | NA | 8 | 1 | 0 | Low-quality | Genome-fragment | 5.09 | AAI-based (high-confidence) | 0 |
| S12_extracted_k141_98943 | 3067 | No | NA | 4 | 0 | 1 | Low-quality | Genome-fragment | 5.36 | AAI-based (medium-confidence) | 0 |
| S16_vs_160776\|\|full | 3024 | No | NA | 4 | 0 | 3 | Not-determined | Genome-fragment | NA | NA | 0 |
| S16_vs_160831\|\|full | 12605 | No | NA | 11 | 0 | 3 | Not-determined | Genome-fragment | NA | NA | 0 |
| S16_vs_160860\|\|full | 2780 | No | NA | 5 | 0 | 1 | Not-determined | Genome-fragment | NA | NA | 0 |
| S12_extracted_k141_93306 | 3067 | No | NA | 8 | 0 | 0 | Low-quality | Genome-fragment | 5.47 | AAI-based (medium-confidence) | 0 |
| S17_vs_k141_1156\|\|full | 3068 | No | NA | 4 | 0 | 2 | Low-quality | Genome-fragment | 10.26 | AAI-based (high-confidence) | 0 |
| S17_vs_k141_109312\|\|full | 3068 | Yes | 2325 | 8 | 1 | 1 | Low-quality | Genome-fragment | 1 | HMM-based (lower-bound) | 24.22 |
| S16_vs_161018\|\|full | 4585 | No | NA | 3 | 0 | 1 | Not-determined | Genome-fragment | NA | NA | 0 |
| S1_extracted_k141_150732 | 3068 | No | NA | 5 | 5 | 0 | Low-quality | Genome-fragment | 4.57 | AAI-based (high-confidence) | 0 |
| S5_extracted_k141_122558 | 3068 | No | NA | 9 | 1 | 0 | Low-quality | Genome-fragment | 5.77 | AAI-based (high-confidence) | 0 |
| S16_vs_161247\|\|full | 5030 | No | NA | 4 | 0 | 1 | Not-determined | Genome-fragment | NA | NA | 0 |
| S16_vs_161310\|\|full | 2639 | No | NA | 3 | 0 | 0 | Not-determined | Genome-fragment | NA | NA | 0 |
| S16_vs_161425\|\|full | 2868 | No | NA | 5 | 0 | 0 | Not-determined | Genome-fragment | NA | NA | 0 |
| S5_extracted_k141_35172 | 3068 | No | NA | 4 | 3 | 0 | Low-quality | Genome-fragment | 10.11 | AAI-based (high-confidence) | 0 |
| S7_extracted_132886 | 3068 | No | NA | 1 | 0 | 0 | Low-quality | Genome-fragment | 2.78 | AAI-based (medium-confidence) | 0 |
| S12_vs_k141_1964\|\|full | 3068 | No | NA | 2 | 0 | 0 | Low-quality | Genome-fragment | 4.04 | AAI-based (high-confidence) | 0 |
| S9_extracted_k141_37999 | 3069 | No | NA | 5 | 1 | 0 | Low-quality | Genome-fragment | 1.99 | AAI-based (medium-confidence) | 0 |
| S9_vs_k141_48095\|\|full | 3069 | No | NA | 7 | 1 | 0 | Low-quality | Genome-fragment | 5.9 | AAI-based (medium-confidence) | 0 |
| S16_vs_161982\|\|full | 1176 | No | NA | 2 | 0 | 0 | Not-determined | Genome-fragment | NA | NA | 0 |
| S16_vs_162034\|\|full | 3771 | No | NA | 4 | 0 | 2 | Not-determined | Genome-fragment | NA | NA | 0 |
| S1_vs_k141_116093\|\|full | 3069 | No | NA | 7 | 2 | 0 | Low-quality | Genome-fragment | 8.99 | AAI-based (high-confidence) | 0 |
| S16_vs_162256\|\|full | 1239 | No | NA | 2 | 0 | 1 | Not-determined | Genome-fragment | NA | NA | 0 |
| S5_vs_k141_122916\|\|full | 3069 | No | NA | 1 | 1 | 0 | Low-quality | Genome-fragment | 5.33 | AAI-based (medium-confidence) | 0 |
| S16_vs_162328\|\|full | 7293 | No | NA | 5 | 0 | 1 | Not-determined | Genome-fragment | NA | NA | 0 |
| S9_extracted_k141_10506 | 3070 | No | NA | 8 | 2 | 0 | Low-quality | Genome-fragment | 4.93 | AAI-based (high-confidence) | 0 |
| S16_vs_162466\|\|full | 6753 | No | NA | 9 | 0 | 1 | Not-determined | Genome-fragment | NA | NA | 0 |
| S16_vs_162586\|\|full | 2123 | No | NA | 3 | 0 | 2 | Not-determined | Genome-fragment | NA | NA | 0 |
| S9_vs_k141_3398\|\|full | 3071 | No | NA | 8 | 0 | 0 | Low-quality | Genome-fragment | 6.55 | AAI-based (medium-confidence) | 0 |
| S16_vs_162644\|\|full | 3731 | No | NA | 7 | 0 | 4 | Not-determined | Genome-fragment | NA | NA | 0 |
| S11_extracted_k141_29292 | 3071 | No | NA | 8 | 2 | 0 | Low-quality | Genome-fragment | 4.75 | AAI-based (high-confidence) | 0 |
| S16_vs_162809\|\|full | 1838 | No | NA | 3 | 0 | 1 | Not-determined | Genome-fragment | NA | NA | 0 |
| S16_vs_162916\|\|full | 1713 | No | NA | 3 | 0 | 0 | Not-determined | Genome-fragment | NA | NA | 0 |
| S16_vs_163041\|\|full | 1434 | No | NA | 2 | 0 | 0 | Not-determined | Genome-fragment | NA | NA | 0 |
| S3_extracted_k141_116913 | 3071 | No | NA | 10 | 1 | 0 | Low-quality | Genome-fragment | 4.19 | AAI-based (high-confidence) | 0 |
| S16_vs_163163\|\|full | 1303 | No | NA | 2 | 0 | 1 | Not-determined | Genome-fragment | NA | NA | 0 |
| S16_vs_163243\|\|full | 1594 | No | NA | 3 | 0 | 1 | Not-determined | Genome-fragment | NA | NA | 0 |
| S16_vs_163339\|\|full | 10261 | No | NA | 17 | 0 | 0 | Not-determined | Genome-fragment | NA | NA | 0 |
| S5_vs_k141_3215\|\|full | 3071 | No | NA | 4 | 0 | 2 | Low-quality | Genome-fragment | 5.09 | AAI-based (medium-confidence) | 0 |
| S16_vs_163404\|\|full | 1704 | No | NA | 3 | 0 | 1 | Not-determined | Genome-fragment | NA | NA | 0 |
| S16_vs_163505\|\|full | 1896 | No | NA | 2 | 0 | 1 | Not-determined | Genome-fragment | NA | NA | 0 |
| S7_extracted_51873 | 3071 | No | NA | 2 | 0 | 0 | Low-quality | Genome-fragment | 1.75 | AAI-based (high-confidence) | 0 |
| S16_vs_163663\|\|full | 1225 | No | NA | 3 | 0 | 0 | Not-determined | Genome-fragment | NA | NA | 0 |
| S16_vs_163672\|\|full | 5553 | No | NA | 10 | 0 | 0 | Not-determined | Genome-fragment | NA | NA | 0 |
| S16_vs_163742\|\|full | 1719 | No | NA | 3 | 0 | 0 | Not-determined | Genome-fragment | NA | NA | 0 |
| S17_vs_k141_152043\|\|full | 3072 | No | NA | 8 | 0 | 0 | Low-quality | Genome-fragment | 8.09 | AAI-based (medium-confidence) | 0 |
| S16_vs_163753\|\|full | 3040 | No | NA | 4 | 0 | 0 | Not-determined | Genome-fragment | NA | NA | 0 |
| S9_extracted_k141_74899 | 3072 | No | NA | 3 | 2 | 0 | Low-quality | Genome-fragment | 2.14 | AAI-based (medium-confidence) | 0 |
| S16_vs_164083\|\|full | 3310 | No | NA | 5 | 0 | 4 | Not-determined | Genome-fragment | NA | NA | 0 |
| S16_vs_164131\|\|full | 5224 | No | NA | 3 | 0 | 3 | Not-determined | Genome-fragment | NA | NA | 0 |
| S16_vs_164192\|\|full | 2606 | No | NA | 2 | 0 | 2 | Not-determined | Genome-fragment | NA | NA | 0 |
| S11_vs_k141_22260\|\|full | 3072 | No | NA | 4 | 3 | 0 | Low-quality | Genome-fragment | 7.59 | AAI-based (medium-confidence) | 0 |
| S5_extracted_k141_940 | 3072 | No | NA | 7 | 0 | 0 | Low-quality | Genome-fragment | 8.62 | AAI-based (high-confidence) | 0 |
| S16_vs_164339\|\|full | 1576 | No | NA | 3 | 0 | 2 | Not-determined | Genome-fragment | NA | NA | 0 |
| S7_vs_82430\|\|full | 3072 | No | NA | 4 | 3 | 0 | Low-quality | Genome-fragment | 5.19 | HMM-based (lower-bound) | 0 |
| S12_extracted_k141_100108 | 3072 | No | NA | 4 | 0 | 0 | Low-quality | Genome-fragment | 5.61 | AAI-based (high-confidence) | 0 |
| S16_vs_164392\|\|full | 2122 | No | NA | 3 | 0 | 2 | Not-determined | Genome-fragment | NA | NA | 0 |
| S11_extracted_k141_2717 | 3073 | No | NA | 3 | 0 | 0 | Low-quality | Genome-fragment | 5.02 | AAI-based (high-confidence) | 0 |
| S14_vs_k141_110744\|\|full | 3073 | No | NA | 6 | 1 | 0 | Low-quality | Genome-fragment | 7.38 | AAI-based (high-confidence) | 0 |
| S16_vs_164521\|\|full | 1943 | No | NA | 3 | 0 | 1 | Not-determined | Genome-fragment | NA | NA | 0 |
| S14_vs_k141_97710\|\|full | 3073 | No | NA | 3 | 0 | 0 | Low-quality | Genome-fragment | 3.83 | AAI-based (medium-confidence) | 0 |
| S5_vs_k141_139135\|\|full | 3073 | No | NA | 4 | 0 | 0 | Low-quality | Genome-fragment | 1.72 | AAI-based (medium-confidence) | 0 |
| S16_vs_164798\|\|full | 1972 | No | NA | 2 | 0 | 0 | Not-determined | Genome-fragment | NA | NA | 0 |
| S16_vs_164858\|\|full | 1900 | No | NA | 6 | 0 | 1 | Not-determined | Genome-fragment | NA | NA | 0 |
| S11_extracted_k141_9677 | 3074 | No | NA | 4 | 2 | 0 | Low-quality | Genome-fragment | 7.41 | AAI-based (high-confidence) | 0 |
| S16_vs_164981\|\|full | 2062 | No | NA | 2 | 0 | 1 | Not-determined | Genome-fragment | NA | NA | 0 |
| S1_vs_k141_51252\|\|full | 3074 | No | NA | 4 | 2 | 0 | Low-quality | Genome-fragment | 8.04 | AAI-based (high-confidence) | 0 |
| S16_vs_165156\|\|full | 5916 | No | NA | 9 | 0 | 0 | Not-determined | Genome-fragment | NA | NA | 0 |
| S16_vs_165198\|\|full | 2001 | No | NA | 3 | 0 | 2 | Not-determined | Genome-fragment | NA | NA | 0 |
| S16_vs_165200\|\|full | 2938 | No | NA | 2 | 0 | 1 | Not-determined | Genome-fragment | NA | NA | 0 |
| S17_vs_k141_23234\|\|full | 3075 | No | NA | 6 | 0 | 0 | Low-quality | Genome-fragment | 5.03 | AAI-based (medium-confidence) | 0 |
| S16_vs_165283\|\|full | 4786 | No | NA | 7 | 0 | 5 | Not-determined | Genome-fragment | NA | NA | 0 |
| S16_vs_165326\|\|full | 1743 | No | NA | 3 | 0 | 2 | Not-determined | Genome-fragment | NA | NA | 0 |
| S3_vs_k141_41175\|\|lt2gene | 3075 | No | NA | 1 | 0 | 0 | Low-quality | Genome-fragment | 5.65 | AAI-based (high-confidence) | 0 |
| S16_extracted_9471 | 3075 | No | NA | 3 | 0 | 1 | Low-quality | Genome-fragment | 6.07 | AAI-based (medium-confidence) | 0 |
| S16_vs_165495\|\|full | 3529 | No | NA | 4 | 0 | 3 | Not-determined | Genome-fragment | NA | NA | 0 |
| S16_vs_165571\|\|full | 4401 | No | NA | 2 | 0 | 1 | Not-determined | Genome-fragment | NA | NA | 0 |
| S16_vs_165655\|\|full | 1919 | No | NA | 3 | 0 | 2 | Not-determined | Genome-fragment | NA | NA | 0 |
| S16_vs_165662\|\|full | 21289 | No | NA | 30 | 13 | 0 | Medium-quality | Genome-fragment | 68.38 | AAI-based (high-confidence) | 0 |
| S6_vs_k141_18645\|\|full | 3075 | No | NA | 4 | 2 | 0 | Low-quality | Genome-fragment | 5.02 | HMM-based (lower-bound) | 0 |
| S16_vs_165688\|\|full | 1224 | No | NA | 2 | 0 | 1 | Not-determined | Genome-fragment | NA | NA | 0 |
| S8_vs_k141_2050\|\|full | 3075 | No | NA | 3 | 0 | 2 | Low-quality | Genome-fragment | 2.74 | AAI-based (medium-confidence) | 0 |
| S16_vs_165781\|\|full | 7210 | No | NA | 12 | 0 | 0 | Not-determined | Genome-fragment | NA | NA | 0 |
| S17_vs_k141_67581\|\|full | 3076 | No | NA | 5 | 2 | 0 | Low-quality | Genome-fragment | 4.66 | AAI-based (high-confidence) | 0 |
| S16_vs_166106\|\|full | 3452 | No | NA | 5 | 0 | 0 | Not-determined | Genome-fragment | NA | NA | 0 |
| S17_vs_k141_5960\|\|full | 3076 | No | NA | 6 | 6 | 0 | Low-quality | Genome-fragment | 6.6 | AAI-based (high-confidence) | 0 |
| S14_extracted_k141_55312 | 3076 | No | NA | 3 | 0 | 1 | Low-quality | Genome-fragment | 2.32 | AAI-based (medium-confidence) | 0 |
| S3_vs_k141_120086\|\|full | 3076 | No | NA | 3 | 0 | 2 | Low-quality | Genome-fragment | 1.38 | AAI-based (medium-confidence) | 0 |
| S7_vs_28099\|\|full | 3076 | No | NA | 3 | 3 | 0 | Low-quality | Genome-fragment | 8.74 | AAI-based (medium-confidence) | 0 |
| S16_vs_166242\|\|full | 1837 | No | NA | 2 | 0 | 1 | Not-determined | Genome-fragment | NA | NA | 0 |
| S16_vs_166246\|\|full | 5041 | No | NA | 4 | 0 | 3 | Not-determined | Genome-fragment | NA | NA | 0 |
| S16_vs_166309\|\|full | 3182 | No | NA | 6 | 0 | 0 | Not-determined | Genome-fragment | NA | NA | 0 |
| S8_extracted_k141_5007 | 3076 | No | NA | 5 | 0 | 0 | Low-quality | Genome-fragment | 4.84 | AAI-based (medium-confidence) | 0 |
| S16_vs_166540\|\|full | 2278 | No | NA | 3 | 0 | 2 | Not-determined | Genome-fragment | NA | NA | 0 |
| S16_vs_166604\|\|full | 2022 | No | NA | 4 | 0 | 0 | Not-determined | Genome-fragment | NA | NA | 0 |
| S8_extracted_k141_165740 | 3077 | No | NA | 7 | 5 | 0 | Low-quality | Genome-fragment | 5.11 | AAI-based (medium-confidence) | 0 |
| S16_vs_166852\|\|full | 1148 | No | NA | 2 | 0 | 0 | Not-determined | Genome-fragment | NA | NA | 0 |
| S16_vs_166938\|\|full | 1160 | No | NA | 2 | 0 | 2 | Not-determined | Genome-fragment | NA | NA | 0 |
| S16_vs_166972\|\|full | 1429 | No | NA | 2 | 0 | 1 | Not-determined | Genome-fragment | NA | NA | 0 |
| S16_vs_166989\|\|full | 1262 | No | NA | 2 | 0 | 0 | Not-determined | Genome-fragment | NA | NA | 0 |
| S16_vs_166997\|\|full | 1471 | No | NA | 3 | 0 | 1 | Not-determined | Genome-fragment | NA | NA | 0 |
| S9_extracted_k141_842 | 3079 | No | NA | 8 | 1 | 0 | Low-quality | Genome-fragment | 2.2 | HMM-based (lower-bound) | 0 |
| S3_vs_k141_28985\|\|full | 3079 | No | NA | 4 | 0 | 0 | Low-quality | Genome-fragment | 1.51 | AAI-based (high-confidence) | 0 |
| S10_extracted_85921 | 3079 | No | NA | 5 | 0 | 1 | Low-quality | Genome-fragment | 5.03 | AAI-based (medium-confidence) | 0 |
| S16_vs_167206\|\|full | 1906 | No | NA | 4 | 0 | 2 | Not-determined | Genome-fragment | NA | NA | 0 |
| S17_extracted_k141_13030 | 3080 | No | NA | 5 | 0 | 1 | Low-quality | Genome-fragment | 4.14 | AAI-based (high-confidence) | 0 |
| S5_extracted_k141_4281 | 3080 | No | NA | 5 | 0 | 4 | Low-quality | Genome-fragment | 1.28 | AAI-based (medium-confidence) | 0 |
| S16_vs_167285\|\|full | 1725 | No | NA | 3 | 0 | 2 | Not-determined | Genome-fragment | NA | NA | 0 |
| S16_vs_167324\|\|full | 2647 | No | NA | 5 | 0 | 1 | Not-determined | Genome-fragment | NA | NA | 0 |
| S7_vs_24806\|\|full | 3080 | No | NA | 4 | 0 | 2 | Low-quality | Genome-fragment | 4.73 | AAI-based (medium-confidence) | 0 |
| S16_vs_167532\|\|full | 8891 | No | NA | 8 | 0 | 5 | Not-determined | Genome-fragment | NA | NA | 0 |
| S16_vs_160861\|\|full | 3080 | No | NA | 4 | 0 | 0 | Low-quality | Genome-fragment | 3.84 | AAI-based (high-confidence) | 0 |
| S16_vs_167605\|\|full | 6539 | No | NA | 6 | 0 | 4 | Not-determined | Genome-fragment | NA | NA | 0 |
| S16_vs_167607\|\|full | 1918 | No | NA | 3 | 0 | 1 | Not-determined | Genome-fragment | NA | NA | 0 |
| S16_vs_167641\|\|full | 1142 | No | NA | 2 | 0 | 1 | Not-determined | Genome-fragment | NA | NA | 0 |
| S10_extracted_47106 | 3080 | No | NA | 9 | 1 | 0 | Low-quality | Genome-fragment | 1.8 | AAI-based (medium-confidence) | 0 |
| S2_extracted_k141_7800 | 3080 | No | NA | 5 | 0 | 1 | Low-quality | Genome-fragment | 5.34 | AAI-based (medium-confidence) | 0 |
| S16_vs_167836\|\|full | 1895 | No | NA | 3 | 0 | 2 | Not-determined | Genome-fragment | NA | NA | 0 |
| S16_vs_167894\|\|full | 1757 | No | NA | 3 | 0 | 2 | Not-determined | Genome-fragment | NA | NA | 0 |
| S16_vs_167905\|\|full | 3224 | No | NA | 2 | 0 | 1 | Not-determined | Genome-fragment | NA | NA | 0 |
| S16_vs_168040\|\|full | 3373 | No | NA | 2 | 0 | 1 | Not-determined | Genome-fragment | NA | NA | 0 |
| S16_vs_168068\|\|full | 1405 | No | NA | 3 | 0 | 0 | Not-determined | Genome-fragment | NA | NA | 0 |
| S16_vs_168087\|\|full | 2186 | No | NA | 3 | 0 | 0 | Not-determined | Genome-fragment | NA | NA | 0 |
| S16_vs_168114\|\|full | 1964 | No | NA | 4 | 0 | 2 | Not-determined | Genome-fragment | NA | NA | 0 |
| S16_vs_168256\|\|full | 1106 | No | NA | 3 | 0 | 1 | Not-determined | Genome-fragment | NA | NA | 0 |
| S16_vs_168377\|\|full | 3133 | No | NA | 3 | 0 | 3 | Not-determined | Genome-fragment | NA | NA | 0 |
| S16_vs_168662\|\|full | 3544 | No | NA | 3 | 0 | 0 | Not-determined | Genome-fragment | NA | NA | 0 |
| S16_vs_168682\|\|full | 4351 | No | NA | 4 | 0 | 3 | Not-determined | Genome-fragment | NA | NA | 0 |
| S16_vs_168757\|\|full | 2907 | No | NA | 3 | 0 | 1 | Not-determined | Genome-fragment | NA | NA | 0 |
| S17_vs_k141_54964\|\|full | 3081 | No | NA | 8 | 1 | 0 | Low-quality | Genome-fragment | 6.41 | AAI-based (high-confidence) | 0 |
| S16_vs_168829\|\|full | 6152 | No | NA | 4 | 0 | 4 | Not-determined | Genome-fragment | NA | NA | 0 |
| S7_vs_73173\|\|full | 3081 | No | NA | 6 | 4 | 0 | Low-quality | Genome-fragment | 9.13 | AAI-based (high-confidence) | 0 |
| S16_vs_169121\|\|full | 5117 | No | NA | 4 | 0 | 4 | Not-determined | Genome-fragment | NA | NA | 0 |
| S16_vs_169202\|\|full | 5757 | No | NA | 5 | 0 | 1 | Not-determined | Genome-fragment | NA | NA | 0 |
| S16_vs_169212\|\|full | 1784 | No | NA | 3 | 0 | 2 | Not-determined | Genome-fragment | NA | NA | 0 |
| S16_vs_169228\|\|full | 2266 | No | NA | 5 | 0 | 2 | Not-determined | Genome-fragment | NA | NA | 0 |
| S16_vs_169273\|\|full | 4156 | No | NA | 6 | 0 | 4 | Not-determined | Genome-fragment | NA | NA | 0 |
| S16_vs_169320\|\|full | 1745 | No | NA | 2 | 0 | 0 | Not-determined | Genome-fragment | NA | NA | 0 |
| S16_vs_169361\|\|full | 1489 | No | NA | 2 | 0 | 2 | Not-determined | Genome-fragment | NA | NA | 0 |
| S12_extracted_k141_41456 | 3082 | No | NA | 4 | 0 | 2 | Low-quality | Genome-fragment | 1.2 | AAI-based (medium-confidence) | 0 |
| S15_extracted_53136 | 3082 | No | NA | 3 | 3 | 0 | Low-quality | Genome-fragment | 7.23 | AAI-based (high-confidence) | 0 |
| S5_vs_k141_123223\|\|full | 3083 | No | NA | 6 | 4 | 0 | Low-quality | Genome-fragment | 8.39 | AAI-based (high-confidence) | 0 |
| S16_vs_169591\|\|full | 2222 | No | NA | 3 | 0 | 2 | Not-determined | Genome-fragment | NA | NA | 0 |
| S16_vs_169650\|\|full | 1175 | No | NA | 3 | 0 | 0 | Not-determined | Genome-fragment | NA | NA | 0 |
| S7_vs_2075\|\|full | 3084 | Yes | 2235 | 6 | 1 | 1 | Low-quality | Genome-fragment | 2.17 | HMM-based (lower-bound) | 27.53 |
| S8_vs_k141_34640\|\|full | 3084 | No | NA | 5 | 0 | 0 | Low-quality | Genome-fragment | 7.4 | AAI-based (medium-confidence) | 0 |
| S17_vs_k141_98515\|\|full | 3085 | No | NA | 4 | 0 | 2 | Low-quality | Genome-fragment | 3.23 | AAI-based (medium-confidence) | 0 |
| S9_extracted_k141_62174 | 3086 | No | NA | 6 | 0 | 1 | Low-quality | Genome-fragment | 2.62 | AAI-based (high-confidence) | 0 |
| S3_extracted_k141_31760 | 3086 | No | NA | 4 | 0 | 0 | Low-quality | Genome-fragment | 7.79 | AAI-based (high-confidence) | 0 |
| S7_extracted_21097 | 3086 | No | NA | 2 | 0 | 2 | Low-quality | Genome-fragment | 1.54 | AAI-based (high-confidence) | 0 |
| S14_extracted_k141_91325 | 3087 | No | NA | 3 | 3 | 0 | Low-quality | Genome-fragment | 10.17 | AAI-based (high-confidence) | 0 |
| S16_vs_170009\|\|full | 1249 | No | NA | 2 | 0 | 0 | Not-determined | Genome-fragment | NA | NA | 0 |
| S16_vs_170040\|\|full | 1768 | No | NA | 3 | 0 | 2 | Not-determined | Genome-fragment | NA | NA | 0 |
| S1_vs_k141_57583\|\|full | 3087 | No | NA | 3 | 1 | 0 | Low-quality | Genome-fragment | 6.5 | AAI-based (high-confidence) | 0 |
| S3_extracted_k141_59054 | 3087 | No | NA | 9 | 0 | 0 | Low-quality | Genome-fragment | 4.98 | AAI-based (high-confidence) | 0 |
| S5_vs_k141_8702\|\|full | 3087 | No | NA | 6 | 1 | 0 | Low-quality | Genome-fragment | 7.75 | AAI-based (high-confidence) | 0 |
| S16_extracted_50555 | 3087 | No | NA | 2 | 2 | 0 | Low-quality | Genome-fragment | 5.1 | AAI-based (high-confidence) | 0 |
| S4_extracted_7639 | 3087 | No | NA | 2 | 0 | 1 | Low-quality | Genome-fragment | 4.88 | AAI-based (high-confidence) | 0 |
| S17_extracted_k141_112120 | 3089 | No | NA | 3 | 0 | 1 | Low-quality | Genome-fragment | 2.25 | AAI-based (medium-confidence) | 0 |
| S16_vs_170254\|\|full | 10419 | No | NA | 8 | 0 | 7 | Not-determined | Genome-fragment | NA | NA | 0 |
| S6_vs_k141_15916\|\|full | 3089 | No | NA | 3 | 0 | 1 | Low-quality | Genome-fragment | 1.34 | AAI-based (medium-confidence) | 0 |
| S16_vs_170356\|\|full | 3097 | No | NA | 2 | 0 | 1 | Not-determined | Genome-fragment | NA | NA | 0 |
| S16_vs_170425\|\|full | 2064 | No | NA | 2 | 0 | 1 | Not-determined | Genome-fragment | NA | NA | 0 |
| S9_vs_k141_80792\|\|full | 3090 | No | NA | 3 | 0 | 2 | Low-quality | Genome-fragment | 2.87 | AAI-based (medium-confidence) | 0 |
| S16_vs_170504\|\|full | 1109 | No | NA | 3 | 0 | 1 | Not-determined | Genome-fragment | NA | NA | 0 |
| S16_vs_170516\|\|full | 1211 | No | NA | 4 | 0 | 0 | Not-determined | Genome-fragment | NA | NA | 0 |
| S16_vs_170520\|\|full | 6392 | No | NA | 11 | 0 | 2 | Not-determined | Genome-fragment | NA | NA | 0 |
| S14_vs_k141_102833\|\|full | 3090 | No | NA | 3 | 3 | 0 | Low-quality | Genome-fragment | 8.27 | HMM-based (lower-bound) | 0 |
| S16_vs_170537\|\|full | 1299 | No | NA | 4 | 0 | 1 | Not-determined | Genome-fragment | NA | NA | 0 |
| S7_vs_94036\|\|full | 3090 | No | NA | 8 | 0 | 0 | Low-quality | Genome-fragment | 8.86 | AAI-based (high-confidence) | 0 |
| S10_vs_88098\|\|full | 3090 | No | NA | 4 | 0 | 1 | Low-quality | Genome-fragment | 7.67 | AAI-based (medium-confidence) | 0 |
| S16_vs_170737\|\|full | 1208 | No | NA | 2 | 0 | 0 | Not-determined | Genome-fragment | NA | NA | 0 |
| S16_vs_170738\|\|full | 1661 | No | NA | 2 | 0 | 0 | Not-determined | Genome-fragment | NA | NA | 0 |
| S16_vs_170800\|\|full | 2626 | No | NA | 4 | 0 | 1 | Not-determined | Genome-fragment | NA | NA | 0 |
| S12_vs_k141_176752\|\|full | 3090 | No | NA | 4 | 1 | 0 | Low-quality | Genome-fragment | 4.68 | HMM-based (lower-bound) | 0 |
| S16_vs_170856\|\|full | 5072 | No | NA | 6 | 0 | 0 | Not-determined | Genome-fragment | NA | NA | 0 |
| S16_vs_170900\|\|full | 1573 | No | NA | 3 | 0 | 1 | Not-determined | Genome-fragment | NA | NA | 0 |
| S16_vs_170988\|\|full | 1653 | No | NA | 3 | 0 | 2 | Not-determined | Genome-fragment | NA | NA | 0 |
| S1_extracted_k141_110365 | 3091 | No | NA | 2 | 0 | 2 | Low-quality | Genome-fragment | 6.19 | AAI-based (medium-confidence) | 0 |
| S3_extracted_k141_113139 | 3091 | Yes | 1501 | 6 | 3 | 1 | Low-quality | Genome-fragment | 2.35 | AAI-based (high-confidence) | 51.44 |
| S16_vs_171129\|\|full | 4147 | No | NA | 4 | 0 | 2 | Not-determined | Genome-fragment | NA | NA | 0 |
| S16_vs_171192\|\|full | 1903 | No | NA | 2 | 0 | 1 | Not-determined | Genome-fragment | NA | NA | 0 |
| S7_vs_544\|\|full | 3091 | No | NA | 4 | 0 | 4 | Low-quality | Genome-fragment | 1.45 | AAI-based (high-confidence) | 0 |
| S16_vs_171243\|\|full | 2583 | No | NA | 4 | 0 | 2 | Not-determined | Genome-fragment | NA | NA | 0 |
| S16_vs_171249\|\|full | 1774 | No | NA | 3 | 0 | 0 | Not-determined | Genome-fragment | NA | NA | 0 |
| S12_vs_k141_42082\|\|full | 3091 | No | NA | 3 | 0 | 3 | Low-quality | Genome-fragment | 3.15 | AAI-based (high-confidence) | 0 |
| S16_vs_171431\|\|full | 2053 | No | NA | 4 | 0 | 0 | Not-determined | Genome-fragment | NA | NA | 0 |
| S16_vs_171598\|\|full | 3153 | No | NA | 4 | 0 | 1 | Not-determined | Genome-fragment | NA | NA | 0 |
| S16_vs_171664\|\|full | 5782 | No | NA | 12 | 0 | 0 | Not-determined | Genome-fragment | NA | NA | 0 |
| S6_extracted_k141_51868 | 3091 | No | NA | 5 | 0 | 3 | Low-quality | Genome-fragment | 1.42 | AAI-based (medium-confidence) | 0 |
| S16_vs_171726\|\|full | 2760 | No | NA | 3 | 0 | 1 | Not-determined | Genome-fragment | NA | NA | 0 |
| S16_vs_171745\|\|full | 2722 | No | NA | 3 | 0 | 1 | Not-determined | Genome-fragment | NA | NA | 0 |
| S16_vs_171944\|\|full | 1719 | No | NA | 4 | 0 | 1 | Not-determined | Genome-fragment | NA | NA | 0 |
| S16_vs_171945\|\|full | 1047 | No | NA | 2 | 0 | 2 | Not-determined | Genome-fragment | NA | NA | 0 |
| S6_vs_k141_26518\|\|full | 3092 | No | NA | 5 | 0 | 0 | Low-quality | Genome-fragment | 1.73 | AAI-based (high-confidence) | 0 |
| S16_vs_172167\|\|full | 1887 | No | NA | 4 | 0 | 3 | Not-determined | Genome-fragment | NA | NA | 0 |
| S14_vs_k141_113447\|\|full | 3093 | No | NA | 2 | 0 | 1 | Low-quality | Genome-fragment | 1.48 | AAI-based (medium-confidence) | 0 |
| S16_vs_172262\|\|full | 4502 | No | NA | 8 | 0 | 1 | Not-determined | Genome-fragment | NA | NA | 0 |
| S12_extracted_k141_104962 | 3093 | No | NA | 3 | 1 | 0 | Low-quality | Genome-fragment | 5.2 | HMM-based (lower-bound) | 0 |
| S7_vs_55433\|\|full | 3094 | No | NA | 8 | 1 | 1 | Low-quality | Genome-fragment | 8.15 | AAI-based (high-confidence) | 0 |
| S8_vs_k141_139322\|\|full | 3094 | No | NA | 4 | 0 | 2 | Low-quality | Genome-fragment | 3.58 | AAI-based (medium-confidence) | 0 |
| S17_vs_k141_25690\|\|full | 3095 | No | NA | 7 | 0 | 0 | Low-quality | Genome-fragment | 3.83 | AAI-based (high-confidence) | 0 |
| S3_extracted_k141_92464 | 3095 | No | NA | 7 | 2 | 0 | Low-quality | Genome-fragment | 1.94 | AAI-based (high-confidence) | 0 |
| S12_vs_k141_40748\|\|0_partial | 3095 | No | NA | 4 | 1 | 0 | Low-quality | Genome-fragment | 37.33 | AAI-based (medium-confidence) | 0 |
| S16_vs_172412\|\|full | 9934 | No | NA | 11 | 0 | 2 | Not-determined | Genome-fragment | NA | NA | 0 |
| S17_vs_k141_160147\|\|full | 3096 | No | NA | 2 | 0 | 1 | Low-quality | Genome-fragment | 1.03 | AAI-based (medium-confidence) | 0 |
| S16_vs_172443\|\|full | 5000 | No | NA | 4 | 0 | 4 | Not-determined | Genome-fragment | NA | NA | 0 |
| S16_vs_172573\|\|full | 2337 | No | NA | 5 | 0 | 0 | Not-determined | Genome-fragment | NA | NA | 0 |
| S16_vs_172677\|\|full | 1087 | No | NA | 2 | 0 | 2 | Not-determined | Genome-fragment | NA | NA | 0 |
| S9_extracted_k141_76409 | 3096 | No | NA | 11 | 1 | 0 | Low-quality | Genome-fragment | 7.65 | AAI-based (high-confidence) | 0 |
| S9_vs_k141_21456\|\|full | 3096 | No | NA | 6 | 1 | 0 | Low-quality | Genome-fragment | 6.85 | AAI-based (medium-confidence) | 0 |
| S16_vs_172769\|\|full | 2425 | No | NA | 2 | 0 | 0 | Not-determined | Genome-fragment | NA | NA | 0 |
| S16_vs_172796\|\|full | 1786 | No | NA | 3 | 0 | 0 | Not-determined | Genome-fragment | NA | NA | 0 |
| S16_vs_172884\|\|full | 4279 | No | NA | 8 | 0 | 0 | Not-determined | Genome-fragment | NA | NA | 0 |
| S16_vs_172885\|\|full | 2121 | No | NA | 3 | 0 | 2 | Not-determined | Genome-fragment | NA | NA | 0 |
| S16_vs_172949\|\|full | 5140 | No | NA | 4 | 0 | 2 | Not-determined | Genome-fragment | NA | NA | 0 |
| S16_vs_172968\|\|full | 1643 | No | NA | 3 | 0 | 0 | Not-determined | Genome-fragment | NA | NA | 0 |
| S3_extracted_k141_96533 | 3096 | Yes | 1088 | 3 | 1 | 1 | Low-quality | Genome-fragment | 0.77 | HMM-based (lower-bound) | 64.86 |
| S16_vs_173080\|\|full | 2286 | No | NA | 4 | 0 | 1 | Not-determined | Genome-fragment | NA | NA | 0 |
| S16_vs_14307\|\|0_partial | 15458 | No | NA | 16 | 0 | 6 | Not-determined | Genome-fragment | NA | NA | 0 |
| S5_vs_k141_36742\|\|full | 3096 | No | NA | 3 | 0 | 1 | Low-quality | Genome-fragment | 2.16 | AAI-based (medium-confidence) | 0 |
| S16_vs_15775\|\|0_partial | 16592 | No | NA | 14 | 0 | 9 | Not-determined | Genome-fragment | NA | NA | 0 |
| S12_extracted_k141_104690 | 3096 | No | NA | 3 | 0 | 2 | Low-quality | Genome-fragment | 0.39 | AAI-based (medium-confidence) | 0 |
| S16_vs_32077\|\|0_partial | 13026 | No | NA | 18 | 0 | 9 | Not-determined | Genome-fragment | NA | NA | 0 |
| S16_vs_34591\|\|0_partial | 11880 | No | NA | 14 | 0 | 7 | Not-determined | Genome-fragment | NA | NA | 0 |
| S16_vs_36988\|\|0_partial | 14227 | No | NA | 10 | 0 | 6 | Not-determined | Genome-fragment | NA | NA | 0 |
| S16_vs_60695\|\|0_partial | 16135 | No | NA | 17 | 0 | 13 | Not-determined | Genome-fragment | NA | NA | 0 |
| S16_vs_88202\|\|0_partial | 17609 | No | NA | 14 | 0 | 4 | Not-determined | Genome-fragment | NA | NA | 0 |
| S16_vs_88268\|\|0_partial | 18597 | No | NA | 12 | 0 | 8 | Not-determined | Genome-fragment | NA | NA | 0 |
| S16_vs_92916\|\|0_partial | 14316 | No | NA | 7 | 0 | 6 | Not-determined | Genome-fragment | NA | NA | 0 |
| S12_vs_k141_18570\|\|full | 3096 | No | NA | 6 | 0 | 1 | Low-quality | Genome-fragment | 5.52 | AAI-based (medium-confidence) | 0 |
| S12_vs_k141_184701\|\|full | 3096 | No | NA | 4 | 0 | 0 | Low-quality | Genome-fragment | 2.03 | AAI-based (high-confidence) | 0 |
| S6_vs_k141_26152\|\|full | 3098 | No | NA | 4 | 1 | 0 | Low-quality | Genome-fragment | 7.61 | AAI-based (high-confidence) | 0 |
| S16_vs_116081\|\|0_partial | 19074 | No | NA | 24 | 0 | 9 | Not-determined | Genome-fragment | NA | NA | 0 |
| S16_vs_140776\|\|0_partial | 12693 | No | NA | 12 | 0 | 8 | Not-determined | Genome-fragment | NA | NA | 0 |
| S12_extracted_k141_151399 | 3099 | No | NA | 3 | 2 | 0 | Low-quality | Genome-fragment | 6.52 | AAI-based (high-confidence) | 0 |
| S16_vs_147914\|\|0_partial | 10060 | No | NA | 15 | 0 | 5 | Not-determined | Genome-fragment | NA | NA | 0 |
| S9_extracted_k141_17730 | 3100 | No | NA | 3 | 2 | 0 | Low-quality | Genome-fragment | 3.87 | HMM-based (lower-bound) | 0 |
| S1_extracted_k141_144972 | 3100 | No | NA | 2 | 0 | 2 | Low-quality | Genome-fragment | 0.91 | AAI-based (medium-confidence) | 0 |
| S14_extracted_k141_69057 | 3101 | No | NA | 4 | 2 | 0 | Low-quality | Genome-fragment | 5.14 | HMM-based (lower-bound) | 0 |
| S1_extracted_k141_140536 | 3101 | No | NA | 7 | 4 | 0 | Low-quality | Genome-fragment | 8.82 | AAI-based (high-confidence) | 0 |
| S3_extracted_k141_79918 | 3101 | No | NA | 6 | 1 | 0 | Low-quality | Genome-fragment | 7.5 | AAI-based (medium-confidence) | 0 |
| S16_vs_3353\|\|lt2gene | 1024 | No | NA | 1 | 0 | 0 | Not-determined | Genome-fragment | NA | NA | 0 |
| S6_extracted_k141_40869 | 3101 | No | NA | 4 | 0 | 1 | Low-quality | Genome-fragment | 2.9 | AAI-based (medium-confidence) | 0 |
| S9_extracted_k141_8839 | 3102 | No | NA | 5 | 4 | 0 | Low-quality | Genome-fragment | 9.76 | AAI-based (high-confidence) | 0 |
| S16_vs_5757\|\|lt2gene | 1029 | No | NA | 1 | 0 | 0 | Not-determined | Genome-fragment | NA | NA | 0 |
| S14_extracted_k141_60559 | 3102 | No | NA | 7 | 5 | 0 | Low-quality | Genome-fragment | 10.23 | AAI-based (high-confidence) | 0 |
| S16_vs_7304\|\|lt2gene | 1536 | No | NA | 2 | 0 | 0 | Not-determined | Genome-fragment | NA | NA | 0 |
| S16_vs_7692\|\|lt2gene | 1077 | No | NA | 2 | 0 | 0 | Not-determined | Genome-fragment | NA | NA | 0 |
| S14_vs_k141_102047\|\|full | 3102 | No | NA | 3 | 3 | 0 | Low-quality | Genome-fragment | 5.17 | AAI-based (high-confidence) | 0 |
| S1_vs_k141_51684\|\|full | 3103 | No | NA | 4 | 1 | 0 | Low-quality | Genome-fragment | 9.33 | AAI-based (high-confidence) | 0 |
| S10_vs_73566\|\|full | 3103 | Yes | 2726 | 8 | 2 | 1 | Low-quality | Genome-fragment | 1.91 | HMM-based (lower-bound) | 12.15 |
| S16_vs_14059\|\|lt2gene | 1083 | No | NA | 2 | 0 | 0 | Not-determined | Genome-fragment | NA | NA | 0 |
| S16_vs_14138\|\|lt2gene | 1247 | No | NA | 2 | 0 | 0 | Not-determined | Genome-fragment | NA | NA | 0 |
| S12_vs_k141_94249\|\|full | 3103 | No | NA | 4 | 0 | 2 | Low-quality | Genome-fragment | 2.75 | AAI-based (medium-confidence) | 0 |
| S16_vs_17310\|\|lt2gene | 1834 | No | NA | 2 | 0 | 0 | Not-determined | Genome-fragment | NA | NA | 0 |
| S11_vs_k141_66353\|\|full | 3105 | Yes | 2748 | 4 | 1 | 1 | Low-quality | Genome-fragment | 1.56 | AAI-based (medium-confidence) | 11.5 |
| S6_extracted_k141_40397 | 3105 | No | NA | 6 | 3 | 0 | Low-quality | Genome-fragment | 6.81 | AAI-based (high-confidence) | 0 |
| S14_extracted_k141_45370 | 3106 | No | NA | 4 | 0 | 1 | Low-quality | Genome-fragment | 6.59 | AAI-based (medium-confidence) | 0 |
| S1_vs_k141_96582\|\|full | 3106 | No | NA | 6 | 4 | 0 | Low-quality | Genome-fragment | 5.85 | HMM-based (lower-bound) | 0 |
| S16_vs_30491\|\|lt2gene | 1918 | No | NA | 1 | 0 | 1 | Not-determined | Genome-fragment | NA | NA | 0 |
| S16_vs_30660\|\|lt2gene | 1490 | No | NA | 1 | 0 | 0 | Not-determined | Genome-fragment | NA | NA | 0 |
| S3_extracted_k141_101860 | 3106 | No | NA | 4 | 1 | 0 | Low-quality | Genome-fragment | 2.03 | AAI-based (high-confidence) | 0 |
| S12_vs_k141_135427\|\|full | 3106 | No | NA | 3 | 0 | 0 | Low-quality | Genome-fragment | 3.51 | AAI-based (high-confidence) | 0 |
| S12_vs_k141_161061\|\|lt2gene | 3106 | No | NA | 1 | 1 | 0 | Low-quality | Genome-fragment | 1.68 | HMM-based (lower-bound) | 0 |
| S8_vs_k141_31882\|\|full | 3106 | No | NA | 3 | 0 | 3 | Low-quality | Genome-fragment | 9.34 | AAI-based (medium-confidence) | 0 |
| S13_vs_1910\|\|full | 3107 | No | NA | 5 | 0 | 1 | Low-quality | Genome-fragment | 5.82 | AAI-based (medium-confidence) | 0 |
| S16_vs_36732\|\|lt2gene | 1280 | No | NA | 2 | 0 | 1 | Not-determined | Genome-fragment | NA | NA | 0 |
| S14_vs_k141_111060\|\|full | 3107 | No | NA | 7 | 5 | 0 | Low-quality | Genome-fragment | 6.52 | AAI-based (high-confidence) | 0 |
| S14_vs_k141_56938\|\|full | 3108 | No | NA | 7 | 1 | 0 | Low-quality | Genome-fragment | 4.34 | AAI-based (high-confidence) | 0 |
| S3_extracted_k141_125354 | 3108 | No | NA | 5 | 0 | 5 | Low-quality | Genome-fragment | 7.93 | AAI-based (medium-confidence) | 0 |
| S3_vs_k141_89336\|\|full | 3108 | No | NA | 3 | 0 | 0 | Low-quality | Genome-fragment | 7.09 | AAI-based (high-confidence) | 0 |
| S2_extracted_k141_3893 | 3108 | No | NA | 9 | 1 | 0 | Low-quality | Genome-fragment | 7.91 | AAI-based (medium-confidence) | 0 |

| | | | | | | | | | | | |
|---|---|---|---|---|---|---|---|---|---|---|---|
| S1_vs_k141_75397\|\|full | 3109 | No | NA | 5 | 3 | 0 | Low-quality | Genome-fragment | 6.35 | AAI-based (high-confidence) | 0 |
| S16_vs_54375\|\|lt2gene | 1043 | No | NA | 2 | 0 | 0 | Not-determined | Genome-fragment | NA | NA | 0 |
| S16_vs_54618\|\|lt2gene | 1803 | No | NA | 2 | 0 | 1 | Not-determined | Genome-fragment | NA | NA | 0 |
| S16_vs_55598\|\|lt2gene | 3528 | No | NA | 2 | 0 | 1 | Not-determined | Genome-fragment | NA | NA | 0 |
| S5_vs_k141_91620\|\|full | 3109 | No | NA | 3 | 2 | 0 | Low-quality | Genome-fragment | 6.55 | AAI-based (high-confidence) | 0 |
| S16_vs_57117\|\|lt2gene | 1291 | No | NA | 2 | 0 | 1 | Not-determined | Genome-fragment | NA | NA | 0 |
| S14_vs_k141_102943\|\|full | 3111 | No | NA | 5 | 2 | 0 | Low-quality | Genome-fragment | 2.33 | HMM-based (lower-bound) | 0 |
| S3_vs_k141_104263\|\|full | 3111 | No | NA | 4 | 0 | 1 | Low-quality | Genome-fragment | 7.57 | AAI-based (medium-confidence) | 0 |
| S5_extracted_k141_42610 | 3111 | No | NA | 4 | 4 | 0 | Low-quality | Genome-fragment | 10.25 | AAI-based (high-confidence) | 0 |
| S12_extracted_k141_161230 | 3111 | No | NA | 1 | 1 | 0 | Low-quality | Genome-fragment | 2.13 | HMM-based (lower-bound) | 0 |
| S16_vs_69662\|\|lt2gene | 1373 | No | NA | 1 | 0 | 0 | Not-determined | Genome-fragment | NA | NA | 0 |
| S16_vs_72979\|\|lt2gene | 1812 | No | NA | 2 | 0 | 0 | Not-determined | Genome-fragment | NA | NA | 0 |
| S16_vs_73775\|\|lt2gene | 2590 | No | NA | 2 | 0 | 1 | Not-determined | Genome-fragment | NA | NA | 0 |
| S16_vs_43805\|\|full | 3112 | No | NA | 4 | 0 | 0 | Low-quality | Genome-fragment | 7.85 | AAI-based (medium-confidence) | 0 |
| S10_extracted_33781 | 3112 | No | NA | 9 | 2 | 0 | Low-quality | Genome-fragment | 1.61 | HMM-based (lower-bound) | 0 |
| S8_vs_k141_5481\|\|full | 3112 | No | NA | 4 | 0 | 1 | Low-quality | Genome-fragment | 11.83 | AAI-based (high-confidence) | 0 |
| S16_vs_78302\|\|lt2gene | 1576 | No | NA | 1 | 0 | 0 | Not-determined | Genome-fragment | NA | NA | 0 |
| S16_vs_79241\|\|lt2gene | 1958 | No | NA | 2 | 0 | 0 | Not-determined | Genome-fragment | NA | NA | 0 |
| S17_vs_k141_131474\|\|full | 3113 | No | NA | 7 | 1 | 0 | Low-quality | Genome-fragment | 1.81 | AAI-based (high-confidence) | 0 |
| S8_extracted_k141_177896 | 3113 | No | NA | 3 | 0 | 1 | Low-quality | Genome-fragment | 7.61 | AAI-based (medium-confidence) | 0 |
| S8_extracted_k141_12792 | 3113 | No | NA | 6 | 0 | 0 | Low-quality | Genome-fragment | 6.61 | AAI-based (high-confidence) | 0 |
| S16_vs_81036\|\|full | 3114 | No | NA | 3 | 0 | 0 | Low-quality | Genome-fragment | 7.53 | AAI-based (medium-confidence) | 0 |
| S8_vs_k141_3305\|\|full | 3114 | No | NA | 4 | 0 | 3 | Low-quality | Genome-fragment | 0.73 | AAI-based (medium-confidence) | 0 |
| S16_vs_87418\|\|lt2gene | 1012 | No | NA | 1 | 0 | 1 | Not-determined | Genome-fragment | NA | NA | 0 |
| S16_vs_87993\|\|lt2gene | 2439 | No | NA | 2 | 0 | 0 | Not-determined | Genome-fragment | NA | NA | 0 |
| S9_extracted_k141_23188 | 3115 | No | NA | 4 | 0 | 3 | Low-quality | Genome-fragment | 7.32 | AAI-based (medium-confidence) | 0 |
| S3_vs_k141_45354\|\|full | 3115 | No | NA | 3 | 0 | 1 | Low-quality | Genome-fragment | 2.25 | AAI-based (high-confidence) | 0 |
| S17_extracted_k141_33904 | 3116 | No | NA | 2 | 0 | 2 | Low-quality | Genome-fragment | 3.7 | AAI-based (medium-confidence) | 0 |
| S16_vs_92374\|\|lt2gene | 2390 | No | NA | 2 | 0 | 2 | Not-determined | Genome-fragment | NA | NA | 0 |
| S11_vs_k141_48810\|\|full | 3116 | No | NA | 5 | 1 | 0 | Low-quality | Genome-fragment | 6.57 | HMM-based (lower-bound) | 0 |
| S13_vs_18873\|\|full | 3116 | No | NA | 7 | 0 | 0 | Low-quality | Genome-fragment | 3.47 | AAI-based (medium-confidence) | 0 |
| S16_vs_93572\|\|lt2gene | 1218 | No | NA | 1 | 0 | 0 | Not-determined | Genome-fragment | NA | NA | 0 |
| S16_vs_95893\|\|lt2gene | 1301 | No | NA | 1 | 0 | 0 | Not-determined | Genome-fragment | NA | NA | 0 |
| S16_vs_96351\|\|lt2gene | 1408 | No | NA | 1 | 0 | 0 | Not-determined | Genome-fragment | NA | NA | 0 |
| S1_extracted_k141_62449 | 3116 | No | NA | 4 | 2 | 0 | Low-quality | Genome-fragment | 2.85 | AAI-based (high-confidence) | 0 |
| S13_vs_41063\|\|full | 3117 | No | NA | 7 | 1 | 0 | Low-quality | Genome-fragment | 5.86 | AAI-based (medium-confidence) | 0 |
| S16_vs_100710\|\|lt2gene | 1375 | No | NA | 2 | 0 | 0 | Not-determined | Genome-fragment | NA | NA | 0 |
| S16_vs_103007\|\|lt2gene | 1145 | No | NA | 1 | 0 | 1 | Not-determined | Genome-fragment | NA | NA | 0 |
| S5_vs_k141_103869\|\|full | 3117 | No | NA | 5 | 1 | 0 | Low-quality | Genome-fragment | 8.42 | AAI-based (medium-confidence) | 0 |
| S6_vs_k141_46310\|\|full | 3117 | No | NA | 5 | 0 | 1 | Low-quality | Genome-fragment | 0.85 | AAI-based (high-confidence) | 0 |
| S14_vs_k141_72019\|\|full | 3118 | No | NA | 8 | 2 | 0 | Low-quality | Genome-fragment | 7.48 | AAI-based (high-confidence) | 0 |
| S11_vs_k141_47699\|\|full | 3119 | No | NA | 3 | 0 | 0 | Low-quality | Genome-fragment | 2.63 | AAI-based (medium-confidence) | 0 |
| S5_vs_k141_76704\|\|full | 3119 | No | NA | 4 | 0 | 0 | Low-quality | Genome-fragment | 6.22 | AAI-based (medium-confidence) | 0 |
| S16_vs_108378\|\|lt2gene | 1478 | No | NA | 1 | 0 | 0 | Not-determined | Genome-fragment | NA | NA | 0 |
| S16_vs_171295\|\|full | 3119 | No | NA | 10 | 2 | 0 | Low-quality | Genome-fragment | 5.38 | HMM-based (lower-bound) | 0 |
| S12_extracted_k141_106336 | 3119 | No | NA | 5 | 1 | 0 | Low-quality | Genome-fragment | 6.8 | AAI-based (medium-confidence) | 0 |
| S1_extracted_k141_30705 | 3120 | No | NA | 6 | 0 | 0 | Low-quality | Genome-fragment | 5.34 | AAI-based (high-confidence) | 0 |
| S16_vs_112893\|\|lt2gene | 1308 | No | NA | 2 | 0 | 1 | Not-determined | Genome-fragment | NA | NA | 0 |
| S3_extracted_k141_107200 | 3120 | No | NA | 5 | 0 | 0 | Low-quality | Genome-fragment | 8.35 | AAI-based (high-confidence) | 0 |
| S7_extracted_4409 | 3120 | No | NA | 5 | 0 | 0 | Low-quality | Genome-fragment | 6.35 | AAI-based (high-confidence) | 0 |
| S16_vs_114291\|\|lt2gene | 2611 | No | NA | 1 | 0 | 1 | Not-determined | Genome-fragment | NA | NA | 0 |
| S7_extracted_12382 | 3120 | No | NA | 1 | 0 | 1 | Low-quality | Genome-fragment | 0.28 | AAI-based (medium-confidence) | 0 |
| S5_vs_k141_117395\|\|full | 3122 | No | NA | 4 | 0 | 2 | Low-quality | Genome-fragment | 3 | AAI-based (medium-confidence) | 0 |
| S16_vs_115341\|\|lt2gene | 1557 | No | NA | 2 | 0 | 0 | Not-determined | Genome-fragment | NA | NA | 0 |
| S16_vs_118905\|\|lt2gene | 1269 | No | NA | 2 | 0 | 1 | Not-determined | Genome-fragment | NA | NA | 0 |
| S7_vs_20867\|\|full | 3122 | No | NA | 6 | 2 | 0 | Low-quality | Genome-fragment | 1.33 | HMM-based (lower-bound) | 0 |
| S8_extracted_k141_116717 | 3122 | No | NA | 7 | 2 | 0 | Low-quality | Genome-fragment | 6.86 | AAI-based (high-confidence) | 0 |
| S5_vs_k141_44935\|\|full | 3123 | No | NA | 6 | 1 | 0 | Low-quality | Genome-fragment | 5.14 | AAI-based (high-confidence) | 0 |
| S11_extracted_k141_44224 | 3124 | No | NA | 5 | 0 | 1 | Low-quality | Genome-fragment | 5.33 | AAI-based (high-confidence) | 0 |
| S10_extracted_54274 | 3124 | No | NA | 3 | 0 | 1 | Low-quality | Genome-fragment | 2.18 | AAI-based (medium-confidence) | 0 |
| S17_extracted_k141_17204 | 3125 | No | NA | 9 | 9 | 0 | Low-quality | Genome-fragment | 10.29 | AAI-based (high-confidence) | 0 |
| S14_extracted_k141_17573 | 3125 | No | NA | 9 | 9 | 0 | Low-quality | Genome-fragment | 10.28 | AAI-based (high-confidence) | 0 |
| S6_vs_k141_22840\|\|full | 3125 | Yes | 1429 | 3 | 1 | 1 | Low-quality | Genome-fragment | 1.77 | HMM-based (lower-bound) | 54.27 |
| S16_vs_126303\|\|lt2gene | 1037 | No | NA | 1 | 0 | 1 | Not-determined | Genome-fragment | NA | NA | 0 |
| S8_extracted_k141_81290 | 3125 | No | NA | 3 | 0 | 2 | Low-quality | Genome-fragment | 0.44 | AAI-based (medium-confidence) | 0 |
| S13_extracted_93390 | 3126 | No | NA | 1 | 1 | 0 | Low-quality | Genome-fragment | 5.12 | AAI-based (high-confidence) | 0 |
| S16_vs_129068\|\|lt2gene | 2063 | No | NA | 2 | 0 | 1 | Not-determined | Genome-fragment | NA | NA | 0 |
| S7_vs_35425\|\|full | 3126 | No | NA | 3 | 1 | 0 | Low-quality | Genome-fragment | 4.7 | HMM-based (lower-bound) | 0 |
| S4_extracted_15930 | 3127 | No | NA | 5 | 0 | 0 | Low-quality | Genome-fragment | 6.86 | AAI-based (medium-confidence) | 0 |
| S7_vs_94509\|\|full | 3128 | No | NA | 6 | 2 | 0 | Low-quality | Genome-fragment | 8.75 | AAI-based (high-confidence) | 0 |
| S12_extracted_k141_165147 | 3128 | No | NA | 4 | 0 | 0 | Low-quality | Genome-fragment | 2.88 | AAI-based (high-confidence) | 0 |
| S16_vs_17481\|\|full | 3129 | No | NA | 8 | 0 | 0 | Low-quality | Genome-fragment | 5.15 | AAI-based (high-confidence) | 0 |
| S10_extracted_78653 | 3129 | No | NA | 8 | 0 | 0 | Low-quality | Genome-fragment | 8.31 | AAI-based (medium-confidence) | 0 |
| S16_vs_132662\|\|lt2gene | 1281 | No | NA | 1 | 0 | 1 | Not-determined | Genome-fragment | NA | NA | 0 |
| S17_vs_k141_146620\|\|full | 3130 | No | NA | 4 | 0 | 1 | Low-quality | Genome-fragment | 4.82 | AAI-based (medium-confidence) | 0 |
| S16_vs_137453\|\|lt2gene | 4715 | No | NA | 1 | 0 | 1 | Not-determined | Genome-fragment | NA | NA | 0 |
| S9_extracted_k141_69799 | 3130 | No | NA | 3 | 2 | 0 | Low-quality | Genome-fragment | 8.29 | AAI-based (high-confidence) | 0 |
| S3_vs_k141_20957\|\|full | 3130 | No | NA | 5 | 0 | 0 | Low-quality | Genome-fragment | 6.13 | AAI-based (medium-confidence) | 0 |
| S5_vs_k141_120239\|\|full | 3130 | No | NA | 5 | 3 | 0 | Low-quality | Genome-fragment | 6.83 | AAI-based (high-confidence) | 0 |
| S10_vs_7527\|\|full | 3130 | No | NA | 10 | 2 | 0 | Low-quality | Genome-fragment | 4.88 | AAI-based (high-confidence) | 0 |
| S9_vs_k141_87855\|\|full | 3131 | No | NA | 5 | 0 | 0 | Low-quality | Genome-fragment | 7.5 | AAI-based (high-confidence) | 0 |
| S10_vs_89595\|\|full | 3131 | No | NA | 3 | 0 | 0 | Low-quality | Genome-fragment | 7.81 | AAI-based (high-confidence) | 0 |
| S16_vs_141335\|\|lt2gene | 1149 | No | NA | 1 | 0 | 1 | Not-determined | Genome-fragment | NA | NA | 0 |
| S6_extracted_k141_10240 | 3131 | No | NA | 9 | 1 | 0 | Low-quality | Genome-fragment | 2.27 | AAI-based (high-confidence) | 0 |
| S8_extracted_k141_44911 | 3131 | No | NA | 4 | 2 | 0 | Low-quality | Genome-fragment | 16.02 | AAI-based (high-confidence) | 0 |
| S1_extracted_k141_154303 | 3132 | No | NA | 6 | 1 | 0 | Low-quality | Genome-fragment | 7.08 | AAI-based (high-confidence) | 0 |
| S16_vs_144083\|\|lt2gene | 1090 | No | NA | 1 | 0 | 0 | Not-determined | Genome-fragment | NA | NA | 0 |
| S16_vs_144100\|\|lt2gene | 1099 | No | NA | 1 | 0 | 0 | Not-determined | Genome-fragment | NA | NA | 0 |
| S3_vs_k141_111419\|\|full | 3133 | No | NA | 5 | 1 | 0 | Low-quality | Genome-fragment | 1.53 | AAI-based (high-confidence) | 0 |
| S16_vs_146534\|\|lt2gene | 2902 | No | NA | 1 | 0 | 0 | Not-determined | Genome-fragment | NA | NA | 0 |
| S16_vs_147046\|\|lt2gene | 2225 | No | NA | 2 | 0 | 0 | Not-determined | Genome-fragment | NA | NA | 0 |
| S16_vs_147637\|\|lt2gene | 1920 | No | NA | 2 | 0 | 0 | Not-determined | Genome-fragment | NA | NA | 0 |
| S7_vs_122897\|\|full | 3133 | No | NA | 4 | 0 | 0 | Low-quality | Genome-fragment | 6.16 | AAI-based (medium-confidence) | 0 |
| S10_vs_2230\|\|full | 3133 | No | NA | 4 | 1 | 0 | Low-quality | Genome-fragment | 3.83 | HMM-based (lower-bound) | 0 |
| S8_extracted_k141_73169 | 3133 | Yes | 400 | 4 | 1 | 1 | Low-quality | Genome-fragment | 0.18 | HMM-based (lower-bound) | 87.23 |
| S9_vs_k141_10086\|\|full | 3134 | No | NA | 5 | 5 | 0 | Low-quality | Genome-fragment | 9.27 | AAI-based (high-confidence) | 0 |
| S5_vs_k141_43416\|\|full | 3135 | No | NA | 2 | 1 | 0 | Low-quality | Genome-fragment | 9.72 | AAI-based (high-confidence) | 0 |
| S5_vs_k141_50041\|\|full | 3135 | No | NA | 2 | 1 | 0 | Low-quality | Genome-fragment | 9.71 | AAI-based (high-confidence) | 0 |
| S16_vs_95823\|\|full | 3135 | No | NA | 7 | 0 | 0 | Low-quality | Genome-fragment | 7.5 | AAI-based (high-confidence) | 0 |
| S5_extracted_k141_65199 | 3136 | No | NA | 1 | 0 | 1 | Low-quality | Genome-fragment | 6.75 | AAI-based (high-confidence) | 0 |
| S16_vs_154750\|\|lt2gene | 1362 | No | NA | 1 | 0 | 1 | Not-determined | Genome-fragment | NA | NA | 0 |
| S7_vs_66639\|\|full | 3136 | No | NA | 8 | 2 | 0 | Low-quality | Genome-fragment | 8.62 | AAI-based (medium-confidence) | 0 |
| S12_vs_k141_13846\|\|full | 3136 | No | NA | 4 | 0 | 3 | Low-quality | Genome-fragment | 3.1 | AAI-based (medium-confidence) | 0 |
| S1_extracted_k141_43599 | 3137 | No | NA | 4 | 2 | 0 | Low-quality | Genome-fragment | 5.4 | AAI-based (high-confidence) | 0 |
| S16_vs_157004\|\|lt2gene | 1065 | No | NA | 1 | 0 | 0 | Not-determined | Genome-fragment | NA | NA | 0 |
| S5_extracted_k141_101702 | 3137 | No | NA | 5 | 4 | 0 | Low-quality | Genome-fragment | 14.42 | AAI-based (high-confidence) | 0 |
| S5_vs_k141_61918\|\|full | 3137 | No | NA | 6 | 3 | 0 | Low-quality | Genome-fragment | 3.95 | HMM-based (lower-bound) | 0 |
| S16_vs_78935\|\|full | 3137 | No | NA | 3 | 0 | 2 | Low-quality | Genome-fragment | 1.46 | AAI-based (medium-confidence) | 0 |
| S5_extracted_k141_139065 | 3138 | No | NA | 2 | 2 | 0 | Low-quality | Genome-fragment | 8.24 | AAI-based (high-confidence) | 0 |
| S5_vs_k141_143723\|\|full | 3138 | No | NA | 3 | 2 | 0 | Low-quality | Genome-fragment | 3.61 | HMM-based (lower-bound) | 0 |
| S7_extracted_67748 | 3138 | No | NA | 5 | 0 | 2 | Low-quality | Genome-fragment | 4.06 | AAI-based (medium-confidence) | 0 |
| S12_extracted_k141_166519 | 3138 | No | NA | 5 | 0 | 2 | Low-quality | Genome-fragment | 2.92 | AAI-based (high-confidence) | 0 |
| S5_extracted_k141_138490 | 3139 | No | NA | 4 | 3 | 0 | Low-quality | Genome-fragment | 6.83 | AAI-based (high-confidence) | 0 |
| S10_extracted_28 | 1118 | No | NA | 1 | 0 | 0 | Not-determined | Genome-fragment | NA | NA | 0 |
| S10_extracted_49 | 1730 | No | NA | 2 | 0 | 1 | Not-determined | Genome-fragment | NA | NA | 0 |
| S10_extracted_183 | 3626 | No | NA | 2 | 0 | 1 | Not-determined | Genome-fragment | NA | NA | 0 |
| S10_extracted_295 | 1676 | No | NA | 3 | 0 | 1 | Not-determined | Genome-fragment | NA | NA | 0 |
| S10_extracted_312 | 1407 | No | NA | 2 | 0 | 1 | Not-determined | Genome-fragment | NA | NA | 0 |
| S5_vs_k141_132109\|\|full | 3139 | No | NA | 3 | 2 | 0 | Low-quality | Genome-fragment | 5.12 | HMM-based (lower-bound) | 0 |
| S10_extracted_572 | 1503 | No | NA | 2 | 0 | 2 | Not-determined | Genome-fragment | NA | NA | 0 |
| S5_vs_k141_30292\|\|full | 3140 | No | NA | 2 | 0 | 2 | Low-quality | Genome-fragment | 4.52 | AAI-based (medium-confidence) | 0 |
| S10_extracted_836 | 1103 | No | NA | 2 | 0 | 2 | Not-determined | Genome-fragment | NA | NA | 0 |
| S10_extracted_930 | 1770 | No | NA | 1 | 0 | 1 | Not-determined | Genome-fragment | NA | NA | 0 |
| S10_extracted_66338 | 3140 | No | NA | 3 | 0 | 1 | Low-quality | Genome-fragment | 6.2 | AAI-based (medium-confidence) | 0 |
| S10_extracted_1109 | 3218 | No | NA | 4 | 0 | 2 | Not-determined | Genome-fragment | NA | NA | 0 |
| S8_extracted_k141_43036 | 3140 | No | NA | 3 | 0 | 0 | Low-quality | Genome-fragment | 6.22 | AAI-based (high-confidence) | 0 |
| S14_vs_k141_19209\|\|full | 3141 | No | NA | 6 | 0 | 0 | Low-quality | Genome-fragment | 6.6 | AAI-based (high-confidence) | 0 |
| S10_extracted_1321 | 2975 | No | NA | 1 | 0 | 1 | Not-determined | Genome-fragment | NA | NA | 0 |
| S10_extracted_1347 | 1231 | No | NA | 1 | 0 | 1 | Not-determined | Genome-fragment | NA | NA | 0 |
| S5_extracted_k141_98001 | 3141 | No | NA | 3 | 0 | 1 | Low-quality | Genome-fragment | 6.57 | AAI-based (high-confidence) | 0 |
| S10_extracted_1374 | 2039 | No | NA | 2 | 0 | 0 | Not-determined | Genome-fragment | NA | NA | 0 |
| S17_vs_k141_156913\|\|full | 3142 | No | NA | 4 | 0 | 1 | Low-quality | Genome-fragment | 1.51 | AAI-based (medium-confidence) | 0 |
| S10_extracted_1729 | 1003 | No | NA | 1 | 0 | 1 | Not-determined | Genome-fragment | NA | NA | 0 |
| S10_extracted_1892 | 1475 | No | NA | 1 | 0 | 1 | Not-determined | Genome-fragment | NA | NA | 0 |
| S13_vs_3242\|\|full | 3142 | No | NA | 8 | 0 | 0 | Low-quality | Genome-fragment | 2.88 | AAI-based (high-confidence) | 0 |
| S10_extracted_1958 | 1053 | No | NA | 3 | 0 | 0 | Not-determined | Genome-fragment | NA | NA | 0 |
| S10_extracted_1977 | 2239 | No | NA | 4 | 0 | 3 | Not-determined | Genome-fragment | NA | NA | 0 |
| S10_extracted_2016 | 1050 | No | NA | 2 | 0 | 2 | Not-determined | Genome-fragment | NA | NA | 0 |
| S5_vs_k141_26889\|\|full | 3142 | No | NA | 6 | 3 | 0 | Low-quality | Genome-fragment | 8.34 | AAI-based (medium-confidence) | 0 |
| S10_extracted_2157 | 1695 | No | NA | 2 | 0 | 1 | Not-determined | Genome-fragment | NA | NA | 0 |
| S10_vs_52662\|\|full | 3142 | No | NA | 7 | 1 | 0 | Low-quality | Genome-fragment | 7.47 | AAI-based (high-confidence) | 0 |
| S10_extracted_2259 | 1780 | No | NA | 1 | 0 | 1 | Not-determined | Genome-fragment | NA | NA | 0 |
| S10_extracted_2601 | 1074 | No | NA | 2 | 0 | 1 | Not-determined | Genome-fragment | NA | NA | 0 |
| S15_extracted_61334 | 3143 | No | NA | 7 | 2 | 0 | Low-quality | Genome-fragment | 6.91 | AAI-based (high-confidence) | 0 |
| S9_extracted_k141_73489 | 3144 | Yes | 2447 | 5 | 1 | 1 | Low-quality | Genome-fragment | 2.93 | AAI-based (high-confidence) | 22.17 |
| S14_extracted_k141_17386 | 3144 | No | NA | 5 | 3 | 0 | Low-quality | Genome-fragment | 6.65 | AAI-based (high-confidence) | 0 |
| S10_extracted_2972 | 3161 | No | NA | 5 | 0 | 4 | Not-determined | Genome-fragment | NA | NA | 0 |
| S10_extracted_2973 | 1150 | No | NA | 2 | 0 | 1 | Not-determined | Genome-fragment | NA | NA | 0 |
| S16_vs_83829\|\|full | 3144 | No | NA | 4 | 0 | 4 | Low-quality | Genome-fragment | 4.27 | AAI-based (medium-confidence) | 0 |
| S10_extracted_3093 | 1406 | No | NA | 1 | 0 | 1 | Not-determined | Genome-fragment | NA | NA | 0 |
| S9_extracted_k141_32069 | 3145 | No | NA | 7 | 0 | 1 | Low-quality | Genome-fragment | 4.98 | AAI-based (medium-confidence) | 0 |
| S10_extracted_3178 | 1257 | No | NA | 3 | 0 | 0 | Not-determined | Genome-fragment | NA | NA | 0 |
| S10_extracted_3287 | 3577 | No | NA | 3 | 0 | 2 | Not-determined | Genome-fragment | NA | NA | 0 |
| S5_vs_k141_69675\|\|full | 3146 | No | NA | 4 | 0 | 1 | Low-quality | Genome-fragment | 5.5 | AAI-based (medium-confidence) | 0 |
| S10_extracted_3309 | 1468 | No | NA | 3 | 0 | 3 | Not-determined | Genome-fragment | NA | NA | 0 |
| S10_extracted_3439 | 2526 | No | NA | 3 | 0 | 1 | Not-determined | Genome-fragment | NA | NA | 0 |
| S10_vs_1265\|\|full | 3147 | No | NA | 8 | 0 | 0 | Low-quality | Genome-fragment | 7.35 | AAI-based (high-confidence) | 0 |
| S10_extracted_3588 | 10293 | No | NA | 6 | 0 | 3 | Not-determined | Genome-fragment | NA | NA | 0 |
| S10_extracted_3644 | 103067 | No | NA | 92 | 16 | 1 | High-quality | High-quality | 100 | AAI-based (high-confidence) | 0 |
| S10_extracted_3672 | 2046 | No | NA | 2 | 0 | 1 | Not-determined | Genome-fragment | NA | NA | 0 |
| S10_extracted_3811 | 2559 | No | NA | 3 | 0 | 2 | Not-determined | Genome-fragment | NA | NA | 0 |
| S10_extracted_3914 | 2769 | No | NA | 3 | 0 | 3 | Not-determined | Genome-fragment | NA | NA | 0 |
| S10_extracted_4049 | 5097 | No | NA | 5 | 0 | 4 | Not-determined | Genome-fragment | NA | NA | 0 |
| S10_extracted_4087 | 2084 | No | NA | 3 | 0 | 2 | Not-determined | Genome-fragment | NA | NA | 0 |
| S10_extracted_4183 | 3351 | No | NA | 2 | 0 | 1 | Not-determined | Genome-fragment | NA | NA | 0 |
| S10_extracted_4229 | 6376 | No | NA | 3 | 0 | 2 | Not-determined | Genome-fragment | NA | NA | 0 |
| S10_extracted_4319 | 1227 | No | NA | 2 | 0 | 2 | Not-determined | Genome-fragment | NA | NA | 0 |
| S8_vs_k141_21850\|\|full | 3147 | No | NA | 6 | 0 | 0 | Low-quality | Genome-fragment | 0.76 | AAI-based (medium-confidence) | 0 |
| S10_extracted_4381 | 1392 | No | NA | 2 | 0 | 1 | Not-determined | Genome-fragment | NA | NA | 0 |
| S1_extracted_k141_10970 | 3148 | No | NA | 2 | 0 | 2 | Low-quality | Genome-fragment | 0.46 | AAI-based (medium-confidence) | 0 |
| S10_extracted_4473 | 1234 | No | NA | 3 | 0 | 2 | Not-determined | Genome-fragment | NA | NA | 0 |
| S3_vs_k141_98955\|\|full | 3148 | No | NA | 7 | 1 | 0 | Low-quality | Genome-fragment | 9.56 | AAI-based (high-confidence) | 0 |
| S10_extracted_4621 | 1729 | No | NA | 2 | 0 | 2 | Not-determined | Genome-fragment | NA | NA | 0 |
| S10_extracted_4967 | 1299 | No | NA | 1 | 0 | 0 | Not-determined | Genome-fragment | NA | NA | 0 |
| S10_extracted_4969 | 1192 | No | NA | 1 | 0 | 1 | Not-determined | Genome-fragment | NA | NA | 0 |
| S10_extracted_5005 | 2249 | No | NA | 6 | 0 | 0 | Not-determined | Genome-fragment | NA | NA | 0 |
| S10_extracted_5017 | 1347 | No | NA | 2 | 0 | 1 | Not-determined | Genome-fragment | NA | NA | 0 |
| S10_extracted_5056 | 1700 | No | NA | 2 | 0 | 1 | Not-determined | Genome-fragment | NA | NA | 0 |
| S10_extracted_5122 | 5275 | No | NA | 6 | 0 | 5 | Not-determined | Genome-fragment | NA | NA | 0 |
| S10_extracted_5129 | 4375 | No | NA | 4 | 0 | 2 | Not-determined | Genome-fragment | NA | NA | 0 |
| S10_extracted_5149 | 1161 | No | NA | 2 | 0 | 0 | Not-determined | Genome-fragment | NA | NA | 0 |
| S10_extracted_5199 | 1044 | No | NA | 1 | 0 | 1 | Not-determined | Genome-fragment | NA | NA | 0 |
| S10_extracted_5207 | 7388 | No | NA | 10 | 0 | 7 | Not-determined | Genome-fragment | NA | NA | 0 |
| S10_extracted_5208 | 8663 | No | NA | 6 | 0 | 3 | Not-determined | Genome-fragment | NA | NA | 0 |
| S10_extracted_5222 | 2807 | No | NA | 3 | 0 | 0 | Not-determined | Genome-fragment | NA | NA | 0 |
| S10_extracted_5352 | 1285 | No | NA | 1 | 0 | 1 | Not-determined | Genome-fragment | NA | NA | 0 |
| S10_extracted_5394 | 3444 | No | NA | 2 | 0 | 2 | Not-determined | Genome-fragment | NA | NA | 0 |
| S10_extracted_5401 | 1110 | No | NA | 2 | 0 | 2 | Not-determined | Genome-fragment | NA | NA | 0 |
| S10_extracted_5403 | 7616 | No | NA | 4 | 0 | 4 | Not-determined | Genome-fragment | NA | NA | 0 |
| S10_extracted_5411 | 5802 | No | NA | 7 | 0 | 4 | Not-determined | Genome-fragment | NA | NA | 0 |
| S7_vs_93096\|\|full | 3148 | No | NA | 3 | 3 | 0 | Low-quality | Genome-fragment | 4.84 | AAI-based (high-confidence) | 0 |
| S16_extracted_93048 | 3148 | No | NA | 8 | 2 | 0 | Low-quality | Genome-fragment | 7.56 | AAI-based (high-confidence) | 0 |
| S10_extracted_5506 | 3036 | No | NA | 5 | 0 | 1 | Not-determined | Genome-fragment | NA | NA | 0 |
| S16_extracted_96513 | 3148 | No | NA | 3 | 0 | 3 | Low-quality | Genome-fragment | 1.16 | AAI-based (medium-confidence) | 0 |
| S8_vs_k141_20267\|\|full | 3148 | No | NA | 4 | 0 | 0 | Low-quality | Genome-fragment | 8.78 | AAI-based (high-confidence) | 0 |
| S10_extracted_5667 | 1790 | No | NA | 2 | 0 | 2 | Not-determined | Genome-fragment | NA | NA | 0 |
| S10_extracted_5727 | 1092 | No | NA | 2 | 0 | 1 | Not-determined | Genome-fragment | NA | NA | 0 |
| S10_extracted_5762 | 5404 | No | NA | 4 | 0 | 4 | Not-determined | Genome-fragment | NA | NA | 0 |
| S10_extracted_5777 | 1281 | No | NA | 1 | 0 | 0 | Not-determined | Genome-fragment | NA | NA | 0 |
| S10_extracted_5877 | 3370 | No | NA | 3 | 0 | 1 | Not-determined | Genome-fragment | NA | NA | 0 |
| S10_extracted_5920 | 3343 | No | NA | 3 | 0 | 1 | Not-determined | Genome-fragment | NA | NA | 0 |
| S10_extracted_5938 | 1321 | No | NA | 4 | 0 | 0 | Not-determined | Genome-fragment | NA | NA | 0 |
| S10_extracted_6050 | 3063 | No | NA | 2 | 0 | 1 | Not-determined | Genome-fragment | NA | NA | 0 |
| S10_extracted_6231 | 2408 | No | NA | 4 | 0 | 0 | Not-determined | Genome-fragment | NA | NA | 0 |
| S9_extracted_k141_66769 | 3149 | No | NA | 5 | 0 | 0 | Low-quality | Genome-fragment | 1.86 | AAI-based (high-confidence) | 0 |
| S14_vs_k141_73896\|\|full | 3149 | No | NA | 7 | 3 | 0 | Low-quality | Genome-fragment | 7.59 | AAI-based (high-confidence) | 0 |
| S7_extracted_136128 | 3149 | No | NA | 4 | 0 | 1 | Low-quality | Genome-fragment | 6.38 | AAI-based (high-confidence) | 0 |
| S10_extracted_6430 | 1069 | No | NA | 1 | 0 | 0 | Not-determined | Genome-fragment | NA | NA | 0 |
| S10_vs_20544\|\|full | 3149 | No | NA | 7 | 1 | 0 | Low-quality | Genome-fragment | 6.31 | HMM-based (lower-bound) | 0 |
| S10_extracted_6491 | 3187 | No | NA | 9 | 0 | 1 | Not-determined | Genome-fragment | NA | NA | 0 |
| S10_extracted_6498 | 3444 | No | NA | 3 | 0 | 2 | Not-determined | Genome-fragment | NA | NA | 0 |
| S10_extracted_6603 | 3728 | No | NA | 2 | 0 | 2 | Not-determined | Genome-fragment | NA | NA | 0 |
| S10_extracted_6758 | 6259 | No | NA | 4 | 0 | 4 | Not-determined | Genome-fragment | NA | NA | 0 |
| S10_extracted_6765 | 1727 | No | NA | 2 | 0 | 0 | Not-determined | Genome-fragment | NA | NA | 0 |
| S12_extracted_k141_34743 | 3149 | No | NA | 7 | 0 | 4 | Low-quality | Genome-fragment | 4.59 | AAI-based (medium-confidence) | 0 |
| S10_extracted_6851 | 7908 | No | NA | 9 | 0 | 2 | Not-determined | Genome-fragment | NA | NA | 0 |
| S10_extracted_6886 | 1639 | No | NA | 4 | 0 | 3 | Not-determined | Genome-fragment | NA | NA | 0 |
| S10_extracted_7024 | 1495 | No | NA | 1 | 0 | 0 | Not-determined | Genome-fragment | NA | NA | 0 |
| S10_extracted_7056 | 1437 | No | NA | 2 | 0 | 1 | Not-determined | Genome-fragment | NA | NA | 0 |
| S10_extracted_7269 | 1062 | No | NA | 1 | 0 | 0 | Not-determined | Genome-fragment | NA | NA | 0 |
| S10_extracted_7302 | 1408 | No | NA | 3 | 0 | 1 | Not-determined | Genome-fragment | NA | NA | 0 |
| S10_extracted_7329 | 2003 | No | NA | 4 | 0 | 1 | Not-determined | Genome-fragment | NA | NA | 0 |
| S10_extracted_7336 | 1092 | No | NA | 1 | 0 | 0 | Not-determined | Genome-fragment | NA | NA | 0 |
| S11_vs_k141_69339\|\|full | 3150 | No | NA | 10 | 1 | 0 | Low-quality | Genome-fragment | 3.44 | HMM-based (lower-bound) | 0 |
| S5_vs_k141_70288\|\|full | 3150 | No | NA | 8 | 2 | 0 | Low-quality | Genome-fragment | 7.49 | AAI-based (medium-confidence) | 0 |
| S8_extracted_k141_201134 | 3150 | No | NA | 3 | 0 | 1 | Low-quality | Genome-fragment | 0.88 | AAI-based (high-confidence) | 0 |
| S10_extracted_7509 | 4619 | No | NA | 7 | 0 | 2 | Not-determined | Genome-fragment | NA | NA | 0 |
| S10_extracted_7644 | 1001 | No | NA | 1 | 0 | 1 | Not-determined | Genome-fragment | NA | NA | 0 |
| S10_extracted_7665 | 4415 | No | NA | 5 | 0 | 2 | Not-determined | Genome-fragment | NA | NA | 0 |
| S10_extracted_7782 | 10421 | No | NA | 8 | 0 | 5 | Not-determined | Genome-fragment | NA | NA | 0 |
| S10_extracted_8118 | 3169 | No | NA | 3 | 0 | 1 | Not-determined | Genome-fragment | NA | NA | 0 |
| S10_vs_25453\|\|full | 3151 | No | NA | 4 | 2 | 0 | Low-quality | Genome-fragment | 7.95 | AAI-based (high-confidence) | 0 |
| S10_extracted_8318 | 1008 | No | NA | 1 | 0 | 1 | Not-determined | Genome-fragment | NA | NA | 0 |
| S8_extracted_k141_182540 | 3151 | No | NA | 8 | 0 | 0 | Low-quality | Genome-fragment | 3.51 | AAI-based (high-confidence) | 0 |
| S10_extracted_8552 | 1046 | No | NA | 2 | 0 | 1 | Not-determined | Genome-fragment | NA | NA | 0 |
| S1_extracted_k141_89033 | 3152 | No | NA | 3 | 0 | 2 | Low-quality | Genome-fragment | 5.5 | AAI-based (high-confidence) | 0 |
| S10_extracted_8579 | 1875 | No | NA | 2 | 0 | 0 | Not-determined | Genome-fragment | NA | NA | 0 |
| S1_vs_k141_17797\|\|full | 3152 | No | NA | 4 | 3 | 0 | Low-quality | Genome-fragment | 7.38 | AAI-based (high-confidence) | 0 |
| S15_extracted_7265 | 3152 | No | NA | 3 | 1 | 0 | Low-quality | Genome-fragment | 2.31 | AAI-based (medium-confidence) | 0 |
| S10_extracted_8799 | 5133 | No | NA | 6 | 0 | 0 | Not-determined | Genome-fragment | NA | NA | 0 |
| S10_extracted_8833 | 1957 | No | NA | 2 | 0 | 0 | Not-determined | Genome-fragment | NA | NA | 0 |
| S10_extracted_8860 | 3278 | No | NA | 3 | 0 | 0 | Not-determined | Genome-fragment | NA | NA | 0 |
| S8_vs_k141_52292\|\|full | 3152 | No | NA | 4 | 0 | 0 | Low-quality | Genome-fragment | 5.97 | AAI-based (medium-confidence) | 0 |
| S16_extracted_136639 | 3153 | No | NA | 2 | 0 | 2 | Low-quality | Genome-fragment | 8.89 | AAI-based (medium-confidence) | 0 |
| S10_extracted_28259 | 3153 | No | NA | 3 | 0 | 3 | Low-quality | Genome-fragment | 0.85 | AAI-based (medium-confidence) | 0 |
| S10_extracted_9046 | 1081 | No | NA | 1 | 0 | 1 | Not-determined | Genome-fragment | NA | NA | 0 |
| S10_extracted_9105 | 1086 | No | NA | 2 | 0 | 0 | Not-determined | Genome-fragment | NA | NA | 0 |
| S10_extracted_9144 | 1171 | No | NA | 1 | 0 | 0 | Not-determined | Genome-fragment | NA | NA | 0 |
| S11_extracted_k141_48868 | 3154 | No | NA | 8 | 2 | 0 | Low-quality | Genome-fragment | 6.47 | AAI-based (high-confidence) | 0 |
| S3_vs_k141_19857\|\|full | 3155 | No | NA | 5 | 1 | 0 | Low-quality | Genome-fragment | 2.06 | AAI-based (high-confidence) | 0 |
| S5_vs_k141_36135\|\|full | 3155 | No | NA | 4 | 0 | 2 | Low-quality | Genome-fragment | 2.85 | AAI-based (medium-confidence) | 0 |
| S10_extracted_9452 | 1042 | No | NA | 2 | 0 | 1 | Not-determined | Genome-fragment | NA | NA | 0 |
| S10_extracted_9592 | 4273 | No | NA | 7 | 0 | 0 | Not-determined | Genome-fragment | NA | NA | 0 |
| S10_extracted_9613 | 1078 | No | NA | 1 | 0 | 0 | Not-determined | Genome-fragment | NA | NA | 0 |
| S16_vs_170123\|\|full | 3155 | No | NA | 5 | 0 | 3 | Low-quality | Genome-fragment | 3.98 | AAI-based (medium-confidence) | 0 |
| S10_extracted_9670 | 3862 | No | NA | 3 | 0 | 2 | Not-determined | Genome-fragment | NA | NA | 0 |
| S10_extracted_9703 | 1410 | No | NA | 2 | 0 | 1 | Not-determined | Genome-fragment | NA | NA | 0 |
| S10_extracted_9709 | 1072 | No | NA | 1 | 0 | 1 | Not-determined | Genome-fragment | NA | NA | 0 |
| S10_extracted_9780 | 3103 | No | NA | 3 | 0 | 0 | Not-determined | Genome-fragment | NA | NA | 0 |
| S1_extracted_k141_98135 | 3156 | No | NA | 5 | 1 | 0 | Low-quality | Genome-fragment | 5.43 | AAI-based (high-confidence) | 0 |
| S10_extracted_9898 | 1149 | No | NA | 2 | 0 | 1 | Not-determined | Genome-fragment | NA | NA | 0 |
| S10_extracted_10031 | 2068 | No | NA | 3 | 0 | 1 | Not-determined | Genome-fragment | NA | NA | 0 |
| S10_extracted_10138 | 1786 | No | NA | 1 | 0 | 0 | Not-determined | Genome-fragment | NA | NA | 0 |
| S10_extracted_10243 | 1234 | No | NA | 1 | 0 | 0 | Not-determined | Genome-fragment | NA | NA | 0 |
| S10_extracted_10401 | 1259 | No | NA | 2 | 0 | 0 | Not-determined | Genome-fragment | NA | NA | 0 |
| S10_extracted_10417 | 3138 | No | NA | 1 | 0 | 0 | Not-determined | Genome-fragment | NA | NA | 0 |
| S10_extracted_10503 | 3368 | No | NA | 3 | 0 | 3 | Not-determined | Genome-fragment | NA | NA | 0 |
| S10_extracted_10518 | 1448 | No | NA | 1 | 0 | 1 | Not-determined | Genome-fragment | NA | NA | 0 |
| S10_extracted_10521 | 2495 | No | NA | 4 | 0 | 3 | Not-determined | Genome-fragment | NA | NA | 0 |
| S10_extracted_10656 | 1212 | No | NA | 2 | 0 | 0 | Not-determined | Genome-fragment | NA | NA | 0 |
| S3_vs_k141_90791\|\|full | 3156 | No | NA | 4 | 0 | 1 | Low-quality | Genome-fragment | 2.26 | AAI-based (high-confidence) | 0 |
| S10_extracted_10879 | 2439 | No | NA | 2 | 0 | 0 | Not-determined | Genome-fragment | NA | NA | 0 |
| S10_extracted_10895 | 1233 | No | NA | 1 | 0 | 1 | Not-determined | Genome-fragment | NA | NA | 0 |
| S10_extracted_10904 | 1094 | No | NA | 2 | 0 | 2 | Not-determined | Genome-fragment | NA | NA | 0 |
| S10_extracted_11076 | 2353 | No | NA | 3 | 0 | 3 | Not-determined | Genome-fragment | NA | NA | 0 |
| S10_extracted_11128 | 2219 | No | NA | 2 | 0 | 1 | Not-determined | Genome-fragment | NA | NA | 0 |
| S10_extracted_11232 | 1231 | No | NA | 1 | 0 | 1 | Not-determined | Genome-fragment | NA | NA | 0 |
| S10_extracted_11283 | 1477 | No | NA | 1 | 0 | 1 | Not-determined | Genome-fragment | NA | NA | 0 |
| S12_extracted_k141_56430 | 3156 | No | NA | 6 | 0 | 6 | Low-quality | Genome-fragment | 1.02 | AAI-based (medium-confidence) | 0 |
| S10_extracted_11342 | 1141 | No | NA | 2 | 0 | 2 | Not-determined | Genome-fragment | NA | NA | 0 |
| S10_extracted_11357 | 4068 | No | NA | 6 | 0 | 6 | Not-determined | Genome-fragment | NA | NA | 0 |
| S10_extracted_11375 | 1315 | No | NA | 2 | 0 | 1 | Not-determined | Genome-fragment | NA | NA | 0 |
| S8_extracted_k141_17217 | 3156 | No | NA | 4 | 0 | 1 | Low-quality | Genome-fragment | 1.71 | AAI-based (medium-confidence) | 0 |
| S7_extracted_76819 | 3157 | No | NA | 4 | 1 | 0 | Low-quality | Genome-fragment | 2.03 | AAI-based (medium-confidence) | 0 |
| S10_extracted_11550 | 1003 | No | NA | 1 | 0 | 1 | Not-determined | Genome-fragment | NA | NA | 0 |
| S10_extracted_11627 | 1332 | No | NA | 1 | 0 | 0 | Not-determined | Genome-fragment | NA | NA | 0 |
| S10_extracted_11715 | 2369 | No | NA | 4 | 0 | 4 | Not-determined | Genome-fragment | NA | NA | 0 |
| S7_vs_21208\|\|full | 3157 | No | NA | 3 | 0 | 0 | Low-quality | Genome-fragment | 8.65 | AAI-based (medium-confidence) | 0 |
| S10_extracted_11815 | 3331 | No | NA | 5 | 0 | 3 | Not-determined | Genome-fragment | NA | NA | 0 |
| S10_extracted_11901 | 1843 | No | NA | 2 | 0 | 1 | Not-determined | Genome-fragment | NA | NA | 0 |
| S7_vs_29349\|\|full | 3157 | No | NA | 9 | 2 | 0 | Low-quality | Genome-fragment | 7.74 | AAI-based (medium-confidence) | 0 |
| S10_extracted_12002 | 1477 | No | NA | 2 | 0 | 1 | Not-determined | Genome-fragment | NA | NA | 0 |
| S10_extracted_12047 | 1845 | No | NA | 3 | 0 | 3 | Not-determined | Genome-fragment | NA | NA | 0 |
| S10_extracted_12092 | 1049 | No | NA | 2 | 0 | 1 | Not-determined | Genome-fragment | NA | NA | 0 |
| S12_extracted_k141_179654 | 3157 | No | NA | 8 | 2 | 0 | Low-quality | Genome-fragment | 2.9 | AAI-based (high-confidence) | 0 |
| S10_extracted_12142 | 8250 | No | NA | 6 | 0 | 4 | Not-determined | Genome-fragment | NA | NA | 0 |
| S12_vs_k141_38004\|\|full | 3157 | No | NA | 7 | 3 | 0 | Low-quality | Genome-fragment | 7.24 | AAI-based (medium-confidence) | 0 |
| S10_extracted_12348 | 4294 | No | NA | 4 | 0 | 3 | Not-determined | Genome-fragment | NA | NA | 0 |
| S10_extracted_12418 | 6518 | No | NA | 9 | 0 | 0 | Not-determined | Genome-fragment | NA | NA | 0 |
| S10_extracted_12520 | 1273 | No | NA | 2 | 0 | 1 | Not-determined | Genome-fragment | NA | NA | 0 |
| S10_extracted_12536 | 1062 | No | NA | 1 | 0 | 1 | Not-determined | Genome-fragment | NA | NA | 0 |
| S10_extracted_12710 | 8543 | No | NA | 5 | 0 | 4 | Not-determined | Genome-fragment | NA | NA | 0 |
| S10_extracted_12739 | 1081 | No | NA | 1 | 0 | 0 | Not-determined | Genome-fragment | NA | NA | 0 |
| S8_vs_k141_132076\|\|full | 3158 | No | NA | 6 | 1 | 0 | Low-quality | Genome-fragment | 6.93 | AAI-based (high-confidence) | 0 |
| S10_extracted_12954 | 5611 | No | NA | 11 | 0 | 10 | Not-determined | Genome-fragment | NA | NA | 0 |
| S10_extracted_13043 | 1881 | No | NA | 2 | 0 | 2 | Not-determined | Genome-fragment | NA | NA | 0 |
| S9_vs_k141_81015\|\|full | 3160 | No | NA | 3 | 3 | 0 | Low-quality | Genome-fragment | 8.3 | AAI-based (high-confidence) | 0 |
| S14_vs_k141_109138\|\|full | 3160 | Yes | 2815 | 3 | 1 | 1 | Low-quality | Genome-fragment | 1.65 | AAI-based (high-confidence) | 10.92 |
| S16_extracted_16822 | 3160 | No | NA | 3 | 0 | 0 | Low-quality | Genome-fragment | 4.76 | AAI-based (high-confidence) | 0 |
| S16_extracted_151733 | 3160 | No | NA | 4 | 1 | 0 | Low-quality | Genome-fragment | 7.98 | AAI-based (high-confidence) | 0 |
| S3_extracted_k141_113747 | 3161 | No | NA | 7 | 1 | 0 | Low-quality | Genome-fragment | 5.52 | AAI-based (high-confidence) | 0 |
| S16_extracted_8304 | 3161 | No | NA | 8 | 0 | 0 | Low-quality | Genome-fragment | 2.9 | AAI-based (high-confidence) | 0 |
| S10_extracted_13229 | 1830 | No | NA | 4 | 0 | 2 | Not-determined | Genome-fragment | NA | NA | 0 |
| S10_extracted_13233 | 1589 | No | NA | 2 | 0 | 0 | Not-determined | Genome-fragment | NA | NA | 0 |
| S10_extracted_13328 | 1207 | No | NA | 2 | 0 | 1 | Not-determined | Genome-fragment | NA | NA | 0 |
| S16_extracted_104479 | 3161 | No | NA | 3 | 0 | 3 | Low-quality | Genome-fragment | 0.96 | AAI-based (medium-confidence) | 0 |
| S16_vs_110252\|\|full | 3161 | No | NA | 7 | 3 | 0 | Low-quality | Genome-fragment | 6.33 | AAI-based (medium-confidence) | 0 |
| S10_extracted_13466 | 1004 | No | NA | 1 | 0 | 1 | Not-determined | Genome-fragment | NA | NA | 0 |
| S10_extracted_13516 | 1725 | No | NA | 2 | 0 | 2 | Not-determined | Genome-fragment | NA | NA | 0 |
| S10_extracted_13578 | 10102 | No | NA | 12 | 0 | 5 | Not-determined | Genome-fragment | NA | NA | 0 |
| S10_extracted_13657 | 6039 | No | NA | 4 | 0 | 2 | Not-determined | Genome-fragment | NA | NA | 0 |
| S10_extracted_13677 | 4163 | No | NA | 4 | 0 | 2 | Not-determined | Genome-fragment | NA | NA | 0 |
| S10_extracted_13760 | 1324 | No | NA | 2 | 0 | 1 | Not-determined | Genome-fragment | NA | NA | 0 |
| S10_extracted_13812 | 1113 | No | NA | 3 | 0 | 1 | Not-determined | Genome-fragment | NA | NA | 0 |
| S10_extracted_13913 | 1595 | No | NA | 2 | 0 | 2 | Not-determined | Genome-fragment | NA | NA | 0 |
| S5_vs_k141_80514\|\|full | 3162 | No | NA | 4 | 0 | 2 | Low-quality | Genome-fragment | 1.65 | AAI-based (medium-confidence) | 0 |
| S14_vs_k141_46048\|\|full | 3164 | No | NA | 6 | 0 | 4 | Low-quality | Genome-fragment | 0.84 | AAI-based (medium-confidence) | 0 |
| S10_extracted_14049 | 1368 | No | NA | 2 | 0 | 0 | Not-determined | Genome-fragment | NA | NA | 0 |
| S10_extracted_14072 | 2674 | No | NA | 1 | 0 | 1 | Not-determined | Genome-fragment | NA | NA | 0 |
| S10_extracted_14330 | 2418 | No | NA | 1 | 0 | 1 | Not-determined | Genome-fragment | NA | NA | 0 |
| S10_extracted_14359 | 1903 | No | NA | 2 | 0 | 1 | Not-determined | Genome-fragment | NA | NA | 0 |
| S10_extracted_14464 | 5046 | No | NA | 7 | 0 | 2 | Not-determined | Genome-fragment | NA | NA | 0 |
| S14_vs_k141_11132\|\|full | 3164 | No | NA | 5 | 0 | 1 | Low-quality | Genome-fragment | 11.19 | AAI-based (medium-confidence) | 0 |
| S10_extracted_14481 | 5969 | No | NA | 3 | 0 | 3 | Not-determined | Genome-fragment | NA | NA | 0 |
| S10_extracted_14670 | 1278 | No | NA | 2 | 0 | 2 | Not-determined | Genome-fragment | NA | NA | 0 |
| S1_vs_k141_86314\|\|full | 3164 | No | NA | 3 | 2 | 0 | Low-quality | Genome-fragment | 7.63 | AAI-based (high-confidence) | 0 |
| S10_extracted_14820 | 1228 | No | NA | 2 | 0 | 2 | Not-determined | Genome-fragment | NA | NA | 0 |
| S10_extracted_14882 | 2807 | No | NA | 3 | 0 | 1 | Not-determined | Genome-fragment | NA | NA | 0 |
| S10_extracted_15147 | 1022 | No | NA | 1 | 0 | 0 | Not-determined | Genome-fragment | NA | NA | 0 |
| S10_extracted_15166 | 3956 | No | NA | 8 | 0 | 7 | Not-determined | Genome-fragment | NA | NA | 0 |
| S10_extracted_15309 | 1082 | No | NA | 1 | 0 | 0 | Not-determined | Genome-fragment | NA | NA | 0 |
| S7_extracted_18708 | 3164 | No | NA | 6 | 0 | 0 | Low-quality | Genome-fragment | 6.69 | AAI-based (medium-confidence) | 0 |
| S10_extracted_15412 | 1514 | No | NA | 2 | 0 | 0 | Not-determined | Genome-fragment | NA | NA | 0 |
| S16_extracted_146127 | 3164 | No | NA | 4 | 0 | 2 | Low-quality | Genome-fragment | 7.33 | AAI-based (medium-confidence) | 0 |
| S10_extracted_15525 | 1320 | No | NA | 1 | 0 | 0 | Not-determined | Genome-fragment | NA | NA | 0 |
| S10_extracted_15558 | 2463 | No | NA | 3 | 0 | 2 | Not-determined | Genome-fragment | NA | NA | 0 |
| S10_extracted_15695 | 2155 | No | NA | 2 | 0 | 1 | Not-determined | Genome-fragment | NA | NA | 0 |
| S8_extracted_k141_84852 | 3164 | No | NA | 7 | 0 | 0 | Low-quality | Genome-fragment | 5.75 | AAI-based (high-confidence) | 0 |
| S10_extracted_15882 | 1081 | No | NA | 1 | 0 | 1 | Not-determined | Genome-fragment | NA | NA | 0 |
| S5_vs_k141_91708\|\|full | 3165 | No | NA | 3 | 2 | 0 | Low-quality | Genome-fragment | 5.55 | HMM-based (lower-bound) | 0 |
| S16_vs_150782\|\|full | 3165 | No | NA | 3 | 0 | 2 | Low-quality | Genome-fragment | 2.62 | AAI-based (medium-confidence) | 0 |
| S10_extracted_15997 | 1289 | No | NA | 2 | 0 | 0 | Not-determined | Genome-fragment | NA | NA | 0 |
| S6_vs_k141_32313\|\|full | 3165 | No | NA | 11 | 2 | 0 | Low-quality | Genome-fragment | 7.05 | AAI-based (medium-confidence) | 0 |
| S10_extracted_16164 | 1099 | No | NA | 1 | 0 | 0 | Not-determined | Genome-fragment | NA | NA | 0 |
| S10_extracted_16314 | 1027 | No | NA | 2 | 0 | 0 | Not-determined | Genome-fragment | NA | NA | 0 |
| S11_vs_k141_79453\|\|full | 3166 | No | NA | 5 | 2 | 0 | Low-quality | Genome-fragment | 4.63 | AAI-based (medium-confidence) | 0 |
| S8_vs_k141_81043\|\|full | 3166 | Yes | 1033 | 3 | 1 | 1 | Low-quality | Genome-fragment | 1.61 | HMM-based (lower-bound) | 67.37 |
| S10_extracted_16652 | 1047 | No | NA | 1 | 0 | 0 | Not-determined | Genome-fragment | NA | NA | 0 |
| S11_vs_k141_30770\|\|full | 3167 | No | NA | 4 | 0 | 0 | Low-quality | Genome-fragment | 6.27 | AAI-based (medium-confidence) | 0 |
| S7_vs_11701\|\|full | 3167 | No | NA | 6 | 1 | 0 | Low-quality | Genome-fragment | 2.27 | HMM-based (lower-bound) | 0 |
| S10_extracted_16871 | 4350 | No | NA | 3 | 0 | 3 | Not-determined | Genome-fragment | NA | NA | 0 |
| S9_extracted_k141_81005 | 3169 | No | NA | 7 | 1 | 0 | Low-quality | Genome-fragment | 7.86 | AAI-based (high-confidence) | 0 |
| S10_extracted_17020 | 5441 | No | NA | 3 | 0 | 2 | Not-determined | Genome-fragment | NA | NA | 0 |
| S10_extracted_17108 | 1367 | No | NA | 2 | 0 | 0 | Not-determined | Genome-fragment | NA | NA | 0 |
| S4_extracted_13223 | 3169 | No | NA | 8 | 0 | 0 | Low-quality | Genome-fragment | 1.74 | AAI-based (medium-confidence) | 0 |
| S10_extracted_17274 | 2368 | No | NA | 1 | 0 | 1 | Not-determined | Genome-fragment | NA | NA | 0 |
| S10_extracted_17465 | 3124 | No | NA | 5 | 0 | 1 | Not-determined | Genome-fragment | NA | NA | 0 |
| S10_extracted_17493 | 6843 | No | NA | 7 | 0 | 3 | Not-determined | Genome-fragment | NA | NA | 0 |
| S10_extracted_17543 | 1549 | No | NA | 1 | 0 | 0 | Not-determined | Genome-fragment | NA | NA | 0 |
| S10_extracted_17634 | 1458 | No | NA | 1 | 0 | 1 | Not-determined | Genome-fragment | NA | NA | 0 |
| S13_vs_113527\|\|full | 3171 | No | NA | 3 | 0 | 2 | Low-quality | Genome-fragment | 2.91 | AAI-based (medium-confidence) | 0 |
| S10_extracted_17722 | 1882 | No | NA | 5 | 0 | 2 | Not-determined | Genome-fragment | NA | NA | 0 |
| S10_extracted_17756 | 1916 | No | NA | 2 | 0 | 0 | Not-determined | Genome-fragment | NA | NA | 0 |
| S5_extracted_k141_129061 | 3172 | No | NA | 2 | 0 | 2 | Low-quality | Genome-fragment | 1.01 | AAI-based (medium-confidence) | 0 |
| S10_extracted_17858 | 3115 | No | NA | 1 | 0 | 1 | Not-determined | Genome-fragment | NA | NA | 0 |
| S10_extracted_17941 | 1077 | No | NA | 3 | 0 | 2 | Not-determined | Genome-fragment | NA | NA | 0 |
| S10_extracted_18024 | 3481 | No | NA | 3 | 0 | 2 | Not-determined | Genome-fragment | NA | NA | 0 |
| S10_vs_19260\|\|full | 3172 | No | NA | 3 | 0 | 1 | Low-quality | Genome-fragment | 0.54 | AAI-based (medium-confidence) | 0 |
| S10_extracted_18168 | 1074 | No | NA | 1 | 0 | 0 | Not-determined | Genome-fragment | NA | NA | 0 |
| S10_extracted_18193 | 1276 | No | NA | 2 | 0 | 1 | Not-determined | Genome-fragment | NA | NA | 0 |
| S10_extracted_18237 | 1897 | No | NA | 2 | 0 | 2 | Not-determined | Genome-fragment | NA | NA | 0 |
| S10_extracted_18472 | 1013 | No | NA | 1 | 0 | 0 | Not-determined | Genome-fragment | NA | NA | 0 |
| S10_extracted_18502 | 1084 | No | NA | 1 | 0 | 1 | Not-determined | Genome-fragment | NA | NA | 0 |
| S10_extracted_18513 | 3473 | No | NA | 1 | 0 | 0 | Not-determined | Genome-fragment | NA | NA | 0 |
| S10_extracted_18579 | 9215 | No | NA | 13 | 0 | 3 | Not-determined | Genome-fragment | NA | NA | 0 |
| S10_extracted_18621 | 1894 | No | NA | 3 | 0 | 2 | Not-determined | Genome-fragment | NA | NA | 0 |
| S10_extracted_18671 | 44897 | No | NA | 59 | 12 | 1 | Complete | High-quality | 100 | DTR (high-confidence) | 0 |
| S12_extracted_k141_142216 | 3172 | No | NA | 2 | 0 | 2 | Low-quality | Genome-fragment | 1.4 | AAI-based (high-confidence) | 0 |
| S10_extracted_18692 | 1681 | No | NA | 2 | 0 | 0 | Not-determined | Genome-fragment | NA | NA | 0 |
| S12_vs_k141_143629\|\|full | 3173 | Yes | 1374 | 3 | 1 | 1 | Low-quality | Genome-fragment | 0.94 | HMM-based (lower-bound) | 56.7 |
| S10_extracted_18767 | 3357 | No | NA | 2 | 0 | 2 | Not-determined | Genome-fragment | NA | NA | 0 |
| S10_extracted_18782 | 1347 | No | NA | 2 | 0 | 2 | Not-determined | Genome-fragment | NA | NA | 0 |
| S10_extracted_18810 | 3628 | No | NA | 6 | 0 | 1 | Not-determined | Genome-fragment | NA | NA | 0 |
| S10_extracted_18909 | 1486 | No | NA | 2 | 0 | 2 | Not-determined | Genome-fragment | NA | NA | 0 |
| S6_extracted_k141_18954 | 3173 | No | NA | 4 | 2 | 0 | Low-quality | Genome-fragment | 3.41 | AAI-based (high-confidence) | 0 |
| S10_extracted_18945 | 5668 | No | NA | 4 | 0 | 4 | Not-determined | Genome-fragment | NA | NA | 0 |
| S6_extracted_k141_45095 | 3173 | No | NA | 3 | 0 | 0 | Low-quality | Genome-fragment | 4.79 | AAI-based (medium-confidence) | 0 |
| S10_extracted_19039 | 1227 | No | NA | 3 | 0 | 0 | Not-determined | Genome-fragment | NA | NA | 0 |
| S10_extracted_19062 | 1051 | No | NA | 3 | 0 | 0 | Not-determined | Genome-fragment | NA | NA | 0 |
| S10_extracted_19201 | 1214 | No | NA | 2 | 0 | 2 | Not-determined | Genome-fragment | NA | NA | 0 |
| S10_extracted_19225 | 1222 | No | NA | 1 | 0 | 0 | Not-determined | Genome-fragment | NA | NA | 0 |
| S7_extracted_28931 | 3174 | No | NA | 3 | 0 | 1 | Low-quality | Genome-fragment | 8.13 | AAI-based (high-confidence) | 0 |
| S10_extracted_19299 | 3696 | No | NA | 4 | 0 | 1 | Not-determined | Genome-fragment | NA | NA | 0 |
| S10_extracted_19319 | 1831 | No | NA | 2 | 0 | 0 | Not-determined | Genome-fragment | NA | NA | 0 |
| S16_vs_16690\|\|full | 3174 | No | NA | 4 | 2 | 0 | Low-quality | Genome-fragment | 8.77 | AAI-based (high-confidence) | 0 |
| S10_extracted_19564 | 2262 | No | NA | 1 | 0 | 0 | Not-determined | Genome-fragment | NA | NA | 0 |
| S10_extracted_19623 | 1521 | No | NA | 3 | 0 | 3 | Not-determined | Genome-fragment | NA | NA | 0 |
| S10_extracted_19698 | 3350 | No | NA | 3 | 0 | 2 | Not-determined | Genome-fragment | NA | NA | 0 |
| S10_extracted_19720 | 1632 | No | NA | 2 | 0 | 0 | Not-determined | Genome-fragment | NA | NA | 0 |
| S10_extracted_19762 | 1664 | No | NA | 3 | 0 | 3 | Not-determined | Genome-fragment | NA | NA | 0 |
| S10_extracted_19764 | 1016 | No | NA | 2 | 0 | 0 | Not-determined | Genome-fragment | NA | NA | 0 |
| S7_extracted_25773 | 3175 | No | NA | 6 | 1 | 0 | Low-quality | Genome-fragment | 6.33 | AAI-based (high-confidence) | 0 |
| S16_extracted_13072 | 3175 | No | NA | 4 | 0 | 4 | Low-quality | Genome-fragment | 5.87 | AAI-based (medium-confidence) | 0 |

| | | | | | | | | | | | |
|---|---|---|---|---|---|---|---|---|---|---|---|
| S10_extracted_76265 | 3175 | No | NA | 6 | 0 | 0 | Low-quality | Genome-fragment | 3.95 | AAI-based (high-confidence) | 0 |
| S10_extracted_19910 | 1185 | No | NA | 2 | 0 | 1 | Not-determined | Genome-fragment | NA | NA | 0 |
| S16_extracted_24960 | 3176 | No | NA | 2 | 0 | 1 | Low-quality | Genome-fragment | 6.57 | AAI-based (high-confidence) | 0 |
| S10_extracted_20136 | 1329 | No | NA | 1 | 0 | 0 | Not-determined | Genome-fragment | NA | NA | 0 |
| S10_extracted_20319 | 1292 | No | NA | 1 | 0 | 0 | Not-determined | Genome-fragment | NA | NA | 0 |
| S10_extracted_20321 | 1414 | No | NA | 2 | 0 | 2 | Not-determined | Genome-fragment | NA | NA | 0 |
| S10_extracted_20333 | 1025 | No | NA | 2 | 0 | 0 | Not-determined | Genome-fragment | NA | NA | 0 |
| S10_extracted_20347 | 1010 | No | NA | 2 | 0 | 1 | Not-determined | Genome-fragment | NA | NA | 0 |
| S10_extracted_20353 | 6284 | No | NA | 3 | 0 | 2 | Not-determined | Genome-fragment | NA | NA | 0 |
| S10_extracted_20563 | 6958 | No | NA | 7 | 0 | 2 | Not-determined | Genome-fragment | NA | NA | 0 |
| S10_extracted_20622 | 2415 | No | NA | 2 | 0 | 2 | Not-determined | Genome-fragment | NA | NA | 0 |
| S12_vs_k141_110381\|\|full | 3176 | No | NA | 5 | 1 | 0 | Low-quality | Genome-fragment | 7.73 | AAI-based (medium-confidence) | 0 |
| S10_extracted_20714 | 3225 | No | NA | 3 | 0 | 2 | Not-determined | Genome-fragment | NA | NA | 0 |
| S8_extracted_k141_11477 | 3176 | No | NA | 3 | 0 | 1 | Low-quality | Genome-fragment | 6.39 | AAI-based (high-confidence) | 0 |
| S8_vs_k141_109296\|\|full | 3176 | No | NA | 7 | 1 | 0 | Low-quality | Genome-fragment | 5.39 | AAI-based (high-confidence) | 0 |
| S17_vs_k141_16453\|\|full | 3178 | No | NA | 5 | 0 | 1 | Low-quality | Genome-fragment | 3.4 | AAI-based (medium-confidence) | 0 |
| S11_vs_k141_9311\|\|full | 3178 | No | NA | 8 | 2 | 0 | Low-quality | Genome-fragment | 8.25 | AAI-based (high-confidence) | 0 |
| S10_extracted_21096 | 3198 | No | NA | 3 | 0 | 3 | Not-determined | Genome-fragment | NA | NA | 0 |
| S10_extracted_21097 | 1430 | No | NA | 1 | 0 | 1 | Not-determined | Genome-fragment | NA | NA | 0 |
| S10_extracted_21272 | 1695 | No | NA | 2 | 0 | 2 | Not-determined | Genome-fragment | NA | NA | 0 |
| S16_vs_164617\|\|full | 3178 | No | NA | 9 | 0 | 0 | Low-quality | Genome-fragment | 9.24 | AAI-based (high-confidence) | 0 |
| S3_vs_k141_107529\|\|full | 3179 | No | NA | 7 | 7 | 0 | Low-quality | Genome-fragment | 7.99 | AAI-based (high-confidence) | 0 |
| S5_extracted_k141_23628 | 3179 | No | NA | 4 | 0 | 0 | Low-quality | Genome-fragment | 7.68 | AAI-based (high-confidence) | 0 |
| S11_extracted_k141_92784 | 3180 | No | NA | 1 | 1 | 0 | Low-quality | Genome-fragment | 5.6 | AAI-based (medium-confidence) | 0 |
| S10_extracted_21921 | 1203 | No | NA | 2 | 0 | 1 | Not-determined | Genome-fragment | NA | NA | 0 |
| S10_extracted_22058 | 1369 | No | NA | 2 | 0 | 2 | Not-determined | Genome-fragment | NA | NA | 0 |
| S3_extracted_k141_97958 | 3180 | No | NA | 4 | 0 | 2 | Low-quality | Genome-fragment | 5.67 | AAI-based (medium-confidence) | 0 |
| S5_extracted_k141_65156 | 3180 | No | NA | 3 | 3 | 0 | Low-quality | Genome-fragment | 7.89 | AAI-based (high-confidence) | 0 |
| S10_extracted_22190 | 3676 | No | NA | 4 | 0 | 2 | Not-determined | Genome-fragment | NA | NA | 0 |
| S12_vs_k141_171746\|\|full | 3180 | No | NA | 3 | 0 | 1 | Low-quality | Genome-fragment | 1.05 | AAI-based (medium-confidence) | 0 |
| S10_extracted_22215 | 4745 | No | NA | 4 | 0 | 4 | Not-determined | Genome-fragment | NA | NA | 0 |
| S17_extracted_k141_7201 | 3181 | No | NA | 8 | 2 | 0 | Low-quality | Genome-fragment | 4.9 | HMM-based (lower-bound) | 0 |
| S10_extracted_22231 | 1243 | No | NA | 1 | 0 | 0 | Not-determined | Genome-fragment | NA | NA | 0 |
| S14_extracted_k141_71287 | 3181 | No | NA | 6 | 1 | 0 | Low-quality | Genome-fragment | 1.85 | AAI-based (medium-confidence) | 0 |
| S10_extracted_22329 | 3279 | No | NA | 4 | 0 | 4 | Not-determined | Genome-fragment | NA | NA | 0 |
| S10_extracted_22442 | 2218 | No | NA | 2 | 0 | 1 | Not-determined | Genome-fragment | NA | NA | 0 |
| S7_extracted_101455 | 3181 | No | NA | 9 | 1 | 0 | Low-quality | Genome-fragment | 2.15 | AAI-based (medium-confidence) | 0 |
| S8_vs_k141_194173\|\|full | 3181 | No | NA | 4 | 0 | 1 | Low-quality | Genome-fragment | 4.33 | AAI-based (medium-confidence) | 0 |
| S10_extracted_22565 | 3266 | No | NA | 3 | 0 | 2 | Not-determined | Genome-fragment | NA | NA | 0 |
| S10_extracted_22650 | 1189 | No | NA | 1 | 0 | 1 | Not-determined | Genome-fragment | NA | NA | 0 |
| S10_extracted_22719 | 3211 | No | NA | 3 | 0 | 0 | Not-determined | Genome-fragment | NA | NA | 0 |
| S10_extracted_22740 | 1345 | No | NA | 2 | 0 | 0 | Not-determined | Genome-fragment | NA | NA | 0 |
| S10_extracted_22906 | 1894 | No | NA | 2 | 0 | 1 | Not-determined | Genome-fragment | NA | NA | 0 |
| S10_extracted_22918 | 1828 | No | NA | 2 | 0 | 2 | Not-determined | Genome-fragment | NA | NA | 0 |
| S10_extracted_22952 | 1377 | No | NA | 2 | 0 | 1 | Not-determined | Genome-fragment | NA | NA | 0 |
| S10_extracted_23212 | 1094 | No | NA | 1 | 0 | 0 | Not-determined | Genome-fragment | NA | NA | 0 |
| S10_extracted_23225 | 4036 | No | NA | 3 | 0 | 3 | Not-determined | Genome-fragment | NA | NA | 0 |
| S16_vs_94911\|\|full | 3182 | No | NA | 4 | 3 | 0 | Low-quality | Genome-fragment | 7.8 | AAI-based (high-confidence) | 0 |
| S10_extracted_23274 | 4888 | No | NA | 4 | 0 | 3 | Not-determined | Genome-fragment | NA | NA | 0 |
| S10_vs_64149\|\|full | 3182 | No | NA | 4 | 0 | 0 | Low-quality | Genome-fragment | 4.81 | AAI-based (medium-confidence) | 0 |
| S10_extracted_23352 | 1608 | No | NA | 2 | 0 | 1 | Not-determined | Genome-fragment | NA | NA | 0 |
| S10_extracted_23515 | 1575 | No | NA | 2 | 0 | 1 | Not-determined | Genome-fragment | NA | NA | 0 |
| S10_extracted_23581 | 2266 | No | NA | 2 | 0 | 1 | Not-determined | Genome-fragment | NA | NA | 0 |
| S10_extracted_23595 | 5956 | No | NA | 3 | 0 | 3 | Not-determined | Genome-fragment | NA | NA | 0 |
| S9_vs_k141_48212\|\|full | 3183 | No | NA | 3 | 1 | 0 | Low-quality | Genome-fragment | 1.36 | HMM-based (lower-bound) | 0 |
| S1_extracted_k141_109688 | 3183 | No | NA | 2 | 0 | 0 | Low-quality | Genome-fragment | 2.07 | AAI-based (high-confidence) | 0 |
| S12_vs_k141_78885\|\|full | 3183 | No | NA | 4 | 0 | 1 | Low-quality | Genome-fragment | 0.87 | AAI-based (medium-confidence) | 0 |
| S10_extracted_23909 | 1188 | No | NA | 2 | 0 | 1 | Not-determined | Genome-fragment | NA | NA | 0 |
| S7_vs_109168\|\|lt2gene | 3185 | No | NA | 1 | 1 | 0 | Low-quality | Genome-fragment | 6.07 | AAI-based (medium-confidence) | 0 |
| S10_extracted_24022 | 3516 | No | NA | 7 | 0 | 0 | Not-determined | Genome-fragment | NA | NA | 0 |
| S10_extracted_24048 | 1190 | No | NA | 1 | 0 | 0 | Not-determined | Genome-fragment | NA | NA | 0 |
| S10_extracted_24098 | 3106 | No | NA | 11 | 0 | 0 | Not-determined | Genome-fragment | NA | NA | 0 |
| S10_extracted_24315 | 6161 | No | NA | 3 | 0 | 3 | Not-determined | Genome-fragment | NA | NA | 0 |
| S10_extracted_24356 | 1075 | No | NA | 2 | 0 | 1 | Not-determined | Genome-fragment | NA | NA | 0 |
| S4_extracted_19106 | 3186 | No | NA | 7 | 2 | 0 | Low-quality | Genome-fragment | 5.82 | AAI-based (high-confidence) | 0 |
| S10_extracted_24573 | 5179 | No | NA | 10 | 0 | 9 | Not-determined | Genome-fragment | NA | NA | 0 |
| S10_extracted_24668 | 3624 | No | NA | 6 | 0 | 0 | Not-determined | Genome-fragment | NA | NA | 0 |
| S10_extracted_24751 | 3170 | No | NA | 5 | 0 | 2 | Not-determined | Genome-fragment | NA | NA | 0 |
| S10_extracted_24791 | 44832 | No | NA | 71 | 23 | 0 | High-quality | High-quality | 91.98 | AAI-based (high-confidence) | 0 |
| S10_extracted_25039 | 1382 | No | NA | 2 | 0 | 0 | Not-determined | Genome-fragment | NA | NA | 0 |
| S10_extracted_25142 | 1144 | No | NA | 5 | 0 | 0 | Not-determined | Genome-fragment | NA | NA | 0 |
| S10_extracted_25158 | 3508 | No | NA | 2 | 0 | 2 | Not-determined | Genome-fragment | NA | NA | 0 |
| S10_extracted_25241 | 7209 | No | NA | 16 | 0 | 0 | Medium-quality | Genome-fragment | 60.65 | AAI-based (high-confidence) | 0 |
| S8_vs_k141_35004\|\|full | 3186 | No | NA | 6 | 0 | 0 | Low-quality | Genome-fragment | 7.74 | AAI-based (high-confidence) | 0 |
| S10_extracted_25432 | 2966 | No | NA | 4 | 0 | 4 | Not-determined | Genome-fragment | NA | NA | 0 |
| S10_extracted_25511 | 1902 | No | NA | 2 | 0 | 1 | Not-determined | Genome-fragment | NA | NA | 0 |
| S7_extracted_36808 | 3187 | No | NA | 5 | 0 | 1 | Low-quality | Genome-fragment | 7.5 | AAI-based (high-confidence) | 0 |
| S10_extracted_25565 | 1015 | No | NA | 1 | 0 | 1 | Not-determined | Genome-fragment | NA | NA | 0 |
| S10_extracted_25585 | 1250 | No | NA | 2 | 0 | 1 | Not-determined | Genome-fragment | NA | NA | 0 |
| S16_vs_9258\|\|full | 3188 | No | NA | 5 | 0 | 0 | Low-quality | Genome-fragment | 5.57 | AAI-based (medium-confidence) | 0 |
| S15_extracted_55175 | 3188 | No | NA | 4 | 2 | 0 | Low-quality | Genome-fragment | 9.43 | HMM-based (lower-bound) | 0 |
| S10_extracted_25790 | 1169 | No | NA | 1 | 0 | 1 | Not-determined | Genome-fragment | NA | NA | 0 |
| S10_extracted_25823 | 1456 | No | NA | 2 | 0 | 1 | Not-determined | Genome-fragment | NA | NA | 0 |
| S14_extracted_k141_1196 | 3189 | Yes | 1544 | 4 | 1 | 1 | Low-quality | Genome-fragment | 1.41 | HMM-based (lower-bound) | 51.58 |
| S10_extracted_25967 | 1354 | No | NA | 1 | 0 | 1 | Not-determined | Genome-fragment | NA | NA | 0 |
| S3_vs_k141_120298\|\|full | 3189 | No | NA | 5 | 0 | 0 | Low-quality | Genome-fragment | 5.12 | AAI-based (medium-confidence) | 0 |
| S10_extracted_26022 | 5528 | No | NA | 3 | 0 | 2 | Not-determined | Genome-fragment | NA | NA | 0 |
| S10_extracted_26095 | 1673 | No | NA | 1 | 0 | 1 | Not-determined | Genome-fragment | NA | NA | 0 |
| S10_extracted_26125 | 1971 | No | NA | 3 | 0 | 0 | Not-determined | Genome-fragment | NA | NA | 0 |
| S10_extracted_26185 | 3711 | No | NA | 6 | 0 | 0 | Not-determined | Genome-fragment | NA | NA | 0 |
| S10_extracted_26188 | 1254 | No | NA | 3 | 0 | 0 | Not-determined | Genome-fragment | NA | NA | 0 |
| S16_vs_79403\|\|full | 3189 | No | NA | 4 | 0 | 3 | Low-quality | Genome-fragment | 2.57 | AAI-based (high-confidence) | 0 |
| S10_extracted_26200 | 5737 | No | NA | 5 | 0 | 3 | Not-determined | Genome-fragment | NA | NA | 0 |
| S10_extracted_26280 | 1804 | No | NA | 2 | 0 | 2 | Not-determined | Genome-fragment | NA | NA | 0 |
| S10_extracted_26474 | 3356 | No | NA | 5 | 0 | 1 | Not-determined | Genome-fragment | NA | NA | 0 |
| S10_extracted_26563 | 1258 | No | NA | 2 | 0 | 1 | Not-determined | Genome-fragment | NA | NA | 0 |
| S10_extracted_26635 | 1141 | No | NA | 1 | 0 | 1 | Not-determined | Genome-fragment | NA | NA | 0 |
| S16_extracted_96143 | 3190 | No | NA | 3 | 0 | 2 | Low-quality | Genome-fragment | 1.73 | AAI-based (medium-confidence) | 0 |
| S12_extracted_k141_169126 | 3190 | No | NA | 5 | 0 | 0 | Low-quality | Genome-fragment | 1.92 | AAI-based (high-confidence) | 0 |
| S10_extracted_26795 | 1313 | No | NA | 2 | 0 | 0 | Not-determined | Genome-fragment | NA | NA | 0 |
| S3_vs_k141_111065\|\|full | 3191 | No | NA | 9 | 0 | 0 | Low-quality | Genome-fragment | 4.86 | AAI-based (high-confidence) | 0 |
| S10_extracted_26808 | 1199 | No | NA | 2 | 0 | 2 | Not-determined | Genome-fragment | NA | NA | 0 |
| S10_extracted_26854 | 1346 | No | NA | 2 | 0 | 0 | Not-determined | Genome-fragment | NA | NA | 0 |
| S10_extracted_110208 | 3191 | No | NA | 7 | 0 | 0 | Low-quality | Genome-fragment | 6.33 | AAI-based (high-confidence) | 0 |
| S10_extracted_27056 | 4164 | No | NA | 4 | 0 | 0 | Not-determined | Genome-fragment | NA | NA | 0 |
| S10_extracted_27065 | 1755 | No | NA | 1 | 0 | 0 | Not-determined | Genome-fragment | NA | NA | 0 |
| S10_extracted_27155 | 2183 | No | NA | 2 | 0 | 2 | Not-determined | Genome-fragment | NA | NA | 0 |
| S8_vs_k141_50751\|\|full | 3191 | No | NA | 6 | 0 | 2 | Low-quality | Genome-fragment | 3 | AAI-based (medium-confidence) | 0 |
| S17_vs_k141_77515\|\|full | 3192 | No | NA | 4 | 0 | 3 | Low-quality | Genome-fragment | 1.76 | AAI-based (medium-confidence) | 0 |
| S13_vs_66544\|\|full | 3192 | No | NA | 4 | 1 | 2 | Low-quality | Genome-fragment | 2.23 | AAI-based (high-confidence) | 0 |
| S10_extracted_27632 | 1012 | No | NA | 1 | 0 | 0 | Not-determined | Genome-fragment | NA | NA | 0 |
| S10_extracted_27687 | 1105 | No | NA | 2 | 0 | 1 | Not-determined | Genome-fragment | NA | NA | 0 |
| S5_extracted_k141_50142 | 3192 | No | NA | 6 | 0 | 0 | Low-quality | Genome-fragment | 7.46 | AAI-based (high-confidence) | 0 |
| S10_extracted_28211 | 3864 | No | NA | 4 | 0 | 2 | Not-determined | Genome-fragment | NA | NA | 0 |
| S10_extracted_28228 | 1052 | No | NA | 1 | 0 | 1 | Not-determined | Genome-fragment | NA | NA | 0 |
| S10_extracted_28238 | 2000 | No | NA | 3 | 0 | 2 | Not-determined | Genome-fragment | NA | NA | 0 |
| S10_extracted_28328 | 1116 | No | NA | 1 | 0 | 1 | Not-determined | Genome-fragment | NA | NA | 0 |
| S10_extracted_28486 | 1218 | No | NA | 2 | 0 | 1 | Not-determined | Genome-fragment | NA | NA | 0 |
| S10_extracted_28490 | 2751 | No | NA | 5 | 0 | 1 | Not-determined | Genome-fragment | NA | NA | 0 |
| S10_extracted_28612 | 1035 | No | NA | 1 | 0 | 1 | Not-determined | Genome-fragment | NA | NA | 0 |
| S10_extracted_28623 | 1866 | No | NA | 2 | 0 | 2 | Not-determined | Genome-fragment | NA | NA | 0 |
| S10_extracted_28727 | 4073 | No | NA | 4 | 0 | 3 | Not-determined | Genome-fragment | NA | NA | 0 |
| S10_extracted_28761 | 4818 | No | NA | 2 | 0 | 2 | Not-determined | Genome-fragment | NA | NA | 0 |
| S10_extracted_28930 | 1276 | No | NA | 1 | 0 | 0 | Not-determined | Genome-fragment | NA | NA | 0 |
| S10_extracted_28972 | 1829 | No | NA | 1 | 0 | 1 | Not-determined | Genome-fragment | NA | NA | 0 |
| S10_extracted_29216 | 1484 | No | NA | 4 | 0 | 1 | Not-determined | Genome-fragment | NA | NA | 0 |
| S10_extracted_29425 | 2479 | No | NA | 5 | 0 | 4 | Not-determined | Genome-fragment | NA | NA | 0 |
| S10_extracted_29499 | 1407 | No | NA | 2 | 0 | 0 | Not-determined | Genome-fragment | NA | NA | 0 |
| S11_extracted_k141_42320 | 3193 | No | NA | 4 | 0 | 3 | Low-quality | Genome-fragment | 2.36 | AAI-based (medium-confidence) | 0 |
| S14_vs_k141_50811\|\|full | 3193 | No | NA | 6 | 0 | 0 | Low-quality | Genome-fragment | 5.89 | AAI-based (medium-confidence) | 0 |
| S10_extracted_29724 | 6976 | No | NA | 4 | 0 | 3 | Not-determined | Genome-fragment | NA | NA | 0 |
| S7_vs_48070\|\|full | 3193 | No | NA | 4 | 2 | 0 | Low-quality | Genome-fragment | 7.32 | AAI-based (high-confidence) | 0 |
| S10_extracted_29804 | 1200 | No | NA | 1 | 0 | 1 | Not-determined | Genome-fragment | NA | NA | 0 |
| S10_extracted_29866 | 1327 | No | NA | 2 | 0 | 0 | Not-determined | Genome-fragment | NA | NA | 0 |
| S10_extracted_29874 | 2629 | No | NA | 2 | 0 | 2 | Not-determined | Genome-fragment | NA | NA | 0 |
| S10_extracted_29963 | 1724 | No | NA | 3 | 0 | 3 | Not-determined | Genome-fragment | NA | NA | 0 |
| S10_extracted_30047 | 1083 | No | NA | 1 | 0 | 0 | Not-determined | Genome-fragment | NA | NA | 0 |
| S10_extracted_30057 | 1018 | No | NA | 1 | 0 | 1 | Not-determined | Genome-fragment | NA | NA | 0 |
| S10_extracted_30198 | 8061 | No | NA | 6 | 0 | 5 | Not-determined | Genome-fragment | NA | NA | 0 |
| S14_vs_k141_44155\|\|full | 3194 | No | NA | 2 | 2 | 0 | Low-quality | Genome-fragment | 5.61 | HMM-based (lower-bound) | 0 |
| S14_vs_k141_115481\|\|full | 3194 | No | NA | 7 | 2 | 1 | Low-quality | Genome-fragment | 4.18 | AAI-based (high-confidence) | 0 |
| S1_extracted_k141_102489 | 3194 | No | NA | 6 | 2 | 0 | Low-quality | Genome-fragment | 3.97 | AAI-based (high-confidence) | 0 |
| S10_extracted_30351 | 1345 | No | NA | 3 | 0 | 1 | Not-determined | Genome-fragment | NA | NA | 0 |
| S16_vs_2166\|\|lt2gene | 3194 | No | NA | 2 | 1 | 0 | Low-quality | Genome-fragment | 2.21 | HMM-based (lower-bound) | 0 |
| S11_extracted_k141_52347 | 3195 | No | NA | 6 | 0 | 0 | Low-quality | Genome-fragment | 3.76 | AAI-based (medium-confidence) | 0 |
| S10_extracted_30521 | 1562 | No | NA | 2 | 0 | 2 | Not-determined | Genome-fragment | NA | NA | 0 |
| S10_extracted_30532 | 1615 | No | NA | 1 | 0 | 1 | Not-determined | Genome-fragment | NA | NA | 0 |
| S10_extracted_30552 | 1505 | No | NA | 2 | 0 | 0 | Not-determined | Genome-fragment | NA | NA | 0 |
| S10_extracted_30559 | 1258 | No | NA | 1 | 0 | 1 | Not-determined | Genome-fragment | NA | NA | 0 |
| S10_extracted_30885 | 1571 | No | NA | 1 | 0 | 0 | Not-determined | Genome-fragment | NA | NA | 0 |
| S10_extracted_31056 | 2061 | No | NA | 2 | 0 | 0 | Not-determined | Genome-fragment | NA | NA | 0 |
| S10_extracted_31161 | 10672 | No | NA | 10 | 0 | 3 | Not-determined | Genome-fragment | NA | NA | 0 |
| S10_extracted_31304 | 11795 | No | NA | 9 | 0 | 6 | Not-determined | Genome-fragment | NA | NA | 0 |
| S10_extracted_31364 | 1411 | No | NA | 6 | 0 | 0 | Not-determined | Genome-fragment | NA | NA | 0 |
| S10_extracted_31415 | 5389 | No | NA | 4 | 0 | 1 | Not-determined | Genome-fragment | NA | NA | 0 |
| S10_extracted_31429 | 1617 | No | NA | 1 | 0 | 1 | Not-determined | Genome-fragment | NA | NA | 0 |
| S10_extracted_31487 | 1007 | No | NA | 1 | 0 | 1 | Not-determined | Genome-fragment | NA | NA | 0 |
| S10_extracted_31491 | 1317 | No | NA | 3 | 0 | 0 | Not-determined | Genome-fragment | NA | NA | 0 |
| S10_extracted_31548 | 1315 | No | NA | 2 | 0 | 0 | Not-determined | Genome-fragment | NA | NA | 0 |
| S7_extracted_81431 | 3195 | No | NA | 6 | 0 | 0 | Low-quality | Genome-fragment | 8.55 | AAI-based (high-confidence) | 0 |
| S10_extracted_104279 | 3196 | No | NA | 3 | 2 | 0 | Low-quality | Genome-fragment | 10.18 | AAI-based (high-confidence) | 0 |
| S10_extracted_31728 | 1488 | No | NA | 3 | 0 | 0 | Not-determined | Genome-fragment | NA | NA | 0 |
| S10_extracted_31775 | 2092 | No | NA | 3 | 0 | 3 | Not-determined | Genome-fragment | NA | NA | 0 |
| S10_extracted_31812 | 1151 | No | NA | 4 | 0 | 3 | Not-determined | Genome-fragment | NA | NA | 0 |
| S10_extracted_31827 | 1010 | No | NA | 2 | 0 | 0 | Not-determined | Genome-fragment | NA | NA | 0 |
| S10_extracted_31872 | 1179 | No | NA | 2 | 0 | 0 | Not-determined | Genome-fragment | NA | NA | 0 |
| S8_vs_k141_22718\|\|full | 3196 | No | NA | 4 | 0 | 0 | Low-quality | Genome-fragment | 5.08 | AAI-based (high-confidence) | 0 |
| S10_extracted_32153 | 1006 | No | NA | 2 | 0 | 1 | Not-determined | Genome-fragment | NA | NA | 0 |
| S17_vs_k141_47579\|\|full | 3197 | No | NA | 3 | 0 | 3 | Low-quality | Genome-fragment | 1.74 | AAI-based (medium-confidence) | 0 |
| S14_vs_k141_67780\|\|full | 3198 | No | NA | 3 | 1 | 0 | Low-quality | Genome-fragment | 2.3 | HMM-based (lower-bound) | 0 |
| S5_extracted_k141_57372 | 3198 | No | NA | 3 | 0 | 0 | Low-quality | Genome-fragment | 4.86 | AAI-based (medium-confidence) | 0 |
| S10_extracted_32725 | 1120 | No | NA | 2 | 0 | 2 | Not-determined | Genome-fragment | NA | NA | 0 |
| S10_extracted_8512 | 3198 | No | NA | 3 | 0 | 1 | Low-quality | Genome-fragment | 6.73 | AAI-based (medium-confidence) | 0 |
| S12_extracted_k141_138277 | 3198 | No | NA | 5 | 1 | 0 | Low-quality | Genome-fragment | 2.21 | AAI-based (high-confidence) | 0 |
| S10_extracted_32829 | 1064 | No | NA | 2 | 0 | 2 | Not-determined | Genome-fragment | NA | NA | 0 |
| S1_vs_k141_76867\|\|full | 3199 | No | NA | 4 | 0 | 0 | Low-quality | Genome-fragment | 4.68 | AAI-based (medium-confidence) | 0 |
| S10_extracted_32892 | 1651 | No | NA | 2 | 0 | 2 | Not-determined | Genome-fragment | NA | NA | 0 |
| S10_extracted_32897 | 5621 | No | NA | 6 | 0 | 4 | Not-determined | Genome-fragment | NA | NA | 0 |
| S10_extracted_32953 | 6755 | No | NA | 6 | 0 | 1 | Not-determined | Genome-fragment | NA | NA | 0 |
| S10_extracted_33007 | 1800 | No | NA | 2 | 0 | 2 | Not-determined | Genome-fragment | NA | NA | 0 |
| S10_extracted_33072 | 1319 | No | NA | 2 | 0 | 1 | Not-determined | Genome-fragment | NA | NA | 0 |
| S10_extracted_33100 | 2586 | No | NA | 1 | 0 | 1 | Not-determined | Genome-fragment | NA | NA | 0 |
| S3_extracted_k141_105903 | 3199 | No | NA | 5 | 0 | 1 | Low-quality | Genome-fragment | 6.45 | AAI-based (high-confidence) | 0 |
| S10_extracted_33471 | 1273 | No | NA | 1 | 0 | 1 | Not-determined | Genome-fragment | NA | NA | 0 |
| S10_extracted_33584 | 1461 | No | NA | 2 | 0 | 0 | Not-determined | Genome-fragment | NA | NA | 0 |
| S10_extracted_33637 | 1650 | No | NA | 4 | 0 | 3 | Not-determined | Genome-fragment | NA | NA | 0 |
| S10_extracted_33649 | 1715 | No | NA | 2 | 0 | 1 | Not-determined | Genome-fragment | NA | NA | 0 |
| S10_extracted_33740 | 1763 | No | NA | 3 | 0 | 2 | Not-determined | Genome-fragment | NA | NA | 0 |
| S16_vs_14582\|\|full | 3199 | No | NA | 5 | 2 | 0 | Low-quality | Genome-fragment | 1.26 | HMM-based (lower-bound) | 0 |
| S10_extracted_33809 | 1114 | No | NA | 1 | 0 | 0 | Not-determined | Genome-fragment | NA | NA | 0 |
| S10_extracted_33881 | 3254 | No | NA | 5 | 0 | 3 | Not-determined | Genome-fragment | NA | NA | 0 |
| S10_vs_10268\|\|full | 3199 | No | NA | 4 | 0 | 1 | Low-quality | Genome-fragment | 19.86 | AAI-based (medium-confidence) | 0 |
| S10_extracted_34021 | 3999 | No | NA | 3 | 0 | 3 | Not-determined | Genome-fragment | NA | NA | 0 |
| S10_extracted_34039 | 2971 | No | NA | 3 | 0 | 1 | Not-determined | Genome-fragment | NA | NA | 0 |
| S10_extracted_34056 | 1603 | No | NA | 2 | 0 | 0 | Not-determined | Genome-fragment | NA | NA | 0 |
| S10_extracted_34233 | 2395 | No | NA | 2 | 0 | 1 | Not-determined | Genome-fragment | NA | NA | 0 |
| S3_extracted_k141_72258 | 3200 | No | NA | 1 | 1 | 0 | Low-quality | Genome-fragment | 3.98 | HMM-based (lower-bound) | 0 |
| S10_extracted_34351 | 1667 | No | NA | 3 | 0 | 1 | Not-determined | Genome-fragment | NA | NA | 0 |
| S10_extracted_34395 | 1121 | No | NA | 1 | 0 | 0 | Not-determined | Genome-fragment | NA | NA | 0 |
| S10_extracted_34414 | 1164 | No | NA | 1 | 0 | 0 | Not-determined | Genome-fragment | NA | NA | 0 |
| S13_extracted_57018 | 3201 | No | NA | 2 | 0 | 1 | Low-quality | Genome-fragment | 1.89 | AAI-based (medium-confidence) | 0 |
| S10_extracted_34541 | 2370 | No | NA | 2 | 0 | 1 | Not-determined | Genome-fragment | NA | NA | 0 |
| S10_extracted_34574 | 2157 | No | NA | 4 | 0 | 4 | Not-determined | Genome-fragment | NA | NA | 0 |
| S3_vs_k141_57764\|\|full | 3201 | No | NA | 6 | 2 | 0 | Low-quality | Genome-fragment | 5.99 | AAI-based (high-confidence) | 0 |
| S10_extracted_34817 | 1392 | No | NA | 2 | 0 | 1 | Not-determined | Genome-fragment | NA | NA | 0 |
| S3_vs_k141_109702\|\|full | 3201 | No | NA | 5 | 0 | 1 | Low-quality | Genome-fragment | 7.38 | AAI-based (high-confidence) | 0 |
| S1_vs_k141_63550\|\|full | 3202 | No | NA | 6 | 1 | 0 | Low-quality | Genome-fragment | 6.77 | AAI-based (high-confidence) | 0 |
| S10_extracted_34949 | 3379 | No | NA | 2 | 0 | 2 | Not-determined | Genome-fragment | NA | NA | 0 |
| S10_extracted_34960 | 1389 | No | NA | 1 | 0 | 1 | Not-determined | Genome-fragment | NA | NA | 0 |
| S10_extracted_35226 | 1203 | No | NA | 1 | 0 | 0 | Not-determined | Genome-fragment | NA | NA | 0 |
| S3_vs_k141_121834\|\|full | 3202 | No | NA | 4 | 0 | 0 | Low-quality | Genome-fragment | 6.2 | AAI-based (high-confidence) | 0 |
| S10_extracted_35311 | 1313 | No | NA | 1 | 0 | 1 | Not-determined | Genome-fragment | NA | NA | 0 |
| S10_extracted_35381 | 1288 | No | NA | 1 | 0 | 1 | Not-determined | Genome-fragment | NA | NA | 0 |
| S8_extracted_k141_177909 | 3202 | No | NA | 3 | 0 | 1 | Low-quality | Genome-fragment | 0.84 | AAI-based (medium-confidence) | 0 |
| S10_extracted_35584 | 1151 | No | NA | 2 | 0 | 1 | Not-determined | Genome-fragment | NA | NA | 0 |
| S10_extracted_35669 | 1026 | No | NA | 1 | 0 | 1 | Not-determined | Genome-fragment | NA | NA | 0 |
| S10_extracted_35693 | 1526 | No | NA | 2 | 0 | 0 | Not-determined | Genome-fragment | NA | NA | 0 |
| S10_extracted_35754 | 4639 | No | NA | 4 | 0 | 3 | Not-determined | Genome-fragment | NA | NA | 0 |
| S10_extracted_35857 | 2421 | No | NA | 2 | 0 | 1 | Not-determined | Genome-fragment | NA | NA | 0 |
| S10_extracted_35911 | 1242 | No | NA | 1 | 0 | 1 | Not-determined | Genome-fragment | NA | NA | 0 |
| S10_extracted_35938 | 1550 | No | NA | 2 | 0 | 2 | Not-determined | Genome-fragment | NA | NA | 0 |
| S10_extracted_36012 | 1732 | No | NA | 3 | 0 | 2 | Not-determined | Genome-fragment | NA | NA | 0 |
| S10_extracted_36035 | 1425 | No | NA | 2 | 0 | 2 | Not-determined | Genome-fragment | NA | NA | 0 |
| S10_extracted_36044 | 1090 | No | NA | 2 | 0 | 1 | Not-determined | Genome-fragment | NA | NA | 0 |
| S17_vs_k141_85688\|\|full | 3203 | No | NA | 5 | 0 | 0 | Low-quality | Genome-fragment | 3.41 | AAI-based (high-confidence) | 0 |
| S3_extracted_k141_94821 | 3203 | No | NA | 3 | 0 | 2 | Low-quality | Genome-fragment | 4.23 | AAI-based (medium-confidence) | 0 |
| S10_extracted_36543 | 1122 | No | NA | 2 | 0 | 0 | Not-determined | Genome-fragment | NA | NA | 0 |
| S10_extracted_36606 | 1532 | No | NA | 2 | 0 | 1 | Not-determined | Genome-fragment | NA | NA | 0 |
| S5_vs_k141_127918\|\|full | 3204 | No | NA | 7 | 1 | 0 | Low-quality | Genome-fragment | 7.29 | AAI-based (medium-confidence) | 0 |
| S10_extracted_36729 | 11130 | No | NA | 14 | 0 | 1 | Not-determined | Genome-fragment | NA | NA | 0 |
| S10_extracted_36742 | 4500 | No | NA | 5 | 0 | 2 | Not-determined | Genome-fragment | NA | NA | 0 |
| S10_extracted_36744 | 3871 | No | NA | 5 | 0 | 2 | Not-determined | Genome-fragment | NA | NA | 0 |
| S10_extracted_36836 | 10017 | No | NA | 7 | 0 | 5 | Not-determined | Genome-fragment | NA | NA | 0 |
| S10_extracted_36858 | 3505 | No | NA | 5 | 0 | 1 | Not-determined | Genome-fragment | NA | NA | 0 |
| S10_extracted_36880 | 93310 | No | NA | 91 | 10 | 0 | Complete | High-quality | 100 | DTR (high-confidence) | 0 |
| S10_extracted_37001 | 3569 | No | NA | 3 | 0 | 1 | Not-determined | Genome-fragment | NA | NA | 0 |
| S10_extracted_37052 | 1787 | No | NA | 1 | 0 | 1 | Not-determined | Genome-fragment | NA | NA | 0 |
| S10_extracted_37198 | 5247 | No | NA | 4 | 0 | 0 | Not-determined | Genome-fragment | NA | NA | 0 |
| S10_extracted_37284 | 1440 | No | NA | 1 | 0 | 1 | Not-determined | Genome-fragment | NA | NA | 0 |
| S10_extracted_37739 | 1908 | No | NA | 4 | 0 | 4 | Not-determined | Genome-fragment | NA | NA | 0 |
| S10_extracted_37873 | 1143 | No | NA | 1 | 0 | 0 | Not-determined | Genome-fragment | NA | NA | 0 |
| S12_vs_k141_171260\|\|full | 3204 | No | NA | 2 | 0 | 2 | Low-quality | Genome-fragment | 1.54 | AAI-based (medium-confidence) | 0 |
| S10_extracted_38166 | 4091 | No | NA | 2 | 0 | 2 | Not-determined | Genome-fragment | NA | NA | 0 |
| S11_vs_k141_101322\|\|full | 3205 | No | NA | 4 | 0 | 0 | Low-quality | Genome-fragment | 2.67 | AAI-based (high-confidence) | 0 |
| S10_extracted_38273 | 1157 | No | NA | 1 | 0 | 1 | Not-determined | Genome-fragment | NA | NA | 0 |
| S10_extracted_38390 | 3343 | No | NA | 4 | 0 | 0 | Not-determined | Genome-fragment | NA | NA | 0 |
| S10_extracted_38483 | 1638 | No | NA | 2 | 0 | 0 | Not-determined | Genome-fragment | NA | NA | 0 |
| S10_extracted_38486 | 2203 | No | NA | 3 | 0 | 3 | Not-determined | Genome-fragment | NA | NA | 0 |
| S10_extracted_38541 | 3347 | No | NA | 7 | 0 | 5 | Not-determined | Genome-fragment | NA | NA | 0 |
| S3_vs_k141_125224\|\|full | 3205 | No | NA | 5 | 2 | 0 | Low-quality | Genome-fragment | 7.44 | AAI-based (medium-confidence) | 0 |
| S10_extracted_38643 | 1237 | No | NA | 2 | 0 | 1 | Not-determined | Genome-fragment | NA | NA | 0 |
| S10_extracted_38649 | 1023 | No | NA | 1 | 0 | 0 | Not-determined | Genome-fragment | NA | NA | 0 |
| S7_extracted_109126 | 3205 | No | NA | 10 | 1 | 0 | Low-quality | Genome-fragment | 3.5 | HMM-based (lower-bound) | 0 |
| S10_extracted_38832 | 7532 | No | NA | 9 | 0 | 6 | Not-determined | Genome-fragment | NA | NA | 0 |
| S10_extracted_38934 | 1097 | No | NA | 1 | 0 | 0 | Not-determined | Genome-fragment | NA | NA | 0 |
| S10_extracted_38950 | 1452 | No | NA | 3 | 0 | 1 | Not-determined | Genome-fragment | NA | NA | 0 |
| S10_extracted_38952 | 1095 | No | NA | 3 | 0 | 1 | Not-determined | Genome-fragment | NA | NA | 0 |
| S10_extracted_38974 | 1132 | No | NA | 1 | 0 | 1 | Not-determined | Genome-fragment | NA | NA | 0 |
| S10_extracted_39066 | 1140 | No | NA | 2 | 0 | 0 | Not-determined | Genome-fragment | NA | NA | 0 |
| S10_extracted_39105 | 3982 | No | NA | 2 | 0 | 2 | Not-determined | Genome-fragment | NA | NA | 0 |
| S16_extracted_4161 | 3205 | No | NA | 9 | 1 | 0 | Low-quality | Genome-fragment | 6.53 | AAI-based (high-confidence) | 0 |
| S10_extracted_39250 | 1203 | No | NA | 2 | 0 | 0 | Not-determined | Genome-fragment | NA | NA | 0 |
| S10_extracted_39447 | 3424 | No | NA | 2 | 0 | 1 | Not-determined | Genome-fragment | NA | NA | 0 |
| S10_extracted_39453 | 2571 | No | NA | 4 | 0 | 3 | Not-determined | Genome-fragment | NA | NA | 0 |
| S10_extracted_39549 | 5895 | No | NA | 4 | 0 | 4 | Not-determined | Genome-fragment | NA | NA | 0 |
| S10_extracted_39779 | 1287 | No | NA | 1 | 0 | 0 | Not-determined | Genome-fragment | NA | NA | 0 |
| S16_vs_95879\|\|full | 3205 | No | NA | 2 | 1 | 0 | Low-quality | Genome-fragment | 9.77 | AAI-based (high-confidence) | 0 |
| S11_extracted_k141_25656 | 3206 | No | NA | 7 | 6 | 0 | Low-quality | Genome-fragment | 7.92 | AAI-based (high-confidence) | 0 |
| S10_extracted_40073 | 1622 | No | NA | 2 | 0 | 2 | Not-determined | Genome-fragment | NA | NA | 0 |
| S3_extracted_k141_19282 | 3206 | No | NA | 4 | 2 | 0 | Low-quality | Genome-fragment | 14.31 | AAI-based (high-confidence) | 0 |
| S10_extracted_40134 | 8936 | No | NA | 8 | 0 | 3 | Not-determined | Genome-fragment | NA | NA | 0 |
| S10_extracted_40251 | 1126 | No | NA | 3 | 0 | 0 | Not-determined | Genome-fragment | NA | NA | 0 |
| S10_extracted_40324 | 1218 | No | NA | 2 | 0 | 0 | Not-determined | Genome-fragment | NA | NA | 0 |
| S10_extracted_40360 | 1159 | No | NA | 1 | 0 | 0 | Not-determined | Genome-fragment | NA | NA | 0 |
| S13_extracted_124963 | 3207 | No | NA | 7 | 0 | 0 | Low-quality | Genome-fragment | 7.36 | AAI-based (medium-confidence) | 0 |
| S10_extracted_40555 | 1041 | No | NA | 1 | 0 | 0 | Not-determined | Genome-fragment | NA | NA | 0 |
| S5_vs_k141_9317\|\|full | 3207 | No | NA | 8 | 3 | 0 | Low-quality | Genome-fragment | 5.29 | AAI-based (high-confidence) | 0 |
| S10_extracted_40689 | 1004 | No | NA | 1 | 0 | 1 | Not-determined | Genome-fragment | NA | NA | 0 |
| S10_extracted_40799 | 1374 | No | NA | 3 | 0 | 0 | Not-determined | Genome-fragment | NA | NA | 0 |
| S16_vs_15904\|\|full | 3207 | No | NA | 4 | 0 | 2 | Low-quality | Genome-fragment | 1.84 | AAI-based (medium-confidence) | 0 |
| S10_extracted_41305 | 3303 | No | NA | 7 | 0 | 6 | Not-determined | Genome-fragment | NA | NA | 0 |
| S10_extracted_41380 | 5198 | No | NA | 4 | 0 | 4 | Not-determined | Genome-fragment | NA | NA | 0 |
| S10_extracted_41427 | 1399 | No | NA | 1 | 0 | 1 | Not-determined | Genome-fragment | NA | NA | 0 |
| S10_extracted_41456 | 1182 | No | NA | 1 | 0 | 1 | Not-determined | Genome-fragment | NA | NA | 0 |
| S10_extracted_41481 | 2857 | No | NA | 1 | 0 | 0 | Not-determined | Genome-fragment | NA | NA | 0 |
| S10_extracted_41527 | 1009 | No | NA | 1 | 0 | 0 | Not-determined | Genome-fragment | NA | NA | 0 |
| S10_extracted_41664 | 4698 | No | NA | 5 | 0 | 4 | Not-determined | Genome-fragment | NA | NA | 0 |
| S6_vs_k141_8009\|\|full | 3207 | No | NA | 2 | 0 | 1 | Low-quality | Genome-fragment | 1.01 | AAI-based (medium-confidence) | 0 |
| S10_extracted_41934 | 1148 | No | NA | 2 | 0 | 1 | Not-determined | Genome-fragment | NA | NA | 0 |
| S10_extracted_41954 | 1015 | No | NA | 2 | 0 | 0 | Not-determined | Genome-fragment | NA | NA | 0 |
| S10_extracted_42022 | 1230 | No | NA | 2 | 0 | 1 | Not-determined | Genome-fragment | NA | NA | 0 |
| S10_extracted_42239 | 1158 | No | NA | 3 | 0 | 3 | Not-determined | Genome-fragment | NA | NA | 0 |
| S10_extracted_42292 | 1293 | No | NA | 1 | 0 | 1 | Not-determined | Genome-fragment | NA | NA | 0 |
| S11_vs_k141_22187\|\|full | 3208 | No | NA | 3 | 0 | 1 | Low-quality | Genome-fragment | 1.1 | AAI-based (medium-confidence) | 0 |
| S10_extracted_42378 | 2394 | No | NA | 5 | 0 | 0 | Not-determined | Genome-fragment | NA | NA | 0 |
| S8_vs_k141_182847\|\|full | 3208 | No | NA | 4 | 2 | 0 | Low-quality | Genome-fragment | 8.33 | AAI-based (high-confidence) | 0 |
| S10_extracted_42420 | 1057 | No | NA | 2 | 0 | 1 | Not-determined | Genome-fragment | NA | NA | 0 |
| S10_extracted_42435 | 1095 | No | NA | 1 | 0 | 1 | Not-determined | Genome-fragment | NA | NA | 0 |
| S10_extracted_42460 | 5371 | No | NA | 7 | 0 | 0 | Not-determined | Genome-fragment | NA | NA | 0 |
| S8_vs_k141_83570\|\|full | 3208 | No | NA | 2 | 0 | 1 | Low-quality | Genome-fragment | 8.4 | AAI-based (high-confidence) | 0 |
| S10_extracted_42843 | 1309 | No | NA | 1 | 0 | 0 | Not-determined | Genome-fragment | NA | NA | 0 |
| S10_extracted_42978 | 1925 | No | NA | 4 | 0 | 0 | Not-determined | Genome-fragment | NA | NA | 0 |
| S10_extracted_43016 | 2619 | No | NA | 2 | 0 | 1 | Not-determined | Genome-fragment | NA | NA | 0 |
| S10_extracted_43093 | 1490 | No | NA | 1 | 0 | 1 | Not-determined | Genome-fragment | NA | NA | 0 |
| S17_vs_k141_5228\|\|full | 3209 | No | NA | 8 | 2 | 0 | Low-quality | Genome-fragment | 4.85 | AAI-based (high-confidence) | 0 |
| S7_vs_16226\|\|full | 3209 | No | NA | 4 | 0 | 1 | Low-quality | Genome-fragment | 1.14 | AAI-based (high-confidence) | 0 |
| S16_extracted_151353 | 3209 | No | NA | 3 | 2 | 0 | Low-quality | Genome-fragment | 1.91 | HMM-based (lower-bound) | 0 |
| S6_vs_k141_20680\|\|full | 3209 | No | NA | 3 | 0 | 1 | Low-quality | Genome-fragment | 4.37 | AAI-based (high-confidence) | 0 |
| S11_extracted_k141_78402 | 3210 | No | NA | 4 | 1 | 0 | Low-quality | Genome-fragment | 1.11 | HMM-based (lower-bound) | 0 |
| S10_extracted_43569 | 1051 | No | NA | 2 | 0 | 0 | Not-determined | Genome-fragment | NA | NA | 0 |
| S10_extracted_43623 | 5440 | No | NA | 6 | 0 | 3 | Not-determined | Genome-fragment | NA | NA | 0 |
| S10_extracted_43764 | 2695 | No | NA | 4 | 0 | 2 | Not-determined | Genome-fragment | NA | NA | 0 |
| S10_extracted_43964 | 93939 | No | NA | 143 | 15 | 0 | Complete | High-quality | 100 | DTR (high-confidence) | 0 |
| S1_extracted_k141_113056 | 3210 | No | NA | 4 | 0 | 4 | Low-quality | Genome-fragment | 0.93 | AAI-based (medium-confidence) | 0 |
| S10_extracted_43999 | 1346 | No | NA | 1 | 0 | 1 | Not-determined | Genome-fragment | NA | NA | 0 |
| S3_vs_k141_97452\|\|full | 3210 | No | NA | 2 | 0 | 2 | Low-quality | Genome-fragment | 2.23 | AAI-based (medium-confidence) | 0 |
| S16_vs_2336\|\|full | 3210 | No | NA | 7 | 2 | 0 | Low-quality | Genome-fragment | 8.06 | AAI-based (high-confidence) | 0 |
| S10_extracted_44161 | 2481 | No | NA | 2 | 0 | 2 | Not-determined | Genome-fragment | NA | NA | 0 |
| S10_extracted_44316 | 2646 | No | NA | 3 | 0 | 0 | Not-determined | Genome-fragment | NA | NA | 0 |
| S10_extracted_44332 | 1122 | No | NA | 2 | 0 | 2 | Not-determined | Genome-fragment | NA | NA | 0 |
| S10_extracted_44474 | 1337 | No | NA | 2 | 0 | 1 | Not-determined | Genome-fragment | NA | NA | 0 |
| S17_extracted_k141_123439 | 3211 | No | NA | 2 | 1 | 0 | Low-quality | Genome-fragment | 4.86 | HMM-based (lower-bound) | 0 |
| S11_vs_k141_83651\|\|full | 3211 | No | NA | 7 | 1 | 0 | Low-quality | Genome-fragment | 3.38 | HMM-based (lower-bound) | 0 |
| S10_extracted_44637 | 3331 | No | NA | 4 | 0 | 0 | Not-determined | Genome-fragment | NA | NA | 0 |
| S10_extracted_44750 | 4858 | No | NA | 7 | 0 | 5 | Not-determined | Genome-fragment | NA | NA | 0 |
| S10_extracted_44821 | 1152 | No | NA | 1 | 0 | 1 | Not-determined | Genome-fragment | NA | NA | 0 |
| S10_extracted_44859 | 1040 | No | NA | 1 | 0 | 0 | Not-determined | Genome-fragment | NA | NA | 0 |
| S5_vs_k141_40547\|\|full | 3211 | No | NA | 3 | 3 | 0 | Low-quality | Genome-fragment | 9.2 | AAI-based (high-confidence) | 0 |
| S10_extracted_44949 | 1044 | No | NA | 1 | 0 | 1 | Not-determined | Genome-fragment | NA | NA | 0 |
| S10_extracted_45003 | 1110 | No | NA | 2 | 0 | 1 | Not-determined | Genome-fragment | NA | NA | 0 |
| S10_extracted_45004 | 5021 | No | NA | 5 | 0 | 3 | Not-determined | Genome-fragment | NA | NA | 0 |
| S10_extracted_45117 | 3256 | No | NA | 3 | 0 | 3 | Not-determined | Genome-fragment | NA | NA | 0 |
| S10_extracted_45152 | 2009 | No | NA | 1 | 0 | 0 | Not-determined | Genome-fragment | NA | NA | 0 |
| S10_extracted_45157 | 1150 | No | NA | 1 | 0 | 1 | Not-determined | Genome-fragment | NA | NA | 0 |
| S10_extracted_45352 | 3281 | No | NA | 4 | 0 | 1 | Not-determined | Genome-fragment | NA | NA | 0 |
| S10_extracted_45362 | 3010 | No | NA | 2 | 0 | 2 | Not-determined | Genome-fragment | NA | NA | 0 |
| S10_extracted_45580 | 1455 | No | NA | 3 | 0 | 0 | Not-determined | Genome-fragment | NA | NA | 0 |
| S10_extracted_45638 | 2933 | No | NA | 4 | 0 | 0 | Not-determined | Genome-fragment | NA | NA | 0 |
| S10_extracted_45668 | 1006 | No | NA | 1 | 0 | 1 | Not-determined | Genome-fragment | NA | NA | 0 |
| S10_extracted_45714 | 1328 | No | NA | 3 | 0 | 1 | Not-determined | Genome-fragment | NA | NA | 0 |
| S10_extracted_45758 | 1331 | No | NA | 3 | 0 | 0 | Not-determined | Genome-fragment | NA | NA | 0 |
| S7_vs_79919\|\|full | 3211 | No | NA | 5 | 0 | 2 | Low-quality | Genome-fragment | 3.36 | AAI-based (medium-confidence) | 0 |
| S10_extracted_45948 | 1503 | No | NA | 2 | 0 | 1 | Not-determined | Genome-fragment | NA | NA | 0 |
| S10_extracted_45958 | 1078 | No | NA | 2 | 0 | 1 | Not-determined | Genome-fragment | NA | NA | 0 |
| S10_vs_3128\|\|full | 3211 | No | NA | 4 | 0 | 0 | Low-quality | Genome-fragment | 2.22 | AAI-based (high-confidence) | 0 |
| S10_extracted_46090 | 1052 | No | NA | 1 | 0 | 1 | Not-determined | Genome-fragment | NA | NA | 0 |
| S10_extracted_46139 | 1078 | No | NA | 1 | 0 | 1 | Not-determined | Genome-fragment | NA | NA | 0 |
| S12_vs_k141_12310\|\|full | 3211 | No | NA | 3 | 3 | 0 | Low-quality | Genome-fragment | 5.61 | HMM-based (lower-bound) | 0 |
| S10_extracted_46257 | 3619 | No | NA | 2 | 0 | 1 | Not-determined | Genome-fragment | NA | NA | 0 |
| S10_extracted_46281 | 1002 | No | NA | 2 | 0 | 2 | Not-determined | Genome-fragment | NA | NA | 0 |
| S10_extracted_46285 | 2786 | No | NA | 4 | 0 | 4 | Not-determined | Genome-fragment | NA | NA | 0 |
| S10_extracted_46302 | 1071 | No | NA | 1 | 0 | 1 | Not-determined | Genome-fragment | NA | NA | 0 |
| S10_extracted_46307 | 3909 | No | NA | 4 | 0 | 3 | Not-determined | Genome-fragment | NA | NA | 0 |
| S10_extracted_46328 | 24210 | No | NA | 32 | 12 | 0 | Medium-quality | Genome-fragment | 55.73 | AAI-based (high-confidence) | 0 |
| S10_extracted_46444 | 4061 | No | NA | 5 | 0 | 4 | Not-determined | Genome-fragment | NA | NA | 0 |
| S10_extracted_46522 | 1067 | No | NA | 1 | 0 | 0 | Not-determined | Genome-fragment | NA | NA | 0 |
| S10_extracted_46732 | 3435 | No | NA | 2 | 0 | 2 | Not-determined | Genome-fragment | NA | NA | 0 |
| S10_extracted_46760 | 1656 | No | NA | 2 | 0 | 2 | Not-determined | Genome-fragment | NA | NA | 0 |
| S16_extracted_115061 | 3212 | No | NA | 5 | 4 | 0 | Low-quality | Genome-fragment | 5.34 | AAI-based (high-confidence) | 0 |
| S10_extracted_46824 | 6076 | No | NA | 2 | 0 | 0 | Not-determined | Genome-fragment | NA | NA | 0 |
| S10_extracted_46828 | 1398 | No | NA | 1 | 0 | 0 | Not-determined | Genome-fragment | NA | NA | 0 |
| S10_extracted_46862 | 1801 | No | NA | 4 | 0 | 3 | Not-determined | Genome-fragment | NA | NA | 0 |
| S15_vs_57898\|\|full | 3212 | No | NA | 8 | 0 | 0 | Low-quality | Genome-fragment | 5.24 | AAI-based (high-confidence) | 0 |
| S10_extracted_47170 | 6477 | No | NA | 6 | 1 | 0 | Medium-quality | Genome-fragment | 54.49 | AAI-based (medium-confidence) | 0 |
| S9_extracted_k141_6107 | 3213 | No | NA | 6 | 1 | 0 | Low-quality | Genome-fragment | 7.35 | AAI-based (high-confidence) | 0 |
| S10_extracted_47511 | 1115 | No | NA | 1 | 0 | 1 | Not-determined | Genome-fragment | NA | NA | 0 |
| S10_extracted_47615 | 3039 | No | NA | 5 | 0 | 1 | Not-determined | Genome-fragment | NA | NA | 0 |
| S10_extracted_47792 | 1091 | No | NA | 1 | 0 | 1 | Not-determined | Genome-fragment | NA | NA | 0 |
| S10_extracted_47816 | 1229 | No | NA | 2 | 0 | 0 | Not-determined | Genome-fragment | NA | NA | 0 |
| S10_extracted_47859 | 1135 | No | NA | 1 | 0 | 0 | Not-determined | Genome-fragment | NA | NA | 0 |
| S10_extracted_47872 | 1171 | No | NA | 1 | 0 | 1 | Not-determined | Genome-fragment | NA | NA | 0 |
| S10_extracted_47912 | 1934 | No | NA | 2 | 0 | 1 | Not-determined | Genome-fragment | NA | NA | 0 |
| S10_extracted_47913 | 1532 | No | NA | 2 | 0 | 1 | Not-determined | Genome-fragment | NA | NA | 0 |
| S10_extracted_47948 | 1502 | No | NA | 1 | 0 | 0 | Not-determined | Genome-fragment | NA | NA | 0 |
| S10_extracted_48109 | 1252 | No | NA | 1 | 0 | 0 | Not-determined | Genome-fragment | NA | NA | 0 |
| S10_extracted_48112 | 1622 | No | NA | 3 | 0 | 2 | Not-determined | Genome-fragment | NA | NA | 0 |
| S10_extracted_48194 | 8916 | No | NA | 7 | 0 | 4 | Not-determined | Genome-fragment | NA | NA | 0 |
| S10_extracted_48230 | 3657 | No | NA | 5 | 0 | 4 | Not-determined | Genome-fragment | NA | NA | 0 |
| S10_extracted_48275 | 1217 | No | NA | 3 | 0 | 3 | Not-determined | Genome-fragment | NA | NA | 0 |
| S3_extracted_k141_50700 | 3213 | No | NA | 2 | 0 | 1 | Low-quality | Genome-fragment | 1.62 | AAI-based (high-confidence) | 0 |
| S3_vs_k141_24967\|\|full | 3213 | No | NA | 10 | 2 | 0 | Low-quality | Genome-fragment | 8.12 | AAI-based (high-confidence) | 0 |
| S10_extracted_48738 | 20494 | No | NA | 10 | 0 | 6 | Not-determined | Genome-fragment | NA | NA | 0 |
| S10_extracted_48751 | 1661 | No | NA | 2 | 0 | 0 | Not-determined | Genome-fragment | NA | NA | 0 |
| S5_vs_k141_37441\|\|full | 3213 | No | NA | 4 | 0 | 2 | Low-quality | Genome-fragment | 0.79 | AAI-based (medium-confidence) | 0 |
| S5_vs_k141_86118\|\|full | 3213 | No | NA | 3 | 0 | 0 | Low-quality | Genome-fragment | 7.28 | AAI-based (high-confidence) | 0 |
| S10_extracted_4462 | 3213 | No | NA | 4 | 2 | 0 | Low-quality | Genome-fragment | 7.15 | AAI-based (medium-confidence) | 0 |
| S10_extracted_48908 | 1241 | No | NA | 2 | 0 | 1 | Not-determined | Genome-fragment | NA | NA | 0 |
| S10_extracted_48938 | 1244 | No | NA | 3 | 0 | 1 | Not-determined | Genome-fragment | NA | NA | 0 |
| S11_vs_k141_77815\|\|full | 3214 | No | NA | 6 | 1 | 0 | Low-quality | Genome-fragment | 2.02 | HMM-based (lower-bound) | 0 |
| S10_extracted_49180 | 4238 | No | NA | 2 | 0 | 2 | Not-determined | Genome-fragment | NA | NA | 0 |
| S10_extracted_49322 | 4275 | No | NA | 2 | 0 | 2 | Not-determined | Genome-fragment | NA | NA | 0 |
| S10_extracted_49401 | 2955 | No | NA | 3 | 0 | 2 | Not-determined | Genome-fragment | NA | NA | 0 |
| S10_extracted_49468 | 1067 | No | NA | 1 | 0 | 1 | Not-determined | Genome-fragment | NA | NA | 0 |
| S16_extracted_10021 | 3214 | No | NA | 8 | 3 | 0 | Low-quality | Genome-fragment | 6.35 | AAI-based (high-confidence) | 0 |
| S10_extracted_49532 | 1012 | No | NA | 1 | 0 | 0 | Not-determined | Genome-fragment | NA | NA | 0 |
| S10_extracted_49552 | 1135 | No | NA | 2 | 0 | 0 | Not-determined | Genome-fragment | NA | NA | 0 |
| S10_extracted_49703 | 1369 | No | NA | 3 | 0 | 1 | Not-determined | Genome-fragment | NA | NA | 0 |
| S10_extracted_49837 | 2021 | No | NA | 4 | 0 | 4 | Not-determined | Genome-fragment | NA | NA | 0 |
| S10_extracted_49851 | 15276 | No | NA | 14 | 0 | 4 | Not-determined | Genome-fragment | NA | NA | 0 |
| S10_extracted_49914 | 1290 | No | NA | 1 | 0 | 1 | Not-determined | Genome-fragment | NA | NA | 0 |
| S10_extracted_49994 | 1109 | No | NA | 2 | 0 | 2 | Not-determined | Genome-fragment | NA | NA | 0 |
| S10_extracted_49996 | 1820 | No | NA | 3 | 0 | 1 | Not-determined | Genome-fragment | NA | NA | 0 |
| S10_extracted_50014 | 1685 | No | NA | 2 | 0 | 2 | Not-determined | Genome-fragment | NA | NA | 0 |
| S5_vs_k141_28221\|\|full | 3215 | No | NA | 9 | 3 | 0 | Low-quality | Genome-fragment | 2.64 | HMM-based (lower-bound) | 0 |
| S7_extracted_5479 | 3215 | No | NA | 7 | 1 | 0 | Low-quality | Genome-fragment | 8.97 | AAI-based (high-confidence) | 0 |
| S7_vs_110952\|\|full | 3215 | No | NA | 4 | 3 | 0 | Low-quality | Genome-fragment | 6.74 | AAI-based (high-confidence) | 0 |
| S10_extracted_50397 | 1456 | No | NA | 2 | 0 | 1 | Not-determined | Genome-fragment | NA | NA | 0 |
| S10_extracted_50703 | 3010 | No | NA | 3 | 0 | 2 | Not-determined | Genome-fragment | NA | NA | 0 |
| S10_extracted_50797 | 2043 | No | NA | 2 | 0 | 1 | Not-determined | Genome-fragment | NA | NA | 0 |
| S14_vs_k141_80705\|\|full | 3217 | No | NA | 3 | 2 | 0 | Low-quality | Genome-fragment | 10.41 | AAI-based (high-confidence) | 0 |
| S5_vs_k141_136889\|\|full | 3217 | No | NA | 8 | 0 | 0 | Low-quality | Genome-fragment | 8.63 | AAI-based (medium-confidence) | 0 |
| S7_extracted_34561 | 3217 | No | NA | 5 | 1 | 0 | Low-quality | Genome-fragment | 1.85 | AAI-based (high-confidence) | 0 |
| S8_vs_k141_202206\|\|full | 3217 | No | NA | 3 | 1 | 0 | Low-quality | Genome-fragment | 7.33 | AAI-based (medium-confidence) | 0 |

| | | | | | | | | | | | |
|---|---|---|---|---|---|---|---|---|---|---|---|
| S10_extracted_51103 | 1087 | No | NA | 1 | 0 | 1 | Not-determined | Genome-fragment | NA | NA | 0 |
| S10_extracted_51116 | 3001 | No | NA | 4 | 0 | 0 | Not-determined | Genome-fragment | NA | NA | 0 |
| S11_vs_k141_51980\|\|full | 3218 | No | NA | 7 | 0 | 0 | Low-quality | Genome-fragment | 6.08 | AAI-based (high-confidence) | 0 |
| S10_extracted_51288 | 7984 | No | NA | 6 | 0 | 4 | Not-determined | Genome-fragment | NA | NA | 0 |
| S8_vs_k141_62450\|\|full | 3218 | No | NA | 4 | 0 | 2 | Low-quality | Genome-fragment | 1.85 | AAI-based (high-confidence) | 0 |
| S10_extracted_51379 | 1521 | No | NA | 3 | 0 | 2 | Not-determined | Genome-fragment | NA | NA | 0 |
| S10_extracted_51418 | 1555 | No | NA | 3 | 0 | 0 | Not-determined | Genome-fragment | NA | NA | 0 |
| S10_extracted_51430 | 1344 | No | NA | 2 | 0 | 1 | Not-determined | Genome-fragment | NA | NA | 0 |
| S14_vs_k141_111897\|\|full | 3219 | No | NA | 3 | 0 | 2 | Low-quality | Genome-fragment | 1.85 | AAI-based (medium-confidence) | 0 |
| S10_extracted_51672 | 1506 | No | NA | 1 | 0 | 0 | Not-determined | Genome-fragment | NA | NA | 0 |
| S10_extracted_51742 | 6600 | No | NA | 7 | 0 | 4 | Not-determined | Genome-fragment | NA | NA | 0 |
| S10_extracted_51860 | 2518 | No | NA | 2 | 0 | 2 | Not-determined | Genome-fragment | NA | NA | 0 |
| S3_extracted_k141_54558 | 3219 | No | NA | 7 | 2 | 0 | Low-quality | Genome-fragment | 9.5 | AAI-based (high-confidence) | 0 |
| S10_extracted_51914 | 3205 | No | NA | 4 | 0 | 3 | Not-determined | Genome-fragment | NA | NA | 0 |
| S10_extracted_52136 | 1592 | No | NA | 1 | 0 | 1 | Not-determined | Genome-fragment | NA | NA | 0 |
| S7_extracted_48019 | 3219 | No | NA | 7 | 1 | 0 | Low-quality | Genome-fragment | 5.55 | HMM-based (lower-bound) | 0 |
| S10_extracted_52169 | 4660 | No | NA | 4 | 0 | 3 | Not-determined | Genome-fragment | NA | NA | 0 |
| S9_vs_k141_87940\|\|full | 3220 | No | NA | 7 | 2 | 0 | Low-quality | Genome-fragment | 7.89 | AAI-based (high-confidence) | 0 |
| S14_extracted_k141_95782 | 3220 | No | NA | 8 | 1 | 0 | Low-quality | Genome-fragment | 2.26 | AAI-based (medium-confidence) | 0 |
| S10_extracted_52399 | 1728 | No | NA | 1 | 0 | 1 | Not-determined | Genome-fragment | NA | NA | 0 |
| S10_extracted_52430 | 1344 | No | NA | 1 | 0 | 1 | Not-determined | Genome-fragment | NA | NA | 0 |
| S10_extracted_52510 | 1048 | No | NA | 1 | 0 | 1 | Not-determined | Genome-fragment | NA | NA | 0 |
| S10_extracted_52511 | 1604 | No | NA | 2 | 0 | 1 | Not-determined | Genome-fragment | NA | NA | 0 |
| S10_extracted_52553 | 1058 | No | NA | 1 | 0 | 1 | Not-determined | Genome-fragment | NA | NA | 0 |
| S10_extracted_52577 | 1221 | No | NA | 2 | 0 | 0 | Not-determined | Genome-fragment | NA | NA | 0 |
| S10_extracted_52628 | 1040 | No | NA | 3 | 0 | 1 | Not-determined | Genome-fragment | NA | NA | 0 |
| S10_extracted_52674 | 2997 | No | NA | 1 | 0 | 1 | Not-determined | Genome-fragment | NA | NA | 0 |
| S10_extracted_52893 | 1028 | No | NA | 3 | 0 | 2 | Not-determined | Genome-fragment | NA | NA | 0 |
| S10_extracted_52953 | 1283 | No | NA | 1 | 0 | 0 | Not-determined | Genome-fragment | NA | NA | 0 |
| S10_extracted_53052 | 2046 | No | NA | 2 | 0 | 1 | Not-determined | Genome-fragment | NA | NA | 0 |
| S10_extracted_53064 | 1202 | No | NA | 3 | 0 | 0 | Not-determined | Genome-fragment | NA | NA | 0 |
| S10_extracted_53254 | 1077 | No | NA | 2 | 0 | 1 | Not-determined | Genome-fragment | NA | NA | 0 |
| S10_extracted_53288 | 1741 | No | NA | 2 | 0 | 0 | Not-determined | Genome-fragment | NA | NA | 0 |
| S7_vs_522\|\|full | 3220 | No | NA | 6 | 2 | 0 | Low-quality | Genome-fragment | 7.54 | AAI-based (high-confidence) | 0 |
| S10_extracted_53439 | 5770 | No | NA | 4 | 0 | 2 | Not-determined | Genome-fragment | NA | NA | 0 |
| S10_extracted_65088 | 3220 | No | NA | 4 | 0 | 3 | Low-quality | Genome-fragment | 2.28 | AAI-based (medium-confidence) | 0 |
| S5_vs_k141_114105\|\|full | 3222 | No | NA | 7 | 0 | 0 | Low-quality | Genome-fragment | 7.49 | AAI-based (medium-confidence) | 0 |
| S10_extracted_53604 | 1256 | No | NA | 2 | 0 | 1 | Not-determined | Genome-fragment | NA | NA | 0 |
| S10_extracted_53637 | 2968 | No | NA | 3 | 0 | 3 | Not-determined | Genome-fragment | NA | NA | 0 |
| S10_extracted_53676 | 1142 | No | NA | 1 | 0 | 1 | Not-determined | Genome-fragment | NA | NA | 0 |
| S8_vs_k141_18364\|\|full | 3222 | No | NA | 2 | 0 | 1 | Low-quality | Genome-fragment | 3.84 | AAI-based (medium-confidence) | 0 |
| S10_extracted_53778 | 1149 | No | NA | 1 | 0 | 0 | Not-determined | Genome-fragment | NA | NA | 0 |
| S10_extracted_53816 | 1983 | No | NA | 6 | 0 | 0 | Not-determined | Genome-fragment | NA | NA | 0 |
| S10_extracted_53927 | 1608 | No | NA | 2 | 0 | 0 | Not-determined | Genome-fragment | NA | NA | 0 |
| S11_vs_k141_63162\|\|full | 3223 | No | NA | 7 | 0 | 1 | Low-quality | Genome-fragment | 9.4 | AAI-based (high-confidence) | 0 |
| S12_vs_k141_84188\|\|full | 3223 | No | NA | 3 | 3 | 0 | Low-quality | Genome-fragment | 9.06 | AAI-based (medium-confidence) | 0 |
| S10_extracted_54150 | 1120 | No | NA | 2 | 0 | 0 | Not-determined | Genome-fragment | NA | NA | 0 |
| S4_extracted_4719 | 3223 | No | NA | 4 | 3 | 0 | Low-quality | Genome-fragment | 10.62 | AAI-based (high-confidence) | 0 |
| S10_extracted_54341 | 3386 | No | NA | 4 | 0 | 0 | Not-determined | Genome-fragment | NA | NA | 0 |
| S10_extracted_54367 | 1309 | No | NA | 2 | 0 | 1 | Not-determined | Genome-fragment | NA | NA | 0 |
| S10_extracted_54390 | 4462 | No | NA | 7 | 0 | 0 | Not-determined | Genome-fragment | NA | NA | 0 |
| S10_extracted_54461 | 1022 | No | NA | 2 | 0 | 0 | Not-determined | Genome-fragment | NA | NA | 0 |
| S10_extracted_54515 | 4261 | No | NA | 2 | 0 | 2 | Not-determined | Genome-fragment | NA | NA | 0 |
| S10_extracted_54591 | 1113 | No | NA | 1 | 0 | 1 | Not-determined | Genome-fragment | NA | NA | 0 |
| S10_extracted_54600 | 1089 | No | NA | 1 | 0 | 1 | Not-determined | Genome-fragment | NA | NA | 0 |
| S10_extracted_54601 | 5846 | No | NA | 3 | 0 | 0 | Not-determined | Genome-fragment | NA | NA | 0 |
| S9_vs_k141_4742\|\|lt2gene | 3225 | No | NA | 1 | 1 | 0 | Low-quality | Genome-fragment | 4.71 | HMM-based (lower-bound) | 0 |
| S10_extracted_54750 | 1459 | No | NA | 1 | 0 | 1 | Not-determined | Genome-fragment | NA | NA | 0 |
| S10_extracted_54763 | 5994 | No | NA | 6 | 0 | 6 | Not-determined | Genome-fragment | NA | NA | 0 |
| S14_extracted_k141_7399 | 3225 | No | NA | 6 | 0 | 1 | Low-quality | Genome-fragment | 5.91 | AAI-based (medium-confidence) | 0 |
| S10_extracted_54894 | 1062 | No | NA | 2 | 0 | 0 | Not-determined | Genome-fragment | NA | NA | 0 |
| S10_extracted_55011 | 1258 | No | NA | 2 | 0 | 1 | Not-determined | Genome-fragment | NA | NA | 0 |
| S10_extracted_55184 | 1252 | No | NA | 2 | 0 | 1 | Not-determined | Genome-fragment | NA | NA | 0 |
| S10_extracted_55249 | 1391 | No | NA | 2 | 0 | 0 | Not-determined | Genome-fragment | NA | NA | 0 |
| S10_extracted_55258 | 1170 | No | NA | 1 | 0 | 0 | Not-determined | Genome-fragment | NA | NA | 0 |
| S12_vs_k141_32186\|\|full | 3225 | No | NA | 3 | 0 | 0 | Low-quality | Genome-fragment | 7.42 | AAI-based (medium-confidence) | 0 |
| S10_extracted_55409 | 1015 | No | NA | 2 | 0 | 0 | Not-determined | Genome-fragment | NA | NA | 0 |
| S10_extracted_55534 | 1502 | No | NA | 4 | 0 | 0 | Not-determined | Genome-fragment | NA | NA | 0 |
| S10_extracted_55550 | 3300 | No | NA | 2 | 0 | 2 | Not-determined | Genome-fragment | NA | NA | 0 |
| S10_extracted_55844 | 4857 | No | NA | 4 | 0 | 2 | Not-determined | Genome-fragment | NA | NA | 0 |
| S14_vs_k141_75254\|\|full | 3226 | No | NA | 5 | 1 | 0 | Low-quality | Genome-fragment | 6.9 | AAI-based (high-confidence) | 0 |
| S16_extracted_119730 | 3226 | No | NA | 6 | 1 | 0 | Low-quality | Genome-fragment | 8.36 | AAI-based (high-confidence) | 0 |
| S10_extracted_56081 | 1061 | No | NA | 2 | 0 | 0 | Not-determined | Genome-fragment | NA | NA | 0 |
| S16_extracted_11094 | 3227 | No | NA | 4 | 0 | 3 | Low-quality | Genome-fragment | 0.94 | AAI-based (medium-confidence) | 0 |
| S10_extracted_56154 | 3875 | No | NA | 2 | 0 | 2 | Not-determined | Genome-fragment | NA | NA | 0 |
| S10_extracted_56176 | 1856 | No | NA | 3 | 0 | 2 | Not-determined | Genome-fragment | NA | NA | 0 |
| S10_extracted_56221 | 9084 | No | NA | 4 | 0 | 1 | Not-determined | Genome-fragment | NA | NA | 0 |
| S10_extracted_56358 | 1674 | No | NA | 1 | 0 | 0 | Not-determined | Genome-fragment | NA | NA | 0 |
| S10_extracted_56448 | 3356 | No | NA | 4 | 0 | 0 | Not-determined | Genome-fragment | NA | NA | 0 |
| S10_extracted_56518 | 1336 | No | NA | 2 | 0 | 0 | Not-determined | Genome-fragment | NA | NA | 0 |
| S10_extracted_56570 | 6394 | No | NA | 4 | 0 | 4 | Not-determined | Genome-fragment | NA | NA | 0 |
| S10_extracted_56614 | 5151 | No | NA | 7 | 0 | 1 | Not-determined | Genome-fragment | NA | NA | 0 |
| S10_extracted_56624 | 2113 | No | NA | 1 | 0 | 1 | Not-determined | Genome-fragment | NA | NA | 0 |
| S10_extracted_56706 | 1185 | No | NA | 1 | 0 | 1 | Not-determined | Genome-fragment | NA | NA | 0 |
| S10_extracted_56836 | 2124 | No | NA | 1 | 0 | 1 | Not-determined | Genome-fragment | NA | NA | 0 |
| S16_vs_49200\|\|full | 3227 | Yes | 1788 | 4 | 1 | 1 | Low-quality | Genome-fragment | 2.71 | HMM-based (lower-bound) | 44.59 |
| S10_extracted_57007 | 1278 | No | NA | 2 | 0 | 0 | Not-determined | Genome-fragment | NA | NA | 0 |
| S13_extracted_33147 | 3228 | No | NA | 4 | 0 | 1 | Low-quality | Genome-fragment | 2.23 | AAI-based (high-confidence) | 0 |
| S10_extracted_57109 | 1698 | No | NA | 2 | 0 | 0 | Not-determined | Genome-fragment | NA | NA | 0 |
| S16_extracted_157781 | 3228 | No | NA | 3 | 2 | 0 | Low-quality | Genome-fragment | 7.05 | AAI-based (high-confidence) | 0 |
| S12_vs_k141_95051\|\|full | 3228 | No | NA | 7 | 1 | 0 | Low-quality | Genome-fragment | 5.43 | HMM-based (lower-bound) | 0 |
| S10_extracted_57233 | 1048 | No | NA | 3 | 0 | 3 | Not-determined | Genome-fragment | NA | NA | 0 |
| S17_vs_k141_92978\|\|full | 3229 | No | NA | 3 | 0 | 2 | Low-quality | Genome-fragment | 1.09 | AAI-based (medium-confidence) | 0 |
| S10_extracted_57358 | 1339 | No | NA | 1 | 0 | 1 | Not-determined | Genome-fragment | NA | NA | 0 |
| S10_extracted_57437 | 1165 | No | NA | 2 | 0 | 2 | Not-determined | Genome-fragment | NA | NA | 0 |
| S10_extracted_57447 | 1006 | No | NA | 1 | 0 | 1 | Not-determined | Genome-fragment | NA | NA | 0 |
| S14_extracted_k141_27469 | 3229 | No | NA | 7 | 7 | 0 | Low-quality | Genome-fragment | 10.64 | AAI-based (high-confidence) | 0 |
| S16_vs_16406\|\|full | 3229 | No | NA | 4 | 3 | 0 | Low-quality | Genome-fragment | 9.53 | AAI-based (high-confidence) | 0 |
| S10_extracted_57593 | 5158 | No | NA | 4 | 0 | 4 | Not-determined | Genome-fragment | NA | NA | 0 |
| S10_extracted_57644 | 1336 | No | NA | 4 | 0 | 2 | Not-determined | Genome-fragment | NA | NA | 0 |
| S10_extracted_57705 | 13928 | No | NA | 18 | 6 | 0 | Medium-quality | Genome-fragment | 62.54 | AAI-based (medium-confidence) | 0 |
| S6_vs_k141_22193\|\|full | 3229 | No | NA | 6 | 0 | 1 | Low-quality | Genome-fragment | 7.55 | AAI-based (high-confidence) | 0 |
| S10_extracted_57880 | 1855 | No | NA | 1 | 0 | 1 | Not-determined | Genome-fragment | NA | NA | 0 |
| S10_extracted_57903 | 6837 | No | NA | 4 | 0 | 2 | Not-determined | Genome-fragment | NA | NA | 0 |
| S14_extracted_k141_75012 | 3230 | No | NA | 4 | 3 | 0 | Low-quality | Genome-fragment | 5.58 | AAI-based (high-confidence) | 0 |
| S14_extracted_k141_98128 | 3230 | No | NA | 8 | 6 | 0 | Low-quality | Genome-fragment | 10.44 | AAI-based (high-confidence) | 0 |
| S10_extracted_58408 | 1675 | No | NA | 1 | 0 | 1 | Not-determined | Genome-fragment | NA | NA | 0 |
| S10_extracted_58538 | 4242 | No | NA | 6 | 0 | 6 | Not-determined | Genome-fragment | NA | NA | 0 |
| S10_extracted_58540 | 2973 | No | NA | 1 | 0 | 1 | Not-determined | Genome-fragment | NA | NA | 0 |
| S14_vs_k141_95839\|\|full | 3230 | No | NA | 6 | 0 | 0 | Low-quality | Genome-fragment | 9.77 | AAI-based (high-confidence) | 0 |
| S10_extracted_58665 | 1084 | No | NA | 1 | 0 | 1 | Not-determined | Genome-fragment | NA | NA | 0 |
| S10_extracted_58679 | 2398 | No | NA | 3 | 0 | 2 | Not-determined | Genome-fragment | NA | NA | 0 |
| S10_extracted_58687 | 1630 | No | NA | 1 | 0 | 0 | Not-determined | Genome-fragment | NA | NA | 0 |
| S14_vs_k141_100157\|\|full | 3230 | No | NA | 3 | 2 | 0 | Low-quality | Genome-fragment | 5.16 | AAI-based (high-confidence) | 0 |
| S8_vs_k141_106194\|\|full | 3230 | No | NA | 4 | 1 | 0 | Low-quality | Genome-fragment | 1.5 | HMM-based (lower-bound) | 0 |
| S12_extracted_k141_152816 | 3231 | No | NA | 6 | 1 | 0 | Low-quality | Genome-fragment | 4.16 | HMM-based (lower-bound) | 0 |
| S10_extracted_58835 | 3089 | No | NA | 5 | 0 | 4 | Not-determined | Genome-fragment | NA | NA | 0 |
| S10_extracted_59015 | 1175 | No | NA | 1 | 0 | 1 | Not-determined | Genome-fragment | NA | NA | 0 |
| S10_extracted_59019 | 1035 | No | NA | 2 | 0 | 1 | Not-determined | Genome-fragment | NA | NA | 0 |
| S10_extracted_59020 | 1323 | No | NA | 3 | 0 | 2 | Not-determined | Genome-fragment | NA | NA | 0 |
| S10_extracted_59260 | 1481 | No | NA | 2 | 0 | 0 | Not-determined | Genome-fragment | NA | NA | 0 |
| S10_extracted_59270 | 2404 | No | NA | 5 | 0 | 2 | Not-determined | Genome-fragment | NA | NA | 0 |
| S10_extracted_59414 | 2153 | No | NA | 2 | 0 | 1 | Not-determined | Genome-fragment | NA | NA | 0 |
| S10_extracted_59575 | 5301 | No | NA | 4 | 0 | 1 | Not-determined | Genome-fragment | NA | NA | 0 |
| S10_extracted_59712 | 2664 | No | NA | 2 | 0 | 2 | Not-determined | Genome-fragment | NA | NA | 0 |
| S10_extracted_59732 | 3423 | No | NA | 5 | 0 | 4 | Not-determined | Genome-fragment | NA | NA | 0 |
| S10_extracted_59772 | 1022 | No | NA | 2 | 0 | 0 | Not-determined | Genome-fragment | NA | NA | 0 |
| S10_extracted_59785 | 1286 | No | NA | 1 | 0 | 1 | Not-determined | Genome-fragment | NA | NA | 0 |
| S10_extracted_59839 | 1003 | No | NA | 2 | 0 | 2 | Not-determined | Genome-fragment | NA | NA | 0 |
| S10_extracted_59906 | 1071 | No | NA | 2 | 0 | 1 | Not-determined | Genome-fragment | NA | NA | 0 |
| S10_extracted_59974 | 1005 | No | NA | 4 | 0 | 0 | Not-determined | Genome-fragment | NA | NA | 0 |
| S12_vs_k141_42864\|\|full | 3231 | No | NA | 4 | 3 | 0 | Low-quality | Genome-fragment | 9 | AAI-based (medium-confidence) | 0 |
| S3_vs_k141_78597\|\|full | 3232 | No | NA | 8 | 0 | 0 | Low-quality | Genome-fragment | 8.48 | AAI-based (high-confidence) | 0 |
| S10_extracted_60173 | 2315 | No | NA | 2 | 0 | 0 | Not-determined | Genome-fragment | NA | NA | 0 |
| S12_vs_k141_17968\|\|full | 3232 | No | NA | 3 | 0 | 0 | Low-quality | Genome-fragment | 5.49 | AAI-based (medium-confidence) | 0 |
| S10_extracted_60305 | 6711 | No | NA | 4 | 0 | 3 | Not-determined | Genome-fragment | NA | NA | 0 |
| S12_extracted_k141_188238 | 3234 | No | NA | 4 | 0 | 2 | Low-quality | Genome-fragment | 0.78 | AAI-based (high-confidence) | 0 |
| S10_extracted_60396 | 3184 | No | NA | 3 | 0 | 2 | Not-determined | Genome-fragment | NA | NA | 0 |
| S10_extracted_60458 | 3136 | No | NA | 4 | 0 | 1 | Not-determined | Genome-fragment | NA | NA | 0 |
| S15_extracted_3962 | 3234 | No | NA | 4 | 0 | 2 | Low-quality | Genome-fragment | 1.95 | AAI-based (high-confidence) | 0 |
| S10_extracted_60698 | 1024 | No | NA | 1 | 0 | 1 | Not-determined | Genome-fragment | NA | NA | 0 |
| S10_extracted_60868 | 1164 | No | NA | 1 | 0 | 1 | Not-determined | Genome-fragment | NA | NA | 0 |
| S10_extracted_60897 | 1236 | No | NA | 1 | 0 | 0 | Not-determined | Genome-fragment | NA | NA | 0 |
| S10_extracted_61048 | 1274 | No | NA | 4 | 0 | 1 | Not-determined | Genome-fragment | NA | NA | 0 |
| S10_extracted_61283 | 1465 | No | NA | 2 | 0 | 1 | Not-determined | Genome-fragment | NA | NA | 0 |
| S15_extracted_27285 | 3234 | No | NA | 7 | 0 | 0 | Low-quality | Genome-fragment | 5.02 | AAI-based (medium-confidence) | 0 |
| S10_extracted_61547 | 4579 | No | NA | 4 | 0 | 3 | Not-determined | Genome-fragment | NA | NA | 0 |
| S7_extracted_122964 | 3235 | No | NA | 6 | 5 | 0 | Low-quality | Genome-fragment | 8.13 | AAI-based (high-confidence) | 0 |
| S12_vs_k141_91335\|\|full | 3235 | No | NA | 6 | 0 | 1 | Low-quality | Genome-fragment | 5.23 | AAI-based (medium-confidence) | 0 |
| S17_extracted_k141_39001 | 3236 | No | NA | 6 | 0 | 0 | Low-quality | Genome-fragment | 3.92 | AAI-based (medium-confidence) | 0 |
| S10_extracted_62075 | 1122 | No | NA | 1 | 0 | 0 | Not-determined | Genome-fragment | NA | NA | 0 |
| S10_extracted_62152 | 1871 | No | NA | 1 | 0 | 1 | Not-determined | Genome-fragment | NA | NA | 0 |
| S10_extracted_62230 | 23520 | No | NA | 28 | 23 | 0 | Medium-quality | Genome-fragment | 58.57 | AAI-based (medium-confidence) | 0 |
| S10_extracted_62255 | 1605 | No | NA | 1 | 0 | 1 | Not-determined | Genome-fragment | NA | NA | 0 |
| S10_extracted_62270 | 3909 | No | NA | 2 | 0 | 1 | Not-determined | Genome-fragment | NA | NA | 0 |
| S16_extracted_20071 | 3236 | No | NA | 3 | 0 | 1 | Low-quality | Genome-fragment | 0.37 | AAI-based (medium-confidence) | 0 |
| S10_extracted_62358 | 1282 | No | NA | 1 | 0 | 1 | Not-determined | Genome-fragment | NA | NA | 0 |
| S10_extracted_62362 | 2144 | No | NA | 2 | 0 | 1 | Not-determined | Genome-fragment | NA | NA | 0 |
| S10_extracted_62375 | 1302 | No | NA | 2 | 0 | 0 | Not-determined | Genome-fragment | NA | NA | 0 |
| S10_extracted_62442 | 1467 | No | NA | 2 | 0 | 0 | Not-determined | Genome-fragment | NA | NA | 0 |
| S10_extracted_62508 | 2243 | No | NA | 2 | 0 | 2 | Not-determined | Genome-fragment | NA | NA | 0 |
| S16_extracted_92207 | 3236 | No | NA | 4 | 1 | 0 | Low-quality | Genome-fragment | 5.46 | AAI-based (high-confidence) | 0 |
| S10_extracted_62582 | 2125 | No | NA | 1 | 0 | 1 | Not-determined | Genome-fragment | NA | NA | 0 |
| S10_extracted_62597 | 3740 | No | NA | 2 | 0 | 1 | Not-determined | Genome-fragment | NA | NA | 0 |
| S16_vs_116844\|\|full | 3236 | No | NA | 3 | 0 | 2 | Low-quality | Genome-fragment | 0.67 | AAI-based (medium-confidence) | 0 |
| S10_extracted_62910 | 2170 | No | NA | 2 | 0 | 1 | Not-determined | Genome-fragment | NA | NA | 0 |
| S10_extracted_63060 | 1148 | No | NA | 2 | 0 | 1 | Not-determined | Genome-fragment | NA | NA | 0 |
| S10_extracted_63096 | 1478 | No | NA | 2 | 0 | 0 | Not-determined | Genome-fragment | NA | NA | 0 |
| S10_extracted_63105 | 3544 | No | NA | 4 | 0 | 2 | Not-determined | Genome-fragment | NA | NA | 0 |
| S10_vs_105157\|\|full | 3236 | No | NA | 8 | 1 | 0 | Low-quality | Genome-fragment | 2.26 | HMM-based (lower-bound) | 0 |
| S10_extracted_63175 | 12157 | No | NA | 11 | 0 | 9 | Not-determined | Genome-fragment | NA | NA | 0 |
| S10_extracted_63221 | 3116 | No | NA | 3 | 0 | 3 | Not-determined | Genome-fragment | NA | NA | 0 |
| S10_extracted_63412 | 1168 | No | NA | 1 | 0 | 1 | Not-determined | Genome-fragment | NA | NA | 0 |
| S10_extracted_63526 | 2276 | No | NA | 2 | 0 | 1 | Not-determined | Genome-fragment | NA | NA | 0 |
| S12_vs_k141_59018\|\|full | 3236 | No | NA | 4 | 1 | 0 | Low-quality | Genome-fragment | 7.5 | AAI-based (medium-confidence) | 0 |
| S17_vs_k141_99512\|\|full | 3237 | No | NA | 6 | 0 | 0 | Low-quality | Genome-fragment | 6.98 | AAI-based (high-confidence) | 0 |
| S10_extracted_63934 | 7025 | No | NA | 9 | 0 | 6 | Not-determined | Genome-fragment | NA | NA | 0 |
| S10_extracted_64020 | 1130 | No | NA | 1 | 0 | 0 | Not-determined | Genome-fragment | NA | NA | 0 |
| S10_extracted_64037 | 1013 | No | NA | 3 | 0 | 0 | Not-determined | Genome-fragment | NA | NA | 0 |
| S10_extracted_64062 | 7518 | No | NA | 7 | 0 | 5 | Not-determined | Genome-fragment | NA | NA | 0 |
| S5_extracted_k141_85468 | 3237 | No | NA | 8 | 0 | 0 | Low-quality | Genome-fragment | 7.53 | AAI-based (high-confidence) | 0 |
| S10_extracted_64226 | 7762 | No | NA | 4 | 0 | 4 | Not-determined | Genome-fragment | NA | NA | 0 |
| S10_extracted_64234 | 4748 | No | NA | 12 | 0 | 12 | Not-determined | Genome-fragment | NA | NA | 0 |
| S5_extracted_k141_62196 | 3237 | No | NA | 3 | 0 | 3 | Low-quality | Genome-fragment | 4.41 | AAI-based (medium-confidence) | 0 |
| S10_extracted_64371 | 59185 | No | NA | 76 | 9 | 0 | Complete | High-quality | 100 | DTR (high-confidence) | 0 |
| S10_extracted_64422 | 1055 | No | NA | 1 | 0 | 0 | Not-determined | Genome-fragment | NA | NA | 0 |
| S10_extracted_64458 | 1402 | No | NA | 1 | 0 | 1 | Not-determined | Genome-fragment | NA | NA | 0 |
| S10_extracted_64478 | 1876 | No | NA | 2 | 0 | 1 | Not-determined | Genome-fragment | NA | NA | 0 |
| S10_extracted_64549 | 5150 | No | NA | 7 | 0 | 1 | Not-determined | Genome-fragment | NA | NA | 0 |
| S10_extracted_64576 | 1026 | No | NA | 2 | 0 | 0 | Not-determined | Genome-fragment | NA | NA | 0 |
| S10_extracted_64615 | 14379 | No | NA | 18 | 0 | 14 | Not-determined | Genome-fragment | NA | NA | 0 |
| S10_extracted_64662 | 4253 | No | NA | 4 | 0 | 2 | Not-determined | Genome-fragment | NA | NA | 0 |
| S10_extracted_64719 | 1070 | No | NA | 1 | 0 | 0 | Not-determined | Genome-fragment | NA | NA | 0 |
| S10_extracted_64763 | 1394 | No | NA | 1 | 0 | 1 | Not-determined | Genome-fragment | NA | NA | 0 |
| S17_vs_k141_22382\|\|full | 3238 | No | NA | 6 | 1 | 0 | Low-quality | Genome-fragment | 5.05 | AAI-based (high-confidence) | 0 |
| S10_extracted_64849 | 1158 | No | NA | 2 | 0 | 2 | Not-determined | Genome-fragment | NA | NA | 0 |
| S10_extracted_64884 | 1352 | No | NA | 1 | 0 | 1 | Not-determined | Genome-fragment | NA | NA | 0 |
| S10_extracted_64917 | 1230 | No | NA | 3 | 0 | 0 | Not-determined | Genome-fragment | NA | NA | 0 |
| S10_extracted_64932 | 4495 | No | NA | 4 | 0 | 2 | Not-determined | Genome-fragment | NA | NA | 0 |
| S10_extracted_64933 | 1312 | No | NA | 1 | 0 | 1 | Not-determined | Genome-fragment | NA | NA | 0 |
| S14_extracted_k141_103383 | 3238 | No | NA | 3 | 1 | 0 | Low-quality | Genome-fragment | 6.6 | AAI-based (high-confidence) | 0 |
| S10_extracted_65093 | 1297 | No | NA | 1 | 0 | 0 | Not-determined | Genome-fragment | NA | NA | 0 |
| S10_extracted_65116 | 1085 | No | NA | 2 | 0 | 0 | Not-determined | Genome-fragment | NA | NA | 0 |
| S10_extracted_65207 | 1022 | No | NA | 2 | 0 | 0 | Not-determined | Genome-fragment | NA | NA | 0 |
| S10_extracted_65216 | 1733 | No | NA | 1 | 0 | 1 | Not-determined | Genome-fragment | NA | NA | 0 |
| S10_extracted_65262 | 1022 | No | NA | 2 | 0 | 0 | Not-determined | Genome-fragment | NA | NA | 0 |
| S10_extracted_65311 | 6216 | No | NA | 4 | 0 | 1 | Not-determined | Genome-fragment | NA | NA | 0 |
| S16_extracted_143564 | 3238 | No | NA | 10 | 1 | 0 | Low-quality | Genome-fragment | 6.63 | AAI-based (high-confidence) | 0 |
| S16_vs_120846\|\|full | 3238 | No | NA | 6 | 1 | 0 | Low-quality | Genome-fragment | 1.33 | AAI-based (medium-confidence) | 0 |
| S10_extracted_65801 | 1016 | No | NA | 1 | 0 | 0 | Not-determined | Genome-fragment | NA | NA | 0 |
| S10_extracted_66249 | 1016 | No | NA | 1 | 0 | 0 | Not-determined | Genome-fragment | NA | NA | 0 |
| S10_extracted_66257 | 1160 | No | NA | 1 | 0 | 0 | Not-determined | Genome-fragment | NA | NA | 0 |
| S6_vs_k141_35873\|\|full | 3238 | No | NA | 3 | 0 | 2 | Low-quality | Genome-fragment | 2.28 | AAI-based (medium-confidence) | 0 |
| S10_extracted_66520 | 2119 | No | NA | 3 | 0 | 1 | Not-determined | Genome-fragment | NA | NA | 0 |
| S7_vs_12035\|\|full | 3239 | No | NA | 8 | 2 | 0 | Low-quality | Genome-fragment | 8.01 | AAI-based (high-confidence) | 0 |
| S12_extracted_k141_110420 | 3239 | No | NA | 4 | 0 | 0 | Low-quality | Genome-fragment | 10.13 | AAI-based (medium-confidence) | 0 |
| S10_extracted_66753 | 1884 | No | NA | 3 | 0 | 1 | Not-determined | Genome-fragment | NA | NA | 0 |
| S10_extracted_66757 | 1047 | No | NA | 2 | 0 | 0 | Not-determined | Genome-fragment | NA | NA | 0 |
| S10_extracted_66827 | 4137 | No | NA | 4 | 0 | 2 | Not-determined | Genome-fragment | NA | NA | 0 |
| S10_extracted_66926 | 3954 | No | NA | 5 | 0 | 0 | Not-determined | Genome-fragment | NA | NA | 0 |
| S10_extracted_66962 | 1551 | No | NA | 2 | 0 | 1 | Not-determined | Genome-fragment | NA | NA | 0 |
| S14_extracted_k141_24998 | 3240 | No | NA | 5 | 0 | 1 | Low-quality | Genome-fragment | 1.76 | AAI-based (medium-confidence) | 0 |
| S14_vs_k141_108288\|\|full | 3240 | Yes | 2573 | 10 | 1 | 1 | Low-quality | Genome-fragment | 3.45 | AAI-based (high-confidence) | 20.59 |
| S10_extracted_67042 | 1065 | No | NA | 1 | 0 | 1 | Not-determined | Genome-fragment | NA | NA | 0 |
| S10_extracted_67058 | 1870 | No | NA | 6 | 0 | 0 | Not-determined | Genome-fragment | NA | NA | 0 |
| S10_extracted_67078 | 1665 | No | NA | 3 | 0 | 1 | Not-determined | Genome-fragment | NA | NA | 0 |
| S10_extracted_67236 | 1733 | No | NA | 2 | 0 | 1 | Not-determined | Genome-fragment | NA | NA | 0 |
| S1_extracted_k141_136901 | 3240 | No | NA | 7 | 1 | 0 | Low-quality | Genome-fragment | 7.08 | AAI-based (high-confidence) | 0 |
| S9_vs_k141_12010\|\|full | 3241 | No | NA | 4 | 1 | 0 | Low-quality | Genome-fragment | 5.55 | AAI-based (high-confidence) | 0 |
| S16_extracted_14201 | 3241 | No | NA | 6 | 0 | 0 | Low-quality | Genome-fragment | 4.11 | AAI-based (high-confidence) | 0 |
| S10_extracted_67685 | 1241 | No | NA | 1 | 0 | 1 | Not-determined | Genome-fragment | NA | NA | 0 |
| S7_extracted_18924 | 3242 | No | NA | 6 | 1 | 0 | Low-quality | Genome-fragment | 5.9 | AAI-based (high-confidence) | 0 |
| S10_extracted_67710 | 3550 | No | NA | 4 | 0 | 2 | Not-determined | Genome-fragment | NA | NA | 0 |
| S10_extracted_67751 | 4562 | No | NA | 6 | 0 | 1 | Not-determined | Genome-fragment | NA | NA | 0 |
| S10_extracted_67788 | 1019 | No | NA | 2 | 0 | 1 | Not-determined | Genome-fragment | NA | NA | 0 |
| S10_extracted_67943 | 2040 | No | NA | 2 | 0 | 1 | Not-determined | Genome-fragment | NA | NA | 0 |
| S10_extracted_67993 | 1828 | No | NA | 1 | 0 | 1 | Not-determined | Genome-fragment | NA | NA | 0 |
| S10_extracted_68233 | 3593 | No | NA | 4 | 0 | 3 | Not-determined | Genome-fragment | NA | NA | 0 |
| S10_extracted_68415 | 1316 | No | NA | 1 | 0 | 1 | Not-determined | Genome-fragment | NA | NA | 0 |
| S12_vs_k141_148915\|\|full | 3242 | No | NA | 4 | 0 | 2 | Low-quality | Genome-fragment | 4.68 | AAI-based (medium-confidence) | 0 |
| S10_extracted_68571 | 1305 | No | NA | 2 | 0 | 0 | Not-determined | Genome-fragment | NA | NA | 0 |
| S10_extracted_68638 | 1119 | No | NA | 2 | 0 | 0 | Not-determined | Genome-fragment | NA | NA | 0 |
| S10_extracted_68690 | 8431 | No | NA | 6 | 0 | 4 | Not-determined | Genome-fragment | NA | NA | 0 |
| S10_extracted_68706 | 6144 | No | NA | 4 | 0 | 1 | Not-determined | Genome-fragment | NA | NA | 0 |
| S10_extracted_68726 | 2179 | No | NA | 2 | 0 | 1 | Not-determined | Genome-fragment | NA | NA | 0 |
| S10_extracted_68775 | 1586 | No | NA | 1 | 0 | 0 | Not-determined | Genome-fragment | NA | NA | 0 |
| S10_extracted_69028 | 1292 | No | NA | 1 | 0 | 0 | Not-determined | Genome-fragment | NA | NA | 0 |
| S14_extracted_k141_84722 | 3243 | No | NA | 4 | 1 | 0 | Low-quality | Genome-fragment | 2.18 | HMM-based (lower-bound) | 0 |
| S7_vs_23971\|\|full | 3244 | No | NA | 4 | 0 | 0 | Low-quality | Genome-fragment | 2.43 | AAI-based (medium-confidence) | 0 |
| S16_extracted_124123 | 3244 | No | NA | 3 | 0 | 2 | Low-quality | Genome-fragment | 1.56 | AAI-based (high-confidence) | 0 |
| S10_extracted_69315 | 1082 | No | NA | 1 | 0 | 1 | Not-determined | Genome-fragment | NA | NA | 0 |
| S10_extracted_69372 | 1388 | No | NA | 3 | 0 | 0 | Not-determined | Genome-fragment | NA | NA | 0 |
| S10_extracted_69517 | 10461 | No | NA | 9 | 0 | 2 | Not-determined | Genome-fragment | NA | NA | 0 |
| S10_extracted_69569 | 1336 | No | NA | 1 | 0 | 0 | Not-determined | Genome-fragment | NA | NA | 0 |
| S10_extracted_69805 | 1057 | No | NA | 2 | 0 | 1 | Not-determined | Genome-fragment | NA | NA | 0 |
| S10_extracted_69850 | 1224 | No | NA | 1 | 0 | 1 | Not-determined | Genome-fragment | NA | NA | 0 |
| S10_extracted_70023 | 1980 | No | NA | 6 | 0 | 6 | Not-determined | Genome-fragment | NA | NA | 0 |
| S10_extracted_70176 | 1082 | No | NA | 2 | 0 | 2 | Not-determined | Genome-fragment | NA | NA | 0 |
| S16_vs_38191\|\|full | 3244 | Yes | 2266 | 4 | 1 | 1 | Low-quality | Genome-fragment | 5.03 | AAI-based (medium-confidence) | 30.15 |
| S12_vs_k141_150078\|\|full | 3244 | No | NA | 6 | 1 | 0 | Low-quality | Genome-fragment | 1.4 | HMM-based (lower-bound) | 0 |
| S10_extracted_70612 | 1820 | No | NA | 2 | 0 | 1 | Not-determined | Genome-fragment | NA | NA | 0 |
| S10_extracted_70668 | 1067 | No | NA | 1 | 0 | 0 | Not-determined | Genome-fragment | NA | NA | 0 |
| S12_vs_k141_138100\|\|full | 3244 | No | NA | 5 | 0 | 2 | Low-quality | Genome-fragment | 4.61 | AAI-based (medium-confidence) | 0 |
| S10_extracted_70865 | 1394 | No | NA | 1 | 0 | 1 | Not-determined | Genome-fragment | NA | NA | 0 |
| S10_extracted_70888 | 3490 | No | NA | 1 | 0 | 1 | Not-determined | Genome-fragment | NA | NA | 0 |
| S10_extracted_70889 | 1461 | No | NA | 2 | 0 | 2 | Not-determined | Genome-fragment | NA | NA | 0 |
| S10_extracted_71003 | 5413 | No | NA | 3 | 0 | 1 | Not-determined | Genome-fragment | NA | NA | 0 |
| S10_extracted_71071 | 6478 | No | NA | 5 | 0 | 5 | Not-determined | Genome-fragment | NA | NA | 0 |
| S10_extracted_71104 | 1223 | No | NA | 2 | 0 | 0 | Not-determined | Genome-fragment | NA | NA | 0 |
| S10_extracted_71158 | 1472 | No | NA | 4 | 0 | 0 | Not-determined | Genome-fragment | NA | NA | 0 |
| S17_vs_k141_86053\|\|full | 3245 | No | NA | 5 | 0 | 0 | Low-quality | Genome-fragment | 4.12 | AAI-based (high-confidence) | 0 |
| S10_extracted_71329 | 1180 | No | NA | 2 | 0 | 2 | Not-determined | Genome-fragment | NA | NA | 0 |
| S9_extracted_k141_83498 | 3245 | No | NA | 4 | 3 | 0 | Low-quality | Genome-fragment | 6.9 | AAI-based (high-confidence) | 0 |
| S14_vs_k141_31202\|\|full | 3245 | No | NA | 6 | 3 | 0 | Low-quality | Genome-fragment | 9.56 | AAI-based (high-confidence) | 0 |
| S10_extracted_71403 | 1626 | No | NA | 2 | 0 | 1 | Not-determined | Genome-fragment | NA | NA | 0 |
| S10_extracted_71405 | 4731 | No | NA | 4 | 0 | 1 | Not-determined | Genome-fragment | NA | NA | 0 |
| S10_extracted_71478 | 3355 | No | NA | 1 | 0 | 1 | Not-determined | Genome-fragment | NA | NA | 0 |
| S10_extracted_71513 | 3356 | No | NA | 4 | 0 | 3 | Not-determined | Genome-fragment | NA | NA | 0 |
| S10_extracted_71553 | 4007 | No | NA | 4 | 0 | 2 | Not-determined | Genome-fragment | NA | NA | 0 |
| S10_extracted_71687 | 1075 | No | NA | 2 | 0 | 1 | Not-determined | Genome-fragment | NA | NA | 0 |
| S1_vs_k141_94617\|\|full | 3245 | No | NA | 5 | 0 | 0 | Low-quality | Genome-fragment | 5.83 | AAI-based (high-confidence) | 0 |
| S10_extracted_71825 | 1209 | No | NA | 3 | 0 | 2 | Not-determined | Genome-fragment | NA | NA | 0 |
| S7_vs_126001\|\|full | 3245 | No | NA | 3 | 0 | 2 | Low-quality | Genome-fragment | 2.62 | AAI-based (high-confidence) | 0 |
| S10_extracted_71927 | 1980 | No | NA | 1 | 0 | 1 | Not-determined | Genome-fragment | NA | NA | 0 |
| S10_extracted_72038 | 2371 | No | NA | 4 | 0 | 1 | Not-determined | Genome-fragment | NA | NA | 0 |
| S10_extracted_72107 | 2653 | No | NA | 2 | 0 | 1 | Not-determined | Genome-fragment | NA | NA | 0 |
| S2_vs_k141_9281\|\|full | 3245 | No | NA | 8 | 1 | 0 | Low-quality | Genome-fragment | 6.79 | AAI-based (high-confidence) | 0 |
| S10_extracted_72226 | 1106 | No | NA | 5 | 0 | 3 | Not-determined | Genome-fragment | NA | NA | 0 |
| S10_extracted_72284 | 1067 | No | NA | 2 | 0 | 0 | Not-determined | Genome-fragment | NA | NA | 0 |
| S10_extracted_72604 | 1620 | No | NA | 2 | 0 | 1 | Not-determined | Genome-fragment | NA | NA | 0 |
| S8_vs_k141_32425\|\|full | 3245 | No | NA | 4 | 3 | 0 | Low-quality | Genome-fragment | 10.08 | AAI-based (high-confidence) | 0 |
| S10_extracted_72943 | 1438 | No | NA | 3 | 0 | 0 | Not-determined | Genome-fragment | NA | NA | 0 |
| S10_extracted_73047 | 1240 | No | NA | 3 | 0 | 0 | Not-determined | Genome-fragment | NA | NA | 0 |
| S8_vs_k141_138282\|\|full | 3245 | No | NA | 9 | 1 | 0 | Low-quality | Genome-fragment | 5.28 | AAI-based (high-confidence) | 0 |
| S1_vs_k141_62867\|\|full | 3246 | No | NA | 4 | 0 | 0 | Low-quality | Genome-fragment | 7.76 | AAI-based (medium-confidence) | 0 |
| S10_extracted_73147 | 1349 | No | NA | 1 | 0 | 1 | Not-determined | Genome-fragment | NA | NA | 0 |
| S10_extracted_73280 | 1236 | No | NA | 2 | 0 | 1 | Not-determined | Genome-fragment | NA | NA | 0 |
| S10_extracted_73423 | 1064 | No | NA | 2 | 0 | 0 | Not-determined | Genome-fragment | NA | NA | 0 |
| S10_extracted_73441 | 1342 | No | NA | 1 | 0 | 0 | Not-determined | Genome-fragment | NA | NA | 0 |
| S10_extracted_73452 | 1921 | No | NA | 3 | 0 | 2 | Not-determined | Genome-fragment | NA | NA | 0 |
| S10_extracted_73569 | 2648 | No | NA | 1 | 0 | 1 | Not-determined | Genome-fragment | NA | NA | 0 |
| S3_vs_k141_124995\|\|full | 3247 | No | NA | 2 | 0 | 1 | Low-quality | Genome-fragment | 1.31 | AAI-based (medium-confidence) | 0 |
| S5_vs_k141_115563\|\|full | 3247 | No | NA | 6 | 1 | 0 | Low-quality | Genome-fragment | 7.31 | AAI-based (medium-confidence) | 0 |
| S10_extracted_74069 | 7177 | No | NA | 8 | 0 | 1 | Not-determined | Genome-fragment | NA | NA | 0 |
| S10_extracted_74222 | 1535 | No | NA | 3 | 0 | 1 | Not-determined | Genome-fragment | NA | NA | 0 |
| S10_extracted_39886 | 3247 | No | NA | 3 | 0 | 0 | Low-quality | Genome-fragment | 5.03 | AAI-based (high-confidence) | 0 |
| S10_vs_55034\|\|full | 3247 | No | NA | 6 | 0 | 0 | Low-quality | Genome-fragment | 8.25 | AAI-based (medium-confidence) | 0 |
| S10_extracted_74459 | 1031 | No | NA | 1 | 0 | 0 | Not-determined | Genome-fragment | NA | NA | 0 |
| S10_extracted_74574 | 2924 | No | NA | 5 | 0 | 4 | Not-determined | Genome-fragment | NA | NA | 0 |
| S10_extracted_74602 | 1216 | No | NA | 3 | 0 | 1 | Not-determined | Genome-fragment | NA | NA | 0 |
| S10_extracted_74671 | 4834 | No | NA | 5 | 0 | 3 | Not-determined | Genome-fragment | NA | NA | 0 |
| S10_extracted_74761 | 1154 | No | NA | 3 | 0 | 0 | Not-determined | Genome-fragment | NA | NA | 0 |
| S10_extracted_74965 | 1010 | No | NA | 2 | 0 | 0 | Not-determined | Genome-fragment | NA | NA | 0 |
| S10_extracted_74986 | 2029 | No | NA | 1 | 0 | 1 | Not-determined | Genome-fragment | NA | NA | 0 |
| S10_extracted_75077 | 1501 | No | NA | 2 | 0 | 2 | Not-determined | Genome-fragment | NA | NA | 0 |
| S12_extracted_k141_152778 | 3247 | No | NA | 4 | 0 | 2 | Low-quality | Genome-fragment | 5.41 | AAI-based (medium-confidence) | 0 |
| S3_vs_k141_93660\|\|full | 3248 | No | NA | 3 | 1 | 0 | Low-quality | Genome-fragment | 7.02 | AAI-based (medium-confidence) | 0 |
| S10_extracted_75432 | 1016 | No | NA | 1 | 0 | 1 | Not-determined | Genome-fragment | NA | NA | 0 |
| S10_extracted_75507 | 3051 | No | NA | 3 | 0 | 1 | Not-determined | Genome-fragment | NA | NA | 0 |
| S7_vs_85628\|\|full | 3248 | No | NA | 7 | 0 | 1 | Low-quality | Genome-fragment | 4.82 | AAI-based (high-confidence) | 0 |
| S10_extracted_75917 | 1140 | No | NA | 2 | 0 | 1 | Not-determined | Genome-fragment | NA | NA | 0 |
| S10_extracted_75919 | 1939 | No | NA | 2 | 0 | 1 | Not-determined | Genome-fragment | NA | NA | 0 |
| S10_extracted_75950 | 5317 | No | NA | 5 | 0 | 2 | Not-determined | Genome-fragment | NA | NA | 0 |
| S10_extracted_76003 | 1256 | No | NA | 2 | 0 | 1 | Not-determined | Genome-fragment | NA | NA | 0 |
| S8_extracted_k141_123038 | 3249 | No | NA | 5 | 1 | 0 | Low-quality | Genome-fragment | 3.23 | AAI-based (high-confidence) | 0 |
| S10_extracted_76148 | 1097 | No | NA | 2 | 0 | 1 | Not-determined | Genome-fragment | NA | NA | 0 |
| S10_extracted_76187 | 1808 | No | NA | 3 | 0 | 1 | Not-determined | Genome-fragment | NA | NA | 0 |
| S10_extracted_76224 | 1328 | No | NA | 1 | 0 | 0 | Not-determined | Genome-fragment | NA | NA | 0 |
| S17_vs_k141_95284\|\|full | 3250 | No | NA | 6 | 0 | 2 | Low-quality | Genome-fragment | 3.82 | AAI-based (medium-confidence) | 0 |
| S10_extracted_76338 | 1345 | No | NA | 1 | 0 | 1 | Not-determined | Genome-fragment | NA | NA | 0 |
| S10_extracted_76389 | 1585 | No | NA | 1 | 0 | 1 | Not-determined | Genome-fragment | NA | NA | 0 |
| S10_extracted_76647 | 1017 | No | NA | 4 | 0 | 0 | Not-determined | Genome-fragment | NA | NA | 0 |
| S10_extracted_76696 | 1076 | No | NA | 2 | 0 | 1 | Not-determined | Genome-fragment | NA | NA | 0 |
| S1_extracted_k141_10098 | 3250 | No | NA | 2 | 0 | 2 | Low-quality | Genome-fragment | 2.35 | AAI-based (medium-confidence) | 0 |
| S10_extracted_76766 | 4049 | No | NA | 4 | 0 | 4 | Not-determined | Genome-fragment | NA | NA | 0 |
| S5_vs_k141_46230\|\|full | 3250 | No | NA | 4 | 2 | 0 | Low-quality | Genome-fragment | 4.39 | HMM-based (lower-bound) | 0 |
| S10_extracted_84891 | 3250 | No | NA | 6 | 3 | 0 | Low-quality | Genome-fragment | 7.61 | AAI-based (high-confidence) | 0 |
| S10_extracted_76913 | 6208 | No | NA | 7 | 0 | 1 | Not-determined | Genome-fragment | NA | NA | 0 |
| S10_extracted_76927 | 1255 | No | NA | 1 | 0 | 1 | Not-determined | Genome-fragment | NA | NA | 0 |
| S3_vs_k141_27570\|\|full | 3251 | No | NA | 7 | 0 | 0 | Low-quality | Genome-fragment | 6.56 | AAI-based (high-confidence) | 0 |
| S10_extracted_76951 | 1042 | No | NA | 1 | 0 | 1 | Not-determined | Genome-fragment | NA | NA | 0 |
| S10_extracted_77274 | 1617 | No | NA | 4 | 0 | 0 | Not-determined | Genome-fragment | NA | NA | 0 |
| S10_extracted_77286 | 1185 | No | NA | 2 | 0 | 1 | Not-determined | Genome-fragment | NA | NA | 0 |
| S10_extracted_77346 | 1635 | No | NA | 3 | 0 | 3 | Not-determined | Genome-fragment | NA | NA | 0 |
| S10_extracted_22266 | 3251 | No | NA | 2 | 1 | 0 | Low-quality | Genome-fragment | 6.31 | AAI-based (high-confidence) | 0 |
| S10_extracted_77572 | 4006 | No | NA | 7 | 0 | 0 | Not-determined | Genome-fragment | NA | NA | 0 |
| S10_extracted_77668 | 3422 | No | NA | 4 | 0 | 1 | Not-determined | Genome-fragment | NA | NA | 0 |
| S10_extracted_77696 | 2090 | No | NA | 2 | 0 | 2 | Not-determined | Genome-fragment | NA | NA | 0 |
| S10_extracted_77722 | 1100 | No | NA | 2 | 0 | 1 | Not-determined | Genome-fragment | NA | NA | 0 |
| S10_extracted_77779 | 2472 | No | NA | 4 | 0 | 1 | Not-determined | Genome-fragment | NA | NA | 0 |
| S10_extracted_77785 | 1043 | No | NA | 1 | 0 | 0 | Not-determined | Genome-fragment | NA | NA | 0 |
| S10_extracted_77820 | 4290 | No | NA | 4 | 0 | 3 | Not-determined | Genome-fragment | NA | NA | 0 |
| S10_extracted_77901 | 3669 | No | NA | 4 | 0 | 2 | Not-determined | Genome-fragment | NA | NA | 0 |
| S10_extracted_78149 | 1157 | No | NA | 1 | 0 | 1 | Not-determined | Genome-fragment | NA | NA | 0 |
| S10_extracted_78250 | 2322 | No | NA | 2 | 0 | 2 | Not-determined | Genome-fragment | NA | NA | 0 |
| S10_extracted_78261 | 1096 | No | NA | 1 | 0 | 0 | Not-determined | Genome-fragment | NA | NA | 0 |
| S6_extracted_k141_16328 | 3251 | No | NA | 5 | 1 | 0 | Low-quality | Genome-fragment | 1.95 | AAI-based (medium-confidence) | 0 |
| S10_extracted_78289 | 2120 | No | NA | 3 | 0 | 0 | Not-determined | Genome-fragment | NA | NA | 0 |
| S10_extracted_78415 | 3877 | No | NA | 4 | 0 | 2 | Not-determined | Genome-fragment | NA | NA | 0 |
| S10_extracted_78426 | 1000 | No | NA | 2 | 0 | 0 | Not-determined | Genome-fragment | NA | NA | 0 |
| S14_vs_k141_12187\|\|full | 3252 | No | NA | 4 | 0 | 0 | Low-quality | Genome-fragment | 3.62 | AAI-based (medium-confidence) | 0 |
| S1_extracted_k141_41641 | 3253 | No | NA | 6 | 1 | 0 | Low-quality | Genome-fragment | 9.17 | AAI-based (high-confidence) | 0 |
| S10_extracted_78593 | 1312 | No | NA | 2 | 0 | 1 | Not-determined | Genome-fragment | NA | NA | 0 |
| S10_extracted_78620 | 1780 | No | NA | 2 | 0 | 1 | Not-determined | Genome-fragment | NA | NA | 0 |
| S10_extracted_78637 | 1934 | No | NA | 2 | 0 | 2 | Not-determined | Genome-fragment | NA | NA | 0 |
| S9_extracted_k141_15972 | 3254 | No | NA | 10 | 0 | 0 | Low-quality | Genome-fragment | 8.56 | AAI-based (medium-confidence) | 0 |
| S10_extracted_78816 | 1190 | No | NA | 1 | 0 | 1 | Not-determined | Genome-fragment | NA | NA | 0 |
| S9_vs_k141_37637\|\|full | 3254 | No | NA | 4 | 3 | 0 | Low-quality | Genome-fragment | 7 | AAI-based (medium-confidence) | 0 |
| S10_extracted_78859 | 1057 | No | NA | 1 | 0 | 1 | Not-determined | Genome-fragment | NA | NA | 0 |
| S14_vs_k141_17936\|\|full | 3254 | No | NA | 5 | 2 | 0 | Low-quality | Genome-fragment | 7.96 | AAI-based (high-confidence) | 0 |
| S10_extracted_78997 | 3606 | No | NA | 2 | 0 | 0 | Not-determined | Genome-fragment | NA | NA | 0 |
| S3_extracted_k141_94521 | 3254 | No | NA | 5 | 1 | 1 | Low-quality | Genome-fragment | 1.98 | AAI-based (high-confidence) | 0 |
| S7_vs_11519\|\|full | 3254 | No | NA | 6 | 3 | 0 | Low-quality | Genome-fragment | 6.82 | AAI-based (high-confidence) | 0 |
| S16_extracted_123060 | 3254 | No | NA | 6 | 2 | 0 | Low-quality | Genome-fragment | 8.32 | AAI-based (high-confidence) | 0 |
| S12_vs_k141_146656\|\|full | 3254 | No | NA | 4 | 0 | 0 | Low-quality | Genome-fragment | 4.61 | AAI-based (medium-confidence) | 0 |
| S10_extracted_79417 | 1516 | No | NA | 3 | 0 | 1 | Not-determined | Genome-fragment | NA | NA | 0 |
| S10_extracted_79450 | 1796 | No | NA | 2 | 0 | 0 | Not-determined | Genome-fragment | NA | NA | 0 |
| S5_vs_k141_126117\|\|full | 3255 | No | NA | 6 | 3 | 0 | Low-quality | Genome-fragment | 9.39 | AAI-based (high-confidence) | 0 |
| S10_extracted_79495 | 1550 | No | NA | 1 | 0 | 1 | Not-determined | Genome-fragment | NA | NA | 0 |
| S10_extracted_79509 | 1086 | No | NA | 2 | 0 | 0 | Not-determined | Genome-fragment | NA | NA | 0 |
| S10_extracted_79534 | 2147 | No | NA | 3 | 0 | 0 | Not-determined | Genome-fragment | NA | NA | 0 |
| S10_extracted_79584 | 10202 | No | NA | 9 | 0 | 4 | Not-determined | Genome-fragment | NA | NA | 0 |
| S10_extracted_79659 | 1656 | No | NA | 1 | 0 | 0 | Not-determined | Genome-fragment | NA | NA | 0 |
| S10_extracted_79875 | 2086 | No | NA | 2 | 0 | 2 | Not-determined | Genome-fragment | NA | NA | 0 |
| S10_extracted_79917 | 1165 | No | NA | 1 | 0 | 1 | Not-determined | Genome-fragment | NA | NA | 0 |
| S10_extracted_79953 | 1021 | No | NA | 2 | 0 | 0 | Not-determined | Genome-fragment | NA | NA | 0 |
| S15_vs_53914\|\|full | 3256 | No | NA | 7 | 1 | 0 | Low-quality | Genome-fragment | 4.45 | AAI-based (high-confidence) | 0 |
| S10_extracted_80022 | 3761 | No | NA | 5 | 0 | 1 | Not-determined | Genome-fragment | NA | NA | 0 |
| S10_extracted_80076 | 4041 | No | NA | 5 | 0 | 2 | Not-determined | Genome-fragment | NA | NA | 0 |
| S10_extracted_80103 | 1731 | No | NA | 3 | 0 | 1 | Not-determined | Genome-fragment | NA | NA | 0 |
| S3_extracted_k141_113348 | 3257 | No | NA | 3 | 3 | 0 | Low-quality | Genome-fragment | 3.7 | AAI-based (medium-confidence) | 0 |
| S12_extracted_k141_103803 | 3257 | Yes | 653 | 5 | 1 | 1 | Low-quality | Genome-fragment | 0.7 | AAI-based (high-confidence) | 79.95 |
| S10_extracted_80308 | 2659 | No | NA | 3 | 0 | 3 | Not-determined | Genome-fragment | NA | NA | 0 |
| S10_extracted_80317 | 2511 | No | NA | 2 | 0 | 1 | Not-determined | Genome-fragment | NA | NA | 0 |
| S10_extracted_80498 | 1836 | No | NA | 6 | 0 | 6 | Not-determined | Genome-fragment | NA | NA | 0 |
| S10_extracted_80504 | 2946 | No | NA | 2 | 0 | 1 | Not-determined | Genome-fragment | NA | NA | 0 |
| S5_vs_k141_33105\|\|full | 3258 | No | NA | 8 | 3 | 0 | Low-quality | Genome-fragment | 7.44 | AAI-based (high-confidence) | 0 |
| S10_extracted_80712 | 116981 | No | NA | 141 | 27 | 0 | Medium-quality | Genome-fragment | 72.67 | AAI-based (high-confidence) | 0 |
| S5_vs_k141_5892\|\|lt2gene | 3259 | No | NA | 2 | 2 | 0 | Low-quality | Genome-fragment | 10.74 | AAI-based (high-confidence) | 0 |
| S10_extracted_80964 | 1413 | No | NA | 2 | 0 | 2 | Not-determined | Genome-fragment | NA | NA | 0 |
| S10_extracted_81208 | 1619 | No | NA | 5 | 0 | 2 | Not-determined | Genome-fragment | NA | NA | 0 |
| S10_extracted_81214 | 1784 | No | NA | 3 | 0 | 1 | Not-determined | Genome-fragment | NA | NA | 0 |
| S10_extracted_81238 | 5219 | No | NA | 12 | 0 | 12 | Not-determined | Genome-fragment | NA | NA | 0 |
| S10_extracted_81253 | 2831 | No | NA | 3 | 0 | 2 | Not-determined | Genome-fragment | NA | NA | 0 |
| S10_extracted_81353 | 2214 | No | NA | 1 | 0 | 1 | Not-determined | Genome-fragment | NA | NA | 0 |
| S10_extracted_81368 | 1010 | No | NA | 1 | 0 | 0 | Not-determined | Genome-fragment | NA | NA | 0 |
| S12_vs_k141_150484\|\|full | 3259 | No | NA | 3 | 1 | 0 | Low-quality | Genome-fragment | 8.77 | AAI-based (high-confidence) | 0 |
| S10_extracted_81697 | 1424 | No | NA | 2 | 0 | 2 | Not-determined | Genome-fragment | NA | NA | 0 |
| S10_extracted_81719 | 6376 | No | NA | 11 | 0 | 10 | Not-determined | Genome-fragment | NA | NA | 0 |
| S2_vs_k141_16679\|\|full | 3259 | No | NA | 3 | 2 | 0 | Low-quality | Genome-fragment | 8.29 | AAI-based (high-confidence) | 0 |
| S10_extracted_81876 | 5312 | No | NA | 6 | 0 | 4 | Not-determined | Genome-fragment | NA | NA | 0 |
| S10_extracted_82145 | 1086 | No | NA | 2 | 0 | 1 | Not-determined | Genome-fragment | NA | NA | 0 |
| S14_vs_k141_28536\|\|full | 3260 | No | NA | 6 | 3 | 0 | Low-quality | Genome-fragment | 6 | HMM-based (lower-bound) | 0 |

| | | | | | | | | | | | |
|---|---|---|---|---|---|---|---|---|---|---|---|
| S10_extracted_82315 | 1633 | No | NA | 1 | 0 | 1 | Not-determined | Genome-fragment | NA | NA | 0 |
| S1_vs_k141_110116\|\|full | 3260 | No | NA | 4 | 0 | 3 | Low-quality | Genome-fragment | 2.27 | AAI-based (medium-confidence) | 0 |
| S8_vs_k141_122025\|\|full | 3260 | No | NA | 6 | 0 | 1 | Low-quality | Genome-fragment | 1.9 | AAI-based (medium-confidence) | 0 |
| S10_extracted_82598 | 1275 | No | NA | 1 | 0 | 0 | Not-determined | Genome-fragment | NA | NA | 0 |
| S10_extracted_82603 | 1410 | No | NA | 1 | 0 | 0 | Not-determined | Genome-fragment | NA | NA | 0 |
| S10_extracted_82662 | 6839 | No | NA | 7 | 0 | 0 | Not-determined | Genome-fragment | NA | NA | 0 |
| S10_extracted_82669 | 1187 | No | NA | 2 | 0 | 0 | Not-determined | Genome-fragment | NA | NA | 0 |
| S10_extracted_82680 | 1482 | No | NA | 2 | 0 | 2 | Not-determined | Genome-fragment | NA | NA | 0 |
| S10_extracted_82698 | 10790 | No | NA | 11 | 0 | 3 | Not-determined | Genome-fragment | NA | NA | 0 |
| S10_extracted_82794 | 3979 | No | NA | 6 | 0 | 1 | Not-determined | Genome-fragment | NA | NA | 0 |
| S10_extracted_82809 | 1184 | No | NA | 1 | 0 | 1 | Not-determined | Genome-fragment | NA | NA | 0 |
| S10_extracted_82821 | 2005 | No | NA | 2 | 0 | 0 | Not-determined | Genome-fragment | NA | NA | 0 |
| S10_extracted_82928 | 3506 | No | NA | 5 | 0 | 3 | Not-determined | Genome-fragment | NA | NA | 0 |
| S9_extracted_k141_65310 | 3261 | No | NA | 9 | 1 | 0 | Low-quality | Genome-fragment | 7.81 | AAI-based (high-confidence) | 0 |
| S10_extracted_83134 | 1232 | No | NA | 3 | 0 | 3 | Not-determined | Genome-fragment | NA | NA | 0 |
| S10_extracted_83181 | 3726 | No | NA | 4 | 0 | 2 | Not-determined | Genome-fragment | NA | NA | 0 |
| S16_vs_166165\|\|full | 3261 | No | NA | 4 | 0 | 1 | Low-quality | Genome-fragment | 0.76 | AAI-based (medium-confidence) | 0 |
| S10_vs_34228\|\|full | 3261 | No | NA | 3 | 0 | 3 | Low-quality | Genome-fragment | 1.66 | AAI-based (medium-confidence) | 0 |
| S1_vs_k141_88417\|\|full | 3262 | No | NA | 6 | 2 | 0 | Low-quality | Genome-fragment | 4.25 | AAI-based (medium-confidence) | 0 |
| S10_extracted_83423 | 1737 | No | NA | 3 | 0 | 0 | Not-determined | Genome-fragment | NA | NA | 0 |
| S10_extracted_83428 | 1137 | No | NA | 2 | 0 | 1 | Not-determined | Genome-fragment | NA | NA | 0 |
| S10_extracted_83497 | 1444 | No | NA | 2 | 0 | 1 | Not-determined | Genome-fragment | NA | NA | 0 |
| S10_extracted_83757 | 2544 | No | NA | 4 | 0 | 3 | Not-determined | Genome-fragment | NA | NA | 0 |
| S10_extracted_84064 | 7153 | No | NA | 7 | 0 | 5 | Not-determined | Genome-fragment | NA | NA | 0 |
| S10_extracted_84133 | 1184 | No | NA | 2 | 0 | 1 | Not-determined | Genome-fragment | NA | NA | 0 |
| S10_extracted_84261 | 1443 | No | NA | 2 | 0 | 1 | Not-determined | Genome-fragment | NA | NA | 0 |
| S10_extracted_84347 | 1760 | No | NA | 4 | 0 | 0 | Not-determined | Genome-fragment | NA | NA | 0 |
| S10_extracted_84517 | 2033 | No | NA | 3 | 0 | 1 | Not-determined | Genome-fragment | NA | NA | 0 |
| S10_extracted_84814 | 1136 | No | NA | 1 | 0 | 1 | Not-determined | Genome-fragment | NA | NA | 0 |
| S10_extracted_83277 | 3262 | No | NA | 3 | 0 | 3 | Low-quality | Genome-fragment | 0.76 | AAI-based (medium-confidence) | 0 |
| S12_vs_k141_150723\|\|full | 3262 | No | NA | 4 | 0 | 2 | Low-quality | Genome-fragment | 6.28 | AAI-based (medium-confidence) | 0 |
| S17_vs_k141_45650\|\|full | 3265 | No | NA | 4 | 0 | 1 | Low-quality | Genome-fragment | 3.54 | AAI-based (medium-confidence) | 0 |
| S10_extracted_85111 | 1003 | No | NA | 1 | 0 | 1 | Not-determined | Genome-fragment | NA | NA | 0 |
| S9_vs_k141_60749\|\|full | 3265 | No | NA | 5 | 3 | 0 | Low-quality | Genome-fragment | 9.25 | AAI-based (high-confidence) | 0 |
| S10_extracted_85243 | 1538 | No | NA | 3 | 0 | 0 | Not-determined | Genome-fragment | NA | NA | 0 |
| S10_extracted_85253 | 1511 | No | NA | 2 | 0 | 0 | Not-determined | Genome-fragment | NA | NA | 0 |
| S10_extracted_85334 | 6372 | No | NA | 6 | 0 | 5 | Not-determined | Genome-fragment | NA | NA | 0 |
| S7_extracted_16361 | 3265 | Yes | 2733 | 7 | 1 | 1 | Low-quality | Genome-fragment | 4.72 | AAI-based (medium-confidence) | 16.29 |
| S10_extracted_85372 | 1227 | No | NA | 2 | 0 | 1 | Not-determined | Genome-fragment | NA | NA | 0 |
| S8_extracted_k141_43233 | 3265 | No | NA | 3 | 0 | 3 | Low-quality | Genome-fragment | 1.76 | AAI-based (medium-confidence) | 0 |
| S10_extracted_85510 | 1023 | No | NA | 1 | 0 | 1 | Not-determined | Genome-fragment | NA | NA | 0 |
| S10_extracted_85533 | 1008 | No | NA | 1 | 0 | 1 | Not-determined | Genome-fragment | NA | NA | 0 |
| S5_vs_k141_37592\|\|full | 3267 | No | NA | 6 | 3 | 0 | Low-quality | Genome-fragment | 10.99 | AAI-based (high-confidence) | 0 |
| S10_extracted_25956 | 3267 | No | NA | 3 | 0 | 3 | Low-quality | Genome-fragment | 0.5 | AAI-based (medium-confidence) | 0 |
| S10_extracted_85797 | 3739 | No | NA | 6 | 0 | 5 | Not-determined | Genome-fragment | NA | NA | 0 |
| S10_vs_23666\|\|full | 3268 | No | NA | 5 | 2 | 0 | Low-quality | Genome-fragment | 6.78 | AAI-based (medium-confidence) | 0 |
| S10_extracted_86016 | 1650 | No | NA | 2 | 0 | 2 | Not-determined | Genome-fragment | NA | NA | 0 |
| S13_extracted_142127 | 3269 | No | NA | 4 | 1 | 0 | Low-quality | Genome-fragment | 9.78 | AAI-based (high-confidence) | 0 |
| S10_extracted_86108 | 1095 | No | NA | 2 | 0 | 1 | Not-determined | Genome-fragment | NA | NA | 0 |
| S10_extracted_86129 | 1243 | No | NA | 2 | 0 | 1 | Not-determined | Genome-fragment | NA | NA | 0 |
| S10_extracted_86255 | 6309 | No | NA | 4 | 0 | 3 | Not-determined | Genome-fragment | NA | NA | 0 |
| S10_extracted_86260 | 1303 | No | NA | 1 | 0 | 0 | Not-determined | Genome-fragment | NA | NA | 0 |
| S13_vs_94331\|\|full | 3269 | No | NA | 5 | 1 | 0 | Low-quality | Genome-fragment | 2 | HMM-based (lower-bound) | 0 |
| S10_extracted_86392 | 1131 | No | NA | 2 | 0 | 1 | Not-determined | Genome-fragment | NA | NA | 0 |
| S10_extracted_86404 | 1289 | No | NA | 1 | 0 | 1 | Not-determined | Genome-fragment | NA | NA | 0 |
| S10_extracted_86428 | 4372 | No | NA | 6 | 0 | 0 | Not-determined | Genome-fragment | NA | NA | 0 |
| S10_extracted_86439 | 1167 | No | NA | 2 | 0 | 1 | Not-determined | Genome-fragment | NA | NA | 0 |
| S10_extracted_86451 | 1054 | No | NA | 1 | 0 | 0 | Not-determined | Genome-fragment | NA | NA | 0 |
| S10_extracted_86460 | 2407 | No | NA | 5 | 0 | 3 | Not-determined | Genome-fragment | NA | NA | 0 |
| S5_vs_k141_68580\|\|lt2gene | 3271 | No | NA | 1 | 1 | 0 | Low-quality | Genome-fragment | 5.4 | AAI-based (medium-confidence) | 0 |
| S7_vs_35094\|\|full | 3271 | Yes | 1704 | 4 | 2 | 1 | Low-quality | Genome-fragment | 3.88 | AAI-based (medium-confidence) | 47.91 |
| S10_extracted_86570 | 1074 | No | NA | 1 | 0 | 0 | Not-determined | Genome-fragment | NA | NA | 0 |
| S10_extracted_86653 | 2762 | No | NA | 2 | 0 | 1 | Not-determined | Genome-fragment | NA | NA | 0 |
| S10_extracted_86684 | 1694 | No | NA | 2 | 0 | 0 | Not-determined | Genome-fragment | NA | NA | 0 |
| S7_vs_62824\|\|full | 3271 | No | NA | 3 | 0 | 0 | Low-quality | Genome-fragment | 2.34 | AAI-based (medium-confidence) | 0 |
| S10_extracted_86800 | 5278 | No | NA | 4 | 0 | 3 | Not-determined | Genome-fragment | NA | NA | 0 |
| S10_extracted_86930 | 8895 | No | NA | 5 | 0 | 4 | Not-determined | Genome-fragment | NA | NA | 0 |
| S10_extracted_86999 | 1171 | No | NA | 2 | 0 | 0 | Not-determined | Genome-fragment | NA | NA | 0 |
| S10_extracted_87114 | 1203 | No | NA | 4 | 0 | 4 | Not-determined | Genome-fragment | NA | NA | 0 |
| S10_extracted_87244 | 2175 | No | NA | 3 | 0 | 0 | Not-determined | Genome-fragment | NA | NA | 0 |
| S10_extracted_87267 | 2154 | No | NA | 1 | 0 | 0 | Not-determined | Genome-fragment | NA | NA | 0 |
| S10_extracted_87411 | 1841 | No | NA | 2 | 0 | 2 | Not-determined | Genome-fragment | NA | NA | 0 |
| S10_extracted_116721 | 3271 | No | NA | 3 | 0 | 3 | Low-quality | Genome-fragment | 9.42 | AAI-based (medium-confidence) | 0 |
| S10_extracted_87475 | 1182 | No | NA | 1 | 0 | 1 | Not-determined | Genome-fragment | NA | NA | 0 |
| S10_extracted_87500 | 1597 | No | NA | 1 | 0 | 1 | Not-determined | Genome-fragment | NA | NA | 0 |
| S10_extracted_87558 | 1115 | No | NA | 1 | 0 | 1 | Not-determined | Genome-fragment | NA | NA | 0 |
| S10_extracted_87685 | 1075 | No | NA | 2 | 0 | 1 | Not-determined | Genome-fragment | NA | NA | 0 |
| S10_extracted_87827 | 1487 | No | NA | 2 | 0 | 1 | Not-determined | Genome-fragment | NA | NA | 0 |
| S1_extracted_k141_77622 | 3273 | No | NA | 12 | 2 | 0 | Low-quality | Genome-fragment | 5.52 | AAI-based (high-confidence) | 0 |
| S10_extracted_88371 | 1119 | No | NA | 3 | 0 | 1 | Not-determined | Genome-fragment | NA | NA | 0 |
| S10_extracted_88591 | 3807 | No | NA | 5 | 0 | 2 | Not-determined | Genome-fragment | NA | NA | 0 |
| S10_extracted_88636 | 1124 | No | NA | 2 | 0 | 0 | Not-determined | Genome-fragment | NA | NA | 0 |
| S10_extracted_88878 | 1034 | No | NA | 1 | 0 | 1 | Not-determined | Genome-fragment | NA | NA | 0 |
| S10_extracted_88973 | 1039 | No | NA | 1 | 0 | 0 | Not-determined | Genome-fragment | NA | NA | 0 |
| S10_extracted_88985 | 1847 | No | NA | 1 | 0 | 0 | Not-determined | Genome-fragment | NA | NA | 0 |
| S10_extracted_79492 | 3274 | No | NA | 3 | 0 | 0 | Low-quality | Genome-fragment | 8.33 | AAI-based (high-confidence) | 0 |
| S10_extracted_89187 | 1891 | No | NA | 2 | 0 | 2 | Not-determined | Genome-fragment | NA | NA | 0 |
| S10_extracted_89258 | 1518 | No | NA | 2 | 0 | 0 | Not-determined | Genome-fragment | NA | NA | 0 |
| S10_extracted_89425 | 4198 | No | NA | 5 | 0 | 3 | Not-determined | Genome-fragment | NA | NA | 0 |
| S10_extracted_89431 | 1126 | No | NA | 3 | 0 | 0 | Not-determined | Genome-fragment | NA | NA | 0 |
| S10_extracted_89450 | 1079 | No | NA | 1 | 0 | 1 | Not-determined | Genome-fragment | NA | NA | 0 |
| S10_extracted_89514 | 1213 | No | NA | 1 | 0 | 1 | Not-determined | Genome-fragment | NA | NA | 0 |
| S10_extracted_89604 | 2257 | No | NA | 2 | 0 | 2 | Not-determined | Genome-fragment | NA | NA | 0 |
| S10_extracted_89683 | 1714 | No | NA | 2 | 0 | 1 | Not-determined | Genome-fragment | NA | NA | 0 |
| S10_extracted_89724 | 1198 | No | NA | 1 | 0 | 1 | Not-determined | Genome-fragment | NA | NA | 0 |
| S10_extracted_90046 | 1709 | No | NA | 1 | 0 | 1 | Not-determined | Genome-fragment | NA | NA | 0 |
| S6_extracted_k141_18649 | 3274 | No | NA | 7 | 2 | 0 | Low-quality | Genome-fragment | 2.51 | HMM-based (lower-bound) | 0 |
| S10_extracted_90159 | 6811 | No | NA | 3 | 0 | 2 | Not-determined | Genome-fragment | NA | NA | 0 |
| S10_extracted_90170 | 2209 | No | NA | 3 | 0 | 1 | Not-determined | Genome-fragment | NA | NA | 0 |
| S17_vs_k141_126005\|\|full | 3276 | No | NA | 5 | 1 | 0 | Low-quality | Genome-fragment | 1.9 | AAI-based (high-confidence) | 0 |
| S10_extracted_90248 | 1421 | No | NA | 1 | 0 | 1 | Not-determined | Genome-fragment | NA | NA | 0 |
| S10_extracted_90318 | 3211 | No | NA | 3 | 0 | 3 | Not-determined | Genome-fragment | NA | NA | 0 |
| S10_extracted_90351 | 1612 | No | NA | 2 | 0 | 1 | Not-determined | Genome-fragment | NA | NA | 0 |
| S10_extracted_90391 | 8205 | No | NA | 7 | 0 | 0 | Not-determined | Genome-fragment | NA | NA | 0 |
| S10_extracted_90410 | 3381 | No | NA | 2 | 0 | 0 | Not-determined | Genome-fragment | NA | NA | 0 |
| S17_vs_k141_5772\|\|full | 3276 | No | NA | 4 | 0 | 1 | Low-quality | Genome-fragment | 3.81 | AAI-based (medium-confidence) | 0 |
| S10_extracted_90458 | 2001 | No | NA | 2 | 0 | 1 | Not-determined | Genome-fragment | NA | NA | 0 |
| S11_vs_k141_51676\|\|full | 3276 | No | NA | 5 | 3 | 0 | Low-quality | Genome-fragment | 23.96 | AAI-based (high-confidence) | 0 |
| S10_extracted_90489 | 3044 | No | NA | 2 | 0 | 2 | Not-determined | Genome-fragment | NA | NA | 0 |
| S10_extracted_90521 | 1081 | No | NA | 2 | 0 | 1 | Not-determined | Genome-fragment | NA | NA | 0 |
| S10_extracted_90565 | 1180 | No | NA | 3 | 0 | 0 | Not-determined | Genome-fragment | NA | NA | 0 |
| S3_extracted_k141_124605 | 3276 | No | NA | 13 | 1 | 0 | Low-quality | Genome-fragment | 9.45 | AAI-based (high-confidence) | 0 |
| S10_extracted_90647 | 1188 | No | NA | 2 | 0 | 0 | Not-determined | Genome-fragment | NA | NA | 0 |
| S10_extracted_90649 | 1630 | No | NA | 3 | 0 | 1 | Not-determined | Genome-fragment | NA | NA | 0 |
| S7_extracted_80788 | 3276 | No | NA | 1 | 0 | 0 | Low-quality | Genome-fragment | 3.07 | AAI-based (medium-confidence) | 0 |
| S17_extracted_k141_128138 | 3277 | No | NA | 5 | 0 | 1 | Low-quality | Genome-fragment | 6.96 | AAI-based (high-confidence) | 0 |
| S10_extracted_90701 | 6390 | No | NA | 6 | 0 | 6 | Not-determined | Genome-fragment | NA | NA | 0 |
| S10_extracted_90753 | 1991 | No | NA | 1 | 0 | 1 | Not-determined | Genome-fragment | NA | NA | 0 |
| S10_extracted_90781 | 1278 | No | NA | 3 | 0 | 0 | Not-determined | Genome-fragment | NA | NA | 0 |
| S10_extracted_90855 | 3338 | No | NA | 6 | 0 | 4 | Not-determined | Genome-fragment | NA | NA | 0 |
| S10_extracted_91060 | 1540 | No | NA | 2 | 0 | 2 | Not-determined | Genome-fragment | NA | NA | 0 |
| S16_vs_86568\|\|full | 3277 | No | NA | 4 | 0 | 0 | Low-quality | Genome-fragment | 6.62 | AAI-based (high-confidence) | 0 |
| S10_extracted_91103 | 1025 | No | NA | 1 | 0 | 0 | Not-determined | Genome-fragment | NA | NA | 0 |
| S16_vs_132052\|\|full | 3277 | No | NA | 3 | 3 | 0 | Low-quality | Genome-fragment | 8.63 | AAI-based (high-confidence) | 0 |
| S8_vs_k141_207881\|\|full | 3277 | No | NA | 6 | 6 | 0 | Low-quality | Genome-fragment | 7.53 | AAI-based (high-confidence) | 0 |
| S10_extracted_91197 | 2380 | No | NA | 2 | 0 | 1 | Not-determined | Genome-fragment | NA | NA | 0 |
| S10_extracted_91242 | 12024 | No | NA | 6 | 0 | 3 | Not-determined | Genome-fragment | NA | NA | 0 |
| S10_extracted_91300 | 2140 | No | NA | 2 | 0 | 2 | Not-determined | Genome-fragment | NA | NA | 0 |
| S10_extracted_91323 | 3053 | No | NA | 2 | 0 | 1 | Not-determined | Genome-fragment | NA | NA | 0 |
| S10_extracted_91544 | 3848 | No | NA | 5 | 0 | 0 | Not-determined | Genome-fragment | NA | NA | 0 |
| S10_extracted_91651 | 1110 | No | NA | 1 | 0 | 0 | Not-determined | Genome-fragment | NA | NA | 0 |
| S10_extracted_91767 | 1370 | No | NA | 2 | 0 | 1 | Not-determined | Genome-fragment | NA | NA | 0 |
| S10_extracted_91798 | 9829 | No | NA | 9 | 0 | 5 | Not-determined | Genome-fragment | NA | NA | 0 |
| S10_extracted_91862 | 5523 | No | NA | 6 | 0 | 4 | Not-determined | Genome-fragment | NA | NA | 0 |
| S10_extracted_91912 | 1075 | No | NA | 2 | 0 | 0 | Not-determined | Genome-fragment | NA | NA | 0 |
| S10_extracted_92009 | 1580 | No | NA | 2 | 0 | 1 | Not-determined | Genome-fragment | NA | NA | 0 |
| S10_extracted_92075 | 1018 | No | NA | 1 | 0 | 0 | Not-determined | Genome-fragment | NA | NA | 0 |
| S10_extracted_92107 | 1043 | No | NA | 2 | 0 | 1 | Not-determined | Genome-fragment | NA | NA | 0 |
| S10_extracted_92172 | 3040 | No | NA | 5 | 0 | 2 | Not-determined | Genome-fragment | NA | NA | 0 |
| S10_extracted_92184 | 1574 | No | NA | 3 | 0 | 1 | Not-determined | Genome-fragment | NA | NA | 0 |
| S10_extracted_92255 | 5457 | No | NA | 4 | 0 | 4 | Not-determined | Genome-fragment | NA | NA | 0 |
| S10_extracted_92301 | 1448 | No | NA | 3 | 0 | 2 | Not-determined | Genome-fragment | NA | NA | 0 |
| S10_extracted_92328 | 1728 | No | NA | 3 | 0 | 3 | Not-determined | Genome-fragment | NA | NA | 0 |
| S10_extracted_92516 | 2421 | No | NA | 2 | 0 | 1 | Not-determined | Genome-fragment | NA | NA | 0 |
| S12_vs_k141_97901\|\|full | 3278 | No | NA | 4 | 2 | 0 | Low-quality | Genome-fragment | 8.69 | AAI-based (high-confidence) | 0 |
| S10_extracted_92554 | 3115 | No | NA | 3 | 0 | 3 | Not-determined | Genome-fragment | NA | NA | 0 |
| S7_vs_35244\|\|full | 3279 | No | NA | 4 | 0 | 3 | Low-quality | Genome-fragment | 0.98 | AAI-based (medium-confidence) | 0 |
| S10_extracted_92804 | 5672 | No | NA | 5 | 0 | 2 | Not-determined | Genome-fragment | NA | NA | 0 |
| S10_extracted_92886 | 7656 | No | NA | 9 | 0 | 6 | Not-determined | Genome-fragment | NA | NA | 0 |
| S10_extracted_92931 | 1259 | No | NA | 1 | 0 | 1 | Not-determined | Genome-fragment | NA | NA | 0 |
| S10_extracted_93070 | 1483 | No | NA | 1 | 0 | 0 | Not-determined | Genome-fragment | NA | NA | 0 |
| S3_extracted_k141_92916 | 3281 | No | NA | 7 | 1 | 0 | Low-quality | Genome-fragment | 1.41 | HMM-based (lower-bound) | 0 |
| S10_extracted_93127 | 1064 | No | NA | 2 | 0 | 2 | Not-determined | Genome-fragment | NA | NA | 0 |
| S10_extracted_93170 | 1550 | No | NA | 2 | 0 | 2 | Not-determined | Genome-fragment | NA | NA | 0 |
| S10_extracted_93218 | 1445 | No | NA | 1 | 0 | 0 | Not-determined | Genome-fragment | NA | NA | 0 |
| S10_extracted_93244 | 1860 | No | NA | 4 | 0 | 1 | Not-determined | Genome-fragment | NA | NA | 0 |
| S10_extracted_93266 | 1012 | No | NA | 3 | 0 | 0 | Not-determined | Genome-fragment | NA | NA | 0 |
| S10_extracted_93295 | 1242 | No | NA | 1 | 0 | 1 | Not-determined | Genome-fragment | NA | NA | 0 |
| S10_extracted_93365 | 2739 | No | NA | 1 | 0 | 1 | Not-determined | Genome-fragment | NA | NA | 0 |
| S10_extracted_93378 | 1508 | No | NA | 1 | 0 | 0 | Not-determined | Genome-fragment | NA | NA | 0 |
| S10_extracted_93459 | 1308 | No | NA | 1 | 0 | 1 | Not-determined | Genome-fragment | NA | NA | 0 |
| S10_extracted_93534 | 10887 | No | NA | 15 | 0 | 0 | Not-determined | Genome-fragment | NA | NA | 0 |
| S10_extracted_93764 | 4290 | No | NA | 5 | 0 | 2 | Not-determined | Genome-fragment | NA | NA | 0 |
| S10_extracted_93833 | 1129 | No | NA | 2 | 0 | 0 | Not-determined | Genome-fragment | NA | NA | 0 |
| S16_vs_74799\|\|full | 3281 | No | NA | 4 | 2 | 0 | Low-quality | Genome-fragment | 7.91 | AAI-based (medium-confidence) | 0 |
| S10_extracted_93912 | 4340 | No | NA | 5 | 0 | 0 | Not-determined | Genome-fragment | NA | NA | 0 |
| S10_extracted_94029 | 1573 | No | NA | 1 | 0 | 1 | Not-determined | Genome-fragment | NA | NA | 0 |
| S10_extracted_94108 | 1174 | No | NA | 1 | 0 | 1 | Not-determined | Genome-fragment | NA | NA | 0 |
| S10_extracted_94127 | 1721 | No | NA | 1 | 0 | 0 | Not-determined | Genome-fragment | NA | NA | 0 |
| S10_extracted_94215 | 8402 | No | NA | 6 | 0 | 3 | Not-determined | Genome-fragment | NA | NA | 0 |
| S10_extracted_94333 | 2277 | No | NA | 2 | 0 | 2 | Not-determined | Genome-fragment | NA | NA | 0 |
| S13_vs_41103\|\|full | 3282 | No | NA | 3 | 0 | 1 | Low-quality | Genome-fragment | 2.65 | AAI-based (medium-confidence) | 0 |
| S10_extracted_94399 | 4179 | No | NA | 3 | 0 | 0 | Not-determined | Genome-fragment | NA | NA | 0 |
| S10_extracted_94465 | 15606 | No | NA | 13 | 0 | 3 | Not-determined | Genome-fragment | NA | NA | 0 |
| S10_extracted_94526 | 2027 | No | NA | 1 | 0 | 1 | Not-determined | Genome-fragment | NA | NA | 0 |
| S10_extracted_94538 | 1445 | No | NA | 2 | 0 | 0 | Not-determined | Genome-fragment | NA | NA | 0 |
| S5_extracted_k141_36451 | 3282 | No | NA | 3 | 0 | 3 | Low-quality | Genome-fragment | 0.96 | AAI-based (high-confidence) | 0 |
| S16_vs_56843\|\|full | 3282 | No | NA | 4 | 2 | 0 | Low-quality | Genome-fragment | 4.56 | HMM-based (lower-bound) | 0 |
| S10_extracted_94596 | 3183 | No | NA | 5 | 0 | 0 | Not-determined | Genome-fragment | NA | NA | 0 |
| S10_extracted_94660 | 2905 | No | NA | 2 | 0 | 1 | Not-determined | Genome-fragment | NA | NA | 0 |
| S10_extracted_94663 | 1203 | No | NA | 2 | 0 | 1 | Not-determined | Genome-fragment | NA | NA | 0 |
| S10_extracted_94873 | 1329 | No | NA | 3 | 0 | 2 | Not-determined | Genome-fragment | NA | NA | 0 |
| S5_vs_k141_43137\|\|full | 3283 | No | NA | 3 | 2 | 0 | Low-quality | Genome-fragment | 4.76 | AAI-based (medium-confidence) | 0 |
| S10_extracted_95093 | 5116 | No | NA | 2 | 0 | 1 | Not-determined | Genome-fragment | NA | NA | 0 |
| S10_extracted_95116 | 9461 | No | NA | 6 | 0 | 4 | Not-determined | Genome-fragment | NA | NA | 0 |
| S10_extracted_95135 | 10056 | No | NA | 10 | 0 | 10 | Not-determined | Genome-fragment | NA | NA | 0 |
| S16_vs_140005\|\|full | 3283 | No | NA | 4 | 0 | 0 | Low-quality | Genome-fragment | 5.11 | AAI-based (medium-confidence) | 0 |
| S10_extracted_95272 | 2005 | No | NA | 6 | 0 | 5 | Not-determined | Genome-fragment | NA | NA | 0 |
| S10_extracted_95352 | 1786 | No | NA | 1 | 0 | 1 | Not-determined | Genome-fragment | NA | NA | 0 |
| S6_extracted_k141_4800 | 3284 | No | NA | 3 | 0 | 1 | Low-quality | Genome-fragment | 6.7 | AAI-based (high-confidence) | 0 |
| S4_extracted_21808 | 3285 | No | NA | 6 | 4 | 0 | Low-quality | Genome-fragment | 5.36 | AAI-based (high-confidence) | 0 |
| S7_vs_28478\|\|full | 3286 | No | NA | 6 | 2 | 0 | Low-quality | Genome-fragment | 2.9 | HMM-based (lower-bound) | 0 |
| S10_extracted_95755 | 1674 | No | NA | 1 | 0 | 0 | Not-determined | Genome-fragment | NA | NA | 0 |
| S10_extracted_95766 | 5494 | No | NA | 6 | 0 | 4 | Not-determined | Genome-fragment | NA | NA | 0 |
| S10_extracted_95823 | 6643 | No | NA | 8 | 0 | 7 | Not-determined | Genome-fragment | NA | NA | 0 |
| S10_extracted_95973 | 1082 | No | NA | 2 | 0 | 1 | Not-determined | Genome-fragment | NA | NA | 0 |
| S10_extracted_96013 | 2144 | No | NA | 1 | 0 | 1 | Not-determined | Genome-fragment | NA | NA | 0 |
| S10_extracted_96056 | 1217 | No | NA | 2 | 0 | 1 | Not-determined | Genome-fragment | NA | NA | 0 |
| S10_extracted_96491 | 1122 | No | NA | 3 | 0 | 1 | Not-determined | Genome-fragment | NA | NA | 0 |
| S10_extracted_96604 | 1535 | No | NA | 4 | 0 | 2 | Not-determined | Genome-fragment | NA | NA | 0 |
| S10_extracted_96690 | 1735 | No | NA | 1 | 0 | 1 | Not-determined | Genome-fragment | NA | NA | 0 |
| S10_extracted_96769 | 1009 | No | NA | 2 | 0 | 1 | Not-determined | Genome-fragment | NA | NA | 0 |
| S10_extracted_96794 | 2309 | No | NA | 2 | 0 | 2 | Not-determined | Genome-fragment | NA | NA | 0 |
| S10_extracted_96908 | 1046 | No | NA | 2 | 0 | 2 | Not-determined | Genome-fragment | NA | NA | 0 |
| S10_extracted_97003 | 8548 | No | NA | 4 | 0 | 3 | Not-determined | Genome-fragment | NA | NA | 0 |
| S10_extracted_97163 | 2601 | No | NA | 3 | 0 | 0 | Not-determined | Genome-fragment | NA | NA | 0 |
| S10_extracted_97242 | 1288 | No | NA | 1 | 0 | 1 | Not-determined | Genome-fragment | NA | NA | 0 |
| S9_vs_k141_29540\|\|full | 3287 | No | NA | 6 | 0 | 0 | Low-quality | Genome-fragment | 7.75 | AAI-based (medium-confidence) | 0 |
| S7_vs_106315\|\|full | 3287 | No | NA | 4 | 0 | 1 | Low-quality | Genome-fragment | 5.46 | AAI-based (high-confidence) | 0 |
| S10_extracted_97370 | 1115 | No | NA | 2 | 0 | 1 | Not-determined | Genome-fragment | NA | NA | 0 |
| S10_extracted_97482 | 1901 | No | NA | 4 | 0 | 2 | Not-determined | Genome-fragment | NA | NA | 0 |
| S10_extracted_97490 | 1135 | No | NA | 2 | 0 | 2 | Not-determined | Genome-fragment | NA | NA | 0 |
| S10_extracted_97549 | 4977 | No | NA | 7 | 0 | 3 | Not-determined | Genome-fragment | NA | NA | 0 |
| S12_vs_k141_176604\|\|full | 3287 | No | NA | 3 | 0 | 1 | Low-quality | Genome-fragment | 5.2 | AAI-based (medium-confidence) | 0 |
| S10_extracted_97676 | 2631 | No | NA | 3 | 0 | 1 | Not-determined | Genome-fragment | NA | NA | 0 |
| S10_extracted_97757 | 5463 | No | NA | 7 | 0 | 2 | Not-determined | Genome-fragment | NA | NA | 0 |
| S10_extracted_97778 | 1266 | No | NA | 3 | 0 | 1 | Not-determined | Genome-fragment | NA | NA | 0 |
| S10_extracted_97805 | 3884 | No | NA | 2 | 0 | 2 | Not-determined | Genome-fragment | NA | NA | 0 |
| S9_extracted_k141_14508 | 3288 | No | NA | 7 | 2 | 0 | Low-quality | Genome-fragment | 7.6 | AAI-based (medium-confidence) | 0 |
| S10_extracted_97867 | 1199 | No | NA | 1 | 0 | 1 | Not-determined | Genome-fragment | NA | NA | 0 |
| S10_extracted_98029 | 1231 | No | NA | 4 | 0 | 0 | Not-determined | Genome-fragment | NA | NA | 0 |
| S10_extracted_98164 | 7442 | No | NA | 5 | 0 | 2 | Not-determined | Genome-fragment | NA | NA | 0 |
| S10_extracted_98187 | 1183 | No | NA | 2 | 0 | 2 | Not-determined | Genome-fragment | NA | NA | 0 |
| S13_vs_90596\|\|full | 3288 | No | NA | 7 | 1 | 0 | Low-quality | Genome-fragment | 7.78 | AAI-based (high-confidence) | 0 |
| S10_extracted_98345 | 1156 | No | NA | 1 | 0 | 0 | Not-determined | Genome-fragment | NA | NA | 0 |
| S16_extracted_26005 | 3288 | No | NA | 8 | 2 | 0 | Low-quality | Genome-fragment | 7.83 | AAI-based (medium-confidence) | 0 |
| S17_vs_k141_69314\|\|full | 3289 | No | NA | 6 | 0 | 1 | Low-quality | Genome-fragment | 6.94 | AAI-based (medium-confidence) | 0 |
| S10_extracted_98666 | 1510 | No | NA | 2 | 0 | 0 | Not-determined | Genome-fragment | NA | NA | 0 |
| S11_extracted_k141_29036 | 3289 | No | NA | 10 | 1 | 0 | Low-quality | Genome-fragment | 3.69 | AAI-based (high-confidence) | 0 |
| S10_extracted_98737 | 7968 | No | NA | 6 | 0 | 1 | Not-determined | Genome-fragment | NA | NA | 0 |
| S10_extracted_98867 | 3125 | No | NA | 2 | 0 | 0 | Not-determined | Genome-fragment | NA | NA | 0 |
| S16_vs_15081\|\|full | 3289 | No | NA | 10 | 1 | 0 | Low-quality | Genome-fragment | 6.69 | AAI-based (high-confidence) | 0 |
| S10_extracted_99055 | 1163 | No | NA | 2 | 0 | 1 | Not-determined | Genome-fragment | NA | NA | 0 |
| S6_vs_k141_32397\|\|full | 3289 | No | NA | 4 | 0 | 3 | Low-quality | Genome-fragment | 6.09 | AAI-based (medium-confidence) | 0 |
| S10_extracted_99199 | 1164 | No | NA | 3 | 0 | 3 | Not-determined | Genome-fragment | NA | NA | 0 |
| S1_extracted_k141_86371 | 3290 | No | NA | 2 | 0 | 2 | Low-quality | Genome-fragment | 0.49 | AAI-based (medium-confidence) | 0 |
| S10_extracted_99513 | 1123 | No | NA | 1 | 0 | 1 | Not-determined | Genome-fragment | NA | NA | 0 |
| S10_extracted_99607 | 1525 | No | NA | 2 | 0 | 1 | Not-determined | Genome-fragment | NA | NA | 0 |
| S16_vs_125136\|\|full | 3290 | No | NA | 12 | 1 | 0 | Low-quality | Genome-fragment | 3.96 | AAI-based (high-confidence) | 0 |
| S10_extracted_99675 | 1361 | No | NA | 3 | 0 | 0 | Not-determined | Genome-fragment | NA | NA | 0 |
| S10_extracted_99756 | 1600 | No | NA | 2 | 0 | 1 | Not-determined | Genome-fragment | NA | NA | 0 |
| S10_extracted_99783 | 3612 | No | NA | 4 | 0 | 4 | Not-determined | Genome-fragment | NA | NA | 0 |
| S10_extracted_99827 | 1047 | No | NA | 2 | 0 | 0 | Not-determined | Genome-fragment | NA | NA | 0 |
| S10_extracted_100197 | 3684 | No | NA | 3 | 0 | 3 | Not-determined | Genome-fragment | NA | NA | 0 |
| S1_vs_k141_140583\|\|full | 3291 | No | NA | 3 | 0 | 1 | Low-quality | Genome-fragment | 1.45 | AAI-based (medium-confidence) | 0 |
| S10_extracted_100755 | 1517 | No | NA | 3 | 0 | 1 | Not-determined | Genome-fragment | NA | NA | 0 |
| S10_extracted_100911 | 1786 | No | NA | 1 | 0 | 0 | Not-determined | Genome-fragment | NA | NA | 0 |
| S12_extracted_k141_54717 | 3291 | No | NA | 4 | 0 | 2 | Low-quality | Genome-fragment | 1.96 | AAI-based (medium-confidence) | 0 |
| S10_extracted_101007 | 3247 | No | NA | 3 | 0 | 2 | Not-determined | Genome-fragment | NA | NA | 0 |
| S10_extracted_101103 | 3048 | No | NA | 4 | 0 | 4 | Not-determined | Genome-fragment | NA | NA | 0 |
| S10_extracted_101129 | 5523 | No | NA | 5 | 0 | 5 | Not-determined | Genome-fragment | NA | NA | 0 |
| S10_extracted_101235 | 1095 | No | NA | 1 | 0 | 0 | Not-determined | Genome-fragment | NA | NA | 0 |
| S10_extracted_101350 | 1019 | No | NA | 1 | 0 | 1 | Not-determined | Genome-fragment | NA | NA | 0 |
| S10_extracted_101396 | 1045 | No | NA | 1 | 0 | 0 | Not-determined | Genome-fragment | NA | NA | 0 |
| S10_extracted_101533 | 1933 | No | NA | 2 | 0 | 2 | Not-determined | Genome-fragment | NA | NA | 0 |
| S10_extracted_101601 | 1846 | No | NA | 1 | 0 | 0 | Not-determined | Genome-fragment | NA | NA | 0 |
| S10_extracted_101655 | 1026 | No | NA | 1 | 0 | 0 | Not-determined | Genome-fragment | NA | NA | 0 |
| S14_extracted_k141_80530 | 3292 | No | NA | 4 | 0 | 3 | Low-quality | Genome-fragment | 2.51 | AAI-based (medium-confidence) | 0 |
| S10_extracted_101765 | 1043 | No | NA | 1 | 0 | 0 | Not-determined | Genome-fragment | NA | NA | 0 |
| S7_extracted_93093 | 3292 | No | NA | 2 | 1 | 0 | Low-quality | Genome-fragment | 2.5 | AAI-based (high-confidence) | 0 |
| S16_vs_36873\|\|full | 3292 | No | NA | 5 | 0 | 1 | Low-quality | Genome-fragment | 2.49 | AAI-based (high-confidence) | 0 |
| S10_extracted_102040 | 1024 | No | NA | 1 | 0 | 1 | Not-determined | Genome-fragment | NA | NA | 0 |
| S10_extracted_102048 | 1548 | No | NA | 2 | 0 | 1 | Not-determined | Genome-fragment | NA | NA | 0 |
| S10_extracted_102098 | 3455 | No | NA | 7 | 0 | 4 | Not-determined | Genome-fragment | NA | NA | 0 |
| S10_extracted_102184 | 1106 | No | NA | 1 | 0 | 0 | Not-determined | Genome-fragment | NA | NA | 0 |
| S10_extracted_102204 | 2039 | No | NA | 1 | 0 | 1 | Not-determined | Genome-fragment | NA | NA | 0 |
| S10_vs_112942\|\|full | 3292 | Yes | 1691 | 6 | 1 | 1 | Low-quality | Genome-fragment | 4.61 | AAI-based (high-confidence) | 48.63 |
| S10_extracted_102415 | 2110 | No | NA | 1 | 0 | 1 | Not-determined | Genome-fragment | NA | NA | 0 |
| S17_extracted_k141_111536 | 3293 | No | NA | 2 | 0 | 2 | Low-quality | Genome-fragment | 1.03 | AAI-based (medium-confidence) | 0 |
| S10_extracted_102596 | 5520 | No | NA | 4 | 0 | 4 | Not-determined | Genome-fragment | NA | NA | 0 |
| S10_extracted_102688 | 3140 | No | NA | 6 | 0 | 6 | Not-determined | Genome-fragment | NA | NA | 0 |
| S10_extracted_102739 | 5273 | No | NA | 4 | 0 | 2 | Not-determined | Genome-fragment | NA | NA | 0 |
| S10_extracted_102887 | 1806 | No | NA | 3 | 0 | 2 | Not-determined | Genome-fragment | NA | NA | 0 |
| S5_vs_k141_81198\|\|full | 3293 | No | NA | 3 | 3 | 0 | Low-quality | Genome-fragment | 2.42 | AAI-based (medium-confidence) | 0 |
| S10_extracted_102944 | 1111 | No | NA | 2 | 0 | 1 | Not-determined | Genome-fragment | NA | NA | 0 |
| S10_extracted_102995 | 1202 | No | NA | 1 | 0 | 1 | Not-determined | Genome-fragment | NA | NA | 0 |
| S10_extracted_103243 | 2154 | No | NA | 3 | 0 | 2 | Not-determined | Genome-fragment | NA | NA | 0 |
| S10_extracted_103307 | 1098 | No | NA | 0 | 0 | 0 | Not-determined | Genome-fragment | NA | NA | 0 |
| S10_extracted_103318 | 1339 | No | NA | 1 | 0 | 1 | Not-determined | Genome-fragment | NA | NA | 0 |
| S10_extracted_103386 | 1065 | No | NA | 2 | 0 | 1 | Not-determined | Genome-fragment | NA | NA | 0 |
| S10_extracted_103438 | 2366 | No | NA | 3 | 0 | 1 | Not-determined | Genome-fragment | NA | NA | 0 |
| S10_extracted_103466 | 1310 | No | NA | 1 | 0 | 1 | Not-determined | Genome-fragment | NA | NA | 0 |
| S10_extracted_103579 | 1510 | No | NA | 3 | 0 | 0 | Not-determined | Genome-fragment | NA | NA | 0 |
| S10_extracted_103616 | 1841 | No | NA | 2 | 0 | 2 | Not-determined | Genome-fragment | NA | NA | 0 |
| S10_extracted_103673 | 5443 | No | NA | 5 | 0 | 4 | Not-determined | Genome-fragment | NA | NA | 0 |
| S10_extracted_103811 | 1278 | No | NA | 2 | 0 | 1 | Not-determined | Genome-fragment | NA | NA | 0 |
| S10_extracted_103873 | 1671 | No | NA | 1 | 0 | 0 | Not-determined | Genome-fragment | NA | NA | 0 |
| S10_extracted_103925 | 1047 | No | NA | 1 | 0 | 1 | Not-determined | Genome-fragment | NA | NA | 0 |
| S10_extracted_103943 | 1450 | No | NA | 1 | 0 | 0 | Not-determined | Genome-fragment | NA | NA | 0 |
| S10_extracted_20828 | 3293 | No | NA | 4 | 0 | 4 | Low-quality | Genome-fragment | 1.36 | AAI-based (high-confidence) | 0 |
| S10_extracted_104095 | 1098 | No | NA | 2 | 0 | 0 | Not-determined | Genome-fragment | NA | NA | 0 |
| S10_extracted_104103 | 1276 | No | NA | 2 | 0 | 2 | Not-determined | Genome-fragment | NA | NA | 0 |
| S10_extracted_104190 | 1097 | No | NA | 3 | 0 | 3 | Not-determined | Genome-fragment | NA | NA | 0 |
| S8_vs_k141_15271\|\|full | 3293 | No | NA | 10 | 0 | 0 | Low-quality | Genome-fragment | 5.99 | AAI-based (medium-confidence) | 0 |
| S5_vs_k141_70133\|\|full | 3294 | No | NA | 4 | 0 | 4 | Low-quality | Genome-fragment | 1.52 | AAI-based (medium-confidence) | 0 |
| S10_extracted_104536 | 2501 | No | NA | 3 | 0 | 1 | Not-determined | Genome-fragment | NA | NA | 0 |
| S16_vs_62521\|\|full | 3295 | No | NA | 11 | 0 | 0 | Low-quality | Genome-fragment | 10.29 | AAI-based (medium-confidence) | 0 |
| S10_extracted_118639 | 3295 | No | NA | 2 | 1 | 0 | Low-quality | Genome-fragment | 1.28 | AAI-based (medium-confidence) | 0 |
| S10_extracted_104718 | 1142 | No | NA | 2 | 0 | 0 | Not-determined | Genome-fragment | NA | NA | 0 |
| S10_extracted_104813 | 1244 | No | NA | 2 | 0 | 1 | Not-determined | Genome-fragment | NA | NA | 0 |
| S10_extracted_104838 | 1218 | No | NA | 2 | 0 | 0 | Not-determined | Genome-fragment | NA | NA | 0 |
| S10_extracted_104933 | 1078 | No | NA | 1 | 0 | 1 | Not-determined | Genome-fragment | NA | NA | 0 |
| S12_vs_k141_182969\|\|full | 3295 | No | NA | 5 | 0 | 1 | Low-quality | Genome-fragment | 7.55 | AAI-based (high-confidence) | 0 |
| S10_extracted_105030 | 1225 | No | NA | 2 | 0 | 0 | Not-determined | Genome-fragment | NA | NA | 0 |
| S10_extracted_105037 | 7951 | No | NA | 15 | 0 | 15 | Not-determined | Genome-fragment | NA | NA | 0 |
| S4_extracted_21846 | 3296 | No | NA | 8 | 2 | 0 | Low-quality | Genome-fragment | 7.83 | AAI-based (high-confidence) | 0 |
| S10_extracted_105107 | 1227 | No | NA | 2 | 0 | 0 | Not-determined | Genome-fragment | NA | NA | 0 |
| S10_extracted_105111 | 2348 | No | NA | 2 | 0 | 1 | Not-determined | Genome-fragment | NA | NA | 0 |
| S10_extracted_105203 | 1367 | No | NA | 1 | 0 | 1 | Not-determined | Genome-fragment | NA | NA | 0 |
| S6_vs_k141_44884\|\|full | 3296 | No | NA | 8 | 1 | 0 | Low-quality | Genome-fragment | 7.66 | AAI-based (high-confidence) | 0 |
| S10_extracted_105261 | 3483 | No | NA | 5 | 0 | 2 | Not-determined | Genome-fragment | NA | NA | 0 |
| S10_extracted_105329 | 7894 | No | NA | 7 | 0 | 6 | Not-determined | Genome-fragment | NA | NA | 0 |
| S8_vs_k141_161480\|\|full | 3296 | No | NA | 4 | 0 | 0 | Low-quality | Genome-fragment | 6.44 | AAI-based (high-confidence) | 0 |
| S10_extracted_105335 | 6000 | No | NA | 5 | 0 | 4 | Not-determined | Genome-fragment | NA | NA | 0 |
| S10_extracted_105336 | 1018 | No | NA | 1 | 0 | 1 | Not-determined | Genome-fragment | NA | NA | 0 |
| S10_extracted_105375 | 1238 | No | NA | 2 | 0 | 1 | Not-determined | Genome-fragment | NA | NA | 0 |
| S1_vs_k141_142243\|\|full | 3298 | No | NA | 3 | 0 | 1 | Low-quality | Genome-fragment | 1.5 | AAI-based (medium-confidence) | 0 |
| S10_extracted_105434 | 1777 | No | NA | 2 | 0 | 1 | Not-determined | Genome-fragment | NA | NA | 0 |
| S10_extracted_105498 | 1006 | No | NA | 1 | 0 | 1 | Not-determined | Genome-fragment | NA | NA | 0 |
| S10_extracted_105631 | 1401 | No | NA | 2 | 0 | 2 | Not-determined | Genome-fragment | NA | NA | 0 |
| S10_extracted_105689 | 2593 | No | NA | 2 | 0 | 2 | Not-determined | Genome-fragment | NA | NA | 0 |
| S10_extracted_105808 | 1320 | No | NA | 1 | 0 | 1 | Not-determined | Genome-fragment | NA | NA | 0 |
| S10_extracted_105938 | 11198 | No | NA | 13 | 0 | 3 | Not-determined | Genome-fragment | NA | NA | 0 |
| S10_extracted_105998 | 1113 | No | NA | 1 | 0 | 1 | Not-determined | Genome-fragment | NA | NA | 0 |
| S10_extracted_106035 | 6743 | No | NA | 5 | 0 | 3 | Not-determined | Genome-fragment | NA | NA | 0 |
| S10_extracted_106116 | 1183 | No | NA | 1 | 0 | 1 | Not-determined | Genome-fragment | NA | NA | 0 |
| S14_extracted_k141_87421 | 3299 | No | NA | 3 | 0 | 1 | Low-quality | Genome-fragment | 7.01 | AAI-based (medium-confidence) | 0 |
| S5_extracted_k141_89579 | 3299 | No | NA | 6 | 5 | 0 | Low-quality | Genome-fragment | 7.18 | AAI-based (medium-confidence) | 0 |
| S10_extracted_48870 | 3299 | No | NA | 4 | 0 | 0 | Low-quality | Genome-fragment | 7.09 | AAI-based (high-confidence) | 0 |
| S10_extracted_106337 | 1649 | No | NA | 1 | 0 | 1 | Not-determined | Genome-fragment | NA | NA | 0 |
| S10_extracted_106341 | 5379 | No | NA | 5 | 0 | 1 | Not-determined | Genome-fragment | NA | NA | 0 |
| S10_extracted_106543 | 3506 | No | NA | 4 | 0 | 1 | Not-determined | Genome-fragment | NA | NA | 0 |
| S10_extracted_106597 | 1551 | No | NA | 2 | 0 | 2 | Not-determined | Genome-fragment | NA | NA | 0 |
| S10_extracted_106791 | 3145 | No | NA | 3 | 0 | 1 | Not-determined | Genome-fragment | NA | NA | 0 |
| S11_vs_k141_93990\|\|full | 3300 | No | NA | 2 | 0 | 2 | Low-quality | Genome-fragment | 1.68 | AAI-based (medium-confidence) | 0 |
| S10_extracted_107021 | 1496 | No | NA | 1 | 0 | 1 | Not-determined | Genome-fragment | NA | NA | 0 |
| S10_extracted_107061 | 3938 | No | NA | 2 | 0 | 2 | Not-determined | Genome-fragment | NA | NA | 0 |
| S16_extracted_18919 | 3300 | No | NA | 6 | 0 | 0 | Low-quality | Genome-fragment | 5.1 | AAI-based (high-confidence) | 0 |
| S10_extracted_107422 | 7151 | No | NA | 4 | 0 | 2 | Not-determined | Genome-fragment | NA | NA | 0 |
| S16_vs_151593\|\|full | 3301 | No | NA | 3 | 0 | 1 | Low-quality | Genome-fragment | 1.4 | AAI-based (medium-confidence) | 0 |
| S10_extracted_107680 | 2398 | No | NA | 5 | 0 | 0 | Not-determined | Genome-fragment | NA | NA | 0 |
| S10_extracted_107915 | 1546 | No | NA | 1 | 0 | 1 | Not-determined | Genome-fragment | NA | NA | 0 |
| S8_extracted_k141_132012 | 3301 | No | NA | 7 | 0 | 0 | Low-quality | Genome-fragment | 4.55 | AAI-based (high-confidence) | 0 |
| S10_extracted_108011 | 8728 | No | NA | 4 | 0 | 4 | Not-determined | Genome-fragment | NA | NA | 0 |
| S10_extracted_108525 | 5159 | No | NA | 7 | 0 | 2 | Not-determined | Genome-fragment | NA | NA | 0 |
| S10_extracted_108651 | 1295 | No | NA | 2 | 0 | 0 | Not-determined | Genome-fragment | NA | NA | 0 |
| S10_extracted_108671 | 1692 | No | NA | 2 | 0 | 1 | Not-determined | Genome-fragment | NA | NA | 0 |
| S8_vs_k141_199922\|\|full | 3301 | No | NA | 4 | 0 | 2 | Low-quality | Genome-fragment | 8.49 | AAI-based (medium-confidence) | 0 |
| S10_extracted_108753 | 1408 | No | NA | 2 | 0 | 1 | Not-determined | Genome-fragment | NA | NA | 0 |
| S10_extracted_108823 | 3661 | No | NA | 4 | 0 | 3 | Not-determined | Genome-fragment | NA | NA | 0 |
| S11_vs_k141_20307\|\|full | 3302 | No | NA | 8 | 2 | 0 | Low-quality | Genome-fragment | 6.12 | AAI-based (medium-confidence) | 0 |
| S10_extracted_108888 | 3480 | No | NA | 5 | 0 | 4 | Not-determined | Genome-fragment | NA | NA | 0 |
| S10_extracted_109023 | 3141 | No | NA | 4 | 0 | 0 | Not-determined | Genome-fragment | NA | NA | 0 |
| S10_extracted_109028 | 3123 | No | NA | 4 | 0 | 3 | Not-determined | Genome-fragment | NA | NA | 0 |
| S10_extracted_109035 | 1439 | No | NA | 1 | 0 | 0 | Not-determined | Genome-fragment | NA | NA | 0 |
| S10_extracted_109147 | 1516 | No | NA | 2 | 0 | 2 | Not-determined | Genome-fragment | NA | NA | 0 |
| S16_extracted_11619 | 3303 | No | NA | 10 | 1 | 0 | Low-quality | Genome-fragment | 5.35 | AAI-based (high-confidence) | 0 |
| S4_extracted_28678 | 3303 | No | NA | 7 | 0 | 0 | Low-quality | Genome-fragment | 6.17 | AAI-based (high-confidence) | 0 |
| S10_extracted_109438 | 1311 | No | NA | 2 | 0 | 1 | Not-determined | Genome-fragment | NA | NA | 0 |
| S10_extracted_109453 | 1332 | No | NA | 2 | 0 | 2 | Not-determined | Genome-fragment | NA | NA | 0 |
| S3_vs_k141_63235\|\|full | 3304 | No | NA | 5 | 1 | 0 | Low-quality | Genome-fragment | 7.27 | AAI-based (medium-confidence) | 0 |
| S10_extracted_109642 | 1210 | No | NA | 1 | 0 | 1 | Not-determined | Genome-fragment | NA | NA | 0 |
| S16_vs_112375\|\|full | 3304 | No | NA | 4 | 0 | 0 | Low-quality | Genome-fragment | 2.89 | AAI-based (medium-confidence) | 0 |
| S10_extracted_109832 | 1534 | No | NA | 1 | 0 | 1 | Not-determined | Genome-fragment | NA | NA | 0 |
| S12_extracted_k141_120498 | 3304 | No | NA | 4 | 0 | 3 | Low-quality | Genome-fragment | 1.21 | AAI-based (medium-confidence) | 0 |
| S10_extracted_110295 | 2081 | No | NA | 2 | 0 | 2 | Not-determined | Genome-fragment | NA | NA | 0 |
| S10_extracted_110334 | 9353 | No | NA | 8 | 0 | 7 | Not-determined | Genome-fragment | NA | NA | 0 |
| S10_extracted_110500 | 1247 | No | NA | 2 | 0 | 1 | Not-determined | Genome-fragment | NA | NA | 0 |
| S10_extracted_110592 | 1062 | No | NA | 1 | 0 | 1 | Not-determined | Genome-fragment | NA | NA | 0 |
| S2_extracted_k141_18029 | 3304 | No | NA | 10 | 8 | 0 | Low-quality | Genome-fragment | 10.9 | AAI-based (high-confidence) | 0 |
| S8_vs_k141_52769\|\|full | 3304 | No | NA | 6 | 5 | 0 | Low-quality | Genome-fragment | 7.49 | AAI-based (high-confidence) | 0 |
| S10_extracted_111093 | 1008 | No | NA | 1 | 0 | 1 | Not-determined | Genome-fragment | NA | NA | 0 |
| S10_extracted_111100 | 1208 | No | NA | 1 | 0 | 1 | Not-determined | Genome-fragment | NA | NA | 0 |
| S10_extracted_111432 | 1036 | No | NA | 1 | 0 | 1 | Not-determined | Genome-fragment | NA | NA | 0 |
| S10_extracted_111462 | 1159 | No | NA | 1 | 0 | 0 | Not-determined | Genome-fragment | NA | NA | 0 |
| S10_extracted_111785 | 1763 | No | NA | 2 | 0 | 2 | Not-determined | Genome-fragment | NA | NA | 0 |
| S10_extracted_111907 | 1273 | No | NA | 2 | 0 | 2 | Not-determined | Genome-fragment | NA | NA | 0 |
| S10_extracted_111930 | 1526 | No | NA | 2 | 0 | 2 | Not-determined | Genome-fragment | NA | NA | 0 |
| S10_extracted_111933 | 1162 | No | NA | 1 | 0 | 0 | Not-determined | Genome-fragment | NA | NA | 0 |
| S10_extracted_111959 | 1324 | No | NA | 3 | 0 | 0 | Not-determined | Genome-fragment | NA | NA | 0 |
| S10_extracted_111967 | 1009 | No | NA | 1 | 0 | 0 | Not-determined | Genome-fragment | NA | NA | 0 |
| S10_extracted_111969 | 1114 | No | NA | 2 | 0 | 1 | Not-determined | Genome-fragment | NA | NA | 0 |
| S10_extracted_112016 | 1689 | No | NA | 2 | 0 | 2 | Not-determined | Genome-fragment | NA | NA | 0 |
| S10_extracted_112151 | 1777 | No | NA | 2 | 0 | 0 | Not-determined | Genome-fragment | NA | NA | 0 |
| S8_extracted_k141_116183 | 3305 | No | NA | 4 | 0 | 4 | Low-quality | Genome-fragment | 4.01 | AAI-based (medium-confidence) | 0 |
| S10_extracted_112333 | 1480 | No | NA | 5 | 0 | 4 | Not-determined | Genome-fragment | NA | NA | 0 |
| S10_extracted_112529 | 2539 | No | NA | 2 | 0 | 1 | Not-determined | Genome-fragment | NA | NA | 0 |
| S10_extracted_112600 | 1199 | No | NA | 1 | 0 | 1 | Not-determined | Genome-fragment | NA | NA | 0 |
| S10_extracted_112681 | 9629 | No | NA | 5 | 0 | 4 | Not-determined | Genome-fragment | NA | NA | 0 |
| S14_extracted_k141_113499 | 3306 | No | NA | 2 | 0 | 0 | Low-quality | Genome-fragment | 7.15 | AAI-based (high-confidence) | 0 |
| S10_extracted_112957 | 1503 | No | NA | 1 | 0 | 1 | Not-determined | Genome-fragment | NA | NA | 0 |
| S10_extracted_113060 | 1181 | No | NA | 2 | 0 | 0 | Not-determined | Genome-fragment | NA | NA | 0 |
| S10_extracted_113133 | 1730 | No | NA | 1 | 0 | 1 | Not-determined | Genome-fragment | NA | NA | 0 |
| S3_extracted_k141_106166 | 3306 | No | NA | 9 | 2 | 0 | Low-quality | Genome-fragment | 7.92 | AAI-based (high-confidence) | 0 |
| S10_extracted_113305 | 1000 | No | NA | 2 | 0 | 1 | Not-determined | Genome-fragment | NA | NA | 0 |
| S10_extracted_113489 | 27189 | No | NA | 44 | 10 | 0 | Medium-quality | Genome-fragment | 58.41 | AAI-based (high-confidence) | 0 |
| S10_extracted_113528 | 1522 | No | NA | 2 | 0 | 0 | Not-determined | Genome-fragment | NA | NA | 0 |
| S10_extracted_113589 | 3197 | No | NA | 3 | 0 | 2 | Not-determined | Genome-fragment | NA | NA | 0 |
| S10_extracted_113678 | 2384 | No | NA | 3 | 0 | 2 | Not-determined | Genome-fragment | NA | NA | 0 |
| S10_extracted_113840 | 1149 | No | NA | 2 | 0 | 2 | Not-determined | Genome-fragment | NA | NA | 0 |
| S12_extracted_k141_17684 | 3306 | No | NA | 2 | 0 | 1 | Low-quality | Genome-fragment | 3.83 | AAI-based (medium-confidence) | 0 |
| S10_extracted_113876 | 1080 | No | NA | 1 | 0 | 1 | Not-determined | Genome-fragment | NA | NA | 0 |
| S14_extracted_k141_37290 | 3307 | No | NA | 9 | 8 | 0 | Low-quality | Genome-fragment | 10.91 | AAI-based (high-confidence) | 0 |
| S10_extracted_114077 | 2928 | No | NA | 3 | 0 | 3 | Not-determined | Genome-fragment | NA | NA | 0 |
| S3_vs_k141_89373\|\|full | 3307 | No | NA | 5 | 0 | 2 | Low-quality | Genome-fragment | 4.97 | AAI-based (medium-confidence) | 0 |
| S5_vs_k141_61612\|\|full | 3307 | No | NA | 3 | 0 | 2 | Low-quality | Genome-fragment | 1.1 | AAI-based (medium-confidence) | 0 |
| S10_extracted_114260 | 1217 | No | NA | 3 | 0 | 2 | Not-determined | Genome-fragment | NA | NA | 0 |
| S10_extracted_114338 | 4411 | No | NA | 3 | 0 | 3 | Not-determined | Genome-fragment | NA | NA | 0 |
| S16_vs_12768\|\|full | 3307 | No | NA | 4 | 0 | 2 | Low-quality | Genome-fragment | 4.93 | AAI-based (high-confidence) | 0 |
| S17_vs_k141_53727\|\|full | 3309 | Yes | 1973 | 5 | 1 | 1 | Low-quality | Genome-fragment | 1.37 | HMM-based (lower-bound) | 40.37 |
| S10_extracted_114488 | 3436 | No | NA | 5 | 0 | 5 | Not-determined | Genome-fragment | NA | NA | 0 |
| S10_extracted_114528 | 1252 | No | NA | 1 | 0 | 1 | Not-determined | Genome-fragment | NA | NA | 0 |
| S10_extracted_114536 | 1784 | No | NA | 2 | 0 | 1 | Not-determined | Genome-fragment | NA | NA | 0 |

| | | | | | | | | | | | |
|---|---|---|---|---|---|---|---|---|---|---|---|
| S10_extracted_114569 | 1004 | No | NA | 2 | 0 | 2 | Not-determined | Genome-fragment | NA | NA | 0 |
| S9_extracted_k141_53738 | 3309 | No | NA | 9 | 1 | 0 | Low-quality | Genome-fragment | 7.69 | AAI-based (high-confidence) | 0 |
| S10_extracted_114600 | 3811 | No | NA | 7 | 0 | 4 | Not-determined | Genome-fragment | NA | NA | 0 |
| S10_extracted_114656 | 11344 | No | NA | 5 | 0 | 2 | Not-determined | Genome-fragment | NA | NA | 0 |
| S9_extracted_k141_89369 | 3309 | No | NA | 4 | 0 | 0 | Low-quality | Genome-fragment | 1.97 | AAI-based (high-confidence) | 0 |
| S10_extracted_115003 | 1328 | No | NA | 4 | 0 | 0 | Not-determined | Genome-fragment | NA | NA | 0 |
| S10_extracted_115231 | 3542 | No | NA | 3 | 0 | 1 | Not-determined | Genome-fragment | NA | NA | 0 |
| S10_extracted_115387 | 1122 | No | NA | 1 | 0 | 1 | Not-determined | Genome-fragment | NA | NA | 0 |
| S13_vs_45161\|\|0_partial | 3309 | Yes | 1948 | 4 | 1 | 1 | Low-quality | Genome-fragment | 5.31 | AAI-based (medium-confidence) | 41.13 |
| S10_extracted_115481 | 4035 | No | NA | 3 | 0 | 3 | Not-determined | Genome-fragment | NA | NA | 0 |
| S10_extracted_115517 | 1153 | No | NA | 1 | 0 | 0 | Not-determined | Genome-fragment | NA | NA | 0 |
| S10_extracted_115537 | 2716 | No | NA | 4 | 0 | 0 | Not-determined | Genome-fragment | NA | NA | 0 |
| S5_extracted_k141_17917 | 3309 | No | NA | 4 | 0 | 3 | Low-quality | Genome-fragment | 2.11 | AAI-based (medium-confidence) | 0 |
| S10_extracted_115645 | 6900 | No | NA | 4 | 0 | 2 | Not-determined | Genome-fragment | NA | NA | 0 |
| S10_extracted_115674 | 3333 | No | NA | 5 | 0 | 2 | Not-determined | Genome-fragment | NA | NA | 0 |
| S10_extracted_115689 | 1105 | No | NA | 1 | 0 | 1 | Not-determined | Genome-fragment | NA | NA | 0 |
| S10_extracted_115772 | 7141 | No | NA | 7 | 0 | 7 | Not-determined | Genome-fragment | NA | NA | 0 |
| S10_extracted_115855 | 1042 | No | NA | 1 | 0 | 0 | Not-determined | Genome-fragment | NA | NA | 0 |
| S10_extracted_116023 | 4285 | No | NA | 1 | 0 | 1 | Not-determined | Genome-fragment | NA | NA | 0 |
| S10_extracted_116092 | 1225 | No | NA | 3 | 0 | 1 | Not-determined | Genome-fragment | NA | NA | 0 |
| S17_vs_k141_29323\|\|full | 3310 | No | NA | 3 | 0 | 0 | Low-quality | Genome-fragment | 1.42 | AAI-based (medium-confidence) | 0 |
| S10_extracted_116405 | 1230 | No | NA | 2 | 0 | 1 | Not-determined | Genome-fragment | NA | NA | 0 |
| S10_extracted_116448 | 3652 | No | NA | 8 | 0 | 7 | Not-determined | Genome-fragment | NA | NA | 0 |
| S10_extracted_116516 | 3045 | No | NA | 2 | 0 | 0 | Not-determined | Genome-fragment | NA | NA | 0 |
| S10_extracted_116551 | 1804 | No | NA | 3 | 0 | 0 | Not-determined | Genome-fragment | NA | NA | 0 |
| S10_extracted_116709 | 1146 | No | NA | 2 | 0 | 2 | Not-determined | Genome-fragment | NA | NA | 0 |
| S3_vs_k141_17207\|\|full | 3310 | No | NA | 6 | 1 | 0 | Low-quality | Genome-fragment | 1.74 | HMM-based (lower-bound) | 0 |
| S10_extracted_116734 | 1361 | No | NA | 1 | 0 | 1 | Not-determined | Genome-fragment | NA | NA | 0 |
| S10_extracted_116743 | 1151 | No | NA | 2 | 0 | 1 | Not-determined | Genome-fragment | NA | NA | 0 |
| S10_extracted_116762 | 1763 | No | NA | 2 | 0 | 1 | Not-determined | Genome-fragment | NA | NA | 0 |
| S10_extracted_86085 | 3310 | No | NA | 9 | 1 | 0 | Low-quality | Genome-fragment | 4.47 | AAI-based (medium-confidence) | 0 |
| S10_extracted_116962 | 1914 | No | NA | 1 | 0 | 0 | Not-determined | Genome-fragment | NA | NA | 0 |
| S17_vs_k141_71094\|\|full | 3311 | No | NA | 4 | 0 | 0 | Low-quality | Genome-fragment | 1.93 | AAI-based (medium-confidence) | 0 |
| S9_vs_k141_77567\|\|full | 3311 | No | NA | 4 | 3 | 0 | Low-quality | Genome-fragment | 2.16 | HMM-based (lower-bound) | 0 |
| S11_extracted_k141_67252 | 3311 | No | NA | 11 | 1 | 0 | Low-quality | Genome-fragment | 5.79 | AAI-based (high-confidence) | 0 |
| S10_extracted_117807 | 1715 | No | NA | 1 | 0 | 0 | Not-determined | Genome-fragment | NA | NA | 0 |
| S10_extracted_117825 | 2984 | No | NA | 2 | 0 | 2 | Not-determined | Genome-fragment | NA | NA | 0 |
| S10_extracted_117831 | 1537 | No | NA | 3 | 0 | 2 | Not-determined | Genome-fragment | NA | NA | 0 |
| S10_extracted_117904 | 1060 | No | NA | 1 | 0 | 0 | Not-determined | Genome-fragment | NA | NA | 0 |
| S10_extracted_118195 | 2180 | No | NA | 2 | 0 | 2 | Not-determined | Genome-fragment | NA | NA | 0 |
| S10_extracted_118215 | 1040 | No | NA | 1 | 0 | 1 | Not-determined | Genome-fragment | NA | NA | 0 |
| S13_vs_45045\|\|full | 3311 | No | NA | 4 | 0 | 0 | Low-quality | Genome-fragment | 3.55 | AAI-based (high-confidence) | 0 |
| S10_extracted_118363 | 1305 | No | NA | 2 | 0 | 2 | Not-determined | Genome-fragment | NA | NA | 0 |
| S10_extracted_118366 | 1198 | No | NA | 1 | 0 | 1 | Not-determined | Genome-fragment | NA | NA | 0 |
| S10_extracted_118419 | 1032 | No | NA | 1 | 0 | 0 | Not-determined | Genome-fragment | NA | NA | 0 |
| S16_vs_15963\|\|full | 3311 | No | NA | 5 | 3 | 0 | Low-quality | Genome-fragment | 9.21 | AAI-based (high-confidence) | 0 |
| S10_extracted_118779 | 1430 | No | NA | 2 | 0 | 1 | Not-determined | Genome-fragment | NA | NA | 0 |
| S12_vs_k141_73808\|\|full | 3312 | No | NA | 6 | 0 | 0 | Low-quality | Genome-fragment | 6.4 | AAI-based (medium-confidence) | 0 |
| S10_extracted_119164 | 2125 | No | NA | 2 | 0 | 2 | Not-determined | Genome-fragment | NA | NA | 0 |
| S4_extracted_10510 | 3312 | No | NA | 4 | 3 | 0 | Low-quality | Genome-fragment | 11.68 | HMM-based (lower-bound) | 0 |
| S10_extracted_119394 | 1105 | No | NA | 1 | 0 | 1 | Not-determined | Genome-fragment | NA | NA | 0 |
| S10_extracted_119507 | 1144 | No | NA | 1 | 0 | 0 | Not-determined | Genome-fragment | NA | NA | 0 |
| S10_extracted_119529 | 4243 | No | NA | 3 | 0 | 3 | Not-determined | Genome-fragment | NA | NA | 0 |
| S10_extracted_119585 | 1341 | No | NA | 1 | 0 | 1 | Not-determined | Genome-fragment | NA | NA | 0 |
| S10_extracted_119713 | 3456 | No | NA | 6 | 0 | 0 | Not-determined | Genome-fragment | NA | NA | 0 |
| S10_extracted_119842 | 1746 | No | NA | 1 | 0 | 1 | Not-determined | Genome-fragment | NA | NA | 0 |
| S10_extracted_119857 | 1061 | No | NA | 2 | 0 | 0 | Not-determined | Genome-fragment | NA | NA | 0 |
| S10_extracted_119924 | 1347 | No | NA | 2 | 0 | 1 | Not-determined | Genome-fragment | NA | NA | 0 |
| S10_extracted_119943 | 1424 | No | NA | 1 | 0 | 1 | Not-determined | Genome-fragment | NA | NA | 0 |
| S10_extracted_120055 | 1515 | No | NA | 2 | 0 | 1 | Not-determined | Genome-fragment | NA | NA | 0 |
| S10_extracted_120362 | 1097 | No | NA | 3 | 0 | 2 | Not-determined | Genome-fragment | NA | NA | 0 |
| S10_extracted_120454 | 2290 | No | NA | 4 | 0 | 0 | Not-determined | Genome-fragment | NA | NA | 0 |
| S10_extracted_120457 | 1028 | No | NA | 2 | 0 | 2 | Not-determined | Genome-fragment | NA | NA | 0 |
| S10_extracted_120558 | 1090 | No | NA | 2 | 0 | 1 | Not-determined | Genome-fragment | NA | NA | 0 |
| S3_extracted_k141_66876 | 3313 | No | NA | 8 | 2 | 0 | Low-quality | Genome-fragment | 22.99 | AAI-based (high-confidence) | 0 |
| S10_extracted_120620 | 3103 | No | NA | 3 | 0 | 0 | Not-determined | Genome-fragment | NA | NA | 0 |
| S10_extracted_120683 | 2041 | No | NA | 1 | 0 | 0 | Not-determined | Genome-fragment | NA | NA | 0 |
| S10_extracted_120738 | 138855 | No | NA | 182 | 37 | 6 | Complete | High-quality | 100 | DTR (high-confidence) | 0 |
| S10_extracted_120762 | 3616 | No | NA | 5 | 0 | 1 | Not-determined | Genome-fragment | NA | NA | 0 |
| S10_extracted_120913 | 1025 | No | NA | 2 | 0 | 2 | Not-determined | Genome-fragment | NA | NA | 0 |
| S10_extracted_120918 | 1020 | No | NA | 2 | 0 | 0 | Not-determined | Genome-fragment | NA | NA | 0 |
| S10_extracted_120958 | 1088 | No | NA | 2 | 0 | 1 | Not-determined | Genome-fragment | NA | NA | 0 |
| S10_extracted_120981 | 1070 | No | NA | 1 | 0 | 1 | Not-determined | Genome-fragment | NA | NA | 0 |
| S5_extracted_k141_135643 | 3313 | No | NA | 3 | 0 | 3 | Low-quality | Genome-fragment | 0.94 | AAI-based (medium-confidence) | 0 |
| S10_extracted_121015 | 3071 | No | NA | 2 | 0 | 1 | Not-determined | Genome-fragment | NA | NA | 0 |
| S10_extracted_121047 | 1137 | No | NA | 2 | 0 | 1 | Not-determined | Genome-fragment | NA | NA | 0 |
| S5_vs_k141_27838\|\|full | 3313 | No | NA | 8 | 4 | 0 | Low-quality | Genome-fragment | 7.57 | AAI-based (high-confidence) | 0 |
| S1_extracted_k141_118736 | 3314 | No | NA | 6 | 0 | 0 | Low-quality | Genome-fragment | 5.26 | AAI-based (medium-confidence) | 0 |
| S10_vs_23835\|\|full | 4326 | No | NA | 6 | 0 | 0 | Not-determined | Genome-fragment | NA | NA | 0 |
| S10_vs_43473\|\|full | 5985 | No | NA | 8 | 4 | 0 | High-quality | High-quality | 98.05 | AAI-based (high-confidence) | 0 |
| S10_vs_120489\|\|full | 34012 | No | NA | 51 | 8 | 1 | High-quality | High-quality | 93.7 | AAI-based (high-confidence) | 0 |
| S10_vs_120541\|\|full | 39544 | No | NA | 73 | 21 | 1 | High-quality | High-quality | 99.08 | AAI-based (high-confidence) | 0 |
| S10_vs_120570\|\|full | 35054 | No | NA | 49 | 20 | 0 | High-quality | High-quality | 99.04 | AAI-based (high-confidence) | 0 |
| S16_vs_46344\|\|full | 3315 | No | NA | 6 | 1 | 0 | Low-quality | Genome-fragment | 7.69 | AAI-based (medium-confidence) | 0 |
| S6_vs_k141_52005\|\|full | 3315 | No | NA | 4 | 0 | 0 | Low-quality | Genome-fragment | 5.53 | AAI-based (medium-confidence) | 0 |
| S8_extracted_k141_92929 | 3315 | No | NA | 3 | 1 | 0 | Low-quality | Genome-fragment | 5.27 | AAI-based (high-confidence) | 0 |
| S10_vs_55\|\|full | 1772 | No | NA | 2 | 0 | 0 | Not-determined | Genome-fragment | NA | NA | 0 |
| S10_vs_206\|\|full | 2676 | No | NA | 4 | 0 | 3 | Not-determined | Genome-fragment | NA | NA | 0 |
| S10_vs_231\|\|full | 2739 | No | NA | 7 | 0 | 0 | Not-determined | Genome-fragment | NA | NA | 0 |
| S10_vs_358\|\|full | 6180 | No | NA | 5 | 0 | 4 | Not-determined | Genome-fragment | NA | NA | 0 |
| S10_vs_379\|\|full | 1724 | No | NA | 3 | 0 | 2 | Not-determined | Genome-fragment | NA | NA | 0 |
| S17_vs_k141_51851\|\|full | 3316 | No | NA | 3 | 0 | 3 | Low-quality | Genome-fragment | 0.91 | AAI-based (medium-confidence) | 0 |
| S16_extracted_1844 | 3316 | No | NA | 3 | 0 | 0 | Low-quality | Genome-fragment | 2.12 | AAI-based (high-confidence) | 0 |
| S10_vs_18423\|\|full | 3317 | No | NA | 4 | 1 | 0 | Low-quality | Genome-fragment | 6.64 | HMM-based (lower-bound) | 0 |
| S10_vs_967\|\|full | 14615 | No | NA | 11 | 0 | 8 | Not-determined | Genome-fragment | NA | NA | 0 |
| S3_extracted_k141_97244 | 3318 | No | NA | 2 | 0 | 0 | Low-quality | Genome-fragment | 11.5 | AAI-based (high-confidence) | 0 |
| S5_extracted_k141_53363 | 3318 | No | NA | 5 | 3 | 0 | Low-quality | Genome-fragment | 6.91 | AAI-based (high-confidence) | 0 |
| S7_vs_134053\|\|full | 3318 | No | NA | 4 | 0 | 3 | Low-quality | Genome-fragment | 1.6 | AAI-based (medium-confidence) | 0 |
| S8_extracted_k141_201967 | 3318 | No | NA | 10 | 0 | 0 | Low-quality | Genome-fragment | 2.06 | AAI-based (high-confidence) | 0 |
| S13_extracted_15296 | 3319 | No | NA | 5 | 1 | 0 | Low-quality | Genome-fragment | 4.84 | HMM-based (lower-bound) | 0 |
| S10_vs_1441\|\|full | 3194 | No | NA | 4 | 0 | 3 | Not-determined | Genome-fragment | NA | NA | 0 |
| S10_vs_1468\|\|full | 1645 | No | NA | 4 | 0 | 0 | Not-determined | Genome-fragment | NA | NA | 0 |
| S10_vs_1476\|\|full | 2297 | No | NA | 3 | 0 | 2 | Not-determined | Genome-fragment | NA | NA | 0 |
| S10_vs_1493\|\|full | 2112 | No | NA | 4 | 0 | 2 | Not-determined | Genome-fragment | NA | NA | 0 |
| S10_vs_1496\|\|full | 2956 | No | NA | 5 | 0 | 2 | Not-determined | Genome-fragment | NA | NA | 0 |
| S10_vs_1583\|\|full | 3591 | No | NA | 6 | 0 | 1 | Not-determined | Genome-fragment | NA | NA | 0 |
| S5_extracted_k141_20987 | 3319 | No | NA | 4 | 0 | 0 | Low-quality | Genome-fragment | 2.34 | AAI-based (high-confidence) | 0 |
| S7_vs_121520\|\|full | 3319 | No | NA | 3 | 1 | 0 | Low-quality | Genome-fragment | 5.68 | AAI-based (high-confidence) | 0 |
| S10_extracted_21865 | 3319 | No | NA | 3 | 0 | 2 | Low-quality | Genome-fragment | 4 | AAI-based (medium-confidence) | 0 |
| S5_extracted_k141_63722 | 3320 | No | NA | 7 | 2 | 0 | Low-quality | Genome-fragment | 7.4 | AAI-based (high-confidence) | 0 |
| S10_vs_1963\|\|full | 2426 | No | NA | 3 | 0 | 3 | Not-determined | Genome-fragment | NA | NA | 0 |
| S16_extracted_152346 | 3320 | No | NA | 4 | 0 | 2 | Low-quality | Genome-fragment | 1.03 | AAI-based (high-confidence) | 0 |
| S10_vs_2071\|\|full | 2381 | No | NA | 4 | 0 | 1 | Not-determined | Genome-fragment | NA | NA | 0 |
| S7_vs_113813\|\|full | 3321 | No | NA | 3 | 2 | 0 | Low-quality | Genome-fragment | 8.17 | AAI-based (medium-confidence) | 0 |
| S10_vs_2115\|\|full | 6220 | No | NA | 5 | 0 | 5 | Not-determined | Genome-fragment | NA | NA | 0 |
| S16_extracted_95246 | 3321 | No | NA | 8 | 0 | 1 | Low-quality | Genome-fragment | 7.78 | AAI-based (high-confidence) | 0 |
| S8_vs_k141_187927\|\|full | 3322 | Yes | 1324 | 8 | 1 | 2 | Low-quality | Genome-fragment | 2.65 | HMM-based (lower-bound) | 60.14 |
| S10_vs_2434\|\|full | 9043 | No | NA | 9 | 0 | 1 | Not-determined | Genome-fragment | NA | NA | 0 |
| S16_extracted_88367 | 3323 | No | NA | 4 | 3 | 0 | Low-quality | Genome-fragment | 10.33 | AAI-based (high-confidence) | 0 |
| S10_vs_2499\|\|full | 7612 | No | NA | 7 | 0 | 0 | Not-determined | Genome-fragment | NA | NA | 0 |
| S10_vs_2538\|\|full | 32212 | No | NA | 64 | 19 | 1 | High-quality | High-quality | 93.27 | AAI-based (high-confidence) | 0 |
| S6_vs_k141_18483\|\|full | 3323 | No | NA | 6 | 2 | 0 | Low-quality | Genome-fragment | 6.95 | AAI-based (medium-confidence) | 0 |
| S11_vs_k141_22406\|\|full | 3324 | Yes | 1607 | 5 | 1 | 1 | Low-quality | Genome-fragment | 1.81 | AAI-based (medium-confidence) | 51.65 |
| S11_vs_k141_56732\|\|full | 3324 | Yes | 2549 | 4 | 2 | 1 | Low-quality | Genome-fragment | 1.58 | AAI-based (medium-confidence) | 23.32 |
| S7_vs_89621\|\|full | 3324 | No | NA | 5 | 0 | 0 | Low-quality | Genome-fragment | 6.15 | AAI-based (medium-confidence) | 0 |
| S10_vs_2961\|\|full | 2513 | No | NA | 3 | 0 | 2 | Not-determined | Genome-fragment | NA | NA | 0 |
| S11_vs_k141_6809\|\|full | 3325 | No | NA | 4 | 0 | 0 | Low-quality | Genome-fragment | 7.71 | AAI-based (high-confidence) | 0 |
| S1_vs_k141_25971\|\|full | 3325 | No | NA | 7 | 0 | 0 | Low-quality | Genome-fragment | 9.95 | AAI-based (high-confidence) | 0 |
| S14_vs_k141_104887\|\|full | 3326 | No | NA | 9 | 2 | 0 | Low-quality | Genome-fragment | 5.35 | AAI-based (high-confidence) | 0 |
| S10_vs_3447\|\|full | 2154 | No | NA | 3 | 0 | 3 | Not-determined | Genome-fragment | NA | NA | 0 |
| S10_vs_3456\|\|full | 2887 | No | NA | 3 | 0 | 0 | Not-determined | Genome-fragment | NA | NA | 0 |
| S10_vs_3473\|\|full | 3172 | No | NA | 4 | 0 | 1 | Not-determined | Genome-fragment | NA | NA | 0 |
| S16_extracted_33751 | 3326 | No | NA | 11 | 2 | 0 | Low-quality | Genome-fragment | 9.59 | AAI-based (high-confidence) | 0 |
| S10_vs_3629\|\|full | 2647 | No | NA | 3 | 0 | 3 | Not-determined | Genome-fragment | NA | NA | 0 |
| S10_vs_3666\|\|full | 6555 | No | NA | 8 | 0 | 3 | Not-determined | Genome-fragment | NA | NA | 0 |
| S10_vs_3691\|\|full | 13880 | No | NA | 18 | 0 | 2 | Not-determined | Genome-fragment | NA | NA | 0 |
| S10_vs_3702\|\|full | 2076 | No | NA | 4 | 0 | 0 | Not-determined | Genome-fragment | NA | NA | 0 |
| S16_extracted_142010 | 3326 | No | NA | 3 | 0 | 2 | Low-quality | Genome-fragment | 1.53 | AAI-based (high-confidence) | 0 |
| S10_vs_3751\|\|full | 1211 | No | NA | 2 | 0 | 1 | Not-determined | Genome-fragment | NA | NA | 0 |
| S11_extracted_k141_4077 | 3327 | No | NA | 8 | 7 | 0 | Low-quality | Genome-fragment | 5.47 | AAI-based (high-confidence) | 0 |
| S7_extracted_121167 | 3327 | No | NA | 7 | 3 | 0 | Low-quality | Genome-fragment | 5.65 | AAI-based (high-confidence) | 0 |
| S17_extracted_k141_102361 | 3328 | No | NA | 4 | 0 | 4 | Low-quality | Genome-fragment | 8.49 | AAI-based (medium-confidence) | 0 |
| S11_vs_k141_1176\|\|full | 3328 | No | NA | 3 | 0 | 2 | Low-quality | Genome-fragment | 4.44 | AAI-based (medium-confidence) | 0 |
| S1_vs_k141_106959\|\|lt2gene | 3328 | No | NA | 1 | 1 | 0 | Low-quality | Genome-fragment | 6.23 | AAI-based (high-confidence) | 0 |
| S5_vs_k141_122368\|\|full | 3328 | No | NA | 9 | 1 | 0 | Low-quality | Genome-fragment | 5.72 | AAI-based (medium-confidence) | 0 |
| S10_vs_4114\|\|full | 14764 | No | NA | 19 | 0 | 3 | Not-determined | Genome-fragment | NA | NA | 0 |
| S10_vs_4123\|\|full | 8886 | No | NA | 9 | 0 | 2 | Not-determined | Genome-fragment | NA | NA | 0 |
| S12_extracted_k141_150001 | 3328 | No | NA | 3 | 1 | 0 | Low-quality | Genome-fragment | 5.36 | AAI-based (high-confidence) | 0 |
| S1_extracted_k141_63765 | 3329 | No | NA | 5 | 2 | 0 | Low-quality | Genome-fragment | 7.28 | AAI-based (high-confidence) | 0 |
| S17_extracted_k141_77861 | 3330 | No | NA | 3 | 0 | 0 | Low-quality | Genome-fragment | 7.14 | AAI-based (high-confidence) | 0 |
| S10_vs_4624\|\|full | 8629 | No | NA | 7 | 0 | 5 | Not-determined | Genome-fragment | NA | NA | 0 |
| S10_vs_4627\|\|full | 2673 | No | NA | 9 | 0 | 0 | Not-determined | Genome-fragment | NA | NA | 0 |
| S6_vs_k141_17723\|\|full | 3330 | No | NA | 3 | 0 | 1 | Low-quality | Genome-fragment | 1.1 | AAI-based (medium-confidence) | 0 |
| S9_extracted_k141_78000 | 3331 | No | NA | 2 | 0 | 1 | Low-quality | Genome-fragment | 1.38 | AAI-based (medium-confidence) | 0 |
| S13_vs_45953\|\|full | 3331 | No | NA | 5 | 0 | 1 | Low-quality | Genome-fragment | 4.3 | AAI-based (medium-confidence) | 0 |
| S10_vs_4790\|\|full | 1415 | No | NA | 3 | 0 | 1 | Not-determined | Genome-fragment | NA | NA | 0 |
| S10_vs_4812\|\|full | 1028 | No | NA | 2 | 0 | 1 | Not-determined | Genome-fragment | NA | NA | 0 |
| S10_vs_5173\|\|full | 1586 | No | NA | 2 | 0 | 2 | Not-determined | Genome-fragment | NA | NA | 0 |
| S10_vs_5177\|\|full | 1885 | No | NA | 4 | 0 | 0 | Not-determined | Genome-fragment | NA | NA | 0 |
| S1_extracted_k141_19352 | 3331 | No | NA | 5 | 0 | 0 | Low-quality | Genome-fragment | 3.37 | AAI-based (high-confidence) | 0 |
| S7_extracted_1266 | 3331 | Yes | 2212 | 8 | 2 | 1 | Low-quality | Genome-fragment | 5.98 | AAI-based (high-confidence) | 33.59 |
| S12_vs_k141_14373\|\|full | 3331 | No | NA | 4 | 0 | 3 | Low-quality | Genome-fragment | 4.52 | AAI-based (medium-confidence) | 0 |
| S17_vs_k141_33745\|\|full | 3332 | No | NA | 3 | 0 | 2 | Low-quality | Genome-fragment | 1.72 | AAI-based (high-confidence) | 0 |
| S10_vs_5608\|\|full | 1723 | No | NA | 3 | 0 | 2 | Not-determined | Genome-fragment | NA | NA | 0 |
| S10_vs_5632\|\|full | 4635 | No | NA | 7 | 0 | 0 | Not-determined | Genome-fragment | NA | NA | 0 |
| S10_vs_5825\|\|full | 1777 | No | NA | 3 | 0 | 1 | Not-determined | Genome-fragment | NA | NA | 0 |
| S10_vs_5924\|\|full | 2543 | No | NA | 4 | 0 | 1 | Not-determined | Genome-fragment | NA | NA | 0 |
| S10_vs_5982\|\|full | 3676 | No | NA | 3 | 0 | 2 | Not-determined | Genome-fragment | NA | NA | 0 |
| S3_extracted_k141_115965 | 3332 | No | NA | 9 | 0 | 0 | Low-quality | Genome-fragment | 5.83 | AAI-based (high-confidence) | 0 |
| S10_vs_6015\|\|full | 2556 | No | NA | 3 | 0 | 1 | Not-determined | Genome-fragment | NA | NA | 0 |
| S12_extracted_k141_49926 | 3332 | No | NA | 4 | 3 | 0 | Low-quality | Genome-fragment | 9.55 | HMM-based (lower-bound) | 0 |
| S10_vs_6557\|\|full | 2184 | No | NA | 3 | 0 | 2 | Not-determined | Genome-fragment | NA | NA | 0 |
| S8_vs_k141_125930\|\|full | 3332 | No | NA | 4 | 0 | 3 | Low-quality | Genome-fragment | 2.38 | AAI-based (medium-confidence) | 0 |
| S10_vs_6613\|\|full | 2106 | No | NA | 2 | 0 | 1 | Not-determined | Genome-fragment | NA | NA | 0 |
| S10_vs_6660\|\|full | 15085 | No | NA | 20 | 0 | 2 | Not-determined | Genome-fragment | NA | NA | 0 |
| S7_vs_112910\|\|full | 3333 | No | NA | 7 | 1 | 0 | Low-quality | Genome-fragment | 8.29 | AAI-based (high-confidence) | 0 |
| S10_vs_6867\|\|full | 2110 | No | NA | 4 | 0 | 0 | Not-determined | Genome-fragment | NA | NA | 0 |
| S10_vs_7020\|\|full | 1715 | No | NA | 2 | 0 | 2 | Not-determined | Genome-fragment | NA | NA | 0 |
| S16_extracted_121258 | 3333 | No | NA | 4 | 0 | 2 | Low-quality | Genome-fragment | 1.57 | AAI-based (medium-confidence) | 0 |
| S10_vs_7050\|\|full | 1625 | No | NA | 3 | 0 | 0 | Not-determined | Genome-fragment | NA | NA | 0 |
| S10_vs_7218\|\|full | 2286 | No | NA | 2 | 0 | 1 | Not-determined | Genome-fragment | NA | NA | 0 |
| S16_vs_56871\|\|full | 3334 | No | NA | 7 | 1 | 0 | Low-quality | Genome-fragment | 5.16 | AAI-based (high-confidence) | 0 |
| S10_vs_7326\|\|full | 3633 | No | NA | 4 | 0 | 0 | Not-determined | Genome-fragment | NA | NA | 0 |
| S10_vs_7367\|\|full | 1579 | No | NA | 3 | 0 | 0 | Not-determined | Genome-fragment | NA | NA | 0 |
| S10_vs_7393\|\|full | 2554 | No | NA | 4 | 0 | 1 | Not-determined | Genome-fragment | NA | NA | 0 |
| S8_extracted_k141_156322 | 3334 | No | NA | 3 | 1 | 0 | Low-quality | Genome-fragment | 7.65 | AAI-based (high-confidence) | 0 |
| S17_vs_k141_64629\|\|full | 3335 | No | NA | 3 | 0 | 1 | Low-quality | Genome-fragment | 1.84 | AAI-based (medium-confidence) | 0 |
| S10_vs_7579\|\|full | 2829 | No | NA | 6 | 0 | 0 | Not-determined | Genome-fragment | NA | NA | 0 |
| S9_vs_k141_68666\|\|full | 3335 | No | NA | 3 | 0 | 3 | Low-quality | Genome-fragment | 1.14 | AAI-based (medium-confidence) | 0 |
| S11_extracted_k141_15108 | 3335 | No | NA | 5 | 5 | 0 | Low-quality | Genome-fragment | 7.62 | AAI-based (high-confidence) | 0 |
| S1_extracted_k141_66810 | 3335 | No | NA | 4 | 2 | 0 | Low-quality | Genome-fragment | 7.33 | AAI-based (high-confidence) | 0 |
| S5_vs_k141_2017\|\|full | 3335 | No | NA | 6 | 5 | 0 | Low-quality | Genome-fragment | 5.5 | AAI-based (medium-confidence) | 0 |
| S16_vs_57064\|\|full | 3335 | No | NA | 6 | 0 | 0 | Low-quality | Genome-fragment | 10.34 | AAI-based (high-confidence) | 0 |
| S10_vs_7850\|\|full | 4701 | No | NA | 4 | 0 | 2 | Not-determined | Genome-fragment | NA | NA | 0 |
| S10_vs_7901\|\|full | 1253 | No | NA | 2 | 0 | 0 | Not-determined | Genome-fragment | NA | NA | 0 |
| S12_vs_k141_39092\|\|full | 3335 | No | NA | 4 | 0 | 2 | Low-quality | Genome-fragment | 2.42 | AAI-based (medium-confidence) | 0 |
| S5_extracted_k141_27872 | 3336 | No | NA | 5 | 0 | 0 | Low-quality | Genome-fragment | 6.08 | AAI-based (high-confidence) | 0 |
| S10_vs_8106\|\|full | 2258 | No | NA | 4 | 0 | 1 | Not-determined | Genome-fragment | NA | NA | 0 |
| S10_vs_8199\|\|full | 1075 | No | NA | 2 | 0 | 0 | Not-determined | Genome-fragment | NA | NA | 0 |
| S10_vs_27151\|\|full | 3336 | No | NA | 6 | 0 | 0 | Low-quality | Genome-fragment | 4.18 | AAI-based (high-confidence) | 0 |
| S4_extracted_2966 | 3336 | No | NA | 8 | 1 | 0 | Low-quality | Genome-fragment | 7.37 | AAI-based (high-confidence) | 0 |
| S10_vs_8616\|\|full | 1254 | No | NA | 2 | 0 | 0 | Not-determined | Genome-fragment | NA | NA | 0 |
| S10_vs_8722\|\|full | 2419 | No | NA | 4 | 0 | 1 | Not-determined | Genome-fragment | NA | NA | 0 |
| S11_extracted_k141_21575 | 3337 | No | NA | 3 | 0 | 2 | Low-quality | Genome-fragment | 0.5 | AAI-based (medium-confidence) | 0 |
| S3_vs_k141_108657\|\|full | 3337 | No | NA | 7 | 2 | 0 | Low-quality | Genome-fragment | 2.18 | AAI-based (high-confidence) | 0 |
| S3_vs_k141_5031\|\|full | 3337 | No | NA | 5 | 2 | 0 | Low-quality | Genome-fragment | 8.05 | AAI-based (high-confidence) | 0 |
| S10_vs_8855\|\|full | 1458 | No | NA | 3 | 0 | 1 | Not-determined | Genome-fragment | NA | NA | 0 |
| S14_vs_k141_58983\|\|full | 3338 | No | NA | 6 | 1 | 0 | Low-quality | Genome-fragment | 5.59 | AAI-based (high-confidence) | 0 |
| S12_vs_k141_27490\|\|full | 3338 | No | NA | 5 | 2 | 0 | Low-quality | Genome-fragment | 7.67 | AAI-based (medium-confidence) | 0 |
| S3_vs_k141_23454\|\|full | 3339 | No | NA | 3 | 0 | 3 | Low-quality | Genome-fragment | 1.85 | AAI-based (medium-confidence) | 0 |
| S10_vs_9432\|\|full | 2458 | No | NA | 5 | 0 | 1 | Not-determined | Genome-fragment | NA | NA | 0 |
| S16_vs_25573\|\|full | 3339 | No | NA | 5 | 1 | 0 | Low-quality | Genome-fragment | 9.2 | AAI-based (high-confidence) | 0 |
| S10_vs_9728\|\|full | 3076 | No | NA | 3 | 0 | 2 | Not-determined | Genome-fragment | NA | NA | 0 |
| S12_vs_k141_83439\|\|full | 3339 | No | NA | 9 | 1 | 0 | Low-quality | Genome-fragment | 10.91 | AAI-based (high-confidence) | 0 |
| S6_extracted_k141_11532 | 3339 | No | NA | 3 | 3 | 0 | Low-quality | Genome-fragment | 3.7 | HMM-based (lower-bound) | 0 |
| S11_extracted_k141_88923 | 3340 | No | NA | 3 | 0 | 1 | Low-quality | Genome-fragment | 6.63 | AAI-based (medium-confidence) | 0 |
| S3_vs_k141_30581\|\|full | 3340 | No | NA | 4 | 2 | 0 | Low-quality | Genome-fragment | 8.1 | AAI-based (medium-confidence) | 0 |
| S16_vs_164060\|\|full | 3340 | No | NA | 10 | 4 | 0 | Low-quality | Genome-fragment | 8.26 | AAI-based (medium-confidence) | 0 |
| S10_vs_10721\|\|full | 2147 | No | NA | 3 | 0 | 0 | Not-determined | Genome-fragment | NA | NA | 0 |
| S10_vs_10778\|\|full | 1565 | No | NA | 4 | 0 | 1 | Not-determined | Genome-fragment | NA | NA | 0 |
| S10_vs_10806\|\|full | 1471 | No | NA | 3 | 0 | 2 | Not-determined | Genome-fragment | NA | NA | 0 |
| S13_extracted_95411 | 3341 | No | NA | 7 | 0 | 0 | Low-quality | Genome-fragment | 6.4 | AAI-based (high-confidence) | 0 |
| S14_vs_k141_65255\|\|full | 3341 | No | NA | 6 | 2 | 0 | Low-quality | Genome-fragment | 7.89 | AAI-based (high-confidence) | 0 |
| S8_extracted_k141_173449 | 3341 | No | NA | 4 | 0 | 1 | Low-quality | Genome-fragment | 5.05 | AAI-based (medium-confidence) | 0 |
| S10_vs_11143\|\|full | 2020 | No | NA | 4 | 0 | 3 | Not-determined | Genome-fragment | NA | NA | 0 |
| S10_vs_11289\|\|full | 1896 | No | NA | 3 | 0 | 0 | Not-determined | Genome-fragment | NA | NA | 0 |
| S14_vs_k141_16412\|\|full | 3342 | No | NA | 4 | 1 | 0 | Low-quality | Genome-fragment | 7.94 | AAI-based (high-confidence) | 0 |
| S5_vs_k141_28099\|\|full | 3342 | No | NA | 4 | 0 | 4 | Low-quality | Genome-fragment | 2.88 | AAI-based (medium-confidence) | 0 |
| S9_extracted_k141_13010 | 3343 | No | NA | 2 | 0 | 2 | Low-quality | Genome-fragment | 1.19 | AAI-based (medium-confidence) | 0 |
| S10_vs_12255\|\|full | 1648 | No | NA | 2 | 0 | 0 | Not-determined | Genome-fragment | NA | NA | 0 |
| S10_vs_12515\|\|full | 1729 | No | NA | 3 | 0 | 2 | Not-determined | Genome-fragment | NA | NA | 0 |
| S7_extracted_36226 | 3343 | No | NA | 4 | 3 | 0 | Low-quality | Genome-fragment | 8.96 | AAI-based (high-confidence) | 0 |
| S12_vs_k141_173674\|\|full | 3343 | No | NA | 3 | 2 | 0 | Low-quality | Genome-fragment | 8.94 | AAI-based (medium-confidence) | 0 |
| S10_vs_12795\|\|full | 6086 | No | NA | 16 | 0 | 0 | Not-determined | Genome-fragment | NA | NA | 0 |
| S10_vs_12830\|\|full | 2152 | No | NA | 3 | 0 | 1 | Not-determined | Genome-fragment | NA | NA | 0 |
| S10_vs_12868\|\|full | 1520 | No | NA | 2 | 0 | 0 | Not-determined | Genome-fragment | NA | NA | 0 |
| S5_vs_k141_79112\|\|full | 3344 | No | NA | 10 | 1 | 0 | Low-quality | Genome-fragment | 9.34 | AAI-based (medium-confidence) | 0 |
| S10_vs_12980\|\|full | 1079 | No | NA | 2 | 0 | 1 | Not-determined | Genome-fragment | NA | NA | 0 |
| S10_vs_13140\|\|full | 4771 | No | NA | 7 | 0 | 0 | Not-determined | Genome-fragment | NA | NA | 0 |
| S16_vs_20027\|\|full | 3345 | No | NA | 5 | 1 | 0 | Low-quality | Genome-fragment | 5.13 | HMM-based (lower-bound) | 0 |
| S10_vs_13251\|\|full | 1299 | No | NA | 3 | 0 | 0 | Not-determined | Genome-fragment | NA | NA | 0 |
| S9_vs_k141_78509\|\|full | 3346 | No | NA | 4 | 0 | 2 | Low-quality | Genome-fragment | 3.93 | AAI-based (medium-confidence) | 0 |
| S10_vs_13486\|\|full | 9103 | No | NA | 15 | 0 | 2 | Not-determined | Genome-fragment | NA | NA | 0 |
| S10_vs_13511\|\|full | 1901 | No | NA | 3 | 0 | 1 | Not-determined | Genome-fragment | NA | NA | 0 |
| S10_vs_13567\|\|full | 2113 | No | NA | 2 | 0 | 2 | Not-determined | Genome-fragment | NA | NA | 0 |
| S5_vs_k141_86624\|\|full | 3346 | No | NA | 7 | 1 | 0 | Low-quality | Genome-fragment | 4.04 | HMM-based (lower-bound) | 0 |
| S12_vs_k141_48402\|\|full | 3346 | No | NA | 4 | 0 | 0 | Low-quality | Genome-fragment | 3.07 | AAI-based (high-confidence) | 0 |
| S10_vs_13705\|\|full | 2742 | No | NA | 3 | 0 | 2 | Not-determined | Genome-fragment | NA | NA | 0 |
| S10_vs_13744\|\|full | 7347 | No | NA | 3 | 0 | 3 | Not-determined | Genome-fragment | NA | NA | 0 |
| S1_vs_k141_68222\|\|full | 3348 | No | NA | 9 | 2 | 0 | Low-quality | Genome-fragment | 7.37 | AAI-based (high-confidence) | 0 |
| S12_extracted_k141_4125 | 3348 | No | NA | 4 | 0 | 0 | Low-quality | Genome-fragment | 5.48 | AAI-based (medium-confidence) | 0 |
| S10_vs_13828\|\|full | 2123 | No | NA | 4 | 0 | 1 | Not-determined | Genome-fragment | NA | NA | 0 |
| S10_vs_14001\|\|full | 10480 | No | NA | 6 | 0 | 4 | Not-determined | Genome-fragment | NA | NA | 0 |
| S10_vs_14042\|\|full | 2883 | No | NA | 3 | 0 | 2 | Not-determined | Genome-fragment | NA | NA | 0 |
| S12_vs_k141_46092\|\|full | 3348 | No | NA | 6 | 1 | 0 | Low-quality | Genome-fragment | 4.32 | HMM-based (lower-bound) | 0 |
| S10_vs_14332\|\|full | 1187 | No | NA | 3 | 0 | 1 | Not-determined | Genome-fragment | NA | NA | 0 |
| S10_vs_14557\|\|full | 93657 | Yes | 41728 | 121 | 4 | 43 | Complete | High-quality | 100 | Provirus (medium-confidence) | 55.45 |
| S5_extracted_k141_47984 | 3349 | No | NA | 6 | 0 | 1 | Low-quality | Genome-fragment | 2.09 | AAI-based (medium-confidence) | 0 |
| S16_vs_114793\|\|full | 3349 | No | NA | 4 | 0 | 0 | Low-quality | Genome-fragment | 9.58 | AAI-based (high-confidence) | 0 |
| S10_vs_64376\|\|full | 3349 | No | NA | 6 | 0 | 0 | Low-quality | Genome-fragment | 5.29 | AAI-based (high-confidence) | 0 |
| S12_vs_k141_135641\|\|full | 3349 | No | NA | 6 | 1 | 0 | Low-quality | Genome-fragment | 9.27 | AAI-based (high-confidence) | 0 |
| S11_vs_k141_37589\|\|full | 3350 | No | NA | 7 | 0 | 1 | Low-quality | Genome-fragment | 2.42 | AAI-based (high-confidence) | 0 |
| S11_extracted_k141_58239 | 3352 | No | NA | 3 | 0 | 2 | Low-quality | Genome-fragment | 0.92 | AAI-based (high-confidence) | 0 |
| S10_vs_14949\|\|full | 1058 | No | NA | 3 | 0 | 0 | Not-determined | Genome-fragment | NA | NA | 0 |
| S10_vs_14988\|\|full | 8039 | No | NA | 7 | 0 | 3 | Not-determined | Genome-fragment | NA | NA | 0 |
| S10_vs_14991\|\|full | 1885 | No | NA | 5 | 0 | 0 | Not-determined | Genome-fragment | NA | NA | 0 |
| S8_vs_k141_199706\|\|full | 3352 | No | NA | 7 | 0 | 3 | Low-quality | Genome-fragment | 8.88 | AAI-based (medium-confidence) | 0 |
| S17_extracted_k141_59027 | 3353 | No | NA | 4 | 1 | 0 | Low-quality | Genome-fragment | 5.03 | AAI-based (high-confidence) | 0 |
| S10_vs_15248\|\|full | 2541 | No | NA | 3 | 0 | 1 | Not-determined | Genome-fragment | NA | NA | 0 |
| S16_vs_139805\|\|full | 3353 | No | NA | 4 | 0 | 2 | Low-quality | Genome-fragment | 1.56 | AAI-based (medium-confidence) | 0 |
| S10_vs_15433\|\|full | 2416 | No | NA | 3 | 0 | 1 | Not-determined | Genome-fragment | NA | NA | 0 |
| S8_extracted_k141_27760 | 3353 | No | NA | 9 | 0 | 0 | Low-quality | Genome-fragment | 2.09 | AAI-based (high-confidence) | 0 |
| S8_vs_k141_41920\|\|full | 3353 | No | NA | 9 | 0 | 0 | Low-quality | Genome-fragment | 9.59 | AAI-based (medium-confidence) | 0 |
| S9_vs_k141_23532\|\|full | 3354 | No | NA | 10 | 0 | 0 | Low-quality | Genome-fragment | 5.41 | AAI-based (high-confidence) | 0 |
| S10_vs_15639\|\|full | 1672 | No | NA | 3 | 0 | 1 | Not-determined | Genome-fragment | NA | NA | 0 |
| S10_vs_15642\|\|full | 3949 | No | NA | 4 | 0 | 2 | Not-determined | Genome-fragment | NA | NA | 0 |
| S1_vs_k141_54351\|\|full | 3354 | No | NA | 5 | 0 | 0 | Low-quality | Genome-fragment | 8.71 | AAI-based (high-confidence) | 0 |
| S10_vs_15764\|\|full | 9250 | No | NA | 12 | 0 | 5 | Not-determined | Genome-fragment | NA | NA | 0 |
| S1_vs_k141_45760\|\|full | 3354 | No | NA | 6 | 2 | 0 | Low-quality | Genome-fragment | 5.11 | HMM-based (lower-bound) | 0 |
| S10_vs_15960\|\|full | 1090 | No | NA | 2 | 0 | 1 | Not-determined | Genome-fragment | NA | NA | 0 |
| S4_extracted_30878 | 3355 | No | NA | 11 | 1 | 0 | Low-quality | Genome-fragment | 7.47 | AAI-based (high-confidence) | 0 |
| S10_vs_16182\|\|full | 1230 | No | NA | 3 | 0 | 1 | Not-determined | Genome-fragment | NA | NA | 0 |
| S10_vs_16185\|\|full | 1762 | No | NA | 2 | 0 | 2 | Not-determined | Genome-fragment | NA | NA | 0 |
| S10_vs_16225\|\|full | 2503 | No | NA | 5 | 0 | 2 | Not-determined | Genome-fragment | NA | NA | 0 |
| S10_vs_16263\|\|full | 2112 | No | NA | 3 | 0 | 0 | Not-determined | Genome-fragment | NA | NA | 0 |
| S3_extracted_k141_112657 | 3357 | No | NA | 8 | 1 | 1 | Low-quality | Genome-fragment | 9.19 | AAI-based (high-confidence) | 0 |
| S10_vs_16380\|\|full | 1286 | No | NA | 3 | 0 | 0 | Not-determined | Genome-fragment | NA | NA | 0 |
| S10_vs_16519\|\|full | 2218 | No | NA | 3 | 0 | 1 | Not-determined | Genome-fragment | NA | NA | 0 |
| S10_vs_16572\|\|full | 1642 | No | NA | 3 | 0 | 0 | Not-determined | Genome-fragment | NA | NA | 0 |
| S10_vs_16632\|\|full | 1873 | No | NA | 5 | 0 | 0 | Not-determined | Genome-fragment | NA | NA | 0 |
| S10_vs_16658\|\|full | 3619 | No | NA | 4 | 0 | 2 | Not-determined | Genome-fragment | NA | NA | 0 |
| S5_vs_k141_102935\|\|full | 3357 | No | NA | 7 | 1 | 0 | Low-quality | Genome-fragment | 4.93 | HMM-based (lower-bound) | 0 |
| S10_vs_17019\|\|full | 6403 | No | NA | 5 | 0 | 5 | Not-determined | Genome-fragment | NA | NA | 0 |
| S3_vs_k141_125954\|\|full | 3358 | No | NA | 4 | 2 | 0 | Low-quality | Genome-fragment | 7.48 | AAI-based (high-confidence) | 0 |
| S10_vs_17363\|\|full | 5051 | No | NA | 7 | 0 | 4 | Not-determined | Genome-fragment | NA | NA | 0 |
| S10_vs_17395\|\|full | 4953 | No | NA | 5 | 0 | 4 | Not-determined | Genome-fragment | NA | NA | 0 |
| S10_vs_17402\|\|full | 8288 | No | NA | 4 | 0 | 3 | Not-determined | Genome-fragment | NA | NA | 0 |
| S5_vs_k141_1335\|\|full | 3358 | No | NA | 8 | 1 | 0 | Low-quality | Genome-fragment | 2.03 | HMM-based (lower-bound) | 0 |
| S10_vs_17525\|\|full | 3162 | No | NA | 6 | 0 | 1 | Not-determined | Genome-fragment | NA | NA | 0 |
| S12_vs_k141_79632\|\|full | 3358 | No | NA | 8 | 5 | 0 | Low-quality | Genome-fragment | 9.2 | AAI-based (high-confidence) | 0 |
| S8_extracted_k141_151823 | 3358 | No | NA | 3 | 1 | 0 | Low-quality | Genome-fragment | 3.5 | HMM-based (lower-bound) | 0 |
| S10_vs_17750\|\|full | 1302 | No | NA | 3 | 0 | 1 | Not-determined | Genome-fragment | NA | NA | 0 |
| S3_extracted_k141_76372 | 3359 | No | NA | 6 | 0 | 1 | Low-quality | Genome-fragment | 6.18 | AAI-based (medium-confidence) | 0 |
| S10_vs_18131\|\|full | 1462 | No | NA | 1 | 0 | 1 | Not-determined | Genome-fragment | NA | NA | 0 |
| S10_vs_18157\|\|full | 1071 | No | NA | 2 | 0 | 1 | Not-determined | Genome-fragment | NA | NA | 0 |
| S14_vs_k141_83178\|\|full | 3360 | No | NA | 7 | 0 | 0 | Low-quality | Genome-fragment | 8.36 | AAI-based (medium-confidence) | 0 |
| S10_vs_18488\|\|full | 1004 | No | NA | 3 | 0 | 0 | Not-determined | Genome-fragment | NA | NA | 0 |
| S10_vs_18537\|\|full | 1572 | No | NA | 2 | 0 | 0 | Not-determined | Genome-fragment | NA | NA | 0 |
| S9_vs_k141_18017\|\|full | 3361 | No | NA | 5 | 0 | 0 | Low-quality | Genome-fragment | 5.7 | AAI-based (medium-confidence) | 0 |
| S10_vs_18623\|\|full | 1665 | No | NA | 3 | 0 | 0 | Not-determined | Genome-fragment | NA | NA | 0 |
| S10_vs_18864\|\|full | 4822 | No | NA | 5 | 0 | 5 | Not-determined | Genome-fragment | NA | NA | 0 |
| S10_vs_18901\|\|full | 42132 | No | NA | 59 | 11 | 0 | High-quality | High-quality | 100 | AAI-based (medium-confidence) | 0 |
| S10_vs_18931\|\|full | 1477 | No | NA | 3 | 0 | 1 | Not-determined | Genome-fragment | NA | NA | 0 |
| S10_vs_18988\|\|full | 7098 | No | NA | 8 | 0 | 5 | Not-determined | Genome-fragment | NA | NA | 0 |
| S13_vs_11739\|\|full | 3362 | No | NA | 3 | 3 | 0 | Low-quality | Genome-fragment | 5.63 | AAI-based (high-confidence) | 0 |
| S10_vs_19079\|\|full | 2133 | No | NA | 3 | 0 | 0 | Not-determined | Genome-fragment | NA | NA | 0 |
| S14_vs_k141_98473\|\|full | 3362 | No | NA | 7 | 1 | 0 | Low-quality | Genome-fragment | 5.94 | AAI-based (medium-confidence) | 0 |
| S10_vs_19339\|\|full | 1636 | No | NA | 3 | 0 | 1 | Not-determined | Genome-fragment | NA | NA | 0 |
| S10_vs_19435\|\|full | 1470 | No | NA | 3 | 0 | 2 | Not-determined | Genome-fragment | NA | NA | 0 |
| S10_vs_19448\|\|full | 1509 | No | NA | 3 | 0 | 2 | Not-determined | Genome-fragment | NA | NA | 0 |
| S9_extracted_k141_56986 | 3363 | No | NA | 8 | 1 | 0 | Low-quality | Genome-fragment | 7.53 | AAI-based (high-confidence) | 0 |
| S10_vs_19466\|\|full | 9270 | No | NA | 5 | 0 | 3 | Not-determined | Genome-fragment | NA | NA | 0 |
| S10_vs_19597\|\|full | 1540 | No | NA | 2 | 0 | 0 | Not-determined | Genome-fragment | NA | NA | 0 |
| S10_vs_19691\|\|full | 1326 | No | NA | 2 | 0 | 1 | Not-determined | Genome-fragment | NA | NA | 0 |
| S10_vs_19734\|\|full | 1407 | No | NA | 2 | 0 | 0 | Not-determined | Genome-fragment | NA | NA | 0 |
| S10_vs_19810\|\|full | 1227 | No | NA | 2 | 0 | 0 | Not-determined | Genome-fragment | NA | NA | 0 |
| S1_extracted_k141_129286 | 3364 | No | NA | 3 | 1 | 0 | Low-quality | Genome-fragment | 3.42 | AAI-based (high-confidence) | 0 |
| S10_vs_19976\|\|full | 1294 | No | NA | 2 | 0 | 2 | Not-determined | Genome-fragment | NA | NA | 0 |
| S10_vs_20000\|\|full | 6219 | No | NA | 8 | 0 | 1 | Not-determined | Genome-fragment | NA | NA | 0 |
| S10_vs_20057\|\|full | 6568 | No | NA | 5 | 0 | 4 | Not-determined | Genome-fragment | NA | NA | 0 |
| S5_vs_k141_51866\|\|full | 3364 | No | NA | 9 | 2 | 0 | Low-quality | Genome-fragment | 2.91 | AAI-based (medium-confidence) | 0 |
| S10_vs_20140\|\|full | 2088 | No | NA | 3 | 0 | 0 | Not-determined | Genome-fragment | NA | NA | 0 |
| S7_extracted_6670 | 3364 | No | NA | 7 | 0 | 0 | Low-quality | Genome-fragment | 6.65 | AAI-based (high-confidence) | 0 |
| S16_vs_75497\|\|full | 3364 | Yes | 1645 | 6 | 1 | 1 | Low-quality | Genome-fragment | 1.39 | AAI-based (high-confidence) | 51.1 |
| S10_vs_20338\|\|full | 6564 | No | NA | 11 | 0 | 1 | Not-determined | Genome-fragment | NA | NA | 0 |
| S8_extracted_k141_48959 | 3364 | Yes | 1545 | 5 | 2 | 2 | Low-quality | Genome-fragment | 3.13 | AAI-based (medium-confidence) | 54.07 |
| S10_vs_20400\|\|full | 20725 | No | NA | 33 | 11 | 0 | Medium-quality | Genome-fragment | 51.99 | AAI-based (high-confidence) | 0 |
| S3_extracted_k141_5508 | 3365 | No | NA | 3 | 0 | 3 | Low-quality | Genome-fragment | 1.81 | AAI-based (medium-confidence) | 0 |
| S7_extracted_112431 | 3366 | No | NA | 6 | 0 | 1 | Low-quality | Genome-fragment | 2.5 | AAI-based (medium-confidence) | 0 |
| S17_extracted_k141_59946 | 3367 | No | NA | 6 | 1 | 0 | Low-quality | Genome-fragment | 7.16 | AAI-based (high-confidence) | 0 |
| S14_extracted_k141_84151 | 3367 | No | NA | 6 | 3 | 0 | Low-quality | Genome-fragment | 7.15 | AAI-based (high-confidence) | 0 |
| S10_vs_20668\|\|full | 1246 | No | NA | 2 | 0 | 2 | Not-determined | Genome-fragment | NA | NA | 0 |
| S10_vs_20768\|\|full | 3109 | No | NA | 5 | 0 | 1 | Not-determined | Genome-fragment | NA | NA | 0 |
| S10_vs_20802\|\|full | 2859 | No | NA | 5 | 0 | 0 | Not-determined | Genome-fragment | NA | NA | 0 |
| S10_vs_20930\|\|full | 2825 | No | NA | 6 | 0 | 1 | Not-determined | Genome-fragment | NA | NA | 0 |
| S10_vs_20993\|\|full | 1769 | No | NA | 3 | 0 | 1 | Not-determined | Genome-fragment | NA | NA | 0 |
| S11_extracted_k141_53861 | 3368 | No | NA | 3 | 0 | 2 | Low-quality | Genome-fragment | 2.4 | AAI-based (medium-confidence) | 0 |
| S16_vs_63809\|\|full | 3368 | No | NA | 3 | 0 | 2 | Low-quality | Genome-fragment | 1.1 | AAI-based (medium-confidence) | 0 |
| S6_vs_k141_24009\|\|full | 3368 | No | NA | 6 | 1 | 0 | Low-quality | Genome-fragment | 7.5 | AAI-based (high-confidence) | 0 |
| S4_extracted_12574 | 3369 | No | NA | 4 | 1 | 0 | Low-quality | Genome-fragment | 2.08 | HMM-based (lower-bound) | 0 |
| S10_vs_21370\|\|full | 1471 | No | NA | 3 | 0 | 1 | Not-determined | Genome-fragment | NA | NA | 0 |
| S17_vs_k141_64489\|\|full | 3370 | No | NA | 5 | 3 | 0 | Low-quality | Genome-fragment | 7.7 | AAI-based (high-confidence) | 0 |
| S10_vs_21613\|\|full | 2754 | No | NA | 5 | 0 | 1 | Not-determined | Genome-fragment | NA | NA | 0 |
| S10_vs_21718\|\|full | 1004 | No | NA | 2 | 0 | 1 | Not-determined | Genome-fragment | NA | NA | 0 |
| S10_vs_21946\|\|full | 3518 | No | NA | 6 | 0 | 0 | Not-determined | Genome-fragment | NA | NA | 0 |
| S5_vs_k141_15189\|\|full | 3370 | No | NA | 7 | 5 | 0 | Low-quality | Genome-fragment | 7.9 | AAI-based (high-confidence) | 0 |
| S10_vs_22502\|\|full | 3513 | No | NA | 8 | 0 | 8 | Not-determined | Genome-fragment | NA | NA | 0 |
| S10_vs_22607\|\|full | 3082 | No | NA | 4 | 0 | 2 | Not-determined | Genome-fragment | NA | NA | 0 |
| S10_vs_22676\|\|full | 3660 | No | NA | 6 | 0 | 2 | Not-determined | Genome-fragment | NA | NA | 0 |
| S10_vs_22681\|\|full | 1680 | No | NA | 3 | 0 | 0 | Not-determined | Genome-fragment | NA | NA | 0 |
| S10_vs_22728\|\|full | 4584 | No | NA | 4 | 0 | 2 | Not-determined | Genome-fragment | NA | NA | 0 |
| S4_extracted_29231 | 3370 | No | NA | 7 | 1 | 0 | Low-quality | Genome-fragment | 4.14 | AAI-based (high-confidence) | 0 |
| S10_vs_23059\|\|full | 1990 | No | NA | 4 | 0 | 0 | Not-determined | Genome-fragment | NA | NA | 0 |
| S9_vs_k141_34504\|\|full | 3371 | No | NA | 8 | 0 | 0 | Low-quality | Genome-fragment | 5.37 | AAI-based (high-confidence) | 0 |
| S10_vs_23120\|\|full | 2181 | No | NA | 3 | 0 | 0 | Not-determined | Genome-fragment | NA | NA | 0 |
| S10_vs_23207\|\|full | 6672 | No | NA | 5 | 0 | 3 | Not-determined | Genome-fragment | NA | NA | 0 |
| S10_vs_23344\|\|full | 1955 | No | NA | 5 | 0 | 0 | Not-determined | Genome-fragment | NA | NA | 0 |
| S10_vs_23360\|\|full | 2174 | No | NA | 7 | 0 | 0 | Not-determined | Genome-fragment | NA | NA | 0 |
| S10_vs_23378\|\|full | 3015 | No | NA | 3 | 0 | 1 | Not-determined | Genome-fragment | NA | NA | 0 |
| S17_extracted_k141_143656 | 3372 | No | NA | 6 | 4 | 0 | Low-quality | Genome-fragment | 5.72 | AAI-based (medium-confidence) | 0 |
| S10_vs_23423\|\|full | 43891 | No | NA | 42 | 0 | 3 | Not-determined | Genome-fragment | NA | NA | 0 |
| S11_extracted_k141_103203 | 3372 | No | NA | 5 | 1 | 0 | Low-quality | Genome-fragment | 1.88 | HMM-based (lower-bound) | 0 |
| S9_extracted_k141_14333 | 3373 | No | NA | 4 | 0 | 3 | Low-quality | Genome-fragment | 1.26 | AAI-based (high-confidence) | 0 |
| S10_vs_23697\|\|full | 3206 | No | NA | 5 | 0 | 0 | Not-determined | Genome-fragment | NA | NA | 0 |
| S10_vs_23771\|\|full | 1356 | No | NA | 3 | 0 | 0 | Not-determined | Genome-fragment | NA | NA | 0 |
| S10_vs_23853\|\|full | 1802 | No | NA | 3 | 0 | 2 | Not-determined | Genome-fragment | NA | NA | 0 |
| S10_vs_51706\|\|full | 3373 | No | NA | 6 | 3 | 0 | Low-quality | Genome-fragment | 9.65 | AAI-based (high-confidence) | 0 |
| S10_vs_23900\|\|full | 4856 | No | NA | 6 | 0 | 2 | Not-determined | Genome-fragment | NA | NA | 0 |
| S10_vs_24159\|\|full | 1953 | No | NA | 4 | 0 | 0 | Not-determined | Genome-fragment | NA | NA | 0 |
| S12_extracted_k141_98680 | 3373 | No | NA | 4 | 4 | 0 | Low-quality | Genome-fragment | 5.11 | HMM-based (lower-bound) | 0 |
| S8_vs_k141_36876\|\|full | 3373 | Yes | 2008 | 5 | 2 | 1 | Low-quality | Genome-fragment | 2.31 | HMM-based (lower-bound) | 40.47 |
| S10_vs_24229\|\|full | 3750 | No | NA | 6 | 0 | 0 | Not-determined | Genome-fragment | NA | NA | 0 |
| S5_extracted_k141_54818 | 3374 | No | NA | 5 | 0 | 0 | Low-quality | Genome-fragment | 8.27 | AAI-based (high-confidence) | 0 |
| S10_vs_24298\|\|full | 4541 | No | NA | 6 | 0 | 3 | Not-determined | Genome-fragment | NA | NA | 0 |
| S2_vs_k141_97\|\|full | 3374 | No | NA | 8 | 2 | 0 | Low-quality | Genome-fragment | 8.21 | AAI-based (high-confidence) | 0 |
| S17_extracted_k141_31419 | 3375 | Yes | 2069 | 3 | 1 | 1 | Low-quality | Genome-fragment | 0.96 | AAI-based (medium-confidence) | 38.7 |
| S10_vs_24543\|\|full | 1881 | No | NA | 3 | 0 | 0 | Not-determined | Genome-fragment | NA | NA | 0 |
| S9_vs_k141_16927\|\|full | 3375 | No | NA | 8 | 0 | 0 | Low-quality | Genome-fragment | 8.35 | AAI-based (high-confidence) | 0 |
| S10_vs_24781\|\|full | 1103 | No | NA | 2 | 0 | 1 | Not-determined | Genome-fragment | NA | NA | 0 |
| S10_vs_24863\|\|full | 1001 | No | NA | 2 | 0 | 0 | Not-determined | Genome-fragment | NA | NA | 0 |
| S17_extracted_k141_93008 | 3376 | No | NA | 3 | 0 | 0 | Low-quality | Genome-fragment | 3.24 | AAI-based (medium-confidence) | 0 |
| S9_vs_k141_46031\|\|full | 3376 | No | NA | 4 | 0 | 0 | Low-quality | Genome-fragment | 4.98 | AAI-based (medium-confidence) | 0 |
| S7_vs_71356\|\|full | 3376 | No | NA | 8 | 0 | 1 | Low-quality | Genome-fragment | 9.16 | AAI-based (high-confidence) | 0 |
| S10_vs_25468\|\|full | 3913 | No | NA | 4 | 0 | 0 | Not-determined | Genome-fragment | NA | NA | 0 |
| S9_extracted_k141_36314 | 3377 | No | NA | 6 | 3 | 0 | Low-quality | Genome-fragment | 1.94 | AAI-based (high-confidence) | 0 |
| S10_vs_25580\|\|full | 1811 | No | NA | 4 | 0 | 3 | Not-determined | Genome-fragment | NA | NA | 0 |
| S1_vs_k141_86061\|\|full | 3377 | No | NA | 3 | 0 | 2 | Low-quality | Genome-fragment | 1.61 | AAI-based (high-confidence) | 0 |
| S16_vs_167278\|\|full | 3378 | No | NA | 4 | 0 | 0 | Low-quality | Genome-fragment | 10.64 | AAI-based (high-confidence) | 0 |
| S10_vs_25971\|\|full | 1360 | No | NA | 2 | 0 | 1 | Not-determined | Genome-fragment | NA | NA | 0 |
| S10_vs_25992\|\|full | 2693 | No | NA | 2 | 0 | 1 | Not-determined | Genome-fragment | NA | NA | 0 |
| S10_vs_26093\|\|full | 1821 | No | NA | 2 | 0 | 2 | Not-determined | Genome-fragment | NA | NA | 0 |
| S10_vs_26251\|\|full | 3169 | No | NA | 3 | 0 | 2 | Not-determined | Genome-fragment | NA | NA | 0 |
| S10_vs_26343\|\|full | 5221 | No | NA | 7 | 0 | 4 | Not-determined | Genome-fragment | NA | NA | 0 |
| S10_vs_26365\|\|full | 58279 | No | NA | 82 | 32 | 1 | High-quality | High-quality | 98.08 | AAI-based (high-confidence) | 0 |
| S10_vs_26402\|\|full | 1009 | No | NA | 2 | 0 | 2 | Not-determined | Genome-fragment | NA | NA | 0 |
| S10_vs_26583\|\|full | 3303 | No | NA | 4 | 0 | 2 | Not-determined | Genome-fragment | NA | NA | 0 |
| S15_extracted_54823 | 3378 | No | NA | 4 | 1 | 0 | Low-quality | Genome-fragment | 2.08 | HMM-based (lower-bound) | 0 |
| S10_vs_26707\|\|full | 1095 | No | NA | 1 | 0 | 0 | Not-determined | Genome-fragment | NA | NA | 0 |
| S11_vs_k141_39485\|\|full | 3379 | No | NA | 6 | 2 | 0 | Low-quality | Genome-fragment | 8.5 | AAI-based (high-confidence) | 0 |
| S11_vs_k141_65072\|\|full | 3379 | No | NA | 5 | 0 | 0 | Low-quality | Genome-fragment | 8.36 | AAI-based (high-confidence) | 0 |
| S10_vs_27116\|\|full | 1360 | No | NA | 2 | 0 | 1 | Not-determined | Genome-fragment | NA | NA | 0 |
| S1_extracted_k141_151760 | 3379 | No | NA | 7 | 1 | 0 | Low-quality | Genome-fragment | 5.85 | AAI-based (high-confidence) | 0 |
| S10_vs_27223\|\|full | 3706 | No | NA | 3 | 0 | 0 | Not-determined | Genome-fragment | NA | NA | 0 |

| | | | | | | | | | | | | | |
|---|---|---|---|---|---|---|---|---|---|---|---|---|---|
| S10_vs_27586\|\|full | 4105 | No | NA | | 5 | 0 | 4 | Not-determined | Genome-fragment | NA | | NA | 0 |
| S7_extracted_97885 | 3379 | No | NA | | 6 | 1 | 0 | Low-quality | Genome-fragment | | 7.53 | AAI-based (high-confidence) | 0 |
| S16_extracted_79511 | 3379 | No | NA | | 3 | 1 | 0 | Low-quality | Genome-fragment | | 6.26 | AAI-based (medium-confidence) | 0 |
| S10_vs_27802\|\|full | 13834 | No | NA | | 6 | 0 | 4 | Not-determined | Genome-fragment | NA | | NA | 0 |
| S12_extracted_k141_177760 | 3379 | No | NA | | 4 | 0 | 1 | Low-quality | Genome-fragment | | 1.69 | AAI-based (medium-confidence) | 0 |
| S10_vs_27865\|\|full | 2878 | No | NA | | 2 | 0 | 2 | Not-determined | Genome-fragment | NA | | NA | 0 |
| S15_vs_22807\|\|full | 3379 | No | NA | | 3 | 1 | 0 | Low-quality | Genome-fragment | | 1.82 | AAI-based (high-confidence) | 0 |
| S8_extracted_k141_194507 | 3379 | No | NA | | 2 | 0 | 1 | Low-quality | Genome-fragment | | 4.07 | AAI-based (medium-confidence) | 0 |
| S8_vs_k141_166191\|\|full | 3380 | No | NA | | 4 | 1 | 2 | Low-quality | Genome-fragment | | 1.83 | HMM-based (lower-bound) | 0 |
| S10_vs_28484\|\|full | 2178 | No | NA | | 3 | 0 | 0 | Not-determined | Genome-fragment | NA | | NA | 0 |
| S10_vs_28646\|\|full | 8521 | No | NA | | 7 | 0 | 3 | Not-determined | Genome-fragment | NA | | NA | 0 |
| S10_vs_28658\|\|full | 3043 | No | NA | | 3 | 0 | 1 | Not-determined | Genome-fragment | NA | | NA | 0 |
| S17_extracted_k141_89103 | 3381 | No | NA | | 3 | 1 | 0 | Low-quality | Genome-fragment | | 4.2 | AAI-based (high-confidence) | 0 |
| S10_vs_28824\|\|full | 1474 | No | NA | | 3 | 0 | 2 | Not-determined | Genome-fragment | NA | | NA | 0 |
| S10_vs_28916\|\|full | 8385 | No | NA | | 16 | 0 | 0 | Not-determined | Genome-fragment | NA | | NA | 0 |
| S10_vs_29103\|\|full | 2789 | No | NA | | 4 | 0 | 2 | Not-determined | Genome-fragment | NA | | NA | 0 |
| S10_vs_29200\|\|full | 2183 | No | NA | | 3 | 0 | 1 | Not-determined | Genome-fragment | NA | | NA | 0 |
| S10_vs_29280\|\|full | 2604 | No | NA | | 3 | 0 | 1 | Not-determined | Genome-fragment | NA | | NA | 0 |
| S10_vs_29450\|\|full | 6128 | No | NA | | 6 | 0 | 4 | Not-determined | Genome-fragment | NA | | NA | 0 |
| S12_extracted_k141_49369 | 3381 | No | NA | | 5 | 0 | 1 | Low-quality | Genome-fragment | | 2.33 | AAI-based (high-confidence) | 0 |
| S10_vs_29651\|\|full | 1610 | No | NA | | 4 | 0 | 1 | Not-determined | Genome-fragment | NA | | NA | 0 |
| S10_vs_29704\|\|full | 2837 | No | NA | | 7 | 0 | 0 | Not-determined | Genome-fragment | NA | | NA | 0 |
| S3_extracted_k141_103466 | 3383 | No | NA | | 7 | 1 | 0 | Low-quality | Genome-fragment | | 8.98 | AAI-based (high-confidence) | 0 |
| S7_extracted_71925 | 3383 | No | NA | | 4 | 0 | 0 | Low-quality | Genome-fragment | | 11.05 | AAI-based (high-confidence) | 0 |
| S7_extracted_121861 | 3383 | No | NA | | 5 | 0 | 0 | Low-quality | Genome-fragment | | 3.52 | AAI-based (high-confidence) | 0 |
| S7_vs_33819\|\|full | 3383 | No | NA | | 5 | 0 | 0 | Low-quality | Genome-fragment | | 6.47 | AAI-based (high-confidence) | 0 |
| S2_vs_k141_18943\|\|full | 3383 | No | NA | | 4 | 0 | 3 | Low-quality | Genome-fragment | | 2.02 | AAI-based (medium-confidence) | 0 |
| S10_vs_30031\|\|full | 2108 | No | NA | | 4 | 0 | 0 | Not-determined | Genome-fragment | NA | | NA | 0 |
| S10_vs_30148\|\|full | 3593 | No | NA | | 6 | 0 | 2 | Not-determined | Genome-fragment | NA | | NA | 0 |
| S5_extracted_k141_72962 | 3384 | No | NA | | 4 | 2 | 0 | Low-quality | Genome-fragment | | 1.61 | AAI-based (medium-confidence) | 0 |
| S10_vs_30355\|\|full | 2188 | No | NA | | 4 | 0 | 0 | Not-determined | Genome-fragment | NA | | NA | 0 |
| S17_vs_k141_26212\|\|full | 3385 | No | NA | | 5 | 0 | 1 | Low-quality | Genome-fragment | | 3.44 | AAI-based (medium-confidence) | 0 |
| S10_vs_30468\|\|full | 2469 | No | NA | | 6 | 0 | 1 | Not-determined | Genome-fragment | NA | | NA | 0 |
| S3_vs_k141_69256\|\|full | 3386 | Yes | | 2993 | 7 | 2 | 1 | Low-quality | Genome-fragment | | 5.26 | AAI-based (medium-confidence) | 11.61 |
| S16_extracted_48282 | 3386 | No | NA | | 1 | 1 | 0 | Low-quality | Genome-fragment | | 5.58 | AAI-based (high-confidence) | 0 |
| S10_vs_30801\|\|full | 2434 | No | NA | | 3 | 0 | 1 | Not-determined | Genome-fragment | NA | | NA | 0 |
| S10_vs_30931\|\|full | 10466 | No | NA | | 10 | 0 | 6 | Not-determined | Genome-fragment | NA | | NA | 0 |
| S16_vs_54971\|\|full | 3386 | No | NA | | 6 | 2 | 0 | Low-quality | Genome-fragment | | 7.46 | AAI-based (high-confidence) | 0 |
| S10_vs_31238\|\|full | 2555 | No | NA | | 4 | 0 | 2 | Not-determined | Genome-fragment | NA | | NA | 0 |
| S16_vs_78031\|\|full | 3386 | No | NA | | 8 | 0 | 0 | Low-quality | Genome-fragment | | 7.54 | AAI-based (high-confidence) | 0 |
| S10_vs_31731\|\|full | 1787 | No | NA | | 5 | 0 | 0 | Not-determined | Genome-fragment | NA | | NA | 0 |
| S11_vs_k141_9262\|\|full | 3388 | No | NA | | 7 | 0 | 0 | Low-quality | Genome-fragment | | 7.6 | AAI-based (medium-confidence) | 0 |
| S10_vs_31855\|\|full | 1965 | No | NA | | 2 | 0 | 1 | Not-determined | Genome-fragment | NA | | NA | 0 |
| S1_extracted_k141_67814 | 3388 | No | NA | | 3 | 3 | 0 | Low-quality | Genome-fragment | | 8.91 | AAI-based (high-confidence) | 0 |
| S10_vs_31895\|\|full | 1091 | No | NA | | 3 | 0 | 1 | Not-determined | Genome-fragment | NA | | NA | 0 |
| S10_vs_31974\|\|full | 2636 | No | NA | | 2 | 0 | 1 | Not-determined | Genome-fragment | NA | | NA | 0 |
| S10_vs_32008\|\|full | 4249 | No | NA | | 3 | 0 | 2 | Not-determined | Genome-fragment | NA | | NA | 0 |
| S10_vs_32076\|\|full | 2697 | No | NA | | 2 | 0 | 1 | Not-determined | Genome-fragment | NA | | NA | 0 |
| S12_extracted_k141_164102 | 3388 | No | NA | | 4 | 1 | 0 | Low-quality | Genome-fragment | | 8.57 | AAI-based (high-confidence) | 0 |
| S8_vs_k141_179521\|\|full | 3389 | No | NA | | 8 | 0 | 0 | Low-quality | Genome-fragment | | 2.09 | AAI-based (high-confidence) | 0 |
| S10_vs_32643\|\|full | 2077 | No | NA | | 3 | 0 | 1 | Not-determined | Genome-fragment | NA | | NA | 0 |
| S5_extracted_k141_101608 | 3390 | No | NA | | 2 | 2 | 0 | Low-quality | Genome-fragment | | 2.01 | AAI-based (medium-confidence) | 0 |
| S3_vs_k141_1471\|\|full | 3391 | No | NA | | 5 | 0 | 0 | Low-quality | Genome-fragment | | 5.02 | AAI-based (medium-confidence) | 0 |
| S12_extracted_k141_81567 | 3391 | No | NA | | 5 | 0 | 0 | Low-quality | Genome-fragment | | 5.08 | AAI-based (medium-confidence) | 0 |
| S1_vs_k141_64277\|\|full | 3392 | No | NA | | 7 | 0 | 1 | Low-quality | Genome-fragment | | 5.76 | AAI-based (high-confidence) | 0 |
| S10_vs_33009\|\|full | 1379 | No | NA | | 3 | 0 | 0 | Not-determined | Genome-fragment | NA | | NA | 0 |
| S10_vs_33122\|\|full | 1140 | No | NA | | 2 | 0 | 2 | Not-determined | Genome-fragment | NA | | NA | 0 |
| S10_vs_33270\|\|full | 2988 | No | NA | | 3 | 0 | 2 | Not-determined | Genome-fragment | NA | | NA | 0 |
| S5_vs_k141_43543\|\|full | 3392 | No | NA | | 5 | 0 | 3 | Low-quality | Genome-fragment | | 2.25 | AAI-based (medium-confidence) | 0 |
| S10_vs_96121\|\|full | 3393 | No | NA | | 6 | 0 | 0 | Low-quality | Genome-fragment | | 6.23 | AAI-based (high-confidence) | 0 |
| S10_vs_33366\|\|full | 1122 | No | NA | | 2 | 0 | 1 | Not-determined | Genome-fragment | NA | | NA | 0 |
| S10_vs_33473\|\|full | 1396 | No | NA | | 3 | 0 | 2 | Not-determined | Genome-fragment | NA | | NA | 0 |
| S10_vs_33592\|\|full | 2476 | No | NA | | 3 | 0 | 2 | Not-determined | Genome-fragment | NA | | NA | 0 |
| S10_vs_33670\|\|full | 2987 | No | NA | | 3 | 0 | 3 | Not-determined | Genome-fragment | NA | | NA | 0 |
| S9_extracted_k141_87242 | 3394 | No | NA | | 4 | 0 | 1 | Low-quality | Genome-fragment | | 7.56 | AAI-based (high-confidence) | 0 |
| S10_extracted_14700 | 3394 | No | NA | | 5 | 4 | 0 | Low-quality | Genome-fragment | | 8.96 | AAI-based (high-confidence) | 0 |
| S10_vs_33950\|\|full | 2873 | No | NA | | 4 | 0 | 1 | Not-determined | Genome-fragment | NA | | NA | 0 |
| S6_vs_k141_38573\|\|full | 3394 | No | NA | | 5 | 0 | 0 | Low-quality | Genome-fragment | | 10.31 | AAI-based (high-confidence) | 0 |
| S10_vs_34117\|\|full | 1067 | No | NA | | 2 | 0 | 2 | Not-determined | Genome-fragment | NA | | NA | 0 |
| S16_extracted_15847 | 3395 | No | NA | | 2 | 0 | 2 | Low-quality | Genome-fragment | | 0.32 | AAI-based (medium-confidence) | 0 |
| S8_extracted_k141_154199 | 3395 | No | NA | | 5 | 1 | 0 | Low-quality | Genome-fragment | | 3.46 | AAI-based (high-confidence) | 0 |
| S10_extracted_113914 | 3396 | No | NA | | 4 | 0 | 0 | Low-quality | Genome-fragment | | 5.28 | AAI-based (high-confidence) | 0 |
| S10_extracted_57308 | 3398 | No | NA | | 4 | 1 | 0 | Low-quality | Genome-fragment | | 5.31 | HMM-based (lower-bound) | 0 |
| S12_vs_k141_162114\|\|full | 3398 | No | NA | | 4 | 0 | 3 | Low-quality | Genome-fragment | | 6.39 | AAI-based (medium-confidence) | 0 |
| S10_vs_34624\|\|full | 1327 | No | NA | | 3 | 0 | 2 | Not-determined | Genome-fragment | NA | | NA | 0 |
| S10_vs_34627\|\|full | 1609 | No | NA | | 5 | 0 | 0 | Not-determined | Genome-fragment | NA | | NA | 0 |
| S8_extracted_k141_58380 | 3398 | No | NA | | 2 | 0 | 0 | Low-quality | Genome-fragment | | 3.33 | AAI-based (high-confidence) | 0 |
| S11_extracted_k141_76680 | 3400 | No | NA | | 5 | 0 | 0 | Low-quality | Genome-fragment | | 7.18 | AAI-based (medium-confidence) | 0 |
| S10_vs_34833\|\|full | 2094 | No | NA | | 4 | 0 | 0 | Not-determined | Genome-fragment | NA | | NA | 0 |
| S10_vs_34923\|\|full | 1207 | No | NA | | 3 | 0 | 0 | Not-determined | Genome-fragment | NA | | NA | 0 |
| S7_extracted_129669 | 3400 | No | NA | | 3 | 0 | 2 | Low-quality | Genome-fragment | | 9.59 | AAI-based (medium-confidence) | 0 |
| S8_vs_k141_170008\|\|full | 3400 | No | NA | | 6 | 1 | 0 | Low-quality | Genome-fragment | | 8.4 | AAI-based (high-confidence) | 0 |
| S7_vs_4269\|\|full | 3401 | No | NA | | 9 | 1 | 0 | Low-quality | Genome-fragment | | 6.82 | AAI-based (medium-confidence) | 0 |
| S12_extracted_k141_33416 | 3401 | No | NA | | 9 | 4 | 0 | Low-quality | Genome-fragment | | 9.06 | AAI-based (high-confidence) | 0 |
| S15_extracted_33484 | 3401 | No | NA | | 4 | 0 | 0 | Low-quality | Genome-fragment | | 2.39 | AAI-based (medium-confidence) | 0 |
| S10_vs_35308\|\|full | 5948 | No | NA | | 6 | 0 | 5 | Not-determined | Genome-fragment | NA | | NA | 0 |
| S17_extracted_k141_76842 | 3402 | No | NA | | 6 | 0 | 0 | Low-quality | Genome-fragment | | 4.6 | AAI-based (high-confidence) | 0 |
| S10_vs_35401\|\|full | 1046 | No | NA | | 5 | 0 | 0 | Not-determined | Genome-fragment | NA | | NA | 0 |
| S14_extracted_k141_112902 | 3402 | No | NA | | 6 | 1 | 0 | Low-quality | Genome-fragment | | 4.22 | AAI-based (medium-confidence) | 0 |
| S10_vs_35479\|\|full | 8897 | No | NA | | 9 | 0 | 7 | Not-determined | Genome-fragment | NA | | NA | 0 |
| S10_vs_35553\|\|full | 1364 | No | NA | | 2 | 0 | 2 | Not-determined | Genome-fragment | NA | | NA | 0 |
| S16_vs_145758\|\|full | 3402 | No | NA | | 3 | 0 | 3 | Low-quality | Genome-fragment | | 2.14 | AAI-based (medium-confidence) | 0 |
| S10_vs_35828\|\|full | 5867 | No | NA | | 6 | 0 | 2 | Not-determined | Genome-fragment | NA | | NA | 0 |
| S14_extracted_k141_66405 | 3403 | No | NA | | 11 | 10 | 0 | Low-quality | Genome-fragment | | 11.25 | AAI-based (high-confidence) | 0 |
| S10_vs_35854\|\|full | 51045 | No | NA | | 95 | 9 | 0 | Medium-quality | Genome-fragment | | 75.43 | AAI-based (high-confidence) | 0 |
| S14_vs_k141_743\|\|full | 3404 | No | NA | | 3 | 1 | 0 | Low-quality | Genome-fragment | | 2.1 | HMM-based (lower-bound) | 0 |
| S10_vs_35893\|\|full | 1715 | No | NA | | 3 | 0 | 2 | Not-determined | Genome-fragment | NA | | NA | 0 |
| S16_extracted_113833 | 3404 | No | NA | | 3 | 0 | 3 | Low-quality | Genome-fragment | | 0.96 | AAI-based (medium-confidence) | 0 |
| S14_extracted_k141_92270 | 3405 | Yes | | 2612 | 6 | 1 | 1 | Low-quality | Genome-fragment | | 6.72 | AAI-based (high-confidence) | 23.29 |
| S5_extracted_k141_51017 | 3405 | No | NA | | 4 | 0 | 1 | Low-quality | Genome-fragment | | 6.81 | AAI-based (medium-confidence) | 0 |
| S8_extracted_k141_72759 | 3405 | No | NA | | 8 | 3 | 0 | Low-quality | Genome-fragment | | 8.51 | AAI-based (medium-confidence) | 0 |
| S13_extracted_117058 | 3406 | No | NA | | 4 | 0 | 0 | Low-quality | Genome-fragment | | 8.91 | AAI-based (medium-confidence) | 0 |
| S10_vs_36296\|\|full | 1845 | No | NA | | 3 | 0 | 1 | Not-determined | Genome-fragment | NA | | NA | 0 |
| S8_extracted_k141_66569 | 3406 | No | NA | | 3 | 1 | 0 | Low-quality | Genome-fragment | | 12.15 | HMM-based (lower-bound) | 0 |
| S10_vs_36407\|\|full | 24518 | No | NA | | 45 | 9 | 1 | Medium-quality | Genome-fragment | | 58.94 | AAI-based (medium-confidence) | 0 |
| S10_vs_36457\|\|full | 3389 | No | NA | | 7 | 0 | 0 | Not-determined | Genome-fragment | NA | | NA | 0 |
| S10_vs_36473\|\|full | 6773 | No | NA | | 8 | 0 | 4 | Not-determined | Genome-fragment | NA | | NA | 0 |
| S10_vs_36489\|\|full | 1518 | No | NA | | 2 | 0 | 1 | Not-determined | Genome-fragment | NA | | NA | 0 |
| S10_vs_36501\|\|full | 2072 | No | NA | | 3 | 0 | 2 | Not-determined | Genome-fragment | NA | | NA | 0 |
| S10_vs_36598\|\|full | 29836 | No | NA | | 35 | 11 | 2 | Medium-quality | Genome-fragment | | 50.18 | AAI-based (high-confidence) | 0 |
| S3_vs_k141_123051\|\|full | 3408 | No | NA | | 6 | 0 | 0 | Low-quality | Genome-fragment | | 9 | AAI-based (high-confidence) | 0 |
| S16_extracted_110986 | 3408 | No | NA | | 4 | 2 | 0 | Low-quality | Genome-fragment | | 3.17 | AAI-based (high-confidence) | 0 |
| S6_extracted_k141_42013 | 3408 | No | NA | | 1 | 0 | 1 | Low-quality | Genome-fragment | | 1.46 | AAI-based (medium-confidence) | 0 |
| S8_vs_k141_126577\|\|full | 3409 | No | NA | | 3 | 0 | 1 | Low-quality | Genome-fragment | | 7.58 | AAI-based (high-confidence) | 0 |
| S10_vs_36861\|\|full | 1034 | No | NA | | 2 | 0 | 1 | Not-determined | Genome-fragment | NA | | NA | 0 |
| S14_extracted_k141_405 | 3410 | No | NA | | 8 | 4 | 0 | Low-quality | Genome-fragment | | 11.26 | AAI-based (high-confidence) | 0 |
| S10_extracted_9293 | 3410 | No | NA | | 4 | 0 | 0 | Low-quality | Genome-fragment | | 11.8 | AAI-based (medium-confidence) | 0 |
| S10_vs_37155\|\|full | 3250 | No | NA | | 5 | 0 | 4 | Not-determined | Genome-fragment | NA | | NA | 0 |
| S8_extracted_k141_204570 | 3410 | No | NA | | 4 | 1 | 0 | Low-quality | Genome-fragment | | 5.81 | AAI-based (high-confidence) | 0 |
| S7_extracted_36776 | 3411 | No | NA | | 9 | 4 | 0 | Low-quality | Genome-fragment | | 8.35 | AAI-based (high-confidence) | 0 |
| S10_vs_37379\|\|full | 109791 | No | NA | | 151 | 17 | 1 | Medium-quality | Genome-fragment | | 71.38 | AAI-based (high-confidence) | 0 |
| S10_vs_37393\|\|full | 2284 | No | NA | | 3 | 0 | 2 | Not-determined | Genome-fragment | NA | | NA | 0 |
| S4_extracted_2783 | 3411 | No | NA | | 6 | 0 | 0 | Low-quality | Genome-fragment | | 3.6 | AAI-based (high-confidence) | 0 |
| S10_vs_37410\|\|full | 1240 | No | NA | | 3 | 0 | 1 | Not-determined | Genome-fragment | NA | | NA | 0 |
| S10_vs_37537\|\|full | 1061 | No | NA | | 3 | 0 | 1 | Not-determined | Genome-fragment | NA | | NA | 0 |
| S10_vs_37694\|\|full | 27829 | No | NA | | 33 | 4 | 0 | Medium-quality | Genome-fragment | | 63.54 | AAI-based (high-confidence) | 0 |
| S9_extracted_k141_55200 | 3412 | No | NA | | 8 | 0 | 0 | Low-quality | Genome-fragment | | 1.94 | AAI-based (high-confidence) | 0 |
| S10_vs_37766\|\|full | 1045 | No | NA | | 2 | 0 | 0 | Not-determined | Genome-fragment | NA | | NA | 0 |
| S10_vs_37796\|\|full | 2378 | No | NA | | 4 | 0 | 3 | Not-determined | Genome-fragment | NA | | NA | 0 |
| S10_vs_37845\|\|full | 1624 | No | NA | | 3 | 0 | 2 | Not-determined | Genome-fragment | NA | | NA | 0 |
| S9_vs_k141_7192\|\|full | 3412 | No | NA | | 11 | 0 | 0 | Low-quality | Genome-fragment | | 7.83 | AAI-based (high-confidence) | 0 |
| S10_vs_38110\|\|full | 1066 | No | NA | | 2 | 0 | 1 | Not-determined | Genome-fragment | NA | | NA | 0 |
| S14_vs_k141_60867\|\|full | 3412 | No | NA | | 7 | 0 | 0 | Low-quality | Genome-fragment | | 8.5 | AAI-based (high-confidence) | 0 |
| S12_vs_k141_33193\|\|full | 3412 | No | NA | | 3 | 0 | 2 | Low-quality | Genome-fragment | | 1.15 | AAI-based (medium-confidence) | 0 |
| S4_extracted_6763 | 3412 | No | NA | | 9 | 9 | 0 | Low-quality | Genome-fragment | | 11.24 | AAI-based (high-confidence) | 0 |
| S10_vs_38427\|\|full | 3487 | No | NA | | 3 | 0 | 3 | Not-determined | Genome-fragment | NA | | NA | 0 |
| S14_extracted_k141_60978 | 3413 | No | NA | | 2 | 0 | 2 | Low-quality | Genome-fragment | | 1.64 | AAI-based (medium-confidence) | 0 |
| S8_vs_k141_123028\|\|full | 3413 | No | NA | | 7 | 0 | 0 | Low-quality | Genome-fragment | | 7.97 | AAI-based (high-confidence) | 0 |
| S16_extracted_104463 | 3414 | No | NA | | 4 | 0 | 3 | Low-quality | Genome-fragment | | 1.72 | AAI-based (medium-confidence) | 0 |
| S10_vs_38554\|\|full | 1031 | No | NA | | 2 | 0 | 1 | Not-determined | Genome-fragment | NA | | NA | 0 |
| S10_vs_38567\|\|full | 2411 | No | NA | | 3 | 0 | 1 | Not-determined | Genome-fragment | NA | | NA | 0 |
| S16_vs_19336\|\|full | 3414 | No | NA | | 5 | 5 | 0 | Low-quality | Genome-fragment | | 8.53 | AAI-based (medium-confidence) | 0 |
| S10_vs_38772\|\|full | 4240 | No | NA | | 4 | 0 | 2 | Not-determined | Genome-fragment | NA | | NA | 0 |
| S17_vs_k141_118790\|\|full | 3415 | No | NA | | 6 | 3 | 0 | Low-quality | Genome-fragment | | 7.33 | AAI-based (high-confidence) | 0 |
| S10_vs_38957\|\|full | 2074 | No | NA | | 3 | 0 | 1 | Not-determined | Genome-fragment | NA | | NA | 0 |
| S10_vs_39026\|\|full | 1348 | No | NA | | 2 | 0 | 1 | Not-determined | Genome-fragment | NA | | NA | 0 |
| S7_vs_29067\|\|full | 3415 | No | NA | | 4 | 1 | 0 | Low-quality | Genome-fragment | | 2.03 | AAI-based (medium-confidence) | 0 |
| S10_vs_39329\|\|full | 1279 | No | NA | | 2 | 0 | 0 | Not-determined | Genome-fragment | NA | | NA | 0 |
| S10_vs_39400\|\|full | 1475 | No | NA | | 3 | 0 | 0 | Not-determined | Genome-fragment | NA | | NA | 0 |
| S10_vs_39407\|\|full | 1916 | No | NA | | 4 | 0 | 2 | Not-determined | Genome-fragment | NA | | NA | 0 |
| S17_vs_k141_61740\|\|lt2gene | 3416 | No | NA | | 2 | 0 | 1 | Low-quality | Genome-fragment | | 1.76 | AAI-based (medium-confidence) | 0 |
| S14_extracted_k141_223 | 3416 | No | NA | | 2 | 2 | 0 | Low-quality | Genome-fragment | | 11.21 | AAI-based (high-confidence) | 0 |
| S12_vs_k141_27194\|\|full | 3416 | Yes | | 2678 | 3 | 1 | 1 | Low-quality | Genome-fragment | | 6.07 | AAI-based (high-confidence) | 21.6 |
| S10_vs_39617\|\|full | 2712 | No | NA | | 3 | 0 | 2 | Not-determined | Genome-fragment | NA | | NA | 0 |
| S10_vs_39717\|\|full | 1202 | No | NA | | 2 | 0 | 1 | Not-determined | Genome-fragment | NA | | NA | 0 |
| S10_vs_39754\|\|full | 2603 | No | NA | | 6 | 0 | 1 | Not-determined | Genome-fragment | NA | | NA | 0 |
| S10_vs_39919\|\|full | 2283 | No | NA | | 6 | 0 | 0 | Not-determined | Genome-fragment | NA | | NA | 0 |
| S8_vs_k141_17041\|\|full | 3416 | No | NA | | 6 | 1 | 0 | Low-quality | Genome-fragment | | 5.43 | AAI-based (medium-confidence) | 0 |
| S10_vs_40381\|\|full | 4480 | No | NA | | 3 | 0 | 1 | Not-determined | Genome-fragment | NA | | NA | 0 |
| S10_vs_40557\|\|full | 1108 | No | NA | | 2 | 0 | 1 | Not-determined | Genome-fragment | NA | | NA | 0 |
| S1_extracted_k141_55963 | 3417 | No | NA | | 5 | 0 | 1 | Low-quality | Genome-fragment | | 3.41 | AAI-based (high-confidence) | 0 |
| S10_vs_40676\|\|full | 1184 | No | NA | | 2 | 0 | 0 | Not-determined | Genome-fragment | NA | | NA | 0 |
| S1_extracted_k141_35761 | 3417 | No | NA | | 8 | 1 | 0 | Low-quality | Genome-fragment | | 7.99 | AAI-based (high-confidence) | 0 |
| S12_vs_k141_87624\|\|full | 3417 | No | NA | | 7 | 4 | 0 | Low-quality | Genome-fragment | | 9.83 | AAI-based (high-confidence) | 0 |
| S10_vs_40792\|\|full | 1101 | No | NA | | 3 | 0 | 0 | Not-determined | Genome-fragment | NA | | NA | 0 |
| S10_vs_40909\|\|full | 4900 | No | NA | | 8 | 0 | 0 | Not-determined | Genome-fragment | NA | | NA | 0 |
| S6_extracted_k141_19118 | 3417 | No | NA | | 4 | 0 | 4 | Low-quality | Genome-fragment | | 4.19 | AAI-based (medium-confidence) | 0 |
| S10_vs_41044\|\|full | 3255 | No | NA | | 4 | 0 | 2 | Not-determined | Genome-fragment | NA | | NA | 0 |
| S10_vs_41049\|\|full | 2905 | No | NA | | 5 | 0 | 0 | Not-determined | Genome-fragment | NA | | NA | 0 |
| S10_vs_41055\|\|full | 4680 | No | NA | | 4 | 0 | 0 | Not-determined | Genome-fragment | NA | | NA | 0 |
| S10_vs_41105\|\|full | 1760 | No | NA | | 3 | 0 | 2 | Not-determined | Genome-fragment | NA | | NA | 0 |
| S13_extracted_53639 | 3418 | No | NA | | 3 | 3 | 0 | Low-quality | Genome-fragment | | 5.77 | AAI-based (high-confidence) | 0 |
| S12_vs_k141_3136\|\|full | 3418 | No | NA | | 5 | 0 | 0 | Low-quality | Genome-fragment | | 1.43 | AAI-based (medium-confidence) | 0 |
| S4_vs_VIRSorter_4640-cat_2 | 3418 | No | NA | | 9 | 5 | 0 | Low-quality | Genome-fragment | | 8.09 | AAI-based (high-confidence) | 0 |
| S10_vs_41243\|\|full | 3089 | No | NA | | 4 | 0 | 1 | Not-determined | Genome-fragment | NA | | NA | 0 |
| S14_vs_k141_9613\|\|full | 3419 | No | NA | | 7 | 4 | 0 | Low-quality | Genome-fragment | | 8.05 | AAI-based (medium-confidence) | 0 |
| S7_extracted_3893 | 3419 | No | NA | | 6 | 0 | 0 | Low-quality | Genome-fragment | | 2.23 | AAI-based (high-confidence) | 0 |
| S10_vs_41706\|\|full | 3181 | No | NA | | 3 | 0 | 3 | Not-determined | Genome-fragment | NA | | NA | 0 |
| S16_vs_101575\|\|full | 3420 | No | NA | | 3 | 0 | 1 | Low-quality | Genome-fragment | | 6.45 | AAI-based (medium-confidence) | 0 |
| S10_vs_42089\|\|full | 9093 | No | NA | | 6 | 0 | 6 | Not-determined | Genome-fragment | NA | | NA | 0 |
| S2_extracted_k141_6795 | 3420 | No | NA | | 9 | 0 | 1 | Low-quality | Genome-fragment | | 6.89 | AAI-based (medium-confidence) | 0 |
| S10_vs_42300\|\|full | 1950 | No | NA | | 3 | 0 | 2 | Not-determined | Genome-fragment | NA | | NA | 0 |
| S10_extracted_65347 | 3421 | No | NA | | 10 | 0 | 0 | Low-quality | Genome-fragment | | 5.43 | AAI-based (high-confidence) | 0 |
| S10_vs_42417\|\|full | 1742 | No | NA | | 3 | 0 | 0 | Not-determined | Genome-fragment | NA | | NA | 0 |
| S10_vs_42463\|\|full | 2157 | No | NA | | 3 | 0 | 2 | Not-determined | Genome-fragment | NA | | NA | 0 |
| S10_vs_42477\|\|full | 10436 | No | NA | | 13 | 0 | 1 | Not-determined | Genome-fragment | NA | | NA | 0 |
| S10_vs_42578\|\|full | 2478 | No | NA | | 4 | 0 | 1 | Not-determined | Genome-fragment | NA | | NA | 0 |
| S10_extracted_72879 | 3421 | No | NA | | 15 | 0 | 1 | Low-quality | Genome-fragment | | 5.21 | AAI-based (medium-confidence) | 0 |
| S16_extracted_151239 | 3422 | No | NA | | 3 | 0 | 2 | Low-quality | Genome-fragment | | 2.4 | AAI-based (medium-confidence) | 0 |
| S7_vs_118167\|\|full | 3424 | No | NA | | 3 | 0 | 0 | Low-quality | Genome-fragment | | 7.25 | AAI-based (medium-confidence) | 0 |
| S6_extracted_k141_14128 | 3424 | No | NA | | 6 | 4 | 0 | Low-quality | Genome-fragment | | 9.17 | AAI-based (high-confidence) | 0 |
| S8_extracted_k141_131936 | 3424 | No | NA | | 4 | 3 | 0 | Low-quality | Genome-fragment | | 2.34 | AAI-based (high-confidence) | 0 |
| S10_vs_43009\|\|full | 1583 | No | NA | | 3 | 0 | 1 | Not-determined | Genome-fragment | NA | | NA | 0 |
| S10_vs_43025\|\|full | 3910 | No | NA | | 10 | 0 | 0 | Not-determined | Genome-fragment | NA | | NA | 0 |
| S10_vs_43089\|\|full | 1947 | No | NA | | 2 | 0 | 2 | Not-determined | Genome-fragment | NA | | NA | 0 |
| S8_vs_k141_76406\|\|full | 3424 | No | NA | | 4 | 3 | 0 | Low-quality | Genome-fragment | | 9.98 | AAI-based (high-confidence) | 0 |
| S8_vs_k141_50708\|\|full | 3425 | No | NA | | 7 | 1 | 0 | Low-quality | Genome-fragment | | 6.04 | AAI-based (high-confidence) | 0 |
| S10_vs_43402\|\|full | 1882 | No | NA | | 3 | 0 | 1 | Not-determined | Genome-fragment | NA | | NA | 0 |
| S10_vs_43435\|\|full | 1048 | No | NA | | 2 | 0 | 0 | Not-determined | Genome-fragment | NA | | NA | 0 |
| S17_vs_k141_88736\|\|full | 3426 | No | NA | | 2 | 0 | 1 | Low-quality | Genome-fragment | | 4.38 | AAI-based (medium-confidence) | 0 |
| S10_vs_43771\|\|full | 1591 | No | NA | | 2 | 0 | 0 | Not-determined | Genome-fragment | NA | | NA | 0 |
| S10_vs_65536\|\|full | 3426 | No | NA | | 3 | 0 | 3 | Low-quality | Genome-fragment | | 1.88 | AAI-based (medium-confidence) | 0 |
| S14_extracted_k141_112330 | 3427 | No | NA | | 9 | 1 | 0 | Low-quality | Genome-fragment | | 5.87 | AAI-based (high-confidence) | 0 |
| S12_extracted_k141_115632 | 3427 | No | NA | | 4 | 0 | 1 | Low-quality | Genome-fragment | | 1.12 | AAI-based (medium-confidence) | 0 |
| S10_vs_44076\|\|full | 2734 | No | NA | | 4 | 0 | 1 | Not-determined | Genome-fragment | NA | | NA | 0 |
| S10_vs_44128\|\|full | 5728 | No | NA | | 6 | 0 | 1 | Not-determined | Genome-fragment | NA | | NA | 0 |
| S10_vs_44319\|\|full | 9535 | No | NA | | 10 | 0 | 4 | Not-determined | Genome-fragment | NA | | NA | 0 |
| S10_vs_44412\|\|full | 1777 | No | NA | | 2 | 0 | 1 | Not-determined | Genome-fragment | NA | | NA | 0 |
| S17_extracted_k141_110462 | 3428 | No | NA | | 2 | 0 | 1 | Low-quality | Genome-fragment | | 7.38 | AAI-based (high-confidence) | 0 |
| S17_vs_k141_94642\|\|full | 3429 | No | NA | | 8 | 3 | 0 | Low-quality | Genome-fragment | | 8.01 | AAI-based (high-confidence) | 0 |
| S10_vs_44551\|\|full | 1742 | No | NA | | 3 | 0 | 0 | Not-determined | Genome-fragment | NA | | NA | 0 |
| S14_extracted_k141_8718 | 3429 | No | NA | | 5 | 1 | 0 | Low-quality | Genome-fragment | | 8.07 | AAI-based (high-confidence) | 0 |
| S10_vs_44684\|\|full | 4624 | No | NA | | 5 | 0 | 4 | Not-determined | Genome-fragment | NA | | NA | 0 |
| S10_vs_44739\|\|full | 1410 | No | NA | | 2 | 0 | 0 | Not-determined | Genome-fragment | NA | | NA | 0 |
| S7_extracted_134662 | 3430 | No | NA | | 2 | 0 | 2 | Low-quality | Genome-fragment | | 0.5 | AAI-based (medium-confidence) | 0 |
| S10_vs_44833\|\|full | 8432 | No | NA | | 3 | 0 | 3 | Not-determined | Genome-fragment | NA | | NA | 0 |
| S10_vs_44847\|\|full | 4636 | No | NA | | 4 | 0 | 1 | Not-determined | Genome-fragment | NA | | NA | 0 |
| S10_vs_44891\|\|full | 5002 | No | NA | | 6 | 0 | 2 | Not-determined | Genome-fragment | NA | | NA | 0 |
| S10_vs_44963\|\|full | 3069 | No | NA | | 6 | 0 | 4 | Not-determined | Genome-fragment | NA | | NA | 0 |
| S10_vs_45039\|\|full | 1198 | No | NA | | 3 | 0 | 0 | Not-determined | Genome-fragment | NA | | NA | 0 |
| S4_extracted_4557 | 3430 | No | NA | | 11 | 1 | 0 | Low-quality | Genome-fragment | | 7.56 | AAI-based (high-confidence) | 0 |
| S10_vs_45345\|\|full | 1494 | No | NA | | 4 | 0 | 0 | Not-determined | Genome-fragment | NA | | NA | 0 |
| S10_vs_45424\|\|full | 1488 | No | NA | | 3 | 0 | 0 | Not-determined | Genome-fragment | NA | | NA | 0 |
| S11_vs_k141_18499\|\|full | 3431 | No | NA | | 5 | 2 | 0 | Low-quality | Genome-fragment | | 5.13 | AAI-based (high-confidence) | 0 |
| S10_vs_45629\|\|full | 92747 | No | NA | | 98 | 0 | 18 | Not-determined | Genome-fragment | NA | | NA | 0 |
| S10_extracted_100732 | 3431 | No | NA | | 4 | 0 | 0 | Low-quality | Genome-fragment | | 1.4 | AAI-based (medium-confidence) | 0 |
| S12_vs_k141_86417\|\|full | 3431 | No | NA | | 5 | 0 | 2 | Low-quality | Genome-fragment | | 2.54 | AAI-based (high-confidence) | 0 |
| S10_vs_45832\|\|full | 1337 | No | NA | | 4 | 0 | 0 | Not-determined | Genome-fragment | NA | | NA | 0 |
| S8_extracted_k141_55168 | 3432 | No | NA | | 4 | 0 | 2 | Low-quality | Genome-fragment | | 1.8 | AAI-based (medium-confidence) | 0 |
| S10_vs_46014\|\|full | 2759 | No | NA | | 3 | 0 | 2 | Not-determined | Genome-fragment | NA | | NA | 0 |
| S10_vs_46096\|\|full | 1565 | No | NA | | 3 | 0 | 1 | Not-determined | Genome-fragment | NA | | NA | 0 |
| S11_extracted_k141_2608 | 3433 | No | NA | | 7 | 1 | 0 | Low-quality | Genome-fragment | | 7.99 | AAI-based (high-confidence) | 0 |
| S10_vs_29645\|\|full | 3433 | No | NA | | 3 | 0 | 2 | Low-quality | Genome-fragment | | 1.77 | AAI-based (medium-confidence) | 0 |
| S10_vs_46228\|\|full | 1252 | No | NA | | 2 | 0 | 0 | Not-determined | Genome-fragment | NA | | NA | 0 |
| S10_vs_46265\|\|full | 5600 | No | NA | | 7 | 0 | 2 | Not-determined | Genome-fragment | NA | | NA | 0 |
| S16_extracted_124438 | 3434 | No | NA | | 4 | 3 | 0 | Low-quality | Genome-fragment | | 9.9 | AAI-based (high-confidence) | 0 |
| S10_vs_46508\|\|full | 4642 | No | NA | | 5 | 0 | 2 | Not-determined | Genome-fragment | NA | | NA | 0 |
| S17_extracted_k141_170317 | 3435 | No | NA | | 2 | 0 | 1 | Low-quality | Genome-fragment | | 12.4 | AAI-based (medium-confidence) | 0 |
| S5_vs_k141_1792\|\|full | 3435 | No | NA | | 3 | 0 | 2 | Low-quality | Genome-fragment | | 3.64 | AAI-based (medium-confidence) | 0 |
| S10_vs_46632\|\|full | 1233 | No | NA | | 2 | 0 | 1 | Not-determined | Genome-fragment | NA | | NA | 0 |
| S10_vs_46663\|\|full | 1189 | No | NA | | 3 | 0 | 1 | Not-determined | Genome-fragment | NA | | NA | 0 |
| S10_vs_46819\|\|full | 34565 | No | NA | | 51 | 19 | 1 | High-quality | High-quality | | 100 | AAI-based (high-confidence) | 0 |
| S12_vs_k141_102604\|\|lt2gene | 3436 | No | NA | | 2 | 0 | 0 | Low-quality | Genome-fragment | | 7.65 | AAI-based (medium-confidence) | 0 |
| S10_vs_46952\|\|full | 2897 | No | NA | | 3 | 0 | 3 | Not-determined | Genome-fragment | NA | | NA | 0 |
| S10_vs_47199\|\|full | 6444 | No | NA | | 6 | 0 | 4 | Not-determined | Genome-fragment | NA | | NA | 0 |
| S10_vs_47432\|\|full | 1681 | No | NA | | 2 | 0 | 2 | Not-determined | Genome-fragment | NA | | NA | 0 |
| S10_vs_47722\|\|full | 1525 | No | NA | | 4 | 0 | 0 | Not-determined | Genome-fragment | NA | | NA | 0 |
| S10_vs_47957\|\|full | 2851 | No | NA | | 3 | 0 | 1 | Not-determined | Genome-fragment | NA | | NA | 0 |
| S10_vs_48024\|\|full | 1748 | No | NA | | 4 | 0 | 0 | Not-determined | Genome-fragment | NA | | NA | 0 |
| S8_extracted_k141_44417 | 3436 | No | NA | | 3 | 0 | 2 | Low-quality | Genome-fragment | | 4.46 | AAI-based (medium-confidence) | 0 |
| S8_extracted_k141_157799 | 3436 | No | NA | | 2 | 0 | 1 | Low-quality | Genome-fragment | | 0.3 | AAI-based (medium-confidence) | 0 |
| S3_extracted_k141_59922 | 3437 | No | NA | | 3 | 1 | 0 | Low-quality | Genome-fragment | | 3.9 | AAI-based (high-confidence) | 0 |
| S10_vs_48141\|\|full | 7663 | No | NA | | 9 | 0 | 3 | Not-determined | Genome-fragment | NA | | NA | 0 |
| S8_extracted_k141_57513 | 3437 | No | NA | | 5 | 0 | 0 | Low-quality | Genome-fragment | | 12.23 | AAI-based (medium-confidence) | 0 |
| S3_vs_k141_53713\|\|full | 3438 | No | NA | | 4 | 3 | 0 | Low-quality | Genome-fragment | | 11.25 | AAI-based (high-confidence) | 0 |
| S10_extracted_117206 | 3438 | No | NA | | 6 | 3 | 0 | Low-quality | Genome-fragment | | 4.62 | HMM-based (lower-bound) | 0 |
| S10_vs_48333\|\|full | 1248 | No | NA | | 3 | 0 | 0 | Not-determined | Genome-fragment | NA | | NA | 0 |
| S6_vs_k141_34848\|\|full | 3438 | No | NA | | 4 | 1 | 0 | Low-quality | Genome-fragment | | 3.22 | HMM-based (lower-bound) | 0 |
| S10_vs_48360\|\|full | 2033 | No | NA | | 3 | 0 | 1 | Not-determined | Genome-fragment | NA | | NA | 0 |
| S8_vs_k141_135617\|\|full | 3438 | No | NA | | 6 | 1 | 0 | Low-quality | Genome-fragment | | 4.08 | HMM-based (lower-bound) | 0 |
| S10_vs_48393\|\|full | 2602 | No | NA | | 2 | 0 | 1 | Not-determined | Genome-fragment | NA | | NA | 0 |
| S10_vs_48576\|\|full | 3721 | No | NA | | 5 | 0 | 1 | Not-determined | Genome-fragment | NA | | NA | 0 |
| S10_vs_48654\|\|full | 1314 | No | NA | | 3 | 0 | 1 | Not-determined | Genome-fragment | NA | | NA | 0 |
| S3_extracted_k141_32570 | 3439 | Yes | | 1757 | 6 | 1 | 1 | Low-quality | Genome-fragment | | 0.94 | HMM-based (lower-bound) | 48.91 |
| S5_vs_k141_106183\|\|full | 3439 | No | NA | | 5 | 0 | 0 | Low-quality | Genome-fragment | | 3 | AAI-based (high-confidence) | 0 |
| S11_vs_k141_22687\|\|full | 3441 | No | NA | | 7 | 3 | 0 | Low-quality | Genome-fragment | | 9.29 | AAI-based (medium-confidence) | 0 |
| S10_vs_48993\|\|full | 6521 | No | NA | | 7 | 0 | 5 | Not-determined | Genome-fragment | NA | | NA | 0 |
| S10_vs_49001\|\|full | 2887 | No | NA | | 4 | 0 | 1 | Not-determined | Genome-fragment | NA | | NA | 0 |
| S10_vs_49124\|\|full | 2286 | No | NA | | 3 | 0 | 2 | Not-determined | Genome-fragment | NA | | NA | 0 |
| S10_vs_49136\|\|full | 2447 | No | NA | | 3 | 0 | 2 | Not-determined | Genome-fragment | NA | | NA | 0 |
| S3_vs_k141_69161\|\|full | 3441 | No | NA | | 8 | 3 | 0 | Low-quality | Genome-fragment | | 10.69 | AAI-based (high-confidence) | 0 |
| S10_vs_49243\|\|full | 2236 | No | NA | | 3 | 0 | 2 | Not-determined | Genome-fragment | NA | | NA | 0 |
| S12_vs_k141_99363\|\|full | 3441 | No | NA | | 5 | 3 | 0 | Low-quality | Genome-fragment | | 8.72 | AAI-based (high-confidence) | 0 |
| S10_vs_49290\|\|full | 5735 | No | NA | | 10 | 0 | 0 | Not-determined | Genome-fragment | NA | | NA | 0 |
| S8_vs_k141_40352\|\|full | 3441 | No | NA | | 7 | 0 | 0 | Low-quality | Genome-fragment | | 8.61 | AAI-based (medium-confidence) | 0 |
| S3_extracted_k141_34737 | 3442 | No | NA | | 10 | 1 | 0 | Low-quality | Genome-fragment | | 9.43 | AAI-based (high-confidence) | 0 |
| S5_extracted_k141_14024 | 3442 | No | NA | | 3 | 2 | 0 | Low-quality | Genome-fragment | | 8.45 | AAI-based (high-confidence) | 0 |
| S14_vs_k141_52421\|\|full | 3445 | No | NA | | 9 | 4 | 0 | Low-quality | Genome-fragment | | 8.89 | AAI-based (high-confidence) | 0 |
| S10_vs_49394\|\|full | 3237 | No | NA | | 4 | 0 | 2 | Not-determined | Genome-fragment | NA | | NA | 0 |
| S10_vs_49429\|\|full | 1323 | No | NA | | 3 | 0 | 1 | Not-determined | Genome-fragment | NA | | NA | 0 |
| S14_vs_k141_58913\|\|full | 3446 | No | NA | | 4 | 0 | 0 | Low-quality | Genome-fragment | | 3.3 | AAI-based (high-confidence) | 0 |
| S10_vs_49473\|\|full | 1013 | No | NA | | 2 | 0 | 0 | Not-determined | Genome-fragment | NA | | NA | 0 |
| S10_vs_49540\|\|full | 1353 | No | NA | | 2 | 0 | 1 | Not-determined | Genome-fragment | NA | | NA | 0 |
| S10_vs_49599\|\|full | 1114 | No | NA | | 3 | 0 | 1 | Not-determined | Genome-fragment | NA | | NA | 0 |
| S1_extracted_k141_25667 | 3446 | No | NA | | 3 | 3 | 0 | Low-quality | Genome-fragment | | 10.19 | AAI-based (high-confidence) | 0 |
| S10_vs_49627\|\|full | 1404 | No | NA | | 2 | 0 | 2 | Not-determined | Genome-fragment | NA | | NA | 0 |
| S10_vs_49694\|\|full | 4416 | No | NA | | 5 | 0 | 0 | Not-determined | Genome-fragment | NA | | NA | 0 |
| S10_vs_49716\|\|full | 3531 | No | NA | | 4 | 0 | 3 | Not-determined | Genome-fragment | NA | | NA | 0 |
| S10_vs_49726\|\|full | 4574 | No | NA | | 2 | 0 | 2 | Not-determined | Genome-fragment | NA | | NA | 0 |
| S10_vs_49985\|\|full | 1309 | No | NA | | 3 | 0 | 1 | Not-determined | Genome-fragment | NA | | NA | 0 |
| S16_extracted_17902 | 3446 | No | NA | | 7 | 1 | 0 | Low-quality | Genome-fragment | | 8.28 | AAI-based (high-confidence) | 0 |
| S16_extracted_75364 | 3446 | No | NA | | 1 | 0 | 1 | Low-quality | Genome-fragment | | 0.87 | AAI-based (high-confidence) | 0 |
| S10_vs_50130\|\|full | 1440 | No | NA | | 3 | 0 | 0 | Not-determined | Genome-fragment | NA | | NA | 0 |
| S10_vs_50319\|\|full | 2425 | No | NA | | 2 | 0 | 2 | Not-determined | Genome-fragment | NA | | NA | 0 |
| S10_vs_50350\|\|full | 1574 | No | NA | | 3 | 0 | 1 | Not-determined | Genome-fragment | NA | | NA | 0 |
| S6_vs_k141_40320\|\|full | 3446 | No | NA | | 7 | 0 | 0 | Low-quality | Genome-fragment | | 8.2 | AAI-based (high-confidence) | 0 |
| S10_vs_50459\|\|full | 37645 | No | NA | | 49 | 0 | 7 | Not-determined | Genome-fragment | NA | | NA | 0 |
| S1_extracted_k141_10199 | 3448 | No | NA | | 5 | 0 | 0 | Low-quality | Genome-fragment | | 8.42 | AAI-based (high-confidence) | 0 |
| S11_vs_k141_48160\|\|full | 3449 | No | NA | | 5 | 0 | 0 | Low-quality | Genome-fragment | | 5.17 | AAI-based (medium-confidence) | 0 |
| S16_vs_53303\|\|full | 3450 | No | NA | | 5 | 0 | 1 | Low-quality | Genome-fragment | | 25.58 | AAI-based (medium-confidence) | 0 |
| S10_vs_50796\|\|full | 5951 | No | NA | | 6 | 0 | 3 | Not-determined | Genome-fragment | NA | | NA | 0 |
| S6_extracted_k141_11284 | 3450 | No | NA | | 2 | 0 | 0 | Low-quality | Genome-fragment | | 2.44 | AAI-based (medium-confidence) | 0 |
| S5_extracted_k141_31772 | 3451 | No | NA | | 4 | 0 | 1 | Low-quality | Genome-fragment | | 5.76 | AAI-based (medium-confidence) | 0 |
| S16_vs_18767\|\|full | 3451 | No | NA | | 4 | 0 | 3 | Low-quality | Genome-fragment | | 11.02 | AAI-based (medium-confidence) | 0 |
| S8_extracted_k141_63599 | 3451 | No | NA | | 3 | 0 | 0 | Low-quality | Genome-fragment | | 1.51 | AAI-based (medium-confidence) | 0 |
| S9_extracted_k141_81998 | 3452 | No | NA | | 7 | 5 | 0 | Low-quality | Genome-fragment | | 11.38 | AAI-based (high-confidence) | 0 |
| S10_vs_51321\|\|full | 4623 | No | NA | | 6 | 0 | 4 | Not-determined | Genome-fragment | NA | | NA | 0 |
| S3_vs_k141_42766\|\|full | 3452 | No | NA | | 7 | 2 | 0 | Low-quality | Genome-fragment | | 10.15 | AAI-based (high-confidence) | 0 |
| S10_vs_51371\|\|full | 2005 | No | NA | | 4 | 0 | 0 | Not-determined | Genome-fragment | NA | | NA | 0 |
| S10_vs_51412\|\|full | 2846 | No | NA | | 2 | 0 | 1 | Not-determined | Genome-fragment | NA | | NA | 0 |
| S16_extracted_107945 | 3452 | No | NA | | 4 | 3 | 0 | Low-quality | Genome-fragment | | 9.53 | AAI-based (high-confidence) | 0 |
| S10_vs_51490\|\|full | 2912 | No | NA | | 3 | 0 | 2 | Not-determined | Genome-fragment | NA | | NA | 0 |
| S10_vs_51537\|\|full | 5247 | No | NA | | 5 | 0 | 4 | Not-determined | Genome-fragment | NA | | NA | 0 |
| S8_vs_k141_162198\|\|full | 3453 | No | NA | | 3 | 0 | 1 | Low-quality | Genome-fragment | | 5.48 | AAI-based (medium-confidence) | 0 |
| S10_vs_51588\|\|full | 2844 | No | NA | | 5 | 0 | 3 | Not-determined | Genome-fragment | NA | | NA | 0 |
| S3_vs_k141_13666\|\|full | 3454 | No | NA | | 5 | 1 | 0 | Low-quality | Genome-fragment | | 4.22 | HMM-based (lower-bound) | 0 |
| S15_extracted_33110 | 3455 | No | NA | | 5 | 0 | 1 | Low-quality | Genome-fragment | | 4.96 | AAI-based (high-confidence) | 0 |
| S10_vs_51653\|\|full | 1552 | No | NA | | 3 | 0 | 1 | Not-determined | Genome-fragment | NA | | NA | 0 |
| S12_extracted_k141_186699 | 3456 | No | NA | | 5 | 1 | 0 | Low-quality | Genome-fragment | | 8.47 | AAI-based (high-confidence) | 0 |
| S15_vs_8490\|\|full | 3456 | No | NA | | 3 | 0 | 3 | Low-quality | Genome-fragment | | 7.43 | AAI-based (medium-confidence) | 0 |
| S10_vs_51752\|\|full | 5949 | No | NA | | 6 | 0 | 5 | Not-determined | Genome-fragment | NA | | NA | 0 |
| S10_vs_51754\|\|full | 1291 | No | NA | | 2 | 0 | 0 | Not-determined | Genome-fragment | NA | | NA | 0 |
| S11_extracted_k141_34695 | 3457 | No | NA | | 4 | 0 | 2 | Low-quality | Genome-fragment | | 1.76 | AAI-based (medium-confidence) | 0 |
| S16_extracted_168494 | 3457 | No | NA | | 3 | 2 | 0 | Low-quality | Genome-fragment | | 2.16 | AAI-based (medium-confidence) | 0 |
| S6_vs_k141_8708\|\|full | 3457 | No | NA | | 5 | 3 | 0 | Low-quality | Genome-fragment | | 5.7 | AAI-based (high-confidence) | 0 |
| S10_vs_51962\|\|full | 3640 | No | NA | | 4 | 0 | 1 | Not-determined | Genome-fragment | NA | | NA | 0 |
| S10_vs_52115\|\|full | 1451 | No | NA | | 2 | 0 | 0 | Not-determined | Genome-fragment | NA | | NA | 0 |
| S17_vs_k141_154968\|\|full | 3458 | No | NA | | 8 | 0 | 0 | Low-quality | Genome-fragment | | 2.01 | AAI-based (high-confidence) | 0 |
| S16_vs_137734\|\|full | 3458 | No | NA | | 3 | 0 | 0 | Low-quality | Genome-fragment | | 5.42 | AAI-based (medium-confidence) | 0 |
| S10_vs_52394\|\|full | 4290 | No | NA | | 6 | 0 | 0 | Not-determined | Genome-fragment | NA | | NA | 0 |
| S6_extracted_k141_29925 | 3458 | No | NA | | 4 | 2 | 0 | Low-quality | Genome-fragment | | 15.72 | AAI-based (high-confidence) | 0 |
| S5_extracted_k141_96152 | 3459 | No | NA | | 4 | 4 | 0 | Low-quality | Genome-fragment | | 8.71 | AAI-based (high-confidence) | 0 |
| S17_extracted_k141_112140 | 3460 | No | NA | | 10 | 1 | 0 | Low-quality | Genome-fragment | | 2.28 | HMM-based (lower-bound) | 0 |
| S11_vs_k141_54594\|\|full | 3460 | No | NA | | 6 | 0 | 0 | Low-quality | Genome-fragment | | 9.67 | AAI-based (medium-confidence) | 0 |
| S10_vs_52692\|\|full | 1314 | No | NA | | 3 | 0 | 2 | Not-determined | Genome-fragment | NA | | NA | 0 |
| S3_vs_k141_57298\|\|full | 3460 | No | NA | | 6 | 1 | 0 | Low-quality | Genome-fragment | | 7.47 | AAI-based (medium-confidence) | 0 |
| S10_vs_52990\|\|full | 1885 | No | NA | | 3 | 0 | 1 | Not-determined | Genome-fragment | NA | | NA | 0 |
| S10_extracted_43103 | 3460 | No | NA | | 4 | 0 | 4 | Low-quality | Genome-fragment | | 3.75 | AAI-based (medium-confidence) | 0 |
| S12_extracted_k141_93521 | 3460 | No | NA | | 1 | 0 | 1 | Low-quality | Genome-fragment | | 8.22 | AAI-based (medium-confidence) | 0 |
| S10_vs_53553\|\|full | 5374 | No | NA | | 9 | 0 | 4 | Not-determined | Genome-fragment | NA | | NA | 0 |
| S13_vs_69068\|\|full | 3461 | No | NA | | 3 | 1 | 0 | Low-quality | Genome-fragment | | 1.88 | HMM-based (lower-bound) | 0 |
| S10_vs_53687\|\|full | 2751 | No | NA | | 3 | 0 | 1 | Not-determined | Genome-fragment | NA | | NA | 0 |
| S10_vs_53715\|\|full | 1119 | No | NA | | 3 | 0 | 0 | Not-determined | Genome-fragment | NA | | NA | 0 |
| S10_vs_53909\|\|full | 1914 | No | NA | | 3 | 0 | 3 | Not-determined | Genome-fragment | NA | | NA | 0 |
| S5_extracted_k141_128050 | 3463 | No | NA | | 2 | 1 | 0 | Low-quality | Genome-fragment | | 4.53 | AAI-based (medium-confidence) | 0 |
| S1_vs_k141_136102\|\|full | 3464 | No | NA | | 2 | 0 | 2 | Low-quality | Genome-fragment | | 2.24 | AAI-based (medium-confidence) | 0 |
| S10_vs_54164\|\|full | 1930 | No | NA | | 3 | 0 | 0 | Not-determined | Genome-fragment | NA | | NA | 0 |
| S10_vs_54202\|\|full | 2668 | No | NA | | 4 | 0 | 0 | Not-determined | Genome-fragment | NA | | NA | 0 |
| S16_vs_10924\|\|full | 3464 | No | NA | | 6 | 1 | 0 | Low-quality | Genome-fragment | | 6.17 | HMM-based (lower-bound) | 0 |
| S10_vs_54333\|\|full | 1729 | No | NA | | 3 | 0 | 0 | Not-determined | Genome-fragment | NA | | NA | 0 |
| S10_vs_54408\|\|full | 1572 | No | NA | | 4 | 0 | 0 | Not-determined | Genome-fragment | NA | | NA | 0 |
| S11_extracted_k141_87540 | 3465 | No | NA | | 6 | 1 | 0 | Low-quality | Genome-fragment | | 2.24 | HMM-based (lower-bound) | 0 |
| S3_vs_k141_45007\|\|full | 3465 | No | NA | | 7 | 1 | 0 | Low-quality | Genome-fragment | | 5.63 | AAI-based (high-confidence) | 0 |
| S5_vs_k141_48861\|\|full | 3465 | No | NA | | 7 | 1 | 0 | Low-quality | Genome-fragment | | 5.8 | AAI-based (high-confidence) | 0 |
| S10_vs_54627\|\|full | 2772 | No | NA | | 3 | 0 | 2 | Not-determined | Genome-fragment | NA | | NA | 0 |
| S10_vs_54640\|\|full | 7436 | No | NA | | 9 | 0 | 5 | Not-determined | Genome-fragment | NA | | NA | 0 |
| S10_vs_54659\|\|full | 2214 | No | NA | | 3 | 0 | 2 | Not-determined | Genome-fragment | NA | | NA | 0 |
| S10_vs_54670\|\|full | 1733 | No | NA | | 3 | 0 | 1 | Not-determined | Genome-fragment | NA | | NA | 0 |
| S10_vs_54861\|\|full | 1107 | No | NA | | 2 | 0 | 0 | Not-determined | Genome-fragment | NA | | NA | 0 |
| S8_extracted_k141_142919 | 3465 | No | NA | | 5 | 0 | 2 | Low-quality | Genome-fragment | | 0.99 | AAI-based (high-confidence) | 0 |
| S1_vs_k141_69364\|\|full | 3466 | No | NA | | 5 | 1 | 0 | Low-quality | Genome-fragment | | 9.61 | AAI-based (medium-confidence) | 0 |
| S7_vs_1102\|\|full | 3466 | No | NA | | 4 | 0 | 0 | Low-quality | Genome-fragment | | 6.76 | AAI-based (high-confidence) | 0 |
| S10_vs_55112\|\|full | 1473 | No | NA | | 3 | 0 | 1 | Not-determined | Genome-fragment | NA | | NA | 0 |
| S14_extracted_k141_103792 | 3467 | No | NA | | 3 | 0 | 3 | Low-quality | Genome-fragment | | 1.62 | AAI-based (medium-confidence) | 0 |
| S10_vs_55276\|\|full | 1063 | No | NA | | 3 | 0 | 0 | Not-determined | Genome-fragment | NA | | NA | 0 |
| S10_vs_55361\|\|full | 1433 | No | NA | | 2 | 0 | 0 | Not-determined | Genome-fragment | NA | | NA | 0 |
| S10_vs_55497\|\|full | 2529 | No | NA | | 3 | 0 | 3 | Not-determined | Genome-fragment | NA | | NA | 0 |
| S10_vs_55725\|\|full | 4549 | No | NA | | 2 | 0 | 1 | Not-determined | Genome-fragment | NA | | NA | 0 |
| S10_extracted_95732 | 3467 | No | NA | | 8 | 2 | 1 | Low-quality | Genome-fragment | | 1.99 | AAI-based (high-confidence) | 0 |
| S8_extracted_k141_31360 | 3467 | No | NA | | 3 | 0 | 3 | Low-quality | Genome-fragment | | 1.07 | AAI-based (high-confidence) | 0 |
| S10_vs_55783\|\|full | 2201 | No | NA | | 2 | 0 | 0 | Not-determined | Genome-fragment | NA | | NA | 0 |
| S10_vs_55790\|\|full | 1362 | No | NA | | 3 | 0 | 1 | Not-determined | Genome-fragment | NA | | NA | 0 |
| S10_vs_55799\|\|full | 1420 | No | NA | | 3 | 0 | 2 | Not-determined | Genome-fragment | NA | | NA | 0 |
| S5_vs_k141_29406\|\|full | 3468 | No | NA | | 5 | 1 | 0 | Low-quality | Genome-fragment | | 7.29 | AAI-based (high-confidence) | 0 |
| S8_extracted_k141_73786 | 3468 | No | NA | | 5 | 0 | 3 | Low-quality | Genome-fragment | | 1.33 | AAI-based (medium-confidence) | 0 |
| S10_vs_55895\|\|full | 1498 | No | NA | | 4 | 0 | 2 | Not-determined | Genome-fragment | NA | | NA | 0 |
| S10_vs_55925\|\|full | 1574 | No | NA | | 3 | 0 | 1 | Not-determined | Genome-fragment | NA | | NA | 0 |
| S7_vs_29311\|\|full | 3469 | Yes | | 2215 | 5 | 1 | 1 | Low-quality | Genome-fragment | | 3.75 | AAI-based (medium-confidence) | 36.15 |
| S10_vs_56000\|\|full | 3040 | No | NA | | 4 | 0 | 0 | Not-determined | Genome-fragment | NA | | NA | 0 |
| S10_vs_56007\|\|full | 1939 | No | NA | | 3 | 0 | 1 | Not-determined | Genome-fragment | NA | | NA | 0 |
| S10_extracted_76746 | 3469 | No | NA | | 2 | 0 | 1 | Low-quality | Genome-fragment | | 3.62 | AAI-based (high-confidence) | 0 |
| S6_vs_k141_29828\|\|full | 3469 | No | NA | | 6 | 1 | 0 | Low-quality | Genome-fragment | | 5.05 | AAI-based (high-confidence) | 0 |
| S17_vs_k141_170340\|\|full | 3470 | No | NA | | 8 | 0 | 0 | Low-quality | Genome-fragment | | 8.1 | AAI-based (high-confidence) | 0 |
| S11_extracted_k141_79867 | 3470 | No | NA | | 7 | 0 | 1 | Low-quality | Genome-fragment | | 3.21 | AAI-based (medium-confidence) | 0 |
| S11_extracted_k141_63087 | 3470 | No | NA | | 9 | 3 | 0 | Low-quality | Genome-fragment | | 5.9 | AAI-based (medium-confidence) | 0 |
| S10_vs_56387\|\|full | 1296 | No | NA | | 3 | 0 | 0 | Not-determined | Genome-fragment | NA | | NA | 0 |
| S1_vs_k141_150556\|\|full | 3470 | No | NA | | 9 | 1 | 0 | Low-quality | Genome-fragment | | 8.18 | AAI-based (medium-confidence) | 0 |
| S10_vs_56421\|\|full | 4036 | No | NA | | 6 | 0 | 1 | Not-determined | Genome-fragment | NA | | NA | 0 |
| S12_vs_k141_52432\|\|full | 3470 | No | NA | | 5 | 0 | 1 | Low-quality | Genome-fragment | | 9.96 | AAI-based (high-confidence) | 0 |
| S17_extracted_k141_15349 | 3471 | Yes | | 1470 | 4 | 1 | 1 | Low-quality | Genome-fragment | | 1.08 | HMM-based (lower-bound) | 57.65 |
| S3_vs_k141_112598\|\|full | 3471 | No | NA | | 9 | 3 | 0 | Low-quality | Genome-fragment | | 2.05 | AAI-based (high-confidence) | 0 |
| S10_vs_56780\|\|full | 1428 | No | NA | | 2 | 0 | 2 | Not-determined | Genome-fragment | NA | | NA | 0 |
| S15_extracted_63325 | 3471 | No | NA | | 4 | 1 | 0 | Low-quality | Genome-fragment | | 9.85 | AAI-based (high-confidence) | 0 |
| S10_vs_57099\|\|full | 3328 | No | NA | | 7 | 0 | 1 | Not-determined | Genome-fragment | NA | | NA | 0 |
| S10_vs_57101\|\|full | 3247 | No | NA | | 3 | 0 | 2 | Not-determined | Genome-fragment | NA | | NA | 0 |
| S10_vs_57120\|\|full | 13222 | No | NA | | 14 | 0 | 4 | Not-determined | Genome-fragment | NA | | NA | 0 |
| S9_vs_k141_90813\|\|full | 3472 | No | NA | | 3 | 0 | 2 | Low-quality | Genome-fragment | | 2.77 | AAI-based (medium-confidence) | 0 |
| S3_extracted_k141_28381 | 3472 | No | NA | | 9 | 5 | 0 | Low-quality | Genome-fragment | | 15.84 | AAI-based (high-confidence) | 0 |
| S6_vs_k141_45209\|\|full | 3472 | No | NA | | 8 | 7 | 0 | Low-quality | Genome-fragment | | 8.67 | AAI-based (high-confidence) | 0 |
| S8_extracted_k141_147133 | 3472 | No | NA | | 5 | 2 | 0 | Low-quality | Genome-fragment | | 9.29 | AAI-based (high-confidence) | 0 |
| S10_vs_57436\|\|full | 4129 | No | NA | | 4 | 0 | 1 | Not-determined | Genome-fragment | NA | | NA | 0 |
| S10_vs_57517\|\|full | 5670 | No | NA | | 7 | 0 | 3 | Not-determined | Genome-fragment | NA | | NA | 0 |
| S14_extracted_k141_57286 | 3473 | No | NA | | 4 | 3 | 0 | Low-quality | Genome-fragment | | 8.37 | AAI-based (high-confidence) | 0 |
| S7_vs_60786\|\|full | 3473 | No | NA | | 9 | 4 | 0 | Low-quality | Genome-fragment | | 8.07 | AAI-based (medium-confidence) | 0 |
| S10_vs_57686\|\|full | 2237 | No | NA | | 3 | 0 | 2 | Not-determined | Genome-fragment | NA | | NA | 0 |
| S10_vs_58009\|\|full | 7401 | No | NA | | 7 | 0 | 4 | Not-determined | Genome-fragment | NA | | NA | 0 |
| S10_vs_58014\|\|full | 35404 | No | NA | | 30 | 6 | 2 | Medium-quality | Genome-fragment | | 82.66 | AAI-based (high-confidence) | 0 |
| S10_vs_58048\|\|full | 1404 | No | NA | | 2 | 0 | 0 | Not-determined | Genome-fragment | NA | | NA | 0 |
| S7_vs_116551\|\|full | 3473 | No | NA | | 4 | 0 | 1 | Low-quality | Genome-fragment | | 7.87 | AAI-based (high-confidence) | 0 |
| S10_vs_58204\|\|full | 2305 | No | NA | | 3 | 0 | 0 | Not-determined | Genome-fragment | NA | | NA | 0 |
| S10_vs_58223\|\|full | 2135 | No | NA | | 2 | 0 | 1 | Not-determined | Genome-fragment | NA | | NA | 0 |
| S11_vs_k141_6538\|\|full | 3474 | No | NA | | 6 | 4 | 0 | Low-quality | Genome-fragment | | 8.56 | AAI-based (high-confidence) | 0 |
| S10_vs_38694\|\|full | 3474 | No | NA | | 4 | 0 | 2 | Low-quality | Genome-fragment | | 2.21 | AAI-based (medium-confidence) | 0 |

| | | | | | | | | | | | |
|---|---|---|---|---|---|---|---|---|---|---|---|
| S10_vs_58505\|\|full | 12794 | No | NA | 11 | 0 | 11 | Not-determined | Genome-fragment | NA | NA | 0 |
| S10_vs_58567\|\|full | 3495 | No | NA | 3 | 0 | 1 | Not-determined | Genome-fragment | NA | NA | 0 |
| S9_vs_k141_44394\|\|full | 3476 | No | NA | 4 | 0 | 1 | Low-quality | Genome-fragment | 1.93 | AAI-based (medium-confidence) | 0 |
| S10_vs_58722\|\|full | 1657 | No | NA | 3 | 0 | 0 | Not-determined | Genome-fragment | NA | NA | 0 |
| S10_vs_58745\|\|full | 2889 | No | NA | 5 | 0 | 2 | Not-determined | Genome-fragment | NA | NA | 0 |
| S14_extracted_k141_105641 | 3476 | No | NA | 9 | 9 | 0 | Low-quality | Genome-fragment | 11.47 | AAI-based (high-confidence) | 0 |
| S10_vs_58891\|\|full | 1179 | No | NA | 3 | 0 | 1 | Not-determined | Genome-fragment | NA | NA | 0 |
| S17_vs_k141_170070\|\|full | 3477 | No | NA | 4 | 0 | 3 | Low-quality | Genome-fragment | 11.1 | AAI-based (medium-confidence) | 0 |
| S1_vs_k141_35417\|\|full | 3477 | No | NA | 3 | 3 | 0 | Low-quality | Genome-fragment | 9.28 | AAI-based (high-confidence) | 0 |
| S10_vs_59004\|\|full | 1475 | No | NA | 2 | 0 | 0 | Not-determined | Genome-fragment | NA | NA | 0 |
| S7_vs_130081\|\|full | 3477 | No | NA | 7 | 3 | 0 | Low-quality | Genome-fragment | 7.89 | AAI-based (high-confidence) | 0 |
| S10_vs_59188\|\|full | 2127 | No | NA | 2 | 0 | 2 | Not-determined | Genome-fragment | NA | NA | 0 |
| S10_vs_59230\|\|full | 2335 | No | NA | 2 | 0 | 0 | Not-determined | Genome-fragment | NA | NA | 0 |
| S10_vs_59256\|\|full | 5648 | No | NA | 2 | 0 | 2 | Not-determined | Genome-fragment | NA | NA | 0 |
| S10_vs_59302\|\|full | 1227 | No | NA | 2 | 0 | 1 | Not-determined | Genome-fragment | NA | NA | 0 |
| S10_vs_59487\|\|full | 1344 | No | NA | 2 | 0 | 1 | Not-determined | Genome-fragment | NA | NA | 0 |
| S10_vs_6705\|\|full | 3477 | No | NA | 3 | 1 | 0 | Low-quality | Genome-fragment | 1.76 | HMM-based (lower-bound) | 0 |
| S10_vs_59929\|\|full | 1529 | No | NA | 2 | 0 | 2 | Not-determined | Genome-fragment | NA | NA | 0 |
| S11_vs_k141_2159\|\|full | 3478 | No | NA | 5 | 0 | 0 | Low-quality | Genome-fragment | 8.12 | AAI-based (medium-confidence) | 0 |
| S10_vs_60276\|\|full | 5662 | No | NA | 6 | 0 | 1 | Not-determined | Genome-fragment | NA | NA | 0 |
| S10_vs_60294\|\|full | 2976 | No | NA | 4 | 0 | 3 | Not-determined | Genome-fragment | NA | NA | 0 |
| S10_vs_60316\|\|full | 1068 | No | NA | 3 | 0 | 0 | Not-determined | Genome-fragment | NA | NA | 0 |
| S12_extracted_k141_735 | 3479 | No | NA | 4 | 2 | 0 | Low-quality | Genome-fragment | 5.29 | HMM-based (lower-bound) | 0 |
| S10_vs_60404\|\|full | 5126 | No | NA | 3 | 0 | 2 | Not-determined | Genome-fragment | NA | NA | 0 |
| S12_extracted_k141_70305 | 3479 | No | NA | 4 | 1 | 0 | Low-quality | Genome-fragment | 5.43 | HMM-based (lower-bound) | 0 |
| S12_vs_k141_46870\|\|full | 3479 | No | NA | 4 | 0 | 2 | Low-quality | Genome-fragment | 1.34 | AAI-based (medium-confidence) | 0 |
| S10_vs_60621\|\|full | 1232 | No | NA | 2 | 0 | 0 | Not-determined | Genome-fragment | NA | NA | 0 |
| S10_vs_60675\|\|full | 11671 | No | NA | 19 | 0 | 1 | Not-determined | Genome-fragment | NA | NA | 0 |
| S10_vs_60677\|\|full | 5372 | No | NA | 4 | 0 | 2 | Not-determined | Genome-fragment | NA | NA | 0 |
| S10_vs_60742\|\|full | 2252 | No | NA | 1 | 0 | 1 | Not-determined | Genome-fragment | NA | NA | 0 |
| S13_vs_56415\|\|full | 3480 | No | NA | 8 | 0 | 1 | Low-quality | Genome-fragment | 8.12 | AAI-based (high-confidence) | 0 |
| S10_vs_60810\|\|full | 3105 | No | NA | 5 | 0 | 0 | Not-determined | Genome-fragment | NA | NA | 0 |
| S10_vs_60937\|\|full | 1108 | No | NA | 3 | 0 | 1 | Not-determined | Genome-fragment | NA | NA | 0 |
| S14_extracted_k141_66839 | 3480 | No | NA | 6 | 5 | 0 | Low-quality | Genome-fragment | 11.48 | AAI-based (high-confidence) | 0 |
| S9_extracted_k141_81478 | 3481 | No | NA | 7 | 2 | 0 | Low-quality | Genome-fragment | 6.91 | AAI-based (high-confidence) | 0 |
| S10_vs_61200\|\|full | 2832 | No | NA | 3 | 0 | 2 | Not-determined | Genome-fragment | NA | NA | 0 |
| S14_vs_k141_69976\|\|full | 3481 | No | NA | 2 | 1 | 0 | Low-quality | Genome-fragment | 6.49 | AAI-based (medium-confidence) | 0 |
| S14_extracted_k141_37700 | 3482 | No | NA | 2 | 0 | 0 | Low-quality | Genome-fragment | 8.42 | AAI-based (medium-confidence) | 0 |
| S10_vs_61658\|\|full | 3935 | No | NA | 6 | 0 | 1 | Not-determined | Genome-fragment | NA | NA | 0 |
| S3_extracted_k141_50738 | 3482 | No | NA | 5 | 0 | 0 | Low-quality | Genome-fragment | 1.6 | AAI-based (high-confidence) | 0 |
| S10_vs_61869\|\|full | 5675 | No | NA | 5 | 0 | 3 | Not-determined | Genome-fragment | NA | NA | 0 |
| S10_vs_61888\|\|full | 4737 | No | NA | 4 | 0 | 3 | Not-determined | Genome-fragment | NA | NA | 0 |
| S16_extracted_96283 | 3482 | No | NA | 5 | 0 | 1 | Low-quality | Genome-fragment | 2.32 | AAI-based (medium-confidence) | 0 |
| S10_vs_120955\|\|full | 3482 | No | NA | 7 | 0 | 0 | Low-quality | Genome-fragment | 6.55 | AAI-based (high-confidence) | 0 |
| S6_vs_k141_27953\|\|full | 3482 | No | NA | 9 | 0 | 0 | Low-quality | Genome-fragment | 5.74 | AAI-based (high-confidence) | 0 |
| S10_vs_62142\|\|full | 8089 | No | NA | 7 | 0 | 5 | Not-determined | Genome-fragment | NA | NA | 0 |
| S8_extracted_k141_177530 | 3482 | No | NA | 5 | 1 | 0 | Low-quality | Genome-fragment | 2.54 | AAI-based (high-confidence) | 0 |
| S5_vs_k141_50928\|\|full | 3483 | No | NA | 5 | 1 | 0 | Low-quality | Genome-fragment | 8.04 | AAI-based (high-confidence) | 0 |
| S5_vs_k141_125288\|\|lt2gene | 3483 | No | NA | 2 | 1 | 0 | Low-quality | Genome-fragment | 2.23 | HMM-based (lower-bound) | 0 |
| S10_vs_62438\|\|full | 11690 | No | NA | 7 | 0 | 4 | Not-determined | Genome-fragment | NA | NA | 0 |
| S10_vs_62496\|\|full | 1576 | No | NA | 2 | 0 | 1 | Not-determined | Genome-fragment | NA | NA | 0 |
| S16_vs_160696\|\|full | 3483 | No | NA | 7 | 1 | 0 | Low-quality | Genome-fragment | 5.24 | AAI-based (medium-confidence) | 0 |
| S10_vs_62629\|\|full | 2323 | No | NA | 4 | 0 | 1 | Not-determined | Genome-fragment | NA | NA | 0 |
| S10_extracted_26727 | 3484 | No | NA | 5 | 0 | 0 | Low-quality | Genome-fragment | 7.49 | AAI-based (high-confidence) | 0 |
| S10_vs_62776\|\|full | 63036 | No | NA | 91 | 13 | 2 | High-quality | High-quality | 90.32 | AAI-based (high-confidence) | 0 |
| S10_vs_62866\|\|full | 17922 | No | NA | 12 | 0 | 3 | Not-determined | Genome-fragment | NA | NA | 0 |
| S10_vs_63001\|\|full | 1092 | No | NA | 2 | 0 | 1 | Not-determined | Genome-fragment | NA | NA | 0 |
| S17_vs_k141_25516\|\|full | 3486 | No | NA | 5 | 0 | 1 | Low-quality | Genome-fragment | 7.43 | AAI-based (high-confidence) | 0 |
| S10_vs_63045\|\|full | 2788 | No | NA | 2 | 0 | 1 | Not-determined | Genome-fragment | NA | NA | 0 |
| S10_vs_63246\|\|full | 4267 | No | NA | 4 | 0 | 4 | Not-determined | Genome-fragment | NA | NA | 0 |
| S10_vs_63272\|\|full | 1746 | No | NA | 3 | 0 | 1 | Not-determined | Genome-fragment | NA | NA | 0 |
| S16_extracted_139136 | 3486 | No | NA | 5 | 0 | 3 | Low-quality | Genome-fragment | 11.63 | AAI-based (medium-confidence) | 0 |
| S11_vs_k141_1117\|\|full | 3487 | No | NA | 8 | 3 | 0 | Low-quality | Genome-fragment | 5.82 | AAI-based (medium-confidence) | 0 |
| S3_extracted_k141_28654 | 3487 | No | NA | 2 | 0 | 2 | Low-quality | Genome-fragment | 5.69 | AAI-based (medium-confidence) | 0 |
| S10_vs_63344\|\|full | 1271 | No | NA | 3 | 0 | 1 | Not-determined | Genome-fragment | NA | NA | 0 |
| S10_vs_63373\|\|full | 6825 | No | NA | 7 | 0 | 3 | Not-determined | Genome-fragment | NA | NA | 0 |
| S12_extracted_k141_45987 | 3487 | No | NA | 4 | 2 | 0 | Low-quality | Genome-fragment | 15.62 | AAI-based (high-confidence) | 0 |
| S6_vs_k141_37276\|\|full | 3487 | No | NA | 4 | 0 | 4 | Low-quality | Genome-fragment | 2.88 | AAI-based (medium-confidence) | 0 |
| S10_vs_63632\|\|full | 3603 | No | NA | 4 | 0 | 3 | Not-determined | Genome-fragment | NA | NA | 0 |
| S10_vs_63681\|\|full | 2947 | No | NA | 3 | 0 | 1 | Not-determined | Genome-fragment | NA | NA | 0 |
| S10_vs_63704\|\|full | 4099 | No | NA | 5 | 0 | 3 | Not-determined | Genome-fragment | NA | NA | 0 |
| S17_vs_k141_50138\|\|full | 3488 | No | NA | 4 | 0 | 3 | Low-quality | Genome-fragment | 2.84 | AAI-based (medium-confidence) | 0 |
| S10_vs_63979\|\|full | 2623 | No | NA | 6 | 0 | 0 | Not-determined | Genome-fragment | NA | NA | 0 |
| S10_vs_64065\|\|full | 5013 | No | NA | 6 | 0 | 1 | Not-determined | Genome-fragment | NA | NA | 0 |
| S5_extracted_k141_4457 | 3489 | No | NA | 9 | 3 | 0 | Low-quality | Genome-fragment | 8.34 | AAI-based (high-confidence) | 0 |
| S10_vs_64239\|\|full | 1834 | No | NA | 2 | 0 | 2 | Not-determined | Genome-fragment | NA | NA | 0 |
| S14_vs_k141_77114\|\|full | 3490 | No | NA | 3 | 0 | 0 | Low-quality | Genome-fragment | 2.23 | AAI-based (high-confidence) | 0 |
| S7_vs_62750\|\|full | 3491 | No | NA | 6 | 0 | 0 | Low-quality | Genome-fragment | 7.24 | AAI-based (medium-confidence) | 0 |
| S10_vs_64492\|\|full | 2288 | No | NA | 2 | 0 | 1 | Not-determined | Genome-fragment | NA | NA | 0 |
| S10_vs_64510\|\|full | 3290 | No | NA | 2 | 0 | 0 | Not-determined | Genome-fragment | NA | NA | 0 |
| S10_vs_64737\|\|full | 2444 | No | NA | 3 | 0 | 1 | Not-determined | Genome-fragment | NA | NA | 0 |
| S8_vs_k141_18244\|\|full | 3491 | No | NA | 4 | 0 | 0 | Low-quality | Genome-fragment | 6.09 | AAI-based (medium-confidence) | 0 |
| S17_extracted_k141_59945 | 3492 | No | NA | 5 | 2 | 0 | Low-quality | Genome-fragment | 5.25 | AAI-based (high-confidence) | 0 |
| S10_vs_65196\|\|full | 1312 | No | NA | 1 | 0 | 0 | Not-determined | Genome-fragment | NA | NA | 0 |
| S10_vs_65278\|\|full | 8428 | No | NA | 10 | 0 | 4 | Not-determined | Genome-fragment | NA | NA | 0 |
| S10_vs_65281\|\|full | 3466 | No | NA | 4 | 0 | 2 | Not-determined | Genome-fragment | NA | NA | 0 |
| S16_vs_84819\|\|full | 3492 | No | NA | 3 | 0 | 1 | Low-quality | Genome-fragment | 5.59 | AAI-based (medium-confidence) | 0 |
| S14_extracted_k141_56125 | 3493 | No | NA | 4 | 1 | 0 | Low-quality | Genome-fragment | 1.76 | HMM-based (lower-bound) | 0 |
| S10_vs_65718\|\|full | 1661 | No | NA | 3 | 0 | 1 | Not-determined | Genome-fragment | NA | NA | 0 |
| S10_vs_65915\|\|full | 5794 | No | NA | 6 | 0 | 2 | Not-determined | Genome-fragment | NA | NA | 0 |
| S10_vs_65979\|\|full | 3655 | No | NA | 7 | 0 | 3 | Not-determined | Genome-fragment | NA | NA | 0 |
| S16_extracted_19035 | 3493 | No | NA | 6 | 1 | 0 | Low-quality | Genome-fragment | 6.87 | AAI-based (high-confidence) | 0 |
| S10_vs_66153\|\|full | 1148 | No | NA | 2 | 0 | 0 | Not-determined | Genome-fragment | NA | NA | 0 |
| S10_vs_66208\|\|full | 1932 | No | NA | 3 | 0 | 1 | Not-determined | Genome-fragment | NA | NA | 0 |
| S16_vs_13864\|\|full | 3493 | No | NA | 3 | 0 | 1 | Low-quality | Genome-fragment | 1.88 | AAI-based (medium-confidence) | 0 |
| S10_vs_66404\|\|full | 1545 | No | NA | 3 | 0 | 2 | Not-determined | Genome-fragment | NA | NA | 0 |
| S10_vs_66452\|\|full | 2489 | No | NA | 3 | 0 | 0 | Not-determined | Genome-fragment | NA | NA | 0 |
| S10_vs_104775\|\|full | 3493 | No | NA | 10 | 0 | 0 | Low-quality | Genome-fragment | 5.92 | AAI-based (high-confidence) | 0 |
| S17_extracted_k141_74783 | 3494 | No | NA | 5 | 0 | 1 | Low-quality | Genome-fragment | 1.83 | AAI-based (medium-confidence) | 0 |
| S11_vs_k141_20983\|\|lt2gene | 3494 | No | NA | 2 | 0 | 0 | Low-quality | Genome-fragment | 5.84 | AAI-based (medium-confidence) | 0 |
| S10_vs_66709\|\|full | 1953 | No | NA | 3 | 0 | 1 | Not-determined | Genome-fragment | NA | NA | 0 |
| S5_extracted_k141_139943 | 3494 | No | NA | 7 | 7 | 0 | Low-quality | Genome-fragment | 11.52 | AAI-based (high-confidence) | 0 |
| S10_vs_67069\|\|full | 1318 | No | NA | 3 | 0 | 2 | Not-determined | Genome-fragment | NA | NA | 0 |
| S7_vs_52584\|\|full | 3495 | No | NA | 3 | 0 | 1 | Low-quality | Genome-fragment | 7.12 | AAI-based (medium-confidence) | 0 |
| S10_vs_67135\|\|full | 5043 | No | NA | 6 | 0 | 1 | Not-determined | Genome-fragment | NA | NA | 0 |
| S12_vs_k141_185553\|\|full | 3495 | No | NA | 4 | 3 | 0 | Low-quality | Genome-fragment | 4.47 | HMM-based (lower-bound) | 0 |
| S10_vs_67247\|\|full | 3090 | No | NA | 5 | 0 | 0 | Not-determined | Genome-fragment | NA | NA | 0 |
| S9_vs_k141_1408\|\|full | 3496 | No | NA | 6 | 0 | 0 | Low-quality | Genome-fragment | 3.06 | AAI-based (medium-confidence) | 0 |
| S10_vs_67319\|\|full | 1469 | No | NA | 3 | 0 | 1 | Not-determined | Genome-fragment | NA | NA | 0 |
| S3_extracted_k141_109089 | 3496 | No | NA | 3 | 1 | 0 | Low-quality | Genome-fragment | 5.54 | AAI-based (high-confidence) | 0 |
| S17_extracted_k141_126765 | 3497 | No | NA | 6 | 0 | 0 | Low-quality | Genome-fragment | 9.92 | AAI-based (medium-confidence) | 0 |
| S10_vs_67471\|\|full | 38042 | No | NA | 48 | 9 | 0 | Medium-quality | Genome-fragment | 76.5 | AAI-based (high-confidence) | 0 |
| S5_vs_k141_104104\|\|full | 3497 | Yes | 1814 | 6 | 1 | 1 | Low-quality | Genome-fragment | 3.23 | HMM-based (lower-bound) | 48.13 |
| S10_vs_67588\|\|full | 1781 | No | NA | 3 | 0 | 2 | Not-determined | Genome-fragment | NA | NA | 0 |
| S10_vs_67607\|\|full | 2012 | No | NA | 5 | 0 | 0 | Not-determined | Genome-fragment | NA | NA | 0 |
| S17_extracted_k141_98644 | 3498 | No | NA | 6 | 6 | 0 | Low-quality | Genome-fragment | 5.88 | AAI-based (medium-confidence) | 0 |
| S10_vs_67624\|\|full | 2156 | No | NA | 3 | 0 | 1 | Not-determined | Genome-fragment | NA | NA | 0 |
| S10_vs_67626\|\|full | 1590 | No | NA | 2 | 0 | 0 | Not-determined | Genome-fragment | NA | NA | 0 |
| S10_vs_67643\|\|full | 2041 | No | NA | 3 | 0 | 0 | Not-determined | Genome-fragment | NA | NA | 0 |
| S10_vs_67664\|\|full | 1632 | No | NA | 3 | 0 | 0 | Not-determined | Genome-fragment | NA | NA | 0 |
| S14_vs_k141_96625\|\|full | 3499 | No | NA | 7 | 4 | 0 | Low-quality | Genome-fragment | 6.73 | AAI-based (high-confidence) | 0 |
| S10_vs_67682\|\|full | 2122 | No | NA | 3 | 0 | 2 | Not-determined | Genome-fragment | NA | NA | 0 |
| S1_vs_k141_97725\|\|full | 3501 | No | NA | 4 | 2 | 0 | Low-quality | Genome-fragment | 7.91 | AAI-based (medium-confidence) | 0 |
| S10_vs_67770\|\|full | 2993 | No | NA | 4 | 0 | 0 | Not-determined | Genome-fragment | NA | NA | 0 |
| S10_vs_67772\|\|full | 1951 | No | NA | 3 | 0 | 0 | Not-determined | Genome-fragment | NA | NA | 0 |
| S16_extracted_12418 | 3501 | No | NA | 4 | 0 | 2 | Low-quality | Genome-fragment | 5.08 | AAI-based (medium-confidence) | 0 |
| S10_vs_67795\|\|full | 1860 | No | NA | 3 | 0 | 2 | Not-determined | Genome-fragment | NA | NA | 0 |
| S10_vs_67983\|\|full | 2319 | No | NA | 6 | 0 | 1 | Not-determined | Genome-fragment | NA | NA | 0 |
| S10_vs_68036\|\|full | 3438 | No | NA | 6 | 0 | 4 | Not-determined | Genome-fragment | NA | NA | 0 |
| S16_extracted_35486 | 3502 | Yes | 1102 | 6 | 1 | 1 | Low-quality | Genome-fragment | 1.97 | AAI-based (high-confidence) | 68.53 |
| S16_vs_104817\|\|full | 3502 | No | NA | 7 | 2 | 0 | Low-quality | Genome-fragment | 5.56 | AAI-based (high-confidence) | 0 |
| S12_vs_k141_4267\|\|full | 3502 | No | NA | 9 | 1 | 0 | Low-quality | Genome-fragment | 8.81 | AAI-based (high-confidence) | 0 |
| S17_vs_k141_71371\|\|full | 3503 | No | NA | 6 | 2 | 0 | Low-quality | Genome-fragment | 2.03 | AAI-based (high-confidence) | 0 |
| S9_extracted_k141_43544 | 3503 | No | NA | 3 | 0 | 2 | Low-quality | Genome-fragment | 1.39 | AAI-based (medium-confidence) | 0 |
| S10_vs_68658\|\|full | 1108 | No | NA | 2 | 0 | 2 | Not-determined | Genome-fragment | NA | NA | 0 |
| S14_vs_k141_782\|\|full | 3503 | No | NA | 4 | 0 | 2 | Low-quality | Genome-fragment | 1 | AAI-based (high-confidence) | 0 |
| S10_vs_68746\|\|full | 1899 | No | NA | 3 | 0 | 2 | Not-determined | Genome-fragment | NA | NA | 0 |
| S10_vs_68799\|\|full | 1798 | No | NA | 2 | 0 | 1 | Not-determined | Genome-fragment | NA | NA | 0 |
| S1_vs_k141_99935\|\|full | 3503 | No | NA | 3 | 0 | 0 | Low-quality | Genome-fragment | 24.49 | AAI-based (high-confidence) | 0 |
| S16_extracted_166549 | 3503 | No | NA | 4 | 0 | 2 | Low-quality | Genome-fragment | 3.57 | AAI-based (medium-confidence) | 0 |
| S8_extracted_k141_86008 | 3504 | No | NA | 9 | 1 | 0 | Low-quality | Genome-fragment | 12.01 | HMM-based (lower-bound) | 0 |
| S11_extracted_k141_13066 | 3505 | No | NA | 3 | 0 | 3 | Low-quality | Genome-fragment | 1.89 | AAI-based (medium-confidence) | 0 |
| S1_vs_k141_107455\|\|full | 3505 | No | NA | 5 | 1 | 0 | Low-quality | Genome-fragment | 6.88 | AAI-based (high-confidence) | 0 |
| S7_vs_54761\|\|full | 3505 | No | NA | 6 | 2 | 0 | Low-quality | Genome-fragment | 8.54 | AAI-based (medium-confidence) | 0 |
| S12_vs_k141_128778\|\|full | 3505 | No | NA | 3 | 0 | 1 | Low-quality | Genome-fragment | 1.77 | AAI-based (medium-confidence) | 0 |
| S8_extracted_k141_88170 | 3505 | No | NA | 3 | 0 | 3 | Low-quality | Genome-fragment | 4.22 | AAI-based (high-confidence) | 0 |
| S10_vs_69294\|\|full | 7718 | No | NA | 5 | 0 | 4 | Not-determined | Genome-fragment | NA | NA | 0 |
| S10_vs_69561\|\|full | 3687 | No | NA | 6 | 0 | 1 | Not-determined | Genome-fragment | NA | NA | 0 |
| S10_vs_69671\|\|full | 6078 | No | NA | 8 | 0 | 4 | Not-determined | Genome-fragment | NA | NA | 0 |
| S10_vs_69675\|\|full | 1327 | No | NA | 2 | 0 | 2 | Not-determined | Genome-fragment | NA | NA | 0 |
| S1_extracted_k141_108051 | 3506 | No | NA | 5 | 1 | 0 | Low-quality | Genome-fragment | 9.1 | AAI-based (medium-confidence) | 0 |
| S10_vs_69801\|\|full | 1527 | No | NA | 2 | 0 | 0 | Not-determined | Genome-fragment | NA | NA | 0 |
| S10_vs_69845\|\|full | 2354 | No | NA | 3 | 0 | 3 | Not-determined | Genome-fragment | NA | NA | 0 |
| S10_vs_70151\|\|full | 6958 | No | NA | 6 | 0 | 2 | Not-determined | Genome-fragment | NA | NA | 0 |
| S10_vs_70156\|\|full | 5900 | No | NA | 10 | 0 | 1 | Not-determined | Genome-fragment | NA | NA | 0 |
| S10_vs_70194\|\|full | 4460 | No | NA | 3 | 0 | 0 | Not-determined | Genome-fragment | NA | NA | 0 |
| S10_vs_70222\|\|full | 12962 | No | NA | 11 | 0 | 3 | Not-determined | Genome-fragment | NA | NA | 0 |
| S10_vs_70247\|\|full | 1143 | No | NA | 2 | 0 | 0 | Not-determined | Genome-fragment | NA | NA | 0 |
| S15_extracted_34368 | 3506 | No | NA | 4 | 3 | 0 | Low-quality | Genome-fragment | 18.2 | AAI-based (medium-confidence) | 0 |
| S10_vs_36344\|\|full | 3507 | No | NA | 4 | 0 | 2 | Low-quality | Genome-fragment | 1.23 | AAI-based (medium-confidence) | 0 |
| S10_vs_116257\|\|full | 3507 | No | NA | 3 | 0 | 1 | Low-quality | Genome-fragment | 2.76 | AAI-based (medium-confidence) | 0 |
| S8_extracted_k141_157670 | 3507 | No | NA | 1 | 0 | 0 | Low-quality | Genome-fragment | 3.53 | AAI-based (high-confidence) | 0 |
| S7_extracted_105981 | 3508 | No | NA | 2 | 0 | 2 | Low-quality | Genome-fragment | 0.89 | AAI-based (high-confidence) | 0 |
| S10_vs_71091\|\|full | 36895 | No | NA | 28 | 0 | 21 | Not-determined | Genome-fragment | NA | NA | 0 |
| S7_vs_13698\|\|full | 3509 | No | NA | 3 | 0 | 0 | Low-quality | Genome-fragment | 4.64 | AAI-based (medium-confidence) | 0 |
| S10_vs_71584\|\|full | 4736 | No | NA | 3 | 0 | 2 | Not-determined | Genome-fragment | NA | NA | 0 |
| S10_vs_71731\|\|full | 7791 | No | NA | 9 | 0 | 7 | Not-determined | Genome-fragment | NA | NA | 0 |
| S10_vs_71739\|\|full | 3720 | No | NA | 5 | 0 | 2 | Not-determined | Genome-fragment | NA | NA | 0 |
| S5_vs_k141_102735\|\|full | 3510 | No | NA | 3 | 0 | 1 | Low-quality | Genome-fragment | 8.05 | AAI-based (medium-confidence) | 0 |
| S10_vs_71900\|\|full | 26708 | No | NA | 27 | 2 | 0 | Medium-quality | Genome-fragment | 60.38 | AAI-based (high-confidence) | 0 |
| S12_vs_k141_84063\|\|full | 3510 | No | NA | 5 | 0 | 3 | Low-quality | Genome-fragment | 3.2 | AAI-based (medium-confidence) | 0 |
| S4_extracted_10059 | 3510 | No | NA | 2 | 0 | 1 | Low-quality | Genome-fragment | 6.32 | AAI-based (medium-confidence) | 0 |
| S10_vs_72061\|\|full | 4345 | No | NA | 7 | 0 | 1 | Not-determined | Genome-fragment | NA | NA | 0 |
| S10_vs_72075\|\|full | 2688 | No | NA | 6 | 0 | 4 | Not-determined | Genome-fragment | NA | NA | 0 |
| S10_vs_72116\|\|full | 1624 | No | NA | 3 | 0 | 0 | Not-determined | Genome-fragment | NA | NA | 0 |
| S11_extracted_k141_8464 | 3511 | No | NA | 4 | 0 | 2 | Low-quality | Genome-fragment | 6.79 | AAI-based (medium-confidence) | 0 |
| S10_vs_72263\|\|full | 1556 | No | NA | 3 | 0 | 2 | Not-determined | Genome-fragment | NA | NA | 0 |
| S10_vs_72483\|\|full | 1239 | No | NA | 5 | 0 | 0 | Not-determined | Genome-fragment | NA | NA | 0 |
| S1_extracted_k141_121243 | 3511 | No | NA | 6 | 1 | 0 | Low-quality | Genome-fragment | 6.71 | AAI-based (high-confidence) | 0 |
| S10_vs_72981\|\|full | 5903 | No | NA | 7 | 0 | 3 | Not-determined | Genome-fragment | NA | NA | 0 |
| S10_vs_34730\|\|full | 3511 | No | NA | 3 | 0 | 1 | Low-quality | Genome-fragment | 13.48 | AAI-based (medium-confidence) | 0 |
| S12_vs_k141_164355\|\|full | 3511 | Yes | 2487 | 6 | 1 | 1 | Low-quality | Genome-fragment | 1.68 | HMM-based (lower-bound) | 29.17 |
| S10_vs_73238\|\|full | 1499 | No | NA | 4 | 0 | 0 | Not-determined | Genome-fragment | NA | NA | 0 |
| S8_vs_k141_62016\|\|full | 3511 | No | NA | 3 | 0 | 2 | Low-quality | Genome-fragment | 2.33 | AAI-based (medium-confidence) | 0 |
| S10_vs_73521\|\|full | 2152 | No | NA | 5 | 0 | 1 | Not-determined | Genome-fragment | NA | NA | 0 |
| S11_vs_k141_69823\|\|full | 3512 | No | NA | 4 | 0 | 0 | Low-quality | Genome-fragment | 3.04 | AAI-based (medium-confidence) | 0 |
| S10_vs_73688\|\|full | 6164 | No | NA | 7 | 0 | 1 | Not-determined | Genome-fragment | NA | NA | 0 |
| S13_extracted_93230 | 3512 | No | NA | 4 | 1 | 0 | Low-quality | Genome-fragment | 3.49 | HMM-based (lower-bound) | 0 |
| S10_vs_73794\|\|full | 5985 | No | NA | 10 | 0 | 8 | Not-determined | Genome-fragment | NA | NA | 0 |
| S10_vs_73878\|\|full | 2226 | No | NA | 3 | 0 | 0 | Not-determined | Genome-fragment | NA | NA | 0 |
| S10_vs_73979\|\|full | 11934 | No | NA | 17 | 0 | 2 | Not-determined | Genome-fragment | NA | NA | 0 |
| S10_vs_74019\|\|full | 14211 | No | NA | 13 | 0 | 9 | Not-determined | Genome-fragment | NA | NA | 0 |
| S10_vs_74028\|\|full | 1914 | No | NA | 3 | 0 | 2 | Not-determined | Genome-fragment | NA | NA | 0 |
| S14_vs_k141_85422\|\|full | 3512 | No | NA | 6 | 0 | 3 | Low-quality | Genome-fragment | 2.4 | AAI-based (medium-confidence) | 0 |
| S10_vs_74110\|\|full | 2224 | No | NA | 8 | 0 | 1 | Not-determined | Genome-fragment | NA | NA | 0 |
| S10_vs_74216\|\|full | 1658 | No | NA | 2 | 0 | 2 | Not-determined | Genome-fragment | NA | NA | 0 |
| S3_extracted_k141_81731 | 3512 | No | NA | 9 | 7 | 0 | Low-quality | Genome-fragment | 11.58 | AAI-based (high-confidence) | 0 |
| S10_vs_74463\|\|full | 1324 | No | NA | 2 | 0 | 0 | Not-determined | Genome-fragment | NA | NA | 0 |
| S3_vs_k141_17932\|\|full | 3514 | No | NA | 7 | 3 | 0 | Low-quality | Genome-fragment | 7.56 | AAI-based (high-confidence) | 0 |
| S5_vs_k141_92039\|\|full | 3514 | No | NA | 5 | 0 | 0 | Low-quality | Genome-fragment | 2.5 | AAI-based (high-confidence) | 0 |
| S10_vs_74899\|\|full | 2137 | No | NA | 4 | 0 | 0 | Not-determined | Genome-fragment | NA | NA | 0 |
| S10_vs_75035\|\|full | 9003 | No | NA | 8 | 0 | 2 | Not-determined | Genome-fragment | NA | NA | 0 |
| S10_vs_75163\|\|full | 5536 | No | NA | 5 | 0 | 1 | Not-determined | Genome-fragment | NA | NA | 0 |
| S12_vs_k141_79219\|\|full | 3514 | No | NA | 4 | 0 | 2 | Low-quality | Genome-fragment | 5.95 | AAI-based (high-confidence) | 0 |
| S5_vs_k141_10868\|\|full | 3515 | No | NA | 8 | 4 | 0 | Low-quality | Genome-fragment | 5.68 | AAI-based (high-confidence) | 0 |
| S7_vs_94334\|\|full | 3515 | Yes | 2495 | 7 | 2 | 1 | Low-quality | Genome-fragment | 4.07 | AAI-based (high-confidence) | 29.02 |
| S4_extracted_952 | 3515 | No | NA | 3 | 0 | 0 | Low-quality | Genome-fragment | 4 | AAI-based (high-confidence) | 0 |
| S10_vs_75682\|\|full | 2436 | No | NA | 3 | 0 | 1 | Not-determined | Genome-fragment | NA | NA | 0 |
| S10_vs_75757\|\|full | 3725 | No | NA | 5 | 0 | 4 | Not-determined | Genome-fragment | NA | NA | 0 |
| S1_vs_k141_42188\|\|full | 3516 | No | NA | 4 | 1 | 0 | Low-quality | Genome-fragment | 1.91 | HMM-based (lower-bound) | 0 |
| S10_vs_75931\|\|full | 1443 | No | NA | 2 | 0 | 0 | Not-determined | Genome-fragment | NA | NA | 0 |
| S10_vs_75935\|\|full | 2218 | No | NA | 4 | 0 | 1 | Not-determined | Genome-fragment | NA | NA | 0 |
| S10_vs_75970\|\|full | 1531 | No | NA | 3 | 0 | 0 | Not-determined | Genome-fragment | NA | NA | 0 |
| S7_vs_28911\|\|full | 3518 | No | NA | 4 | 0 | 3 | Low-quality | Genome-fragment | 1.64 | AAI-based (high-confidence) | 0 |
| S16_vs_161763\|\|full | 3518 | No | NA | 2 | 0 | 2 | Low-quality | Genome-fragment | 3.55 | AAI-based (medium-confidence) | 0 |
| S10_vs_76048\|\|full | 1266 | No | NA | 3 | 0 | 0 | Not-determined | Genome-fragment | NA | NA | 0 |
| S10_vs_76213\|\|full | 2629 | No | NA | 4 | 0 | 1 | Not-determined | Genome-fragment | NA | NA | 0 |
| S11_vs_k141_92463\|\|full | 3519 | No | NA | 4 | 0 | 0 | Low-quality | Genome-fragment | 5.83 | AAI-based (medium-confidence) | 0 |
| S16_extracted_145160 | 3519 | No | NA | 8 | 0 | 0 | Low-quality | Genome-fragment | 8.42 | AAI-based (high-confidence) | 0 |
| S9_vs_k141_66858\|\|full | 3520 | No | NA | 7 | 4 | 0 | Low-quality | Genome-fragment | 8 | AAI-based (high-confidence) | 0 |
| S13_vs_47059\|\|full | 3520 | Yes | 1129 | 5 | 1 | 3 | Low-quality | Genome-fragment | 0.61 | HMM-based (lower-bound) | 67.93 |
| S7_extracted_74678 | 3520 | No | NA | 4 | 0 | 2 | Low-quality | Genome-fragment | 2.1 | AAI-based (medium-confidence) | 0 |
| S10_vs_76660\|\|full | 3617 | No | NA | 4 | 0 | 1 | Not-determined | Genome-fragment | NA | NA | 0 |
| S10_vs_76664\|\|full | 3041 | No | NA | 3 | 0 | 3 | Not-determined | Genome-fragment | NA | NA | 0 |
| S5_extracted_k141_20685 | 3521 | No | NA | 2 | 0 | 2 | Low-quality | Genome-fragment | 1.59 | AAI-based (high-confidence) | 0 |
| S10_vs_76854\|\|full | 2295 | No | NA | 4 | 0 | 3 | Not-determined | Genome-fragment | NA | NA | 0 |
| S5_vs_k141_116062\|\|full | 3521 | Yes | 2210 | 5 | 2 | 1 | Low-quality | Genome-fragment | 4.68 | AAI-based (high-confidence) | 37.23 |
| S10_vs_77004\|\|full | 35621 | No | NA | 49 | 16 | 1 | High-quality | High-quality | 97.19 | AAI-based (medium-confidence) | 0 |
| S11_vs_k141_62458\|\|full | 3522 | No | NA | 3 | 2 | 0 | Low-quality | Genome-fragment | 9.4 | AAI-based (high-confidence) | 0 |
| S10_vs_77228\|\|full | 6978 | No | NA | 9 | 0 | 5 | Not-determined | Genome-fragment | NA | NA | 0 |
| S5_vs_k141_46797\|\|full | 3522 | No | NA | 5 | 4 | 0 | Low-quality | Genome-fragment | 5.27 | HMM-based (lower-bound) | 0 |
| S10_vs_77403\|\|full | 2233 | No | NA | 3 | 0 | 1 | Not-determined | Genome-fragment | NA | NA | 0 |
| S10_vs_77554\|\|full | 1986 | No | NA | 3 | 0 | 3 | Not-determined | Genome-fragment | NA | NA | 0 |
| S10_vs_77656\|\|full | 2366 | No | NA | 2 | 0 | 2 | Not-determined | Genome-fragment | NA | NA | 0 |
| S12_vs_k141_137132\|\|full | 3522 | No | NA | 5 | 0 | 0 | Low-quality | Genome-fragment | 6.26 | AAI-based (medium-confidence) | 0 |
| S6_extracted_k141_8128 | 3522 | No | NA | 1 | 0 | 1 | Low-quality | Genome-fragment | 3.21 | AAI-based (high-confidence) | 0 |
| S6_vs_k141_3522\|\|full | 3522 | No | NA | 5 | 0 | 0 | Low-quality | Genome-fragment | 6 | AAI-based (medium-confidence) | 0 |
| S10_vs_77969\|\|full | 3152 | No | NA | 4 | 0 | 0 | Not-determined | Genome-fragment | NA | NA | 0 |
| S10_vs_78080\|\|full | 2029 | No | NA | 4 | 0 | 1 | Not-determined | Genome-fragment | NA | NA | 0 |
| S10_vs_78098\|\|full | 14912 | No | NA | 15 | 0 | 2 | Not-determined | Genome-fragment | NA | NA | 0 |
| S10_vs_78217\|\|full | 3567 | No | NA | 3 | 0 | 3 | Not-determined | Genome-fragment | NA | NA | 0 |
| S17_extracted_k141_68541 | 3524 | No | NA | 5 | 1 | 0 | Low-quality | Genome-fragment | 3.22 | AAI-based (high-confidence) | 0 |
| S17_vs_k141_175357\|\|full | 3524 | Yes | 2223 | 3 | 1 | 2 | Low-quality | Genome-fragment | 0.65 | HMM-based (lower-bound) | 36.92 |
| S10_vs_78490\|\|full | 1920 | No | NA | 3 | 0 | 0 | Not-determined | Genome-fragment | NA | NA | 0 |
| S10_vs_78539\|\|full | 3833 | No | NA | 3 | 0 | 3 | Not-determined | Genome-fragment | NA | NA | 0 |
| S10_vs_78565\|\|full | 1903 | No | NA | 2 | 0 | 1 | Not-determined | Genome-fragment | NA | NA | 0 |
| S11_vs_k141_13548\|\|full | 3524 | No | NA | 4 | 1 | 0 | Low-quality | Genome-fragment | 2.67 | AAI-based (high-confidence) | 0 |
| S10_vs_78657\|\|full | 2658 | No | NA | 3 | 0 | 3 | Not-determined | Genome-fragment | NA | NA | 0 |
| S10_vs_44590\|\|full | 3524 | No | NA | 5 | 0 | 1 | Low-quality | Genome-fragment | 5.55 | AAI-based (medium-confidence) | 0 |
| S4_extracted_6662 | 3524 | No | NA | 5 | 0 | 0 | Low-quality | Genome-fragment | 3.75 | AAI-based (high-confidence) | 0 |
| S13_extracted_111355 | 3525 | No | NA | 3 | 0 | 3 | Low-quality | Genome-fragment | 1.61 | AAI-based (high-confidence) | 0 |
| S10_vs_79237\|\|full | 3470 | No | NA | 5 | 0 | 4 | Not-determined | Genome-fragment | NA | NA | 0 |
| S14_vs_k141_90506\|\|full | 3525 | No | NA | 10 | 2 | 0 | Low-quality | Genome-fragment | 5.12 | HMM-based (lower-bound) | 0 |
| S10_vs_79418\|\|full | 1113 | No | NA | 3 | 0 | 0 | Not-determined | Genome-fragment | NA | NA | 0 |
| S10_vs_79518\|\|full | 2518 | No | NA | 2 | 0 | 1 | Not-determined | Genome-fragment | NA | NA | 0 |
| S14_vs_k141_47256\|\|full | 3525 | No | NA | 6 | 5 | 0 | Low-quality | Genome-fragment | 7.52 | AAI-based (medium-confidence) | 0 |
| S7_vs_92020\|\|full | 3525 | No | NA | 2 | 0 | 1 | Low-quality | Genome-fragment | 1.77 | AAI-based (medium-confidence) | 0 |
| S9_vs_k141_76046\|\|full | 3526 | No | NA | 5 | 0 | 1 | Low-quality | Genome-fragment | 6.81 | AAI-based (medium-confidence) | 0 |
| S5_vs_k141_47106\|\|full | 3526 | No | NA | 7 | 0 | 0 | Low-quality | Genome-fragment | 5.84 | AAI-based (medium-confidence) | 0 |
| S16_extracted_47381 | 3526 | No | NA | 7 | 2 | 0 | Low-quality | Genome-fragment | 7.29 | AAI-based (high-confidence) | 0 |
| S10_vs_80216\|\|full | 4378 | No | NA | 12 | 0 | 0 | Not-determined | Genome-fragment | NA | NA | 0 |
| S16_extracted_171437 | 3526 | No | NA | 3 | 0 | 3 | Low-quality | Genome-fragment | 0.5 | AAI-based (medium-confidence) | 0 |
| S12_extracted_k141_9917 | 3526 | No | NA | 4 | 3 | 0 | Low-quality | Genome-fragment | 8.77 | AAI-based (high-confidence) | 0 |
| S10_vs_80381\|\|full | 1902 | No | NA | 4 | 0 | 1 | Not-determined | Genome-fragment | NA | NA | 0 |
| S10_vs_80409\|\|full | 6728 | No | NA | 5 | 0 | 3 | Not-determined | Genome-fragment | NA | NA | 0 |
| S5_vs_k141_127298\|\|full | 3527 | No | NA | 3 | 0 | 2 | Low-quality | Genome-fragment | 4.29 | AAI-based (medium-confidence) | 0 |
| S10_vs_80427\|\|full | 2872 | No | NA | 3 | 0 | 1 | Not-determined | Genome-fragment | NA | NA | 0 |
| S7_extracted_52197 | 3528 | No | NA | 7 | 3 | 0 | Low-quality | Genome-fragment | 2.43 | AAI-based (high-confidence) | 0 |
| S10_vs_80512\|\|full | 7144 | No | NA | 9 | 0 | 0 | Not-determined | Genome-fragment | NA | NA | 0 |
| S16_extracted_3897 | 3528 | No | NA | 2 | 0 | 2 | Low-quality | Genome-fragment | 0.32 | AAI-based (medium-confidence) | 0 |
| S10_vs_80556\|\|full | 2953 | No | NA | 4 | 0 | 3 | Not-determined | Genome-fragment | NA | NA | 0 |
| S10_vs_80559\|\|full | 1172 | No | NA | 2 | 0 | 1 | Not-determined | Genome-fragment | NA | NA | 0 |
| S10_vs_80580\|\|full | 3037 | No | NA | 5 | 0 | 0 | Not-determined | Genome-fragment | NA | NA | 0 |
| S10_vs_1915\|\|full | 3528 | No | NA | 6 | 3 | 0 | Low-quality | Genome-fragment | 5.86 | AAI-based (high-confidence) | 0 |
| S10_vs_80595\|\|full | 1116 | No | NA | 2 | 0 | 1 | Not-determined | Genome-fragment | NA | NA | 0 |
| S10_vs_80923\|\|full | 1269 | No | NA | 2 | 0 | 0 | Not-determined | Genome-fragment | NA | NA | 0 |
| S10_vs_80967\|\|full | 2216 | No | NA | 3 | 0 | 2 | Not-determined | Genome-fragment | NA | NA | 0 |
| S13_vs_47217\|\|full | 3529 | No | NA | 5 | 0 | 3 | Low-quality | Genome-fragment | 8.32 | AAI-based (medium-confidence) | 0 |
| S14_vs_k141_5599\|\|full | 3529 | No | NA | 4 | 4 | 0 | Low-quality | Genome-fragment | 9.76 | AAI-based (high-confidence) | 0 |
| S5_vs_k141_75315\|\|full | 3529 | No | NA | 2 | 0 | 2 | Low-quality | Genome-fragment | 1.13 | AAI-based (medium-confidence) | 0 |
| S10_vs_81393\|\|full | 1469 | No | NA | 2 | 0 | 0 | Not-determined | Genome-fragment | NA | NA | 0 |
| S10_vs_81456\|\|full | 3634 | No | NA | 8 | 0 | 0 | Not-determined | Genome-fragment | NA | NA | 0 |
| S10_vs_81457\|\|full | 4089 | No | NA | 9 | 0 | 0 | Not-determined | Genome-fragment | NA | NA | 0 |
| S16_vs_44833\|\|full | 3529 | No | NA | 3 | 0 | 1 | Low-quality | Genome-fragment | 7.09 | AAI-based (medium-confidence) | 0 |
| S10_vs_81562\|\|full | 1415 | No | NA | 3 | 0 | 2 | Not-determined | Genome-fragment | NA | NA | 0 |
| S8_vs_k141_113739\|\|full | 3529 | No | NA | 5 | 3 | 0 | Low-quality | Genome-fragment | 9.25 | AAI-based (high-confidence) | 0 |
| S10_vs_81614\|\|full | 4319 | No | NA | 5 | 0 | 1 | Not-determined | Genome-fragment | NA | NA | 0 |
| S12_vs_k141_172315\|\|full | 3530 | No | NA | 8 | 5 | 0 | Low-quality | Genome-fragment | 7.34 | AAI-based (high-confidence) | 0 |
| S5_extracted_k141_63233 | 3532 | No | NA | 4 | 3 | 0 | Low-quality | Genome-fragment | 9.93 | AAI-based (high-confidence) | 0 |
| S10_vs_81933\|\|full | 1906 | No | NA | 3 | 0 | 0 | Not-determined | Genome-fragment | NA | NA | 0 |
| S10_vs_82011\|\|full | 3528 | No | NA | 4 | 0 | 0 | Not-determined | Genome-fragment | NA | NA | 0 |
| S10_vs_57366\|\|full | 3532 | No | NA | 7 | 3 | 0 | Low-quality | Genome-fragment | 7.59 | AAI-based (high-confidence) | 0 |
| S10_vs_82119\|\|full | 1205 | No | NA | 2 | 0 | 2 | Not-determined | Genome-fragment | NA | NA | 0 |
| S10_vs_82208\|\|full | 1877 | No | NA | 3 | 0 | 2 | Not-determined | Genome-fragment | NA | NA | 0 |
| S10_vs_82232\|\|full | 1278 | No | NA | 3 | 0 | 2 | Not-determined | Genome-fragment | NA | NA | 0 |
| S1_vs_k141_119857\|\|full | 3533 | No | NA | 10 | 0 | 0 | Low-quality | Genome-fragment | 7.61 | AAI-based (high-confidence) | 0 |
| S10_vs_41130\|\|full | 3533 | No | NA | 6 | 0 | 3 | Low-quality | Genome-fragment | 1.7 | AAI-based (high-confidence) | 0 |
| S17_vs_k141_62365\|\|full | 3534 | No | NA | 5 | 0 | 0 | Low-quality | Genome-fragment | 6.53 | AAI-based (medium-confidence) | 0 |
| S7_vs_37373\|\|full | 3534 | No | NA | 9 | 1 | 0 | Low-quality | Genome-fragment | 9.44 | AAI-based (high-confidence) | 0 |
| S10_vs_82418\|\|full | 1611 | No | NA | 2 | 0 | 2 | Not-determined | Genome-fragment | NA | NA | 0 |
| S10_vs_82512\|\|full | 1901 | No | NA | 2 | 0 | 1 | Not-determined | Genome-fragment | NA | NA | 0 |
| S10_vs_82571\|\|full | 4126 | No | NA | 5 | 0 | 2 | Not-determined | Genome-fragment | NA | NA | 0 |
| S10_vs_82667\|\|full | 2884 | No | NA | 4 | 0 | 1 | Not-determined | Genome-fragment | NA | NA | 0 |
| S10_vs_82963\|\|full | 1827 | No | NA | 3 | 0 | 2 | Not-determined | Genome-fragment | NA | NA | 0 |
| S11_vs_k141_19895\|\|full | 3535 | No | NA | 5 | 1 | 0 | Low-quality | Genome-fragment | 7.45 | AAI-based (high-confidence) | 0 |
| S10_vs_83040\|\|full | 4644 | No | NA | 6 | 0 | 1 | Not-determined | Genome-fragment | NA | NA | 0 |
| S10_vs_83052\|\|full | 3556 | No | NA | 5 | 0 | 4 | Not-determined | Genome-fragment | NA | NA | 0 |
| S5_vs_k141_25165\|\|full | 3535 | No | NA | 5 | 0 | 2 | Low-quality | Genome-fragment | 2.58 | AAI-based (high-confidence) | 0 |
| S10_vs_83289\|\|full | 1768 | No | NA | 2 | 0 | 2 | Not-determined | Genome-fragment | NA | NA | 0 |
| S10_vs_83342\|\|full | 2645 | No | NA | 3 | 0 | 2 | Not-determined | Genome-fragment | NA | NA | 0 |
| S10_vs_83366\|\|full | 2208 | No | NA | 3 | 0 | 2 | Not-determined | Genome-fragment | NA | NA | 0 |
| S10_vs_83462\|\|full | 2099 | No | NA | 3 | 0 | 2 | Not-determined | Genome-fragment | NA | NA | 0 |
| S7_vs_115285\|\|full | 3535 | No | NA | 5 | 4 | 0 | Low-quality | Genome-fragment | 9.15 | AAI-based (high-confidence) | 0 |
| S11_vs_k141_63390\|\|full | 3536 | No | NA | 3 | 0 | 0 | Low-quality | Genome-fragment | 8.76 | AAI-based (high-confidence) | 0 |
| S10_vs_110709\|\|full | 3536 | No | NA | 3 | 0 | 0 | Low-quality | Genome-fragment | 6.8 | AAI-based (medium-confidence) | 0 |
| S17_vs_k141_163064\|\|full | 3538 | No | NA | 7 | 1 | 0 | Low-quality | Genome-fragment | 9.95 | AAI-based (high-confidence) | 0 |
| S10_vs_83748\|\|full | 1459 | No | NA | 2 | 0 | 0 | Not-determined | Genome-fragment | NA | NA | 0 |
| S10_vs_83796\|\|full | 5319 | No | NA | 3 | 0 | 2 | Not-determined | Genome-fragment | NA | NA | 0 |
| S7_vs_33339\|\|full | 3538 | No | NA | 9 | 0 | 1 | Low-quality | Genome-fragment | 9.59 | AAI-based (high-confidence) | 0 |
| S16_extracted_55934 | 3539 | No | NA | 2 | 0 | 1 | Low-quality | Genome-fragment | 5.82 | AAI-based (medium-confidence) | 0 |
| S10_vs_83918\|\|full | 1691 | No | NA | 3 | 0 | 0 | Not-determined | Genome-fragment | NA | NA | 0 |
| S9_extracted_k141_38540 | 3540 | No | NA | 6 | 5 | 0 | Low-quality | Genome-fragment | 5.9 | AAI-based (high-confidence) | 0 |
| S10_vs_84051\|\|full | 2209 | No | NA | 4 | 0 | 2 | Not-determined | Genome-fragment | NA | NA | 0 |
| S10_vs_84124\|\|full | 2746 | No | NA | 3 | 0 | 2 | Not-determined | Genome-fragment | NA | NA | 0 |
| S10_vs_84150\|\|full | 19446 | No | NA | 22 | 0 | 6 | Not-determined | Genome-fragment | NA | NA | 0 |
| S7_vs_85186\|\|full | 3540 | No | NA | 5 | 0 | 3 | Low-quality | Genome-fragment | 4.49 | AAI-based (medium-confidence) | 0 |
| S10_extracted_15854 | 3540 | No | NA | 4 | 2 | 0 | Low-quality | Genome-fragment | 8.86 | AAI-based (high-confidence) | 0 |
| S10_vs_84686\|\|full | 2211 | No | NA | 4 | 0 | 0 | Not-determined | Genome-fragment | NA | NA | 0 |
| S8_vs_k141_141645\|\|full | 3540 | No | NA | 4 | 0 | 0 | Low-quality | Genome-fragment | 5.52 | AAI-based (medium-confidence) | 0 |
| S10_vs_84694\|\|full | 1999 | No | NA | 3 | 0 | 2 | Not-determined | Genome-fragment | NA | NA | 0 |
| S10_vs_84713\|\|full | 1765 | No | NA | 2 | 0 | 1 | Not-determined | Genome-fragment | NA | NA | 0 |
| S8_vs_k141_25709\|\|full | 3540 | No | NA | 5 | 0 | 1 | Low-quality | Genome-fragment | 5.45 | AAI-based (medium-confidence) | 0 |
| S10_vs_85083\|\|full | 2217 | No | NA | 4 | 0 | 2 | Not-determined | Genome-fragment | NA | NA | 0 |
| S10_vs_85085\|\|full | 1903 | No | NA | 3 | 0 | 1 | Not-determined | Genome-fragment | NA | NA | 0 |
| S10_vs_85090\|\|full | 2083 | No | NA | 3 | 0 | 0 | Not-determined | Genome-fragment | NA | NA | 0 |
| S10_vs_85347\|\|full | 1389 | No | NA | 2 | 0 | 0 | Not-determined | Genome-fragment | NA | NA | 0 |
| S14_extracted_k141_106915 | 3541 | No | NA | 3 | 1 | 0 | Low-quality | Genome-fragment | 1.41 | HMM-based (lower-bound) | 0 |
| S10_vs_85387\|\|full | 1456 | No | NA | 2 | 0 | 2 | Not-determined | Genome-fragment | NA | NA | 0 |
| S1_vs_k141_132998\|\|full | 3541 | No | NA | 6 | 6 | 0 | Low-quality | Genome-fragment | 9.76 | AAI-based (high-confidence) | 0 |
| S7_vs_48803\|\|full | 3541 | No | NA | 7 | 0 | 1 | Low-quality | Genome-fragment | 6.02 | AAI-based (medium-confidence) | 0 |
| S10_vs_83907\|\|full | 3541 | No | NA | 4 | 0 | 0 | Low-quality | Genome-fragment | 6.58 | AAI-based (medium-confidence) | 0 |
| S10_vs_85768\|\|full | 2739 | No | NA | 3 | 0 | 1 | Not-determined | Genome-fragment | NA | NA | 0 |
| S10_vs_85782\|\|full | 2026 | No | NA | 4 | 0 | 1 | Not-determined | Genome-fragment | NA | NA | 0 |
| S3_vs_k141_52422\|\|full | 3542 | No | NA | 3 | 0 | 0 | Low-quality | Genome-fragment | 2.33 | AAI-based (high-confidence) | 0 |
| S10_vs_85834\|\|full | 1106 | No | NA | 3 | 0 | 0 | Not-determined | Genome-fragment | NA | NA | 0 |
| S5_vs_k141_91652\|\|full | 3542 | No | NA | 5 | 0 | 0 | Low-quality | Genome-fragment | 7.96 | AAI-based (high-confidence) | 0 |
| S7_vs_37218\|\|full | 3542 | No | NA | 3 | 0 | 1 | Low-quality | Genome-fragment | 5.61 | AAI-based (medium-confidence) | 0 |
| S17_vs_k141_83134\|\|full | 3544 | No | NA | 3 | 0 | 0 | Low-quality | Genome-fragment | 6.09 | AAI-based (medium-confidence) | 0 |
| S7_extracted_121224 | 3544 | No | NA | 7 | 0 | 0 | Low-quality | Genome-fragment | 3.62 | AAI-based (medium-confidence) | 0 |
| S17_extracted_k141_102816 | 3545 | No | NA | 4 | 2 | 0 | Low-quality | Genome-fragment | 16.24 | AAI-based (high-confidence) | 0 |
| S14_vs_k141_56710\|\|full | 3545 | No | NA | 7 | 3 | 0 | Low-quality | Genome-fragment | 7.91 | AAI-based (medium-confidence) | 0 |
| S14_vs_k141_63959\|\|full | 3545 | No | NA | 9 | 0 | 0 | Low-quality | Genome-fragment | 7.05 | AAI-based (high-confidence) | 0 |
| S10_vs_86300\|\|full | 2148 | No | NA | 3 | 0 | 1 | Not-determined | Genome-fragment | NA | NA | 0 |
| S10_vs_86458\|\|full | 2013 | No | NA | 3 | 0 | 3 | Not-determined | Genome-fragment | NA | NA | 0 |
| S10_vs_86604\|\|full | 1107 | No | NA | 3 | 0 | 1 | Not-determined | Genome-fragment | NA | NA | 0 |
| S10_vs_86673\|\|full | 3029 | No | NA | 6 | 0 | 4 | Not-determined | Genome-fragment | NA | NA | 0 |
| S10_vs_86711\|\|full | 1652 | No | NA | 4 | 0 | 1 | Not-determined | Genome-fragment | NA | NA | 0 |
| S1_vs_k141_93741\|\|full | 3546 | No | NA | 3 | 2 | 0 | Low-quality | Genome-fragment | 10.37 | AAI-based (high-confidence) | 0 |
| S10_vs_86745\|\|full | 2016 | No | NA | 3 | 0 | 0 | Not-determined | Genome-fragment | NA | NA | 0 |
| S10_vs_86852\|\|full | 5751 | No | NA | 5 | 0 | 5 | Not-determined | Genome-fragment | NA | NA | 0 |
| S10_vs_86864\|\|full | 5274 | No | NA | 5 | 0 | 3 | Not-determined | Genome-fragment | NA | NA | 0 |
| S7_vs_16113\|\|full | 3546 | No | NA | 9 | 2 | 0 | Low-quality | Genome-fragment | 7.49 | AAI-based (high-confidence) | 0 |
| S10_vs_87051\|\|full | 1195 | No | NA | 4 | 0 | 1 | Not-determined | Genome-fragment | NA | NA | 0 |
| S10_vs_87162\|\|full | 4294 | No | NA | 4 | 0 | 3 | Not-determined | Genome-fragment | NA | NA | 0 |
| S10_vs_87250\|\|full | 1488 | No | NA | 2 | 0 | 1 | Not-determined | Genome-fragment | NA | NA | 0 |
| S16_vs_104423\|\|full | 3546 | No | NA | 3 | 0 | 1 | Low-quality | Genome-fragment | 6.81 | AAI-based (medium-confidence) | 0 |
| S10_vs_87378\|\|full | 3714 | No | NA | 4 | 0 | 2 | Not-determined | Genome-fragment | NA | NA | 0 |
| S10_vs_87561\|\|full | 1239 | No | NA | 3 | 0 | 1 | Not-determined | Genome-fragment | NA | NA | 0 |
| S10_vs_87600\|\|full | 2282 | No | NA | 3 | 0 | 3 | Not-determined | Genome-fragment | NA | NA | 0 |
| S10_extracted_38796 | 3546 | No | NA | 6 | 0 | 0 | Low-quality | Genome-fragment | 7.35 | AAI-based (high-confidence) | 0 |
| S10_vs_87837\|\|full | 1591 | No | NA | 2 | 0 | 0 | Not-determined | Genome-fragment | NA | NA | 0 |
| S8_extracted_k141_159406 | 3546 | No | NA | 7 | 1 | 0 | Low-quality | Genome-fragment | 4.1 | HMM-based (lower-bound) | 0 |
| S10_vs_87933\|\|full | 3182 | No | NA | 3 | 0 | 1 | Not-determined | Genome-fragment | NA | NA | 0 |
| S5_extracted_k141_138657 | 3547 | No | NA | 9 | 4 | 0 | Low-quality | Genome-fragment | 7.34 | AAI-based (high-confidence) | 0 |
| S1_vs_k141_96955\|\|full | 3548 | No | NA | 2 | 0 | 2 | Low-quality | Genome-fragment | 2.32 | AAI-based (medium-confidence) | 0 |
| S1_vs_k141_62356\|\|full | 3548 | No | NA | 7 | 4 | 0 | Low-quality | Genome-fragment | 6.59 | AAI-based (medium-confidence) | 0 |
| S10_vs_88120\|\|full | 1789 | No | NA | 3 | 0 | 1 | Not-determined | Genome-fragment | NA | NA | 0 |
| S3_extracted_k141_42121 | 3548 | No | NA | 3 | 0 | 1 | Low-quality | Genome-fragment | 5.82 | AAI-based (medium-confidence) | 0 |
| S10_vs_88397\|\|full | 20563 | No | NA | 25 | 0 | 2 | Not-determined | Genome-fragment | NA | NA | 0 |
| S10_vs_88484\|\|full | 1582 | No | NA | 5 | 0 | 0 | Not-determined | Genome-fragment | NA | NA | 0 |
| S10_vs_88546\|\|full | 1441 | No | NA | 4 | 0 | 1 | Not-determined | Genome-fragment | NA | NA | 0 |
| S5_vs_k141_3327\|\|full | 3548 | No | NA | 4 | 2 | 0 | Low-quality | Genome-fragment | 4.36 | HMM-based (lower-bound) | 0 |
| S10_vs_88872\|\|full | 1177 | No | NA | 2 | 0 | 2 | Not-determined | Genome-fragment | NA | NA | 0 |
| S10_vs_89108\|\|full | 4614 | No | NA | 4 | 0 | 3 | Not-determined | Genome-fragment | NA | NA | 0 |
| S14_vs_k141_40553\|\|full | 3549 | No | NA | 5 | 0 | 3 | Low-quality | Genome-fragment | 5.99 | AAI-based (high-confidence) | 0 |
| S1_vs_k141_24729\|\|full | 3549 | No | NA | 7 | 3 | 0 | Low-quality | Genome-fragment | 8.52 | AAI-based (medium-confidence) | 0 |
| S10_vs_89399\|\|full | 2580 | No | NA | 4 | 0 | 0 | Not-determined | Genome-fragment | NA | NA | 0 |
| S10_vs_89462\|\|full | 1663 | No | NA | 3 | 0 | 1 | Not-determined | Genome-fragment | NA | NA | 0 |
| S16_extracted_57565 | 3550 | No | NA | 3 | 0 | 0 | Low-quality | Genome-fragment | 3.88 | AAI-based (medium-confidence) | 0 |
| S11_vs_k141_84792\|\|full | 3551 | No | NA | 3 | 1 | 0 | Low-quality | Genome-fragment | 3.13 | AAI-based (high-confidence) | 0 |
| S14_extracted_k141_75344 | 3551 | No | NA | 6 | 0 | 0 | Low-quality | Genome-fragment | 5.57 | AAI-based (high-confidence) | 0 |
| S10_vs_89690\|\|full | 1347 | No | NA | 3 | 0 | 1 | Not-determined | Genome-fragment | NA | NA | 0 |
| S16_vs_148675\|\|full | 3551 | No | NA | 7 | 1 | 0 | Low-quality | Genome-fragment | 3.01 | AAI-based (high-confidence) | 0 |
| S10_vs_89767\|\|full | 6481 | No | NA | 3 | 0 | 3 | Not-determined | Genome-fragment | NA | NA | 0 |
| S10_extracted_106193 | 3551 | No | NA | 3 | 0 | 0 | Low-quality | Genome-fragment | 2.11 | AAI-based (high-confidence) | 0 |
| S12_vs_k141_99415\|\|full | 3551 | No | NA | 6 | 3 | 0 | Low-quality | Genome-fragment | 8.32 | AAI-based (high-confidence) | 0 |
| S10_vs_90084\|\|full | 8671 | No | NA | 9 | 0 | 6 | Not-determined | Genome-fragment | NA | NA | 0 |
| S10_vs_90209\|\|full | 13243 | No | NA | 11 | 0 | 9 | Not-determined | Genome-fragment | NA | NA | 0 |
| S10_vs_90264\|\|full | 6927 | No | NA | 11 | 0 | 2 | Not-determined | Genome-fragment | NA | NA | 0 |
| S8_extracted_k141_174518 | 3551 | No | NA | 3 | 0 | 1 | Low-quality | Genome-fragment | 0.31 | AAI-based (medium-confidence) | 0 |
| S10_vs_90365\|\|full | 4926 | No | NA | 5 | 0 | 1 | Not-determined | Genome-fragment | NA | NA | 0 |
| S10_vs_90452\|\|full | 4404 | No | NA | 3 | 0 | 1 | Not-determined | Genome-fragment | NA | NA | 0 |
| S10_vs_90497\|\|full | 3746 | No | NA | 3 | 0 | 3 | Not-determined | Genome-fragment | NA | NA | 0 |
| S9_vs_k141_42276\|\|full | 3552 | No | NA | 5 | 0 | 0 | Low-quality | Genome-fragment | 2.04 | AAI-based (high-confidence) | 0 |
| S10_vs_90702\|\|full | 4748 | No | NA | 5 | 0 | 4 | Not-determined | Genome-fragment | NA | NA | 0 |
| S5_extracted_k141_99370 | 3552 | No | NA | 6 | 2 | 0 | Low-quality | Genome-fragment | 2.17 | AAI-based (high-confidence) | 0 |
| S10_vs_91346\|\|full | 3286 | No | NA | 4 | 0 | 3 | Not-determined | Genome-fragment | NA | NA | 0 |
| S8_extracted_k141_139525 | 3552 | No | NA | 3 | 0 | 2 | Low-quality | Genome-fragment | 1.68 | AAI-based (high-confidence) | 0 |
| S7_extracted_68404 | 3553 | No | NA | 9 | 0 | 0 | Low-quality | Genome-fragment | 5.6 | AAI-based (high-confidence) | 0 |
| S17_extracted_k141_39576 | 3554 | No | NA | 1 | 0 | 1 | Low-quality | Genome-fragment | 2.03 | AAI-based (medium-confidence) | 0 |
| S7_extracted_18164 | 3554 | No | NA | 6 | 0 | 0 | Low-quality | Genome-fragment | 7.19 | AAI-based (high-confidence) | 0 |
| S8_vs_k141_164500\|\|full | 3554 | No | NA | 4 | 0 | 3 | Low-quality | Genome-fragment | 1.7 | AAI-based (medium-confidence) | 0 |
| S9_extracted_k141_20591 | 3555 | No | NA | 4 | 0 | 1 | Low-quality | Genome-fragment | 8.24 | AAI-based (medium-confidence) | 0 |

| | | | | | | | | | | | |
|---|---|---|---|---|---|---|---|---|---|---|---|
| S16_vs_9027\|\|full | 3555 | Yes | 2052 | 4 | 2 | 1 | Low-quality | Genome-fragment | 5.47 | AAI-based (medium-confidence) | 42.28 |
| S8_extracted_k141_18611 | 3555 | No | NA | 6 | 0 | 2 | Low-quality | Genome-fragment | 2.52 | AAI-based (medium-confidence) | 0 |
| S11_vs_k141_11326\|\|full | 3556 | No | NA | 4 | 0 | 0 | Low-quality | Genome-fragment | 8.62 | AAI-based (high-confidence) | 0 |
| S10_vs_92351\|\|full | 2142 | No | NA | 2 | 0 | 2 | Not-determined | Genome-fragment | NA | NA | 0 |
| S16_vs_77186\|\|full | 3556 | No | NA | 5 | 1 | 2 | Low-quality | Genome-fragment | 1.57 | HMM-based (lower-bound) | 0 |
| S16_vs_165124\|\|full | 3556 | No | NA | 2 | 0 | 1 | Low-quality | Genome-fragment | 1.47 | AAI-based (medium-confidence) | 0 |
| S5_extracted_k141_63767 | 3557 | No | NA | 4 | 0 | 0 | Low-quality | Genome-fragment | 12.21 | AAI-based (medium-confidence) | 0 |
| S10_vs_92770\|\|full | 2346 | No | NA | 3 | 0 | 2 | Not-determined | Genome-fragment | NA | NA | 0 |
| S5_extracted_k141_21354 | 3558 | No | NA | 2 | 0 | 0 | Low-quality | Genome-fragment | 5.9 | AAI-based (medium-confidence) | 0 |
| S5_vs_k141_3883\|\|full | 3558 | No | NA | 11 | 2 | 0 | Low-quality | Genome-fragment | 9.68 | AAI-based (high-confidence) | 0 |
| S10_vs_93084\|\|full | 1684 | No | NA | 3 | 0 | 0 | Not-determined | Genome-fragment | NA | NA | 0 |
| S16_vs_9727\|\|full | 3558 | No | NA | 4 | 2 | 0 | Low-quality | Genome-fragment | 9.48 | AAI-based (medium-confidence) | 0 |
| S10_vs_93431\|\|full | 9656 | No | NA | 6 | 0 | 4 | Not-determined | Genome-fragment | NA | NA | 0 |
| S10_vs_93479\|\|full | 2285 | No | NA | 3 | 0 | 0 | Not-determined | Genome-fragment | NA | NA | 0 |
| S10_vs_93578\|\|full | 3018 | No | NA | 4 | 0 | 4 | Not-determined | Genome-fragment | NA | NA | 0 |
| S10_vs_93603\|\|full | 2083 | No | NA | 3 | 0 | 2 | Not-determined | Genome-fragment | NA | NA | 0 |
| S10_vs_93697\|\|full | 1779 | No | NA | 3 | 0 | 1 | Not-determined | Genome-fragment | NA | NA | 0 |
| S17_extracted_k141_85774 | 3559 | No | NA | 4 | 0 | 0 | Low-quality | Genome-fragment | 5.18 | AAI-based (high-confidence) | 0 |
| S10_vs_93763\|\|full | 2239 | No | NA | 3 | 0 | 3 | Not-determined | Genome-fragment | NA | NA | 0 |
| S1_vs_k141_51040\|\|full | 3559 | No | NA | 5 | 0 | 2 | Low-quality | Genome-fragment | 8.31 | AAI-based (high-confidence) | 0 |
| S5_vs_k141_113319\|\|full | 3559 | Yes | 1566 | 6 | 1 | 1 | Low-quality | Genome-fragment | 1.67 | AAI-based (medium-confidence) | 56 |
| S16_extracted_130566 | 3559 | No | NA | 6 | 1 | 0 | Low-quality | Genome-fragment | 6.23 | AAI-based (high-confidence) | 0 |
| S10_vs_94138\|\|full | 6964 | No | NA | 11 | 0 | 4 | Not-determined | Genome-fragment | NA | NA | 0 |
| S10_vs_94170\|\|full | 1530 | No | NA | 2 | 0 | 1 | Not-determined | Genome-fragment | NA | NA | 0 |
| S10_vs_94317\|\|full | 2304 | No | NA | 3 | 0 | 2 | Not-determined | Genome-fragment | NA | NA | 0 |
| S10_vs_94363\|\|full | 1581 | No | NA | 3 | 0 | 1 | Not-determined | Genome-fragment | NA | NA | 0 |
| S17_vs_k141_107567\|\|full | 3560 | No | NA | 5 | 0 | 0 | Low-quality | Genome-fragment | 5.54 | AAI-based (high-confidence) | 0 |
| S14_extracted_k141_52026 | 3560 | No | NA | 4 | 3 | 0 | Low-quality | Genome-fragment | 11.73 | AAI-based (high-confidence) | 0 |
| S3_extracted_k141_12807 | 3560 | No | NA | 8 | 0 | 0 | Low-quality | Genome-fragment | 4.38 | AAI-based (medium-confidence) | 0 |
| S10_vs_95358\|\|full | 4256 | No | NA | 5 | 0 | 2 | Not-determined | Genome-fragment | NA | NA | 0 |
| S10_vs_95452\|\|full | 3689 | No | NA | 3 | 0 | 2 | Not-determined | Genome-fragment | NA | NA | 0 |
| S3_vs_k141_76269\|\|full | 3560 | No | NA | 5 | 2 | 0 | Low-quality | Genome-fragment | 6.15 | HMM-based (lower-bound) | 0 |
| S1_vs_k141_37669\|\|full | 3561 | No | NA | 4 | 0 | 0 | Low-quality | Genome-fragment | 4.66 | AAI-based (high-confidence) | 0 |
| S10_vs_95773\|\|full | 1345 | No | NA | 2 | 0 | 0 | Not-determined | Genome-fragment | NA | NA | 0 |
| S10_vs_48335\|\|full | 3561 | No | NA | 4 | 0 | 3 | Low-quality | Genome-fragment | 3.78 | AAI-based (medium-confidence) | 0 |
| S10_vs_96018\|\|full | 4215 | No | NA | 6 | 0 | 4 | Not-determined | Genome-fragment | NA | NA | 0 |
| S10_vs_96081\|\|full | 3169 | No | NA | 5 | 0 | 2 | Not-determined | Genome-fragment | NA | NA | 0 |
| S12_extracted_k141_98361 | 3561 | No | NA | 5 | 0 | 0 | Low-quality | Genome-fragment | 5.98 | AAI-based (medium-confidence) | 0 |
| S7_extracted_28322 | 3562 | No | NA | 3 | 0 | 1 | Low-quality | Genome-fragment | 5.21 | AAI-based (high-confidence) | 0 |
| S8_vs_k141_116256\|\|full | 3562 | No | NA | 3 | 0 | 2 | Low-quality | Genome-fragment | 1.79 | AAI-based (medium-confidence) | 0 |
| S10_vs_96277\|\|full | 8605 | No | NA | 9 | 0 | 2 | Not-determined | Genome-fragment | NA | NA | 0 |
| S10_vs_96340\|\|full | 17176 | No | NA | 18 | 0 | 11 | Not-determined | Genome-fragment | NA | NA | 0 |
| S16_extracted_128131 | 3563 | No | NA | 7 | 0 | 0 | Low-quality | Genome-fragment | 6.14 | AAI-based (high-confidence) | 0 |
| S10_vs_96422\|\|full | 1219 | No | NA | 4 | 0 | 0 | Not-determined | Genome-fragment | NA | NA | 0 |
| S10_vs_96462\|\|full | 1289 | No | NA | 3 | 0 | 2 | Not-determined | Genome-fragment | NA | NA | 0 |
| S10_extracted_32418 | 3563 | No | NA | 6 | 1 | 0 | Low-quality | Genome-fragment | 9.13 | AAI-based (high-confidence) | 0 |
| S10_vs_80447\|\|full | 3563 | No | NA | 5 | 0 | 1 | Low-quality | Genome-fragment | 8.32 | AAI-based (high-confidence) | 0 |
| S10_vs_96678\|\|full | 1869 | No | NA | 3 | 0 | 1 | Not-determined | Genome-fragment | NA | NA | 0 |
| S10_vs_96773\|\|full | 1517 | No | NA | 2 | 0 | 2 | Not-determined | Genome-fragment | NA | NA | 0 |
| S3_vs_k141_91859\|\|full | 3564 | No | NA | 3 | 1 | 0 | Low-quality | Genome-fragment | 3.72 | HMM-based (lower-bound) | 0 |
| S10_vs_96824\|\|full | 1046 | No | NA | 4 | 0 | 0 | Not-determined | Genome-fragment | NA | NA | 0 |
| S17_vs_k141_175826\|\|full | 3565 | No | NA | 5 | 0 | 3 | Low-quality | Genome-fragment | 2.05 | AAI-based (medium-confidence) | 0 |
| S10_vs_97143\|\|full | 1394 | No | NA | 2 | 0 | 1 | Not-determined | Genome-fragment | NA | NA | 0 |
| S16_extracted_172330 | 3565 | No | NA | 7 | 3 | 0 | Low-quality | Genome-fragment | 3.7 | AAI-based (high-confidence) | 0 |
| S10_vs_97320\|\|full | 1810 | No | NA | 2 | 0 | 1 | Not-determined | Genome-fragment | NA | NA | 0 |
| S10_vs_97366\|\|full | 3445 | No | NA | 4 | 0 | 1 | Not-determined | Genome-fragment | NA | NA | 0 |
| S10_vs_97420\|\|full | 1819 | No | NA | 3 | 0 | 1 | Not-determined | Genome-fragment | NA | NA | 0 |
| S10_vs_97425\|\|full | 1760 | No | NA | 4 | 0 | 1 | Not-determined | Genome-fragment | NA | NA | 0 |
| S10_vs_97712\|\|full | 1584 | No | NA | 2 | 0 | 1 | Not-determined | Genome-fragment | NA | NA | 0 |
| S13_extracted_94499 | 3566 | No | NA | 6 | 1 | 0 | Low-quality | Genome-fragment | 1.43 | HMM-based (lower-bound) | 0 |
| S10_vs_97823\|\|full | 2610 | No | NA | 2 | 0 | 1 | Not-determined | Genome-fragment | NA | NA | 0 |
| S10_vs_97996\|\|full | 2413 | No | NA | 5 | 0 | 0 | Not-determined | Genome-fragment | NA | NA | 0 |
| S10_vs_98006\|\|full | 2655 | No | NA | 3 | 0 | 0 | Not-determined | Genome-fragment | NA | NA | 0 |
| S10_vs_98317\|\|full | 1550 | No | NA | 2 | 0 | 2 | Not-determined | Genome-fragment | NA | NA | 0 |
| S10_vs_98325\|\|full | 1171 | No | NA | 3 | 0 | 1 | Not-determined | Genome-fragment | NA | NA | 0 |
| S10_vs_98340\|\|full | 3566 | No | NA | 3 | 0 | 2 | Not-determined | Genome-fragment | NA | NA | 0 |
| S10_vs_98407\|\|full | 2396 | No | NA | 4 | 0 | 2 | Not-determined | Genome-fragment | NA | NA | 0 |
| S5_extracted_k141_40839 | 3566 | No | NA | 13 | 1 | 0 | Low-quality | Genome-fragment | 2.28 | HMM-based (lower-bound) | 0 |
| S10_vs_98527\|\|full | 5110 | No | NA | 5 | 0 | 2 | Not-determined | Genome-fragment | NA | NA | 0 |
| S3_vs_k141_2557\|\|full | 3567 | No | NA | 3 | 3 | 0 | Low-quality | Genome-fragment | 5.94 | AAI-based (high-confidence) | 0 |
| S8_vs_k141_11928\|\|full | 3567 | No | NA | 4 | 0 | 0 | Low-quality | Genome-fragment | 7.01 | AAI-based (high-confidence) | 0 |
| S10_vs_98905\|\|full | 3529 | No | NA | 5 | 0 | 2 | Not-determined | Genome-fragment | NA | NA | 0 |
| S7_vs_5299\|\|full | 3568 | Yes | 666 | 3 | 1 | 1 | Low-quality | Genome-fragment | 1.14 | AAI-based (high-confidence) | 81.33 |
| S10_vs_98959\|\|full | 5448 | No | NA | 6 | 0 | 3 | Not-determined | Genome-fragment | NA | NA | 0 |
| S10_vs_99163\|\|full | 3066 | No | NA | 3 | 0 | 3 | Not-determined | Genome-fragment | NA | NA | 0 |
| S10_vs_99193\|\|full | 2101 | No | NA | 3 | 0 | 3 | Not-determined | Genome-fragment | NA | NA | 0 |
| S10_vs_99208\|\|full | 22537 | No | NA | 10 | 0 | 7 | Not-determined | Genome-fragment | NA | NA | 0 |
| S10_vs_99250\|\|full | 4846 | No | NA | 4 | 0 | 4 | Not-determined | Genome-fragment | NA | NA | 0 |
| S10_vs_99307\|\|full | 1712 | No | NA | 3 | 0 | 2 | Not-determined | Genome-fragment | NA | NA | 0 |
| S16_extracted_97679 | 3568 | No | NA | 6 | 2 | 0 | Low-quality | Genome-fragment | 6.71 | AAI-based (high-confidence) | 0 |
| S10_vs_99482\|\|full | 1777 | No | NA | 3 | 0 | 1 | Not-determined | Genome-fragment | NA | NA | 0 |
| S10_vs_99609\|\|full | 1196 | No | NA | 3 | 0 | 0 | Not-determined | Genome-fragment | NA | NA | 0 |
| S12_vs_k141_137880\|\|full | 3568 | Yes | 2800 | 10 | 2 | 1 | Low-quality | Genome-fragment | 7.72 | AAI-based (high-confidence) | 21.52 |
| S10_vs_99919\|\|full | 11506 | No | NA | 6 | 0 | 3 | Not-determined | Genome-fragment | NA | NA | 0 |
| S10_vs_100090\|\|full | 2059 | No | NA | 2 | 0 | 0 | Not-determined | Genome-fragment | NA | NA | 0 |
| S10_vs_100131\|\|full | 1923 | No | NA | 2 | 0 | 0 | Not-determined | Genome-fragment | NA | NA | 0 |
| S10_vs_100313\|\|full | 7027 | No | NA | 8 | 0 | 3 | Not-determined | Genome-fragment | NA | NA | 0 |
| S10_vs_100391\|\|full | 1947 | No | NA | 2 | 0 | 2 | Not-determined | Genome-fragment | NA | NA | 0 |
| S16_extracted_5322 | 3569 | No | NA | 5 | 0 | 0 | Low-quality | Genome-fragment | 12.28 | AAI-based (medium-confidence) | 0 |
| S10_vs_100742\|\|full | 1516 | No | NA | 3 | 0 | 2 | Not-determined | Genome-fragment | NA | NA | 0 |
| S16_extracted_135679 | 3569 | No | NA | 6 | 0 | 0 | Low-quality | Genome-fragment | 6.41 | AAI-based (medium-confidence) | 0 |
| S16_vs_79346\|\|full | 3569 | No | NA | 4 | 2 | 0 | Low-quality | Genome-fragment | 8.46 | AAI-based (high-confidence) | 0 |
| S16_vs_172203\|\|full | 3569 | Yes | 1514 | 5 | 1 | 2 | Low-quality | Genome-fragment | 2.88 | HMM-based (lower-bound) | 57.58 |
| S8_extracted_k141_146501 | 3569 | No | NA | 2 | 0 | 1 | Low-quality | Genome-fragment | 1.83 | AAI-based (medium-confidence) | 0 |
| S8_vs_k141_160745\|\|full | 3569 | No | NA | 4 | 0 | 4 | Low-quality | Genome-fragment | 1.77 | AAI-based (medium-confidence) | 0 |
| S10_vs_101479\|\|full | 1093 | No | NA | 2 | 0 | 0 | Not-determined | Genome-fragment | NA | NA | 0 |
| S8_extracted_k141_45848 | 3570 | No | NA | 5 | 2 | 0 | Low-quality | Genome-fragment | 17.57 | AAI-based (medium-confidence) | 0 |
| S10_vs_101535\|\|full | 1560 | No | NA | 3 | 0 | 0 | Not-determined | Genome-fragment | NA | NA | 0 |
| S8_vs_k141_179386\|\|full | 3570 | No | NA | 4 | 0 | 1 | Low-quality | Genome-fragment | 1.14 | AAI-based (medium-confidence) | 0 |
| S14_extracted_k141_81094 | 3571 | No | NA | 4 | 0 | 1 | Low-quality | Genome-fragment | 6.1 | AAI-based (medium-confidence) | 0 |
| S7_extracted_83194 | 3571 | No | NA | 9 | 2 | 0 | Low-quality | Genome-fragment | 7.82 | AAI-based (high-confidence) | 0 |
| S10_vs_101749\|\|full | 1129 | No | NA | 2 | 0 | 0 | Not-determined | Genome-fragment | NA | NA | 0 |
| S10_vs_101843\|\|full | 1588 | No | NA | 3 | 0 | 0 | Not-determined | Genome-fragment | NA | NA | 0 |
| S12_extracted_k141_154497 | 3571 | No | NA | 3 | 1 | 0 | Low-quality | Genome-fragment | 2.06 | HMM-based (lower-bound) | 0 |
| S16_vs_112197\|\|full | 3572 | No | NA | 3 | 0 | 1 | Low-quality | Genome-fragment | 1.02 | AAI-based (medium-confidence) | 0 |
| S10_vs_102127\|\|full | 1003 | No | NA | 2 | 0 | 0 | Not-determined | Genome-fragment | NA | NA | 0 |
| S10_vs_102178\|\|full | 5772 | No | NA | 5 | 0 | 4 | Not-determined | Genome-fragment | NA | NA | 0 |
| S10_vs_102253\|\|full | 1683 | No | NA | 4 | 0 | 1 | Not-determined | Genome-fragment | NA | NA | 0 |
| S10_extracted_25625 | 3572 | No | NA | 2 | 0 | 1 | Low-quality | Genome-fragment | 1.64 | AAI-based (medium-confidence) | 0 |
| S10_vs_102339\|\|full | 2916 | No | NA | 3 | 0 | 2 | Not-determined | Genome-fragment | NA | NA | 0 |
| S10_vs_102355\|\|full | 1761 | No | NA | 3 | 0 | 0 | Not-determined | Genome-fragment | NA | NA | 0 |
| S10_vs_102368\|\|full | 1428 | No | NA | 3 | 0 | 1 | Not-determined | Genome-fragment | NA | NA | 0 |
| S10_vs_102389\|\|full | 4545 | No | NA | 12 | 0 | 2 | Not-determined | Genome-fragment | NA | NA | 0 |
| S10_vs_102447\|\|full | 2438 | No | NA | 2 | 0 | 1 | Not-determined | Genome-fragment | NA | NA | 0 |
| S11_extracted_k141_98220 | 3573 | No | NA | 5 | 0 | 1 | Low-quality | Genome-fragment | 6.3 | AAI-based (medium-confidence) | 0 |
| S3_extracted_k141_116852 | 3573 | No | NA | 3 | 0 | 3 | Low-quality | Genome-fragment | 6.03 | AAI-based (high-confidence) | 0 |
| S10_vs_102633\|\|full | 1436 | No | NA | 2 | 0 | 1 | Not-determined | Genome-fragment | NA | NA | 0 |
| S7_vs_62603\|\|full | 3573 | No | NA | 7 | 2 | 0 | Low-quality | Genome-fragment | 2.37 | AAI-based (medium-confidence) | 0 |
| S10_vs_102745\|\|full | 2100 | No | NA | 3 | 0 | 1 | Not-determined | Genome-fragment | NA | NA | 0 |
| S16_vs_168795\|\|full | 3574 | No | NA | 4 | 0 | 1 | Low-quality | Genome-fragment | 9.33 | AAI-based (medium-confidence) | 0 |
| S10_vs_102965\|\|full | 7822 | No | NA | 7 | 0 | 5 | Not-determined | Genome-fragment | NA | NA | 0 |
| S8_vs_k141_55421\|\|full | 3574 | No | NA | 3 | 0 | 1 | Low-quality | Genome-fragment | 7.88 | AAI-based (medium-confidence) | 0 |
| S10_vs_103085\|\|full | 1301 | No | NA | 3 | 0 | 1 | Not-determined | Genome-fragment | NA | NA | 0 |
| S3_extracted_k141_27079 | 3575 | No | NA | 6 | 0 | 0 | Low-quality | Genome-fragment | 1.97 | AAI-based (high-confidence) | 0 |
| S10_vs_103266\|\|full | 1133 | No | NA | 3 | 0 | 1 | Not-determined | Genome-fragment | NA | NA | 0 |
| S12_vs_k141_55075\|\|full | 3575 | No | NA | 4 | 0 | 2 | Low-quality | Genome-fragment | 5.07 | AAI-based (medium-confidence) | 0 |
| S7_extracted_133557 | 3576 | No | NA | 4 | 4 | 0 | Low-quality | Genome-fragment | 10.34 | AAI-based (high-confidence) | 0 |
| S16_extracted_115012 | 3576 | No | NA | 3 | 0 | 2 | Low-quality | Genome-fragment | 1.45 | AAI-based (medium-confidence) | 0 |
| S10_vs_103581\|\|full | 2635 | No | NA | 3 | 0 | 2 | Not-determined | Genome-fragment | NA | NA | 0 |
| S10_vs_103709\|\|full | 2703 | No | NA | 6 | 0 | 0 | Not-determined | Genome-fragment | NA | NA | 0 |
| S12_extracted_k141_115064 | 3576 | No | NA | 6 | 0 | 5 | Low-quality | Genome-fragment | 2.37 | AAI-based (high-confidence) | 0 |
| S4_extracted_13731 | 3576 | No | NA | 4 | 0 | 0 | Low-quality | Genome-fragment | 9.16 | AAI-based (high-confidence) | 0 |
| S6_extracted_k141_1246 | 3576 | No | NA | 8 | 0 | 0 | Low-quality | Genome-fragment | 8.23 | AAI-based (high-confidence) | 0 |
| S9_extracted_k141_47507 | 3577 | No | NA | 8 | 4 | 0 | Low-quality | Genome-fragment | 10.06 | AAI-based (medium-confidence) | 0 |
| S11_extracted_k141_52533 | 3577 | No | NA | 6 | 3 | 0 | Low-quality | Genome-fragment | 6.5 | AAI-based (medium-confidence) | 0 |
| S3_vs_k141_54770\|\|full | 3577 | No | NA | 5 | 2 | 0 | Low-quality | Genome-fragment | 6.24 | AAI-based (high-confidence) | 0 |
| S7_extracted_24505 | 3577 | No | NA | 5 | 0 | 1 | Low-quality | Genome-fragment | 6.39 | AAI-based (medium-confidence) | 0 |
| S10_vs_96118\|\|full | 3577 | No | NA | 5 | 2 | 0 | Low-quality | Genome-fragment | 5.96 | HMM-based (lower-bound) | 0 |
| S10_vs_104113\|\|full | 1256 | No | NA | 2 | 0 | 0 | Not-determined | Genome-fragment | NA | NA | 0 |
| S1_extracted_k141_27321 | 3578 | No | NA | 4 | 0 | 3 | Low-quality | Genome-fragment | 1.64 | AAI-based (medium-confidence) | 0 |
| S17_extracted_k141_158609 | 3579 | No | NA | 4 | 0 | 3 | Low-quality | Genome-fragment | 3.12 | AAI-based (medium-confidence) | 0 |
| S10_vs_104245\|\|full | 6210 | No | NA | 6 | 0 | 3 | Not-determined | Genome-fragment | NA | NA | 0 |
| S10_vs_104304\|\|full | 1464 | No | NA | 3 | 0 | 1 | Not-determined | Genome-fragment | NA | NA | 0 |
| S10_vs_104342\|\|full | 5048 | No | NA | 5 | 0 | 4 | Not-determined | Genome-fragment | NA | NA | 0 |
| S12_vs_k141_11785\|\|full | 3581 | No | NA | 4 | 0 | 2 | Low-quality | Genome-fragment | 1.66 | AAI-based (medium-confidence) | 0 |
| S8_extracted_k141_35490 | 3581 | No | NA | 9 | 3 | 0 | Low-quality | Genome-fragment | 5.52 | HMM-based (lower-bound) | 0 |
| S10_vs_104550\|\|full | 3100 | No | NA | 4 | 0 | 0 | Not-determined | Genome-fragment | NA | NA | 0 |
| S3_extracted_k141_69422 | 3582 | No | NA | 4 | 0 | 1 | Low-quality | Genome-fragment | 7.61 | AAI-based (medium-confidence) | 0 |
| S1_vs_k141_155160\|\|full | 3583 | No | NA | 4 | 0 | 3 | Low-quality | Genome-fragment | 2.82 | AAI-based (high-confidence) | 0 |
| S9_vs_k141_77141\|\|full | 3584 | Yes | 1961 | 4 | 1 | 1 | Low-quality | Genome-fragment | 1.42 | AAI-based (high-confidence) | 45.28 |
| S10_vs_104909\|\|full | 6859 | No | NA | 5 | 0 | 4 | Not-determined | Genome-fragment | NA | NA | 0 |
| S10_vs_104922\|\|full | 2037 | No | NA | 3 | 0 | 0 | Not-determined | Genome-fragment | NA | NA | 0 |
| S3_vs_k141_80406\|\|full | 3584 | No | NA | 5 | 2 | 0 | Low-quality | Genome-fragment | 2.04 | AAI-based (high-confidence) | 0 |
| S10_extracted_75543 | 3584 | No | NA | 5 | 0 | 0 | Low-quality | Genome-fragment | 3.64 | AAI-based (medium-confidence) | 0 |
| S8_extracted_k141_64354 | 3584 | No | NA | 2 | 0 | 1 | Low-quality | Genome-fragment | 0.99 | AAI-based (medium-confidence) | 0 |
| S10_vs_105164\|\|full | 9110 | No | NA | 11 | 0 | 5 | Not-determined | Genome-fragment | NA | NA | 0 |
| S10_vs_105183\|\|full | 2591 | No | NA | 6 | 0 | 1 | Not-determined | Genome-fragment | NA | NA | 0 |
| S16_extracted_19338 | 3585 | No | NA | 3 | 0 | 0 | Low-quality | Genome-fragment | 3.37 | AAI-based (medium-confidence) | 0 |
| S6_vs_k141_41217\|\|full | 3585 | No | NA | 4 | 0 | 2 | Low-quality | Genome-fragment | 3.01 | AAI-based (medium-confidence) | 0 |
| S10_vs_105399\|\|full | 3020 | No | NA | 4 | 0 | 4 | Not-determined | Genome-fragment | NA | NA | 0 |
| S10_vs_105415\|\|full | 2505 | No | NA | 3 | 0 | 1 | Not-determined | Genome-fragment | NA | NA | 0 |
| S10_vs_105547\|\|full | 6502 | No | NA | 5 | 0 | 3 | Not-determined | Genome-fragment | NA | NA | 0 |
| S10_vs_105597\|\|full | 2631 | No | NA | 4 | 0 | 2 | Not-determined | Genome-fragment | NA | NA | 0 |
| S10_vs_105832\|\|full | 2041 | No | NA | 3 | 0 | 3 | Not-determined | Genome-fragment | NA | NA | 0 |
| S7_vs_43928\|\|full | 3586 | No | NA | 7 | 2 | 0 | Low-quality | Genome-fragment | 8.18 | AAI-based (medium-confidence) | 0 |
| S10_vs_106090\|\|full | 3169 | No | NA | 5 | 0 | 3 | Not-determined | Genome-fragment | NA | NA | 0 |
| S10_vs_106107\|\|full | 1510 | No | NA | 2 | 0 | 2 | Not-determined | Genome-fragment | NA | NA | 0 |
| S10_vs_106197\|\|full | 3439 | No | NA | 5 | 0 | 3 | Not-determined | Genome-fragment | NA | NA | 0 |
| S12_extracted_k141_154528 | 3586 | No | NA | 2 | 0 | 1 | Low-quality | Genome-fragment | 0.32 | AAI-based (medium-confidence) | 0 |
| S1_extracted_k141_15457 | 3588 | No | NA | 5 | 2 | 0 | Low-quality | Genome-fragment | 7.54 | AAI-based (high-confidence) | 0 |
| S10_vs_106373\|\|full | 4454 | No | NA | 5 | 0 | 0 | Not-determined | Genome-fragment | NA | NA | 0 |
| S14_vs_k141_108745\|\|full | 3589 | No | NA | 3 | 0 | 2 | Low-quality | Genome-fragment | 12.53 | AAI-based (medium-confidence) | 0 |
| S12_vs_k141_120887\|\|full | 3589 | No | NA | 3 | 1 | 0 | Low-quality | Genome-fragment | 9.77 | AAI-based (medium-confidence) | 0 |
| S10_vs_106591\|\|full | 2342 | No | NA | 4 | 0 | 0 | Not-determined | Genome-fragment | NA | NA | 0 |
| S1_vs_k141_149643\|\|full | 3590 | No | NA | 2 | 0 | 2 | Low-quality | Genome-fragment | 2.03 | AAI-based (medium-confidence) | 0 |
| S12_extracted_k141_185663 | 3590 | No | NA | 2 | 0 | 2 | Low-quality | Genome-fragment | 0.62 | AAI-based (medium-confidence) | 0 |
| S10_vs_106690\|\|full | 2092 | No | NA | 4 | 0 | 1 | Not-determined | Genome-fragment | NA | NA | 0 |
| S8_vs_k141_178125\|\|full | 3591 | No | NA | 1 | 1 | 0 | Low-quality | Genome-fragment | 6.16 | AAI-based (high-confidence) | 0 |
| S17_vs_k141_62978\|\|full | 3592 | No | NA | 4 | 0 | 2 | Low-quality | Genome-fragment | 3.01 | AAI-based (medium-confidence) | 0 |
| S10_vs_106825\|\|full | 4988 | No | NA | 3 | 0 | 2 | Not-determined | Genome-fragment | NA | NA | 0 |
| S10_vs_106834\|\|full | 31000 | No | NA | 48 | 12 | 2 | Medium-quality | Genome-fragment | 55.18 | AAI-based (high-confidence) | 0 |
| S3_extracted_k141_109475 | 3592 | No | NA | 4 | 0 | 4 | Low-quality | Genome-fragment | 2.36 | AAI-based (high-confidence) | 0 |
| S10_vs_107131\|\|full | 2901 | No | NA | 6 | 0 | 2 | Not-determined | Genome-fragment | NA | NA | 0 |
| S8_extracted_k141_123324 | 3592 | No | NA | 4 | 0 | 4 | Low-quality | Genome-fragment | 1.35 | AAI-based (high-confidence) | 0 |
| S17_extracted_k141_16031 | 3593 | No | NA | 4 | 0 | 2 | Low-quality | Genome-fragment | 8.34 | AAI-based (medium-confidence) | 0 |
| S1_extracted_k141_32458 | 3593 | No | NA | 7 | 4 | 0 | Low-quality | Genome-fragment | 7.63 | AAI-based (high-confidence) | 0 |
| S1_vs_k141_119109\|\|full | 3593 | No | NA | 2 | 0 | 2 | Low-quality | Genome-fragment | 6.34 | AAI-based (medium-confidence) | 0 |
| S10_vs_107402\|\|full | 7361 | No | NA | 10 | 0 | 4 | Not-determined | Genome-fragment | NA | NA | 0 |
| S10_vs_107423\|\|full | 2861 | No | NA | 3 | 0 | 1 | Not-determined | Genome-fragment | NA | NA | 0 |
| S10_vs_107424\|\|full | 1368 | No | NA | 4 | 0 | 1 | Not-determined | Genome-fragment | NA | NA | 0 |
| S10_vs_107441\|\|full | 6540 | No | NA | 5 | 0 | 4 | Not-determined | Genome-fragment | NA | NA | 0 |
| S10_vs_107477\|\|full | 1175 | No | NA | 3 | 0 | 0 | Not-determined | Genome-fragment | NA | NA | 0 |
| S9_extracted_k141_20519 | 3594 | No | NA | 10 | 1 | 0 | Low-quality | Genome-fragment | 6.7 | AAI-based (high-confidence) | 0 |
| S10_vs_107577\|\|full | 1373 | No | NA | 2 | 0 | 1 | Not-determined | Genome-fragment | NA | NA | 0 |
| S9_extracted_k141_14822 | 3594 | No | NA | 4 | 0 | 2 | Low-quality | Genome-fragment | 5.03 | AAI-based (medium-confidence) | 0 |
| S16_extracted_4654 | 3594 | No | NA | 2 | 0 | 2 | Low-quality | Genome-fragment | 8.55 | AAI-based (high-confidence) | 0 |
| S10_vs_107890\|\|full | 1318 | No | NA | 3 | 0 | 1 | Not-determined | Genome-fragment | NA | NA | 0 |
| S12_vs_k141_16879\|\|full | 3595 | No | NA | 4 | 0 | 1 | Low-quality | Genome-fragment | 1.69 | AAI-based (medium-confidence) | 0 |
| S3_vs_k141_107865\|\|full | 3596 | No | NA | 5 | 4 | 0 | Low-quality | Genome-fragment | 9.68 | AAI-based (high-confidence) | 0 |
| S8_vs_k141_90455\|\|full | 3597 | No | NA | 3 | 0 | 2 | Low-quality | Genome-fragment | 1.62 | AAI-based (medium-confidence) | 0 |
| S10_vs_108208\|\|full | 3107 | No | NA | 6 | 0 | 6 | Not-determined | Genome-fragment | NA | NA | 0 |
| S10_vs_108439\|\|full | 1278 | No | NA | 3 | 0 | 0 | Not-determined | Genome-fragment | NA | NA | 0 |
| S10_vs_108532\|\|full | 2289 | No | NA | 2 | 0 | 2 | Not-determined | Genome-fragment | NA | NA | 0 |
| S10_vs_108652\|\|full | 1584 | No | NA | 2 | 0 | 1 | Not-determined | Genome-fragment | NA | NA | 0 |
| S10_vs_108708\|\|full | 1645 | No | NA | 4 | 0 | 0 | Not-determined | Genome-fragment | NA | NA | 0 |
| S17_extracted_k141_144246 | 3598 | No | NA | 6 | 0 | 0 | Low-quality | Genome-fragment | 5.58 | AAI-based (medium-confidence) | 0 |
| S10_vs_108840\|\|full | 3722 | No | NA | 3 | 0 | 1 | Not-determined | Genome-fragment | NA | NA | 0 |
| S7_vs_52515\|\|full | 3598 | No | NA | 5 | 2 | 0 | Low-quality | Genome-fragment | 8.93 | AAI-based (medium-confidence) | 0 |
| S7_extracted_24438 | 3599 | No | NA | 5 | 3 | 0 | Low-quality | Genome-fragment | 6.63 | AAI-based (medium-confidence) | 0 |
| S16_extracted_96107 | 3599 | No | NA | 3 | 0 | 2 | Low-quality | Genome-fragment | 6.23 | AAI-based (medium-confidence) | 0 |
| S10_vs_109119\|\|full | 1930 | No | NA | 4 | 0 | 0 | Not-determined | Genome-fragment | NA | NA | 0 |
| S14_extracted_k141_107109 | 3600 | No | NA | 5 | 3 | 0 | Low-quality | Genome-fragment | 3.2 | HMM-based (lower-bound) | 0 |
| S10_vs_109169\|\|full | 1412 | No | NA | 2 | 0 | 1 | Not-determined | Genome-fragment | NA | NA | 0 |
| S10_vs_109209\|\|full | 1681 | No | NA | 3 | 0 | 2 | Not-determined | Genome-fragment | NA | NA | 0 |
| S15_vs_47781\|\|full | 3600 | No | NA | 4 | 0 | 0 | Low-quality | Genome-fragment | 6.75 | AAI-based (high-confidence) | 0 |
| S10_vs_109398\|\|full | 2042 | No | NA | 4 | 0 | 0 | Not-determined | Genome-fragment | NA | NA | 0 |
| S9_extracted_k141_21975 | 3601 | No | NA | 2 | 0 | 1 | Low-quality | Genome-fragment | 7.65 | AAI-based (medium-confidence) | 0 |
| S10_vs_109468\|\|full | 1284 | No | NA | 3 | 0 | 0 | Not-determined | Genome-fragment | NA | NA | 0 |
| S16_vs_117145\|\|full | 3601 | No | NA | 2 | 0 | 2 | Low-quality | Genome-fragment | 4.85 | AAI-based (medium-confidence) | 0 |
| S10_vs_109686\|\|full | 4848 | No | NA | 5 | 0 | 4 | Not-determined | Genome-fragment | NA | NA | 0 |
| S10_vs_109731\|\|full | 2410 | No | NA | 2 | 0 | 2 | Not-determined | Genome-fragment | NA | NA | 0 |
| S12_vs_k141_24423\|\|full | 3601 | Yes | 1504 | 5 | 1 | 1 | Low-quality | Genome-fragment | 2.14 | HMM-based (lower-bound) | 58.23 |
| S10_vs_109773\|\|full | 1895 | No | NA | 2 | 0 | 0 | Not-determined | Genome-fragment | NA | NA | 0 |
| S10_vs_109867\|\|full | 2513 | No | NA | 4 | 0 | 0 | Not-determined | Genome-fragment | NA | NA | 0 |
| S8_vs_k141_136935\|\|full | 3601 | No | NA | 2 | 0 | 2 | Low-quality | Genome-fragment | 6.49 | AAI-based (medium-confidence) | 0 |
| S10_vs_110099\|\|full | 1812 | No | NA | 2 | 0 | 1 | Not-determined | Genome-fragment | NA | NA | 0 |
| S14_vs_k141_53651\|\|full | 3602 | No | NA | 4 | 0 | 0 | Low-quality | Genome-fragment | 8.93 | AAI-based (medium-confidence) | 0 |
| S10_vs_110167\|\|full | 6130 | No | NA | 3 | 0 | 3 | Not-determined | Genome-fragment | NA | NA | 0 |
| S17_vs_k141_90993\|\|full | 3603 | No | NA | 3 | 0 | 2 | Low-quality | Genome-fragment | 12.04 | AAI-based (high-confidence) | 0 |
| S10_vs_110437\|\|full | 2607 | No | NA | 3 | 0 | 0 | Not-determined | Genome-fragment | NA | NA | 0 |
| S16_extracted_10025 | 3603 | No | NA | 7 | 1 | 0 | Low-quality | Genome-fragment | 6.75 | AAI-based (high-confidence) | 0 |
| S10_vs_110553\|\|full | 3111 | No | NA | 3 | 0 | 2 | Not-determined | Genome-fragment | NA | NA | 0 |
| S10_vs_110569\|\|full | 1017 | No | NA | 2 | 0 | 1 | Not-determined | Genome-fragment | NA | NA | 0 |
| S10_vs_110658\|\|full | 2149 | No | NA | 2 | 0 | 1 | Not-determined | Genome-fragment | NA | NA | 0 |
| S16_extracted_15550 | 3603 | No | NA | 1 | 1 | 0 | Low-quality | Genome-fragment | 2.49 | HMM-based (lower-bound) | 0 |
| S10_vs_110835\|\|full | 1314 | No | NA | 4 | 0 | 0 | Not-determined | Genome-fragment | NA | NA | 0 |
| S9_vs_k141_58973\|\|full | 3604 | No | NA | 4 | 2 | 0 | Low-quality | Genome-fragment | 9.01 | AAI-based (high-confidence) | 0 |
| S8_extracted_k141_119197 | 3605 | No | NA | 3 | 0 | 3 | Low-quality | Genome-fragment | 6.23 | AAI-based (medium-confidence) | 0 |
| S5_extracted_k141_106361 | 3606 | No | NA | 5 | 2 | 0 | Low-quality | Genome-fragment | 8.67 | AAI-based (medium-confidence) | 0 |
| S5_vs_k141_63818\|\|full | 3606 | No | NA | 11 | 2 | 0 | Low-quality | Genome-fragment | 7.42 | AAI-based (high-confidence) | 0 |
| S10_vs_111603\|\|full | 5636 | No | NA | 4 | 0 | 2 | Not-determined | Genome-fragment | NA | NA | 0 |
| S9_vs_k141_42534\|\|full | 3608 | Yes | 1560 | 6 | 2 | 1 | Low-quality | Genome-fragment | 2.81 | HMM-based (lower-bound) | 56.76 |
| S10_vs_112023\|\|full | 6941 | No | NA | 5 | 0 | 3 | Not-determined | Genome-fragment | NA | NA | 0 |
| S5_vs_k141_17613\|\|full | 3608 | No | NA | 6 | 0 | 3 | Low-quality | Genome-fragment | 5.02 | AAI-based (medium-confidence) | 0 |
| S10_vs_112224\|\|full | 1136 | No | NA | 3 | 0 | 0 | Not-determined | Genome-fragment | NA | NA | 0 |
| S16_vs_150391\|\|full | 3608 | No | NA | 4 | 0 | 3 | Low-quality | Genome-fragment | 0.91 | AAI-based (medium-confidence) | 0 |
| S11_vs_k141_9951\|\|full | 3609 | No | NA | 4 | 0 | 0 | Low-quality | Genome-fragment | 4.2 | AAI-based (medium-confidence) | 0 |
| S10_vs_112496\|\|full | 1780 | No | NA | 4 | 0 | 0 | Not-determined | Genome-fragment | NA | NA | 0 |
| S10_vs_112497\|\|full | 1222 | No | NA | 2 | 0 | 1 | Not-determined | Genome-fragment | NA | NA | 0 |
| S10_vs_112524\|\|full | 1115 | No | NA | 3 | 0 | 0 | Not-determined | Genome-fragment | NA | NA | 0 |
| S10_vs_112525\|\|full | 1087 | No | NA | 2 | 0 | 2 | Not-determined | Genome-fragment | NA | NA | 0 |
| S14_vs_k141_57927\|\|full | 3609 | No | NA | 3 | 0 | 2 | Low-quality | Genome-fragment | 1.67 | AAI-based (medium-confidence) | 0 |
| S1_extracted_k141_10733 | 3609 | No | NA | 7 | 0 | 0 | Low-quality | Genome-fragment | 9.32 | AAI-based (medium-confidence) | 0 |
| S5_vs_k141_112715\|\|full | 3609 | No | NA | 11 | 1 | 0 | Low-quality | Genome-fragment | 6.84 | AAI-based (medium-confidence) | 0 |
| S10_vs_112861\|\|full | 1552 | No | NA | 2 | 0 | 0 | Not-determined | Genome-fragment | NA | NA | 0 |
| S12_extracted_k141_79497 | 3609 | No | NA | 6 | 3 | 0 | Low-quality | Genome-fragment | 8.16 | AAI-based (high-confidence) | 0 |
| S10_vs_113218\|\|full | 2536 | No | NA | 3 | 0 | 3 | Not-determined | Genome-fragment | NA | NA | 0 |
| S10_vs_113241\|\|full | 2298 | No | NA | 3 | 0 | 1 | Not-determined | Genome-fragment | NA | NA | 0 |
| S12_extracted_k141_111240 | 3609 | No | NA | 6 | 1 | 0 | Low-quality | Genome-fragment | 6.97 | AAI-based (high-confidence) | 0 |
| S10_vs_113364\|\|full | 1450 | No | NA | 3 | 0 | 0 | Not-determined | Genome-fragment | NA | NA | 0 |
| S14_extracted_k141_37191 | 3610 | No | NA | 4 | 3 | 0 | Low-quality | Genome-fragment | 11.79 | AAI-based (high-confidence) | 0 |
| S10_vs_113730\|\|full | 4577 | No | NA | 4 | 0 | 0 | Not-determined | Genome-fragment | NA | NA | 0 |
| S10_vs_113732\|\|full | 6059 | No | NA | 9 | 0 | 1 | Not-determined | Genome-fragment | NA | NA | 0 |
| S5_vs_k141_56156\|\|full | 3610 | No | NA | 3 | 0 | 1 | Low-quality | Genome-fragment | 1.54 | AAI-based (medium-confidence) | 0 |
| S1_vs_k141_26132\|\|full | 3611 | No | NA | 3 | 0 | 1 | Low-quality | Genome-fragment | 5.78 | AAI-based (medium-confidence) | 0 |
| S10_vs_114217\|\|full | 2245 | No | NA | 4 | 0 | 3 | Not-determined | Genome-fragment | NA | NA | 0 |
| S7_extracted_117395 | 3611 | No | NA | 2 | 2 | 0 | Low-quality | Genome-fragment | 8.79 | AAI-based (medium-confidence) | 0 |
| S12_vs_k141_167163\|\|full | 3612 | No | NA | 9 | 1 | 0 | Low-quality | Genome-fragment | 7.71 | AAI-based (high-confidence) | 0 |
| S10_vs_114388\|\|full | 4573 | No | NA | 7 | 0 | 1 | Not-determined | Genome-fragment | NA | NA | 0 |
| S12_vs_k141_161958\|\|full | 3612 | No | NA | 7 | 1 | 0 | Low-quality | Genome-fragment | 4.76 | HMM-based (lower-bound) | 0 |
| S10_vs_114418\|\|full | 4842 | No | NA | 6 | 0 | 4 | Not-determined | Genome-fragment | NA | NA | 0 |
| S10_vs_114583\|\|full | 1618 | No | NA | 3 | 0 | 1 | Not-determined | Genome-fragment | NA | NA | 0 |
| S10_vs_114606\|\|full | 2462 | No | NA | 2 | 0 | 2 | Not-determined | Genome-fragment | NA | NA | 0 |
| S10_vs_114673\|\|full | 9652 | No | NA | 5 | 0 | 3 | Not-determined | Genome-fragment | NA | NA | 0 |
| S6_vs_k141_32370\|\|full | 3612 | No | NA | 2 | 0 | 1 | Low-quality | Genome-fragment | 1.46 | AAI-based (medium-confidence) | 0 |
| S10_vs_114966\|\|full | 1793 | No | NA | 2 | 0 | 2 | Not-determined | Genome-fragment | NA | NA | 0 |
| S10_vs_115015\|\|full | 7218 | No | NA | 12 | 0 | 2 | Not-determined | Genome-fragment | NA | NA | 0 |
| S10_vs_115033\|\|full | 1684 | No | NA | 2 | 0 | 1 | Not-determined | Genome-fragment | NA | NA | 0 |
| S1_vs_k141_108350\|\|full | 3613 | No | NA | 5 | 0 | 2 | Low-quality | Genome-fragment | 1.58 | AAI-based (medium-confidence) | 0 |
| S10_vs_115060\|\|full | 3262 | No | NA | 4 | 0 | 3 | Not-determined | Genome-fragment | NA | NA | 0 |
| S10_vs_115155\|\|full | 1706 | No | NA | 3 | 0 | 1 | Not-determined | Genome-fragment | NA | NA | 0 |
| S10_vs_115253\|\|full | 1737 | No | NA | 4 | 0 | 0 | Not-determined | Genome-fragment | NA | NA | 0 |
| S10_vs_115314\|\|full | 1763 | No | NA | 3 | 0 | 2 | Not-determined | Genome-fragment | NA | NA | 0 |
| S10_vs_115361\|\|full | 2344 | No | NA | 2 | 0 | 1 | Not-determined | Genome-fragment | NA | NA | 0 |
| S5_extracted_k141_582 | 3614 | No | NA | 8 | 5 | 0 | Low-quality | Genome-fragment | 16.48 | AAI-based (high-confidence) | 0 |
| S10_vs_115501\|\|full | 2616 | No | NA | 4 | 0 | 2 | Not-determined | Genome-fragment | NA | NA | 0 |
| S10_vs_115722\|\|full | 3712 | No | NA | 4 | 0 | 1 | Not-determined | Genome-fragment | NA | NA | 0 |
| S12_extracted_k141_152653 | 3614 | No | NA | 11 | 3 | 0 | Low-quality | Genome-fragment | 3.7 | AAI-based (medium-confidence) | 0 |
| S10_vs_115877\|\|full | 5426 | No | NA | 6 | 0 | 3 | Not-determined | Genome-fragment | NA | NA | 0 |
| S10_vs_115903\|\|full | 7556 | No | NA | 9 | 0 | 4 | Not-determined | Genome-fragment | NA | NA | 0 |
| S17_vs_k141_115013\|\|full | 3615 | No | NA | 7 | 1 | 0 | Low-quality | Genome-fragment | 5.2 | AAI-based (high-confidence) | 0 |
| S10_vs_116137\|\|full | 1410 | No | NA | 2 | 0 | 1 | Not-determined | Genome-fragment | NA | NA | 0 |
| S1_vs_k141_31703\|\|full | 3615 | No | NA | 6 | 3 | 0 | Low-quality | Genome-fragment | 8.78 | AAI-based (high-confidence) | 0 |
| S5_extracted_k141_79931 | 3615 | No | NA | 5 | 0 | 0 | Low-quality | Genome-fragment | 7.35 | AAI-based (high-confidence) | 0 |
| S10_vs_116287\|\|full | 3714 | No | NA | 3 | 0 | 2 | Not-determined | Genome-fragment | NA | NA | 0 |
| S10_vs_116297\|\|full | 1174 | No | NA | 2 | 0 | 1 | Not-determined | Genome-fragment | NA | NA | 0 |
| S10_vs_116426\|\|full | 5415 | No | NA | 5 | 0 | 4 | Not-determined | Genome-fragment | NA | NA | 0 |
| S7_vs_83797\|\|full | 3615 | No | NA | 5 | 2 | 0 | Low-quality | Genome-fragment | 10.15 | AAI-based (medium-confidence) | 0 |
| S8_extracted_k141_168032 | 3616 | No | NA | 7 | 1 | 0 | Low-quality | Genome-fragment | 7.21 | AAI-based (medium-confidence) | 0 |
| S10_vs_116636\|\|full | 1591 | No | NA | 2 | 0 | 0 | Not-determined | Genome-fragment | NA | NA | 0 |
| S10_vs_116670\|\|full | 1323 | No | NA | 3 | 0 | 0 | Not-determined | Genome-fragment | NA | NA | 0 |
| S7_extracted_107889 | 3618 | No | NA | 5 | 1 | 0 | Low-quality | Genome-fragment | 7.57 | AAI-based (high-confidence) | 0 |
| S10_vs_116779\|\|full | 2172 | No | NA | 3 | 0 | 2 | Not-determined | Genome-fragment | NA | NA | 0 |
| S16_vs_142334\|\|full | 3618 | Yes | 2496 | 7 | 1 | 1 | Low-quality | Genome-fragment | 3 | HMM-based (lower-bound) | 31.01 |
| S10_vs_116966\|\|full | 1862 | No | NA | 3 | 0 | 0 | Not-determined | Genome-fragment | NA | NA | 0 |
| S16_vs_143270\|\|full | 3618 | No | NA | 2 | 0 | 2 | Low-quality | Genome-fragment | 0.42 | AAI-based (medium-confidence) | 0 |
| S12_extracted_k141_53762 | 3618 | No | NA | 2 | 0 | 2 | Low-quality | Genome-fragment | 1.22 | AAI-based (high-confidence) | 0 |
| S10_vs_117150\|\|full | 1836 | No | NA | 4 | 0 | 0 | Not-determined | Genome-fragment | NA | NA | 0 |
| S17_extracted_k141_59740 | 3619 | No | NA | 3 | 0 | 2 | Low-quality | Genome-fragment | 7.79 | AAI-based (medium-confidence) | 0 |
| S17_extracted_k141_18988 | 3619 | No | NA | 3 | 0 | 1 | Low-quality | Genome-fragment | 4.17 | AAI-based (medium-confidence) | 0 |
| S10_vs_117383\|\|full | 2090 | No | NA | 3 | 0 | 1 | Not-determined | Genome-fragment | NA | NA | 0 |
| S14_vs_k141_111887\|\|full | 3619 | No | NA | 8 | 1 | 0 | Low-quality | Genome-fragment | 8.36 | AAI-based (high-confidence) | 0 |
| S12_extracted_k141_177994 | 3619 | No | NA | 2 | 0 | 1 | Low-quality | Genome-fragment | 1.73 | AAI-based (medium-confidence) | 0 |
| S10_vs_117699\|\|full | 1056 | No | NA | 2 | 0 | 0 | Not-determined | Genome-fragment | NA | NA | 0 |
| S10_vs_117836\|\|full | 1124 | No | NA | 2 | 0 | 0 | Not-determined | Genome-fragment | NA | NA | 0 |
| S10_vs_117921\|\|full | 3126 | No | NA | 3 | 0 | 2 | Not-determined | Genome-fragment | NA | NA | 0 |
| S10_vs_117945\|\|full | 2451 | No | NA | 4 | 0 | 3 | Not-determined | Genome-fragment | NA | NA | 0 |
| S10_vs_118090\|\|full | 3822 | No | NA | 7 | 0 | 2 | Not-determined | Genome-fragment | NA | NA | 0 |
| S3_extracted_k141_91234 | 3620 | No | NA | 5 | 0 | 1 | Low-quality | Genome-fragment | 4.68 | AAI-based (medium-confidence) | 0 |
| S10_vs_118184\|\|full | 1099 | No | NA | 3 | 0 | 1 | Not-determined | Genome-fragment | NA | NA | 0 |
| S10_vs_118189\|\|full | 3188 | No | NA | 4 | 0 | 0 | Not-determined | Genome-fragment | NA | NA | 0 |
| S8_vs_k141_81208\|\|full | 3620 | No | NA | 4 | 0 | 0 | Low-quality | Genome-fragment | 5.8 | AAI-based (medium-confidence) | 0 |
| S10_vs_118254\|\|full | 1368 | No | NA | 3 | 0 | 2 | Not-determined | Genome-fragment | NA | NA | 0 |
| S10_vs_118320\|\|full | 2825 | No | NA | 3 | 0 | 1 | Not-determined | Genome-fragment | NA | NA | 0 |
| S10_vs_118493\|\|full | 2229 | No | NA | 3 | 0 | 2 | Not-determined | Genome-fragment | NA | NA | 0 |
| S10_vs_118575\|\|full | 2329 | No | NA | 3 | 0 | 2 | Not-determined | Genome-fragment | NA | NA | 0 |
| S6_extracted_k141_15586 | 3621 | No | NA | 4 | 1 | 0 | Low-quality | Genome-fragment | 2.68 | AAI-based (high-confidence) | 0 |
| S10_vs_118776\|\|full | 1405 | No | NA | 3 | 0 | 0 | Not-determined | Genome-fragment | NA | NA | 0 |
| S5_vs_k141_55692\|\|full | 3622 | No | NA | 4 | 0 | 1 | Low-quality | Genome-fragment | 2 | AAI-based (medium-confidence) | 0 |
| S7_vs_57920\|\|full | 3622 | No | NA | 7 | 0 | 0 | Low-quality | Genome-fragment | 8.16 | AAI-based (medium-confidence) | 0 |
| S3_extracted_k141_21290 | 3623 | No | NA | 4 | 0 | 0 | Low-quality | Genome-fragment | 8.34 | AAI-based (high-confidence) | 0 |
| S10_vs_119035\|\|full | 1215 | No | NA | 3 | 0 | 0 | Not-determined | Genome-fragment | NA | NA | 0 |
| S10_vs_119245\|\|full | 8694 | No | NA | 12 | 0 | 1 | Not-determined | Genome-fragment | NA | NA | 0 |
| S5_extracted_k141_768 | 3624 | No | NA | 6 | 0 | 6 | Low-quality | Genome-fragment | 2.37 | AAI-based (high-confidence) | 0 |
| S10_vs_119298\|\|full | 49258 | No | NA | 66 | 14 | 1 | High-quality | High-quality | 97.09 | AAI-based (high-confidence) | 0 |
| S16_vs_113174\|\|full | 3625 | No | NA | 3 | 0 | 1 | Low-quality | Genome-fragment | 2.16 | AAI-based (medium-confidence) | 0 |
| S10_vs_119317\|\|full | 4703 | No | NA | 4 | 0 | 3 | Not-determined | Genome-fragment | NA | NA | 0 |
| S10_vs_119376\|\|full | 5273 | No | NA | 4 | 0 | 4 | Not-determined | Genome-fragment | NA | NA | 0 |
| S10_vs_119389\|\|full | 1438 | No | NA | 3 | 0 | 1 | Not-determined | Genome-fragment | NA | NA | 0 |
| S10_vs_119523\|\|full | 1039 | No | NA | 2 | 0 | 1 | Not-determined | Genome-fragment | NA | NA | 0 |
| S9_vs_k141_27246\|\|full | 3626 | No | NA | 5 | 0 | 0 | Low-quality | Genome-fragment | 2.25 | AAI-based (high-confidence) | 0 |
| S10_vs_119628\|\|full | 4045 | No | NA | 5 | 0 | 2 | Not-determined | Genome-fragment | NA | NA | 0 |
| S3_vs_k141_2303\|\|full | 3626 | No | NA | 9 | 3 | 0 | Low-quality | Genome-fragment | 7.28 | AAI-based (medium-confidence) | 0 |
| S7_extracted_52192 | 3626 | No | NA | 4 | 0 | 0 | Low-quality | Genome-fragment | 4.43 | AAI-based (medium-confidence) | 0 |
| S10_vs_119833\|\|full | 3136 | No | NA | 4 | 0 | 0 | Not-determined | Genome-fragment | NA | NA | 0 |
| S8_vs_k141_184965\|\|full | 3626 | No | NA | 5 | 0 | 0 | Low-quality | Genome-fragment | 7.01 | AAI-based (medium-confidence) | 0 |
| S9_vs_k141_6318\|\|full | 3627 | No | NA | 7 | 0 | 0 | Low-quality | Genome-fragment | 6.76 | AAI-based (high-confidence) | 0 |
| S10_vs_119947\|\|full | 2763 | No | NA | 5 | 0 | 3 | Not-determined | Genome-fragment | NA | NA | 0 |
| S10_vs_119953\|\|full | 2423 | No | NA | 2 | 0 | 2 | Not-determined | Genome-fragment | NA | NA | 0 |
| S10_extracted_60390 | 3628 | Yes | 1334 | 6 | 1 | 3 | Low-quality | Genome-fragment | 0.87 | AAI-based (medium-confidence) | 63.23 |
| S10_vs_120214\|\|full | 1084 | No | NA | 2 | 0 | 0 | Not-determined | Genome-fragment | NA | NA | 0 |
| S1_vs_k141_100828\|\|full | 3629 | No | NA | 7 | 0 | 0 | Low-quality | Genome-fragment | 2.04 | AAI-based (medium-confidence) | 0 |
| S7_extracted_47853 | 3629 | No | NA | 12 | 1 | 0 | Low-quality | Genome-fragment | 2.23 | AAI-based (medium-confidence) | 0 |
| S10_vs_120366\|\|full | 1744 | No | NA | 3 | 0 | 2 | Not-determined | Genome-fragment | NA | NA | 0 |
| S10_vs_120367\|\|full | 1006 | No | NA | 2 | 0 | 2 | Not-determined | Genome-fragment | NA | NA | 0 |
| S10_vs_120410\|\|full | 5827 | No | NA | 7 | 0 | 5 | Not-determined | Genome-fragment | NA | NA | 0 |
| S14_vs_k141_901\|\|full | 3630 | No | NA | 5 | 1 | 0 | Low-quality | Genome-fragment | 9.61 | AAI-based (high-confidence) | 0 |
| S5_vs_k141_47687\|\|full | 3630 | No | NA | 11 | 1 | 1 | Low-quality | Genome-fragment | 4.88 | HMM-based (lower-bound) | 0 |
| S7_vs_104346\|\|full | 3630 | No | NA | 4 | 0 | 0 | Low-quality | Genome-fragment | 8.24 | AAI-based (high-confidence) | 0 |
| S10_vs_120624\|\|full | 2281 | No | NA | 4 | 0 | 3 | Not-determined | Genome-fragment | NA | NA | 0 |
| S10_vs_120734\|\|full | 1425 | No | NA | 2 | 0 | 1 | Not-determined | Genome-fragment | NA | NA | 0 |
| S10_vs_120749\|\|full | 2475 | No | NA | 3 | 0 | 0 | Not-determined | Genome-fragment | NA | NA | 0 |
| S10_vs_120799\|\|full | 3800 | No | NA | 3 | 0 | 2 | Not-determined | Genome-fragment | NA | NA | 0 |
| S11_vs_k141_42909\|\|full | 3631 | No | NA | 11 | 0 | 0 | Low-quality | Genome-fragment | 7.34 | AAI-based (medium-confidence) | 0 |
| S10_vs_121014\|\|full | 2033 | No | NA | 4 | 0 | 0 | Not-determined | Genome-fragment | NA | NA | 0 |
| S3_extracted_k141_104522 | 3631 | No | NA | 2 | 0 | 1 | Low-quality | Genome-fragment | 0.56 | AAI-based (medium-confidence) | 0 |
| S9_vs_k141_63271\|\|full | 3632 | No | NA | 4 | 0 | 1 | Low-quality | Genome-fragment | 2.71 | AAI-based (medium-confidence) | 0 |
| S10_vs_13905\|\|0_partial | 14078 | No | NA | 15 | 0 | 5 | Not-determined | Genome-fragment | NA | NA | 0 |
| S4_extracted_8752 | 3632 | No | NA | 9 | 8 | 0 | Low-quality | Genome-fragment | 11.95 | AAI-based (high-confidence) | 0 |
| S10_vs_20642\|\|0_partial | 12407 | No | NA | 11 | 0 | 9 | Not-determined | Genome-fragment | NA | NA | 0 |
| S10_vs_42162\|\|0_partial | 29461 | No | NA | 32 | 0 | 7 | Not-determined | Genome-fragment | NA | NA | 0 |
| S3_extracted_k141_67437 | 3633 | No | NA | 3 | 3 | 0 | Low-quality | Genome-fragment | 9.7 | AAI-based (high-confidence) | 0 |
| S15_extracted_29249 | 3633 | No | NA | 4 | 0 | 1 | Low-quality | Genome-fragment | 2.79 | AAI-based (medium-confidence) | 0 |
| S10_vs_62668\|\|1_partial | 27745 | No | NA | 20 | 0 | 16 | Not-determined | Genome-fragment | NA | NA | 0 |
| S7_vs_82411\|\|full | 3634 | No | NA | 4 | 4 | 0 | Low-quality | Genome-fragment | 8.35 | AAI-based (high-confidence) | 0 |
| S10_vs_70031\|\|0_partial | 21280 | No | NA | 15 | 0 | 8 | Not-determined | Genome-fragment | NA | NA | 0 |
| S10_vs_83125\|\|0_partial | 15212 | No | NA | 8 | 0 | 3 | Not-determined | Genome-fragment | NA | NA | 0 |
| S4_extracted_20835 | 3634 | No | NA | 9 | 9 | 0 | Low-quality | Genome-fragment | 11.96 | AAI-based (high-confidence) | 0 |
| S16_vs_161792\|\|full | 3635 | No | NA | 4 | 1 | 0 | Low-quality | Genome-fragment | 2.27 | AAI-based (high-confidence) | 0 |
| S11_extracted_k141_22012 | 3637 | No | NA | 4 | 1 | 0 | Low-quality | Genome-fragment | 2.36 | AAI-based (high-confidence) | 0 |
| S13_extracted_121227 | 3637 | No | NA | 4 | 0 | 0 | Low-quality | Genome-fragment | 7.61 | AAI-based (medium-confidence) | 0 |
| S16_extracted_42901 | 3637 | No | NA | 6 | 4 | 0 | Low-quality | Genome-fragment | 2.11 | AAI-based (high-confidence) | 0 |
| S10_vs_3871\|\|lt2gene | 1645 | No | NA | 1 | 0 | 1 | Not-determined | Genome-fragment | NA | NA | 0 |
| S12_vs_k141_105345\|\|full | 3637 | No | NA | 5 | 1 | 0 | Low-quality | Genome-fragment | 3.64 | AAI-based (medium-confidence) | 0 |
| S8_vs_k141_4704\|\|full | 3639 | Yes | 2096 | 5 | 1 | 2 | Low-quality | Genome-fragment | 1.24 | HMM-based (lower-bound) | 42.4 |
| S17_vs_k141_53256\|\|full | 3640 | No | NA | 2 | 0 | 1 | Low-quality | Genome-fragment | 1.71 | AAI-based (medium-confidence) | 0 |
| S10_vs_9342\|\|lt2gene | 1006 | No | NA | 2 | 0 | 1 | Not-determined | Genome-fragment | NA | NA | 0 |
| S7_vs_65620\|\|full | 3640 | No | NA | 3 | 0 | 2 | Low-quality | Genome-fragment | 2.18 | AAI-based (medium-confidence) | 0 |
| S10_vs_31792\|\|full | 3640 | No | NA | 5 | 0 | 4 | Low-quality | Genome-fragment | 1.53 | AAI-based (high-confidence) | 0 |
| S17_extracted_k141_40337 | 3641 | No | NA | 3 | 2 | 0 | Low-quality | Genome-fragment | 6.09 | AAI-based (high-confidence) | 0 |
| S11_vs_k141_89010\|\|full | 3641 | No | NA | 3 | 0 | 1 | Low-quality | Genome-fragment | 5.95 | AAI-based (medium-confidence) | 0 |
| S5_extracted_k141_140059 | 3641 | No | NA | 7 | 1 | 0 | Low-quality | Genome-fragment | 8.43 | AAI-based (high-confidence) | 0 |
| S7_vs_76685\|\|lt2gene | 3641 | No | NA | 2 | 2 | 0 | Low-quality | Genome-fragment | 7.47 | AAI-based (high-confidence) | 0 |
| S16_vs_128076\|\|full | 3642 | No | NA | 5 | 3 | 0 | Low-quality | Genome-fragment | 8.99 | AAI-based (high-confidence) | 0 |
| S10_vs_62567\|\|full | 3643 | No | NA | 12 | 0 | 0 | Low-quality | Genome-fragment | 7.38 | AAI-based (high-confidence) | 0 |
| S11_vs_k141_15327\|\|full | 3644 | No | NA | 7 | 1 | 0 | Low-quality | Genome-fragment | 5.84 | HMM-based (lower-bound) | 0 |
| S10_vs_26559\|\|lt2gene | 2214 | No | NA | 2 | 0 | 1 | Not-determined | Genome-fragment | NA | NA | 0 |
| S7_vs_57495\|\|full | 3644 | No | NA | 6 | 2 | 0 | Low-quality | Genome-fragment | 11.86 | AAI-based (high-confidence) | 0 |
| S16_vs_37613\|\|full | 3644 | No | NA | 2 | 0 | 2 | Low-quality | Genome-fragment | 1.91 | AAI-based (medium-confidence) | 0 |
| S16_vs_172431\|\|full | 3644 | No | NA | 2 | 0 | 2 | Low-quality | Genome-fragment | 0.43 | AAI-based (medium-confidence) | 0 |
| S4_extracted_30563 | 3644 | No | NA | 3 | 0 | 0 | Low-quality | Genome-fragment | 6.33 | AAI-based (high-confidence) | 0 |
| S8_extracted_k141_142365 | 3644 | No | NA | 5 | 0 | 0 | Low-quality | Genome-fragment | 9.18 | AAI-based (medium-confidence) | 0 |
| S8_vs_k141_71550\|\|full | 3644 | No | NA | 3 | 0 | 0 | Low-quality | Genome-fragment | 2.42 | AAI-based (medium-confidence) | 0 |
| S13_extracted_87494 | 3645 | No | NA | 4 | 0 | 4 | Low-quality | Genome-fragment | 1.13 | AAI-based (high-confidence) | 0 |
| S5_extracted_k141_137232 | 3645 | No | NA | 3 | 0 | 1 | Low-quality | Genome-fragment | 3.32 | AAI-based (medium-confidence) | 0 |
| S6_extracted_k141_41519 | 3645 | No | NA | 1 | 0 | 1 | Low-quality | Genome-fragment | 4.62 | AAI-based (high-confidence) | 0 |
| S8_extracted_k141_119530 | 3645 | No | NA | 2 | 0 | 2 | Low-quality | Genome-fragment | 1.86 | AAI-based (medium-confidence) | 0 |
| S8_vs_k141_30194\|\|full | 3645 | No | NA | 3 | 0 | 3 | Low-quality | Genome-fragment | 2.48 | AAI-based (medium-confidence) | 0 |
| S5_extracted_k141_123162 | 3646 | No | NA | 8 | 2 | 0 | Low-quality | Genome-fragment | 7.34 | AAI-based (high-confidence) | 0 |
| S8_extracted_k141_108292 | 3646 | No | NA | 3 | 0 | 2 | Low-quality | Genome-fragment | 0.67 | AAI-based (medium-confidence) | 0 |
| S12_extracted_k141_66484 | 3647 | No | NA | 5 | 0 | 5 | Low-quality | Genome-fragment | 4.26 | AAI-based (medium-confidence) | 0 |
| S11_extracted_k141_32203 | 3648 | No | NA | 5 | 1 | 0 | Low-quality | Genome-fragment | 6.32 | AAI-based (high-confidence) | 0 |
| S5_extracted_k141_68566 | 3648 | No | NA | 7 | 0 | 0 | Low-quality | Genome-fragment | 3.91 | AAI-based (high-confidence) | 0 |

| | | | | | | | | | | | |
|---|---|---|---|---|---|---|---|---|---|---|---|
| S7_extracted_62250 | 3648 | No | NA | 4 | 0 | 1 | Low-quality | Genome-fragment | 1.54 | AAI-based (medium-confidence) | 0 |
| S6_extracted_k141_35391 | 3648 | No | NA | 5 | 0 | 3 | Low-quality | Genome-fragment | 7.02 | AAI-based (medium-confidence) | 0 |
| S14_extracted_k141_104226 | 3649 | No | NA | 4 | 4 | 0 | Low-quality | Genome-fragment | 11.99 | AAI-based (high-confidence) | 0 |
| S15_extracted_74611 | 3649 | No | NA | 6 | 2 | 0 | Low-quality | Genome-fragment | 8.07 | AAI-based (high-confidence) | 0 |
| S14_extracted_k141_94441 | 3650 | No | NA | 14 | 11 | 0 | Low-quality | Genome-fragment | 12.04 | AAI-based (high-confidence) | 0 |
| S10_vs_56201\|\|lt2gene | 1153 | No | NA | 1 | 0 | 0 | Not-determined | Genome-fragment | NA | NA | 0 |
| S11_vs_k141_85911\|\|full | 3651 | No | NA | 4 | 3 | 0 | Low-quality | Genome-fragment | 9.37 | AAI-based (high-confidence) | 0 |
| S3_extracted_k141_38527 | 3651 | No | NA | 2 | 0 | 2 | Low-quality | Genome-fragment | 0.43 | AAI-based (medium-confidence) | 0 |
| S10_vs_66016\|\|lt2gene | 1828 | No | NA | 1 | 0 | 0 | Not-determined | Genome-fragment | NA | NA | 0 |
| S7_vs_47415\|\|full | 3651 | No | NA | 8 | 1 | 0 | Low-quality | Genome-fragment | 6.73 | AAI-based (medium-confidence) | 0 |
| S7_vs_4495\|\|full | 3652 | No | NA | 5 | 1 | 0 | Low-quality | Genome-fragment | 8.75 | AAI-based (medium-confidence) | 0 |
| S7_vs_67435\|\|full | 3652 | No | NA | 9 | 2 | 0 | Low-quality | Genome-fragment | 5.67 | AAI-based (high-confidence) | 0 |
| S10_extracted_50293 | 3652 | No | NA | 8 | 1 | 0 | Low-quality | Genome-fragment | 2.45 | HMM-based (lower-bound) | 0 |
| S3_vs_k141_90375\|\|full | 3653 | No | NA | 10 | 0 | 0 | Low-quality | Genome-fragment | 2.16 | AAI-based (high-confidence) | 0 |
| S12_vs_k141_185763\|\|full | 3653 | No | NA | 3 | 0 | 2 | Low-quality | Genome-fragment | 2.26 | AAI-based (medium-confidence) | 0 |
| S10_extracted_50941 | 3654 | No | NA | 3 | 0 | 1 | Low-quality | Genome-fragment | 1.88 | AAI-based (medium-confidence) | 0 |
| S14_extracted_k141_2658 | 3655 | No | NA | 10 | 2 | 0 | Low-quality | Genome-fragment | 7.56 | AAI-based (high-confidence) | 0 |
| S1_vs_k141_32624\|\|full | 3655 | No | NA | 6 | 0 | 1 | Low-quality | Genome-fragment | 1.69 | AAI-based (medium-confidence) | 0 |
| S5_vs_k141_113074\|\|full | 3655 | No | NA | 3 | 0 | 3 | Low-quality | Genome-fragment | 2.44 | AAI-based (medium-confidence) | 0 |
| S11_extracted_k141_61550 | 3656 | No | NA | 5 | 1 | 0 | Low-quality | Genome-fragment | 1.42 | AAI-based (medium-confidence) | 0 |
| S11_vs_k141_13132\|\|full | 3656 | No | NA | 4 | 1 | 0 | Low-quality | Genome-fragment | 9.22 | AAI-based (high-confidence) | 0 |
| S3_vs_k141_90000\|\|full | 3656 | No | NA | 5 | 4 | 0 | Low-quality | Genome-fragment | 10.49 | AAI-based (high-confidence) | 0 |
| S10_vs_83862\|\|lt2gene | 1415 | No | NA | 1 | 0 | 0 | Not-determined | Genome-fragment | NA | NA | 0 |
| S5_vs_k141_140239\|\|lt2gene | 3656 | No | NA | 1 | 1 | 0 | Low-quality | Genome-fragment | 7.58 | AAI-based (high-confidence) | 0 |
| S7_extracted_135518 | 3656 | No | NA | 2 | 0 | 1 | Low-quality | Genome-fragment | 0.67 | AAI-based (medium-confidence) | 0 |
| S8_vs_k141_207900\|\|full | 3656 | No | NA | 9 | 4 | 0 | Low-quality | Genome-fragment | 6.12 | HMM-based (lower-bound) | 0 |
| S1_extracted_k141_149005 | 3657 | No | NA | 2 | 1 | 0 | Low-quality | Genome-fragment | 7.23 | HMM-based (lower-bound) | 0 |
| S7_vs_79463\|\|full | 3657 | No | NA | 6 | 2 | 0 | Low-quality | Genome-fragment | 8.86 | AAI-based (high-confidence) | 0 |
| S10_vs_106737\|\|full | 3657 | No | NA | 11 | 0 | 0 | Low-quality | Genome-fragment | 8.62 | AAI-based (high-confidence) | 0 |
| S10_vs_89567\|\|lt2gene | 1333 | No | NA | 2 | 0 | 1 | Not-determined | Genome-fragment | NA | NA | 0 |
| S17_vs_k141_40675\|\|full | 3658 | No | NA | 4 | 0 | 1 | Low-quality | Genome-fragment | 10.45 | AAI-based (medium-confidence) | 0 |
| S11_extracted_k141_85675 | 3658 | No | NA | 4 | 0 | 1 | Low-quality | Genome-fragment | 9.05 | AAI-based (medium-confidence) | 0 |
| S14_extracted_k141_15548 | 3658 | No | NA | 9 | 4 | 0 | Low-quality | Genome-fragment | 9.09 | AAI-based (high-confidence) | 0 |
| S12_vs_k141_163811\|\|full | 3658 | No | NA | 6 | 0 | 0 | Low-quality | Genome-fragment | 8.5 | AAI-based (medium-confidence) | 0 |
| S7_extracted_104969 | 3659 | No | NA | 6 | 1 | 0 | Low-quality | Genome-fragment | 2.19 | AAI-based (medium-confidence) | 0 |
| S7_vs_65992\|\|full | 3659 | No | NA | 4 | 2 | 0 | Low-quality | Genome-fragment | 8.04 | AAI-based (high-confidence) | 0 |
| S17_extracted_k141_70841 | 3660 | No | NA | 9 | 0 | 0 | Low-quality | Genome-fragment | 2.12 | AAI-based (high-confidence) | 0 |
| S10_vs_113010\|\|lt2gene | 1207 | No | NA | 1 | 0 | 0 | Not-determined | Genome-fragment | NA | NA | 0 |
| S10_vs_112125\|\|full | 3660 | No | NA | 4 | 0 | 2 | Low-quality | Genome-fragment | 1.64 | AAI-based (medium-confidence) | 0 |
| S8_extracted_k141_178148 | 3660 | No | NA | 7 | 1 | 0 | Low-quality | Genome-fragment | 7.31 | AAI-based (high-confidence) | 0 |
| S10_vs_114193\|\|lt2gene | 1476 | No | NA | 1 | 0 | 0 | Not-determined | Genome-fragment | NA | NA | 0 |
| S7_extracted_21297 | 3661 | No | NA | 2 | 1 | 0 | Low-quality | Genome-fragment | 3.78 | AAI-based (high-confidence) | 0 |
| S10_vs_118871\|\|lt2gene | 1025 | No | NA | 1 | 0 | 1 | Not-determined | Genome-fragment | NA | NA | 0 |
| S12_extracted_k141_117520 | 3661 | No | NA | 4 | 0 | 0 | Low-quality | Genome-fragment | 8.39 | AAI-based (high-confidence) | 0 |
| S12_extracted_k141_94490 | 1098 | No | NA | 2 | 0 | 0 | Not-determined | Genome-fragment | NA | NA | 0 |
| S12_extracted_k141_125990 | 1053 | No | NA | 2 | 0 | 0 | Not-determined | Genome-fragment | NA | NA | 0 |
| S12_extracted_k141_157478 | 1271 | No | NA | 1 | 0 | 1 | Not-determined | Genome-fragment | NA | NA | 0 |
| S12_extracted_k141_78749 | 1157 | No | NA | 1 | 0 | 0 | Not-determined | Genome-fragment | NA | NA | 0 |
| S12_extracted_k141_125996 | 1004 | No | NA | 2 | 0 | 0 | Not-determined | Genome-fragment | NA | NA | 0 |
| S12_extracted_k141_181113 | 1995 | No | NA | 1 | 0 | 1 | Not-determined | Genome-fragment | NA | NA | 0 |
| S8_extracted_k141_104607 | 3661 | No | NA | 6 | 0 | 0 | Low-quality | Genome-fragment | 3.27 | AAI-based (high-confidence) | 0 |
| S12_extracted_k141_86637 | 1651 | No | NA | 1 | 0 | 1 | Not-determined | Genome-fragment | NA | NA | 0 |
| S12_extracted_k141_86641 | 2008 | No | NA | 4 | 1 | 0 | Not-determined | Genome-fragment | NA | NA | 0 |
| S12_extracted_k141_23651 | 1413 | No | NA | 3 | 0 | 2 | Not-determined | Genome-fragment | NA | NA | 0 |
| S12_extracted_k141_78773 | 2127 | No | NA | 2 | 0 | 1 | Not-determined | Genome-fragment | NA | NA | 0 |
| S12_extracted_k141_31523 | 3136 | No | NA | 4 | 0 | 0 | Not-determined | Genome-fragment | NA | NA | 0 |
| S12_extracted_k141_133899 | 1698 | No | NA | 2 | 0 | 1 | Not-determined | Genome-fragment | NA | NA | 0 |
| S8_vs_k141_82978\|\|full | 3661 | No | NA | 5 | 0 | 0 | Low-quality | Genome-fragment | 8.23 | AAI-based (medium-confidence) | 0 |
| S12_extracted_k141_110267 | 1439 | No | NA | 4 | 0 | 1 | Not-determined | Genome-fragment | NA | NA | 0 |
| S12_extracted_k141_31552 | 1200 | No | NA | 1 | 0 | 0 | Not-determined | Genome-fragment | NA | NA | 0 |
| S3_vs_k141_22841\|\|full | 3662 | No | NA | 5 | 0 | 0 | Low-quality | Genome-fragment | 8.47 | AAI-based (medium-confidence) | 0 |
| S12_extracted_k141_110274 | 1713 | No | NA | 2 | 0 | 2 | Not-determined | Genome-fragment | NA | NA | 0 |
| S16_vs_8209\|\|full | 3663 | No | NA | 2 | 0 | 1 | Low-quality | Genome-fragment | 2.06 | AAI-based (medium-confidence) | 0 |
| S12_extracted_k141_94549 | 1033 | No | NA | 1 | 0 | 1 | Not-determined | Genome-fragment | NA | NA | 0 |
| S10_extracted_43493 | 3663 | No | NA | 3 | 1 | 0 | Low-quality | Genome-fragment | 7.97 | AAI-based (high-confidence) | 0 |
| S12_extracted_k141_126060 | 2044 | No | NA | 1 | 0 | 1 | Not-determined | Genome-fragment | NA | NA | 0 |
| S7_vs_125527\|\|full | 3664 | No | NA | 3 | 0 | 2 | Low-quality | Genome-fragment | 8.47 | AAI-based (medium-confidence) | 0 |
| S12_extracted_k141_102437 | 1558 | No | NA | 2 | 0 | 2 | Not-determined | Genome-fragment | NA | NA | 0 |
| S8_extracted_k141_132581 | 3664 | No | NA | 4 | 0 | 4 | Low-quality | Genome-fragment | 1.37 | AAI-based (medium-confidence) | 0 |
| S12_extracted_k141_110299 | 1508 | No | NA | 1 | 0 | 1 | Not-determined | Genome-fragment | NA | NA | 0 |
| S12_extracted_k141_149682 | 1633 | No | NA | 2 | 0 | 0 | Not-determined | Genome-fragment | NA | NA | 0 |
| S12_extracted_k141_110303 | 1454 | No | NA | 2 | 0 | 2 | Not-determined | Genome-fragment | NA | NA | 0 |
| S7_extracted_67494 | 3665 | No | NA | 10 | 0 | 0 | Low-quality | Genome-fragment | 8.15 | AAI-based (high-confidence) | 0 |
| S12_extracted_k141_173301 | 1197 | No | NA | 2 | 0 | 2 | Not-determined | Genome-fragment | NA | NA | 0 |
| S12_extracted_k141_23695 | 12745 | No | NA | 13 | 0 | 3 | Not-determined | Genome-fragment | NA | NA | 0 |
| S12_extracted_k141_70950 | 1530 | No | NA | 2 | 0 | 0 | Not-determined | Genome-fragment | NA | NA | 0 |
| S12_extracted_k141_173306 | 1179 | No | NA | 2 | 0 | 0 | Not-determined | Genome-fragment | NA | NA | 0 |
| S12_extracted_k141_118161 | 1357 | No | NA | 2 | 0 | 2 | Not-determined | Genome-fragment | NA | NA | 0 |
| S14_vs_k141_104246\|\|full | 3666 | No | NA | 5 | 0 | 1 | Low-quality | Genome-fragment | 6.44 | AAI-based (medium-confidence) | 0 |
| S12_extracted_k141_181182 | 1067 | No | NA | 2 | 0 | 1 | Not-determined | Genome-fragment | NA | NA | 0 |
| S12_extracted_k141_15807 | 2174 | No | NA | 2 | 0 | 2 | Not-determined | Genome-fragment | NA | NA | 0 |
| S12_extracted_k141_149698 | 1258 | No | NA | 1 | 0 | 1 | Not-determined | Genome-fragment | NA | NA | 0 |
| S12_extracted_k141_23726 | 1003 | No | NA | 1 | 0 | 1 | Not-determined | Genome-fragment | NA | NA | 0 |
| S14_vs_k141_83504\|\|full | 3666 | No | NA | 3 | 0 | 3 | Low-quality | Genome-fragment | 3.17 | AAI-based (medium-confidence) | 0 |
| S14_vs_k141_84912\|\|full | 3666 | No | NA | 7 | 4 | 0 | Low-quality | Genome-fragment | 8.86 | AAI-based (high-confidence) | 0 |
| S12_extracted_k141_149725 | 1416 | No | NA | 1 | 0 | 0 | Not-determined | Genome-fragment | NA | NA | 0 |
| S12_extracted_k141_39446 | 1961 | No | NA | 3 | 0 | 2 | Not-determined | Genome-fragment | NA | NA | 0 |
| S12_extracted_k141_165495 | 5685 | No | NA | 4 | 0 | 4 | Not-determined | Genome-fragment | NA | NA | 0 |
| S12_extracted_k141_71007 | 1207 | No | NA | 1 | 0 | 0 | Not-determined | Genome-fragment | NA | NA | 0 |
| S12_extracted_k141_173355 | 1604 | No | NA | 2 | 0 | 1 | Not-determined | Genome-fragment | NA | NA | 0 |
| S12_extracted_k141_102488 | 2887 | No | NA | 2 | 0 | 1 | Not-determined | Genome-fragment | NA | NA | 0 |
| S12_extracted_k141_126149 | 8840 | No | NA | 10 | 0 | 3 | Not-determined | Genome-fragment | NA | NA | 0 |
| S12_extracted_k141_118218 | 1101 | No | NA | 2 | 0 | 1 | Not-determined | Genome-fragment | NA | NA | 0 |
| S12_extracted_k141_55175 | 42177 | No | NA | 46 | 9 | 1 | Complete | High-quality | 100 | DTR (high-confidence) | 0 |
| S12_extracted_k141_8027 | 3974 | No | NA | 2 | 0 | 0 | Not-determined | Genome-fragment | NA | NA | 0 |
| S12_extracted_k141_63120 | 1179 | No | NA | 2 | 0 | 0 | Not-determined | Genome-fragment | NA | NA | 0 |
| S5_vs_k141_18062\|\|full | 3666 | Yes | 1457 | 4 | 1 | 1 | Low-quality | Genome-fragment | 0.12 | HMM-based (lower-bound) | 60.26 |
| S12_extracted_k141_165536 | 1006 | No | NA | 1 | 0 | 1 | Not-determined | Genome-fragment | NA | NA | 0 |
| S12_extracted_k141_47333 | 1250 | No | NA | 1 | 0 | 0 | Not-determined | Genome-fragment | NA | NA | 0 |
| S12_extracted_k141_124936 | 3666 | No | NA | 3 | 0 | 2 | Low-quality | Genome-fragment | 1.8 | AAI-based (medium-confidence) | 0 |
| S12_extracted_k141_173391 | 1157 | No | NA | 2 | 0 | 1 | Not-determined | Genome-fragment | NA | NA | 0 |
| S12_vs_k141_155899\|\|full | 3666 | No | NA | 4 | 0 | 2 | Low-quality | Genome-fragment | 1.03 | AAI-based (high-confidence) | 0 |
| S12_extracted_k141_71053 | 4117 | No | NA | 4 | 0 | 2 | Not-determined | Genome-fragment | NA | NA | 0 |
| S12_extracted_k141_165552 | 1867 | No | NA | 1 | 0 | 1 | Not-determined | Genome-fragment | NA | NA | 0 |
| S12_extracted_k141_254 | 2212 | No | NA | 1 | 0 | 0 | Not-determined | Genome-fragment | NA | NA | 0 |
| S12_extracted_k141_78907 | 1664 | No | NA | 2 | 0 | 1 | Not-determined | Genome-fragment | NA | NA | 0 |
| S11_extracted_k141_21073 | 3667 | No | NA | 8 | 1 | 0 | Low-quality | Genome-fragment | 8.08 | HMM-based (lower-bound) | 0 |
| S12_extracted_k141_173404 | 1043 | No | NA | 1 | 0 | 1 | Not-determined | Genome-fragment | NA | NA | 0 |
| S5_vs_k141_12990\|\|full | 3667 | No | NA | 9 | 4 | 0 | Low-quality | Genome-fragment | 7.3 | HMM-based (lower-bound) | 0 |
| S12_extracted_k141_47376 | 1118 | No | NA | 2 | 0 | 2 | Not-determined | Genome-fragment | NA | NA | 0 |
| S12_extracted_k141_134025 | 7535 | No | NA | 12 | 0 | 4 | Not-determined | Genome-fragment | NA | NA | 0 |
| S12_extracted_k141_47384 | 1091 | No | NA | 1 | 0 | 0 | Not-determined | Genome-fragment | NA | NA | 0 |
| S12_extracted_k141_102561 | 1258 | No | NA | 1 | 0 | 0 | Not-determined | Genome-fragment | NA | NA | 0 |
| S11_extracted_k141_31936 | 3668 | No | NA | 3 | 0 | 1 | Low-quality | Genome-fragment | 3.56 | AAI-based (medium-confidence) | 0 |
| S12_extracted_k141_110410 | 1413 | No | NA | 3 | 0 | 3 | Not-determined | Genome-fragment | NA | NA | 0 |
| S12_extracted_k141_94695 | 2044 | No | NA | 3 | 0 | 3 | Not-determined | Genome-fragment | NA | NA | 0 |
| S12_extracted_k141_134049 | 3221 | No | NA | 5 | 0 | 3 | Not-determined | Genome-fragment | NA | NA | 0 |
| S7_extracted_16274 | 3669 | No | NA | 3 | 2 | 0 | Low-quality | Genome-fragment | 16.73 | AAI-based (high-confidence) | 0 |
| S16_extracted_18718 | 3669 | No | NA | 11 | 0 | 0 | Low-quality | Genome-fragment | 7.42 | AAI-based (high-confidence) | 0 |
| S12_extracted_k141_78956 | 2161 | No | NA | 3 | 0 | 1 | Not-determined | Genome-fragment | NA | NA | 0 |
| S12_extracted_k141_181320 | 2058 | No | NA | 1 | 0 | 1 | Not-determined | Genome-fragment | NA | NA | 0 |
| S16_extracted_165517 | 3669 | No | NA | 4 | 0 | 2 | Low-quality | Genome-fragment | 7.79 | AAI-based (medium-confidence) | 0 |
| S12_extracted_k141_134064 | 2061 | No | NA | 3 | 0 | 0 | Not-determined | Genome-fragment | NA | NA | 0 |
| S12_extracted_k141_15870 | 3251 | No | NA | 3 | 0 | 1 | Not-determined | Genome-fragment | NA | NA | 0 |
| S12_extracted_k141_63217 | 1615 | No | NA | 2 | 0 | 1 | Not-determined | Genome-fragment | NA | NA | 0 |
| S12_extracted_k141_181331 | 3940 | No | NA | 2 | 0 | 2 | Not-determined | Genome-fragment | NA | NA | 0 |
| S12_extracted_k141_327 | 1878 | No | NA | 3 | 0 | 1 | Not-determined | Genome-fragment | NA | NA | 0 |
| S12_extracted_k141_173493 | 1426 | No | NA | 2 | 0 | 0 | Not-determined | Genome-fragment | NA | NA | 0 |
| S12_extracted_k141_149821 | 1284 | No | NA | 2 | 0 | 1 | Not-determined | Genome-fragment | NA | NA | 0 |
| S12_extracted_k141_39548 | 1575 | No | NA | 3 | 0 | 1 | Not-determined | Genome-fragment | NA | NA | 0 |
| S12_extracted_k141_332 | 1714 | No | NA | 1 | 0 | 0 | Not-determined | Genome-fragment | NA | NA | 0 |
| S12_extracted_k141_126243 | 1631 | No | NA | 2 | 0 | 1 | Not-determined | Genome-fragment | NA | NA | 0 |
| S12_extracted_k141_173502 | 1397 | No | NA | 1 | 0 | 0 | Not-determined | Genome-fragment | NA | NA | 0 |
| S12_extracted_k141_141924 | 1397 | No | NA | 2 | 0 | 0 | Not-determined | Genome-fragment | NA | NA | 0 |
| S12_extracted_k141_39558 | 1041 | No | NA | 1 | 0 | 1 | Not-determined | Genome-fragment | NA | NA | 0 |
| S12_extracted_k141_110482 | 1262 | No | NA | 2 | 0 | 0 | Not-determined | Genome-fragment | NA | NA | 0 |
| S12_extracted_k141_63249 | 2217 | No | NA | 2 | 0 | 0 | Not-determined | Genome-fragment | NA | NA | 0 |
| S12_extracted_k141_343 | 1700 | No | NA | 2 | 0 | 1 | Not-determined | Genome-fragment | NA | NA | 0 |
| S12_extracted_k141_126258 | 6004 | No | NA | 5 | 0 | 1 | Not-determined | Genome-fragment | NA | NA | 0 |
| S12_extracted_k141_39591 | 1033 | No | NA | 1 | 0 | 0 | Not-determined | Genome-fragment | NA | NA | 0 |
| S15_vs_53388\|\|full | 3669 | No | NA | 4 | 3 | 0 | Low-quality | Genome-fragment | 17.17 | AAI-based (medium-confidence) | 0 |
| S10_extracted_112316 | 3670 | No | NA | 3 | 0 | 0 | Low-quality | Genome-fragment | 5.26 | AAI-based (medium-confidence) | 0 |
| S12_extracted_k141_15897 | 1819 | No | NA | 1 | 0 | 0 | Not-determined | Genome-fragment | NA | NA | 0 |
| S12_extracted_k141_94811 | 1000 | No | NA | 1 | 0 | 0 | Not-determined | Genome-fragment | NA | NA | 0 |
| S12_vs_k141_114984\|\|full | 3670 | No | NA | 3 | 0 | 3 | Low-quality | Genome-fragment | 2.4 | AAI-based (medium-confidence) | 0 |
| S12_extracted_k141_110532 | 1015 | No | NA | 2 | 0 | 0 | Not-determined | Genome-fragment | NA | NA | 0 |
| S17_vs_k141_23998\|\|full | 3671 | No | NA | 4 | 0 | 0 | Low-quality | Genome-fragment | 8.92 | AAI-based (medium-confidence) | 0 |
| S12_extracted_k141_134142 | 6648 | No | NA | 7 | 0 | 3 | Not-determined | Genome-fragment | NA | NA | 0 |
| S12_extracted_k141_141993 | 3037 | No | NA | 4 | 0 | 2 | Not-determined | Genome-fragment | NA | NA | 0 |
| S12_extracted_k141_71198 | 1387 | No | NA | 1 | 0 | 0 | Not-determined | Genome-fragment | NA | NA | 0 |
| S12_extracted_k141_86960 | 4553 | No | NA | 3 | 0 | 1 | Not-determined | Genome-fragment | NA | NA | 0 |
| S12_extracted_k141_8173 | 13877 | No | NA | 17 | 7 | 0 | Medium-quality | Genome-fragment | 62.34 | AAI-based (medium-confidence) | 0 |
| S12_extracted_k141_427 | 1883 | No | NA | 2 | 0 | 0 | Not-determined | Genome-fragment | NA | NA | 0 |
| S12_extracted_k141_431 | 2663 | No | NA | 4 | 0 | 1 | Not-determined | Genome-fragment | NA | NA | 0 |
| S12_extracted_k141_165678 | 1127 | No | NA | 2 | 0 | 2 | Not-determined | Genome-fragment | NA | NA | 0 |
| S12_extracted_k141_86996 | 1690 | No | NA | 2 | 0 | 0 | Not-determined | Genome-fragment | NA | NA | 0 |
| S12_extracted_k141_63327 | 1201 | No | NA | 2 | 0 | 2 | Not-determined | Genome-fragment | NA | NA | 0 |
| S12_extracted_k141_63329 | 1395 | No | NA | 1 | 0 | 1 | Not-determined | Genome-fragment | NA | NA | 0 |
| S12_extracted_k141_15959 | 3694 | No | NA | 3 | 0 | 1 | Not-determined | Genome-fragment | NA | NA | 0 |
| S11_vs_k141_5369\|\|full | 3671 | No | NA | 5 | 0 | 0 | Low-quality | Genome-fragment | 7.29 | AAI-based (high-confidence) | 0 |
| S12_extracted_k141_55382 | 1021 | No | NA | 2 | 0 | 0 | Not-determined | Genome-fragment | NA | NA | 0 |
| S13_extracted_10522 | 3672 | No | NA | 3 | 0 | 1 | Low-quality | Genome-fragment | 6.57 | AAI-based (medium-confidence) | 0 |
| S12_extracted_k141_55390 | 9834 | No | NA | 12 | 0 | 3 | Not-determined | Genome-fragment | NA | NA | 0 |
| S5_vs_k141_110519\|\|full | 3672 | No | NA | 3 | 0 | 1 | Low-quality | Genome-fragment | 5.66 | AAI-based (medium-confidence) | 0 |
| S12_extracted_k141_87038 | 3783 | No | NA | 4 | 0 | 3 | Not-determined | Genome-fragment | NA | NA | 0 |
| S12_extracted_k141_55404 | 1322 | No | NA | 1 | 0 | 1 | Not-determined | Genome-fragment | NA | NA | 0 |
| S5_vs_k141_102365\|\|full | 3672 | No | NA | 5 | 0 | 1 | Low-quality | Genome-fragment | 5.3 | AAI-based (medium-confidence) | 0 |
| S12_extracted_k141_142082 | 4271 | No | NA | 4 | 0 | 3 | Not-determined | Genome-fragment | NA | NA | 0 |
| S12_extracted_k141_149892 | 2096 | No | NA | 5 | 0 | 0 | Not-determined | Genome-fragment | NA | NA | 0 |
| S12_extracted_k141_15984 | 56306 | No | NA | 30 | 0 | 24 | Not-determined | Genome-fragment | NA | NA | 0 |
| S12_extracted_k141_39754 | 4849 | No | NA | 4 | 0 | 4 | Not-determined | Genome-fragment | NA | NA | 0 |
| S12_extracted_k141_181409 | 1679 | No | NA | 3 | 0 | 1 | Not-determined | Genome-fragment | NA | NA | 0 |
| S12_extracted_k141_55442 | 1383 | No | NA | 3 | 0 | 1 | Not-determined | Genome-fragment | NA | NA | 0 |
| S12_extracted_k141_79130 | 3570 | No | NA | 4 | 0 | 2 | Not-determined | Genome-fragment | NA | NA | 0 |
| S12_extracted_k141_94904 | 1592 | No | NA | 1 | 0 | 1 | Not-determined | Genome-fragment | NA | NA | 0 |
| S12_extracted_k141_509 | 7064 | No | NA | 5 | 0 | 1 | Not-determined | Genome-fragment | NA | NA | 0 |
| S16_extracted_81079 | 3672 | No | NA | 4 | 0 | 2 | Low-quality | Genome-fragment | 1.45 | AAI-based (medium-confidence) | 0 |
| S12_extracted_k141_79131 | 1794 | No | NA | 2 | 0 | 0 | Not-determined | Genome-fragment | NA | NA | 0 |
| S12_extracted_k141_79133 | 1530 | No | NA | 1 | 0 | 1 | Not-determined | Genome-fragment | NA | NA | 0 |
| S12_extracted_k141_181422 | 1226 | No | NA | 2 | 0 | 0 | Not-determined | Genome-fragment | NA | NA | 0 |
| S12_extracted_k141_31767 | 1101 | No | NA | 1 | 0 | 1 | Not-determined | Genome-fragment | NA | NA | 0 |
| S12_extracted_k141_142154 | 5957 | No | NA | 5 | 0 | 1 | Not-determined | Genome-fragment | NA | NA | 0 |
| S12_extracted_k141_181430 | 2825 | No | NA | 2 | 0 | 1 | Not-determined | Genome-fragment | NA | NA | 0 |
| S12_extracted_k141_181433 | 1292 | No | NA | 2 | 0 | 0 | Not-determined | Genome-fragment | NA | NA | 0 |
| S12_extracted_k141_23813 | 1488 | No | NA | 2 | 0 | 2 | Not-determined | Genome-fragment | NA | NA | 0 |
| S12_extracted_k141_63420 | 1661 | No | NA | 2 | 0 | 2 | Not-determined | Genome-fragment | NA | NA | 0 |
| S8_extracted_k141_63270 | 3672 | No | NA | 2 | 0 | 1 | Low-quality | Genome-fragment | 1.52 | AAI-based (medium-confidence) | 0 |
| S12_extracted_k141_142183 | 1333 | No | NA | 1 | 0 | 1 | Not-determined | Genome-fragment | NA | NA | 0 |
| S12_extracted_k141_31815 | 1023 | No | NA | 2 | 0 | 1 | Not-determined | Genome-fragment | NA | NA | 0 |
| S9_vs_k141_82632\|\|full | 3674 | No | NA | 4 | 0 | 1 | Low-quality | Genome-fragment | 2.32 | AAI-based (medium-confidence) | 0 |
| S12_extracted_k141_87092 | 1025 | No | NA | 1 | 0 | 1 | Not-determined | Genome-fragment | NA | NA | 0 |
| S12_extracted_k141_157881 | 1851 | No | NA | 4 | 0 | 4 | Not-determined | Genome-fragment | NA | NA | 0 |
| S12_extracted_k141_79189 | 5230 | No | NA | 7 | 0 | 0 | Not-determined | Genome-fragment | NA | NA | 0 |
| S12_extracted_k141_142195 | 3477 | No | NA | 7 | 0 | 3 | Not-determined | Genome-fragment | NA | NA | 0 |
| S3_vs_k141_30480\|\|full | 3674 | No | NA | 3 | 0 | 3 | Low-quality | Genome-fragment | 2.48 | AAI-based (medium-confidence) | 0 |
| S16_extracted_62385 | 3674 | No | NA | 2 | 0 | 2 | Low-quality | Genome-fragment | 3.6 | AAI-based (high-confidence) | 0 |
| S12_extracted_k141_63453 | 1441 | No | NA | 1 | 0 | 1 | Not-determined | Genome-fragment | NA | NA | 0 |
| S12_extracted_k141_16049 | 5462 | No | NA | 6 | 0 | 2 | Not-determined | Genome-fragment | NA | NA | 0 |
| S17_extracted_k141_67470 | 3675 | No | NA | 2 | 0 | 2 | Low-quality | Genome-fragment | 0.32 | AAI-based (medium-confidence) | 0 |
| S12_extracted_k141_586 | 1336 | No | NA | 2 | 0 | 1 | Not-determined | Genome-fragment | NA | NA | 0 |
| S12_extracted_k141_126512 | 1000 | No | NA | 2 | 0 | 2 | Not-determined | Genome-fragment | NA | NA | 0 |
| S12_extracted_k141_142217 | 5625 | No | NA | 4 | 0 | 4 | Not-determined | Genome-fragment | NA | NA | 0 |
| S12_extracted_k141_142219 | 1255 | No | NA | 3 | 0 | 2 | Not-determined | Genome-fragment | NA | NA | 0 |
| S12_extracted_k141_134163 | 1982 | No | NA | 3 | 0 | 1 | Not-determined | Genome-fragment | NA | NA | 0 |
| S12_extracted_k141_87123 | 1826 | No | NA | 2 | 0 | 1 | Not-determined | Genome-fragment | NA | NA | 0 |
| S11_vs_k141_28038\|\|full | 3676 | Yes | 2571 | 4 | 1 | 1 | Low-quality | Genome-fragment | 5.46 | AAI-based (high-confidence) | 30.06 |
| S12_extracted_k141_31868 | 2308 | No | NA | 2 | 0 | 1 | Not-determined | Genome-fragment | NA | NA | 0 |
| S12_extracted_k141_16075 | 2021 | No | NA | 1 | 0 | 1 | Not-determined | Genome-fragment | NA | NA | 0 |
| S12_extracted_k141_118583 | 1117 | No | NA | 1 | 0 | 1 | Not-determined | Genome-fragment | NA | NA | 0 |
| S12_extracted_k141_655 | 2204 | No | NA | 2 | 0 | 2 | Not-determined | Genome-fragment | NA | NA | 0 |
| S1_extracted_k141_145217 | 3676 | No | NA | 8 | 0 | 0 | Low-quality | Genome-fragment | 3.94 | AAI-based (high-confidence) | 0 |
| S12_extracted_k141_659 | 4580 | No | NA | 8 | 0 | 2 | Not-determined | Genome-fragment | NA | NA | 0 |
| S12_extracted_k141_181533 | 5844 | No | NA | 7 | 0 | 2 | Not-determined | Genome-fragment | NA | NA | 0 |
| S12_extracted_k141_55556 | 2756 | No | NA | 2 | 0 | 0 | Not-determined | Genome-fragment | NA | NA | 0 |
| S5_vs_k141_117606\|\|full | 3676 | No | NA | 3 | 0 | 0 | Low-quality | Genome-fragment | 3.37 | AAI-based (medium-confidence) | 0 |
| S12_extracted_k141_87173 | 1089 | No | NA | 1 | 0 | 0 | Not-determined | Genome-fragment | NA | NA | 0 |
| S12_extracted_k141_87174 | 1623 | No | NA | 2 | 0 | 1 | Not-determined | Genome-fragment | NA | NA | 0 |
| S5_extracted_k141_38349 | 3677 | No | NA | 7 | 0 | 0 | Low-quality | Genome-fragment | 2.06 | AAI-based (high-confidence) | 0 |
| S12_extracted_k141_165840 | 1081 | No | NA | 1 | 0 | 1 | Not-determined | Genome-fragment | NA | NA | 0 |
| S12_extracted_k141_71306 | 1017 | No | NA | 1 | 0 | 1 | Not-determined | Genome-fragment | NA | NA | 0 |
| S11_extracted_k141_98419 | 3679 | No | NA | 5 | 0 | 1 | Low-quality | Genome-fragment | 12.51 | AAI-based (medium-confidence) | 0 |
| S12_extracted_k141_697 | 1905 | No | NA | 1 | 0 | 0 | Not-determined | Genome-fragment | NA | NA | 0 |
| S12_extracted_k141_8284 | 1064 | No | NA | 2 | 0 | 1 | Not-determined | Genome-fragment | NA | NA | 0 |
| S12_extracted_k141_79284 | 5446 | No | NA | 6 | 0 | 5 | Not-determined | Genome-fragment | NA | NA | 0 |
| S7_vs_4804\|\|full | 3679 | No | NA | 5 | 0 | 1 | Low-quality | Genome-fragment | 5.96 | AAI-based (high-confidence) | 0 |
| S12_extracted_k141_181571 | 6963 | No | NA | 11 | 0 | 2 | Not-determined | Genome-fragment | NA | NA | 0 |
| S12_extracted_k141_150059 | 1482 | No | NA | 1 | 0 | 1 | Not-determined | Genome-fragment | NA | NA | 0 |
| S12_extracted_k141_142342 | 1896 | No | NA | 1 | 0 | 1 | Not-determined | Genome-fragment | NA | NA | 0 |
| S16_vs_145504\|\|full | 3679 | No | NA | 7 | 1 | 0 | Low-quality | Genome-fragment | 6.76 | AAI-based (high-confidence) | 0 |
| S12_extracted_k141_31898 | 5387 | No | NA | 4 | 0 | 4 | Not-determined | Genome-fragment | NA | NA | 0 |
| S12_extracted_k141_31904 | 2527 | No | NA | 3 | 0 | 3 | Not-determined | Genome-fragment | NA | NA | 0 |
| S12_extracted_k141_94996 | 1542 | No | NA | 1 | 0 | 1 | Not-determined | Genome-fragment | NA | NA | 0 |
| S12_extracted_k141_31909 | 1591 | No | NA | 2 | 0 | 1 | Not-determined | Genome-fragment | NA | NA | 0 |
| S12_extracted_k141_95000 | 1359 | No | NA | 2 | 0 | 0 | Not-determined | Genome-fragment | NA | NA | 0 |
| S12_extracted_k141_31917 | 1260 | No | NA | 1 | 0 | 0 | Not-determined | Genome-fragment | NA | NA | 0 |
| S12_extracted_k141_732 | 1024 | No | NA | 2 | 0 | 0 | Not-determined | Genome-fragment | NA | NA | 0 |
| S12_extracted_k141_102828 | 1069 | No | NA | 1 | 0 | 0 | Not-determined | Genome-fragment | NA | NA | 0 |
| S8_extracted_k141_74119 | 3680 | No | NA | 2 | 1 | 0 | Low-quality | Genome-fragment | 13.13 | HMM-based (lower-bound) | 0 |
| S12_extracted_k141_8321 | 1022 | No | NA | 2 | 0 | 1 | Not-determined | Genome-fragment | NA | NA | 0 |
| S12_extracted_k141_747 | 3039 | No | NA | 3 | 0 | 1 | Not-determined | Genome-fragment | NA | NA | 0 |
| S12_extracted_k141_39885 | 1293 | No | NA | 2 | 0 | 1 | Not-determined | Genome-fragment | NA | NA | 0 |
| S12_extracted_k141_8334 | 1216 | No | NA | 1 | 0 | 1 | Not-determined | Genome-fragment | NA | NA | 0 |
| S8_vs_k141_129837\|\|full | 3680 | No | NA | 5 | 0 | 0 | Low-quality | Genome-fragment | 4.56 | AAI-based (medium-confidence) | 0 |
| S12_extracted_k141_150132 | 1029 | No | NA | 2 | 0 | 1 | Not-determined | Genome-fragment | NA | NA | 0 |
| S12_extracted_k141_102879 | 2293 | No | NA | 3 | 0 | 2 | Not-determined | Genome-fragment | NA | NA | 0 |
| S12_extracted_k141_118744 | 1025 | No | NA | 2 | 0 | 0 | Not-determined | Genome-fragment | NA | NA | 0 |
| S12_extracted_k141_786 | 1514 | No | NA | 1 | 0 | 0 | Not-determined | Genome-fragment | NA | NA | 0 |
| S17_vs_k141_105050\|\|full | 3681 | No | NA | 7 | 3 | 0 | Low-quality | Genome-fragment | 8.56 | AAI-based (high-confidence) | 0 |
| S3_extracted_k141_86879 | 3681 | No | NA | 9 | 0 | 0 | Low-quality | Genome-fragment | 2.03 | AAI-based (high-confidence) | 0 |
| S16_extracted_36472 | 3681 | No | NA | 5 | 0 | 4 | Low-quality | Genome-fragment | 1.38 | AAI-based (high-confidence) | 0 |
| S12_extracted_k141_158081 | 2289 | No | NA | 2 | 0 | 0 | Not-determined | Genome-fragment | NA | NA | 0 |
| S12_extracted_k141_142419 | 3351 | No | NA | 8 | 0 | 7 | Not-determined | Genome-fragment | NA | NA | 0 |
| S14_vs_k141_78435\|\|full | 3682 | No | NA | 7 | 1 | 0 | Low-quality | Genome-fragment | 6.32 | AAI-based (medium-confidence) | 0 |
| S12_extracted_k141_63712 | 3449 | No | NA | 3 | 0 | 1 | Not-determined | Genome-fragment | NA | NA | 0 |
| S15_vs_39960\|\|full | 3682 | No | NA | 8 | 2 | 0 | Low-quality | Genome-fragment | 7.61 | AAI-based (high-confidence) | 0 |
| S12_extracted_k141_47605 | 1622 | No | NA | 3 | 0 | 2 | Not-determined | Genome-fragment | NA | NA | 0 |
| S12_extracted_k141_16124 | 3767 | No | NA | 3 | 0 | 0 | Not-determined | Genome-fragment | NA | NA | 0 |
| S12_extracted_k141_134302 | 7964 | No | NA | 6 | 0 | 2 | Not-determined | Genome-fragment | NA | NA | 0 |
| S8_vs_k141_130274\|\|full | 3682 | No | NA | 5 | 0 | 3 | Low-quality | Genome-fragment | 0.91 | AAI-based (medium-confidence) | 0 |
| S12_extracted_k141_32023 | 2220 | No | NA | 4 | 0 | 4 | Not-determined | Genome-fragment | NA | NA | 0 |
| S12_extracted_k141_39956 | 10001 | No | NA | 6 | 0 | 3 | Not-determined | Genome-fragment | NA | NA | 0 |
| S16_vs_97655\|\|full | 3683 | No | NA | 5 | 0 | 0 | Low-quality | Genome-fragment | 2.17 | AAI-based (high-confidence) | 0 |
| S12_extracted_k141_118797 | 11596 | No | NA | 5 | 0 | 3 | Not-determined | Genome-fragment | NA | NA | 0 |
| S12_extracted_k141_16163 | 1265 | No | NA | 2 | 0 | 2 | Not-determined | Genome-fragment | NA | NA | 0 |
| S12_extracted_k141_134346 | 1226 | No | NA | 2 | 0 | 0 | Not-determined | Genome-fragment | NA | NA | 0 |
| S12_extracted_k141_95164 | 1013 | No | NA | 1 | 0 | 0 | Not-determined | Genome-fragment | NA | NA | 0 |
| S12_extracted_k141_173666 | 30650 | No | NA | 44 | 18 | 1 | High-quality | High-quality | 95.84 | AAI-based (high-confidence) | 0 |
| S12_extracted_k141_23671 | 3683 | No | NA | 8 | 1 | 0 | Low-quality | Genome-fragment | 4.7 | AAI-based (high-confidence) | 0 |
| S12_extracted_k141_32056 | 8543 | No | NA | 9 | 0 | 2 | Not-determined | Genome-fragment | NA | NA | 0 |
| S12_extracted_k141_55709 | 3012 | No | NA | 3 | 0 | 1 | Not-determined | Genome-fragment | NA | NA | 0 |
| S12_extracted_k141_126656 | 3749 | No | NA | 5 | 0 | 5 | Not-determined | Genome-fragment | NA | NA | 0 |
| S12_extracted_k141_95180 | 4665 | No | NA | 7 | 0 | 3 | Not-determined | Genome-fragment | NA | NA | 0 |
| S12_extracted_k141_32061 | 7946 | No | NA | 4 | 0 | 2 | Not-determined | Genome-fragment | NA | NA | 0 |
| S12_extracted_k141_150230 | 1684 | No | NA | 1 | 0 | 1 | Not-determined | Genome-fragment | NA | NA | 0 |
| S8_vs_k141_126511\|\|full | 3683 | No | NA | 3 | 0 | 2 | Low-quality | Genome-fragment | 1.15 | AAI-based (medium-confidence) | 0 |
| S12_extracted_k141_173675 | 2312 | No | NA | 2 | 0 | 1 | Not-determined | Genome-fragment | NA | NA | 0 |
| S3_vs_k141_1950\|\|full | 3684 | No | NA | 1 | 0 | 1 | Low-quality | Genome-fragment | 2.04 | AAI-based (medium-confidence) | 0 |
| S12_extracted_k141_11605 | 3684 | No | NA | 6 | 1 | 0 | Low-quality | Genome-fragment | 8.9 | AAI-based (medium-confidence) | 0 |
| S11_extracted_k141_21258 | 3685 | No | NA | 5 | 1 | 0 | Low-quality | Genome-fragment | 9.1 | AAI-based (medium-confidence) | 0 |
| S12_extracted_k141_24209 | 1212 | No | NA | 1 | 0 | 1 | Not-determined | Genome-fragment | NA | NA | 0 |
| S14_extracted_k141_51535 | 3685 | No | NA | 9 | 9 | 0 | Low-quality | Genome-fragment | 12.12 | AAI-based (high-confidence) | 0 |
| S12_extracted_k141_24211 | 1127 | No | NA | 2 | 0 | 0 | Not-determined | Genome-fragment | NA | NA | 0 |
| S12_vs_k141_111272\|\|full | 3685 | Yes | 1802 | 6 | 1 | 2 | Low-quality | Genome-fragment | 0.94 | HMM-based (lower-bound) | 51.1 |
| S12_vs_k141_116875\|\|full | 3685 | No | NA | 4 | 0 | 2 | Low-quality | Genome-fragment | 2.53 | AAI-based (high-confidence) | 0 |
| S16_extracted_34778 | 3686 | No | NA | 8 | 1 | 0 | Low-quality | Genome-fragment | 8.55 | AAI-based (high-confidence) | 0 |
| S12_extracted_k141_87347 | 3137 | No | NA | 4 | 0 | 0 | Not-determined | Genome-fragment | NA | NA | 0 |
| S12_extracted_k141_166066 | 1099 | No | NA | 2 | 0 | 1 | Not-determined | Genome-fragment | NA | NA | 0 |
| S13_vs_57457\|\|full | 3687 | No | NA | 5 | 0 | 1 | Low-quality | Genome-fragment | 8.79 | AAI-based (medium-confidence) | 0 |
| S12_extracted_k141_55765 | 1105 | No | NA | 1 | 0 | 0 | Not-determined | Genome-fragment | NA | NA | 0 |
| S12_extracted_k141_40072 | 1006 | No | NA | 1 | 0 | 0 | Not-determined | Genome-fragment | NA | NA | 0 |
| S12_extracted_k141_55789 | 1058 | No | NA | 1 | 0 | 0 | Not-determined | Genome-fragment | NA | NA | 0 |
| S12_extracted_k141_158163 | 3730 | No | NA | 5 | 0 | 4 | Not-determined | Genome-fragment | NA | NA | 0 |
| S12_extracted_k141_8552 | 1153 | No | NA | 2 | 0 | 1 | Not-determined | Genome-fragment | NA | NA | 0 |
| S12_extracted_k141_150299 | 3310 | No | NA | 3 | 0 | 0 | Not-determined | Genome-fragment | NA | NA | 0 |
| S12_extracted_k141_16280 | 9800 | No | NA | 12 | 0 | 3 | Not-determined | Genome-fragment | NA | NA | 0 |
| S12_extracted_k141_150302 | 1104 | No | NA | 3 | 0 | 2 | Not-determined | Genome-fragment | NA | NA | 0 |
| S12_extracted_k141_103119 | 5124 | No | NA | 5 | 0 | 3 | Not-determined | Genome-fragment | NA | NA | 0 |
| S12_extracted_k141_158195 | 1141 | No | NA | 2 | 0 | 0 | Not-determined | Genome-fragment | NA | NA | 0 |
| S12_extracted_k141_940 | 3385 | No | NA | 7 | 0 | 1 | Not-determined | Genome-fragment | NA | NA | 0 |
| S12_extracted_k141_55816 | 3617 | No | NA | 3 | 0 | 2 | Not-determined | Genome-fragment | NA | NA | 0 |
| S12_extracted_k141_82703 | 3687 | No | NA | 1 | 0 | 1 | Low-quality | Genome-fragment | 1.68 | AAI-based (high-confidence) | 0 |
| S12_extracted_k141_150340 | 1961 | No | NA | 2 | 0 | 2 | Not-determined | Genome-fragment | NA | NA | 0 |
| S12_extracted_k141_40134 | 1206 | No | NA | 2 | 0 | 1 | Not-determined | Genome-fragment | NA | NA | 0 |
| S16_vs_48149\|\|full | 3688 | No | NA | 4 | 0 | 1 | Low-quality | Genome-fragment | 4.24 | AAI-based (medium-confidence) | 0 |
| S12_extracted_k141_118887 | 1458 | No | NA | 1 | 0 | 0 | Not-determined | Genome-fragment | NA | NA | 0 |
| S12_extracted_k141_150345 | 7119 | No | NA | 8 | 0 | 1 | Not-determined | Genome-fragment | NA | NA | 0 |
| S12_extracted_k141_166105 | 5704 | No | NA | 6 | 0 | 1 | Not-determined | Genome-fragment | NA | NA | 0 |
| S17_vs_k141_90439\|\|full | 3689 | No | NA | 5 | 3 | 0 | Low-quality | Genome-fragment | 8.64 | AAI-based (high-confidence) | 0 |
| S12_extracted_k141_983 | 1070 | No | NA | 1 | 0 | 0 | Not-determined | Genome-fragment | NA | NA | 0 |
| S12_extracted_k141_173827 | 1032 | No | NA | 1 | 0 | 0 | Not-determined | Genome-fragment | NA | NA | 0 |
| S11_extracted_k141_9924 | 3689 | No | NA | 8 | 0 | 1 | Low-quality | Genome-fragment | 26.24 | AAI-based (high-confidence) | 0 |
| S12_extracted_k141_118902 | 12196 | No | NA | 15 | 0 | 2 | Not-determined | Genome-fragment | NA | NA | 0 |
| S12_extracted_k141_40148 | 6232 | No | NA | 6 | 0 | 5 | Not-determined | Genome-fragment | NA | NA | 0 |
| S12_extracted_k141_24339 | 5705 | No | NA | 6 | 0 | 5 | Not-determined | Genome-fragment | NA | NA | 0 |
| S12_extracted_k141_173832 | 1211 | No | NA | 2 | 0 | 0 | Not-determined | Genome-fragment | NA | NA | 0 |
| S12_extracted_k141_40154 | 1325 | No | NA | 3 | 0 | 2 | Not-determined | Genome-fragment | NA | NA | 0 |
| S12_extracted_k141_55881 | 1055 | No | NA | 1 | 0 | 1 | Not-determined | Genome-fragment | NA | NA | 0 |
| S12_extracted_k141_71649 | 1453 | No | NA | 2 | 0 | 1 | Not-determined | Genome-fragment | NA | NA | 0 |
| S13_vs_15438\|\|full | 3689 | No | NA | 6 | 2 | 0 | Low-quality | Genome-fragment | 8.59 | AAI-based (high-confidence) | 0 |
| S13_vs_101199\|\|full | 3689 | No | NA | 3 | 0 | 1 | Low-quality | Genome-fragment | 1.99 | AAI-based (medium-confidence) | 0 |
| S12_extracted_k141_24355 | 1426 | No | NA | 1 | 0 | 0 | Not-determined | Genome-fragment | NA | NA | 0 |
| S12_extracted_k141_32231 | 1563 | No | NA | 3 | 0 | 0 | Not-determined | Genome-fragment | NA | NA | 0 |
| S12_extracted_k141_103230 | 1260 | No | NA | 1 | 0 | 0 | Not-determined | Genome-fragment | NA | NA | 0 |
| S8_vs_k141_206735\|\|full | 3689 | No | NA | 5 | 0 | 3 | Low-quality | Genome-fragment | 2.12 | AAI-based (high-confidence) | 0 |
| S12_extracted_k141_150389 | 4473 | No | NA | 3 | 0 | 2 | Not-determined | Genome-fragment | NA | NA | 0 |
| S12_extracted_k141_166141 | 1274 | No | NA | 2 | 0 | 2 | Not-determined | Genome-fragment | NA | NA | 0 |
| S12_extracted_k141_173869 | 2631 | No | NA | 5 | 0 | 4 | Not-determined | Genome-fragment | NA | NA | 0 |
| S12_extracted_k141_32240 | 1946 | No | NA | 2 | 0 | 1 | Not-determined | Genome-fragment | NA | NA | 0 |
| S12_extracted_k141_55910 | 1229 | No | NA | 1 | 0 | 0 | Not-determined | Genome-fragment | NA | NA | 0 |
| S12_extracted_k141_47746 | 1508 | No | NA | 3 | 0 | 3 | Not-determined | Genome-fragment | NA | NA | 0 |
| S12_extracted_k141_103252 | 1959 | No | NA | 2 | 0 | 1 | Not-determined | Genome-fragment | NA | NA | 0 |
| S12_extracted_k141_95273 | 1288 | No | NA | 3 | 0 | 0 | Not-determined | Genome-fragment | NA | NA | 0 |
| S12_extracted_k141_150411 | 4520 | No | NA | 2 | 0 | 2 | Not-determined | Genome-fragment | NA | NA | 0 |
| S11_vs_k141_90813\|\|full | 3690 | No | NA | 8 | 5 | 0 | Low-quality | Genome-fragment | 5.67 | AAI-based (high-confidence) | 0 |
| S12_extracted_k141_63874 | 3195 | No | NA | 5 | 0 | 5 | Not-determined | Genome-fragment | NA | NA | 0 |
| S12_extracted_k141_95277 | 1492 | No | NA | 1 | 0 | 0 | Not-determined | Genome-fragment | NA | NA | 0 |
| S12_extracted_k141_134487 | 3193 | No | NA | 4 | 0 | 0 | Not-determined | Genome-fragment | NA | NA | 0 |
| S12_extracted_k141_47766 | 1226 | No | NA | 4 | 0 | 2 | Not-determined | Genome-fragment | NA | NA | 0 |
| S16_extracted_114224 | 3690 | No | NA | 3 | 1 | 0 | Low-quality | Genome-fragment | 5.36 | AAI-based (high-confidence) | 0 |
| S12_extracted_k141_71673 | 1295 | No | NA | 1 | 0 | 1 | Not-determined | Genome-fragment | NA | NA | 0 |
| S12_extracted_k141_173913 | 1098 | No | NA | 2 | 0 | 1 | Not-determined | Genome-fragment | NA | NA | 0 |
| S12_extracted_k141_24409 | 1667 | No | NA | 3 | 0 | 1 | Not-determined | Genome-fragment | NA | NA | 0 |
| S12_extracted_k141_32306 | 1305 | No | NA | 2 | 0 | 0 | Not-determined | Genome-fragment | NA | NA | 0 |
| S12_extracted_k141_16374 | 59768 | Yes | 53251 | 80 | 13 | 4 | High-quality | High-quality | 91.23 | AAI-based (high-confidence) | 10.9 |
| S12_extracted_k141_24424 | 1068 | No | NA | 1 | 0 | 1 | Not-determined | Genome-fragment | NA | NA | 0 |
| S12_extracted_k141_79601 | 7141 | No | NA | 8 | 0 | 2 | Not-determined | Genome-fragment | NA | NA | 0 |
| S10_vs_63281\|\|full | 3690 | No | NA | 7 | 4 | 0 | Low-quality | Genome-fragment | 8.81 | AAI-based (high-confidence) | 0 |
| S12_extracted_k141_103320 | 1548 | No | NA | 2 | 0 | 2 | Not-determined | Genome-fragment | NA | NA | 0 |
| S12_extracted_k141_126791 | 1002 | No | NA | 1 | 0 | 1 | Not-determined | Genome-fragment | NA | NA | 0 |
| S12_extracted_k141_142659 | 5393 | No | NA | 4 | 0 | 1 | Not-determined | Genome-fragment | NA | NA | 0 |
| S12_extracted_k141_181942 | 2691 | No | NA | 3 | 0 | 1 | Not-determined | Genome-fragment | NA | NA | 0 |
| S12_extracted_k141_134544 | 1646 | No | NA | 2 | 0 | 0 | Not-determined | Genome-fragment | NA | NA | 0 |
| S12_extracted_k141_79621 | 1166 | No | NA | 2 | 0 | 2 | Not-determined | Genome-fragment | NA | NA | 0 |
| S12_extracted_k141_71696 | 3025 | No | NA | 3 | 0 | 1 | Not-determined | Genome-fragment | NA | NA | 0 |
| S12_extracted_k141_166251 | 3206 | No | NA | 4 | 0 | 3 | Not-determined | Genome-fragment | NA | NA | 0 |
| S12_extracted_k141_16409 | 4950 | No | NA | 4 | 0 | 1 | Not-determined | Genome-fragment | NA | NA | 0 |
| S12_extracted_k141_142684 | 1169 | No | NA | 2 | 0 | 0 | Not-determined | Genome-fragment | NA | NA | 0 |
| S12_vs_k141_165940\|\|full | 3690 | No | NA | 6 | 1 | 0 | Low-quality | Genome-fragment | 9.71 | AAI-based (high-confidence) | 0 |
| S12_extracted_k141_173937 | 2689 | No | NA | 2 | 0 | 2 | Not-determined | Genome-fragment | NA | NA | 0 |
| S12_extracted_k141_134588 | 1408 | No | NA | 2 | 0 | 1 | Not-determined | Genome-fragment | NA | NA | 0 |
| S12_extracted_k141_95326 | 1418 | No | NA | 2 | 0 | 0 | Not-determined | Genome-fragment | NA | NA | 0 |
| S12_extracted_k141_16421 | 2041 | No | NA | 1 | 0 | 0 | Not-determined | Genome-fragment | NA | NA | 0 |
| S12_extracted_k141_47878 | 1083 | No | NA | 2 | 0 | 0 | Not-determined | Genome-fragment | NA | NA | 0 |
| S12_extracted_k141_95328 | 2310 | No | NA | 2 | 0 | 0 | Not-determined | Genome-fragment | NA | NA | 0 |
| S12_extracted_k141_1072 | 4153 | No | NA | 3 | 0 | 3 | Not-determined | Genome-fragment | NA | NA | 0 |
| S12_extracted_k141_173940 | 3712 | No | NA | 6 | 0 | 5 | Not-determined | Genome-fragment | NA | NA | 0 |
| S12_extracted_k141_63983 | 1044 | No | NA | 1 | 0 | 1 | Not-determined | Genome-fragment | NA | NA | 0 |
| S12_extracted_k141_134603 | 1069 | No | NA | 2 | 0 | 0 | Not-determined | Genome-fragment | NA | NA | 0 |
| S12_extracted_k141_166287 | 5491 | No | NA | 4 | 0 | 3 | Not-determined | Genome-fragment | NA | NA | 0 |
| S12_extracted_k141_1096 | 4162 | No | NA | 4 | 0 | 3 | Not-determined | Genome-fragment | NA | NA | 0 |
| S12_extracted_k141_119036 | 1288 | No | NA | 2 | 0 | 1 | Not-determined | Genome-fragment | NA | NA | 0 |
| S12_extracted_k141_95369 | 1899 | No | NA | 3 | 0 | 1 | Not-determined | Genome-fragment | NA | NA | 0 |
| S12_extracted_k141_40287 | 1154 | No | NA | 2 | 0 | 1 | Not-determined | Genome-fragment | NA | NA | 0 |
| S12_extracted_k141_126856 | 1395 | No | NA | 1 | 0 | 0 | Not-determined | Genome-fragment | NA | NA | 0 |
| S12_extracted_k141_95371 | 1330 | No | NA | 2 | 0 | 2 | Not-determined | Genome-fragment | NA | NA | 0 |
| S12_extracted_k141_40292 | 1086 | No | NA | 1 | 0 | 1 | Not-determined | Genome-fragment | NA | NA | 0 |
| S12_extracted_k141_126872 | 1064 | No | NA | 1 | 0 | 1 | Not-determined | Genome-fragment | NA | NA | 0 |
| S17_extracted_k141_16761 | 3693 | No | NA | 4 | 0 | 2 | Low-quality | Genome-fragment | 0.39 | AAI-based (medium-confidence) | 0 |
| S5_extracted_k141_71464 | 3693 | No | NA | 4 | 0 | 0 | Low-quality | Genome-fragment | 9.38 | AAI-based (medium-confidence) | 0 |
| S5_vs_k141_102199\|\|full | 3693 | No | NA | 4 | 1 | 0 | Low-quality | Genome-fragment | 7.98 | AAI-based (high-confidence) | 0 |
| S12_extracted_k141_158427 | 2055 | No | NA | 3 | 0 | 1 | Not-determined | Genome-fragment | NA | NA | 0 |
| S9_vs_k141_77619\|\|full | 3694 | No | NA | 6 | 0 | 0 | Low-quality | Genome-fragment | 1.87 | AAI-based (high-confidence) | 0 |
| S14_vs_k141_45640\|\|full | 3694 | No | NA | 6 | 0 | 2 | Low-quality | Genome-fragment | 2.36 | AAI-based (medium-confidence) | 0 |
| S12_extracted_k141_16489 | 1485 | No | NA | 2 | 0 | 2 | Not-determined | Genome-fragment | NA | NA | 0 |
| S12_extracted_k141_56082 | 10917 | No | NA | 7 | 0 | 6 | Not-determined | Genome-fragment | NA | NA | 0 |
| S12_extracted_k141_32385 | 4720 | No | NA | 4 | 0 | 3 | Not-determined | Genome-fragment | NA | NA | 0 |
| S12_extracted_k141_110845 | 11911 | No | NA | 7 | 0 | 5 | Not-determined | Genome-fragment | NA | NA | 0 |
| S12_extracted_k141_103444 | 3058 | No | NA | 2 | 0 | 0 | Not-determined | Genome-fragment | NA | NA | 0 |
| S12_extracted_k141_103450 | 1102 | No | NA | 2 | 0 | 1 | Not-determined | Genome-fragment | NA | NA | 0 |
| S5_vs_k141_68569\|\|full | 3694 | No | NA | 9 | 1 | 0 | Low-quality | Genome-fragment | 7.82 | AAI-based (medium-confidence) | 0 |
| S12_extracted_k141_166366 | 3084 | No | NA | 3 | 0 | 2 | Not-determined | Genome-fragment | NA | NA | 0 |
| S11_vs_k141_50010\|\|full | 3695 | No | NA | 7 | 0 | 0 | Low-quality | Genome-fragment | 7.59 | AAI-based (high-confidence) | 0 |
| S12_extracted_k141_16533 | 1516 | No | NA | 2 | 0 | 1 | Not-determined | Genome-fragment | NA | NA | 0 |
| S12_extracted_k141_182140 | 1214 | No | NA | 1 | 0 | 1 | Not-determined | Genome-fragment | NA | NA | 0 |
| S12_extracted_k141_40386 | 1113 | No | NA | 3 | 0 | 0 | Not-determined | Genome-fragment | NA | NA | 0 |
| S12_extracted_k141_48027 | 1071 | No | NA | 2 | 0 | 1 | Not-determined | Genome-fragment | NA | NA | 0 |
| S12_extracted_k141_110917 | 1251 | No | NA | 2 | 0 | 2 | Not-determined | Genome-fragment | NA | NA | 0 |
| S13_extracted_13140 | 3695 | No | NA | 4 | 0 | 0 | Low-quality | Genome-fragment | 3.37 | AAI-based (medium-confidence) | 0 |
| S12_extracted_k141_103494 | 2215 | No | NA | 1 | 0 | 1 | Not-determined | Genome-fragment | NA | NA | 0 |
| S12_extracted_k141_119095 | 1259 | No | NA | 1 | 0 | 1 | Not-determined | Genome-fragment | NA | NA | 0 |
| S12_extracted_k141_182204 | 1320 | No | NA | 4 | 0 | 0 | Not-determined | Genome-fragment | NA | NA | 0 |
| S12_extracted_k141_16576 | 1684 | No | NA | 2 | 0 | 1 | Not-determined | Genome-fragment | NA | NA | 0 |
| S12_extracted_k141_150569 | 1546 | No | NA | 1 | 0 | 1 | Not-determined | Genome-fragment | NA | NA | 0 |
| S12_extracted_k141_95518 | 6188 | No | NA | 4 | 0 | 4 | Not-determined | Genome-fragment | NA | NA | 0 |
| S12_extracted_k141_110943 | 1805 | No | NA | 3 | 0 | 3 | Not-determined | Genome-fragment | NA | NA | 0 |
| S12_extracted_k141_174052 | 1005 | No | NA | 2 | 0 | 1 | Not-determined | Genome-fragment | NA | NA | 0 |
| S12_extracted_k141_174065 | 1646 | No | NA | 3 | 0 | 0 | Not-determined | Genome-fragment | NA | NA | 0 |
| S12_extracted_k141_24640 | 1027 | No | NA | 1 | 0 | 1 | Not-determined | Genome-fragment | NA | NA | 0 |
| S12_extracted_k141_79650 | 1192 | No | NA | 1 | 0 | 0 | Not-determined | Genome-fragment | NA | NA | 0 |
| S12_extracted_k141_182242 | 3264 | No | NA | 7 | 0 | 1 | Not-determined | Genome-fragment | NA | NA | 0 |
| S12_extracted_k141_71835 | 11874 | No | NA | 8 | 0 | 6 | Not-determined | Genome-fragment | NA | NA | 0 |
| S12_extracted_k141_134674 | 5734 | No | NA | 6 | 0 | 1 | Not-determined | Genome-fragment | NA | NA | 0 |
| S12_extracted_k141_56227 | 1181 | No | NA | 2 | 0 | 1 | Not-determined | Genome-fragment | NA | NA | 0 |

| | | | | | | | | | | | |
|---|---|---|---|---|---|---|---|---|---|---|---|
| S12_extracted_k141_64153 | 3320 | No | NA | 3 | 0 | 3 | Not-determined | Genome-fragment | NA | NA | 0 |
| S14_vs_k141_71859\|\|full | 3695 | No | NA | 10 | 0 | 0 | Low-quality | Genome-fragment | 10.86 | AAI-based (high-confidence) | 0 |
| S12_extracted_k141_119125 | 1301 | No | NA | 2 | 0 | 1 | Not-determined | Genome-fragment | NA | NA | 0 |
| S16_vs_103252\|\|full | 3695 | No | NA | 3 | 0 | 3 | Low-quality | Genome-fragment | 2.61 | AAI-based (medium-confidence) | 0 |
| S8_extracted_k141_105323 | 3695 | No | NA | 2 | 0 | 1 | Low-quality | Genome-fragment | 1.99 | AAI-based (medium-confidence) | 0 |
| S12_extracted_k141_8787 | 1593 | No | NA | 2 | 0 | 1 | Not-determined | Genome-fragment | NA | NA | 0 |
| S12_extracted_k141_182271 | 1082 | No | NA | 1 | 0 | 0 | Not-determined | Genome-fragment | NA | NA | 0 |
| S12_extracted_k141_8788 | 3535 | No | NA | 2 | 0 | 2 | Not-determined | Genome-fragment | NA | NA | 0 |
| S12_extracted_k141_142872 | 2883 | No | NA | 4 | 0 | 2 | Not-determined | Genome-fragment | NA | NA | 0 |
| S12_extracted_k141_134681 | 2045 | No | NA | 3 | 0 | 0 | Not-determined | Genome-fragment | NA | NA | 0 |
| S12_extracted_k141_87658 | 1001 | No | NA | 1 | 0 | 1 | Not-determined | Genome-fragment | NA | NA | 0 |
| S12_extracted_k141_40481 | 1134 | No | NA | 1 | 0 | 0 | Not-determined | Genome-fragment | NA | NA | 0 |
| S12_extracted_k141_71902 | 1871 | No | NA | 1 | 0 | 1 | Not-determined | Genome-fragment | NA | NA | 0 |
| S12_extracted_k141_174120 | 2210 | No | NA | 2 | 0 | 2 | Not-determined | Genome-fragment | NA | NA | 0 |
| S12_extracted_k141_48083 | 1094 | No | NA | 2 | 0 | 2 | Not-determined | Genome-fragment | NA | NA | 0 |
| S12_extracted_k141_79666 | 46218 | No | NA | 68 | 13 | 1 | High-quality | High-quality | 100 | AAI-based (high-confidence) | 0 |
| S12_extracted_k141_87684 | 1439 | No | NA | 2 | 0 | 2 | Not-determined | Genome-fragment | NA | NA | 0 |
| S12_extracted_k141_79676 | 5590 | No | NA | 5 | 0 | 1 | Not-determined | Genome-fragment | NA | NA | 0 |
| S13_extracted_82574 | 3696 | No | NA | 5 | 2 | 0 | Low-quality | Genome-fragment | 8.53 | AAI-based (medium-confidence) | 0 |
| S12_extracted_k141_134695 | 1674 | No | NA | 2 | 0 | 0 | Not-determined | Genome-fragment | NA | NA | 0 |
| S12_extracted_k141_56295 | 1033 | No | NA | 1 | 0 | 0 | Not-determined | Genome-fragment | NA | NA | 0 |
| S12_extracted_k141_24751 | 1117 | No | NA | 2 | 0 | 1 | Not-determined | Genome-fragment | NA | NA | 0 |
| S5_vs_k141_5161\|\|full | 3696 | No | NA | 4 | 0 | 1 | Low-quality | Genome-fragment | 9.18 | AAI-based (high-confidence) | 0 |
| S7_vs_136538\|\|full | 3696 | No | NA | 3 | 0 | 3 | Low-quality | Genome-fragment | 1.64 | AAI-based (medium-confidence) | 0 |
| S12_extracted_k141_119240 | 2925 | No | NA | 3 | 0 | 1 | Not-determined | Genome-fragment | NA | NA | 0 |
| S12_extracted_k141_56321 | 1547 | No | NA | 1 | 0 | 1 | Not-determined | Genome-fragment | NA | NA | 0 |
| S12_extracted_k141_40561 | 1750 | No | NA | 2 | 0 | 1 | Not-determined | Genome-fragment | NA | NA | 0 |
| S16_vs_90571\|\|full | 3696 | No | NA | 3 | 0 | 2 | Low-quality | Genome-fragment | 2.12 | AAI-based (medium-confidence) | 0 |
| S12_extracted_k141_79717 | 2331 | No | NA | 5 | 0 | 4 | Not-determined | Genome-fragment | NA | NA | 0 |
| S17_vs_k141_32429\|\|full | 3698 | No | NA | 8 | 2 | 0 | Low-quality | Genome-fragment | 6.88 | HMM-based (lower-bound) | 0 |
| S16_extracted_18532 | 3698 | No | NA | 3 | 0 | 1 | Low-quality | Genome-fragment | 6.07 | AAI-based (medium-confidence) | 0 |
| S12_extracted_k141_32489 | 1191 | No | NA | 1 | 0 | 1 | Not-determined | Genome-fragment | NA | NA | 0 |
| S10_extracted_64783 | 3698 | No | NA | 3 | 0 | 3 | Low-quality | Genome-fragment | 1.26 | AAI-based (medium-confidence) | 0 |
| S12_extracted_k141_142891 | 1281 | No | NA | 1 | 0 | 0 | Not-determined | Genome-fragment | NA | NA | 0 |
| S12_extracted_k141_79726 | 5253 | No | NA | 5 | 0 | 3 | Not-determined | Genome-fragment | NA | NA | 0 |
| S12_extracted_k141_79739 | 3879 | No | NA | 4 | 0 | 3 | Not-determined | Genome-fragment | NA | NA | 0 |
| S12_extracted_k141_71960 | 30634 | No | NA | 34 | 0 | 4 | Not-determined | Genome-fragment | NA | NA | 0 |
| S12_extracted_k141_111130 | 1320 | No | NA | 1 | 0 | 1 | Not-determined | Genome-fragment | NA | NA | 0 |
| S12_extracted_k141_142916 | 2959 | No | NA | 3 | 0 | 2 | Not-determined | Genome-fragment | NA | NA | 0 |
| S12_extracted_k141_174220 | 2173 | No | NA | 3 | 0 | 0 | Not-determined | Genome-fragment | NA | NA | 0 |
| S12_extracted_k141_79772 | 1130 | No | NA | 2 | 0 | 1 | Not-determined | Genome-fragment | NA | NA | 0 |
| S17_extracted_k141_53580 | 3699 | No | NA | 4 | 0 | 1 | Low-quality | Genome-fragment | 2.06 | AAI-based (medium-confidence) | 0 |
| S12_extracted_k141_95758 | 3453 | No | NA | 3 | 0 | 2 | Not-determined | Genome-fragment | NA | NA | 0 |
| S5_extracted_k141_40500 | 3699 | No | NA | 2 | 0 | 2 | Low-quality | Genome-fragment | 1 | AAI-based (high-confidence) | 0 |
| S12_extracted_k141_111153 | 1320 | No | NA | 1 | 0 | 1 | Not-determined | Genome-fragment | NA | NA | 0 |
| S12_extracted_k141_127112 | 3092 | No | NA | 3 | 0 | 2 | Not-determined | Genome-fragment | NA | NA | 0 |
| S12_extracted_k141_87835 | 1099 | No | NA | 1 | 0 | 0 | Not-determined | Genome-fragment | NA | NA | 0 |
| S12_extracted_k141_166592 | 1680 | No | NA | 1 | 0 | 1 | Not-determined | Genome-fragment | NA | NA | 0 |
| S12_extracted_k141_87851 | 1063 | No | NA | 1 | 0 | 0 | Not-determined | Genome-fragment | NA | NA | 0 |
| S17_extracted_k141_10183 | 3700 | No | NA | 7 | 3 | 0 | Low-quality | Genome-fragment | 11.67 | AAI-based (high-confidence) | 0 |
| S12_extracted_k141_119347 | 1678 | No | NA | 1 | 0 | 0 | Not-determined | Genome-fragment | NA | NA | 0 |
| S12_extracted_k141_166616 | 1267 | No | NA | 1 | 0 | 1 | Not-determined | Genome-fragment | NA | NA | 0 |
| S10_extracted_13192 | 3701 | No | NA | 2 | 1 | 0 | Low-quality | Genome-fragment | 2.27 | AAI-based (high-confidence) | 0 |
| S12_extracted_k141_150671 | 1038 | No | NA | 1 | 0 | 1 | Not-determined | Genome-fragment | NA | NA | 0 |
| S17_extracted_k141_97824 | 3702 | No | NA | 4 | 1 | 0 | Low-quality | Genome-fragment | 5.55 | AAI-based (high-confidence) | 0 |
| S12_extracted_k141_40674 | 1099 | No | NA | 1 | 0 | 1 | Not-determined | Genome-fragment | NA | NA | 0 |
| S12_extracted_k141_8967 | 1468 | No | NA | 3 | 0 | 2 | Not-determined | Genome-fragment | NA | NA | 0 |
| S12_extracted_k141_64363 | 1086 | No | NA | 1 | 0 | 1 | Not-determined | Genome-fragment | NA | NA | 0 |
| S7_extracted_89534 | 3703 | No | NA | 4 | 4 | 0 | Low-quality | Genome-fragment | 2.5 | AAI-based (high-confidence) | 0 |
| S12_vs_k141_91009\|\|full | 3703 | No | NA | 4 | 0 | 0 | Low-quality | Genome-fragment | 6.62 | AAI-based (high-confidence) | 0 |
| S12_vs_k141_147958\|\|full | 3703 | No | NA | 3 | 0 | 2 | Low-quality | Genome-fragment | 3.48 | AAI-based (medium-confidence) | 0 |
| S12_extracted_k141_111244 | 3892 | No | NA | 4 | 0 | 4 | Not-determined | Genome-fragment | NA | NA | 0 |
| S12_extracted_k141_40685 | 2921 | No | NA | 1 | 0 | 0 | Not-determined | Genome-fragment | NA | NA | 0 |
| S12_extracted_k141_48278 | 42164 | No | NA | 55 | 24 | 1 | Complete | High-quality | 100 | DTR (high-confidence) | 0 |
| S3_extracted_k141_96001 | 3704 | No | NA | 2 | 0 | 1 | Low-quality | Genome-fragment | 3.42 | AAI-based (medium-confidence) | 0 |
| S12_extracted_k141_166709 | 1328 | No | NA | 2 | 0 | 1 | Not-determined | Genome-fragment | NA | NA | 0 |
| S12_extracted_k141_56479 | 2854 | No | NA | 3 | 0 | 2 | Not-determined | Genome-fragment | NA | NA | 0 |
| S12_extracted_k141_111282 | 1140 | No | NA | 4 | 0 | 0 | Not-determined | Genome-fragment | NA | NA | 0 |
| S12_extracted_k141_127263 | 1301 | No | NA | 1 | 0 | 1 | Not-determined | Genome-fragment | NA | NA | 0 |
| S12_extracted_k141_1467 | 1197 | No | NA | 1 | 0 | 1 | Not-determined | Genome-fragment | NA | NA | 0 |
| S12_extracted_k141_9022 | 1419 | No | NA | 2 | 0 | 0 | Not-determined | Genome-fragment | NA | NA | 0 |
| S12_extracted_k141_127269 | 1104 | No | NA | 1 | 0 | 0 | Not-determined | Genome-fragment | NA | NA | 0 |
| S12_extracted_k141_56508 | 1057 | No | NA | 1 | 0 | 0 | Not-determined | Genome-fragment | NA | NA | 0 |
| S12_extracted_k141_16922 | 2853 | No | NA | 2 | 0 | 2 | Not-determined | Genome-fragment | NA | NA | 0 |
| S12_extracted_k141_174321 | 1795 | No | NA | 2 | 0 | 0 | Not-determined | Genome-fragment | NA | NA | 0 |
| S12_extracted_k141_182498 | 1593 | No | NA | 2 | 0 | 1 | Not-determined | Genome-fragment | NA | NA | 0 |
| S12_extracted_k141_127302 | 1015 | No | NA | 1 | 0 | 1 | Not-determined | Genome-fragment | NA | NA | 0 |
| S3_vs_k141_52117\|\|full | 3704 | No | NA | 8 | 0 | 0 | Low-quality | Genome-fragment | 9.79 | AAI-based (medium-confidence) | 0 |
| S12_extracted_k141_64434 | 1004 | No | NA | 1 | 0 | 0 | Not-determined | Genome-fragment | NA | NA | 0 |
| S12_extracted_k141_79891 | 2479 | No | NA | 2 | 0 | 2 | Not-determined | Genome-fragment | NA | NA | 0 |
| S12_extracted_k141_24995 | 1051 | No | NA | 1 | 0 | 1 | Not-determined | Genome-fragment | NA | NA | 0 |
| S12_extracted_k141_9068 | 1307 | No | NA | 1 | 0 | 1 | Not-determined | Genome-fragment | NA | NA | 0 |
| S8_vs_k141_18723\|\|full | 3704 | No | NA | 3 | 0 | 2 | Low-quality | Genome-fragment | 2.57 | AAI-based (medium-confidence) | 0 |
| S12_extracted_k141_56549 | 1787 | No | NA | 2 | 0 | 1 | Not-determined | Genome-fragment | NA | NA | 0 |
| S12_extracted_k141_24998 | 4151 | No | NA | 3 | 0 | 2 | Not-determined | Genome-fragment | NA | NA | 0 |
| S12_extracted_k141_158655 | 1196 | No | NA | 1 | 0 | 1 | Not-determined | Genome-fragment | NA | NA | 0 |
| S12_extracted_k141_9103 | 1153 | No | NA | 3 | 0 | 2 | Not-determined | Genome-fragment | NA | NA | 0 |
| S12_extracted_k141_95925 | 2032 | No | NA | 1 | 0 | 1 | Not-determined | Genome-fragment | NA | NA | 0 |
| S12_extracted_k141_119466 | 2217 | No | NA | 2 | 0 | 1 | Not-determined | Genome-fragment | NA | NA | 0 |
| S12_extracted_k141_135038 | 1153 | No | NA | 1 | 0 | 1 | Not-determined | Genome-fragment | NA | NA | 0 |
| S12_extracted_k141_64462 | 2177 | No | NA | 5 | 0 | 1 | Not-determined | Genome-fragment | NA | NA | 0 |
| S12_extracted_k141_150744 | 4123 | No | NA | 4 | 0 | 1 | Not-determined | Genome-fragment | NA | NA | 0 |
| S12_extracted_k141_88073 | 1194 | No | NA | 1 | 0 | 1 | Not-determined | Genome-fragment | NA | NA | 0 |
| S12_extracted_k141_16989 | 5583 | No | NA | 2 | 0 | 2 | Not-determined | Genome-fragment | NA | NA | 0 |
| S12_extracted_k141_64475 | 3999 | No | NA | 2 | 0 | 1 | Not-determined | Genome-fragment | NA | NA | 0 |
| S12_extracted_k141_88083 | 5132 | No | NA | 3 | 0 | 1 | Not-determined | Genome-fragment | NA | NA | 0 |
| S12_extracted_k141_150767 | 1074 | No | NA | 2 | 0 | 1 | Not-determined | Genome-fragment | NA | NA | 0 |
| S12_extracted_k141_17008 | 1396 | No | NA | 2 | 0 | 2 | Not-determined | Genome-fragment | NA | NA | 0 |
| S12_extracted_k141_103747 | 1909 | No | NA | 3 | 0 | 0 | Not-determined | Genome-fragment | NA | NA | 0 |
| S12_extracted_k141_119512 | 1540 | No | NA | 1 | 0 | 0 | Not-determined | Genome-fragment | NA | NA | 0 |
| S13_extracted_21287 | 3705 | No | NA | 10 | 2 | 0 | Low-quality | Genome-fragment | 2.06 | HMM-based (lower-bound) | 0 |
| S12_extracted_k141_32615 | 174902 | No | NA | 191 | 32 | 4 | Complete | High-quality | 100 | DTR (high-confidence) | 0 |
| S12_extracted_k141_103754 | 1681 | No | NA | 1 | 0 | 0 | Not-determined | Genome-fragment | NA | NA | 0 |
| S12_extracted_k141_95971 | 3099 | No | NA | 1 | 0 | 1 | Not-determined | Genome-fragment | NA | NA | 0 |
| S12_extracted_k141_95972 | 1143 | No | NA | 2 | 0 | 0 | Not-determined | Genome-fragment | NA | NA | 0 |
| S12_extracted_k141_79932 | 2477 | No | NA | 3 | 0 | 2 | Not-determined | Genome-fragment | NA | NA | 0 |
| S14_vs_k141_5836\|\|full | 3705 | No | NA | 3 | 0 | 0 | Low-quality | Genome-fragment | 7.42 | AAI-based (high-confidence) | 0 |
| S12_extracted_k141_103760 | 1304 | No | NA | 2 | 0 | 2 | Not-determined | Genome-fragment | NA | NA | 0 |
| S12_extracted_k141_135070 | 2513 | No | NA | 2 | 0 | 2 | Not-determined | Genome-fragment | NA | NA | 0 |
| S12_extracted_k141_135074 | 3498 | No | NA | 4 | 0 | 2 | Not-determined | Genome-fragment | NA | NA | 0 |
| S12_extracted_k141_103763 | 1602 | No | NA | 2 | 0 | 1 | Not-determined | Genome-fragment | NA | NA | 0 |
| S12_extracted_k141_9125 | 10677 | No | NA | 16 | 0 | 2 | Not-determined | Genome-fragment | NA | NA | 0 |
| S12_extracted_k141_25100 | 1354 | No | NA | 1 | 0 | 1 | Not-determined | Genome-fragment | NA | NA | 0 |
| S12_extracted_k141_1517 | 3104 | No | NA | 4 | 0 | 3 | Not-determined | Genome-fragment | NA | NA | 0 |
| S12_extracted_k141_174365 | 1439 | No | NA | 1 | 0 | 0 | Not-determined | Genome-fragment | NA | NA | 0 |
| S12_extracted_k141_119567 | 1153 | No | NA | 1 | 0 | 1 | Not-determined | Genome-fragment | NA | NA | 0 |
| S12_extracted_k141_72322 | 2039 | No | NA | 2 | 0 | 2 | Not-determined | Genome-fragment | NA | NA | 0 |
| S12_extracted_k141_56669 | 4389 | No | NA | 6 | 0 | 0 | Not-determined | Genome-fragment | NA | NA | 0 |
| S12_extracted_k141_64527 | 1012 | No | NA | 2 | 0 | 1 | Not-determined | Genome-fragment | NA | NA | 0 |
| S12_extracted_k141_119584 | 1098 | No | NA | 1 | 0 | 0 | Not-determined | Genome-fragment | NA | NA | 0 |
| S1_extracted_k141_2724 | 3706 | No | NA | 2 | 0 | 0 | Low-quality | Genome-fragment | 5.36 | AAI-based (high-confidence) | 0 |
| S12_extracted_k141_1552 | 1512 | No | NA | 2 | 0 | 1 | Not-determined | Genome-fragment | NA | NA | 0 |
| S12_extracted_k141_96055 | 2215 | No | NA | 4 | 0 | 0 | Not-determined | Genome-fragment | NA | NA | 0 |
| S12_extracted_k141_9188 | 1331 | No | NA | 2 | 0 | 1 | Not-determined | Genome-fragment | NA | NA | 0 |
| S12_extracted_k141_56695 | 1104 | No | NA | 2 | 0 | 2 | Not-determined | Genome-fragment | NA | NA | 0 |
| S12_extracted_k141_119607 | 2538 | No | NA | 2 | 0 | 2 | Not-determined | Genome-fragment | NA | NA | 0 |
| S12_extracted_k141_103800 | 1272 | No | NA | 1 | 0 | 1 | Not-determined | Genome-fragment | NA | NA | 0 |
| S10_extracted_16938 | 3706 | No | NA | 3 | 2 | 0 | Low-quality | Genome-fragment | 1.62 | AAI-based (high-confidence) | 0 |
| S12_extracted_k141_135143 | 1400 | No | NA | 2 | 0 | 2 | Not-determined | Genome-fragment | NA | NA | 0 |
| S15_vs_2662\|\|full | 3706 | No | NA | 10 | 0 | 0 | Low-quality | Genome-fragment | 4.04 | AAI-based (high-confidence) | 0 |
| S12_extracted_k141_72377 | 1343 | No | NA | 1 | 0 | 0 | Not-determined | Genome-fragment | NA | NA | 0 |
| S12_vs_k141_131164\|\|full | 3707 | No | NA | 5 | 0 | 1 | Low-quality | Genome-fragment | 7.8 | AAI-based (high-confidence) | 0 |
| S12_extracted_k141_174374 | 40225 | No | NA | 54 | 8 | 1 | High-quality | High-quality | 99.22 | AAI-based (high-confidence) | 0 |
| S12_extracted_k141_166923 | 1014 | No | NA | 1 | 0 | 1 | Not-determined | Genome-fragment | NA | NA | 0 |
| S12_extracted_k141_25179 | 1049 | No | NA | 2 | 0 | 1 | Not-determined | Genome-fragment | NA | NA | 0 |
| S12_extracted_k141_96066 | 3717 | No | NA | 5 | 0 | 4 | Not-determined | Genome-fragment | NA | NA | 0 |
| S12_extracted_k141_166936 | 1660 | No | NA | 2 | 0 | 0 | Not-determined | Genome-fragment | NA | NA | 0 |
| S12_extracted_k141_143212 | 1766 | No | NA | 2 | 0 | 0 | Not-determined | Genome-fragment | NA | NA | 0 |
| S12_extracted_k141_103833 | 3450 | No | NA | 6 | 0 | 0 | Not-determined | Genome-fragment | NA | NA | 0 |
| S12_extracted_k141_166952 | 1924 | No | NA | 2 | 0 | 0 | Not-determined | Genome-fragment | NA | NA | 0 |
| S12_extracted_k141_182634 | 7102 | No | NA | 8 | 0 | 0 | Not-determined | Genome-fragment | NA | NA | 0 |
| S12_extracted_k141_9241 | 1572 | No | NA | 5 | 0 | 5 | Not-determined | Genome-fragment | NA | NA | 0 |
| S14_extracted_k141_65882 | 3708 | No | NA | 5 | 2 | 0 | Low-quality | Genome-fragment | 3.16 | AAI-based (high-confidence) | 0 |
| S12_extracted_k141_166967 | 1265 | No | NA | 2 | 0 | 0 | Not-determined | Genome-fragment | NA | NA | 0 |
| S5_vs_k141_88477\|\|full | 3708 | No | NA | 9 | 3 | 0 | Low-quality | Genome-fragment | 8.62 | AAI-based (medium-confidence) | 0 |
| S12_extracted_k141_182688 | 1819 | No | NA | 2 | 0 | 2 | Not-determined | Genome-fragment | NA | NA | 0 |
| S16_vs_4049\|\|full | 3708 | No | NA | 4 | 0 | 1 | Low-quality | Genome-fragment | 2.16 | AAI-based (medium-confidence) | 0 |
| S12_extracted_k141_56768 | 11819 | No | NA | 14 | 0 | 10 | Not-determined | Genome-fragment | NA | NA | 0 |
| S12_extracted_k141_40766 | 1894 | No | NA | 2 | 0 | 0 | Not-determined | Genome-fragment | NA | NA | 0 |
| S12_extracted_k141_127480 | 1062 | No | NA | 2 | 0 | 2 | Not-determined | Genome-fragment | NA | NA | 0 |
| S16_vs_63699\|\|full | 3708 | No | NA | 4 | 0 | 3 | Low-quality | Genome-fragment | 4.19 | AAI-based (medium-confidence) | 0 |
| S12_extracted_k141_150943 | 1306 | No | NA | 2 | 0 | 0 | Not-determined | Genome-fragment | NA | NA | 0 |
| S12_extracted_k141_48308 | 8941 | No | NA | 4 | 0 | 1 | Not-determined | Genome-fragment | NA | NA | 0 |
| S3_vs_k141_51342\|\|full | 3709 | No | NA | 4 | 0 | 1 | Low-quality | Genome-fragment | 5.93 | AAI-based (medium-confidence) | 0 |
| S12_extracted_k141_127501 | 3508 | No | NA | 4 | 0 | 3 | Not-determined | Genome-fragment | NA | NA | 0 |
| S12_extracted_k141_127504 | 1202 | No | NA | 4 | 0 | 0 | Not-determined | Genome-fragment | NA | NA | 0 |
| S11_vs_k141_52102\|\|full | 3710 | No | NA | 5 | 0 | 4 | Low-quality | Genome-fragment | 8.57 | AAI-based (medium-confidence) | 0 |
| S12_extracted_k141_64682 | 1173 | No | NA | 2 | 0 | 1 | Not-determined | Genome-fragment | NA | NA | 0 |
| S12_extracted_k141_111455 | 1445 | No | NA | 2 | 0 | 1 | Not-determined | Genome-fragment | NA | NA | 0 |
| S11_vs_k141_26189\|\|full | 3710 | No | NA | 4 | 4 | 0 | Low-quality | Genome-fragment | 10.02 | AAI-based (high-confidence) | 0 |
| S12_extracted_k141_143307 | 1368 | No | NA | 1 | 0 | 0 | Not-determined | Genome-fragment | NA | NA | 0 |
| S12_extracted_k141_96242 | 1006 | No | NA | 1 | 0 | 0 | Not-determined | Genome-fragment | NA | NA | 0 |
| S12_extracted_k141_25242 | 1070 | No | NA | 2 | 0 | 2 | Not-determined | Genome-fragment | NA | NA | 0 |
| S12_extracted_k141_64714 | 5148 | No | NA | 8 | 0 | 8 | Not-determined | Genome-fragment | NA | NA | 0 |
| S12_extracted_k141_103957 | 1220 | No | NA | 1 | 0 | 1 | Not-determined | Genome-fragment | NA | NA | 0 |
| S12_extracted_k141_40840 | 1278 | No | NA | 2 | 0 | 2 | Not-determined | Genome-fragment | NA | NA | 0 |
| S3_extracted_k141_85809 | 3710 | No | NA | 3 | 0 | 0 | Low-quality | Genome-fragment | 3.27 | AAI-based (medium-confidence) | 0 |
| S12_extracted_k141_25272 | 2023 | No | NA | 3 | 0 | 1 | Not-determined | Genome-fragment | NA | NA | 0 |
| S16_extracted_165882 | 3710 | No | NA | 5 | 0 | 4 | Low-quality | Genome-fragment | 3.32 | AAI-based (medium-confidence) | 0 |
| S12_extracted_k141_25282 | 1247 | No | NA | 3 | 0 | 1 | Not-determined | Genome-fragment | NA | NA | 0 |
| S1_extracted_k141_33164 | 3711 | No | NA | 4 | 0 | 1 | Low-quality | Genome-fragment | 2.62 | AAI-based (high-confidence) | 0 |
| S12_extracted_k141_17196 | 1306 | No | NA | 1 | 0 | 1 | Not-determined | Genome-fragment | NA | NA | 0 |
| S12_extracted_k141_127594 | 1038 | No | NA | 1 | 0 | 1 | Not-determined | Genome-fragment | NA | NA | 0 |
| S12_extracted_k141_96301 | 11490 | No | NA | 7 | 0 | 6 | Not-determined | Genome-fragment | NA | NA | 0 |
| S3_extracted_k141_97177 | 3711 | No | NA | 6 | 2 | 0 | Low-quality | Genome-fragment | 2.42 | AAI-based (high-confidence) | 0 |
| S12_extracted_k141_111510 | 1333 | No | NA | 1 | 0 | 1 | Not-determined | Genome-fragment | NA | NA | 0 |
| S12_extracted_k141_119893 | 1175 | No | NA | 2 | 0 | 1 | Not-determined | Genome-fragment | NA | NA | 0 |
| S12_extracted_k141_104023 | 3548 | No | NA | 3 | 0 | 1 | Not-determined | Genome-fragment | NA | NA | 0 |
| S12_extracted_k141_25327 | 4423 | No | NA | 6 | 0 | 0 | Not-determined | Genome-fragment | NA | NA | 0 |
| S12_extracted_k141_96358 | 4023 | No | NA | 4 | 0 | 1 | Not-determined | Genome-fragment | NA | NA | 0 |
| S12_extracted_k141_1828 | 2786 | No | NA | 3 | 0 | 1 | Not-determined | Genome-fragment | NA | NA | 0 |
| S12_extracted_k141_119932 | 3299 | No | NA | 2 | 0 | 0 | Not-determined | Genome-fragment | NA | NA | 0 |
| S5_extracted_k141_51765 | 3711 | No | NA | 2 | 0 | 0 | Low-quality | Genome-fragment | 9.33 | AAI-based (high-confidence) | 0 |
| S12_extracted_k141_1837 | 1536 | No | NA | 3 | 0 | 0 | Not-determined | Genome-fragment | NA | NA | 0 |
| S12_extracted_k141_40908 | 3067 | No | NA | 3 | 0 | 2 | Not-determined | Genome-fragment | NA | NA | 0 |
| S12_extracted_k141_182780 | 1294 | No | NA | 2 | 0 | 2 | Not-determined | Genome-fragment | NA | NA | 0 |
| S16_extracted_48214 | 3711 | Yes | 814 | 3 | 1 | 2 | Low-quality | Genome-fragment | 0.47 | HMM-based (lower-bound) | 78.07 |
| S12_extracted_k141_56986 | 4814 | No | NA | 6 | 0 | 4 | Not-determined | Genome-fragment | NA | NA | 0 |
| S12_extracted_k141_135319 | 1207 | No | NA | 1 | 0 | 1 | Not-determined | Genome-fragment | NA | NA | 0 |
| S6_vs_k141_20252\|\|lt2gene | 3711 | No | NA | 1 | 1 | 0 | Low-quality | Genome-fragment | 6.38 | AAI-based (medium-confidence) | 0 |
| S5_extracted_k141_67757 | 3712 | No | NA | 4 | 0 | 1 | Low-quality | Genome-fragment | 6.11 | AAI-based (medium-confidence) | 0 |
| S5_vs_k141_72867\|\|full | 3712 | No | NA | 5 | 0 | 2 | Low-quality | Genome-fragment | 5.04 | AAI-based (medium-confidence) | 0 |
| S12_extracted_k141_40935 | 1665 | No | NA | 3 | 0 | 0 | Not-determined | Genome-fragment | NA | NA | 0 |
| S12_extracted_k141_48458 | 1942 | No | NA | 1 | 0 | 1 | Not-determined | Genome-fragment | NA | NA | 0 |
| S12_extracted_k141_104114 | 2412 | No | NA | 2 | 0 | 2 | Not-determined | Genome-fragment | NA | NA | 0 |
| S12_extracted_k141_88301 | 2392 | No | NA | 1 | 0 | 1 | Not-determined | Genome-fragment | NA | NA | 0 |
| S12_extracted_k141_120019 | 1263 | No | NA | 2 | 0 | 1 | Not-determined | Genome-fragment | NA | NA | 0 |
| S12_extracted_k141_182811 | 1043 | No | NA | 1 | 0 | 1 | Not-determined | Genome-fragment | NA | NA | 0 |
| S12_extracted_k141_88316 | 1776 | No | NA | 2 | 0 | 1 | Not-determined | Genome-fragment | NA | NA | 0 |
| S12_extracted_k141_17274 | 22207 | No | NA | 18 | 0 | 12 | Not-determined | Genome-fragment | NA | NA | 0 |
| S7_extracted_17417 | 3712 | No | NA | 2 | 0 | 1 | Low-quality | Genome-fragment | 3.37 | AAI-based (high-confidence) | 0 |
| S12_extracted_k141_1873 | 4451 | No | NA | 3 | 0 | 2 | Not-determined | Genome-fragment | NA | NA | 0 |
| S16_extracted_23842 | 3714 | No | NA | 8 | 2 | 0 | Low-quality | Genome-fragment | 6.4 | AAI-based (high-confidence) | 0 |
| S12_extracted_k141_182826 | 3230 | No | NA | 2 | 0 | 2 | Not-determined | Genome-fragment | NA | NA | 0 |
| S12_extracted_k141_80097 | 1554 | No | NA | 1 | 0 | 1 | Not-determined | Genome-fragment | NA | NA | 0 |
| S12_extracted_k141_41010 | 2196 | No | NA | 2 | 0 | 0 | Not-determined | Genome-fragment | NA | NA | 0 |
| S16_extracted_26985 | 3714 | No | NA | 13 | 1 | 0 | Low-quality | Genome-fragment | 7.82 | AAI-based (high-confidence) | 0 |
| S12_extracted_k141_48480 | 4566 | No | NA | 5 | 0 | 1 | Not-determined | Genome-fragment | NA | NA | 0 |
| S12_extracted_k141_120071 | 1015 | No | NA | 1 | 0 | 1 | Not-determined | Genome-fragment | NA | NA | 0 |
| S12_extracted_k141_127656 | 2081 | No | NA | 2 | 0 | 0 | Not-determined | Genome-fragment | NA | NA | 0 |
| S12_extracted_k141_104174 | 1024 | No | NA | 2 | 0 | 2 | Not-determined | Genome-fragment | NA | NA | 0 |
| S12_extracted_k141_1901 | 3248 | No | NA | 4 | 0 | 2 | Not-determined | Genome-fragment | NA | NA | 0 |
| S12_vs_k141_21475\|\|full | 3714 | No | NA | 4 | 0 | 2 | Low-quality | Genome-fragment | 0.64 | AAI-based (medium-confidence) | 0 |
| S12_extracted_k141_151037 | 1428 | No | NA | 2 | 0 | 2 | Not-determined | Genome-fragment | NA | NA | 0 |
| S12_extracted_k141_104182 | 1536 | No | NA | 1 | 0 | 1 | Not-determined | Genome-fragment | NA | NA | 0 |
| S12_extracted_k141_88382 | 1477 | No | NA | 2 | 0 | 1 | Not-determined | Genome-fragment | NA | NA | 0 |
| S12_extracted_k141_111676 | 2408 | No | NA | 1 | 0 | 0 | Not-determined | Genome-fragment | NA | NA | 0 |
| S17_extracted_k141_116855 | 3716 | No | NA | 8 | 3 | 0 | Low-quality | Genome-fragment | 2.16 | AAI-based (high-confidence) | 0 |
| S12_extracted_k141_104192 | 3712 | No | NA | 4 | 0 | 3 | Not-determined | Genome-fragment | NA | NA | 0 |
| S3_vs_k141_106330\|\|full | 3717 | No | NA | 8 | 1 | 0 | Low-quality | Genome-fragment | 2.21 | AAI-based (high-confidence) | 0 |
| S12_extracted_k141_32751 | 1138 | No | NA | 1 | 0 | 0 | Not-determined | Genome-fragment | NA | NA | 0 |
| S12_extracted_k141_104207 | 1052 | No | NA | 2 | 0 | 1 | Not-determined | Genome-fragment | NA | NA | 0 |
| S12_extracted_k141_182844 | 35312 | No | NA | 32 | 0 | 20 | Not-determined | Genome-fragment | NA | NA | 0 |
| S12_extracted_k141_57111 | 1207 | No | NA | 2 | 0 | 1 | Not-determined | Genome-fragment | NA | NA | 0 |
| S12_extracted_k141_1939 | 3804 | No | NA | 7 | 0 | 2 | Not-determined | Genome-fragment | NA | NA | 0 |
| S12_extracted_k141_41075 | 6427 | No | NA | 6 | 0 | 1 | Not-determined | Genome-fragment | NA | NA | 0 |
| S12_extracted_k141_88397 | 1158 | No | NA | 4 | 0 | 4 | Not-determined | Genome-fragment | NA | NA | 0 |
| S12_extracted_k141_9403 | 1304 | No | NA | 2 | 0 | 2 | Not-determined | Genome-fragment | NA | NA | 0 |
| S3_extracted_k141_87808 | 3719 | No | NA | 7 | 2 | 0 | Low-quality | Genome-fragment | 6.2 | AAI-based (high-confidence) | 0 |
| S12_extracted_k141_127707 | 1627 | No | NA | 2 | 0 | 2 | Not-determined | Genome-fragment | NA | NA | 0 |
| S12_extracted_k141_1958 | 5980 | No | NA | 8 | 0 | 0 | Not-determined | Genome-fragment | NA | NA | 0 |
| S12_extracted_k141_88432 | 1209 | No | NA | 2 | 0 | 1 | Not-determined | Genome-fragment | NA | NA | 0 |
| S9_vs_k141_45910\|\|full | 3720 | No | NA | 6 | 5 | 0 | Low-quality | Genome-fragment | 7.72 | AAI-based (medium-confidence) | 0 |
| S12_extracted_k141_88436 | 3319 | No | NA | 5 | 0 | 2 | Not-determined | Genome-fragment | NA | NA | 0 |
| S12_extracted_k141_64877 | 3532 | No | NA | 7 | 0 | 6 | Not-determined | Genome-fragment | NA | NA | 0 |
| S3_extracted_k141_39981 | 3720 | No | NA | 6 | 0 | 0 | Low-quality | Genome-fragment | 2.44 | AAI-based (high-confidence) | 0 |
| S12_extracted_k141_96602 | 3565 | No | NA | 5 | 0 | 0 | Not-determined | Genome-fragment | NA | NA | 0 |
| S12_extracted_k141_88481 | 3138 | No | NA | 5 | 0 | 0 | Not-determined | Genome-fragment | NA | NA | 0 |
| S12_extracted_k141_151123 | 3462 | No | NA | 5 | 0 | 1 | Not-determined | Genome-fragment | NA | NA | 0 |
| S12_vs_k141_130679\|\|full | 3720 | No | NA | 4 | 1 | 0 | Low-quality | Genome-fragment | 7.75 | AAI-based (high-confidence) | 0 |
| S12_extracted_k141_41151 | 1286 | No | NA | 2 | 0 | 2 | Not-determined | Genome-fragment | NA | NA | 0 |
| S12_extracted_k141_151132 | 1025 | No | NA | 1 | 0 | 0 | Not-determined | Genome-fragment | NA | NA | 0 |
| S12_extracted_k141_88495 | 1319 | No | NA | 2 | 0 | 0 | Not-determined | Genome-fragment | NA | NA | 0 |
| S12_extracted_k141_182935 | 1358 | No | NA | 1 | 0 | 1 | Not-determined | Genome-fragment | NA | NA | 0 |
| S12_extracted_k141_48574 | 3417 | No | NA | 3 | 0 | 2 | Not-determined | Genome-fragment | NA | NA | 0 |
| S12_extracted_k141_88511 | 1119 | No | NA | 1 | 0 | 1 | Not-determined | Genome-fragment | NA | NA | 0 |
| S12_extracted_k141_174532 | 1070 | No | NA | 2 | 0 | 1 | Not-determined | Genome-fragment | NA | NA | 0 |
| S14_extracted_k141_73673 | 3721 | No | NA | 7 | 3 | 0 | Low-quality | Genome-fragment | 18.52 | AAI-based (medium-confidence) | 0 |
| S12_extracted_k141_48590 | 1400 | No | NA | 1 | 0 | 1 | Not-determined | Genome-fragment | NA | NA | 0 |
| S12_extracted_k141_41200 | 1014 | No | NA | 1 | 0 | 1 | Not-determined | Genome-fragment | NA | NA | 0 |
| S12_extracted_k141_1994 | 1502 | No | NA | 1 | 0 | 0 | Not-determined | Genome-fragment | NA | NA | 0 |
| S12_extracted_k141_2005 | 1020 | No | NA | 1 | 0 | 0 | Not-determined | Genome-fragment | NA | NA | 0 |
| S12_extracted_k141_48615 | 1115 | No | NA | 1 | 0 | 0 | Not-determined | Genome-fragment | NA | NA | 0 |
| S12_extracted_k141_25599 | 1881 | No | NA | 2 | 0 | 1 | Not-determined | Genome-fragment | NA | NA | 0 |
| S12_extracted_k141_32808 | 1226 | No | NA | 1 | 0 | 1 | Not-determined | Genome-fragment | NA | NA | 0 |
| S12_extracted_k141_80191 | 1229 | No | NA | 2 | 0 | 0 | Not-determined | Genome-fragment | NA | NA | 0 |
| S12_extracted_k141_167299 | 1757 | No | NA | 1 | 0 | 1 | Not-determined | Genome-fragment | NA | NA | 0 |
| S12_extracted_k141_41223 | 7559 | No | NA | 9 | 0 | 1 | Not-determined | Genome-fragment | NA | NA | 0 |
| S12_extracted_k141_48636 | 1338 | No | NA | 1 | 0 | 0 | Not-determined | Genome-fragment | NA | NA | 0 |
| S12_extracted_k141_88565 | 1267 | No | NA | 1 | 0 | 0 | Not-determined | Genome-fragment | NA | NA | 0 |
| S12_extracted_k141_32832 | 1019 | No | NA | 2 | 0 | 0 | Not-determined | Genome-fragment | NA | NA | 0 |
| S12_extracted_k141_64973 | 1031 | No | NA | 1 | 0 | 1 | Not-determined | Genome-fragment | NA | NA | 0 |
| S12_extracted_k141_41244 | 1033 | No | NA | 2 | 0 | 1 | Not-determined | Genome-fragment | NA | NA | 0 |
| S12_extracted_k141_104361 | 3004 | No | NA | 3 | 0 | 3 | Not-determined | Genome-fragment | NA | NA | 0 |
| S12_extracted_k141_88594 | 1514 | No | NA | 2 | 0 | 1 | Not-determined | Genome-fragment | NA | NA | 0 |
| S12_extracted_k141_183022 | 1215 | No | NA | 1 | 0 | 1 | Not-determined | Genome-fragment | NA | NA | 0 |
| S12_extracted_k141_135518 | 1285 | No | NA | 2 | 0 | 1 | Not-determined | Genome-fragment | NA | NA | 0 |
| S12_extracted_k141_135520 | 6185 | No | NA | 4 | 0 | 3 | Not-determined | Genome-fragment | NA | NA | 0 |
| S12_extracted_k141_32875 | 1217 | No | NA | 1 | 0 | 0 | Not-determined | Genome-fragment | NA | NA | 0 |
| S12_extracted_k141_120229 | 1024 | No | NA | 1 | 0 | 1 | Not-determined | Genome-fragment | NA | NA | 0 |
| S14_extracted_k141_92662 | 3721 | No | NA | 4 | 4 | 0 | Low-quality | Genome-fragment | 7.99 | AAI-based (medium-confidence) | 0 |
| S12_extracted_k141_183030 | 1521 | No | NA | 2 | 0 | 2 | Not-determined | Genome-fragment | NA | NA | 0 |
| S12_extracted_k141_120234 | 2302 | No | NA | 1 | 0 | 1 | Not-determined | Genome-fragment | NA | NA | 0 |
| S12_extracted_k141_48685 | 13299 | No | NA | 19 | 0 | 2 | Not-determined | Genome-fragment | NA | NA | 0 |
| S12_extracted_k141_143726 | 3345 | No | NA | 3 | 0 | 2 | Not-determined | Genome-fragment | NA | NA | 0 |
| S12_extracted_k141_96736 | 1150 | No | NA | 2 | 0 | 2 | Not-determined | Genome-fragment | NA | NA | 0 |
| S12_extracted_k141_25662 | 4859 | No | NA | 5 | 0 | 1 | Not-determined | Genome-fragment | NA | NA | 0 |
| S12_extracted_k141_135546 | 1241 | No | NA | 1 | 0 | 0 | Not-determined | Genome-fragment | NA | NA | 0 |
| S12_extracted_k141_41337 | 1192 | No | NA | 1 | 0 | 1 | Not-determined | Genome-fragment | NA | NA | 0 |
| S12_extracted_k141_80304 | 1491 | No | NA | 1 | 0 | 1 | Not-determined | Genome-fragment | NA | NA | 0 |
| S12_extracted_k141_143737 | 1006 | No | NA | 1 | 0 | 0 | Not-determined | Genome-fragment | NA | NA | 0 |
| S12_extracted_k141_48717 | 1910 | No | NA | 2 | 0 | 2 | Not-determined | Genome-fragment | NA | NA | 0 |
| S12_extracted_k141_143745 | 1801 | No | NA | 1 | 0 | 1 | Not-determined | Genome-fragment | NA | NA | 0 |
| S12_extracted_k141_183097 | 1368 | No | NA | 2 | 0 | 1 | Not-determined | Genome-fragment | NA | NA | 0 |
| S12_extracted_k141_41353 | 5452 | No | NA | 11 | 0 | 0 | Not-determined | Genome-fragment | NA | NA | 0 |
| S12_extracted_k141_104413 | 1145 | No | NA | 1 | 0 | 0 | Not-determined | Genome-fragment | NA | NA | 0 |
| S12_extracted_k141_174625 | 2445 | No | NA | 4 | 0 | 0 | Not-determined | Genome-fragment | NA | NA | 0 |
| S12_extracted_k141_159030 | 2674 | No | NA | 3 | 0 | 1 | Not-determined | Genome-fragment | NA | NA | 0 |
| S5_extracted_k141_139506 | 3721 | No | NA | 2 | 2 | 0 | Low-quality | Genome-fragment | 9.2 | AAI-based (high-confidence) | 0 |
| S12_extracted_k141_80342 | 3811 | No | NA | 7 | 0 | 5 | Not-determined | Genome-fragment | NA | NA | 0 |
| S12_extracted_k141_174664 | 1145 | No | NA | 1 | 0 | 0 | Not-determined | Genome-fragment | NA | NA | 0 |
| S12_extracted_k141_25689 | 3703 | No | NA | 6 | 0 | 2 | Not-determined | Genome-fragment | NA | NA | 0 |
| S12_extracted_k141_167366 | 1041 | No | NA | 2 | 0 | 1 | Not-determined | Genome-fragment | NA | NA | 0 |
| S12_extracted_k141_143780 | 1125 | No | NA | 2 | 0 | 1 | Not-determined | Genome-fragment | NA | NA | 0 |
| S12_extracted_k141_135573 | 1196 | No | NA | 1 | 0 | 0 | Not-determined | Genome-fragment | NA | NA | 0 |
| S12_extracted_k141_9636 | 1241 | No | NA | 1 | 0 | 0 | Not-determined | Genome-fragment | NA | NA | 0 |
| S12_extracted_k141_104451 | 1137 | No | NA | 1 | 0 | 1 | Not-determined | Genome-fragment | NA | NA | 0 |
| S12_extracted_k141_159075 | 1054 | No | NA | 2 | 0 | 0 | Not-determined | Genome-fragment | NA | NA | 0 |
| S12_extracted_k141_41371 | 3230 | No | NA | 4 | 0 | 1 | Not-determined | Genome-fragment | NA | NA | 0 |
| S12_extracted_k141_104455 | 2192 | No | NA | 1 | 0 | 1 | Not-determined | Genome-fragment | NA | NA | 0 |
| S7_extracted_44611 | 3721 | No | NA | 6 | 3 | 0 | Low-quality | Genome-fragment | 7.44 | AAI-based (high-confidence) | 0 |
| S12_extracted_k141_118255 | 3721 | No | NA | 3 | 0 | 1 | Low-quality | Genome-fragment | 5.52 | AAI-based (medium-confidence) | 0 |
| S12_extracted_k141_151182 | 1020 | No | NA | 2 | 0 | 0 | Not-determined | Genome-fragment | NA | NA | 0 |
| S12_extracted_k141_57368 | 6850 | No | NA | 7 | 0 | 3 | Not-determined | Genome-fragment | NA | NA | 0 |
| S12_extracted_k141_41383 | 3682 | No | NA | 3 | 0 | 1 | Not-determined | Genome-fragment | NA | NA | 0 |
| S12_extracted_k141_151184 | 3794 | No | NA | 4 | 0 | 1 | Not-determined | Genome-fragment | NA | NA | 0 |
| S12_extracted_k141_174684 | 2565 | No | NA | 2 | 0 | 1 | Not-determined | Genome-fragment | NA | NA | 0 |
| S16_extracted_118250 | 3722 | No | NA | 3 | 0 | 1 | Low-quality | Genome-fragment | 2.81 | AAI-based (medium-confidence) | 0 |
| S12_vs_k141_1592\|\|full | 3722 | No | NA | 7 | 0 | 1 | Low-quality | Genome-fragment | 6.42 | AAI-based (medium-confidence) | 0 |
| S12_extracted_k141_151186 | 1303 | No | NA | 1 | 0 | 0 | Not-determined | Genome-fragment | NA | NA | 0 |
| S8_vs_k141_46211\|\|full | 3722 | No | NA | 5 | 0 | 3 | Low-quality | Genome-fragment | 2.5 | AAI-based (high-confidence) | 0 |
| S12_extracted_k141_104484 | 1114 | No | NA | 2 | 0 | 1 | Not-determined | Genome-fragment | NA | NA | 0 |
| S12_extracted_k141_80403 | 1265 | No | NA | 2 | 0 | 2 | Not-determined | Genome-fragment | NA | NA | 0 |
| S12_extracted_k141_33017 | 1164 | No | NA | 1 | 0 | 1 | Not-determined | Genome-fragment | NA | NA | 0 |
| S12_extracted_k141_80409 | 1926 | No | NA | 2 | 0 | 1 | Not-determined | Genome-fragment | NA | NA | 0 |
| S12_extracted_k141_57387 | 1913 | No | NA | 1 | 0 | 1 | Not-determined | Genome-fragment | NA | NA | 0 |
| S12_extracted_k141_183164 | 3362 | No | NA | 4 | 0 | 2 | Not-determined | Genome-fragment | NA | NA | 0 |
| S11_extracted_k141_3287 | 3723 | No | NA | 6 | 2 | 0 | Low-quality | Genome-fragment | 8.61 | AAI-based (high-confidence) | 0 |
| S12_extracted_k141_104490 | 1350 | No | NA | 2 | 0 | 0 | Not-determined | Genome-fragment | NA | NA | 0 |
| S12_extracted_k141_127894 | 1061 | No | NA | 1 | 0 | 0 | Not-determined | Genome-fragment | NA | NA | 0 |
| S12_extracted_k141_159123 | 1378 | No | NA | 1 | 0 | 0 | Not-determined | Genome-fragment | NA | NA | 0 |
| S12_extracted_k141_183179 | 1932 | No | NA | 1 | 0 | 0 | Not-determined | Genome-fragment | NA | NA | 0 |
| S11_vs_k141_59002\|\|full | 3723 | No | NA | 3 | 1 | 0 | Low-quality | Genome-fragment | 9.82 | AAI-based (high-confidence) | 0 |
| S8_extracted_k141_84049 | 3723 | No | NA | 4 | 0 | 1 | Low-quality | Genome-fragment | 2.36 | AAI-based (medium-confidence) | 0 |
| S12_extracted_k141_174730 | 1250 | No | NA | 1 | 0 | 1 | Not-determined | Genome-fragment | NA | NA | 0 |
| S12_extracted_k141_65117 | 21588 | No | NA | 19 | 0 | 6 | Not-determined | Genome-fragment | NA | NA | 0 |
| S12_extracted_k141_111774 | 1342 | No | NA | 2 | 0 | 2 | Not-determined | Genome-fragment | NA | NA | 0 |
| S12_extracted_k141_183190 | 1208 | No | NA | 1 | 0 | 1 | Not-determined | Genome-fragment | NA | NA | 0 |
| S12_extracted_k141_167430 | 1809 | No | NA | 2 | 0 | 2 | Not-determined | Genome-fragment | NA | NA | 0 |
| S12_extracted_k141_33049 | 2573 | No | NA | 2 | 0 | 1 | Not-determined | Genome-fragment | NA | NA | 0 |
| S11_extracted_k141_88781 | 3724 | No | NA | 4 | 0 | 3 | Low-quality | Genome-fragment | 2.04 | AAI-based (medium-confidence) | 0 |
| S7_vs_102855\|\|full | 3724 | No | NA | 7 | 3 | 0 | Low-quality | Genome-fragment | 7.81 | AAI-based (high-confidence) | 0 |
| S6_extracted_k141_38174 | 3724 | No | NA | 5 | 0 | 0 | Low-quality | Genome-fragment | 5.11 | AAI-based (high-confidence) | 0 |
| S12_extracted_k141_2196 | 1808 | No | NA | 2 | 0 | 1 | Not-determined | Genome-fragment | NA | NA | 0 |
| S12_extracted_k141_111787 | 6989 | No | NA | 6 | 0 | 0 | Not-determined | Genome-fragment | NA | NA | 0 |
| S12_extracted_k141_159158 | 1467 | No | NA | 1 | 0 | 1 | Not-determined | Genome-fragment | NA | NA | 0 |
| S12_extracted_k141_120366 | 1126 | No | NA | 2 | 0 | 1 | Not-determined | Genome-fragment | NA | NA | 0 |
| S12_extracted_k141_33083 | 1314 | No | NA | 2 | 0 | 2 | Not-determined | Genome-fragment | NA | NA | 0 |
| S12_extracted_k141_174782 | 1127 | No | NA | 1 | 0 | 0 | Not-determined | Genome-fragment | NA | NA | 0 |
| S12_extracted_k141_25792 | 1303 | No | NA | 3 | 0 | 2 | Not-determined | Genome-fragment | NA | NA | 0 |
| S12_extracted_k141_65163 | 3413 | No | NA | 4 | 0 | 2 | Not-determined | Genome-fragment | NA | NA | 0 |
| S12_extracted_k141_57430 | 2598 | No | NA | 3 | 0 | 1 | Not-determined | Genome-fragment | NA | NA | 0 |
| S5_vs_k141_19665\|\|full | 3725 | No | NA | 5 | 2 | 0 | Low-quality | Genome-fragment | 11.37 | AAI-based (medium-confidence) | 0 |
| S12_extracted_k141_2216 | 7736 | No | NA | 4 | 0 | 3 | Not-determined | Genome-fragment | NA | NA | 0 |
| S11_vs_k141_30648\|\|full | 3727 | No | NA | 4 | 0 | 1 | Low-quality | Genome-fragment | 4.84 | AAI-based (medium-confidence) | 0 |
| S12_extracted_k141_135712 | 3480 | No | NA | 2 | 0 | 2 | Not-determined | Genome-fragment | NA | NA | 0 |
| S12_extracted_k141_41490 | 3289 | No | NA | 2 | 0 | 2 | Not-determined | Genome-fragment | NA | NA | 0 |
| S6_extracted_k141_19425 | 3728 | No | NA | 3 | 0 | 3 | Low-quality | Genome-fragment | 0.75 | AAI-based (high-confidence) | 0 |
| S12_extracted_k141_88806 | 1333 | No | NA | 3 | 0 | 0 | Not-determined | Genome-fragment | NA | NA | 0 |
| S12_extracted_k141_127951 | 1044 | No | NA | 1 | 0 | 0 | Not-determined | Genome-fragment | NA | NA | 0 |
| S12_extracted_k141_143878 | 4344 | No | NA | 4 | 0 | 3 | Not-determined | Genome-fragment | NA | NA | 0 |
| S12_extracted_k141_104518 | 1069 | No | NA | 2 | 0 | 2 | Not-determined | Genome-fragment | NA | NA | 0 |
| S12_extracted_k141_48872 | 1333 | No | NA | 1 | 0 | 1 | Not-determined | Genome-fragment | NA | NA | 0 |
| S12_extracted_k141_96945 | 1037 | No | NA | 1 | 0 | 0 | Not-determined | Genome-fragment | NA | NA | 0 |
| S12_extracted_k141_57475 | 2729 | No | NA | 3 | 0 | 1 | Not-determined | Genome-fragment | NA | NA | 0 |
| S3_extracted_k141_57905 | 3729 | No | NA | 8 | 8 | 0 | Low-quality | Genome-fragment | 8.03 | AAI-based (medium-confidence) | 0 |
| S12_extracted_k141_65219 | 1419 | No | NA | 1 | 0 | 1 | Not-determined | Genome-fragment | NA | NA | 0 |
| S12_extracted_k141_25850 | 1443 | No | NA | 1 | 0 | 1 | Not-determined | Genome-fragment | NA | NA | 0 |
| S12_extracted_k141_33190 | 1001 | No | NA | 1 | 0 | 1 | Not-determined | Genome-fragment | NA | NA | 0 |
| S12_extracted_k141_33191 | 4593 | No | NA | 4 | 0 | 0 | Not-determined | Genome-fragment | NA | NA | 0 |
| S5_vs_k141_7958\|\|full | 3729 | No | NA | 4 | 3 | 0 | Low-quality | Genome-fragment | 6.08 | HMM-based (lower-bound) | 0 |
| S12_extracted_k141_80590 | 1212 | No | NA | 1 | 0 | 1 | Not-determined | Genome-fragment | NA | NA | 0 |
| S12_extracted_k141_2234 | 1457 | No | NA | 3 | 0 | 0 | Not-determined | Genome-fragment | NA | NA | 0 |
| S12_extracted_k141_135757 | 1055 | No | NA | 1 | 0 | 1 | Not-determined | Genome-fragment | NA | NA | 0 |
| S12_extracted_k141_41595 | 1036 | No | NA | 1 | 0 | 0 | Not-determined | Genome-fragment | NA | NA | 0 |
| S12_extracted_k141_88901 | 3624 | No | NA | 2 | 0 | 2 | Not-determined | Genome-fragment | NA | NA | 0 |
| S12_extracted_k141_17835 | 1364 | No | NA | 2 | 0 | 1 | Not-determined | Genome-fragment | NA | NA | 0 |
| S12_extracted_k141_88909 | 1450 | No | NA | 2 | 0 | 1 | Not-determined | Genome-fragment | NA | NA | 0 |
| S7_vs_21358\|\|full | 3729 | No | NA | 4 | 3 | 0 | Low-quality | Genome-fragment | 6.29 | AAI-based (high-confidence) | 0 |
| S7_vs_38164\|\|full | 3729 | No | NA | 4 | 0 | 0 | Low-quality | Genome-fragment | 3.14 | AAI-based (medium-confidence) | 0 |
| S12_extracted_k141_151345 | 1746 | No | NA | 3 | 0 | 2 | Not-determined | Genome-fragment | NA | NA | 0 |
| S12_extracted_k141_128039 | 1216 | No | NA | 1 | 0 | 0 | Not-determined | Genome-fragment | NA | NA | 0 |
| S12_extracted_k141_17856 | 5128 | No | NA | 11 | 0 | 1 | Not-determined | Genome-fragment | NA | NA | 0 |
| S12_extracted_k141_80663 | 1035 | No | NA | 1 | 0 | 0 | Not-determined | Genome-fragment | NA | NA | 0 |
| S12_extracted_k141_25898 | 1107 | No | NA | 1 | 0 | 1 | Not-determined | Genome-fragment | NA | NA | 0 |
| S12_extracted_k141_159255 | 1036 | No | NA | 2 | 0 | 0 | Not-determined | Genome-fragment | NA | NA | 0 |
| S8_vs_k141_119273\|\|full | 3729 | No | NA | 9 | 2 | 0 | Low-quality | Genome-fragment | 4.58 | HMM-based (lower-bound) | 0 |
| S12_extracted_k141_2278 | 1155 | No | NA | 1 | 0 | 0 | Not-determined | Genome-fragment | NA | NA | 0 |
| S17_extracted_k141_41005 | 3730 | No | NA | 2 | 0 | 2 | Low-quality | Genome-fragment | 1.56 | AAI-based (medium-confidence) | 0 |
| S12_extracted_k141_88933 | 2383 | No | NA | 1 | 0 | 1 | Not-determined | Genome-fragment | NA | NA | 0 |
| S13_extracted_89982 | 3730 | No | NA | 2 | 0 | 1 | Low-quality | Genome-fragment | 3.41 | AAI-based (medium-confidence) | 0 |
| S12_extracted_k141_183302 | 3311 | No | NA | 2 | 0 | 2 | Not-determined | Genome-fragment | NA | NA | 0 |
| S12_extracted_k141_80707 | 2365 | No | NA | 4 | 0 | 2 | Not-determined | Genome-fragment | NA | NA | 0 |
| S12_extracted_k141_111897 | 4285 | No | NA | 2 | 0 | 2 | Not-determined | Genome-fragment | NA | NA | 0 |
| S12_extracted_k141_88943 | 1146 | No | NA | 2 | 0 | 1 | Not-determined | Genome-fragment | NA | NA | 0 |
| S12_extracted_k141_104656 | 3229 | No | NA | 4 | 0 | 0 | Not-determined | Genome-fragment | NA | NA | 0 |
| S13_vs_66616\|\|full | 3731 | No | NA | 4 | 0 | 0 | Low-quality | Genome-fragment | 1.55 | AAI-based (medium-confidence) | 0 |
| S7_vs_24542\|\|full | 3731 | No | NA | 8 | 1 | 0 | Low-quality | Genome-fragment | 9.52 | AAI-based (medium-confidence) | 0 |
| S12_extracted_k141_120564 | 4202 | No | NA | 4 | 0 | 1 | Not-determined | Genome-fragment | NA | NA | 0 |
| S12_extracted_k141_33263 | 1027 | No | NA | 2 | 0 | 0 | Not-determined | Genome-fragment | NA | NA | 0 |
| S12_extracted_k141_88968 | 1117 | No | NA | 1 | 0 | 0 | Not-determined | Genome-fragment | NA | NA | 0 |
| S12_extracted_k141_33272 | 3249 | No | NA | 3 | 0 | 2 | Not-determined | Genome-fragment | NA | NA | 0 |
| S12_extracted_k141_135864 | 2773 | No | NA | 3 | 0 | 0 | Not-determined | Genome-fragment | NA | NA | 0 |
| S12_extracted_k141_88986 | 1804 | No | NA | 1 | 0 | 0 | Not-determined | Genome-fragment | NA | NA | 0 |
| S12_extracted_k141_144003 | 1298 | No | NA | 3 | 0 | 0 | Not-determined | Genome-fragment | NA | NA | 0 |
| S12_extracted_k141_25924 | 1069 | No | NA | 3 | 0 | 0 | Not-determined | Genome-fragment | NA | NA | 0 |
| S12_extracted_k141_151440 | 2227 | No | NA | 1 | 0 | 0 | Not-determined | Genome-fragment | NA | NA | 0 |
| S12_extracted_k141_183343 | 1005 | No | NA | 2 | 0 | 0 | Not-determined | Genome-fragment | NA | NA | 0 |
| S12_extracted_k141_151444 | 1367 | No | NA | 2 | 0 | 1 | Not-determined | Genome-fragment | NA | NA | 0 |
| S16_vs_143849\|\|full | 3731 | No | NA | 9 | 1 | 0 | Low-quality | Genome-fragment | 8.02 | AAI-based (high-confidence) | 0 |
| S12_extracted_k141_144023 | 1204 | No | NA | 1 | 0 | 1 | Not-determined | Genome-fragment | NA | NA | 0 |
| S12_extracted_k141_111968 | 1118 | No | NA | 2 | 0 | 0 | Not-determined | Genome-fragment | NA | NA | 0 |
| S12_extracted_k141_65369 | 1124 | No | NA | 1 | 0 | 1 | Not-determined | Genome-fragment | NA | NA | 0 |
| S12_extracted_k141_111972 | 1063 | No | NA | 1 | 0 | 1 | Not-determined | Genome-fragment | NA | NA | 0 |
| S12_extracted_k141_104684 | 3747 | No | NA | 3 | 0 | 2 | Not-determined | Genome-fragment | NA | NA | 0 |
| S12_extracted_k141_89002 | 1494 | No | NA | 2 | 0 | 1 | Not-determined | Genome-fragment | NA | NA | 0 |
| S11_vs_k141_25300\|\|full | 3733 | No | NA | 5 | 0 | 0 | Low-quality | Genome-fragment | 2.29 | AAI-based (high-confidence) | 0 |
| S12_extracted_k141_17924 | 3244 | No | NA | 2 | 0 | 0 | Not-determined | Genome-fragment | NA | NA | 0 |
| S12_extracted_k141_128158 | 3594 | No | NA | 8 | 0 | 8 | Not-determined | Genome-fragment | NA | NA | 0 |
| S12_extracted_k141_25978 | 1173 | No | NA | 1 | 0 | 1 | Not-determined | Genome-fragment | NA | NA | 0 |
| S12_extracted_k141_159298 | 1408 | No | NA | 2 | 0 | 2 | Not-determined | Genome-fragment | NA | NA | 0 |
| S7_vs_118402\|\|full | 3733 | No | NA | 9 | 2 | 0 | Low-quality | Genome-fragment | 10.04 | AAI-based (high-confidence) | 0 |
| S12_extracted_k141_144075 | 6761 | No | NA | 8 | 0 | 5 | Not-determined | Genome-fragment | NA | NA | 0 |

| | | | | | | | | | | | |
|---|---|---|---|---|---|---|---|---|---|---|---|
| S12_extracted_k141_128179 | 5005 | No | NA | 4 | 0 | 3 | Not-determined | Genome-fragment | NA | NA | 0 |
| S12_extracted_k141_89050 | 1151 | No | NA | 2 | 0 | 0 | Not-determined | Genome-fragment | NA | NA | 0 |
| S12_extracted_k141_112021 | 1938 | No | NA | 1 | 0 | 0 | Not-determined | Genome-fragment | NA | NA | 0 |
| S12_extracted_k141_26015 | 1228 | No | NA | 2 | 0 | 2 | Not-determined | Genome-fragment | NA | NA | 0 |
| S16_extracted_131116 | 3733 | No | NA | 6 | 2 | 0 | Low-quality | Genome-fragment | 3.42 | AAI-based (high-confidence) | 0 |
| S11_vs_k141_74555\|\|full | 3734 | Yes | 2816 | 6 | 1 | 1 | Low-quality | Genome-fragment | 5.17 | HMM-based (lower-bound) | 24.58 |
| S12_extracted_k141_97188 | 1096 | No | NA | 1 | 0 | 1 | Not-determined | Genome-fragment | NA | NA | 0 |
| S12_extracted_k141_33361 | 1315 | No | NA | 2 | 0 | 1 | Not-determined | Genome-fragment | NA | NA | 0 |
| S13_extracted_65916 | 3734 | No | NA | 2 | 0 | 1 | Low-quality | Genome-fragment | 3.37 | AAI-based (medium-confidence) | 0 |
| S12_extracted_k141_104774 | 3762 | No | NA | 3 | 0 | 2 | Not-determined | Genome-fragment | NA | NA | 0 |
| S12_vs_k141_99179\|\|full | 3734 | No | NA | 8 | 0 | 0 | Low-quality | Genome-fragment | 5.5 | AAI-based (high-confidence) | 0 |
| S12_extracted_k141_120619 | 1363 | No | NA | 1 | 0 | 1 | Not-determined | Genome-fragment | NA | NA | 0 |
| S12_extracted_k141_120626 | 1087 | No | NA | 1 | 0 | 0 | Not-determined | Genome-fragment | NA | NA | 0 |
| S12_extracted_k141_183439 | 2297 | No | NA | 3 | 0 | 0 | Not-determined | Genome-fragment | NA | NA | 0 |
| S12_extracted_k141_41840 | 6530 | No | NA | 5 | 0 | 2 | Not-determined | Genome-fragment | NA | NA | 0 |
| S12_vs_k141_170214\|\|full | 3734 | No | NA | 7 | 2 | 0 | Low-quality | Genome-fragment | 7.76 | AAI-based (high-confidence) | 0 |
| S12_extracted_k141_57713 | 1560 | No | NA | 1 | 0 | 1 | Not-determined | Genome-fragment | NA | NA | 0 |
| S12_extracted_k141_151544 | 1062 | No | NA | 3 | 0 | 2 | Not-determined | Genome-fragment | NA | NA | 0 |
| S12_extracted_k141_17979 | 2540 | No | NA | 1 | 0 | 0 | Not-determined | Genome-fragment | NA | NA | 0 |
| S12_extracted_k141_57718 | 1064 | No | NA | 1 | 0 | 0 | Not-determined | Genome-fragment | NA | NA | 0 |
| S12_extracted_k141_183451 | 1479 | No | NA | 2 | 0 | 0 | Not-determined | Genome-fragment | NA | NA | 0 |
| S12_extracted_k141_151566 | 1560 | No | NA | 4 | 0 | 4 | Not-determined | Genome-fragment | NA | NA | 0 |
| S12_extracted_k141_89140 | 1237 | No | NA | 1 | 0 | 0 | Not-determined | Genome-fragment | NA | NA | 0 |
| S12_extracted_k141_65487 | 3019 | No | NA | 2 | 0 | 2 | Not-determined | Genome-fragment | NA | NA | 0 |
| S12_extracted_k141_2434 | 1050 | No | NA | 1 | 0 | 1 | Not-determined | Genome-fragment | NA | NA | 0 |
| S12_extracted_k141_167467 | 2157 | No | NA | 1 | 0 | 1 | Not-determined | Genome-fragment | NA | NA | 0 |
| S12_extracted_k141_49130 | 1360 | No | NA | 2 | 0 | 2 | Not-determined | Genome-fragment | NA | NA | 0 |
| S7_extracted_45639 | 3736 | No | NA | 2 | 0 | 1 | Low-quality | Genome-fragment | 0.48 | AAI-based (medium-confidence) | 0 |
| S12_extracted_k141_159358 | 7318 | No | NA | 4 | 0 | 2 | Not-determined | Genome-fragment | NA | NA | 0 |
| S12_extracted_k141_26062 | 1337 | No | NA | 3 | 0 | 0 | Not-determined | Genome-fragment | NA | NA | 0 |
| S7_vs_74783\|\|full | 3736 | No | NA | 6 | 1 | 0 | Low-quality | Genome-fragment | 8.54 | AAI-based (medium-confidence) | 0 |
| S12_extracted_k141_104825 | 1232 | No | NA | 2 | 0 | 2 | Not-determined | Genome-fragment | NA | NA | 0 |
| S12_extracted_k141_104827 | 1393 | No | NA | 2 | 0 | 2 | Not-determined | Genome-fragment | NA | NA | 0 |
| S12_extracted_k141_72726 | 2559 | No | NA | 3 | 0 | 2 | Not-determined | Genome-fragment | NA | NA | 0 |
| S8_extracted_k141_45170 | 3736 | No | NA | 6 | 2 | 0 | Low-quality | Genome-fragment | 11.59 | AAI-based (high-confidence) | 0 |
| S12_extracted_k141_97318 | 1049 | No | NA | 1 | 0 | 0 | Not-determined | Genome-fragment | NA | NA | 0 |
| S12_extracted_k141_41916 | 1454 | No | NA | 1 | 0 | 0 | Not-determined | Genome-fragment | NA | NA | 0 |
| S12_extracted_k141_89215 | 1113 | No | NA | 1 | 0 | 1 | Not-determined | Genome-fragment | NA | NA | 0 |
| S12_extracted_k141_112151 | 2737 | No | NA | 5 | 0 | 2 | Not-determined | Genome-fragment | NA | NA | 0 |
| S12_extracted_k141_159391 | 3740 | No | NA | 4 | 0 | 1 | Not-determined | Genome-fragment | NA | NA | 0 |
| S12_extracted_k141_72753 | 1341 | No | NA | 1 | 0 | 0 | Not-determined | Genome-fragment | NA | NA | 0 |
| S12_extracted_k141_18051 | 1648 | No | NA | 3 | 0 | 1 | Not-determined | Genome-fragment | NA | NA | 0 |
| S12_extracted_k141_159397 | 3397 | No | NA | 3 | 0 | 0 | Not-determined | Genome-fragment | NA | NA | 0 |
| S12_extracted_k141_167487 | 2206 | No | NA | 1 | 0 | 1 | Not-determined | Genome-fragment | NA | NA | 0 |
| S1_vs_k141_54476\|\|full | 3737 | No | NA | 2 | 1 | 0 | Low-quality | Genome-fragment | 7.07 | AAI-based (high-confidence) | 0 |
| S16_vs_128358\|\|full | 3739 | No | NA | 9 | 0 | 0 | Low-quality | Genome-fragment | 8.64 | AAI-based (medium-confidence) | 0 |
| S12_extracted_k141_183528 | 1266 | No | NA | 1 | 0 | 0 | Not-determined | Genome-fragment | NA | NA | 0 |
| S12_extracted_k141_112165 | 4447 | No | NA | 1 | 0 | 1 | Not-determined | Genome-fragment | NA | NA | 0 |
| S12_extracted_k141_57794 | 1186 | No | NA | 2 | 0 | 2 | Not-determined | Genome-fragment | NA | NA | 0 |
| S12_extracted_k141_104836 | 3050 | No | NA | 3 | 0 | 0 | Not-determined | Genome-fragment | NA | NA | 0 |
| S12_extracted_k141_130533 | 3739 | No | NA | 4 | 0 | 3 | Low-quality | Genome-fragment | 1.34 | AAI-based (medium-confidence) | 0 |
| S14_vs_k141_55952\|\|full | 3741 | No | NA | 3 | 0 | 1 | Low-quality | Genome-fragment | 1.89 | AAI-based (medium-confidence) | 0 |
| S12_extracted_k141_18105 | 1615 | No | NA | 1 | 0 | 1 | Not-determined | Genome-fragment | NA | NA | 0 |
| S12_extracted_k141_72800 | 1968 | No | NA | 1 | 0 | 0 | Not-determined | Genome-fragment | NA | NA | 0 |
| S12_extracted_k141_159474 | 2525 | No | NA | 2 | 0 | 2 | Not-determined | Genome-fragment | NA | NA | 0 |
| S12_extracted_k141_136143 | 1001 | No | NA | 2 | 0 | 2 | Not-determined | Genome-fragment | NA | NA | 0 |
| S12_extracted_k141_42009 | 1189 | No | NA | 2 | 0 | 1 | Not-determined | Genome-fragment | NA | NA | 0 |
| S7_vs_92287\|\|full | 3742 | No | NA | 2 | 2 | 0 | Low-quality | Genome-fragment | 9.13 | AAI-based (high-confidence) | 0 |
| S12_extracted_k141_167553 | 1372 | No | NA | 1 | 0 | 1 | Not-determined | Genome-fragment | NA | NA | 0 |
| S12_extracted_k141_72818 | 1334 | No | NA | 2 | 0 | 0 | Not-determined | Genome-fragment | NA | NA | 0 |
| S12_extracted_k141_144260 | 1472 | No | NA | 2 | 0 | 2 | Not-determined | Genome-fragment | NA | NA | 0 |
| S12_extracted_k141_57815 | 1434 | No | NA | 3 | 0 | 3 | Not-determined | Genome-fragment | NA | NA | 0 |
| S12_extracted_k141_97434 | 1909 | No | NA | 2 | 0 | 2 | Not-determined | Genome-fragment | NA | NA | 0 |
| S12_extracted_k141_183585 | 1116 | No | NA | 1 | 0 | 1 | Not-determined | Genome-fragment | NA | NA | 0 |
| S4_extracted_22202 | 3742 | No | NA | 4 | 0 | 0 | Low-quality | Genome-fragment | 11.15 | AAI-based (medium-confidence) | 0 |
| S16_vs_11043\|\|full | 3744 | No | NA | 3 | 0 | 2 | Low-quality | Genome-fragment | 5.65 | AAI-based (medium-confidence) | 0 |
| S12_extracted_k141_57847 | 1045 | No | NA | 2 | 0 | 1 | Not-determined | Genome-fragment | NA | NA | 0 |
| S12_extracted_k141_26159 | 1868 | No | NA | 2 | 0 | 0 | Not-determined | Genome-fragment | NA | NA | 0 |
| S12_extracted_k141_144279 | 11171 | No | NA | 14 | 0 | 10 | Not-determined | Genome-fragment | NA | NA | 0 |
| S12_extracted_k141_89332 | 1160 | No | NA | 1 | 0 | 0 | Not-determined | Genome-fragment | NA | NA | 0 |
| S12_extracted_k141_33506 | 6905 | No | NA | 6 | 0 | 5 | Not-determined | Genome-fragment | NA | NA | 0 |
| S9_vs_k141_82514\|\|full | 3745 | No | NA | 5 | 0 | 0 | Low-quality | Genome-fragment | 6.19 | AAI-based (high-confidence) | 0 |
| S13_vs_95131\|\|full | 3745 | No | NA | 3 | 2 | 0 | Low-quality | Genome-fragment | 9 | AAI-based (high-confidence) | 0 |
| S12_extracted_k141_26187 | 1623 | No | NA | 3 | 0 | 2 | Not-determined | Genome-fragment | NA | NA | 0 |
| S12_extracted_k141_104920 | 2499 | No | NA | 4 | 0 | 2 | Not-determined | Genome-fragment | NA | NA | 0 |
| S6_vs_k141_22003\|\|full | 3745 | No | NA | 9 | 0 | 0 | Low-quality | Genome-fragment | 2.72 | AAI-based (high-confidence) | 0 |
| S12_extracted_k141_33512 | 7649 | No | NA | 9 | 0 | 2 | Not-determined | Genome-fragment | NA | NA | 0 |
| S12_extracted_k141_57897 | 1354 | No | NA | 1 | 0 | 1 | Not-determined | Genome-fragment | NA | NA | 0 |
| S12_extracted_k141_151641 | 40715 | No | NA | 46 | 0 | 29 | Not-determined | Genome-fragment | NA | NA | 0 |
| S12_extracted_k141_65692 | 3257 | No | NA | 2 | 0 | 1 | Not-determined | Genome-fragment | NA | NA | 0 |
| S12_extracted_k141_72865 | 2948 | No | NA | 4 | 0 | 1 | Not-determined | Genome-fragment | NA | NA | 0 |
| S1_vs_k141_33136\|\|full | 3746 | No | NA | 2 | 0 | 2 | Low-quality | Genome-fragment | 4.36 | AAI-based (medium-confidence) | 0 |
| S5_vs_k141_98383\|\|full | 3746 | No | NA | 7 | 4 | 0 | Low-quality | Genome-fragment | 9.21 | AAI-based (high-confidence) | 0 |
| S10_vs_92084\|\|full | 3746 | No | NA | 5 | 0 | 3 | Low-quality | Genome-fragment | 10.23 | AAI-based (medium-confidence) | 0 |
| S9_vs_k141_12408\|\|full | 3748 | No | NA | 10 | 3 | 0 | Low-quality | Genome-fragment | 9.5 | AAI-based (high-confidence) | 0 |
| S12_extracted_k141_42104 | 3669 | No | NA | 5 | 0 | 0 | Not-determined | Genome-fragment | NA | NA | 0 |
| S12_extracted_k141_2675 | 1490 | No | NA | 2 | 0 | 0 | Not-determined | Genome-fragment | NA | NA | 0 |
| S5_extracted_k141_141098 | 3748 | No | NA | 11 | 2 | 0 | Low-quality | Genome-fragment | 10.2 | AAI-based (high-confidence) | 0 |
| S12_extracted_k141_9912 | 1037 | No | NA | 2 | 0 | 0 | Not-determined | Genome-fragment | NA | NA | 0 |
| S7_vs_108580\|\|full | 3748 | No | NA | 4 | 2 | 0 | Low-quality | Genome-fragment | 10.47 | AAI-based (high-confidence) | 0 |
| S12_extracted_k141_144345 | 1025 | No | NA | 2 | 0 | 0 | Not-determined | Genome-fragment | NA | NA | 0 |
| S5_extracted_k141_93101 | 3749 | No | NA | 9 | 3 | 0 | Low-quality | Genome-fragment | 26.85 | AAI-based (high-confidence) | 0 |
| S12_extracted_k141_183666 | 1063 | No | NA | 1 | 0 | 1 | Not-determined | Genome-fragment | NA | NA | 0 |
| S12_extracted_k141_49229 | 1031 | No | NA | 1 | 0 | 1 | Not-determined | Genome-fragment | NA | NA | 0 |
| S12_extracted_k141_183693 | 1209 | No | NA | 1 | 0 | 1 | Not-determined | Genome-fragment | NA | NA | 0 |
| S12_extracted_k141_33628 | 3033 | No | NA | 3 | 0 | 1 | Not-determined | Genome-fragment | NA | NA | 0 |
| S5_extracted_k141_27147 | 3749 | No | NA | 4 | 3 | 0 | Low-quality | Genome-fragment | 8.62 | AAI-based (high-confidence) | 0 |
| S13_vs_24733\|\|full | 3751 | No | NA | 4 | 0 | 0 | Low-quality | Genome-fragment | 8.95 | AAI-based (high-confidence) | 0 |
| S12_extracted_k141_72921 | 1028 | No | NA | 2 | 0 | 2 | Not-determined | Genome-fragment | NA | NA | 0 |
| S5_vs_k141_78053\|\|full | 3752 | No | NA | 3 | 0 | 3 | Low-quality | Genome-fragment | 1.99 | AAI-based (medium-confidence) | 0 |
| S12_extracted_k141_112298 | 1343 | No | NA | 1 | 0 | 1 | Not-determined | Genome-fragment | NA | NA | 0 |
| S10_vs_118666\|\|full | 3753 | No | NA | 8 | 0 | 0 | Low-quality | Genome-fragment | 5.57 | AAI-based (high-confidence) | 0 |
| S12_extracted_k141_49259 | 1742 | No | NA | 3 | 0 | 1 | Not-determined | Genome-fragment | NA | NA | 0 |
| S12_extracted_k141_136324 | 1418 | No | NA | 2 | 0 | 0 | Not-determined | Genome-fragment | NA | NA | 0 |
| S12_extracted_k141_105056 | 2471 | No | NA | 2 | 0 | 0 | Not-determined | Genome-fragment | NA | NA | 0 |
| S12_extracted_k141_128375 | 1287 | No | NA | 1 | 0 | 1 | Not-determined | Genome-fragment | NA | NA | 0 |
| S7_extracted_88248 | 3754 | No | NA | 2 | 0 | 2 | Low-quality | Genome-fragment | 3.78 | AAI-based (medium-confidence) | 0 |
| S12_extracted_k141_144425 | 6825 | No | NA | 9 | 0 | 3 | Not-determined | Genome-fragment | NA | NA | 0 |
| S7_vs_136204\|\|lt2gene | 3754 | No | NA | 1 | 1 | 0 | Low-quality | Genome-fragment | 7.59 | AAI-based (medium-confidence) | 0 |
| S12_extracted_k141_18299 | 1025 | No | NA | 2 | 0 | 2 | Not-determined | Genome-fragment | NA | NA | 0 |
| S12_extracted_k141_18303 | 1229 | No | NA | 2 | 0 | 0 | Not-determined | Genome-fragment | NA | NA | 0 |
| S12_extracted_k141_175323 | 1109 | No | NA | 2 | 0 | 0 | Not-determined | Genome-fragment | NA | NA | 0 |
| S12_extracted_k141_128390 | 3822 | No | NA | 3 | 0 | 2 | Not-determined | Genome-fragment | NA | NA | 0 |
| S12_extracted_k141_183748 | 4949 | No | NA | 3 | 0 | 2 | Not-determined | Genome-fragment | NA | NA | 0 |
| S12_extracted_k141_18312 | 1165 | No | NA | 1 | 0 | 0 | Not-determined | Genome-fragment | NA | NA | 0 |
| S12_extracted_k141_175330 | 1719 | No | NA | 1 | 0 | 1 | Not-determined | Genome-fragment | NA | NA | 0 |
| S12_extracted_k141_42237 | 3038 | No | NA | 4 | 0 | 2 | Not-determined | Genome-fragment | NA | NA | 0 |
| S12_extracted_k141_167743 | 1265 | No | NA | 2 | 0 | 0 | Not-determined | Genome-fragment | NA | NA | 0 |
| S12_extracted_k141_18325 | 1259 | No | NA | 3 | 0 | 0 | Not-determined | Genome-fragment | NA | NA | 0 |
| S12_extracted_k141_10018 | 1008 | No | NA | 1 | 0 | 1 | Not-determined | Genome-fragment | NA | NA | 0 |
| S12_extracted_k141_58041 | 2167 | No | NA | 1 | 0 | 0 | Not-determined | Genome-fragment | NA | NA | 0 |
| S12_extracted_k141_175372 | 1199 | No | NA | 1 | 0 | 1 | Not-determined | Genome-fragment | NA | NA | 0 |
| S12_extracted_k141_97535 | 1527 | No | NA | 2 | 0 | 2 | Not-determined | Genome-fragment | NA | NA | 0 |
| S4_vs_VIRSorter_26379-cat_2 | 3756 | No | NA | 5 | 0 | 0 | Low-quality | Genome-fragment | 8.26 | AAI-based (high-confidence) | 0 |
| S5_vs_k141_71846\|\|full | 3758 | No | NA | 2 | 1 | 0 | Low-quality | Genome-fragment | 2.2 | AAI-based (medium-confidence) | 0 |
| S8_vs_k141_65078\|\|full | 3758 | No | NA | 6 | 1 | 0 | Low-quality | Genome-fragment | 5.46 | AAI-based (medium-confidence) | 0 |
| S12_extracted_k141_89550 | 3570 | No | NA | 2 | 0 | 1 | Not-determined | Genome-fragment | NA | NA | 0 |
| S12_extracted_k141_58079 | 3218 | No | NA | 2 | 0 | 0 | Not-determined | Genome-fragment | NA | NA | 0 |
| S12_extracted_k141_2800 | 2700 | No | NA | 3 | 0 | 2 | Not-determined | Genome-fragment | NA | NA | 0 |
| S12_extracted_k141_89557 | 1166 | No | NA | 2 | 0 | 1 | Not-determined | Genome-fragment | NA | NA | 0 |
| S12_extracted_k141_58095 | 1331 | No | NA | 1 | 0 | 1 | Not-determined | Genome-fragment | NA | NA | 0 |
| S17_extracted_k141_59453 | 3759 | No | NA | 12 | 1 | 0 | Low-quality | Genome-fragment | 4.33 | AAI-based (high-confidence) | 0 |
| S12_extracted_k141_2819 | 1665 | No | NA | 3 | 0 | 2 | Not-determined | Genome-fragment | NA | NA | 0 |
| S1_vs_k141_46091\|\|full | 3759 | No | NA | 7 | 3 | 0 | Low-quality | Genome-fragment | 11.24 | AAI-based (high-confidence) | 0 |
| S12_extracted_k141_73029 | 1060 | No | NA | 2 | 0 | 1 | Not-determined | Genome-fragment | NA | NA | 0 |
| S17_vs_k141_167392\|\|full | 3760 | No | NA | 6 | 3 | 0 | Low-quality | Genome-fragment | 5.6 | AAI-based (high-confidence) | 0 |
| S13_vs_123006\|\|full | 3760 | No | NA | 6 | 0 | 1 | Low-quality | Genome-fragment | 2.67 | AAI-based (high-confidence) | 0 |
| S3_vs_k141_73440\|\|full | 3760 | No | NA | 3 | 0 | 3 | Low-quality | Genome-fragment | 2.11 | AAI-based (medium-confidence) | 0 |
| S12_extracted_k141_97614 | 1434 | No | NA | 3 | 0 | 0 | Not-determined | Genome-fragment | NA | NA | 0 |
| S12_extracted_k141_73055 | 1014 | No | NA | 1 | 0 | 1 | Not-determined | Genome-fragment | NA | NA | 0 |
| S7_vs_131831\|\|full | 3760 | No | NA | 3 | 0 | 1 | Low-quality | Genome-fragment | 2.12 | AAI-based (medium-confidence) | 0 |
| S12_extracted_k141_18425 | 1236 | No | NA | 2 | 0 | 1 | Not-determined | Genome-fragment | NA | NA | 0 |
| S16_vs_11769\|\|full | 3760 | No | NA | 3 | 2 | 0 | Low-quality | Genome-fragment | 10.28 | AAI-based (high-confidence) | 0 |
| S12_extracted_k141_42365 | 4933 | No | NA | 3 | 0 | 3 | Not-determined | Genome-fragment | NA | NA | 0 |
| S12_extracted_k141_167857 | 3027 | No | NA | 3 | 0 | 0 | Not-determined | Genome-fragment | NA | NA | 0 |
| S13_vs_8368\|\|full | 3761 | No | NA | 8 | 1 | 0 | Low-quality | Genome-fragment | 9.67 | AAI-based (high-confidence) | 0 |
| S5_vs_k141_108965\|\|full | 3761 | No | NA | 5 | 0 | 2 | Low-quality | Genome-fragment | 0.53 | AAI-based (medium-confidence) | 0 |
| S12_extracted_k141_159709 | 1277 | No | NA | 3 | 0 | 2 | Not-determined | Genome-fragment | NA | NA | 0 |
| S7_vs_5723\|\|full | 3761 | No | NA | 7 | 4 | 0 | Low-quality | Genome-fragment | 10 | AAI-based (high-confidence) | 0 |
| S10_extracted_48452 | 3761 | No | NA | 4 | 1 | 0 | Low-quality | Genome-fragment | 1.6 | AAI-based (high-confidence) | 0 |
| S12_extracted_k141_58158 | 2512 | No | NA | 4 | 0 | 1 | Not-determined | Genome-fragment | NA | NA | 0 |
| S12_extracted_k141_2902 | 3688 | No | NA | 2 | 0 | 2 | Not-determined | Genome-fragment | NA | NA | 0 |
| S12_extracted_k141_105237 | 1361 | No | NA | 2 | 0 | 1 | Not-determined | Genome-fragment | NA | NA | 0 |
| S12_extracted_k141_159755 | 8053 | No | NA | 5 | 0 | 4 | Not-determined | Genome-fragment | NA | NA | 0 |
| S12_extracted_k141_105238 | 1224 | No | NA | 1 | 0 | 1 | Not-determined | Genome-fragment | NA | NA | 0 |
| S12_extracted_k141_144431 | 2302 | No | NA | 3 | 0 | 2 | Not-determined | Genome-fragment | NA | NA | 0 |
| S12_extracted_k141_65914 | 1292 | No | NA | 2 | 0 | 2 | Not-determined | Genome-fragment | NA | NA | 0 |
| S12_extracted_k141_105271 | 1898 | No | NA | 2 | 0 | 1 | Not-determined | Genome-fragment | NA | NA | 0 |
| S12_extracted_k141_58203 | 1022 | No | NA | 2 | 0 | 1 | Not-determined | Genome-fragment | NA | NA | 0 |
| S12_extracted_k141_65939 | 2378 | No | NA | 2 | 0 | 0 | Not-determined | Genome-fragment | NA | NA | 0 |
| S12_extracted_k141_97680 | 3949 | No | NA | 3 | 0 | 0 | Not-determined | Genome-fragment | NA | NA | 0 |
| S12_extracted_k141_159777 | 3539 | No | NA | 4 | 0 | 4 | Not-determined | Genome-fragment | NA | NA | 0 |
| S12_extracted_k141_42424 | 3446 | No | NA | 4 | 0 | 0 | Not-determined | Genome-fragment | NA | NA | 0 |
| S12_extracted_k141_105303 | 1374 | No | NA | 3 | 0 | 0 | Not-determined | Genome-fragment | NA | NA | 0 |
| S12_extracted_k141_97713 | 1281 | No | NA | 3 | 0 | 0 | Not-determined | Genome-fragment | NA | NA | 0 |
| S12_extracted_k141_120717 | 1196 | No | NA | 1 | 0 | 1 | Not-determined | Genome-fragment | NA | NA | 0 |
| S12_extracted_k141_175418 | 6252 | No | NA | 8 | 0 | 2 | Not-determined | Genome-fragment | NA | NA | 0 |
| S12_extracted_k141_65984 | 1956 | No | NA | 3 | 0 | 2 | Not-determined | Genome-fragment | NA | NA | 0 |
| S12_extracted_k141_97739 | 1032 | No | NA | 2 | 0 | 1 | Not-determined | Genome-fragment | NA | NA | 0 |
| S12_extracted_k141_33828 | 1308 | No | NA | 1 | 0 | 0 | Not-determined | Genome-fragment | NA | NA | 0 |
| S16_extracted_110134 | 3762 | No | NA | 4 | 0 | 4 | Low-quality | Genome-fragment | 2.21 | AAI-based (high-confidence) | 0 |
| S12_extracted_k141_58248 | 6084 | No | NA | 5 | 0 | 3 | Not-determined | Genome-fragment | NA | NA | 0 |
| S12_extracted_k141_120758 | 1214 | No | NA | 1 | 0 | 0 | Not-determined | Genome-fragment | NA | NA | 0 |
| S12_extracted_k141_168030 | 1454 | No | NA | 1 | 0 | 1 | Not-determined | Genome-fragment | NA | NA | 0 |
| S12_extracted_k141_58271 | 1757 | No | NA | 1 | 0 | 1 | Not-determined | Genome-fragment | NA | NA | 0 |
| S12_extracted_k141_183970 | 1120 | No | NA | 1 | 0 | 0 | Not-determined | Genome-fragment | NA | NA | 0 |
| S12_extracted_k141_18645 | 1308 | No | NA | 2 | 0 | 2 | Not-determined | Genome-fragment | NA | NA | 0 |
| S12_extracted_k141_105342 | 1742 | No | NA | 1 | 0 | 1 | Not-determined | Genome-fragment | NA | NA | 0 |
| S12_extracted_k141_151979 | 1971 | No | NA | 2 | 0 | 2 | Not-determined | Genome-fragment | NA | NA | 0 |
| S12_extracted_k141_66007 | 8300 | No | NA | 7 | 0 | 6 | Not-determined | Genome-fragment | NA | NA | 0 |
| S12_extracted_k141_105355 | 1348 | No | NA | 1 | 0 | 1 | Not-determined | Genome-fragment | NA | NA | 0 |
| S12_extracted_k141_175501 | 2215 | No | NA | 3 | 0 | 2 | Not-determined | Genome-fragment | NA | NA | 0 |
| S3_extracted_k141_127414 | 3763 | No | NA | 5 | 0 | 0 | Low-quality | Genome-fragment | 4.81 | AAI-based (medium-confidence) | 0 |
| S12_extracted_k141_81413 | 3391 | No | NA | 4 | 0 | 3 | Not-determined | Genome-fragment | NA | NA | 0 |
| S12_extracted_k141_112624 | 2552 | No | NA | 3 | 0 | 3 | Not-determined | Genome-fragment | NA | NA | 0 |
| S12_extracted_k141_120769 | 2015 | No | NA | 1 | 0 | 0 | Not-determined | Genome-fragment | NA | NA | 0 |
| S3_vs_k141_119266\|\|full | 3763 | No | NA | 9 | 1 | 0 | Low-quality | Genome-fragment | 5.77 | AAI-based (medium-confidence) | 0 |
| S12_extracted_k141_112629 | 1238 | No | NA | 1 | 0 | 0 | Not-determined | Genome-fragment | NA | NA | 0 |
| S12_extracted_k141_184005 | 1073 | No | NA | 3 | 0 | 2 | Not-determined | Genome-fragment | NA | NA | 0 |
| S12_extracted_k141_144526 | 1103 | No | NA | 1 | 0 | 1 | Not-determined | Genome-fragment | NA | NA | 0 |
| S11_vs_k141_101836\|\|full | 3764 | No | NA | 6 | 1 | 0 | Low-quality | Genome-fragment | 1.51 | HMM-based (lower-bound) | 0 |
| S12_extracted_k141_94716 | 3765 | No | NA | 4 | 0 | 1 | Low-quality | Genome-fragment | 1.18 | AAI-based (medium-confidence) | 0 |
| S12_extracted_k141_27916 | 3767 | Yes | 3138 | 6 | 1 | 1 | Low-quality | Genome-fragment | 2.65 | AAI-based (high-confidence) | 16.7 |
| S12_extracted_k141_175541 | 1514 | No | NA | 3 | 0 | 0 | Not-determined | Genome-fragment | NA | NA | 0 |
| S12_extracted_k141_26449 | 1046 | No | NA | 2 | 0 | 0 | Not-determined | Genome-fragment | NA | NA | 0 |
| S12_extracted_k141_184015 | 1122 | No | NA | 1 | 0 | 0 | Not-determined | Genome-fragment | NA | NA | 0 |
| S12_extracted_k141_184017 | 2971 | No | NA | 2 | 0 | 2 | Not-determined | Genome-fragment | NA | NA | 0 |
| S12_extracted_k141_58332 | 1171 | No | NA | 2 | 0 | 1 | Not-determined | Genome-fragment | NA | NA | 0 |
| S12_extracted_k141_26471 | 1762 | No | NA | 2 | 0 | 1 | Not-determined | Genome-fragment | NA | NA | 0 |
| S12_extracted_k141_89776 | 3745 | No | NA | 4 | 0 | 1 | Not-determined | Genome-fragment | NA | NA | 0 |
| S12_extracted_k141_112681 | 1047 | No | NA | 2 | 0 | 2 | Not-determined | Genome-fragment | NA | NA | 0 |
| S12_extracted_k141_49640 | 1008 | No | NA | 1 | 0 | 1 | Not-determined | Genome-fragment | NA | NA | 0 |
| S12_extracted_k141_89811 | 1111 | No | NA | 1 | 0 | 0 | Not-determined | Genome-fragment | NA | NA | 0 |
| S12_extracted_k141_73242 | 1306 | No | NA | 3 | 0 | 0 | Not-determined | Genome-fragment | NA | NA | 0 |
| S12_extracted_k141_18744 | 1018 | No | NA | 2 | 0 | 2 | Not-determined | Genome-fragment | NA | NA | 0 |
| S12_extracted_k141_3106 | 1008 | No | NA | 1 | 0 | 1 | Not-determined | Genome-fragment | NA | NA | 0 |
| S11_vs_k141_68763\|\|full | 3768 | No | NA | 3 | 2 | 0 | Low-quality | Genome-fragment | 2.05 | HMM-based (lower-bound) | 0 |
| S12_extracted_k141_81478 | 1492 | No | NA | 2 | 0 | 1 | Not-determined | Genome-fragment | NA | NA | 0 |
| S12_vs_k141_105244\|\|full | 3768 | No | NA | 5 | 0 | 4 | Low-quality | Genome-fragment | 4.83 | AAI-based (medium-confidence) | 0 |
| S3_vs_k141_25892\|\|full | 3769 | No | NA | 5 | 0 | 0 | Low-quality | Genome-fragment | 8.65 | AAI-based (medium-confidence) | 0 |
| S5_vs_k141_82478\|\|full | 3770 | No | NA | 5 | 1 | 0 | Low-quality | Genome-fragment | 9.52 | AAI-based (high-confidence) | 0 |
| S12_extracted_k141_49749 | 1175 | No | NA | 2 | 0 | 0 | Not-determined | Genome-fragment | NA | NA | 0 |
| S12_extracted_k141_112754 | 1214 | No | NA | 1 | 0 | 1 | Not-determined | Genome-fragment | NA | NA | 0 |
| S12_extracted_k141_112756 | 1019 | No | NA | 2 | 0 | 0 | Not-determined | Genome-fragment | NA | NA | 0 |
| S12_extracted_k141_152101 | 1053 | No | NA | 1 | 0 | 1 | Not-determined | Genome-fragment | NA | NA | 0 |
| S12_extracted_k141_89886 | 1383 | No | NA | 1 | 0 | 1 | Not-determined | Genome-fragment | NA | NA | 0 |
| S12_extracted_k141_81530 | 3195 | No | NA | 3 | 0 | 3 | Not-determined | Genome-fragment | NA | NA | 0 |
| S12_extracted_k141_144641 | 2289 | No | NA | 2 | 0 | 1 | Not-determined | Genome-fragment | NA | NA | 0 |
| S12_extracted_k141_97974 | 3563 | No | NA | 7 | 0 | 3 | Not-determined | Genome-fragment | NA | NA | 0 |
| S12_extracted_k141_184099 | 2476 | No | NA | 3 | 0 | 2 | Not-determined | Genome-fragment | NA | NA | 0 |
| S12_extracted_k141_66080 | 1302 | No | NA | 1 | 0 | 0 | Not-determined | Genome-fragment | NA | NA | 0 |
| S11_vs_k141_4853\|\|full | 3771 | No | NA | 6 | 0 | 0 | Low-quality | Genome-fragment | 8.78 | AAI-based (high-confidence) | 0 |
| S5_extracted_k141_6629 | 3771 | No | NA | 9 | 3 | 0 | Low-quality | Genome-fragment | 11.58 | AAI-based (high-confidence) | 0 |
| S3_vs_k141_66154\|\|full | 3772 | Yes | 2410 | 5 | 1 | 1 | Low-quality | Genome-fragment | 1.55 | HMM-based (lower-bound) | 36.11 |
| S5_vs_k141_41600\|\|full | 3772 | No | NA | 4 | 0 | 0 | Low-quality | Genome-fragment | 1.67 | AAI-based (medium-confidence) | 0 |
| S16_extracted_27004 | 3772 | No | NA | 5 | 0 | 0 | Low-quality | Genome-fragment | 7.58 | AAI-based (medium-confidence) | 0 |
| S12_extracted_k141_66093 | 3262 | No | NA | 5 | 0 | 5 | Not-determined | Genome-fragment | NA | NA | 0 |
| S12_extracted_k141_3197 | 4314 | No | NA | 4 | 0 | 2 | Not-determined | Genome-fragment | NA | NA | 0 |
| S12_extracted_k141_168167 | 2221 | No | NA | 4 | 0 | 2 | Not-determined | Genome-fragment | NA | NA | 0 |
| S12_extracted_k141_98030 | 1613 | No | NA | 3 | 0 | 1 | Not-determined | Genome-fragment | NA | NA | 0 |
| S12_extracted_k141_168169 | 1068 | No | NA | 2 | 0 | 0 | Not-determined | Genome-fragment | NA | NA | 0 |
| S12_extracted_k141_144666 | 3111 | No | NA | 4 | 0 | 0 | Not-determined | Genome-fragment | NA | NA | 0 |
| S17_extracted_k141_39091 | 3774 | No | NA | 7 | 3 | 0 | Low-quality | Genome-fragment | 8.02 | AAI-based (high-confidence) | 0 |
| S12_extracted_k141_184120 | 1153 | No | NA | 3 | 0 | 0 | Not-determined | Genome-fragment | NA | NA | 0 |
| S12_extracted_k141_136779 | 1461 | No | NA | 1 | 0 | 0 | Not-determined | Genome-fragment | NA | NA | 0 |
| S12_extracted_k141_89952 | 2430 | No | NA | 4 | 0 | 2 | Not-determined | Genome-fragment | NA | NA | 0 |
| S12_extracted_k141_144679 | 2232 | No | NA | 1 | 0 | 1 | Not-determined | Genome-fragment | NA | NA | 0 |
| S12_extracted_k141_3232 | 1299 | No | NA | 1 | 0 | 1 | Not-determined | Genome-fragment | NA | NA | 0 |
| S12_extracted_k141_81577 | 1442 | No | NA | 2 | 0 | 0 | Not-determined | Genome-fragment | NA | NA | 0 |
| S12_extracted_k141_73345 | 7950 | No | NA | 5 | 0 | 2 | Not-determined | Genome-fragment | NA | NA | 0 |
| S12_extracted_k141_168210 | 1484 | No | NA | 1 | 0 | 0 | Not-determined | Genome-fragment | NA | NA | 0 |
| S12_extracted_k141_152165 | 3191 | No | NA | 3 | 0 | 0 | Not-determined | Genome-fragment | NA | NA | 0 |
| S12_extracted_k141_26627 | 1334 | No | NA | 1 | 0 | 1 | Not-determined | Genome-fragment | NA | NA | 0 |
| S12_extracted_k141_136817 | 12456 | No | NA | 9 | 0 | 2 | Not-determined | Genome-fragment | NA | NA | 0 |
| S12_extracted_k141_152187 | 1001 | No | NA | 2 | 0 | 0 | Not-determined | Genome-fragment | NA | NA | 0 |
| S12_extracted_k141_18858 | 1081 | No | NA | 2 | 0 | 1 | Not-determined | Genome-fragment | NA | NA | 0 |
| S12_extracted_k141_144734 | 5509 | No | NA | 3 | 0 | 3 | Not-determined | Genome-fragment | NA | NA | 0 |
| S12_extracted_k141_105542 | 43702 | Yes | 33068 | 62 | 18 | 6 | High-quality | High-quality | 94.07 | AAI-based (high-confidence) | 24.33 |
| S12_extracted_k141_144744 | 1313 | No | NA | 1 | 0 | 0 | Not-determined | Genome-fragment | NA | NA | 0 |
| S12_extracted_k141_89998 | 1321 | No | NA | 2 | 0 | 0 | Not-determined | Genome-fragment | NA | NA | 0 |
| S12_extracted_k141_26669 | 1079 | No | NA | 2 | 0 | 1 | Not-determined | Genome-fragment | NA | NA | 0 |
| S12_extracted_k141_128647 | 1112 | No | NA | 3 | 0 | 2 | Not-determined | Genome-fragment | NA | NA | 0 |
| S12_extracted_k141_152212 | 1083 | No | NA | 2 | 0 | 0 | Not-determined | Genome-fragment | NA | NA | 0 |
| S12_extracted_k141_18892 | 4687 | No | NA | 6 | 0 | 3 | Not-determined | Genome-fragment | NA | NA | 0 |
| S10_extracted_60618 | 3774 | No | NA | 4 | 0 | 0 | Low-quality | Genome-fragment | 1.65 | AAI-based (high-confidence) | 0 |
| S11_vs_k141_28574\|\|full | 3775 | No | NA | 5 | 1 | 0 | Low-quality | Genome-fragment | 6.38 | AAI-based (high-confidence) | 0 |
| S12_extracted_k141_105569 | 3410 | No | NA | 4 | 0 | 0 | Not-determined | Genome-fragment | NA | NA | 0 |
| S12_extracted_k141_90038 | 1086 | No | NA | 3 | 0 | 0 | Not-determined | Genome-fragment | NA | NA | 0 |
| S11_vs_k141_77883\|\|full | 3776 | No | NA | 5 | 2 | 0 | Low-quality | Genome-fragment | 9.55 | AAI-based (high-confidence) | 0 |
| S5_vs_k141_126110\|\|full | 3776 | No | NA | 2 | 0 | 2 | Low-quality | Genome-fragment | 1.04 | AAI-based (medium-confidence) | 0 |
| S8_vs_k141_58463\|\|full | 3776 | No | NA | 4 | 0 | 1 | Low-quality | Genome-fragment | 4.22 | AAI-based (high-confidence) | 0 |
| S12_extracted_k141_168324 | 1023 | No | NA | 1 | 0 | 1 | Not-determined | Genome-fragment | NA | NA | 0 |
| S3_vs_k141_102310\|\|full | 3777 | No | NA | 5 | 0 | 2 | Low-quality | Genome-fragment | 2.24 | AAI-based (medium-confidence) | 0 |
| S12_extracted_k141_81702 | 4183 | No | NA | 4 | 0 | 3 | Not-determined | Genome-fragment | NA | NA | 0 |
| S16_vs_118024\|\|full | 3777 | No | NA | 4 | 1 | 0 | Low-quality | Genome-fragment | 6.38 | AAI-based (medium-confidence) | 0 |
| S10_vs_116237\|\|full | 3777 | No | NA | 5 | 0 | 1 | Low-quality | Genome-fragment | 7.86 | AAI-based (medium-confidence) | 0 |
| S12_extracted_k141_66206 | 4340 | No | NA | 5 | 0 | 3 | Not-determined | Genome-fragment | NA | NA | 0 |
| S12_extracted_k141_73470 | 1155 | No | NA | 1 | 0 | 1 | Not-determined | Genome-fragment | NA | NA | 0 |
| S12_extracted_k141_184259 | 1264 | No | NA | 2 | 0 | 1 | Not-determined | Genome-fragment | NA | NA | 0 |
| S12_extracted_k141_160138 | 1602 | No | NA | 1 | 0 | 1 | Not-determined | Genome-fragment | NA | NA | 0 |
| S12_extracted_k141_184265 | 1101 | No | NA | 2 | 0 | 0 | Not-determined | Genome-fragment | NA | NA | 0 |
| S12_extracted_k141_128722 | 4549 | No | NA | 7 | 0 | 1 | Not-determined | Genome-fragment | NA | NA | 0 |
| S12_extracted_k141_26746 | 4094 | No | NA | 6 | 0 | 0 | Not-determined | Genome-fragment | NA | NA | 0 |
| S12_extracted_k141_160161 | 1124 | No | NA | 1 | 0 | 1 | Not-determined | Genome-fragment | NA | NA | 0 |
| S9_extracted_k141_29049 | 3778 | No | NA | 6 | 3 | 0 | Low-quality | Genome-fragment | 11.02 | AAI-based (medium-confidence) | 0 |
| S12_extracted_k141_136897 | 1806 | No | NA | 2 | 0 | 2 | Not-determined | Genome-fragment | NA | NA | 0 |
| S12_extracted_k141_90134 | 3976 | No | NA | 2 | 0 | 2 | Not-determined | Genome-fragment | NA | NA | 0 |
| S12_extracted_k141_175776 | 1012 | No | NA | 2 | 0 | 0 | Not-determined | Genome-fragment | NA | NA | 0 |
| S12_extracted_k141_90138 | 2260 | No | NA | 2 | 0 | 1 | Not-determined | Genome-fragment | NA | NA | 0 |
| S12_extracted_k141_81744 | 1283 | No | NA | 2 | 0 | 2 | Not-determined | Genome-fragment | NA | NA | 0 |
| S7_vs_89439\|\|full | 3778 | Yes | 2706 | 7 | 1 | 1 | Low-quality | Genome-fragment | 7.18 | AAI-based (high-confidence) | 28.37 |
| S17_vs_k141_112799\|\|full | 3779 | No | NA | 2 | 0 | 0 | Low-quality | Genome-fragment | 1.52 | AAI-based (high-confidence) | 0 |
| S12_extracted_k141_10294 | 2484 | No | NA | 1 | 0 | 1 | Not-determined | Genome-fragment | NA | NA | 0 |
| S3_vs_k141_119252\|\|full | 3779 | No | NA | 9 | 1 | 0 | Low-quality | Genome-fragment | 5.36 | AAI-based (high-confidence) | 0 |
| S12_extracted_k141_66263 | 1482 | No | NA | 2 | 0 | 2 | Not-determined | Genome-fragment | NA | NA | 0 |
| S8_vs_k141_107454\|\|full | 3780 | No | NA | 3 | 0 | 2 | Low-quality | Genome-fragment | 0.73 | AAI-based (medium-confidence) | 0 |
| S12_extracted_k141_58561 | 1141 | No | NA | 3 | 0 | 1 | Not-determined | Genome-fragment | NA | NA | 0 |
| S8_vs_k141_11992\|\|full | 3780 | No | NA | 3 | 1 | 0 | Low-quality | Genome-fragment | 8.07 | AAI-based (high-confidence) | 0 |
| S12_extracted_k141_113021 | 1050 | No | NA | 1 | 0 | 0 | Not-determined | Genome-fragment | NA | NA | 0 |
| S12_extracted_k141_73551 | 1223 | No | NA | 2 | 0 | 1 | Not-determined | Genome-fragment | NA | NA | 0 |
| S12_extracted_k141_105741 | 1061 | No | NA | 1 | 0 | 1 | Not-determined | Genome-fragment | NA | NA | 0 |
| S12_extracted_k141_175830 | 3936 | No | NA | 7 | 0 | 4 | Not-determined | Genome-fragment | NA | NA | 0 |
| S12_extracted_k141_18902 | 117200 | No | NA | 148 | 34 | 4 | Complete | High-quality | 100 | DTR (high-confidence) | 0 |
| S12_extracted_k141_90225 | 1075 | No | NA | 3 | 0 | 0 | Not-determined | Genome-fragment | NA | NA | 0 |
| S9_extracted_k141_62599 | 3781 | No | NA | 2 | 0 | 1 | Low-quality | Genome-fragment | 1.88 | AAI-based (medium-confidence) | 0 |
| S12_extracted_k141_105773 | 1044 | No | NA | 1 | 0 | 0 | Not-determined | Genome-fragment | NA | NA | 0 |
| S12_extracted_k141_42976 | 2188 | No | NA | 2 | 0 | 1 | Not-determined | Genome-fragment | NA | NA | 0 |
| S12_extracted_k141_26799 | 1264 | No | NA | 3 | 0 | 2 | Not-determined | Genome-fragment | NA | NA | 0 |
| S12_extracted_k141_128794 | 45090 | No | NA | 67 | 14 | 0 | Complete | High-quality | 100 | DTR (high-confidence) | 0 |
| S12_extracted_k141_144963 | 1236 | No | NA | 2 | 0 | 0 | Not-determined | Genome-fragment | NA | NA | 0 |
| S12_extracted_k141_136996 | 2155 | No | NA | 2 | 0 | 1 | Not-determined | Genome-fragment | NA | NA | 0 |
| S11_extracted_k141_6861 | 3781 | No | NA | 4 | 0 | 3 | Low-quality | Genome-fragment | 0.42 | AAI-based (medium-confidence) | 0 |
| S12_extracted_k141_144973 | 1213 | No | NA | 1 | 0 | 0 | Not-determined | Genome-fragment | NA | NA | 0 |
| S12_extracted_k141_128803 | 1774 | No | NA | 3 | 0 | 2 | Not-determined | Genome-fragment | NA | NA | 0 |
| S11_vs_k141_84866\|\|full | 3781 | No | NA | 5 | 0 | 2 | Low-quality | Genome-fragment | 4.95 | AAI-based (high-confidence) | 0 |
| S12_extracted_k141_144983 | 7977 | No | NA | 5 | 0 | 5 | Not-determined | Genome-fragment | NA | NA | 0 |
| S12_extracted_k141_26819 | 9165 | No | NA | 9 | 0 | 0 | Not-determined | Genome-fragment | NA | NA | 0 |
| S12_extracted_k141_152358 | 1001 | No | NA | 1 | 0 | 1 | Not-determined | Genome-fragment | NA | NA | 0 |
| S12_extracted_k141_34186 | 2133 | No | NA | 3 | 0 | 0 | Not-determined | Genome-fragment | NA | NA | 0 |
| S12_extracted_k141_144994 | 4062 | No | NA | 4 | 0 | 4 | Not-determined | Genome-fragment | NA | NA | 0 |
| S12_extracted_k141_90278 | 1008 | No | NA | 1 | 0 | 0 | Not-determined | Genome-fragment | NA | NA | 0 |
| S12_extracted_k141_26842 | 1431 | No | NA | 2 | 0 | 1 | Not-determined | Genome-fragment | NA | NA | 0 |
| S9_extracted_k141_49320 | 3782 | No | NA | 3 | 0 | 2 | Low-quality | Genome-fragment | 8.13 | AAI-based (medium-confidence) | 0 |
| S12_extracted_k141_58662 | 1031 | No | NA | 1 | 0 | 1 | Not-determined | Genome-fragment | NA | NA | 0 |
| S14_vs_k141_87059\|\|full | 3782 | No | NA | 3 | 0 | 1 | Low-quality | Genome-fragment | 3.71 | AAI-based (medium-confidence) | 0 |
| S12_extracted_k141_168535 | 1043 | No | NA | 1 | 0 | 0 | Not-determined | Genome-fragment | NA | NA | 0 |
| S12_extracted_k141_175899 | 1706 | No | NA | 2 | 0 | 1 | Not-determined | Genome-fragment | NA | NA | 0 |
| S12_extracted_k141_73605 | 4090 | No | NA | 2 | 0 | 2 | Not-determined | Genome-fragment | NA | NA | 0 |
| S12_extracted_k141_66322 | 3121 | No | NA | 3 | 0 | 1 | Not-determined | Genome-fragment | NA | NA | 0 |
| S12_extracted_k141_168557 | 1952 | No | NA | 3 | 0 | 2 | Not-determined | Genome-fragment | NA | NA | 0 |
| S12_extracted_k141_168562 | 1097 | No | NA | 2 | 0 | 0 | Not-determined | Genome-fragment | NA | NA | 0 |
| S8_extracted_k141_98884 | 3783 | No | NA | 4 | 0 | 1 | Low-quality | Genome-fragment | 4.8 | AAI-based (medium-confidence) | 0 |
| S12_extracted_k141_18943 | 3098 | No | NA | 4 | 0 | 1 | Not-determined | Genome-fragment | NA | NA | 0 |
| S8_vs_k141_168114\|\|full | 3783 | No | NA | 3 | 0 | 3 | Low-quality | Genome-fragment | 1.61 | AAI-based (medium-confidence) | 0 |
| S12_extracted_k141_90298 | 1065 | No | NA | 2 | 0 | 1 | Not-determined | Genome-fragment | NA | NA | 0 |
| S12_extracted_k141_34251 | 1422 | No | NA | 1 | 0 | 0 | Not-determined | Genome-fragment | NA | NA | 0 |
| S12_extracted_k141_34253 | 1829 | No | NA | 1 | 0 | 1 | Not-determined | Genome-fragment | NA | NA | 0 |
| S17_vs_k141_93877\|\|full | 3784 | No | NA | 4 | 0 | 3 | Low-quality | Genome-fragment | 2.31 | AAI-based (medium-confidence) | 0 |
| S12_extracted_k141_66361 | 1268 | No | NA | 3 | 0 | 2 | Not-determined | Genome-fragment | NA | NA | 0 |
| S12_extracted_k141_34269 | 3493 | No | NA | 3 | 0 | 2 | Not-determined | Genome-fragment | NA | NA | 0 |
| S12_extracted_k141_43101 | 3960 | No | NA | 3 | 0 | 2 | Not-determined | Genome-fragment | NA | NA | 0 |
| S14_extracted_k141_74615 | 3784 | No | NA | 4 | 2 | 0 | Low-quality | Genome-fragment | 10.14 | AAI-based (high-confidence) | 0 |
| S12_extracted_k141_128933 | 1084 | No | NA | 1 | 0 | 0 | Not-determined | Genome-fragment | NA | NA | 0 |
| S3_extracted_k141_44497 | 3784 | No | NA | 4 | 0 | 0 | Low-quality | Genome-fragment | 5.71 | AAI-based (high-confidence) | 0 |
| S12_extracted_k141_3548 | 2505 | No | NA | 1 | 0 | 1 | Not-determined | Genome-fragment | NA | NA | 0 |
| S17_extracted_k141_166532 | 3785 | No | NA | 7 | 1 | 0 | Low-quality | Genome-fragment | 5.44 | AAI-based (high-confidence) | 0 |
| S1_extracted_k141_130373 | 3785 | No | NA | 4 | 0 | 0 | Low-quality | Genome-fragment | 8.33 | AAI-based (high-confidence) | 0 |
| S12_extracted_k141_152493 | 1390 | No | NA | 1 | 0 | 1 | Not-determined | Genome-fragment | NA | NA | 0 |
| S12_extracted_k141_90343 | 1457 | No | NA | 3 | 0 | 0 | Not-determined | Genome-fragment | NA | NA | 0 |
| S12_extracted_k141_3597 | 1116 | No | NA | 1 | 0 | 1 | Not-determined | Genome-fragment | NA | NA | 0 |
| S12_extracted_k141_3638 | 1012 | No | NA | 1 | 0 | 0 | Not-determined | Genome-fragment | NA | NA | 0 |
| S12_extracted_k141_19031 | 1960 | No | NA | 4 | 0 | 2 | Not-determined | Genome-fragment | NA | NA | 0 |
| S12_extracted_k141_3641 | 1317 | No | NA | 3 | 0 | 1 | Not-determined | Genome-fragment | NA | NA | 0 |
| S12_extracted_k141_43179 | 1277 | No | NA | 1 | 0 | 0 | Not-determined | Genome-fragment | NA | NA | 0 |
| S1_vs_k141_95842\|\|full | 3785 | No | NA | 6 | 1 | 0 | Low-quality | Genome-fragment | 8.43 | AAI-based (high-confidence) | 0 |
| S5_extracted_k141_138119 | 3785 | No | NA | 2 | 0 | 2 | Low-quality | Genome-fragment | 1.04 | AAI-based (medium-confidence) | 0 |
| S16_vs_100221\|\|full | 3785 | No | NA | 4 | 0 | 2 | Low-quality | Genome-fragment | 1.63 | AAI-based (medium-confidence) | 0 |
| S16_vs_134834\|\|full | 3785 | No | NA | 4 | 0 | 0 | Low-quality | Genome-fragment | 3.06 | AAI-based (medium-confidence) | 0 |
| S12_extracted_k141_26919 | 1379 | No | NA | 3 | 0 | 2 | Not-determined | Genome-fragment | NA | NA | 0 |
| S12_extracted_k141_160466 | 2475 | No | NA | 3 | 0 | 2 | Not-determined | Genome-fragment | NA | NA | 0 |
| S12_extracted_k141_50114 | 1029 | No | NA | 1 | 0 | 1 | Not-determined | Genome-fragment | NA | NA | 0 |
| S12_extracted_k141_10519 | 1000 | No | NA | 1 | 0 | 0 | Not-determined | Genome-fragment | NA | NA | 0 |
| S12_extracted_k141_10545 | 1398 | No | NA | 2 | 0 | 1 | Not-determined | Genome-fragment | NA | NA | 0 |
| S12_extracted_k141_152547 | 4227 | No | NA | 5 | 0 | 1 | Not-determined | Genome-fragment | NA | NA | 0 |
| S12_extracted_k141_3728 | 1049 | No | NA | 2 | 0 | 1 | Not-determined | Genome-fragment | NA | NA | 0 |
| S12_extracted_k141_113217 | 1144 | No | NA | 2 | 0 | 1 | Not-determined | Genome-fragment | NA | NA | 0 |
| S12_extracted_k141_111044 | 3785 | No | NA | 4 | 0 | 4 | Low-quality | Genome-fragment | 3.55 | AAI-based (medium-confidence) | 0 |
| S12_extracted_k141_145044 | 168494 | No | NA | 191 | 32 | 2 | High-quality | High-quality | 100 | AAI-based (high-confidence) | 0 |
| S12_extracted_k141_26969 | 5354 | No | NA | 9 | 0 | 0 | Not-determined | Genome-fragment | NA | NA | 0 |
| S12_extracted_k141_43295 | 1132 | No | NA | 1 | 0 | 0 | Not-determined | Genome-fragment | NA | NA | 0 |
| S12_extracted_k141_82109 | 1990 | No | NA | 2 | 0 | 1 | Not-determined | Genome-fragment | NA | NA | 0 |
| S12_extracted_k141_121149 | 1250 | No | NA | 1 | 0 | 0 | Not-determined | Genome-fragment | NA | NA | 0 |
| S12_extracted_k141_73843 | 1782 | No | NA | 2 | 0 | 2 | Not-determined | Genome-fragment | NA | NA | 0 |
| S6_extracted_k141_44861 | 3785 | No | NA | 3 | 0 | 0 | Low-quality | Genome-fragment | 6.22 | AAI-based (high-confidence) | 0 |
| S6_vs_k141_18586\|\|full | 3785 | No | NA | 4 | 0 | 0 | Low-quality | Genome-fragment | 2.48 | AAI-based (high-confidence) | 0 |
| S12_extracted_k141_43331 | 1033 | No | NA | 3 | 0 | 1 | Not-determined | Genome-fragment | NA | NA | 0 |
| S6_vs_k141_6865\|\|full | 3785 | No | NA | 3 | 0 | 0 | Low-quality | Genome-fragment | 2.55 | AAI-based (high-confidence) | 0 |
| S12_extracted_k141_73867 | 4653 | No | NA | 3 | 0 | 2 | Not-determined | Genome-fragment | NA | NA | 0 |
| S12_extracted_k141_121168 | 1120 | No | NA | 1 | 0 | 1 | Not-determined | Genome-fragment | NA | NA | 0 |
| S12_extracted_k141_176168 | 3031 | No | NA | 2 | 0 | 0 | Not-determined | Genome-fragment | NA | NA | 0 |
| S12_extracted_k141_152618 | 1276 | No | NA | 2 | 0 | 2 | Not-determined | Genome-fragment | NA | NA | 0 |
| S5_vs_k141_43650\|\|full | 3786 | No | NA | 5 | 0 | 2 | Low-quality | Genome-fragment | 8.12 | AAI-based (high-confidence) | 0 |
| S12_extracted_k141_160594 | 4276 | No | NA | 1 | 0 | 1 | Not-determined | Genome-fragment | NA | NA | 0 |
| S12_extracted_k141_145111 | 1110 | No | NA | 1 | 0 | 0 | Not-determined | Genome-fragment | NA | NA | 0 |
| S12_extracted_k141_98580 | 4454 | No | NA | 7 | 0 | 2 | Not-determined | Genome-fragment | NA | NA | 0 |
| S12_extracted_k141_10643 | 1036 | No | NA | 1 | 0 | 0 | Not-determined | Genome-fragment | NA | NA | 0 |
| S12_extracted_k141_106137 | 2958 | No | NA | 2 | 0 | 0 | Not-determined | Genome-fragment | NA | NA | 0 |
| S4_extracted_23012 | 3786 | No | NA | 2 | 1 | 0 | Low-quality | Genome-fragment | 9.56 | AAI-based (high-confidence) | 0 |
| S12_extracted_k141_137275 | 1031 | No | NA | 2 | 0 | 0 | Not-determined | Genome-fragment | NA | NA | 0 |
| S12_extracted_k141_137280 | 1261 | No | NA | 2 | 0 | 0 | Not-determined | Genome-fragment | NA | NA | 0 |
| S12_extracted_k141_90587 | 1023 | No | NA | 1 | 0 | 1 | Not-determined | Genome-fragment | NA | NA | 0 |
| S12_extracted_k141_90589 | 2614 | No | NA | 3 | 0 | 1 | Not-determined | Genome-fragment | NA | NA | 0 |
| S12_extracted_k141_176207 | 5719 | No | NA | 7 | 0 | 0 | Not-determined | Genome-fragment | NA | NA | 0 |
| S12_extracted_k141_145156 | 2688 | No | NA | 2 | 0 | 0 | Not-determined | Genome-fragment | NA | NA | 0 |
| S12_extracted_k141_43358 | 1560 | No | NA | 3 | 0 | 1 | Not-determined | Genome-fragment | NA | NA | 0 |
| S12_extracted_k141_121231 | 1139 | No | NA | 1 | 0 | 1 | Not-determined | Genome-fragment | NA | NA | 0 |
| S12_extracted_k141_10681 | 1950 | No | NA | 2 | 0 | 0 | Not-determined | Genome-fragment | NA | NA | 0 |
| S12_extracted_k141_184710 | 1248 | No | NA | 1 | 0 | 0 | Not-determined | Genome-fragment | NA | NA | 0 |
| S12_extracted_k141_3890 | 1079 | No | NA | 2 | 0 | 1 | Not-determined | Genome-fragment | NA | NA | 0 |
| S12_extracted_k141_98634 | 1088 | No | NA | 1 | 0 | 0 | Not-determined | Genome-fragment | NA | NA | 0 |
| S12_extracted_k141_160630 | 1241 | No | NA | 3 | 0 | 1 | Not-determined | Genome-fragment | NA | NA | 0 |
| S12_extracted_k141_50289 | 1353 | No | NA | 2 | 0 | 2 | Not-determined | Genome-fragment | NA | NA | 0 |
| S8_extracted_k141_33915 | 3786 | No | NA | 4 | 1 | 0 | Low-quality | Genome-fragment | 2.18 | HMM-based (lower-bound) | 0 |
| S12_extracted_k141_152662 | 1744 | No | NA | 3 | 0 | 1 | Not-determined | Genome-fragment | NA | NA | 0 |
| S8_extracted_k141_163748 | 3786 | No | NA | 3 | 0 | 2 | Low-quality | Genome-fragment | 3.27 | AAI-based (medium-confidence) | 0 |
| S12_extracted_k141_129062 | 1081 | No | NA | 2 | 0 | 1 | Not-determined | Genome-fragment | NA | NA | 0 |
| S12_extracted_k141_129063 | 1036 | No | NA | 3 | 0 | 0 | Not-determined | Genome-fragment | NA | NA | 0 |
| S11_vs_k141_85787\|\|full | 3788 | No | NA | 4 | 0 | 1 | Low-quality | Genome-fragment | 4.73 | AAI-based (high-confidence) | 0 |
| S12_extracted_k141_27059 | 1063 | No | NA | 3 | 0 | 1 | Not-determined | Genome-fragment | NA | NA | 0 |
| S13_vs_23340\|\|full | 3789 | No | NA | 8 | 1 | 0 | Low-quality | Genome-fragment | 9.05 | AAI-based (medium-confidence) | 0 |
| S12_extracted_k141_176261 | 5109 | No | NA | 5 | 0 | 4 | Not-determined | Genome-fragment | NA | NA | 0 |
| S12_extracted_k141_137360 | 3245 | No | NA | 3 | 0 | 1 | Not-determined | Genome-fragment | NA | NA | 0 |
| S12_extracted_k141_176271 | 2078 | No | NA | 2 | 0 | 0 | Not-determined | Genome-fragment | NA | NA | 0 |
| S12_extracted_k141_145176 | 1459 | No | NA | 2 | 0 | 2 | Not-determined | Genome-fragment | NA | NA | 0 |
| S12_extracted_k141_176273 | 3125 | No | NA | 2 | 0 | 1 | Not-determined | Genome-fragment | NA | NA | 0 |
| S12_extracted_k141_129103 | 1024 | No | NA | 1 | 0 | 1 | Not-determined | Genome-fragment | NA | NA | 0 |
| S12_extracted_k141_34473 | 1565 | No | NA | 2 | 0 | 0 | Not-determined | Genome-fragment | NA | NA | 0 |
| S12_extracted_k141_184788 | 1724 | No | NA | 2 | 0 | 1 | Not-determined | Genome-fragment | NA | NA | 0 |
| S12_extracted_k141_145209 | 1243 | No | NA | 2 | 0 | 1 | Not-determined | Genome-fragment | NA | NA | 0 |
| S12_extracted_k141_152736 | 1641 | No | NA | 3 | 0 | 3 | Not-determined | Genome-fragment | NA | NA | 0 |
| S12_extracted_k141_27101 | 1078 | No | NA | 3 | 0 | 2 | Not-determined | Genome-fragment | NA | NA | 0 |
| S10_vs_66618\|\|full | 3789 | Yes | 1785 | 8 | 1 | 1 | Low-quality | Genome-fragment | 1.28 | HMM-based (lower-bound) | 52.89 |
| S12_extracted_k141_121294 | 5372 | No | NA | 6 | 0 | 5 | Not-determined | Genome-fragment | NA | NA | 0 |

| | | | | | | | | | | | |
|---|---|---|---|---|---|---|---|---|---|---|---|
| S12_extracted_k141_168748 | 1709 | No | NA | 2 | 0 | 1 | Not-determined | Genome-fragment | NA | NA | 0 |
| S16_extracted_140447 | 3792 | No | NA | 4 | 0 | 4 | Low-quality | Genome-fragment | 1.4 | AAI-based (high-confidence) | 0 |
| S3_vs_k141_53640\|\|full | 3794 | No | NA | 3 | 0 | 2 | Low-quality | Genome-fragment | 2.83 | AAI-based (medium-confidence) | 0 |
| S12_extracted_k141_160679 | 1165 | No | NA | 2 | 0 | 0 | Not-determined | Genome-fragment | NA | NA | 0 |
| S12_extracted_k141_74026 | 2303 | No | NA | 2 | 0 | 1 | Not-determined | Genome-fragment | NA | NA | 0 |
| S12_extracted_k141_113435 | 1154 | No | NA | 1 | 0 | 1 | Not-determined | Genome-fragment | NA | NA | 0 |
| S12_extracted_k141_27191 | 1129 | No | NA | 2 | 0 | 0 | Not-determined | Genome-fragment | NA | NA | 0 |
| S12_extracted_k141_152796 | 2143 | No | NA | 3 | 0 | 0 | Not-determined | Genome-fragment | NA | NA | 0 |
| S12_extracted_k141_10824 | 1847 | No | NA | 3 | 0 | 1 | Not-determined | Genome-fragment | NA | NA | 0 |
| S12_extracted_k141_113440 | 1043 | No | NA | 2 | 0 | 0 | Not-determined | Genome-fragment | NA | NA | 0 |
| S12_extracted_k141_137450 | 1143 | No | NA | 3 | 0 | 1 | Not-determined | Genome-fragment | NA | NA | 0 |
| S12_extracted_k141_106270 | 1185 | No | NA | 2 | 0 | 0 | Not-determined | Genome-fragment | NA | NA | 0 |
| S12_extracted_k141_50428 | 5092 | No | NA | 6 | 0 | 5 | Not-determined | Genome-fragment | NA | NA | 0 |
| S12_extracted_k141_113461 | 1117 | No | NA | 1 | 0 | 0 | Not-determined | Genome-fragment | NA | NA | 0 |
| S3_extracted_k141_34756 | 3795 | No | NA | 6 | 4 | 0 | Low-quality | Genome-fragment | 9.47 | AAI-based (high-confidence) | 0 |
| S12_extracted_k141_58940 | 1216 | No | NA | 1 | 0 | 1 | Not-determined | Genome-fragment | NA | NA | 0 |
| S12_extracted_k141_66575 | 1672 | No | NA | 2 | 0 | 0 | Not-determined | Genome-fragment | NA | NA | 0 |
| S12_extracted_k141_121356 | 5648 | No | NA | 6 | 0 | 4 | Not-determined | Genome-fragment | NA | NA | 0 |
| S12_extracted_k141_58953 | 4246 | No | NA | 5 | 0 | 0 | Not-determined | Genome-fragment | NA | NA | 0 |
| S12_extracted_k141_50477 | 3327 | No | NA | 5 | 0 | 0 | Not-determined | Genome-fragment | NA | NA | 0 |
| S12_extracted_k141_145329 | 1574 | No | NA | 4 | 0 | 1 | Not-determined | Genome-fragment | NA | NA | 0 |
| S7_extracted_127724 | 3796 | No | NA | 4 | 0 | 1 | Low-quality | Genome-fragment | 7.46 | AAI-based (medium-confidence) | 0 |
| S12_extracted_k141_82289 | 1389 | No | NA | 1 | 0 | 0 | Not-determined | Genome-fragment | NA | NA | 0 |
| S12_extracted_k141_98856 | 1293 | No | NA | 2 | 0 | 0 | Not-determined | Genome-fragment | NA | NA | 0 |
| S12_extracted_k141_50499 | 1070 | No | NA | 2 | 0 | 1 | Not-determined | Genome-fragment | NA | NA | 0 |
| S16_vs_58354\|\|full | 3796 | No | NA | 5 | 0 | 4 | Low-quality | Genome-fragment | 4.61 | AAI-based (medium-confidence) | 0 |
| S12_extracted_k141_66614 | 1340 | No | NA | 1 | 0 | 0 | Not-determined | Genome-fragment | NA | NA | 0 |
| S15_extracted_72938 | 3796 | No | NA | 5 | 0 | 4 | Low-quality | Genome-fragment | 1.4 | AAI-based (high-confidence) | 0 |
| S17_vs_k141_57026\|\|full | 3797 | No | NA | 3 | 0 | 0 | Low-quality | Genome-fragment | 7.28 | AAI-based (high-confidence) | 0 |
| S5_vs_k141_24974\|\|full | 3797 | No | NA | 4 | 0 | 3 | Low-quality | Genome-fragment | 4.04 | AAI-based (medium-confidence) | 0 |
| S12_extracted_k141_19333 | 2481 | No | NA | 4 | 0 | 4 | Not-determined | Genome-fragment | NA | NA | 0 |
| S12_extracted_k141_184907 | 4682 | No | NA | 3 | 0 | 2 | Not-determined | Genome-fragment | NA | NA | 0 |
| S12_extracted_k141_27317 | 1972 | No | NA | 1 | 0 | 0 | Not-determined | Genome-fragment | NA | NA | 0 |
| S12_extracted_k141_66633 | 1126 | No | NA | 1 | 0 | 1 | Not-determined | Genome-fragment | NA | NA | 0 |
| S12_extracted_k141_66645 | 1375 | No | NA | 2 | 0 | 2 | Not-determined | Genome-fragment | NA | NA | 0 |
| S12_extracted_k141_129346 | 1226 | No | NA | 3 | 0 | 1 | Not-determined | Genome-fragment | NA | NA | 0 |
| S12_extracted_k141_66649 | 1102 | No | NA | 2 | 0 | 2 | Not-determined | Genome-fragment | NA | NA | 0 |
| S9_extracted_k141_41658 | 3798 | No | NA | 6 | 2 | 0 | Low-quality | Genome-fragment | 5.33 | AAI-based (high-confidence) | 0 |
| S12_extracted_k141_50568 | 2702 | No | NA | 4 | 0 | 1 | Not-determined | Genome-fragment | NA | NA | 0 |
| S7_extracted_48644 | 3798 | No | NA | 3 | 1 | 0 | Low-quality | Genome-fragment | 4.15 | HMM-based (lower-bound) | 0 |
| S12_extracted_k141_10887 | 65312 | No | NA | 92 | 19 | 0 | Complete | High-quality | 100 | DTR (high-confidence) | 0 |
| S12_extracted_k141_82363 | 1126 | No | NA | 1 | 0 | 1 | Not-determined | Genome-fragment | NA | NA | 0 |
| S12_extracted_k141_121439 | 2528 | No | NA | 3 | 0 | 1 | Not-determined | Genome-fragment | NA | NA | 0 |
| S12_extracted_k141_121440 | 3054 | No | NA | 2 | 0 | 1 | Not-determined | Genome-fragment | NA | NA | 0 |
| S12_extracted_k141_66676 | 3719 | No | NA | 3 | 0 | 2 | Not-determined | Genome-fragment | NA | NA | 0 |
| S12_extracted_k141_129374 | 2200 | No | NA | 3 | 0 | 2 | Not-determined | Genome-fragment | NA | NA | 0 |
| S12_extracted_k141_184951 | 1576 | No | NA | 1 | 0 | 0 | Not-determined | Genome-fragment | NA | NA | 0 |
| S12_extracted_k141_50589 | 1197 | No | NA | 2 | 0 | 1 | Not-determined | Genome-fragment | NA | NA | 0 |
| S12_extracted_k141_74178 | 1137 | No | NA | 1 | 0 | 1 | Not-determined | Genome-fragment | NA | NA | 0 |
| S12_extracted_k141_34633 | 2669 | No | NA | 4 | 0 | 0 | Not-determined | Genome-fragment | NA | NA | 0 |
| S16_extracted_168955 | 3799 | No | NA | 6 | 0 | 1 | Low-quality | Genome-fragment | 4.13 | AAI-based (medium-confidence) | 0 |
| S12_extracted_k141_121463 | 1088 | No | NA | 1 | 0 | 0 | Not-determined | Genome-fragment | NA | NA | 0 |
| S12_extracted_k141_168879 | 1201 | No | NA | 2 | 0 | 1 | Not-determined | Genome-fragment | NA | NA | 0 |
| S12_extracted_k141_176437 | 15756 | No | NA | 21 | 7 | 0 | Medium-quality | Genome-fragment | 67.39 | AAI-based (high-confidence) | 0 |
| S8_vs_k141_189040\|\|full | 3799 | No | NA | 4 | 0 | 2 | Low-quality | Genome-fragment | 4.96 | AAI-based (medium-confidence) | 0 |
| S17_extracted_k141_81882 | 3800 | No | NA | 2 | 0 | 2 | Low-quality | Genome-fragment | 1.49 | AAI-based (medium-confidence) | 0 |
| S12_extracted_k141_10955 | 2052 | No | NA | 2 | 0 | 2 | Not-determined | Genome-fragment | NA | NA | 0 |
| S12_extracted_k141_145453 | 1268 | No | NA | 2 | 0 | 2 | Not-determined | Genome-fragment | NA | NA | 0 |
| S12_extracted_k141_185015 | 1519 | No | NA | 1 | 0 | 1 | Not-determined | Genome-fragment | NA | NA | 0 |
| S12_extracted_k141_34682 | 2954 | No | NA | 3 | 0 | 0 | Not-determined | Genome-fragment | NA | NA | 0 |
| S12_extracted_k141_34687 | 1103 | No | NA | 1 | 0 | 1 | Not-determined | Genome-fragment | NA | NA | 0 |
| S12_extracted_k141_121503 | 1144 | No | NA | 1 | 0 | 1 | Not-determined | Genome-fragment | NA | NA | 0 |
| S14_extracted_k141_103718 | 3800 | No | NA | 9 | 2 | 0 | Low-quality | Genome-fragment | 5.56 | HMM-based (lower-bound) | 0 |
| S12_extracted_k141_145478 | 4675 | No | NA | 5 | 0 | 1 | Not-determined | Genome-fragment | NA | NA | 0 |
| S12_extracted_k141_50649 | 3619 | No | NA | 3 | 0 | 2 | Not-determined | Genome-fragment | NA | NA | 0 |
| S12_extracted_k141_168950 | 2892 | No | NA | 4 | 0 | 0 | Not-determined | Genome-fragment | NA | NA | 0 |
| S12_extracted_k141_168955 | 3027 | No | NA | 3 | 0 | 3 | Not-determined | Genome-fragment | NA | NA | 0 |
| S12_extracted_k141_59091 | 3662 | No | NA | 4 | 0 | 2 | Not-determined | Genome-fragment | NA | NA | 0 |
| S12_extracted_k141_4069 | 3794 | No | NA | 3 | 0 | 1 | Not-determined | Genome-fragment | NA | NA | 0 |
| S12_extracted_k141_27463 | 3160 | No | NA | 4 | 0 | 2 | Not-determined | Genome-fragment | NA | NA | 0 |
| S14_extracted_k141_20579 | 3800 | No | NA | 6 | 0 | 0 | Low-quality | Genome-fragment | 8.81 | AAI-based (high-confidence) | 0 |
| S12_extracted_k141_106443 | 1529 | No | NA | 3 | 0 | 1 | Not-determined | Genome-fragment | NA | NA | 0 |
| S1_vs_k141_107771\|\|full | 3800 | No | NA | 5 | 0 | 0 | Low-quality | Genome-fragment | 6.88 | AAI-based (medium-confidence) | 0 |
| S12_extracted_k141_106466 | 1387 | No | NA | 1 | 0 | 1 | Not-determined | Genome-fragment | NA | NA | 0 |
| S12_extracted_k141_145526 | 1035 | No | NA | 2 | 0 | 1 | Not-determined | Genome-fragment | NA | NA | 0 |
| S12_extracted_k141_66825 | 1808 | No | NA | 2 | 0 | 1 | Not-determined | Genome-fragment | NA | NA | 0 |
| S12_extracted_k141_27501 | 1717 | No | NA | 4 | 0 | 3 | Not-determined | Genome-fragment | NA | NA | 0 |
| S3_extracted_k141_27104 | 3800 | No | NA | 9 | 1 | 0 | Low-quality | Genome-fragment | 6.63 | AAI-based (high-confidence) | 0 |
| S12_extracted_k141_59144 | 7532 | No | NA | 8 | 0 | 2 | Not-determined | Genome-fragment | NA | NA | 0 |
| S12_extracted_k141_43778 | 1009 | No | NA | 2 | 0 | 1 | Not-determined | Genome-fragment | NA | NA | 0 |
| S12_extracted_k141_99047 | 1301 | No | NA | 2 | 0 | 1 | Not-determined | Genome-fragment | NA | NA | 0 |
| S12_extracted_k141_113644 | 2236 | No | NA | 5 | 0 | 0 | Not-determined | Genome-fragment | NA | NA | 0 |
| S12_extracted_k141_50729 | 1431 | No | NA | 2 | 0 | 0 | Not-determined | Genome-fragment | NA | NA | 0 |
| S5_vs_k141_4387\|\|full | 3800 | No | NA | 5 | 0 | 0 | Low-quality | Genome-fragment | 9.18 | AAI-based (medium-confidence) | 0 |
| S12_extracted_k141_137746 | 1438 | No | NA | 1 | 0 | 1 | Not-determined | Genome-fragment | NA | NA | 0 |
| S12_extracted_k141_121630 | 1033 | No | NA | 1 | 0 | 1 | Not-determined | Genome-fragment | NA | NA | 0 |
| S12_extracted_k141_137759 | 1760 | No | NA | 2 | 0 | 1 | Not-determined | Genome-fragment | NA | NA | 0 |
| S12_extracted_k141_137761 | 1012 | No | NA | 2 | 0 | 2 | Not-determined | Genome-fragment | NA | NA | 0 |
| S12_extracted_k141_160882 | 6367 | No | NA | 7 | 0 | 5 | Not-determined | Genome-fragment | NA | NA | 0 |
| S12_extracted_k141_106511 | 1336 | No | NA | 2 | 0 | 0 | Not-determined | Genome-fragment | NA | NA | 0 |
| S12_extracted_k141_113656 | 1100 | No | NA | 2 | 0 | 2 | Not-determined | Genome-fragment | NA | NA | 0 |
| S3_extracted_k141_38023 | 3801 | No | NA | 6 | 1 | 0 | Low-quality | Genome-fragment | 2.48 | AAI-based (high-confidence) | 0 |
| S16_vs_91126\|\|full | 3801 | No | NA | 6 | 1 | 0 | Low-quality | Genome-fragment | 2.41 | HMM-based (lower-bound) | 0 |
| S12_extracted_k141_11014 | 1104 | No | NA | 1 | 0 | 0 | Not-determined | Genome-fragment | NA | NA | 0 |
| S12_extracted_k141_106520 | 1126 | No | NA | 2 | 0 | 0 | Not-determined | Genome-fragment | NA | NA | 0 |
| S12_extracted_k141_34816 | 1281 | No | NA | 1 | 0 | 0 | Not-determined | Genome-fragment | NA | NA | 0 |
| S12_extracted_k141_59212 | 2860 | No | NA | 6 | 0 | 4 | Not-determined | Genome-fragment | NA | NA | 0 |
| S12_extracted_k141_27571 | 4528 | No | NA | 6 | 0 | 0 | Not-determined | Genome-fragment | NA | NA | 0 |
| S7_vs_118133\|\|full | 3802 | No | NA | 3 | 0 | 0 | Low-quality | Genome-fragment | 3.79 | AAI-based (medium-confidence) | 0 |
| S12_extracted_k141_153110 | 8392 | No | NA | 6 | 0 | 4 | Not-determined | Genome-fragment | NA | NA | 0 |
| S12_extracted_k141_50786 | 1243 | No | NA | 3 | 0 | 2 | Not-determined | Genome-fragment | NA | NA | 0 |
| S12_extracted_k141_82512 | 9321 | No | NA | 6 | 0 | 0 | Not-determined | Genome-fragment | NA | NA | 0 |
| S12_extracted_k141_129528 | 1349 | No | NA | 2 | 0 | 0 | Not-determined | Genome-fragment | NA | NA | 0 |
| S12_extracted_k141_90885 | 8229 | No | NA | 7 | 0 | 5 | Not-determined | Genome-fragment | NA | NA | 0 |
| S12_extracted_k141_160946 | 1586 | No | NA | 3 | 0 | 3 | Not-determined | Genome-fragment | NA | NA | 0 |
| S12_extracted_k141_50794 | 3284 | No | NA | 3 | 0 | 3 | Not-determined | Genome-fragment | NA | NA | 0 |
| S12_extracted_k141_169106 | 1005 | No | NA | 1 | 0 | 1 | Not-determined | Genome-fragment | NA | NA | 0 |
| S12_extracted_k141_34854 | 5630 | No | NA | 4 | 0 | 4 | Not-determined | Genome-fragment | NA | NA | 0 |
| S12_extracted_k141_27597 | 1159 | No | NA | 2 | 0 | 0 | Not-determined | Genome-fragment | NA | NA | 0 |
| S12_extracted_k141_145647 | 1236 | No | NA | 1 | 0 | 0 | Not-determined | Genome-fragment | NA | NA | 0 |
| S12_extracted_k141_185216 | 1184 | No | NA | 1 | 0 | 1 | Not-determined | Genome-fragment | NA | NA | 0 |
| S12_extracted_k141_145654 | 1160 | No | NA | 1 | 0 | 1 | Not-determined | Genome-fragment | NA | NA | 0 |
| S12_extracted_k141_50804 | 4361 | No | NA | 5 | 0 | 2 | Not-determined | Genome-fragment | NA | NA | 0 |
| S12_extracted_k141_59239 | 3385 | No | NA | 4 | 0 | 2 | Not-determined | Genome-fragment | NA | NA | 0 |
| S1_extracted_k141_6346 | 3803 | No | NA | 4 | 0 | 1 | Low-quality | Genome-fragment | 1.95 | AAI-based (medium-confidence) | 0 |
| S12_extracted_k141_19576 | 2335 | No | NA | 2 | 0 | 2 | Not-determined | Genome-fragment | NA | NA | 0 |
| S12_extracted_k141_66872 | 1333 | No | NA | 2 | 0 | 0 | Not-determined | Genome-fragment | NA | NA | 0 |
| S5_vs_k141_88573\|\|full | 3803 | No | NA | 6 | 0 | 0 | Low-quality | Genome-fragment | 9.38 | AAI-based (high-confidence) | 0 |
| S12_vs_k141_58254\|\|full | 3803 | No | NA | 4 | 0 | 3 | Low-quality | Genome-fragment | 0.71 | AAI-based (high-confidence) | 0 |
| S12_extracted_k141_59260 | 1052 | No | NA | 2 | 0 | 2 | Not-determined | Genome-fragment | NA | NA | 0 |
| S12_extracted_k141_99127 | 9427 | No | NA | 9 | 0 | 4 | Not-determined | Genome-fragment | NA | NA | 0 |
| S12_extracted_k141_169149 | 3376 | No | NA | 4 | 0 | 3 | Not-determined | Genome-fragment | NA | NA | 0 |
| S12_extracted_k141_106613 | 1312 | No | NA | 1 | 0 | 1 | Not-determined | Genome-fragment | NA | NA | 0 |
| S12_extracted_k141_145696 | 3794 | No | NA | 3 | 0 | 2 | Not-determined | Genome-fragment | NA | NA | 0 |
| S14_vs_k141_100254\|\|full | 3804 | No | NA | 10 | 0 | 0 | Low-quality | Genome-fragment | 9.51 | AAI-based (high-confidence) | 0 |
| S12_extracted_k141_169167 | 1398 | No | NA | 2 | 0 | 2 | Not-determined | Genome-fragment | NA | NA | 0 |
| S1_extracted_k141_127183 | 3804 | No | NA | 8 | 4 | 0 | Low-quality | Genome-fragment | 6.27 | AAI-based (high-confidence) | 0 |
| S4_extracted_6742 | 3804 | No | NA | 6 | 0 | 0 | Low-quality | Genome-fragment | 4.08 | AAI-based (high-confidence) | 0 |
| S12_extracted_k141_11122 | 1158 | No | NA | 1 | 0 | 1 | Not-determined | Genome-fragment | NA | NA | 0 |
| S12_extracted_k141_4231 | 3599 | No | NA | 5 | 0 | 4 | Not-determined | Genome-fragment | NA | NA | 0 |
| S12_extracted_k141_129552 | 7481 | No | NA | 8 | 0 | 1 | Not-determined | Genome-fragment | NA | NA | 0 |
| S12_extracted_k141_50845 | 1151 | No | NA | 1 | 0 | 0 | Not-determined | Genome-fragment | NA | NA | 0 |
| S12_extracted_k141_137893 | 1600 | No | NA | 3 | 0 | 0 | Not-determined | Genome-fragment | NA | NA | 0 |
| S13_vs_50405\|\|full | 3805 | No | NA | 4 | 4 | 0 | Low-quality | Genome-fragment | 10.02 | AAI-based (high-confidence) | 0 |
| S12_extracted_k141_153248 | 1050 | No | NA | 1 | 0 | 0 | Not-determined | Genome-fragment | NA | NA | 0 |
| S12_extracted_k141_90991 | 1060 | No | NA | 2 | 0 | 1 | Not-determined | Genome-fragment | NA | NA | 0 |
| S12_extracted_k141_145723 | 1779 | No | NA | 2 | 0 | 1 | Not-determined | Genome-fragment | NA | NA | 0 |
| S12_extracted_k141_50868 | 1294 | No | NA | 2 | 0 | 2 | Not-determined | Genome-fragment | NA | NA | 0 |
| S12_extracted_k141_74435 | 1054 | No | NA | 1 | 0 | 1 | Not-determined | Genome-fragment | NA | NA | 0 |
| S12_extracted_k141_66941 | 2127 | No | NA | 2 | 0 | 0 | Not-determined | Genome-fragment | NA | NA | 0 |
| S12_extracted_k141_34957 | 3090 | No | NA | 4 | 0 | 2 | Not-determined | Genome-fragment | NA | NA | 0 |
| S12_extracted_k141_27720 | 1131 | No | NA | 2 | 0 | 2 | Not-determined | Genome-fragment | NA | NA | 0 |
| S12_extracted_k141_27723 | 1213 | No | NA | 1 | 0 | 1 | Not-determined | Genome-fragment | NA | NA | 0 |
| S12_extracted_k141_4298 | 1046 | No | NA | 1 | 0 | 1 | Not-determined | Genome-fragment | NA | NA | 0 |
| S12_extracted_k141_137943 | 1245 | No | NA | 1 | 0 | 0 | Not-determined | Genome-fragment | NA | NA | 0 |
| S12_extracted_k141_169234 | 1006 | No | NA | 1 | 0 | 1 | Not-determined | Genome-fragment | NA | NA | 0 |
| S12_extracted_k141_145764 | 1020 | No | NA | 2 | 0 | 0 | Not-determined | Genome-fragment | NA | NA | 0 |
| S12_extracted_k141_50916 | 3216 | No | NA | 3 | 0 | 2 | Not-determined | Genome-fragment | NA | NA | 0 |
| S12_extracted_k141_4332 | 1862 | No | NA | 1 | 0 | 0 | Not-determined | Genome-fragment | NA | NA | 0 |
| S12_extracted_k141_137972 | 1219 | No | NA | 2 | 0 | 2 | Not-determined | Genome-fragment | NA | NA | 0 |
| S14_vs_k141_36046\|\|full | 3805 | No | NA | 4 | 0 | 1 | Low-quality | Genome-fragment | 6.55 | AAI-based (high-confidence) | 0 |
| S12_extracted_k141_82707 | 1191 | No | NA | 2 | 0 | 2 | Not-determined | Genome-fragment | NA | NA | 0 |
| S12_extracted_k141_50951 | 1240 | No | NA | 1 | 0 | 1 | Not-determined | Genome-fragment | NA | NA | 0 |
| S12_extracted_k141_74476 | 1588 | No | NA | 3 | 0 | 2 | Not-determined | Genome-fragment | NA | NA | 0 |
| S12_extracted_k141_4366 | 1014 | No | NA | 1 | 0 | 0 | Not-determined | Genome-fragment | NA | NA | 0 |
| S10_extracted_62557 | 3805 | No | NA | 6 | 0 | 6 | Low-quality | Genome-fragment | 1.76 | AAI-based (medium-confidence) | 0 |
| S12_extracted_k141_91071 | 1293 | No | NA | 2 | 0 | 1 | Not-determined | Genome-fragment | NA | NA | 0 |
| S1_extracted_k141_148762 | 3806 | No | NA | 7 | 0 | 0 | Low-quality | Genome-fragment | 2.48 | AAI-based (high-confidence) | 0 |
| S12_extracted_k141_44003 | 11256 | No | NA | 12 | 0 | 4 | Not-determined | Genome-fragment | NA | NA | 0 |
| S12_extracted_k141_82746 | 2305 | No | NA | 1 | 0 | 0 | Not-determined | Genome-fragment | NA | NA | 0 |
| S12_extracted_k141_169249 | 3239 | No | NA | 4 | 0 | 1 | Not-determined | Genome-fragment | NA | NA | 0 |
| S17_extracted_k141_77708 | 3807 | No | NA | 3 | 0 | 2 | Low-quality | Genome-fragment | 2.11 | AAI-based (medium-confidence) | 0 |
| S12_extracted_k141_113815 | 1292 | No | NA | 1 | 0 | 0 | Not-determined | Genome-fragment | NA | NA | 0 |
| S12_extracted_k141_161266 | 1821 | No | NA | 2 | 0 | 2 | Not-determined | Genome-fragment | NA | NA | 0 |
| S12_extracted_k141_185401 | 1884 | No | NA | 2 | 0 | 0 | Not-determined | Genome-fragment | NA | NA | 0 |
| S12_extracted_k141_145884 | 1198 | No | NA | 1 | 0 | 1 | Not-determined | Genome-fragment | NA | NA | 0 |
| S17_vs_k141_134052\|\|full | 3807 | No | NA | 3 | 0 | 2 | Low-quality | Genome-fragment | 4.38 | AAI-based (medium-confidence) | 0 |
| S11_vs_k141_81441\|\|full | 3808 | No | NA | 9 | 0 | 1 | Low-quality | Genome-fragment | 2.46 | AAI-based (high-confidence) | 0 |
| S12_extracted_k141_51012 | 1465 | No | NA | 2 | 0 | 1 | Not-determined | Genome-fragment | NA | NA | 0 |
| S12_extracted_k141_145905 | 1350 | No | NA | 3 | 0 | 0 | Not-determined | Genome-fragment | NA | NA | 0 |
| S16_extracted_90739 | 3808 | No | NA | 3 | 0 | 0 | Low-quality | Genome-fragment | 6.42 | AAI-based (medium-confidence) | 0 |
| S12_extracted_k141_67111 | 1303 | No | NA | 2 | 0 | 2 | Not-determined | Genome-fragment | NA | NA | 0 |
| S7_vs_76409\|\|full | 3809 | No | NA | 8 | 1 | 0 | Low-quality | Genome-fragment | 6.8 | AAI-based (high-confidence) | 0 |
| S16_vs_135297\|\|full | 3809 | No | NA | 3 | 0 | 2 | Low-quality | Genome-fragment | 1.1 | AAI-based (medium-confidence) | 0 |
| S12_extracted_k141_113848 | 1046 | No | NA | 1 | 0 | 0 | Not-determined | Genome-fragment | NA | NA | 0 |
| S12_extracted_k141_176501 | 1015 | No | NA | 1 | 0 | 1 | Not-determined | Genome-fragment | NA | NA | 0 |
| S12_extracted_k141_145943 | 3108 | No | NA | 4 | 0 | 2 | Not-determined | Genome-fragment | NA | NA | 0 |
| S12_extracted_k141_169316 | 1052 | No | NA | 1 | 0 | 1 | Not-determined | Genome-fragment | NA | NA | 0 |
| S8_vs_k141_5778\|\|full | 3809 | No | NA | 3 | 0 | 0 | Low-quality | Genome-fragment | 9.76 | AAI-based (medium-confidence) | 0 |
| S5_extracted_k141_88508 | 3810 | No | NA | 7 | 1 | 0 | Low-quality | Genome-fragment | 8.47 | HMM-based (lower-bound) | 0 |
| S12_extracted_k141_161309 | 5803 | No | NA | 4 | 0 | 3 | Not-determined | Genome-fragment | NA | NA | 0 |
| S12_extracted_k141_176515 | 1568 | No | NA | 3 | 0 | 3 | Not-determined | Genome-fragment | NA | NA | 0 |
| S12_extracted_k141_153358 | 1205 | No | NA | 2 | 0 | 1 | Not-determined | Genome-fragment | NA | NA | 0 |
| S12_extracted_k141_51065 | 3280 | No | NA | 3 | 0 | 2 | Not-determined | Genome-fragment | NA | NA | 0 |
| S12_extracted_k141_74639 | 1221 | No | NA | 2 | 0 | 2 | Not-determined | Genome-fragment | NA | NA | 0 |
| S7_vs_62641\|\|full | 3810 | No | NA | 7 | 1 | 0 | Low-quality | Genome-fragment | 2.4 | HMM-based (lower-bound) | 0 |
| S12_extracted_k141_59496 | 1144 | No | NA | 2 | 0 | 1 | Not-determined | Genome-fragment | NA | NA | 0 |
| S12_extracted_k141_161349 | 1001 | No | NA | 1 | 0 | 1 | Not-determined | Genome-fragment | NA | NA | 0 |
| S12_extracted_k141_27841 | 1052 | No | NA | 3 | 0 | 3 | Not-determined | Genome-fragment | NA | NA | 0 |
| S12_extracted_k141_99293 | 1124 | No | NA | 1 | 0 | 0 | Not-determined | Genome-fragment | NA | NA | 0 |
| S12_extracted_k141_185510 | 2442 | No | NA | 2 | 0 | 2 | Not-determined | Genome-fragment | NA | NA | 0 |
| S9_extracted_k141_86386 | 3811 | No | NA | 4 | 0 | 0 | Low-quality | Genome-fragment | 9.43 | AAI-based (medium-confidence) | 0 |
| S12_extracted_k141_11260 | 1620 | No | NA | 1 | 0 | 1 | Not-determined | Genome-fragment | NA | NA | 0 |
| S12_extracted_k141_11264 | 1785 | No | NA | 2 | 0 | 0 | Not-determined | Genome-fragment | NA | NA | 0 |
| S12_extracted_k141_35167 | 1176 | No | NA | 1 | 0 | 0 | Not-determined | Genome-fragment | NA | NA | 0 |
| S5_vs_k141_110714\|\|full | 3811 | No | NA | 7 | 3 | 0 | Low-quality | Genome-fragment | 10.65 | AAI-based (high-confidence) | 0 |
| S7_vs_10586\|\|full | 3811 | No | NA | 3 | 2 | 0 | Low-quality | Genome-fragment | 8.13 | AAI-based (high-confidence) | 0 |
| S12_extracted_k141_106834 | 1644 | No | NA | 2 | 0 | 1 | Not-determined | Genome-fragment | NA | NA | 0 |
| S12_extracted_k141_146019 | 3627 | No | NA | 4 | 0 | 1 | Not-determined | Genome-fragment | NA | NA | 0 |
| S12_extracted_k141_44178 | 1551 | No | NA | 2 | 0 | 0 | Not-determined | Genome-fragment | NA | NA | 0 |
| S12_extracted_k141_19667 | 3229 | No | NA | 6 | 0 | 5 | Not-determined | Genome-fragment | NA | NA | 0 |
| S12_extracted_k141_59577 | 1703 | No | NA | 2 | 0 | 1 | Not-determined | Genome-fragment | NA | NA | 0 |
| S12_extracted_k141_153456 | 1745 | No | NA | 2 | 0 | 2 | Not-determined | Genome-fragment | NA | NA | 0 |
| S12_extracted_k141_161413 | 1022 | No | NA | 1 | 0 | 0 | Not-determined | Genome-fragment | NA | NA | 0 |
| S12_extracted_k141_169406 | 3184 | No | NA | 2 | 0 | 1 | Not-determined | Genome-fragment | NA | NA | 0 |
| S12_extracted_k141_27899 | 1359 | No | NA | 3 | 0 | 2 | Not-determined | Genome-fragment | NA | NA | 0 |
| S12_extracted_k141_138177 | 1340 | No | NA | 2 | 0 | 1 | Not-determined | Genome-fragment | NA | NA | 0 |
| S12_extracted_k141_161416 | 2194 | No | NA | 2 | 0 | 1 | Not-determined | Genome-fragment | NA | NA | 0 |
| S16_vs_6539\|\|full | 3811 | No | NA | 5 | 0 | 1 | Low-quality | Genome-fragment | 7.36 | AAI-based (high-confidence) | 0 |
| S12_extracted_k141_121773 | 2772 | No | NA | 1 | 0 | 1 | Not-determined | Genome-fragment | NA | NA | 0 |
| S12_extracted_k141_67188 | 57094 | No | NA | 56 | 0 | 49 | Not-determined | Genome-fragment | NA | NA | 0 |
| S12_extracted_k141_91300 | 1120 | No | NA | 2 | 0 | 1 | Not-determined | Genome-fragment | NA | NA | 0 |
| S16_vs_98647\|\|full | 3811 | No | NA | 4 | 0 | 0 | Low-quality | Genome-fragment | 3.59 | AAI-based (medium-confidence) | 0 |
| S11_vs_k141_81213\|\|full | 3812 | No | NA | 8 | 0 | 1 | Low-quality | Genome-fragment | 9.46 | AAI-based (medium-confidence) | 0 |
| S12_extracted_k141_99334 | 1580 | No | NA | 2 | 0 | 0 | Not-determined | Genome-fragment | NA | NA | 0 |
| S12_extracted_k141_176618 | 1924 | No | NA | 2 | 0 | 0 | Not-determined | Genome-fragment | NA | NA | 0 |
| S12_extracted_k141_82875 | 1039 | No | NA | 1 | 0 | 0 | Not-determined | Genome-fragment | NA | NA | 0 |
| S12_extracted_k141_153512 | 1803 | No | NA | 2 | 0 | 0 | Not-determined | Genome-fragment | NA | NA | 0 |
| S1_vs_k141_29621\|\|full | 3812 | No | NA | 5 | 0 | 2 | Low-quality | Genome-fragment | 2.38 | AAI-based (medium-confidence) | 0 |
| S12_extracted_k141_153531 | 1889 | No | NA | 3 | 0 | 2 | Not-determined | Genome-fragment | NA | NA | 0 |
| S5_vs_k141_30063\|\|full | 3812 | No | NA | 4 | 0 | 4 | Low-quality | Genome-fragment | 1.32 | AAI-based (medium-confidence) | 0 |
| S12_extracted_k141_59650 | 1293 | No | NA | 1 | 0 | 1 | Not-determined | Genome-fragment | NA | NA | 0 |
| S12_extracted_k141_91347 | 1023 | No | NA | 1 | 0 | 1 | Not-determined | Genome-fragment | NA | NA | 0 |
| S12_extracted_k141_67263 | 1141 | No | NA | 1 | 0 | 1 | Not-determined | Genome-fragment | NA | NA | 0 |
| S3_vs_k141_92168\|\|full | 3813 | No | NA | 5 | 3 | 0 | Low-quality | Genome-fragment | 10.03 | AAI-based (high-confidence) | 0 |
| S12_extracted_k141_106962 | 1233 | No | NA | 2 | 0 | 1 | Not-determined | Genome-fragment | NA | NA | 0 |
| S11_extracted_k141_49521 | 3814 | No | NA | 5 | 1 | 0 | Low-quality | Genome-fragment | 10.78 | AAI-based (high-confidence) | 0 |
| S12_extracted_k141_4635 | 3578 | No | NA | 3 | 0 | 1 | Not-determined | Genome-fragment | NA | NA | 0 |
| S12_extracted_k141_114074 | 1699 | No | NA | 4 | 0 | 4 | Not-determined | Genome-fragment | NA | NA | 0 |
| S12_extracted_k141_44242 | 1038 | No | NA | 1 | 0 | 0 | Not-determined | Genome-fragment | NA | NA | 0 |
| S12_extracted_k141_19715 | 3979 | No | NA | 5 | 0 | 0 | Not-determined | Genome-fragment | NA | NA | 0 |
| S12_extracted_k141_4642 | 6514 | No | NA | 6 | 0 | 3 | Not-determined | Genome-fragment | NA | NA | 0 |
| S11_vs_k141_39532\|\|full | 3814 | Yes | 1447 | 4 | 1 | 1 | Low-quality | Genome-fragment | 2.38 | AAI-based (medium-confidence) | 62.06 |
| S12_extracted_k141_129738 | 3143 | No | NA | 1 | 0 | 0 | Not-determined | Genome-fragment | NA | NA | 0 |
| S7_vs_71126\|\|full | 3816 | No | NA | 10 | 2 | 0 | Low-quality | Genome-fragment | 9.04 | AAI-based (medium-confidence) | 0 |
| S12_extracted_k141_106989 | 2921 | No | NA | 1 | 0 | 1 | Not-determined | Genome-fragment | NA | NA | 0 |
| S12_extracted_k141_176690 | 3596 | No | NA | 10 | 0 | 0 | Not-determined | Genome-fragment | NA | NA | 0 |
| S12_extracted_k141_129741 | 2408 | No | NA | 2 | 0 | 0 | Not-determined | Genome-fragment | NA | NA | 0 |
| S12_extracted_k141_27982 | 6332 | No | NA | 6 | 0 | 2 | Not-determined | Genome-fragment | NA | NA | 0 |
| S12_extracted_k141_176694 | 3070 | No | NA | 4 | 0 | 1 | Not-determined | Genome-fragment | NA | NA | 0 |
| S12_extracted_k141_91362 | 1467 | No | NA | 3 | 0 | 2 | Not-determined | Genome-fragment | NA | NA | 0 |
| S4_extracted_19807 | 3816 | No | NA | 7 | 1 | 0 | Low-quality | Genome-fragment | 9.11 | AAI-based (high-confidence) | 0 |
| S12_extracted_k141_146121 | 2806 | No | NA | 2 | 0 | 2 | Not-determined | Genome-fragment | NA | NA | 0 |
| S12_extracted_k141_51150 | 1181 | No | NA | 2 | 0 | 1 | Not-determined | Genome-fragment | NA | NA | 0 |
| S12_extracted_k141_44290 | 1129 | No | NA | 1 | 0 | 0 | Not-determined | Genome-fragment | NA | NA | 0 |
| S12_extracted_k141_185718 | 1842 | No | NA | 1 | 0 | 1 | Not-determined | Genome-fragment | NA | NA | 0 |
| S12_extracted_k141_114135 | 1032 | No | NA | 4 | 0 | 0 | Not-determined | Genome-fragment | NA | NA | 0 |
| S12_extracted_k141_161533 | 3088 | No | NA | 2 | 0 | 2 | Not-determined | Genome-fragment | NA | NA | 0 |
| S12_extracted_k141_185723 | 2317 | No | NA | 3 | 0 | 0 | Not-determined | Genome-fragment | NA | NA | 0 |
| S12_extracted_k141_99426 | 6108 | No | NA | 2 | 0 | 1 | Not-determined | Genome-fragment | NA | NA | 0 |
| S12_extracted_k141_146158 | 3367 | No | NA | 4 | 0 | 4 | Not-determined | Genome-fragment | NA | NA | 0 |
| S12_extracted_k141_161558 | 1242 | No | NA | 2 | 0 | 0 | Not-determined | Genome-fragment | NA | NA | 0 |
| S12_extracted_k141_153643 | 1844 | No | NA | 2 | 0 | 1 | Not-determined | Genome-fragment | NA | NA | 0 |
| S12_extracted_k141_44325 | 1031 | No | NA | 1 | 0 | 1 | Not-determined | Genome-fragment | NA | NA | 0 |
| S17_vs_k141_79095\|\|full | 3819 | No | NA | 3 | 0 | 2 | Low-quality | Genome-fragment | 0.34 | AAI-based (medium-confidence) | 0 |
| S12_extracted_k141_176735 | 2162 | No | NA | 4 | 0 | 2 | Not-determined | Genome-fragment | NA | NA | 0 |
| S5_extracted_k141_72320 | 3819 | No | NA | 5 | 0 | 0 | Low-quality | Genome-fragment | 3.48 | AAI-based (high-confidence) | 0 |
| S12_extracted_k141_146193 | 1790 | No | NA | 2 | 0 | 2 | Not-determined | Genome-fragment | NA | NA | 0 |
| S12_extracted_k141_91452 | 1372 | No | NA | 2 | 0 | 1 | Not-determined | Genome-fragment | NA | NA | 0 |
| S12_extracted_k141_146196 | 1965 | No | NA | 2 | 0 | 2 | Not-determined | Genome-fragment | NA | NA | 0 |
| S12_extracted_k141_11433 | 1154 | No | NA | 2 | 0 | 0 | Not-determined | Genome-fragment | NA | NA | 0 |
| S12_extracted_k141_51195 | 1121 | No | NA | 1 | 0 | 0 | Not-determined | Genome-fragment | NA | NA | 0 |
| S10_extracted_107991 | 3819 | No | NA | 5 | 4 | 0 | Low-quality | Genome-fragment | 11.96 | AAI-based (high-confidence) | 0 |
| S7_vs_72952\|\|full | 3820 | No | NA | 5 | 2 | 0 | Low-quality | Genome-fragment | 9.28 | AAI-based (high-confidence) | 0 |
| S1_vs_k141_123509\|\|full | 3821 | No | NA | 3 | 2 | 0 | Low-quality | Genome-fragment | 10.4 | AAI-based (high-confidence) | 0 |
| S12_extracted_k141_28053 | 4346 | No | NA | 2 | 0 | 0 | Not-determined | Genome-fragment | NA | NA | 0 |
| S12_extracted_k141_51213 | 11731 | No | NA | 19 | 0 | 6 | Not-determined | Genome-fragment | NA | NA | 0 |
| S17_extracted_k141_174136 | 3822 | No | NA | 6 | 1 | 0 | Low-quality | Genome-fragment | 8.17 | HMM-based (lower-bound) | 0 |
| S12_extracted_k141_51228 | 7645 | No | NA | 9 | 0 | 3 | Not-determined | Genome-fragment | NA | NA | 0 |
| S12_extracted_k141_59808 | 2538 | No | NA | 2 | 0 | 2 | Not-determined | Genome-fragment | NA | NA | 0 |
| S12_extracted_k141_4727 | 1243 | No | NA | 1 | 0 | 1 | Not-determined | Genome-fragment | NA | NA | 0 |
| S12_extracted_k141_161683 | 1258 | No | NA | 1 | 0 | 1 | Not-determined | Genome-fragment | NA | NA | 0 |
| S12_extracted_k141_11499 | 1094 | No | NA | 1 | 0 | 0 | Not-determined | Genome-fragment | NA | NA | 0 |
| S12_extracted_k141_185825 | 1119 | No | NA | 1 | 0 | 1 | Not-determined | Genome-fragment | NA | NA | 0 |
| S12_extracted_k141_176798 | 2179 | No | NA | 2 | 0 | 1 | Not-determined | Genome-fragment | NA | NA | 0 |
| S16_vs_131115\|\|full | 3822 | No | NA | 4 | 0 | 2 | Low-quality | Genome-fragment | 5.5 | AAI-based (medium-confidence) | 0 |
| S12_extracted_k141_161697 | 1641 | No | NA | 1 | 0 | 1 | Not-determined | Genome-fragment | NA | NA | 0 |
| S12_extracted_k141_185838 | 1976 | No | NA | 1 | 0 | 1 | Not-determined | Genome-fragment | NA | NA | 0 |
| S12_extracted_k141_74951 | 1287 | No | NA | 2 | 0 | 0 | Not-determined | Genome-fragment | NA | NA | 0 |
| S12_extracted_k141_99557 | 1921 | No | NA | 3 | 0 | 2 | Not-determined | Genome-fragment | NA | NA | 0 |
| S12_extracted_k141_67438 | 1608 | No | NA | 2 | 0 | 1 | Not-determined | Genome-fragment | NA | NA | 0 |
| S3_extracted_k141_93527 | 3823 | Yes | 1133 | 5 | 1 | 1 | Low-quality | Genome-fragment | 0.61 | AAI-based (high-confidence) | 70.36 |
| S12_extracted_k141_161714 | 1014 | No | NA | 2 | 0 | 1 | Not-determined | Genome-fragment | NA | NA | 0 |
| S12_extracted_k141_176817 | 1483 | No | NA | 2 | 0 | 2 | Not-determined | Genome-fragment | NA | NA | 0 |
| S12_extracted_k141_107192 | 1029 | No | NA | 2 | 0 | 1 | Not-determined | Genome-fragment | NA | NA | 0 |
| S12_extracted_k141_67477 | 2186 | No | NA | 2 | 0 | 1 | Not-determined | Genome-fragment | NA | NA | 0 |
| S12_vs_k141_164543\|\|full | 3824 | No | NA | 5 | 0 | 3 | Low-quality | Genome-fragment | 2.07 | AAI-based (medium-confidence) | 0 |
| S15_vs_14667\|\|full | 3824 | No | NA | 6 | 0 | 1 | Low-quality | Genome-fragment | 8.93 | AAI-based (high-confidence) | 0 |
| S12_extracted_k141_35403 | 2253 | No | NA | 2 | 0 | 2 | Not-determined | Genome-fragment | NA | NA | 0 |
| S17_extracted_k141_170755 | 3825 | No | NA | 11 | 10 | 0 | Low-quality | Genome-fragment | 12.62 | AAI-based (high-confidence) | 0 |
| S12_extracted_k141_153733 | 1328 | No | NA | 3 | 0 | 0 | Not-determined | Genome-fragment | NA | NA | 0 |
| S12_extracted_k141_107207 | 1124 | No | NA | 1 | 0 | 0 | Not-determined | Genome-fragment | NA | NA | 0 |
| S3_vs_k141_27676\|\|full | 3825 | No | NA | 4 | 1 | 0 | Low-quality | Genome-fragment | 9.02 | AAI-based (high-confidence) | 0 |
| S12_extracted_k141_176842 | 2383 | No | NA | 4 | 0 | 2 | Not-determined | Genome-fragment | NA | NA | 0 |
| S12_extracted_k141_146278 | 7993 | No | NA | 15 | 0 | 14 | Not-determined | Genome-fragment | NA | NA | 0 |
| S12_extracted_k141_4795 | 1610 | No | NA | 2 | 0 | 0 | Not-determined | Genome-fragment | NA | NA | 0 |
| S12_extracted_k141_67515 | 4854 | No | NA | 6 | 0 | 2 | Not-determined | Genome-fragment | NA | NA | 0 |
| S12_extracted_k141_129971 | 4487 | No | NA | 5 | 0 | 1 | Not-determined | Genome-fragment | NA | NA | 0 |
| S16_extracted_16138 | 3825 | No | NA | 5 | 3 | 0 | Low-quality | Genome-fragment | 8.12 | AAI-based (high-confidence) | 0 |
| S14_extracted_k141_66513 | 3826 | No | NA | 6 | 5 | 0 | Low-quality | Genome-fragment | 12.63 | AAI-based (high-confidence) | 0 |
| S16_extracted_161726 | 3826 | No | NA | 4 | 0 | 0 | Low-quality | Genome-fragment | 6.45 | AAI-based (high-confidence) | 0 |
| S16_vs_68845\|\|full | 3826 | No | NA | 6 | 0 | 2 | Low-quality | Genome-fragment | 6.96 | AAI-based (high-confidence) | 0 |
| S12_extracted_k141_185902 | 1475 | No | NA | 1 | 0 | 1 | Not-determined | Genome-fragment | NA | NA | 0 |
| S5_vs_k141_4509\|\|full | 3827 | No | NA | 8 | 0 | 0 | Low-quality | Genome-fragment | 11.91 | AAI-based (high-confidence) | 0 |
| S12_extracted_k141_51336 | 1750 | No | NA | 2 | 0 | 2 | Not-determined | Genome-fragment | NA | NA | 0 |
| S12_extracted_k141_130016 | 7790 | No | NA | 5 | 0 | 2 | Not-determined | Genome-fragment | NA | NA | 0 |
| S12_extracted_k141_161812 | 1346 | No | NA | 3 | 0 | 2 | Not-determined | Genome-fragment | NA | NA | 0 |
| S12_extracted_k141_107283 | 3426 | No | NA | 4 | 0 | 3 | Not-determined | Genome-fragment | NA | NA | 0 |
| S12_extracted_k141_91618 | 1771 | No | NA | 1 | 0 | 1 | Not-determined | Genome-fragment | NA | NA | 0 |
| S12_extracted_k141_176937 | 3782 | No | NA | 2 | 0 | 1 | Not-determined | Genome-fragment | NA | NA | 0 |
| S12_extracted_k141_107300 | 1026 | No | NA | 1 | 0 | 1 | Not-determined | Genome-fragment | NA | NA | 0 |
| S12_extracted_k141_169686 | 1956 | No | NA | 2 | 0 | 1 | Not-determined | Genome-fragment | NA | NA | 0 |
| S10_vs_96505\|\|full | 3827 | No | NA | 5 | 0 | 2 | Low-quality | Genome-fragment | 1.69 | AAI-based (medium-confidence) | 0 |
| S12_extracted_k141_51357 | 4497 | No | NA | 3 | 0 | 1 | Not-determined | Genome-fragment | NA | NA | 0 |
| S12_extracted_k141_44580 | 1095 | No | NA | 1 | 0 | 1 | Not-determined | Genome-fragment | NA | NA | 0 |
| S12_extracted_k141_75027 | 2546 | No | NA | 2 | 0 | 0 | Not-determined | Genome-fragment | NA | NA | 0 |
| S12_extracted_k141_169691 | 1066 | No | NA | 1 | 0 | 0 | Not-determined | Genome-fragment | NA | NA | 0 |
| S12_extracted_k141_122004 | 1610 | No | NA | 2 | 0 | 1 | Not-determined | Genome-fragment | NA | NA | 0 |
| S12_extracted_k141_169724 | 1214 | No | NA | 1 | 0 | 0 | Not-determined | Genome-fragment | NA | NA | 0 |
| S12_extracted_k141_75034 | 3521 | No | NA | 3 | 0 | 3 | Not-determined | Genome-fragment | NA | NA | 0 |
| S12_extracted_k141_122007 | 1048 | No | NA | 1 | 0 | 0 | Not-determined | Genome-fragment | NA | NA | 0 |
| S17_vs_k141_166163\|\|full | 3828 | No | NA | 9 | 6 | 0 | Low-quality | Genome-fragment | 8.8 | AAI-based (high-confidence) | 0 |
| S12_extracted_k141_138520 | 1045 | No | NA | 1 | 0 | 1 | Not-determined | Genome-fragment | NA | NA | 0 |
| S12_extracted_k141_44626 | 3281 | No | NA | 6 | 0 | 0 | Not-determined | Genome-fragment | NA | NA | 0 |
| S12_extracted_k141_83204 | 2892 | No | NA | 3 | 0 | 0 | Not-determined | Genome-fragment | NA | NA | 0 |
| S12_extracted_k141_169753 | 1667 | No | NA | 2 | 0 | 0 | Not-determined | Genome-fragment | NA | NA | 0 |
| S4_extracted_15625 | 3828 | No | NA | 7 | 6 | 0 | Low-quality | Genome-fragment | 12.63 | AAI-based (high-confidence) | 0 |
| S12_extracted_k141_28282 | 1042 | No | NA | 1 | 0 | 1 | Not-determined | Genome-fragment | NA | NA | 0 |
| S12_extracted_k141_20053 | 1139 | No | NA | 2 | 0 | 0 | Not-determined | Genome-fragment | NA | NA | 0 |
| S7_extracted_90275 | 3829 | No | NA | 3 | 0 | 0 | Low-quality | Genome-fragment | 6.4 | AAI-based (high-confidence) | 0 |
| S12_extracted_k141_28293 | 2021 | No | NA | 3 | 0 | 1 | Not-determined | Genome-fragment | NA | NA | 0 |
| S12_extracted_k141_153837 | 1363 | No | NA | 4 | 0 | 3 | Not-determined | Genome-fragment | NA | NA | 0 |
| S11_extracted_k141_64321 | 3830 | No | NA | 6 | 0 | 0 | Low-quality | Genome-fragment | 2.96 | AAI-based (high-confidence) | 0 |
| S12_extracted_k141_153844 | 1440 | No | NA | 1 | 0 | 0 | Not-determined | Genome-fragment | NA | NA | 0 |
| S12_extracted_k141_107391 | 1177 | No | NA | 2 | 0 | 2 | Not-determined | Genome-fragment | NA | NA | 0 |
| S3_vs_k141_104112\|\|full | 3831 | No | NA | 8 | 1 | 0 | Low-quality | Genome-fragment | 7.03 | AAI-based (high-confidence) | 0 |
| S12_extracted_k141_35570 | 1502 | No | NA | 2 | 0 | 0 | Not-determined | Genome-fragment | NA | NA | 0 |
| S16_extracted_139231 | 3831 | No | NA | 5 | 0 | 1 | Low-quality | Genome-fragment | 5.93 | AAI-based (high-confidence) | 0 |
| S12_extracted_k141_186065 | 2722 | No | NA | 4 | 0 | 0 | Not-determined | Genome-fragment | NA | NA | 0 |
| S12_extracted_k141_130135 | 1711 | No | NA | 1 | 0 | 0 | Not-determined | Genome-fragment | NA | NA | 0 |
| S12_extracted_k141_99747 | 3741 | No | NA | 2 | 0 | 0 | Not-determined | Genome-fragment | NA | NA | 0 |
| S8_extracted_k141_84032 | 3831 | No | NA | 4 | 0 | 4 | Low-quality | Genome-fragment | 9.66 | AAI-based (medium-confidence) | 0 |
| S12_extracted_k141_51444 | 6578 | No | NA | 10 | 0 | 6 | Not-determined | Genome-fragment | NA | NA | 0 |
| S12_extracted_k141_130151 | 1272 | No | NA | 3 | 0 | 0 | Not-determined | Genome-fragment | NA | NA | 0 |
| S11_extracted_k141_98899 | 3832 | No | NA | 4 | 2 | 0 | Low-quality | Genome-fragment | 9.48 | AAI-based (high-confidence) | 0 |
| S12_extracted_k141_186086 | 9281 | No | NA | 10 | 0 | 3 | Not-determined | Genome-fragment | NA | NA | 0 |
| S4_extracted_23741 | 3832 | No | NA | 5 | 0 | 1 | Low-quality | Genome-fragment | 9.84 | AAI-based (medium-confidence) | 0 |
| S12_extracted_k141_99768 | 1313 | No | NA | 2 | 0 | 2 | Not-determined | Genome-fragment | NA | NA | 0 |
| S12_extracted_k141_186092 | 1169 | No | NA | 2 | 0 | 1 | Not-determined | Genome-fragment | NA | NA | 0 |
| S17_vs_k141_175432\|\|full | 3833 | No | NA | 4 | 0 | 1 | Low-quality | Genome-fragment | 1.08 | AAI-based (high-confidence) | 0 |
| S12_extracted_k141_146360 | 1377 | No | NA | 1 | 0 | 1 | Not-determined | Genome-fragment | NA | NA | 0 |
| S12_extracted_k141_67630 | 3109 | No | NA | 5 | 0 | 1 | Not-determined | Genome-fragment | NA | NA | 0 |
| S12_extracted_k141_44753 | 1567 | No | NA | 3 | 0 | 2 | Not-determined | Genome-fragment | NA | NA | 0 |
| S12_extracted_k141_153951 | 1019 | No | NA | 1 | 0 | 1 | Not-determined | Genome-fragment | NA | NA | 0 |
| S12_extracted_k141_153963 | 1634 | No | NA | 1 | 0 | 1 | Not-determined | Genome-fragment | NA | NA | 0 |
| S13_extracted_10920 | 3833 | No | NA | 10 | 2 | 0 | Low-quality | Genome-fragment | 2.42 | AAI-based (high-confidence) | 0 |
| S12_extracted_k141_44762 | 1002 | No | NA | 2 | 0 | 1 | Not-determined | Genome-fragment | NA | NA | 0 |
| S12_extracted_k141_20169 | 1067 | No | NA | 1 | 0 | 1 | Not-determined | Genome-fragment | NA | NA | 0 |
| S12_extracted_k141_177088 | 1027 | No | NA | 1 | 0 | 0 | Not-determined | Genome-fragment | NA | NA | 0 |
| S12_extracted_k141_186159 | 1035 | No | NA | 2 | 0 | 0 | Not-determined | Genome-fragment | NA | NA | 0 |
| S8_vs_k141_34655\|\|full | 3833 | Yes | 2409 | 7 | 1 | 1 | Low-quality | Genome-fragment | 4.42 | HMM-based (lower-bound) | 37.15 |
| S17_vs_k141_134830\|\|full | 3834 | No | NA | 3 | 0 | 2 | Low-quality | Genome-fragment | 1.94 | AAI-based (medium-confidence) | 0 |
| S12_extracted_k141_99819 | 1573 | No | NA | 4 | 0 | 4 | Not-determined | Genome-fragment | NA | NA | 0 |
| S12_extracted_k141_186174 | 2640 | No | NA | 2 | 0 | 2 | Not-determined | Genome-fragment | NA | NA | 0 |
| S13_vs_68257\|\|full | 3834 | No | NA | 3 | 0 | 2 | Low-quality | Genome-fragment | 1.96 | AAI-based (medium-confidence) | 0 |
| S12_extracted_k141_83290 | 5552 | No | NA | 3 | 0 | 3 | Not-determined | Genome-fragment | NA | NA | 0 |
| S12_extracted_k141_107520 | 1427 | No | NA | 3 | 0 | 0 | Not-determined | Genome-fragment | NA | NA | 0 |
| S12_extracted_k141_44809 | 1024 | No | NA | 2 | 0 | 0 | Not-determined | Genome-fragment | NA | NA | 0 |
| S12_extracted_k141_11641 | 1134 | No | NA | 3 | 0 | 0 | Not-determined | Genome-fragment | NA | NA | 0 |
| S12_extracted_k141_130247 | 1769 | No | NA | 1 | 0 | 1 | Not-determined | Genome-fragment | NA | NA | 0 |
| S12_extracted_k141_162049 | 3453 | No | NA | 3 | 0 | 1 | Not-determined | Genome-fragment | NA | NA | 0 |
| S12_extracted_k141_75166 | 1208 | No | NA | 2 | 0 | 0 | Not-determined | Genome-fragment | NA | NA | 0 |
| S3_vs_k141_12245\|\|full | 3834 | No | NA | 6 | 0 | 3 | Low-quality | Genome-fragment | 2.21 | AAI-based (medium-confidence) | 0 |
| S7_extracted_28007 | 3834 | No | NA | 6 | 3 | 0 | Low-quality | Genome-fragment | 7.35 | AAI-based (high-confidence) | 0 |
| S7_extracted_55030 | 3834 | Yes | 1017 | 7 | 1 | 1 | Low-quality | Genome-fragment | 2.15 | AAI-based (high-confidence) | 73.47 |
| S12_extracted_k141_20215 | 1334 | No | NA | 1 | 0 | 1 | Not-determined | Genome-fragment | NA | NA | 0 |
| S12_extracted_k141_44835 | 1088 | No | NA | 1 | 0 | 0 | Not-determined | Genome-fragment | NA | NA | 0 |
| S12_extracted_k141_162072 | 1511 | No | NA | 2 | 0 | 0 | Not-determined | Genome-fragment | NA | NA | 0 |
| S12_extracted_k141_177164 | 1258 | No | NA | 2 | 0 | 0 | Not-determined | Genome-fragment | NA | NA | 0 |
| S12_extracted_k141_154022 | 3349 | No | NA | 4 | 0 | 2 | Not-determined | Genome-fragment | NA | NA | 0 |
| S12_extracted_k141_162088 | 2772 | No | NA | 3 | 0 | 1 | Not-determined | Genome-fragment | NA | NA | 0 |
| S12_extracted_k141_11709 | 1014 | No | NA | 1 | 0 | 1 | Not-determined | Genome-fragment | NA | NA | 0 |
| S12_extracted_k141_28421 | 1196 | No | NA | 1 | 0 | 1 | Not-determined | Genome-fragment | NA | NA | 0 |
| S12_extracted_k141_169796 | 130696 | No | NA | 180 | 36 | 2 | High-quality | High-quality | 98.19 | AAI-based (high-confidence) | 0 |
| S10_vs_101981\|\|full | 3834 | No | NA | 3 | 0 | 1 | Low-quality | Genome-fragment | 0.44 | AAI-based (medium-confidence) | 0 |
| S12_extracted_k141_51592 | 1194 | No | NA | 2 | 0 | 0 | Not-determined | Genome-fragment | NA | NA | 0 |
| S12_extracted_k141_146469 | 1209 | No | NA | 1 | 0 | 0 | Not-determined | Genome-fragment | NA | NA | 0 |
| S12_extracted_k141_51593 | 1034 | No | NA | 2 | 0 | 1 | Not-determined | Genome-fragment | NA | NA | 0 |
| S12_extracted_k141_11717 | 2366 | No | NA | 3 | 0 | 0 | Not-determined | Genome-fragment | NA | NA | 0 |
| S15_extracted_73119 | 3834 | No | NA | 5 | 0 | 0 | Low-quality | Genome-fragment | 5.18 | AAI-based (medium-confidence) | 0 |
| S12_extracted_k141_146474 | 1772 | No | NA | 1 | 0 | 0 | Not-determined | Genome-fragment | NA | NA | 0 |
| S12_extracted_k141_75228 | 1411 | No | NA | 1 | 0 | 0 | Not-determined | Genome-fragment | NA | NA | 0 |
| S12_extracted_k141_35753 | 7615 | No | NA | 5 | 0 | 3 | Not-determined | Genome-fragment | NA | NA | 0 |
| S12_extracted_k141_186290 | 1524 | No | NA | 2 | 0 | 0 | Not-determined | Genome-fragment | NA | NA | 0 |
| S9_vs_k141_76178\|\|full | 3835 | No | NA | 4 | 4 | 0 | Low-quality | Genome-fragment | 6.37 | AAI-based (high-confidence) | 0 |
| S12_extracted_k141_107597 | 3691 | No | NA | 4 | 0 | 2 | Not-determined | Genome-fragment | NA | NA | 0 |
| S12_extracted_k141_122194 | 2538 | No | NA | 1 | 0 | 0 | Not-determined | Genome-fragment | NA | NA | 0 |
| S12_extracted_k141_75238 | 1542 | No | NA | 2 | 0 | 1 | Not-determined | Genome-fragment | NA | NA | 0 |
| S12_extracted_k141_11731 | 3710 | No | NA | 1 | 0 | 1 | Not-determined | Genome-fragment | NA | NA | 0 |
| S12_extracted_k141_75252 | 1701 | No | NA | 4 | 0 | 2 | Not-determined | Genome-fragment | NA | NA | 0 |
| S12_extracted_k141_107614 | 2904 | No | NA | 1 | 0 | 1 | Not-determined | Genome-fragment | NA | NA | 0 |
| S12_vs_k141_143646\|\|full | 3835 | No | NA | 2 | 1 | 0 | Low-quality | Genome-fragment | 1.77 | AAI-based (high-confidence) | 0 |
| S12_extracted_k141_114662 | 1166 | No | NA | 1 | 0 | 1 | Not-determined | Genome-fragment | NA | NA | 0 |
| S6_vs_k141_24951\|\|full | 3836 | No | NA | 5 | 0 | 1 | Low-quality | Genome-fragment | 2.88 | AAI-based (high-confidence) | 0 |
| S12_extracted_k141_99917 | 1114 | No | NA | 2 | 0 | 2 | Not-determined | Genome-fragment | NA | NA | 0 |
| S12_extracted_k141_177257 | 1120 | No | NA | 3 | 0 | 2 | Not-determined | Genome-fragment | NA | NA | 0 |
| S12_extracted_k141_35805 | 1245 | No | NA | 1 | 0 | 1 | Not-determined | Genome-fragment | NA | NA | 0 |
| S12_extracted_k141_75283 | 1609 | No | NA | 2 | 0 | 0 | Not-determined | Genome-fragment | NA | NA | 0 |
| S12_extracted_k141_122244 | 1859 | No | NA | 2 | 0 | 2 | Not-determined | Genome-fragment | NA | NA | 0 |
| S12_extracted_k141_75299 | 1633 | No | NA | 1 | 0 | 0 | Not-determined | Genome-fragment | NA | NA | 0 |
| S12_extracted_k141_154046 | 1009 | No | NA | 1 | 0 | 0 | Not-determined | Genome-fragment | NA | NA | 0 |
| S12_extracted_k141_51676 | 1110 | No | NA | 2 | 0 | 1 | Not-determined | Genome-fragment | NA | NA | 0 |
| S12_extracted_k141_28522 | 1144 | No | NA | 4 | 0 | 0 | Not-determined | Genome-fragment | NA | NA | 0 |

| | | | | | | | | | | | |
|---|---|---|---|---|---|---|---|---|---|---|---|
| S1_extracted_k141_82202 | 3837 | Yes | 2421 | 6 | 2 | 1 | Low-quality | Genome-fragment | 4.99 | AAI-based (medium-confidence) | 36.9 |
| S12_extracted_k141_44919 | 1364 | No | NA | 2 | 0 | 1 | Not-determined | Genome-fragment | NA | NA | 0 |
| S12_extracted_k141_177299 | 1561 | No | NA | 2 | 0 | 0 | Not-determined | Genome-fragment | NA | NA | 0 |
| S12_extracted_k141_75322 | 1341 | No | NA | 1 | 0 | 0 | Not-determined | Genome-fragment | NA | NA | 0 |
| S12_extracted_k141_28543 | 1118 | No | NA | 1 | 0 | 0 | Not-determined | Genome-fragment | NA | NA | 0 |
| S7_extracted_105053 | 3838 | No | NA | 7 | 1 | 0 | Low-quality | Genome-fragment | 10.2 | AAI-based (medium-confidence) | 0 |
| S12_extracted_k141_107695 | 1059 | No | NA | 2 | 0 | 0 | Not-determined | Genome-fragment | NA | NA | 0 |
| S12_extracted_k141_83437 | 1651 | No | NA | 4 | 0 | 4 | Not-determined | Genome-fragment | NA | NA | 0 |
| S12_extracted_k141_107703 | 4915 | No | NA | 5 | 0 | 3 | Not-determined | Genome-fragment | NA | NA | 0 |
| S12_extracted_k141_186359 | 1373 | No | NA | 1 | 0 | 1 | Not-determined | Genome-fragment | NA | NA | 0 |
| S12_extracted_k141_146580 | 2799 | No | NA | 3 | 0 | 0 | Not-determined | Genome-fragment | NA | NA | 0 |
| S12_extracted_k141_100025 | 1382 | No | NA | 1 | 0 | 1 | Not-determined | Genome-fragment | NA | NA | 0 |
| S12_extracted_k141_100034 | 1166 | No | NA | 3 | 0 | 0 | Not-determined | Genome-fragment | NA | NA | 0 |
| S12_extracted_k141_186378 | 2286 | No | NA | 2 | 0 | 1 | Not-determined | Genome-fragment | NA | NA | 0 |
| S12_extracted_k141_146607 | 1824 | No | NA | 1 | 0 | 1 | Not-determined | Genome-fragment | NA | NA | 0 |
| S5_extracted_k141_71699 | 3839 | No | NA | 4 | 0 | 0 | Low-quality | Genome-fragment | 6.82 | AAI-based (medium-confidence) | 0 |
| S13_extracted_38126 | 3840 | No | NA | 6 | 0 | 4 | Low-quality | Genome-fragment | 3.72 | AAI-based (medium-confidence) | 0 |
| S12_extracted_k141_130404 | 1194 | No | NA | 1 | 0 | 1 | Not-determined | Genome-fragment | NA | NA | 0 |
| S12_extracted_k141_146626 | 1425 | No | NA | 2 | 0 | 0 | Not-determined | Genome-fragment | NA | NA | 0 |
| S12_extracted_k141_75424 | 1665 | No | NA | 2 | 0 | 1 | Not-determined | Genome-fragment | NA | NA | 0 |
| S12_extracted_k141_114830 | 2556 | No | NA | 2 | 0 | 1 | Not-determined | Genome-fragment | NA | NA | 0 |
| S12_vs_k141_83511\|\|full | 3840 | No | NA | 3 | 0 | 1 | Low-quality | Genome-fragment | 5.82 | AAI-based (medium-confidence) | 0 |
| S12_extracted_k141_107723 | 1511 | No | NA | 2 | 0 | 2 | Not-determined | Genome-fragment | NA | NA | 0 |
| S12_extracted_k141_75435 | 3850 | No | NA | 3 | 0 | 2 | Not-determined | Genome-fragment | NA | NA | 0 |
| S7_vs_110511\|\|full | 3841 | No | NA | 2 | 0 | 2 | Low-quality | Genome-fragment | 2.54 | AAI-based (medium-confidence) | 0 |
| S12_extracted_k141_162291 | 1473 | No | NA | 1 | 0 | 1 | Not-determined | Genome-fragment | NA | NA | 0 |
| S12_extracted_k141_138972 | 1155 | No | NA | 1 | 0 | 0 | Not-determined | Genome-fragment | NA | NA | 0 |
| S12_extracted_k141_100103 | 7586 | No | NA | 9 | 0 | 5 | Not-determined | Genome-fragment | NA | NA | 0 |
| S12_extracted_k141_83496 | 6854 | No | NA | 4 | 0 | 3 | Not-determined | Genome-fragment | NA | NA | 0 |
| S12_extracted_k141_138980 | 1886 | No | NA | 2 | 0 | 1 | Not-determined | Genome-fragment | NA | NA | 0 |
| S16_extracted_156501 | 3841 | No | NA | 5 | 3 | 0 | Low-quality | Genome-fragment | 4.25 | HMM-based (lower-bound) | 0 |
| S16_vs_36292\|\|full | 3841 | No | NA | 3 | 0 | 2 | Low-quality | Genome-fragment | 2.59 | AAI-based (medium-confidence) | 0 |
| S12_extracted_k141_114862 | 1061 | No | NA | 2 | 0 | 0 | Not-determined | Genome-fragment | NA | NA | 0 |
| S12_extracted_k141_162318 | 1053 | No | NA | 2 | 0 | 1 | Not-determined | Genome-fragment | NA | NA | 0 |
| S12_extracted_k141_75493 | 1159 | No | NA | 1 | 0 | 0 | Not-determined | Genome-fragment | NA | NA | 0 |
| S12_extracted_k141_114876 | 1288 | No | NA | 1 | 0 | 1 | Not-determined | Genome-fragment | NA | NA | 0 |
| S12_extracted_k141_83515 | 1458 | No | NA | 1 | 0 | 1 | Not-determined | Genome-fragment | NA | NA | 0 |
| S12_extracted_k141_20541 | 2086 | No | NA | 3 | 0 | 0 | Not-determined | Genome-fragment | NA | NA | 0 |
| S10_extracted_86346 | 3841 | No | NA | 9 | 0 | 0 | Low-quality | Genome-fragment | 7.27 | AAI-based (high-confidence) | 0 |
| S12_extracted_k141_107773 | 3692 | No | NA | 8 | 0 | 7 | Not-determined | Genome-fragment | NA | NA | 0 |
| S12_extracted_k141_92096 | 1058 | No | NA | 2 | 0 | 2 | Not-determined | Genome-fragment | NA | NA | 0 |
| S12_extracted_k141_146720 | 1980 | No | NA | 3 | 0 | 2 | Not-determined | Genome-fragment | NA | NA | 0 |
| S12_extracted_k141_92100 | 1061 | No | NA | 2 | 0 | 1 | Not-determined | Genome-fragment | NA | NA | 0 |
| S8_extracted_k141_153165 | 3841 | No | NA | 7 | 1 | 0 | Low-quality | Genome-fragment | 2.49 | HMM-based (lower-bound) | 0 |
| S12_extracted_k141_67878 | 1202 | No | NA | 2 | 0 | 0 | Not-determined | Genome-fragment | NA | NA | 0 |
| S16_extracted_27857 | 3843 | No | NA | 4 | 1 | 0 | Low-quality | Genome-fragment | 5.81 | HMM-based (lower-bound) | 0 |
| S12_extracted_k141_20563 | 1304 | No | NA | 5 | 0 | 0 | Not-determined | Genome-fragment | NA | NA | 0 |
| S12_extracted_k141_146727 | 1142 | No | NA | 1 | 0 | 1 | Not-determined | Genome-fragment | NA | NA | 0 |
| S12_extracted_k141_83530 | 1056 | No | NA | 3 | 0 | 1 | Not-determined | Genome-fragment | NA | NA | 0 |
| S12_extracted_k141_67882 | 1286 | No | NA | 2 | 0 | 1 | Not-determined | Genome-fragment | NA | NA | 0 |
| S12_extracted_k141_170651 | 3843 | No | NA | 4 | 0 | 0 | Low-quality | Genome-fragment | 6.5 | AAI-based (high-confidence) | 0 |
| S12_extracted_k141_92131 | 1210 | No | NA | 1 | 0 | 1 | Not-determined | Genome-fragment | NA | NA | 0 |
| S12_extracted_k141_154251 | 1006 | No | NA | 2 | 0 | 1 | Not-determined | Genome-fragment | NA | NA | 0 |
| S12_extracted_k141_5314 | 1131 | No | NA | 2 | 0 | 1 | Not-determined | Genome-fragment | NA | NA | 0 |
| S12_extracted_k141_75539 | 3346 | No | NA | 1 | 0 | 0 | Not-determined | Genome-fragment | NA | NA | 0 |
| S12_extracted_k141_146739 | 1312 | No | NA | 1 | 0 | 1 | Not-determined | Genome-fragment | NA | NA | 0 |
| S12_extracted_k141_5329 | 1400 | No | NA | 2 | 0 | 0 | Not-determined | Genome-fragment | NA | NA | 0 |
| S12_extracted_k141_177376 | 2577 | No | NA | 2 | 0 | 0 | Not-determined | Genome-fragment | NA | NA | 0 |
| S12_extracted_k141_35948 | 1137 | No | NA | 1 | 0 | 1 | Not-determined | Genome-fragment | NA | NA | 0 |
| S12_extracted_k141_139094 | 1118 | No | NA | 1 | 0 | 1 | Not-determined | Genome-fragment | NA | NA | 0 |
| S12_extracted_k141_20599 | 1054 | No | NA | 2 | 0 | 2 | Not-determined | Genome-fragment | NA | NA | 0 |
| S4_extracted_2038 | 3843 | No | NA | 10 | 1 | 0 | Low-quality | Genome-fragment | 5.68 | AAI-based (high-confidence) | 0 |
| S12_extracted_k141_130613 | 1524 | No | NA | 2 | 0 | 1 | Not-determined | Genome-fragment | NA | NA | 0 |
| S12_extracted_k141_162476 | 1603 | No | NA | 1 | 0 | 1 | Not-determined | Genome-fragment | NA | NA | 0 |
| S12_extracted_k141_162478 | 1110 | No | NA | 2 | 0 | 1 | Not-determined | Genome-fragment | NA | NA | 0 |
| S1_vs_k141_96818\|\|full | 3844 | No | NA | 2 | 0 | 1 | Low-quality | Genome-fragment | 1.87 | AAI-based (medium-confidence) | 0 |
| S16_extracted_149727 | 3844 | No | NA | 6 | 5 | 0 | Low-quality | Genome-fragment | 2.26 | AAI-based (medium-confidence) | 0 |
| S12_extracted_k141_170103 | 1060 | No | NA | 2 | 0 | 1 | Not-determined | Genome-fragment | NA | NA | 0 |
| S12_extracted_k141_5399 | 1267 | No | NA | 1 | 0 | 0 | Not-determined | Genome-fragment | NA | NA | 0 |
| S12_extracted_k141_20631 | 1006 | No | NA | 1 | 0 | 0 | Not-determined | Genome-fragment | NA | NA | 0 |
| S10_vs_42933\|\|full | 3844 | No | NA | 4 | 0 | 3 | Low-quality | Genome-fragment | 4.29 | AAI-based (medium-confidence) | 0 |
| S12_extracted_k141_20644 | 3014 | No | NA | 3 | 0 | 1 | Not-determined | Genome-fragment | NA | NA | 0 |
| S12_extracted_k141_100273 | 8099 | No | NA | 11 | 0 | 6 | Not-determined | Genome-fragment | NA | NA | 0 |
| S14_extracted_k141_88697 | 3846 | No | NA | 7 | 0 | 1 | Low-quality | Genome-fragment | 2.18 | AAI-based (medium-confidence) | 0 |
| S12_extracted_k141_170135 | 1323 | No | NA | 2 | 0 | 2 | Not-determined | Genome-fragment | NA | NA | 0 |
| S12_extracted_k141_20653 | 2099 | No | NA | 2 | 0 | 0 | Not-determined | Genome-fragment | NA | NA | 0 |
| S12_extracted_k141_83567 | 1036 | No | NA | 2 | 0 | 1 | Not-determined | Genome-fragment | NA | NA | 0 |
| S12_extracted_k141_100301 | 2193 | No | NA | 1 | 0 | 1 | Not-determined | Genome-fragment | NA | NA | 0 |
| S12_extracted_k141_45082 | 1708 | No | NA | 3 | 0 | 0 | Not-determined | Genome-fragment | NA | NA | 0 |
| S12_extracted_k141_186582 | 1418 | No | NA | 3 | 0 | 1 | Not-determined | Genome-fragment | NA | NA | 0 |
| S8_vs_k141_150192\|\|full | 3846 | No | NA | 7 | 4 | 0 | Low-quality | Genome-fragment | 9.71 | AAI-based (high-confidence) | 0 |
| S12_extracted_k141_177536 | 1221 | No | NA | 2 | 0 | 1 | Not-determined | Genome-fragment | NA | NA | 0 |
| S12_extracted_k141_67909 | 116083 | No | NA | 139 | 31 | 5 | Medium-quality | Genome-fragment | 79.42 | AAI-based (medium-confidence) | 0 |
| S12_extracted_k141_60224 | 54526 | Yes | 47561 | 72 | 11 | 5 | Complete | High-quality | 100 | Provirus (high-confidence) | 12.77 |
| S12_extracted_k141_12081 | 1566 | No | NA | 2 | 0 | 1 | Not-determined | Genome-fragment | NA | NA | 0 |
| S12_extracted_k141_73424 | 3847 | No | NA | 4 | 1 | 0 | Low-quality | Genome-fragment | 8.58 | AAI-based (medium-confidence) | 0 |
| S12_extracted_k141_20723 | 1280 | No | NA | 3 | 0 | 0 | Not-determined | Genome-fragment | NA | NA | 0 |
| S12_extracted_k141_100364 | 1137 | No | NA | 1 | 0 | 1 | Not-determined | Genome-fragment | NA | NA | 0 |
| S12_extracted_k141_92206 | 8785 | No | NA | 8 | 0 | 4 | Not-determined | Genome-fragment | NA | NA | 0 |
| S12_extracted_k141_20726 | 3366 | No | NA | 3 | 0 | 1 | Not-determined | Genome-fragment | NA | NA | 0 |
| S11_vs_k141_81494\|\|full | 3848 | No | NA | 7 | 2 | 0 | Low-quality | Genome-fragment | 10.27 | AAI-based (high-confidence) | 0 |
| S12_extracted_k141_177542 | 1124 | No | NA | 3 | 0 | 1 | Not-determined | Genome-fragment | NA | NA | 0 |
| S10_extracted_2163 | 3848 | No | NA | 8 | 5 | 0 | Low-quality | Genome-fragment | 9.37 | AAI-based (high-confidence) | 0 |
| S12_extracted_k141_45132 | 3155 | No | NA | 2 | 0 | 1 | Not-determined | Genome-fragment | NA | NA | 0 |
| S12_extracted_k141_114985 | 5097 | No | NA | 5 | 0 | 4 | Not-determined | Genome-fragment | NA | NA | 0 |
| S8_extracted_k141_183537 | 3850 | No | NA | 4 | 4 | 0 | Low-quality | Genome-fragment | 10.85 | AAI-based (high-confidence) | 0 |
| S12_extracted_k141_186624 | 1154 | No | NA | 1 | 0 | 0 | Not-determined | Genome-fragment | NA | NA | 0 |
| S12_extracted_k141_45143 | 6614 | No | NA | 4 | 0 | 4 | Not-determined | Genome-fragment | NA | NA | 0 |
| S12_extracted_k141_100392 | 1811 | No | NA | 2 | 0 | 1 | Not-determined | Genome-fragment | NA | NA | 0 |
| S12_extracted_k141_130641 | 3271 | No | NA | 2 | 0 | 2 | Not-determined | Genome-fragment | NA | NA | 0 |
| S12_extracted_k141_100405 | 1201 | No | NA | 1 | 0 | 0 | Not-determined | Genome-fragment | NA | NA | 0 |
| S12_extracted_k141_154404 | 1295 | No | NA | 1 | 0 | 1 | Not-determined | Genome-fragment | NA | NA | 0 |
| S12_extracted_k141_139260 | 1039 | No | NA | 2 | 0 | 1 | Not-determined | Genome-fragment | NA | NA | 0 |
| S12_extracted_k141_67999 | 1077 | No | NA | 1 | 0 | 1 | Not-determined | Genome-fragment | NA | NA | 0 |
| S3_extracted_k141_48864 | 3851 | No | NA | 8 | 0 | 0 | Low-quality | Genome-fragment | 6.09 | AAI-based (high-confidence) | 0 |
| S12_extracted_k141_100433 | 1119 | No | NA | 1 | 0 | 1 | Not-determined | Genome-fragment | NA | NA | 0 |
| S12_extracted_k141_36112 | 1179 | No | NA | 2 | 0 | 0 | Not-determined | Genome-fragment | NA | NA | 0 |
| S12_extracted_k141_28784 | 2166 | No | NA | 1 | 0 | 1 | Not-determined | Genome-fragment | NA | NA | 0 |
| S12_extracted_k141_154449 | 1453 | No | NA | 2 | 0 | 1 | Not-determined | Genome-fragment | NA | NA | 0 |
| S12_extracted_k141_107877 | 1107 | No | NA | 2 | 0 | 0 | Not-determined | Genome-fragment | NA | NA | 0 |
| S8_extracted_k141_165267 | 3851 | No | NA | 5 | 0 | 1 | Low-quality | Genome-fragment | 8.25 | AAI-based (medium-confidence) | 0 |
| S12_extracted_k141_107884 | 8114 | No | NA | 5 | 0 | 3 | Not-determined | Genome-fragment | NA | NA | 0 |
| S12_extracted_k141_186691 | 6265 | No | NA | 6 | 0 | 4 | Not-determined | Genome-fragment | NA | NA | 0 |
| S12_extracted_k141_45208 | 1326 | No | NA | 1 | 0 | 0 | Not-determined | Genome-fragment | NA | NA | 0 |
| S12_extracted_k141_177649 | 1035 | No | NA | 2 | 0 | 2 | Not-determined | Genome-fragment | NA | NA | 0 |
| S1_vs_k141_120965\|\|full | 3853 | No | NA | 4 | 3 | 0 | Low-quality | Genome-fragment | 5.74 | HMM-based (lower-bound) | 0 |
| S12_vs_k141_93896\|\|full | 3854 | No | NA | 4 | 2 | 0 | Low-quality | Genome-fragment | 7.16 | HMM-based (lower-bound) | 0 |
| S12_extracted_k141_60240 | 3363 | No | NA | 4 | 0 | 3 | Not-determined | Genome-fragment | NA | NA | 0 |
| S12_extracted_k141_45217 | 1154 | No | NA | 1 | 0 | 1 | Not-determined | Genome-fragment | NA | NA | 0 |
| S12_extracted_k141_68041 | 1118 | No | NA | 1 | 0 | 0 | Not-determined | Genome-fragment | NA | NA | 0 |
| S17_extracted_k141_56346 | 3856 | No | NA | 3 | 0 | 0 | Low-quality | Genome-fragment | 8.65 | AAI-based (high-confidence) | 0 |
| S7_extracted_30211 | 3856 | No | NA | 6 | 0 | 3 | Low-quality | Genome-fragment | 4.67 | AAI-based (medium-confidence) | 0 |
| S7_extracted_41107 | 3856 | No | NA | 4 | 0 | 4 | Low-quality | Genome-fragment | 0.35 | AAI-based (medium-confidence) | 0 |
| S7_vs_118871\|\|full | 3856 | No | NA | 4 | 0 | 2 | Low-quality | Genome-fragment | 2.91 | AAI-based (medium-confidence) | 0 |
| S12_extracted_k141_107916 | 1207 | No | NA | 1 | 0 | 0 | Not-determined | Genome-fragment | NA | NA | 0 |
| S12_extracted_k141_45237 | 1652 | No | NA | 1 | 0 | 1 | Not-determined | Genome-fragment | NA | NA | 0 |
| S12_extracted_k141_68074 | 1201 | No | NA | 3 | 0 | 1 | Not-determined | Genome-fragment | NA | NA | 0 |
| S12_extracted_k141_5614 | 1122 | No | NA | 2 | 0 | 1 | Not-determined | Genome-fragment | NA | NA | 0 |
| S12_extracted_k141_5616 | 1581 | No | NA | 2 | 0 | 2 | Not-determined | Genome-fragment | NA | NA | 0 |
| S11_extracted_k141_100581 | 3857 | No | NA | 5 | 0 | 1 | Low-quality | Genome-fragment | 7.61 | AAI-based (medium-confidence) | 0 |
| S12_extracted_k141_28843 | 4666 | No | NA | 4 | 0 | 1 | Not-determined | Genome-fragment | NA | NA | 0 |
| S12_extracted_k141_130774 | 1425 | No | NA | 1 | 0 | 1 | Not-determined | Genome-fragment | NA | NA | 0 |
| S16_vs_36853\|\|full | 3857 | No | NA | 2 | 1 | 0 | Low-quality | Genome-fragment | 1.14 | HMM-based (lower-bound) | 0 |
| S12_extracted_k141_5638 | 1291 | No | NA | 2 | 0 | 1 | Not-determined | Genome-fragment | NA | NA | 0 |
| S17_vs_k141_110349\|\|full | 3858 | No | NA | 8 | 3 | 0 | Low-quality | Genome-fragment | 10.78 | AAI-based (high-confidence) | 0 |
| S11_vs_k141_89736\|\|full | 3858 | No | NA | 5 | 1 | 0 | Low-quality | Genome-fragment | 9.14 | AAI-based (high-confidence) | 0 |
| S12_extracted_k141_130797 | 1534 | No | NA | 3 | 0 | 3 | Not-determined | Genome-fragment | NA | NA | 0 |
| S12_extracted_k141_154548 | 1202 | No | NA | 1 | 0 | 0 | Not-determined | Genome-fragment | NA | NA | 0 |
| S12_extracted_k141_12210 | 1439 | No | NA | 3 | 0 | 1 | Not-determined | Genome-fragment | NA | NA | 0 |
| S12_extracted_k141_170393 | 4468 | No | NA | 5 | 0 | 1 | Not-determined | Genome-fragment | NA | NA | 0 |
| S12_extracted_k141_12214 | 1128 | No | NA | 1 | 0 | 1 | Not-determined | Genome-fragment | NA | NA | 0 |
| S12_extracted_k141_177709 | 3639 | No | NA | 7 | 0 | 0 | Not-determined | Genome-fragment | NA | NA | 0 |
| S12_extracted_k141_45283 | 1188 | No | NA | 1 | 0 | 1 | Not-determined | Genome-fragment | NA | NA | 0 |
| S12_extracted_k141_130815 | 1407 | No | NA | 2 | 0 | 1 | Not-determined | Genome-fragment | NA | NA | 0 |
| S12_extracted_k141_68109 | 8382 | No | NA | 7 | 0 | 5 | Not-determined | Genome-fragment | NA | NA | 0 |
| S12_extracted_k141_107984 | 1364 | No | NA | 1 | 0 | 1 | Not-determined | Genome-fragment | NA | NA | 0 |
| S12_extracted_k141_12232 | 1427 | No | NA | 1 | 0 | 1 | Not-determined | Genome-fragment | NA | NA | 0 |
| S12_extracted_k141_75885 | 4003 | No | NA | 4 | 0 | 2 | Not-determined | Genome-fragment | NA | NA | 0 |
| S12_extracted_k141_186820 | 1024 | No | NA | 1 | 0 | 1 | Not-determined | Genome-fragment | NA | NA | 0 |
| S12_extracted_k141_12248 | 1254 | No | NA | 2 | 0 | 1 | Not-determined | Genome-fragment | NA | NA | 0 |
| S12_extracted_k141_20769 | 1622 | No | NA | 2 | 0 | 2 | Not-determined | Genome-fragment | NA | NA | 0 |
| S12_extracted_k141_139387 | 1010 | No | NA | 3 | 0 | 0 | Not-determined | Genome-fragment | NA | NA | 0 |
| S5_vs_k141_23856\|\|full | 3858 | No | NA | 5 | 1 | 0 | Low-quality | Genome-fragment | 5.69 | AAI-based (high-confidence) | 0 |
| S12_extracted_k141_12261 | 1270 | No | NA | 1 | 0 | 1 | Not-determined | Genome-fragment | NA | NA | 0 |
| S12_extracted_k141_12270 | 1447 | No | NA | 2 | 0 | 1 | Not-determined | Genome-fragment | NA | NA | 0 |
| S12_extracted_k141_20779 | 5350 | No | NA | 7 | 0 | 3 | Not-determined | Genome-fragment | NA | NA | 0 |
| S16_vs_139891\|\|full | 3858 | No | NA | 8 | 1 | 0 | Low-quality | Genome-fragment | 5.79 | AAI-based (high-confidence) | 0 |
| S12_extracted_k141_12277 | 1063 | No | NA | 1 | 0 | 0 | Not-determined | Genome-fragment | NA | NA | 0 |
| S12_extracted_k141_177745 | 2194 | No | NA | 1 | 0 | 1 | Not-determined | Genome-fragment | NA | NA | 0 |
| S12_extracted_k141_170427 | 1382 | No | NA | 3 | 0 | 1 | Not-determined | Genome-fragment | NA | NA | 0 |
| S12_extracted_k141_60386 | 3326 | No | NA | 1 | 0 | 0 | Not-determined | Genome-fragment | NA | NA | 0 |
| S12_vs_k141_105126\|\|full | 3858 | No | NA | 3 | 0 | 1 | Low-quality | Genome-fragment | 2.03 | AAI-based (medium-confidence) | 0 |
| S12_extracted_k141_154629 | 1103 | No | NA | 1 | 0 | 1 | Not-determined | Genome-fragment | NA | NA | 0 |
| S12_extracted_k141_92453 | 1286 | No | NA | 3 | 0 | 0 | Not-determined | Genome-fragment | NA | NA | 0 |
| S12_extracted_k141_115098 | 1236 | No | NA | 2 | 0 | 1 | Not-determined | Genome-fragment | NA | NA | 0 |
| S12_extracted_k141_68191 | 1274 | No | NA | 2 | 0 | 1 | Not-determined | Genome-fragment | NA | NA | 0 |
| S12_extracted_k141_83695 | 1076 | No | NA | 2 | 0 | 0 | Not-determined | Genome-fragment | NA | NA | 0 |
| S12_extracted_k141_139416 | 27145 | No | NA | 25 | 0 | 8 | Not-determined | Genome-fragment | NA | NA | 0 |
| S16_vs_67864\|\|full | 3859 | No | NA | 3 | 0 | 2 | Low-quality | Genome-fragment | 2.03 | AAI-based (medium-confidence) | 0 |
| S12_extracted_k141_115120 | 1201 | No | NA | 1 | 0 | 1 | Not-determined | Genome-fragment | NA | NA | 0 |
| S12_vs_k141_134758\|\|full | 3859 | No | NA | 7 | 3 | 0 | Low-quality | Genome-fragment | 7.56 | AAI-based (medium-confidence) | 0 |
| S12_extracted_k141_12340 | 1235 | No | NA | 2 | 0 | 1 | Not-determined | Genome-fragment | NA | NA | 0 |
| S12_extracted_k141_130889 | 1206 | No | NA | 2 | 0 | 0 | Not-determined | Genome-fragment | NA | NA | 0 |
| S12_extracted_k141_177826 | 1486 | No | NA | 1 | 0 | 1 | Not-determined | Genome-fragment | NA | NA | 0 |
| S11_vs_k141_50018\|\|full | 3860 | Yes | 2828 | 8 | 1 | 1 | Low-quality | Genome-fragment | 1.82 | HMM-based (lower-bound) | 26.74 |
| S12_extracted_k141_115154 | 1021 | No | NA | 2 | 0 | 1 | Not-determined | Genome-fragment | NA | NA | 0 |
| S12_extracted_k141_12371 | 3857 | No | NA | 4 | 0 | 2 | Not-determined | Genome-fragment | NA | NA | 0 |
| S13_extracted_136853 | 3860 | No | NA | 2 | 0 | 1 | Low-quality | Genome-fragment | 1.92 | AAI-based (medium-confidence) | 0 |
| S12_extracted_k141_100595 | 3558 | No | NA | 4 | 0 | 2 | Not-determined | Genome-fragment | NA | NA | 0 |
| S12_extracted_k141_186966 | 1888 | No | NA | 4 | 0 | 0 | Not-determined | Genome-fragment | NA | NA | 0 |
| S12_extracted_k141_177898 | 1215 | No | NA | 1 | 0 | 1 | Not-determined | Genome-fragment | NA | NA | 0 |
| S12_extracted_k141_100613 | 1090 | No | NA | 2 | 0 | 0 | Not-determined | Genome-fragment | NA | NA | 0 |
| S12_extracted_k141_5834 | 1782 | No | NA | 1 | 0 | 0 | Not-determined | Genome-fragment | NA | NA | 0 |
| S10_vs_113717\|\|full | 3862 | No | NA | 6 | 4 | 0 | Low-quality | Genome-fragment | 8.69 | AAI-based (high-confidence) | 0 |
| S12_extracted_k141_147166 | 1205 | No | NA | 2 | 0 | 1 | Not-determined | Genome-fragment | NA | NA | 0 |
| S12_extracted_k141_20924 | 1284 | No | NA | 2 | 0 | 2 | Not-determined | Genome-fragment | NA | NA | 0 |
| S12_extracted_k141_186986 | 4679 | No | NA | 2 | 0 | 1 | Not-determined | Genome-fragment | NA | NA | 0 |
| S12_extracted_k141_130967 | 1182 | No | NA | 2 | 0 | 2 | Not-determined | Genome-fragment | NA | NA | 0 |
| S3_vs_k141_75870\|\|full | 3863 | No | NA | 3 | 0 | 0 | Low-quality | Genome-fragment | 1.96 | AAI-based (high-confidence) | 0 |
| S12_extracted_k141_36298 | 1101 | No | NA | 2 | 0 | 2 | Not-determined | Genome-fragment | NA | NA | 0 |
| S12_extracted_k141_154742 | 2296 | No | NA | 3 | 0 | 2 | Not-determined | Genome-fragment | NA | NA | 0 |
| S16_vs_163397\|\|full | 3863 | No | NA | 3 | 0 | 0 | Low-quality | Genome-fragment | 9.99 | AAI-based (medium-confidence) | 0 |
| S6_vs_k141_28481\|\|full | 3863 | No | NA | 3 | 0 | 1 | Low-quality | Genome-fragment | 1.65 | AAI-based (medium-confidence) | 0 |
| S17_vs_k141_172029\|\|full | 3864 | No | NA | 3 | 0 | 1 | Low-quality | Genome-fragment | 12.34 | AAI-based (high-confidence) | 0 |
| S14_vs_k141_8365\|\|full | 3864 | No | NA | 3 | 0 | 1 | Low-quality | Genome-fragment | 6.05 | AAI-based (medium-confidence) | 0 |
| S12_extracted_k141_52402 | 1155 | No | NA | 1 | 0 | 0 | Not-determined | Genome-fragment | NA | NA | 0 |
| S12_extracted_k141_122889 | 2309 | No | NA | 2 | 0 | 0 | Not-determined | Genome-fragment | NA | NA | 0 |
| S12_extracted_k141_12449 | 3249 | No | NA | 5 | 0 | 1 | Not-determined | Genome-fragment | NA | NA | 0 |
| S6_extracted_k141_10220 | 3864 | No | NA | 1 | 0 | 1 | Low-quality | Genome-fragment | 1.63 | AAI-based (medium-confidence) | 0 |
| S12_extracted_k141_115260 | 1120 | No | NA | 2 | 0 | 1 | Not-determined | Genome-fragment | NA | NA | 0 |
| S12_extracted_k141_187039 | 1211 | No | NA | 2 | 0 | 2 | Not-determined | Genome-fragment | NA | NA | 0 |
| S12_extracted_k141_177972 | 1296 | No | NA | 1 | 0 | 0 | Not-determined | Genome-fragment | NA | NA | 0 |
| S12_extracted_k141_52418 | 1143 | No | NA | 1 | 0 | 0 | Not-determined | Genome-fragment | NA | NA | 0 |
| S12_extracted_k141_177974 | 1152 | No | NA | 2 | 0 | 0 | Not-determined | Genome-fragment | NA | NA | 0 |
| S12_extracted_k141_68300 | 1286 | No | NA | 2 | 0 | 2 | Not-determined | Genome-fragment | NA | NA | 0 |
| S12_extracted_k141_20975 | 1486 | No | NA | 2 | 0 | 0 | Not-determined | Genome-fragment | NA | NA | 0 |
| S12_extracted_k141_122917 | 1293 | No | NA | 2 | 0 | 0 | Not-determined | Genome-fragment | NA | NA | 0 |
| S8_vs_k141_167559\|\|full | 3864 | No | NA | 4 | 0 | 2 | Low-quality | Genome-fragment | 1.28 | AAI-based (medium-confidence) | 0 |
| S12_extracted_k141_20978 | 2412 | No | NA | 2 | 0 | 1 | Not-determined | Genome-fragment | NA | NA | 0 |
| S17_vs_k141_10926\|\|full | 3865 | No | NA | 3 | 0 | 3 | Low-quality | Genome-fragment | 3.32 | AAI-based (medium-confidence) | 0 |
| S12_extracted_k141_76082 | 1058 | No | NA | 2 | 0 | 2 | Not-determined | Genome-fragment | NA | NA | 0 |
| S12_extracted_k141_60558 | 1270 | No | NA | 2 | 0 | 2 | Not-determined | Genome-fragment | NA | NA | 0 |
| S12_extracted_k141_52440 | 1721 | No | NA | 2 | 0 | 0 | Not-determined | Genome-fragment | NA | NA | 0 |
| S12_extracted_k141_178008 | 1013 | No | NA | 1 | 0 | 0 | Not-determined | Genome-fragment | NA | NA | 0 |
| S12_extracted_k141_29059 | 3256 | No | NA | 2 | 0 | 2 | Not-determined | Genome-fragment | NA | NA | 0 |
| S12_extracted_k141_12487 | 1023 | No | NA | 2 | 0 | 2 | Not-determined | Genome-fragment | NA | NA | 0 |
| S12_extracted_k141_83802 | 3533 | No | NA | 4 | 0 | 4 | Not-determined | Genome-fragment | NA | NA | 0 |
| S12_extracted_k141_83804 | 1095 | No | NA | 2 | 0 | 1 | Not-determined | Genome-fragment | NA | NA | 0 |
| S12_extracted_k141_52468 | 1125 | No | NA | 1 | 0 | 1 | Not-determined | Genome-fragment | NA | NA | 0 |
| S12_extracted_k141_60597 | 1105 | No | NA | 2 | 0 | 0 | Not-determined | Genome-fragment | NA | NA | 0 |
| S3_extracted_k141_19949 | 3867 | No | NA | 5 | 4 | 0 | Low-quality | Genome-fragment | 17.64 | AAI-based (high-confidence) | 0 |
| S12_extracted_k141_178036 | 1122 | No | NA | 2 | 0 | 1 | Not-determined | Genome-fragment | NA | NA | 0 |
| S12_extracted_k141_29092 | 1134 | No | NA | 2 | 0 | 1 | Not-determined | Genome-fragment | NA | NA | 0 |
| S12_extracted_k141_178041 | 1416 | No | NA | 2 | 0 | 2 | Not-determined | Genome-fragment | NA | NA | 0 |
| S5_vs_k141_130959\|\|full | 3867 | No | NA | 7 | 6 | 0 | Low-quality | Genome-fragment | 8.55 | AAI-based (medium-confidence) | 0 |
| S12_extracted_k141_21013 | 1042 | No | NA | 1 | 0 | 0 | Not-determined | Genome-fragment | NA | NA | 0 |
| S12_extracted_k141_162718 | 1012 | No | NA | 2 | 0 | 1 | Not-determined | Genome-fragment | NA | NA | 0 |
| S12_extracted_k141_154818 | 1353 | No | NA | 2 | 0 | 1 | Not-determined | Genome-fragment | NA | NA | 0 |
| S12_extracted_k141_147248 | 1384 | No | NA | 1 | 0 | 0 | Not-determined | Genome-fragment | NA | NA | 0 |
| S12_extracted_k141_83827 | 2492 | No | NA | 2 | 0 | 1 | Not-determined | Genome-fragment | NA | NA | 0 |
| S7_extracted_113530 | 3867 | No | NA | 4 | 0 | 4 | Low-quality | Genome-fragment | 2.54 | AAI-based (medium-confidence) | 0 |
| S12_extracted_k141_36376 | 1396 | No | NA | 2 | 0 | 0 | Not-determined | Genome-fragment | NA | NA | 0 |
| S12_extracted_k141_76169 | 1075 | No | NA | 1 | 0 | 0 | Not-determined | Genome-fragment | NA | NA | 0 |
| S11_extracted_k141_58954 | 3868 | No | NA | 4 | 0 | 1 | Low-quality | Genome-fragment | 3.65 | AAI-based (medium-confidence) | 0 |
| S12_extracted_k141_108222 | 1300 | No | NA | 2 | 0 | 1 | Not-determined | Genome-fragment | NA | NA | 0 |
| S16_extracted_18926 | 3869 | No | NA | 5 | 2 | 0 | Low-quality | Genome-fragment | 7.39 | AAI-based (high-confidence) | 0 |
| S12_extracted_k141_68336 | 16859 | No | NA | 13 | 0 | 10 | Not-determined | Genome-fragment | NA | NA | 0 |
| S7_extracted_30054 | 3870 | No | NA | 7 | 0 | 0 | Low-quality | Genome-fragment | 3.24 | AAI-based (medium-confidence) | 0 |
| S16_extracted_131242 | 3870 | No | NA | 3 | 0 | 3 | Low-quality | Genome-fragment | 0.35 | AAI-based (medium-confidence) | 0 |
| S10_vs_39612\|\|full | 3870 | No | NA | 7 | 0 | 2 | Low-quality | Genome-fragment | 7.41 | AAI-based (high-confidence) | 0 |
| S12_extracted_k141_115354 | 1062 | No | NA | 1 | 0 | 1 | Not-determined | Genome-fragment | NA | NA | 0 |
| S8_extracted_k141_43769 | 3870 | No | NA | 4 | 1 | 0 | Low-quality | Genome-fragment | 9.77 | AAI-based (high-confidence) | 0 |
| S12_extracted_k141_108259 | 1920 | No | NA | 3 | 0 | 2 | Not-determined | Genome-fragment | NA | NA | 0 |
| S11_extracted_k141_81522 | 3871 | No | NA | 4 | 1 | 0 | Low-quality | Genome-fragment | 2.51 | AAI-based (high-confidence) | 0 |
| S12_extracted_k141_100790 | 1134 | No | NA | 2 | 0 | 1 | Not-determined | Genome-fragment | NA | NA | 0 |
| S12_extracted_k141_170594 | 1002 | No | NA | 1 | 0 | 0 | Not-determined | Genome-fragment | NA | NA | 0 |
| S7_vs_122594\|\|full | 3871 | No | NA | 5 | 4 | 0 | Low-quality | Genome-fragment | 10.37 | AAI-based (high-confidence) | 0 |
| S12_extracted_k141_170597 | 2219 | No | NA | 2 | 0 | 0 | Not-determined | Genome-fragment | NA | NA | 0 |
| S12_extracted_k141_76222 | 1072 | No | NA | 1 | 0 | 0 | Not-determined | Genome-fragment | NA | NA | 0 |
| S12_extracted_k141_100804 | 1821 | No | NA | 1 | 0 | 1 | Not-determined | Genome-fragment | NA | NA | 0 |
| S12_extracted_k141_36416 | 1404 | No | NA | 2 | 0 | 0 | Not-determined | Genome-fragment | NA | NA | 0 |
| S12_extracted_k141_187177 | 1066 | No | NA | 3 | 0 | 1 | Not-determined | Genome-fragment | NA | NA | 0 |
| S12_extracted_k141_139686 | 1227 | No | NA | 2 | 0 | 0 | Not-determined | Genome-fragment | NA | NA | 0 |
| S12_extracted_k141_123041 | 1483 | No | NA | 2 | 0 | 1 | Not-determined | Genome-fragment | NA | NA | 0 |
| S12_extracted_k141_147265 | 1943 | No | NA | 1 | 0 | 1 | Not-determined | Genome-fragment | NA | NA | 0 |
| S12_extracted_k141_170613 | 3545 | No | NA | 5 | 0 | 2 | Not-determined | Genome-fragment | NA | NA | 0 |
| S12_vs_k141_181528\|\|full | 3871 | No | NA | 5 | 3 | 0 | Low-quality | Genome-fragment | 7.24 | AAI-based (medium-confidence) | 0 |
| S12_extracted_k141_60665 | 2874 | No | NA | 1 | 0 | 1 | Not-determined | Genome-fragment | NA | NA | 0 |
| S12_extracted_k141_139714 | 1646 | No | NA | 1 | 0 | 0 | Not-determined | Genome-fragment | NA | NA | 0 |
| S12_extracted_k141_68411 | 3588 | No | NA | 1 | 0 | 0 | Not-determined | Genome-fragment | NA | NA | 0 |
| S12_extracted_k141_170614 | 16108 | No | NA | 20 | 6 | 0 | Medium-quality | Genome-fragment | 69.63 | AAI-based (medium-confidence) | 0 |
| S12_extracted_k141_45681 | 1210 | No | NA | 3 | 0 | 0 | Not-determined | Genome-fragment | NA | NA | 0 |
| S12_extracted_k141_170628 | 1995 | No | NA | 2 | 0 | 2 | Not-determined | Genome-fragment | NA | NA | 0 |
| S12_extracted_k141_12613 | 2662 | No | NA | 2 | 0 | 1 | Not-determined | Genome-fragment | NA | NA | 0 |
| S12_extracted_k141_21094 | 1465 | No | NA | 1 | 0 | 1 | Not-determined | Genome-fragment | NA | NA | 0 |
| S12_extracted_k141_139751 | 8484 | No | NA | 6 | 0 | 4 | Not-determined | Genome-fragment | NA | NA | 0 |
| S8_vs_k141_26447\|\|full | 3871 | No | NA | 7 | 0 | 1 | Low-quality | Genome-fragment | 7.43 | AAI-based (high-confidence) | 0 |
| S12_extracted_k141_139757 | 1494 | No | NA | 1 | 0 | 1 | Not-determined | Genome-fragment | NA | NA | 0 |
| S12_extracted_k141_68457 | 1015 | No | NA | 1 | 0 | 1 | Not-determined | Genome-fragment | NA | NA | 0 |
| S12_extracted_k141_92700 | 2758 | No | NA | 2 | 0 | 1 | Not-determined | Genome-fragment | NA | NA | 0 |
| S5_vs_k141_32305\|\|full | 3872 | No | NA | 4 | 0 | 0 | Low-quality | Genome-fragment | 5.26 | AAI-based (medium-confidence) | 0 |
| S5_vs_k141_73522\|\|lt2gene | 3872 | No | NA | 2 | 1 | 0 | Low-quality | Genome-fragment | 3.46 | AAI-based (medium-confidence) | 0 |
| S12_extracted_k141_115452 | 1235 | No | NA | 1 | 0 | 1 | Not-determined | Genome-fragment | NA | NA | 0 |
| S12_extracted_k141_162852 | 1233 | No | NA | 1 | 0 | 1 | Not-determined | Genome-fragment | NA | NA | 0 |
| S12_extracted_k141_162853 | 1051 | No | NA | 2 | 0 | 0 | Not-determined | Genome-fragment | NA | NA | 0 |
| S12_extracted_k141_21153 | 1002 | No | NA | 2 | 0 | 2 | Not-determined | Genome-fragment | NA | NA | 0 |
| S12_extracted_k141_115454 | 6843 | No | NA | 9 | 0 | 6 | Not-determined | Genome-fragment | NA | NA | 0 |
| S12_extracted_k141_21157 | 2921 | No | NA | 1 | 0 | 1 | Not-determined | Genome-fragment | NA | NA | 0 |
| S12_extracted_k141_108398 | 1127 | No | NA | 3 | 0 | 1 | Not-determined | Genome-fragment | NA | NA | 0 |
| S12_extracted_k141_100946 | 1408 | No | NA | 1 | 0 | 1 | Not-determined | Genome-fragment | NA | NA | 0 |
| S12_extracted_k141_68524 | 1204 | No | NA | 2 | 0 | 0 | Not-determined | Genome-fragment | NA | NA | 0 |
| S12_extracted_k141_115498 | 1543 | No | NA | 3 | 0 | 0 | Not-determined | Genome-fragment | NA | NA | 0 |
| S12_extracted_k141_12719 | 1101 | No | NA | 2 | 0 | 1 | Not-determined | Genome-fragment | NA | NA | 0 |
| S12_extracted_k141_178293 | 2654 | No | NA | 4 | 0 | 4 | Not-determined | Genome-fragment | NA | NA | 0 |
| S12_extracted_k141_115513 | 1639 | No | NA | 2 | 0 | 1 | Not-determined | Genome-fragment | NA | NA | 0 |
| S12_extracted_k141_170744 | 1555 | No | NA | 1 | 0 | 0 | Not-determined | Genome-fragment | NA | NA | 0 |
| S12_extracted_k141_92779 | 1428 | No | NA | 2 | 0 | 0 | Not-determined | Genome-fragment | NA | NA | 0 |
| S1_extracted_k141_5274 | 3874 | No | NA | 8 | 7 | 0 | Low-quality | Genome-fragment | 8.84 | AAI-based (high-confidence) | 0 |
| S12_extracted_k141_92782 | 1898 | No | NA | 1 | 0 | 0 | Not-determined | Genome-fragment | NA | NA | 0 |
| S12_extracted_k141_100959 | 34914 | No | NA | 34 | 0 | 5 | Not-determined | Genome-fragment | NA | NA | 0 |
| S12_extracted_k141_68585 | 1868 | No | NA | 3 | 0 | 2 | Not-determined | Genome-fragment | NA | NA | 0 |
| S12_extracted_k141_84033 | 1028 | No | NA | 2 | 0 | 1 | Not-determined | Genome-fragment | NA | NA | 0 |
| S12_extracted_k141_139853 | 1128 | No | NA | 1 | 0 | 0 | Not-determined | Genome-fragment | NA | NA | 0 |
| S12_extracted_k141_12737 | 5060 | No | NA | 2 | 0 | 1 | Not-determined | Genome-fragment | NA | NA | 0 |
| S12_extracted_k141_123219 | 3344 | No | NA | 4 | 0 | 2 | Not-determined | Genome-fragment | NA | NA | 0 |
| S12_extracted_k141_84051 | 1658 | No | NA | 2 | 0 | 0 | Not-determined | Genome-fragment | NA | NA | 0 |
| S12_extracted_k141_92837 | 3651 | No | NA | 2 | 0 | 2 | Not-determined | Genome-fragment | NA | NA | 0 |
| S14_extracted_k141_111095 | 3875 | No | NA | 10 | 9 | 0 | Low-quality | Genome-fragment | 12.73 | AAI-based (high-confidence) | 0 |
| S12_extracted_k141_147507 | 1050 | No | NA | 3 | 0 | 0 | Not-determined | Genome-fragment | NA | NA | 0 |
| S12_extracted_k141_21296 | 1385 | No | NA | 1 | 0 | 1 | Not-determined | Genome-fragment | NA | NA | 0 |
| S12_extracted_k141_29196 | 3709 | No | NA | 5 | 0 | 4 | Not-determined | Genome-fragment | NA | NA | 0 |
| S12_extracted_k141_92852 | 1114 | No | NA | 2 | 0 | 2 | Not-determined | Genome-fragment | NA | NA | 0 |
| S12_extracted_k141_29201 | 4875 | No | NA | 3 | 0 | 0 | Not-determined | Genome-fragment | NA | NA | 0 |
| S12_extracted_k141_92858 | 3696 | No | NA | 4 | 0 | 1 | Not-determined | Genome-fragment | NA | NA | 0 |
| S7_extracted_32294 | 3876 | No | NA | 3 | 1 | 0 | Low-quality | Genome-fragment | 3.04 | HMM-based (lower-bound) | 0 |
| S12_extracted_k141_101048 | 5393 | No | NA | 4 | 0 | 2 | Not-determined | Genome-fragment | NA | NA | 0 |
| S12_extracted_k141_178367 | 1024 | No | NA | 1 | 0 | 1 | Not-determined | Genome-fragment | NA | NA | 0 |
| S12_extracted_k141_6155 | 3292 | No | NA | 2 | 0 | 0 | Not-determined | Genome-fragment | NA | NA | 0 |
| S12_extracted_k141_29224 | 1026 | No | NA | 2 | 0 | 1 | Not-determined | Genome-fragment | NA | NA | 0 |
| S12_extracted_k141_36536 | 5964 | No | NA | 14 | 0 | 1 | Not-determined | Genome-fragment | NA | NA | 0 |
| S12_extracted_k141_131284 | 1162 | No | NA | 1 | 0 | 1 | Not-determined | Genome-fragment | NA | NA | 0 |
| S12_extracted_k141_163008 | 1079 | No | NA | 1 | 0 | 1 | Not-determined | Genome-fragment | NA | NA | 0 |
| S12_extracted_k141_21339 | 1063 | No | NA | 2 | 0 | 2 | Not-determined | Genome-fragment | NA | NA | 0 |
| S12_extracted_k141_60909 | 1910 | No | NA | 2 | 0 | 0 | Not-determined | Genome-fragment | NA | NA | 0 |
| S12_extracted_k141_187333 | 10188 | No | NA | 5 | 0 | 1 | Not-determined | Genome-fragment | NA | NA | 0 |
| S12_extracted_k141_147552 | 3304 | No | NA | 5 | 0 | 5 | Not-determined | Genome-fragment | NA | NA | 0 |
| S12_extracted_k141_101088 | 1683 | No | NA | 1 | 0 | 0 | Not-determined | Genome-fragment | NA | NA | 0 |
| S12_extracted_k141_178396 | 1018 | No | NA | 1 | 0 | 1 | Not-determined | Genome-fragment | NA | NA | 0 |
| S12_extracted_k141_147564 | 1042 | No | NA | 2 | 0 | 1 | Not-determined | Genome-fragment | NA | NA | 0 |
| S12_extracted_k141_108511 | 3450 | No | NA | 6 | 0 | 1 | Not-determined | Genome-fragment | NA | NA | 0 |
| S12_extracted_k141_29270 | 1079 | No | NA | 1 | 0 | 0 | Not-determined | Genome-fragment | NA | NA | 0 |
| S12_extracted_k141_6194 | 3107 | No | NA | 2 | 0 | 0 | Not-determined | Genome-fragment | NA | NA | 0 |
| S12_extracted_k141_36587 | 1329 | No | NA | 2 | 0 | 0 | Not-determined | Genome-fragment | NA | NA | 0 |
| S12_extracted_k141_147589 | 1491 | No | NA | 1 | 0 | 0 | Not-determined | Genome-fragment | NA | NA | 0 |
| S12_extracted_k141_155072 | 1188 | No | NA | 1 | 0 | 0 | Not-determined | Genome-fragment | NA | NA | 0 |
| S12_extracted_k141_123365 | 5424 | No | NA | 2 | 0 | 2 | Not-determined | Genome-fragment | NA | NA | 0 |
| S12_extracted_k141_6226 | 1283 | No | NA | 1 | 0 | 1 | Not-determined | Genome-fragment | NA | NA | 0 |
| S12_extracted_k141_139993 | 1830 | No | NA | 2 | 0 | 2 | Not-determined | Genome-fragment | NA | NA | 0 |
| S12_extracted_k141_131336 | 9058 | No | NA | 8 | 0 | 4 | Not-determined | Genome-fragment | NA | NA | 0 |
| S12_extracted_k141_155074 | 5835 | No | NA | 6 | 0 | 4 | Not-determined | Genome-fragment | NA | NA | 0 |
| S17_extracted_k141_69254 | 3877 | No | NA | 2 | 0 | 0 | Low-quality | Genome-fragment | 3.92 | AAI-based (high-confidence) | 0 |
| S12_extracted_k141_178430 | 1156 | No | NA | 2 | 0 | 2 | Not-determined | Genome-fragment | NA | NA | 0 |
| S12_extracted_k141_84144 | 4198 | No | NA | 3 | 0 | 0 | Not-determined | Genome-fragment | NA | NA | 0 |
| S12_extracted_k141_68684 | 1199 | No | NA | 3 | 0 | 0 | Not-determined | Genome-fragment | NA | NA | 0 |
| S5_extracted_k141_101052 | 3877 | No | NA | 7 | 0 | 0 | Low-quality | Genome-fragment | 2.28 | AAI-based (medium-confidence) | 0 |
| S16_extracted_57494 | 3877 | No | NA | 4 | 0 | 3 | Low-quality | Genome-fragment | 1.68 | AAI-based (medium-confidence) | 0 |
| S1_vs_k141_22143\|\|full | 3878 | No | NA | 8 | 1 | 0 | Low-quality | Genome-fragment | 5.51 | HMM-based (lower-bound) | 0 |
| S12_extracted_k141_45899 | 1973 | No | NA | 4 | 0 | 0 | Not-determined | Genome-fragment | NA | NA | 0 |
| S12_extracted_k141_155105 | 1145 | No | NA | 2 | 0 | 1 | Not-determined | Genome-fragment | NA | NA | 0 |
| S10_vs_51649\|\|full | 3879 | No | NA | 6 | 0 | 1 | Low-quality | Genome-fragment | 8.25 | AAI-based (medium-confidence) | 0 |
| S12_extracted_k141_123411 | 1195 | No | NA | 2 | 0 | 0 | Not-determined | Genome-fragment | NA | NA | 0 |
| S17_extracted_k141_111237 | 3880 | Yes | 2909 | 3 | 1 | 1 | Low-quality | Genome-fragment | 3.55 | AAI-based (high-confidence) | 25.03 |
| S12_extracted_k141_84195 | 1236 | No | NA | 2 | 0 | 0 | Not-determined | Genome-fragment | NA | NA | 0 |
| S12_extracted_k141_140045 | 1140 | No | NA | 1 | 0 | 0 | Not-determined | Genome-fragment | NA | NA | 0 |
| S12_extracted_k141_108596 | 1307 | No | NA | 2 | 0 | 1 | Not-determined | Genome-fragment | NA | NA | 0 |
| S11_extracted_k141_38346 | 3880 | No | NA | 6 | 0 | 1 | Low-quality | Genome-fragment | 7.56 | AAI-based (medium-confidence) | 0 |
| S12_extracted_k141_108612 | 1278 | No | NA | 2 | 0 | 1 | Not-determined | Genome-fragment | NA | NA | 0 |
| S12_extracted_k141_6269 | 1140 | No | NA | 1 | 0 | 0 | Not-determined | Genome-fragment | NA | NA | 0 |
| S12_extracted_k141_163119 | 1627 | No | NA | 4 | 0 | 0 | Not-determined | Genome-fragment | NA | NA | 0 |
| S12_extracted_k141_36652 | 5424 | No | NA | 6 | 0 | 1 | Not-determined | Genome-fragment | NA | NA | 0 |
| S12_extracted_k141_84235 | 1135 | No | NA | 1 | 0 | 1 | Not-determined | Genome-fragment | NA | NA | 0 |
| S12_extracted_k141_76495 | 1127 | No | NA | 1 | 0 | 1 | Not-determined | Genome-fragment | NA | NA | 0 |
| S12_extracted_k141_101195 | 1018 | No | NA | 3 | 0 | 0 | Not-determined | Genome-fragment | NA | NA | 0 |
| S12_extracted_k141_178512 | 1043 | No | NA | 2 | 0 | 1 | Not-determined | Genome-fragment | NA | NA | 0 |
| S12_extracted_k141_178521 | 1264 | No | NA | 1 | 0 | 1 | Not-determined | Genome-fragment | NA | NA | 0 |
| S12_extracted_k141_147652 | 1463 | No | NA | 2 | 0 | 1 | Not-determined | Genome-fragment | NA | NA | 0 |
| S11_vs_k141_47286\|\|full | 3881 | No | NA | 7 | 0 | 0 | Low-quality | Genome-fragment | 5.5 | AAI-based (high-confidence) | 0 |
| S12_extracted_k141_178524 | 1339 | No | NA | 2 | 0 | 2 | Not-determined | Genome-fragment | NA | NA | 0 |
| S6_extracted_k141_25312 | 3881 | No | NA | 3 | 0 | 1 | Low-quality | Genome-fragment | 4.01 | AAI-based (medium-confidence) | 0 |
| S12_extracted_k141_178529 | 1001 | No | NA | 1 | 0 | 0 | Not-determined | Genome-fragment | NA | NA | 0 |
| S12_extracted_k141_68765 | 3941 | No | NA | 5 | 0 | 5 | Not-determined | Genome-fragment | NA | NA | 0 |
| S12_extracted_k141_21438 | 2596 | No | NA | 4 | 0 | 1 | Not-determined | Genome-fragment | NA | NA | 0 |
| S12_extracted_k141_36685 | 2202 | No | NA | 5 | 0 | 1 | Not-determined | Genome-fragment | NA | NA | 0 |
| S13_vs_51473\|\|full | 3882 | No | NA | 3 | 0 | 1 | Low-quality | Genome-fragment | 1.82 | AAI-based (medium-confidence) | 0 |
| S5_vs_k141_8684\|\|full | 3882 | Yes | 2372 | 7 | 1 | 1 | Low-quality | Genome-fragment | 3.19 | HMM-based (lower-bound) | 38.9 |
| S12_extracted_k141_115636 | 1183 | No | NA | 2 | 0 | 2 | Not-determined | Genome-fragment | NA | NA | 0 |
| S12_extracted_k141_115644 | 1294 | No | NA | 1 | 0 | 1 | Not-determined | Genome-fragment | NA | NA | 0 |
| S12_extracted_k141_155170 | 4172 | No | NA | 3 | 0 | 3 | Not-determined | Genome-fragment | NA | NA | 0 |
| S12_extracted_k141_101243 | 1325 | No | NA | 1 | 0 | 0 | Not-determined | Genome-fragment | NA | NA | 0 |
| S12_extracted_k141_61068 | 1200 | No | NA | 2 | 0 | 1 | Not-determined | Genome-fragment | NA | NA | 0 |
| S12_extracted_k141_115650 | 1527 | No | NA | 1 | 0 | 0 | Not-determined | Genome-fragment | NA | NA | 0 |
| S12_extracted_k141_155175 | 6407 | No | NA | 5 | 0 | 1 | Not-determined | Genome-fragment | NA | NA | 0 |
| S12_extracted_k141_108667 | 1009 | No | NA | 2 | 0 | 0 | Not-determined | Genome-fragment | NA | NA | 0 |
| S12_extracted_k141_36711 | 1108 | No | NA | 2 | 0 | 1 | Not-determined | Genome-fragment | NA | NA | 0 |
| S12_extracted_k141_12895 | 1578 | No | NA | 2 | 0 | 0 | Not-determined | Genome-fragment | NA | NA | 0 |
| S12_extracted_k141_36715 | 1092 | No | NA | 2 | 0 | 1 | Not-determined | Genome-fragment | NA | NA | 0 |
| S12_extracted_k141_108674 | 5413 | No | NA | 4 | 0 | 1 | Not-determined | Genome-fragment | NA | NA | 0 |
| S12_extracted_k141_170988 | 1316 | No | NA | 2 | 0 | 2 | Not-determined | Genome-fragment | NA | NA | 0 |
| S7_extracted_40304 | 3884 | No | NA | 9 | 2 | 0 | Low-quality | Genome-fragment | 8.38 | AAI-based (high-confidence) | 0 |
| S12_extracted_k141_68788 | 25886 | No | NA | 19 | 0 | 17 | Not-determined | Genome-fragment | NA | NA | 0 |
| S12_extracted_k141_29446 | 1215 | No | NA | 3 | 0 | 1 | Not-determined | Genome-fragment | NA | NA | 0 |
| S12_extracted_k141_155194 | 5081 | No | NA | 1 | 0 | 0 | Not-determined | Genome-fragment | NA | NA | 0 |
| S12_extracted_k141_123542 | 1201 | No | NA | 1 | 0 | 0 | Not-determined | Genome-fragment | NA | NA | 0 |
| S12_extracted_k141_171014 | 1242 | No | NA | 3 | 0 | 0 | Not-determined | Genome-fragment | NA | NA | 0 |
| S12_extracted_k141_147719 | 1554 | No | NA | 1 | 0 | 0 | Not-determined | Genome-fragment | NA | NA | 0 |
| S12_extracted_k141_147723 | 1076 | No | NA | 2 | 0 | 2 | Not-determined | Genome-fragment | NA | NA | 0 |
| S8_vs_k141_55870\|\|full | 3885 | No | NA | 5 | 3 | 0 | Low-quality | Genome-fragment | 9.15 | AAI-based (medium-confidence) | 0 |
| S12_extracted_k141_84348 | 1376 | No | NA | 2 | 0 | 0 | Not-determined | Genome-fragment | NA | NA | 0 |
| S12_extracted_k141_76585 | 1001 | No | NA | 1 | 0 | 1 | Not-determined | Genome-fragment | NA | NA | 0 |
| S5_vs_k141_3802\|\|full | 3886 | No | NA | 4 | 3 | 0 | Low-quality | Genome-fragment | 9.56 | AAI-based (medium-confidence) | 0 |
| S12_extracted_k141_46088 | 10684 | No | NA | 14 | 0 | 1 | Not-determined | Genome-fragment | NA | NA | 0 |
| S12_extracted_k141_101353 | 1534 | No | NA | 2 | 0 | 1 | Not-determined | Genome-fragment | NA | NA | 0 |
| S12_extracted_k141_52848 | 1142 | No | NA | 1 | 0 | 1 | Not-determined | Genome-fragment | NA | NA | 0 |
| S12_extracted_k141_92893 | 1322 | No | NA | 1 | 0 | 1 | Not-determined | Genome-fragment | NA | NA | 0 |
| S5_vs_k141_28341\|\|full | 3886 | No | NA | 5 | 2 | 0 | Low-quality | Genome-fragment | 9.73 | AAI-based (medium-confidence) | 0 |
| S12_extracted_k141_101359 | 2160 | No | NA | 3 | 0 | 0 | Not-determined | Genome-fragment | NA | NA | 0 |
| S5_extracted_k141_106869 | 3887 | No | NA | 6 | 4 | 0 | Low-quality | Genome-fragment | 9.73 | AAI-based (high-confidence) | 0 |
| S12_extracted_k141_140235 | 1127 | No | NA | 3 | 0 | 1 | Not-determined | Genome-fragment | NA | NA | 0 |
| S5_extracted_k141_54478 | 3887 | No | NA | 9 | 5 | 0 | Low-quality | Genome-fragment | 9.74 | AAI-based (high-confidence) | 0 |
| S12_extracted_k141_21536 | 3161 | No | NA | 3 | 0 | 3 | Not-determined | Genome-fragment | NA | NA | 0 |
| S5_vs_k141_141830\|\|full | 3887 | No | NA | 4 | 0 | 3 | Low-quality | Genome-fragment | 1.96 | AAI-based (medium-confidence) | 0 |
| S12_extracted_k141_52867 | 1274 | No | NA | 2 | 0 | 0 | Not-determined | Genome-fragment | NA | NA | 0 |
| S12_extracted_k141_68846 | 1716 | No | NA | 2 | 0 | 2 | Not-determined | Genome-fragment | NA | NA | 0 |
| S12_extracted_k141_61138 | 1052 | No | NA | 2 | 0 | 2 | Not-determined | Genome-fragment | NA | NA | 0 |
| S12_extracted_k141_147781 | 3891 | No | NA | 5 | 0 | 3 | Not-determined | Genome-fragment | NA | NA | 0 |
| S12_extracted_k141_68859 | 3198 | No | NA | 2 | 0 | 2 | Not-determined | Genome-fragment | NA | NA | 0 |
| S8_vs_k141_74390\|\|full | 3887 | No | NA | 7 | 1 | 0 | Low-quality | Genome-fragment | 11.41 | AAI-based (high-confidence) | 0 |
| S12_extracted_k141_46145 | 3140 | No | NA | 3 | 0 | 1 | Not-determined | Genome-fragment | NA | NA | 0 |
| S12_extracted_k141_61158 | 1173 | No | NA | 2 | 0 | 1 | Not-determined | Genome-fragment | NA | NA | 0 |
| S12_extracted_k141_140291 | 1127 | No | NA | 1 | 0 | 1 | Not-determined | Genome-fragment | NA | NA | 0 |
| S12_extracted_k141_68878 | 1378 | No | NA | 2 | 0 | 1 | Not-determined | Genome-fragment | NA | NA | 0 |
| S12_extracted_k141_68882 | 1558 | No | NA | 2 | 0 | 2 | Not-determined | Genome-fragment | NA | NA | 0 |
| S7_vs_78243\|\|full | 3888 | No | NA | 4 | 0 | 4 | Low-quality | Genome-fragment | 6.1 | AAI-based (medium-confidence) | 0 |

| | | | | | | | | | | | |
|---|---|---|---|---|---|---|---|---|---|---|---|
| S12_extracted_k141_123604 | 2096 | No | NA | 2 | 0 | 1 | Not-determined | Genome-fragment | NA | NA | 0 |
| S12_extracted_k141_13037 | 1135 | No | NA | 2 | 0 | 0 | Not-determined | Genome-fragment | NA | NA | 0 |
| S3_vs_k141_53905\|\|full | 3889 | No | NA | 8 | 2 | 0 | Low-quality | Genome-fragment | 10.58 | AAI-based (high-confidence) | 0 |
| S12_extracted_k141_187579 | 2185 | No | NA | 1 | 0 | 0 | Not-determined | Genome-fragment | NA | NA | 0 |
| S12_extracted_k141_36866 | 4101 | No | NA | 3 | 0 | 3 | Not-determined | Genome-fragment | NA | NA | 0 |
| S12_extracted_k141_13055 | 1147 | No | NA | 2 | 0 | 1 | Not-determined | Genome-fragment | NA | NA | 0 |
| S16_vs_34766\|\|full | 3889 | No | NA | 3 | 0 | 2 | Low-quality | Genome-fragment | 2.04 | AAI-based (medium-confidence) | 0 |
| S12_extracted_k141_36877 | 1222 | No | NA | 1 | 0 | 0 | Not-determined | Genome-fragment | NA | NA | 0 |
| S12_extracted_k141_36881 | 1007 | No | NA | 1 | 0 | 1 | Not-determined | Genome-fragment | NA | NA | 0 |
| S12_extracted_k141_178682 | 1049 | No | NA | 1 | 0 | 1 | Not-determined | Genome-fragment | NA | NA | 0 |
| S12_extracted_k141_68904 | 1060 | No | NA | 2 | 0 | 1 | Not-determined | Genome-fragment | NA | NA | 0 |
| S12_extracted_k141_46173 | 1193 | No | NA | 1 | 0 | 0 | Not-determined | Genome-fragment | NA | NA | 0 |
| S12_extracted_k141_61178 | 16204 | No | NA | 31 | 0 | 30 | Not-determined | Genome-fragment | NA | NA | 0 |
| S16_vs_143328\|\|full | 3889 | No | NA | 3 | 0 | 2 | Low-quality | Genome-fragment | 2.52 | AAI-based (medium-confidence) | 0 |
| S12_extracted_k141_178705 | 1506 | No | NA | 2 | 0 | 2 | Not-determined | Genome-fragment | NA | NA | 0 |
| S12_extracted_k141_84436 | 1777 | No | NA | 2 | 0 | 0 | Not-determined | Genome-fragment | NA | NA | 0 |
| S12_extracted_k141_171138 | 2628 | No | NA | 2 | 0 | 2 | Not-determined | Genome-fragment | NA | NA | 0 |
| S12_extracted_k141_101495 | 4604 | No | NA | 4 | 0 | 4 | Not-determined | Genome-fragment | NA | NA | 0 |
| S12_extracted_k141_115833 | 1074 | No | NA | 1 | 0 | 1 | Not-determined | Genome-fragment | NA | NA | 0 |
| S11_extracted_k141_69553 | 3890 | No | NA | 6 | 1 | 0 | Low-quality | Genome-fragment | 1.34 | HMM-based (lower-bound) | 0 |
| S12_extracted_k141_13146 | 4823 | No | NA | 3 | 0 | 2 | Not-determined | Genome-fragment | NA | NA | 0 |
| S12_extracted_k141_21582 | 3047 | No | NA | 3 | 0 | 0 | Not-determined | Genome-fragment | NA | NA | 0 |
| S12_extracted_k141_115848 | 1514 | No | NA | 3 | 0 | 3 | Not-determined | Genome-fragment | NA | NA | 0 |
| S12_extracted_k141_163374 | 1053 | No | NA | 2 | 0 | 1 | Not-determined | Genome-fragment | NA | NA | 0 |
| S12_extracted_k141_76757 | 1464 | No | NA | 1 | 0 | 1 | Not-determined | Genome-fragment | NA | NA | 0 |
| S12_extracted_k141_76760 | 1417 | No | NA | 2 | 0 | 0 | Not-determined | Genome-fragment | NA | NA | 0 |
| S12_extracted_k141_13200 | 1571 | No | NA | 1 | 0 | 1 | Not-determined | Genome-fragment | NA | NA | 0 |
| S12_extracted_k141_163405 | 4017 | No | NA | 3 | 0 | 3 | Not-determined | Genome-fragment | NA | NA | 0 |
| S13_extracted_84830 | 3890 | No | NA | 3 | 0 | 1 | Low-quality | Genome-fragment | 6.41 | AAI-based (medium-confidence) | 0 |
| S12_extracted_k141_53001 | 2285 | No | NA | 2 | 0 | 0 | Not-determined | Genome-fragment | NA | NA | 0 |
| S12_extracted_k141_140444 | 5682 | No | NA | 3 | 0 | 0 | Not-determined | Genome-fragment | NA | NA | 0 |
| S12_extracted_k141_13225 | 1858 | No | NA | 1 | 0 | 1 | Not-determined | Genome-fragment | NA | NA | 0 |
| S12_extracted_k141_37010 | 2845 | No | NA | 2 | 0 | 2 | Not-determined | Genome-fragment | NA | NA | 0 |
| S12_extracted_k141_147924 | 1070 | No | NA | 1 | 0 | 1 | Not-determined | Genome-fragment | NA | NA | 0 |
| S12_extracted_k141_69025 | 1728 | No | NA | 1 | 0 | 0 | Not-determined | Genome-fragment | NA | NA | 0 |
| S12_extracted_k141_131732 | 1331 | No | NA | 2 | 0 | 1 | Not-determined | Genome-fragment | NA | NA | 0 |
| S12_extracted_k141_76781 | 1088 | No | NA | 3 | 0 | 1 | Not-determined | Genome-fragment | NA | NA | 0 |
| S12_extracted_k141_76787 | 1025 | No | NA | 1 | 0 | 0 | Not-determined | Genome-fragment | NA | NA | 0 |
| S12_extracted_k141_29652 | 2694 | No | NA | 3 | 0 | 0 | Not-determined | Genome-fragment | NA | NA | 0 |
| S14_vs_k141_97846\|\|full | 3890 | No | NA | 6 | 0 | 0 | Low-quality | Genome-fragment | 2.64 | AAI-based (high-confidence) | 0 |
| S12_extracted_k141_131748 | 1097 | No | NA | 2 | 0 | 1 | Not-determined | Genome-fragment | NA | NA | 0 |
| S12_extracted_k141_53039 | 1024 | No | NA | 2 | 0 | 0 | Not-determined | Genome-fragment | NA | NA | 0 |
| S12_extracted_k141_140475 | 1778 | No | NA | 1 | 0 | 1 | Not-determined | Genome-fragment | NA | NA | 0 |
| S12_extracted_k141_29664 | 1877 | No | NA | 4 | 0 | 0 | Not-determined | Genome-fragment | NA | NA | 0 |
| S12_extracted_k141_6528 | 1040 | No | NA | 2 | 0 | 2 | Not-determined | Genome-fragment | NA | NA | 0 |
| S7_extracted_91298 | 3891 | No | NA | 12 | 0 | 0 | Low-quality | Genome-fragment | 9.09 | AAI-based (high-confidence) | 0 |
| S12_extracted_k141_61299 | 16337 | No | NA | 24 | 6 | 0 | Medium-quality | Genome-fragment | 69.2 | AAI-based (high-confidence) | 0 |
| S12_extracted_k141_115915 | 2618 | No | NA | 3 | 0 | 0 | Not-determined | Genome-fragment | NA | NA | 0 |
| S12_extracted_k141_140486 | 2340 | No | NA | 3 | 0 | 3 | Not-determined | Genome-fragment | NA | NA | 0 |
| S12_extracted_k141_13270 | 1246 | No | NA | 1 | 0 | 1 | Not-determined | Genome-fragment | NA | NA | 0 |
| S12_extracted_k141_108917 | 1001 | No | NA | 1 | 0 | 0 | Not-determined | Genome-fragment | NA | NA | 0 |
| S12_extracted_k141_131772 | 1185 | No | NA | 2 | 0 | 1 | Not-determined | Genome-fragment | NA | NA | 0 |
| S12_extracted_k141_61325 | 3163 | No | NA | 2 | 0 | 2 | Not-determined | Genome-fragment | NA | NA | 0 |
| S17_vs_k141_844\|\|full | 3892 | No | NA | 8 | 2 | 0 | Low-quality | Genome-fragment | 9.51 | AAI-based (high-confidence) | 0 |
| S7_vs_30678\|\|full | 3892 | No | NA | 5 | 3 | 0 | Low-quality | Genome-fragment | 11.37 | AAI-based (high-confidence) | 0 |
| S12_extracted_k141_61331 | 1202 | No | NA | 2 | 0 | 0 | Not-determined | Genome-fragment | NA | NA | 0 |
| S12_extracted_k141_37082 | 3586 | No | NA | 3 | 0 | 1 | Not-determined | Genome-fragment | NA | NA | 0 |
| S12_extracted_k141_6558 | 1099 | No | NA | 2 | 0 | 2 | Not-determined | Genome-fragment | NA | NA | 0 |
| S12_extracted_k141_76840 | 1229 | No | NA | 1 | 0 | 0 | Not-determined | Genome-fragment | NA | NA | 0 |
| S12_extracted_k141_76859 | 1085 | No | NA | 2 | 0 | 2 | Not-determined | Genome-fragment | NA | NA | 0 |
| S12_extracted_k141_163473 | 2585 | No | NA | 1 | 0 | 1 | Not-determined | Genome-fragment | NA | NA | 0 |
| S12_extracted_k141_53102 | 1142 | No | NA | 2 | 0 | 1 | Not-determined | Genome-fragment | NA | NA | 0 |
| S12_extracted_k141_61369 | 9076 | No | NA | 6 | 0 | 4 | Not-determined | Genome-fragment | NA | NA | 0 |
| S12_extracted_k141_53111 | 1269 | No | NA | 1 | 0 | 0 | Not-determined | Genome-fragment | NA | NA | 0 |
| S12_extracted_k141_46327 | 4125 | No | NA | 6 | 0 | 3 | Not-determined | Genome-fragment | NA | NA | 0 |
| S6_vs_k141_37696\|\|full | 3892 | No | NA | 3 | 0 | 3 | Low-quality | Genome-fragment | 2.52 | AAI-based (medium-confidence) | 0 |
| S12_extracted_k141_93253 | 1098 | No | NA | 1 | 0 | 1 | Not-determined | Genome-fragment | NA | NA | 0 |
| S12_extracted_k141_187842 | 1752 | No | NA | 2 | 0 | 0 | Not-determined | Genome-fragment | NA | NA | 0 |
| S12_extracted_k141_109014 | 3202 | No | NA | 1 | 0 | 0 | Not-determined | Genome-fragment | NA | NA | 0 |
| S14_vs_k141_89000\|\|full | 3895 | No | NA | 8 | 2 | 0 | Low-quality | Genome-fragment | 7.26 | AAI-based (high-confidence) | 0 |
| S12_extracted_k141_37132 | 1190 | No | NA | 2 | 0 | 1 | Not-determined | Genome-fragment | NA | NA | 0 |
| S12_extracted_k141_101688 | 2353 | No | NA | 3 | 0 | 2 | Not-determined | Genome-fragment | NA | NA | 0 |
| S12_extracted_k141_76908 | 1227 | No | NA | 1 | 0 | 1 | Not-determined | Genome-fragment | NA | NA | 0 |
| S12_extracted_k141_76912 | 2604 | No | NA | 1 | 0 | 1 | Not-determined | Genome-fragment | NA | NA | 0 |
| S12_extracted_k141_46356 | 7618 | No | NA | 7 | 0 | 5 | Not-determined | Genome-fragment | NA | NA | 0 |
| S7_vs_30429\|\|full | 3895 | No | NA | 3 | 0 | 2 | Low-quality | Genome-fragment | 4.14 | AAI-based (medium-confidence) | 0 |
| S7_vs_76693\|\|full | 3895 | No | NA | 5 | 0 | 0 | Low-quality | Genome-fragment | 2.65 | AAI-based (high-confidence) | 0 |
| S12_extracted_k141_6624 | 1910 | No | NA | 2 | 0 | 0 | Not-determined | Genome-fragment | NA | NA | 0 |
| S12_extracted_k141_109045 | 1243 | No | NA | 2 | 0 | 1 | Not-determined | Genome-fragment | NA | NA | 0 |
| S7_vs_80101\|\|full | 3897 | Yes | 2598 | 10 | 5 | 1 | Low-quality | Genome-fragment | 3.99 | AAI-based (high-confidence) | 33.33 |
| S12_extracted_k141_131897 | 1309 | No | NA | 1 | 0 | 0 | Not-determined | Genome-fragment | NA | NA | 0 |
| S16_extracted_30313 | 3897 | No | NA | 6 | 1 | 0 | Low-quality | Genome-fragment | 7.5 | AAI-based (medium-confidence) | 0 |
| S12_extracted_k141_69136 | 32310 | No | NA | 36 | 8 | 0 | Medium-quality | Genome-fragment | 63.65 | AAI-based (high-confidence) | 0 |
| S12_extracted_k141_6639 | 1062 | No | NA | 1 | 0 | 1 | Not-determined | Genome-fragment | NA | NA | 0 |
| S7_extracted_112582 | 3898 | No | NA | 7 | 3 | 0 | Low-quality | Genome-fragment | 9.09 | AAI-based (high-confidence) | 0 |
| S12_extracted_k141_171357 | 1195 | No | NA | 2 | 0 | 1 | Not-determined | Genome-fragment | NA | NA | 0 |
| S12_extracted_k141_13366 | 1127 | No | NA | 1 | 0 | 1 | Not-determined | Genome-fragment | NA | NA | 0 |
| S12_extracted_k141_101751 | 1398 | No | NA | 2 | 0 | 0 | Not-determined | Genome-fragment | NA | NA | 0 |
| S12_extracted_k141_76991 | 1123 | No | NA | 1 | 0 | 1 | Not-determined | Genome-fragment | NA | NA | 0 |
| S7_extracted_128697 | 3898 | No | NA | 4 | 3 | 0 | Low-quality | Genome-fragment | 10.66 | AAI-based (medium-confidence) | 0 |
| S12_extracted_k141_123821 | 1689 | No | NA | 2 | 0 | 2 | Not-determined | Genome-fragment | NA | NA | 0 |
| S12_extracted_k141_85472 | 3898 | No | NA | 7 | 0 | 0 | Low-quality | Genome-fragment | 10.2 | AAI-based (high-confidence) | 0 |
| S12_extracted_k141_140600 | 1519 | No | NA | 2 | 0 | 0 | Not-determined | Genome-fragment | NA | NA | 0 |
| S12_extracted_k141_61511 | 1865 | No | NA | 2 | 0 | 1 | Not-determined | Genome-fragment | NA | NA | 0 |
| S12_extracted_k141_77020 | 1008 | No | NA | 2 | 0 | 0 | Not-determined | Genome-fragment | NA | NA | 0 |
| S12_extracted_k141_116072 | 1251 | No | NA | 2 | 0 | 0 | Not-determined | Genome-fragment | NA | NA | 0 |
| S12_extracted_k141_93337 | 1609 | No | NA | 2 | 0 | 0 | Not-determined | Genome-fragment | NA | NA | 0 |
| S12_extracted_k141_29881 | 1338 | No | NA | 1 | 0 | 0 | Not-determined | Genome-fragment | NA | NA | 0 |
| S12_extracted_k141_163652 | 3365 | No | NA | 4 | 0 | 1 | Not-determined | Genome-fragment | NA | NA | 0 |
| S12_extracted_k141_13423 | 1486 | No | NA | 2 | 0 | 2 | Not-determined | Genome-fragment | NA | NA | 0 |
| S12_extracted_k141_140609 | 16931 | No | NA | 25 | 0 | 4 | Not-determined | Genome-fragment | NA | NA | 0 |
| S12_extracted_k141_61542 | 1060 | No | NA | 3 | 0 | 0 | Not-determined | Genome-fragment | NA | NA | 0 |
| S12_extracted_k141_6708 | 1423 | No | NA | 2 | 0 | 1 | Not-determined | Genome-fragment | NA | NA | 0 |
| S12_extracted_k141_13438 | 1862 | No | NA | 3 | 0 | 0 | Not-determined | Genome-fragment | NA | NA | 0 |
| S12_extracted_k141_163674 | 4776 | No | NA | 3 | 0 | 3 | Not-determined | Genome-fragment | NA | NA | 0 |
| S12_extracted_k141_163678 | 1897 | No | NA | 2 | 0 | 2 | Not-determined | Genome-fragment | NA | NA | 0 |
| S12_extracted_k141_101788 | 5090 | No | NA | 5 | 0 | 2 | Not-determined | Genome-fragment | NA | NA | 0 |
| S12_extracted_k141_37270 | 3010 | No | NA | 2 | 0 | 2 | Not-determined | Genome-fragment | NA | NA | 0 |
| S12_extracted_k141_155553 | 1000 | No | NA | 3 | 0 | 0 | Not-determined | Genome-fragment | NA | NA | 0 |
| S12_extracted_k141_61582 | 4054 | No | NA | 3 | 0 | 3 | Not-determined | Genome-fragment | NA | NA | 0 |
| S12_extracted_k141_84728 | 1125 | No | NA | 1 | 0 | 0 | Not-determined | Genome-fragment | NA | NA | 0 |
| S12_extracted_k141_21724 | 2211 | No | NA | 2 | 0 | 1 | Not-determined | Genome-fragment | NA | NA | 0 |
| S12_extracted_k141_155571 | 1684 | No | NA | 2 | 0 | 2 | Not-determined | Genome-fragment | NA | NA | 0 |
| S12_extracted_k141_178929 | 3570 | No | NA | 3 | 0 | 2 | Not-determined | Genome-fragment | NA | NA | 0 |
| S12_extracted_k141_37272 | 1954 | No | NA | 1 | 0 | 1 | Not-determined | Genome-fragment | NA | NA | 0 |
| S12_extracted_k141_29946 | 1122 | No | NA | 2 | 0 | 0 | Not-determined | Genome-fragment | NA | NA | 0 |
| S12_extracted_k141_187941 | 56466 | No | NA | 87 | 10 | 1 | Medium-quality | Genome-fragment | 74.11 | AAI-based (medium-confidence) | 0 |
| S12_extracted_k141_69265 | 1570 | No | NA | 3 | 0 | 0 | Not-determined | Genome-fragment | NA | NA | 0 |
| S12_extracted_k141_187950 | 3155 | No | NA | 3 | 0 | 2 | Not-determined | Genome-fragment | NA | NA | 0 |
| S14_extracted_k141_107071 | 3901 | No | NA | 6 | 0 | 1 | Low-quality | Genome-fragment | 5.1 | AAI-based (high-confidence) | 0 |
| S12_extracted_k141_61616 | 3064 | No | NA | 3 | 0 | 2 | Not-determined | Genome-fragment | NA | NA | 0 |
| S12_extracted_k141_123926 | 1019 | No | NA | 1 | 0 | 0 | Not-determined | Genome-fragment | NA | NA | 0 |
| S16_vs_70753\|\|full | 3901 | No | NA | 5 | 0 | 3 | Low-quality | Genome-fragment | 2.61 | AAI-based (medium-confidence) | 0 |
| S12_extracted_k141_155617 | 1518 | No | NA | 2 | 0 | 2 | Not-determined | Genome-fragment | NA | NA | 0 |
| S12_extracted_k141_46510 | 5517 | No | NA | 4 | 0 | 3 | Not-determined | Genome-fragment | NA | NA | 0 |
| S12_extracted_k141_69302 | 4575 | No | NA | 3 | 0 | 1 | Not-determined | Genome-fragment | NA | NA | 0 |
| S12_extracted_k141_140717 | 1413 | No | NA | 1 | 0 | 0 | Not-determined | Genome-fragment | NA | NA | 0 |
| S1_vs_k141_130754\|\|full | 3902 | No | NA | 3 | 1 | 0 | Low-quality | Genome-fragment | 4.07 | HMM-based (lower-bound) | 0 |
| S16_vs_2645\|\|full | 3905 | No | NA | 3 | 0 | 0 | Low-quality | Genome-fragment | 7.18 | AAI-based (medium-confidence) | 0 |
| S12_extracted_k141_53382 | 1023 | No | NA | 2 | 0 | 1 | Not-determined | Genome-fragment | NA | NA | 0 |
| S10_extracted_85384 | 3906 | No | NA | 8 | 4 | 0 | Low-quality | Genome-fragment | 9.18 | AAI-based (medium-confidence) | 0 |
| S12_extracted_k141_30030 | 1242 | No | NA | 2 | 0 | 1 | Not-determined | Genome-fragment | NA | NA | 0 |
| S12_extracted_k141_132053 | 1244 | No | NA | 2 | 0 | 0 | Not-determined | Genome-fragment | NA | NA | 0 |
| S6_extracted_k141_22671 | 3906 | No | NA | 3 | 0 | 1 | Low-quality | Genome-fragment | 1.58 | AAI-based (medium-confidence) | 0 |
| S8_extracted_k141_87789 | 3906 | No | NA | 6 | 0 | 1 | Low-quality | Genome-fragment | 4.17 | AAI-based (high-confidence) | 0 |
| S12_extracted_k141_101862 | 1057 | No | NA | 1 | 0 | 0 | Not-determined | Genome-fragment | NA | NA | 0 |
| S8_vs_k141_175483\|\|full | 3906 | No | NA | 5 | 0 | 4 | Low-quality | Genome-fragment | 2.93 | AAI-based (medium-confidence) | 0 |
| S17_extracted_k141_105369 | 3908 | No | NA | 2 | 0 | 1 | Low-quality | Genome-fragment | 1.28 | AAI-based (high-confidence) | 0 |
| S12_extracted_k141_163795 | 2792 | No | NA | 5 | 0 | 5 | Not-determined | Genome-fragment | NA | NA | 0 |
| S12_extracted_k141_69366 | 1077 | No | NA | 2 | 0 | 1 | Not-determined | Genome-fragment | NA | NA | 0 |
| S12_extracted_k141_179029 | 1302 | No | NA | 2 | 0 | 1 | Not-determined | Genome-fragment | NA | NA | 0 |
| S12_extracted_k141_30108 | 1143 | No | NA | 2 | 0 | 1 | Not-determined | Genome-fragment | NA | NA | 0 |
| S12_extracted_k141_132104 | 1395 | No | NA | 1 | 0 | 1 | Not-determined | Genome-fragment | NA | NA | 0 |
| S3_vs_k141_46826\|\|full | 3909 | No | NA | 12 | 0 | 0 | Low-quality | Genome-fragment | 2.49 | AAI-based (high-confidence) | 0 |
| S5_vs_k141_15288\|\|lt2gene | 3909 | No | NA | 1 | 1 | 0 | Low-quality | Genome-fragment | 10.28 | AAI-based (high-confidence) | 0 |
| S12_extracted_k141_77223 | 3175 | No | NA | 3 | 0 | 1 | Not-determined | Genome-fragment | NA | NA | 0 |
| S12_extracted_k141_21856 | 1287 | No | NA | 1 | 0 | 1 | Not-determined | Genome-fragment | NA | NA | 0 |
| S12_extracted_k141_21857 | 1429 | No | NA | 1 | 0 | 1 | Not-determined | Genome-fragment | NA | NA | 0 |
| S12_extracted_k141_13635 | 2506 | No | NA | 3 | 0 | 2 | Not-determined | Genome-fragment | NA | NA | 0 |
| S16_extracted_52692 | 3909 | No | NA | 5 | 4 | 0 | Low-quality | Genome-fragment | 10.77 | AAI-based (high-confidence) | 0 |
| S12_extracted_k141_116266 | 1450 | No | NA | 1 | 0 | 0 | Not-determined | Genome-fragment | NA | NA | 0 |
| S10_extracted_85569 | 3909 | No | NA | 7 | 5 | 0 | Low-quality | Genome-fragment | 7.44 | AAI-based (medium-confidence) | 0 |
| S12_extracted_k141_69415 | 1340 | No | NA | 2 | 0 | 2 | Not-determined | Genome-fragment | NA | NA | 0 |
| S12_extracted_k141_46644 | 1329 | No | NA | 1 | 0 | 0 | Not-determined | Genome-fragment | NA | NA | 0 |
| S12_extracted_k141_6874 | 1285 | No | NA | 1 | 0 | 0 | Not-determined | Genome-fragment | NA | NA | 0 |
| S12_extracted_k141_140817 | 3127 | No | NA | 1 | 0 | 1 | Not-determined | Genome-fragment | NA | NA | 0 |
| S13_vs_144433\|\|full | 3910 | No | NA | 4 | 0 | 3 | Low-quality | Genome-fragment | 1.42 | AAI-based (medium-confidence) | 0 |
| S12_extracted_k141_61689 | 1601 | No | NA | 2 | 0 | 2 | Not-determined | Genome-fragment | NA | NA | 0 |
| S12_extracted_k141_84911 | 1125 | No | NA | 1 | 0 | 1 | Not-determined | Genome-fragment | NA | NA | 0 |
| S16_vs_166208\|\|full | 3910 | No | NA | 6 | 0 | 0 | Low-quality | Genome-fragment | 5.44 | AAI-based (medium-confidence) | 0 |
| S12_extracted_k141_30198 | 1655 | No | NA | 3 | 0 | 2 | Not-determined | Genome-fragment | NA | NA | 0 |
| S12_extracted_k141_6901 | 1299 | No | NA | 2 | 0 | 0 | Not-determined | Genome-fragment | NA | NA | 0 |
| S12_extracted_k141_148353 | 1234 | No | NA | 2 | 0 | 1 | Not-determined | Genome-fragment | NA | NA | 0 |
| S12_extracted_k141_30235 | 2985 | No | NA | 3 | 0 | 0 | Not-determined | Genome-fragment | NA | NA | 0 |
| S12_extracted_k141_93586 | 1925 | No | NA | 2 | 0 | 2 | Not-determined | Genome-fragment | NA | NA | 0 |
| S8_vs_k141_107869\|\|full | 3910 | No | NA | 3 | 0 | 0 | Low-quality | Genome-fragment | 8.33 | AAI-based (high-confidence) | 0 |
| S11_vs_k141_75210\|\|full | 3911 | No | NA | 6 | 1 | 0 | Low-quality | Genome-fragment | 2.54 | AAI-based (high-confidence) | 0 |
| S12_extracted_k141_132229 | 1120 | No | NA | 2 | 0 | 1 | Not-determined | Genome-fragment | NA | NA | 0 |
| S12_extracted_k141_132230 | 1013 | No | NA | 1 | 0 | 0 | Not-determined | Genome-fragment | NA | NA | 0 |
| S6_vs_k141_48211\|\|full | 3911 | No | NA | 4 | 0 | 3 | Low-quality | Genome-fragment | 3.95 | AAI-based (medium-confidence) | 0 |
| S12_extracted_k141_124121 | 1874 | No | NA | 2 | 0 | 2 | Not-determined | Genome-fragment | NA | NA | 0 |
| S12_extracted_k141_6967 | 1578 | No | NA | 3 | 0 | 1 | Not-determined | Genome-fragment | NA | NA | 0 |
| S12_extracted_k141_109413 | 1194 | No | NA | 1 | 0 | 1 | Not-determined | Genome-fragment | NA | NA | 0 |
| S12_extracted_k141_53615 | 1211 | No | NA | 2 | 0 | 2 | Not-determined | Genome-fragment | NA | NA | 0 |
| S12_extracted_k141_171564 | 1133 | No | NA | 2 | 0 | 0 | Not-determined | Genome-fragment | NA | NA | 0 |
| S7_extracted_56484 | 3912 | No | NA | 3 | 0 | 1 | Low-quality | Genome-fragment | 5.68 | AAI-based (medium-confidence) | 0 |
| S12_extracted_k141_69581 | 1140 | No | NA | 1 | 0 | 1 | Not-determined | Genome-fragment | NA | NA | 0 |
| S12_extracted_k141_13724 | 1179 | No | NA | 2 | 0 | 0 | Not-determined | Genome-fragment | NA | NA | 0 |
| S12_extracted_k141_140910 | 3522 | No | NA | 2 | 0 | 1 | Not-determined | Genome-fragment | NA | NA | 0 |
| S12_extracted_k141_61774 | 1098 | No | NA | 1 | 0 | 1 | Not-determined | Genome-fragment | NA | NA | 0 |
| S13_extracted_131614 | 3913 | No | NA | 2 | 0 | 1 | Low-quality | Genome-fragment | 2.01 | AAI-based (medium-confidence) | 0 |
| S12_extracted_k141_171586 | 1181 | No | NA | 1 | 0 | 0 | Not-determined | Genome-fragment | NA | NA | 0 |
| S12_extracted_k141_188092 | 2206 | No | NA | 3 | 0 | 1 | Not-determined | Genome-fragment | NA | NA | 0 |
| S12_extracted_k141_21998 | 2259 | No | NA | 2 | 0 | 0 | Not-determined | Genome-fragment | NA | NA | 0 |
| S12_extracted_k141_61789 | 1693 | No | NA | 3 | 0 | 1 | Not-determined | Genome-fragment | NA | NA | 0 |
| S12_extracted_k141_140937 | 4942 | No | NA | 9 | 0 | 8 | Not-determined | Genome-fragment | NA | NA | 0 |
| S12_extracted_k141_85067 | 2521 | No | NA | 3 | 0 | 0 | Not-determined | Genome-fragment | NA | NA | 0 |
| S12_extracted_k141_93682 | 1338 | No | NA | 2 | 0 | 0 | Not-determined | Genome-fragment | NA | NA | 0 |
| S12_extracted_k141_53675 | 1074 | No | NA | 1 | 0 | 1 | Not-determined | Genome-fragment | NA | NA | 0 |
| S12_extracted_k141_171622 | 1074 | No | NA | 3 | 0 | 0 | Not-determined | Genome-fragment | NA | NA | 0 |
| S12_extracted_k141_85081 | 3125 | No | NA | 6 | 0 | 1 | Not-determined | Genome-fragment | NA | NA | 0 |
| S12_extracted_k141_53688 | 1313 | No | NA | 2 | 0 | 1 | Not-determined | Genome-fragment | NA | NA | 0 |
| S12_extracted_k141_77393 | 1014 | No | NA | 2 | 0 | 1 | Not-determined | Genome-fragment | NA | NA | 0 |
| S17_vs_k141_55841\|\|full | 3914 | No | NA | 4 | 0 | 0 | Low-quality | Genome-fragment | 6.4 | AAI-based (high-confidence) | 0 |
| S12_extracted_k141_30310 | 1073 | No | NA | 2 | 0 | 1 | Not-determined | Genome-fragment | NA | NA | 0 |
| S12_extracted_k141_93707 | 1241 | No | NA | 1 | 0 | 1 | Not-determined | Genome-fragment | NA | NA | 0 |
| S11_extracted_k141_71779 | 3914 | No | NA | 7 | 0 | 0 | Low-quality | Genome-fragment | 9.11 | AAI-based (medium-confidence) | 0 |
| S14_extracted_k141_93057 | 3914 | No | NA | 3 | 3 | 0 | Low-quality | Genome-fragment | 6.54 | AAI-based (high-confidence) | 0 |
| S12_extracted_k141_53713 | 1002 | No | NA | 2 | 0 | 0 | Not-determined | Genome-fragment | NA | NA | 0 |
| S12_extracted_k141_30325 | 1646 | No | NA | 4 | 0 | 2 | Not-determined | Genome-fragment | NA | NA | 0 |
| S12_vs_k141_147927\|\|full | 3916 | No | NA | 3 | 0 | 2 | Low-quality | Genome-fragment | 1.77 | AAI-based (medium-confidence) | 0 |
| S12_extracted_k141_102089 | 3405 | No | NA | 1 | 0 | 0 | Not-determined | Genome-fragment | NA | NA | 0 |
| S12_extracted_k141_22063 | 2052 | No | NA | 2 | 0 | 2 | Not-determined | Genome-fragment | NA | NA | 0 |
| S12_extracted_k141_124203 | 1017 | No | NA | 1 | 0 | 1 | Not-determined | Genome-fragment | NA | NA | 0 |
| S12_extracted_k141_109543 | 1122 | No | NA | 2 | 0 | 2 | Not-determined | Genome-fragment | NA | NA | 0 |
| S12_extracted_k141_179181 | 3521 | No | NA | 7 | 0 | 3 | Not-determined | Genome-fragment | NA | NA | 0 |
| S5_vs_k141_79879\|\|full | 3918 | No | NA | 6 | 0 | 2 | Low-quality | Genome-fragment | 7.91 | AAI-based (high-confidence) | 0 |
| S12_extracted_k141_53721 | 1471 | No | NA | 1 | 0 | 1 | Not-determined | Genome-fragment | NA | NA | 0 |
| S12_extracted_k141_13818 | 1394 | No | NA | 1 | 0 | 1 | Not-determined | Genome-fragment | NA | NA | 0 |
| S12_extracted_k141_46810 | 1487 | No | NA | 1 | 0 | 0 | Not-determined | Genome-fragment | NA | NA | 0 |
| S12_extracted_k141_53743 | 1268 | No | NA | 2 | 0 | 1 | Not-determined | Genome-fragment | NA | NA | 0 |
| S12_extracted_k141_155906 | 4344 | No | NA | 4 | 0 | 2 | Not-determined | Genome-fragment | NA | NA | 0 |
| S16_extracted_64686 | 3918 | No | NA | 11 | 0 | 0 | Low-quality | Genome-fragment | 6.71 | AAI-based (medium-confidence) | 0 |
| S16_vs_66295\|\|full | 3918 | No | NA | 7 | 1 | 0 | Low-quality | Genome-fragment | 7.41 | AAI-based (high-confidence) | 0 |
| S4_vs_VIRSorter_10386-cat_1 | 3918 | No | NA | 6 | 3 | 0 | Low-quality | Genome-fragment | 9.76 | AAI-based (high-confidence) | 0 |
| S12_extracted_k141_141019 | 1084 | No | NA | 2 | 0 | 1 | Not-determined | Genome-fragment | NA | NA | 0 |
| S12_extracted_k141_148504 | 1026 | No | NA | 1 | 0 | 1 | Not-determined | Genome-fragment | NA | NA | 0 |
| S14_vs_k141_25712\|\|full | 3920 | No | NA | 9 | 1 | 0 | Low-quality | Genome-fragment | 4.28 | HMM-based (lower-bound) | 0 |
| S7_extracted_68737 | 3920 | No | NA | 6 | 1 | 0 | Low-quality | Genome-fragment | 2.88 | HMM-based (lower-bound) | 0 |
| S7_vs_116070\|\|full | 3920 | No | NA | 6 | 1 | 0 | Low-quality | Genome-fragment | 9.83 | AAI-based (high-confidence) | 0 |
| S12_extracted_k141_188179 | 7621 | No | NA | 7 | 0 | 0 | Not-determined | Genome-fragment | NA | NA | 0 |
| S12_extracted_k141_188180 | 1822 | No | NA | 1 | 0 | 1 | Not-determined | Genome-fragment | NA | NA | 0 |
| S12_extracted_k141_188186 | 1058 | No | NA | 2 | 0 | 2 | Not-determined | Genome-fragment | NA | NA | 0 |
| S12_extracted_k141_85167 | 1686 | No | NA | 3 | 0 | 0 | Not-determined | Genome-fragment | NA | NA | 0 |
| S12_extracted_k141_164025 | 1270 | No | NA | 2 | 0 | 2 | Not-determined | Genome-fragment | NA | NA | 0 |
| S16_extracted_39053 | 3920 | No | NA | 8 | 0 | 0 | Low-quality | Genome-fragment | 8.3 | AAI-based (medium-confidence) | 0 |
| S12_extracted_k141_102142 | 1242 | No | NA | 2 | 0 | 0 | Not-determined | Genome-fragment | NA | NA | 0 |
| S12_vs_k141_134164\|\|lt2gene | 3921 | No | NA | 2 | 1 | 0 | Low-quality | Genome-fragment | 7.52 | AAI-based (medium-confidence) | 0 |
| S12_extracted_k141_124244 | 1557 | No | NA | 1 | 0 | 1 | Not-determined | Genome-fragment | NA | NA | 0 |
| S12_extracted_k141_53793 | 1178 | No | NA | 1 | 0 | 1 | Not-determined | Genome-fragment | NA | NA | 0 |
| S12_extracted_k141_61937 | 1395 | No | NA | 2 | 0 | 1 | Not-determined | Genome-fragment | NA | NA | 0 |
| S12_extracted_k141_109606 | 1034 | No | NA | 1 | 0 | 1 | Not-determined | Genome-fragment | NA | NA | 0 |
| S12_extracted_k141_109608 | 1669 | No | NA | 1 | 0 | 0 | Not-determined | Genome-fragment | NA | NA | 0 |
| S12_extracted_k141_132405 | 1386 | No | NA | 1 | 0 | 1 | Not-determined | Genome-fragment | NA | NA | 0 |
| S12_extracted_k141_188225 | 2652 | No | NA | 1 | 0 | 1 | Not-determined | Genome-fragment | NA | NA | 0 |
| S12_extracted_k141_93831 | 1247 | No | NA | 1 | 0 | 1 | Not-determined | Genome-fragment | NA | NA | 0 |
| S12_extracted_k141_93839 | 1455 | No | NA | 1 | 0 | 0 | Not-determined | Genome-fragment | NA | NA | 0 |
| S12_extracted_k141_124280 | 1054 | No | NA | 1 | 0 | 0 | Not-determined | Genome-fragment | NA | NA | 0 |
| S12_extracted_k141_141088 | 1539 | No | NA | 1 | 0 | 1 | Not-determined | Genome-fragment | NA | NA | 0 |
| S12_extracted_k141_179266 | 1048 | No | NA | 1 | 0 | 0 | Not-determined | Genome-fragment | NA | NA | 0 |
| S12_extracted_k141_141089 | 3259 | No | NA | 5 | 0 | 3 | Not-determined | Genome-fragment | NA | NA | 0 |
| S12_extracted_k141_179269 | 3354 | No | NA | 4 | 0 | 0 | Not-determined | Genome-fragment | NA | NA | 0 |
| S12_extracted_k141_69798 | 6044 | No | NA | 5 | 0 | 4 | Not-determined | Genome-fragment | NA | NA | 0 |
| S3_vs_k141_46736\|\|full | 3922 | No | NA | 7 | 0 | 0 | Low-quality | Genome-fragment | 4.61 | AAI-based (high-confidence) | 0 |
| S12_extracted_k141_132470 | 1029 | No | NA | 1 | 0 | 1 | Not-determined | Genome-fragment | NA | NA | 0 |
| S11_vs_k141_68606\|\|full | 3923 | No | NA | 4 | 0 | 3 | Low-quality | Genome-fragment | 3.21 | AAI-based (medium-confidence) | 0 |
| S12_extracted_k141_69807 | 3317 | No | NA | 2 | 0 | 2 | Not-determined | Genome-fragment | NA | NA | 0 |
| S12_extracted_k141_155958 | 2318 | No | NA | 3 | 0 | 0 | Not-determined | Genome-fragment | NA | NA | 0 |
| S9_vs_k141_10071\|\|full | 3924 | No | NA | 6 | 1 | 0 | Low-quality | Genome-fragment | 8.71 | AAI-based (high-confidence) | 0 |
| S12_extracted_k141_46896 | 1222 | No | NA | 3 | 0 | 2 | Not-determined | Genome-fragment | NA | NA | 0 |
| S3_vs_k141_25632\|\|full | 3924 | No | NA | 4 | 0 | 0 | Low-quality | Genome-fragment | 4.5 | AAI-based (medium-confidence) | 0 |
| S12_extracted_k141_188256 | 1956 | No | NA | 2 | 0 | 1 | Not-determined | Genome-fragment | NA | NA | 0 |
| S11_vs_k141_85120\|\|full | 3925 | No | NA | 6 | 1 | 0 | Low-quality | Genome-fragment | 6.41 | HMM-based (lower-bound) | 0 |
| S12_extracted_k141_69829 | 1147 | No | NA | 1 | 0 | 1 | Not-determined | Genome-fragment | NA | NA | 0 |
| S12_extracted_k141_46917 | 1006 | No | NA | 1 | 0 | 0 | Not-determined | Genome-fragment | NA | NA | 0 |
| S12_extracted_k141_30488 | 3126 | No | NA | 2 | 0 | 1 | Not-determined | Genome-fragment | NA | NA | 0 |
| S14_extracted_k141_38216 | 3926 | No | NA | 4 | 3 | 0 | Low-quality | Genome-fragment | 9.74 | AAI-based (medium-confidence) | 0 |
| S12_extracted_k141_102206 | 1895 | No | NA | 3 | 0 | 2 | Not-determined | Genome-fragment | NA | NA | 0 |
| S7_extracted_46630 | 3926 | Yes | 3068 | 10 | 1 | 1 | Low-quality | Genome-fragment | 7.88 | AAI-based (high-confidence) | 21.85 |
| S16_extracted_152773 | 3927 | No | NA | 7 | 0 | 0 | Low-quality | Genome-fragment | 12.2 | AAI-based (high-confidence) | 0 |
| S12_extracted_k141_62045 | 1438 | No | NA | 1 | 0 | 0 | Not-determined | Genome-fragment | NA | NA | 0 |
| S12_extracted_k141_46947 | 1740 | No | NA | 1 | 0 | 1 | Not-determined | Genome-fragment | NA | NA | 0 |
| S12_extracted_k141_22289 | 1042 | No | NA | 2 | 0 | 1 | Not-determined | Genome-fragment | NA | NA | 0 |
| S12_extracted_k141_141166 | 7796 | No | NA | 6 | 0 | 3 | Not-determined | Genome-fragment | NA | NA | 0 |
| S12_extracted_k141_171831 | 4694 | No | NA | 2 | 0 | 1 | Not-determined | Genome-fragment | NA | NA | 0 |
| S12_extracted_k141_85305 | 1506 | No | NA | 1 | 0 | 1 | Not-determined | Genome-fragment | NA | NA | 0 |
| S12_extracted_k141_62086 | 2001 | No | NA | 2 | 0 | 2 | Not-determined | Genome-fragment | NA | NA | 0 |
| S12_extracted_k141_69895 | 2196 | No | NA | 1 | 0 | 0 | Not-determined | Genome-fragment | NA | NA | 0 |
| S12_extracted_k141_116604 | 1131 | No | NA | 1 | 0 | 1 | Not-determined | Genome-fragment | NA | NA | 0 |
| S12_extracted_k141_108128 | 3927 | No | NA | 8 | 6 | 0 | Low-quality | Genome-fragment | 8.93 | HMM-based (lower-bound) | 0 |
| S12_extracted_k141_148742 | 2903 | No | NA | 3 | 0 | 0 | Not-determined | Genome-fragment | NA | NA | 0 |
| S11_extracted_k141_38164 | 3929 | Yes | 853 | 6 | 1 | 1 | Low-quality | Genome-fragment | 1.99 | AAI-based (medium-confidence) | 78.29 |
| S12_extracted_k141_171850 | 1012 | No | NA | 1 | 0 | 1 | Not-determined | Genome-fragment | NA | NA | 0 |
| S12_extracted_k141_188364 | 1370 | No | NA | 1 | 0 | 1 | Not-determined | Genome-fragment | NA | NA | 0 |
| S12_extracted_k141_164231 | 1233 | No | NA | 1 | 0 | 0 | Not-determined | Genome-fragment | NA | NA | 0 |
| S3_extracted_k141_87626 | 3929 | No | NA | 2 | 0 | 1 | Low-quality | Genome-fragment | 1.67 | AAI-based (medium-confidence) | 0 |
| S16_vs_85480\|\|full | 3929 | Yes | 1899 | 5 | 1 | 1 | Low-quality | Genome-fragment | 3.91 | AAI-based (medium-confidence) | 51.67 |
| S12_extracted_k141_22341 | 1182 | No | NA | 2 | 0 | 0 | Not-determined | Genome-fragment | NA | NA | 0 |
| S8_vs_k141_63897\|\|full | 3929 | No | NA | 4 | 0 | 1 | Low-quality | Genome-fragment | 7.06 | AAI-based (medium-confidence) | 0 |
| S13_vs_98172\|\|full | 3930 | No | NA | 5 | 1 | 0 | Low-quality | Genome-fragment | 9.6 | AAI-based (high-confidence) | 0 |
| S3_extracted_k141_101702 | 3931 | No | NA | 4 | 1 | 0 | Low-quality | Genome-fragment | 9.81 | AAI-based (medium-confidence) | 0 |
| S12_extracted_k141_30555 | 4868 | No | NA | 5 | 0 | 1 | Not-determined | Genome-fragment | NA | NA | 0 |
| S5_vs_k141_74706\|\|full | 3931 | No | NA | 8 | 4 | 0 | Low-quality | Genome-fragment | 6.96 | AAI-based (high-confidence) | 0 |
| S12_extracted_k141_188407 | 1038 | No | NA | 1 | 0 | 0 | Not-determined | Genome-fragment | NA | NA | 0 |
| S12_extracted_k141_188420 | 1278 | No | NA | 2 | 0 | 2 | Not-determined | Genome-fragment | NA | NA | 0 |
| S12_extracted_k141_37796 | 1487 | No | NA | 2 | 0 | 2 | Not-determined | Genome-fragment | NA | NA | 0 |
| S12_extracted_k141_77650 | 1658 | No | NA | 2 | 0 | 0 | Not-determined | Genome-fragment | NA | NA | 0 |
| S12_extracted_k141_93953 | 1731 | No | NA | 2 | 0 | 1 | Not-determined | Genome-fragment | NA | NA | 0 |
| S12_extracted_k141_70746 | 3931 | No | NA | 5 | 2 | 0 | Low-quality | Genome-fragment | 3.59 | AAI-based (high-confidence) | 0 |
| S12_extracted_k141_22418 | 1525 | No | NA | 3 | 0 | 2 | Not-determined | Genome-fragment | NA | NA | 0 |
| S15_extracted_59262 | 3931 | No | NA | 9 | 4 | 0 | Low-quality | Genome-fragment | 8.8 | AAI-based (high-confidence) | 0 |
| S12_extracted_k141_93968 | 3004 | No | NA | 3 | 0 | 3 | Not-determined | Genome-fragment | NA | NA | 0 |
| S12_extracted_k141_62210 | 1291 | No | NA | 2 | 0 | 2 | Not-determined | Genome-fragment | NA | NA | 0 |
| S12_extracted_k141_124457 | 1524 | No | NA | 1 | 0 | 1 | Not-determined | Genome-fragment | NA | NA | 0 |
| S12_extracted_k141_132681 | 1052 | No | NA | 1 | 0 | 1 | Not-determined | Genome-fragment | NA | NA | 0 |
| S12_extracted_k141_179457 | 1921 | No | NA | 2 | 0 | 0 | Not-determined | Genome-fragment | NA | NA | 0 |
| S8_vs_k141_23458\|\|full | 3931 | Yes | 2176 | 4 | 2 | 1 | Low-quality | Genome-fragment | 4.64 | AAI-based (high-confidence) | 44.65 |
| S12_extracted_k141_14079 | 2772 | No | NA | 2 | 0 | 2 | Not-determined | Genome-fragment | NA | NA | 0 |
| S8_vs_k141_147694\|\|full | 3932 | No | NA | 8 | 0 | 1 | Low-quality | Genome-fragment | 9.28 | AAI-based (medium-confidence) | 0 |
| S12_extracted_k141_188483 | 1184 | No | NA | 3 | 0 | 2 | Not-determined | Genome-fragment | NA | NA | 0 |
| S1_vs_k141_9762\|\|full | 3933 | No | NA | 4 | 4 | 0 | Low-quality | Genome-fragment | 10.34 | AAI-based (high-confidence) | 0 |
| S12_extracted_k141_7403 | 1377 | No | NA | 2 | 0 | 0 | Not-determined | Genome-fragment | NA | NA | 0 |
| S12_extracted_k141_148877 | 1301 | No | NA | 2 | 0 | 0 | Not-determined | Genome-fragment | NA | NA | 0 |
| S12_extracted_k141_109796 | 1973 | No | NA | 3 | 0 | 1 | Not-determined | Genome-fragment | NA | NA | 0 |
| S12_extracted_k141_124530 | 1457 | No | NA | 3 | 0 | 1 | Not-determined | Genome-fragment | NA | NA | 0 |
| S12_extracted_k141_132703 | 1440 | No | NA | 1 | 0 | 1 | Not-determined | Genome-fragment | NA | NA | 0 |
| S5_vs_k141_79110\|\|full | 3933 | No | NA | 3 | 3 | 0 | Low-quality | Genome-fragment | 6.27 | AAI-based (high-confidence) | 0 |
| S12_extracted_k141_124546 | 1293 | No | NA | 1 | 0 | 0 | Not-determined | Genome-fragment | NA | NA | 0 |
| S12_extracted_k141_109820 | 1052 | No | NA | 2 | 0 | 0 | Not-determined | Genome-fragment | NA | NA | 0 |
| S12_extracted_k141_62297 | 2681 | No | NA | 2 | 0 | 1 | Not-determined | Genome-fragment | NA | NA | 0 |
| S12_extracted_k141_141343 | 1736 | No | NA | 2 | 0 | 1 | Not-determined | Genome-fragment | NA | NA | 0 |
| S8_vs_k141_19778\|\|full | 3933 | No | NA | 4 | 0 | 3 | Low-quality | Genome-fragment | 2.38 | AAI-based (medium-confidence) | 0 |
| S1_vs_k141_132221\|\|full | 3934 | No | NA | 3 | 3 | 0 | Low-quality | Genome-fragment | 8.11 | AAI-based (medium-confidence) | 0 |
| S12_extracted_k141_62308 | 2184 | No | NA | 2 | 0 | 1 | Not-determined | Genome-fragment | NA | NA | 0 |
| S12_extracted_k141_70088 | 3315 | No | NA | 4 | 0 | 2 | Not-determined | Genome-fragment | NA | NA | 0 |
| S12_extracted_k141_132730 | 4707 | No | NA | 7 | 0 | 4 | Not-determined | Genome-fragment | NA | NA | 0 |
| S12_extracted_k141_7461 | 1001 | No | NA | 1 | 0 | 0 | Not-determined | Genome-fragment | NA | NA | 0 |
| S12_extracted_k141_148941 | 1098 | No | NA | 2 | 0 | 2 | Not-determined | Genome-fragment | NA | NA | 0 |
| S12_extracted_k141_148957 | 3292 | No | NA | 3 | 0 | 2 | Not-determined | Genome-fragment | NA | NA | 0 |
| S12_extracted_k141_70128 | 1121 | No | NA | 1 | 0 | 1 | Not-determined | Genome-fragment | NA | NA | 0 |
| S12_extracted_k141_164345 | 16057 | No | NA | 28 | 0 | 26 | Not-determined | Genome-fragment | NA | NA | 0 |
| S12_extracted_k141_54109 | 1426 | No | NA | 2 | 0 | 0 | Not-determined | Genome-fragment | NA | NA | 0 |
| S7_extracted_54085 | 3934 | No | NA | 4 | 3 | 0 | Low-quality | Genome-fragment | 10.57 | AAI-based (high-confidence) | 0 |
| S12_extracted_k141_47204 | 4646 | No | NA | 7 | 0 | 4 | Not-determined | Genome-fragment | NA | NA | 0 |
| S12_extracted_k141_188555 | 2290 | No | NA | 3 | 0 | 2 | Not-determined | Genome-fragment | NA | NA | 0 |
| S12_extracted_k141_7521 | 2594 | No | NA | 3 | 0 | 2 | Not-determined | Genome-fragment | NA | NA | 0 |
| S12_extracted_k141_77849 | 2126 | No | NA | 4 | 0 | 1 | Not-determined | Genome-fragment | NA | NA | 0 |
| S12_extracted_k141_156025 | 1691 | No | NA | 2 | 0 | 1 | Not-determined | Genome-fragment | NA | NA | 0 |
| S7_vs_122522\|\|full | 3934 | No | NA | 5 | 4 | 0 | Low-quality | Genome-fragment | 10.84 | AAI-based (high-confidence) | 0 |
| S12_extracted_k141_179637 | 2343 | No | NA | 2 | 0 | 2 | Not-determined | Genome-fragment | NA | NA | 0 |
| S12_extracted_k141_37972 | 1101 | No | NA | 3 | 0 | 0 | Not-determined | Genome-fragment | NA | NA | 0 |
| S12_extracted_k141_149023 | 2461 | No | NA | 3 | 0 | 3 | Not-determined | Genome-fragment | NA | NA | 0 |
| S12_extracted_k141_62409 | 1165 | No | NA | 1 | 0 | 0 | Not-determined | Genome-fragment | NA | NA | 0 |
| S6_vs_k141_16365\|\|full | 3934 | No | NA | 9 | 0 | 0 | Low-quality | Genome-fragment | 9.49 | AAI-based (medium-confidence) | 0 |
| S12_extracted_k141_70190 | 1432 | No | NA | 1 | 0 | 1 | Not-determined | Genome-fragment | NA | NA | 0 |
| S17_vs_k141_23524\|\|full | 3935 | No | NA | 6 | 0 | 0 | Low-quality | Genome-fragment | 9.19 | AAI-based (high-confidence) | 0 |
| S12_extracted_k141_124675 | 1057 | No | NA | 2 | 0 | 1 | Not-determined | Genome-fragment | NA | NA | 0 |
| S12_extracted_k141_124680 | 1266 | No | NA | 1 | 0 | 1 | Not-determined | Genome-fragment | NA | NA | 0 |
| S12_extracted_k141_7540 | 4322 | No | NA | 2 | 0 | 1 | Not-determined | Genome-fragment | NA | NA | 0 |
| S12_extracted_k141_149075 | 2142 | No | NA | 3 | 0 | 1 | Not-determined | Genome-fragment | NA | NA | 0 |
| S12_extracted_k141_85646 | 1135 | No | NA | 1 | 0 | 0 | Not-determined | Genome-fragment | NA | NA | 0 |
| S12_extracted_k141_109960 | 1005 | No | NA | 2 | 0 | 1 | Not-determined | Genome-fragment | NA | NA | 0 |
| S12_extracted_k141_132880 | 1468 | No | NA | 1 | 0 | 0 | Not-determined | Genome-fragment | NA | NA | 0 |
| S12_extracted_k141_164457 | 5238 | No | NA | 7 | 0 | 0 | Not-determined | Genome-fragment | NA | NA | 0 |
| S12_extracted_k141_54218 | 3358 | No | NA | 2 | 0 | 1 | Not-determined | Genome-fragment | NA | NA | 0 |
| S12_extracted_k141_109981 | 1168 | No | NA | 1 | 0 | 0 | Not-determined | Genome-fragment | NA | NA | 0 |
| S17_vs_k141_86543\|\|full | 3935 | No | NA | 4 | 0 | 3 | Low-quality | Genome-fragment | 2.57 | AAI-based (medium-confidence) | 0 |
| S12_extracted_k141_85657 | 2229 | No | NA | 2 | 0 | 1 | Not-determined | Genome-fragment | NA | NA | 0 |
| S12_extracted_k141_54240 | 1259 | No | NA | 2 | 0 | 1 | Not-determined | Genome-fragment | NA | NA | 0 |
| S12_extracted_k141_116884 | 1001 | No | NA | 2 | 0 | 1 | Not-determined | Genome-fragment | NA | NA | 0 |
| S12_extracted_k141_179716 | 1290 | No | NA | 3 | 0 | 0 | Not-determined | Genome-fragment | NA | NA | 0 |
| S12_extracted_k141_85664 | 4166 | No | NA | 5 | 0 | 3 | Not-determined | Genome-fragment | NA | NA | 0 |
| S12_extracted_k141_7563 | 1245 | No | NA | 1 | 0 | 1 | Not-determined | Genome-fragment | NA | NA | 0 |
| S12_extracted_k141_62479 | 4227 | No | NA | 3 | 0 | 3 | Not-determined | Genome-fragment | NA | NA | 0 |
| S12_extracted_k141_22560 | 2477 | No | NA | 2 | 0 | 2 | Not-determined | Genome-fragment | NA | NA | 0 |
| S12_extracted_k141_110025 | 1337 | No | NA | 2 | 0 | 0 | Not-determined | Genome-fragment | NA | NA | 0 |
| S12_extracted_k141_62496 | 3273 | No | NA | 2 | 0 | 0 | Not-determined | Genome-fragment | NA | NA | 0 |
| S11_extracted_k141_45112 | 3935 | No | NA | 11 | 2 | 1 | Low-quality | Genome-fragment | 6.71 | AAI-based (medium-confidence) | 0 |
| S12_extracted_k141_14371 | 1211 | No | NA | 1 | 0 | 0 | Not-determined | Genome-fragment | NA | NA | 0 |
| S12_extracted_k141_22594 | 1317 | No | NA | 2 | 0 | 0 | Not-determined | Genome-fragment | NA | NA | 0 |
| S5_extracted_k141_120460 | 3935 | No | NA | 2 | 0 | 2 | Low-quality | Genome-fragment | 0.45 | AAI-based (medium-confidence) | 0 |
| S12_extracted_k141_22607 | 1103 | No | NA | 1 | 0 | 0 | Not-determined | Genome-fragment | NA | NA | 0 |
| S12_extracted_k141_7632 | 1275 | No | NA | 2 | 0 | 0 | Not-determined | Genome-fragment | NA | NA | 0 |
| S12_extracted_k141_85728 | 4035 | No | NA | 4 | 0 | 1 | Not-determined | Genome-fragment | NA | NA | 0 |
| S16_vs_103769\|\|full | 3935 | No | NA | 5 | 1 | 0 | Low-quality | Genome-fragment | 6.14 | HMM-based (lower-bound) | 0 |
| S12_extracted_k141_85734 | 1210 | No | NA | 1 | 0 | 0 | Not-determined | Genome-fragment | NA | NA | 0 |
| S12_extracted_k141_62535 | 1003 | No | NA | 1 | 0 | 1 | Not-determined | Genome-fragment | NA | NA | 0 |
| S12_extracted_k141_110059 | 1067 | No | NA | 1 | 0 | 0 | Not-determined | Genome-fragment | NA | NA | 0 |
| S12_extracted_k141_149171 | 1273 | No | NA | 1 | 0 | 1 | Not-determined | Genome-fragment | NA | NA | 0 |
| S12_extracted_k141_62540 | 1339 | No | NA | 2 | 0 | 0 | Not-determined | Genome-fragment | NA | NA | 0 |
| S1_extracted_k141_126806 | 3936 | No | NA | 6 | 0 | 1 | Low-quality | Genome-fragment | 4.9 | AAI-based (high-confidence) | 0 |
| S12_extracted_k141_54302 | 3734 | No | NA | 2 | 0 | 1 | Not-determined | Genome-fragment | NA | NA | 0 |
| S12_extracted_k141_141520 | 2228 | No | NA | 2 | 0 | 1 | Not-determined | Genome-fragment | NA | NA | 0 |
| S12_extracted_k141_164564 | 1440 | No | NA | 1 | 0 | 1 | Not-determined | Genome-fragment | NA | NA | 0 |
| S12_extracted_k141_85756 | 1285 | No | NA | 1 | 0 | 1 | Not-determined | Genome-fragment | NA | NA | 0 |
| S12_extracted_k141_179828 | 1671 | No | NA | 4 | 0 | 1 | Not-determined | Genome-fragment | NA | NA | 0 |
| S12_extracted_k141_94099 | 5733 | No | NA | 6 | 0 | 1 | Not-determined | Genome-fragment | NA | NA | 0 |
| S16_extracted_25558 | 3936 | No | NA | 4 | 3 | 0 | Low-quality | Genome-fragment | 11.38 | AAI-based (high-confidence) | 0 |
| S16_vs_24288\|\|full | 3936 | No | NA | 4 | 0 | 1 | Low-quality | Genome-fragment | 7.65 | AAI-based (medium-confidence) | 0 |
| S12_extracted_k141_94113 | 1312 | No | NA | 1 | 0 | 1 | Not-determined | Genome-fragment | NA | NA | 0 |
| S12_extracted_k141_94115 | 1204 | No | NA | 2 | 0 | 2 | Not-determined | Genome-fragment | NA | NA | 0 |
| S12_extracted_k141_156108 | 1525 | No | NA | 2 | 0 | 2 | Not-determined | Genome-fragment | NA | NA | 0 |
| S12_extracted_k141_110111 | 1787 | No | NA | 2 | 0 | 2 | Not-determined | Genome-fragment | NA | NA | 0 |
| S12_extracted_k141_62573 | 2867 | No | NA | 2 | 0 | 1 | Not-determined | Genome-fragment | NA | NA | 0 |
| S12_extracted_k141_149214 | 4433 | No | NA | 2 | 0 | 2 | Not-determined | Genome-fragment | NA | NA | 0 |
| S11_extracted_k141_54541 | 3937 | No | NA | 3 | 1 | 0 | Low-quality | Genome-fragment | 7.01 | AAI-based (high-confidence) | 0 |
| S12_extracted_k141_70913 | 3937 | No | NA | 2 | 0 | 2 | Low-quality | Genome-fragment | 0.78 | AAI-based (medium-confidence) | 0 |
| S12_extracted_k141_149221 | 1861 | No | NA | 1 | 0 | 1 | Not-determined | Genome-fragment | NA | NA | 0 |
| S12_extracted_k141_70384 | 1159 | No | NA | 2 | 0 | 2 | Not-determined | Genome-fragment | NA | NA | 0 |
| S12_extracted_k141_116968 | 1598 | No | NA | 2 | 0 | 0 | Not-determined | Genome-fragment | NA | NA | 0 |
| S12_extracted_k141_116975 | 1914 | No | NA | 2 | 0 | 1 | Not-determined | Genome-fragment | NA | NA | 0 |
| S12_extracted_k141_149225 | 1404 | No | NA | 1 | 0 | 0 | Not-determined | Genome-fragment | NA | NA | 0 |
| S12_extracted_k141_116978 | 1681 | No | NA | 2 | 0 | 1 | Not-determined | Genome-fragment | NA | NA | 0 |
| S12_extracted_k141_22725 | 1011 | No | NA | 3 | 0 | 0 | Not-determined | Genome-fragment | NA | NA | 0 |
| S16_extracted_22200 | 3938 | No | NA | 3 | 0 | 1 | Low-quality | Genome-fragment | 5.92 | AAI-based (medium-confidence) | 0 |
| S12_extracted_k141_7712 | 1717 | No | NA | 2 | 0 | 0 | Not-determined | Genome-fragment | NA | NA | 0 |
| S12_extracted_k141_171953 | 1014 | No | NA | 2 | 0 | 0 | Not-determined | Genome-fragment | NA | NA | 0 |
| S12_extracted_k141_7716 | 1588 | No | NA | 1 | 0 | 0 | Not-determined | Genome-fragment | NA | NA | 0 |
| S16_extracted_101221 | 3938 | No | NA | 9 | 0 | 0 | Low-quality | Genome-fragment | 8.62 | AAI-based (high-confidence) | 0 |
| S7_extracted_38115 | 3941 | No | NA | 5 | 0 | 3 | Low-quality | Genome-fragment | 1.68 | AAI-based (medium-confidence) | 0 |
| S12_extracted_k141_22785 | 1192 | No | NA | 2 | 0 | 1 | Not-determined | Genome-fragment | NA | NA | 0 |
| S12_extracted_k141_38135 | 1546 | No | NA | 2 | 0 | 0 | Not-determined | Genome-fragment | NA | NA | 0 |
| S12_extracted_k141_22786 | 1242 | No | NA | 1 | 0 | 0 | Not-determined | Genome-fragment | NA | NA | 0 |
| S12_extracted_k141_179914 | 1151 | No | NA | 1 | 0 | 0 | Not-determined | Genome-fragment | NA | NA | 0 |
| S12_extracted_k141_70462 | 2372 | No | NA | 3 | 0 | 1 | Not-determined | Genome-fragment | NA | NA | 0 |
| S12_extracted_k141_124777 | 5257 | No | NA | 6 | 0 | 2 | Not-determined | Genome-fragment | NA | NA | 0 |
| S12_extracted_k141_38155 | 4622 | No | NA | 2 | 0 | 2 | Not-determined | Genome-fragment | NA | NA | 0 |
| S12_extracted_k141_179936 | 1271 | No | NA | 1 | 0 | 1 | Not-determined | Genome-fragment | NA | NA | 0 |
| S12_extracted_k141_117024 | 2245 | No | NA | 3 | 0 | 2 | Not-determined | Genome-fragment | NA | NA | 0 |
| S12_extracted_k141_94153 | 1296 | No | NA | 1 | 0 | 1 | Not-determined | Genome-fragment | NA | NA | 0 |
| S16_extracted_42100 | 3942 | No | NA | 4 | 0 | 2 | Low-quality | Genome-fragment | 1.92 | AAI-based (medium-confidence) | 0 |
| S12_extracted_k141_62650 | 1064 | No | NA | 1 | 0 | 1 | Not-determined | Genome-fragment | NA | NA | 0 |
| S12_extracted_k141_38162 | 1259 | No | NA | 1 | 0 | 1 | Not-determined | Genome-fragment | NA | NA | 0 |

| | | | | | | | | | | | |
|---|---|---|---|---|---|---|---|---|---|---|---|
| S12_extracted_k141_171999 | 1039 | No | NA | 1 | 0 | 0 | Not-determined | Genome-fragment | NA | NA | 0 |
| S12_extracted_k141_141694 | 1495 | No | NA | 1 | 0 | 1 | Not-determined | Genome-fragment | NA | NA | 0 |
| S12_extracted_k141_14527 | 5924 | No | NA | 5 | 0 | 2 | Not-determined | Genome-fragment | NA | NA | 0 |
| S12_extracted_k141_14529 | 1941 | No | NA | 2 | 0 | 0 | Not-determined | Genome-fragment | NA | NA | 0 |
| S12_extracted_k141_156148 | 2069 | No | NA | 2 | 0 | 0 | Not-determined | Genome-fragment | NA | NA | 0 |
| S12_extracted_k141_78155 | 1808 | No | NA | 2 | 0 | 2 | Not-determined | Genome-fragment | NA | NA | 0 |
| S12_extracted_k141_117027 | 1197 | No | NA | 1 | 0 | 1 | Not-determined | Genome-fragment | NA | NA | 0 |
| S12_extracted_k141_156159 | 1048 | No | NA | 1 | 0 | 1 | Not-determined | Genome-fragment | NA | NA | 0 |
| S12_extracted_k141_85843 | 1531 | No | NA | 2 | 0 | 2 | Not-determined | Genome-fragment | NA | NA | 0 |
| S12_extracted_k141_31005 | 1949 | No | NA | 4 | 0 | 4 | Not-determined | Genome-fragment | NA | NA | 0 |
| S12_extracted_k141_172055 | 1002 | No | NA | 2 | 0 | 1 | Not-determined | Genome-fragment | NA | NA | 0 |
| S12_extracted_k141_78194 | 3037 | No | NA | 2 | 0 | 2 | Not-determined | Genome-fragment | NA | NA | 0 |
| S12_extracted_k141_124841 | 1159 | No | NA | 1 | 0 | 0 | Not-determined | Genome-fragment | NA | NA | 0 |
| S12_extracted_k141_7794 | 1014 | No | NA | 2 | 0 | 2 | Not-determined | Genome-fragment | NA | NA | 0 |
| S12_extracted_k141_164666 | 6240 | No | NA | 5 | 0 | 2 | Not-determined | Genome-fragment | NA | NA | 0 |
| S12_extracted_k141_172072 | 2475 | No | NA | 3 | 0 | 2 | Not-determined | Genome-fragment | NA | NA | 0 |
| S12_extracted_k141_62720 | 8243 | No | NA | 9 | 0 | 5 | Not-determined | Genome-fragment | NA | NA | 0 |
| S16_vs_156089\|\|full | 3942 | No | NA | 8 | 2 | 0 | Low-quality | Genome-fragment | 7.32 | AAI-based (high-confidence) | 0 |
| S13_extracted_124879 | 3943 | No | NA | 7 | 0 | 1 | Low-quality | Genome-fragment | 5.61 | AAI-based (medium-confidence) | 0 |
| S12_extracted_k141_180007 | 1068 | No | NA | 1 | 0 | 1 | Not-determined | Genome-fragment | NA | NA | 0 |
| S16_vs_169500\|\|full | 3943 | No | NA | 4 | 0 | 0 | Low-quality | Genome-fragment | 10.37 | AAI-based (medium-confidence) | 0 |
| S12_extracted_k141_62738 | 1077 | No | NA | 1 | 0 | 0 | Not-determined | Genome-fragment | NA | NA | 0 |
| S9_vs_k141_29746\|\|full | 3944 | No | NA | 5 | 0 | 0 | Low-quality | Genome-fragment | 8.93 | AAI-based (high-confidence) | 0 |
| S12_extracted_k141_156218 | 4604 | No | NA | 5 | 0 | 1 | Not-determined | Genome-fragment | NA | NA | 0 |
| S16_vs_58738\|\|full | 3944 | No | NA | 6 | 5 | 0 | Low-quality | Genome-fragment | 2.4 | AAI-based (medium-confidence) | 0 |
| S12_extracted_k141_22856 | 1362 | No | NA | 1 | 0 | 1 | Not-determined | Genome-fragment | NA | NA | 0 |
| S12_extracted_k141_124894 | 1448 | No | NA | 2 | 0 | 0 | Not-determined | Genome-fragment | NA | NA | 0 |
| S12_extracted_k141_180021 | 1328 | No | NA | 1 | 0 | 0 | Not-determined | Genome-fragment | NA | NA | 0 |
| S13_extracted_79510 | 3945 | No | NA | 3 | 0 | 1 | Low-quality | Genome-fragment | 3.84 | AAI-based (medium-confidence) | 0 |
| S12_extracted_k141_117104 | 2267 | No | NA | 1 | 0 | 1 | Not-determined | Genome-fragment | NA | NA | 0 |
| S11_vs_k141_101308\|\|full | 3947 | No | NA | 6 | 1 | 0 | Low-quality | Genome-fragment | 7.36 | AAI-based (high-confidence) | 0 |
| S12_extracted_k141_31025 | 1626 | No | NA | 2 | 0 | 1 | Not-determined | Genome-fragment | NA | NA | 0 |
| S12_extracted_k141_149396 | 10108 | No | NA | 11 | 0 | 5 | Not-determined | Genome-fragment | NA | NA | 0 |
| S1_vs_k141_30800\|\|full | 3947 | No | NA | 7 | 1 | 0 | Low-quality | Genome-fragment | 8.97 | AAI-based (high-confidence) | 0 |
| S12_extracted_k141_164743 | 1656 | No | NA | 5 | 0 | 3 | Not-determined | Genome-fragment | NA | NA | 0 |
| S12_extracted_k141_117134 | 1425 | No | NA | 2 | 0 | 0 | Not-determined | Genome-fragment | NA | NA | 0 |
| S12_extracted_k141_85934 | 1477 | No | NA | 2 | 0 | 1 | Not-determined | Genome-fragment | NA | NA | 0 |
| S12_extracted_k141_38264 | 1042 | No | NA | 1 | 0 | 0 | Not-determined | Genome-fragment | NA | NA | 0 |
| S12_extracted_k141_14665 | 1507 | No | NA | 1 | 0 | 1 | Not-determined | Genome-fragment | NA | NA | 0 |
| S12_extracted_k141_188745 | 4611 | No | NA | 3 | 0 | 2 | Not-determined | Genome-fragment | NA | NA | 0 |
| S12_extracted_k141_164762 | 1262 | No | NA | 1 | 0 | 1 | Not-determined | Genome-fragment | NA | NA | 0 |
| S3_vs_k141_92724\|\|full | 3947 | No | NA | 9 | 0 | 0 | Low-quality | Genome-fragment | 7.99 | AAI-based (high-confidence) | 0 |
| S12_extracted_k141_164782 | 1241 | No | NA | 1 | 0 | 1 | Not-determined | Genome-fragment | NA | NA | 0 |
| S12_extracted_k141_78289 | 1021 | No | NA | 2 | 0 | 1 | Not-determined | Genome-fragment | NA | NA | 0 |
| S5_extracted_k141_61703 | 3947 | No | NA | 2 | 0 | 1 | Low-quality | Genome-fragment | 1.65 | AAI-based (medium-confidence) | 0 |
| S12_extracted_k141_85991 | 1489 | No | NA | 1 | 0 | 0 | Not-determined | Genome-fragment | NA | NA | 0 |
| S12_extracted_k141_156296 | 1361 | No | NA | 1 | 0 | 1 | Not-determined | Genome-fragment | NA | NA | 0 |
| S12_extracted_k141_54562 | 2783 | No | NA | 2 | 0 | 1 | Not-determined | Genome-fragment | NA | NA | 0 |
| S12_extracted_k141_149439 | 1058 | No | NA | 1 | 0 | 1 | Not-determined | Genome-fragment | NA | NA | 0 |
| S12_extracted_k141_156311 | 1009 | No | NA | 2 | 0 | 1 | Not-determined | Genome-fragment | NA | NA | 0 |
| S12_extracted_k141_124997 | 1100 | No | NA | 1 | 0 | 1 | Not-determined | Genome-fragment | NA | NA | 0 |
| S12_extracted_k141_164802 | 2829 | No | NA | 3 | 0 | 0 | Not-determined | Genome-fragment | NA | NA | 0 |
| S12_extracted_k141_86008 | 1686 | No | NA | 3 | 0 | 2 | Not-determined | Genome-fragment | NA | NA | 0 |
| S12_extracted_k141_31120 | 1058 | No | NA | 2 | 0 | 0 | Not-determined | Genome-fragment | NA | NA | 0 |
| S12_extracted_k141_70629 | 1752 | No | NA | 2 | 0 | 1 | Not-determined | Genome-fragment | NA | NA | 0 |
| S12_extracted_k141_86019 | 3016 | No | NA | 3 | 0 | 1 | Not-determined | Genome-fragment | NA | NA | 0 |
| S12_extracted_k141_70631 | 1523 | No | NA | 3 | 0 | 0 | Not-determined | Genome-fragment | NA | NA | 0 |
| S12_extracted_k141_31135 | 1047 | No | NA | 2 | 0 | 2 | Not-determined | Genome-fragment | NA | NA | 0 |
| S12_extracted_k141_164820 | 4031 | No | NA | 3 | 0 | 3 | Not-determined | Genome-fragment | NA | NA | 0 |
| S10_vs_98883\|\|full | 3947 | No | NA | 4 | 0 | 4 | Low-quality | Genome-fragment | 1.09 | AAI-based (medium-confidence) | 0 |
| S12_extracted_k141_164829 | 1493 | No | NA | 1 | 0 | 1 | Not-determined | Genome-fragment | NA | NA | 0 |
| S12_extracted_k141_38336 | 1077 | No | NA | 2 | 0 | 0 | Not-determined | Genome-fragment | NA | NA | 0 |
| S12_extracted_k141_54619 | 1084 | No | NA | 1 | 0 | 0 | Not-determined | Genome-fragment | NA | NA | 0 |
| S12_extracted_k141_156377 | 1775 | No | NA | 2 | 0 | 1 | Not-determined | Genome-fragment | NA | NA | 0 |
| S12_extracted_k141_54635 | 1378 | No | NA | 2 | 0 | 2 | Not-determined | Genome-fragment | NA | NA | 0 |
| S8_extracted_k141_20361 | 3947 | No | NA | 4 | 0 | 0 | Low-quality | Genome-fragment | 4.49 | AAI-based (medium-confidence) | 0 |
| S12_extracted_k141_172240 | 1528 | No | NA | 3 | 0 | 0 | Not-determined | Genome-fragment | NA | NA | 0 |
| S12_extracted_k141_156394 | 1508 | No | NA | 1 | 0 | 1 | Not-determined | Genome-fragment | NA | NA | 0 |
| S12_extracted_k141_94376 | 3629 | No | NA | 4 | 0 | 4 | Not-determined | Genome-fragment | NA | NA | 0 |
| S12_extracted_k141_78410 | 1185 | No | NA | 1 | 0 | 0 | Not-determined | Genome-fragment | NA | NA | 0 |
| S12_extracted_k141_14786 | 1257 | No | NA | 3 | 0 | 3 | Not-determined | Genome-fragment | NA | NA | 0 |
| S12_extracted_k141_172263 | 1069 | No | NA | 2 | 0 | 0 | Not-determined | Genome-fragment | NA | NA | 0 |
| S12_extracted_k141_23069 | 1201 | No | NA | 1 | 0 | 0 | Not-determined | Genome-fragment | NA | NA | 0 |
| S12_extracted_k141_86107 | 1279 | No | NA | 1 | 0 | 1 | Not-determined | Genome-fragment | NA | NA | 0 |
| S12_extracted_k141_180137 | 2038 | No | NA | 2 | 0 | 0 | Not-determined | Genome-fragment | NA | NA | 0 |
| S12_extracted_k141_172298 | 5421 | No | NA | 5 | 0 | 3 | Not-determined | Genome-fragment | NA | NA | 0 |
| S12_extracted_k141_172300 | 1271 | No | NA | 1 | 0 | 1 | Not-determined | Genome-fragment | NA | NA | 0 |
| S12_extracted_k141_14832 | 1362 | No | NA | 2 | 0 | 0 | Not-determined | Genome-fragment | NA | NA | 0 |
| S12_extracted_k141_149583 | 1991 | No | NA | 2 | 0 | 2 | Not-determined | Genome-fragment | NA | NA | 0 |
| S12_extracted_k141_78471 | 1149 | No | NA | 3 | 0 | 1 | Not-determined | Genome-fragment | NA | NA | 0 |
| S12_extracted_k141_78479 | 2251 | No | NA | 2 | 0 | 1 | Not-determined | Genome-fragment | NA | NA | 0 |
| S12_extracted_k141_38436 | 1295 | No | NA | 2 | 0 | 1 | Not-determined | Genome-fragment | NA | NA | 0 |
| S12_extracted_k141_54711 | 2606 | No | NA | 1 | 0 | 0 | Not-determined | Genome-fragment | NA | NA | 0 |
| S3_extracted_k141_89471 | 3949 | No | NA | 12 | 0 | 0 | Low-quality | Genome-fragment | 1.96 | AAI-based (high-confidence) | 0 |
| S16_extracted_109474 | 3949 | No | NA | 7 | 0 | 5 | Low-quality | Genome-fragment | 2.54 | AAI-based (medium-confidence) | 0 |
| S12_extracted_k141_133260 | 1092 | No | NA | 2 | 0 | 0 | Not-determined | Genome-fragment | NA | NA | 0 |
| S12_extracted_k141_27548 | 3950 | Yes | 1995 | 3 | 1 | 1 | Low-quality | Genome-fragment | 4.51 | AAI-based (medium-confidence) | 49.49 |
| S12_vs_k141_76256\|\|full | 3950 | No | NA | 9 | 4 | 1 | Low-quality | Genome-fragment | 10.68 | AAI-based (high-confidence) | 0 |
| S10_vs_68378\|\|full | 3951 | No | NA | 10 | 2 | 0 | Low-quality | Genome-fragment | 7.43 | AAI-based (medium-confidence) | 0 |
| S12_extracted_k141_78509 | 1597 | No | NA | 2 | 0 | 1 | Not-determined | Genome-fragment | NA | NA | 0 |
| S9_vs_k141_3357\|\|full | 3952 | No | NA | 3 | 1 | 0 | Low-quality | Genome-fragment | 9.13 | AAI-based (high-confidence) | 0 |
| S12_extracted_k141_125109 | 3316 | No | NA | 4 | 0 | 2 | Not-determined | Genome-fragment | NA | NA | 0 |
| S12_extracted_k141_133294 | 1579 | No | NA | 3 | 0 | 0 | Not-determined | Genome-fragment | NA | NA | 0 |
| S12_extracted_k141_70775 | 1258 | No | NA | 3 | 0 | 0 | Not-determined | Genome-fragment | NA | NA | 0 |
| S12_extracted_k141_180208 | 1076 | No | NA | 1 | 0 | 0 | Not-determined | Genome-fragment | NA | NA | 0 |
| S12_extracted_k141_125123 | 1089 | No | NA | 1 | 0 | 1 | Not-determined | Genome-fragment | NA | NA | 0 |
| S12_extracted_k141_180227 | 1062 | No | NA | 2 | 0 | 0 | Not-determined | Genome-fragment | NA | NA | 0 |
| S12_extracted_k141_23184 | 3271 | No | NA | 3 | 0 | 3 | Not-determined | Genome-fragment | NA | NA | 0 |
| S7_vs_129183\|\|full | 3952 | No | NA | 3 | 1 | 0 | Low-quality | Genome-fragment | 1.93 | HMM-based (lower-bound) | 0 |
| S12_extracted_k141_14882 | 1661 | No | NA | 3 | 0 | 1 | Not-determined | Genome-fragment | NA | NA | 0 |
| S12_extracted_k141_164947 | 3958 | No | NA | 5 | 0 | 1 | Not-determined | Genome-fragment | NA | NA | 0 |
| S12_extracted_k141_70835 | 1120 | No | NA | 1 | 0 | 0 | Not-determined | Genome-fragment | NA | NA | 0 |
| S12_extracted_k141_86297 | 3755 | No | NA | 4 | 0 | 3 | Not-determined | Genome-fragment | NA | NA | 0 |
| S12_extracted_k141_14893 | 1071 | No | NA | 1 | 0 | 0 | Not-determined | Genome-fragment | NA | NA | 0 |
| S12_extracted_k141_54806 | 2040 | No | NA | 2 | 0 | 1 | Not-determined | Genome-fragment | NA | NA | 0 |
| S12_extracted_k141_188819 | 216219 | No | NA | 281 | 42 | 6 | Complete | High-quality | 100 | DTR (high-confidence) | 0 |
| S12_extracted_k141_54827 | 6226 | No | NA | 5 | 0 | 1 | Not-determined | Genome-fragment | NA | NA | 0 |
| S12_extracted_k141_133372 | 1589 | No | NA | 2 | 0 | 2 | Not-determined | Genome-fragment | NA | NA | 0 |
| S12_extracted_k141_38605 | 1139 | No | NA | 1 | 0 | 1 | Not-determined | Genome-fragment | NA | NA | 0 |
| S12_extracted_k141_31398 | 1342 | No | NA | 2 | 0 | 0 | Not-determined | Genome-fragment | NA | NA | 0 |
| S12_extracted_k141_78667 | 4455 | No | NA | 4 | 0 | 3 | Not-determined | Genome-fragment | NA | NA | 0 |
| S12_extracted_k141_133382 | 1232 | No | NA | 3 | 0 | 1 | Not-determined | Genome-fragment | NA | NA | 0 |
| S12_extracted_k141_125259 | 2189 | No | NA | 3 | 0 | 1 | Not-determined | Genome-fragment | NA | NA | 0 |
| S12_extracted_k141_86365 | 1133 | No | NA | 1 | 0 | 1 | Not-determined | Genome-fragment | NA | NA | 0 |
| S12_extracted_k141_117465 | 1441 | No | NA | 1 | 0 | 1 | Not-determined | Genome-fragment | NA | NA | 0 |
| S12_extracted_k141_180306 | 1680 | No | NA | 2 | 0 | 1 | Not-determined | Genome-fragment | NA | NA | 0 |
| S15_extracted_41022 | 3952 | No | NA | 4 | 1 | 0 | Low-quality | Genome-fragment | 0.99 | AAI-based (high-confidence) | 0 |
| S12_extracted_k141_117468 | 1115 | No | NA | 2 | 0 | 0 | Not-determined | Genome-fragment | NA | NA | 0 |
| S12_extracted_k141_125274 | 1346 | No | NA | 2 | 0 | 0 | Not-determined | Genome-fragment | NA | NA | 0 |
| S12_extracted_k141_180315 | 1397 | No | NA | 1 | 0 | 1 | Not-determined | Genome-fragment | NA | NA | 0 |
| S6_extracted_k141_17150 | 3952 | No | NA | 1 | 0 | 1 | Low-quality | Genome-fragment | 2.38 | AAI-based (medium-confidence) | 0 |
| S12_extracted_k141_156594 | 8650 | No | NA | 7 | 0 | 2 | Not-determined | Genome-fragment | NA | NA | 0 |
| S12_extracted_k141_38676 | 1100 | No | NA | 2 | 0 | 0 | Not-determined | Genome-fragment | NA | NA | 0 |
| S12_extracted_k141_180353 | 1150 | No | NA | 3 | 0 | 3 | Not-determined | Genome-fragment | NA | NA | 0 |
| S12_extracted_k141_23331 | 1367 | No | NA | 1 | 0 | 1 | Not-determined | Genome-fragment | NA | NA | 0 |
| S12_extracted_k141_165069 | 1046 | No | NA | 1 | 0 | 0 | Not-determined | Genome-fragment | NA | NA | 0 |
| S12_extracted_k141_133464 | 2229 | No | NA | 2 | 0 | 2 | Not-determined | Genome-fragment | NA | NA | 0 |
| S12_extracted_k141_180370 | 1339 | No | NA | 1 | 0 | 1 | Not-determined | Genome-fragment | NA | NA | 0 |
| S12_extracted_k141_23356 | 1465 | No | NA | 2 | 0 | 0 | Not-determined | Genome-fragment | NA | NA | 0 |
| S12_extracted_k141_133486 | 3278 | No | NA | 3 | 0 | 3 | Not-determined | Genome-fragment | NA | NA | 0 |
| S12_extracted_k141_54933 | 3109 | No | NA | 3 | 0 | 2 | Not-determined | Genome-fragment | NA | NA | 0 |
| S12_extracted_k141_156635 | 8913 | No | NA | 7 | 0 | 5 | Not-determined | Genome-fragment | NA | NA | 0 |
| S12_extracted_k141_117514 | 1389 | No | NA | 1 | 0 | 1 | Not-determined | Genome-fragment | NA | NA | 0 |
| S14_extracted_k141_73984 | 3953 | No | NA | 10 | 5 | 0 | Low-quality | Genome-fragment | 5.5 | AAI-based (high-confidence) | 0 |
| S12_extracted_k141_172461 | 1092 | No | NA | 1 | 0 | 1 | Not-determined | Genome-fragment | NA | NA | 0 |
| S12_extracted_k141_55000 | 1332 | No | NA | 1 | 0 | 1 | Not-determined | Genome-fragment | NA | NA | 0 |
| S12_extracted_k141_38739 | 1084 | No | NA | 2 | 0 | 0 | Not-determined | Genome-fragment | NA | NA | 0 |
| S12_extracted_k141_156689 | 1211 | No | NA | 2 | 0 | 1 | Not-determined | Genome-fragment | NA | NA | 0 |
| S10_vs_93126\|\|full | 3953 | Yes | 2018 | 4 | 1 | 1 | Low-quality | Genome-fragment | 1.27 | HMM-based (lower-bound) | 48.95 |
| S12_extracted_k141_172512 | 1356 | No | NA | 1 | 0 | 1 | Not-determined | Genome-fragment | NA | NA | 0 |
| S12_extracted_k141_125388 | 1455 | No | NA | 3 | 0 | 2 | Not-determined | Genome-fragment | NA | NA | 0 |
| S12_extracted_k141_188822 | 60827 | No | NA | 77 | 16 | 0 | Complete | High-quality | 100 | DTR (high-confidence) | 0 |
| S12_vs_k141_135400\|\|full | 3954 | No | NA | 9 | 4 | 0 | Low-quality | Genome-fragment | 10.63 | AAI-based (high-confidence) | 0 |
| S12_extracted_k141_38776 | 1439 | No | NA | 1 | 0 | 0 | Not-determined | Genome-fragment | NA | NA | 0 |
| S5_extracted_k141_126998 | 3955 | No | NA | 6 | 0 | 0 | Low-quality | Genome-fragment | 6.11 | AAI-based (medium-confidence) | 0 |
| S12_extracted_k141_86534 | 1064 | No | NA | 2 | 0 | 1 | Not-determined | Genome-fragment | NA | NA | 0 |
| S16_extracted_61103 | 3956 | No | NA | 6 | 0 | 1 | Low-quality | Genome-fragment | 7.93 | AAI-based (medium-confidence) | 0 |
| S12_extracted_k141_172546 | 3046 | No | NA | 5 | 0 | 3 | Not-determined | Genome-fragment | NA | NA | 0 |
| S12_extracted_k141_38795 | 1078 | No | NA | 2 | 0 | 2 | Not-determined | Genome-fragment | NA | NA | 0 |
| S12_extracted_k141_117539 | 2085 | No | NA | 2 | 0 | 1 | Not-determined | Genome-fragment | NA | NA | 0 |
| S14_extracted_k141_116060 | 3957 | No | NA | 3 | 0 | 3 | Low-quality | Genome-fragment | 0.98 | AAI-based (medium-confidence) | 0 |
| S12_extracted_k141_86555 | 1579 | No | NA | 2 | 0 | 1 | Not-determined | Genome-fragment | NA | NA | 0 |
| S9_extracted_k141_9453 | 3958 | No | NA | 8 | 4 | 0 | Low-quality | Genome-fragment | 9.89 | AAI-based (high-confidence) | 0 |
| S12_extracted_k141_172587 | 1537 | No | NA | 2 | 0 | 0 | Not-determined | Genome-fragment | NA | NA | 0 |
| S12_extracted_k141_86577 | 2321 | No | NA | 4 | 0 | 0 | Not-determined | Genome-fragment | NA | NA | 0 |
| S3_extracted_k141_55391 | 3958 | No | NA | 3 | 1 | 0 | Low-quality | Genome-fragment | 6.76 | AAI-based (high-confidence) | 0 |
| S12_extracted_k141_172643 | 1544 | No | NA | 1 | 0 | 1 | Not-determined | Genome-fragment | NA | NA | 0 |
| S13_extracted_110068 | 3959 | No | NA | 4 | 0 | 0 | Low-quality | Genome-fragment | 6.19 | AAI-based (high-confidence) | 0 |
| S12_extracted_k141_180588 | 4779 | No | NA | 7 | 0 | 7 | Not-determined | Genome-fragment | NA | NA | 0 |
| S12_extracted_k141_180600 | 1146 | No | NA | 2 | 0 | 1 | Not-determined | Genome-fragment | NA | NA | 0 |
| S7_vs_107691\|\|full | 3959 | No | NA | 6 | 3 | 0 | Low-quality | Genome-fragment | 6.41 | AAI-based (high-confidence) | 0 |
| S12_extracted_k141_38915 | 1107 | No | NA | 3 | 0 | 0 | Not-determined | Genome-fragment | NA | NA | 0 |
| S12_extracted_k141_156827 | 1399 | No | NA | 1 | 0 | 1 | Not-determined | Genome-fragment | NA | NA | 0 |
| S12_extracted_k141_55080 | 4468 | No | NA | 6 | 0 | 2 | Not-determined | Genome-fragment | NA | NA | 0 |
| S12_extracted_k141_188824 | 87909 | No | NA | 112 | 16 | 4 | Complete | High-quality | 100 | DTR (high-confidence) | 0 |
| S12_extracted_k141_156850 | 1987 | No | NA | 2 | 0 | 1 | Not-determined | Genome-fragment | NA | NA | 0 |
| S12_extracted_k141_117695 | 1325 | No | NA | 1 | 0 | 1 | Not-determined | Genome-fragment | NA | NA | 0 |
| S15_vs_60479\|\|full | 3959 | No | NA | 4 | 0 | 1 | Low-quality | Genome-fragment | 4.18 | AAI-based (medium-confidence) | 0 |
| S9_vs_k141_54643\|\|full | 3960 | No | NA | 8 | 1 | 0 | Low-quality | Genome-fragment | 11.28 | AAI-based (high-confidence) | 0 |
| S12_extracted_k141_38950 | 1725 | No | NA | 1 | 0 | 0 | Not-determined | Genome-fragment | NA | NA | 0 |
| S12_extracted_k141_133760 | 1601 | No | NA | 1 | 0 | 0 | Not-determined | Genome-fragment | NA | NA | 0 |
| S12_extracted_k141_15162 | 5237 | No | NA | 5 | 0 | 4 | Not-determined | Genome-fragment | NA | NA | 0 |
| S5_extracted_k141_60700 | 3960 | No | NA | 5 | 5 | 0 | Low-quality | Genome-fragment | 9.71 | AAI-based (high-confidence) | 0 |
| S12_extracted_k141_117783 | 1045 | No | NA | 1 | 0 | 1 | Not-determined | Genome-fragment | NA | NA | 0 |
| S12_extracted_k141_125486 | 1003 | No | NA | 1 | 0 | 1 | Not-determined | Genome-fragment | NA | NA | 0 |
| S12_extracted_k141_125501 | 1438 | No | NA | 2 | 0 | 0 | Not-determined | Genome-fragment | NA | NA | 0 |
| S12_extracted_k141_188827 | 2601 | No | NA | 5 | 2 | 0 | Complete | High-quality | 100 | DTR (high-confidence) | 0 |
| S14_vs_k141_73958\|\|full | 3961 | Yes | 2626 | 10 | 2 | 2 | Low-quality | Genome-fragment | 6.13 | AAI-based (high-confidence) | 33.7 |
| S12_extracted_k141_23607 | 1526 | No | NA | 1 | 0 | 1 | Not-determined | Genome-fragment | NA | NA | 0 |
| S12_extracted_k141_125554 | 1023 | No | NA | 2 | 0 | 0 | Not-determined | Genome-fragment | NA | NA | 0 |
| S12_extracted_k141_156981 | 1191 | No | NA | 2 | 0 | 1 | Not-determined | Genome-fragment | NA | NA | 0 |
| S12_vs_k141_72153\|\|full | 3962 | No | NA | 3 | 0 | 2 | Low-quality | Genome-fragment | 3.2 | AAI-based (high-confidence) | 0 |
| S12_extracted_k141_180634 | 1572 | No | NA | 2 | 0 | 1 | Not-determined | Genome-fragment | NA | NA | 0 |
| S3_vs_k141_120497\|\|full | 3963 | No | NA | 6 | 0 | 1 | Low-quality | Genome-fragment | 6.2 | AAI-based (medium-confidence) | 0 |
| S12_extracted_k141_39123 | 1440 | No | NA | 3 | 0 | 1 | Not-determined | Genome-fragment | NA | NA | 0 |
| S12_extracted_k141_188830 | 68192 | No | NA | 80 | 5 | 0 | Complete | High-quality | 100 | DTR (high-confidence) | 0 |
| S12_extracted_k141_172971 | 1651 | No | NA | 2 | 0 | 1 | Not-determined | Genome-fragment | NA | NA | 0 |
| S12_extracted_k141_15387 | 1214 | No | NA | 1 | 0 | 1 | Not-determined | Genome-fragment | NA | NA | 0 |
| S12_extracted_k141_39141 | 1983 | No | NA | 2 | 0 | 2 | Not-determined | Genome-fragment | NA | NA | 0 |
| S12_extracted_k141_15400 | 3332 | No | NA | 3 | 0 | 1 | Not-determined | Genome-fragment | NA | NA | 0 |
| S12_extracted_k141_157067 | 1041 | No | NA | 2 | 0 | 0 | Not-determined | Genome-fragment | NA | NA | 0 |
| S16_extracted_155644 | 3963 | No | NA | 4 | 4 | 0 | Low-quality | Genome-fragment | 11.26 | AAI-based (high-confidence) | 0 |
| S10_vs_86963\|\|full | 3963 | No | NA | 8 | 1 | 0 | Low-quality | Genome-fragment | 1.94 | HMM-based (lower-bound) | 0 |
| S12_extracted_k141_117970 | 1502 | No | NA | 2 | 0 | 2 | Not-determined | Genome-fragment | NA | NA | 0 |
| S2_vs_k141_12272\|\|full | 3963 | No | NA | 7 | 0 | 0 | Low-quality | Genome-fragment | 6.65 | AAI-based (high-confidence) | 0 |
| S12_extracted_k141_188838 | 5233 | No | NA | 9 | 0 | 0 | Complete | High-quality | 100 | DTR (high-confidence) | 0 |
| S12_extracted_k141_39211 | 1189 | No | NA | 1 | 0 | 0 | Not-determined | Genome-fragment | NA | NA | 0 |
| S8_vs_k141_40442\|\|full | 3963 | No | NA | 9 | 4 | 0 | Low-quality | Genome-fragment | 11.02 | AAI-based (high-confidence) | 0 |
| S8_extracted_k141_98307 | 3964 | No | NA | 3 | 0 | 2 | Low-quality | Genome-fragment | 2.19 | AAI-based (high-confidence) | 0 |
| S12_extracted_k141_173071 | 1217 | No | NA | 1 | 0 | 1 | Not-determined | Genome-fragment | NA | NA | 0 |
| S1_extracted_k141_150959 | 3965 | No | NA | 7 | 1 | 0 | Low-quality | Genome-fragment | 6.94 | AAI-based (high-confidence) | 0 |
| S12_extracted_k141_157151 | 1016 | No | NA | 2 | 0 | 0 | Not-determined | Genome-fragment | NA | NA | 0 |
| S12_extracted_k141_188846 | 5661 | No | NA | 8 | 1 | 0 | Complete | High-quality | 100 | DTR (high-confidence) | 0 |
| S12_extracted_k141_39250 | 1282 | No | NA | 1 | 0 | 1 | Not-determined | Genome-fragment | NA | NA | 0 |
| S12_extracted_k141_188865 | 3605 | No | NA | 5 | 0 | 0 | Not-determined | Genome-fragment | NA | NA | 0 |
| S12_extracted_k141_118009 | 30980 | No | NA | 30 | 0 | 20 | Not-determined | Genome-fragment | NA | NA | 0 |
| S12_extracted_k141_118011 | 1381 | No | NA | 2 | 0 | 1 | Not-determined | Genome-fragment | NA | NA | 0 |
| S8_vs_k141_12223\|\|full | 3965 | No | NA | 11 | 2 | 0 | Low-quality | Genome-fragment | 12.21 | AAI-based (high-confidence) | 0 |
| S12_extracted_k141_157205 | 1120 | No | NA | 2 | 0 | 2 | Not-determined | Genome-fragment | NA | NA | 0 |
| S12_extracted_k141_125738 | 1262 | No | NA | 3 | 0 | 1 | Not-determined | Genome-fragment | NA | NA | 0 |
| S12_extracted_k141_157209 | 1164 | No | NA | 1 | 0 | 1 | Not-determined | Genome-fragment | NA | NA | 0 |
| S12_extracted_k141_39303 | 2152 | No | NA | 2 | 0 | 1 | Not-determined | Genome-fragment | NA | NA | 0 |
| S12_extracted_k141_15568 | 2097 | No | NA | 2 | 0 | 2 | Not-determined | Genome-fragment | NA | NA | 0 |
| S12_extracted_k141_180797 | 3403 | No | NA | 3 | 0 | 1 | Not-determined | Genome-fragment | NA | NA | 0 |
| S12_extracted_k141_39365 | 2944 | No | NA | 2 | 0 | 2 | Not-determined | Genome-fragment | NA | NA | 0 |
| S12_extracted_k141_180856 | 1571 | No | NA | 2 | 0 | 2 | Not-determined | Genome-fragment | NA | NA | 0 |
| S3_extracted_k141_24018 | 3967 | No | NA | 6 | 6 | 0 | Low-quality | Genome-fragment | 18.1 | AAI-based (high-confidence) | 0 |
| S12_extracted_k141_180872 | 1172 | No | NA | 1 | 0 | 1 | Not-determined | Genome-fragment | NA | NA | 0 |
| S12_extracted_k141_157302 | 1264 | No | NA | 2 | 0 | 0 | Not-determined | Genome-fragment | NA | NA | 0 |
| S5_vs_k141_92106\|\|full | 3967 | No | NA | 7 | 0 | 0 | Low-quality | Genome-fragment | 7.48 | AAI-based (medium-confidence) | 0 |
| S12_extracted_k141_157350 | 2162 | No | NA | 4 | 0 | 0 | Not-determined | Genome-fragment | NA | NA | 0 |
| S12_extracted_k141_15721 | 4048 | No | NA | 5 | 0 | 2 | Not-determined | Genome-fragment | NA | NA | 0 |
| S12_extracted_k141_173208 | 18073 | No | NA | 35 | 3 | 0 | Medium-quality | Genome-fragment | 56.07 | AAI-based (high-confidence) | 0 |
| S12_extracted_k141_180995 | 5066 | No | NA | 3 | 0 | 2 | Not-determined | Genome-fragment | NA | NA | 0 |
| S12_extracted_k141_125938 | 1642 | No | NA | 2 | 0 | 0 | Not-determined | Genome-fragment | NA | NA | 0 |
| S12_extracted_k141_181024 | 71259 | No | NA | 85 | 22 | 1 | Medium-quality | Genome-fragment | 65.59 | AAI-based (high-confidence) | 0 |
| S12_vs_k141_55201\|\|full | 4212 | No | NA | 5 | 0 | 1 | Not-determined | Genome-fragment | NA | NA | 0 |
| S12_vs_k141_86928\|\|full | 41629 | No | NA | 55 | 6 | 2 | Medium-quality | Genome-fragment | 54.85 | HMM-based (lower-bound) | 0 |
| S12_vs_k141_87088\|\|full | 39892 | No | NA | 50 | 23 | 0 | High-quality | High-quality | 96.05 | AAI-based (high-confidence) | 0 |
| S12_vs_k141_79785\|\|full | 44275 | No | NA | 68 | 32 | 0 | High-quality | High-quality | 97.36 | AAI-based (high-confidence) | 0 |
| S16_extracted_71269 | 3968 | No | NA | 6 | 1 | 1 | Low-quality | Genome-fragment | 2.53 | AAI-based (high-confidence) | 0 |
| S12_vs_k141_167155\|\|full | 42619 | No | NA | 66 | 13 | 0 | High-quality | High-quality | 97.15 | AAI-based (high-confidence) | 0 |
| S12_vs_k141_1980\|\|full | 47205 | No | NA | 62 | 5 | 0 | High-quality | High-quality | 100 | AAI-based (high-confidence) | 0 |
| S12_vs_k141_111815\|\|full | 53432 | No | NA | 78 | 12 | 2 | High-quality | High-quality | 99.42 | AAI-based (medium-confidence) | 0 |
| S12_vs_k141_183891\|\|full | 5176 | No | NA | 5 | 1 | 0 | High-quality | High-quality | 94.21 | AAI-based (high-confidence) | 0 |
| S12_vs_k141_105523\|\|full | 9799 | No | NA | 11 | 0 | 0 | Not-determined | Genome-fragment | NA | NA | 0 |
| S12_vs_k141_137069\|\|full | 10908 | No | NA | 14 | 0 | 1 | Medium-quality | Genome-fragment | 74.66 | AAI-based (high-confidence) | 0 |
| S8_vs_k141_150226\|\|full | 3968 | No | NA | 12 | 1 | 0 | Low-quality | Genome-fragment | 3.9 | HMM-based (lower-bound) | 0 |
| S16_extracted_77177 | 3969 | No | NA | 8 | 3 | 0 | Low-quality | Genome-fragment | 5.32 | HMM-based (lower-bound) | 0 |
| S12_vs_k141_188820\|\|full | 52666 | No | NA | 87 | 14 | 0 | High-quality | High-quality | 100 | AAI-based (high-confidence) | 0 |
| S12_vs_k141_188821\|\|full | 59341 | No | NA | 84 | 35 | 2 | High-quality | High-quality | 98.95 | AAI-based (high-confidence) | 0 |
| S12_vs_k141_188823\|\|full | 32687 | No | NA | 56 | 9 | 2 | High-quality | High-quality | 100 | AAI-based (high-confidence) | 0 |
| S12_vs_k141_188825\|\|full | 41036 | No | NA | 55 | 11 | 0 | Medium-quality | Genome-fragment | 51.07 | HMM-based (lower-bound) | 0 |
| S12_vs_k141_188828\|\|full | 34951 | No | NA | 54 | 13 | 0 | Medium-quality | Genome-fragment | 84.99 | AAI-based (medium-confidence) | 0 |
| S12_vs_k141_188829\|\|full | 6511 | No | NA | 15 | 0 | 0 | Not-determined | Genome-fragment | NA | NA | 0 |
| S12_vs_k141_188832\|\|full | 4680 | No | NA | 8 | 4 | 0 | High-quality | High-quality | 96.7 | AAI-based (medium-confidence) | 0 |
| S12_vs_k141_188833\|\|full | 32121 | No | NA | 42 | 14 | 1 | High-quality | High-quality | 98.29 | AAI-based (high-confidence) | 0 |
| S12_vs_k141_188836\|\|full | 5325 | No | NA | 9 | 0 | 1 | Not-determined | Genome-fragment | NA | NA | 0 |
| S12_vs_k141_188841\|\|full | 3152 | No | NA | 5 | 0 | 0 | Not-determined | Genome-fragment | NA | NA | 0 |
| S12_vs_k141_188863\|\|full | 2229 | No | NA | 3 | 0 | 0 | Not-determined | Genome-fragment | NA | NA | 0 |
| S12_vs_k141_188879\|\|full | 1799 | No | NA | 2 | 0 | 0 | Not-determined | Genome-fragment | NA | NA | 0 |
| S16_vs_170430\|\|full | 3969 | No | NA | 3 | 3 | 0 | Low-quality | Genome-fragment | 9.97 | AAI-based (high-confidence) | 0 |
| S12_vs_k141_165365\|\|full | 2607 | No | NA | 3 | 0 | 2 | Not-determined | Genome-fragment | NA | NA | 0 |
| S14_vs_k141_4912\|\|full | 3971 | No | NA | 7 | 4 | 0 | Low-quality | Genome-fragment | 10.29 | AAI-based (high-confidence) | 0 |
| S3_vs_k141_115485\|\|full | 3971 | No | NA | 7 | 0 | 1 | Low-quality | Genome-fragment | 9.53 | AAI-based (high-confidence) | 0 |
| S12_vs_k141_110262\|\|full | 3039 | No | NA | 3 | 0 | 1 | Not-determined | Genome-fragment | NA | NA | 0 |
| S12_vs_k141_181122\|\|full | 3795 | No | NA | 8 | 0 | 8 | Not-determined | Genome-fragment | NA | NA | 0 |
| S7_vs_58318\|\|full | 3972 | No | NA | 7 | 3 | 0 | Low-quality | Genome-fragment | 8.6 | AAI-based (high-confidence) | 0 |
| S7_vs_136635\|\|full | 3972 | Yes | 327 | 3 | 1 | 2 | Low-quality | Genome-fragment | 0.53 | HMM-based (lower-bound) | 91.77 |
| S12_vs_k141_126025\|\|full | 1450 | No | NA | 3 | 0 | 1 | Not-determined | Genome-fragment | NA | NA | 0 |
| S12_vs_k141_55\|\|full | 2207 | No | NA | 4 | 0 | 0 | Not-determined | Genome-fragment | NA | NA | 0 |
| S7_vs_5148\|\|full | 3973 | No | NA | 4 | 0 | 2 | Low-quality | Genome-fragment | 2.28 | AAI-based (medium-confidence) | 0 |
| S12_vs_k141_165419\|\|full | 1991 | No | NA | 2 | 0 | 2 | Not-determined | Genome-fragment | NA | NA | 0 |
| S12_vs_k141_7908\|\|full | 4824 | No | NA | 6 | 0 | 3 | Not-determined | Genome-fragment | NA | NA | 0 |
| S12_vs_k141_94550\|\|full | 1650 | No | NA | 3 | 0 | 1 | Not-determined | Genome-fragment | NA | NA | 0 |
| S12_vs_k141_70934\|\|full | 2037 | No | NA | 3 | 0 | 1 | Not-determined | Genome-fragment | NA | NA | 0 |
| S12_vs_k141_152646\|\|full | 3974 | No | NA | 5 | 1 | 0 | Low-quality | Genome-fragment | 2.89 | AAI-based (high-confidence) | 0 |
| S12_vs_k141_39403\|\|full | 1694 | No | NA | 3 | 0 | 1 | Not-determined | Genome-fragment | NA | NA | 0 |
| S11_extracted_k141_76785 | 3975 | No | NA | 5 | 4 | 0 | Low-quality | Genome-fragment | 11.56 | AAI-based (high-confidence) | 0 |
| S11_vs_k141_43684\|\|full | 3975 | No | NA | 5 | 0 | 0 | Low-quality | Genome-fragment | 9.61 | AAI-based (medium-confidence) | 0 |
| S12_vs_k141_70953\|\|full | 1718 | No | NA | 3 | 0 | 2 | Not-determined | Genome-fragment | NA | NA | 0 |
| S12_vs_k141_118154\|\|full | 30177 | No | NA | 45 | 19 | 0 | High-quality | High-quality | 100 | AAI-based (high-confidence) | 0 |
| S10_extracted_104966 | 3975 | No | NA | 4 | 0 | 4 | Low-quality | Genome-fragment | 6.24 | AAI-based (high-confidence) | 0 |
| S12_vs_k141_157570\|\|full | 3338 | No | NA | 4 | 0 | 2 | Not-determined | Genome-fragment | NA | NA | 0 |
| S12_vs_k141_70957\|\|full | 3150 | No | NA | 3 | 0 | 1 | Not-determined | Genome-fragment | NA | NA | 0 |
| S16_extracted_74011 | 3977 | No | NA | 7 | 4 | 0 | Low-quality | Genome-fragment | 5.93 | AAI-based (high-confidence) | 0 |
| S12_vs_k141_118171\|\|full | 1641 | No | NA | 2 | 0 | 1 | Not-determined | Genome-fragment | NA | NA | 0 |
| S10_vs_49298\|\|full | 3977 | No | NA | 8 | 1 | 0 | Low-quality | Genome-fragment | 4.34 | AAI-based (high-confidence) | 0 |
| S9_vs_k141_49761\|\|full | 3978 | No | NA | 4 | 3 | 0 | Low-quality | Genome-fragment | 8.81 | AAI-based (high-confidence) | 0 |
| S7_vs_10102\|\|full | 3979 | No | NA | 8 | 4 | 0 | Low-quality | Genome-fragment | 7.21 | AAI-based (high-confidence) | 0 |
| S16_extracted_40651 | 3980 | No | NA | 3 | 0 | 2 | Low-quality | Genome-fragment | 6.6 | AAI-based (medium-confidence) | 0 |
| S12_vs_k141_86751\|\|full | 2634 | No | NA | 3 | 0 | 0 | Not-determined | Genome-fragment | NA | NA | 0 |
| S12_vs_k141_110353\|\|full | 1214 | No | NA | 3 | 0 | 1 | Not-determined | Genome-fragment | NA | NA | 0 |
| S13_vs_42476\|\|full | 3981 | No | NA | 5 | 0 | 0 | Low-quality | Genome-fragment | 4.28 | AAI-based (high-confidence) | 0 |
| S12_vs_k141_133979\|\|full | 3834 | No | NA | 4 | 0 | 1 | Not-determined | Genome-fragment | NA | NA | 0 |
| S12_vs_k141_63121\|\|full | 1154 | No | NA | 2 | 0 | 1 | Not-determined | Genome-fragment | NA | NA | 0 |
| S12_vs_k141_204\|\|full | 1877 | No | NA | 2 | 0 | 0 | Not-determined | Genome-fragment | NA | NA | 0 |
| S12_vs_k141_118247\|\|full | 1290 | No | NA | 2 | 0 | 1 | Not-determined | Genome-fragment | NA | NA | 0 |
| S16_extracted_36055 | 3981 | No | NA | 8 | 7 | 0 | Low-quality | Genome-fragment | 13.11 | AAI-based (high-confidence) | 0 |
| S12_vs_k141_165539\|\|full | 1499 | No | NA | 3 | 0 | 2 | Not-determined | Genome-fragment | NA | NA | 0 |
| S12_vs_k141_165542\|\|full | 1888 | No | NA | 4 | 0 | 0 | Not-determined | Genome-fragment | NA | NA | 0 |
| S12_vs_k141_141848\|\|full | 2571 | No | NA | 3 | 0 | 3 | Not-determined | Genome-fragment | NA | NA | 0 |
| S8_vs_k141_80084\|\|full | 3981 | No | NA | 5 | 0 | 1 | Low-quality | Genome-fragment | 4.63 | AAI-based (medium-confidence) | 0 |
| S12_vs_k141_78888\|\|full | 5523 | No | NA | 8 | 0 | 5 | Not-determined | Genome-fragment | NA | NA | 0 |
| S11_vs_k141_27836\|\|full | 3983 | No | NA | 3 | 0 | 0 | Low-quality | Genome-fragment | 4.73 | AAI-based (high-confidence) | 0 |
| S11_vs_k141_68306\|\|full | 3984 | No | NA | 10 | 0 | 0 | Low-quality | Genome-fragment | 4.47 | AAI-based (high-confidence) | 0 |
| S12_vs_k141_232\|\|full | 2999 | No | NA | 5 | 0 | 1 | Not-determined | Genome-fragment | NA | NA | 0 |
| S12_vs_k141_134002\|\|full | 4199 | No | NA | 5 | 0 | 3 | Not-determined | Genome-fragment | NA | NA | 0 |
| S11_vs_k141_89441\|\|full | 3984 | No | NA | 3 | 0 | 0 | Low-quality | Genome-fragment | 7.83 | AAI-based (high-confidence) | 0 |
| S13_vs_143428\|\|full | 3984 | No | NA | 10 | 2 | 0 | Low-quality | Genome-fragment | 11.03 | AAI-based (high-confidence) | 0 |
| S1_vs_k141_127610\|\|full | 3985 | No | NA | 4 | 0 | 2 | Low-quality | Genome-fragment | 1.87 | AAI-based (high-confidence) | 0 |
| S12_vs_k141_173419\|\|full | 1372 | No | NA | 3 | 0 | 0 | Not-determined | Genome-fragment | NA | NA | 0 |
| S17_vs_k141_9483\|\|full | 3986 | No | NA | 3 | 0 | 3 | Low-quality | Genome-fragment | 2.22 | AAI-based (medium-confidence) | 0 |
| S12_vs_k141_181297\|\|full | 9880 | No | NA | 7 | 0 | 3 | Not-determined | Genome-fragment | NA | NA | 0 |
| S12_vs_k141_102587\|\|full | 2305 | No | NA | 3 | 0 | 3 | Not-determined | Genome-fragment | NA | NA | 0 |
| S12_vs_k141_63178\|\|full | 1687 | No | NA | 3 | 0 | 1 | Not-determined | Genome-fragment | NA | NA | 0 |
| S17_extracted_k141_165399 | 3987 | No | NA | 3 | 3 | 0 | Low-quality | Genome-fragment | 13.14 | AAI-based (high-confidence) | 0 |
| S12_vs_k141_181304\|\|full | 5181 | No | NA | 6 | 0 | 3 | Not-determined | Genome-fragment | NA | NA | 0 |
| S5_extracted_k141_145619 | 3988 | No | NA | 6 | 0 | 0 | Low-quality | Genome-fragment | 25.31 | AAI-based (medium-confidence) | 0 |
| S12_vs_k141_94742\|\|full | 3483 | No | NA | 3 | 0 | 1 | Not-determined | Genome-fragment | NA | NA | 0 |
| S8_vs_k141_88879\|\|full | 3988 | No | NA | 5 | 0 | 0 | Low-quality | Genome-fragment | 6.72 | AAI-based (medium-confidence) | 0 |
| S3_extracted_k141_65746 | 3989 | No | NA | 2 | 0 | 2 | Low-quality | Genome-fragment | 0.35 | AAI-based (medium-confidence) | 0 |
| S16_extracted_36102 | 3990 | No | NA | 7 | 1 | 0 | Low-quality | Genome-fragment | 7.84 | AAI-based (high-confidence) | 0 |
| S5_vs_k141_87153\|\|full | 3991 | No | NA | 4 | 0 | 1 | Low-quality | Genome-fragment | 1.44 | AAI-based (medium-confidence) | 0 |
| S16_vs_440\|\|full | 3991 | No | NA | 8 | 0 | 0 | Low-quality | Genome-fragment | 6.39 | AAI-based (medium-confidence) | 0 |
| S16_vs_172721\|\|full | 3991 | Yes | 1337 | 3 | 1 | 1 | Low-quality | Genome-fragment | 3.03 | AAI-based (medium-confidence) | 66.5 |
| S12_vs_k141_94769\|\|full | 1526 | No | NA | 3 | 0 | 0 | Not-determined | Genome-fragment | NA | NA | 0 |
| S12_vs_k141_8107\|\|full | 4711 | No | NA | 3 | 0 | 2 | Not-determined | Genome-fragment | NA | NA | 0 |
| S8_extracted_k141_6781 | 3991 | No | NA | 3 | 0 | 2 | Low-quality | Genome-fragment | 1.1 | AAI-based (medium-confidence) | 0 |
| S12_vs_k141_55273\|\|full | 1142 | No | NA | 3 | 0 | 0 | Not-determined | Genome-fragment | NA | NA | 0 |
| S12_vs_k141_55277\|\|full | 2854 | No | NA | 3 | 0 | 1 | Not-determined | Genome-fragment | NA | NA | 0 |
| S12_vs_k141_102658\|\|full | 1186 | No | NA | 3 | 0 | 0 | Not-determined | Genome-fragment | NA | NA | 0 |
| S8_vs_k141_9367\|\|full | 3991 | No | NA | 9 | 2 | 0 | Low-quality | Genome-fragment | 4.91 | HMM-based (lower-bound) | 0 |
| S12_vs_k141_102677\|\|full | 5365 | No | NA | 3 | 0 | 0 | Not-determined | Genome-fragment | NA | NA | 0 |
| S5_vs_k141_104786\|\|full | 3993 | No | NA | 7 | 0 | 3 | Low-quality | Genome-fragment | 4.58 | AAI-based (medium-confidence) | 0 |
| S8_extracted_k141_27258 | 3994 | No | NA | 4 | 0 | 4 | Low-quality | Genome-fragment | 3.09 | AAI-based (medium-confidence) | 0 |
| S12_vs_k141_47414\|\|full | 2562 | No | NA | 3 | 0 | 1 | Not-determined | Genome-fragment | NA | NA | 0 |
| S14_vs_k141_107244\|\|full | 3995 | Yes | 2704 | 6 | 1 | 1 | Low-quality | Genome-fragment | 7.41 | AAI-based (high-confidence) | 32.32 |
| S12_vs_k141_94805\|\|full | 4087 | No | NA | 6 | 0 | 0 | Not-determined | Genome-fragment | NA | NA | 0 |
| S8_extracted_k141_10790 | 3996 | No | NA | 4 | 0 | 1 | Low-quality | Genome-fragment | 6.8 | AAI-based (medium-confidence) | 0 |
| S12_vs_k141_102692\|\|full | 1554 | No | NA | 3 | 0 | 2 | Not-determined | Genome-fragment | NA | NA | 0 |
| S12_vs_k141_94813\|\|full | 1331 | No | NA | 3 | 0 | 3 | Not-determined | Genome-fragment | NA | NA | 0 |
| S12_vs_k141_8146\|\|full | 1503 | No | NA | 5 | 0 | 0 | Not-determined | Genome-fragment | NA | NA | 0 |
| S12_vs_k141_71175\|\|full | 1584 | No | NA | 2 | 0 | 1 | Not-determined | Genome-fragment | NA | NA | 0 |
| S12_vs_k141_79010\|\|full | 1301 | No | NA | 3 | 0 | 2 | Not-determined | Genome-fragment | NA | NA | 0 |
| S12_vs_k141_383\|\|full | 2955 | No | NA | 3 | 0 | 0 | Not-determined | Genome-fragment | NA | NA | 0 |
| S12_vs_k141_86956\|\|full | 4794 | No | NA | 3 | 0 | 3 | Not-determined | Genome-fragment | NA | NA | 0 |
| S12_vs_k141_15932\|\|full | 1796 | No | NA | 3 | 0 | 2 | Not-determined | Genome-fragment | NA | NA | 0 |
| S9_extracted_k141_34058 | 3997 | No | NA | 9 | 4 | 0 | Low-quality | Genome-fragment | 10.06 | AAI-based (high-confidence) | 0 |
| S12_vs_k141_71191\|\|full | 1635 | No | NA | 3 | 0 | 0 | Not-determined | Genome-fragment | NA | NA | 0 |
| S8_extracted_k141_100158 | 3997 | No | NA | 3 | 0 | 1 | Low-quality | Genome-fragment | 8.62 | AAI-based (high-confidence) | 0 |
| S14_vs_k141_44564\|\|full | 3998 | No | NA | 6 | 0 | 0 | Low-quality | Genome-fragment | 16.18 | AAI-based (medium-confidence) | 0 |
| S12_vs_k141_409\|\|full | 1831 | No | NA | 3 | 0 | 1 | Not-determined | Genome-fragment | NA | NA | 0 |
| S17_extracted_k141_131629 | 3999 | No | NA | 10 | 1 | 0 | Low-quality | Genome-fragment | 2.09 | HMM-based (lower-bound) | 0 |
| S17_vs_k141_25199\|\|full | 3999 | No | NA | 3 | 0 | 0 | Low-quality | Genome-fragment | 5.23 | AAI-based (high-confidence) | 0 |
| S12_vs_k141_55350\|\|full | 1365 | No | NA | 2 | 0 | 0 | Not-determined | Genome-fragment | NA | NA | 0 |
| S16_vs_38528\|\|full | 3999 | No | NA | 7 | 1 | 0 | Low-quality | Genome-fragment | 2.69 | HMM-based (lower-bound) | 0 |
| S3_extracted_k141_18408 | 4000 | No | NA | 7 | 0 | 0 | Low-quality | Genome-fragment | 9.12 | AAI-based (high-confidence) | 0 |
| S10_extracted_73108 | 4000 | No | NA | 6 | 0 | 0 | Low-quality | Genome-fragment | 9.69 | AAI-based (high-confidence) | 0 |
| S8_extracted_k141_110817 | 4000 | No | NA | 5 | 2 | 0 | Low-quality | Genome-fragment | 9.69 | AAI-based (high-confidence) | 0 |
| S16_extracted_45652 | 4001 | No | NA | 4 | 0 | 3 | Low-quality | Genome-fragment | 2.31 | AAI-based (medium-confidence) | 0 |
| S12_vs_k141_442\|\|full | 2065 | No | NA | 4 | 0 | 1 | Not-determined | Genome-fragment | NA | NA | 0 |
| S16_vs_4504\|\|full | 4001 | No | NA | 4 | 1 | 0 | Low-quality | Genome-fragment | 9.82 | AAI-based (high-confidence) | 0 |
| S12_vs_k141_149889\|\|full | 5083 | No | NA | 5 | 0 | 1 | Not-determined | Genome-fragment | NA | NA | 0 |
| S12_vs_k141_87057\|\|full | 2248 | No | NA | 3 | 0 | 2 | Not-determined | Genome-fragment | NA | NA | 0 |
| S12_vs_k141_149891\|\|full | 3350 | No | NA | 5 | 0 | 4 | Not-determined | Genome-fragment | NA | NA | 0 |
| S12_vs_k141_55429\|\|full | 2441 | No | NA | 3 | 0 | 0 | Not-determined | Genome-fragment | NA | NA | 0 |
| S11_extracted_k141_5418 | 4002 | No | NA | 3 | 0 | 3 | Low-quality | Genome-fragment | 2.56 | AAI-based (medium-confidence) | 0 |
| S16_extracted_62713 | 4002 | Yes | 3336 | 5 | 1 | 1 | Low-quality | Genome-fragment | 5.89 | AAI-based (high-confidence) | 16.64 |
| S12_vs_k141_31749\|\|full | 1316 | No | NA | 2 | 0 | 1 | Not-determined | Genome-fragment | NA | NA | 0 |
| S16_vs_22901\|\|full | 4002 | No | NA | 5 | 0 | 0 | Low-quality | Genome-fragment | 7.1 | AAI-based (medium-confidence) | 0 |
| S6_vs_k141_16693\|\|full | 4002 | Yes | 2220 | 4 | 1 | 1 | Low-quality | Genome-fragment | 5.14 | AAI-based (high-confidence) | 44.53 |
| S9_vs_k141_44257\|\|full | 4004 | No | NA | 10 | 9 | 0 | Low-quality | Genome-fragment | 9.2 | AAI-based (high-confidence) | 0 |
| S3_vs_k141_122253\|\|full | 4005 | No | NA | 4 | 0 | 1 | Low-quality | Genome-fragment | 4.83 | AAI-based (high-confidence) | 0 |
| S12_vs_k141_15994\|\|full | 16763 | No | NA | 16 | 0 | 4 | Not-determined | Genome-fragment | NA | NA | 0 |
| S5_vs_k141_109830\|\|full | 4005 | No | NA | 6 | 1 | 0 | Low-quality | Genome-fragment | 8.93 | AAI-based (medium-confidence) | 0 |
| S12_vs_k141_110617\|\|full | 10268 | No | NA | 11 | 0 | 4 | Not-determined | Genome-fragment | NA | NA | 0 |
| S16_extracted_113913 | 4005 | No | NA | 4 | 0 | 4 | Low-quality | Genome-fragment | 2.2 | AAI-based (medium-confidence) | 0 |
| S13_extracted_65574 | 4006 | No | NA | 5 | 0 | 1 | Low-quality | Genome-fragment | 7.69 | AAI-based (medium-confidence) | 0 |
| S8_extracted_k141_155448 | 4006 | No | NA | 4 | 0 | 3 | Low-quality | Genome-fragment | 2.76 | AAI-based (medium-confidence) | 0 |
| S12_vs_k141_118511\|\|full | 4455 | No | NA | 7 | 0 | 2 | Not-determined | Genome-fragment | NA | NA | 0 |
| S12_vs_k141_63442\|\|full | 1216 | No | NA | 2 | 0 | 2 | Not-determined | Genome-fragment | NA | NA | 0 |
| S3_extracted_k141_98057 | 4007 | No | NA | 4 | 0 | 4 | Low-quality | Genome-fragment | 1.25 | AAI-based (medium-confidence) | 0 |
| S12_vs_k141_16045\|\|full | 8275 | No | NA | 12 | 0 | 1 | Not-determined | Genome-fragment | NA | NA | 0 |
| S16_extracted_100765 | 4007 | No | NA | 4 | 0 | 4 | Low-quality | Genome-fragment | 0.67 | AAI-based (high-confidence) | 0 |
| S3_vs_k141_67643\|\|full | 4008 | No | NA | 6 | 0 | 0 | Low-quality | Genome-fragment | 6.67 | AAI-based (medium-confidence) | 0 |
| S12_vs_k141_118540\|\|full | 2587 | No | NA | 4 | 0 | 2 | Not-determined | Genome-fragment | NA | NA | 0 |
| S12_vs_k141_8204\|\|full | 155395 | No | NA | 252 | 28 | 9 | Medium-quality | Genome-fragment | 86.93 | AAI-based (high-confidence) | 0 |
| S12_vs_k141_63482\|\|full | 3767 | No | NA | 4 | 0 | 2 | Not-determined | Genome-fragment | NA | NA | 0 |
| S7_extracted_37789 | 4008 | No | NA | 2 | 0 | 2 | Low-quality | Genome-fragment | 1.26 | AAI-based (medium-confidence) | 0 |
| S8_vs_k141_205418\|\|full | 4008 | No | NA | 6 | 1 | 0 | Low-quality | Genome-fragment | 8.79 | AAI-based (high-confidence) | 0 |
| S12_vs_k141_157916\|\|full | 5007 | No | NA | 6 | 0 | 4 | Not-determined | Genome-fragment | NA | NA | 0 |
| S5_extracted_k141_116577 | 4009 | No | NA | 4 | 0 | 0 | Low-quality | Genome-fragment | 9.36 | AAI-based (medium-confidence) | 0 |
| S16_extracted_101437 | 4010 | No | NA | 5 | 4 | 0 | Low-quality | Genome-fragment | 6.59 | AAI-based (high-confidence) | 0 |
| S1_extracted_k141_55161 | 4011 | No | NA | 5 | 1 | 0 | Low-quality | Genome-fragment | 9.63 | AAI-based (high-confidence) | 0 |
| S3_vs_k141_95592\|\|full | 4012 | No | NA | 5 | 4 | 0 | Low-quality | Genome-fragment | 9.65 | AAI-based (high-confidence) | 0 |
| S5_vs_k141_43805\|\|full | 4012 | No | NA | 4 | 0 | 3 | Low-quality | Genome-fragment | 1.26 | AAI-based (medium-confidence) | 0 |
| S12_vs_k141_47504\|\|full | 1977 | No | NA | 3 | 0 | 1 | Not-determined | Genome-fragment | NA | NA | 0 |
| S12_vs_k141_118567\|\|full | 1217 | No | NA | 3 | 0 | 1 | Not-determined | Genome-fragment | NA | NA | 0 |
| S12_vs_k141_23923\|\|full | 5830 | No | NA | 6 | 0 | 4 | Not-determined | Genome-fragment | NA | NA | 0 |
| S11_vs_k141_83060\|\|full | 4013 | No | NA | 9 | 2 | 0 | Low-quality | Genome-fragment | 7.04 | AAI-based (high-confidence) | 0 |
| S7_extracted_65369 | 4013 | No | NA | 9 | 3 | 0 | Low-quality | Genome-fragment | 8.8 | AAI-based (high-confidence) | 0 |
| S3_extracted_k141_121441 | 4014 | No | NA | 2 | 2 | 0 | Low-quality | Genome-fragment | 9.94 | AAI-based (high-confidence) | 0 |
| S12_vs_k141_118581\|\|full | 1674 | No | NA | 3 | 0 | 2 | Not-determined | Genome-fragment | NA | NA | 0 |
| S3_extracted_k141_101297 | 4014 | No | NA | 3 | 0 | 1 | Low-quality | Genome-fragment | 2.69 | AAI-based (high-confidence) | 0 |
| S16_vs_157702\|\|full | 4014 | No | NA | 10 | 0 | 1 | Low-quality | Genome-fragment | 6.13 | AAI-based (medium-confidence) | 0 |
| S12_vs_k141_150003\|\|full | 2495 | No | NA | 2 | 0 | 0 | Not-determined | Genome-fragment | NA | NA | 0 |
| S12_vs_k141_23960\|\|full | 2362 | No | NA | 3 | 0 | 3 | Not-determined | Genome-fragment | NA | NA | 0 |
| S12_vs_k141_663\|\|full | 1402 | No | NA | 2 | 0 | 2 | Not-determined | Genome-fragment | NA | NA | 0 |
| S13_extracted_136487 | 4015 | No | NA | 13 | 0 | 0 | Low-quality | Genome-fragment | 10.13 | AAI-based (high-confidence) | 0 |
| S12_vs_k141_55559\|\|full | 5921 | No | NA | 4 | 0 | 2 | Not-determined | Genome-fragment | NA | NA | 0 |
| S14_vs_k141_111506\|\|full | 4015 | No | NA | 6 | 0 | 0 | Low-quality | Genome-fragment | 7.04 | AAI-based (high-confidence) | 0 |
| S1_extracted_k141_28124 | 4015 | No | NA | 4 | 0 | 4 | Low-quality | Genome-fragment | 0.51 | AAI-based (medium-confidence) | 0 |
| S1_extracted_k141_3717 | 4015 | No | NA | 3 | 0 | 2 | Low-quality | Genome-fragment | 1.45 | AAI-based (medium-confidence) | 0 |
| S14_vs_k141_54891\|\|full | 4016 | No | NA | 5 | 4 | 0 | Low-quality | Genome-fragment | 10.53 | AAI-based (high-confidence) | 0 |
| S3_extracted_k141_85496 | 4016 | No | NA | 3 | 0 | 3 | Low-quality | Genome-fragment | 0.35 | AAI-based (medium-confidence) | 0 |
| S5_extracted_k141_115779 | 4016 | No | NA | 9 | 1 | 0 | Low-quality | Genome-fragment | 2.24 | HMM-based (lower-bound) | 0 |
| S5_vs_k141_102030\|\|full | 4016 | No | NA | 11 | 1 | 0 | Low-quality | Genome-fragment | 6.6 | AAI-based (high-confidence) | 0 |
| S12_vs_k141_713\|\|full | 1243 | No | NA | 3 | 0 | 1 | Not-determined | Genome-fragment | NA | NA | 0 |
| S12_vs_k141_150066\|\|full | 1215 | No | NA | 3 | 0 | 2 | Not-determined | Genome-fragment | NA | NA | 0 |
| S12_vs_k141_158006\|\|full | 4347 | No | NA | 5 | 0 | 4 | Not-determined | Genome-fragment | NA | NA | 0 |
| S12_vs_k141_86937\|\|full | 4016 | No | NA | 2 | 0 | 2 | Low-quality | Genome-fragment | 1.98 | AAI-based (medium-confidence) | 0 |
| S12_vs_k141_181590\|\|full | 1109 | No | NA | 2 | 0 | 2 | Not-determined | Genome-fragment | NA | NA | 0 |

| | | | | | | | | | | | |
|---|---|---|---|---|---|---|---|---|---|---|---|
| S12_vs_k141_68510\|\|full | 4016 | No | NA | 3 | 0 | 1 | Low-quality | Genome-fragment | 2.16 | AAI-based (medium-confidence) | 0 |
| S11_vs_k141_104077\|\|full | 4019 | No | NA | 9 | 0 | 1 | Low-quality | Genome-fragment | 22.71 | AAI-based (high-confidence) | 0 |
| S12_vs_k141_126597\|\|full | 2137 | No | NA | 2 | 0 | 2 | Not-determined | Genome-fragment | NA | NA | 0 |
| S13_extracted_147136 | 4020 | No | NA | 6 | 0 | 0 | Low-quality | Genome-fragment | 9.75 | AAI-based (high-confidence) | 0 |
| S8_extracted_k141_183954 | 4020 | No | NA | 4 | 1 | 2 | Low-quality | Genome-fragment | 2.18 | HMM-based (lower-bound) | 0 |
| S12_vs_k141_79326\|\|full | 1925 | No | NA | 2 | 0 | 1 | Not-determined | Genome-fragment | NA | NA | 0 |
| S12_vs_k141_71375\|\|full | 4521 | No | NA | 4 | 0 | 2 | Not-determined | Genome-fragment | NA | NA | 0 |
| S12_vs_k141_8307\|\|full | 2031 | No | NA | 3 | 0 | 1 | Not-determined | Genome-fragment | NA | NA | 0 |
| S12_vs_k141_63599\|\|full | 3992 | No | NA | 5 | 0 | 4 | Not-determined | Genome-fragment | NA | NA | 0 |
| S3_extracted_k141_91613 | 4021 | No | NA | 6 | 4 | 0 | Low-quality | Genome-fragment | 18.3 | AAI-based (high-confidence) | 0 |
| S12_vs_k141_150103\|\|full | 6616 | No | NA | 9 | 0 | 3 | Not-determined | Genome-fragment | NA | NA | 0 |
| S12_vs_k141_63636\|\|full | 1894 | No | NA | 2 | 0 | 2 | Not-determined | Genome-fragment | NA | NA | 0 |
| S12_vs_k141_8347\|\|full | 2383 | No | NA | 3 | 0 | 1 | Not-determined | Genome-fragment | NA | NA | 0 |
| S12_vs_k141_19504\|\|full | 4021 | No | NA | 8 | 0 | 0 | Low-quality | Genome-fragment | 9.44 | AAI-based (high-confidence) | 0 |
| S12_vs_k141_71424\|\|full | 7115 | No | NA | 8 | 0 | 4 | Not-determined | Genome-fragment | NA | NA | 0 |
| S12_vs_k141_71434\|\|full | 1712 | No | NA | 4 | 0 | 1 | Not-determined | Genome-fragment | NA | NA | 0 |
| S12_vs_k141_16103\|\|full | 1146 | No | NA | 3 | 0 | 1 | Not-determined | Genome-fragment | NA | NA | 0 |
| S16_extracted_48629 | 4022 | No | NA | 4 | 0 | 0 | Low-quality | Genome-fragment | 3.55 | AAI-based (medium-confidence) | 0 |
| S12_vs_k141_142415\|\|full | 2830 | No | NA | 4 | 0 | 0 | Not-determined | Genome-fragment | NA | NA | 0 |
| S12_vs_k141_110766\|\|full | 1006 | No | NA | 2 | 0 | 1 | Not-determined | Genome-fragment | NA | NA | 0 |
| S16_extracted_101335 | 4022 | No | NA | 2 | 0 | 1 | Low-quality | Genome-fragment | 0.35 | AAI-based (medium-confidence) | 0 |
| S12_vs_k141_181707\|\|full | 1300 | No | NA | 3 | 0 | 0 | Not-determined | Genome-fragment | NA | NA | 0 |
| S12_vs_k141_79378\|\|full | 1387 | No | NA | 2 | 0 | 1 | Not-determined | Genome-fragment | NA | NA | 0 |
| S7_vs_106875\|\|full | 4023 | No | NA | 5 | 4 | 0 | Low-quality | Genome-fragment | 11.74 | AAI-based (high-confidence) | 0 |
| S12_vs_k141_39952\|\|full | 1472 | No | NA | 2 | 0 | 2 | Not-determined | Genome-fragment | NA | NA | 0 |
| S12_vs_k141_87226\|\|full | 3249 | No | NA | 2 | 0 | 1 | Not-determined | Genome-fragment | NA | NA | 0 |
| S9_vs_k141_79894\|\|full | 4025 | No | NA | 6 | 0 | 3 | Low-quality | Genome-fragment | 2.84 | AAI-based (high-confidence) | 0 |
| S5_vs_k141_88442\|\|full | 4029 | Yes | 2373 | 5 | 2 | 1 | Low-quality | Genome-fragment | 4.4 | AAI-based (high-confidence) | 41.1 |
| S10_vs_92244\|\|lt2gene | 4030 | No | NA | 1 | 1 | 0 | Low-quality | Genome-fragment | 1.7 | HMM-based (lower-bound) | 0 |
| S16_extracted_84564 | 4031 | No | NA | 6 | 0 | 4 | Low-quality | Genome-fragment | 5.85 | AAI-based (medium-confidence) | 0 |
| S10_vs_60578\|\|full | 4031 | No | NA | 8 | 2 | 0 | Low-quality | Genome-fragment | 6.5 | AAI-based (high-confidence) | 0 |
| S12_vs_k141_165960\|\|full | 1667 | No | NA | 2 | 0 | 0 | Not-determined | Genome-fragment | NA | NA | 0 |
| S15_extracted_59253 | 4031 | No | NA | 7 | 0 | 0 | Low-quality | Genome-fragment | 2.18 | AAI-based (high-confidence) | 0 |
| S8_vs_k141_76152\|\|full | 4034 | No | NA | 4 | 0 | 2 | Low-quality | Genome-fragment | 1.96 | AAI-based (medium-confidence) | 0 |
| S12_vs_k141_830\|\|full | 1472 | No | NA | 2 | 0 | 1 | Not-determined | Genome-fragment | NA | NA | 0 |
| S12_vs_k141_833\|\|full | 3077 | No | NA | 2 | 0 | 1 | Not-determined | Genome-fragment | NA | NA | 0 |
| S12_vs_k141_8455\|\|full | 1093 | No | NA | 3 | 0 | 1 | Not-determined | Genome-fragment | NA | NA | 0 |
| S12_vs_k141_103006\|\|full | 2513 | No | NA | 3 | 0 | 3 | Not-determined | Genome-fragment | NA | NA | 0 |
| S14_vs_k141_73382\|\|full | 4036 | No | NA | 6 | 4 | 0 | Low-quality | Genome-fragment | 8.58 | AAI-based (high-confidence) | 0 |
| S12_vs_k141_126658\|\|full | 2507 | No | NA | 3 | 0 | 1 | Not-determined | Genome-fragment | NA | NA | 0 |
| S8_extracted_k141_149348 | 4036 | No | NA | 5 | 0 | 0 | Low-quality | Genome-fragment | 7.45 | AAI-based (high-confidence) | 0 |
| S17_extracted_k141_87647 | 4037 | No | NA | 5 | 0 | 3 | Low-quality | Genome-fragment | 2.31 | AAI-based (medium-confidence) | 0 |
| S12_vs_k141_181776\|\|full | 2180 | No | NA | 3 | 0 | 3 | Not-determined | Genome-fragment | NA | NA | 0 |
| S13_extracted_9391 | 4037 | No | NA | 8 | 1 | 0 | Low-quality | Genome-fragment | 6.31 | AAI-based (high-confidence) | 0 |
| S12_vs_k141_16231\|\|full | 1489 | No | NA | 2 | 0 | 1 | Not-determined | Genome-fragment | NA | NA | 0 |
| S12_vs_k141_71525\|\|full | 31779 | No | NA | 39 | 4 | 1 | Medium-quality | Genome-fragment | 56.31 | HMM-based (lower-bound) | 0 |
| S13_extracted_17442 | 4037 | No | NA | 5 | 0 | 1 | Low-quality | Genome-fragment | 9.15 | AAI-based (medium-confidence) | 0 |
| S7_vs_18588\|\|full | 4038 | No | NA | 5 | 0 | 0 | Low-quality | Genome-fragment | 10.53 | AAI-based (high-confidence) | 0 |
| S7_vs_31554\|\|full | 4040 | No | NA | 5 | 2 | 0 | Low-quality | Genome-fragment | 4.93 | HMM-based (lower-bound) | 0 |
| S12_extracted_k141_132275 | 4040 | No | NA | 4 | 0 | 2 | Low-quality | Genome-fragment | 11.84 | AAI-based (medium-confidence) | 0 |
| S12_vs_k141_47647\|\|full | 4035 | No | NA | 7 | 0 | 1 | Not-determined | Genome-fragment | NA | NA | 0 |
| S12_extracted_k141_133274 | 4040 | Yes | 1754 | 4 | 1 | 1 | Low-quality | Genome-fragment | 4.99 | AAI-based (high-confidence) | 56.58 |
| S14_vs_k141_59445\|\|full | 4041 | No | NA | 6 | 2 | 0 | Low-quality | Genome-fragment | 8.73 | AAI-based (high-confidence) | 0 |
| S12_vs_k141_55760\|\|full | 4131 | No | NA | 3 | 0 | 1 | Not-determined | Genome-fragment | NA | NA | 0 |
| S12_vs_k141_24242\|\|full | 1737 | No | NA | 2 | 0 | 0 | Not-determined | Genome-fragment | NA | NA | 0 |
| S1_vs_k141_105145\|\|full | 4041 | No | NA | 5 | 1 | 0 | Low-quality | Genome-fragment | 10.46 | AAI-based (medium-confidence) | 0 |
| S12_vs_k141_55764\|\|full | 1523 | No | NA | 3 | 0 | 0 | Not-determined | Genome-fragment | NA | NA | 0 |
| S12_vs_k141_166072\|\|full | 1835 | No | NA | 3 | 0 | 0 | Not-determined | Genome-fragment | NA | NA | 0 |
| S8_extracted_k141_49342 | 4041 | No | NA | 9 | 1 | 0 | Low-quality | Genome-fragment | 3.52 | AAI-based (high-confidence) | 0 |
| S12_vs_k141_24251\|\|full | 2738 | No | NA | 2 | 0 | 0 | Not-determined | Genome-fragment | NA | NA | 0 |
| S14_extracted_k141_3332 | 4042 | No | NA | 3 | 0 | 0 | Low-quality | Genome-fragment | 6.88 | AAI-based (high-confidence) | 0 |
| S12_vs_k141_150279\|\|full | 11444 | No | NA | 15 | 0 | 0 | Not-determined | Genome-fragment | NA | NA | 0 |
| S12_vs_k141_24252\|\|full | 3111 | No | NA | 2 | 0 | 2 | Not-determined | Genome-fragment | NA | NA | 0 |
| S3_extracted_k141_111546 | 4042 | No | NA | 7 | 1 | 0 | Low-quality | Genome-fragment | 5.4 | AAI-based (high-confidence) | 0 |
| S12_vs_k141_87373\|\|full | 1533 | No | NA | 2 | 0 | 2 | Not-determined | Genome-fragment | NA | NA | 0 |
| S12_vs_k141_47672\|\|full | 1655 | No | NA | 3 | 0 | 1 | Not-determined | Genome-fragment | NA | NA | 0 |
| S12_vs_k141_40101\|\|full | 2937 | No | NA | 3 | 0 | 1 | Not-determined | Genome-fragment | NA | NA | 0 |
| S12_vs_k141_79473\|\|full | 1359 | No | NA | 2 | 0 | 0 | Not-determined | Genome-fragment | NA | NA | 0 |
| S12_vs_k141_71583\|\|full | 1892 | No | NA | 3 | 0 | 1 | Not-determined | Genome-fragment | NA | NA | 0 |
| S12_vs_k141_24272\|\|full | 2205 | No | NA | 2 | 0 | 1 | Not-determined | Genome-fragment | NA | NA | 0 |
| S12_vs_k141_126719\|\|full | 1196 | No | NA | 3 | 0 | 1 | Not-determined | Genome-fragment | NA | NA | 0 |
| S7_extracted_91970 | 4042 | No | NA | 5 | 2 | 0 | Low-quality | Genome-fragment | 6.55 | AAI-based (high-confidence) | 0 |
| S12_vs_k141_71596\|\|full | 2409 | No | NA | 3 | 0 | 2 | Not-determined | Genome-fragment | NA | NA | 0 |
| S17_extracted_k141_139176 | 4043 | No | NA | 8 | 0 | 2 | Low-quality | Genome-fragment | 8.66 | AAI-based (medium-confidence) | 0 |
| S12_vs_k141_150344\|\|full | 1122 | No | NA | 3 | 0 | 0 | Not-determined | Genome-fragment | NA | NA | 0 |
| S1_vs_k141_52811\|\|full | 4043 | No | NA | 4 | 0 | 0 | Low-quality | Genome-fragment | 9.86 | AAI-based (high-confidence) | 0 |
| S8_vs_k141_187294\|\|full | 4043 | No | NA | 3 | 0 | 1 | Low-quality | Genome-fragment | 3.8 | AAI-based (medium-confidence) | 0 |
| S11_vs_k141_13922\|\|full | 4045 | No | NA | 3 | 2 | 0 | Low-quality | Genome-fragment | 3.65 | AAI-based (medium-confidence) | 0 |
| S12_vs_k141_63824\|\|full | 4547 | No | NA | 3 | 0 | 2 | Not-determined | Genome-fragment | NA | NA | 0 |
| S14_vs_k141_105024\|\|full | 4045 | No | NA | 5 | 0 | 0 | Low-quality | Genome-fragment | 9.54 | AAI-based (high-confidence) | 0 |
| S12_vs_k141_977\|\|full | 1713 | No | NA | 3 | 0 | 1 | Not-determined | Genome-fragment | NA | NA | 0 |
| S12_vs_k141_24324\|\|full | 3456 | No | NA | 5 | 0 | 1 | Not-determined | Genome-fragment | NA | NA | 0 |
| S3_vs_k141_85254\|\|full | 4045 | No | NA | 6 | 0 | 2 | Low-quality | Genome-fragment | 2.29 | AAI-based (high-confidence) | 0 |
| S12_vs_k141_156201\|\|full | 4045 | No | NA | 5 | 4 | 0 | Low-quality | Genome-fragment | 12.13 | AAI-based (high-confidence) | 0 |
| S5_extracted_k141_118632 | 4046 | No | NA | 8 | 0 | 0 | Low-quality | Genome-fragment | 6.76 | AAI-based (high-confidence) | 0 |
| S12_vs_k141_40166\|\|full | 1238 | No | NA | 2 | 0 | 0 | Not-determined | Genome-fragment | NA | NA | 0 |
| S12_vs_k141_126755\|\|full | 2834 | No | NA | 4 | 0 | 0 | Not-determined | Genome-fragment | NA | NA | 0 |
| S12_vs_k141_32226\|\|full | 15546 | No | NA | 14 | 0 | 6 | Not-determined | Genome-fragment | NA | NA | 0 |
| S10_vs_19074\|\|full | 4048 | No | NA | 7 | 1 | 0 | Low-quality | Genome-fragment | 11.79 | AAI-based (high-confidence) | 0 |
| S12_vs_k141_134465\|\|full | 1326 | No | NA | 2 | 0 | 0 | Not-determined | Genome-fragment | NA | NA | 0 |
| S3_extracted_k141_31868 | 4049 | No | NA | 4 | 0 | 0 | Low-quality | Genome-fragment | 3.57 | AAI-based (high-confidence) | 0 |
| S7_vs_73653\|\|full | 4049 | No | NA | 8 | 0 | 1 | Low-quality | Genome-fragment | 7 | AAI-based (high-confidence) | 0 |
| S14_vs_k141_75227\|\|full | 4050 | No | NA | 3 | 0 | 3 | Low-quality | Genome-fragment | 1.78 | AAI-based (medium-confidence) | 0 |
| S12_vs_k141_79569\|\|full | 1687 | No | NA | 3 | 0 | 0 | Not-determined | Genome-fragment | NA | NA | 0 |
| S8_extracted_k141_43192 | 4050 | No | NA | 3 | 0 | 2 | Low-quality | Genome-fragment | 0.57 | AAI-based (medium-confidence) | 0 |
| S12_vs_k141_40190\|\|full | 1056 | No | NA | 2 | 0 | 1 | Not-determined | Genome-fragment | NA | NA | 0 |
| S14_extracted_k141_15657 | 4052 | No | NA | 8 | 7 | 0 | Low-quality | Genome-fragment | 13.38 | AAI-based (high-confidence) | 0 |
| S16_extracted_95602 | 4052 | No | NA | 8 | 2 | 0 | Low-quality | Genome-fragment | 2.79 | AAI-based (high-confidence) | 0 |
| S12_vs_k141_142625\|\|full | 80692 | Yes | 41808 | 78 | 5 | 23 | Complete | High-quality | 100 | Provirus (medium-confidence) | 48.19 |
| S12_vs_k141_87420\|\|full | 1508 | No | NA | 3 | 0 | 0 | Not-determined | Genome-fragment | NA | NA | 0 |
| S12_vs_k141_134493\|\|full | 1421 | No | NA | 3 | 0 | 1 | Not-determined | Genome-fragment | NA | NA | 0 |
| S12_vs_k141_95283\|\|full | 3346 | No | NA | 3 | 0 | 2 | Not-determined | Genome-fragment | NA | NA | 0 |
| S12_vs_k141_158327\|\|full | 3920 | No | NA | 6 | 0 | 4 | Not-determined | Genome-fragment | NA | NA | 0 |
| S12_vs_k141_1008\|\|full | 1760 | No | NA | 3 | 0 | 1 | Not-determined | Genome-fragment | NA | NA | 0 |
| S16_vs_97466\|\|full | 4052 | No | NA | 4 | 0 | 1 | Low-quality | Genome-fragment | 2.56 | AAI-based (medium-confidence) | 0 |
| S12_vs_k141_134525\|\|full | 5832 | No | NA | 5 | 0 | 4 | Not-determined | Genome-fragment | NA | NA | 0 |
| S12_vs_k141_75135\|\|full | 4054 | No | NA | 10 | 3 | 0 | Low-quality | Genome-fragment | 10.86 | AAI-based (high-confidence) | 0 |
| S12_vs_k141_47819\|\|full | 8732 | No | NA | 7 | 0 | 2 | Not-determined | Genome-fragment | NA | NA | 0 |
| S12_vs_k141_142660\|\|full | 1756 | No | NA | 3 | 0 | 2 | Not-determined | Genome-fragment | NA | NA | 0 |
| S12_vs_k141_129855\|\|full | 4055 | No | NA | 7 | 2 | 0 | Low-quality | Genome-fragment | 9.55 | AAI-based (high-confidence) | 0 |
| S6_vs_k141_4604\|\|full | 4055 | No | NA | 4 | 0 | 2 | Low-quality | Genome-fragment | 8.11 | AAI-based (medium-confidence) | 0 |
| S12_vs_k141_166250\|\|full | 1471 | No | NA | 3 | 0 | 0 | Not-determined | Genome-fragment | NA | NA | 0 |
| S12_vs_k141_16406\|\|full | 1637 | No | NA | 3 | 0 | 2 | Not-determined | Genome-fragment | NA | NA | 0 |
| S12_vs_k141_16412\|\|full | 1465 | No | NA | 2 | 0 | 1 | Not-determined | Genome-fragment | NA | NA | 0 |
| S12_vs_k141_181967\|\|full | 1395 | No | NA | 2 | 0 | 0 | Not-determined | Genome-fragment | NA | NA | 0 |
| S8_vs_k141_132824\|\|full | 4055 | No | NA | 5 | 0 | 1 | Low-quality | Genome-fragment | 3.39 | AAI-based (medium-confidence) | 0 |
| S12_vs_k141_181984\|\|full | 2142 | No | NA | 3 | 0 | 1 | Not-determined | Genome-fragment | NA | NA | 0 |
| S11_extracted_k141_78751 | 4056 | No | NA | 2 | 0 | 0 | Low-quality | Genome-fragment | 4.5 | AAI-based (high-confidence) | 0 |
| S14_vs_k141_2765\|\|full | 4058 | No | NA | 9 | 2 | 0 | Low-quality | Genome-fragment | 4.48 | HMM-based (lower-bound) | 0 |
| S12_vs_k141_142729\|\|full | 2308 | No | NA | 2 | 0 | 1 | Not-determined | Genome-fragment | NA | NA | 0 |
| S1_vs_k141_155286\|\|full | 4058 | No | NA | 4 | 0 | 2 | Low-quality | Genome-fragment | 1.21 | AAI-based (medium-confidence) | 0 |
| S3_extracted_k141_58167 | 4058 | No | NA | 3 | 0 | 0 | Low-quality | Genome-fragment | 6.03 | AAI-based (high-confidence) | 0 |
| S12_vs_k141_150465\|\|full | 2709 | No | NA | 4 | 0 | 0 | Not-determined | Genome-fragment | NA | NA | 0 |
| S12_vs_k141_126867\|\|full | 1893 | No | NA | 4 | 0 | 1 | Not-determined | Genome-fragment | NA | NA | 0 |
| S12_vs_k141_8646\|\|full | 3812 | No | NA | 4 | 0 | 0 | Not-determined | Genome-fragment | NA | NA | 0 |
| S5_vs_k141_90404\|\|full | 4059 | No | NA | 7 | 2 | 1 | Low-quality | Genome-fragment | 9.78 | AAI-based (medium-confidence) | 0 |
| S12_vs_k141_79642\|\|full | 6344 | No | NA | 8 | 0 | 1 | Not-determined | Genome-fragment | NA | NA | 0 |
| S8_vs_k141_7097\|\|full | 4059 | No | NA | 8 | 0 | 0 | Low-quality | Genome-fragment | 9.47 | AAI-based (medium-confidence) | 0 |
| S12_vs_k141_95388\|\|full | 2070 | No | NA | 5 | 0 | 1 | Not-determined | Genome-fragment | NA | NA | 0 |
| S12_vs_k141_16483\|\|full | 2857 | No | NA | 4 | 0 | 3 | Not-determined | Genome-fragment | NA | NA | 0 |
| S13_vs_113225\|\|full | 4060 | No | NA | 5 | 1 | 1 | Low-quality | Genome-fragment | 6.52 | AAI-based (medium-confidence) | 0 |
| S12_vs_k141_71723\|\|full | 1750 | No | NA | 2 | 0 | 1 | Not-determined | Genome-fragment | NA | NA | 0 |
| S5_extracted_k141_59144 | 4060 | No | NA | 10 | 3 | 0 | Low-quality | Genome-fragment | 4.05 | HMM-based (lower-bound) | 0 |
| S16_extracted_14737 | 4060 | No | NA | 4 | 0 | 1 | Low-quality | Genome-fragment | 6.98 | AAI-based (medium-confidence) | 0 |
| S12_vs_k141_173959\|\|full | 3043 | No | NA | 3 | 0 | 3 | Not-determined | Genome-fragment | NA | NA | 0 |
| S8_vs_k141_137023\|\|full | 4060 | No | NA | 5 | 0 | 3 | Low-quality | Genome-fragment | 6.13 | AAI-based (medium-confidence) | 0 |
| S14_extracted_k141_62123 | 4061 | No | NA | 4 | 4 | 0 | Low-quality | Genome-fragment | 13.4 | AAI-based (high-confidence) | 0 |
| S8_extracted_k141_113273 | 4061 | No | NA | 4 | 0 | 3 | Low-quality | Genome-fragment | 0.82 | AAI-based (medium-confidence) | 0 |
| S5_vs_k141_42837\|\|full | 4062 | No | NA | 8 | 2 | 0 | Low-quality | Genome-fragment | 10.07 | AAI-based (high-confidence) | 0 |
| S12_vs_k141_103454\|\|full | 1099 | No | NA | 3 | 0 | 1 | Not-determined | Genome-fragment | NA | NA | 0 |
| S12_vs_k141_119074\|\|full | 1748 | No | NA | 3 | 0 | 0 | Not-determined | Genome-fragment | NA | NA | 0 |
| S10_extracted_7499 | 4063 | No | NA | 3 | 0 | 2 | Low-quality | Genome-fragment | 6.17 | AAI-based (high-confidence) | 0 |
| S12_vs_k141_95460\|\|full | 1790 | No | NA | 3 | 0 | 0 | Not-determined | Genome-fragment | NA | NA | 0 |
| S12_vs_k141_56117\|\|full | 1312 | No | NA | 3 | 0 | 1 | Not-determined | Genome-fragment | NA | NA | 0 |
| S12_vs_k141_16520\|\|full | 1422 | No | NA | 2 | 0 | 1 | Not-determined | Genome-fragment | NA | NA | 0 |
| S10_vs_32950\|\|full | 4063 | No | NA | 4 | 0 | 1 | Low-quality | Genome-fragment | 8.72 | AAI-based (medium-confidence) | 0 |
| S12_vs_k141_119079\|\|full | 2455 | No | NA | 3 | 0 | 2 | Not-determined | Genome-fragment | NA | NA | 0 |
| S1_extracted_k141_66977 | 4064 | No | NA | 1 | 0 | 1 | Low-quality | Genome-fragment | 2.1 | AAI-based (medium-confidence) | 0 |
| S12_vs_k141_103456\|\|full | 16298 | No | NA | 14 | 0 | 10 | Not-determined | Genome-fragment | NA | NA | 0 |
| S5_vs_k141_140346\|\|full | 4065 | No | NA | 6 | 4 | 0 | Low-quality | Genome-fragment | 10.6 | AAI-based (medium-confidence) | 0 |
| S12_vs_k141_95474\|\|full | 1186 | No | NA | 3 | 0 | 1 | Not-determined | Genome-fragment | NA | NA | 0 |
| S12_vs_k141_71789\|\|full | 5566 | No | NA | 4 | 0 | 4 | Not-determined | Genome-fragment | NA | NA | 0 |
| S12_vs_k141_166406\|\|full | 2613 | No | NA | 2 | 0 | 1 | Not-determined | Genome-fragment | NA | NA | 0 |
| S5_vs_k141_42162\|\|full | 4065 | No | NA | 3 | 1 | 0 | Low-quality | Genome-fragment | 6.93 | AAI-based (medium-confidence) | 0 |
| S17_vs_k141_139429\|\|full | 4066 | No | NA | 10 | 1 | 0 | Low-quality | Genome-fragment | 2.41 | AAI-based (high-confidence) | 0 |
| S8_extracted_k141_61370 | 4066 | No | NA | 14 | 0 | 0 | Low-quality | Genome-fragment | 2.53 | AAI-based (high-confidence) | 0 |
| S8_extracted_k141_206798 | 4066 | No | NA | 4 | 0 | 2 | Low-quality | Genome-fragment | 12.99 | AAI-based (high-confidence) | 0 |
| S1_extracted_k141_58012 | 4068 | No | NA | 9 | 0 | 0 | Low-quality | Genome-fragment | 9.38 | AAI-based (high-confidence) | 0 |
| S1_vs_k141_98050\|\|full | 4068 | No | NA | 6 | 1 | 0 | Low-quality | Genome-fragment | 9.97 | AAI-based (high-confidence) | 0 |
| S14_vs_k141_45459\|\|full | 4069 | No | NA | 6 | 0 | 0 | Low-quality | Genome-fragment | 10.47 | AAI-based (medium-confidence) | 0 |
| S3_extracted_k141_29647 | 4069 | No | NA | 2 | 2 | 0 | Low-quality | Genome-fragment | 10.43 | AAI-based (high-confidence) | 0 |
| S12_vs_k141_56209\|\|full | 1003 | No | NA | 2 | 0 | 0 | Not-determined | Genome-fragment | NA | NA | 0 |
| S12_vs_k141_103529\|\|full | 1135 | No | NA | 2 | 0 | 1 | Not-determined | Genome-fragment | NA | NA | 0 |
| S5_vs_k141_43603\|\|full | 4069 | No | NA | 5 | 0 | 1 | Low-quality | Genome-fragment | 4.15 | AAI-based (high-confidence) | 0 |
| S12_vs_k141_40425\|\|full | 3783 | No | NA | 3 | 0 | 1 | Not-determined | Genome-fragment | NA | NA | 0 |
| S12_vs_k141_119118\|\|full | 1568 | No | NA | 3 | 0 | 0 | Not-determined | Genome-fragment | NA | NA | 0 |
| S12_vs_k141_56241\|\|full | 1720 | No | NA | 2 | 0 | 0 | Not-determined | Genome-fragment | NA | NA | 0 |
| S12_vs_k141_48045\|\|full | 32825 | No | NA | 48 | 23 | 0 | Medium-quality | Genome-fragment | 64.36 | AAI-based (high-confidence) | 0 |
| S7_vs_98161\|\|full | 4071 | No | NA | 5 | 1 | 0 | Low-quality | Genome-fragment | 9.81 | AAI-based (medium-confidence) | 0 |
| S16_vs_136809\|\|full | 4071 | No | NA | 6 | 1 | 0 | Low-quality | Genome-fragment | 9.35 | AAI-based (high-confidence) | 0 |
| S12_vs_k141_95581\|\|full | 2307 | No | NA | 3 | 0 | 2 | Not-determined | Genome-fragment | NA | NA | 0 |
| S12_vs_k141_16647\|\|full | 1839 | No | NA | 3 | 0 | 2 | Not-determined | Genome-fragment | NA | NA | 0 |
| S15_vs_52680\|\|full | 4071 | No | NA | 5 | 1 | 0 | Low-quality | Genome-fragment | 2.92 | AAI-based (high-confidence) | 0 |
| S17_vs_k141_103356\|\|full | 4072 | No | NA | 4 | 0 | 3 | Low-quality | Genome-fragment | 2.21 | AAI-based (medium-confidence) | 0 |
| S12_vs_k141_174107\|\|full | 3604 | No | NA | 8 | 0 | 0 | Not-determined | Genome-fragment | NA | NA | 0 |
| S7_vs_111896\|\|full | 4072 | No | NA | 3 | 2 | 0 | Low-quality | Genome-fragment | 2.74 | AAI-based (high-confidence) | 0 |
| S9_extracted_k141_18431 | 4073 | No | NA | 3 | 0 | 0 | Low-quality | Genome-fragment | 2.92 | AAI-based (medium-confidence) | 0 |
| S12_vs_k141_87660\|\|full | 5504 | No | NA | 5 | 0 | 4 | Not-determined | Genome-fragment | NA | NA | 0 |
| S12_vs_k141_127010\|\|full | 5329 | No | NA | 5 | 0 | 0 | Not-determined | Genome-fragment | NA | NA | 0 |
| S12_vs_k141_48091\|\|full | 1313 | No | NA | 3 | 0 | 0 | Not-determined | Genome-fragment | NA | NA | 0 |
| S12_vs_k141_174133\|\|full | 1750 | No | NA | 2 | 0 | 1 | Not-determined | Genome-fragment | NA | NA | 0 |
| S12_vs_k141_134689\|\|full | 6043 | No | NA | 6 | 0 | 6 | Not-determined | Genome-fragment | NA | NA | 0 |
| S12_vs_k141_64193\|\|full | 5736 | No | NA | 6 | 0 | 2 | Not-determined | Genome-fragment | NA | NA | 0 |
| S16_vs_150569\|\|full | 4073 | No | NA | 4 | 0 | 3 | Low-quality | Genome-fragment | 1.96 | AAI-based (medium-confidence) | 0 |
| S8_vs_k141_140381\|\|full | 4073 | No | NA | 4 | 2 | 0 | Low-quality | Genome-fragment | 4.78 | HMM-based (lower-bound) | 0 |
| S12_vs_k141_24750\|\|full | 1212 | No | NA | 2 | 0 | 0 | Not-determined | Genome-fragment | NA | NA | 0 |
| S12_vs_k141_40530\|\|full | 2032 | No | NA | 2 | 0 | 0 | Not-determined | Genome-fragment | NA | NA | 0 |
| S17_vs_k141_74304\|\|full | 4074 | No | NA | 4 | 0 | 3 | Low-quality | Genome-fragment | 0.99 | AAI-based (medium-confidence) | 0 |
| S14_extracted_k141_21405 | 4074 | No | NA | 10 | 8 | 0 | Low-quality | Genome-fragment | 13.44 | AAI-based (high-confidence) | 0 |
| S3_extracted_k141_37210 | 4074 | No | NA | 5 | 0 | 0 | Low-quality | Genome-fragment | 7.68 | AAI-based (medium-confidence) | 0 |
| S12_vs_k141_119234\|\|full | 8392 | No | NA | 4 | 0 | 3 | Not-determined | Genome-fragment | NA | NA | 0 |
| S12_vs_k141_127035\|\|full | 1994 | No | NA | 1 | 0 | 1 | Not-determined | Genome-fragment | NA | NA | 0 |
| S12_vs_k141_127036\|\|full | 5450 | No | NA | 7 | 0 | 3 | Not-determined | Genome-fragment | NA | NA | 0 |
| S15_vs_40405\|\|full | 4074 | No | NA | 5 | 3 | 0 | Low-quality | Genome-fragment | 2.71 | AAI-based (high-confidence) | 0 |
| S5_extracted_k141_11163 | 4076 | No | NA | 3 | 2 | 0 | Low-quality | Genome-fragment | 6.39 | AAI-based (high-confidence) | 0 |
| S12_vs_k141_119249\|\|full | 1524 | No | NA | 3 | 0 | 1 | Not-determined | Genome-fragment | NA | NA | 0 |
| S12_vs_k141_150640\|\|full | 1351 | No | NA | 2 | 0 | 1 | Not-determined | Genome-fragment | NA | NA | 0 |
| S4_extracted_3973 | 4077 | No | NA | 5 | 1 | 0 | Low-quality | Genome-fragment | 20.37 | AAI-based (medium-confidence) | 0 |
| S8_vs_k141_167226\|\|full | 4078 | No | NA | 10 | 0 | 0 | Low-quality | Genome-fragment | 10.11 | AAI-based (medium-confidence) | 0 |
| S12_vs_k141_150649\|\|full | 3986 | No | NA | 4 | 0 | 1 | Not-determined | Genome-fragment | NA | NA | 0 |
| S1_extracted_k141_134471 | 4079 | No | NA | 6 | 2 | 0 | Low-quality | Genome-fragment | 8.34 | AAI-based (high-confidence) | 0 |
| S16_vs_143262\|\|full | 4079 | No | NA | 7 | 2 | 0 | Low-quality | Genome-fragment | 11.97 | AAI-based (high-confidence) | 0 |
| S9_vs_k141_72693\|\|full | 4080 | No | NA | 6 | 1 | 0 | Low-quality | Genome-fragment | 9.02 | AAI-based (high-confidence) | 0 |
| S14_extracted_k141_22587 | 4080 | No | NA | 5 | 5 | 0 | Low-quality | Genome-fragment | 13.31 | AAI-based (high-confidence) | 0 |
| S12_vs_k141_25766\|\|full | 4081 | No | NA | 8 | 0 | 0 | Low-quality | Genome-fragment | 7.07 | AAI-based (medium-confidence) | 0 |
| S8_extracted_k141_96843 | 4081 | Yes | 3117 | 7 | 1 | 1 | Low-quality | Genome-fragment | 2.17 | HMM-based (lower-bound) | 23.62 |
| S11_vs_k141_6877\|\|full | 4083 | No | NA | 11 | 2 | 0 | Low-quality | Genome-fragment | 3.12 | HMM-based (lower-bound) | 0 |
| S14_vs_k141_82652\|\|full | 4083 | No | NA | 5 | 0 | 1 | Low-quality | Genome-fragment | 6.22 | AAI-based (medium-confidence) | 0 |
| S12_vs_k141_119284\|\|full | 6137 | No | NA | 5 | 0 | 2 | Not-determined | Genome-fragment | NA | NA | 0 |
| S8_extracted_k141_85167 | 4083 | No | NA | 6 | 0 | 1 | Low-quality | Genome-fragment | 9.61 | AAI-based (high-confidence) | 0 |
| S12_vs_k141_16730\|\|full | 3426 | No | NA | 3 | 0 | 2 | Not-determined | Genome-fragment | NA | NA | 0 |
| S12_vs_k141_182371\|\|full | 1482 | No | NA | 3 | 0 | 1 | Not-determined | Genome-fragment | NA | NA | 0 |
| S12_vs_k141_111143\|\|full | 1668 | No | NA | 3 | 0 | 2 | Not-determined | Genome-fragment | NA | NA | 0 |
| S12_vs_k141_8888\|\|full | 6874 | No | NA | 8 | 0 | 6 | Not-determined | Genome-fragment | NA | NA | 0 |
| S9_vs_k141_3443\|\|full | 4084 | No | NA | 7 | 3 | 0 | Low-quality | Genome-fragment | 10.04 | AAI-based (medium-confidence) | 0 |
| S12_vs_k141_119292\|\|full | 7488 | No | NA | 7 | 0 | 1 | Not-determined | Genome-fragment | NA | NA | 0 |
| S12_vs_k141_166562\|\|full | 1067 | No | NA | 2 | 0 | 1 | Not-determined | Genome-fragment | NA | NA | 0 |
| S16_vs_52345\|\|full | 4085 | No | NA | 4 | 0 | 2 | Low-quality | Genome-fragment | 2.47 | AAI-based (medium-confidence) | 0 |
| S12_vs_k141_24837\|\|full | 2333 | No | NA | 4 | 0 | 1 | Not-determined | Genome-fragment | NA | NA | 0 |
| S12_vs_k141_111167\|\|full | 3015 | No | NA | 3 | 0 | 2 | Not-determined | Genome-fragment | NA | NA | 0 |
| S16_vs_108334\|\|full | 4085 | No | NA | 6 | 0 | 0 | Low-quality | Genome-fragment | 6.75 | AAI-based (medium-confidence) | 0 |
| S12_vs_k141_1311\|\|full | 10090 | No | NA | 16 | 0 | 2 | Not-determined | Genome-fragment | NA | NA | 0 |
| S13_vs_2577\|\|full | 4086 | No | NA | 9 | 1 | 0 | Low-quality | Genome-fragment | 3.44 | AAI-based (medium-confidence) | 0 |
| S3_vs_k141_37550\|\|full | 4087 | Yes | 1864 | 10 | 1 | 1 | Low-quality | Genome-fragment | 3.91 | AAI-based (medium-confidence) | 54.39 |
| S12_vs_k141_134785\|\|full | 3626 | No | NA | 3 | 0 | 1 | Not-determined | Genome-fragment | NA | NA | 0 |
| S12_vs_k141_56414\|\|full | 1742 | No | NA | 2 | 0 | 0 | Not-determined | Genome-fragment | NA | NA | 0 |
| S12_vs_k141_24886\|\|full | 1908 | No | NA | 4 | 0 | 4 | Not-determined | Genome-fragment | NA | NA | 0 |
| S12_vs_k141_119349\|\|full | 6436 | No | NA | 5 | 0 | 4 | Not-determined | Genome-fragment | NA | NA | 0 |
| S10_vs_101684\|\|full | 4087 | No | NA | 4 | 0 | 4 | Low-quality | Genome-fragment | 1.63 | AAI-based (medium-confidence) | 0 |
| S12_vs_k141_134817\|\|full | 1307 | No | NA | 2 | 0 | 0 | Not-determined | Genome-fragment | NA | NA | 0 |
| S8_vs_k141_92331\|\|full | 4087 | No | NA | 5 | 0 | 2 | Low-quality | Genome-fragment | 1.51 | AAI-based (medium-confidence) | 0 |
| S9_vs_k141_917\|\|full | 4088 | No | NA | 5 | 0 | 1 | Low-quality | Genome-fragment | 8.38 | AAI-based (high-confidence) | 0 |
| S12_vs_k141_182401\|\|full | 1529 | No | NA | 3 | 0 | 0 | Not-determined | Genome-fragment | NA | NA | 0 |
| S12_vs_k141_56452\|\|full | 1030 | No | NA | 3 | 0 | 0 | Not-determined | Genome-fragment | NA | NA | 0 |
| S13_extracted_23202 | 4088 | No | NA | 3 | 0 | 2 | Low-quality | Genome-fragment | 5.52 | AAI-based (high-confidence) | 0 |
| S4_extracted_10991 | 4088 | No | NA | 6 | 0 | 1 | Low-quality | Genome-fragment | 4.47 | AAI-based (medium-confidence) | 0 |
| S12_vs_k141_1401\|\|full | 2531 | No | NA | 2 | 0 | 0 | Not-determined | Genome-fragment | NA | NA | 0 |
| S5_vs_k141_106346\|\|full | 4090 | No | NA | 8 | 8 | 0 | Low-quality | Genome-fragment | 8.82 | AAI-based (medium-confidence) | 0 |
| S4_extracted_29911 | 4090 | No | NA | 5 | 0 | 1 | Low-quality | Genome-fragment | 7.59 | AAI-based (medium-confidence) | 0 |
| S1_vs_k141_27803\|\|full | 4091 | No | NA | 3 | 0 | 1 | Low-quality | Genome-fragment | 2.76 | AAI-based (high-confidence) | 0 |
| S12_vs_k141_182450\|\|full | 2205 | No | NA | 3 | 0 | 0 | Not-determined | Genome-fragment | NA | NA | 0 |
| S12_vs_k141_158568\|\|full | 2003 | No | NA | 2 | 0 | 0 | Not-determined | Genome-fragment | NA | NA | 0 |
| S3_vs_k141_94098\|\|full | 4091 | No | NA | 3 | 0 | 1 | Low-quality | Genome-fragment | 3.57 | AAI-based (medium-confidence) | 0 |
| S12_vs_k141_8983\|\|full | 2195 | No | NA | 4 | 0 | 0 | Not-determined | Genome-fragment | NA | NA | 0 |
| S12_vs_k141_40710\|\|full | 2344 | No | NA | 4 | 0 | 2 | Not-determined | Genome-fragment | NA | NA | 0 |
| S5_extracted_k141_11938 | 4091 | No | NA | 6 | 3 | 0 | Low-quality | Genome-fragment | 9.76 | AAI-based (medium-confidence) | 0 |
| S12_vs_k141_2583\|\|full | 4091 | No | NA | 4 | 0 | 2 | Low-quality | Genome-fragment | 1.99 | AAI-based (medium-confidence) | 0 |
| S12_vs_k141_16893\|\|full | 3010 | No | NA | 4 | 0 | 3 | Not-determined | Genome-fragment | NA | NA | 0 |
| S15_vs_75971\|\|full | 4091 | No | NA | 2 | 0 | 0 | Low-quality | Genome-fragment | 7.14 | AAI-based (high-confidence) | 0 |
| S12_vs_k141_134909\|\|full | 1269 | No | NA | 3 | 0 | 0 | Not-determined | Genome-fragment | NA | NA | 0 |
| S1_vs_k141_65763\|\|full | 4092 | No | NA | 3 | 0 | 0 | Low-quality | Genome-fragment | 2.66 | AAI-based (high-confidence) | 0 |
| S12_vs_k141_24974\|\|full | 1717 | No | NA | 2 | 0 | 1 | Not-determined | Genome-fragment | NA | NA | 0 |
| S14_extracted_k141_59379 | 4093 | No | NA | 8 | 5 | 0 | Low-quality | Genome-fragment | 9.9 | AAI-based (high-confidence) | 0 |
| S5_vs_k141_134204\|\|full | 4093 | No | NA | 5 | 1 | 0 | Low-quality | Genome-fragment | 9.35 | AAI-based (medium-confidence) | 0 |
| S12_vs_k141_158632\|\|full | 2141 | No | NA | 4 | 0 | 1 | Not-determined | Genome-fragment | NA | NA | 0 |
| S12_vs_k141_24984\|\|full | 4239 | No | NA | 6 | 0 | 0 | Not-determined | Genome-fragment | NA | NA | 0 |
| S12_vs_k141_24985\|\|full | 2349 | No | NA | 3 | 0 | 2 | Not-determined | Genome-fragment | NA | NA | 0 |
| S12_vs_k141_182512\|\|full | 3242 | No | NA | 3 | 0 | 2 | Not-determined | Genome-fragment | NA | NA | 0 |
| S12_vs_k141_9062\|\|full | 3610 | No | NA | 4 | 0 | 3 | Not-determined | Genome-fragment | NA | NA | 0 |
| S16_extracted_156512 | 4093 | No | NA | 7 | 0 | 0 | Low-quality | Genome-fragment | 3.76 | AAI-based (high-confidence) | 0 |
| S12_vs_k141_88026\|\|full | 2166 | No | NA | 3 | 0 | 1 | Not-determined | Genome-fragment | NA | NA | 0 |
| S12_vs_k141_119431\|\|full | 2526 | No | NA | 5 | 0 | 4 | Not-determined | Genome-fragment | NA | NA | 0 |
| S17_vs_k141_65027\|\|full | 4094 | No | NA | 4 | 0 | 2 | Low-quality | Genome-fragment | 0.47 | AAI-based (medium-confidence) | 0 |
| S11_extracted_k141_3076 | 4094 | No | NA | 7 | 2 | 0 | Low-quality | Genome-fragment | 11.2 | AAI-based (high-confidence) | 0 |
| S12_vs_k141_111286\|\|full | 65591 | No | NA | 77 | 13 | 4 | High-quality | High-quality | 100 | AAI-based (high-confidence) | 0 |
| S12_vs_k141_88061\|\|full | 7670 | No | NA | 9 | 0 | 2 | Not-determined | Genome-fragment | NA | NA | 0 |
| S12_vs_k141_53638\|\|full | 4094 | No | NA | 5 | 4 | 0 | Low-quality | Genome-fragment | 11.92 | AAI-based (high-confidence) | 0 |
| S5_extracted_k141_18326 | 4095 | No | NA | 2 | 0 | 0 | Low-quality | Genome-fragment | 6.79 | AAI-based (medium-confidence) | 0 |
| S12_vs_k141_16995\|\|full | 1796 | No | NA | 4 | 0 | 1 | Not-determined | Genome-fragment | NA | NA | 0 |
| S12_vs_k141_135057\|\|full | 2103 | No | NA | 4 | 0 | 1 | Not-determined | Genome-fragment | NA | NA | 0 |
| S12_vs_k141_56610\|\|full | 6891 | No | NA | 10 | 0 | 0 | Not-determined | Genome-fragment | NA | NA | 0 |
| S12_vs_k141_25064\|\|full | 2248 | No | NA | 2 | 0 | 1 | Not-determined | Genome-fragment | NA | NA | 0 |
| S12_vs_k141_9123\|\|full | 3610 | No | NA | 4 | 0 | 3 | Not-determined | Genome-fragment | NA | NA | 0 |
| S12_vs_k141_79933\|\|full | 1328 | No | NA | 3 | 0 | 1 | Not-determined | Genome-fragment | NA | NA | 0 |
| S12_vs_k141_32622\|\|full | 2170 | No | NA | 2 | 0 | 2 | Not-determined | Genome-fragment | NA | NA | 0 |
| S10_vs_85687\|\|full | 4095 | No | NA | 6 | 2 | 0 | Low-quality | Genome-fragment | 9.96 | AAI-based (high-confidence) | 0 |
| S12_vs_k141_150794\|\|full | 2835 | No | NA | 4 | 0 | 0 | Not-determined | Genome-fragment | NA | NA | 0 |
| S3_extracted_k141_98049 | 4096 | No | NA | 3 | 0 | 2 | Low-quality | Genome-fragment | 1.56 | AAI-based (medium-confidence) | 0 |
| S12_vs_k141_166853\|\|full | 1145 | No | NA | 2 | 0 | 1 | Not-determined | Genome-fragment | NA | NA | 0 |
| S12_vs_k141_79975\|\|full | 1016 | No | NA | 2 | 0 | 1 | Not-determined | Genome-fragment | NA | NA | 0 |
| S10_vs_120773\|\|full | 4097 | No | NA | 4 | 0 | 0 | Low-quality | Genome-fragment | 4.29 | AAI-based (medium-confidence) | 0 |
| S12_vs_k141_25113\|\|full | 2825 | No | NA | 3 | 0 | 3 | Not-determined | Genome-fragment | NA | NA | 0 |
| S12_extracted_k141_95101 | 4097 | No | NA | 3 | 0 | 0 | Low-quality | Genome-fragment | 5.23 | AAI-based (medium-confidence) | 0 |
| S9_vs_k141_27479\|\|full | 4098 | Yes | 1951 | 5 | 1 | 1 | Low-quality | Genome-fragment | 4.38 | AAI-based (medium-confidence) | 52.39 |
| S12_vs_k141_9171\|\|full | 1381 | No | NA | 2 | 0 | 1 | Not-determined | Genome-fragment | NA | NA | 0 |
| S12_vs_k141_150812\|\|full | 37545 | No | NA | 47 | 31 | 0 | Medium-quality | Genome-fragment | 82.79 | AAI-based (high-confidence) | 0 |
| S12_vs_k141_25148\|\|full | 1339 | No | NA | 2 | 0 | 0 | Not-determined | Genome-fragment | NA | NA | 0 |
| S12_vs_k141_135135\|\|full | 1270 | No | NA | 3 | 0 | 0 | Not-determined | Genome-fragment | NA | NA | 0 |
| S12_vs_k141_17054\|\|full | 2779 | No | NA | 4 | 0 | 1 | Not-determined | Genome-fragment | NA | NA | 0 |
| S12_vs_k141_9198\|\|full | 1541 | No | NA | 2 | 0 | 0 | Not-determined | Genome-fragment | NA | NA | 0 |
| S12_extracted_k141_42786 | 4098 | No | NA | 4 | 0 | 0 | Low-quality | Genome-fragment | 3.71 | AAI-based (medium-confidence) | 0 |
| S12_vs_k141_56722\|\|full | 1825 | No | NA | 3 | 0 | 1 | Not-determined | Genome-fragment | NA | NA | 0 |
| S12_vs_k141_182624\|\|full | 1766 | No | NA | 3 | 0 | 3 | Not-determined | Genome-fragment | NA | NA | 0 |
| S12_vs_k141_80022\|\|full | 1635 | No | NA | 2 | 0 | 0 | Not-determined | Genome-fragment | NA | NA | 0 |
| S12_vs_k141_1590\|\|full | 1402 | No | NA | 4 | 0 | 0 | Not-determined | Genome-fragment | NA | NA | 0 |
| S12_vs_k141_174401\|\|full | 2292 | No | NA | 4 | 0 | 1 | Not-determined | Genome-fragment | NA | NA | 0 |
| S8_vs_k141_41060\|\|full | 4098 | No | NA | 5 | 0 | 4 | Low-quality | Genome-fragment | 4.42 | AAI-based (medium-confidence) | 0 |
| S12_vs_k141_143202\|\|full | 1239 | No | NA | 2 | 0 | 1 | Not-determined | Genome-fragment | NA | NA | 0 |
| S17_vs_k141_103952\|\|full | 4099 | No | NA | 7 | 2 | 0 | Low-quality | Genome-fragment | 8.22 | AAI-based (medium-confidence) | 0 |
| S14_extracted_k141_87818 | 4099 | Yes | 1521 | 5 | 1 | 2 | Low-quality | Genome-fragment | 1.88 | HMM-based (lower-bound) | 62.89 |
| S12_vs_k141_96080\|\|full | 1619 | No | NA | 3 | 0 | 2 | Not-determined | Genome-fragment | NA | NA | 0 |
| S8_vs_k141_59060\|\|full | 4099 | No | NA | 10 | 4 | 0 | Low-quality | Genome-fragment | 9.84 | AAI-based (high-confidence) | 0 |
| S3_vs_k141_60942\|\|full | 4100 | No | NA | 3 | 0 | 0 | Low-quality | Genome-fragment | 5.21 | AAI-based (medium-confidence) | 0 |
| S9_extracted_k141_39647 | 4101 | No | NA | 3 | 2 | 0 | Low-quality | Genome-fragment | 6.85 | AAI-based (medium-confidence) | 0 |
| S12_vs_k141_96092\|\|full | 4018 | No | NA | 5 | 0 | 0 | Not-determined | Genome-fragment | NA | NA | 0 |
| S14_vs_k141_98469\|\|full | 4101 | No | NA | 9 | 8 | 0 | Low-quality | Genome-fragment | 12.33 | AAI-based (high-confidence) | 0 |
| S3_vs_k141_54800\|\|full | 4101 | No | NA | 3 | 0 | 2 | Low-quality | Genome-fragment | 1.57 | AAI-based (medium-confidence) | 0 |
| S11_extracted_k141_78850 | 4103 | No | NA | 6 | 1 | 0 | Low-quality | Genome-fragment | 11.46 | AAI-based (medium-confidence) | 0 |
| S12_vs_k141_182671\|\|full | 4006 | No | NA | 5 | 0 | 3 | Not-determined | Genome-fragment | NA | NA | 0 |
| S12_vs_k141_150896\|\|full | 1436 | No | NA | 2 | 0 | 1 | Not-determined | Genome-fragment | NA | NA | 0 |
| S12_vs_k141_174449\|\|full | 1814 | No | NA | 4 | 0 | 0 | Not-determined | Genome-fragment | NA | NA | 0 |
| S12_vs_k141_127443\|\|full | 1439 | No | NA | 4 | 0 | 1 | Not-determined | Genome-fragment | NA | NA | 0 |
| S12_vs_k141_150918\|\|full | 6181 | No | NA | 7 | 0 | 0 | Not-determined | Genome-fragment | NA | NA | 0 |
| S13_vs_126847\|\|full | 4103 | No | NA | 3 | 0 | 2 | Low-quality | Genome-fragment | 5.68 | AAI-based (medium-confidence) | 0 |
| S12_vs_k141_64638\|\|full | 2196 | No | NA | 2 | 0 | 0 | Not-determined | Genome-fragment | NA | NA | 0 |
| S12_vs_k141_1668\|\|full | 1549 | No | NA | 2 | 0 | 1 | Not-determined | Genome-fragment | NA | NA | 0 |
| S12_vs_k141_119760\|\|full | 1442 | No | NA | 1 | 0 | 0 | Not-determined | Genome-fragment | NA | NA | 0 |
| S12_vs_k141_166978\|\|full | 5259 | No | NA | 3 | 0 | 3 | Not-determined | Genome-fragment | NA | NA | 0 |
| S12_extracted_k141_124606 | 4103 | No | NA | 4 | 0 | 4 | Low-quality | Genome-fragment | 1.53 | AAI-based (high-confidence) | 0 |
| S16_vs_111276\|\|full | 4105 | No | NA | 4 | 0 | 1 | Low-quality | Genome-fragment | 0.47 | AAI-based (medium-confidence) | 0 |
| S6_extracted_k141_51289 | 4105 | No | NA | 4 | 0 | 0 | Low-quality | Genome-fragment | 2.6 | AAI-based (high-confidence) | 0 |
| S12_vs_k141_96198\|\|full | 1009 | No | NA | 3 | 0 | 1 | Not-determined | Genome-fragment | NA | NA | 0 |
| S10_vs_32553\|\|full | 4107 | No | NA | 8 | 4 | 0 | Low-quality | Genome-fragment | 6.76 | AAI-based (medium-confidence) | 0 |
| S5_vs_k141_134182\|\|full | 4108 | No | NA | 3 | 0 | 2 | Low-quality | Genome-fragment | 3.77 | AAI-based (medium-confidence) | 0 |
| S16_extracted_103774 | 4108 | No | NA | 3 | 0 | 3 | Low-quality | Genome-fragment | 1.16 | AAI-based (medium-confidence) | 0 |
| S14_vs_k141_100305\|\|full | 4109 | No | NA | 2 | 0 | 2 | Low-quality | Genome-fragment | 1.43 | AAI-based (medium-confidence) | 0 |
| S12_vs_k141_111464\|\|full | 6132 | No | NA | 5 | 0 | 4 | Not-determined | Genome-fragment | NA | NA | 0 |
| S10_vs_91758\|\|full | 4111 | No | NA | 10 | 1 | 0 | Low-quality | Genome-fragment | 12.62 | AAI-based (high-confidence) | 0 |
| S12_vs_k141_127541\|\|full | 2457 | No | NA | 3 | 0 | 0 | Not-determined | Genome-fragment | NA | NA | 0 |
| S12_vs_k141_143317\|\|full | 2382 | No | NA | 4 | 0 | 3 | Not-determined | Genome-fragment | NA | NA | 0 |
| S9_extracted_k141_34840 | 4112 | No | NA | 8 | 3 | 0 | Low-quality | Genome-fragment | 2.43 | AAI-based (high-confidence) | 0 |
| S9_extracted_k141_73618 | 4113 | No | NA | 7 | 4 | 0 | Low-quality | Genome-fragment | 10.39 | AAI-based (high-confidence) | 0 |
| S3_vs_k141_70464\|\|full | 4114 | No | NA | 4 | 0 | 1 | Low-quality | Genome-fragment | 2.51 | AAI-based (high-confidence) | 0 |
| S12_vs_k141_143337\|\|full | 1358 | No | NA | 3 | 0 | 2 | Not-determined | Genome-fragment | NA | NA | 0 |
| S12_vs_k141_64723\|\|full | 1937 | No | NA | 2 | 0 | 2 | Not-determined | Genome-fragment | NA | NA | 0 |
| S12_vs_k141_48358\|\|full | 1862 | No | NA | 5 | 0 | 2 | Not-determined | Genome-fragment | NA | NA | 0 |
| S12_vs_k141_64742\|\|full | 2365 | No | NA | 3 | 0 | 2 | Not-determined | Genome-fragment | NA | NA | 0 |
| S5_extracted_k141_8396 | 4114 | No | NA | 2 | 2 | 0 | Low-quality | Genome-fragment | 3.67 | AAI-based (medium-confidence) | 0 |
| S12_vs_k141_56909\|\|full | 2245 | No | NA | 3 | 0 | 0 | Not-determined | Genome-fragment | NA | NA | 0 |
| S12_vs_k141_1767\|\|full | 4319 | No | NA | 6 | 0 | 5 | Not-determined | Genome-fragment | NA | NA | 0 |
| S12_vs_k141_25299\|\|full | 1037 | No | NA | 2 | 0 | 2 | Not-determined | Genome-fragment | NA | NA | 0 |
| S8_extracted_k141_179878 | 4114 | No | NA | 4 | 0 | 0 | Low-quality | Genome-fragment | 9.64 | AAI-based (medium-confidence) | 0 |
| S12_vs_k141_56955\|\|full | 2072 | No | NA | 3 | 0 | 1 | Not-determined | Genome-fragment | NA | NA | 0 |
| S12_vs_k141_25319\|\|full | 4474 | No | NA | 3 | 0 | 1 | Not-determined | Genome-fragment | NA | NA | 0 |
| S9_vs_k141_7829\|\|full | 4115 | No | NA | 8 | 0 | 0 | Low-quality | Genome-fragment | 10.34 | AAI-based (high-confidence) | 0 |
| S12_vs_k141_56956\|\|full | 7097 | No | NA | 5 | 0 | 2 | Not-determined | Genome-fragment | NA | NA | 0 |
| S12_vs_k141_119927\|\|full | 2754 | No | NA | 4 | 0 | 1 | Not-determined | Genome-fragment | NA | NA | 0 |
| S12_vs_k141_121220\|\|full | 4115 | No | NA | 4 | 0 | 2 | Low-quality | Genome-fragment | 2.23 | AAI-based (medium-confidence) | 0 |
| S12_vs_k141_25337\|\|full | 1862 | No | NA | 4 | 0 | 0 | Not-determined | Genome-fragment | NA | NA | 0 |
| S12_vs_k141_150997\|\|full | 2342 | No | NA | 4 | 0 | 1 | Not-determined | Genome-fragment | NA | NA | 0 |
| S12_vs_k141_135320\|\|full | 1944 | No | NA | 3 | 0 | 2 | Not-determined | Genome-fragment | NA | NA | 0 |
| S12_vs_k141_40923\|\|full | 2439 | No | NA | 3 | 0 | 2 | Not-determined | Genome-fragment | NA | NA | 0 |
| S12_vs_k141_48453\|\|full | 2698 | No | NA | 4 | 0 | 1 | Not-determined | Genome-fragment | NA | NA | 0 |
| S7_extracted_125481 | 4116 | No | NA | 7 | 0 | 0 | Low-quality | Genome-fragment | 6.32 | AAI-based (high-confidence) | 0 |
| S7_vs_67090\|\|full | 4116 | No | NA | 7 | 0 | 0 | Low-quality | Genome-fragment | 7.69 | AAI-based (medium-confidence) | 0 |
| S14_vs_k141_73061\|\|full | 4118 | No | NA | 4 | 0 | 1 | Low-quality | Genome-fragment | 7.67 | AAI-based (medium-confidence) | 0 |
| S1_vs_k141_37371\|\|full | 4118 | No | NA | 4 | 2 | 0 | Low-quality | Genome-fragment | 9.83 | AAI-based (high-confidence) | 0 |
| S5_vs_k141_63421\|\|full | 4119 | No | NA | 4 | 0 | 2 | Low-quality | Genome-fragment | 5.43 | AAI-based (medium-confidence) | 0 |
| S12_vs_k141_57040\|\|full | 1300 | No | NA | 3 | 0 | 2 | Not-determined | Genome-fragment | NA | NA | 0 |
| S8_vs_k141_156459\|\|full | 4119 | No | NA | 7 | 1 | 0 | Low-quality | Genome-fragment | 8.71 | AAI-based (high-confidence) | 0 |
| S12_vs_k141_182800\|\|full | 1453 | No | NA | 2 | 0 | 1 | Not-determined | Genome-fragment | NA | NA | 0 |
| S12_vs_k141_104137\|\|full | 1130 | No | NA | 2 | 0 | 1 | Not-determined | Genome-fragment | NA | NA | 0 |
| S12_vs_k141_104156\|\|full | 2164 | No | NA | 2 | 0 | 1 | Not-determined | Genome-fragment | NA | NA | 0 |
| S12_vs_k141_80101\|\|full | 2117 | No | NA | 3 | 0 | 2 | Not-determined | Genome-fragment | NA | NA | 0 |
| S12_vs_k141_64781\|\|full | 3436 | No | NA | 5 | 0 | 5 | Not-determined | Genome-fragment | NA | NA | 0 |
| S11_vs_k141_23271\|\|full | 4120 | No | NA | 7 | 3 | 0 | Low-quality | Genome-fragment | 11.2 | AAI-based (medium-confidence) | 0 |
| S12_vs_k141_104163\|\|full | 2381 | No | NA | 3 | 0 | 0 | Not-determined | Genome-fragment | NA | NA | 0 |
| S10_extracted_3140 | 4120 | No | NA | 7 | 0 | 6 | Low-quality | Genome-fragment | 2.71 | AAI-based (high-confidence) | 0 |
| S5_extracted_k141_29068 | 4121 | No | NA | 3 | 0 | 1 | Low-quality | Genome-fragment | 4.87 | AAI-based (medium-confidence) | 0 |
| S5_vs_k141_68903\|\|full | 4121 | No | NA | 6 | 0 | 1 | Low-quality | Genome-fragment | 3.14 | AAI-based (high-confidence) | 0 |
| S12_vs_k141_64812\|\|full | 5454 | No | NA | 6 | 0 | 1 | Not-determined | Genome-fragment | NA | NA | 0 |
| S12_vs_k141_17317\|\|full | 1921 | No | NA | 3 | 0 | 1 | Not-determined | Genome-fragment | NA | NA | 0 |
| S7_extracted_52041 | 4121 | No | NA | 5 | 0 | 1 | Low-quality | Genome-fragment | 6.58 | AAI-based (medium-confidence) | 0 |
| S12_vs_k141_41069\|\|full | 1400 | No | NA | 3 | 0 | 1 | Not-determined | Genome-fragment | NA | NA | 0 |
| S12_vs_k141_1934\|\|full | 3156 | No | NA | 2 | 0 | 2 | Not-determined | Genome-fragment | NA | NA | 0 |
| S12_vs_k141_48507\|\|full | 2103 | No | NA | 8 | 0 | 0 | Not-determined | Genome-fragment | NA | NA | 0 |
| S7_vs_71971\|\|full | 4121 | No | NA | 4 | 0 | 0 | Low-quality | Genome-fragment | 13.46 | AAI-based (high-confidence) | 0 |
| S12_vs_k141_182862\|\|full | 1982 | No | NA | 2 | 0 | 0 | Not-determined | Genome-fragment | NA | NA | 0 |
| S12_vs_k141_64845\|\|full | 1858 | No | NA | 2 | 0 | 2 | Not-determined | Genome-fragment | NA | NA | 0 |
| S12_vs_k141_135376\|\|full | 3620 | No | NA | 5 | 0 | 3 | Not-determined | Genome-fragment | NA | NA | 0 |
| S12_vs_k141_127703\|\|full | 1997 | No | NA | 3 | 0 | 1 | Not-determined | Genome-fragment | NA | NA | 0 |
| S3_vs_k141_97445\|\|full | 4122 | No | NA | 4 | 4 | 0 | Low-quality | Genome-fragment | 8.41 | AAI-based (high-confidence) | 0 |
| S17_extracted_k141_123852 | 4123 | No | NA | 5 | 0 | 1 | Low-quality | Genome-fragment | 5.13 | AAI-based (medium-confidence) | 0 |
| S16_extracted_60270 | 4128 | No | NA | 3 | 0 | 1 | Low-quality | Genome-fragment | 8.88 | AAI-based (medium-confidence) | 0 |
| S11_vs_k141_24666\|\|full | 4129 | No | NA | 7 | 0 | 0 | Low-quality | Genome-fragment | 6.88 | AAI-based (high-confidence) | 0 |
| S11_vs_k141_65242\|\|full | 4130 | No | NA | 8 | 1 | 0 | Low-quality | Genome-fragment | 6.47 | AAI-based (medium-confidence) | 0 |
| S12_vs_k141_9465\|\|full | 4269 | No | NA | 3 | 0 | 1 | Not-determined | Genome-fragment | NA | NA | 0 |
| S7_extracted_7946 | 4130 | No | NA | 3 | 0 | 1 | Low-quality | Genome-fragment | 1.67 | AAI-based (medium-confidence) | 0 |
| S11_vs_k141_62599\|\|full | 4131 | No | NA | 7 | 2 | 0 | Low-quality | Genome-fragment | 9.07 | AAI-based (high-confidence) | 0 |
| S12_vs_k141_95760\|\|full | 4132 | No | NA | 5 | 0 | 2 | Low-quality | Genome-fragment | 7.83 | AAI-based (medium-confidence) | 0 |
| S12_vs_k141_88485\|\|full | 2733 | No | NA | 3 | 0 | 3 | Not-determined | Genome-fragment | NA | NA | 0 |
| S4_extracted_12254 | 4132 | No | NA | 4 | 2 | 0 | Low-quality | Genome-fragment | 7.71 | AAI-based (high-confidence) | 0 |
| S12_vs_k141_17432\|\|full | 7127 | No | NA | 8 | 0 | 0 | Not-determined | Genome-fragment | NA | NA | 0 |
| S17_vs_k141_160491\|\|full | 4133 | No | NA | 7 | 0 | 0 | Low-quality | Genome-fragment | 6.49 | AAI-based (high-confidence) | 0 |
| S12_vs_k141_48575\|\|full | 1012 | No | NA | 2 | 0 | 0 | Not-determined | Genome-fragment | NA | NA | 0 |
| S12_vs_k141_57221\|\|full | 2046 | No | NA | 2 | 0 | 0 | Not-determined | Genome-fragment | NA | NA | 0 |
| S14_extracted_k141_59107 | 4133 | No | NA | 7 | 1 | 0 | Low-quality | Genome-fragment | 2.84 | AAI-based (medium-confidence) | 0 |
| S17_extracted_k141_164187 | 4134 | No | NA | 7 | 2 | 0 | Low-quality | Genome-fragment | 10.78 | AAI-based (high-confidence) | 0 |
| S10_extracted_3009 | 4134 | No | NA | 9 | 3 | 0 | Low-quality | Genome-fragment | 7.9 | AAI-based (high-confidence) | 0 |
| S12_vs_k141_17457\|\|full | 2026 | No | NA | 7 | 0 | 0 | Not-determined | Genome-fragment | NA | NA | 0 |
| S12_vs_k141_52376\|\|full | 4134 | No | NA | 10 | 3 | 0 | Low-quality | Genome-fragment | 8.17 | AAI-based (high-confidence) | 0 |
| S14_extracted_k141_15370 | 4135 | No | NA | 11 | 10 | 0 | Low-quality | Genome-fragment | 13.65 | AAI-based (high-confidence) | 0 |
| S12_vs_k141_57232\|\|full | 6740 | No | NA | 7 | 0 | 3 | Not-determined | Genome-fragment | NA | NA | 0 |
| S16_extracted_166255 | 4136 | No | NA | 2 | 0 | 0 | Low-quality | Genome-fragment | 6.43 | AAI-based (medium-confidence) | 0 |
| S12_vs_k141_32800\|\|full | 3614 | No | NA | 4 | 0 | 0 | Not-determined | Genome-fragment | NA | NA | 0 |
| S12_vs_k141_167286\|\|full | 1836 | No | NA | 2 | 0 | 2 | Not-determined | Genome-fragment | NA | NA | 0 |
| S12_vs_k141_64964\|\|full | 1341 | No | NA | 3 | 0 | 0 | Not-determined | Genome-fragment | NA | NA | 0 |
| S8_vs_k141_77748\|\|full | 4136 | Yes | 3455 | 7 | 3 | 1 | Low-quality | Genome-fragment | 8.73 | AAI-based (high-confidence) | 16.47 |
| S11_extracted_k141_17135 | 4138 | No | NA | 6 | 2 | 0 | Low-quality | Genome-fragment | 1.6 | AAI-based (medium-confidence) | 0 |
| S12_vs_k141_135495\|\|full | 1988 | No | NA | 3 | 0 | 2 | Not-determined | Genome-fragment | NA | NA | 0 |
| S12_vs_k141_57257\|\|full | 3524 | No | NA | 2 | 0 | 1 | Not-determined | Genome-fragment | NA | NA | 0 |
| S7_extracted_108199 | 4138 | Yes | 1006 | 3 | 1 | 1 | Low-quality | Genome-fragment | 2.11 | AAI-based (medium-confidence) | 75.69 |
| S12_vs_k141_88577\|\|full | 2618 | No | NA | 3 | 0 | 1 | Not-determined | Genome-fragment | NA | NA | 0 |

| | | | | | | | | | | | |
|---|---|---|---|---|---|---|---|---|---|---|---|
| S7_vs_92219\|\|full | 4139 | No | NA | 3 | 0 | 1 | Low-quality | Genome-fragment | 9.83 | AAI-based (high-confidence) | 0 |
| S12_vs_k141_183002\|\|full | 1001 | No | NA | 2 | 0 | 0 | Not-determined | Genome-fragment | NA | NA | 0 |
| S8_extracted_k141_6562 | 4139 | No | NA | 3 | 0 | 1 | Low-quality | Genome-fragment | 6.86 | AAI-based (medium-confidence) | 0 |
| S17_vs_k141_164661\|\|full | 4140 | Yes | 2266 | 4 | 1 | 2 | Low-quality | Genome-fragment | 3.46 | AAI-based (medium-confidence) | 45.27 |
| S12_vs_k141_104373\|\|full | 4241 | No | NA | 4 | 0 | 2 | Not-determined | Genome-fragment | NA | NA | 0 |
| S14_vs_k141_64518\|\|full | 4141 | No | NA | 4 | 0 | 2 | Low-quality | Genome-fragment | 3.36 | AAI-based (medium-confidence) | 0 |
| S12_vs_k141_65000\|\|full | 2680 | No | NA | 3 | 0 | 0 | Not-determined | Genome-fragment | NA | NA | 0 |
| S12_vs_k141_174583\|\|full | 1922 | No | NA | 3 | 0 | 2 | Not-determined | Genome-fragment | NA | NA | 0 |
| S3_extracted_k141_91744 | 4141 | No | NA | 7 | 4 | 0 | Low-quality | Genome-fragment | 6.18 | AAI-based (high-confidence) | 0 |
| S12_vs_k141_48677\|\|full | 1658 | No | NA | 3 | 0 | 0 | Not-determined | Genome-fragment | NA | NA | 0 |
| S8_extracted_k141_95743 | 4141 | No | NA | 4 | 0 | 4 | Low-quality | Genome-fragment | 2.03 | AAI-based (medium-confidence) | 0 |
| S12_vs_k141_17553\|\|full | 2405 | No | NA | 2 | 0 | 1 | Not-determined | Genome-fragment | NA | NA | 0 |
| S8_vs_k141_91273\|\|full | 4141 | No | NA | 4 | 0 | 1 | Low-quality | Genome-fragment | 3.27 | AAI-based (medium-confidence) | 0 |
| S12_vs_k141_80271\|\|full | 3232 | No | NA | 2 | 0 | 1 | Not-determined | Genome-fragment | NA | NA | 0 |
| S12_vs_k141_158970\|\|full | 4041 | No | NA | 5 | 0 | 1 | Not-determined | Genome-fragment | NA | NA | 0 |
| S12_vs_k141_41320\|\|full | 5883 | No | NA | 12 | 0 | 0 | Not-determined | Genome-fragment | NA | NA | 0 |
| S12_vs_k141_57341\|\|full | 8974 | No | NA | 10 | 0 | 1 | Not-determined | Genome-fragment | NA | NA | 0 |
| S12_vs_k141_183054\|\|full | 3055 | No | NA | 3 | 0 | 2 | Not-determined | Genome-fragment | NA | NA | 0 |
| S1_extracted_k141_34185 | 4142 | No | NA | 5 | 1 | 0 | Low-quality | Genome-fragment | 12.08 | AAI-based (high-confidence) | 0 |
| S12_vs_k141_32914\|\|full | 2124 | No | NA | 4 | 0 | 1 | Not-determined | Genome-fragment | NA | NA | 0 |
| S4_extracted_23853 | 4143 | No | NA | 5 | 0 | 2 | Low-quality | Genome-fragment | 3.14 | AAI-based (medium-confidence) | 0 |
| S7_vs_77365\|\|full | 4144 | Yes | 2569 | 7 | 2 | 1 | Low-quality | Genome-fragment | 5.91 | HMM-based (lower-bound) | 38.01 |
| S12_vs_k141_96741\|\|full | 1339 | No | NA | 3 | 0 | 0 | Not-determined | Genome-fragment | NA | NA | 0 |
| S12_vs_k141_41346\|\|full | 1002 | No | NA | 3 | 0 | 0 | Not-determined | Genome-fragment | NA | NA | 0 |
| S12_vs_k141_80307\|\|full | 1525 | No | NA | 4 | 0 | 0 | Not-determined | Genome-fragment | NA | NA | 0 |
| S12_vs_k141_41349\|\|full | 1317 | No | NA | 3 | 0 | 0 | Not-determined | Genome-fragment | NA | NA | 0 |
| S6_extracted_k141_28278 | 4144 | No | NA | 8 | 7 | 0 | Low-quality | Genome-fragment | 13.67 | AAI-based (high-confidence) | 0 |
| S14_extracted_k141_58273 | 4146 | No | NA | 6 | 4 | 0 | Low-quality | Genome-fragment | 9.11 | AAI-based (high-confidence) | 0 |
| S1_vs_k141_79099\|\|full | 4146 | No | NA | 6 | 3 | 0 | Low-quality | Genome-fragment | 7.04 | AAI-based (high-confidence) | 0 |
| S12_vs_k141_2086\|\|full | 1480 | No | NA | 3 | 0 | 0 | Not-determined | Genome-fragment | NA | NA | 0 |
| S16_extracted_144864 | 4146 | No | NA | 3 | 0 | 2 | Low-quality | Genome-fragment | 5.59 | AAI-based (medium-confidence) | 0 |
| S12_vs_k141_183104\|\|full | 1232 | No | NA | 3 | 0 | 0 | Not-determined | Genome-fragment | NA | NA | 0 |
| S13_vs_108581\|\|full | 4147 | No | NA | 5 | 0 | 2 | Low-quality | Genome-fragment | 2.23 | AAI-based (high-confidence) | 0 |
| S12_vs_k141_167340\|\|full | 5315 | No | NA | 12 | 0 | 1 | Not-determined | Genome-fragment | NA | NA | 0 |
| S12_vs_k141_17611\|\|full | 2721 | No | NA | 3 | 0 | 1 | Not-determined | Genome-fragment | NA | NA | 0 |
| S14_extracted_k141_60185 | 4147 | No | NA | 11 | 10 | 0 | Low-quality | Genome-fragment | 13.59 | AAI-based (high-confidence) | 0 |
| S7_extracted_9312 | 4147 | No | NA | 4 | 1 | 0 | Low-quality | Genome-fragment | 5.41 | AAI-based (medium-confidence) | 0 |
| S8_extracted_k141_78769 | 4147 | No | NA | 5 | 0 | 3 | Low-quality | Genome-fragment | 6.87 | AAI-based (high-confidence) | 0 |
| S16_vs_9560\|\|full | 4148 | No | NA | 8 | 0 | 0 | Low-quality | Genome-fragment | 7.15 | AAI-based (high-confidence) | 0 |
| S12_vs_k141_80365\|\|full | 4621 | No | NA | 4 | 0 | 3 | Not-determined | Genome-fragment | NA | NA | 0 |
| S12_vs_k141_25697\|\|full | 5568 | No | NA | 6 | 0 | 6 | Not-determined | Genome-fragment | NA | NA | 0 |
| S12_vs_k141_17640\|\|full | 1250 | No | NA | 2 | 0 | 1 | Not-determined | Genome-fragment | NA | NA | 0 |
| S17_extracted_k141_33607 | 4150 | No | NA | 5 | 4 | 0 | Low-quality | Genome-fragment | 13.69 | AAI-based (high-confidence) | 0 |
| S12_vs_k141_88660\|\|full | 2184 | No | NA | 2 | 0 | 2 | Not-determined | Genome-fragment | NA | NA | 0 |
| S12_vs_k141_151177\|\|full | 1743 | No | NA | 2 | 0 | 0 | Not-determined | Genome-fragment | NA | NA | 0 |
| S17_extracted_k141_124889 | 4150 | No | NA | 3 | 0 | 0 | Low-quality | Genome-fragment | 3.53 | AAI-based (medium-confidence) | 0 |
| S12_vs_k141_33007\|\|full | 2690 | No | NA | 4 | 0 | 1 | Not-determined | Genome-fragment | NA | NA | 0 |
| S1_vs_k141_158536\|\|full | 4150 | No | NA | 4 | 1 | 0 | Low-quality | Genome-fragment | 9.31 | AAI-based (high-confidence) | 0 |
| S7_vs_12606\|\|full | 4150 | No | NA | 7 | 3 | 0 | Low-quality | Genome-fragment | 9.55 | AAI-based (medium-confidence) | 0 |
| S12_vs_k141_2154\|\|full | 3995 | No | NA | 3 | 0 | 1 | Not-determined | Genome-fragment | NA | NA | 0 |
| S12_vs_k141_41425\|\|full | 2993 | No | NA | 3 | 0 | 0 | Not-determined | Genome-fragment | NA | NA | 0 |
| S11_vs_k141_49252\|\|full | 4152 | No | NA | 7 | 1 | 3 | Low-quality | Genome-fragment | 8.32 | HMM-based (lower-bound) | 0 |
| S12_vs_k141_135645\|\|full | 2428 | No | NA | 4 | 0 | 0 | Not-determined | Genome-fragment | NA | NA | 0 |
| S13_extracted_77420 | 4152 | Yes | 2753 | 3 | 2 | 1 | Low-quality | Genome-fragment | 1.64 | HMM-based (lower-bound) | 33.69 |
| S12_vs_k141_127909\|\|full | 6876 | No | NA | 6 | 0 | 3 | Not-determined | Genome-fragment | NA | NA | 0 |
| S12_vs_k141_33055\|\|full | 2591 | No | NA | 5 | 0 | 2 | Not-determined | Genome-fragment | NA | NA | 0 |
| S5_vs_k141_3629\|\|full | 4152 | No | NA | 4 | 0 | 1 | Low-quality | Genome-fragment | 7.82 | AAI-based (medium-confidence) | 0 |
| S10_vs_85757\|\|full | 4152 | No | NA | 9 | 0 | 1 | Low-quality | Genome-fragment | 9.71 | AAI-based (high-confidence) | 0 |
| S12_vs_k141_111791\|\|full | 1689 | No | NA | 2 | 0 | 1 | Not-determined | Genome-fragment | NA | NA | 0 |
| S12_vs_k141_143839\|\|full | 5862 | No | NA | 6 | 0 | 3 | Not-determined | Genome-fragment | NA | NA | 0 |
| S9_vs_k141_23687\|\|full | 4153 | No | NA | 6 | 1 | 0 | Low-quality | Genome-fragment | 2.37 | AAI-based (high-confidence) | 0 |
| S11_extracted_k141_78449 | 4153 | Yes | 3813 | 7 | 2 | 1 | Low-quality | Genome-fragment | 2.32 | AAI-based (medium-confidence) | 8.19 |
| S12_extracted_k141_60045 | 4153 | No | NA | 4 | 0 | 0 | Low-quality | Genome-fragment | 9.53 | AAI-based (medium-confidence) | 0 |
| S1_extracted_k141_20576 | 4154 | No | NA | 6 | 2 | 0 | Low-quality | Genome-fragment | 11.58 | AAI-based (high-confidence) | 0 |
| S2_extracted_k141_18540 | 4154 | No | NA | 2 | 1 | 0 | Low-quality | Genome-fragment | 3.95 | AAI-based (high-confidence) | 0 |
| S5_vs_k141_119537\|\|full | 4156 | No | NA | 5 | 0 | 0 | Low-quality | Genome-fragment | 3.65 | AAI-based (medium-confidence) | 0 |
| S12_vs_k141_135693\|\|full | 1128 | No | NA | 3 | 0 | 0 | Not-determined | Genome-fragment | NA | NA | 0 |
| S12_vs_k141_48837\|\|full | 2911 | No | NA | 5 | 0 | 0 | Not-determined | Genome-fragment | NA | NA | 0 |
| S12_extracted_k141_159399 | 4156 | No | NA | 1 | 0 | 0 | Low-quality | Genome-fragment | 3.86 | AAI-based (high-confidence) | 0 |
| S9_vs_k141_37462\|\|full | 4157 | No | NA | 4 | 3 | 0 | Low-quality | Genome-fragment | 11.71 | AAI-based (high-confidence) | 0 |
| S12_vs_k141_127958\|\|full | 2193 | No | NA | 3 | 0 | 2 | Not-determined | Genome-fragment | NA | NA | 0 |
| S12_vs_k141_33142\|\|full | 2605 | No | NA | 2 | 0 | 0 | Not-determined | Genome-fragment | NA | NA | 0 |
| S16_extracted_7575 | 4157 | No | NA | 4 | 0 | 2 | Low-quality | Genome-fragment | 1.18 | AAI-based (high-confidence) | 0 |
| S12_vs_k141_159213\|\|full | 1386 | No | NA | 3 | 0 | 1 | Not-determined | Genome-fragment | NA | NA | 0 |
| S12_vs_k141_80553\|\|full | 1360 | No | NA | 3 | 0 | 0 | Not-determined | Genome-fragment | NA | NA | 0 |
| S17_extracted_k141_29656 | 4158 | No | NA | 3 | 0 | 0 | Low-quality | Genome-fragment | 10.43 | AAI-based (medium-confidence) | 0 |
| S12_vs_k141_17789\|\|full | 2171 | No | NA | 2 | 0 | 2 | Not-determined | Genome-fragment | NA | NA | 0 |
| S12_vs_k141_120489\|\|full | 1672 | No | NA | 3 | 0 | 0 | Not-determined | Genome-fragment | NA | NA | 0 |
| S12_vs_k141_151292\|\|full | 63916 | Yes | 57412 | 90 | 11 | 4 | Medium-quality | Genome-fragment | 81.3 | AAI-based (high-confidence) | 10.18 |
| S3_extracted_k141_41440 | 4158 | No | NA | 5 | 0 | 0 | Low-quality | Genome-fragment | 14.52 | AAI-based (medium-confidence) | 0 |
| S17_extracted_k141_174340 | 4159 | No | NA | 3 | 1 | 0 | Low-quality | Genome-fragment | 7.38 | AAI-based (high-confidence) | 0 |
| S8_extracted_k141_7626 | 4159 | No | NA | 2 | 2 | 0 | Low-quality | Genome-fragment | 4.12 | AAI-based (high-confidence) | 0 |
| S12_vs_k141_104573\|\|full | 1926 | No | NA | 3 | 0 | 2 | Not-determined | Genome-fragment | NA | NA | 0 |
| S12_vs_k141_135756\|\|full | 1203 | No | NA | 2 | 0 | 1 | Not-determined | Genome-fragment | NA | NA | 0 |
| S11_vs_k141_3170\|\|full | 4163 | No | NA | 6 | 2 | 0 | Low-quality | Genome-fragment | 7.41 | HMM-based (lower-bound) | 0 |
| S12_vs_k141_183254\|\|full | 1669 | No | NA | 3 | 0 | 0 | Not-determined | Genome-fragment | NA | NA | 0 |
| S13_extracted_93908 | 4163 | No | NA | 8 | 1 | 1 | Low-quality | Genome-fragment | 6.5 | HMM-based (lower-bound) | 0 |
| S3_extracted_k141_67998 | 4163 | No | NA | 4 | 0 | 3 | Low-quality | Genome-fragment | 2.69 | AAI-based (medium-confidence) | 0 |
| S12_extracted_k141_76898 | 4163 | No | NA | 6 | 0 | 2 | Low-quality | Genome-fragment | 6.85 | AAI-based (medium-confidence) | 0 |
| S12_vs_k141_41604\|\|full | 1436 | No | NA | 3 | 0 | 1 | Not-determined | Genome-fragment | NA | NA | 0 |
| S12_vs_k141_120530\|\|full | 3119 | No | NA | 5 | 0 | 2 | Not-determined | Genome-fragment | NA | NA | 0 |
| S5_vs_k141_16565\|\|full | 4164 | No | NA | 10 | 0 | 0 | Low-quality | Genome-fragment | 3.14 | AAI-based (high-confidence) | 0 |
| S12_vs_k141_65286\|\|full | 2177 | No | NA | 3 | 0 | 1 | Not-determined | Genome-fragment | NA | NA | 0 |
| S12_vs_k141_57563\|\|full | 1063 | No | NA | 2 | 0 | 0 | Not-determined | Genome-fragment | NA | NA | 0 |
| S15_extracted_26462 | 4164 | No | NA | 8 | 2 | 0 | Low-quality | Genome-fragment | 6.74 | AAI-based (high-confidence) | 0 |
| S12_vs_k141_151357\|\|full | 1575 | No | NA | 4 | 0 | 0 | Not-determined | Genome-fragment | NA | NA | 0 |
| S12_vs_k141_159248\|\|full | 4479 | No | NA | 4 | 0 | 3 | Not-determined | Genome-fragment | NA | NA | 0 |
| S12_vs_k141_65310\|\|full | 1932 | No | NA | 2 | 0 | 2 | Not-determined | Genome-fragment | NA | NA | 0 |
| S8_vs_k141_161777\|\|full | 4164 | No | NA | 4 | 3 | 0 | Low-quality | Genome-fragment | 21.31 | AAI-based (medium-confidence) | 0 |
| S7_vs_100805\|\|full | 4165 | No | NA | 8 | 4 | 0 | Low-quality | Genome-fragment | 8.66 | AAI-based (high-confidence) | 0 |
| S17_vs_k141_51493\|\|full | 4167 | No | NA | 3 | 0 | 0 | Low-quality | Genome-fragment | 1.36 | AAI-based (medium-confidence) | 0 |
| S12_vs_k141_159284\|\|full | 1915 | No | NA | 2 | 0 | 1 | Not-determined | Genome-fragment | NA | NA | 0 |
| S3_extracted_k141_107758 | 4167 | No | NA | 5 | 1 | 0 | Low-quality | Genome-fragment | 2.24 | HMM-based (lower-bound) | 0 |
| S7_extracted_72720 | 4167 | No | NA | 5 | 4 | 0 | Low-quality | Genome-fragment | 2.56 | AAI-based (medium-confidence) | 0 |
| S12_vs_k141_97096\|\|full | 3268 | No | NA | 5 | 0 | 0 | Not-determined | Genome-fragment | NA | NA | 0 |
| S17_extracted_k141_51781 | 4168 | No | NA | 8 | 2 | 0 | Low-quality | Genome-fragment | 6.41 | AAI-based (high-confidence) | 0 |
| S12_vs_k141_111923\|\|full | 3172 | No | NA | 5 | 0 | 0 | Not-determined | Genome-fragment | NA | NA | 0 |
| S16_vs_158726\|\|full | 4168 | No | NA | 7 | 6 | 0 | Low-quality | Genome-fragment | 5.13 | AAI-based (high-confidence) | 0 |
| S12_vs_k141_65345\|\|full | 1776 | No | NA | 5 | 0 | 0 | Not-determined | Genome-fragment | NA | NA | 0 |
| S12_vs_k141_57611\|\|full | 2011 | No | NA | 3 | 0 | 0 | Not-determined | Genome-fragment | NA | NA | 0 |
| S13_extracted_58029 | 4169 | No | NA | 5 | 0 | 4 | Low-quality | Genome-fragment | 1.76 | AAI-based (high-confidence) | 0 |
| S7_extracted_30532 | 4169 | No | NA | 5 | 3 | 0 | Low-quality | Genome-fragment | 14.71 | HMM-based (lower-bound) | 0 |
| S12_vs_k141_111979\|\|full | 1580 | No | NA | 3 | 0 | 0 | Not-determined | Genome-fragment | NA | NA | 0 |
| S12_vs_k141_17931\|\|full | 1412 | No | NA | 3 | 0 | 1 | Not-determined | Genome-fragment | NA | NA | 0 |
| S12_vs_k141_151472\|\|full | 1267 | No | NA | 2 | 0 | 1 | Not-determined | Genome-fragment | NA | NA | 0 |
| S12_vs_k141_148575\|\|full | 4169 | No | NA | 5 | 0 | 2 | Low-quality | Genome-fragment | 2.08 | AAI-based (medium-confidence) | 0 |
| S12_vs_k141_26017\|\|full | 1998 | No | NA | 2 | 0 | 1 | Not-determined | Genome-fragment | NA | NA | 0 |
| S12_vs_k141_144087\|\|full | 1119 | No | NA | 2 | 0 | 1 | Not-determined | Genome-fragment | NA | NA | 0 |
| S6_extracted_k141_8711 | 4169 | No | NA | 4 | 0 | 3 | Low-quality | Genome-fragment | 1.56 | AAI-based (high-confidence) | 0 |
| S12_vs_k141_175045\|\|full | 2999 | No | NA | 3 | 0 | 1 | Not-determined | Genome-fragment | NA | NA | 0 |
| S12_vs_k141_49088\|\|full | 1897 | No | NA | 2 | 0 | 0 | Not-determined | Genome-fragment | NA | NA | 0 |
| S12_vs_k141_151523\|\|full | 3108 | No | NA | 4 | 0 | 2 | Not-determined | Genome-fragment | NA | NA | 0 |
| S8_extracted_k141_148387 | 4169 | No | NA | 5 | 5 | 0 | Low-quality | Genome-fragment | 13.74 | AAI-based (high-confidence) | 0 |
| S11_extracted_k141_49267 | 4170 | No | NA | 9 | 7 | 0 | Low-quality | Genome-fragment | 10.02 | AAI-based (high-confidence) | 0 |
| S16_extracted_162381 | 4171 | No | NA | 2 | 0 | 2 | Low-quality | Genome-fragment | 4.42 | AAI-based (medium-confidence) | 0 |
| S12_vs_k141_49108\|\|full | 3887 | No | NA | 5 | 0 | 4 | Not-determined | Genome-fragment | NA | NA | 0 |
| S12_vs_k141_112076\|\|full | 1358 | No | NA | 2 | 0 | 2 | Not-determined | Genome-fragment | NA | NA | 0 |
| S12_vs_k141_104778\|\|full | 2601 | No | NA | 4 | 0 | 0 | Not-determined | Genome-fragment | NA | NA | 0 |
| S12_extracted_k141_30241 | 4172 | No | NA | 3 | 0 | 3 | Low-quality | Genome-fragment | 1.72 | AAI-based (medium-confidence) | 0 |
| S12_vs_k141_65473\|\|full | 1408 | No | NA | 3 | 0 | 1 | Not-determined | Genome-fragment | NA | NA | 0 |
| S12_extracted_k141_156729 | 4172 | No | NA | 5 | 0 | 0 | Low-quality | Genome-fragment | 6.07 | AAI-based (medium-confidence) | 0 |
| S3_extracted_k141_4527 | 4173 | No | NA | 12 | 1 | 0 | Low-quality | Genome-fragment | 8.31 | AAI-based (high-confidence) | 0 |
| S14_vs_k141_74556\|\|full | 4174 | No | NA | 5 | 0 | 0 | Low-quality | Genome-fragment | 3.3 | AAI-based (medium-confidence) | 0 |
| S12_vs_k141_25019\|\|full | 4174 | No | NA | 6 | 0 | 2 | Low-quality | Genome-fragment | 2.39 | AAI-based (medium-confidence) | 0 |
| S12_extracted_k141_59179 | 4175 | No | NA | 4 | 0 | 1 | Low-quality | Genome-fragment | 6.89 | AAI-based (medium-confidence) | 0 |
| S12_vs_k141_65504\|\|full | 1668 | No | NA | 3 | 0 | 2 | Not-determined | Genome-fragment | NA | NA | 0 |
| S12_vs_k141_72714\|\|full | 2102 | No | NA | 3 | 0 | 1 | Not-determined | Genome-fragment | NA | NA | 0 |
| S14_extracted_k141_80694 | 4176 | No | NA | 9 | 4 | 0 | Low-quality | Genome-fragment | 10.55 | AAI-based (medium-confidence) | 0 |
| S12_vs_k141_151609\|\|full | 4238 | No | NA | 5 | 0 | 5 | Not-determined | Genome-fragment | NA | NA | 0 |
| S12_vs_k141_159362\|\|full | 4153 | No | NA | 4 | 0 | 4 | Not-determined | Genome-fragment | NA | NA | 0 |
| S12_vs_k141_57763\|\|full | 2567 | No | NA | 3 | 0 | 0 | Not-determined | Genome-fragment | NA | NA | 0 |
| S1_vs_k141_54003\|\|full | 4176 | No | NA | 5 | 2 | 0 | Low-quality | Genome-fragment | 8.74 | AAI-based (high-confidence) | 0 |
| S12_vs_k141_65548\|\|full | 1073 | No | NA | 3 | 0 | 1 | Not-determined | Genome-fragment | NA | NA | 0 |
| S12_vs_k141_80932\|\|full | 1051 | No | NA | 3 | 0 | 1 | Not-determined | Genome-fragment | NA | NA | 0 |
| S1_vs_k141_17596\|\|full | 4176 | No | NA | 6 | 0 | 0 | Low-quality | Genome-fragment | 6.75 | AAI-based (medium-confidence) | 0 |
| S7_vs_42270\|\|full | 4176 | No | NA | 8 | 2 | 1 | Low-quality | Genome-fragment | 8.79 | AAI-based (high-confidence) | 0 |
| S8_vs_k141_78578\|\|full | 4180 | No | NA | 3 | 0 | 3 | Low-quality | Genome-fragment | 2.56 | AAI-based (medium-confidence) | 0 |
| S14_extracted_k141_87991 | 4181 | No | NA | 9 | 3 | 1 | Low-quality | Genome-fragment | 10.2 | AAI-based (medium-confidence) | 0 |
| S8_vs_k141_183987\|\|full | 4181 | No | NA | 4 | 0 | 0 | Low-quality | Genome-fragment | 7.7 | AAI-based (medium-confidence) | 0 |
| S1_extracted_k141_127374 | 4182 | No | NA | 5 | 0 | 0 | Low-quality | Genome-fragment | 6.65 | AAI-based (high-confidence) | 0 |
| S12_vs_k141_89237\|\|full | 3240 | No | NA | 3 | 0 | 1 | Not-determined | Genome-fragment | NA | NA | 0 |
| S7_vs_62955\|\|full | 4182 | No | NA | 4 | 0 | 0 | Low-quality | Genome-fragment | 7.22 | AAI-based (medium-confidence) | 0 |
| S12_vs_k141_120692\|\|full | 2446 | No | NA | 3 | 0 | 1 | Not-determined | Genome-fragment | NA | NA | 0 |
| S10_vs_107190\|\|full | 4182 | No | NA | 7 | 2 | 0 | Low-quality | Genome-fragment | 10.01 | AAI-based (high-confidence) | 0 |
| S12_vs_k141_183539\|\|full | 4463 | No | NA | 4 | 0 | 1 | Not-determined | Genome-fragment | NA | NA | 0 |
| S8_extracted_k141_107925 | 4182 | No | NA | 7 | 1 | 0 | Low-quality | Genome-fragment | 13.14 | AAI-based (high-confidence) | 0 |
| S14_vs_k141_9372\|\|full | 4183 | No | NA | 8 | 0 | 0 | Low-quality | Genome-fragment | 10.9 | AAI-based (medium-confidence) | 0 |
| S12_vs_k141_104847\|\|full | 1780 | No | NA | 4 | 0 | 2 | Not-determined | Genome-fragment | NA | NA | 0 |
| S12_vs_k141_97408\|\|full | 7407 | No | NA | 7 | 0 | 4 | Not-determined | Genome-fragment | NA | NA | 0 |
| S16_vs_94990\|\|full | 4183 | No | NA | 9 | 0 | 0 | Low-quality | Genome-fragment | 5.01 | AAI-based (high-confidence) | 0 |
| S6_extracted_k141_29619 | 4183 | No | NA | 4 | 0 | 3 | Low-quality | Genome-fragment | 7.48 | AAI-based (medium-confidence) | 0 |
| S12_vs_k141_80974\|\|full | 2404 | No | NA | 2 | 0 | 0 | Not-determined | Genome-fragment | NA | NA | 0 |
| S12_vs_k141_41995\|\|full | 3107 | No | NA | 2 | 0 | 1 | Not-determined | Genome-fragment | NA | NA | 0 |
| S12_vs_k141_80976\|\|full | 1427 | No | NA | 3 | 0 | 1 | Not-determined | Genome-fragment | NA | NA | 0 |
| S12_vs_k141_144251\|\|full | 2546 | No | NA | 4 | 0 | 1 | Not-determined | Genome-fragment | NA | NA | 0 |
| S8_vs_k141_166494\|\|full | 4183 | No | NA | 2 | 0 | 2 | Low-quality | Genome-fragment | 1.32 | AAI-based (medium-confidence) | 0 |
| S12_vs_k141_26122\|\|full | 1455 | No | NA | 3 | 0 | 2 | Not-determined | Genome-fragment | NA | NA | 0 |
| S11_vs_k141_43117\|\|full | 4184 | No | NA | 3 | 0 | 0 | Low-quality | Genome-fragment | 6.94 | AAI-based (medium-confidence) | 0 |
| S12_vs_k141_80992\|\|full | 1670 | No | NA | 2 | 0 | 1 | Not-determined | Genome-fragment | NA | NA | 0 |
| S12_vs_k141_26133\|\|full | 1388 | No | NA | 3 | 0 | 2 | Not-determined | Genome-fragment | NA | NA | 0 |
| S12_vs_k141_89309\|\|full | 6505 | No | NA | 6 | 0 | 6 | Not-determined | Genome-fragment | NA | NA | 0 |
| S7_vs_113356\|\|full | 4184 | No | NA | 7 | 0 | 3 | Low-quality | Genome-fragment | 5.68 | AAI-based (high-confidence) | 0 |
| S16_extracted_7084 | 4184 | No | NA | 5 | 1 | 0 | Low-quality | Genome-fragment | 8.58 | HMM-based (lower-bound) | 0 |
| S12_vs_k141_104888\|\|full | 2411 | No | NA | 3 | 0 | 2 | Not-determined | Genome-fragment | NA | NA | 0 |
| S14_vs_k141_5988\|\|full | 4185 | No | NA | 8 | 2 | 0 | Low-quality | Genome-fragment | 12.54 | AAI-based (high-confidence) | 0 |
| S12_vs_k141_183609\|\|full | 2338 | No | NA | 4 | 0 | 4 | Not-determined | Genome-fragment | NA | NA | 0 |
| S12_vs_k141_65658\|\|full | 2606 | No | NA | 3 | 0 | 2 | Not-determined | Genome-fragment | NA | NA | 0 |
| S12_vs_k141_89349\|\|full | 1682 | No | NA | 2 | 0 | 2 | Not-determined | Genome-fragment | NA | NA | 0 |
| S3_vs_k141_55769\|\|full | 4186 | No | NA | 3 | 3 | 0 | Low-quality | Genome-fragment | 6.99 | AAI-based (medium-confidence) | 0 |
| S5_vs_k141_80491\|\|full | 4186 | No | NA | 8 | 3 | 0 | Low-quality | Genome-fragment | 10.4 | AAI-based (high-confidence) | 0 |
| S12_vs_k141_2628\|\|full | 1427 | No | NA | 3 | 0 | 2 | Not-determined | Genome-fragment | NA | NA | 0 |
| S12_vs_k141_26203\|\|full | 1259 | No | NA | 2 | 0 | 0 | Not-determined | Genome-fragment | NA | NA | 0 |
| S12_vs_k141_159545\|\|full | 1714 | No | NA | 2 | 0 | 1 | Not-determined | Genome-fragment | NA | NA | 0 |
| S16_extracted_117064 | 4186 | No | NA | 4 | 1 | 0 | Low-quality | Genome-fragment | 7.42 | AAI-based (medium-confidence) | 0 |
| S12_vs_k141_9887\|\|full | 2339 | No | NA | 3 | 0 | 2 | Not-determined | Genome-fragment | NA | NA | 0 |
| S8_extracted_k141_58618 | 4186 | No | NA | 3 | 0 | 3 | Low-quality | Genome-fragment | 1.64 | AAI-based (medium-confidence) | 0 |
| S12_vs_k141_42084\|\|full | 1871 | No | NA | 2 | 0 | 1 | Not-determined | Genome-fragment | NA | NA | 0 |
| S12_vs_k141_159546\|\|full | 2580 | No | NA | 3 | 0 | 0 | Not-determined | Genome-fragment | NA | NA | 0 |
| S5_extracted_k141_119568 | 4187 | No | NA | 3 | 0 | 1 | Low-quality | Genome-fragment | 3.15 | AAI-based (medium-confidence) | 0 |
| S12_vs_k141_65708\|\|full | 2135 | No | NA | 4 | 0 | 2 | Not-determined | Genome-fragment | NA | NA | 0 |
| S14_vs_k141_87175\|\|full | 4188 | No | NA | 11 | 2 | 0 | Low-quality | Genome-fragment | 13.01 | AAI-based (high-confidence) | 0 |
| S17_extracted_k141_166770 | 4189 | No | NA | 7 | 0 | 1 | Low-quality | Genome-fragment | 7.7 | AAI-based (high-confidence) | 0 |
| S17_extracted_k141_18240 | 4191 | No | NA | 4 | 0 | 4 | Low-quality | Genome-fragment | 1.13 | AAI-based (medium-confidence) | 0 |
| S12_extracted_k141_73676 | 4191 | No | NA | 5 | 1 | 0 | Low-quality | Genome-fragment | 3.96 | AAI-based (high-confidence) | 0 |
| S12_vs_k141_136247\|\|full | 2251 | No | NA | 2 | 0 | 0 | Not-determined | Genome-fragment | NA | NA | 0 |
| S3_vs_k141_9492\|\|full | 4192 | No | NA | 8 | 3 | 1 | Low-quality | Genome-fragment | 2.04 | AAI-based (high-confidence) | 0 |
| S12_vs_k141_183653\|\|full | 4133 | No | NA | 4 | 0 | 2 | Not-determined | Genome-fragment | NA | NA | 0 |
| S12_vs_k141_18221\|\|full | 1065 | No | NA | 2 | 0 | 0 | Not-determined | Genome-fragment | NA | NA | 0 |
| S8_extracted_k141_57951 | 4192 | No | NA | 4 | 0 | 4 | Low-quality | Genome-fragment | 3.37 | AAI-based (medium-confidence) | 0 |
| S12_vs_k141_175309\|\|full | 6165 | No | NA | 8 | 0 | 3 | Not-determined | Genome-fragment | NA | NA | 0 |
| S12_vs_k141_136269\|\|full | 7029 | No | NA | 8 | 0 | 2 | Not-determined | Genome-fragment | NA | NA | 0 |
| S12_vs_k141_81141\|\|full | 2608 | No | NA | 4 | 0 | 1 | Not-determined | Genome-fragment | NA | NA | 0 |
| S1_extracted_k141_128184 | 4193 | No | NA | 7 | 1 | 1 | Low-quality | Genome-fragment | 5.21 | AAI-based (high-confidence) | 0 |
| S12_vs_k141_57957\|\|full | 4874 | No | NA | 6 | 0 | 4 | Not-determined | Genome-fragment | NA | NA | 0 |
| S16_extracted_136664 | 4193 | No | NA | 4 | 0 | 3 | Low-quality | Genome-fragment | 2.51 | AAI-based (medium-confidence) | 0 |
| S12_vs_k141_81165\|\|full | 3294 | No | NA | 4 | 0 | 1 | Not-determined | Genome-fragment | NA | NA | 0 |
| S12_vs_k141_128344\|\|full | 3241 | No | NA | 5 | 0 | 0 | Not-determined | Genome-fragment | NA | NA | 0 |
| S12_vs_k141_81180\|\|full | 4724 | No | NA | 6 | 0 | 3 | Not-determined | Genome-fragment | NA | NA | 0 |
| S12_vs_k141_183719\|\|full | 3348 | No | NA | 3 | 0 | 0 | Not-determined | Genome-fragment | NA | NA | 0 |
| S12_vs_k141_65767\|\|full | 2302 | No | NA | 3 | 0 | 2 | Not-determined | Genome-fragment | NA | NA | 0 |
| S12_vs_k141_2701\|\|full | 5185 | No | NA | 8 | 0 | 3 | Not-determined | Genome-fragment | NA | NA | 0 |
| S12_vs_k141_49267\|\|full | 1067 | No | NA | 2 | 0 | 0 | Not-determined | Genome-fragment | NA | NA | 0 |
| S12_vs_k141_57999\|\|full | 1717 | No | NA | 3 | 0 | 2 | Not-determined | Genome-fragment | NA | NA | 0 |
| S10_vs_54117\|\|full | 4195 | No | NA | 6 | 0 | 3 | Low-quality | Genome-fragment | 15.31 | AAI-based (medium-confidence) | 0 |
| S12_vs_k141_97508\|\|full | 6216 | No | NA | 8 | 0 | 6 | Not-determined | Genome-fragment | NA | NA | 0 |
| S5_vs_k141_73732\|\|full | 4196 | No | NA | 6 | 4 | 0 | Low-quality | Genome-fragment | 12.22 | AAI-based (high-confidence) | 0 |
| S12_vs_k141_183751\|\|full | 2797 | No | NA | 6 | 0 | 0 | Not-determined | Genome-fragment | NA | NA | 0 |
| S12_vs_k141_81239\|\|full | 1104 | No | NA | 2 | 0 | 1 | Not-determined | Genome-fragment | NA | NA | 0 |
| S12_vs_k141_128399\|\|full | 2148 | No | NA | 4 | 0 | 1 | Not-determined | Genome-fragment | NA | NA | 0 |
| S12_vs_k141_128409\|\|full | 1726 | No | NA | 2 | 0 | 0 | Not-determined | Genome-fragment | NA | NA | 0 |
| S12_vs_k141_183752\|\|full | 14400 | No | NA | 19 | 7 | 0 | Medium-quality | Genome-fragment | 61.3 | AAI-based (medium-confidence) | 0 |
| S9_extracted_k141_85492 | 4197 | No | NA | 5 | 0 | 1 | Low-quality | Genome-fragment | 3.95 | AAI-based (medium-confidence) | 0 |
| S7_vs_26320\|\|full | 4198 | No | NA | 6 | 0 | 3 | Low-quality | Genome-fragment | 3.2 | AAI-based (high-confidence) | 0 |
| S3_extracted_k141_108642 | 4200 | No | NA | 4 | 0 | 0 | Low-quality | Genome-fragment | 2.3 | AAI-based (high-confidence) | 0 |
| S14_vs_k141_7083\|\|full | 4202 | No | NA | 5 | 0 | 4 | Low-quality | Genome-fragment | 12.64 | AAI-based (medium-confidence) | 0 |
| S10_vs_9484\|\|full | 4202 | No | NA | 7 | 0 | 0 | Low-quality | Genome-fragment | 9.81 | AAI-based (high-confidence) | 0 |
| S12_vs_k141_81263\|\|full | 3979 | No | NA | 3 | 0 | 3 | Not-determined | Genome-fragment | NA | NA | 0 |
| S12_vs_k141_183796\|\|full | 2318 | No | NA | 4 | 0 | 0 | Not-determined | Genome-fragment | NA | NA | 0 |
| S12_vs_k141_26305\|\|full | 15381 | No | NA | 20 | 0 | 4 | Not-determined | Genome-fragment | NA | NA | 0 |
| S13_vs_29604\|\|full | 4204 | No | NA | 5 | 0 | 4 | Low-quality | Genome-fragment | 3.46 | AAI-based (medium-confidence) | 0 |
| S1_vs_k141_82197\|\|full | 4204 | No | NA | 2 | 0 | 1 | Low-quality | Genome-fragment | 3.72 | AAI-based (medium-confidence) | 0 |
| S12_vs_k141_183808\|\|full | 8792 | No | NA | 7 | 0 | 5 | Not-determined | Genome-fragment | NA | NA | 0 |
| S12_vs_k141_97571\|\|full | 1334 | No | NA | 2 | 0 | 0 | Not-determined | Genome-fragment | NA | NA | 0 |
| S12_vs_k141_73008\|\|full | 1599 | No | NA | 2 | 0 | 2 | Not-determined | Genome-fragment | NA | NA | 0 |
| S11_extracted_k141_79683 | 4205 | No | NA | 3 | 0 | 1 | Low-quality | Genome-fragment | 5.48 | AAI-based (medium-confidence) | 0 |
| S12_vs_k141_42320\|\|full | 3640 | No | NA | 4 | 0 | 2 | Not-determined | Genome-fragment | NA | NA | 0 |
| S16_extracted_61646 | 4205 | No | NA | 5 | 1 | 0 | Low-quality | Genome-fragment | 10.75 | AAI-based (medium-confidence) | 0 |
| S8_vs_k141_54804\|\|full | 4205 | No | NA | 3 | 0 | 2 | Low-quality | Genome-fragment | 13.43 | AAI-based (medium-confidence) | 0 |
| S12_vs_k141_49387\|\|full | 1976 | No | NA | 3 | 0 | 0 | Not-determined | Genome-fragment | NA | NA | 0 |
| S12_vs_k141_136394\|\|full | 1577 | No | NA | 2 | 0 | 2 | Not-determined | Genome-fragment | NA | NA | 0 |
| S5_vs_k141_44158\|\|full | 4206 | No | NA | 4 | 3 | 0 | Low-quality | Genome-fragment | 10.26 | AAI-based (high-confidence) | 0 |
| S7_extracted_117429 | 4206 | No | NA | 5 | 3 | 0 | Low-quality | Genome-fragment | 12.16 | AAI-based (high-confidence) | 0 |
| S12_vs_k141_136412\|\|full | 2296 | No | NA | 4 | 0 | 0 | Not-determined | Genome-fragment | NA | NA | 0 |
| S12_vs_k141_81347\|\|full | 1285 | No | NA | 2 | 0 | 1 | Not-determined | Genome-fragment | NA | NA | 0 |
| S12_vs_k141_183852\|\|full | 10083 | No | NA | 12 | 0 | 2 | Not-determined | Genome-fragment | NA | NA | 0 |
| S16_extracted_94285 | 4207 | No | NA | 4 | 0 | 3 | Low-quality | Genome-fragment | 2.09 | AAI-based (medium-confidence) | 0 |
| S8_vs_k141_193788\|\|full | 4208 | No | NA | 4 | 0 | 1 | Low-quality | Genome-fragment | 9.84 | AAI-based (medium-confidence) | 0 |
| S12_vs_k141_105197\|\|full | 1651 | No | NA | 3 | 0 | 1 | Not-determined | Genome-fragment | NA | NA | 0 |
| S16_vs_32311\|\|full | 4211 | No | NA | 9 | 5 | 0 | Low-quality | Genome-fragment | 9.5 | AAI-based (high-confidence) | 0 |
| S12_vs_k141_143109\|\|full | 4211 | Yes | 2808 | 10 | 1 | 1 | Low-quality | Genome-fragment | 7.01 | AAI-based (high-confidence) | 33.32 |
| S12_vs_k141_49413\|\|full | 2486 | No | NA | 4 | 0 | 2 | Not-determined | Genome-fragment | NA | NA | 0 |
| S12_vs_k141_42393\|\|full | 7909 | No | NA | 8 | 0 | 3 | Not-determined | Genome-fragment | NA | NA | 0 |
| S12_vs_k141_58148\|\|full | 23751 | No | NA | 30 | 19 | 2 | Medium-quality | Genome-fragment | 68.77 | AAI-based (high-confidence) | 0 |
| S12_vs_k141_159730\|\|full | 1461 | No | NA | 2 | 0 | 1 | Not-determined | Genome-fragment | NA | NA | 0 |
| S12_vs_k141_26336\|\|full | 67092 | Yes | 48897 | 50 | 5 | 11 | Medium-quality | Genome-fragment | 80.77 | AAI-based (medium-confidence) | 27.12 |
| S12_vs_k141_18450\|\|full | 1390 | No | NA | 3 | 0 | 0 | Not-determined | Genome-fragment | NA | NA | 0 |
| S12_vs_k141_2892\|\|full | 1348 | No | NA | 3 | 0 | 0 | Not-determined | Genome-fragment | NA | NA | 0 |
| S8_extracted_k141_49890 | 4211 | No | NA | 3 | 0 | 2 | Low-quality | Genome-fragment | 4.83 | AAI-based (medium-confidence) | 0 |
| S11_vs_k141_32155\|\|full | 4214 | No | NA | 4 | 2 | 0 | Low-quality | Genome-fragment | 12.86 | AAI-based (medium-confidence) | 0 |
| S12_vs_k141_18456\|\|full | 1341 | No | NA | 4 | 0 | 1 | Not-determined | Genome-fragment | NA | NA | 0 |
| S12_vs_k141_26343\|\|full | 2837 | No | NA | 3 | 0 | 2 | Not-determined | Genome-fragment | NA | NA | 0 |
| S12_vs_k141_151857\|\|full | 3503 | No | NA | 5 | 0 | 1 | Not-determined | Genome-fragment | NA | NA | 0 |
| S1_vs_k141_55634\|\|full | 4214 | No | NA | 8 | 1 | 0 | Low-quality | Genome-fragment | 9.18 | AAI-based (high-confidence) | 0 |
| S5_vs_k141_1372\|\|full | 4215 | No | NA | 7 | 0 | 1 | Low-quality | Genome-fragment | 3.84 | AAI-based (medium-confidence) | 0 |
| S16_vs_116049\|\|full | 4215 | No | NA | 5 | 1 | 0 | Low-quality | Genome-fragment | 10.28 | AAI-based (high-confidence) | 0 |
| S10_extracted_20657 | 4215 | No | NA | 3 | 0 | 2 | Low-quality | Genome-fragment | 2.04 | AAI-based (medium-confidence) | 0 |
| S3_extracted_k141_80887 | 4216 | No | NA | 9 | 1 | 0 | Low-quality | Genome-fragment | 7.33 | AAI-based (high-confidence) | 0 |
| S12_vs_k141_49476\|\|full | 1317 | No | NA | 2 | 0 | 0 | Not-determined | Genome-fragment | NA | NA | 0 |
| S12_vs_k141_151863\|\|full | 7812 | No | NA | 12 | 0 | 0 | Not-determined | Genome-fragment | NA | NA | 0 |
| S12_vs_k141_89684\|\|full | 1111 | No | NA | 3 | 0 | 2 | Not-determined | Genome-fragment | NA | NA | 0 |
| S16_vs_145262\|\|full | 4216 | No | NA | 3 | 0 | 1 | Low-quality | Genome-fragment | 5.15 | AAI-based (high-confidence) | 0 |
| S12_vs_k141_136466\|\|full | 1402 | No | NA | 2 | 0 | 2 | Not-determined | Genome-fragment | NA | NA | 0 |
| S13_extracted_44922 | 4217 | No | NA | 6 | 0 | 5 | Low-quality | Genome-fragment | 1.56 | AAI-based (medium-confidence) | 0 |
| S12_vs_k141_33781\|\|full | 2473 | No | NA | 3 | 0 | 2 | Not-determined | Genome-fragment | NA | NA | 0 |
| S12_vs_k141_42411\|\|full | 6826 | No | NA | 12 | 0 | 0 | Not-determined | Genome-fragment | NA | NA | 0 |
| S12_vs_k141_18552\|\|full | 1100 | No | NA | 2 | 0 | 1 | Not-determined | Genome-fragment | NA | NA | 0 |
| S12_vs_k141_2953\|\|full | 2201 | No | NA | 3 | 0 | 2 | Not-determined | Genome-fragment | NA | NA | 0 |
| S12_vs_k141_33793\|\|full | 5064 | No | NA | 7 | 0 | 0 | Not-determined | Genome-fragment | NA | NA | 0 |
| S12_vs_k141_81382\|\|full | 6477 | No | NA | 5 | 0 | 4 | Not-determined | Genome-fragment | NA | NA | 0 |
| S5_vs_k141_60162\|\|full | 4217 | No | NA | 6 | 3 | 0 | Low-quality | Genome-fragment | 11.21 | AAI-based (high-confidence) | 0 |
| S16_vs_152565\|\|full | 4218 | No | NA | 5 | 3 | 0 | Low-quality | Genome-fragment | 2.26 | HMM-based (lower-bound) | 0 |
| S1_extracted_k141_141947 | 4220 | No | NA | 4 | 0 | 3 | Low-quality | Genome-fragment | 0.77 | AAI-based (medium-confidence) | 0 |
| S12_vs_k141_26417\|\|full | 2599 | No | NA | 3 | 0 | 2 | Not-determined | Genome-fragment | NA | NA | 0 |
| S14_extracted_k141_108818 | 4221 | No | NA | 4 | 2 | 0 | Low-quality | Genome-fragment | 6.57 | AAI-based (high-confidence) | 0 |
| S7_vs_10149\|\|full | 4222 | No | NA | 3 | 0 | 0 | Low-quality | Genome-fragment | 2.77 | AAI-based (high-confidence) | 0 |
| S12_vs_k141_18597\|\|full | 6055 | No | NA | 9 | 0 | 2 | Not-determined | Genome-fragment | NA | NA | 0 |
| S12_vs_k141_112603\|\|full | 1165 | No | NA | 3 | 0 | 1 | Not-determined | Genome-fragment | NA | NA | 0 |
| S7_vs_130086\|\|full | 4222 | No | NA | 8 | 3 | 0 | Low-quality | Genome-fragment | 6.62 | AAI-based (medium-confidence) | 0 |
| S12_vs_k141_168022\|\|full | 9295 | No | NA | 6 | 0 | 5 | Not-determined | Genome-fragment | NA | NA | 0 |
| S12_vs_k141_97773\|\|full | 2363 | No | NA | 2 | 0 | 1 | Not-determined | Genome-fragment | NA | NA | 0 |
| S12_vs_k141_97781\|\|full | 3453 | No | NA | 3 | 0 | 2 | Not-determined | Genome-fragment | NA | NA | 0 |
| S12_vs_k141_42524\|\|full | 2530 | No | NA | 6 | 0 | 1 | Not-determined | Genome-fragment | NA | NA | 0 |
| S12_vs_k141_159881\|\|full | 1371 | No | NA | 3 | 0 | 0 | Not-determined | Genome-fragment | NA | NA | 0 |
| S5_vs_k141_68008\|\|full | 4223 | No | NA | 4 | 0 | 2 | Low-quality | Genome-fragment | 3.14 | AAI-based (high-confidence) | 0 |
| S12_vs_k141_97814\|\|full | 1322 | No | NA | 2 | 0 | 2 | Not-determined | Genome-fragment | NA | NA | 0 |
| S12_vs_k141_89716\|\|full | 1645 | No | NA | 2 | 0 | 0 | Not-determined | Genome-fragment | NA | NA | 0 |
| S12_vs_k141_97827\|\|full | 2512 | No | NA | 4 | 0 | 3 | Not-determined | Genome-fragment | NA | NA | 0 |
| S12_vs_k141_58308\|\|full | 2708 | No | NA | 5 | 0 | 3 | Not-determined | Genome-fragment | NA | NA | 0 |
| S12_vs_k141_184001\|\|full | 13560 | No | NA | 14 | 0 | 1 | Not-determined | Genome-fragment | NA | NA | 0 |
| S12_extracted_k141_85346 | 4224 | No | NA | 5 | 0 | 3 | Low-quality | Genome-fragment | 2.03 | AAI-based (high-confidence) | 0 |
| S4_extracted_16438 | 4224 | No | NA | 7 | 0 | 0 | Low-quality | Genome-fragment | 10.42 | AAI-based (high-confidence) | 0 |
| S12_vs_k141_184011\|\|full | 1558 | No | NA | 3 | 0 | 0 | Not-determined | Genome-fragment | NA | NA | 0 |
| S12_vs_k141_184016\|\|full | 1184 | No | NA | 2 | 0 | 0 | Not-determined | Genome-fragment | NA | NA | 0 |
| S3_extracted_k141_80615 | 4225 | No | NA | 4 | 0 | 0 | Low-quality | Genome-fragment | 11.16 | AAI-based (medium-confidence) | 0 |
| S5_vs_k141_48002\|\|full | 4225 | No | NA | 4 | 4 | 0 | Low-quality | Genome-fragment | 8.55 | AAI-based (high-confidence) | 0 |
| S12_vs_k141_3042\|\|full | 2462 | No | NA | 2 | 0 | 2 | Not-determined | Genome-fragment | NA | NA | 0 |
| S7_vs_120190\|\|full | 4225 | No | NA | 3 | 0 | 3 | Low-quality | Genome-fragment | 2.29 | AAI-based (medium-confidence) | 0 |
| S12_vs_k141_136598\|\|full | 1169 | No | NA | 2 | 0 | 0 | Not-determined | Genome-fragment | NA | NA | 0 |
| S12_vs_k141_168103\|\|full | 1152 | No | NA | 3 | 0 | 2 | Not-determined | Genome-fragment | NA | NA | 0 |
| S12_vs_k141_152043\|\|full | 8198 | No | NA | 11 | 0 | 3 | Not-determined | Genome-fragment | NA | NA | 0 |
| S5_vs_k141_106979\|\|full | 4226 | No | NA | 3 | 0 | 3 | Low-quality | Genome-fragment | 4.11 | AAI-based (medium-confidence) | 0 |
| S12_vs_k141_112685\|\|full | 1557 | No | NA | 2 | 0 | 1 | Not-determined | Genome-fragment | NA | NA | 0 |
| S12_vs_k141_97878\|\|full | 2037 | No | NA | 3 | 0 | 0 | Not-determined | Genome-fragment | NA | NA | 0 |
| S12_vs_k141_112686\|\|full | 1130 | No | NA | 3 | 0 | 1 | Not-determined | Genome-fragment | NA | NA | 0 |
| S12_vs_k141_159959\|\|full | 2304 | No | NA | 4 | 0 | 2 | Not-determined | Genome-fragment | NA | NA | 0 |
| S9_vs_k141_47897\|\|full | 4229 | No | NA | 3 | 3 | 0 | Low-quality | Genome-fragment | 10.52 | AAI-based (high-confidence) | 0 |
| S12_vs_k141_138513\|\|full | 4229 | No | NA | 4 | 3 | 0 | Low-quality | Genome-fragment | 3.89 | AAI-based (high-confidence) | 0 |
| S12_vs_k141_136621\|\|full | 4720 | No | NA | 3 | 0 | 3 | Not-determined | Genome-fragment | NA | NA | 0 |
| S12_vs_k141_18742\|\|full | 4194 | No | NA | 5 | 0 | 0 | Not-determined | Genome-fragment | NA | NA | 0 |
| S17_vs_k141_36879\|\|full | 4230 | No | NA | 2 | 0 | 0 | Low-quality | Genome-fragment | 6.19 | AAI-based (high-confidence) | 0 |
| S12_vs_k141_175588\|\|full | 2423 | No | NA | 6 | 0 | 0 | Not-determined | Genome-fragment | NA | NA | 0 |
| S12_vs_k141_3107\|\|full | 8328 | No | NA | 8 | 0 | 8 | Not-determined | Genome-fragment | NA | NA | 0 |
| S14_vs_k141_88437\|\|full | 4230 | No | NA | 8 | 0 | 0 | Low-quality | Genome-fragment | 25.23 | AAI-based (high-confidence) | 0 |
| S12_vs_k141_175597\|\|full | 3660 | No | NA | 6 | 0 | 5 | Not-determined | Genome-fragment | NA | NA | 0 |
| S10_vs_46587\|\|full | 4231 | No | NA | 3 | 0 | 2 | Low-quality | Genome-fragment | 9.6 | AAI-based (medium-confidence) | 0 |
| S12_vs_k141_42630\|\|full | 2092 | No | NA | 3 | 0 | 2 | Not-determined | Genome-fragment | NA | NA | 0 |
| S12_vs_k141_144603\|\|full | 2637 | No | NA | 5 | 0 | 2 | Not-determined | Genome-fragment | NA | NA | 0 |
| S12_vs_k141_73279\|\|full | 3277 | No | NA | 3 | 0 | 2 | Not-determined | Genome-fragment | NA | NA | 0 |
| S12_vs_k141_184078\|\|full | 1161 | No | NA | 2 | 0 | 1 | Not-determined | Genome-fragment | NA | NA | 0 |
| S12_vs_k141_120910\|\|full | 2594 | No | NA | 2 | 0 | 2 | Not-determined | Genome-fragment | NA | NA | 0 |
| S12_vs_k141_175635\|\|full | 1107 | No | NA | 2 | 0 | 0 | Not-determined | Genome-fragment | NA | NA | 0 |
| S5_vs_k141_140866\|\|full | 4232 | No | NA | 2 | 0 | 1 | Low-quality | Genome-fragment | 5.94 | AAI-based (medium-confidence) | 0 |
| S5_vs_k141_130609\|\|full | 4234 | No | NA | 9 | 4 | 0 | Low-quality | Genome-fragment | 9.3 | AAI-based (medium-confidence) | 0 |
| S12_vs_k141_81527\|\|full | 1713 | No | NA | 3 | 0 | 2 | Not-determined | Genome-fragment | NA | NA | 0 |
| S17_extracted_k141_92588 | 4235 | No | NA | 4 | 0 | 2 | Low-quality | Genome-fragment | 9.3 | AAI-based (medium-confidence) | 0 |
| S7_vs_132489\|\|full | 4235 | No | NA | 7 | 5 | 0 | Low-quality | Genome-fragment | 10.98 | AAI-based (high-confidence) | 0 |
| S12_vs_k141_168134\|\|full | 1012 | No | NA | 3 | 0 | 2 | Not-determined | Genome-fragment | NA | NA | 0 |
| S16_extracted_126388 | 4235 | No | NA | 2 | 0 | 0 | Low-quality | Genome-fragment | 3.87 | AAI-based (high-confidence) | 0 |
| S12_vs_k141_26545\|\|full | 1376 | No | NA | 2 | 0 | 0 | Not-determined | Genome-fragment | NA | NA | 0 |
| S16_vs_44990\|\|full | 4237 | No | NA | 4 | 0 | 2 | Low-quality | Genome-fragment | 0.69 | AAI-based (high-confidence) | 0 |
| S12_vs_k141_120932\|\|full | 2449 | No | NA | 3 | 0 | 0 | Not-determined | Genome-fragment | NA | NA | 0 |
| S17_vs_k141_95962\|\|full | 4238 | No | NA | 6 | 0 | 0 | Low-quality | Genome-fragment | 2.92 | AAI-based (medium-confidence) | 0 |
| S9_vs_k141_27295\|\|full | 4238 | No | NA | 6 | 1 | 0 | Low-quality | Genome-fragment | 6.41 | HMM-based (lower-bound) | 0 |
| S12_vs_k141_81552\|\|full | 12353 | No | NA | 6 | 0 | 5 | Not-determined | Genome-fragment | NA | NA | 0 |
| S16_vs_87426\|\|full | 4238 | No | NA | 3 | 0 | 3 | Low-quality | Genome-fragment | 2.38 | AAI-based (medium-confidence) | 0 |
| S12_vs_k141_152131\|\|full | 3172 | No | NA | 4 | 0 | 2 | Not-determined | Genome-fragment | NA | NA | 0 |
| S12_extracted_k141_152289 | 4239 | No | NA | 11 | 0 | 0 | Low-quality | Genome-fragment | 6.31 | AAI-based (high-confidence) | 0 |
| S13_vs_86823\|\|full | 4241 | No | NA | 6 | 3 | 0 | Low-quality | Genome-fragment | 10.26 | AAI-based (high-confidence) | 0 |
| S3_extracted_k141_33055 | 4242 | No | NA | 7 | 4 | 0 | Low-quality | Genome-fragment | 8.91 | AAI-based (medium-confidence) | 0 |
| S12_vs_k141_112821\|\|full | 1232 | No | NA | 3 | 0 | 2 | Not-determined | Genome-fragment | NA | NA | 0 |
| S3_vs_k141_114877\|\|full | 4242 | No | NA | 6 | 0 | 0 | Low-quality | Genome-fragment | 7.97 | AAI-based (high-confidence) | 0 |
| S12_extracted_k141_55336 | 4242 | No | NA | 4 | 0 | 4 | Low-quality | Genome-fragment | 1.83 | AAI-based (high-confidence) | 0 |
| S12_vs_k141_128614\|\|full | 7435 | No | NA | 4 | 0 | 4 | Not-determined | Genome-fragment | NA | NA | 0 |
| S15_vs_65116\|\|full | 4242 | No | NA | 6 | 0 | 0 | Low-quality | Genome-fragment | 4.15 | AAI-based (high-confidence) | 0 |
| S12_vs_k141_34023\|\|full | 2063 | No | NA | 2 | 0 | 0 | Not-determined | Genome-fragment | NA | NA | 0 |
| S8_vs_k141_156073\|\|full | 4242 | No | NA | 3 | 0 | 3 | Low-quality | Genome-fragment | 5.86 | AAI-based (medium-confidence) | 0 |
| S12_vs_k141_73392\|\|full | 1527 | No | NA | 3 | 0 | 0 | Not-determined | Genome-fragment | NA | NA | 0 |
| S12_vs_k141_66122\|\|full | 7983 | No | NA | 3 | 0 | 2 | Not-determined | Genome-fragment | NA | NA | 0 |
| S12_vs_k141_168241\|\|full | 2629 | No | NA | 4 | 0 | 2 | Not-determined | Genome-fragment | NA | NA | 0 |
| S12_vs_k141_98088\|\|full | 2148 | No | NA | 3 | 0 | 1 | Not-determined | Genome-fragment | NA | NA | 0 |
| S12_vs_k141_34026\|\|full | 4844 | No | NA | 4 | 0 | 3 | Not-determined | Genome-fragment | NA | NA | 0 |
| S12_vs_k141_73398\|\|full | 1086 | No | NA | 2 | 0 | 0 | Not-determined | Genome-fragment | NA | NA | 0 |
| S12_vs_k141_10222\|\|full | 2219 | No | NA | 4 | 0 | 1 | Not-determined | Genome-fragment | NA | NA | 0 |
| S12_vs_k141_152205\|\|full | 1249 | No | NA | 3 | 0 | 1 | Not-determined | Genome-fragment | NA | NA | 0 |
| S9_extracted_k141_75999 | 4243 | No | NA | 7 | 1 | 0 | Low-quality | Genome-fragment | 2.58 | HMM-based (lower-bound) | 0 |
| S3_vs_k141_48131\|\|full | 4244 | No | NA | 2 | 1 | 0 | Low-quality | Genome-fragment | 6.32 | AAI-based (high-confidence) | 0 |
| S12_vs_k141_81648\|\|full | 1966 | No | NA | 3 | 0 | 1 | Not-determined | Genome-fragment | NA | NA | 0 |
| S12_vs_k141_90023\|\|full | 1931 | No | NA | 2 | 0 | 2 | Not-determined | Genome-fragment | NA | NA | 0 |
| S12_vs_k141_42828\|\|full | 5618 | No | NA | 7 | 0 | 1 | Not-determined | Genome-fragment | NA | NA | 0 |
| S12_vs_k141_66173\|\|full | 1837 | No | NA | 3 | 0 | 3 | Not-determined | Genome-fragment | NA | NA | 0 |
| S12_vs_k141_105607\|\|full | 1459 | No | NA | 3 | 0 | 0 | Not-determined | Genome-fragment | NA | NA | 0 |
| S12_vs_k141_3290\|\|full | 2108 | No | NA | 2 | 0 | 1 | Not-determined | Genome-fragment | NA | NA | 0 |
| S5_vs_k141_61131\|\|full | 4244 | No | NA | 7 | 4 | 0 | Low-quality | Genome-fragment | 8.58 | AAI-based (high-confidence) | 0 |
| S16_extracted_97337 | 4244 | No | NA | 5 | 0 | 0 | Low-quality | Genome-fragment | 11.86 | AAI-based (medium-confidence) | 0 |
| S5_vs_k141_127468\|\|full | 4246 | No | NA | 4 | 0 | 3 | Low-quality | Genome-fragment | 4.3 | AAI-based (medium-confidence) | 0 |
| S12_vs_k141_112901\|\|full | 7855 | No | NA | 7 | 0 | 4 | Not-determined | Genome-fragment | NA | NA | 0 |
| S8_vs_k141_146044\|\|full | 4246 | No | NA | 8 | 0 | 1 | Low-quality | Genome-fragment | 9.91 | AAI-based (high-confidence) | 0 |
| S12_vs_k141_136864\|\|full | 5289 | No | NA | 6 | 0 | 1 | Not-determined | Genome-fragment | NA | NA | 0 |
| S12_vs_k141_42854\|\|full | 1010 | No | NA | 3 | 0 | 0 | Not-determined | Genome-fragment | NA | NA | 0 |
| S14_vs_k141_75539\|\|full | 4248 | No | NA | 5 | 4 | 0 | Low-quality | Genome-fragment | 10.17 | AAI-based (high-confidence) | 0 |
| S7_vs_2688\|\|full | 4248 | No | NA | 6 | 2 | 0 | Low-quality | Genome-fragment | 9.44 | AAI-based (high-confidence) | 0 |
| S17_vs_k141_90647\|\|full | 4250 | No | NA | 5 | 0 | 0 | Low-quality | Genome-fragment | 9.96 | AAI-based (medium-confidence) | 0 |
| S12_vs_k141_168339\|\|full | 14598 | No | NA | 11 | 0 | 5 | Not-determined | Genome-fragment | NA | NA | 0 |
| S5_vs_k141_28941\|\|full | 4251 | No | NA | 6 | 1 | 0 | Low-quality | Genome-fragment | 7.4 | HMM-based (lower-bound) | 0 |
| S12_vs_k141_121036\|\|full | 1719 | No | NA | 3 | 0 | 0 | Not-determined | Genome-fragment | NA | NA | 0 |
| S12_vs_k141_160150\|\|full | 2080 | No | NA | 3 | 0 | 0 | Not-determined | Genome-fragment | NA | NA | 0 |
| S7_extracted_16835 | 4251 | No | NA | 7 | 0 | 3 | Low-quality | Genome-fragment | 1 | AAI-based (medium-confidence) | 0 |
| S12_vs_k141_160166\|\|full | 1597 | No | NA | 4 | 0 | 0 | Not-determined | Genome-fragment | NA | NA | 0 |
| S17_vs_k141_173348\|\|full | 4253 | No | NA | 7 | 1 | 0 | Low-quality | Genome-fragment | 7.72 | AAI-based (high-confidence) | 0 |
| S12_vs_k141_81729\|\|full | 37456 | No | NA | 62 | 11 | 1 | High-quality | High-quality | 91.57 | AAI-based (high-confidence) | 0 |
| S10_extracted_67024 | 4253 | No | NA | 3 | 0 | 3 | Low-quality | Genome-fragment | 0.54 | AAI-based (medium-confidence) | 0 |
| S12_vs_k141_168382\|\|full | 1610 | No | NA | 2 | 0 | 1 | Not-determined | Genome-fragment | NA | NA | 0 |
| S12_vs_k141_3363\|\|full | 2696 | No | NA | 3 | 0 | 0 | Not-determined | Genome-fragment | NA | NA | 0 |
| S12_vs_k141_98175\|\|full | 22925 | No | NA | 34 | 14 | 0 | Medium-quality | Genome-fragment | 61.01 | AAI-based (high-confidence) | 0 |
| S12_vs_k141_81739\|\|full | 1648 | No | NA | 3 | 0 | 0 | Not-determined | Genome-fragment | NA | NA | 0 |
| S12_vs_k141_168391\|\|full | 2443 | No | NA | 2 | 0 | 1 | Not-determined | Genome-fragment | NA | NA | 0 |
| S13_vs_53406\|\|full | 4254 | No | NA | 7 | 1 | 0 | Low-quality | Genome-fragment | 9.84 | AAI-based (medium-confidence) | 0 |
| S12_vs_k141_112991\|\|full | 2825 | No | NA | 4 | 0 | 3 | Not-determined | Genome-fragment | NA | NA | 0 |
| S12_vs_k141_81761\|\|full | 1134 | No | NA | 3 | 0 | 0 | Not-determined | Genome-fragment | NA | NA | 0 |
| S1_vs_k141_86066\|\|full | 4254 | No | NA | 5 | 2 | 0 | Low-quality | Genome-fragment | 13.04 | AAI-based (high-confidence) | 0 |
| S12_vs_k141_73528\|\|full | 1619 | No | NA | 2 | 0 | 0 | Not-determined | Genome-fragment | NA | NA | 0 |
| S14_extracted_k141_71437 | 4255 | No | NA | 5 | 3 | 0 | Low-quality | Genome-fragment | 10.66 | AAI-based (high-confidence) | 0 |
| S12_vs_k141_66254\|\|full | 9812 | No | NA | 8 | 0 | 7 | Not-determined | Genome-fragment | NA | NA | 0 |
| S12_vs_k141_90175\|\|full | 2778 | No | NA | 3 | 0 | 1 | Not-determined | Genome-fragment | NA | NA | 0 |
| S12_vs_k141_113008\|\|full | 1799 | No | NA | 2 | 0 | 0 | Not-determined | Genome-fragment | NA | NA | 0 |
| S8_extracted_k141_171793 | 4257 | No | NA | 9 | 0 | 1 | Low-quality | Genome-fragment | 2.76 | AAI-based (high-confidence) | 0 |
| S12_vs_k141_58560\|\|full | 43412 | No | NA | 47 | 26 | 0 | Medium-quality | Genome-fragment | 71.57 | AAI-based (medium-confidence) | 0 |
| S12_vs_k141_152317\|\|full | 1479 | No | NA | 3 | 0 | 0 | Not-determined | Genome-fragment | NA | NA | 0 |
| S17_vs_k141_11527\|\|full | 4258 | No | NA | 5 | 0 | 3 | Low-quality | Genome-fragment | 3.57 | AAI-based (medium-confidence) | 0 |
| S12_vs_k141_144936\|\|full | 2208 | No | NA | 4 | 0 | 1 | Not-determined | Genome-fragment | NA | NA | 0 |
| S12_vs_k141_168432\|\|full | 3636 | No | NA | 10 | 0 | 1 | Not-determined | Genome-fragment | NA | NA | 0 |
| S9_vs_k141_21171\|\|full | 4258 | No | NA | 8 | 7 | 0 | Low-quality | Genome-fragment | 9.79 | AAI-based (high-confidence) | 0 |
| S12_vs_k141_113035\|\|full | 6835 | No | NA | 6 | 0 | 5 | Not-determined | Genome-fragment | NA | NA | 0 |

| | | | | | | | | | | | |
|---|---|---|---|---|---|---|---|---|---|---|---|
| S12_vs_k141_144955\|\|full | 1556 | No | NA | 3 | 0 | 1 | Not-determined | Genome-fragment | NA | NA | 0 |
| S4_extracted_7153 | 4258 | No | NA | 3 | 0 | 2 | Low-quality | Genome-fragment | 0.37 | AAI-based (medium-confidence) | 0 |
| S12_extracted_k141_36388 | 4259 | No | NA | 3 | 0 | 3 | Low-quality | Genome-fragment | 0.66 | AAI-based (high-confidence) | 0 |
| S7_extracted_46658 | 4260 | No | NA | 9 | 1 | 0 | Low-quality | Genome-fragment | 10.84 | AAI-based (high-confidence) | 0 |
| S12_vs_k141_137004\|\|full | 2785 | No | NA | 3 | 0 | 3 | Not-determined | Genome-fragment | NA | NA | 0 |
| S12_vs_k141_168490\|\|full | 1819 | No | NA | 3 | 0 | 3 | Not-determined | Genome-fragment | NA | NA | 0 |
| S16_vs_48005\|\|full | 4261 | No | NA | 2 | 0 | 0 | Low-quality | Genome-fragment | 10.13 | AAI-based (high-confidence) | 0 |
| S12_vs_k141_137018\|\|full | 3479 | No | NA | 5 | 0 | 5 | Not-determined | Genome-fragment | NA | NA | 0 |
| S12_vs_k141_137019\|\|full | 2913 | No | NA | 3 | 0 | 1 | Not-determined | Genome-fragment | NA | NA | 0 |
| S12_vs_k141_187243\|\|full | 4261 | No | NA | 6 | 0 | 3 | Low-quality | Genome-fragment | 3.64 | AAI-based (medium-confidence) | 0 |
| S12_vs_k141_43017\|\|full | 1079 | No | NA | 2 | 0 | 1 | Not-determined | Genome-fragment | NA | NA | 0 |
| S5_vs_k141_126284\|\|full | 4263 | No | NA | 5 | 2 | 0 | Low-quality | Genome-fragment | 11.6 | AAI-based (high-confidence) | 0 |
| S12_vs_k141_105830\|\|full | 1258 | No | NA | 3 | 0 | 0 | Not-determined | Genome-fragment | NA | NA | 0 |
| S13_extracted_132056 | 4264 | No | NA | 4 | 0 | 3 | Low-quality | Genome-fragment | 5.16 | AAI-based (medium-confidence) | 0 |
| S12_vs_k141_3483\|\|full | 3238 | No | NA | 2 | 0 | 2 | Not-determined | Genome-fragment | NA | NA | 0 |
| S12_vs_k141_98292\|\|full | 6607 | No | NA | 6 | 0 | 4 | Not-determined | Genome-fragment | NA | NA | 0 |
| S12_vs_k141_81875\|\|full | 1068 | No | NA | 2 | 0 | 1 | Not-determined | Genome-fragment | NA | NA | 0 |
| S14_extracted_k141_98929 | 4264 | No | NA | 3 | 2 | 0 | Low-quality | Genome-fragment | 6.13 | HMM-based (lower-bound) | 0 |
| S12_vs_k141_3498\|\|full | 2763 | No | NA | 4 | 0 | 0 | Not-determined | Genome-fragment | NA | NA | 0 |
| S7_extracted_33780 | 4264 | No | NA | 3 | 0 | 2 | Low-quality | Genome-fragment | 2.16 | AAI-based (medium-confidence) | 0 |
| S16_extracted_148345 | 4264 | No | NA | 2 | 0 | 1 | Low-quality | Genome-fragment | 5.05 | AAI-based (medium-confidence) | 0 |
| S12_vs_k141_73618\|\|full | 3698 | No | NA | 4 | 0 | 3 | Not-determined | Genome-fragment | NA | NA | 0 |
| S3_vs_k141_84764\|\|full | 4265 | No | NA | 11 | 1 | 0 | Low-quality | Genome-fragment | 6.39 | AAI-based (high-confidence) | 0 |
| S7_vs_76301\|\|full | 4265 | No | NA | 5 | 3 | 0 | Low-quality | Genome-fragment | 11.71 | AAI-based (high-confidence) | 0 |
| S12_vs_k141_3510\|\|full | 1745 | No | NA | 3 | 0 | 1 | Not-determined | Genome-fragment | NA | NA | 0 |
| S12_vs_k141_26847\|\|full | 29815 | No | NA | 36 | 12 | 0 | Medium-quality | Genome-fragment | 50.69 | AAI-based (high-confidence) | 0 |
| S7_vs_78907\|\|full | 4265 | Yes | 1459 | 6 | 2 | 3 | Low-quality | Genome-fragment | 1.75 | HMM-based (lower-bound) | 65.79 |
| S16_extracted_116580 | 4265 | Yes | 3427 | 7 | 2 | 1 | Low-quality | Genome-fragment | 6.16 | AAI-based (high-confidence) | 19.65 |
| S12_vs_k141_128907\|\|full | 1092 | No | NA | 2 | 0 | 0 | Not-determined | Genome-fragment | NA | NA | 0 |
| S8_extracted_k141_67169 | 4265 | No | NA | 5 | 2 | 0 | Low-quality | Genome-fragment | 7.27 | AAI-based (high-confidence) | 0 |
| S11_extracted_k141_69923 | 4267 | No | NA | 10 | 1 | 1 | Low-quality | Genome-fragment | 2.31 | HMM-based (lower-bound) | 0 |
| S12_vs_k141_43099\|\|full | 1199 | No | NA | 3 | 0 | 0 | Not-determined | Genome-fragment | NA | NA | 0 |
| S12_vs_k141_113123\|\|full | 2501 | No | NA | 4 | 0 | 1 | Not-determined | Genome-fragment | NA | NA | 0 |
| S12_vs_k141_113131\|\|full | 2364 | No | NA | 3 | 0 | 1 | Not-determined | Genome-fragment | NA | NA | 0 |
| S12_vs_k141_73672\|\|full | 1407 | No | NA | 3 | 0 | 0 | Not-determined | Genome-fragment | NA | NA | 0 |
| S7_vs_13949\|\|full | 4267 | No | NA | 6 | 1 | 0 | Low-quality | Genome-fragment | 8.99 | AAI-based (high-confidence) | 0 |
| S12_vs_k141_105898\|\|full | 1964 | No | NA | 4 | 0 | 2 | Not-determined | Genome-fragment | NA | NA | 0 |
| S12_vs_k141_105901\|\|full | 1317 | No | NA | 2 | 0 | 0 | Not-determined | Genome-fragment | NA | NA | 0 |
| S13_vs_113046\|\|full | 4269 | No | NA | 8 | 1 | 0 | Low-quality | Genome-fragment | 10.13 | AAI-based (medium-confidence) | 0 |
| S1_vs_k141_121371\|\|full | 4269 | No | NA | 6 | 3 | 0 | Low-quality | Genome-fragment | 6.12 | AAI-based (medium-confidence) | 0 |
| S7_vs_52283\|\|full | 4270 | No | NA | 4 | 1 | 0 | Low-quality | Genome-fragment | 11.41 | AAI-based (medium-confidence) | 0 |
| S11_vs_k141_37922\|\|full | 4271 | No | NA | 12 | 2 | 0 | Low-quality | Genome-fragment | 9.47 | AAI-based (high-confidence) | 0 |
| S12_vs_k141_90351\|\|full | 2383 | No | NA | 4 | 0 | 0 | Not-determined | Genome-fragment | NA | NA | 0 |
| S12_vs_k141_66441\|\|full | 2661 | No | NA | 5 | 0 | 2 | Not-determined | Genome-fragment | NA | NA | 0 |
| S12_vs_k141_105933\|\|full | 1430 | No | NA | 3 | 0 | 1 | Not-determined | Genome-fragment | NA | NA | 0 |
| S12_extracted_k141_39181 | 4272 | No | NA | 7 | 3 | 0 | Low-quality | Genome-fragment | 5.14 | HMM-based (lower-bound) | 0 |
| S16_extracted_21406 | 4273 | No | NA | 7 | 0 | 0 | Low-quality | Genome-fragment | 2.44 | AAI-based (high-confidence) | 0 |
| S12_vs_k141_50061\|\|full | 1488 | No | NA | 2 | 0 | 0 | Not-determined | Genome-fragment | NA | NA | 0 |
| S12_vs_k141_82010\|\|full | 1178 | No | NA | 2 | 0 | 1 | Not-determined | Genome-fragment | NA | NA | 0 |
| S12_vs_k141_168628\|\|full | 1266 | No | NA | 2 | 0 | 0 | Not-determined | Genome-fragment | NA | NA | 0 |
| S12_extracted_k141_167473 | 4273 | No | NA | 6 | 0 | 1 | Low-quality | Genome-fragment | 12.17 | AAI-based (medium-confidence) | 0 |
| S13_extracted_84980 | 4274 | No | NA | 5 | 0 | 1 | Low-quality | Genome-fragment | 6.37 | AAI-based (medium-confidence) | 0 |
| S12_vs_k141_10498\|\|full | 5114 | No | NA | 5 | 0 | 2 | Not-determined | Genome-fragment | NA | NA | 0 |
| S12_vs_k141_137123\|\|full | 1132 | No | NA | 3 | 0 | 2 | Not-determined | Genome-fragment | NA | NA | 0 |
| S12_vs_k141_50094\|\|full | 3467 | No | NA | 5 | 0 | 3 | Not-determined | Genome-fragment | NA | NA | 0 |
| S16_extracted_166348 | 4274 | No | NA | 4 | 0 | 2 | Low-quality | Genome-fragment | 7.3 | AAI-based (high-confidence) | 0 |
| S16_extracted_117690 | 4275 | No | NA | 3 | 0 | 3 | Low-quality | Genome-fragment | 3.85 | AAI-based (medium-confidence) | 0 |
| S12_vs_k141_26914\|\|full | 1398 | No | NA | 3 | 0 | 0 | Not-determined | Genome-fragment | NA | NA | 0 |
| S12_vs_k141_26921\|\|full | 1448 | No | NA | 4 | 0 | 0 | Not-determined | Genome-fragment | NA | NA | 0 |
| S1_extracted_k141_96567 | 4276 | No | NA | 3 | 0 | 3 | Low-quality | Genome-fragment | 1.63 | AAI-based (medium-confidence) | 0 |
| S12_vs_k141_34383\|\|full | 3067 | No | NA | 8 | 0 | 1 | Not-determined | Genome-fragment | NA | NA | 0 |
| S12_vs_k141_34385\|\|full | 3344 | No | NA | 3 | 0 | 1 | Not-determined | Genome-fragment | NA | NA | 0 |
| S3_extracted_k141_107801 | 4276 | No | NA | 5 | 1 | 0 | Low-quality | Genome-fragment | 2.52 | AAI-based (high-confidence) | 0 |
| S17_extracted_k141_24713 | 4277 | No | NA | 5 | 0 | 4 | Low-quality | Genome-fragment | 2.24 | AAI-based (medium-confidence) | 0 |
| S14_extracted_k141_78558 | 4277 | No | NA | 3 | 0 | 2 | Low-quality | Genome-fragment | 1.07 | AAI-based (high-confidence) | 0 |
| S12_vs_k141_90470\|\|full | 3961 | No | NA | 3 | 0 | 2 | Not-determined | Genome-fragment | NA | NA | 0 |
| S10_vs_635\|\|full | 4278 | No | NA | 4 | 1 | 0 | Low-quality | Genome-fragment | 3.96 | AAI-based (medium-confidence) | 0 |
| S12_vs_k141_152579\|\|full | 1534 | No | NA | 2 | 0 | 0 | Not-determined | Genome-fragment | NA | NA | 0 |
| S4_extracted_859 | 4278 | No | NA | 3 | 0 | 0 | Low-quality | Genome-fragment | 4.44 | AAI-based (high-confidence) | 0 |
| S12_vs_k141_26965\|\|full | 2566 | No | NA | 4 | 0 | 1 | Not-determined | Genome-fragment | NA | NA | 0 |
| S13_vs_18931\|\|full | 4279 | No | NA | 8 | 0 | 0 | Low-quality | Genome-fragment | 4.77 | AAI-based (medium-confidence) | 0 |
| S3_extracted_k141_120402 | 4279 | No | NA | 3 | 0 | 2 | Low-quality | Genome-fragment | 3.19 | AAI-based (medium-confidence) | 0 |
| S3_vs_k141_57\|\|full | 4279 | No | NA | 4 | 0 | 2 | Low-quality | Genome-fragment | 2.88 | AAI-based (medium-confidence) | 0 |
| S12_extracted_k141_98334 | 4279 | Yes | 1682 | 5 | 1 | 2 | Low-quality | Genome-fragment | 1.53 | HMM-based (lower-bound) | 60.69 |
| S14_vs_k141_852\|\|full | 4280 | No | NA | 8 | 4 | 0 | Low-quality | Genome-fragment | 5.28 | HMM-based (lower-bound) | 0 |
| S12_vs_k141_50241\|\|full | 3335 | No | NA | 6 | 0 | 5 | Not-determined | Genome-fragment | NA | NA | 0 |
| S12_vs_k141_160583\|\|full | 1132 | No | NA | 2 | 0 | 1 | Not-determined | Genome-fragment | NA | NA | 0 |
| S5_vs_k141_82787\|\|full | 4280 | No | NA | 4 | 0 | 3 | Low-quality | Genome-fragment | 2.62 | AAI-based (medium-confidence) | 0 |
| S16_vs_94319\|\|full | 4280 | No | NA | 3 | 1 | 0 | Low-quality | Genome-fragment | 4.33 | AAI-based (medium-confidence) | 0 |
| S3_vs_k141_81810\|\|full | 4281 | No | NA | 9 | 0 | 0 | Low-quality | Genome-fragment | 9.52 | AAI-based (high-confidence) | 0 |
| S7_extracted_98579 | 4281 | No | NA | 11 | 2 | 0 | Low-quality | Genome-fragment | 9.89 | AAI-based (high-confidence) | 0 |
| S12_vs_k141_152623\|\|full | 4079 | No | NA | 5 | 0 | 2 | Not-determined | Genome-fragment | NA | NA | 0 |
| S12_vs_k141_145118\|\|full | 2741 | No | NA | 3 | 0 | 0 | Not-determined | Genome-fragment | NA | NA | 0 |
| S12_vs_k141_145131\|\|full | 3865 | No | NA | 3 | 0 | 3 | Not-determined | Genome-fragment | NA | NA | 0 |
| S16_vs_106061\|\|full | 4282 | No | NA | 8 | 0 | 2 | Low-quality | Genome-fragment | 5.82 | AAI-based (medium-confidence) | 0 |
| S5_extracted_k141_126567 | 4283 | No | NA | 9 | 1 | 0 | Low-quality | Genome-fragment | 2.32 | HMM-based (lower-bound) | 0 |
| S17_extracted_k141_159784 | 4285 | No | NA | 5 | 0 | 2 | Low-quality | Genome-fragment | 2.73 | AAI-based (high-confidence) | 0 |
| S12_vs_k141_98623\|\|full | 1423 | No | NA | 3 | 0 | 1 | Not-determined | Genome-fragment | NA | NA | 0 |
| S13_extracted_50848 | 4285 | No | NA | 3 | 0 | 0 | Low-quality | Genome-fragment | 4.43 | AAI-based (high-confidence) | 0 |
| S12_vs_k141_129036\|\|full | 2161 | No | NA | 4 | 0 | 3 | Not-determined | Genome-fragment | NA | NA | 0 |
| S5_extracted_k141_81813 | 4285 | Yes | 2901 | 5 | 1 | 1 | Low-quality | Genome-fragment | 3.59 | HMM-based (lower-bound) | 32.3 |
| S9_vs_k141_7821\|\|full | 4286 | No | NA | 5 | 0 | 0 | Low-quality | Genome-fragment | 9.47 | AAI-based (medium-confidence) | 0 |
| S7_extracted_68352 | 4286 | No | NA | 3 | 0 | 1 | Low-quality | Genome-fragment | 9.23 | AAI-based (high-confidence) | 0 |
| S12_vs_k141_152663\|\|full | 1461 | No | NA | 3 | 0 | 0 | Not-determined | Genome-fragment | NA | NA | 0 |
| S11_vs_k141_87914\|\|full | 4287 | No | NA | 6 | 1 | 0 | Low-quality | Genome-fragment | 5.31 | HMM-based (lower-bound) | 0 |
| S16_vs_37549\|\|full | 4287 | No | NA | 3 | 0 | 2 | Low-quality | Genome-fragment | 1.34 | AAI-based (medium-confidence) | 0 |
| S17_vs_k141_150099\|\|full | 4288 | No | NA | 4 | 0 | 1 | Low-quality | Genome-fragment | 1.73 | AAI-based (medium-confidence) | 0 |
| S13_extracted_64503 | 4288 | No | NA | 10 | 3 | 0 | Low-quality | Genome-fragment | 6.29 | AAI-based (high-confidence) | 0 |
| S12_vs_k141_27075\|\|full | 1101 | No | NA | 2 | 0 | 1 | Not-determined | Genome-fragment | NA | NA | 0 |
| S12_vs_k141_129093\|\|full | 1766 | No | NA | 3 | 0 | 0 | Not-determined | Genome-fragment | NA | NA | 0 |
| S12_vs_k141_184774\|\|full | 1779 | No | NA | 2 | 0 | 2 | Not-determined | Genome-fragment | NA | NA | 0 |
| S12_vs_k141_27078\|\|full | 2519 | No | NA | 4 | 0 | 4 | Not-determined | Genome-fragment | NA | NA | 0 |
| S12_vs_k141_176283\|\|full | 2669 | No | NA | 2 | 0 | 1 | Not-determined | Genome-fragment | NA | NA | 0 |
| S9_extracted_k141_37154 | 4292 | No | NA | 3 | 0 | 3 | Low-quality | Genome-fragment | 14.26 | AAI-based (high-confidence) | 0 |
| S11_vs_k141_71849\|\|full | 4292 | No | NA | 11 | 1 | 0 | Low-quality | Genome-fragment | 2.79 | AAI-based (high-confidence) | 0 |
| S12_vs_k141_50362\|\|full | 10271 | No | NA | 9 | 0 | 5 | Not-determined | Genome-fragment | NA | NA | 0 |
| S12_vs_k141_129122\|\|full | 15232 | No | NA | 10 | 0 | 3 | Not-determined | Genome-fragment | NA | NA | 0 |
| S3_extracted_k141_56735 | 4292 | No | NA | 8 | 3 | 0 | Low-quality | Genome-fragment | 12.05 | AAI-based (high-confidence) | 0 |
| S12_vs_k141_27123\|\|full | 1900 | No | NA | 3 | 0 | 2 | Not-determined | Genome-fragment | NA | NA | 0 |
| S5_extracted_k141_35598 | 4292 | No | NA | 10 | 2 | 0 | Low-quality | Genome-fragment | 9.17 | AAI-based (high-confidence) | 0 |
| S12_vs_k141_145269\|\|full | 1506 | Yes | 516 | 2 | 1 | 1 | Not-determined | Genome-fragment | NA | NA | 65.74 |
| S12_vs_k141_27137\|\|full | 1504 | No | NA | 2 | 0 | 0 | Not-determined | Genome-fragment | NA | NA | 0 |
| S12_vs_k141_113402\|\|full | 1090 | No | NA | 3 | 0 | 2 | Not-determined | Genome-fragment | NA | NA | 0 |
| S12_extracted_k141_100277 | 4293 | No | NA | 4 | 0 | 4 | Low-quality | Genome-fragment | 1.79 | AAI-based (medium-confidence) | 0 |
| S12_vs_k141_106215\|\|full | 1297 | No | NA | 2 | 0 | 0 | Not-determined | Genome-fragment | NA | NA | 0 |
| S12_vs_k141_176370\|\|full | 1543 | No | NA | 3 | 0 | 1 | Not-determined | Genome-fragment | NA | NA | 0 |
| S4_extracted_8310 | 4293 | No | NA | 9 | 8 | 0 | Low-quality | Genome-fragment | 14.16 | AAI-based (high-confidence) | 0 |
| S17_extracted_k141_33416 | 4294 | No | NA | 12 | 0 | 0 | Low-quality | Genome-fragment | 5.14 | AAI-based (high-confidence) | 0 |
| S15_extracted_41697 | 4294 | No | NA | 8 | 3 | 0 | Low-quality | Genome-fragment | 9.55 | AAI-based (high-confidence) | 0 |
| S12_vs_k141_82242\|\|full | 1363 | No | NA | 2 | 0 | 1 | Not-determined | Genome-fragment | NA | NA | 0 |
| S12_vs_k141_176390\|\|full | 3636 | No | NA | 3 | 0 | 1 | Not-determined | Genome-fragment | NA | NA | 0 |
| S17_vs_k141_49253\|\|full | 4295 | Yes | 1628 | 5 | 1 | 1 | Low-quality | Genome-fragment | 3.34 | HMM-based (lower-bound) | 62.1 |
| S10_vs_25371\|\|full | 4295 | No | NA | 4 | 0 | 1 | Low-quality | Genome-fragment | 5.8 | AAI-based (high-confidence) | 0 |
| S12_vs_k141_145279\|\|full | 1070 | No | NA | 2 | 0 | 0 | Not-determined | Genome-fragment | NA | NA | 0 |
| S12_vs_k141_66565\|\|full | 2197 | No | NA | 2 | 0 | 2 | Not-determined | Genome-fragment | NA | NA | 0 |
| S8_extracted_k141_42131 | 4295 | No | NA | 6 | 0 | 0 | Low-quality | Genome-fragment | 13.66 | AAI-based (high-confidence) | 0 |
| S12_vs_k141_106281\|\|full | 1634 | No | NA | 3 | 0 | 1 | Not-determined | Genome-fragment | NA | NA | 0 |
| S12_vs_k141_50448\|\|full | 1564 | No | NA | 2 | 0 | 0 | Not-determined | Genome-fragment | NA | NA | 0 |
| S12_vs_k141_184850\|\|full | 1278 | No | NA | 2 | 0 | 0 | Not-determined | Genome-fragment | NA | NA | 0 |
| S5_vs_k141_20959\|\|full | 4296 | No | NA | 8 | 0 | 0 | Low-quality | Genome-fragment | 10.23 | AAI-based (medium-confidence) | 0 |
| S6_vs_k141_43033\|\|full | 4296 | No | NA | 4 | 0 | 3 | Low-quality | Genome-fragment | 2.05 | AAI-based (high-confidence) | 0 |
| S12_vs_k141_184860\|\|full | 1240 | No | NA | 2 | 0 | 0 | Not-determined | Genome-fragment | NA | NA | 0 |
| S5_vs_k141_54061\|\|full | 4297 | No | NA | 3 | 0 | 1 | Low-quality | Genome-fragment | 2.19 | AAI-based (medium-confidence) | 0 |
| S12_vs_k141_4007\|\|full | 3890 | No | NA | 3 | 0 | 1 | Not-determined | Genome-fragment | NA | NA | 0 |
| S12_vs_k141_98841\|\|full | 1217 | No | NA | 2 | 0 | 1 | Not-determined | Genome-fragment | NA | NA | 0 |
| S12_vs_k141_82267\|\|full | 6816 | No | NA | 7 | 0 | 6 | Not-determined | Genome-fragment | NA | NA | 0 |
| S12_vs_k141_145328\|\|full | 2321 | No | NA | 2 | 0 | 1 | Not-determined | Genome-fragment | NA | NA | 0 |
| S11_extracted_k141_1851 | 4298 | No | NA | 5 | 2 | 0 | Low-quality | Genome-fragment | 1.66 | AAI-based (medium-confidence) | 0 |
| S3_extracted_k141_84621 | 4298 | No | NA | 5 | 0 | 0 | Low-quality | Genome-fragment | 2.81 | AAI-based (high-confidence) | 0 |
| S12_vs_k141_4023\|\|full | 2016 | No | NA | 4 | 0 | 0 | Not-determined | Genome-fragment | NA | NA | 0 |
| S12_vs_k141_90618\|\|full | 1849 | No | NA | 4 | 0 | 0 | Not-determined | Genome-fragment | NA | NA | 0 |
| S5_vs_k141_35653\|\|full | 4299 | No | NA | 5 | 0 | 2 | Low-quality | Genome-fragment | 5.07 | AAI-based (medium-confidence) | 0 |
| S12_vs_k141_168807\|\|full | 2255 | No | NA | 3 | 0 | 1 | Not-determined | Genome-fragment | NA | NA | 0 |
| S17_vs_k141_129567\|\|full | 4301 | No | NA | 4 | 0 | 3 | Low-quality | Genome-fragment | 1.34 | AAI-based (medium-confidence) | 0 |
| S12_vs_k141_43549\|\|full | 1940 | No | NA | 3 | 0 | 0 | Not-determined | Genome-fragment | NA | NA | 0 |
| S12_vs_k141_152861\|\|full | 2111 | No | NA | 2 | 0 | 0 | Not-determined | Genome-fragment | NA | NA | 0 |
| S10_extracted_13065 | 4301 | No | NA | 7 | 3 | 0 | Low-quality | Genome-fragment | 3.01 | HMM-based (lower-bound) | 0 |
| S12_vs_k141_66620\|\|full | 6996 | No | NA | 10 | 0 | 0 | Not-determined | Genome-fragment | NA | NA | 0 |
| S12_vs_k141_184904\|\|full | 2870 | No | NA | 5 | 0 | 1 | Not-determined | Genome-fragment | NA | NA | 0 |
| S12_vs_k141_50523\|\|full | 5583 | No | NA | 4 | 0 | 4 | Not-determined | Genome-fragment | NA | NA | 0 |
| S9_extracted_k141_3819 | 4302 | No | NA | 5 | 3 | 0 | Low-quality | Genome-fragment | 10.01 | AAI-based (high-confidence) | 0 |
| S12_vs_k141_98905\|\|full | 3123 | No | NA | 4 | 0 | 1 | Not-determined | Genome-fragment | NA | NA | 0 |
| S12_vs_k141_27315\|\|full | 1155 | No | NA | 3 | 0 | 0 | Not-determined | Genome-fragment | NA | NA | 0 |
| S12_vs_k141_58997\|\|full | 2795 | No | NA | 7 | 0 | 0 | Not-determined | Genome-fragment | NA | NA | 0 |
| S12_vs_k141_106329\|\|full | 26070 | No | NA | 40 | 13 | 1 | Medium-quality | Genome-fragment | 76.82 | AAI-based (medium-confidence) | 0 |
| S5_extracted_k141_30928 | 4302 | No | NA | 1 | 0 | 0 | Low-quality | Genome-fragment | 3.96 | AAI-based (high-confidence) | 0 |
| S8_extracted_k141_128872 | 4302 | No | NA | 5 | 1 | 0 | Low-quality | Genome-fragment | 2.64 | AAI-based (medium-confidence) | 0 |
| S12_vs_k141_82348\|\|full | 1170 | No | NA | 3 | 0 | 0 | Not-determined | Genome-fragment | NA | NA | 0 |
| S17_vs_k141_11951\|\|full | 4303 | No | NA | 12 | 2 | 0 | Low-quality | Genome-fragment | 10 | AAI-based (high-confidence) | 0 |
| S12_vs_k141_82360\|\|full | 1049 | No | NA | 2 | 0 | 0 | Not-determined | Genome-fragment | NA | NA | 0 |
| S3_extracted_k141_89862 | 4303 | No | NA | 4 | 1 | 0 | Low-quality | Genome-fragment | 2.78 | HMM-based (lower-bound) | 0 |
| S13_vs_49536\|\|full | 4305 | No | NA | 4 | 0 | 4 | Low-quality | Genome-fragment | 9.95 | AAI-based (medium-confidence) | 0 |
| S12_vs_k141_27353\|\|full | 3401 | No | NA | 2 | 0 | 2 | Not-determined | Genome-fragment | NA | NA | 0 |
| S12_vs_k141_129369\|\|full | 1148 | No | NA | 2 | 0 | 1 | Not-determined | Genome-fragment | NA | NA | 0 |
| S12_vs_k141_176426\|\|full | 2128 | No | NA | 3 | 0 | 2 | Not-determined | Genome-fragment | NA | NA | 0 |
| S7_vs_94722\|\|full | 4305 | No | NA | 4 | 0 | 2 | Low-quality | Genome-fragment | 8.15 | AAI-based (medium-confidence) | 0 |
| S12_vs_k141_98946\|\|full | 2270 | No | NA | 3 | 0 | 2 | Not-determined | Genome-fragment | NA | NA | 0 |
| S12_vs_k141_176428\|\|full | 5763 | No | NA | 2 | 0 | 2 | Not-determined | Genome-fragment | NA | NA | 0 |
| S3_vs_k141_6739\|\|full | 4306 | No | NA | 5 | 2 | 0 | Low-quality | Genome-fragment | 9.16 | AAI-based (high-confidence) | 0 |
| S12_vs_k141_129392\|\|full | 1407 | No | NA | 2 | 0 | 0 | Not-determined | Genome-fragment | NA | NA | 0 |
| S12_vs_k141_129407\|\|full | 1143 | No | NA | 3 | 0 | 0 | Not-determined | Genome-fragment | NA | NA | 0 |
| S12_vs_k141_66734\|\|full | 3543 | No | NA | 4 | 0 | 2 | Not-determined | Genome-fragment | NA | NA | 0 |
| S12_vs_k141_89575\|\|full | 4306 | No | NA | 5 | 0 | 3 | Low-quality | Genome-fragment | 2.25 | AAI-based (medium-confidence) | 0 |
| S12_vs_k141_121486\|\|full | 6167 | No | NA | 9 | 0 | 2 | Not-determined | Genome-fragment | NA | NA | 0 |
| S12_vs_k141_82412\|\|full | 1025 | No | NA | 3 | 0 | 0 | Not-determined | Genome-fragment | NA | NA | 0 |
| S9_vs_k141_8199\|\|full | 4307 | No | NA | 10 | 3 | 0 | Low-quality | Genome-fragment | 9.55 | AAI-based (high-confidence) | 0 |
| S14_vs_k141_113987\|\|full | 4308 | No | NA | 3 | 0 | 3 | Low-quality | Genome-fragment | 2.41 | AAI-based (medium-confidence) | 0 |
| S5_extracted_k141_69851 | 4308 | No | NA | 5 | 0 | 1 | Low-quality | Genome-fragment | 8.54 | AAI-based (medium-confidence) | 0 |
| S10_extracted_2794 | 4308 | No | NA | 3 | 0 | 1 | Low-quality | Genome-fragment | 2.21 | AAI-based (medium-confidence) | 0 |
| S12_vs_k141_50639\|\|full | 6335 | No | NA | 4 | 0 | 3 | Not-determined | Genome-fragment | NA | NA | 0 |
| S16_extracted_77703 | 4309 | No | NA | 4 | 3 | 0 | Low-quality | Genome-fragment | 11.28 | AAI-based (high-confidence) | 0 |
| S7_vs_5257\|\|full | 4310 | No | NA | 2 | 0 | 1 | Low-quality | Genome-fragment | 3.84 | AAI-based (medium-confidence) | 0 |
| S10_vs_25172\|\|full | 4310 | No | NA | 3 | 0 | 2 | Low-quality | Genome-fragment | 2.23 | AAI-based (medium-confidence) | 0 |
| S17_extracted_k141_174844 | 4311 | No | NA | 5 | 1 | 3 | Low-quality | Genome-fragment | 1.27 | HMM-based (lower-bound) | 0 |
| S12_vs_k141_185040\|\|full | 3199 | No | NA | 4 | 0 | 2 | Not-determined | Genome-fragment | NA | NA | 0 |
| S12_vs_k141_90746\|\|full | 3846 | No | NA | 5 | 0 | 1 | Not-determined | Genome-fragment | NA | NA | 0 |
| S12_vs_k141_32938\|\|full | 4312 | Yes | 3643 | 6 | 3 | 1 | Low-quality | Genome-fragment | 2.05 | AAI-based (medium-confidence) | 15.51 |
| S6_vs_k141_50192\|\|full | 4312 | No | NA | 4 | 0 | 4 | Low-quality | Genome-fragment | 9.04 | AAI-based (medium-confidence) | 0 |
| S3_vs_k141_15695\|\|full | 4313 | No | NA | 6 | 1 | 0 | Low-quality | Genome-fragment | 9.72 | AAI-based (high-confidence) | 0 |
| S12_vs_k141_82464\|\|full | 1809 | No | NA | 2 | 0 | 1 | Not-determined | Genome-fragment | NA | NA | 0 |
| S5_vs_k141_21318\|\|full | 4314 | Yes | 3844 | 6 | 1 | 1 | Low-quality | Genome-fragment | 4.36 | HMM-based (lower-bound) | 10.89 |
| S12_vs_k141_106429\|\|full | 1039 | No | NA | 2 | 0 | 1 | Not-determined | Genome-fragment | NA | NA | 0 |
| S12_vs_k141_99023\|\|full | 2102 | No | NA | 3 | 0 | 1 | Not-determined | Genome-fragment | NA | NA | 0 |
| S12_vs_k141_90793\|\|full | 2297 | No | NA | 3 | 0 | 0 | Not-determined | Genome-fragment | NA | NA | 0 |
| S12_vs_k141_74280\|\|full | 14091 | No | NA | 15 | 3 | 0 | Medium-quality | Genome-fragment | 53.58 | AAI-based (high-confidence) | 0 |
| S12_vs_k141_4094\|\|full | 1007 | No | NA | 3 | 0 | 1 | Not-determined | Genome-fragment | NA | NA | 0 |
| S12_vs_k141_137711\|\|full | 3473 | No | NA | 3 | 0 | 1 | Not-determined | Genome-fragment | NA | NA | 0 |
| S7_vs_112968\|\|full | 4314 | No | NA | 10 | 2 | 0 | Low-quality | Genome-fragment | 8.94 | AAI-based (medium-confidence) | 0 |
| S12_vs_k141_74291\|\|full | 4113 | No | NA | 3 | 0 | 2 | Not-determined | Genome-fragment | NA | NA | 0 |
| S10_vs_67758\|\|full | 4314 | No | NA | 3 | 0 | 2 | Low-quality | Genome-fragment | 1.35 | AAI-based (medium-confidence) | 0 |
| S4_extracted_25728 | 4314 | No | NA | 3 | 3 | 0 | Low-quality | Genome-fragment | 14.21 | AAI-based (high-confidence) | 0 |
| S8_vs_k141_143946\|\|full | 4314 | No | NA | 13 | 0 | 0 | Low-quality | Genome-fragment | 11.03 | AAI-based (medium-confidence) | 0 |
| S12_vs_k141_27519\|\|full | 3305 | No | NA | 3 | 0 | 2 | Not-determined | Genome-fragment | NA | NA | 0 |
| S13_vs_93818\|\|full | 4315 | No | NA | 5 | 0 | 2 | Low-quality | Genome-fragment | 12.01 | AAI-based (medium-confidence) | 0 |
| S6_extracted_k141_36239 | 4315 | No | NA | 9 | 9 | 0 | Low-quality | Genome-fragment | 14.19 | AAI-based (high-confidence) | 0 |
| S12_vs_k141_121614\|\|full | 1527 | No | NA | 3 | 0 | 0 | Not-determined | Genome-fragment | NA | NA | 0 |
| S11_vs_k141_19386\|\|full | 4316 | No | NA | 7 | 1 | 0 | Low-quality | Genome-fragment | 10.15 | AAI-based (high-confidence) | 0 |
| S12_vs_k141_82471\|\|full | 1756 | No | NA | 2 | 0 | 1 | Not-determined | Genome-fragment | NA | NA | 0 |
| S12_vs_k141_160894\|\|full | 1403 | No | NA | 2 | 0 | 0 | Not-determined | Genome-fragment | NA | NA | 0 |
| S12_vs_k141_113662\|\|full | 2459 | No | NA | 5 | 0 | 2 | Not-determined | Genome-fragment | NA | NA | 0 |
| S12_vs_k141_99077\|\|full | 2678 | No | NA | 3 | 0 | 2 | Not-determined | Genome-fragment | NA | NA | 0 |
| S12_vs_k141_106518\|\|full | 1966 | No | NA | 3 | 0 | 2 | Not-determined | Genome-fragment | NA | NA | 0 |
| S7_vs_40189\|\|full | 4316 | No | NA | 5 | 5 | 0 | Low-quality | Genome-fragment | 12.5 | AAI-based (high-confidence) | 0 |
| S12_vs_k141_106521\|\|full | 4943 | No | NA | 3 | 0 | 3 | Not-determined | Genome-fragment | NA | NA | 0 |
| S12_vs_k141_82500\|\|full | 1383 | No | NA | 3 | 0 | 1 | Not-determined | Genome-fragment | NA | NA | 0 |
| S16_vs_145382\|\|full | 4316 | No | NA | 6 | 0 | 0 | Low-quality | Genome-fragment | 10.91 | AAI-based (medium-confidence) | 0 |
| S12_vs_k141_11026\|\|full | 5196 | No | NA | 6 | 0 | 1 | Not-determined | Genome-fragment | NA | NA | 0 |
| S12_vs_k141_11033\|\|full | 5897 | No | NA | 5 | 0 | 4 | Not-determined | Genome-fragment | NA | NA | 0 |
| S12_vs_k141_19549\|\|full | 2060 | No | NA | 3 | 0 | 2 | Not-determined | Genome-fragment | NA | NA | 0 |
| S5_extracted_k141_127408 | 4318 | No | NA | 8 | 0 | 1 | Low-quality | Genome-fragment | 5.53 | AAI-based (medium-confidence) | 0 |
| S12_vs_k141_99092\|\|full | 35261 | No | NA | 49 | 20 | 1 | High-quality | High-quality | 100 | AAI-based (high-confidence) | 0 |
| S2_extracted_k141_5360 | 4319 | No | NA | 11 | 2 | 0 | Low-quality | Genome-fragment | 11.1 | AAI-based (high-confidence) | 0 |
| S12_vs_k141_82534\|\|full | 2108 | No | NA | 3 | 0 | 2 | Not-determined | Genome-fragment | NA | NA | 0 |
| S12_vs_k141_59238\|\|full | 19380 | No | NA | 20 | 0 | 12 | Not-determined | Genome-fragment | NA | NA | 0 |
| S12_vs_k141_27606\|\|full | 6639 | No | NA | 10 | 0 | 4 | Not-determined | Genome-fragment | NA | NA | 0 |
| S16_vs_123087\|\|full | 4320 | No | NA | 5 | 0 | 3 | Low-quality | Genome-fragment | 1.22 | AAI-based (medium-confidence) | 0 |
| S3_extracted_k141_9339 | 4321 | No | NA | 3 | 3 | 0 | Low-quality | Genome-fragment | 11.78 | AAI-based (high-confidence) | 0 |
| S12_vs_k141_145672\|\|full | 5403 | No | NA | 2 | 0 | 1 | Not-determined | Genome-fragment | NA | NA | 0 |
| S12_vs_k141_145681\|\|full | 1358 | No | NA | 2 | 0 | 0 | Not-determined | Genome-fragment | NA | NA | 0 |
| S12_vs_k141_160991\|\|full | 15714 | No | NA | 9 | 0 | 3 | Not-determined | Genome-fragment | NA | NA | 0 |
| S4_extracted_18174 | 4321 | No | NA | 3 | 3 | 0 | Low-quality | Genome-fragment | 14.17 | AAI-based (high-confidence) | 0 |
| S12_vs_k141_27654\|\|full | 2513 | No | NA | 3 | 0 | 2 | Not-determined | Genome-fragment | NA | NA | 0 |
| S12_vs_k141_74370\|\|full | 1275 | No | NA | 2 | 0 | 1 | Not-determined | Genome-fragment | NA | NA | 0 |
| S12_extracted_k141_36374 | 4322 | No | NA | 4 | 1 | 0 | Low-quality | Genome-fragment | 7.78 | AAI-based (medium-confidence) | 0 |
| S1_vs_k141_93486\|\|full | 4324 | No | NA | 8 | 0 | 0 | Low-quality | Genome-fragment | 7.84 | AAI-based (high-confidence) | 0 |
| S12_vs_k141_71627\|\|full | 4325 | No | NA | 6 | 1 | 0 | Low-quality | Genome-fragment | 8.56 | AAI-based (medium-confidence) | 0 |
| S12_vs_k141_43884\|\|full | 8411 | No | NA | 12 | 0 | 11 | Not-determined | Genome-fragment | NA | NA | 0 |
| S12_vs_k141_59298\|\|full | 1371 | No | NA | 2 | 0 | 0 | Not-determined | Genome-fragment | NA | NA | 0 |
| S10_extracted_66591 | 4326 | No | NA | 9 | 0 | 0 | Low-quality | Genome-fragment | 3.94 | AAI-based (high-confidence) | 0 |
| S3_vs_k141_3726\|\|full | 4327 | No | NA | 9 | 0 | 0 | Low-quality | Genome-fragment | 12.27 | AAI-based (high-confidence) | 0 |
| S14_vs_k141_71398\|\|full | 4328 | No | NA | 4 | 0 | 0 | Low-quality | Genome-fragment | 3.57 | AAI-based (medium-confidence) | 0 |
| S5_extracted_k141_69579 | 4328 | No | NA | 3 | 2 | 0 | Low-quality | Genome-fragment | 15.75 | HMM-based (lower-bound) | 0 |
| S12_vs_k141_27676\|\|full | 3047 | No | NA | 3 | 0 | 0 | Not-determined | Genome-fragment | NA | NA | 0 |
| S12_vs_k141_27683\|\|full | 1496 | No | NA | 3 | 0 | 0 | Not-determined | Genome-fragment | NA | NA | 0 |
| S16_vs_114372\|\|full | 4328 | Yes | 2861 | 7 | 1 | 1 | Low-quality | Genome-fragment | 1.48 | HMM-based (lower-bound) | 33.9 |
| S8_vs_k141_150122\|\|full | 4328 | No | NA | 4 | 2 | 0 | Low-quality | Genome-fragment | 7.17 | AAI-based (high-confidence) | 0 |
| S12_vs_k141_34952\|\|full | 21836 | No | NA | 14 | 0 | 1 | Not-determined | Genome-fragment | NA | NA | 0 |
| S12_vs_k141_82633\|\|full | 1171 | No | NA | 4 | 0 | 0 | Not-determined | Genome-fragment | NA | NA | 0 |
| S8_extracted_k141_62937 | 4329 | No | NA | 1 | 0 | 1 | Low-quality | Genome-fragment | 2.01 | AAI-based (medium-confidence) | 0 |
| S9_vs_k141_32695\|\|full | 4330 | No | NA | 4 | 2 | 0 | Low-quality | Genome-fragment | 8.74 | AAI-based (medium-confidence) | 0 |
| S12_vs_k141_91000\|\|full | 5808 | No | NA | 4 | 0 | 2 | Not-determined | Genome-fragment | NA | NA | 0 |
| S12_vs_k141_27728\|\|full | 1816 | No | NA | 2 | 0 | 2 | Not-determined | Genome-fragment | NA | NA | 0 |
| S16_extracted_114271 | 4331 | No | NA | 7 | 2 | 0 | Low-quality | Genome-fragment | 10.19 | AAI-based (medium-confidence) | 0 |
| S12_vs_k141_82666\|\|full | 1255 | No | NA | 3 | 0 | 2 | Not-determined | Genome-fragment | NA | NA | 0 |
| S8_vs_k141_183209\|\|lt2gene | 4331 | No | NA | 2 | 0 | 0 | Low-quality | Genome-fragment | 4.38 | AAI-based (high-confidence) | 0 |
| S16_extracted_13499 | 4332 | No | NA | 5 | 1 | 0 | Low-quality | Genome-fragment | 5.42 | AAI-based (high-confidence) | 0 |
| S10_vs_42387\|\|full | 4332 | No | NA | 7 | 0 | 0 | Low-quality | Genome-fragment | 10.04 | AAI-based (medium-confidence) | 0 |
| S8_vs_k141_9463\|\|full | 4332 | No | NA | 3 | 0 | 1 | Low-quality | Genome-fragment | 2.18 | AAI-based (medium-confidence) | 0 |
| S12_vs_k141_185361\|\|full | 2613 | No | NA | 4 | 0 | 1 | Not-determined | Genome-fragment | NA | NA | 0 |
| S12_vs_k141_82719\|\|full | 4538 | No | NA | 4 | 0 | 0 | Not-determined | Genome-fragment | NA | NA | 0 |
| S7_vs_37152\|\|full | 4333 | No | NA | 3 | 2 | 0 | Low-quality | Genome-fragment | 2.98 | AAI-based (medium-confidence) | 0 |
| S12_vs_k141_99219\|\|full | 65968 | No | NA | 63 | 6 | 3 | High-quality | High-quality | 100 | AAI-based (high-confidence) | 0 |
| S12_vs_k141_113789\|\|full | 1473 | No | NA | 1 | 0 | 1 | Not-determined | Genome-fragment | NA | NA | 0 |
| S12_extracted_k141_125820 | 4333 | No | NA | 6 | 0 | 0 | Low-quality | Genome-fragment | 10.26 | AAI-based (high-confidence) | 0 |
| S12_vs_k141_74496\|\|full | 3120 | No | NA | 3 | 0 | 1 | Not-determined | Genome-fragment | NA | NA | 0 |
| S12_vs_k141_59420\|\|full | 2333 | No | NA | 4 | 0 | 0 | Not-determined | Genome-fragment | NA | NA | 0 |
| S5_vs_k141_28804\|\|full | 4335 | No | NA | 13 | 0 | 0 | Low-quality | Genome-fragment | 10.97 | AAI-based (high-confidence) | 0 |
| S11_vs_k141_31250\|\|full | 4337 | No | NA | 5 | 0 | 0 | Low-quality | Genome-fragment | 3.97 | AAI-based (medium-confidence) | 0 |
| S8_extracted_k141_53777 | 4337 | No | NA | 8 | 6 | 0 | Low-quality | Genome-fragment | 11.99 | AAI-based (high-confidence) | 0 |
| S12_vs_k141_35073\|\|full | 1185 | No | NA | 4 | 0 | 0 | Not-determined | Genome-fragment | NA | NA | 0 |
| S8_vs_k141_39030\|\|full | 4337 | No | NA | 6 | 0 | 4 | Low-quality | Genome-fragment | 1.82 | AAI-based (medium-confidence) | 0 |
| S3_extracted_k141_81409 | 4338 | No | NA | 9 | 0 | 0 | Low-quality | Genome-fragment | 9.96 | AAI-based (high-confidence) | 0 |
| S12_vs_k141_74553\|\|full | 7305 | No | NA | 9 | 0 | 4 | Not-determined | Genome-fragment | NA | NA | 0 |
| S12_vs_k141_169276\|\|full | 7392 | No | NA | 8 | 0 | 4 | Not-determined | Genome-fragment | NA | NA | 0 |
| S12_vs_k141_185423\|\|full | 2274 | No | NA | 2 | 0 | 2 | Not-determined | Genome-fragment | NA | NA | 0 |
| S12_vs_k141_59462\|\|full | 1930 | No | NA | 5 | 0 | 0 | Not-determined | Genome-fragment | NA | NA | 0 |
| S8_extracted_k141_49575 | 4339 | No | NA | 5 | 3 | 0 | Low-quality | Genome-fragment | 10.39 | AAI-based (high-confidence) | 0 |
| S12_vs_k141_106727\|\|full | 2397 | No | NA | 4 | 0 | 0 | Not-determined | Genome-fragment | NA | NA | 0 |
| S12_vs_k141_44101\|\|full | 3551 | No | NA | 2 | 0 | 0 | Not-determined | Genome-fragment | NA | NA | 0 |
| S11_vs_k141_90540\|\|full | 4340 | No | NA | 11 | 3 | 0 | Low-quality | Genome-fragment | 6.09 | AAI-based (high-confidence) | 0 |
| S12_vs_k141_74619\|\|full | 2147 | No | NA | 4 | 0 | 1 | Not-determined | Genome-fragment | NA | NA | 0 |
| S12_vs_k141_113857\|\|full | 2298 | No | NA | 4 | 0 | 1 | Not-determined | Genome-fragment | NA | NA | 0 |
| S3_vs_k141_89051\|\|full | 4340 | No | NA | 4 | 0 | 0 | Low-quality | Genome-fragment | 10.12 | AAI-based (high-confidence) | 0 |
| S12_vs_k141_23383\|\|full | 4341 | No | NA | 5 | 0 | 0 | Low-quality | Genome-fragment | 4.08 | AAI-based (high-confidence) | 0 |
| S12_vs_k141_161328\|\|full | 2143 | No | NA | 4 | 0 | 3 | Not-determined | Genome-fragment | NA | NA | 0 |
| S12_vs_k141_67169\|\|full | 1306 | No | NA | 4 | 0 | 2 | Not-determined | Genome-fragment | NA | NA | 0 |
| S1_vs_k141_69546\|\|full | 4342 | No | NA | 8 | 4 | 0 | Low-quality | Genome-fragment | 9.21 | AAI-based (medium-confidence) | 0 |
| S12_vs_k141_99281\|\|full | 1960 | No | NA | 5 | 0 | 3 | Not-determined | Genome-fragment | NA | NA | 0 |
| S16_extracted_170196 | 4343 | No | NA | 7 | 4 | 0 | Low-quality | Genome-fragment | 8.99 | AAI-based (high-confidence) | 0 |
| S16_vs_133428\|\|full | 4343 | No | NA | 7 | 1 | 0 | Low-quality | Genome-fragment | 8.77 | AAI-based (high-confidence) | 0 |
| S12_vs_k141_35158\|\|full | 3546 | No | NA | 5 | 0 | 1 | Not-determined | Genome-fragment | NA | NA | 0 |
| S12_vs_k141_91239\|\|full | 1206 | No | NA | 4 | 0 | 1 | Not-determined | Genome-fragment | NA | NA | 0 |
| S12_vs_k141_59519\|\|full | 1837 | No | NA | 3 | 0 | 3 | Not-determined | Genome-fragment | NA | NA | 0 |
| S10_vs_2656\|\|full | 4343 | No | NA | 5 | 0 | 0 | Low-quality | Genome-fragment | 6.81 | AAI-based (medium-confidence) | 0 |
| S12_vs_k141_113928\|\|full | 1960 | No | NA | 3 | 0 | 0 | Not-determined | Genome-fragment | NA | NA | 0 |
| S13_extracted_30791 | 4345 | No | NA | 1 | 0 | 1 | Low-quality | Genome-fragment | 7.79 | AAI-based (high-confidence) | 0 |
| S12_vs_k141_138125\|\|full | 1688 | No | NA | 3 | 0 | 0 | Not-determined | Genome-fragment | NA | NA | 0 |
| S1_extracted_k141_128808 | 4345 | No | NA | 11 | 1 | 0 | Low-quality | Genome-fragment | 10.51 | AAI-based (high-confidence) | 0 |
| S10_extracted_9420 | 4345 | No | NA | 3 | 0 | 1 | Low-quality | Genome-fragment | 3.95 | AAI-based (medium-confidence) | 0 |
| S12_vs_k141_169385\|\|full | 4096 | No | NA | 8 | 0 | 0 | Not-determined | Genome-fragment | NA | NA | 0 |
| S8_vs_k141_57381\|\|full | 4345 | No | NA | 4 | 3 | 0 | Low-quality | Genome-fragment | 4.9 | HMM-based (lower-bound) | 0 |
| S5_vs_k141_95495\|\|full | 4347 | No | NA | 11 | 1 | 0 | Low-quality | Genome-fragment | 8.18 | AAI-based (high-confidence) | 0 |
| S12_vs_k141_176594\|\|full | 5017 | No | NA | 7 | 0 | 1 | Not-determined | Genome-fragment | NA | NA | 0 |
| S12_vs_k141_121792\|\|full | 1164 | No | NA | 2 | 0 | 1 | Not-determined | Genome-fragment | NA | NA | 0 |
| S17_vs_k141_169063\|\|full | 4348 | No | NA | 5 | 0 | 3 | Low-quality | Genome-fragment | 2.29 | AAI-based (medium-confidence) | 0 |
| S12_vs_k141_138208\|\|full | 1685 | No | NA | 3 | 0 | 0 | Not-determined | Genome-fragment | NA | NA | 0 |
| S12_vs_k141_67197\|\|full | 3454 | No | NA | 7 | 0 | 7 | Not-determined | Genome-fragment | NA | NA | 0 |
| S9_vs_k141_83562\|\|full | 4349 | No | NA | 10 | 2 | 0 | Low-quality | Genome-fragment | 12.25 | AAI-based (high-confidence) | 0 |
| S3_vs_k141_49410\|\|full | 4349 | No | NA | 7 | 2 | 0 | Low-quality | Genome-fragment | 2.66 | AAI-based (high-confidence) | 0 |
| S12_vs_k141_67205\|\|full | 1217 | No | NA | 3 | 0 | 0 | Not-determined | Genome-fragment | NA | NA | 0 |
| S16_vs_134363\|\|full | 4349 | No | NA | 4 | 0 | 1 | Low-quality | Genome-fragment | 4.41 | AAI-based (medium-confidence) | 0 |
| S12_vs_k141_109482\|\|full | 4349 | No | NA | 7 | 0 | 0 | Low-quality | Genome-fragment | 8.56 | AAI-based (medium-confidence) | 0 |
| S12_vs_k141_185618\|\|full | 2035 | No | NA | 2 | 0 | 2 | Not-determined | Genome-fragment | NA | NA | 0 |
| S12_vs_k141_153509\|\|full | 4228 | No | NA | 5 | 0 | 4 | Not-determined | Genome-fragment | NA | NA | 0 |
| S12_vs_k141_114017\|\|full | 2701 | No | NA | 3 | 0 | 1 | Not-determined | Genome-fragment | NA | NA | 0 |
| S12_vs_k141_121808\|\|full | 5870 | No | NA | 4 | 0 | 2 | Not-determined | Genome-fragment | NA | NA | 0 |
| S3_vs_k141_13489\|\|full | 4350 | Yes | 876 | 4 | 1 | 3 | Low-quality | Genome-fragment | 0.41 | HMM-based (lower-bound) | 79.86 |
| S16_extracted_78339 | 4351 | No | NA | 5 | 0 | 2 | Low-quality | Genome-fragment | 5.52 | AAI-based (medium-confidence) | 0 |
| S16_vs_166914\|\|lt2gene | 4351 | No | NA | 2 | 1 | 0 | Low-quality | Genome-fragment | 6.71 | AAI-based (high-confidence) | 0 |
| S10_extracted_27926 | 4352 | No | NA | 7 | 2 | 0 | Low-quality | Genome-fragment | 7.31 | AAI-based (high-confidence) | 0 |
| S11_vs_k141_44591\|\|full | 4354 | No | NA | 10 | 1 | 0 | Low-quality | Genome-fragment | 6.98 | HMM-based (lower-bound) | 0 |
| S12_vs_k141_82922\|\|full | 1983 | No | NA | 2 | 0 | 1 | Not-determined | Genome-fragment | NA | NA | 0 |
| S8_extracted_k141_149183 | 4354 | No | NA | 7 | 0 | 1 | Low-quality | Genome-fragment | 2.93 | AAI-based (medium-confidence) | 0 |
| S12_vs_k141_35241\|\|full | 1489 | No | NA | 2 | 0 | 1 | Not-determined | Genome-fragment | NA | NA | 0 |
| S8_vs_k141_75749\|\|full | 4355 | No | NA | 9 | 0 | 0 | Low-quality | Genome-fragment | 9.22 | AAI-based (high-confidence) | 0 |
| S8_vs_k141_6943\|\|full | 4356 | No | NA | 9 | 4 | 0 | Low-quality | Genome-fragment | 11.66 | AAI-based (high-confidence) | 0 |
| S12_vs_k141_74814\|\|full | 1618 | No | NA | 3 | 0 | 1 | Not-determined | Genome-fragment | NA | NA | 0 |
| S10_extracted_27308 | 4357 | No | NA | 6 | 4 | 0 | Low-quality | Genome-fragment | 15.1 | AAI-based (high-confidence) | 0 |
| S12_vs_k141_176687\|\|full | 1145 | No | NA | 2 | 0 | 1 | Not-determined | Genome-fragment | NA | NA | 0 |
| S12_vs_k141_185664\|\|full | 1973 | No | NA | 2 | 0 | 1 | Not-determined | Genome-fragment | NA | NA | 0 |
| S13_extracted_128360 | 4358 | No | NA | 1 | 0 | 0 | Low-quality | Genome-fragment | 4.64 | AAI-based (medium-confidence) | 0 |
| S12_vs_k141_161495\|\|full | 8199 | No | NA | 8 | 0 | 7 | Not-determined | Genome-fragment | NA | NA | 0 |
| S13_vs_127846\|\|full | 4358 | Yes | 927 | 4 | 1 | 1 | Low-quality | Genome-fragment | 0.5 | HMM-based (lower-bound) | 78.73 |
| S12_vs_k141_188268\|\|full | 4358 | No | NA | 7 | 1 | 0 | Low-quality | Genome-fragment | 3.5 | HMM-based (lower-bound) | 0 |
| S12_vs_k141_185689\|\|full | 2967 | No | NA | 4 | 0 | 0 | Not-determined | Genome-fragment | NA | NA | 0 |
| S3_vs_k141_66308\|\|full | 4359 | No | NA | 8 | 1 | 0 | Low-quality | Genome-fragment | 11.54 | AAI-based (high-confidence) | 0 |
| S12_vs_k141_91371\|\|full | 1106 | No | NA | 2 | 0 | 0 | Not-determined | Genome-fragment | NA | NA | 0 |
| S12_vs_k141_107027\|\|full | 1606 | No | NA | 3 | 0 | 1 | Not-determined | Genome-fragment | NA | NA | 0 |
| S10_vs_92487\|\|full | 4359 | No | NA | 4 | 0 | 2 | Low-quality | Genome-fragment | 5.52 | AAI-based (medium-confidence) | 0 |
| S12_vs_k141_59687\|\|full | 3915 | No | NA | 5 | 0 | 4 | Not-determined | Genome-fragment | NA | NA | 0 |
| S12_extracted_k141_47325 | 4359 | No | NA | 4 | 0 | 4 | Low-quality | Genome-fragment | 2.01 | AAI-based (high-confidence) | 0 |
| S17_vs_k141_16296\|\|full | 4360 | No | NA | 3 | 0 | 1 | Low-quality | Genome-fragment | 5.66 | AAI-based (medium-confidence) | 0 |
| S12_vs_k141_176722\|\|full | 14031 | No | NA | 14 | 0 | 6 | Not-determined | Genome-fragment | NA | NA | 0 |
| S9_vs_k141_69251\|\|full | 4360 | No | NA | 5 | 1 | 0 | Low-quality | Genome-fragment | 12.22 | AAI-based (high-confidence) | 0 |
| S12_vs_k141_138261\|\|full | 63040 | No | NA | 68 | 12 | 2 | High-quality | High-quality | 99.25 | AAI-based (high-confidence) | 0 |
| S12_vs_k141_161554\|\|full | 1581 | No | NA | 2 | 0 | 2 | Not-determined | Genome-fragment | NA | NA | 0 |
| S12_vs_k141_121897\|\|full | 1878 | No | NA | 3 | 0 | 2 | Not-determined | Genome-fragment | NA | NA | 0 |
| S12_vs_k141_185727\|\|full | 28635 | No | NA | 30 | 0 | 17 | Not-determined | Genome-fragment | NA | NA | 0 |
| S12_vs_k141_82989\|\|full | 2115 | No | NA | 3 | 0 | 0 | Not-determined | Genome-fragment | NA | NA | 0 |
| S12_vs_k141_185732\|\|full | 2558 | No | NA | 2 | 0 | 2 | Not-determined | Genome-fragment | NA | NA | 0 |
| S12_vs_k141_44348\|\|full | 2929 | No | NA | 3 | 0 | 1 | Not-determined | Genome-fragment | NA | NA | 0 |
| S12_vs_k141_114181\|\|full | 2029 | No | NA | 4 | 0 | 0 | Not-determined | Genome-fragment | NA | NA | 0 |
| S6_vs_k141_14261\|\|full | 4360 | No | NA | 4 | 0 | 0 | Low-quality | Genome-fragment | 2.61 | AAI-based (medium-confidence) | 0 |
| S1_vs_k141_75170\|\|full | 4361 | No | NA | 3 | 3 | 0 | Low-quality | Genome-fragment | 10.61 | AAI-based (high-confidence) | 0 |
| S1_vs_k141_97051\|\|full | 4361 | No | NA | 6 | 0 | 6 | Low-quality | Genome-fragment | 3.64 | AAI-based (medium-confidence) | 0 |
| S16_vs_6556\|\|full | 4363 | Yes | 3092 | 6 | 1 | 1 | Low-quality | Genome-fragment | 6.12 | AAI-based (high-confidence) | 29.13 |
| S12_vs_k141_114207\|\|full | 1168 | No | NA | 2 | 0 | 2 | Not-determined | Genome-fragment | NA | NA | 0 |
| S14_extracted_k141_105970 | 4365 | No | NA | 6 | 1 | 0 | Low-quality | Genome-fragment | 9.63 | AAI-based (high-confidence) | 0 |
| S7_extracted_136327 | 4365 | No | NA | 9 | 2 | 0 | Low-quality | Genome-fragment | 7.33 | AAI-based (high-confidence) | 0 |
| S16_vs_12939\|\|full | 4365 | No | NA | 5 | 0 | 3 | Low-quality | Genome-fragment | 12.01 | AAI-based (medium-confidence) | 0 |
| S6_extracted_k141_495 | 4366 | No | NA | 9 | 3 | 0 | Low-quality | Genome-fragment | 9.13 | AAI-based (high-confidence) | 0 |
| S12_vs_k141_169548\|\|full | 1403 | No | NA | 2 | 0 | 0 | Not-determined | Genome-fragment | NA | NA | 0 |
| S8_vs_k141_56700\|\|full | 4366 | No | NA | 4 | 0 | 1 | Low-quality | Genome-fragment | 1.3 | AAI-based (medium-confidence) | 0 |
| S12_vs_k141_74927\|\|full | 4165 | No | NA | 3 | 0 | 0 | Not-determined | Genome-fragment | NA | NA | 0 |
| S12_vs_k141_83019\|\|full | 4035 | No | NA | 7 | 0 | 2 | Not-determined | Genome-fragment | NA | NA | 0 |
| S12_vs_k141_59815\|\|full | 2296 | No | NA | 2 | 0 | 1 | Not-determined | Genome-fragment | NA | NA | 0 |
| S8_vs_k141_91332\|\|full | 4366 | No | NA | 5 | 0 | 0 | Low-quality | Genome-fragment | 2.82 | AAI-based (high-confidence) | 0 |
| S3_extracted_k141_52137 | 4367 | No | NA | 6 | 1 | 0 | Low-quality | Genome-fragment | 2.67 | AAI-based (high-confidence) | 0 |
| S1_vs_k141_22155\|\|full | 4368 | No | NA | 3 | 0 | 1 | Low-quality | Genome-fragment | 3.45 | AAI-based (medium-confidence) | 0 |
| S12_vs_k141_4764\|\|full | 1744 | No | NA | 3 | 0 | 1 | Not-determined | Genome-fragment | NA | NA | 0 |
| S12_vs_k141_99560\|\|full | 5504 | No | NA | 10 | 0 | 0 | Not-determined | Genome-fragment | NA | NA | 0 |
| S12_vs_k141_176810\|\|full | 7516 | No | NA | 8 | 0 | 2 | Not-determined | Genome-fragment | NA | NA | 0 |
| S7_vs_79279\|\|full | 4368 | No | NA | 4 | 0 | 0 | Low-quality | Genome-fragment | 10.11 | AAI-based (high-confidence) | 0 |
| S12_vs_k141_74958\|\|full | 6830 | No | NA | 13 | 0 | 0 | Not-determined | Genome-fragment | NA | NA | 0 |
| S16_vs_56933\|\|full | 4368 | No | NA | 3 | 0 | 3 | Low-quality | Genome-fragment | 2.19 | AAI-based (medium-confidence) | 0 |
| S8_vs_k141_156613\|\|full | 4368 | No | NA | 4 | 0 | 2 | Low-quality | Genome-fragment | 5.17 | AAI-based (medium-confidence) | 0 |
| S12_vs_k141_51284\|\|full | 2460 | No | NA | 4 | 0 | 1 | Not-determined | Genome-fragment | NA | NA | 0 |
| S12_vs_k141_35392\|\|full | 1951 | No | NA | 4 | 0 | 0 | Not-determined | Genome-fragment | NA | NA | 0 |
| S12_vs_k141_138393\|\|full | 1370 | No | NA | 2 | 0 | 0 | Not-determined | Genome-fragment | NA | NA | 0 |
| S13_vs_68275\|\|full | 4369 | No | NA | 4 | 0 | 3 | Low-quality | Genome-fragment | 0.77 | AAI-based (medium-confidence) | 0 |
| S5_extracted_k141_104144 | 4369 | No | NA | 6 | 5 | 0 | Low-quality | Genome-fragment | 11.72 | AAI-based (high-confidence) | 0 |
| S7_vs_102763\|\|full | 4369 | Yes | 2412 | 8 | 3 | 2 | Low-quality | Genome-fragment | 2.33 | HMM-based (lower-bound) | 44.79 |
| S12_extracted_k141_32678 | 4369 | No | NA | 9 | 0 | 1 | Low-quality | Genome-fragment | 10.03 | AAI-based (medium-confidence) | 0 |
| S12_vs_k141_121913\|\|full | 131670 | Yes | 74298 | 133 | 11 | 24 | Medium-quality | Genome-fragment | 66.92 | AAI-based (medium-confidence) | 43.57 |
| S12_vs_k141_153776\|\|full | 1990 | No | NA | 4 | 0 | 1 | Not-determined | Genome-fragment | NA | NA | 0 |
| S12_vs_k141_91581\|\|full | 5132 | No | NA | 5 | 0 | 5 | Not-determined | Genome-fragment | NA | NA | 0 |
| S12_vs_k141_91585\|\|full | 1367 | No | NA | 3 | 0 | 1 | Not-determined | Genome-fragment | NA | NA | 0 |
| S12_vs_k141_185883\|\|full | 2459 | No | NA | 4 | 0 | 1 | Not-determined | Genome-fragment | NA | NA | 0 |
| S12_vs_k141_176899\|\|full | 1855 | No | NA | 2 | 0 | 1 | Not-determined | Genome-fragment | NA | NA | 0 |
| S12_vs_k141_114326\|\|full | 1338 | No | NA | 3 | 0 | 0 | Not-determined | Genome-fragment | NA | NA | 0 |
| S8_extracted_k141_125664 | 4369 | No | NA | 6 | 0 | 2 | Low-quality | Genome-fragment | 5.21 | AAI-based (high-confidence) | 0 |
| S12_vs_k141_176904\|\|full | 1595 | No | NA | 2 | 0 | 1 | Not-determined | Genome-fragment | NA | NA | 0 |
| S4_extracted_17208 | 4370 | No | NA | 12 | 10 | 0 | Low-quality | Genome-fragment | 14.38 | AAI-based (high-confidence) | 0 |
| S7_vs_137429\|\|full | 4371 | No | NA | 4 | 3 | 1 | Low-quality | Genome-fragment | 8.16 | AAI-based (high-confidence) | 0 |
| S10_vs_3836\|\|full | 4371 | No | NA | 7 | 1 | 0 | Low-quality | Genome-fragment | 5.41 | HMM-based (lower-bound) | 0 |
| S14_vs_k141_47799\|\|full | 4372 | No | NA | 5 | 0 | 1 | Low-quality | Genome-fragment | 2.23 | AAI-based (medium-confidence) | 0 |
| S12_vs_k141_59936\|\|full | 1514 | No | NA | 3 | 0 | 0 | Not-determined | Genome-fragment | NA | NA | 0 |
| S3_vs_k141_124183\|\|full | 4372 | No | NA | 4 | 0 | 2 | Low-quality | Genome-fragment | 3.31 | AAI-based (high-confidence) | 0 |
| S12_vs_k141_161825\|\|full | 3559 | No | NA | 5 | 0 | 1 | Not-determined | Genome-fragment | NA | NA | 0 |
| S11_extracted_k141_87390 | 4373 | No | NA | 11 | 1 | 0 | Low-quality | Genome-fragment | 11.17 | AAI-based (high-confidence) | 0 |
| S12_vs_k141_91627\|\|full | 2739 | No | NA | 7 | 0 | 0 | Not-determined | Genome-fragment | NA | NA | 0 |
| S8_vs_k141_198740\|\|full | 4373 | No | NA | 3 | 0 | 3 | Low-quality | Genome-fragment | 11.42 | AAI-based (medium-confidence) | 0 |
| S12_vs_k141_185943\|\|full | 2579 | No | NA | 3 | 0 | 1 | Not-determined | Genome-fragment | NA | NA | 0 |
| S10_vs_20536\|\|full | 4374 | No | NA | 5 | 0 | 5 | Low-quality | Genome-fragment | 0.54 | AAI-based (medium-confidence) | 0 |
| S12_vs_k141_121986\|\|full | 8155 | No | NA | 7 | 0 | 0 | Not-determined | Genome-fragment | NA | NA | 0 |
| S12_vs_k141_35510\|\|full | 1494 | No | NA | 3 | 0 | 2 | Not-determined | Genome-fragment | NA | NA | 0 |
| S8_extracted_k141_194946 | 4374 | No | NA | 7 | 2 | 0 | Low-quality | Genome-fragment | 9.25 | AAI-based (high-confidence) | 0 |
| S12_vs_k141_153780\|\|full | 61281 | No | NA | 72 | 20 | 7 | High-quality | High-quality | 97.63 | HMM-based (lower-bound) | 0 |
| S12_vs_k141_4851\|\|full | 9918 | No | NA | 10 | 0 | 1 | Not-determined | Genome-fragment | NA | NA | 0 |
| S12_vs_k141_185967\|\|full | 2569 | No | NA | 4 | 0 | 0 | Not-determined | Genome-fragment | NA | NA | 0 |
| S12_vs_k141_91659\|\|full | 2358 | No | NA | 3 | 0 | 0 | Not-determined | Genome-fragment | NA | NA | 0 |
| S8_extracted_k141_195379 | 4374 | No | NA | 4 | 3 | 0 | Low-quality | Genome-fragment | 15.43 | HMM-based (lower-bound) | 0 |
| S5_vs_k141_45972\|\|full | 4375 | No | NA | 6 | 4 | 0 | Low-quality | Genome-fragment | 12.33 | AAI-based (medium-confidence) | 0 |

| | | | | | | | | | | | |
|---|---|---|---|---|---|---|---|---|---|---|---|
| S12_vs_k141_72282\|\|full | 4376 | No | NA | 5 | 2 | 0 | Low-quality | Genome-fragment | 8.85 | AAI-based (medium-confidence) | 0 |
| S17_vs_k141_50195\|\|full | 4377 | Yes | 3353 | 8 | 1 | 1 | Low-quality | Genome-fragment | 1.8 | HMM-based (lower-bound) | 23.4 |
| S4_extracted_30560 | 4377 | No | NA | 4 | 4 | 0 | Low-quality | Genome-fragment | 9.62 | AAI-based (high-confidence) | 0 |
| S12_vs_k141_91680\|\|full | 3208 | No | NA | 4 | 0 | 2 | Not-determined | Genome-fragment | NA | NA | 0 |
| S12_vs_k141_114444\|\|full | 2794 | No | NA | 4 | 0 | 3 | Not-determined | Genome-fragment | NA | NA | 0 |
| S12_vs_k141_75086\|\|full | 1782 | No | NA | 3 | 0 | 0 | Not-determined | Genome-fragment | NA | NA | 0 |
| S8_extracted_k141_81743 | 4378 | No | NA | 4 | 0 | 1 | Low-quality | Genome-fragment | 1.81 | AAI-based (medium-confidence) | 0 |
| S12_vs_k141_138564\|\|full | 1747 | No | NA | 3 | 0 | 3 | Not-determined | Genome-fragment | NA | NA | 0 |
| S12_vs_k141_130106\|\|full | 2001 | No | NA | 4 | 0 | 2 | Not-determined | Genome-fragment | NA | NA | 0 |
| S12_vs_k141_28297\|\|full | 5787 | No | NA | 6 | 0 | 0 | Not-determined | Genome-fragment | NA | NA | 0 |
| S5_vs_k141_35482\|\|full | 4379 | No | NA | 5 | 0 | 2 | Low-quality | Genome-fragment | 3.25 | AAI-based (medium-confidence) | 0 |
| S12_vs_k141_51419\|\|full | 1524 | No | NA | 2 | 0 | 0 | Not-determined | Genome-fragment | NA | NA | 0 |
| S12_vs_k141_146325\|\|full | 3674 | No | NA | 5 | 0 | 1 | Not-determined | Genome-fragment | NA | NA | 0 |
| S12_vs_k141_44680\|\|full | 12066 | No | NA | 8 | 0 | 1 | Not-determined | Genome-fragment | NA | NA | 0 |
| S12_vs_k141_4925\|\|full | 1381 | No | NA | 2 | 0 | 1 | Not-determined | Genome-fragment | NA | NA | 0 |
| S12_vs_k141_138600\|\|full | 3676 | No | NA | 4 | 0 | 0 | Not-determined | Genome-fragment | NA | NA | 0 |
| S14_vs_k141_25168\|\|full | 4380 | No | NA | 4 | 3 | 0 | Low-quality | Genome-fragment | 12.3 | AAI-based (high-confidence) | 0 |
| S8_extracted_k141_185807 | 4380 | No | NA | 6 | 4 | 0 | Low-quality | Genome-fragment | 10.97 | HMM-based (lower-bound) | 0 |
| S7_extracted_98473 | 4381 | No | NA | 2 | 0 | 1 | Low-quality | Genome-fragment | 9.45 | AAI-based (medium-confidence) | 0 |
| S12_vs_k141_130159\|\|full | 2888 | No | NA | 4 | 0 | 0 | Not-determined | Genome-fragment | NA | NA | 0 |
| S8_vs_k141_108160\|\|full | 4382 | No | NA | 3 | 0 | 3 | Low-quality | Genome-fragment | 0.77 | AAI-based (medium-confidence) | 0 |
| S12_vs_k141_75098\|\|full | 41466 | Yes | 28858 | 56 | 14 | 4 | Medium-quality | Genome-fragment | 76.14 | AAI-based (high-confidence) | 30.41 |
| S5_extracted_k141_22771 | 4383 | No | NA | 4 | 3 | 0 | Low-quality | Genome-fragment | 11.63 | AAI-based (high-confidence) | 0 |
| S12_vs_k141_114507\|\|full | 2072 | No | NA | 4 | 0 | 1 | Not-determined | Genome-fragment | NA | NA | 0 |
| S12_vs_k141_122070\|\|full | 1549 | No | NA | 2 | 0 | 0 | Not-determined | Genome-fragment | NA | NA | 0 |
| S3_extracted_k141_108490 | 4385 | No | NA | 7 | 0 | 0 | Low-quality | Genome-fragment | 5.56 | AAI-based (high-confidence) | 0 |
| S12_vs_k141_67632\|\|full | 3684 | No | NA | 3 | 0 | 1 | Not-determined | Genome-fragment | NA | NA | 0 |
| S16_vs_112933\|\|full | 4385 | No | NA | 9 | 2 | 0 | Low-quality | Genome-fragment | 10.17 | AAI-based (high-confidence) | 0 |
| S12_vs_k141_108433\|\|full | 4388 | No | NA | 5 | 0 | 0 | Low-quality | Genome-fragment | 11.65 | AAI-based (high-confidence) | 0 |
| S16_vs_104148\|\|full | 4390 | No | NA | 10 | 1 | 0 | Low-quality | Genome-fragment | 5.11 | AAI-based (medium-confidence) | 0 |
| S12_vs_k141_130239\|\|full | 3943 | No | NA | 3 | 0 | 0 | Not-determined | Genome-fragment | NA | NA | 0 |
| S7_vs_92596\|\|full | 4391 | No | NA | 10 | 1 | 0 | Low-quality | Genome-fragment | 5.71 | HMM-based (lower-bound) | 0 |
| S12_vs_k141_4999\|\|full | 1007 | No | NA | 2 | 0 | 2 | Not-determined | Genome-fragment | NA | NA | 0 |
| S12_vs_k141_177142\|\|full | 2290 | No | NA | 3 | 0 | 1 | Not-determined | Genome-fragment | NA | NA | 0 |
| S10_vs_34205\|\|full | 4392 | No | NA | 5 | 0 | 2 | Low-quality | Genome-fragment | 4.04 | AAI-based (medium-confidence) | 0 |
| S12_vs_k141_20197\|\|full | 2250 | No | NA | 4 | 0 | 0 | Not-determined | Genome-fragment | NA | NA | 0 |
| S12_vs_k141_138718\|\|full | 1292 | No | NA | 4 | 0 | 0 | Not-determined | Genome-fragment | NA | NA | 0 |
| S1_vs_k141_108029\|\|full | 4393 | No | NA | 8 | 0 | 0 | Low-quality | Genome-fragment | 11.32 | AAI-based (high-confidence) | 0 |
| S7_vs_58186\|\|full | 4394 | No | NA | 9 | 1 | 0 | Low-quality | Genome-fragment | 11.28 | AAI-based (high-confidence) | 0 |
| S12_extracted_k141_127107 | 4394 | No | NA | 2 | 0 | 0 | Low-quality | Genome-fragment | 5.91 | AAI-based (medium-confidence) | 0 |
| S1_vs_k141_129976\|\|full | 4395 | No | NA | 5 | 0 | 4 | Low-quality | Genome-fragment | 2.03 | AAI-based (medium-confidence) | 0 |
| S5_extracted_k141_75820 | 4395 | No | NA | 13 | 2 | 0 | Low-quality | Genome-fragment | 6.18 | AAI-based (high-confidence) | 0 |
| S5_vs_k141_4792\|\|full | 4395 | No | NA | 9 | 2 | 0 | Low-quality | Genome-fragment | 11.16 | AAI-based (high-confidence) | 0 |
| S11_vs_k141_19227\|\|full | 4396 | No | NA | 6 | 0 | 0 | Low-quality | Genome-fragment | 3.05 | AAI-based (medium-confidence) | 0 |
| S12_vs_k141_35747\|\|full | 6041 | No | NA | 4 | 0 | 4 | Not-determined | Genome-fragment | NA | NA | 0 |
| S11_vs_k141_83284\|\|full | 4397 | No | NA | 9 | 1 | 0 | Low-quality | Genome-fragment | 9.72 | AAI-based (high-confidence) | 0 |
| S14_vs_k141_105033\|\|full | 4397 | No | NA | 3 | 0 | 3 | Low-quality | Genome-fragment | 1.52 | AAI-based (medium-confidence) | 0 |
| S12_vs_k141_122170\|\|full | 1078 | No | NA | 3 | 0 | 1 | Not-determined | Genome-fragment | NA | NA | 0 |
| S16_vs_132417\|\|full | 4398 | No | NA | 3 | 0 | 2 | Low-quality | Genome-fragment | 1.56 | AAI-based (medium-confidence) | 0 |
| S12_vs_k141_51610\|\|full | 2490 | No | NA | 6 | 0 | 0 | Not-determined | Genome-fragment | NA | NA | 0 |
| S12_vs_k141_177220\|\|full | 4143 | No | NA | 3 | 0 | 1 | Not-determined | Genome-fragment | NA | NA | 0 |
| S13_vs_5992\|\|full | 4399 | No | NA | 10 | 1 | 0 | Low-quality | Genome-fragment | 6.99 | AAI-based (high-confidence) | 0 |
| S12_vs_k141_110014\|\|full | 4399 | Yes | 3044 | 8 | 2 | 1 | Low-quality | Genome-fragment | 2.2 | AAI-based (high-confidence) | 30.8 |
| S12_vs_k141_107620\|\|full | 2239 | No | NA | 3 | 0 | 1 | Not-determined | Genome-fragment | NA | NA | 0 |
| S12_vs_k141_20267\|\|full | 3723 | No | NA | 3 | 0 | 1 | Not-determined | Genome-fragment | NA | NA | 0 |
| S5_extracted_k141_75695 | 4400 | No | NA | 4 | 4 | 0 | Low-quality | Genome-fragment | 12.36 | AAI-based (high-confidence) | 0 |
| S10_extracted_62320 | 4400 | No | NA | 5 | 0 | 1 | Low-quality | Genome-fragment | 4.79 | AAI-based (medium-confidence) | 0 |
| S12_vs_k141_146501\|\|full | 3665 | No | NA | 6 | 0 | 1 | Not-determined | Genome-fragment | NA | NA | 0 |
| S12_vs_k141_67768\|\|full | 3867 | No | NA | 4 | 0 | 0 | Not-determined | Genome-fragment | NA | NA | 0 |
| S12_vs_k141_130335\|\|full | 1857 | No | NA | 2 | 0 | 1 | Not-determined | Genome-fragment | NA | NA | 0 |
| S12_vs_k141_114475\|\|full | 4400 | No | NA | 5 | 0 | 1 | Low-quality | Genome-fragment | 0.51 | AAI-based (medium-confidence) | 0 |
| S12_vs_k141_138819\|\|full | 7351 | No | NA | 9 | 0 | 9 | Not-determined | Genome-fragment | NA | NA | 0 |
| S12_vs_k141_138821\|\|full | 1998 | No | NA | 3 | 0 | 1 | Not-determined | Genome-fragment | NA | NA | 0 |
| S12_vs_k141_59986\|\|full | 1767 | No | NA | 4 | 0 | 1 | Not-determined | Genome-fragment | NA | NA | 0 |
| S11_vs_k141_60435\|\|full | 4405 | No | NA | 6 | 4 | 0 | Low-quality | Genome-fragment | 12.05 | AAI-based (high-confidence) | 0 |
| S12_vs_k141_44909\|\|full | 13508 | No | NA | 20 | 0 | 3 | Not-determined | Genome-fragment | NA | NA | 0 |
| S12_vs_k141_99975\|\|full | 1226 | No | NA | 3 | 0 | 1 | Not-determined | Genome-fragment | NA | NA | 0 |
| S12_vs_k141_162150\|\|full | 12636 | No | NA | 6 | 0 | 6 | Not-determined | Genome-fragment | NA | NA | 0 |
| S5_vs_k141_83419\|\|full | 4405 | No | NA | 10 | 1 | 0 | Low-quality | Genome-fragment | 12.88 | AAI-based (high-confidence) | 0 |
| S9_vs_k141_1466\|\|full | 4406 | No | NA | 9 | 0 | 0 | Low-quality | Genome-fragment | 6.51 | AAI-based (high-confidence) | 0 |
| S9_extracted_k141_29118 | 4407 | No | NA | 4 | 0 | 2 | Low-quality | Genome-fragment | 7.42 | AAI-based (medium-confidence) | 0 |
| S12_vs_k141_44952\|\|full | 3962 | No | NA | 6 | 0 | 0 | Not-determined | Genome-fragment | NA | NA | 0 |
| S12_vs_k141_114726\|\|full | 3925 | No | NA | 3 | 0 | 1 | Not-determined | Genome-fragment | NA | NA | 0 |
| S14_vs_k141_19041\|\|full | 4407 | No | NA | 3 | 0 | 2 | Low-quality | Genome-fragment | 1.54 | AAI-based (medium-confidence) | 0 |
| S16_vs_128573\|\|full | 4407 | Yes | 3999 | 3 | 1 | 1 | Low-quality | Genome-fragment | 2.3 | HMM-based (lower-bound) | 9.26 |
| S12_vs_k141_11833\|\|full | 1104 | No | NA | 3 | 0 | 0 | Not-determined | Genome-fragment | NA | NA | 0 |
| S16_vs_11668\|\|full | 4408 | No | NA | 11 | 0 | 1 | Low-quality | Genome-fragment | 7.61 | AAI-based (medium-confidence) | 0 |
| S12_vs_k141_83481\|\|full | 1174 | No | NA | 2 | 0 | 0 | Not-determined | Genome-fragment | NA | NA | 0 |
| S12_vs_k141_5176\|\|full | 1054 | No | NA | 3 | 0 | 0 | Not-determined | Genome-fragment | NA | NA | 0 |
| S16_vs_93234\|\|full | 4409 | No | NA | 10 | 0 | 0 | Low-quality | Genome-fragment | 7.27 | AAI-based (high-confidence) | 0 |
| S5_vs_k141_104409\|\|full | 4410 | No | NA | 7 | 4 | 0 | Low-quality | Genome-fragment | 11.21 | AAI-based (high-confidence) | 0 |
| S12_vs_k141_162230\|\|full | 1194 | No | NA | 3 | 0 | 0 | Not-determined | Genome-fragment | NA | NA | 0 |
| S12_vs_k141_44965\|\|full | 4350 | No | NA | 4 | 0 | 1 | Not-determined | Genome-fragment | NA | NA | 0 |
| S12_vs_k141_75404\|\|full | 3531 | No | NA | 4 | 0 | 0 | Not-determined | Genome-fragment | NA | NA | 0 |
| S12_vs_k141_83489\|\|full | 2357 | No | NA | 3 | 0 | 1 | Not-determined | Genome-fragment | NA | NA | 0 |
| S13_extracted_39506 | 4411 | No | NA | 5 | 0 | 4 | Low-quality | Genome-fragment | 1.85 | AAI-based (high-confidence) | 0 |
| S14_extracted_k141_27733 | 4411 | No | NA | 6 | 3 | 0 | Low-quality | Genome-fragment | 10.77 | AAI-based (high-confidence) | 0 |
| S12_vs_k141_186400\|\|full | 2796 | No | NA | 4 | 0 | 1 | Not-determined | Genome-fragment | NA | NA | 0 |
| S12_vs_k141_44477\|\|full | 4411 | No | NA | 5 | 3 | 0 | Low-quality | Genome-fragment | 17.41 | AAI-based (high-confidence) | 0 |
| S11_vs_k141_14160\|\|full | 4412 | No | NA | 9 | 1 | 0 | Low-quality | Genome-fragment | 11.02 | AAI-based (high-confidence) | 0 |
| S12_vs_k141_107716\|\|full | 26434 | No | NA | 25 | 0 | 13 | Not-determined | Genome-fragment | NA | NA | 0 |
| S12_vs_k141_5205\|\|full | 1684 | No | NA | 6 | 0 | 0 | Not-determined | Genome-fragment | NA | NA | 0 |
| S14_vs_k141_6520\|\|full | 4413 | No | NA | 5 | 5 | 0 | Low-quality | Genome-fragment | 14.4 | AAI-based (high-confidence) | 0 |
| S5_vs_k141_18378\|\|full | 4414 | No | NA | 10 | 6 | 0 | Low-quality | Genome-fragment | 11.18 | AAI-based (high-confidence) | 0 |
| S8_vs_k141_78199\|\|full | 4415 | No | NA | 13 | 0 | 0 | Low-quality | Genome-fragment | 11.96 | AAI-based (high-confidence) | 0 |
| S12_vs_k141_75444\|\|full | 1794 | No | NA | 3 | 0 | 1 | Not-determined | Genome-fragment | NA | NA | 0 |
| S14_extracted_k141_100006 | 4416 | No | NA | 5 | 1 | 1 | Low-quality | Genome-fragment | 1.16 | HMM-based (lower-bound) | 0 |
| S8_extracted_k141_117718 | 4416 | No | NA | 11 | 4 | 0 | Low-quality | Genome-fragment | 10.05 | AAI-based (high-confidence) | 0 |
| S12_vs_k141_45041\|\|full | 5283 | No | NA | 2 | 0 | 2 | Not-determined | Genome-fragment | NA | NA | 0 |
| S12_vs_k141_186456\|\|full | 1166 | No | NA | 3 | 0 | 1 | Not-determined | Genome-fragment | NA | NA | 0 |
| S13_extracted_27160 | 4417 | No | NA | 5 | 5 | 0 | Low-quality | Genome-fragment | 13.67 | AAI-based (high-confidence) | 0 |
| S12_vs_k141_5236\|\|full | 2253 | No | NA | 3 | 0 | 1 | Not-determined | Genome-fragment | NA | NA | 0 |
| S12_vs_k141_20488\|\|full | 2706 | No | NA | 3 | 0 | 0 | Not-determined | Genome-fragment | NA | NA | 0 |
| S12_vs_k141_45053\|\|full | 7962 | No | NA | 10 | 0 | 4 | Not-determined | Genome-fragment | NA | NA | 0 |
| S12_vs_k141_67831\|\|full | 1668 | No | NA | 2 | 0 | 2 | Not-determined | Genome-fragment | NA | NA | 0 |
| S12_vs_k141_130475\|\|full | 5981 | No | NA | 7 | 0 | 1 | Not-determined | Genome-fragment | NA | NA | 0 |
| S12_vs_k141_51760\|\|full | 1666 | No | NA | 3 | 0 | 1 | Not-determined | Genome-fragment | NA | NA | 0 |
| S12_vs_k141_5248\|\|full | 10905 | No | NA | 10 | 0 | 7 | Not-determined | Genome-fragment | NA | NA | 0 |
| S3_extracted_k141_125083 | 4417 | No | NA | 9 | 3 | 0 | Low-quality | Genome-fragment | 9.61 | AAI-based (medium-confidence) | 0 |
| S10_extracted_53424 | 4417 | No | NA | 5 | 0 | 3 | Low-quality | Genome-fragment | 2.14 | AAI-based (medium-confidence) | 0 |
| S12_vs_k141_139004\|\|full | 1873 | No | NA | 2 | 0 | 1 | Not-determined | Genome-fragment | NA | NA | 0 |
| S12_vs_k141_28661\|\|full | 27374 | No | NA | 33 | 12 | 0 | Medium-quality | Genome-fragment | 76.45 | AAI-based (medium-confidence) | 0 |
| S3_extracted_k141_69728 | 4418 | No | NA | 5 | 0 | 0 | Low-quality | Genome-fragment | 2.38 | AAI-based (medium-confidence) | 0 |
| S10_extracted_51876 | 4418 | No | NA | 4 | 0 | 3 | Low-quality | Genome-fragment | 4.97 | AAI-based (medium-confidence) | 0 |
| S5_extracted_k141_143796 | 4419 | No | NA | 5 | 1 | 0 | Low-quality | Genome-fragment | 4.29 | AAI-based (high-confidence) | 0 |
| S12_vs_k141_67857\|\|full | 2961 | No | NA | 7 | 0 | 0 | Not-determined | Genome-fragment | NA | NA | 0 |
| S12_vs_k141_139017\|\|full | 4379 | No | NA | 4 | 0 | 0 | Not-determined | Genome-fragment | NA | NA | 0 |
| S12_vs_k141_11934\|\|full | 1654 | No | NA | 2 | 0 | 1 | Not-determined | Genome-fragment | NA | NA | 0 |
| S12_vs_k141_130523\|\|full | 1850 | No | NA | 2 | 0 | 0 | Not-determined | Genome-fragment | NA | NA | 0 |
| S12_vs_k141_75510\|\|full | 7013 | No | NA | 6 | 0 | 5 | Not-determined | Genome-fragment | NA | NA | 0 |
| S12_vs_k141_92135\|\|full | 1133 | No | NA | 3 | 0 | 0 | Not-determined | Genome-fragment | NA | NA | 0 |
| S12_vs_k141_20584\|\|full | 1033 | No | NA | 2 | 0 | 0 | Not-determined | Genome-fragment | NA | NA | 0 |
| S12_vs_k141_5303\|\|full | 1675 | No | NA | 3 | 0 | 0 | Not-determined | Genome-fragment | NA | NA | 0 |
| S6_vs_k141_8634\|\|full | 4419 | No | NA | 2 | 0 | 2 | Low-quality | Genome-fragment | 2.08 | AAI-based (medium-confidence) | 0 |
| S12_vs_k141_146747\|\|full | 1540 | No | NA | 2 | 0 | 1 | Not-determined | Genome-fragment | NA | NA | 0 |
| S12_vs_k141_100196\|\|full | 2390 | No | NA | 3 | 0 | 2 | Not-determined | Genome-fragment | NA | NA | 0 |
| S12_vs_k141_162428\|\|full | 2650 | No | NA | 3 | 0 | 2 | Not-determined | Genome-fragment | NA | NA | 0 |
| S12_vs_k141_170099\|\|full | 3827 | No | NA | 6 | 0 | 3 | Not-determined | Genome-fragment | NA | NA | 0 |
| S12_vs_k141_177444\|\|full | 1107 | No | NA | 2 | 0 | 0 | Not-determined | Genome-fragment | NA | NA | 0 |
| S14_extracted_k141_52573 | 4421 | No | NA | 4 | 0 | 1 | Low-quality | Genome-fragment | 6.48 | AAI-based (high-confidence) | 0 |
| S5_extracted_k141_4465 | 4421 | No | NA | 10 | 0 | 0 | Low-quality | Genome-fragment | 13.25 | AAI-based (high-confidence) | 0 |
| S12_vs_k141_177488\|\|full | 2011 | No | NA | 5 | 0 | 1 | Not-determined | Genome-fragment | NA | NA | 0 |
| S12_vs_k141_162517\|\|full | 7925 | No | NA | 6 | 0 | 6 | Not-determined | Genome-fragment | NA | NA | 0 |
| S10_extracted_4364 | 4421 | No | NA | 8 | 1 | 0 | Low-quality | Genome-fragment | 6.46 | AAI-based (high-confidence) | 0 |
| S12_vs_k141_183618\|\|full | 4421 | No | NA | 5 | 1 | 0 | Low-quality | Genome-fragment | 3.23 | AAI-based (high-confidence) | 0 |
| S11_extracted_k141_7662 | 4425 | No | NA | 10 | 3 | 0 | Low-quality | Genome-fragment | 11.67 | AAI-based (high-confidence) | 0 |
| S12_vs_k141_154330\|\|full | 7253 | No | NA | 8 | 0 | 0 | Not-determined | Genome-fragment | NA | NA | 0 |
| S12_vs_k141_162547\|\|full | 1118 | No | NA | 2 | 0 | 1 | Not-determined | Genome-fragment | NA | NA | 0 |
| S11_extracted_k141_77795 | 4425 | No | NA | 6 | 0 | 1 | Low-quality | Genome-fragment | 2.88 | AAI-based (medium-confidence) | 0 |
| S1_vs_k141_45023\|\|full | 4426 | No | NA | 5 | 1 | 0 | Low-quality | Genome-fragment | 11 | AAI-based (high-confidence) | 0 |
| S12_vs_k141_92202\|\|full | 1853 | No | NA | 2 | 0 | 2 | Not-determined | Genome-fragment | NA | NA | 0 |
| S5_extracted_k141_127132 | 4426 | No | NA | 11 | 1 | 0 | Low-quality | Genome-fragment | 11.69 | AAI-based (high-confidence) | 0 |
| S12_vs_k141_114961\|\|full | 6291 | No | NA | 7 | 0 | 0 | Not-determined | Genome-fragment | NA | NA | 0 |
| S4_extracted_24680 | 4426 | No | NA | 7 | 0 | 0 | Low-quality | Genome-fragment | 11.26 | AAI-based (high-confidence) | 0 |
| S3_vs_k141_45832\|\|full | 4427 | No | NA | 6 | 0 | 3 | Low-quality | Genome-fragment | 7.54 | AAI-based (medium-confidence) | 0 |
| S12_vs_k141_186610\|\|full | 7526 | No | NA | 13 | 0 | 0 | Not-determined | Genome-fragment | NA | NA | 0 |
| S17_extracted_k141_49816 | 4428 | No | NA | 12 | 1 | 0 | Low-quality | Genome-fragment | 10.26 | AAI-based (high-confidence) | 0 |
| S9_extracted_k141_21796 | 4428 | No | NA | 1 | 0 | 1 | Low-quality | Genome-fragment | 4.01 | AAI-based (high-confidence) | 0 |
| S12_vs_k141_28713\|\|full | 1298 | No | NA | 2 | 0 | 1 | Not-determined | Genome-fragment | NA | NA | 0 |
| S12_vs_k141_186621\|\|full | 1722 | No | NA | 3 | 0 | 1 | Not-determined | Genome-fragment | NA | NA | 0 |
| S11_vs_k141_18219\|\|full | 4428 | No | NA | 9 | 4 | 0 | Low-quality | Genome-fragment | 12.27 | AAI-based (high-confidence) | 0 |
| S16_extracted_141677 | 4430 | No | NA | 4 | 0 | 3 | Low-quality | Genome-fragment | 1.74 | AAI-based (medium-confidence) | 0 |
| S12_vs_k141_154382\|\|full | 1062 | No | NA | 3 | 0 | 0 | Not-determined | Genome-fragment | NA | NA | 0 |
| S12_vs_k141_67976\|\|full | 3740 | No | NA | 5 | 0 | 4 | Not-determined | Genome-fragment | NA | NA | 0 |
| S12_vs_k141_100408\|\|full | 1098 | No | NA | 3 | 0 | 0 | Not-determined | Genome-fragment | NA | NA | 0 |
| S10_extracted_49487 | 4430 | No | NA | 3 | 2 | 0 | Low-quality | Genome-fragment | 8.72 | AAI-based (high-confidence) | 0 |
| S12_vs_k141_130677\|\|full | 1640 | No | NA | 2 | 0 | 1 | Not-determined | Genome-fragment | NA | NA | 0 |
| S12_vs_k141_115009\|\|full | 2111 | No | NA | 3 | 0 | 2 | Not-determined | Genome-fragment | NA | NA | 0 |
| S12_vs_k141_186664\|\|full | 3748 | No | NA | 5 | 0 | 3 | Not-determined | Genome-fragment | NA | NA | 0 |
| S12_vs_k141_107844\|\|full | 2282 | No | NA | 4 | 0 | 1 | Not-determined | Genome-fragment | NA | NA | 0 |
| S17_vs_k141_123814\|\|full | 4431 | No | NA | 8 | 5 | 0 | Low-quality | Genome-fragment | 10.98 | AAI-based (high-confidence) | 0 |
| S12_vs_k141_68002\|\|full | 3430 | No | NA | 5 | 0 | 1 | Not-determined | Genome-fragment | NA | NA | 0 |
| S12_vs_k141_36111\|\|full | 2346 | No | NA | 3 | 0 | 3 | Not-determined | Genome-fragment | NA | NA | 0 |
| S12_vs_k141_146945\|\|full | 2191 | No | NA | 3 | 0 | 1 | Not-determined | Genome-fragment | NA | NA | 0 |
| S12_vs_k141_28796\|\|full | 2022 | No | NA | 2 | 0 | 1 | Not-determined | Genome-fragment | NA | NA | 0 |
| S14_vs_k141_78337\|\|full | 4432 | No | NA | 7 | 1 | 0 | Low-quality | Genome-fragment | 10.66 | AAI-based (medium-confidence) | 0 |
| S5_extracted_k141_32430 | 4432 | No | NA | 2 | 0 | 1 | Low-quality | Genome-fragment | 4.09 | AAI-based (medium-confidence) | 0 |
| S7_vs_82576\|\|full | 4432 | No | NA | 9 | 3 | 0 | Low-quality | Genome-fragment | 2.64 | AAI-based (high-confidence) | 0 |
| S6_extracted_k141_824 | 4432 | No | NA | 8 | 3 | 0 | Low-quality | Genome-fragment | 9.87 | AAI-based (high-confidence) | 0 |
| S12_vs_k141_122675\|\|full | 3058 | No | NA | 4 | 0 | 2 | Not-determined | Genome-fragment | NA | NA | 0 |
| S17_extracted_k141_32916 | 4434 | No | NA | 4 | 3 | 0 | Low-quality | Genome-fragment | 11.86 | AAI-based (high-confidence) | 0 |
| S11_vs_k141_37026\|\|full | 4434 | No | NA | 3 | 0 | 1 | Low-quality | Genome-fragment | 5.19 | AAI-based (medium-confidence) | 0 |
| S12_vs_k141_52078\|\|full | 9064 | No | NA | 7 | 0 | 6 | Not-determined | Genome-fragment | NA | NA | 0 |
| S11_vs_k141_67951\|\|full | 4435 | No | NA | 11 | 0 | 0 | Low-quality | Genome-fragment | 6.94 | AAI-based (high-confidence) | 0 |
| S12_vs_k141_107939\|\|full | 1295 | No | NA | 3 | 0 | 1 | Not-determined | Genome-fragment | NA | NA | 0 |
| S1_vs_k141_84946\|\|full | 4435 | No | NA | 6 | 0 | 4 | Low-quality | Genome-fragment | 2.06 | AAI-based (medium-confidence) | 0 |
| S12_vs_k141_122706\|\|full | 5997 | No | NA | 7 | 0 | 3 | Not-determined | Genome-fragment | NA | NA | 0 |
| S12_vs_k141_186769\|\|full | 2586 | No | NA | 4 | 0 | 1 | Not-determined | Genome-fragment | NA | NA | 0 |
| S3_extracted_k141_81110 | 4435 | No | NA | 8 | 1 | 0 | Low-quality | Genome-fragment | 10.12 | AAI-based (high-confidence) | 0 |
| S12_vs_k141_92330\|\|full | 1384 | No | NA | 3 | 0 | 1 | Not-determined | Genome-fragment | NA | NA | 0 |
| S12_vs_k141_12199\|\|full | 4053 | No | NA | 4 | 0 | 0 | Not-determined | Genome-fragment | NA | NA | 0 |
| S7_vs_131753\|\|full | 4436 | No | NA | 7 | 0 | 0 | Low-quality | Genome-fragment | 10.3 | AAI-based (high-confidence) | 0 |
| S12_vs_k141_36122\|\|full | 9960 | No | NA | 7 | 0 | 2 | Not-determined | Genome-fragment | NA | NA | 0 |
| S12_vs_k141_20746\|\|full | 2510 | No | NA | 4 | 0 | 2 | Not-determined | Genome-fragment | NA | NA | 0 |
| S12_extracted_k141_138320 | 4436 | No | NA | 5 | 2 | 0 | Low-quality | Genome-fragment | 9.41 | AAI-based (high-confidence) | 0 |
| S14_extracted_k141_55120 | 4437 | No | NA | 5 | 2 | 0 | Low-quality | Genome-fragment | 13.47 | AAI-based (medium-confidence) | 0 |
| S10_vs_53581\|\|full | 4437 | No | NA | 5 | 0 | 3 | Low-quality | Genome-fragment | 3.68 | AAI-based (medium-confidence) | 0 |
| S12_vs_k141_75886\|\|full | 2742 | No | NA | 3 | 0 | 1 | Not-determined | Genome-fragment | NA | NA | 0 |
| S11_extracted_k141_92270 | 4438 | No | NA | 7 | 0 | 0 | Low-quality | Genome-fragment | 8.96 | AAI-based (medium-confidence) | 0 |
| S12_vs_k141_68114\|\|full | 6178 | No | NA | 7 | 0 | 2 | Not-determined | Genome-fragment | NA | NA | 0 |
| S12_vs_k141_5688\|\|full | 1668 | No | NA | 3 | 0 | 1 | Not-determined | Genome-fragment | NA | NA | 0 |
| S12_vs_k141_5689\|\|full | 1272 | No | NA | 3 | 0 | 1 | Not-determined | Genome-fragment | NA | NA | 0 |
| S3_vs_k141_119155\|\|full | 4438 | No | NA | 5 | 2 | 0 | Low-quality | Genome-fragment | 8.5 | HMM-based (lower-bound) | 0 |
| S12_vs_k141_52191\|\|full | 1257 | No | NA | 3 | 0 | 1 | Not-determined | Genome-fragment | NA | NA | 0 |
| S12_vs_k141_92380\|\|full | 2842 | No | NA | 3 | 0 | 0 | Not-determined | Genome-fragment | NA | NA | 0 |
| S17_vs_k141_10278\|\|full | 4441 | No | NA | 7 | 1 | 0 | Low-quality | Genome-fragment | 5.59 | AAI-based (medium-confidence) | 0 |
| S12_vs_k141_52193\|\|full | 1733 | No | NA | 3 | 0 | 0 | Not-determined | Genome-fragment | NA | NA | 0 |
| S10_extracted_84851 | 4441 | No | NA | 3 | 0 | 2 | Low-quality | Genome-fragment | 4.41 | AAI-based (high-confidence) | 0 |
| S10_extracted_52188 | 4442 | No | NA | 4 | 0 | 3 | Low-quality | Genome-fragment | 2.15 | AAI-based (medium-confidence) | 0 |
| S11_extracted_k141_73798 | 4444 | No | NA | 8 | 2 | 0 | Low-quality | Genome-fragment | 10.91 | AAI-based (high-confidence) | 0 |
| S12_vs_k141_5721\|\|full | 1916 | No | NA | 2 | 0 | 1 | Not-determined | Genome-fragment | NA | NA | 0 |
| S15_extracted_18146 | 4444 | No | NA | 2 | 2 | 0 | Low-quality | Genome-fragment | 2.36 | AAI-based (high-confidence) | 0 |
| S10_vs_15161\|\|full | 4446 | No | NA | 5 | 5 | 0 | Low-quality | Genome-fragment | 11.67 | AAI-based (high-confidence) | 0 |
| S6_extracted_k141_46369 | 4446 | No | NA | 9 | 3 | 1 | Low-quality | Genome-fragment | 7.37 | AAI-based (high-confidence) | 0 |
| S14_extracted_k141_60279 | 4448 | No | NA | 6 | 5 | 0 | Low-quality | Genome-fragment | 14.69 | AAI-based (high-confidence) | 0 |
| S12_vs_k141_186888\|\|full | 3677 | No | NA | 6 | 0 | 3 | Not-determined | Genome-fragment | NA | NA | 0 |
| S12_vs_k141_177789\|\|full | 1185 | No | NA | 2 | 0 | 1 | Not-determined | Genome-fragment | NA | NA | 0 |
| S5_extracted_k141_113667 | 4449 | No | NA | 5 | 4 | 0 | Low-quality | Genome-fragment | 10.77 | AAI-based (high-confidence) | 0 |
| S11_vs_k141_33063\|\|full | 4450 | No | NA | 4 | 0 | 0 | Low-quality | Genome-fragment | 3.89 | AAI-based (medium-confidence) | 0 |
| S12_vs_k141_100538\|\|full | 1203 | No | NA | 2 | 0 | 0 | Not-determined | Genome-fragment | NA | NA | 0 |
| S5_vs_k141_62608\|\|full | 4450 | No | NA | 4 | 0 | 2 | Low-quality | Genome-fragment | 2.33 | AAI-based (high-confidence) | 0 |
| S12_vs_k141_36252\|\|full | 2845 | No | NA | 3 | 0 | 0 | Not-determined | Genome-fragment | NA | NA | 0 |
| S7_extracted_63415 | 4452 | No | NA | 6 | 1 | 0 | Low-quality | Genome-fragment | 5.39 | AAI-based (medium-confidence) | 0 |
| S12_vs_k141_139432\|\|full | 2193 | No | NA | 3 | 0 | 2 | Not-determined | Genome-fragment | NA | NA | 0 |
| S7_vs_107211\|\|full | 4452 | No | NA | 7 | 0 | 0 | Low-quality | Genome-fragment | 10.33 | AAI-based (high-confidence) | 0 |
| S12_vs_k141_147120\|\|full | 1542 | No | NA | 2 | 0 | 2 | Not-determined | Genome-fragment | NA | NA | 0 |
| S12_vs_k141_100570\|\|full | 4891 | No | NA | 6 | 0 | 4 | Not-determined | Genome-fragment | NA | NA | 0 |
| S10_vs_57643\|\|full | 4452 | Yes | 3576 | 4 | 1 | 1 | Low-quality | Genome-fragment | 2.4 | HMM-based (lower-bound) | 19.68 |
| S12_vs_k141_122829\|\|full | 2145 | No | NA | 5 | 0 | 2 | Not-determined | Genome-fragment | NA | NA | 0 |
| S17_vs_k141_81041\|\|full | 4454 | No | NA | 6 | 0 | 4 | Low-quality | Genome-fragment | 7.94 | AAI-based (medium-confidence) | 0 |
| S5_vs_k141_25730\|\|full | 4454 | No | NA | 5 | 4 | 0 | Low-quality | Genome-fragment | 9.26 | AAI-based (high-confidence) | 0 |
| S12_vs_k141_60443\|\|full | 1937 | No | NA | 3 | 0 | 1 | Not-determined | Genome-fragment | NA | NA | 0 |
| S12_vs_k141_76045\|\|full | 2404 | No | NA | 5 | 0 | 2 | Not-determined | Genome-fragment | NA | NA | 0 |
| S12_vs_k141_108125\|\|full | 3070 | No | NA | 4 | 0 | 1 | Not-determined | Genome-fragment | NA | NA | 0 |
| S12_vs_k141_68257\|\|full | 2201 | No | NA | 3 | 0 | 2 | Not-determined | Genome-fragment | NA | NA | 0 |
| S12_vs_k141_36268\|\|full | 3515 | No | NA | 6 | 0 | 2 | Not-determined | Genome-fragment | NA | NA | 0 |
| S10_vs_85791\|\|full | 4454 | No | NA | 7 | 0 | 2 | Low-quality | Genome-fragment | 3.97 | AAI-based (high-confidence) | 0 |
| S8_extracted_k141_103518 | 4455 | No | NA | 9 | 1 | 0 | Low-quality | Genome-fragment | 8.06 | AAI-based (medium-confidence) | 0 |
| S17_vs_k141_102902\|\|full | 4458 | No | NA | 3 | 0 | 1 | Low-quality | Genome-fragment | 1.54 | AAI-based (medium-confidence) | 0 |
| S17_vs_k141_70561\|\|full | 4459 | Yes | 2133 | 10 | 1 | 1 | Low-quality | Genome-fragment | 3.21 | AAI-based (high-confidence) | 52.16 |
| S11_vs_k141_38847\|\|full | 4459 | No | NA | 3 | 0 | 3 | Low-quality | Genome-fragment | 2.4 | AAI-based (medium-confidence) | 0 |
| S16_vs_90373\|\|full | 4459 | Yes | 789 | 4 | 1 | 1 | Low-quality | Genome-fragment | 1.58 | HMM-based (lower-bound) | 82.31 |
| S12_vs_k141_130977\|\|full | 1962 | No | NA | 3 | 0 | 2 | Not-determined | Genome-fragment | NA | NA | 0 |
| S12_vs_k141_5306\|\|full | 4460 | No | NA | 6 | 3 | 0 | Low-quality | Genome-fragment | 11.22 | AAI-based (medium-confidence) | 0 |
| S12_vs_k141_130988\|\|full | 4129 | No | NA | 6 | 0 | 3 | Not-determined | Genome-fragment | NA | NA | 0 |
| S7_vs_24923\|\|full | 4461 | No | NA | 8 | 1 | 0 | Low-quality | Genome-fragment | 7.65 | AAI-based (medium-confidence) | 0 |
| S12_vs_k141_20963\|\|full | 2185 | No | NA | 5 | 0 | 0 | Not-determined | Genome-fragment | NA | NA | 0 |
| S12_vs_k141_187025\|\|full | 1132 | No | NA | 2 | 0 | 0 | Not-determined | Genome-fragment | NA | NA | 0 |
| S14_extracted_k141_70279 | 4462 | No | NA | 6 | 4 | 0 | Low-quality | Genome-fragment | 11.66 | AAI-based (high-confidence) | 0 |
| S12_vs_k141_29036\|\|full | 3806 | No | NA | 3 | 0 | 3 | Not-determined | Genome-fragment | NA | NA | 0 |
| S3_vs_k141_80441\|\|full | 4463 | No | NA | 5 | 3 | 0 | Low-quality | Genome-fragment | 11.17 | AAI-based (high-confidence) | 0 |
| S12_vs_k141_100677\|\|full | 5170 | No | NA | 8 | 0 | 8 | Not-determined | Genome-fragment | NA | NA | 0 |
| S3_vs_k141_114435\|\|full | 4463 | No | NA | 3 | 0 | 2 | Low-quality | Genome-fragment | 4.47 | AAI-based (medium-confidence) | 0 |
| S8_vs_k141_130243\|\|full | 4463 | No | NA | 4 | 0 | 2 | Low-quality | Genome-fragment | 2.34 | AAI-based (medium-confidence) | 0 |
| S12_vs_k141_68305\|\|full | 2606 | No | NA | 3 | 0 | 2 | Not-determined | Genome-fragment | NA | NA | 0 |
| S12_vs_k141_131038\|\|full | 1113 | No | NA | 2 | 0 | 1 | Not-determined | Genome-fragment | NA | NA | 0 |
| S17_extracted_k141_117245 | 4464 | No | NA | 13 | 1 | 0 | Low-quality | Genome-fragment | 6.33 | AAI-based (high-confidence) | 0 |
| S12_vs_k141_5857\|\|full | 37204 | No | NA | 53 | 14 | 0 | Medium-quality | Genome-fragment | 85.69 | AAI-based (medium-confidence) | 0 |
| S3_vs_k141_67722\|\|full | 4464 | No | NA | 8 | 1 | 0 | Low-quality | Genome-fragment | 2.87 | HMM-based (lower-bound) | 0 |
| S5_extracted_k141_103542 | 4464 | No | NA | 4 | 0 | 0 | Low-quality | Genome-fragment | 2.62 | AAI-based (high-confidence) | 0 |
| S12_vs_k141_20998\|\|full | 2068 | No | NA | 2 | 0 | 2 | Not-determined | Genome-fragment | NA | NA | 0 |
| S12_vs_k141_100709\|\|full | 4054 | No | NA | 5 | 0 | 2 | Not-determined | Genome-fragment | NA | NA | 0 |
| S12_vs_k141_52473\|\|full | 4879 | No | NA | 4 | 0 | 2 | Not-determined | Genome-fragment | NA | NA | 0 |
| S12_vs_k141_92553\|\|full | 1563 | No | NA | 3 | 0 | 0 | Not-determined | Genome-fragment | NA | NA | 0 |
| S16_extracted_118610 | 4464 | No | NA | 5 | 0 | 0 | Low-quality | Genome-fragment | 9.91 | AAI-based (high-confidence) | 0 |
| S7_vs_61501\|\|full | 4465 | No | NA | 5 | 3 | 0 | Low-quality | Genome-fragment | 11.4 | AAI-based (medium-confidence) | 0 |
| S12_vs_k141_52479\|\|full | 2546 | No | NA | 3 | 0 | 3 | Not-determined | Genome-fragment | NA | NA | 0 |
| S12_vs_k141_11461\|\|full | 4465 | No | NA | 6 | 0 | 0 | Low-quality | Genome-fragment | 7.62 | AAI-based (high-confidence) | 0 |
| S12_vs_k141_76148\|\|full | 2973 | No | NA | 5 | 0 | 3 | Not-determined | Genome-fragment | NA | NA | 0 |
| S12_vs_k141_147241\|\|full | 1824 | No | NA | 3 | 0 | 1 | Not-determined | Genome-fragment | NA | NA | 0 |
| S12_vs_k141_5895\|\|full | 2080 | No | NA | 4 | 0 | 0 | Not-determined | Genome-fragment | NA | NA | 0 |
| S11_vs_k141_20499\|\|full | 4467 | No | NA | 4 | 1 | 0 | Low-quality | Genome-fragment | 13.93 | AAI-based (medium-confidence) | 0 |
| S12_vs_k141_92563\|\|full | 2987 | No | NA | 4 | 0 | 1 | Not-determined | Genome-fragment | NA | NA | 0 |
| S12_vs_k141_131083\|\|full | 1528 | No | NA | 3 | 0 | 1 | Not-determined | Genome-fragment | NA | NA | 0 |
| S12_vs_k141_122992\|\|full | 3211 | No | NA | 3 | 0 | 3 | Not-determined | Genome-fragment | NA | NA | 0 |
| S8_vs_k141_163225\|\|full | 4467 | No | NA | 6 | 4 | 0 | Low-quality | Genome-fragment | 10.96 | AAI-based (high-confidence) | 0 |
| S14_extracted_k141_24010 | 4470 | No | NA | 10 | 9 | 0 | Low-quality | Genome-fragment | 6.22 | AAI-based (high-confidence) | 0 |
| S8_extracted_k141_11974 | 4470 | No | NA | 4 | 0 | 4 | Low-quality | Genome-fragment | 1.55 | AAI-based (medium-confidence) | 0 |
| S12_vs_k141_162744\|\|full | 9024 | No | NA | 7 | 0 | 4 | Not-determined | Genome-fragment | NA | NA | 0 |
| S12_vs_k141_147256\|\|full | 1440 | No | NA | 3 | 0 | 1 | Not-determined | Genome-fragment | NA | NA | 0 |
| S3_extracted_k141_113302 | 4471 | No | NA | 9 | 3 | 0 | Low-quality | Genome-fragment | 9.5 | AAI-based (high-confidence) | 0 |
| S12_vs_k141_76209\|\|full | 2054 | No | NA | 5 | 0 | 1 | Not-determined | Genome-fragment | NA | NA | 0 |
| S12_vs_k141_5954\|\|full | 3642 | No | NA | 4 | 0 | 2 | Not-determined | Genome-fragment | NA | NA | 0 |
| S5_extracted_k141_111316 | 4471 | No | NA | 6 | 3 | 0 | Low-quality | Genome-fragment | 12.32 | AAI-based (high-confidence) | 0 |
| S16_extracted_146005 | 4471 | No | NA | 10 | 0 | 1 | Low-quality | Genome-fragment | 7.57 | AAI-based (medium-confidence) | 0 |
| S12_vs_k141_108264\|\|full | 6167 | No | NA | 6 | 0 | 3 | Not-determined | Genome-fragment | NA | NA | 0 |
| S12_vs_k141_154885\|\|full | 7317 | No | NA | 9 | 0 | 4 | Not-determined | Genome-fragment | NA | NA | 0 |
| S13_extracted_18624 | 4472 | No | NA | 7 | 0 | 0 | Low-quality | Genome-fragment | 7.81 | AAI-based (high-confidence) | 0 |
| S12_vs_k141_83887\|\|full | 2359 | No | NA | 3 | 0 | 0 | Not-determined | Genome-fragment | NA | NA | 0 |
| S12_extracted_k141_117806 | 4472 | No | NA | 7 | 1 | 0 | Low-quality | Genome-fragment | 9.85 | AAI-based (medium-confidence) | 0 |
| S8_vs_k141_148098\|\|full | 4473 | No | NA | 8 | 6 | 0 | Low-quality | Genome-fragment | 9.53 | AAI-based (medium-confidence) | 0 |
| S12_vs_k141_115418\|\|full | 3997 | No | NA | 7 | 0 | 0 | Not-determined | Genome-fragment | NA | NA | 0 |
| S12_vs_k141_108308\|\|full | 4258 | No | NA | 5 | 0 | 1 | Not-determined | Genome-fragment | NA | NA | 0 |
| S9_vs_k141_60000\|\|full | 4474 | No | NA | 8 | 0 | 0 | Low-quality | Genome-fragment | 3.24 | AAI-based (high-confidence) | 0 |
| S12_vs_k141_162817\|\|full | 2912 | No | NA | 3 | 0 | 0 | Not-determined | Genome-fragment | NA | NA | 0 |
| S13_vs_56727\|\|full | 4474 | No | NA | 5 | 1 | 0 | Low-quality | Genome-fragment | 6.77 | HMM-based (lower-bound) | 0 |
| S12_vs_k141_60689\|\|full | 7218 | No | NA | 6 | 0 | 3 | Not-determined | Genome-fragment | NA | NA | 0 |
| S12_vs_k141_100876\|\|full | 1464 | No | NA | 3 | 0 | 0 | Not-determined | Genome-fragment | NA | NA | 0 |
| S6_extracted_k141_53811 | 4475 | No | NA | 5 | 4 | 0 | Low-quality | Genome-fragment | 11.12 | AAI-based (high-confidence) | 0 |
| S12_vs_k141_139754\|\|full | 2960 | No | NA | 3 | 0 | 2 | Not-determined | Genome-fragment | NA | NA | 0 |
| S1_vs_k141_19055\|\|full | 4476 | No | NA | 3 | 0 | 3 | Low-quality | Genome-fragment | 2.59 | AAI-based (medium-confidence) | 0 |
| S12_vs_k141_170669\|\|full | 1931 | No | NA | 3 | 0 | 1 | Not-determined | Genome-fragment | NA | NA | 0 |
| S12_vs_k141_21148\|\|full | 2121 | No | NA | 5 | 0 | 2 | Not-determined | Genome-fragment | NA | NA | 0 |
| S12_vs_k141_91329\|\|full | 4476 | No | NA | 6 | 1 | 0 | Low-quality | Genome-fragment | 10.45 | AAI-based (high-confidence) | 0 |
| S11_vs_k141_65787\|\|full | 4478 | No | NA | 6 | 3 | 0 | Low-quality | Genome-fragment | 11.16 | AAI-based (high-confidence) | 0 |
| S10_vs_119308\|\|full | 4478 | No | NA | 4 | 0 | 3 | Low-quality | Genome-fragment | 0.78 | AAI-based (high-confidence) | 0 |
| S3_vs_k141_47400\|\|full | 4481 | No | NA | 4 | 0 | 4 | Low-quality | Genome-fragment | 3.59 | AAI-based (high-confidence) | 0 |
| S16_extracted_8663 | 4481 | No | NA | 6 | 1 | 0 | Low-quality | Genome-fragment | 14.13 | AAI-based (high-confidence) | 0 |
| S16_vs_96906\|\|full | 4481 | No | NA | 7 | 0 | 1 | Low-quality | Genome-fragment | 6.89 | AAI-based (high-confidence) | 0 |
| S12_vs_k141_92743\|\|full | 4693 | No | NA | 9 | 0 | 3 | Not-determined | Genome-fragment | NA | NA | 0 |
| S8_vs_k141_134529\|\|full | 4481 | No | NA | 9 | 1 | 0 | Low-quality | Genome-fragment | 8.2 | AAI-based (high-confidence) | 0 |
| S14_vs_k141_89021\|\|full | 4484 | No | NA | 9 | 4 | 0 | Low-quality | Genome-fragment | 11.87 | AAI-based (medium-confidence) | 0 |
| S16_extracted_110440 | 4484 | No | NA | 7 | 3 | 0 | Low-quality | Genome-fragment | 10.05 | AAI-based (high-confidence) | 0 |
| S13_vs_62053\|\|full | 4487 | No | NA | 6 | 1 | 0 | Low-quality | Genome-fragment | 8.99 | HMM-based (lower-bound) | 0 |
| S17_extracted_k141_20219 | 4488 | No | NA | 3 | 0 | 2 | Low-quality | Genome-fragment | 0.39 | AAI-based (medium-confidence) | 0 |
| S12_vs_k141_154981\|\|full | 4332 | No | NA | 5 | 0 | 0 | Not-determined | Genome-fragment | NA | NA | 0 |
| S16_vs_134077\|\|full | 4488 | No | NA | 11 | 1 | 0 | Low-quality | Genome-fragment | 3.02 | HMM-based (lower-bound) | 0 |
| S12_extracted_k141_129592 | 4488 | No | NA | 5 | 1 | 0 | Low-quality | Genome-fragment | 4.31 | AAI-based (high-confidence) | 0 |
| S12_vs_k141_131212\|\|full | 1269 | No | NA | 3 | 0 | 0 | Not-determined | Genome-fragment | NA | NA | 0 |
| S12_vs_k141_137639\|\|full | 4488 | No | NA | 10 | 1 | 0 | Low-quality | Genome-fragment | 2.25 | HMM-based (lower-bound) | 0 |
| S14_vs_k141_87715\|\|full | 4489 | No | NA | 8 | 0 | 1 | Low-quality | Genome-fragment | 10.43 | AAI-based (medium-confidence) | 0 |
| S12_vs_k141_84012\|\|full | 1208 | No | NA | 2 | 0 | 0 | Not-determined | Genome-fragment | NA | NA | 0 |
| S12_vs_k141_162905\|\|full | 17615 | No | NA | 16 | 0 | 3 | Not-determined | Genome-fragment | NA | NA | 0 |
| S5_vs_k141_118716\|\|full | 4489 | No | NA | 5 | 5 | 0 | Low-quality | Genome-fragment | 9.38 | AAI-based (medium-confidence) | 0 |
| S12_vs_k141_108460\|\|full | 3239 | No | NA | 5 | 0 | 3 | Not-determined | Genome-fragment | NA | NA | 0 |
| S7_extracted_124441 | 4490 | No | NA | 5 | 0 | 1 | Low-quality | Genome-fragment | 2.82 | AAI-based (medium-confidence) | 0 |
| S12_vs_k141_84045\|\|full | 5070 | No | NA | 5 | 0 | 3 | Not-determined | Genome-fragment | NA | NA | 0 |
| S11_vs_k141_70085\|\|full | 4491 | No | NA | 5 | 0 | 0 | Low-quality | Genome-fragment | 3.19 | AAI-based (medium-confidence) | 0 |
| S12_vs_k141_6103\|\|full | 1100 | No | NA | 4 | 0 | 3 | Not-determined | Genome-fragment | NA | NA | 0 |
| S6_extracted_k141_20825 | 4492 | No | NA | 8 | 2 | 0 | Low-quality | Genome-fragment | 12.66 | AAI-based (high-confidence) | 0 |
| S12_vs_k141_139884\|\|full | 3488 | No | NA | 3 | 0 | 2 | Not-determined | Genome-fragment | NA | NA | 0 |
| S12_vs_k141_139886\|\|full | 5211 | No | NA | 5 | 0 | 1 | Not-determined | Genome-fragment | NA | NA | 0 |
| S12_vs_k141_21292\|\|full | 1786 | No | NA | 2 | 0 | 1 | Not-determined | Genome-fragment | NA | NA | 0 |
| S8_extracted_k141_44527 | 4492 | No | NA | 5 | 0 | 5 | Low-quality | Genome-fragment | 14.35 | AAI-based (high-confidence) | 0 |
| S5_vs_k141_14519\|\|full | 4493 | No | NA | 4 | 0 | 2 | Low-quality | Genome-fragment | 2.23 | AAI-based (medium-confidence) | 0 |
| S16_vs_139807\|\|full | 4493 | No | NA | 5 | 0 | 3 | Low-quality | Genome-fragment | 4.43 | AAI-based (medium-confidence) | 0 |
| S16_extracted_24834 | 4494 | No | NA | 5 | 0 | 1 | Low-quality | Genome-fragment | 6.73 | AAI-based (medium-confidence) | 0 |
| S12_vs_k141_187304\|\|full | 2320 | No | NA | 3 | 0 | 1 | Not-determined | Genome-fragment | NA | NA | 0 |
| S8_extracted_k141_19748 | 4494 | No | NA | 4 | 0 | 2 | Low-quality | Genome-fragment | 2.42 | AAI-based (medium-confidence) | 0 |
| S11_vs_k141_47395\|\|full | 4496 | No | NA | 5 | 0 | 1 | Low-quality | Genome-fragment | 11.32 | AAI-based (high-confidence) | 0 |
| S16_extracted_117750 | 4496 | No | NA | 4 | 0 | 0 | Low-quality | Genome-fragment | 7.5 | AAI-based (medium-confidence) | 0 |
| S6_vs_k141_2755\|\|full | 4496 | No | NA | 8 | 0 | 0 | Low-quality | Genome-fragment | 8.15 | AAI-based (high-confidence) | 0 |
| S12_vs_k141_187315\|\|full | 7281 | No | NA | 9 | 0 | 5 | Not-determined | Genome-fragment | NA | NA | 0 |
| S3_vs_k141_49499\|\|full | 4498 | No | NA | 8 | 0 | 0 | Low-quality | Genome-fragment | 11.39 | AAI-based (medium-confidence) | 0 |
| S12_vs_k141_170850\|\|full | 24495 | No | NA | 29 | 0 | 15 | Not-determined | Genome-fragment | NA | NA | 0 |
| S12_vs_k141_147547\|\|full | 1768 | No | NA | 3 | 0 | 0 | Not-determined | Genome-fragment | NA | NA | 0 |
| S12_vs_k141_45830\|\|full | 2008 | No | NA | 3 | 0 | 0 | Not-determined | Genome-fragment | NA | NA | 0 |
| S9_extracted_k141_39888 | 4500 | No | NA | 3 | 0 | 3 | Low-quality | Genome-fragment | 2.26 | AAI-based (medium-confidence) | 0 |
| S12_vs_k141_147549\|\|full | 5080 | No | NA | 6 | 0 | 0 | Not-determined | Genome-fragment | NA | NA | 0 |
| S12_vs_k141_68641\|\|full | 2634 | No | NA | 4 | 0 | 3 | Not-determined | Genome-fragment | NA | NA | 0 |
| S12_vs_k141_131296\|\|full | 3596 | No | NA | 8 | 0 | 2 | Not-determined | Genome-fragment | NA | NA | 0 |
| S3_extracted_k141_33909 | 4501 | No | NA | 5 | 4 | 0 | Low-quality | Genome-fragment | 11.96 | AAI-based (high-confidence) | 0 |
| S13_extracted_43532 | 4502 | No | NA | 6 | 0 | 2 | Low-quality | Genome-fragment | 3.27 | AAI-based (medium-confidence) | 0 |
| S12_vs_k141_60925\|\|full | 1385 | No | NA | 4 | 0 | 1 | Not-determined | Genome-fragment | NA | NA | 0 |
| S12_vs_k141_131326\|\|full | 1319 | No | NA | 3 | 0 | 0 | Not-determined | Genome-fragment | NA | NA | 0 |
| S12_vs_k141_108514\|\|full | 5024 | No | NA | 3 | 0 | 3 | Not-determined | Genome-fragment | NA | NA | 0 |
| S12_vs_k141_29277\|\|full | 3015 | No | NA | 6 | 0 | 2 | Not-determined | Genome-fragment | NA | NA | 0 |
| S12_vs_k141_52648\|\|full | 1770 | No | NA | 2 | 0 | 2 | Not-determined | Genome-fragment | NA | NA | 0 |
| S7_vs_3094\|\|full | 4503 | No | NA | 12 | 0 | 0 | Low-quality | Genome-fragment | 10.03 | AAI-based (high-confidence) | 0 |
| S16_vs_130129\|\|full | 4503 | No | NA | 4 | 0 | 2 | Low-quality | Genome-fragment | 4.74 | AAI-based (medium-confidence) | 0 |
| S12_vs_k141_29295\|\|full | 13014 | No | NA | 12 | 0 | 4 | Not-determined | Genome-fragment | NA | NA | 0 |
| S12_vs_k141_178445\|\|full | 1209 | No | NA | 2 | 0 | 1 | Not-determined | Genome-fragment | NA | NA | 0 |
| S12_vs_k141_123408\|\|full | 1595 | No | NA | 2 | 0 | 1 | Not-determined | Genome-fragment | NA | NA | 0 |
| S12_vs_k141_108568\|\|full | 2628 | No | NA | 3 | 0 | 0 | Not-determined | Genome-fragment | NA | NA | 0 |
| S8_vs_k141_2931\|\|full | 4503 | No | NA | 5 | 0 | 2 | Low-quality | Genome-fragment | 4.24 | AAI-based (high-confidence) | 0 |
| S12_vs_k141_92868\|\|full | 85010 | No | NA | 128 | 40 | 1 | High-quality | High-quality | 100 | AAI-based (high-confidence) | 0 |
| S5_vs_k141_49083\|\|full | 4505 | No | NA | 5 | 0 | 1 | Low-quality | Genome-fragment | 5.5 | AAI-based (medium-confidence) | 0 |
| S6_vs_k141_11979\|\|full | 4506 | No | NA | 3 | 0 | 1 | Low-quality | Genome-fragment | 6.85 | AAI-based (medium-confidence) | 0 |
| S12_vs_k141_21408\|\|full | 1157 | No | NA | 2 | 0 | 1 | Not-determined | Genome-fragment | NA | NA | 0 |
| S12_vs_k141_52723\|\|full | 11783 | No | NA | 12 | 0 | 8 | Not-determined | Genome-fragment | NA | NA | 0 |
| S12_vs_k141_147635\|\|full | 1513 | No | NA | 3 | 0 | 0 | Not-determined | Genome-fragment | NA | NA | 0 |
| S12_vs_k141_131389\|\|full | 3716 | No | NA | 2 | 0 | 2 | Not-determined | Genome-fragment | NA | NA | 0 |
| S12_vs_k141_147636\|\|full | 1787 | No | NA | 3 | 0 | 1 | Not-determined | Genome-fragment | NA | NA | 0 |
| S12_vs_k141_84233\|\|full | 4084 | No | NA | 3 | 0 | 1 | Not-determined | Genome-fragment | NA | NA | 0 |
| S12_vs_k141_21425\|\|full | 1930 | No | NA | 2 | 0 | 2 | Not-determined | Genome-fragment | NA | NA | 0 |
| S7_vs_110505\|\|full | 4507 | No | NA | 4 | 0 | 1 | Low-quality | Genome-fragment | 2.26 | AAI-based (medium-confidence) | 0 |
| S12_vs_k141_108638\|\|full | 1883 | No | NA | 3 | 0 | 0 | Not-determined | Genome-fragment | NA | NA | 0 |
| S17_extracted_k141_77162 | 4508 | No | NA | 7 | 1 | 0 | Low-quality | Genome-fragment | 6.82 | HMM-based (lower-bound) | 0 |
| S17_vs_k141_9952\|\|full | 4508 | No | NA | 7 | 0 | 1 | Low-quality | Genome-fragment | 3.99 | AAI-based (high-confidence) | 0 |
| S12_vs_k141_68758\|\|full | 10045 | No | NA | 9 | 0 | 3 | Not-determined | Genome-fragment | NA | NA | 0 |
| S12_vs_k141_76514\|\|full | 2873 | No | NA | 4 | 0 | 2 | Not-determined | Genome-fragment | NA | NA | 0 |
| S7_vs_70139\|\|full | 4508 | No | NA | 13 | 2 | 0 | Low-quality | Genome-fragment | 10.2 | AAI-based (high-confidence) | 0 |
| S12_vs_k141_84248\|\|full | 5378 | No | NA | 5 | 0 | 3 | Not-determined | Genome-fragment | NA | NA | 0 |
| S16_extracted_114064 | 4509 | No | NA | 3 | 0 | 2 | Low-quality | Genome-fragment | 1.67 | AAI-based (high-confidence) | 0 |
| S8_extracted_k141_122896 | 4510 | No | NA | 6 | 1 | 2 | Low-quality | Genome-fragment | 5.58 | HMM-based (lower-bound) | 0 |
| S11_vs_k141_42813\|\|full | 4512 | No | NA | 7 | 1 | 0 | Low-quality | Genome-fragment | 12.55 | AAI-based (medium-confidence) | 0 |
| S1_extracted_k141_97989 | 4512 | No | NA | 7 | 1 | 0 | Low-quality | Genome-fragment | 9.87 | AAI-based (high-confidence) | 0 |
| S12_vs_k141_36694\|\|full | 2293 | No | NA | 2 | 0 | 0 | Not-determined | Genome-fragment | NA | NA | 0 |
| S12_vs_k141_123499\|\|full | 11604 | No | NA | 13 | 0 | 3 | Not-determined | Genome-fragment | NA | NA | 0 |
| S12_vs_k141_140147\|\|full | 1689 | No | NA | 4 | 0 | 0 | Not-determined | Genome-fragment | NA | NA | 0 |
| S12_vs_k141_46037\|\|full | 1346 | No | NA | 3 | 0 | 0 | Not-determined | Genome-fragment | NA | NA | 0 |
| S3_extracted_k141_102406 | 4512 | No | NA | 4 | 0 | 0 | Low-quality | Genome-fragment | 3.16 | AAI-based (medium-confidence) | 0 |
| S3_vs_k141_2383\|\|full | 4513 | No | NA | 6 | 1 | 0 | Low-quality | Genome-fragment | 2.95 | AAI-based (high-confidence) | 0 |
| S12_vs_k141_163185\|\|full | 6111 | No | NA | 3 | 0 | 2 | Not-determined | Genome-fragment | NA | NA | 0 |
| S12_vs_k141_178570\|\|full | 2706 | No | NA | 3 | 0 | 2 | Not-determined | Genome-fragment | NA | NA | 0 |
| S16_extracted_90582 | 4514 | No | NA | 7 | 0 | 1 | Low-quality | Genome-fragment | 3.57 | AAI-based (high-confidence) | 0 |
| S12_vs_k141_46058\|\|full | 2247 | No | NA | 2 | 0 | 1 | Not-determined | Genome-fragment | NA | NA | 0 |
| S8_extracted_k141_62951 | 4514 | No | NA | 8 | 5 | 0 | Low-quality | Genome-fragment | 20.59 | AAI-based (high-confidence) | 0 |
| S8_vs_k141_199856\|\|full | 4514 | No | NA | 5 | 0 | 3 | Low-quality | Genome-fragment | 2.07 | AAI-based (medium-confidence) | 0 |
| S12_vs_k141_36755\|\|full | 1583 | No | NA | 2 | 0 | 2 | Not-determined | Genome-fragment | NA | NA | 0 |
| S7_vs_67782\|\|full | 4515 | No | NA | 4 | 3 | 0 | Low-quality | Genome-fragment | 26.43 | AAI-based (high-confidence) | 0 |
| S12_vs_k141_12957\|\|full | 1744 | No | NA | 2 | 0 | 0 | Not-determined | Genome-fragment | NA | NA | 0 |
| S1_extracted_k141_88025 | 4516 | No | NA | 7 | 2 | 0 | Low-quality | Genome-fragment | 16.44 | HMM-based (lower-bound) | 0 |
| S5_vs_k141_99024\|\|full | 4516 | No | NA | 7 | 1 | 0 | Low-quality | Genome-fragment | 9.42 | AAI-based (medium-confidence) | 0 |
| S12_vs_k141_12974\|\|full | 5344 | No | NA | 5 | 0 | 4 | Not-determined | Genome-fragment | NA | NA | 0 |
| S11_vs_k141_47993\|\|full | 4519 | No | NA | 10 | 1 | 0 | Low-quality | Genome-fragment | 7.93 | AAI-based (high-confidence) | 0 |
| S12_vs_k141_21534\|\|full | 4089 | No | NA | 4 | 0 | 1 | Not-determined | Genome-fragment | NA | NA | 0 |
| S12_vs_k141_6353\|\|full | 2482 | No | NA | 3 | 0 | 0 | Not-determined | Genome-fragment | NA | NA | 0 |
| S17_vs_k141_79771\|\|full | 4522 | No | NA | 6 | 1 | 2 | Low-quality | Genome-fragment | 2.22 | HMM-based (lower-bound) | 0 |
| S12_vs_k141_6357\|\|full | 1329 | No | NA | 2 | 0 | 0 | Not-determined | Genome-fragment | NA | NA | 0 |
| S11_vs_k141_14404\|\|full | 4523 | No | NA | 9 | 2 | 0 | Low-quality | Genome-fragment | 8.73 | HMM-based (lower-bound) | 0 |
| S12_vs_k141_76639\|\|full | 1659 | No | NA | 4 | 0 | 0 | Not-determined | Genome-fragment | NA | NA | 0 |
| S12_vs_k141_61142\|\|full | 5296 | No | NA | 6 | 0 | 1 | Not-determined | Genome-fragment | NA | NA | 0 |
| S13_extracted_47971 | 4524 | No | NA | 4 | 3 | 0 | Low-quality | Genome-fragment | 11.2 | HMM-based (lower-bound) | 0 |
| S3_vs_k141_106130\|\|full | 4524 | No | NA | 5 | 0 | 4 | Low-quality | Genome-fragment | 3.04 | AAI-based (medium-confidence) | 0 |
| S12_vs_k141_76648\|\|full | 6476 | No | NA | 6 | 0 | 5 | Not-determined | Genome-fragment | NA | NA | 0 |
| S12_vs_k141_163295\|\|full | 4335 | No | NA | 8 | 0 | 0 | Not-determined | Genome-fragment | NA | NA | 0 |
| S16_vs_93915\|\|full | 4525 | No | NA | 4 | 0 | 3 | Low-quality | Genome-fragment | 2.97 | AAI-based (medium-confidence) | 0 |
| S12_vs_k141_115750\|\|full | 1685 | No | NA | 2 | 0 | 0 | Not-determined | Genome-fragment | NA | NA | 0 |
| S12_vs_k141_92969\|\|full | 3172 | No | NA | 4 | 0 | 3 | Not-determined | Genome-fragment | NA | NA | 0 |
| S12_vs_k141_163319\|\|full | 1738 | No | NA | 2 | 0 | 1 | Not-determined | Genome-fragment | NA | NA | 0 |
| S12_vs_k141_84405\|\|full | 1007 | No | NA | 2 | 0 | 0 | Not-determined | Genome-fragment | NA | NA | 0 |
| S17_vs_k141_40728\|\|full | 4526 | No | NA | 5 | 0 | 0 | Low-quality | Genome-fragment | 2.49 | AAI-based (medium-confidence) | 0 |

| | | | | | | | | | | | |
|---|---|---|---|---|---|---|---|---|---|---|---|
| S12_vs_k141_140341\|\|full | 1504 | No | NA | 2 | 0 | 0 | Not-determined | Genome-fragment | NA | NA | 0 |
| S12_vs_k141_187611\|\|full | 4484 | No | NA | 7 | 0 | 0 | Not-determined | Genome-fragment | NA | NA | 0 |
| S12_vs_k141_115780\|\|full | 1385 | No | NA | 2 | 0 | 0 | Not-determined | Genome-fragment | NA | NA | 0 |
| S12_vs_k141_178696\|\|full | 2212 | No | NA | 3 | 0 | 0 | Not-determined | Genome-fragment | NA | NA | 0 |
| S12_vs_k141_36907\|\|full | 8898 | No | NA | 13 | 0 | 2 | Not-determined | Genome-fragment | NA | NA | 0 |
| S7_vs_9171\|\|full | 4526 | No | NA | 5 | 4 | 0 | Low-quality | Genome-fragment | 11.78 | AAI-based (high-confidence) | 0 |
| S12_vs_k141_187645\|\|full | 1439 | No | NA | 3 | 0 | 2 | Not-determined | Genome-fragment | NA | NA | 0 |
| S17_extracted_k141_76815 | 4528 | Yes | 3237 | 5 | 2 | 1 | Low-quality | Genome-fragment | 5.29 | AAI-based (high-confidence) | 28.51 |
| S17_extracted_k141_144443 | 4528 | No | NA | 6 | 0 | 0 | Low-quality | Genome-fragment | 7.84 | AAI-based (medium-confidence) | 0 |
| S17_vs_k141_29249\|\|full | 4528 | No | NA | 4 | 0 | 3 | Low-quality | Genome-fragment | 0.77 | AAI-based (high-confidence) | 0 |
| S12_vs_k141_21575\|\|full | 1312 | No | NA | 3 | 0 | 0 | Not-determined | Genome-fragment | NA | NA | 0 |
| S14_extracted_k141_66022 | 4528 | No | NA | 12 | 9 | 0 | Low-quality | Genome-fragment | 14.9 | AAI-based (high-confidence) | 0 |
| S12_vs_k141_76741\|\|full | 3490 | No | NA | 4 | 0 | 2 | Not-determined | Genome-fragment | NA | NA | 0 |
| S12_vs_k141_93050\|\|full | 1485 | No | NA | 2 | 0 | 0 | Not-determined | Genome-fragment | NA | NA | 0 |
| S3_vs_k141_20871\|\|full | 4529 | No | NA | 6 | 0 | 1 | Low-quality | Genome-fragment | 5.74 | AAI-based (high-confidence) | 0 |
| S12_vs_k141_171179\|\|full | 1756 | No | NA | 3 | 0 | 2 | Not-determined | Genome-fragment | NA | NA | 0 |
| S12_vs_k141_13170\|\|full | 2056 | No | NA | 3 | 0 | 1 | Not-determined | Genome-fragment | NA | NA | 0 |
| S16_extracted_141648 | 4529 | No | NA | 5 | 0 | 0 | Low-quality | Genome-fragment | 3.67 | AAI-based (high-confidence) | 0 |
| S12_vs_k141_84472\|\|full | 8000 | No | NA | 6 | 0 | 2 | Not-determined | Genome-fragment | NA | NA | 0 |
| S12_vs_k141_187703\|\|full | 3198 | No | NA | 4 | 0 | 2 | Not-determined | Genome-fragment | NA | NA | 0 |
| S9_extracted_k141_1924 | 4530 | No | NA | 4 | 4 | 0 | Low-quality | Genome-fragment | 14.92 | AAI-based (high-confidence) | 0 |
| S12_vs_k141_101547\|\|full | 2831 | No | NA | 3 | 0 | 1 | Not-determined | Genome-fragment | NA | NA | 0 |
| S12_vs_k141_69021\|\|full | 7168 | No | NA | 5 | 0 | 1 | Not-determined | Genome-fragment | NA | NA | 0 |
| S5_vs_k141_21211\|\|full | 4530 | No | NA | 11 | 3 | 0 | Low-quality | Genome-fragment | 11.2 | AAI-based (high-confidence) | 0 |
| S7_extracted_11517 | 4530 | No | NA | 9 | 1 | 0 | Low-quality | Genome-fragment | 3.14 | AAI-based (high-confidence) | 0 |
| S12_vs_k141_61278\|\|full | 1103 | No | NA | 4 | 0 | 0 | Not-determined | Genome-fragment | NA | NA | 0 |
| S8_vs_k141_98681\|\|full | 4530 | Yes | 1485 | 7 | 1 | 1 | Low-quality | Genome-fragment | 3.32 | AAI-based (medium-confidence) | 67.22 |
| S12_vs_k141_123620\|\|full | 115852 | Yes | 32587 | 145 | 13 | 45 | Complete | High-quality | 100 | Provirus (medium-confidence) | 71.87 |
| S16_extracted_6327 | 4533 | No | NA | 9 | 0 | 9 | Low-quality | Genome-fragment | 2.99 | AAI-based (high-confidence) | 0 |
| S16_extracted_94896 | 4533 | No | NA | 5 | 0 | 0 | Low-quality | Genome-fragment | 1.13 | AAI-based (high-confidence) | 0 |
| S5_vs_k141_51016\|\|full | 4536 | No | NA | 3 | 0 | 3 | Low-quality | Genome-fragment | 1.94 | AAI-based (medium-confidence) | 0 |
| S12_vs_k141_29667\|\|full | 2071 | No | NA | 3 | 0 | 1 | Not-determined | Genome-fragment | NA | NA | 0 |
| S12_vs_k141_37046\|\|full | 5370 | No | NA | 4 | 0 | 0 | Not-determined | Genome-fragment | NA | NA | 0 |
| S8_vs_k141_89009\|\|full | 4536 | No | NA | 8 | 1 | 0 | Low-quality | Genome-fragment | 9.86 | AAI-based (medium-confidence) | 0 |
| S8_vs_k141_206904\|\|full | 4536 | No | NA | 8 | 1 | 0 | Low-quality | Genome-fragment | 9.91 | AAI-based (medium-confidence) | 0 |
| S12_vs_k141_108921\|\|full | 3272 | No | NA | 6 | 0 | 0 | Not-determined | Genome-fragment | NA | NA | 0 |
| S12_vs_k141_46280\|\|full | 1343 | No | NA | 3 | 0 | 0 | Not-determined | Genome-fragment | NA | NA | 0 |
| S7_vs_70047\|\|full | 4539 | Yes | 2806 | 8 | 1 | 1 | Low-quality | Genome-fragment | 6.6 | AAI-based (medium-confidence) | 38.18 |
| S12_extracted_k141_8481 | 4539 | No | NA | 4 | 0 | 3 | Low-quality | Genome-fragment | 0.77 | AAI-based (medium-confidence) | 0 |
| S12_vs_k141_101606\|\|full | 3470 | No | NA | 5 | 0 | 3 | Not-determined | Genome-fragment | NA | NA | 0 |
| S17_extracted_k141_140830 | 4542 | No | NA | 3 | 2 | 0 | Low-quality | Genome-fragment | 7.76 | AAI-based (medium-confidence) | 0 |
| S9_vs_k141_25178\|\|full | 4542 | No | NA | 10 | 3 | 0 | Low-quality | Genome-fragment | 3.01 | AAI-based (high-confidence) | 0 |
| S12_vs_k141_46314\|\|full | 3327 | No | NA | 4 | 0 | 0 | Not-determined | Genome-fragment | NA | NA | 0 |
| S13_vs_18568\|\|full | 4542 | No | NA | 10 | 0 | 0 | Low-quality | Genome-fragment | 9.06 | AAI-based (high-confidence) | 0 |
| S14_vs_k141_39894\|\|full | 4543 | No | NA | 5 | 0 | 1 | Low-quality | Genome-fragment | 7.62 | AAI-based (medium-confidence) | 0 |
| S5_vs_k141_145551\|\|full | 4543 | No | NA | 5 | 0 | 1 | Low-quality | Genome-fragment | 7.51 | AAI-based (medium-confidence) | 0 |
| S12_vs_k141_76839\|\|full | 2909 | No | NA | 2 | 0 | 2 | Not-determined | Genome-fragment | NA | NA | 0 |
| S12_vs_k141_46321\|\|full | 1373 | No | NA | 4 | 0 | 0 | Not-determined | Genome-fragment | NA | NA | 0 |
| S11_vs_k141_30533\|\|full | 4544 | No | NA | 8 | 1 | 0 | Low-quality | Genome-fragment | 6.74 | HMM-based (lower-bound) | 0 |
| S12_vs_k141_115967\|\|full | 6903 | No | NA | 10 | 0 | 3 | Not-determined | Genome-fragment | NA | NA | 0 |
| S12_vs_k141_187796\|\|full | 3752 | No | NA | 4 | 0 | 2 | Not-determined | Genome-fragment | NA | NA | 0 |
| S12_vs_k141_84544\|\|full | 1206 | No | NA | 2 | 0 | 1 | Not-determined | Genome-fragment | NA | NA | 0 |
| S1_vs_k141_20078\|\|full | 4545 | No | NA | 9 | 5 | 0 | Low-quality | Genome-fragment | 10.05 | AAI-based (high-confidence) | 0 |
| S12_vs_k141_101649\|\|full | 2353 | No | NA | 3 | 0 | 0 | Not-determined | Genome-fragment | NA | NA | 0 |
| S3_extracted_k141_2410 | 4546 | No | NA | 6 | 1 | 0 | Low-quality | Genome-fragment | 20.83 | AAI-based (high-confidence) | 0 |
| S12_vs_k141_84546\|\|full | 5258 | No | NA | 4 | 0 | 0 | Not-determined | Genome-fragment | NA | NA | 0 |
| S12_vs_k141_163479\|\|full | 2351 | No | NA | 3 | 0 | 2 | Not-determined | Genome-fragment | NA | NA | 0 |
| S12_vs_k141_76879\|\|full | 2717 | No | NA | 2 | 0 | 0 | Not-determined | Genome-fragment | NA | NA | 0 |
| S12_vs_k141_101670\|\|full | 1318 | No | NA | 3 | 0 | 0 | Not-determined | Genome-fragment | NA | NA | 0 |
| S12_vs_k141_148030\|\|full | 1129 | No | NA | 2 | 0 | 0 | Not-determined | Genome-fragment | NA | NA | 0 |
| S8_vs_k141_124470\|\|full | 4546 | No | NA | 6 | 1 | 0 | Low-quality | Genome-fragment | 10.41 | AAI-based (medium-confidence) | 0 |
| S12_vs_k141_155493\|\|full | 1549 | No | NA | 3 | 0 | 1 | Not-determined | Genome-fragment | NA | NA | 0 |
| S12_vs_k141_37129\|\|full | 1859 | No | NA | 2 | 0 | 1 | Not-determined | Genome-fragment | NA | NA | 0 |
| S17_vs_k141_14110\|\|full | 4548 | No | NA | 2 | 1 | 0 | Low-quality | Genome-fragment | 6.55 | AAI-based (high-confidence) | 0 |
| S5_vs_k141_26241\|\|full | 4548 | No | NA | 14 | 4 | 0 | Low-quality | Genome-fragment | 8.44 | AAI-based (high-confidence) | 0 |
| S12_extracted_k141_17889 | 4548 | No | NA | 6 | 0 | 0 | Low-quality | Genome-fragment | 3.83 | AAI-based (high-confidence) | 0 |
| S12_vs_k141_84619\|\|full | 1116 | No | NA | 3 | 0 | 0 | Not-determined | Genome-fragment | NA | NA | 0 |
| S12_vs_k141_29853\|\|full | 2474 | No | NA | 3 | 0 | 2 | Not-determined | Genome-fragment | NA | NA | 0 |
| S12_vs_k141_6625\|\|full | 12656 | No | NA | 7 | 1 | 1 | Not-determined | Genome-fragment | NA | NA | 0 |
| S12_vs_k141_13337\|\|full | 2876 | No | NA | 2 | 0 | 2 | Not-determined | Genome-fragment | NA | NA | 0 |
| S11_vs_k141_26376\|\|full | 4549 | No | NA | 5 | 4 | 0 | Low-quality | Genome-fragment | 9.68 | AAI-based (high-confidence) | 0 |
| S3_extracted_k141_6997 | 4549 | No | NA | 5 | 1 | 0 | Low-quality | Genome-fragment | 2.97 | AAI-based (high-confidence) | 0 |
| S12_vs_k141_131912\|\|full | 1807 | No | NA | 2 | 0 | 0 | Not-determined | Genome-fragment | NA | NA | 0 |
| S12_vs_k141_93320\|\|full | 2184 | No | NA | 3 | 0 | 1 | Not-determined | Genome-fragment | NA | NA | 0 |
| S12_vs_k141_69185\|\|full | 1720 | No | NA | 3 | 0 | 1 | Not-determined | Genome-fragment | NA | NA | 0 |
| S12_vs_k141_172888\|\|full | 4549 | No | NA | 7 | 5 | 0 | Low-quality | Genome-fragment | 11.96 | AAI-based (medium-confidence) | 0 |
| S12_vs_k141_131937\|\|full | 2429 | No | NA | 2 | 0 | 2 | Not-determined | Genome-fragment | NA | NA | 0 |
| S12_vs_k141_69199\|\|full | 1940 | No | NA | 2 | 0 | 2 | Not-determined | Genome-fragment | NA | NA | 0 |
| S12_vs_k141_53255\|\|full | 3428 | No | NA | 4 | 0 | 0 | Not-determined | Genome-fragment | NA | NA | 0 |
| S8_vs_k141_49793\|\|full | 4553 | No | NA | 6 | 1 | 0 | Low-quality | Genome-fragment | 10.95 | AAI-based (high-confidence) | 0 |
| S5_vs_k141_27534\|\|full | 4555 | No | NA | 4 | 1 | 0 | Low-quality | Genome-fragment | 10.75 | AAI-based (medium-confidence) | 0 |
| S7_vs_70065\|\|full | 4555 | No | NA | 6 | 0 | 2 | Low-quality | Genome-fragment | 8.17 | AAI-based (high-confidence) | 0 |
| S12_vs_k141_61570\|\|full | 2215 | No | NA | 5 | 0 | 0 | Not-determined | Genome-fragment | NA | NA | 0 |
| S12_vs_k141_148112\|\|full | 2432 | No | NA | 4 | 0 | 1 | Not-determined | Genome-fragment | NA | NA | 0 |
| S12_extracted_k141_136171 | 4556 | No | NA | 4 | 0 | 3 | Low-quality | Genome-fragment | 1.39 | AAI-based (medium-confidence) | 0 |
| S12_vs_k141_109169\|\|full | 1048 | No | NA | 2 | 0 | 1 | Not-determined | Genome-fragment | NA | NA | 0 |
| S14_vs_k141_99756\|\|full | 4557 | No | NA | 5 | 0 | 0 | Low-quality | Genome-fragment | 4.97 | AAI-based (medium-confidence) | 0 |
| S10_vs_112285\|\|full | 4557 | No | NA | 5 | 1 | 0 | Low-quality | Genome-fragment | 12.01 | AAI-based (medium-confidence) | 0 |
| S14_vs_k141_10853\|\|full | 4558 | No | NA | 4 | 0 | 0 | Low-quality | Genome-fragment | 4.77 | AAI-based (medium-confidence) | 0 |
| S12_vs_k141_123892\|\|full | 1937 | No | NA | 3 | 0 | 1 | Not-determined | Genome-fragment | NA | NA | 0 |
| S12_vs_k141_21723\|\|full | 10845 | No | NA | 9 | 0 | 6 | Not-determined | Genome-fragment | NA | NA | 0 |
| S16_vs_32040\|\|full | 4558 | No | NA | 5 | 0 | 2 | Low-quality | Genome-fragment | 5.94 | AAI-based (medium-confidence) | 0 |
| S12_vs_k141_46452\|\|full | 3757 | No | NA | 4 | 0 | 3 | Not-determined | Genome-fragment | NA | NA | 0 |
| S12_vs_k141_93357\|\|full | 40010 | No | NA | 55 | 24 | 0 | Medium-quality | Genome-fragment | 80.94 | AAI-based (high-confidence) | 0 |
| S12_vs_k141_140673\|\|full | 6155 | No | NA | 7 | 0 | 4 | Not-determined | Genome-fragment | NA | NA | 0 |
| S6_vs_k141_17533\|\|full | 4559 | No | NA | 8 | 1 | 0 | Low-quality | Genome-fragment | 11.19 | AAI-based (high-confidence) | 0 |
| S12_vs_k141_21730\|\|full | 1358 | No | NA | 3 | 0 | 1 | Not-determined | Genome-fragment | NA | NA | 0 |
| S8_vs_k141_147893\|\|full | 4561 | No | NA | 4 | 0 | 1 | Low-quality | Genome-fragment | 3.25 | AAI-based (high-confidence) | 0 |
| S12_vs_k141_61602\|\|full | 7718 | No | NA | 7 | 0 | 3 | Not-determined | Genome-fragment | NA | NA | 0 |
| S12_vs_k141_148141\|\|full | 2695 | No | NA | 4 | 0 | 3 | Not-determined | Genome-fragment | NA | NA | 0 |
| S3_extracted_k141_24424 | 4562 | No | NA | 6 | 0 | 1 | Low-quality | Genome-fragment | 8.97 | AAI-based (medium-confidence) | 0 |
| S8_vs_k141_142489\|\|full | 4562 | No | NA | 9 | 3 | 0 | Low-quality | Genome-fragment | 10.25 | AAI-based (high-confidence) | 0 |
| S7_vs_15549\|\|full | 4563 | No | NA | 7 | 0 | 1 | Low-quality | Genome-fragment | 10.4 | AAI-based (high-confidence) | 0 |
| S12_vs_k141_77088\|\|full | 1159 | No | NA | 3 | 0 | 0 | Not-determined | Genome-fragment | NA | NA | 0 |
| S12_vs_k141_17541\|\|full | 4564 | No | NA | 7 | 2 | 0 | Low-quality | Genome-fragment | 8.46 | AAI-based (high-confidence) | 0 |
| S12_vs_k141_123950\|\|full | 2674 | No | NA | 2 | 0 | 1 | Not-determined | Genome-fragment | NA | NA | 0 |
| S8_extracted_k141_168311 | 4564 | Yes | 3500 | 6 | 1 | 1 | Low-quality | Genome-fragment | 4.73 | HMM-based (lower-bound) | 23.31 |
| S12_vs_k141_109242\|\|full | 1046 | No | NA | 3 | 0 | 1 | Not-determined | Genome-fragment | NA | NA | 0 |
| S17_extracted_k141_152959 | 4568 | No | NA | 3 | 0 | 3 | Low-quality | Genome-fragment | 1.27 | AAI-based (medium-confidence) | 0 |
| S12_vs_k141_116216\|\|full | 2245 | No | NA | 4 | 0 | 0 | Not-determined | Genome-fragment | NA | NA | 0 |
| S12_vs_k141_30058\|\|full | 4262 | No | NA | 4 | 0 | 3 | Not-determined | Genome-fragment | NA | NA | 0 |
| S12_vs_k141_116219\|\|full | 1116 | No | NA | 2 | 0 | 2 | Not-determined | Genome-fragment | NA | NA | 0 |
| S12_vs_k141_188046\|\|full | 1271 | No | NA | 3 | 0 | 0 | Not-determined | Genome-fragment | NA | NA | 0 |
| S12_vs_k141_6829\|\|full | 1897 | No | NA | 4 | 0 | 0 | Not-determined | Genome-fragment | NA | NA | 0 |
| S12_vs_k141_77167\|\|full | 4648 | No | NA | 6 | 0 | 0 | Not-determined | Genome-fragment | NA | NA | 0 |
| S12_vs_k141_132078\|\|full | 1984 | No | NA | 3 | 0 | 2 | Not-determined | Genome-fragment | NA | NA | 0 |
| S12_vs_k141_124018\|\|full | 1037 | No | NA | 3 | 0 | 0 | Not-determined | Genome-fragment | NA | NA | 0 |
| S12_vs_k141_93456\|\|full | 5039 | No | NA | 4 | 0 | 4 | Not-determined | Genome-fragment | NA | NA | 0 |
| S3_extracted_k141_60166 | 4570 | No | NA | 3 | 0 | 3 | Low-quality | Genome-fragment | 2.61 | AAI-based (medium-confidence) | 0 |
| S17_extracted_k141_96364 | 4572 | No | NA | 12 | 11 | 0 | Low-quality | Genome-fragment | 15.05 | AAI-based (high-confidence) | 0 |
| S12_vs_k141_69359\|\|full | 1675 | No | NA | 2 | 0 | 1 | Not-determined | Genome-fragment | NA | NA | 0 |
| S12_vs_k141_21828\|\|full | 8379 | No | NA | 9 | 0 | 5 | Not-determined | Genome-fragment | NA | NA | 0 |
| S12_vs_k141_179025\|\|full | 4258 | No | NA | 5 | 0 | 1 | Not-determined | Genome-fragment | NA | NA | 0 |
| S7_vs_26253\|\|full | 4572 | No | NA | 7 | 4 | 0 | Low-quality | Genome-fragment | 9.71 | AAI-based (high-confidence) | 0 |
| S8_vs_k141_149646\|\|full | 4572 | No | NA | 3 | 0 | 2 | Low-quality | Genome-fragment | 3.36 | AAI-based (medium-confidence) | 0 |
| S12_vs_k141_124041\|\|full | 1110 | No | NA | 1 | 0 | 1 | Not-determined | Genome-fragment | NA | NA | 0 |
| S12_vs_k141_179042\|\|full | 1206 | No | NA | 4 | 0 | 0 | Not-determined | Genome-fragment | NA | NA | 0 |
| S12_vs_k141_6836\|\|full | 26571 | No | NA | 42 | 19 | 1 | Medium-quality | Genome-fragment | 56 | AAI-based (high-confidence) | 0 |
| S12_vs_k141_13627\|\|full | 1033 | No | NA | 2 | 0 | 1 | Not-determined | Genome-fragment | NA | NA | 0 |
| S8_vs_k141_40961\|\|full | 4572 | No | NA | 3 | 0 | 2 | Low-quality | Genome-fragment | 2.32 | AAI-based (medium-confidence) | 0 |
| S8_extracted_k141_6941 | 4573 | No | NA | 3 | 0 | 2 | Low-quality | Genome-fragment | 2.27 | AAI-based (medium-confidence) | 0 |
| S7_extracted_91903 | 4575 | No | NA | 13 | 1 | 0 | Low-quality | Genome-fragment | 3.49 | HMM-based (lower-bound) | 0 |
| S12_vs_k141_132137\|\|full | 1263 | No | NA | 1 | 0 | 0 | Not-determined | Genome-fragment | NA | NA | 0 |
| S12_extracted_k141_72090 | 4575 | No | NA | 7 | 0 | 3 | Low-quality | Genome-fragment | 9.8 | AAI-based (high-confidence) | 0 |
| S12_vs_k141_46654\|\|full | 3797 | No | NA | 7 | 0 | 0 | Not-determined | Genome-fragment | NA | NA | 0 |
| S12_vs_k141_132153\|\|full | 4367 | No | NA | 7 | 0 | 2 | Not-determined | Genome-fragment | NA | NA | 0 |
| S3_extracted_k141_121136 | 4578 | No | NA | 11 | 3 | 1 | Low-quality | Genome-fragment | 5.84 | AAI-based (high-confidence) | 0 |
| S16_extracted_94186 | 4578 | Yes | 2715 | 10 | 1 | 1 | Low-quality | Genome-fragment | 3.81 | HMM-based (lower-bound) | 40.69 |
| S12_vs_k141_30193\|\|full | 3137 | No | NA | 4 | 0 | 1 | Not-determined | Genome-fragment | NA | NA | 0 |
| S12_vs_k141_155743\|\|full | 3206 | No | NA | 4 | 0 | 1 | Not-determined | Genome-fragment | NA | NA | 0 |
| S12_vs_k141_93547\|\|full | 1923 | No | NA | 3 | 0 | 1 | Not-determined | Genome-fragment | NA | NA | 0 |
| S12_vs_k141_163908\|\|full | 3579 | No | NA | 3 | 0 | 1 | Not-determined | Genome-fragment | NA | NA | 0 |
| S12_vs_k141_140854\|\|full | 1553 | No | NA | 3 | 0 | 0 | Not-determined | Genome-fragment | NA | NA | 0 |
| S8_vs_k141_143008\|\|full | 4582 | No | NA | 5 | 1 | 0 | Low-quality | Genome-fragment | 1.28 | HMM-based (lower-bound) | 0 |
| S12_vs_k141_171545\|\|full | 1680 | No | NA | 2 | 0 | 0 | Not-determined | Genome-fragment | NA | NA | 0 |
| S12_vs_k141_124088\|\|full | 2716 | No | NA | 3 | 0 | 0 | Not-determined | Genome-fragment | NA | NA | 0 |
| S7_vs_102386\|\|full | 4583 | No | NA | 5 | 0 | 4 | Low-quality | Genome-fragment | 1.84 | AAI-based (medium-confidence) | 0 |
| S12_vs_k141_132204\|\|full | 1402 | No | NA | 3 | 0 | 0 | Not-determined | Genome-fragment | NA | NA | 0 |
| S12_vs_k141_155784\|\|full | 1177 | No | NA | 3 | 0 | 1 | Not-determined | Genome-fragment | NA | NA | 0 |
| S12_vs_k141_37479\|\|full | 1786 | No | NA | 3 | 0 | 1 | Not-determined | Genome-fragment | NA | NA | 0 |
| S12_vs_k141_46695\|\|full | 1336 | No | NA | 3 | 0 | 2 | Not-determined | Genome-fragment | NA | NA | 0 |
| S12_vs_k141_46702\|\|full | 1150 | No | NA | 5 | 0 | 0 | Not-determined | Genome-fragment | NA | NA | 0 |
| S17_vs_k141_12240\|\|full | 4584 | No | NA | 8 | 0 | 1 | Low-quality | Genome-fragment | 4.88 | AAI-based (medium-confidence) | 0 |
| S14_vs_k141_26194\|\|full | 4585 | No | NA | 7 | 0 | 0 | Low-quality | Genome-fragment | 12.81 | AAI-based (high-confidence) | 0 |
| S5_extracted_k141_92798 | 4585 | No | NA | 8 | 7 | 0 | Low-quality | Genome-fragment | 15.13 | AAI-based (high-confidence) | 0 |
| S17_extracted_k141_165852 | 4586 | No | NA | 5 | 0 | 4 | Low-quality | Genome-fragment | 3.45 | AAI-based (medium-confidence) | 0 |
| S12_vs_k141_93614\|\|full | 1019 | No | NA | 2 | 0 | 2 | Not-determined | Genome-fragment | NA | NA | 0 |
| S17_extracted_k141_42369 | 4586 | No | NA | 8 | 5 | 0 | Low-quality | Genome-fragment | 7.53 | AAI-based (high-confidence) | 0 |
| S14_vs_k141_84360\|\|full | 4587 | Yes | 1608 | 7 | 1 | 4 | Low-quality | Genome-fragment | 0.65 | HMM-based (lower-bound) | 64.94 |
| S1_extracted_k141_45980 | 4587 | No | NA | 4 | 2 | 0 | Low-quality | Genome-fragment | 7.85 | AAI-based (high-confidence) | 0 |
| S3_vs_k141_118345\|\|full | 4588 | No | NA | 8 | 2 | 0 | Low-quality | Genome-fragment | 3.08 | AAI-based (high-confidence) | 0 |
| S12_vs_k141_85019\|\|full | 6861 | No | NA | 7 | 0 | 3 | Not-determined | Genome-fragment | NA | NA | 0 |
| S7_extracted_249 | 4589 | No | NA | 5 | 4 | 0 | Low-quality | Genome-fragment | 7.67 | AAI-based (high-confidence) | 0 |
| S10_extracted_27396 | 4589 | No | NA | 4 | 0 | 3 | Low-quality | Genome-fragment | 4.2 | AAI-based (medium-confidence) | 0 |
| S1_vs_k141_56589\|\|full | 4590 | No | NA | 3 | 0 | 0 | Low-quality | Genome-fragment | 11.71 | AAI-based (high-confidence) | 0 |
| S12_vs_k141_53651\|\|full | 1057 | No | NA | 3 | 0 | 2 | Not-determined | Genome-fragment | NA | NA | 0 |
| S12_vs_k141_109463\|\|full | 1370 | No | NA | 1 | 0 | 0 | Not-determined | Genome-fragment | NA | NA | 0 |
| S17_extracted_k141_105551 | 4591 | No | NA | 1 | 0 | 0 | Low-quality | Genome-fragment | 4.51 | AAI-based (high-confidence) | 0 |
| S12_vs_k141_116391\|\|full | 1020 | No | NA | 1 | 0 | 0 | Not-determined | Genome-fragment | NA | NA | 0 |
| S12_vs_k141_179149\|\|full | 2276 | No | NA | 2 | 0 | 0 | Not-determined | Genome-fragment | NA | NA | 0 |
| S12_vs_k141_116395\|\|full | 1620 | No | NA | 3 | 0 | 1 | Not-determined | Genome-fragment | NA | NA | 0 |
| S12_vs_k141_53681\|\|full | 2080 | No | NA | 4 | 0 | 0 | Not-determined | Genome-fragment | NA | NA | 0 |
| S12_vs_k141_179155\|\|full | 3188 | No | NA | 2 | 0 | 2 | Not-determined | Genome-fragment | NA | NA | 0 |
| S12_vs_k141_85077\|\|full | 3013 | No | NA | 3 | 0 | 2 | Not-determined | Genome-fragment | NA | NA | 0 |
| S14_vs_k141_36298\|\|full | 4591 | No | NA | 6 | 0 | 1 | Low-quality | Genome-fragment | 9.99 | AAI-based (high-confidence) | 0 |
| S12_vs_k141_188132\|\|full | 7034 | No | NA | 6 | 0 | 1 | Not-determined | Genome-fragment | NA | NA | 0 |
| S12_vs_k141_22016\|\|full | 2590 | No | NA | 3 | 0 | 2 | Not-determined | Genome-fragment | NA | NA | 0 |
| S12_vs_k141_148450\|\|full | 3234 | No | NA | 5 | 0 | 0 | Not-determined | Genome-fragment | NA | NA | 0 |
| S12_vs_k141_155857\|\|full | 2585 | No | NA | 4 | 0 | 4 | Not-determined | Genome-fragment | NA | NA | 0 |
| S16_extracted_65041 | 4591 | No | NA | 9 | 3 | 0 | Low-quality | Genome-fragment | 3.7 | AAI-based (high-confidence) | 0 |
| S5_extracted_k141_30182 | 4592 | No | NA | 6 | 0 | 0 | Low-quality | Genome-fragment | 2.26 | AAI-based (high-confidence) | 0 |
| S8_vs_k141_69482\|\|full | 4593 | No | NA | 7 | 2 | 0 | Low-quality | Genome-fragment | 10.91 | AAI-based (high-confidence) | 0 |
| S12_vs_k141_30314\|\|full | 2829 | No | NA | 3 | 0 | 1 | Not-determined | Genome-fragment | NA | NA | 0 |
| S12_vs_k141_30322\|\|full | 1445 | No | NA | 2 | 0 | 0 | Not-determined | Genome-fragment | NA | NA | 0 |
| S7_extracted_7741 | 4594 | No | NA | 3 | 0 | 2 | Low-quality | Genome-fragment | 2.45 | AAI-based (medium-confidence) | 0 |
| S10_extracted_34536 | 4595 | No | NA | 11 | 0 | 0 | Low-quality | Genome-fragment | 7.62 | AAI-based (high-confidence) | 0 |
| S12_vs_k141_102090\|\|full | 1020 | No | NA | 2 | 0 | 1 | Not-determined | Genome-fragment | NA | NA | 0 |
| S3_vs_k141_50756\|\|full | 4597 | No | NA | 10 | 2 | 0 | Low-quality | Genome-fragment | 10.3 | AAI-based (high-confidence) | 0 |
| S12_vs_k141_96251\|\|full | 4597 | No | NA | 2 | 0 | 1 | Low-quality | Genome-fragment | 4.2 | AAI-based (medium-confidence) | 0 |
| S12_vs_k141_163990\|\|full | 61244 | No | NA | 81 | 31 | 1 | Medium-quality | Genome-fragment | 81.13 | AAI-based (high-confidence) | 0 |
| S12_vs_k141_77438\|\|full | 1830 | No | NA | 2 | 0 | 1 | Not-determined | Genome-fragment | NA | NA | 0 |
| S12_vs_k141_37619\|\|full | 1973 | No | NA | 3 | 0 | 2 | Not-determined | Genome-fragment | NA | NA | 0 |
| S14_vs_k141_59486\|\|full | 4599 | No | NA | 3 | 0 | 1 | Low-quality | Genome-fragment | 5.09 | AAI-based (medium-confidence) | 0 |
| S12_vs_k141_148493\|\|full | 2743 | No | NA | 2 | 0 | 1 | Not-determined | Genome-fragment | NA | NA | 0 |
| S5_extracted_k141_10607 | 4600 | No | NA | 7 | 1 | 0 | Low-quality | Genome-fragment | 7.23 | AAI-based (high-confidence) | 0 |
| S12_vs_k141_102112\|\|full | 3178 | No | NA | 3 | 0 | 1 | Not-determined | Genome-fragment | NA | NA | 0 |
| S12_vs_k141_109557\|\|full | 1775 | No | NA | 3 | 0 | 1 | Not-determined | Genome-fragment | NA | NA | 0 |
| S12_vs_k141_148498\|\|full | 3593 | No | NA | 3 | 0 | 3 | Not-determined | Genome-fragment | NA | NA | 0 |
| S12_vs_k141_109567\|\|full | 3036 | No | NA | 2 | 0 | 1 | Not-determined | Genome-fragment | NA | NA | 0 |
| S12_vs_k141_53763\|\|full | 1318 | No | NA | 4 | 0 | 2 | Not-determined | Genome-fragment | NA | NA | 0 |
| S12_vs_k141_22107\|\|full | 3259 | No | NA | 3 | 0 | 1 | Not-determined | Genome-fragment | NA | NA | 0 |
| S3_vs_k141_91927\|\|full | 4601 | No | NA | 4 | 0 | 0 | Low-quality | Genome-fragment | 10.69 | AAI-based (medium-confidence) | 0 |
| S12_vs_k141_85162\|\|full | 1318 | No | NA | 3 | 0 | 1 | Not-determined | Genome-fragment | NA | NA | 0 |
| S16_extracted_90717 | 4602 | No | NA | 2 | 1 | 0 | Low-quality | Genome-fragment | 2.5 | HMM-based (lower-bound) | 0 |
| S12_vs_k141_188195\|\|full | 3196 | No | NA | 3 | 0 | 3 | Not-determined | Genome-fragment | NA | NA | 0 |
| S12_vs_k141_171719\|\|full | 1074 | No | NA | 2 | 0 | 0 | Not-determined | Genome-fragment | NA | NA | 0 |
| S16_vs_15448\|\|full | 4602 | No | NA | 3 | 0 | 2 | Low-quality | Genome-fragment | 2.3 | AAI-based (medium-confidence) | 0 |
| S12_vs_k141_77520\|\|full | 1614 | No | NA | 3 | 0 | 0 | Not-determined | Genome-fragment | NA | NA | 0 |
| S12_vs_k141_148550\|\|full | 1586 | No | NA | 3 | 0 | 1 | Not-determined | Genome-fragment | NA | NA | 0 |
| S12_vs_k141_116498\|\|full | 9077 | No | NA | 9 | 0 | 6 | Not-determined | Genome-fragment | NA | NA | 0 |
| S8_vs_k141_88552\|\|full | 4604 | No | NA | 8 | 0 | 0 | Low-quality | Genome-fragment | 8.39 | AAI-based (high-confidence) | 0 |
| S17_vs_k141_16758\|\|full | 4607 | No | NA | 5 | 2 | 0 | Low-quality | Genome-fragment | 10.9 | AAI-based (high-confidence) | 0 |
| S12_vs_k141_164056\|\|full | 6664 | No | NA | 9 | 1 | 1 | High-quality | High-quality | 100 | HMM-based (lower-bound) | 0 |
| S1_vs_k141_49948\|\|full | 4607 | No | NA | 7 | 0 | 3 | Low-quality | Genome-fragment | 1.96 | AAI-based (medium-confidence) | 0 |
| S3_extracted_k141_25384 | 4607 | Yes | 2103 | 8 | 1 | 2 | Low-quality | Genome-fragment | 1.33 | HMM-based (lower-bound) | 54.35 |
| S8_extracted_k141_65542 | 4607 | No | NA | 6 | 0 | 0 | Low-quality | Genome-fragment | 11.39 | AAI-based (high-confidence) | 0 |
| S12_vs_k141_22179\|\|full | 1079 | No | NA | 2 | 0 | 1 | Not-determined | Genome-fragment | NA | NA | 0 |
| S3_extracted_k141_17705 | 4608 | No | NA | 11 | 3 | 0 | Low-quality | Genome-fragment | 8.02 | AAI-based (high-confidence) | 0 |
| S14_vs_k141_13382\|\|full | 4609 | No | NA | 7 | 1 | 0 | Low-quality | Genome-fragment | 3.1 | HMM-based (lower-bound) | 0 |
| S16_vs_31644\|\|full | 4609 | No | NA | 8 | 3 | 0 | Low-quality | Genome-fragment | 7.27 | AAI-based (high-confidence) | 0 |
| S12_vs_k141_171770\|\|full | 3036 | No | NA | 4 | 0 | 2 | Not-determined | Genome-fragment | NA | NA | 0 |
| S12_vs_k141_53836\|\|full | 3290 | No | NA | 3 | 0 | 1 | Not-determined | Genome-fragment | NA | NA | 0 |
| S13_extracted_72183 | 4612 | No | NA | 4 | 0 | 2 | Low-quality | Genome-fragment | 2.28 | AAI-based (medium-confidence) | 0 |
| S10_vs_66649\|\|full | 4612 | No | NA | 5 | 0 | 1 | Low-quality | Genome-fragment | 14.25 | AAI-based (medium-confidence) | 0 |
| S12_vs_k141_179272\|\|full | 5271 | No | NA | 5 | 0 | 1 | Not-determined | Genome-fragment | NA | NA | 0 |
| S8_extracted_k141_107099 | 4612 | No | NA | 4 | 0 | 0 | Low-quality | Genome-fragment | 7.86 | AAI-based (medium-confidence) | 0 |
| S10_extracted_43527 | 4613 | No | NA | 6 | 0 | 2 | Low-quality | Genome-fragment | 7.71 | AAI-based (medium-confidence) | 0 |
| S12_vs_k141_124311\|\|full | 4862 | No | NA | 6 | 0 | 4 | Not-determined | Genome-fragment | NA | NA | 0 |
| S12_vs_k141_116530\|\|full | 5395 | No | NA | 7 | 0 | 3 | Not-determined | Genome-fragment | NA | NA | 0 |
| S12_vs_k141_93884\|\|full | 2723 | No | NA | 4 | 0 | 2 | Not-determined | Genome-fragment | NA | NA | 0 |
| S13_vs_118655\|\|full | 4614 | No | NA | 4 | 0 | 2 | Low-quality | Genome-fragment | 3.03 | AAI-based (medium-confidence) | 0 |
| S5_vs_k141_83320\|\|full | 4614 | No | NA | 7 | 5 | 0 | Low-quality | Genome-fragment | 6.79 | HMM-based (lower-bound) | 0 |
| S11_vs_k141_59206\|\|full | 4616 | No | NA | 5 | 1 | 0 | Low-quality | Genome-fragment | 10.96 | AAI-based (medium-confidence) | 0 |
| S3_vs_k141_74782\|\|full | 4616 | No | NA | 12 | 2 | 0 | Low-quality | Genome-fragment | 5.99 | HMM-based (lower-bound) | 0 |
| S3_vs_k141_16524\|\|full | 4617 | No | NA | 4 | 0 | 2 | Low-quality | Genome-fragment | 3.32 | AAI-based (medium-confidence) | 0 |
| S12_vs_k141_188269\|\|full | 1300 | No | NA | 3 | 0 | 0 | Not-determined | Genome-fragment | NA | NA | 0 |
| S5_vs_k141_124313\|\|full | 4618 | No | NA | 3 | 0 | 1 | Low-quality | Genome-fragment | 1.54 | AAI-based (medium-confidence) | 0 |
| S8_extracted_k141_122599 | 4618 | No | NA | 7 | 0 | 0 | Low-quality | Genome-fragment | 4.92 | AAI-based (high-confidence) | 0 |
| S1_vs_k141_68800\|\|full | 4621 | No | NA | 8 | 0 | 3 | Low-quality | Genome-fragment | 0.42 | AAI-based (medium-confidence) | 0 |
| S12_vs_k141_188292\|\|full | 1448 | No | NA | 3 | 0 | 0 | Not-determined | Genome-fragment | NA | NA | 0 |
| S12_vs_k141_7207\|\|full | 1000 | No | NA | 2 | 0 | 0 | Not-determined | Genome-fragment | NA | NA | 0 |
| S16_extracted_154791 | 4622 | No | NA | 2 | 0 | 2 | Low-quality | Genome-fragment | 2.35 | AAI-based (medium-confidence) | 0 |
| S12_vs_k141_7213\|\|full | 1457 | No | NA | 2 | 0 | 2 | Not-determined | Genome-fragment | NA | NA | 0 |
| S16_extracted_3461 | 4623 | Yes | 3718 | 4 | 1 | 1 | Low-quality | Genome-fragment | 6.24 | HMM-based (lower-bound) | 19.58 |
| S12_vs_k141_148717\|\|full | 1057 | No | NA | 3 | 0 | 1 | Not-determined | Genome-fragment | NA | NA | 0 |
| S12_vs_k141_62087\|\|full | 1204 | No | NA | 4 | 0 | 0 | Not-determined | Genome-fragment | NA | NA | 0 |
| S16_vs_10095\|\|full | 4627 | No | NA | 6 | 0 | 1 | Low-quality | Genome-fragment | 7.31 | AAI-based (high-confidence) | 0 |
| S12_vs_k141_53884\|\|full | 1818 | No | NA | 3 | 0 | 1 | Not-determined | Genome-fragment | NA | NA | 0 |
| S10_vs_56760\|\|full | 4631 | No | NA | 4 | 3 | 0 | Low-quality | Genome-fragment | 10.98 | AAI-based (high-confidence) | 0 |
| S12_vs_k141_124397\|\|full | 2725 | No | NA | 3 | 0 | 1 | Not-determined | Genome-fragment | NA | NA | 0 |
| S16_vs_158574\|\|full | 4635 | No | NA | 3 | 0 | 1 | Low-quality | Genome-fragment | 2.62 | AAI-based (medium-confidence) | 0 |
| S12_vs_k141_179375\|\|full | 1250 | No | NA | 2 | 0 | 1 | Not-determined | Genome-fragment | NA | NA | 0 |
| S14_extracted_k141_84608 | 4636 | No | NA | 12 | 1 | 0 | Low-quality | Genome-fragment | 5.7 | AAI-based (medium-confidence) | 0 |
| S16_extracted_7865 | 4636 | No | NA | 7 | 1 | 0 | Low-quality | Genome-fragment | 7.64 | AAI-based (high-confidence) | 0 |
| S12_vs_k141_13980\|\|full | 3837 | No | NA | 6 | 0 | 2 | Not-determined | Genome-fragment | NA | NA | 0 |
| S12_vs_k141_179377\|\|full | 11977 | No | NA | 10 | 0 | 6 | Not-determined | Genome-fragment | NA | NA | 0 |
| S12_vs_k141_13985\|\|full | 1545 | No | NA | 3 | 0 | 0 | Not-determined | Genome-fragment | NA | NA | 0 |
| S12_vs_k141_188387\|\|full | 1926 | No | NA | 2 | 0 | 0 | Not-determined | Genome-fragment | NA | NA | 0 |
| S12_vs_k141_85380\|\|full | 1890 | No | NA | 2 | 0 | 1 | Not-determined | Genome-fragment | NA | NA | 0 |
| S8_extracted_k141_27923 | 4636 | No | NA | 6 | 0 | 0 | Low-quality | Genome-fragment | 4.65 | AAI-based (high-confidence) | 0 |
| S12_extracted_k141_109817 | 4637 | No | NA | 9 | 1 | 0 | Low-quality | Genome-fragment | 10.13 | AAI-based (high-confidence) | 0 |
| S12_vs_k141_77639\|\|full | 3557 | No | NA | 3 | 0 | 2 | Not-determined | Genome-fragment | NA | NA | 0 |
| S12_vs_k141_53920\|\|full | 3768 | No | NA | 4 | 0 | 2 | Not-determined | Genome-fragment | NA | NA | 0 |
| S12_vs_k141_164267\|\|full | 5295 | No | NA | 5 | 0 | 5 | Not-determined | Genome-fragment | NA | NA | 0 |
| S6_vs_k141_5336\|\|full | 4637 | No | NA | 14 | 0 | 0 | Low-quality | Genome-fragment | 13.44 | AAI-based (high-confidence) | 0 |
| S10_vs_71470\|\|full | 4638 | Yes | 1115 | 6 | 1 | 3 | Low-quality | Genome-fragment | 1.73 | HMM-based (lower-bound) | 75.96 |
| S12_vs_k141_14038\|\|full | 1479 | No | NA | 2 | 0 | 0 | Not-determined | Genome-fragment | NA | NA | 0 |
| S12_vs_k141_141290\|\|full | 2414 | No | NA | 3 | 0 | 1 | Not-determined | Genome-fragment | NA | NA | 0 |
| S12_vs_k141_7364\|\|full | 1992 | No | NA | 3 | 0 | 1 | Not-determined | Genome-fragment | NA | NA | 0 |
| S8_extracted_k141_7083 | 4638 | No | NA | 9 | 0 | 0 | Low-quality | Genome-fragment | 4.63 | AAI-based (high-confidence) | 0 |
| S14_extracted_k141_74765 | 4639 | No | NA | 9 | 3 | 0 | Low-quality | Genome-fragment | 12.43 | AAI-based (high-confidence) | 0 |
| S12_vs_k141_85444\|\|full | 1556 | No | NA | 2 | 0 | 0 | Not-determined | Genome-fragment | NA | NA | 0 |
| S12_vs_k141_37857\|\|full | 1175 | No | NA | 2 | 0 | 1 | Not-determined | Genome-fragment | NA | NA | 0 |
| S7_vs_100741\|\|full | 4640 | No | NA | 7 | 0 | 0 | Low-quality | Genome-fragment | 7.86 | AAI-based (high-confidence) | 0 |
| S7_vs_85761\|\|full | 4641 | No | NA | 4 | 0 | 1 | Low-quality | Genome-fragment | 3.37 | AAI-based (high-confidence) | 0 |
| S12_vs_k141_102306\|\|full | 3219 | No | NA | 4 | 0 | 2 | Not-determined | Genome-fragment | NA | NA | 0 |
| S12_vs_k141_124497\|\|full | 1581 | No | NA | 3 | 0 | 0 | Not-determined | Genome-fragment | NA | NA | 0 |
| S7_vs_91984\|\|full | 4641 | No | NA | 11 | 4 | 0 | Low-quality | Genome-fragment | 6.24 | AAI-based (high-confidence) | 0 |
| S11_vs_k141_5790\|\|full | 4642 | No | NA | 6 | 1 | 0 | Low-quality | Genome-fragment | 10.33 | HMM-based (lower-bound) | 0 |
| S12_vs_k141_47141\|\|full | 3211 | No | NA | 3 | 0 | 2 | Not-determined | Genome-fragment | NA | NA | 0 |
| S12_vs_k141_22488\|\|full | 2073 | No | NA | 3 | 0 | 1 | Not-determined | Genome-fragment | NA | NA | 0 |
| S12_vs_k141_179532\|\|full | 1646 | No | NA | 3 | 0 | 0 | Not-determined | Genome-fragment | NA | NA | 0 |
| S12_vs_k141_62295\|\|full | 1235 | No | NA | 3 | 0 | 0 | Not-determined | Genome-fragment | NA | NA | 0 |
| S12_vs_k141_132710\|\|full | 2736 | No | NA | 4 | 0 | 2 | Not-determined | Genome-fragment | NA | NA | 0 |
| S17_extracted_k141_96596 | 4643 | No | NA | 4 | 0 | 2 | Low-quality | Genome-fragment | 4.3 | AAI-based (high-confidence) | 0 |
| S12_vs_k141_141337\|\|full | 44369 | No | NA | 44 | 5 | 3 | High-quality | High-quality | 100 | AAI-based (high-confidence) | 0 |
| S5_extracted_k141_78230 | 4643 | No | NA | 5 | 3 | 0 | Low-quality | Genome-fragment | 10.35 | AAI-based (high-confidence) | 0 |
| S12_vs_k141_14156\|\|full | 1646 | No | NA | 4 | 0 | 0 | Not-determined | Genome-fragment | NA | NA | 0 |
| S11_vs_k141_64285\|\|full | 4644 | No | NA | 7 | 0 | 1 | Low-quality | Genome-fragment | 10.84 | AAI-based (medium-confidence) | 0 |
| S13_extracted_21664 | 4644 | No | NA | 2 | 0 | 2 | Low-quality | Genome-fragment | 2.92 | AAI-based (medium-confidence) | 0 |
| S1_vs_k141_103317\|\|full | 4645 | No | NA | 9 | 0 | 0 | Low-quality | Genome-fragment | 11.83 | AAI-based (high-confidence) | 0 |
| S8_vs_k141_12407\|\|full | 4645 | No | NA | 9 | 2 | 0 | Low-quality | Genome-fragment | 12.69 | AAI-based (high-confidence) | 0 |
| S17_extracted_k141_115971 | 4646 | No | NA | 10 | 2 | 0 | Low-quality | Genome-fragment | 7.63 | AAI-based (high-confidence) | 0 |
| S12_vs_k141_85526\|\|full | 1785 | No | NA | 3 | 0 | 1 | Not-determined | Genome-fragment | NA | NA | 0 |
| S13_extracted_26796 | 4646 | No | NA | 7 | 0 | 0 | Low-quality | Genome-fragment | 1.16 | AAI-based (high-confidence) | 0 |
| S9_extracted_k141_23159 | 4647 | No | NA | 6 | 0 | 0 | Low-quality | Genome-fragment | 10.61 | AAI-based (high-confidence) | 0 |
| S12_vs_k141_54107\|\|full | 1969 | No | NA | 4 | 0 | 0 | Not-determined | Genome-fragment | NA | NA | 0 |
| S16_vs_90930\|\|full | 4647 | No | NA | 3 | 1 | 0 | Low-quality | Genome-fragment | 2.52 | HMM-based (lower-bound) | 0 |
| S5_vs_k141_62983\|\|full | 4648 | No | NA | 9 | 1 | 0 | Low-quality | Genome-fragment | 31.02 | AAI-based (high-confidence) | 0 |
| S9_vs_k141_44478\|\|full | 4651 | No | NA | 7 | 4 | 0 | Low-quality | Genome-fragment | 9.77 | AAI-based (high-confidence) | 0 |
| S12_vs_k141_7519\|\|full | 1694 | No | NA | 4 | 0 | 3 | Not-determined | Genome-fragment | NA | NA | 0 |
| S16_vs_24510\|\|full | 4652 | No | NA | 5 | 0 | 0 | Low-quality | Genome-fragment | 9.16 | AAI-based (medium-confidence) | 0 |
| S17_vs_k141_127706\|\|full | 4654 | No | NA | 7 | 2 | 0 | Low-quality | Genome-fragment | 10.39 | HMM-based (lower-bound) | 0 |
| S12_vs_k141_14210\|\|full | 3327 | No | NA | 4 | 0 | 3 | Not-determined | Genome-fragment | NA | NA | 0 |
| S12_vs_k141_62367\|\|full | 6326 | No | NA | 8 | 0 | 7 | Not-determined | Genome-fragment | NA | NA | 0 |
| S16_vs_158974\|\|full | 4656 | No | NA | 4 | 0 | 2 | Low-quality | Genome-fragment | 3.72 | AAI-based (medium-confidence) | 0 |
| S12_vs_k141_188580\|\|full | 1567 | No | NA | 2 | 0 | 0 | Not-determined | Genome-fragment | NA | NA | 0 |
| S12_vs_k141_132821\|\|full | 2148 | No | NA | 4 | 0 | 0 | Not-determined | Genome-fragment | NA | NA | 0 |
| S14_extracted_k141_112131 | 4657 | No | NA | 5 | 4 | 0 | Low-quality | Genome-fragment | 15.2 | AAI-based (high-confidence) | 0 |
| S12_vs_k141_109944\|\|full | 2440 | No | NA | 3 | 0 | 2 | Not-determined | Genome-fragment | NA | NA | 0 |
| S12_vs_k141_156078\|\|full | 1899 | No | NA | 2 | 0 | 2 | Not-determined | Genome-fragment | NA | NA | 0 |
| S12_vs_k141_62418\|\|full | 1403 | No | NA | 2 | 0 | 0 | Not-determined | Genome-fragment | NA | NA | 0 |
| S12_vs_k141_132855\|\|full | 5137 | No | NA | 4 | 0 | 3 | Not-determined | Genome-fragment | NA | NA | 0 |
| S10_vs_13436\|\|full | 4657 | No | NA | 4 | 0 | 0 | Low-quality | Genome-fragment | 2.1 | AAI-based (high-confidence) | 0 |
| S8_extracted_k141_35204 | 4657 | No | NA | 2 | 0 | 2 | Low-quality | Genome-fragment | 7.68 | AAI-based (high-confidence) | 0 |
| S14_vs_k141_45844\|\|full | 4659 | Yes | 1971 | 3 | 1 | 1 | Low-quality | Genome-fragment | 0.17 | HMM-based (lower-bound) | 57.69 |
| S16_extracted_9055 | 4662 | No | NA | 3 | 0 | 2 | Low-quality | Genome-fragment | 2.18 | AAI-based (medium-confidence) | 0 |
| S12_vs_k141_188651\|\|full | 4730 | No | NA | 6 | 0 | 1 | Not-determined | Genome-fragment | NA | NA | 0 |
| S17_extracted_k141_15848 | 4664 | No | NA | 5 | 0 | 0 | Low-quality | Genome-fragment | 5.2 | AAI-based (high-confidence) | 0 |
| S17_extracted_k141_41995 | 4664 | No | NA | 5 | 0 | 0 | Low-quality | Genome-fragment | 2.74 | AAI-based (high-confidence) | 0 |
| S12_vs_k141_54231\|\|full | 1105 | No | NA | 2 | 0 | 1 | Not-determined | Genome-fragment | NA | NA | 0 |
| S2_vs_k141_18631\|\|full | 4664 | No | NA | 12 | 1 | 0 | Low-quality | Genome-fragment | 10.86 | AAI-based (high-confidence) | 0 |
| S12_vs_k141_30768\|\|full | 16489 | No | NA | 10 | 0 | 8 | Not-determined | Genome-fragment | NA | NA | 0 |
| S14_vs_k141_12266\|\|full | 4665 | No | NA | 10 | 3 | 0 | Low-quality | Genome-fragment | 8.86 | AAI-based (medium-confidence) | 0 |
| S12_vs_k141_70241\|\|full | 3052 | No | NA | 4 | 1 | 0 | Medium-quality | Genome-fragment | 73.34 | HMM-based (lower-bound) | 0 |
| S12_vs_k141_7562\|\|full | 1399 | No | NA | 3 | 0 | 1 | Not-determined | Genome-fragment | NA | NA | 0 |
| S12_vs_k141_149119\|\|full | 3335 | No | NA | 3 | 0 | 1 | Not-determined | Genome-fragment | NA | NA | 0 |
| S12_vs_k141_110007\|\|full | 3406 | No | NA | 6 | 0 | 2 | Not-determined | Genome-fragment | NA | NA | 0 |
| S12_vs_k141_37671\|\|full | 4665 | No | NA | 8 | 7 | 0 | Low-quality | Genome-fragment | 12.58 | AAI-based (high-confidence) | 0 |
| S12_vs_k141_62484\|\|full | 1243 | No | NA | 2 | 0 | 2 | Not-determined | Genome-fragment | NA | NA | 0 |
| S12_vs_k141_164490\|\|full | 5092 | No | NA | 5 | 0 | 3 | Not-determined | Genome-fragment | NA | NA | 0 |
| S12_vs_k141_14370\|\|full | 8213 | No | NA | 5 | 0 | 3 | Not-determined | Genome-fragment | NA | NA | 0 |
| S3_vs_k141_84001\|\|full | 4666 | No | NA | 5 | 0 | 1 | Low-quality | Genome-fragment | 6.3 | AAI-based (high-confidence) | 0 |
| S5_extracted_k141_127504 | 4666 | No | NA | 2 | 0 | 2 | Low-quality | Genome-fragment | 7.93 | AAI-based (high-confidence) | 0 |
| S10_extracted_53727 | 4666 | No | NA | 7 | 0 | 0 | Low-quality | Genome-fragment | 13.04 | AAI-based (medium-confidence) | 0 |
| S1_vs_k141_87165\|\|full | 4668 | No | NA | 7 | 2 | 0 | Low-quality | Genome-fragment | 12.49 | AAI-based (high-confidence) | 0 |
| S11_extracted_k141_37855 | 4669 | No | NA | 8 | 1 | 0 | Low-quality | Genome-fragment | 8.3 | AAI-based (medium-confidence) | 0 |
| S12_vs_k141_64127\|\|full | 4670 | No | NA | 12 | 0 | 0 | Low-quality | Genome-fragment | 12.59 | AAI-based (high-confidence) | 0 |
| S12_vs_k141_94042\|\|full | 39033 | Yes | 36811 | 56 | 12 | 3 | Medium-quality | Genome-fragment | 72.42 | AAI-based (high-confidence) | 5.69 |
| S12_vs_k141_124718\|\|full | 1458 | No | NA | 3 | 0 | 1 | Not-determined | Genome-fragment | NA | NA | 0 |
| S6_extracted_k141_40729 | 4670 | No | NA | 4 | 0 | 4 | Low-quality | Genome-fragment | 3.74 | AAI-based (high-confidence) | 0 |
| S9_vs_k141_36233\|\|full | 4671 | No | NA | 12 | 3 | 0 | Low-quality | Genome-fragment | 6.71 | AAI-based (high-confidence) | 0 |
| S12_vs_k141_124722\|\|full | 1097 | No | NA | 3 | 0 | 0 | Not-determined | Genome-fragment | NA | NA | 0 |
| S16_vs_43012\|\|full | 4671 | Yes | 1022 | 4 | 1 | 2 | Low-quality | Genome-fragment | 1.45 | AAI-based (medium-confidence) | 78.12 |
| S12_extracted_k141_141741 | 4672 | No | NA | 5 | 3 | 0 | Low-quality | Genome-fragment | 11.55 | AAI-based (high-confidence) | 0 |
| S12_vs_k141_30845\|\|full | 1118 | No | NA | 2 | 0 | 1 | Not-determined | Genome-fragment | NA | NA | 0 |
| S12_vs_k141_78016\|\|full | 7357 | No | NA | 6 | 0 | 5 | Not-determined | Genome-fragment | NA | NA | 0 |
| S16_extracted_173108 | 4673 | No | NA | 5 | 0 | 0 | Low-quality | Genome-fragment | 5.74 | AAI-based (high-confidence) | 0 |
| S12_vs_k141_141514\|\|full | 3695 | No | NA | 5 | 0 | 0 | Not-determined | Genome-fragment | NA | NA | 0 |
| S12_vs_k141_22640\|\|full | 1119 | No | NA | 3 | 0 | 0 | Not-determined | Genome-fragment | NA | NA | 0 |
| S12_vs_k141_149183\|\|full | 2528 | No | NA | 3 | 0 | 1 | Not-determined | Genome-fragment | NA | NA | 0 |
| S12_vs_k141_94092\|\|full | 1043 | No | NA | 1 | 0 | 0 | Not-determined | Genome-fragment | NA | NA | 0 |
| S12_vs_k141_94097\|\|full | 1523 | No | NA | 2 | 0 | 2 | Not-determined | Genome-fragment | NA | NA | 0 |
| S12_vs_k141_116933\|\|full | 3198 | No | NA | 2 | 0 | 2 | Not-determined | Genome-fragment | NA | NA | 0 |
| S16_vs_39321\|\|full | 4673 | No | NA | 7 | 0 | 0 | Low-quality | Genome-fragment | 10.25 | AAI-based (medium-confidence) | 0 |
| S12_vs_k141_141550\|\|full | 5422 | No | NA | 5 | 0 | 4 | Not-determined | Genome-fragment | NA | NA | 0 |
| S12_vs_k141_179851\|\|full | 4109 | No | NA | 4 | 0 | 2 | Not-determined | Genome-fragment | NA | NA | 0 |
| S12_vs_k141_38081\|\|full | 3821 | No | NA | 3 | 0 | 3 | Not-determined | Genome-fragment | NA | NA | 0 |
| S12_vs_k141_133022\|\|full | 2289 | No | NA | 3 | 0 | 2 | Not-determined | Genome-fragment | NA | NA | 0 |
| S12_vs_k141_179864\|\|full | 1216 | No | NA | 2 | 0 | 0 | Not-determined | Genome-fragment | NA | NA | 0 |
| S9_extracted_k141_13843 | 4674 | No | NA | 1 | 1 | 0 | Low-quality | Genome-fragment | 0.38 | HMM-based (lower-bound) | 0 |
| S12_vs_k141_110143\|\|full | 1741 | No | NA | 3 | 0 | 0 | Not-determined | Genome-fragment | NA | NA | 0 |
| S12_vs_k141_14467\|\|full | 19385 | No | NA | 35 | 3 | 3 | Medium-quality | Genome-fragment | 50.18 | AAI-based (medium-confidence) | 0 |
| S5_extracted_k141_102751 | 4674 | No | NA | 7 | 0 | 0 | Low-quality | Genome-fragment | 13.01 | AAI-based (high-confidence) | 0 |
| S7_extracted_119828 | 4674 | No | NA | 2 | 0 | 1 | Low-quality | Genome-fragment | 6.99 | AAI-based (medium-confidence) | 0 |
| S12_vs_k141_124772\|\|full | 4311 | No | NA | 3 | 0 | 1 | Not-determined | Genome-fragment | NA | NA | 0 |
| S8_extracted_k141_124354 | 4674 | No | NA | 10 | 0 | 1 | Low-quality | Genome-fragment | 2.69 | AAI-based (high-confidence) | 0 |
| S12_vs_k141_133065\|\|full | 2223 | No | NA | 3 | 0 | 0 | Not-determined | Genome-fragment | NA | NA | 0 |
| S12_vs_k141_85806\|\|full | 1901 | No | NA | 2 | 0 | 0 | Not-determined | Genome-fragment | NA | NA | 0 |
| S12_vs_k141_62616\|\|full | 1796 | No | NA | 2 | 0 | 1 | Not-determined | Genome-fragment | NA | NA | 0 |
| S12_vs_k141_30963\|\|full | 3503 | No | NA | 5 | 0 | 4 | Not-determined | Genome-fragment | NA | NA | 0 |
| S12_vs_k141_38145\|\|full | 1142 | No | NA | 3 | 0 | 1 | Not-determined | Genome-fragment | NA | NA | 0 |
| S12_vs_k141_164626\|\|full | 1301 | No | NA | 2 | 0 | 1 | Not-determined | Genome-fragment | NA | NA | 0 |
| S8_vs_k141_9347\|\|full | 4674 | No | NA | 10 | 5 | 0 | Low-quality | Genome-fragment | 13.07 | AAI-based (medium-confidence) | 0 |
| S16_extracted_9385 | 4675 | No | NA | 5 | 3 | 0 | Low-quality | Genome-fragment | 10.42 | AAI-based (medium-confidence) | 0 |
| S5_extracted_k141_9281 | 4677 | No | NA | 9 | 6 | 0 | Low-quality | Genome-fragment | 14.82 | AAI-based (high-confidence) | 0 |
| S8_extracted_k141_132898 | 4677 | No | NA | 4 | 0 | 1 | Low-quality | Genome-fragment | 0.4 | AAI-based (medium-confidence) | 0 |
| S12_vs_k141_179951\|\|full | 2424 | No | NA | 4 | 0 | 1 | Not-determined | Genome-fragment | NA | NA | 0 |
| S13_extracted_126814 | 4678 | No | NA | 4 | 0 | 4 | Low-quality | Genome-fragment | 2.14 | AAI-based (high-confidence) | 0 |
| S16_extracted_60838 | 4680 | No | NA | 2 | 0 | 1 | Low-quality | Genome-fragment | 5.07 | AAI-based (medium-confidence) | 0 |
| S12_vs_k141_179969\|\|full | 2307 | No | NA | 2 | 0 | 0 | Not-determined | Genome-fragment | NA | NA | 0 |
| S12_vs_k141_14548\|\|full | 1711 | No | NA | 3 | 0 | 2 | Not-determined | Genome-fragment | NA | NA | 0 |
| S12_vs_k141_94181\|\|full | 12565 | No | NA | 8 | 0 | 6 | Not-determined | Genome-fragment | NA | NA | 0 |
| S12_extracted_k141_137860 | 4680 | No | NA | 7 | 3 | 0 | Low-quality | Genome-fragment | 4.49 | AAI-based (high-confidence) | 0 |
| S12_vs_k141_156179\|\|full | 1393 | No | NA | 3 | 0 | 0 | Not-determined | Genome-fragment | NA | NA | 0 |
| S1_vs_k141_154416\|\|full | 4681 | No | NA | 3 | 0 | 2 | Low-quality | Genome-fragment | 2.39 | AAI-based (medium-confidence) | 0 |
| S12_vs_k141_70486\|\|full | 3340 | No | NA | 4 | 0 | 0 | Not-determined | Genome-fragment | NA | NA | 0 |
| S5_extracted_k141_117457 | 4682 | No | NA | 9 | 1 | 0 | Low-quality | Genome-fragment | 10.16 | AAI-based (medium-confidence) | 0 |
| S12_vs_k141_141721\|\|full | 3508 | No | NA | 3 | 0 | 1 | Not-determined | Genome-fragment | NA | NA | 0 |
| S12_vs_k141_54476\|\|full | 1415 | No | NA | 3 | 0 | 1 | Not-determined | Genome-fragment | NA | NA | 0 |
| S10_extracted_97360 | 4682 | No | NA | 10 | 1 | 0 | Low-quality | Genome-fragment | 9.32 | AAI-based (high-confidence) | 0 |
| S12_vs_k141_124867\|\|full | 1828 | No | NA | 2 | 0 | 0 | Not-determined | Genome-fragment | NA | NA | 0 |
| S17_extracted_k141_126234 | 4684 | No | NA | 7 | 6 | 0 | Low-quality | Genome-fragment | 15.44 | AAI-based (high-confidence) | 0 |
| S12_vs_k141_22853\|\|full | 30882 | No | NA | 47 | 27 | 1 | Medium-quality | Genome-fragment | 89.68 | AAI-based (medium-confidence) | 0 |
| S5_extracted_k141_38563 | 4684 | No | NA | 9 | 8 | 0 | Low-quality | Genome-fragment | 15.45 | AAI-based (high-confidence) | 0 |
| S5_extracted_k141_93783 | 4685 | No | NA | 4 | 2 | 0 | Low-quality | Genome-fragment | 11.03 | AAI-based (high-confidence) | 0 |
| S12_vs_k141_149383\|\|full | 3617 | No | NA | 4 | 0 | 1 | Not-determined | Genome-fragment | NA | NA | 0 |
| S12_vs_k141_78233\|\|full | 1442 | No | NA | 2 | 0 | 1 | Not-determined | Genome-fragment | NA | NA | 0 |
| S12_vs_k141_149389\|\|full | 1924 | No | NA | 3 | 0 | 0 | Not-determined | Genome-fragment | NA | NA | 0 |
| S12_vs_k141_18006\|\|full | 4685 | No | NA | 10 | 1 | 0 | Low-quality | Genome-fragment | 12.03 | AAI-based (high-confidence) | 0 |
| S17_extracted_k141_38194 | 4686 | No | NA | 1 | 0 | 1 | Low-quality | Genome-fragment | 7.09 | AAI-based (high-confidence) | 0 |
| S5_extracted_k141_85751 | 4688 | No | NA | 9 | 6 | 0 | Low-quality | Genome-fragment | 14.44 | AAI-based (high-confidence) | 0 |
| S12_vs_k141_164716\|\|full | 1583 | No | NA | 2 | 0 | 1 | Not-determined | Genome-fragment | NA | NA | 0 |
| S10_vs_72019\|\|full | 4688 | No | NA | 8 | 1 | 0 | Low-quality | Genome-fragment | 10.96 | AAI-based (high-confidence) | 0 |
| S12_vs_k141_188718\|\|full | 1595 | No | NA | 3 | 0 | 0 | Not-determined | Genome-fragment | NA | NA | 0 |
| S12_vs_k141_124923\|\|full | 3022 | No | NA | 2 | 0 | 2 | Not-determined | Genome-fragment | NA | NA | 0 |
| S11_extracted_k141_68209 | 4689 | No | NA | 5 | 1 | 0 | Low-quality | Genome-fragment | 12.36 | AAI-based (high-confidence) | 0 |
| S12_vs_k141_156236\|\|full | 3603 | No | NA | 8 | 0 | 0 | Not-determined | Genome-fragment | NA | NA | 0 |
| S12_vs_k141_94266\|\|full | 1878 | No | NA | 3 | 0 | 0 | Not-determined | Genome-fragment | NA | NA | 0 |
| S12_vs_k141_149405\|\|full | 1844 | No | NA | 2 | 0 | 2 | Not-determined | Genome-fragment | NA | NA | 0 |
| S12_vs_k141_133070\|\|full | 132222 | Yes | 76271 | 145 | 17 | 40 | Medium-quality | Genome-fragment | 56.81 | AAI-based (high-confidence) | 42.32 |
| S14_vs_k141_54205\|\|full | 4690 | No | NA | 5 | 3 | 0 | Low-quality | Genome-fragment | 13.58 | AAI-based (high-confidence) | 0 |
| S12_vs_k141_156264\|\|full | 1255 | No | NA | 4 | 0 | 0 | Not-determined | Genome-fragment | NA | NA | 0 |

| | | | | | | | | | | | | | |
|---|---|---|---|---|---|---|---|---|---|---|---|---|---|
| S5_vs_k141_66359\|\|full | 4690 | No | NA | | 14 | 1 | 0 | Low-quality | Genome-fragment | | 9.88 | AAI-based (high-confidence) | 0 |
| S12_vs_k141_164760\|\|full | 1569 | No | NA | | 4 | 0 | 1 | Not-determined | Genome-fragment | NA | | NA | 0 |
| S12_vs_k141_77629\|\|full | 4690 | No | NA | | 7 | 1 | 0 | Low-quality | Genome-fragment | | 3 | HMM-based (lower-bound) | 0 |
| S12_vs_k141_70589\|\|full | 9339 | No | NA | | 10 | 0 | 7 | Not-determined | Genome-fragment | NA | | NA | 0 |
| S8_extracted_k141_31469 | 4690 | No | NA | | 6 | 3 | 0 | Low-quality | Genome-fragment | | 14.03 | AAI-based (high-confidence) | 0 |
| S5_vs_k141_53253\|\|full | 4692 | No | NA | | 7 | 3 | 0 | Low-quality | Genome-fragment | | 12.04 | AAI-based (high-confidence) | 0 |
| S12_vs_k141_20005\|\|full | 4692 | No | NA | | 6 | 0 | 3 | Low-quality | Genome-fragment | | 0.41 | AAI-based (medium-confidence) | 0 |
| S12_vs_k141_117193\|\|full | 2400 | No | NA | | 4 | 0 | 4 | Not-determined | Genome-fragment | NA | | NA | 0 |
| S5_extracted_k141_112336 | 4693 | No | NA | | 6 | 0 | 1 | Low-quality | Genome-fragment | | 5.74 | AAI-based (medium-confidence) | 0 |
| S12_vs_k141_62855\|\|full | 1159 | No | NA | | 3 | 0 | 2 | Not-determined | Genome-fragment | NA | | NA | 0 |
| S12_vs_k141_125006\|\|full | 1263 | No | NA | | 2 | 0 | 1 | Not-determined | Genome-fragment | NA | | NA | 0 |
| S12_vs_k141_117202\|\|full | 4723 | No | NA | | 4 | 0 | 2 | Not-determined | Genome-fragment | NA | | NA | 0 |
| S1_extracted_k141_54550 | 4698 | No | NA | | 6 | 2 | 0 | Low-quality | Genome-fragment | | 11.21 | AAI-based (high-confidence) | 0 |
| S3_vs_k141_87469\|\|full | 4698 | No | NA | | 4 | 0 | 0 | Low-quality | Genome-fragment | | 12.08 | AAI-based (high-confidence) | 0 |
| S11_vs_k141_36152\|\|full | 4699 | No | NA | | 5 | 0 | 0 | Low-quality | Genome-fragment | | 10.57 | AAI-based (high-confidence) | 0 |
| S12_vs_k141_117231\|\|full | 2287 | No | NA | | 3 | 0 | 2 | Not-determined | Genome-fragment | NA | | NA | 0 |
| S12_vs_k141_180092\|\|full | 1213 | No | NA | | 2 | 0 | 0 | Not-determined | Genome-fragment | NA | | NA | 0 |
| S12_vs_k141_62922\|\|full | 5795 | No | NA | | 6 | 0 | 6 | Not-determined | Genome-fragment | NA | | NA | 0 |
| S12_vs_k141_14777\|\|full | 1723 | No | NA | | 4 | 0 | 0 | Not-determined | Genome-fragment | NA | | NA | 0 |
| S12_vs_k141_149527\|\|full | 2682 | No | NA | | 4 | 0 | 3 | Not-determined | Genome-fragment | NA | | NA | 0 |
| S12_vs_k141_180112\|\|full | 1029 | No | NA | | 3 | 0 | 0 | Not-determined | Genome-fragment | NA | | NA | 0 |
| S12_extracted_k141_97043 | 4700 | No | NA | | 4 | 0 | 2 | Low-quality | Genome-fragment | | 0.81 | AAI-based (medium-confidence) | 0 |
| S12_vs_k141_62933\|\|full | 2306 | No | NA | | 2 | 0 | 0 | Not-determined | Genome-fragment | NA | | NA | 0 |
| S12_vs_k141_149539\|\|full | 1510 | No | NA | | 3 | 0 | 0 | Not-determined | Genome-fragment | NA | | NA | 0 |
| S12_vs_k141_54649\|\|full | 3846 | No | NA | | 3 | 0 | 0 | Not-determined | Genome-fragment | NA | | NA | 0 |
| S8_vs_k141_118945\|\|full | 4700 | No | NA | | 5 | 0 | 2 | Low-quality | Genome-fragment | | 3.43 | AAI-based (high-confidence) | 0 |
| S12_vs_k141_38383\|\|full | 5841 | No | NA | | 5 | 0 | 3 | Not-determined | Genome-fragment | NA | | NA | 0 |
| S12_vs_k141_180136\|\|full | 1214 | No | NA | | 2 | 0 | 0 | Not-determined | Genome-fragment | NA | | NA | 0 |
| S13_extracted_28415 | 4702 | No | NA | | 4 | 0 | 4 | Low-quality | Genome-fragment | | 1.25 | AAI-based (medium-confidence) | 0 |
| S12_vs_k141_54670\|\|full | 1403 | No | NA | | 2 | 0 | 0 | Not-determined | Genome-fragment | NA | | NA | 0 |
| S12_vs_k141_31179\|\|full | 1672 | No | NA | | 2 | 0 | 2 | Not-determined | Genome-fragment | NA | | NA | 0 |
| S12_vs_k141_70709\|\|full | 1646 | No | NA | | 3 | 0 | 2 | Not-determined | Genome-fragment | NA | | NA | 0 |
| S12_vs_k141_31182\|\|full | 2768 | No | NA | | 7 | 0 | 0 | Not-determined | Genome-fragment | NA | | NA | 0 |
| S7_vs_81880\|\|full | 4703 | No | NA | | 15 | 0 | 0 | Low-quality | Genome-fragment | | 3.25 | AAI-based (high-confidence) | 0 |
| S8_extracted_k141_164611 | 4703 | No | NA | | 4 | 3 | 0 | Low-quality | Genome-fragment | | 7.83 | AAI-based (high-confidence) | 0 |
| S12_vs_k141_133246\|\|full | 5942 | No | NA | | 7 | 0 | 5 | Not-determined | Genome-fragment | NA | | NA | 0 |
| S3_extracted_k141_123523 | 4704 | No | NA | | 5 | 1 | 0 | Low-quality | Genome-fragment | | 2.95 | AAI-based (high-confidence) | 0 |
| S3_vs_k141_68946\|\|full | 4704 | No | NA | | 5 | 0 | 4 | Low-quality | Genome-fragment | | 2.24 | AAI-based (medium-confidence) | 0 |
| S15_extracted_2801 | 4704 | No | NA | | 4 | 0 | 3 | Low-quality | Genome-fragment | | 0.83 | AAI-based (medium-confidence) | 0 |
| S12_vs_k141_133271\|\|full | 1578 | No | NA | | 3 | 0 | 1 | Not-determined | Genome-fragment | NA | | NA | 0 |
| S12_vs_k141_164910\|\|full | 3071 | No | NA | | 4 | 0 | 2 | Not-determined | Genome-fragment | NA | | NA | 0 |
| S12_vs_k141_38481\|\|full | 2757 | No | NA | | 3 | 0 | 2 | Not-determined | Genome-fragment | NA | | NA | 0 |
| S12_vs_k141_78533\|\|full | 3400 | No | NA | | 4 | 0 | 2 | Not-determined | Genome-fragment | NA | | NA | 0 |
| S6_vs_k141_55255\|\|full | 4704 | No | NA | | 4 | 4 | 0 | Low-quality | Genome-fragment | | 12.42 | AAI-based (high-confidence) | 0 |
| S12_vs_k141_86259\|\|full | 2115 | No | NA | | 4 | 0 | 0 | Not-determined | Genome-fragment | NA | | NA | 0 |
| S12_vs_k141_54789\|\|full | 1990 | No | NA | | 2 | 0 | 1 | Not-determined | Genome-fragment | NA | | NA | 0 |
| S17_extracted_k141_70361 | 4705 | No | NA | | 4 | 0 | 2 | Low-quality | Genome-fragment | | 1.7 | AAI-based (medium-confidence) | 0 |
| S5_vs_k141_45047\|\|full | 4705 | No | NA | | 10 | 1 | 0 | Low-quality | Genome-fragment | | 2.85 | HMM-based (lower-bound) | 0 |
| S13_extracted_149804 | 4706 | No | NA | | 9 | 4 | 0 | Low-quality | Genome-fragment | | 7.54 | AAI-based (high-confidence) | 0 |
| S16_extracted_9866 | 4706 | No | NA | | 6 | 1 | 0 | Low-quality | Genome-fragment | | 8.95 | HMM-based (lower-bound) | 0 |
| S12_vs_k141_31380\|\|full | 2649 | No | NA | | 5 | 0 | 1 | Not-determined | Genome-fragment | NA | | NA | 0 |
| S12_vs_k141_35552\|\|full | 4706 | No | NA | | 4 | 0 | 3 | Low-quality | Genome-fragment | | 4.82 | AAI-based (medium-confidence) | 0 |
| S12_vs_k141_23244\|\|full | 1066 | No | NA | | 3 | 0 | 0 | Not-determined | Genome-fragment | NA | | NA | 0 |
| S1_vs_k141_75503\|\|full | 4707 | No | NA | | 7 | 5 | 0 | Low-quality | Genome-fragment | | 11.72 | AAI-based (high-confidence) | 0 |
| S12_vs_k141_180296\|\|full | 1565 | No | NA | | 2 | 0 | 0 | Not-determined | Genome-fragment | NA | | NA | 0 |
| S12_vs_k141_133385\|\|full | 1076 | No | NA | | 3 | 0 | 0 | Not-determined | Genome-fragment | NA | | NA | 0 |
| S3_vs_k141_125650\|\|full | 4707 | No | NA | | 7 | 0 | 3 | Low-quality | Genome-fragment | | 9.56 | AAI-based (medium-confidence) | 0 |
| S5_vs_k141_29470\|\|full | 4707 | No | NA | | 7 | 4 | 0 | Low-quality | Genome-fragment | | 12.36 | AAI-based (high-confidence) | 0 |
| S12_vs_k141_172379\|\|full | 170104 | Yes | | 31748 | 189 | 15 | 82 | Medium-quality | Genome-fragment | | 69.36 | HMM-based (lower-bound) | 81.34 |
| S12_vs_k141_156630\|\|full | 2226 | No | NA | | 4 | 0 | 1 | Not-determined | Genome-fragment | NA | | NA | 0 |
| S12_vs_k141_172383\|\|full | 2512 | No | NA | | 3 | 0 | 0 | Not-determined | Genome-fragment | NA | | NA | 0 |
| S12_vs_k141_172418\|\|full | 1271 | No | NA | | 3 | 0 | 0 | Not-determined | Genome-fragment | NA | | NA | 0 |
| S8_extracted_k141_50764 | 4707 | No | NA | | 5 | 0 | 1 | Low-quality | Genome-fragment | | 10.82 | AAI-based (medium-confidence) | 0 |
| S12_vs_k141_133511\|\|full | 4275 | No | NA | | 6 | 0 | 1 | Not-determined | Genome-fragment | NA | | NA | 0 |
| S12_vs_k141_14927\|\|full | 86511 | No | NA | | 79 | 6 | 0 | High-quality | High-quality | | 94.18 | AAI-based (high-confidence) | 0 |
| S12_vs_k141_14928\|\|full | 2272 | No | NA | | 4 | 0 | 2 | Not-determined | Genome-fragment | NA | | NA | 0 |
| S3_vs_k141_102831\|\|full | 4708 | No | NA | | 6 | 0 | 3 | Low-quality | Genome-fragment | | 1.57 | AAI-based (medium-confidence) | 0 |
| S5_vs_k141_62592\|\|full | 4708 | No | NA | | 3 | 0 | 3 | Low-quality | Genome-fragment | | 1.86 | AAI-based (medium-confidence) | 0 |
| S12_vs_k141_165128\|\|full | 4879 | No | NA | | 2 | 0 | 1 | Not-determined | Genome-fragment | NA | | NA | 0 |
| S16_extracted_8845 | 4708 | No | NA | | 5 | 5 | 0 | Low-quality | Genome-fragment | | 12.86 | AAI-based (high-confidence) | 0 |
| S12_vs_k141_117518\|\|full | 4403 | No | NA | | 5 | 0 | 2 | Not-determined | Genome-fragment | NA | | NA | 0 |
| S12_vs_k141_180475\|\|full | 1226 | No | NA | | 1 | 0 | 0 | Not-determined | Genome-fragment | NA | | NA | 0 |
| S12_vs_k141_14981\|\|full | 4393 | No | NA | | 4 | 0 | 3 | Not-determined | Genome-fragment | NA | | NA | 0 |
| S14_extracted_k141_59496 | 4710 | No | NA | | 2 | 0 | 2 | Low-quality | Genome-fragment | | 1.86 | AAI-based (medium-confidence) | 0 |
| S3_extracted_k141_99390 | 4711 | No | NA | | 2 | 0 | 1 | Low-quality | Genome-fragment | | 7.76 | AAI-based (medium-confidence) | 0 |
| S12_vs_k141_86569\|\|full | 1198 | No | NA | | 5 | 0 | 0 | Not-determined | Genome-fragment | NA | | NA | 0 |
| S12_vs_k141_65898\|\|full | 4713 | No | NA | | 4 | 0 | 3 | Low-quality | Genome-fragment | | 2.76 | AAI-based (medium-confidence) | 0 |
| S12_vs_k141_86580\|\|full | 7213 | No | NA | | 7 | 0 | 5 | Not-determined | Genome-fragment | NA | | NA | 0 |
| S1_vs_k141_89772\|\|full | 4714 | No | NA | | 7 | 0 | 6 | Low-quality | Genome-fragment | | 1.4 | AAI-based (high-confidence) | 0 |
| S12_vs_k141_86604\|\|full | 1358 | No | NA | | 3 | 0 | 1 | Not-determined | Genome-fragment | NA | | NA | 0 |
| S5_extracted_k141_101936 | 4714 | No | NA | | 2 | 0 | 2 | Low-quality | Genome-fragment | | 4.34 | AAI-based (medium-confidence) | 0 |
| S4_extracted_22159 | 4717 | No | NA | | 5 | 3 | 0 | Low-quality | Genome-fragment | | 11.76 | AAI-based (high-confidence) | 0 |
| S11_extracted_k141_76887 | 4719 | No | NA | | 5 | 1 | 0 | Low-quality | Genome-fragment | | 8.47 | AAI-based (medium-confidence) | 0 |
| S12_vs_k141_38939\|\|full | 2195 | No | NA | | 3 | 0 | 2 | Not-determined | Genome-fragment | NA | | NA | 0 |
| S15_vs_46889\|\|full | 4719 | No | NA | | 4 | 0 | 3 | Low-quality | Genome-fragment | | 3.88 | AAI-based (high-confidence) | 0 |
| S12_vs_k141_156896\|\|full | 2727 | No | NA | | 2 | 0 | 2 | Not-determined | Genome-fragment | NA | | NA | 0 |
| S12_vs_k141_23574\|\|full | 1257 | No | NA | | 2 | 0 | 1 | Not-determined | Genome-fragment | NA | | NA | 0 |
| S11_extracted_k141_103543 | 4721 | No | NA | | 6 | 1 | 0 | Low-quality | Genome-fragment | | 4.26 | HMM-based (lower-bound) | 0 |
| S12_vs_k141_117759\|\|full | 1621 | No | NA | | 3 | 0 | 2 | Not-determined | Genome-fragment | NA | | NA | 0 |
| S12_vs_k141_15201\|\|full | 1825 | No | NA | | 3 | 0 | 2 | Not-determined | Genome-fragment | NA | | NA | 0 |
| S5_vs_k141_93891\|\|full | 4721 | No | NA | | 5 | 1 | 0 | Low-quality | Genome-fragment | | 7.68 | AAI-based (medium-confidence) | 0 |
| S7_vs_123614\|\|full | 4722 | No | NA | | 8 | 0 | 0 | Low-quality | Genome-fragment | | 13.42 | AAI-based (high-confidence) | 0 |
| S7_extracted_98313 | 4723 | No | NA | | 5 | 4 | 0 | Low-quality | Genome-fragment | | 14.23 | AAI-based (high-confidence) | 0 |
| S11_vs_k141_60579\|\|full | 4724 | No | NA | | 5 | 0 | 0 | Low-quality | Genome-fragment | | 10.41 | AAI-based (high-confidence) | 0 |
| S12_vs_k141_133843\|\|full | 6741 | No | NA | | 7 | 0 | 5 | Not-determined | Genome-fragment | NA | | NA | 0 |
| S5_vs_k141_28654\|\|full | 4724 | No | NA | | 13 | 1 | 0 | Low-quality | Genome-fragment | | 10.07 | AAI-based (medium-confidence) | 0 |
| S12_vs_k141_125520\|\|full | 1126 | No | NA | | 3 | 0 | 0 | Not-determined | Genome-fragment | NA | | NA | 0 |
| S1_vs_k141_36427\|\|full | 4725 | No | NA | | 3 | 0 | 1 | Low-quality | Genome-fragment | | 3.16 | AAI-based (medium-confidence) | 0 |
| S12_vs_k141_125527\|\|full | 1857 | No | NA | | 3 | 0 | 1 | Not-determined | Genome-fragment | NA | | NA | 0 |
| S12_vs_k141_39054\|\|full | 2326 | No | NA | | 3 | 0 | 2 | Not-determined | Genome-fragment | NA | | NA | 0 |
| S3_vs_k141_51793\|\|full | 4725 | No | NA | | 3 | 0 | 1 | Low-quality | Genome-fragment | | 7.61 | AAI-based (medium-confidence) | 0 |
| S12_vs_k141_157003\|\|full | 1214 | No | NA | | 2 | 0 | 1 | Not-determined | Genome-fragment | NA | | NA | 0 |
| S12_vs_k141_151316\|\|full | 4725 | No | NA | | 7 | 0 | 0 | Low-quality | Genome-fragment | | 8.06 | AAI-based (high-confidence) | 0 |
| S12_vs_k141_15339\|\|full | 1733 | No | NA | | 2 | 0 | 2 | Not-determined | Genome-fragment | NA | | NA | 0 |
| S17_vs_k141_159630\|\|full | 4726 | No | NA | | 7 | 1 | 1 | Low-quality | Genome-fragment | | 2.74 | AAI-based (high-confidence) | 0 |
| S1_extracted_k141_19851 | 4727 | No | NA | | 6 | 1 | 0 | Low-quality | Genome-fragment | | 7.07 | HMM-based (lower-bound) | 0 |
| S12_vs_k141_117948\|\|full | 2495 | No | NA | | 4 | 0 | 2 | Not-determined | Genome-fragment | NA | | NA | 0 |
| S12_vs_k141_125659\|\|full | 1314 | No | NA | | 2 | 0 | 0 | Not-determined | Genome-fragment | NA | | NA | 0 |
| S10_vs_79807\|\|full | 4727 | No | NA | | 3 | 0 | 0 | Low-quality | Genome-fragment | | 5.89 | AAI-based (medium-confidence) | 0 |
| S12_vs_k141_102047\|\|full | 4728 | Yes | | 3569 | 7 | 1 | 2 | Low-quality | Genome-fragment | | 7.2 | AAI-based (medium-confidence) | 24.51 |
| S12_vs_k141_173025\|\|full | 2584 | No | NA | | 3 | 0 | 2 | Not-determined | Genome-fragment | NA | | NA | 0 |
| S12_vs_k141_121866\|\|full | 4729 | No | NA | | 9 | 2 | 0 | Low-quality | Genome-fragment | | 11.69 | AAI-based (high-confidence) | 0 |
| S12_vs_k141_180692\|\|full | 2950 | No | NA | | 4 | 0 | 0 | Not-determined | Genome-fragment | NA | | NA | 0 |
| S5_vs_k141_43235\|\|full | 4731 | No | NA | | 5 | 0 | 0 | Low-quality | Genome-fragment | | 9.13 | AAI-based (high-confidence) | 0 |
| S12_vs_k141_125702\|\|full | 1701 | No | NA | | 3 | 0 | 0 | Not-determined | Genome-fragment | NA | | NA | 0 |
| S8_extracted_k141_108083 | 4732 | No | NA | | 4 | 0 | 3 | Low-quality | Genome-fragment | | 2.6 | AAI-based (medium-confidence) | 0 |
| S17_extracted_k141_70526 | 4733 | No | NA | | 6 | 4 | 0 | Low-quality | Genome-fragment | | 7.52 | HMM-based (lower-bound) | 0 |
| S12_vs_k141_39260\|\|full | 1512 | No | NA | | 3 | 0 | 0 | Not-determined | Genome-fragment | NA | | NA | 0 |
| S12_vs_k141_173132\|\|full | 1589 | No | NA | | 4 | 0 | 3 | Not-determined | Genome-fragment | NA | | NA | 0 |
| S12_vs_k141_125714\|\|full | 9069 | No | NA | | 7 | 0 | 4 | Not-determined | Genome-fragment | NA | | NA | 0 |
| S16_extracted_112920 | 4733 | No | NA | | 3 | 3 | 0 | Low-quality | Genome-fragment | | 11.64 | AAI-based (high-confidence) | 0 |
| S8_extracted_k141_46640 | 4735 | No | NA | | 6 | 3 | 0 | Low-quality | Genome-fragment | | 24.23 | AAI-based (high-confidence) | 0 |
| S10_vs_106627\|\|full | 4736 | No | NA | | 6 | 0 | 0 | Low-quality | Genome-fragment | | 8.56 | AAI-based (high-confidence) | 0 |
| S12_vs_k141_15596\|\|full | 1798 | No | NA | | 2 | 0 | 0 | Not-determined | Genome-fragment | NA | | NA | 0 |
| S12_vs_k141_15621\|\|full | 1667 | No | NA | | 3 | 0 | 0 | Not-determined | Genome-fragment | NA | | NA | 0 |
| S8_extracted_k141_95041 | 4737 | No | NA | | 9 | 0 | 0 | Low-quality | Genome-fragment | | 10.68 | AAI-based (high-confidence) | 0 |
| S11_vs_k141_55201\|\|full | 4738 | No | NA | | 9 | 0 | 0 | Low-quality | Genome-fragment | | 3.8 | AAI-based (high-confidence) | 0 |
| S13_vs_46653\|\|full | 4738 | No | NA | | 8 | 1 | 0 | Low-quality | Genome-fragment | | 11.84 | AAI-based (high-confidence) | 0 |
| S12_vs_k141_180878\|\|full | 2075 | No | NA | | 2 | 0 | 0 | Not-determined | Genome-fragment | NA | | NA | 0 |
| S12_vs_k141_180915\|\|full | 1634 | No | NA | | 3 | 0 | 0 | Not-determined | Genome-fragment | NA | | NA | 0 |
| S12_vs_k141_180930\|\|full | 1801 | No | NA | | 3 | 0 | 1 | Not-determined | Genome-fragment | NA | | NA | 0 |
| S12_vs_k141_23796\|\|0_partial | 63354 | Yes | | 38428 | 55 | 9 | 13 | Medium-quality | Genome-fragment | | 89.72 | AAI-based (medium-confidence) | 39.34 |
| S16_extracted_130867 | 4738 | No | NA | | 7 | 1 | 0 | Low-quality | Genome-fragment | | 10.57 | AAI-based (high-confidence) | 0 |
| S8_extracted_k141_71739 | 4738 | No | NA | | 4 | 1 | 0 | Low-quality | Genome-fragment | | 9.56 | AAI-based (high-confidence) | 0 |
| S12_vs_k141_158058\|\|0_partial | 13533 | No | NA | | 19 | 0 | 10 | Not-determined | Genome-fragment | NA | | NA | 0 |
| S8_vs_k141_64587\|\|full | 4738 | No | NA | | 8 | 0 | 0 | Low-quality | Genome-fragment | | 6.78 | AAI-based (medium-confidence) | 0 |
| S3_vs_k141_50252\|\|full | 4739 | No | NA | | 4 | 1 | 0 | Low-quality | Genome-fragment | | 2.8 | AAI-based (medium-confidence) | 0 |
| S12_vs_k141_103765\|\|0_partial | 25201 | No | NA | | 12 | 0 | 5 | Not-determined | Genome-fragment | NA | | NA | 0 |
| S8_vs_k141_74906\|\|full | 4739 | No | NA | | 6 | 0 | 3 | Low-quality | Genome-fragment | | 13.98 | AAI-based (high-confidence) | 0 |
| S5_extracted_k141_135638 | 4740 | No | NA | | 4 | 4 | 0 | Low-quality | Genome-fragment | | 11.66 | AAI-based (high-confidence) | 0 |
| S3_extracted_k141_93103 | 4741 | No | NA | | 4 | 0 | 4 | Low-quality | Genome-fragment | | 4.77 | AAI-based (medium-confidence) | 0 |
| S12_vs_k141_32669\|\|0_partial | 24358 | No | NA | | 25 | 0 | 7 | Medium-quality | Genome-fragment | | 69.41 | AAI-based (medium-confidence) | 0 |
| S12_extracted_k141_44092 | 4741 | No | NA | | 6 | 0 | 3 | Low-quality | Genome-fragment | | 1.86 | AAI-based (medium-confidence) | 0 |
| S12_vs_k141_25672\|\|0_partial | 13729 | No | NA | | 14 | 0 | 2 | Not-determined | Genome-fragment | NA | | NA | 0 |
| S17_extracted_k141_149665 | 4744 | No | NA | | 6 | 3 | 0 | Low-quality | Genome-fragment | | 11.63 | AAI-based (medium-confidence) | 0 |
| S12_vs_k141_183908\|\|0_partial | 15965 | No | NA | | 17 | 0 | 11 | Not-determined | Genome-fragment | NA | | NA | 0 |
| S1_vs_k141_104833\|\|full | 4744 | No | NA | | 9 | 0 | 0 | Low-quality | Genome-fragment | | 9.11 | AAI-based (high-confidence) | 0 |
| S14_extracted_k141_75528 | 4745 | No | NA | | 6 | 5 | 0 | Low-quality | Genome-fragment | | 15.62 | AAI-based (high-confidence) | 0 |
| S12_vs_k141_58711\|\|0_partial | 16022 | No | NA | | 11 | 0 | 8 | Not-determined | Genome-fragment | NA | | NA | 0 |
| S12_vs_k141_19581\|\|0_partial | 25769 | No | NA | | 20 | 0 | 9 | Not-determined | Genome-fragment | NA | | NA | 0 |
| S12_vs_k141_106720\|\|0_partial | 11750 | No | NA | | 11 | 0 | 8 | Not-determined | Genome-fragment | NA | | NA | 0 |
| S8_vs_k141_163821\|\|full | 4746 | No | NA | | 3 | 0 | 0 | Low-quality | Genome-fragment | | 5.17 | AAI-based (medium-confidence) | 0 |
| S16_extracted_87713 | 4747 | No | NA | | 2 | 2 | 0 | Low-quality | Genome-fragment | | 12.78 | AAI-based (high-confidence) | 0 |
| S10_vs_11637\|\|full | 4747 | No | NA | | 11 | 0 | 0 | Low-quality | Genome-fragment | | 11.66 | AAI-based (high-confidence) | 0 |
| S7_extracted_113393 | 4749 | No | NA | | 7 | 4 | 0 | Low-quality | Genome-fragment | | 11.78 | AAI-based (high-confidence) | 0 |
| S12_vs_k141_6860\|\|0_partial | 12553 | No | NA | | 5 | 0 | 2 | Not-determined | Genome-fragment | NA | | NA | 0 |
| S12_vs_k141_164274\|\|0_partial | 11758 | No | NA | | 7 | 0 | 4 | Not-determined | Genome-fragment | NA | | NA | 0 |
| S12_vs_k141_171917\|\|0_partial | 11728 | No | NA | | 12 | 0 | 3 | Not-determined | Genome-fragment | NA | | NA | 0 |
| S12_vs_k141_125063\|\|0_partial | 14010 | No | NA | | 11 | 0 | 1 | Not-determined | Genome-fragment | NA | | NA | 0 |
| S16_extracted_11869 | 4750 | No | NA | | 5 | 2 | 0 | Low-quality | Genome-fragment | | 5.62 | AAI-based (high-confidence) | 0 |
| S12_vs_k141_165294\|\|0_partial | 16135 | No | NA | | 23 | 0 | 5 | Not-determined | Genome-fragment | NA | | NA | 0 |
| S7_extracted_44613 | 4751 | No | NA | | 9 | 1 | 0 | Low-quality | Genome-fragment | | 9.78 | AAI-based (high-confidence) | 0 |
| S16_vs_1858\|\|full | 4751 | No | NA | | 13 | 2 | 0 | Low-quality | Genome-fragment | | 12.18 | AAI-based (medium-confidence) | 0 |
| S15_vs_50996\|\|full | 4752 | No | NA | | 5 | 0 | 0 | Low-quality | Genome-fragment | | 8.51 | AAI-based (medium-confidence) | 0 |
| S5_extracted_k141_47093 | 4754 | No | NA | | 4 | 3 | 0 | Low-quality | Genome-fragment | | 14.75 | AAI-based (high-confidence) | 0 |
| S10_extracted_55920 | 4754 | No | NA | | 3 | 0 | 1 | Low-quality | Genome-fragment | | 7.94 | AAI-based (high-confidence) | 0 |
| S8_vs_k141_156368\|\|full | 4754 | No | NA | | 6 | 0 | 0 | Low-quality | Genome-fragment | | 10.91 | AAI-based (medium-confidence) | 0 |
| S12_vs_k141_39995\|\|lt2gene | 1537 | No | NA | | 1 | 0 | 0 | Not-determined | Genome-fragment | NA | | NA | 0 |
| S8_vs_k141_137807\|\|full | 4754 | No | NA | | 6 | 0 | 1 | Low-quality | Genome-fragment | | 5.75 | AAI-based (medium-confidence) | 0 |
| S6_vs_k141_31756\|\|full | 4755 | No | NA | | 8 | 0 | 1 | Low-quality | Genome-fragment | | 6.63 | AAI-based (medium-confidence) | 0 |
| S8_vs_k141_165738\|\|full | 4755 | No | NA | | 3 | 1 | 0 | Low-quality | Genome-fragment | | 1.62 | HMM-based (lower-bound) | 0 |
| S8_extracted_k141_139810 | 4756 | No | NA | | 9 | 0 | 0 | Low-quality | Genome-fragment | | 8.35 | AAI-based (medium-confidence) | 0 |
| S4_extracted_30434 | 4757 | No | NA | | 8 | 3 | 0 | Low-quality | Genome-fragment | | 8.63 | AAI-based (high-confidence) | 0 |
| S3_extracted_k141_103653 | 4758 | No | NA | | 5 | 0 | 3 | Low-quality | Genome-fragment | | 3.04 | AAI-based (high-confidence) | 0 |
| S16_vs_6144\|\|full | 4758 | No | NA | | 5 | 2 | 0 | Low-quality | Genome-fragment | | 1.67 | HMM-based (lower-bound) | 0 |
| S10_extracted_36515 | 4759 | No | NA | | 6 | 4 | 0 | Low-quality | Genome-fragment | | 11.2 | AAI-based (high-confidence) | 0 |
| S9_vs_k141_3414\|\|full | 4761 | No | NA | | 7 | 1 | 0 | Low-quality | Genome-fragment | | 10.93 | AAI-based (high-confidence) | 0 |
| S12_vs_k141_79765\|\|lt2gene | 1109 | No | NA | | 1 | 0 | 1 | Not-determined | Genome-fragment | NA | | NA | 0 |
| S14_vs_k141_55180\|\|full | 4761 | No | NA | | 7 | 3 | 0 | Low-quality | Genome-fragment | | 12.13 | AAI-based (medium-confidence) | 0 |
| S1_vs_k141_45899\|\|full | 4761 | No | NA | | 7 | 6 | 0 | Low-quality | Genome-fragment | | 3.27 | AAI-based (high-confidence) | 0 |
| S5_vs_k141_122413\|\|full | 4761 | No | NA | | 6 | 0 | 0 | Low-quality | Genome-fragment | | 10.03 | AAI-based (medium-confidence) | 0 |
| S12_vs_k141_56852\|\|lt2gene | 1169 | No | NA | | 2 | 0 | 0 | Not-determined | Genome-fragment | NA | | NA | 0 |
| S12_vs_k141_103977\|\|lt2gene | 1703 | No | NA | | 1 | 0 | 0 | Not-determined | Genome-fragment | NA | | NA | 0 |
| S12_vs_k141_104056\|\|lt2gene | 1555 | No | NA | | 2 | 0 | 0 | Not-determined | Genome-fragment | NA | | NA | 0 |
| S10_extracted_313 | 4761 | No | NA | | 8 | 0 | 0 | Low-quality | Genome-fragment | | 10.34 | AAI-based (high-confidence) | 0 |
| S12_extracted_k141_28239 | 4762 | Yes | | 3821 | 4 | 1 | 1 | Low-quality | Genome-fragment | | 7.15 | HMM-based (lower-bound) | 19.76 |
| S8_vs_k141_45421\|\|full | 4762 | No | NA | | 5 | 0 | 2 | Low-quality | Genome-fragment | | 2.45 | AAI-based (medium-confidence) | 0 |
| S12_vs_k141_2262\|\|lt2gene | 1348 | No | NA | | 2 | 0 | 2 | Not-determined | Genome-fragment | NA | | NA | 0 |
| S8_extracted_k141_55094 | 4766 | No | NA | | 3 | 1 | 0 | Low-quality | Genome-fragment | | 9.42 | AAI-based (high-confidence) | 0 |
| S8_extracted_k141_15348 | 4767 | No | NA | | 5 | 0 | 2 | Low-quality | Genome-fragment | | 0.68 | AAI-based (medium-confidence) | 0 |
| S7_vs_54712\|\|full | 4769 | No | NA | | 9 | 3 | 0 | Low-quality | Genome-fragment | | 10.17 | AAI-based (high-confidence) | 0 |
| S12_vs_k141_97486\|\|lt2gene | 1604 | No | NA | | 2 | 0 | 0 | Not-determined | Genome-fragment | NA | | NA | 0 |
| S6_extracted_k141_24820 | 4769 | Yes | | 4024 | 7 | 1 | 1 | Low-quality | Genome-fragment | | 10.63 | AAI-based (high-confidence) | 15.62 |
| S8_extracted_k141_173718 | 4769 | No | NA | | 4 | 0 | 1 | Low-quality | Genome-fragment | | 0.41 | AAI-based (medium-confidence) | 0 |
| S9_vs_k141_1879\|\|full | 4770 | No | NA | | 3 | 1 | 0 | Low-quality | Genome-fragment | | 7.41 | AAI-based (high-confidence) | 0 |
| S17_vs_k141_94121\|\|full | 4772 | Yes | | 2339 | 5 | 1 | 2 | Low-quality | Genome-fragment | | 4.1 | HMM-based (lower-bound) | 50.98 |
| S12_vs_k141_58293\|\|lt2gene | 1476 | No | NA | | 1 | 0 | 0 | Not-determined | Genome-fragment | NA | | NA | 0 |
| S12_vs_k141_152021\|\|lt2gene | 1078 | No | NA | | 1 | 0 | 1 | Not-determined | Genome-fragment | NA | | NA | 0 |
| S14_vs_k141_98573\|\|full | 4772 | No | NA | | 4 | 0 | 2 | Low-quality | Genome-fragment | | 6.76 | AAI-based (medium-confidence) | 0 |
| S12_vs_k141_66115\|\|lt2gene | 1596 | No | NA | | 2 | 0 | 0 | Not-determined | Genome-fragment | NA | | NA | 0 |
| S10_vs_50624\|\|full | 4772 | No | NA | | 6 | 4 | 0 | Low-quality | Genome-fragment | | 10.05 | AAI-based (high-confidence) | 0 |
| S12_vs_k141_136966\|\|lt2gene | 1060 | No | NA | | 1 | 0 | 0 | Not-determined | Genome-fragment | NA | | NA | 0 |
| S12_vs_k141_136988\|\|lt2gene | 1012 | No | NA | | 2 | 0 | 2 | Not-determined | Genome-fragment | NA | | NA | 0 |
| S12_vs_k141_58634\|\|lt2gene | 1725 | No | NA | | 2 | 0 | 1 | Not-determined | Genome-fragment | NA | | NA | 0 |
| S12_vs_k141_3591\|\|lt2gene | 1096 | No | NA | | 2 | 0 | 2 | Not-determined | Genome-fragment | NA | | NA | 0 |
| S11_vs_k141_54203\|\|full | 4773 | No | NA | | 9 | 3 | 0 | Low-quality | Genome-fragment | | 9.5 | AAI-based (high-confidence) | 0 |
| S5_vs_k141_44025\|\|0_partial | 4774 | Yes | | 2266 | 6 | 1 | 1 | Low-quality | Genome-fragment | | 1.99 | AAI-based (high-confidence) | 52.53 |
| S5_vs_k141_79015\|\|full | 4775 | No | NA | | 3 | 0 | 2 | Low-quality | Genome-fragment | | 4.7 | AAI-based (medium-confidence) | 0 |
| S3_vs_k141_1158\|\|full | 4776 | No | NA | | 5 | 0 | 1 | Low-quality | Genome-fragment | | 8.15 | AAI-based (medium-confidence) | 0 |
| S7_vs_16363\|\|full | 4776 | No | NA | | 8 | 2 | 0 | Low-quality | Genome-fragment | | 10.99 | AAI-based (medium-confidence) | 0 |
| S12_vs_k141_106113\|\|lt2gene | 1456 | No | NA | | 1 | 0 | 1 | Not-determined | Genome-fragment | NA | | NA | 0 |
| S12_vs_k141_50243\|\|lt2gene | 1092 | No | NA | | 1 | 0 | 0 | Not-determined | Genome-fragment | NA | | NA | 0 |
| S12_vs_k141_43407\|\|lt2gene | 1590 | No | NA | | 2 | 0 | 0 | Not-determined | Genome-fragment | NA | | NA | 0 |
| S11_vs_k141_47545\|\|full | 4777 | No | NA | | 5 | 0 | 0 | Low-quality | Genome-fragment | | 8.07 | AAI-based (medium-confidence) | 0 |
| S7_extracted_115769 | 4778 | No | NA | | 4 | 0 | 3 | Low-quality | Genome-fragment | | 1.89 | AAI-based (medium-confidence) | 0 |
| S15_vs_55853\|\|full | 4778 | No | NA | | 4 | 0 | 4 | Low-quality | Genome-fragment | | 2.73 | AAI-based (medium-confidence) | 0 |
| S9_vs_k141_61361\|\|full | 4780 | No | NA | | 10 | 5 | 0 | Low-quality | Genome-fragment | | 9.65 | AAI-based (medium-confidence) | 0 |
| S7_vs_87651\|\|full | 4780 | No | NA | | 4 | 2 | 0 | Low-quality | Genome-fragment | | 8.92 | AAI-based (medium-confidence) | 0 |
| S16_extracted_111786 | 4781 | No | NA | | 4 | 0 | 1 | Low-quality | Genome-fragment | | 4.96 | AAI-based (medium-confidence) | 0 |
| S3_vs_k141_93751\|\|full | 4782 | No | NA | | 6 | 0 | 1 | Low-quality | Genome-fragment | | 12.32 | AAI-based (high-confidence) | 0 |
| S7_vs_14959\|\|full | 4785 | No | NA | | 8 | 0 | 1 | Low-quality | Genome-fragment | | 8.91 | AAI-based (medium-confidence) | 0 |
| S15_vs_36094\|\|full | 4785 | No | NA | | 3 | 0 | 2 | Low-quality | Genome-fragment | | 4 | AAI-based (medium-confidence) | 0 |
| S14_extracted_k141_51782 | 4788 | No | NA | | 5 | 5 | 0 | Low-quality | Genome-fragment | | 15.8 | AAI-based (high-confidence) | 0 |
| S3_vs_k141_49227\|\|full | 4789 | No | NA | | 6 | 0 | 0 | Low-quality | Genome-fragment | | 8.11 | AAI-based (high-confidence) | 0 |
| S11_vs_k141_11680\|\|full | 4790 | No | NA | | 7 | 2 | 0 | Low-quality | Genome-fragment | | 3.77 | HMM-based (lower-bound) | 0 |
| S1_vs_k141_137406\|\|full | 4790 | No | NA | | 10 | 0 | 0 | Low-quality | Genome-fragment | | 3.12 | AAI-based (high-confidence) | 0 |
| S12_vs_k141_129652\|\|lt2gene | 1521 | No | NA | | 2 | 0 | 0 | Not-determined | Genome-fragment | NA | | NA | 0 |
| S2_extracted_k141_437 | 4790 | No | NA | | 6 | 4 | 0 | Low-quality | Genome-fragment | | 7.82 | AAI-based (high-confidence) | 0 |
| S12_extracted_k141_137626 | 4792 | No | NA | | 7 | 0 | 1 | Low-quality | Genome-fragment | | 12.21 | AAI-based (high-confidence) | 0 |
| S10_extracted_12096 | 4793 | No | NA | | 4 | 0 | 0 | Low-quality | Genome-fragment | | 9.39 | AAI-based (medium-confidence) | 0 |
| S12_vs_k141_130023\|\|lt2gene | 2507 | No | NA | | 1 | 0 | 1 | Not-determined | Genome-fragment | NA | | NA | 0 |
| S4_extracted_2663 | 4794 | No | NA | | 9 | 1 | 0 | Low-quality | Genome-fragment | | 13.29 | AAI-based (high-confidence) | 0 |
| S12_vs_k141_161925\|\|lt2gene | 2078 | No | NA | | 2 | 0 | 0 | Not-determined | Genome-fragment | NA | | NA | 0 |
| S12_vs_k141_67633\|\|lt2gene | 1032 | No | NA | | 1 | 0 | 0 | Not-determined | Genome-fragment | NA | | NA | 0 |
| S12_vs_k141_28441\|\|lt2gene | 1003 | No | NA | | 1 | 0 | 1 | Not-determined | Genome-fragment | NA | | NA | 0 |
| S12_vs_k141_186340\|\|lt2gene | 1167 | No | NA | | 1 | 0 | 0 | Not-determined | Genome-fragment | NA | | NA | 0 |
| S16_vs_76010\|\|full | 4795 | No | NA | | 12 | 1 | 0 | Low-quality | Genome-fragment | | 8.86 | AAI-based (high-confidence) | 0 |
| S1_vs_k141_98028\|\|full | 4798 | No | NA | | 6 | 2 | 0 | Low-quality | Genome-fragment | | 17.47 | HMM-based (lower-bound) | 0 |
| S11_vs_k141_32401\|\|full | 4799 | No | NA | | 4 | 0 | 4 | Low-quality | Genome-fragment | | 8.17 | AAI-based (medium-confidence) | 0 |
| S6_vs_k141_42274\|\|full | 4801 | No | NA | | 9 | 1 | 0 | Low-quality | Genome-fragment | | 8.2 | AAI-based (high-confidence) | 0 |
| S8_extracted_k141_208083 | 4801 | No | NA | | 7 | 2 | 0 | Low-quality | Genome-fragment | | 12.21 | AAI-based (medium-confidence) | 0 |
| S12_vs_k141_29068\|\|lt2gene | 1647 | No | NA | | 2 | 0 | 0 | Not-determined | Genome-fragment | NA | | NA | 0 |
| S11_vs_k141_57675\|\|full | 4803 | No | NA | | 8 | 0 | 0 | Low-quality | Genome-fragment | | 12.84 | AAI-based (high-confidence) | 0 |
| S2_extracted_k141_17460 | 4804 | No | NA | | 7 | 4 | 0 | Low-quality | Genome-fragment | | 24.77 | AAI-based (medium-confidence) | 0 |
| S1_vs_k141_104803\|\|full | 4805 | No | NA | | 12 | 1 | 0 | Low-quality | Genome-fragment | | 13.13 | AAI-based (high-confidence) | 0 |
| S16_extracted_151668 | 4805 | No | NA | | 9 | 3 | 0 | Low-quality | Genome-fragment | | 5.23 | HMM-based (lower-bound) | 0 |
| S12_vs_k141_187277\|\|lt2gene | 1717 | No | NA | | 1 | 0 | 0 | Not-determined | Genome-fragment | NA | | NA | 0 |
| S9_extracted_k141_23240 | 4806 | No | NA | | 7 | 1 | 0 | Low-quality | Genome-fragment | | 10.71 | AAI-based (high-confidence) | 0 |
| S16_vs_165365\|\|full | 4806 | No | NA | | 8 | 0 | 0 | Low-quality | Genome-fragment | | 11.22 | AAI-based (medium-confidence) | 0 |
| S10_vs_2904\|\|full | 4806 | No | NA | | 3 | 0 | 2 | Low-quality | Genome-fragment | | 3.11 | AAI-based (high-confidence) | 0 |
| S17_vs_k141_136462\|\|full | 4807 | No | NA | | 8 | 7 | 0 | Low-quality | Genome-fragment | | 7.77 | AAI-based (high-confidence) | 0 |
| S17_vs_k141_168026\|\|full | 4809 | No | NA | | 5 | 0 | 3 | Low-quality | Genome-fragment | | 2.64 | AAI-based (medium-confidence) | 0 |
| S10_vs_43165\|\|full | 4809 | No | NA | | 6 | 3 | 0 | Low-quality | Genome-fragment | | 11.63 | AAI-based (medium-confidence) | 0 |
| S14_vs_k141_25370\|\|full | 4810 | No | NA | | 7 | 1 | 0 | Low-quality | Genome-fragment | | 9.49 | HMM-based (lower-bound) | 0 |
| S15_extracted_44153 | 4810 | No | NA | | 7 | 1 | 0 | Low-quality | Genome-fragment | | 5.39 | HMM-based (lower-bound) | 0 |
| S12_vs_k141_62047\|\|lt2gene | 1218 | No | NA | | 2 | 0 | 0 | Not-determined | Genome-fragment | NA | | NA | 0 |
| S12_vs_k141_46946\|\|lt2gene | 1093 | No | NA | | 1 | 0 | 0 | Not-determined | Genome-fragment | NA | | NA | 0 |
| S1_extracted_k141_99185 | 4811 | No | NA | | 6 | 2 | 0 | Low-quality | Genome-fragment | | 15.88 | HMM-based (lower-bound) | 0 |
| S1_extracted_k141_112408 | 4811 | No | NA | | 8 | 3 | 0 | Low-quality | Genome-fragment | | 10.32 | AAI-based (high-confidence) | 0 |
| S7_vs_55853\|\|full | 4811 | No | NA | | 9 | 0 | 0 | Low-quality | Genome-fragment | | 11.4 | AAI-based (medium-confidence) | 0 |
| S3_extracted_k141_2120 | 4812 | No | NA | | 19 | 0 | 0 | Low-quality | Genome-fragment | | 2.41 | AAI-based (high-confidence) | 0 |
| S9_vs_k141_35095\|\|full | 4813 | No | NA | | 8 | 1 | 0 | Low-quality | Genome-fragment | | 12.74 | AAI-based (high-confidence) | 0 |
| S7_vs_8054\|\|full | 4813 | No | NA | | 3 | 0 | 0 | Low-quality | Genome-fragment | | 12.67 | AAI-based (medium-confidence) | 0 |
| S3_vs_k141_36613\|\|full | 4814 | No | NA | | 7 | 0 | 3 | Low-quality | Genome-fragment | | 2.98 | AAI-based (medium-confidence) | 0 |
| S12_vs_k141_107726\|\|full | 4814 | No | NA | | 4 | 0 | 1 | Low-quality | Genome-fragment | | 2.99 | AAI-based (high-confidence) | 0 |
| S1_extracted_k141_108007 | 4815 | No | NA | | 7 | 3 | 0 | Low-quality | Genome-fragment | | 10.55 | AAI-based (high-confidence) | 0 |
| S12_vs_k141_173024\|\|lt2gene | 1244 | No | NA | | 1 | 0 | 0 | Not-determined | Genome-fragment | NA | | NA | 0 |
| S15_extracted_122 | 1367 | No | NA | | 3 | 0 | 2 | Not-determined | Genome-fragment | NA | | NA | 0 |
| S15_extracted_574 | 1168 | No | NA | | 2 | 0 | 1 | Not-determined | Genome-fragment | NA | | NA | 0 |
| S16_vs_162316\|\|full | 4815 | Yes | | 2954 | 9 | 1 | 1 | Low-quality | Genome-fragment | | 3.5 | HMM-based (lower-bound) | 38.65 |
| S15_extracted_717 | 3200 | No | NA | | 4 | 0 | 1 | Not-determined | Genome-fragment | NA | | NA | 0 |
| S15_extracted_737 | 12556 | No | NA | | 15 | 0 | 3 | Not-determined | Genome-fragment | NA | | NA | 0 |
| S10_vs_35960\|\|full | 4817 | No | NA | | 4 | 0 | 3 | Low-quality | Genome-fragment | | 2.36 | AAI-based (medium-confidence) | 0 |
| S9_vs_k141_87586\|\|full | 4819 | No | NA | | 5 | 4 | 0 | Low-quality | Genome-fragment | | 12.67 | AAI-based (high-confidence) | 0 |
| S15_extracted_1092 | 2123 | No | NA | | 2 | 0 | 2 | Not-determined | Genome-fragment | NA | | NA | 0 |
| S15_extracted_1190 | 2261 | No | NA | | 1 | 0 | 1 | Not-determined | Genome-fragment | NA | | NA | 0 |
| S1_vs_k141_69623\|\|full | 4819 | No | NA | | 12 | 2 | 0 | Low-quality | Genome-fragment | | 12.2 | AAI-based (high-confidence) | 0 |
| S15_extracted_1310 | 1194 | No | NA | | 3 | 0 | 0 | Not-determined | Genome-fragment | NA | | NA | 0 |
| S15_extracted_1582 | 3255 | No | NA | | 3 | 0 | 2 | Not-determined | Genome-fragment | NA | | NA | 0 |
| S15_extracted_1856 | 2048 | No | NA | | 2 | 0 | 0 | Not-determined | Genome-fragment | NA | | NA | 0 |
| S15_extracted_2032 | 68670 | No | NA | | 106 | 18 | 0 | Complete | High-quality | | 100 | DTR (high-confidence) | 0 |
| S15_extracted_2087 | 1230 | No | NA | | 2 | 0 | 0 | Not-determined | Genome-fragment | NA | | NA | 0 |
| S15_extracted_2109 | 1126 | No | NA | | 3 | 0 | 1 | Not-determined | Genome-fragment | NA | | NA | 0 |
| S15_extracted_2305 | 1066 | No | NA | | 1 | 0 | 1 | Not-determined | Genome-fragment | NA | | NA | 0 |
| S15_extracted_2351 | 3842 | No | NA | | 4 | 0 | 4 | Not-determined | Genome-fragment | NA | | NA | 0 |
| S15_extracted_2372 | 5298 | No | NA | | 5 | 0 | 3 | Not-determined | Genome-fragment | NA | | NA | 0 |
| S15_extracted_2382 | 1574 | No | NA | | 2 | 0 | 2 | Not-determined | Genome-fragment | NA | | NA | 0 |
| S15_extracted_2405 | 1059 | No | NA | | 2 | 0 | 0 | Not-determined | Genome-fragment | NA | | NA | 0 |
| S15_extracted_2513 | 57304 | No | NA | | 92 | 15 | 0 | High-quality | High-quality | | 100 | AAI-based (high-confidence) | 0 |
| S11_extracted_k141_70294 | 4820 | No | NA | | 7 | 1 | 0 | Low-quality | Genome-fragment | | 13.08 | AAI-based (high-confidence) | 0 |
| S3_vs_k141_11555\|\|full | 4820 | No | NA | | 6 | 3 | 0 | Low-quality | Genome-fragment | | 12.82 | AAI-based (high-confidence) | 0 |
| S16_vs_112306\|\|full | 4820 | No | NA | | 5 | 0 | 0 | Low-quality | Genome-fragment | | 2.82 | AAI-based (medium-confidence) | 0 |
| S15_extracted_2934 | 7630 | No | NA | | 4 | 0 | 2 | Not-determined | Genome-fragment | NA | | NA | 0 |
| S15_extracted_3006 | 1437 | No | NA | | 2 | 0 | 2 | Not-determined | Genome-fragment | NA | | NA | 0 |
| S5_vs_k141_5159\|\|full | 4823 | No | NA | | 7 | 2 | 0 | Low-quality | Genome-fragment | | 12.69 | AAI-based (high-confidence) | 0 |
| S15_extracted_3251 | 1098 | No | NA | | 1 | 0 | 0 | Not-determined | Genome-fragment | NA | | NA | 0 |
| S3_vs_k141_37457\|\|full | 4824 | No | NA | | 5 | 3 | 0 | Low-quality | Genome-fragment | | 3.3 | AAI-based (medium-confidence) | 0 |
| S15_extracted_3462 | 1184 | No | NA | | 2 | 0 | 2 | Not-determined | Genome-fragment | NA | | NA | 0 |
| S15_extracted_3506 | 4267 | No | NA | | 3 | 0 | 3 | Not-determined | Genome-fragment | NA | | NA | 0 |
| S15_extracted_3757 | 3182 | No | NA | | 3 | 0 | 0 | Not-determined | Genome-fragment | NA | | NA | 0 |
| S15_extracted_3831 | 13430 | No | NA | | 17 | 0 | 0 | Not-determined | Genome-fragment | NA | | NA | 0 |
| S15_extracted_3887 | 1137 | No | NA | | 1 | 0 | 0 | Not-determined | Genome-fragment | NA | | NA | 0 |
| S13_extracted_30032 | 4825 | No | NA | | 7 | 0 | 2 | Low-quality | Genome-fragment | | 5.11 | AAI-based (high-confidence) | 0 |
| S15_extracted_4196 | 1921 | No | NA | | 1 | 0 | 1 | Not-determined | Genome-fragment | NA | | NA | 0 |
| S15_extracted_4257 | 1587 | No | NA | | 1 | 0 | 1 | Not-determined | Genome-fragment | NA | | NA | 0 |
| S15_extracted_4490 | 3013 | No | NA | | 4 | 0 | 1 | Not-determined | Genome-fragment | NA | | NA | 0 |
| S15_extracted_4601 | 1074 | No | NA | | 1 | 0 | 0 | Not-determined | Genome-fragment | NA | | NA | 0 |
| S15_extracted_4680 | 3914 | No | NA | | 4 | 0 | 4 | Not-determined | Genome-fragment | NA | | NA | 0 |
| S15_extracted_4941 | 1369 | No | NA | | 2 | 0 | 2 | Not-determined | Genome-fragment | NA | | NA | 0 |
| S15_extracted_5029 | 6015 | No | NA | | 9 | 0 | 1 | Not-determined | Genome-fragment | NA | | NA | 0 |
| S15_extracted_5199 | 1768 | No | NA | | 1 | 0 | 1 | Not-determined | Genome-fragment | NA | | NA | 0 |
| S15_extracted_5498 | 2467 | No | NA | | 3 | 0 | 3 | Not-determined | Genome-fragment | NA | | NA | 0 |
| S9_vs_k141_59377\|\|full | 4827 | No | NA | | 4 | 0 | 1 | Low-quality | Genome-fragment | | 8.11 | AAI-based (medium-confidence) | 0 |
| S15_extracted_5756 | 1827 | No | NA | | 1 | 0 | 1 | Not-determined | Genome-fragment | NA | | NA | 0 |
| S15_extracted_5759 | 1071 | No | NA | | 1 | 0 | 1 | Not-determined | Genome-fragment | NA | | NA | 0 |
| S15_extracted_5820 | 1140 | No | NA | | 1 | 0 | 0 | Not-determined | Genome-fragment | NA | | NA | 0 |
| S15_extracted_6013 | 1070 | No | NA | | 1 | 0 | 1 | Not-determined | Genome-fragment | NA | | NA | 0 |
| S15_extracted_6026 | 1955 | No | NA | | 2 | 0 | 0 | Not-determined | Genome-fragment | NA | | NA | 0 |
| S15_extracted_6140 | 1458 | No | NA | | 2 | 0 | 1 | Not-determined | Genome-fragment | NA | | NA | 0 |
| S15_extracted_6318 | 1457 | No | NA | | 2 | 0 | 1 | Not-determined | Genome-fragment | NA | | NA | 0 |
| S15_extracted_6420 | 7131 | No | NA | | 7 | 0 | 3 | Not-determined | Genome-fragment | NA | | NA | 0 |
| S8_extracted_k141_115480 | 4828 | No | NA | | 5 | 0 | 0 | Low-quality | Genome-fragment | | 2.21 | AAI-based (medium-confidence) | 0 |
| S15_extracted_6515 | 1179 | No | NA | | 2 | 0 | 0 | Not-determined | Genome-fragment | NA | | NA | 0 |
| S8_extracted_k141_200280 | 4830 | No | NA | | 8 | 0 | 7 | Low-quality | Genome-fragment | | 3.17 | AAI-based (high-confidence) | 0 |
| S15_extracted_6850 | 3200 | No | NA | | 3 | 0 | 1 | Not-determined | Genome-fragment | NA | | NA | 0 |
| S7_extracted_16529 | 4831 | No | NA | | 7 | 2 | 0 | Low-quality | Genome-fragment | | 3.15 | HMM-based (lower-bound) | 0 |
| S15_extracted_7165 | 1021 | No | NA | | 1 | 0 | 0 | Not-determined | Genome-fragment | NA | | NA | 0 |
| S15_extracted_7200 | 6990 | No | NA | | 9 | 0 | 2 | Not-determined | Genome-fragment | NA | | NA | 0 |
| S10_vs_22040\|\|full | 4831 | No | NA | | 5 | 0 | 4 | Low-quality | Genome-fragment | | 2.21 | AAI-based (medium-confidence) | 0 |
| S6_extracted_k141_18125 | 4831 | No | NA | | 4 | 2 | 0 | Low-quality | Genome-fragment | | 2.88 | AAI-based (high-confidence) | 0 |
| S15_extracted_7539 | 1846 | No | NA | | 4 | 0 | 0 | Not-determined | Genome-fragment | NA | | NA | 0 |
| S15_extracted_7594 | 1770 | No | NA | | 2 | 0 | 2 | Not-determined | Genome-fragment | NA | | NA | 0 |
| S15_extracted_7731 | 1241 | No | NA | | 2 | 0 | 1 | Not-determined | Genome-fragment | NA | | NA | 0 |
| S15_extracted_7738 | 4940 | No | NA | | 4 | 0 | 2 | Not-determined | Genome-fragment | NA | | NA | 0 |
| S16_extracted_19846 | 4832 | No | NA | | 5 | 3 | 0 | Low-quality | Genome-fragment | | 13.47 | AAI-based (high-confidence) | 0 |
| S15_extracted_7910 | 1189 | No | NA | | 1 | 0 | 1 | Not-determined | Genome-fragment | NA | | NA | 0 |
| S15_extracted_64881 | 4832 | No | NA | | 3 | 0 | 0 | Low-quality | Genome-fragment | | 5 | AAI-based (medium-confidence) | 0 |
| S15_extracted_8095 | 1241 | No | NA | | 2 | 0 | 1 | Not-determined | Genome-fragment | NA | | NA | 0 |
| S15_extracted_8112 | 1037 | No | NA | | 1 | 0 | 0 | Not-determined | Genome-fragment | NA | | NA | 0 |
| S15_extracted_8398 | 2071 | No | NA | | 2 | 0 | 1 | Not-determined | Genome-fragment | NA | | NA | 0 |
| S5_vs_k141_86586\|\|full | 4833 | No | NA | | 6 | 3 | 0 | Low-quality | Genome-fragment | | 13.01 | AAI-based (high-confidence) | 0 |
| S15_extracted_8548 | 4759 | No | NA | | 6 | 0 | 1 | Not-determined | Genome-fragment | NA | | NA | 0 |
| S15_extracted_8609 | 1371 | No | NA | | 3 | 0 | 1 | Not-determined | Genome-fragment | NA | | NA | 0 |
| S15_extracted_8846 | 3593 | No | NA | | 4 | 0 | 2 | Not-determined | Genome-fragment | NA | | NA | 0 |
| S15_extracted_8939 | 1695 | No | NA | | 3 | 0 | 3 | Not-determined | Genome-fragment | NA | | NA | 0 |
| S15_extracted_8946 | 5739 | No | NA | | 8 | 0 | 0 | Not-determined | Genome-fragment | NA | | NA | 0 |
| S15_extracted_8987 | 1632 | No | NA | | 3 | 0 | 1 | Not-determined | Genome-fragment | NA | | NA | 0 |
| S10_extracted_78919 | 4834 | No | NA | | 7 | 5 | 0 | Low-quality | Genome-fragment | | 11.11 | AAI-based (high-confidence) | 0 |
| S15_extracted_9416 | 1212 | No | NA | | 2 | 0 | 2 | Not-determined | Genome-fragment | NA | | NA | 0 |
| S12_extracted_k141_2130 | 4834 | No | NA | | 7 | 2 | 0 | Low-quality | Genome-fragment | | 10.67 | AAI-based (high-confidence) | 0 |
| S15_extracted_9555 | 3896 | No | NA | | 13 | 0 | 0 | Not-determined | Genome-fragment | NA | | NA | 0 |
| S2_extracted_k141_10854 | 4835 | No | NA | | 7 | 4 | 0 | Low-quality | Genome-fragment | | 8.17 | AAI-based (high-confidence) | 0 |
| S15_extracted_9726 | 1449 | No | NA | | 3 | 0 | 0 | Not-determined | Genome-fragment | NA | | NA | 0 |
| S15_extracted_9757 | 1092 | No | NA | | 2 | 0 | 0 | Not-determined | Genome-fragment | NA | | NA | 0 |
| S11_extracted_k141_57217 | 4836 | No | NA | | 8 | 0 | 1 | Low-quality | Genome-fragment | | 8.23 | AAI-based (medium-confidence) | 0 |
| S15_extracted_9793 | 1509 | No | NA | | 2 | 0 | 0 | Not-determined | Genome-fragment | NA | | NA | 0 |
| S15_extracted_9853 | 1106 | No | NA | | 1 | 0 | 1 | Not-determined | Genome-fragment | NA | | NA | 0 |
| S15_extracted_9996 | 1883 | No | NA | | 1 | 0 | 0 | Not-determined | Genome-fragment | NA | | NA | 0 |
| S15_extracted_10140 | 1011 | No | NA | | 2 | 0 | 0 | Not-determined | Genome-fragment | NA | | NA | 0 |
| S15_extracted_10245 | 4273 | No | NA | | 3 | 0 | 0 | Not-determined | Genome-fragment | NA | | NA | 0 |
| S15_extracted_10792 | 1078 | No | NA | | 2 | 0 | 2 | Not-determined | Genome-fragment | NA | | NA | 0 |
| S15_extracted_10850 | 1010 | No | NA | | 3 | 0 | 2 | Not-determined | Genome-fragment | NA | | NA | 0 |
| S15_extracted_10878 | 3054 | No | NA | | 4 | 0 | 1 | Not-determined | Genome-fragment | NA | | NA | 0 |
| S15_extracted_11038 | 1222 | No | NA | | 1 | 0 | 1 | Not-determined | Genome-fragment | NA | | NA | 0 |
| S15_extracted_11206 | 1219 | No | NA | | 5 | 0 | 0 | Not-determined | Genome-fragment | NA | | NA | 0 |
| S3_extracted_k141_84742 | 4837 | No | NA | | 9 | 1 | 1 | Low-quality | Genome-fragment | | 8.88 | AAI-based (high-confidence) | 0 |
| S8_vs_k141_58555\|\|full | 4838 | No | NA | | 12 | 2 | 0 | Low-quality | Genome-fragment | | 12.36 | AAI-based (medium-confidence) | 0 |
| S15_extracted_11617 | 1037 | No | NA | | 1 | 0 | 0 | Not-determined | Genome-fragment | NA | | NA | 0 |
| S16_extracted_160954 | 4842 | No | NA | | 6 | 1 | 0 | Low-quality | Genome-fragment | | 11.78 | HMM-based (lower-bound) | 0 |
| S15_extracted_11843 | 1202 | No | NA | | 1 | 0 | 1 | Not-determined | Genome-fragment | NA | | NA | 0 |
| S15_extracted_11943 | 1329 | No | NA | | 1 | 0 | 0 | Not-determined | Genome-fragment | NA | | NA | 0 |
| S15_extracted_12153 | 1126 | No | NA | | 2 | 0 | 1 | Not-determined | Genome-fragment | NA | | NA | 0 |
| S15_extracted_12169 | 1692 | No | NA | | 2 | 0 | 2 | Not-determined | Genome-fragment | NA | | NA | 0 |
| S4_extracted_26647 | 4842 | No | NA | | 6 | 3 | 0 | Low-quality | Genome-fragment | | 10.46 | AAI-based (high-confidence) | 0 |
| S15_extracted_12353 | 4109 | No | NA | | 3 | 0 | 1 | Not-determined | Genome-fragment | NA | | NA | 0 |
| S3_vs_k141_111819\|\|full | 4844 | No | NA | | 8 | 1 | 0 | Low-quality | Genome-fragment | | 11.36 | AAI-based (high-confidence) | 0 |
| S15_extracted_12566 | 1220 | No | NA | | 1 | 0 | 1 | Not-determined | Genome-fragment | NA | | NA | 0 |
| S15_extracted_13089 | 1683 | No | NA | | 1 | 0 | 1 | Not-determined | Genome-fragment | NA | | NA | 0 |
| S5_vs_k141_43645\|\|full | 4844 | No | NA | | 7 | 0 | 0 | Low-quality | Genome-fragment | | 8.3 | AAI-based (medium-confidence) | 0 |
| S15_extracted_13238 | 1101 | No | NA | | 1 | 0 | 0 | Not-determined | Genome-fragment | NA | | NA | 0 |
| S15_extracted_13362 | 1045 | No | NA | | 1 | 0 | 1 | Not-determined | Genome-fragment | NA | | NA | 0 |
| S4_extracted_17164 | 4844 | No | NA | | 7 | 2 | 0 | Low-quality | Genome-fragment | | 10.75 | AAI-based (high-confidence) | 0 |
| S15_extracted_13647 | 3325 | No | NA | | 4 | 0 | 2 | Not-determined | Genome-fragment | NA | | NA | 0 |
| S15_extracted_13761 | 1035 | No | NA | | 3 | 0 | 0 | Not-determined | Genome-fragment | NA | | NA | 0 |
| S15_extracted_13767 | 1581 | No | NA | | 2 | 0 | 2 | Not-determined | Genome-fragment | NA | | NA | 0 |
| S6_vs_k141_25324\|\|full | 4845 | Yes | | 3834 | 7 | 1 | 1 | Low-quality | Genome-fragment | | 7.17 | HMM-based (lower-bound) | 20.87 |
| S15_extracted_13818 | 1123 | No | NA | | 1 | 0 | 0 | Not-determined | Genome-fragment | NA | | NA | 0 |
| S8_extracted_k141_139315 | 4845 | No | NA | | 9 | 1 | 0 | Low-quality | Genome-fragment | | 10.94 | AAI-based (high-confidence) | 0 |
| S15_extracted_13994 | 1084 | No | NA | | 2 | 0 | 1 | Not-determined | Genome-fragment | NA | | NA | 0 |
| S15_extracted_14042 | 11217 | No | NA | | 11 | 0 | 11 | Not-determined | Genome-fragment | NA | | NA | 0 |
| S15_extracted_14080 | 1379 | No | NA | | 2 | 0 | 2 | Not-determined | Genome-fragment | NA | | NA | 0 |
| S14_vs_k141_52977\|\|full | 4846 | No | NA | | 6 | 0 | 1 | Low-quality | Genome-fragment | | 8.21 | AAI-based (medium-confidence) | 0 |
| S14_vs_k141_98404\|\|full | 4846 | No | NA | | 7 | 1 | 0 | Low-quality | Genome-fragment | | 3.05 | HMM-based (lower-bound) | 0 |
| S15_extracted_14350 | 1222 | No | NA | | 2 | 0 | 1 | Not-determined | Genome-fragment | NA | | NA | 0 |
| S15_extracted_14407 | 1022 | No | NA | | 1 | 0 | 0 | Not-determined | Genome-fragment | NA | | NA | 0 |
| S15_extracted_14419 | 1631 | No | NA | | 1 | 0 | 1 | Not-determined | Genome-fragment | NA | | NA | 0 |
| S3_extracted_k141_61114 | 4848 | No | NA | | 6 | 0 | 1 | Low-quality | Genome-fragment | | 40.78 | AAI-based (high-confidence) | 0 |
| S15_extracted_14746 | 1747 | No | NA | | 1 | 0 | 1 | Not-determined | Genome-fragment | NA | | NA | 0 |
| S15_extracted_14766 | 1042 | No | NA | | 3 | 0 | 0 | Not-determined | Genome-fragment | NA | | NA | 0 |
| S15_extracted_14998 | 1168 | No | NA | | 1 | 0 | 1 | Not-determined | Genome-fragment | NA | | NA | 0 |
| S15_extracted_15270 | 1148 | No | NA | | 2 | 0 | 1 | Not-determined | Genome-fragment | NA | | NA | 0 |
| S15_extracted_15644 | 4664 | No | NA | | 2 | 0 | 2 | Not-determined | Genome-fragment | NA | | NA | 0 |
| S7_extracted_83653 | 4849 | No | NA | | 11 | 1 | 0 | Low-quality | Genome-fragment | | 4.58 | AAI-based (high-confidence) | 0 |
| S13_vs_120032\|\|full | 4851 | No | NA | | 5 | 0 | 4 | Low-quality | Genome-fragment | | 2.61 | AAI-based (medium-confidence) | 0 |
| S15_extracted_16212 | 1016 | No | NA | | 1 | 0 | 1 | Not-determined | Genome-fragment | NA | | NA | 0 |
| S7_extracted_87258 | 4852 | No | NA | | 7 | 0 | 1 | Low-quality | Genome-fragment | | 11.19 | AAI-based (medium-confidence) | 0 |
| S15_extracted_16282 | 5048 | No | NA | | 4 | 0 | 2 | Not-determined | Genome-fragment | NA | | NA | 0 |
| S15_extracted_16448 | 1103 | No | NA | | 2 | 0 | 0 | Not-determined | Genome-fragment | NA | | NA | 0 |
| S15_extracted_16457 | 1171 | No | NA | | 1 | 0 | 0 | Not-determined | Genome-fragment | NA | | NA | 0 |
| S15_extracted_16600 | 1483 | No | NA | | 2 | 0 | 1 | Not-determined | Genome-fragment | NA | | NA | 0 |
| S15_extracted_16691 | 1102 | No | NA | | 1 | 0 | 0 | Not-determined | Genome-fragment | NA | | NA | 0 |
| S15_extracted_16878 | 1065 | No | NA | | 3 | 0 | 2 | Not-determined | Genome-fragment | NA | | NA | 0 |
| S15_extracted_17226 | 1805 | No | NA | | 2 | 0 | 1 | Not-determined | Genome-fragment | NA | | NA | 0 |

| | | | | | | | | | | | |
|---|---|---|---|---|---|---|---|---|---|---|---|
| S15_extracted_17232 | 5186 | No | NA | 6 | 0 | 3 | Not-determined | Genome-fragment | NA | NA | 0 |
| S15_extracted_17385 | 1076 | No | NA | 1 | 0 | 0 | Not-determined | Genome-fragment | NA | NA | 0 |
| S12_vs_k141_25682\|\|full | 4852 | No | NA | 4 | 2 | 1 | Low-quality | Genome-fragment | 4.49 | AAI-based (high-confidence) | 0 |
| S11_vs_k141_93547\|\|full | 4853 | No | NA | 9 | 1 | 0 | Low-quality | Genome-fragment | 8.45 | AAI-based (high-confidence) | 0 |
| S3_vs_k141_106599\|\|full | 4853 | No | NA | 3 | 0 | 1 | Low-quality | Genome-fragment | 1.51 | AAI-based (medium-confidence) | 0 |
| S15_extracted_18038 | 1573 | No | NA | 2 | 0 | 1 | Not-determined | Genome-fragment | NA | NA | 0 |
| S10_extracted_5660 | 4853 | No | NA | 7 | 0 | 3 | Low-quality | Genome-fragment | 6.8 | AAI-based (medium-confidence) | 0 |
| S15_extracted_18129 | 1330 | No | NA | 3 | 0 | 1 | Not-determined | Genome-fragment | NA | NA | 0 |
| S4_extracted_10831 | 4855 | No | NA | 5 | 1 | 0 | Low-quality | Genome-fragment | 8.39 | AAI-based (medium-confidence) | 0 |
| S15_extracted_18428 | 1232 | No | NA | 1 | 0 | 1 | Not-determined | Genome-fragment | NA | NA | 0 |
| S15_extracted_18435 | 1151 | No | NA | 2 | 0 | 0 | Not-determined | Genome-fragment | NA | NA | 0 |
| S4_extracted_13917 | 4855 | No | NA | 3 | 0 | 0 | Low-quality | Genome-fragment | 5.07 | AAI-based (medium-confidence) | 0 |
| S15_extracted_18805 | 1403 | No | NA | 3 | 0 | 2 | Not-determined | Genome-fragment | NA | NA | 0 |
| S15_extracted_18870 | 1009 | No | NA | 1 | 0 | 1 | Not-determined | Genome-fragment | NA | NA | 0 |
| S16_vs_158057\|\|full | 4857 | No | NA | 6 | 0 | 0 | Low-quality | Genome-fragment | 3.29 | AAI-based (medium-confidence) | 0 |
| S15_extracted_19283 | 1167 | No | NA | 2 | 0 | 1 | Not-determined | Genome-fragment | NA | NA | 0 |
| S15_extracted_19287 | 1029 | No | NA | 1 | 0 | 1 | Not-determined | Genome-fragment | NA | NA | 0 |
| S15_extracted_19367 | 1078 | No | NA | 1 | 0 | 1 | Not-determined | Genome-fragment | NA | NA | 0 |
| S15_extracted_19421 | 4626 | No | NA | 7 | 0 | 0 | Not-determined | Genome-fragment | NA | NA | 0 |
| S15_extracted_19523 | 1009 | No | NA | 1 | 0 | 1 | Not-determined | Genome-fragment | NA | NA | 0 |
| S15_extracted_19609 | 1184 | No | NA | 1 | 0 | 1 | Not-determined | Genome-fragment | NA | NA | 0 |
| S15_extracted_20382 | 5410 | No | NA | 5 | 0 | 4 | Not-determined | Genome-fragment | NA | NA | 0 |
| S15_extracted_20644 | 77306 | No | NA | 100 | 12 | 0 | High-quality | High-quality | 98.07 | AAI-based (high-confidence) | 0 |
| S15_extracted_20676 | 2559 | No | NA | 3 | 0 | 0 | Not-determined | Genome-fragment | NA | NA | 0 |
| S15_extracted_20743 | 1030 | No | NA | 2 | 0 | 0 | Not-determined | Genome-fragment | NA | NA | 0 |
| S15_extracted_21275 | 14533 | No | NA | 16 | 0 | 4 | Not-determined | Genome-fragment | NA | NA | 0 |
| S15_extracted_21343 | 1383 | No | NA | 2 | 0 | 1 | Not-determined | Genome-fragment | NA | NA | 0 |
| S15_extracted_21708 | 1022 | No | NA | 2 | 0 | 0 | Not-determined | Genome-fragment | NA | NA | 0 |
| S15_extracted_21873 | 4198 | No | NA | 4 | 0 | 0 | Not-determined | Genome-fragment | NA | NA | 0 |
| S15_extracted_21889 | 1993 | No | NA | 2 | 0 | 1 | Not-determined | Genome-fragment | NA | NA | 0 |
| S15_extracted_21957 | 2308 | No | NA | 2 | 0 | 2 | Not-determined | Genome-fragment | NA | NA | 0 |
| S15_extracted_22101 | 5915 | No | NA | 4 | 0 | 2 | Not-determined | Genome-fragment | NA | NA | 0 |
| S15_extracted_22117 | 1075 | No | NA | 1 | 0 | 1 | Not-determined | Genome-fragment | NA | NA | 0 |
| S15_extracted_22228 | 2778 | No | NA | 3 | 0 | 1 | Not-determined | Genome-fragment | NA | NA | 0 |
| S17_vs_k141_57230\|\|full | 4859 | No | NA | 7 | 0 | 0 | Low-quality | Genome-fragment | 9.75 | AAI-based (medium-confidence) | 0 |
| S1_extracted_k141_131846 | 4861 | No | NA | 7 | 2 | 0 | Low-quality | Genome-fragment | 34.02 | AAI-based (medium-confidence) | 0 |
| S15_extracted_22453 | 1010 | No | NA | 2 | 0 | 1 | Not-determined | Genome-fragment | NA | NA | 0 |
| S15_extracted_22456 | 5352 | No | NA | 5 | 0 | 2 | Not-determined | Genome-fragment | NA | NA | 0 |
| S7_vs_37855\|\|full | 4861 | No | NA | 6 | 0 | 3 | Low-quality | Genome-fragment | 4.7 | AAI-based (medium-confidence) | 0 |
| S15_extracted_22510 | 1409 | No | NA | 2 | 0 | 2 | Not-determined | Genome-fragment | NA | NA | 0 |
| S15_extracted_22581 | 3061 | No | NA | 3 | 0 | 2 | Not-determined | Genome-fragment | NA | NA | 0 |
| S15_extracted_22654 | 1093 | No | NA | 1 | 0 | 1 | Not-determined | Genome-fragment | NA | NA | 0 |
| S15_extracted_22659 | 1409 | No | NA | 2 | 0 | 0 | Not-determined | Genome-fragment | NA | NA | 0 |
| S15_extracted_22930 | 1051 | No | NA | 1 | 0 | 1 | Not-determined | Genome-fragment | NA | NA | 0 |
| S15_extracted_22951 | 3096 | No | NA | 3 | 0 | 2 | Not-determined | Genome-fragment | NA | NA | 0 |
| S15_extracted_23049 | 1047 | No | NA | 1 | 0 | 1 | Not-determined | Genome-fragment | NA | NA | 0 |
| S15_extracted_23059 | 6843 | No | NA | 10 | 0 | 1 | Not-determined | Genome-fragment | NA | NA | 0 |
| S15_extracted_23080 | 1252 | No | NA | 1 | 0 | 1 | Not-determined | Genome-fragment | NA | NA | 0 |
| S15_extracted_23087 | 1335 | No | NA | 1 | 0 | 1 | Not-determined | Genome-fragment | NA | NA | 0 |
| S6_vs_k141_17095\|\|full | 4861 | No | NA | 4 | 0 | 3 | Low-quality | Genome-fragment | 1.52 | AAI-based (medium-confidence) | 0 |
| S15_extracted_23563 | 1112 | No | NA | 2 | 0 | 1 | Not-determined | Genome-fragment | NA | NA | 0 |
| S15_extracted_23603 | 1332 | No | NA | 2 | 0 | 1 | Not-determined | Genome-fragment | NA | NA | 0 |
| S15_extracted_23684 | 4085 | No | NA | 4 | 0 | 3 | Not-determined | Genome-fragment | NA | NA | 0 |
| S8_vs_k141_196500\|\|full | 4862 | No | NA | 3 | 0 | 3 | Low-quality | Genome-fragment | 5.65 | AAI-based (medium-confidence) | 0 |
| S17_vs_k141_160991\|\|full | 4864 | No | NA | 3 | 0 | 3 | Low-quality | Genome-fragment | 2.95 | AAI-based (medium-confidence) | 0 |
| S15_extracted_24139 | 1261 | No | NA | 1 | 0 | 1 | Not-determined | Genome-fragment | NA | NA | 0 |
| S15_extracted_24299 | 1580 | No | NA | 2 | 0 | 2 | Not-determined | Genome-fragment | NA | NA | 0 |
| S15_extracted_24400 | 3565 | No | NA | 3 | 0 | 3 | Not-determined | Genome-fragment | NA | NA | 0 |
| S14_vs_k141_7962\|\|full | 4864 | No | NA | 4 | 3 | 0 | Low-quality | Genome-fragment | 14.35 | AAI-based (high-confidence) | 0 |
| S15_extracted_24829 | 1386 | No | NA | 3 | 0 | 2 | Not-determined | Genome-fragment | NA | NA | 0 |
| S10_vs_84284\|\|full | 4865 | No | NA | 10 | 0 | 3 | Low-quality | Genome-fragment | 10.47 | AAI-based (high-confidence) | 0 |
| S12_extracted_k141_168367 | 4865 | No | NA | 8 | 2 | 0 | Low-quality | Genome-fragment | 12.84 | AAI-based (medium-confidence) | 0 |
| S15_extracted_25490 | 1434 | No | NA | 1 | 0 | 1 | Not-determined | Genome-fragment | NA | NA | 0 |
| S8_extracted_k141_63628 | 4866 | No | NA | 7 | 5 | 0 | Low-quality | Genome-fragment | 8.13 | AAI-based (high-confidence) | 0 |
| S15_extracted_25573 | 1216 | No | NA | 2 | 0 | 2 | Not-determined | Genome-fragment | NA | NA | 0 |
| S11_extracted_k141_28106 | 4868 | No | NA | 5 | 5 | 0 | Low-quality | Genome-fragment | 13.99 | AAI-based (high-confidence) | 0 |
| S15_extracted_26460 | 4611 | No | NA | 8 | 0 | 1 | Not-determined | Genome-fragment | NA | NA | 0 |
| S13_extracted_10602 | 4868 | No | NA | 16 | 5 | 0 | Low-quality | Genome-fragment | 10.52 | AAI-based (high-confidence) | 0 |
| S15_extracted_26800 | 3863 | No | NA | 4 | 0 | 0 | Not-determined | Genome-fragment | NA | NA | 0 |
| S15_extracted_26848 | 1070 | No | NA | 1 | 0 | 0 | Not-determined | Genome-fragment | NA | NA | 0 |
| S15_extracted_27002 | 17881 | No | NA | 13 | 0 | 2 | Not-determined | Genome-fragment | NA | NA | 0 |
| S15_extracted_27062 | 1431 | No | NA | 3 | 0 | 1 | Not-determined | Genome-fragment | NA | NA | 0 |
| S15_extracted_27117 | 1289 | No | NA | 2 | 0 | 1 | Not-determined | Genome-fragment | NA | NA | 0 |
| S15_extracted_27146 | 4466 | No | NA | 5 | 0 | 4 | Not-determined | Genome-fragment | NA | NA | 0 |
| S8_extracted_k141_198830 | 4868 | No | NA | 4 | 0 | 2 | Low-quality | Genome-fragment | 2.26 | AAI-based (high-confidence) | 0 |
| S15_extracted_27298 | 1526 | No | NA | 3 | 0 | 2 | Not-determined | Genome-fragment | NA | NA | 0 |
| S15_extracted_27339 | 1064 | No | NA | 4 | 0 | 0 | Not-determined | Genome-fragment | NA | NA | 0 |
| S15_extracted_27465 | 1789 | No | NA | 1 | 0 | 0 | Not-determined | Genome-fragment | NA | NA | 0 |
| S15_extracted_27535 | 1967 | No | NA | 3 | 0 | 2 | Not-determined | Genome-fragment | NA | NA | 0 |
| S15_extracted_27973 | 3297 | No | NA | 5 | 0 | 0 | Not-determined | Genome-fragment | NA | NA | 0 |
| S15_extracted_28168 | 3194 | No | NA | 5 | 0 | 1 | Not-determined | Genome-fragment | NA | NA | 0 |
| S17_extracted_k141_122512 | 4870 | No | NA | 8 | 1 | 0 | Low-quality | Genome-fragment | 10.35 | AAI-based (high-confidence) | 0 |
| S15_extracted_28860 | 1322 | No | NA | 1 | 0 | 1 | Not-determined | Genome-fragment | NA | NA | 0 |
| S5_vs_k141_24639\|\|full | 4870 | No | NA | 5 | 1 | 0 | Low-quality | Genome-fragment | 10.63 | AAI-based (medium-confidence) | 0 |
| S7_vs_88046\|\|full | 4874 | No | NA | 11 | 1 | 0 | Low-quality | Genome-fragment | 2.98 | HMM-based (lower-bound) | 0 |
| S10_extracted_90211 | 4874 | No | NA | 11 | 2 | 0 | Low-quality | Genome-fragment | 10.67 | AAI-based (high-confidence) | 0 |
| S10_vs_10935\|\|full | 4874 | No | NA | 10 | 2 | 0 | Low-quality | Genome-fragment | 9.77 | AAI-based (high-confidence) | 0 |
| S15_extracted_29642 | 1010 | No | NA | 1 | 0 | 0 | Not-determined | Genome-fragment | NA | NA | 0 |
| S15_extracted_29839 | 1311 | No | NA | 3 | 0 | 0 | Not-determined | Genome-fragment | NA | NA | 0 |
| S15_extracted_29849 | 1752 | No | NA | 2 | 0 | 2 | Not-determined | Genome-fragment | NA | NA | 0 |
| S15_extracted_30264 | 1213 | No | NA | 1 | 0 | 1 | Not-determined | Genome-fragment | NA | NA | 0 |
| S8_vs_k141_200984\|\|full | 4874 | No | NA | 3 | 2 | 0 | Low-quality | Genome-fragment | 13.04 | AAI-based (medium-confidence) | 0 |
| S15_extracted_30351 | 1297 | No | NA | 2 | 0 | 1 | Not-determined | Genome-fragment | NA | NA | 0 |
| S15_extracted_30496 | 1397 | No | NA | 3 | 0 | 0 | Not-determined | Genome-fragment | NA | NA | 0 |
| S11_extracted_k141_36063 | 4875 | No | NA | 6 | 1 | 0 | Low-quality | Genome-fragment | 1.25 | HMM-based (lower-bound) | 0 |
| S15_extracted_30577 | 2618 | No | NA | 3 | 0 | 3 | Not-determined | Genome-fragment | NA | NA | 0 |
| S15_extracted_30676 | 4214 | No | NA | 2 | 0 | 0 | Not-determined | Genome-fragment | NA | NA | 0 |
| S15_extracted_30989 | 1132 | No | NA | 1 | 0 | 0 | Not-determined | Genome-fragment | NA | NA | 0 |
| S15_extracted_31096 | 1544 | No | NA | 2 | 0 | 0 | Not-determined | Genome-fragment | NA | NA | 0 |
| S15_extracted_31260 | 2552 | No | NA | 4 | 0 | 2 | Not-determined | Genome-fragment | NA | NA | 0 |
| S3_extracted_k141_18842 | 4875 | No | NA | 11 | 0 | 0 | Low-quality | Genome-fragment | 3.12 | AAI-based (medium-confidence) | 0 |
| S15_extracted_32052 | 1494 | No | NA | 1 | 0 | 1 | Not-determined | Genome-fragment | NA | NA | 0 |
| S15_extracted_32123 | 1047 | No | NA | 1 | 0 | 1 | Not-determined | Genome-fragment | NA | NA | 0 |
| S15_extracted_32402 | 5603 | No | NA | 2 | 0 | 0 | Not-determined | Genome-fragment | NA | NA | 0 |
| S15_extracted_32406 | 1327 | No | NA | 2 | 0 | 2 | Not-determined | Genome-fragment | NA | NA | 0 |
| S15_extracted_32646 | 2224 | No | NA | 2 | 0 | 1 | Not-determined | Genome-fragment | NA | NA | 0 |
| S15_extracted_32681 | 5100 | No | NA | 2 | 0 | 2 | Not-determined | Genome-fragment | NA | NA | 0 |
| S13_vs_7448\|\|full | 4876 | No | NA | 7 | 2 | 0 | Low-quality | Genome-fragment | 33.48 | AAI-based (medium-confidence) | 0 |
| S15_extracted_32884 | 1683 | No | NA | 2 | 0 | 0 | Not-determined | Genome-fragment | NA | NA | 0 |
| S16_extracted_44264 | 4876 | No | NA | 4 | 4 | 0 | Low-quality | Genome-fragment | 8.09 | AAI-based (high-confidence) | 0 |
| S15_extracted_33178 | 1267 | No | NA | 2 | 0 | 1 | Not-determined | Genome-fragment | NA | NA | 0 |
| S15_extracted_33201 | 3203 | No | NA | 2 | 0 | 2 | Not-determined | Genome-fragment | NA | NA | 0 |
| S15_extracted_33256 | 1783 | No | NA | 2 | 0 | 0 | Not-determined | Genome-fragment | NA | NA | 0 |
| S17_extracted_k141_164667 | 4877 | No | NA | 8 | 1 | 0 | Low-quality | Genome-fragment | 7.56 | AAI-based (high-confidence) | 0 |
| S11_vs_k141_29773\|\|full | 4877 | Yes | 2540 | 7 | 1 | 1 | Low-quality | Genome-fragment | 4.03 | AAI-based (high-confidence) | 47.92 |
| S15_extracted_33505 | 174703 | No | NA | 192 | 27 | 4 | Complete | High-quality | 100 | DTR (high-confidence) | 0 |
| S15_extracted_33560 | 2075 | No | NA | 4 | 0 | 2 | Not-determined | Genome-fragment | NA | NA | 0 |
| S7_vs_48009\|\|full | 4877 | No | NA | 4 | 0 | 4 | Low-quality | Genome-fragment | 2.62 | AAI-based (medium-confidence) | 0 |
| S15_extracted_33683 | 1128 | No | NA | 2 | 0 | 2 | Not-determined | Genome-fragment | NA | NA | 0 |
| S15_extracted_33973 | 4470 | No | NA | 8 | 0 | 0 | Not-determined | Genome-fragment | NA | NA | 0 |
| S15_extracted_34027 | 1212 | No | NA | 2 | 0 | 2 | Not-determined | Genome-fragment | NA | NA | 0 |
| S15_extracted_34325 | 1373 | No | NA | 1 | 0 | 0 | Not-determined | Genome-fragment | NA | NA | 0 |
| S12_vs_k141_64716\|\|full | 4877 | No | NA | 10 | 2 | 0 | Low-quality | Genome-fragment | 10.18 | AAI-based (medium-confidence) | 0 |
| S15_extracted_34492 | 1022 | No | NA | 1 | 0 | 0 | Not-determined | Genome-fragment | NA | NA | 0 |
| S7_extracted_36455 | 4878 | No | NA | 10 | 1 | 1 | Low-quality | Genome-fragment | 8.35 | AAI-based (high-confidence) | 0 |
| S15_extracted_34567 | 1456 | No | NA | 4 | 0 | 2 | Not-determined | Genome-fragment | NA | NA | 0 |
| S15_extracted_34577 | 1286 | No | NA | 2 | 0 | 0 | Not-determined | Genome-fragment | NA | NA | 0 |
| S15_extracted_34860 | 2769 | No | NA | 3 | 0 | 1 | Not-determined | Genome-fragment | NA | NA | 0 |
| S16_extracted_45751 | 4878 | No | NA | 6 | 0 | 1 | Low-quality | Genome-fragment | 7.68 | AAI-based (medium-confidence) | 0 |
| S15_extracted_35004 | 1182 | No | NA | 1 | 0 | 0 | Not-determined | Genome-fragment | NA | NA | 0 |
| S3_extracted_k141_36439 | 4879 | No | NA | 7 | 5 | 0 | Low-quality | Genome-fragment | 22.23 | AAI-based (high-confidence) | 0 |
| S15_extracted_35168 | 3169 | No | NA | 3 | 0 | 1 | Not-determined | Genome-fragment | NA | NA | 0 |
| S15_extracted_35632 | 1458 | No | NA | 2 | 0 | 1 | Not-determined | Genome-fragment | NA | NA | 0 |
| S16_extracted_125248 | 4879 | No | NA | 6 | 0 | 0 | Low-quality | Genome-fragment | 4.59 | AAI-based (high-confidence) | 0 |
| S1_vs_k141_132183\|\|full | 4880 | No | NA | 12 | 3 | 0 | Low-quality | Genome-fragment | 11.46 | AAI-based (high-confidence) | 0 |
| S15_extracted_36065 | 1009 | No | NA | 1 | 0 | 1 | Not-determined | Genome-fragment | NA | NA | 0 |
| S15_extracted_36110 | 1996 | No | NA | 3 | 0 | 2 | Not-determined | Genome-fragment | NA | NA | 0 |
| S15_extracted_36165 | 3091 | No | NA | 2 | 0 | 2 | Not-determined | Genome-fragment | NA | NA | 0 |
| S15_extracted_36274 | 1008 | No | NA | 1 | 0 | 1 | Not-determined | Genome-fragment | NA | NA | 0 |
| S15_extracted_36291 | 1132 | No | NA | 3 | 0 | 1 | Not-determined | Genome-fragment | NA | NA | 0 |
| S15_extracted_36361 | 1439 | No | NA | 2 | 0 | 1 | Not-determined | Genome-fragment | NA | NA | 0 |
| S15_extracted_36411 | 1018 | No | NA | 2 | 0 | 0 | Not-determined | Genome-fragment | NA | NA | 0 |
| S15_extracted_36474 | 1027 | No | NA | 2 | 0 | 2 | Not-determined | Genome-fragment | NA | NA | 0 |
| S3_extracted_k141_49584 | 4881 | No | NA | 11 | 3 | 0 | Low-quality | Genome-fragment | 5.48 | AAI-based (high-confidence) | 0 |
| S11_vs_k141_15676\|\|full | 4882 | No | NA | 10 | 4 | 0 | Low-quality | Genome-fragment | 12.42 | AAI-based (high-confidence) | 0 |
| S15_extracted_37666 | 4882 | No | NA | 9 | 0 | 0 | Low-quality | Genome-fragment | 3.25 | AAI-based (high-confidence) | 0 |
| S15_extracted_37593 | 1315 | No | NA | 2 | 0 | 2 | Not-determined | Genome-fragment | NA | NA | 0 |
| S8_vs_k141_39082\|\|full | 4886 | No | NA | 4 | 0 | 2 | Low-quality | Genome-fragment | 10.91 | AAI-based (medium-confidence) | 0 |
| S10_vs_8836\|\|full | 4887 | No | NA | 6 | 0 | 1 | Low-quality | Genome-fragment | 4.09 | AAI-based (medium-confidence) | 0 |
| S15_extracted_37765 | 2019 | No | NA | 3 | 0 | 1 | Not-determined | Genome-fragment | NA | NA | 0 |
| S15_extracted_37884 | 1322 | No | NA | 2 | 0 | 1 | Not-determined | Genome-fragment | NA | NA | 0 |
| S8_vs_k141_202535\|\|full | 4887 | No | NA | 3 | 0 | 3 | Low-quality | Genome-fragment | 5.02 | AAI-based (medium-confidence) | 0 |
| S15_extracted_37959 | 1245 | No | NA | 2 | 0 | 2 | Not-determined | Genome-fragment | NA | NA | 0 |
| S15_extracted_38023 | 3040 | No | NA | 3 | 0 | 2 | Not-determined | Genome-fragment | NA | NA | 0 |
| S15_extracted_38035 | 1152 | No | NA | 1 | 0 | 0 | Not-determined | Genome-fragment | NA | NA | 0 |
| S15_extracted_38146 | 1160 | No | NA | 2 | 0 | 2 | Not-determined | Genome-fragment | NA | NA | 0 |
| S15_extracted_38296 | 3251 | No | NA | 2 | 0 | 0 | Not-determined | Genome-fragment | NA | NA | 0 |
| S5_vs_k141_73198\|\|full | 4890 | No | NA | 3 | 0 | 2 | Low-quality | Genome-fragment | 4.51 | AAI-based (medium-confidence) | 0 |
| S11_vs_k141_39064\|\|full | 4891 | No | NA | 7 | 1 | 1 | Low-quality | Genome-fragment | 13.95 | AAI-based (medium-confidence) | 0 |
| S15_extracted_39091 | 1950 | No | NA | 1 | 0 | 1 | Not-determined | Genome-fragment | NA | NA | 0 |
| S15_extracted_39156 | 1346 | No | NA | 1 | 0 | 0 | Not-determined | Genome-fragment | NA | NA | 0 |
| S15_extracted_39317 | 1416 | No | NA | 1 | 0 | 1 | Not-determined | Genome-fragment | NA | NA | 0 |
| S15_extracted_39775 | 1215 | No | NA | 1 | 0 | 1 | Not-determined | Genome-fragment | NA | NA | 0 |
| S15_extracted_40060 | 3719 | No | NA | 4 | 0 | 4 | Not-determined | Genome-fragment | NA | NA | 0 |
| S10_vs_120745\|\|full | 4892 | No | NA | 8 | 1 | 1 | Low-quality | Genome-fragment | 6.87 | HMM-based (lower-bound) | 0 |
| S15_extracted_40417 | 1189 | No | NA | 2 | 0 | 2 | Not-determined | Genome-fragment | NA | NA | 0 |
| S15_extracted_41014 | 1335 | No | NA | 1 | 0 | 1 | Not-determined | Genome-fragment | NA | NA | 0 |
| S12_extracted_k141_48469 | 4892 | No | NA | 14 | 2 | 0 | Low-quality | Genome-fragment | 8.99 | AAI-based (high-confidence) | 0 |
| S15_extracted_41023 | 2009 | No | NA | 2 | 0 | 0 | Not-determined | Genome-fragment | NA | NA | 0 |
| S15_extracted_41054 | 2279 | No | NA | 3 | 0 | 3 | Not-determined | Genome-fragment | NA | NA | 0 |
| S15_extracted_41201 | 3013 | No | NA | 4 | 0 | 1 | Not-determined | Genome-fragment | NA | NA | 0 |
| S15_extracted_41202 | 1212 | No | NA | 2 | 0 | 1 | Not-determined | Genome-fragment | NA | NA | 0 |
| S14_extracted_k141_7634 | 4895 | No | NA | 4 | 2 | 0 | Low-quality | Genome-fragment | 10.78 | AAI-based (high-confidence) | 0 |
| S5_vs_k141_7566\|\|full | 4895 | No | NA | 10 | 2 | 0 | Low-quality | Genome-fragment | 10.41 | AAI-based (medium-confidence) | 0 |
| S15_extracted_41525 | 1056 | No | NA | 3 | 0 | 2 | Not-determined | Genome-fragment | NA | NA | 0 |
| S13_extracted_18981 | 4896 | No | NA | 3 | 0 | 2 | Low-quality | Genome-fragment | 7.69 | AAI-based (medium-confidence) | 0 |
| S12_vs_k141_12801\|\|full | 4898 | Yes | 1598 | 5 | 1 | 1 | Low-quality | Genome-fragment | 2.57 | AAI-based (medium-confidence) | 67.37 |
| S14_vs_k141_23823\|\|full | 4900 | No | NA | 6 | 0 | 4 | Low-quality | Genome-fragment | 1.45 | AAI-based (medium-confidence) | 0 |
| S3_vs_k141_2528\|\|full | 4900 | No | NA | 9 | 1 | 0 | Low-quality | Genome-fragment | 11.33 | AAI-based (high-confidence) | 0 |
| S15_extracted_41830 | 1156 | No | NA | 2 | 0 | 1 | Not-determined | Genome-fragment | NA | NA | 0 |
| S5_vs_k141_124784\|\|full | 4900 | No | NA | 10 | 1 | 0 | Low-quality | Genome-fragment | 3.72 | HMM-based (lower-bound) | 0 |
| S9_vs_k141_3484\|\|full | 4901 | No | NA | 6 | 0 | 1 | Low-quality | Genome-fragment | 7.34 | AAI-based (medium-confidence) | 0 |
| S15_extracted_42168 | 1180 | No | NA | 1 | 0 | 1 | Not-determined | Genome-fragment | NA | NA | 0 |
| S15_extracted_42193 | 1247 | No | NA | 1 | 0 | 1 | Not-determined | Genome-fragment | NA | NA | 0 |
| S15_extracted_42323 | 7078 | No | NA | 3 | 0 | 2 | Not-determined | Genome-fragment | NA | NA | 0 |
| S7_vs_104033\|\|full | 4901 | No | NA | 5 | 5 | 0 | Low-quality | Genome-fragment | 10.46 | AAI-based (high-confidence) | 0 |
| S15_extracted_42401 | 2418 | No | NA | 3 | 0 | 1 | Not-determined | Genome-fragment | NA | NA | 0 |
| S15_extracted_42659 | 4050 | No | NA | 4 | 0 | 2 | Not-determined | Genome-fragment | NA | NA | 0 |
| S15_extracted_43075 | 4200 | No | NA | 5 | 0 | 3 | Not-determined | Genome-fragment | NA | NA | 0 |
| S15_extracted_43212 | 5504 | No | NA | 6 | 0 | 0 | Not-determined | Genome-fragment | NA | NA | 0 |
| S15_extracted_43372 | 1493 | No | NA | 2 | 0 | 1 | Not-determined | Genome-fragment | NA | NA | 0 |
| S2_vs_k141_12861\|\|full | 4901 | No | NA | 6 | 1 | 0 | Low-quality | Genome-fragment | 8.4 | AAI-based (high-confidence) | 0 |
| S15_extracted_43574 | 1325 | No | NA | 2 | 0 | 2 | Not-determined | Genome-fragment | NA | NA | 0 |
| S15_extracted_43690 | 1379 | No | NA | 6 | 0 | 4 | Not-determined | Genome-fragment | NA | NA | 0 |
| S15_extracted_43791 | 5531 | No | NA | 4 | 0 | 3 | Not-determined | Genome-fragment | NA | NA | 0 |
| S15_extracted_43990 | 18327 | No | NA | 14 | 0 | 12 | Not-determined | Genome-fragment | NA | NA | 0 |
| S15_extracted_44075 | 1189 | No | NA | 1 | 0 | 1 | Not-determined | Genome-fragment | NA | NA | 0 |
| S15_extracted_44123 | 1162 | No | NA | 2 | 0 | 2 | Not-determined | Genome-fragment | NA | NA | 0 |
| S15_extracted_44151 | 3862 | No | NA | 3 | 0 | 3 | Not-determined | Genome-fragment | NA | NA | 0 |
| S9_extracted_k141_36059 | 4904 | No | NA | 5 | 3 | 0 | Low-quality | Genome-fragment | 13.23 | AAI-based (high-confidence) | 0 |
| S15_extracted_44221 | 4384 | No | NA | 4 | 0 | 2 | Not-determined | Genome-fragment | NA | NA | 0 |
| S5_vs_k141_76731\|\|full | 4905 | No | NA | 8 | 6 | 1 | Low-quality | Genome-fragment | 12.49 | AAI-based (high-confidence) | 0 |
| S8_vs_k141_37694\|\|full | 4905 | No | NA | 5 | 0 | 2 | Low-quality | Genome-fragment | 3.65 | AAI-based (high-confidence) | 0 |
| S15_extracted_44495 | 1489 | No | NA | 3 | 0 | 0 | Not-determined | Genome-fragment | NA | NA | 0 |
| S15_extracted_44800 | 1244 | No | NA | 2 | 0 | 1 | Not-determined | Genome-fragment | NA | NA | 0 |
| S3_extracted_k141_36391 | 4906 | No | NA | 10 | 3 | 0 | Low-quality | Genome-fragment | 8.12 | AAI-based (high-confidence) | 0 |
| S15_extracted_45152 | 1110 | No | NA | 1 | 0 | 1 | Not-determined | Genome-fragment | NA | NA | 0 |
| S15_extracted_45174 | 1142 | No | NA | 3 | 0 | 0 | Not-determined | Genome-fragment | NA | NA | 0 |
| S16_vs_24616\|\|full | 4908 | No | NA | 10 | 3 | 0 | Low-quality | Genome-fragment | 13.58 | AAI-based (high-confidence) | 0 |
| S17_extracted_k141_172874 | 4909 | No | NA | 5 | 0 | 3 | Low-quality | Genome-fragment | 0.82 | AAI-based (medium-confidence) | 0 |
| S15_extracted_45443 | 1027 | No | NA | 1 | 0 | 1 | Not-determined | Genome-fragment | NA | NA | 0 |
| S15_extracted_45497 | 1063 | No | NA | 2 | 0 | 1 | Not-determined | Genome-fragment | NA | NA | 0 |
| S15_extracted_45615 | 1276 | No | NA | 2 | 0 | 0 | Not-determined | Genome-fragment | NA | NA | 0 |
| S15_extracted_45691 | 1112 | No | NA | 1 | 0 | 0 | Not-determined | Genome-fragment | NA | NA | 0 |
| S15_extracted_45780 | 1229 | No | NA | 2 | 0 | 0 | Not-determined | Genome-fragment | NA | NA | 0 |
| S15_extracted_45879 | 9588 | No | NA | 9 | 0 | 1 | Not-determined | Genome-fragment | NA | NA | 0 |
| S15_extracted_45918 | 1087 | No | NA | 1 | 0 | 1 | Not-determined | Genome-fragment | NA | NA | 0 |
| S1_vs_k141_91914\|\|full | 4909 | No | NA | 7 | 3 | 0 | Low-quality | Genome-fragment | 13.25 | AAI-based (high-confidence) | 0 |
| S15_extracted_45957 | 2158 | No | NA | 3 | 0 | 1 | Not-determined | Genome-fragment | NA | NA | 0 |
| S15_extracted_46090 | 2916 | No | NA | 3 | 0 | 1 | Not-determined | Genome-fragment | NA | NA | 0 |
| S15_extracted_46182 | 4551 | No | NA | 4 | 0 | 3 | Not-determined | Genome-fragment | NA | NA | 0 |
| S7_vs_102327\|\|full | 4909 | No | NA | 7 | 1 | 0 | Low-quality | Genome-fragment | 12.56 | AAI-based (medium-confidence) | 0 |
| S16_vs_10318\|\|full | 4909 | No | NA | 11 | 0 | 0 | Low-quality | Genome-fragment | 7.74 | AAI-based (high-confidence) | 0 |
| S15_extracted_46566 | 1145 | No | NA | 2 | 0 | 1 | Not-determined | Genome-fragment | NA | NA | 0 |
| S15_extracted_46717 | 1050 | No | NA | 3 | 0 | 0 | Not-determined | Genome-fragment | NA | NA | 0 |
| S11_extracted_k141_69162 | 4910 | No | NA | 8 | 4 | 0 | Low-quality | Genome-fragment | 8.59 | AAI-based (high-confidence) | 0 |
| S15_extracted_47255 | 1400 | No | NA | 1 | 0 | 0 | Not-determined | Genome-fragment | NA | NA | 0 |
| S15_extracted_47313 | 1151 | No | NA | 1 | 0 | 0 | Not-determined | Genome-fragment | NA | NA | 0 |
| S5_vs_k141_44072\|\|full | 4910 | No | NA | 7 | 1 | 0 | Low-quality | Genome-fragment | 12.29 | AAI-based (high-confidence) | 0 |
| S15_extracted_47421 | 3213 | No | NA | 4 | 0 | 4 | Not-determined | Genome-fragment | NA | NA | 0 |
| S15_extracted_47940 | 3520 | No | NA | 2 | 0 | 2 | Not-determined | Genome-fragment | NA | NA | 0 |
| S15_extracted_47960 | 1209 | No | NA | 1 | 0 | 1 | Not-determined | Genome-fragment | NA | NA | 0 |
| S15_extracted_47963 | 1279 | No | NA | 2 | 0 | 1 | Not-determined | Genome-fragment | NA | NA | 0 |
| S15_extracted_48368 | 1688 | No | NA | 1 | 0 | 1 | Not-determined | Genome-fragment | NA | NA | 0 |
| S7_vs_100242\|\|full | 4910 | No | NA | 5 | 1 | 0 | Low-quality | Genome-fragment | 12.03 | AAI-based (high-confidence) | 0 |
| S15_extracted_48544 | 1292 | No | NA | 1 | 0 | 0 | Not-determined | Genome-fragment | NA | NA | 0 |
| S15_extracted_48585 | 1109 | No | NA | 1 | 0 | 0 | Not-determined | Genome-fragment | NA | NA | 0 |
| S15_extracted_48586 | 1792 | No | NA | 2 | 0 | 1 | Not-determined | Genome-fragment | NA | NA | 0 |
| S11_vs_k141_102250\|\|full | 4911 | No | NA | 6 | 1 | 0 | Low-quality | Genome-fragment | 12.87 | AAI-based (high-confidence) | 0 |
| S15_extracted_49180 | 4393 | No | NA | 7 | 0 | 1 | Not-determined | Genome-fragment | NA | NA | 0 |
| S15_extracted_49210 | 1077 | No | NA | 2 | 0 | 0 | Not-determined | Genome-fragment | NA | NA | 0 |
| S15_extracted_49267 | 1034 | No | NA | 2 | 0 | 1 | Not-determined | Genome-fragment | NA | NA | 0 |
| S15_extracted_49507 | 1133 | No | NA | 1 | 0 | 1 | Not-determined | Genome-fragment | NA | NA | 0 |
| S15_extracted_49639 | 4694 | No | NA | 2 | 0 | 2 | Not-determined | Genome-fragment | NA | NA | 0 |
| S15_extracted_49812 | 9340 | No | NA | 8 | 0 | 7 | Not-determined | Genome-fragment | NA | NA | 0 |
| S15_extracted_50315 | 1008 | No | NA | 1 | 0 | 1 | Not-determined | Genome-fragment | NA | NA | 0 |
| S15_extracted_50357 | 1320 | No | NA | 1 | 0 | 0 | Not-determined | Genome-fragment | NA | NA | 0 |
| S15_extracted_50471 | 1157 | No | NA | 1 | 0 | 1 | Not-determined | Genome-fragment | NA | NA | 0 |
| S16_vs_77252\|\|full | 4911 | No | NA | 5 | 0 | 3 | Low-quality | Genome-fragment | 2.48 | AAI-based (medium-confidence) | 0 |
| S17_extracted_k141_99553 | 4912 | No | NA | 5 | 0 | 0 | Low-quality | Genome-fragment | 8.13 | AAI-based (medium-confidence) | 0 |
| S15_extracted_51163 | 1034 | No | NA | 2 | 0 | 0 | Not-determined | Genome-fragment | NA | NA | 0 |
| S14_extracted_k141_33714 | 4912 | No | NA | 8 | 2 | 0 | Low-quality | Genome-fragment | 3.68 | HMM-based (lower-bound) | 0 |
| S12_vs_k141_22241\|\|full | 4912 | No | NA | 5 | 0 | 3 | Low-quality | Genome-fragment | 2.13 | AAI-based (medium-confidence) | 0 |
| S15_extracted_51766 | 1241 | No | NA | 1 | 0 | 1 | Not-determined | Genome-fragment | NA | NA | 0 |
| S15_extracted_51879 | 1759 | No | NA | 2 | 0 | 0 | Not-determined | Genome-fragment | NA | NA | 0 |
| S15_extracted_51920 | 2345 | No | NA | 2 | 0 | 2 | Not-determined | Genome-fragment | NA | NA | 0 |
| S15_extracted_51935 | 1011 | No | NA | 1 | 0 | 1 | Not-determined | Genome-fragment | NA | NA | 0 |
| S11_extracted_k141_46510 | 4913 | No | NA | 17 | 1 | 0 | Low-quality | Genome-fragment | 8.17 | AAI-based (medium-confidence) | 0 |
| S15_extracted_52118 | 3360 | No | NA | 2 | 0 | 2 | Not-determined | Genome-fragment | NA | NA | 0 |
| S11_extracted_k141_29177 | 4913 | No | NA | 12 | 1 | 0 | Low-quality | Genome-fragment | 2.92 | HMM-based (lower-bound) | 0 |
| S15_extracted_52231 | 1247 | No | NA | 2 | 0 | 2 | Not-determined | Genome-fragment | NA | NA | 0 |
| S15_extracted_52240 | 3200 | No | NA | 2 | 0 | 0 | Not-determined | Genome-fragment | NA | NA | 0 |
| S5_vs_k141_109835\|\|full | 4918 | No | NA | 5 | 4 | 0 | Low-quality | Genome-fragment | 13.37 | AAI-based (high-confidence) | 0 |
| S15_extracted_52369 | 1131 | No | NA | 3 | 0 | 0 | Not-determined | Genome-fragment | NA | NA | 0 |
| S15_extracted_52525 | 1042 | No | NA | 1 | 0 | 0 | Not-determined | Genome-fragment | NA | NA | 0 |
| S15_extracted_52779 | 1339 | No | NA | 1 | 0 | 1 | Not-determined | Genome-fragment | NA | NA | 0 |
| S11_vs_k141_8588\|\|full | 4921 | No | NA | 5 | 4 | 0 | Low-quality | Genome-fragment | 11.62 | AAI-based (high-confidence) | 0 |
| S15_extracted_53233 | 6881 | No | NA | 12 | 0 | 0 | Not-determined | Genome-fragment | NA | NA | 0 |
| S15_extracted_53578 | 3306 | No | NA | 3 | 0 | 1 | Not-determined | Genome-fragment | NA | NA | 0 |
| S1_extracted_k141_90663 | 4921 | No | NA | 5 | 1 | 0 | Low-quality | Genome-fragment | 5.83 | AAI-based (high-confidence) | 0 |
| S4_extracted_2435 | 4921 | No | NA | 13 | 12 | 0 | Low-quality | Genome-fragment | 16.24 | AAI-based (high-confidence) | 0 |
| S15_extracted_54164 | 4201 | No | NA | 7 | 0 | 0 | Not-determined | Genome-fragment | NA | NA | 0 |
| S15_extracted_54579 | 1249 | No | NA | 1 | 0 | 1 | Not-determined | Genome-fragment | NA | NA | 0 |
| S15_extracted_54590 | 1074 | No | NA | 1 | 0 | 0 | Not-determined | Genome-fragment | NA | NA | 0 |
| S15_extracted_54733 | 1172 | No | NA | 1 | 0 | 1 | Not-determined | Genome-fragment | NA | NA | 0 |
| S3_extracted_k141_109177 | 4922 | No | NA | 8 | 1 | 0 | Low-quality | Genome-fragment | 2.77 | AAI-based (medium-confidence) | 0 |
| S15_vs_6747\|\|full | 4922 | No | NA | 3 | 0 | 3 | Low-quality | Genome-fragment | 2.71 | AAI-based (medium-confidence) | 0 |
| S15_extracted_55041 | 1178 | No | NA | 3 | 0 | 0 | Not-determined | Genome-fragment | NA | NA | 0 |
| S8_extracted_k141_42647 | 4922 | No | NA | 5 | 4 | 0 | Low-quality | Genome-fragment | 15.76 | AAI-based (high-confidence) | 0 |
| S15_extracted_55287 | 4497 | No | NA | 3 | 0 | 1 | Not-determined | Genome-fragment | NA | NA | 0 |
| S15_extracted_55400 | 1182 | No | NA | 1 | 0 | 0 | Not-determined | Genome-fragment | NA | NA | 0 |
| S15_extracted_55639 | 13570 | No | NA | 19 | 6 | 1 | High-quality | High-quality | 98.5 | AAI-based (high-confidence) | 0 |
| S15_extracted_55741 | 1243 | No | NA | 1 | 0 | 1 | Not-determined | Genome-fragment | NA | NA | 0 |
| S15_extracted_55747 | 3396 | No | NA | 2 | 0 | 2 | Not-determined | Genome-fragment | NA | NA | 0 |
| S17_extracted_k141_171223 | 4924 | No | NA | 5 | 5 | 0 | Low-quality | Genome-fragment | 16.23 | AAI-based (high-confidence) | 0 |
| S15_extracted_56048 | 1054 | No | NA | 1 | 0 | 1 | Not-determined | Genome-fragment | NA | NA | 0 |
| S15_extracted_56104 | 2150 | No | NA | 2 | 0 | 0 | Not-determined | Genome-fragment | NA | NA | 0 |
| S3_extracted_k141_126627 | 4926 | No | NA | 15 | 2 | 0 | Low-quality | Genome-fragment | 9.62 | AAI-based (high-confidence) | 0 |
| S5_vs_k141_71294\|\|full | 4928 | No | NA | 3 | 0 | 2 | Low-quality | Genome-fragment | 2.65 | AAI-based (high-confidence) | 0 |
| S12_vs_k141_105785\|\|full | 4928 | No | NA | 5 | 0 | 1 | Low-quality | Genome-fragment | 2.01 | AAI-based (medium-confidence) | 0 |
| S15_extracted_56246 | 1055 | No | NA | 2 | 0 | 2 | Not-determined | Genome-fragment | NA | NA | 0 |
| S15_extracted_56285 | 2584 | No | NA | 4 | 0 | 0 | Not-determined | Genome-fragment | NA | NA | 0 |
| S16_vs_135877\|\|full | 4929 | No | NA | 3 | 0 | 3 | Low-quality | Genome-fragment | 4.42 | AAI-based (medium-confidence) | 0 |
| S15_extracted_56503 | 1177 | No | NA | 1 | 0 | 0 | Not-determined | Genome-fragment | NA | NA | 0 |
| S15_extracted_56602 | 4425 | No | NA | 2 | 0 | 2 | Not-determined | Genome-fragment | NA | NA | 0 |
| S15_extracted_56685 | 1007 | No | NA | 2 | 0 | 1 | Not-determined | Genome-fragment | NA | NA | 0 |
| S9_vs_k141_15536\|\|full | 4930 | No | NA | 4 | 0 | 0 | Low-quality | Genome-fragment | 10.88 | AAI-based (high-confidence) | 0 |
| S15_extracted_56983 | 1881 | No | NA | 3 | 0 | 1 | Not-determined | Genome-fragment | NA | NA | 0 |
| S15_extracted_57097 | 1899 | No | NA | 1 | 0 | 1 | Not-determined | Genome-fragment | NA | NA | 0 |
| S15_extracted_57135 | 2168 | No | NA | 2 | 0 | 1 | Not-determined | Genome-fragment | NA | NA | 0 |
| S11_vs_k141_64430\|\|full | 4930 | Yes | 4171 | 7 | 1 | 1 | Low-quality | Genome-fragment | 1.36 | HMM-based (lower-bound) | 15.4 |
| S15_extracted_57286 | 1070 | No | NA | 1 | 0 | 1 | Not-determined | Genome-fragment | NA | NA | 0 |
| S3_extracted_k141_110422 | 4930 | No | NA | 8 | 2 | 0 | Low-quality | Genome-fragment | 11.42 | AAI-based (high-confidence) | 0 |
| S15_extracted_57401 | 1116 | No | NA | 1 | 0 | 0 | Not-determined | Genome-fragment | NA | NA | 0 |
| S15_extracted_57660 | 1591 | No | NA | 2 | 0 | 2 | Not-determined | Genome-fragment | NA | NA | 0 |
| S15_extracted_57911 | 1039 | No | NA | 2 | 0 | 1 | Not-determined | Genome-fragment | NA | NA | 0 |
| S3_vs_k141_33333\|\|full | 4930 | No | NA | 3 | 2 | 0 | Low-quality | Genome-fragment | 2.97 | AAI-based (medium-confidence) | 0 |
| S15_extracted_58117 | 5963 | No | NA | 7 | 0 | 3 | Not-determined | Genome-fragment | NA | NA | 0 |
| S3_vs_k141_47602\|\|full | 4933 | No | NA | 4 | 0 | 3 | Low-quality | Genome-fragment | 9.99 | AAI-based (medium-confidence) | 0 |
| S10_vs_6393\|\|full | 4934 | No | NA | 9 | 4 | 0 | Low-quality | Genome-fragment | 10.86 | AAI-based (high-confidence) | 0 |
| S15_extracted_58201 | 3173 | No | NA | 4 | 0 | 1 | Not-determined | Genome-fragment | NA | NA | 0 |
| S14_extracted_k141_38132 | 4935 | No | NA | 8 | 4 | 0 | Low-quality | Genome-fragment | 2.85 | HMM-based (lower-bound) | 0 |
| S15_extracted_58259 | 1943 | No | NA | 2 | 0 | 2 | Not-determined | Genome-fragment | NA | NA | 0 |
| S15_extracted_58329 | 1045 | No | NA | 3 | 0 | 0 | Not-determined | Genome-fragment | NA | NA | 0 |
| S15_extracted_58467 | 91453 | No | NA | 94 | 24 | 0 | High-quality | High-quality | 99.98 | AAI-based (high-confidence) | 0 |
| S15_extracted_58948 | 1244 | No | NA | 1 | 0 | 1 | Not-determined | Genome-fragment | NA | NA | 0 |
| S15_extracted_59049 | 27951 | No | NA | 34 | 0 | 8 | Not-determined | Genome-fragment | NA | NA | 0 |
| S15_extracted_59106 | 4461 | No | NA | 3 | 0 | 1 | Not-determined | Genome-fragment | NA | NA | 0 |
| S12_extracted_k141_1631 | 4937 | No | NA | 4 | 3 | 0 | Low-quality | Genome-fragment | 13.54 | AAI-based (high-confidence) | 0 |
| S11_vs_k141_92274\|\|full | 4942 | No | NA | 9 | 1 | 0 | Low-quality | Genome-fragment | 2.98 | AAI-based (high-confidence) | 0 |
| S15_extracted_59334 | 1086 | No | NA | 1 | 0 | 1 | Not-determined | Genome-fragment | NA | NA | 0 |
| S15_extracted_59350 | 12904 | No | NA | 17 | 0 | 3 | Not-determined | Genome-fragment | NA | NA | 0 |
| S16_vs_16152\|\|full | 4942 | No | NA | 9 | 5 | 0 | Low-quality | Genome-fragment | 14.14 | AAI-based (high-confidence) | 0 |
| S15_extracted_59427 | 1513 | No | NA | 3 | 0 | 2 | Not-determined | Genome-fragment | NA | NA | 0 |
| S15_extracted_59593 | 1142 | No | NA | 2 | 0 | 0 | Not-determined | Genome-fragment | NA | NA | 0 |
| S15_extracted_59653 | 1078 | No | NA | 2 | 0 | 1 | Not-determined | Genome-fragment | NA | NA | 0 |
| S15_extracted_59801 | 1014 | No | NA | 3 | 0 | 2 | Not-determined | Genome-fragment | NA | NA | 0 |
| S15_extracted_59841 | 1075 | No | NA | 1 | 0 | 1 | Not-determined | Genome-fragment | NA | NA | 0 |
| S5_extracted_k141_71789 | 4943 | No | NA | 9 | 4 | 0 | Low-quality | Genome-fragment | 10.93 | AAI-based (high-confidence) | 0 |
| S15_extracted_59911 | 1178 | No | NA | 3 | 0 | 0 | Not-determined | Genome-fragment | NA | NA | 0 |
| S15_extracted_59968 | 1565 | No | NA | 2 | 0 | 1 | Not-determined | Genome-fragment | NA | NA | 0 |
| S15_extracted_60180 | 1952 | No | NA | 1 | 0 | 1 | Not-determined | Genome-fragment | NA | NA | 0 |
| S15_extracted_60417 | 1121 | No | NA | 2 | 0 | 0 | Not-determined | Genome-fragment | NA | NA | 0 |
| S10_vs_85380\|\|full | 4943 | No | NA | 13 | 3 | 0 | Low-quality | Genome-fragment | 13.38 | AAI-based (high-confidence) | 0 |
| S11_vs_k141_37298\|\|full | 4945 | No | NA | 5 | 2 | 0 | Low-quality | Genome-fragment | 11.8 | AAI-based (high-confidence) | 0 |
| S15_extracted_60951 | 1121 | No | NA | 1 | 0 | 1 | Not-determined | Genome-fragment | NA | NA | 0 |
| S15_extracted_61111 | 1068 | No | NA | 1 | 0 | 1 | Not-determined | Genome-fragment | NA | NA | 0 |
| S13_vs_97225\|\|full | 4945 | No | NA | 3 | 0 | 0 | Low-quality | Genome-fragment | 5.83 | AAI-based (medium-confidence) | 0 |
| S15_extracted_61404 | 1205 | No | NA | 1 | 0 | 0 | Not-determined | Genome-fragment | NA | NA | 0 |
| S15_extracted_61564 | 1211 | No | NA | 2 | 0 | 2 | Not-determined | Genome-fragment | NA | NA | 0 |
| S15_extracted_61952 | 1184 | No | NA | 1 | 0 | 0 | Not-determined | Genome-fragment | NA | NA | 0 |
| S14_extracted_k141_41253 | 4945 | No | NA | 6 | 0 | 0 | Low-quality | Genome-fragment | 4.78 | AAI-based (high-confidence) | 0 |
| S15_extracted_62084 | 1010 | No | NA | 1 | 0 | 1 | Not-determined | Genome-fragment | NA | NA | 0 |
| S15_extracted_62564 | 3722 | No | NA | 6 | 0 | 1 | Not-determined | Genome-fragment | NA | NA | 0 |
| S15_extracted_62625 | 2510 | No | NA | 3 | 0 | 1 | Not-determined | Genome-fragment | NA | NA | 0 |
| S15_extracted_62657 | 1238 | No | NA | 1 | 0 | 0 | Not-determined | Genome-fragment | NA | NA | 0 |
| S15_extracted_62660 | 2519 | No | NA | 4 | 0 | 1 | Not-determined | Genome-fragment | NA | NA | 0 |
| S15_extracted_62791 | 1900 | No | NA | 1 | 0 | 1 | Not-determined | Genome-fragment | NA | NA | 0 |
| S3_vs_k141_86285\|\|full | 4946 | No | NA | 12 | 2 | 0 | Low-quality | Genome-fragment | 9.05 | HMM-based (lower-bound) | 0 |
| S15_extracted_63389 | 1238 | No | NA | 2 | 0 | 0 | Not-determined | Genome-fragment | NA | NA | 0 |
| S15_extracted_63617 | 1062 | No | NA | 2 | 0 | 1 | Not-determined | Genome-fragment | NA | NA | 0 |
| S11_vs_k141_16415\|\|full | 4948 | No | NA | 5 | 0 | 1 | Low-quality | Genome-fragment | 2.84 | AAI-based (medium-confidence) | 0 |
| S15_extracted_63774 | 1158 | No | NA | 1 | 0 | 0 | Not-determined | Genome-fragment | NA | NA | 0 |
| S11_vs_k141_40201\|\|full | 4950 | No | NA | 6 | 0 | 0 | Low-quality | Genome-fragment | 13.46 | AAI-based (medium-confidence) | 0 |
| S8_extracted_k141_182096 | 4950 | No | NA | 3 | 0 | 1 | Low-quality | Genome-fragment | 4.82 | AAI-based (high-confidence) | 0 |
| S15_extracted_63976 | 1275 | No | NA | 1 | 0 | 1 | Not-determined | Genome-fragment | NA | NA | 0 |
| S15_extracted_64018 | 1912 | No | NA | 3 | 0 | 1 | Not-determined | Genome-fragment | NA | NA | 0 |
| S8_extracted_k141_78432 | 4951 | No | NA | 5 | 3 | 0 | Low-quality | Genome-fragment | 22.59 | AAI-based (high-confidence) | 0 |
| S15_extracted_64054 | 1477 | No | NA | 2 | 0 | 1 | Not-determined | Genome-fragment | NA | NA | 0 |
| S15_extracted_64190 | 1052 | No | NA | 3 | 0 | 0 | Not-determined | Genome-fragment | NA | NA | 0 |
| S15_extracted_64224 | 5458 | No | NA | 4 | 0 | 2 | Not-determined | Genome-fragment | NA | NA | 0 |
| S15_extracted_64267 | 2413 | No | NA | 2 | 0 | 2 | Not-determined | Genome-fragment | NA | NA | 0 |
| S15_extracted_64340 | 1069 | No | NA | 1 | 0 | 1 | Not-determined | Genome-fragment | NA | NA | 0 |
| S15_extracted_64613 | 1298 | No | NA | 2 | 0 | 0 | Not-determined | Genome-fragment | NA | NA | 0 |
| S15_extracted_64686 | 1104 | No | NA | 2 | 0 | 2 | Not-determined | Genome-fragment | NA | NA | 0 |
| S14_extracted_k141_89714 | 4952 | No | NA | 11 | 11 | 0 | Low-quality | Genome-fragment | 16.34 | AAI-based (high-confidence) | 0 |
| S14_extracted_k141_82021 | 4955 | No | NA | 6 | 2 | 0 | Low-quality | Genome-fragment | 12.93 | AAI-based (high-confidence) | 0 |
| S15_extracted_65421 | 1577 | No | NA | 1 | 0 | 0 | Not-determined | Genome-fragment | NA | NA | 0 |
| S15_extracted_65481 | 1579 | No | NA | 2 | 0 | 2 | Not-determined | Genome-fragment | NA | NA | 0 |
| S15_extracted_65538 | 1465 | No | NA | 1 | 0 | 1 | Not-determined | Genome-fragment | NA | NA | 0 |
| S15_extracted_65710 | 1137 | No | NA | 1 | 0 | 0 | Not-determined | Genome-fragment | NA | NA | 0 |
| S17_extracted_k141_2819 | 4959 | No | NA | 6 | 4 | 0 | Low-quality | Genome-fragment | 7.43 | AAI-based (high-confidence) | 0 |
| S15_extracted_65910 | 1200 | No | NA | 4 | 0 | 0 | Not-determined | Genome-fragment | NA | NA | 0 |
| S15_extracted_65925 | 1048 | No | NA | 1 | 0 | 1 | Not-determined | Genome-fragment | NA | NA | 0 |
| S15_extracted_65970 | 4284 | No | NA | 6 | 0 | 0 | Not-determined | Genome-fragment | NA | NA | 0 |
| S15_vs_21557\|\|full | 4959 | No | NA | 10 | 5 | 0 | Low-quality | Genome-fragment | 2.63 | AAI-based (high-confidence) | 0 |
| S7_vs_112026\|\|full | 4960 | No | NA | 9 | 0 | 1 | Low-quality | Genome-fragment | 14.04 | AAI-based (high-confidence) | 0 |
| S15_extracted_66531 | 1157 | No | NA | 2 | 0 | 1 | Not-determined | Genome-fragment | NA | NA | 0 |
| S15_extracted_66620 | 1494 | No | NA | 1 | 0 | 1 | Not-determined | Genome-fragment | NA | NA | 0 |
| S3_vs_k141_114816\|\|full | 4963 | No | NA | 11 | 0 | 0 | Low-quality | Genome-fragment | 10.02 | AAI-based (high-confidence) | 0 |
| S15_extracted_66818 | 1230 | No | NA | 2 | 0 | 2 | Not-determined | Genome-fragment | NA | NA | 0 |
| S14_vs_k141_27056\|\|full | 4964 | No | NA | 8 | 1 | 0 | Low-quality | Genome-fragment | 10.55 | AAI-based (high-confidence) | 0 |
| S15_extracted_66970 | 1127 | No | NA | 1 | 0 | 1 | Not-determined | Genome-fragment | NA | NA | 0 |
| S15_extracted_67293 | 3158 | No | NA | 2 | 0 | 2 | Not-determined | Genome-fragment | NA | NA | 0 |
| S15_extracted_67297 | 1163 | No | NA | 2 | 0 | 2 | Not-determined | Genome-fragment | NA | NA | 0 |
| S15_extracted_67427 | 4276 | No | NA | 4 | 0 | 3 | Not-determined | Genome-fragment | NA | NA | 0 |
| S2_extracted_k141_6427 | 4964 | No | NA | 9 | 7 | 0 | Low-quality | Genome-fragment | 15.17 | AAI-based (high-confidence) | 0 |
| S15_extracted_67626 | 1201 | No | NA | 3 | 0 | 2 | Not-determined | Genome-fragment | NA | NA | 0 |
| S15_extracted_67942 | 1928 | No | NA | 4 | 0 | 1 | Not-determined | Genome-fragment | NA | NA | 0 |
| S15_extracted_68042 | 1151 | No | NA | 2 | 0 | 2 | Not-determined | Genome-fragment | NA | NA | 0 |
| S15_extracted_68280 | 1231 | No | NA | 2 | 0 | 1 | Not-determined | Genome-fragment | NA | NA | 0 |
| S8_extracted_k141_55621 | 4964 | No | NA | 10 | 1 | 0 | Low-quality | Genome-fragment | 9.03 | AAI-based (high-confidence) | 0 |
| S15_extracted_68507 | 1254 | No | NA | 2 | 0 | 0 | Not-determined | Genome-fragment | NA | NA | 0 |
| S5_extracted_k141_100075 | 4967 | No | NA | 5 | 1 | 0 | Low-quality | Genome-fragment | 6.03 | AAI-based (high-confidence) | 0 |
| S15_extracted_68961 | 1094 | No | NA | 2 | 0 | 2 | Not-determined | Genome-fragment | NA | NA | 0 |
| S15_extracted_69205 | 1445 | No | NA | 2 | 0 | 0 | Not-determined | Genome-fragment | NA | NA | 0 |
| S3_extracted_k141_126958 | 4968 | No | NA | 9 | 0 | 0 | Low-quality | Genome-fragment | 2.58 | AAI-based (high-confidence) | 0 |
| S15_extracted_69644 | 1954 | No | NA | 3 | 0 | 2 | Not-determined | Genome-fragment | NA | NA | 0 |
| S15_extracted_69747 | 2009 | No | NA | 1 | 0 | 1 | Not-determined | Genome-fragment | NA | NA | 0 |
| S15_extracted_69786 | 1008 | No | NA | 1 | 0 | 0 | Not-determined | Genome-fragment | NA | NA | 0 |
| S14_extracted_k141_60395 | 4969 | No | NA | 9 | 1 | 0 | Low-quality | Genome-fragment | 33.45 | AAI-based (high-confidence) | 0 |
| S15_extracted_70068 | 1794 | No | NA | 2 | 0 | 2 | Not-determined | Genome-fragment | NA | NA | 0 |
| S5_vs_k141_62346\|\|full | 4972 | No | NA | 9 | 1 | 0 | Low-quality | Genome-fragment | 12.03 | AAI-based (high-confidence) | 0 |
| S8_extracted_k141_128232 | 4972 | No | NA | 11 | 0 | 0 | Low-quality | Genome-fragment | 8.43 | AAI-based (high-confidence) | 0 |
| S14_vs_k141_40211\|\|lt2gene | 4973 | No | NA | 2 | 2 | 0 | Low-quality | Genome-fragment | 8.19 | AAI-based (high-confidence) | 0 |
| S7_extracted_65867 | 4973 | No | NA | 8 | 0 | 0 | Low-quality | Genome-fragment | 7.04 | AAI-based (high-confidence) | 0 |
| S15_extracted_70773 | 1179 | No | NA | 1 | 0 | 0 | Not-determined | Genome-fragment | NA | NA | 0 |
| S15_extracted_71204 | 1246 | No | NA | 1 | 0 | 1 | Not-determined | Genome-fragment | NA | NA | 0 |
| S15_extracted_71296 | 1378 | No | NA | 1 | 0 | 1 | Not-determined | Genome-fragment | NA | NA | 0 |
| S15_extracted_71530 | 1042 | No | NA | 1 | 0 | 1 | Not-determined | Genome-fragment | NA | NA | 0 |
| S15_extracted_71583 | 1098 | No | NA | 1 | 0 | 1 | Not-determined | Genome-fragment | NA | NA | 0 |
| S12_extracted_k141_117018 | 4976 | No | NA | 5 | 5 | 0 | Low-quality | Genome-fragment | 14.52 | AAI-based (high-confidence) | 0 |
| S15_extracted_71616 | 13826 | No | NA | 17 | 7 | 0 | Medium-quality | Genome-fragment | 62.04 | AAI-based (medium-confidence) | 0 |

| | | | | | | | | | | | |
|---|---|---|---|---|---|---|---|---|---|---|---|
| S15_extracted_71676 | 1734 | No | NA | 2 | 0 | 0 | Not-determined | Genome-fragment | NA | NA | 0 |
| S14_extracted_k141_46499 | 4978 | No | NA | 6 | 0 | 0 | Low-quality | Genome-fragment | 17.21 | AAI-based (medium-confidence) | 0 |
| S12_vs_k141_185376\|\|full | 4979 | No | NA | 8 | 0 | 1 | Low-quality | Genome-fragment | 9.03 | AAI-based (high-confidence) | 0 |
| S15_extracted_72459 | 1091 | No | NA | 2 | 0 | 1 | Not-determined | Genome-fragment | NA | NA | 0 |
| S15_extracted_72533 | 1137 | No | NA | 2 | 0 | 1 | Not-determined | Genome-fragment | NA | NA | 0 |
| S15_extracted_72737 | 1564 | No | NA | 2 | 0 | 1 | Not-determined | Genome-fragment | NA | NA | 0 |
| S5_vs_k141_79191\|\|full | 4980 | No | NA | 8 | 1 | 1 | Low-quality | Genome-fragment | 11.33 | AAI-based (high-confidence) | 0 |
| S15_vs_5102\|\|full | 4981 | No | NA | 3 | 0 | 1 | Low-quality | Genome-fragment | 3.71 | AAI-based (high-confidence) | 0 |
| S3_vs_k141_92235\|\|full | 4982 | No | NA | 9 | 6 | 0 | Low-quality | Genome-fragment | 13.9 | AAI-based (medium-confidence) | 0 |
| S12_extracted_k141_153847 | 4982 | No | NA | 2 | 0 | 2 | Low-quality | Genome-fragment | 2.95 | AAI-based (medium-confidence) | 0 |
| S3_vs_k141_57052\|\|full | 4983 | No | NA | 5 | 0 | 4 | Low-quality | Genome-fragment | 9.97 | AAI-based (medium-confidence) | 0 |
| S16_vs_131665\|\|full | 4984 | No | NA | 4 | 0 | 1 | Low-quality | Genome-fragment | 4.4 | AAI-based (medium-confidence) | 0 |
| S12_extracted_k141_14525 | 4984 | No | NA | 6 | 1 | 0 | Low-quality | Genome-fragment | 12.4 | AAI-based (high-confidence) | 0 |
| S15_extracted_73424 | 3511 | No | NA | 2 | 0 | 0 | Not-determined | Genome-fragment | NA | NA | 0 |
| S15_extracted_73449 | 3224 | No | NA | 1 | 0 | 0 | Not-determined | Genome-fragment | NA | NA | 0 |
| S11_vs_k141_35162\|\|full | 4985 | No | NA | 9 | 2 | 0 | Low-quality | Genome-fragment | 3.09 | HMM-based (lower-bound) | 0 |
| S15_extracted_74024 | 1626 | No | NA | 1 | 0 | 0 | Not-determined | Genome-fragment | NA | NA | 0 |
| S15_extracted_74148 | 1452 | No | NA | 2 | 0 | 0 | Not-determined | Genome-fragment | NA | NA | 0 |
| S15_extracted_74228 | 1039 | No | NA | 2 | 0 | 1 | Not-determined | Genome-fragment | NA | NA | 0 |
| S15_extracted_74351 | 1321 | No | NA | 1 | 0 | 0 | Not-determined | Genome-fragment | NA | NA | 0 |
| S15_extracted_74405 | 1135 | No | NA | 1 | 0 | 1 | Not-determined | Genome-fragment | NA | NA | 0 |
| S15_extracted_74449 | 1050 | No | NA | 1 | 0 | 0 | Not-determined | Genome-fragment | NA | NA | 0 |
| S5_extracted_k141_129451 | 4985 | No | NA | 5 | 0 | 0 | Low-quality | Genome-fragment | 7.84 | AAI-based (high-confidence) | 0 |
| S15_extracted_74586 | 84900 | No | NA | 112 | 12 | 4 | Complete | High-quality | 100 | DTR (high-confidence) | 0 |
| S5_extracted_k141_99061 | 4985 | No | NA | 4 | 0 | 4 | Low-quality | Genome-fragment | 0.73 | AAI-based (medium-confidence) | 0 |
| S15_extracted_74719 | 1433 | No | NA | 2 | 0 | 2 | Not-determined | Genome-fragment | NA | NA | 0 |
| S15_extracted_74903 | 1114 | No | NA | 1 | 0 | 0 | Not-determined | Genome-fragment | NA | NA | 0 |
| S15_extracted_75077 | 1191 | No | NA | 2 | 0 | 1 | Not-determined | Genome-fragment | NA | NA | 0 |
| S15_extracted_75386 | 1356 | No | NA | 3 | 0 | 2 | Not-determined | Genome-fragment | NA | NA | 0 |
| S15_extracted_75433 | 1048 | No | NA | 1 | 0 | 0 | Not-determined | Genome-fragment | NA | NA | 0 |
| S15_extracted_75739 | 1108 | No | NA | 2 | 0 | 1 | Not-determined | Genome-fragment | NA | NA | 0 |
| S15_extracted_75914 | 1071 | No | NA | 1 | 0 | 0 | Not-determined | Genome-fragment | NA | NA | 0 |
| S15_extracted_75917 | 5130 | No | NA | 10 | 0 | 0 | Not-determined | Genome-fragment | NA | NA | 0 |
| S15_extracted_75924 | 185145 | No | NA | 232 | 26 | 3 | Complete | High-quality | 100 | DTR (high-confidence) | 0 |
| S15_extracted_76063 | 1088 | No | NA | 1 | 0 | 1 | Not-determined | Genome-fragment | NA | NA | 0 |
| S3_vs_k141_96852\|\|full | 4986 | No | NA | 7 | 0 | 0 | Low-quality | Genome-fragment | 11.59 | AAI-based (high-confidence) | 0 |
| S8_vs_k141_9547\|\|full | 4987 | No | NA | 8 | 0 | 3 | Low-quality | Genome-fragment | 8.81 | AAI-based (high-confidence) | 0 |
| S15_extracted_76787 | 1346 | No | NA | 4 | 0 | 0 | Not-determined | Genome-fragment | NA | NA | 0 |
| S15_extracted_76884 | 7142 | No | NA | 10 | 0 | 7 | Not-determined | Genome-fragment | NA | NA | 0 |
| S15_extracted_76997 | 1208 | No | NA | 1 | 0 | 1 | Not-determined | Genome-fragment | NA | NA | 0 |
| S10_extracted_95374 | 4988 | No | NA | 5 | 0 | 1 | Low-quality | Genome-fragment | 10.73 | AAI-based (high-confidence) | 0 |
| S15_extracted_77240 | 1860 | No | NA | 2 | 0 | 2 | Not-determined | Genome-fragment | NA | NA | 0 |
| S14_vs_k141_54262\|\|full | 4990 | No | NA | 6 | 1 | 0 | Low-quality | Genome-fragment | 11.83 | AAI-based (high-confidence) | 0 |
| S5_extracted_k141_109591 | 4990 | No | NA | 11 | 3 | 0 | Low-quality | Genome-fragment | 12.05 | AAI-based (high-confidence) | 0 |
| S15_extracted_77515 | 1040 | No | NA | 1 | 0 | 1 | Not-determined | Genome-fragment | NA | NA | 0 |
| S15_vs_1407\|\|full | 13007 | No | NA | 20 | 7 | 0 | High-quality | High-quality | 98.04 | AAI-based (high-confidence) | 0 |
| S15_vs_7103\|\|full | 40282 | No | NA | 77 | 17 | 2 | High-quality | High-quality | 93.75 | AAI-based (high-confidence) | 0 |
| S15_vs_18274\|\|full | 35785 | No | NA | 59 | 9 | 0 | High-quality | High-quality | 98.64 | AAI-based (high-confidence) | 0 |
| S15_vs_76390\|\|full | 22356 | No | NA | 47 | 4 | 0 | High-quality | High-quality | 97.63 | AAI-based (high-confidence) | 0 |
| S8_extracted_k141_195322 | 4990 | No | NA | 6 | 0 | 1 | Low-quality | Genome-fragment | 7.02 | AAI-based (medium-confidence) | 0 |
| S15_vs_76435\|\|full | 4149 | No | NA | 9 | 0 | 2 | Not-determined | Genome-fragment | NA | NA | 0 |
| S15_vs_76554\|\|full | 6740 | No | NA | 13 | 0 | 1 | Not-determined | Genome-fragment | NA | NA | 0 |
| S15_vs_151\|\|full | 2271 | No | NA | 5 | 0 | 1 | Not-determined | Genome-fragment | NA | NA | 0 |
| S14_extracted_k141_50245 | 4991 | No | NA | 8 | 2 | 0 | Low-quality | Genome-fragment | 14 | AAI-based (high-confidence) | 0 |
| S15_vs_360\|\|full | 2963 | No | NA | 3 | 0 | 3 | Not-determined | Genome-fragment | NA | NA | 0 |
| S5_vs_k141_141156\|\|full | 4991 | Yes | 4266 | 5 | 1 | 1 | Low-quality | Genome-fragment | 2.68 | HMM-based (lower-bound) | 14.53 |
| S12_vs_k141_179354\|\|full | 4991 | No | NA | 3 | 0 | 0 | Low-quality | Genome-fragment | 4.76 | AAI-based (high-confidence) | 0 |
| S15_vs_1590\|\|full | 1454 | No | NA | 3 | 0 | 2 | Not-determined | Genome-fragment | NA | NA | 0 |
| S8_extracted_k141_68847 | 4991 | Yes | 3665 | 9 | 3 | 1 | Low-quality | Genome-fragment | 4.34 | HMM-based (lower-bound) | 26.57 |
| S15_vs_1734\|\|full | 3041 | No | NA | 3 | 0 | 1 | Not-determined | Genome-fragment | NA | NA | 0 |
| S15_vs_1978\|\|full | 3174 | No | NA | 2 | 0 | 1 | Not-determined | Genome-fragment | NA | NA | 0 |
| S15_vs_2067\|\|full | 1811 | No | NA | 2 | 0 | 1 | Not-determined | Genome-fragment | NA | NA | 0 |
| S14_extracted_k141_4629 | 4994 | No | NA | 4 | 3 | 0 | Low-quality | Genome-fragment | 13.38 | AAI-based (high-confidence) | 0 |
| S7_extracted_2050 | 4994 | No | NA | 13 | 0 | 0 | Low-quality | Genome-fragment | 8.17 | AAI-based (high-confidence) | 0 |
| S7_vs_121375\|\|full | 4994 | No | NA | 11 | 0 | 1 | Low-quality | Genome-fragment | 16.93 | AAI-based (medium-confidence) | 0 |
| S7_vs_129275\|\|full | 4994 | No | NA | 5 | 0 | 3 | Low-quality | Genome-fragment | 7.51 | AAI-based (medium-confidence) | 0 |
| S15_vs_3868\|\|full | 57618 | Yes | 53922 | 80 | 13 | 3 | High-quality | High-quality | 95.21 | AAI-based (high-confidence) | 6.41 |
| S15_vs_3997\|\|full | 3077 | No | NA | 6 | 0 | 2 | Not-determined | Genome-fragment | NA | NA | 0 |
| S10_extracted_23837 | 4996 | No | NA | 16 | 2 | 1 | Low-quality | Genome-fragment | 7.24 | HMM-based (lower-bound) | 0 |
| S15_vs_4199\|\|full | 4040 | No | NA | 3 | 0 | 1 | Not-determined | Genome-fragment | NA | NA | 0 |
| S15_vs_4606\|\|full | 9736 | No | NA | 7 | 0 | 5 | Not-determined | Genome-fragment | NA | NA | 0 |
| S8_extracted_k141_183152 | 4996 | No | NA | 3 | 0 | 0 | Low-quality | Genome-fragment | 5.04 | AAI-based (high-confidence) | 0 |
| S5_vs_k141_55427\|\|full | 4998 | Yes | 777 | 7 | 1 | 5 | Low-quality | Genome-fragment | 0.73 | AAI-based (medium-confidence) | 84.45 |
| S16_extracted_116746 | 4998 | Yes | 1940 | 5 | 2 | 2 | Low-quality | Genome-fragment | 5.34 | AAI-based (high-confidence) | 61.18 |
| S12_vs_k141_169549\|\|full | 4998 | No | NA | 3 | 1 | 0 | Low-quality | Genome-fragment | 8.39 | HMM-based (lower-bound) | 0 |
| S6_vs_k141_13046\|\|full | 4998 | No | NA | 8 | 4 | 0 | Low-quality | Genome-fragment | 12.39 | AAI-based (high-confidence) | 0 |
| S15_vs_5369\|\|full | 1265 | No | NA | 3 | 0 | 1 | Not-determined | Genome-fragment | NA | NA | 0 |
| S7_vs_18417\|\|full | 4999 | No | NA | 5 | 3 | 0 | Low-quality | Genome-fragment | 11.64 | AAI-based (high-confidence) | 0 |
| S15_vs_5510\|\|full | 1995 | No | NA | 3 | 0 | 2 | Not-determined | Genome-fragment | NA | NA | 0 |
| S8_vs_k141_4483\|\|full | 5001 | No | NA | 4 | 0 | 4 | Low-quality | Genome-fragment | 7.3 | AAI-based (medium-confidence) | 0 |
| S15_vs_5634\|\|full | 1221 | No | NA | 4 | 0 | 0 | Not-determined | Genome-fragment | NA | NA | 0 |
| S15_vs_5636\|\|full | 5256 | No | NA | 3 | 0 | 1 | Not-determined | Genome-fragment | NA | NA | 0 |
| S11_vs_k141_48783\|\|full | 5002 | Yes | 1280 | 5 | 1 | 1 | Low-quality | Genome-fragment | 2.39 | AAI-based (high-confidence) | 74.41 |
| S8_extracted_k141_135163 | 5003 | No | NA | 6 | 0 | 6 | Low-quality | Genome-fragment | 5.61 | AAI-based (high-confidence) | 0 |
| S15_vs_6079\|\|full | 1867 | No | NA | 3 | 0 | 3 | Not-determined | Genome-fragment | NA | NA | 0 |
| S15_vs_6132\|\|full | 5258 | No | NA | 7 | 0 | 5 | Not-determined | Genome-fragment | NA | NA | 0 |
| S17_extracted_k141_65943 | 5004 | No | NA | 6 | 2 | 0 | Low-quality | Genome-fragment | 4.06 | HMM-based (lower-bound) | 0 |
| S15_vs_6454\|\|full | 1028 | No | NA | 2 | 0 | 1 | Not-determined | Genome-fragment | NA | NA | 0 |
| S15_vs_6743\|\|full | 2041 | No | NA | 3 | 0 | 1 | Not-determined | Genome-fragment | NA | NA | 0 |
| S7_extracted_120229 | 5004 | No | NA | 5 | 2 | 0 | Low-quality | Genome-fragment | 8.59 | AAI-based (high-confidence) | 0 |
| S15_vs_6766\|\|full | 5075 | No | NA | 7 | 0 | 3 | Not-determined | Genome-fragment | NA | NA | 0 |
| S15_vs_6787\|\|full | 2431 | No | NA | 4 | 0 | 0 | Not-determined | Genome-fragment | NA | NA | 0 |
| S15_vs_8446\|\|full | 1072 | No | NA | 3 | 0 | 0 | Not-determined | Genome-fragment | NA | NA | 0 |
| S11_vs_k141_35975\|\|full | 5005 | No | NA | 4 | 3 | 0 | Low-quality | Genome-fragment | 9.82 | AAI-based (medium-confidence) | 0 |
| S2_vs_k141_3808\|\|full | 5009 | No | NA | 9 | 0 | 0 | Low-quality | Genome-fragment | 12.17 | AAI-based (high-confidence) | 0 |
| S15_vs_8929\|\|full | 1540 | No | NA | 4 | 0 | 0 | Not-determined | Genome-fragment | NA | NA | 0 |
| S16_vs_137322\|\|full | 5011 | No | NA | 4 | 0 | 2 | Low-quality | Genome-fragment | 5.01 | AAI-based (medium-confidence) | 0 |
| S15_vs_9198\|\|full | 2484 | No | NA | 3 | 0 | 1 | Not-determined | Genome-fragment | NA | NA | 0 |
| S15_vs_9449\|\|full | 2699 | No | NA | 4 | 0 | 4 | Not-determined | Genome-fragment | NA | NA | 0 |
| S14_vs_k141_54006\|\|full | 5014 | No | NA | 6 | 0 | 0 | Low-quality | Genome-fragment | 7.5 | AAI-based (medium-confidence) | 0 |
| S15_vs_9660\|\|full | 2006 | No | NA | 3 | 0 | 1 | Not-determined | Genome-fragment | NA | NA | 0 |
| S15_vs_9776\|\|full | 1178 | No | NA | 3 | 0 | 1 | Not-determined | Genome-fragment | NA | NA | 0 |
| S3_extracted_k141_50803 | 5014 | No | NA | 6 | 1 | 0 | Low-quality | Genome-fragment | 3.85 | AAI-based (high-confidence) | 0 |
| S15_vs_9934\|\|full | 1524 | No | NA | 3 | 0 | 2 | Not-determined | Genome-fragment | NA | NA | 0 |
| S13_vs_72005\|\|full | 5017 | No | NA | 9 | 2 | 0 | Low-quality | Genome-fragment | 12.44 | AAI-based (high-confidence) | 0 |
| S12_extracted_k141_112106 | 5017 | No | NA | 3 | 0 | 2 | Low-quality | Genome-fragment | 16.68 | AAI-based (high-confidence) | 0 |
| S15_vs_10239\|\|full | 3313 | No | NA | 3 | 0 | 3 | Not-determined | Genome-fragment | NA | NA | 0 |
| S15_vs_10411\|\|full | 1033 | No | NA | 1 | 0 | 0 | Not-determined | Genome-fragment | NA | NA | 0 |
| S15_vs_10521\|\|full | 1863 | No | NA | 3 | 0 | 2 | Not-determined | Genome-fragment | NA | NA | 0 |
| S15_vs_10532\|\|full | 2365 | No | NA | 4 | 0 | 0 | Not-determined | Genome-fragment | NA | NA | 0 |
| S3_vs_k141_32086\|\|full | 5018 | No | NA | 12 | 0 | 0 | Low-quality | Genome-fragment | 12.21 | AAI-based (high-confidence) | 0 |
| S12_vs_k141_110666\|\|full | 5018 | No | NA | 9 | 6 | 0 | Low-quality | Genome-fragment | 13.79 | AAI-based (medium-confidence) | 0 |
| S8_vs_k141_134557\|\|full | 5019 | No | NA | 13 | 3 | 0 | Low-quality | Genome-fragment | 7.69 | AAI-based (high-confidence) | 0 |
| S15_vs_11207\|\|full | 1741 | No | NA | 2 | 0 | 1 | Not-determined | Genome-fragment | NA | NA | 0 |
| S3_vs_k141_43311\|\|full | 5020 | No | NA | 7 | 0 | 0 | Low-quality | Genome-fragment | 9.19 | AAI-based (medium-confidence) | 0 |
| S3_vs_k141_100691\|\|full | 5022 | No | NA | 4 | 0 | 3 | Low-quality | Genome-fragment | 5.88 | AAI-based (medium-confidence) | 0 |
| S15_vs_11435\|\|full | 1218 | No | NA | 3 | 0 | 1 | Not-determined | Genome-fragment | NA | NA | 0 |
| S15_vs_11546\|\|full | 4776 | No | NA | 5 | 0 | 3 | Not-determined | Genome-fragment | NA | NA | 0 |
| S7_extracted_13828 | 5023 | No | NA | 6 | 0 | 4 | Low-quality | Genome-fragment | 3.42 | AAI-based (medium-confidence) | 0 |
| S15_vs_11876\|\|full | 6691 | No | NA | 8 | 0 | 4 | Not-determined | Genome-fragment | NA | NA | 0 |
| S5_vs_k141_82184\|\|full | 5026 | No | NA | 6 | 0 | 1 | Low-quality | Genome-fragment | 8.16 | AAI-based (medium-confidence) | 0 |
| S8_vs_k141_127109\|\|full | 5027 | No | NA | 8 | 2 | 1 | Low-quality | Genome-fragment | 6.99 | HMM-based (lower-bound) | 0 |
| S15_vs_12174\|\|full | 1337 | No | NA | 4 | 0 | 1 | Not-determined | Genome-fragment | NA | NA | 0 |
| S11_extracted_k141_87172 | 5031 | No | NA | 8 | 4 | 0 | Low-quality | Genome-fragment | 11.31 | AAI-based (high-confidence) | 0 |
| S15_vs_12622\|\|full | 1992 | No | NA | 2 | 0 | 2 | Not-determined | Genome-fragment | NA | NA | 0 |
| S15_vs_12858\|\|full | 2920 | No | NA | 5 | 0 | 0 | Not-determined | Genome-fragment | NA | NA | 0 |
| S15_vs_13285\|\|full | 4424 | No | NA | 6 | 0 | 1 | Not-determined | Genome-fragment | NA | NA | 0 |
| S15_vs_13914\|\|full | 3506 | No | NA | 4 | 0 | 3 | Not-determined | Genome-fragment | NA | NA | 0 |
| S15_vs_13967\|\|full | 2475 | No | NA | 2 | 0 | 1 | Not-determined | Genome-fragment | NA | NA | 0 |
| S15_vs_13970\|\|full | 8966 | No | NA | 9 | 0 | 8 | Not-determined | Genome-fragment | NA | NA | 0 |
| S15_vs_14110\|\|full | 3801 | No | NA | 4 | 0 | 1 | Not-determined | Genome-fragment | NA | NA | 0 |
| S15_vs_14252\|\|full | 1553 | No | NA | 4 | 0 | 1 | Not-determined | Genome-fragment | NA | NA | 0 |
| S3_vs_k141_40007\|\|full | 5032 | No | NA | 7 | 1 | 0 | Low-quality | Genome-fragment | 3.51 | HMM-based (lower-bound) | 0 |
| S15_vs_14866\|\|full | 3588 | No | NA | 4 | 0 | 1 | Not-determined | Genome-fragment | NA | NA | 0 |
| S15_vs_14882\|\|full | 1435 | No | NA | 3 | 0 | 0 | Not-determined | Genome-fragment | NA | NA | 0 |
| S15_vs_15117\|\|full | 2141 | No | NA | 6 | 0 | 1 | Not-determined | Genome-fragment | NA | NA | 0 |
| S15_vs_15196\|\|full | 3974 | No | NA | 3 | 0 | 2 | Not-determined | Genome-fragment | NA | NA | 0 |
| S3_vs_k141_88493\|\|full | 5032 | No | NA | 12 | 3 | 0 | Low-quality | Genome-fragment | 2.58 | AAI-based (high-confidence) | 0 |
| S15_vs_15949\|\|full | 1275 | No | NA | 2 | 0 | 2 | Not-determined | Genome-fragment | NA | NA | 0 |
| S15_vs_16134\|\|full | 2199 | No | NA | 3 | 0 | 2 | Not-determined | Genome-fragment | NA | NA | 0 |
| S5_extracted_k141_141726 | 5032 | No | NA | 7 | 4 | 0 | Low-quality | Genome-fragment | 14.56 | AAI-based (medium-confidence) | 0 |
| S15_vs_16199\|\|full | 1339 | No | NA | 3 | 0 | 0 | Not-determined | Genome-fragment | NA | NA | 0 |
| S15_vs_16231\|\|full | 1496 | No | NA | 3 | 0 | 0 | Not-determined | Genome-fragment | NA | NA | 0 |
| S7_extracted_41060 | 5032 | No | NA | 5 | 0 | 0 | Low-quality | Genome-fragment | 8.13 | AAI-based (medium-confidence) | 0 |
| S15_vs_16363\|\|full | 1317 | No | NA | 3 | 0 | 1 | Not-determined | Genome-fragment | NA | NA | 0 |
| S14_vs_k141_21363\|\|full | 5033 | No | NA | 12 | 3 | 1 | Low-quality | Genome-fragment | 7.11 | AAI-based (high-confidence) | 0 |
| S15_vs_16619\|\|full | 2715 | No | NA | 4 | 0 | 2 | Not-determined | Genome-fragment | NA | NA | 0 |
| S15_vs_17079\|\|full | 2713 | No | NA | 5 | 0 | 0 | Not-determined | Genome-fragment | NA | NA | 0 |
| S15_vs_17114\|\|full | 15295 | No | NA | 10 | 0 | 5 | Not-determined | Genome-fragment | NA | NA | 0 |
| S7_vs_37137\|\|full | 5035 | No | NA | 10 | 5 | 0 | Low-quality | Genome-fragment | 11.7 | AAI-based (high-confidence) | 0 |
| S15_vs_17238\|\|full | 29383 | No | NA | 32 | 0 | 22 | Not-determined | Genome-fragment | NA | NA | 0 |
| S15_vs_17361\|\|full | 6667 | No | NA | 9 | 0 | 6 | Not-determined | Genome-fragment | NA | NA | 0 |
| S11_vs_k141_13551\|\|full | 5036 | No | NA | 5 | 1 | 0 | Low-quality | Genome-fragment | 13.37 | AAI-based (high-confidence) | 0 |
| S15_vs_17471\|\|full | 1448 | No | NA | 4 | 0 | 0 | Not-determined | Genome-fragment | NA | NA | 0 |
| S15_vs_17594\|\|full | 6008 | No | NA | 5 | 0 | 5 | Not-determined | Genome-fragment | NA | NA | 0 |
| S15_vs_17664\|\|full | 2837 | No | NA | 3 | 0 | 1 | Not-determined | Genome-fragment | NA | NA | 0 |
| S5_vs_k141_145595\|\|full | 5036 | No | NA | 7 | 0 | 2 | Low-quality | Genome-fragment | 7.61 | AAI-based (medium-confidence) | 0 |
| S12_extracted_k141_32454 | 5042 | No | NA | 10 | 2 | 0 | Low-quality | Genome-fragment | 9.02 | AAI-based (high-confidence) | 0 |
| S3_vs_k141_122973\|\|full | 5043 | No | NA | 7 | 4 | 0 | Low-quality | Genome-fragment | 12.44 | AAI-based (medium-confidence) | 0 |
| S3_vs_k141_8414\|\|full | 5044 | No | NA | 10 | 2 | 0 | Low-quality | Genome-fragment | 12.72 | AAI-based (high-confidence) | 0 |
| S15_vs_18522\|\|full | 1349 | No | NA | 2 | 0 | 1 | Not-determined | Genome-fragment | NA | NA | 0 |
| S15_vs_18587\|\|full | 5035 | No | NA | 5 | 0 | 1 | Not-determined | Genome-fragment | NA | NA | 0 |
| S8_vs_k141_103428\|\|full | 5047 | No | NA | 9 | 0 | 0 | Low-quality | Genome-fragment | 11.71 | AAI-based (medium-confidence) | 0 |
| S15_vs_18789\|\|full | 2706 | No | NA | 4 | 0 | 0 | Not-determined | Genome-fragment | NA | NA | 0 |
| S15_vs_18949\|\|full | 5125 | No | NA | 3 | 0 | 1 | Not-determined | Genome-fragment | NA | NA | 0 |
| S12_vs_k141_22825\|\|full | 5048 | No | NA | 13 | 3 | 0 | Low-quality | Genome-fragment | 5.59 | HMM-based (lower-bound) | 0 |
| S3_vs_k141_53674\|\|full | 5050 | No | NA | 4 | 0 | 3 | Low-quality | Genome-fragment | 1.68 | AAI-based (medium-confidence) | 0 |
| S13_vs_85028\|\|full | 5051 | No | NA | 4 | 0 | 0 | Low-quality | Genome-fragment | 10.97 | AAI-based (high-confidence) | 0 |
| S15_vs_19964\|\|full | 1041 | No | NA | 2 | 0 | 1 | Not-determined | Genome-fragment | NA | NA | 0 |
| S11_extracted_k141_87601 | 5052 | No | NA | 7 | 2 | 0 | Low-quality | Genome-fragment | 11.05 | AAI-based (high-confidence) | 0 |
| S15_vs_20445\|\|full | 9095 | No | NA | 12 | 0 | 0 | Not-determined | Genome-fragment | NA | NA | 0 |
| S15_vs_20606\|\|full | 1183 | No | NA | 2 | 0 | 0 | Not-determined | Genome-fragment | NA | NA | 0 |
| S15_vs_21078\|\|full | 1885 | No | NA | 3 | 0 | 2 | Not-determined | Genome-fragment | NA | NA | 0 |
| S15_vs_21122\|\|full | 1442 | No | NA | 4 | 0 | 0 | Not-determined | Genome-fragment | NA | NA | 0 |
| S11_extracted_k141_32223 | 5052 | No | NA | 6 | 0 | 2 | Low-quality | Genome-fragment | 7.82 | AAI-based (medium-confidence) | 0 |
| S11_vs_k141_45177\|\|full | 5054 | No | NA | 3 | 3 | 0 | Low-quality | Genome-fragment | 12.02 | AAI-based (medium-confidence) | 0 |
| S15_vs_21519\|\|full | 2496 | No | NA | 4 | 0 | 0 | Not-determined | Genome-fragment | NA | NA | 0 |
| S3_extracted_k141_119377 | 5055 | No | NA | 14 | 1 | 0 | Low-quality | Genome-fragment | 3.12 | AAI-based (high-confidence) | 0 |
| S6_extracted_k141_30192 | 5055 | No | NA | 10 | 3 | 0 | Low-quality | Genome-fragment | 7.48 | AAI-based (high-confidence) | 0 |
| S7_extracted_1945 | 5057 | No | NA | 6 | 2 | 0 | Low-quality | Genome-fragment | 23.11 | AAI-based (high-confidence) | 0 |
| S9_extracted_k141_33193 | 5058 | No | NA | 11 | 6 | 0 | Low-quality | Genome-fragment | 11.69 | AAI-based (medium-confidence) | 0 |
| S15_vs_22299\|\|full | 2152 | No | NA | 3 | 0 | 2 | Not-determined | Genome-fragment | NA | NA | 0 |
| S7_extracted_22121 | 5058 | No | NA | 14 | 3 | 0 | Low-quality | Genome-fragment | 9.17 | AAI-based (high-confidence) | 0 |
| S15_vs_54176\|\|full | 5058 | No | NA | 6 | 1 | 0 | Low-quality | Genome-fragment | 6.36 | AAI-based (medium-confidence) | 0 |
| S15_vs_23250\|\|full | 2319 | No | NA | 3 | 0 | 3 | Not-determined | Genome-fragment | NA | NA | 0 |
| S4_vs_VIRSorter_1584-cat_2 | 5059 | Yes | 4096 | 9 | 5 | 1 | Low-quality | Genome-fragment | 11.07 | AAI-based (high-confidence) | 19.04 |
| S15_vs_24480\|\|full | 6320 | No | NA | 7 | 0 | 4 | Not-determined | Genome-fragment | NA | NA | 0 |
| S15_vs_24792\|\|full | 1151 | No | NA | 3 | 0 | 0 | Not-determined | Genome-fragment | NA | NA | 0 |
| S15_vs_25104\|\|full | 4315 | No | NA | 3 | 0 | 3 | Not-determined | Genome-fragment | NA | NA | 0 |
| S15_vs_25195\|\|full | 2720 | No | NA | 3 | 0 | 2 | Not-determined | Genome-fragment | NA | NA | 0 |
| S15_vs_25291\|\|full | 1235 | No | NA | 2 | 0 | 1 | Not-determined | Genome-fragment | NA | NA | 0 |
| S15_vs_25493\|\|full | 1259 | No | NA | 3 | 0 | 1 | Not-determined | Genome-fragment | NA | NA | 0 |
| S9_vs_k141_42784\|\|full | 5060 | No | NA | 6 | 0 | 2 | Low-quality | Genome-fragment | 1.8 | AAI-based (medium-confidence) | 0 |
| S15_vs_25606\|\|full | 2255 | No | NA | 4 | 0 | 0 | Not-determined | Genome-fragment | NA | NA | 0 |
| S15_vs_25673\|\|full | 9464 | No | NA | 7 | 0 | 5 | Not-determined | Genome-fragment | NA | NA | 0 |
| S15_vs_26015\|\|full | 4600 | No | NA | 7 | 0 | 2 | Not-determined | Genome-fragment | NA | NA | 0 |
| S11_extracted_k141_97842 | 5067 | No | NA | 13 | 1 | 0 | Low-quality | Genome-fragment | 8.08 | AAI-based (high-confidence) | 0 |
| S16_extracted_144340 | 5067 | No | NA | 4 | 0 | 1 | Low-quality | Genome-fragment | 0.44 | AAI-based (medium-confidence) | 0 |
| S15_vs_26463\|\|full | 1503 | No | NA | 3 | 0 | 2 | Not-determined | Genome-fragment | NA | NA | 0 |
| S14_extracted_k141_99108 | 5070 | No | NA | 10 | 1 | 0 | Low-quality | Genome-fragment | 11.98 | AAI-based (high-confidence) | 0 |
| S5_extracted_k141_140377 | 5070 | No | NA | 10 | 2 | 0 | Low-quality | Genome-fragment | 8.45 | AAI-based (high-confidence) | 0 |
| S15_vs_27291\|\|full | 2413 | No | NA | 4 | 0 | 2 | Not-determined | Genome-fragment | NA | NA | 0 |
| S15_vs_27390\|\|full | 3456 | No | NA | 3 | 0 | 0 | Not-determined | Genome-fragment | NA | NA | 0 |
| S7_extracted_18402 | 5070 | No | NA | 11 | 4 | 0 | Low-quality | Genome-fragment | 3.05 | AAI-based (medium-confidence) | 0 |
| S15_vs_27510\|\|full | 57438 | No | NA | 86 | 16 | 2 | High-quality | High-quality | 92.69 | AAI-based (high-confidence) | 0 |
| S15_vs_27530\|\|full | 1471 | No | NA | 3 | 0 | 0 | Not-determined | Genome-fragment | NA | NA | 0 |
| S15_vs_27682\|\|full | 1088 | No | NA | 2 | 0 | 2 | Not-determined | Genome-fragment | NA | NA | 0 |
| S15_vs_27813\|\|full | 2305 | No | NA | 3 | 0 | 1 | Not-determined | Genome-fragment | NA | NA | 0 |
| S15_vs_27904\|\|full | 1916 | No | NA | 3 | 0 | 0 | Not-determined | Genome-fragment | NA | NA | 0 |
| S14_vs_k141_32896\|\|full | 5071 | No | NA | 8 | 0 | 1 | Low-quality | Genome-fragment | 8.9 | AAI-based (medium-confidence) | 0 |
| S14_vs_k141_98849\|\|full | 5074 | No | NA | 6 | 2 | 0 | Low-quality | Genome-fragment | 11.13 | AAI-based (high-confidence) | 0 |
| S12_vs_k141_57429\|\|full | 5074 | No | NA | 7 | 0 | 5 | Low-quality | Genome-fragment | 10.03 | AAI-based (medium-confidence) | 0 |
| S15_vs_28329\|\|full | 1197 | No | NA | 3 | 0 | 1 | Not-determined | Genome-fragment | NA | NA | 0 |
| S17_vs_k141_156145\|\|full | 5075 | No | NA | 7 | 0 | 5 | Low-quality | Genome-fragment | 16.21 | AAI-based (medium-confidence) | 0 |
| S15_vs_29001\|\|full | 3367 | No | NA | 4 | 0 | 3 | Not-determined | Genome-fragment | NA | NA | 0 |
| S15_vs_29487\|\|full | 1865 | No | NA | 4 | 0 | 0 | Not-determined | Genome-fragment | NA | NA | 0 |
| S3_extracted_k141_9999 | 5075 | No | NA | 7 | 0 | 0 | Low-quality | Genome-fragment | 11.65 | AAI-based (medium-confidence) | 0 |
| S15_vs_29615\|\|full | 2386 | No | NA | 4 | 0 | 4 | Not-determined | Genome-fragment | NA | NA | 0 |
| S15_vs_29720\|\|full | 9771 | No | NA | 10 | 0 | 8 | Not-determined | Genome-fragment | NA | NA | 0 |
| S15_vs_29831\|\|full | 20459 | No | NA | 18 | 0 | 13 | Not-determined | Genome-fragment | NA | NA | 0 |
| S15_vs_30158\|\|full | 10952 | No | NA | 11 | 0 | 5 | Not-determined | Genome-fragment | NA | NA | 0 |
| S15_vs_30558\|\|full | 3605 | No | NA | 3 | 0 | 3 | Not-determined | Genome-fragment | NA | NA | 0 |
| S15_vs_30967\|\|full | 3436 | No | NA | 4 | 0 | 3 | Not-determined | Genome-fragment | NA | NA | 0 |
| S1_vs_k141_126659\|\|full | 5076 | No | NA | 7 | 3 | 0 | Low-quality | Genome-fragment | 12.43 | AAI-based (high-confidence) | 0 |
| S15_vs_31155\|\|full | 2655 | No | NA | 2 | 0 | 1 | Not-determined | Genome-fragment | NA | NA | 0 |
| S15_vs_31246\|\|full | 3305 | No | NA | 4 | 0 | 0 | Not-determined | Genome-fragment | NA | NA | 0 |
| S15_vs_31318\|\|full | 2372 | No | NA | 3 | 0 | 2 | Not-determined | Genome-fragment | NA | NA | 0 |
| S15_vs_31536\|\|full | 5902 | No | NA | 4 | 0 | 2 | Not-determined | Genome-fragment | NA | NA | 0 |
| S17_vs_k141_55043\|\|full | 5078 | No | NA | 7 | 5 | 0 | Low-quality | Genome-fragment | 15.39 | AAI-based (high-confidence) | 0 |
| S14_vs_k141_84366\|\|full | 5080 | No | NA | 4 | 2 | 0 | Low-quality | Genome-fragment | 13.18 | AAI-based (high-confidence) | 0 |
| S1_vs_k141_17087\|\|full | 5080 | No | NA | 10 | 2 | 0 | Low-quality | Genome-fragment | 10.44 | AAI-based (high-confidence) | 0 |
| S9_extracted_k141_24484 | 5081 | No | NA | 15 | 0 | 0 | Low-quality | Genome-fragment | 3.11 | AAI-based (high-confidence) | 0 |
| S15_vs_31946\|\|full | 8133 | No | NA | 6 | 0 | 1 | Not-determined | Genome-fragment | NA | NA | 0 |
| S15_vs_32044\|\|full | 1478 | No | NA | 3 | 0 | 0 | Not-determined | Genome-fragment | NA | NA | 0 |
| S1_vs_k141_153070\|\|full | 5081 | No | NA | 7 | 0 | 3 | Low-quality | Genome-fragment | 5.84 | AAI-based (high-confidence) | 0 |
| S15_vs_32136\|\|full | 5682 | No | NA | 5 | 0 | 5 | Not-determined | Genome-fragment | NA | NA | 0 |
| S15_vs_32204\|\|full | 1438 | No | NA | 2 | 0 | 2 | Not-determined | Genome-fragment | NA | NA | 0 |
| S14_extracted_k141_39637 | 5082 | No | NA | 5 | 0 | 3 | Low-quality | Genome-fragment | 16.84 | AAI-based (high-confidence) | 0 |
| S15_vs_32803\|\|full | 14911 | No | NA | 12 | 0 | 10 | Not-determined | Genome-fragment | NA | NA | 0 |
| S15_vs_32834\|\|full | 2313 | No | NA | 3 | 0 | 2 | Not-determined | Genome-fragment | NA | NA | 0 |
| S15_vs_33087\|\|full | 3070 | No | NA | 3 | 0 | 0 | Not-determined | Genome-fragment | NA | NA | 0 |
| S15_vs_33158\|\|full | 2974 | No | NA | 4 | 0 | 1 | Not-determined | Genome-fragment | NA | NA | 0 |
| S15_vs_33648\|\|full | 7897 | No | NA | 5 | 0 | 4 | Not-determined | Genome-fragment | NA | NA | 0 |
| S5_vs_k141_1064\|\|full | 5083 | No | NA | 9 | 0 | 0 | Low-quality | Genome-fragment | 11.78 | AAI-based (high-confidence) | 0 |
| S15_vs_34108\|\|full | 1948 | No | NA | 2 | 0 | 2 | Not-determined | Genome-fragment | NA | NA | 0 |
| S15_vs_34193\|\|full | 2221 | No | NA | 3 | 0 | 1 | Not-determined | Genome-fragment | NA | NA | 0 |
| S15_vs_34581\|\|full | 3516 | No | NA | 5 | 0 | 0 | Not-determined | Genome-fragment | NA | NA | 0 |
| S16_vs_121710\|\|full | 5085 | No | NA | 7 | 1 | 0 | Low-quality | Genome-fragment | 3.42 | HMM-based (lower-bound) | 0 |
| S15_vs_35445\|\|full | 41163 | No | NA | 24 | 1 | 13 | Medium-quality | Genome-fragment | 61.58 | HMM-based (lower-bound) | 0 |
| S5_vs_k141_71009\|\|full | 5086 | No | NA | 12 | 2 | 1 | Low-quality | Genome-fragment | 14.52 | AAI-based (high-confidence) | 0 |
| S15_vs_35473\|\|full | 3000 | No | NA | 4 | 0 | 1 | Not-determined | Genome-fragment | NA | NA | 0 |
| S15_vs_35544\|\|full | 2199 | No | NA | 3 | 0 | 0 | Not-determined | Genome-fragment | NA | NA | 0 |
| S14_vs_k141_27630\|\|full | 5089 | No | NA | 6 | 4 | 0 | Low-quality | Genome-fragment | 11.93 | AAI-based (high-confidence) | 0 |
| S12_vs_k141_44833\|\|full | 5089 | No | NA | 6 | 4 | 0 | Low-quality | Genome-fragment | 14.03 | AAI-based (high-confidence) | 0 |
| S15_vs_35861\|\|full | 2383 | No | NA | 2 | 0 | 0 | Not-determined | Genome-fragment | NA | NA | 0 |
| S15_vs_36010\|\|full | 1904 | No | NA | 3 | 0 | 1 | Not-determined | Genome-fragment | NA | NA | 0 |
| S14_vs_k141_77569\|\|full | 5090 | No | NA | 3 | 1 | 0 | Low-quality | Genome-fragment | 8.83 | HMM-based (lower-bound) | 0 |
| S15_vs_36194\|\|full | 3261 | No | NA | 3 | 0 | 1 | Not-determined | Genome-fragment | NA | NA | 0 |
| S9_vs_k141_35724\|\|full | 5092 | No | NA | 7 | 5 | 0 | Low-quality | Genome-fragment | 11.78 | AAI-based (high-confidence) | 0 |
| S16_extracted_56196 | 5092 | No | NA | 6 | 1 | 0 | Low-quality | Genome-fragment | 10.37 | AAI-based (medium-confidence) | 0 |
| S15_vs_36583\|\|full | 1814 | No | NA | 3 | 0 | 1 | Not-determined | Genome-fragment | NA | NA | 0 |
| S15_vs_36967\|\|full | 1080 | No | NA | 2 | 0 | 0 | Not-determined | Genome-fragment | NA | NA | 0 |
| S15_vs_37523\|\|full | 3851 | No | NA | 6 | 0 | 1 | Not-determined | Genome-fragment | NA | NA | 0 |
| S15_vs_37530\|\|full | 8944 | No | NA | 7 | 0 | 6 | Not-determined | Genome-fragment | NA | NA | 0 |
| S6_vs_k141_44965\|\|full | 5094 | No | NA | 4 | 0 | 1 | Low-quality | Genome-fragment | 5.46 | AAI-based (high-confidence) | 0 |
| S15_vs_38280\|\|full | 2890 | No | NA | 2 | 0 | 0 | Not-determined | Genome-fragment | NA | NA | 0 |
| S12_extracted_k141_85794 | 5095 | No | NA | 3 | 2 | 0 | Low-quality | Genome-fragment | 8.01 | AAI-based (medium-confidence) | 0 |
| S15_vs_38471\|\|full | 2361 | No | NA | 3 | 0 | 1 | Not-determined | Genome-fragment | NA | NA | 0 |
| S15_vs_38614\|\|full | 2738 | No | NA | 5 | 0 | 1 | Not-determined | Genome-fragment | NA | NA | 0 |
| S15_vs_38710\|\|full | 4235 | No | NA | 3 | 0 | 3 | Not-determined | Genome-fragment | NA | NA | 0 |
| S15_vs_38874\|\|full | 1952 | No | NA | 3 | 0 | 2 | Not-determined | Genome-fragment | NA | NA | 0 |
| S15_vs_39804\|\|full | 32952 | No | NA | 44 | 17 | 0 | Medium-quality | Genome-fragment | 80.83 | AAI-based (high-confidence) | 0 |
| S14_vs_k141_73711\|\|full | 5096 | No | NA | 7 | 2 | 0 | Low-quality | Genome-fragment | 12.15 | AAI-based (high-confidence) | 0 |
| S15_vs_40277\|\|full | 2971 | No | NA | 3 | 0 | 1 | Not-determined | Genome-fragment | NA | NA | 0 |
| S14_vs_k141_113904\|\|full | 5099 | No | NA | 10 | 1 | 0 | Low-quality | Genome-fragment | 11.63 | AAI-based (high-confidence) | 0 |
| S5_vs_k141_35206\|\|full | 5099 | No | NA | 7 | 2 | 0 | Low-quality | Genome-fragment | 12.31 | AAI-based (high-confidence) | 0 |
| S16_vs_157549\|\|full | 5102 | No | NA | 4 | 0 | 3 | Low-quality | Genome-fragment | 3.08 | AAI-based (medium-confidence) | 0 |
| S15_vs_40498\|\|full | 2347 | No | NA | 3 | 0 | 3 | Not-determined | Genome-fragment | NA | NA | 0 |
| S15_vs_40775\|\|full | 2255 | No | NA | 3 | 0 | 1 | Not-determined | Genome-fragment | NA | NA | 0 |
| S15_vs_41013\|\|full | 3914 | No | NA | 4 | 0 | 1 | Not-determined | Genome-fragment | NA | NA | 0 |
| S15_vs_41475\|\|full | 2050 | No | NA | 2 | 0 | 1 | Not-determined | Genome-fragment | NA | NA | 0 |
| S7_extracted_5404 | 5108 | No | NA | 6 | 0 | 1 | Low-quality | Genome-fragment | 12.19 | AAI-based (medium-confidence) | 0 |
| S7_extracted_12682 | 5108 | No | NA | 4 | 0 | 2 | Low-quality | Genome-fragment | 5.04 | AAI-based (medium-confidence) | 0 |
| S16_extracted_158538 | 5108 | No | NA | 4 | 0 | 1 | Low-quality | Genome-fragment | 7.43 | AAI-based (medium-confidence) | 0 |
| S15_vs_42043\|\|full | 193486 | Yes | 60217 | 197 | 15 | 84 | High-quality | High-quality | 99.53 | AAI-based (medium-confidence) | 68.88 |
| S15_vs_42109\|\|full | 11602 | No | NA | 11 | 0 | 6 | Not-determined | Genome-fragment | NA | NA | 0 |
| S10_vs_82248\|\|full | 5108 | No | NA | 4 | 0 | 3 | Low-quality | Genome-fragment | 7.4 | AAI-based (medium-confidence) | 0 |
| S14_vs_k141_30764\|\|full | 5110 | No | NA | 8 | 1 | 0 | Low-quality | Genome-fragment | 8.62 | HMM-based (lower-bound) | 0 |
| S15_vs_42644\|\|full | 7479 | No | NA | 5 | 0 | 4 | Not-determined | Genome-fragment | NA | NA | 0 |
| S15_vs_42680\|\|full | 2299 | No | NA | 2 | 0 | 0 | Not-determined | Genome-fragment | NA | NA | 0 |
| S15_vs_42725\|\|full | 4959 | No | NA | 5 | 0 | 0 | Not-determined | Genome-fragment | NA | NA | 0 |
| S15_vs_42805\|\|full | 1280 | No | NA | 3 | 0 | 1 | Not-determined | Genome-fragment | NA | NA | 0 |
| S15_vs_42872\|\|full | 8695 | No | NA | 13 | 0 | 2 | Not-determined | Genome-fragment | NA | NA | 0 |
| S15_vs_43017\|\|full | 11115 | No | NA | 10 | 0 | 4 | Not-determined | Genome-fragment | NA | NA | 0 |
| S15_vs_43454\|\|full | 1841 | No | NA | 4 | 0 | 0 | Not-determined | Genome-fragment | NA | NA | 0 |
| S5_vs_k141_79668\|\|full | 5111 | No | NA | 4 | 0 | 3 | Low-quality | Genome-fragment | 4.94 | AAI-based (medium-confidence) | 0 |
| S15_vs_43709\|\|full | 2521 | No | NA | 3 | 0 | 2 | Not-determined | Genome-fragment | NA | NA | 0 |
| S15_vs_43804\|\|full | 1320 | No | NA | 3 | 0 | 0 | Not-determined | Genome-fragment | NA | NA | 0 |
| S15_vs_43913\|\|full | 44889 | Yes | 34075 | 45 | 5 | 5 | Medium-quality | Genome-fragment | 58.36 | AAI-based (high-confidence) | 24.09 |
| S15_vs_44308\|\|full | 4003 | No | NA | 4 | 0 | 2 | Not-determined | Genome-fragment | NA | NA | 0 |
| S16_extracted_135627 | 5111 | No | NA | 3 | 0 | 2 | Low-quality | Genome-fragment | 2.29 | AAI-based (medium-confidence) | 0 |
| S10_extracted_75424 | 5111 | No | NA | 9 | 2 | 0 | Low-quality | Genome-fragment | 8.08 | AAI-based (high-confidence) | 0 |
| S3_extracted_k141_55879 | 5112 | No | NA | 7 | 5 | 0 | Low-quality | Genome-fragment | 12.85 | AAI-based (high-confidence) | 0 |
| S8_extracted_k141_142151 | 5112 | No | NA | 5 | 4 | 0 | Low-quality | Genome-fragment | 16.24 | AAI-based (high-confidence) | 0 |
| S15_vs_45380\|\|full | 3086 | No | NA | 4 | 0 | 2 | Not-determined | Genome-fragment | NA | NA | 0 |
| S17_extracted_k141_6808 | 5114 | No | NA | 3 | 0 | 2 | Low-quality | Genome-fragment | 2.75 | AAI-based (high-confidence) | 0 |
| S1_vs_k141_2905\|\|full | 5115 | No | NA | 8 | 3 | 0 | Low-quality | Genome-fragment | 11.47 | AAI-based (high-confidence) | 0 |
| S5_vs_k141_117140\|\|full | 5117 | Yes | 4665 | 7 | 1 | 1 | Low-quality | Genome-fragment | 7.06 | HMM-based (lower-bound) | 8.83 |
| S15_vs_46128\|\|full | 4832 | No | NA | 7 | 0 | 0 | Not-determined | Genome-fragment | NA | NA | 0 |
| S15_extracted_62013 | 5119 | No | NA | 8 | 1 | 0 | Low-quality | Genome-fragment | 14.57 | AAI-based (high-confidence) | 0 |
| S15_vs_46331\|\|full | 1405 | No | NA | 2 | 0 | 1 | Not-determined | Genome-fragment | NA | NA | 0 |
| S15_vs_46336\|\|full | 1582 | No | NA | 3 | 0 | 2 | Not-determined | Genome-fragment | NA | NA | 0 |
| S15_vs_46355\|\|full | 7793 | No | NA | 9 | 0 | 7 | Not-determined | Genome-fragment | NA | NA | 0 |
| S15_vs_46478\|\|full | 1053 | No | NA | 3 | 0 | 0 | Not-determined | Genome-fragment | NA | NA | 0 |
| S15_vs_46838\|\|full | 1514 | No | NA | 2 | 0 | 1 | Not-determined | Genome-fragment | NA | NA | 0 |
| S17_vs_k141_174807\|\|full | 5120 | No | NA | 7 | 0 | 0 | Low-quality | Genome-fragment | 7.69 | AAI-based (medium-confidence) | 0 |
| S15_vs_47316\|\|full | 2221 | No | NA | 5 | 0 | 0 | Not-determined | Genome-fragment | NA | NA | 0 |
| S15_vs_47402\|\|full | 2578 | No | NA | 2 | 0 | 2 | Not-determined | Genome-fragment | NA | NA | 0 |
| S14_extracted_k141_116486 | 5120 | No | NA | 6 | 0 | 1 | Low-quality | Genome-fragment | 2.41 | AAI-based (medium-confidence) | 0 |
| S15_vs_48332\|\|full | 1175 | No | NA | 3 | 0 | 3 | Not-determined | Genome-fragment | NA | NA | 0 |
| S10_extracted_16526 | 5120 | No | NA | 4 | 0 | 1 | Low-quality | Genome-fragment | 4.16 | AAI-based (high-confidence) | 0 |
| S15_vs_48613\|\|full | 1751 | No | NA | 2 | 0 | 0 | Not-determined | Genome-fragment | NA | NA | 0 |
| S15_vs_48667\|\|full | 4095 | No | NA | 3 | 0 | 1 | Not-determined | Genome-fragment | NA | NA | 0 |
| S15_vs_48930\|\|full | 1823 | No | NA | 3 | 0 | 1 | Not-determined | Genome-fragment | NA | NA | 0 |
| S8_extracted_k141_1564 | 5120 | Yes | 4347 | 9 | 4 | 1 | Low-quality | Genome-fragment | 10.21 | AAI-based (high-confidence) | 15.1 |
| S15_vs_49004\|\|full | 1326 | No | NA | 3 | 0 | 0 | Not-determined | Genome-fragment | NA | NA | 0 |
| S13_vs_150686\|\|full | 5121 | Yes | 3129 | 9 | 1 | 1 | Low-quality | Genome-fragment | 2.1 | HMM-based (lower-bound) | 38.9 |
| S15_vs_49993\|\|full | 1139 | No | NA | 2 | 0 | 0 | Not-determined | Genome-fragment | NA | NA | 0 |
| S1_extracted_k141_149054 | 5121 | No | NA | 12 | 2 | 1 | Low-quality | Genome-fragment | 10.48 | AAI-based (medium-confidence) | 0 |
| S8_extracted_k141_102305 | 5121 | No | NA | 3 | 0 | 2 | Low-quality | Genome-fragment | 2.18 | AAI-based (medium-confidence) | 0 |
| S15_vs_50904\|\|full | 3399 | No | NA | 3 | 0 | 2 | Not-determined | Genome-fragment | NA | NA | 0 |
| S15_vs_50947\|\|full | 1570 | No | NA | 3 | 0 | 2 | Not-determined | Genome-fragment | NA | NA | 0 |
| S4_extracted_10730 | 5122 | No | NA | 6 | 0 | 4 | Low-quality | Genome-fragment | 1.57 | AAI-based (medium-confidence) | 0 |
| S15_vs_51052\|\|full | 6012 | No | NA | 7 | 0 | 1 | Not-determined | Genome-fragment | NA | NA | 0 |
| S15_vs_51250\|\|full | 1976 | No | NA | 2 | 0 | 0 | Not-determined | Genome-fragment | NA | NA | 0 |
| S5_vs_k141_129408\|\|full | 5123 | No | NA | 9 | 3 | 0 | Low-quality | Genome-fragment | 12.86 | AAI-based (high-confidence) | 0 |
| S5_vs_k141_5389\|\|full | 5124 | No | NA | 9 | 1 | 1 | Low-quality | Genome-fragment | 3.32 | HMM-based (lower-bound) | 0 |
| S15_vs_51378\|\|full | 4413 | No | NA | 5 | 0 | 1 | Not-determined | Genome-fragment | NA | NA | 0 |
| S15_vs_51780\|\|full | 2128 | No | NA | 3 | 0 | 1 | Not-determined | Genome-fragment | NA | NA | 0 |
| S15_vs_51798\|\|full | 1895 | No | NA | 3 | 0 | 1 | Not-determined | Genome-fragment | NA | NA | 0 |
| S8_extracted_k141_81003 | 5128 | No | NA | 5 | 0 | 1 | Low-quality | Genome-fragment | 10.89 | AAI-based (medium-confidence) | 0 |
| S7_vs_128966\|\|full | 5129 | No | NA | 5 | 1 | 0 | Low-quality | Genome-fragment | 11.42 | AAI-based (medium-confidence) | 0 |
| S15_vs_52011\|\|full | 4832 | No | NA | 8 | 0 | 1 | Not-determined | Genome-fragment | NA | NA | 0 |
| S16_extracted_63429 | 5129 | No | NA | 16 | 3 | 0 | Low-quality | Genome-fragment | 9.67 | AAI-based (high-confidence) | 0 |
| S15_vs_52311\|\|full | 6981 | No | NA | 6 | 0 | 5 | Not-determined | Genome-fragment | NA | NA | 0 |
| S15_vs_52405\|\|full | 1107 | No | NA | 2 | 0 | 2 | Not-determined | Genome-fragment | NA | NA | 0 |
| S15_vs_52653\|\|full | 5480 | No | NA | 6 | 0 | 1 | Not-determined | Genome-fragment | NA | NA | 0 |
| S12_vs_k141_171981\|\|full | 5129 | No | NA | 8 | 0 | 0 | Low-quality | Genome-fragment | 8.01 | AAI-based (medium-confidence) | 0 |
| S14_vs_k141_52540\|\|full | 5130 | No | NA | 10 | 0 | 1 | Low-quality | Genome-fragment | 13.22 | AAI-based (medium-confidence) | 0 |
| S15_vs_53211\|\|full | 7418 | No | NA | 6 | 0 | 3 | Not-determined | Genome-fragment | NA | NA | 0 |
| S1_vs_k141_9965\|\|full | 5131 | Yes | 1591 | 4 | 1 | 1 | Low-quality | Genome-fragment | 2.45 | HMM-based (lower-bound) | 68.99 |
| S16_extracted_79835 | 5135 | Yes | 2209 | 6 | 2 | 1 | Low-quality | Genome-fragment | 3.55 | AAI-based (high-confidence) | 56.98 |
| S3_extracted_k141_81004 | 5136 | No | NA | 7 | 2 | 0 | Low-quality | Genome-fragment | 1.57 | HMM-based (lower-bound) | 0 |
| S15_vs_54049\|\|full | 2112 | No | NA | 2 | 0 | 0 | Not-determined | Genome-fragment | NA | NA | 0 |
| S4_extracted_6330 | 5139 | No | NA | 9 | 7 | 0 | Low-quality | Genome-fragment | 13.83 | AAI-based (medium-confidence) | 0 |
| S15_vs_54675\|\|full | 3242 | No | NA | 4 | 0 | 3 | Not-determined | Genome-fragment | NA | NA | 0 |
| S15_vs_54767\|\|full | 2094 | No | NA | 4 | 0 | 1 | Not-determined | Genome-fragment | NA | NA | 0 |
| S16_vs_6657\|\|full | 5142 | No | NA | 6 | 0 | 1 | Low-quality | Genome-fragment | 9.17 | AAI-based (high-confidence) | 0 |
| S15_vs_55062\|\|full | 5182 | No | NA | 4 | 0 | 3 | Not-determined | Genome-fragment | NA | NA | 0 |
| S15_vs_55071\|\|full | 1726 | No | NA | 2 | 0 | 1 | Not-determined | Genome-fragment | NA | NA | 0 |
| S15_vs_55079\|\|full | 1809 | No | NA | 2 | 0 | 1 | Not-determined | Genome-fragment | NA | NA | 0 |
| S15_vs_55196\|\|full | 1432 | No | NA | 3 | 0 | 0 | Not-determined | Genome-fragment | NA | NA | 0 |
| S10_extracted_19843 | 5143 | No | NA | 9 | 4 | 0 | Low-quality | Genome-fragment | 11.67 | AAI-based (high-confidence) | 0 |
| S11_vs_k141_64953\|\|full | 5145 | No | NA | 4 | 4 | 0 | Low-quality | Genome-fragment | 12.88 | AAI-based (medium-confidence) | 0 |
| S15_vs_56112\|\|full | 6611 | No | NA | 3 | 0 | 3 | Not-determined | Genome-fragment | NA | NA | 0 |
| S9_vs_k141_25243\|\|full | 5147 | No | NA | 6 | 5 | 0 | Low-quality | Genome-fragment | 11.43 | AAI-based (medium-confidence) | 0 |
| S15_vs_56272\|\|full | 2727 | No | NA | 2 | 0 | 2 | Not-determined | Genome-fragment | NA | NA | 0 |
| S15_vs_56961\|\|full | 1330 | No | NA | 3 | 0 | 0 | Not-determined | Genome-fragment | NA | NA | 0 |
| S7_extracted_91589 | 5147 | No | NA | 17 | 2 | 0 | Low-quality | Genome-fragment | 6.83 | HMM-based (lower-bound) | 0 |
| S15_vs_57018\|\|full | 1204 | No | NA | 3 | 0 | 1 | Not-determined | Genome-fragment | NA | NA | 0 |
| S15_vs_57353\|\|full | 3407 | No | NA | 4 | 0 | 1 | Not-determined | Genome-fragment | NA | NA | 0 |
| S6_extracted_k141_48299 | 5147 | No | NA | 5 | 2 | 0 | Low-quality | Genome-fragment | 5.44 | AAI-based (medium-confidence) | 0 |
| S11_extracted_k141_9926 | 5148 | No | NA | 3 | 1 | 0 | Low-quality | Genome-fragment | 8.99 | AAI-based (high-confidence) | 0 |
| S15_vs_57572\|\|full | 1505 | No | NA | 2 | 0 | 1 | Not-determined | Genome-fragment | NA | NA | 0 |
| S15_vs_57893\|\|full | 6048 | No | NA | 6 | 0 | 5 | Not-determined | Genome-fragment | NA | NA | 0 |
| S16_extracted_138835 | 5148 | No | NA | 2 | 0 | 2 | Low-quality | Genome-fragment | 0.44 | AAI-based (medium-confidence) | 0 |
| S13_vs_104405\|\|full | 5150 | No | NA | 9 | 0 | 1 | Low-quality | Genome-fragment | 10.42 | AAI-based (high-confidence) | 0 |
| S15_vs_58529\|\|full | 1458 | No | NA | 2 | 0 | 1 | Not-determined | Genome-fragment | NA | NA | 0 |
| S15_vs_58638\|\|full | 1734 | No | NA | 4 | 0 | 1 | Not-determined | Genome-fragment | NA | NA | 0 |
| S15_vs_58836\|\|full | 3926 | No | NA | 3 | 0 | 2 | Not-determined | Genome-fragment | NA | NA | 0 |
| S15_vs_58917\|\|full | 1354 | No | NA | 3 | 0 | 0 | Not-determined | Genome-fragment | NA | NA | 0 |
| S10_extracted_118253 | 5150 | No | NA | 8 | 4 | 0 | Low-quality | Genome-fragment | 12.23 | AAI-based (medium-confidence) | 0 |
| S15_vs_59488\|\|full | 3384 | No | NA | 6 | 0 | 2 | Not-determined | Genome-fragment | NA | NA | 0 |
| S5_vs_k141_104397\|\|full | 5153 | No | NA | 4 | 4 | 0 | Low-quality | Genome-fragment | 11.27 | AAI-based (high-confidence) | 0 |
| S15_vs_59704\|\|full | 3285 | No | NA | 4 | 0 | 1 | Not-determined | Genome-fragment | NA | NA | 0 |
| S15_vs_60001\|\|full | 1372 | No | NA | 2 | 0 | 2 | Not-determined | Genome-fragment | NA | NA | 0 |
| S1_vs_k141_27384\|\|full | 5154 | No | NA | 3 | 0 | 2 | Low-quality | Genome-fragment | 2.83 | AAI-based (medium-confidence) | 0 |
| S15_vs_60832\|\|full | 3312 | No | NA | 2 | 0 | 2 | Not-determined | Genome-fragment | NA | NA | 0 |
| S3_extracted_k141_47333 | 5159 | No | NA | 9 | 5 | 0 | Low-quality | Genome-fragment | 11.98 | AAI-based (high-confidence) | 0 |
| S15_vs_61337\|\|full | 2980 | No | NA | 6 | 0 | 0 | Not-determined | Genome-fragment | NA | NA | 0 |
| S15_extracted_77162 | 5161 | No | NA | 5 | 0 | 2 | Low-quality | Genome-fragment | 4.82 | AAI-based (medium-confidence) | 0 |
| S9_extracted_k141_48100 | 5162 | No | NA | 4 | 3 | 0 | Low-quality | Genome-fragment | 11.81 | AAI-based (high-confidence) | 0 |
| S5_vs_k141_112515\|\|full | 5166 | No | NA | 3 | 0 | 0 | Low-quality | Genome-fragment | 8.63 | AAI-based (high-confidence) | 0 |
| S16_extracted_38959 | 5169 | No | NA | 5 | 0 | 4 | Low-quality | Genome-fragment | 1.43 | AAI-based (medium-confidence) | 0 |
| S3_extracted_k141_122054 | 5171 | No | NA | 5 | 0 | 2 | Low-quality | Genome-fragment | 8.86 | AAI-based (medium-confidence) | 0 |
| S15_vs_62039\|\|full | 9563 | No | NA | 8 | 0 | 2 | Not-determined | Genome-fragment | NA | NA | 0 |
| S14_vs_k141_5101\|\|full | 5173 | No | NA | 4 | 0 | 3 | Low-quality | Genome-fragment | 4.19 | AAI-based (medium-confidence) | 0 |
| S15_vs_62178\|\|full | 2164 | No | NA | 3 | 0 | 2 | Not-determined | Genome-fragment | NA | NA | 0 |
| S15_vs_62354\|\|full | 1594 | No | NA | 3 | 0 | 0 | Not-determined | Genome-fragment | NA | NA | 0 |
| S15_vs_62638\|\|full | 3414 | No | NA | 2 | 0 | 2 | Not-determined | Genome-fragment | NA | NA | 0 |
| S4_extracted_8060 | 5173 | No | NA | 7 | 0 | 0 | Low-quality | Genome-fragment | 12.08 | AAI-based (high-confidence) | 0 |
| S15_vs_63053\|\|full | 1128 | No | NA | 3 | 0 | 1 | Not-determined | Genome-fragment | NA | NA | 0 |
| S5_vs_k141_31587\|\|full | 5174 | No | NA | 7 | 1 | 0 | Low-quality | Genome-fragment | 10.39 | AAI-based (high-confidence) | 0 |
| S15_vs_63568\|\|full | 1651 | No | NA | 4 | 0 | 2 | Not-determined | Genome-fragment | NA | NA | 0 |
| S15_extracted_2543 | 5175 | No | NA | 7 | 5 | 0 | Low-quality | Genome-fragment | 3.09 | AAI-based (high-confidence) | 0 |
| S15_vs_63917\|\|full | 1106 | No | NA | 3 | 0 | 1 | Not-determined | Genome-fragment | NA | NA | 0 |
| S5_vs_k141_9615\|\|full | 5176 | No | NA | 14 | 3 | 1 | Low-quality | Genome-fragment | 14.89 | AAI-based (high-confidence) | 0 |
| S15_vs_63999\|\|full | 2062 | No | NA | 3 | 0 | 1 | Not-determined | Genome-fragment | NA | NA | 0 |
| S14_vs_k141_54113\|\|full | 5178 | No | NA | 3 | 0 | 1 | Low-quality | Genome-fragment | 5.45 | AAI-based (medium-confidence) | 0 |
| S3_vs_k141_5540\|\|full | 5179 | Yes | 1127 | 5 | 1 | 1 | Low-quality | Genome-fragment | 1.07 | AAI-based (medium-confidence) | 78.24 |
| S15_vs_64043\|\|full | 1568 | No | NA | 2 | 0 | 0 | Not-determined | Genome-fragment | NA | NA | 0 |
| S15_vs_64066\|\|full | 5779 | No | NA | 4 | 0 | 2 | Not-determined | Genome-fragment | NA | NA | 0 |
| S15_vs_64236\|\|full | 1925 | No | NA | 3 | 0 | 0 | Not-determined | Genome-fragment | NA | NA | 0 |

| | | | | | | | | | | | |
|---|---|---|---|---|---|---|---|---|---|---|---|
| S15_vs_64417\|\|full | 7990 | No | NA | 7 | 0 | 2 | Not-determined | Genome-fragment | NA | NA | 0 |
| S15_vs_64520\|\|full | 1341 | No | NA | 4 | 0 | 0 | Not-determined | Genome-fragment | NA | NA | 0 |
| S15_vs_64714\|\|full | 3099 | No | NA | 3 | 0 | 0 | Not-determined | Genome-fragment | NA | NA | 0 |
| S8_vs_k141_58007\|\|full | 5180 | No | NA | 10 | 2 | 0 | Low-quality | Genome-fragment | 8.34 | AAI-based (high-confidence) | 0 |
| S5_extracted_k141_42494 | 5182 | No | NA | 7 | 3 | 0 | Low-quality | Genome-fragment | 8.12 | AAI-based (high-confidence) | 0 |
| S15_vs_65248\|\|full | 10971 | No | NA | 18 | 0 | 1 | Not-determined | Genome-fragment | NA | NA | 0 |
| S16_vs_145283\|\|full | 5183 | No | NA | 7 | 0 | 4 | Low-quality | Genome-fragment | 4.87 | AAI-based (medium-confidence) | 0 |
| S15_vs_65428\|\|full | 1118 | No | NA | 3 | 0 | 1 | Not-determined | Genome-fragment | NA | NA | 0 |
| S17_vs_k141_10162\|\|full | 5185 | No | NA | 9 | 0 | 0 | Low-quality | Genome-fragment | 9.45 | AAI-based (medium-confidence) | 0 |
| S15_vs_65948\|\|full | 1362 | No | NA | 3 | 0 | 0 | Not-determined | Genome-fragment | NA | NA | 0 |
| S15_vs_65949\|\|full | 3168 | No | NA | 3 | 0 | 1 | Not-determined | Genome-fragment | NA | NA | 0 |
| S11_extracted_k141_15698 | 5185 | No | NA | 5 | 0 | 0 | Low-quality | Genome-fragment | 8.51 | AAI-based (medium-confidence) | 0 |
| S15_vs_66077\|\|full | 1356 | No | NA | 3 | 0 | 0 | Not-determined | Genome-fragment | NA | NA | 0 |
| S15_vs_66269\|\|full | 1311 | No | NA | 2 | 0 | 0 | Not-determined | Genome-fragment | NA | NA | 0 |
| S15_vs_66794\|\|full | 1486 | No | NA | 2 | 0 | 0 | Not-determined | Genome-fragment | NA | NA | 0 |
| S15_vs_67178\|\|full | 2423 | No | NA | 3 | 0 | 1 | Not-determined | Genome-fragment | NA | NA | 0 |
| S14_vs_k141_93820\|\|full | 5186 | No | NA | 12 | 3 | 0 | Low-quality | Genome-fragment | 10.11 | AAI-based (high-confidence) | 0 |
| S4_extracted_24258 | 5186 | No | NA | 13 | 2 | 0 | Low-quality | Genome-fragment | 12.46 | AAI-based (high-confidence) | 0 |
| S15_vs_68418\|\|full | 2076 | No | NA | 2 | 0 | 1 | Not-determined | Genome-fragment | NA | NA | 0 |
| S15_vs_68444\|\|full | 1652 | No | NA | 3 | 0 | 0 | Not-determined | Genome-fragment | NA | NA | 0 |
| S15_vs_68490\|\|full | 1416 | No | NA | 2 | 0 | 0 | Not-determined | Genome-fragment | NA | NA | 0 |
| S3_vs_k141_82863\|\|full | 5192 | No | NA | 7 | 5 | 0 | Low-quality | Genome-fragment | 10.87 | AAI-based (high-confidence) | 0 |
| S3_extracted_k141_81464 | 5195 | No | NA | 14 | 8 | 0 | Low-quality | Genome-fragment | 23.71 | AAI-based (high-confidence) | 0 |
| S3_extracted_k141_95202 | 5196 | No | NA | 5 | 0 | 0 | Low-quality | Genome-fragment | 8.27 | AAI-based (medium-confidence) | 0 |
| S15_extracted_8089 | 5196 | No | NA | 12 | 2 | 0 | Low-quality | Genome-fragment | 3.13 | AAI-based (medium-confidence) | 0 |
| S15_vs_69553\|\|full | 6564 | No | NA | 6 | 0 | 5 | Not-determined | Genome-fragment | NA | NA | 0 |
| S15_vs_69708\|\|full | 1696 | No | NA | 2 | 0 | 1 | Not-determined | Genome-fragment | NA | NA | 0 |
| S6_vs_k141_12125\|\|full | 5196 | No | NA | 10 | 5 | 0 | Low-quality | Genome-fragment | 11.16 | AAI-based (high-confidence) | 0 |
| S15_vs_69902\|\|full | 3738 | No | NA | 3 | 0 | 0 | Not-determined | Genome-fragment | NA | NA | 0 |
| S15_vs_69950\|\|full | 1320 | No | NA | 2 | 0 | 1 | Not-determined | Genome-fragment | NA | NA | 0 |
| S8_vs_k141_67498\|\|full | 5196 | No | NA | 8 | 5 | 0 | Low-quality | Genome-fragment | 11.43 | AAI-based (medium-confidence) | 0 |
| S12_vs_k141_74982\|\|full | 5198 | No | NA | 7 | 1 | 0 | Low-quality | Genome-fragment | 8.73 | AAI-based (high-confidence) | 0 |
| S5_vs_k141_74039\|\|full | 5200 | No | NA | 9 | 1 | 0 | Low-quality | Genome-fragment | 13.04 | HMM-based (lower-bound) | 0 |
| S8_extracted_k141_6754 | 5202 | No | NA | 2 | 0 | 0 | Low-quality | Genome-fragment | 5.6 | AAI-based (medium-confidence) | 0 |
| S5_extracted_k141_19313 | 5204 | No | NA | 10 | 1 | 0 | Low-quality | Genome-fragment | 2.82 | HMM-based (lower-bound) | 0 |
| S7_vs_39632\|\|full | 5205 | No | NA | 4 | 0 | 3 | Low-quality | Genome-fragment | 4.28 | AAI-based (high-confidence) | 0 |
| S15_vs_70744\|\|full | 2000 | No | NA | 3 | 0 | 2 | Not-determined | Genome-fragment | NA | NA | 0 |
| S15_vs_70783\|\|full | 1254 | No | NA | 3 | 0 | 2 | Not-determined | Genome-fragment | NA | NA | 0 |
| S1_extracted_k141_87910 | 5210 | No | NA | 7 | 3 | 0 | Low-quality | Genome-fragment | 16.47 | HMM-based (lower-bound) | 0 |
| S12_vs_k141_7260\|\|full | 5210 | No | NA | 15 | 1 | 0 | Low-quality | Genome-fragment | 11.92 | AAI-based (high-confidence) | 0 |
| S8_extracted_k141_100513 | 5210 | No | NA | 6 | 5 | 0 | Low-quality | Genome-fragment | 23.76 | AAI-based (high-confidence) | 0 |
| S5_extracted_k141_119366 | 5211 | No | NA | 7 | 1 | 0 | Low-quality | Genome-fragment | 2.91 | HMM-based (lower-bound) | 0 |
| S15_vs_71734\|\|full | 3032 | No | NA | 6 | 0 | 4 | Not-determined | Genome-fragment | NA | NA | 0 |
| S15_vs_71787\|\|full | 1257 | No | NA | 2 | 0 | 1 | Not-determined | Genome-fragment | NA | NA | 0 |
| S14_vs_k141_9685\|\|full | 5214 | No | NA | 6 | 1 | 0 | Low-quality | Genome-fragment | 13.31 | AAI-based (high-confidence) | 0 |
| S15_vs_71968\|\|full | 39396 | Yes | 19842 | 41 | 11 | 9 | Medium-quality | Genome-fragment | 53.86 | AAI-based (high-confidence) | 49.63 |
| S15_vs_72230\|\|full | 1896 | No | NA | 2 | 0 | 0 | Not-determined | Genome-fragment | NA | NA | 0 |
| S15_vs_72648\|\|full | 1047 | No | NA | 3 | 0 | 2 | Not-determined | Genome-fragment | NA | NA | 0 |
| S15_vs_72793\|\|full | 1893 | No | NA | 3 | 0 | 2 | Not-determined | Genome-fragment | NA | NA | 0 |
| S15_vs_73475\|\|full | 1943 | No | NA | 3 | 0 | 0 | Not-determined | Genome-fragment | NA | NA | 0 |
| S15_vs_73556\|\|full | 1033 | No | NA | 2 | 0 | 0 | Not-determined | Genome-fragment | NA | NA | 0 |
| S16_vs_73593\|\|full | 5214 | No | NA | 9 | 2 | 0 | Low-quality | Genome-fragment | 8.64 | AAI-based (high-confidence) | 0 |
| S15_vs_73789\|\|full | 1304 | No | NA | 4 | 0 | 0 | Not-determined | Genome-fragment | NA | NA | 0 |
| S11_vs_k141_72671\|\|full | 5216 | No | NA | 3 | 3 | 0 | Low-quality | Genome-fragment | 8.87 | AAI-based (high-confidence) | 0 |
| S15_vs_74327\|\|full | 2600 | No | NA | 2 | 0 | 1 | Not-determined | Genome-fragment | NA | NA | 0 |
| S1_extracted_k141_154205 | 5216 | No | NA | 9 | 4 | 0 | Low-quality | Genome-fragment | 11.75 | AAI-based (high-confidence) | 0 |
| S3_extracted_k141_68297 | 5216 | No | NA | 10 | 0 | 0 | Low-quality | Genome-fragment | 3.4 | AAI-based (high-confidence) | 0 |
| S15_vs_74463\|\|full | 3083 | No | NA | 2 | 0 | 0 | Not-determined | Genome-fragment | NA | NA | 0 |
| S15_vs_74477\|\|full | 1465 | No | NA | 3 | 0 | 2 | Not-determined | Genome-fragment | NA | NA | 0 |
| S8_vs_k141_172950\|\|full | 5217 | No | NA | 14 | 0 | 0 | Low-quality | Genome-fragment | 13.44 | AAI-based (high-confidence) | 0 |
| S7_vs_66017\|\|full | 5218 | Yes | 3319 | 9 | 1 | 2 | Low-quality | Genome-fragment | 9.41 | AAI-based (high-confidence) | 36.39 |
| S7_vs_19007\|\|full | 5221 | No | NA | 3 | 3 | 0 | Low-quality | Genome-fragment | 11.78 | AAI-based (high-confidence) | 0 |
| S15_vs_75474\|\|full | 12193 | No | NA | 11 | 0 | 2 | Not-determined | Genome-fragment | NA | NA | 0 |
| S15_vs_75507\|\|full | 7610 | No | NA | 10 | 0 | 0 | Not-determined | Genome-fragment | NA | NA | 0 |
| S11_extracted_k141_39340 | 5222 | No | NA | 12 | 0 | 0 | Low-quality | Genome-fragment | 5.56 | AAI-based (high-confidence) | 0 |
| S7_extracted_1744 | 5223 | No | NA | 12 | 1 | 0 | Low-quality | Genome-fragment | 10.95 | AAI-based (medium-confidence) | 0 |
| S1_extracted_k141_43216 | 5224 | No | NA | 5 | 0 | 3 | Low-quality | Genome-fragment | 2.56 | AAI-based (medium-confidence) | 0 |
| S7_extracted_29891 | 5225 | No | NA | 16 | 2 | 0 | Low-quality | Genome-fragment | 8.47 | AAI-based (high-confidence) | 0 |
| S16_vs_82779\|\|full | 5228 | No | NA | 6 | 0 | 2 | Low-quality | Genome-fragment | 7.87 | AAI-based (medium-confidence) | 0 |
| S12_vs_k141_162275\|\|full | 5229 | No | NA | 4 | 0 | 0 | Low-quality | Genome-fragment | 12.05 | AAI-based (high-confidence) | 0 |
| S16_vs_165240\|\|full | 5233 | No | NA | 4 | 0 | 4 | Low-quality | Genome-fragment | 1.23 | AAI-based (medium-confidence) | 0 |
| S8_extracted_k141_198154 | 5233 | Yes | 1470 | 5 | 1 | 3 | Low-quality | Genome-fragment | 0.58 | HMM-based (lower-bound) | 71.91 |
| S15_vs_77065\|\|full | 1009 | No | NA | 2 | 0 | 0 | Not-determined | Genome-fragment | NA | NA | 0 |
| S15_vs_77074\|\|full | 1310 | No | NA | 3 | 0 | 0 | Not-determined | Genome-fragment | NA | NA | 0 |
| S15_vs_77154\|\|full | 18949 | No | NA | 14 | 0 | 3 | Not-determined | Genome-fragment | NA | NA | 0 |
| S15_vs_77171\|\|full | 10593 | No | NA | 10 | 0 | 3 | Not-determined | Genome-fragment | NA | NA | 0 |
| S15_vs_77255\|\|full | 1502 | No | NA | 5 | 0 | 3 | Not-determined | Genome-fragment | NA | NA | 0 |
| S15_vs_14765\|\|0_partial | 11939 | No | NA | 11 | 0 | 8 | Not-determined | Genome-fragment | NA | NA | 0 |
| S14_vs_k141_105404\|\|full | 5236 | No | NA | 11 | 5 | 0 | Low-quality | Genome-fragment | 11.88 | AAI-based (high-confidence) | 0 |
| S15_vs_42979\|\|0_partial | 21026 | No | NA | 23 | 0 | 7 | Not-determined | Genome-fragment | NA | NA | 0 |
| S16_extracted_1044 | 5236 | No | NA | 7 | 1 | 0 | Low-quality | Genome-fragment | 6.49 | AAI-based (high-confidence) | 0 |
| S10_extracted_78527 | 5237 | No | NA | 5 | 2 | 0 | Low-quality | Genome-fragment | 2.35 | AAI-based (high-confidence) | 0 |
| S15_vs_4970\|\|lt2gene | 1033 | No | NA | 1 | 0 | 0 | Not-determined | Genome-fragment | NA | NA | 0 |
| S5_vs_k141_119666\|\|full | 5240 | No | NA | 9 | 0 | 0 | Low-quality | Genome-fragment | 14.49 | AAI-based (medium-confidence) | 0 |
| S15_vs_7112\|\|lt2gene | 1014 | No | NA | 1 | 0 | 0 | Not-determined | Genome-fragment | NA | NA | 0 |
| S3_extracted_k141_29633 | 5241 | No | NA | 4 | 0 | 1 | Low-quality | Genome-fragment | 6.64 | AAI-based (high-confidence) | 0 |
| S1_extracted_k141_114042 | 5243 | No | NA | 4 | 0 | 2 | Low-quality | Genome-fragment | 3.81 | AAI-based (medium-confidence) | 0 |
| S5_vs_k141_48686\|\|full | 5245 | No | NA | 9 | 8 | 0 | Low-quality | Genome-fragment | 11.33 | AAI-based (medium-confidence) | 0 |
| S15_vs_16676\|\|lt2gene | 1346 | No | NA | 1 | 0 | 0 | Not-determined | Genome-fragment | NA | NA | 0 |
| S10_extracted_61854 | 5246 | No | NA | 15 | 1 | 0 | Low-quality | Genome-fragment | 8.07 | AAI-based (medium-confidence) | 0 |
| S15_vs_26294\|\|lt2gene | 2955 | No | NA | 2 | 0 | 0 | Not-determined | Genome-fragment | NA | NA | 0 |
| S1_vs_k141_46480\|\|full | 5248 | No | NA | 9 | 4 | 0 | Low-quality | Genome-fragment | 12.53 | AAI-based (high-confidence) | 0 |
| S16_extracted_45174 | 5248 | No | NA | 5 | 0 | 1 | Low-quality | Genome-fragment | 2.34 | AAI-based (medium-confidence) | 0 |
| S15_vs_34181\|\|lt2gene | 1198 | No | NA | 1 | 0 | 1 | Not-determined | Genome-fragment | NA | NA | 0 |
| S16_extracted_27890 | 5250 | No | NA | 9 | 2 | 0 | Low-quality | Genome-fragment | 9.11 | AAI-based (high-confidence) | 0 |
| S11_vs_k141_9300\|\|full | 5253 | No | NA | 7 | 0 | 4 | Low-quality | Genome-fragment | 8.13 | AAI-based (high-confidence) | 0 |
| S15_vs_41955\|\|lt2gene | 1600 | No | NA | 2 | 0 | 2 | Not-determined | Genome-fragment | NA | NA | 0 |
| S9_vs_k141_17112\|\|full | 5258 | No | NA | 8 | 5 | 0 | Low-quality | Genome-fragment | 12.86 | AAI-based (high-confidence) | 0 |
| S15_vs_42837\|\|lt2gene | 1032 | No | NA | 2 | 0 | 0 | Not-determined | Genome-fragment | NA | NA | 0 |
| S2_extracted_k141_4721 | 5258 | No | NA | 12 | 1 | 0 | Low-quality | Genome-fragment | 8.7 | AAI-based (high-confidence) | 0 |
| S8_vs_k141_63552\|\|full | 5258 | Yes | 3773 | 10 | 4 | 2 | Low-quality | Genome-fragment | 8.34 | AAI-based (high-confidence) | 28.24 |
| S17_vs_k141_92886\|\|full | 5259 | No | NA | 13 | 0 | 0 | Low-quality | Genome-fragment | 6.66 | AAI-based (high-confidence) | 0 |
| S3_vs_k141_31993\|\|full | 5260 | Yes | 1059 | 7 | 1 | 3 | Low-quality | Genome-fragment | 0.43 | HMM-based (lower-bound) | 79.87 |
| S15_vs_55414\|\|lt2gene | 1768 | No | NA | 2 | 0 | 1 | Not-determined | Genome-fragment | NA | NA | 0 |
| S7_vs_71101\|\|full | 5260 | No | NA | 5 | 0 | 4 | Low-quality | Genome-fragment | 3.77 | AAI-based (medium-confidence) | 0 |
| S12_vs_k141_138042\|\|full | 5261 | No | NA | 10 | 3 | 0 | Low-quality | Genome-fragment | 10.03 | AAI-based (medium-confidence) | 0 |
| S15_vs_62741\|\|lt2gene | 3627 | No | NA | 2 | 0 | 1 | Not-determined | Genome-fragment | NA | NA | 0 |
| S14_vs_k141_38076\|\|full | 5262 | No | NA | 11 | 3 | 0 | Low-quality | Genome-fragment | 17.39 | AAI-based (high-confidence) | 0 |
| S1_extracted_k141_52303 | 5263 | No | NA | 2 | 0 | 0 | Low-quality | Genome-fragment | 12.53 | AAI-based (high-confidence) | 0 |
| S1_extracted_k141_130676 | 5266 | Yes | 4832 | 2 | 1 | 1 | Low-quality | Genome-fragment | 7.63 | AAI-based (high-confidence) | 8.24 |
| S11_extracted_k141_82062 | 5268 | No | NA | 6 | 4 | 0 | Low-quality | Genome-fragment | 13.45 | AAI-based (high-confidence) | 0 |
| S2_extracted_k141_9472 | 1074 | No | NA | 2 | 0 | 1 | Not-determined | Genome-fragment | NA | NA | 0 |
| S2_extracted_k141_21 | 6073 | No | NA | 7 | 0 | 3 | Not-determined | Genome-fragment | NA | NA | 0 |
| S7_vs_134069\|\|full | 5268 | Yes | 2642 | 7 | 1 | 1 | Low-quality | Genome-fragment | 3.27 | HMM-based (lower-bound) | 49.85 |
| S2_extracted_k141_2742 | 1139 | No | NA | 2 | 0 | 2 | Not-determined | Genome-fragment | NA | NA | 0 |
| S2_extracted_k141_12202 | 1145 | No | NA | 2 | 0 | 0 | Not-determined | Genome-fragment | NA | NA | 0 |
| S2_extracted_k141_17612 | 1359 | No | NA | 2 | 0 | 2 | Not-determined | Genome-fragment | NA | NA | 0 |
| S2_extracted_k141_6793 | 1511 | No | NA | 3 | 0 | 0 | Not-determined | Genome-fragment | NA | NA | 0 |
| S11_vs_k141_41243\|\|full | 5273 | No | NA | 9 | 3 | 0 | Low-quality | Genome-fragment | 5.91 | AAI-based (high-confidence) | 0 |
| S3_vs_k141_90456\|\|full | 5273 | No | NA | 9 | 3 | 0 | Low-quality | Genome-fragment | 12.29 | AAI-based (high-confidence) | 0 |
| S2_extracted_k141_6802 | 1037 | No | NA | 2 | 0 | 0 | Not-determined | Genome-fragment | NA | NA | 0 |
| S16_vs_13161\|\|full | 5273 | No | NA | 4 | 0 | 3 | Low-quality | Genome-fragment | 6.25 | AAI-based (medium-confidence) | 0 |
| S2_extracted_k141_17635 | 1141 | No | NA | 2 | 0 | 1 | Not-determined | Genome-fragment | NA | NA | 0 |
| S2_extracted_k141_8176 | 2049 | No | NA | 2 | 0 | 1 | Not-determined | Genome-fragment | NA | NA | 0 |
| S2_extracted_k141_60 | 6498 | No | NA | 5 | 0 | 4 | Not-determined | Genome-fragment | NA | NA | 0 |
| S14_extracted_k141_107699 | 5276 | Yes | 2485 | 6 | 1 | 2 | Low-quality | Genome-fragment | 3.76 | HMM-based (lower-bound) | 52.9 |
| S16_vs_48702\|\|full | 5276 | No | NA | 8 | 4 | 0 | Low-quality | Genome-fragment | 17.86 | AAI-based (high-confidence) | 0 |
| S11_vs_k141_39184\|\|full | 5278 | No | NA | 3 | 2 | 0 | Low-quality | Genome-fragment | 7.97 | AAI-based (high-confidence) | 0 |
| S2_extracted_k141_20395 | 3195 | No | NA | 3 | 0 | 1 | Not-determined | Genome-fragment | NA | NA | 0 |
| S2_extracted_k141_20398 | 1223 | No | NA | 1 | 0 | 1 | Not-determined | Genome-fragment | NA | NA | 0 |
| S2_extracted_k141_17688 | 1034 | No | NA | 2 | 0 | 2 | Not-determined | Genome-fragment | NA | NA | 0 |
| S2_extracted_k141_5478 | 1297 | No | NA | 3 | 0 | 1 | Not-determined | Genome-fragment | NA | NA | 0 |
| S5_vs_k141_102560\|\|full | 5281 | No | NA | 7 | 2 | 0 | Low-quality | Genome-fragment | 3.56 | HMM-based (lower-bound) | 0 |
| S2_extracted_k141_10891 | 1801 | No | NA | 2 | 0 | 1 | Not-determined | Genome-fragment | NA | NA | 0 |
| S2_extracted_k141_2836 | 1005 | No | NA | 2 | 0 | 1 | Not-determined | Genome-fragment | NA | NA | 0 |
| S3_extracted_k141_98239 | 5283 | No | NA | 3 | 0 | 0 | Low-quality | Genome-fragment | 8.95 | AAI-based (medium-confidence) | 0 |
| S2_extracted_k141_6878 | 1536 | No | NA | 2 | 0 | 1 | Not-determined | Genome-fragment | NA | NA | 0 |
| S2_extracted_k141_4130 | 100094 | No | NA | 100 | 17 | 1 | High-quality | High-quality | 100 | AAI-based (high-confidence) | 0 |
| S11_extracted_k141_25071 | 5286 | No | NA | 10 | 4 | 0 | Low-quality | Genome-fragment | 8.18 | AAI-based (high-confidence) | 0 |
| S5_vs_k141_126677\|\|full | 5286 | No | NA | 10 | 2 | 0 | Low-quality | Genome-fragment | 8.42 | AAI-based (medium-confidence) | 0 |
| S2_extracted_k141_19073 | 1351 | No | NA | 2 | 0 | 2 | Not-determined | Genome-fragment | NA | NA | 0 |
| S2_extracted_k141_15028 | 1221 | No | NA | 1 | 0 | 0 | Not-determined | Genome-fragment | NA | NA | 0 |
| S2_extracted_k141_16360 | 1620 | No | NA | 2 | 0 | 1 | Not-determined | Genome-fragment | NA | NA | 0 |
| S5_vs_k141_6720\|\|full | 5292 | No | NA | 2 | 0 | 2 | Low-quality | Genome-fragment | 0.52 | AAI-based (medium-confidence) | 0 |
| S2_extracted_k141_163 | 1066 | No | NA | 1 | 0 | 0 | Not-determined | Genome-fragment | NA | NA | 0 |
| S2_extracted_k141_1403 | 1277 | No | NA | 2 | 0 | 1 | Not-determined | Genome-fragment | NA | NA | 0 |
| S2_extracted_k141_19099 | 1177 | No | NA | 1 | 0 | 1 | Not-determined | Genome-fragment | NA | NA | 0 |
| S2_extracted_k141_19102 | 1357 | No | NA | 1 | 0 | 0 | Not-determined | Genome-fragment | NA | NA | 0 |
| S2_extracted_k141_20497 | 1266 | No | NA | 3 | 0 | 1 | Not-determined | Genome-fragment | NA | NA | 0 |
| S2_extracted_k141_13720 | 1411 | No | NA | 1 | 0 | 0 | Not-determined | Genome-fragment | NA | NA | 0 |
| S10_extracted_60225 | 5294 | No | NA | 5 | 0 | 2 | Low-quality | Genome-fragment | 2.4 | AAI-based (medium-confidence) | 0 |
| S2_extracted_k141_16420 | 1051 | No | NA | 1 | 0 | 0 | Not-determined | Genome-fragment | NA | NA | 0 |
| S2_extracted_k141_6950 | 1188 | No | NA | 1 | 0 | 1 | Not-determined | Genome-fragment | NA | NA | 0 |
| S2_extracted_k141_13790 | 1008 | No | NA | 1 | 0 | 0 | Not-determined | Genome-fragment | NA | NA | 0 |
| S2_extracted_k141_9683 | 1203 | No | NA | 1 | 0 | 1 | Not-determined | Genome-fragment | NA | NA | 0 |
| S8_vs_k141_70513\|\|full | 5295 | No | NA | 3 | 0 | 2 | Low-quality | Genome-fragment | 2.69 | AAI-based (medium-confidence) | 0 |
| S2_extracted_k141_5623 | 1326 | No | NA | 1 | 0 | 1 | Not-determined | Genome-fragment | NA | NA | 0 |
| S2_extracted_k141_12373 | 2105 | No | NA | 4 | 0 | 0 | Not-determined | Genome-fragment | NA | NA | 0 |
| S2_extracted_k141_9726 | 4231 | No | NA | 3 | 0 | 3 | Not-determined | Genome-fragment | NA | NA | 0 |
| S2_extracted_k141_19203 | 7125 | No | NA | 6 | 0 | 2 | Not-determined | Genome-fragment | NA | NA | 0 |
| S3_extracted_k141_32717 | 5297 | No | NA | 9 | 3 | 0 | Low-quality | Genome-fragment | 12.96 | AAI-based (high-confidence) | 0 |
| S7_extracted_28907 | 5297 | No | NA | 5 | 0 | 0 | Low-quality | Genome-fragment | 6.14 | AAI-based (medium-confidence) | 0 |
| S2_extracted_k141_9748 | 1006 | No | NA | 1 | 0 | 1 | Not-determined | Genome-fragment | NA | NA | 0 |
| S17_vs_k141_68732\|\|full | 5298 | No | NA | 3 | 3 | 0 | Low-quality | Genome-fragment | 8.73 | AAI-based (high-confidence) | 0 |
| S2_extracted_k141_9754 | 1678 | No | NA | 4 | 0 | 1 | Not-determined | Genome-fragment | NA | NA | 0 |
| S2_extracted_k141_17850 | 15371 | No | NA | 21 | 7 | 0 | Medium-quality | Genome-fragment | 65.08 | AAI-based (high-confidence) | 0 |
| S2_extracted_k141_230 | 1222 | No | NA | 1 | 0 | 1 | Not-determined | Genome-fragment | NA | NA | 0 |
| S2_extracted_k141_11116 | 1250 | No | NA | 2 | 0 | 2 | Not-determined | Genome-fragment | NA | NA | 0 |
| S2_extracted_k141_12457 | 1408 | No | NA | 2 | 0 | 2 | Not-determined | Genome-fragment | NA | NA | 0 |
| S13_vs_67227\|\|full | 5298 | No | NA | 8 | 0 | 2 | Low-quality | Genome-fragment | 8.07 | AAI-based (medium-confidence) | 0 |
| S2_extracted_k141_9797 | 1037 | No | NA | 1 | 0 | 0 | Not-determined | Genome-fragment | NA | NA | 0 |
| S2_extracted_k141_16616 | 3886 | No | NA | 3 | 0 | 2 | Not-determined | Genome-fragment | NA | NA | 0 |
| S2_extracted_k141_7132 | 3924 | No | NA | 8 | 0 | 0 | Not-determined | Genome-fragment | NA | NA | 0 |
| S2_extracted_k141_9830 | 4453 | No | NA | 5 | 0 | 2 | Not-determined | Genome-fragment | NA | NA | 0 |
| S2_extracted_k141_7167 | 1345 | No | NA | 1 | 0 | 0 | Not-determined | Genome-fragment | NA | NA | 0 |
| S2_extracted_k141_11195 | 1808 | No | NA | 2 | 0 | 1 | Not-determined | Genome-fragment | NA | NA | 0 |
| S16_vs_36599\|\|full | 5298 | No | NA | 4 | 0 | 1 | Low-quality | Genome-fragment | 2.67 | AAI-based (medium-confidence) | 0 |
| S2_extracted_k141_9847 | 1661 | No | NA | 3 | 0 | 2 | Not-determined | Genome-fragment | NA | NA | 0 |
| S17_vs_k141_50352\|\|full | 5300 | No | NA | 6 | 5 | 0 | Low-quality | Genome-fragment | 8.64 | AAI-based (high-confidence) | 0 |
| S2_extracted_k141_15382 | 2853 | No | NA | 3 | 0 | 0 | Not-determined | Genome-fragment | NA | NA | 0 |
| S2_extracted_k141_17926 | 3497 | No | NA | 3 | 0 | 2 | Not-determined | Genome-fragment | NA | NA | 0 |
| S5_vs_k141_78076\|\|full | 5300 | No | NA | 7 | 1 | 0 | Low-quality | Genome-fragment | 11.76 | HMM-based (lower-bound) | 0 |
| S14_extracted_k141_1795 | 5301 | No | NA | 6 | 6 | 0 | Low-quality | Genome-fragment | 17.49 | AAI-based (high-confidence) | 0 |
| S2_extracted_k141_7242 | 3072 | No | NA | 4 | 0 | 2 | Not-determined | Genome-fragment | NA | NA | 0 |
| S7_vs_134248\|\|full | 5303 | No | NA | 5 | 3 | 0 | Low-quality | Genome-fragment | 12.77 | AAI-based (high-confidence) | 0 |
| S2_extracted_k141_20722 | 1225 | No | NA | 1 | 0 | 1 | Not-determined | Genome-fragment | NA | NA | 0 |
| S2_extracted_k141_1615 | 1127 | No | NA | 2 | 0 | 0 | Not-determined | Genome-fragment | NA | NA | 0 |
| S16_vs_121088\|\|full | 5306 | No | NA | 5 | 0 | 3 | Low-quality | Genome-fragment | 6.86 | AAI-based (high-confidence) | 0 |
| S2_extracted_k141_16802 | 1278 | No | NA | 4 | 0 | 0 | Not-determined | Genome-fragment | NA | NA | 0 |
| S2_extracted_k141_18052 | 1044 | No | NA | 1 | 0 | 1 | Not-determined | Genome-fragment | NA | NA | 0 |
| S2_extracted_k141_18075 | 1274 | No | NA | 2 | 0 | 0 | Not-determined | Genome-fragment | NA | NA | 0 |
| S2_extracted_k141_19534 | 1558 | No | NA | 1 | 0 | 1 | Not-determined | Genome-fragment | NA | NA | 0 |
| S2_extracted_k141_4555 | 3071 | No | NA | 5 | 0 | 0 | Not-determined | Genome-fragment | NA | NA | 0 |
| S2_extracted_k141_12776 | 1473 | No | NA | 4 | 0 | 2 | Not-determined | Genome-fragment | NA | NA | 0 |
| S5_vs_k141_107913\|\|full | 5307 | No | NA | 7 | 4 | 0 | Low-quality | Genome-fragment | 13.01 | AAI-based (high-confidence) | 0 |
| S2_extracted_k141_16829 | 17189 | No | NA | 25 | 7 | 0 | Medium-quality | Genome-fragment | 88.25 | AAI-based (medium-confidence) | 0 |
| S8_vs_k141_101994\|\|full | 5308 | No | NA | 4 | 0 | 3 | Low-quality | Genome-fragment | 6.57 | AAI-based (medium-confidence) | 0 |
| S17_vs_k141_34493\|\|full | 5309 | No | NA | 9 | 2 | 0 | Low-quality | Genome-fragment | 15.7 | HMM-based (lower-bound) | 0 |
| S2_extracted_k141_14096 | 2510 | No | NA | 1 | 0 | 0 | Not-determined | Genome-fragment | NA | NA | 0 |
| S2_extracted_k141_20770 | 1284 | No | NA | 1 | 0 | 1 | Not-determined | Genome-fragment | NA | NA | 0 |
| S2_extracted_k141_1689 | 1003 | No | NA | 2 | 0 | 0 | Not-determined | Genome-fragment | NA | NA | 0 |
| S3_vs_k141_97756\|\|full | 5309 | No | NA | 7 | 1 | 0 | Low-quality | Genome-fragment | 12.22 | AAI-based (high-confidence) | 0 |
| S3_extracted_k141_83786 | 5310 | No | NA | 8 | 5 | 0 | Low-quality | Genome-fragment | 8.63 | AAI-based (high-confidence) | 0 |
| S5_vs_k141_79339\|\|full | 5310 | No | NA | 7 | 4 | 0 | Low-quality | Genome-fragment | 5.04 | HMM-based (lower-bound) | 0 |
| S2_extracted_k141_4648 | 1000 | No | NA | 2 | 0 | 1 | Not-determined | Genome-fragment | NA | NA | 0 |
| S2_extracted_k141_413 | 1724 | No | NA | 2 | 0 | 2 | Not-determined | Genome-fragment | NA | NA | 0 |
| S2_extracted_k141_3236 | 7967 | No | NA | 6 | 0 | 1 | Not-determined | Genome-fragment | NA | NA | 0 |
| S9_vs_k141_21671\|\|full | 5311 | No | NA | 9 | 1 | 0 | Low-quality | Genome-fragment | 14.38 | AAI-based (high-confidence) | 0 |
| S2_extracted_k141_10079 | 1548 | No | NA | 2 | 0 | 0 | Not-determined | Genome-fragment | NA | NA | 0 |
| S2_extracted_k141_12937 | 1655 | No | NA | 2 | 0 | 1 | Not-determined | Genome-fragment | NA | NA | 0 |
| S2_extracted_k141_7367 | 4980 | No | NA | 5 | 0 | 2 | Not-determined | Genome-fragment | NA | NA | 0 |
| S2_extracted_k141_20779 | 1096 | No | NA | 1 | 0 | 0 | Not-determined | Genome-fragment | NA | NA | 0 |
| S16_vs_33819\|\|full | 5313 | Yes | 3357 | 9 | 2 | 1 | Low-quality | Genome-fragment | 9.27 | AAI-based (high-confidence) | 36.82 |
| S2_extracted_k141_12953 | 1738 | No | NA | 2 | 0 | 2 | Not-determined | Genome-fragment | NA | NA | 0 |
| S2_extracted_k141_5918 | 1412 | No | NA | 1 | 0 | 1 | Not-determined | Genome-fragment | NA | NA | 0 |
| S2_extracted_k141_11431 | 1015 | No | NA | 1 | 0 | 0 | Not-determined | Genome-fragment | NA | NA | 0 |
| S12_vs_k141_42059\|\|full | 5313 | No | NA | 9 | 2 | 1 | Low-quality | Genome-fragment | 14.52 | AAI-based (medium-confidence) | 0 |
| S13_vs_55896\|\|full | 5314 | No | NA | 4 | 0 | 2 | Low-quality | Genome-fragment | 12.56 | AAI-based (medium-confidence) | 0 |
| S2_extracted_k141_7378 | 8001 | No | NA | 6 | 0 | 3 | Not-determined | Genome-fragment | NA | NA | 0 |
| S2_extracted_k141_14229 | 1637 | No | NA | 1 | 0 | 0 | Not-determined | Genome-fragment | NA | NA | 0 |
| S2_extracted_k141_1745 | 1935 | No | NA | 1 | 0 | 1 | Not-determined | Genome-fragment | NA | NA | 0 |
| S2_extracted_k141_15667 | 2386 | No | NA | 2 | 0 | 1 | Not-determined | Genome-fragment | NA | NA | 0 |
| S2_extracted_k141_8647 | 1351 | No | NA | 1 | 0 | 1 | Not-determined | Genome-fragment | NA | NA | 0 |
| S2_extracted_k141_8650 | 1402 | No | NA | 2 | 0 | 1 | Not-determined | Genome-fragment | NA | NA | 0 |
| S2_extracted_k141_18295 | 1297 | No | NA | 2 | 0 | 2 | Not-determined | Genome-fragment | NA | NA | 0 |
| S1_vs_k141_10865\|\|full | 5315 | No | NA | 12 | 1 | 0 | Low-quality | Genome-fragment | 11.69 | AAI-based (high-confidence) | 0 |
| S2_extracted_k141_5970 | 2793 | No | NA | 2 | 0 | 1 | Not-determined | Genome-fragment | NA | NA | 0 |
| S2_extracted_k141_8660 | 1320 | No | NA | 1 | 0 | 1 | Not-determined | Genome-fragment | NA | NA | 0 |
| S12_extracted_k141_168017 | 5316 | No | NA | 14 | 1 | 0 | Low-quality | Genome-fragment | 8.48 | AAI-based (high-confidence) | 0 |
| S2_extracted_k141_3305 | 1139 | No | NA | 5 | 0 | 0 | Not-determined | Genome-fragment | NA | NA | 0 |
| S2_extracted_k141_18310 | 7448 | No | NA | 7 | 0 | 3 | Not-determined | Genome-fragment | NA | NA | 0 |
| S2_extracted_k141_16940 | 1081 | No | NA | 1 | 0 | 0 | Not-determined | Genome-fragment | NA | NA | 0 |
| S2_extracted_k141_4739 | 1065 | No | NA | 2 | 0 | 1 | Not-determined | Genome-fragment | NA | NA | 0 |
| S1_vs_k141_145174\|\|full | 5317 | No | NA | 7 | 3 | 0 | Low-quality | Genome-fragment | 12.98 | AAI-based (high-confidence) | 0 |
| S2_extracted_k141_8703 | 2136 | No | NA | 2 | 0 | 1 | Not-determined | Genome-fragment | NA | NA | 0 |
| S2_extracted_k141_7420 | 3066 | No | NA | 3 | 0 | 2 | Not-determined | Genome-fragment | NA | NA | 0 |
| S17_vs_k141_135583\|\|full | 5318 | No | NA | 5 | 0 | 2 | Low-quality | Genome-fragment | 6.92 | AAI-based (medium-confidence) | 0 |
| S2_extracted_k141_20811 | 1039 | No | NA | 1 | 0 | 0 | Not-determined | Genome-fragment | NA | NA | 0 |
| S2_extracted_k141_20815 | 1078 | No | NA | 2 | 0 | 0 | Not-determined | Genome-fragment | NA | NA | 0 |
| S9_vs_k141_7379\|\|full | 5318 | No | NA | 8 | 2 | 0 | Low-quality | Genome-fragment | 12.68 | AAI-based (high-confidence) | 0 |
| S2_extracted_k141_15747 | 1184 | No | NA | 2 | 0 | 0 | Not-determined | Genome-fragment | NA | NA | 0 |
| S2_extracted_k141_19752 | 1149 | No | NA | 2 | 0 | 2 | Not-determined | Genome-fragment | NA | NA | 0 |
| S2_extracted_k141_8758 | 1374 | No | NA | 2 | 0 | 2 | Not-determined | Genome-fragment | NA | NA | 0 |
| S2_extracted_k141_4769 | 5667 | No | NA | 5 | 0 | 3 | Not-determined | Genome-fragment | NA | NA | 0 |
| S2_extracted_k141_3356 | 13599 | No | NA | 17 | 3 | 0 | Complete | High-quality | 100 | ITR (high-confidence) | 0 |
| S2_extracted_k141_18390 | 2095 | No | NA | 2 | 0 | 1 | Not-determined | Genome-fragment | NA | NA | 0 |
| S2_extracted_k141_14311 | 1103 | No | NA | 2 | 0 | 2 | Not-determined | Genome-fragment | NA | NA | 0 |
| S6_vs_k141_18399\|\|full | 5318 | No | NA | 7 | 1 | 0 | Low-quality | Genome-fragment | 14.42 | AAI-based (medium-confidence) | 0 |
| S17_vs_k141_8430\|\|full | 5319 | No | NA | 10 | 9 | 0 | Low-quality | Genome-fragment | 15.98 | AAI-based (high-confidence) | 0 |
| S2_extracted_k141_14323 | 1110 | No | NA | 2 | 0 | 2 | Not-determined | Genome-fragment | NA | NA | 0 |
| S2_extracted_k141_8837 | 1549 | No | NA | 2 | 0 | 1 | Not-determined | Genome-fragment | NA | NA | 0 |
| S16_extracted_12345 | 5320 | No | NA | 4 | 0 | 4 | Low-quality | Genome-fragment | 0.46 | AAI-based (medium-confidence) | 0 |
| S12_vs_k141_181158\|\|full | 5320 | No | NA | 5 | 0 | 1 | Low-quality | Genome-fragment | 2.47 | AAI-based (medium-confidence) | 0 |
| S7_vs_70225\|\|full | 5321 | No | NA | 11 | 2 | 0 | Low-quality | Genome-fragment | 12.92 | AAI-based (high-confidence) | 0 |
| S16_extracted_91874 | 5321 | No | NA | 4 | 0 | 2 | Low-quality | Genome-fragment | 1.43 | AAI-based (high-confidence) | 0 |
| S2_extracted_k141_6194 | 1368 | No | NA | 2 | 0 | 0 | Not-determined | Genome-fragment | NA | NA | 0 |
| S12_extracted_k141_118289 | 5324 | No | NA | 4 | 0 | 1 | Low-quality | Genome-fragment | 5.96 | AAI-based (medium-confidence) | 0 |
| S2_extracted_k141_18478 | 1038 | No | NA | 1 | 0 | 0 | Not-determined | Genome-fragment | NA | NA | 0 |
| S2_extracted_k141_20910 | 15926 | No | NA | 22 | 7 | 0 | Medium-quality | Genome-fragment | 52.63 | HMM-based (lower-bound) | 0 |
| S1_vs_k141_101263\|\|full | 5325 | No | NA | 4 | 0 | 3 | Low-quality | Genome-fragment | 4.16 | AAI-based (medium-confidence) | 0 |
| S2_extracted_k141_20917 | 1255 | No | NA | 1 | 0 | 1 | Not-determined | Genome-fragment | NA | NA | 0 |
| S2_extracted_k141_20919 | 1279 | No | NA | 1 | 0 | 0 | Not-determined | Genome-fragment | NA | NA | 0 |
| S2_extracted_k141_17072 | 1529 | No | NA | 1 | 0 | 1 | Not-determined | Genome-fragment | NA | NA | 0 |
| S2_extracted_k141_20929 | 1365 | No | NA | 1 | 0 | 1 | Not-determined | Genome-fragment | NA | NA | 0 |
| S2_extracted_k141_14377 | 13044 | No | NA | 20 | 0 | 8 | Not-determined | Genome-fragment | NA | NA | 0 |
| S2_extracted_k141_8938 | 1387 | No | NA | 2 | 0 | 1 | Not-determined | Genome-fragment | NA | NA | 0 |
| S5_vs_k141_27012\|\|full | 5325 | No | NA | 7 | 4 | 0 | Low-quality | Genome-fragment | 8.87 | AAI-based (high-confidence) | 0 |
| S5_vs_k141_86201\|\|full | 5326 | No | NA | 13 | 1 | 0 | Low-quality | Genome-fragment | 3.43 | HMM-based (lower-bound) | 0 |
| S2_extracted_k141_680 | 1223 | No | NA | 2 | 0 | 1 | Not-determined | Genome-fragment | NA | NA | 0 |
| S2_extracted_k141_17117 | 1164 | No | NA | 1 | 0 | 1 | Not-determined | Genome-fragment | NA | NA | 0 |
| S2_extracted_k141_15893 | 1053 | No | NA | 2 | 0 | 2 | Not-determined | Genome-fragment | NA | NA | 0 |
| S2_extracted_k141_4910 | 17143 | No | NA | 23 | 8 | 0 | Medium-quality | Genome-fragment | 80.99 | AAI-based (medium-confidence) | 0 |
| S16_extracted_47809 | 5326 | No | NA | 15 | 1 | 0 | Low-quality | Genome-fragment | 3.51 | HMM-based (lower-bound) | 0 |
| S2_extracted_k141_699 | 1227 | No | NA | 2 | 0 | 1 | Not-determined | Genome-fragment | NA | NA | 0 |
| S2_extracted_k141_702 | 1030 | No | NA | 1 | 0 | 1 | Not-determined | Genome-fragment | NA | NA | 0 |
| S12_vs_k141_178087\|\|full | 5328 | No | NA | 4 | 0 | 3 | Low-quality | Genome-fragment | 3.54 | AAI-based (medium-confidence) | 0 |
| S2_extracted_k141_21018 | 1818 | No | NA | 2 | 0 | 1 | Not-determined | Genome-fragment | NA | NA | 0 |
| S2_extracted_k141_4933 | 1323 | No | NA | 2 | 0 | 1 | Not-determined | Genome-fragment | NA | NA | 0 |
| S10_vs_63587\|\|full | 5330 | No | NA | 10 | 1 | 0 | Low-quality | Genome-fragment | 2.98 | AAI-based (high-confidence) | 0 |
| S2_extracted_k141_3529 | 1519 | No | NA | 2 | 0 | 1 | Not-determined | Genome-fragment | NA | NA | 0 |
| S9_extracted_k141_11573 | 5332 | No | NA | 9 | 4 | 0 | Low-quality | Genome-fragment | 11.29 | AAI-based (high-confidence) | 0 |
| S2_extracted_k141_3549 | 1128 | No | NA | 3 | 0 | 2 | Not-determined | Genome-fragment | NA | NA | 0 |
| S15_extracted_11450 | 5332 | No | NA | 7 | 0 | 0 | Low-quality | Genome-fragment | 2.83 | AAI-based (high-confidence) | 0 |
| S2_extracted_k141_791 | 1207 | No | NA | 1 | 0 | 1 | Not-determined | Genome-fragment | NA | NA | 0 |
| S2_extracted_k141_9043 | 1053 | No | NA | 1 | 0 | 1 | Not-determined | Genome-fragment | NA | NA | 0 |
| S2_extracted_k141_14509 | 5819 | No | NA | 2 | 0 | 2 | Not-determined | Genome-fragment | NA | NA | 0 |
| S2_extracted_k141_20000 | 1338 | No | NA | 1 | 0 | 1 | Not-determined | Genome-fragment | NA | NA | 0 |
| S11_vs_k141_60165\|\|full | 5335 | No | NA | 8 | 1 | 0 | Low-quality | Genome-fragment | 12.74 | AAI-based (medium-confidence) | 0 |
| S1_vs_k141_88317\|\|full | 5335 | No | NA | 6 | 2 | 0 | Low-quality | Genome-fragment | 8.97 | AAI-based (medium-confidence) | 0 |
| S7_vs_94023\|\|full | 5335 | No | NA | 4 | 0 | 2 | Low-quality | Genome-fragment | 2.93 | AAI-based (medium-confidence) | 0 |
| S16_extracted_89067 | 5336 | No | NA | 10 | 1 | 0 | Low-quality | Genome-fragment | 2.14 | HMM-based (lower-bound) | 0 |
| S5_extracted_k141_85848 | 5337 | No | NA | 9 | 2 | 0 | Low-quality | Genome-fragment | 12.41 | HMM-based (lower-bound) | 0 |
| S2_extracted_k141_869 | 1049 | No | NA | 2 | 0 | 0 | Not-determined | Genome-fragment | NA | NA | 0 |
| S16_extracted_71601 | 5337 | No | NA | 5 | 1 | 0 | Low-quality | Genome-fragment | 6.96 | HMM-based (lower-bound) | 0 |
| S2_extracted_k141_20040 | 2627 | No | NA | 2 | 0 | 0 | Not-determined | Genome-fragment | NA | NA | 0 |
| S2_extracted_k141_17240 | 1912 | No | NA | 1 | 0 | 1 | Not-determined | Genome-fragment | NA | NA | 0 |
| S2_extracted_k141_897 | 1185 | No | NA | 2 | 0 | 1 | Not-determined | Genome-fragment | NA | NA | 0 |
| S2_extracted_k141_21122 | 7141 | No | NA | 22 | 0 | 0 | Not-determined | Genome-fragment | NA | NA | 0 |
| S8_vs_k141_26692\|\|full | 5337 | No | NA | 4 | 0 | 1 | Low-quality | Genome-fragment | 2.69 | AAI-based (medium-confidence) | 0 |
| S16_extracted_106925 | 5338 | No | NA | 3 | 0 | 2 | Low-quality | Genome-fragment | 4.82 | AAI-based (medium-confidence) | 0 |
| S12_vs_k141_89685\|\|full | 5338 | No | NA | 6 | 3 | 0 | Low-quality | Genome-fragment | 6.26 | HMM-based (lower-bound) | 0 |
| S2_extracted_k141_3700 | 1604 | No | NA | 1 | 0 | 1 | Not-determined | Genome-fragment | NA | NA | 0 |
| S2_extracted_k141_3701 | 1003 | No | NA | 2 | 0 | 0 | Not-determined | Genome-fragment | NA | NA | 0 |
| S2_extracted_k141_21162 | 1027 | No | NA | 1 | 0 | 0 | Not-determined | Genome-fragment | NA | NA | 0 |
| S2_extracted_k141_928 | 3907 | No | NA | 5 | 0 | 4 | Not-determined | Genome-fragment | NA | NA | 0 |
| S2_extracted_k141_21184 | 1268 | No | NA | 2 | 0 | 1 | Not-determined | Genome-fragment | NA | NA | 0 |
| S14_extracted_k141_76582 | 5339 | No | NA | 14 | 1 | 0 | Low-quality | Genome-fragment | 8.12 | HMM-based (lower-bound) | 0 |
| S2_extracted_k141_955 | 1171 | No | NA | 2 | 0 | 1 | Not-determined | Genome-fragment | NA | NA | 0 |
| S2_extracted_k141_3780 | 1257 | No | NA | 2 | 0 | 2 | Not-determined | Genome-fragment | NA | NA | 0 |
| S2_extracted_k141_2218 | 1180 | No | NA | 1 | 0 | 0 | Not-determined | Genome-fragment | NA | NA | 0 |
| S3_vs_k141_97834\|\|full | 5339 | No | NA | 12 | 5 | 0 | Low-quality | Genome-fragment | 13.26 | AAI-based (medium-confidence) | 0 |
| S2_extracted_k141_2252 | 2564 | No | NA | 2 | 0 | 2 | Not-determined | Genome-fragment | NA | NA | 0 |
| S2_extracted_k141_2254 | 2452 | No | NA | 2 | 0 | 0 | Not-determined | Genome-fragment | NA | NA | 0 |
| S2_extracted_k141_2263 | 1186 | No | NA | 2 | 0 | 1 | Not-determined | Genome-fragment | NA | NA | 0 |
| S7_vs_27741\|\|full | 5342 | No | NA | 8 | 1 | 0 | Low-quality | Genome-fragment | 8.59 | HMM-based (lower-bound) | 0 |
| S2_extracted_k141_5142 | 1150 | No | NA | 2 | 0 | 1 | Not-determined | Genome-fragment | NA | NA | 0 |
| S2_extracted_k141_17346 | 1272 | No | NA | 1 | 0 | 1 | Not-determined | Genome-fragment | NA | NA | 0 |
| S2_extracted_k141_16112 | 1584 | No | NA | 3 | 0 | 0 | Not-determined | Genome-fragment | NA | NA | 0 |
| S2_extracted_k141_10330 | 2830 | No | NA | 3 | 0 | 2 | Not-determined | Genome-fragment | NA | NA | 0 |
| S2_extracted_k141_20256 | 1164 | No | NA | 1 | 0 | 1 | Not-determined | Genome-fragment | NA | NA | 0 |
| S2_extracted_k141_14693 | 1012 | No | NA | 2 | 0 | 0 | Not-determined | Genome-fragment | NA | NA | 0 |
| S15_vs_31929\|\|full | 5343 | No | NA | 7 | 0 | 2 | Low-quality | Genome-fragment | 2.44 | AAI-based (medium-confidence) | 0 |
| S3_extracted_k141_64623 | 5345 | No | NA | 6 | 2 | 0 | Low-quality | Genome-fragment | 24.25 | AAI-based (high-confidence) | 0 |
| S2_extracted_k141_6350 | 2029 | No | NA | 2 | 0 | 1 | Not-determined | Genome-fragment | NA | NA | 0 |
| S10_vs_17561\|\|full | 5347 | No | NA | 10 | 1 | 0 | Low-quality | Genome-fragment | 13.28 | AAI-based (high-confidence) | 0 |
| S2_extracted_k141_9392 | 1344 | No | NA | 2 | 0 | 1 | Not-determined | Genome-fragment | NA | NA | 0 |
| S9_vs_k141_73408\|\|full | 5352 | No | NA | 7 | 0 | 2 | Low-quality | Genome-fragment | 9.61 | AAI-based (medium-confidence) | 0 |
| S2_extracted_k141_2305 | 1331 | No | NA | 2 | 0 | 2 | Not-determined | Genome-fragment | NA | NA | 0 |
| S2_extracted_k141_12106 | 1742 | No | NA | 2 | 0 | 2 | Not-determined | Genome-fragment | NA | NA | 0 |
| S2_extracted_k141_5235 | 1908 | No | NA | 3 | 0 | 2 | Not-determined | Genome-fragment | NA | NA | 0 |
| S2_extracted_k141_6413 | 1157 | No | NA | 1 | 0 | 0 | Not-determined | Genome-fragment | NA | NA | 0 |
| S11_vs_k141_34204\|\|full | 5354 | No | NA | 8 | 0 | 1 | Low-quality | Genome-fragment | 4.5 | AAI-based (high-confidence) | 0 |
| S2_extracted_k141_17420 | 1476 | No | NA | 3 | 0 | 1 | Not-determined | Genome-fragment | NA | NA | 0 |
| S2_extracted_k141_18900 | 4044 | No | NA | 3 | 0 | 1 | Not-determined | Genome-fragment | NA | NA | 0 |
| S2_extracted_k141_14808 | 1505 | No | NA | 1 | 0 | 1 | Not-determined | Genome-fragment | NA | NA | 0 |
| S12_extracted_k141_72662 | 5354 | No | NA | 9 | 5 | 0 | Low-quality | Genome-fragment | 15.16 | AAI-based (high-confidence) | 0 |
| S2_extracted_k141_1151 | 1352 | No | NA | 2 | 0 | 0 | Not-determined | Genome-fragment | NA | NA | 0 |
| S9_extracted_k141_71054 | 5356 | No | NA | 11 | 1 | 0 | Low-quality | Genome-fragment | 3.26 | AAI-based (high-confidence) | 0 |
| S2_extracted_k141_3915 | 1175 | No | NA | 2 | 0 | 0 | Not-determined | Genome-fragment | NA | NA | 0 |
| S10_extracted_25972 | 5356 | No | NA | 9 | 6 | 0 | Low-quality | Genome-fragment | 8.9 | AAI-based (high-confidence) | 0 |
| S2_extracted_k141_1212 | 1049 | No | NA | 1 | 0 | 1 | Not-determined | Genome-fragment | NA | NA | 0 |
| S2_extracted_k141_1213 | 1192 | No | NA | 1 | 0 | 1 | Not-determined | Genome-fragment | NA | NA | 0 |
| S2_extracted_k141_1215 | 1461 | No | NA | 1 | 0 | 0 | Not-determined | Genome-fragment | NA | NA | 0 |
| S2_extracted_k141_1217 | 1540 | No | NA | 1 | 0 | 1 | Not-determined | Genome-fragment | NA | NA | 0 |
| S2_extracted_k141_1226 | 1287 | No | NA | 1 | 0 | 0 | Not-determined | Genome-fragment | NA | NA | 0 |
| S7_vs_18067\|\|full | 5357 | Yes | 1264 | 6 | 1 | 3 | Low-quality | Genome-fragment | 2.53 | HMM-based (lower-bound) | 76.4 |
| S7_vs_53357\|\|full | 5358 | No | NA | 7 | 1 | 0 | Low-quality | Genome-fragment | 6.01 | HMM-based (lower-bound) | 0 |
| S16_vs_42144\|\|full | 5358 | No | NA | 4 | 3 | 0 | Low-quality | Genome-fragment | 3.17 | AAI-based (high-confidence) | 0 |
| S2_extracted_k141_4043 | 1301 | No | NA | 3 | 0 | 1 | Not-determined | Genome-fragment | NA | NA | 0 |
| S1_extracted_k141_92636 | 5360 | No | NA | 3 | 0 | 1 | Low-quality | Genome-fragment | 13.66 | AAI-based (medium-confidence) | 0 |
| S2_extracted_k141_17583 | 2777 | No | NA | 3 | 0 | 3 | Not-determined | Genome-fragment | NA | NA | 0 |
| S8_vs_k141_47315\|\|full | 5360 | No | NA | 10 | 0 | 0 | Low-quality | Genome-fragment | 10.12 | AAI-based (high-confidence) | 0 |
| S2_extracted_k141_1352 | 1235 | No | NA | 2 | 0 | 0 | Not-determined | Genome-fragment | NA | NA | 0 |
| S2_extracted_k141_21511 | 64083 | No | NA | 95 | 28 | 0 | High-quality | High-quality | 97.85 | AAI-based (high-confidence) | 0 |
| S3_extracted_k141_13245 | 5364 | No | NA | 4 | 0 | 2 | Low-quality | Genome-fragment | 2.67 | AAI-based (medium-confidence) | 0 |
| S2_extracted_k141_8072 | 1297 | No | NA | 2 | 0 | 2 | Not-determined | Genome-fragment | NA | NA | 0 |
| S2_extracted_k141_11781 | 5366 | No | NA | 13 | 1 | 1 | Low-quality | Genome-fragment | 9.83 | AAI-based (high-confidence) | 0 |
| S7_vs_31553\|\|full | 5367 | Yes | 1182 | 5 | 1 | 3 | Low-quality | Genome-fragment | 1.22 | AAI-based (medium-confidence) | 77.98 |
| S7_vs_74327\|\|full | 5368 | No | NA | 4 | 0 | 0 | Low-quality | Genome-fragment | 10.82 | AAI-based (medium-confidence) | 0 |
| S2_extracted_k141_10584 | 1139 | No | NA | 2 | 0 | 1 | Not-determined | Genome-fragment | NA | NA | 0 |
| S2_extracted_k141_6673 | 1833 | No | NA | 2 | 0 | 2 | Not-determined | Genome-fragment | NA | NA | 0 |
| S16_extracted_43067 | 5368 | No | NA | 5 | 2 | 0 | Low-quality | Genome-fragment | 12.9 | AAI-based (high-confidence) | 0 |
| S2_extracted_k141_10750 | 1229 | No | NA | 1 | 0 | 0 | Not-determined | Genome-fragment | NA | NA | 0 |
| S2_extracted_k141_21630 | 48927 | No | NA | 72 | 24 | 1 | Complete | High-quality | 100 | DTR (high-confidence) | 0 |
| S2_vs_k141_7306\|\|full | 46830 | No | NA | 61 | 15 | 1 | High-quality | High-quality | 100 | AAI-based (high-confidence) | 0 |
| S2_vs_k141_10280\|\|full | 2749 | No | NA | 3 | 0 | 0 | Not-determined | Genome-fragment | NA | NA | 0 |
| S2_vs_k141_21629\|\|full | 2859 | No | NA | 4 | 0 | 0 | Not-determined | Genome-fragment | NA | NA | 0 |
| S16_vs_125494\|\|full | 5368 | No | NA | 11 | 5 | 0 | Low-quality | Genome-fragment | 14.3 | AAI-based (high-confidence) | 0 |
| S2_vs_k141_13530\|\|full | 1956 | No | NA | 4 | 0 | 2 | Not-determined | Genome-fragment | NA | NA | 0 |
| S9_extracted_k141_51706 | 5369 | No | NA | 10 | 1 | 0 | Low-quality | Genome-fragment | 11.81 | AAI-based (high-confidence) | 0 |
| S2_vs_k141_16265\|\|full | 2148 | No | NA | 3 | 0 | 0 | Not-determined | Genome-fragment | NA | NA | 0 |
| S17_extracted_k141_2566 | 5371 | No | NA | 13 | 12 | 0 | Low-quality | Genome-fragment | 17.65 | AAI-based (high-confidence) | 0 |
| S16_vs_4310\|\|full | 5372 | No | NA | 7 | 0 | 1 | Low-quality | Genome-fragment | 3.62 | AAI-based (medium-confidence) | 0 |
| S2_vs_k141_6839\|\|full | 3236 | No | NA | 2 | 0 | 1 | Not-determined | Genome-fragment | NA | NA | 0 |
| S2_vs_k141_2849\|\|full | 1828 | No | NA | 3 | 0 | 0 | Not-determined | Genome-fragment | NA | NA | 0 |
| S2_vs_k141_10914\|\|full | 1349 | No | NA | 5 | 0 | 4 | Not-determined | Genome-fragment | NA | NA | 0 |
| S2_vs_k141_118\|\|full | 2720 | No | NA | 8 | 0 | 0 | Not-determined | Genome-fragment | NA | NA | 0 |
| S8_extracted_k141_56657 | 5374 | No | NA | 6 | 0 | 2 | Low-quality | Genome-fragment | 7.08 | AAI-based (medium-confidence) | 0 |
| S14_extracted_k141_60583 | 5375 | No | NA | 6 | 0 | 3 | Low-quality | Genome-fragment | 8.98 | AAI-based (high-confidence) | 0 |
| S5_vs_k141_89270\|\|full | 5377 | No | NA | 2 | 0 | 2 | Low-quality | Genome-fragment | 5 | AAI-based (medium-confidence) | 0 |
| S7_vs_93577\|\|full | 5377 | No | NA | 9 | 1 | 0 | Low-quality | Genome-fragment | 8.14 | HMM-based (lower-bound) | 0 |
| S10_extracted_26194 | 5377 | No | NA | 6 | 0 | 0 | Low-quality | Genome-fragment | 11.75 | AAI-based (medium-confidence) | 0 |
| S11_vs_k141_25436\|\|full | 5379 | No | NA | 6 | 3 | 0 | Low-quality | Genome-fragment | 15.52 | AAI-based (medium-confidence) | 0 |
| S14_extracted_k141_97255 | 5380 | No | NA | 7 | 5 | 0 | Low-quality | Genome-fragment | 20.02 | AAI-based (high-confidence) | 0 |
| S12_vs_k141_86993\|\|full | 5381 | No | NA | 7 | 1 | 0 | Low-quality | Genome-fragment | 9.02 | AAI-based (high-confidence) | 0 |
| S17_extracted_k141_61626 | 5382 | No | NA | 5 | 0 | 4 | Low-quality | Genome-fragment | 3.58 | AAI-based (medium-confidence) | 0 |
| S2_vs_k141_11046\|\|full | 2450 | No | NA | 4 | 0 | 3 | Not-determined | Genome-fragment | NA | NA | 0 |
| S16_vs_142202\|\|full | 5383 | No | NA | 2 | 0 | 1 | Low-quality | Genome-fragment | 6.48 | AAI-based (high-confidence) | 0 |
| S10_extracted_97865 | 5383 | No | NA | 6 | 1 | 0 | Low-quality | Genome-fragment | 2.2 | AAI-based (high-confidence) | 0 |
| S12_extracted_k141_37706 | 5387 | No | NA | 3 | 0 | 0 | Low-quality | Genome-fragment | 9.33 | AAI-based (high-confidence) | 0 |
| S10_extracted_1160 | 5392 | No | NA | 4 | 4 | 0 | Low-quality | Genome-fragment | 8.95 | AAI-based (high-confidence) | 0 |
| S8_vs_k141_185018\|\|full | 5396 | No | NA | 7 | 4 | 0 | Low-quality | Genome-fragment | 11.96 | AAI-based (high-confidence) | 0 |
| S2_vs_k141_11088\|\|full | 5116 | No | NA | 5 | 0 | 0 | Not-determined | Genome-fragment | NA | NA | 0 |
| S14_extracted_k141_51729 | 5398 | No | NA | 11 | 1 | 0 | Low-quality | Genome-fragment | 9.05 | AAI-based (high-confidence) | 0 |
| S8_extracted_k141_46846 | 5400 | No | NA | 6 | 6 | 0 | Low-quality | Genome-fragment | 9.06 | AAI-based (medium-confidence) | 0 |
| S2_vs_k141_20597\|\|full | 3423 | No | NA | 6 | 0 | 3 | Not-determined | Genome-fragment | NA | NA | 0 |
| S12_vs_k141_96093\|\|full | 5401 | No | NA | 5 | 0 | 2 | Low-quality | Genome-fragment | 3.72 | AAI-based (medium-confidence) | 0 |
| S2_vs_k141_4343\|\|full | 2155 | No | NA | 3 | 0 | 1 | Not-determined | Genome-fragment | NA | NA | 0 |
| S2_vs_k141_9815\|\|full | 1124 | No | NA | 3 | 0 | 0 | Not-determined | Genome-fragment | NA | NA | 0 |
| S8_extracted_k141_144561 | 5402 | Yes | 1186 | 5 | 1 | 3 | Low-quality | Genome-fragment | 1.54 | HMM-based (lower-bound) | 78.05 |
| S2_vs_k141_4368\|\|full | 20983 | No | NA | 36 | 8 | 0 | Medium-quality | Genome-fragment | 69.27 | AAI-based (high-confidence) | 0 |
| S7_extracted_53039 | 5408 | No | NA | 8 | 0 | 1 | Low-quality | Genome-fragment | 9.03 | AAI-based (high-confidence) | 0 |
| S12_vs_k141_34975\|\|full | 5410 | No | NA | 9 | 1 | 0 | Low-quality | Genome-fragment | 9.72 | AAI-based (high-confidence) | 0 |
| S7_vs_35941\|\|full | 5411 | No | NA | 11 | 2 | 0 | Low-quality | Genome-fragment | 12.69 | AAI-based (medium-confidence) | 0 |
| S5_extracted_k141_29860 | 5413 | No | NA | 7 | 2 | 0 | Low-quality | Genome-fragment | 3.29 | AAI-based (high-confidence) | 0 |
| S16_vs_36527\|\|full | 5413 | No | NA | 10 | 9 | 0 | Low-quality | Genome-fragment | 15.73 | AAI-based (high-confidence) | 0 |
| S16_vs_129215\|\|full | 5415 | No | NA | 5 | 0 | 4 | Low-quality | Genome-fragment | 2.15 | AAI-based (medium-confidence) | 0 |
| S9_vs_k141_17471\|\|full | 5418 | No | NA | 12 | 1 | 0 | Low-quality | Genome-fragment | 12.94 | AAI-based (high-confidence) | 0 |
| S2_vs_k141_7229\|\|full | 1206 | No | NA | 3 | 0 | 2 | Not-determined | Genome-fragment | NA | NA | 0 |
| S7_vs_71327\|\|full | 5419 | No | NA | 4 | 1 | 0 | Low-quality | Genome-fragment | 16.31 | AAI-based (high-confidence) | 0 |
| S2_vs_k141_15413\|\|full | 2915 | No | NA | 4 | 0 | 0 | Not-determined | Genome-fragment | NA | NA | 0 |
| S2_vs_k141_14049\|\|full | 3290 | No | NA | 4 | 0 | 3 | Not-determined | Genome-fragment | NA | NA | 0 |
| S14_extracted_k141_69617 | 5421 | No | NA | 4 | 0 | 4 | Low-quality | Genome-fragment | 3.22 | AAI-based (medium-confidence) | 0 |

| | | | | | | | | | | | |
|---|---|---|---|---|---|---|---|---|---|---|---|
| S16_extracted_67634 | 5421 | No | NA | 8 | 3 | 0 | Low-quality | Genome-fragment | 9.02 | AAI-based (high-confidence) | 0 |
| S2_vs_k141_4528\|\|full | 6857 | No | NA | 6 | 0 | 4 | Not-determined | Genome-fragment | NA | NA | 0 |
| S14_extracted_k141_83755 | 5425 | No | NA | 10 | 2 | 0 | Low-quality | Genome-fragment | 12.01 | AAI-based (medium-confidence) | 0 |
| S3_extracted_k141_32609 | 5425 | No | NA | 6 | 0 | 1 | Low-quality | Genome-fragment | 9.93 | AAI-based (high-confidence) | 0 |
| S7_vs_9256\|\|full | 5426 | No | NA | 3 | 0 | 0 | Low-quality | Genome-fragment | 13.95 | AAI-based (high-confidence) | 0 |
| S12_vs_k141_105854\|\|full | 5426 | No | NA | 5 | 0 | 3 | Low-quality | Genome-fragment | 3.84 | AAI-based (medium-confidence) | 0 |
| S9_vs_k141_41408\|\|full | 5429 | No | NA | 11 | 3 | 0 | Low-quality | Genome-fragment | 14.1 | AAI-based (high-confidence) | 0 |
| S11_vs_k141_7346\|\|full | 5430 | No | NA | 9 | 2 | 1 | Low-quality | Genome-fragment | 8.45 | HMM-based (lower-bound) | 0 |
| S11_vs_k141_90408\|\|full | 5430 | No | NA | 10 | 1 | 0 | Low-quality | Genome-fragment | 3.33 | AAI-based (high-confidence) | 0 |
| S2_vs_k141_18201\|\|full | 1880 | No | NA | 3 | 0 | 3 | Not-determined | Genome-fragment | NA | NA | 0 |
| S5_extracted_k141_115658 | 5430 | No | NA | 5 | 5 | 0 | Low-quality | Genome-fragment | 17.9 | AAI-based (high-confidence) | 0 |
| S5_vs_k141_91175\|\|full | 5430 | No | NA | 6 | 1 | 0 | Low-quality | Genome-fragment | 13.04 | AAI-based (medium-confidence) | 0 |
| S5_vs_k141_143893\|\|full | 5430 | Yes | 3844 | 10 | 1 | 1 | Low-quality | Genome-fragment | 9.75 | AAI-based (medium-confidence) | 29.21 |
| S2_vs_k141_18252\|\|full | 4668 | No | NA | 4 | 0 | 1 | Not-determined | Genome-fragment | NA | NA | 0 |
| S2_vs_k141_7375\|\|full | 2183 | No | NA | 3 | 0 | 1 | Not-determined | Genome-fragment | NA | NA | 0 |
| S3_extracted_k141_57934 | 5431 | No | NA | 7 | 1 | 0 | Low-quality | Genome-fragment | 4.06 | AAI-based (high-confidence) | 0 |
| S2_vs_k141_11462\|\|full | 2096 | No | NA | 3 | 0 | 0 | Not-determined | Genome-fragment | NA | NA | 0 |
| S8_vs_k141_13184\|\|full | 5433 | No | NA | 7 | 0 | 1 | Low-quality | Genome-fragment | 10.35 | AAI-based (medium-confidence) | 0 |
| S3_vs_k141_64034\|\|full | 5435 | No | NA | 6 | 0 | 2 | Low-quality | Genome-fragment | 4.24 | AAI-based (high-confidence) | 0 |
| S8_vs_k141_140415\|\|full | 5435 | No | NA | 6 | 0 | 4 | Low-quality | Genome-fragment | 2.81 | AAI-based (medium-confidence) | 0 |
| S17_extracted_k141_7897 | 5436 | No | NA | 12 | 5 | 0 | Low-quality | Genome-fragment | 6.92 | HMM-based (lower-bound) | 0 |
| S4_vs_VIRSorter_2992-cat_2 | 5436 | No | NA | 4 | 0 | 0 | Low-quality | Genome-fragment | 11.93 | AAI-based (high-confidence) | 0 |
| S8_extracted_k141_23657 | 5436 | No | NA | 12 | 4 | 0 | Low-quality | Genome-fragment | 12.16 | AAI-based (medium-confidence) | 0 |
| S2_vs_k141_14285\|\|full | 1492 | No | NA | 3 | 0 | 1 | Not-determined | Genome-fragment | NA | NA | 0 |
| S3_extracted_k141_89210 | 5438 | No | NA | 9 | 8 | 0 | Low-quality | Genome-fragment | 17.95 | AAI-based (high-confidence) | 0 |
| S2_vs_k141_8783\|\|full | 1703 | No | NA | 3 | 0 | 2 | Not-determined | Genome-fragment | NA | NA | 0 |
| S12_vs_k141_167667\|\|full | 5438 | No | NA | 10 | 2 | 1 | Low-quality | Genome-fragment | 4.22 | AAI-based (medium-confidence) | 0 |
| S8_extracted_k141_143330 | 5440 | No | NA | 1 | 0 | 0 | Low-quality | Genome-fragment | 6.55 | AAI-based (medium-confidence) | 0 |
| S13_vs_41176\|\|full | 5441 | No | NA | 9 | 0 | 0 | Low-quality | Genome-fragment | 10.32 | AAI-based (high-confidence) | 0 |
| S12_extracted_k141_176017 | 5441 | No | NA | 6 | 1 | 0 | Low-quality | Genome-fragment | 11.21 | AAI-based (medium-confidence) | 0 |
| S5_vs_k141_41734\|\|full | 5443 | No | NA | 2 | 0 | 2 | Low-quality | Genome-fragment | 5.2 | AAI-based (medium-confidence) | 0 |
| S2_vs_k141_14331\|\|full | 8429 | No | NA | 5 | 0 | 2 | Not-determined | Genome-fragment | NA | NA | 0 |
| S16_extracted_69016 | 5444 | No | NA | 8 | 3 | 0 | Low-quality | Genome-fragment | 10.7 | AAI-based (medium-confidence) | 0 |
| S3_extracted_k141_111357 | 5445 | No | NA | 7 | 0 | 0 | Low-quality | Genome-fragment | 37.88 | AAI-based (high-confidence) | 0 |
| S2_vs_k141_8886\|\|full | 2138 | No | NA | 4 | 0 | 0 | Not-determined | Genome-fragment | NA | NA | 0 |
| S2_vs_k141_4847\|\|full | 1988 | No | NA | 3 | 0 | 1 | Not-determined | Genome-fragment | NA | NA | 0 |
| S14_extracted_k141_22336 | 5446 | No | NA | 5 | 1 | 0 | Low-quality | Genome-fragment | 3.83 | AAI-based (medium-confidence) | 0 |
| S14_vs_k141_15215\|\|full | 5447 | No | NA | 8 | 0 | 0 | Low-quality | Genome-fragment | 13.85 | AAI-based (medium-confidence) | 0 |
| S11_vs_k141_20987\|\|full | 5448 | No | NA | 14 | 2 | 0 | Low-quality | Genome-fragment | 13.55 | AAI-based (medium-confidence) | 0 |
| S2_vs_k141_17146\|\|full | 2320 | No | NA | 3 | 0 | 1 | Not-determined | Genome-fragment | NA | NA | 0 |
| S2_vs_k141_11747\|\|full | 1458 | No | NA | 3 | 0 | 0 | Not-determined | Genome-fragment | NA | NA | 0 |
| S3_extracted_k141_27138 | 5449 | No | NA | 4 | 0 | 0 | Low-quality | Genome-fragment | 9.5 | AAI-based (high-confidence) | 0 |
| S2_vs_k141_755\|\|full | 1063 | No | NA | 2 | 0 | 1 | Not-determined | Genome-fragment | NA | NA | 0 |
| S2_vs_k141_10256\|\|full | 1368 | No | NA | 6 | 0 | 0 | Not-determined | Genome-fragment | NA | NA | 0 |
| S3_extracted_k141_14778 | 5450 | No | NA | 13 | 3 | 0 | Low-quality | Genome-fragment | 9.58 | AAI-based (high-confidence) | 0 |
| S13_vs_100903\|\|full | 5454 | No | NA | 3 | 0 | 2 | Low-quality | Genome-fragment | 2.68 | AAI-based (high-confidence) | 0 |
| S16_vs_144384\|\|full | 5455 | No | NA | 4 | 0 | 2 | Low-quality | Genome-fragment | 3.67 | AAI-based (high-confidence) | 0 |
| S8_vs_k141_139117\|\|full | 5456 | No | NA | 7 | 2 | 0 | Low-quality | Genome-fragment | 10.74 | AAI-based (high-confidence) | 0 |
| S15_extracted_48414 | 5457 | No | NA | 3 | 0 | 0 | Low-quality | Genome-fragment | 5.49 | AAI-based (high-confidence) | 0 |
| S5_vs_k141_135677\|\|full | 5461 | No | NA | 8 | 3 | 0 | Low-quality | Genome-fragment | 4.91 | HMM-based (lower-bound) | 0 |
| S2_vs_k141_17173\|\|full | 2658 | No | NA | 3 | 0 | 1 | Not-determined | Genome-fragment | NA | NA | 0 |
| S7_vs_27501\|\|full | 5463 | No | NA | 7 | 0 | 1 | Low-quality | Genome-fragment | 3.9 | AAI-based (high-confidence) | 0 |
| S2_vs_k141_5003\|\|full | 2745 | No | NA | 3 | 0 | 2 | Not-determined | Genome-fragment | NA | NA | 0 |
| S8_vs_k141_86258\|\|full | 5463 | No | NA | 5 | 0 | 0 | Low-quality | Genome-fragment | 12.96 | AAI-based (high-confidence) | 0 |
| S7_vs_121942\|\|full | 5464 | No | NA | 11 | 1 | 0 | Low-quality | Genome-fragment | 3.67 | HMM-based (lower-bound) | 0 |
| S2_vs_k141_10289\|\|full | 46594 | Yes | 30604 | 49 | 6 | 8 | Medium-quality | Genome-fragment | 56.92 | AAI-based (high-confidence) | 34.32 |
| S3_extracted_k141_6126 | 5467 | No | NA | 8 | 4 | 0 | Low-quality | Genome-fragment | 24.93 | AAI-based (high-confidence) | 0 |
| S2_vs_k141_16020\|\|full | 6517 | No | NA | 11 | 0 | 0 | Not-determined | Genome-fragment | NA | NA | 0 |
| S2_vs_k141_9210\|\|full | 2417 | No | NA | 2 | 0 | 2 | Not-determined | Genome-fragment | NA | NA | 0 |
| S2_vs_k141_10306\|\|full | 12183 | No | NA | 13 | 0 | 8 | Not-determined | Genome-fragment | NA | NA | 0 |
| S10_extracted_41714 | 5468 | No | NA | 13 | 4 | 0 | Low-quality | Genome-fragment | 13.11 | AAI-based (high-confidence) | 0 |
| S2_vs_k141_9277\|\|full | 2038 | No | NA | 3 | 0 | 2 | Not-determined | Genome-fragment | NA | NA | 0 |
| S12_vs_k141_160949\|\|full | 5468 | No | NA | 11 | 2 | 0 | Low-quality | Genome-fragment | 21.75 | AAI-based (high-confidence) | 0 |
| S5_extracted_k141_121956 | 5473 | No | NA | 14 | 3 | 0 | Low-quality | Genome-fragment | 15.91 | AAI-based (high-confidence) | 0 |
| S2_vs_k141_991\|\|full | 3017 | No | NA | 3 | 0 | 2 | Not-determined | Genome-fragment | NA | NA | 0 |
| S8_extracted_k141_97923 | 5474 | No | NA | 6 | 1 | 0 | Low-quality | Genome-fragment | 8.46 | AAI-based (high-confidence) | 0 |
| S8_vs_k141_68516\|\|full | 5474 | No | NA | 13 | 4 | 0 | Low-quality | Genome-fragment | 12.95 | AAI-based (high-confidence) | 0 |
| S2_vs_k141_17330\|\|full | 3457 | No | NA | 2 | 0 | 1 | Not-determined | Genome-fragment | NA | NA | 0 |
| S13_vs_112051\|\|full | 5477 | No | NA | 6 | 1 | 0 | Low-quality | Genome-fragment | 41 | AAI-based (medium-confidence) | 0 |
| S4_extracted_26672 | 5477 | No | NA | 10 | 0 | 0 | Low-quality | Genome-fragment | 13.28 | AAI-based (high-confidence) | 0 |
| S2_vs_k141_6303\|\|full | 6083 | No | NA | 6 | 0 | 5 | Not-determined | Genome-fragment | NA | NA | 0 |
| S2_vs_k141_5166\|\|full | 1216 | No | NA | 3 | 0 | 1 | Not-determined | Genome-fragment | NA | NA | 0 |
| S7_vs_26817\|\|full | 5478 | No | NA | 5 | 2 | 0 | Low-quality | Genome-fragment | 7.07 | HMM-based (lower-bound) | 0 |
| S1_vs_k141_70348\|\|full | 5480 | No | NA | 7 | 5 | 0 | Low-quality | Genome-fragment | 17.05 | AAI-based (high-confidence) | 0 |
| S2_vs_k141_3890\|\|full | 7550 | No | NA | 10 | 0 | 4 | Not-determined | Genome-fragment | NA | NA | 0 |
| S17_vs_k141_83354\|\|full | 5481 | No | NA | 13 | 1 | 0 | Low-quality | Genome-fragment | 5.39 | HMM-based (lower-bound) | 0 |
| S11_vs_k141_43875\|\|full | 5483 | No | NA | 6 | 4 | 0 | Low-quality | Genome-fragment | 15.38 | AAI-based (high-confidence) | 0 |
| S12_vs_k141_123192\|\|full | 5483 | No | NA | 8 | 0 | 7 | Low-quality | Genome-fragment | 3.92 | AAI-based (medium-confidence) | 0 |
| S3_vs_k141_113523\|\|full | 5484 | No | NA | 7 | 1 | 0 | Low-quality | Genome-fragment | 7.26 | HMM-based (lower-bound) | 0 |
| S7_vs_45987\|\|full | 5485 | No | NA | 13 | 4 | 0 | Low-quality | Genome-fragment | 11.99 | AAI-based (medium-confidence) | 0 |
| S2_vs_k141_6514\|\|full | 1298 | No | NA | 2 | 0 | 1 | Not-determined | Genome-fragment | NA | NA | 0 |
| S2_vs_k141_1207\|\|full | 1082 | No | NA | 4 | 0 | 0 | Not-determined | Genome-fragment | NA | NA | 0 |
| S11_vs_k141_89074\|\|full | 5486 | No | NA | 13 | 2 | 0 | Low-quality | Genome-fragment | 6.79 | HMM-based (lower-bound) | 0 |
| S8_vs_k141_69509\|\|full | 5486 | No | NA | 9 | 2 | 1 | Low-quality | Genome-fragment | 12.99 | AAI-based (high-confidence) | 0 |
| S11_vs_k141_46119\|\|full | 5487 | No | NA | 9 | 7 | 0 | Low-quality | Genome-fragment | 13.72 | AAI-based (medium-confidence) | 0 |
| S17_vs_k141_56351\|\|full | 5490 | Yes | 2991 | 4 | 1 | 1 | Low-quality | Genome-fragment | 1.39 | HMM-based (lower-bound) | 45.52 |
| S2_vs_k141_6602\|\|full | 2093 | No | NA | 3 | 0 | 2 | Not-determined | Genome-fragment | NA | NA | 0 |
| S2_vs_k141_10519\|\|full | 1782 | No | NA | 2 | 0 | 0 | Not-determined | Genome-fragment | NA | NA | 0 |
| S2_vs_k141_6628\|\|full | 8489 | No | NA | 8 | 0 | 6 | Not-determined | Genome-fragment | NA | NA | 0 |
| S9_extracted_k141_54278 | 5491 | No | NA | 7 | 0 | 0 | Low-quality | Genome-fragment | 3.16 | AAI-based (high-confidence) | 0 |
| S11_extracted_k141_7178 | 5491 | No | NA | 12 | 5 | 0 | Low-quality | Genome-fragment | 12.16 | AAI-based (high-confidence) | 0 |
| S8_vs_k141_44439\|\|full | 5491 | No | NA | 5 | 0 | 0 | Low-quality | Genome-fragment | 7.31 | AAI-based (medium-confidence) | 0 |
| S2_vs_k141_20583\|\|lt2gene | 1007 | No | NA | 2 | 0 | 0 | Not-determined | Genome-fragment | NA | NA | 0 |
| S13_vs_108990\|\|full | 5493 | No | NA | 3 | 0 | 3 | Low-quality | Genome-fragment | 4.72 | AAI-based (medium-confidence) | 0 |
| S15_extracted_76253 | 5493 | No | NA | 15 | 6 | 0 | Low-quality | Genome-fragment | 9.15 | AAI-based (high-confidence) | 0 |
| S17_extracted_k141_82261 | 5495 | No | NA | 8 | 1 | 0 | Low-quality | Genome-fragment | 6.83 | AAI-based (high-confidence) | 0 |
| S9_vs_k141_29751\|\|full | 5496 | No | NA | 10 | 4 | 1 | Low-quality | Genome-fragment | 11.54 | AAI-based (high-confidence) | 0 |
| S4_extracted_87 | 2416 | No | NA | 2 | 0 | 0 | Not-determined | Genome-fragment | NA | NA | 0 |
| S4_extracted_93 | 1144 | No | NA | 1 | 0 | 1 | Not-determined | Genome-fragment | NA | NA | 0 |
| S16_vs_80694\|\|full | 5498 | No | NA | 12 | 0 | 0 | Low-quality | Genome-fragment | 13.08 | AAI-based (high-confidence) | 0 |
| S4_extracted_159 | 1254 | No | NA | 2 | 0 | 1 | Not-determined | Genome-fragment | NA | NA | 0 |
| S4_extracted_305 | 1003 | No | NA | 1 | 0 | 1 | Not-determined | Genome-fragment | NA | NA | 0 |
| S4_extracted_320 | 10416 | No | NA | 8 | 0 | 7 | Not-determined | Genome-fragment | NA | NA | 0 |
| S4_extracted_508 | 1540 | No | NA | 2 | 0 | 2 | Not-determined | Genome-fragment | NA | NA | 0 |
| S13_extracted_74508 | 5501 | No | NA | 7 | 0 | 2 | Low-quality | Genome-fragment | 6.82 | AAI-based (medium-confidence) | 0 |
| S4_extracted_568 | 6401 | No | NA | 4 | 0 | 2 | Not-determined | Genome-fragment | NA | NA | 0 |
| S3_extracted_k141_89687 | 5503 | No | NA | 5 | 0 | 1 | Low-quality | Genome-fragment | 3.46 | AAI-based (high-confidence) | 0 |
| S4_extracted_651 | 1144 | No | NA | 1 | 0 | 1 | Not-determined | Genome-fragment | NA | NA | 0 |
| S12_vs_k141_138066\|\|full | 5506 | No | NA | 7 | 1 | 0 | Low-quality | Genome-fragment | 5.5 | HMM-based (lower-bound) | 0 |
| S8_extracted_k141_45367 | 5506 | No | NA | 6 | 0 | 5 | Low-quality | Genome-fragment | 16.56 | AAI-based (medium-confidence) | 0 |
| S4_extracted_813 | 6838 | No | NA | 3 | 0 | 1 | Not-determined | Genome-fragment | NA | NA | 0 |
| S9_vs_k141_61245\|\|full | 5510 | Yes | 3611 | 5 | 1 | 1 | Low-quality | Genome-fragment | 7.22 | AAI-based (medium-confidence) | 34.46 |
| S4_extracted_900 | 3238 | No | NA | 2 | 0 | 2 | Not-determined | Genome-fragment | NA | NA | 0 |
| S4_extracted_931 | 7331 | No | NA | 7 | 0 | 6 | Not-determined | Genome-fragment | NA | NA | 0 |
| S4_extracted_934 | 1494 | No | NA | 1 | 0 | 1 | Not-determined | Genome-fragment | NA | NA | 0 |
| S4_extracted_938 | 6237 | No | NA | 9 | 0 | 5 | Not-determined | Genome-fragment | NA | NA | 0 |
| S11_vs_k141_79976\|\|full | 5510 | No | NA | 5 | 0 | 3 | Low-quality | Genome-fragment | 3.97 | AAI-based (medium-confidence) | 0 |
| S3_vs_k141_15749\|\|full | 5510 | No | NA | 6 | 4 | 0 | Low-quality | Genome-fragment | 3.27 | AAI-based (high-confidence) | 0 |
| S4_extracted_1280 | 3962 | No | NA | 4 | 0 | 0 | Not-determined | Genome-fragment | NA | NA | 0 |
| S4_extracted_1283 | 2449 | No | NA | 2 | 0 | 1 | Not-determined | Genome-fragment | NA | NA | 0 |
| S17_extracted_k141_2850 | 5512 | No | NA | 7 | 0 | 1 | Low-quality | Genome-fragment | 10.65 | AAI-based (high-confidence) | 0 |
| S4_extracted_1290 | 3900 | No | NA | 4 | 0 | 2 | Not-determined | Genome-fragment | NA | NA | 0 |
| S4_extracted_1431 | 9168 | No | NA | 6 | 0 | 4 | Not-determined | Genome-fragment | NA | NA | 0 |
| S4_extracted_1445 | 1394 | No | NA | 2 | 0 | 0 | Not-determined | Genome-fragment | NA | NA | 0 |
| S11_vs_k141_64012\|\|full | 5513 | No | NA | 10 | 0 | 0 | Low-quality | Genome-fragment | 16.17 | AAI-based (high-confidence) | 0 |
| S4_extracted_1489 | 1019 | No | NA | 1 | 0 | 0 | Not-determined | Genome-fragment | NA | NA | 0 |
| S4_extracted_1583 | 1933 | No | NA | 4 | 0 | 1 | Not-determined | Genome-fragment | NA | NA | 0 |
| S4_extracted_1587 | 1288 | No | NA | 2 | 0 | 1 | Not-determined | Genome-fragment | NA | NA | 0 |
| S4_extracted_1759 | 1660 | No | NA | 2 | 0 | 2 | Not-determined | Genome-fragment | NA | NA | 0 |
| S4_extracted_1784 | 9674 | No | NA | 8 | 0 | 3 | Not-determined | Genome-fragment | NA | NA | 0 |
| S16_vs_137094\|\|full | 5513 | No | NA | 4 | 0 | 4 | Low-quality | Genome-fragment | 4.56 | AAI-based (medium-confidence) | 0 |
| S7_vs_2696\|\|full | 5515 | No | NA | 9 | 0 | 1 | Low-quality | Genome-fragment | 13.47 | AAI-based (high-confidence) | 0 |
| S12_vs_k141_65068\|\|full | 5515 | No | NA | 14 | 5 | 1 | Low-quality | Genome-fragment | 13.89 | AAI-based (high-confidence) | 0 |
| S5_vs_k141_115835\|\|full | 5516 | No | NA | 6 | 4 | 0 | Low-quality | Genome-fragment | 12.8 | AAI-based (medium-confidence) | 0 |
| S16_extracted_36692 | 5516 | No | NA | 5 | 0 | 1 | Low-quality | Genome-fragment | 5.82 | AAI-based (medium-confidence) | 0 |
| S14_vs_k141_15168\|\|full | 5519 | No | NA | 13 | 1 | 0 | Low-quality | Genome-fragment | 10.13 | AAI-based (medium-confidence) | 0 |
| S4_extracted_2091 | 1009 | No | NA | 2 | 0 | 0 | Not-determined | Genome-fragment | NA | NA | 0 |
| S5_vs_k141_67693\|\|full | 5519 | No | NA | 7 | 1 | 0 | Low-quality | Genome-fragment | 14.08 | AAI-based (high-confidence) | 0 |
| S4_extracted_2357 | 2115 | No | NA | 2 | 0 | 0 | Not-determined | Genome-fragment | NA | NA | 0 |
| S4_extracted_2388 | 4765 | No | NA | 4 | 0 | 1 | Not-determined | Genome-fragment | NA | NA | 0 |
| S4_extracted_3490 | 5519 | No | NA | 5 | 5 | 0 | Low-quality | Genome-fragment | 18.2 | AAI-based (high-confidence) | 0 |
| S4_extracted_2475 | 1328 | No | NA | 1 | 0 | 0 | Not-determined | Genome-fragment | NA | NA | 0 |
| S9_vs_k141_69366\|\|full | 5521 | No | NA | 9 | 7 | 0 | Low-quality | Genome-fragment | 14.43 | AAI-based (high-confidence) | 0 |
| S4_extracted_2524 | 13131 | No | NA | 20 | 7 | 0 | High-quality | High-quality | 99.04 | AAI-based (high-confidence) | 0 |
| S11_vs_k141_9718\|\|full | 5521 | No | NA | 7 | 2 | 0 | Low-quality | Genome-fragment | 4.25 | AAI-based (medium-confidence) | 0 |
| S4_extracted_2604 | 1289 | No | NA | 2 | 0 | 1 | Not-determined | Genome-fragment | NA | NA | 0 |
| S13_vs_30532\|\|full | 5521 | No | NA | 7 | 1 | 0 | Low-quality | Genome-fragment | 13.05 | AAI-based (high-confidence) | 0 |
| S12_vs_k141_146\|\|full | 5521 | No | NA | 8 | 1 | 0 | Low-quality | Genome-fragment | 9.39 | AAI-based (medium-confidence) | 0 |
| S4_extracted_2692 | 4985 | No | NA | 5 | 0 | 3 | Not-determined | Genome-fragment | NA | NA | 0 |
| S2_extracted_k141_12112 | 5522 | No | NA | 9 | 0 | 0 | Low-quality | Genome-fragment | 9.96 | AAI-based (high-confidence) | 0 |
| S4_extracted_2926 | 5733 | No | NA | 6 | 0 | 3 | Not-determined | Genome-fragment | NA | NA | 0 |
| S4_extracted_2947 | 1224 | No | NA | 3 | 0 | 0 | Not-determined | Genome-fragment | NA | NA | 0 |
| S7_vs_12385\|\|full | 5524 | No | NA | 6 | 0 | 1 | Low-quality | Genome-fragment | 12.9 | AAI-based (high-confidence) | 0 |
| S15_vs_45266\|\|full | 5524 | No | NA | 9 | 0 | 1 | Low-quality | Genome-fragment | 7.1 | AAI-based (high-confidence) | 0 |
| S4_extracted_3093 | 5455 | No | NA | 7 | 0 | 5 | Not-determined | Genome-fragment | NA | NA | 0 |
| S4_extracted_3109 | 4026 | No | NA | 5 | 0 | 0 | Not-determined | Genome-fragment | NA | NA | 0 |
| S4_extracted_3168 | 4673 | No | NA | 2 | 0 | 1 | Not-determined | Genome-fragment | NA | NA | 0 |
| S4_extracted_3246 | 4290 | No | NA | 6 | 0 | 5 | Not-determined | Genome-fragment | NA | NA | 0 |
| S10_vs_35850\|\|full | 5528 | No | NA | 5 | 3 | 0 | Low-quality | Genome-fragment | 11.09 | AAI-based (high-confidence) | 0 |
| S4_extracted_3272 | 3324 | No | NA | 3 | 0 | 3 | Not-determined | Genome-fragment | NA | NA | 0 |
| S4_extracted_26363 | 5528 | No | NA | 3 | 3 | 0 | Low-quality | Genome-fragment | 18.22 | AAI-based (high-confidence) | 0 |
| S13_vs_55264\|\|full | 5532 | No | NA | 6 | 0 | 1 | Low-quality | Genome-fragment | 10.71 | AAI-based (medium-confidence) | 0 |
| S5_vs_k141_126525\|\|full | 5532 | No | NA | 7 | 1 | 0 | Low-quality | Genome-fragment | 10.67 | AAI-based (high-confidence) | 0 |
| S4_extracted_3600 | 1020 | No | NA | 1 | 0 | 1 | Not-determined | Genome-fragment | NA | NA | 0 |
| S10_extracted_99083 | 5532 | No | NA | 6 | 1 | 0 | Low-quality | Genome-fragment | 3.39 | AAI-based (high-confidence) | 0 |
| S16_vs_33661\|\|full | 5533 | No | NA | 12 | 0 | 1 | Low-quality | Genome-fragment | 12.09 | AAI-based (medium-confidence) | 0 |
| S12_vs_k141_8194\|\|full | 5533 | No | NA | 6 | 2 | 0 | Low-quality | Genome-fragment | 22.01 | AAI-based (high-confidence) | 0 |
| S4_extracted_3743 | 12812 | No | NA | 17 | 0 | 6 | Not-determined | Genome-fragment | NA | NA | 0 |
| S16_vs_49701\|\|full | 5537 | No | NA | 7 | 2 | 0 | Low-quality | Genome-fragment | 13.51 | AAI-based (high-confidence) | 0 |
| S4_extracted_3876 | 8073 | No | NA | 4 | 0 | 3 | Not-determined | Genome-fragment | NA | NA | 0 |
| S4_extracted_3906 | 1079 | No | NA | 1 | 0 | 1 | Not-determined | Genome-fragment | NA | NA | 0 |
| S4_extracted_3960 | 1054 | No | NA | 1 | 0 | 0 | Not-determined | Genome-fragment | NA | NA | 0 |
| S16_extracted_105179 | 5539 | No | NA | 4 | 0 | 3 | Low-quality | Genome-fragment | 1.53 | AAI-based (medium-confidence) | 0 |
| S11_extracted_k141_97323 | 5540 | No | NA | 3 | 0 | 0 | Low-quality | Genome-fragment | 5.01 | AAI-based (medium-confidence) | 0 |
| S4_extracted_4037 | 1205 | No | NA | 1 | 0 | 1 | Not-determined | Genome-fragment | NA | NA | 0 |
| S14_vs_k141_75052\|\|full | 5540 | No | NA | 8 | 4 | 0 | Low-quality | Genome-fragment | 12.35 | AAI-based (high-confidence) | 0 |
| S13_extracted_97619 | 5541 | No | NA | 5 | 0 | 3 | Low-quality | Genome-fragment | 4.9 | AAI-based (medium-confidence) | 0 |
| S12_vs_k141_167435\|\|full | 5543 | No | NA | 10 | 0 | 0 | Low-quality | Genome-fragment | 12.92 | AAI-based (medium-confidence) | 0 |
| S9_vs_k141_21657\|\|full | 5545 | No | NA | 8 | 0 | 0 | Low-quality | Genome-fragment | 13.11 | AAI-based (high-confidence) | 0 |
| S4_extracted_4244 | 6019 | No | NA | 8 | 0 | 3 | Not-determined | Genome-fragment | NA | NA | 0 |
| S16_vs_142354\|\|full | 5546 | No | NA | 15 | 1 | 0 | Low-quality | Genome-fragment | 9.19 | AAI-based (high-confidence) | 0 |
| S4_extracted_4456 | 7906 | No | NA | 3 | 0 | 2 | Not-determined | Genome-fragment | NA | NA | 0 |
| S14_vs_k141_24171\|\|full | 5547 | No | NA | 10 | 2 | 0 | Low-quality | Genome-fragment | 12.93 | AAI-based (high-confidence) | 0 |
| S4_extracted_4533 | 2592 | No | NA | 3 | 0 | 1 | Not-determined | Genome-fragment | NA | NA | 0 |
| S14_extracted_k141_110263 | 5548 | No | NA | 16 | 14 | 0 | Low-quality | Genome-fragment | 18.25 | AAI-based (high-confidence) | 0 |
| S5_extracted_k141_130187 | 5549 | No | NA | 16 | 15 | 0 | Low-quality | Genome-fragment | 18.26 | AAI-based (high-confidence) | 0 |
| S4_extracted_4769 | 1069 | No | NA | 2 | 0 | 1 | Not-determined | Genome-fragment | NA | NA | 0 |
| S10_extracted_116234 | 5549 | No | NA | 8 | 2 | 0 | Low-quality | Genome-fragment | 10.9 | AAI-based (high-confidence) | 0 |
| S4_extracted_4838 | 99642 | No | NA | 100 | 12 | 1 | Complete | High-quality | 100 | DTR (high-confidence) | 0 |
| S4_extracted_4887 | 12974 | No | NA | 6 | 0 | 5 | Not-determined | Genome-fragment | NA | NA | 0 |
| S4_extracted_4891 | 43130 | No | NA | 71 | 11 | 1 | Medium-quality | Genome-fragment | 64.14 | AAI-based (high-confidence) | 0 |
| S17_extracted_k141_116050 | 5550 | No | NA | 12 | 2 | 1 | Low-quality | Genome-fragment | 9.14 | AAI-based (high-confidence) | 0 |
| S4_extracted_5021 | 15316 | No | NA | 25 | 8 | 0 | Medium-quality | Genome-fragment | 66.65 | AAI-based (medium-confidence) | 0 |
| S4_extracted_5070 | 1266 | No | NA | 1 | 0 | 0 | Not-determined | Genome-fragment | NA | NA | 0 |
| S4_extracted_5135 | 1560 | No | NA | 2 | 0 | 0 | Not-determined | Genome-fragment | NA | NA | 0 |
| S4_extracted_5223 | 3507 | No | NA | 3 | 0 | 1 | Not-determined | Genome-fragment | NA | NA | 0 |
| S4_extracted_5315 | 1252 | No | NA | 1 | 0 | 1 | Not-determined | Genome-fragment | NA | NA | 0 |
| S5_extracted_k141_59064 | 5550 | Yes | 2044 | 8 | 1 | 2 | Low-quality | Genome-fragment | 2.94 | HMM-based (lower-bound) | 63.17 |
| S4_extracted_5426 | 1014 | No | NA | 3 | 0 | 0 | Not-determined | Genome-fragment | NA | NA | 0 |
| S12_extracted_k141_8249 | 5550 | No | NA | 10 | 0 | 0 | Low-quality | Genome-fragment | 3.05 | AAI-based (high-confidence) | 0 |
| S4_extracted_5533 | 1362 | No | NA | 2 | 0 | 2 | Not-determined | Genome-fragment | NA | NA | 0 |
| S1_extracted_k141_51670 | 5551 | No | NA | 4 | 0 | 4 | Low-quality | Genome-fragment | 2.99 | AAI-based (medium-confidence) | 0 |
| S13_extracted_14458 | 5553 | No | NA | 4 | 0 | 3 | Low-quality | Genome-fragment | 8.45 | AAI-based (medium-confidence) | 0 |
| S4_extracted_5695 | 5127 | No | NA | 7 | 0 | 0 | Not-determined | Genome-fragment | NA | NA | 0 |
| S4_extracted_5704 | 1006 | No | NA | 1 | 0 | 0 | Not-determined | Genome-fragment | NA | NA | 0 |
| S8_extracted_k141_168545 | 5553 | No | NA | 4 | 0 | 2 | Low-quality | Genome-fragment | 4.8 | AAI-based (medium-confidence) | 0 |
| S11_vs_k141_72783\|\|full | 5555 | No | NA | 8 | 0 | 0 | Low-quality | Genome-fragment | 3.61 | AAI-based (high-confidence) | 0 |
| S5_extracted_k141_83224 | 5555 | No | NA | 9 | 1 | 0 | Low-quality | Genome-fragment | 3.61 | AAI-based (medium-confidence) | 0 |
| S4_extracted_6256 | 1463 | No | NA | 2 | 0 | 1 | Not-determined | Genome-fragment | NA | NA | 0 |
| S7_extracted_16073 | 5557 | No | NA | 8 | 3 | 0 | Low-quality | Genome-fragment | 9 | AAI-based (high-confidence) | 0 |
| S13_extracted_23010 | 5559 | No | NA | 11 | 4 | 0 | Low-quality | Genome-fragment | 12.28 | AAI-based (high-confidence) | 0 |
| S13_vs_10400\|\|full | 5559 | No | NA | 10 | 0 | 0 | Low-quality | Genome-fragment | 10.05 | AAI-based (medium-confidence) | 0 |
| S4_extracted_6616 | 1360 | No | NA | 1 | 0 | 1 | Not-determined | Genome-fragment | NA | NA | 0 |
| S1_vs_k141_31601\|\|full | 5560 | No | NA | 9 | 4 | 0 | Low-quality | Genome-fragment | 14.92 | AAI-based (high-confidence) | 0 |
| S16_vs_27679\|\|full | 5560 | No | NA | 3 | 0 | 2 | Low-quality | Genome-fragment | 4.15 | AAI-based (medium-confidence) | 0 |
| S16_vs_116757\|\|full | 5560 | No | NA | 6 | 2 | 0 | Low-quality | Genome-fragment | 13.4 | AAI-based (high-confidence) | 0 |
| S4_extracted_6713 | 1423 | No | NA | 3 | 0 | 0 | Not-determined | Genome-fragment | NA | NA | 0 |
| S12_vs_k141_111478\|\|full | 5560 | Yes | 4142 | 7 | 2 | 2 | Low-quality | Genome-fragment | 9.1 | AAI-based (high-confidence) | 25.5 |
| S13_extracted_101278 | 5561 | No | NA | 9 | 1 | 0 | Low-quality | Genome-fragment | 11.92 | AAI-based (medium-confidence) | 0 |
| S4_extracted_6854 | 1963 | No | NA | 1 | 0 | 1 | Not-determined | Genome-fragment | NA | NA | 0 |
| S4_extracted_6899 | 2083 | No | NA | 2 | 0 | 0 | Not-determined | Genome-fragment | NA | NA | 0 |
| S4_extracted_6904 | 1007 | No | NA | 2 | 0 | 2 | Not-determined | Genome-fragment | NA | NA | 0 |
| S12_vs_k141_28168\|\|full | 5561 | Yes | 3104 | 10 | 1 | 1 | Low-quality | Genome-fragment | 8.1 | AAI-based (high-confidence) | 44.18 |
| S10_vs_68814\|\|full | 5563 | No | NA | 4 | 1 | 0 | Low-quality | Genome-fragment | 11.11 | AAI-based (medium-confidence) | 0 |
| S4_extracted_7129 | 1001 | No | NA | 1 | 0 | 0 | Not-determined | Genome-fragment | NA | NA | 0 |
| S8_extracted_k141_95536 | 5563 | No | NA | 5 | 0 | 4 | Low-quality | Genome-fragment | 4.95 | AAI-based (medium-confidence) | 0 |
| S16_extracted_16094 | 5564 | No | NA | 9 | 8 | 0 | Low-quality | Genome-fragment | 18.37 | AAI-based (high-confidence) | 0 |
| S16_extracted_149543 | 5564 | No | NA | 5 | 0 | 4 | Low-quality | Genome-fragment | 14.53 | AAI-based (medium-confidence) | 0 |
| S3_vs_k141_48682\|\|full | 5565 | Yes | 1426 | 10 | 2 | 2 | Low-quality | Genome-fragment | 2.37 | AAI-based (high-confidence) | 74.38 |
| S13_vs_129220\|\|full | 5566 | No | NA | 14 | 0 | 0 | Low-quality | Genome-fragment | 12.41 | AAI-based (high-confidence) | 0 |
| S1_extracted_k141_80925 | 5567 | No | NA | 7 | 0 | 2 | Low-quality | Genome-fragment | 10.58 | AAI-based (high-confidence) | 0 |
| S4_extracted_7541 | 3185 | No | NA | 3 | 0 | 0 | Not-determined | Genome-fragment | NA | NA | 0 |
| S4_extracted_7629 | 1114 | No | NA | 1 | 0 | 0 | Not-determined | Genome-fragment | NA | NA | 0 |
| S8_extracted_k141_1292 | 5568 | Yes | 4536 | 8 | 1 | 2 | Low-quality | Genome-fragment | 8.46 | HMM-based (lower-bound) | 18.53 |
| S14_vs_k141_54431\|\|full | 5571 | No | NA | 11 | 1 | 0 | Low-quality | Genome-fragment | 10.18 | AAI-based (high-confidence) | 0 |
| S4_extracted_7734 | 1396 | No | NA | 2 | 0 | 1 | Not-determined | Genome-fragment | NA | NA | 0 |
| S4_extracted_7828 | 1031 | No | NA | 2 | 0 | 0 | Not-determined | Genome-fragment | NA | NA | 0 |
| S1_vs_k141_114341\|\|full | 5572 | No | NA | 4 | 0 | 3 | Low-quality | Genome-fragment | 2.63 | AAI-based (medium-confidence) | 0 |
| S15_extracted_35135 | 5574 | No | NA | 6 | 2 | 0 | Low-quality | Genome-fragment | 11.54 | AAI-based (high-confidence) | 0 |
| S9_vs_k141_28505\|\|full | 5576 | No | NA | 10 | 6 | 0 | Low-quality | Genome-fragment | 14.73 | AAI-based (medium-confidence) | 0 |
| S8_extracted_k141_18750 | 5577 | No | NA | 8 | 6 | 0 | Low-quality | Genome-fragment | 17.18 | AAI-based (high-confidence) | 0 |
| S3_extracted_k141_21194 | 5578 | No | NA | 5 | 0 | 5 | Low-quality | Genome-fragment | 2.53 | AAI-based (high-confidence) | 0 |
| S4_extracted_7965 | 1203 | No | NA | 2 | 0 | 0 | Not-determined | Genome-fragment | NA | NA | 0 |
| S4_extracted_7996 | 1210 | No | NA | 2 | 0 | 2 | Not-determined | Genome-fragment | NA | NA | 0 |
| S4_extracted_8018 | 3920 | No | NA | 4 | 0 | 3 | Not-determined | Genome-fragment | NA | NA | 0 |
| S3_vs_k141_13616\|\|full | 5578 | Yes | 2378 | 9 | 2 | 1 | Low-quality | Genome-fragment | 7.15 | AAI-based (high-confidence) | 57.37 |
| S5_vs_k141_143919\|\|full | 5578 | Yes | 4936 | 5 | 1 | 1 | Low-quality | Genome-fragment | 14.15 | AAI-based (high-confidence) | 11.51 |
| S4_extracted_8231 | 1247 | No | NA | 3 | 0 | 1 | Not-determined | Genome-fragment | NA | NA | 0 |
| S4_extracted_8239 | 1060 | No | NA | 2 | 0 | 1 | Not-determined | Genome-fragment | NA | NA | 0 |
| S5_extracted_k141_52531 | 5579 | No | NA | 13 | 1 | 0 | Low-quality | Genome-fragment | 13.23 | AAI-based (high-confidence) | 0 |
| S8_extracted_k141_154123 | 5580 | No | NA | 13 | 1 | 1 | Low-quality | Genome-fragment | 17.53 | AAI-based (high-confidence) | 0 |
| S14_extracted_k141_102870 | 5581 | No | NA | 8 | 3 | 0 | Low-quality | Genome-fragment | 15.02 | AAI-based (high-confidence) | 0 |
| S3_extracted_k141_109213 | 5581 | No | NA | 8 | 1 | 0 | Low-quality | Genome-fragment | 4.17 | AAI-based (high-confidence) | 0 |
| S4_extracted_8448 | 3494 | No | NA | 5 | 0 | 4 | Not-determined | Genome-fragment | NA | NA | 0 |
| S4_extracted_8474 | 11599 | No | NA | 9 | 0 | 6 | Not-determined | Genome-fragment | NA | NA | 0 |
| S17_extracted_k141_103703 | 5583 | No | NA | 9 | 0 | 1 | Low-quality | Genome-fragment | 3.05 | AAI-based (high-confidence) | 0 |
| S1_vs_k141_58904\|\|full | 5583 | No | NA | 14 | 3 | 0 | Low-quality | Genome-fragment | 9.12 | AAI-based (high-confidence) | 0 |
| S4_extracted_8497 | 2180 | No | NA | 2 | 0 | 1 | Not-determined | Genome-fragment | NA | NA | 0 |
| S12_vs_k141_87914\|\|full | 5583 | No | NA | 5 | 0 | 3 | Low-quality | Genome-fragment | 7.63 | AAI-based (medium-confidence) | 0 |
| S8_extracted_k141_1164 | 5584 | No | NA | 7 | 1 | 0 | Low-quality | Genome-fragment | 5.59 | AAI-based (high-confidence) | 0 |
| S5_extracted_k141_17967 | 5585 | No | NA | 15 | 8 | 0 | Low-quality | Genome-fragment | 25.48 | AAI-based (high-confidence) | 0 |
| S7_extracted_38339 | 5586 | No | NA | 7 | 0 | 0 | Low-quality | Genome-fragment | 8.51 | AAI-based (medium-confidence) | 0 |
| S3_extracted_k141_101039 | 5587 | No | NA | 1 | 0 | 0 | Low-quality | Genome-fragment | 6.27 | AAI-based (high-confidence) | 0 |
| S8_extracted_k141_104224 | 5587 | No | NA | 8 | 0 | 1 | Low-quality | Genome-fragment | 13.78 | AAI-based (high-confidence) | 0 |
| S4_extracted_8998 | 2009 | No | NA | 1 | 0 | 1 | Not-determined | Genome-fragment | NA | NA | 0 |
| S4_extracted_9037 | 31165 | No | NA | 14 | 0 | 12 | Not-determined | Genome-fragment | NA | NA | 0 |
| S4_extracted_9039 | 1146 | No | NA | 3 | 0 | 0 | Not-determined | Genome-fragment | NA | NA | 0 |
| S11_extracted_k141_44538 | 5594 | No | NA | 12 | 1 | 0 | Low-quality | Genome-fragment | 8.97 | HMM-based (lower-bound) | 0 |
| S16_vs_29959\|\|full | 5594 | Yes | 2152 | 8 | 1 | 3 | Low-quality | Genome-fragment | 2.49 | HMM-based (lower-bound) | 61.53 |
| S11_extracted_k141_48072 | 5598 | No | NA | 11 | 0 | 0 | Low-quality | Genome-fragment | 7.32 | AAI-based (high-confidence) | 0 |
| S5_extracted_k141_112989 | 5600 | No | NA | 6 | 0 | 3 | Low-quality | Genome-fragment | 3.93 | AAI-based (medium-confidence) | 0 |
| S16_extracted_77848 | 5601 | No | NA | 10 | 4 | 0 | Low-quality | Genome-fragment | 5.1 | AAI-based (high-confidence) | 0 |
| S4_extracted_9530 | 1867 | No | NA | 2 | 0 | 0 | Not-determined | Genome-fragment | NA | NA | 0 |
| S4_extracted_9637 | 1497 | No | NA | 1 | 0 | 1 | Not-determined | Genome-fragment | NA | NA | 0 |
| S4_extracted_9651 | 2264 | No | NA | 3 | 0 | 1 | Not-determined | Genome-fragment | NA | NA | 0 |
| S14_extracted_k141_41367 | 5602 | No | NA | 7 | 0 | 0 | Low-quality | Genome-fragment | 3.92 | AAI-based (medium-confidence) | 0 |
| S10_extracted_104654 | 5604 | No | NA | 11 | 4 | 0 | Low-quality | Genome-fragment | 13.91 | AAI-based (medium-confidence) | 0 |
| S4_extracted_15842 | 5606 | No | NA | 13 | 2 | 0 | Low-quality | Genome-fragment | 14.01 | AAI-based (high-confidence) | 0 |
| S4_extracted_9994 | 1582 | No | NA | 2 | 0 | 2 | Not-determined | Genome-fragment | NA | NA | 0 |
| S3_vs_k141_115502\|\|full | 5608 | No | NA | 5 | 0 | 4 | Low-quality | Genome-fragment | 12.72 | AAI-based (medium-confidence) | 0 |
| S4_extracted_10200 | 2315 | No | NA | 3 | 0 | 0 | Not-determined | Genome-fragment | NA | NA | 0 |
| S4_extracted_10252 | 1620 | No | NA | 2 | 0 | 1 | Not-determined | Genome-fragment | NA | NA | 0 |
| S7_extracted_11278 | 5611 | No | NA | 7 | 4 | 0 | Low-quality | Genome-fragment | 11.91 | AAI-based (high-confidence) | 0 |
| S4_extracted_10378 | 1971 | No | NA | 2 | 0 | 0 | Not-determined | Genome-fragment | NA | NA | 0 |
| S12_extracted_k141_151019 | 5617 | No | NA | 6 | 0 | 1 | Low-quality | Genome-fragment | 11.73 | AAI-based (medium-confidence) | 0 |
| S5_vs_k141_134648\|\|full | 5618 | No | NA | 16 | 0 | 0 | Low-quality | Genome-fragment | 12.66 | AAI-based (high-confidence) | 0 |
| S4_extracted_10445 | 4270 | No | NA | 5 | 0 | 5 | Not-determined | Genome-fragment | NA | NA | 0 |
| S4_extracted_10506 | 50427 | No | NA | 69 | 16 | 0 | Complete | High-quality | 100 | DTR (high-confidence) | 0 |
| S16_extracted_43986 | 5621 | No | NA | 3 | 0 | 2 | Low-quality | Genome-fragment | 0.5 | AAI-based (medium-confidence) | 0 |
| S12_vs_k141_155672\|\|full | 5622 | No | NA | 7 | 0 | 1 | Low-quality | Genome-fragment | 13.05 | AAI-based (medium-confidence) | 0 |
| S10_extracted_115577 | 5626 | No | NA | 6 | 1 | 0 | Low-quality | Genome-fragment | 10.3 | AAI-based (high-confidence) | 0 |
| S4_extracted_10667 | 1782 | No | NA | 2 | 0 | 2 | Not-determined | Genome-fragment | NA | NA | 0 |
| S17_extracted_k141_70947 | 5628 | No | NA | 4 | 0 | 3 | Low-quality | Genome-fragment | 1.5 | AAI-based (high-confidence) | 0 |
| S4_extracted_10780 | 6554 | No | NA | 5 | 0 | 0 | Not-determined | Genome-fragment | NA | NA | 0 |
| S13_vs_132092\|\|full | 5633 | No | NA | 7 | 0 | 2 | Low-quality | Genome-fragment | 14.12 | AAI-based (medium-confidence) | 0 |
| S4_extracted_10843 | 1533 | No | NA | 1 | 0 | 1 | Not-determined | Genome-fragment | NA | NA | 0 |
| S4_vs_VIRSorter_10786-cat_1 | 5633 | No | NA | 5 | 4 | 0 | Low-quality | Genome-fragment | 15.69 | AAI-based (high-confidence) | 0 |
| S6_vs_k141_48906\|\|full | 5633 | No | NA | 7 | 4 | 0 | Low-quality | Genome-fragment | 10.94 | AAI-based (high-confidence) | 0 |
| S11_vs_k141_58905\|\|full | 5637 | No | NA | 8 | 0 | 1 | Low-quality | Genome-fragment | 12.82 | AAI-based (high-confidence) | 0 |
| S3_extracted_k141_96314 | 5637 | No | NA | 11 | 0 | 0 | Low-quality | Genome-fragment | 7.14 | AAI-based (high-confidence) | 0 |
| S6_vs_k141_43074\|\|full | 5638 | No | NA | 7 | 0 | 2 | Low-quality | Genome-fragment | 5.92 | AAI-based (high-confidence) | 0 |
| S4_extracted_11093 | 4004 | No | NA | 3 | 0 | 1 | Not-determined | Genome-fragment | NA | NA | 0 |
| S4_extracted_11097 | 17775 | No | NA | 8 | 0 | 7 | Not-determined | Genome-fragment | NA | NA | 0 |
| S16_extracted_47990 | 5640 | No | NA | 7 | 0 | 0 | Low-quality | Genome-fragment | 6.85 | AAI-based (high-confidence) | 0 |
| S4_extracted_11300 | 1208 | No | NA | 3 | 0 | 0 | Not-determined | Genome-fragment | NA | NA | 0 |
| S6_vs_k141_50459\|\|full | 5640 | Yes | 4747 | 8 | 1 | 1 | Low-quality | Genome-fragment | 3.65 | HMM-based (lower-bound) | 15.83 |
| S4_extracted_11308 | 32786 | No | NA | 43 | 19 | 0 | High-quality | High-quality | 92.35 | AAI-based (high-confidence) | 0 |
| S4_extracted_11311 | 3242 | No | NA | 3 | 0 | 0 | Not-determined | Genome-fragment | NA | NA | 0 |
| S16_extracted_4685 | 5641 | No | NA | 4 | 0 | 1 | Low-quality | Genome-fragment | 9.21 | AAI-based (medium-confidence) | 0 |
| S4_extracted_11438 | 1909 | No | NA | 2 | 0 | 2 | Not-determined | Genome-fragment | NA | NA | 0 |
| S4_extracted_11447 | 1701 | No | NA | 2 | 0 | 1 | Not-determined | Genome-fragment | NA | NA | 0 |
| S4_extracted_11523 | 4573 | No | NA | 8 | 0 | 2 | Not-determined | Genome-fragment | NA | NA | 0 |
| S17_vs_k141_50464\|\|full | 5643 | No | NA | 7 | 5 | 0 | Low-quality | Genome-fragment | 12.89 | AAI-based (high-confidence) | 0 |
| S4_extracted_11642 | 1936 | No | NA | 2 | 0 | 2 | Not-determined | Genome-fragment | NA | NA | 0 |
| S3_vs_k141_16256\|\|full | 5643 | No | NA | 6 | 1 | 0 | Low-quality | Genome-fragment | 12.13 | AAI-based (high-confidence) | 0 |
| S4_extracted_11736 | 2256 | No | NA | 2 | 0 | 1 | Not-determined | Genome-fragment | NA | NA | 0 |
| S4_extracted_11757 | 1020 | No | NA | 3 | 0 | 1 | Not-determined | Genome-fragment | NA | NA | 0 |
| S4_extracted_11784 | 1411 | No | NA | 1 | 0 | 0 | Not-determined | Genome-fragment | NA | NA | 0 |
| S4_extracted_11813 | 1110 | No | NA | 1 | 0 | 1 | Not-determined | Genome-fragment | NA | NA | 0 |
| S17_vs_k141_103864\|\|full | 5644 | No | NA | 6 | 0 | 0 | Low-quality | Genome-fragment | 8.59 | AAI-based (medium-confidence) | 0 |
| S4_extracted_12142 | 1072 | No | NA | 1 | 0 | 0 | Not-determined | Genome-fragment | NA | NA | 0 |
| S5_vs_k141_86027\|\|full | 5645 | No | NA | 12 | 3 | 0 | Low-quality | Genome-fragment | 10.89 | AAI-based (high-confidence) | 0 |
| S4_extracted_12386 | 2112 | No | NA | 1 | 0 | 0 | Not-determined | Genome-fragment | NA | NA | 0 |
| S8_vs_k141_199669\|\|full | 5645 | No | NA | 8 | 2 | 0 | Low-quality | Genome-fragment | 14.29 | AAI-based (high-confidence) | 0 |
| S12_vs_k141_109994\|\|full | 5649 | No | NA | 13 | 0 | 0 | Low-quality | Genome-fragment | 6.99 | AAI-based (high-confidence) | 0 |
| S1_extracted_k141_18168 | 5650 | No | NA | 13 | 2 | 0 | Low-quality | Genome-fragment | 9.93 | AAI-based (high-confidence) | 0 |
| S7_extracted_101942 | 5652 | No | NA | 11 | 1 | 0 | Low-quality | Genome-fragment | 8.78 | HMM-based (lower-bound) | 0 |
| S4_extracted_12600 | 1862 | No | NA | 3 | 0 | 3 | Not-determined | Genome-fragment | NA | NA | 0 |
| S4_extracted_12614 | 44016 | No | NA | 81 | 13 | 3 | Medium-quality | Genome-fragment | 65 | AAI-based (high-confidence) | 0 |
| S4_extracted_12624 | 16529 | No | NA | 25 | 7 | 0 | High-quality | High-quality | 96.9 | AAI-based (high-confidence) | 0 |
| S16_extracted_119685 | 5652 | No | NA | 11 | 1 | 0 | Low-quality | Genome-fragment | 2.77 | HMM-based (lower-bound) | 0 |
| S4_extracted_12936 | 1113 | No | NA | 3 | 0 | 0 | Not-determined | Genome-fragment | NA | NA | 0 |
| S4_extracted_13014 | 7448 | No | NA | 5 | 0 | 4 | Not-determined | Genome-fragment | NA | NA | 0 |
| S7_extracted_9392 | 5653 | No | NA | 3 | 0 | 0 | Low-quality | Genome-fragment | 5.42 | AAI-based (medium-confidence) | 0 |
| S4_extracted_13195 | 1212 | No | NA | 2 | 0 | 0 | Not-determined | Genome-fragment | NA | NA | 0 |
| S3_extracted_k141_67596 | 5655 | Yes | 4899 | 9 | 2 | 1 | Low-quality | Genome-fragment | 3.2 | AAI-based (high-confidence) | 13.37 |
| S3_vs_k141_20683\|\|full | 5656 | No | NA | 7 | 1 | 0 | Low-quality | Genome-fragment | 11.51 | AAI-based (medium-confidence) | 0 |
| S4_extracted_13353 | 49534 | No | NA | 79 | 25 | 0 | High-quality | High-quality | 100 | AAI-based (high-confidence) | 0 |
| S11_vs_k141_65200\|\|full | 5658 | No | NA | 7 | 0 | 0 | Low-quality | Genome-fragment | 10.84 | AAI-based (medium-confidence) | 0 |
| S4_extracted_13395 | 21440 | No | NA | 16 | 0 | 9 | Not-determined | Genome-fragment | NA | NA | 0 |
| S4_extracted_13444 | 1788 | No | NA | 4 | 0 | 2 | Not-determined | Genome-fragment | NA | NA | 0 |
| S10_vs_15567\|\|full | 5658 | No | NA | 5 | 0 | 2 | Low-quality | Genome-fragment | 1.51 | AAI-based (medium-confidence) | 0 |
| S4_extracted_13612 | 3392 | No | NA | 3 | 0 | 3 | Not-determined | Genome-fragment | NA | NA | 0 |
| S4_extracted_13627 | 16749 | No | NA | 20 | 8 | 0 | Medium-quality | Genome-fragment | 74.12 | AAI-based (medium-confidence) | 0 |
| S4_extracted_13631 | 1187 | No | NA | 1 | 0 | 0 | Not-determined | Genome-fragment | NA | NA | 0 |
| S4_extracted_13682 | 1517 | No | NA | 2 | 0 | 0 | Not-determined | Genome-fragment | NA | NA | 0 |
| S5_extracted_k141_21892 | 5659 | No | NA | 10 | 0 | 2 | Low-quality | Genome-fragment | 6.31 | AAI-based (medium-confidence) | 0 |
| S4_extracted_13738 | 1136 | No | NA | 2 | 0 | 2 | Not-determined | Genome-fragment | NA | NA | 0 |
| S4_extracted_13759 | 21084 | No | NA | 28 | 15 | 0 | Medium-quality | Genome-fragment | 53.32 | AAI-based (high-confidence) | 0 |
| S4_extracted_13763 | 10813 | No | NA | 4 | 0 | 3 | Not-determined | Genome-fragment | NA | NA | 0 |
| S4_extracted_13783 | 46158 | No | NA | 54 | 0 | 51 | Not-determined | Genome-fragment | NA | NA | 0 |
| S4_extracted_13831 | 16727 | No | NA | 25 | 6 | 0 | Medium-quality | Genome-fragment | 56.3 | HMM-based (lower-bound) | 0 |
| S7_extracted_49257 | 5659 | No | NA | 7 | 4 | 0 | Low-quality | Genome-fragment | 12.12 | AAI-based (high-confidence) | 0 |
| S16_extracted_112304 | 5660 | No | NA | 6 | 0 | 5 | Low-quality | Genome-fragment | 7.95 | AAI-based (medium-confidence) | 0 |
| S4_extracted_13941 | 1749 | No | NA | 2 | 0 | 1 | Not-determined | Genome-fragment | NA | NA | 0 |
| S14_extracted_k141_103264 | 5661 | No | NA | 12 | 5 | 0 | Low-quality | Genome-fragment | 12.99 | AAI-based (high-confidence) | 0 |
| S7_vs_75657\|\|full | 5662 | No | NA | 5 | 0 | 5 | Low-quality | Genome-fragment | 12.85 | AAI-based (medium-confidence) | 0 |
| S4_extracted_14013 | 6368 | No | NA | 5 | 0 | 5 | Not-determined | Genome-fragment | NA | NA | 0 |
| S4_extracted_14033 | 1470 | No | NA | 2 | 0 | 1 | Not-determined | Genome-fragment | NA | NA | 0 |
| S4_extracted_14053 | 1229 | No | NA | 1 | 0 | 0 | Not-determined | Genome-fragment | NA | NA | 0 |
| S10_vs_48928\|\|full | 5662 | No | NA | 10 | 0 | 1 | Low-quality | Genome-fragment | 8.73 | AAI-based (high-confidence) | 0 |
| S8_extracted_k141_182098 | 5664 | No | NA | 10 | 7 | 0 | Low-quality | Genome-fragment | 8.35 | HMM-based (lower-bound) | 0 |
| S4_extracted_14146 | 4821 | No | NA | 6 | 0 | 1 | Not-determined | Genome-fragment | NA | NA | 0 |
| S17_vs_k141_118152\|\|full | 5666 | No | NA | 7 | 1 | 0 | Low-quality | Genome-fragment | 8.57 | HMM-based (lower-bound) | 0 |
| S4_extracted_14229 | 2005 | No | NA | 3 | 0 | 1 | Not-determined | Genome-fragment | NA | NA | 0 |
| S4_extracted_14255 | 1121 | No | NA | 2 | 0 | 1 | Not-determined | Genome-fragment | NA | NA | 0 |
| S4_extracted_14340 | 2887 | No | NA | 4 | 0 | 0 | Not-determined | Genome-fragment | NA | NA | 0 |
| S7_vs_132201\|\|full | 5668 | No | NA | 5 | 0 | 4 | Low-quality | Genome-fragment | 9.61 | AAI-based (medium-confidence) | 0 |
| S12_vs_k141_57462\|\|full | 5668 | No | NA | 8 | 4 | 0 | Low-quality | Genome-fragment | 4.46 | AAI-based (high-confidence) | 0 |
| S12_extracted_k141_143169 | 5669 | No | NA | 10 | 0 | 1 | Low-quality | Genome-fragment | 11.85 | AAI-based (medium-confidence) | 0 |
| S5_extracted_k141_140196 | 5670 | No | NA | 8 | 1 | 0 | Low-quality | Genome-fragment | 3.08 | HMM-based (lower-bound) | 0 |
| S4_extracted_14623 | 1067 | No | NA | 1 | 0 | 0 | Not-determined | Genome-fragment | NA | NA | 0 |
| S6_extracted_k141_51287 | 5672 | No | NA | 12 | 4 | 1 | Low-quality | Genome-fragment | 13.44 | AAI-based (high-confidence) | 0 |
| S4_extracted_14772 | 7354 | No | NA | 4 | 0 | 4 | Not-determined | Genome-fragment | NA | NA | 0 |
| S4_extracted_14806 | 1023 | No | NA | 3 | 0 | 1 | Not-determined | Genome-fragment | NA | NA | 0 |
| S4_extracted_14876 | 1886 | No | NA | 1 | 0 | 1 | Not-determined | Genome-fragment | NA | NA | 0 |
| S4_extracted_14924 | 1823 | No | NA | 2 | 0 | 0 | Not-determined | Genome-fragment | NA | NA | 0 |
| S3_vs_k141_33225\|\|full | 5674 | No | NA | 6 | 2 | 0 | Low-quality | Genome-fragment | 12.56 | AAI-based (medium-confidence) | 0 |
| S4_extracted_15016 | 1975 | No | NA | 3 | 0 | 1 | Not-determined | Genome-fragment | NA | NA | 0 |
| S4_extracted_15031 | 4345 | No | NA | 4 | 0 | 0 | Not-determined | Genome-fragment | NA | NA | 0 |
| S4_extracted_15054 | 1398 | No | NA | 2 | 0 | 2 | Not-determined | Genome-fragment | NA | NA | 0 |
| S7_vs_49537\|\|full | 5675 | No | NA | 12 | 2 | 1 | Low-quality | Genome-fragment | 5.4 | HMM-based (lower-bound) | 0 |
| S4_extracted_15141 | 1224 | No | NA | 2 | 0 | 1 | Not-determined | Genome-fragment | NA | NA | 0 |
| S4_extracted_15186 | 8353 | No | NA | 4 | 0 | 4 | Not-determined | Genome-fragment | NA | NA | 0 |
| S3_vs_k141_85864\|\|full | 5676 | No | NA | 14 | 0 | 0 | Low-quality | Genome-fragment | 3 | AAI-based (high-confidence) | 0 |
| S8_extracted_k141_99704 | 5676 | No | NA | 7 | 4 | 0 | Low-quality | Genome-fragment | 17.51 | AAI-based (high-confidence) | 0 |
| S8_vs_k141_26319\|\|full | 5676 | No | NA | 12 | 4 | 0 | Low-quality | Genome-fragment | 12.16 | AAI-based (high-confidence) | 0 |
| S16_vs_70662\|\|full | 5678 | No | NA | 4 | 0 | 4 | Low-quality | Genome-fragment | 1.32 | AAI-based (medium-confidence) | 0 |
| S12_vs_k141_136959\|\|full | 5678 | No | NA | 4 | 3 | 0 | Low-quality | Genome-fragment | 15.46 | AAI-based (high-confidence) | 0 |
| S10_vs_120255\|\|full | 5679 | No | NA | 13 | 1 | 0 | Low-quality | Genome-fragment | 11.6 | AAI-based (high-confidence) | 0 |
| S10_extracted_63153 | 5680 | No | NA | 12 | 1 | 0 | Low-quality | Genome-fragment | 13.48 | AAI-based (high-confidence) | 0 |
| S8_vs_k141_183547\|\|full | 5680 | No | NA | 8 | 3 | 0 | Low-quality | Genome-fragment | 7.6 | HMM-based (lower-bound) | 0 |
| S4_extracted_16093 | 5001 | No | NA | 3 | 0 | 3 | Not-determined | Genome-fragment | NA | NA | 0 |
| S4_extracted_16167 | 2190 | No | NA | 2 | 0 | 0 | Not-determined | Genome-fragment | NA | NA | 0 |
| S3_vs_k141_85361\|\|full | 5683 | No | NA | 7 | 3 | 0 | Low-quality | Genome-fragment | 3.46 | AAI-based (high-confidence) | 0 |
| S4_extracted_16226 | 1589 | No | NA | 2 | 0 | 1 | Not-determined | Genome-fragment | NA | NA | 0 |
| S4_extracted_16239 | 1045 | No | NA | 1 | 0 | 1 | Not-determined | Genome-fragment | NA | NA | 0 |
| S2_vs_k141_15731\|\|full | 5683 | No | NA | 5 | 0 | 2 | Low-quality | Genome-fragment | 4.8 | AAI-based (high-confidence) | 0 |
| S17_extracted_k141_66803 | 5685 | No | NA | 9 | 1 | 0 | Low-quality | Genome-fragment | 12.06 | AAI-based (high-confidence) | 0 |
| S4_extracted_16411 | 1185 | No | NA | 1 | 0 | 0 | Not-determined | Genome-fragment | NA | NA | 0 |
| S10_vs_63296\|\|full | 5685 | No | NA | 3 | 0 | 0 | Low-quality | Genome-fragment | 11.75 | AAI-based (high-confidence) | 0 |
| S4_extracted_16485 | 12964 | No | NA | 19 | 4 | 0 | High-quality | High-quality | 94.97 | AAI-based (medium-confidence) | 0 |
| S4_extracted_16518 | 1496 | No | NA | 1 | 0 | 0 | Not-determined | Genome-fragment | NA | NA | 0 |
| S4_extracted_16607 | 2735 | No | NA | 2 | 0 | 1 | Not-determined | Genome-fragment | NA | NA | 0 |
| S4_extracted_16707 | 1158 | No | NA | 2 | 0 | 0 | Not-determined | Genome-fragment | NA | NA | 0 |
| S4_extracted_16751 | 3765 | No | NA | 6 | 0 | 0 | Not-determined | Genome-fragment | NA | NA | 0 |
| S4_extracted_16770 | 6612 | No | NA | 5 | 0 | 5 | Not-determined | Genome-fragment | NA | NA | 0 |
| S4_extracted_16926 | 1005 | No | NA | 1 | 0 | 1 | Not-determined | Genome-fragment | NA | NA | 0 |
| S4_extracted_16943 | 1693 | No | NA | 3 | 0 | 2 | Not-determined | Genome-fragment | NA | NA | 0 |
| S4_extracted_17150 | 2027 | No | NA | 1 | 0 | 0 | Not-determined | Genome-fragment | NA | NA | 0 |
| S3_vs_k141_4752\|\|full | 5686 | No | NA | 7 | 5 | 0 | Low-quality | Genome-fragment | 14.88 | AAI-based (medium-confidence) | 0 |
| S4_extracted_17180 | 1139 | No | NA | 2 | 0 | 2 | Not-determined | Genome-fragment | NA | NA | 0 |
| S7_vs_110926\|\|full | 5688 | No | NA | 6 | 0 | 0 | Low-quality | Genome-fragment | 16.63 | AAI-based (high-confidence) | 0 |

| | | | | | | | | | | | | |
|---|---|---|---|---|---|---|---|---|---|---|---|---|
| S5_vs_k141_64019\|\|full | 5689 | No | NA | | 12 | 6 | 0 | Low-quality | Genome-fragment | 16.28 | AAI-based (high-confidence) | 0 |
| S10_vs_50036\|\|full | 5692 | No | NA | | 9 | 6 | 0 | Low-quality | Genome-fragment | 15.23 | AAI-based (high-confidence) | 0 |
| S4_extracted_17711 | 3842 | No | NA | | 3 | 0 | 2 | Not-determined | Genome-fragment | NA | NA | 0 |
| S4_extracted_14419 | 5693 | No | NA | | 12 | 2 | 0 | Low-quality | Genome-fragment | 11.91 | AAI-based (high-confidence) | 0 |
| S4_extracted_17948 | 6590 | No | NA | | 7 | 3 | 0 | High-quality | High-quality | 96.57 | AAI-based (high-confidence) | 0 |
| S7_extracted_119052 | 5698 | No | NA | | 7 | 0 | 1 | Low-quality | Genome-fragment | 7.04 | AAI-based (medium-confidence) | 0 |
| S4_extracted_18022 | 1415 | No | NA | | 3 | 0 | 2 | Not-determined | Genome-fragment | NA | NA | 0 |
| S4_extracted_18094 | 1574 | No | NA | | 1 | 0 | 0 | Not-determined | Genome-fragment | NA | NA | 0 |
| S12_extracted_k141_92952 | 5698 | No | NA | | 2 | 1 | 0 | Low-quality | Genome-fragment | 8.96 | AAI-based (high-confidence) | 0 |
| S8_extracted_k141_177316 | 5700 | No | NA | | 8 | 7 | 0 | Low-quality | Genome-fragment | 18.8 | AAI-based (high-confidence) | 0 |
| S2_extracted_k141_6520 | 5701 | No | NA | | 8 | 1 | 0 | Low-quality | Genome-fragment | 5.94 | AAI-based (high-confidence) | 0 |
| S3_extracted_k141_69384 | 5702 | No | NA | | 10 | 4 | 0 | Low-quality | Genome-fragment | 4.75 | AAI-based (high-confidence) | 0 |
| S4_extracted_18184 | 1113 | No | NA | | 1 | 0 | 1 | Not-determined | Genome-fragment | NA | NA | 0 |
| S4_extracted_18266 | 10177 | No | NA | | 6 | 0 | 6 | Not-determined | Genome-fragment | NA | NA | 0 |
| S16_extracted_8862 | 5702 | No | NA | | 4 | 0 | 2 | Low-quality | Genome-fragment | 2.85 | AAI-based (medium-confidence) | 0 |
| S16_vs_32716\|\|full | 5702 | No | NA | | 6 | 6 | 0 | Low-quality | Genome-fragment | 11.16 | AAI-based (high-confidence) | 0 |
| S4_extracted_18392 | 1422 | No | NA | | 3 | 0 | 0 | Not-determined | Genome-fragment | NA | NA | 0 |
| S4_extracted_18437 | 1244 | No | NA | | 2 | 0 | 2 | Not-determined | Genome-fragment | NA | NA | 0 |
| S14_extracted_k141_819 | 5703 | No | NA | | 8 | 8 | 0 | Low-quality | Genome-fragment | 18.81 | AAI-based (high-confidence) | 0 |
| S4_extracted_18512 | 1485 | No | NA | | 1 | 0 | 0 | Not-determined | Genome-fragment | NA | NA | 0 |
| S15_extracted_37417 | 5703 | No | NA | | 2 | 0 | 0 | Low-quality | Genome-fragment | 5.62 | AAI-based (high-confidence) | 0 |
| S7_vs_78496\|\|full | 5704 | No | NA | | 7 | 0 | 3 | Low-quality | Genome-fragment | 4.77 | AAI-based (medium-confidence) | 0 |
| S4_extracted_18569 | 4402 | No | NA | | 3 | 0 | 0 | Not-determined | Genome-fragment | NA | NA | 0 |
| S4_extracted_18772 | 1592 | No | NA | | 1 | 0 | 0 | Not-determined | Genome-fragment | NA | NA | 0 |
| S4_extracted_18789 | 1381 | No | NA | | 1 | 0 | 0 | Not-determined | Genome-fragment | NA | NA | 0 |
| S16_vs_144798\|\|full | 5705 | No | NA | | 9 | 5 | 0 | Low-quality | Genome-fragment | 15.38 | AAI-based (high-confidence) | 0 |
| S4_extracted_18803 | 28435 | No | NA | | 28 | 13 | 0 | Medium-quality | Genome-fragment | 69.23 | AAI-based (high-confidence) | 0 |
| S7_extracted_15547 | 5708 | Yes | | 3727 | 9 | 1 | 1 | Low-quality | Genome-fragment | 3.37 | AAI-based (high-confidence) | 34.71 |
| S4_extracted_19014 | 1161 | No | NA | | 1 | 0 | 0 | Not-determined | Genome-fragment | NA | NA | 0 |
| S8_vs_k141_45192\|\|full | 5708 | Yes | | 3377 | 10 | 1 | 1 | Low-quality | Genome-fragment | 9.85 | AAI-based (high-confidence) | 40.84 |
| S4_extracted_19248 | 1898 | No | NA | | 4 | 0 | 2 | Not-determined | Genome-fragment | NA | NA | 0 |
| S4_extracted_19253 | 1249 | No | NA | | 1 | 0 | 0 | Not-determined | Genome-fragment | NA | NA | 0 |
| S7_vs_100021\|\|full | 5710 | No | NA | | 15 | 5 | 0 | Low-quality | Genome-fragment | 15.74 | AAI-based (high-confidence) | 0 |
| S4_extracted_19282 | 1848 | No | NA | | 2 | 0 | 2 | Not-determined | Genome-fragment | NA | NA | 0 |
| S15_vs_56979\|\|full | 5710 | No | NA | | 10 | 1 | 1 | Low-quality | Genome-fragment | 10.92 | AAI-based (high-confidence) | 0 |
| S1_vs_k141_72311\|\|full | 5716 | No | NA | | 10 | 3 | 0 | Low-quality | Genome-fragment | 13.96 | AAI-based (medium-confidence) | 0 |
| S3_vs_k141_36098\|\|full | 5717 | No | NA | | 14 | 1 | 0 | Low-quality | Genome-fragment | 13.34 | AAI-based (high-confidence) | 0 |
| S9_vs_k141_89695\|\|full | 5718 | No | NA | | 6 | 3 | 0 | Low-quality | Genome-fragment | 14.1 | AAI-based (high-confidence) | 0 |
| S4_extracted_19785 | 2124 | No | NA | | 3 | 0 | 2 | Not-determined | Genome-fragment | NA | NA | 0 |
| S12_vs_k141_176170\|\|full | 5718 | No | NA | | 14 | 0 | 0 | Low-quality | Genome-fragment | 13.31 | AAI-based (high-confidence) | 0 |
| S11_vs_k141_45782\|\|full | 5720 | No | NA | | 4 | 1 | 0 | Low-quality | Genome-fragment | 2.95 | HMM-based (lower-bound) | 0 |
| S4_extracted_20038 | 3356 | No | NA | | 4 | 0 | 3 | Not-determined | Genome-fragment | NA | NA | 0 |
| S5_vs_k141_117601\|\|full | 5722 | No | NA | | 9 | 0 | 2 | Low-quality | Genome-fragment | 9.27 | AAI-based (high-confidence) | 0 |
| S12_extracted_k141_89572 | 5722 | No | NA | | 11 | 1 | 1 | Low-quality | Genome-fragment | 4.92 | HMM-based (lower-bound) | 0 |
| S4_extracted_20323 | 1092 | No | NA | | 1 | 0 | 0 | Not-determined | Genome-fragment | NA | NA | 0 |
| S11_vs_k141_76094\|\|full | 5723 | No | NA | | 12 | 1 | 0 | Low-quality | Genome-fragment | 9.32 | AAI-based (medium-confidence) | 0 |
| S4_extracted_20440 | 63073 | No | NA | | 94 | 16 | 3 | Medium-quality | Genome-fragment | 83.01 | AAI-based (high-confidence) | 0 |
| S4_extracted_20603 | 1931 | No | NA | | 2 | 0 | 2 | Not-determined | Genome-fragment | NA | NA | 0 |
| S17_vs_k141_91118\|\|full | 5725 | No | NA | | 8 | 2 | 0 | Low-quality | Genome-fragment | 7.1 | AAI-based (high-confidence) | 0 |
| S7_vs_115418\|\|full | 5726 | No | NA | | 7 | 1 | 0 | Low-quality | Genome-fragment | 15.15 | AAI-based (high-confidence) | 0 |
| S14_extracted_k141_113773 | 5728 | No | NA | | 6 | 4 | 0 | Low-quality | Genome-fragment | 12.7 | AAI-based (high-confidence) | 0 |
| S4_extracted_20965 | 7434 | No | NA | | 6 | 0 | 5 | Not-determined | Genome-fragment | NA | NA | 0 |
| S4_extracted_20981 | 1979 | No | NA | | 2 | 0 | 1 | Not-determined | Genome-fragment | NA | NA | 0 |
| S5_vs_k141_77757\|\|full | 5729 | No | NA | | 9 | 1 | 1 | Low-quality | Genome-fragment | 3.6 | HMM-based (lower-bound) | 0 |
| S7_vs_11070\|\|full | 5729 | No | NA | | 12 | 3 | 0 | Low-quality | Genome-fragment | 3.66 | HMM-based (lower-bound) | 0 |
| S15_extracted_77345 | 5731 | No | NA | | 11 | 1 | 0 | Low-quality | Genome-fragment | 14.03 | AAI-based (high-confidence) | 0 |
| S4_extracted_21586 | 4178 | No | NA | | 6 | 0 | 1 | Not-determined | Genome-fragment | NA | NA | 0 |
| S4_extracted_21744 | 20906 | No | NA | | 9 | 0 | 8 | Not-determined | Genome-fragment | NA | NA | 0 |
| S4_extracted_21762 | 3113 | No | NA | | 3 | 0 | 2 | Not-determined | Genome-fragment | NA | NA | 0 |
| S4_extracted_21768 | 1783 | No | NA | | 3 | 0 | 0 | Not-determined | Genome-fragment | NA | NA | 0 |
| S8_extracted_k141_63886 | 5732 | No | NA | | 9 | 0 | 0 | Low-quality | Genome-fragment | 15.3 | AAI-based (medium-confidence) | 0 |
| S5_vs_k141_94328\|\|full | 5738 | No | NA | | 13 | 1 | 0 | Low-quality | Genome-fragment | 11.39 | AAI-based (medium-confidence) | 0 |
| S6_vs_k141_41579\|\|full | 5741 | No | NA | | 11 | 2 | 0 | Low-quality | Genome-fragment | 3.64 | HMM-based (lower-bound) | 0 |
| S10_vs_50469\|\|full | 5742 | Yes | | 3124 | 9 | 1 | 3 | Low-quality | Genome-fragment | 1.96 | HMM-based (lower-bound) | 45.59 |
| S4_extracted_22153 | 1099 | No | NA | | 1 | 0 | 1 | Not-determined | Genome-fragment | NA | NA | 0 |
| S11_vs_k141_15792\|\|full | 5744 | No | NA | | 6 | 0 | 5 | Low-quality | Genome-fragment | 3.75 | AAI-based (medium-confidence) | 0 |
| S13_vs_76515\|\|full | 5744 | No | NA | | 3 | 0 | 3 | Low-quality | Genome-fragment | 5.33 | AAI-based (medium-confidence) | 0 |
| S16_vs_169872\|\|full | 5744 | No | NA | | 3 | 0 | 1 | Low-quality | Genome-fragment | 5.72 | AAI-based (medium-confidence) | 0 |
| S4_extracted_22219 | 84867 | No | NA | | 108 | 24 | 8 | Complete | High-quality | 100 | DTR (high-confidence) | 0 |
| S16_vs_145523\|\|full | 5745 | No | NA | | 9 | 6 | 0 | Low-quality | Genome-fragment | 15.93 | AAI-based (medium-confidence) | 0 |
| S4_extracted_22303 | 3165 | No | NA | | 4 | 0 | 1 | Not-determined | Genome-fragment | NA | NA | 0 |
| S7_vs_27895\|\|full | 5746 | No | NA | | 11 | 9 | 0 | Low-quality | Genome-fragment | 19.68 | AAI-based (high-confidence) | 0 |
| S4_extracted_22447 | 1208 | No | NA | | 3 | 0 | 1 | Not-determined | Genome-fragment | NA | NA | 0 |
| S4_extracted_22489 | 7733 | No | NA | | 5 | 0 | 4 | Not-determined | Genome-fragment | NA | NA | 0 |
| S4_extracted_22580 | 6003 | No | NA | | 5 | 0 | 3 | Not-determined | Genome-fragment | NA | NA | 0 |
| S4_extracted_22612 | 3223 | No | NA | | 2 | 0 | 2 | Not-determined | Genome-fragment | NA | NA | 0 |
| S4_extracted_22669 | 1199 | No | NA | | 1 | 0 | 1 | Not-determined | Genome-fragment | NA | NA | 0 |
| S11_vs_k141_22583\|\|full | 5747 | No | NA | | 8 | 1 | 0 | Low-quality | Genome-fragment | 3.47 | AAI-based (high-confidence) | 0 |
| S4_extracted_22707 | 3106 | No | NA | | 3 | 0 | 0 | Not-determined | Genome-fragment | NA | NA | 0 |
| S4_extracted_22760 | 3056 | No | NA | | 1 | 0 | 1 | Not-determined | Genome-fragment | NA | NA | 0 |
| S4_extracted_22860 | 16230 | No | NA | | 8 | 0 | 7 | Not-determined | Genome-fragment | NA | NA | 0 |
| S4_extracted_22952 | 4568 | No | NA | | 5 | 0 | 0 | Not-determined | Genome-fragment | NA | NA | 0 |
| S14_vs_k141_44819\|\|full | 5748 | No | NA | | 12 | 1 | 0 | Low-quality | Genome-fragment | 8.95 | HMM-based (lower-bound) | 0 |
| S5_extracted_k141_48880 | 5748 | No | NA | | 6 | 3 | 0 | Low-quality | Genome-fragment | 26.23 | AAI-based (high-confidence) | 0 |
| S4_extracted_23078 | 2836 | No | NA | | 4 | 0 | 1 | Not-determined | Genome-fragment | NA | NA | 0 |
| S4_extracted_23104 | 2522 | No | NA | | 10 | 0 | 0 | Not-determined | Genome-fragment | NA | NA | 0 |
| S3_extracted_k141_56753 | 5749 | No | NA | | 8 | 6 | 0 | Low-quality | Genome-fragment | 14.92 | AAI-based (high-confidence) | 0 |
| S4_extracted_23222 | 4887 | No | NA | | 6 | 0 | 5 | Not-determined | Genome-fragment | NA | NA | 0 |
| S4_extracted_23307 | 4861 | No | NA | | 6 | 0 | 1 | Not-determined | Genome-fragment | NA | NA | 0 |
| S8_vs_k141_184253\|\|full | 5749 | No | NA | | 6 | 6 | 0 | Low-quality | Genome-fragment | 16.64 | AAI-based (high-confidence) | 0 |
| S4_extracted_23356 | 4622 | No | NA | | 13 | 0 | 0 | Not-determined | Genome-fragment | NA | NA | 0 |
| S14_vs_k141_18319\|\|full | 5750 | No | NA | | 12 | 3 | 0 | Low-quality | Genome-fragment | 9.17 | AAI-based (medium-confidence) | 0 |
| S4_extracted_23576 | 12765 | No | NA | | 10 | 0 | 7 | Not-determined | Genome-fragment | NA | NA | 0 |
| S4_extracted_23657 | 2096 | No | NA | | 4 | 0 | 1 | Not-determined | Genome-fragment | NA | NA | 0 |
| S4_extracted_23691 | 1161 | No | NA | | 1 | 0 | 1 | Not-determined | Genome-fragment | NA | NA | 0 |
| S1_extracted_k141_26096 | 5750 | No | NA | | 2 | 0 | 0 | Low-quality | Genome-fragment | 4.15 | AAI-based (high-confidence) | 0 |
| S6_extracted_k141_10561 | 5751 | No | NA | | 6 | 4 | 0 | Low-quality | Genome-fragment | 9.51 | AAI-based (high-confidence) | 0 |
| S6_vs_k141_35183\|\|full | 5751 | No | NA | | 14 | 2 | 0 | Low-quality | Genome-fragment | 4.2 | AAI-based (high-confidence) | 0 |
| S16_vs_160733\|\|full | 5752 | No | NA | | 9 | 1 | 0 | Low-quality | Genome-fragment | 7.02 | AAI-based (high-confidence) | 0 |
| S12_vs_k141_161472\|\|full | 5755 | No | NA | | 4 | 0 | 2 | Low-quality | Genome-fragment | 13.06 | AAI-based (medium-confidence) | 0 |
| S9_vs_k141_52161\|\|full | 5759 | No | NA | | 10 | 0 | 0 | Low-quality | Genome-fragment | 14.68 | AAI-based (high-confidence) | 0 |
| S4_extracted_24145 | 1221 | No | NA | | 1 | 0 | 1 | Not-determined | Genome-fragment | NA | NA | 0 |
| S7_vs_114308\|\|full | 5760 | No | NA | | 8 | 2 | 0 | Low-quality | Genome-fragment | 15.41 | AAI-based (medium-confidence) | 0 |
| S4_extracted_24283 | 1046 | No | NA | | 2 | 0 | 1 | Not-determined | Genome-fragment | NA | NA | 0 |
| S4_extracted_24286 | 3231 | No | NA | | 4 | 0 | 0 | Not-determined | Genome-fragment | NA | NA | 0 |
| S4_extracted_24329 | 2000 | No | NA | | 2 | 0 | 1 | Not-determined | Genome-fragment | NA | NA | 0 |
| S4_extracted_24335 | 1098 | No | NA | | 2 | 0 | 2 | Not-determined | Genome-fragment | NA | NA | 0 |
| S4_extracted_24380 | 1594 | No | NA | | 2 | 0 | 0 | Not-determined | Genome-fragment | NA | NA | 0 |
| S4_extracted_24408 | 1007 | No | NA | | 2 | 0 | 2 | Not-determined | Genome-fragment | NA | NA | 0 |
| S11_vs_k141_96979\|\|full | 5761 | No | NA | | 13 | 2 | 0 | Low-quality | Genome-fragment | 3.93 | AAI-based (high-confidence) | 0 |
| S4_extracted_24845 | 1102 | No | NA | | 1 | 0 | 0 | Not-determined | Genome-fragment | NA | NA | 0 |
| S12_vs_k141_4004\|\|full | 5761 | No | NA | | 7 | 2 | 1 | Low-quality | Genome-fragment | 9.74 | AAI-based (high-confidence) | 0 |
| S10_vs_103285\|\|full | 5764 | No | NA | | 8 | 1 | 0 | Low-quality | Genome-fragment | 15.38 | AAI-based (high-confidence) | 0 |
| S4_extracted_24972 | 1989 | No | NA | | 2 | 0 | 1 | Not-determined | Genome-fragment | NA | NA | 0 |
| S4_extracted_25009 | 1043 | No | NA | | 1 | 0 | 0 | Not-determined | Genome-fragment | NA | NA | 0 |
| S17_vs_k141_93978\|\|full | 5767 | No | NA | | 13 | 5 | 0 | Low-quality | Genome-fragment | 14.12 | AAI-based (medium-confidence) | 0 |
| S4_extracted_25092 | 4178 | No | NA | | 5 | 0 | 4 | Not-determined | Genome-fragment | NA | NA | 0 |
| S14_extracted_k141_44049 | 5769 | No | NA | | 6 | 3 | 0 | Low-quality | Genome-fragment | 9.89 | AAI-based (high-confidence) | 0 |
| S8_vs_k141_198181\|\|full | 5769 | No | NA | | 8 | 0 | 4 | Low-quality | Genome-fragment | 18.42 | AAI-based (high-confidence) | 0 |
| S17_extracted_k141_76836 | 5771 | No | NA | | 3 | 0 | 2 | Low-quality | Genome-fragment | 0.94 | AAI-based (high-confidence) | 0 |
| S4_extracted_25304 | 2517 | No | NA | | 2 | 0 | 0 | Not-determined | Genome-fragment | NA | NA | 0 |
| S4_extracted_25491 | 3147 | No | NA | | 5 | 0 | 1 | Not-determined | Genome-fragment | NA | NA | 0 |
| S8_vs_k141_80836\|\|full | 5772 | No | NA | | 5 | 0 | 4 | Low-quality | Genome-fragment | 1.66 | AAI-based (medium-confidence) | 0 |
| S4_extracted_25575 | 1589 | No | NA | | 1 | 0 | 1 | Not-determined | Genome-fragment | NA | NA | 0 |
| S16_vs_16070\|\|full | 5773 | No | NA | | 15 | 2 | 0 | Low-quality | Genome-fragment | 27.39 | AAI-based (medium-confidence) | 0 |
| S4_extracted_26026 | 1543 | No | NA | | 0 | 0 | 0 | Not-determined | Genome-fragment | NA | NA | 0 |
| S4_extracted_26027 | 1438 | No | NA | | 1 | 0 | 0 | Not-determined | Genome-fragment | NA | NA | 0 |
| S4_extracted_26065 | 3670 | No | NA | | 4 | 0 | 2 | Not-determined | Genome-fragment | NA | NA | 0 |
| S14_vs_k141_44755\|\|full | 5774 | No | NA | | 12 | 3 | 0 | Low-quality | Genome-fragment | 14.83 | AAI-based (medium-confidence) | 0 |
| S4_extracted_26208 | 1037 | No | NA | | 1 | 0 | 1 | Not-determined | Genome-fragment | NA | NA | 0 |
| S6_extracted_k141_36789 | 5775 | No | NA | | 3 | 0 | 2 | Low-quality | Genome-fragment | 5.71 | AAI-based (medium-confidence) | 0 |
| S4_extracted_26353 | 1056 | No | NA | | 1 | 0 | 1 | Not-determined | Genome-fragment | NA | NA | 0 |
| S13_extracted_59201 | 5776 | Yes | | 4199 | 10 | 1 | 1 | Low-quality | Genome-fragment | 8.26 | AAI-based (medium-confidence) | 27.3 |
| S4_extracted_26417 | 83882 | No | NA | | 84 | 9 | 0 | High-quality | High-quality | 99.57 | AAI-based (high-confidence) | 0 |
| S4_extracted_26472 | 6555 | No | NA | | 4 | 0 | 3 | Not-determined | Genome-fragment | NA | NA | 0 |
| S17_vs_k141_157485\|\|full | 5777 | No | NA | | 9 | 0 | 1 | Low-quality | Genome-fragment | 5.52 | AAI-based (medium-confidence) | 0 |
| S4_extracted_26533 | 12406 | No | NA | | 8 | 0 | 1 | Not-determined | Genome-fragment | NA | NA | 0 |
| S5_vs_k141_95332\|\|full | 5777 | Yes | | 5092 | 10 | 1 | 1 | Low-quality | Genome-fragment | 13.73 | AAI-based (high-confidence) | 11.86 |
| S16_extracted_122713 | 5779 | No | NA | | 4 | 0 | 2 | Low-quality | Genome-fragment | 3.45 | AAI-based (medium-confidence) | 0 |
| S4_extracted_26686 | 12981 | No | NA | | 18 | 0 | 2 | Not-determined | Genome-fragment | NA | NA | 0 |
| S4_extracted_26717 | 2393 | No | NA | | 2 | 0 | 0 | Not-determined | Genome-fragment | NA | NA | 0 |
| S3_extracted_k141_15778 | 5782 | Yes | | 2954 | 8 | 2 | 2 | Low-quality | Genome-fragment | 7.39 | AAI-based (high-confidence) | 48.91 |
| S4_extracted_26854 | 1241 | No | NA | | 2 | 0 | 1 | Not-determined | Genome-fragment | NA | NA | 0 |
| S8_extracted_k141_78283 | 5782 | No | NA | | 8 | 5 | 0 | Low-quality | Genome-fragment | 9.33 | AAI-based (high-confidence) | 0 |
| S4_extracted_26907 | 2951 | No | NA | | 2 | 0 | 0 | Not-determined | Genome-fragment | NA | NA | 0 |
| S7_vs_102067\|\|full | 5783 | No | NA | | 14 | 7 | 0 | Low-quality | Genome-fragment | 6.72 | HMM-based (lower-bound) | 0 |
| S1_extracted_k141_22244 | 5784 | No | NA | | 9 | 4 | 1 | Low-quality | Genome-fragment | 10.11 | AAI-based (high-confidence) | 0 |
| S14_extracted_k141_74728 | 5788 | No | NA | | 7 | 7 | 0 | Low-quality | Genome-fragment | 19.09 | AAI-based (high-confidence) | 0 |
| S4_extracted_27861 | 87384 | No | NA | | 117 | 15 | 2 | High-quality | High-quality | 100 | AAI-based (high-confidence) | 0 |
| S17_extracted_k141_62733 | 5789 | No | NA | | 4 | 0 | 3 | Low-quality | Genome-fragment | 1.87 | AAI-based (high-confidence) | 0 |
| S4_extracted_28072 | 1098 | No | NA | | 3 | 0 | 0 | Not-determined | Genome-fragment | NA | NA | 0 |
| S4_extracted_28126 | 3268 | No | NA | | 8 | 0 | 3 | Not-determined | Genome-fragment | NA | NA | 0 |
| S5_vs_k141_115448\|\|full | 5789 | No | NA | | 11 | 6 | 1 | Low-quality | Genome-fragment | 15.06 | AAI-based (high-confidence) | 0 |
| S4_extracted_28429 | 1589 | No | NA | | 3 | 0 | 0 | Not-determined | Genome-fragment | NA | NA | 0 |
| S4_extracted_28601 | 38342 | No | NA | | 54 | 22 | 1 | High-quality | High-quality | 100 | AAI-based (medium-confidence) | 0 |
| S12_vs_k141_901\|\|full | 5789 | No | NA | | 2 | 0 | 0 | Low-quality | Genome-fragment | 6.05 | AAI-based (medium-confidence) | 0 |
| S4_extracted_28710 | 18383 | No | NA | | 9 | 0 | 4 | Not-determined | Genome-fragment | NA | NA | 0 |
| S4_extracted_28737 | 1571 | No | NA | | 3 | 0 | 0 | Not-determined | Genome-fragment | NA | NA | 0 |
| S8_vs_k141_94034\|\|full | 5789 | No | NA | | 5 | 0 | 4 | Low-quality | Genome-fragment | 5.34 | AAI-based (medium-confidence) | 0 |
| S12_extracted_k141_13262 | 5790 | No | NA | | 9 | 2 | 0 | Low-quality | Genome-fragment | 9.99 | HMM-based (lower-bound) | 0 |
| S4_extracted_28900 | 1630 | No | NA | | 2 | 0 | 1 | Not-determined | Genome-fragment | NA | NA | 0 |
| S4_extracted_28930 | 1055 | No | NA | | 1 | 0 | 1 | Not-determined | Genome-fragment | NA | NA | 0 |
| S17_extracted_k141_43614 | 5792 | No | NA | | 4 | 0 | 2 | Low-quality | Genome-fragment | 0.5 | AAI-based (medium-confidence) | 0 |
| S4_extracted_29066 | 1134 | No | NA | | 1 | 0 | 0 | Not-determined | Genome-fragment | NA | NA | 0 |
| S14_extracted_k141_24676 | 5792 | No | NA | | 12 | 0 | 1 | Low-quality | Genome-fragment | 10.63 | AAI-based (high-confidence) | 0 |
| S5_vs_k141_144059\|\|full | 5793 | No | NA | | 10 | 8 | 0 | Low-quality | Genome-fragment | 11.19 | AAI-based (high-confidence) | 0 |
| S4_extracted_29380 | 3357 | No | NA | | 4 | 0 | 3 | Not-determined | Genome-fragment | NA | NA | 0 |
| S16_vs_140522\|\|full | 5793 | No | NA | | 5 | 1 | 0 | Low-quality | Genome-fragment | 7.56 | HMM-based (lower-bound) | 0 |
| S4_extracted_29507 | 8608 | No | NA | | 9 | 0 | 3 | Not-determined | Genome-fragment | NA | NA | 0 |
| S4_extracted_29537 | 1031 | No | NA | | 2 | 0 | 1 | Not-determined | Genome-fragment | NA | NA | 0 |
| S4_extracted_29770 | 1011 | No | NA | | 1 | 0 | 0 | Not-determined | Genome-fragment | NA | NA | 0 |
| S16_vs_33726\|\|full | 5799 | No | NA | | 8 | 0 | 0 | Low-quality | Genome-fragment | 7.04 | AAI-based (medium-confidence) | 0 |
| S4_extracted_29964 | 8011 | No | NA | | 8 | 0 | 1 | Not-determined | Genome-fragment | NA | NA | 0 |
| S4_extracted_30103 | 15716 | No | NA | | 18 | 4 | 0 | Medium-quality | Genome-fragment | 52.71 | HMM-based (lower-bound) | 0 |
| S4_extracted_30153 | 2494 | No | NA | | 2 | 0 | 1 | Not-determined | Genome-fragment | NA | NA | 0 |
| S4_extracted_30177 | 1610 | No | NA | | 2 | 0 | 2 | Not-determined | Genome-fragment | NA | NA | 0 |
| S16_vs_160975\|\|full | 5799 | No | NA | | 15 | 4 | 0 | Low-quality | Genome-fragment | 3.94 | AAI-based (medium-confidence) | 0 |
| S3_extracted_k141_43032 | 5800 | No | NA | | 7 | 2 | 0 | Low-quality | Genome-fragment | 4.88 | AAI-based (high-confidence) | 0 |
| S4_extracted_3252 | 5805 | No | NA | | 9 | 8 | 0 | Low-quality | Genome-fragment | 19.13 | AAI-based (high-confidence) | 0 |
| S3_vs_k141_26522\|\|full | 5807 | No | NA | | 3 | 0 | 0 | Low-quality | Genome-fragment | 3.48 | AAI-based (high-confidence) | 0 |
| S4_extracted_30619 | 4567 | No | NA | | 4 | 0 | 2 | Not-determined | Genome-fragment | NA | NA | 0 |
| S5_extracted_k141_97018 | 5808 | No | NA | | 9 | 4 | 0 | Low-quality | Genome-fragment | 13.91 | AAI-based (medium-confidence) | 0 |
| S4_extracted_30700 | 3331 | No | NA | | 6 | 0 | 4 | Not-determined | Genome-fragment | NA | NA | 0 |
| S17_vs_k141_64628\|\|full | 5809 | No | NA | | 6 | 0 | 2 | Low-quality | Genome-fragment | 2.72 | AAI-based (medium-confidence) | 0 |
| S4_extracted_30824 | 1232 | No | NA | | 2 | 0 | 0 | Not-determined | Genome-fragment | NA | NA | 0 |
| S1_vs_k141_124148\|\|full | 5809 | No | NA | | 5 | 0 | 3 | Low-quality | Genome-fragment | 1.82 | AAI-based (medium-confidence) | 0 |
| S4_extracted_30851 | 1167 | No | NA | | 1 | 0 | 1 | Not-determined | Genome-fragment | NA | NA | 0 |
| S7_extracted_16161 | 5811 | No | NA | | 16 | 1 | 0 | Low-quality | Genome-fragment | 10.98 | AAI-based (medium-confidence) | 0 |
| S8_vs_k141_61776\|\|full | 5813 | No | NA | | 7 | 1 | 2 | Low-quality | Genome-fragment | 8.8 | HMM-based (lower-bound) | 0 |
| S5_vs_k141_52987\|\|full | 5814 | No | NA | | 7 | 1 | 0 | Low-quality | Genome-fragment | 9.49 | HMM-based (lower-bound) | 0 |
| S10_vs_35904\|\|full | 5815 | No | NA | | 13 | 3 | 1 | Low-quality | Genome-fragment | 11.4 | AAI-based (high-confidence) | 0 |
| S12_vs_k141_101552\|\|full | 5820 | No | NA | | 5 | 0 | 0 | Low-quality | Genome-fragment | 10.45 | AAI-based (high-confidence) | 0 |
| S4_extracted_31359 | 1331 | No | NA | | 4 | 0 | 3 | Not-determined | Genome-fragment | NA | NA | 0 |
| S6_extracted_k141_27142 | 5821 | No | NA | | 3 | 0 | 3 | Low-quality | Genome-fragment | 12.37 | AAI-based (medium-confidence) | 0 |
| S9_extracted_k141_64310 | 5826 | No | NA | | 10 | 2 | 0 | Low-quality | Genome-fragment | 2.62 | HMM-based (lower-bound) | 0 |
| S4_extracted_31568 | 1127 | No | NA | | 1 | 0 | 1 | Not-determined | Genome-fragment | NA | NA | 0 |
| S4_extracted_31625 | 6270 | No | NA | | 10 | 0 | 0 | Not-determined | Genome-fragment | NA | NA | 0 |
| S4_extracted_31682 | 37597 | No | NA | | 50 | 22 | 0 | Complete | High-quality | 100 | DTR (high-confidence) | 0 |
| S3_extracted_k141_55170 | 5826 | No | NA | | 9 | 0 | 1 | Low-quality | Genome-fragment | 12.52 | AAI-based (high-confidence) | 0 |
| S4_extracted_31825 | 98149 | No | NA | | 97 | 14 | 1 | Complete | High-quality | 100 | DTR (high-confidence) | 0 |
| S4_extracted_31841 | 13611 | No | NA | | 7 | 0 | 4 | Not-determined | Genome-fragment | NA | NA | 0 |
| S4_extracted_31929 | 4879 | No | NA | | 5 | 0 | 5 | Not-determined | Genome-fragment | NA | NA | 0 |
| S4_extracted_31933 | 5510 | No | NA | | 2 | 0 | 2 | Not-determined | Genome-fragment | NA | NA | 0 |
| S4_extracted_31973 | 118464 | No | NA | | 166 | 30 | 2 | Complete | High-quality | 100 | DTR (high-confidence) | 0 |
| S4_extracted_31978 | 56513 | No | NA | | 97 | 20 | 1 | Complete | High-quality | 100 | DTR (high-confidence) | 0 |
| S4_extracted_31979 | 42371 | No | NA | | 70 | 17 | 0 | Complete | High-quality | 100 | DTR (high-confidence) | 0 |
| S4_extracted_31981 | 91583 | No | NA | | 96 | 10 | 1 | Complete | High-quality | 100 | DTR (high-confidence) | 0 |
| S4_extracted_31982 | 33157 | No | NA | | 53 | 23 | 0 | Complete | High-quality | 100 | DTR (high-confidence) | 0 |
| S14_extracted_k141_5593 | 5830 | No | NA | | 6 | 0 | 3 | Low-quality | Genome-fragment | 20.21 | AAI-based (medium-confidence) | 0 |
| S4_vs_VIRSorter_31975-circular-cat_1 | 69179 | No | NA | | 105 | 15 | 5 | Medium-quality | Genome-fragment | 88.2 | AAI-based (high-confidence) | 0 |
| S4_vs_VIRSorter_6022-circular-cat_1 | 43563 | No | NA | | 58 | 10 | 0 | High-quality | High-quality | 96.49 | AAI-based (high-confidence) | 0 |
| S4_vs_VIRSorter_31916-circular-cat_1 | 41577 | No | NA | | 58 | 19 | 1 | High-quality | High-quality | 100 | AAI-based (medium-confidence) | 0 |
| S4_vs_VIRSorter_3006-cat_1 | 62793 | No | NA | | 94 | 35 | 2 | High-quality | High-quality | 100 | AAI-based (high-confidence) | 0 |
| S4_vs_VIRSorter_9913-cat_1 | 25002 | No | NA | | 40 | 9 | 0 | Medium-quality | Genome-fragment | 55.32 | AAI-based (high-confidence) | 0 |
| S8_vs_k141_145905\|\|full | 5831 | No | NA | | 11 | 4 | 0 | Low-quality | Genome-fragment | 9.32 | AAI-based (high-confidence) | 0 |
| S1_vs_k141_11122\|\|full | 5833 | No | NA | | 7 | 1 | 0 | Low-quality | Genome-fragment | 11.67 | HMM-based (lower-bound) | 0 |
| S13_vs_23410\|\|full | 5837 | Yes | | 4167 | 8 | 1 | 1 | Low-quality | Genome-fragment | 6.31 | HMM-based (lower-bound) | 28.61 |
| S15_extracted_50876 | 5839 | No | NA | | 4 | 1 | 0 | Low-quality | Genome-fragment | 6.14 | AAI-based (high-confidence) | 0 |
| S3_extracted_k141_12593 | 5840 | No | NA | | 9 | 1 | 0 | Low-quality | Genome-fragment | 3.85 | AAI-based (high-confidence) | 0 |
| S11_vs_k141_68133\|\|full | 5849 | No | NA | | 10 | 0 | 1 | Low-quality | Genome-fragment | 13.17 | AAI-based (medium-confidence) | 0 |
| S4_vs_VIRSorter_12831-circular-cat_2 | 37678 | No | NA | | 61 | 13 | 3 | High-quality | High-quality | 100 | AAI-based (high-confidence) | 0 |
| S4_vs_VIRSorter_31196-circular-cat_2 | 44132 | No | NA | | 75 | 14 | 0 | High-quality | High-quality | 99.71 | AAI-based (high-confidence) | 0 |
| S4_vs_VIRSorter_31560-circular-cat_2 | 48517 | No | NA | | 77 | 21 | 1 | High-quality | High-quality | 100 | AAI-based (high-confidence) | 0 |
| S4_vs_VIRSorter_31977-circular-cat_2 | 39384 | No | NA | | 60 | 19 | 2 | High-quality | High-quality | 95.55 | AAI-based (medium-confidence) | 0 |
| S4_vs_VIRSorter_7386-circular-cat_2 | 44589 | No | NA | | 69 | 16 | 1 | High-quality | High-quality | 99.34 | AAI-based (high-confidence) | 0 |
| S4_vs_VIRSorter_31171-circular-cat_2 | 30681 | No | NA | | 49 | 15 | 0 | Medium-quality | Genome-fragment | 89.39 | AAI-based (high-confidence) | 0 |
| S4_vs_VIRSorter_31972-circular-cat_2 | 37842 | No | NA | | 56 | 19 | 0 | High-quality | High-quality | 100 | AAI-based (high-confidence) | 0 |
| S4_vs_VIRSorter_31976-circular-cat_2 | 58467 | No | NA | | 81 | 18 | 1 | High-quality | High-quality | 97.59 | AAI-based (high-confidence) | 0 |
| S4_vs_VIRSorter_31980-circular-cat_2 | 42928 | No | NA | | 65 | 23 | 0 | High-quality | High-quality | 93.59 | AAI-based (medium-confidence) | 0 |
| S4_vs_VIRSorter_13190-cat_2 | 98394 | No | NA | | 87 | 11 | 2 | Medium-quality | Genome-fragment | 65.21 | AAI-based (high-confidence) | 0 |
| S4_vs_VIRSorter_30222-cat_2 | 87349 | No | NA | | 132 | 38 | 1 | High-quality | High-quality | 100 | AAI-based (high-confidence) | 0 |
| S4_vs_VIRSorter_27316-cat_2 | 69191 | No | NA | | 111 | 41 | 3 | High-quality | High-quality | 100 | AAI-based (high-confidence) | 0 |
| S4_vs_VIRSorter_4839-cat_2 | 61401 | No | NA | | 79 | 30 | 1 | High-quality | High-quality | 98.93 | AAI-based (high-confidence) | 0 |
| S4_vs_VIRSorter_8622-cat_2 | 43660 | No | NA | | 67 | 18 | 0 | High-quality | High-quality | 92.6 | AAI-based (high-confidence) | 0 |
| S4_vs_VIRSorter_15837-cat_2 | 42134 | No | NA | | 60 | 30 | 0 | High-quality | High-quality | 97.63 | AAI-based (high-confidence) | 0 |
| S4_vs_VIRSorter_16493-cat_2 | 40455 | No | NA | | 59 | 19 | 0 | High-quality | High-quality | 99.66 | AAI-based (high-confidence) | 0 |
| S4_vs_VIRSorter_21643-cat_2 | 39545 | No | NA | | 58 | 25 | 0 | High-quality | High-quality | 100 | AAI-based (high-confidence) | 0 |
| S4_vs_VIRSorter_5698-cat_2 | 39508 | No | NA | | 61 | 18 | 0 | High-quality | High-quality | 100 | AAI-based (high-confidence) | 0 |
| S4_vs_VIRSorter_19324-cat_2 | 37383 | No | NA | | 51 | 19 | 0 | High-quality | High-quality | 100 | AAI-based (high-confidence) | 0 |
| S5_vs_k141_38249\|\|full | 5855 | No | NA | | 6 | 1 | 0 | Low-quality | Genome-fragment | 6.57 | HMM-based (lower-bound) | 0 |
| S4_vs_VIRSorter_1135-cat_2 | 36647 | No | NA | | 55 | 25 | 1 | Medium-quality | Genome-fragment | 81 | AAI-based (medium-confidence) | 0 |
| S4_vs_VIRSorter_4959-cat_2 | 36586 | No | NA | | 50 | 19 | 1 | Medium-quality | Genome-fragment | 63.54 | AAI-based (high-confidence) | 0 |
| S4_vs_VIRSorter_16803-cat_2 | 33696 | No | NA | | 51 | 17 | 1 | High-quality | High-quality | 97.4 | AAI-based (medium-confidence) | 0 |
| S4_vs_VIRSorter_13488-cat_2 | 32441 | No | NA | | 49 | 15 | 2 | High-quality | High-quality | 94.84 | AAI-based (high-confidence) | 0 |
| S4_vs_VIRSorter_15112-cat_2 | 31917 | No | NA | | 46 | 15 | 0 | Medium-quality | Genome-fragment | 85.6 | AAI-based (high-confidence) | 0 |
| S4_vs_VIRSorter_29332-cat_2 | 21229 | No | NA | | 29 | 10 | 1 | Medium-quality | Genome-fragment | 52.71 | AAI-based (high-confidence) | 0 |
| S11_vs_k141_53210\|\|full | 5858 | No | NA | | 6 | 2 | 0 | Low-quality | Genome-fragment | 2.27 | AAI-based (medium-confidence) | 0 |
| S14_extracted_k141_75688 | 5859 | No | NA | | 9 | 2 | 0 | Low-quality | Genome-fragment | 9.78 | AAI-based (high-confidence) | 0 |
| S4_vs_VIRSorter_4746-cat_2 | 15960 | No | NA | | 22 | 7 | 0 | Medium-quality | Genome-fragment | 70.75 | AAI-based (high-confidence) | 0 |
| S3_vs_k141_73188\|\|full | 5864 | No | NA | | 6 | 0 | 3 | Low-quality | Genome-fragment | 3.86 | AAI-based (high-confidence) | 0 |
| S16_vs_5784\|\|full | 5867 | No | NA | | 6 | 0 | 0 | Low-quality | Genome-fragment | 10.68 | AAI-based (medium-confidence) | 0 |
| S12_vs_k141_57724\|\|full | 5867 | No | NA | | 5 | 0 | 3 | Low-quality | Genome-fragment | 5.24 | AAI-based (medium-confidence) | 0 |
| S7_vs_1626\|\|full | 5868 | No | NA | | 11 | 9 | 0 | Low-quality | Genome-fragment | 9.02 | AAI-based (high-confidence) | 0 |
| S12_vs_k141_154682\|\|full | 5872 | No | NA | | 9 | 2 | 0 | Low-quality | Genome-fragment | 13.31 | AAI-based (high-confidence) | 0 |
| S16_vs_49204\|\|full | 5873 | No | NA | | 5 | 1 | 0 | Low-quality | Genome-fragment | 3.33 | HMM-based (lower-bound) | 0 |
| S8_extracted_k141_80365 | 5873 | No | NA | | 14 | 14 | 0 | Low-quality | Genome-fragment | 19.31 | AAI-based (high-confidence) | 0 |
| S13_vs_12359\|\|full | 5880 | No | NA | | 7 | 0 | 2 | Low-quality | Genome-fragment | 3.51 | AAI-based (medium-confidence) | 0 |
| S15_extracted_36504 | 5883 | No | NA | | 6 | 1 | 0 | Low-quality | Genome-fragment | 12.97 | AAI-based (high-confidence) | 0 |
| S8_vs_k141_157835\|\|full | 5884 | No | NA | | 9 | 1 | 0 | Low-quality | Genome-fragment | 5.04 | AAI-based (high-confidence) | 0 |
| S11_extracted_k141_38684 | 5886 | No | NA | | 4 | 0 | 3 | Low-quality | Genome-fragment | 3.1 | AAI-based (medium-confidence) | 0 |
| S10_vs_39409\|\|full | 5888 | No | NA | | 11 | 1 | 0 | Low-quality | Genome-fragment | 12.24 | AAI-based (medium-confidence) | 0 |
| S7_vs_77904\|\|full | 5889 | No | NA | | 12 | 1 | 0 | Low-quality | Genome-fragment | 13.41 | AAI-based (medium-confidence) | 0 |
| S10_extracted_38642 | 5890 | No | NA | | 4 | 0 | 3 | Low-quality | Genome-fragment | 5.22 | AAI-based (medium-confidence) | 0 |
| S12_vs_k141_147809\|\|full | 5890 | Yes | | 1669 | 7 | 1 | 1 | Low-quality | Genome-fragment | 1.49 | AAI-based (medium-confidence) | 71.66 |
| S9_vs_k141_76383\|\|full | 5891 | Yes | | 4133 | 8 | 2 | 1 | Low-quality | Genome-fragment | 12.2 | AAI-based (high-confidence) | 29.84 |
| S4_vs_VIRSorter_2744-cat_3 | 20584 | No | NA | | 41 | 8 | 1 | Medium-quality | Genome-fragment | 51.54 | AAI-based (high-confidence) | 0 |
| S8_vs_k141_61253\|\|full | 5893 | No | NA | | 10 | 1 | 0 | Low-quality | Genome-fragment | 7.51 | HMM-based (lower-bound) | 0 |
| S6_extracted_k141_37703 | 3537 | No | NA | | 5 | 0 | 3 | Not-determined | Genome-fragment | NA | NA | 0 |
| S3_vs_k141_102322\|\|full | 5894 | No | NA | | 6 | 4 | 0 | Low-quality | Genome-fragment | 16.87 | AAI-based (high-confidence) | 0 |
| S6_extracted_k141_18872 | 1171 | No | NA | | 4 | 0 | 2 | Not-determined | Genome-fragment | NA | NA | 0 |
| S6_extracted_k141_16527 | 1203 | No | NA | | 3 | 0 | 3 | Not-determined | Genome-fragment | NA | NA | 0 |
| S6_extracted_k141_44786 | 1264 | No | NA | | 1 | 0 | 0 | Not-determined | Genome-fragment | NA | NA | 0 |
| S6_extracted_k141_33017 | 1066 | No | NA | | 2 | 0 | 2 | Not-determined | Genome-fragment | NA | NA | 0 |
| S16_extracted_118214 | 5895 | No | NA | | 2 | 1 | 0 | Low-quality | Genome-fragment | 10.16 | AAI-based (high-confidence) | 0 |
| S6_extracted_k141_49513 | 2045 | No | NA | | 2 | 0 | 2 | Not-determined | Genome-fragment | NA | NA | 0 |
| S16_vs_144376\|\|full | 5895 | No | NA | | 10 | 5 | 0 | Low-quality | Genome-fragment | 16.8 | AAI-based (medium-confidence) | 0 |
| S6_extracted_k141_47166 | 3360 | No | NA | | 4 | 0 | 2 | Not-determined | Genome-fragment | NA | NA | 0 |
| S10_vs_106735\|\|full | 5895 | No | NA | | 11 | 4 | 0 | Low-quality | Genome-fragment | 7.5 | HMM-based (lower-bound) | 0 |
| S6_extracted_k141_47170 | 1053 | No | NA | | 2 | 0 | 0 | Not-determined | Genome-fragment | NA | NA | 0 |
| S6_extracted_k141_47172 | 1306 | No | NA | | 1 | 0 | 0 | Not-determined | Genome-fragment | NA | NA | 0 |
| S6_extracted_k141_30673 | 1218 | No | NA | | 2 | 0 | 2 | Not-determined | Genome-fragment | NA | NA | 0 |
| S6_extracted_k141_49544 | 1118 | No | NA | | 2 | 0 | 1 | Not-determined | Genome-fragment | NA | NA | 0 |
| S6_extracted_k141_54238 | 1048 | No | NA | | 1 | 0 | 0 | Not-determined | Genome-fragment | NA | NA | 0 |
| S6_extracted_k141_30688 | 1682 | No | NA | | 2 | 0 | 1 | Not-determined | Genome-fragment | NA | NA | 0 |
| S6_extracted_k141_42509 | 1851 | No | NA | | 1 | 0 | 0 | Not-determined | Genome-fragment | NA | NA | 0 |
| S6_extracted_k141_28305 | 1136 | No | NA | | 1 | 0 | 0 | Not-determined | Genome-fragment | NA | NA | 0 |
| S6_extracted_k141_42532 | 2471 | No | NA | | 2 | 0 | 2 | Not-determined | Genome-fragment | NA | NA | 0 |
| S3_extracted_k141_113350 | 5898 | No | NA | | 5 | 3 | 0 | Low-quality | Genome-fragment | 15.12 | AAI-based (high-confidence) | 0 |
| S6_extracted_k141_33079 | 2438 | No | NA | | 2 | 0 | 0 | Not-determined | Genome-fragment | NA | NA | 0 |
| S10_vs_15287\|\|full | 5898 | No | NA | | 7 | 0 | 1 | Low-quality | Genome-fragment | 10.95 | AAI-based (medium-confidence) | 0 |
| S14_vs_k141_87481\|\|full | 5899 | No | NA | | 13 | 4 | 0 | Low-quality | Genome-fragment | 4.59 | HMM-based (lower-bound) | 0 |
| S6_extracted_k141_16637 | 1074 | No | NA | | 1 | 0 | 1 | Not-determined | Genome-fragment | NA | NA | 0 |
| S6_extracted_k141_16641 | 1131 | No | NA | | 1 | 0 | 0 | Not-determined | Genome-fragment | NA | NA | 0 |
| S7_extracted_75453 | 5904 | No | NA | | 5 | 0 | 2 | Low-quality | Genome-fragment | 2.94 | AAI-based (medium-confidence) | 0 |
| S3_extracted_k141_89817 | 5906 | No | NA | | 4 | 1 | 0 | Low-quality | Genome-fragment | 4.42 | AAI-based (high-confidence) | 0 |
| S17_vs_k141_68587\|\|full | 5907 | No | NA | | 12 | 0 | 0 | Low-quality | Genome-fragment | 9.32 | AAI-based (high-confidence) | 0 |
| S6_extracted_k141_49621 | 3291 | No | NA | | 1 | 0 | 0 | Not-determined | Genome-fragment | NA | NA | 0 |
| S6_extracted_k141_9515 | 3915 | No | NA | | 6 | 0 | 0 | Not-determined | Genome-fragment | NA | NA | 0 |
| S6_extracted_k141_44894 | 1337 | No | NA | | 1 | 0 | 0 | Not-determined | Genome-fragment | NA | NA | 0 |
| S7_vs_93715\|\|full | 5911 | No | NA | | 8 | 4 | 0 | Low-quality | Genome-fragment | 12.58 | AAI-based (high-confidence) | 0 |
| S6_extracted_k141_28414 | 1157 | No | NA | | 3 | 0 | 0 | Not-determined | Genome-fragment | NA | NA | 0 |
| S6_extracted_k141_49629 | 1260 | No | NA | | 1 | 0 | 0 | Not-determined | Genome-fragment | NA | NA | 0 |
| S6_extracted_k141_42635 | 3666 | No | NA | | 6 | 0 | 1 | Not-determined | Genome-fragment | NA | NA | 0 |
| S6_extracted_k141_14320 | 1383 | No | NA | | 3 | 0 | 2 | Not-determined | Genome-fragment | NA | NA | 0 |
| S6_extracted_k141_44926 | 1073 | No | NA | | 1 | 0 | 1 | Not-determined | Genome-fragment | NA | NA | 0 |
| S8_vs_k141_129737\|\|full | 5912 | No | NA | | 6 | 0 | 4 | Low-quality | Genome-fragment | 2.75 | AAI-based (medium-confidence) | 0 |
| S6_extracted_k141_230 | 4269 | No | NA | | 5 | 0 | 1 | Not-determined | Genome-fragment | NA | NA | 0 |
| S16_vs_63890\|\|full | 5914 | No | NA | | 9 | 3 | 0 | Low-quality | Genome-fragment | 7.66 | HMM-based (lower-bound) | 0 |
| S6_extracted_k141_235 | 1750 | No | NA | | 2 | 0 | 0 | Not-determined | Genome-fragment | NA | NA | 0 |
| S6_extracted_k141_44954 | 2478 | No | NA | | 4 | 0 | 3 | Not-determined | Genome-fragment | NA | NA | 0 |
| S6_extracted_k141_252 | 1018 | No | NA | | 1 | 0 | 0 | Not-determined | Genome-fragment | NA | NA | 0 |
| S6_extracted_k141_33200 | 1425 | No | NA | | 2 | 0 | 1 | Not-determined | Genome-fragment | NA | NA | 0 |
| S6_extracted_k141_16699 | 1053 | No | NA | | 1 | 0 | 1 | Not-determined | Genome-fragment | NA | NA | 0 |
| S6_extracted_k141_33204 | 2151 | No | NA | | 4 | 0 | 4 | Not-determined | Genome-fragment | NA | NA | 0 |
| S6_extracted_k141_26045 | 1640 | No | NA | | 4 | 0 | 3 | Not-determined | Genome-fragment | NA | NA | 0 |
| S2_extracted_k141_8899 | 5914 | No | NA | | 9 | 8 | 0 | Low-quality | Genome-fragment | 14.52 | AAI-based (high-confidence) | 0 |
| S6_extracted_k141_4848 | 5478 | No | NA | | 7 | 0 | 0 | Not-determined | Genome-fragment | NA | NA | 0 |
| S12_vs_k141_183218\|\|full | 5915 | No | NA | | 8 | 6 | 0 | Low-quality | Genome-fragment | 14.69 | AAI-based (high-confidence) | 0 |
| S6_extracted_k141_306 | 1184 | No | NA | | 1 | 0 | 1 | Not-determined | Genome-fragment | NA | NA | 0 |
| S6_extracted_k141_40241 | 1603 | No | NA | | 2 | 0 | 1 | Not-determined | Genome-fragment | NA | NA | 0 |
| S6_extracted_k141_2514 | 1021 | No | NA | | 1 | 0 | 1 | Not-determined | Genome-fragment | NA | NA | 0 |
| S6_extracted_k141_40250 | 3304 | No | NA | | 2 | 0 | 0 | Not-determined | Genome-fragment | NA | NA | 0 |
| S6_extracted_k141_26098 | 1664 | No | NA | | 1 | 0 | 0 | Not-determined | Genome-fragment | NA | NA | 0 |
| S6_extracted_k141_7191 | 1361 | No | NA | | 1 | 0 | 0 | Not-determined | Genome-fragment | NA | NA | 0 |
| S6_extracted_k141_376 | 1943 | No | NA | | 2 | 0 | 1 | Not-determined | Genome-fragment | NA | NA | 0 |
| S6_extracted_k141_52138 | 2147 | No | NA | | 1 | 0 | 0 | Not-determined | Genome-fragment | NA | NA | 0 |
| S6_extracted_k141_33283 | 5785 | No | NA | | 6 | 0 | 5 | Not-determined | Genome-fragment | NA | NA | 0 |
| S6_extracted_k141_45042 | 1804 | No | NA | | 3 | 0 | 3 | Not-determined | Genome-fragment | NA | NA | 0 |
| S14_vs_k141_55569\|\|full | 5916 | No | NA | | 8 | 4 | 0 | Low-quality | Genome-fragment | 6.1 | HMM-based (lower-bound) | 0 |
| S6_extracted_k141_33296 | 1479 | No | NA | | 2 | 0 | 0 | Not-determined | Genome-fragment | NA | NA | 0 |
| S1_vs_k141_44611\|\|full | 5916 | No | NA | | 8 | 4 | 1 | Low-quality | Genome-fragment | 13.69 | AAI-based (medium-confidence) | 0 |
| S6_extracted_k141_2640 | 1415 | No | NA | | 2 | 0 | 2 | Not-determined | Genome-fragment | NA | NA | 0 |
| S6_extracted_k141_16758 | 1492 | No | NA | | 1 | 0 | 1 | Not-determined | Genome-fragment | NA | NA | 0 |
| S6_extracted_k141_4896 | 2484 | No | NA | | 1 | 0 | 1 | Not-determined | Genome-fragment | NA | NA | 0 |
| S6_extracted_k141_45073 | 1388 | No | NA | | 2 | 0 | 1 | Not-determined | Genome-fragment | NA | NA | 0 |
| S6_extracted_k141_45077 | 1627 | No | NA | | 1 | 0 | 1 | Not-determined | Genome-fragment | NA | NA | 0 |
| S6_extracted_k141_38138 | 2470 | No | NA | | 3 | 0 | 1 | Not-determined | Genome-fragment | NA | NA | 0 |
| S6_extracted_k141_47238 | 12508 | No | NA | | 16 | 5 | 0 | High-quality | High-quality | 100 | AAI-based (high-confidence) | 0 |
| S12_vs_k141_11280\|\|full | 5921 | No | NA | | 6 | 6 | 0 | Low-quality | Genome-fragment | 9.87 | AAI-based (high-confidence) | 0 |
| S16_vs_126993\|\|full | 5926 | No | NA | | 6 | 0 | 1 | Low-quality | Genome-fragment | 10.64 | AAI-based (high-confidence) | 0 |
| S8_vs_k141_21460\|\|full | 5928 | No | NA | | 6 | 1 | 5 | Low-quality | Genome-fragment | 2.99 | HMM-based (lower-bound) | 0 |
| S6_extracted_k141_2700 | 1834 | No | NA | | 1 | 0 | 1 | Not-determined | Genome-fragment | NA | NA | 0 |
| S6_extracted_k141_9648 | 2354 | No | NA | | 3 | 0 | 1 | Not-determined | Genome-fragment | NA | NA | 0 |
| S6_extracted_k141_16788 | 1370 | No | NA | | 1 | 0 | 1 | Not-determined | Genome-fragment | NA | NA | 0 |
| S6_extracted_k141_26122 | 1167 | No | NA | | 2 | 0 | 0 | Not-determined | Genome-fragment | NA | NA | 0 |
| S6_extracted_k141_26128 | 1255 | No | NA | | 3 | 0 | 3 | Not-determined | Genome-fragment | NA | NA | 0 |
| S6_extracted_k141_9681 | 1111 | No | NA | | 1 | 0 | 1 | Not-determined | Genome-fragment | NA | NA | 0 |
| S6_extracted_k141_33391 | 1641 | No | NA | | 2 | 0 | 2 | Not-determined | Genome-fragment | NA | NA | 0 |
| S6_extracted_k141_23842 | 1523 | No | NA | | 2 | 0 | 2 | Not-determined | Genome-fragment | NA | NA | 0 |
| S16_vs_42643\|\|full | 5930 | Yes | | 858 | 4 | 1 | 3 | Low-quality | Genome-fragment | 0.49 | HMM-based (lower-bound) | 85.53 |
| S6_extracted_k141_9692 | 1770 | No | NA | | 1 | 0 | 1 | Not-determined | Genome-fragment | NA | NA | 0 |
| S6_extracted_k141_38192 | 1152 | No | NA | | 2 | 0 | 0 | Not-determined | Genome-fragment | NA | NA | 0 |
| S17_extracted_k141_79290 | 5933 | No | NA | | 10 | 3 | 0 | Low-quality | Genome-fragment | 13.26 | AAI-based (high-confidence) | 0 |
| S6_extracted_k141_487 | 2882 | No | NA | | 2 | 0 | 0 | Not-determined | Genome-fragment | NA | NA | 0 |
| S3_vs_k141_98024\|\|full | 5933 | No | NA | | 16 | 2 | 0 | Low-quality | Genome-fragment | 4.44 | AAI-based (high-confidence) | 0 |
| S17_vs_k141_87629\|\|full | 5936 | No | NA | | 8 | 3 | 0 | Low-quality | Genome-fragment | 14.65 | AAI-based (high-confidence) | 0 |
| S6_extracted_k141_52229 | 2074 | No | NA | | 1 | 0 | 0 | Not-determined | Genome-fragment | NA | NA | 0 |
| S6_extracted_k141_47296 | 3423 | No | NA | | 1 | 0 | 0 | Not-determined | Genome-fragment | NA | NA | 0 |
| S6_extracted_k141_26146 | 3220 | No | NA | | 4 | 0 | 3 | Not-determined | Genome-fragment | NA | NA | 0 |
| S14_vs_k141_4775\|\|full | 5936 | No | NA | | 5 | 2 | 0 | Low-quality | Genome-fragment | 4.02 | HMM-based (lower-bound) | 0 |
| S17_extracted_k141_105335 | 5941 | No | NA | | 7 | 7 | 0 | Low-quality | Genome-fragment | 19.58 | AAI-based (high-confidence) | 0 |
| S6_extracted_k141_19031 | 1073 | No | NA | | 1 | 0 | 0 | Not-determined | Genome-fragment | NA | NA | 0 |
| S10_extracted_744 | 5943 | No | NA | | 12 | 1 | 1 | Low-quality | Genome-fragment | 3.22 | HMM-based (lower-bound) | 0 |
| S5_vs_k141_110620\|\|full | 5944 | No | NA | | 8 | 2 | 0 | Low-quality | Genome-fragment | 3.5 | HMM-based (lower-bound) | 0 |
| S6_extracted_k141_16858 | 3925 | No | NA | | 3 | 0 | 3 | Not-determined | Genome-fragment | NA | NA | 0 |
| S6_extracted_k141_52304 | 1074 | No | NA | | 1 | 0 | 1 | Not-determined | Genome-fragment | NA | NA | 0 |
| S6_extracted_k141_42848 | 3413 | No | NA | | 4 | 0 | 3 | Not-determined | Genome-fragment | NA | NA | 0 |
| S6_extracted_k141_2875 | 1395 | No | NA | | 2 | 0 | 1 | Not-determined | Genome-fragment | NA | NA | 0 |
| S6_extracted_k141_7275 | 2273 | No | NA | | 3 | 0 | 3 | Not-determined | Genome-fragment | NA | NA | 0 |
| S6_extracted_k141_4960 | 3934 | No | NA | | 7 | 0 | 1 | Not-determined | Genome-fragment | NA | NA | 0 |
| S6_extracted_k141_47314 | 1087 | No | NA | | 1 | 0 | 1 | Not-determined | Genome-fragment | NA | NA | 0 |
| S12_vs_k141_13467\|\|full | 5944 | Yes | | 2398 | 7 | 2 | 1 | Low-quality | Genome-fragment | 5.85 | AAI-based (medium-confidence) | 59.66 |
| S6_extracted_k141_19066 | 1093 | No | NA | | 3 | 0 | 1 | Not-determined | Genome-fragment | NA | NA | 0 |
| S6_extracted_k141_52410 | 1768 | No | NA | | 2 | 0 | 2 | Not-determined | Genome-fragment | NA | NA | 0 |
| S6_extracted_k141_12217 | 1521 | No | NA | | 1 | 0 | 0 | Not-determined | Genome-fragment | NA | NA | 0 |
| S6_extracted_k141_45179 | 1228 | No | NA | | 2 | 0 | 2 | Not-determined | Genome-fragment | NA | NA | 0 |
| S3_extracted_k141_15559 | 5947 | No | NA | | 6 | 2 | 0 | Low-quality | Genome-fragment | 6.67 | AAI-based (high-confidence) | 0 |
| S6_extracted_k141_544 | 1756 | No | NA | | 3 | 0 | 1 | Not-determined | Genome-fragment | NA | NA | 0 |
| S6_extracted_k141_7330 | 5279 | No | NA | | 3 | 0 | 3 | Not-determined | Genome-fragment | NA | NA | 0 |
| S6_extracted_k141_16923 | 2193 | No | NA | | 3 | 0 | 1 | Not-determined | Genome-fragment | NA | NA | 0 |
| S9_vs_k141_85757\|\|full | 5952 | No | NA | | 5 | 1 | 0 | Low-quality | Genome-fragment | 3.32 | HMM-based (lower-bound) | 0 |
| S6_extracted_k141_33488 | 1372 | No | NA | | 2 | 0 | 1 | Not-determined | Genome-fragment | NA | NA | 0 |
| S1_extracted_k141_65025 | 5954 | No | NA | | 12 | 4 | 0 | Low-quality | Genome-fragment | 9.84 | AAI-based (high-confidence) | 0 |
| S6_extracted_k141_12270 | 1446 | No | NA | | 2 | 0 | 2 | Not-determined | Genome-fragment | NA | NA | 0 |
| S6_extracted_k141_16964 | 1225 | No | NA | | 1 | 0 | 1 | Not-determined | Genome-fragment | NA | NA | 0 |
| S16_extracted_95382 | 5954 | No | NA | | 8 | 3 | 0 | Low-quality | Genome-fragment | 9.12 | AAI-based (high-confidence) | 0 |
| S6_extracted_k141_2990 | 2057 | No | NA | | 1 | 0 | 0 | Not-determined | Genome-fragment | NA | NA | 0 |
| S6_extracted_k141_47395 | 1317 | No | NA | | 1 | 0 | 1 | Not-determined | Genome-fragment | NA | NA | 0 |
| S6_extracted_k141_28708 | 10820 | No | NA | | 8 | 0 | 4 | Not-determined | Genome-fragment | NA | NA | 0 |
| S6_extracted_k141_45228 | 15593 | No | NA | | 21 | 7 | 0 | Medium-quality | Genome-fragment | 72.68 | AAI-based (medium-confidence) | 0 |
| S6_extracted_k141_28721 | 1149 | No | NA | | 1 | 0 | 0 | Not-determined | Genome-fragment | NA | NA | 0 |
| S6_extracted_k141_628 | 1679 | No | NA | | 1 | 0 | 0 | Not-determined | Genome-fragment | NA | NA | 0 |
| S6_extracted_k141_28722 | 1147 | No | NA | | 2 | 0 | 2 | Not-determined | Genome-fragment | NA | NA | 0 |
| S6_extracted_k141_52560 | 1185 | No | NA | | 2 | 0 | 1 | Not-determined | Genome-fragment | NA | NA | 0 |
| S11_vs_k141_57351\|\|full | 5958 | No | NA | | 12 | 0 | 0 | Low-quality | Genome-fragment | 3.87 | AAI-based (high-confidence) | 0 |
| S6_extracted_k141_3011 | 1415 | No | NA | | 2 | 0 | 2 | Not-determined | Genome-fragment | NA | NA | 0 |
| S6_extracted_k141_30881 | 1415 | No | NA | | 2 | 0 | 0 | Not-determined | Genome-fragment | NA | NA | 0 |
| S7_vs_108544\|\|full | 5958 | No | NA | | 10 | 1 | 0 | Low-quality | Genome-fragment | 5.4 | HMM-based (lower-bound) | 0 |
| S6_extracted_k141_54296 | 1152 | No | NA | | 1 | 0 | 0 | Not-determined | Genome-fragment | NA | NA | 0 |
| S6_extracted_k141_30888 | 1513 | No | NA | | 3 | 0 | 1 | Not-determined | Genome-fragment | NA | NA | 0 |
| S6_extracted_k141_40602 | 1197 | No | NA | | 2 | 0 | 2 | Not-determined | Genome-fragment | NA | NA | 0 |
| S6_extracted_k141_26292 | 8247 | No | NA | | 11 | 0 | 1 | Not-determined | Genome-fragment | NA | NA | 0 |
| S3_extracted_k141_16365 | 5962 | No | NA | | 15 | 7 | 0 | Low-quality | Genome-fragment | 17.2 | AAI-based (high-confidence) | 0 |
| S6_extracted_k141_54312 | 3032 | No | NA | | 3 | 0 | 3 | Not-determined | Genome-fragment | NA | NA | 0 |
| S6_extracted_k141_21399 | 5618 | No | NA | | 3 | 0 | 1 | Not-determined | Genome-fragment | NA | NA | 0 |
| S6_extracted_k141_30915 | 1397 | No | NA | | 2 | 0 | 1 | Not-determined | Genome-fragment | NA | NA | 0 |
| S6_extracted_k141_54319 | 1989 | No | NA | | 1 | 0 | 1 | Not-determined | Genome-fragment | NA | NA | 0 |
| S6_extracted_k141_3048 | 1169 | No | NA | | 2 | 0 | 2 | Not-determined | Genome-fragment | NA | NA | 0 |
| S6_extracted_k141_38309 | 1053 | No | NA | | 1 | 0 | 0 | Not-determined | Genome-fragment | NA | NA | 0 |
| S6_extracted_k141_28756 | 1110 | No | NA | | 1 | 0 | 1 | Not-determined | Genome-fragment | NA | NA | 0 |
| S8_vs_k141_194138\|\|full | 5964 | Yes | | 4862 | 8 | 2 | 1 | Low-quality | Genome-fragment | 10.17 | AAI-based (high-confidence) | 18.48 |
| S6_extracted_k141_24103 | 1004 | No | NA | | 1 | 0 | 1 | Not-determined | Genome-fragment | NA | NA | 0 |
| S6_extracted_k141_38337 | 2125 | No | NA | | 4 | 0 | 3 | Not-determined | Genome-fragment | NA | NA | 0 |

| | | | | | | | | | | | |
|---|---|---|---|---|---|---|---|---|---|---|---|
| S6_extracted_k141_38342 | 1603 | No | NA | 2 | 0 | 2 | Not-determined | Genome-fragment | NA | NA | 0 |
| S6_extracted_k141_9893 | 1236 | No | NA | 1 | 0 | 1 | Not-determined | Genome-fragment | NA | NA | 0 |
| S6_extracted_k141_12448 | 1061 | No | NA | 2 | 0 | 1 | Not-determined | Genome-fragment | NA | NA | 0 |
| S2_extracted_k141_824 | 5965 | No | NA | 6 | 3 | 0 | Low-quality | Genome-fragment | 21.04 | HMM-based (lower-bound) | 0 |
| S3_extracted_k141_51814 | 5967 | No | NA | 15 | 3 | 0 | Low-quality | Genome-fragment | 3.46 | AAI-based (high-confidence) | 0 |
| S6_extracted_k141_26350 | 7110 | No | NA | 15 | 0 | 14 | Not-determined | Genome-fragment | NA | NA | 0 |
| S15_extracted_72758 | 5970 | No | NA | 2 | 0 | 1 | Low-quality | Genome-fragment | 3.6 | AAI-based (high-confidence) | 0 |
| S6_extracted_k141_12498 | 1215 | No | NA | 1 | 0 | 0 | Not-determined | Genome-fragment | NA | NA | 0 |
| S3_extracted_k141_40028 | 5974 | No | NA | 12 | 2 | 0 | Low-quality | Genome-fragment | 12.09 | AAI-based (high-confidence) | 0 |
| S3_vs_k141_96674\|\|full | 5974 | No | NA | 3 | 1 | 0 | Low-quality | Genome-fragment | 13.24 | AAI-based (high-confidence) | 0 |
| S10_vs_106525\|\|full | 5976 | No | NA | 9 | 6 | 0 | Low-quality | Genome-fragment | 12.97 | AAI-based (high-confidence) | 0 |
| S6_extracted_k141_14763 | 1171 | No | NA | 1 | 0 | 0 | Not-determined | Genome-fragment | NA | NA | 0 |
| S6_extracted_k141_26395 | 1590 | No | NA | 3 | 0 | 1 | Not-determined | Genome-fragment | NA | NA | 0 |
| S6_extracted_k141_21468 | 1116 | No | NA | 1 | 0 | 0 | Not-determined | Genome-fragment | NA | NA | 0 |
| S6_extracted_k141_35831 | 1407 | No | NA | 1 | 0 | 1 | Not-determined | Genome-fragment | NA | NA | 0 |
| S6_extracted_k141_24177 | 1810 | No | NA | 2 | 0 | 2 | Not-determined | Genome-fragment | NA | NA | 0 |
| S6_extracted_k141_24178 | 1506 | No | NA | 2 | 0 | 1 | Not-determined | Genome-fragment | NA | NA | 0 |
| S6_extracted_k141_24218 | 1027 | No | NA | 3 | 0 | 2 | Not-determined | Genome-fragment | NA | NA | 0 |
| S6_extracted_k141_33668 | 1351 | No | NA | 1 | 0 | 0 | Not-determined | Genome-fragment | NA | NA | 0 |
| S3_extracted_k141_24004 | 5977 | No | NA | 4 | 0 | 0 | Low-quality | Genome-fragment | 5.52 | AAI-based (high-confidence) | 0 |
| S6_extracted_k141_26436 | 1148 | No | NA | 1 | 0 | 1 | Not-determined | Genome-fragment | NA | NA | 0 |
| S6_extracted_k141_24236 | 3140 | No | NA | 2 | 0 | 0 | Not-determined | Genome-fragment | NA | NA | 0 |
| S8_vs_k141_148178\|\|full | 5977 | No | NA | 12 | 1 | 1 | Low-quality | Genome-fragment | 13.69 | AAI-based (high-confidence) | 0 |
| S6_extracted_k141_21516 | 1048 | No | NA | 1 | 0 | 1 | Not-determined | Genome-fragment | NA | NA | 0 |
| S3_extracted_k141_122204 | 5980 | No | NA | 14 | 3 | 0 | Low-quality | Genome-fragment | 12.07 | AAI-based (high-confidence) | 0 |
| S6_extracted_k141_49948 | 1204 | No | NA | 1 | 0 | 1 | Not-determined | Genome-fragment | NA | NA | 0 |
| S6_extracted_k141_5157 | 1868 | No | NA | 1 | 0 | 0 | Not-determined | Genome-fragment | NA | NA | 0 |
| S6_extracted_k141_7542 | 4023 | No | NA | 1 | 0 | 1 | Not-determined | Genome-fragment | NA | NA | 0 |
| S6_extracted_k141_24299 | 3791 | No | NA | 3 | 0 | 1 | Not-determined | Genome-fragment | NA | NA | 0 |
| S11_vs_k141_39795\|\|full | 5981 | No | NA | 4 | 0 | 0 | Low-quality | Genome-fragment | 3.67 | AAI-based (high-confidence) | 0 |
| S6_extracted_k141_24303 | 1089 | No | NA | 1 | 0 | 1 | Not-determined | Genome-fragment | NA | NA | 0 |
| S6_extracted_k141_38404 | 1036 | No | NA | 1 | 0 | 1 | Not-determined | Genome-fragment | NA | NA | 0 |
| S6_extracted_k141_45434 | 2631 | No | NA | 2 | 0 | 2 | Not-determined | Genome-fragment | NA | NA | 0 |
| S6_extracted_k141_35894 | 1019 | No | NA | 2 | 0 | 1 | Not-determined | Genome-fragment | NA | NA | 0 |
| S6_extracted_k141_50028 | 1536 | No | NA | 1 | 0 | 1 | Not-determined | Genome-fragment | NA | NA | 0 |
| S7_vs_89980\|\|full | 5981 | No | NA | 5 | 5 | 0 | Low-quality | Genome-fragment | 14.61 | AAI-based (high-confidence) | 0 |
| S6_extracted_k141_24320 | 3103 | No | NA | 2 | 0 | 1 | Not-determined | Genome-fragment | NA | NA | 0 |
| S6_extracted_k141_5212 | 1219 | No | NA | 2 | 0 | 1 | Not-determined | Genome-fragment | NA | NA | 0 |
| S6_extracted_k141_9993 | 2559 | No | NA | 2 | 0 | 0 | Not-determined | Genome-fragment | NA | NA | 0 |
| S6_extracted_k141_35943 | 1154 | No | NA | 1 | 0 | 0 | Not-determined | Genome-fragment | NA | NA | 0 |
| S6_extracted_k141_7585 | 1902 | No | NA | 1 | 0 | 0 | Not-determined | Genome-fragment | NA | NA | 0 |
| S1_extracted_k141_106234 | 5984 | No | NA | 2 | 0 | 2 | Low-quality | Genome-fragment | 3.38 | AAI-based (medium-confidence) | 0 |
| S6_extracted_k141_47625 | 1007 | No | NA | 2 | 0 | 0 | Not-determined | Genome-fragment | NA | NA | 0 |
| S4_vs_VIRSorter_29122-cat_2 | 5986 | No | NA | 12 | 2 | 0 | Low-quality | Genome-fragment | 6.85 | AAI-based (high-confidence) | 0 |
| S6_extracted_k141_31131 | 2008 | No | NA | 1 | 0 | 1 | Not-determined | Genome-fragment | NA | NA | 0 |
| S6_extracted_k141_43077 | 2478 | No | NA | 3 | 0 | 2 | Not-determined | Genome-fragment | NA | NA | 0 |
| S17_extracted_k141_27809 | 5989 | No | NA | 10 | 2 | 0 | Low-quality | Genome-fragment | 8.28 | AAI-based (high-confidence) | 0 |
| S6_extracted_k141_35976 | 1146 | No | NA | 1 | 0 | 0 | Not-determined | Genome-fragment | NA | NA | 0 |
| S6_extracted_k141_43086 | 1113 | No | NA | 2 | 0 | 1 | Not-determined | Genome-fragment | NA | NA | 0 |
| S6_extracted_k141_47661 | 1626 | No | NA | 3 | 0 | 2 | Not-determined | Genome-fragment | NA | NA | 0 |
| S6_extracted_k141_31202 | 1314 | No | NA | 2 | 0 | 2 | Not-determined | Genome-fragment | NA | NA | 0 |
| S6_extracted_k141_5317 | 1100 | No | NA | 4 | 0 | 0 | Not-determined | Genome-fragment | NA | NA | 0 |
| S17_vs_k141_52892\|\|full | 5991 | Yes | 656 | 7 | 1 | 4 | Low-quality | Genome-fragment | 0.45 | HMM-based (lower-bound) | 89.05 |
| S8_extracted_k141_160095 | 5994 | No | NA | 11 | 1 | 0 | Low-quality | Genome-fragment | 4.03 | HMM-based (lower-bound) | 0 |
| S7_extracted_23873 | 5995 | No | NA | 7 | 0 | 0 | Low-quality | Genome-fragment | 6.82 | AAI-based (medium-confidence) | 0 |
| S6_extracted_k141_47725 | 2125 | No | NA | 1 | 0 | 1 | Not-determined | Genome-fragment | NA | NA | 0 |
| S6_extracted_k141_33819 | 1040 | No | NA | 2 | 0 | 1 | Not-determined | Genome-fragment | NA | NA | 0 |
| S8_vs_k141_148924\|\|full | 5995 | No | NA | 9 | 5 | 0 | Low-quality | Genome-fragment | 14.17 | AAI-based (high-confidence) | 0 |
| S6_extracted_k141_33845 | 1049 | No | NA | 2 | 0 | 0 | Not-determined | Genome-fragment | NA | NA | 0 |
| S6_extracted_k141_17337 | 1581 | No | NA | 2 | 0 | 2 | Not-determined | Genome-fragment | NA | NA | 0 |
| S6_extracted_k141_45618 | 44283 | No | NA | 82 | 15 | 2 | Complete | High-quality | 100 | DTR (high-confidence) | 0 |
| S6_extracted_k141_40796 | 1767 | No | NA | 2 | 0 | 0 | Not-determined | Genome-fragment | NA | NA | 0 |
| S6_extracted_k141_40804 | 1228 | No | NA | 2 | 0 | 1 | Not-determined | Genome-fragment | NA | NA | 0 |
| S6_extracted_k141_45624 | 6637 | No | NA | 8 | 0 | 2 | Not-determined | Genome-fragment | NA | NA | 0 |
| S6_extracted_k141_38485 | 1162 | No | NA | 1 | 0 | 1 | Not-determined | Genome-fragment | NA | NA | 0 |
| S6_extracted_k141_3289 | 1296 | No | NA | 2 | 0 | 2 | Not-determined | Genome-fragment | NA | NA | 0 |
| S6_extracted_k141_43093 | 3239 | No | NA | 6 | 0 | 6 | Not-determined | Genome-fragment | NA | NA | 0 |
| S6_extracted_k141_26482 | 1563 | No | NA | 1 | 0 | 1 | Not-determined | Genome-fragment | NA | NA | 0 |
| S6_extracted_k141_28992 | 1075 | No | NA | 2 | 0 | 2 | Not-determined | Genome-fragment | NA | NA | 0 |
| S8_vs_k141_150993\|\|full | 5995 | No | NA | 6 | 0 | 3 | Low-quality | Genome-fragment | 8.5 | AAI-based (medium-confidence) | 0 |
| S6_extracted_k141_14882 | 1900 | No | NA | 4 | 0 | 2 | Not-determined | Genome-fragment | NA | NA | 0 |
| S6_extracted_k141_12738 | 1364 | No | NA | 1 | 0 | 0 | Not-determined | Genome-fragment | NA | NA | 0 |
| S6_extracted_k141_24553 | 16941 | No | NA | 25 | 8 | 0 | Medium-quality | Genome-fragment | 79.21 | AAI-based (medium-confidence) | 0 |
| S6_extracted_k141_45673 | 1036 | No | NA | 2 | 0 | 2 | Not-determined | Genome-fragment | NA | NA | 0 |
| S6_extracted_k141_52844 | 1105 | No | NA | 1 | 0 | 0 | Not-determined | Genome-fragment | NA | NA | 0 |
| S6_extracted_k141_33955 | 1007 | No | NA | 2 | 0 | 1 | Not-determined | Genome-fragment | NA | NA | 0 |
| S6_extracted_k141_24598 | 1261 | No | NA | 1 | 0 | 0 | Not-determined | Genome-fragment | NA | NA | 0 |
| S6_extracted_k141_7802 | 1126 | No | NA | 1 | 0 | 1 | Not-determined | Genome-fragment | NA | NA | 0 |
| S3_extracted_k141_76586 | 6003 | No | NA | 5 | 0 | 1 | Low-quality | Genome-fragment | 4.74 | AAI-based (medium-confidence) | 0 |
| S6_extracted_k141_24602 | 1607 | No | NA | 3 | 0 | 0 | Not-determined | Genome-fragment | NA | NA | 0 |
| S3_extracted_k141_3969 | 6004 | No | NA | 8 | 0 | 0 | Low-quality | Genome-fragment | 3.67 | AAI-based (high-confidence) | 0 |
| S3_vs_k141_17044\|\|full | 6004 | No | NA | 8 | 5 | 0 | Low-quality | Genome-fragment | 13.96 | AAI-based (high-confidence) | 0 |
| S6_extracted_k141_36078 | 1714 | No | NA | 4 | 0 | 0 | Not-determined | Genome-fragment | NA | NA | 0 |
| S6_extracted_k141_19264 | 1811 | No | NA | 1 | 0 | 0 | Not-determined | Genome-fragment | NA | NA | 0 |
| S6_extracted_k141_30153 | 6005 | No | NA | 12 | 4 | 0 | Low-quality | Genome-fragment | 13.24 | AAI-based (high-confidence) | 0 |
| S6_extracted_k141_10267 | 1242 | No | NA | 2 | 0 | 2 | Not-determined | Genome-fragment | NA | NA | 0 |
| S6_extracted_k141_24665 | 1093 | No | NA | 1 | 0 | 1 | Not-determined | Genome-fragment | NA | NA | 0 |
| S6_extracted_k141_45809 | 1151 | No | NA | 3 | 0 | 0 | Not-determined | Genome-fragment | NA | NA | 0 |
| S6_vs_k141_17081\|\|full | 6006 | No | NA | 7 | 1 | 0 | Low-quality | Genome-fragment | 16.25 | AAI-based (high-confidence) | 0 |
| S6_extracted_k141_1002 | 1589 | No | NA | 1 | 0 | 1 | Not-determined | Genome-fragment | NA | NA | 0 |
| S6_extracted_k141_10277 | 1144 | No | NA | 2 | 0 | 0 | Not-determined | Genome-fragment | NA | NA | 0 |
| S6_extracted_k141_31372 | 1117 | No | NA | 1 | 0 | 1 | Not-determined | Genome-fragment | NA | NA | 0 |
| S6_extracted_k141_34030 | 1259 | No | NA | 3 | 0 | 0 | Not-determined | Genome-fragment | NA | NA | 0 |
| S6_extracted_k141_3435 | 12140 | No | NA | 23 | 14 | 0 | Medium-quality | Genome-fragment | 55.39 | AAI-based (high-confidence) | 0 |
| S16_vs_75306\|\|full | 6011 | No | NA | 8 | 0 | 0 | Low-quality | Genome-fragment | 9.62 | AAI-based (medium-confidence) | 0 |
| S6_extracted_k141_5476 | 1335 | No | NA | 3 | 0 | 0 | Not-determined | Genome-fragment | NA | NA | 0 |
| S6_extracted_k141_3443 | 1208 | No | NA | 2 | 0 | 0 | Not-determined | Genome-fragment | NA | NA | 0 |
| S6_extracted_k141_38631 | 5027 | No | NA | 9 | 0 | 0 | Not-determined | Genome-fragment | NA | NA | 0 |
| S8_vs_k141_189803\|\|full | 6012 | No | NA | 13 | 0 | 0 | Low-quality | Genome-fragment | 17.64 | AAI-based (high-confidence) | 0 |
| S6_extracted_k141_29134 | 2798 | No | NA | 1 | 0 | 0 | Not-determined | Genome-fragment | NA | NA | 0 |
| S8_vs_k141_25758\|\|full | 6014 | No | NA | 15 | 1 | 0 | Low-quality | Genome-fragment | 14.05 | AAI-based (high-confidence) | 0 |
| S14_vs_k141_10572\|\|full | 6016 | No | NA | 9 | 5 | 0 | Low-quality | Genome-fragment | 15.04 | AAI-based (medium-confidence) | 0 |
| S6_extracted_k141_38642 | 1800 | No | NA | 4 | 0 | 1 | Not-determined | Genome-fragment | NA | NA | 0 |
| S6_extracted_k141_40924 | 3045 | No | NA | 3 | 0 | 3 | Not-determined | Genome-fragment | NA | NA | 0 |
| S16_vs_45001\|\|full | 6016 | No | NA | 14 | 2 | 0 | Low-quality | Genome-fragment | 17.32 | AAI-based (high-confidence) | 0 |
| S6_extracted_k141_5504 | 1298 | No | NA | 1 | 0 | 1 | Not-determined | Genome-fragment | NA | NA | 0 |
| S6_extracted_k141_36185 | 1160 | No | NA | 3 | 0 | 0 | Not-determined | Genome-fragment | NA | NA | 0 |
| S6_extracted_k141_12801 | 1124 | No | NA | 1 | 0 | 1 | Not-determined | Genome-fragment | NA | NA | 0 |
| S8_extracted_k141_4488 | 6018 | No | NA | 7 | 1 | 0 | Low-quality | Genome-fragment | 3.56 | AAI-based (high-confidence) | 0 |
| S6_extracted_k141_34138 | 1025 | No | NA | 1 | 0 | 0 | Not-determined | Genome-fragment | NA | NA | 0 |
| S6_extracted_k141_12818 | 1042 | No | NA | 1 | 0 | 1 | Not-determined | Genome-fragment | NA | NA | 0 |
| S6_extracted_k141_19300 | 3397 | No | NA | 4 | 0 | 1 | Not-determined | Genome-fragment | NA | NA | 0 |
| S6_extracted_k141_19301 | 1046 | No | NA | 1 | 0 | 0 | Not-determined | Genome-fragment | NA | NA | 0 |
| S6_extracted_k141_21797 | 1565 | No | NA | 2 | 0 | 2 | Not-determined | Genome-fragment | NA | NA | 0 |
| S6_extracted_k141_50231 | 1034 | No | NA | 1 | 0 | 1 | Not-determined | Genome-fragment | NA | NA | 0 |
| S6_extracted_k141_24744 | 1060 | No | NA | 1 | 0 | 1 | Not-determined | Genome-fragment | NA | NA | 0 |
| S6_extracted_k141_19321 | 1420 | No | NA | 1 | 0 | 1 | Not-determined | Genome-fragment | NA | NA | 0 |
| S14_extracted_k141_76835 | 6019 | No | NA | 8 | 1 | 0 | Low-quality | Genome-fragment | 14.37 | AAI-based (medium-confidence) | 0 |
| S6_extracted_k141_54689 | 1685 | No | NA | 1 | 0 | 0 | Not-determined | Genome-fragment | NA | NA | 0 |
| S6_extracted_k141_15122 | 1512 | No | NA | 2 | 0 | 1 | Not-determined | Genome-fragment | NA | NA | 0 |
| S6_extracted_k141_36274 | 6160 | No | NA | 8 | 0 | 2 | Not-determined | Genome-fragment | NA | NA | 0 |
| S5_extracted_k141_56403 | 6019 | No | NA | 9 | 2 | 0 | Low-quality | Genome-fragment | 43.34 | AAI-based (high-confidence) | 0 |
| S4_extracted_11320 | 6019 | No | NA | 7 | 0 | 0 | Low-quality | Genome-fragment | 8.39 | AAI-based (high-confidence) | 0 |
| S6_extracted_k141_34208 | 7172 | No | NA | 7 | 0 | 3 | Not-determined | Genome-fragment | NA | NA | 0 |
| S6_extracted_k141_54747 | 1851 | No | NA | 2 | 0 | 2 | Not-determined | Genome-fragment | NA | NA | 0 |
| S6_extracted_k141_47996 | 2169 | No | NA | 2 | 0 | 2 | Not-determined | Genome-fragment | NA | NA | 0 |
| S6_extracted_k141_38772 | 3128 | No | NA | 2 | 0 | 0 | Not-determined | Genome-fragment | NA | NA | 0 |
| S6_extracted_k141_10318 | 1331 | No | NA | 1 | 0 | 1 | Not-determined | Genome-fragment | NA | NA | 0 |
| S6_extracted_k141_50309 | 1176 | No | NA | 1 | 0 | 0 | Not-determined | Genome-fragment | NA | NA | 0 |
| S6_extracted_k141_26613 | 2103 | No | NA | 2 | 0 | 0 | Not-determined | Genome-fragment | NA | NA | 0 |
| S6_extracted_k141_26615 | 1611 | No | NA | 3 | 0 | 3 | Not-determined | Genome-fragment | NA | NA | 0 |
| S6_extracted_k141_54781 | 1027 | No | NA | 2 | 0 | 1 | Not-determined | Genome-fragment | NA | NA | 0 |
| S6_extracted_k141_31501 | 1378 | No | NA | 1 | 0 | 1 | Not-determined | Genome-fragment | NA | NA | 0 |
| S6_extracted_k141_31502 | 1237 | No | NA | 1 | 0 | 0 | Not-determined | Genome-fragment | NA | NA | 0 |
| S16_vs_104991\|\|full | 6020 | No | NA | 7 | 0 | 3 | Low-quality | Genome-fragment | 9.24 | AAI-based (medium-confidence) | 0 |
| S6_extracted_k141_34257 | 3930 | No | NA | 3 | 0 | 1 | Not-determined | Genome-fragment | NA | NA | 0 |
| S6_extracted_k141_10366 | 2103 | No | NA | 1 | 0 | 1 | Not-determined | Genome-fragment | NA | NA | 0 |
| S6_extracted_k141_5658 | 1868 | No | NA | 3 | 0 | 1 | Not-determined | Genome-fragment | NA | NA | 0 |
| S11_vs_k141_56577\|\|full | 6021 | No | NA | 8 | 5 | 0 | Low-quality | Genome-fragment | 13.31 | AAI-based (high-confidence) | 0 |
| S6_extracted_k141_10384 | 1549 | No | NA | 1 | 0 | 0 | Not-determined | Genome-fragment | NA | NA | 0 |
| S6_extracted_k141_41015 | 1244 | No | NA | 2 | 0 | 1 | Not-determined | Genome-fragment | NA | NA | 0 |
| S6_extracted_k141_21904 | 1035 | No | NA | 1 | 0 | 0 | Not-determined | Genome-fragment | NA | NA | 0 |
| S6_extracted_k141_10396 | 1091 | No | NA | 1 | 0 | 0 | Not-determined | Genome-fragment | NA | NA | 0 |
| S6_extracted_k141_19401 | 1226 | No | NA | 2 | 0 | 0 | Not-determined | Genome-fragment | NA | NA | 0 |
| S6_extracted_k141_38839 | 1136 | No | NA | 2 | 0 | 1 | Not-determined | Genome-fragment | NA | NA | 0 |
| S6_extracted_k141_17685 | 1291 | No | NA | 1 | 0 | 1 | Not-determined | Genome-fragment | NA | NA | 0 |
| S6_extracted_k141_15201 | 1033 | No | NA | 1 | 0 | 1 | Not-determined | Genome-fragment | NA | NA | 0 |
| S6_extracted_k141_3684 | 1015 | No | NA | 2 | 0 | 1 | Not-determined | Genome-fragment | NA | NA | 0 |
| S6_extracted_k141_10469 | 1023 | No | NA | 2 | 0 | 1 | Not-determined | Genome-fragment | NA | NA | 0 |
| S6_extracted_k141_10499 | 1105 | No | NA | 2 | 0 | 2 | Not-determined | Genome-fragment | NA | NA | 0 |
| S6_extracted_k141_15248 | 1459 | No | NA | 2 | 0 | 2 | Not-determined | Genome-fragment | NA | NA | 0 |
| S6_extracted_k141_17722 | 1831 | No | NA | 3 | 0 | 3 | Not-determined | Genome-fragment | NA | NA | 0 |
| S16_vs_44217\|\|full | 6021 | No | NA | 7 | 0 | 0 | Low-quality | Genome-fragment | 13.94 | AAI-based (medium-confidence) | 0 |
| S3_extracted_k141_109599 | 6022 | No | NA | 19 | 10 | 0 | Low-quality | Genome-fragment | 27.47 | AAI-based (high-confidence) | 0 |
| S6_extracted_k141_36344 | 1062 | No | NA | 2 | 0 | 0 | Not-determined | Genome-fragment | NA | NA | 0 |
| S6_extracted_k141_38921 | 1076 | No | NA | 1 | 0 | 1 | Not-determined | Genome-fragment | NA | NA | 0 |
| S6_extracted_k141_17763 | 1763 | No | NA | 2 | 0 | 1 | Not-determined | Genome-fragment | NA | NA | 0 |
| S6_extracted_k141_29431 | 1007 | No | NA | 3 | 0 | 0 | Not-determined | Genome-fragment | NA | NA | 0 |
| S6_extracted_k141_8149 | 2759 | No | NA | 2 | 0 | 2 | Not-determined | Genome-fragment | NA | NA | 0 |
| S6_extracted_k141_17774 | 1170 | No | NA | 1 | 0 | 0 | Not-determined | Genome-fragment | NA | NA | 0 |
| S5_vs_k141_105929\|\|full | 6022 | No | NA | 4 | 2 | 0 | Low-quality | Genome-fragment | 12.21 | AAI-based (high-confidence) | 0 |
| S6_extracted_k141_15314 | 1145 | No | NA | 2 | 0 | 1 | Not-determined | Genome-fragment | NA | NA | 0 |
| S14_vs_k141_89604\|\|full | 6023 | Yes | 2007 | 6 | 1 | 2 | Low-quality | Genome-fragment | 0.93 | HMM-based (lower-bound) | 66.68 |
| S1_vs_k141_120728\|\|full | 6024 | No | NA | 8 | 0 | 0 | Low-quality | Genome-fragment | 18.18 | AAI-based (high-confidence) | 0 |
| S6_extracted_k141_36378 | 1117 | No | NA | 2 | 0 | 0 | Not-determined | Genome-fragment | NA | NA | 0 |
| S6_extracted_k141_19464 | 1139 | No | NA | 1 | 0 | 1 | Not-determined | Genome-fragment | NA | NA | 0 |
| S6_extracted_k141_36382 | 1759 | No | NA | 1 | 0 | 1 | Not-determined | Genome-fragment | NA | NA | 0 |
| S6_extracted_k141_29534 | 1418 | No | NA | 1 | 0 | 0 | Not-determined | Genome-fragment | NA | NA | 0 |
| S6_extracted_k141_17848 | 1256 | No | NA | 3 | 0 | 0 | Not-determined | Genome-fragment | NA | NA | 0 |
| S6_extracted_k141_31741 | 1053 | No | NA | 3 | 0 | 3 | Not-determined | Genome-fragment | NA | NA | 0 |
| S17_vs_k141_85173\|\|full | 6026 | No | NA | 12 | 1 | 0 | Low-quality | Genome-fragment | 7.63 | AAI-based (high-confidence) | 0 |
| S1_extracted_k141_135007 | 6027 | No | NA | 11 | 2 | 0 | Low-quality | Genome-fragment | 44.08 | AAI-based (medium-confidence) | 0 |
| S6_extracted_k141_29555 | 3161 | No | NA | 1 | 0 | 0 | Not-determined | Genome-fragment | NA | NA | 0 |
| S6_extracted_k141_29563 | 1982 | No | NA | 3 | 0 | 1 | Not-determined | Genome-fragment | NA | NA | 0 |
| S6_extracted_k141_29564 | 1216 | No | NA | 2 | 0 | 2 | Not-determined | Genome-fragment | NA | NA | 0 |
| S6_extracted_k141_5867 | 3090 | No | NA | 4 | 0 | 0 | Not-determined | Genome-fragment | NA | NA | 0 |
| S6_extracted_k141_5868 | 2998 | No | NA | 2 | 0 | 1 | Not-determined | Genome-fragment | NA | NA | 0 |
| S6_extracted_k141_50480 | 1100 | No | NA | 3 | 0 | 1 | Not-determined | Genome-fragment | NA | NA | 0 |
| S6_extracted_k141_22105 | 1153 | No | NA | 2 | 0 | 0 | Not-determined | Genome-fragment | NA | NA | 0 |
| S8_vs_k141_25938\|\|full | 6028 | No | NA | 5 | 1 | 1 | Low-quality | Genome-fragment | 3.27 | HMM-based (lower-bound) | 0 |
| S17_extracted_k141_74152 | 6034 | No | NA | 4 | 0 | 2 | Low-quality | Genome-fragment | 6.36 | AAI-based (medium-confidence) | 0 |
| S6_extracted_k141_41160 | 1367 | No | NA | 2 | 0 | 1 | Not-determined | Genome-fragment | NA | NA | 0 |
| S6_extracted_k141_13152 | 1359 | No | NA | 1 | 0 | 0 | Not-determined | Genome-fragment | NA | NA | 0 |
| S8_extracted_k141_142067 | 6034 | No | NA | 6 | 6 | 0 | Low-quality | Genome-fragment | 19.17 | AAI-based (high-confidence) | 0 |
| S6_extracted_k141_1248 | 1080 | No | NA | 2 | 0 | 2 | Not-determined | Genome-fragment | NA | NA | 0 |
| S6_extracted_k141_29658 | 1743 | No | NA | 2 | 0 | 1 | Not-determined | Genome-fragment | NA | NA | 0 |
| S6_extracted_k141_36511 | 1357 | No | NA | 1 | 0 | 1 | Not-determined | Genome-fragment | NA | NA | 0 |
| S6_extracted_k141_46079 | 1134 | No | NA | 1 | 0 | 1 | Not-determined | Genome-fragment | NA | NA | 0 |
| S6_extracted_k141_10736 | 2175 | No | NA | 3 | 0 | 2 | Not-determined | Genome-fragment | NA | NA | 0 |
| S6_extracted_k141_31819 | 3031 | No | NA | 2 | 0 | 1 | Not-determined | Genome-fragment | NA | NA | 0 |
| S6_extracted_k141_13254 | 3170 | No | NA | 5 | 0 | 2 | Not-determined | Genome-fragment | NA | NA | 0 |
| S9_vs_k141_52264\|\|full | 6035 | No | NA | 8 | 6 | 0 | Low-quality | Genome-fragment | 17.6 | AAI-based (high-confidence) | 0 |
| S6_extracted_k141_29717 | 1288 | No | NA | 1 | 0 | 1 | Not-determined | Genome-fragment | NA | NA | 0 |
| S6_extracted_k141_54948 | 1100 | No | NA | 2 | 0 | 2 | Not-determined | Genome-fragment | NA | NA | 0 |
| S6_extracted_k141_36552 | 1193 | No | NA | 2 | 0 | 0 | Not-determined | Genome-fragment | NA | NA | 0 |
| S6_extracted_k141_36555 | 1101 | No | NA | 2 | 0 | 0 | Not-determined | Genome-fragment | NA | NA | 0 |
| S6_extracted_k141_25092 | 1392 | No | NA | 2 | 0 | 1 | Not-determined | Genome-fragment | NA | NA | 0 |
| S6_extracted_k141_34568 | 2040 | No | NA | 2 | 0 | 1 | Not-determined | Genome-fragment | NA | NA | 0 |
| S6_extracted_k141_41251 | 1123 | No | NA | 1 | 0 | 1 | Not-determined | Genome-fragment | NA | NA | 0 |
| S17_vs_k141_124057\|\|full | 6037 | No | NA | 12 | 3 | 0 | Low-quality | Genome-fragment | 17.33 | AAI-based (high-confidence) | 0 |
| S8_vs_k141_167681\|\|full | 6037 | No | NA | 5 | 0 | 3 | Low-quality | Genome-fragment | 3.26 | AAI-based (medium-confidence) | 0 |
| S6_extracted_k141_34579 | 1552 | No | NA | 2 | 0 | 1 | Not-determined | Genome-fragment | NA | NA | 0 |
| S6_extracted_k141_1341 | 1029 | No | NA | 1 | 0 | 1 | Not-determined | Genome-fragment | NA | NA | 0 |
| S16_vs_83258\|\|full | 6038 | Yes | 5288 | 8 | 1 | 1 | Low-quality | Genome-fragment | 3.37 | HMM-based (lower-bound) | 12.42 |
| S12_extracted_k141_84301 | 6041 | No | NA | 5 | 0 | 0 | Low-quality | Genome-fragment | 13.93 | AAI-based (medium-confidence) | 0 |
| S6_extracted_k141_1356 | 1377 | No | NA | 3 | 0 | 1 | Not-determined | Genome-fragment | NA | NA | 0 |
| S3_extracted_k141_28197 | 6042 | No | NA | 6 | 5 | 0 | Low-quality | Genome-fragment | 10.09 | AAI-based (high-confidence) | 0 |
| S6_extracted_k141_34672 | 1207 | No | NA | 1 | 0 | 0 | Not-determined | Genome-fragment | NA | NA | 0 |
| S8_extracted_k141_158113 | 6043 | No | NA | 3 | 1 | 0 | Low-quality | Genome-fragment | 6.02 | AAI-based (high-confidence) | 0 |
| S11_extracted_k141_15619 | 6044 | No | NA | 12 | 1 | 0 | Low-quality | Genome-fragment | 8.56 | AAI-based (high-confidence) | 0 |
| S17_extracted_k141_101487 | 6046 | No | NA | 16 | 1 | 0 | Low-quality | Genome-fragment | 9.06 | AAI-based (high-confidence) | 0 |
| S7_vs_103336\|\|full | 6047 | No | NA | 5 | 3 | 0 | Low-quality | Genome-fragment | 9.33 | HMM-based (lower-bound) | 0 |
| S6_extracted_k141_8387 | 1482 | No | NA | 2 | 0 | 2 | Not-determined | Genome-fragment | NA | NA | 0 |
| S6_extracted_k141_46221 | 1016 | No | NA | 2 | 0 | 0 | Not-determined | Genome-fragment | NA | NA | 0 |
| S6_extracted_k141_4092 | 1355 | No | NA | 2 | 0 | 0 | Not-determined | Genome-fragment | NA | NA | 0 |
| S7_vs_125910\|\|full | 6052 | No | NA | 7 | 0 | 0 | Low-quality | Genome-fragment | 10.36 | AAI-based (high-confidence) | 0 |
| S6_extracted_k141_18154 | 1124 | No | NA | 2 | 0 | 2 | Not-determined | Genome-fragment | NA | NA | 0 |
| S6_extracted_k141_34713 | 1130 | No | NA | 2 | 0 | 0 | Not-determined | Genome-fragment | NA | NA | 0 |
| S6_extracted_k141_41420 | 1398 | No | NA | 4 | 0 | 0 | Not-determined | Genome-fragment | NA | NA | 0 |
| S6_extracted_k141_19634 | 1211 | No | NA | 1 | 0 | 0 | Not-determined | Genome-fragment | NA | NA | 0 |
| S14_vs_k141_43042\|\|full | 6053 | No | NA | 12 | 5 | 0 | Low-quality | Genome-fragment | 14.12 | AAI-based (high-confidence) | 0 |
| S9_vs_k141_19806\|\|full | 6055 | No | NA | 8 | 0 | 0 | Low-quality | Genome-fragment | 14.3 | AAI-based (high-confidence) | 0 |
| S14_vs_k141_68202\|\|full | 6058 | No | NA | 7 | 5 | 0 | Low-quality | Genome-fragment | 16.29 | AAI-based (high-confidence) | 0 |
| S1_vs_k141_121394\|\|full | 6063 | No | NA | 6 | 6 | 0 | Low-quality | Genome-fragment | 12.93 | AAI-based (high-confidence) | 0 |
| S6_extracted_k141_13418 | 1662 | No | NA | 1 | 0 | 1 | Not-determined | Genome-fragment | NA | NA | 0 |
| S17_extracted_k141_17948 | 6064 | No | NA | 8 | 3 | 0 | Low-quality | Genome-fragment | 9.09 | AAI-based (high-confidence) | 0 |
| S6_extracted_k141_25268 | 1184 | No | NA | 3 | 0 | 0 | Not-determined | Genome-fragment | NA | NA | 0 |
| S6_extracted_k141_13448 | 1779 | No | NA | 1 | 0 | 0 | Not-determined | Genome-fragment | NA | NA | 0 |
| S6_extracted_k141_41487 | 1312 | No | NA | 1 | 0 | 0 | Not-determined | Genome-fragment | NA | NA | 0 |
| S6_extracted_k141_8537 | 1886 | No | NA | 3 | 0 | 2 | Not-determined | Genome-fragment | NA | NA | 0 |
| S6_extracted_k141_8539 | 3012 | No | NA | 5 | 0 | 3 | Not-determined | Genome-fragment | NA | NA | 0 |
| S6_extracted_k141_8542 | 1966 | No | NA | 3 | 0 | 1 | Not-determined | Genome-fragment | NA | NA | 0 |
| S6_extracted_k141_13474 | 1277 | No | NA | 2 | 0 | 1 | Not-determined | Genome-fragment | NA | NA | 0 |
| S1_extracted_k141_37186 | 6066 | No | NA | 8 | 6 | 0 | Low-quality | Genome-fragment | 18.18 | AAI-based (high-confidence) | 0 |
| S6_extracted_k141_48360 | 1075 | No | NA | 2 | 0 | 0 | Not-determined | Genome-fragment | NA | NA | 0 |
| S6_extracted_k141_15680 | 1010 | No | NA | 2 | 0 | 1 | Not-determined | Genome-fragment | NA | NA | 0 |
| S6_extracted_k141_56499 | 6067 | No | NA | 8 | 0 | 0 | Low-quality | Genome-fragment | 5.1 | AAI-based (high-confidence) | 0 |
| S17_extracted_k141_3772 | 6071 | Yes | 5720 | 8 | 1 | 1 | Low-quality | Genome-fragment | 10.25 | AAI-based (high-confidence) | 5.78 |
| S6_extracted_k141_1404 | 1615 | No | NA | 1 | 0 | 1 | Not-determined | Genome-fragment | NA | NA | 0 |
| S6_extracted_k141_4214 | 1385 | No | NA | 1 | 0 | 1 | Not-determined | Genome-fragment | NA | NA | 0 |
| S6_extracted_k141_4229 | 1139 | No | NA | 1 | 0 | 1 | Not-determined | Genome-fragment | NA | NA | 0 |
| S6_extracted_k141_46331 | 1043 | No | NA | 1 | 0 | 0 | Not-determined | Genome-fragment | NA | NA | 0 |
| S5_extracted_k141_76418 | 6075 | No | NA | 5 | 0 | 4 | Low-quality | Genome-fragment | 5.66 | AAI-based (medium-confidence) | 0 |
| S6_extracted_k141_25329 | 1200 | No | NA | 2 | 0 | 2 | Not-determined | Genome-fragment | NA | NA | 0 |
| S6_extracted_k141_43699 | 1365 | No | NA | 1 | 0 | 0 | Not-determined | Genome-fragment | NA | NA | 0 |
| S6_extracted_k141_19801 | 4610 | No | NA | 9 | 0 | 1 | Not-determined | Genome-fragment | NA | NA | 0 |
| S6_extracted_k141_13536 | 2112 | No | NA | 1 | 0 | 0 | Not-determined | Genome-fragment | NA | NA | 0 |
| S5_vs_k141_3835\|\|full | 6075 | No | NA | 7 | 5 | 0 | Low-quality | Genome-fragment | 15.99 | AAI-based (high-confidence) | 0 |
| S6_extracted_k141_46370 | 2245 | No | NA | 1 | 0 | 1 | Not-determined | Genome-fragment | NA | NA | 0 |
| S5_vs_k141_61891\|\|full | 6076 | Yes | 2142 | 6 | 2 | 3 | Low-quality | Genome-fragment | 4.98 | AAI-based (high-confidence) | 64.75 |
| S6_extracted_k141_1427 | 4440 | No | NA | 6 | 0 | 5 | Not-determined | Genome-fragment | NA | NA | 0 |
| S14_vs_k141_102322\|\|full | 6079 | No | NA | 3 | 0 | 1 | Low-quality | Genome-fragment | 12.18 | AAI-based (medium-confidence) | 0 |
| S16_vs_121972\|\|full | 6079 | No | NA | 11 | 1 | 2 | Low-quality | Genome-fragment | 4.09 | HMM-based (lower-bound) | 0 |
| S6_extracted_k141_29869 | 1641 | No | NA | 5 | 0 | 5 | Not-determined | Genome-fragment | NA | NA | 0 |
| S6_extracted_k141_29872 | 1177 | No | NA | 2 | 0 | 2 | Not-determined | Genome-fragment | NA | NA | 0 |
| S6_extracted_k141_15756 | 3686 | No | NA | 3 | 0 | 0 | Not-determined | Genome-fragment | NA | NA | 0 |
| S6_extracted_k141_43718 | 2840 | No | NA | 3 | 0 | 2 | Not-determined | Genome-fragment | NA | NA | 0 |
| S6_extracted_k141_13569 | 1073 | No | NA | 2 | 0 | 1 | Not-determined | Genome-fragment | NA | NA | 0 |
| S6_extracted_k141_1461 | 1202 | No | NA | 3 | 0 | 1 | Not-determined | Genome-fragment | NA | NA | 0 |
| S6_extracted_k141_22379 | 2389 | No | NA | 3 | 0 | 3 | Not-determined | Genome-fragment | NA | NA | 0 |
| S6_extracted_k141_6158 | 3048 | No | NA | 3 | 0 | 3 | Not-determined | Genome-fragment | NA | NA | 0 |
| S17_vs_k141_66040\|\|full | 6080 | No | NA | 12 | 7 | 0 | Low-quality | Genome-fragment | 8.66 | AAI-based (high-confidence) | 0 |
| S6_extracted_k141_53413 | 2329 | No | NA | 1 | 0 | 0 | Not-determined | Genome-fragment | NA | NA | 0 |
| S5_extracted_k141_126748 | 6080 | No | NA | 13 | 3 | 0 | Low-quality | Genome-fragment | 14.19 | AAI-based (medium-confidence) | 0 |
| S6_extracted_k141_34881 | 1057 | No | NA | 1 | 0 | 1 | Not-determined | Genome-fragment | NA | NA | 0 |
| S6_extracted_k141_22411 | 1092 | No | NA | 1 | 0 | 1 | Not-determined | Genome-fragment | NA | NA | 0 |
| S6_extracted_k141_55108 | 1064 | No | NA | 1 | 0 | 1 | Not-determined | Genome-fragment | NA | NA | 0 |
| S16_extracted_49495 | 6082 | No | NA | 9 | 0 | 0 | Low-quality | Genome-fragment | 14.74 | AAI-based (high-confidence) | 0 |
| S6_extracted_k141_29905 | 3933 | No | NA | 2 | 0 | 1 | Not-determined | Genome-fragment | NA | NA | 0 |
| S12_extracted_k141_10517 | 6084 | No | NA | 10 | 3 | 0 | Low-quality | Genome-fragment | 9.4 | AAI-based (high-confidence) | 0 |
| S8_extracted_k141_182108 | 6084 | No | NA | 13 | 1 | 0 | Low-quality | Genome-fragment | 5.55 | HMM-based (lower-bound) | 0 |
| S1_extracted_k141_23678 | 6085 | No | NA | 13 | 2 | 1 | Low-quality | Genome-fragment | 7.55 | AAI-based (high-confidence) | 0 |
| S6_extracted_k141_27214 | 1008 | No | NA | 2 | 0 | 2 | Not-determined | Genome-fragment | NA | NA | 0 |
| S6_extracted_k141_18334 | 1839 | No | NA | 2 | 0 | 1 | Not-determined | Genome-fragment | NA | NA | 0 |
| S8_vs_k141_4161\|\|full | 6085 | No | NA | 7 | 0 | 1 | Low-quality | Genome-fragment | 8.28 | AAI-based (high-confidence) | 0 |
| S6_extracted_k141_41749 | 1701 | No | NA | 1 | 0 | 1 | Not-determined | Genome-fragment | NA | NA | 0 |
| S5_vs_k141_14440\|\|full | 6088 | No | NA | 8 | 7 | 0 | Low-quality | Genome-fragment | 14.34 | AAI-based (high-confidence) | 0 |
| S6_extracted_k141_27250 | 1092 | No | NA | 1 | 0 | 0 | Not-determined | Genome-fragment | NA | NA | 0 |
| S11_vs_k141_34567\|\|full | 6090 | No | NA | 8 | 4 | 0 | Low-quality | Genome-fragment | 14.25 | AAI-based (medium-confidence) | 0 |
| S11_vs_k141_54904\|\|full | 6090 | No | NA | 6 | 0 | 6 | Low-quality | Genome-fragment | 3.03 | AAI-based (medium-confidence) | 0 |
| S6_extracted_k141_46477 | 1749 | No | NA | 2 | 0 | 2 | Not-determined | Genome-fragment | NA | NA | 0 |
| S6_extracted_k141_30051 | 5045 | No | NA | 5 | 0 | 1 | Not-determined | Genome-fragment | NA | NA | 0 |
| S6_extracted_k141_6314 | 7600 | No | NA | 7 | 4 | 0 | Medium-quality | Genome-fragment | 54.72 | AAI-based (high-confidence) | 0 |
| S6_extracted_k141_53575 | 1094 | No | NA | 4 | 0 | 0 | Not-determined | Genome-fragment | NA | NA | 0 |
| S6_extracted_k141_43789 | 1533 | No | NA | 2 | 0 | 1 | Not-determined | Genome-fragment | NA | NA | 0 |
| S15_extracted_66789 | 6091 | No | NA | 3 | 0 | 1 | Low-quality | Genome-fragment | 5.61 | AAI-based (high-confidence) | 0 |
| S6_extracted_k141_22536 | 1071 | No | NA | 2 | 0 | 1 | Not-determined | Genome-fragment | NA | NA | 0 |
| S6_extracted_k141_32194 | 1890 | No | NA | 1 | 0 | 0 | Not-determined | Genome-fragment | NA | NA | 0 |
| S6_extracted_k141_55287 | 1269 | No | NA | 1 | 0 | 1 | Not-determined | Genome-fragment | NA | NA | 0 |
| S6_extracted_k141_39121 | 2079 | No | NA | 3 | 0 | 3 | Not-determined | Genome-fragment | NA | NA | 0 |
| S6_extracted_k141_32210 | 1017 | No | NA | 2 | 0 | 0 | Not-determined | Genome-fragment | NA | NA | 0 |
| S16_extracted_137562 | 6095 | No | NA | 8 | 5 | 0 | Low-quality | Genome-fragment | 17.14 | AAI-based (high-confidence) | 0 |
| S6_extracted_k141_22561 | 1345 | No | NA | 2 | 0 | 1 | Not-determined | Genome-fragment | NA | NA | 0 |
| S6_extracted_k141_55309 | 1171 | No | NA | 1 | 0 | 1 | Not-determined | Genome-fragment | NA | NA | 0 |
| S6_extracted_k141_55313 | 1999 | No | NA | 1 | 0 | 0 | Not-determined | Genome-fragment | NA | NA | 0 |
| S6_extracted_k141_18408 | 1314 | No | NA | 2 | 0 | 2 | Not-determined | Genome-fragment | NA | NA | 0 |
| S16_vs_28348\|\|full | 6096 | No | NA | 9 | 1 | 0 | Low-quality | Genome-fragment | 17.62 | AAI-based (high-confidence) | 0 |
| S6_vs_k141_28260\|\|full | 6096 | No | NA | 8 | 6 | 0 | Low-quality | Genome-fragment | 17.5 | AAI-based (high-confidence) | 0 |
| S6_extracted_k141_15971 | 3932 | No | NA | 3 | 0 | 1 | Not-determined | Genome-fragment | NA | NA | 0 |
| S6_extracted_k141_19901 | 1145 | No | NA | 1 | 0 | 0 | Not-determined | Genome-fragment | NA | NA | 0 |
| S6_extracted_k141_35001 | 1319 | No | NA | 1 | 0 | 0 | Not-determined | Genome-fragment | NA | NA | 0 |
| S10_vs_80063\|\|full | 6098 | No | NA | 10 | 2 | 1 | Low-quality | Genome-fragment | 13.99 | AAI-based (high-confidence) | 0 |
| S6_extracted_k141_35003 | 1172 | No | NA | 2 | 0 | 1 | Not-determined | Genome-fragment | NA | NA | 0 |
| S6_extracted_k141_6425 | 1006 | No | NA | 2 | 0 | 0 | Not-determined | Genome-fragment | NA | NA | 0 |
| S6_extracted_k141_1558 | 1849 | No | NA | 2 | 0 | 1 | Not-determined | Genome-fragment | NA | NA | 0 |
| S6_extracted_k141_36840 | 1489 | No | NA | 2 | 0 | 0 | Not-determined | Genome-fragment | NA | NA | 0 |
| S6_extracted_k141_27317 | 1384 | No | NA | 1 | 0 | 1 | Not-determined | Genome-fragment | NA | NA | 0 |
| S12_vs_k141_59826\|\|full | 6100 | No | NA | 12 | 4 | 0 | Low-quality | Genome-fragment | 8.43 | HMM-based (lower-bound) | 0 |
| S8_vs_k141_100049\|\|full | 6101 | Yes | 3215 | 7 | 2 | 1 | Low-quality | Genome-fragment | 7.63 | AAI-based (medium-confidence) | 47.3 |
| S11_extracted_k141_79468 | 6105 | No | NA | 6 | 0 | 1 | Low-quality | Genome-fragment | 13.13 | AAI-based (medium-confidence) | 0 |
| S6_extracted_k141_6466 | 1503 | No | NA | 2 | 0 | 1 | Not-determined | Genome-fragment | NA | NA | 0 |
| S6_extracted_k141_41951 | 1011 | No | NA | 1 | 0 | 1 | Not-determined | Genome-fragment | NA | NA | 0 |
| S6_extracted_k141_32336 | 1260 | No | NA | 2 | 0 | 1 | Not-determined | Genome-fragment | NA | NA | 0 |
| S6_extracted_k141_16011 | 1772 | No | NA | 1 | 0 | 1 | Not-determined | Genome-fragment | NA | NA | 0 |
| S3_extracted_k141_1370 | 6105 | No | NA | 9 | 5 | 0 | Low-quality | Genome-fragment | 3.72 | AAI-based (high-confidence) | 0 |
| S6_extracted_k141_19943 | 2773 | No | NA | 1 | 0 | 1 | Not-determined | Genome-fragment | NA | NA | 0 |
| S16_vs_59032\|\|full | 6106 | No | NA | 12 | 2 | 0 | Low-quality | Genome-fragment | 14.53 | HMM-based (lower-bound) | 0 |
| S6_extracted_k141_46642 | 1230 | No | NA | 1 | 0 | 0 | Not-determined | Genome-fragment | NA | NA | 0 |
| S16_vs_31259\|\|full | 6109 | No | NA | 7 | 3 | 0 | Low-quality | Genome-fragment | 46.76 | AAI-based (medium-confidence) | 0 |
| S6_extracted_k141_27389 | 1747 | No | NA | 2 | 0 | 2 | Not-determined | Genome-fragment | NA | NA | 0 |
| S6_extracted_k141_18511 | 1034 | No | NA | 1 | 0 | 0 | Not-determined | Genome-fragment | NA | NA | 0 |
| S6_extracted_k141_27400 | 1626 | No | NA | 1 | 0 | 1 | Not-determined | Genome-fragment | NA | NA | 0 |
| S6_extracted_k141_18533 | 1088 | No | NA | 2 | 0 | 0 | Not-determined | Genome-fragment | NA | NA | 0 |
| S12_extracted_k141_132600 | 6112 | No | NA | 7 | 1 | 0 | Low-quality | Genome-fragment | 9.91 | AAI-based (high-confidence) | 0 |
| S6_extracted_k141_13855 | 1273 | No | NA | 2 | 0 | 0 | Not-determined | Genome-fragment | NA | NA | 0 |
| S6_extracted_k141_4540 | 1034 | No | NA | 1 | 0 | 1 | Not-determined | Genome-fragment | NA | NA | 0 |
| S6_extracted_k141_27438 | 2476 | No | NA | 4 | 0 | 2 | Not-determined | Genome-fragment | NA | NA | 0 |
| S6_extracted_k141_27447 | 1082 | No | NA | 1 | 0 | 1 | Not-determined | Genome-fragment | NA | NA | 0 |
| S6_extracted_k141_27460 | 1363 | No | NA | 1 | 0 | 0 | Not-determined | Genome-fragment | NA | NA | 0 |
| S6_extracted_k141_32402 | 1688 | No | NA | 5 | 0 | 1 | Not-determined | Genome-fragment | NA | NA | 0 |
| S6_extracted_k141_48448 | 8330 | No | NA | 8 | 0 | 5 | Not-determined | Genome-fragment | NA | NA | 0 |
| S3_extracted_k141_52075 | 6113 | No | NA | 15 | 4 | 0 | Low-quality | Genome-fragment | 15.03 | AAI-based (high-confidence) | 0 |
| S6_extracted_k141_42096 | 1020 | No | NA | 1 | 0 | 1 | Not-determined | Genome-fragment | NA | NA | 0 |
| S7_vs_14606\|\|full | 6113 | No | NA | 8 | 0 | 0 | Low-quality | Genome-fragment | 13.6 | AAI-based (high-confidence) | 0 |
| S7_vs_64421\|\|full | 6113 | No | NA | 6 | 1 | 0 | Low-quality | Genome-fragment | 3.41 | HMM-based (lower-bound) | 0 |
| S6_extracted_k141_27510 | 10720 | No | NA | 15 | 0 | 3 | Not-determined | Genome-fragment | NA | NA | 0 |
| S6_extracted_k141_27513 | 1172 | No | NA | 3 | 0 | 2 | Not-determined | Genome-fragment | NA | NA | 0 |
| S16_extracted_105180 | 6113 | No | NA | 4 | 0 | 4 | Low-quality | Genome-fragment | 1.99 | AAI-based (medium-confidence) | 0 |
| S6_extracted_k141_13990 | 1047 | No | NA | 2 | 0 | 2 | Not-determined | Genome-fragment | NA | NA | 0 |
| S6_extracted_k141_8845 | 2818 | No | NA | 5 | 0 | 0 | Not-determined | Genome-fragment | NA | NA | 0 |
| S5_vs_k141_62229\|\|full | 6114 | No | NA | 10 | 5 | 0 | Low-quality | Genome-fragment | 16.76 | AAI-based (high-confidence) | 0 |
| S16_vs_24307\|\|full | 6115 | No | NA | 11 | 3 | 0 | Low-quality | Genome-fragment | 4.41 | AAI-based (high-confidence) | 0 |
| S6_extracted_k141_42169 | 1065 | No | NA | 2 | 0 | 1 | Not-determined | Genome-fragment | NA | NA | 0 |
| S6_extracted_k141_53795 | 1077 | No | NA | 2 | 0 | 0 | Not-determined | Genome-fragment | NA | NA | 0 |
| S6_extracted_k141_20043 | 1195 | No | NA | 4 | 0 | 0 | Not-determined | Genome-fragment | NA | NA | 0 |
| S6_extracted_k141_22855 | 1002 | No | NA | 2 | 0 | 0 | Not-determined | Genome-fragment | NA | NA | 0 |
| S6_extracted_k141_39322 | 1506 | No | NA | 1 | 0 | 1 | Not-determined | Genome-fragment | NA | NA | 0 |
| S3_vs_k141_69221\|\|full | 6118 | No | NA | 12 | 2 | 0 | Low-quality | Genome-fragment | 11.33 | AAI-based (high-confidence) | 0 |
| S6_extracted_k141_39333 | 1315 | No | NA | 2 | 0 | 1 | Not-determined | Genome-fragment | NA | NA | 0 |
| S6_extracted_k141_25492 | 1657 | No | NA | 1 | 0 | 0 | Not-determined | Genome-fragment | NA | NA | 0 |
| S6_extracted_k141_32592 | 1770 | No | NA | 2 | 0 | 1 | Not-determined | Genome-fragment | NA | NA | 0 |
| S6_extracted_k141_22923 | 1009 | No | NA | 1 | 0 | 1 | Not-determined | Genome-fragment | NA | NA | 0 |
| S6_extracted_k141_52412 | 6125 | No | NA | 1 | 0 | 0 | Low-quality | Genome-fragment | 7.76 | AAI-based (medium-confidence) | 0 |
| S6_extracted_k141_14045 | 1660 | No | NA | 2 | 0 | 2 | Not-determined | Genome-fragment | NA | NA | 0 |
| S6_extracted_k141_30308 | 1021 | No | NA | 2 | 0 | 0 | Not-determined | Genome-fragment | NA | NA | 0 |
| S14_vs_k141_59822\|\|full | 6127 | No | NA | 9 | 2 | 3 | Low-quality | Genome-fragment | 10.34 | HMM-based (lower-bound) | 0 |
| S6_extracted_k141_1705 | 1070 | No | NA | 2 | 0 | 1 | Not-determined | Genome-fragment | NA | NA | 0 |
| S14_extracted_k141_81601 | 6128 | No | NA | 9 | 9 | 0 | Low-quality | Genome-fragment | 20.19 | AAI-based (high-confidence) | 0 |
| S6_extracted_k141_20128 | 1281 | No | NA | 2 | 0 | 1 | Not-determined | Genome-fragment | NA | NA | 0 |
| S6_extracted_k141_1715 | 1643 | No | NA | 4 | 0 | 4 | Not-determined | Genome-fragment | NA | NA | 0 |
| S6_extracted_k141_32658 | 1071 | No | NA | 2 | 0 | 2 | Not-determined | Genome-fragment | NA | NA | 0 |
| S17_extracted_k141_157041 | 6130 | No | NA | 10 | 0 | 1 | Low-quality | Genome-fragment | 10.24 | AAI-based (high-confidence) | 0 |
| S6_extracted_k141_39427 | 3197 | No | NA | 4 | 0 | 2 | Not-determined | Genome-fragment | NA | NA | 0 |
| S6_extracted_k141_32665 | 1016 | No | NA | 1 | 0 | 1 | Not-determined | Genome-fragment | NA | NA | 0 |
| S6_extracted_k141_14061 | 2939 | No | NA | 2 | 0 | 1 | Not-determined | Genome-fragment | NA | NA | 0 |
| S6_extracted_k141_53871 | 1296 | No | NA | 2 | 0 | 0 | Not-determined | Genome-fragment | NA | NA | 0 |
| S6_extracted_k141_14076 | 1276 | No | NA | 2 | 0 | 2 | Not-determined | Genome-fragment | NA | NA | 0 |
| S6_extracted_k141_48584 | 5154 | No | NA | 4 | 0 | 0 | Not-determined | Genome-fragment | NA | NA | 0 |
| S6_extracted_k141_23005 | 2650 | No | NA | 1 | 0 | 0 | Not-determined | Genome-fragment | NA | NA | 0 |
| S6_extracted_k141_43968 | 1002 | No | NA | 2 | 0 | 1 | Not-determined | Genome-fragment | NA | NA | 0 |
| S14_vs_k141_97012\|\|full | 6135 | No | NA | 6 | 1 | 0 | Low-quality | Genome-fragment | 15.51 | AAI-based (high-confidence) | 0 |
| S6_extracted_k141_43987 | 1037 | No | NA | 2 | 0 | 2 | Not-determined | Genome-fragment | NA | NA | 0 |
| S6_extracted_k141_16217 | 1899 | No | NA | 4 | 0 | 2 | Not-determined | Genome-fragment | NA | NA | 0 |
| S6_extracted_k141_16220 | 3739 | No | NA | 3 | 0 | 3 | Not-determined | Genome-fragment | NA | NA | 0 |
| S6_extracted_k141_44016 | 1666 | No | NA | 3 | 0 | 0 | Not-determined | Genome-fragment | NA | NA | 0 |
| S6_extracted_k141_32763 | 1438 | No | NA | 2 | 0 | 1 | Not-determined | Genome-fragment | NA | NA | 0 |
| S6_extracted_k141_18726 | 3232 | No | NA | 4 | 0 | 1 | Not-determined | Genome-fragment | NA | NA | 0 |
| S6_extracted_k141_53908 | 1052 | No | NA | 2 | 0 | 0 | Not-determined | Genome-fragment | NA | NA | 0 |
| S6_extracted_k141_55545 | 1527 | No | NA | 4 | 0 | 1 | Not-determined | Genome-fragment | NA | NA | 0 |
| S6_extracted_k141_53946 | 1063 | No | NA | 1 | 0 | 1 | Not-determined | Genome-fragment | NA | NA | 0 |
| S6_extracted_k141_35667 | 6136 | No | NA | 9 | 0 | 2 | Low-quality | Genome-fragment | 12.2 | AAI-based (high-confidence) | 0 |
| S6_extracted_k141_27567 | 1646 | No | NA | 3 | 0 | 1 | Not-determined | Genome-fragment | NA | NA | 0 |
| S6_extracted_k141_36882 | 1360 | No | NA | 2 | 0 | 0 | Not-determined | Genome-fragment | NA | NA | 0 |
| S6_extracted_k141_8888 | 19917 | No | NA | 17 | 0 | 3 | Not-determined | Genome-fragment | NA | NA | 0 |
| S6_extracted_k141_44089 | 3965 | No | NA | 1 | 0 | 0 | Not-determined | Genome-fragment | NA | NA | 0 |
| S6_extracted_k141_11367 | 1359 | No | NA | 2 | 0 | 1 | Not-determined | Genome-fragment | NA | NA | 0 |
| S6_extracted_k141_23160 | 1028 | No | NA | 2 | 0 | 1 | Not-determined | Genome-fragment | NA | NA | 0 |
| S3_vs_k141_60011\|\|full | 6143 | No | NA | 3 | 0 | 3 | Low-quality | Genome-fragment | 3.31 | AAI-based (medium-confidence) | 0 |
| S6_extracted_k141_54014 | 1740 | No | NA | 3 | 0 | 1 | Not-determined | Genome-fragment | NA | NA | 0 |
| S5_vs_k141_31206\|\|full | 6144 | No | NA | 13 | 3 | 1 | Low-quality | Genome-fragment | 9.15 | AAI-based (high-confidence) | 0 |
| S6_extracted_k141_39550 | 1425 | No | NA | 2 | 0 | 0 | Not-determined | Genome-fragment | NA | NA | 0 |
| S6_extracted_k141_39565 | 1939 | No | NA | 1 | 0 | 0 | Not-determined | Genome-fragment | NA | NA | 0 |
| S6_extracted_k141_35305 | 4759 | No | NA | 3 | 0 | 2 | Not-determined | Genome-fragment | NA | NA | 0 |
| S6_extracted_k141_36954 | 1296 | No | NA | 2 | 0 | 1 | Not-determined | Genome-fragment | NA | NA | 0 |
| S9_vs_k141_33485\|\|full | 6146 | No | NA | 15 | 1 | 2 | Low-quality | Genome-fragment | 3.24 | HMM-based (lower-bound) | 0 |
| S7_extracted_102256 | 6147 | No | NA | 7 | 4 | 0 | Low-quality | Genome-fragment | 21.53 | HMM-based (lower-bound) | 0 |
| S6_extracted_k141_20342 | 1215 | No | NA | 1 | 0 | 1 | Not-determined | Genome-fragment | NA | NA | 0 |
| S6_extracted_k141_46795 | 7359 | No | NA | 5 | 0 | 3 | Not-determined | Genome-fragment | NA | NA | 0 |
| S6_extracted_k141_1808 | 1592 | No | NA | 2 | 0 | 0 | Not-determined | Genome-fragment | NA | NA | 0 |
| S14_extracted_k141_112874 | 6149 | No | NA | 9 | 0 | 0 | Low-quality | Genome-fragment | 13.82 | AAI-based (high-confidence) | 0 |
| S6_extracted_k141_36992 | 1126 | No | NA | 3 | 0 | 0 | Not-determined | Genome-fragment | NA | NA | 0 |
| S6_extracted_k141_25680 | 4727 | No | NA | 6 | 0 | 2 | Not-determined | Genome-fragment | NA | NA | 0 |
| S6_extracted_k141_27662 | 1530 | No | NA | 1 | 0 | 1 | Not-determined | Genome-fragment | NA | NA | 0 |
| S6_extracted_k141_46849 | 8502 | No | NA | 10 | 0 | 4 | Not-determined | Genome-fragment | NA | NA | 0 |
| S6_extracted_k141_25696 | 1461 | No | NA | 2 | 0 | 1 | Not-determined | Genome-fragment | NA | NA | 0 |
| S6_extracted_k141_54152 | 1494 | No | NA | 2 | 0 | 1 | Not-determined | Genome-fragment | NA | NA | 0 |
| S16_extracted_160394 | 6151 | No | NA | 11 | 0 | 3 | Low-quality | Genome-fragment | 14.48 | AAI-based (high-confidence) | 0 |
| S6_extracted_k141_25710 | 41050 | No | NA | 51 | 6 | 0 | High-quality | High-quality | 100 | AAI-based (high-confidence) | 0 |
| S6_extracted_k141_16352 | 1283 | No | NA | 1 | 0 | 1 | Not-determined | Genome-fragment | NA | NA | 0 |
| S6_vs_k141_3652\|\|full | 6151 | No | NA | 8 | 0 | 1 | Low-quality | Genome-fragment | 9.86 | AAI-based (high-confidence) | 0 |
| S6_extracted_k141_39655 | 2543 | No | NA | 5 | 0 | 2 | Not-determined | Genome-fragment | NA | NA | 0 |
| S6_extracted_k141_23401 | 6183 | No | NA | 7 | 0 | 7 | Not-determined | Genome-fragment | NA | NA | 0 |
| S3_vs_k141_120071\|\|full | 6153 | No | NA | 9 | 1 | 0 | Low-quality | Genome-fragment | 14.85 | AAI-based (high-confidence) | 0 |

| | | | | | | | | | | | |
|---|---|---|---|---|---|---|---|---|---|---|---|
| S6_extracted_k141_30538 | 1014 | No | NA | 1 | 0 | 1 | Not-determined | Genome-fragment | NA | NA | 0 |
| S6_extracted_k141_48930 | 1253 | No | NA | 2 | 0 | 2 | Not-determined | Genome-fragment | NA | NA | 0 |
| S6_extracted_k141_39710 | 1447 | No | NA | 1 | 0 | 0 | Not-determined | Genome-fragment | NA | NA | 0 |
| S6_extracted_k141_55663 | 1093 | No | NA | 1 | 0 | 0 | Not-determined | Genome-fragment | NA | NA | 0 |
| S6_extracted_k141_25800 | 1091 | No | NA | 2 | 0 | 2 | Not-determined | Genome-fragment | NA | NA | 0 |
| S6_extracted_k141_55668 | 1161 | No | NA | 3 | 0 | 2 | Not-determined | Genome-fragment | NA | NA | 0 |
| S6_extracted_k141_44262 | 11802 | No | NA | 14 | 0 | 1 | Not-determined | Genome-fragment | NA | NA | 0 |
| S12_vs_k141_27528\|\|full | 6153 | Yes | 3561 | 7 | 1 | 1 | Low-quality | Genome-fragment | 2.18 | HMM-based (lower-bound) | 42.13 |
| S6_extracted_k141_55690 | 6525 | No | NA | 7 | 0 | 4 | Not-determined | Genome-fragment | NA | NA | 0 |
| S6_extracted_k141_25838 | 1305 | No | NA | 2 | 0 | 0 | Not-determined | Genome-fragment | NA | NA | 0 |
| S6_extracted_k141_48991 | 1146 | No | NA | 1 | 0 | 1 | Not-determined | Genome-fragment | NA | NA | 0 |
| S6_extracted_k141_25844 | 7701 | No | NA | 3 | 0 | 3 | Not-determined | Genome-fragment | NA | NA | 0 |
| S11_extracted_k141_78700 | 6154 | No | NA | 10 | 3 | 0 | Low-quality | Genome-fragment | 17.71 | AAI-based (medium-confidence) | 0 |
| S12_vs_k141_186406\|\|full | 6154 | No | NA | 8 | 0 | 1 | Low-quality | Genome-fragment | 11.66 | AAI-based (medium-confidence) | 0 |
| S6_extracted_k141_23519 | 1465 | No | NA | 2 | 0 | 2 | Not-determined | Genome-fragment | NA | NA | 0 |
| S6_extracted_k141_55753 | 1332 | No | NA | 3 | 0 | 2 | Not-determined | Genome-fragment | NA | NA | 0 |
| S6_extracted_k141_16426 | 1554 | No | NA | 1 | 0 | 0 | Not-determined | Genome-fragment | NA | NA | 0 |
| S6_extracted_k141_20447 | 1271 | No | NA | 2 | 0 | 2 | Not-determined | Genome-fragment | NA | NA | 0 |
| S6_extracted_k141_16436 | 1242 | No | NA | 1 | 0 | 1 | Not-determined | Genome-fragment | NA | NA | 0 |
| S6_extracted_k141_8985 | 1172 | No | NA | 1 | 0 | 1 | Not-determined | Genome-fragment | NA | NA | 0 |
| S6_extracted_k141_1936 | 1308 | No | NA | 5 | 0 | 0 | Not-determined | Genome-fragment | NA | NA | 0 |
| S6_extracted_k141_9057 | 1052 | No | NA | 1 | 0 | 0 | Not-determined | Genome-fragment | NA | NA | 0 |
| S6_extracted_k141_37216 | 1106 | No | NA | 3 | 0 | 3 | Not-determined | Genome-fragment | NA | NA | 0 |
| S6_extracted_k141_39912 | 1311 | No | NA | 3 | 0 | 0 | Not-determined | Genome-fragment | NA | NA | 0 |
| S7_vs_32626\|\|full | 6156 | No | NA | 4 | 0 | 2 | Low-quality | Genome-fragment | 14.37 | AAI-based (high-confidence) | 0 |
| S6_extracted_k141_39930 | 1332 | No | NA | 2 | 0 | 0 | Not-determined | Genome-fragment | NA | NA | 0 |
| S6_extracted_k141_9076 | 4578 | No | NA | 5 | 0 | 4 | Not-determined | Genome-fragment | NA | NA | 0 |
| S7_vs_69678\|\|full | 6157 | No | NA | 9 | 5 | 0 | Low-quality | Genome-fragment | 17.1 | AAI-based (medium-confidence) | 0 |
| S6_extracted_k141_9090 | 1318 | No | NA | 1 | 0 | 1 | Not-determined | Genome-fragment | NA | NA | 0 |
| S16_vs_55718\|\|full | 6159 | No | NA | 7 | 0 | 6 | Low-quality | Genome-fragment | 4.24 | AAI-based (medium-confidence) | 0 |
| S6_extracted_k141_9144 | 1154 | No | NA | 2 | 0 | 0 | Not-determined | Genome-fragment | NA | NA | 0 |
| S12_vs_k141_151923\|\|full | 6160 | No | NA | 5 | 0 | 2 | Low-quality | Genome-fragment | 1.76 | AAI-based (high-confidence) | 0 |
| S6_extracted_k141_55825 | 1614 | No | NA | 2 | 0 | 1 | Not-determined | Genome-fragment | NA | NA | 0 |
| S10_vs_51623\|\|full | 6164 | No | NA | 6 | 3 | 0 | Low-quality | Genome-fragment | 10.7 | AAI-based (high-confidence) | 0 |
| S6_extracted_k141_55834 | 1334 | No | NA | 1 | 0 | 1 | Not-determined | Genome-fragment | NA | NA | 0 |
| S6_extracted_k141_9178 | 1233 | No | NA | 1 | 0 | 1 | Not-determined | Genome-fragment | NA | NA | 0 |
| S6_extracted_k141_2059 | 2954 | No | NA | 2 | 0 | 2 | Not-determined | Genome-fragment | NA | NA | 0 |
| S1_vs_k141_39109\|\|full | 6167 | No | NA | 4 | 2 | 0 | Low-quality | Genome-fragment | 10.72 | AAI-based (high-confidence) | 0 |
| S6_extracted_k141_49197 | 1145 | No | NA | 1 | 0 | 0 | Not-determined | Genome-fragment | NA | NA | 0 |
| S6_extracted_k141_27700 | 3026 | No | NA | 5 | 0 | 1 | Not-determined | Genome-fragment | NA | NA | 0 |
| S6_extracted_k141_27708 | 3319 | No | NA | 1 | 0 | 0 | Not-determined | Genome-fragment | NA | NA | 0 |
| S6_extracted_k141_55924 | 1714 | No | NA | 3 | 0 | 2 | Not-determined | Genome-fragment | NA | NA | 0 |
| S6_vs_k141_24590\|\|full | 6169 | Yes | 3937 | 8 | 4 | 1 | Low-quality | Genome-fragment | 8.77 | AAI-based (high-confidence) | 36.18 |
| S6_extracted_k141_55967 | 1557 | No | NA | 1 | 0 | 0 | Not-determined | Genome-fragment | NA | NA | 0 |
| S7_vs_72610\|\|full | 6171 | No | NA | 17 | 3 | 0 | Low-quality | Genome-fragment | 17.11 | AAI-based (high-confidence) | 0 |
| S6_extracted_k141_51177 | 1002 | No | NA | 2 | 0 | 0 | Not-determined | Genome-fragment | NA | NA | 0 |
| S6_extracted_k141_49303 | 1710 | No | NA | 2 | 0 | 2 | Not-determined | Genome-fragment | NA | NA | 0 |
| S6_extracted_k141_49304 | 2018 | No | NA | 2 | 0 | 0 | Not-determined | Genome-fragment | NA | NA | 0 |
| S16_extracted_93732 | 6171 | Yes | 4553 | 10 | 2 | 1 | Low-quality | Genome-fragment | 2.4 | HMM-based (lower-bound) | 26.22 |
| S6_extracted_k141_37391 | 1152 | No | NA | 1 | 0 | 0 | Not-determined | Genome-fragment | NA | NA | 0 |
| S15_extracted_24508 | 6173 | No | NA | 7 | 3 | 0 | Low-quality | Genome-fragment | 6.19 | AAI-based (high-confidence) | 0 |
| S6_extracted_k141_56026 | 1114 | No | NA | 1 | 0 | 1 | Not-determined | Genome-fragment | NA | NA | 0 |
| S8_extracted_k141_43823 | 6173 | No | NA | 14 | 0 | 0 | Low-quality | Genome-fragment | 6.18 | AAI-based (high-confidence) | 0 |
| S11_vs_k141_67735\|\|full | 6174 | No | NA | 12 | 2 | 0 | Low-quality | Genome-fragment | 10.67 | HMM-based (lower-bound) | 0 |
| S11_extracted_k141_18830 | 6176 | No | NA | 7 | 0 | 4 | Low-quality | Genome-fragment | 4.94 | AAI-based (medium-confidence) | 0 |
| S6_extracted_k141_40036 | 1394 | No | NA | 3 | 0 | 1 | Not-determined | Genome-fragment | NA | NA | 0 |
| S5_vs_k141_17315\|\|full | 6177 | No | NA | 11 | 2 | 0 | Low-quality | Genome-fragment | 13.92 | AAI-based (high-confidence) | 0 |
| S6_extracted_k141_9372 | 2376 | No | NA | 2 | 0 | 0 | Not-determined | Genome-fragment | NA | NA | 0 |
| S6_extracted_k141_20676 | 1164 | No | NA | 1 | 0 | 1 | Not-determined | Genome-fragment | NA | NA | 0 |
| S6_extracted_k141_51334 | 1320 | No | NA | 2 | 0 | 0 | Not-determined | Genome-fragment | NA | NA | 0 |
| S6_extracted_k141_51429 | 1197 | No | NA | 2 | 0 | 2 | Not-determined | Genome-fragment | NA | NA | 0 |
| S6_extracted_k141_37591 | 1092 | No | NA | 1 | 0 | 1 | Not-determined | Genome-fragment | NA | NA | 0 |
| S6_extracted_k141_51496 | 1233 | No | NA | 1 | 0 | 1 | Not-determined | Genome-fragment | NA | NA | 0 |
| S6_extracted_k141_37630 | 2945 | No | NA | 5 | 0 | 5 | Not-determined | Genome-fragment | NA | NA | 0 |
| S6_extracted_k141_56237 | 2674 | No | NA | 7 | 0 | 6 | Not-determined | Genome-fragment | NA | NA | 0 |
| S6_extracted_k141_56240 | 7147 | No | NA | 5 | 0 | 4 | Not-determined | Genome-fragment | NA | NA | 0 |
| S6_extracted_k141_56246 | 5315 | No | NA | 3 | 0 | 2 | Not-determined | Genome-fragment | NA | NA | 0 |
| S6_extracted_k141_37654 | 2497 | No | NA | 2 | 0 | 0 | Not-determined | Genome-fragment | NA | NA | 0 |
| S6_extracted_k141_37665 | 1299 | No | NA | 1 | 0 | 1 | Not-determined | Genome-fragment | NA | NA | 0 |
| S8_vs_k141_140930\|\|full | 6177 | No | NA | 13 | 2 | 0 | Low-quality | Genome-fragment | 7.34 | HMM-based (lower-bound) | 0 |
| S6_extracted_k141_20834 | 2277 | No | NA | 3 | 0 | 2 | Not-determined | Genome-fragment | NA | NA | 0 |
| S6_extracted_k141_51615 | 1412 | No | NA | 1 | 0 | 1 | Not-determined | Genome-fragment | NA | NA | 0 |
| S6_extracted_k141_27930 | 1019 | No | NA | 1 | 0 | 1 | Not-determined | Genome-fragment | NA | NA | 0 |
| S1_vs_k141_73309\|\|full | 6179 | No | NA | 8 | 4 | 0 | Low-quality | Genome-fragment | 16.06 | AAI-based (medium-confidence) | 0 |
| S14_vs_k141_104144\|\|full | 6182 | No | NA | 11 | 1 | 1 | Low-quality | Genome-fragment | 17.71 | AAI-based (high-confidence) | 0 |
| S6_extracted_k141_51669 | 1433 | No | NA | 2 | 0 | 0 | Not-determined | Genome-fragment | NA | NA | 0 |
| S6_extracted_k141_56356 | 1068 | No | NA | 1 | 0 | 0 | Not-determined | Genome-fragment | NA | NA | 0 |
| S6_extracted_k141_21017 | 1149 | No | NA | 2 | 0 | 0 | Not-determined | Genome-fragment | NA | NA | 0 |
| S3_vs_k141_66500\|\|full | 6182 | No | NA | 15 | 0 | 1 | Low-quality | Genome-fragment | 4.15 | AAI-based (high-confidence) | 0 |
| S6_extracted_k141_56450 | 4601 | No | NA | 5 | 0 | 3 | Not-determined | Genome-fragment | NA | NA | 0 |
| S5_vs_k141_62217\|\|full | 6184 | No | NA | 12 | 0 | 0 | Low-quality | Genome-fragment | 17.02 | AAI-based (medium-confidence) | 0 |
| S6_extracted_k141_28243 | 1128 | No | NA | 1 | 0 | 1 | Not-determined | Genome-fragment | NA | NA | 0 |
| S6_extracted_k141_56484 | 47845 | No | NA | 63 | 27 | 1 | Complete | High-quality | 100 | DTR (high-confidence) | 0 |
| S13_vs_94817\|\|full | 6186 | No | NA | 13 | 1 | 0 | Low-quality | Genome-fragment | 6.73 | AAI-based (high-confidence) | 0 |
| S6_extracted_k141_56486 | 107433 | No | NA | 147 | 18 | 4 | Complete | High-quality | 100 | DTR (high-confidence) | 0 |
| S7_extracted_99125 | 6186 | No | NA | 10 | 6 | 0 | Low-quality | Genome-fragment | 3.53 | AAI-based (high-confidence) | 0 |
| S6_vs_k141_32886\|\|full | 38613 | No | NA | 60 | 10 | 1 | High-quality | High-quality | 100 | AAI-based (medium-confidence) | 0 |
| S6_vs_k141_56485\|\|full | 40038 | No | NA | 59 | 22 | 0 | High-quality | High-quality | 95.95 | AAI-based (high-confidence) | 0 |
| S6_vs_k141_56490\|\|full | 35374 | No | NA | 63 | 24 | 2 | High-quality | High-quality | 100 | AAI-based (high-confidence) | 0 |
| S6_vs_k141_56491\|\|full | 46642 | No | NA | 77 | 26 | 0 | High-quality | High-quality | 100 | AAI-based (medium-confidence) | 0 |
| S6_vs_k141_56492\|\|full | 45676 | No | NA | 77 | 13 | 0 | High-quality | High-quality | 93.65 | AAI-based (high-confidence) | 0 |
| S6_vs_k141_56493\|\|full | 13326 | No | NA | 23 | 2 | 1 | High-quality | High-quality | 91.72 | AAI-based (high-confidence) | 0 |
| S6_vs_k141_56496\|\|full | 49540 | No | NA | 75 | 20 | 0 | High-quality | High-quality | 100 | AAI-based (high-confidence) | 0 |
| S6_vs_k141_56498\|\|full | 33635 | No | NA | 52 | 15 | 1 | High-quality | High-quality | 96.19 | AAI-based (high-confidence) | 0 |
| S6_vs_k141_51844\|\|full | 1681 | No | NA | 3 | 0 | 0 | Not-determined | Genome-fragment | NA | NA | 0 |
| S6_vs_k141_51846\|\|full | 2974 | No | NA | 3 | 0 | 2 | Not-determined | Genome-fragment | NA | NA | 0 |
| S5_extracted_k141_124473 | 6191 | No | NA | 9 | 5 | 0 | Low-quality | Genome-fragment | 14.42 | AAI-based (medium-confidence) | 0 |
| S17_vs_k141_149224\|\|full | 6192 | No | NA | 8 | 0 | 0 | Low-quality | Genome-fragment | 8.68 | AAI-based (high-confidence) | 0 |
| S6_vs_k141_23612\|\|full | 1461 | No | NA | 3 | 0 | 2 | Not-determined | Genome-fragment | NA | NA | 0 |
| S12_vs_k141_447\|\|full | 6194 | No | NA | 9 | 3 | 0 | Low-quality | Genome-fragment | 14.09 | AAI-based (high-confidence) | 0 |
| S3_vs_k141_46149\|\|full | 6196 | No | NA | 19 | 4 | 0 | Low-quality | Genome-fragment | 10.26 | HMM-based (lower-bound) | 0 |
| S6_vs_k141_30651\|\|full | 1014 | No | NA | 4 | 0 | 0 | Not-determined | Genome-fragment | NA | NA | 0 |
| S2_vs_k141_14056\|\|full | 6199 | No | NA | 13 | 1 | 0 | Low-quality | Genome-fragment | 15.08 | AAI-based (high-confidence) | 0 |
| S6_vs_k141_38\|\|full | 1105 | No | NA | 2 | 0 | 1 | Not-determined | Genome-fragment | NA | NA | 0 |
| S13_vs_7689\|\|full | 6201 | No | NA | 7 | 0 | 2 | Low-quality | Genome-fragment | 10.95 | AAI-based (high-confidence) | 0 |
| S6_vs_k141_54241\|\|full | 4170 | No | NA | 4 | 0 | 2 | Not-determined | Genome-fragment | NA | NA | 0 |
| S6_vs_k141_47189\|\|full | 2308 | No | NA | 3 | 0 | 1 | Not-determined | Genome-fragment | NA | NA | 0 |
| S6_vs_k141_42505\|\|full | 2561 | No | NA | 3 | 0 | 0 | Not-determined | Genome-fragment | NA | NA | 0 |
| S6_vs_k141_18917\|\|full | 11205 | No | NA | 13 | 0 | 10 | Not-determined | Genome-fragment | NA | NA | 0 |
| S6_vs_k141_30736\|\|full | 4179 | No | NA | 5 | 0 | 3 | Not-determined | Genome-fragment | NA | NA | 0 |
| S8_vs_k141_190074\|\|full | 6201 | No | NA | 12 | 0 | 0 | Low-quality | Genome-fragment | 3.53 | AAI-based (high-confidence) | 0 |
| S17_vs_k141_119624\|\|full | 6206 | No | NA | 8 | 0 | 2 | Low-quality | Genome-fragment | 10.74 | AAI-based (medium-confidence) | 0 |
| S6_vs_k141_25973\|\|full | 1149 | No | NA | 3 | 0 | 1 | Not-determined | Genome-fragment | NA | NA | 0 |
| S16_vs_128475\|\|full | 6207 | No | NA | 10 | 7 | 0 | Low-quality | Genome-fragment | 14.9 | AAI-based (high-confidence) | 0 |
| S10_extracted_90462 | 6207 | No | NA | 11 | 3 | 0 | Low-quality | Genome-fragment | 8.5 | AAI-based (high-confidence) | 0 |
| S6_vs_k141_4781\|\|full | 1267 | No | NA | 2 | 0 | 1 | Not-determined | Genome-fragment | NA | NA | 0 |
| S8_extracted_k141_67533 | 6213 | No | NA | 11 | 0 | 0 | Low-quality | Genome-fragment | 3.89 | AAI-based (high-confidence) | 0 |
| S6_vs_k141_14253\|\|full | 1064 | No | NA | 2 | 0 | 0 | Not-determined | Genome-fragment | NA | NA | 0 |
| S10_extracted_110943 | 6215 | No | NA | 8 | 2 | 0 | Low-quality | Genome-fragment | 14.05 | AAI-based (high-confidence) | 0 |
| S9_vs_k141_34875\|\|full | 6216 | No | NA | 13 | 3 | 0 | Low-quality | Genome-fragment | 3.7 | AAI-based (high-confidence) | 0 |
| S16_vs_166014\|\|full | 6216 | No | NA | 14 | 3 | 0 | Low-quality | Genome-fragment | 15.2 | AAI-based (medium-confidence) | 0 |
| S7_vs_74244\|\|full | 6217 | No | NA | 14 | 0 | 1 | Low-quality | Genome-fragment | 3.85 | AAI-based (medium-confidence) | 0 |
| S6_vs_k141_37812\|\|full | 32138 | No | NA | 42 | 13 | 2 | Medium-quality | Genome-fragment | 63.18 | AAI-based (medium-confidence) | 0 |
| S10_vs_62084\|\|full | 6220 | Yes | 4776 | 10 | 2 | 1 | Low-quality | Genome-fragment | 2.37 | HMM-based (lower-bound) | 23.22 |
| S6_extracted_k141_12055 | 6222 | No | NA | 7 | 5 | 0 | Low-quality | Genome-fragment | 28.4 | AAI-based (high-confidence) | 0 |
| S6_vs_k141_33151\|\|full | 4400 | No | NA | 6 | 0 | 2 | Not-determined | Genome-fragment | NA | NA | 0 |
| S1_vs_k141_110922\|\|full | 6224 | No | NA | 5 | 5 | 0 | Low-quality | Genome-fragment | 13.8 | AAI-based (high-confidence) | 0 |
| S5_vs_k141_137599\|\|full | 6224 | No | NA | 13 | 2 | 0 | Low-quality | Genome-fragment | 9.44 | HMM-based (lower-bound) | 0 |
| S6_vs_k141_14288\|\|full | 1362 | No | NA | 3 | 0 | 0 | Not-determined | Genome-fragment | NA | NA | 0 |
| S6_vs_k141_52042\|\|full | 3451 | No | NA | 4 | 0 | 0 | Not-determined | Genome-fragment | NA | NA | 0 |
| S6_vs_k141_9524\|\|full | 2217 | No | NA | 4 | 0 | 2 | Not-determined | Genome-fragment | NA | NA | 0 |
| S6_vs_k141_44900\|\|full | 1292 | No | NA | 3 | 0 | 0 | Not-determined | Genome-fragment | NA | NA | 0 |
| S6_vs_k141_44915\|\|full | 1202 | No | NA | 3 | 0 | 2 | Not-determined | Genome-fragment | NA | NA | 0 |
| S3_extracted_k141_4050 | 6228 | No | NA | 9 | 0 | 0 | Low-quality | Genome-fragment | 3.03 | AAI-based (high-confidence) | 0 |
| S16_extracted_82028 | 6228 | No | NA | 7 | 0 | 2 | Low-quality | Genome-fragment | 3.57 | AAI-based (high-confidence) | 0 |
| S7_vs_67487\|\|full | 6232 | No | NA | 9 | 1 | 0 | Low-quality | Genome-fragment | 9.85 | AAI-based (high-confidence) | 0 |
| S16_vs_20134\|\|full | 6235 | No | NA | 5 | 0 | 0 | Low-quality | Genome-fragment | 6.32 | AAI-based (medium-confidence) | 0 |
| S6_vs_k141_16671\|\|full | 1963 | No | NA | 2 | 0 | 2 | Not-determined | Genome-fragment | NA | NA | 0 |
| S16_vs_145745\|\|full | 6235 | No | NA | 8 | 0 | 0 | Low-quality | Genome-fragment | 16.43 | AAI-based (high-confidence) | 0 |
| S6_vs_k141_23659\|\|full | 1168 | No | NA | 2 | 0 | 1 | Not-determined | Genome-fragment | NA | NA | 0 |
| S6_vs_k141_244\|\|full | 5109 | No | NA | 8 | 0 | 3 | Not-determined | Genome-fragment | NA | NA | 0 |
| S6_vs_k141_37949\|\|full | 1930 | No | NA | 4 | 0 | 1 | Not-determined | Genome-fragment | NA | NA | 0 |
| S6_vs_k141_23667\|\|full | 1474 | No | NA | 3 | 0 | 0 | Not-determined | Genome-fragment | NA | NA | 0 |
| S6_vs_k141_26013\|\|full | 5361 | No | NA | 5 | 0 | 4 | Not-determined | Genome-fragment | NA | NA | 0 |
| S14_extracted_k141_106866 | 6236 | No | NA | 6 | 6 | 0 | Low-quality | Genome-fragment | 3.99 | AAI-based (medium-confidence) | 0 |
| S6_vs_k141_37967\|\|full | 2100 | No | NA | 3 | 0 | 2 | Not-determined | Genome-fragment | NA | NA | 0 |
| S3_vs_k141_6525\|\|full | 6237 | No | NA | 12 | 1 | 0 | Low-quality | Genome-fragment | 45.28 | AAI-based (high-confidence) | 0 |
| S6_vs_k141_40228\|\|full | 5067 | No | NA | 4 | 0 | 3 | Not-determined | Genome-fragment | NA | NA | 0 |
| S6_vs_k141_11900\|\|full | 4802 | No | NA | 8 | 0 | 1 | Not-determined | Genome-fragment | NA | NA | 0 |
| S10_vs_55098\|\|full | 6237 | No | NA | 9 | 1 | 0 | Low-quality | Genome-fragment | 4.04 | HMM-based (lower-bound) | 0 |
| S8_vs_k141_97484\|\|full | 6241 | No | NA | 9 | 2 | 1 | Low-quality | Genome-fragment | 10.35 | AAI-based (high-confidence) | 0 |
| S6_vs_k141_2492\|\|full | 2675 | No | NA | 3 | 0 | 0 | Not-determined | Genome-fragment | NA | NA | 0 |
| S6_vs_k141_286\|\|full | 1549 | No | NA | 3 | 0 | 1 | Not-determined | Genome-fragment | NA | NA | 0 |
| S6_vs_k141_37986\|\|full | 3238 | No | NA | 2 | 0 | 1 | Not-determined | Genome-fragment | NA | NA | 0 |
| S6_vs_k141_37989\|\|full | 2286 | No | NA | 4 | 0 | 0 | Not-determined | Genome-fragment | NA | NA | 0 |
| S9_vs_k141_54163\|\|full | 6243 | No | NA | 10 | 0 | 1 | Low-quality | Genome-fragment | 11.34 | AAI-based (medium-confidence) | 0 |
| S16_extracted_14128 | 6243 | No | NA | 7 | 0 | 2 | Low-quality | Genome-fragment | 21.18 | AAI-based (medium-confidence) | 0 |
| S5_extracted_k141_69260 | 6244 | Yes | 5189 | 8 | 1 | 2 | Low-quality | Genome-fragment | 1.98 | HMM-based (lower-bound) | 16.9 |
| S11_vs_k141_11386\|\|full | 6245 | No | NA | 12 | 3 | 0 | Low-quality | Genome-fragment | 17.78 | AAI-based (high-confidence) | 0 |
| S17_extracted_k141_70935 | 6246 | No | NA | 10 | 1 | 0 | Low-quality | Genome-fragment | 3.39 | HMM-based (lower-bound) | 0 |
| S12_extracted_k141_159642 | 6246 | No | NA | 13 | 2 | 0 | Low-quality | Genome-fragment | 10.41 | AAI-based (high-confidence) | 0 |
| S6_vs_k141_34700\|\|full | 6246 | No | NA | 11 | 1 | 0 | Low-quality | Genome-fragment | 12.19 | AAI-based (high-confidence) | 0 |
| S3_extracted_k141_85523 | 6248 | Yes | 4864 | 6 | 1 | 1 | Low-quality | Genome-fragment | 12.51 | AAI-based (medium-confidence) | 22.15 |
| S6_vs_k141_21312\|\|full | 1731 | No | NA | 3 | 0 | 1 | Not-determined | Genome-fragment | NA | NA | 0 |
| S6_vs_k141_45011\|\|full | 4236 | No | NA | 4 | 0 | 3 | Not-determined | Genome-fragment | NA | NA | 0 |
| S17_extracted_k141_30738 | 6258 | No | NA | 8 | 2 | 0 | Low-quality | Genome-fragment | 10.32 | AAI-based (high-confidence) | 0 |
| S3_extracted_k141_28514 | 6267 | No | NA | 5 | 0 | 5 | Low-quality | Genome-fragment | 3.25 | AAI-based (high-confidence) | 0 |
| S17_vs_k141_11440\|\|full | 6273 | Yes | 2990 | 5 | 1 | 2 | Low-quality | Genome-fragment | 1.39 | HMM-based (lower-bound) | 52.34 |
| S6_vs_k141_33255\|\|full | 3657 | No | NA | 5 | 0 | 2 | Not-determined | Genome-fragment | NA | NA | 0 |
| S6_vs_k141_9609\|\|full | 2228 | No | NA | 2 | 0 | 0 | Not-determined | Genome-fragment | NA | NA | 0 |
| S6_vs_k141_40295\|\|full | 2441 | No | NA | 3 | 0 | 2 | Not-determined | Genome-fragment | NA | NA | 0 |
| S6_vs_k141_402\|\|full | 1436 | No | NA | 2 | 0 | 1 | Not-determined | Genome-fragment | NA | NA | 0 |
| S6_vs_k141_33315\|\|full | 1526 | No | NA | 2 | 0 | 0 | Not-determined | Genome-fragment | NA | NA | 0 |
| S6_vs_k141_47236\|\|full | 2032 | No | NA | 3 | 0 | 2 | Not-determined | Genome-fragment | NA | NA | 0 |
| S7_extracted_101918 | 6273 | No | NA | 6 | 0 | 1 | Low-quality | Genome-fragment | 9.91 | AAI-based (medium-confidence) | 0 |
| S9_vs_k141_63884\|\|full | 6274 | No | NA | 8 | 7 | 0 | Low-quality | Genome-fragment | 17.14 | AAI-based (high-confidence) | 0 |
| S6_vs_k141_12058\|\|full | 3552 | No | NA | 4 | 0 | 4 | Not-determined | Genome-fragment | NA | NA | 0 |
| S12_vs_k141_134831\|\|full | 6277 | No | NA | 8 | 0 | 4 | Low-quality | Genome-fragment | 2.93 | AAI-based (medium-confidence) | 0 |
| S6_vs_k141_33333\|\|full | 2303 | No | NA | 3 | 0 | 1 | Not-determined | Genome-fragment | NA | NA | 0 |
| S6_vs_k141_23792\|\|full | 1466 | No | NA | 2 | 0 | 1 | Not-determined | Genome-fragment | NA | NA | 0 |
| S6_vs_k141_12068\|\|full | 4471 | No | NA | 6 | 0 | 1 | Not-determined | Genome-fragment | NA | NA | 0 |
| S12_vs_k141_45029\|\|full | 6277 | No | NA | 13 | 3 | 0 | Low-quality | Genome-fragment | 15.87 | AAI-based (medium-confidence) | 0 |
| S6_vs_k141_4930\|\|full | 7202 | No | NA | 8 | 0 | 3 | Not-determined | Genome-fragment | NA | NA | 0 |
| S6_vs_k141_12078\|\|full | 1380 | No | NA | 2 | 0 | 2 | Not-determined | Genome-fragment | NA | NA | 0 |
| S6_vs_k141_28543\|\|full | 3329 | No | NA | 5 | 0 | 4 | Not-determined | Genome-fragment | NA | NA | 0 |
| S11_extracted_k141_70347 | 6278 | No | NA | 3 | 0 | 2 | Low-quality | Genome-fragment | 5.68 | AAI-based (medium-confidence) | 0 |
| S6_vs_k141_35580\|\|full | 2802 | No | NA | 4 | 0 | 3 | Not-determined | Genome-fragment | NA | NA | 0 |
| S6_vs_k141_7247\|\|full | 2962 | No | NA | 4 | 0 | 1 | Not-determined | Genome-fragment | NA | NA | 0 |
| S6_vs_k141_47282\|\|full | 1499 | No | NA | 4 | 0 | 1 | Not-determined | Genome-fragment | NA | NA | 0 |
| S7_vs_10778\|\|full | 6282 | No | NA | 8 | 0 | 0 | Low-quality | Genome-fragment | 15.72 | AAI-based (medium-confidence) | 0 |
| S8_vs_k141_40312\|\|full | 6286 | No | NA | 9 | 6 | 0 | Low-quality | Genome-fragment | 19.09 | AAI-based (high-confidence) | 0 |
| S11_vs_k141_13439\|\|full | 6288 | No | NA | 7 | 6 | 0 | Low-quality | Genome-fragment | 13.59 | AAI-based (medium-confidence) | 0 |
| S7_vs_61686\|\|full | 6291 | No | NA | 11 | 2 | 0 | Low-quality | Genome-fragment | 10.76 | AAI-based (high-confidence) | 0 |
| S11_vs_k141_5641\|\|full | 6293 | No | NA | 13 | 1 | 1 | Low-quality | Genome-fragment | 15.1 | AAI-based (medium-confidence) | 0 |
| S3_extracted_k141_66761 | 6294 | No | NA | 5 | 0 | 0 | Low-quality | Genome-fragment | 4.11 | AAI-based (high-confidence) | 0 |
| S16_extracted_159246 | 6294 | No | NA | 8 | 6 | 0 | Low-quality | Genome-fragment | 17.09 | AAI-based (medium-confidence) | 0 |
| S6_vs_k141_2759\|\|full | 1216 | No | NA | 3 | 0 | 1 | Not-determined | Genome-fragment | NA | NA | 0 |
| S6_vs_k141_52275\|\|full | 10082 | No | NA | 10 | 0 | 3 | Not-determined | Genome-fragment | NA | NA | 0 |
| S4_vs_VIRSorter_24249-cat_2 | 6294 | No | NA | 8 | 1 | 0 | Low-quality | Genome-fragment | 14.39 | AAI-based (high-confidence) | 0 |
| S6_vs_k141_33438\|\|full | 2797 | No | NA | 5 | 0 | 3 | Not-determined | Genome-fragment | NA | NA | 0 |
| S5_extracted_k141_99671 | 6295 | No | NA | 14 | 2 | 0 | Low-quality | Genome-fragment | 8.15 | AAI-based (high-confidence) | 0 |
| S8_extracted_k141_5343 | 6297 | No | NA | 17 | 1 | 1 | Low-quality | Genome-fragment | 19.3 | AAI-based (high-confidence) | 0 |
| S7_vs_77781\|\|full | 6299 | No | NA | 15 | 2 | 0 | Low-quality | Genome-fragment | 18.62 | AAI-based (high-confidence) | 0 |
| S6_vs_k141_38220\|\|full | 5353 | No | NA | 5 | 0 | 2 | Not-determined | Genome-fragment | NA | NA | 0 |
| S6_vs_k141_45134\|\|full | 3256 | No | NA | 5 | 0 | 0 | Not-determined | Genome-fragment | NA | NA | 0 |
| S16_vs_154361\|\|full | 6299 | No | NA | 7 | 5 | 0 | Low-quality | Genome-fragment | 16.18 | AAI-based (high-confidence) | 0 |
| S3_extracted_k141_24799 | 6301 | No | NA | 4 | 0 | 1 | Low-quality | Genome-fragment | 6.1 | AAI-based (high-confidence) | 0 |
| S6_vs_k141_7282\|\|full | 1089 | No | NA | 3 | 0 | 1 | Not-determined | Genome-fragment | NA | NA | 0 |
| S16_extracted_128446 | 6303 | No | NA | 5 | 0 | 1 | Low-quality | Genome-fragment | 11.49 | AAI-based (medium-confidence) | 0 |
| S6_vs_k141_16898\|\|full | 1807 | No | NA | 4 | 0 | 0 | Not-determined | Genome-fragment | NA | NA | 0 |
| S6_vs_k141_14586\|\|full | 1217 | No | NA | 2 | 0 | 0 | Not-determined | Genome-fragment | NA | NA | 0 |
| S1_extracted_k141_120978 | 6304 | No | NA | 3 | 0 | 0 | Low-quality | Genome-fragment | 6.26 | AAI-based (medium-confidence) | 0 |
| S8_extracted_k141_86789 | 6305 | No | NA | 7 | 0 | 0 | Low-quality | Genome-fragment | 3.63 | AAI-based (high-confidence) | 0 |
| S9_vs_k141_70570\|\|full | 6311 | No | NA | 15 | 4 | 0 | Low-quality | Genome-fragment | 14.49 | AAI-based (high-confidence) | 0 |
| S9_vs_k141_56196\|\|full | 6311 | Yes | 5678 | 5 | 1 | 1 | Low-quality | Genome-fragment | 6.37 | HMM-based (lower-bound) | 10.03 |
| S1_extracted_k141_27563 | 6312 | No | NA | 7 | 3 | 0 | Low-quality | Genome-fragment | 10.99 | AAI-based (high-confidence) | 0 |
| S3_extracted_k141_115700 | 6312 | No | NA | 13 | 7 | 0 | Low-quality | Genome-fragment | 3.67 | AAI-based (high-confidence) | 0 |
| S6_vs_k141_38250\|\|full | 23419 | No | NA | 33 | 14 | 0 | Medium-quality | Genome-fragment | 62.87 | AAI-based (medium-confidence) | 0 |
| S6_vs_k141_47349\|\|full | 4622 | No | NA | 4 | 0 | 3 | Not-determined | Genome-fragment | NA | NA | 0 |
| S3_vs_k141_111921\|\|full | 6313 | No | NA | 10 | 4 | 0 | Low-quality | Genome-fragment | 13.2 | AAI-based (high-confidence) | 0 |
| S7_vs_28338\|\|full | 6313 | No | NA | 11 | 1 | 1 | Low-quality | Genome-fragment | 5.76 | AAI-based (medium-confidence) | 0 |
| S6_vs_k141_2967\|\|full | 1306 | No | NA | 3 | 0 | 1 | Not-determined | Genome-fragment | NA | NA | 0 |
| S3_extracted_k141_103601 | 6316 | No | NA | 7 | 0 | 3 | Low-quality | Genome-fragment | 20.17 | AAI-based (high-confidence) | 0 |
| S6_vs_k141_9784\|\|full | 59903 | Yes | 35541 | 70 | 13 | 16 | Complete | High-quality | 100 | Provirus (medium-confidence) | 40.67 |
| S9_vs_k141_51225\|\|full | 6320 | No | NA | 10 | 5 | 0 | Low-quality | Genome-fragment | 10.35 | AAI-based (high-confidence) | 0 |
| S5_vs_k141_93553\|\|full | 6322 | Yes | 4096 | 7 | 1 | 3 | Low-quality | Genome-fragment | 9.24 | AAI-based (medium-confidence) | 35.21 |
| S6_vs_k141_42909\|\|full | 1110 | No | NA | 3 | 0 | 0 | Not-determined | Genome-fragment | NA | NA | 0 |
| S6_vs_k141_7357\|\|full | 2536 | No | NA | 3 | 0 | 1 | Not-determined | Genome-fragment | NA | NA | 0 |
| S6_vs_k141_2993\|\|full | 2657 | No | NA | 3 | 0 | 1 | Not-determined | Genome-fragment | NA | NA | 0 |
| S6_vs_k141_38276\|\|full | 1051 | No | NA | 2 | 0 | 1 | Not-determined | Genome-fragment | NA | NA | 0 |
| S12_vs_k141_6615\|\|full | 6324 | No | NA | 10 | 0 | 1 | Low-quality | Genome-fragment | 14.2 | AAI-based (high-confidence) | 0 |
| S6_vs_k141_33534\|\|full | 5242 | No | NA | 6 | 0 | 2 | Not-determined | Genome-fragment | NA | NA | 0 |
| S6_vs_k141_7394\|\|full | 1100 | No | NA | 2 | 0 | 0 | Not-determined | Genome-fragment | NA | NA | 0 |
| S6_vs_k141_40592\|\|full | 2966 | No | NA | 2 | 0 | 2 | Not-determined | Genome-fragment | NA | NA | 0 |
| S6_vs_k141_26287\|\|full | 5178 | No | NA | 3 | 0 | 2 | Not-determined | Genome-fragment | NA | NA | 0 |
| S12_vs_k141_146000\|\|full | 6328 | No | NA | 9 | 0 | 1 | Low-quality | Genome-fragment | 8.23 | AAI-based (medium-confidence) | 0 |
| S6_vs_k141_38290\|\|full | 1400 | No | NA | 3 | 0 | 2 | Not-determined | Genome-fragment | NA | NA | 0 |
| S6_vs_k141_52596\|\|full | 2606 | No | NA | 3 | 0 | 0 | Not-determined | Genome-fragment | NA | NA | 0 |
| S6_vs_k141_26294\|\|full | 2562 | No | NA | 4 | 0 | 0 | Not-determined | Genome-fragment | NA | NA | 0 |
| S6_vs_k141_33588\|\|full | 1079 | No | NA | 3 | 0 | 1 | Not-determined | Genome-fragment | NA | NA | 0 |
| S17_extracted_k141_155720 | 6330 | No | NA | 7 | 0 | 0 | Low-quality | Genome-fragment | 11.89 | AAI-based (medium-confidence) | 0 |
| S6_vs_k141_12395\|\|full | 4557 | No | NA | 3 | 0 | 1 | Not-determined | Genome-fragment | NA | NA | 0 |
| S10_extracted_67691 | 6332 | No | NA | 12 | 2 | 0 | Low-quality | Genome-fragment | 9.84 | AAI-based (high-confidence) | 0 |
| S6_vs_k141_33604\|\|full | 3016 | No | NA | 4 | 0 | 2 | Not-determined | Genome-fragment | NA | NA | 0 |
| S8_vs_k141_164439\|\|full | 6333 | No | NA | 18 | 1 | 0 | Low-quality | Genome-fragment | 8.24 | HMM-based (lower-bound) | 0 |
| S13_extracted_4234 | 6338 | No | NA | 12 | 3 | 1 | Low-quality | Genome-fragment | 5.81 | AAI-based (high-confidence) | 0 |
| S15_extracted_46834 | 6339 | No | NA | 12 | 1 | 1 | Low-quality | Genome-fragment | 3.44 | HMM-based (lower-bound) | 0 |
| S6_vs_k141_38334\|\|full | 3630 | No | NA | 5 | 0 | 4 | Not-determined | Genome-fragment | NA | NA | 0 |
| S1_vs_k141_76731\|\|full | 6340 | No | NA | 3 | 0 | 3 | Low-quality | Genome-fragment | 3.94 | AAI-based (medium-confidence) | 0 |
| S8_vs_k141_21547\|\|full | 6342 | No | NA | 6 | 0 | 4 | Low-quality | Genome-fragment | 5.58 | AAI-based (medium-confidence) | 0 |
| S8_extracted_k141_202914 | 6343 | No | NA | 5 | 0 | 0 | Low-quality | Genome-fragment | 20.07 | AAI-based (medium-confidence) | 0 |
| S10_extracted_22563 | 6344 | No | NA | 7 | 0 | 5 | Low-quality | Genome-fragment | 2.54 | AAI-based (medium-confidence) | 0 |
| S2_extracted_k141_9105 | 6346 | No | NA | 15 | 2 | 0 | Low-quality | Genome-fragment | 15.17 | AAI-based (high-confidence) | 0 |
| S11_extracted_k141_5967 | 6348 | No | NA | 8 | 3 | 0 | Low-quality | Genome-fragment | 16.16 | AAI-based (medium-confidence) | 0 |
| S6_vs_k141_5079\|\|full | 1951 | No | NA | 5 | 0 | 0 | Not-determined | Genome-fragment | NA | NA | 0 |
| S11_vs_k141_18241\|\|full | 6349 | No | NA | 7 | 0 | 3 | Low-quality | Genome-fragment | 4.67 | AAI-based (medium-confidence) | 0 |
| S13_extracted_48588 | 6352 | No | NA | 13 | 2 | 0 | Low-quality | Genome-fragment | 10.66 | AAI-based (high-confidence) | 0 |
| S8_extracted_k141_82754 | 6352 | No | NA | 5 | 0 | 5 | Low-quality | Genome-fragment | 0.75 | AAI-based (medium-confidence) | 0 |
| S6_vs_k141_38360\|\|full | 6968 | No | NA | 9 | 0 | 1 | Not-determined | Genome-fragment | NA | NA | 0 |
| S1_vs_k141_79911\|\|full | 6358 | No | NA | 8 | 6 | 0 | Low-quality | Genome-fragment | 13.88 | AAI-based (high-confidence) | 0 |
| S10_extracted_81757 | 6359 | No | NA | 11 | 0 | 1 | Low-quality | Genome-fragment | 10.69 | AAI-based (medium-confidence) | 0 |
| S6_vs_k141_54409\|\|full | 1184 | No | NA | 2 | 0 | 1 | Not-determined | Genome-fragment | NA | NA | 0 |
| S6_vs_k141_26351\|\|full | 3142 | No | NA | 3 | 0 | 3 | Not-determined | Genome-fragment | NA | NA | 0 |
| S10_extracted_92547 | 6362 | No | NA | 16 | 1 | 0 | Low-quality | Genome-fragment | 14.31 | AAI-based (high-confidence) | 0 |
| S6_vs_k141_7433\|\|full | 2134 | No | NA | 4 | 0 | 1 | Not-determined | Genome-fragment | NA | NA | 0 |
| S5_vs_k141_127813\|\|full | 6365 | No | NA | 8 | 6 | 0 | Low-quality | Genome-fragment | 13.9 | AAI-based (high-confidence) | 0 |
| S8_extracted_k141_155879 | 6366 | No | NA | 2 | 0 | 1 | Low-quality | Genome-fragment | 9.66 | AAI-based (high-confidence) | 0 |
| S6_vs_k141_43001\|\|full | 2081 | No | NA | 3 | 0 | 1 | Not-determined | Genome-fragment | NA | NA | 0 |
| S6_vs_k141_26370\|\|full | 2932 | No | NA | 6 | 0 | 0 | Not-determined | Genome-fragment | NA | NA | 0 |
| S6_vs_k141_21458\|\|full | 2485 | No | NA | 5 | 0 | 0 | Not-determined | Genome-fragment | NA | NA | 0 |
| S3_extracted_k141_110730 | 6372 | No | NA | 3 | 0 | 2 | Low-quality | Genome-fragment | 5.99 | AAI-based (high-confidence) | 0 |
| S5_vs_k141_17317\|\|full | 6372 | No | NA | 16 | 2 | 0 | Low-quality | Genome-fragment | 14.7 | AAI-based (high-confidence) | 0 |
| S7_vs_37877\|\|full | 6372 | No | NA | 9 | 0 | 1 | Low-quality | Genome-fragment | 11.78 | AAI-based (medium-confidence) | 0 |
| S5_vs_k141_35559\|\|full | 6373 | No | NA | 9 | 1 | 1 | Low-quality | Genome-fragment | 9.15 | HMM-based (lower-bound) | 0 |
| S12_vs_k141_20993\|\|full | 6375 | No | NA | 9 | 0 | 1 | Low-quality | Genome-fragment | 11.99 | AAI-based (high-confidence) | 0 |
| S6_vs_k141_31025\|\|full | 1550 | No | NA | 3 | 0 | 1 | Not-determined | Genome-fragment | NA | NA | 0 |
| S1_vs_k141_84010\|\|full | 6376 | No | NA | 12 | 5 | 0 | Low-quality | Genome-fragment | 11.07 | AAI-based (high-confidence) | 0 |
| S8_vs_k141_121996\|\|full | 6380 | No | NA | 18 | 1 | 1 | Low-quality | Genome-fragment | 3.71 | HMM-based (lower-bound) | 0 |
| S6_vs_k141_19112\|\|full | 6480 | No | NA | 9 | 0 | 0 | Not-determined | Genome-fragment | NA | NA | 0 |
| S17_vs_k141_125825\|\|full | 6383 | No | NA | 16 | 2 | 0 | Low-quality | Genome-fragment | 10.11 | AAI-based (high-confidence) | 0 |
| S6_vs_k141_49924\|\|full | 9779 | No | NA | 2 | 0 | 1 | Not-determined | Genome-fragment | NA | NA | 0 |
| S8_vs_k141_42881\|\|full | 6384 | No | NA | 5 | 0 | 1 | Low-quality | Genome-fragment | 1.2 | AAI-based (medium-confidence) | 0 |
| S6_vs_k141_31071\|\|full | 1628 | No | NA | 4 | 0 | 1 | Not-determined | Genome-fragment | NA | NA | 0 |
| S6_vs_k141_7504\|\|full | 1914 | No | NA | 4 | 0 | 2 | Not-determined | Genome-fragment | NA | NA | 0 |
| S17_extracted_k141_6210 | 6386 | No | NA | 6 | 1 | 0 | Low-quality | Genome-fragment | 5.71 | AAI-based (high-confidence) | 0 |
| S1_vs_k141_152196\|\|full | 6386 | No | NA | 7 | 2 | 0 | Low-quality | Genome-fragment | 14.38 | AAI-based (high-confidence) | 0 |
| S5_vs_k141_139083\|\|full | 6386 | No | NA | 8 | 0 | 3 | Low-quality | Genome-fragment | 7.53 | AAI-based (medium-confidence) | 0 |
| S3_extracted_k141_98003 | 6388 | No | NA | 9 | 5 | 0 | Low-quality | Genome-fragment | 15.08 | AAI-based (high-confidence) | 0 |
| S6_vs_k141_17161\|\|full | 1207 | No | NA | 3 | 0 | 3 | Not-determined | Genome-fragment | NA | NA | 0 |
| S6_vs_k141_21574\|\|full | 1301 | No | NA | 2 | 0 | 1 | Not-determined | Genome-fragment | NA | NA | 0 |
| S6_vs_k141_28818\|\|full | 2633 | No | NA | 4 | 0 | 0 | Not-determined | Genome-fragment | NA | NA | 0 |
| S6_vs_k141_35907\|\|full | 3600 | No | NA | 4 | 0 | 3 | Not-determined | Genome-fragment | NA | NA | 0 |
| S8_vs_k141_122270\|\|full | 6388 | No | NA | 11 | 0 | 0 | Low-quality | Genome-fragment | 6.86 | AAI-based (high-confidence) | 0 |
| S16_vs_19453\|\|full | 6391 | No | NA | 8 | 0 | 4 | Low-quality | Genome-fragment | 3.58 | AAI-based (medium-confidence) | 0 |
| S12_extracted_k141_183866 | 6392 | No | NA | 14 | 2 | 0 | Low-quality | Genome-fragment | 13.03 | AAI-based (high-confidence) | 0 |
| S6_vs_k141_45479\|\|full | 7361 | No | NA | 11 | 0 | 0 | Not-determined | Genome-fragment | NA | NA | 0 |
| S6_vs_k141_17220\|\|full | 1453 | No | NA | 3 | 0 | 0 | Not-determined | Genome-fragment | NA | NA | 0 |
| S16_vs_136127\|\|full | 6394 | No | NA | 13 | 1 | 0 | Low-quality | Genome-fragment | 10.08 | AAI-based (high-confidence) | 0 |
| S6_vs_k141_10009\|\|full | 1774 | No | NA | 3 | 0 | 2 | Not-determined | Genome-fragment | NA | NA | 0 |
| S14_extracted_k141_108331 | 6395 | No | NA | 15 | 1 | 0 | Low-quality | Genome-fragment | 13.92 | AAI-based (high-confidence) | 0 |
| S14_vs_k141_16335\|\|full | 6399 | No | NA | 10 | 0 | 0 | Low-quality | Genome-fragment | 15.6 | AAI-based (medium-confidence) | 0 |
| S7_vs_56267\|\|full | 6399 | No | NA | 6 | 0 | 1 | Low-quality | Genome-fragment | 6.5 | AAI-based (medium-confidence) | 0 |
| S12_extracted_k141_127659 | 6399 | No | NA | 8 | 1 | 0 | Low-quality | Genome-fragment | 4.74 | AAI-based (medium-confidence) | 0 |
| S11_extracted_k141_91611 | 6401 | No | NA | 14 | 3 | 0 | Low-quality | Genome-fragment | 14.09 | AAI-based (high-confidence) | 0 |
| S6_vs_k141_33711\|\|full | 36137 | No | NA | 56 | 13 | 1 | High-quality | High-quality | 96.73 | AAI-based (medium-confidence) | 0 |
| S6_vs_k141_14816\|\|full | 2082 | No | NA | 4 | 0 | 2 | Not-determined | Genome-fragment | NA | NA | 0 |
| S3_extracted_k141_66269 | 6404 | No | NA | 8 | 2 | 0 | Low-quality | Genome-fragment | 4.79 | AAI-based (high-confidence) | 0 |
| S4_extracted_13541 | 6406 | No | NA | 4 | 1 | 0 | Low-quality | Genome-fragment | 10.41 | AAI-based (high-confidence) | 0 |
| S6_vs_k141_43082\|\|full | 1955 | No | NA | 2 | 0 | 1 | Not-determined | Genome-fragment | NA | NA | 0 |
| S6_extracted_k141_4313 | 6407 | No | NA | 6 | 0 | 2 | Low-quality | Genome-fragment | 2.55 | AAI-based (medium-confidence) | 0 |
| S3_extracted_k141_26896 | 6408 | No | NA | 13 | 1 | 0 | Low-quality | Genome-fragment | 9.94 | AAI-based (high-confidence) | 0 |
| S6_vs_k141_12605\|\|full | 7882 | No | NA | 8 | 0 | 6 | Not-determined | Genome-fragment | NA | NA | 0 |
| S5_vs_k141_22996\|\|full | 6412 | No | NA | 13 | 2 | 0 | Low-quality | Genome-fragment | 16.32 | AAI-based (medium-confidence) | 0 |
| S11_extracted_k141_75658 | 6414 | No | NA | 10 | 3 | 0 | Low-quality | Genome-fragment | 8.23 | AAI-based (high-confidence) | 0 |
| S6_vs_k141_40762\|\|full | 2677 | No | NA | 3 | 0 | 2 | Not-determined | Genome-fragment | NA | NA | 0 |
| S6_vs_k141_47700\|\|full | 1275 | No | NA | 3 | 0 | 0 | Not-determined | Genome-fragment | NA | NA | 0 |
| S16_extracted_55714 | 6414 | No | NA | 9 | 1 | 1 | Low-quality | Genome-fragment | 7.2 | HMM-based (lower-bound) | 0 |
| S6_vs_k141_7694\|\|full | 2267 | No | NA | 4 | 0 | 1 | Not-determined | Genome-fragment | NA | NA | 0 |
| S16_extracted_99183 | 6414 | No | NA | 15 | 7 | 0 | Low-quality | Genome-fragment | 8.18 | AAI-based (high-confidence) | 0 |
| S13_vs_93602\|\|full | 6417 | No | NA | 9 | 0 | 1 | Low-quality | Genome-fragment | 11.95 | AAI-based (high-confidence) | 0 |
| S11_vs_k141_23412\|\|full | 6423 | No | NA | 10 | 3 | 0 | Low-quality | Genome-fragment | 2.49 | AAI-based (medium-confidence) | 0 |
| S5_vs_k141_27705\|\|full | 6424 | No | NA | 7 | 0 | 2 | Low-quality | Genome-fragment | 13.01 | AAI-based (medium-confidence) | 0 |
| S6_vs_k141_33807\|\|full | 2111 | No | NA | 5 | 0 | 1 | Not-determined | Genome-fragment | NA | NA | 0 |
| S6_vs_k141_50154\|\|full | 2085 | No | NA | 2 | 0 | 2 | Not-determined | Genome-fragment | NA | NA | 0 |
| S3_extracted_k141_85694 | 6426 | No | NA | 17 | 1 | 0 | Low-quality | Genome-fragment | 3.64 | AAI-based (high-confidence) | 0 |
| S12_vs_k141_108194\|\|full | 6426 | No | NA | 9 | 1 | 0 | Low-quality | Genome-fragment | 4.09 | HMM-based (lower-bound) | 0 |
| S17_vs_k141_171795\|\|full | 6428 | Yes | 5441 | 7 | 2 | 1 | Low-quality | Genome-fragment | 7.36 | AAI-based (high-confidence) | 15.35 |
| S3_extracted_k141_125073 | 6428 | No | NA | 13 | 5 | 0 | Low-quality | Genome-fragment | 3.82 | AAI-based (high-confidence) | 0 |
| S6_vs_k141_40805\|\|full | 10774 | No | NA | 10 | 0 | 3 | Not-determined | Genome-fragment | NA | NA | 0 |
| S6_vs_k141_43097\|\|full | 1354 | No | NA | 2 | 0 | 1 | Not-determined | Genome-fragment | NA | NA | 0 |
| S6_vs_k141_14877\|\|full | 6595 | No | NA | 8 | 0 | 5 | Not-determined | Genome-fragment | NA | NA | 0 |
| S6_vs_k141_12711\|\|full | 2354 | No | NA | 3 | 0 | 3 | Not-determined | Genome-fragment | NA | NA | 0 |
| S11_vs_k141_78980\|\|full | 6433 | No | NA | 8 | 5 | 0 | Low-quality | Genome-fragment | 15.13 | AAI-based (high-confidence) | 0 |
| S10_extracted_31665 | 6436 | No | NA | 4 | 0 | 3 | Low-quality | Genome-fragment | 1.61 | AAI-based (medium-confidence) | 0 |
| S16_vs_162227\|\|full | 6438 | No | NA | 5 | 0 | 5 | Low-quality | Genome-fragment | 1.64 | AAI-based (medium-confidence) | 0 |
| S7_vs_77051\|\|full | 6440 | No | NA | 15 | 6 | 0 | Low-quality | Genome-fragment | 8.67 | HMM-based (lower-bound) | 0 |
| S3_vs_k141_73743\|\|full | 6444 | No | NA | 9 | 4 | 0 | Low-quality | Genome-fragment | 15.55 | AAI-based (high-confidence) | 0 |
| S6_vs_k141_45725\|\|full | 2357 | No | NA | 3 | 0 | 2 | Not-determined | Genome-fragment | NA | NA | 0 |
| S16_vs_36260\|\|full | 6444 | No | NA | 12 | 0 | 0 | Low-quality | Genome-fragment | 16.13 | AAI-based (medium-confidence) | 0 |
| S4_vs_VIRSorter_13524-cat_2 | 6446 | No | NA | 10 | 4 | 0 | Low-quality | Genome-fragment | 13.64 | AAI-based (medium-confidence) | 0 |
| S6_vs_k141_24600\|\|full | 3641 | No | NA | 4 | 0 | 2 | Not-determined | Genome-fragment | NA | NA | 0 |
| S6_vs_k141_19233\|\|full | 1459 | No | NA | 2 | 0 | 0 | Not-determined | Genome-fragment | NA | NA | 0 |
| S6_vs_k141_29057\|\|full | 1891 | No | NA | 3 | 0 | 3 | Not-determined | Genome-fragment | NA | NA | 0 |
| S5_vs_k141_66720\|\|full | 6447 | No | NA | 11 | 4 | 0 | Low-quality | Genome-fragment | 15.56 | AAI-based (high-confidence) | 0 |
| S16_extracted_80006 | 6450 | No | NA | 6 | 0 | 2 | Low-quality | Genome-fragment | 2.91 | AAI-based (medium-confidence) | 0 |
| S9_vs_k141_21800\|\|full | 6452 | No | NA | 9 | 0 | 0 | Low-quality | Genome-fragment | 15.05 | AAI-based (high-confidence) | 0 |
| S16_vs_164370\|\|full | 6452 | No | NA | 17 | 2 | 0 | Low-quality | Genome-fragment | 16.65 | AAI-based (high-confidence) | 0 |
| S6_vs_k141_31317\|\|full | 2156 | No | NA | 3 | 0 | 1 | Not-determined | Genome-fragment | NA | NA | 0 |
| S5_extracted_k141_16078 | 6455 | Yes | 5860 | 10 | 4 | 1 | Low-quality | Genome-fragment | 12.28 | AAI-based (high-confidence) | 9.22 |
| S3_extracted_k141_86762 | 6459 | No | NA | 5 | 4 | 0 | Low-quality | Genome-fragment | 22.62 | HMM-based (lower-bound) | 0 |
| S16_extracted_107782 | 6467 | No | NA | 12 | 4 | 0 | Low-quality | Genome-fragment | 14.37 | AAI-based (high-confidence) | 0 |
| S6_vs_k141_45806\|\|full | 2959 | No | NA | 4 | 0 | 1 | Not-determined | Genome-fragment | NA | NA | 0 |
| S16_vs_23157\|\|full | 6467 | No | NA | 4 | 2 | 0 | Low-quality | Genome-fragment | 7.85 | HMM-based (lower-bound) | 0 |
| S8_extracted_k141_65709 | 6467 | No | NA | 8 | 0 | 4 | Low-quality | Genome-fragment | 10.07 | AAI-based (medium-confidence) | 0 |
| S16_vs_116702\|\|full | 6469 | No | NA | 7 | 0 | 0 | Low-quality | Genome-fragment | 1.61 | AAI-based (high-confidence) | 0 |
| S6_vs_k141_1000\|\|full | 2839 | No | NA | 3 | 0 | 2 | Not-determined | Genome-fragment | NA | NA | 0 |
| S12_extracted_k141_44086 | 6469 | No | NA | 7 | 4 | 0 | Low-quality | Genome-fragment | 16.98 | AAI-based (medium-confidence) | 0 |
| S6_vs_k141_31373\|\|full | 3734 | No | NA | 9 | 0 | 0 | Not-determined | Genome-fragment | NA | NA | 0 |
| S8_vs_k141_144251\|\|full | 6469 | No | NA | 15 | 4 | 0 | Low-quality | Genome-fragment | 12.75 | AAI-based (high-confidence) | 0 |
| S6_vs_k141_7945\|\|full | 2329 | No | NA | 2 | 0 | 1 | Not-determined | Genome-fragment | NA | NA | 0 |
| S6_vs_k141_15002\|\|full | 1187 | No | NA | 3 | 0 | 1 | Not-determined | Genome-fragment | NA | NA | 0 |
| S14_extracted_k141_110845 | 6473 | No | NA | 8 | 8 | 0 | Low-quality | Genome-fragment | 21.33 | AAI-based (high-confidence) | 0 |
| S12_vs_k141_107952\|\|full | 6475 | No | NA | 7 | 1 | 0 | Low-quality | Genome-fragment | 1.82 | HMM-based (lower-bound) | 0 |
| S6_vs_k141_40882\|\|full | 2858 | No | NA | 3 | 0 | 0 | Not-determined | Genome-fragment | NA | NA | 0 |
| S6_vs_k141_43247\|\|full | 6927 | No | NA | 6 | 0 | 3 | Not-determined | Genome-fragment | NA | NA | 0 |
| S9_extracted_k141_57739 | 6478 | No | NA | 18 | 2 | 0 | Low-quality | Genome-fragment | 13.01 | AAI-based (high-confidence) | 0 |
| S3_extracted_k141_46338 | 6479 | No | NA | 11 | 0 | 0 | Low-quality | Genome-fragment | 14.71 | AAI-based (medium-confidence) | 0 |
| S6_vs_k141_54606\|\|full | 65938 | Yes | 21709 | 84 | 11 | 28 | Medium-quality | Genome-fragment | 62.05 | AAI-based (high-confidence) | 67.08 |
| S6_vs_k141_43261\|\|full | 1529 | No | NA | 3 | 0 | 2 | Not-determined | Genome-fragment | NA | NA | 0 |
| S3_vs_k141_113839\|\|full | 6481 | Yes | 3910 | 3 | 1 | 1 | Low-quality | Genome-fragment | 7.84 | AAI-based (high-confidence) | 39.67 |
| S17_extracted_k141_27767 | 6483 | No | NA | 5 | 0 | 1 | Low-quality | Genome-fragment | 6.27 | AAI-based (medium-confidence) | 0 |
| S7_extracted_93551 | 6485 | No | NA | 7 | 0 | 6 | Low-quality | Genome-fragment | 2.68 | AAI-based (high-confidence) | 0 |
| S6_vs_k141_10309\|\|full | 1777 | No | NA | 3 | 0 | 2 | Not-determined | Genome-fragment | NA | NA | 0 |
| S10_vs_81521\|\|full | 6485 | Yes | 4129 | 7 | 1 | 2 | Low-quality | Genome-fragment | 1.92 | HMM-based (lower-bound) | 36.33 |
| S6_vs_k141_54645\|\|full | 11708 | No | NA | 8 | 0 | 6 | Not-determined | Genome-fragment | NA | NA | 0 |
| S5_vs_k141_88697\|\|full | 6491 | No | NA | 3 | 0 | 3 | Low-quality | Genome-fragment | 2.51 | AAI-based (medium-confidence) | 0 |
| S16_extracted_104690 | 6499 | No | NA | 7 | 1 | 0 | Low-quality | Genome-fragment | 4.24 | AAI-based (high-confidence) | 0 |
| S6_vs_k141_21739\|\|full | 1667 | No | NA | 2 | 0 | 1 | Not-determined | Genome-fragment | NA | NA | 0 |
| S6_vs_k141_38734\|\|full | 1287 | No | NA | 5 | 0 | 0 | Not-determined | Genome-fragment | NA | NA | 0 |
| S17_extracted_k141_155563 | 6507 | No | NA | 11 | 4 | 0 | Low-quality | Genome-fragment | 14.39 | AAI-based (high-confidence) | 0 |
| S6_vs_k141_29152\|\|full | 71717 | Yes | 53494 | 104 | 24 | 13 | Complete | High-quality | 100 | Provirus (high-confidence) | 25.41 |
| S3_extracted_k141_51642 | 6507 | No | NA | 11 | 1 | 2 | Low-quality | Genome-fragment | 8.38 | HMM-based (lower-bound) | 0 |
| S6_vs_k141_5557\|\|full | 2627 | No | NA | 3 | 0 | 2 | Not-determined | Genome-fragment | NA | NA | 0 |
| S6_vs_k141_54688\|\|full | 23930 | No | NA | 12 | 0 | 7 | Not-determined | Genome-fragment | NA | NA | 0 |
| S6_vs_k141_15121\|\|full | 1653 | No | NA | 3 | 0 | 1 | Not-determined | Genome-fragment | NA | NA | 0 |
| S3_extracted_k141_51422 | 6509 | No | NA | 11 | 5 | 0 | Low-quality | Genome-fragment | 16.74 | AAI-based (medium-confidence) | 0 |
| S16_vs_86524\|\|full | 6515 | No | NA | 9 | 0 | 0 | Low-quality | Genome-fragment | 21.89 | AAI-based (high-confidence) | 0 |
| S16_extracted_34821 | 6516 | No | NA | 5 | 0 | 0 | Low-quality | Genome-fragment | 10.25 | AAI-based (high-confidence) | 0 |
| S8_extracted_k141_81554 | 6517 | No | NA | 4 | 0 | 1 | Low-quality | Genome-fragment | 5.96 | AAI-based (medium-confidence) | 0 |
| S4_extracted_4142 | 6519 | No | NA | 19 | 2 | 0 | Low-quality | Genome-fragment | 14.45 | AAI-based (high-confidence) | 0 |
| S6_vs_k141_24821\|\|full | 3809 | No | NA | 7 | 0 | 1 | Not-determined | Genome-fragment | NA | NA | 0 |
| S6_vs_k141_34204\|\|full | 1954 | No | NA | 3 | 0 | 2 | Not-determined | Genome-fragment | NA | NA | 0 |
| S6_vs_k141_17609\|\|full | 1800 | No | NA | 3 | 0 | 1 | Not-determined | Genome-fragment | NA | NA | 0 |
| S6_vs_k141_10314\|\|full | 83752 | Yes | 28882 | 100 | 14 | 37 | Complete | High-quality | 100 | Provirus (medium-confidence) | 65.51 |
| S9_vs_k141_54996\|\|full | 6520 | No | NA | 16 | 2 | 0 | Low-quality | Genome-fragment | 5.26 | AAI-based (high-confidence) | 0 |
| S6_vs_k141_43376\|\|full | 1293 | No | NA | 3 | 0 | 0 | Not-determined | Genome-fragment | NA | NA | 0 |
| S6_vs_k141_53051\|\|full | 3028 | No | NA | 4 | 0 | 4 | Not-determined | Genome-fragment | NA | NA | 0 |
| S6_vs_k141_34227\|\|full | 1898 | No | NA | 3 | 0 | 1 | Not-determined | Genome-fragment | NA | NA | 0 |
| S9_extracted_k141_14013 | 6522 | No | NA | 10 | 2 | 0 | Low-quality | Genome-fragment | 4.64 | AAI-based (high-confidence) | 0 |
| S6_vs_k141_40987\|\|full | 1931 | No | NA | 3 | 0 | 1 | Not-determined | Genome-fragment | NA | NA | 0 |
| S6_vs_k141_5632\|\|full | 1882 | No | NA | 3 | 0 | 0 | Not-determined | Genome-fragment | NA | NA | 0 |
| S6_vs_k141_40992\|\|full | 1759 | No | NA | 3 | 0 | 1 | Not-determined | Genome-fragment | NA | NA | 0 |
| S5_vs_k141_74436\|\|full | 6522 | No | NA | 3 | 1 | 0 | Low-quality | Genome-fragment | 14.46 | AAI-based (high-confidence) | 0 |
| S6_extracted_k141_8490 | 6524 | No | NA | 15 | 13 | 0 | Low-quality | Genome-fragment | 21.5 | AAI-based (high-confidence) | 0 |
| S6_vs_k141_3592\|\|full | 2802 | No | NA | 2 | 0 | 2 | Not-determined | Genome-fragment | NA | NA | 0 |
| S11_vs_k141_42220\|\|full | 6527 | No | NA | 10 | 1 | 0 | Low-quality | Genome-fragment | 6.12 | HMM-based (lower-bound) | 0 |
| S6_vs_k141_31515\|\|full | 2002 | No | NA | 3 | 0 | 2 | Not-determined | Genome-fragment | NA | NA | 0 |
| S6_vs_k141_3602\|\|full | 3318 | No | NA | 4 | 0 | 0 | Not-determined | Genome-fragment | NA | NA | 0 |
| S12_vs_k141_40479\|\|full | 6527 | No | NA | 8 | 0 | 0 | Low-quality | Genome-fragment | 9.62 | AAI-based (high-confidence) | 0 |
| S12_extracted_k141_65799 | 6529 | No | NA | 7 | 0 | 1 | Low-quality | Genome-fragment | 8.7 | AAI-based (medium-confidence) | 0 |
| S17_extracted_k141_119559 | 6530 | No | NA | 9 | 1 | 0 | Low-quality | Genome-fragment | 8.29 | AAI-based (high-confidence) | 0 |
| S6_vs_k141_54831\|\|full | 11314 | No | NA | 13 | 0 | 11 | Not-determined | Genome-fragment | NA | NA | 0 |
| S6_vs_k141_5678\|\|full | 1617 | No | NA | 3 | 0 | 0 | Not-determined | Genome-fragment | NA | NA | 0 |

| | | | | | | | | | | | |
|---|---|---|---|---|---|---|---|---|---|---|---|
| S16_vs_59500\|\|full | 6538 | No | NA | 11 | 3 | 0 | Low-quality | Genome-fragment | 6.03 | HMM-based (lower-bound) | 0 |
| S11_vs_k141_45935\|\|full | 6540 | No | NA | 6 | 0 | 2 | Low-quality | Genome-fragment | 11.73 | AAI-based (medium-confidence) | 0 |
| S7_extracted_117718 | 6540 | No | NA | 7 | 0 | 0 | Low-quality | Genome-fragment | 11.64 | AAI-based (medium-confidence) | 0 |
| S6_vs_k141_41024\|\|full | 1730 | No | NA | 2 | 0 | 2 | Not-determined | Genome-fragment | NA | NA | 0 |
| S6_vs_k141_1146\|\|full | 53780 | Yes | 34975 | 72 | 20 | 12 | High-quality | High-quality | 100 | AAI-based (medium-confidence) | 34.97 |
| S6_vs_k141_31569\|\|full | 1183 | No | NA | 2 | 0 | 2 | Not-determined | Genome-fragment | NA | NA | 0 |
| S17_extracted_k141_2636 | 6542 | No | NA | 12 | 3 | 0 | Low-quality | Genome-fragment | 14.5 | AAI-based (high-confidence) | 0 |
| S6_vs_k141_21940\|\|full | 1327 | No | NA | 2 | 0 | 1 | Not-determined | Genome-fragment | NA | NA | 0 |
| S10_vs_120490\|\|full | 6544 | No | NA | 11 | 1 | 0 | Low-quality | Genome-fragment | 17.26 | AAI-based (high-confidence) | 0 |
| S12_extracted_k141_146777 | 6544 | No | NA | 12 | 1 | 0 | Low-quality | Genome-fragment | 5.48 | AAI-based (high-confidence) | 0 |
| S17_extracted_k141_2179 | 6545 | Yes | 6057 | 7 | 1 | 1 | Low-quality | Genome-fragment | 4.44 | HMM-based (lower-bound) | 7.46 |
| S17_vs_k141_38706\|\|full | 6547 | No | NA | 6 | 0 | 4 | Low-quality | Genome-fragment | 2.15 | AAI-based (medium-confidence) | 0 |
| S6_vs_k141_46039\|\|full | 111911 | No | NA | 121 | 2 | 23 | Medium-quality | Genome-fragment | 69.39 | HMM-based (lower-bound) | 0 |
| S6_vs_k141_8092\|\|full | 3462 | No | NA | 5 | 0 | 1 | Not-determined | Genome-fragment | NA | NA | 0 |
| S6_vs_k141_53137\|\|full | 1404 | No | NA | 3 | 0 | 0 | Not-determined | Genome-fragment | NA | NA | 0 |
| S6_vs_k141_12945\|\|full | 39137 | No | NA | 58 | 17 | 1 | Medium-quality | Genome-fragment | 68.58 | HMM-based (lower-bound) | 0 |
| S4_extracted_4102 | 6549 | No | NA | 12 | 11 | 0 | Low-quality | Genome-fragment | 21.6 | AAI-based (high-confidence) | 0 |
| S14_vs_k141_51749\|\|full | 6554 | No | NA | 8 | 4 | 0 | Low-quality | Genome-fragment | 13.88 | AAI-based (high-confidence) | 0 |
| S6_vs_k141_15254\|\|full | 1593 | No | NA | 3 | 0 | 0 | Not-determined | Genome-fragment | NA | NA | 0 |
| S6_vs_k141_29391\|\|full | 1104 | No | NA | 3 | 0 | 0 | Not-determined | Genome-fragment | NA | NA | 0 |
| S8_extracted_k141_103084 | 6556 | No | NA | 9 | 8 | 0 | Low-quality | Genome-fragment | 20.39 | AAI-based (high-confidence) | 0 |
| S8_extracted_k141_38480 | 6560 | No | NA | 10 | 3 | 0 | Low-quality | Genome-fragment | 47.23 | AAI-based (high-confidence) | 0 |
| S6_vs_k141_15263\|\|full | 1938 | No | NA | 5 | 0 | 3 | Not-determined | Genome-fragment | NA | NA | 0 |
| S6_vs_k141_5743\|\|full | 4823 | No | NA | 4 | 0 | 3 | Not-determined | Genome-fragment | NA | NA | 0 |
| S6_vs_k141_8119\|\|full | 1288 | No | NA | 2 | 0 | 0 | Not-determined | Genome-fragment | NA | NA | 0 |
| S8_vs_k141_188764\|\|full | 6560 | Yes | 5738 | 10 | 1 | 1 | Low-quality | Genome-fragment | 3.86 | HMM-based (lower-bound) | 12.53 |
| S6_vs_k141_31610\|\|full | 2577 | No | NA | 4 | 0 | 2 | Not-determined | Genome-fragment | NA | NA | 0 |
| S6_vs_k141_29424\|\|full | 3109 | No | NA | 3 | 0 | 2 | Not-determined | Genome-fragment | NA | NA | 0 |
| S6_vs_k141_5762\|\|full | 1423 | No | NA | 2 | 0 | 1 | Not-determined | Genome-fragment | NA | NA | 0 |
| S6_vs_k141_36343\|\|full | 3902 | No | NA | 5 | 0 | 4 | Not-determined | Genome-fragment | NA | NA | 0 |
| S5_vs_k141_70933\|\|full | 6562 | Yes | 5551 | 8 | 2 | 2 | Low-quality | Genome-fragment | 13.14 | AAI-based (medium-confidence) | 15.41 |
| S12_vs_k141_35916\|\|full | 6562 | No | NA | 12 | 1 | 0 | Low-quality | Genome-fragment | 13.42 | AAI-based (high-confidence) | 0 |
| S6_vs_k141_38924\|\|full | 3653 | No | NA | 4 | 0 | 3 | Not-determined | Genome-fragment | NA | NA | 0 |
| S4_extracted_23882 | 6562 | No | NA | 10 | 0 | 0 | Low-quality | Genome-fragment | 14.54 | AAI-based (high-confidence) | 0 |
| S6_vs_k141_21981\|\|full | 5169 | No | NA | 4 | 0 | 3 | Not-determined | Genome-fragment | NA | NA | 0 |
| S1_vs_k141_107851\|\|full | 6567 | No | NA | 7 | 4 | 0 | Low-quality | Genome-fragment | 11.5 | AAI-based (high-confidence) | 0 |
| S13_extracted_117599 | 6568 | No | NA | 10 | 2 | 1 | Low-quality | Genome-fragment | 11.02 | AAI-based (high-confidence) | 0 |
| S6_vs_k141_5812\|\|full | 1379 | No | NA | 3 | 0 | 1 | Not-determined | Genome-fragment | NA | NA | 0 |
| S6_vs_k141_22005\|\|full | 2619 | No | NA | 3 | 0 | 3 | Not-determined | Genome-fragment | NA | NA | 0 |
| S1_vs_k141_20142\|\|full | 6570 | No | NA | 4 | 4 | 0 | Low-quality | Genome-fragment | 10.9 | AAI-based (high-confidence) | 0 |
| S1_extracted_k141_34884 | 6573 | No | NA | 7 | 2 | 0 | Low-quality | Genome-fragment | 17.66 | AAI-based (high-confidence) | 0 |
| S6_vs_k141_26725\|\|full | 2112 | No | NA | 3 | 0 | 0 | Not-determined | Genome-fragment | NA | NA | 0 |
| S6_vs_k141_10604\|\|full | 1258 | No | NA | 3 | 0 | 1 | Not-determined | Genome-fragment | NA | NA | 0 |
| S16_vs_42365\|\|full | 6573 | No | NA | 6 | 0 | 1 | Low-quality | Genome-fragment | 4.11 | AAI-based (medium-confidence) | 0 |
| S11_vs_k141_96797\|\|full | 6580 | No | NA | 8 | 0 | 0 | Low-quality | Genome-fragment | 14.57 | AAI-based (high-confidence) | 0 |
| S4_extracted_11014 | 6580 | No | NA | 11 | 3 | 0 | Low-quality | Genome-fragment | 12.26 | AAI-based (high-confidence) | 0 |
| S1_vs_k141_109646\|\|full | 6582 | No | NA | 14 | 3 | 0 | Low-quality | Genome-fragment | 8.97 | HMM-based (lower-bound) | 0 |
| S6_vs_k141_34453\|\|full | 65621 | Yes | 61847 | 93 | 31 | 6 | High-quality | High-quality | 100 | AAI-based (high-confidence) | 5.75 |
| S1_vs_k141_113952\|\|full | 6584 | No | NA | 13 | 1 | 1 | Low-quality | Genome-fragment | 1.73 | HMM-based (lower-bound) | 0 |
| S6_vs_k141_50471\|\|full | 1139 | No | NA | 3 | 0 | 0 | Not-determined | Genome-fragment | NA | NA | 0 |
| S16_extracted_24844 | 6586 | No | NA | 4 | 0 | 3 | Low-quality | Genome-fragment | 5.92 | AAI-based (medium-confidence) | 0 |
| S6_vs_k141_15390\|\|full | 42083 | No | NA | 50 | 12 | 1 | Medium-quality | Genome-fragment | 82.88 | AAI-based (high-confidence) | 0 |
| S16_extracted_66903 | 6592 | No | NA | 15 | 0 | 0 | Low-quality | Genome-fragment | 6.1 | AAI-based (high-confidence) | 0 |
| S5_extracted_k141_41688 | 6595 | No | NA | 5 | 0 | 4 | Low-quality | Genome-fragment | 14.2 | AAI-based (high-confidence) | 0 |
| S9_vs_k141_6746\|\|full | 6596 | No | NA | 7 | 1 | 0 | Low-quality | Genome-fragment | 15.69 | AAI-based (medium-confidence) | 0 |
| S14_vs_k141_11476\|\|full | 6596 | No | NA | 14 | 0 | 0 | Low-quality | Genome-fragment | 18.97 | AAI-based (high-confidence) | 0 |
| S6_vs_k141_41115\|\|full | 2222 | No | NA | 3 | 0 | 1 | Not-determined | Genome-fragment | NA | NA | 0 |
| S7_vs_134429\|\|full | 6596 | No | NA | 12 | 4 | 0 | Low-quality | Genome-fragment | 15.84 | AAI-based (medium-confidence) | 0 |
| S6_vs_k141_15407\|\|full | 2234 | No | NA | 2 | 0 | 2 | Not-determined | Genome-fragment | NA | NA | 0 |
| S7_vs_26047\|\|full | 6600 | No | NA | 8 | 0 | 0 | Low-quality | Genome-fragment | 22.5 | AAI-based (medium-confidence) | 0 |
| S3_extracted_k141_3823 | 6601 | No | NA | 6 | 2 | 0 | Low-quality | Genome-fragment | 3.59 | HMM-based (lower-bound) | 0 |
| S8_vs_k141_54945\|\|full | 6601 | No | NA | 9 | 7 | 0 | Low-quality | Genome-fragment | 19.43 | AAI-based (high-confidence) | 0 |
| S6_vs_k141_17938\|\|full | 1398 | No | NA | 3 | 0 | 1 | Not-determined | Genome-fragment | NA | NA | 0 |
| S11_vs_k141_72365\|\|full | 6605 | No | NA | 5 | 1 | 0 | Low-quality | Genome-fragment | 2.56 | AAI-based (medium-confidence) | 0 |
| S11_vs_k141_18633\|\|full | 6606 | No | NA | 11 | 2 | 1 | Low-quality | Genome-fragment | 14.58 | AAI-based (high-confidence) | 0 |
| S3_extracted_k141_114991 | 6609 | Yes | 4287 | 9 | 1 | 1 | Low-quality | Genome-fragment | 5.43 | AAI-based (high-confidence) | 35.13 |
| S6_vs_k141_41193\|\|full | 6619 | No | NA | 5 | 0 | 3 | Not-determined | Genome-fragment | NA | NA | 0 |
| S6_vs_k141_39025\|\|full | 57387 | Yes | 32460 | 75 | 20 | 15 | Medium-quality | Genome-fragment | 54.9 | HMM-based (lower-bound) | 43.44 |
| S5_vs_k141_127840\|\|full | 6610 | No | NA | 7 | 3 | 0 | Low-quality | Genome-fragment | 14.25 | AAI-based (high-confidence) | 0 |
| S6_vs_k141_34519\|\|full | 1205 | No | NA | 3 | 0 | 0 | Not-determined | Genome-fragment | NA | NA | 0 |
| S6_vs_k141_17995\|\|full | 1472 | No | NA | 4 | 0 | 0 | Not-determined | Genome-fragment | NA | NA | 0 |
| S8_extracted_k141_26652 | 6610 | Yes | 5518 | 7 | 1 | 1 | Low-quality | Genome-fragment | 3.18 | HMM-based (lower-bound) | 16.52 |
| S6_vs_k141_29695\|\|full | 2696 | No | NA | 3 | 0 | 2 | Not-determined | Genome-fragment | NA | NA | 0 |
| S9_vs_k141_89924\|\|full | 6612 | No | NA | 16 | 2 | 0 | Low-quality | Genome-fragment | 19.1 | AAI-based (high-confidence) | 0 |
| S7_vs_41520\|\|full | 6612 | No | NA | 9 | 1 | 0 | Low-quality | Genome-fragment | 17.04 | AAI-based (medium-confidence) | 0 |
| S6_vs_k141_41212\|\|full | 1100 | No | NA | 2 | 0 | 1 | Not-determined | Genome-fragment | NA | NA | 0 |
| S5_vs_k141_106139\|\|full | 6613 | No | NA | 11 | 1 | 4 | Low-quality | Genome-fragment | 7.42 | HMM-based (lower-bound) | 0 |
| S15_extracted_25551 | 6613 | No | NA | 11 | 1 | 0 | Low-quality | Genome-fragment | 7.2 | AAI-based (high-confidence) | 0 |
| S6_vs_k141_46091\|\|full | 30750 | No | NA | 49 | 21 | 2 | High-quality | High-quality | 93.45 | AAI-based (high-confidence) | 0 |
| S6_extracted_k141_5145 | 6613 | No | NA | 12 | 4 | 0 | Low-quality | Genome-fragment | 14.73 | AAI-based (high-confidence) | 0 |
| S3_extracted_k141_76108 | 6615 | No | NA | 6 | 1 | 0 | Low-quality | Genome-fragment | 4.95 | AAI-based (high-confidence) | 0 |
| S6_vs_k141_50580\|\|full | 1357 | No | NA | 3 | 0 | 0 | Not-determined | Genome-fragment | NA | NA | 0 |
| S17_vs_k141_133847\|\|full | 6616 | No | NA | 18 | 3 | 0 | Low-quality | Genome-fragment | 9.8 | AAI-based (high-confidence) | 0 |
| S4_extracted_29314 | 6620 | No | NA | 4 | 0 | 1 | Low-quality | Genome-fragment | 6.98 | AAI-based (medium-confidence) | 0 |
| S8_extracted_k141_178383 | 6620 | No | NA | 8 | 0 | 5 | Low-quality | Genome-fragment | 4.65 | AAI-based (medium-confidence) | 0 |
| S6_vs_k141_18063\|\|full | 3561 | No | NA | 3 | 0 | 2 | Not-determined | Genome-fragment | NA | NA | 0 |
| S7_vs_69544\|\|full | 6621 | No | NA | 15 | 1 | 0 | Low-quality | Genome-fragment | 18.11 | AAI-based (medium-confidence) | 0 |
| S5_extracted_k141_4415 | 6623 | No | NA | 9 | 5 | 0 | Low-quality | Genome-fragment | 15.4 | AAI-based (medium-confidence) | 0 |
| S6_vs_k141_18067\|\|full | 1281 | No | NA | 3 | 0 | 0 | Not-determined | Genome-fragment | NA | NA | 0 |
| S6_vs_k141_41234\|\|full | 2393 | No | NA | 2 | 0 | 2 | Not-determined | Genome-fragment | NA | NA | 0 |
| S16_vs_110229\|\|full | 6643 | No | NA | 5 | 1 | 0 | Low-quality | Genome-fragment | 1.75 | HMM-based (lower-bound) | 0 |
| S6_vs_k141_41272\|\|full | 1649 | No | NA | 3 | 0 | 1 | Not-determined | Genome-fragment | NA | NA | 0 |
| S11_extracted_k141_24940 | 6647 | No | NA | 6 | 3 | 0 | Low-quality | Genome-fragment | 10.55 | AAI-based (high-confidence) | 0 |
| S17_vs_k141_3592\|\|full | 6649 | No | NA | 12 | 1 | 0 | Low-quality | Genome-fragment | 1.82 | HMM-based (lower-bound) | 0 |
| S3_vs_k141_121135\|\|full | 6649 | No | NA | 8 | 4 | 0 | Low-quality | Genome-fragment | 15.85 | AAI-based (high-confidence) | 0 |
| S6_vs_k141_18087\|\|full | 1930 | No | NA | 3 | 0 | 1 | Not-determined | Genome-fragment | NA | NA | 0 |
| S6_vs_k141_53340\|\|full | 2988 | No | NA | 2 | 0 | 2 | Not-determined | Genome-fragment | NA | NA | 0 |
| S6_vs_k141_10826\|\|full | 2554 | No | NA | 3 | 0 | 0 | Not-determined | Genome-fragment | NA | NA | 0 |
| S6_vs_k141_41333\|\|full | 1521 | No | NA | 3 | 0 | 0 | Not-determined | Genome-fragment | NA | NA | 0 |
| S9_vs_k141_73892\|\|full | 6650 | No | NA | 4 | 1 | 0 | Low-quality | Genome-fragment | 16.85 | AAI-based (high-confidence) | 0 |
| S6_vs_k141_15514\|\|full | 2994 | No | NA | 3 | 0 | 1 | Not-determined | Genome-fragment | NA | NA | 0 |
| S6_vs_k141_18102\|\|full | 3729 | No | NA | 3 | 0 | 2 | Not-determined | Genome-fragment | NA | NA | 0 |
| S13_extracted_107744 | 6653 | No | NA | 7 | 0 | 0 | Low-quality | Genome-fragment | 16.82 | AAI-based (high-confidence) | 0 |
| S6_vs_k141_55044\|\|full | 1247 | No | NA | 3 | 0 | 1 | Not-determined | Genome-fragment | NA | NA | 0 |
| S6_vs_k141_13362\|\|full | 2226 | No | NA | 3 | 0 | 1 | Not-determined | Genome-fragment | NA | NA | 0 |
| S14_vs_k141_32020\|\|full | 6667 | No | NA | 18 | 2 | 0 | Low-quality | Genome-fragment | 17.1 | AAI-based (high-confidence) | 0 |
| S5_vs_k141_140564\|\|full | 6667 | No | NA | 10 | 9 | 0 | Low-quality | Genome-fragment | 8.77 | HMM-based (lower-bound) | 0 |
| S15_vs_49694\|\|full | 6669 | No | NA | 4 | 0 | 3 | Low-quality | Genome-fragment | 2.21 | AAI-based (medium-confidence) | 0 |
| S6_vs_k141_48279\|\|full | 13097 | No | NA | 13 | 0 | 1 | Not-determined | Genome-fragment | NA | NA | 0 |
| S6_vs_k141_8401\|\|full | 4419 | No | NA | 6 | 0 | 4 | Not-determined | Genome-fragment | NA | NA | 0 |
| S8_extracted_k141_119795 | 6670 | No | NA | 6 | 0 | 1 | Low-quality | Genome-fragment | 15.19 | AAI-based (medium-confidence) | 0 |
| S6_vs_k141_41397\|\|full | 1679 | No | NA | 2 | 0 | 1 | Not-determined | Genome-fragment | NA | NA | 0 |
| S6_vs_k141_41400\|\|full | 1434 | No | NA | 3 | 0 | 1 | Not-determined | Genome-fragment | NA | NA | 0 |
| S8_vs_k141_60337\|\|full | 6670 | No | NA | 12 | 2 | 0 | Low-quality | Genome-fragment | 6.93 | HMM-based (lower-bound) | 0 |
| S6_vs_k141_13404\|\|full | 1853 | No | NA | 2 | 0 | 1 | Not-determined | Genome-fragment | NA | NA | 0 |
| S6_vs_k141_18191\|\|full | 1173 | No | NA | 3 | 0 | 1 | Not-determined | Genome-fragment | NA | NA | 0 |
| S6_vs_k141_27010\|\|full | 1401 | No | NA | 3 | 0 | 0 | Not-determined | Genome-fragment | NA | NA | 0 |
| S6_vs_k141_25219\|\|full | 1773 | No | NA | 2 | 0 | 0 | Not-determined | Genome-fragment | NA | NA | 0 |
| S6_vs_k141_22283\|\|full | 2505 | No | NA | 5 | 0 | 3 | Not-determined | Genome-fragment | NA | NA | 0 |
| S6_vs_k141_8476\|\|full | 1063 | No | NA | 3 | 0 | 1 | Not-determined | Genome-fragment | NA | NA | 0 |
| S8_extracted_k141_47493 | 6671 | No | NA | 8 | 0 | 4 | Low-quality | Genome-fragment | 15.87 | AAI-based (high-confidence) | 0 |
| S6_vs_k141_34765\|\|full | 1598 | No | NA | 3 | 0 | 2 | Not-determined | Genome-fragment | NA | NA | 0 |
| S6_vs_k141_25242\|\|full | 2022 | No | NA | 4 | 0 | 2 | Not-determined | Genome-fragment | NA | NA | 0 |
| S5_extracted_k141_38629 | 6673 | No | NA | 8 | 0 | 0 | Low-quality | Genome-fragment | 10.54 | AAI-based (high-confidence) | 0 |
| S12_extracted_k141_38990 | 6677 | No | NA | 7 | 2 | 0 | Low-quality | Genome-fragment | 9.88 | HMM-based (lower-bound) | 0 |
| S6_vs_k141_15649\|\|full | 1204 | No | NA | 3 | 0 | 0 | Not-determined | Genome-fragment | NA | NA | 0 |
| S12_vs_k141_102393\|\|full | 6677 | No | NA | 4 | 2 | 1 | Low-quality | Genome-fragment | 7.84 | HMM-based (lower-bound) | 0 |
| S6_vs_k141_34831\|\|full | 1187 | No | NA | 3 | 0 | 1 | Not-determined | Genome-fragment | NA | NA | 0 |
| S6_vs_k141_25302\|\|full | 4153 | No | NA | 5 | 0 | 3 | Not-determined | Genome-fragment | NA | NA | 0 |
| S6_vs_k141_29795\|\|full | 2990 | No | NA | 4 | 0 | 1 | Not-determined | Genome-fragment | NA | NA | 0 |
| S6_vs_k141_1395\|\|full | 2000 | No | NA | 2 | 0 | 2 | Not-determined | Genome-fragment | NA | NA | 0 |
| S8_extracted_k141_141158 | 6681 | No | NA | 12 | 1 | 0 | Low-quality | Genome-fragment | 5.31 | AAI-based (high-confidence) | 0 |
| S6_vs_k141_8587\|\|full | 1642 | No | NA | 3 | 0 | 0 | Not-determined | Genome-fragment | NA | NA | 0 |
| S6_vs_k141_29819\|\|full | 2200 | No | NA | 2 | 0 | 2 | Not-determined | Genome-fragment | NA | NA | 0 |
| S7_extracted_129544 | 6682 | No | NA | 7 | 1 | 0 | Low-quality | Genome-fragment | 4.99 | AAI-based (medium-confidence) | 0 |
| S6_vs_k141_41546\|\|full | 1664 | No | NA | 3 | 0 | 1 | Not-determined | Genome-fragment | NA | NA | 0 |
| S14_extracted_k141_11733 | 6685 | No | NA | 5 | 0 | 4 | Low-quality | Genome-fragment | 2.98 | AAI-based (medium-confidence) | 0 |
| S8_extracted_k141_83720 | 6690 | No | NA | 10 | 7 | 0 | Low-quality | Genome-fragment | 10.41 | HMM-based (lower-bound) | 0 |
| S6_vs_k141_15729\|\|full | 1348 | No | NA | 2 | 0 | 1 | Not-determined | Genome-fragment | NA | NA | 0 |
| S5_extracted_k141_15259 | 6695 | No | NA | 11 | 1 | 0 | Low-quality | Genome-fragment | 12 | AAI-based (medium-confidence) | 0 |
| S6_vs_k141_8625\|\|full | 1804 | No | NA | 3 | 0 | 0 | Not-determined | Genome-fragment | NA | NA | 0 |
| S8_vs_k141_120944\|\|full | 6695 | No | NA | 7 | 6 | 0 | Low-quality | Genome-fragment | 11.61 | AAI-based (high-confidence) | 0 |
| S6_vs_k141_10944\|\|full | 1278 | No | NA | 3 | 0 | 1 | Not-determined | Genome-fragment | NA | NA | 0 |
| S6_vs_k141_4264\|\|full | 1915 | No | NA | 4 | 0 | 0 | Not-determined | Genome-fragment | NA | NA | 0 |
| S6_vs_k141_13550\|\|full | 1408 | No | NA | 3 | 0 | 1 | Not-determined | Genome-fragment | NA | NA | 0 |
| S9_extracted_k141_44651 | 6696 | No | NA | 12 | 3 | 0 | Low-quality | Genome-fragment | 3.99 | AAI-based (high-confidence) | 0 |
| S12_extracted_k141_172135 | 6699 | No | NA | 12 | 1 | 0 | Low-quality | Genome-fragment | 6.15 | AAI-based (high-confidence) | 0 |
| S6_vs_k141_4310\|\|full | 1355 | No | NA | 2 | 0 | 0 | Not-determined | Genome-fragment | NA | NA | 0 |
| S11_vs_k141_78020\|\|full | 6700 | No | NA | 2 | 0 | 0 | Low-quality | Genome-fragment | 4.11 | AAI-based (high-confidence) | 0 |
| S3_vs_k141_107719\|\|full | 6702 | No | NA | 9 | 2 | 0 | Low-quality | Genome-fragment | 14.18 | AAI-based (high-confidence) | 0 |
| S6_vs_k141_4319\|\|full | 3760 | No | NA | 4 | 0 | 1 | Not-determined | Genome-fragment | NA | NA | 0 |
| S6_vs_k141_22403\|\|full | 1585 | No | NA | 3 | 0 | 2 | Not-determined | Genome-fragment | NA | NA | 0 |
| S6_vs_k141_18206\|\|full | 1006 | No | NA | 2 | 0 | 1 | Not-determined | Genome-fragment | NA | NA | 0 |
| S6_vs_k141_4329\|\|full | 1487 | No | NA | 3 | 0 | 1 | Not-determined | Genome-fragment | NA | NA | 0 |
| S6_vs_k141_55096\|\|full | 1731 | No | NA | 3 | 0 | 1 | Not-determined | Genome-fragment | NA | NA | 0 |
| S6_vs_k141_8688\|\|full | 5695 | No | NA | 5 | 0 | 0 | Not-determined | Genome-fragment | NA | NA | 0 |
| S3_extracted_k141_112353 | 6704 | No | NA | 7 | 3 | 0 | Low-quality | Genome-fragment | 13.87 | AAI-based (high-confidence) | 0 |
| S3_extracted_k141_17637 | 6704 | No | NA | 9 | 4 | 0 | Low-quality | Genome-fragment | 16.72 | AAI-based (high-confidence) | 0 |
| S6_vs_k141_41636\|\|full | 38964 | No | NA | 27 | 0 | 20 | Not-determined | Genome-fragment | NA | NA | 0 |
| S3_extracted_k141_43654 | 6704 | No | NA | 7 | 5 | 0 | Low-quality | Genome-fragment | 30.58 | AAI-based (high-confidence) | 0 |
| S17_vs_k141_60795\|\|full | 6709 | No | NA | 12 | 1 | 0 | Low-quality | Genome-fragment | 15.93 | AAI-based (medium-confidence) | 0 |
| S6_vs_k141_36704\|\|full | 1884 | No | NA | 4 | 0 | 1 | Not-determined | Genome-fragment | NA | NA | 0 |
| S6_vs_k141_29934\|\|full | 2931 | No | NA | 4 | 0 | 4 | Not-determined | Genome-fragment | NA | NA | 0 |
| S16_extracted_107025 | 6711 | Yes | 4767 | 8 | 2 | 1 | Low-quality | Genome-fragment | 2.57 | AAI-based (high-confidence) | 28.97 |
| S8_extracted_k141_108334 | 6714 | No | NA | 9 | 5 | 0 | Low-quality | Genome-fragment | 10.31 | HMM-based (lower-bound) | 0 |
| S3_extracted_k141_47483 | 6716 | No | NA | 10 | 0 | 9 | Low-quality | Genome-fragment | 17.14 | AAI-based (medium-confidence) | 0 |
| S5_extracted_k141_52365 | 6717 | Yes | 5231 | 8 | 1 | 1 | Low-quality | Genome-fragment | 37.65 | AAI-based (high-confidence) | 22.12 |
| S7_extracted_132857 | 6718 | No | NA | 7 | 4 | 0 | Low-quality | Genome-fragment | 30.62 | AAI-based (high-confidence) | 0 |
| S6_vs_k141_15813\|\|full | 46227 | No | NA | 77 | 17 | 0 | High-quality | High-quality | 100 | AAI-based (high-confidence) | 0 |
| S11_extracted_k141_61147 | 6719 | No | NA | 14 | 1 | 0 | Low-quality | Genome-fragment | 4.36 | AAI-based (high-confidence) | 0 |
| S6_vs_k141_44288\|\|full | 6722 | No | NA | 9 | 0 | 0 | Low-quality | Genome-fragment | 10.77 | AAI-based (high-confidence) | 0 |
| S7_vs_48687\|\|full | 6723 | No | NA | 8 | 5 | 0 | Low-quality | Genome-fragment | 12.75 | AAI-based (medium-confidence) | 0 |
| S4_extracted_14515 | 6723 | No | NA | 9 | 1 | 3 | Low-quality | Genome-fragment | 16.99 | HMM-based (lower-bound) | 0 |
| S3_vs_k141_95102\|\|full | 6728 | No | NA | 8 | 6 | 0 | Low-quality | Genome-fragment | 19.56 | AAI-based (high-confidence) | 0 |
| S7_vs_91153\|\|full | 6728 | No | NA | 12 | 1 | 0 | Low-quality | Genome-fragment | 15.66 | AAI-based (medium-confidence) | 0 |
| S6_vs_k141_50271\|\|full | 6731 | No | NA | 5 | 0 | 3 | Low-quality | Genome-fragment | 3.62 | AAI-based (medium-confidence) | 0 |
| S1_extracted_k141_41381 | 6732 | No | NA | 3 | 0 | 3 | Low-quality | Genome-fragment | 2.53 | AAI-based (medium-confidence) | 0 |
| S5_vs_k141_50361\|\|full | 6732 | No | NA | 17 | 2 | 0 | Low-quality | Genome-fragment | 14.57 | AAI-based (medium-confidence) | 0 |
| S8_extracted_k141_139332 | 6734 | No | NA | 5 | 1 | 0 | Low-quality | Genome-fragment | 4.39 | AAI-based (high-confidence) | 0 |
| S6_vs_k141_53546\|\|full | 3036 | No | NA | 4 | 0 | 3 | Not-determined | Genome-fragment | NA | NA | 0 |
| S6_vs_k141_11031\|\|full | 1315 | No | NA | 3 | 0 | 1 | Not-determined | Genome-fragment | NA | NA | 0 |
| S17_vs_k141_1383\|\|full | 6735 | No | NA | 12 | 5 | 0 | Low-quality | Genome-fragment | 9.5 | AAI-based (high-confidence) | 0 |
| S6_vs_k141_55233\|\|full | 2558 | No | NA | 3 | 0 | 2 | Not-determined | Genome-fragment | NA | NA | 0 |
| S12_vs_k141_70145\|\|full | 6736 | No | NA | 5 | 0 | 4 | Low-quality | Genome-fragment | 11.69 | AAI-based (high-confidence) | 0 |
| S6_vs_k141_53564\|\|full | 1558 | No | NA | 3 | 0 | 1 | Not-determined | Genome-fragment | NA | NA | 0 |
| S3_vs_k141_32960\|\|full | 6740 | No | NA | 10 | 1 | 0 | Low-quality | Genome-fragment | 4.15 | AAI-based (medium-confidence) | 0 |
| S6_vs_k141_30022\|\|full | 1250 | No | NA | 2 | 0 | 0 | Not-determined | Genome-fragment | NA | NA | 0 |
| S5_vs_k141_82682\|\|full | 6740 | No | NA | 11 | 3 | 0 | Low-quality | Genome-fragment | 14.98 | AAI-based (high-confidence) | 0 |
| S14_vs_k141_47266\|\|full | 6744 | No | NA | 9 | 4 | 0 | Low-quality | Genome-fragment | 17.7 | AAI-based (high-confidence) | 0 |
| S6_vs_k141_32162\|\|full | 2648 | No | NA | 3 | 0 | 3 | Not-determined | Genome-fragment | NA | NA | 0 |
| S11_vs_k141_10313\|\|full | 6745 | No | NA | 15 | 1 | 0 | Low-quality | Genome-fragment | 13.35 | HMM-based (lower-bound) | 0 |
| S16_vs_8173\|\|full | 6746 | No | NA | 12 | 2 | 0 | Low-quality | Genome-fragment | 13.8 | AAI-based (high-confidence) | 0 |
| S14_vs_k141_67908\|\|full | 6747 | No | NA | 6 | 1 | 2 | Low-quality | Genome-fragment | 7.8 | HMM-based (lower-bound) | 0 |
| S6_vs_k141_34951\|\|full | 1587 | No | NA | 3 | 0 | 1 | Not-determined | Genome-fragment | NA | NA | 0 |
| S6_vs_k141_30077\|\|full | 1968 | No | NA | 3 | 0 | 0 | Not-determined | Genome-fragment | NA | NA | 0 |
| S17_extracted_k141_82838 | 6750 | No | NA | 4 | 0 | 0 | Low-quality | Genome-fragment | 3.91 | AAI-based (high-confidence) | 0 |
| S5_vs_k141_29562\|\|full | 6751 | No | NA | 13 | 3 | 0 | Low-quality | Genome-fragment | 16.1 | AAI-based (high-confidence) | 0 |
| S6_vs_k141_15919\|\|full | 1545 | No | NA | 3 | 0 | 1 | Not-determined | Genome-fragment | NA | NA | 0 |
| S7_extracted_81843 | 6752 | No | NA | 10 | 1 | 0 | Low-quality | Genome-fragment | 3.9 | AAI-based (medium-confidence) | 0 |
| S15_extracted_42026 | 6752 | No | NA | 14 | 2 | 0 | Low-quality | Genome-fragment | 14.92 | AAI-based (high-confidence) | 0 |
| S7_extracted_785 | 6753 | No | NA | 6 | 1 | 0 | Low-quality | Genome-fragment | 16.65 | AAI-based (medium-confidence) | 0 |
| S12_extracted_k141_97913 | 6754 | No | NA | 6 | 0 | 0 | Low-quality | Genome-fragment | 5.87 | AAI-based (high-confidence) | 0 |
| S6_vs_k141_46556\|\|full | 1725 | No | NA | 3 | 0 | 1 | Not-determined | Genome-fragment | NA | NA | 0 |
| S6_vs_k141_53605\|\|full | 2459 | No | NA | 3 | 0 | 3 | Not-determined | Genome-fragment | NA | NA | 0 |
| S8_vs_k141_129561\|\|full | 6757 | No | NA | 8 | 2 | 0 | Low-quality | Genome-fragment | 16.01 | AAI-based (high-confidence) | 0 |
| S6_vs_k141_1546\|\|full | 2787 | No | NA | 3 | 0 | 0 | Not-determined | Genome-fragment | NA | NA | 0 |
| S6_vs_k141_55372\|\|full | 1479 | No | NA | 3 | 0 | 1 | Not-determined | Genome-fragment | NA | NA | 0 |
| S4_vs_VIRSorter_31146-cat_1 | 6759 | No | NA | 11 | 9 | 0 | Low-quality | Genome-fragment | 19.32 | AAI-based (high-confidence) | 0 |
| S9_vs_k141_69040\|\|full | 6761 | No | NA | 9 | 3 | 0 | Low-quality | Genome-fragment | 18.91 | AAI-based (high-confidence) | 0 |
| S5_vs_k141_92164\|\|full | 6761 | No | NA | 7 | 0 | 1 | Low-quality | Genome-fragment | 9.05 | AAI-based (medium-confidence) | 0 |
| S9_extracted_k141_68579 | 6762 | No | NA | 7 | 1 | 0 | Low-quality | Genome-fragment | 17.47 | AAI-based (high-confidence) | 0 |
| S12_vs_k141_9557\|\|full | 6763 | No | NA | 11 | 1 | 2 | Low-quality | Genome-fragment | 4.71 | HMM-based (lower-bound) | 0 |
| S16_extracted_33333 | 6764 | No | NA | 10 | 0 | 3 | Low-quality | Genome-fragment | 9.13 | AAI-based (high-confidence) | 0 |
| S7_vs_33055\|\|full | 6767 | No | NA | 8 | 3 | 0 | Low-quality | Genome-fragment | 15.72 | AAI-based (high-confidence) | 0 |
| S6_vs_k141_18422\|\|full | 3024 | No | NA | 3 | 0 | 2 | Not-determined | Genome-fragment | NA | NA | 0 |
| S6_vs_k141_22637\|\|full | 29837 | No | NA | 41 | 15 | 0 | Medium-quality | Genome-fragment | 72.51 | AAI-based (medium-confidence) | 0 |
| S3_vs_k141_256\|\|full | 6779 | No | NA | 7 | 2 | 0 | Low-quality | Genome-fragment | 17.15 | AAI-based (medium-confidence) | 0 |
| S13_vs_111001\|\|full | 6784 | No | NA | 9 | 4 | 0 | Low-quality | Genome-fragment | 19.78 | HMM-based (lower-bound) | 0 |
| S16_vs_139762\|\|full | 6786 | No | NA | 3 | 1 | 0 | Low-quality | Genome-fragment | 16.62 | AAI-based (medium-confidence) | 0 |
| S6_vs_k141_53693\|\|full | 1528 | No | NA | 3 | 0 | 1 | Not-determined | Genome-fragment | NA | NA | 0 |
| S8_vs_k141_174412\|\|full | 6789 | No | NA | 16 | 5 | 0 | Low-quality | Genome-fragment | 14.42 | AAI-based (medium-confidence) | 0 |
| S12_vs_k141_109781\|\|full | 6790 | No | NA | 7 | 0 | 6 | Low-quality | Genome-fragment | 6.92 | AAI-based (medium-confidence) | 0 |
| S6_vs_k141_25824\|\|full | 6790 | No | NA | 7 | 4 | 0 | Low-quality | Genome-fragment | 14.93 | AAI-based (high-confidence) | 0 |
| S13_vs_25099\|\|full | 6792 | No | NA | 6 | 0 | 3 | Low-quality | Genome-fragment | 6.4 | AAI-based (medium-confidence) | 0 |
| S8_vs_k141_109346\|\|full | 6793 | No | NA | 9 | 3 | 0 | Low-quality | Genome-fragment | 18.38 | AAI-based (high-confidence) | 0 |
| S6_vs_k141_19935\|\|full | 2844 | No | NA | 3 | 0 | 3 | Not-determined | Genome-fragment | NA | NA | 0 |
| S6_vs_k141_50923\|\|full | 1752 | No | NA | 2 | 0 | 2 | Not-determined | Genome-fragment | NA | NA | 0 |
| S6_vs_k141_18481\|\|full | 3485 | No | NA | 4 | 0 | 2 | Not-determined | Genome-fragment | NA | NA | 0 |
| S6_vs_k141_6483\|\|full | 3282 | No | NA | 4 | 0 | 2 | Not-determined | Genome-fragment | NA | NA | 0 |
| S8_extracted_k141_90868 | 6796 | Yes | 4853 | 10 | 1 | 2 | Low-quality | Genome-fragment | 7.56 | HMM-based (lower-bound) | 28.59 |
| S8_vs_k141_26362\|\|full | 6796 | No | NA | 6 | 0 | 4 | Low-quality | Genome-fragment | 4.83 | AAI-based (medium-confidence) | 0 |
| S6_vs_k141_18496\|\|full | 1709 | No | NA | 3 | 0 | 0 | Not-determined | Genome-fragment | NA | NA | 0 |
| S17_extracted_k141_1714 | 6798 | Yes | 5240 | 8 | 1 | 1 | Low-quality | Genome-fragment | 8.29 | AAI-based (high-confidence) | 22.92 |
| S16_vs_125298\|\|full | 6798 | No | NA | 5 | 1 | 0 | Low-quality | Genome-fragment | 10.83 | HMM-based (lower-bound) | 0 |
| S5_extracted_k141_138355 | 6805 | No | NA | 8 | 0 | 5 | Low-quality | Genome-fragment | 3.11 | AAI-based (medium-confidence) | 0 |
| S17_extracted_k141_172674 | 6810 | No | NA | 4 | 0 | 2 | Low-quality | Genome-fragment | 4.37 | AAI-based (high-confidence) | 0 |
| S5_vs_k141_106501\|\|full | 6814 | No | NA | 13 | 6 | 0 | Low-quality | Genome-fragment | 18.37 | AAI-based (high-confidence) | 0 |
| S7_vs_131946\|\|full | 6819 | No | NA | 11 | 0 | 0 | Low-quality | Genome-fragment | 15.42 | AAI-based (medium-confidence) | 0 |
| S8_extracted_k141_86192 | 6822 | No | NA | 5 | 0 | 0 | Low-quality | Genome-fragment | 9.34 | AAI-based (high-confidence) | 0 |
| S11_extracted_k141_42862 | 6824 | No | NA | 6 | 4 | 0 | Low-quality | Genome-fragment | 4.35 | AAI-based (medium-confidence) | 0 |
| S6_vs_k141_18547\|\|full | 3047 | No | NA | 3 | 0 | 0 | Not-determined | Genome-fragment | NA | NA | 0 |
| S6_vs_k141_39237\|\|full | 1725 | No | NA | 3 | 0 | 0 | Not-determined | Genome-fragment | NA | NA | 0 |
| S6_vs_k141_46664\|\|full | 1645 | No | NA | 3 | 0 | 1 | Not-determined | Genome-fragment | NA | NA | 0 |
| S6_vs_k141_48444\|\|full | 1043 | No | NA | 3 | 0 | 1 | Not-determined | Genome-fragment | NA | NA | 0 |
| S8_vs_k141_81454\|\|full | 6824 | No | NA | 7 | 0 | 0 | Low-quality | Genome-fragment | 14.94 | AAI-based (medium-confidence) | 0 |
| S7_vs_40202\|\|full | 6828 | No | NA | 13 | 2 | 0 | Low-quality | Genome-fragment | 16.7 | AAI-based (medium-confidence) | 0 |
| S17_extracted_k141_45462 | 6829 | No | NA | 8 | 1 | 0 | Low-quality | Genome-fragment | 8.34 | AAI-based (medium-confidence) | 0 |
| S14_extracted_k141_88586 | 6831 | No | NA | 20 | 4 | 0 | Low-quality | Genome-fragment | 16.5 | AAI-based (high-confidence) | 0 |
| S12_vs_k141_55445\|\|full | 6832 | No | NA | 4 | 0 | 0 | Low-quality | Genome-fragment | 12.27 | AAI-based (high-confidence) | 0 |
| S6_vs_k141_35184\|\|full | 3420 | No | NA | 5 | 0 | 2 | Not-determined | Genome-fragment | NA | NA | 0 |
| S6_vs_k141_42073\|\|full | 3368 | No | NA | 3 | 0 | 2 | Not-determined | Genome-fragment | NA | NA | 0 |
| S3_extracted_k141_72942 | 6833 | No | NA | 18 | 4 | 0 | Low-quality | Genome-fragment | 3.86 | AAI-based (high-confidence) | 0 |
| S13_extracted_153734 | 6846 | No | NA | 4 | 1 | 0 | Low-quality | Genome-fragment | 7.26 | AAI-based (medium-confidence) | 0 |
| S3_extracted_k141_88388 | 6847 | No | NA | 11 | 2 | 1 | Low-quality | Genome-fragment | 4.22 | HMM-based (lower-bound) | 0 |
| S16_vs_134504\|\|full | 6847 | Yes | 5786 | 5 | 1 | 1 | Low-quality | Genome-fragment | 9.1 | AAI-based (high-confidence) | 15.5 |
| S15_extracted_9296 | 6848 | No | NA | 6 | 2 | 0 | Low-quality | Genome-fragment | 11.22 | AAI-based (high-confidence) | 0 |
| S6_vs_k141_46739\|\|full | 2599 | No | NA | 6 | 0 | 0 | Not-determined | Genome-fragment | NA | NA | 0 |
| S6_vs_k141_32519\|\|full | 2086 | No | NA | 7 | 0 | 0 | Not-determined | Genome-fragment | NA | NA | 0 |
| S15_vs_66020\|\|full | 6849 | Yes | 6612 | 9 | 1 | 1 | Low-quality | Genome-fragment | 5.35 | AAI-based (medium-confidence) | 3.46 |
| S6_vs_k141_53789\|\|full | 1187 | No | NA | 3 | 0 | 0 | Not-determined | Genome-fragment | NA | NA | 0 |
| S6_vs_k141_4670\|\|full | 2047 | No | NA | 4 | 0 | 0 | Not-determined | Genome-fragment | NA | NA | 0 |
| S6_vs_k141_42198\|\|full | 1653 | No | NA | 2 | 0 | 2 | Not-determined | Genome-fragment | NA | NA | 0 |
| S6_vs_k141_43900\|\|full | 2789 | No | NA | 3 | 0 | 1 | Not-determined | Genome-fragment | NA | NA | 0 |
| S13_vs_83238\|\|full | 6850 | No | NA | 9 | 4 | 0 | Low-quality | Genome-fragment | 14.63 | AAI-based (high-confidence) | 0 |
| S3_extracted_k141_20202 | 6852 | No | NA | 13 | 4 | 0 | Low-quality | Genome-fragment | 9.36 | AAI-based (high-confidence) | 0 |
| S6_vs_k141_4681\|\|full | 43497 | No | NA | 41 | 0 | 14 | Not-determined | Genome-fragment | NA | NA | 0 |
| S6_vs_k141_13587\|\|full | 6853 | No | NA | 8 | 0 | 3 | Low-quality | Genome-fragment | 5.23 | AAI-based (medium-confidence) | 0 |
| S6_vs_k141_25201\|\|full | 6858 | No | NA | 6 | 0 | 1 | Low-quality | Genome-fragment | 6.2 | AAI-based (medium-confidence) | 0 |
| S6_vs_k141_30301\|\|full | 1676 | No | NA | 3 | 0 | 0 | Not-determined | Genome-fragment | NA | NA | 0 |
| S6_vs_k141_53852\|\|full | 2520 | No | NA | 3 | 0 | 3 | Not-determined | Genome-fragment | NA | NA | 0 |
| S16_vs_169426\|\|full | 6861 | No | NA | 9 | 1 | 0 | Low-quality | Genome-fragment | 4.27 | HMM-based (lower-bound) | 0 |
| S3_extracted_k141_72522 | 6864 | No | NA | 2 | 2 | 0 | Low-quality | Genome-fragment | 7.08 | AAI-based (high-confidence) | 0 |
| S6_vs_k141_50126\|\|full | 6864 | No | NA | 16 | 4 | 0 | Low-quality | Genome-fragment | 12.02 | AAI-based (medium-confidence) | 0 |
| S16_vs_38804\|\|full | 6865 | No | NA | 13 | 6 | 0 | Low-quality | Genome-fragment | 20.5 | AAI-based (high-confidence) | 0 |
| S10_vs_13792\|\|full | 6872 | No | NA | 11 | 5 | 0 | Low-quality | Genome-fragment | 13.13 | AAI-based (medium-confidence) | 0 |
| S8_vs_k141_166104\|\|full | 6872 | No | NA | 11 | 2 | 0 | Low-quality | Genome-fragment | 15.79 | AAI-based (high-confidence) | 0 |
| S6_vs_k141_22992\|\|full | 1516 | No | NA | 4 | 0 | 1 | Not-determined | Genome-fragment | NA | NA | 0 |
| S1_extracted_k141_93977 | 6875 | No | NA | 8 | 7 | 0 | Low-quality | Genome-fragment | 11.17 | AAI-based (high-confidence) | 0 |
| S6_vs_k141_11300\|\|full | 1274 | No | NA | 2 | 0 | 1 | Not-determined | Genome-fragment | NA | NA | 0 |
| S17_vs_k141_119727\|\|full | 6876 | No | NA | 14 | 2 | 0 | Low-quality | Genome-fragment | 12.58 | AAI-based (high-confidence) | 0 |
| S1_vs_k141_29023\|\|full | 6876 | No | NA | 8 | 6 | 0 | Low-quality | Genome-fragment | 16.07 | AAI-based (high-confidence) | 0 |
| S6_vs_k141_16196\|\|full | 1171 | No | NA | 2 | 0 | 1 | Not-determined | Genome-fragment | NA | NA | 0 |
| S6_vs_k141_32710\|\|full | 1226 | No | NA | 2 | 0 | 0 | Not-determined | Genome-fragment | NA | NA | 0 |
| S6_vs_k141_1784\|\|full | 2723 | No | NA | 3 | 0 | 1 | Not-determined | Genome-fragment | NA | NA | 0 |
| S6_vs_k141_48617\|\|full | 3445 | No | NA | 4 | 0 | 2 | Not-determined | Genome-fragment | NA | NA | 0 |
| S8_extracted_k141_28813 | 6882 | No | NA | 8 | 4 | 0 | Low-quality | Genome-fragment | 11.5 | AAI-based (medium-confidence) | 0 |
| S11_extracted_k141_95929 | 6883 | No | NA | 13 | 1 | 1 | Low-quality | Genome-fragment | 12.15 | AAI-based (high-confidence) | 0 |
| S6_vs_k141_44032\|\|full | 3548 | No | NA | 6 | 0 | 3 | Not-determined | Genome-fragment | NA | NA | 0 |
| S16_extracted_45537 | 6883 | No | NA | 3 | 0 | 0 | Low-quality | Genome-fragment | 10.65 | AAI-based (high-confidence) | 0 |
| S6_vs_k141_55535\|\|full | 4451 | No | NA | 4 | 0 | 3 | Not-determined | Genome-fragment | NA | NA | 0 |
| S6_vs_k141_27548\|\|full | 1300 | No | NA | 3 | 0 | 0 | Not-determined | Genome-fragment | NA | NA | 0 |
| S8_extracted_k141_131854 | 6896 | No | NA | 9 | 4 | 0 | Low-quality | Genome-fragment | 13.83 | AAI-based (medium-confidence) | 0 |
| S8_vs_k141_161774\|\|full | 6900 | No | NA | 8 | 6 | 0 | Low-quality | Genome-fragment | 11.51 | AAI-based (high-confidence) | 0 |
| S7_vs_89489\|\|full | 6901 | No | NA | 13 | 2 | 0 | Low-quality | Genome-fragment | 9.64 | HMM-based (lower-bound) | 0 |
| S9_vs_k141_31931\|\|full | 6902 | No | NA | 10 | 0 | 2 | Low-quality | Genome-fragment | 12.75 | AAI-based (medium-confidence) | 0 |
| S6_vs_k141_42412\|\|full | 6682 | No | NA | 9 | 0 | 7 | Not-determined | Genome-fragment | NA | NA | 0 |
| S6_vs_k141_44070\|\|full | 18695 | No | NA | 23 | 11 | 0 | Medium-quality | Genome-fragment | 51.55 | AAI-based (high-confidence) | 0 |
| S6_vs_k141_48702\|\|full | 2809 | No | NA | 2 | 0 | 1 | Not-determined | Genome-fragment | NA | NA | 0 |
| S6_vs_k141_36886\|\|full | 2463 | No | NA | 7 | 0 | 1 | Not-determined | Genome-fragment | NA | NA | 0 |
| S5_vs_k141_59871\|\|full | 6907 | No | NA | 8 | 0 | 5 | Low-quality | Genome-fragment | 5.6 | AAI-based (medium-confidence) | 0 |
| S16_extracted_39867 | 6909 | No | NA | 10 | 5 | 0 | Low-quality | Genome-fragment | 16.28 | AAI-based (medium-confidence) | 0 |
| S6_vs_k141_53966\|\|full | 28962 | Yes | 24133 | 44 | 14 | 2 | Medium-quality | Genome-fragment | 68.8 | AAI-based (high-confidence) | 16.67 |
| S6_vs_k141_32856\|\|full | 1492 | No | NA | 3 | 0 | 0 | Not-determined | Genome-fragment | NA | NA | 0 |
| S6_vs_k141_30418\|\|full | 2638 | No | NA | 4 | 0 | 1 | Not-determined | Genome-fragment | NA | NA | 0 |
| S6_vs_k141_44161\|\|full | 1165 | No | NA | 3 | 0 | 1 | Not-determined | Genome-fragment | NA | NA | 0 |
| S6_vs_k141_8926\|\|full | 3578 | No | NA | 3 | 0 | 0 | Not-determined | Genome-fragment | NA | NA | 0 |
| S7_vs_78677\|\|full | 6910 | Yes | 4947 | 8 | 1 | 1 | Low-quality | Genome-fragment | 11.34 | AAI-based (medium-confidence) | 28.41 |
| S16_vs_54822\|\|full | 6918 | No | NA | 4 | 0 | 0 | Low-quality | Genome-fragment | 6.28 | AAI-based (high-confidence) | 0 |
| S6_vs_k141_49756\|\|full | 6921 | No | NA | 9 | 5 | 0 | Low-quality | Genome-fragment | 13.45 | AAI-based (high-confidence) | 0 |
| S8_extracted_k141_63908 | 6925 | No | NA | 3 | 1 | 0 | Low-quality | Genome-fragment | 11.62 | HMM-based (lower-bound) | 0 |
| S5_extracted_k141_47398 | 6931 | No | NA | 8 | 2 | 0 | Low-quality | Genome-fragment | 10.86 | AAI-based (high-confidence) | 0 |
| S14_extracted_k141_60174 | 6932 | No | NA | 8 | 6 | 0 | Low-quality | Genome-fragment | 15.58 | AAI-based (high-confidence) | 0 |
| S6_vs_k141_23211\|\|full | 2238 | No | NA | 2 | 0 | 0 | Not-determined | Genome-fragment | NA | NA | 0 |
| S6_vs_k141_35318\|\|full | 1103 | No | NA | 1 | 0 | 1 | Not-determined | Genome-fragment | NA | NA | 0 |
| S6_vs_k141_27633\|\|full | 1843 | No | NA | 2 | 0 | 1 | Not-determined | Genome-fragment | NA | NA | 0 |
| S6_vs_k141_20357\|\|full | 4350 | No | NA | 4 | 0 | 3 | Not-determined | Genome-fragment | NA | NA | 0 |
| S6_vs_k141_48762\|\|full | 3519 | No | NA | 4 | 0 | 1 | Not-determined | Genome-fragment | NA | NA | 0 |
| S5_extracted_k141_93625 | 6933 | No | NA | 13 | 1 | 1 | Low-quality | Genome-fragment | 3.76 | HMM-based (lower-bound) | 0 |
| S7_extracted_47191 | 6934 | No | NA | 15 | 2 | 0 | Low-quality | Genome-fragment | 10.91 | AAI-based (high-confidence) | 0 |
| S2_extracted_k141_2125 | 6938 | No | NA | 12 | 2 | 0 | Low-quality | Genome-fragment | 20.52 | HMM-based (lower-bound) | 0 |
| S7_extracted_67566 | 6939 | No | NA | 13 | 4 | 0 | Low-quality | Genome-fragment | 18.26 | AAI-based (high-confidence) | 0 |
| S6_vs_k141_35337\|\|full | 1828 | No | NA | 2 | 0 | 2 | Not-determined | Genome-fragment | NA | NA | 0 |
| S16_extracted_111702 | 6939 | No | NA | 12 | 3 | 0 | Low-quality | Genome-fragment | 6.9 | HMM-based (lower-bound) | 0 |
| S3_extracted_k141_98315 | 6943 | No | NA | 12 | 2 | 0 | Low-quality | Genome-fragment | 15.37 | HMM-based (lower-bound) | 0 |
| S10_vs_92185\|\|full | 6947 | No | NA | 11 | 3 | 0 | Low-quality | Genome-fragment | 12.56 | AAI-based (high-confidence) | 0 |
| S12_vs_k141_22875\|\|full | 6953 | No | NA | 13 | 5 | 0 | Low-quality | Genome-fragment | 15.45 | AAI-based (high-confidence) | 0 |
| S6_vs_k141_23315\|\|full | 4005 | No | NA | 6 | 0 | 4 | Not-determined | Genome-fragment | NA | NA | 0 |
| S6_vs_k141_27679\|\|full | 1195 | No | NA | 3 | 0 | 0 | Not-determined | Genome-fragment | NA | NA | 0 |
| S6_vs_k141_23319\|\|full | 1744 | No | NA | 2 | 0 | 2 | Not-determined | Genome-fragment | NA | NA | 0 |
| S6_vs_k141_11539\|\|full | 1463 | No | NA | 3 | 0 | 1 | Not-determined | Genome-fragment | NA | NA | 0 |
| S6_vs_k141_25727\|\|full | 2397 | No | NA | 3 | 0 | 2 | Not-determined | Genome-fragment | NA | NA | 0 |
| S17_vs_k141_94000\|\|full | 6954 | No | NA | 13 | 0 | 0 | Low-quality | Genome-fragment | 17.24 | AAI-based (medium-confidence) | 0 |
| S4_vs_VIRSorter_23346-cat_1 | 6958 | No | NA | 12 | 6 | 0 | Low-quality | Genome-fragment | 17.29 | AAI-based (high-confidence) | 0 |
| S6_vs_k141_11588\|\|full | 1108 | No | NA | 2 | 0 | 0 | Not-determined | Genome-fragment | NA | NA | 0 |
| S3_vs_k141_94290\|\|full | 6966 | No | NA | 5 | 3 | 0 | Low-quality | Genome-fragment | 4.93 | AAI-based (high-confidence) | 0 |
| S4_extracted_17822 | 6968 | No | NA | 17 | 16 | 0 | Low-quality | Genome-fragment | 22.92 | AAI-based (high-confidence) | 0 |
| S6_vs_k141_11636\|\|full | 1740 | No | NA | 3 | 0 | 0 | Not-determined | Genome-fragment | NA | NA | 0 |
| S7_vs_136273\|\|full | 6969 | No | NA | 7 | 0 | 0 | Low-quality | Genome-fragment | 11.93 | AAI-based (high-confidence) | 0 |
| S6_vs_k141_25781\|\|full | 1107 | No | NA | 2 | 0 | 1 | Not-determined | Genome-fragment | NA | NA | 0 |
| S16_extracted_66609 | 6970 | No | NA | 7 | 0 | 4 | Low-quality | Genome-fragment | 1.03 | AAI-based (medium-confidence) | 0 |
| S7_vs_80530\|\|full | 6971 | No | NA | 7 | 3 | 0 | Low-quality | Genome-fragment | 16.79 | AAI-based (high-confidence) | 0 |
| S6_vs_k141_23448\|\|full | 6095 | No | NA | 6 | 0 | 1 | Not-determined | Genome-fragment | NA | NA | 0 |
| S6_vs_k141_23454\|\|full | 4530 | No | NA | 4 | 0 | 3 | Not-determined | Genome-fragment | NA | NA | 0 |
| S6_vs_k141_39749\|\|full | 1176 | No | NA | 3 | 0 | 0 | Not-determined | Genome-fragment | NA | NA | 0 |
| S12_vs_k141_30761\|\|full | 6971 | No | NA | 8 | 0 | 6 | Low-quality | Genome-fragment | 3.03 | AAI-based (high-confidence) | 0 |
| S14_extracted_k141_46727 | 6975 | No | NA | 17 | 8 | 1 | Low-quality | Genome-fragment | 20.12 | AAI-based (high-confidence) | 0 |
| S14_vs_k141_36999\|\|full | 6975 | No | NA | 19 | 2 | 0 | Low-quality | Genome-fragment | 19.92 | AAI-based (high-confidence) | 0 |
| S3_extracted_k141_107246 | 6976 | No | NA | 9 | 8 | 0 | Low-quality | Genome-fragment | 16.42 | AAI-based (high-confidence) | 0 |
| S6_vs_k141_1832\|\|full | 12039 | No | NA | 12 | 0 | 2 | Not-determined | Genome-fragment | NA | NA | 0 |
| S1_extracted_k141_59422 | 6977 | No | NA | 9 | 0 | 1 | Low-quality | Genome-fragment | 47.74 | AAI-based (high-confidence) | 0 |
| S17_vs_k141_60173\|\|full | 6978 | No | NA | 13 | 3 | 0 | Low-quality | Genome-fragment | 12.78 | AAI-based (high-confidence) | 0 |
| S12_extracted_k141_7278 | 6979 | No | NA | 11 | 1 | 0 | Low-quality | Genome-fragment | 11.91 | AAI-based (high-confidence) | 0 |
| S14_extracted_k141_12634 | 6982 | No | NA | 14 | 1 | 1 | Low-quality | Genome-fragment | 3.79 | HMM-based (lower-bound) | 0 |
| S6_vs_k141_8970\|\|full | 1561 | No | NA | 3 | 0 | 2 | Not-determined | Genome-fragment | NA | NA | 0 |
| S6_vs_k141_23540\|\|full | 2510 | No | NA | 3 | 0 | 2 | Not-determined | Genome-fragment | NA | NA | 0 |
| S5_extracted_k141_143884 | 6983 | No | NA | 13 | 3 | 0 | Low-quality | Genome-fragment | 16.23 | AAI-based (high-confidence) | 0 |
| S4_extracted_9974 | 6983 | No | NA | 15 | 11 | 0 | Low-quality | Genome-fragment | 22.85 | AAI-based (high-confidence) | 0 |
| S14_vs_k141_31040\|\|full | 6995 | No | NA | 9 | 0 | 3 | Low-quality | Genome-fragment | 5.71 | AAI-based (high-confidence) | 0 |
| S6_vs_k141_49065\|\|full | 1049 | No | NA | 2 | 0 | 1 | Not-determined | Genome-fragment | NA | NA | 0 |
| S9_extracted_k141_72175 | 6996 | No | NA | 6 | 0 | 0 | Low-quality | Genome-fragment | 4.03 | AAI-based (high-confidence) | 0 |
| S16_vs_82179\|\|full | 7001 | No | NA | 13 | 4 | 0 | Low-quality | Genome-fragment | 12.07 | AAI-based (high-confidence) | 0 |
| S13_vs_42284\|\|full | 7004 | No | NA | 6 | 2 | 0 | Low-quality | Genome-fragment | 3.14 | HMM-based (lower-bound) | 0 |
| S5_vs_k141_137782\|\|full | 7004 | Yes | 4793 | 9 | 1 | 1 | Low-quality | Genome-fragment | 5.33 | HMM-based (lower-bound) | 31.57 |
| S14_vs_k141_108427\|\|full | 7011 | No | NA | 14 | 1 | 0 | Low-quality | Genome-fragment | 14.23 | AAI-based (high-confidence) | 0 |
| S5_vs_k141_36680\|\|full | 7012 | No | NA | 9 | 4 | 0 | Low-quality | Genome-fragment | 17.52 | AAI-based (medium-confidence) | 0 |
| S7_vs_28305\|\|full | 7012 | No | NA | 10 | 0 | 0 | Low-quality | Genome-fragment | 4.6 | AAI-based (high-confidence) | 0 |
| S4_extracted_2064 | 7012 | No | NA | 20 | 18 | 0 | Low-quality | Genome-fragment | 23.13 | AAI-based (high-confidence) | 0 |
| S11_extracted_k141_6303 | 7016 | No | NA | 8 | 7 | 0 | Low-quality | Genome-fragment | 18.62 | AAI-based (high-confidence) | 0 |
| S6_vs_k141_51106\|\|full | 1706 | No | NA | 3 | 0 | 3 | Not-determined | Genome-fragment | NA | NA | 0 |
| S17_extracted_k141_122173 | 7019 | No | NA | 7 | 0 | 4 | Low-quality | Genome-fragment | 7.74 | AAI-based (medium-confidence) | 0 |
| S6_vs_k141_55911\|\|full | 1790 | No | NA | 3 | 0 | 2 | Not-determined | Genome-fragment | NA | NA | 0 |
| S14_extracted_k141_51296 | 7022 | No | NA | 8 | 2 | 1 | Low-quality | Genome-fragment | 11.37 | AAI-based (medium-confidence) | 0 |
| S10_extracted_71229 | 7026 | No | NA | 11 | 5 | 0 | Low-quality | Genome-fragment | 11.43 | AAI-based (high-confidence) | 0 |
| S4_extracted_30826 | 7028 | Yes | 4824 | 10 | 1 | 1 | Low-quality | Genome-fragment | 11.95 | AAI-based (high-confidence) | 31.36 |
| S6_vs_k141_2139\|\|full | 2658 | No | NA | 4 | 0 | 2 | Not-determined | Genome-fragment | NA | NA | 0 |
| S6_vs_k141_9300\|\|full | 2006 | No | NA | 3 | 0 | 2 | Not-determined | Genome-fragment | NA | NA | 0 |
| S7_extracted_21498 | 7030 | No | NA | 8 | 1 | 0 | Low-quality | Genome-fragment | 11.06 | AAI-based (high-confidence) | 0 |
| S6_vs_k141_9349\|\|full | 2849 | No | NA | 2 | 0 | 1 | Not-determined | Genome-fragment | NA | NA | 0 |
| S6_vs_k141_47055\|\|full | 1372 | No | NA | 2 | 0 | 1 | Not-determined | Genome-fragment | NA | NA | 0 |
| S6_vs_k141_20622\|\|full | 4003 | No | NA | 4 | 0 | 1 | Not-determined | Genome-fragment | NA | NA | 0 |
| S7_extracted_108653 | 7031 | No | NA | 3 | 0 | 3 | Low-quality | Genome-fragment | 2.98 | AAI-based (medium-confidence) | 0 |
| S4_extracted_8544 | 7032 | No | NA | 9 | 9 | 0 | Low-quality | Genome-fragment | 23.21 | AAI-based (high-confidence) | 0 |
| S5_vs_k141_6958\|\|full | 7034 | No | NA | 17 | 2 | 0 | Low-quality | Genome-fragment | 20.38 | AAI-based (high-confidence) | 0 |
| S6_vs_k141_44648\|\|full | 1043 | No | NA | 2 | 0 | 1 | Not-determined | Genome-fragment | NA | NA | 0 |
| S6_vs_k141_37485\|\|full | 2921 | No | NA | 2 | 0 | 1 | Not-determined | Genome-fragment | NA | NA | 0 |
| S6_vs_k141_49444\|\|full | 31632 | No | NA | 30 | 0 | 18 | Not-determined | Genome-fragment | NA | NA | 0 |
| S13_extracted_131399 | 7038 | No | NA | 5 | 0 | 4 | Low-quality | Genome-fragment | 2.54 | AAI-based (medium-confidence) | 0 |
| S6_vs_k141_49458\|\|full | 4212 | No | NA | 7 | 0 | 1 | Not-determined | Genome-fragment | NA | NA | 0 |
| S6_vs_k141_51393\|\|full | 4172 | No | NA | 5 | 0 | 2 | Not-determined | Genome-fragment | NA | NA | 0 |
| S1_vs_k141_95079\|\|full | 7038 | No | NA | 11 | 2 | 0 | Low-quality | Genome-fragment | 14.65 | AAI-based (high-confidence) | 0 |
| S6_vs_k141_51492\|\|full | 1758 | No | NA | 2 | 0 | 1 | Not-determined | Genome-fragment | NA | NA | 0 |
| S6_vs_k141_51522\|\|full | 12252 | No | NA | 12 | 0 | 9 | Not-determined | Genome-fragment | NA | NA | 0 |
| S14_vs_k141_70044\|\|full | 7043 | No | NA | 11 | 8 | 0 | Low-quality | Genome-fragment | 17.42 | AAI-based (high-confidence) | 0 |
| S10_vs_98957\|\|full | 7045 | No | NA | 12 | 6 | 0 | Low-quality | Genome-fragment | 11.31 | AAI-based (high-confidence) | 0 |
| S3_vs_k141_63036\|\|full | 7046 | No | NA | 5 | 0 | 3 | Low-quality | Genome-fragment | 1.83 | AAI-based (medium-confidence) | 0 |
| S6_vs_k141_51578\|\|full | 3390 | No | NA | 3 | 0 | 2 | Not-determined | Genome-fragment | NA | NA | 0 |
| S6_vs_k141_51601\|\|full | 1185 | No | NA | 2 | 0 | 2 | Not-determined | Genome-fragment | NA | NA | 0 |
| S6_vs_k141_20952\|\|full | 2085 | No | NA | 3 | 0 | 1 | Not-determined | Genome-fragment | NA | NA | 0 |
| S6_vs_k141_20959\|\|full | 2579 | No | NA | 3 | 0 | 1 | Not-determined | Genome-fragment | NA | NA | 0 |
| S7_vs_88160\|\|full | 7048 | No | NA | 12 | 1 | 2 | Low-quality | Genome-fragment | 3.03 | HMM-based (lower-bound) | 0 |
| S6_extracted_k141_12887 | 7049 | No | NA | 6 | 0 | 2 | Low-quality | Genome-fragment | 6.84 | AAI-based (medium-confidence) | 0 |
| S17_extracted_k141_149447 | 7050 | Yes | 5824 | 6 | 1 | 1 | Low-quality | Genome-fragment | 9.08 | AAI-based (high-confidence) | 17.39 |
| S17_extracted_k141_54007 | 7050 | No | NA | 11 | 3 | 0 | Low-quality | Genome-fragment | 11.02 | AAI-based (high-confidence) | 0 |
| S6_vs_k141_27980\|\|full | 2319 | No | NA | 2 | 0 | 2 | Not-determined | Genome-fragment | NA | NA | 0 |
| S6_vs_k141_56331\|\|full | 2021 | No | NA | 2 | 0 | 0 | Not-determined | Genome-fragment | NA | NA | 0 |
| S6_vs_k141_28077\|\|full | 2370 | No | NA | 3 | 0 | 3 | Not-determined | Genome-fragment | NA | NA | 0 |
| S6_vs_k141_51807\|\|full | 1048 | No | NA | 3 | 0 | 3 | Not-determined | Genome-fragment | NA | NA | 0 |
| S1_vs_k141_27940\|\|full | 7050 | No | NA | 14 | 3 | 0 | Low-quality | Genome-fragment | 16.63 | AAI-based (high-confidence) | 0 |
| S6_vs_k141_28099\|\|full | 2449 | No | NA | 6 | 0 | 1 | Not-determined | Genome-fragment | NA | NA | 0 |
| S6_vs_k141_56469\|\|full | 1045 | No | NA | 3 | 0 | 0 | Not-determined | Genome-fragment | NA | NA | 0 |
| S6_vs_k141_21106\|\|full | 1650 | No | NA | 3 | 0 | 2 | Not-determined | Genome-fragment | NA | NA | 0 |
| S8_extracted_k141_18282 | 7053 | No | NA | 12 | 11 | 0 | Low-quality | Genome-fragment | 16.22 | AAI-based (high-confidence) | 0 |
| S5_vs_k141_8783\|\|full | 7060 | Yes | 4743 | 6 | 1 | 1 | Low-quality | Genome-fragment | 11.94 | AAI-based (high-confidence) | 32.82 |

| | | | | | | | | | | | |
|---|---|---|---|---|---|---|---|---|---|---|---|
| S6_vs_k141_14480||0_partial | 20333 | No | NA | 15 | 0 | 9 | Not-determined | Genome-fragment | NA | NA | 0 |
| S6_vs_k141_25980||0_partial | 12479 | No | NA | 10 | 0 | 6 | Not-determined | Genome-fragment | NA | NA | 0 |
| S3_vs_k141_82501||full | 7061 | No | NA | 9 | 0 | 0 | Low-quality | Genome-fragment | 4.4 | AAI-based (high-confidence) | 0 |
| S6_vs_k141_50185||0_partial | 57623 | No | NA | 72 | 0 | 14 | Not-determined | Genome-fragment | NA | NA | 0 |
| S14_vs_k141_91678||full | 7062 | No | NA | 9 | 0 | 3 | Low-quality | Genome-fragment | 8.48 | AAI-based (high-confidence) | 0 |
| S6_vs_k141_54925||0_partial | 22598 | No | NA | 17 | 0 | 4 | Not-determined | Genome-fragment | NA | NA | 0 |
| S14_vs_k141_103105||full | 7066 | No | NA | 14 | 1 | 1 | Low-quality | Genome-fragment | 19.2 | AAI-based (medium-confidence) | 0 |
| S6_vs_k141_10908||0_partial | 17302 | No | NA | 18 | 0 | 3 | Not-determined | Genome-fragment | NA | NA | 0 |
| S6_vs_k141_55084||0_partial | 21437 | No | NA | 14 | 0 | 7 | Not-determined | Genome-fragment | NA | NA | 0 |
| S6_vs_k141_39065||0_partial | 47509 | Yes | 29537 | 42 | 5 | 8 | Medium-quality | Genome-fragment | 66.32 | AAI-based (medium-confidence) | 37.83 |
| S6_vs_k141_1504||0_partial | 12430 | No | NA | 24 | 0 | 6 | Not-determined | Genome-fragment | NA | NA | 0 |
| S3_vs_k141_40843||full | 7068 | No | NA | 14 | 3 | 0 | Low-quality | Genome-fragment | 11.11 | HMM-based (lower-bound) | 0 |
| S6_vs_k141_39286||0_partial | 15247 | No | NA | 11 | 0 | 6 | Not-determined | Genome-fragment | NA | NA | 0 |
| S6_vs_k141_30297||0_partial | 20627 | No | NA | 15 | 0 | 6 | Not-determined | Genome-fragment | NA | NA | 0 |
| S6_vs_k141_55570||0_partial | 13784 | No | NA | 13 | 0 | 8 | Not-determined | Genome-fragment | NA | NA | 0 |
| S16_extracted_172136 | 7068 | No | NA | 19 | 3 | 0 | Low-quality | Genome-fragment | 15.34 | AAI-based (high-confidence) | 0 |
| S8_vs_k141_77846||full | 7068 | No | NA | 6 | 0 | 6 | Low-quality | Genome-fragment | 1.99 | AAI-based (medium-confidence) | 0 |
| S7_extracted_56712 | 7087 | No | NA | 6 | 0 | 1 | Low-quality | Genome-fragment | 8.98 | AAI-based (medium-confidence) | 0 |
| S10_extracted_119210 | 7088 | No | NA | 6 | 0 | 6 | Low-quality | Genome-fragment | 3.81 | AAI-based (medium-confidence) | 0 |
| S8_extracted_k141_40966 | 7089 | No | NA | 12 | 6 | 0 | Low-quality | Genome-fragment | 16.08 | AAI-based (high-confidence) | 0 |
| S17_vs_k141_118848||full | 7091 | No | NA | 11 | 3 | 0 | Low-quality | Genome-fragment | 19.81 | AAI-based (medium-confidence) | 0 |
| S12_vs_k141_137127||full | 7095 | No | NA | 8 | 3 | 0 | Low-quality | Genome-fragment | 6.82 | HMM-based (lower-bound) | 0 |
| S5_extracted_k141_34419 | 7099 | No | NA | 8 | 0 | 2 | Low-quality | Genome-fragment | 5.71 | AAI-based (high-confidence) | 0 |
| S6_vs_k141_3028||lt2gene | 1312 | No | NA | 1 | 0 | 1 | Not-determined | Genome-fragment | NA | NA | 0 |
| S16_extracted_2704 | 7107 | No | NA | 2 | 0 | 1 | Low-quality | Genome-fragment | 12.65 | AAI-based (medium-confidence) | 0 |
| S6_vs_k141_26340||lt2gene | 1800 | No | NA | 1 | 0 | 0 | Not-determined | Genome-fragment | NA | NA | 0 |
| S6_vs_k141_35841||lt2gene | 1509 | No | NA | 1 | 0 | 1 | Not-determined | Genome-fragment | NA | NA | 0 |
| S10_vs_85685||full | 7108 | No | NA | 14 | 1 | 0 | Low-quality | Genome-fragment | 18.64 | AAI-based (high-confidence) | 0 |
| S15_extracted_7427 | 7113 | No | NA | 19 | 2 | 0 | Low-quality | Genome-fragment | 4.07 | AAI-based (high-confidence) | 0 |
| S6_vs_k141_36110||lt2gene | 1487 | No | NA | 1 | 0 | 1 | Not-determined | Genome-fragment | NA | NA | 0 |
| S6_vs_k141_26554||lt2gene | 1698 | No | NA | 1 | 0 | 0 | Not-determined | Genome-fragment | NA | NA | 0 |
| S6_vs_k141_12835||lt2gene | 1317 | No | NA | 2 | 0 | 0 | Not-determined | Genome-fragment | NA | NA | 0 |
| S9_extracted_k141_56789 | 7116 | No | NA | 9 | 4 | 2 | Low-quality | Genome-fragment | 11.86 | AAI-based (high-confidence) | 0 |
| S6_vs_k141_38872||lt2gene | 1081 | No | NA | 1 | 0 | 0 | Not-determined | Genome-fragment | NA | NA | 0 |
| S3_extracted_k141_54199 | 7119 | No | NA | 8 | 0 | 6 | Low-quality | Genome-fragment | 1.08 | AAI-based (medium-confidence) | 0 |
| S12_extracted_k141_136418 | 7120 | No | NA | 11 | 5 | 0 | Low-quality | Genome-fragment | 18.53 | AAI-based (medium-confidence) | 0 |
| S11_vs_k141_7863||full | 7122 | No | NA | 11 | 2 | 0 | Low-quality | Genome-fragment | 7.57 | AAI-based (medium-confidence) | 0 |
| S3_vs_k141_88628||full | 7123 | No | NA | 12 | 9 | 0 | Low-quality | Genome-fragment | 20.71 | AAI-based (high-confidence) | 0 |
| S6_vs_k141_19599||lt2gene | 2285 | No | NA | 1 | 0 | 0 | Not-determined | Genome-fragment | NA | NA | 0 |
| S3_vs_k141_105108||full | 7123 | No | NA | 7 | 3 | 0 | Low-quality | Genome-fragment | 8.46 | HMM-based (lower-bound) | 0 |
| S16_vs_93494||full | 7124 | No | NA | 12 | 5 | 0 | Low-quality | Genome-fragment | 13.27 | AAI-based (high-confidence) | 0 |
| S6_vs_k141_32050||lt2gene | 1110 | No | NA | 1 | 0 | 1 | Not-determined | Genome-fragment | NA | NA | 0 |
| S6_vs_k141_41674||lt2gene | 1032 | No | NA | 2 | 0 | 0 | Not-determined | Genome-fragment | NA | NA | 0 |
| S10_vs_83642||full | 7131 | No | NA | 13 | 3 | 0 | Low-quality | Genome-fragment | 11.95 | HMM-based (lower-bound) | 0 |
| S1_extracted_k141_141358 | 7138 | No | NA | 11 | 1 | 1 | Low-quality | Genome-fragment | 12.46 | AAI-based (high-confidence) | 0 |
| S2_vs_k141_11805||full | 7144 | No | NA | 5 | 0 | 2 | Low-quality | Genome-fragment | 15.53 | AAI-based (high-confidence) | 0 |
| S2_vs_k141_17520||full | 7144 | No | NA | 7 | 2 | 0 | Low-quality | Genome-fragment | 17.54 | AAI-based (high-confidence) | 0 |
| S11_extracted_k141_1095 | 7150 | No | NA | 11 | 3 | 0 | Low-quality | Genome-fragment | 16.83 | AAI-based (high-confidence) | 0 |
| S6_vs_k141_30284||lt2gene | 1469 | No | NA | 1 | 0 | 1 | Not-determined | Genome-fragment | NA | NA | 0 |
| S17_extracted_k141_4926 | 7152 | No | NA | 10 | 1 | 0 | Low-quality | Genome-fragment | 3.51 | HMM-based (lower-bound) | 0 |
| S16_vs_172744||full | 7152 | No | NA | 20 | 2 | 0 | Low-quality | Genome-fragment | 8.31 | HMM-based (lower-bound) | 0 |
| S3_extracted_k141_127398 | 7153 | No | NA | 14 | 1 | 0 | Low-quality | Genome-fragment | 12.32 | HMM-based (lower-bound) | 0 |
| S14_vs_k141_94258||full | 7164 | No | NA | 10 | 0 | 6 | Low-quality | Genome-fragment | 6.25 | AAI-based (medium-confidence) | 0 |
| S5_vs_k141_15262||full | 7164 | No | NA | 11 | 10 | 0 | Low-quality | Genome-fragment | 19.31 | AAI-based (high-confidence) | 0 |
| S3_extracted_k141_69458 | 7165 | No | NA | 15 | 0 | 0 | Low-quality | Genome-fragment | 5.48 | AAI-based (high-confidence) | 0 |
| S7_vs_5855||full | 7170 | No | NA | 15 | 0 | 0 | Low-quality | Genome-fragment | 15.08 | AAI-based (high-confidence) | 0 |
| S16_vs_3041||full | 7174 | No | NA | 11 | 0 | 0 | Low-quality | Genome-fragment | 8.59 | AAI-based (high-confidence) | 0 |
| S1_extracted_k141_58337 | 7176 | No | NA | 5 | 2 | 0 | Low-quality | Genome-fragment | 20.32 | AAI-based (high-confidence) | 0 |
| S8_vs_k141_174928||full | 7177 | No | NA | 17 | 1 | 0 | Low-quality | Genome-fragment | 11.86 | AAI-based (medium-confidence) | 0 |
| S8_extracted_k141_164873 | 1599 | No | NA | 1 | 0 | 0 | Not-determined | Genome-fragment | NA | NA | 0 |
| S8_extracted_k141_17358 | 1056 | No | NA | 2 | 0 | 0 | Not-determined | Genome-fragment | NA | NA | 0 |
| S8_extracted_k141_60748 | 2232 | No | NA | 3 | 0 | 2 | Not-determined | Genome-fragment | NA | NA | 0 |
| S8_extracted_k141_78109 | 1083 | No | NA | 2 | 0 | 2 | Not-determined | Genome-fragment | NA | NA | 0 |
| S8_extracted_k141_156204 | 1210 | No | NA | 2 | 0 | 0 | Not-determined | Genome-fragment | NA | NA | 0 |
| S17_vs_k141_101004||full | 7179 | No | NA | 16 | 2 | 0 | Low-quality | Genome-fragment | 11.8 | AAI-based (high-confidence) | 0 |
| S8_extracted_k141_17366 | 2959 | No | NA | 1 | 0 | 1 | Not-determined | Genome-fragment | NA | NA | 0 |
| S8_extracted_k141_190921 | 3222 | No | NA | 4 | 0 | 3 | Not-determined | Genome-fragment | NA | NA | 0 |
| S16_vs_122308||full | 7180 | No | NA | 3 | 0 | 2 | Low-quality | Genome-fragment | 2.23 | AAI-based (high-confidence) | 0 |
| S8_extracted_k141_173561 | 2966 | No | NA | 2 | 0 | 0 | Not-determined | Genome-fragment | NA | NA | 0 |
| S8_extracted_k141_156216 | 2914 | No | NA | 4 | 0 | 0 | Not-determined | Genome-fragment | NA | NA | 0 |
| S8_extracted_k141_121506 | 1179 | No | NA | 1 | 0 | 0 | Not-determined | Genome-fragment | NA | NA | 0 |
| S8_extracted_k141_104151 | 7599 | No | NA | 7 | 0 | 3 | Not-determined | Genome-fragment | NA | NA | 0 |
| S10_vs_11101||full | 7181 | No | NA | 9 | 0 | 7 | Low-quality | Genome-fragment | 2.02 | AAI-based (medium-confidence) | 0 |
| S8_extracted_k141_69447 | 1065 | No | NA | 1 | 0 | 1 | Not-determined | Genome-fragment | NA | NA | 0 |
| S8_extracted_k141_86808 | 2538 | No | NA | 2 | 0 | 2 | Not-determined | Genome-fragment | NA | NA | 0 |
| S8_extracted_k141_182253 | 1132 | No | NA | 1 | 0 | 0 | Not-determined | Genome-fragment | NA | NA | 0 |
| S8_extracted_k141_138864 | 2454 | No | NA | 4 | 0 | 1 | Not-determined | Genome-fragment | NA | NA | 0 |
| S8_extracted_k141_43422 | 2003 | No | NA | 2 | 0 | 1 | Not-determined | Genome-fragment | NA | NA | 0 |
| S8_extracted_k141_182262 | 1054 | No | NA | 2 | 0 | 2 | Not-determined | Genome-fragment | NA | NA | 0 |
| S8_extracted_k141_26079 | 5054 | No | NA | 5 | 0 | 0 | Not-determined | Genome-fragment | NA | NA | 0 |
| S8_extracted_k141_78119 | 1044 | No | NA | 2 | 0 | 1 | Not-determined | Genome-fragment | NA | NA | 0 |
| S17_vs_k141_78567||full | 7185 | No | NA | 3 | 0 | 0 | Low-quality | Genome-fragment | 7.12 | AAI-based (medium-confidence) | 0 |
| S8_extracted_k141_43430 | 3260 | No | NA | 3 | 0 | 3 | Not-determined | Genome-fragment | NA | NA | 0 |
| S8_extracted_k141_69462 | 3236 | No | NA | 3 | 0 | 1 | Not-determined | Genome-fragment | NA | NA | 0 |
| S8_extracted_k141_199616 | 1100 | No | NA | 2 | 0 | 1 | Not-determined | Genome-fragment | NA | NA | 0 |
| S3_extracted_k141_16813 | 7185 | No | NA | 10 | 2 | 0 | Low-quality | Genome-fragment | 19.02 | AAI-based (high-confidence) | 0 |
| S8_extracted_k141_60801 | 1108 | No | NA | 1 | 0 | 0 | Not-determined | Genome-fragment | NA | NA | 0 |
| S8_extracted_k141_95516 | 10416 | No | NA | 6 | 0 | 5 | Not-determined | Genome-fragment | NA | NA | 0 |
| S8_extracted_k141_199626 | 1241 | No | NA | 1 | 0 | 0 | Not-determined | Genome-fragment | NA | NA | 0 |
| S8_extracted_k141_26096 | 3273 | No | NA | 3 | 0 | 2 | Not-determined | Genome-fragment | NA | NA | 0 |
| S7_vs_5446||full | 7185 | No | NA | 4 | 1 | 0 | Low-quality | Genome-fragment | 3.65 | AAI-based (high-confidence) | 0 |
| S8_extracted_k141_52120 | 10614 | No | NA | 8 | 0 | 6 | Not-determined | Genome-fragment | NA | NA | 0 |
| S8_extracted_k141_69477 | 1142 | No | NA | 1 | 0 | 0 | Not-determined | Genome-fragment | NA | NA | 0 |
| S8_extracted_k141_86856 | 1009 | No | NA | 1 | 0 | 0 | Not-determined | Genome-fragment | NA | NA | 0 |
| S16_vs_39179||full | 7185 | No | NA | 15 | 2 | 0 | Low-quality | Genome-fragment | 8.1 | HMM-based (lower-bound) | 0 |
| S17_vs_k141_100596||full | 7187 | Yes | 3260 | 7 | 1 | 3 | Low-quality | Genome-fragment | 4.93 | HMM-based (lower-bound) | 54.64 |
| S8_extracted_k141_190982 | 1258 | No | NA | 2 | 0 | 2 | Not-determined | Genome-fragment | NA | NA | 0 |
| S8_extracted_k141_69484 | 1799 | No | NA | 3 | 0 | 1 | Not-determined | Genome-fragment | NA | NA | 0 |
| S8_extracted_k141_60826 | 3259 | No | NA | 4 | 0 | 3 | Not-determined | Genome-fragment | NA | NA | 0 |
| S12_vs_k141_78731||lt2gene | 7187 | No | NA | 2 | 1 | 0 | Low-quality | Genome-fragment | 11.06 | AAI-based (medium-confidence) | 0 |
| S8_extracted_k141_26129 | 1695 | No | NA | 2 | 0 | 2 | Not-determined | Genome-fragment | NA | NA | 0 |
| S8_extracted_k141_110484 | 7187 | No | NA | 4 | 0 | 3 | Low-quality | Genome-fragment | 3.87 | AAI-based (medium-confidence) | 0 |
| S7_vs_69333||full | 7188 | No | NA | 7 | 0 | 4 | Low-quality | Genome-fragment | 1.41 | AAI-based (medium-confidence) | 0 |
| S8_extracted_k141_8772 | 4880 | No | NA | 7 | 0 | 1 | Not-determined | Genome-fragment | NA | NA | 0 |
| S8_extracted_k141_43483 | 7365 | No | NA | 7 | 5 | 0 | Medium-quality | Genome-fragment | 53.03 | AAI-based (high-confidence) | 0 |
| S8_extracted_k141_147623 | 1279 | No | NA | 2 | 0 | 1 | Not-determined | Genome-fragment | NA | NA | 0 |
| S5_vs_k141_134340||full | 7193 | No | NA | 17 | 2 | 0 | Low-quality | Genome-fragment | 17.87 | AAI-based (high-confidence) | 0 |
| S1_vs_k141_89635||full | 7197 | No | NA | 8 | 0 | 3 | Low-quality | Genome-fragment | 12.83 | AAI-based (medium-confidence) | 0 |
| S8_extracted_k141_95539 | 1060 | No | NA | 1 | 0 | 1 | Not-determined | Genome-fragment | NA | NA | 0 |
| S8_extracted_k141_164966 | 1409 | No | NA | 2 | 0 | 2 | Not-determined | Genome-fragment | NA | NA | 0 |
| S8_extracted_k141_78193 | 1305 | No | NA | 2 | 0 | 1 | Not-determined | Genome-fragment | NA | NA | 0 |
| S8_extracted_k141_78194 | 1206 | No | NA | 1 | 0 | 1 | Not-determined | Genome-fragment | NA | NA | 0 |
| S16_extracted_119882 | 7197 | Yes | 5028 | 10 | 3 | 1 | Low-quality | Genome-fragment | 4.06 | AAI-based (high-confidence) | 30.14 |
| S8_extracted_k141_121601 | 1058 | No | NA | 1 | 0 | 1 | Not-determined | Genome-fragment | NA | NA | 0 |
| S10_extracted_52160 | 7197 | No | NA | 12 | 0 | 2 | Low-quality | Genome-fragment | 39.61 | AAI-based (medium-confidence) | 0 |
| S8_extracted_k141_34793 | 1483 | No | NA | 1 | 0 | 0 | Not-determined | Genome-fragment | NA | NA | 0 |
| S8_extracted_k141_121604 | 6195 | No | NA | 5 | 0 | 4 | Not-determined | Genome-fragment | NA | NA | 0 |
| S8_extracted_k141_34802 | 4239 | No | NA | 3 | 0 | 2 | Not-determined | Genome-fragment | NA | NA | 0 |
| S8_extracted_k141_147666 | 1973 | No | NA | 2 | 0 | 0 | Not-determined | Genome-fragment | NA | NA | 0 |
| S8_extracted_k141_104242 | 2342 | No | NA | 3 | 0 | 0 | Not-determined | Genome-fragment | NA | NA | 0 |
| S8_extracted_k141_156341 | 3133 | No | NA | 5 | 0 | 2 | Not-determined | Genome-fragment | NA | NA | 0 |
| S8_extracted_k141_104244 | 2323 | No | NA | 2 | 0 | 0 | Not-determined | Genome-fragment | NA | NA | 0 |
| S8_extracted_k141_8813 | 1126 | No | NA | 1 | 0 | 1 | Not-determined | Genome-fragment | NA | NA | 0 |
| S8_extracted_k141_156347 | 3294 | No | NA | 3 | 0 | 3 | Not-determined | Genome-fragment | NA | NA | 0 |
| S8_extracted_k141_168 | 1395 | No | NA | 1 | 0 | 1 | Not-determined | Genome-fragment | NA | NA | 0 |
| S8_extracted_k141_138976 | 3474 | No | NA | 2 | 0 | 2 | Not-determined | Genome-fragment | NA | NA | 0 |
| S8_extracted_k141_138978 | 1223 | No | NA | 1 | 0 | 0 | Not-determined | Genome-fragment | NA | NA | 0 |
| S8_extracted_k141_121633 | 1304 | No | NA | 1 | 0 | 1 | Not-determined | Genome-fragment | NA | NA | 0 |
| S8_extracted_k141_173660 | 1062 | No | NA | 1 | 0 | 0 | Not-determined | Genome-fragment | NA | NA | 0 |
| S8_extracted_k141_86972 | 1321 | No | NA | 2 | 0 | 0 | Not-determined | Genome-fragment | NA | NA | 0 |
| S8_extracted_k141_52203 | 3373 | No | NA | 3 | 0 | 3 | Not-determined | Genome-fragment | NA | NA | 0 |
| S8_extracted_k141_199710 | 4892 | No | NA | 3 | 0 | 2 | Not-determined | Genome-fragment | NA | NA | 0 |
| S9_vs_k141_29457||full | 7199 | No | NA | 12 | 2 | 0 | Low-quality | Genome-fragment | 15.61 | AAI-based (medium-confidence) | 0 |
| S8_extracted_k141_173667 | 1546 | No | NA | 3 | 0 | 1 | Not-determined | Genome-fragment | NA | NA | 0 |
| S5_vs_k141_2564||full | 7206 | No | NA | 14 | 1 | 0 | Low-quality | Genome-fragment | 17.3 | AAI-based (medium-confidence) | 0 |
| S8_extracted_k141_78258 | 1338 | No | NA | 1 | 0 | 1 | Not-determined | Genome-fragment | NA | NA | 0 |
| S8_extracted_k141_17490 | 2476 | No | NA | 2 | 0 | 2 | Not-determined | Genome-fragment | NA | NA | 0 |
| S8_extracted_k141_26231 | 4241 | No | NA | 3 | 0 | 0 | Not-determined | Genome-fragment | NA | NA | 0 |
| S8_extracted_k141_86997 | 2449 | No | NA | 3 | 0 | 2 | Not-determined | Genome-fragment | NA | NA | 0 |
| S8_extracted_k141_8849 | 2441 | No | NA | 2 | 0 | 0 | Not-determined | Genome-fragment | NA | NA | 0 |
| S12_vs_k141_5781||full | 7208 | No | NA | 11 | 5 | 0 | Low-quality | Genome-fragment | 17.34 | AAI-based (high-confidence) | 0 |
| S11_vs_k141_98490||full | 7211 | No | NA | 5 | 3 | 0 | Low-quality | Genome-fragment | 12.43 | AAI-based (high-confidence) | 0 |
| S8_extracted_k141_112993 | 3356 | No | NA | 5 | 0 | 4 | Not-determined | Genome-fragment | NA | NA | 0 |
| S8_extracted_k141_87017 | 1304 | No | NA | 1 | 0 | 1 | Not-determined | Genome-fragment | NA | NA | 0 |
| S8_extracted_k141_43538 | 3835 | No | NA | 2 | 0 | 1 | Not-determined | Genome-fragment | NA | NA | 0 |
| S8_extracted_k141_52222 | 3779 | No | NA | 5 | 0 | 3 | Not-determined | Genome-fragment | NA | NA | 0 |
| S8_extracted_k141_43546 | 1259 | No | NA | 1 | 0 | 1 | Not-determined | Genome-fragment | NA | NA | 0 |
| S8_extracted_k141_139012 | 1233 | No | NA | 1 | 0 | 1 | Not-determined | Genome-fragment | NA | NA | 0 |
| S8_extracted_k141_34879 | 5091 | No | NA | 6 | 0 | 5 | Not-determined | Genome-fragment | NA | NA | 0 |
| S8_extracted_k141_43551 | 1378 | No | NA | 1 | 0 | 0 | Not-determined | Genome-fragment | NA | NA | 0 |
| S8_extracted_k141_34882 | 1273 | No | NA | 1 | 0 | 0 | Not-determined | Genome-fragment | NA | NA | 0 |
| S8_extracted_k141_231 | 2344 | No | NA | 3 | 0 | 2 | Not-determined | Genome-fragment | NA | NA | 0 |
| S8_extracted_k141_121676 | 1712 | No | NA | 3 | 0 | 1 | Not-determined | Genome-fragment | NA | NA | 0 |
| S14_extracted_k141_27064 | 7219 | No | NA | 13 | 6 | 0 | Low-quality | Genome-fragment | 17.8 | AAI-based (high-confidence) | 0 |
| S8_extracted_k141_87046 | 3365 | No | NA | 4 | 0 | 2 | Not-determined | Genome-fragment | NA | NA | 0 |
| S8_extracted_k141_69550 | 1458 | No | NA | 2 | 0 | 1 | Not-determined | Genome-fragment | NA | NA | 0 |
| S8_extracted_k141_130316 | 1280 | No | NA | 1 | 0 | 1 | Not-determined | Genome-fragment | NA | NA | 0 |
| S1_extracted_k141_133507 | 7225 | No | NA | 9 | 3 | 0 | Low-quality | Genome-fragment | 23.08 | HMM-based (lower-bound) | 0 |
| S8_extracted_k141_78303 | 1034 | No | NA | 2 | 0 | 1 | Not-determined | Genome-fragment | NA | NA | 0 |
| S3_vs_k141_30989||full | 7227 | No | NA | 17 | 4 | 0 | Low-quality | Genome-fragment | 13.72 | AAI-based (medium-confidence) | 0 |
| S8_extracted_k141_60969 | 5559 | No | NA | 2 | 0 | 1 | Not-determined | Genome-fragment | NA | NA | 0 |
| S8_extracted_k141_139052 | 3221 | No | NA | 4 | 0 | 1 | Not-determined | Genome-fragment | NA | NA | 0 |
| S8_extracted_k141_113032 | 1456 | No | NA | 1 | 0 | 1 | Not-determined | Genome-fragment | NA | NA | 0 |
| S8_extracted_k141_43589 | 1342 | No | NA | 2 | 0 | 2 | Not-determined | Genome-fragment | NA | NA | 0 |
| S8_extracted_k141_139063 | 1069 | No | NA | 1 | 0 | 1 | Not-determined | Genome-fragment | NA | NA | 0 |
| S8_extracted_k141_8919 | 3004 | No | NA | 3 | 0 | 2 | Not-determined | Genome-fragment | NA | NA | 0 |
| S12_vs_k141_63998||full | 7227 | No | NA | 6 | 1 | 0 | Low-quality | Genome-fragment | 12.33 | AAI-based (high-confidence) | 0 |
| S6_extracted_k141_8174 | 7228 | No | NA | 9 | 5 | 0 | Low-quality | Genome-fragment | 4.35 | AAI-based (high-confidence) | 0 |
| S8_extracted_k141_52278 | 2397 | No | NA | 3 | 0 | 2 | Not-determined | Genome-fragment | NA | NA | 0 |
| S8_extracted_k141_191158 | 5216 | No | NA | 2 | 0 | 2 | Not-determined | Genome-fragment | NA | NA | 0 |
| S1_extracted_k141_92407 | 7229 | No | NA | 8 | 0 | 2 | Low-quality | Genome-fragment | 10.88 | AAI-based (medium-confidence) | 0 |
| S8_extracted_k141_52283 | 4689 | No | NA | 6 | 0 | 4 | Not-determined | Genome-fragment | NA | NA | 0 |
| S8_extracted_k141_165103 | 1180 | No | NA | 2 | 0 | 1 | Not-determined | Genome-fragment | NA | NA | 0 |
| S8_extracted_k141_121730 | 2933 | No | NA | 2 | 0 | 1 | Not-determined | Genome-fragment | NA | NA | 0 |
| S8_extracted_k141_306 | 1432 | No | NA | 2 | 0 | 1 | Not-determined | Genome-fragment | NA | NA | 0 |
| S5_vs_k141_139897||full | 7233 | No | NA | 9 | 3 | 0 | Low-quality | Genome-fragment | 15.6 | AAI-based (high-confidence) | 0 |
| S8_extracted_k141_191189 | 1296 | No | NA | 2 | 0 | 2 | Not-determined | Genome-fragment | NA | NA | 0 |
| S8_extracted_k141_78349 | 1650 | No | NA | 1 | 0 | 1 | Not-determined | Genome-fragment | NA | NA | 0 |
| S8_extracted_k141_121754 | 2249 | No | NA | 1 | 0 | 1 | Not-determined | Genome-fragment | NA | NA | 0 |
| S8_extracted_k141_130405 | 1131 | No | NA | 2 | 0 | 0 | Not-determined | Genome-fragment | NA | NA | 0 |
| S8_extracted_k141_113079 | 1743 | No | NA | 2 | 0 | 0 | Not-determined | Genome-fragment | NA | NA | 0 |
| S8_extracted_k141_69614 | 1348 | No | NA | 1 | 0 | 1 | Not-determined | Genome-fragment | NA | NA | 0 |
| S8_extracted_k141_121759 | 1222 | No | NA | 1 | 0 | 0 | Not-determined | Genome-fragment | NA | NA | 0 |
| S8_extracted_k141_69618 | 1555 | No | NA | 2 | 0 | 0 | Not-determined | Genome-fragment | NA | NA | 0 |
| S8_extracted_k141_121763 | 1396 | No | NA | 1 | 0 | 1 | Not-determined | Genome-fragment | NA | NA | 0 |
| S8_extracted_k141_328 | 3120 | No | NA | 2 | 0 | 2 | Not-determined | Genome-fragment | NA | NA | 0 |
| S8_extracted_k141_156445 | 6193 | No | NA | 2 | 0 | 2 | Not-determined | Genome-fragment | NA | NA | 0 |
| S8_extracted_k141_130429 | 1606 | No | NA | 2 | 0 | 1 | Not-determined | Genome-fragment | NA | NA | 0 |
| S8_extracted_k141_104328 | 1064 | No | NA | 1 | 0 | 0 | Not-determined | Genome-fragment | NA | NA | 0 |
| S8_extracted_k141_165135 | 3270 | No | NA | 2 | 0 | 1 | Not-determined | Genome-fragment | NA | NA | 0 |
| S8_extracted_k141_69633 | 1215 | No | NA | 1 | 0 | 1 | Not-determined | Genome-fragment | NA | NA | 0 |
| S8_extracted_k141_69638 | 1973 | No | NA | 2 | 0 | 2 | Not-determined | Genome-fragment | NA | NA | 0 |
| S8_extracted_k141_87151 | 10330 | No | NA | 11 | 0 | 4 | Not-determined | Genome-fragment | NA | NA | 0 |
| S8_extracted_k141_182359 | 8343 | No | NA | 5 | 0 | 4 | Not-determined | Genome-fragment | NA | NA | 0 |
| S12_vs_k141_94220||full | 7236 | Yes | 2296 | 7 | 1 | 4 | Low-quality | Genome-fragment | 3.47 | HMM-based (lower-bound) | 68.27 |
| S8_extracted_k141_35019 | 1499 | No | NA | 2 | 0 | 2 | Not-determined | Genome-fragment | NA | NA | 0 |
| S8_extracted_k141_130473 | 1045 | No | NA | 2 | 0 | 2 | Not-determined | Genome-fragment | NA | NA | 0 |
| S8_extracted_k141_69653 | 1638 | No | NA | 2 | 0 | 2 | Not-determined | Genome-fragment | NA | NA | 0 |
| S17_extracted_k141_151981 | 7237 | No | NA | 14 | 0 | 1 | Low-quality | Genome-fragment | 15.06 | AAI-based (high-confidence) | 0 |
| S8_extracted_k141_173742 | 1082 | No | NA | 2 | 0 | 1 | Not-determined | Genome-fragment | NA | NA | 0 |
| S8_extracted_k141_156470 | 1034 | No | NA | 1 | 0 | 0 | Not-determined | Genome-fragment | NA | NA | 0 |
| S9_vs_k141_3803||full | 7239 | No | NA | 11 | 1 | 0 | Low-quality | Genome-fragment | 4.35 | AAI-based (high-confidence) | 0 |
| S8_extracted_k141_61069 | 1169 | No | NA | 1 | 0 | 0 | Not-determined | Genome-fragment | NA | NA | 0 |
| S8_extracted_k141_173751 | 1077 | No | NA | 2 | 0 | 0 | Not-determined | Genome-fragment | NA | NA | 0 |
| S8_extracted_k141_156485 | 1063 | No | NA | 2 | 0 | 0 | Not-determined | Genome-fragment | NA | NA | 0 |
| S8_extracted_k141_139140 | 1099 | No | NA | 1 | 0 | 0 | Not-determined | Genome-fragment | NA | NA | 0 |
| S8_extracted_k141_182414 | 1523 | No | NA | 2 | 0 | 0 | Not-determined | Genome-fragment | NA | NA | 0 |
| S14_vs_k141_100812||full | 7243 | No | NA | 15 | 2 | 3 | Low-quality | Genome-fragment | 11.38 | HMM-based (lower-bound) | 0 |
| S8_extracted_k141_87189 | 1156 | No | NA | 4 | 0 | 0 | Not-determined | Genome-fragment | NA | NA | 0 |
| S7_vs_76284||full | 7244 | No | NA | 10 | 4 | 0 | Low-quality | Genome-fragment | 16.14 | AAI-based (high-confidence) | 0 |
| S8_extracted_k141_52399 | 1169 | No | NA | 2 | 0 | 2 | Not-determined | Genome-fragment | NA | NA | 0 |
| S10_extracted_7339 | 7246 | No | NA | 13 | 1 | 1 | Low-quality | Genome-fragment | 3.93 | HMM-based (lower-bound) | 0 |
| S11_vs_k141_102534||full | 7247 | No | NA | 11 | 1 | 0 | Low-quality | Genome-fragment | 16.59 | AAI-based (medium-confidence) | 0 |
| S8_extracted_k141_43673 | 5084 | No | NA | 7 | 0 | 4 | Not-determined | Genome-fragment | NA | NA | 0 |
| S8_extracted_k141_78437 | 1233 | No | NA | 1 | 0 | 0 | Not-determined | Genome-fragment | NA | NA | 0 |
| S7_vs_352||full | 7251 | No | NA | 11 | 7 | 0 | Low-quality | Genome-fragment | 17.54 | AAI-based (medium-confidence) | 0 |
| S8_extracted_k141_199849 | 4324 | No | NA | 4 | 0 | 1 | Not-determined | Genome-fragment | NA | NA | 0 |
| S8_extracted_k141_173789 | 1917 | No | NA | 1 | 0 | 1 | Not-determined | Genome-fragment | NA | NA | 0 |
| S8_extracted_k141_113160 | 2631 | No | NA | 3 | 0 | 1 | Not-determined | Genome-fragment | NA | NA | 0 |
| S4_extracted_7325 | 7252 | No | NA | 16 | 14 | 0 | Low-quality | Genome-fragment | 23.89 | AAI-based (high-confidence) | 0 |
| S8_extracted_k141_139155 | 2873 | No | NA | 4 | 0 | 1 | Not-determined | Genome-fragment | NA | NA | 0 |
| S8_extracted_k141_26350 | 4837 | No | NA | 4 | 0 | 3 | Not-determined | Genome-fragment | NA | NA | 0 |
| S8_extracted_k141_121828 | 1235 | No | NA | 2 | 0 | 2 | Not-determined | Genome-fragment | NA | NA | 0 |
| S8_extracted_k141_69683 | 1164 | No | NA | 1 | 0 | 0 | Not-determined | Genome-fragment | NA | NA | 0 |
| S8_extracted_k141_99913 | 7252 | No | NA | 6 | 0 | 2 | Low-quality | Genome-fragment | 17.02 | AAI-based (high-confidence) | 0 |
| S8_extracted_k141_182436 | 4600 | No | NA | 2 | 0 | 1 | Not-determined | Genome-fragment | NA | NA | 0 |
| S8_extracted_k141_199858 | 3721 | No | NA | 4 | 0 | 3 | Not-determined | Genome-fragment | NA | NA | 0 |
| S8_extracted_k141_139178 | 4777 | No | NA | 4 | 0 | 3 | Not-determined | Genome-fragment | NA | NA | 0 |
| S8_extracted_k141_191273 | 1233 | No | NA | 2 | 0 | 0 | Not-determined | Genome-fragment | NA | NA | 0 |
| S8_extracted_k141_121839 | 1038 | No | NA | 1 | 0 | 1 | Not-determined | Genome-fragment | NA | NA | 0 |
| S16_extracted_68235 | 7258 | No | NA | 13 | 2 | 0 | Low-quality | Genome-fragment | 12.71 | AAI-based (high-confidence) | 0 |
| S8_extracted_k141_95672 | 3149 | No | NA | 4 | 0 | 3 | Not-determined | Genome-fragment | NA | NA | 0 |
| S8_extracted_k141_104422 | 7103 | No | NA | 4 | 0 | 4 | Not-determined | Genome-fragment | NA | NA | 0 |
| S8_extracted_k141_191296 | 1146 | No | NA | 2 | 0 | 1 | Not-determined | Genome-fragment | NA | NA | 0 |
| S8_extracted_k141_121852 | 7936 | No | NA | 5 | 0 | 3 | Not-determined | Genome-fragment | NA | NA | 0 |
| S12_extracted_k141_167630 | 7258 | No | NA | 9 | 1 | 2 | Low-quality | Genome-fragment | 5.82 | HMM-based (lower-bound) | 0 |
| S8_extracted_k141_147709 | 12425 | No | NA | 5 | 0 | 3 | Not-determined | Genome-fragment | NA | NA | 0 |
| S8_extracted_k141_61120 | 3073 | No | NA | 3 | 0 | 1 | Not-determined | Genome-fragment | NA | NA | 0 |
| S8_extracted_k141_121863 | 8687 | No | NA | 8 | 0 | 4 | Not-determined | Genome-fragment | NA | NA | 0 |
| S8_extracted_k141_156581 | 3856 | No | NA | 6 | 0 | 2 | Not-determined | Genome-fragment | NA | NA | 0 |
| S8_extracted_k141_95708 | 1141 | No | NA | 3 | 0 | 2 | Not-determined | Genome-fragment | NA | NA | 0 |
| S8_extracted_k141_147720 | 1016 | No | NA | 1 | 0 | 1 | Not-determined | Genome-fragment | NA | NA | 0 |
| S11_vs_k141_39674||full | 7261 | No | NA | 14 | 1 | 0 | Low-quality | Genome-fragment | 4.39 | HMM-based (lower-bound) | 0 |
| S8_extracted_k141_17576 | 3132 | No | NA | 2 | 0 | 0 | Not-determined | Genome-fragment | NA | NA | 0 |
| S3_vs_k141_30057||full | 7261 | No | NA | 9 | 7 | 0 | Low-quality | Genome-fragment | 15.46 | AAI-based (high-confidence) | 0 |
| S8_extracted_k141_130565 | 1358 | No | NA | 1 | 0 | 1 | Not-determined | Genome-fragment | NA | NA | 0 |
| S8_extracted_k141_191350 | 2175 | No | NA | 3 | 0 | 1 | Not-determined | Genome-fragment | NA | NA | 0 |
| S8_extracted_k141_87293 | 5400 | No | NA | 2 | 0 | 1 | Not-determined | Genome-fragment | NA | NA | 0 |
| S8_extracted_k141_87299 | 1375 | No | NA | 2 | 0 | 1 | Not-determined | Genome-fragment | NA | NA | 0 |
| S8_extracted_k141_87301 | 1137 | No | NA | 1 | 0 | 0 | Not-determined | Genome-fragment | NA | NA | 0 |
| S8_extracted_k141_35137 | 1053 | No | NA | 1 | 0 | 1 | Not-determined | Genome-fragment | NA | NA | 0 |
| S3_extracted_k141_72510 | 7264 | Yes | 3597 | 7 | 1 | 2 | Low-quality | Genome-fragment | 2.2 | HMM-based (lower-bound) | 50.48 |
| S8_extracted_k141_147750 | 3752 | No | NA | 4 | 0 | 2 | Not-determined | Genome-fragment | NA | NA | 0 |
| S8_extracted_k141_78548 | 4696 | No | NA | 5 | 0 | 3 | Not-determined | Genome-fragment | NA | NA | 0 |
| S8_extracted_k141_191387 | 1904 | No | NA | 3 | 0 | 2 | Not-determined | Genome-fragment | NA | NA | 0 |
| S8_extracted_k141_182535 | 1374 | No | NA | 1 | 0 | 1 | Not-determined | Genome-fragment | NA | NA | 0 |
| S8_extracted_k141_130608 | 1573 | No | NA | 2 | 0 | 1 | Not-determined | Genome-fragment | NA | NA | 0 |
| S8_extracted_k141_78557 | 1378 | No | NA | 1 | 0 | 1 | Not-determined | Genome-fragment | NA | NA | 0 |
| S8_extracted_k141_165311 | 1128 | No | NA | 1 | 0 | 1 | Not-determined | Genome-fragment | NA | NA | 0 |
| S7_extracted_128687 | 7267 | No | NA | 14 | 4 | 0 | Low-quality | Genome-fragment | 11.77 | AAI-based (high-confidence) | 0 |
| S14_vs_k141_41916||full | 7275 | No | NA | 20 | 3 | 0 | Low-quality | Genome-fragment | 5.64 | HMM-based (lower-bound) | 0 |
| S12_extracted_k141_112263 | 7278 | No | NA | 18 | 0 | 0 | Low-quality | Genome-fragment | 6.68 | AAI-based (high-confidence) | 0 |
| S8_extracted_k141_87331 | 3301 | No | NA | 3 | 0 | 1 | Not-determined | Genome-fragment | NA | NA | 0 |
| S3_extracted_k141_38921 | 7283 | No | NA | 8 | 1 | 1 | Low-quality | Genome-fragment | 9.08 | HMM-based (lower-bound) | 0 |
| S8_extracted_k141_104483 | 6389 | No | NA | 5 | 0 | 2 | Not-determined | Genome-fragment | NA | NA | 0 |
| S7_extracted_98448 | 7285 | No | NA | 9 | 6 | 0 | Low-quality | Genome-fragment | 18.94 | AAI-based (high-confidence) | 0 |
| S8_extracted_k141_69723 | 1475 | No | NA | 2 | 0 | 2 | Not-determined | Genome-fragment | NA | NA | 0 |
| S8_extracted_k141_61152 | 1025 | No | NA | 2 | 0 | 1 | Not-determined | Genome-fragment | NA | NA | 0 |
| S7_vs_6652||full | 7286 | No | NA | 11 | 10 | 0 | Low-quality | Genome-fragment | 19.26 | AAI-based (high-confidence) | 0 |
| S14_extracted_k141_18137 | 7287 | No | NA | 10 | 2 | 0 | Low-quality | Genome-fragment | 12.85 | AAI-based (high-confidence) | 0 |
| S8_extracted_k141_199970 | 2715 | No | NA | 2 | 0 | 1 | Not-determined | Genome-fragment | NA | NA | 0 |
| S8_extracted_k141_69732 | 1469 | No | NA | 3 | 0 | 1 | Not-determined | Genome-fragment | NA | NA | 0 |
| S8_extracted_k141_130630 | 1560 | No | NA | 3 | 0 | 2 | Not-determined | Genome-fragment | NA | NA | 0 |
| S8_extracted_k141_78590 | 1140 | No | NA | 2 | 0 | 0 | Not-determined | Genome-fragment | NA | NA | 0 |
| S8_extracted_k141_191429 | 1707 | No | NA | 4 | 0 | 1 | Not-determined | Genome-fragment | NA | NA | 0 |
| S8_extracted_k141_173908 | 1048 | No | NA | 2 | 0 | 0 | Not-determined | Genome-fragment | NA | NA | 0 |
| S8_extracted_k141_87354 | 3839 | No | NA | 5 | 0 | 0 | Not-determined | Genome-fragment | NA | NA | 0 |
| S8_extracted_k141_121934 | 1595 | No | NA | 2 | 0 | 1 | Not-determined | Genome-fragment | NA | NA | 0 |
| S8_extracted_k141_35188 | 4615 | No | NA | 2 | 0 | 2 | Not-determined | Genome-fragment | NA | NA | 0 |
| S8_extracted_k141_165354 | 2068 | No | NA | 2 | 0 | 1 | Not-determined | Genome-fragment | NA | NA | 0 |
| S8_extracted_k141_78600 | 3564 | No | NA | 3 | 0 | 3 | Not-determined | Genome-fragment | NA | NA | 0 |
| S8_extracted_k141_191441 | 4530 | No | NA | 6 | 0 | 6 | Not-determined | Genome-fragment | NA | NA | 0 |
| S8_extracted_k141_87365 | 1141 | No | NA | 2 | 0 | 0 | Not-determined | Genome-fragment | NA | NA | 0 |
| S8_extracted_k141_130640 | 1378 | No | NA | 1 | 0 | 1 | Not-determined | Genome-fragment | NA | NA | 0 |
| S8_extracted_k141_104512 | 12924 | No | NA | 11 | 0 | 9 | Not-determined | Genome-fragment | NA | NA | 0 |
| S8_extracted_k141_498 | 1722 | No | NA | 4 | 0 | 0 | Not-determined | Genome-fragment | NA | NA | 0 |
| S17_extracted_k141_107910 | 7292 | No | NA | 9 | 0 | 0 | Low-quality | Genome-fragment | 10.47 | AAI-based (high-confidence) | 0 |
| S8_extracted_k141_121946 | 2780 | No | NA | 3 | 0 | 1 | Not-determined | Genome-fragment | NA | NA | 0 |
| S17_vs_k141_18107||full | 7293 | No | NA | 12 | 3 | 0 | Low-quality | Genome-fragment | 9.07 | AAI-based (high-confidence) | 0 |
| S8_extracted_k141_113217 | 1129 | No | NA | 2 | 0 | 0 | Not-determined | Genome-fragment | NA | NA | 0 |
| S8_extracted_k141_43815 | 8958 | No | NA | 9 | 0 | 4 | Not-determined | Genome-fragment | NA | NA | 0 |
| S7_vs_84669||full | 7297 | No | NA | 10 | 9 | 0 | Low-quality | Genome-fragment | 21.59 | AAI-based (medium-confidence) | 0 |
| S8_extracted_k141_165383 | 1995 | No | NA | 1 | 0 | 0 | Not-determined | Genome-fragment | NA | NA | 0 |
| S8_extracted_k141_104530 | 4687 | No | NA | 5 | 0 | 3 | Not-determined | Genome-fragment | NA | NA | 0 |
| S8_extracted_k141_61169 | 1691 | No | NA | 1 | 0 | 1 | Not-determined | Genome-fragment | NA | NA | 0 |
| S8_extracted_k141_78638 | 6942 | No | NA | 4 | 0 | 3 | Not-determined | Genome-fragment | NA | NA | 0 |
| S8_extracted_k141_165399 | 1190 | No | NA | 1 | 0 | 1 | Not-determined | Genome-fragment | NA | NA | 0 |
| S8_extracted_k141_61194 | 1597 | No | NA | 3 | 0 | 0 | Not-determined | Genome-fragment | NA | NA | 0 |
| S8_extracted_k141_130678 | 1864 | No | NA | 3 | 0 | 2 | Not-determined | Genome-fragment | NA | NA | 0 |
| S8_extracted_k141_541 | 1566 | No | NA | 1 | 0 | 0 | Not-determined | Genome-fragment | NA | NA | 0 |
| S8_extracted_k141_165403 | 2225 | No | NA | 1 | 0 | 1 | Not-determined | Genome-fragment | NA | NA | 0 |
| S12_vs_k141_4868||full | 7297 | No | NA | 16 | 1 | 0 | Low-quality | Genome-fragment | 17.27 | AAI-based (high-confidence) | 0 |
| S8_extracted_k141_52564 | 1710 | No | NA | 1 | 0 | 1 | Not-determined | Genome-fragment | NA | NA | 0 |
| S8_extracted_k141_139290 | 4314 | No | NA | 3 | 0 | 1 | Not-determined | Genome-fragment | NA | NA | 0 |
| S8_extracted_k141_17656 | 1221 | No | NA | 3 | 0 | 0 | Not-determined | Genome-fragment | NA | NA | 0 |
| S8_extracted_k141_52569 | 1892 | No | NA | 2 | 0 | 2 | Not-determined | Genome-fragment | NA | NA | 0 |
| S8_extracted_k141_147849 | 1128 | No | NA | 2 | 0 | 1 | Not-determined | Genome-fragment | NA | NA | 0 |
| S8_extracted_k141_35230 | 2240 | No | NA | 1 | 0 | 1 | Not-determined | Genome-fragment | NA | NA | 0 |
| S8_extracted_k141_78656 | 1002 | No | NA | 1 | 0 | 1 | Not-determined | Genome-fragment | NA | NA | 0 |
| S8_extracted_k141_130695 | 1318 | No | NA | 2 | 0 | 0 | Not-determined | Genome-fragment | NA | NA | 0 |
| S8_extracted_k141_139314 | 2835 | No | NA | 3 | 0 | 0 | Not-determined | Genome-fragment | NA | NA | 0 |
| S8_extracted_k141_173938 | 1238 | No | NA | 1 | 0 | 0 | Not-determined | Genome-fragment | NA | NA | 0 |
| S8_extracted_k141_35247 | 1138 | No | NA | 2 | 0 | 2 | Not-determined | Genome-fragment | NA | NA | 0 |
| S11_vs_k141_35881||full | 7299 | No | NA | 13 | 10 | 0 | Low-quality | Genome-fragment | 23.99 | AAI-based (high-confidence) | 0 |
| S8_extracted_k141_147871 | 1492 | No | NA | 1 | 0 | 1 | Not-determined | Genome-fragment | NA | NA | 0 |
| S8_extracted_k141_52599 | 1045 | No | NA | 1 | 0 | 1 | Not-determined | Genome-fragment | NA | NA | 0 |
| S16_vs_19445||full | 7299 | No | NA | 11 | 3 | 0 | Low-quality | Genome-fragment | 7.31 | HMM-based (lower-bound) | 0 |
| S5_vs_k141_6814||full | 7312 | No | NA | 16 | 2 | 1 | Low-quality | Genome-fragment | 14.98 | AAI-based (medium-confidence) | 0 |
| S8_extracted_k141_26520 | 1715 | No | NA | 2 | 0 | 0 | Not-determined | Genome-fragment | NA | NA | 0 |
| S15_extracted_41582 | 7317 | No | NA | 13 | 5 | 0 | Low-quality | Genome-fragment | 14.16 | AAI-based (high-confidence) | 0 |
| S8_extracted_k141_61252 | 1284 | No | NA | 1 | 0 | 1 | Not-determined | Genome-fragment | NA | NA | 0 |
| S8_extracted_k141_17680 | 1797 | No | NA | 4 | 0 | 2 | Not-determined | Genome-fragment | NA | NA | 0 |
| S8_extracted_k141_191533 | 3293 | No | NA | 3 | 0 | 2 | Not-determined | Genome-fragment | NA | NA | 0 |
| S8_extracted_k141_182555 | 1045 | No | NA | 1 | 0 | 0 | Not-determined | Genome-fragment | NA | NA | 0 |
| S8_extracted_k141_165457 | 2080 | No | NA | 2 | 0 | 2 | Not-determined | Genome-fragment | NA | NA | 0 |
| S8_extracted_k141_69773 | 1086 | No | NA | 3 | 0 | 0 | Not-determined | Genome-fragment | NA | NA | 0 |
| S8_extracted_k141_122921 | 7317 | No | NA | 7 | 5 | 0 | Low-quality | Genome-fragment | 21.5 | AAI-based (high-confidence) | 0 |
| S8_extracted_k141_61264 | 1044 | No | NA | 3 | 0 | 0 | Not-determined | Genome-fragment | NA | NA | 0 |
| S8_extracted_k141_35281 | 2239 | No | NA | 5 | 0 | 2 | Not-determined | Genome-fragment | NA | NA | 0 |
| S8_extracted_k141_61288 | 1143 | No | NA | 1 | 0 | 1 | Not-determined | Genome-fragment | NA | NA | 0 |
| S3_vs_k141_69090||full | 7319 | No | NA | 15 | 1 | 0 | Low-quality | Genome-fragment | 11.76 | AAI-based (medium-confidence) | 0 |
| S8_extracted_k141_607 | 1308 | No | NA | 3 | 0 | 1 | Not-determined | Genome-fragment | NA | NA | 0 |
| S8_extracted_k141_52631 | 1393 | No | NA | 2 | 0 | 2 | Not-determined | Genome-fragment | NA | NA | 0 |
| S8_extracted_k141_182593 | 4098 | No | NA | 4 | 0 | 3 | Not-determined | Genome-fragment | NA | NA | 0 |
| S8_extracted_k141_122048 | 1267 | No | NA | 1 | 0 | 1 | Not-determined | Genome-fragment | NA | NA | 0 |
| S8_extracted_k141_61297 | 1035 | No | NA | 2 | 0 | 0 | Not-determined | Genome-fragment | NA | NA | 0 |
| S8_extracted_k141_61322 | 1267 | No | NA | 2 | 0 | 2 | Not-determined | Genome-fragment | NA | NA | 0 |
| S8_extracted_k141_35307 | 4418 | No | NA | 4 | 0 | 0 | Not-determined | Genome-fragment | NA | NA | 0 |
| S8_extracted_k141_199989 | 1409 | No | NA | 1 | 0 | 1 | Not-determined | Genome-fragment | NA | NA | 0 |
| S8_extracted_k141_122069 | 3321 | No | NA | 3 | 0 | 1 | Not-determined | Genome-fragment | NA | NA | 0 |
| S8_extracted_k141_165511 | 1042 | No | NA | 1 | 0 | 1 | Not-determined | Genome-fragment | NA | NA | 0 |
| S8_extracted_k141_139374 | 3443 | No | NA | 5 | 0 | 2 | Not-determined | Genome-fragment | NA | NA | 0 |
| S8_extracted_k141_17731 | 1402 | No | NA | 2 | 0 | 2 | Not-determined | Genome-fragment | NA | NA | 0 |
| S8_extracted_k141_130734 | 1173 | No | NA | 2 | 0 | 1 | Not-determined | Genome-fragment | NA | NA | 0 |
| S8_extracted_k141_182612 | 1135 | No | NA | 2 | 0 | 0 | Not-determined | Genome-fragment | NA | NA | 0 |
| S8_extracted_k141_9252 | 11430 | No | NA | 13 | 0 | 2 | Not-determined | Genome-fragment | NA | NA | 0 |
| S8_extracted_k141_165526 | 1179 | No | NA | 2 | 0 | 0 | Not-determined | Genome-fragment | NA | NA | 0 |
| S8_extracted_k141_69835 | 1818 | No | NA | 3 | 0 | 3 | Not-determined | Genome-fragment | NA | NA | 0 |
| S8_extracted_k141_182624 | 1868 | No | NA | 2 | 0 | 1 | Not-determined | Genome-fragment | NA | NA | 0 |
| S8_extracted_k141_78666 | 1052 | No | NA | 1 | 0 | 1 | Not-determined | Genome-fragment | NA | NA | 0 |
| S8_extracted_k141_52694 | 1762 | No | NA | 1 | 0 | 1 | Not-determined | Genome-fragment | NA | NA | 0 |
| S8_extracted_k141_122102 | 3994 | No | NA | 4 | 0 | 0 | Not-determined | Genome-fragment | NA | NA | 0 |
| S8_extracted_k141_69843 | 3650 | No | NA | 8 | 0 | 2 | Not-determined | Genome-fragment | NA | NA | 0 |
| S8_extracted_k141_664 | 1953 | No | NA | 3 | 0 | 3 | Not-determined | Genome-fragment | NA | NA | 0 |
| S5_extracted_k141_6186 | 7320 | No | NA | 12 | 1 | 1 | Low-quality | Genome-fragment | 3.97 | HMM-based (lower-bound) | 0 |
| S8_extracted_k141_52696 | 1095 | No | NA | 1 | 0 | 0 | Not-determined | Genome-fragment | NA | NA | 0 |
| S14_vs_k141_53020||full | 7323 | No | NA | 9 | 2 | 0 | Low-quality | Genome-fragment | 16.56 | AAI-based (high-confidence) | 0 |
| S8_extracted_k141_113284 | 1134 | No | NA | 2 | 0 | 1 | Not-determined | Genome-fragment | NA | NA | 0 |
| S7_extracted_130522 | 7327 | No | NA | 10 | 1 | 0 | Low-quality | Genome-fragment | 15.2 | AAI-based (high-confidence) | 0 |
| S8_vs_k141_42738||full | 7330 | No | NA | 5 | 0 | 4 | Low-quality | Genome-fragment | 5.04 | AAI-based (medium-confidence) | 0 |
| S8_extracted_k141_104706 | 1261 | No | NA | 1 | 0 | 0 | Not-determined | Genome-fragment | NA | NA | 0 |
| S13_vs_104066||full | 7331 | Yes | 1718 | 5 | 1 | 4 | Low-quality | Genome-fragment | 1.06 | HMM-based (lower-bound) | 76.57 |
| S14_extracted_k141_85050 | 7331 | No | NA | 15 | 1 | 0 | Low-quality | Genome-fragment | 13.32 | AAI-based (high-confidence) | 0 |
| S8_extracted_k141_148006 | 1042 | No | NA | 2 | 0 | 1 | Not-determined | Genome-fragment | NA | NA | 0 |
| S8_extracted_k141_26550 | 67068 | No | NA | 81 | 0 | 48 | Not-determined | Genome-fragment | NA | NA | 0 |
| S8_extracted_k141_35310 | 6012 | No | NA | 3 | 0 | 0 | Not-determined | Genome-fragment | NA | NA | 0 |
| S8_extracted_k141_26552 | 1540 | No | NA | 4 | 0 | 1 | Not-determined | Genome-fragment | NA | NA | 0 |
| S8_extracted_k141_35311 | 1180 | No | NA | 1 | 0 | 1 | Not-determined | Genome-fragment | NA | NA | 0 |
| S8_extracted_k141_78679 | 2541 | No | NA | 3 | 0 | 3 | Not-determined | Genome-fragment | NA | NA | 0 |
| S8_extracted_k141_95957 | 1367 | No | NA | 3 | 0 | 2 | Not-determined | Genome-fragment | NA | NA | 0 |
| S8_extracted_k141_165556 | 1399 | No | NA | 2 | 0 | 0 | Not-determined | Genome-fragment | NA | NA | 0 |
| S8_extracted_k141_148023 | 1083 | No | NA | 1 | 0 | 1 | Not-determined | Genome-fragment | NA | NA | 0 |
| S8_extracted_k141_681 | 4498 | No | NA | 5 | 0 | 4 | Not-determined | Genome-fragment | NA | NA | 0 |
| S8_extracted_k141_26560 | 6151 | No | NA | 7 | 0 | 3 | Not-determined | Genome-fragment | NA | NA | 0 |
| S8_extracted_k141_17782 | 7805 | No | NA | 10 | 0 | 9 | Not-determined | Genome-fragment | NA | NA | 0 |
| S8_extracted_k141_113314 | 1103 | No | NA | 2 | 0 | 1 | Not-determined | Genome-fragment | NA | NA | 0 |
| S8_extracted_k141_87522 | 1026 | No | NA | 2 | 0 | 1 | Not-determined | Genome-fragment | NA | NA | 0 |
| S8_extracted_k141_148039 | 1676 | No | NA | 2 | 0 | 2 | Not-determined | Genome-fragment | NA | NA | 0 |
| S8_extracted_k141_122149 | 6080 | No | NA | 6 | 0 | 4 | Not-determined | Genome-fragment | NA | NA | 0 |
| S11_vs_k141_52665||full | 7338 | No | NA | 15 | 1 | 0 | Low-quality | Genome-fragment | 4.77 | AAI-based (high-confidence) | 0 |
| S8_vs_k141_195615||full | 7338 | No | NA | 17 | 1 | 1 | Low-quality | Genome-fragment | 11.65 | AAI-based (high-confidence) | 0 |
| S8_extracted_k141_52744 | 1437 | No | NA | 1 | 0 | 1 | Not-determined | Genome-fragment | NA | NA | 0 |
| S8_extracted_k141_17802 | 1186 | No | NA | 1 | 0 | 0 | Not-determined | Genome-fragment | NA | NA | 0 |
| S8_extracted_k141_26587 | 3280 | No | NA | 3 | 0 | 1 | Not-determined | Genome-fragment | NA | NA | 0 |
| S8_extracted_k141_69911 | 1852 | No | NA | 2 | 0 | 1 | Not-determined | Genome-fragment | NA | NA | 0 |
| S8_extracted_k141_122159 | 4030 | No | NA | 2 | 0 | 2 | Not-determined | Genome-fragment | NA | NA | 0 |
| S8_extracted_k141_165604 | 4976 | No | NA | 3 | 0 | 3 | Not-determined | Genome-fragment | NA | NA | 0 |
| S8_extracted_k141_130838 | 1971 | No | NA | 2 | 0 | 1 | Not-determined | Genome-fragment | NA | NA | 0 |
| S8_extracted_k141_122170 | 2248 | No | NA | 2 | 0 | 1 | Not-determined | Genome-fragment | NA | NA | 0 |
| S8_extracted_k141_200083 | 9165 | No | NA | 11 | 0 | 1 | Not-determined | Genome-fragment | NA | NA | 0 |
| S8_extracted_k141_17820 | 1047 | No | NA | 2 | 0 | 1 | Not-determined | Genome-fragment | NA | NA | 0 |
| S8_extracted_k141_35349 | 5645 | No | NA | 5 | 0 | 4 | Not-determined | Genome-fragment | NA | NA | 0 |
| S8_extracted_k141_130843 | 2225 | No | NA | 1 | 0 | 1 | Not-determined | Genome-fragment | NA | NA | 0 |
| S8_extracted_k141_113355 | 1246 | No | NA | 1 | 0 | 1 | Not-determined | Genome-fragment | NA | NA | 0 |
| S8_extracted_k141_113358 | 1180 | No | NA | 1 | 0 | 1 | Not-determined | Genome-fragment | NA | NA | 0 |
| S8_extracted_k141_96009 | 1307 | No | NA | 2 | 0 | 2 | Not-determined | Genome-fragment | NA | NA | 0 |
| S8_extracted_k141_35358 | 2233 | No | NA | 1 | 0 | 0 | Not-determined | Genome-fragment | NA | NA | 0 |
| S8_extracted_k141_730 | 3833 | No | NA | 5 | 0 | 4 | Not-determined | Genome-fragment | NA | NA | 0 |
| S8_extracted_k141_26606 | 1218 | No | NA | 1 | 0 | 0 | Not-determined | Genome-fragment | NA | NA | 0 |
| S8_extracted_k141_191665 | 1200 | No | NA | 3 | 0 | 0 | Not-determined | Genome-fragment | NA | NA | 0 |
| S8_extracted_k141_139479 | 1086 | No | NA | 2 | 0 | 2 | Not-determined | Genome-fragment | NA | NA | 0 |
| S3_vs_k141_18675||full | 7340 | No | NA | 16 | 4 | 0 | Low-quality | Genome-fragment | 17.21 | AAI-based (high-confidence) | 0 |
| S1_extracted_k141_96449 | 7341 | No | NA | 9 | 3 | 0 | Low-quality | Genome-fragment | 12.82 | AAI-based (high-confidence) | 0 |
| S12_extracted_k141_104489 | 7341 | No | NA | 8 | 0 | 1 | Low-quality | Genome-fragment | 11.74 | AAI-based (medium-confidence) | 0 |
| S8_extracted_k141_156705 | 7354 | No | NA | 11 | 4 | 0 | Low-quality | Genome-fragment | 15.8 | AAI-based (medium-confidence) | 0 |
| S8_extracted_k141_61437 | 4549 | No | NA | 2 | 0 | 1 | Not-determined | Genome-fragment | NA | NA | 0 |
| S8_extracted_k141_26614 | 1777 | No | NA | 1 | 0 | 1 | Not-determined | Genome-fragment | NA | NA | 0 |
| S16_extracted_54256 | 7355 | No | NA | 11 | 4 | 0 | Low-quality | Genome-fragment | 17.94 | AAI-based (high-confidence) | 0 |
| S8_extracted_k141_760 | 1091 | No | NA | 2 | 0 | 2 | Not-determined | Genome-fragment | NA | NA | 0 |
| S8_extracted_k141_96039 | 1576 | No | NA | 2 | 0 | 1 | Not-determined | Genome-fragment | NA | NA | 0 |
| S8_vs_k141_34691||full | 7358 | No | NA | 3 | 2 | 0 | Low-quality | Genome-fragment | 6.19 | HMM-based (lower-bound) | 0 |
| S8_extracted_k141_43949 | 2027 | No | NA | 1 | 0 | 1 | Not-determined | Genome-fragment | NA | NA | 0 |
| S8_extracted_k141_9333 | 1091 | No | NA | 1 | 0 | 0 | Not-determined | Genome-fragment | NA | NA | 0 |

| | | | | | | | | | | | |
|---|---|---|---|---|---|---|---|---|---|---|---|
| S5_vs_k141_75023\|\|full | 7361 | No | NA | 17 | 1 | 0 | Low-quality | Genome-fragment | 9.69 | AAI-based (high-confidence) | 0 |
| S8_extracted_k141_773 | 1242 | No | NA | 1 | 0 | 1 | Not-determined | Genome-fragment | NA | NA | 0 |
| S8_extracted_k141_26641 | 1890 | No | NA | 1 | 0 | 1 | Not-determined | Genome-fragment | NA | NA | 0 |
| S8_extracted_k141_26643 | 2568 | No | NA | 3 | 0 | 0 | Not-determined | Genome-fragment | NA | NA | 0 |
| S8_extracted_k141_43978 | 1368 | No | NA | 2 | 0 | 1 | Not-determined | Genome-fragment | NA | NA | 0 |
| S13_extracted_41163 | 7374 | No | NA | 10 | 3 | 0 | Low-quality | Genome-fragment | 16.29 | AAI-based (high-confidence) | 0 |
| S8_extracted_k141_156746 | 1068 | No | NA | 1 | 0 | 1 | Not-determined | Genome-fragment | NA | NA | 0 |
| S8_extracted_k141_191703 | 2205 | No | NA | 1 | 0 | 1 | Not-determined | Genome-fragment | NA | NA | 0 |
| S3_vs_k141_112884\|\|full | 7377 | Yes | 3942 | 6 | 1 | 1 | Low-quality | Genome-fragment | 5.9 | HMM-based (lower-bound) | 46.56 |
| S8_extracted_k141_52821 | 6201 | No | NA | 5 | 0 | 1 | Not-determined | Genome-fragment | NA | NA | 0 |
| S5_extracted_k141_62218 | 7378 | No | NA | 7 | 0 | 2 | Low-quality | Genome-fragment | 7.8 | AAI-based (high-confidence) | 0 |
| S8_extracted_k141_104769 | 1099 | No | NA | 2 | 0 | 2 | Not-determined | Genome-fragment | NA | NA | 0 |
| S8_extracted_k141_78810 | 1574 | No | NA | 2 | 0 | 2 | Not-determined | Genome-fragment | NA | NA | 0 |
| S8_extracted_k141_122206 | 56408 | Yes | 54016 | 70 | 12 | 4 | High-quality | High-quality | 100 | AAI-based (high-confidence) | 4.24 |
| S8_extracted_k141_139544 | 1391 | No | NA | 2 | 0 | 1 | Not-determined | Genome-fragment | NA | NA | 0 |
| S8_extracted_k141_156773 | 1676 | No | NA | 3 | 0 | 0 | Not-determined | Genome-fragment | NA | NA | 0 |
| S8_extracted_k141_165678 | 1150 | No | NA | 2 | 0 | 2 | Not-determined | Genome-fragment | NA | NA | 0 |
| S8_extracted_k141_26687 | 1791 | No | NA | 1 | 0 | 1 | Not-determined | Genome-fragment | NA | NA | 0 |
| S8_extracted_k141_61503 | 2630 | No | NA | 2 | 0 | 2 | Not-determined | Genome-fragment | NA | NA | 0 |
| S8_extracted_k141_52845 | 1059 | No | NA | 1 | 0 | 0 | Not-determined | Genome-fragment | NA | NA | 0 |
| S8_extracted_k141_182741 | 1380 | No | NA | 1 | 0 | 1 | Not-determined | Genome-fragment | NA | NA | 0 |
| S8_extracted_k141_70005 | 1914 | No | NA | 2 | 0 | 1 | Not-determined | Genome-fragment | NA | NA | 0 |
| S8_extracted_k141_191743 | 1197 | No | NA | 1 | 0 | 1 | Not-determined | Genome-fragment | NA | NA | 0 |
| S8_extracted_k141_44033 | 1210 | No | NA | 2 | 0 | 0 | Not-determined | Genome-fragment | NA | NA | 0 |
| S8_extracted_k141_96073 | 1194 | No | NA | 3 | 0 | 0 | Not-determined | Genome-fragment | NA | NA | 0 |
| S14_vs_k141_48829\|\|full | 7379 | No | NA | 11 | 8 | 0 | Low-quality | Genome-fragment | 19.06 | AAI-based (high-confidence) | 0 |
| S8_extracted_k141_96076 | 1225 | No | NA | 1 | 0 | 1 | Not-determined | Genome-fragment | NA | NA | 0 |
| S8_extracted_k141_44044 | 1128 | No | NA | 2 | 0 | 2 | Not-determined | Genome-fragment | NA | NA | 0 |
| S8_extracted_k141_130963 | 1661 | No | NA | 1 | 0 | 1 | Not-determined | Genome-fragment | NA | NA | 0 |
| S8_extracted_k141_35408 | 1272 | No | NA | 1 | 0 | 0 | Not-determined | Genome-fragment | NA | NA | 0 |
| S8_extracted_k141_17945 | 1465 | No | NA | 2 | 0 | 2 | Not-determined | Genome-fragment | NA | NA | 0 |
| S8_extracted_k141_17950 | 1334 | No | NA | 1 | 0 | 1 | Not-determined | Genome-fragment | NA | NA | 0 |
| S8_extracted_k141_200124 | 7637 | No | NA | 12 | 0 | 0 | Not-determined | Genome-fragment | NA | NA | 0 |
| S8_extracted_k141_139599 | 8774 | No | NA | 9 | 0 | 1 | Not-determined | Genome-fragment | NA | NA | 0 |
| S8_extracted_k141_87698 | 1106 | No | NA | 1 | 0 | 1 | Not-determined | Genome-fragment | NA | NA | 0 |
| S8_extracted_k141_9439 | 4055 | No | NA | 5 | 0 | 2 | Not-determined | Genome-fragment | NA | NA | 0 |
| S11_vs_k141_63764\|\|full | 7381 | No | NA | 9 | 2 | 0 | Low-quality | Genome-fragment | 17.75 | AAI-based (high-confidence) | 0 |
| S8_extracted_k141_70055 | 4117 | No | NA | 3 | 0 | 1 | Not-determined | Genome-fragment | NA | NA | 0 |
| S8_extracted_k141_139610 | 5719 | No | NA | 6 | 0 | 6 | Not-determined | Genome-fragment | NA | NA | 0 |
| S8_extracted_k141_1673 | 7382 | No | NA | 8 | 0 | 6 | Low-quality | Genome-fragment | 5.91 | AAI-based (high-confidence) | 0 |
| S12_extracted_k141_54321 | 7388 | No | NA | 3 | 0 | 0 | Low-quality | Genome-fragment | 4.37 | AAI-based (medium-confidence) | 0 |
| S8_extracted_k141_61521 | 1355 | No | NA | 1 | 0 | 1 | Not-determined | Genome-fragment | NA | NA | 0 |
| S8_extracted_k141_122305 | 3564 | No | NA | 7 | 0 | 2 | Not-determined | Genome-fragment | NA | NA | 0 |
| S8_extracted_k141_52858 | 31668 | No | NA | 18 | 0 | 10 | Not-determined | Genome-fragment | NA | NA | 0 |
| S8_extracted_k141_200148 | 1454 | No | NA | 1 | 0 | 0 | Not-determined | Genome-fragment | NA | NA | 0 |
| S8_extracted_k141_122320 | 2557 | No | NA | 1 | 0 | 1 | Not-determined | Genome-fragment | NA | NA | 0 |
| S8_extracted_k141_78916 | 1400 | No | NA | 2 | 0 | 2 | Not-determined | Genome-fragment | NA | NA | 0 |
| S8_extracted_k141_26782 | 1498 | No | NA | 1 | 0 | 0 | Not-determined | Genome-fragment | NA | NA | 0 |
| S8_extracted_k141_191798 | 3240 | No | NA | 4 | 0 | 0 | Not-determined | Genome-fragment | NA | NA | 0 |
| S8_extracted_k141_61538 | 2563 | No | NA | 4 | 0 | 1 | Not-determined | Genome-fragment | NA | NA | 0 |
| S8_extracted_k141_148201 | 3003 | No | NA | 2 | 0 | 1 | Not-determined | Genome-fragment | NA | NA | 0 |
| S16_vs_64092\|\|full | 7394 | No | NA | 12 | 1 | 0 | Low-quality | Genome-fragment | 6.91 | AAI-based (high-confidence) | 0 |
| S8_extracted_k141_26805 | 2723 | No | NA | 1 | 0 | 1 | Not-determined | Genome-fragment | NA | NA | 0 |
| S8_extracted_k141_52890 | 2522 | No | NA | 1 | 0 | 0 | Not-determined | Genome-fragment | NA | NA | 0 |
| S8_extracted_k141_182814 | 1587 | No | NA | 4 | 0 | 0 | Not-determined | Genome-fragment | NA | NA | 0 |
| S8_extracted_k141_9485 | 1408 | No | NA | 2 | 0 | 1 | Not-determined | Genome-fragment | NA | NA | 0 |
| S8_extracted_k141_44174 | 3038 | No | NA | 2 | 0 | 1 | Not-determined | Genome-fragment | NA | NA | 0 |
| S8_extracted_k141_35506 | 2810 | No | NA | 2 | 0 | 2 | Not-determined | Genome-fragment | NA | NA | 0 |
| S8_extracted_k141_26823 | 3255 | No | NA | 4 | 0 | 1 | Not-determined | Genome-fragment | NA | NA | 0 |
| S11_vs_k141_72602\|\|full | 7395 | No | NA | 12 | 3 | 0 | Low-quality | Genome-fragment | 4.78 | AAI-based (high-confidence) | 0 |
| S8_extracted_k141_131009 | 57372 | No | NA | 94 | 18 | 1 | Complete | High-quality | 100 | DTR (high-confidence) | 0 |
| S3_extracted_k141_119611 | 7395 | No | NA | 9 | 2 | 0 | Low-quality | Genome-fragment | 5.97 | AAI-based (high-confidence) | 0 |
| S8_extracted_k141_96198 | 1227 | No | NA | 2 | 0 | 1 | Not-determined | Genome-fragment | NA | NA | 0 |
| S16_extracted_88409 | 7400 | No | NA | 4 | 0 | 3 | Low-quality | Genome-fragment | 0.64 | AAI-based (medium-confidence) | 0 |
| S8_extracted_k141_44203 | 1576 | No | NA | 2 | 0 | 2 | Not-determined | Genome-fragment | NA | NA | 0 |
| S8_extracted_k141_964 | 1417 | No | NA | 2 | 0 | 2 | Not-determined | Genome-fragment | NA | NA | 0 |
| S8_extracted_k141_156881 | 2037 | No | NA | 1 | 0 | 0 | Not-determined | Genome-fragment | NA | NA | 0 |
| S8_extracted_k141_148244 | 1911 | No | NA | 2 | 0 | 1 | Not-determined | Genome-fragment | NA | NA | 0 |
| S7_vs_30348\|\|full | 7401 | No | NA | 5 | 1 | 0 | Low-quality | Genome-fragment | 17.91 | AAI-based (medium-confidence) | 0 |
| S8_extracted_k141_52902 | 11366 | No | NA | 13 | 0 | 3 | Not-determined | Genome-fragment | NA | NA | 0 |
| S8_extracted_k141_148252 | 1198 | No | NA | 1 | 0 | 0 | Not-determined | Genome-fragment | NA | NA | 0 |
| S8_extracted_k141_104902 | 1024 | No | NA | 1 | 0 | 0 | Not-determined | Genome-fragment | NA | NA | 0 |
| S3_vs_k141_53711\|\|full | 7403 | No | NA | 7 | 4 | 0 | Low-quality | Genome-fragment | 17.65 | AAI-based (high-confidence) | 0 |
| S8_extracted_k141_182853 | 1077 | No | NA | 1 | 0 | 0 | Not-determined | Genome-fragment | NA | NA | 0 |
| S8_extracted_k141_131059 | 1026 | No | NA | 2 | 0 | 0 | Not-determined | Genome-fragment | NA | NA | 0 |
| S11_vs_k141_17254\|\|full | 7404 | No | NA | 14 | 1 | 0 | Low-quality | Genome-fragment | 18.43 | HMM-based (lower-bound) | 0 |
| S8_extracted_k141_104908 | 4360 | No | NA | 6 | 0 | 0 | Not-determined | Genome-fragment | NA | NA | 0 |
| S8_extracted_k141_122408 | 6158 | No | NA | 1 | 0 | 1 | Not-determined | Genome-fragment | NA | NA | 0 |
| S8_extracted_k141_87755 | 54718 | No | NA | 59 | 4 | 0 | Medium-quality | Genome-fragment | 51.78 | AAI-based (high-confidence) | 0 |
| S8_extracted_k141_182864 | 2856 | No | NA | 4 | 0 | 1 | Not-determined | Genome-fragment | NA | NA | 0 |
| S8_extracted_k141_61629 | 4274 | No | NA | 3 | 0 | 1 | Not-determined | Genome-fragment | NA | NA | 0 |
| S8_extracted_k141_52925 | 2032 | No | NA | 3 | 0 | 1 | Not-determined | Genome-fragment | NA | NA | 0 |
| S8_extracted_k141_104920 | 1005 | No | NA | 1 | 0 | 1 | Not-determined | Genome-fragment | NA | NA | 0 |
| S8_extracted_k141_191924 | 3252 | No | NA | 6 | 0 | 0 | Not-determined | Genome-fragment | NA | NA | 0 |
| S8_extracted_k141_9553 | 2044 | No | NA | 1 | 0 | 1 | Not-determined | Genome-fragment | NA | NA | 0 |
| S8_extracted_k141_998 | 1893 | No | NA | 3 | 0 | 0 | Not-determined | Genome-fragment | NA | NA | 0 |
| S8_extracted_k141_131094 | 1069 | No | NA | 1 | 0 | 1 | Not-determined | Genome-fragment | NA | NA | 0 |
| S8_extracted_k141_9562 | 1303 | No | NA | 2 | 0 | 2 | Not-determined | Genome-fragment | NA | NA | 0 |
| S8_extracted_k141_26921 | 1066 | No | NA | 1 | 0 | 1 | Not-determined | Genome-fragment | NA | NA | 0 |
| S8_extracted_k141_200183 | 1057 | No | NA | 1 | 0 | 1 | Not-determined | Genome-fragment | NA | NA | 0 |
| S8_extracted_k141_191946 | 1379 | No | NA | 2 | 0 | 2 | Not-determined | Genome-fragment | NA | NA | 0 |
| S8_extracted_k141_191952 | 1003 | No | NA | 2 | 0 | 1 | Not-determined | Genome-fragment | NA | NA | 0 |
| S8_extracted_k141_200190 | 1010 | No | NA | 1 | 0 | 1 | Not-determined | Genome-fragment | NA | NA | 0 |
| S8_extracted_k141_113578 | 2355 | No | NA | 2 | 0 | 2 | Not-determined | Genome-fragment | NA | NA | 0 |
| S8_extracted_k141_113581 | 1010 | No | NA | 3 | 0 | 0 | Not-determined | Genome-fragment | NA | NA | 0 |
| S8_extracted_k141_35576 | 1159 | No | NA | 3 | 0 | 0 | Not-determined | Genome-fragment | NA | NA | 0 |
| S8_extracted_k141_148317 | 2528 | No | NA | 1 | 0 | 1 | Not-determined | Genome-fragment | NA | NA | 0 |
| S8_extracted_k141_139758 | 1165 | No | NA | 2 | 0 | 2 | Not-determined | Genome-fragment | NA | NA | 0 |
| S8_extracted_k141_44225 | 1446 | No | NA | 1 | 0 | 0 | Not-determined | Genome-fragment | NA | NA | 0 |
| S8_extracted_k141_96289 | 2049 | No | NA | 3 | 0 | 1 | Not-determined | Genome-fragment | NA | NA | 0 |
| S8_extracted_k141_131118 | 10267 | No | NA | 6 | 0 | 3 | Not-determined | Genome-fragment | NA | NA | 0 |
| S9_extracted_k141_18895 | 7405 | No | NA | 9 | 0 | 2 | Low-quality | Genome-fragment | 5.61 | AAI-based (high-confidence) | 0 |
| S8_extracted_k141_156941 | 3569 | No | NA | 1 | 0 | 1 | Not-determined | Genome-fragment | NA | NA | 0 |
| S8_extracted_k141_79057 | 1051 | No | NA | 1 | 0 | 1 | Not-determined | Genome-fragment | NA | NA | 0 |
| S8_extracted_k141_26944 | 8408 | No | NA | 2 | 0 | 2 | Not-determined | Genome-fragment | NA | NA | 0 |
| S17_vs_k141_174131\|\|full | 7409 | No | NA | 16 | 2 | 1 | Low-quality | Genome-fragment | 11.64 | AAI-based (high-confidence) | 0 |
| S8_extracted_k141_156956 | 1704 | No | NA | 2 | 0 | 0 | Not-determined | Genome-fragment | NA | NA | 0 |
| S14_extracted_k141_114920 | 7413 | No | NA | 9 | 9 | 0 | Low-quality | Genome-fragment | 24.45 | AAI-based (high-confidence) | 0 |
| S8_extracted_k141_44238 | 5543 | No | NA | 4 | 0 | 3 | Not-determined | Genome-fragment | NA | NA | 0 |
| S8_extracted_k141_87794 | 1510 | No | NA | 4 | 0 | 1 | Not-determined | Genome-fragment | NA | NA | 0 |
| S3_extracted_k141_91188 | 7416 | No | NA | 13 | 1 | 1 | Low-quality | Genome-fragment | 15.76 | AAI-based (high-confidence) | 0 |
| S8_extracted_k141_113608 | 2435 | No | NA | 2 | 0 | 2 | Not-determined | Genome-fragment | NA | NA | 0 |
| S8_extracted_k141_113611 | 1638 | No | NA | 1 | 0 | 1 | Not-determined | Genome-fragment | NA | NA | 0 |
| S8_extracted_k141_122468 | 1387 | No | NA | 1 | 0 | 0 | Not-determined | Genome-fragment | NA | NA | 0 |
| S8_extracted_k141_104981 | 3288 | No | NA | 6 | 0 | 1 | Not-determined | Genome-fragment | NA | NA | 0 |
| S3_extracted_k141_38857 | 7424 | No | NA | 11 | 6 | 0 | Low-quality | Genome-fragment | 15.77 | AAI-based (high-confidence) | 0 |
| S8_extracted_k141_200200 | 1003 | No | NA | 1 | 0 | 0 | Not-determined | Genome-fragment | NA | NA | 0 |
| S5_extracted_k141_26612 | 7425 | No | NA | 14 | 1 | 1 | Low-quality | Genome-fragment | 4.03 | HMM-based (lower-bound) | 0 |
| S8_extracted_k141_113616 | 8971 | No | NA | 8 | 0 | 6 | Not-determined | Genome-fragment | NA | NA | 0 |
| S8_extracted_k141_131153 | 3245 | No | NA | 3 | 0 | 1 | Not-determined | Genome-fragment | NA | NA | 0 |
| S8_extracted_k141_52991 | 1201 | No | NA | 1 | 0 | 1 | Not-determined | Genome-fragment | NA | NA | 0 |
| S8_extracted_k141_157000 | 1143 | No | NA | 2 | 0 | 0 | Not-determined | Genome-fragment | NA | NA | 0 |
| S8_extracted_k141_104998 | 1423 | No | NA | 2 | 0 | 0 | Not-determined | Genome-fragment | NA | NA | 0 |
| S8_extracted_k141_61716 | 1081 | No | NA | 2 | 0 | 0 | Not-determined | Genome-fragment | NA | NA | 0 |
| S8_extracted_k141_87822 | 1139 | No | NA | 3 | 0 | 0 | Not-determined | Genome-fragment | NA | NA | 0 |
| S6_vs_k141_5449\|\|full | 7432 | No | NA | 10 | 7 | 0 | Low-quality | Genome-fragment | 15.58 | AAI-based (medium-confidence) | 0 |
| S8_extracted_k141_200210 | 9596 | No | NA | 8 | 0 | 3 | Not-determined | Genome-fragment | NA | NA | 0 |
| S8_extracted_k141_165814 | 3151 | No | NA | 3 | 0 | 1 | Not-determined | Genome-fragment | NA | NA | 0 |
| S12_vs_k141_24971\|\|full | 7433 | No | NA | 9 | 1 | 0 | Low-quality | Genome-fragment | 1.37 | HMM-based (lower-bound) | 0 |
| S8_extracted_k141_87827 | 3051 | No | NA | 8 | 0 | 8 | Not-determined | Genome-fragment | NA | NA | 0 |
| S5_vs_k141_48126\|\|full | 7435 | No | NA | 7 | 2 | 0 | Low-quality | Genome-fragment | 15.47 | AAI-based (high-confidence) | 0 |
| S8_extracted_k141_182925 | 1069 | No | NA | 2 | 0 | 0 | Not-determined | Genome-fragment | NA | NA | 0 |
| S8_extracted_k141_44274 | 1193 | No | NA | 1 | 0 | 1 | Not-determined | Genome-fragment | NA | NA | 0 |
| S8_extracted_k141_79102 | 1379 | No | NA | 2 | 0 | 1 | Not-determined | Genome-fragment | NA | NA | 0 |
| S8_extracted_k141_9577 | 2100 | No | NA | 3 | 0 | 2 | Not-determined | Genome-fragment | NA | NA | 0 |
| S17_vs_k141_14999\|\|full | 7438 | No | NA | 12 | 1 | 0 | Low-quality | Genome-fragment | 11.92 | AAI-based (medium-confidence) | 0 |
| S10_vs_57300\|\|full | 7444 | No | NA | 5 | 3 | 1 | Low-quality | Genome-fragment | 3.32 | HMM-based (lower-bound) | 0 |
| S14_vs_k141_7734\|\|full | 7448 | No | NA | 6 | 2 | 0 | Low-quality | Genome-fragment | 12.63 | HMM-based (lower-bound) | 0 |
| S8_extracted_k141_174281 | 2812 | No | NA | 3 | 0 | 1 | Not-determined | Genome-fragment | NA | NA | 0 |
| S8_extracted_k141_148374 | 1039 | No | NA | 1 | 0 | 1 | Not-determined | Genome-fragment | NA | NA | 0 |
| S8_extracted_k141_87866 | 1483 | No | NA | 2 | 0 | 1 | Not-determined | Genome-fragment | NA | NA | 0 |
| S8_extracted_k141_113679 | 1008 | No | NA | 1 | 0 | 1 | Not-determined | Genome-fragment | NA | NA | 0 |
| S3_vs_k141_101038\|\|full | 7449 | No | NA | 6 | 0 | 1 | Low-quality | Genome-fragment | 6.73 | AAI-based (medium-confidence) | 0 |
| S12_vs_k141_152798\|\|full | 7450 | No | NA | 10 | 10 | 0 | Low-quality | Genome-fragment | 21.89 | AAI-based (high-confidence) | 0 |
| S8_extracted_k141_44309 | 3787 | No | NA | 7 | 0 | 0 | Not-determined | Genome-fragment | NA | NA | 0 |
| S8_extracted_k141_113692 | 2831 | No | NA | 5 | 0 | 1 | Not-determined | Genome-fragment | NA | NA | 0 |
| S8_extracted_k141_113698 | 1826 | No | NA | 3 | 0 | 1 | Not-determined | Genome-fragment | NA | NA | 0 |
| S8_extracted_k141_131202 | 4632 | No | NA | 4 | 0 | 4 | Not-determined | Genome-fragment | NA | NA | 0 |
| S8_extracted_k141_79143 | 1184 | No | NA | 2 | 0 | 1 | Not-determined | Genome-fragment | NA | NA | 0 |
| S8_extracted_k141_131204 | 1191 | No | NA | 2 | 0 | 0 | Not-determined | Genome-fragment | NA | NA | 0 |
| S8_extracted_k141_174313 | 1143 | No | NA | 2 | 0 | 2 | Not-determined | Genome-fragment | NA | NA | 0 |
| S8_extracted_k141_70206 | 1099 | No | NA | 1 | 0 | 1 | Not-determined | Genome-fragment | NA | NA | 0 |
| S8_extracted_k141_53038 | 3443 | No | NA | 4 | 0 | 0 | Not-determined | Genome-fragment | NA | NA | 0 |
| S3_extracted_k141_2664 | 7452 | No | NA | 5 | 0 | 4 | Low-quality | Genome-fragment | 4.01 | AAI-based (medium-confidence) | 0 |
| S8_extracted_k141_174322 | 3876 | No | NA | 2 | 0 | 1 | Not-determined | Genome-fragment | NA | NA | 0 |
| S8_extracted_k141_139872 | 6179 | No | NA | 10 | 0 | 3 | Not-determined | Genome-fragment | NA | NA | 0 |
| S8_extracted_k141_18112 | 1078 | No | NA | 2 | 0 | 1 | Not-determined | Genome-fragment | NA | NA | 0 |
| S7_vs_5627\|\|full | 7455 | No | NA | 5 | 2 | 0 | Low-quality | Genome-fragment | 17.93 | AAI-based (high-confidence) | 0 |
| S8_extracted_k141_18122 | 1549 | No | NA | 2 | 0 | 1 | Not-determined | Genome-fragment | NA | NA | 0 |
| S8_extracted_k141_79180 | 1272 | No | NA | 3 | 0 | 1 | Not-determined | Genome-fragment | NA | NA | 0 |
| S8_extracted_k141_105061 | 3679 | No | NA | 2 | 0 | 2 | Not-determined | Genome-fragment | NA | NA | 0 |
| S8_extracted_k141_131217 | 1455 | No | NA | 1 | 0 | 1 | Not-determined | Genome-fragment | NA | NA | 0 |
| S8_extracted_k141_9659 | 3301 | No | NA | 4 | 0 | 2 | Not-determined | Genome-fragment | NA | NA | 0 |
| S8_extracted_k141_87924 | 1217 | No | NA | 1 | 0 | 1 | Not-determined | Genome-fragment | NA | NA | 0 |
| S11_vs_k141_41299\|\|full | 7457 | No | NA | 14 | 1 | 2 | Low-quality | Genome-fragment | 4.61 | HMM-based (lower-bound) | 0 |
| S8_extracted_k141_174351 | 1139 | No | NA | 1 | 0 | 0 | Not-determined | Genome-fragment | NA | NA | 0 |
| S8_extracted_k141_18129 | 3117 | No | NA | 8 | 0 | 3 | Not-determined | Genome-fragment | NA | NA | 0 |
| S8_extracted_k141_96435 | 1146 | No | NA | 2 | 0 | 1 | Not-determined | Genome-fragment | NA | NA | 0 |
| S8_extracted_k141_87931 | 1101 | No | NA | 2 | 0 | 0 | Not-determined | Genome-fragment | NA | NA | 0 |
| S1_vs_k141_138181\|\|full | 7457 | No | NA | 11 | 3 | 0 | Low-quality | Genome-fragment | 25.99 | HMM-based (lower-bound) | 0 |
| S8_extracted_k141_148396 | 1273 | No | NA | 1 | 0 | 0 | Not-determined | Genome-fragment | NA | NA | 0 |
| S8_extracted_k141_35770 | 1035 | No | NA | 2 | 0 | 0 | Not-determined | Genome-fragment | NA | NA | 0 |
| S8_extracted_k141_96439 | 2323 | No | NA | 1 | 0 | 1 | Not-determined | Genome-fragment | NA | NA | 0 |
| S8_extracted_k141_131236 | 1024 | No | NA | 1 | 0 | 0 | Not-determined | Genome-fragment | NA | NA | 0 |
| S8_extracted_k141_105104 | 1017 | No | NA | 2 | 0 | 0 | Not-determined | Genome-fragment | NA | NA | 0 |
| S8_extracted_k141_9684 | 9298 | No | NA | 9 | 0 | 3 | Not-determined | Genome-fragment | NA | NA | 0 |
| S8_extracted_k141_131238 | 1157 | No | NA | 1 | 0 | 1 | Not-determined | Genome-fragment | NA | NA | 0 |
| S8_extracted_k141_27106 | 5313 | No | NA | 4 | 0 | 4 | Not-determined | Genome-fragment | NA | NA | 0 |
| S8_extracted_k141_139925 | 1087 | No | NA | 2 | 0 | 2 | Not-determined | Genome-fragment | NA | NA | 0 |
| S8_extracted_k141_70222 | 28436 | No | NA | 33 | 0 | 23 | Not-determined | Genome-fragment | NA | NA | 0 |
| S8_extracted_k141_105114 | 1040 | No | NA | 1 | 0 | 0 | Not-determined | Genome-fragment | NA | NA | 0 |
| S8_extracted_k141_79215 | 5102 | No | NA | 4 | 0 | 1 | Not-determined | Genome-fragment | NA | NA | 0 |
| S8_extracted_k141_53087 | 2433 | No | NA | 2 | 0 | 2 | Not-determined | Genome-fragment | NA | NA | 0 |
| S8_extracted_k141_105127 | 1342 | No | NA | 1 | 0 | 1 | Not-determined | Genome-fragment | NA | NA | 0 |
| S3_vs_k141_34745\|\|full | 7458 | No | NA | 7 | 1 | 0 | Low-quality | Genome-fragment | 6.58 | HMM-based (lower-bound) | 0 |
| S8_extracted_k141_139933 | 1659 | No | NA | 2 | 0 | 0 | Not-determined | Genome-fragment | NA | NA | 0 |
| S8_vs_k141_81897\|\|full | 7459 | No | NA | 12 | 4 | 0 | Low-quality | Genome-fragment | 15 | AAI-based (high-confidence) | 0 |
| S8_extracted_k141_87956 | 1014 | No | NA | 1 | 0 | 1 | Not-determined | Genome-fragment | NA | NA | 0 |
| S16_vs_1738\|\|full | 7460 | No | NA | 5 | 0 | 4 | Low-quality | Genome-fragment | 9.54 | AAI-based (medium-confidence) | 0 |
| S8_extracted_k141_70240 | 1095 | No | NA | 1 | 0 | 1 | Not-determined | Genome-fragment | NA | NA | 0 |
| S8_extracted_k141_35812 | 1141 | No | NA | 3 | 0 | 0 | Not-determined | Genome-fragment | NA | NA | 0 |
| S8_extracted_k141_131263 | 1046 | No | NA | 1 | 0 | 1 | Not-determined | Genome-fragment | NA | NA | 0 |
| S8_extracted_k141_113769 | 17481 | No | NA | 16 | 0 | 9 | Not-determined | Genome-fragment | NA | NA | 0 |
| S8_extracted_k141_18178 | 2011 | No | NA | 2 | 0 | 0 | Not-determined | Genome-fragment | NA | NA | 0 |
| S8_extracted_k141_174399 | 2511 | No | NA | 3 | 0 | 3 | Not-determined | Genome-fragment | NA | NA | 0 |
| S8_extracted_k141_131275 | 1121 | No | NA | 1 | 0 | 0 | Not-determined | Genome-fragment | NA | NA | 0 |
| S8_extracted_k141_9732 | 1367 | No | NA | 1 | 0 | 0 | Not-determined | Genome-fragment | NA | NA | 0 |
| S8_extracted_k141_35836 | 3220 | No | NA | 2 | 0 | 0 | Not-determined | Genome-fragment | NA | NA | 0 |
| S8_extracted_k141_96497 | 3742 | No | NA | 4 | 0 | 1 | Not-determined | Genome-fragment | NA | NA | 0 |
| S8_extracted_k141_87969 | 14464 | No | NA | 2 | 0 | 1 | Not-determined | Genome-fragment | NA | NA | 0 |
| S8_extracted_k141_96501 | 6723 | No | NA | 7 | 0 | 5 | Not-determined | Genome-fragment | NA | NA | 0 |
| S8_extracted_k141_1124 | 1112 | No | NA | 1 | 0 | 1 | Not-determined | Genome-fragment | NA | NA | 0 |
| S10_extracted_114153 | 7463 | No | NA | 13 | 2 | 2 | Low-quality | Genome-fragment | 18.07 | HMM-based (lower-bound) | 0 |
| S8_extracted_k141_96504 | 1069 | No | NA | 3 | 0 | 0 | Not-determined | Genome-fragment | NA | NA | 0 |
| S8_extracted_k141_18209 | 1063 | No | NA | 1 | 0 | 1 | Not-determined | Genome-fragment | NA | NA | 0 |
| S8_extracted_k141_53134 | 2039 | No | NA | 1 | 0 | 1 | Not-determined | Genome-fragment | NA | NA | 0 |
| S6_extracted_k141_27200 | 7466 | No | NA | 6 | 0 | 3 | Low-quality | Genome-fragment | 5.35 | AAI-based (high-confidence) | 0 |
| S8_extracted_k141_35854 | 4164 | No | NA | 3 | 0 | 2 | Not-determined | Genome-fragment | NA | NA | 0 |
| S8_extracted_k141_1134 | 5361 | No | NA | 3 | 0 | 0 | Not-determined | Genome-fragment | NA | NA | 0 |
| S8_extracted_k141_96509 | 1370 | No | NA | 2 | 0 | 1 | Not-determined | Genome-fragment | NA | NA | 0 |
| S8_extracted_k141_192163 | 3918 | No | NA | 5 | 0 | 0 | Not-determined | Genome-fragment | NA | NA | 0 |
| S8_extracted_k141_1145 | 1393 | No | NA | 1 | 0 | 1 | Not-determined | Genome-fragment | NA | NA | 0 |
| S8_extracted_k141_79288 | 1027 | No | NA | 2 | 0 | 0 | Not-determined | Genome-fragment | NA | NA | 0 |
| S8_extracted_k141_35877 | 1599 | No | NA | 1 | 0 | 1 | Not-determined | Genome-fragment | NA | NA | 0 |
| S8_extracted_k141_105163 | 1971 | No | NA | 2 | 0 | 1 | Not-determined | Genome-fragment | NA | NA | 0 |
| S8_extracted_k141_96539 | 1075 | No | NA | 3 | 0 | 0 | Not-determined | Genome-fragment | NA | NA | 0 |
| S8_extracted_k141_61855 | 1986 | No | NA | 2 | 0 | 2 | Not-determined | Genome-fragment | NA | NA | 0 |
| S8_extracted_k141_157133 | 1140 | No | NA | 2 | 0 | 1 | Not-determined | Genome-fragment | NA | NA | 0 |
| S8_extracted_k141_140002 | 1464 | No | NA | 2 | 0 | 1 | Not-determined | Genome-fragment | NA | NA | 0 |
| S8_vs_k141_95613\|\|full | 7467 | No | NA | 17 | 4 | 0 | Low-quality | Genome-fragment | 17.19 | AAI-based (medium-confidence) | 0 |
| S8_extracted_k141_18243 | 5577 | No | NA | 3 | 0 | 2 | Not-determined | Genome-fragment | NA | NA | 0 |
| S8_extracted_k141_148554 | 2117 | No | NA | 4 | 0 | 1 | Not-determined | Genome-fragment | NA | NA | 0 |
| S8_extracted_k141_122661 | 1129 | No | NA | 1 | 0 | 1 | Not-determined | Genome-fragment | NA | NA | 0 |
| S8_extracted_k141_122669 | 2131 | No | NA | 2 | 0 | 1 | Not-determined | Genome-fragment | NA | NA | 0 |
| S3_extracted_k141_40381 | 7473 | No | NA | 14 | 1 | 0 | Low-quality | Genome-fragment | 5.87 | AAI-based (high-confidence) | 0 |
| S8_extracted_k141_79349 | 3205 | No | NA | 5 | 0 | 2 | Not-determined | Genome-fragment | NA | NA | 0 |
| S8_extracted_k141_200353 | 1198 | No | NA | 1 | 0 | 0 | Not-determined | Genome-fragment | NA | NA | 0 |
| S8_extracted_k141_1205 | 1373 | No | NA | 3 | 0 | 0 | Not-determined | Genome-fragment | NA | NA | 0 |
| S8_extracted_k141_96557 | 1996 | No | NA | 1 | 0 | 1 | Not-determined | Genome-fragment | NA | NA | 0 |
| S8_extracted_k141_122671 | 11403 | No | NA | 5 | 0 | 4 | Not-determined | Genome-fragment | NA | NA | 0 |
| S5_vs_k141_70920\|\|full | 7473 | No | NA | 9 | 3 | 0 | Low-quality | Genome-fragment | 11.83 | HMM-based (lower-bound) | 0 |
| S8_extracted_k141_1215 | 3919 | No | NA | 4 | 0 | 1 | Not-determined | Genome-fragment | NA | NA | 0 |
| S8_extracted_k141_182933 | 2110 | No | NA | 4 | 0 | 3 | Not-determined | Genome-fragment | NA | NA | 0 |
| S8_extracted_k141_131386 | 2047 | No | NA | 3 | 0 | 3 | Not-determined | Genome-fragment | NA | NA | 0 |
| S8_extracted_k141_192232 | 1214 | No | NA | 1 | 0 | 0 | Not-determined | Genome-fragment | NA | NA | 0 |
| S8_extracted_k141_1230 | 1132 | No | NA | 1 | 0 | 1 | Not-determined | Genome-fragment | NA | NA | 0 |
| S8_extracted_k141_165990 | 4650 | No | NA | 2 | 0 | 1 | Not-determined | Genome-fragment | NA | NA | 0 |
| S8_extracted_k141_122689 | 1943 | No | NA | 4 | 0 | 1 | Not-determined | Genome-fragment | NA | NA | 0 |
| S8_extracted_k141_182937 | 1738 | No | NA | 2 | 0 | 0 | Not-determined | Genome-fragment | NA | NA | 0 |
| S8_extracted_k141_148612 | 1574 | No | NA | 1 | 0 | 1 | Not-determined | Genome-fragment | NA | NA | 0 |
| S10_vs_63798\|\|full | 7474 | No | NA | 15 | 3 | 0 | Low-quality | Genome-fragment | 18.3 | AAI-based (high-confidence) | 0 |
| S8_extracted_k141_166003 | 1636 | No | NA | 3 | 0 | 1 | Not-determined | Genome-fragment | NA | NA | 0 |
| S11_extracted_k141_68934 | 7478 | No | NA | 13 | 1 | 1 | Low-quality | Genome-fragment | 4.86 | AAI-based (high-confidence) | 0 |
| S11_vs_k141_78062\|\|full | 7478 | No | NA | 10 | 4 | 0 | Low-quality | Genome-fragment | 20.68 | AAI-based (high-confidence) | 0 |
| S8_extracted_k141_148632 | 2757 | No | NA | 2 | 0 | 0 | Not-determined | Genome-fragment | NA | NA | 0 |
| S8_extracted_k141_113827 | 1192 | No | NA | 1 | 0 | 1 | Not-determined | Genome-fragment | NA | NA | 0 |
| S8_extracted_k141_61937 | 3289 | No | NA | 2 | 0 | 2 | Not-determined | Genome-fragment | NA | NA | 0 |
| S8_extracted_k141_96580 | 1625 | No | NA | 3 | 0 | 1 | Not-determined | Genome-fragment | NA | NA | 0 |
| S3_extracted_k141_110790 | 7483 | No | NA | 7 | 0 | 6 | Low-quality | Genome-fragment | 3.35 | AAI-based (medium-confidence) | 0 |
| S8_extracted_k141_166023 | 2091 | No | NA | 4 | 0 | 1 | Not-determined | Genome-fragment | NA | NA | 0 |
| S8_extracted_k141_174492 | 2433 | No | NA | 4 | 0 | 3 | Not-determined | Genome-fragment | NA | NA | 0 |
| S8_extracted_k141_148644 | 1352 | No | NA | 1 | 0 | 0 | Not-determined | Genome-fragment | NA | NA | 0 |
| S8_extracted_k141_96584 | 2815 | No | NA | 2 | 0 | 0 | Not-determined | Genome-fragment | NA | NA | 0 |
| S12_vs_k141_157523\|\|full | 7486 | No | NA | 12 | 6 | 0 | Low-quality | Genome-fragment | 12.5 | AAI-based (high-confidence) | 0 |
| S8_extracted_k141_18288 | 1164 | No | NA | 2 | 0 | 0 | Not-determined | Genome-fragment | NA | NA | 0 |
| S8_extracted_k141_131428 | 1739 | No | NA | 2 | 0 | 2 | Not-determined | Genome-fragment | NA | NA | 0 |
| S8_extracted_k141_79392 | 5710 | No | NA | 4 | 0 | 2 | Not-determined | Genome-fragment | NA | NA | 0 |
| S8_extracted_k141_131430 | 1070 | No | NA | 1 | 0 | 1 | Not-determined | Genome-fragment | NA | NA | 0 |
| S8_extracted_k141_61944 | 1346 | No | NA | 3 | 0 | 1 | Not-determined | Genome-fragment | NA | NA | 0 |
| S8_extracted_k141_157226 | 5188 | No | NA | 6 | 0 | 3 | Not-determined | Genome-fragment | NA | NA | 0 |
| S8_extracted_k141_122732 | 3376 | No | NA | 3 | 0 | 1 | Not-determined | Genome-fragment | NA | NA | 0 |
| S12_vs_k141_20969\|\|full | 7487 | Yes | 4092 | 8 | 1 | 3 | Low-quality | Genome-fragment | 2.75 | HMM-based (lower-bound) | 45.35 |
| S8_extracted_k141_44395 | 5316 | No | NA | 5 | 0 | 3 | Not-determined | Genome-fragment | NA | NA | 0 |
| S8_extracted_k141_27306 | 1270 | No | NA | 2 | 0 | 1 | Not-determined | Genome-fragment | NA | NA | 0 |
| S8_extracted_k141_18304 | 1473 | No | NA | 2 | 0 | 0 | Not-determined | Genome-fragment | NA | NA | 0 |
| S8_extracted_k141_182973 | 7828 | No | NA | 7 | 0 | 5 | Not-determined | Genome-fragment | NA | NA | 0 |
| S8_extracted_k141_166033 | 2754 | No | NA | 3 | 0 | 1 | Not-determined | Genome-fragment | NA | NA | 0 |
| S8_extracted_k141_113880 | 6449 | No | NA | 4 | 0 | 3 | Not-determined | Genome-fragment | NA | NA | 0 |
| S17_vs_k141_53858\|\|full | 7488 | No | NA | 12 | 2 | 0 | Low-quality | Genome-fragment | 12.17 | AAI-based (high-confidence) | 0 |
| S8_extracted_k141_18333 | 2039 | No | NA | 6 | 0 | 0 | Not-determined | Genome-fragment | NA | NA | 0 |
| S8_extracted_k141_44418 | 3915 | No | NA | 4 | 0 | 0 | Not-determined | Genome-fragment | NA | NA | 0 |
| S8_extracted_k141_192328 | 6890 | No | NA | 11 | 0 | 4 | Not-determined | Genome-fragment | NA | NA | 0 |
| S5_extracted_k141_137732 | 7495 | No | NA | 19 | 6 | 0 | Low-quality | Genome-fragment | 21.29 | AAI-based (high-confidence) | 0 |
| S8_extracted_k141_96636 | 1240 | No | NA | 2 | 0 | 0 | Not-determined | Genome-fragment | NA | NA | 0 |
| S1_extracted_k141_118348 | 7500 | No | NA | 19 | 4 | 0 | Low-quality | Genome-fragment | 17.9 | AAI-based (high-confidence) | 0 |
| S8_extracted_k141_1300 | 2572 | No | NA | 1 | 0 | 0 | Not-determined | Genome-fragment | NA | NA | 0 |
| S5_vs_k141_104149\|\|full | 7503 | No | NA | 17 | 0 | 0 | Low-quality | Genome-fragment | 16.72 | AAI-based (medium-confidence) | 0 |
| S1_extracted_k141_155388 | 7509 | No | NA | 11 | 8 | 0 | Low-quality | Genome-fragment | 19.84 | AAI-based (high-confidence) | 0 |
| S8_extracted_k141_96650 | 1050 | No | NA | 1 | 0 | 1 | Not-determined | Genome-fragment | NA | NA | 0 |
| S8_extracted_k141_192344 | 1849 | No | NA | 3 | 0 | 0 | Not-determined | Genome-fragment | NA | NA | 0 |
| S8_extracted_k141_157296 | 5146 | No | NA | 3 | 0 | 3 | Not-determined | Genome-fragment | NA | NA | 0 |
| S8_extracted_k141_96654 | 6255 | No | NA | 7 | 0 | 4 | Not-determined | Genome-fragment | NA | NA | 0 |
| S8_extracted_k141_62002 | 1167 | No | NA | 1 | 0 | 1 | Not-determined | Genome-fragment | NA | NA | 0 |
| S8_extracted_k141_113910 | 1503 | No | NA | 2 | 0 | 0 | Not-determined | Genome-fragment | NA | NA | 0 |
| S8_extracted_k141_183035 | 2408 | No | NA | 4 | 0 | 3 | Not-determined | Genome-fragment | NA | NA | 0 |
| S8_extracted_k141_140067 | 1292 | No | NA | 2 | 0 | 0 | Not-determined | Genome-fragment | NA | NA | 0 |
| S8_extracted_k141_122793 | 1982 | No | NA | 5 | 0 | 2 | Not-determined | Genome-fragment | NA | NA | 0 |
| S8_extracted_k141_140072 | 1018 | No | NA | 1 | 0 | 0 | Not-determined | Genome-fragment | NA | NA | 0 |
| S8_extracted_k141_105301 | 1445 | No | NA | 2 | 0 | 0 | Not-determined | Genome-fragment | NA | NA | 0 |
| S8_extracted_k141_27412 | 1337 | No | NA | 2 | 0 | 1 | Not-determined | Genome-fragment | NA | NA | 0 |
| S8_extracted_k141_122797 | 3559 | No | NA | 3 | 0 | 3 | Not-determined | Genome-fragment | NA | NA | 0 |
| S8_extracted_k141_96673 | 2085 | No | NA | 4 | 0 | 1 | Not-determined | Genome-fragment | NA | NA | 0 |
| S8_extracted_k141_192380 | 1106 | No | NA | 1 | 0 | 1 | Not-determined | Genome-fragment | NA | NA | 0 |
| S8_extracted_k141_27419 | 1523 | No | NA | 4 | 0 | 0 | Not-determined | Genome-fragment | NA | NA | 0 |
| S8_extracted_k141_192381 | 2137 | No | NA | 3 | 0 | 1 | Not-determined | Genome-fragment | NA | NA | 0 |
| S8_extracted_k141_96679 | 2425 | No | NA | 2 | 0 | 0 | Not-determined | Genome-fragment | NA | NA | 0 |
| S8_extracted_k141_79476 | 1170 | No | NA | 2 | 0 | 2 | Not-determined | Genome-fragment | NA | NA | 0 |
| S8_extracted_k141_1339 | 7717 | No | NA | 5 | 0 | 2 | Not-determined | Genome-fragment | NA | NA | 0 |
| S8_extracted_k141_148724 | 1690 | No | NA | 2 | 0 | 1 | Not-determined | Genome-fragment | NA | NA | 0 |
| S10_vs_118179\|\|full | 7509 | No | NA | 11 | 1 | 0 | Low-quality | Genome-fragment | 19.11 | AAI-based (high-confidence) | 0 |
| S8_extracted_k141_113940 | 1261 | No | NA | 1 | 0 | 1 | Not-determined | Genome-fragment | NA | NA | 0 |
| S8_extracted_k141_88113 | 1509 | No | NA | 2 | 0 | 1 | Not-determined | Genome-fragment | NA | NA | 0 |
| S8_extracted_k141_122824 | 1328 | No | NA | 1 | 0 | 0 | Not-determined | Genome-fragment | NA | NA | 0 |
| S8_extracted_k141_105345 | 1627 | No | NA | 2 | 0 | 0 | Not-determined | Genome-fragment | NA | NA | 0 |
| S8_extracted_k141_174559 | 16202 | No | NA | 17 | 8 | 0 | Medium-quality | Genome-fragment | 79.32 | AAI-based (high-confidence) | 0 |
| S8_extracted_k141_79505 | 1152 | No | NA | 1 | 0 | 1 | Not-determined | Genome-fragment | NA | NA | 0 |
| S8_extracted_k141_105355 | 1643 | No | NA | 3 | 0 | 0 | Not-determined | Genome-fragment | NA | NA | 0 |
| S8_extracted_k141_200514 | 1740 | No | NA | 2 | 0 | 0 | Not-determined | Genome-fragment | NA | NA | 0 |
| S8_extracted_k141_36076 | 1198 | No | NA | 1 | 0 | 0 | Not-determined | Genome-fragment | NA | NA | 0 |
| S4_extracted_7896 | 7509 | No | NA | 12 | 8 | 0 | Low-quality | Genome-fragment | 16.39 | AAI-based (high-confidence) | 0 |
| S8_extracted_k141_27473 | 1761 | No | NA | 3 | 0 | 2 | Not-determined | Genome-fragment | NA | NA | 0 |
| S8_extracted_k141_200531 | 2275 | No | NA | 4 | 0 | 4 | Not-determined | Genome-fragment | NA | NA | 0 |
| S8_extracted_k141_96727 | 2720 | No | NA | 4 | 0 | 0 | Not-determined | Genome-fragment | NA | NA | 0 |
| S8_extracted_k141_183045 | 46717 | No | NA | 58 | 18 | 2 | Complete | High-quality | 100 | DTR (high-confidence) | 0 |
| S8_extracted_k141_9809 | 1030 | No | NA | 2 | 0 | 1 | Not-determined | Genome-fragment | NA | NA | 0 |
| S8_extracted_k141_79512 | 1136 | No | NA | 2 | 0 | 1 | Not-determined | Genome-fragment | NA | NA | 0 |
| S8_extracted_k141_96732 | 1509 | No | NA | 1 | 0 | 0 | Not-determined | Genome-fragment | NA | NA | 0 |
| S10_vs_7669\|\|full | 7511 | No | NA | 15 | 1 | 1 | Low-quality | Genome-fragment | 12.64 | AAI-based (high-confidence) | 0 |
| S16_extracted_149579 | 7512 | No | NA | 17 | 7 | 0 | Low-quality | Genome-fragment | 12.51 | AAI-based (high-confidence) | 0 |
| S8_extracted_k141_96734 | 4303 | No | NA | 9 | 0 | 0 | Not-determined | Genome-fragment | NA | NA | 0 |
| S8_extracted_k141_192412 | 1038 | No | NA | 1 | 0 | 1 | Not-determined | Genome-fragment | NA | NA | 0 |
| S8_extracted_k141_105398 | 1260 | No | NA | 1 | 0 | 1 | Not-determined | Genome-fragment | NA | NA | 0 |
| S8_extracted_k141_18417 | 2270 | No | NA | 3 | 0 | 2 | Not-determined | Genome-fragment | NA | NA | 0 |
| S8_extracted_k141_105403 | 1043 | No | NA | 2 | 0 | 1 | Not-determined | Genome-fragment | NA | NA | 0 |
| S8_extracted_k141_131524 | 1419 | No | NA | 3 | 0 | 1 | Not-determined | Genome-fragment | NA | NA | 0 |
| S8_extracted_k141_27513 | 1679 | No | NA | 1 | 0 | 1 | Not-determined | Genome-fragment | NA | NA | 0 |
| S16_vs_97287\|\|full | 7512 | No | NA | 7 | 0 | 1 | Low-quality | Genome-fragment | 13.54 | AAI-based (medium-confidence) | 0 |
| S8_extracted_k141_200562 | 1122 | No | NA | 1 | 0 | 0 | Not-determined | Genome-fragment | NA | NA | 0 |
| S8_extracted_k141_114008 | 1550 | No | NA | 1 | 0 | 1 | Not-determined | Genome-fragment | NA | NA | 0 |
| S8_extracted_k141_27523 | 2437 | No | NA | 2 | 0 | 1 | Not-determined | Genome-fragment | NA | NA | 0 |
| S8_extracted_k141_88168 | 4642 | No | NA | 6 | 0 | 0 | Not-determined | Genome-fragment | NA | NA | 0 |
| S5_vs_k141_111530\|\|full | 7516 | No | NA | 20 | 4 | 0 | Low-quality | Genome-fragment | 16.21 | AAI-based (medium-confidence) | 0 |
| S8_extracted_k141_192460 | 1097 | No | NA | 2 | 0 | 0 | Not-determined | Genome-fragment | NA | NA | 0 |
| S8_extracted_k141_96769 | 1887 | No | NA | 1 | 0 | 1 | Not-determined | Genome-fragment | NA | NA | 0 |
| S1_vs_k141_82519\|\|full | 7517 | No | NA | 10 | 4 | 0 | Low-quality | Genome-fragment | 17.89 | AAI-based (high-confidence) | 0 |
| S8_extracted_k141_148800 | 2201 | No | NA | 2 | 0 | 0 | Not-determined | Genome-fragment | NA | NA | 0 |
| S12_vs_k141_9502\|\|full | 7519 | No | NA | 11 | 1 | 0 | Low-quality | Genome-fragment | 6.96 | HMM-based (lower-bound) | 0 |
| S8_extracted_k141_70496 | 1315 | No | NA | 2 | 0 | 1 | Not-determined | Genome-fragment | NA | NA | 0 |
| S8_vs_k141_44594\|\|full | 7523 | No | NA | 7 | 0 | 1 | Low-quality | Genome-fragment | 8.89 | AAI-based (medium-confidence) | 0 |
| S8_extracted_k141_140182 | 1036 | No | NA | 1 | 0 | 1 | Not-determined | Genome-fragment | NA | NA | 0 |
| S8_extracted_k141_114043 | 1277 | No | NA | 2 | 0 | 2 | Not-determined | Genome-fragment | NA | NA | 0 |
| S8_extracted_k141_166192 | 1214 | No | NA | 1 | 0 | 1 | Not-determined | Genome-fragment | NA | NA | 0 |
| S8_extracted_k141_192478 | 3307 | No | NA | 3 | 0 | 1 | Not-determined | Genome-fragment | NA | NA | 0 |
| S8_extracted_k141_131536 | 3327 | No | NA | 1 | 0 | 1 | Not-determined | Genome-fragment | NA | NA | 0 |
| S8_extracted_k141_192484 | 1397 | No | NA | 3 | 0 | 0 | Not-determined | Genome-fragment | NA | NA | 0 |
| S8_extracted_k141_53480 | 1298 | No | NA | 3 | 0 | 3 | Not-determined | Genome-fragment | NA | NA | 0 |
| S16_vs_53992\|\|full | 7526 | No | NA | 15 | 0 | 1 | Low-quality | Genome-fragment | 15.16 | AAI-based (medium-confidence) | 0 |
| S8_extracted_k141_140215 | 1164 | No | NA | 2 | 0 | 0 | Not-determined | Genome-fragment | NA | NA | 0 |
| S8_extracted_k141_1477 | 8201 | No | NA | 9 | 0 | 1 | Not-determined | Genome-fragment | NA | NA | 0 |
| S8_extracted_k141_36182 | 4314 | No | NA | 4 | 0 | 2 | Not-determined | Genome-fragment | NA | NA | 0 |
| S8_extracted_k141_105464 | 3218 | No | NA | 2 | 0 | 2 | Not-determined | Genome-fragment | NA | NA | 0 |
| S8_extracted_k141_131548 | 6275 | No | NA | 7 | 0 | 4 | Not-determined | Genome-fragment | NA | NA | 0 |
| S8_extracted_k141_88211 | 1161 | No | NA | 2 | 0 | 0 | Not-determined | Genome-fragment | NA | NA | 0 |
| S8_vs_k141_186088\|\|full | 7527 | No | NA | 12 | 1 | 0 | Low-quality | Genome-fragment | 17.21 | AAI-based (medium-confidence) | 0 |
| S3_vs_k141_47904\|\|full | 7529 | No | NA | 13 | 0 | 0 | Low-quality | Genome-fragment | 19.68 | AAI-based (high-confidence) | 0 |
| S8_extracted_k141_36197 | 3511 | No | NA | 3 | 0 | 2 | Not-determined | Genome-fragment | NA | NA | 0 |
| S8_extracted_k141_36199 | 1371 | No | NA | 2 | 0 | 1 | Not-determined | Genome-fragment | NA | NA | 0 |
| S8_extracted_k141_27591 | 1316 | No | NA | 2 | 0 | 2 | Not-determined | Genome-fragment | NA | NA | 0 |
| S8_extracted_k141_174685 | 1115 | No | NA | 1 | 0 | 0 | Not-determined | Genome-fragment | NA | NA | 0 |
| S8_extracted_k141_36217 | 1466 | No | NA | 1 | 0 | 1 | Not-determined | Genome-fragment | NA | NA | 0 |
| S8_extracted_k141_200590 | 1934 | No | NA | 2 | 0 | 2 | Not-determined | Genome-fragment | NA | NA | 0 |
| S17_vs_k141_74660\|\|full | 7539 | Yes | 5263 | 10 | 1 | 1 | Low-quality | Genome-fragment | 8.21 | AAI-based (medium-confidence) | 30.19 |
| S8_extracted_k141_27607 | 3096 | No | NA | 4 | 0 | 2 | Not-determined | Genome-fragment | NA | NA | 0 |
| S8_extracted_k141_1518 | 1878 | No | NA | 3 | 0 | 2 | Not-determined | Genome-fragment | NA | NA | 0 |
| S8_extracted_k141_174694 | 1147 | No | NA | 3 | 0 | 1 | Not-determined | Genome-fragment | NA | NA | 0 |
| S5_vs_k141_351\|\|full | 7541 | No | NA | 9 | 0 | 1 | Low-quality | Genome-fragment | 12.48 | AAI-based (medium-confidence) | 0 |
| S8_vs_k141_204793\|\|full | 7542 | No | NA | 11 | 2 | 1 | Low-quality | Genome-fragment | 13.28 | AAI-based (medium-confidence) | 0 |
| S3_vs_k141_51330\|\|full | 7543 | No | NA | 20 | 0 | 0 | Low-quality | Genome-fragment | 13.71 | AAI-based (high-confidence) | 0 |
| S8_extracted_k141_96879 | 4498 | No | NA | 3 | 0 | 1 | Not-determined | Genome-fragment | NA | NA | 0 |
| S8_extracted_k141_96880 | 3001 | No | NA | 4 | 0 | 3 | Not-determined | Genome-fragment | NA | NA | 0 |
| S8_extracted_k141_1530 | 2267 | No | NA | 1 | 0 | 0 | Not-determined | Genome-fragment | NA | NA | 0 |
| S8_extracted_k141_36263 | 1353 | No | NA | 2 | 0 | 0 | Not-determined | Genome-fragment | NA | NA | 0 |
| S8_extracted_k141_79597 | 1240 | No | NA | 2 | 0 | 1 | Not-determined | Genome-fragment | NA | NA | 0 |
| S8_extracted_k141_122967 | 1067 | No | NA | 1 | 0 | 1 | Not-determined | Genome-fragment | NA | NA | 0 |
| S8_extracted_k141_143140 | 7544 | No | NA | 7 | 0 | 1 | Low-quality | Genome-fragment | 13.5 | AAI-based (medium-confidence) | 0 |
| S8_extracted_k141_27643 | 2118 | No | NA | 3 | 0 | 0 | Not-determined | Genome-fragment | NA | NA | 0 |
| S8_extracted_k141_122969 | 1320 | No | NA | 2 | 0 | 1 | Not-determined | Genome-fragment | NA | NA | 0 |
| S8_extracted_k141_183170 | 1892 | No | NA | 1 | 0 | 1 | Not-determined | Genome-fragment | NA | NA | 0 |
| S1_vs_k141_90691\|\|full | 7546 | No | NA | 10 | 2 | 0 | Low-quality | Genome-fragment | 18.83 | AAI-based (high-confidence) | 0 |
| S8_extracted_k141_36266 | 11658 | No | NA | 9 | 0 | 4 | Not-determined | Genome-fragment | NA | NA | 0 |
| S5_vs_k141_122381\|\|full | 7548 | No | NA | 12 | 1 | 0 | Low-quality | Genome-fragment | 11.29 | HMM-based (lower-bound) | 0 |
| S5_vs_k141_342\|\|full | 7557 | Yes | 4665 | 10 | 2 | 1 | Low-quality | Genome-fragment | 4.93 | HMM-based (lower-bound) | 38.27 |
| S17_vs_k141_11180\|\|full | 7558 | No | NA | 12 | 4 | 0 | Low-quality | Genome-fragment | 9.57 | AAI-based (high-confidence) | 0 |
| S3_vs_k141_63508\|\|full | 7565 | No | NA | 4 | 0 | 3 | Low-quality | Genome-fragment | 3.26 | AAI-based (medium-confidence) | 0 |
| S8_vs_k141_188175\|\|full | 7569 | No | NA | 14 | 1 | 0 | Low-quality | Genome-fragment | 14.74 | AAI-based (high-confidence) | 0 |
| S8_extracted_k141_183184 | 1349 | No | NA | 2 | 0 | 0 | Not-determined | Genome-fragment | NA | NA | 0 |
| S8_extracted_k141_96906 | 3232 | No | NA | 3 | 0 | 1 | Not-determined | Genome-fragment | NA | NA | 0 |
| S8_extracted_k141_53551 | 3227 | No | NA | 3 | 0 | 2 | Not-determined | Genome-fragment | NA | NA | 0 |
| S8_extracted_k141_140321 | 1062 | No | NA | 3 | 0 | 1 | Not-determined | Genome-fragment | NA | NA | 0 |
| S16_extracted_3348 | 7572 | No | NA | 18 | 1 | 0 | Low-quality | Genome-fragment | 7.05 | AAI-based (high-confidence) | 0 |
| S8_extracted_k141_79621 | 1298 | No | NA | 3 | 0 | 3 | Not-determined | Genome-fragment | NA | NA | 0 |
| S8_extracted_k141_166289 | 1219 | No | NA | 1 | 0 | 0 | Not-determined | Genome-fragment | NA | NA | 0 |
| S8_extracted_k141_174748 | 1256 | No | NA | 2 | 0 | 2 | Not-determined | Genome-fragment | NA | NA | 0 |
| S8_extracted_k141_183191 | 2207 | No | NA | 3 | 0 | 3 | Not-determined | Genome-fragment | NA | NA | 0 |
| S8_extracted_k141_18605 | 2356 | No | NA | 7 | 0 | 0 | Not-determined | Genome-fragment | NA | NA | 0 |
| S8_extracted_k141_183193 | 1467 | No | NA | 2 | 0 | 2 | Not-determined | Genome-fragment | NA | NA | 0 |
| S8_extracted_k141_131569 | 5179 | No | NA | 4 | 0 | 3 | Not-determined | Genome-fragment | NA | NA | 0 |
| S8_extracted_k141_174753 | 3426 | No | NA | 4 | 0 | 2 | Not-determined | Genome-fragment | NA | NA | 0 |
| S11_vs_k141_23558\|\|full | 7573 | No | NA | 12 | 4 | 0 | Low-quality | Genome-fragment | 17.75 | AAI-based (medium-confidence) | 0 |
| S11_extracted_k141_82820 | 7576 | No | NA | 6 | 0 | 3 | Low-quality | Genome-fragment | 4.08 | AAI-based (medium-confidence) | 0 |
| S14_vs_k141_76580\|\|full | 7579 | No | NA | 18 | 4 | 0 | Low-quality | Genome-fragment | 17.84 | AAI-based (medium-confidence) | 0 |
| S8_extracted_k141_157536 | 2498 | No | NA | 3 | 0 | 2 | Not-determined | Genome-fragment | NA | NA | 0 |
| S8_extracted_k141_44619 | 1832 | No | NA | 2 | 0 | 1 | Not-determined | Genome-fragment | NA | NA | 0 |
| S8_extracted_k141_1578 | 1112 | No | NA | 1 | 0 | 0 | Not-determined | Genome-fragment | NA | NA | 0 |
| S8_extracted_k141_70660 | 1198 | No | NA | 1 | 0 | 0 | Not-determined | Genome-fragment | NA | NA | 0 |
| S8_extracted_k141_27674 | 1209 | No | NA | 1 | 0 | 1 | Not-determined | Genome-fragment | NA | NA | 0 |
| S8_extracted_k141_53589 | 1897 | No | NA | 3 | 0 | 1 | Not-determined | Genome-fragment | NA | NA | 0 |
| S8_vs_k141_87506\|\|full | 7583 | No | NA | 5 | 0 | 3 | Low-quality | Genome-fragment | 6.63 | AAI-based (high-confidence) | 0 |
| S8_extracted_k141_131601 | 1523 | No | NA | 2 | 0 | 0 | Not-determined | Genome-fragment | NA | NA | 0 |
| S8_extracted_k141_18630 | 2131 | No | NA | 3 | 0 | 2 | Not-determined | Genome-fragment | NA | NA | 0 |
| S8_extracted_k141_148914 | 1977 | No | NA | 2 | 0 | 1 | Not-determined | Genome-fragment | NA | NA | 0 |
| S8_extracted_k141_122981 | 3181 | No | NA | 1 | 0 | 1 | Not-determined | Genome-fragment | NA | NA | 0 |
| S8_extracted_k141_148915 | 2473 | No | NA | 2 | 0 | 2 | Not-determined | Genome-fragment | NA | NA | 0 |

| | | | | | | | | | | | |
|---|---|---|---|---|---|---|---|---|---|---|---|
| S8_extracted_k141_131609 | 1094 | No | NA | 1 | 0 | 0 | Not-determined | Genome-fragment | NA | NA | 0 |
| S8_extracted_k141_140370 | 1275 | No | NA | 1 | 0 | 1 | Not-determined | Genome-fragment | NA | NA | 0 |
| S17_extracted_k141_115588 | 7585 | No | NA | 7 | 0 | 1 | Low-quality | Genome-fragment | 16.32 | AAI-based (medium-confidence) | 0 |
| S8_extracted_k141_79710 | 1106 | No | NA | 1 | 0 | 1 | Not-determined | Genome-fragment | NA | NA | 0 |
| S8_extracted_k141_49757 | 7585 | No | NA | 11 | 1 | 1 | Low-quality | Genome-fragment | 3.05 | HMM-based (lower-bound) | 0 |
| S8_extracted_k141_62165 | 1073 | No | NA | 3 | 0 | 1 | Not-determined | Genome-fragment | NA | NA | 0 |
| S17_extracted_k141_44546 | 7586 | No | NA | 17 | 2 | 0 | Low-quality | Genome-fragment | 16.99 | AAI-based (high-confidence) | 0 |
| S8_extracted_k141_18650 | 1830 | No | NA | 3 | 0 | 2 | Not-determined | Genome-fragment | NA | NA | 0 |
| S8_extracted_k141_105593 | 1185 | No | NA | 3 | 0 | 2 | Not-determined | Genome-fragment | NA | NA | 0 |
| S8_extracted_k141_44684 | 1442 | No | NA | 2 | 0 | 1 | Not-determined | Genome-fragment | NA | NA | 0 |
| S12_extracted_k141_118610 | 7586 | No | NA | 4 | 0 | 3 | Low-quality | Genome-fragment | 4.55 | AAI-based (medium-confidence) | 0 |
| S8_extracted_k141_180823 | 7586 | No | NA | 20 | 0 | 0 | Low-quality | Genome-fragment | 5.7 | AAI-based (high-confidence) | 0 |
| S8_extracted_k141_131637 | 1494 | No | NA | 1 | 0 | 0 | Not-determined | Genome-fragment | NA | NA | 0 |
| S8_extracted_k141_123017 | 5868 | No | NA | 5 | 0 | 2 | Not-determined | Genome-fragment | NA | NA | 0 |
| S8_extracted_k141_27726 | 1313 | No | NA | 1 | 0 | 1 | Not-determined | Genome-fragment | NA | NA | 0 |
| S8_extracted_k141_105610 | 3647 | No | NA | 4 | 0 | 2 | Not-determined | Genome-fragment | NA | NA | 0 |
| S8_extracted_k141_18673 | 1184 | No | NA | 2 | 0 | 2 | Not-determined | Genome-fragment | NA | NA | 0 |
| S8_extracted_k141_27727 | 9510 | No | NA | 11 | 0 | 4 | Not-determined | Genome-fragment | NA | NA | 0 |
| S8_extracted_k141_166341 | 3453 | No | NA | 3 | 0 | 2 | Not-determined | Genome-fragment | NA | NA | 0 |
| S8_extracted_k141_62202 | 1254 | No | NA | 2 | 0 | 0 | Not-determined | Genome-fragment | NA | NA | 0 |
| S8_extracted_k141_62207 | 1108 | No | NA | 1 | 0 | 0 | Not-determined | Genome-fragment | NA | NA | 0 |
| S8_extracted_k141_36336 | 2500 | No | NA | 1 | 0 | 1 | Not-determined | Genome-fragment | NA | NA | 0 |
| S8_extracted_k141_123031 | 1730 | No | NA | 2 | 0 | 2 | Not-determined | Genome-fragment | NA | NA | 0 |
| S8_extracted_k141_62209 | 3487 | No | NA | 4 | 0 | 2 | Not-determined | Genome-fragment | NA | NA | 0 |
| S9_vs_k141_70958\|\|full | 7587 | No | NA | 22 | 2 | 0 | Low-quality | Genome-fragment | 7.29 | HMM-based (lower-bound) | 0 |
| S8_extracted_k141_79736 | 1189 | No | NA | 1 | 0 | 1 | Not-determined | Genome-fragment | NA | NA | 0 |
| S7_vs_2295\|\|full | 7587 | No | NA | 15 | 0 | 0 | Low-quality | Genome-fragment | 17.31 | AAI-based (high-confidence) | 0 |
| S16_vs_109480\|\|full | 7588 | No | NA | 6 | 0 | 3 | Low-quality | Genome-fragment | 3.8 | AAI-based (medium-confidence) | 0 |
| S8_extracted_k141_1612 | 1884 | No | NA | 1 | 0 | 0 | Not-determined | Genome-fragment | NA | NA | 0 |
| S8_extracted_k141_36353 | 7129 | No | NA | 5 | 0 | 4 | Not-determined | Genome-fragment | NA | NA | 0 |
| S8_extracted_k141_166376 | 1273 | No | NA | 2 | 0 | 2 | Not-determined | Genome-fragment | NA | NA | 0 |
| S8_extracted_k141_114216 | 2110 | No | NA | 3 | 0 | 1 | Not-determined | Genome-fragment | NA | NA | 0 |
| S8_extracted_k141_105647 | 1664 | No | NA | 2 | 0 | 0 | Not-determined | Genome-fragment | NA | NA | 0 |
| S8_extracted_k141_70753 | 1031 | No | NA | 1 | 0 | 1 | Not-determined | Genome-fragment | NA | NA | 0 |
| S8_extracted_k141_174877 | 1555 | No | NA | 3 | 0 | 0 | Not-determined | Genome-fragment | NA | NA | 0 |
| S5_vs_k141_26477\|\|full | 7590 | No | NA | 13 | 2 | 0 | Low-quality | Genome-fragment | 21.67 | AAI-based (high-confidence) | 0 |
| S8_extracted_k141_114247 | 1176 | No | NA | 2 | 0 | 0 | Not-determined | Genome-fragment | NA | NA | 0 |
| S8_extracted_k141_183289 | 1198 | No | NA | 2 | 0 | 1 | Not-determined | Genome-fragment | NA | NA | 0 |
| S8_extracted_k141_166398 | 2559 | No | NA | 2 | 0 | 1 | Not-determined | Genome-fragment | NA | NA | 0 |
| S8_extracted_k141_183299 | 1500 | No | NA | 1 | 0 | 0 | Not-determined | Genome-fragment | NA | NA | 0 |
| S12_extracted_k141_70232 | 7594 | No | NA | 6 | 0 | 4 | Low-quality | Genome-fragment | 3.57 | AAI-based (medium-confidence) | 0 |
| S8_extracted_k141_192614 | 2366 | No | NA | 4 | 0 | 2 | Not-determined | Genome-fragment | NA | NA | 0 |
| S8_extracted_k141_157562 | 1576 | No | NA | 1 | 0 | 1 | Not-determined | Genome-fragment | NA | NA | 0 |
| S8_extracted_k141_105664 | 5868 | No | NA | 1 | 0 | 1 | Not-determined | Genome-fragment | NA | NA | 0 |
| S7_vs_32610\|\|full | 7595 | No | NA | 11 | 3 | 0 | Low-quality | Genome-fragment | 16.49 | AAI-based (high-confidence) | 0 |
| S8_extracted_k141_88262 | 2806 | No | NA | 3 | 0 | 1 | Not-determined | Genome-fragment | NA | NA | 0 |
| S8_extracted_k141_114275 | 1321 | No | NA | 1 | 0 | 1 | Not-determined | Genome-fragment | NA | NA | 0 |
| S8_extracted_k141_97057 | 1066 | No | NA | 1 | 0 | 0 | Not-determined | Genome-fragment | NA | NA | 0 |
| S8_extracted_k141_200796 | 1658 | No | NA | 1 | 0 | 1 | Not-determined | Genome-fragment | NA | NA | 0 |
| S8_extracted_k141_166403 | 28905 | No | NA | 43 | 10 | 0 | Medium-quality | Genome-fragment | 84.65 | AAI-based (high-confidence) | 0 |
| S8_vs_k141_50273\|\|full | 7598 | No | NA | 17 | 0 | 0 | Low-quality | Genome-fragment | 9.27 | AAI-based (high-confidence) | 0 |
| S8_extracted_k141_123120 | 1453 | No | NA | 3 | 0 | 2 | Not-determined | Genome-fragment | NA | NA | 0 |
| S6_vs_k141_50145\|\|full | 7600 | No | NA | 8 | 0 | 0 | Low-quality | Genome-fragment | 8.32 | AAI-based (medium-confidence) | 0 |
| S8_extracted_k141_53683 | 1010 | No | NA | 1 | 0 | 1 | Not-determined | Genome-fragment | NA | NA | 0 |
| S8_extracted_k141_183330 | 2507 | No | NA | 2 | 0 | 2 | Not-determined | Genome-fragment | NA | NA | 0 |
| S8_extracted_k141_36438 | 1013 | No | NA | 1 | 0 | 0 | Not-determined | Genome-fragment | NA | NA | 0 |
| S9_vs_k141_11174\|\|full | 7611 | No | NA | 11 | 6 | 0 | Low-quality | Genome-fragment | 17.75 | HMM-based (lower-bound) | 0 |
| S1_vs_k141_126746\|\|full | 7611 | No | NA | 10 | 2 | 0 | Low-quality | Genome-fragment | 17.41 | AAI-based (high-confidence) | 0 |
| S8_extracted_k141_166422 | 1738 | No | NA | 1 | 0 | 1 | Not-determined | Genome-fragment | NA | NA | 0 |
| S8_extracted_k141_192676 | 1382 | No | NA | 1 | 0 | 0 | Not-determined | Genome-fragment | NA | NA | 0 |
| S8_extracted_k141_157625 | 1193 | No | NA | 1 | 0 | 1 | Not-determined | Genome-fragment | NA | NA | 0 |
| S8_extracted_k141_49104 | 7617 | No | NA | 2 | 0 | 2 | Low-quality | Genome-fragment | 2.94 | AAI-based (high-confidence) | 0 |
| S8_extracted_k141_105724 | 4224 | No | NA | 4 | 0 | 2 | Not-determined | Genome-fragment | NA | NA | 0 |
| S8_extracted_k141_157632 | 1024 | No | NA | 5 | 0 | 0 | Not-determined | Genome-fragment | NA | NA | 0 |
| S8_extracted_k141_79769 | 65808 | No | NA | 78 | 14 | 1 | Complete | High-quality | 100 | DTR (high-confidence) | 0 |
| S8_extracted_k141_166448 | 1312 | No | NA | 1 | 0 | 0 | Not-determined | Genome-fragment | NA | NA | 0 |
| S8_extracted_k141_36483 | 4062 | No | NA | 6 | 0 | 3 | Not-determined | Genome-fragment | NA | NA | 0 |
| S8_extracted_k141_44772 | 1174 | No | NA | 1 | 0 | 0 | Not-determined | Genome-fragment | NA | NA | 0 |
| S8_extracted_k141_88329 | 1038 | No | NA | 1 | 0 | 0 | Not-determined | Genome-fragment | NA | NA | 0 |
| S8_extracted_k141_114335 | 2773 | No | NA | 6 | 0 | 0 | Not-determined | Genome-fragment | NA | NA | 0 |
| S8_extracted_k141_123162 | 1072 | No | NA | 1 | 0 | 1 | Not-determined | Genome-fragment | NA | NA | 0 |
| S8_extracted_k141_88333 | 3237 | No | NA | 1 | 0 | 1 | Not-determined | Genome-fragment | NA | NA | 0 |
| S8_extracted_k141_114342 | 1023 | No | NA | 1 | 0 | 0 | Not-determined | Genome-fragment | NA | NA | 0 |
| S3_vs_k141_91452\|\|full | 7621 | Yes | 4385 | 10 | 2 | 1 | Low-quality | Genome-fragment | 5.04 | AAI-based (medium-confidence) | 42.46 |
| S8_extracted_k141_123168 | 1005 | No | NA | 1 | 0 | 0 | Not-determined | Genome-fragment | NA | NA | 0 |
| S8_extracted_k141_114345 | 6385 | No | NA | 7 | 0 | 5 | Not-determined | Genome-fragment | NA | NA | 0 |
| S8_extracted_k141_79789 | 1781 | No | NA | 2 | 0 | 0 | Not-determined | Genome-fragment | NA | NA | 0 |
| S8_extracted_k141_166482 | 1767 | No | NA | 1 | 0 | 1 | Not-determined | Genome-fragment | NA | NA | 0 |
| S3_extracted_k141_104303 | 7622 | No | NA | 17 | 4 | 0 | Low-quality | Genome-fragment | 16.19 | AAI-based (high-confidence) | 0 |
| S8_extracted_k141_44774 | 2024 | No | NA | 1 | 0 | 1 | Not-determined | Genome-fragment | NA | NA | 0 |
| S8_extracted_k141_88360 | 1045 | No | NA | 2 | 0 | 0 | Not-determined | Genome-fragment | NA | NA | 0 |
| S8_extracted_k141_97152 | 1062 | No | NA | 1 | 0 | 0 | Not-determined | Genome-fragment | NA | NA | 0 |
| S8_extracted_k141_88373 | 1120 | No | NA | 2 | 0 | 0 | Not-determined | Genome-fragment | NA | NA | 0 |
| S8_extracted_k141_166500 | 1228 | No | NA | 1 | 0 | 0 | Not-determined | Genome-fragment | NA | NA | 0 |
| S8_extracted_k141_36519 | 1840 | No | NA | 2 | 0 | 1 | Not-determined | Genome-fragment | NA | NA | 0 |
| S8_extracted_k141_157665 | 3717 | No | NA | 3 | 0 | 2 | Not-determined | Genome-fragment | NA | NA | 0 |
| S8_extracted_k141_119663 | 7623 | No | NA | 10 | 3 | 1 | Low-quality | Genome-fragment | 7.63 | AAI-based (high-confidence) | 0 |
| S8_extracted_k141_27846 | 1413 | No | NA | 2 | 0 | 1 | Not-determined | Genome-fragment | NA | NA | 0 |
| S8_extracted_k141_157667 | 3106 | No | NA | 1 | 0 | 1 | Not-determined | Genome-fragment | NA | NA | 0 |
| S14_extracted_k141_39652 | 7624 | No | NA | 10 | 8 | 0 | Low-quality | Genome-fragment | 16.54 | AAI-based (medium-confidence) | 0 |
| S8_extracted_k141_192724 | 15073 | No | NA | 17 | 5 | 0 | Medium-quality | Genome-fragment | 50.58 | HMM-based (lower-bound) | 0 |
| S8_extracted_k141_114360 | 3671 | No | NA | 2 | 0 | 2 | Not-determined | Genome-fragment | NA | NA | 0 |
| S9_extracted_k141_42726 | 7629 | No | NA | 12 | 1 | 1 | Low-quality | Genome-fragment | 16 | AAI-based (high-confidence) | 0 |
| S8_extracted_k141_36535 | 5061 | No | NA | 3 | 0 | 2 | Not-determined | Genome-fragment | NA | NA | 0 |
| S8_extracted_k141_114363 | 4265 | No | NA | 6 | 0 | 2 | Not-determined | Genome-fragment | NA | NA | 0 |
| S7_vs_46372\|\|full | 7631 | No | NA | 12 | 4 | 0 | Low-quality | Genome-fragment | 16.56 | AAI-based (high-confidence) | 0 |
| S17_extracted_k141_54664 | 7632 | No | NA | 10 | 0 | 0 | Low-quality | Genome-fragment | 26.46 | AAI-based (medium-confidence) | 0 |
| S8_extracted_k141_157700 | 1751 | No | NA | 3 | 0 | 1 | Not-determined | Genome-fragment | NA | NA | 0 |
| S8_extracted_k141_166536 | 1927 | No | NA | 2 | 0 | 2 | Not-determined | Genome-fragment | NA | NA | 0 |
| S8_extracted_k141_140547 | 7530 | No | NA | 7 | 0 | 4 | Not-determined | Genome-fragment | NA | NA | 0 |
| S8_extracted_k141_1767 | 1003 | No | NA | 1 | 0 | 0 | Not-determined | Genome-fragment | NA | NA | 0 |
| S5_extracted_k141_859 | 7636 | No | NA | 6 | 0 | 2 | Low-quality | Genome-fragment | 7.17 | AAI-based (medium-confidence) | 0 |
| S8_extracted_k141_192761 | 1613 | No | NA | 1 | 0 | 1 | Not-determined | Genome-fragment | NA | NA | 0 |
| S3_extracted_k141_84325 | 7637 | No | NA | 13 | 2 | 0 | Low-quality | Genome-fragment | 16.09 | HMM-based (lower-bound) | 0 |
| S8_extracted_k141_175039 | 1035 | No | NA | 2 | 0 | 1 | Not-determined | Genome-fragment | NA | NA | 0 |
| S8_vs_k141_199951\|\|full | 7638 | No | NA | 8 | 3 | 0 | Low-quality | Genome-fragment | 5.5 | AAI-based (high-confidence) | 0 |
| S8_extracted_k141_114399 | 1227 | No | NA | 1 | 0 | 1 | Not-determined | Genome-fragment | NA | NA | 0 |
| S8_extracted_k141_1782 | 1360 | No | NA | 4 | 0 | 0 | Not-determined | Genome-fragment | NA | NA | 0 |
| S8_extracted_k141_36583 | 1402 | No | NA | 2 | 0 | 0 | Not-determined | Genome-fragment | NA | NA | 0 |
| S8_extracted_k141_18885 | 2425 | No | NA | 3 | 0 | 1 | Not-determined | Genome-fragment | NA | NA | 0 |
| S8_extracted_k141_175053 | 4247 | No | NA | 4 | 0 | 4 | Not-determined | Genome-fragment | NA | NA | 0 |
| S8_extracted_k141_114415 | 1788 | No | NA | 2 | 0 | 2 | Not-determined | Genome-fragment | NA | NA | 0 |
| S8_extracted_k141_97201 | 5625 | No | NA | 5 | 0 | 5 | Not-determined | Genome-fragment | NA | NA | 0 |
| S8_extracted_k141_1800 | 1627 | No | NA | 2 | 0 | 0 | Not-determined | Genome-fragment | NA | NA | 0 |
| S8_extracted_k141_175062 | 2673 | No | NA | 1 | 0 | 1 | Not-determined | Genome-fragment | NA | NA | 0 |
| S8_extracted_k141_1802 | 2022 | No | NA | 3 | 0 | 0 | Not-determined | Genome-fragment | NA | NA | 0 |
| S8_extracted_k141_192795 | 5328 | No | NA | 4 | 0 | 1 | Not-determined | Genome-fragment | NA | NA | 0 |
| S8_extracted_k141_18911 | 1410 | No | NA | 1 | 0 | 0 | Not-determined | Genome-fragment | NA | NA | 0 |
| S8_extracted_k141_149012 | 1320 | No | NA | 1 | 0 | 0 | Not-determined | Genome-fragment | NA | NA | 0 |
| S8_extracted_k141_97218 | 1105 | No | NA | 1 | 0 | 0 | Not-determined | Genome-fragment | NA | NA | 0 |
| S8_extracted_k141_157719 | 1619 | No | NA | 2 | 0 | 1 | Not-determined | Genome-fragment | NA | NA | 0 |
| S8_extracted_k141_183402 | 1352 | No | NA | 2 | 0 | 0 | Not-determined | Genome-fragment | NA | NA | 0 |
| S8_extracted_k141_149026 | 1511 | No | NA | 1 | 0 | 0 | Not-determined | Genome-fragment | NA | NA | 0 |
| S8_extracted_k141_53878 | 3795 | No | NA | 3 | 0 | 3 | Not-determined | Genome-fragment | NA | NA | 0 |
| S8_extracted_k141_166603 | 1345 | No | NA | 1 | 0 | 0 | Not-determined | Genome-fragment | NA | NA | 0 |
| S8_extracted_k141_200938 | 3554 | No | NA | 4 | 0 | 1 | Not-determined | Genome-fragment | NA | NA | 0 |
| S8_extracted_k141_183425 | 1276 | No | NA | 1 | 0 | 1 | Not-determined | Genome-fragment | NA | NA | 0 |
| S8_extracted_k141_62370 | 4228 | No | NA | 6 | 0 | 5 | Not-determined | Genome-fragment | NA | NA | 0 |
| S8_extracted_k141_62372 | 1244 | No | NA | 3 | 0 | 1 | Not-determined | Genome-fragment | NA | NA | 0 |
| S8_extracted_k141_123272 | 1116 | No | NA | 2 | 0 | 1 | Not-determined | Genome-fragment | NA | NA | 0 |
| S3_vs_k141_104660\|\|full | 7643 | No | NA | 10 | 2 | 0 | Low-quality | Genome-fragment | 5.22 | HMM-based (lower-bound) | 0 |
| S3_vs_k141_49386\|\|full | 7647 | No | NA | 12 | 2 | 1 | Low-quality | Genome-fragment | 16.24 | AAI-based (high-confidence) | 0 |
| S8_extracted_k141_62380 | 3365 | No | NA | 2 | 0 | 2 | Not-determined | Genome-fragment | NA | NA | 0 |
| S17_vs_k141_55871\|\|full | 7650 | No | NA | 15 | 1 | 1 | Low-quality | Genome-fragment | 9.95 | HMM-based (lower-bound) | 0 |
| S5_vs_k141_142200\|\|full | 7651 | No | NA | 13 | 5 | 1 | Low-quality | Genome-fragment | 15.61 | AAI-based (high-confidence) | 0 |
| S8_extracted_k141_1839 | 1581 | No | NA | 4 | 0 | 0 | Not-determined | Genome-fragment | NA | NA | 0 |
| S8_extracted_k141_149070 | 1253 | No | NA | 1 | 0 | 0 | Not-determined | Genome-fragment | NA | NA | 0 |
| S8_extracted_k141_183442 | 1438 | No | NA | 1 | 0 | 1 | Not-determined | Genome-fragment | NA | NA | 0 |
| S8_extracted_k141_131802 | 99043 | No | NA | 97 | 18 | 1 | Medium-quality | Genome-fragment | 64.45 | HMM-based (lower-bound) | 0 |
| S10_extracted_13386 | 7652 | No | NA | 7 | 0 | 2 | Low-quality | Genome-fragment | 7.08 | AAI-based (medium-confidence) | 0 |
| S8_extracted_k141_70763 | 1094 | No | NA | 1 | 0 | 1 | Not-determined | Genome-fragment | NA | NA | 0 |
| S8_extracted_k141_123292 | 2722 | No | NA | 3 | 0 | 0 | Not-determined | Genome-fragment | NA | NA | 0 |
| S8_extracted_k141_131820 | 1403 | No | NA | 2 | 0 | 1 | Not-determined | Genome-fragment | NA | NA | 0 |
| S8_extracted_k141_97293 | 1654 | No | NA | 2 | 0 | 0 | Not-determined | Genome-fragment | NA | NA | 0 |
| S8_extracted_k141_192846 | 4310 | No | NA | 4 | 0 | 1 | Not-determined | Genome-fragment | NA | NA | 0 |
| S7_extracted_93746 | 7655 | No | NA | 12 | 0 | 1 | Low-quality | Genome-fragment | 16.99 | AAI-based (high-confidence) | 0 |
| S16_extracted_145611 | 7658 | No | NA | 15 | 3 | 1 | Low-quality | Genome-fragment | 11.91 | AAI-based (high-confidence) | 0 |
| S11_extracted_k141_135 | 7659 | No | NA | 6 | 4 | 0 | Low-quality | Genome-fragment | 12.45 | AAI-based (medium-confidence) | 0 |
| S8_extracted_k141_18964 | 1027 | No | NA | 2 | 0 | 0 | Not-determined | Genome-fragment | NA | NA | 0 |
| S8_extracted_k141_131835 | 1134 | No | NA | 2 | 0 | 2 | Not-determined | Genome-fragment | NA | NA | 0 |
| S8_extracted_k141_79892 | 3156 | No | NA | 5 | 0 | 3 | Not-determined | Genome-fragment | NA | NA | 0 |
| S8_extracted_k141_105875 | 3567 | No | NA | 3 | 0 | 2 | Not-determined | Genome-fragment | NA | NA | 0 |
| S8_extracted_k141_10253 | 4009 | No | NA | 1 | 0 | 1 | Not-determined | Genome-fragment | NA | NA | 0 |
| S8_extracted_k141_72161 | 7662 | No | NA | 6 | 0 | 3 | Low-quality | Genome-fragment | 4.53 | AAI-based (high-confidence) | 0 |
| S1_extracted_k141_117448 | 7665 | No | NA | 16 | 3 | 0 | Low-quality | Genome-fragment | 16.98 | AAI-based (high-confidence) | 0 |
| S8_extracted_k141_140679 | 1346 | No | NA | 2 | 0 | 1 | Not-determined | Genome-fragment | NA | NA | 0 |
| S8_extracted_k141_200971 | 2817 | No | NA | 3 | 0 | 2 | Not-determined | Genome-fragment | NA | NA | 0 |
| S8_extracted_k141_10259 | 8038 | No | NA | 6 | 0 | 4 | Not-determined | Genome-fragment | NA | NA | 0 |
| S8_extracted_k141_200975 | 1193 | No | NA | 1 | 0 | 0 | Not-determined | Genome-fragment | NA | NA | 0 |
| S8_extracted_k141_53947 | 8390 | No | NA | 6 | 0 | 3 | Not-determined | Genome-fragment | NA | NA | 0 |
| S12_vs_k141_28329\|\|full | 7666 | No | NA | 8 | 1 | 2 | Low-quality | Genome-fragment | 8.85 | HMM-based (lower-bound) | 0 |
| S8_extracted_k141_27934 | 5549 | No | NA | 6 | 0 | 1 | Not-determined | Genome-fragment | NA | NA | 0 |
| S8_extracted_k141_114516 | 4409 | No | NA | 2 | 0 | 1 | Not-determined | Genome-fragment | NA | NA | 0 |
| S8_extracted_k141_53950 | 1034 | No | NA | 2 | 0 | 0 | Not-determined | Genome-fragment | NA | NA | 0 |
| S8_extracted_k141_131864 | 1168 | No | NA | 1 | 0 | 1 | Not-determined | Genome-fragment | NA | NA | 0 |
| S8_extracted_k141_79919 | 1513 | No | NA | 3 | 0 | 0 | Not-determined | Genome-fragment | NA | NA | 0 |
| S6_extracted_k141_38259 | 7667 | No | NA | 18 | 1 | 0 | Low-quality | Genome-fragment | 5.56 | AAI-based (high-confidence) | 0 |
| S8_extracted_k141_44948 | 3086 | No | NA | 3 | 0 | 3 | Not-determined | Genome-fragment | NA | NA | 0 |
| S8_extracted_k141_18982 | 2971 | No | NA | 4 | 0 | 4 | Not-determined | Genome-fragment | NA | NA | 0 |
| S3_vs_k141_23594\|\|full | 7668 | No | NA | 14 | 2 | 0 | Low-quality | Genome-fragment | 19.39 | AAI-based (high-confidence) | 0 |
| S8_extracted_k141_131877 | 1029 | No | NA | 2 | 0 | 0 | Not-determined | Genome-fragment | NA | NA | 0 |
| S8_extracted_k141_70807 | 1228 | No | NA | 1 | 0 | 0 | Not-determined | Genome-fragment | NA | NA | 0 |
| S8_extracted_k141_88547 | 1212 | No | NA | 1 | 0 | 1 | Not-determined | Genome-fragment | NA | NA | 0 |
| S8_extracted_k141_10280 | 3222 | No | NA | 2 | 0 | 2 | Not-determined | Genome-fragment | NA | NA | 0 |
| S8_extracted_k141_140697 | 3649 | No | NA | 5 | 0 | 3 | Not-determined | Genome-fragment | NA | NA | 0 |
| S8_extracted_k141_53969 | 1210 | No | NA | 1 | 0 | 0 | Not-determined | Genome-fragment | NA | NA | 0 |
| S8_extracted_k141_10284 | 1241 | No | NA | 1 | 0 | 0 | Not-determined | Genome-fragment | NA | NA | 0 |
| S8_extracted_k141_105900 | 6298 | No | NA | 4 | 0 | 4 | Not-determined | Genome-fragment | NA | NA | 0 |
| S8_extracted_k141_201009 | 1378 | No | NA | 2 | 0 | 2 | Not-determined | Genome-fragment | NA | NA | 0 |
| S8_extracted_k141_36708 | 4155 | No | NA | 5 | 0 | 1 | Not-determined | Genome-fragment | NA | NA | 0 |
| S12_extracted_k141_76682 | 7671 | No | NA | 8 | 1 | 1 | Low-quality | Genome-fragment | 5.63 | HMM-based (lower-bound) | 0 |
| S8_extracted_k141_149167 | 1244 | No | NA | 1 | 0 | 0 | Not-determined | Genome-fragment | NA | NA | 0 |
| S8_extracted_k141_70819 | 3037 | No | NA | 2 | 0 | 1 | Not-determined | Genome-fragment | NA | NA | 0 |
| S8_extracted_k141_131924 | 1223 | No | NA | 1 | 0 | 1 | Not-determined | Genome-fragment | NA | NA | 0 |
| S8_extracted_k141_36723 | 1007 | No | NA | 1 | 0 | 1 | Not-determined | Genome-fragment | NA | NA | 0 |
| S8_extracted_k141_36725 | 3608 | No | NA | 3 | 0 | 3 | Not-determined | Genome-fragment | NA | NA | 0 |
| S12_vs_k141_160426\|\|full | 7671 | Yes | 2663 | 8 | 1 | 2 | Low-quality | Genome-fragment | 3.08 | HMM-based (lower-bound) | 65.28 |
| S8_extracted_k141_201041 | 5323 | No | NA | 3 | 0 | 1 | Not-determined | Genome-fragment | NA | NA | 0 |
| S17_extracted_k141_47459 | 7673 | No | NA | 11 | 0 | 0 | Low-quality | Genome-fragment | 12.87 | AAI-based (high-confidence) | 0 |
| S16_extracted_125130 | 7675 | No | NA | 7 | 6 | 0 | Low-quality | Genome-fragment | 20.66 | AAI-based (high-confidence) | 0 |
| S10_vs_106371\|\|full | 7677 | No | NA | 5 | 0 | 5 | Low-quality | Genome-fragment | 0.91 | AAI-based (medium-confidence) | 0 |
| S8_extracted_k141_79988 | 1695 | No | NA | 1 | 0 | 1 | Not-determined | Genome-fragment | NA | NA | 0 |
| S8_extracted_k141_131940 | 1157 | No | NA | 2 | 0 | 0 | Not-determined | Genome-fragment | NA | NA | 0 |
| S8_extracted_k141_70837 | 1849 | No | NA | 3 | 0 | 0 | Not-determined | Genome-fragment | NA | NA | 0 |
| S8_extracted_k141_79991 | 3231 | No | NA | 3 | 0 | 1 | Not-determined | Genome-fragment | NA | NA | 0 |
| S8_extracted_k141_28026 | 2015 | No | NA | 4 | 0 | 0 | Not-determined | Genome-fragment | NA | NA | 0 |
| S8_extracted_k141_123387 | 1016 | No | NA | 1 | 0 | 1 | Not-determined | Genome-fragment | NA | NA | 0 |
| S8_extracted_k141_88589 | 2575 | No | NA | 2 | 0 | 2 | Not-determined | Genome-fragment | NA | NA | 0 |
| S8_extracted_k141_45027 | 3708 | No | NA | 6 | 0 | 0 | Not-determined | Genome-fragment | NA | NA | 0 |
| S8_extracted_k141_183571 | 1118 | No | NA | 2 | 0 | 0 | Not-determined | Genome-fragment | NA | NA | 0 |
| S8_extracted_k141_54044 | 6771 | No | NA | 4 | 0 | 2 | Not-determined | Genome-fragment | NA | NA | 0 |
| S8_extracted_k141_10365 | 1046 | No | NA | 1 | 0 | 1 | Not-determined | Genome-fragment | NA | NA | 0 |
| S8_extracted_k141_80007 | 1064 | No | NA | 1 | 0 | 0 | Not-determined | Genome-fragment | NA | NA | 0 |
| S8_extracted_k141_19050 | 1058 | No | NA | 1 | 0 | 1 | Not-determined | Genome-fragment | NA | NA | 0 |
| S8_extracted_k141_183574 | 1691 | No | NA | 3 | 0 | 0 | Not-determined | Genome-fragment | NA | NA | 0 |
| S8_extracted_k141_192932 | 1046 | No | NA | 1 | 0 | 1 | Not-determined | Genome-fragment | NA | NA | 0 |
| S8_extracted_k141_175211 | 1607 | No | NA | 3 | 0 | 2 | Not-determined | Genome-fragment | NA | NA | 0 |
| S8_extracted_k141_149219 | 1708 | No | NA | 1 | 0 | 1 | Not-determined | Genome-fragment | NA | NA | 0 |
| S8_extracted_k141_10376 | 1143 | No | NA | 1 | 0 | 1 | Not-determined | Genome-fragment | NA | NA | 0 |
| S8_extracted_k141_28065 | 2454 | No | NA | 3 | 0 | 2 | Not-determined | Genome-fragment | NA | NA | 0 |
| S8_extracted_k141_157831 | 1035 | No | NA | 2 | 0 | 1 | Not-determined | Genome-fragment | NA | NA | 0 |
| S8_extracted_k141_62519 | 2043 | No | NA | 1 | 0 | 0 | Not-determined | Genome-fragment | NA | NA | 0 |
| S8_extracted_k141_19061 | 1019 | No | NA | 1 | 0 | 0 | Not-determined | Genome-fragment | NA | NA | 0 |
| S8_extracted_k141_183589 | 2088 | No | NA | 3 | 0 | 2 | Not-determined | Genome-fragment | NA | NA | 0 |
| S8_extracted_k141_140730 | 7514 | No | NA | 6 | 0 | 3 | Not-determined | Genome-fragment | NA | NA | 0 |
| S8_extracted_k141_192953 | 1335 | No | NA | 2 | 0 | 0 | Not-determined | Genome-fragment | NA | NA | 0 |
| S8_extracted_k141_183599 | 1242 | No | NA | 2 | 0 | 1 | Not-determined | Genome-fragment | NA | NA | 0 |
| S8_extracted_k141_105993 | 1543 | No | NA | 1 | 0 | 1 | Not-determined | Genome-fragment | NA | NA | 0 |
| S8_extracted_k141_149271 | 1294 | No | NA | 2 | 0 | 1 | Not-determined | Genome-fragment | NA | NA | 0 |
| S8_extracted_k141_45048 | 1086 | No | NA | 1 | 0 | 0 | Not-determined | Genome-fragment | NA | NA | 0 |
| S8_extracted_k141_149273 | 1053 | No | NA | 2 | 0 | 0 | Not-determined | Genome-fragment | NA | NA | 0 |
| S8_extracted_k141_114603 | 2403 | No | NA | 2 | 0 | 0 | Not-determined | Genome-fragment | NA | NA | 0 |
| S8_extracted_k141_201069 | 3591 | No | NA | 7 | 0 | 3 | Not-determined | Genome-fragment | NA | NA | 0 |
| S8_vs_k141_53035\|\|full | 7678 | No | NA | 15 | 1 | 1 | Low-quality | Genome-fragment | 6.87 | HMM-based (lower-bound) | 0 |
| S8_extracted_k141_88637 | 1323 | No | NA | 2 | 0 | 1 | Not-determined | Genome-fragment | NA | NA | 0 |
| S8_extracted_k141_201080 | 1061 | No | NA | 1 | 0 | 0 | Not-determined | Genome-fragment | NA | NA | 0 |
| S8_extracted_k141_88652 | 1213 | No | NA | 2 | 0 | 2 | Not-determined | Genome-fragment | NA | NA | 0 |
| S8_extracted_k141_19095 | 1245 | No | NA | 3 | 0 | 0 | Not-determined | Genome-fragment | NA | NA | 0 |
| S8_extracted_k141_1915 | 1616 | No | NA | 2 | 0 | 1 | Not-determined | Genome-fragment | NA | NA | 0 |
| S8_extracted_k141_123448 | 1601 | No | NA | 2 | 0 | 1 | Not-determined | Genome-fragment | NA | NA | 0 |
| S8_extracted_k141_166677 | 92899 | No | NA | 91 | 11 | 0 | High-quality | High-quality | 96.85 | AAI-based (high-confidence) | 0 |
| S8_extracted_k141_149297 | 1290 | No | NA | 1 | 0 | 0 | Not-determined | Genome-fragment | NA | NA | 0 |
| S8_extracted_k141_114631 | 1301 | No | NA | 1 | 0 | 1 | Not-determined | Genome-fragment | NA | NA | 0 |
| S8_extracted_k141_140776 | 1252 | No | NA | 1 | 0 | 0 | Not-determined | Genome-fragment | NA | NA | 0 |
| S8_extracted_k141_201091 | 3620 | No | NA | 2 | 0 | 2 | Not-determined | Genome-fragment | NA | NA | 0 |
| S12_vs_k141_157861\|\|full | 7679 | No | NA | 6 | 0 | 2 | Low-quality | Genome-fragment | 3.25 | AAI-based (medium-confidence) | 0 |
| S8_extracted_k141_28131 | 1457 | No | NA | 2 | 0 | 0 | Not-determined | Genome-fragment | NA | NA | 0 |
| S8_extracted_k141_149308 | 1363 | No | NA | 2 | 0 | 1 | Not-determined | Genome-fragment | NA | NA | 0 |
| S11_vs_k141_2696\|\|full | 7683 | No | NA | 17 | 2 | 0 | Low-quality | Genome-fragment | 17.05 | AAI-based (medium-confidence) | 0 |
| S8_extracted_k141_149309 | 1731 | No | NA | 2 | 0 | 0 | Not-determined | Genome-fragment | NA | NA | 0 |
| S8_extracted_k141_149310 | 1013 | No | NA | 1 | 0 | 0 | Not-determined | Genome-fragment | NA | NA | 0 |
| S16_vs_130649\|\|full | 7687 | No | NA | 5 | 0 | 4 | Low-quality | Genome-fragment | 4.06 | AAI-based (medium-confidence) | 0 |
| S8_extracted_k141_149317 | 1638 | No | NA | 1 | 0 | 0 | Not-determined | Genome-fragment | NA | NA | 0 |
| S9_vs_k141_26115\|\|full | 7688 | No | NA | 9 | 6 | 0 | Low-quality | Genome-fragment | 16.71 | AAI-based (medium-confidence) | 0 |
| S8_extracted_k141_192989 | 1626 | No | NA | 2 | 0 | 1 | Not-determined | Genome-fragment | NA | NA | 0 |
| S5_extracted_k141_22241 | 7700 | No | NA | 13 | 7 | 0 | Low-quality | Genome-fragment | 15.37 | AAI-based (medium-confidence) | 0 |
| S8_extracted_k141_203322 | 7700 | No | NA | 8 | 0 | 6 | Low-quality | Genome-fragment | 2.41 | AAI-based (high-confidence) | 0 |
| S8_extracted_k141_183632 | 3369 | No | NA | 2 | 0 | 1 | Not-determined | Genome-fragment | NA | NA | 0 |
| S8_extracted_k141_10446 | 1049 | No | NA | 2 | 0 | 1 | Not-determined | Genome-fragment | NA | NA | 0 |
| S8_extracted_k141_1950 | 1288 | No | NA | 2 | 0 | 1 | Not-determined | Genome-fragment | NA | NA | 0 |
| S8_extracted_k141_19166 | 1079 | No | NA | 1 | 0 | 1 | Not-determined | Genome-fragment | NA | NA | 0 |
| S8_extracted_k141_132028 | 1098 | No | NA | 1 | 0 | 1 | Not-determined | Genome-fragment | NA | NA | 0 |
| S8_extracted_k141_62586 | 2463 | No | NA | 1 | 0 | 1 | Not-determined | Genome-fragment | NA | NA | 0 |
| S8_extracted_k141_80126 | 1282 | No | NA | 4 | 0 | 0 | Not-determined | Genome-fragment | NA | NA | 0 |
| S5_vs_k141_68869\|\|full | 7709 | No | NA | 18 | 3 | 0 | Low-quality | Genome-fragment | 17.38 | AAI-based (medium-confidence) | 0 |
| S8_extracted_k141_183669 | 1211 | No | NA | 2 | 0 | 0 | Not-determined | Genome-fragment | NA | NA | 0 |
| S8_extracted_k141_106039 | 2008 | No | NA | 2 | 0 | 2 | Not-determined | Genome-fragment | NA | NA | 0 |
| S8_extracted_k141_193021 | 7467 | No | NA | 5 | 0 | 4 | Not-determined | Genome-fragment | NA | NA | 0 |
| S8_extracted_k141_1979 | 1667 | No | NA | 1 | 0 | 0 | Not-determined | Genome-fragment | NA | NA | 0 |
| S8_extracted_k141_80129 | 4256 | No | NA | 4 | 0 | 1 | Not-determined | Genome-fragment | NA | NA | 0 |
| S5_vs_k141_51908\|\|full | 7711 | No | NA | 10 | 3 | 0 | Low-quality | Genome-fragment | 18.57 | AAI-based (medium-confidence) | 0 |
| S12_vs_k141_63130\|\|full | 7712 | No | NA | 5 | 0 | 5 | Low-quality | Genome-fragment | 11.54 | AAI-based (medium-confidence) | 0 |
| S15_vs_75680\|\|full | 7712 | No | NA | 12 | 3 | 0 | Low-quality | Genome-fragment | 13.82 | AAI-based (high-confidence) | 0 |
| S8_extracted_k141_166752 | 1759 | No | NA | 1 | 0 | 1 | Not-determined | Genome-fragment | NA | NA | 0 |
| S8_extracted_k141_175263 | 3630 | No | NA | 4 | 0 | 3 | Not-determined | Genome-fragment | NA | NA | 0 |
| S8_extracted_k141_193042 | 1277 | No | NA | 2 | 0 | 1 | Not-determined | Genome-fragment | NA | NA | 0 |
| S8_extracted_k141_157948 | 3858 | No | NA | 2 | 0 | 2 | Not-determined | Genome-fragment | NA | NA | 0 |
| S8_extracted_k141_54157 | 2998 | No | NA | 4 | 0 | 4 | Not-determined | Genome-fragment | NA | NA | 0 |
| S8_extracted_k141_28182 | 2214 | No | NA | 2 | 0 | 0 | Not-determined | Genome-fragment | NA | NA | 0 |
| S17_vs_k141_32462\|\|full | 7717 | No | NA | 8 | 0 | 4 | Low-quality | Genome-fragment | 17.84 | AAI-based (medium-confidence) | 0 |
| S8_extracted_k141_36885 | 6353 | No | NA | 7 | 0 | 7 | Not-determined | Genome-fragment | NA | NA | 0 |
| S11_vs_k141_93775\|\|full | 7721 | No | NA | 21 | 0 | 0 | Low-quality | Genome-fragment | 15.09 | AAI-based (medium-confidence) | 0 |
| S8_extracted_k141_114695 | 5246 | No | NA | 6 | 0 | 3 | Not-determined | Genome-fragment | NA | NA | 0 |
| S8_extracted_k141_45127 | 3099 | No | NA | 2 | 0 | 2 | Not-determined | Genome-fragment | NA | NA | 0 |
| S8_extracted_k141_157950 | 6948 | No | NA | 4 | 0 | 2 | Not-determined | Genome-fragment | NA | NA | 0 |
| S8_extracted_k141_123520 | 2387 | No | NA | 1 | 0 | 1 | Not-determined | Genome-fragment | NA | NA | 0 |
| S3_vs_k141_39841\|\|full | 7721 | No | NA | 8 | 1 | 0 | Low-quality | Genome-fragment | 20.22 | AAI-based (high-confidence) | 0 |
| S7_extracted_99026 | 7725 | No | NA | 19 | 6 | 0 | Low-quality | Genome-fragment | 17.33 | AAI-based (high-confidence) | 0 |
| S8_vs_k141_4328\|\|full | 7726 | No | NA | 9 | 1 | 0 | Low-quality | Genome-fragment | 7.99 | AAI-based (medium-confidence) | 0 |
| S8_extracted_k141_106094 | 1802 | No | NA | 2 | 0 | 1 | Not-determined | Genome-fragment | NA | NA | 0 |
| S8_extracted_k141_114716 | 1234 | No | NA | 2 | 0 | 1 | Not-determined | Genome-fragment | NA | NA | 0 |
| S8_extracted_k141_62660 | 1250 | No | NA | 1 | 0 | 1 | Not-determined | Genome-fragment | NA | NA | 0 |
| S8_extracted_k141_201185 | 1601 | No | NA | 3 | 0 | 3 | Not-determined | Genome-fragment | NA | NA | 0 |
| S8_extracted_k141_36939 | 5807 | No | NA | 8 | 0 | 5 | Not-determined | Genome-fragment | NA | NA | 0 |
| S14_vs_k141_110264\|\|full | 7727 | Yes | 5756 | 10 | 4 | 1 | Low-quality | Genome-fragment | 8.87 | AAI-based (medium-confidence) | 25.51 |
| S8_extracted_k141_175309 | 1052 | No | NA | 1 | 0 | 0 | Not-determined | Genome-fragment | NA | NA | 0 |
| S8_extracted_k141_80179 | 1318 | No | NA | 1 | 0 | 0 | Not-determined | Genome-fragment | NA | NA | 0 |
| S8_extracted_k141_97447 | 1456 | No | NA | 2 | 0 | 2 | Not-determined | Genome-fragment | NA | NA | 0 |
| S8_extracted_k141_166782 | 2628 | No | NA | 2 | 0 | 1 | Not-determined | Genome-fragment | NA | NA | 0 |
| S8_extracted_k141_201191 | 1210 | No | NA | 2 | 0 | 0 | Not-determined | Genome-fragment | NA | NA | 0 |
| S8_extracted_k141_45157 | 1752 | No | NA | 2 | 0 | 2 | Not-determined | Genome-fragment | NA | NA | 0 |
| S12_vs_k141_150241\|\|full | 7727 | No | NA | 7 | 0 | 5 | Low-quality | Genome-fragment | 5.48 | AAI-based (medium-confidence) | 0 |
| S8_extracted_k141_193047 | 7732 | No | NA | 8 | 1 | 0 | Low-quality | Genome-fragment | 7.87 | AAI-based (high-confidence) | 0 |
| S5_extracted_k141_71482 | 7740 | No | NA | 5 | 0 | 3 | Low-quality | Genome-fragment | 16.44 | AAI-based (medium-confidence) | 0 |
| S8_extracted_k141_181690 | 7741 | No | NA | 8 | 1 | 0 | Low-quality | Genome-fragment | 7.71 | AAI-based (high-confidence) | 0 |
| S8_extracted_k141_149410 | 3984 | No | NA | 2 | 0 | 2 | Not-determined | Genome-fragment | NA | NA | 0 |
| S8_extracted_k141_114752 | 1720 | No | NA | 2 | 0 | 0 | Not-determined | Genome-fragment | NA | NA | 0 |
| S5_vs_k141_67397\|\|full | 7742 | No | NA | 10 | 2 | 0 | Low-quality | Genome-fragment | 15.36 | AAI-based (medium-confidence) | 0 |
| S8_extracted_k141_2065 | 1953 | No | NA | 2 | 0 | 2 | Not-determined | Genome-fragment | NA | NA | 0 |
| S5_vs_k141_112920\|\|full | 7747 | No | NA | 13 | 2 | 0 | Low-quality | Genome-fragment | 16.92 | AAI-based (high-confidence) | 0 |
| S8_extracted_k141_114764 | 2602 | No | NA | 2 | 0 | 2 | Not-determined | Genome-fragment | NA | NA | 0 |
| S8_extracted_k141_62715 | 1625 | No | NA | 2 | 0 | 1 | Not-determined | Genome-fragment | NA | NA | 0 |
| S8_extracted_k141_19210 | 1990 | No | NA | 1 | 0 | 0 | Not-determined | Genome-fragment | NA | NA | 0 |
| S8_extracted_k141_2074 | 2385 | No | NA | 1 | 0 | 0 | Not-determined | Genome-fragment | NA | NA | 0 |
| S8_extracted_k141_28240 | 3035 | No | NA | 3 | 0 | 0 | Not-determined | Genome-fragment | NA | NA | 0 |
| S8_extracted_k141_149421 | 2903 | No | NA | 4 | 0 | 2 | Not-determined | Genome-fragment | NA | NA | 0 |
| S8_extracted_k141_183754 | 9936 | No | NA | 5 | 0 | 5 | Not-determined | Genome-fragment | NA | NA | 0 |
| S8_extracted_k141_88839 | 1109 | No | NA | 1 | 0 | 0 | Not-determined | Genome-fragment | NA | NA | 0 |
| S8_extracted_k141_183763 | 1502 | No | NA | 1 | 0 | 0 | Not-determined | Genome-fragment | NA | NA | 0 |
| S4_extracted_20624 | 7748 | No | NA | 13 | 3 | 0 | Low-quality | Genome-fragment | 16.99 | AAI-based (high-confidence) | 0 |
| S8_extracted_k141_140910 | 2362 | No | NA | 3 | 0 | 0 | Not-determined | Genome-fragment | NA | NA | 0 |
| S8_extracted_k141_193132 | 1841 | No | NA | 2 | 0 | 0 | Not-determined | Genome-fragment | NA | NA | 0 |
| S8_extracted_k141_45225 | 1563 | No | NA | 1 | 0 | 1 | Not-determined | Genome-fragment | NA | NA | 0 |
| S8_extracted_k141_45227 | 1044 | No | NA | 2 | 0 | 1 | Not-determined | Genome-fragment | NA | NA | 0 |
| S16_vs_157373\|\|full | 7749 | No | NA | 14 | 0 | 0 | Low-quality | Genome-fragment | 3.99 | AAI-based (high-confidence) | 0 |
| S16_vs_167710\|\|full | 7752 | No | NA | 12 | 2 | 1 | Low-quality | Genome-fragment | 18.25 | AAI-based (high-confidence) | 0 |
| S8_extracted_k141_166842 | 1992 | No | NA | 3 | 0 | 1 | Not-determined | Genome-fragment | NA | NA | 0 |
| S12_vs_k141_104423\|\|full | 7752 | No | NA | 10 | 0 | 2 | Low-quality | Genome-fragment | 1.87 | AAI-based (medium-confidence) | 0 |
| S8_extracted_k141_2120 | 1378 | No | NA | 2 | 0 | 1 | Not-determined | Genome-fragment | NA | NA | 0 |
| S8_extracted_k141_201291 | 1077 | No | NA | 2 | 0 | 1 | Not-determined | Genome-fragment | NA | NA | 0 |
| S14_vs_k141_37006\|\|full | 7757 | No | NA | 17 | 0 | 0 | Low-quality | Genome-fragment | 22.58 | AAI-based (high-confidence) | 0 |
| S8_extracted_k141_201300 | 2467 | No | NA | 1 | 0 | 1 | Not-determined | Genome-fragment | NA | NA | 0 |
| S8_extracted_k141_106210 | 1088 | No | NA | 2 | 0 | 1 | Not-determined | Genome-fragment | NA | NA | 0 |
| S8_extracted_k141_114847 | 1964 | No | NA | 2 | 0 | 1 | Not-determined | Genome-fragment | NA | NA | 0 |
| S8_extracted_k141_62796 | 2708 | No | NA | 3 | 0 | 3 | Not-determined | Genome-fragment | NA | NA | 0 |
| S8_vs_k141_192815\|\|full | 7762 | Yes | 5707 | 5 | 1 | 2 | Low-quality | Genome-fragment | 3.23 | HMM-based (lower-bound) | 26.48 |
| S8_extracted_k141_201321 | 5089 | No | NA | 3 | 0 | 1 | Not-determined | Genome-fragment | NA | NA | 0 |
| S8_extracted_k141_175453 | 1155 | No | NA | 1 | 0 | 1 | Not-determined | Genome-fragment | NA | NA | 0 |
| S8_extracted_k141_158085 | 4044 | No | NA | 5 | 0 | 4 | Not-determined | Genome-fragment | NA | NA | 0 |
| S3_extracted_k141_120468 | 7766 | No | NA | 8 | 1 | 0 | Low-quality | Genome-fragment | 5.31 | AAI-based (high-confidence) | 0 |
| S8_extracted_k141_183815 | 1758 | No | NA | 1 | 0 | 1 | Not-determined | Genome-fragment | NA | NA | 0 |
| S8_extracted_k141_71061 | 1039 | No | NA | 1 | 0 | 0 | Not-determined | Genome-fragment | NA | NA | 0 |
| S8_extracted_k141_140975 | 4378 | No | NA | 4 | 0 | 2 | Not-determined | Genome-fragment | NA | NA | 0 |
| S8_extracted_k141_54322 | 1899 | No | NA | 2 | 0 | 1 | Not-determined | Genome-fragment | NA | NA | 0 |
| S8_extracted_k141_114891 | 1522 | No | NA | 1 | 0 | 1 | Not-determined | Genome-fragment | NA | NA | 0 |
| S8_extracted_k141_201340 | 6486 | No | NA | 4 | 0 | 3 | Not-determined | Genome-fragment | NA | NA | 0 |
| S8_extracted_k141_62819 | 1480 | No | NA | 1 | 0 | 1 | Not-determined | Genome-fragment | NA | NA | 0 |
| S8_extracted_k141_45295 | 1227 | No | NA | 1 | 0 | 1 | Not-determined | Genome-fragment | NA | NA | 0 |
| S16_vs_66649\|\|full | 7767 | No | NA | 10 | 5 | 0 | Low-quality | Genome-fragment | 12.02 | AAI-based (high-confidence) | 0 |
| S8_extracted_k141_88959 | 1146 | No | NA | 1 | 0 | 1 | Not-determined | Genome-fragment | NA | NA | 0 |
| S8_extracted_k141_201345 | 1091 | No | NA | 2 | 0 | 0 | Not-determined | Genome-fragment | NA | NA | 0 |
| S8_extracted_k141_166906 | 8573 | No | NA | 5 | 0 | 4 | Not-determined | Genome-fragment | NA | NA | 0 |
| S8_extracted_k141_132158 | 1603 | No | NA | 3 | 0 | 2 | Not-determined | Genome-fragment | NA | NA | 0 |
| S8_extracted_k141_71092 | 1892 | No | NA | 2 | 0 | 0 | Not-determined | Genome-fragment | NA | NA | 0 |
| S8_extracted_k141_183824 | 1110 | No | NA | 3 | 0 | 2 | Not-determined | Genome-fragment | NA | NA | 0 |
| S8_extracted_k141_2180 | 2326 | No | NA | 4 | 0 | 1 | Not-determined | Genome-fragment | NA | NA | 0 |
| S8_vs_k141_110614\|\|full | 7772 | No | NA | 13 | 3 | 0 | Low-quality | Genome-fragment | 6.27 | HMM-based (lower-bound) | 0 |
| S8_extracted_k141_45318 | 1031 | No | NA | 1 | 0 | 1 | Not-determined | Genome-fragment | NA | NA | 0 |
| S8_extracted_k141_19358 | 3195 | No | NA | 2 | 0 | 2 | Not-determined | Genome-fragment | NA | NA | 0 |
| S8_extracted_k141_19361 | 1276 | No | NA | 1 | 0 | 0 | Not-determined | Genome-fragment | NA | NA | 0 |
| S8_extracted_k141_166921 | 6210 | No | NA | 2 | 0 | 2 | Not-determined | Genome-fragment | NA | NA | 0 |
| S8_extracted_k141_193220 | 2842 | No | NA | 4 | 0 | 2 | Not-determined | Genome-fragment | NA | NA | 0 |
| S8_extracted_k141_37057 | 1116 | No | NA | 1 | 0 | 0 | Not-determined | Genome-fragment | NA | NA | 0 |
| S8_extracted_k141_158132 | 1036 | No | NA | 2 | 0 | 2 | Not-determined | Genome-fragment | NA | NA | 0 |
| S8_extracted_k141_97539 | 2875 | No | NA | 2 | 0 | 0 | Not-determined | Genome-fragment | NA | NA | 0 |
| S8_extracted_k141_54369 | 1582 | No | NA | 2 | 0 | 0 | Not-determined | Genome-fragment | NA | NA | 0 |
| S8_extracted_k141_62864 | 1115 | No | NA | 1 | 0 | 1 | Not-determined | Genome-fragment | NA | NA | 0 |
| S8_extracted_k141_37069 | 1559 | No | NA | 1 | 0 | 1 | Not-determined | Genome-fragment | NA | NA | 0 |
| S8_extracted_k141_149569 | 3277 | No | NA | 1 | 0 | 1 | Not-determined | Genome-fragment | NA | NA | 0 |
| S5_extracted_k141_142395 | 7774 | No | NA | 17 | 4 | 0 | Low-quality | Genome-fragment | 9.72 | AAI-based (high-confidence) | 0 |
| S7_extracted_103425 | 7774 | No | NA | 13 | 5 | 0 | Low-quality | Genome-fragment | 16.34 | AAI-based (high-confidence) | 0 |
| S14_vs_k141_28937\|\|full | 7778 | No | NA | 18 | 3 | 1 | Low-quality | Genome-fragment | 9.99 | AAI-based (high-confidence) | 0 |
| S8_extracted_k141_106320 | 2156 | No | NA | 2 | 0 | 1 | Not-determined | Genome-fragment | NA | NA | 0 |
| S8_extracted_k141_28344 | 3299 | No | NA | 1 | 0 | 1 | Not-determined | Genome-fragment | NA | NA | 0 |
| S8_extracted_k141_71134 | 2062 | No | NA | 3 | 0 | 2 | Not-determined | Genome-fragment | NA | NA | 0 |
| S3_extracted_k141_125153 | 7779 | No | NA | 20 | 2 | 1 | Low-quality | Genome-fragment | 6.26 | AAI-based (high-confidence) | 0 |
| S8_extracted_k141_28346 | 1248 | No | NA | 3 | 0 | 2 | Not-determined | Genome-fragment | NA | NA | 0 |
| S8_extracted_k141_132203 | 17853 | No | NA | 8 | 0 | 7 | Not-determined | Genome-fragment | NA | NA | 0 |
| S8_extracted_k141_97583 | 1354 | No | NA | 2 | 0 | 1 | Not-determined | Genome-fragment | NA | NA | 0 |
| S8_extracted_k141_10570 | 4085 | No | NA | 4 | 0 | 1 | Not-determined | Genome-fragment | NA | NA | 0 |
| S6_extracted_k141_24167 | 7785 | No | NA | 13 | 4 | 1 | Low-quality | Genome-fragment | 22.01 | AAI-based (high-confidence) | 0 |
| S8_extracted_k141_10573 | 1565 | No | NA | 3 | 0 | 2 | Not-determined | Genome-fragment | NA | NA | 0 |
| S8_extracted_k141_89010 | 1240 | No | NA | 2 | 0 | 1 | Not-determined | Genome-fragment | NA | NA | 0 |
| S8_extracted_k141_201398 | 3684 | No | NA | 2 | 0 | 2 | Not-determined | Genome-fragment | NA | NA | 0 |
| S8_vs_k141_152109\|\|full | 7786 | No | NA | 3 | 0 | 3 | Low-quality | Genome-fragment | 0.9 | AAI-based (medium-confidence) | 0 |
| S8_extracted_k141_28367 | 2266 | No | NA | 2 | 0 | 1 | Not-determined | Genome-fragment | NA | NA | 0 |
| S8_extracted_k141_71156 | 5521 | No | NA | 2 | 0 | 2 | Not-determined | Genome-fragment | NA | NA | 0 |
| S8_extracted_k141_2265 | 2235 | No | NA | 1 | 0 | 1 | Not-determined | Genome-fragment | NA | NA | 0 |
| S8_extracted_k141_141009 | 1506 | No | NA | 1 | 0 | 1 | Not-determined | Genome-fragment | NA | NA | 0 |
| S5_vs_k141_105807\|\|full | 7788 | No | NA | 16 | 1 | 1 | Low-quality | Genome-fragment | 17.92 | AAI-based (medium-confidence) | 0 |
| S8_extracted_k141_2267 | 2383 | No | NA | 3 | 0 | 3 | Not-determined | Genome-fragment | NA | NA | 0 |
| S8_extracted_k141_28386 | 1055 | No | NA | 2 | 0 | 1 | Not-determined | Genome-fragment | NA | NA | 0 |
| S1_extracted_k141_132082 | 7790 | No | NA | 7 | 0 | 3 | Low-quality | Genome-fragment | 3.78 | AAI-based (medium-confidence) | 0 |
| S8_extracted_k141_166982 | 3807 | No | NA | 1 | 0 | 0 | Not-determined | Genome-fragment | NA | NA | 0 |
| S8_extracted_k141_123788 | 1208 | No | NA | 1 | 0 | 1 | Not-determined | Genome-fragment | NA | NA | 0 |
| S8_extracted_k141_89039 | 1579 | No | NA | 2 | 0 | 0 | Not-determined | Genome-fragment | NA | NA | 0 |
| S5_extracted_k141_90341 | 7793 | No | NA | 6 | 0 | 5 | Low-quality | Genome-fragment | 15.86 | AAI-based (medium-confidence) | 0 |
| S8_extracted_k141_197851 | 7796 | No | NA | 8 | 0 | 1 | Low-quality | Genome-fragment | 23.17 | AAI-based (medium-confidence) | 0 |
| S14_vs_k141_53969\|\|full | 7801 | No | NA | 12 | 10 | 0 | Low-quality | Genome-fragment | 16.75 | AAI-based (medium-confidence) | 0 |
| S8_extracted_k141_71181 | 1156 | No | NA | 1 | 0 | 1 | Not-determined | Genome-fragment | NA | NA | 0 |
| S8_extracted_k141_54445 | 3613 | No | NA | 4 | 0 | 1 | Not-determined | Genome-fragment | NA | NA | 0 |
| S8_extracted_k141_167006 | 1002 | No | NA | 1 | 0 | 1 | Not-determined | Genome-fragment | NA | NA | 0 |
| S8_extracted_k141_71187 | 1211 | No | NA | 1 | 0 | 0 | Not-determined | Genome-fragment | NA | NA | 0 |
| S8_extracted_k141_158193 | 1065 | No | NA | 4 | 0 | 2 | Not-determined | Genome-fragment | NA | NA | 0 |
| S8_extracted_k141_10597 | 5426 | No | NA | 5 | 0 | 2 | Not-determined | Genome-fragment | NA | NA | 0 |
| S8_extracted_k141_167025 | 1155 | No | NA | 3 | 0 | 0 | Not-determined | Genome-fragment | NA | NA | 0 |
| S8_extracted_k141_149680 | 3521 | No | NA | 2 | 0 | 1 | Not-determined | Genome-fragment | NA | NA | 0 |
| S8_extracted_k141_80353 | 1162 | No | NA | 1 | 0 | 1 | Not-determined | Genome-fragment | NA | NA | 0 |
| S14_vs_k141_62709\|\|full | 7805 | No | NA | 12 | 1 | 3 | Low-quality | Genome-fragment | 12.99 | AAI-based (medium-confidence) | 0 |
| S8_extracted_k141_114974 | 3102 | No | NA | 5 | 0 | 1 | Not-determined | Genome-fragment | NA | NA | 0 |
| S15_extracted_23702 | 7805 | No | NA | 16 | 3 | 1 | Low-quality | Genome-fragment | 11.13 | AAI-based (high-confidence) | 0 |
| S8_extracted_k141_2306 | 1320 | No | NA | 1 | 0 | 1 | Not-determined | Genome-fragment | NA | NA | 0 |
| S7_vs_83390\|\|full | 7808 | No | NA | 6 | 5 | 0 | Low-quality | Genome-fragment | 22.05 | AAI-based (high-confidence) | 0 |
| S8_extracted_k141_97635 | 2208 | No | NA | 4 | 0 | 0 | Not-determined | Genome-fragment | NA | NA | 0 |
| S8_extracted_k141_132275 | 1182 | No | NA | 2 | 0 | 2 | Not-determined | Genome-fragment | NA | NA | 0 |
| S8_extracted_k141_89075 | 3736 | No | NA | 3 | 0 | 0 | Not-determined | Genome-fragment | NA | NA | 0 |
| S10_extracted_60111 | 7809 | No | NA | 6 | 0 | 1 | Low-quality | Genome-fragment | 17.98 | AAI-based (medium-confidence) | 0 |
| S8_extracted_k141_193354 | 1009 | No | NA | 3 | 0 | 0 | Not-determined | Genome-fragment | NA | NA | 0 |
| S8_extracted_k141_193356 | 1371 | No | NA | 1 | 0 | 1 | Not-determined | Genome-fragment | NA | NA | 0 |
| S8_extracted_k141_97656 | 1643 | No | NA | 1 | 0 | 0 | Not-determined | Genome-fragment | NA | NA | 0 |
| S8_extracted_k141_193368 | 1416 | No | NA | 2 | 0 | 1 | Not-determined | Genome-fragment | NA | NA | 0 |
| S8_extracted_k141_2341 | 1418 | No | NA | 2 | 0 | 0 | Not-determined | Genome-fragment | NA | NA | 0 |
| S8_extracted_k141_54488 | 4262 | No | NA | 3 | 0 | 2 | Not-determined | Genome-fragment | NA | NA | 0 |

| | | | | | | | | | | | |
|---|---|---|---|---|---|---|---|---|---|---|---|
| S11_vs_k141_45308\|\|full | 7815 | No | NA | 11 | 9 | 0 | Low-quality | Genome-fragment | 22.56 | AAI-based (high-confidence) | 0 |
| S8_extracted_k141_80413 | 1212 | No | NA | 2 | 0 | 2 | Not-determined | Genome-fragment | NA | NA | 0 |
| S8_extracted_k141_115009 | 7808 | No | NA | 5 | 0 | 4 | Not-determined | Genome-fragment | NA | NA | 0 |
| S8_extracted_k141_19483 | 1293 | No | NA | 1 | 0 | 0 | Not-determined | Genome-fragment | NA | NA | 0 |
| S8_extracted_k141_106479 | 1414 | No | NA | 2 | 0 | 2 | Not-determined | Genome-fragment | NA | NA | 0 |
| S8_extracted_k141_167076 | 4580 | No | NA | 5 | 0 | 0 | Not-determined | Genome-fragment | NA | NA | 0 |
| S8_extracted_k141_97691 | 1181 | No | NA | 1 | 0 | 0 | Not-determined | Genome-fragment | NA | NA | 0 |
| S8_extracted_k141_141091 | 1056 | No | NA | 2 | 0 | 2 | Not-determined | Genome-fragment | NA | NA | 0 |
| S8_extracted_k141_28510 | 1048 | No | NA | 2 | 0 | 2 | Not-determined | Genome-fragment | NA | NA | 0 |
| S8_extracted_k141_106486 | 1215 | No | NA | 1 | 0 | 1 | Not-determined | Genome-fragment | NA | NA | 0 |
| S8_extracted_k141_37168 | 1014 | No | NA | 1 | 0 | 0 | Not-determined | Genome-fragment | NA | NA | 0 |
| S14_vs_k141_55833\|\|full | 7815 | No | NA | 9 | 2 | 0 | Low-quality | Genome-fragment | 18.02 | AAI-based (high-confidence) | 0 |
| S1_extracted_k141_48966 | 7817 | No | NA | 10 | 0 | 0 | Low-quality | Genome-fragment | 19.05 | AAI-based (high-confidence) | 0 |
| S8_extracted_k141_201493 | 1017 | No | NA | 2 | 0 | 0 | Not-determined | Genome-fragment | NA | NA | 0 |
| S8_extracted_k141_80445 | 1558 | No | NA | 1 | 0 | 1 | Not-determined | Genome-fragment | NA | NA | 0 |
| S8_extracted_k141_141104 | 3086 | No | NA | 3 | 0 | 1 | Not-determined | Genome-fragment | NA | NA | 0 |
| S13_extracted_89590 | 7819 | Yes | 3489 | 6 | 1 | 1 | Low-quality | Genome-fragment | 7.38 | AAI-based (medium-confidence) | 55.38 |
| S8_extracted_k141_10701 | 1662 | No | NA | 2 | 0 | 0 | Not-determined | Genome-fragment | NA | NA | 0 |
| S8_extracted_k141_71214 | 23486 | No | NA | 37 | 9 | 0 | Medium-quality | Genome-fragment | 74.7 | AAI-based (high-confidence) | 0 |
| S8_extracted_k141_89136 | 1028 | No | NA | 2 | 0 | 1 | Not-determined | Genome-fragment | NA | NA | 0 |
| S8_extracted_k141_28539 | 1128 | No | NA | 1 | 0 | 0 | Not-determined | Genome-fragment | NA | NA | 0 |
| S8_extracted_k141_37176 | 3649 | No | NA | 4 | 0 | 3 | Not-determined | Genome-fragment | NA | NA | 0 |
| S8_extracted_k141_63041 | 1135 | No | NA | 1 | 0 | 1 | Not-determined | Genome-fragment | NA | NA | 0 |
| S8_extracted_k141_28549 | 4226 | No | NA | 7 | 0 | 0 | Not-determined | Genome-fragment | NA | NA | 0 |
| S8_extracted_k141_97734 | 1814 | No | NA | 2 | 0 | 1 | Not-determined | Genome-fragment | NA | NA | 0 |
| S8_extracted_k141_63052 | 1081 | No | NA | 1 | 0 | 0 | Not-determined | Genome-fragment | NA | NA | 0 |
| S8_extracted_k141_80457 | 4063 | No | NA | 4 | 0 | 3 | Not-determined | Genome-fragment | NA | NA | 0 |
| S8_extracted_k141_80461 | 1765 | No | NA | 2 | 0 | 2 | Not-determined | Genome-fragment | NA | NA | 0 |
| S8_extracted_k141_193420 | 2012 | No | NA | 3 | 0 | 1 | Not-determined | Genome-fragment | NA | NA | 0 |
| S8_extracted_k141_175692 | 1247 | No | NA | 1 | 0 | 0 | Not-determined | Genome-fragment | NA | NA | 0 |
| S8_extracted_k141_10742 | 1152 | No | NA | 1 | 0 | 1 | Not-determined | Genome-fragment | NA | NA | 0 |
| S17_extracted_k141_62525 | 7830 | No | NA | 15 | 4 | 0 | Low-quality | Genome-fragment | 13.59 | AAI-based (high-confidence) | 0 |
| S12_vs_k141_81574\|\|full | 7831 | No | NA | 14 | 1 | 0 | Low-quality | Genome-fragment | 13.9 | AAI-based (medium-confidence) | 0 |
| S8_vs_k141_162341\|\|full | 7834 | No | NA | 19 | 4 | 0 | Low-quality | Genome-fragment | 19.83 | AAI-based (high-confidence) | 0 |
| S9_vs_k141_65960\|\|full | 7837 | No | NA | 8 | 1 | 0 | Low-quality | Genome-fragment | 18.78 | AAI-based (high-confidence) | 0 |
| S8_extracted_k141_158225 | 90855 | No | NA | 88 | 7 | 0 | Complete | High-quality | 100 | DTR (high-confidence) | 0 |
| S8_extracted_k141_183962 | 1168 | No | NA | 1 | 0 | 1 | Not-determined | Genome-fragment | NA | NA | 0 |
| S8_extracted_k141_10767 | 1650 | No | NA | 1 | 0 | 1 | Not-determined | Genome-fragment | NA | NA | 0 |
| S5_vs_k141_40009\|\|full | 7840 | No | NA | 9 | 0 | 0 | Low-quality | Genome-fragment | 17.86 | AAI-based (high-confidence) | 0 |
| S8_extracted_k141_167140 | 1016 | No | NA | 1 | 0 | 1 | Not-determined | Genome-fragment | NA | NA | 0 |
| S8_extracted_k141_183966 | 1590 | No | NA | 2 | 0 | 1 | Not-determined | Genome-fragment | NA | NA | 0 |
| S8_extracted_k141_175707 | 1257 | No | NA | 3 | 0 | 0 | Not-determined | Genome-fragment | NA | NA | 0 |
| S8_extracted_k141_123901 | 1401 | No | NA | 2 | 0 | 1 | Not-determined | Genome-fragment | NA | NA | 0 |
| S8_extracted_k141_106563 | 1172 | No | NA | 2 | 0 | 1 | Not-determined | Genome-fragment | NA | NA | 0 |
| S8_extracted_k141_123904 | 1060 | No | NA | 2 | 0 | 0 | Not-determined | Genome-fragment | NA | NA | 0 |
| S8_extracted_k141_28575 | 1396 | No | NA | 2 | 0 | 2 | Not-determined | Genome-fragment | NA | NA | 0 |
| S8_extracted_k141_54514 | 53614 | No | NA | 99 | 16 | 0 | Complete | High-quality | 100 | DTR (high-confidence) | 0 |
| S8_extracted_k141_28584 | 1673 | No | NA | 1 | 0 | 0 | Not-determined | Genome-fragment | NA | NA | 0 |
| S8_extracted_k141_19605 | 1254 | No | NA | 2 | 0 | 1 | Not-determined | Genome-fragment | NA | NA | 0 |
| S8_extracted_k141_45450 | 6845 | No | NA | 5 | 0 | 2 | Not-determined | Genome-fragment | NA | NA | 0 |
| S8_extracted_k141_106566 | 5254 | No | NA | 5 | 0 | 3 | Not-determined | Genome-fragment | NA | NA | 0 |
| S8_extracted_k141_106567 | 1403 | No | NA | 3 | 0 | 1 | Not-determined | Genome-fragment | NA | NA | 0 |
| S8_extracted_k141_201557 | 1541 | No | NA | 1 | 0 | 1 | Not-determined | Genome-fragment | NA | NA | 0 |
| S8_extracted_k141_63115 | 1531 | No | NA | 2 | 0 | 2 | Not-determined | Genome-fragment | NA | NA | 0 |
| S8_extracted_k141_149716 | 5832 | No | NA | 6 | 0 | 5 | Not-determined | Genome-fragment | NA | NA | 0 |
| S8_extracted_k141_45475 | 1011 | No | NA | 1 | 0 | 0 | Not-determined | Genome-fragment | NA | NA | 0 |
| S8_extracted_k141_184005 | 1140 | No | NA | 1 | 0 | 0 | Not-determined | Genome-fragment | NA | NA | 0 |
| S8_extracted_k141_115089 | 1950 | No | NA | 8 | 0 | 0 | Not-determined | Genome-fragment | NA | NA | 0 |
| S8_extracted_k141_123947 | 1227 | No | NA | 2 | 0 | 1 | Not-determined | Genome-fragment | NA | NA | 0 |
| S3_vs_k141_50979\|\|full | 7850 | No | NA | 14 | 2 | 1 | Low-quality | Genome-fragment | 23.75 | AAI-based (high-confidence) | 0 |
| S8_extracted_k141_80515 | 4248 | No | NA | 3 | 0 | 3 | Not-determined | Genome-fragment | NA | NA | 0 |
| S8_extracted_k141_63167 | 1804 | No | NA | 2 | 0 | 2 | Not-determined | Genome-fragment | NA | NA | 0 |
| S8_extracted_k141_123960 | 1475 | No | NA | 1 | 0 | 1 | Not-determined | Genome-fragment | NA | NA | 0 |
| S8_extracted_k141_28652 | 1102 | No | NA | 1 | 0 | 1 | Not-determined | Genome-fragment | NA | NA | 0 |
| S8_extracted_k141_45483 | 25130 | No | NA | 23 | 0 | 18 | Not-determined | Genome-fragment | NA | NA | 0 |
| S8_extracted_k141_2454 | 1073 | No | NA | 1 | 0 | 1 | Not-determined | Genome-fragment | NA | NA | 0 |
| S14_extracted_k141_42465 | 7852 | No | NA | 7 | 0 | 3 | Low-quality | Genome-fragment | 6.01 | AAI-based (medium-confidence) | 0 |
| S8_extracted_k141_167189 | 1148 | No | NA | 1 | 0 | 1 | Not-determined | Genome-fragment | NA | NA | 0 |
| S8_extracted_k141_2456 | 1118 | No | NA | 3 | 0 | 1 | Not-determined | Genome-fragment | NA | NA | 0 |
| S8_extracted_k141_123978 | 1725 | No | NA | 1 | 0 | 1 | Not-determined | Genome-fragment | NA | NA | 0 |
| S8_extracted_k141_63183 | 2564 | No | NA | 3 | 0 | 2 | Not-determined | Genome-fragment | NA | NA | 0 |
| S8_extracted_k141_19678 | 1926 | No | NA | 2 | 0 | 1 | Not-determined | Genome-fragment | NA | NA | 0 |
| S8_extracted_k141_167197 | 1139 | No | NA | 1 | 0 | 1 | Not-determined | Genome-fragment | NA | NA | 0 |
| S8_extracted_k141_10803 | 15142 | No | NA | 12 | 0 | 7 | Not-determined | Genome-fragment | NA | NA | 0 |
| S8_extracted_k141_37337 | 3848 | No | NA | 5 | 0 | 5 | Not-determined | Genome-fragment | NA | NA | 0 |
| S8_extracted_k141_80559 | 1302 | No | NA | 1 | 0 | 1 | Not-determined | Genome-fragment | NA | NA | 0 |
| S8_extracted_k141_193512 | 1424 | No | NA | 2 | 0 | 1 | Not-determined | Genome-fragment | NA | NA | 0 |
| S8_extracted_k141_28704 | 1087 | No | NA | 1 | 0 | 1 | Not-determined | Genome-fragment | NA | NA | 0 |
| S8_extracted_k141_175797 | 1196 | No | NA | 2 | 0 | 0 | Not-determined | Genome-fragment | NA | NA | 0 |
| S8_extracted_k141_10828 | 1109 | No | NA | 2 | 0 | 0 | Not-determined | Genome-fragment | NA | NA | 0 |
| S9_vs_k141_90168\|\|full | 7853 | No | NA | 17 | 1 | 0 | Low-quality | Genome-fragment | 19.19 | AAI-based (medium-confidence) | 0 |
| S8_extracted_k141_141248 | 1096 | No | NA | 2 | 0 | 0 | Not-determined | Genome-fragment | NA | NA | 0 |
| S12_extracted_k141_128572 | 7858 | No | NA | 10 | 1 | 0 | Low-quality | Genome-fragment | 19.38 | AAI-based (high-confidence) | 0 |
| S8_extracted_k141_124022 | 3713 | No | NA | 2 | 0 | 2 | Not-determined | Genome-fragment | NA | NA | 0 |
| S8_extracted_k141_115110 | 1005 | No | NA | 1 | 0 | 1 | Not-determined | Genome-fragment | NA | NA | 0 |
| S5_extracted_k141_115529 | 7859 | No | NA | 22 | 9 | 0 | Low-quality | Genome-fragment | 35.85 | AAI-based (high-confidence) | 0 |
| S8_extracted_k141_193520 | 3492 | No | NA | 4 | 0 | 3 | Not-determined | Genome-fragment | NA | NA | 0 |
| S8_extracted_k141_19709 | 1146 | No | NA | 1 | 0 | 1 | Not-determined | Genome-fragment | NA | NA | 0 |
| S8_extracted_k141_149792 | 1640 | No | NA | 1 | 0 | 1 | Not-determined | Genome-fragment | NA | NA | 0 |
| S8_extracted_k141_141260 | 1975 | No | NA | 3 | 0 | 1 | Not-determined | Genome-fragment | NA | NA | 0 |
| S8_extracted_k141_141274 | 1571 | No | NA | 1 | 0 | 1 | Not-determined | Genome-fragment | NA | NA | 0 |
| S8_extracted_k141_80603 | 1912 | No | NA | 2 | 0 | 1 | Not-determined | Genome-fragment | NA | NA | 0 |
| S8_extracted_k141_19720 | 3496 | No | NA | 5 | 0 | 3 | Not-determined | Genome-fragment | NA | NA | 0 |
| S8_extracted_k141_10872 | 1368 | No | NA | 2 | 0 | 1 | Not-determined | Genome-fragment | NA | NA | 0 |
| S7_vs_62640\|\|full | 7860 | Yes | 4809 | 6 | 2 | 1 | Low-quality | Genome-fragment | 6.68 | AAI-based (medium-confidence) | 38.82 |
| S8_extracted_k141_71363 | 8695 | No | NA | 9 | 0 | 2 | Not-determined | Genome-fragment | NA | NA | 0 |
| S8_extracted_k141_175853 | 1626 | No | NA | 4 | 0 | 3 | Not-determined | Genome-fragment | NA | NA | 0 |
| S8_extracted_k141_115133 | 6346 | No | NA | 3 | 0 | 2 | Not-determined | Genome-fragment | NA | NA | 0 |
| S17_extracted_k141_59642 | 7861 | No | NA | 15 | 4 | 0 | Low-quality | Genome-fragment | 16.66 | AAI-based (high-confidence) | 0 |
| S8_extracted_k141_54607 | 1993 | No | NA | 1 | 0 | 1 | Not-determined | Genome-fragment | NA | NA | 0 |
| S2_extracted_k141_1808 | 7872 | No | NA | 15 | 1 | 0 | Low-quality | Genome-fragment | 13.66 | AAI-based (medium-confidence) | 0 |
| S8_extracted_k141_54609 | 4469 | No | NA | 5 | 0 | 2 | Not-determined | Genome-fragment | NA | NA | 0 |
| S8_extracted_k141_89180 | 1004 | No | NA | 1 | 0 | 0 | Not-determined | Genome-fragment | NA | NA | 0 |
| S8_extracted_k141_54614 | 1103 | No | NA | 1 | 0 | 0 | Not-determined | Genome-fragment | NA | NA | 0 |
| S8_extracted_k141_10899 | 4332 | No | NA | 4 | 0 | 0 | Not-determined | Genome-fragment | NA | NA | 0 |
| S8_extracted_k141_2510 | 1324 | No | NA | 2 | 0 | 1 | Not-determined | Genome-fragment | NA | NA | 0 |
| S8_extracted_k141_167257 | 2984 | No | NA | 3 | 0 | 3 | Not-determined | Genome-fragment | NA | NA | 0 |
| S8_extracted_k141_201695 | 45510 | No | NA | 76 | 15 | 1 | Medium-quality | Genome-fragment | 83.41 | AAI-based (high-confidence) | 0 |
| S8_extracted_k141_71376 | 4274 | No | NA | 4 | 0 | 4 | Not-determined | Genome-fragment | NA | NA | 0 |
| S8_extracted_k141_80655 | 1002 | No | NA | 1 | 0 | 1 | Not-determined | Genome-fragment | NA | NA | 0 |
| S8_extracted_k141_10908 | 1600 | No | NA | 3 | 0 | 0 | Not-determined | Genome-fragment | NA | NA | 0 |
| S8_extracted_k141_149849 | 1122 | No | NA | 1 | 0 | 1 | Not-determined | Genome-fragment | NA | NA | 0 |
| S8_extracted_k141_124087 | 1079 | No | NA | 3 | 0 | 0 | Not-determined | Genome-fragment | NA | NA | 0 |
| S8_extracted_k141_141310 | 11133 | No | NA | 16 | 0 | 6 | Not-determined | Genome-fragment | NA | NA | 0 |
| S8_extracted_k141_10914 | 1420 | No | NA | 1 | 0 | 0 | Not-determined | Genome-fragment | NA | NA | 0 |
| S8_extracted_k141_89187 | 7868 | No | NA | 2 | 0 | 2 | Not-determined | Genome-fragment | NA | NA | 0 |
| S8_extracted_k141_19753 | 6489 | No | NA | 5 | 0 | 2 | Not-determined | Genome-fragment | NA | NA | 0 |
| S8_extracted_k141_115163 | 3195 | No | NA | 5 | 0 | 5 | Not-determined | Genome-fragment | NA | NA | 0 |
| S16_vs_137317\|\|full | 7873 | No | NA | 9 | 0 | 0 | Low-quality | Genome-fragment | 17.05 | AAI-based (medium-confidence) | 0 |
| S8_extracted_k141_63279 | 1665 | No | NA | 1 | 0 | 1 | Not-determined | Genome-fragment | NA | NA | 0 |
| S8_extracted_k141_193612 | 2975 | No | NA | 2 | 0 | 1 | Not-determined | Genome-fragment | NA | NA | 0 |
| S8_extracted_k141_80674 | 6209 | No | NA | 4 | 0 | 1 | Not-determined | Genome-fragment | NA | NA | 0 |
| S8_extracted_k141_71394 | 1135 | No | NA | 1 | 0 | 0 | Not-determined | Genome-fragment | NA | NA | 0 |
| S8_extracted_k141_141335 | 6991 | No | NA | 9 | 0 | 1 | Not-determined | Genome-fragment | NA | NA | 0 |
| S8_extracted_k141_193614 | 8299 | No | NA | 6 | 0 | 2 | Not-determined | Genome-fragment | NA | NA | 0 |
| S8_extracted_k141_10935 | 1404 | No | NA | 1 | 0 | 1 | Not-determined | Genome-fragment | NA | NA | 0 |
| S8_extracted_k141_158268 | 10955 | No | NA | 10 | 0 | 2 | Not-determined | Genome-fragment | NA | NA | 0 |
| S8_extracted_k141_80686 | 3278 | No | NA | 3 | 0 | 3 | Not-determined | Genome-fragment | NA | NA | 0 |
| S8_vs_k141_129400\|\|full | 7873 | No | NA | 5 | 1 | 0 | Low-quality | Genome-fragment | 3.55 | HMM-based (lower-bound) | 0 |
| S8_extracted_k141_201712 | 7484 | No | NA | 1 | 0 | 0 | Not-determined | Genome-fragment | NA | NA | 0 |
| S8_extracted_k141_124122 | 2969 | No | NA | 3 | 0 | 2 | Not-determined | Genome-fragment | NA | NA | 0 |
| S8_extracted_k141_132527 | 1379 | No | NA | 3 | 0 | 2 | Not-determined | Genome-fragment | NA | NA | 0 |
| S8_extracted_k141_167282 | 15850 | No | NA | 30 | 0 | 28 | Not-determined | Genome-fragment | NA | NA | 0 |
| S8_extracted_k141_19794 | 1229 | No | NA | 3 | 0 | 2 | Not-determined | Genome-fragment | NA | NA | 0 |
| S8_extracted_k141_175931 | 2925 | No | NA | 5 | 0 | 0 | Not-determined | Genome-fragment | NA | NA | 0 |
| S16_vs_76756\|\|full | 7874 | No | NA | 6 | 0 | 4 | Low-quality | Genome-fragment | 17.59 | AAI-based (medium-confidence) | 0 |
| S8_extracted_k141_141351 | 1236 | No | NA | 2 | 0 | 1 | Not-determined | Genome-fragment | NA | NA | 0 |
| S8_extracted_k141_19801 | 2840 | No | NA | 4 | 0 | 3 | Not-determined | Genome-fragment | NA | NA | 0 |
| S8_extracted_k141_28847 | 15245 | No | NA | 14 | 0 | 3 | Not-determined | Genome-fragment | NA | NA | 0 |
| S8_extracted_k141_106638 | 2414 | No | NA | 3 | 0 | 1 | Not-determined | Genome-fragment | NA | NA | 0 |
| S8_extracted_k141_167301 | 4197 | No | NA | 3 | 0 | 2 | Not-determined | Genome-fragment | NA | NA | 0 |
| S16_vs_8929\|\|full | 7876 | No | NA | 8 | 1 | 3 | Low-quality | Genome-fragment | 8.94 | HMM-based (lower-bound) | 0 |
| S8_extracted_k141_115204 | 1037 | No | NA | 1 | 0 | 1 | Not-determined | Genome-fragment | NA | NA | 0 |
| S8_extracted_k141_19821 | 1840 | No | NA | 1 | 0 | 0 | Not-determined | Genome-fragment | NA | NA | 0 |
| S8_extracted_k141_19824 | 1081 | No | NA | 1 | 0 | 1 | Not-determined | Genome-fragment | NA | NA | 0 |
| S8_extracted_k141_89236 | 7952 | No | NA | 6 | 0 | 2 | Not-determined | Genome-fragment | NA | NA | 0 |
| S8_extracted_k141_175972 | 1126 | No | NA | 2 | 0 | 2 | Not-determined | Genome-fragment | NA | NA | 0 |
| S8_extracted_k141_193655 | 1714 | No | NA | 3 | 0 | 2 | Not-determined | Genome-fragment | NA | NA | 0 |
| S12_vs_k141_39820\|\|full | 7878 | No | NA | 5 | 0 | 4 | Low-quality | Genome-fragment | 0.92 | AAI-based (medium-confidence) | 0 |
| S8_extracted_k141_28871 | 1077 | No | NA | 1 | 0 | 1 | Not-determined | Genome-fragment | NA | NA | 0 |
| S8_extracted_k141_141400 | 5989 | No | NA | 8 | 0 | 0 | Not-determined | Genome-fragment | NA | NA | 0 |
| S8_extracted_k141_2620 | 1255 | No | NA | 1 | 0 | 0 | Not-determined | Genome-fragment | NA | NA | 0 |
| S8_extracted_k141_193659 | 1003 | No | NA | 2 | 0 | 0 | Not-determined | Genome-fragment | NA | NA | 0 |
| S16_vs_66459\|\|full | 7879 | No | NA | 4 | 0 | 3 | Low-quality | Genome-fragment | 2.43 | AAI-based (high-confidence) | 0 |
| S8_extracted_k141_45637 | 3813 | No | NA | 2 | 0 | 1 | Not-determined | Genome-fragment | NA | NA | 0 |
| S8_extracted_k141_28879 | 1463 | No | NA | 2 | 0 | 1 | Not-determined | Genome-fragment | NA | NA | 0 |
| S8_extracted_k141_115231 | 1333 | No | NA | 1 | 0 | 1 | Not-determined | Genome-fragment | NA | NA | 0 |
| S5_vs_k141_4558\|\|full | 7881 | No | NA | 10 | 2 | 0 | Low-quality | Genome-fragment | 10.74 | HMM-based (lower-bound) | 0 |
| S8_extracted_k141_106684 | 3268 | No | NA | 3 | 0 | 2 | Not-determined | Genome-fragment | NA | NA | 0 |
| S8_extracted_k141_193677 | 1767 | No | NA | 2 | 0 | 2 | Not-determined | Genome-fragment | NA | NA | 0 |
| S8_extracted_k141_97848 | 1965 | No | NA | 3 | 0 | 0 | Not-determined | Genome-fragment | NA | NA | 0 |
| S8_extracted_k141_45660 | 1610 | No | NA | 2 | 0 | 2 | Not-determined | Genome-fragment | NA | NA | 0 |
| S8_extracted_k141_106688 | 3300 | No | NA | 2 | 0 | 0 | Not-determined | Genome-fragment | NA | NA | 0 |
| S16_vs_84361\|\|full | 7883 | No | NA | 4 | 0 | 2 | Low-quality | Genome-fragment | 7.79 | AAI-based (medium-confidence) | 0 |
| S8_extracted_k141_141440 | 2107 | No | NA | 2 | 0 | 1 | Not-determined | Genome-fragment | NA | NA | 0 |
| S8_extracted_k141_28896 | 1797 | No | NA | 1 | 0 | 0 | Not-determined | Genome-fragment | NA | NA | 0 |
| S8_extracted_k141_71491 | 1353 | No | NA | 3 | 0 | 2 | Not-determined | Genome-fragment | NA | NA | 0 |
| S8_extracted_k141_19872 | 1622 | No | NA | 1 | 0 | 0 | Not-determined | Genome-fragment | NA | NA | 0 |
| S8_extracted_k141_71503 | 1007 | No | NA | 1 | 0 | 0 | Not-determined | Genome-fragment | NA | NA | 0 |
| S8_extracted_k141_141474 | 1504 | No | NA | 2 | 0 | 0 | Not-determined | Genome-fragment | NA | NA | 0 |
| S8_extracted_k141_45693 | 3287 | No | NA | 3 | 0 | 2 | Not-determined | Genome-fragment | NA | NA | 0 |
| S5_extracted_k141_114825 | 7895 | No | NA | 15 | 4 | 0 | Low-quality | Genome-fragment | 21.45 | AAI-based (medium-confidence) | 0 |
| S8_extracted_k141_158346 | 2800 | No | NA | 2 | 0 | 1 | Not-determined | Genome-fragment | NA | NA | 0 |
| S8_extracted_k141_115261 | 4617 | No | NA | 3 | 0 | 3 | Not-determined | Genome-fragment | NA | NA | 0 |
| S8_extracted_k141_141486 | 1588 | No | NA | 2 | 0 | 2 | Not-determined | Genome-fragment | NA | NA | 0 |
| S8_extracted_k141_45701 | 1077 | No | NA | 2 | 0 | 1 | Not-determined | Genome-fragment | NA | NA | 0 |
| S8_extracted_k141_89298 | 2300 | No | NA | 1 | 0 | 0 | Not-determined | Genome-fragment | NA | NA | 0 |
| S8_extracted_k141_54695 | 1412 | No | NA | 1 | 0 | 0 | Not-determined | Genome-fragment | NA | NA | 0 |
| S8_extracted_k141_106723 | 1047 | No | NA | 1 | 0 | 0 | Not-determined | Genome-fragment | NA | NA | 0 |
| S8_extracted_k141_193732 | 1733 | No | NA | 2 | 0 | 1 | Not-determined | Genome-fragment | NA | NA | 0 |
| S8_extracted_k141_89304 | 3013 | No | NA | 1 | 0 | 1 | Not-determined | Genome-fragment | NA | NA | 0 |
| S8_extracted_k141_37412 | 3176 | No | NA | 3 | 0 | 2 | Not-determined | Genome-fragment | NA | NA | 0 |
| S8_extracted_k141_132617 | 2842 | No | NA | 2 | 0 | 2 | Not-determined | Genome-fragment | NA | NA | 0 |
| S17_vs_k141_59377\|\|full | 7899 | No | NA | 11 | 1 | 3 | Low-quality | Genome-fragment | 7.18 | HMM-based (lower-bound) | 0 |
| S8_extracted_k141_115282 | 1056 | No | NA | 2 | 0 | 2 | Not-determined | Genome-fragment | NA | NA | 0 |
| S8_extracted_k141_2662 | 1523 | No | NA | 3 | 0 | 1 | Not-determined | Genome-fragment | NA | NA | 0 |
| S8_extracted_k141_132624 | 2951 | No | NA | 2 | 0 | 1 | Not-determined | Genome-fragment | NA | NA | 0 |
| S14_vs_k141_32417\|\|full | 7900 | No | NA | 15 | 4 | 1 | Low-quality | Genome-fragment | 16.8 | AAI-based (medium-confidence) | 0 |
| S8_extracted_k141_141512 | 3687 | No | NA | 4 | 0 | 2 | Not-determined | Genome-fragment | NA | NA | 0 |
| S8_extracted_k141_63444 | 1515 | No | NA | 2 | 0 | 0 | Not-determined | Genome-fragment | NA | NA | 0 |
| S8_extracted_k141_201763 | 1147 | No | NA | 1 | 0 | 1 | Not-determined | Genome-fragment | NA | NA | 0 |
| S11_extracted_k141_14923 | 7905 | No | NA | 10 | 4 | 0 | Low-quality | Genome-fragment | 12.21 | HMM-based (lower-bound) | 0 |
| S8_extracted_k141_176121 | 2206 | No | NA | 2 | 0 | 2 | Not-determined | Genome-fragment | NA | NA | 0 |
| S8_extracted_k141_71554 | 1898 | No | NA | 2 | 0 | 2 | Not-determined | Genome-fragment | NA | NA | 0 |
| S8_extracted_k141_2677 | 1256 | No | NA | 1 | 0 | 1 | Not-determined | Genome-fragment | NA | NA | 0 |
| S14_vs_k141_95827\|\|full | 7905 | No | NA | 9 | 0 | 5 | Low-quality | Genome-fragment | 3.44 | AAI-based (medium-confidence) | 0 |
| S8_extracted_k141_97925 | 1284 | No | NA | 1 | 0 | 0 | Not-determined | Genome-fragment | NA | NA | 0 |
| S8_extracted_k141_89351 | 4456 | No | NA | 5 | 0 | 2 | Not-determined | Genome-fragment | NA | NA | 0 |
| S8_extracted_k141_71562 | 1560 | No | NA | 2 | 0 | 0 | Not-determined | Genome-fragment | NA | NA | 0 |
| S8_extracted_k141_28937 | 1924 | No | NA | 3 | 0 | 3 | Not-determined | Genome-fragment | NA | NA | 0 |
| S8_extracted_k141_63486 | 1159 | No | NA | 2 | 0 | 0 | Not-determined | Genome-fragment | NA | NA | 0 |
| S8_extracted_k141_89358 | 3626 | No | NA | 3 | 0 | 3 | Not-determined | Genome-fragment | NA | NA | 0 |
| S8_extracted_k141_63496 | 1558 | No | NA | 4 | 0 | 3 | Not-determined | Genome-fragment | NA | NA | 0 |
| S8_extracted_k141_176151 | 1015 | No | NA | 1 | 0 | 1 | Not-determined | Genome-fragment | NA | NA | 0 |
| S8_extracted_k141_71564 | 11380 | No | NA | 4 | 0 | 0 | Not-determined | Genome-fragment | NA | NA | 0 |
| S8_extracted_k141_37469 | 1476 | No | NA | 1 | 0 | 1 | Not-determined | Genome-fragment | NA | NA | 0 |
| S8_extracted_k141_193807 | 1308 | No | NA | 1 | 0 | 0 | Not-determined | Genome-fragment | NA | NA | 0 |
| S8_extracted_k141_132685 | 1090 | No | NA | 1 | 0 | 0 | Not-determined | Genome-fragment | NA | NA | 0 |
| S8_extracted_k141_37476 | 3017 | No | NA | 3 | 0 | 2 | Not-determined | Genome-fragment | NA | NA | 0 |
| S8_extracted_k141_132686 | 1178 | No | NA | 3 | 0 | 0 | Not-determined | Genome-fragment | NA | NA | 0 |
| S8_extracted_k141_63504 | 1086 | No | NA | 1 | 0 | 1 | Not-determined | Genome-fragment | NA | NA | 0 |
| S8_extracted_k141_124318 | 3628 | No | NA | 5 | 0 | 5 | Not-determined | Genome-fragment | NA | NA | 0 |
| S8_extracted_k141_45776 | 36660 | No | NA | 77 | 70 | 0 | High-quality | High-quality | 100 | AAI-based (high-confidence) | 0 |
| S8_extracted_k141_54785 | 1026 | No | NA | 2 | 0 | 0 | Not-determined | Genome-fragment | NA | NA | 0 |
| S8_extracted_k141_124321 | 2817 | No | NA | 1 | 0 | 0 | Not-determined | Genome-fragment | NA | NA | 0 |
| S8_extracted_k141_89397 | 1055 | No | NA | 1 | 0 | 1 | Not-determined | Genome-fragment | NA | NA | 0 |
| S8_extracted_k141_54789 | 1805 | No | NA | 1 | 0 | 1 | Not-determined | Genome-fragment | NA | NA | 0 |
| S8_extracted_k141_124336 | 3376 | No | NA | 2 | 0 | 1 | Not-determined | Genome-fragment | NA | NA | 0 |
| S8_extracted_k141_63528 | 1781 | No | NA | 2 | 0 | 1 | Not-determined | Genome-fragment | NA | NA | 0 |
| S8_extracted_k141_80858 | 1679 | No | NA | 2 | 0 | 1 | Not-determined | Genome-fragment | NA | NA | 0 |
| S8_extracted_k141_28956 | 43413 | Yes | 25419 | 46 | 10 | 9 | Medium-quality | Genome-fragment | 88.11 | AAI-based (high-confidence) | 41.45 |
| S8_extracted_k141_63546 | 2015 | No | NA | 1 | 0 | 1 | Not-determined | Genome-fragment | NA | NA | 0 |
| S8_extracted_k141_106831 | 4691 | No | NA | 4 | 0 | 3 | Not-determined | Genome-fragment | NA | NA | 0 |
| S3_vs_k141_30674\|\|full | 7908 | No | NA | 11 | 4 | 1 | Low-quality | Genome-fragment | 18.44 | AAI-based (medium-confidence) | 0 |
| S15_vs_19939\|\|full | 7910 | No | NA | 14 | 3 | 1 | Low-quality | Genome-fragment | 18.52 | HMM-based (lower-bound) | 0 |
| S8_extracted_k141_193840 | 2423 | No | NA | 4 | 0 | 0 | Not-determined | Genome-fragment | NA | NA | 0 |
| S8_extracted_k141_124362 | 1022 | No | NA | 3 | 0 | 1 | Not-determined | Genome-fragment | NA | NA | 0 |
| S8_extracted_k141_98016 | 1232 | No | NA | 1 | 0 | 1 | Not-determined | Genome-fragment | NA | NA | 0 |
| S8_extracted_k141_167526 | 4459 | No | NA | 6 | 0 | 3 | Not-determined | Genome-fragment | NA | NA | 0 |
| S8_extracted_k141_28989 | 1023 | No | NA | 1 | 0 | 1 | Not-determined | Genome-fragment | NA | NA | 0 |
| S8_extracted_k141_11137 | 1037 | No | NA | 1 | 0 | 1 | Not-determined | Genome-fragment | NA | NA | 0 |
| S8_extracted_k141_80902 | 2256 | No | NA | 1 | 0 | 0 | Not-determined | Genome-fragment | NA | NA | 0 |
| S3_extracted_k141_1131 | 7911 | No | NA | 9 | 0 | 1 | Low-quality | Genome-fragment | 18.06 | AAI-based (medium-confidence) | 0 |
| S8_extracted_k141_98036 | 1893 | No | NA | 2 | 0 | 1 | Not-determined | Genome-fragment | NA | NA | 0 |
| S8_extracted_k141_201904 | 2342 | No | NA | 4 | 0 | 3 | Not-determined | Genome-fragment | NA | NA | 0 |
| S8_extracted_k141_132697 | 59637 | No | NA | 107 | 19 | 0 | Complete | High-quality | 100 | DTR (high-confidence) | 0 |
| S3_vs_k141_76615\|\|full | 7918 | No | NA | 17 | 4 | 0 | Low-quality | Genome-fragment | 19.37 | AAI-based (high-confidence) | 0 |
| S8_extracted_k141_71591 | 4969 | No | NA | 7 | 0 | 0 | Not-determined | Genome-fragment | NA | NA | 0 |
| S10_extracted_114867 | 7924 | No | NA | 6 | 0 | 6 | Low-quality | Genome-fragment | 4.51 | AAI-based (medium-confidence) | 0 |
| S8_extracted_k141_201911 | 3094 | No | NA | 4 | 0 | 1 | Not-determined | Genome-fragment | NA | NA | 0 |
| S8_extracted_k141_80924 | 1439 | No | NA | 2 | 0 | 2 | Not-determined | Genome-fragment | NA | NA | 0 |
| S8_extracted_k141_89497 | 1022 | No | NA | 1 | 0 | 1 | Not-determined | Genome-fragment | NA | NA | 0 |
| S8_extracted_k141_115407 | 1571 | No | NA | 1 | 0 | 0 | Not-determined | Genome-fragment | NA | NA | 0 |
| S8_extracted_k141_54829 | 3344 | No | NA | 3 | 0 | 3 | Not-determined | Genome-fragment | NA | NA | 0 |
| S8_extracted_k141_115412 | 1349 | No | NA | 1 | 0 | 0 | Not-determined | Genome-fragment | NA | NA | 0 |
| S12_vs_k141_10021\|\|full | 7926 | No | NA | 11 | 2 | 0 | Low-quality | Genome-fragment | 20.09 | AAI-based (high-confidence) | 0 |
| S8_extracted_k141_158510 | 3278 | No | NA | 2 | 0 | 2 | Not-determined | Genome-fragment | NA | NA | 0 |
| S8_extracted_k141_11162 | 1242 | No | NA | 2 | 0 | 1 | Not-determined | Genome-fragment | NA | NA | 0 |
| S14_vs_k141_6433\|\|full | 7930 | Yes | 2031 | 6 | 1 | 2 | Low-quality | Genome-fragment | 4.16 | AAI-based (medium-confidence) | 74.39 |
| S8_extracted_k141_193844 | 1822 | No | NA | 1 | 0 | 1 | Not-determined | Genome-fragment | NA | NA | 0 |
| S8_extracted_k141_20076 | 7192 | No | NA | 8 | 0 | 6 | Not-determined | Genome-fragment | NA | NA | 0 |
| S14_vs_k141_73593\|\|full | 7931 | Yes | 5893 | 9 | 1 | 1 | Low-quality | Genome-fragment | 3.96 | HMM-based (lower-bound) | 25.7 |
| S8_extracted_k141_89520 | 1546 | No | NA | 1 | 0 | 0 | Not-determined | Genome-fragment | NA | NA | 0 |
| S8_extracted_k141_158530 | 3537 | No | NA | 3 | 0 | 2 | Not-determined | Genome-fragment | NA | NA | 0 |
| S8_extracted_k141_115439 | 1577 | No | NA | 1 | 0 | 0 | Not-determined | Genome-fragment | NA | NA | 0 |
| S8_extracted_k141_54862 | 1793 | No | NA | 1 | 0 | 0 | Not-determined | Genome-fragment | NA | NA | 0 |
| S8_extracted_k141_80618 | 7933 | No | NA | 5 | 0 | 4 | Low-quality | Genome-fragment | 14.46 | AAI-based (high-confidence) | 0 |
| S9_extracted_k141_31185 | 7940 | No | NA | 19 | 3 | 1 | Low-quality | Genome-fragment | 18.01 | AAI-based (high-confidence) | 0 |
| S8_extracted_k141_89528 | 4839 | No | NA | 5 | 0 | 1 | Not-determined | Genome-fragment | NA | NA | 0 |
| S8_extracted_k141_106925 | 2273 | No | NA | 3 | 0 | 1 | Not-determined | Genome-fragment | NA | NA | 0 |
| S8_extracted_k141_98089 | 2849 | No | NA | 1 | 0 | 1 | Not-determined | Genome-fragment | NA | NA | 0 |
| S8_extracted_k141_158552 | 1277 | No | NA | 1 | 0 | 0 | Not-determined | Genome-fragment | NA | NA | 0 |
| S12_extracted_k141_52192 | 7940 | No | NA | 10 | 2 | 0 | Low-quality | Genome-fragment | 12.47 | AAI-based (high-confidence) | 0 |
| S7_vs_8255\|\|full | 7941 | No | NA | 12 | 8 | 0 | Low-quality | Genome-fragment | 22.89 | AAI-based (high-confidence) | 0 |
| S14_vs_k141_12733\|\|full | 7944 | No | NA | 8 | 0 | 0 | Low-quality | Genome-fragment | 15.42 | AAI-based (medium-confidence) | 0 |
| S8_extracted_k141_132749 | 1030 | No | NA | 1 | 0 | 1 | Not-determined | Genome-fragment | NA | NA | 0 |
| S8_extracted_k141_201950 | 3924 | No | NA | 4 | 0 | 2 | Not-determined | Genome-fragment | NA | NA | 0 |
| S8_extracted_k141_115473 | 2558 | No | NA | 2 | 0 | 2 | Not-determined | Genome-fragment | NA | NA | 0 |
| S8_extracted_k141_54880 | 1655 | No | NA | 2 | 0 | 1 | Not-determined | Genome-fragment | NA | NA | 0 |
| S8_extracted_k141_63642 | 6281 | No | NA | 3 | 0 | 3 | Not-determined | Genome-fragment | NA | NA | 0 |
| S8_extracted_k141_141619 | 1206 | No | NA | 3 | 0 | 0 | Not-determined | Genome-fragment | NA | NA | 0 |
| S8_extracted_k141_124422 | 1201 | No | NA | 2 | 0 | 0 | Not-determined | Genome-fragment | NA | NA | 0 |
| S8_extracted_k141_45892 | 1528 | No | NA | 1 | 0 | 0 | Not-determined | Genome-fragment | NA | NA | 0 |
| S11_vs_k141_50313\|\|full | 7951 | No | NA | 11 | 5 | 0 | Low-quality | Genome-fragment | 19.48 | AAI-based (high-confidence) | 0 |
| S8_extracted_k141_158587 | 2237 | No | NA | 2 | 0 | 1 | Not-determined | Genome-fragment | NA | NA | 0 |
| S8_extracted_k141_54898 | 2288 | No | NA | 2 | 0 | 1 | Not-determined | Genome-fragment | NA | NA | 0 |
| S8_extracted_k141_45899 | 2926 | No | NA | 3 | 0 | 1 | Not-determined | Genome-fragment | NA | NA | 0 |
| S1_vs_k141_78263\|\|full | 7955 | No | NA | 6 | 0 | 4 | Low-quality | Genome-fragment | 3.43 | AAI-based (medium-confidence) | 0 |
| S8_extracted_k141_63659 | 1412 | No | NA | 2 | 0 | 2 | Not-determined | Genome-fragment | NA | NA | 0 |
| S8_extracted_k141_141631 | 1112 | No | NA | 2 | 0 | 2 | Not-determined | Genome-fragment | NA | NA | 0 |
| S8_extracted_k141_63663 | 1753 | No | NA | 2 | 0 | 2 | Not-determined | Genome-fragment | NA | NA | 0 |
| S8_extracted_k141_124428 | 5247 | No | NA | 5 | 0 | 3 | Not-determined | Genome-fragment | NA | NA | 0 |
| S12_extracted_k141_97440 | 7955 | No | NA | 4 | 1 | 0 | Low-quality | Genome-fragment | 6.88 | HMM-based (lower-bound) | 0 |
| S17_extracted_k141_89360 | 7956 | No | NA | 18 | 17 | 0 | Low-quality | Genome-fragment | 26.24 | AAI-based (high-confidence) | 0 |
| S3_extracted_k141_24076 | 7960 | No | NA | 13 | 2 | 0 | Low-quality | Genome-fragment | 16.4 | AAI-based (medium-confidence) | 0 |
| S12_vs_k141_3939\|\|full | 7960 | No | NA | 10 | 6 | 0 | Low-quality | Genome-fragment | 22.2 | AAI-based (high-confidence) | 0 |
| S2_vs_k141_14187\|\|full | 7965 | No | NA | 9 | 2 | 0 | Low-quality | Genome-fragment | 20.23 | AAI-based (high-confidence) | 0 |
| S8_extracted_k141_63677 | 1989 | No | NA | 3 | 0 | 2 | Not-determined | Genome-fragment | NA | NA | 0 |
| S8_extracted_k141_98156 | 1089 | No | NA | 1 | 0 | 1 | Not-determined | Genome-fragment | NA | NA | 0 |
| S8_extracted_k141_45927 | 1704 | No | NA | 4 | 0 | 4 | Not-determined | Genome-fragment | NA | NA | 0 |
| S8_extracted_k141_45932 | 1034 | No | NA | 1 | 0 | 1 | Not-determined | Genome-fragment | NA | NA | 0 |
| S8_extracted_k141_167654 | 1830 | No | NA | 2 | 0 | 0 | Not-determined | Genome-fragment | NA | NA | 0 |
| S8_extracted_k141_63685 | 1138 | No | NA | 1 | 0 | 1 | Not-determined | Genome-fragment | NA | NA | 0 |
| S8_extracted_k141_106946 | 42628 | No | NA | 48 | 27 | 0 | Complete | High-quality | 100 | DTR (high-confidence) | 0 |
| S8_extracted_k141_54917 | 5526 | No | NA | 6 | 0 | 0 | Not-determined | Genome-fragment | NA | NA | 0 |
| S8_extracted_k141_98175 | 2811 | No | NA | 4 | 0 | 0 | Not-determined | Genome-fragment | NA | NA | 0 |
| S8_extracted_k141_89560 | 2450 | No | NA | 2 | 0 | 2 | Not-determined | Genome-fragment | NA | NA | 0 |
| S8_extracted_k141_132787 | 5274 | No | NA | 6 | 0 | 4 | Not-determined | Genome-fragment | NA | NA | 0 |
| S8_extracted_k141_193896 | 1159 | No | NA | 1 | 0 | 1 | Not-determined | Genome-fragment | NA | NA | 0 |
| S8_extracted_k141_202016 | 5358 | No | NA | 9 | 0 | 0 | Not-determined | Genome-fragment | NA | NA | 0 |
| S8_extracted_k141_54941 | 4011 | No | NA | 3 | 0 | 1 | Not-determined | Genome-fragment | NA | NA | 0 |
| S1_extracted_k141_67670 | 7982 | No | NA | 10 | 6 | 0 | Low-quality | Genome-fragment | 14.43 | AAI-based (high-confidence) | 0 |
| S8_extracted_k141_115570 | 1471 | No | NA | 1 | 0 | 1 | Not-determined | Genome-fragment | NA | NA | 0 |
| S11_extracted_k141_53961 | 7983 | No | NA | 10 | 3 | 0 | Low-quality | Genome-fragment | 20.17 | AAI-based (medium-confidence) | 0 |
| S8_extracted_k141_106984 | 1429 | No | NA | 1 | 0 | 1 | Not-determined | Genome-fragment | NA | NA | 0 |
| S8_extracted_k141_141660 | 1965 | No | NA | 5 | 0 | 0 | Not-determined | Genome-fragment | NA | NA | 0 |
| S8_extracted_k141_193903 | 4556 | No | NA | 5 | 0 | 0 | Not-determined | Genome-fragment | NA | NA | 0 |
| S8_extracted_k141_132805 | 1528 | No | NA | 1 | 0 | 1 | Not-determined | Genome-fragment | NA | NA | 0 |
| S8_extracted_k141_132807 | 1641 | No | NA | 2 | 0 | 1 | Not-determined | Genome-fragment | NA | NA | 0 |
| S8_extracted_k141_176394 | 3657 | No | NA | 3 | 0 | 1 | Not-determined | Genome-fragment | NA | NA | 0 |
| S8_extracted_k141_54454 | 7983 | No | NA | 7 | 0 | 1 | Low-quality | Genome-fragment | 10.79 | AAI-based (medium-confidence) | 0 |
| S9_vs_k141_18711\|\|full | 7991 | No | NA | 9 | 0 | 3 | Low-quality | Genome-fragment | 4.51 | AAI-based (high-confidence) | 0 |
| S8_extracted_k141_37655 | 4217 | No | NA | 4 | 0 | 3 | Not-determined | Genome-fragment | NA | NA | 0 |
| S8_extracted_k141_141674 | 1926 | No | NA | 2 | 0 | 2 | Not-determined | Genome-fragment | NA | NA | 0 |
| S8_extracted_k141_98235 | 1277 | No | NA | 2 | 0 | 1 | Not-determined | Genome-fragment | NA | NA | 0 |
| S8_extracted_k141_150163 | 5869 | No | NA | 7 | 0 | 2 | Not-determined | Genome-fragment | NA | NA | 0 |
| S8_extracted_k141_176400 | 1267 | No | NA | 2 | 0 | 1 | Not-determined | Genome-fragment | NA | NA | 0 |
| S8_extracted_k141_115593 | 5106 | No | NA | 4 | 0 | 3 | Not-determined | Genome-fragment | NA | NA | 0 |
| S8_extracted_k141_132822 | 1504 | No | NA | 3 | 0 | 1 | Not-determined | Genome-fragment | NA | NA | 0 |
| S8_extracted_k141_202042 | 1763 | No | NA | 2 | 0 | 1 | Not-determined | Genome-fragment | NA | NA | 0 |
| S8_extracted_k141_81087 | 1242 | No | NA | 1 | 0 | 1 | Not-determined | Genome-fragment | NA | NA | 0 |
| S8_extracted_k141_124509 | 1703 | No | NA | 1 | 0 | 0 | Not-determined | Genome-fragment | NA | NA | 0 |
| S8_extracted_k141_11272 | 3162 | No | NA | 3 | 0 | 2 | Not-determined | Genome-fragment | NA | NA | 0 |
| S8_extracted_k141_45996 | 1185 | No | NA | 2 | 0 | 1 | Not-determined | Genome-fragment | NA | NA | 0 |
| S8_extracted_k141_107024 | 1225 | No | NA | 2 | 0 | 1 | Not-determined | Genome-fragment | NA | NA | 0 |
| S8_extracted_k141_150177 | 2070 | No | NA | 2 | 0 | 2 | Not-determined | Genome-fragment | NA | NA | 0 |
| S1_vs_k141_24784\|\|full | 8004 | No | NA | 9 | 4 | 0 | Low-quality | Genome-fragment | 5.52 | AAI-based (high-confidence) | 0 |
| S8_extracted_k141_71707 | 1001 | No | NA | 2 | 0 | 0 | Not-determined | Genome-fragment | NA | NA | 0 |
| S8_extracted_k141_54988 | 1034 | No | NA | 2 | 0 | 1 | Not-determined | Genome-fragment | NA | NA | 0 |
| S8_extracted_k141_37663 | 1037 | No | NA | 2 | 0 | 0 | Not-determined | Genome-fragment | NA | NA | 0 |
| S8_extracted_k141_167682 | 2626 | No | NA | 3 | 0 | 1 | Not-determined | Genome-fragment | NA | NA | 0 |
| S8_extracted_k141_20212 | 2324 | No | NA | 4 | 0 | 3 | Not-determined | Genome-fragment | NA | NA | 0 |
| S8_extracted_k141_54999 | 1530 | No | NA | 2 | 0 | 0 | Not-determined | Genome-fragment | NA | NA | 0 |
| S8_extracted_k141_107047 | 1274 | No | NA | 2 | 0 | 1 | Not-determined | Genome-fragment | NA | NA | 0 |
| S8_extracted_k141_2856 | 3600 | No | NA | 4 | 0 | 0 | Not-determined | Genome-fragment | NA | NA | 0 |
| S8_extracted_k141_20219 | 1606 | No | NA | 2 | 0 | 1 | Not-determined | Genome-fragment | NA | NA | 0 |
| S8_extracted_k141_55003 | 1697 | No | NA | 4 | 0 | 4 | Not-determined | Genome-fragment | NA | NA | 0 |
| S8_extracted_k141_202075 | 1206 | No | NA | 1 | 0 | 0 | Not-determined | Genome-fragment | NA | NA | 0 |
| S8_extracted_k141_11291 | 1539 | No | NA | 3 | 0 | 3 | Not-determined | Genome-fragment | NA | NA | 0 |
| S8_extracted_k141_55013 | 2055 | No | NA | 3 | 0 | 2 | Not-determined | Genome-fragment | NA | NA | 0 |
| S8_extracted_k141_46020 | 13470 | No | NA | 11 | 0 | 0 | Not-determined | Genome-fragment | NA | NA | 0 |
| S8_extracted_k141_63758 | 1942 | No | NA | 3 | 0 | 2 | Not-determined | Genome-fragment | NA | NA | 0 |
| S8_extracted_k141_55021 | 1021 | No | NA | 3 | 0 | 3 | Not-determined | Genome-fragment | NA | NA | 0 |
| S8_extracted_k141_124539 | 1980 | No | NA | 1 | 0 | 1 | Not-determined | Genome-fragment | NA | NA | 0 |
| S8_extracted_k141_81131 | 2183 | No | NA | 4 | 0 | 0 | Not-determined | Genome-fragment | NA | NA | 0 |
| S8_extracted_k141_89635 | 1228 | No | NA | 3 | 0 | 2 | Not-determined | Genome-fragment | NA | NA | 0 |
| S8_extracted_k141_46044 | 3324 | No | NA | 3 | 0 | 1 | Not-determined | Genome-fragment | NA | NA | 0 |
| S8_extracted_k141_63771 | 2529 | No | NA | 3 | 0 | 2 | Not-determined | Genome-fragment | NA | NA | 0 |
| S8_extracted_k141_11315 | 3956 | No | NA | 5 | 0 | 0 | Not-determined | Genome-fragment | NA | NA | 0 |
| S8_extracted_k141_20275 | 1033 | No | NA | 1 | 0 | 0 | Not-determined | Genome-fragment | NA | NA | 0 |
| S8_extracted_k141_98597 | 8005 | No | NA | 12 | 3 | 0 | Low-quality | Genome-fragment | 18.32 | HMM-based (lower-bound) | 0 |
| S12_vs_k141_21233\|\|full | 8010 | No | NA | 21 | 3 | 0 | Low-quality | Genome-fragment | 22.44 | AAI-based (medium-confidence) | 0 |
| S8_extracted_k141_107081 | 4240 | No | NA | 2 | 0 | 2 | Not-determined | Genome-fragment | NA | NA | 0 |
| S8_extracted_k141_141736 | 1760 | No | NA | 1 | 0 | 0 | Not-determined | Genome-fragment | NA | NA | 0 |
| S8_extracted_k141_98318 | 1484 | No | NA | 1 | 0 | 1 | Not-determined | Genome-fragment | NA | NA | 0 |
| S8_extracted_k141_20287 | 3287 | No | NA | 5 | 0 | 3 | Not-determined | Genome-fragment | NA | NA | 0 |
| S8_vs_k141_125011\|\|full | 8015 | No | NA | 9 | 7 | 0 | Low-quality | Genome-fragment | 12.8 | AAI-based (high-confidence) | 0 |
| S8_extracted_k141_46070 | 3368 | No | NA | 2 | 0 | 2 | Not-determined | Genome-fragment | NA | NA | 0 |
| S11_extracted_k141_30750 | 8018 | No | NA | 7 | 2 | 0 | Low-quality | Genome-fragment | 6.62 | AAI-based (medium-confidence) | 0 |
| S8_extracted_k141_46073 | 1903 | No | NA | 5 | 0 | 0 | Not-determined | Genome-fragment | NA | NA | 0 |
| S10_extracted_99008 | 8018 | Yes | 6530 | 9 | 3 | 1 | Low-quality | Genome-fragment | 12.79 | AAI-based (high-confidence) | 18.56 |
| S8_extracted_k141_46075 | 3274 | No | NA | 4 | 0 | 2 | Not-determined | Genome-fragment | NA | NA | 0 |
| S8_extracted_k141_37733 | 1727 | No | NA | 2 | 0 | 1 | Not-determined | Genome-fragment | NA | NA | 0 |
| S11_vs_k141_36310\|\|full | 8023 | No | NA | 13 | 2 | 0 | Low-quality | Genome-fragment | 17.29 | AAI-based (medium-confidence) | 0 |
| S8_extracted_k141_2900 | 1120 | No | NA | 1 | 0 | 0 | Not-determined | Genome-fragment | NA | NA | 0 |
| S8_extracted_k141_89682 | 1469 | No | NA | 2 | 0 | 2 | Not-determined | Genome-fragment | NA | NA | 0 |
| S8_extracted_k141_71785 | 1017 | No | NA | 1 | 0 | 1 | Not-determined | Genome-fragment | NA | NA | 0 |
| S5_vs_k141_119610\|\|full | 8025 | No | NA | 11 | 0 | 1 | Low-quality | Genome-fragment | 18.48 | AAI-based (medium-confidence) | 0 |
| S8_extracted_k141_29118 | 5720 | No | NA | 4 | 0 | 0 | Not-determined | Genome-fragment | NA | NA | 0 |
| S8_extracted_k141_46093 | 1708 | No | NA | 2 | 0 | 2 | Not-determined | Genome-fragment | NA | NA | 0 |
| S8_extracted_k141_98343 | 1161 | No | NA | 2 | 0 | 0 | Not-determined | Genome-fragment | NA | NA | 0 |
| S8_extracted_k141_29127 | 1025 | No | NA | 2 | 0 | 0 | Not-determined | Genome-fragment | NA | NA | 0 |
| S8_extracted_k141_46096 | 1537 | No | NA | 2 | 0 | 2 | Not-determined | Genome-fragment | NA | NA | 0 |
| S8_extracted_k141_176504 | 3346 | No | NA | 3 | 0 | 2 | Not-determined | Genome-fragment | NA | NA | 0 |
| S8_extracted_k141_98358 | 1546 | No | NA | 2 | 0 | 2 | Not-determined | Genome-fragment | NA | NA | 0 |
| S8_extracted_k141_202145 | 4431 | No | NA | 5 | 0 | 3 | Not-determined | Genome-fragment | NA | NA | 0 |
| S13_extracted_32408 | 8027 | No | NA | 22 | 2 | 0 | Low-quality | Genome-fragment | 17.74 | AAI-based (high-confidence) | 0 |
| S9_extracted_k141_87953 | 8032 | No | NA | 8 | 5 | 0 | Low-quality | Genome-fragment | 5.28 | AAI-based (medium-confidence) | 0 |
| S8_extracted_k141_184510 | 2041 | No | NA | 2 | 0 | 2 | Not-determined | Genome-fragment | NA | NA | 0 |
| S8_extracted_k141_98362 | 2887 | No | NA | 3 | 0 | 1 | Not-determined | Genome-fragment | NA | NA | 0 |
| S8_extracted_k141_176524 | 3591 | No | NA | 3 | 0 | 2 | Not-determined | Genome-fragment | NA | NA | 0 |
| S8_extracted_k141_98368 | 1012 | No | NA | 2 | 0 | 1 | Not-determined | Genome-fragment | NA | NA | 0 |
| S8_extracted_k141_176525 | 1654 | No | NA | 1 | 0 | 1 | Not-determined | Genome-fragment | NA | NA | 0 |
| S8_extracted_k141_55102 | 1384 | No | NA | 1 | 0 | 0 | Not-determined | Genome-fragment | NA | NA | 0 |
| S8_vs_k141_44769\|\|full | 8038 | No | NA | 9 | 0 | 2 | Low-quality | Genome-fragment | 5.58 | AAI-based (medium-confidence) | 0 |
| S8_extracted_k141_2961 | 3237 | No | NA | 2 | 0 | 1 | Not-determined | Genome-fragment | NA | NA | 0 |
| S8_extracted_k141_202155 | 5516 | No | NA | 4 | 0 | 1 | Not-determined | Genome-fragment | NA | NA | 0 |
| S8_extracted_k141_2962 | 4155 | No | NA | 3 | 0 | 1 | Not-determined | Genome-fragment | NA | NA | 0 |
| S17_vs_k141_54375\|\|full | 8040 | No | NA | 8 | 0 | 7 | Low-quality | Genome-fragment | 24.18 | AAI-based (medium-confidence) | 0 |
| S8_extracted_k141_89768 | 1850 | No | NA | 2 | 0 | 2 | Not-determined | Genome-fragment | NA | NA | 0 |
| S8_extracted_k141_37793 | 1059 | No | NA | 1 | 0 | 0 | Not-determined | Genome-fragment | NA | NA | 0 |
| S8_extracted_k141_55128 | 3504 | No | NA | 5 | 0 | 0 | Not-determined | Genome-fragment | NA | NA | 0 |
| S10_vs_2318\|\|full | 8040 | No | NA | 16 | 2 | 1 | Low-quality | Genome-fragment | 5.33 | AAI-based (medium-confidence) | 0 |
| S8_extracted_k141_2984 | 2151 | No | NA | 3 | 0 | 0 | Not-determined | Genome-fragment | NA | NA | 0 |
| S8_extracted_k141_89782 | 2455 | No | NA | 2 | 0 | 1 | Not-determined | Genome-fragment | NA | NA | 0 |
| S8_extracted_k141_107178 | 2480 | No | NA | 2 | 0 | 2 | Not-determined | Genome-fragment | NA | NA | 0 |
| S8_extracted_k141_193988 | 4901 | No | NA | 5 | 0 | 1 | Not-determined | Genome-fragment | NA | NA | 0 |
| S8_extracted_k141_29208 | 2072 | No | NA | 5 | 0 | 5 | Not-determined | Genome-fragment | NA | NA | 0 |
| S8_extracted_k141_71865 | 7771 | No | NA | 5 | 0 | 4 | Not-determined | Genome-fragment | NA | NA | 0 |
| S8_extracted_k141_37815 | 1225 | No | NA | 3 | 0 | 0 | Not-determined | Genome-fragment | NA | NA | 0 |
| S11_vs_k141_58184\|\|full | 8045 | No | NA | 16 | 1 | 0 | Low-quality | Genome-fragment | 17.53 | AAI-based (high-confidence) | 0 |
| S8_extracted_k141_176565 | 1215 | No | NA | 2 | 0 | 1 | Not-determined | Genome-fragment | NA | NA | 0 |
| S11_vs_k141_34149\|\|full | 8047 | Yes | 4897 | 14 | 2 | 2 | Low-quality | Genome-fragment | 8.2 | AAI-based (high-confidence) | 39.15 |
| S8_extracted_k141_71886 | 1340 | No | NA | 2 | 0 | 1 | Not-determined | Genome-fragment | NA | NA | 0 |
| S8_extracted_k141_81206 | 7078 | No | NA | 7 | 0 | 2 | Not-determined | Genome-fragment | NA | NA | 0 |
| S8_extracted_k141_63842 | 1005 | No | NA | 2 | 0 | 0 | Not-determined | Genome-fragment | NA | NA | 0 |
| S8_extracted_k141_176578 | 2038 | No | NA | 2 | 0 | 1 | Not-determined | Genome-fragment | NA | NA | 0 |
| S8_extracted_k141_20375 | 1494 | No | NA | 3 | 0 | 1 | Not-determined | Genome-fragment | NA | NA | 0 |
| S8_extracted_k141_71895 | 2261 | No | NA | 2 | 0 | 1 | Not-determined | Genome-fragment | NA | NA | 0 |
| S8_extracted_k141_141842 | 1061 | No | NA | 2 | 0 | 1 | Not-determined | Genome-fragment | NA | NA | 0 |
| S11_vs_k141_82964\|\|full | 8048 | No | NA | 10 | 0 | 2 | Low-quality | Genome-fragment | 10.29 | AAI-based (medium-confidence) | 0 |
| S13_extracted_123509 | 8050 | No | NA | 18 | 2 | 0 | Low-quality | Genome-fragment | 17.3 | AAI-based (high-confidence) | 0 |
| S5_extracted_k141_79389 | 8050 | No | NA | 12 | 10 | 0 | Low-quality | Genome-fragment | 19.14 | AAI-based (high-confidence) | 0 |

| | | | | | | | | | | | |
|---|---|---|---|---|---|---|---|---|---|---|---|
| S8_extracted_k141_81215 | 1087 | No | NA | 2 | 0 | 1 | Not-determined | Genome-fragment | NA | NA | 0 |
| S16_vs_151035\|\|full | 8052 | No | NA | 8 | 0 | 3 | Low-quality | Genome-fragment | 4.71 | AAI-based (high-confidence) | 0 |
| S8_extracted_k141_171622 | 8053 | No | NA | 11 | 2 | 2 | Low-quality | Genome-fragment | 4.22 | AAI-based (medium-confidence) | 0 |
| S8_extracted_k141_184576 | 8172 | No | NA | 7 | 0 | 7 | Not-determined | Genome-fragment | NA | NA | 0 |
| S8_extracted_k141_107230 | 4078 | No | NA | 4 | 0 | 2 | Not-determined | Genome-fragment | NA | NA | 0 |
| S8_extracted_k141_11447 | 1382 | No | NA | 4 | 0 | 3 | Not-determined | Genome-fragment | NA | NA | 0 |
| S8_extracted_k141_81234 | 1335 | No | NA | 1 | 0 | 0 | Not-determined | Genome-fragment | NA | NA | 0 |
| S8_extracted_k141_124683 | 1024 | No | NA | 2 | 0 | 1 | Not-determined | Genome-fragment | NA | NA | 0 |
| S8_extracted_k141_202228 | 4288 | No | NA | 3 | 0 | 0 | Not-determined | Genome-fragment | NA | NA | 0 |
| S8_extracted_k141_81238 | 3126 | No | NA | 3 | 0 | 2 | Not-determined | Genome-fragment | NA | NA | 0 |
| S14_vs_k141_79138\|\|full | 8056 | No | NA | 12 | 9 | 0 | Low-quality | Genome-fragment | 19.2 | AAI-based (high-confidence) | 0 |
| S8_extracted_k141_89799 | 1138 | No | NA | 0 | 0 | 0 | Not-determined | Genome-fragment | NA | NA | 0 |
| S8_extracted_k141_29259 | 14554 | No | NA | 4 | 0 | 2 | Not-determined | Genome-fragment | NA | NA | 0 |
| S8_extracted_k141_3038 | 1643 | No | NA | 5 | 0 | 0 | Not-determined | Genome-fragment | NA | NA | 0 |
| S8_extracted_k141_141873 | 1249 | No | NA | 1 | 0 | 1 | Not-determined | Genome-fragment | NA | NA | 0 |
| S3_extracted_k141_98756 | 8056 | No | NA | 18 | 2 | 0 | Low-quality | Genome-fragment | 18.27 | AAI-based (high-confidence) | 0 |
| S8_extracted_k141_98470 | 2662 | No | NA | 3 | 0 | 2 | Not-determined | Genome-fragment | NA | NA | 0 |
| S8_extracted_k141_194063 | 1452 | No | NA | 1 | 0 | 0 | Not-determined | Genome-fragment | NA | NA | 0 |
| S8_extracted_k141_20442 | 3017 | No | NA | 7 | 0 | 7 | Not-determined | Genome-fragment | NA | NA | 0 |
| S8_extracted_k141_115799 | 3180 | No | NA | 2 | 0 | 2 | Not-determined | Genome-fragment | NA | NA | 0 |
| S8_extracted_k141_184619 | 1075 | No | NA | 1 | 0 | 1 | Not-determined | Genome-fragment | NA | NA | 0 |
| S8_extracted_k141_202272 | 1538 | No | NA | 1 | 0 | 1 | Not-determined | Genome-fragment | NA | NA | 0 |
| S11_vs_k141_88356\|\|full | 8058 | No | NA | 17 | 0 | 2 | Low-quality | Genome-fragment | 17.07 | AAI-based (medium-confidence) | 0 |
| S8_extracted_k141_3064 | 5864 | No | NA | 6 | 0 | 5 | Not-determined | Genome-fragment | NA | NA | 0 |
| S1_extracted_k141_66259 | 8059 | No | NA | 12 | 1 | 0 | Low-quality | Genome-fragment | 19.08 | AAI-based (high-confidence) | 0 |
| S8_extracted_k141_176611 | 1257 | No | NA | 1 | 0 | 1 | Not-determined | Genome-fragment | NA | NA | 0 |
| S8_extracted_k141_11491 | 1260 | No | NA | 1 | 0 | 1 | Not-determined | Genome-fragment | NA | NA | 0 |
| S8_extracted_k141_3075 | 1523 | No | NA | 2 | 0 | 1 | Not-determined | Genome-fragment | NA | NA | 0 |
| S8_vs_k141_32868\|\|full | 8061 | No | NA | 12 | 1 | 0 | Low-quality | Genome-fragment | 23.64 | AAI-based (high-confidence) | 0 |
| S8_extracted_k141_37884 | 1158 | No | NA | 2 | 0 | 0 | Not-determined | Genome-fragment | NA | NA | 0 |
| S8_extracted_k141_3087 | 1310 | No | NA | 2 | 0 | 1 | Not-determined | Genome-fragment | NA | NA | 0 |
| S8_extracted_k141_124740 | 1008 | No | NA | 1 | 0 | 0 | Not-determined | Genome-fragment | NA | NA | 0 |
| S8_extracted_k141_141921 | 1505 | No | NA | 4 | 0 | 0 | Not-determined | Genome-fragment | NA | NA | 0 |
| S8_extracted_k141_176630 | 1569 | No | NA | 1 | 0 | 1 | Not-determined | Genome-fragment | NA | NA | 0 |
| S8_extracted_k141_194085 | 1759 | No | NA | 1 | 0 | 1 | Not-determined | Genome-fragment | NA | NA | 0 |
| S8_extracted_k141_124756 | 1163 | No | NA | 2 | 0 | 0 | Not-determined | Genome-fragment | NA | NA | 0 |
| S14_vs_k141_90774\|\|full | 8063 | No | NA | 12 | 1 | 1 | Low-quality | Genome-fragment | 17.62 | AAI-based (high-confidence) | 0 |
| S8_extracted_k141_141934 | 3563 | No | NA | 4 | 0 | 3 | Not-determined | Genome-fragment | NA | NA | 0 |
| S8_extracted_k141_194097 | 3308 | No | NA | 3 | 0 | 2 | Not-determined | Genome-fragment | NA | NA | 0 |
| S8_extracted_k141_63920 | 1389 | No | NA | 1 | 0 | 1 | Not-determined | Genome-fragment | NA | NA | 0 |
| S8_extracted_k141_89873 | 1819 | No | NA | 3 | 0 | 0 | Not-determined | Genome-fragment | NA | NA | 0 |
| S8_extracted_k141_133163 | 3707 | No | NA | 4 | 0 | 3 | Not-determined | Genome-fragment | NA | NA | 0 |
| S8_extracted_k141_194119 | 1360 | No | NA | 3 | 0 | 0 | Not-determined | Genome-fragment | NA | NA | 0 |
| S8_extracted_k141_55207 | 6533 | No | NA | 4 | 0 | 2 | Not-determined | Genome-fragment | NA | NA | 0 |
| S8_extracted_k141_3124 | 3245 | No | NA | 4 | 0 | 2 | Not-determined | Genome-fragment | NA | NA | 0 |
| S8_extracted_k141_20474 | 1338 | No | NA | 2 | 0 | 2 | Not-determined | Genome-fragment | NA | NA | 0 |
| S15_extracted_37627 | 8063 | No | NA | 13 | 1 | 0 | Low-quality | Genome-fragment | 7.22 | HMM-based (lower-bound) | 0 |
| S8_extracted_k141_176679 | 3560 | No | NA | 3 | 0 | 2 | Not-determined | Genome-fragment | NA | NA | 0 |
| S8_extracted_k141_20484 | 3032 | No | NA | 5 | 0 | 1 | Not-determined | Genome-fragment | NA | NA | 0 |
| S7_vs_126362\|\|full | 8064 | No | NA | 4 | 0 | 4 | Low-quality | Genome-fragment | 4.34 | AAI-based (medium-confidence) | 0 |
| S8_extracted_k141_115888 | 1133 | No | NA | 2 | 0 | 1 | Not-determined | Genome-fragment | NA | NA | 0 |
| S8_extracted_k141_194139 | 1382 | No | NA | 2 | 0 | 1 | Not-determined | Genome-fragment | NA | NA | 0 |
| S8_extracted_k141_46244 | 5451 | No | NA | 7 | 0 | 1 | Not-determined | Genome-fragment | NA | NA | 0 |
| S7_vs_8146\|\|full | 8065 | No | NA | 13 | 1 | 0 | Low-quality | Genome-fragment | 9.21 | AAI-based (high-confidence) | 0 |
| S8_extracted_k141_194153 | 1258 | No | NA | 3 | 0 | 1 | Not-determined | Genome-fragment | NA | NA | 0 |
| S8_extracted_k141_55235 | 1977 | No | NA | 2 | 0 | 2 | Not-determined | Genome-fragment | NA | NA | 0 |
| S8_extracted_k141_46260 | 2755 | No | NA | 4 | 0 | 4 | Not-determined | Genome-fragment | NA | NA | 0 |
| S8_extracted_k141_167859 | 1932 | No | NA | 2 | 0 | 1 | Not-determined | Genome-fragment | NA | NA | 0 |
| S8_extracted_k141_63972 | 19221 | No | NA | 9 | 0 | 7 | Not-determined | Genome-fragment | NA | NA | 0 |
| S8_extracted_k141_158967 | 1196 | No | NA | 1 | 0 | 1 | Not-determined | Genome-fragment | NA | NA | 0 |
| S8_extracted_k141_3154 | 1104 | No | NA | 1 | 0 | 0 | Not-determined | Genome-fragment | NA | NA | 0 |
| S8_extracted_k141_72007 | 1109 | No | NA | 2 | 0 | 0 | Not-determined | Genome-fragment | NA | NA | 0 |
| S8_extracted_k141_133210 | 3233 | No | NA | 5 | 0 | 3 | Not-determined | Genome-fragment | NA | NA | 0 |
| S8_extracted_k141_184706 | 1620 | No | NA | 2 | 0 | 2 | Not-determined | Genome-fragment | NA | NA | 0 |
| S14_vs_k141_77071\|\|full | 8067 | No | NA | 5 | 2 | 0 | Low-quality | Genome-fragment | 20.9 | AAI-based (medium-confidence) | 0 |
| S8_extracted_k141_55259 | 2608 | No | NA | 2 | 0 | 1 | Not-determined | Genome-fragment | NA | NA | 0 |
| S8_extracted_k141_98580 | 1856 | No | NA | 3 | 0 | 0 | Not-determined | Genome-fragment | NA | NA | 0 |
| S8_extracted_k141_184720 | 1676 | No | NA | 2 | 0 | 0 | Not-determined | Genome-fragment | NA | NA | 0 |
| S8_extracted_k141_11566 | 1414 | No | NA | 1 | 0 | 1 | Not-determined | Genome-fragment | NA | NA | 0 |
| S8_extracted_k141_98584 | 1024 | No | NA | 1 | 0 | 0 | Not-determined | Genome-fragment | NA | NA | 0 |
| S8_extracted_k141_98587 | 1443 | No | NA | 2 | 0 | 2 | Not-determined | Genome-fragment | NA | NA | 0 |
| S8_extracted_k141_124791 | 1242 | No | NA | 1 | 0 | 0 | Not-determined | Genome-fragment | NA | NA | 0 |
| S7_extracted_76029 | 8071 | No | NA | 2 | 1 | 0 | Low-quality | Genome-fragment | 5.53 | HMM-based (lower-bound) | 0 |
| S8_extracted_k141_3179 | 3839 | No | NA | 2 | 0 | 2 | Not-determined | Genome-fragment | NA | NA | 0 |
| S3_vs_k141_123252\|\|full | 8077 | No | NA | 15 | 1 | 0 | Low-quality | Genome-fragment | 5.51 | AAI-based (medium-confidence) | 0 |
| S8_extracted_k141_89908 | 3799 | No | NA | 6 | 0 | 3 | Not-determined | Genome-fragment | NA | NA | 0 |
| S5_vs_k141_93267\|\|full | 8078 | No | NA | 9 | 5 | 0 | Low-quality | Genome-fragment | 16.52 | AAI-based (medium-confidence) | 0 |
| S8_extracted_k141_29453 | 1567 | No | NA | 2 | 0 | 0 | Not-determined | Genome-fragment | NA | NA | 0 |
| S5_vs_k141_18208\|\|full | 8082 | Yes | 5704 | 10 | 1 | 1 | Low-quality | Genome-fragment | 12.67 | AAI-based (medium-confidence) | 29.42 |
| S8_extracted_k141_202388 | 1830 | No | NA | 1 | 0 | 0 | Not-determined | Genome-fragment | NA | NA | 0 |
| S8_extracted_k141_167942 | 1234 | No | NA | 1 | 0 | 1 | Not-determined | Genome-fragment | NA | NA | 0 |
| S10_extracted_78818 | 8083 | No | NA | 12 | 0 | 0 | Low-quality | Genome-fragment | 4.96 | AAI-based (high-confidence) | 0 |
| S8_extracted_k141_124829 | 1095 | No | NA | 1 | 0 | 0 | Not-determined | Genome-fragment | NA | NA | 0 |
| S8_extracted_k141_124830 | 1074 | No | NA | 2 | 0 | 0 | Not-determined | Genome-fragment | NA | NA | 0 |
| S8_extracted_k141_133255 | 4268 | No | NA | 2 | 0 | 0 | Not-determined | Genome-fragment | NA | NA | 0 |
| S8_extracted_k141_202398 | 1034 | No | NA | 1 | 0 | 0 | Not-determined | Genome-fragment | NA | NA | 0 |
| S8_extracted_k141_133264 | 1582 | No | NA | 1 | 0 | 0 | Not-determined | Genome-fragment | NA | NA | 0 |
| S10_extracted_107254 | 8083 | No | NA | 10 | 3 | 0 | Low-quality | Genome-fragment | 15.44 | AAI-based (high-confidence) | 0 |
| S11_extracted_k141_76596 | 8084 | Yes | 6559 | 15 | 2 | 2 | Low-quality | Genome-fragment | 17.8 | AAI-based (medium-confidence) | 18.86 |
| S8_extracted_k141_202407 | 1724 | No | NA | 1 | 0 | 0 | Not-determined | Genome-fragment | NA | NA | 0 |
| S8_extracted_k141_133271 | 1930 | No | NA | 2 | 0 | 0 | Not-determined | Genome-fragment | NA | NA | 0 |
| S8_extracted_k141_150533 | 1221 | No | NA | 3 | 0 | 0 | Not-determined | Genome-fragment | NA | NA | 0 |
| S14_vs_k141_20523\|\|full | 8085 | No | NA | 22 | 3 | 0 | Low-quality | Genome-fragment | 14.2 | AAI-based (high-confidence) | 0 |
| S8_extracted_k141_81323 | 1560 | No | NA | 2 | 0 | 0 | Not-determined | Genome-fragment | NA | NA | 0 |
| S8_extracted_k141_142071 | 3505 | No | NA | 3 | 0 | 2 | Not-determined | Genome-fragment | NA | NA | 0 |
| S8_extracted_k141_64067 | 3669 | No | NA | 4 | 0 | 2 | Not-determined | Genome-fragment | NA | NA | 0 |
| S8_extracted_k141_38049 | 1596 | No | NA | 3 | 0 | 0 | Not-determined | Genome-fragment | NA | NA | 0 |
| S8_extracted_k141_142081 | 1285 | No | NA | 2 | 0 | 2 | Not-determined | Genome-fragment | NA | NA | 0 |
| S8_extracted_k141_55313 | 5389 | No | NA | 5 | 0 | 1 | Not-determined | Genome-fragment | NA | NA | 0 |
| S9_vs_k141_65390\|\|full | 8097 | No | NA | 16 | 6 | 0 | Low-quality | Genome-fragment | 19.4 | AAI-based (high-confidence) | 0 |
| S8_extracted_k141_38061 | 1163 | No | NA | 1 | 0 | 1 | Not-determined | Genome-fragment | NA | NA | 0 |
| S8_extracted_k141_46273 | 2858 | No | NA | 2 | 0 | 2 | Not-determined | Genome-fragment | NA | NA | 0 |
| S8_extracted_k141_98648 | 1002 | No | NA | 1 | 0 | 0 | Not-determined | Genome-fragment | NA | NA | 0 |
| S8_vs_k141_152228\|\|full | 8101 | No | NA | 14 | 0 | 0 | Low-quality | Genome-fragment | 18.6 | AAI-based (medium-confidence) | 0 |
| S8_extracted_k141_202454 | 2920 | No | NA | 2 | 0 | 2 | Not-determined | Genome-fragment | NA | NA | 0 |
| S12_vs_k141_26039\|\|full | 8102 | No | NA | 10 | 1 | 0 | Low-quality | Genome-fragment | 3.97 | HMM-based (lower-bound) | 0 |
| S12_vs_k141_152860\|\|full | 8105 | No | NA | 15 | 5 | 0 | Low-quality | Genome-fragment | 21.22 | AAI-based (high-confidence) | 0 |
| S8_extracted_k141_142095 | 1908 | No | NA | 4 | 0 | 3 | Not-determined | Genome-fragment | NA | NA | 0 |
| S8_extracted_k141_176755 | 6081 | No | NA | 4 | 0 | 2 | Not-determined | Genome-fragment | NA | NA | 0 |
| S8_extracted_k141_89969 | 15126 | No | NA | 18 | 0 | 16 | Not-determined | Genome-fragment | NA | NA | 0 |
| S11_vs_k141_100019\|\|full | 8106 | No | NA | 6 | 0 | 4 | Low-quality | Genome-fragment | 2.45 | AAI-based (medium-confidence) | 0 |
| S8_extracted_k141_72119 | 9055 | No | NA | 8 | 0 | 5 | Not-determined | Genome-fragment | NA | NA | 0 |
| S8_extracted_k141_124878 | 8402 | No | NA | 1 | 0 | 1 | Not-determined | Genome-fragment | NA | NA | 0 |
| S8_extracted_k141_11704 | 1329 | No | NA | 2 | 0 | 0 | Not-determined | Genome-fragment | NA | NA | 0 |
| S8_extracted_k141_72122 | 1347 | No | NA | 2 | 0 | 1 | Not-determined | Genome-fragment | NA | NA | 0 |
| S8_extracted_k141_168074 | 4893 | No | NA | 5 | 0 | 3 | Not-determined | Genome-fragment | NA | NA | 0 |
| S8_extracted_k141_184846 | 1294 | No | NA | 2 | 0 | 2 | Not-determined | Genome-fragment | NA | NA | 0 |
| S8_extracted_k141_20641 | 1566 | No | NA | 2 | 0 | 2 | Not-determined | Genome-fragment | NA | NA | 0 |
| S8_extracted_k141_194272 | 4075 | No | NA | 7 | 0 | 3 | Not-determined | Genome-fragment | NA | NA | 0 |
| S12_extracted_k141_28800 | 8113 | No | NA | 8 | 0 | 7 | Low-quality | Genome-fragment | 6.5 | AAI-based (high-confidence) | 0 |
| S8_extracted_k141_194273 | 1396 | No | NA | 3 | 0 | 3 | Not-determined | Genome-fragment | NA | NA | 0 |
| S3_vs_k141_104733\|\|full | 8117 | No | NA | 6 | 0 | 5 | Low-quality | Genome-fragment | 4.29 | AAI-based (high-confidence) | 0 |
| S8_extracted_k141_3228 | 6041 | No | NA | 7 | 0 | 1 | Not-determined | Genome-fragment | NA | NA | 0 |
| S8_extracted_k141_202522 | 1626 | No | NA | 1 | 0 | 1 | Not-determined | Genome-fragment | NA | NA | 0 |
| S8_extracted_k141_70 | 8130 | No | NA | 14 | 2 | 0 | Low-quality | Genome-fragment | 7.75 | AAI-based (high-confidence) | 0 |
| S8_extracted_k141_184869 | 2183 | No | NA | 2 | 0 | 2 | Not-determined | Genome-fragment | NA | NA | 0 |
| S8_extracted_k141_107499 | 1423 | No | NA | 1 | 0 | 1 | Not-determined | Genome-fragment | NA | NA | 0 |
| S8_extracted_k141_159168 | 1101 | No | NA | 2 | 0 | 0 | Not-determined | Genome-fragment | NA | NA | 0 |
| S8_extracted_k141_89992 | 1029 | No | NA | 2 | 0 | 1 | Not-determined | Genome-fragment | NA | NA | 0 |
| S8_extracted_k141_124920 | 9140 | No | NA | 11 | 0 | 3 | Not-determined | Genome-fragment | NA | NA | 0 |
| S8_extracted_k141_107508 | 3245 | No | NA | 2 | 0 | 1 | Not-determined | Genome-fragment | NA | NA | 0 |
| S8_extracted_k141_20672 | 1213 | No | NA | 2 | 0 | 1 | Not-determined | Genome-fragment | NA | NA | 0 |
| S3_vs_k141_117077\|\|full | 8133 | No | NA | 12 | 4 | 0 | Low-quality | Genome-fragment | 13.51 | AAI-based (high-confidence) | 0 |
| S11_extracted_k141_58836 | 8134 | No | NA | 14 | 2 | 0 | Low-quality | Genome-fragment | 16.99 | AAI-based (high-confidence) | 0 |
| S8_extracted_k141_90004 | 1652 | No | NA | 2 | 0 | 1 | Not-determined | Genome-fragment | NA | NA | 0 |
| S10_vs_48269\|\|full | 8135 | No | NA | 15 | 4 | 0 | Low-quality | Genome-fragment | 13.59 | AAI-based (medium-confidence) | 0 |
| S13_extracted_69320 | 8136 | No | NA | 12 | 3 | 0 | Low-quality | Genome-fragment | 13 | AAI-based (high-confidence) | 0 |
| S8_extracted_k141_159173 | 16137 | No | NA | 23 | 7 | 0 | Medium-quality | Genome-fragment | 74.52 | AAI-based (medium-confidence) | 0 |
| S8_extracted_k141_38171 | 1992 | No | NA | 3 | 0 | 1 | Not-determined | Genome-fragment | NA | NA | 0 |
| S8_extracted_k141_184898 | 1080 | No | NA | 1 | 0 | 0 | Not-determined | Genome-fragment | NA | NA | 0 |
| S8_extracted_k141_107531 | 2974 | No | NA | 4 | 0 | 0 | Not-determined | Genome-fragment | NA | NA | 0 |
| S8_extracted_k141_81453 | 3222 | No | NA | 3 | 0 | 1 | Not-determined | Genome-fragment | NA | NA | 0 |
| S8_extracted_k141_3267 | 1857 | No | NA | 1 | 0 | 1 | Not-determined | Genome-fragment | NA | NA | 0 |
| S8_extracted_k141_11771 | 1034 | No | NA | 1 | 0 | 1 | Not-determined | Genome-fragment | NA | NA | 0 |
| S8_extracted_k141_90005 | 15542 | No | NA | 17 | 0 | 3 | Not-determined | Genome-fragment | NA | NA | 0 |
| S8_extracted_k141_38181 | 1931 | No | NA | 1 | 0 | 0 | Not-determined | Genome-fragment | NA | NA | 0 |
| S8_extracted_k141_194336 | 1899 | No | NA | 1 | 0 | 1 | Not-determined | Genome-fragment | NA | NA | 0 |
| S10_extracted_16154 | 8138 | No | NA | 16 | 1 | 4 | Low-quality | Genome-fragment | 20.92 | AAI-based (high-confidence) | 0 |
| S8_extracted_k141_46362 | 1161 | No | NA | 4 | 0 | 2 | Not-determined | Genome-fragment | NA | NA | 0 |
| S8_extracted_k141_107547 | 4489 | No | NA | 5 | 0 | 0 | Not-determined | Genome-fragment | NA | NA | 0 |
| S8_extracted_k141_184918 | 3205 | No | NA | 4 | 0 | 3 | Not-determined | Genome-fragment | NA | NA | 0 |
| S8_extracted_k141_72206 | 2008 | No | NA | 1 | 0 | 1 | Not-determined | Genome-fragment | NA | NA | 0 |
| S8_extracted_k141_72207 | 2386 | No | NA | 3 | 0 | 1 | Not-determined | Genome-fragment | NA | NA | 0 |
| S8_extracted_k141_194364 | 1814 | No | NA | 2 | 0 | 2 | Not-determined | Genome-fragment | NA | NA | 0 |
| S8_extracted_k141_115975 | 1716 | No | NA | 4 | 0 | 1 | Not-determined | Genome-fragment | NA | NA | 0 |
| S16_vs_38696\|\|full | 8150 | No | NA | 8 | 1 | 0 | Low-quality | Genome-fragment | 4.42 | HMM-based (lower-bound) | 0 |
| S8_extracted_k141_159230 | 1088 | No | NA | 1 | 0 | 0 | Not-determined | Genome-fragment | NA | NA | 0 |
| S8_extracted_k141_194383 | 1599 | No | NA | 2 | 0 | 0 | Not-determined | Genome-fragment | NA | NA | 0 |
| S8_extracted_k141_194390 | 1034 | No | NA | 1 | 0 | 1 | Not-determined | Genome-fragment | NA | NA | 0 |
| S8_extracted_k141_38226 | 1665 | No | NA | 2 | 0 | 1 | Not-determined | Genome-fragment | NA | NA | 0 |
| S8_extracted_k141_194396 | 1564 | No | NA | 2 | 0 | 2 | Not-determined | Genome-fragment | NA | NA | 0 |
| S8_extracted_k141_98749 | 2177 | No | NA | 2 | 0 | 2 | Not-determined | Genome-fragment | NA | NA | 0 |
| S8_extracted_k141_20756 | 1287 | No | NA | 1 | 0 | 1 | Not-determined | Genome-fragment | NA | NA | 0 |
| S8_extracted_k141_194400 | 4850 | No | NA | 5 | 0 | 0 | Not-determined | Genome-fragment | NA | NA | 0 |
| S8_extracted_k141_107578 | 3704 | No | NA | 2 | 0 | 1 | Not-determined | Genome-fragment | NA | NA | 0 |
| S8_extracted_k141_29641 | 2010 | No | NA | 1 | 0 | 1 | Not-determined | Genome-fragment | NA | NA | 0 |
| S8_extracted_k141_3346 | 1247 | No | NA | 4 | 0 | 4 | Not-determined | Genome-fragment | NA | NA | 0 |
| S8_extracted_k141_125014 | 5182 | No | NA | 8 | 0 | 2 | Not-determined | Genome-fragment | NA | NA | 0 |
| S8_extracted_k141_3349 | 1426 | No | NA | 1 | 0 | 0 | Not-determined | Genome-fragment | NA | NA | 0 |
| S9_vs_k141_27783\|\|full | 8154 | No | NA | 10 | 0 | 0 | Low-quality | Genome-fragment | 7.77 | AAI-based (medium-confidence) | 0 |
| S3_extracted_k141_115704 | 8170 | No | NA | 3 | 0 | 3 | Low-quality | Genome-fragment | 3.59 | AAI-based (high-confidence) | 0 |
| S8_extracted_k141_194424 | 1433 | No | NA | 1 | 0 | 1 | Not-determined | Genome-fragment | NA | NA | 0 |
| S8_extracted_k141_125018 | 1000 | No | NA | 2 | 0 | 1 | Not-determined | Genome-fragment | NA | NA | 0 |
| S8_extracted_k141_90076 | 1237 | No | NA | 1 | 0 | 0 | Not-determined | Genome-fragment | NA | NA | 0 |
| S8_extracted_k141_194437 | 2069 | No | NA | 1 | 0 | 1 | Not-determined | Genome-fragment | NA | NA | 0 |
| S8_extracted_k141_142205 | 4721 | No | NA | 1 | 0 | 1 | Not-determined | Genome-fragment | NA | NA | 0 |
| S8_extracted_k141_194444 | 1053 | No | NA | 2 | 0 | 1 | Not-determined | Genome-fragment | NA | NA | 0 |
| S8_extracted_k141_116038 | 1271 | No | NA | 2 | 0 | 0 | Not-determined | Genome-fragment | NA | NA | 0 |
| S8_extracted_k141_98777 | 4755 | No | NA | 5 | 0 | 3 | Not-determined | Genome-fragment | NA | NA | 0 |
| S8_vs_k141_66944\|\|full | 8175 | No | NA | 11 | 10 | 0 | Low-quality | Genome-fragment | 24.41 | AAI-based (high-confidence) | 0 |
| S8_extracted_k141_55476 | 47707 | No | NA | 74 | 20 | 1 | Complete | High-quality | 100 | DTR (high-confidence) | 0 |
| S8_extracted_k141_98789 | 3302 | No | NA | 4 | 0 | 1 | Not-determined | Genome-fragment | NA | NA | 0 |
| S14_vs_k141_13616\|\|full | 8177 | No | NA | 8 | 1 | 0 | Low-quality | Genome-fragment | 24.74 | AAI-based (high-confidence) | 0 |
| S8_extracted_k141_107625 | 7538 | No | NA | 5 | 0 | 3 | Not-determined | Genome-fragment | NA | NA | 0 |
| S8_extracted_k141_38279 | 4357 | No | NA | 4 | 0 | 2 | Not-determined | Genome-fragment | NA | NA | 0 |
| S8_extracted_k141_3410 | 16677 | No | NA | 19 | 8 | 0 | High-quality | High-quality | 97.92 | AAI-based (medium-confidence) | 0 |
| S8_extracted_k141_194457 | 5011 | No | NA | 8 | 0 | 4 | Not-determined | Genome-fragment | NA | NA | 0 |
| S8_extracted_k141_202596 | 1334 | No | NA | 2 | 0 | 0 | Not-determined | Genome-fragment | NA | NA | 0 |
| S8_extracted_k141_202604 | 3067 | No | NA | 6 | 0 | 3 | Not-determined | Genome-fragment | NA | NA | 0 |
| S8_extracted_k141_90147 | 2807 | No | NA | 3 | 0 | 2 | Not-determined | Genome-fragment | NA | NA | 0 |
| S8_extracted_k141_46458 | 1437 | No | NA | 2 | 0 | 1 | Not-determined | Genome-fragment | NA | NA | 0 |
| S17_vs_k141_61263\|\|full | 8179 | No | NA | 15 | 2 | 1 | Low-quality | Genome-fragment | 12.9 | AAI-based (high-confidence) | 0 |
| S8_extracted_k141_29750 | 1007 | No | NA | 1 | 0 | 1 | Not-determined | Genome-fragment | NA | NA | 0 |
| S8_extracted_k141_194470 | 1147 | No | NA | 3 | 0 | 1 | Not-determined | Genome-fragment | NA | NA | 0 |
| S8_extracted_k141_159353 | 1386 | No | NA | 1 | 0 | 1 | Not-determined | Genome-fragment | NA | NA | 0 |
| S8_extracted_k141_107652 | 5872 | No | NA | 4 | 0 | 1 | Not-determined | Genome-fragment | NA | NA | 0 |
| S8_extracted_k141_3438 | 6562 | No | NA | 4 | 0 | 4 | Not-determined | Genome-fragment | NA | NA | 0 |
| S5_vs_k141_32444\|\|full | 8181 | No | NA | 9 | 0 | 9 | Low-quality | Genome-fragment | 3.4 | AAI-based (medium-confidence) | 0 |
| S8_extracted_k141_72329 | 42922 | No | NA | 72 | 19 | 0 | Complete | High-quality | 100 | DTR (high-confidence) | 0 |
| S8_extracted_k141_150659 | 7104 | No | NA | 4 | 0 | 3 | Not-determined | Genome-fragment | NA | NA | 0 |
| S8_extracted_k141_168204 | 1406 | No | NA | 1 | 0 | 1 | Not-determined | Genome-fragment | NA | NA | 0 |
| S12_vs_k141_75643\|\|full | 8183 | Yes | 5489 | 11 | 3 | 2 | Low-quality | Genome-fragment | 15.17 | AAI-based (high-confidence) | 32.92 |
| S8_extracted_k141_98846 | 1355 | No | NA | 1 | 0 | 1 | Not-determined | Genome-fragment | NA | NA | 0 |
| S8_extracted_k141_194493 | 2195 | No | NA | 3 | 0 | 0 | Not-determined | Genome-fragment | NA | NA | 0 |
| S8_extracted_k141_11794 | 133413 | No | NA | 182 | 36 | 4 | Complete | High-quality | 100 | DTR (high-confidence) | 0 |
| S8_extracted_k141_202632 | 1217 | No | NA | 1 | 0 | 0 | Not-determined | Genome-fragment | NA | NA | 0 |
| S8_extracted_k141_81608 | 1356 | No | NA | 1 | 0 | 0 | Not-determined | Genome-fragment | NA | NA | 0 |
| S8_extracted_k141_168226 | 1034 | No | NA | 1 | 0 | 1 | Not-determined | Genome-fragment | NA | NA | 0 |
| S8_extracted_k141_159386 | 3872 | No | NA | 2 | 0 | 1 | Not-determined | Genome-fragment | NA | NA | 0 |
| S8_extracted_k141_168230 | 1208 | No | NA | 2 | 0 | 0 | Not-determined | Genome-fragment | NA | NA | 0 |
| S16_vs_90987\|\|full | 8185 | No | NA | 6 | 0 | 5 | Low-quality | Genome-fragment | 5.84 | AAI-based (medium-confidence) | 0 |
| S5_extracted_k141_10159 | 8189 | No | NA | 4 | 4 | 0 | Low-quality | Genome-fragment | 6.66 | AAI-based (medium-confidence) | 0 |
| S8_extracted_k141_72367 | 1422 | No | NA | 2 | 0 | 2 | Not-determined | Genome-fragment | NA | NA | 0 |
| S8_extracted_k141_55566 | 1120 | No | NA | 1 | 0 | 0 | Not-determined | Genome-fragment | NA | NA | 0 |
| S8_extracted_k141_202644 | 4347 | No | NA | 2 | 0 | 2 | Not-determined | Genome-fragment | NA | NA | 0 |
| S3_extracted_k141_27957 | 8198 | No | NA | 4 | 0 | 1 | Low-quality | Genome-fragment | 4.64 | AAI-based (medium-confidence) | 0 |
| S3_vs_k141_86379\|\|full | 8200 | No | NA | 17 | 3 | 0 | Low-quality | Genome-fragment | 4.8 | AAI-based (medium-confidence) | 0 |
| S8_extracted_k141_107706 | 1094 | No | NA | 2 | 0 | 0 | Not-determined | Genome-fragment | NA | NA | 0 |
| S8_extracted_k141_55577 | 2313 | No | NA | 2 | 0 | 2 | Not-determined | Genome-fragment | NA | NA | 0 |
| S8_extracted_k141_150709 | 1280 | No | NA | 2 | 0 | 0 | Not-determined | Genome-fragment | NA | NA | 0 |
| S8_extracted_k141_202661 | 1469 | No | NA | 2 | 0 | 1 | Not-determined | Genome-fragment | NA | NA | 0 |
| S16_vs_81773\|\|full | 8200 | No | NA | 11 | 1 | 0 | Low-quality | Genome-fragment | 11.9 | AAI-based (high-confidence) | 0 |
| S8_extracted_k141_46540 | 1213 | No | NA | 3 | 0 | 2 | Not-determined | Genome-fragment | NA | NA | 0 |
| S8_extracted_k141_185047 | 1582 | No | NA | 1 | 0 | 1 | Not-determined | Genome-fragment | NA | NA | 0 |
| S8_extracted_k141_3520 | 1275 | No | NA | 1 | 0 | 0 | Not-determined | Genome-fragment | NA | NA | 0 |
| S10_extracted_18916 | 8202 | No | NA | 9 | 0 | 1 | Low-quality | Genome-fragment | 15.64 | AAI-based (high-confidence) | 0 |
| S14_vs_k141_77646\|\|full | 8206 | No | NA | 18 | 8 | 0 | Low-quality | Genome-fragment | 22.97 | AAI-based (high-confidence) | 0 |
| S8_extracted_k141_168260 | 2253 | No | NA | 5 | 0 | 0 | Not-determined | Genome-fragment | NA | NA | 0 |
| S8_extracted_k141_185054 | 1801 | No | NA | 2 | 0 | 1 | Not-determined | Genome-fragment | NA | NA | 0 |
| S8_extracted_k141_98886 | 1256 | No | NA | 1 | 0 | 1 | Not-determined | Genome-fragment | NA | NA | 0 |
| S8_extracted_k141_159427 | 1256 | No | NA | 1 | 0 | 1 | Not-determined | Genome-fragment | NA | NA | 0 |
| S8_extracted_k141_46555 | 1480 | No | NA | 2 | 0 | 0 | Not-determined | Genome-fragment | NA | NA | 0 |
| S8_extracted_k141_159434 | 1234 | No | NA | 2 | 0 | 1 | Not-determined | Genome-fragment | NA | NA | 0 |
| S8_extracted_k141_90215 | 1609 | No | NA | 2 | 0 | 2 | Not-determined | Genome-fragment | NA | NA | 0 |
| S8_extracted_k141_125073 | 5019 | No | NA | 6 | 0 | 3 | Not-determined | Genome-fragment | NA | NA | 0 |
| S8_extracted_k141_81648 | 6957 | No | NA | 5 | 0 | 2 | Not-determined | Genome-fragment | NA | NA | 0 |
| S8_extracted_k141_107761 | 1317 | No | NA | 2 | 0 | 1 | Not-determined | Genome-fragment | NA | NA | 0 |
| S6_extracted_k141_29444 | 8208 | No | NA | 18 | 1 | 1 | Low-quality | Genome-fragment | 25.44 | AAI-based (high-confidence) | 0 |
| S12_extracted_k141_51398 | 8209 | No | NA | 22 | 1 | 0 | Low-quality | Genome-fragment | 14.09 | AAI-based (high-confidence) | 0 |
| S8_extracted_k141_185097 | 1772 | No | NA | 2 | 0 | 1 | Not-determined | Genome-fragment | NA | NA | 0 |
| S8_extracted_k141_142370 | 1116 | No | NA | 1 | 0 | 0 | Not-determined | Genome-fragment | NA | NA | 0 |
| S7_vs_66608\|\|full | 8210 | No | NA | 10 | 2 | 0 | Low-quality | Genome-fragment | 17.4 | AAI-based (medium-confidence) | 0 |
| S8_extracted_k141_142373 | 1054 | No | NA | 2 | 0 | 0 | Not-determined | Genome-fragment | NA | NA | 0 |
| S8_extracted_k141_168308 | 1118 | No | NA | 1 | 0 | 1 | Not-determined | Genome-fragment | NA | NA | 0 |
| S8_extracted_k141_142378 | 1114 | No | NA | 2 | 0 | 2 | Not-determined | Genome-fragment | NA | NA | 0 |
| S8_extracted_k141_159464 | 1447 | No | NA | 2 | 0 | 2 | Not-determined | Genome-fragment | NA | NA | 0 |
| S8_extracted_k141_55637 | 1556 | No | NA | 2 | 0 | 1 | Not-determined | Genome-fragment | NA | NA | 0 |
| S5_extracted_k141_120063 | 8213 | Yes | 5610 | 8 | 2 | 1 | Low-quality | Genome-fragment | 8.89 | AAI-based (high-confidence) | 31.69 |
| S1_extracted_k141_130873 | 8214 | No | NA | 14 | 2 | 1 | Low-quality | Genome-fragment | 14.25 | AAI-based (high-confidence) | 0 |
| S8_vs_k141_137609\|\|full | 8216 | No | NA | 12 | 0 | 2 | Low-quality | Genome-fragment | 9.96 | AAI-based (medium-confidence) | 0 |
| S8_extracted_k141_98951 | 1299 | No | NA | 3 | 0 | 2 | Not-determined | Genome-fragment | NA | NA | 0 |
| S8_extracted_k141_20961 | 2907 | No | NA | 1 | 0 | 1 | Not-determined | Genome-fragment | NA | NA | 0 |
| S8_extracted_k141_81675 | 1856 | No | NA | 2 | 0 | 1 | Not-determined | Genome-fragment | NA | NA | 0 |
| S8_extracted_k141_168321 | 1332 | No | NA | 2 | 0 | 2 | Not-determined | Genome-fragment | NA | NA | 0 |
| S8_extracted_k141_90245 | 1306 | No | NA | 2 | 0 | 1 | Not-determined | Genome-fragment | NA | NA | 0 |
| S8_extracted_k141_20967 | 1037 | No | NA | 2 | 0 | 1 | Not-determined | Genome-fragment | NA | NA | 0 |
| S8_extracted_k141_177092 | 1023 | No | NA | 2 | 0 | 2 | Not-determined | Genome-fragment | NA | NA | 0 |
| S8_extracted_k141_81685 | 2508 | No | NA | 2 | 0 | 1 | Not-determined | Genome-fragment | NA | NA | 0 |
| S8_extracted_k141_98959 | 1911 | No | NA | 2 | 0 | 1 | Not-determined | Genome-fragment | NA | NA | 0 |
| S8_extracted_k141_202758 | 3762 | No | NA | 3 | 0 | 0 | Not-determined | Genome-fragment | NA | NA | 0 |
| S8_extracted_k141_177098 | 1597 | No | NA | 2 | 0 | 0 | Not-determined | Genome-fragment | NA | NA | 0 |
| S17_vs_k141_155625\|\|full | 8220 | No | NA | 9 | 3 | 0 | Low-quality | Genome-fragment | 26.63 | HMM-based (lower-bound) | 0 |
| S8_vs_k141_126790\|\|full | 8224 | No | NA | 7 | 0 | 2 | Low-quality | Genome-fragment | 4.5 | AAI-based (medium-confidence) | 0 |
| S8_extracted_k141_98968 | 1062 | No | NA | 2 | 0 | 0 | Not-determined | Genome-fragment | NA | NA | 0 |
| S8_extracted_k141_107815 | 1151 | No | NA | 1 | 0 | 1 | Not-determined | Genome-fragment | NA | NA | 0 |
| S8_extracted_k141_38310 | 1021 | No | NA | 1 | 0 | 1 | Not-determined | Genome-fragment | NA | NA | 0 |
| S8_extracted_k141_185129 | 13196 | No | NA | 17 | 0 | 2 | Not-determined | Genome-fragment | NA | NA | 0 |
| S8_extracted_k141_3637 | 2134 | No | NA | 2 | 0 | 2 | Not-determined | Genome-fragment | NA | NA | 0 |
| S8_extracted_k141_38313 | 2719 | No | NA | 1 | 0 | 1 | Not-determined | Genome-fragment | NA | NA | 0 |
| S8_extracted_k141_98991 | 1667 | No | NA | 2 | 0 | 1 | Not-determined | Genome-fragment | NA | NA | 0 |
| S8_extracted_k141_116241 | 1321 | No | NA | 1 | 0 | 1 | Not-determined | Genome-fragment | NA | NA | 0 |
| S8_extracted_k141_116242 | 1015 | No | NA | 1 | 0 | 0 | Not-determined | Genome-fragment | NA | NA | 0 |
| S8_extracted_k141_177129 | 1088 | No | NA | 1 | 0 | 1 | Not-determined | Genome-fragment | NA | NA | 0 |
| S13_vs_119802\|\|full | 8226 | No | NA | 19 | 1 | 0 | Low-quality | Genome-fragment | 20.35 | AAI-based (high-confidence) | 0 |
| S8_extracted_k141_168373 | 5817 | No | NA | 4 | 0 | 4 | Not-determined | Genome-fragment | NA | NA | 0 |
| S8_extracted_k141_116248 | 1198 | No | NA | 1 | 0 | 1 | Not-determined | Genome-fragment | NA | NA | 0 |
| S8_extracted_k141_11902 | 1129 | No | NA | 2 | 0 | 2 | Not-determined | Genome-fragment | NA | NA | 0 |
| S8_extracted_k141_20992 | 1980 | No | NA | 4 | 0 | 3 | Not-determined | Genome-fragment | NA | NA | 0 |
| S16_extracted_57276 | 8230 | No | NA | 7 | 0 | 6 | Low-quality | Genome-fragment | 7.5 | AAI-based (medium-confidence) | 0 |
| S8_extracted_k141_125189 | 1227 | No | NA | 2 | 0 | 0 | Not-determined | Genome-fragment | NA | NA | 0 |
| S8_extracted_k141_125190 | 1798 | No | NA | 3 | 0 | 1 | Not-determined | Genome-fragment | NA | NA | 0 |
| S8_vs_k141_59642\|\|full | 8231 | No | NA | 13 | 4 | 0 | Low-quality | Genome-fragment | 20.01 | AAI-based (high-confidence) | 0 |
| S8_extracted_k141_90314 | 5386 | No | NA | 7 | 0 | 4 | Not-determined | Genome-fragment | NA | NA | 0 |
| S8_extracted_k141_55689 | 1307 | No | NA | 4 | 0 | 0 | Not-determined | Genome-fragment | NA | NA | 0 |
| S8_extracted_k141_107861 | 1021 | No | NA | 3 | 0 | 1 | Not-determined | Genome-fragment | NA | NA | 0 |
| S8_extracted_k141_107864 | 1097 | No | NA | 2 | 0 | 1 | Not-determined | Genome-fragment | NA | NA | 0 |
| S8_extracted_k141_142432 | 1257 | No | NA | 2 | 0 | 0 | Not-determined | Genome-fragment | NA | NA | 0 |
| S8_extracted_k141_38329 | 1438 | No | NA | 1 | 0 | 1 | Not-determined | Genome-fragment | NA | NA | 0 |
| S8_extracted_k141_159535 | 4064 | No | NA | 3 | 0 | 1 | Not-determined | Genome-fragment | NA | NA | 0 |
| S8_extracted_k141_3676 | 1900 | No | NA | 2 | 0 | 1 | Not-determined | Genome-fragment | NA | NA | 0 |
| S8_extracted_k141_185160 | 1128 | No | NA | 3 | 0 | 1 | Not-determined | Genome-fragment | NA | NA | 0 |
| S11_extracted_k141_91335 | 8234 | Yes | 6031 | 9 | 1 | 1 | Low-quality | Genome-fragment | 4.32 | HMM-based (lower-bound) | 26.75 |
| S8_extracted_k141_55704 | 2148 | No | NA | 3 | 0 | 1 | Not-determined | Genome-fragment | NA | NA | 0 |
| S8_extracted_k141_150833 | 1070 | No | NA | 2 | 0 | 1 | Not-determined | Genome-fragment | NA | NA | 0 |
| S8_extracted_k141_159546 | 3391 | No | NA | 4 | 0 | 2 | Not-determined | Genome-fragment | NA | NA | 0 |
| S8_extracted_k141_202808 | 1396 | No | NA | 2 | 0 | 1 | Not-determined | Genome-fragment | NA | NA | 0 |
| S8_extracted_k141_125207 | 1351 | No | NA | 1 | 0 | 1 | Not-determined | Genome-fragment | NA | NA | 0 |
| S8_extracted_k141_168415 | 1071 | No | NA | 1 | 0 | 1 | Not-determined | Genome-fragment | NA | NA | 0 |
| S8_extracted_k141_90342 | 1131 | No | NA | 1 | 0 | 1 | Not-determined | Genome-fragment | NA | NA | 0 |
| S8_extracted_k141_194658 | 1401 | No | NA | 2 | 0 | 0 | Not-determined | Genome-fragment | NA | NA | 0 |
| S8_extracted_k141_125211 | 1798 | No | NA | 1 | 0 | 1 | Not-determined | Genome-fragment | NA | NA | 0 |
| S8_extracted_k141_168418 | 3945 | No | NA | 3 | 0 | 0 | Not-determined | Genome-fragment | NA | NA | 0 |
| S8_extracted_k141_168421 | 1059 | No | NA | 2 | 0 | 2 | Not-determined | Genome-fragment | NA | NA | 0 |
| S12_vs_k141_19164\|\|full | 8236 | No | NA | 17 | 4 | 0 | Low-quality | Genome-fragment | 14.94 | AAI-based (high-confidence) | 0 |
| S8_extracted_k141_72526 | 1222 | No | NA | 2 | 0 | 2 | Not-determined | Genome-fragment | NA | NA | 0 |
| S8_extracted_k141_29945 | 1297 | No | NA | 3 | 0 | 0 | Not-determined | Genome-fragment | NA | NA | 0 |
| S8_extracted_k141_185187 | 2555 | No | NA | 3 | 0 | 2 | Not-determined | Genome-fragment | NA | NA | 0 |
| S8_extracted_k141_159556 | 3935 | No | NA | 2 | 0 | 2 | Not-determined | Genome-fragment | NA | NA | 0 |
| S8_extracted_k141_90358 | 2164 | No | NA | 2 | 0 | 0 | Not-determined | Genome-fragment | NA | NA | 0 |
| S8_extracted_k141_107885 | 2460 | No | NA | 6 | 0 | 2 | Not-determined | Genome-fragment | NA | NA | 0 |
| S8_extracted_k141_55734 | 3079 | No | NA | 4 | 0 | 2 | Not-determined | Genome-fragment | NA | NA | 0 |
| S8_extracted_k141_72534 | 1403 | No | NA | 1 | 0 | 1 | Not-determined | Genome-fragment | NA | NA | 0 |
| S8_extracted_k141_116316 | 1301 | No | NA | 1 | 0 | 1 | Not-determined | Genome-fragment | NA | NA | 0 |
| S8_extracted_k141_55743 | 3815 | No | NA | 5 | 0 | 4 | Not-determined | Genome-fragment | NA | NA | 0 |
| S8_extracted_k141_81741 | 1390 | No | NA | 3 | 0 | 3 | Not-determined | Genome-fragment | NA | NA | 0 |
| S8_extracted_k141_3725 | 1022 | No | NA | 1 | 0 | 1 | Not-determined | Genome-fragment | NA | NA | 0 |
| S8_extracted_k141_90379 | 8537 | No | NA | 5 | 0 | 2 | Not-determined | Genome-fragment | NA | NA | 0 |
| S8_extracted_k141_125245 | 3625 | No | NA | 3 | 0 | 0 | Not-determined | Genome-fragment | NA | NA | 0 |
| S17_vs_k141_142027\|\|full | 8241 | Yes | 5832 | 8 | 2 | 1 | Low-quality | Genome-fragment | 5.23 | AAI-based (medium-confidence) | 29.23 |
| S8_extracted_k141_55757 | 4466 | No | NA | 2 | 0 | 2 | Not-determined | Genome-fragment | NA | NA | 0 |
| S8_extracted_k141_72563 | 2095 | No | NA | 3 | 0 | 2 | Not-determined | Genome-fragment | NA | NA | 0 |
| S8_extracted_k141_46705 | 1164 | No | NA | 1 | 0 | 1 | Not-determined | Genome-fragment | NA | NA | 0 |
| S3_vs_k141_36331\|\|full | 8243 | No | NA | 13 | 1 | 1 | Low-quality | Genome-fragment | 15.9 | AAI-based (medium-confidence) | 0 |
| S8_extracted_k141_90393 | 3960 | No | NA | 5 | 0 | 5 | Not-determined | Genome-fragment | NA | NA | 0 |
| S8_extracted_k141_29991 | 4479 | No | NA | 5 | 0 | 2 | Not-determined | Genome-fragment | NA | NA | 0 |
| S8_extracted_k141_116350 | 1016 | No | NA | 2 | 0 | 1 | Not-determined | Genome-fragment | NA | NA | 0 |
| S8_extracted_k141_81762 | 1842 | No | NA | 2 | 0 | 2 | Not-determined | Genome-fragment | NA | NA | 0 |
| S8_extracted_k141_150875 | 1495 | No | NA | 1 | 0 | 0 | Not-determined | Genome-fragment | NA | NA | 0 |
| S8_extracted_k141_81765 | 2269 | No | NA | 3 | 0 | 0 | Not-determined | Genome-fragment | NA | NA | 0 |
| S5_vs_k141_113676\|\|full | 8255 | No | NA | 11 | 8 | 0 | Low-quality | Genome-fragment | 17.84 | AAI-based (medium-confidence) | 0 |
| S8_extracted_k141_72588 | 1118 | No | NA | 1 | 0 | 1 | Not-determined | Genome-fragment | NA | NA | 0 |
| S14_vs_k141_106512\|\|full | 8264 | No | NA | 20 | 4 | 0 | Low-quality | Genome-fragment | 18.27 | AAI-based (high-confidence) | 0 |
| S8_extracted_k141_125267 | 1762 | No | NA | 2 | 0 | 0 | Not-determined | Genome-fragment | NA | NA | 0 |
| S8_vs_k141_22526\|\|full | 8266 | No | NA | 12 | 9 | 0 | Low-quality | Genome-fragment | 13.7 | AAI-based (high-confidence) | 0 |
| S8_extracted_k141_107926 | 3041 | No | NA | 2 | 0 | 2 | Not-determined | Genome-fragment | NA | NA | 0 |
| S8_extracted_k141_168487 | 3203 | No | NA | 5 | 0 | 3 | Not-determined | Genome-fragment | NA | NA | 0 |
| S8_extracted_k141_107939 | 1248 | No | NA | 2 | 0 | 1 | Not-determined | Genome-fragment | NA | NA | 0 |
| S16_extracted_36137 | 8272 | No | NA | 3 | 1 | 0 | Low-quality | Genome-fragment | 21.44 | AAI-based (medium-confidence) | 0 |
| S8_extracted_k141_142516 | 1350 | No | NA | 2 | 0 | 1 | Not-determined | Genome-fragment | NA | NA | 0 |
| S8_extracted_k141_46735 | 2515 | No | NA | 5 | 0 | 3 | Not-determined | Genome-fragment | NA | NA | 0 |
| S8_extracted_k141_46741 | 1511 | No | NA | 1 | 0 | 1 | Not-determined | Genome-fragment | NA | NA | 0 |
| S8_extracted_k141_159634 | 1038 | No | NA | 1 | 0 | 1 | Not-determined | Genome-fragment | NA | NA | 0 |
| S8_extracted_k141_202892 | 1214 | No | NA | 2 | 0 | 2 | Not-determined | Genome-fragment | NA | NA | 0 |
| S7_vs_2354\|\|full | 8277 | Yes | 6430 | 11 | 3 | 2 | Low-quality | Genome-fragment | 8.15 | HMM-based (lower-bound) | 22.31 |
| S8_extracted_k141_116401 | 1208 | No | NA | 2 | 0 | 1 | Not-determined | Genome-fragment | NA | NA | 0 |
| S8_extracted_k141_55811 | 2147 | No | NA | 3 | 0 | 1 | Not-determined | Genome-fragment | NA | NA | 0 |
| S8_extracted_k141_150911 | 1655 | No | NA | 1 | 0 | 1 | Not-determined | Genome-fragment | NA | NA | 0 |
| S8_extracted_k141_55814 | 1495 | No | NA | 2 | 0 | 1 | Not-determined | Genome-fragment | NA | NA | 0 |
| S8_extracted_k141_159652 | 3356 | No | NA | 3 | 0 | 3 | Not-determined | Genome-fragment | NA | NA | 0 |
| S8_extracted_k141_107973 | 9656 | No | NA | 9 | 0 | 4 | Not-determined | Genome-fragment | NA | NA | 0 |
| S8_extracted_k141_168535 | 1073 | No | NA | 3 | 0 | 1 | Not-determined | Genome-fragment | NA | NA | 0 |
| S8_extracted_k141_142561 | 1696 | No | NA | 2 | 0 | 1 | Not-determined | Genome-fragment | NA | NA | 0 |
| S8_extracted_k141_177246 | 7131 | No | NA | 7 | 0 | 6 | Not-determined | Genome-fragment | NA | NA | 0 |
| S8_extracted_k141_107980 | 1234 | No | NA | 3 | 0 | 0 | Not-determined | Genome-fragment | NA | NA | 0 |
| S12_vs_k141_60442\|\|full | 8289 | No | NA | 7 | 0 | 3 | Low-quality | Genome-fragment | 2.67 | AAI-based (medium-confidence) | 0 |
| S4_extracted_12572 | 8296 | No | NA | 15 | 7 | 0 | Low-quality | Genome-fragment | 15.15 | AAI-based (high-confidence) | 0 |
| S8_extracted_k141_46772 | 1102 | No | NA | 2 | 0 | 0 | Not-determined | Genome-fragment | NA | NA | 0 |
| S8_extracted_k141_21082 | 1055 | No | NA | 1 | 0 | 1 | Not-determined | Genome-fragment | NA | NA | 0 |
| S1_extracted_k141_114116 | 8300 | No | NA | 8 | 1 | 0 | Low-quality | Genome-fragment | 6.26 | AAI-based (high-confidence) | 0 |
| S8_extracted_k141_202919 | 1347 | No | NA | 2 | 0 | 1 | Not-determined | Genome-fragment | NA | NA | 0 |
| S8_extracted_k141_90436 | 3074 | No | NA | 4 | 0 | 2 | Not-determined | Genome-fragment | NA | NA | 0 |
| S8_extracted_k141_133616 | 1039 | No | NA | 2 | 0 | 0 | Not-determined | Genome-fragment | NA | NA | 0 |
| S11_vs_k141_4686\|\|full | 8307 | No | NA | 10 | 7 | 0 | Low-quality | Genome-fragment | 18.05 | AAI-based (medium-confidence) | 0 |
| S8_extracted_k141_3825 | 1508 | No | NA | 2 | 0 | 0 | Not-determined | Genome-fragment | NA | NA | 0 |
| S8_extracted_k141_46782 | 1032 | No | NA | 3 | 0 | 2 | Not-determined | Genome-fragment | NA | NA | 0 |
| S8_extracted_k141_55844 | 4278 | No | NA | 6 | 0 | 3 | Not-determined | Genome-fragment | NA | NA | 0 |
| S8_extracted_k141_11939 | 2020 | No | NA | 3 | 0 | 1 | Not-determined | Genome-fragment | NA | NA | 0 |
| S8_extracted_k141_202935 | 1017 | No | NA | 2 | 0 | 0 | Not-determined | Genome-fragment | NA | NA | 0 |
| S8_extracted_k141_30079 | 1077 | No | NA | 2 | 0 | 2 | Not-determined | Genome-fragment | NA | NA | 0 |
| S8_extracted_k141_90456 | 4000 | No | NA | 5 | 0 | 0 | Not-determined | Genome-fragment | NA | NA | 0 |
| S8_extracted_k141_90458 | 1127 | No | NA | 1 | 0 | 1 | Not-determined | Genome-fragment | NA | NA | 0 |
| S8_extracted_k141_125330 | 1371 | No | NA | 1 | 0 | 0 | Not-determined | Genome-fragment | NA | NA | 0 |
| S8_extracted_k141_55867 | 4959 | No | NA | 2 | 0 | 1 | Not-determined | Genome-fragment | NA | NA | 0 |
| S3_extracted_k141_83102 | 8309 | No | NA | 12 | 10 | 0 | Low-quality | Genome-fragment | 27.4 | AAI-based (high-confidence) | 0 |
| S8_extracted_k141_125331 | 1078 | No | NA | 1 | 0 | 1 | Not-determined | Genome-fragment | NA | NA | 0 |
| S8_extracted_k141_30094 | 2513 | No | NA | 1 | 0 | 0 | Not-determined | Genome-fragment | NA | NA | 0 |
| S16_extracted_41589 | 8310 | No | NA | 12 | 4 | 0 | Low-quality | Genome-fragment | 26.45 | HMM-based (lower-bound) | 0 |
| S8_extracted_k141_3869 | 1346 | No | NA | 2 | 0 | 2 | Not-determined | Genome-fragment | NA | NA | 0 |
| S8_extracted_k141_72709 | 1758 | No | NA | 4 | 0 | 0 | Not-determined | Genome-fragment | NA | NA | 0 |
| S8_extracted_k141_108019 | 2674 | No | NA | 3 | 0 | 2 | Not-determined | Genome-fragment | NA | NA | 0 |
| S8_extracted_k141_72710 | 1200 | No | NA | 1 | 0 | 1 | Not-determined | Genome-fragment | NA | NA | 0 |
| S8_extracted_k141_177307 | 1487 | No | NA | 2 | 0 | 1 | Not-determined | Genome-fragment | NA | NA | 0 |
| S8_extracted_k141_177309 | 1036 | No | NA | 1 | 0 | 1 | Not-determined | Genome-fragment | NA | NA | 0 |
| S8_extracted_k141_90478 | 1040 | No | NA | 2 | 0 | 2 | Not-determined | Genome-fragment | NA | NA | 0 |
| S8_extracted_k141_150985 | 3074 | No | NA | 1 | 0 | 1 | Not-determined | Genome-fragment | NA | NA | 0 |
| S8_extracted_k141_202959 | 4060 | No | NA | 3 | 0 | 3 | Not-determined | Genome-fragment | NA | NA | 0 |
| S8_extracted_k141_55892 | 2209 | No | NA | 4 | 0 | 2 | Not-determined | Genome-fragment | NA | NA | 0 |
| S8_extracted_k141_116474 | 1661 | No | NA | 2 | 0 | 2 | Not-determined | Genome-fragment | NA | NA | 0 |
| S8_extracted_k141_90491 | 1148 | No | NA | 1 | 0 | 1 | Not-determined | Genome-fragment | NA | NA | 0 |
| S5_vs_k141_9065\|\|full | 8320 | No | NA | 11 | 1 | 2 | Low-quality | Genome-fragment | 17.51 | AAI-based (medium-confidence) | 0 |
| S1_vs_k141_18950\|\|full | 8322 | No | NA | 5 | 0 | 4 | Low-quality | Genome-fragment | 3.85 | AAI-based (medium-confidence) | 0 |
| S8_extracted_k141_11984 | 1117 | No | NA | 1 | 0 | 1 | Not-determined | Genome-fragment | NA | NA | 0 |
| S8_extracted_k141_90495 | 10373 | No | NA | 10 | 0 | 2 | Not-determined | Genome-fragment | NA | NA | 0 |
| S6_vs_k141_26406\|\|full | 8329 | Yes | 5384 | 6 | 1 | 1 | Low-quality | Genome-fragment | 9.29 | AAI-based (medium-confidence) | 35.36 |
| S8_extracted_k141_168607 | 1233 | No | NA | 1 | 0 | 0 | Not-determined | Genome-fragment | NA | NA | 0 |
| S8_extracted_k141_168608 | 4658 | No | NA | 4 | 0 | 3 | Not-determined | Genome-fragment | NA | NA | 0 |
| S8_extracted_k141_12002 | 1899 | No | NA | 2 | 0 | 2 | Not-determined | Genome-fragment | NA | NA | 0 |
| S8_extracted_k141_12004 | 1357 | No | NA | 1 | 0 | 0 | Not-determined | Genome-fragment | NA | NA | 0 |
| S3_extracted_k141_78956 | 8330 | No | NA | 21 | 0 | 0 | Low-quality | Genome-fragment | 4.06 | AAI-based (high-confidence) | 0 |
| S8_extracted_k141_151038 | 2861 | No | NA | 2 | 0 | 2 | Not-determined | Genome-fragment | NA | NA | 0 |
| S8_extracted_k141_81806 | 2173 | No | NA | 2 | 0 | 0 | Not-determined | Genome-fragment | NA | NA | 0 |
| S8_extracted_k141_202981 | 3209 | No | NA | 1 | 0 | 0 | Not-determined | Genome-fragment | NA | NA | 0 |
| S8_extracted_k141_194813 | 3474 | No | NA | 3 | 0 | 2 | Not-determined | Genome-fragment | NA | NA | 0 |
| S8_extracted_k141_125398 | 1655 | No | NA | 1 | 0 | 1 | Not-determined | Genome-fragment | NA | NA | 0 |
| S8_extracted_k141_99270 | 1983 | No | NA | 2 | 0 | 1 | Not-determined | Genome-fragment | NA | NA | 0 |
| S10_extracted_23135 | 8330 | No | NA | 15 | 5 | 0 | Low-quality | Genome-fragment | 19.71 | AAI-based (high-confidence) | 0 |
| S8_extracted_k141_72766 | 5065 | No | NA | 6 | 0 | 4 | Not-determined | Genome-fragment | NA | NA | 0 |
| S8_extracted_k141_151055 | 3090 | No | NA | 3 | 0 | 1 | Not-determined | Genome-fragment | NA | NA | 0 |
| S8_extracted_k141_21189 | 1019 | No | NA | 2 | 0 | 2 | Not-determined | Genome-fragment | NA | NA | 0 |
| S8_extracted_k141_108076 | 2856 | No | NA | 4 | 0 | 4 | Not-determined | Genome-fragment | NA | NA | 0 |
| S8_extracted_k141_151059 | 1792 | No | NA | 3 | 0 | 3 | Not-determined | Genome-fragment | NA | NA | 0 |
| S8_extracted_k141_72769 | 3908 | No | NA | 2 | 0 | 1 | Not-determined | Genome-fragment | NA | NA | 0 |
| S11_vs_k141_71588\|\|full | 8339 | No | NA | 13 | 4 | 0 | Low-quality | Genome-fragment | 13.66 | AAI-based (high-confidence) | 0 |

| | | | | | | | | | | | |
|---|---|---|---|---|---|---|---|---|---|---|---|
| S8_extracted_k141_116532 | 1622 | No | NA | 3 | 0 | 0 | Not-determined | Genome-fragment | NA | NA | 0 |
| S8_extracted_k141_30217 | 1215 | No | NA | 1 | 0 | 0 | Not-determined | Genome-fragment | NA | NA | 0 |
| S8_extracted_k141_116550 | 1396 | No | NA | 2 | 0 | 1 | Not-determined | Genome-fragment | NA | NA | 0 |
| S8_extracted_k141_194844 | 1223 | No | NA | 2 | 0 | 1 | Not-determined | Genome-fragment | NA | NA | 0 |
| S13_vs_106253\|\|full | 8347 | No | NA | 6 | 0 | 3 | Low-quality | Genome-fragment | 11.37 | AAI-based (medium-confidence) | 0 |
| S8_vs_k141_62577\|\|full | 8350 | Yes | 5390 | 7 | 1 | 2 | Low-quality | Genome-fragment | 3.62 | HMM-based (lower-bound) | 35.45 |
| S15_extracted_18076 | 8355 | No | NA | 5 | 0 | 2 | Low-quality | Genome-fragment | 8.2 | AAI-based (high-confidence) | 0 |
| S8_extracted_k141_142700 | 16231 | No | NA | 18 | 6 | 0 | Medium-quality | Genome-fragment | 54.63 | HMM-based (lower-bound) | 0 |
| S8_extracted_k141_21206 | 8078 | No | NA | 9 | 0 | 7 | Not-determined | Genome-fragment | NA | NA | 0 |
| S8_extracted_k141_116554 | 8118 | No | NA | 6 | 0 | 2 | Not-determined | Genome-fragment | NA | NA | 0 |
| S8_extracted_k141_3911 | 1621 | No | NA | 2 | 0 | 0 | Not-determined | Genome-fragment | NA | NA | 0 |
| S8_extracted_k141_168676 | 1084 | No | NA | 2 | 0 | 2 | Not-determined | Genome-fragment | NA | NA | 0 |
| S8_extracted_k141_159721 | 1082 | No | NA | 2 | 0 | 0 | Not-determined | Genome-fragment | NA | NA | 0 |
| S8_extracted_k141_133801 | 3396 | No | NA | 3 | 0 | 2 | Not-determined | Genome-fragment | NA | NA | 0 |
| S8_extracted_k141_159737 | 1118 | No | NA | 1 | 0 | 0 | Not-determined | Genome-fragment | NA | NA | 0 |
| S8_extracted_k141_64364 | 1048 | No | NA | 2 | 0 | 1 | Not-determined | Genome-fragment | NA | NA | 0 |
| S8_extracted_k141_72773 | 42287 | No | NA | 50 | 11 | 0 | Complete | High-quality | 100 | DTR (high-confidence) | 0 |
| S8_extracted_k141_90600 | 1596 | No | NA | 2 | 0 | 2 | Not-determined | Genome-fragment | NA | NA | 0 |
| S3_extracted_k141_12115 | 8371 | No | NA | 5 | 0 | 4 | Low-quality | Genome-fragment | 8.52 | AAI-based (medium-confidence) | 0 |
| S8_extracted_k141_30257 | 1339 | No | NA | 2 | 0 | 1 | Not-determined | Genome-fragment | NA | NA | 0 |
| S8_extracted_k141_185368 | 2171 | No | NA | 5 | 0 | 0 | Not-determined | Genome-fragment | NA | NA | 0 |
| S8_extracted_k141_38514 | 4230 | No | NA | 4 | 0 | 2 | Not-determined | Genome-fragment | NA | NA | 0 |
| S8_extracted_k141_30264 | 1216 | No | NA | 2 | 0 | 2 | Not-determined | Genome-fragment | NA | NA | 0 |
| S8_extracted_k141_194872 | 1809 | No | NA | 2 | 0 | 1 | Not-determined | Genome-fragment | NA | NA | 0 |
| S7_extracted_56423 | 8372 | No | NA | 12 | 4 | 0 | Low-quality | Genome-fragment | 20.85 | AAI-based (high-confidence) | 0 |
| S8_extracted_k141_203044 | 17395 | No | NA | 22 | 7 | 0 | Medium-quality | Genome-fragment | 81.45 | AAI-based (medium-confidence) | 0 |
| S8_extracted_k141_194873 | 2577 | No | NA | 2 | 0 | 1 | Not-determined | Genome-fragment | NA | NA | 0 |
| S11_extracted_k141_58094 | 8381 | No | NA | 8 | 1 | 0 | Low-quality | Genome-fragment | 5.45 | AAI-based (high-confidence) | 0 |
| S8_extracted_k141_151102 | 7108 | No | NA | 10 | 0 | 4 | Not-determined | Genome-fragment | NA | NA | 0 |
| S8_extracted_k141_185378 | 1151 | No | NA | 1 | 0 | 1 | Not-determined | Genome-fragment | NA | NA | 0 |
| S8_extracted_k141_159773 | 1368 | No | NA | 1 | 0 | 0 | Not-determined | Genome-fragment | NA | NA | 0 |
| S8_extracted_k141_108122 | 1838 | No | NA | 5 | 0 | 0 | Not-determined | Genome-fragment | NA | NA | 0 |
| S8_extracted_k141_12129 | 1279 | No | NA | 1 | 0 | 1 | Not-determined | Genome-fragment | NA | NA | 0 |
| S7_extracted_138217 | 8381 | No | NA | 15 | 2 | 1 | Low-quality | Genome-fragment | 17.83 | HMM-based (lower-bound) | 0 |
| S8_extracted_k141_90634 | 2388 | No | NA | 3 | 0 | 3 | Not-determined | Genome-fragment | NA | NA | 0 |
| S8_extracted_k141_38529 | 2444 | No | NA | 1 | 0 | 1 | Not-determined | Genome-fragment | NA | NA | 0 |
| S8_extracted_k141_159788 | 2179 | No | NA | 4 | 0 | 1 | Not-determined | Genome-fragment | NA | NA | 0 |
| S8_extracted_k141_151129 | 1379 | No | NA | 1 | 0 | 1 | Not-determined | Genome-fragment | NA | NA | 0 |
| S2_extracted_k141_19276 | 8381 | No | NA | 9 | 1 | 0 | Low-quality | Genome-fragment | 9 | AAI-based (high-confidence) | 0 |
| S8_extracted_k141_185408 | 1751 | No | NA | 1 | 0 | 1 | Not-determined | Genome-fragment | NA | NA | 0 |
| S8_extracted_k141_168730 | 6104 | No | NA | 6 | 0 | 3 | Not-determined | Genome-fragment | NA | NA | 0 |
| S9_vs_k141_81453\|\|full | 8388 | No | NA | 13 | 2 | 0 | Low-quality | Genome-fragment | 21.97 | AAI-based (high-confidence) | 0 |
| S8_extracted_k141_194914 | 1322 | No | NA | 1 | 0 | 0 | Not-determined | Genome-fragment | NA | NA | 0 |
| S8_extracted_k141_90660 | 1537 | No | NA | 3 | 0 | 0 | Not-determined | Genome-fragment | NA | NA | 0 |
| S3_extracted_k141_113245 | 8388 | Yes | 2542 | 10 | 1 | 4 | Low-quality | Genome-fragment | 0.71 | HMM-based (lower-bound) | 69.69 |
| S8_extracted_k141_81871 | 3702 | No | NA | 2 | 0 | 2 | Not-determined | Genome-fragment | NA | NA | 0 |
| S3_vs_k141_107181\|\|full | 8389 | No | NA | 13 | 4 | 0 | Low-quality | Genome-fragment | 12.34 | AAI-based (high-confidence) | 0 |
| S8_extracted_k141_114143 | 8389 | No | NA | 5 | 2 | 0 | Low-quality | Genome-fragment | 15.76 | AAI-based (medium-confidence) | 0 |
| S8_extracted_k141_90678 | 1800 | No | NA | 1 | 0 | 1 | Not-determined | Genome-fragment | NA | NA | 0 |
| S8_extracted_k141_72843 | 7351 | No | NA | 7 | 0 | 1 | Not-determined | Genome-fragment | NA | NA | 0 |
| S8_extracted_k141_177465 | 3751 | No | NA | 5 | 0 | 1 | Not-determined | Genome-fragment | NA | NA | 0 |
| S8_extracted_k141_203089 | 1252 | No | NA | 1 | 0 | 1 | Not-determined | Genome-fragment | NA | NA | 0 |
| S8_extracted_k141_125546 | 1926 | No | NA | 3 | 0 | 0 | Not-determined | Genome-fragment | NA | NA | 0 |
| S8_extracted_k141_4036 | 1119 | No | NA | 1 | 0 | 1 | Not-determined | Genome-fragment | NA | NA | 0 |
| S8_extracted_k141_194944 | 2153 | No | NA | 2 | 0 | 2 | Not-determined | Genome-fragment | NA | NA | 0 |
| S8_extracted_k141_185435 | 6726 | No | NA | 7 | 0 | 6 | Not-determined | Genome-fragment | NA | NA | 0 |
| S7_vs_5048\|\|full | 8397 | No | NA | 16 | 3 | 0 | Low-quality | Genome-fragment | 18.62 | AAI-based (high-confidence) | 0 |
| S8_extracted_k141_38575 | 1446 | No | NA | 2 | 0 | 0 | Not-determined | Genome-fragment | NA | NA | 0 |
| S8_extracted_k141_177469 | 1450 | No | NA | 2 | 0 | 2 | Not-determined | Genome-fragment | NA | NA | 0 |
| S8_extracted_k141_72854 | 5775 | No | NA | 4 | 0 | 2 | Not-determined | Genome-fragment | NA | NA | 0 |
| S8_extracted_k141_12197 | 1030 | No | NA | 2 | 0 | 2 | Not-determined | Genome-fragment | NA | NA | 0 |
| S8_extracted_k141_194956 | 1643 | No | NA | 3 | 0 | 3 | Not-determined | Genome-fragment | NA | NA | 0 |
| S5_vs_k141_135172\|\|full | 8404 | No | NA | 6 | 0 | 1 | Low-quality | Genome-fragment | 11.69 | AAI-based (medium-confidence) | 0 |
| S8_extracted_k141_108193 | 1247 | No | NA | 2 | 0 | 0 | Not-determined | Genome-fragment | NA | NA | 0 |
| S8_extracted_k141_81896 | 3798 | No | NA | 5 | 0 | 3 | Not-determined | Genome-fragment | NA | NA | 0 |
| S8_extracted_k141_203099 | 1360 | No | NA | 1 | 0 | 1 | Not-determined | Genome-fragment | NA | NA | 0 |
| S8_extracted_k141_72866 | 6136 | No | NA | 7 | 0 | 4 | Not-determined | Genome-fragment | NA | NA | 0 |
| S8_extracted_k141_125570 | 1311 | No | NA | 1 | 0 | 1 | Not-determined | Genome-fragment | NA | NA | 0 |
| S8_extracted_k141_30380 | 2187 | No | NA | 1 | 0 | 1 | Not-determined | Genome-fragment | NA | NA | 0 |
| S8_extracted_k141_56091 | 3127 | No | NA | 4 | 0 | 4 | Not-determined | Genome-fragment | NA | NA | 0 |
| S8_extracted_k141_151178 | 2485 | No | NA | 3 | 0 | 3 | Not-determined | Genome-fragment | NA | NA | 0 |
| S12_extracted_k141_169360 | 8406 | No | NA | 8 | 0 | 1 | Low-quality | Genome-fragment | 18.08 | AAI-based (medium-confidence) | 0 |
| S8_extracted_k141_21295 | 10360 | No | NA | 11 | 0 | 7 | Not-determined | Genome-fragment | NA | NA | 0 |
| S3_extracted_k141_99881 | 8409 | No | NA | 15 | 5 | 0 | Low-quality | Genome-fragment | 14.42 | AAI-based (high-confidence) | 0 |
| S8_extracted_k141_203132 | 1012 | No | NA | 1 | 0 | 1 | Not-determined | Genome-fragment | NA | NA | 0 |
| S8_extracted_k141_81910 | 4920 | No | NA | 5 | 0 | 3 | Not-determined | Genome-fragment | NA | NA | 0 |
| S8_extracted_k141_46935 | 3116 | No | NA | 3 | 0 | 2 | Not-determined | Genome-fragment | NA | NA | 0 |
| S8_extracted_k141_108214 | 2035 | No | NA | 2 | 0 | 0 | Not-determined | Genome-fragment | NA | NA | 0 |
| S8_extracted_k141_159835 | 1349 | No | NA | 3 | 0 | 3 | Not-determined | Genome-fragment | NA | NA | 0 |
| S8_extracted_k141_125596 | 4625 | No | NA | 2 | 0 | 2 | Not-determined | Genome-fragment | NA | NA | 0 |
| S8_extracted_k141_195016 | 1311 | No | NA | 2 | 0 | 1 | Not-determined | Genome-fragment | NA | NA | 0 |
| S8_extracted_k141_133941 | 1557 | No | NA | 1 | 0 | 0 | Not-determined | Genome-fragment | NA | NA | 0 |
| S8_extracted_k141_64469 | 1897 | No | NA | 1 | 0 | 1 | Not-determined | Genome-fragment | NA | NA | 0 |
| S8_extracted_k141_46956 | 1043 | No | NA | 1 | 0 | 0 | Not-determined | Genome-fragment | NA | NA | 0 |
| S16_vs_25581\|\|full | 8410 | No | NA | 10 | 0 | 0 | Low-quality | Genome-fragment | 19.56 | AAI-based (medium-confidence) | 0 |
| S8_extracted_k141_4051 | 3063 | No | NA | 4 | 0 | 3 | Not-determined | Genome-fragment | NA | NA | 0 |
| S8_extracted_k141_38634 | 1231 | No | NA | 1 | 0 | 1 | Not-determined | Genome-fragment | NA | NA | 0 |
| S3_vs_k141_118813\|\|full | 8412 | No | NA | 6 | 0 | 2 | Low-quality | Genome-fragment | 5.64 | AAI-based (high-confidence) | 0 |
| S8_extracted_k141_21316 | 1272 | No | NA | 2 | 0 | 1 | Not-determined | Genome-fragment | NA | NA | 0 |
| S8_extracted_k141_125612 | 3188 | No | NA | 1 | 0 | 1 | Not-determined | Genome-fragment | NA | NA | 0 |
| S8_extracted_k141_99427 | 2827 | No | NA | 4 | 0 | 2 | Not-determined | Genome-fragment | NA | NA | 0 |
| S8_extracted_k141_125620 | 3399 | No | NA | 2 | 0 | 2 | Not-determined | Genome-fragment | NA | NA | 0 |
| S8_extracted_k141_177551 | 1136 | No | NA | 1 | 0 | 0 | Not-determined | Genome-fragment | NA | NA | 0 |
| S8_extracted_k141_151228 | 1000 | No | NA | 4 | 0 | 0 | Not-determined | Genome-fragment | NA | NA | 0 |
| S8_extracted_k141_12212 | 39177 | Yes | 30236 | 61 | 13 | 6 | Medium-quality | Genome-fragment | 81.54 | AAI-based (high-confidence) | 22.82 |
| S16_vs_144728\|\|full | 8418 | No | NA | 18 | 2 | 0 | Low-quality | Genome-fragment | 23.43 | AAI-based (high-confidence) | 0 |
| S8_extracted_k141_46998 | 2930 | No | NA | 4 | 0 | 0 | Not-determined | Genome-fragment | NA | NA | 0 |
| S8_extracted_k141_12220 | 1766 | No | NA | 2 | 0 | 2 | Not-determined | Genome-fragment | NA | NA | 0 |
| S8_extracted_k141_203197 | 1261 | No | NA | 1 | 0 | 1 | Not-determined | Genome-fragment | NA | NA | 0 |
| S7_vs_15071\|\|full | 8422 | No | NA | 9 | 1 | 2 | Low-quality | Genome-fragment | 5.45 | HMM-based (lower-bound) | 0 |
| S8_extracted_k141_195032 | 20853 | No | NA | 37 | 5 | 0 | Medium-quality | Genome-fragment | 63.24 | AAI-based (medium-confidence) | 0 |
| S10_vs_120576\|\|full | 8424 | No | NA | 11 | 1 | 0 | Low-quality | Genome-fragment | 15.08 | AAI-based (medium-confidence) | 0 |
| S8_extracted_k141_133979 | 4596 | No | NA | 5 | 0 | 2 | Not-determined | Genome-fragment | NA | NA | 0 |
| S11_extracted_k141_36404 | 8429 | No | NA | 15 | 3 | 0 | Low-quality | Genome-fragment | 15.38 | AAI-based (medium-confidence) | 0 |
| S8_extracted_k141_159919 | 4720 | No | NA | 4 | 0 | 2 | Not-determined | Genome-fragment | NA | NA | 0 |
| S8_vs_k141_74454\|\|full | 8430 | No | NA | 12 | 8 | 0 | Low-quality | Genome-fragment | 24.53 | AAI-based (high-confidence) | 0 |
| S8_extracted_k141_12258 | 1387 | No | NA | 2 | 0 | 1 | Not-determined | Genome-fragment | NA | NA | 0 |
| S3_extracted_k141_71813 | 8442 | No | NA | 12 | 6 | 0 | Low-quality | Genome-fragment | 5.78 | AAI-based (high-confidence) | 0 |
| S13_extracted_104210 | 8447 | No | NA | 9 | 0 | 6 | Low-quality | Genome-fragment | 3.91 | AAI-based (high-confidence) | 0 |
| S8_extracted_k141_90847 | 3095 | No | NA | 2 | 0 | 2 | Not-determined | Genome-fragment | NA | NA | 0 |
| S8_extracted_k141_35198 | 8447 | No | NA | 10 | 2 | 0 | Low-quality | Genome-fragment | 20.28 | AAI-based (medium-confidence) | 0 |
| S8_vs_k141_192537\|\|full | 8447 | No | NA | 10 | 0 | 7 | Low-quality | Genome-fragment | 2.37 | AAI-based (medium-confidence) | 0 |
| S8_extracted_k141_99500 | 1180 | No | NA | 2 | 0 | 2 | Not-determined | Genome-fragment | NA | NA | 0 |
| S8_extracted_k141_177617 | 4758 | No | NA | 3 | 0 | 2 | Not-determined | Genome-fragment | NA | NA | 0 |
| S17_vs_k141_55059\|\|full | 8455 | No | NA | 3 | 0 | 1 | Low-quality | Genome-fragment | 8.61 | AAI-based (medium-confidence) | 0 |
| S8_extracted_k141_30486 | 1116 | No | NA | 2 | 0 | 2 | Not-determined | Genome-fragment | NA | NA | 0 |
| S8_extracted_k141_12303 | 2084 | No | NA | 1 | 0 | 1 | Not-determined | Genome-fragment | NA | NA | 0 |
| S8_extracted_k141_134016 | 1066 | No | NA | 1 | 0 | 0 | Not-determined | Genome-fragment | NA | NA | 0 |
| S14_extracted_k141_33985 | 8456 | No | NA | 5 | 4 | 0 | Low-quality | Genome-fragment | 21.84 | AAI-based (high-confidence) | 0 |
| S8_extracted_k141_168832 | 14662 | No | NA | 11 | 0 | 7 | Not-determined | Genome-fragment | NA | NA | 0 |
| S8_extracted_k141_195082 | 8428 | No | NA | 6 | 0 | 2 | Not-determined | Genome-fragment | NA | NA | 0 |
| S8_extracted_k141_203252 | 1120 | No | NA | 2 | 0 | 1 | Not-determined | Genome-fragment | NA | NA | 0 |
| S8_extracted_k141_21419 | 1537 | No | NA | 2 | 0 | 1 | Not-determined | Genome-fragment | NA | NA | 0 |
| S8_extracted_k141_30511 | 1096 | No | NA | 1 | 0 | 1 | Not-determined | Genome-fragment | NA | NA | 0 |
| S8_extracted_k141_12337 | 1001 | No | NA | 2 | 0 | 1 | Not-determined | Genome-fragment | NA | NA | 0 |
| S8_extracted_k141_47058 | 2781 | No | NA | 3 | 0 | 1 | Not-determined | Genome-fragment | NA | NA | 0 |
| S8_extracted_k141_125696 | 1024 | No | NA | 1 | 0 | 0 | Not-determined | Genome-fragment | NA | NA | 0 |
| S8_extracted_k141_82058 | 1496 | No | NA | 1 | 0 | 1 | Not-determined | Genome-fragment | NA | NA | 0 |
| S8_extracted_k141_168869 | 1118 | No | NA | 3 | 0 | 1 | Not-determined | Genome-fragment | NA | NA | 0 |
| S8_extracted_k141_177662 | 1177 | No | NA | 2 | 0 | 0 | Not-determined | Genome-fragment | NA | NA | 0 |
| S8_extracted_k141_134054 | 1180 | No | NA | 2 | 0 | 1 | Not-determined | Genome-fragment | NA | NA | 0 |
| S8_extracted_k141_12368 | 1405 | No | NA | 1 | 0 | 0 | Not-determined | Genome-fragment | NA | NA | 0 |
| S8_extracted_k141_99558 | 1188 | No | NA | 2 | 0 | 0 | Not-determined | Genome-fragment | NA | NA | 0 |
| S8_extracted_k141_134060 | 1014 | No | NA | 2 | 0 | 1 | Not-determined | Genome-fragment | NA | NA | 0 |
| S8_extracted_k141_38768 | 1006 | No | NA | 1 | 0 | 0 | Not-determined | Genome-fragment | NA | NA | 0 |
| S8_extracted_k141_116689 | 1239 | No | NA | 1 | 0 | 1 | Not-determined | Genome-fragment | NA | NA | 0 |
| S10_vs_106838\|\|full | 8460 | No | NA | 13 | 3 | 1 | Low-quality | Genome-fragment | 20.84 | AAI-based (high-confidence) | 0 |
| S8_extracted_k141_151355 | 5025 | No | NA | 7 | 0 | 0 | Not-determined | Genome-fragment | NA | NA | 0 |
| S8_extracted_k141_21445 | 1732 | No | NA | 2 | 0 | 2 | Not-determined | Genome-fragment | NA | NA | 0 |
| S8_extracted_k141_116690 | 5273 | No | NA | 5 | 0 | 3 | Not-determined | Genome-fragment | NA | NA | 0 |
| S8_extracted_k141_116693 | 2774 | No | NA | 3 | 0 | 2 | Not-determined | Genome-fragment | NA | NA | 0 |
| S13_extracted_19481 | 8467 | No | NA | 10 | 1 | 2 | Low-quality | Genome-fragment | 7.95 | AAI-based (medium-confidence) | 0 |
| S8_extracted_k141_134082 | 1287 | No | NA | 1 | 0 | 0 | Not-determined | Genome-fragment | NA | NA | 0 |
| S8_extracted_k141_168901 | 1335 | No | NA | 2 | 0 | 1 | Not-determined | Genome-fragment | NA | NA | 0 |
| S9_extracted_k141_90853 | 8472 | No | NA | 16 | 4 | 0 | Low-quality | Genome-fragment | 16.74 | HMM-based (lower-bound) | 0 |
| S8_extracted_k141_30557 | 1214 | No | NA | 3 | 0 | 0 | Not-determined | Genome-fragment | NA | NA | 0 |
| S8_vs_k141_20991\|\|full | 8472 | No | NA | 16 | 5 | 0 | Low-quality | Genome-fragment | 24.21 | AAI-based (medium-confidence) | 0 |
| S8_extracted_k141_4131 | 1603 | No | NA | 2 | 0 | 1 | Not-determined | Genome-fragment | NA | NA | 0 |
| S12_extracted_k141_10434 | 8480 | No | NA | 16 | 1 | 2 | Low-quality | Genome-fragment | 4.6 | HMM-based (lower-bound) | 0 |
| S5_extracted_k141_13041 | 8482 | No | NA | 17 | 6 | 0 | Low-quality | Genome-fragment | 20.42 | AAI-based (medium-confidence) | 0 |
| S8_extracted_k141_142983 | 4012 | No | NA | 8 | 0 | 2 | Not-determined | Genome-fragment | NA | NA | 0 |
| S8_extracted_k141_108357 | 16136 | No | NA | 8 | 0 | 6 | Not-determined | Genome-fragment | NA | NA | 0 |
| S8_extracted_k141_12437 | 1619 | No | NA | 1 | 0 | 0 | Not-determined | Genome-fragment | NA | NA | 0 |
| S8_extracted_k141_195149 | 4962 | No | NA | 4 | 0 | 2 | Not-determined | Genome-fragment | NA | NA | 0 |
| S8_extracted_k141_21483 | 2007 | No | NA | 1 | 0 | 1 | Not-determined | Genome-fragment | NA | NA | 0 |
| S8_extracted_k141_116736 | 2746 | No | NA | 3 | 0 | 2 | Not-determined | Genome-fragment | NA | NA | 0 |
| S8_extracted_k141_90991 | 1073 | No | NA | 2 | 0 | 0 | Not-determined | Genome-fragment | NA | NA | 0 |
| S8_extracted_k141_143003 | 4017 | No | NA | 5 | 0 | 2 | Not-determined | Genome-fragment | NA | NA | 0 |
| S8_extracted_k141_56301 | 1175 | No | NA | 2 | 0 | 0 | Not-determined | Genome-fragment | NA | NA | 0 |
| S8_extracted_k141_160013 | 1637 | No | NA | 1 | 0 | 1 | Not-determined | Genome-fragment | NA | NA | 0 |
| S8_extracted_k141_143014 | 1581 | No | NA | 2 | 0 | 0 | Not-determined | Genome-fragment | NA | NA | 0 |
| S8_extracted_k141_160018 | 1989 | No | NA | 1 | 0 | 0 | Not-determined | Genome-fragment | NA | NA | 0 |
| S8_extracted_k141_73048 | 1398 | No | NA | 1 | 0 | 1 | Not-determined | Genome-fragment | NA | NA | 0 |
| S8_extracted_k141_21505 | 2834 | No | NA | 3 | 0 | 0 | Not-determined | Genome-fragment | NA | NA | 0 |
| S8_extracted_k141_160019 | 1129 | No | NA | 1 | 0 | 1 | Not-determined | Genome-fragment | NA | NA | 0 |
| S12_vs_k141_138722\|\|full | 8483 | No | NA | 5 | 0 | 5 | Low-quality | Genome-fragment | 9.04 | AAI-based (medium-confidence) | 0 |
| S8_extracted_k141_12477 | 4221 | No | NA | 2 | 0 | 1 | Not-determined | Genome-fragment | NA | NA | 0 |
| S8_extracted_k141_143020 | 1633 | No | NA | 2 | 0 | 0 | Not-determined | Genome-fragment | NA | NA | 0 |
| S16_vs_155321\|\|full | 8488 | No | NA | 12 | 1 | 2 | Low-quality | Genome-fragment | 10.82 | HMM-based (lower-bound) | 0 |
| S8_extracted_k141_21517 | 3975 | No | NA | 3 | 0 | 3 | Not-determined | Genome-fragment | NA | NA | 0 |
| S8_extracted_k141_151384 | 51637 | No | NA | 78 | 22 | 0 | High-quality | High-quality | 100 | AAI-based (medium-confidence) | 0 |
| S8_extracted_k141_21518 | 1468 | No | NA | 3 | 0 | 1 | Not-determined | Genome-fragment | NA | NA | 0 |
| S8_extracted_k141_56341 | 1651 | No | NA | 2 | 0 | 1 | Not-determined | Genome-fragment | NA | NA | 0 |
| S8_extracted_k141_4157 | 4447 | No | NA | 5 | 0 | 2 | Not-determined | Genome-fragment | NA | NA | 0 |
| S8_extracted_k141_56345 | 3982 | No | NA | 5 | 0 | 5 | Not-determined | Genome-fragment | NA | NA | 0 |
| S11_vs_k141_85812\|\|full | 8503 | No | NA | 12 | 12 | 0 | Low-quality | Genome-fragment | 24.49 | AAI-based (high-confidence) | 0 |
| S8_extracted_k141_160064 | 1051 | No | NA | 2 | 0 | 1 | Not-determined | Genome-fragment | NA | NA | 0 |
| S8_extracted_k141_38820 | 10135 | No | NA | 13 | 0 | 3 | Not-determined | Genome-fragment | NA | NA | 0 |
| S8_extracted_k141_160070 | 3201 | No | NA | 2 | 0 | 2 | Not-determined | Genome-fragment | NA | NA | 0 |
| S8_extracted_k141_12517 | 1510 | No | NA | 1 | 0 | 1 | Not-determined | Genome-fragment | NA | NA | 0 |
| S8_extracted_k141_203318 | 1821 | No | NA | 2 | 0 | 2 | Not-determined | Genome-fragment | NA | NA | 0 |
| S8_vs_k141_169154\|\|full | 8506 | No | NA | 13 | 3 | 0 | Low-quality | Genome-fragment | 21.46 | AAI-based (high-confidence) | 0 |
| S7_vs_87668\|\|full | 8512 | No | NA | 8 | 3 | 0 | Low-quality | Genome-fragment | 9.96 | HMM-based (lower-bound) | 0 |
| S8_extracted_k141_99634 | 1026 | No | NA | 1 | 0 | 0 | Not-determined | Genome-fragment | NA | NA | 0 |
| S8_extracted_k141_108443 | 3339 | No | NA | 1 | 0 | 1 | Not-determined | Genome-fragment | NA | NA | 0 |
| S8_extracted_k141_177811 | 1052 | No | NA | 2 | 0 | 2 | Not-determined | Genome-fragment | NA | NA | 0 |
| S16_extracted_145572 | 8514 | No | NA | 5 | 0 | 3 | Low-quality | Genome-fragment | 4.58 | AAI-based (medium-confidence) | 0 |
| S8_extracted_k141_143074 | 1358 | No | NA | 2 | 0 | 1 | Not-determined | Genome-fragment | NA | NA | 0 |
| S7_extracted_62927 | 8521 | No | NA | 4 | 2 | 0 | Low-quality | Genome-fragment | 27.84 | AAI-based (high-confidence) | 0 |
| S8_extracted_k141_168965 | 11463 | No | NA | 8 | 0 | 7 | Not-determined | Genome-fragment | NA | NA | 0 |
| S8_extracted_k141_185656 | 1295 | No | NA | 3 | 0 | 0 | Not-determined | Genome-fragment | NA | NA | 0 |
| S8_extracted_k141_108453 | 1022 | No | NA | 2 | 0 | 1 | Not-determined | Genome-fragment | NA | NA | 0 |
| S8_extracted_k141_151430 | 1322 | No | NA | 1 | 0 | 0 | Not-determined | Genome-fragment | NA | NA | 0 |
| S8_extracted_k141_91033 | 16317 | No | NA | 20 | 6 | 0 | Medium-quality | Genome-fragment | 83.74 | AAI-based (medium-confidence) | 0 |
| S11_vs_k141_26600\|\|full | 8532 | No | NA | 16 | 3 | 0 | Low-quality | Genome-fragment | 15.48 | AAI-based (high-confidence) | 0 |
| S8_extracted_k141_195249 | 4094 | No | NA | 4 | 0 | 4 | Not-determined | Genome-fragment | NA | NA | 0 |
| S8_extracted_k141_91038 | 1620 | No | NA | 4 | 0 | 2 | Not-determined | Genome-fragment | NA | NA | 0 |
| S8_extracted_k141_47189 | 1112 | No | NA | 1 | 0 | 0 | Not-determined | Genome-fragment | NA | NA | 0 |
| S8_extracted_k141_177839 | 7361 | No | NA | 3 | 0 | 3 | Not-determined | Genome-fragment | NA | NA | 0 |
| S8_extracted_k141_56407 | 1013 | No | NA | 1 | 0 | 0 | Not-determined | Genome-fragment | NA | NA | 0 |
| S16_extracted_86294 | 8541 | No | NA | 8 | 0 | 6 | Low-quality | Genome-fragment | 2.68 | AAI-based (medium-confidence) | 0 |
| S8_extracted_k141_177841 | 2335 | No | NA | 4 | 0 | 1 | Not-determined | Genome-fragment | NA | NA | 0 |
| S8_extracted_k141_185683 | 1300 | No | NA | 1 | 0 | 0 | Not-determined | Genome-fragment | NA | NA | 0 |
| S8_extracted_k141_99667 | 8281 | No | NA | 4 | 0 | 3 | Not-determined | Genome-fragment | NA | NA | 0 |
| S8_extracted_k141_82225 | 1137 | No | NA | 3 | 0 | 3 | Not-determined | Genome-fragment | NA | NA | 0 |
| S8_extracted_k141_177843 | 4297 | No | NA | 2 | 0 | 1 | Not-determined | Genome-fragment | NA | NA | 0 |
| S8_extracted_k141_73146 | 3268 | No | NA | 3 | 0 | 0 | Not-determined | Genome-fragment | NA | NA | 0 |
| S8_extracted_k141_99671 | 2162 | No | NA | 4 | 0 | 1 | Not-determined | Genome-fragment | NA | NA | 0 |
| S8_extracted_k141_38878 | 5798 | No | NA | 7 | 0 | 6 | Not-determined | Genome-fragment | NA | NA | 0 |
| S16_vs_9248\|\|full | 8546 | No | NA | 14 | 1 | 4 | Low-quality | Genome-fragment | 12.06 | AAI-based (medium-confidence) | 0 |
| S8_extracted_k141_99694 | 2730 | No | NA | 2 | 0 | 2 | Not-determined | Genome-fragment | NA | NA | 0 |
| S3_vs_k141_85431\|\|full | 8552 | No | NA | 19 | 2 | 0 | Low-quality | Genome-fragment | 5.03 | AAI-based (high-confidence) | 0 |
| S17_vs_k141_95172\|\|full | 8553 | Yes | 4898 | 7 | 1 | 1 | Low-quality | Genome-fragment | 11.13 | AAI-based (medium-confidence) | 42.73 |
| S8_extracted_k141_143126 | 1183 | No | NA | 1 | 0 | 1 | Not-determined | Genome-fragment | NA | NA | 0 |
| S8_extracted_k141_195296 | 1423 | No | NA | 2 | 0 | 0 | Not-determined | Genome-fragment | NA | NA | 0 |
| S8_extracted_k141_169046 | 1780 | No | NA | 2 | 0 | 2 | Not-determined | Genome-fragment | NA | NA | 0 |
| S8_extracted_k141_21629 | 1919 | No | NA | 4 | 0 | 0 | Not-determined | Genome-fragment | NA | NA | 0 |
| S5_extracted_k141_51127 | 8554 | No | NA | 12 | 9 | 0 | Low-quality | Genome-fragment | 22.19 | AAI-based (medium-confidence) | 0 |
| S5_vs_k141_110404\|\|full | 8558 | No | NA | 19 | 3 | 2 | Low-quality | Genome-fragment | 13.6 | AAI-based (high-confidence) | 0 |
| S5_vs_k141_46638\|\|full | 8567 | No | NA | 13 | 7 | 0 | Low-quality | Genome-fragment | 22.37 | AAI-based (medium-confidence) | 0 |
| S13_extracted_17154 | 8569 | No | NA | 5 | 3 | 0 | Low-quality | Genome-fragment | 9.28 | AAI-based (high-confidence) | 0 |
| S8_extracted_k141_56443 | 2752 | No | NA | 2 | 0 | 2 | Not-determined | Genome-fragment | NA | NA | 0 |
| S8_extracted_k141_108532 | 1457 | No | NA | 1 | 0 | 0 | Not-determined | Genome-fragment | NA | NA | 0 |
| S8_extracted_k141_82293 | 2402 | No | NA | 6 | 0 | 0 | Not-determined | Genome-fragment | NA | NA | 0 |
| S8_extracted_k141_134216 | 2092 | No | NA | 2 | 0 | 2 | Not-determined | Genome-fragment | NA | NA | 0 |
| S8_extracted_k141_47246 | 1121 | No | NA | 1 | 0 | 1 | Not-determined | Genome-fragment | NA | NA | 0 |
| S12_extracted_k141_169826 | 8576 | No | NA | 13 | 3 | 0 | Low-quality | Genome-fragment | 16.68 | AAI-based (high-confidence) | 0 |
| S8_extracted_k141_169059 | 1313 | No | NA | 3 | 0 | 1 | Not-determined | Genome-fragment | NA | NA | 0 |
| S8_extracted_k141_108545 | 1047 | No | NA | 2 | 0 | 2 | Not-determined | Genome-fragment | NA | NA | 0 |
| S8_extracted_k141_91114 | 1875 | No | NA | 1 | 0 | 1 | Not-determined | Genome-fragment | NA | NA | 0 |
| S16_vs_34799\|\|full | 8578 | No | NA | 13 | 0 | 1 | Low-quality | Genome-fragment | 13.2 | AAI-based (high-confidence) | 0 |
| S8_extracted_k141_82308 | 1913 | No | NA | 1 | 0 | 1 | Not-determined | Genome-fragment | NA | NA | 0 |
| S8_extracted_k141_125883 | 1131 | No | NA | 2 | 0 | 0 | Not-determined | Genome-fragment | NA | NA | 0 |
| S2_vs_k141_13711\|\|full | 8582 | No | NA | 20 | 2 | 0 | Low-quality | Genome-fragment | 14.3 | AAI-based (high-confidence) | 0 |
| S8_extracted_k141_21653 | 3677 | No | NA | 5 | 0 | 1 | Not-determined | Genome-fragment | NA | NA | 0 |
| S6_vs_k141_14991\|\|full | 8587 | No | NA | 12 | 4 | 1 | Low-quality | Genome-fragment | 25.06 | AAI-based (high-confidence) | 0 |
| S8_extracted_k141_82321 | 1972 | No | NA | 1 | 0 | 1 | Not-determined | Genome-fragment | NA | NA | 0 |
| S8_extracted_k141_73240 | 2085 | No | NA | 2 | 0 | 1 | Not-determined | Genome-fragment | NA | NA | 0 |
| S8_extracted_k141_195344 | 2037 | No | NA | 2 | 0 | 1 | Not-determined | Genome-fragment | NA | NA | 0 |
| S8_extracted_k141_64731 | 1359 | No | NA | 1 | 0 | 1 | Not-determined | Genome-fragment | NA | NA | 0 |
| S8_extracted_k141_82333 | 1146 | No | NA | 1 | 0 | 1 | Not-determined | Genome-fragment | NA | NA | 0 |
| S8_extracted_k141_185741 | 2370 | No | NA | 1 | 0 | 1 | Not-determined | Genome-fragment | NA | NA | 0 |
| S5_vs_k141_140471\|\|full | 8589 | No | NA | 4 | 0 | 3 | Low-quality | Genome-fragment | 2.64 | AAI-based (high-confidence) | 0 |
| S8_extracted_k141_125912 | 1097 | No | NA | 1 | 0 | 0 | Not-determined | Genome-fragment | NA | NA | 0 |
| S8_extracted_k141_116871 | 2564 | No | NA | 4 | 0 | 3 | Not-determined | Genome-fragment | NA | NA | 0 |
| S8_extracted_k141_125922 | 1026 | No | NA | 2 | 0 | 1 | Not-determined | Genome-fragment | NA | NA | 0 |
| S8_extracted_k141_134285 | 2998 | No | NA | 3 | 0 | 2 | Not-determined | Genome-fragment | NA | NA | 0 |
| S8_extracted_k141_169135 | 1680 | No | NA | 2 | 0 | 0 | Not-determined | Genome-fragment | NA | NA | 0 |
| S8_extracted_k141_64757 | 1882 | No | NA | 2 | 0 | 2 | Not-determined | Genome-fragment | NA | NA | 0 |
| S8_extracted_k141_134300 | 1991 | No | NA | 1 | 0 | 1 | Not-determined | Genome-fragment | NA | NA | 0 |
| S8_extracted_k141_21691 | 2272 | No | NA | 2 | 0 | 2 | Not-determined | Genome-fragment | NA | NA | 0 |
| S8_extracted_k141_12632 | 5030 | No | NA | 5 | 0 | 2 | Not-determined | Genome-fragment | NA | NA | 0 |
| S16_vs_17255\|\|full | 8595 | No | NA | 3 | 0 | 2 | Low-quality | Genome-fragment | 10.26 | AAI-based (medium-confidence) | 0 |
| S8_extracted_k141_30618 | 1140 | No | NA | 3 | 0 | 2 | Not-determined | Genome-fragment | NA | NA | 0 |
| S8_extracted_k141_73282 | 1862 | No | NA | 1 | 0 | 1 | Not-determined | Genome-fragment | NA | NA | 0 |
| S8_extracted_k141_21704 | 1749 | No | NA | 2 | 0 | 1 | Not-determined | Genome-fragment | NA | NA | 0 |
| S8_extracted_k141_160116 | 1875 | No | NA | 2 | 0 | 2 | Not-determined | Genome-fragment | NA | NA | 0 |
| S8_extracted_k141_21708 | 1202 | No | NA | 2 | 0 | 1 | Not-determined | Genome-fragment | NA | NA | 0 |
| S8_extracted_k141_116924 | 3058 | No | NA | 1 | 0 | 1 | Not-determined | Genome-fragment | NA | NA | 0 |
| S9_extracted_k141_88157 | 8602 | No | NA | 10 | 1 | 0 | Low-quality | Genome-fragment | 4.83 | AAI-based (high-confidence) | 0 |
| S8_extracted_k141_203511 | 1359 | No | NA | 1 | 0 | 1 | Not-determined | Genome-fragment | NA | NA | 0 |
| S5_vs_k141_131707\|\|full | 8603 | No | NA | 12 | 3 | 0 | Low-quality | Genome-fragment | 19.78 | AAI-based (medium-confidence) | 0 |
| S8_extracted_k141_151587 | 3198 | No | NA | 10 | 0 | 0 | Not-determined | Genome-fragment | NA | NA | 0 |
| S8_extracted_k141_160130 | 4027 | No | NA | 2 | 0 | 1 | Not-determined | Genome-fragment | NA | NA | 0 |
| S9_vs_k141_90865\|\|full | 8612 | No | NA | 14 | 2 | 0 | Low-quality | Genome-fragment | 18.86 | AAI-based (medium-confidence) | 0 |
| S8_extracted_k141_195376 | 5643 | No | NA | 6 | 0 | 1 | Not-determined | Genome-fragment | NA | NA | 0 |
| S5_vs_k141_135866\|\|full | 8620 | No | NA | 9 | 0 | 1 | Low-quality | Genome-fragment | 15.63 | AAI-based (medium-confidence) | 0 |
| S8_extracted_k141_143261 | 1038 | No | NA | 1 | 0 | 1 | Not-determined | Genome-fragment | NA | NA | 0 |
| S3_extracted_k141_17677 | 8622 | No | NA | 12 | 11 | 0 | Low-quality | Genome-fragment | 21.93 | AAI-based (high-confidence) | 0 |
| S8_extracted_k141_169190 | 1310 | No | NA | 1 | 0 | 1 | Not-determined | Genome-fragment | NA | NA | 0 |
| S8_extracted_k141_64796 | 1610 | No | NA | 1 | 0 | 0 | Not-determined | Genome-fragment | NA | NA | 0 |
| S8_extracted_k141_116955 | 5796 | No | NA | 6 | 0 | 3 | Not-determined | Genome-fragment | NA | NA | 0 |
| S8_extracted_k141_47273 | 2222 | No | NA | 3 | 0 | 1 | Not-determined | Genome-fragment | NA | NA | 0 |
| S11_vs_k141_88723\|\|full | 8623 | No | NA | 11 | 2 | 0 | Low-quality | Genome-fragment | 21.14 | AAI-based (medium-confidence) | 0 |
| S8_extracted_k141_56571 | 4804 | No | NA | 2 | 0 | 2 | Not-determined | Genome-fragment | NA | NA | 0 |
| S8_extracted_k141_185792 | 5502 | No | NA | 7 | 0 | 3 | Not-determined | Genome-fragment | NA | NA | 0 |
| S8_extracted_k141_203545 | 4669 | No | NA | 3 | 0 | 2 | Not-determined | Genome-fragment | NA | NA | 0 |
| S8_extracted_k141_108642 | 2804 | No | NA | 2 | 0 | 1 | Not-determined | Genome-fragment | NA | NA | 0 |
| S8_extracted_k141_134361 | 1563 | No | NA | 1 | 0 | 0 | Not-determined | Genome-fragment | NA | NA | 0 |
| S8_extracted_k141_169207 | 4850 | No | NA | 3 | 0 | 2 | Not-determined | Genome-fragment | NA | NA | 0 |
| S8_extracted_k141_47302 | 1222 | No | NA | 1 | 0 | 0 | Not-determined | Genome-fragment | NA | NA | 0 |
| S8_extracted_k141_56590 | 2041 | No | NA | 2 | 0 | 0 | Not-determined | Genome-fragment | NA | NA | 0 |
| S8_extracted_k141_39055 | 1743 | No | NA | 1 | 0 | 0 | Not-determined | Genome-fragment | NA | NA | 0 |
| S8_extracted_k141_169236 | 1178 | No | NA | 3 | 0 | 0 | Not-determined | Genome-fragment | NA | NA | 0 |
| S8_extracted_k141_117018 | 2020 | No | NA | 3 | 0 | 0 | Not-determined | Genome-fragment | NA | NA | 0 |
| S8_extracted_k141_151639 | 2200 | No | NA | 2 | 0 | 0 | Not-determined | Genome-fragment | NA | NA | 0 |
| S8_extracted_k141_178029 | 1223 | No | NA | 2 | 0 | 0 | Not-determined | Genome-fragment | NA | NA | 0 |
| S8_extracted_k141_4464 | 3380 | No | NA | 3 | 0 | 3 | Not-determined | Genome-fragment | NA | NA | 0 |
| S8_extracted_k141_56610 | 3867 | No | NA | 4 | 0 | 2 | Not-determined | Genome-fragment | NA | NA | 0 |
| S8_extracted_k141_39077 | 2544 | No | NA | 1 | 0 | 1 | Not-determined | Genome-fragment | NA | NA | 0 |
| S8_extracted_k141_134394 | 3081 | No | NA | 3 | 0 | 3 | Not-determined | Genome-fragment | NA | NA | 0 |
| S3_vs_k141_33019\|\|full | 8624 | No | NA | 16 | 3 | 0 | Low-quality | Genome-fragment | 24.07 | AAI-based (medium-confidence) | 0 |
| S8_extracted_k141_64862 | 1760 | No | NA | 1 | 0 | 1 | Not-determined | Genome-fragment | NA | NA | 0 |
| S5_extracted_k141_68772 | 8626 | No | NA | 16 | 2 | 0 | Low-quality | Genome-fragment | 19.87 | AAI-based (medium-confidence) | 0 |
| S8_extracted_k141_203610 | 1144 | No | NA | 1 | 0 | 0 | Not-determined | Genome-fragment | NA | NA | 0 |
| S5_vs_k141_39855\|\|full | 8636 | No | NA | 11 | 4 | 0 | Low-quality | Genome-fragment | 19.85 | AAI-based (medium-confidence) | 0 |
| S8_extracted_k141_39089 | 2565 | No | NA | 2 | 0 | 0 | Not-determined | Genome-fragment | NA | NA | 0 |
| S11_vs_k141_83602\|\|full | 8638 | No | NA | 14 | 4 | 0 | Low-quality | Genome-fragment | 19.53 | HMM-based (lower-bound) | 0 |
| S14_extracted_k141_82837 | 8638 | No | NA | 15 | 3 | 0 | Low-quality | Genome-fragment | 5.59 | AAI-based (medium-confidence) | 0 |
| S8_extracted_k141_178060 | 1708 | No | NA | 2 | 0 | 0 | Not-determined | Genome-fragment | NA | NA | 0 |
| S8_extracted_k141_73432 | 2279 | No | NA | 3 | 0 | 2 | Not-determined | Genome-fragment | NA | NA | 0 |
| S8_extracted_k141_185816 | 2906 | No | NA | 5 | 0 | 3 | Not-determined | Genome-fragment | NA | NA | 0 |
| S8_extracted_k141_151695 | 1424 | No | NA | 1 | 0 | 0 | Not-determined | Genome-fragment | NA | NA | 0 |
| S8_extracted_k141_160291 | 3322 | No | NA | 3 | 0 | 2 | Not-determined | Genome-fragment | NA | NA | 0 |
| S8_extracted_k141_99883 | 1896 | No | NA | 1 | 0 | 1 | Not-determined | Genome-fragment | NA | NA | 0 |
| S8_extracted_k141_47385 | 2171 | No | NA | 2 | 0 | 0 | Not-determined | Genome-fragment | NA | NA | 0 |
| S8_extracted_k141_160294 | 1849 | No | NA | 2 | 0 | 1 | Not-determined | Genome-fragment | NA | NA | 0 |
| S3_extracted_k141_49559 | 8639 | No | NA | 17 | 6 | 0 | Low-quality | Genome-fragment | 15.13 | AAI-based (high-confidence) | 0 |
| S8_extracted_k141_82465 | 1053 | No | NA | 2 | 0 | 2 | Not-determined | Genome-fragment | NA | NA | 0 |
| S8_extracted_k141_47386 | 3026 | No | NA | 4 | 0 | 3 | Not-determined | Genome-fragment | NA | NA | 0 |
| S8_extracted_k141_73455 | 2009 | No | NA | 3 | 0 | 2 | Not-determined | Genome-fragment | NA | NA | 0 |
| S8_extracted_k141_12785 | 2527 | No | NA | 1 | 0 | 1 | Not-determined | Genome-fragment | NA | NA | 0 |
| S8_extracted_k141_178081 | 1282 | No | NA | 1 | 0 | 0 | Not-determined | Genome-fragment | NA | NA | 0 |
| S1_vs_k141_44356\|\|full | 8646 | No | NA | 20 | 4 | 0 | Low-quality | Genome-fragment | 19.65 | AAI-based (high-confidence) | 0 |
| S8_extracted_k141_117085 | 1341 | No | NA | 1 | 0 | 1 | Not-determined | Genome-fragment | NA | NA | 0 |
| S8_extracted_k141_73463 | 1036 | No | NA | 2 | 0 | 0 | Not-determined | Genome-fragment | NA | NA | 0 |
| S8_extracted_k141_203657 | 1032 | No | NA | 2 | 0 | 0 | Not-determined | Genome-fragment | NA | NA | 0 |
| S3_vs_k141_31615\|\|full | 8655 | No | NA | 15 | 0 | 3 | Low-quality | Genome-fragment | 20.21 | AAI-based (medium-confidence) | 0 |
| S9_vs_k141_36131\|\|full | 8661 | No | NA | 20 | 3 | 0 | Low-quality | Genome-fragment | 19.69 | AAI-based (high-confidence) | 0 |
| S8_extracted_k141_30803 | 4897 | No | NA | 4 | 0 | 2 | Not-determined | Genome-fragment | NA | NA | 0 |
| S8_extracted_k141_64914 | 3514 | No | NA | 1 | 0 | 1 | Not-determined | Genome-fragment | NA | NA | 0 |
| S8_extracted_k141_47404 | 1297 | No | NA | 2 | 0 | 0 | Not-determined | Genome-fragment | NA | NA | 0 |
| S8_extracted_k141_178087 | 3404 | No | NA | 2 | 0 | 1 | Not-determined | Genome-fragment | NA | NA | 0 |
| S11_vs_k141_60870\|\|full | 8662 | No | NA | 9 | 0 | 0 | Low-quality | Genome-fragment | 3.57 | AAI-based (medium-confidence) | 0 |
| S8_extracted_k141_195518 | 5991 | No | NA | 5 | 0 | 1 | Not-determined | Genome-fragment | NA | NA | 0 |
| S8_extracted_k141_195520 | 1517 | No | NA | 1 | 0 | 0 | Not-determined | Genome-fragment | NA | NA | 0 |
| S8_extracted_k141_185847 | 5809 | No | NA | 3 | 0 | 2 | Not-determined | Genome-fragment | NA | NA | 0 |
| S8_extracted_k141_4564 | 1056 | No | NA | 2 | 0 | 1 | Not-determined | Genome-fragment | NA | NA | 0 |
| S8_extracted_k141_56673 | 1169 | No | NA | 1 | 0 | 1 | Not-determined | Genome-fragment | NA | NA | 0 |
| S8_extracted_k141_47414 | 4291 | No | NA | 3 | 0 | 3 | Not-determined | Genome-fragment | NA | NA | 0 |
| S7_extracted_11308 | 8667 | No | NA | 17 | 5 | 0 | Low-quality | Genome-fragment | 21.62 | HMM-based (lower-bound) | 0 |
| S8_extracted_k141_151775 | 1454 | No | NA | 1 | 0 | 1 | Not-determined | Genome-fragment | NA | NA | 0 |
| S8_extracted_k141_82531 | 1985 | No | NA | 1 | 0 | 1 | Not-determined | Genome-fragment | NA | NA | 0 |
| S8_extracted_k141_91206 | 1421 | No | NA | 3 | 0 | 0 | Not-determined | Genome-fragment | NA | NA | 0 |
| S8_extracted_k141_203713 | 1524 | No | NA | 2 | 0 | 0 | Not-determined | Genome-fragment | NA | NA | 0 |
| S8_extracted_k141_143361 | 1248 | No | NA | 2 | 0 | 2 | Not-determined | Genome-fragment | NA | NA | 0 |
| S8_extracted_k141_39182 | 1104 | No | NA | 2 | 0 | 1 | Not-determined | Genome-fragment | NA | NA | 0 |
| S11_vs_k141_59445\|\|full | 8668 | No | NA | 24 | 0 | 1 | Low-quality | Genome-fragment | 5.59 | AAI-based (high-confidence) | 0 |
| S8_extracted_k141_30845 | 3375 | No | NA | 4 | 0 | 3 | Not-determined | Genome-fragment | NA | NA | 0 |
| S8_extracted_k141_151809 | 1578 | No | NA | 2 | 0 | 1 | Not-determined | Genome-fragment | NA | NA | 0 |
| S8_extracted_k141_134466 | 1803 | No | NA | 2 | 0 | 2 | Not-determined | Genome-fragment | NA | NA | 0 |
| S8_extracted_k141_169389 | 1029 | No | NA | 3 | 0 | 0 | Not-determined | Genome-fragment | NA | NA | 0 |
| S1_extracted_k141_12730 | 8668 | No | NA | 19 | 3 | 0 | Low-quality | Genome-fragment | 14.83 | AAI-based (high-confidence) | 0 |
| S8_extracted_k141_21959 | 1566 | No | NA | 1 | 0 | 0 | Not-determined | Genome-fragment | NA | NA | 0 |
| S8_extracted_k141_91252 | 2475 | No | NA | 2 | 0 | 1 | Not-determined | Genome-fragment | NA | NA | 0 |
| S8_extracted_k141_21963 | 1244 | No | NA | 1 | 0 | 0 | Not-determined | Genome-fragment | NA | NA | 0 |
| S8_extracted_k141_203736 | 1759 | No | NA | 3 | 0 | 2 | Not-determined | Genome-fragment | NA | NA | 0 |
| S7_vs_98863\|\|full | 8670 | No | NA | 13 | 9 | 0 | Low-quality | Genome-fragment | 24.23 | AAI-based (high-confidence) | 0 |
| S8_extracted_k141_65015 | 1200 | No | NA | 3 | 0 | 2 | Not-determined | Genome-fragment | NA | NA | 0 |
| S8_extracted_k141_143406 | 1009 | No | NA | 2 | 0 | 0 | Not-determined | Genome-fragment | NA | NA | 0 |
| S8_extracted_k141_117175 | 6102 | No | NA | 3 | 0 | 2 | Not-determined | Genome-fragment | NA | NA | 0 |
| S15_extracted_52314 | 8674 | No | NA | 9 | 3 | 0 | Low-quality | Genome-fragment | 8.35 | AAI-based (medium-confidence) | 0 |
| S8_extracted_k141_203762 | 1815 | No | NA | 1 | 0 | 1 | Not-determined | Genome-fragment | NA | NA | 0 |
| S8_extracted_k141_203765 | 1594 | No | NA | 3 | 0 | 0 | Not-determined | Genome-fragment | NA | NA | 0 |
| S8_extracted_k141_178230 | 1023 | No | NA | 1 | 0 | 1 | Not-determined | Genome-fragment | NA | NA | 0 |
| S8_extracted_k141_185974 | 3650 | No | NA | 3 | 0 | 1 | Not-determined | Genome-fragment | NA | NA | 0 |
| S8_extracted_k141_203772 | 2335 | No | NA | 5 | 0 | 0 | Not-determined | Genome-fragment | NA | NA | 0 |
| S8_extracted_k141_56731 | 1330 | No | NA | 1 | 0 | 1 | Not-determined | Genome-fragment | NA | NA | 0 |
| S8_extracted_k141_12895 | 1746 | No | NA | 2 | 0 | 1 | Not-determined | Genome-fragment | NA | NA | 0 |
| S8_extracted_k141_47532 | 3355 | No | NA | 5 | 0 | 0 | Not-determined | Genome-fragment | NA | NA | 0 |
| S8_extracted_k141_56737 | 3274 | No | NA | 3 | 0 | 2 | Not-determined | Genome-fragment | NA | NA | 0 |
| S8_extracted_k141_108826 | 2228 | No | NA | 3 | 0 | 0 | Not-determined | Genome-fragment | NA | NA | 0 |
| S8_extracted_k141_22009 | 1616 | No | NA | 2 | 0 | 1 | Not-determined | Genome-fragment | NA | NA | 0 |
| S8_vs_k141_87426\|\|full | 8675 | No | NA | 9 | 0 | 6 | Low-quality | Genome-fragment | 27.7 | AAI-based (high-confidence) | 0 |
| S8_extracted_k141_30927 | 1311 | No | NA | 1 | 0 | 1 | Not-determined | Genome-fragment | NA | NA | 0 |
| S8_extracted_k141_134538 | 1309 | No | NA | 1 | 0 | 1 | Not-determined | Genome-fragment | NA | NA | 0 |
| S8_extracted_k141_56769 | 1150 | No | NA | 1 | 0 | 1 | Not-determined | Genome-fragment | NA | NA | 0 |
| S8_extracted_k141_91318 | 2015 | No | NA | 1 | 0 | 0 | Not-determined | Genome-fragment | NA | NA | 0 |
| S8_extracted_k141_203807 | 1297 | No | NA | 2 | 0 | 2 | Not-determined | Genome-fragment | NA | NA | 0 |
| S8_extracted_k141_185989 | 1144 | No | NA | 2 | 0 | 1 | Not-determined | Genome-fragment | NA | NA | 0 |
| S8_extracted_k141_203813 | 2450 | No | NA | 2 | 0 | 2 | Not-determined | Genome-fragment | NA | NA | 0 |
| S8_extracted_k141_65095 | 1518 | No | NA | 2 | 0 | 2 | Not-determined | Genome-fragment | NA | NA | 0 |
| S8_extracted_k141_117269 | 1459 | No | NA | 3 | 0 | 2 | Not-determined | Genome-fragment | NA | NA | 0 |
| S8_extracted_k141_73570 | 1141 | No | NA | 1 | 0 | 0 | Not-determined | Genome-fragment | NA | NA | 0 |
| S8_extracted_k141_185998 | 1024 | No | NA | 1 | 0 | 1 | Not-determined | Genome-fragment | NA | NA | 0 |
| S14_vs_k141_12212\|\|full | 8676 | No | NA | 10 | 4 | 0 | Low-quality | Genome-fragment | 20.87 | AAI-based (high-confidence) | 0 |
| S8_extracted_k141_22054 | 1625 | No | NA | 3 | 0 | 3 | Not-determined | Genome-fragment | NA | NA | 0 |
| S8_extracted_k141_143458 | 2238 | No | NA | 2 | 0 | 2 | Not-determined | Genome-fragment | NA | NA | 0 |
| S8_extracted_k141_203834 | 1125 | No | NA | 0 | 0 | 0 | Not-determined | Genome-fragment | NA | NA | 0 |
| S8_extracted_k141_126134 | 1444 | No | NA | 2 | 0 | 1 | Not-determined | Genome-fragment | NA | NA | 0 |
| S8_extracted_k141_143463 | 1048 | No | NA | 2 | 0 | 0 | Not-determined | Genome-fragment | NA | NA | 0 |
| S8_extracted_k141_186013 | 1248 | No | NA | 1 | 0 | 0 | Not-determined | Genome-fragment | NA | NA | 0 |
| S8_extracted_k141_126143 | 1036 | No | NA | 2 | 0 | 0 | Not-determined | Genome-fragment | NA | NA | 0 |
| S8_extracted_k141_100069 | 1136 | No | NA | 1 | 0 | 0 | Not-determined | Genome-fragment | NA | NA | 0 |
| S8_extracted_k141_82611 | 1937 | No | NA | 1 | 0 | 1 | Not-determined | Genome-fragment | NA | NA | 0 |
| S8_extracted_k141_22077 | 1089 | No | NA | 1 | 0 | 1 | Not-determined | Genome-fragment | NA | NA | 0 |
| S8_extracted_k141_117302 | 1346 | No | NA | 2 | 0 | 1 | Not-determined | Genome-fragment | NA | NA | 0 |
| S8_extracted_k141_178280 | 1019 | No | NA | 1 | 0 | 0 | Not-determined | Genome-fragment | NA | NA | 0 |
| S8_extracted_k141_82621 | 1213 | No | NA | 1 | 0 | 0 | Not-determined | Genome-fragment | NA | NA | 0 |
| S8_extracted_k141_186042 | 1451 | No | NA | 2 | 0 | 2 | Not-determined | Genome-fragment | NA | NA | 0 |
| S8_extracted_k141_22094 | 2113 | No | NA | 1 | 0 | 1 | Not-determined | Genome-fragment | NA | NA | 0 |
| S8_extracted_k141_117329 | 1241 | No | NA | 2 | 0 | 0 | Not-determined | Genome-fragment | NA | NA | 0 |
| S8_extracted_k141_151910 | 1483 | No | NA | 2 | 0 | 1 | Not-determined | Genome-fragment | NA | NA | 0 |
| S8_extracted_k141_56831 | 9192 | No | NA | 4 | 0 | 4 | Not-determined | Genome-fragment | NA | NA | 0 |
| S13_vs_65924\|\|full | 8688 | No | NA | 12 | 0 | 4 | Low-quality | Genome-fragment | 5.59 | AAI-based (high-confidence) | 0 |
| S8_extracted_k141_151915 | 2269 | No | NA | 2 | 0 | 2 | Not-determined | Genome-fragment | NA | NA | 0 |
| S8_extracted_k141_134590 | 1705 | No | NA | 2 | 0 | 1 | Not-determined | Genome-fragment | NA | NA | 0 |
| S8_extracted_k141_160390 | 2110 | No | NA | 4 | 0 | 0 | Not-determined | Genome-fragment | NA | NA | 0 |
| S8_extracted_k141_143528 | 1324 | No | NA | 1 | 0 | 0 | Not-determined | Genome-fragment | NA | NA | 0 |
| S8_extracted_k141_169502 | 3933 | No | NA | 5 | 0 | 4 | Not-determined | Genome-fragment | NA | NA | 0 |
| S8_extracted_k141_39330 | 3860 | No | NA | 3 | 0 | 2 | Not-determined | Genome-fragment | NA | NA | 0 |
| S8_extracted_k141_143534 | 4823 | No | NA | 5 | 0 | 3 | Not-determined | Genome-fragment | NA | NA | 0 |
| S8_extracted_k141_30980 | 15953 | No | NA | 14 | 0 | 5 | Not-determined | Genome-fragment | NA | NA | 0 |
| S5_extracted_k141_62615 | 8691 | No | NA | 15 | 5 | 0 | Low-quality | Genome-fragment | 22.16 | AAI-based (medium-confidence) | 0 |
| S12_vs_k141_152341\|\|full | 8695 | No | NA | 14 | 2 | 0 | Low-quality | Genome-fragment | 19.71 | AAI-based (medium-confidence) | 0 |
| S8_extracted_k141_134606 | 1736 | No | NA | 1 | 0 | 1 | Not-determined | Genome-fragment | NA | NA | 0 |
| S8_extracted_k141_91420 | 2996 | No | NA | 2 | 0 | 1 | Not-determined | Genome-fragment | NA | NA | 0 |
| S8_extracted_k141_126199 | 4389 | No | NA | 4 | 0 | 1 | Not-determined | Genome-fragment | NA | NA | 0 |
| S8_extracted_k141_160417 | 1416 | No | NA | 1 | 0 | 0 | Not-determined | Genome-fragment | NA | NA | 0 |
| S8_extracted_k141_91431 | 2334 | No | NA | 4 | 0 | 3 | Not-determined | Genome-fragment | NA | NA | 0 |
| S8_extracted_k141_151976 | 1028 | No | NA | 2 | 0 | 1 | Not-determined | Genome-fragment | NA | NA | 0 |
| S8_extracted_k141_160419 | 3315 | No | NA | 2 | 0 | 2 | Not-determined | Genome-fragment | NA | NA | 0 |
| S3_extracted_k141_3320 | 8697 | No | NA | 7 | 0 | 7 | Low-quality | Genome-fragment | 7.95 | AAI-based (medium-confidence) | 0 |

| | | | | | | | | | | | |
|---|---|---|---|---|---|---|---|---|---|---|---|
| S8_extracted_k141_126215 | 1018 | No | NA | 1 | 0 | 0 | Not-determined | Genome-fragment | NA | NA | 0 |
| S8_extracted_k141_134631 | 2501 | No | NA | 2 | 0 | 1 | Not-determined | Genome-fragment | NA | NA | 0 |
| S8_extracted_k141_13003 | 1800 | No | NA | 2 | 0 | 1 | Not-determined | Genome-fragment | NA | NA | 0 |
| S8_extracted_k141_31019 | 1118 | No | NA | 1 | 0 | 1 | Not-determined | Genome-fragment | NA | NA | 0 |
| S8_extracted_k141_151997 | 1033 | No | NA | 2 | 0 | 0 | Not-determined | Genome-fragment | NA | NA | 0 |
| S8_extracted_k141_178349 | 7652 | No | NA | 7 | 0 | 2 | Not-determined | Genome-fragment | NA | NA | 0 |
| S8_extracted_k141_91438 | 11225 | No | NA | 6 | 0 | 0 | Not-determined | Genome-fragment | NA | NA | 0 |
| S14_extracted_k141_62252 | 8703 | No | NA | 24 | 21 | 0 | Low-quality | Genome-fragment | 28.66 | AAI-based (high-confidence) | 0 |
| S8_extracted_k141_169547 | 1173 | No | NA | 2 | 0 | 0 | Not-determined | Genome-fragment | NA | NA | 0 |
| S8_extracted_k141_169548 | 1317 | No | NA | 3 | 0 | 1 | Not-determined | Genome-fragment | NA | NA | 0 |
| S8_extracted_k141_22200 | 3882 | No | NA | 8 | 0 | 8 | Not-determined | Genome-fragment | NA | NA | 0 |
| S8_extracted_k141_100172 | 2134 | No | NA | 2 | 0 | 2 | Not-determined | Genome-fragment | NA | NA | 0 |
| S8_extracted_k141_169561 | 3180 | No | NA | 2 | 0 | 2 | Not-determined | Genome-fragment | NA | NA | 0 |
| S11_vs_k141_104050\|\|full | 8709 | No | NA | 11 | 1 | 0 | Low-quality | Genome-fragment | 22.14 | AAI-based (medium-confidence) | 0 |
| S8_extracted_k141_22211 | 2748 | No | NA | 2 | 0 | 2 | Not-determined | Genome-fragment | NA | NA | 0 |
| S8_extracted_k141_169567 | 1524 | No | NA | 3 | 0 | 0 | Not-determined | Genome-fragment | NA | NA | 0 |
| S8_extracted_k141_195757 | 1557 | No | NA | 2 | 0 | 1 | Not-determined | Genome-fragment | NA | NA | 0 |
| S17_vs_k141_190\|\|full | 8714 | No | NA | 7 | 0 | 0 | Low-quality | Genome-fragment | 20.35 | AAI-based (high-confidence) | 0 |
| S8_extracted_k141_178376 | 2583 | No | NA | 1 | 0 | 0 | Not-determined | Genome-fragment | NA | NA | 0 |
| S8_extracted_k141_169576 | 1517 | No | NA | 5 | 0 | 3 | Not-determined | Genome-fragment | NA | NA | 0 |
| S3_vs_k141_49\|\|full | 8730 | No | NA | 13 | 0 | 1 | Low-quality | Genome-fragment | 13.02 | AAI-based (high-confidence) | 0 |
| S7_extracted_128477 | 8730 | No | NA | 9 | 6 | 0 | Low-quality | Genome-fragment | 22.69 | AAI-based (medium-confidence) | 0 |
| S8_extracted_k141_65218 | 2198 | No | NA | 3 | 0 | 3 | Not-determined | Genome-fragment | NA | NA | 0 |
| S8_extracted_k141_31078 | 1850 | No | NA | 2 | 0 | 0 | Not-determined | Genome-fragment | NA | NA | 0 |
| S8_extracted_k141_56917 | 6946 | No | NA | 8 | 0 | 4 | Not-determined | Genome-fragment | NA | NA | 0 |
| S8_extracted_k141_73708 | 1968 | No | NA | 4 | 0 | 0 | Not-determined | Genome-fragment | NA | NA | 0 |
| S8_extracted_k141_186099 | 3044 | No | NA | 3 | 0 | 2 | Not-determined | Genome-fragment | NA | NA | 0 |
| S17_vs_k141_76458\|\|full | 8732 | No | NA | 16 | 1 | 1 | Low-quality | Genome-fragment | 19.96 | AAI-based (medium-confidence) | 0 |
| S5_vs_k141_59217\|\|full | 8735 | Yes | 3433 | 9 | 2 | 2 | Low-quality | Genome-fragment | 1.82 | HMM-based (lower-bound) | 60.7 |
| S8_extracted_k141_203993 | 1843 | No | NA | 2 | 0 | 1 | Not-determined | Genome-fragment | NA | NA | 0 |
| S8_extracted_k141_56930 | 1031 | No | NA | 2 | 0 | 1 | Not-determined | Genome-fragment | NA | NA | 0 |
| S8_extracted_k141_203995 | 2426 | No | NA | 2 | 0 | 1 | Not-determined | Genome-fragment | NA | NA | 0 |
| S8_extracted_k141_134705 | 2437 | No | NA | 2 | 0 | 0 | Not-determined | Genome-fragment | NA | NA | 0 |
| S8_extracted_k141_169605 | 1309 | No | NA | 1 | 0 | 1 | Not-determined | Genome-fragment | NA | NA | 0 |
| S12_vs_k141_92184\|\|full | 8736 | No | NA | 6 | 0 | 5 | Low-quality | Genome-fragment | 4.51 | AAI-based (medium-confidence) | 0 |
| S8_extracted_k141_13076 | 1146 | No | NA | 1 | 0 | 0 | Not-determined | Genome-fragment | NA | NA | 0 |
| S8_extracted_k141_56934 | 3294 | No | NA | 4 | 0 | 2 | Not-determined | Genome-fragment | NA | NA | 0 |
| S8_extracted_k141_152072 | 5203 | No | NA | 3 | 0 | 2 | Not-determined | Genome-fragment | NA | NA | 0 |
| S8_extracted_k141_4665 | 5369 | No | NA | 10 | 0 | 1 | Not-determined | Genome-fragment | NA | NA | 0 |
| S8_extracted_k141_31104 | 1428 | No | NA | 4 | 0 | 0 | Not-determined | Genome-fragment | NA | NA | 0 |
| S8_extracted_k141_22272 | 3952 | No | NA | 7 | 0 | 1 | Not-determined | Genome-fragment | NA | NA | 0 |
| S8_extracted_k141_31106 | 1159 | No | NA | 1 | 0 | 0 | Not-determined | Genome-fragment | NA | NA | 0 |
| S8_extracted_k141_117449 | 3681 | No | NA | 2 | 0 | 2 | Not-determined | Genome-fragment | NA | NA | 0 |
| S8_extracted_k141_117451 | 1005 | No | NA | 2 | 0 | 1 | Not-determined | Genome-fragment | NA | NA | 0 |
| S5_vs_k141_141538\|\|full | 8740 | No | NA | 14 | 4 | 0 | Low-quality | Genome-fragment | 20.34 | AAI-based (high-confidence) | 0 |
| S1_vs_k141_133958\|\|full | 8744 | No | NA | 16 | 1 | 1 | Low-quality | Genome-fragment | 6.04 | AAI-based (high-confidence) | 0 |
| S8_extracted_k141_4675 | 1814 | No | NA | 3 | 0 | 2 | Not-determined | Genome-fragment | NA | NA | 0 |
| S7_vs_19100\|\|full | 8747 | No | NA | 10 | 3 | 0 | Low-quality | Genome-fragment | 28.57 | AAI-based (high-confidence) | 0 |
| S8_extracted_k141_31123 | 1611 | No | NA | 2 | 0 | 1 | Not-determined | Genome-fragment | NA | NA | 0 |
| S8_extracted_k141_4689 | 1215 | No | NA | 1 | 0 | 0 | Not-determined | Genome-fragment | NA | NA | 0 |
| S8_extracted_k141_47759 | 1034 | No | NA | 1 | 0 | 0 | Not-determined | Genome-fragment | NA | NA | 0 |
| S9_vs_k141_640\|\|full | 8751 | No | NA | 14 | 3 | 1 | Low-quality | Genome-fragment | 15.04 | HMM-based (lower-bound) | 0 |
| S8_extracted_k141_204080 | 4467 | No | NA | 5 | 0 | 2 | Not-determined | Genome-fragment | NA | NA | 0 |
| S8_extracted_k141_56981 | 1269 | No | NA | 2 | 0 | 2 | Not-determined | Genome-fragment | NA | NA | 0 |
| S8_extracted_k141_22334 | 9009 | No | NA | 5 | 0 | 3 | Not-determined | Genome-fragment | NA | NA | 0 |
| S8_extracted_k141_117495 | 1050 | No | NA | 1 | 0 | 0 | Not-determined | Genome-fragment | NA | NA | 0 |
| S8_extracted_k141_39447 | 3452 | No | NA | 5 | 0 | 1 | Not-determined | Genome-fragment | NA | NA | 0 |
| S8_extracted_k141_117496 | 2320 | No | NA | 3 | 0 | 0 | Not-determined | Genome-fragment | NA | NA | 0 |
| S7_extracted_123395 | 8760 | No | NA | 16 | 6 | 1 | Low-quality | Genome-fragment | 5.93 | AAI-based (high-confidence) | 0 |
| S13_vs_131409\|\|full | 8764 | No | NA | 16 | 2 | 0 | Low-quality | Genome-fragment | 15.04 | AAI-based (high-confidence) | 0 |
| S16_extracted_108860 | 8767 | No | NA | 13 | 6 | 0 | Low-quality | Genome-fragment | 15.11 | AAI-based (high-confidence) | 0 |
| S8_extracted_k141_195817 | 1789 | No | NA | 1 | 0 | 1 | Not-determined | Genome-fragment | NA | NA | 0 |
| S17_vs_k141_51213\|\|full | 8769 | No | NA | 17 | 1 | 0 | Low-quality | Genome-fragment | 12.94 | AAI-based (high-confidence) | 0 |
| S8_extracted_k141_143715 | 2086 | No | NA | 3 | 0 | 0 | Not-determined | Genome-fragment | NA | NA | 0 |
| S8_extracted_k141_47804 | 1242 | No | NA | 3 | 0 | 0 | Not-determined | Genome-fragment | NA | NA | 0 |
| S8_extracted_k141_73841 | 1075 | No | NA | 2 | 0 | 1 | Not-determined | Genome-fragment | NA | NA | 0 |
| S8_extracted_k141_109004 | 4152 | No | NA | 4 | 0 | 1 | Not-determined | Genome-fragment | NA | NA | 0 |
| S5_vs_k141_121005\|\|full | 8773 | No | NA | 20 | 3 | 1 | Low-quality | Genome-fragment | 18.2 | AAI-based (high-confidence) | 0 |
| S8_extracted_k141_82752 | 1811 | No | NA | 2 | 0 | 1 | Not-determined | Genome-fragment | NA | NA | 0 |
| S8_extracted_k141_143741 | 2477 | No | NA | 4 | 0 | 1 | Not-determined | Genome-fragment | NA | NA | 0 |
| S8_extracted_k141_169662 | 1265 | No | NA | 1 | 0 | 0 | Not-determined | Genome-fragment | NA | NA | 0 |
| S12_vs_k141_69133\|\|full | 8775 | No | NA | 3 | 0 | 3 | Low-quality | Genome-fragment | 5.17 | AAI-based (high-confidence) | 0 |
| S10_vs_65523\|\|full | 8778 | No | NA | 6 | 0 | 2 | Low-quality | Genome-fragment | 9.57 | AAI-based (medium-confidence) | 0 |
| S16_extracted_43246 | 8794 | No | NA | 15 | 12 | 0 | Low-quality | Genome-fragment | 19.65 | AAI-based (high-confidence) | 0 |
| S8_extracted_k141_20897 | 8797 | No | NA | 8 | 2 | 0 | Low-quality | Genome-fragment | 22.27 | AAI-based (high-confidence) | 0 |
| S8_extracted_k141_47836 | 1587 | No | NA | 4 | 0 | 0 | Not-determined | Genome-fragment | NA | NA | 0 |
| S8_extracted_k141_22390 | 1496 | No | NA | 1 | 0 | 0 | Not-determined | Genome-fragment | NA | NA | 0 |
| S8_extracted_k141_169679 | 1022 | No | NA | 2 | 0 | 0 | Not-determined | Genome-fragment | NA | NA | 0 |
| S8_extracted_k141_100339 | 1630 | No | NA | 2 | 0 | 2 | Not-determined | Genome-fragment | NA | NA | 0 |
| S8_extracted_k141_91524 | 1245 | No | NA | 3 | 0 | 1 | Not-determined | Genome-fragment | NA | NA | 0 |
| S8_extracted_k141_126321 | 1798 | No | NA | 4 | 0 | 2 | Not-determined | Genome-fragment | NA | NA | 0 |
| S8_extracted_k141_143768 | 2532 | No | NA | 2 | 0 | 1 | Not-determined | Genome-fragment | NA | NA | 0 |
| S8_extracted_k141_195877 | 1307 | No | NA | 1 | 0 | 1 | Not-determined | Genome-fragment | NA | NA | 0 |
| S8_extracted_k141_22409 | 1069 | No | NA | 1 | 0 | 1 | Not-determined | Genome-fragment | NA | NA | 0 |
| S8_extracted_k141_39505 | 5953 | No | NA | 6 | 0 | 5 | Not-determined | Genome-fragment | NA | NA | 0 |
| S3_extracted_k141_36390 | 8799 | No | NA | 12 | 4 | 1 | Low-quality | Genome-fragment | 21.79 | AAI-based (medium-confidence) | 0 |
| S8_extracted_k141_31215 | 6943 | No | NA | 4 | 0 | 3 | Not-determined | Genome-fragment | NA | NA | 0 |
| S8_extracted_k141_178430 | 1046 | No | NA | 2 | 0 | 2 | Not-determined | Genome-fragment | NA | NA | 0 |
| S8_extracted_k141_204153 | 2360 | No | NA | 3 | 0 | 0 | Not-determined | Genome-fragment | NA | NA | 0 |
| S8_extracted_k141_39524 | 1047 | No | NA | 2 | 0 | 1 | Not-determined | Genome-fragment | NA | NA | 0 |
| S8_extracted_k141_117606 | 1003 | No | NA | 1 | 0 | 0 | Not-determined | Genome-fragment | NA | NA | 0 |
| S8_extracted_k141_143820 | 1780 | No | NA | 1 | 0 | 0 | Not-determined | Genome-fragment | NA | NA | 0 |
| S8_extracted_k141_134807 | 3695 | No | NA | 2 | 0 | 2 | Not-determined | Genome-fragment | NA | NA | 0 |
| S13_vs_98911\|\|full | 8801 | No | NA | 7 | 0 | 7 | Low-quality | Genome-fragment | 3.04 | AAI-based (medium-confidence) | 0 |
| S6_vs_k141_23658\|\|full | 8809 | No | NA | 21 | 3 | 0 | Low-quality | Genome-fragment | 16.95 | AAI-based (medium-confidence) | 0 |
| S8_extracted_k141_57118 | 1031 | No | NA | 1 | 0 | 1 | Not-determined | Genome-fragment | NA | NA | 0 |
| S8_extracted_k141_22463 | 5809 | No | NA | 3 | 0 | 1 | Not-determined | Genome-fragment | NA | NA | 0 |
| S8_extracted_k141_39553 | 1694 | No | NA | 3 | 0 | 1 | Not-determined | Genome-fragment | NA | NA | 0 |
| S8_extracted_k141_65422 | 1496 | No | NA | 2 | 0 | 0 | Not-determined | Genome-fragment | NA | NA | 0 |
| S8_extracted_k141_204176 | 2339 | No | NA | 3 | 0 | 0 | Not-determined | Genome-fragment | NA | NA | 0 |
| S8_extracted_k141_117624 | 3693 | No | NA | 4 | 0 | 4 | Not-determined | Genome-fragment | NA | NA | 0 |
| S17_extracted_k141_112931 | 8810 | Yes | 4874 | 20 | 3 | 2 | Low-quality | Genome-fragment | 6.05 | AAI-based (high-confidence) | 44.68 |
| S15_vs_33756\|\|full | 8811 | No | NA | 9 | 0 | 7 | Low-quality | Genome-fragment | 4.19 | AAI-based (medium-confidence) | 0 |
| S11_vs_k141_23228\|\|full | 8818 | No | NA | 9 | 1 | 1 | Low-quality | Genome-fragment | 8.52 | AAI-based (high-confidence) | 0 |
| S8_extracted_k141_152225 | 1431 | No | NA | 4 | 0 | 0 | Not-determined | Genome-fragment | NA | NA | 0 |
| S8_extracted_k141_143848 | 4656 | No | NA | 7 | 0 | 6 | Not-determined | Genome-fragment | NA | NA | 0 |
| S12_extracted_k141_103204 | 8819 | No | NA | 15 | 2 | 0 | Low-quality | Genome-fragment | 8.08 | AAI-based (high-confidence) | 0 |
| S8_extracted_k141_91618 | 1139 | No | NA | 1 | 0 | 1 | Not-determined | Genome-fragment | NA | NA | 0 |
| S8_extracted_k141_186148 | 12771 | No | NA | 9 | 0 | 3 | Not-determined | Genome-fragment | NA | NA | 0 |
| S8_extracted_k141_117637 | 5801 | No | NA | 2 | 0 | 2 | Not-determined | Genome-fragment | NA | NA | 0 |
| S8_extracted_k141_152229 | 1020 | No | NA | 1 | 0 | 1 | Not-determined | Genome-fragment | NA | NA | 0 |
| S3_vs_k141_61359\|\|full | 8827 | No | NA | 12 | 7 | 0 | Low-quality | Genome-fragment | 26.09 | AAI-based (high-confidence) | 0 |
| S8_extracted_k141_195977 | 1364 | No | NA | 1 | 0 | 1 | Not-determined | Genome-fragment | NA | NA | 0 |
| S8_extracted_k141_57155 | 1192 | No | NA | 2 | 0 | 0 | Not-determined | Genome-fragment | NA | NA | 0 |
| S8_extracted_k141_204200 | 1205 | No | NA | 2 | 0 | 2 | Not-determined | Genome-fragment | NA | NA | 0 |
| S8_extracted_k141_195984 | 2250 | No | NA | 3 | 0 | 0 | Not-determined | Genome-fragment | NA | NA | 0 |
| S8_extracted_k141_13223 | 1869 | No | NA | 3 | 0 | 0 | Not-determined | Genome-fragment | NA | NA | 0 |
| S8_extracted_k141_65472 | 1517 | No | NA | 2 | 0 | 0 | Not-determined | Genome-fragment | NA | NA | 0 |
| S10_extracted_80002 | 8827 | Yes | 6917 | 6 | 2 | 1 | Low-quality | Genome-fragment | 12.7 | AAI-based (high-confidence) | 21.64 |
| S8_extracted_k141_178493 | 1370 | No | NA | 2 | 0 | 0 | Not-determined | Genome-fragment | NA | NA | 0 |
| S8_extracted_k141_73917 | 1666 | No | NA | 1 | 0 | 1 | Not-determined | Genome-fragment | NA | NA | 0 |
| S8_extracted_k141_109128 | 1183 | No | NA | 1 | 0 | 1 | Not-determined | Genome-fragment | NA | NA | 0 |
| S8_extracted_k141_91654 | 2962 | No | NA | 2 | 0 | 2 | Not-determined | Genome-fragment | NA | NA | 0 |
| S8_extracted_k141_126443 | 1856 | No | NA | 3 | 0 | 2 | Not-determined | Genome-fragment | NA | NA | 0 |
| S8_extracted_k141_13244 | 2296 | No | NA | 1 | 0 | 0 | Not-determined | Genome-fragment | NA | NA | 0 |
| S8_extracted_k141_169819 | 1054 | No | NA | 3 | 0 | 1 | Not-determined | Genome-fragment | NA | NA | 0 |
| S8_extracted_k141_178502 | 1328 | No | NA | 1 | 0 | 1 | Not-determined | Genome-fragment | NA | NA | 0 |
| S7_vs_99784\|\|full | 8836 | No | NA | 6 | 5 | 0 | Low-quality | Genome-fragment | 22.46 | AAI-based (medium-confidence) | 0 |
| S8_extracted_k141_22536 | 1436 | No | NA | 2 | 0 | 1 | Not-determined | Genome-fragment | NA | NA | 0 |
| S8_extracted_k141_82863 | 2897 | No | NA | 2 | 0 | 2 | Not-determined | Genome-fragment | NA | NA | 0 |
| S8_extracted_k141_4870 | 1130 | No | NA | 1 | 0 | 1 | Not-determined | Genome-fragment | NA | NA | 0 |
| S16_vs_164218\|\|full | 8860 | No | NA | 7 | 0 | 1 | Low-quality | Genome-fragment | 25.55 | AAI-based (medium-confidence) | 0 |
| S11_vs_k141_98501\|\|full | 8861 | No | NA | 16 | 1 | 4 | Low-quality | Genome-fragment | 16.59 | AAI-based (high-confidence) | 0 |
| S8_extracted_k141_47908 | 2595 | No | NA | 2 | 0 | 2 | Not-determined | Genome-fragment | NA | NA | 0 |
| S8_extracted_k141_31342 | 3862 | No | NA | 3 | 0 | 0 | Not-determined | Genome-fragment | NA | NA | 0 |
| S8_extracted_k141_160652 | 3311 | No | NA | 4 | 0 | 3 | Not-determined | Genome-fragment | NA | NA | 0 |
| S8_extracted_k141_57226 | 1557 | No | NA | 3 | 0 | 1 | Not-determined | Genome-fragment | NA | NA | 0 |
| S8_extracted_k141_126479 | 1211 | No | NA | 1 | 0 | 0 | Not-determined | Genome-fragment | NA | NA | 0 |
| S8_extracted_k141_82899 | 3389 | No | NA | 4 | 0 | 0 | Not-determined | Genome-fragment | NA | NA | 0 |
| S17_extracted_k141_160143 | 8864 | No | NA | 10 | 0 | 3 | Low-quality | Genome-fragment | 15.66 | AAI-based (medium-confidence) | 0 |
| S8_extracted_k141_82914 | 1311 | No | NA | 2 | 0 | 1 | Not-determined | Genome-fragment | NA | NA | 0 |
| S8_extracted_k141_109209 | 1280 | No | NA | 4 | 0 | 0 | Not-determined | Genome-fragment | NA | NA | 0 |
| S3_vs_k141_31147\|\|full | 8866 | Yes | 7267 | 9 | 1 | 2 | Low-quality | Genome-fragment | 5.24 | HMM-based (lower-bound) | 18.04 |
| S8_extracted_k141_143978 | 1001 | No | NA | 2 | 0 | 0 | Not-determined | Genome-fragment | NA | NA | 0 |
| S8_extracted_k141_178594 | 4369 | No | NA | 7 | 0 | 3 | Not-determined | Genome-fragment | NA | NA | 0 |
| S17_vs_k141_30171\|\|full | 8872 | No | NA | 10 | 0 | 0 | Low-quality | Genome-fragment | 12.19 | AAI-based (medium-confidence) | 0 |
| S8_extracted_k141_91749 | 2888 | No | NA | 3 | 0 | 3 | Not-determined | Genome-fragment | NA | NA | 0 |
| S8_extracted_k141_186231 | 1351 | No | NA | 1 | 0 | 0 | Not-determined | Genome-fragment | NA | NA | 0 |
| S8_extracted_k141_57272 | 2126 | No | NA | 1 | 0 | 1 | Not-determined | Genome-fragment | NA | NA | 0 |
| S8_extracted_k141_178603 | 1036 | No | NA | 1 | 0 | 0 | Not-determined | Genome-fragment | NA | NA | 0 |
| S8_extracted_k141_22567 | 4200 | No | NA | 4 | 0 | 2 | Not-determined | Genome-fragment | NA | NA | 0 |
| S3_vs_k141_104507\|\|full | 8883 | No | NA | 7 | 6 | 0 | Low-quality | Genome-fragment | 4.78 | AAI-based (high-confidence) | 0 |
| S8_extracted_k141_143988 | 2543 | No | NA | 4 | 0 | 3 | Not-determined | Genome-fragment | NA | NA | 0 |
| S8_extracted_k141_65522 | 28993 | No | NA | 36 | 14 | 0 | Medium-quality | Genome-fragment | 87.82 | AAI-based (high-confidence) | 0 |
| S8_extracted_k141_31398 | 1239 | No | NA | 1 | 0 | 0 | Not-determined | Genome-fragment | NA | NA | 0 |
| S8_vs_k141_118210\|\|full | 8885 | No | NA | 11 | 4 | 0 | Low-quality | Genome-fragment | 21.08 | AAI-based (high-confidence) | 0 |
| S8_extracted_k141_109241 | 5251 | No | NA | 5 | 0 | 2 | Not-determined | Genome-fragment | NA | NA | 0 |
| S8_extracted_k141_186264 | 2200 | No | NA | 1 | 0 | 1 | Not-determined | Genome-fragment | NA | NA | 0 |
| S5_vs_k141_9010\|\|full | 8887 | No | NA | 12 | 6 | 0 | Low-quality | Genome-fragment | 18.82 | AAI-based (high-confidence) | 0 |
| S8_extracted_k141_109247 | 1899 | No | NA | 4 | 0 | 3 | Not-determined | Genome-fragment | NA | NA | 0 |
| S8_extracted_k141_179846 | 8890 | No | NA | 12 | 8 | 0 | Low-quality | Genome-fragment | 5.61 | AAI-based (high-confidence) | 0 |
| S8_extracted_k141_47950 | 3830 | No | NA | 5 | 0 | 3 | Not-determined | Genome-fragment | NA | NA | 0 |
| S8_extracted_k141_204315 | 1170 | No | NA | 2 | 0 | 1 | Not-determined | Genome-fragment | NA | NA | 0 |
| S8_extracted_k141_160697 | 1109 | No | NA | 1 | 0 | 1 | Not-determined | Genome-fragment | NA | NA | 0 |
| S6_vs_k141_28377\|\|full | 8896 | No | NA | 18 | 2 | 0 | Low-quality | Genome-fragment | 22.19 | AAI-based (high-confidence) | 0 |
| S8_extracted_k141_73992 | 6457 | No | NA | 6 | 0 | 4 | Not-determined | Genome-fragment | NA | NA | 0 |
| S8_extracted_k141_91814 | 1027 | No | NA | 4 | 0 | 0 | Not-determined | Genome-fragment | NA | NA | 0 |
| S8_extracted_k141_47964 | 3103 | No | NA | 2 | 0 | 0 | Not-determined | Genome-fragment | NA | NA | 0 |
| S8_extracted_k141_178645 | 1034 | No | NA | 1 | 0 | 1 | Not-determined | Genome-fragment | NA | NA | 0 |
| S8_extracted_k141_65553 | 1076 | No | NA | 1 | 0 | 1 | Not-determined | Genome-fragment | NA | NA | 0 |
| S14_vs_k141_3598\|\|full | 8898 | No | NA | 10 | 3 | 0 | Low-quality | Genome-fragment | 14.91 | AAI-based (high-confidence) | 0 |
| S12_vs_k141_2064\|\|full | 8898 | No | NA | 13 | 10 | 0 | Low-quality | Genome-fragment | 24.53 | AAI-based (high-confidence) | 0 |
| S8_extracted_k141_134943 | 64005 | No | NA | 90 | 8 | 0 | High-quality | High-quality | 100 | AAI-based (high-confidence) | 0 |
| S17_vs_k141_152459\|\|full | 8901 | No | NA | 12 | 11 | 0 | Low-quality | Genome-fragment | 26.48 | AAI-based (high-confidence) | 0 |
| S8_extracted_k141_22650 | 1122 | No | NA | 1 | 0 | 1 | Not-determined | Genome-fragment | NA | NA | 0 |
| S8_extracted_k141_47994 | 1008 | No | NA | 1 | 0 | 0 | Not-determined | Genome-fragment | NA | NA | 0 |
| S8_extracted_k141_126538 | 5632 | No | NA | 8 | 0 | 0 | Not-determined | Genome-fragment | NA | NA | 0 |
| S8_extracted_k141_178673 | 1042 | No | NA | 1 | 0 | 0 | Not-determined | Genome-fragment | NA | NA | 0 |
| S8_extracted_k141_22654 | 4736 | No | NA | 6 | 0 | 1 | Not-determined | Genome-fragment | NA | NA | 0 |
| S8_extracted_k141_57339 | 5604 | No | NA | 9 | 0 | 1 | Not-determined | Genome-fragment | NA | NA | 0 |
| S8_extracted_k141_109299 | 1411 | No | NA | 1 | 0 | 1 | Not-determined | Genome-fragment | NA | NA | 0 |
| S8_extracted_k141_152436 | 1658 | No | NA | 1 | 0 | 1 | Not-determined | Genome-fragment | NA | NA | 0 |
| S8_extracted_k141_117796 | 42349 | No | NA | 40 | 9 | 2 | High-quality | High-quality | 98.87 | AAI-based (medium-confidence) | 0 |
| S8_extracted_k141_160734 | 2647 | No | NA | 3 | 0 | 0 | Not-determined | Genome-fragment | NA | NA | 0 |
| S8_extracted_k141_22698 | 1937 | No | NA | 2 | 0 | 1 | Not-determined | Genome-fragment | NA | NA | 0 |
| S8_extracted_k141_74039 | 1007 | No | NA | 2 | 0 | 1 | Not-determined | Genome-fragment | NA | NA | 0 |
| S8_extracted_k141_83047 | 5339 | No | NA | 4 | 0 | 2 | Not-determined | Genome-fragment | NA | NA | 0 |
| S8_extracted_k141_204364 | 1183 | No | NA | 1 | 0 | 0 | Not-determined | Genome-fragment | NA | NA | 0 |
| S8_extracted_k141_196133 | 1640 | No | NA | 3 | 0 | 1 | Not-determined | Genome-fragment | NA | NA | 0 |
| S8_extracted_k141_186399 | 1321 | No | NA | 1 | 0 | 1 | Not-determined | Genome-fragment | NA | NA | 0 |
| S8_extracted_k141_109328 | 1089 | No | NA | 1 | 0 | 1 | Not-determined | Genome-fragment | NA | NA | 0 |
| S8_extracted_k141_178691 | 1055 | No | NA | 2 | 0 | 1 | Not-determined | Genome-fragment | NA | NA | 0 |
| S8_extracted_k141_65633 | 5661 | No | NA | 8 | 0 | 6 | Not-determined | Genome-fragment | NA | NA | 0 |
| S8_extracted_k141_74063 | 1943 | No | NA | 1 | 0 | 1 | Not-determined | Genome-fragment | NA | NA | 0 |
| S8_extracted_k141_65635 | 3151 | No | NA | 1 | 0 | 1 | Not-determined | Genome-fragment | NA | NA | 0 |
| S8_extracted_k141_152494 | 1380 | No | NA | 3 | 0 | 2 | Not-determined | Genome-fragment | NA | NA | 0 |
| S8_vs_k141_148361\|\|full | 8903 | No | NA | 7 | 0 | 5 | Low-quality | Genome-fragment | 8.54 | AAI-based (medium-confidence) | 0 |
| S8_extracted_k141_152498 | 3103 | No | NA | 5 | 0 | 3 | Not-determined | Genome-fragment | NA | NA | 0 |
| S8_extracted_k141_186431 | 2735 | No | NA | 2 | 0 | 2 | Not-determined | Genome-fragment | NA | NA | 0 |
| S8_extracted_k141_57412 | 1729 | No | NA | 1 | 0 | 1 | Not-determined | Genome-fragment | NA | NA | 0 |
| S8_extracted_k141_83106 | 1065 | No | NA | 2 | 0 | 2 | Not-determined | Genome-fragment | NA | NA | 0 |
| S8_extracted_k141_152507 | 1460 | No | NA | 2 | 0 | 1 | Not-determined | Genome-fragment | NA | NA | 0 |
| S8_extracted_k141_48073 | 2123 | No | NA | 4 | 0 | 1 | Not-determined | Genome-fragment | NA | NA | 0 |
| S8_extracted_k141_152508 | 2065 | No | NA | 1 | 0 | 0 | Not-determined | Genome-fragment | NA | NA | 0 |
| S5_vs_k141_100206\|\|full | 8905 | No | NA | 11 | 0 | 2 | Low-quality | Genome-fragment | 4.24 | AAI-based (medium-confidence) | 0 |
| S8_extracted_k141_152512 | 1531 | No | NA | 3 | 0 | 2 | Not-determined | Genome-fragment | NA | NA | 0 |
| S8_extracted_k141_186457 | 1271 | No | NA | 2 | 0 | 1 | Not-determined | Genome-fragment | NA | NA | 0 |
| S8_extracted_k141_117846 | 1489 | No | NA | 3 | 0 | 2 | Not-determined | Genome-fragment | NA | NA | 0 |
| S8_extracted_k141_48094 | 1731 | No | NA | 1 | 0 | 1 | Not-determined | Genome-fragment | NA | NA | 0 |
| S8_extracted_k141_39928 | 1302 | No | NA | 1 | 0 | 0 | Not-determined | Genome-fragment | NA | NA | 0 |
| S8_extracted_k141_152542 | 3689 | No | NA | 4 | 0 | 2 | Not-determined | Genome-fragment | NA | NA | 0 |
| S2_vs_k141_19187\|\|full | 8905 | No | NA | 10 | 3 | 0 | Low-quality | Genome-fragment | 27.1 | HMM-based (lower-bound) | 0 |
| S8_extracted_k141_83131 | 3836 | No | NA | 5 | 0 | 5 | Not-determined | Genome-fragment | NA | NA | 0 |
| S8_extracted_k141_91840 | 3469 | No | NA | 1 | 0 | 0 | Not-determined | Genome-fragment | NA | NA | 0 |
| S8_extracted_k141_39945 | 1981 | No | NA | 1 | 0 | 1 | Not-determined | Genome-fragment | NA | NA | 0 |
| S8_extracted_k141_117884 | 2020 | No | NA | 4 | 0 | 3 | Not-determined | Genome-fragment | NA | NA | 0 |
| S8_extracted_k141_83151 | 1569 | No | NA | 1 | 0 | 1 | Not-determined | Genome-fragment | NA | NA | 0 |
| S8_extracted_k141_13564 | 4058 | No | NA | 4 | 0 | 2 | Not-determined | Genome-fragment | NA | NA | 0 |
| S8_extracted_k141_91867 | 1065 | No | NA | 1 | 0 | 1 | Not-determined | Genome-fragment | NA | NA | 0 |
| S8_extracted_k141_91872 | 1454 | No | NA | 2 | 0 | 2 | Not-determined | Genome-fragment | NA | NA | 0 |
| S11_extracted_k141_2645 | 8910 | No | NA | 11 | 2 | 0 | Low-quality | Genome-fragment | 5.47 | AAI-based (high-confidence) | 0 |
| S8_extracted_k141_22818 | 3102 | No | NA | 4 | 0 | 2 | Not-determined | Genome-fragment | NA | NA | 0 |
| S8_extracted_k141_135048 | 1756 | No | NA | 2 | 0 | 1 | Not-determined | Genome-fragment | NA | NA | 0 |
| S14_vs_k141_64107\|\|full | 8910 | No | NA | 7 | 0 | 5 | Low-quality | Genome-fragment | 5.09 | AAI-based (medium-confidence) | 0 |
| S8_extracted_k141_160859 | 3179 | No | NA | 4 | 0 | 1 | Not-determined | Genome-fragment | NA | NA | 0 |
| S8_extracted_k141_135060 | 3656 | No | NA | 4 | 0 | 3 | Not-determined | Genome-fragment | NA | NA | 0 |
| S8_extracted_k141_40008 | 1492 | No | NA | 2 | 0 | 1 | Not-determined | Genome-fragment | NA | NA | 0 |
| S8_extracted_k141_65734 | 1112 | No | NA | 2 | 0 | 0 | Not-determined | Genome-fragment | NA | NA | 0 |
| S8_extracted_k141_91912 | 4836 | No | NA | 4 | 0 | 2 | Not-determined | Genome-fragment | NA | NA | 0 |
| S13_vs_99506\|\|full | 8920 | No | NA | 12 | 3 | 0 | Low-quality | Genome-fragment | 30.21 | HMM-based (lower-bound) | 0 |
| S8_extracted_k141_135076 | 2969 | No | NA | 4 | 0 | 4 | Not-determined | Genome-fragment | NA | NA | 0 |
| S8_extracted_k141_170178 | 1471 | No | NA | 2 | 0 | 1 | Not-determined | Genome-fragment | NA | NA | 0 |
| S8_extracted_k141_178798 | 1508 | No | NA | 1 | 0 | 1 | Not-determined | Genome-fragment | NA | NA | 0 |
| S8_extracted_k141_91949 | 3069 | No | NA | 4 | 0 | 0 | Not-determined | Genome-fragment | NA | NA | 0 |
| S16_vs_105884\|\|full | 8928 | No | NA | 6 | 0 | 5 | Low-quality | Genome-fragment | 6.05 | AAI-based (medium-confidence) | 0 |
| S8_extracted_k141_170209 | 1117 | No | NA | 2 | 0 | 0 | Not-determined | Genome-fragment | NA | NA | 0 |
| S8_extracted_k141_160918 | 1897 | No | NA | 2 | 0 | 2 | Not-determined | Genome-fragment | NA | NA | 0 |
| S5_extracted_k141_104932 | 8932 | No | NA | 10 | 0 | 9 | Low-quality | Genome-fragment | 22.8 | AAI-based (medium-confidence) | 0 |
| S8_extracted_k141_74118 | 1643 | No | NA | 1 | 0 | 1 | Not-determined | Genome-fragment | NA | NA | 0 |
| S8_extracted_k141_160923 | 2438 | No | NA | 1 | 0 | 1 | Not-determined | Genome-fragment | NA | NA | 0 |
| S8_extracted_k141_57536 | 4987 | No | NA | 5 | 0 | 4 | Not-determined | Genome-fragment | NA | NA | 0 |
| S15_extracted_25823 | 8942 | No | NA | 21 | 0 | 0 | Low-quality | Genome-fragment | 4.74 | AAI-based (high-confidence) | 0 |
| S8_extracted_k141_196281 | 1344 | No | NA | 5 | 0 | 5 | Not-determined | Genome-fragment | NA | NA | 0 |
| S8_extracted_k141_126602 | 7681 | No | NA | 5 | 0 | 4 | Not-determined | Genome-fragment | NA | NA | 0 |
| S8_extracted_k141_22913 | 1042 | No | NA | 2 | 0 | 1 | Not-determined | Genome-fragment | NA | NA | 0 |
| S8_extracted_k141_152644 | 1167 | No | NA | 1 | 0 | 0 | Not-determined | Genome-fragment | NA | NA | 0 |
| S8_extracted_k141_178850 | 3850 | No | NA | 5 | 0 | 3 | Not-determined | Genome-fragment | NA | NA | 0 |
| S3_vs_k141_35785\|\|full | 8947 | No | NA | 17 | 1 | 0 | Low-quality | Genome-fragment | 6.69 | AAI-based (high-confidence) | 0 |
| S8_extracted_k141_186580 | 2498 | No | NA | 2 | 0 | 0 | Not-determined | Genome-fragment | NA | NA | 0 |
| S12_vs_k141_171012\|\|full | 8947 | No | NA | 15 | 6 | 0 | Low-quality | Genome-fragment | 23.61 | AAI-based (high-confidence) | 0 |
| S8_extracted_k141_109494 | 1658 | No | NA | 4 | 0 | 0 | Not-determined | Genome-fragment | NA | NA | 0 |
| S8_extracted_k141_152646 | 1021 | No | NA | 1 | 0 | 1 | Not-determined | Genome-fragment | NA | NA | 0 |
| S8_extracted_k141_40056 | 1192 | No | NA | 2 | 0 | 1 | Not-determined | Genome-fragment | NA | NA | 0 |
| S8_extracted_k141_152652 | 1759 | No | NA | 3 | 0 | 3 | Not-determined | Genome-fragment | NA | NA | 0 |
| S3_vs_k141_79907\|\|full | 8956 | No | NA | 15 | 3 | 0 | Low-quality | Genome-fragment | 4.35 | AAI-based (high-confidence) | 0 |
| S8_extracted_k141_186587 | 8658 | No | NA | 12 | 0 | 2 | Not-determined | Genome-fragment | NA | NA | 0 |
| S8_extracted_k141_178862 | 5719 | No | NA | 6 | 0 | 3 | Not-determined | Genome-fragment | NA | NA | 0 |
| S8_extracted_k141_31476 | 1203 | No | NA | 1 | 0 | 1 | Not-determined | Genome-fragment | NA | NA | 0 |
| S8_extracted_k141_160958 | 2177 | No | NA | 2 | 0 | 2 | Not-determined | Genome-fragment | NA | NA | 0 |
| S8_extracted_k141_126638 | 1778 | No | NA | 2 | 0 | 2 | Not-determined | Genome-fragment | NA | NA | 0 |
| S8_extracted_k141_83295 | 1907 | No | NA | 1 | 0 | 1 | Not-determined | Genome-fragment | NA | NA | 0 |
| S3_vs_k141_109358\|\|full | 8956 | No | NA | 18 | 1 | 0 | Low-quality | Genome-fragment | 16.44 | AAI-based (high-confidence) | 0 |
| S5_vs_k141_35195\|\|full | 8956 | No | NA | 5 | 0 | 1 | Low-quality | Genome-fragment | 16.83 | AAI-based (medium-confidence) | 0 |
| S8_extracted_k141_170266 | 1445 | No | NA | 1 | 0 | 0 | Not-determined | Genome-fragment | NA | NA | 0 |
| S8_extracted_k141_152674 | 1852 | No | NA | 2 | 0 | 0 | Not-determined | Genome-fragment | NA | NA | 0 |
| S7_vs_33361\|\|full | 8966 | No | NA | 9 | 1 | 3 | Low-quality | Genome-fragment | 5.54 | HMM-based (lower-bound) | 0 |
| S8_extracted_k141_160972 | 1040 | No | NA | 1 | 0 | 0 | Not-determined | Genome-fragment | NA | NA | 0 |
| S8_extracted_k141_92006 | 1029 | No | NA | 2 | 0 | 0 | Not-determined | Genome-fragment | NA | NA | 0 |
| S8_extracted_k141_74170 | 1185 | No | NA | 1 | 0 | 0 | Not-determined | Genome-fragment | NA | NA | 0 |
| S8_extracted_k141_152679 | 1318 | No | NA | 1 | 0 | 0 | Not-determined | Genome-fragment | NA | NA | 0 |
| S8_extracted_k141_92007 | 1614 | No | NA | 2 | 0 | 0 | Not-determined | Genome-fragment | NA | NA | 0 |
| S8_extracted_k141_40074 | 1115 | No | NA | 1 | 0 | 1 | Not-determined | Genome-fragment | NA | NA | 0 |
| S8_extracted_k141_170277 | 1172 | No | NA | 1 | 0 | 1 | Not-determined | Genome-fragment | NA | NA | 0 |
| S8_extracted_k141_135179 | 4846 | No | NA | 4 | 0 | 2 | Not-determined | Genome-fragment | NA | NA | 0 |
| S8_extracted_k141_83319 | 1116 | No | NA | 3 | 0 | 3 | Not-determined | Genome-fragment | NA | NA | 0 |
| S8_extracted_k141_204525 | 2886 | No | NA | 3 | 0 | 1 | Not-determined | Genome-fragment | NA | NA | 0 |
| S6_vs_k141_6339\|\|full | 8966 | No | NA | 13 | 9 | 0 | Low-quality | Genome-fragment | 23.68 | AAI-based (high-confidence) | 0 |
| S8_extracted_k141_65817 | 4667 | No | NA | 3 | 0 | 1 | Not-determined | Genome-fragment | NA | NA | 0 |
| S8_extracted_k141_83324 | 3300 | No | NA | 4 | 0 | 4 | Not-determined | Genome-fragment | NA | NA | 0 |
| S5_extracted_k141_93003 | 8967 | No | NA | 15 | 6 | 0 | Low-quality | Genome-fragment | 17.95 | AAI-based (high-confidence) | 0 |
| S4_extracted_4323 | 8968 | No | NA | 22 | 16 | 0 | Low-quality | Genome-fragment | 29.52 | AAI-based (high-confidence) | 0 |
| S8_extracted_k141_126681 | 2267 | No | NA | 2 | 0 | 2 | Not-determined | Genome-fragment | NA | NA | 0 |
| S8_extracted_k141_5187 | 2541 | No | NA | 3 | 0 | 2 | Not-determined | Genome-fragment | NA | NA | 0 |
| S8_extracted_k141_74202 | 3528 | No | NA | 3 | 0 | 1 | Not-determined | Genome-fragment | NA | NA | 0 |
| S8_extracted_k141_48229 | 1520 | No | NA | 2 | 0 | 0 | Not-determined | Genome-fragment | NA | NA | 0 |
| S8_extracted_k141_74227 | 2668 | No | NA | 3 | 0 | 2 | Not-determined | Genome-fragment | NA | NA | 0 |
| S8_extracted_k141_48237 | 5037 | No | NA | 5 | 0 | 4 | Not-determined | Genome-fragment | NA | NA | 0 |
| S8_extracted_k141_170301 | 1588 | No | NA | 1 | 0 | 0 | Not-determined | Genome-fragment | NA | NA | 0 |
| S7_vs_111553\|\|full | 8978 | No | NA | 22 | 7 | 0 | Low-quality | Genome-fragment | 20.21 | AAI-based (medium-confidence) | 0 |
| S7_extracted_9946 | 8980 | No | NA | 17 | 4 | 0 | Low-quality | Genome-fragment | 40.09 | AAI-based (medium-confidence) | 0 |
| S8_extracted_k141_13709 | 36057 | No | NA | 66 | 19 | 1 | Complete | High-quality | 100 | DTR (high-confidence) | 0 |
| S13_vs_13552\|\|full | 8984 | No | NA | 9 | 0 | 2 | Low-quality | Genome-fragment | 7.04 | AAI-based (medium-confidence) | 0 |
| S8_extracted_k141_48244 | 1260 | No | NA | 1 | 0 | 0 | Not-determined | Genome-fragment | NA | NA | 0 |
| S8_extracted_k141_170320 | 1407 | No | NA | 2 | 0 | 0 | Not-determined | Genome-fragment | NA | NA | 0 |
| S8_vs_k141_124130\|\|full | 8987 | No | NA | 10 | 1 | 0 | Low-quality | Genome-fragment | 6.04 | HMM-based (lower-bound) | 0 |
| S17_extracted_k141_129865 | 8988 | No | NA | 17 | 16 | 0 | Low-quality | Genome-fragment | 29.65 | AAI-based (high-confidence) | 0 |
| S8_extracted_k141_31543 | 1384 | No | NA | 1 | 0 | 0 | Not-determined | Genome-fragment | NA | NA | 0 |
| S8_extracted_k141_83372 | 3655 | No | NA | 4 | 0 | 3 | Not-determined | Genome-fragment | NA | NA | 0 |
| S8_extracted_k141_135209 | 1621 | No | NA | 1 | 0 | 0 | Not-determined | Genome-fragment | NA | NA | 0 |
| S16_vs_74925\|\|full | 8989 | No | NA | 11 | 1 | 0 | Low-quality | Genome-fragment | 6.08 | HMM-based (lower-bound) | 0 |
| S8_extracted_k141_135212 | 1939 | No | NA | 2 | 0 | 2 | Not-determined | Genome-fragment | NA | NA | 0 |
| S8_extracted_k141_109586 | 1753 | No | NA | 2 | 0 | 1 | Not-determined | Genome-fragment | NA | NA | 0 |
| S8_extracted_k141_126752 | 3292 | No | NA | 2 | 0 | 1 | Not-determined | Genome-fragment | NA | NA | 0 |
| S8_extracted_k141_170358 | 1694 | No | NA | 2 | 0 | 2 | Not-determined | Genome-fragment | NA | NA | 0 |
| S8_extracted_k141_57676 | 1633 | No | NA | 1 | 0 | 0 | Not-determined | Genome-fragment | NA | NA | 0 |
| S17_vs_k141_98969\|\|full | 8998 | No | NA | 17 | 3 | 0 | Low-quality | Genome-fragment | 10.77 | AAI-based (high-confidence) | 0 |
| S5_extracted_k141_119274 | 9008 | No | NA | 15 | 5 | 0 | Low-quality | Genome-fragment | 21.82 | AAI-based (high-confidence) | 0 |
| S8_extracted_k141_31572 | 3606 | No | NA | 3 | 0 | 0 | Not-determined | Genome-fragment | NA | NA | 0 |
| S9_vs_k141_74068\|\|full | 9014 | No | NA | 12 | 9 | 0 | Low-quality | Genome-fragment | 19.04 | AAI-based (high-confidence) | 0 |
| S13_extracted_21940 | 9014 | Yes | 1942 | 10 | 1 | 2 | Low-quality | Genome-fragment | 1.63 | AAI-based (medium-confidence) | 78.46 |
| S12_vs_k141_18339\|\|full | 9016 | No | NA | 8 | 2 | 1 | Low-quality | Genome-fragment | 5.79 | HMM-based (lower-bound) | 0 |
| S8_extracted_k141_196416 | 1847 | No | NA | 2 | 0 | 2 | Not-determined | Genome-fragment | NA | NA | 0 |
| S11_vs_k141_101603\|\|full | 9029 | No | NA | 8 | 7 | 0 | Low-quality | Genome-fragment | 26.82 | AAI-based (medium-confidence) | 0 |
| S8_extracted_k141_48320 | 4742 | No | NA | 3 | 0 | 2 | Not-determined | Genome-fragment | NA | NA | 0 |
| S8_extracted_k141_170390 | 1279 | No | NA | 2 | 0 | 1 | Not-determined | Genome-fragment | NA | NA | 0 |
| S8_extracted_k141_161124 | 6663 | No | NA | 4 | 0 | 2 | Not-determined | Genome-fragment | NA | NA | 0 |
| S8_extracted_k141_13758 | 20841 | No | NA | 37 | 13 | 0 | Medium-quality | Genome-fragment | 58.98 | AAI-based (medium-confidence) | 0 |
| S8_extracted_k141_144324 | 1098 | No | NA | 1 | 0 | 1 | Not-determined | Genome-fragment | NA | NA | 0 |
| S8_extracted_k141_196435 | 3436 | No | NA | 2 | 0 | 2 | Not-determined | Genome-fragment | NA | NA | 0 |
| S8_extracted_k141_204602 | 1776 | No | NA | 2 | 0 | 0 | Not-determined | Genome-fragment | NA | NA | 0 |
| S8_extracted_k141_126795 | 2071 | No | NA | 2 | 0 | 2 | Not-determined | Genome-fragment | NA | NA | 0 |
| S8_extracted_k141_5269 | 1155 | No | NA | 2 | 0 | 1 | Not-determined | Genome-fragment | NA | NA | 0 |
| S8_extracted_k141_57717 | 1033 | No | NA | 1 | 0 | 0 | Not-determined | Genome-fragment | NA | NA | 0 |
| S8_extracted_k141_83459 | 1450 | No | NA | 1 | 0 | 1 | Not-determined | Genome-fragment | NA | NA | 0 |
| S8_extracted_k141_135295 | 3134 | No | NA | 2 | 0 | 2 | Not-determined | Genome-fragment | NA | NA | 0 |
| S8_extracted_k141_48356 | 1489 | No | NA | 2 | 0 | 2 | Not-determined | Genome-fragment | NA | NA | 0 |
| S8_extracted_k141_74340 | 2569 | No | NA | 3 | 0 | 1 | Not-determined | Genome-fragment | NA | NA | 0 |
| S8_extracted_k141_109646 | 1108 | No | NA | 2 | 0 | 2 | Not-determined | Genome-fragment | NA | NA | 0 |
| S8_extracted_k141_204611 | 2218 | No | NA | 4 | 0 | 3 | Not-determined | Genome-fragment | NA | NA | 0 |
| S8_extracted_k141_196469 | 1629 | No | NA | 2 | 0 | 2 | Not-determined | Genome-fragment | NA | NA | 0 |
| S8_extracted_k141_48361 | 2450 | No | NA | 1 | 0 | 0 | Not-determined | Genome-fragment | NA | NA | 0 |
| S8_extracted_k141_135310 | 1111 | No | NA | 3 | 0 | 2 | Not-determined | Genome-fragment | NA | NA | 0 |
| S8_extracted_k141_83469 | 3152 | No | NA | 4 | 0 | 1 | Not-determined | Genome-fragment | NA | NA | 0 |
| S8_extracted_k141_126821 | 2147 | No | NA | 1 | 0 | 0 | Not-determined | Genome-fragment | NA | NA | 0 |
| S8_extracted_k141_48368 | 3342 | No | NA | 3 | 0 | 3 | Not-determined | Genome-fragment | NA | NA | 0 |
| S8_extracted_k141_144360 | 2167 | No | NA | 3 | 0 | 2 | Not-determined | Genome-fragment | NA | NA | 0 |
| S8_extracted_k141_74354 | 1470 | No | NA | 3 | 0 | 0 | Not-determined | Genome-fragment | NA | NA | 0 |
| S8_extracted_k141_5297 | 4045 | No | NA | 4 | 0 | 3 | Not-determined | Genome-fragment | NA | NA | 0 |
| S8_extracted_k141_83476 | 1315 | No | NA | 4 | 0 | 0 | Not-determined | Genome-fragment | NA | NA | 0 |
| S8_extracted_k141_57752 | 1008 | No | NA | 2 | 0 | 2 | Not-determined | Genome-fragment | NA | NA | 0 |
| S8_extracted_k141_92198 | 1368 | No | NA | 1 | 0 | 1 | Not-determined | Genome-fragment | NA | NA | 0 |
| S8_extracted_k141_144367 | 1291 | No | NA | 3 | 0 | 0 | Not-determined | Genome-fragment | NA | NA | 0 |
| S8_extracted_k141_135324 | 1318 | No | NA | 1 | 0 | 1 | Not-determined | Genome-fragment | NA | NA | 0 |
| S8_extracted_k141_126823 | 1157 | No | NA | 1 | 0 | 0 | Not-determined | Genome-fragment | NA | NA | 0 |
| S8_extracted_k141_126825 | 1777 | No | NA | 2 | 0 | 0 | Not-determined | Genome-fragment | NA | NA | 0 |
| S8_extracted_k141_13774 | 1232 | No | NA | 1 | 0 | 1 | Not-determined | Genome-fragment | NA | NA | 0 |
| S8_extracted_k141_48385 | 1969 | No | NA | 2 | 0 | 1 | Not-determined | Genome-fragment | NA | NA | 0 |
| S8_extracted_k141_144381 | 5955 | No | NA | 5 | 0 | 4 | Not-determined | Genome-fragment | NA | NA | 0 |
| S8_extracted_k141_179092 | 2360 | No | NA | 4 | 0 | 3 | Not-determined | Genome-fragment | NA | NA | 0 |
| S8_extracted_k141_179098 | 2110 | No | NA | 2 | 0 | 2 | Not-determined | Genome-fragment | NA | NA | 0 |
| S8_extracted_k141_83503 | 1628 | No | NA | 3 | 0 | 2 | Not-determined | Genome-fragment | NA | NA | 0 |
| S8_extracted_k141_196511 | 1119 | No | NA | 1 | 0 | 0 | Not-determined | Genome-fragment | NA | NA | 0 |
| S8_vs_k141_20968\|\|full | 9031 | No | NA | 23 | 1 | 1 | Low-quality | Genome-fragment | 17.79 | AAI-based (high-confidence) | 0 |
| S10_vs_32789\|\|full | 9033 | No | NA | 15 | 1 | 2 | Low-quality | Genome-fragment | 13.94 | AAI-based (high-confidence) | 0 |
| S8_extracted_k141_104208 | 9039 | No | NA | 16 | 8 | 0 | Low-quality | Genome-fragment | 24.19 | AAI-based (high-confidence) | 0 |
| S8_extracted_k141_152878 | 4421 | No | NA | 5 | 0 | 3 | Not-determined | Genome-fragment | NA | NA | 0 |
| S8_extracted_k141_92210 | 1238 | No | NA | 1 | 0 | 0 | Not-determined | Genome-fragment | NA | NA | 0 |
| S8_extracted_k141_100423 | 3403 | No | NA | 2 | 0 | 1 | Not-determined | Genome-fragment | NA | NA | 0 |
| S16_vs_42222\|\|full | 9042 | No | NA | 17 | 0 | 2 | Low-quality | Genome-fragment | 15.59 | AAI-based (high-confidence) | 0 |
| S8_extracted_k141_31716 | 2520 | No | NA | 3 | 0 | 2 | Not-determined | Genome-fragment | NA | NA | 0 |
| S8_extracted_k141_100426 | 1905 | No | NA | 5 | 0 | 0 | Not-determined | Genome-fragment | NA | NA | 0 |
| S8_extracted_k141_100427 | 1027 | No | NA | 1 | 0 | 1 | Not-determined | Genome-fragment | NA | NA | 0 |
| S8_extracted_k141_40153 | 1154 | No | NA | 2 | 0 | 1 | Not-determined | Genome-fragment | NA | NA | 0 |
| S8_extracted_k141_57799 | 1473 | No | NA | 1 | 0 | 1 | Not-determined | Genome-fragment | NA | NA | 0 |
| S8_extracted_k141_170430 | 48662 | No | NA | 61 | 13 | 1 | Complete | High-quality | 100 | DTR (high-confidence) | 0 |
| S8_extracted_k141_144397 | 1068 | No | NA | 1 | 0 | 0 | Not-determined | Genome-fragment | NA | NA | 0 |
| S8_extracted_k141_74430 | 2691 | No | NA | 3 | 0 | 1 | Not-determined | Genome-fragment | NA | NA | 0 |
| S8_extracted_k141_57802 | 1924 | No | NA | 2 | 0 | 0 | Not-determined | Genome-fragment | NA | NA | 0 |
| S8_extracted_k141_179125 | 3627 | No | NA | 3 | 0 | 3 | Not-determined | Genome-fragment | NA | NA | 0 |
| S8_extracted_k141_57807 | 1903 | No | NA | 3 | 0 | 2 | Not-determined | Genome-fragment | NA | NA | 0 |
| S8_extracted_k141_100440 | 1312 | No | NA | 2 | 0 | 0 | Not-determined | Genome-fragment | NA | NA | 0 |
| S8_extracted_k141_135370 | 6124 | No | NA | 5 | 0 | 4 | Not-determined | Genome-fragment | NA | NA | 0 |
| S8_extracted_k141_40162 | 3781 | No | NA | 1 | 0 | 0 | Not-determined | Genome-fragment | NA | NA | 0 |
| S8_extracted_k141_13806 | 4596 | No | NA | 7 | 0 | 1 | Not-determined | Genome-fragment | NA | NA | 0 |
| S8_extracted_k141_152899 | 2544 | No | NA | 2 | 0 | 2 | Not-determined | Genome-fragment | NA | NA | 0 |
| S8_extracted_k141_126853 | 1414 | No | NA | 1 | 0 | 1 | Not-determined | Genome-fragment | NA | NA | 0 |
| S8_extracted_k141_92229 | 3857 | No | NA | 2 | 0 | 0 | Not-determined | Genome-fragment | NA | NA | 0 |
| S8_extracted_k141_135391 | 1829 | No | NA | 1 | 0 | 1 | Not-determined | Genome-fragment | NA | NA | 0 |
| S8_extracted_k141_40182 | 3424 | No | NA | 5 | 0 | 1 | Not-determined | Genome-fragment | NA | NA | 0 |
| S8_extracted_k141_204713 | 3823 | No | NA | 4 | 0 | 2 | Not-determined | Genome-fragment | NA | NA | 0 |
| S8_extracted_k141_186771 | 1572 | No | NA | 2 | 0 | 0 | Not-determined | Genome-fragment | NA | NA | 0 |
| S8_extracted_k141_135413 | 1800 | No | NA | 2 | 0 | 0 | Not-determined | Genome-fragment | NA | NA | 0 |
| S8_extracted_k141_13812 | 5032 | No | NA | 7 | 0 | 6 | Not-determined | Genome-fragment | NA | NA | 0 |
| S8_extracted_k141_186777 | 1322 | No | NA | 2 | 0 | 1 | Not-determined | Genome-fragment | NA | NA | 0 |
| S8_extracted_k141_179187 | 1428 | No | NA | 2 | 0 | 1 | Not-determined | Genome-fragment | NA | NA | 0 |
| S8_vs_k141_154178\|\|full | 9046 | No | NA | 13 | 12 | 0 | Low-quality | Genome-fragment | 24.44 | AAI-based (high-confidence) | 0 |
| S5_extracted_k141_54124 | 9052 | No | NA | 14 | 3 | 0 | Low-quality | Genome-fragment | 17.87 | HMM-based (lower-bound) | 0 |
| S8_extracted_k141_186789 | 1063 | No | NA | 3 | 0 | 0 | Not-determined | Genome-fragment | NA | NA | 0 |
| S8_extracted_k141_13821 | 6565 | No | NA | 2 | 0 | 1 | Not-determined | Genome-fragment | NA | NA | 0 |
| S8_extracted_k141_170462 | 4010 | No | NA | 7 | 0 | 3 | Not-determined | Genome-fragment | NA | NA | 0 |
| S8_extracted_k141_152942 | 2392 | No | NA | 1 | 0 | 1 | Not-determined | Genome-fragment | NA | NA | 0 |
| S8_extracted_k141_13825 | 1273 | No | NA | 2 | 0 | 0 | Not-determined | Genome-fragment | NA | NA | 0 |
| S8_extracted_k141_179199 | 2045 | No | NA | 3 | 0 | 2 | Not-determined | Genome-fragment | NA | NA | 0 |
| S8_extracted_k141_57882 | 1394 | No | NA | 2 | 0 | 0 | Not-determined | Genome-fragment | NA | NA | 0 |
| S8_extracted_k141_100526 | 1004 | No | NA | 1 | 0 | 0 | Not-determined | Genome-fragment | NA | NA | 0 |
| S8_extracted_k141_126880 | 1055 | No | NA | 2 | 0 | 0 | Not-determined | Genome-fragment | NA | NA | 0 |
| S8_extracted_k141_40250 | 1146 | No | NA | 1 | 0 | 1 | Not-determined | Genome-fragment | NA | NA | 0 |
| S8_extracted_k141_92298 | 4280 | No | NA | 5 | 0 | 1 | Not-determined | Genome-fragment | NA | NA | 0 |
| S8_extracted_k141_23171 | 1958 | No | NA | 1 | 0 | 1 | Not-determined | Genome-fragment | NA | NA | 0 |
| S8_extracted_k141_31795 | 1068 | No | NA | 2 | 0 | 0 | Not-determined | Genome-fragment | NA | NA | 0 |
| S8_extracted_k141_74497 | 2933 | No | NA | 4 | 0 | 0 | Not-determined | Genome-fragment | NA | NA | 0 |
| S8_extracted_k141_161279 | 6422 | No | NA | 4 | 0 | 1 | Not-determined | Genome-fragment | NA | NA | 0 |
| S8_extracted_k141_118313 | 2562 | No | NA | 3 | 0 | 1 | Not-determined | Genome-fragment | NA | NA | 0 |
| S8_extracted_k141_5435 | 1034 | No | NA | 3 | 0 | 0 | Not-determined | Genome-fragment | NA | NA | 0 |
| S8_extracted_k141_23182 | 3018 | No | NA | 4 | 0 | 0 | Not-determined | Genome-fragment | NA | NA | 0 |
| S8_extracted_k141_13855 | 1394 | No | NA | 2 | 0 | 1 | Not-determined | Genome-fragment | NA | NA | 0 |
| S12_vs_k141_39095\|\|full | 9052 | No | NA | 12 | 1 | 2 | Low-quality | Genome-fragment | 9.74 | HMM-based (lower-bound) | 0 |
| S8_extracted_k141_5437 | 3483 | No | NA | 7 | 0 | 7 | Not-determined | Genome-fragment | NA | NA | 0 |
| S8_extracted_k141_100563 | 1157 | No | NA | 1 | 0 | 1 | Not-determined | Genome-fragment | NA | NA | 0 |
| S17_extracted_k141_15933 | 9054 | No | NA | 14 | 1 | 1 | Low-quality | Genome-fragment | 4.91 | HMM-based (lower-bound) | 0 |
| S8_extracted_k141_196646 | 1634 | No | NA | 2 | 0 | 2 | Not-determined | Genome-fragment | NA | NA | 0 |
| S8_extracted_k141_40274 | 1269 | No | NA | 2 | 0 | 0 | Not-determined | Genome-fragment | NA | NA | 0 |
| S8_extracted_k141_196647 | 5863 | No | NA | 5 | 0 | 1 | Not-determined | Genome-fragment | NA | NA | 0 |

| | | | | | | | | | | | |
|---|---|---|---|---|---|---|---|---|---|---|---|
| S8_extracted_k141_186834 | 3283 | No | NA | 3 | 0 | 3 | Not-determined | Genome-fragment | NA | NA | 0 |
| S8_extracted_k141_118338 | 1135 | No | NA | 1 | 0 | 0 | Not-determined | Genome-fragment | NA | NA | 0 |
| S8_extracted_k141_170513 | 1490 | No | NA | 2 | 0 | 1 | Not-determined | Genome-fragment | NA | NA | 0 |
| S8_extracted_k141_135515 | 1459 | No | NA | 2 | 0 | 0 | Not-determined | Genome-fragment | NA | NA | 0 |
| S8_extracted_k141_66035 | 1023 | No | NA | 1 | 0 | 0 | Not-determined | Genome-fragment | NA | NA | 0 |
| S3_extracted_k141_67657 | 9057 | No | NA | 9 | 2 | 0 | Low-quality | Genome-fragment | 7.37 | AAI-based (medium-confidence) | 0 |
| S8_extracted_k141_5483 | 1021 | No | NA | 2 | 0 | 1 | Not-determined | Genome-fragment | NA | NA | 0 |
| S8_extracted_k141_161333 | 5924 | No | NA | 6 | 0 | 1 | Not-determined | Genome-fragment | NA | NA | 0 |
| S8_extracted_k141_74550 | 1291 | No | NA | 1 | 0 | 0 | Not-determined | Genome-fragment | NA | NA | 0 |
| S5_extracted_k141_94392 | 9059 | No | NA | 14 | 4 | 0 | Low-quality | Genome-fragment | 6.51 | HMM-based (lower-bound) | 0 |
| S8_extracted_k141_126940 | 1173 | No | NA | 2 | 0 | 1 | Not-determined | Genome-fragment | NA | NA | 0 |
| S8_extracted_k141_57964 | 3805 | No | NA | 4 | 0 | 0 | Not-determined | Genome-fragment | NA | NA | 0 |
| S9_vs_k141_49309\|\|full | 9065 | No | NA | 18 | 1 | 1 | Low-quality | Genome-fragment | 20.47 | AAI-based (medium-confidence) | 0 |
| S8_extracted_k141_135549 | 1259 | No | NA | 2 | 0 | 0 | Not-determined | Genome-fragment | NA | NA | 0 |
| S16_extracted_149092 | 9065 | No | NA | 9 | 0 | 5 | Low-quality | Genome-fragment | 2.87 | AAI-based (medium-confidence) | 0 |
| S8_extracted_k141_196670 | 15088 | No | NA | 20 | 7 | 0 | Medium-quality | Genome-fragment | 72.82 | AAI-based (medium-confidence) | 0 |
| S1_vs_k141_102159\|\|full | 9075 | No | NA | 24 | 0 | 0 | Low-quality | Genome-fragment | 28.26 | AAI-based (high-confidence) | 0 |
| S8_extracted_k141_92380 | 4071 | No | NA | 5 | 0 | 0 | Not-determined | Genome-fragment | NA | NA | 0 |
| S8_extracted_k141_153084 | 1225 | No | NA | 2 | 0 | 2 | Not-determined | Genome-fragment | NA | NA | 0 |
| S8_extracted_k141_74590 | 1391 | No | NA | 1 | 0 | 1 | Not-determined | Genome-fragment | NA | NA | 0 |
| S8_extracted_k141_5524 | 1156 | No | NA | 2 | 0 | 1 | Not-determined | Genome-fragment | NA | NA | 0 |
| S8_extracted_k141_109814 | 1009 | No | NA | 1 | 0 | 1 | Not-determined | Genome-fragment | NA | NA | 0 |
| S8_extracted_k141_126977 | 1084 | No | NA | 1 | 0 | 1 | Not-determined | Genome-fragment | NA | NA | 0 |
| S9_vs_k141_83148\|\|full | 9089 | No | NA | 24 | 5 | 1 | Low-quality | Genome-fragment | 22.26 | AAI-based (medium-confidence) | 0 |
| S8_extracted_k141_40343 | 2362 | No | NA | 2 | 0 | 2 | Not-determined | Genome-fragment | NA | NA | 0 |
| S8_extracted_k141_118420 | 1203 | No | NA | 2 | 0 | 2 | Not-determined | Genome-fragment | NA | NA | 0 |
| S8_extracted_k141_153109 | 5235 | No | NA | 4 | 0 | 0 | Not-determined | Genome-fragment | NA | NA | 0 |
| S8_extracted_k141_13928 | 3431 | No | NA | 3 | 0 | 1 | Not-determined | Genome-fragment | NA | NA | 0 |
| S8_extracted_k141_118429 | 1090 | No | NA | 2 | 0 | 0 | Not-determined | Genome-fragment | NA | NA | 0 |
| S8_extracted_k141_13932 | 1922 | No | NA | 2 | 0 | 2 | Not-determined | Genome-fragment | NA | NA | 0 |
| S8_extracted_k141_196697 | 1023 | No | NA | 1 | 0 | 1 | Not-determined | Genome-fragment | NA | NA | 0 |
| S8_extracted_k141_92424 | 1877 | No | NA | 2 | 0 | 0 | Not-determined | Genome-fragment | NA | NA | 0 |
| S8_extracted_k141_21909 | 1883 | No | NA | 2 | 0 | 0 | Not-determined | Genome-fragment | NA | NA | 0 |
| S11_vs_k141_64014\|\|full | 9091 | No | NA | 20 | 1 | 0 | Low-quality | Genome-fragment | 26.58 | AAI-based (high-confidence) | 0 |
| S8_extracted_k141_92436 | 1257 | No | NA | 2 | 0 | 1 | Not-determined | Genome-fragment | NA | NA | 0 |
| S14_vs_k141_39537\|\|full | 9092 | No | NA | 6 | 0 | 4 | Low-quality | Genome-fragment | 10.9 | AAI-based (medium-confidence) | 0 |
| S8_extracted_k141_179212 | 1754 | No | NA | 4 | 0 | 3 | Not-determined | Genome-fragment | NA | NA | 0 |
| S8_extracted_k141_135607 | 1545 | No | NA | 1 | 0 | 1 | Not-determined | Genome-fragment | NA | NA | 0 |
| S8_extracted_k141_13953 | 2251 | No | NA | 4 | 0 | 3 | Not-determined | Genome-fragment | NA | NA | 0 |
| S10_extracted_80527 | 9094 | No | NA | 6 | 2 | 0 | Low-quality | Genome-fragment | 14.33 | AAI-based (medium-confidence) | 0 |
| S8_extracted_k141_135608 | 1538 | No | NA | 2 | 0 | 2 | Not-determined | Genome-fragment | NA | NA | 0 |
| S8_extracted_k141_204816 | 1106 | No | NA | 2 | 0 | 1 | Not-determined | Genome-fragment | NA | NA | 0 |
| S8_extracted_k141_161416 | 1303 | No | NA | 2 | 0 | 2 | Not-determined | Genome-fragment | NA | NA | 0 |
| S8_extracted_k141_118463 | 1037 | No | NA | 1 | 0 | 1 | Not-determined | Genome-fragment | NA | NA | 0 |
| S8_extracted_k141_58023 | 1035 | No | NA | 1 | 0 | 0 | Not-determined | Genome-fragment | NA | NA | 0 |
| S8_extracted_k141_179215 | 3056 | No | NA | 4 | 0 | 3 | Not-determined | Genome-fragment | NA | NA | 0 |
| S8_extracted_k141_144540 | 1308 | No | NA | 1 | 0 | 1 | Not-determined | Genome-fragment | NA | NA | 0 |
| S3_extracted_k141_27305 | 9096 | No | NA | 20 | 1 | 1 | Low-quality | Genome-fragment | 5.53 | AAI-based (high-confidence) | 0 |
| S8_extracted_k141_92463 | 1172 | No | NA | 2 | 0 | 2 | Not-determined | Genome-fragment | NA | NA | 0 |
| S8_extracted_k141_179224 | 2411 | No | NA | 1 | 0 | 0 | Not-determined | Genome-fragment | NA | NA | 0 |
| S8_extracted_k141_186942 | 1223 | No | NA | 1 | 0 | 1 | Not-determined | Genome-fragment | NA | NA | 0 |
| S8_extracted_k141_196738 | 3477 | No | NA | 4 | 0 | 1 | Not-determined | Genome-fragment | NA | NA | 0 |
| S8_extracted_k141_5570 | 1208 | No | NA | 2 | 0 | 2 | Not-determined | Genome-fragment | NA | NA | 0 |
| S8_extracted_k141_13971 | 1759 | No | NA | 1 | 0 | 0 | Not-determined | Genome-fragment | NA | NA | 0 |
| S8_extracted_k141_23234 | 1197 | No | NA | 1 | 0 | 0 | Not-determined | Genome-fragment | NA | NA | 0 |
| S8_extracted_k141_23236 | 1665 | No | NA | 3 | 0 | 0 | Not-determined | Genome-fragment | NA | NA | 0 |
| S4_extracted_9193 | 9101 | No | NA | 16 | 4 | 1 | Low-quality | Genome-fragment | 6.03 | AAI-based (high-confidence) | 0 |
| S8_extracted_k141_161444 | 2452 | No | NA | 2 | 0 | 1 | Not-determined | Genome-fragment | NA | NA | 0 |
| S8_extracted_k141_92477 | 1406 | No | NA | 1 | 0 | 1 | Not-determined | Genome-fragment | NA | NA | 0 |
| S8_extracted_k141_124832 | 9101 | No | NA | 16 | 4 | 0 | Low-quality | Genome-fragment | 16.64 | AAI-based (high-confidence) | 0 |
| S8_extracted_k141_196749 | 4797 | No | NA | 3 | 0 | 0 | Not-determined | Genome-fragment | NA | NA | 0 |
| S8_extracted_k141_186965 | 1160 | No | NA | 2 | 0 | 0 | Not-determined | Genome-fragment | NA | NA | 0 |
| S8_extracted_k141_161446 | 2305 | No | NA | 1 | 0 | 1 | Not-determined | Genome-fragment | NA | NA | 0 |
| S17_vs_k141_24905\|\|full | 9104 | No | NA | 14 | 1 | 0 | Low-quality | Genome-fragment | 12.31 | AAI-based (high-confidence) | 0 |
| S7_vs_59924\|\|full | 9105 | Yes | 5347 | 6 | 2 | 1 | Low-quality | Genome-fragment | 8.82 | AAI-based (high-confidence) | 41.27 |
| S8_extracted_k141_66112 | 1215 | No | NA | 1 | 0 | 1 | Not-determined | Genome-fragment | NA | NA | 0 |
| S7_extracted_49357 | 9110 | No | NA | 13 | 0 | 0 | Low-quality | Genome-fragment | 20.28 | AAI-based (medium-confidence) | 0 |
| S8_extracted_k141_127051 | 2076 | No | NA | 3 | 0 | 1 | Not-determined | Genome-fragment | NA | NA | 0 |
| S8_extracted_k141_161474 | 3942 | No | NA | 2 | 0 | 2 | Not-determined | Genome-fragment | NA | NA | 0 |
| S8_extracted_k141_100681 | 1187 | No | NA | 1 | 0 | 1 | Not-determined | Genome-fragment | NA | NA | 0 |
| S8_vs_k141_25614\|\|full | 9112 | No | NA | 20 | 2 | 0 | Low-quality | Genome-fragment | 21.19 | AAI-based (high-confidence) | 0 |
| S8_extracted_k141_58068 | 4546 | No | NA | 2 | 0 | 2 | Not-determined | Genome-fragment | NA | NA | 0 |
| S8_extracted_k141_179286 | 3597 | No | NA | 4 | 0 | 3 | Not-determined | Genome-fragment | NA | NA | 0 |
| S8_extracted_k141_109903 | 1160 | No | NA | 2 | 0 | 1 | Not-determined | Genome-fragment | NA | NA | 0 |
| S8_extracted_k141_170665 | 3333 | No | NA | 2 | 0 | 2 | Not-determined | Genome-fragment | NA | NA | 0 |
| S8_extracted_k141_48709 | 1057 | No | NA | 1 | 0 | 1 | Not-determined | Genome-fragment | NA | NA | 0 |
| S8_extracted_k141_118544 | 1583 | No | NA | 2 | 0 | 2 | Not-determined | Genome-fragment | NA | NA | 0 |
| S8_extracted_k141_23286 | 1388 | No | NA | 2 | 0 | 0 | Not-determined | Genome-fragment | NA | NA | 0 |
| S3_vs_k141_113247\|\|full | 9115 | No | NA | 12 | 0 | 1 | Low-quality | Genome-fragment | 34.66 | AAI-based (high-confidence) | 0 |
| S8_extracted_k141_135693 | 1106 | No | NA | 1 | 0 | 0 | Not-determined | Genome-fragment | NA | NA | 0 |
| S8_extracted_k141_23293 | 3328 | No | NA | 4 | 0 | 2 | Not-determined | Genome-fragment | NA | NA | 0 |
| S11_vs_k141_69967\|\|full | 9125 | No | NA | 11 | 2 | 0 | Low-quality | Genome-fragment | 23.84 | AAI-based (high-confidence) | 0 |
| S8_extracted_k141_135704 | 2734 | No | NA | 3 | 0 | 2 | Not-determined | Genome-fragment | NA | NA | 0 |
| S8_extracted_k141_100726 | 1065 | No | NA | 3 | 0 | 0 | Not-determined | Genome-fragment | NA | NA | 0 |
| S8_extracted_k141_127085 | 1735 | No | NA | 2 | 0 | 1 | Not-determined | Genome-fragment | NA | NA | 0 |
| S8_extracted_k141_100728 | 1086 | No | NA | 2 | 0 | 0 | Not-determined | Genome-fragment | NA | NA | 0 |
| S8_extracted_k141_40477 | 2014 | No | NA | 3 | 0 | 2 | Not-determined | Genome-fragment | NA | NA | 0 |
| S8_extracted_k141_92529 | 1221 | No | NA | 1 | 0 | 1 | Not-determined | Genome-fragment | NA | NA | 0 |
| S8_extracted_k141_100749 | 1010 | No | NA | 1 | 0 | 0 | Not-determined | Genome-fragment | NA | NA | 0 |
| S8_extracted_k141_161528 | 2924 | No | NA | 3 | 0 | 2 | Not-determined | Genome-fragment | NA | NA | 0 |
| S8_extracted_k141_40491 | 1858 | No | NA | 1 | 0 | 1 | Not-determined | Genome-fragment | NA | NA | 0 |
| S8_extracted_k141_32014 | 1465 | No | NA | 1 | 0 | 1 | Not-determined | Genome-fragment | NA | NA | 0 |
| S8_extracted_k141_48746 | 1040 | No | NA | 1 | 0 | 1 | Not-determined | Genome-fragment | NA | NA | 0 |
| S8_extracted_k141_109947 | 1987 | No | NA | 1 | 0 | 1 | Not-determined | Genome-fragment | NA | NA | 0 |
| S8_extracted_k141_179359 | 3404 | No | NA | 2 | 0 | 2 | Not-determined | Genome-fragment | NA | NA | 0 |
| S8_extracted_k141_118581 | 7013 | No | NA | 5 | 0 | 4 | Not-determined | Genome-fragment | NA | NA | 0 |
| S8_extracted_k141_48750 | 1527 | No | NA | 3 | 0 | 1 | Not-determined | Genome-fragment | NA | NA | 0 |
| S8_extracted_k141_161541 | 4440 | No | NA | 2 | 0 | 1 | Not-determined | Genome-fragment | NA | NA | 0 |
| S8_extracted_k141_5674 | 3585 | No | NA | 5 | 0 | 2 | Not-determined | Genome-fragment | NA | NA | 0 |
| S8_extracted_k141_14095 | 2571 | No | NA | 1 | 0 | 1 | Not-determined | Genome-fragment | NA | NA | 0 |
| S3_vs_k141_733\|\|full | 9128 | No | NA | 10 | 0 | 5 | Low-quality | Genome-fragment | 16.47 | AAI-based (medium-confidence) | 0 |
| S8_extracted_k141_74733 | 1081 | No | NA | 2 | 0 | 1 | Not-determined | Genome-fragment | NA | NA | 0 |
| S8_extracted_k141_161549 | 1873 | No | NA | 3 | 0 | 2 | Not-determined | Genome-fragment | NA | NA | 0 |
| S3_vs_k141_26383\|\|full | 9129 | No | NA | 22 | 4 | 0 | Low-quality | Genome-fragment | 14.88 | AAI-based (high-confidence) | 0 |
| S8_extracted_k141_23367 | 3324 | No | NA | 4 | 0 | 1 | Not-determined | Genome-fragment | NA | NA | 0 |
| S17_extracted_k141_102593 | 9132 | No | NA | 14 | 5 | 0 | Low-quality | Genome-fragment | 23.3 | AAI-based (high-confidence) | 0 |
| S8_extracted_k141_153243 | 4789 | No | NA | 6 | 0 | 4 | Not-determined | Genome-fragment | NA | NA | 0 |
| S10_vs_32658\|\|full | 9136 | No | NA | 15 | 2 | 0 | Low-quality | Genome-fragment | 5.59 | AAI-based (high-confidence) | 0 |
| S8_extracted_k141_161586 | 3128 | No | NA | 4 | 0 | 2 | Not-determined | Genome-fragment | NA | NA | 0 |
| S8_extracted_k141_40562 | 1077 | No | NA | 1 | 0 | 0 | Not-determined | Genome-fragment | NA | NA | 0 |
| S8_extracted_k141_153249 | 5287 | No | NA | 10 | 0 | 9 | Not-determined | Genome-fragment | NA | NA | 0 |
| S8_extracted_k141_100819 | 1836 | No | NA | 3 | 0 | 2 | Not-determined | Genome-fragment | NA | NA | 0 |
| S17_vs_k141_70522\|\|full | 9138 | No | NA | 9 | 0 | 1 | Low-quality | Genome-fragment | 7.7 | AAI-based (medium-confidence) | 0 |
| S8_extracted_k141_74799 | 1051 | No | NA | 2 | 0 | 0 | Not-determined | Genome-fragment | NA | NA | 0 |
| S8_extracted_k141_83689 | 1717 | No | NA | 6 | 0 | 0 | Not-determined | Genome-fragment | NA | NA | 0 |
| S8_extracted_k141_179395 | 7558 | No | NA | 9 | 0 | 6 | Not-determined | Genome-fragment | NA | NA | 0 |
| S8_extracted_k141_58167 | 1096 | No | NA | 2 | 0 | 1 | Not-determined | Genome-fragment | NA | NA | 0 |
| S14_extracted_k141_89650 | 9141 | No | NA | 23 | 20 | 0 | Low-quality | Genome-fragment | 30.04 | AAI-based (high-confidence) | 0 |
| S8_extracted_k141_179399 | 1707 | No | NA | 2 | 0 | 2 | Not-determined | Genome-fragment | NA | NA | 0 |
| S8_extracted_k141_179403 | 1178 | No | NA | 1 | 0 | 1 | Not-determined | Genome-fragment | NA | NA | 0 |
| S8_extracted_k141_205024 | 3411 | No | NA | 3 | 0 | 1 | Not-determined | Genome-fragment | NA | NA | 0 |
| S8_extracted_k141_14142 | 1301 | No | NA | 2 | 0 | 1 | Not-determined | Genome-fragment | NA | NA | 0 |
| S8_extracted_k141_187073 | 1141 | No | NA | 1 | 0 | 1 | Not-determined | Genome-fragment | NA | NA | 0 |
| S8_vs_k141_92211\|\|full | 9145 | No | NA | 19 | 2 | 1 | Low-quality | Genome-fragment | 23.03 | AAI-based (high-confidence) | 0 |
| S8_extracted_k141_205036 | 1606 | No | NA | 2 | 0 | 1 | Not-determined | Genome-fragment | NA | NA | 0 |
| S8_extracted_k141_110040 | 1616 | No | NA | 2 | 0 | 0 | Not-determined | Genome-fragment | NA | NA | 0 |
| S8_extracted_k141_74826 | 4281 | No | NA | 4 | 0 | 2 | Not-determined | Genome-fragment | NA | NA | 0 |
| S8_extracted_k141_144724 | 1093 | No | NA | 2 | 0 | 1 | Not-determined | Genome-fragment | NA | NA | 0 |
| S8_extracted_k141_161635 | 1115 | No | NA | 1 | 0 | 0 | Not-determined | Genome-fragment | NA | NA | 0 |
| S8_extracted_k141_127138 | 1161 | No | NA | 1 | 0 | 0 | Not-determined | Genome-fragment | NA | NA | 0 |
| S8_extracted_k141_153266 | 12670 | No | NA | 10 | 0 | 4 | Not-determined | Genome-fragment | NA | NA | 0 |
| S10_extracted_67261 | 9153 | No | NA | 4 | 0 | 4 | Low-quality | Genome-fragment | 3.47 | AAI-based (high-confidence) | 0 |
| S8_extracted_k141_179419 | 3502 | No | NA | 5 | 0 | 4 | Not-determined | Genome-fragment | NA | NA | 0 |
| S5_vs_k141_67935\|\|full | 9156 | No | NA | 13 | 3 | 1 | Low-quality | Genome-fragment | 20.83 | AAI-based (high-confidence) | 0 |
| S8_extracted_k141_161649 | 1309 | No | NA | 2 | 0 | 1 | Not-determined | Genome-fragment | NA | NA | 0 |
| S8_extracted_k141_144760 | 1554 | No | NA | 3 | 0 | 2 | Not-determined | Genome-fragment | NA | NA | 0 |
| S8_extracted_k141_5791 | 1278 | No | NA | 2 | 0 | 2 | Not-determined | Genome-fragment | NA | NA | 0 |
| S8_extracted_k141_74861 | 6412 | No | NA | 5 | 0 | 3 | Not-determined | Genome-fragment | NA | NA | 0 |
| S8_extracted_k141_144764 | 1330 | No | NA | 1 | 0 | 1 | Not-determined | Genome-fragment | NA | NA | 0 |
| S8_extracted_k141_110089 | 1418 | No | NA | 2 | 0 | 2 | Not-determined | Genome-fragment | NA | NA | 0 |
| S8_extracted_k141_74868 | 1003 | No | NA | 1 | 0 | 1 | Not-determined | Genome-fragment | NA | NA | 0 |
| S8_extracted_k141_161660 | 2160 | No | NA | 2 | 0 | 2 | Not-determined | Genome-fragment | NA | NA | 0 |
| S8_extracted_k141_32127 | 1203 | No | NA | 3 | 0 | 0 | Not-determined | Genome-fragment | NA | NA | 0 |
| S8_extracted_k141_153294 | 3436 | No | NA | 3 | 0 | 3 | Not-determined | Genome-fragment | NA | NA | 0 |
| S4_extracted_23013 | 9156 | No | NA | 21 | 3 | 0 | Low-quality | Genome-fragment | 4.76 | HMM-based (lower-bound) | 0 |
| S8_extracted_k141_66318 | 5034 | No | NA | 7 | 0 | 4 | Not-determined | Genome-fragment | NA | NA | 0 |
| S8_extracted_k141_196935 | 2338 | No | NA | 1 | 0 | 1 | Not-determined | Genome-fragment | NA | NA | 0 |
| S8_extracted_k141_161675 | 1398 | No | NA | 2 | 0 | 1 | Not-determined | Genome-fragment | NA | NA | 0 |
| S8_extracted_k141_66321 | 1149 | No | NA | 1 | 0 | 0 | Not-determined | Genome-fragment | NA | NA | 0 |
| S8_extracted_k141_40628 | 6138 | No | NA | 7 | 0 | 3 | Not-determined | Genome-fragment | NA | NA | 0 |
| S8_extracted_k141_5812 | 2870 | No | NA | 3 | 0 | 0 | Not-determined | Genome-fragment | NA | NA | 0 |
| S3_vs_k141_46764\|\|full | 9159 | No | NA | 15 | 2 | 1 | Low-quality | Genome-fragment | 21.64 | AAI-based (medium-confidence) | 0 |
| S8_extracted_k141_74900 | 1775 | No | NA | 2 | 0 | 1 | Not-determined | Genome-fragment | NA | NA | 0 |
| S8_extracted_k141_144796 | 1241 | No | NA | 1 | 0 | 1 | Not-determined | Genome-fragment | NA | NA | 0 |
| S8_extracted_k141_153309 | 3987 | No | NA | 5 | 0 | 0 | Not-determined | Genome-fragment | NA | NA | 0 |
| S8_extracted_k141_48883 | 3404 | No | NA | 2 | 0 | 2 | Not-determined | Genome-fragment | NA | NA | 0 |
| S8_extracted_k141_110125 | 1076 | No | NA | 3 | 0 | 0 | Not-determined | Genome-fragment | NA | NA | 0 |
| S8_extracted_k141_179473 | 1989 | No | NA | 2 | 0 | 1 | Not-determined | Genome-fragment | NA | NA | 0 |
| S8_extracted_k141_74913 | 1136 | No | NA | 2 | 0 | 0 | Not-determined | Genome-fragment | NA | NA | 0 |
| S7_vs_104348\|\|full | 9159 | No | NA | 14 | 9 | 0 | Low-quality | Genome-fragment | 25.28 | AAI-based (medium-confidence) | 0 |
| S5_vs_k141_63595\|\|full | 9166 | No | NA | 9 | 2 | 0 | Low-quality | Genome-fragment | 24.88 | AAI-based (medium-confidence) | 0 |
| S8_extracted_k141_170743 | 23022 | No | NA | 5 | 0 | 4 | Not-determined | Genome-fragment | NA | NA | 0 |
| S8_extracted_k141_144828 | 1671 | No | NA | 3 | 0 | 1 | Not-determined | Genome-fragment | NA | NA | 0 |
| S8_extracted_k141_170744 | 1428 | No | NA | 2 | 0 | 2 | Not-determined | Genome-fragment | NA | NA | 0 |
| S8_extracted_k141_205133 | 1231 | No | NA | 1 | 0 | 1 | Not-determined | Genome-fragment | NA | NA | 0 |
| S11_extracted_k141_70784 | 9167 | No | NA | 10 | 1 | 0 | Low-quality | Genome-fragment | 3.67 | AAI-based (medium-confidence) | 0 |
| S8_extracted_k141_58242 | 1146 | No | NA | 1 | 0 | 0 | Not-determined | Genome-fragment | NA | NA | 0 |
| S16_extracted_149503 | 9169 | No | NA | 19 | 2 | 0 | Low-quality | Genome-fragment | 20.84 | AAI-based (medium-confidence) | 0 |
| S7_extracted_109603 | 9173 | No | NA | 6 | 3 | 0 | Low-quality | Genome-fragment | 17.92 | AAI-based (medium-confidence) | 0 |
| S8_extracted_k141_100952 | 3719 | No | NA | 2 | 0 | 1 | Not-determined | Genome-fragment | NA | NA | 0 |
| S8_extracted_k141_23479 | 7860 | No | NA | 9 | 0 | 6 | Not-determined | Genome-fragment | NA | NA | 0 |
| S8_extracted_k141_5882 | 1397 | No | NA | 2 | 0 | 0 | Not-determined | Genome-fragment | NA | NA | 0 |
| S8_extracted_k141_187203 | 2836 | No | NA | 5 | 0 | 3 | Not-determined | Genome-fragment | NA | NA | 0 |
| S8_extracted_k141_127270 | 2718 | No | NA | 4 | 0 | 2 | Not-determined | Genome-fragment | NA | NA | 0 |
| S8_extracted_k141_118755 | 15190 | No | NA | 13 | 0 | 8 | Not-determined | Genome-fragment | NA | NA | 0 |
| S8_extracted_k141_179522 | 1041 | No | NA | 2 | 0 | 2 | Not-determined | Genome-fragment | NA | NA | 0 |
| S8_extracted_k141_66395 | 1220 | No | NA | 2 | 0 | 1 | Not-determined | Genome-fragment | NA | NA | 0 |
| S8_extracted_k141_205175 | 1621 | No | NA | 3 | 0 | 2 | Not-determined | Genome-fragment | NA | NA | 0 |
| S8_extracted_k141_100977 | 2503 | No | NA | 3 | 0 | 1 | Not-determined | Genome-fragment | NA | NA | 0 |
| S8_extracted_k141_74961 | 1021 | No | NA | 1 | 0 | 1 | Not-determined | Genome-fragment | NA | NA | 0 |
| S17_extracted_k141_169135 | 9180 | No | NA | 16 | 2 | 0 | Low-quality | Genome-fragment | 8.21 | AAI-based (high-confidence) | 0 |
| S8_extracted_k141_127281 | 4264 | No | NA | 6 | 0 | 3 | Not-determined | Genome-fragment | NA | NA | 0 |
| S13_vs_127622\|\|full | 9183 | No | NA | 19 | 4 | 2 | Low-quality | Genome-fragment | 18.26 | AAI-based (medium-confidence) | 0 |
| S8_extracted_k141_40717 | 1874 | No | NA | 2 | 0 | 2 | Not-determined | Genome-fragment | NA | NA | 0 |
| S8_extracted_k141_170802 | 1267 | No | NA | 1 | 0 | 0 | Not-determined | Genome-fragment | NA | NA | 0 |
| S8_extracted_k141_48977 | 1273 | No | NA | 2 | 0 | 2 | Not-determined | Genome-fragment | NA | NA | 0 |
| S8_extracted_k141_48980 | 1157 | No | NA | 1 | 0 | 1 | Not-determined | Genome-fragment | NA | NA | 0 |
| S12_extracted_k141_187095 | 9188 | Yes | 7887 | 7 | 1 | 1 | Low-quality | Genome-fragment | 0.63 | HMM-based (lower-bound) | 14.16 |
| S8_extracted_k141_205206 | 2557 | No | NA | 5 | 0 | 1 | Not-determined | Genome-fragment | NA | NA | 0 |
| S11_vs_k141_60551\|\|full | 9196 | No | NA | 4 | 0 | 0 | Low-quality | Genome-fragment | 5.98 | AAI-based (high-confidence) | 0 |
| S14_vs_k141_39502\|\|full | 9206 | No | NA | 5 | 0 | 3 | Low-quality | Genome-fragment | 5.43 | AAI-based (medium-confidence) | 0 |
| S8_extracted_k141_135740 | 1009 | No | NA | 1 | 0 | 1 | Not-determined | Genome-fragment | NA | NA | 0 |
| S8_extracted_k141_5916 | 3079 | No | NA | 8 | 0 | 8 | Not-determined | Genome-fragment | NA | NA | 0 |
| S8_extracted_k141_66427 | 1825 | No | NA | 2 | 0 | 0 | Not-determined | Genome-fragment | NA | NA | 0 |
| S8_extracted_k141_92693 | 1882 | No | NA | 1 | 0 | 0 | Not-determined | Genome-fragment | NA | NA | 0 |
| S8_extracted_k141_194464 | 9208 | No | NA | 8 | 0 | 6 | Low-quality | Genome-fragment | 1.22 | AAI-based (medium-confidence) | 0 |
| S8_extracted_k141_110212 | 1424 | No | NA | 4 | 0 | 3 | Not-determined | Genome-fragment | NA | NA | 0 |
| S8_extracted_k141_40768 | 1498 | No | NA | 1 | 0 | 1 | Not-determined | Genome-fragment | NA | NA | 0 |
| S8_extracted_k141_66444 | 2167 | No | NA | 5 | 0 | 1 | Not-determined | Genome-fragment | NA | NA | 0 |
| S8_extracted_k141_127330 | 1015 | No | NA | 2 | 0 | 1 | Not-determined | Genome-fragment | NA | NA | 0 |
| S8_extracted_k141_40776 | 1191 | No | NA | 1 | 0 | 1 | Not-determined | Genome-fragment | NA | NA | 0 |
| S8_extracted_k141_197033 | 3239 | No | NA | 3 | 0 | 1 | Not-determined | Genome-fragment | NA | NA | 0 |
| S8_extracted_k141_58323 | 1608 | No | NA | 2 | 0 | 0 | Not-determined | Genome-fragment | NA | NA | 0 |
| S8_extracted_k141_135777 | 1359 | No | NA | 1 | 0 | 1 | Not-determined | Genome-fragment | NA | NA | 0 |
| S14_vs_k141_102697\|\|full | 9210 | No | NA | 19 | 2 | 1 | Low-quality | Genome-fragment | 23.31 | AAI-based (high-confidence) | 0 |
| S8_extracted_k141_170857 | 3521 | No | NA | 6 | 0 | 6 | Not-determined | Genome-fragment | NA | NA | 0 |
| S8_extracted_k141_40789 | 2291 | No | NA | 2 | 0 | 0 | Not-determined | Genome-fragment | NA | NA | 0 |
| S8_extracted_k141_144955 | 4101 | No | NA | 4 | 0 | 3 | Not-determined | Genome-fragment | NA | NA | 0 |
| S8_extracted_k141_92699 | 1380 | No | NA | 1 | 0 | 1 | Not-determined | Genome-fragment | NA | NA | 0 |
| S8_extracted_k141_153378 | 2763 | No | NA | 1 | 0 | 1 | Not-determined | Genome-fragment | NA | NA | 0 |
| S16_extracted_12235 | 9214 | No | NA | 6 | 0 | 2 | Low-quality | Genome-fragment | 7.77 | AAI-based (medium-confidence) | 0 |
| S8_extracted_k141_161703 | 102913 | No | NA | 135 | 25 | 3 | High-quality | High-quality | 100 | AAI-based (high-confidence) | 0 |
| S8_extracted_k141_153383 | 1189 | No | NA | 1 | 0 | 1 | Not-determined | Genome-fragment | NA | NA | 0 |
| S8_extracted_k141_14325 | 1051 | No | NA | 1 | 0 | 1 | Not-determined | Genome-fragment | NA | NA | 0 |
| S8_extracted_k141_127364 | 2398 | No | NA | 2 | 0 | 0 | Not-determined | Genome-fragment | NA | NA | 0 |
| S8_extracted_k141_144972 | 1059 | No | NA | 1 | 0 | 1 | Not-determined | Genome-fragment | NA | NA | 0 |
| S8_extracted_k141_127369 | 1407 | No | NA | 2 | 0 | 1 | Not-determined | Genome-fragment | NA | NA | 0 |
| S8_extracted_k141_135815 | 1058 | No | NA | 2 | 0 | 2 | Not-determined | Genome-fragment | NA | NA | 0 |
| S8_extracted_k141_14341 | 2599 | No | NA | 2 | 0 | 1 | Not-determined | Genome-fragment | NA | NA | 0 |
| S8_extracted_k141_23640 | 2355 | No | NA | 3 | 0 | 1 | Not-determined | Genome-fragment | NA | NA | 0 |
| S8_extracted_k141_6007 | 1113 | No | NA | 1 | 0 | 1 | Not-determined | Genome-fragment | NA | NA | 0 |
| S8_extracted_k141_110265 | 3090 | No | NA | 3 | 0 | 1 | Not-determined | Genome-fragment | NA | NA | 0 |
| S9_vs_k141_35910\|\|full | 9227 | No | NA | 25 | 3 | 1 | Low-quality | Genome-fragment | 22.25 | AAI-based (high-confidence) | 0 |
| S8_extracted_k141_40815 | 4754 | No | NA | 3 | 0 | 0 | Not-determined | Genome-fragment | NA | NA | 0 |
| S8_extracted_k141_66450 | 1357 | No | NA | 2 | 0 | 0 | Not-determined | Genome-fragment | NA | NA | 0 |
| S8_extracted_k141_6015 | 1031 | No | NA | 1 | 0 | 1 | Not-determined | Genome-fragment | NA | NA | 0 |
| S8_extracted_k141_75044 | 1628 | No | NA | 1 | 0 | 1 | Not-determined | Genome-fragment | NA | NA | 0 |
| S8_extracted_k141_6022 | 2044 | No | NA | 2 | 0 | 2 | Not-determined | Genome-fragment | NA | NA | 0 |
| S11_vs_k141_7049\|\|full | 9227 | No | NA | 20 | 4 | 1 | Low-quality | Genome-fragment | 11.95 | HMM-based (lower-bound) | 0 |
| S5_vs_k141_36054\|\|full | 9231 | No | NA | 18 | 3 | 0 | Low-quality | Genome-fragment | 22.58 | AAI-based (high-confidence) | 0 |
| S8_extracted_k141_187348 | 5736 | No | NA | 3 | 0 | 0 | Not-determined | Genome-fragment | NA | NA | 0 |
| S11_extracted_k141_60476 | 9235 | No | NA | 10 | 5 | 0 | Low-quality | Genome-fragment | 18.32 | AAI-based (high-confidence) | 0 |
| S8_extracted_k141_14359 | 1516 | No | NA | 2 | 0 | 0 | Not-determined | Genome-fragment | NA | NA | 0 |
| S8_extracted_k141_197061 | 39543 | No | NA | 67 | 24 | 0 | Complete | High-quality | 100 | ITR (high-confidence) | 0 |
| S8_extracted_k141_118869 | 1278 | No | NA | 2 | 0 | 2 | Not-determined | Genome-fragment | NA | NA | 0 |
| S3_vs_k141_32689\|\|full | 9242 | No | NA | 11 | 11 | 0 | Low-quality | Genome-fragment | 26.75 | AAI-based (high-confidence) | 0 |
| S8_extracted_k141_40842 | 3319 | No | NA | 3 | 0 | 2 | Not-determined | Genome-fragment | NA | NA | 0 |
| S8_extracted_k141_135863 | 3251 | No | NA | 2 | 0 | 2 | Not-determined | Genome-fragment | NA | NA | 0 |
| S8_extracted_k141_197063 | 2746 | No | NA | 2 | 0 | 1 | Not-determined | Genome-fragment | NA | NA | 0 |
| S8_extracted_k141_127427 | 1041 | No | NA | 2 | 0 | 1 | Not-determined | Genome-fragment | NA | NA | 0 |
| S11_vs_k141_60677\|\|full | 9256 | No | NA | 16 | 6 | 0 | Low-quality | Genome-fragment | 23.01 | AAI-based (high-confidence) | 0 |
| S8_extracted_k141_32196 | 1839 | No | NA | 2 | 0 | 2 | Not-determined | Genome-fragment | NA | NA | 0 |
| S8_extracted_k141_170878 | 60026 | Yes | 51809 | 83 | 19 | 4 | High-quality | High-quality | 100 | AAI-based (high-confidence) | 13.69 |
| S8_extracted_k141_58423 | 1148 | No | NA | 1 | 0 | 0 | Not-determined | Genome-fragment | NA | NA | 0 |
| S3_extracted_k141_84522 | 9258 | No | NA | 8 | 3 | 0 | Low-quality | Genome-fragment | 10.76 | AAI-based (high-confidence) | 0 |
| S8_extracted_k141_32207 | 1035 | No | NA | 1 | 0 | 0 | Not-determined | Genome-fragment | NA | NA | 0 |
| S8_extracted_k141_14379 | 5432 | No | NA | 4 | 0 | 3 | Not-determined | Genome-fragment | NA | NA | 0 |
| S5_vs_k141_95239\|\|full | 9259 | No | NA | 8 | 0 | 2 | Low-quality | Genome-fragment | 8.42 | AAI-based (medium-confidence) | 0 |
| S8_extracted_k141_135873 | 3361 | No | NA | 2 | 0 | 1 | Not-determined | Genome-fragment | NA | NA | 0 |
| S8_extracted_k141_58429 | 1671 | No | NA | 2 | 0 | 1 | Not-determined | Genome-fragment | NA | NA | 0 |
| S8_extracted_k141_66494 | 5407 | No | NA | 4 | 0 | 3 | Not-determined | Genome-fragment | NA | NA | 0 |
| S8_extracted_k141_75097 | 1393 | No | NA | 2 | 0 | 2 | Not-determined | Genome-fragment | NA | NA | 0 |
| S8_extracted_k141_83890 | 1137 | No | NA | 1 | 0 | 1 | Not-determined | Genome-fragment | NA | NA | 0 |
| S8_extracted_k141_101124 | 1535 | No | NA | 4 | 0 | 0 | Not-determined | Genome-fragment | NA | NA | 0 |
| S8_extracted_k141_6076 | 1210 | No | NA | 2 | 0 | 2 | Not-determined | Genome-fragment | NA | NA | 0 |
| S14_vs_k141_74563\|\|full | 9262 | No | NA | 14 | 3 | 2 | Low-quality | Genome-fragment | 22.34 | AAI-based (high-confidence) | 0 |
| S8_extracted_k141_66505 | 3131 | No | NA | 5 | 0 | 0 | Not-determined | Genome-fragment | NA | NA | 0 |
| S8_extracted_k141_153529 | 1932 | No | NA | 2 | 0 | 2 | Not-determined | Genome-fragment | NA | NA | 0 |
| S8_extracted_k141_127456 | 1900 | No | NA | 1 | 0 | 1 | Not-determined | Genome-fragment | NA | NA | 0 |
| S11_extracted_k141_76717 | 9272 | No | NA | 14 | 10 | 0 | Low-quality | Genome-fragment | 19.55 | AAI-based (high-confidence) | 0 |
| S14_extracted_k141_31059 | 9279 | No | NA | 10 | 0 | 1 | Low-quality | Genome-fragment | 12.6 | AAI-based (medium-confidence) | 0 |
| S8_extracted_k141_187436 | 1190 | No | NA | 1 | 0 | 0 | Not-determined | Genome-fragment | NA | NA | 0 |
| S13_vs_66919\|\|full | 9280 | No | NA | 11 | 1 | 1 | Low-quality | Genome-fragment | 10.42 | HMM-based (lower-bound) | 0 |
| S8_extracted_k141_23756 | 1536 | No | NA | 1 | 0 | 1 | Not-determined | Genome-fragment | NA | NA | 0 |
| S8_extracted_k141_14434 | 1713 | No | NA | 1 | 0 | 0 | Not-determined | Genome-fragment | NA | NA | 0 |
| S8_extracted_k141_75130 | 4154 | No | NA | 5 | 0 | 2 | Not-determined | Genome-fragment | NA | NA | 0 |
| S8_extracted_k141_83925 | 1393 | No | NA | 1 | 0 | 1 | Not-determined | Genome-fragment | NA | NA | 0 |
| S8_extracted_k141_135909 | 1058 | No | NA | 1 | 0 | 0 | Not-determined | Genome-fragment | NA | NA | 0 |
| S8_extracted_k141_83928 | 1575 | No | NA | 2 | 0 | 1 | Not-determined | Genome-fragment | NA | NA | 0 |
| S12_extracted_k141_168154 | 9292 | No | NA | 10 | 2 | 0 | Low-quality | Genome-fragment | 21.81 | AAI-based (high-confidence) | 0 |
| S8_extracted_k141_75137 | 1354 | No | NA | 2 | 0 | 2 | Not-determined | Genome-fragment | NA | NA | 0 |
| S8_extracted_k141_153568 | 5134 | No | NA | 5 | 0 | 4 | Not-determined | Genome-fragment | NA | NA | 0 |
| S8_extracted_k141_40930 | 1725 | No | NA | 4 | 0 | 4 | Not-determined | Genome-fragment | NA | NA | 0 |
| S3_vs_k141_9500\|\|full | 9300 | No | NA | 7 | 0 | 6 | Low-quality | Genome-fragment | 5.01 | AAI-based (medium-confidence) | 0 |
| S3_extracted_k141_44565 | 9302 | Yes | 4835 | 10 | 1 | 1 | Low-quality | Genome-fragment | 3.12 | AAI-based (high-confidence) | 48.02 |
| S8_extracted_k141_179674 | 2942 | No | NA | 2 | 0 | 1 | Not-determined | Genome-fragment | NA | NA | 0 |
| S8_extracted_k141_110609 | 1233 | No | NA | 3 | 0 | 1 | Not-determined | Genome-fragment | NA | NA | 0 |
| S8_extracted_k141_49094 | 7272 | No | NA | 9 | 0 | 4 | Not-determined | Genome-fragment | NA | NA | 0 |
| S10_vs_89977\|\|full | 9303 | No | NA | 11 | 8 | 0 | Low-quality | Genome-fragment | 26.94 | AAI-based (medium-confidence) | 0 |
| S8_extracted_k141_58491 | 1778 | No | NA | 1 | 0 | 1 | Not-determined | Genome-fragment | NA | NA | 0 |
| S8_extracted_k141_32285 | 1075 | No | NA | 2 | 0 | 0 | Not-determined | Genome-fragment | NA | NA | 0 |
| S8_vs_k141_199613\|\|full | 9303 | Yes | 827 | 10 | 1 | 7 | Low-quality | Genome-fragment | 0.56 | HMM-based (lower-bound) | 91.11 |
| S8_vs_k141_206944\|\|full | 9316 | No | NA | 7 | 0 | 6 | Low-quality | Genome-fragment | 3.14 | AAI-based (medium-confidence) | 0 |
| S8_extracted_k141_197123 | 1408 | No | NA | 1 | 0 | 1 | Not-determined | Genome-fragment | NA | NA | 0 |
| S12_vs_k141_98769\|\|full | 9320 | No | NA | 12 | 7 | 0 | Low-quality | Genome-fragment | 20.01 | AAI-based (high-confidence) | 0 |
| S6_vs_k141_14233\|\|full | 9324 | No | NA | 14 | 6 | 0 | Low-quality | Genome-fragment | 20.27 | AAI-based (medium-confidence) | 0 |
| S8_extracted_k141_23791 | 3724 | No | NA | 4 | 0 | 2 | Not-determined | Genome-fragment | NA | NA | 0 |
| S5_extracted_k141_135439 | 9327 | No | NA | 13 | 9 | 0 | Low-quality | Genome-fragment | 19.5 | AAI-based (high-confidence) | 0 |
| S8_extracted_k141_23794 | 1023 | No | NA | 1 | 0 | 0 | Not-determined | Genome-fragment | NA | NA | 0 |
| S11_vs_k141_59395\|\|full | 9335 | No | NA | 13 | 1 | 0 | Low-quality | Genome-fragment | 6.05 | AAI-based (high-confidence) | 0 |
| S8_extracted_k141_118977 | 1997 | No | NA | 1 | 0 | 0 | Not-determined | Genome-fragment | NA | NA | 0 |
| S8_extracted_k141_101204 | 1561 | No | NA | 2 | 0 | 2 | Not-determined | Genome-fragment | NA | NA | 0 |
| S8_extracted_k141_153610 | 1410 | No | NA | 2 | 0 | 0 | Not-determined | Genome-fragment | NA | NA | 0 |
| S17_extracted_k141_41935 | 9340 | No | NA | 16 | 0 | 2 | Low-quality | Genome-fragment | 7.3 | AAI-based (medium-confidence) | 0 |
| S8_extracted_k141_110467 | 1118 | No | NA | 1 | 0 | 0 | Not-determined | Genome-fragment | NA | NA | 0 |
| S14_vs_k141_45012\|\|full | 9343 | No | NA | 15 | 7 | 0 | Low-quality | Genome-fragment | 24.39 | AAI-based (high-confidence) | 0 |
| S8_extracted_k141_153612 | 1491 | No | NA | 2 | 0 | 1 | Not-determined | Genome-fragment | NA | NA | 0 |
| S7_vs_87122\|\|full | 9351 | No | NA | 18 | 2 | 0 | Low-quality | Genome-fragment | 16.3 | AAI-based (high-confidence) | 0 |
| S7_vs_9413\|\|full | 9355 | No | NA | 12 | 1 | 7 | Low-quality | Genome-fragment | 3.23 | HMM-based (lower-bound) | 0 |
| S8_extracted_k141_58530 | 1133 | No | NA | 1 | 0 | 1 | Not-determined | Genome-fragment | NA | NA | 0 |
| S8_extracted_k141_118993 | 1068 | No | NA | 2 | 0 | 2 | Not-determined | Genome-fragment | NA | NA | 0 |
| S3_vs_k141_99719\|\|full | 9358 | No | NA | 12 | 0 | 7 | Low-quality | Genome-fragment | 15.96 | AAI-based (medium-confidence) | 0 |
| S8_extracted_k141_197167 | 2019 | No | NA | 2 | 0 | 0 | Not-determined | Genome-fragment | NA | NA | 0 |
| S8_extracted_k141_6186 | 2039 | No | NA | 3 | 0 | 1 | Not-determined | Genome-fragment | NA | NA | 0 |
| S3_extracted_k141_111291 | 9363 | No | NA | 14 | 0 | 0 | Low-quality | Genome-fragment | 16.52 | AAI-based (high-confidence) | 0 |
| S8_extracted_k141_127601 | 1382 | No | NA | 3 | 0 | 2 | Not-determined | Genome-fragment | NA | NA | 0 |
| S8_extracted_k141_58547 | 1029 | No | NA | 1 | 0 | 1 | Not-determined | Genome-fragment | NA | NA | 0 |
| S8_extracted_k141_23839 | 1038 | No | NA | 2 | 0 | 0 | Not-determined | Genome-fragment | NA | NA | 0 |
| S8_extracted_k141_83986 | 1030 | No | NA | 2 | 0 | 1 | Not-determined | Genome-fragment | NA | NA | 0 |
| S8_extracted_k141_171020 | 1156 | No | NA | 1 | 0 | 1 | Not-determined | Genome-fragment | NA | NA | 0 |
| S8_extracted_k141_145090 | 1358 | No | NA | 1 | 0 | 1 | Not-determined | Genome-fragment | NA | NA | 0 |
| S8_extracted_k141_66601 | 1428 | No | NA | 2 | 0 | 2 | Not-determined | Genome-fragment | NA | NA | 0 |
| S8_extracted_k141_14534 | 1197 | No | NA | 2 | 0 | 1 | Not-determined | Genome-fragment | NA | NA | 0 |
| S8_extracted_k141_161937 | 1502 | No | NA | 2 | 0 | 0 | Not-determined | Genome-fragment | NA | NA | 0 |
| S8_extracted_k141_49147 | 2442 | No | NA | 1 | 0 | 0 | Not-determined | Genome-fragment | NA | NA | 0 |
| S8_extracted_k141_119032 | 1001 | No | NA | 2 | 0 | 1 | Not-determined | Genome-fragment | NA | NA | 0 |
| S12_vs_k141_84494\|\|full | 9364 | No | NA | 13 | 4 | 0 | Low-quality | Genome-fragment | 27.61 | AAI-based (medium-confidence) | 0 |
| S8_extracted_k141_92828 | 1335 | No | NA | 4 | 0 | 0 | Not-determined | Genome-fragment | NA | NA | 0 |
| S8_extracted_k141_187548 | 1214 | No | NA | 2 | 0 | 0 | Not-determined | Genome-fragment | NA | NA | 0 |
| S8_extracted_k141_171051 | 1512 | No | NA | 2 | 0 | 1 | Not-determined | Genome-fragment | NA | NA | 0 |
| S8_extracted_k141_187552 | 1041 | No | NA | 2 | 0 | 1 | Not-determined | Genome-fragment | NA | NA | 0 |
| S8_extracted_k141_66630 | 1646 | No | NA | 2 | 0 | 1 | Not-determined | Genome-fragment | NA | NA | 0 |
| S8_extracted_k141_179754 | 1649 | No | NA | 3 | 0 | 1 | Not-determined | Genome-fragment | NA | NA | 0 |
| S7_vs_100647\|\|full | 9376 | No | NA | 10 | 3 | 0 | Low-quality | Genome-fragment | 17.21 | AAI-based (high-confidence) | 0 |
| S8_extracted_k141_145119 | 9768 | No | NA | 12 | 0 | 1 | Not-determined | Genome-fragment | NA | NA | 0 |
| S3_extracted_k141_6037 | 9379 | No | NA | 8 | 4 | 0 | Low-quality | Genome-fragment | 22.31 | AAI-based (high-confidence) | 0 |
| S8_extracted_k141_127641 | 3741 | No | NA | 2 | 0 | 2 | Not-determined | Genome-fragment | NA | NA | 0 |
| S8_extracted_k141_187557 | 1287 | No | NA | 3 | 0 | 1 | Not-determined | Genome-fragment | NA | NA | 0 |
| S8_extracted_k141_171057 | 6881 | No | NA | 6 | 0 | 1 | Not-determined | Genome-fragment | NA | NA | 0 |
| S8_extracted_k141_136054 | 1974 | No | NA | 4 | 0 | 2 | Not-determined | Genome-fragment | NA | NA | 0 |
| S8_extracted_k141_14569 | 2232 | No | NA | 3 | 0 | 3 | Not-determined | Genome-fragment | NA | NA | 0 |
| S5_vs_k141_107441\|\|full | 9385 | No | NA | 16 | 3 | 0 | Low-quality | Genome-fragment | 37.8 | AAI-based (medium-confidence) | 0 |
| S17_extracted_k141_133886 | 9387 | No | NA | 14 | 2 | 0 | Low-quality | Genome-fragment | 19.17 | AAI-based (high-confidence) | 0 |
| S8_extracted_k141_32353 | 5183 | No | NA | 11 | 0 | 0 | Not-determined | Genome-fragment | NA | NA | 0 |
| S8_extracted_k141_23872 | 7461 | No | NA | 4 | 0 | 3 | Not-determined | Genome-fragment | NA | NA | 0 |
| S8_extracted_k141_187580 | 1250 | No | NA | 2 | 0 | 1 | Not-determined | Genome-fragment | NA | NA | 0 |
| S15_extracted_713 | 9389 | No | NA | 10 | 0 | 2 | Low-quality | Genome-fragment | 7.93 | AAI-based (medium-confidence) | 0 |
| S8_extracted_k141_153696 | 16486 | No | NA | 22 | 6 | 0 | Medium-quality | Genome-fragment | 74.36 | AAI-based (medium-confidence) | 0 |
| S8_extracted_k141_136082 | 4452 | No | NA | 2 | 0 | 0 | Not-determined | Genome-fragment | NA | NA | 0 |
| S8_extracted_k141_179781 | 1865 | No | NA | 3 | 0 | 2 | Not-determined | Genome-fragment | NA | NA | 0 |
| S8_extracted_k141_197247 | 1203 | No | NA | 1 | 0 | 0 | Not-determined | Genome-fragment | NA | NA | 0 |
| S8_extracted_k141_110529 | 1376 | No | NA | 1 | 0 | 0 | Not-determined | Genome-fragment | NA | NA | 0 |
| S8_extracted_k141_101283 | 32234 | No | NA | 40 | 17 | 0 | Medium-quality | Genome-fragment | 58.69 | AAI-based (high-confidence) | 0 |
| S8_extracted_k141_14586 | 1167 | No | NA | 2 | 0 | 1 | Not-determined | Genome-fragment | NA | NA | 0 |
| S8_extracted_k141_161985 | 1551 | No | NA | 3 | 0 | 0 | Not-determined | Genome-fragment | NA | NA | 0 |
| S8_extracted_k141_127664 | 5749 | No | NA | 5 | 0 | 5 | Not-determined | Genome-fragment | NA | NA | 0 |
| S1_vs_k141_56304\|\|full | 9390 | No | NA | 11 | 3 | 0 | Low-quality | Genome-fragment | 15.7 | AAI-based (high-confidence) | 0 |
| S8_extracted_k141_162002 | 1135 | No | NA | 2 | 0 | 1 | Not-determined | Genome-fragment | NA | NA | 0 |
| S8_extracted_k141_205393 | 8146 | No | NA | 4 | 0 | 3 | Not-determined | Genome-fragment | NA | NA | 0 |
| S8_extracted_k141_23914 | 2948 | No | NA | 4 | 0 | 0 | Not-determined | Genome-fragment | NA | NA | 0 |
| S8_extracted_k141_197286 | 1084 | No | NA | 1 | 0 | 1 | Not-determined | Genome-fragment | NA | NA | 0 |
| S8_extracted_k141_197288 | 2409 | No | NA | 1 | 0 | 1 | Not-determined | Genome-fragment | NA | NA | 0 |
| S8_extracted_k141_197297 | 1699 | No | NA | 3 | 0 | 1 | Not-determined | Genome-fragment | NA | NA | 0 |
| S8_extracted_k141_136125 | 5750 | No | NA | 5 | 0 | 3 | Not-determined | Genome-fragment | NA | NA | 0 |
| S12_extracted_k141_34286 | 9390 | Yes | 7689 | 3 | 1 | 1 | Low-quality | Genome-fragment | 19.42 | AAI-based (medium-confidence) | 18.12 |
| S8_extracted_k141_14641 | 1210 | No | NA | 2 | 0 | 1 | Not-determined | Genome-fragment | NA | NA | 0 |
| S8_extracted_k141_110589 | 3068 | No | NA | 1 | 0 | 0 | Not-determined | Genome-fragment | NA | NA | 0 |
| S8_extracted_k141_6312 | 1414 | No | NA | 2 | 0 | 1 | Not-determined | Genome-fragment | NA | NA | 0 |
| S8_extracted_k141_179845 | 1032 | No | NA | 1 | 0 | 1 | Not-determined | Genome-fragment | NA | NA | 0 |
| S13_vs_56005\|\|full | 9399 | No | NA | 9 | 0 | 2 | Low-quality | Genome-fragment | 8.51 | AAI-based (medium-confidence) | 0 |
| S13_extracted_79387 | 9408 | No | NA | 14 | 3 | 0 | Low-quality | Genome-fragment | 17.2 | AAI-based (high-confidence) | 0 |
| S16_vs_172343\|\|full | 9408 | No | NA | 7 | 0 | 3 | Low-quality | Genome-fragment | 6.82 | AAI-based (high-confidence) | 0 |
| S8_extracted_k141_101339 | 3421 | No | NA | 5 | 0 | 3 | Not-determined | Genome-fragment | NA | NA | 0 |
| S8_extracted_k141_136136 | 1045 | No | NA | 2 | 0 | 0 | Not-determined | Genome-fragment | NA | NA | 0 |
| S8_extracted_k141_75405 | 12035 | No | NA | 4 | 0 | 2 | Not-determined | Genome-fragment | NA | NA | 0 |
| S8_extracted_k141_179858 | 1028 | No | NA | 1 | 0 | 1 | Not-determined | Genome-fragment | NA | NA | 0 |
| S8_extracted_k141_6342 | 1750 | No | NA | 2 | 0 | 2 | Not-determined | Genome-fragment | NA | NA | 0 |
| S8_extracted_k141_162063 | 1091 | No | NA | 1 | 0 | 0 | Not-determined | Genome-fragment | NA | NA | 0 |
| S8_extracted_k141_171178 | 1235 | No | NA | 2 | 0 | 1 | Not-determined | Genome-fragment | NA | NA | 0 |
| S8_extracted_k141_187644 | 1794 | No | NA | 2 | 0 | 0 | Not-determined | Genome-fragment | NA | NA | 0 |
| S16_vs_12812\|\|full | 9411 | No | NA | 15 | 4 | 0 | Low-quality | Genome-fragment | 21.41 | AAI-based (high-confidence) | 0 |
| S8_extracted_k141_110624 | 1377 | No | NA | 1 | 0 | 1 | Not-determined | Genome-fragment | NA | NA | 0 |
| S8_extracted_k141_136148 | 3017 | No | NA | 4 | 0 | 2 | Not-determined | Genome-fragment | NA | NA | 0 |
| S5_vs_k141_1215\|\|full | 9412 | No | NA | 11 | 2 | 0 | Low-quality | Genome-fragment | 7.35 | AAI-based (medium-confidence) | 0 |
| S4_extracted_18330 | 9418 | No | NA | 18 | 0 | 0 | Low-quality | Genome-fragment | 10.1 | AAI-based (high-confidence) | 0 |
| S8_extracted_k141_58742 | 1153 | No | NA | 2 | 0 | 1 | Not-determined | Genome-fragment | NA | NA | 0 |
| S8_extracted_k141_205464 | 4179 | No | NA | 2 | 0 | 2 | Not-determined | Genome-fragment | NA | NA | 0 |
| S8_extracted_k141_58758 | 3044 | No | NA | 4 | 0 | 3 | Not-determined | Genome-fragment | NA | NA | 0 |
| S8_extracted_k141_119156 | 1013 | No | NA | 2 | 0 | 2 | Not-determined | Genome-fragment | NA | NA | 0 |
| S8_extracted_k141_93002 | 1160 | No | NA | 1 | 0 | 0 | Not-determined | Genome-fragment | NA | NA | 0 |
| S17_extracted_k141_94002 | 9419 | No | NA | 10 | 1 | 1 | Low-quality | Genome-fragment | 11.61 | HMM-based (lower-bound) | 0 |
| S8_extracted_k141_179922 | 1021 | No | NA | 2 | 0 | 0 | Not-determined | Genome-fragment | NA | NA | 0 |
| S8_extracted_k141_93015 | 1290 | No | NA | 1 | 0 | 1 | Not-determined | Genome-fragment | NA | NA | 0 |
| S8_extracted_k141_162087 | 1109 | No | NA | 2 | 0 | 1 | Not-determined | Genome-fragment | NA | NA | 0 |
| S8_extracted_k141_14708 | 2536 | No | NA | 3 | 0 | 1 | Not-determined | Genome-fragment | NA | NA | 0 |
| S8_extracted_k141_6405 | 1229 | No | NA | 2 | 0 | 2 | Not-determined | Genome-fragment | NA | NA | 0 |
| S8_extracted_k141_119179 | 1110 | No | NA | 2 | 0 | 0 | Not-determined | Genome-fragment | NA | NA | 0 |
| S8_extracted_k141_66753 | 1280 | No | NA | 2 | 0 | 2 | Not-determined | Genome-fragment | NA | NA | 0 |
| S8_extracted_k141_162100 | 1156 | No | NA | 2 | 0 | 2 | Not-determined | Genome-fragment | NA | NA | 0 |
| S8_extracted_k141_153861 | 1295 | No | NA | 1 | 0 | 1 | Not-determined | Genome-fragment | NA | NA | 0 |
| S8_extracted_k141_14720 | 3959 | No | NA | 5 | 0 | 5 | Not-determined | Genome-fragment | NA | NA | 0 |
| S8_extracted_k141_58798 | 3671 | No | NA | 2 | 0 | 1 | Not-determined | Genome-fragment | NA | NA | 0 |
| S8_extracted_k141_153871 | 3168 | No | NA | 5 | 0 | 0 | Not-determined | Genome-fragment | NA | NA | 0 |
| S8_extracted_k141_32532 | 1331 | No | NA | 1 | 0 | 0 | Not-determined | Genome-fragment | NA | NA | 0 |
| S5_vs_k141_145627\|\|full | 9421 | No | NA | 14 | 0 | 0 | Low-quality | Genome-fragment | 25.05 | AAI-based (medium-confidence) | 0 |
| S8_extracted_k141_153873 | 3287 | No | NA | 4 | 0 | 1 | Not-determined | Genome-fragment | NA | NA | 0 |
| S8_extracted_k141_197423 | 1941 | No | NA | 1 | 0 | 1 | Not-determined | Genome-fragment | NA | NA | 0 |
| S8_extracted_k141_41109 | 2243 | No | NA | 3 | 0 | 2 | Not-determined | Genome-fragment | NA | NA | 0 |
| S8_extracted_k141_187712 | 4709 | No | NA | 1 | 0 | 1 | Not-determined | Genome-fragment | NA | NA | 0 |
| S8_extracted_k141_153895 | 1237 | No | NA | 2 | 0 | 2 | Not-determined | Genome-fragment | NA | NA | 0 |
| S8_extracted_k141_14746 | 8606 | No | NA | 3 | 0 | 1 | Not-determined | Genome-fragment | NA | NA | 0 |
| S8_extracted_k141_49191 | 1006 | No | NA | 1 | 0 | 1 | Not-determined | Genome-fragment | NA | NA | 0 |
| S8_extracted_k141_119221 | 2013 | No | NA | 5 | 0 | 1 | Not-determined | Genome-fragment | NA | NA | 0 |
| S8_extracted_k141_32571 | 1045 | No | NA | 3 | 0 | 0 | Not-determined | Genome-fragment | NA | NA | 0 |
| S8_extracted_k141_6455 | 3735 | No | NA | 3 | 0 | 2 | Not-determined | Genome-fragment | NA | NA | 0 |
| S7_vs_23473\|\|full | 9424 | No | NA | 14 | 1 | 0 | Low-quality | Genome-fragment | 21.89 | AAI-based (medium-confidence) | 0 |
| S8_extracted_k141_180003 | 1862 | No | NA | 3 | 0 | 1 | Not-determined | Genome-fragment | NA | NA | 0 |
| S14_vs_k141_117156\|\|full | 9430 | No | NA | 17 | 3 | 0 | Low-quality | Genome-fragment | 20.96 | AAI-based (medium-confidence) | 0 |
| S8_extracted_k141_101476 | 1137 | No | NA | 2 | 0 | 1 | Not-determined | Genome-fragment | NA | NA | 0 |
| S8_extracted_k141_49207 | 4817 | No | NA | 2 | 0 | 1 | Not-determined | Genome-fragment | NA | NA | 0 |
| S12_vs_k141_184362\|\|full | 9431 | Yes | 2476 | 7 | 1 | 2 | Low-quality | Genome-fragment | 3.51 | HMM-based (lower-bound) | 73.75 |
| S8_extracted_k141_41144 | 3087 | No | NA | 4 | 0 | 0 | Not-determined | Genome-fragment | NA | NA | 0 |
| S8_extracted_k141_32585 | 1741 | No | NA | 1 | 0 | 1 | Not-determined | Genome-fragment | NA | NA | 0 |

| | | | | | | | | | | | | | |
|---|---|---|---|---|---|---|---|---|---|---|---|---|---|
| S8_extracted_k141_75495 | 1546 | No | NA | | 2 | 0 | 1 | Not-determined | Genome-fragment | NA | | NA | 0 |
| S8_extracted_k141_162178 | 1126 | No | NA | | 2 | 0 | 2 | Not-determined | Genome-fragment | NA | | NA | 0 |
| S8_extracted_k141_205542 | 1508 | No | NA | | 3 | 0 | 1 | Not-determined | Genome-fragment | NA | | NA | 0 |
| S8_extracted_k141_23955 | 92207 | No | NA | | 159 | 42 | 0 | High-quality | High-quality | | 99.85 | AAI-based (high-confidence) | 0 |
| S8_extracted_k141_145277 | 9079 | No | NA | | 6 | 0 | 4 | Not-determined | Genome-fragment | NA | | NA | 0 |
| S8_extracted_k141_136235 | 15891 | No | NA | | 17 | 7 | 0 | Medium-quality | Genome-fragment | | 54.83 | HMM-based (lower-bound) | 0 |
| S7_extracted_43809 | 9434 | No | NA | | 6 | 2 | 0 | Low-quality | Genome-fragment | | 13.81 | AAI-based (medium-confidence) | 0 |
| S8_extracted_k141_119254 | 2548 | No | NA | | 2 | 0 | 1 | Not-determined | Genome-fragment | NA | | NA | 0 |
| S8_extracted_k141_93103 | 2125 | No | NA | | 2 | 0 | 2 | Not-determined | Genome-fragment | NA | | NA | 0 |
| S8_extracted_k141_180036 | 2066 | No | NA | | 2 | 0 | 1 | Not-determined | Genome-fragment | NA | | NA | 0 |
| S8_extracted_k141_110741 | 8633 | No | NA | | 2 | 0 | 1 | Not-determined | Genome-fragment | NA | | NA | 0 |
| S8_extracted_k141_58907 | 1060 | No | NA | | 2 | 0 | 0 | Not-determined | Genome-fragment | NA | | NA | 0 |
| S8_extracted_k141_41193 | 2135 | No | NA | | 2 | 0 | 2 | Not-determined | Genome-fragment | NA | | NA | 0 |
| S8_extracted_k141_136250 | 7585 | No | NA | | 5 | 0 | 3 | Not-determined | Genome-fragment | NA | | NA | 0 |
| S8_extracted_k141_119261 | 1206 | No | NA | | 2 | 0 | 2 | Not-determined | Genome-fragment | NA | | NA | 0 |
| S10_extracted_44931 | 9436 | No | NA | | 5 | 0 | 0 | Low-quality | Genome-fragment | | 5.78 | AAI-based (high-confidence) | 0 |
| S3_extracted_k141_53325 | 9442 | No | NA | | 17 | 9 | 0 | Low-quality | Genome-fragment | | 23.44 | AAI-based (high-confidence) | 0 |
| S5_vs_k141_85031\|\|full | 9445 | No | NA | | 13 | 6 | 0 | Low-quality | Genome-fragment | | 10.96 | AAI-based (high-confidence) | 0 |
| S8_extracted_k141_14814 | 1200 | No | NA | | 2 | 0 | 2 | Not-determined | Genome-fragment | NA | | NA | 0 |
| S8_extracted_k141_75549 | 1015 | No | NA | | 2 | 0 | 2 | Not-determined | Genome-fragment | NA | | NA | 0 |
| S8_extracted_k141_145312 | 1076 | No | NA | | 1 | 0 | 1 | Not-determined | Genome-fragment | NA | | NA | 0 |
| S16_vs_87197\|\|full | 9446 | No | NA | | 11 | 6 | 1 | Low-quality | Genome-fragment | | 11.72 | AAI-based (high-confidence) | 0 |
| S8_extracted_k141_136274 | 1362 | No | NA | | 2 | 0 | 0 | Not-determined | Genome-fragment | NA | | NA | 0 |
| S8_extracted_k141_14834 | 3463 | No | NA | | 3 | 0 | 1 | Not-determined | Genome-fragment | NA | | NA | 0 |
| S8_extracted_k141_75566 | 1301 | No | NA | | 2 | 0 | 0 | Not-determined | Genome-fragment | NA | | NA | 0 |
| S8_extracted_k141_84290 | 2127 | No | NA | | 2 | 0 | 1 | Not-determined | Genome-fragment | NA | | NA | 0 |
| S8_extracted_k141_66861 | 6204 | No | NA | | 8 | 0 | 1 | Not-determined | Genome-fragment | NA | | NA | 0 |
| S8_extracted_k141_14838 | 4844 | No | NA | | 3 | 0 | 2 | Not-determined | Genome-fragment | NA | | NA | 0 |
| S8_extracted_k141_14844 | 1609 | No | NA | | 2 | 0 | 2 | Not-determined | Genome-fragment | NA | | NA | 0 |
| S8_extracted_k141_119319 | 1549 | No | NA | | 2 | 0 | 0 | Not-determined | Genome-fragment | NA | | NA | 0 |
| S8_extracted_k141_24017 | 1126 | No | NA | | 1 | 0 | 1 | Not-determined | Genome-fragment | NA | | NA | 0 |
| S8_extracted_k141_205627 | 4117 | No | NA | | 2 | 0 | 2 | Not-determined | Genome-fragment | NA | | NA | 0 |
| S14_vs_k141_77511\|\|full | 9447 | No | NA | | 22 | 2 | 0 | Low-quality | Genome-fragment | | 27.59 | AAI-based (high-confidence) | 0 |
| S8_extracted_k141_127927 | 4626 | No | NA | | 3 | 0 | 3 | Not-determined | Genome-fragment | NA | | NA | 0 |
| S8_extracted_k141_24027 | 3790 | No | NA | | 4 | 0 | 1 | Not-determined | Genome-fragment | NA | | NA | 0 |
| S8_extracted_k141_101596 | 1186 | No | NA | | 2 | 0 | 1 | Not-determined | Genome-fragment | NA | | NA | 0 |
| S8_extracted_k141_145350 | 3301 | No | NA | | 3 | 0 | 1 | Not-determined | Genome-fragment | NA | | NA | 0 |
| S8_extracted_k141_136302 | 1350 | No | NA | | 2 | 0 | 0 | Not-determined | Genome-fragment | NA | | NA | 0 |
| S8_extracted_k141_153993 | 1035 | No | NA | | 1 | 0 | 1 | Not-determined | Genome-fragment | NA | | NA | 0 |
| S8_extracted_k141_110831 | 1011 | No | NA | | 2 | 0 | 0 | Not-determined | Genome-fragment | NA | | NA | 0 |
| S7_vs_65641\|\|full | 9451 | No | NA | | 12 | 9 | 0 | Low-quality | Genome-fragment | | 27.04 | AAI-based (high-confidence) | 0 |
| S8_extracted_k141_93193 | 1932 | No | NA | | 2 | 0 | 1 | Not-determined | Genome-fragment | NA | | NA | 0 |
| S8_extracted_k141_93197 | 1285 | No | NA | | 1 | 0 | 1 | Not-determined | Genome-fragment | NA | | NA | 0 |
| S8_extracted_k141_14914 | 3172 | No | NA | | 3 | 0 | 2 | Not-determined | Genome-fragment | NA | | NA | 0 |
| S7_vs_116644\|\|full | 9453 | No | NA | | 10 | 3 | 0 | Low-quality | Genome-fragment | | 22.76 | AAI-based (high-confidence) | 0 |
| S8_extracted_k141_101627 | 1420 | No | NA | | 1 | 0 | 1 | Not-determined | Genome-fragment | NA | | NA | 0 |
| S8_extracted_k141_136342 | 2548 | No | NA | | 2 | 0 | 2 | Not-determined | Genome-fragment | NA | | NA | 0 |
| S8_extracted_k141_24074 | 1048 | No | NA | | 2 | 0 | 1 | Not-determined | Genome-fragment | NA | | NA | 0 |
| S8_extracted_k141_41252 | 1624 | No | NA | | 3 | 0 | 2 | Not-determined | Genome-fragment | NA | | NA | 0 |
| S8_extracted_k141_2232 | 9453 | No | NA | | 5 | 0 | 3 | Low-quality | Genome-fragment | | 6.03 | AAI-based (high-confidence) | 0 |
| S8_extracted_k141_24080 | 1337 | No | NA | | 2 | 0 | 2 | Not-determined | Genome-fragment | NA | | NA | 0 |
| S8_extracted_k141_187742 | 126096 | No | NA | | 181 | 27 | 3 | High-quality | High-quality | | 94.41 | AAI-based (high-confidence) | 0 |
| S8_extracted_k141_93222 | 1291 | No | NA | | 1 | 0 | 1 | Not-determined | Genome-fragment | NA | | NA | 0 |
| S8_extracted_k141_154040 | 1054 | No | NA | | 2 | 0 | 2 | Not-determined | Genome-fragment | NA | | NA | 0 |
| S8_extracted_k141_24088 | 1373 | No | NA | | 1 | 0 | 1 | Not-determined | Genome-fragment | NA | | NA | 0 |
| S8_extracted_k141_6596 | 3024 | No | NA | | 3 | 0 | 0 | Not-determined | Genome-fragment | NA | | NA | 0 |
| S8_extracted_k141_162247 | 1081 | No | NA | | 1 | 0 | 1 | Not-determined | Genome-fragment | NA | | NA | 0 |
| S8_extracted_k141_154051 | 1671 | No | NA | | 4 | 0 | 3 | Not-determined | Genome-fragment | NA | | NA | 0 |
| S8_extracted_k141_136354 | 1617 | No | NA | | 2 | 0 | 2 | Not-determined | Genome-fragment | NA | | NA | 0 |
| S8_extracted_k141_136362 | 1020 | No | NA | | 1 | 0 | 0 | Not-determined | Genome-fragment | NA | | NA | 0 |
| S8_extracted_k141_59009 | 1248 | No | NA | | 1 | 0 | 0 | Not-determined | Genome-fragment | NA | | NA | 0 |
| S8_extracted_k141_75674 | 1036 | No | NA | | 1 | 0 | 1 | Not-determined | Genome-fragment | NA | | NA | 0 |
| S8_extracted_k141_197626 | 10436 | No | NA | | 9 | 0 | 6 | Not-determined | Genome-fragment | NA | | NA | 0 |
| S8_extracted_k141_180170 | 1359 | No | NA | | 1 | 0 | 0 | Not-determined | Genome-fragment | NA | | NA | 0 |
| S8_extracted_k141_49282 | 99212 | No | NA | | 94 | 14 | 1 | High-quality | High-quality | | 99.74 | AAI-based (high-confidence) | 0 |
| S8_extracted_k141_32714 | 1208 | No | NA | | 3 | 0 | 1 | Not-determined | Genome-fragment | NA | | NA | 0 |
| S8_extracted_k141_162252 | 1207 | No | NA | | 1 | 0 | 0 | Not-determined | Genome-fragment | NA | | NA | 0 |
| S14_vs_k141_5965\|\|full | 9455 | No | NA | | 11 | 1 | 2 | Low-quality | Genome-fragment | | 6.81 | HMM-based (lower-bound) | 0 |
| S8_extracted_k141_24124 | 3453 | No | NA | | 5 | 0 | 3 | Not-determined | Genome-fragment | NA | | NA | 0 |
| S8_extracted_k141_14984 | 9931 | No | NA | | 9 | 0 | 6 | Not-determined | Genome-fragment | NA | | NA | 0 |
| S7_extracted_47374 | 9455 | No | NA | | 6 | 0 | 6 | Low-quality | Genome-fragment | | 3.98 | AAI-based (medium-confidence) | 0 |
| S8_extracted_k141_171484 | 1270 | No | NA | | 2 | 0 | 1 | Not-determined | Genome-fragment | NA | | NA | 0 |
| S8_extracted_k141_136418 | 1609 | No | NA | | 2 | 0 | 0 | Not-determined | Genome-fragment | NA | | NA | 0 |
| S8_extracted_k141_136424 | 2586 | No | NA | | 2 | 0 | 1 | Not-determined | Genome-fragment | NA | | NA | 0 |
| S8_extracted_k141_162292 | 4718 | No | NA | | 5 | 0 | 5 | Not-determined | Genome-fragment | NA | | NA | 0 |
| S8_extracted_k141_101730 | 1132 | No | NA | | 2 | 0 | 2 | Not-determined | Genome-fragment | NA | | NA | 0 |
| S8_extracted_k141_6664 | 5127 | No | NA | | 6 | 0 | 5 | Not-determined | Genome-fragment | NA | | NA | 0 |
| S8_extracted_k141_180247 | 3271 | No | NA | | 3 | 0 | 1 | Not-determined | Genome-fragment | NA | | NA | 0 |
| S3_vs_k141_42463\|\|full | 9460 | No | NA | | 18 | 2 | 0 | Low-quality | Genome-fragment | | 19.66 | AAI-based (high-confidence) | 0 |
| S5_vs_k141_416\|\|full | 9463 | No | NA | | 12 | 1 | 1 | Low-quality | Genome-fragment | | 5.95 | HMM-based (lower-bound) | 0 |
| S8_extracted_k141_101737 | 1262 | No | NA | | 2 | 0 | 1 | Not-determined | Genome-fragment | NA | | NA | 0 |
| S16_extracted_36476 | 9465 | No | NA | | 10 | 0 | 4 | Low-quality | Genome-fragment | | 2.92 | AAI-based (medium-confidence) | 0 |
| S8_extracted_k141_67052 | 2502 | No | NA | | 2 | 0 | 2 | Not-determined | Genome-fragment | NA | | NA | 0 |
| S8_extracted_k141_205790 | 3575 | No | NA | | 4 | 0 | 2 | Not-determined | Genome-fragment | NA | | NA | 0 |
| S8_extracted_k141_59087 | 1497 | No | NA | | 1 | 0 | 0 | Not-determined | Genome-fragment | NA | | NA | 0 |
| S8_extracted_k141_93271 | 1652 | No | NA | | 2 | 0 | 1 | Not-determined | Genome-fragment | NA | | NA | 0 |
| S8_extracted_k141_75760 | 1140 | No | NA | | 1 | 0 | 0 | Not-determined | Genome-fragment | NA | | NA | 0 |
| S8_extracted_k141_197700 | 1163 | No | NA | | 1 | 0 | 0 | Not-determined | Genome-fragment | NA | | NA | 0 |
| S8_extracted_k141_32781 | 1210 | No | NA | | 1 | 0 | 1 | Not-determined | Genome-fragment | NA | | NA | 0 |
| S8_extracted_k141_75762 | 1078 | No | NA | | 1 | 0 | 0 | Not-determined | Genome-fragment | NA | | NA | 0 |
| S14_extracted_k141_105385 | 9477 | No | NA | | 11 | 2 | 0 | Low-quality | Genome-fragment | | 12 | AAI-based (high-confidence) | 0 |
| S8_extracted_k141_24177 | 8473 | No | NA | | 8 | 0 | 4 | Not-determined | Genome-fragment | NA | | NA | 0 |
| S8_extracted_k141_75773 | 1706 | No | NA | | 2 | 0 | 0 | Not-determined | Genome-fragment | NA | | NA | 0 |
| S8_extracted_k141_93279 | 5643 | No | NA | | 4 | 0 | 1 | Not-determined | Genome-fragment | NA | | NA | 0 |
| S8_extracted_k141_84489 | 2771 | No | NA | | 2 | 0 | 2 | Not-determined | Genome-fragment | NA | | NA | 0 |
| S6_vs_k141_25900\|\|full | 9480 | No | NA | | 13 | 0 | 1 | Low-quality | Genome-fragment | | 33.93 | AAI-based (medium-confidence) | 0 |
| S8_extracted_k141_65406 | 9480 | No | NA | | 22 | 6 | 0 | Low-quality | Genome-fragment | | 9.2 | HMM-based (lower-bound) | 0 |
| S8_extracted_k141_67084 | 2062 | No | NA | | 1 | 0 | 1 | Not-determined | Genome-fragment | NA | | NA | 0 |
| S8_extracted_k141_180301 | 3063 | No | NA | | 1 | 0 | 1 | Not-determined | Genome-fragment | NA | | NA | 0 |
| S8_extracted_k141_24201 | 2373 | No | NA | | 2 | 0 | 2 | Not-determined | Genome-fragment | NA | | NA | 0 |
| S8_extracted_k141_171574 | 1108 | No | NA | | 1 | 0 | 1 | Not-determined | Genome-fragment | NA | | NA | 0 |
| S8_extracted_k141_110854 | 1011 | No | NA | | 2 | 0 | 1 | Not-determined | Genome-fragment | NA | | NA | 0 |
| S8_extracted_k141_75801 | 8652 | No | NA | | 6 | 0 | 5 | Not-determined | Genome-fragment | NA | | NA | 0 |
| S8_extracted_k141_24205 | 1174 | No | NA | | 1 | 0 | 1 | Not-determined | Genome-fragment | NA | | NA | 0 |
| S8_extracted_k141_128091 | 10082 | No | NA | | 7 | 0 | 1 | Not-determined | Genome-fragment | NA | | NA | 0 |
| S8_extracted_k141_187838 | 1629 | No | NA | | 2 | 0 | 1 | Not-determined | Genome-fragment | NA | | NA | 0 |
| S8_extracted_k141_187846 | 1608 | No | NA | | 2 | 0 | 0 | Not-determined | Genome-fragment | NA | | NA | 0 |
| S17_vs_k141_140244\|\|full | 9492 | No | NA | | 17 | 0 | 1 | Low-quality | Genome-fragment | | 2.37 | AAI-based (high-confidence) | 0 |
| S8_extracted_k141_187847 | 1169 | No | NA | | 1 | 0 | 1 | Not-determined | Genome-fragment | NA | | NA | 0 |
| S11_vs_k141_20790\|\|full | 9493 | Yes | | 5809 | 9 | 2 | 1 | Low-quality | Genome-fragment | | 6.11 | AAI-based (medium-confidence) | 38.81 |
| S8_extracted_k141_6732 | 2188 | No | NA | | 1 | 0 | 1 | Not-determined | Genome-fragment | NA | | NA | 0 |
| S8_extracted_k141_145565 | 1364 | No | NA | | 2 | 0 | 1 | Not-determined | Genome-fragment | NA | | NA | 0 |
| S8_extracted_k141_24226 | 1047 | No | NA | | 1 | 0 | 1 | Not-determined | Genome-fragment | NA | | NA | 0 |
| S8_extracted_k141_110886 | 1149 | No | NA | | 2 | 0 | 2 | Not-determined | Genome-fragment | NA | | NA | 0 |
| S3_extracted_k141_2158 | 9505 | No | NA | | 18 | 3 | 0 | Low-quality | Genome-fragment | | 5.65 | AAI-based (medium-confidence) | 0 |
| S16_vs_45851\|\|full | 9507 | No | NA | | 13 | 1 | 2 | Low-quality | Genome-fragment | | 12.26 | AAI-based (medium-confidence) | 0 |
| S8_extracted_k141_67128 | 1297 | No | NA | | 1 | 0 | 1 | Not-determined | Genome-fragment | NA | | NA | 0 |
| S8_extracted_k141_171629 | 3345 | No | NA | | 3 | 0 | 3 | Not-determined | Genome-fragment | NA | | NA | 0 |
| S4_extracted_22016 | 9508 | No | NA | | 9 | 0 | 4 | Low-quality | Genome-fragment | | 5.97 | AAI-based (medium-confidence) | 0 |
| S8_extracted_k141_154211 | 1684 | No | NA | | 1 | 0 | 0 | Not-determined | Genome-fragment | NA | | NA | 0 |
| S8_extracted_k141_49410 | 6195 | No | NA | | 6 | 0 | 3 | Not-determined | Genome-fragment | NA | | NA | 0 |
| S8_extracted_k141_119454 | 23223 | No | NA | | 19 | 0 | 4 | Not-determined | Genome-fragment | NA | | NA | 0 |
| S8_extracted_k141_49411 | 3989 | No | NA | | 4 | 0 | 1 | Not-determined | Genome-fragment | NA | | NA | 0 |
| S8_extracted_k141_187888 | 1470 | No | NA | | 1 | 0 | 0 | Not-determined | Genome-fragment | NA | | NA | 0 |
| S8_extracted_k141_24262 | 1376 | No | NA | | 1 | 0 | 1 | Not-determined | Genome-fragment | NA | | NA | 0 |
| S8_extracted_k141_15042 | 1402 | No | NA | | 1 | 0 | 1 | Not-determined | Genome-fragment | NA | | NA | 0 |
| S8_extracted_k141_59184 | 1654 | No | NA | | 1 | 0 | 1 | Not-determined | Genome-fragment | NA | | NA | 0 |
| S16_vs_50088\|\|full | 9512 | Yes | | 3976 | 10 | 2 | 2 | Low-quality | Genome-fragment | | 2.72 | AAI-based (medium-confidence) | 58.2 |
| S8_extracted_k141_136530 | 3464 | No | NA | | 4 | 0 | 2 | Not-determined | Genome-fragment | NA | | NA | 0 |
| S8_extracted_k141_6770 | 1120 | No | NA | | 2 | 0 | 0 | Not-determined | Genome-fragment | NA | | NA | 0 |
| S8_extracted_k141_197795 | 1020 | No | NA | | 1 | 0 | 0 | Not-determined | Genome-fragment | NA | | NA | 0 |
| S8_extracted_k141_128144 | 2148 | No | NA | | 2 | 0 | 2 | Not-determined | Genome-fragment | NA | | NA | 0 |
| S8_extracted_k141_136537 | 1494 | No | NA | | 2 | 0 | 2 | Not-determined | Genome-fragment | NA | | NA | 0 |
| S12_vs_k141_89306\|\|full | 9518 | No | NA | | 9 | 0 | 3 | Low-quality | Genome-fragment | | 19.64 | AAI-based (medium-confidence) | 0 |
| S8_extracted_k141_101900 | 1399 | No | NA | | 1 | 0 | 1 | Not-determined | Genome-fragment | NA | | NA | 0 |
| S13_vs_44758\|\|full | 9526 | No | NA | | 11 | 0 | 8 | Low-quality | Genome-fragment | | 4.52 | AAI-based (medium-confidence) | 0 |
| S8_extracted_k141_59230 | 1147 | No | NA | | 2 | 0 | 2 | Not-determined | Genome-fragment | NA | | NA | 0 |
| S8_extracted_k141_101927 | 1299 | No | NA | | 1 | 0 | 1 | Not-determined | Genome-fragment | NA | | NA | 0 |
| S10_vs_23073\|\|full | 9530 | No | NA | | 13 | 0 | 2 | Low-quality | Genome-fragment | | 18.43 | AAI-based (medium-confidence) | 0 |
| S8_extracted_k141_145637 | 4338 | No | NA | | 4 | 0 | 4 | Not-determined | Genome-fragment | NA | | NA | 0 |
| S8_extracted_k141_128183 | 1140 | No | NA | | 1 | 0 | 1 | Not-determined | Genome-fragment | NA | | NA | 0 |
| S8_extracted_k141_101930 | 1109 | No | NA | | 2 | 0 | 2 | Not-determined | Genome-fragment | NA | | NA | 0 |
| S8_extracted_k141_119486 | 9177 | No | NA | | 6 | 0 | 3 | Not-determined | Genome-fragment | NA | | NA | 0 |
| S8_extracted_k141_49438 | 1161 | No | NA | | 1 | 0 | 1 | Not-determined | Genome-fragment | NA | | NA | 0 |
| S8_extracted_k141_119495 | 1071 | No | NA | | 1 | 0 | 1 | Not-determined | Genome-fragment | NA | | NA | 0 |
| S8_extracted_k141_171711 | 1058 | No | NA | | 1 | 0 | 0 | Not-determined | Genome-fragment | NA | | NA | 0 |
| S8_extracted_k141_6823 | 1901 | No | NA | | 3 | 0 | 1 | Not-determined | Genome-fragment | NA | | NA | 0 |
| S8_extracted_k141_24330 | 1717 | No | NA | | 3 | 0 | 1 | Not-determined | Genome-fragment | NA | | NA | 0 |
| S16_extracted_2323 | 9548 | No | NA | | 16 | 4 | 0 | Low-quality | Genome-fragment | | 25.02 | AAI-based (high-confidence) | 0 |
| S8_extracted_k141_171726 | 2688 | No | NA | | 4 | 0 | 2 | Not-determined | Genome-fragment | NA | | NA | 0 |
| S8_vs_k141_74331\|\|full | 9554 | No | NA | | 15 | 1 | 2 | Low-quality | Genome-fragment | | 23.57 | AAI-based (high-confidence) | 0 |
| S8_extracted_k141_136585 | 1095 | No | NA | | 1 | 0 | 1 | Not-determined | Genome-fragment | NA | | NA | 0 |
| S11_vs_k141_30223\|\|full | 9555 | No | NA | | 20 | 1 | 0 | Low-quality | Genome-fragment | | 23.44 | AAI-based (medium-confidence) | 0 |
| S8_extracted_k141_110935 | 1752 | No | NA | | 2 | 0 | 0 | Not-determined | Genome-fragment | NA | | NA | 0 |
| S8_extracted_k141_101957 | 2042 | No | NA | | 3 | 0 | 0 | Not-determined | Genome-fragment | NA | | NA | 0 |
| S8_extracted_k141_162491 | 7472 | No | NA | | 8 | 0 | 4 | Not-determined | Genome-fragment | NA | | NA | 0 |
| S8_extracted_k141_187955 | 1550 | No | NA | | 5 | 0 | 3 | Not-determined | Genome-fragment | NA | | NA | 0 |
| S8_extracted_k141_128225 | 1330 | No | NA | | 1 | 0 | 1 | Not-determined | Genome-fragment | NA | | NA | 0 |
| S8_extracted_k141_15140 | 1502 | No | NA | | 2 | 0 | 0 | Not-determined | Genome-fragment | NA | | NA | 0 |
| S8_extracted_k141_24345 | 1069 | No | NA | | 2 | 0 | 0 | Not-determined | Genome-fragment | NA | | NA | 0 |
| S8_extracted_k141_49493 | 1087 | No | NA | | 1 | 0 | 0 | Not-determined | Genome-fragment | NA | | NA | 0 |
| S8_extracted_k141_171752 | 3384 | No | NA | | 2 | 0 | 2 | Not-determined | Genome-fragment | NA | | NA | 0 |
| S8_extracted_k141_197865 | 4193 | No | NA | | 5 | 0 | 1 | Not-determined | Genome-fragment | NA | | NA | 0 |
| S7_vs_134886\|\|full | 9560 | No | NA | | 16 | 1 | 0 | Low-quality | Genome-fragment | | 6.15 | AAI-based (high-confidence) | 0 |
| S8_extracted_k141_6866 | 2180 | No | NA | | 4 | 0 | 1 | Not-determined | Genome-fragment | NA | | NA | 0 |
| S8_extracted_k141_93389 | 1275 | No | NA | | 2 | 0 | 2 | Not-determined | Genome-fragment | NA | | NA | 0 |
| S5_extracted_k141_110423 | 9567 | No | NA | | 14 | 9 | 0 | Low-quality | Genome-fragment | | 27.9 | AAI-based (high-confidence) | 0 |
| S8_extracted_k141_136616 | 6380 | No | NA | | 7 | 0 | 5 | Not-determined | Genome-fragment | NA | | NA | 0 |
| S16_extracted_131949 | 9568 | No | NA | | 21 | 5 | 1 | Low-quality | Genome-fragment | | 16.73 | AAI-based (high-confidence) | 0 |
| S8_extracted_k141_128257 | 4883 | No | NA | | 1 | 0 | 0 | Not-determined | Genome-fragment | NA | | NA | 0 |
| S8_extracted_k141_145706 | 1896 | No | NA | | 2 | 0 | 2 | Not-determined | Genome-fragment | NA | | NA | 0 |
| S8_extracted_k141_59333 | 1282 | No | NA | | 1 | 0 | 1 | Not-determined | Genome-fragment | NA | | NA | 0 |
| S7_vs_12720\|\|full | 9570 | Yes | | 7485 | 13 | 5 | 2 | Low-quality | Genome-fragment | | 16.68 | AAI-based (high-confidence) | 21.79 |
| S11_vs_k141_104051\|\|full | 9571 | No | NA | | 17 | 2 | 1 | Low-quality | Genome-fragment | | 12.33 | HMM-based (lower-bound) | 0 |
| S8_extracted_k141_197902 | 6407 | No | NA | | 4 | 0 | 3 | Not-determined | Genome-fragment | NA | | NA | 0 |
| S8_extracted_k141_110998 | 1132 | No | NA | | 1 | 0 | 0 | Not-determined | Genome-fragment | NA | | NA | 0 |
| S2_vs_k141_8551\|\|full | 9573 | No | NA | | 18 | 2 | 1 | Low-quality | Genome-fragment | | 15.13 | AAI-based (high-confidence) | 0 |
| S8_extracted_k141_49552 | 1032 | No | NA | | 2 | 0 | 0 | Not-determined | Genome-fragment | NA | | NA | 0 |
| S8_extracted_k141_102010 | 3044 | No | NA | | 2 | 0 | 1 | Not-determined | Genome-fragment | NA | | NA | 0 |
| S8_extracted_k141_15215 | 1070 | No | NA | | 1 | 0 | 1 | Not-determined | Genome-fragment | NA | | NA | 0 |
| S8_extracted_k141_93447 | 6921 | No | NA | | 5 | 0 | 5 | Not-determined | Genome-fragment | NA | | NA | 0 |
| S8_extracted_k141_197913 | 1717 | No | NA | | 1 | 0 | 1 | Not-determined | Genome-fragment | NA | | NA | 0 |
| S8_extracted_k141_15218 | 3140 | No | NA | | 2 | 0 | 1 | Not-determined | Genome-fragment | NA | | NA | 0 |
| S14_vs_k141_53444\|\|full | 9575 | No | NA | | 17 | 7 | 0 | Low-quality | Genome-fragment | | 25.65 | AAI-based (high-confidence) | 0 |
| S8_extracted_k141_84791 | 2121 | No | NA | | 3 | 0 | 2 | Not-determined | Genome-fragment | NA | | NA | 0 |
| S8_extracted_k141_41376 | 61698 | No | NA | | 90 | 31 | 1 | High-quality | High-quality | | 99.72 | AAI-based (high-confidence) | 0 |
| S8_extracted_k141_67235 | 2819 | No | NA | | 3 | 0 | 1 | Not-determined | Genome-fragment | NA | | NA | 0 |
| S5_vs_k141_69346\|\|full | 9582 | No | NA | | 13 | 11 | 0 | Low-quality | Genome-fragment | | 32.02 | AAI-based (high-confidence) | 0 |
| S8_extracted_k141_93454 | 2475 | No | NA | | 3 | 0 | 2 | Not-determined | Genome-fragment | NA | | NA | 0 |
| S5_vs_k141_30484\|\|full | 9584 | Yes | | 1957 | 9 | 1 | 5 | Low-quality | Genome-fragment | | 2.96 | HMM-based (lower-bound) | 79.58 |
| S8_extracted_k141_67246 | 1498 | No | NA | | 2 | 0 | 2 | Not-determined | Genome-fragment | NA | | NA | 0 |
| S8_extracted_k141_188050 | 1840 | No | NA | | 3 | 0 | 3 | Not-determined | Genome-fragment | NA | | NA | 0 |
| S8_vs_k141_58336\|\|full | 9601 | Yes | | 6729 | 8 | 1 | 3 | Low-quality | Genome-fragment | | 4.11 | AAI-based (medium-confidence) | 29.91 |
| S8_extracted_k141_102030 | 6491 | No | NA | | 4 | 0 | 2 | Not-determined | Genome-fragment | NA | | NA | 0 |
| S8_extracted_k141_93470 | 1471 | No | NA | | 2 | 0 | 2 | Not-determined | Genome-fragment | NA | | NA | 0 |
| S8_extracted_k141_162572 | 1535 | No | NA | | 3 | 0 | 2 | Not-determined | Genome-fragment | NA | | NA | 0 |
| S8_extracted_k141_205918 | 1266 | No | NA | | 2 | 0 | 0 | Not-determined | Genome-fragment | NA | | NA | 0 |
| S8_extracted_k141_32957 | 1044 | No | NA | | 1 | 0 | 1 | Not-determined | Genome-fragment | NA | | NA | 0 |
| S8_extracted_k141_205925 | 1123 | No | NA | | 2 | 0 | 1 | Not-determined | Genome-fragment | NA | | NA | 0 |
| S8_extracted_k141_128310 | 6301 | No | NA | | 7 | 0 | 5 | Not-determined | Genome-fragment | NA | | NA | 0 |
| S8_extracted_k141_136698 | 13264 | No | NA | | 9 | 0 | 7 | Not-determined | Genome-fragment | NA | | NA | 0 |
| S8_extracted_k141_197947 | 4462 | No | NA | | 4 | 0 | 3 | Not-determined | Genome-fragment | NA | | NA | 0 |
| S17_vs_k141_14017\|\|full | 9602 | No | NA | | 12 | 3 | 0 | Low-quality | Genome-fragment | | 13.8 | AAI-based (high-confidence) | 0 |
| S8_extracted_k141_188081 | 1706 | No | NA | | 1 | 0 | 1 | Not-determined | Genome-fragment | NA | | NA | 0 |
| S8_extracted_k141_188089 | 1706 | No | NA | | 2 | 0 | 1 | Not-determined | Genome-fragment | NA | | NA | 0 |
| S8_extracted_k141_171856 | 1067 | No | NA | | 2 | 0 | 0 | Not-determined | Genome-fragment | NA | | NA | 0 |
| S8_extracted_k141_111089 | 1329 | No | NA | | 2 | 0 | 2 | Not-determined | Genome-fragment | NA | | NA | 0 |
| S8_extracted_k141_41439 | 2249 | No | NA | | 3 | 0 | 1 | Not-determined | Genome-fragment | NA | | NA | 0 |
| S8_extracted_k141_102090 | 1073 | No | NA | | 2 | 0 | 0 | Not-determined | Genome-fragment | NA | | NA | 0 |
| S8_extracted_k141_171866 | 3173 | No | NA | | 5 | 0 | 4 | Not-determined | Genome-fragment | NA | | NA | 0 |
| S16_extracted_45965 | 9615 | No | NA | | 15 | 9 | 0 | Low-quality | Genome-fragment | | 15.93 | AAI-based (high-confidence) | 0 |
| S8_extracted_k141_93520 | 1197 | No | NA | | 2 | 0 | 0 | Not-determined | Genome-fragment | NA | | NA | 0 |
| S8_extracted_k141_76083 | 1762 | No | NA | | 1 | 0 | 0 | Not-determined | Genome-fragment | NA | | NA | 0 |
| S8_extracted_k141_41456 | 1857 | No | NA | | 2 | 0 | 1 | Not-determined | Genome-fragment | NA | | NA | 0 |
| S8_extracted_k141_180549 | 7584 | No | NA | | 7 | 0 | 5 | Not-determined | Genome-fragment | NA | | NA | 0 |
| S1_extracted_k141_138247 | 9627 | No | NA | | 18 | 1 | 1 | Low-quality | Genome-fragment | | 16.47 | AAI-based (high-confidence) | 0 |
| S12_extracted_k141_126532 | 9635 | No | NA | | 8 | 1 | 0 | Low-quality | Genome-fragment | | 3.43 | HMM-based (lower-bound) | 0 |
| S8_extracted_k141_59482 | 1429 | No | NA | | 2 | 0 | 0 | Not-determined | Genome-fragment | NA | | NA | 0 |
| S8_extracted_k141_33001 | 5967 | No | NA | | 7 | 0 | 1 | Not-determined | Genome-fragment | NA | | NA | 0 |
| S8_extracted_k141_49662 | 2300 | No | NA | | 2 | 0 | 2 | Not-determined | Genome-fragment | NA | | NA | 0 |
| S8_extracted_k141_33009 | 1392 | No | NA | | 2 | 0 | 0 | Not-determined | Genome-fragment | NA | | NA | 0 |
| S8_extracted_k141_145851 | 1039 | No | NA | | 2 | 0 | 1 | Not-determined | Genome-fragment | NA | | NA | 0 |
| S8_vs_k141_111707\|\|full | 9642 | No | NA | | 10 | 3 | 0 | Low-quality | Genome-fragment | | 23.18 | AAI-based (medium-confidence) | 0 |
| S8_extracted_k141_136775 | 1285 | No | NA | | 2 | 0 | 2 | Not-determined | Genome-fragment | NA | | NA | 0 |
| S8_extracted_k141_67328 | 3482 | No | NA | | 4 | 0 | 0 | Not-determined | Genome-fragment | NA | | NA | 0 |
| S8_extracted_k141_136777 | 1464 | No | NA | | 1 | 0 | 1 | Not-determined | Genome-fragment | NA | | NA | 0 |
| S8_extracted_k141_180583 | 6864 | No | NA | | 3 | 0 | 2 | Not-determined | Genome-fragment | NA | | NA | 0 |
| S8_extracted_k141_15251 | 1791 | No | NA | | 3 | 0 | 2 | Not-determined | Genome-fragment | NA | | NA | 0 |
| S8_extracted_k141_206009 | 1105 | No | NA | | 2 | 0 | 1 | Not-determined | Genome-fragment | NA | | NA | 0 |
| S8_extracted_k141_198048 | 1273 | No | NA | | 1 | 0 | 1 | Not-determined | Genome-fragment | NA | | NA | 0 |
| S8_vs_k141_148699\|\|full | 9644 | No | NA | | 22 | 3 | 0 | Low-quality | Genome-fragment | | 12.52 | HMM-based (lower-bound) | 0 |
| S8_extracted_k141_180589 | 1450 | No | NA | | 2 | 0 | 1 | Not-determined | Genome-fragment | NA | | NA | 0 |
| S8_extracted_k141_76149 | 1896 | No | NA | | 2 | 0 | 0 | Not-determined | Genome-fragment | NA | | NA | 0 |
| S8_extracted_k141_145878 | 1567 | No | NA | | 1 | 0 | 1 | Not-determined | Genome-fragment | NA | | NA | 0 |
| S8_extracted_k141_136820 | 1103 | No | NA | | 2 | 0 | 2 | Not-determined | Genome-fragment | NA | | NA | 0 |
| S8_extracted_k141_180606 | 1389 | No | NA | | 1 | 0 | 1 | Not-determined | Genome-fragment | NA | | NA | 0 |
| S8_extracted_k141_76153 | 1255 | No | NA | | 1 | 0 | 1 | Not-determined | Genome-fragment | NA | | NA | 0 |
| S8_extracted_k141_41463 | 6905 | No | NA | | 4 | 0 | 3 | Not-determined | Genome-fragment | NA | | NA | 0 |
| S8_extracted_k141_119639 | 1000 | No | NA | | 2 | 0 | 1 | Not-determined | Genome-fragment | NA | | NA | 0 |
| S16_extracted_3326 | 9646 | No | NA | | 12 | 0 | 2 | Low-quality | Genome-fragment | | 15.6 | AAI-based (medium-confidence) | 0 |
| S8_extracted_k141_198062 | 2270 | No | NA | | 2 | 0 | 2 | Not-determined | Genome-fragment | NA | | NA | 0 |
| S16_extracted_114153 | 9652 | No | NA | | 6 | 0 | 3 | Low-quality | Genome-fragment | | 11.37 | AAI-based (medium-confidence) | 0 |
| S8_extracted_k141_119649 | 2054 | No | NA | | 2 | 0 | 0 | Not-determined | Genome-fragment | NA | | NA | 0 |
| S8_extracted_k141_162699 | 2288 | No | NA | | 2 | 0 | 2 | Not-determined | Genome-fragment | NA | | NA | 0 |
| S5_vs_k141_61861\|\|full | 9653 | No | NA | | 7 | 3 | 0 | Low-quality | Genome-fragment | | 24.91 | AAI-based (high-confidence) | 0 |
| S8_extracted_k141_6981 | 4229 | No | NA | | 1 | 0 | 1 | Not-determined | Genome-fragment | NA | | NA | 0 |
| S8_extracted_k141_119657 | 5853 | No | NA | | 6 | 0 | 4 | Not-determined | Genome-fragment | NA | | NA | 0 |
| S8_extracted_k141_102166 | 1631 | No | NA | | 3 | 0 | 1 | Not-determined | Genome-fragment | NA | | NA | 0 |
| S8_extracted_k141_24502 | 1166 | No | NA | | 1 | 0 | 0 | Not-determined | Genome-fragment | NA | | NA | 0 |
| S8_extracted_k141_162713 | 4904 | No | NA | | 4 | 0 | 0 | Not-determined | Genome-fragment | NA | | NA | 0 |
| S8_extracted_k141_49734 | 3316 | No | NA | | 4 | 0 | 1 | Not-determined | Genome-fragment | NA | | NA | 0 |
| S8_extracted_k141_84863 | 1114 | No | NA | | 2 | 0 | 1 | Not-determined | Genome-fragment | NA | | NA | 0 |
| S8_extracted_k141_6987 | 1044 | No | NA | | 1 | 0 | 1 | Not-determined | Genome-fragment | NA | | NA | 0 |
| S8_extracted_k141_102173 | 3263 | No | NA | | 1 | 0 | 0 | Not-determined | Genome-fragment | NA | | NA | 0 |
| S3_extracted_k141_95986 | 9657 | No | NA | | 11 | 3 | 0 | Low-quality | Genome-fragment | | 8.1 | AAI-based (high-confidence) | 0 |
| S8_extracted_k141_93589 | 8697 | No | NA | | 7 | 0 | 0 | Not-determined | Genome-fragment | NA | | NA | 0 |
| S8_extracted_k141_6989 | 2692 | No | NA | | 2 | 0 | 2 | Not-determined | Genome-fragment | NA | | NA | 0 |
| S6_vs_k141_11740\|\|full | 9657 | No | NA | | 15 | 12 | 0 | Low-quality | Genome-fragment | | 16.06 | AAI-based (high-confidence) | 0 |
| S8_extracted_k141_136885 | 1030 | No | NA | | 1 | 0 | 1 | Not-determined | Genome-fragment | NA | | NA | 0 |
| S16_vs_67587\|\|full | 9663 | No | NA | | 8 | 1 | 0 | Low-quality | Genome-fragment | | 8.8 | HMM-based (lower-bound) | 0 |
| S8_extracted_k141_172030 | 1079 | No | NA | | 2 | 0 | 0 | Not-determined | Genome-fragment | NA | | NA | 0 |
| S8_extracted_k141_84906 | 2563 | No | NA | | 1 | 0 | 0 | Not-determined | Genome-fragment | NA | | NA | 0 |
| S5_vs_k141_57006\|\|full | 9672 | No | NA | | 11 | 1 | 0 | Low-quality | Genome-fragment | | 18.41 | HMM-based (lower-bound) | 0 |
| S8_extracted_k141_145945 | 1745 | No | NA | | 3 | 0 | 0 | Not-determined | Genome-fragment | NA | | NA | 0 |
| S8_extracted_k141_102202 | 1294 | No | NA | | 2 | 0 | 1 | Not-determined | Genome-fragment | NA | | NA | 0 |
| S3_extracted_k141_125124 | 9673 | No | NA | | 12 | 6 | 1 | Low-quality | Genome-fragment | | 5.68 | AAI-based (high-confidence) | 0 |
| S8_extracted_k141_93633 | 1313 | No | NA | | 2 | 0 | 0 | Not-determined | Genome-fragment | NA | | NA | 0 |
| S8_extracted_k141_119697 | 3287 | No | NA | | 2 | 0 | 1 | Not-determined | Genome-fragment | NA | | NA | 0 |
| S8_extracted_k141_7016 | 3066 | No | NA | | 4 | 0 | 0 | Not-determined | Genome-fragment | NA | | NA | 0 |
| S8_extracted_k141_172034 | 1746 | No | NA | | 3 | 0 | 1 | Not-determined | Genome-fragment | NA | | NA | 0 |
| S8_extracted_k141_102211 | 1050 | No | NA | | 1 | 0 | 1 | Not-determined | Genome-fragment | NA | | NA | 0 |
| S8_extracted_k141_7017 | 2237 | No | NA | | 1 | 0 | 1 | Not-determined | Genome-fragment | NA | | NA | 0 |
| S8_extracted_k141_49769 | 1878 | No | NA | | 2 | 0 | 1 | Not-determined | Genome-fragment | NA | | NA | 0 |
| S8_extracted_k141_15356 | 1300 | No | NA | | 2 | 0 | 1 | Not-determined | Genome-fragment | NA | | NA | 0 |
| S8_extracted_k141_15360 | 1582 | No | NA | | 1 | 0 | 0 | Not-determined | Genome-fragment | NA | | NA | 0 |
| S8_extracted_k141_162780 | 1264 | No | NA | | 1 | 0 | 0 | Not-determined | Genome-fragment | NA | | NA | 0 |
| S8_extracted_k141_93655 | 4416 | No | NA | | 6 | 0 | 4 | Not-determined | Genome-fragment | NA | | NA | 0 |
| S8_extracted_k141_41561 | 5333 | No | NA | | 4 | 0 | 3 | Not-determined | Genome-fragment | NA | | NA | 0 |
| S8_extracted_k141_172057 | 1071 | No | NA | | 3 | 0 | 3 | Not-determined | Genome-fragment | NA | | NA | 0 |
| S8_extracted_k141_162799 | 3485 | No | NA | | 2 | 0 | 0 | Not-determined | Genome-fragment | NA | | NA | 0 |
| S8_extracted_k141_162811 | 1178 | No | NA | | 1 | 0 | 1 | Not-determined | Genome-fragment | NA | | NA | 0 |
| S8_extracted_k141_136917 | 1207 | No | NA | | 2 | 0 | 2 | Not-determined | Genome-fragment | NA | | NA | 0 |
| S8_extracted_k141_162814 | 1118 | No | NA | | 1 | 0 | 1 | Not-determined | Genome-fragment | NA | | NA | 0 |
| S8_extracted_k141_93676 | 1053 | No | NA | | 2 | 0 | 1 | Not-determined | Genome-fragment | NA | | NA | 0 |
| S8_extracted_k141_41585 | 1146 | No | NA | | 2 | 0 | 0 | Not-determined | Genome-fragment | NA | | NA | 0 |
| S8_extracted_k141_198110 | 1500 | No | NA | | 1 | 0 | 0 | Not-determined | Genome-fragment | NA | | NA | 0 |
| S8_extracted_k141_111269 | 1252 | No | NA | | 2 | 0 | 0 | Not-determined | Genome-fragment | NA | | NA | 0 |
| S8_extracted_k141_15394 | 1274 | No | NA | | 2 | 0 | 1 | Not-determined | Genome-fragment | NA | | NA | 0 |
| S8_extracted_k141_24581 | 2150 | No | NA | | 2 | 0 | 0 | Not-determined | Genome-fragment | NA | | NA | 0 |
| S8_extracted_k141_198122 | 1195 | No | NA | | 1 | 0 | 1 | Not-determined | Genome-fragment | NA | | NA | 0 |
| S8_extracted_k141_33138 | 1503 | No | NA | | 3 | 0 | 1 | Not-determined | Genome-fragment | NA | | NA | 0 |
| S8_extracted_k141_67424 | 1182 | No | NA | | 2 | 0 | 2 | Not-determined | Genome-fragment | NA | | NA | 0 |
| S8_vs_k141_195076\|\|full | 9683 | No | NA | | 14 | 0 | 2 | Low-quality | Genome-fragment | | 22.53 | AAI-based (medium-confidence) | 0 |
| S8_extracted_k141_7069 | 1512 | No | NA | | 1 | 0 | 0 | Not-determined | Genome-fragment | NA | | NA | 0 |
| S8_extracted_k141_67439 | 1151 | No | NA | | 1 | 0 | 1 | Not-determined | Genome-fragment | NA | | NA | 0 |
| S8_extracted_k141_188302 | 1349 | No | NA | | 2 | 0 | 2 | Not-determined | Genome-fragment | NA | | NA | 0 |
| S8_extracted_k141_33149 | 5846 | No | NA | | 4 | 0 | 2 | Not-determined | Genome-fragment | NA | | NA | 0 |
| S8_extracted_k141_198140 | 3757 | No | NA | | 5 | 0 | 3 | Not-determined | Genome-fragment | NA | | NA | 0 |
| S8_extracted_k141_206114 | 1574 | No | NA | | 1 | 0 | 1 | Not-determined | Genome-fragment | NA | | NA | 0 |
| S8_extracted_k141_33150 | 1121 | No | NA | | 1 | 0 | 0 | Not-determined | Genome-fragment | NA | | NA | 0 |
| S8_extracted_k141_15415 | 1846 | No | NA | | 1 | 0 | 1 | Not-determined | Genome-fragment | NA | | NA | 0 |
| S8_extracted_k141_172090 | 1056 | No | NA | | 1 | 0 | 1 | Not-determined | Genome-fragment | NA | | NA | 0 |
| S8_extracted_k141_162865 | 2308 | No | NA | | 2 | 0 | 2 | Not-determined | Genome-fragment | NA | | NA | 0 |
| S8_extracted_k141_146035 | 2065 | No | NA | | 3 | 0 | 2 | Not-determined | Genome-fragment | NA | | NA | 0 |
| S8_extracted_k141_111330 | 1657 | No | NA | | 2 | 0 | 1 | Not-determined | Genome-fragment | NA | | NA | 0 |
| S8_extracted_k141_172096 | 1410 | No | NA | | 1 | 0 | 0 | Not-determined | Genome-fragment | NA | | NA | 0 |
| S4_extracted_23943 | 9684 | No | NA | | 7 | 2 | 0 | Low-quality | Genome-fragment | | 25.15 | AAI-based (medium-confidence) | 0 |
| S3_vs_k141_2492\|\|full | 9694 | No | NA | | 11 | 4 | 0 | Low-quality | Genome-fragment | | 19.81 | AAI-based (medium-confidence) | 0 |
| S1_extracted_k141_72774 | 9701 | No | NA | | 21 | 5 | 0 | Low-quality | Genome-fragment | | 12.07 | AAI-based (high-confidence) | 0 |
| S8_vs_k141_125372\|\|full | 9702 | No | NA | | 11 | 0 | 5 | Low-quality | Genome-fragment | | 10.71 | AAI-based (medium-confidence) | 0 |
| S15_extracted_56371 | 9707 | No | NA | | 15 | 7 | 0 | Low-quality | Genome-fragment | | 21.6 | AAI-based (high-confidence) | 0 |
| S8_extracted_k141_180766 | 1609 | No | NA | | 2 | 0 | 2 | Not-determined | Genome-fragment | NA | | NA | 0 |
| S8_extracted_k141_49836 | 1092 | No | NA | | 1 | 0 | 1 | Not-determined | Genome-fragment | NA | | NA | 0 |
| S8_extracted_k141_59571 | 1600 | No | NA | | 2 | 0 | 2 | Not-determined | Genome-fragment | NA | | NA | 0 |
| S11_extracted_k141_17995 | 9716 | No | NA | | 15 | 2 | 0 | Low-quality | Genome-fragment | | 16.36 | AAI-based (high-confidence) | 0 |
| S8_extracted_k141_49837 | 1590 | No | NA | | 1 | 0 | 1 | Not-determined | Genome-fragment | NA | | NA | 0 |
| S11_vs_k141_16887\|\|full | 9721 | No | NA | | 14 | 4 | 0 | Low-quality | Genome-fragment | | 17 | AAI-based (high-confidence) | 0 |
| S5_vs_k141_135673\|\|full | 9725 | No | NA | | 23 | 4 | 1 | Low-quality | Genome-fragment | | 24.37 | AAI-based (high-confidence) | 0 |
| S8_extracted_k141_162886 | 1047 | No | NA | | 2 | 0 | 1 | Not-determined | Genome-fragment | NA | | NA | 0 |
| S8_extracted_k141_111342 | 3816 | No | NA | | 4 | 0 | 2 | Not-determined | Genome-fragment | NA | | NA | 0 |
| S8_extracted_k141_49844 | 1347 | No | NA | | 1 | 0 | 1 | Not-determined | Genome-fragment | NA | | NA | 0 |
| S8_extracted_k141_128462 | 1276 | No | NA | | 2 | 0 | 1 | Not-determined | Genome-fragment | NA | | NA | 0 |
| S8_extracted_k141_85021 | 1693 | No | NA | | 1 | 0 | 0 | Not-determined | Genome-fragment | NA | | NA | 0 |
| S4_extracted_9103 | 9741 | Yes | | 1066 | 9 | 1 | 3 | Low-quality | Genome-fragment | | 1.05 | AAI-based (medium-confidence) | 89.06 |
| S8_extracted_k141_41650 | 4564 | No | NA | | 4 | 0 | 2 | Not-determined | Genome-fragment | NA | | NA | 0 |
| S8_extracted_k141_128472 | 1136 | No | NA | | 1 | 0 | 1 | Not-determined | Genome-fragment | NA | | NA | 0 |
| S8_extracted_k141_172129 | 1306 | No | NA | | 3 | 0 | 2 | Not-determined | Genome-fragment | NA | | NA | 0 |
| S8_extracted_k141_24651 | 1026 | No | NA | | 2 | 0 | 0 | Not-determined | Genome-fragment | NA | | NA | 0 |
| S3_vs_k141_24359\|\|full | 9748 | No | NA | | 20 | 3 | 0 | Low-quality | Genome-fragment | | 30.23 | AAI-based (high-confidence) | 0 |
| S8_extracted_k141_33212 | 1753 | No | NA | | 1 | 0 | 0 | Not-determined | Genome-fragment | NA | | NA | 0 |
| S8_extracted_k141_85039 | 1614 | No | NA | | 2 | 0 | 2 | Not-determined | Genome-fragment | NA | | NA | 0 |
| S8_extracted_k141_172137 | 1494 | No | NA | | 2 | 0 | 2 | Not-determined | Genome-fragment | NA | | NA | 0 |
| S8_extracted_k141_162915 | 1230 | No | NA | | 1 | 0 | 0 | Not-determined | Genome-fragment | NA | | NA | 0 |
| S14_vs_k141_43936\|\|full | 9752 | No | NA | | 17 | 3 | 0 | Low-quality | Genome-fragment | | 12.23 | AAI-based (high-confidence) | 0 |
| S8_extracted_k141_15465 | 7184 | No | NA | | 8 | 0 | 7 | Not-determined | Genome-fragment | NA | | NA | 0 |
| S8_extracted_k141_206153 | 4311 | No | NA | | 4 | 0 | 3 | Not-determined | Genome-fragment | NA | | NA | 0 |
| S8_extracted_k141_136966 | 2512 | No | NA | | 2 | 0 | 2 | Not-determined | Genome-fragment | NA | | NA | 0 |
| S8_extracted_k141_102347 | 3320 | No | NA | | 3 | 0 | 1 | Not-determined | Genome-fragment | NA | | NA | 0 |
| S8_extracted_k141_172165 | 1508 | No | NA | | 2 | 0 | 2 | Not-determined | Genome-fragment | NA | | NA | 0 |
| S11_vs_k141_87673\|\|full | 9753 | No | NA | | 14 | 1 | 0 | Low-quality | Genome-fragment | | 22.27 | AAI-based (high-confidence) | 0 |
| S8_extracted_k141_206162 | 2088 | No | NA | | 5 | 0 | 3 | Not-determined | Genome-fragment | NA | | NA | 0 |
| S8_extracted_k141_128512 | 1585 | No | NA | | 2 | 0 | 1 | Not-determined | Genome-fragment | NA | | NA | 0 |
| S8_vs_k141_78409\|\|full | 9754 | No | NA | | 11 | 1 | 2 | Low-quality | Genome-fragment | | 18.26 | AAI-based (medium-confidence) | 0 |
| S8_extracted_k141_45588 | 9763 | No | NA | | 19 | 2 | 1 | Low-quality | Genome-fragment | | 15.73 | AAI-based (high-confidence) | 0 |
| S8_extracted_k141_41694 | 1043 | No | NA | | 2 | 0 | 1 | Not-determined | Genome-fragment | NA | | NA | 0 |
| S14_extracted_k141_84879 | 9771 | No | NA | | 5 | 0 | 2 | Low-quality | Genome-fragment | | 28.18 | AAI-based (high-confidence) | 0 |
| S8_extracted_k141_188351 | 4199 | No | NA | | 4 | 0 | 4 | Not-determined | Genome-fragment | NA | | NA | 0 |
| S8_extracted_k141_180828 | 1593 | No | NA | | 3 | 0 | 3 | Not-determined | Genome-fragment | NA | | NA | 0 |
| S8_extracted_k141_24692 | 3043 | No | NA | | 3 | 0 | 3 | Not-determined | Genome-fragment | NA | | NA | 0 |
| S8_extracted_k141_15485 | 1127 | No | NA | | 1 | 0 | 1 | Not-determined | Genome-fragment | NA | | NA | 0 |
| S8_extracted_k141_180839 | 1049 | No | NA | | 1 | 0 | 0 | Not-determined | Genome-fragment | NA | | NA | 0 |
| S8_extracted_k141_24695 | 3111 | No | NA | | 4 | 0 | 0 | Not-determined | Genome-fragment | NA | | NA | 0 |
| S8_extracted_k141_180844 | 1441 | No | NA | | 2 | 0 | 0 | Not-determined | Genome-fragment | NA | | NA | 0 |
| S8_extracted_k141_188374 | 3419 | No | NA | | 3 | 0 | 0 | Not-determined | Genome-fragment | NA | | NA | 0 |
| S8_extracted_k141_67476 | 1314 | No | NA | | 1 | 0 | 0 | Not-determined | Genome-fragment | NA | | NA | 0 |
| S8_extracted_k141_162964 | 1021 | No | NA | | 0 | 0 | 0 | Not-determined | Genome-fragment | NA | | NA | 0 |
| S8_extracted_k141_102384 | 1751 | No | NA | | 2 | 0 | 2 | Not-determined | Genome-fragment | NA | | NA | 0 |
| S8_extracted_k141_41727 | 3480 | No | NA | | 1 | 0 | 0 | Not-determined | Genome-fragment | NA | | NA | 0 |
| S8_extracted_k141_7165 | 1449 | No | NA | | 3 | 0 | 1 | Not-determined | Genome-fragment | NA | | NA | 0 |
| S8_extracted_k141_93797 | 1311 | No | NA | | 2 | 0 | 1 | Not-determined | Genome-fragment | NA | | NA | 0 |
| S8_extracted_k141_15514 | 3611 | No | NA | | 3 | 0 | 2 | Not-determined | Genome-fragment | NA | | NA | 0 |
| S8_extracted_k141_102395 | 1938 | No | NA | | 2 | 0 | 0 | Not-determined | Genome-fragment | NA | | NA | 0 |
| S8_extracted_k141_180870 | 1803 | No | NA | | 2 | 0 | 1 | Not-determined | Genome-fragment | NA | | NA | 0 |
| S8_extracted_k141_24733 | 1710 | No | NA | | 1 | 0 | 0 | Not-determined | Genome-fragment | NA | | NA | 0 |
| S8_extracted_k141_198270 | 1013 | No | NA | | 1 | 0 | 1 | Not-determined | Genome-fragment | NA | | NA | 0 |
| S8_extracted_k141_93814 | 1736 | No | NA | | 2 | 0 | 0 | Not-determined | Genome-fragment | NA | | NA | 0 |
| S9_extracted_k141_55700 | 9773 | No | NA | | 11 | 1 | 0 | Low-quality | Genome-fragment | | 5.46 | AAI-based (high-confidence) | 0 |
| S8_extracted_k141_198276 | 1919 | No | NA | | 1 | 0 | 1 | Not-determined | Genome-fragment | NA | | NA | 0 |
| S8_extracted_k141_41754 | 3254 | No | NA | | 4 | 0 | 4 | Not-determined | Genome-fragment | NA | | NA | 0 |
| S8_extracted_k141_119826 | 1048 | No | NA | | 2 | 0 | 0 | Not-determined | Genome-fragment | NA | | NA | 0 |
| S8_extracted_k141_24743 | 1953 | No | NA | | 3 | 0 | 3 | Not-determined | Genome-fragment | NA | | NA | 0 |
| S8_extracted_k141_119832 | 1155 | No | NA | | 1 | 0 | 1 | Not-determined | Genome-fragment | NA | | NA | 0 |
| S8_extracted_k141_119835 | 1405 | No | NA | | 4 | 0 | 3 | Not-determined | Genome-fragment | NA | | NA | 0 |
| S8_extracted_k141_162995 | 2040 | No | NA | | 1 | 0 | 0 | Not-determined | Genome-fragment | NA | | NA | 0 |
| S8_extracted_k141_15542 | 1152 | No | NA | | 1 | 0 | 1 | Not-determined | Genome-fragment | NA | | NA | 0 |
| S8_extracted_k141_172186 | 43104 | No | NA | | 39 | 0 | 26 | Not-determined | Genome-fragment | NA | | NA | 0 |
| S8_extracted_k141_33285 | 4468 | No | NA | | 5 | 0 | 5 | Not-determined | Genome-fragment | NA | | NA | 0 |
| S8_extracted_k141_172194 | 1578 | No | NA | | 2 | 0 | 2 | Not-determined | Genome-fragment | NA | | NA | 0 |
| S14_vs_k141_11324\|\|full | 9778 | No | NA | | 15 | 1 | 0 | Low-quality | Genome-fragment | | 29.58 | AAI-based (high-confidence) | 0 |
| S8_extracted_k141_24754 | 7430 | No | NA | | 5 | 0 | 2 | Not-determined | Genome-fragment | NA | | NA | 0 |
| S12_vs_k141_22377\|\|full | 9782 | No | NA | | 15 | 1 | 0 | Low-quality | Genome-fragment | | 6.04 | HMM-based (lower-bound) | 0 |
| S15_extracted_14213 | 9787 | No | NA | | 1 | 0 | 0 | Low-quality | Genome-fragment | | 9.87 | AAI-based (medium-confidence) | 0 |
| S8_extracted_k141_206231 | 4063 | No | NA | | 4 | 0 | 0 | Not-determined | Genome-fragment | NA | | NA | 0 |
| S8_extracted_k141_146168 | 1573 | No | NA | | 2 | 0 | 2 | Not-determined | Genome-fragment | NA | | NA | 0 |
| S3_vs_k141_127264\|\|full | 9789 | No | NA | | 15 | 3 | 0 | Low-quality | Genome-fragment | | 16.13 | AAI-based (high-confidence) | 0 |
| S8_extracted_k141_24774 | 6263 | No | NA | | 4 | 0 | 3 | Not-determined | Genome-fragment | NA | | NA | 0 |
| S8_extracted_k141_163039 | 7666 | No | NA | | 6 | 0 | 5 | Not-determined | Genome-fragment | NA | | NA | 0 |
| S8_extracted_k141_41807 | 1135 | No | NA | | 2 | 0 | 0 | Not-determined | Genome-fragment | NA | | NA | 0 |
| S14_vs_k141_56398\|\|full | 9792 | No | NA | | 25 | 4 | 0 | Low-quality | Genome-fragment | | 22.81 | AAI-based (high-confidence) | 0 |
| S8_extracted_k141_137089 | 3168 | No | NA | | 4 | 0 | 3 | Not-determined | Genome-fragment | NA | | NA | 0 |
| S3_extracted_k141_72562 | 9794 | No | NA | | 18 | 2 | 0 | Low-quality | Genome-fragment | | 6.36 | AAI-based (high-confidence) | 0 |
| S8_extracted_k141_24805 | 1257 | No | NA | | 2 | 0 | 0 | Not-determined | Genome-fragment | NA | | NA | 0 |
| S8_extracted_k141_188480 | 1480 | No | NA | | 1 | 0 | 1 | Not-determined | Genome-fragment | NA | | NA | 0 |
| S8_extracted_k141_163069 | 1098 | No | NA | | 2 | 0 | 0 | Not-determined | Genome-fragment | NA | | NA | 0 |
| S8_extracted_k141_163073 | 1254 | No | NA | | 2 | 0 | 0 | Not-determined | Genome-fragment | NA | | NA | 0 |
| S8_extracted_k141_15605 | 3163 | No | NA | | 2 | 0 | 2 | Not-determined | Genome-fragment | NA | | NA | 0 |
| S8_extracted_k141_119889 | 3862 | No | NA | | 6 | 0 | 1 | Not-determined | Genome-fragment | NA | | NA | 0 |
| S8_extracted_k141_128610 | 1321 | No | NA | | 1 | 0 | 1 | Not-determined | Genome-fragment | NA | | NA | 0 |
| S16_vs_84173\|\|full | 9794 | No | NA | | 6 | 0 | 0 | Low-quality | Genome-fragment | | 12.04 | AAI-based (medium-confidence) | 0 |
| S7_extracted_55205 | 9802 | No | NA | | 12 | 3 | 0 | Low-quality | Genome-fragment | | 15.43 | AAI-based (high-confidence) | 0 |
| S8_extracted_k141_172276 | 1597 | No | NA | | 2 | 0 | 2 | Not-determined | Genome-fragment | NA | | NA | 0 |
| S8_extracted_k141_180949 | 1092 | No | NA | | 1 | 0 | 0 | Not-determined | Genome-fragment | NA | | NA | 0 |
| S8_extracted_k141_198365 | 1842 | No | NA | | 2 | 0 | 0 | Not-determined | Genome-fragment | NA | | NA | 0 |
| S8_extracted_k141_7243 | 40403 | No | NA | | 44 | 18 | 0 | Medium-quality | Genome-fragment | | 81.63 | AAI-based (high-confidence) | 0 |
| S8_extracted_k141_67567 | 1363 | No | NA | | 3 | 0 | 0 | Not-determined | Genome-fragment | NA | | NA | 0 |
| S5_extracted_k141_142033 | 9809 | No | NA | | 21 | 5 | 0 | Low-quality | Genome-fragment | | 12.79 | AAI-based (medium-confidence) | 0 |
| S8_extracted_k141_102461 | 1198 | No | NA | | 1 | 0 | 0 | Not-determined | Genome-fragment | NA | | NA | 0 |
| S8_extracted_k141_163102 | 1262 | No | NA | | 3 | 0 | 1 | Not-determined | Genome-fragment | NA | | NA | 0 |
| S6_vs_k141_42271\|\|full | 9811 | No | NA | | 13 | 3 | 0 | Low-quality | Genome-fragment | | 23.6 | AAI-based (high-confidence) | 0 |
| S8_extracted_k141_180957 | 3791 | No | NA | | 3 | 0 | 1 | Not-determined | Genome-fragment | NA | | NA | 0 |
| S8_extracted_k141_76375 | 2543 | No | NA | | 5 | 0 | 2 | Not-determined | Genome-fragment | NA | | NA | 0 |
| S8_extracted_k141_7255 | 3119 | No | NA | | 4 | 0 | 1 | Not-determined | Genome-fragment | NA | | NA | 0 |
| S8_extracted_k141_50017 | 1480 | No | NA | | 1 | 0 | 1 | Not-determined | Genome-fragment | NA | | NA | 0 |

| | | | | | | | | | | | |
|---|---|---|---|---|---|---|---|---|---|---|---|
| S8_extracted_k141_15634 | 3053 | No | NA | 2 | 0 | 2 | Not-determined | Genome-fragment | NA | NA | 0 |
| S8_extracted_k141_128629 | 3102 | No | NA | 4 | 0 | 1 | Not-determined | Genome-fragment | NA | NA | 0 |
| S8_extracted_k141_198386 | 1554 | No | NA | 1 | 0 | 1 | Not-determined | Genome-fragment | NA | NA | 0 |
| S8_extracted_k141_146240 | 1767 | No | NA | 2 | 0 | 0 | Not-determined | Genome-fragment | NA | NA | 0 |
| S8_extracted_k141_59729 | 1404 | No | NA | 1 | 0 | 1 | Not-determined | Genome-fragment | NA | NA | 0 |
| S8_extracted_k141_198391 | 1077 | No | NA | 2 | 0 | 1 | Not-determined | Genome-fragment | NA | NA | 0 |
| S6_vs_k141_30079\|\|full | 9812 | No | NA | 17 | 3 | 1 | Low-quality | Genome-fragment | 26.37 | AAI-based (high-confidence) | 0 |
| S8_extracted_k141_7281 | 4001 | No | NA | 7 | 0 | 6 | Not-determined | Genome-fragment | NA | NA | 0 |
| S8_extracted_k141_33417 | 1848 | No | NA | 2 | 0 | 1 | Not-determined | Genome-fragment | NA | NA | 0 |
| S8_extracted_k141_93956 | 1952 | No | NA | 3 | 0 | 1 | Not-determined | Genome-fragment | NA | NA | 0 |
| S8_extracted_k141_154322 | 1096 | No | NA | 2 | 0 | 1 | Not-determined | Genome-fragment | NA | NA | 0 |
| S8_extracted_k141_102505 | 2114 | No | NA | 2 | 0 | 2 | Not-determined | Genome-fragment | NA | NA | 0 |
| S8_extracted_k141_172338 | 2920 | No | NA | 3 | 0 | 2 | Not-determined | Genome-fragment | NA | NA | 0 |
| S8_extracted_k141_180992 | 1042 | No | NA | 1 | 0 | 1 | Not-determined | Genome-fragment | NA | NA | 0 |
| S8_extracted_k141_7293 | 3207 | No | NA | 2 | 0 | 2 | Not-determined | Genome-fragment | NA | NA | 0 |
| S8_extracted_k141_67623 | 1224 | No | NA | 1 | 0 | 0 | Not-determined | Genome-fragment | NA | NA | 0 |
| S8_extracted_k141_206356 | 1393 | No | NA | 1 | 0 | 1 | Not-determined | Genome-fragment | NA | NA | 0 |
| S8_extracted_k141_119979 | 1402 | No | NA | 2 | 0 | 1 | Not-determined | Genome-fragment | NA | NA | 0 |
| S8_extracted_k141_7310 | 1441 | No | NA | 4 | 0 | 1 | Not-determined | Genome-fragment | NA | NA | 0 |
| S8_extracted_k141_7318 | 1784 | No | NA | 3 | 0 | 3 | Not-determined | Genome-fragment | NA | NA | 0 |
| S8_extracted_k141_119995 | 2238 | No | NA | 2 | 0 | 1 | Not-determined | Genome-fragment | NA | NA | 0 |
| S8_extracted_k141_15685 | 1436 | No | NA | 3 | 0 | 0 | Not-determined | Genome-fragment | NA | NA | 0 |
| S5_vs_k141_80389\|\|full | 9813 | No | NA | 13 | 8 | 0 | Low-quality | Genome-fragment | 23.95 | AAI-based (high-confidence) | 0 |
| S8_extracted_k141_76442 | 1431 | No | NA | 2 | 0 | 1 | Not-determined | Genome-fragment | NA | NA | 0 |
| S8_extracted_k141_120007 | 1311 | No | NA | 1 | 0 | 1 | Not-determined | Genome-fragment | NA | NA | 0 |
| S8_extracted_k141_102508 | 36360 | No | NA | 56 | 21 | 0 | High-quality | High-quality | 100 | AAI-based (high-confidence) | 0 |
| S8_extracted_k141_188531 | 3651 | No | NA | 2 | 0 | 1 | Not-determined | Genome-fragment | NA | NA | 0 |
| S12_vs_k141_78262\|\|full | 9821 | Yes | 671 | 10 | 1 | 3 | Low-quality | Genome-fragment | 0.77 | HMM-based (lower-bound) | 93.17 |
| S8_extracted_k141_198439 | 2159 | No | NA | 2 | 0 | 1 | Not-determined | Genome-fragment | NA | NA | 0 |
| S8_extracted_k141_128701 | 2658 | No | NA | 4 | 0 | 1 | Not-determined | Genome-fragment | NA | NA | 0 |
| S8_extracted_k141_33468 | 3066 | No | NA | 2 | 0 | 2 | Not-determined | Genome-fragment | NA | NA | 0 |
| S8_extracted_k141_154366 | 1450 | No | NA | 2 | 0 | 1 | Not-determined | Genome-fragment | NA | NA | 0 |
| S8_extracted_k141_102520 | 4409 | No | NA | 5 | 0 | 4 | Not-determined | Genome-fragment | NA | NA | 0 |
| S8_extracted_k141_50105 | 1131 | No | NA | 1 | 0 | 1 | Not-determined | Genome-fragment | NA | NA | 0 |
| S8_extracted_k141_120024 | 4156 | No | NA | 2 | 0 | 1 | Not-determined | Genome-fragment | NA | NA | 0 |
| S8_extracted_k141_154380 | 2298 | No | NA | 1 | 0 | 1 | Not-determined | Genome-fragment | NA | NA | 0 |
| S8_extracted_k141_128733 | 1055 | No | NA | 1 | 0 | 1 | Not-determined | Genome-fragment | NA | NA | 0 |
| S8_extracted_k141_206403 | 3535 | No | NA | 4 | 0 | 2 | Not-determined | Genome-fragment | NA | NA | 0 |
| S8_extracted_k141_93976 | 26682 | No | NA | 28 | 5 | 0 | Medium-quality | Genome-fragment | 72.63 | AAI-based (high-confidence) | 0 |
| S8_extracted_k141_37518 | 9822 | No | NA | 8 | 0 | 2 | Low-quality | Genome-fragment | 10.85 | AAI-based (medium-confidence) | 0 |
| S8_extracted_k141_206418 | 1186 | No | NA | 1 | 0 | 0 | Not-determined | Genome-fragment | NA | NA | 0 |
| S8_extracted_k141_172428 | 1136 | No | NA | 1 | 0 | 0 | Not-determined | Genome-fragment | NA | NA | 0 |
| S8_extracted_k141_24837 | 3176 | No | NA | 4 | 0 | 1 | Not-determined | Genome-fragment | NA | NA | 0 |
| S8_extracted_k141_102554 | 1823 | No | NA | 2 | 0 | 0 | Not-determined | Genome-fragment | NA | NA | 0 |
| S8_extracted_k141_33513 | 1136 | No | NA | 2 | 0 | 2 | Not-determined | Genome-fragment | NA | NA | 0 |
| S8_extracted_k141_111657 | 1404 | No | NA | 1 | 0 | 0 | Not-determined | Genome-fragment | NA | NA | 0 |
| S8_extracted_k141_181079 | 1029 | No | NA | 1 | 0 | 1 | Not-determined | Genome-fragment | NA | NA | 0 |
| S8_extracted_k141_137245 | 1215 | No | NA | 2 | 0 | 1 | Not-determined | Genome-fragment | NA | NA | 0 |
| S8_extracted_k141_198506 | 3236 | No | NA | 3 | 0 | 1 | Not-determined | Genome-fragment | NA | NA | 0 |
| S5_extracted_k141_20344 | 9831 | No | NA | 8 | 3 | 0 | Low-quality | Genome-fragment | 6.32 | AAI-based (high-confidence) | 0 |
| S8_extracted_k141_128782 | 1336 | No | NA | 1 | 0 | 0 | Not-determined | Genome-fragment | NA | NA | 0 |
| S8_extracted_k141_172457 | 2540 | No | NA | 3 | 0 | 0 | Not-determined | Genome-fragment | NA | NA | 0 |
| S8_extracted_k141_102600 | 1128 | No | NA | 2 | 0 | 0 | Not-determined | Genome-fragment | NA | NA | 0 |
| S8_extracted_k141_198516 | 1198 | No | NA | 1 | 0 | 1 | Not-determined | Genome-fragment | NA | NA | 0 |
| S8_extracted_k141_154451 | 1268 | No | NA | 1 | 0 | 0 | Not-determined | Genome-fragment | NA | NA | 0 |
| S8_extracted_k141_120068 | 22254 | No | NA | 18 | 0 | 18 | Not-determined | Genome-fragment | NA | NA | 0 |
| S8_extracted_k141_198550 | 1083 | No | NA | 1 | 0 | 0 | Not-determined | Genome-fragment | NA | NA | 0 |
| S8_extracted_k141_206496 | 7874 | No | NA | 7 | 0 | 2 | Not-determined | Genome-fragment | NA | NA | 0 |
| S8_extracted_k141_188604 | 1349 | No | NA | 2 | 0 | 2 | Not-determined | Genome-fragment | NA | NA | 0 |
| S8_extracted_k141_67763 | 8587 | No | NA | 10 | 0 | 0 | Not-determined | Genome-fragment | NA | NA | 0 |
| S8_extracted_k141_76569 | 3273 | No | NA | 3 | 0 | 2 | Not-determined | Genome-fragment | NA | NA | 0 |
| S8_extracted_k141_137312 | 2327 | No | NA | 5 | 0 | 0 | Not-determined | Genome-fragment | NA | NA | 0 |
| S8_extracted_k141_33564 | 3215 | No | NA | 3 | 0 | 1 | Not-determined | Genome-fragment | NA | NA | 0 |
| S8_extracted_k141_50219 | 1373 | No | NA | 3 | 0 | 0 | Not-determined | Genome-fragment | NA | NA | 0 |
| S8_extracted_k141_24842 | 46642 | No | NA | 8 | 0 | 6 | Not-determined | Genome-fragment | NA | NA | 0 |
| S8_extracted_k141_154503 | 1730 | No | NA | 2 | 0 | 1 | Not-determined | Genome-fragment | NA | NA | 0 |
| S8_extracted_k141_111739 | 1417 | No | NA | 1 | 0 | 1 | Not-determined | Genome-fragment | NA | NA | 0 |
| S8_extracted_k141_120114 | 1212 | No | NA | 1 | 0 | 1 | Not-determined | Genome-fragment | NA | NA | 0 |
| S8_extracted_k141_206513 | 1213 | No | NA | 1 | 0 | 1 | Not-determined | Genome-fragment | NA | NA | 0 |
| S8_extracted_k141_128846 | 2500 | No | NA | 2 | 0 | 1 | Not-determined | Genome-fragment | NA | NA | 0 |
| S8_extracted_k141_137350 | 4164 | No | NA | 5 | 0 | 3 | Not-determined | Genome-fragment | NA | NA | 0 |
| S8_extracted_k141_172509 | 1123 | No | NA | 2 | 0 | 1 | Not-determined | Genome-fragment | NA | NA | 0 |
| S8_extracted_k141_154543 | 1898 | No | NA | 5 | 0 | 3 | Not-determined | Genome-fragment | NA | NA | 0 |
| S8_extracted_k141_188665 | 5716 | No | NA | 3 | 0 | 2 | Not-determined | Genome-fragment | NA | NA | 0 |
| S8_extracted_k141_76623 | 1048 | No | NA | 2 | 0 | 0 | Not-determined | Genome-fragment | NA | NA | 0 |
| S8_extracted_k141_111765 | 1030 | No | NA | 1 | 0 | 0 | Not-determined | Genome-fragment | NA | NA | 0 |
| S8_extracted_k141_188666 | 1908 | No | NA | 1 | 0 | 1 | Not-determined | Genome-fragment | NA | NA | 0 |
| S8_extracted_k141_163246 | 3406 | No | NA | 3 | 0 | 3 | Not-determined | Genome-fragment | NA | NA | 0 |
| S8_extracted_k141_76632 | 1096 | No | NA | 1 | 0 | 0 | Not-determined | Genome-fragment | NA | NA | 0 |
| S8_extracted_k141_128860 | 1534 | No | NA | 2 | 0 | 0 | Not-determined | Genome-fragment | NA | NA | 0 |
| S8_extracted_k141_198599 | 1084 | No | NA | 2 | 0 | 1 | Not-determined | Genome-fragment | NA | NA | 0 |
| S8_extracted_k141_24884 | 1293 | No | NA | 2 | 0 | 2 | Not-determined | Genome-fragment | NA | NA | 0 |
| S8_extracted_k141_146302 | 1260 | No | NA | 1 | 0 | 1 | Not-determined | Genome-fragment | NA | NA | 0 |
| S8_extracted_k141_76653 | 1375 | No | NA | 1 | 0 | 0 | Not-determined | Genome-fragment | NA | NA | 0 |
| S8_extracted_k141_154558 | 3222 | No | NA | 8 | 0 | 8 | Not-determined | Genome-fragment | NA | NA | 0 |
| S15_vs_28129\|\|full | 9831 | No | NA | 13 | 1 | 2 | Low-quality | Genome-fragment | 2.54 | AAI-based (medium-confidence) | 0 |
| S8_extracted_k141_76661 | 1919 | No | NA | 2 | 0 | 1 | Not-determined | Genome-fragment | NA | NA | 0 |
| S8_extracted_k141_94125 | 3003 | No | NA | 3 | 0 | 2 | Not-determined | Genome-fragment | NA | NA | 0 |
| S16_vs_30441\|\|full | 9833 | No | NA | 9 | 2 | 0 | Low-quality | Genome-fragment | 4.39 | HMM-based (lower-bound) | 0 |
| S8_extracted_k141_128888 | 1492 | No | NA | 2 | 0 | 0 | Not-determined | Genome-fragment | NA | NA | 0 |
| S8_extracted_k141_76670 | 1112 | No | NA | 1 | 0 | 1 | Not-determined | Genome-fragment | NA | NA | 0 |
| S8_extracted_k141_85396 | 1115 | No | NA | 1 | 0 | 0 | Not-determined | Genome-fragment | NA | NA | 0 |
| S8_extracted_k141_188699 | 2168 | No | NA | 2 | 0 | 1 | Not-determined | Genome-fragment | NA | NA | 0 |
| S8_extracted_k141_42082 | 3214 | No | NA | 3 | 0 | 3 | Not-determined | Genome-fragment | NA | NA | 0 |
| S8_extracted_k141_146339 | 1276 | No | NA | 2 | 0 | 0 | Not-determined | Genome-fragment | NA | NA | 0 |
| S8_extracted_k141_120161 | 1048 | No | NA | 2 | 0 | 1 | Not-determined | Genome-fragment | NA | NA | 0 |
| S8_extracted_k141_198653 | 1570 | No | NA | 2 | 0 | 1 | Not-determined | Genome-fragment | NA | NA | 0 |
| S8_extracted_k141_24935 | 1080 | No | NA | 1 | 0 | 1 | Not-determined | Genome-fragment | NA | NA | 0 |
| S8_extracted_k141_33682 | 1199 | No | NA | 2 | 0 | 1 | Not-determined | Genome-fragment | NA | NA | 0 |
| S8_extracted_k141_206536 | 1508 | No | NA | 3 | 0 | 2 | Not-determined | Genome-fragment | NA | NA | 0 |
| S8_extracted_k141_111790 | 25942 | No | NA | 41 | 8 | 0 | Medium-quality | Genome-fragment | 60.63 | AAI-based (high-confidence) | 0 |
| S8_extracted_k141_102704 | 4629 | No | NA | 2 | 0 | 2 | Not-determined | Genome-fragment | NA | NA | 0 |
| S8_extracted_k141_120169 | 3920 | No | NA | 5 | 0 | 0 | Not-determined | Genome-fragment | NA | NA | 0 |
| S8_extracted_k141_188715 | 1270 | No | NA | 1 | 0 | 1 | Not-determined | Genome-fragment | NA | NA | 0 |
| S5_vs_k141_105596\|\|full | 9845 | No | NA | 11 | 0 | 9 | Low-quality | Genome-fragment | 23.43 | AAI-based (medium-confidence) | 0 |
| S8_extracted_k141_198664 | 5039 | No | NA | 5 | 0 | 3 | Not-determined | Genome-fragment | NA | NA | 0 |
| S8_extracted_k141_50324 | 2372 | No | NA | 4 | 0 | 2 | Not-determined | Genome-fragment | NA | NA | 0 |
| S8_extracted_k141_154614 | 3138 | No | NA | 2 | 0 | 0 | Not-determined | Genome-fragment | NA | NA | 0 |
| S8_extracted_k141_76683 | 1489 | No | NA | 2 | 0 | 2 | Not-determined | Genome-fragment | NA | NA | 0 |
| S8_extracted_k141_42111 | 2707 | No | NA | 2 | 0 | 1 | Not-determined | Genome-fragment | NA | NA | 0 |
| S8_extracted_k141_154615 | 1408 | No | NA | 1 | 0 | 1 | Not-determined | Genome-fragment | NA | NA | 0 |
| S8_extracted_k141_111803 | 2079 | No | NA | 2 | 0 | 1 | Not-determined | Genome-fragment | NA | NA | 0 |
| S17_extracted_k141_155089 | 9847 | No | NA | 3 | 1 | 0 | Low-quality | Genome-fragment | 10.34 | AAI-based (high-confidence) | 0 |
| S17_extracted_k141_59125 | 9848 | No | NA | 25 | 22 | 0 | Low-quality | Genome-fragment | 32.46 | AAI-based (high-confidence) | 0 |
| S13_extracted_116107 | 9859 | No | NA | 18 | 10 | 0 | Low-quality | Genome-fragment | 23.16 | AAI-based (high-confidence) | 0 |
| S12_vs_k141_26844\|\|full | 9859 | No | NA | 16 | 1 | 0 | Low-quality | Genome-fragment | 5.52 | HMM-based (lower-bound) | 0 |
| S11_vs_k141_83803\|\|full | 9863 | No | NA | 13 | 1 | 0 | Low-quality | Genome-fragment | 17.89 | AAI-based (high-confidence) | 0 |
| S8_extracted_k141_42136 | 1041 | No | NA | 1 | 0 | 0 | Not-determined | Genome-fragment | NA | NA | 0 |
| S8_extracted_k141_50341 | 4513 | No | NA | 6 | 0 | 2 | Not-determined | Genome-fragment | NA | NA | 0 |
| S8_extracted_k141_120210 | 6816 | No | NA | 7 | 0 | 4 | Not-determined | Genome-fragment | NA | NA | 0 |
| S8_extracted_k141_42139 | 1548 | No | NA | 2 | 0 | 0 | Not-determined | Genome-fragment | NA | NA | 0 |
| S8_extracted_k141_89901 | 9876 | No | NA | 25 | 10 | 0 | Low-quality | Genome-fragment | 26.88 | AAI-based (high-confidence) | 0 |
| S8_extracted_k141_59806 | 1884 | No | NA | 1 | 0 | 1 | Not-determined | Genome-fragment | NA | NA | 0 |
| S8_extracted_k141_128990 | 1117 | No | NA | 1 | 0 | 1 | Not-determined | Genome-fragment | NA | NA | 0 |
| S8_extracted_k141_128999 | 1323 | No | NA | 1 | 0 | 1 | Not-determined | Genome-fragment | NA | NA | 0 |
| S8_extracted_k141_50365 | 1117 | No | NA | 1 | 0 | 0 | Not-determined | Genome-fragment | NA | NA | 0 |
| S8_extracted_k141_146432 | 1479 | No | NA | 2 | 0 | 0 | Not-determined | Genome-fragment | NA | NA | 0 |
| S8_extracted_k141_154660 | 1922 | No | NA | 3 | 0 | 1 | Not-determined | Genome-fragment | NA | NA | 0 |
| S8_extracted_k141_50371 | 1752 | No | NA | 3 | 0 | 0 | Not-determined | Genome-fragment | NA | NA | 0 |
| S10_vs_15568\|\|full | 9878 | No | NA | 24 | 4 | 0 | Low-quality | Genome-fragment | 25.91 | AAI-based (high-confidence) | 0 |
| S8_extracted_k141_85475 | 1699 | No | NA | 2 | 0 | 2 | Not-determined | Genome-fragment | NA | NA | 0 |
| S8_extracted_k141_50381 | 5636 | No | NA | 3 | 0 | 1 | Not-determined | Genome-fragment | NA | NA | 0 |
| S8_extracted_k141_85477 | 9647 | No | NA | 7 | 0 | 6 | Not-determined | Genome-fragment | NA | NA | 0 |
| S1_extracted_k141_89949 | 9882 | No | NA | 23 | 6 | 0 | Low-quality | Genome-fragment | 24.64 | AAI-based (high-confidence) | 0 |
| S1_extracted_k141_4698 | 9898 | No | NA | 15 | 6 | 0 | Low-quality | Genome-fragment | 24.34 | AAI-based (high-confidence) | 0 |
| S8_extracted_k141_15818 | 5203 | No | NA | 4 | 0 | 0 | Not-determined | Genome-fragment | NA | NA | 0 |
| S11_extracted_k141_80701 | 9904 | No | NA | 17 | 1 | 0 | Low-quality | Genome-fragment | 6.07 | AAI-based (high-confidence) | 0 |
| S8_extracted_k141_85487 | 1145 | No | NA | 1 | 0 | 1 | Not-determined | Genome-fragment | NA | NA | 0 |
| S8_extracted_k141_198771 | 5553 | No | NA | 3 | 0 | 3 | Not-determined | Genome-fragment | NA | NA | 0 |
| S8_extracted_k141_198780 | 2125 | No | NA | 2 | 0 | 1 | Not-determined | Genome-fragment | NA | NA | 0 |
| S8_extracted_k141_154686 | 1085 | No | NA | 1 | 0 | 1 | Not-determined | Genome-fragment | NA | NA | 0 |
| S8_extracted_k141_42193 | 3245 | No | NA | 4 | 0 | 2 | Not-determined | Genome-fragment | NA | NA | 0 |
| S8_extracted_k141_146462 | 1894 | No | NA | 1 | 0 | 1 | Not-determined | Genome-fragment | NA | NA | 0 |
| S8_extracted_k141_206618 | 1237 | No | NA | 2 | 0 | 1 | Not-determined | Genome-fragment | NA | NA | 0 |
| S8_extracted_k141_172677 | 2433 | No | NA | 4 | 0 | 2 | Not-determined | Genome-fragment | NA | NA | 0 |
| S8_extracted_k141_67958 | 2485 | No | NA | 3 | 0 | 2 | Not-determined | Genome-fragment | NA | NA | 0 |
| S14_extracted_k141_13793 | 9910 | No | NA | 13 | 5 | 0 | Low-quality | Genome-fragment | 17.02 | AAI-based (high-confidence) | 0 |
| S8_extracted_k141_7627 | 1127 | No | NA | 1 | 0 | 1 | Not-determined | Genome-fragment | NA | NA | 0 |
| S12_extracted_k141_126691 | 9914 | No | NA | 21 | 7 | 0 | Low-quality | Genome-fragment | 25.38 | AAI-based (medium-confidence) | 0 |
| S8_extracted_k141_59884 | 1331 | No | NA | 1 | 0 | 1 | Not-determined | Genome-fragment | NA | NA | 0 |
| S8_extracted_k141_33834 | 1736 | No | NA | 2 | 0 | 1 | Not-determined | Genome-fragment | NA | NA | 0 |
| S8_extracted_k141_94249 | 8460 | No | NA | 10 | 0 | 5 | Not-determined | Genome-fragment | NA | NA | 0 |
| S8_extracted_k141_33843 | 1798 | No | NA | 1 | 0 | 1 | Not-determined | Genome-fragment | NA | NA | 0 |
| S8_extracted_k141_67975 | 1073 | No | NA | 1 | 0 | 1 | Not-determined | Genome-fragment | NA | NA | 0 |
| S8_extracted_k141_181318 | 2108 | No | NA | 2 | 0 | 2 | Not-determined | Genome-fragment | NA | NA | 0 |
| S8_extracted_k141_146479 | 1917 | No | NA | 2 | 0 | 2 | Not-determined | Genome-fragment | NA | NA | 0 |
| S8_extracted_k141_67978 | 4557 | No | NA | 2 | 0 | 2 | Not-determined | Genome-fragment | NA | NA | 0 |
| S3_vs_k141_20769\|\|full | 9924 | No | NA | 26 | 2 | 0 | Low-quality | Genome-fragment | 20.7 | AAI-based (medium-confidence) | 0 |
| S8_extracted_k141_198835 | 1127 | No | NA | 2 | 0 | 1 | Not-determined | Genome-fragment | NA | NA | 0 |
| S8_extracted_k141_137512 | 1184 | No | NA | 1 | 0 | 0 | Not-determined | Genome-fragment | NA | NA | 0 |
| S8_extracted_k141_102730 | 1886 | No | NA | 1 | 0 | 1 | Not-determined | Genome-fragment | NA | NA | 0 |
| S4_vs_VIRSorter_2932-cat_1 | 9924 | No | NA | 12 | 8 | 0 | Low-quality | Genome-fragment | 24.04 | AAI-based (medium-confidence) | 0 |
| S3_vs_k141_38528\|\|full | 9925 | Yes | 7893 | 11 | 2 | 3 | Low-quality | Genome-fragment | 17.25 | HMM-based (lower-bound) | 20.47 |
| S8_extracted_k141_33863 | 1164 | No | NA | 1 | 0 | 1 | Not-determined | Genome-fragment | NA | NA | 0 |
| S8_extracted_k141_76858 | 1425 | No | NA | 4 | 0 | 0 | Not-determined | Genome-fragment | NA | NA | 0 |
| S8_extracted_k141_102756 | 1303 | No | NA | 1 | 0 | 1 | Not-determined | Genome-fragment | NA | NA | 0 |
| S8_extracted_k141_172748 | 1137 | No | NA | 1 | 0 | 0 | Not-determined | Genome-fragment | NA | NA | 0 |
| S8_extracted_k141_85562 | 2580 | No | NA | 1 | 0 | 1 | Not-determined | Genome-fragment | NA | NA | 0 |
| S8_extracted_k141_76872 | 2240 | No | NA | 3 | 0 | 0 | Not-determined | Genome-fragment | NA | NA | 0 |
| S8_extracted_k141_94292 | 4638 | No | NA | 6 | 0 | 4 | Not-determined | Genome-fragment | NA | NA | 0 |
| S8_extracted_k141_146522 | 1133 | No | NA | 1 | 0 | 0 | Not-determined | Genome-fragment | NA | NA | 0 |
| S8_extracted_k141_102764 | 1056 | No | NA | 2 | 0 | 2 | Not-determined | Genome-fragment | NA | NA | 0 |
| S8_extracted_k141_94299 | 1174 | No | NA | 1 | 0 | 0 | Not-determined | Genome-fragment | NA | NA | 0 |
| S8_extracted_k141_120403 | 1143 | No | NA | 2 | 0 | 2 | Not-determined | Genome-fragment | NA | NA | 0 |
| S8_extracted_k141_85568 | 2930 | No | NA | 3 | 0 | 2 | Not-determined | Genome-fragment | NA | NA | 0 |
| S8_extracted_k141_120406 | 1049 | No | NA | 1 | 0 | 0 | Not-determined | Genome-fragment | NA | NA | 0 |
| S12_vs_k141_116337\|\|full | 9927 | No | NA | 13 | 1 | 0 | Low-quality | Genome-fragment | 14.42 | AAI-based (high-confidence) | 0 |
| S8_extracted_k141_68027 | 1117 | No | NA | 1 | 0 | 1 | Not-determined | Genome-fragment | NA | NA | 0 |
| S8_extracted_k141_188806 | 1577 | No | NA | 3 | 0 | 3 | Not-determined | Genome-fragment | NA | NA | 0 |
| S8_extracted_k141_7669 | 1073 | No | NA | 1 | 0 | 0 | Not-determined | Genome-fragment | NA | NA | 0 |
| S8_extracted_k141_85576 | 1692 | No | NA | 2 | 0 | 2 | Not-determined | Genome-fragment | NA | NA | 0 |
| S8_extracted_k141_59951 | 1246 | No | NA | 3 | 0 | 0 | Not-determined | Genome-fragment | NA | NA | 0 |
| S8_extracted_k141_68035 | 4149 | No | NA | 4 | 0 | 2 | Not-determined | Genome-fragment | NA | NA | 0 |
| S8_extracted_k141_59954 | 1836 | No | NA | 3 | 0 | 2 | Not-determined | Genome-fragment | NA | NA | 0 |
| S8_extracted_k141_111872 | 2243 | No | NA | 2 | 0 | 2 | Not-determined | Genome-fragment | NA | NA | 0 |
| S8_extracted_k141_59966 | 1528 | No | NA | 1 | 0 | 1 | Not-determined | Genome-fragment | NA | NA | 0 |
| S7_extracted_95873 | 9929 | No | NA | 14 | 10 | 0 | Low-quality | Genome-fragment | 16.09 | AAI-based (high-confidence) | 0 |
| S12_vs_k141_64139\|\|full | 9930 | No | NA | 21 | 1 | 1 | Low-quality | Genome-fragment | 29.09 | AAI-based (high-confidence) | 0 |
| S8_extracted_k141_33918 | 1191 | No | NA | 2 | 0 | 1 | Not-determined | Genome-fragment | NA | NA | 0 |
| S8_extracted_k141_94332 | 2182 | No | NA | 1 | 0 | 1 | Not-determined | Genome-fragment | NA | NA | 0 |
| S8_extracted_k141_94339 | 3416 | No | NA | 3 | 0 | 0 | Not-determined | Genome-fragment | NA | NA | 0 |
| S8_extracted_k141_42278 | 2019 | No | NA | 2 | 0 | 2 | Not-determined | Genome-fragment | NA | NA | 0 |
| S3_extracted_k141_104768 | 9933 | No | NA | 18 | 3 | 0 | Low-quality | Genome-fragment | 5.56 | AAI-based (high-confidence) | 0 |
| S8_extracted_k141_137605 | 1327 | No | NA | 2 | 0 | 1 | Not-determined | Genome-fragment | NA | NA | 0 |
| S8_extracted_k141_188843 | 1110 | No | NA | 1 | 0 | 0 | Not-determined | Genome-fragment | NA | NA | 0 |
| S8_extracted_k141_42298 | 1178 | No | NA | 1 | 0 | 1 | Not-determined | Genome-fragment | NA | NA | 0 |
| S8_extracted_k141_76966 | 1059 | No | NA | 2 | 0 | 2 | Not-determined | Genome-fragment | NA | NA | 0 |
| S8_extracted_k141_146605 | 2891 | No | NA | 3 | 0 | 0 | Not-determined | Genome-fragment | NA | NA | 0 |
| S8_extracted_k141_25119 | 1120 | No | NA | 1 | 0 | 1 | Not-determined | Genome-fragment | NA | NA | 0 |
| S8_extracted_k141_146608 | 1117 | No | NA | 1 | 0 | 0 | Not-determined | Genome-fragment | NA | NA | 0 |
| S8_extracted_k141_76974 | 1185 | No | NA | 2 | 0 | 2 | Not-determined | Genome-fragment | NA | NA | 0 |
| S8_extracted_k141_146616 | 1151 | No | NA | 2 | 0 | 2 | Not-determined | Genome-fragment | NA | NA | 0 |
| S8_extracted_k141_94361 | 3494 | No | NA | 6 | 5 | 0 | Medium-quality | Genome-fragment | 69.02 | AAI-based (high-confidence) | 0 |
| S8_extracted_k141_60028 | 4063 | No | NA | 3 | 0 | 3 | Not-determined | Genome-fragment | NA | NA | 0 |
| S8_extracted_k141_76977 | 5855 | No | NA | 6 | 0 | 1 | Not-determined | Genome-fragment | NA | NA | 0 |
| S8_extracted_k141_94364 | 1361 | No | NA | 1 | 0 | 1 | Not-determined | Genome-fragment | NA | NA | 0 |
| S8_extracted_k141_68096 | 1376 | No | NA | 1 | 0 | 1 | Not-determined | Genome-fragment | NA | NA | 0 |
| S8_extracted_k141_94372 | 1173 | No | NA | 2 | 0 | 1 | Not-determined | Genome-fragment | NA | NA | 0 |
| S8_extracted_k141_85673 | 2196 | No | NA | 1 | 0 | 0 | Not-determined | Genome-fragment | NA | NA | 0 |
| S8_extracted_k141_129226 | 1708 | No | NA | 1 | 0 | 1 | Not-determined | Genome-fragment | NA | NA | 0 |
| S8_extracted_k141_34002 | 1838 | No | NA | 2 | 0 | 1 | Not-determined | Genome-fragment | NA | NA | 0 |
| S8_extracted_k141_181470 | 3165 | No | NA | 3 | 0 | 3 | Not-determined | Genome-fragment | NA | NA | 0 |
| S8_extracted_k141_154830 | 1492 | No | NA | 3 | 0 | 0 | Not-determined | Genome-fragment | NA | NA | 0 |
| S8_extracted_k141_15958 | 1391 | No | NA | 2 | 0 | 2 | Not-determined | Genome-fragment | NA | NA | 0 |
| S16_vs_140379\|\|full | 9934 | No | NA | 13 | 1 | 1 | Low-quality | Genome-fragment | 15.33 | AAI-based (high-confidence) | 0 |
| S8_extracted_k141_146640 | 1332 | No | NA | 3 | 0 | 2 | Not-determined | Genome-fragment | NA | NA | 0 |
| S13_extracted_14131 | 9936 | No | NA | 20 | 2 | 1 | Low-quality | Genome-fragment | 13.43 | AAI-based (high-confidence) | 0 |
| S3_vs_k141_104360\|\|full | 9936 | No | NA | 13 | 1 | 0 | Low-quality | Genome-fragment | 22.85 | AAI-based (medium-confidence) | 0 |
| S8_extracted_k141_85701 | 1278 | No | NA | 1 | 0 | 1 | Not-determined | Genome-fragment | NA | NA | 0 |
| S16_extracted_110287 | 9938 | No | NA | 14 | 14 | 0 | Low-quality | Genome-fragment | 26.68 | AAI-based (high-confidence) | 0 |
| S8_extracted_k141_102920 | 4095 | No | NA | 1 | 0 | 1 | Not-determined | Genome-fragment | NA | NA | 0 |
| S8_extracted_k141_129244 | 2474 | No | NA | 1 | 0 | 1 | Not-determined | Genome-fragment | NA | NA | 0 |
| S8_extracted_k141_42350 | 2451 | No | NA | 2 | 0 | 2 | Not-determined | Genome-fragment | NA | NA | 0 |
| S8_extracted_k141_25190 | 3075 | No | NA | 2 | 0 | 2 | Not-determined | Genome-fragment | NA | NA | 0 |
| S8_extracted_k141_42359 | 1252 | No | NA | 1 | 0 | 1 | Not-determined | Genome-fragment | NA | NA | 0 |
| S8_extracted_k141_102937 | 1719 | No | NA | 2 | 0 | 1 | Not-determined | Genome-fragment | NA | NA | 0 |
| S8_extracted_k141_34056 | 2999 | No | NA | 2 | 0 | 1 | Not-determined | Genome-fragment | NA | NA | 0 |
| S3_vs_k141_112539\|\|full | 9946 | No | NA | 9 | 0 | 7 | Low-quality | Genome-fragment | 4.95 | AAI-based (medium-confidence) | 0 |
| S8_extracted_k141_112017 | 2939 | No | NA | 1 | 0 | 1 | Not-determined | Genome-fragment | NA | NA | 0 |
| S8_extracted_k141_206853 | 1003 | No | NA | 1 | 0 | 0 | Not-determined | Genome-fragment | NA | NA | 0 |
| S8_extracted_k141_7839 | 4057 | No | NA | 3 | 0 | 2 | Not-determined | Genome-fragment | NA | NA | 0 |
| S8_extracted_k141_60131 | 1309 | No | NA | 2 | 0 | 2 | Not-determined | Genome-fragment | NA | NA | 0 |
| S8_extracted_k141_206863 | 1011 | No | NA | 2 | 0 | 1 | Not-determined | Genome-fragment | NA | NA | 0 |
| S8_extracted_k141_94472 | 1082 | No | NA | 2 | 0 | 0 | Not-determined | Genome-fragment | NA | NA | 0 |
| S8_extracted_k141_188947 | 1255 | No | NA | 2 | 0 | 1 | Not-determined | Genome-fragment | NA | NA | 0 |
| S8_extracted_k141_199047 | 2442 | No | NA | 1 | 0 | 1 | Not-determined | Genome-fragment | NA | NA | 0 |
| S8_extracted_k141_163553 | 1419 | No | NA | 1 | 0 | 0 | Not-determined | Genome-fragment | NA | NA | 0 |
| S8_extracted_k141_181542 | 1440 | No | NA | 2 | 0 | 0 | Not-determined | Genome-fragment | NA | NA | 0 |
| S8_extracted_k141_199050 | 1609 | No | NA | 1 | 0 | 1 | Not-determined | Genome-fragment | NA | NA | 0 |
| S8_extracted_k141_7880 | 2041 | No | NA | 2 | 0 | 2 | Not-determined | Genome-fragment | NA | NA | 0 |
| S8_extracted_k141_34097 | 1238 | No | NA | 1 | 0 | 1 | Not-determined | Genome-fragment | NA | NA | 0 |
| S8_extracted_k141_137768 | 2192 | No | NA | 3 | 0 | 2 | Not-determined | Genome-fragment | NA | NA | 0 |
| S8_extracted_k141_85767 | 2329 | No | NA | 3 | 0 | 2 | Not-determined | Genome-fragment | NA | NA | 0 |
| S12_vs_k141_149430\|\|full | 9949 | No | NA | 14 | 2 | 1 | Low-quality | Genome-fragment | 20.2 | AAI-based (medium-confidence) | 0 |
| S8_extracted_k141_199052 | 6867 | No | NA | 6 | 0 | 3 | Not-determined | Genome-fragment | NA | NA | 0 |
| S8_extracted_k141_154920 | 1174 | No | NA | 2 | 0 | 1 | Not-determined | Genome-fragment | NA | NA | 0 |
| S8_extracted_k141_60177 | 4389 | No | NA | 3 | 0 | 2 | Not-determined | Genome-fragment | NA | NA | 0 |
| S8_extracted_k141_85777 | 1139 | No | NA | 1 | 0 | 0 | Not-determined | Genome-fragment | NA | NA | 0 |
| S8_extracted_k141_154923 | 1733 | No | NA | 1 | 0 | 1 | Not-determined | Genome-fragment | NA | NA | 0 |
| S16_vs_18824\|\|full | 9950 | No | NA | 22 | 5 | 0 | Low-quality | Genome-fragment | 20.95 | AAI-based (high-confidence) | 0 |
| S8_extracted_k141_42401 | 20059 | No | NA | 23 | 0 | 6 | Not-determined | Genome-fragment | NA | NA | 0 |
| S8_extracted_k141_112084 | 1886 | No | NA | 2 | 0 | 2 | Not-determined | Genome-fragment | NA | NA | 0 |
| S8_extracted_k141_77103 | 1942 | No | NA | 1 | 0 | 1 | Not-determined | Genome-fragment | NA | NA | 0 |
| S8_extracted_k141_199079 | 1414 | No | NA | 1 | 0 | 0 | Not-determined | Genome-fragment | NA | NA | 0 |
| S8_extracted_k141_25266 | 1014 | No | NA | 2 | 0 | 2 | Not-determined | Genome-fragment | NA | NA | 0 |
| S8_extracted_k141_34138 | 1145 | No | NA | 1 | 0 | 1 | Not-determined | Genome-fragment | NA | NA | 0 |
| S3_vs_k141_58148\|\|full | 9956 | No | NA | 14 | 2 | 0 | Low-quality | Genome-fragment | 13.95 | AAI-based (high-confidence) | 0 |
| S8_extracted_k141_103033 | 1540 | No | NA | 1 | 0 | 1 | Not-determined | Genome-fragment | NA | NA | 0 |
| S8_extracted_k141_103042 | 1303 | No | NA | 1 | 0 | 0 | Not-determined | Genome-fragment | NA | NA | 0 |
| S8_extracted_k141_172882 | 2728 | No | NA | 3 | 0 | 2 | Not-determined | Genome-fragment | NA | NA | 0 |
| S8_extracted_k141_16103 | 1263 | No | NA | 2 | 0 | 0 | Not-determined | Genome-fragment | NA | NA | 0 |
| S8_extracted_k141_163569 | 1252 | No | NA | 1 | 0 | 0 | Not-determined | Genome-fragment | NA | NA | 0 |
| S8_extracted_k141_189029 | 2148 | No | NA | 5 | 0 | 4 | Not-determined | Genome-fragment | NA | NA | 0 |
| S8_extracted_k141_137834 | 1222 | No | NA | 2 | 0 | 0 | Not-determined | Genome-fragment | NA | NA | 0 |
| S8_extracted_k141_154985 | 2079 | No | NA | 4 | 0 | 4 | Not-determined | Genome-fragment | NA | NA | 0 |
| S8_extracted_k141_94590 | 2574 | No | NA | 3 | 0 | 3 | Not-determined | Genome-fragment | NA | NA | 0 |
| S3_extracted_k141_23204 | 9969 | No | NA | 21 | 0 | 1 | Low-quality | Genome-fragment | 4.86 | AAI-based (high-confidence) | 0 |
| S8_extracted_k141_77159 | 1085 | No | NA | 2 | 0 | 1 | Not-determined | Genome-fragment | NA | NA | 0 |
| S8_extracted_k141_68245 | 2025 | No | NA | 2 | 0 | 1 | Not-determined | Genome-fragment | NA | NA | 0 |
| S13_extracted_133737 | 9974 | No | NA | 9 | 0 | 5 | Low-quality | Genome-fragment | 9.73 | AAI-based (medium-confidence) | 0 |
| S8_extracted_k141_42467 | 1103 | No | NA | 1 | 0 | 0 | Not-determined | Genome-fragment | NA | NA | 0 |
| S8_extracted_k141_206949 | 2104 | No | NA | 4 | 0 | 2 | Not-determined | Genome-fragment | NA | NA | 0 |
| S8_extracted_k141_129331 | 1196 | No | NA | 2 | 0 | 2 | Not-determined | Genome-fragment | NA | NA | 0 |
| S8_extracted_k141_60273 | 4961 | No | NA | 3 | 0 | 2 | Not-determined | Genome-fragment | NA | NA | 0 |
| S8_extracted_k141_85889 | 1145 | No | NA | 2 | 0 | 1 | Not-determined | Genome-fragment | NA | NA | 0 |
| S8_extracted_k141_206958 | 1305 | No | NA | 3 | 0 | 0 | Not-determined | Genome-fragment | NA | NA | 0 |
| S8_extracted_k141_199159 | 1131 | No | NA | 1 | 0 | 1 | Not-determined | Genome-fragment | NA | NA | 0 |
| S8_extracted_k141_181658 | 1307 | No | NA | 2 | 0 | 0 | Not-determined | Genome-fragment | NA | NA | 0 |
| S8_extracted_k141_129341 | 1366 | No | NA | 2 | 0 | 2 | Not-determined | Genome-fragment | NA | NA | 0 |
| S8_extracted_k141_172914 | 1101 | No | NA | 2 | 0 | 2 | Not-determined | Genome-fragment | NA | NA | 0 |
| S8_extracted_k141_199160 | 3064 | No | NA | 3 | 0 | 3 | Not-determined | Genome-fragment | NA | NA | 0 |
| S8_extracted_k141_103111 | 1212 | No | NA | 1 | 0 | 1 | Not-determined | Genome-fragment | NA | NA | 0 |
| S8_extracted_k141_77193 | 1019 | No | NA | 2 | 0 | 1 | Not-determined | Genome-fragment | NA | NA | 0 |
| S8_extracted_k141_103123 | 1201 | No | NA | 2 | 0 | 0 | Not-determined | Genome-fragment | NA | NA | 0 |
| S8_extracted_k141_137856 | 1767 | No | NA | 2 | 0 | 0 | Not-determined | Genome-fragment | NA | NA | 0 |
| S1_vs_k141_41700\|\|full | 9975 | No | NA | 13 | 7 | 0 | Low-quality | Genome-fragment | 25.07 | AAI-based (medium-confidence) | 0 |
| S8_extracted_k141_85923 | 2849 | No | NA | 3 | 0 | 1 | Not-determined | Genome-fragment | NA | NA | 0 |
| S8_extracted_k141_206971 | 1193 | No | NA | 2 | 0 | 0 | Not-determined | Genome-fragment | NA | NA | 0 |
| S8_extracted_k141_25409 | 1155 | No | NA | 2 | 0 | 0 | Not-determined | Genome-fragment | NA | NA | 0 |
| S8_extracted_k141_181699 | 2224 | No | NA | 2 | 0 | 1 | Not-determined | Genome-fragment | NA | NA | 0 |
| S8_extracted_k141_60328 | 1427 | No | NA | 2 | 0 | 0 | Not-determined | Genome-fragment | NA | NA | 0 |
| S8_extracted_k141_189091 | 3864 | No | NA | 3 | 0 | 0 | Not-determined | Genome-fragment | NA | NA | 0 |
| S8_extracted_k141_146867 | 1012 | No | NA | 1 | 0 | 0 | Not-determined | Genome-fragment | NA | NA | 0 |
| S8_extracted_k141_77226 | 1409 | No | NA | 2 | 0 | 2 | Not-determined | Genome-fragment | NA | NA | 0 |
| S8_extracted_k141_25416 | 1855 | No | NA | 1 | 0 | 1 | Not-determined | Genome-fragment | NA | NA | 0 |
| S8_extracted_k141_85958 | 1255 | No | NA | 3 | 0 | 0 | Not-determined | Genome-fragment | NA | NA | 0 |
| S8_extracted_k141_16172 | 37446 | No | NA | 26 | 2 | 0 | Medium-quality | Genome-fragment | 58.96 | AAI-based (high-confidence) | 0 |
| S1_extracted_k141_101189 | 9982 | No | NA | 19 | 5 | 0 | Low-quality | Genome-fragment | 17.07 | AAI-based (high-confidence) | 0 |
| S8_extracted_k141_189106 | 8235 | No | NA | 5 | 0 | 2 | Not-determined | Genome-fragment | NA | NA | 0 |
| S8_extracted_k141_47223 | 9982 | No | NA | 17 | 4 | 0 | Low-quality | Genome-fragment | 13.09 | AAI-based (high-confidence) | 0 |
| S8_extracted_k141_181716 | 3166 | No | NA | 3 | 0 | 1 | Not-determined | Genome-fragment | NA | NA | 0 |
| S8_extracted_k141_206996 | 1020 | No | NA | 1 | 0 | 1 | Not-determined | Genome-fragment | NA | NA | 0 |
| S7_vs_83289\|\|full | 9984 | No | NA | 7 | 0 | 7 | Low-quality | Genome-fragment | 4.98 | AAI-based (medium-confidence) | 0 |
| S8_extracted_k141_78168 | 9989 | No | NA | 11 | 1 | 0 | Low-quality | Genome-fragment | 6.71 | HMM-based (lower-bound) | 0 |
| S8_extracted_k141_112208 | 2631 | No | NA | 4 | 0 | 4 | Not-determined | Genome-fragment | NA | NA | 0 |
| S7_vs_126141\|\|full | 9996 | No | NA | 16 | 2 | 0 | Low-quality | Genome-fragment | 23.54 | AAI-based (high-confidence) | 0 |
| S8_extracted_k141_199232 | 1195 | No | NA | 1 | 0 | 0 | Not-determined | Genome-fragment | NA | NA | 0 |
| S6_extracted_k141_35457 | 9997 | No | NA | 13 | 2 | 0 | Low-quality | Genome-fragment | 3.06 | HMM-based (lower-bound) | 0 |
| S5_vs_k141_68958\|\|full | 10005 | No | NA | 10 | 0 | 2 | Low-quality | Genome-fragment | 24.14 | AAI-based (medium-confidence) | 0 |
| S8_extracted_k141_181763 | 1037 | No | NA | 1 | 0 | 0 | Not-determined | Genome-fragment | NA | NA | 0 |
| S8_extracted_k141_189156 | 1252 | No | NA | 2 | 0 | 0 | Not-determined | Genome-fragment | NA | NA | 0 |
| S8_extracted_k141_25481 | 1084 | No | NA | 2 | 0 | 0 | Not-determined | Genome-fragment | NA | NA | 0 |
| S8_extracted_k141_189157 | 1230 | No | NA | 1 | 0 | 0 | Not-determined | Genome-fragment | NA | NA | 0 |
| S8_extracted_k141_163721 | 1236 | No | NA | 2 | 0 | 0 | Not-determined | Genome-fragment | NA | NA | 0 |
| S16_extracted_134775 | 10005 | No | NA | 13 | 3 | 0 | Low-quality | Genome-fragment | 18.45 | AAI-based (medium-confidence) | 0 |
| S16_vs_66234\|\|full | 10005 | No | NA | 10 | 1 | 1 | Low-quality | Genome-fragment | 7.34 | HMM-based (lower-bound) | 0 |
| S8_extracted_k141_16227 | 1753 | No | NA | 2 | 0 | 2 | Not-determined | Genome-fragment | NA | NA | 0 |
| S8_extracted_k141_146931 | 2092 | No | NA | 1 | 0 | 1 | Not-determined | Genome-fragment | NA | NA | 0 |
| S8_extracted_k141_120683 | 1872 | No | NA | 1 | 0 | 1 | Not-determined | Genome-fragment | NA | NA | 0 |
| S8_extracted_k141_60393 | 2379 | No | NA | 2 | 0 | 0 | Not-determined | Genome-fragment | NA | NA | 0 |
| S8_extracted_k141_189162 | 3599 | No | NA | 2 | 0 | 2 | Not-determined | Genome-fragment | NA | NA | 0 |
| S8_extracted_k141_137956 | 1028 | No | NA | 1 | 0 | 0 | Not-determined | Genome-fragment | NA | NA | 0 |
| S9_vs_k141_395\|\|full | 10007 | No | NA | 20 | 2 | 1 | Low-quality | Genome-fragment | 24.11 | AAI-based (high-confidence) | 0 |
| S8_extracted_k141_146950 | 1920 | No | NA | 2 | 0 | 2 | Not-determined | Genome-fragment | NA | NA | 0 |
| S8_extracted_k141_129423 | 27408 | No | NA | 39 | 18 | 0 | High-quality | High-quality | 95.08 | AAI-based (high-confidence) | 0 |
| S8_extracted_k141_120707 | 1634 | No | NA | 1 | 0 | 1 | Not-determined | Genome-fragment | NA | NA | 0 |
| S11_extracted_k141_47288 | 10014 | No | NA | 20 | 5 | 0 | Low-quality | Genome-fragment | 22.05 | AAI-based (high-confidence) | 0 |
| S12_extracted_k141_185588 | 10014 | No | NA | 15 | 5 | 1 | Low-quality | Genome-fragment | 23.25 | AAI-based (high-confidence) | 0 |
| S5_vs_k141_43074\|\|full | 10028 | Yes | 4377 | 10 | 1 | 5 | Low-quality | Genome-fragment | 0.2 | HMM-based (lower-bound) | 56.35 |
| S8_extracted_k141_42565 | 1943 | No | NA | 2 | 0 | 2 | Not-determined | Genome-fragment | NA | NA | 0 |
| S8_extracted_k141_68395 | 6135 | No | NA | 10 | 0 | 0 | Not-determined | Genome-fragment | NA | NA | 0 |
| S8_extracted_k141_189196 | 2110 | No | NA | 3 | 0 | 3 | Not-determined | Genome-fragment | NA | NA | 0 |
| S8_extracted_k141_42569 | 1091 | No | NA | 1 | 0 | 1 | Not-determined | Genome-fragment | NA | NA | 0 |
| S8_extracted_k141_16254 | 1359 | No | NA | 1 | 0 | 1 | Not-determined | Genome-fragment | NA | NA | 0 |
| S8_extracted_k141_16257 | 1511 | No | NA | 1 | 0 | 0 | Not-determined | Genome-fragment | NA | NA | 0 |
| S8_extracted_k141_77353 | 2890 | No | NA | 4 | 0 | 4 | Not-determined | Genome-fragment | NA | NA | 0 |
| S8_extracted_k141_189199 | 4937 | No | NA | 2 | 0 | 2 | Not-determined | Genome-fragment | NA | NA | 0 |
| S8_extracted_k141_112291 | 2895 | No | NA | 5 | 0 | 1 | Not-determined | Genome-fragment | NA | NA | 0 |
| S5_extracted_k141_62924 | 10032 | No | NA | 18 | 4 | 0 | Low-quality | Genome-fragment | 21.63 | AAI-based (high-confidence) | 0 |
| S8_extracted_k141_25515 | 2564 | No | NA | 2 | 0 | 1 | Not-determined | Genome-fragment | NA | NA | 0 |
| S8_extracted_k141_207066 | 1365 | No | NA | 2 | 0 | 2 | Not-determined | Genome-fragment | NA | NA | 0 |
| S8_extracted_k141_25523 | 1294 | No | NA | 1 | 0 | 1 | Not-determined | Genome-fragment | NA | NA | 0 |
| S8_extracted_k141_77369 | 2726 | No | NA | 2 | 0 | 2 | Not-determined | Genome-fragment | NA | NA | 0 |
| S8_extracted_k141_173050 | 1252 | No | NA | 1 | 0 | 1 | Not-determined | Genome-fragment | NA | NA | 0 |
| S8_extracted_k141_42598 | 1204 | No | NA | 2 | 0 | 1 | Not-determined | Genome-fragment | NA | NA | 0 |
| S12_vs_k141_93871\|\|full | 10040 | No | NA | 11 | 1 | 5 | Low-quality | Genome-fragment | 5.38 | HMM-based (lower-bound) | 0 |
| S8_extracted_k141_103161 | 3327 | No | NA | 3 | 0 | 0 | Not-determined | Genome-fragment | NA | NA | 0 |
| S5_vs_k141_69010\|\|full | 10050 | No | NA | 12 | 3 | 0 | Low-quality | Genome-fragment | 20.94 | AAI-based (high-confidence) | 0 |
| S3_vs_k141_17492\|\|full | 10052 | No | NA | 17 | 2 | 0 | Low-quality | Genome-fragment | 23.11 | AAI-based (medium-confidence) | 0 |
| S8_extracted_k141_60457 | 1671 | No | NA | 1 | 0 | 1 | Not-determined | Genome-fragment | NA | NA | 0 |
| S8_extracted_k141_181837 | 2733 | No | NA | 3 | 0 | 1 | Not-determined | Genome-fragment | NA | NA | 0 |
| S8_extracted_k141_42608 | 3560 | No | NA | 2 | 0 | 1 | Not-determined | Genome-fragment | NA | NA | 0 |
| S8_extracted_k141_60470 | 1504 | No | NA | 2 | 0 | 1 | Not-determined | Genome-fragment | NA | NA | 0 |
| S8_extracted_k141_112354 | 1284 | No | NA | 1 | 0 | 1 | Not-determined | Genome-fragment | NA | NA | 0 |
| S8_extracted_k141_147008 | 3746 | No | NA | 4 | 0 | 1 | Not-determined | Genome-fragment | NA | NA | 0 |
| S8_extracted_k141_199278 | 1514 | No | NA | 4 | 0 | 2 | Not-determined | Genome-fragment | NA | NA | 0 |
| S8_extracted_k141_94686 | 3926 | No | NA | 3 | 0 | 2 | Not-determined | Genome-fragment | NA | NA | 0 |
| S10_vs_10068\|\|full | 10056 | No | NA | 12 | 2 | 1 | Low-quality | Genome-fragment | 3.29 | HMM-based (lower-bound) | 0 |
| S8_extracted_k141_16322 | 2310 | No | NA | 1 | 0 | 1 | Not-determined | Genome-fragment | NA | NA | 0 |
| S8_extracted_k141_86067 | 2510 | No | NA | 3 | 0 | 3 | Not-determined | Genome-fragment | NA | NA | 0 |
| S8_extracted_k141_189245 | 5781 | No | NA | 7 | 0 | 5 | Not-determined | Genome-fragment | NA | NA | 0 |
| S8_extracted_k141_34315 | 2345 | No | NA | 2 | 0 | 1 | Not-determined | Genome-fragment | NA | NA | 0 |
| S8_extracted_k141_155175 | 1176 | No | NA | 1 | 0 | 1 | Not-determined | Genome-fragment | NA | NA | 0 |
| S8_extracted_k141_181858 | 11424 | No | NA | 11 | 0 | 6 | Not-determined | Genome-fragment | NA | NA | 0 |
| S8_extracted_k141_77420 | 1909 | No | NA | 1 | 0 | 0 | Not-determined | Genome-fragment | NA | NA | 0 |
| S3_vs_k141_91100\|\|full | 10063 | No | NA | 16 | 5 | 2 | Low-quality | Genome-fragment | 25.71 | AAI-based (high-confidence) | 0 |
| S5_extracted_k141_30691 | 10076 | No | NA | 16 | 4 | 0 | Low-quality | Genome-fragment | 22.17 | AAI-based (high-confidence) | 0 |
| S8_extracted_k141_173101 | 1040 | No | NA | 3 | 0 | 1 | Not-determined | Genome-fragment | NA | NA | 0 |
| S8_extracted_k141_94715 | 1224 | No | NA | 1 | 0 | 1 | Not-determined | Genome-fragment | NA | NA | 0 |
| S8_extracted_k141_77436 | 4447 | No | NA | 4 | 0 | 1 | Not-determined | Genome-fragment | NA | NA | 0 |
| S8_extracted_k141_16356 | 1690 | No | NA | 1 | 0 | 0 | Not-determined | Genome-fragment | NA | NA | 0 |
| S3_vs_k141_76836\|\|full | 10081 | No | NA | 20 | 3 | 0 | Low-quality | Genome-fragment | 12.77 | AAI-based (high-confidence) | 0 |
| S8_extracted_k141_147048 | 1417 | No | NA | 1 | 0 | 0 | Not-determined | Genome-fragment | NA | NA | 0 |
| S8_extracted_k141_155219 | 1814 | No | NA | 1 | 0 | 0 | Not-determined | Genome-fragment | NA | NA | 0 |
| S8_extracted_k141_207147 | 2470 | No | NA | 3 | 0 | 2 | Not-determined | Genome-fragment | NA | NA | 0 |
| S8_extracted_k141_163760 | 63437 | No | NA | 95 | 33 | 0 | Complete | High-quality | 100 | DTR (high-confidence) | 0 |
| S8_extracted_k141_17294 | 10081 | No | NA | 20 | 19 | 0 | Low-quality | Genome-fragment | 33.19 | AAI-based (high-confidence) | 0 |
| S8_extracted_k141_34352 | 2610 | No | NA | 4 | 0 | 1 | Not-determined | Genome-fragment | NA | NA | 0 |
| S11_vs_k141_16721\|\|full | 10084 | No | NA | 9 | 0 | 1 | Low-quality | Genome-fragment | 22.63 | AAI-based (medium-confidence) | 0 |
| S8_extracted_k141_25576 | 1140 | No | NA | 1 | 0 | 0 | Not-determined | Genome-fragment | NA | NA | 0 |
| S8_extracted_k141_77468 | 1068 | No | NA | 2 | 0 | 2 | Not-determined | Genome-fragment | NA | NA | 0 |
| S8_extracted_k141_199341 | 1236 | No | NA | 2 | 0 | 1 | Not-determined | Genome-fragment | NA | NA | 0 |
| S8_extracted_k141_94767 | 1217 | No | NA | 2 | 0 | 1 | Not-determined | Genome-fragment | NA | NA | 0 |
| S8_extracted_k141_16403 | 1574 | No | NA | 1 | 0 | 0 | Not-determined | Genome-fragment | NA | NA | 0 |
| S8_extracted_k141_60530 | 1967 | No | NA | 3 | 0 | 1 | Not-determined | Genome-fragment | NA | NA | 0 |
| S8_extracted_k141_50652 | 1563 | No | NA | 2 | 0 | 1 | Not-determined | Genome-fragment | NA | NA | 0 |
| S8_extracted_k141_163781 | 4904 | No | NA | 4 | 0 | 2 | Not-determined | Genome-fragment | NA | NA | 0 |
| S8_extracted_k141_207175 | 3345 | No | NA | 4 | 0 | 1 | Not-determined | Genome-fragment | NA | NA | 0 |
| S8_extracted_k141_155244 | 2237 | No | NA | 1 | 0 | 0 | Not-determined | Genome-fragment | NA | NA | 0 |
| S8_extracted_k141_94776 | 1123 | No | NA | 2 | 0 | 0 | Not-determined | Genome-fragment | NA | NA | 0 |
| S8_extracted_k141_16413 | 1082 | No | NA | 2 | 0 | 1 | Not-determined | Genome-fragment | NA | NA | 0 |
| S8_extracted_k141_68538 | 2537 | No | NA | 2 | 0 | 1 | Not-determined | Genome-fragment | NA | NA | 0 |
| S8_extracted_k141_155258 | 3213 | No | NA | 1 | 0 | 1 | Not-determined | Genome-fragment | NA | NA | 0 |
| S8_extracted_k141_138122 | 1109 | No | NA | 2 | 0 | 0 | Not-determined | Genome-fragment | NA | NA | 0 |
| S8_extracted_k141_207193 | 1306 | No | NA | 3 | 0 | 0 | Not-determined | Genome-fragment | NA | NA | 0 |
| S8_extracted_k141_163811 | 1651 | No | NA | 1 | 0 | 1 | Not-determined | Genome-fragment | NA | NA | 0 |
| S10_extracted_77432 | 10084 | No | NA | 14 | 2 | 0 | Low-quality | Genome-fragment | 7.41 | AAI-based (medium-confidence) | 0 |
| S16_vs_156623\|\|full | 10086 | No | NA | 26 | 1 | 0 | Low-quality | Genome-fragment | 29.72 | AAI-based (medium-confidence) | 0 |
| S16_vs_75445\|\|full | 10094 | No | NA | 13 | 6 | 0 | Low-quality | Genome-fragment | 28.58 | AAI-based (high-confidence) | 0 |
| S8_extracted_k141_25637 | 2676 | No | NA | 3 | 0 | 2 | Not-determined | Genome-fragment | NA | NA | 0 |
| S8_extracted_k141_42733 | 3781 | No | NA | 3 | 0 | 1 | Not-determined | Genome-fragment | NA | NA | 0 |
| S8_extracted_k141_77542 | 1449 | No | NA | 2 | 0 | 2 | Not-determined | Genome-fragment | NA | NA | 0 |
| S5_vs_k141_25113\|\|full | 10100 | No | NA | 10 | 5 | 0 | Low-quality | Genome-fragment | 22.92 | AAI-based (medium-confidence) | 0 |
| S8_extracted_k141_129589 | 3301 | No | NA | 5 | 0 | 1 | Not-determined | Genome-fragment | NA | NA | 0 |
| S8_extracted_k141_86152 | 3422 | No | NA | 6 | 0 | 2 | Not-determined | Genome-fragment | NA | NA | 0 |
| S8_extracted_k141_173196 | 6980 | No | NA | 7 | 0 | 0 | Not-determined | Genome-fragment | NA | NA | 0 |
| S8_extracted_k141_86159 | 2719 | No | NA | 3 | 0 | 2 | Not-determined | Genome-fragment | NA | NA | 0 |
| S8_extracted_k141_199397 | 1124 | No | NA | 1 | 0 | 1 | Not-determined | Genome-fragment | NA | NA | 0 |
| S8_extracted_k141_173206 | 1472 | No | NA | 1 | 0 | 1 | Not-determined | Genome-fragment | NA | NA | 0 |
| S5_vs_k141_23242\|\|0_partial | 10104 | Yes | 8984 | 18 | 2 | 2 | Low-quality | Genome-fragment | 27.69 | AAI-based (high-confidence) | 11.08 |
| S8_extracted_k141_199405 | 1568 | No | NA | 3 | 0 | 3 | Not-determined | Genome-fragment | NA | NA | 0 |
| S8_extracted_k141_50699 | 7961 | No | NA | 10 | 0 | 1 | Not-determined | Genome-fragment | NA | NA | 0 |
| S8_extracted_k141_138184 | 1304 | No | NA | 2 | 0 | 1 | Not-determined | Genome-fragment | NA | NA | 0 |

| | | | | | | | | | | | |
|---|---|---|---|---|---|---|---|---|---|---|---|
| S8_extracted_k141_138186 | 1161 | No | NA | 3 | 0 | 0 | Not-determined | Genome-fragment | NA | NA | 0 |
| S8_extracted_k141_155323 | 1207 | No | NA | 3 | 0 | 1 | Not-determined | Genome-fragment | NA | NA | 0 |
| S17_vs_k141_113946\|\|full | 10114 | No | NA | 10 | 1 | 1 | Low-quality | Genome-fragment | 5.67 | HMM-based (lower-bound) | 0 |
| S8_extracted_k141_16487 | 2326 | No | NA | 3 | 0 | 2 | Not-determined | Genome-fragment | NA | NA | 0 |
| S5_vs_k141_12596\|\|full | 10115 | No | NA | 18 | 2 | 1 | Low-quality | Genome-fragment | 26.87 | AAI-based (medium-confidence) | 0 |
| S8_extracted_k141_138189 | 2070 | No | NA | 3 | 0 | 1 | Not-determined | Genome-fragment | NA | NA | 0 |
| S8_extracted_k141_34462 | 1867 | No | NA | 2 | 0 | 2 | Not-determined | Genome-fragment | NA | NA | 0 |
| S8_extracted_k141_68611 | 1163 | No | NA | 2 | 0 | 1 | Not-determined | Genome-fragment | NA | NA | 0 |
| S8_extracted_k141_112495 | 2708 | No | NA | 2 | 0 | 2 | Not-determined | Genome-fragment | NA | NA | 0 |
| S8_extracted_k141_181987 | 1403 | No | NA | 1 | 0 | 1 | Not-determined | Genome-fragment | NA | NA | 0 |
| S8_extracted_k141_129614 | 2761 | No | NA | 1 | 0 | 0 | Not-determined | Genome-fragment | NA | NA | 0 |
| S2_extracted_k141_4058 | 10119 | No | NA | 20 | 4 | 0 | Low-quality | Genome-fragment | 19.24 | AAI-based (high-confidence) | 0 |
| S8_extracted_k141_120791 | 1771 | No | NA | 1 | 0 | 0 | Not-determined | Genome-fragment | NA | NA | 0 |
| S8_extracted_k141_199422 | 4690 | No | NA | 6 | 0 | 2 | Not-determined | Genome-fragment | NA | NA | 0 |
| S8_extracted_k141_129624 | 1353 | No | NA | 3 | 0 | 2 | Not-determined | Genome-fragment | NA | NA | 0 |
| S8_extracted_k141_77596 | 1341 | No | NA | 2 | 0 | 0 | Not-determined | Genome-fragment | NA | NA | 0 |
| S8_extracted_k141_68637 | 1130 | No | NA | 2 | 0 | 0 | Not-determined | Genome-fragment | NA | NA | 0 |
| S8_extracted_k141_199433 | 1029 | No | NA | 2 | 0 | 2 | Not-determined | Genome-fragment | NA | NA | 0 |
| S8_extracted_k141_42771 | 1719 | No | NA | 2 | 0 | 2 | Not-determined | Genome-fragment | NA | NA | 0 |
| S8_extracted_k141_163893 | 2858 | No | NA | 1 | 0 | 1 | Not-determined | Genome-fragment | NA | NA | 0 |
| S8_extracted_k141_16511 | 1185 | No | NA | 1 | 0 | 1 | Not-determined | Genome-fragment | NA | NA | 0 |
| S8_extracted_k141_120806 | 4502 | No | NA | 4 | 0 | 1 | Not-determined | Genome-fragment | NA | NA | 0 |
| S8_extracted_k141_68646 | 1421 | No | NA | 4 | 0 | 1 | Not-determined | Genome-fragment | NA | NA | 0 |
| S12_extracted_k141_52398 | 10127 | No | NA | 24 | 3 | 0 | Low-quality | Genome-fragment | 7.35 | AAI-based (high-confidence) | 0 |
| S8_extracted_k141_199442 | 1170 | No | NA | 1 | 0 | 1 | Not-determined | Genome-fragment | NA | NA | 0 |
| S8_extracted_k141_16515 | 3489 | No | NA | 3 | 0 | 1 | Not-determined | Genome-fragment | NA | NA | 0 |
| S8_extracted_k141_155348 | 1016 | No | NA | 1 | 0 | 0 | Not-determined | Genome-fragment | NA | NA | 0 |
| S8_extracted_k141_25708 | 2392 | No | NA | 3 | 0 | 2 | Not-determined | Genome-fragment | NA | NA | 0 |
| S11_vs_k141_81735\|\|full | 10132 | No | NA | 18 | 0 | 0 | Low-quality | Genome-fragment | 15.52 | AAI-based (high-confidence) | 0 |
| S8_extracted_k141_112528 | 7128 | No | NA | 7 | 0 | 4 | Not-determined | Genome-fragment | NA | NA | 0 |
| S8_extracted_k141_199454 | 1833 | No | NA | 1 | 0 | 1 | Not-determined | Genome-fragment | NA | NA | 0 |
| S8_extracted_k141_138234 | 5099 | No | NA | 4 | 0 | 2 | Not-determined | Genome-fragment | NA | NA | 0 |
| S17_vs_k141_102465\|\|full | 10139 | No | NA | 14 | 4 | 0 | Low-quality | Genome-fragment | 14.9 | AAI-based (high-confidence) | 0 |
| S8_extracted_k141_112544 | 2255 | No | NA | 3 | 0 | 2 | Not-determined | Genome-fragment | NA | NA | 0 |
| S8_extracted_k141_42797 | 1020 | No | NA | 2 | 0 | 1 | Not-determined | Genome-fragment | NA | NA | 0 |
| S8_extracted_k141_138246 | 1904 | No | NA | 1 | 0 | 1 | Not-determined | Genome-fragment | NA | NA | 0 |
| S8_extracted_k141_120852 | 1891 | No | NA | 1 | 0 | 1 | Not-determined | Genome-fragment | NA | NA | 0 |
| S8_extracted_k141_120857 | 1008 | No | NA | 1 | 0 | 1 | Not-determined | Genome-fragment | NA | NA | 0 |
| S14_extracted_k141_36162 | 10148 | No | NA | 14 | 4 | 0 | Low-quality | Genome-fragment | 24.85 | AAI-based (high-confidence) | 0 |
| S8_extracted_k141_147128 | 81855 | No | NA | 107 | 14 | 1 | High-quality | High-quality | 99.52 | AAI-based (high-confidence) | 0 |
| S8_extracted_k141_207308 | 4009 | No | NA | 4 | 0 | 1 | Not-determined | Genome-fragment | NA | NA | 0 |
| S8_extracted_k141_120863 | 1233 | No | NA | 2 | 0 | 1 | Not-determined | Genome-fragment | NA | NA | 0 |
| S8_extracted_k141_112562 | 1248 | No | NA | 2 | 0 | 0 | Not-determined | Genome-fragment | NA | NA | 0 |
| S8_extracted_k141_199482 | 3615 | No | NA | 4 | 0 | 3 | Not-determined | Genome-fragment | NA | NA | 0 |
| S8_extracted_k141_163953 | 1218 | No | NA | 1 | 0 | 1 | Not-determined | Genome-fragment | NA | NA | 0 |
| S8_extracted_k141_86248 | 1106 | No | NA | 2 | 0 | 0 | Not-determined | Genome-fragment | NA | NA | 0 |
| S8_vs_k141_80060\|\|full | 10153 | Yes | 1135 | 10 | 1 | 4 | Low-quality | Genome-fragment | 0.76 | HMM-based (lower-bound) | 88.82 |
| S8_extracted_k141_8310 | 4116 | No | NA | 4 | 0 | 3 | Not-determined | Genome-fragment | NA | NA | 0 |
| S10_vs_40746\|\|full | 10166 | No | NA | 16 | 0 | 0 | Low-quality | Genome-fragment | 25.13 | AAI-based (medium-confidence) | 0 |
| S8_extracted_k141_163969 | 1284 | No | NA | 1 | 0 | 1 | Not-determined | Genome-fragment | NA | NA | 0 |
| S7_extracted_100649 | 10171 | No | NA | 11 | 1 | 0 | Low-quality | Genome-fragment | 27.28 | AAI-based (high-confidence) | 0 |
| S8_extracted_k141_163974 | 1500 | No | NA | 1 | 0 | 1 | Not-determined | Genome-fragment | NA | NA | 0 |
| S8_extracted_k141_77698 | 6343 | No | NA | 7 | 0 | 6 | Not-determined | Genome-fragment | NA | NA | 0 |
| S8_extracted_k141_42830 | 1030 | No | NA | 1 | 0 | 0 | Not-determined | Genome-fragment | NA | NA | 0 |
| S17_vs_k141_35788\|\|full | 10182 | No | NA | 28 | 5 | 0 | Low-quality | Genome-fragment | 22.75 | AAI-based (high-confidence) | 0 |
| S8_extracted_k141_120906 | 1527 | No | NA | 1 | 0 | 1 | Not-determined | Genome-fragment | NA | NA | 0 |
| S7_vs_32030\|\|full | 10183 | No | NA | 8 | 3 | 0 | Low-quality | Genome-fragment | 17.28 | AAI-based (high-confidence) | 0 |
| S13_vs_51840\|\|full | 10187 | No | NA | 8 | 0 | 3 | Low-quality | Genome-fragment | 6.89 | AAI-based (medium-confidence) | 0 |
| S8_extracted_k141_163980 | 1005 | No | NA | 1 | 0 | 1 | Not-determined | Genome-fragment | NA | NA | 0 |
| S8_extracted_k141_120909 | 2483 | No | NA | 1 | 0 | 1 | Not-determined | Genome-fragment | NA | NA | 0 |
| S8_extracted_k141_129713 | 2028 | No | NA | 2 | 0 | 2 | Not-determined | Genome-fragment | NA | NA | 0 |
| S8_extracted_k141_86284 | 2412 | No | NA | 3 | 0 | 1 | Not-determined | Genome-fragment | NA | NA | 0 |
| S8_extracted_k141_199528 | 1574 | No | NA | 3 | 0 | 1 | Not-determined | Genome-fragment | NA | NA | 0 |
| S8_extracted_k141_199533 | 1549 | No | NA | 3 | 0 | 1 | Not-determined | Genome-fragment | NA | NA | 0 |
| S8_extracted_k141_60628 | 3568 | No | NA | 2 | 0 | 1 | Not-determined | Genome-fragment | NA | NA | 0 |
| S14_vs_k141_89890\|\|0_partial | 10201 | No | NA | 13 | 3 | 0 | Low-quality | Genome-fragment | 35.56 | AAI-based (high-confidence) | 0 |
| S8_extracted_k141_173225 | 98174 | No | NA | 123 | 29 | 2 | High-quality | High-quality | 94.9 | AAI-based (high-confidence) | 0 |
| S8_extracted_k141_182116 | 1941 | No | NA | 2 | 0 | 2 | Not-determined | Genome-fragment | NA | NA | 0 |
| S8_extracted_k141_207357 | 1147 | No | NA | 1 | 0 | 0 | Not-determined | Genome-fragment | NA | NA | 0 |
| S10_extracted_13213 | 10217 | No | NA | 17 | 4 | 0 | Low-quality | Genome-fragment | 14.86 | AAI-based (high-confidence) | 0 |
| S8_extracted_k141_68713 | 1061 | No | NA | 1 | 0 | 0 | Not-determined | Genome-fragment | NA | NA | 0 |
| S8_extracted_k141_8377 | 3287 | No | NA | 1 | 0 | 1 | Not-determined | Genome-fragment | NA | NA | 0 |
| S8_extracted_k141_155424 | 1074 | No | NA | 1 | 0 | 0 | Not-determined | Genome-fragment | NA | NA | 0 |
| S8_extracted_k141_8379 | 3394 | No | NA | 4 | 0 | 3 | Not-determined | Genome-fragment | NA | NA | 0 |
| S8_extracted_k141_8380 | 1723 | No | NA | 2 | 0 | 1 | Not-determined | Genome-fragment | NA | NA | 0 |
| S8_extracted_k141_60656 | 1464 | No | NA | 1 | 0 | 0 | Not-determined | Genome-fragment | NA | NA | 0 |
| S8_extracted_k141_16634 | 1897 | No | NA | 2 | 0 | 2 | Not-determined | Genome-fragment | NA | NA | 0 |
| S8_extracted_k141_50778 | 3833 | No | NA | 3 | 0 | 1 | Not-determined | Genome-fragment | NA | NA | 0 |
| S8_extracted_k141_155438 | 2093 | No | NA | 3 | 0 | 2 | Not-determined | Genome-fragment | NA | NA | 0 |
| S8_extracted_k141_103472 | 3499 | No | NA | 2 | 0 | 2 | Not-determined | Genome-fragment | NA | NA | 0 |
| S8_extracted_k141_50784 | 1200 | No | NA | 1 | 0 | 0 | Not-determined | Genome-fragment | NA | NA | 0 |
| S8_extracted_k141_103475 | 1501 | No | NA | 3 | 0 | 2 | Not-determined | Genome-fragment | NA | NA | 0 |
| S12_vs_k141_57765\|\|full | 10218 | No | NA | 13 | 5 | 1 | Low-quality | Genome-fragment | 22.77 | AAI-based (high-confidence) | 0 |
| S8_vs_k141_47554\|\|full | 10219 | No | NA | 12 | 4 | 0 | Low-quality | Genome-fragment | 25.5 | AAI-based (high-confidence) | 0 |
| S8_extracted_k141_155451 | 1082 | No | NA | 2 | 0 | 1 | Not-determined | Genome-fragment | NA | NA | 0 |
| S8_extracted_k141_164022 | 4086 | No | NA | 4 | 0 | 0 | Not-determined | Genome-fragment | NA | NA | 0 |
| S8_extracted_k141_129764 | 3964 | No | NA | 6 | 0 | 1 | Not-determined | Genome-fragment | NA | NA | 0 |
| S8_extracted_k141_77771 | 1229 | No | NA | 1 | 0 | 1 | Not-determined | Genome-fragment | NA | NA | 0 |
| S8_extracted_k141_155460 | 1533 | No | NA | 3 | 0 | 2 | Not-determined | Genome-fragment | NA | NA | 0 |
| S8_extracted_k141_164029 | 5219 | No | NA | 7 | 0 | 4 | Not-determined | Genome-fragment | NA | NA | 0 |
| S8_extracted_k141_86360 | 3281 | No | NA | 5 | 0 | 4 | Not-determined | Genome-fragment | NA | NA | 0 |
| S8_extracted_k141_16674 | 1573 | No | NA | 3 | 0 | 0 | Not-determined | Genome-fragment | NA | NA | 0 |
| S8_extracted_k141_120961 | 4502 | No | NA | 4 | 0 | 4 | Not-determined | Genome-fragment | NA | NA | 0 |
| S8_extracted_k141_16675 | 1876 | No | NA | 2 | 0 | 0 | Not-determined | Genome-fragment | NA | NA | 0 |
| S8_extracted_k141_120971 | 1198 | No | NA | 2 | 0 | 2 | Not-determined | Genome-fragment | NA | NA | 0 |
| S3_extracted_k141_108724 | 10233 | No | NA | 12 | 1 | 0 | Low-quality | Genome-fragment | 27.72 | AAI-based (high-confidence) | 0 |
| S8_extracted_k141_60715 | 1326 | No | NA | 2 | 0 | 0 | Not-determined | Genome-fragment | NA | NA | 0 |
| S8_extracted_k141_86377 | 1117 | No | NA | 2 | 0 | 2 | Not-determined | Genome-fragment | NA | NA | 0 |
| S8_extracted_k141_8439 | 1007 | No | NA | 1 | 0 | 0 | Not-determined | Genome-fragment | NA | NA | 0 |
| S8_extracted_k141_155485 | 1201 | No | NA | 1 | 0 | 1 | Not-determined | Genome-fragment | NA | NA | 0 |
| S8_extracted_k141_34610 | 3638 | No | NA | 4 | 0 | 1 | Not-determined | Genome-fragment | NA | NA | 0 |
| S8_extracted_k141_155487 | 1911 | No | NA | 3 | 0 | 2 | Not-determined | Genome-fragment | NA | NA | 0 |
| S8_extracted_k141_164045 | 1883 | No | NA | 2 | 0 | 0 | Not-determined | Genome-fragment | NA | NA | 0 |
| S8_extracted_k141_120986 | 1342 | No | NA | 1 | 0 | 1 | Not-determined | Genome-fragment | NA | NA | 0 |
| S8_extracted_k141_147247 | 1388 | No | NA | 2 | 0 | 0 | Not-determined | Genome-fragment | NA | NA | 0 |
| S16_vs_113020\|\|0_partial | 10240 | No | NA | 15 | 1 | 7 | Low-quality | Genome-fragment | 6.63 | HMM-based (lower-bound) | 0 |
| S8_extracted_k141_68788 | 6213 | No | NA | 10 | 0 | 5 | Not-determined | Genome-fragment | NA | NA | 0 |
| S8_extracted_k141_182198 | 3614 | No | NA | 2 | 0 | 0 | Not-determined | Genome-fragment | NA | NA | 0 |
| S8_extracted_k141_173275 | 1261 | No | NA | 1 | 0 | 1 | Not-determined | Genome-fragment | NA | NA | 0 |
| S8_extracted_k141_8457 | 1802 | No | NA | 1 | 0 | 1 | Not-determined | Genome-fragment | NA | NA | 0 |
| S8_extracted_k141_138365 | 1482 | No | NA | 3 | 0 | 0 | Not-determined | Genome-fragment | NA | NA | 0 |
| S8_extracted_k141_8458 | 1369 | No | NA | 1 | 0 | 0 | Not-determined | Genome-fragment | NA | NA | 0 |
| S8_extracted_k141_164062 | 4228 | No | NA | 3 | 0 | 3 | Not-determined | Genome-fragment | NA | NA | 0 |
| S8_extracted_k141_42896 | 1420 | No | NA | 1 | 0 | 1 | Not-determined | Genome-fragment | NA | NA | 0 |
| S8_extracted_k141_164066 | 1028 | No | NA | 2 | 0 | 1 | Not-determined | Genome-fragment | NA | NA | 0 |
| S8_extracted_k141_147256 | 3845 | No | NA | 8 | 0 | 0 | Not-determined | Genome-fragment | NA | NA | 0 |
| S8_extracted_k141_16699 | 1183 | No | NA | 1 | 0 | 1 | Not-determined | Genome-fragment | NA | NA | 0 |
| S8_extracted_k141_173282 | 1075 | No | NA | 3 | 0 | 0 | Not-determined | Genome-fragment | NA | NA | 0 |
| S8_extracted_k141_164067 | 3297 | No | NA | 4 | 0 | 3 | Not-determined | Genome-fragment | NA | NA | 0 |
| S10_extracted_85124 | 10242 | No | NA | 12 | 1 | 1 | Low-quality | Genome-fragment | 5.56 | HMM-based (lower-bound) | 0 |
| S11_vs_k141_88248\|\|full | 10251 | No | NA | 9 | 0 | 3 | Low-quality | Genome-fragment | 2.75 | AAI-based (medium-confidence) | 0 |
| S8_extracted_k141_8462 | 6187 | No | NA | 5 | 0 | 1 | Not-determined | Genome-fragment | NA | NA | 0 |
| S8_extracted_k141_16709 | 2342 | No | NA | 2 | 0 | 0 | Not-determined | Genome-fragment | NA | NA | 0 |
| S8_extracted_k141_86423 | 1264 | No | NA | 1 | 0 | 1 | Not-determined | Genome-fragment | NA | NA | 0 |
| S8_extracted_k141_147271 | 4199 | No | NA | 5 | 0 | 2 | Not-determined | Genome-fragment | NA | NA | 0 |
| S8_extracted_k141_207446 | 1604 | No | NA | 3 | 0 | 1 | Not-determined | Genome-fragment | NA | NA | 0 |
| S8_extracted_k141_173296 | 4397 | No | NA | 7 | 0 | 6 | Not-determined | Genome-fragment | NA | NA | 0 |
| S8_extracted_k141_77843 | 1458 | No | NA | 2 | 0 | 2 | Not-determined | Genome-fragment | NA | NA | 0 |
| S8_extracted_k141_189690 | 1067 | No | NA | 1 | 0 | 0 | Not-determined | Genome-fragment | NA | NA | 0 |
| S8_extracted_k141_121044 | 3118 | No | NA | 4 | 0 | 2 | Not-determined | Genome-fragment | NA | NA | 0 |
| S8_extracted_k141_8494 | 2165 | No | NA | 2 | 0 | 1 | Not-determined | Genome-fragment | NA | NA | 0 |
| S8_extracted_k141_8495 | 1621 | No | NA | 2 | 0 | 2 | Not-determined | Genome-fragment | NA | NA | 0 |
| S8_extracted_k141_207472 | 3625 | No | NA | 6 | 0 | 0 | Not-determined | Genome-fragment | NA | NA | 0 |
| S7_extracted_7174 | 10257 | Yes | 6001 | 18 | 3 | 3 | Low-quality | Genome-fragment | 10.41 | HMM-based (lower-bound) | 41.49 |
| S3_extracted_k141_121700 | 10260 | No | NA | 17 | 3 | 0 | Low-quality | Genome-fragment | 11.04 | AAI-based (medium-confidence) | 0 |
| S8_extracted_k141_68853 | 3116 | No | NA | 3 | 0 | 3 | Not-determined | Genome-fragment | NA | NA | 0 |
| S8_extracted_k141_3727 | 10260 | No | NA | 16 | 6 | 0 | Low-quality | Genome-fragment | 17.13 | AAI-based (medium-confidence) | 0 |
| S8_extracted_k141_155568 | 4178 | No | NA | 3 | 0 | 2 | Not-determined | Genome-fragment | NA | NA | 0 |
| S11_extracted_k141_13962 | 10265 | No | NA | 21 | 7 | 0 | Low-quality | Genome-fragment | 21.4 | AAI-based (high-confidence) | 0 |
| S8_extracted_k141_173325 | 1487 | No | NA | 2 | 0 | 1 | Not-determined | Genome-fragment | NA | NA | 0 |
| S8_extracted_k141_103616 | 1053 | No | NA | 2 | 0 | 2 | Not-determined | Genome-fragment | NA | NA | 0 |
| S8_extracted_k141_68877 | 1784 | No | NA | 2 | 0 | 1 | Not-determined | Genome-fragment | NA | NA | 0 |
| S8_extracted_k141_164116 | 3424 | No | NA | 4 | 0 | 2 | Not-determined | Genome-fragment | NA | NA | 0 |
| S8_extracted_k141_103621 | 1116 | No | NA | 1 | 0 | 1 | Not-determined | Genome-fragment | NA | NA | 0 |
| S8_extracted_k141_68880 | 1618 | No | NA | 4 | 0 | 3 | Not-determined | Genome-fragment | NA | NA | 0 |
| S8_extracted_k141_189742 | 4474 | No | NA | 3 | 0 | 2 | Not-determined | Genome-fragment | NA | NA | 0 |
| S8_extracted_k141_121106 | 1473 | No | NA | 1 | 0 | 0 | Not-determined | Genome-fragment | NA | NA | 0 |
| S8_extracted_k141_121112 | 1143 | No | NA | 1 | 0 | 1 | Not-determined | Genome-fragment | NA | NA | 0 |
| S8_extracted_k141_86527 | 1238 | No | NA | 2 | 0 | 0 | Not-determined | Genome-fragment | NA | NA | 0 |
| S8_extracted_k141_129937 | 1161 | No | NA | 2 | 0 | 1 | Not-determined | Genome-fragment | NA | NA | 0 |
| S8_extracted_k141_8561 | 2766 | No | NA | 1 | 0 | 1 | Not-determined | Genome-fragment | NA | NA | 0 |
| S8_extracted_k141_138483 | 1455 | No | NA | 1 | 0 | 0 | Not-determined | Genome-fragment | NA | NA | 0 |
| S11_vs_k141_8230\|\|full | 10282 | No | NA | 8 | 0 | 4 | Low-quality | Genome-fragment | 18.32 | AAI-based (medium-confidence) | 0 |
| S5_vs_k141_11114\|\|full | 10292 | No | NA | 21 | 3 | 0 | Low-quality | Genome-fragment | 10.36 | HMM-based (lower-bound) | 0 |
| S8_extracted_k141_147338 | 34542 | No | NA | 40 | 17 | 0 | Medium-quality | Genome-fragment | 52.69 | AAI-based (high-confidence) | 0 |
| S8_extracted_k141_207527 | 4080 | No | NA | 6 | 0 | 3 | Not-determined | Genome-fragment | NA | NA | 0 |
| S8_extracted_k141_155617 | 1711 | No | NA | 3 | 0 | 0 | Not-determined | Genome-fragment | NA | NA | 0 |
| S5_vs_k141_144329\|\|full | 10312 | No | NA | 12 | 0 | 1 | Low-quality | Genome-fragment | 13.19 | AAI-based (medium-confidence) | 0 |
| S10_vs_78757\|\|full | 10314 | No | NA | 16 | 2 | 1 | Low-quality | Genome-fragment | 27.05 | AAI-based (high-confidence) | 0 |
| S8_extracted_k141_86568 | 1192 | No | NA | 2 | 0 | 1 | Not-determined | Genome-fragment | NA | NA | 0 |
| S3_extracted_k141_79427 | 10321 | No | NA | 13 | 4 | 0 | Low-quality | Genome-fragment | 5.88 | AAI-based (medium-confidence) | 0 |
| S8_extracted_k141_103678 | 2430 | No | NA | 3 | 0 | 1 | Not-determined | Genome-fragment | NA | NA | 0 |
| S8_extracted_k141_43046 | 1045 | No | NA | 2 | 0 | 0 | Not-determined | Genome-fragment | NA | NA | 0 |
| S8_extracted_k141_173363 | 1166 | No | NA | 1 | 0 | 0 | Not-determined | Genome-fragment | NA | NA | 0 |
| S8_extracted_k141_207546 | 2004 | No | NA | 2 | 0 | 1 | Not-determined | Genome-fragment | NA | NA | 0 |
| S12_vs_k141_31501\|\|full | 10324 | No | NA | 15 | 0 | 0 | Low-quality | Genome-fragment | 6.49 | AAI-based (medium-confidence) | 0 |
| S8_extracted_k141_50910 | 1009 | No | NA | 1 | 0 | 1 | Not-determined | Genome-fragment | NA | NA | 0 |
| S8_extracted_k141_207553 | 1339 | No | NA | 3 | 0 | 1 | Not-determined | Genome-fragment | NA | NA | 0 |
| S8_extracted_k141_112613 | 8135 | No | NA | 6 | 0 | 3 | Not-determined | Genome-fragment | NA | NA | 0 |
| S8_extracted_k141_8617 | 1031 | No | NA | 1 | 0 | 0 | Not-determined | Genome-fragment | NA | NA | 0 |
| S8_extracted_k141_155643 | 11633 | No | NA | 6 | 0 | 0 | Not-determined | Genome-fragment | NA | NA | 0 |
| S15_vs_35452\|\|full | 10325 | Yes | 2710 | 10 | 2 | 5 | Low-quality | Genome-fragment | 1.2 | HMM-based (lower-bound) | 73.75 |
| S8_extracted_k141_121205 | 1212 | No | NA | 1 | 0 | 1 | Not-determined | Genome-fragment | NA | NA | 0 |
| S8_extracted_k141_189771 | 2136 | No | NA | 1 | 0 | 1 | Not-determined | Genome-fragment | NA | NA | 0 |
| S8_extracted_k141_207572 | 1178 | No | NA | 3 | 0 | 3 | Not-determined | Genome-fragment | NA | NA | 0 |
| S8_extracted_k141_112636 | 1115 | No | NA | 3 | 0 | 1 | Not-determined | Genome-fragment | NA | NA | 0 |
| S8_extracted_k141_147384 | 1243 | No | NA | 2 | 0 | 1 | Not-determined | Genome-fragment | NA | NA | 0 |
| S8_extracted_k141_8658 | 3713 | No | NA | 4 | 0 | 4 | Not-determined | Genome-fragment | NA | NA | 0 |
| S8_extracted_k141_16872 | 5921 | No | NA | 11 | 0 | 0 | Not-determined | Genome-fragment | NA | NA | 0 |
| S8_extracted_k141_68982 | 3542 | No | NA | 3 | 0 | 3 | Not-determined | Genome-fragment | NA | NA | 0 |
| S8_extracted_k141_43080 | 1493 | No | NA | 2 | 0 | 0 | Not-determined | Genome-fragment | NA | NA | 0 |
| S16_vs_10896\|\|full | 10327 | No | NA | 14 | 1 | 0 | Low-quality | Genome-fragment | 23.46 | AAI-based (medium-confidence) | 0 |
| S8_vs_k141_97333\|\|full | 10329 | No | NA | 15 | 3 | 1 | Low-quality | Genome-fragment | 26.58 | AAI-based (medium-confidence) | 0 |
| S8_extracted_k141_69005 | 2811 | No | NA | 2 | 0 | 2 | Not-determined | Genome-fragment | NA | NA | 0 |
| S8_extracted_k141_103757 | 1208 | No | NA | 1 | 0 | 1 | Not-determined | Genome-fragment | NA | NA | 0 |
| S8_extracted_k141_207603 | 1780 | No | NA | 1 | 0 | 1 | Not-determined | Genome-fragment | NA | NA | 0 |
| S8_extracted_k141_147427 | 1334 | No | NA | 2 | 0 | 0 | Not-determined | Genome-fragment | NA | NA | 0 |
| S8_extracted_k141_69017 | 1222 | No | NA | 1 | 0 | 1 | Not-determined | Genome-fragment | NA | NA | 0 |
| S8_extracted_k141_164243 | 1222 | No | NA | 3 | 0 | 0 | Not-determined | Genome-fragment | NA | NA | 0 |
| S8_extracted_k141_25989 | 2009 | No | NA | 4 | 0 | 3 | Not-determined | Genome-fragment | NA | NA | 0 |
| S8_extracted_k141_16903 | 1540 | No | NA | 1 | 0 | 1 | Not-determined | Genome-fragment | NA | NA | 0 |
| S8_extracted_k141_25991 | 1060 | No | NA | 2 | 0 | 0 | Not-determined | Genome-fragment | NA | NA | 0 |
| S16_extracted_111664 | 10332 | No | NA | 21 | 4 | 0 | Low-quality | Genome-fragment | 26.08 | AAI-based (high-confidence) | 0 |
| S13_vs_56827\|\|full | 10337 | No | NA | 10 | 1 | 0 | Low-quality | Genome-fragment | 26.33 | AAI-based (high-confidence) | 0 |
| S8_extracted_k141_207614 | 1076 | No | NA | 1 | 0 | 1 | Not-determined | Genome-fragment | NA | NA | 0 |
| S8_extracted_k141_51001 | 1544 | No | NA | 1 | 0 | 1 | Not-determined | Genome-fragment | NA | NA | 0 |
| S8_extracted_k141_78037 | 1473 | No | NA | 3 | 0 | 3 | Not-determined | Genome-fragment | NA | NA | 0 |
| S8_extracted_k141_138613 | 1233 | No | NA | 1 | 0 | 1 | Not-determined | Genome-fragment | NA | NA | 0 |
| S8_extracted_k141_78039 | 1288 | No | NA | 2 | 0 | 0 | Not-determined | Genome-fragment | NA | NA | 0 |
| S8_extracted_k141_130092 | 2447 | No | NA | 3 | 0 | 3 | Not-determined | Genome-fragment | NA | NA | 0 |
| S8_extracted_k141_189816 | 4670 | No | NA | 5 | 0 | 5 | Not-determined | Genome-fragment | NA | NA | 0 |
| S8_extracted_k141_16921 | 1153 | No | NA | 1 | 0 | 1 | Not-determined | Genome-fragment | NA | NA | 0 |
| S8_extracted_k141_78056 | 1364 | No | NA | 1 | 0 | 1 | Not-determined | Genome-fragment | NA | NA | 0 |
| S8_extracted_k141_78058 | 1052 | No | NA | 2 | 0 | 1 | Not-determined | Genome-fragment | NA | NA | 0 |
| S8_extracted_k141_16930 | 2513 | No | NA | 1 | 0 | 1 | Not-determined | Genome-fragment | NA | NA | 0 |
| S11_extracted_k141_58871 | 10345 | No | NA | 23 | 5 | 0 | Low-quality | Genome-fragment | 17.76 | AAI-based (high-confidence) | 0 |
| S8_extracted_k141_51033 | 1277 | No | NA | 3 | 0 | 1 | Not-determined | Genome-fragment | NA | NA | 0 |
| S1_vs_k141_88178\|\|full | 10351 | No | NA | 19 | 2 | 0 | Low-quality | Genome-fragment | 25.49 | AAI-based (high-confidence) | 0 |
| S9_extracted_k141_50386 | 10358 | No | NA | 16 | 1 | 0 | Low-quality | Genome-fragment | 6.66 | AAI-based (medium-confidence) | 0 |
| S5_extracted_k141_31181 | 10358 | No | NA | 13 | 1 | 0 | Low-quality | Genome-fragment | 9.41 | HMM-based (lower-bound) | 0 |
| S8_extracted_k141_51034 | 4829 | No | NA | 5 | 0 | 2 | Not-determined | Genome-fragment | NA | NA | 0 |
| S8_extracted_k141_26032 | 1317 | No | NA | 1 | 0 | 1 | Not-determined | Genome-fragment | NA | NA | 0 |
| S8_extracted_k141_207631 | 2162 | No | NA | 3 | 0 | 2 | Not-determined | Genome-fragment | NA | NA | 0 |
| S8_extracted_k141_43147 | 4130 | No | NA | 6 | 0 | 1 | Not-determined | Genome-fragment | NA | NA | 0 |
| S8_extracted_k141_164308 | 1050 | No | NA | 2 | 0 | 1 | Not-determined | Genome-fragment | NA | NA | 0 |
| S8_extracted_k141_78095 | 4360 | No | NA | 4 | 0 | 1 | Not-determined | Genome-fragment | NA | NA | 0 |
| S8_extracted_k141_130116 | 1119 | No | NA | 1 | 0 | 1 | Not-determined | Genome-fragment | NA | NA | 0 |
| S8_extracted_k141_189872 | 1020 | No | NA | 1 | 0 | 0 | Not-determined | Genome-fragment | NA | NA | 0 |
| S8_extracted_k141_173473 | 1251 | No | NA | 1 | 0 | 0 | Not-determined | Genome-fragment | NA | NA | 0 |
| S8_extracted_k141_155748 | 1173 | No | NA | 1 | 0 | 1 | Not-determined | Genome-fragment | NA | NA | 0 |
| S8_extracted_k141_43163 | 1167 | No | NA | 2 | 0 | 0 | Not-determined | Genome-fragment | NA | NA | 0 |
| S8_extracted_k141_207661 | 1517 | No | NA | 4 | 0 | 4 | Not-determined | Genome-fragment | NA | NA | 0 |
| S8_extracted_k141_16985 | 5747 | No | NA | 6 | 0 | 6 | Not-determined | Genome-fragment | NA | NA | 0 |
| S8_extracted_k141_95304 | 6270 | No | NA | 4 | 0 | 4 | Not-determined | Genome-fragment | NA | NA | 0 |
| S8_extracted_k141_86692 | 1422 | No | NA | 2 | 0 | 0 | Not-determined | Genome-fragment | NA | NA | 0 |
| S8_extracted_k141_51062 | 2735 | No | NA | 4 | 0 | 3 | Not-determined | Genome-fragment | NA | NA | 0 |
| S3_extracted_k141_41435 | 10361 | No | NA | 17 | 4 | 0 | Low-quality | Genome-fragment | 18.53 | AAI-based (high-confidence) | 0 |
| S8_extracted_k141_112778 | 1428 | No | NA | 1 | 0 | 0 | Not-determined | Genome-fragment | NA | NA | 0 |
| S8_extracted_k141_43195 | 1306 | No | NA | 3 | 0 | 2 | Not-determined | Genome-fragment | NA | NA | 0 |
| S8_extracted_k141_112786 | 1313 | No | NA | 1 | 0 | 0 | Not-determined | Genome-fragment | NA | NA | 0 |
| S8_extracted_k141_189900 | 1041 | No | NA | 2 | 0 | 1 | Not-determined | Genome-fragment | NA | NA | 0 |
| S8_extracted_k141_121350 | 2316 | No | NA | 2 | 0 | 2 | Not-determined | Genome-fragment | NA | NA | 0 |
| S8_extracted_k141_173508 | 1223 | No | NA | 1 | 0 | 1 | Not-determined | Genome-fragment | NA | NA | 0 |
| S8_extracted_k141_69134 | 1378 | No | NA | 2 | 0 | 1 | Not-determined | Genome-fragment | NA | NA | 0 |
| S8_extracted_k141_95350 | 1089 | No | NA | 2 | 0 | 0 | Not-determined | Genome-fragment | NA | NA | 0 |
| S8_extracted_k141_207692 | 2801 | No | NA | 1 | 0 | 1 | Not-determined | Genome-fragment | NA | NA | 0 |
| S8_extracted_k141_95353 | 1582 | No | NA | 3 | 0 | 2 | Not-determined | Genome-fragment | NA | NA | 0 |
| S8_extracted_k141_121391 | 1261 | No | NA | 2 | 0 | 1 | Not-determined | Genome-fragment | NA | NA | 0 |
| S8_extracted_k141_164378 | 1061 | No | NA | 1 | 0 | 0 | Not-determined | Genome-fragment | NA | NA | 0 |
| S8_extracted_k141_173533 | 1567 | No | NA | 1 | 0 | 1 | Not-determined | Genome-fragment | NA | NA | 0 |
| S8_extracted_k141_121397 | 3063 | No | NA | 2 | 0 | 2 | Not-determined | Genome-fragment | NA | NA | 0 |
| S8_extracted_k141_95371 | 3122 | No | NA | 3 | 0 | 3 | Not-determined | Genome-fragment | NA | NA | 0 |
| S8_extracted_k141_164385 | 5740 | No | NA | 8 | 0 | 0 | Not-determined | Genome-fragment | NA | NA | 0 |
| S10_extracted_99263 | 10361 | No | NA | 28 | 4 | 0 | Low-quality | Genome-fragment | 23.37 | AAI-based (high-confidence) | 0 |
| S8_vs_k141_142696\|\|full | 10363 | No | NA | 13 | 1 | 6 | Low-quality | Genome-fragment | 3.73 | HMM-based (lower-bound) | 0 |
| S8_extracted_k141_95391 | 6182 | No | NA | 7 | 0 | 4 | Not-determined | Genome-fragment | NA | NA | 0 |
| S13_extracted_61695 | 10365 | No | NA | 16 | 4 | 0 | Low-quality | Genome-fragment | 18.67 | AAI-based (high-confidence) | 0 |
| S8_extracted_k141_155842 | 1790 | No | NA | 1 | 0 | 1 | Not-determined | Genome-fragment | NA | NA | 0 |
| S8_extracted_k141_95412 | 1296 | No | NA | 1 | 0 | 1 | Not-determined | Genome-fragment | NA | NA | 0 |
| S8_extracted_k141_207740 | 1115 | No | NA | 1 | 0 | 0 | Not-determined | Genome-fragment | NA | NA | 0 |
| S8_extracted_k141_138650 | 3275 | No | NA | 3 | 0 | 3 | Not-determined | Genome-fragment | NA | NA | 0 |
| S8_extracted_k141_103893 | 4574 | No | NA | 4 | 0 | 0 | Not-determined | Genome-fragment | NA | NA | 0 |
| S8_extracted_k141_95419 | 1241 | No | NA | 2 | 0 | 0 | Not-determined | Genome-fragment | NA | NA | 0 |
| S8_extracted_k141_121460 | 2807 | No | NA | 2 | 0 | 1 | Not-determined | Genome-fragment | NA | NA | 0 |
| S8_extracted_k141_121471 | 1092 | No | NA | 2 | 0 | 1 | Not-determined | Genome-fragment | NA | NA | 0 |
| S16_vs_109309\|\|full | 10365 | No | NA | 19 | 3 | 0 | Low-quality | Genome-fragment | 18.8 | HMM-based (lower-bound) | 0 |
| S17_vs_k141_1115\|\|full | 10380 | No | NA | 8 | 0 | 0 | Low-quality | Genome-fragment | 14.47 | AAI-based (medium-confidence) | 0 |
| S3_extracted_k141_99253 | 10385 | No | NA | 11 | 1 | 0 | Low-quality | Genome-fragment | 6.44 | AAI-based (high-confidence) | 0 |
| S8_extracted_k141_190038 | 1074 | No | NA | 1 | 0 | 1 | Not-determined | Genome-fragment | NA | NA | 0 |
| S8_extracted_k141_31493 | 10390 | No | NA | 11 | 9 | 0 | Low-quality | Genome-fragment | 17.37 | AAI-based (medium-confidence) | 0 |
| S8_extracted_k141_164452 | 2710 | No | NA | 3 | 0 | 2 | Not-determined | Genome-fragment | NA | NA | 0 |
| S11_vs_k141_8051\|\|full | 10396 | No | NA | 12 | 10 | 0 | Low-quality | Genome-fragment | 32.26 | AAI-based (high-confidence) | 0 |
| S8_vs_k141_45780\|\|full | 10402 | Yes | 5866 | 9 | 1 | 2 | Low-quality | Genome-fragment | 12.34 | HMM-based (lower-bound) | 43.61 |
| S8_extracted_k141_164475 | 1147 | No | NA | 1 | 0 | 1 | Not-determined | Genome-fragment | NA | NA | 0 |
| S8_extracted_k141_155912 | 1483 | No | NA | 2 | 0 | 2 | Not-determined | Genome-fragment | NA | NA | 0 |
| S8_extracted_k141_69324 | 1095 | No | NA | 3 | 0 | 0 | Not-determined | Genome-fragment | NA | NA | 0 |
| S8_extracted_k141_17093 | 1381 | No | NA | 1 | 0 | 0 | Not-determined | Genome-fragment | NA | NA | 0 |
| S8_extracted_k141_190102 | 1571 | No | NA | 2 | 0 | 2 | Not-determined | Genome-fragment | NA | NA | 0 |
| S8_extracted_k141_190107 | 2098 | No | NA | 1 | 0 | 1 | Not-determined | Genome-fragment | NA | NA | 0 |
| S11_vs_k141_8421\|\|full | 10403 | No | NA | 10 | 1 | 5 | Low-quality | Genome-fragment | 7.12 | HMM-based (lower-bound) | 0 |
| S8_extracted_k141_164519 | 1059 | No | NA | 2 | 0 | 0 | Not-determined | Genome-fragment | NA | NA | 0 |
| S8_extracted_k141_43315 | 1047 | No | NA | 1 | 0 | 0 | Not-determined | Genome-fragment | NA | NA | 0 |
| S8_extracted_k141_155952 | 2885 | No | NA | 3 | 0 | 0 | Not-determined | Genome-fragment | NA | NA | 0 |
| S8_extracted_k141_69383 | 2980 | No | NA | 1 | 0 | 0 | Not-determined | Genome-fragment | NA | NA | 0 |
| S8_extracted_k141_164535 | 1941 | No | NA | 3 | 0 | 3 | Not-determined | Genome-fragment | NA | NA | 0 |
| S8_extracted_k141_155965 | 1409 | No | NA | 2 | 0 | 1 | Not-determined | Genome-fragment | NA | NA | 0 |
| S16_vs_18320\|\|full | 10408 | No | NA | 12 | 1 | 0 | Low-quality | Genome-fragment | 19.27 | AAI-based (high-confidence) | 0 |
| S8_extracted_k141_207760 | 1878 | No | NA | 2 | 0 | 0 | Not-determined | Genome-fragment | NA | NA | 0 |
| S8_vs_k141_35561\|\|0_partial | 10408 | No | NA | 7 | 0 | 1 | Low-quality | Genome-fragment | 10.87 | AAI-based (medium-confidence) | 0 |
| S8_extracted_k141_190177 | 2122 | No | NA | 2 | 0 | 1 | Not-determined | Genome-fragment | NA | NA | 0 |
| S8_extracted_k141_190191 | 1070 | No | NA | 3 | 0 | 2 | Not-determined | Genome-fragment | NA | NA | 0 |
| S8_extracted_k141_190207 | 1127 | No | NA | 2 | 0 | 1 | Not-determined | Genome-fragment | NA | NA | 0 |
| S8_extracted_k141_51324 | 4623 | No | NA | 4 | 0 | 2 | Not-determined | Genome-fragment | NA | NA | 0 |
| S14_extracted_k141_94371 | 10410 | No | NA | 12 | 9 | 0 | Low-quality | Genome-fragment | 6.89 | AAI-based (medium-confidence) | 0 |
| S8_extracted_k141_51334 | 2393 | No | NA | 2 | 0 | 2 | Not-determined | Genome-fragment | NA | NA | 0 |
| S8_extracted_k141_207806 | 1783 | No | NA | 3 | 0 | 0 | Not-determined | Genome-fragment | NA | NA | 0 |
| S8_extracted_k141_51337 | 1435 | No | NA | 1 | 0 | 1 | Not-determined | Genome-fragment | NA | NA | 0 |
| S17_vs_k141_161362\|\|full | 10417 | No | NA | 19 | 1 | 4 | Low-quality | Genome-fragment | 17.88 | AAI-based (high-confidence) | 0 |
| S8_extracted_k141_51358 | 1265 | No | NA | 1 | 0 | 1 | Not-determined | Genome-fragment | NA | NA | 0 |
| S8_extracted_k141_103975 | 5198 | No | NA | 5 | 0 | 3 | Not-determined | Genome-fragment | NA | NA | 0 |
| S8_extracted_k141_103988 | 1771 | No | NA | 3 | 0 | 0 | Not-determined | Genome-fragment | NA | NA | 0 |
| S8_extracted_k141_207812 | 1076 | No | NA | 1 | 0 | 1 | Not-determined | Genome-fragment | NA | NA | 0 |
| S8_extracted_k141_17122 | 2254 | No | NA | 1 | 0 | 1 | Not-determined | Genome-fragment | NA | NA | 0 |
| S8_extracted_k141_190294 | 1401 | No | NA | 2 | 0 | 0 | Not-determined | Genome-fragment | NA | NA | 0 |
| S8_extracted_k141_207867 | 1076 | No | NA | 1 | 0 | 1 | Not-determined | Genome-fragment | NA | NA | 0 |
| S8_extracted_k141_156086 | 2025 | No | NA | 2 | 0 | 1 | Not-determined | Genome-fragment | NA | NA | 0 |
| S8_extracted_k141_17169 | 2739 | No | NA | 3 | 0 | 3 | Not-determined | Genome-fragment | NA | NA | 0 |
| S8_extracted_k141_104047 | 1148 | No | NA | 1 | 0 | 1 | Not-determined | Genome-fragment | NA | NA | 0 |
| S8_extracted_k141_156097 | 1054 | No | NA | 1 | 0 | 1 | Not-determined | Genome-fragment | NA | NA | 0 |
| S8_extracted_k141_51381 | 4302 | No | NA | 4 | 0 | 4 | Not-determined | Genome-fragment | NA | NA | 0 |
| S8_extracted_k141_207892 | 1235 | No | NA | 2 | 0 | 1 | Not-determined | Genome-fragment | NA | NA | 0 |
| S8_extracted_k141_207896 | 1510 | No | NA | 1 | 0 | 0 | Not-determined | Genome-fragment | NA | NA | 0 |
| S8_extracted_k141_207897 | 1017 | No | NA | 1 | 0 | 1 | Not-determined | Genome-fragment | NA | NA | 0 |
| S8_extracted_k141_156131 | 1090 | No | NA | 2 | 0 | 1 | Not-determined | Genome-fragment | NA | NA | 0 |
| S8_extracted_k141_17199 | 1494 | No | NA | 3 | 0 | 0 | Not-determined | Genome-fragment | NA | NA | 0 |
| S11_vs_k141_70095\|\|full | 10418 | Yes | 2171 | 10 | 1 | 3 | Low-quality | Genome-fragment | 1.93 | HMM-based (lower-bound) | 79.16 |
| S8_extracted_k141_164763 | 3067 | No | NA | 3 | 0 | 2 | Not-determined | Genome-fragment | NA | NA | 0 |
| S8_extracted_k141_104131 | 1118 | No | NA | 1 | 0 | 0 | Not-determined | Genome-fragment | NA | NA | 0 |
| S8_extracted_k141_51474 | 4117 | No | NA | 3 | 0 | 3 | Not-determined | Genome-fragment | NA | NA | 0 |
| S8_extracted_k141_51490 | 1318 | No | NA | 1 | 0 | 1 | Not-determined | Genome-fragment | NA | NA | 0 |
| S8_extracted_k141_17281 | 1404 | No | NA | 1 | 0 | 1 | Not-determined | Genome-fragment | NA | NA | 0 |
| S8_extracted_k141_208018 | 2170 | No | NA | 2 | 0 | 2 | Not-determined | Genome-fragment | NA | NA | 0 |
| S8_extracted_k141_17288 | 1692 | No | NA | 2 | 0 | 0 | Not-determined | Genome-fragment | NA | NA | 0 |
| S8_extracted_k141_208043 | 1071 | No | NA | 1 | 0 | 1 | Not-determined | Genome-fragment | NA | NA | 0 |
| S13_extracted_106280 | 10418 | No | NA | 16 | 5 | 0 | Low-quality | Genome-fragment | 32.08 | HMM-based (lower-bound) | 0 |
| S3_vs_k141_23586\|\|full | 10429 | No | NA | 13 | 6 | 0 | Low-quality | Genome-fragment | 25.68 | AAI-based (high-confidence) | 0 |
| S8_extracted_k141_208071 | 1444 | No | NA | 2 | 0 | 2 | Not-determined | Genome-fragment | NA | NA | 0 |
| S8_extracted_k141_190455 | 1146 | No | NA | 2 | 0 | 2 | Not-determined | Genome-fragment | NA | NA | 0 |
| S3_extracted_k141_24135 | 10443 | No | NA | 20 | 5 | 0 | Low-quality | Genome-fragment | 30.33 | AAI-based (high-confidence) | 0 |
| S8_extracted_k141_51525 | 1394 | No | NA | 1 | 0 | 1 | Not-determined | Genome-fragment | NA | NA | 0 |
| S7_vs_123018\|\|full | 10452 | No | NA | 4 | 0 | 3 | Low-quality | Genome-fragment | 4.34 | AAI-based (high-confidence) | 0 |
| S8_extracted_k141_17331 | 1394 | No | NA | 1 | 0 | 0 | Not-determined | Genome-fragment | NA | NA | 0 |
| S8_extracted_k141_190538 | 1901 | No | NA | 2 | 0 | 1 | Not-determined | Genome-fragment | NA | NA | 0 |
| S13_vs_83609\|\|full | 10460 | Yes | 7064 | 10 | 1 | 3 | Low-quality | Genome-fragment | 10.69 | HMM-based (lower-bound) | 32.47 |
| S8_extracted_k141_208102 | 51637 | No | NA | 60 | 10 | 0 | Complete | High-quality | 100 | DTR (high-confidence) | 0 |
| S8_extracted_k141_51585 | 1091 | No | NA | 1 | 0 | 0 | Not-determined | Genome-fragment | NA | NA | 0 |
| S8_extracted_k141_51589 | 2397 | No | NA | 2 | 0 | 1 | Not-determined | Genome-fragment | NA | NA | 0 |
| S8_extracted_k141_190558 | 1128 | No | NA | 1 | 0 | 1 | Not-determined | Genome-fragment | NA | NA | 0 |
| S8_extracted_k141_51609 | 1120 | No | NA | 3 | 0 | 1 | Not-determined | Genome-fragment | NA | NA | 0 |
| S16_vs_131334\|\|full | 10461 | Yes | 6358 | 15 | 6 | 3 | Low-quality | Genome-fragment | 15.76 | AAI-based (high-confidence) | 39.22 |
| S8_vs_k141_10628\|\|full | 10464 | No | NA | 15 | 8 | 0 | Low-quality | Genome-fragment | 26.65 | AAI-based (high-confidence) | 0 |
| S8_extracted_k141_51633 | 4625 | No | NA | 1 | 0 | 0 | Not-determined | Genome-fragment | NA | NA | 0 |
| S8_extracted_k141_190597 | 1003 | No | NA | 1 | 0 | 1 | Not-determined | Genome-fragment | NA | NA | 0 |
| S8_extracted_k141_190613 | 1272 | No | NA | 2 | 0 | 1 | Not-determined | Genome-fragment | NA | NA | 0 |
| S8_extracted_k141_51671 | 1754 | No | NA | 1 | 0 | 1 | Not-determined | Genome-fragment | NA | NA | 0 |
| S8_extracted_k141_51672 | 1809 | No | NA | 3 | 0 | 0 | Not-determined | Genome-fragment | NA | NA | 0 |
| S11_vs_k141_82797\|\|full | 10469 | No | NA | 19 | 2 | 1 | Low-quality | Genome-fragment | 26.91 | AAI-based (medium-confidence) | 0 |
| S8_extracted_k141_51703 | 1250 | No | NA | 3 | 0 | 2 | Not-determined | Genome-fragment | NA | NA | 0 |
| S8_extracted_k141_190749 | 1586 | No | NA | 2 | 0 | 1 | Not-determined | Genome-fragment | NA | NA | 0 |
| S8_extracted_k141_190766 | 1019 | No | NA | 2 | 0 | 0 | Not-determined | Genome-fragment | NA | NA | 0 |
| S8_extracted_k141_190776 | 1218 | No | NA | 1 | 0 | 0 | Not-determined | Genome-fragment | NA | NA | 0 |
| S8_extracted_k141_51829 | 2450 | No | NA | 3 | 0 | 1 | Not-determined | Genome-fragment | NA | NA | 0 |
| S11_extracted_k141_64640 | 10475 | No | NA | 26 | 1 | 0 | Low-quality | Genome-fragment | 15.2 | AAI-based (high-confidence) | 0 |
| S8_extracted_k141_51871 | 1083 | No | NA | 1 | 0 | 0 | Not-determined | Genome-fragment | NA | NA | 0 |
| S8_extracted_k141_190869 | 4398 | No | NA | 3 | 0 | 1 | Not-determined | Genome-fragment | NA | NA | 0 |
| S12_vs_k141_152583\|\|full | 10475 | No | NA | 16 | 3 | 0 | Low-quality | Genome-fragment | 7.92 | AAI-based (medium-confidence) | 0 |
| S8_extracted_k141_208107 | 52072 | No | NA | 61 | 7 | 0 | Complete | High-quality | 100 | DTR (high-confidence) | 0 |
| S8_extracted_k141_52002 | 1667 | No | NA | 1 | 0 | 1 | Not-determined | Genome-fragment | NA | NA | 0 |
| S8_extracted_k141_52007 | 2816 | No | NA | 1 | 0 | 1 | Not-determined | Genome-fragment | NA | NA | 0 |
| S8_extracted_k141_52050 | 17085 | No | NA | 23 | 8 | 0 | Medium-quality | Genome-fragment | 74.96 | AAI-based (medium-confidence) | 0 |
| S8_extracted_k141_208110 | 137476 | No | NA | 190 | 38 | 4 | Complete | High-quality | 100 | DTR (high-confidence) | 0 |
| S8_extracted_k141_208111 | 62668 | No | NA | 78 | 19 | 0 | Complete | High-quality | 100 | DTR (high-confidence) | 0 |
| S8_extracted_k141_208115 | 5662 | No | NA | 8 | 2 | 0 | Complete | High-quality | 100 | DTR (high-confidence) | 0 |
| S8_extracted_k141_208116 | 5362 | No | NA | 6 | 0 | 0 | Not-determined | Genome-fragment | NA | NA | 0 |
| S8_extracted_k141_208119 | 5827 | No | NA | 8 | 4 | 0 | Complete | High-quality | 100 | DTR (high-confidence) | 0 |
| S8_extracted_k141_208120 | 52201 | No | NA | 70 | 8 | 2 | Complete | High-quality | 100 | DTR (high-confidence) | 0 |
| S8_extracted_k141_208126 | 43499 | No | NA | 59 | 31 | 0 | Complete | High-quality | 100 | DTR (high-confidence) | 0 |
| S8_extracted_k141_208145 | 1953 | No | NA | 3 | 0 | 0 | Not-determined | Genome-fragment | NA | NA | 0 |
| S8_extracted_k141_208146 | 5198 | No | NA | 7 | 5 | 0 | Complete | High-quality | 100 | DTR (high-confidence) | 0 |
| S8_extracted_k141_208152 | 7234 | No | NA | 15 | 0 | 0 | Not-determined | Genome-fragment | NA | NA | 0 |
| S8_vs_k141_156354\|\|full | 13913 | No | NA | 23 | 0 | 6 | Not-determined | Genome-fragment | NA | NA | 0 |
| S8_vs_k141_182288\|\|full | 154466 | No | NA | 183 | 17 | 2 | High-quality | High-quality | 100 | AAI-based (high-confidence) | 0 |
| S8_vs_k141_9114\|\|full | 40264 | No | NA | 59 | 9 | 2 | High-quality | High-quality | 96.69 | AAI-based (high-confidence) | 0 |
| S8_vs_k141_35308\|\|full | 41153 | No | NA | 78 | 17 | 0 | High-quality | High-quality | 100 | AAI-based (high-confidence) | 0 |
| S8_vs_k141_156883\|\|full | 2723 | No | NA | 6 | 0 | 1 | Not-determined | Genome-fragment | NA | NA | 0 |
| S8_vs_k141_87809\|\|full | 9706 | No | NA | 12 | 0 | 0 | Not-determined | Genome-fragment | NA | NA | 0 |
| S8_vs_k141_88013\|\|full | 42463 | No | NA | 75 | 16 | 0 | High-quality | High-quality | 100 | AAI-based (high-confidence) | 0 |
| S8_vs_k141_9987\|\|full | 58206 | No | NA | 84 | 12 | 4 | High-quality | High-quality | 99.97 | AAI-based (high-confidence) | 0 |
| S8_vs_k141_71212\|\|full | 40180 | No | NA | 68 | 7 | 1 | High-quality | High-quality | 100 | AAI-based (high-confidence) | 0 |
| S8_vs_k141_149700\|\|full | 69469 | Yes | 46061 | 90 | 25 | 16 | Medium-quality | Genome-fragment | 83.13 | AAI-based (high-confidence) | 33.7 |
| S8_vs_k141_132359\|\|full | 45715 | No | NA | 58 | 11 | 2 | High-quality | High-quality | 92.4 | AAI-based (medium-confidence) | 0 |
| S8_vs_k141_193843\|\|full | 36636 | No | NA | 55 | 12 | 1 | High-quality | High-quality | 100 | AAI-based (high-confidence) | 0 |
| S8_vs_k141_72117\|\|full | 33104 | No | NA | 48 | 13 | 1 | High-quality | High-quality | 97.22 | AAI-based (high-confidence) | 0 |
| S8_vs_k141_3621\|\|full | 3020 | No | NA | 3 | 0 | 0 | Not-determined | Genome-fragment | NA | NA | 0 |
| S8_vs_k141_40014\|\|full | 33856 | No | NA | 50 | 12 | 0 | High-quality | High-quality | 95.81 | AAI-based (high-confidence) | 0 |
| S8_vs_k141_66480\|\|full | 10486 | No | NA | 20 | 0 | 5 | Low-quality | Genome-fragment | 23.68 | AAI-based (medium-confidence) | 0 |
| S8_vs_k141_162053\|\|full | 15437 | No | NA | 17 | 6 | 0 | Medium-quality | Genome-fragment | 66.12 | AAI-based (high-confidence) | 0 |
| S8_vs_k141_59736\|\|full | 196423 | No | NA | 315 | 42 | 6 | High-quality | High-quality | 100 | AAI-based (high-confidence) | 0 |
| S8_vs_k141_51368\|\|full | 43860 | No | NA | 84 | 13 | 2 | High-quality | High-quality | 98.79 | AAI-based (high-confidence) | 0 |
| S8_vs_k141_208103\|\|full | 99692 | No | NA | 120 | 17 | 1 | High-quality | High-quality | 98.44 | AAI-based (high-confidence) | 0 |
| S8_vs_k141_208108\|\|full | 1986 | No | NA | 4 | 0 | 0 | Not-determined | Genome-fragment | NA | NA | 0 |
| S8_vs_k141_208113\|\|full | 41644 | No | NA | 50 | 10 | 0 | High-quality | High-quality | 100 | AAI-based (high-confidence) | 0 |
| S8_vs_k141_208114\|\|full | 44238 | No | NA | 60 | 31 | 0 | High-quality | High-quality | 98.07 | AAI-based (high-confidence) | 0 |
| S8_vs_k141_208118\|\|full | 44313 | No | NA | 73 | 27 | 0 | High-quality | High-quality | 100 | AAI-based (high-confidence) | 0 |
| S8_vs_k141_208127\|\|full | 48002 | No | NA | 69 | 19 | 2 | High-quality | High-quality | 94.22 | AAI-based (high-confidence) | 0 |
| S8_vs_k141_208129\|\|full | 22668 | No | NA | 50 | 5 | 0 | High-quality | High-quality | 97.57 | AAI-based (high-confidence) | 0 |
| S8_vs_k141_208147\|\|full | 1720 | No | NA | 2 | 0 | 1 | Not-determined | Genome-fragment | NA | NA | 0 |
| S17_extracted_k141_61673 | 10490 | No | NA | 15 | 0 | 2 | Low-quality | Genome-fragment | 25.89 | AAI-based (medium-confidence) | 0 |
| S8_vs_k141_208157\|\|full | 4290 | No | NA | 4 | 0 | 0 | Not-determined | Genome-fragment | NA | NA | 0 |
| S1_vs_k141_47669\|\|full | 10493 | No | NA | 7 | 1 | 0 | Low-quality | Genome-fragment | 6.82 | AAI-based (high-confidence) | 0 |
| S11_vs_k141_93847\|\|full | 10498 | No | NA | 16 | 3 | 0 | Low-quality | Genome-fragment | 20.98 | AAI-based (medium-confidence) | 0 |
| S11_vs_k141_9572\|\|full | 10501 | No | NA | 7 | 1 | 0 | Low-quality | Genome-fragment | 26.08 | AAI-based (medium-confidence) | 0 |
| S12_vs_k141_28418\|\|full | 10504 | No | NA | 19 | 1 | 0 | Low-quality | Genome-fragment | 7.96 | HMM-based (lower-bound) | 0 |
| S8_vs_k141_182234\|\|full | 2776 | No | NA | 5 | 0 | 1 | Not-determined | Genome-fragment | NA | NA | 0 |
| S8_vs_k141_17371\|\|full | 1667 | No | NA | 2 | 0 | 1 | Not-determined | Genome-fragment | NA | NA | 0 |
| S8_vs_k141_190930\|\|full | 2194 | No | NA | 4 | 0 | 2 | Not-determined | Genome-fragment | NA | NA | 0 |
| S8_vs_k141_121509\|\|full | 1338 | No | NA | 2 | 0 | 0 | Not-determined | Genome-fragment | NA | NA | 0 |
| S4_vs_VIRSorter_3742-cat_2 | 10511 | No | NA | 16 | 10 | 0 | Low-quality | Genome-fragment | 22.62 | AAI-based (high-confidence) | 0 |
| S8_vs_k141_34\|\|full | 9073 | No | NA | 8 | 0 | 6 | Not-determined | Genome-fragment | NA | NA | 0 |
| S5_vs_k141_111228\|\|full | 10528 | No | NA | 11 | 1 | 6 | Low-quality | Genome-fragment | 7.21 | HMM-based (lower-bound) | 0 |
| S8_vs_k141_78120\|\|full | 1098 | No | NA | 3 | 0 | 1 | Not-determined | Genome-fragment | NA | NA | 0 |
| S8_vs_k141_26081\|\|full | 2294 | No | NA | 3 | 0 | 0 | Not-determined | Genome-fragment | NA | NA | 0 |
| S1_vs_k141_92098\|\|full | 10536 | No | NA | 11 | 3 | 0 | Low-quality | Genome-fragment | 21.58 | AAI-based (high-confidence) | 0 |
| S8_vs_k141_34765\|\|full | 2650 | No | NA | 3 | 0 | 2 | Not-determined | Genome-fragment | NA | NA | 0 |
| S8_vs_k141_43432\|\|full | 1028 | No | NA | 3 | 0 | 1 | Not-determined | Genome-fragment | NA | NA | 0 |
| S8_vs_k141_199618\|\|full | 1986 | No | NA | 2 | 0 | 1 | Not-determined | Genome-fragment | NA | NA | 0 |
| S16_vs_126310\|\|full | 10537 | No | NA | 26 | 1 | 1 | Low-quality | Genome-fragment | 16.42 | AAI-based (medium-confidence) | 0 |
| S3_extracted_k141_122722 | 10549 | No | NA | 13 | 1 | 4 | Low-quality | Genome-fragment | 6.42 | HMM-based (lower-bound) | 0 |
| S8_vs_k141_26097\|\|full | 1989 | No | NA | 3 | 0 | 0 | Not-determined | Genome-fragment | NA | NA | 0 |
| S17_extracted_k141_24516 | 10553 | No | NA | 19 | 3 | 0 | Low-quality | Genome-fragment | 32.82 | HMM-based (lower-bound) | 0 |
| S5_vs_k141_125899\|\|full | 10554 | No | NA | 20 | 2 | 1 | Low-quality | Genome-fragment | 24.93 | HMM-based (lower-bound) | 0 |
| S8_vs_k141_17421\|\|full | 3635 | No | NA | 9 | 0 | 0 | Not-determined | Genome-fragment | NA | NA | 0 |
| S8_vs_k141_199641\|\|full | 1370 | No | NA | 2 | 0 | 1 | Not-determined | Genome-fragment | NA | NA | 0 |

| | | | | | | | | | | | | | |
|---|---|---|---|---|---|---|---|---|---|---|---|---|---|
| S4_extracted_15245 | 10554 | No | NA | | 16 | 6 | 0 | Low-quality | Genome-fragment | | 19.8 | AAI-based (high-confidence) | 0 |
| S8_vs_k141_52143\|\|full | 1983 | No | NA | | 3 | 0 | 1 | Not-determined | Genome-fragment | NA | | NA | 0 |
| S15_vs_31611\|\|full | 10563 | No | NA | | 15 | 13 | 0 | Low-quality | Genome-fragment | | 30.36 | AAI-based (high-confidence) | 0 |
| S11_vs_k141_47250\|\|full | 10571 | No | NA | | 18 | 5 | 0 | Low-quality | Genome-fragment | | 7.36 | AAI-based (medium-confidence) | 0 |
| S8_vs_k141_111\|\|full | 2222 | No | NA | | 4 | 0 | 1 | Not-determined | Genome-fragment | NA | | NA | 0 |
| S7_vs_71911\|\|full | 10573 | No | NA | | 15 | 6 | 0 | Low-quality | Genome-fragment | | 28.22 | AAI-based (high-confidence) | 0 |
| S8_vs_k141_147621\|\|full | 4658 | No | NA | | 4 | 0 | 4 | Not-determined | Genome-fragment | NA | | NA | 0 |
| S1_vs_k141_150573\|\|full | 10589 | No | NA | | 13 | 1 | 1 | Low-quality | Genome-fragment | | 24.72 | AAI-based (high-confidence) | 0 |
| S8_vs_k141_60850\|\|full | 6813 | No | NA | | 4 | 0 | 4 | Not-determined | Genome-fragment | NA | | NA | 0 |
| S16_extracted_69256 | 10593 | No | NA | | 24 | 3 | 0 | Low-quality | Genome-fragment | | 19.63 | AAI-based (high-confidence) | 0 |
| S12_vs_k141_16106\|\|full | 10594 | No | NA | | 8 | 0 | 5 | Low-quality | Genome-fragment | | 6.92 | AAI-based (medium-confidence) | 0 |
| S3_vs_k141_67026\|\|full | 10596 | No | NA | | 25 | 3 | 0 | Low-quality | Genome-fragment | | 17.51 | AAI-based (high-confidence) | 0 |
| S8_vs_k141_86933\|\|full | 2975 | No | NA | | 3 | 0 | 3 | Not-determined | Genome-fragment | NA | | NA | 0 |
| S8_vs_k141_52167\|\|full | 3280 | No | NA | | 2 | 0 | 0 | Not-determined | Genome-fragment | NA | | NA | 0 |
| S16_vs_67735\|\|full | 10597 | No | NA | | 16 | 1 | 0 | Low-quality | Genome-fragment | | 6.75 | HMM-based (lower-bound) | 0 |
| S8_vs_k141_60866\|\|full | 5113 | No | NA | | 6 | 0 | 0 | Not-determined | Genome-fragment | NA | | NA | 0 |
| S8_vs_k141_78226\|\|full | 1587 | No | NA | | 2 | 0 | 2 | Not-determined | Genome-fragment | NA | | NA | 0 |
| S8_vs_k141_52186\|\|full | 1188 | No | NA | | 3 | 0 | 1 | Not-determined | Genome-fragment | NA | | NA | 0 |
| S8_vs_k141_199697\|\|full | 5572 | No | NA | | 9 | 0 | 3 | Not-determined | Genome-fragment | NA | | NA | 0 |
| S14_vs_k141_6971\|\|full | 10606 | No | NA | | 15 | 2 | 0 | Low-quality | Genome-fragment | | 17.2 | AAI-based (high-confidence) | 0 |
| S8_vs_k141_8820\|\|full | 4942 | No | NA | | 3 | 0 | 0 | Not-determined | Genome-fragment | NA | | NA | 0 |
| S13_extracted_31235 | 10612 | No | NA | | 17 | 2 | 0 | Low-quality | Genome-fragment | | 6.68 | AAI-based (high-confidence) | 0 |
| S8_vs_k141_112971\|\|full | 1304 | No | NA | | 3 | 0 | 1 | Not-determined | Genome-fragment | NA | | NA | 0 |
| S8_vs_k141_147670\|\|full | 23940 | No | NA | | 32 | 12 | 0 | Medium-quality | Genome-fragment | | 62.53 | AAI-based (medium-confidence) | 0 |
| S14_vs_k141_101062\|\|full | 10616 | No | NA | | 17 | 6 | 0 | Low-quality | Genome-fragment | | 25.19 | AAI-based (medium-confidence) | 0 |
| S4_extracted_31985 | 10618 | No | NA | | 11 | 1 | 0 | Low-quality | Genome-fragment | | 0.96 | HMM-based (lower-bound) | 0 |
| S8_vs_k141_103136\|\|full | 10624 | No | NA | | 11 | 1 | 2 | Low-quality | Genome-fragment | | 9.3 | HMM-based (lower-bound) | 0 |
| S8_vs_k141_173668\|\|full | 1248 | No | NA | | 3 | 0 | 0 | Not-determined | Genome-fragment | NA | | NA | 0 |
| S8_vs_k141_86996\|\|full | 1391 | No | NA | | 2 | 0 | 1 | Not-determined | Genome-fragment | NA | | NA | 0 |
| S8_vs_k141_191094\|\|full | 4460 | No | NA | | 6 | 0 | 4 | Not-determined | Genome-fragment | NA | | NA | 0 |
| S1_extracted_k141_15210 | 10629 | No | NA | | 20 | 5 | 0 | Low-quality | Genome-fragment | | 15.72 | AAI-based (high-confidence) | 0 |
| S8_vs_k141_138999\|\|full | 6531 | No | NA | | 8 | 0 | 5 | Not-determined | Genome-fragment | NA | | NA | 0 |
| S8_vs_k141_130283\|\|full | 6290 | No | NA | | 9 | 0 | 2 | Not-determined | Genome-fragment | NA | | NA | 0 |
| S8_vs_k141_121667\|\|full | 1847 | No | NA | | 4 | 0 | 0 | Not-determined | Genome-fragment | NA | | NA | 0 |
| S9_vs_k141_42454\|\|full | 10632 | No | NA | | 21 | 4 | 0 | Low-quality | Genome-fragment | | 24.27 | AAI-based (medium-confidence) | 0 |
| S3_extracted_k141_82845 | 10634 | Yes | | 8598 | 14 | 4 | 2 | Low-quality | Genome-fragment | | 19.69 | AAI-based (medium-confidence) | 19.15 |
| S8_vs_k141_191124\|\|full | 1353 | No | NA | | 2 | 0 | 0 | Not-determined | Genome-fragment | NA | | NA | 0 |
| S8_vs_k141_165070\|\|full | 1845 | No | NA | | 2 | 0 | 2 | Not-determined | Genome-fragment | NA | | NA | 0 |
| S8_vs_k141_199735\|\|full | 3876 | No | NA | | 4 | 0 | 3 | Not-determined | Genome-fragment | NA | | NA | 0 |
| S10_vs_36734\|\|full | 10638 | No | NA | | 8 | 0 | 2 | Low-quality | Genome-fragment | | 7.27 | AAI-based (medium-confidence) | 0 |
| S16_vs_48614\|\|full | 10639 | No | NA | | 13 | 1 | 0 | Low-quality | Genome-fragment | | 4.45 | AAI-based (medium-confidence) | 0 |
| S8_vs_k141_60944\|\|full | 2183 | No | NA | | 5 | 0 | 2 | Not-determined | Genome-fragment | NA | | NA | 0 |
| S8_vs_k141_160978\|\|full | 10643 | No | NA | | 19 | 0 | 0 | Low-quality | Genome-fragment | | 20.18 | AAI-based (high-confidence) | 0 |
| S13_vs_146829\|\|full | 10645 | No | NA | | 15 | 0 | 1 | Low-quality | Genome-fragment | | 12.38 | AAI-based (high-confidence) | 0 |
| S8_vs_k141_60961\|\|full | 1778 | No | NA | | 3 | 0 | 0 | Not-determined | Genome-fragment | NA | | NA | 0 |
| S14_vs_k141_59243\|\|full | 10646 | No | NA | | 18 | 2 | 2 | Low-quality | Genome-fragment | | 17.24 | AAI-based (high-confidence) | 0 |
| S17_extracted_k141_105581 | 10649 | No | NA | | 11 | 0 | 1 | Low-quality | Genome-fragment | | 2.66 | AAI-based (high-confidence) | 0 |
| S5_vs_k141_128231\|\|full | 10655 | No | NA | | 12 | 2 | 1 | Low-quality | Genome-fragment | | 22.02 | AAI-based (medium-confidence) | 0 |
| S8_vs_k141_87074\|\|full | 2816 | No | NA | | 4 | 0 | 0 | Not-determined | Genome-fragment | NA | | NA | 0 |
| S5_vs_k141_101465\|\|full | 10662 | No | NA | | 18 | 12 | 0 | Low-quality | Genome-fragment | | 12.95 | AAI-based (high-confidence) | 0 |
| S8_vs_k141_52270\|\|full | 3358 | No | NA | | 2 | 0 | 0 | Not-determined | Genome-fragment | NA | | NA | 0 |
| S8_vs_k141_113044\|\|full | 2460 | No | NA | | 3 | 0 | 1 | Not-determined | Genome-fragment | NA | | NA | 0 |
| S8_vs_k141_69592\|\|full | 1425 | No | NA | | 5 | 0 | 0 | Not-determined | Genome-fragment | NA | | NA | 0 |
| S8_vs_k141_165095\|\|full | 5228 | No | NA | | 6 | 0 | 3 | Not-determined | Genome-fragment | NA | | NA | 0 |
| S8_vs_k141_26259\|\|full | 3469 | No | NA | | 3 | 0 | 2 | Not-determined | Genome-fragment | NA | | NA | 0 |
| S8_vs_k141_43615\|\|full | 1396 | No | NA | | 3 | 0 | 0 | Not-determined | Genome-fragment | NA | | NA | 0 |
| S8_vs_k141_26264\|\|full | 2959 | No | NA | | 3 | 0 | 3 | Not-determined | Genome-fragment | NA | | NA | 0 |
| S8_vs_k141_113059\|\|full | 3703 | No | NA | | 5 | 4 | 0 | Medium-quality | Genome-fragment | | 72.98 | AAI-based (high-confidence) | 0 |
| S2_vs_k141_21238\|\|full | 10673 | No | NA | | 14 | 2 | 0 | Low-quality | Genome-fragment | | 17.87 | AAI-based (high-confidence) | 0 |
| S8_vs_k141_104282\|\|full | 1605 | No | NA | | 3 | 0 | 1 | Not-determined | Genome-fragment | NA | | NA | 0 |
| S12_vs_k141_40810\|\|full | 10678 | No | NA | | 17 | 1 | 2 | Low-quality | Genome-fragment | | 9.56 | HMM-based (lower-bound) | 0 |
| S8_vs_k141_147689\|\|full | 5160 | No | NA | | 5 | 0 | 5 | Not-determined | Genome-fragment | NA | | NA | 0 |
| S8_vs_k141_8970\|\|full | 1456 | No | NA | | 3 | 0 | 0 | Not-determined | Genome-fragment | NA | | NA | 0 |
| S8_vs_k141_52319\|\|full | 2357 | No | NA | | 3 | 0 | 1 | Not-determined | Genome-fragment | NA | | NA | 0 |
| S8_vs_k141_182331\|\|full | 22065 | No | NA | | 35 | 15 | 0 | Medium-quality | Genome-fragment | | 53.63 | AAI-based (high-confidence) | 0 |
| S6_vs_k141_38694\|\|full | 10679 | No | NA | | 20 | 5 | 0 | Low-quality | Genome-fragment | | 29.37 | AAI-based (medium-confidence) | 0 |
| S8_vs_k141_8998\|\|full | 1427 | No | NA | | 2 | 0 | 0 | Not-determined | Genome-fragment | NA | | NA | 0 |
| S2_vs_k141_374\|\|full | 10682 | No | NA | | 19 | 4 | 2 | Low-quality | Genome-fragment | | 17.74 | AAI-based (high-confidence) | 0 |
| S8_vs_k141_156447\|\|full | 1803 | No | NA | | 3 | 0 | 2 | Not-determined | Genome-fragment | NA | | NA | 0 |
| S3_vs_k141_795\|\|full | 10687 | No | NA | | 14 | 4 | 1 | Low-quality | Genome-fragment | | 13.87 | AAI-based (high-confidence) | 0 |
| S8_vs_k141_112399\|\|full | 10689 | No | NA | | 20 | 3 | 0 | Low-quality | Genome-fragment | | 27.28 | AAI-based (high-confidence) | 0 |
| S8_vs_k141_156449\|\|full | 1990 | No | NA | | 3 | 0 | 2 | Not-determined | Genome-fragment | NA | | NA | 0 |
| S8_vs_k141_52334\|\|full | 1998 | No | NA | | 2 | 0 | 1 | Not-determined | Genome-fragment | NA | | NA | 0 |
| S7_extracted_95619 | 10717 | No | NA | | 14 | 3 | 1 | Low-quality | Genome-fragment | | 22.04 | AAI-based (high-confidence) | 0 |
| S14_vs_k141_16610\|\|full | 10732 | No | NA | | 10 | 1 | 6 | Low-quality | Genome-fragment | | 4.97 | HMM-based (lower-bound) | 0 |
| S9_vs_k141_56161\|\|full | 10733 | No | NA | | 24 | 3 | 1 | Low-quality | Genome-fragment | | 33.12 | AAI-based (high-confidence) | 0 |
| S9_extracted_k141_63973 | 10734 | No | NA | | 23 | 6 | 0 | Low-quality | Genome-fragment | | 27.23 | AAI-based (high-confidence) | 0 |
| S8_vs_k141_35031\|\|full | 6504 | No | NA | | 8 | 0 | 3 | Not-determined | Genome-fragment | NA | | NA | 0 |
| S8_vs_k141_182386\|\|full | 1930 | No | NA | | 3 | 0 | 2 | Not-determined | Genome-fragment | NA | | NA | 0 |
| S16_extracted_44547 | 10743 | No | NA | | 3 | 0 | 3 | Low-quality | Genome-fragment | | 3.74 | AAI-based (medium-confidence) | 0 |
| S9_vs_k141_17785\|\|full | 10746 | No | NA | | 16 | 1 | 1 | Low-quality | Genome-fragment | | 23.95 | AAI-based (medium-confidence) | 0 |
| S16_vs_82722\|\|full | 10758 | No | NA | | 17 | 3 | 2 | Low-quality | Genome-fragment | | 16.65 | HMM-based (lower-bound) | 0 |
| S8_vs_k141_199828\|\|full | 4289 | No | NA | | 3 | 0 | 3 | Not-determined | Genome-fragment | NA | | NA | 0 |
| S8_extracted_k141_66652 | 10758 | Yes | | 2104 | 9 | 1 | 2 | Low-quality | Genome-fragment | | 2.68 | HMM-based (lower-bound) | 80.44 |
| S4_extracted_8400 | 10768 | No | NA | | 21 | 5 | 0 | Low-quality | Genome-fragment | | 23.68 | AAI-based (high-confidence) | 0 |
| S7_extracted_63165 | 10776 | No | NA | | 6 | 0 | 6 | Low-quality | Genome-fragment | | 5.34 | AAI-based (medium-confidence) | 0 |
| S8_vs_k141_17549\|\|full | 2788 | No | NA | | 5 | 0 | 4 | Not-determined | Genome-fragment | NA | | NA | 0 |
| S8_vs_k141_43686\|\|full | 4512 | No | NA | | 5 | 0 | 4 | Not-determined | Genome-fragment | NA | | NA | 0 |
| S8_vs_k141_147697\|\|full | 55352 | No | NA | | 60 | 3 | 9 | Medium-quality | Genome-fragment | | 62.99 | HMM-based (lower-bound) | 0 |
| S8_vs_k141_130541\|\|full | 1150 | No | NA | | 2 | 0 | 1 | Not-determined | Genome-fragment | NA | | NA | 0 |
| S5_vs_k141_105800\|\|full | 10791 | No | NA | | 22 | 4 | 0 | Low-quality | Genome-fragment | | 20.44 | AAI-based (medium-confidence) | 0 |
| S8_vs_k141_78469\|\|full | 1408 | No | NA | | 3 | 0 | 0 | Not-determined | Genome-fragment | NA | | NA | 0 |
| S8_vs_k141_26370\|\|full | 4003 | No | NA | | 4 | 0 | 2 | Not-determined | Genome-fragment | NA | | NA | 0 |
| S12_vs_k141_71385\|\|full | 10792 | No | NA | | 10 | 0 | 9 | Low-quality | Genome-fragment | | 4.13 | AAI-based (medium-confidence) | 0 |
| S8_vs_k141_139195\|\|full | 6838 | No | NA | | 7 | 0 | 2 | Not-determined | Genome-fragment | NA | | NA | 0 |
| S8_vs_k141_95685\|\|full | 2774 | No | NA | | 3 | 0 | 1 | Not-determined | Genome-fragment | NA | | NA | 0 |
| S8_vs_k141_121864\|\|full | 2279 | No | NA | | 3 | 0 | 1 | Not-determined | Genome-fragment | NA | | NA | 0 |
| S8_vs_k141_26406\|\|full | 2616 | No | NA | | 2 | 0 | 1 | Not-determined | Genome-fragment | NA | | NA | 0 |
| S8_vs_k141_104442\|\|full | 2663 | No | NA | | 3 | 0 | 0 | Not-determined | Genome-fragment | NA | | NA | 0 |
| S14_vs_k141_75166\|\|full | 10805 | No | NA | | 13 | 0 | 0 | Low-quality | Genome-fragment | | 24.57 | AAI-based (high-confidence) | 0 |
| S8_vs_k141_113199\|\|full | 1188 | No | NA | | 3 | 0 | 1 | Not-determined | Genome-fragment | NA | | NA | 0 |
| S8_vs_k141_432\|\|full | 8419 | No | NA | | 14 | 0 | 2 | Not-determined | Genome-fragment | NA | | NA | 0 |
| S8_vs_k141_56772\|\|full | 10809 | No | NA | | 17 | 13 | 0 | Low-quality | Genome-fragment | | 17.98 | AAI-based (high-confidence) | 0 |
| S8_vs_k141_87297\|\|full | 1418 | No | NA | | 2 | 0 | 1 | Not-determined | Genome-fragment | NA | | NA | 0 |
| S13_vs_56845\|\|full | 10815 | No | NA | | 13 | 0 | 5 | Low-quality | Genome-fragment | | 25.25 | AAI-based (high-confidence) | 0 |
| S8_extracted_k141_13058 | 10818 | No | NA | | 8 | 0 | 5 | Low-quality | Genome-fragment | | 28.83 | AAI-based (medium-confidence) | 0 |
| S8_vs_k141_35156\|\|full | 2210 | No | NA | | 3 | 0 | 2 | Not-determined | Genome-fragment | NA | | NA | 0 |
| S16_vs_14307\|\|1_partial | 10820 | No | NA | | 11 | 1 | 1 | Low-quality | Genome-fragment | | 26.41 | AAI-based (medium-confidence) | 0 |
| S16_extracted_167270 | 10823 | No | NA | | 14 | 0 | 0 | Low-quality | Genome-fragment | | 4.18 | AAI-based (medium-confidence) | 0 |
| S8_vs_k141_163459\|\|full | 10823 | No | NA | | 16 | 2 | 0 | Low-quality | Genome-fragment | | 13.19 | HMM-based (lower-bound) | 0 |
| S7_extracted_135907 | 10828 | No | NA | | 9 | 1 | 0 | Low-quality | Genome-fragment | | 23.95 | AAI-based (high-confidence) | 0 |
| S12_extracted_k141_125693 | 10838 | No | NA | | 16 | 1 | 4 | Low-quality | Genome-fragment | | 16.4 | HMM-based (lower-bound) | 0 |
| S3_vs_k141_7878\|\|full | 10843 | No | NA | | 15 | 5 | 0 | Low-quality | Genome-fragment | | 18.05 | AAI-based (high-confidence) | 0 |
| S8_vs_k141_104511\|\|full | 2821 | No | NA | | 3 | 0 | 1 | Not-determined | Genome-fragment | NA | | NA | 0 |
| S8_vs_k141_487\|\|full | 3553 | No | NA | | 7 | 0 | 1 | Not-determined | Genome-fragment | NA | | NA | 0 |
| S8_vs_k141_95769\|\|full | 11950 | No | NA | | 13 | 0 | 3 | Not-determined | Genome-fragment | NA | | NA | 0 |
| S8_vs_k141_52513\|\|full | 2118 | No | NA | | 4 | 0 | 3 | Not-determined | Genome-fragment | NA | | NA | 0 |
| S5_vs_k141_61338\|\|full | 10853 | No | NA | | 14 | 1 | 2 | Low-quality | Genome-fragment | | 18.7 | HMM-based (lower-bound) | 0 |
| S8_vs_k141_191448\|\|full | 1893 | No | NA | | 2 | 0 | 0 | Not-determined | Genome-fragment | NA | | NA | 0 |
| S8_vs_k141_87388\|\|full | 1362 | No | NA | | 3 | 0 | 0 | Not-determined | Genome-fragment | NA | | NA | 0 |
| S8_vs_k141_121968\|\|full | 1193 | No | NA | | 2 | 0 | 0 | Not-determined | Genome-fragment | NA | | NA | 0 |
| S5_extracted_k141_41377 | 10858 | No | NA | | 14 | 1 | 0 | Low-quality | Genome-fragment | | 19.43 | AAI-based (medium-confidence) | 0 |
| S8_vs_k141_9215\|\|full | 2705 | No | NA | | 4 | 0 | 1 | Not-determined | Genome-fragment | NA | | NA | 0 |
| S8_vs_k141_130683\|\|full | 1771 | No | NA | | 3 | 0 | 1 | Not-determined | Genome-fragment | NA | | NA | 0 |
| S16_vs_60663\|\|full | 10861 | No | NA | | 19 | 14 | 0 | Low-quality | Genome-fragment | | 30.53 | AAI-based (high-confidence) | 0 |
| S8_vs_k141_35220\|\|full | 14290 | No | NA | | 16 | 0 | 3 | Not-determined | Genome-fragment | NA | | NA | 0 |
| S6_vs_k141_37013\|\|full | 10864 | No | NA | | 13 | 0 | 0 | Low-quality | Genome-fragment | | 19.48 | AAI-based (high-confidence) | 0 |
| S8_vs_k141_191510\|\|full | 1103 | No | NA | | 3 | 0 | 1 | Not-determined | Genome-fragment | NA | | NA | 0 |
| S8_vs_k141_43853\|\|full | 1435 | No | NA | | 3 | 0 | 0 | Not-determined | Genome-fragment | NA | | NA | 0 |
| S8_vs_k141_52581\|\|full | 1946 | No | NA | | 2 | 0 | 2 | Not-determined | Genome-fragment | NA | | NA | 0 |
| S16_extracted_77987 | 10865 | No | NA | | 9 | 0 | 6 | Low-quality | Genome-fragment | | 8.73 | AAI-based (high-confidence) | 0 |
| S8_vs_k141_191513\|\|full | 1602 | No | NA | | 3 | 0 | 1 | Not-determined | Genome-fragment | NA | | NA | 0 |
| S8_extracted_k141_101387 | 10865 | No | NA | | 20 | 5 | 1 | Low-quality | Genome-fragment | | 16.18 | AAI-based (high-confidence) | 0 |
| S10_vs_102644\|\|full | 10883 | No | NA | | 8 | 2 | 0 | Low-quality | Genome-fragment | | 27.47 | AAI-based (high-confidence) | 0 |
| S8_vs_k141_35242\|\|full | 1392 | No | NA | | 2 | 0 | 1 | Not-determined | Genome-fragment | NA | | NA | 0 |
| S17_vs_k141_35486\|\|full | 10885 | No | NA | | 19 | 1 | 0 | Low-quality | Genome-fragment | | 26.03 | AAI-based (high-confidence) | 0 |
| S15_vs_28139\|\|0_partial | 10887 | Yes | | 7506 | 10 | 1 | 1 | Low-quality | Genome-fragment | | 7.34 | AAI-based (medium-confidence) | 31.06 |
| S8_extracted_k141_73994 | 10889 | No | NA | | 23 | 3 | 0 | Low-quality | Genome-fragment | | 22.59 | AAI-based (high-confidence) | 0 |
| S8_vs_k141_173946\|\|full | 2134 | No | NA | | 3 | 0 | 2 | Not-determined | Genome-fragment | NA | | NA | 0 |
| S8_vs_k141_104605\|\|full | 1006 | No | NA | | 3 | 0 | 0 | Not-determined | Genome-fragment | NA | | NA | 0 |
| S8_vs_k141_173948\|\|full | 1005 | No | NA | | 2 | 0 | 2 | Not-determined | Genome-fragment | NA | | NA | 0 |
| S9_vs_k141_70135\|\|full | 10896 | No | NA | | 11 | 0 | 6 | Low-quality | Genome-fragment | | 7.46 | AAI-based (high-confidence) | 0 |
| S8_vs_k141_35268\|\|full | 1779 | No | NA | | 3 | 0 | 0 | Not-determined | Genome-fragment | NA | | NA | 0 |
| S10_extracted_11730 | 10899 | No | NA | | 16 | 4 | 0 | Low-quality | Genome-fragment | | 6.17 | AAI-based (high-confidence) | 0 |
| S8_vs_k141_120725\|\|full | 10908 | No | NA | | 15 | 1 | 1 | Low-quality | Genome-fragment | | 2.99 | HMM-based (lower-bound) | 0 |
| S3_vs_k141_86620\|\|full | 10911 | No | NA | | 4 | 0 | 3 | Low-quality | Genome-fragment | | 3.43 | AAI-based (medium-confidence) | 0 |
| S10_vs_24700\|\|full | 10914 | Yes | | 5880 | 12 | 2 | 2 | Low-quality | Genome-fragment | | 12.85 | HMM-based (lower-bound) | 46.12 |
| S8_vs_k141_26532\|\|full | 4329 | No | NA | | 4 | 0 | 3 | Not-determined | Genome-fragment | NA | | NA | 0 |
| S3_vs_k141_42038\|\|full | 10924 | No | NA | | 9 | 0 | 5 | Low-quality | Genome-fragment | | 9.35 | AAI-based (medium-confidence) | 0 |
| S3_extracted_k141_69210 | 10927 | No | NA | | 17 | 2 | 0 | Low-quality | Genome-fragment | | 8.17 | AAI-based (high-confidence) | 0 |
| S3_vs_k141_22468\|\|full | 10929 | No | NA | | 9 | 0 | 4 | Low-quality | Genome-fragment | | 19.86 | AAI-based (medium-confidence) | 0 |
| S8_vs_k141_139342\|\|full | 1559 | No | NA | | 3 | 0 | 1 | Not-determined | Genome-fragment | NA | | NA | 0 |
| S8_vs_k141_95869\|\|full | 3092 | No | NA | | 3 | 0 | 2 | Not-determined | Genome-fragment | NA | | NA | 0 |
| S3_extracted_k141_43274 | 10933 | No | NA | | 24 | 12 | 0 | Low-quality | Genome-fragment | | 49.91 | AAI-based (high-confidence) | 0 |
| S8_vs_k141_52634\|\|full | 2691 | No | NA | | 2 | 0 | 0 | Not-determined | Genome-fragment | NA | | NA | 0 |
| S8_vs_k141_95896\|\|full | 2094 | No | NA | | 2 | 0 | 0 | Not-determined | Genome-fragment | NA | | NA | 0 |
| S8_vs_k141_35296\|\|full | 6807 | No | NA | | 3 | 0 | 1 | Not-determined | Genome-fragment | NA | | NA | 0 |
| S16_extracted_131450 | 10933 | No | NA | | 13 | 8 | 0 | Low-quality | Genome-fragment | | 28.36 | AAI-based (high-confidence) | 0 |
| S8_vs_k141_147922\|\|full | 5302 | No | NA | | 5 | 0 | 2 | Not-determined | Genome-fragment | NA | | NA | 0 |
| S16_vs_4284\|\|full | 10946 | No | NA | | 14 | 3 | 0 | Low-quality | Genome-fragment | | 18.07 | HMM-based (lower-bound) | 0 |
| S8_vs_k141_173961\|\|full | 37216 | No | NA | | 62 | 20 | 0 | Medium-quality | Genome-fragment | | 88.33 | AAI-based (high-confidence) | 0 |
| S8_vs_k141_87460\|\|full | 2883 | No | NA | | 4 | 0 | 0 | Not-determined | Genome-fragment | NA | | NA | 0 |
| S8_vs_k141_165513\|\|full | 2050 | No | NA | | 3 | 0 | 1 | Not-determined | Genome-fragment | NA | | NA | 0 |
| S13_vs_60808\|\|full | 10949 | No | NA | | 4 | 1 | 0 | Low-quality | Genome-fragment | | 27.94 | AAI-based (high-confidence) | 0 |
| S8_vs_k141_17739\|\|full | 1395 | No | NA | | 4 | 0 | 1 | Not-determined | Genome-fragment | NA | | NA | 0 |
| S8_vs_k141_175167\|\|0_partial | 10950 | No | NA | | 9 | 0 | 7 | Low-quality | Genome-fragment | | 10.01 | AAI-based (medium-confidence) | 0 |
| S8_vs_k141_9254\|\|full | 2288 | No | NA | | 4 | 0 | 0 | Not-determined | Genome-fragment | NA | | NA | 0 |
| S5_vs_k141_71228\|\|full | 10959 | No | NA | | 18 | 12 | 0 | Low-quality | Genome-fragment | | 29.3 | AAI-based (high-confidence) | 0 |
| S8_vs_k141_147994\|\|full | 2582 | No | NA | | 4 | 0 | 2 | Not-determined | Genome-fragment | NA | | NA | 0 |
| S8_vs_k141_139391\|\|full | 26317 | No | NA | | 42 | 12 | 0 | Medium-quality | Genome-fragment | | 64.31 | AAI-based (high-confidence) | 0 |
| S8_vs_k141_122122\|\|full | 2280 | No | NA | | 3 | 0 | 1 | Not-determined | Genome-fragment | NA | | NA | 0 |
| S8_vs_k141_191609\|\|full | 1005 | No | NA | | 2 | 0 | 1 | Not-determined | Genome-fragment | NA | | NA | 0 |
| S15_extracted_30336 | 10962 | No | NA | | 14 | 0 | 4 | Low-quality | Genome-fragment | | 10.97 | AAI-based (medium-confidence) | 0 |
| S8_vs_k141_113313\|\|full | 2740 | No | NA | | 3 | 0 | 1 | Not-determined | Genome-fragment | NA | | NA | 0 |
| S8_vs_k141_35325\|\|full | 3003 | No | NA | | 3 | 0 | 2 | Not-determined | Genome-fragment | NA | | NA | 0 |
| S16_vs_64018\|\|full | 10965 | No | NA | | 15 | 3 | 0 | Low-quality | Genome-fragment | | 5.92 | AAI-based (high-confidence) | 0 |
| S8_vs_k141_122141\|\|full | 3689 | No | NA | | 3 | 0 | 2 | Not-determined | Genome-fragment | NA | | NA | 0 |
| S8_vs_k141_43870\|\|full | 1389 | No | NA | | 3 | 0 | 1 | Not-determined | Genome-fragment | NA | | NA | 0 |
| S8_vs_k141_139418\|\|full | 2102 | No | NA | | 2 | 0 | 2 | Not-determined | Genome-fragment | NA | | NA | 0 |
| S8_vs_k141_174042\|\|full | 5494 | No | NA | | 2 | 0 | 2 | Not-determined | Genome-fragment | NA | | NA | 0 |
| S8_vs_k141_78664\|\|0_partial | 10968 | No | NA | | 12 | 2 | 2 | Low-quality | Genome-fragment | | 13.52 | AAI-based (medium-confidence) | 0 |
| S8_vs_k141_139433\|\|full | 2014 | No | NA | | 3 | 0 | 1 | Not-determined | Genome-fragment | NA | | NA | 0 |
| S8_vs_k141_122152\|\|full | 1127 | No | NA | | 3 | 0 | 2 | Not-determined | Genome-fragment | NA | | NA | 0 |
| S13_extracted_148235 | 10970 | No | NA | | 17 | 12 | 0 | Low-quality | Genome-fragment | | 17.67 | AAI-based (high-confidence) | 0 |
| S3_extracted_k141_45216 | 10971 | No | NA | | 12 | 0 | 8 | Low-quality | Genome-fragment | | 3.07 | AAI-based (high-confidence) | 0 |
| S8_vs_k141_148070\|\|full | 2764 | No | NA | | 3 | 0 | 1 | Not-determined | Genome-fragment | NA | | NA | 0 |
| S8_vs_k141_169202\|\|full | 10971 | No | NA | | 25 | 3 | 2 | Low-quality | Genome-fragment | | 31.29 | AAI-based (high-confidence) | 0 |
| S8_vs_k141_61411\|\|full | 2208 | No | NA | | 4 | 0 | 0 | Not-determined | Genome-fragment | NA | | NA | 0 |
| S7_vs_120623\|\|0_partial | 10980 | No | NA | | 11 | 1 | 6 | Low-quality | Genome-fragment | | 15.43 | HMM-based (lower-bound) | 0 |
| S8_vs_k141_43909\|\|full | 1793 | No | NA | | 5 | 0 | 2 | Not-determined | Genome-fragment | NA | | NA | 0 |
| S5_vs_k141_62502\|\|full | 10986 | No | NA | | 15 | 3 | 0 | Low-quality | Genome-fragment | | 28.01 | AAI-based (medium-confidence) | 0 |
| S8_vs_k141_133228\|\|full | 10996 | No | NA | | 10 | 1 | 3 | Low-quality | Genome-fragment | | 6.5 | HMM-based (lower-bound) | 0 |
| S8_vs_k141_165630\|\|full | 1189 | No | NA | | 2 | 0 | 1 | Not-determined | Genome-fragment | NA | | NA | 0 |
| S11_vs_k141_46825\|\|full | 11001 | No | NA | | 13 | 0 | 5 | Low-quality | Genome-fragment | | 27.13 | AAI-based (high-confidence) | 0 |
| S8_vs_k141_745\|\|full | 2318 | No | NA | | 3 | 0 | 2 | Not-determined | Genome-fragment | NA | | NA | 0 |
| S8_vs_k141_104737\|\|full | 1216 | No | NA | | 2 | 0 | 1 | Not-determined | Genome-fragment | NA | | NA | 0 |
| S8_vs_k141_52784\|\|full | 1478 | No | NA | | 2 | 0 | 1 | Not-determined | Genome-fragment | NA | | NA | 0 |
| S8_vs_k141_139497\|\|full | 11407 | No | NA | | 11 | 0 | 9 | Not-determined | Genome-fragment | NA | | NA | 0 |
| S8_vs_k141_78784\|\|full | 1054 | No | NA | | 2 | 0 | 1 | Not-determined | Genome-fragment | NA | | NA | 0 |
| S5_extracted_k141_74316 | 11001 | No | NA | | 15 | 13 | 0 | Low-quality | Genome-fragment | | 24.11 | AAI-based (high-confidence) | 0 |
| S10_vs_17005\|\|full | 11001 | No | NA | | 17 | 2 | 1 | Low-quality | Genome-fragment | | 22.55 | HMM-based (lower-bound) | 0 |
| S14_vs_k141_30159\|\|full | 11002 | No | NA | | 8 | 0 | 1 | Low-quality | Genome-fragment | | 11.51 | AAI-based (medium-confidence) | 0 |
| S8_vs_k141_87627\|\|full | 2399 | No | NA | | 3 | 0 | 1 | Not-determined | Genome-fragment | NA | | NA | 0 |
| S8_vs_k141_139541\|\|full | 2621 | No | NA | | 4 | 0 | 0 | Not-determined | Genome-fragment | NA | | NA | 0 |
| S14_vs_k141_117149\|\|full | 11005 | No | NA | | 20 | 1 | 0 | Low-quality | Genome-fragment | | 14.32 | HMM-based (lower-bound) | 0 |
| S17_vs_k141_59730\|\|full | 11008 | No | NA | | 18 | 2 | 0 | Low-quality | Genome-fragment | | 24.47 | AAI-based (high-confidence) | 0 |
| S9_vs_k141_90875\|\|full | 11011 | No | NA | | 17 | 1 | 0 | Low-quality | Genome-fragment | | 17.25 | HMM-based (lower-bound) | 0 |
| S8_vs_k141_139557\|\|full | 2236 | No | NA | | 3 | 0 | 2 | Not-determined | Genome-fragment | NA | | NA | 0 |
| S8_vs_k141_156783\|\|full | 2068 | No | NA | | 3 | 0 | 0 | Not-determined | Genome-fragment | NA | | NA | 0 |
| S8_vs_k141_104800\|\|full | 5193 | No | NA | | 4 | 0 | 4 | Not-determined | Genome-fragment | NA | | NA | 0 |
| S8_vs_k141_824\|\|full | 3165 | No | NA | | 4 | 0 | 1 | Not-determined | Genome-fragment | NA | | NA | 0 |
| S8_vs_k141_78846\|\|full | 2287 | No | NA | | 5 | 0 | 1 | Not-determined | Genome-fragment | NA | | NA | 0 |
| S12_vs_k141_159933\|\|full | 11013 | No | NA | | 25 | 3 | 0 | Low-quality | Genome-fragment | | 19.65 | AAI-based (high-confidence) | 0 |
| S8_vs_k141_139579\|\|full | 3097 | No | NA | | 3 | 0 | 2 | Not-determined | Genome-fragment | NA | | NA | 0 |
| S12_vs_k141_27884\|\|full | 11016 | No | NA | | 10 | 1 | 0 | Low-quality | Genome-fragment | | 11.49 | HMM-based (lower-bound) | 0 |
| S8_vs_k141_35423\|\|full | 1786 | No | NA | | 2 | 0 | 1 | Not-determined | Genome-fragment | NA | | NA | 0 |
| S8_vs_k141_70035\|\|full | 2644 | No | NA | | 4 | 0 | 2 | Not-determined | Genome-fragment | NA | | NA | 0 |
| S5_vs_k141_112902\|\|full | 11029 | No | NA | | 18 | 8 | 0 | Low-quality | Genome-fragment | | 24.6 | AAI-based (high-confidence) | 0 |
| S8_vs_k141_853\|\|full | 1378 | No | NA | | 3 | 0 | 1 | Not-determined | Genome-fragment | NA | | NA | 0 |
| S5_vs_k141_42343\|\|full | 11033 | No | NA | | 19 | 3 | 2 | Low-quality | Genome-fragment | | 18.24 | HMM-based (lower-bound) | 0 |
| S1_extracted_k141_46477 | 11041 | No | NA | | 18 | 3 | 0 | Low-quality | Genome-fragment | | 24.33 | AAI-based (high-confidence) | 0 |
| S8_vs_k141_78870\|\|full | 1917 | No | NA | | 3 | 0 | 2 | Not-determined | Genome-fragment | NA | | NA | 0 |
| S5_vs_k141_114802\|\|full | 11070 | No | NA | | 15 | 1 | 2 | Low-quality | Genome-fragment | | 12.43 | HMM-based (lower-bound) | 0 |
| S8_vs_k141_131000\|\|full | 4957 | No | NA | | 7 | 0 | 1 | Not-determined | Genome-fragment | NA | | NA | 0 |
| S3_vs_k141_6985\|\|0_partial | 11075 | No | NA | | 14 | 1 | 7 | Low-quality | Genome-fragment | | 3.97 | HMM-based (lower-bound) | 0 |
| S8_vs_k141_26758\|\|full | 1692 | No | NA | | 3 | 0 | 1 | Not-determined | Genome-fragment | NA | | NA | 0 |
| S8_vs_k141_122296\|\|full | 1435 | No | NA | | 3 | 0 | 0 | Not-determined | Genome-fragment | NA | | NA | 0 |
| S11_extracted_k141_8405 | 11078 | No | NA | | 5 | 0 | 0 | Low-quality | Genome-fragment | | 6.78 | AAI-based (high-confidence) | 0 |
| S16_vs_73059\|\|full | 11079 | No | NA | | 9 | 3 | 0 | Low-quality | Genome-fragment | | 24.57 | AAI-based (high-confidence) | 0 |
| S1_extracted_k141_117795 | 11082 | Yes | | 6085 | 14 | 5 | 2 | Low-quality | Genome-fragment | | 10.41 | AAI-based (high-confidence) | 45.09 |
| S7_vs_61546\|\|full | 11107 | No | NA | | 23 | 1 | 0 | Low-quality | Genome-fragment | | 23.58 | AAI-based (high-confidence) | 0 |
| S8_vs_k141_35469\|\|full | 1191 | No | NA | | 2 | 0 | 1 | Not-determined | Genome-fragment | NA | | NA | 0 |
| S16_extracted_68689 | 11113 | No | NA | | 11 | 2 | 0 | Low-quality | Genome-fragment | | 14.11 | AAI-based (high-confidence) | 0 |
| S16_vs_28256\|\|full | 11122 | No | NA | | 17 | 13 | 0 | Low-quality | Genome-fragment | | 25.59 | AAI-based (high-confidence) | 0 |
| S8_vs_k141_893\|\|full | 2248 | No | NA | | 3 | 0 | 2 | Not-determined | Genome-fragment | NA | | NA | 0 |
| S8_extracted_k141_6026 | 11126 | No | NA | | 16 | 11 | 0 | Low-quality | Genome-fragment | | 16.46 | HMM-based (lower-bound) | 0 |
| S8_vs_k141_9460\|\|full | 6240 | No | NA | | 7 | 0 | 1 | Not-determined | Genome-fragment | NA | | NA | 0 |
| S8_vs_k141_63567\|\|full | 11142 | No | NA | | 14 | 2 | 0 | Low-quality | Genome-fragment | | 13.59 | AAI-based (high-confidence) | 0 |
| S8_vs_k141_44132\|\|full | 1149 | No | NA | | 2 | 0 | 0 | Not-determined | Genome-fragment | NA | | NA | 0 |
| S8_vs_k141_79568\|\|full | 11152 | No | NA | | 6 | 4 | 0 | Low-quality | Genome-fragment | | 18.93 | AAI-based (high-confidence) | 0 |
| S16_extracted_7504 | 11158 | No | NA | | 6 | 0 | 5 | Low-quality | Genome-fragment | | 7.23 | AAI-based (medium-confidence) | 0 |
| S8_vs_k141_35483\|\|full | 8672 | No | NA | | 9 | 0 | 1 | Not-determined | Genome-fragment | NA | | NA | 0 |
| S12_vs_k141_43013\|\|full | 11158 | No | NA | | 13 | 5 | 0 | Low-quality | Genome-fragment | | 21.67 | AAI-based (high-confidence) | 0 |
| S8_vs_k141_148209\|\|full | 3212 | No | NA | | 3 | 0 | 2 | Not-determined | Genome-fragment | NA | | NA | 0 |
| S8_vs_k141_156843\|\|full | 2876 | No | NA | | 4 | 0 | 0 | Not-determined | Genome-fragment | NA | | NA | 0 |
| S8_vs_k141_113473\|\|full | 2704 | No | NA | | 3 | 0 | 2 | Not-determined | Genome-fragment | NA | | NA | 0 |
| S8_vs_k141_61568\|\|full | 4386 | No | NA | | 3 | 0 | 1 | Not-determined | Genome-fragment | NA | | NA | 0 |
| S8_vs_k141_9496\|\|full | 6447 | No | NA | | 5 | 0 | 3 | Not-determined | Genome-fragment | NA | | NA | 0 |
| S8_vs_k141_35522\|\|full | 2144 | No | NA | | 2 | 0 | 1 | Not-determined | Genome-fragment | NA | | NA | 0 |
| S8_vs_k141_18013\|\|full | 3810 | No | NA | | 5 | 0 | 5 | Not-determined | Genome-fragment | NA | | NA | 0 |
| S14_vs_k141_71087\|\|full | 11166 | No | NA | | 27 | 2 | 0 | Low-quality | Genome-fragment | | 28.76 | AAI-based (high-confidence) | 0 |
| S1_extracted_k141_85219 | 11189 | No | NA | | 22 | 4 | 0 | Low-quality | Genome-fragment | | 24.25 | AAI-based (high-confidence) | 0 |
| S8_vs_k141_35539\|\|full | 2206 | No | NA | | 5 | 0 | 0 | Not-determined | Genome-fragment | NA | | NA | 0 |
| S8_vs_k141_44206\|\|full | 3297 | No | NA | | 3 | 0 | 2 | Not-determined | Genome-fragment | NA | | NA | 0 |
| S8_vs_k141_131037\|\|full | 1032 | No | NA | | 2 | 0 | 2 | Not-determined | Genome-fragment | NA | | NA | 0 |
| S12_vs_k141_40246\|\|full | 11213 | No | NA | | 13 | 3 | 0 | Low-quality | Genome-fragment | | 26.95 | AAI-based (medium-confidence) | 0 |
| S3_vs_k141_65147\|\|full | 11219 | Yes | | 6433 | 15 | 3 | 2 | Low-quality | Genome-fragment | | 11.47 | AAI-based (high-confidence) | 42.66 |
| S8_vs_k141_122420\|\|full | 1042 | No | NA | | 2 | 0 | 1 | Not-determined | Genome-fragment | NA | | NA | 0 |
| S5_vs_k141_133803\|\|full | 11219 | No | NA | | 14 | 12 | 0 | Low-quality | Genome-fragment | | 24.94 | AAI-based (medium-confidence) | 0 |
| S17_vs_k141_135182\|\|full | 11230 | No | NA | | 19 | 1 | 2 | Low-quality | Genome-fragment | | 6.67 | HMM-based (lower-bound) | 0 |
| S8_vs_k141_156903\|\|full | 7663 | No | NA | | 9 | 0 | 2 | Not-determined | Genome-fragment | NA | | NA | 0 |
| S8_vs_k141_26904\|\|full | 1082 | No | NA | | 1 | 0 | 0 | Not-determined | Genome-fragment | NA | | NA | 0 |
| S16_vs_53013\|\|full | 11234 | No | NA | | 15 | 1 | 0 | Low-quality | Genome-fragment | | 14.87 | HMM-based (lower-bound) | 0 |
| S8_vs_k141_87771\|\|full | 3052 | No | NA | | 3 | 0 | 2 | Not-determined | Genome-fragment | NA | | NA | 0 |
| S4_extracted_14454 | 11235 | No | NA | | 17 | 5 | 0 | Low-quality | Genome-fragment | | 10.28 | AAI-based (medium-confidence) | 0 |
| S17_vs_k141_9201\|\|full | 11236 | No | NA | | 22 | 2 | 0 | Low-quality | Genome-fragment | | 16.9 | AAI-based (medium-confidence) | 0 |
| S8_vs_k141_44236\|\|full | 4007 | No | NA | | 3 | 0 | 3 | Not-determined | Genome-fragment | NA | | NA | 0 |
| S7_extracted_17856 | 11254 | No | NA | | 15 | 4 | 1 | Low-quality | Genome-fragment | | 22.43 | AAI-based (high-confidence) | 0 |
| S3_vs_k141_94404\|\|full | 11268 | No | NA | | 13 | 0 | 3 | Low-quality | Genome-fragment | | 26.31 | AAI-based (medium-confidence) | 0 |
| S8_vs_k141_52980\|\|full | 6048 | No | NA | | 6 | 0 | 3 | Not-determined | Genome-fragment | NA | | NA | 0 |
| S8_vs_k141_44246\|\|full | 5793 | No | NA | | 5 | 0 | 5 | Not-determined | Genome-fragment | NA | | NA | 0 |
| S17_vs_k141_152433\|\|full | 11272 | No | NA | | 15 | 10 | 0 | Low-quality | Genome-fragment | | 30.21 | AAI-based (high-confidence) | 0 |
| S5_vs_k141_72732\|\|full | 11276 | No | NA | | 15 | 13 | 0 | Low-quality | Genome-fragment | | 22.84 | AAI-based (high-confidence) | 0 |
| S8_vs_k141_52985\|\|full | 7434 | No | NA | | 10 | 0 | 5 | Not-determined | Genome-fragment | NA | | NA | 0 |
| S8_vs_k141_44261\|\|full | 2911 | No | NA | | 3 | 0 | 2 | Not-determined | Genome-fragment | NA | | NA | 0 |
| S7_extracted_77212 | 11276 | No | NA | | 9 | 0 | 3 | Low-quality | Genome-fragment | | 7.35 | AAI-based (medium-confidence) | 0 |
| S1_vs_k141_153593\|\|full | 11277 | No | NA | | 15 | 9 | 0 | Low-quality | Genome-fragment | | 29.76 | AAI-based (high-confidence) | 0 |
| S8_vs_k141_35657\|\|full | 1817 | No | NA | | 3 | 0 | 1 | Not-determined | Genome-fragment | NA | | NA | 0 |
| S7_extracted_134929 | 11277 | Yes | | 3032 | 6 | 1 | 1 | Low-quality | Genome-fragment | | 1.22 | HMM-based (lower-bound) | 73.11 |
| S8_vs_k141_44267\|\|full | 9025 | No | NA | | 15 | 0 | 2 | Not-determined | Genome-fragment | NA | | NA | 0 |
| S8_vs_k141_200213\|\|full | 4242 | No | NA | | 7 | 0 | 5 | Not-determined | Genome-fragment | NA | | NA | 0 |
| S8_vs_k141_131710\|\|full | 11279 | No | NA | | 15 | 4 | 1 | Low-quality | Genome-fragment | | 15.39 | HMM-based (lower-bound) | 0 |
| S1_extracted_k141_17249 | 11282 | No | NA | | 16 | 6 | 0 | Low-quality | Genome-fragment | | 19.58 | AAI-based (high-confidence) | 0 |
| S16_vs_144307\|\|full | 11291 | No | NA | | 4 | 0 | 0 | Low-quality | Genome-fragment | | 27.91 | AAI-based (medium-confidence) | 0 |
| S8_vs_k141_18095\|\|full | 33715 | No | NA | | 28 | 0 | 18 | Not-determined | Genome-fragment | NA | | NA | 0 |
| S14_vs_k141_27035\|\|full | 11293 | No | NA | | 27 | 2 | 0 | Low-quality | Genome-fragment | | 15.56 | AAI-based (high-confidence) | 0 |
| S8_vs_k141_35688\|\|full | 2281 | No | NA | | 3 | 0 | 3 | Not-determined | Genome-fragment | NA | | NA | 0 |
| S8_vs_k141_139828\|\|full | 1215 | No | NA | | 3 | 0 | 0 | Not-determined | Genome-fragment | NA | | NA | 0 |
| S8_vs_k141_165839\|\|full | 1789 | No | NA | | 3 | 0 | 2 | Not-determined | Genome-fragment | NA | | NA | 0 |
| S8_vs_k141_192026\|\|full | 4785 | No | NA | | 7 | 0 | 0 | Not-determined | Genome-fragment | NA | | NA | 0 |
| S8_vs_k141_131184\|\|full | 1897 | No | NA | | 2 | 0 | 2 | Not-determined | Genome-fragment | NA | | NA | 0 |
| S8_vs_k141_9599\|\|full | 1862 | No | NA | | 2 | 0 | 1 | Not-determined | Genome-fragment | NA | | NA | 0 |
| S8_vs_k141_79128\|\|full | 3222 | No | NA | | 4 | 0 | 3 | Not-determined | Genome-fragment | NA | | NA | 0 |
| S8_vs_k141_139840\|\|full | 2099 | No | NA | | 2 | 0 | 1 | Not-determined | Genome-fragment | NA | | NA | 0 |
| S8_vs_k141_87870\|\|full | 1762 | No | NA | | 3 | 0 | 0 | Not-determined | Genome-fragment | NA | | NA | 0 |
| S8_vs_k141_200258\|\|full | 1187 | No | NA | | 2 | 0 | 0 | Not-determined | Genome-fragment | NA | | NA | 0 |
| S8_vs_k141_79136\|\|full | 5140 | No | NA | | 5 | 0 | 2 | Not-determined | Genome-fragment | NA | | NA | 0 |
| S10_vs_103945\|\|full | 11321 | No | NA | | 15 | 3 | 0 | Low-quality | Genome-fragment | | 10.2 | AAI-based (medium-confidence) | 0 |
| S7_vs_19058\|\|full | 11331 | No | NA | | 11 | 6 | 0 | Low-quality | Genome-fragment | | 23.77 | AAI-based (medium-confidence) | 0 |
| S11_vs_k141_52845\|\|full | 11348 | No | NA | | 10 | 3 | 1 | Low-quality | Genome-fragment | | 16.4 | AAI-based (medium-confidence) | 0 |
| S8_vs_k141_35747\|\|full | 1601 | No | NA | | 3 | 0 | 0 | Not-determined | Genome-fragment | NA | | NA | 0 |
| S8_vs_k141_27043\|\|full | 2504 | No | NA | | 3 | 0 | 0 | Not-determined | Genome-fragment | NA | | NA | 0 |
| S8_extracted_k141_78480 | 11359 | No | NA | | 21 | 2 | 0 | Low-quality | Genome-fragment | | 26.53 | AAI-based (high-confidence) | 0 |
| S8_vs_k141_96427\|\|full | 4188 | No | NA | | 4 | 0 | 2 | Not-determined | Genome-fragment | NA | | NA | 0 |
| S15_extracted_45192 | 11361 | No | NA | | 16 | 1 | 0 | Low-quality | Genome-fragment | | 12.37 | AAI-based (high-confidence) | 0 |
| S8_vs_k141_148389\|\|full | 37731 | No | NA | | 44 | 3 | 1 | Medium-quality | Genome-fragment | | 83.4 | AAI-based (high-confidence) | 0 |
| S15_vs_28020\|\|full | 11363 | No | NA | | 10 | 1 | 2 | Low-quality | Genome-fragment | | 23.29 | HMM-based (lower-bound) | 0 |
| S8_vs_k141_113758\|\|full | 3075 | No | NA | | 3 | 0 | 3 | Not-determined | Genome-fragment | NA | | NA | 0 |
| S5_vs_k141_8988\|\|0_partial | 11365 | No | NA | | 11 | 0 | 9 | Low-quality | Genome-fragment | | 7.37 | AAI-based (medium-confidence) | 0 |
| S8_vs_k141_165920\|\|full | 2964 | No | NA | | 5 | 0 | 1 | Not-determined | Genome-fragment | NA | | NA | 0 |
| S8_vs_k141_192120\|\|full | 3034 | No | NA | | 5 | 0 | 5 | Not-determined | Genome-fragment | NA | | NA | 0 |
| S8_vs_k141_192127\|\|full | 4166 | No | NA | | 5 | 0 | 3 | Not-determined | Genome-fragment | NA | | NA | 0 |
| S3_extracted_k141_20391 | 11374 | No | NA | | 13 | 6 | 0 | Low-quality | Genome-fragment | | 38.28 | HMM-based (lower-bound) | 0 |
| S8_vs_k141_27136\|\|full | 1235 | No | NA | | 3 | 0 | 0 | Not-determined | Genome-fragment | NA | | NA | 0 |
| S8_vs_k141_61806\|\|full | 1665 | No | NA | | 3 | 0 | 0 | Not-determined | Genome-fragment | NA | | NA | 0 |
| S8_vs_k141_96485\|\|full | 5637 | No | NA | | 3 | 0 | 2 | Not-determined | Genome-fragment | NA | | NA | 0 |
| S7_vs_122171\|\|full | 11378 | No | NA | | 14 | 6 | 0 | Low-quality | Genome-fragment | | 22.06 | AAI-based (high-confidence) | 0 |
| S5_extracted_k141_138039 | 11387 | No | NA | | 15 | 12 | 0 | Low-quality | Genome-fragment | | 32.76 | AAI-based (high-confidence) | 0 |
| S8_vs_k141_53123\|\|full | 1043 | No | NA | | 2 | 0 | 1 | Not-determined | Genome-fragment | NA | | NA | 0 |
| S8_vs_k141_157115\|\|full | 3226 | No | NA | | 2 | 0 | 1 | Not-determined | Genome-fragment | NA | | NA | 0 |
| S12_extracted_k141_31564 | 11388 | No | NA | | 16 | 1 | 0 | Low-quality | Genome-fragment | | 4.9 | HMM-based (lower-bound) | 0 |
| S8_vs_k141_122625\|\|full | 3748 | No | NA | | 7 | 0 | 3 | Not-determined | Genome-fragment | NA | | NA | 0 |
| S8_vs_k141_174426\|\|full | 2603 | No | NA | | 2 | 0 | 1 | Not-determined | Genome-fragment | NA | | NA | 0 |
| S8_vs_k141_192175\|\|full | 1163 | No | NA | | 2 | 0 | 0 | Not-determined | Genome-fragment | NA | | NA | 0 |
| S8_vs_k141_27191\|\|full | 3163 | No | NA | | 3 | 0 | 2 | Not-determined | Genome-fragment | NA | | NA | 0 |
| S8_vs_k141_79314\|\|full | 1888 | No | NA | | 3 | 0 | 0 | Not-determined | Genome-fragment | NA | | NA | 0 |
| S8_vs_k141_9747\|\|full | 12429 | No | NA | | 11 | 0 | 7 | Not-determined | Genome-fragment | NA | | NA | 0 |
| S8_extracted_k141_165924 | 11388 | No | NA | | 14 | 12 | 0 | Low-quality | Genome-fragment | | 38.77 | AAI-based (high-confidence) | 0 |
| S8_vs_k141_44320\|\|full | 7635 | No | NA | | 4 | 0 | 2 | Not-determined | Genome-fragment | NA | | NA | 0 |
| S12_vs_k141_165729\|\|0_partial | 11405 | No | NA | | 11 | 1 | 1 | Low-quality | Genome-fragment | | 6.45 | HMM-based (lower-bound) | 0 |
| S16_extracted_27789 | 11413 | No | NA | | 18 | 11 | 0 | Low-quality | Genome-fragment | | 17.67 | AAI-based (high-confidence) | 0 |
| S8_vs_k141_200340\|\|full | 9373 | No | NA | | 13 | 0 | 11 | Not-determined | Genome-fragment | NA | | NA | 0 |
| S8_vs_k141_174451\|\|full | 1436 | No | NA | | 2 | 0 | 1 | Not-determined | Genome-fragment | NA | | NA | 0 |
| S16_extracted_23710 | 11422 | No | NA | | 13 | 10 | 0 | Low-quality | Genome-fragment | | 35.53 | AAI-based (high-confidence) | 0 |
| S11_vs_k141_43031\|\|full | 11428 | No | NA | | 10 | 0 | 5 | Low-quality | Genome-fragment | | 3.55 | AAI-based (medium-confidence) | 0 |
| S12_extracted_k141_8749 | 11450 | Yes | | 8941 | 8 | 1 | 1 | Low-quality | Genome-fragment | | 10.71 | AAI-based (medium-confidence) | 21.91 |
| S16_extracted_46894 | 11454 | No | NA | | 18 | 3 | 1 | Low-quality | Genome-fragment | | 30.25 | AAI-based (high-confidence) | 0 |
| S8_vs_k141_35975\|\|full | 4151 | No | NA | | 7 | 0 | 0 | Not-determined | Genome-fragment | NA | | NA | 0 |
| S8_vs_k141_166011\|\|full | 2455 | No | NA | | 3 | 0 | 2 | Not-determined | Genome-fragment | NA | | NA | 0 |
| S8_vs_k141_44380\|\|full | 1749 | No | NA | | 3 | 0 | 3 | Not-determined | Genome-fragment | NA | | NA | 0 |
| S5_vs_k141_87728\|\|full | 11461 | No | NA | | 21 | 4 | 0 | Low-quality | Genome-fragment | | 18.54 | AAI-based (medium-confidence) | 0 |
| S8_vs_k141_105210\|\|full | 1227 | No | NA | | 2 | 0 | 0 | Not-determined | Genome-fragment | NA | | NA | 0 |
| S16_vs_26037\|\|full | 11467 | No | NA | | 13 | 4 | 0 | Low-quality | Genome-fragment | | 27.54 | AAI-based (high-confidence) | 0 |
| S8_vs_k141_148647\|\|full | 1972 | No | NA | | 4 | 0 | 0 | Not-determined | Genome-fragment | NA | | NA | 0 |
| S8_vs_k141_27303\|\|full | 4622 | No | NA | | 4 | 0 | 4 | Not-determined | Genome-fragment | NA | | NA | 0 |
| S16_vs_28240\|\|full | 11471 | No | NA | | 18 | 0 | 1 | Low-quality | Genome-fragment | | 26.13 | AAI-based (medium-confidence) | 0 |
| S8_vs_k141_105236\|\|full | 2008 | No | NA | | 2 | 0 | 1 | Not-determined | Genome-fragment | NA | | NA | 0 |
| S8_vs_k141_27313\|\|full | 1982 | No | NA | | 3 | 0 | 1 | Not-determined | Genome-fragment | NA | | NA | 0 |
| S8_vs_k141_148658\|\|full | 3582 | No | NA | | 4 | 0 | 2 | Not-determined | Genome-fragment | NA | | NA | 0 |
| S8_vs_k141_79415\|\|full | 2342 | No | NA | | 4 | 0 | 0 | Not-determined | Genome-fragment | NA | | NA | 0 |
| S16_vs_77473\|\|full | 11499 | No | NA | | 14 | 0 | 0 | Low-quality | Genome-fragment | | 6.4 | AAI-based (high-confidence) | 0 |
| S8_vs_k141_105269\|\|full | 6706 | No | NA | | 4 | 0 | 4 | Not-determined | Genome-fragment | NA | | NA | 0 |
| S8_vs_k141_166057\|\|full | 2049 | No | NA | | 2 | 0 | 1 | Not-determined | Genome-fragment | NA | | NA | 0 |
| S8_vs_k141_192341\|\|full | 3134 | No | NA | | 5 | 0 | 3 | Not-determined | Genome-fragment | NA | | NA | 0 |
| S3_vs_k141_86775\|\|full | 11501 | No | NA | | 25 | 3 | 0 | Low-quality | Genome-fragment | | 15.61 | AAI-based (medium-confidence) | 0 |
| S16_extracted_73955 | 11511 | No | NA | | 22 | 5 | 1 | Low-quality | Genome-fragment | | 19.71 | AAI-based (high-confidence) | 0 |
| S5_vs_k141_35298\|\|full | 11517 | No | NA | | 20 | 4 | 3 | Low-quality | Genome-fragment | | 27.56 | AAI-based (medium-confidence) | 0 |
| S8_vs_k141_79466\|\|full | 2147 | No | NA | | 2 | 0 | 1 | Not-determined | Genome-fragment | NA | | NA | 0 |
| S3_vs_k141_116740\|\|full | 11525 | No | NA | | 12 | 1 | 1 | Low-quality | Genome-fragment | | 5.62 | AAI-based (high-confidence) | 0 |
| S8_extracted_k141_201948 | 11549 | No | NA | | 10 | 0 | 7 | Low-quality | Genome-fragment | | 2 | AAI-based (medium-confidence) | 0 |
| S8_vs_k141_174530\|\|full | 1872 | No | NA | | 3 | 0 | 0 | Not-determined | Genome-fragment | NA | | NA | 0 |
| S7_vs_68451\|\|full | 11554 | No | NA | | 31 | 3 | 1 | Low-quality | Genome-fragment | | 33.98 | AAI-based (high-confidence) | 0 |
| S8_vs_k141_174536\|\|full | 2833 | No | NA | | 3 | 0 | 3 | Not-determined | Genome-fragment | NA | | NA | 0 |
| S10_vs_89729\|\|full | 11573 | No | NA | | 16 | 3 | 0 | Low-quality | Genome-fragment | | 7.1 | AAI-based (high-confidence) | 0 |
| S5_extracted_k141_54598 | 11574 | No | NA | | 24 | 7 | 1 | Low-quality | Genome-fragment | | 20.17 | AAI-based (high-confidence) | 0 |
| S12_extracted_k141_143865 | 11582 | No | NA | | 17 | 2 | 1 | Low-quality | Genome-fragment | | 10.28 | HMM-based (lower-bound) | 0 |
| S8_vs_k141_44451\|\|full | 1489 | No | NA | | 4 | 0 | 1 | Not-determined | Genome-fragment | NA | | NA | 0 |
| S11_extracted_k141_96649 | 11583 | No | NA | | 6 | 0 | 0 | Low-quality | Genome-fragment | | 7.11 | AAI-based (high-confidence) | 0 |
| S8_vs_k141_88124\|\|full | 6581 | No | NA | | 7 | 0 | 0 | Not-determined | Genome-fragment | NA | | NA | 0 |
| S8_vs_k141_200508\|\|full | 5702 | No | NA | | 2 | 0 | 0 | Not-determined | Genome-fragment | NA | | NA | 0 |
| S16_vs_119370\|\|full | 11596 | No | NA | | 16 | 6 | 0 | Low-quality | Genome-fragment | | 29.62 | AAI-based (high-confidence) | 0 |
| S8_vs_k141_96713\|\|full | 1131 | No | NA | | 2 | 0 | 1 | Not-determined | Genome-fragment | NA | | NA | 0 |
| S5_vs_k141_130158\|\|full | 11604 | No | NA | | 17 | 16 | 0 | Low-quality | Genome-fragment | | 33.52 | AAI-based (medium-confidence) | 0 |
| S3_extracted_k141_111315 | 11608 | No | NA | | 8 | 4 | 0 | Low-quality | Genome-fragment | | 25.39 | AAI-based (medium-confidence) | 0 |
| S8_vs_k141_131482\|\|full | 12701 | No | NA | | 17 | 0 | 9 | Not-determined | Genome-fragment | NA | | NA | 0 |
| S8_vs_k141_122848\|\|full | 3022 | No | NA | | 4 | 0 | 2 | Not-determined | Genome-fragment | NA | | NA | 0 |
| S8_vs_k141_148768\|\|full | 2954 | No | NA | | 3 | 0 | 3 | Not-determined | Genome-fragment | NA | | NA | 0 |
| S11_vs_k141_45044\|\|full | 11615 | No | NA | | 19 | 7 | 0 | Low-quality | Genome-fragment | | 25.72 | AAI-based (high-confidence) | 0 |
| S8_vs_k141_174591\|\|full | 2006 | No | NA | | 2 | 0 | 1 | Not-determined | Genome-fragment | NA | | NA | 0 |
| S16_vs_171302\|\|0_partial | 11615 | No | NA | | 13 | 1 | 7 | Low-quality | Genome-fragment | | 20.08 | AAI-based (medium-confidence) | 0 |
| S1_extracted_k141_54653 | 11617 | No | NA | | 24 | 3 | 1 | Low-quality | Genome-fragment | | 30.24 | AAI-based (high-confidence) | 0 |
| S8_vs_k141_174595\|\|full | 2562 | No | NA | | 3 | 0 | 1 | Not-determined | Genome-fragment | NA | | NA | 0 |
| S8_vs_k141_62067\|\|full | 2031 | No | NA | | 4 | 0 | 0 | Not-determined | Genome-fragment | NA | | NA | 0 |
| S8_vs_k141_96735\|\|full | 2111 | No | NA | | 5 | 0 | 1 | Not-determined | Genome-fragment | NA | | NA | 0 |
| S12_extracted_k141_120553 | 11618 | No | NA | | 14 | 5 | 0 | Low-quality | Genome-fragment | | 38.67 | HMM-based (lower-bound) | 0 |
| S8_vs_k141_114003\|\|full | 1747 | No | NA | | 3 | 0 | 0 | Not-determined | Genome-fragment | NA | | NA | 0 |
| S7_extracted_22953 | 11619 | No | NA | | 11 | 1 | 6 | Low-quality | Genome-fragment | | 19.01 | HMM-based (lower-bound) | 0 |
| S16_vs_30449\|\|full | 11623 | No | NA | | 15 | 1 | 1 | Low-quality | Genome-fragment | | 28.14 | AAI-based (high-confidence) | 0 |
| S6_vs_k141_43653\|\|0_partial | 11631 | Yes | | 2316 | 10 | 1 | 6 | Low-quality | Genome-fragment | | 5 | AAI-based (medium-confidence) | 80.09 |
| S12_vs_k141_31878\|\|0_partial | 11663 | Yes | | 10877 | 9 | 1 | 1 | Low-quality | Genome-fragment | | 17 | AAI-based (medium-confidence) | 6.74 |
| S8_vs_k141_192444\|\|full | 6901 | No | NA | | 8 | 0 | 2 | Not-determined | Genome-fragment | NA | | NA | 0 |
| S8_vs_k141_157378\|\|full | 4186 | No | NA | | 2 | 0 | 2 | Not-determined | Genome-fragment | NA | | NA | 0 |
| S8_vs_k141_27531\|\|full | 2291 | No | NA | | 3 | 0 | 0 | Not-determined | Genome-fragment | NA | | NA | 0 |
| S16_vs_62081\|\|full | 11667 | No | NA | | 15 | 2 | 2 | Low-quality | Genome-fragment | | 36.75 | AAI-based (high-confidence) | 0 |
| S8_vs_k141_1434\|\|full | 1514 | No | NA | | 3 | 0 | 0 | Not-determined | Genome-fragment | NA | | NA | 0 |
| S8_vs_k141_166183\|\|full | 1138 | No | NA | | 3 | 0 | 1 | Not-determined | Genome-fragment | NA | | NA | 0 |
| S8_vs_k141_140175\|\|full | 6459 | No | NA | | 6 | 0 | 2 | Not-determined | Genome-fragment | NA | | NA | 0 |
| S8_vs_k141_122904\|\|full | 1143 | No | NA | | 2 | 0 | 1 | Not-determined | Genome-fragment | NA | | NA | 0 |
| S5_vs_k141_74507\|\|full | 11674 | No | NA | | 27 | 1 | 0 | Low-quality | Genome-fragment | | 26.93 | AAI-based (medium-confidence) | 0 |
| S14_extracted_k141_96127 | 11675 | No | NA | | 12 | 2 | 0 | Low-quality | Genome-fragment | | 29.55 | AAI-based (high-confidence) | 0 |
| S8_vs_k141_1471\|\|full | 4928 | No | NA | | 3 | 0 | 2 | Not-determined | Genome-fragment | NA | | NA | 0 |
| S8_vs_k141_53482\|\|full | 1431 | No | NA | | 3 | 0 | 1 | Not-determined | Genome-fragment | NA | | NA | 0 |
| S7_vs_62791\|\|full | 11675 | No | NA | | 23 | 6 | 1 | Low-quality | Genome-fragment | | 23.11 | AAI-based (medium-confidence) | 0 |
| S8_vs_k141_140231\|\|full | 2446 | No | NA | | 3 | 0 | 1 | Not-determined | Genome-fragment | NA | | NA | 0 |
| S14_vs_k141_90303\|\|full | 11687 | No | NA | | 18 | 9 | 1 | Low-quality | Genome-fragment | | 29.58 | AAI-based (high-confidence) | 0 |
| S14_vs_k141_45422\|\|full | 11688 | No | NA | | 13 | 1 | 0 | Low-quality | Genome-fragment | | 26.95 | AAI-based (medium-confidence) | 0 |
| S8_vs_k141_44581\|\|full | 3145 | No | NA | | 4 | 0 | 1 | Not-determined | Genome-fragment | NA | | NA | 0 |
| S8_vs_k141_122933\|\|full | 3368 | No | NA | | 4 | 0 | 3 | Not-determined | Genome-fragment | NA | | NA | 0 |
| S8_vs_k141_53501\|\|full | 1822 | No | NA | | 2 | 0 | 0 | Not-determined | Genome-fragment | NA | | NA | 0 |
| S1_vs_k141_76130\|\|full | 11692 | No | NA | | 8 | 0 | 3 | Low-quality | Genome-fragment | | 3.65 | AAI-based (medium-confidence) | 0 |
| S8_vs_k141_27602\|\|full | 3716 | No | NA | | 6 | 0 | 1 | Not-determined | Genome-fragment | NA | | NA | 0 |
| S3_vs_k141_113081\|\|full | 11702 | No | NA | | 7 | 1 | 0 | Low-quality | Genome-fragment | | 43.05 | HMM-based (lower-bound) | 0 |
| S8_vs_k141_9959\|\|full | 1697 | No | NA | | 2 | 0 | 1 | Not-determined | Genome-fragment | NA | | NA | 0 |

| | | | | | | | | | | | |
|---|---|---|---|---|---|---|---|---|---|---|---|
| S5_extracted_k141_51810 | 11719 | No | NA | 17 | 12 | 0 | Low-quality | Genome-fragment | 29.6 | AAI-based (medium-confidence) | 0 |
| S8_vs_k141_44813\|\|full | 11720 | No | NA | 24 | 4 | 0 | Low-quality | Genome-fragment | 36.47 | AAI-based (high-confidence) | 0 |
| S12_vs_k141_51104\|\|0_partial | 11723 | Yes | 2047 | 8 | 1 | 5 | Low-quality | Genome-fragment | 4.49 | AAI-based (medium-confidence) | 82.54 |
| S3_vs_k141_79379\|\|full | 11730 | No | NA | 13 | 4 | 1 | Low-quality | Genome-fragment | 31.77 | AAI-based (high-confidence) | 0 |
| S14_extracted_k141_103234 | 11731 | No | NA | 20 | 5 | 0 | Low-quality | Genome-fragment | 30.06 | AAI-based (high-confidence) | 0 |
| S8_vs_k141_70599\|\|full | 2107 | No | NA | 2 | 0 | 0 | Not-determined | Genome-fragment | NA | NA | 0 |
| S16_vs_76001\|\|full | 11746 | No | NA | 11 | 3 | 0 | Low-quality | Genome-fragment | 31.57 | AAI-based (high-confidence) | 0 |
| S8_vs_k141_27641\|\|full | 3727 | No | NA | 2 | 0 | 1 | Not-determined | Genome-fragment | NA | NA | 0 |
| S8_vs_k141_140296\|\|full | 1473 | No | NA | 3 | 0 | 0 | Not-determined | Genome-fragment | NA | NA | 0 |
| S5_vs_k141_117798\|\|full | 11749 | No | NA | 13 | 7 | 0 | Low-quality | Genome-fragment | 24.91 | AAI-based (high-confidence) | 0 |
| S1_vs_k141_27585\|\|full | 11753 | No | NA | 24 | 2 | 0 | Low-quality | Genome-fragment | 27.53 | AAI-based (high-confidence) | 0 |
| S11_vs_k141_66678\|\|full | 11759 | Yes | 7780 | 18 | 2 | 2 | Low-quality | Genome-fragment | 13.04 | HMM-based (lower-bound) | 33.84 |
| S8_vs_k141_27669\|\|full | 1228 | No | NA | 3 | 0 | 2 | Not-determined | Genome-fragment | NA | NA | 0 |
| S8_vs_k141_105544\|\|full | 3129 | No | NA | 4 | 0 | 0 | Not-determined | Genome-fragment | NA | NA | 0 |
| S8_vs_k141_157543\|\|full | 5217 | No | NA | 10 | 0 | 0 | Not-determined | Genome-fragment | NA | NA | 0 |
| S8_vs_k141_44633\|\|full | 1390 | No | NA | 3 | 0 | 1 | Not-determined | Genome-fragment | NA | NA | 0 |
| S8_vs_k141_114505\|\|0_partial | 11762 | Yes | 4667 | 8 | 1 | 4 | Low-quality | Genome-fragment | 22.37 | AAI-based (high-confidence) | 60.32 |
| S8_vs_k141_27683\|\|full | 1218 | No | NA | 2 | 0 | 1 | Not-determined | Genome-fragment | NA | NA | 0 |
| S3_extracted_k141_4403 | 11768 | No | NA | 21 | 1 | 0 | Low-quality | Genome-fragment | 27.47 | AAI-based (high-confidence) | 0 |
| S8_vs_k141_166299\|\|full | 3694 | No | NA | 5 | 0 | 2 | Not-determined | Genome-fragment | NA | NA | 0 |
| S3_vs_k141_42933\|\|full | 11781 | No | NA | 17 | 1 | 2 | Low-quality | Genome-fragment | 5.94 | AAI-based (high-confidence) | 0 |
| S5_vs_k141_43334\|\|full | 11781 | No | NA | 11 | 5 | 0 | Low-quality | Genome-fragment | 28.84 | AAI-based (medium-confidence) | 0 |
| S16_vs_142513\|\|full | 11782 | No | NA | 13 | 1 | 3 | Low-quality | Genome-fragment | 10.31 | HMM-based (lower-bound) | 0 |
| S10_vs_68606\|\|0_partial | 11782 | No | NA | 10 | 0 | 8 | Low-quality | Genome-fragment | 10.77 | AAI-based (medium-confidence) | 0 |
| S5_vs_k141_86243\|\|0_partial | 11794 | Yes | 8283 | 13 | 2 | 4 | Low-quality | Genome-fragment | 5.6 | AAI-based (medium-confidence) | 29.77 |
| S8_vs_k141_166324\|\|full | 5291 | No | NA | 6 | 0 | 3 | Not-determined | Genome-fragment | NA | NA | 0 |
| S7_vs_30140\|\|full | 11799 | No | NA | 14 | 9 | 0 | Low-quality | Genome-fragment | 41.07 | AAI-based (high-confidence) | 0 |
| S10_extracted_76938 | 11805 | No | NA | 15 | 8 | 0 | Low-quality | Genome-fragment | 35.03 | HMM-based (lower-bound) | 0 |
| S8_vs_k141_62199\|\|full | 1582 | No | NA | 4 | 0 | 0 | Not-determined | Genome-fragment | NA | NA | 0 |
| S1_vs_k141_103310\|\|full | 11809 | No | NA | 21 | 3 | 0 | Low-quality | Genome-fragment | 29.11 | AAI-based (high-confidence) | 0 |
| S8_vs_k141_79731\|\|full | 1598 | No | NA | 2 | 0 | 1 | Not-determined | Genome-fragment | NA | NA | 0 |
| S8_vs_k141_27736\|\|full | 1704 | No | NA | 4 | 0 | 0 | Not-determined | Genome-fragment | NA | NA | 0 |
| S12_vs_k141_127678\|\|full | 11811 | No | NA | 7 | 1 | 6 | Low-quality | Genome-fragment | 6.35 | HMM-based (lower-bound) | 0 |
| S12_vs_k141_174415\|\|full | 11820 | No | NA | 18 | 8 | 1 | Low-quality | Genome-fragment | 29.04 | AAI-based (high-confidence) | 0 |
| S14_vs_k141_21408\|\|full | 11824 | No | NA | 13 | 8 | 0 | Low-quality | Genome-fragment | 19.81 | AAI-based (high-confidence) | 0 |
| S8_vs_k141_1630\|\|full | 2456 | No | NA | 3 | 0 | 0 | Not-determined | Genome-fragment | NA | NA | 0 |
| S8_vs_k141_18695\|\|full | 2510 | No | NA | 4 | 0 | 2 | Not-determined | Genome-fragment | NA | NA | 0 |
| S8_vs_k141_27753\|\|full | 4627 | No | NA | 7 | 0 | 5 | Not-determined | Genome-fragment | NA | NA | 0 |
| S8_vs_k141_88226\|\|full | 1903 | No | NA | 2 | 0 | 0 | Not-determined | Genome-fragment | NA | NA | 0 |
| S12_extracted_k141_140093 | 11831 | No | NA | 15 | 7 | 0 | Low-quality | Genome-fragment | 32.22 | HMM-based (lower-bound) | 0 |
| S8_vs_k141_123086\|\|full | 3272 | No | NA | 2 | 0 | 1 | Not-determined | Genome-fragment | NA | NA | 0 |
| S5_vs_k141_144003\|\|full | 11835 | No | NA | 18 | 13 | 0 | Low-quality | Genome-fragment | 40.05 | AAI-based (high-confidence) | 0 |
| S17_vs_k141_163150\|\|full | 11851 | No | NA | 13 | 4 | 1 | Low-quality | Genome-fragment | 26.96 | AAI-based (high-confidence) | 0 |
| S8_vs_k141_62264\|\|full | 9661 | No | NA | 12 | 0 | 1 | Not-determined | Genome-fragment | NA | NA | 0 |
| S8_vs_k141_157571\|\|full | 3371 | No | NA | 3 | 0 | 0 | Not-determined | Genome-fragment | NA | NA | 0 |
| S8_vs_k141_97048\|\|full | 2722 | No | NA | 4 | 0 | 0 | Not-determined | Genome-fragment | NA | NA | 0 |
| S5_vs_k141_73268\|\|full | 11854 | Yes | 7001 | 18 | 4 | 4 | Low-quality | Genome-fragment | 18.07 | AAI-based (high-confidence) | 40.94 |
| S8_vs_k141_10068\|\|full | 1492 | No | NA | 3 | 0 | 0 | Not-determined | Genome-fragment | NA | NA | 0 |
| S8_vs_k141_53663\|\|full | 1372 | No | NA | 4 | 0 | 0 | Not-determined | Genome-fragment | NA | NA | 0 |
| S8_vs_k141_183315\|\|full | 1622 | No | NA | 2 | 0 | 0 | Not-determined | Genome-fragment | NA | NA | 0 |
| S8_vs_k141_149336\|\|0_partial | 11861 | No | NA | 11 | 1 | 2 | Low-quality | Genome-fragment | 6.64 | HMM-based (lower-bound) | 0 |
| S8_vs_k141_114287\|\|full | 2896 | No | NA | 5 | 0 | 4 | Not-determined | Genome-fragment | NA | NA | 0 |
| S8_extracted_k141_100986 | 11874 | No | NA | 15 | 12 | 0 | Low-quality | Genome-fragment | 36.01 | AAI-based (medium-confidence) | 0 |
| S1_vs_k141_119745\|\|full | 11875 | No | NA | 8 | 2 | 0 | Low-quality | Genome-fragment | 28.33 | AAI-based (medium-confidence) | 0 |
| S12_vs_k141_109862\|\|full | 11876 | No | NA | 14 | 3 | 0 | Low-quality | Genome-fragment | 19.03 | HMM-based (lower-bound) | 0 |
| S8_vs_k141_157598\|\|full | 2074 | No | NA | 4 | 0 | 1 | Not-determined | Genome-fragment | NA | NA | 0 |
| S8_vs_k141_36448\|\|full | 1903 | No | NA | 4 | 0 | 3 | Not-determined | Genome-fragment | NA | NA | 0 |
| S17_vs_k141_60196\|\|full | 11880 | No | NA | 18 | 5 | 0 | Low-quality | Genome-fragment | 19.76 | AAI-based (high-confidence) | 0 |
| S8_vs_k141_97090\|\|full | 1550 | No | NA | 2 | 0 | 0 | Not-determined | Genome-fragment | NA | NA | 0 |
| S8_vs_k141_166439\|\|full | 2140 | No | NA | 3 | 0 | 1 | Not-determined | Genome-fragment | NA | NA | 0 |
| S12_extracted_k141_138733 | 11900 | No | NA | 20 | 5 | 0 | Low-quality | Genome-fragment | 39.61 | HMM-based (lower-bound) | 0 |
| S8_vs_k141_44771\|\|full | 3859 | No | NA | 3 | 0 | 3 | Not-determined | Genome-fragment | NA | NA | 0 |
| S12_vs_k141_188648\|\|full | 11901 | No | NA | 19 | 6 | 0 | Low-quality | Genome-fragment | 40.06 | HMM-based (lower-bound) | 0 |
| S8_vs_k141_157650\|\|full | 1003 | No | NA | 2 | 0 | 1 | Not-determined | Genome-fragment | NA | NA | 0 |
| S6_vs_k141_38813\|\|full | 11903 | No | NA | 22 | 10 | 0 | Low-quality | Genome-fragment | 31.75 | AAI-based (high-confidence) | 0 |
| S8_vs_k141_183347\|\|full | 2796 | No | NA | 4 | 0 | 0 | Not-determined | Genome-fragment | NA | NA | 0 |
| S16_vs_165292\|\|1_partial | 11907 | No | NA | 13 | 1 | 2 | Low-quality | Genome-fragment | 17.81 | HMM-based (lower-bound) | 0 |
| S8_vs_k141_36512\|\|full | 2212 | No | NA | 2 | 0 | 2 | Not-determined | Genome-fragment | NA | NA | 0 |
| S12_vs_k141_24899\|\|0_partial | 11907 | Yes | 6051 | 10 | 1 | 4 | Low-quality | Genome-fragment | 3.34 | HMM-based (lower-bound) | 49.18 |
| S1_vs_k141_143956\|\|full | 11916 | No | NA | 19 | 3 | 0 | Low-quality | Genome-fragment | 31.06 | AAI-based (high-confidence) | 0 |
| S8_vs_k141_10166\|\|full | 2444 | No | NA | 2 | 0 | 2 | Not-determined | Genome-fragment | NA | NA | 0 |
| S14_extracted_k141_14147 | 11928 | No | NA | 15 | 3 | 0 | Low-quality | Genome-fragment | 20.34 | AAI-based (high-confidence) | 0 |
| S8_vs_k141_18852\|\|full | 3392 | No | NA | 3 | 0 | 2 | Not-determined | Genome-fragment | NA | NA | 0 |
| S7_vs_36054\|\|full | 11939 | No | NA | 17 | 15 | 0 | Low-quality | Genome-fragment | 26.04 | AAI-based (high-confidence) | 0 |
| S8_vs_k141_97156\|\|full | 27934 | No | NA | 27 | 0 | 24 | Not-determined | Genome-fragment | NA | NA | 0 |
| S5_vs_k141_114653\|\|full | 11946 | No | NA | 24 | 4 | 0 | Low-quality | Genome-fragment | 17.81 | HMM-based (lower-bound) | 0 |
| S8_vs_k141_1766\|\|full | 2221 | No | NA | 3 | 0 | 2 | Not-determined | Genome-fragment | NA | NA | 0 |
| S8_vs_k141_166566\|\|full | 1897 | No | NA | 3 | 0 | 0 | Not-determined | Genome-fragment | NA | NA | 0 |
| S11_vs_k141_52974\|\|full | 11958 | No | NA | 26 | 5 | 0 | Low-quality | Genome-fragment | 14.49 | AAI-based (high-confidence) | 0 |
| S8_vs_k141_36585\|\|full | 4749 | No | NA | 4 | 0 | 3 | Not-determined | Genome-fragment | NA | NA | 0 |
| S8_vs_k141_88440\|\|full | 3716 | No | NA | 3 | 0 | 0 | Not-determined | Genome-fragment | NA | NA | 0 |
| S8_vs_k141_123244\|\|full | 7311 | No | NA | 6 | 0 | 3 | Not-determined | Genome-fragment | NA | NA | 0 |
| S8_vs_k141_149011\|\|full | 3477 | No | NA | 7 | 0 | 3 | Not-determined | Genome-fragment | NA | NA | 0 |
| S8_vs_k141_97215\|\|full | 1326 | No | NA | 2 | 0 | 0 | Not-determined | Genome-fragment | NA | NA | 0 |
| S8_vs_k141_149017\|\|full | 1383 | No | NA | 3 | 0 | 1 | Not-determined | Genome-fragment | NA | NA | 0 |
| S8_vs_k141_200915\|\|full | 1253 | No | NA | 2 | 0 | 0 | Not-determined | Genome-fragment | NA | NA | 0 |
| S8_vs_k141_88453\|\|full | 4791 | No | NA | 5 | 0 | 2 | Not-determined | Genome-fragment | NA | NA | 0 |
| S8_vs_k141_1811\|\|full | 10717 | No | NA | 7 | 0 | 3 | Not-determined | Genome-fragment | NA | NA | 0 |
| S8_vs_k141_157730\|\|full | 1743 | No | NA | 3 | 0 | 0 | Not-determined | Genome-fragment | NA | NA | 0 |
| S10_vs_68062\|\|full | 11988 | No | NA | 15 | 3 | 0 | Low-quality | Genome-fragment | 29.52 | AAI-based (high-confidence) | 0 |
| S8_vs_k141_140607\|\|full | 5159 | No | NA | 7 | 0 | 4 | Not-determined | Genome-fragment | NA | NA | 0 |
| S8_vs_k141_53888\|\|full | 1405 | No | NA | 2 | 0 | 0 | Not-determined | Genome-fragment | NA | NA | 0 |
| S5_vs_k141_38536\|\|full | 11991 | No | NA | 17 | 9 | 0 | Low-quality | Genome-fragment | 25.42 | AAI-based (high-confidence) | 0 |
| S8_vs_k141_140617\|\|full | 2873 | No | NA | 3 | 0 | 1 | Not-determined | Genome-fragment | NA | NA | 0 |
| S3_extracted_k141_86827 | 11997 | No | NA | 23 | 1 | 0 | Low-quality | Genome-fragment | 37.04 | AAI-based (high-confidence) | 0 |
| S7_vs_53141\|\|full | 12004 | No | NA | 13 | 5 | 0 | Low-quality | Genome-fragment | 34.48 | AAI-based (high-confidence) | 0 |
| S8_vs_k141_123284\|\|full | 2444 | No | NA | 3 | 0 | 1 | Not-determined | Genome-fragment | NA | NA | 0 |
| S10_vs_67781\|\|full | 12005 | Yes | 5887 | 6 | 2 | 1 | Low-quality | Genome-fragment | 9.03 | AAI-based (medium-confidence) | 50.96 |
| S8_vs_k141_200964\|\|full | 5677 | No | NA | 5 | 0 | 0 | Not-determined | Genome-fragment | NA | NA | 0 |
| S8_vs_k141_131817\|\|full | 2044 | No | NA | 4 | 0 | 1 | Not-determined | Genome-fragment | NA | NA | 0 |
| S8_vs_k141_105865\|\|full | 1907 | No | NA | 2 | 0 | 1 | Not-determined | Genome-fragment | NA | NA | 0 |
| S8_vs_k141_183460\|\|full | 1424 | No | NA | 3 | 0 | 0 | Not-determined | Genome-fragment | NA | NA | 0 |
| S7_vs_70467\|\|full | 12008 | No | NA | 17 | 4 | 0 | Low-quality | Genome-fragment | 26.13 | AAI-based (high-confidence) | 0 |
| S8_vs_k141_10246\|\|full | 3343 | No | NA | 3 | 0 | 3 | Not-determined | Genome-fragment | NA | NA | 0 |
| S8_vs_k141_62384\|\|full | 2226 | No | NA | 5 | 0 | 0 | Not-determined | Genome-fragment | NA | NA | 0 |
| S8_vs_k141_53932\|\|full | 4699 | No | NA | 4 | 0 | 1 | Not-determined | Genome-fragment | NA | NA | 0 |
| S8_vs_k141_123323\|\|full | 2182 | No | NA | 2 | 0 | 2 | Not-determined | Genome-fragment | NA | NA | 0 |
| S8_vs_k141_149108\|\|full | 10796 | No | NA | 13 | 0 | 6 | Not-determined | Genome-fragment | NA | NA | 0 |
| S8_vs_k141_140677\|\|full | 2092 | No | NA | 3 | 0 | 0 | Not-determined | Genome-fragment | NA | NA | 0 |
| S8_vs_k141_183493\|\|full | 1945 | No | NA | 3 | 0 | 0 | Not-determined | Genome-fragment | NA | NA | 0 |
| S8_vs_k141_166645\|\|full | 2788 | No | NA | 6 | 0 | 0 | Not-determined | Genome-fragment | NA | NA | 0 |
| S14_vs_k141_25912\|\|full | 12018 | No | NA | 22 | 3 | 0 | Low-quality | Genome-fragment | 15.63 | HMM-based (lower-bound) | 0 |
| S5_vs_k141_36890\|\|0_partial | 12025 | No | NA | 8 | 0 | 7 | Low-quality | Genome-fragment | 15.88 | AAI-based (medium-confidence) | 0 |
| S16_vs_80089\|\|full | 12026 | No | NA | 17 | 2 | 0 | Low-quality | Genome-fragment | 21.13 | AAI-based (high-confidence) | 0 |
| S8_vs_k141_44951\|\|full | 1015 | No | NA | 2 | 0 | 0 | Not-determined | Genome-fragment | NA | NA | 0 |
| S8_vs_k141_114520\|\|full | 4137 | No | NA | 4 | 0 | 2 | Not-determined | Genome-fragment | NA | NA | 0 |
| S8_vs_k141_10281\|\|full | 1043 | No | NA | 3 | 0 | 1 | Not-determined | Genome-fragment | NA | NA | 0 |
| S8_vs_k141_53973\|\|full | 1775 | No | NA | 4 | 0 | 0 | Not-determined | Genome-fragment | NA | NA | 0 |
| S4_extracted_1282 | 12030 | No | NA | 17 | 1 | 1 | Low-quality | Genome-fragment | 13.76 | AAI-based (high-confidence) | 0 |
| S8_vs_k141_105907\|\|full | 2244 | No | NA | 2 | 0 | 1 | Not-determined | Genome-fragment | NA | NA | 0 |
| S8_vs_k141_44975\|\|full | 2870 | No | NA | 3 | 0 | 0 | Not-determined | Genome-fragment | NA | NA | 0 |
| S13_extracted_141294 | 12036 | No | NA | 15 | 0 | 7 | Low-quality | Genome-fragment | 5.48 | AAI-based (medium-confidence) | 0 |
| S1_vs_k141_25266\|\|full | 12037 | No | NA | 4 | 0 | 0 | Low-quality | Genome-fragment | 19 | AAI-based (medium-confidence) | 0 |
| S8_vs_k141_114548\|\|full | 2662 | No | NA | 3 | 0 | 0 | Not-determined | Genome-fragment | NA | NA | 0 |
| S12_extracted_k141_148276 | 12038 | No | NA | 11 | 1 | 1 | Low-quality | Genome-fragment | 30.97 | AAI-based (medium-confidence) | 0 |
| S16_extracted_167852 | 12041 | No | NA | 11 | 0 | 4 | Low-quality | Genome-fragment | 16.71 | AAI-based (medium-confidence) | 0 |
| S3_extracted_k141_60732 | 12051 | No | NA | 10 | 3 | 0 | Low-quality | Genome-fragment | 6.95 | AAI-based (high-confidence) | 0 |
| S14_vs_k141_35119\|\|0_partial | 12060 | No | NA | 14 | 1 | 2 | Low-quality | Genome-fragment | 29.91 | AAI-based (high-confidence) | 0 |
| S8_vs_k141_45019\|\|full | 4045 | No | NA | 7 | 0 | 1 | Not-determined | Genome-fragment | NA | NA | 0 |
| S8_vs_k141_131956\|\|full | 2476 | No | NA | 3 | 0 | 2 | Not-determined | Genome-fragment | NA | NA | 0 |
| S8_vs_k141_157809\|\|full | 2212 | No | NA | 3 | 0 | 1 | Not-determined | Genome-fragment | NA | NA | 0 |
| S8_vs_k141_131963\|\|full | 6917 | No | NA | 9 | 0 | 7 | Not-determined | Genome-fragment | NA | NA | 0 |
| S8_vs_k141_80019\|\|full | 2404 | No | NA | 3 | 0 | 0 | Not-determined | Genome-fragment | NA | NA | 0 |
| S15_extracted_63789 | 12066 | No | NA | 20 | 2 | 0 | Low-quality | Genome-fragment | 13.14 | AAI-based (high-confidence) | 0 |
| S8_vs_k141_62518\|\|full | 1295 | No | NA | 3 | 0 | 1 | Not-determined | Genome-fragment | NA | NA | 0 |
| S15_vs_70800\|\|full | 12074 | No | NA | 23 | 4 | 1 | Low-quality | Genome-fragment | 7.18 | AAI-based (high-confidence) | 0 |
| S8_vs_k141_36777\|\|full | 3292 | No | NA | 3 | 0 | 2 | Not-determined | Genome-fragment | NA | NA | 0 |
| S12_vs_k141_26775\|\|full | 12079 | No | NA | 24 | 2 | 1 | Low-quality | Genome-fragment | 30.9 | AAI-based (medium-confidence) | 0 |
| S8_vs_k141_36785\|\|full | 3420 | No | NA | 4 | 0 | 4 | Not-determined | Genome-fragment | NA | NA | 0 |
| S11_vs_k141_88836\|\|full | 12084 | Yes | 10648 | 27 | 4 | 3 | Low-quality | Genome-fragment | 18.01 | AAI-based (high-confidence) | 11.88 |
| S8_vs_k141_106001\|\|full | 1790 | No | NA | 2 | 0 | 0 | Not-determined | Genome-fragment | NA | NA | 0 |
| S8_vs_k141_192969\|\|full | 3994 | No | NA | 5 | 0 | 1 | Not-determined | Genome-fragment | NA | NA | 0 |
| S8_vs_k141_206121\|\|full | 12089 | No | NA | 18 | 1 | 2 | Low-quality | Genome-fragment | 13.95 | HMM-based (lower-bound) | 0 |
| S5_extracted_k141_112288 | 12126 | No | NA | 18 | 2 | 0 | Low-quality | Genome-fragment | 14.73 | AAI-based (high-confidence) | 0 |
| S8_vs_k141_149279\|\|full | 3594 | No | NA | 5 | 0 | 0 | Not-determined | Genome-fragment | NA | NA | 0 |
| S8_vs_k141_183608\|\|full | 7767 | No | NA | 3 | 0 | 2 | Not-determined | Genome-fragment | NA | NA | 0 |
| S8_vs_k141_28112\|\|full | 1817 | No | NA | 4 | 0 | 2 | Not-determined | Genome-fragment | NA | NA | 0 |
| S8_vs_k141_54094\|\|full | 2282 | No | NA | 5 | 0 | 0 | Not-determined | Genome-fragment | NA | NA | 0 |
| S8_vs_k141_131988\|\|full | 2698 | No | NA | 3 | 0 | 2 | Not-determined | Genome-fragment | NA | NA | 0 |
| S8_vs_k141_10416\|\|full | 11236 | No | NA | 11 | 0 | 3 | Not-determined | Genome-fragment | NA | NA | 0 |
| S13_extracted_11943 | 12127 | No | NA | 17 | 7 | 0 | Low-quality | Genome-fragment | 34.43 | HMM-based (lower-bound) | 0 |
| S8_vs_k141_88658\|\|full | 4653 | No | NA | 5 | 0 | 1 | Not-determined | Genome-fragment | NA | NA | 0 |
| S8_vs_k141_149299\|\|full | 1699 | No | NA | 3 | 0 | 1 | Not-determined | Genome-fragment | NA | NA | 0 |
| S8_vs_k141_175235\|\|full | 36143 | No | NA | 62 | 11 | 0 | High-quality | High-quality | 96.8 | AAI-based (high-confidence) | 0 |
| S8_vs_k141_123460\|\|full | 1451 | No | NA | 2 | 0 | 1 | Not-determined | Genome-fragment | NA | NA | 0 |
| S6_vs_k141_4457\|\|0_partial | 12131 | Yes | 5187 | 12 | 3 | 2 | Low-quality | Genome-fragment | 18.08 | AAI-based (high-confidence) | 57.24 |
| S1_vs_k141_128148\|\|full | 12132 | No | NA | 27 | 5 | 0 | Low-quality | Genome-fragment | 25.98 | AAI-based (high-confidence) | 0 |
| S16_extracted_86911 | 12133 | No | NA | 17 | 14 | 0 | Low-quality | Genome-fragment | 20.08 | AAI-based (high-confidence) | 0 |
| S2_extracted_k141_17727 | 12137 | No | NA | 27 | 2 | 0 | Low-quality | Genome-fragment | 23.73 | AAI-based (high-confidence) | 0 |
| S8_vs_k141_183637\|\|full | 2594 | No | NA | 3 | 0 | 0 | Not-determined | Genome-fragment | NA | NA | 0 |
| S8_vs_k141_28154\|\|full | 2234 | No | NA | 4 | 0 | 3 | Not-determined | Genome-fragment | NA | NA | 0 |
| S12_vs_k141_135479\|\|full | 12145 | No | NA | 13 | 1 | 2 | Low-quality | Genome-fragment | 10.97 | HMM-based (lower-bound) | 0 |
| S7_vs_65238\|\|full | 12146 | No | NA | 15 | 4 | 0 | Low-quality | Genome-fragment | 35.99 | HMM-based (lower-bound) | 0 |
| S8_vs_k141_98336\|\|full | 12183 | No | NA | 17 | 8 | 0 | Low-quality | Genome-fragment | 28.29 | AAI-based (high-confidence) | 0 |
| S8_vs_k141_114668\|\|full | 1366 | No | NA | 5 | 0 | 0 | Not-determined | Genome-fragment | NA | NA | 0 |
| S8_vs_k141_54121\|\|full | 12113 | No | NA | 8 | 0 | 6 | Not-determined | Genome-fragment | NA | NA | 0 |
| S8_vs_k141_157919\|\|full | 1159 | No | NA | 2 | 0 | 0 | Not-determined | Genome-fragment | NA | NA | 0 |
| S10_vs_21207\|\|full | 12185 | No | NA | 15 | 1 | 5 | Low-quality | Genome-fragment | 21.02 | AAI-based (high-confidence) | 0 |
| S8_vs_k141_54136\|\|full | 3095 | No | NA | 3 | 0 | 1 | Not-determined | Genome-fragment | NA | NA | 0 |
| S8_vs_k141_1976\|\|full | 1460 | No | NA | 3 | 0 | 0 | Not-determined | Genome-fragment | NA | NA | 0 |
| S8_vs_k141_123492\|\|full | 9783 | No | NA | 9 | 0 | 8 | Not-determined | Genome-fragment | NA | NA | 0 |
| S8_vs_k141_193029\|\|full | 1461 | No | NA | 2 | 0 | 0 | Not-determined | Genome-fragment | NA | NA | 0 |
| S17_extracted_k141_147828 | 12186 | No | NA | 31 | 5 | 0 | Low-quality | Genome-fragment | 15.13 | AAI-based (high-confidence) | 0 |
| S7_vs_4529\|\|full | 12187 | No | NA | 30 | 7 | 0 | Low-quality | Genome-fragment | 32.95 | AAI-based (high-confidence) | 0 |
| S1_vs_k141_92342\|\|full | 12193 | No | NA | 14 | 7 | 0 | Low-quality | Genome-fragment | 32.24 | AAI-based (high-confidence) | 0 |
| S8_vs_k141_88731\|\|full | 5335 | No | NA | 6 | 0 | 0 | Not-determined | Genome-fragment | NA | NA | 0 |
| S8_vs_k141_1987\|\|full | 4752 | No | NA | 7 | 0 | 1 | Not-determined | Genome-fragment | NA | NA | 0 |
| S8_vs_k141_193044\|\|full | 1896 | No | NA | 3 | 0 | 1 | Not-determined | Genome-fragment | NA | NA | 0 |
| S8_vs_k141_132053\|\|full | 1752 | No | NA | 2 | 0 | 0 | Not-determined | Genome-fragment | NA | NA | 0 |
| S8_vs_k141_80155\|\|full | 4738 | No | NA | 6 | 0 | 0 | Not-determined | Genome-fragment | NA | NA | 0 |
| S8_vs_k141_193046\|\|full | 3516 | No | NA | 5 | 0 | 0 | Not-determined | Genome-fragment | NA | NA | 0 |
| S14_extracted_k141_30438 | 12223 | No | NA | 15 | 6 | 0 | Low-quality | Genome-fragment | 20.49 | AAI-based (high-confidence) | 0 |
| S3_vs_k141_51886\|\|full | 12235 | Yes | 11083 | 9 | 2 | 1 | Low-quality | Genome-fragment | 25.31 | AAI-based (medium-confidence) | 9.42 |
| S17_vs_k141_92618\|\|full | 12243 | Yes | 1127 | 8 | 1 | 2 | Low-quality | Genome-fragment | 0.71 | HMM-based (lower-bound) | 90.79 |
| S8_vs_k141_157962\|\|full | 1759 | No | NA | 2 | 0 | 2 | Not-determined | Genome-fragment | NA | NA | 0 |
| S13_extracted_100646 | 12243 | No | NA | 12 | 0 | 5 | Low-quality | Genome-fragment | 2.14 | AAI-based (medium-confidence) | 0 |
| S3_vs_k141_25953\|\|full | 12243 | No | NA | 26 | 1 | 0 | Low-quality | Genome-fragment | 32.4 | AAI-based (high-confidence) | 0 |
| S8_vs_k141_157974\|\|full | 2143 | No | NA | 2 | 0 | 2 | Not-determined | Genome-fragment | NA | NA | 0 |
| S17_extracted_k141_31368 | 12245 | No | NA | 7 | 0 | 2 | Low-quality | Genome-fragment | 10.68 | AAI-based (medium-confidence) | 0 |
| S8_vs_k141_80185\|\|full | 1393 | No | NA | 2 | 0 | 0 | Not-determined | Genome-fragment | NA | NA | 0 |
| S8_vs_k141_201202\|\|full | 1544 | No | NA | 3 | 0 | 0 | Not-determined | Genome-fragment | NA | NA | 0 |
| S3_extracted_k141_112575 | 12247 | No | NA | 12 | 5 | 0 | Low-quality | Genome-fragment | 30.42 | AAI-based (high-confidence) | 0 |
| S8_vs_k141_106145\|\|full | 2780 | No | NA | 5 | 0 | 3 | Not-determined | Genome-fragment | NA | NA | 0 |
| S8_vs_k141_23926\|\|0_partial | 12251 | No | NA | 19 | 3 | 1 | Low-quality | Genome-fragment | 22.16 | AAI-based (high-confidence) | 0 |
| S7_extracted_89110 | 12256 | No | NA | 34 | 6 | 0 | Low-quality | Genome-fragment | 29.81 | AAI-based (medium-confidence) | 0 |
| S3_extracted_k141_41491 | 12262 | No | NA | 20 | 14 | 0 | Low-quality | Genome-fragment | 32.92 | AAI-based (high-confidence) | 0 |
| S5_extracted_k141_128166 | 12269 | No | NA | 12 | 0 | 1 | Low-quality | Genome-fragment | 6.97 | AAI-based (high-confidence) | 0 |
| S8_vs_k141_54183\|\|full | 43572 | No | NA | 78 | 13 | 1 | High-quality | High-quality | 100 | AAI-based (high-confidence) | 0 |
| S8_vs_k141_80222\|\|full | 1842 | No | NA | 3 | 0 | 1 | Not-determined | Genome-fragment | NA | NA | 0 |
| S14_vs_k141_83720\|\|full | 12275 | No | NA | 22 | 1 | 6 | Low-quality | Genome-fragment | 14.73 | HMM-based (lower-bound) | 0 |
| S8_vs_k141_123603\|\|full | 1550 | No | NA | 2 | 0 | 2 | Not-determined | Genome-fragment | NA | NA | 0 |
| S3_vs_k141_78481\|\|full | 12291 | No | NA | 25 | 3 | 0 | Low-quality | Genome-fragment | 8.4 | AAI-based (high-confidence) | 0 |
| S8_vs_k141_54212\|\|full | 1323 | No | NA | 3 | 0 | 1 | Not-determined | Genome-fragment | NA | NA | 0 |
| S8_vs_k141_62736\|\|full | 2533 | No | NA | 2 | 0 | 0 | Not-determined | Genome-fragment | NA | NA | 0 |
| S8_extracted_k141_195370 | 12301 | No | NA | 16 | 5 | 0 | Low-quality | Genome-fragment | 37.36 | HMM-based (lower-bound) | 0 |
| S8_vs_k141_77838\|\|full | 12313 | No | NA | 23 | 3 | 1 | Low-quality | Genome-fragment | 34.07 | AAI-based (high-confidence) | 0 |
| S1_extracted_k141_103657 | 12318 | No | NA | 31 | 4 | 2 | Low-quality | Genome-fragment | 21.33 | AAI-based (high-confidence) | 0 |
| S8_vs_k141_114809\|\|full | 1003 | No | NA | 3 | 0 | 1 | Not-determined | Genome-fragment | NA | NA | 0 |
| S5_extracted_k141_11723 | 12326 | No | NA | 20 | 17 | 0 | Low-quality | Genome-fragment | 25.77 | AAI-based (high-confidence) | 0 |
| S5_extracted_k141_68549 | 12335 | No | NA | 18 | 0 | 5 | Low-quality | Genome-fragment | 6.91 | AAI-based (medium-confidence) | 0 |
| S14_extracted_k141_51440 | 12370 | No | NA | 15 | 11 | 0 | Low-quality | Genome-fragment | 32.35 | AAI-based (medium-confidence) | 0 |
| S13_extracted_144316 | 12400 | No | NA | 26 | 2 | 1 | Low-quality | Genome-fragment | 12.9 | AAI-based (high-confidence) | 0 |
| S8_vs_k141_28288\|\|full | 1701 | No | NA | 3 | 0 | 2 | Not-determined | Genome-fragment | NA | NA | 0 |
| S8_vs_k141_106220\|\|full | 4770 | No | NA | 3 | 0 | 3 | Not-determined | Genome-fragment | NA | NA | 0 |
| S8_vs_k141_19307\|\|full | 2352 | No | NA | 3 | 0 | 0 | Not-determined | Genome-fragment | NA | NA | 0 |
| S8_vs_k141_54315\|\|full | 1787 | No | NA | 4 | 0 | 2 | Not-determined | Genome-fragment | NA | NA | 0 |
| S15_vs_2862\|\|full | 12404 | No | NA | 14 | 1 | 2 | Low-quality | Genome-fragment | 7.65 | HMM-based (lower-bound) | 0 |
| S8_vs_k141_19318\|\|full | 3487 | No | NA | 5 | 0 | 2 | Not-determined | Genome-fragment | NA | NA | 0 |
| S8_vs_k141_79829\|\|0_partial | 12405 | No | NA | 11 | 1 | 8 | Low-quality | Genome-fragment | 14.34 | HMM-based (lower-bound) | 0 |
| S8_vs_k141_45301\|\|full | 2487 | No | NA | 1 | 0 | 1 | Not-determined | Genome-fragment | NA | NA | 0 |
| S8_vs_k141_88976\|\|full | 1545 | No | NA | 3 | 0 | 2 | Not-determined | Genome-fragment | NA | NA | 0 |
| S5_extracted_k141_121030 | 12412 | No | NA | 14 | 1 | 0 | Low-quality | Genome-fragment | 22.44 | AAI-based (high-confidence) | 0 |
| S3_vs_k141_1238\|\|full | 12414 | No | NA | 15 | 1 | 1 | Low-quality | Genome-fragment | 22.52 | AAI-based (high-confidence) | 0 |
| S8_vs_k141_175506\|\|full | 1297 | No | NA | 3 | 0 | 0 | Not-determined | Genome-fragment | NA | NA | 0 |
| S8_vs_k141_62858\|\|full | 1251 | No | NA | 2 | 0 | 0 | Not-determined | Genome-fragment | NA | NA | 0 |
| S8_vs_k141_28311\|\|full | 45747 | No | NA | 65 | 4 | 2 | High-quality | High-quality | 100 | AAI-based (medium-confidence) | 0 |
| S5_vs_k141_134223\|\|full | 12422 | No | NA | 19 | 9 | 0 | Low-quality | Genome-fragment | 30.36 | AAI-based (high-confidence) | 0 |
| S8_vs_k141_106302\|\|full | 1704 | No | NA | 2 | 0 | 0 | Not-determined | Genome-fragment | NA | NA | 0 |
| S8_vs_k141_45338\|\|full | 8002 | No | NA | 6 | 0 | 4 | Not-determined | Genome-fragment | NA | NA | 0 |
| S5_vs_k141_22266\|\|full | 12424 | No | NA | 21 | 9 | 0 | Low-quality | Genome-fragment | 29.6 | AAI-based (high-confidence) | 0 |
| S8_vs_k141_28326\|\|full | 1297 | No | NA | 4 | 0 | 0 | Not-determined | Genome-fragment | NA | NA | 0 |
| S16_extracted_24816 | 12431 | No | NA | 18 | 4 | 0 | Low-quality | Genome-fragment | 41.69 | HMM-based (lower-bound) | 0 |
| S8_vs_k141_123737\|\|full | 1547 | No | NA | 6 | 0 | 0 | Not-determined | Genome-fragment | NA | NA | 0 |
| S6_extracted_k141_10815 | 12435 | No | NA | 8 | 0 | 7 | Low-quality | Genome-fragment | 5.38 | AAI-based (medium-confidence) | 0 |
| S12_extracted_k141_62222 | 12436 | No | NA | 18 | 3 | 0 | Low-quality | Genome-fragment | 36.78 | HMM-based (lower-bound) | 0 |
| S8_vs_k141_149617\|\|full | 5494 | No | NA | 10 | 0 | 0 | Not-determined | Genome-fragment | NA | NA | 0 |
| S8_vs_k141_10585\|\|full | 2175 | No | NA | 3 | 0 | 2 | Not-determined | Genome-fragment | NA | NA | 0 |
| S12_vs_k141_152393\|\|full | 12438 | No | NA | 12 | 1 | 3 | Low-quality | Genome-fragment | 19.03 | AAI-based (medium-confidence) | 0 |
| S3_extracted_k141_85359 | 12447 | No | NA | 24 | 9 | 0 | Low-quality | Genome-fragment | 8.68 | AAI-based (high-confidence) | 0 |
| S8_vs_k141_106353\|\|full | 1560 | No | NA | 2 | 0 | 2 | Not-determined | Genome-fragment | NA | NA | 0 |
| S8_vs_k141_132230\|\|full | 3970 | No | NA | 4 | 0 | 4 | Not-determined | Genome-fragment | NA | NA | 0 |
| S14_vs_k141_73426\|\|full | 12449 | No | NA | 14 | 4 | 0 | Low-quality | Genome-fragment | 28.69 | AAI-based (high-confidence) | 0 |
| S3_vs_k141_36654\|\|full | 12460 | No | NA | 21 | 5 | 1 | Low-quality | Genome-fragment | 20.06 | AAI-based (high-confidence) | 0 |
| S11_extracted_k141_56160 | 12464 | No | NA | 17 | 2 | 0 | Low-quality | Genome-fragment | 28.8 | AAI-based (high-confidence) | 0 |
| S1_extracted_k141_46184 | 12465 | No | NA | 26 | 3 | 0 | Low-quality | Genome-fragment | 24.88 | AAI-based (high-confidence) | 0 |
| S8_vs_k141_175593\|\|full | 3140 | No | NA | 4 | 0 | 2 | Not-determined | Genome-fragment | NA | NA | 0 |
| S8_vs_k141_201431\|\|full | 1279 | No | NA | 3 | 0 | 2 | Not-determined | Genome-fragment | NA | NA | 0 |
| S8_vs_k141_125838\|\|full | 12466 | No | NA | 30 | 7 | 1 | Low-quality | Genome-fragment | 23.77 | AAI-based (high-confidence) | 0 |
| S8_vs_k141_114965\|\|full | 1350 | No | NA | 1 | 0 | 0 | Not-determined | Genome-fragment | NA | NA | 0 |
| S8_vs_k141_114966\|\|full | 2668 | No | NA | 6 | 0 | 5 | Not-determined | Genome-fragment | NA | NA | 0 |
| S8_vs_k141_114968\|\|full | 1292 | No | NA | 3 | 0 | 0 | Not-determined | Genome-fragment | NA | NA | 0 |
| S8_vs_k141_193318\|\|full | 8865 | No | NA | 4 | 0 | 4 | Not-determined | Genome-fragment | NA | NA | 0 |
| S8_vs_k141_97633\|\|full | 1932 | No | NA | 2 | 0 | 2 | Not-determined | Genome-fragment | NA | NA | 0 |
| S8_vs_k141_37125\|\|full | 1180 | No | NA | 3 | 0 | 0 | Not-determined | Genome-fragment | NA | NA | 0 |
| S8_vs_k141_175646\|\|full | 2081 | No | NA | 2 | 0 | 2 | Not-determined | Genome-fragment | NA | NA | 0 |
| S3_vs_k141_96960\|\|full | 12468 | No | NA | 12 | 1 | 6 | Low-quality | Genome-fragment | 11.18 | HMM-based (lower-bound) | 0 |
| S8_vs_k141_149685\|\|full | 3572 | No | NA | 6 | 0 | 0 | Not-determined | Genome-fragment | NA | NA | 0 |
| S8_vs_k141_2336\|\|full | 5559 | No | NA | 5 | 0 | 2 | Not-determined | Genome-fragment | NA | NA | 0 |
| S17_extracted_k141_70416 | 12487 | No | NA | 11 | 0 | 4 | Low-quality | Genome-fragment | 26.86 | AAI-based (high-confidence) | 0 |
| S8_vs_k141_97663\|\|full | 6257 | No | NA | 8 | 0 | 7 | Not-determined | Genome-fragment | NA | NA | 0 |
| S1_extracted_k141_38301 | 12495 | No | NA | 6 | 0 | 0 | Low-quality | Genome-fragment | 13.35 | AAI-based (medium-confidence) | 0 |
| S8_vs_k141_19472\|\|full | 4833 | No | NA | 3 | 0 | 3 | Not-determined | Genome-fragment | NA | NA | 0 |
| S7_vs_23373\|\|0_partial | 12501 | No | NA | 13 | 1 | 3 | Low-quality | Genome-fragment | 8.4 | HMM-based (lower-bound) | 0 |
| S16_vs_62907\|\|full | 12504 | No | NA | 13 | 2 | 0 | Low-quality | Genome-fragment | 31.33 | AAI-based (high-confidence) | 0 |
| S8_vs_k141_19529\|\|full | 2122 | No | NA | 4 | 0 | 0 | Not-determined | Genome-fragment | NA | NA | 0 |
| S17_vs_k141_2095\|\|full | 12517 | No | NA | 14 | 1 | 4 | Low-quality | Genome-fragment | 8.06 | HMM-based (lower-bound) | 0 |
| S3_vs_k141_127403\|\|full | 12537 | No | NA | 17 | 0 | 4 | Low-quality | Genome-fragment | 10.49 | AAI-based (medium-confidence) | 0 |
| S8_vs_k141_134591\|\|full | 12541 | No | NA | 14 | 6 | 0 | Low-quality | Genome-fragment | 30.04 | HMM-based (lower-bound) | 0 |
| S8_vs_k141_37198\|\|full | 1308 | No | NA | 3 | 0 | 2 | Not-determined | Genome-fragment | NA | NA | 0 |
| S8_vs_k141_63058\|\|full | 1024 | No | NA | 2 | 0 | 1 | Not-determined | Genome-fragment | NA | NA | 0 |
| S14_extracted_k141_28606 | 12546 | No | NA | 19 | 7 | 1 | Low-quality | Genome-fragment | 28.79 | AAI-based (high-confidence) | 0 |
| S17_extracted_k141_116256 | 12551 | No | NA | 20 | 0 | 4 | Low-quality | Genome-fragment | 38.63 | AAI-based (medium-confidence) | 0 |
| S8_vs_k141_193428\|\|full | 8971 | No | NA | 10 | 0 | 6 | Not-determined | Genome-fragment | NA | NA | 0 |
| S8_vs_k141_106543\|\|full | 1395 | No | NA | 3 | 0 | 0 | Not-determined | Genome-fragment | NA | NA | 0 |
| S8_vs_k141_123900\|\|full | 3059 | No | NA | 3 | 0 | 2 | Not-determined | Genome-fragment | NA | NA | 0 |
| S8_vs_k141_71262\|\|full | 2227 | No | NA | 3 | 0 | 1 | Not-determined | Genome-fragment | NA | NA | 0 |
| S8_vs_k141_175709\|\|full | 1063 | No | NA | 2 | 0 | 0 | Not-determined | Genome-fragment | NA | NA | 0 |
| S8_vs_k141_193451\|\|full | 2705 | No | NA | 4 | 0 | 1 | Not-determined | Genome-fragment | NA | NA | 0 |
| S8_vs_k141_147166\|\|full | 12554 | No | NA | 21 | 10 | 0 | Low-quality | Genome-fragment | 28.93 | AAI-based (high-confidence) | 0 |
| S17_extracted_k141_634 | 12562 | Yes | 9270 | 13 | 2 | 3 | Low-quality | Genome-fragment | 3.7 | HMM-based (lower-bound) | 26.21 |
| S8_vs_k141_115062\|\|full | 2957 | No | NA | 4 | 0 | 1 | Not-determined | Genome-fragment | NA | NA | 0 |
| S8_vs_k141_175746\|\|full | 2426 | No | NA | 3 | 0 | 0 | Not-determined | Genome-fragment | NA | NA | 0 |
| S8_vs_k141_2417\|\|full | 4174 | No | NA | 6 | 0 | 3 | Not-determined | Genome-fragment | NA | NA | 0 |
| S8_extracted_k141_86050 | 12576 | No | NA | 26 | 4 | 0 | Low-quality | Genome-fragment | 30.5 | AAI-based (high-confidence) | 0 |
| S6_vs_k141_39568\|\|full | 12583 | No | NA | 18 | 4 | 0 | Low-quality | Genome-fragment | 30.24 | AAI-based (medium-confidence) | 0 |
| S8_vs_k141_141170\|\|full | 1571 | No | NA | 2 | 0 | 2 | Not-determined | Genome-fragment | NA | NA | 0 |
| S7_vs_49009\|\|full | 12593 | No | NA | 19 | 7 | 0 | Low-quality | Genome-fragment | 42.39 | HMM-based (lower-bound) | 0 |
| S8_vs_k141_167184\|\|full | 4437 | No | NA | 7 | 0 | 3 | Not-determined | Genome-fragment | NA | NA | 0 |
| S8_vs_k141_19676\|\|full | 1940 | No | NA | 3 | 0 | 3 | Not-determined | Genome-fragment | NA | NA | 0 |
| S8_vs_k141_138275\|\|full | 12620 | No | NA | 14 | 7 | 0 | Low-quality | Genome-fragment | 35.27 | AAI-based (high-confidence) | 0 |
| S8_vs_k141_184059\|\|full | 5004 | No | NA | 6 | 0 | 1 | Not-determined | Genome-fragment | NA | NA | 0 |
| S12_vs_k141_170396\|\|full | 12628 | No | NA | 26 | 5 | 0 | Low-quality | Genome-fragment | 29.83 | AAI-based (high-confidence) | 0 |
| S8_vs_k141_149765\|\|full | 2354 | No | NA | 3 | 0 | 2 | Not-determined | Genome-fragment | NA | NA | 0 |
| S8_vs_k141_141244\|\|full | 1026 | No | NA | 3 | 0 | 1 | Not-determined | Genome-fragment | NA | NA | 0 |
| S8_vs_k141_193515\|\|full | 1422 | No | NA | 3 | 0 | 2 | Not-determined | Genome-fragment | NA | NA | 0 |
| S16_vs_19989\|\|full | 12631 | No | NA | 22 | 3 | 1 | Low-quality | Genome-fragment | 18.24 | AAI-based (high-confidence) | 0 |
| S1_extracted_k141_104407 | 12642 | No | NA | 34 | 5 | 0 | Low-quality | Genome-fragment | 28.45 | AAI-based (high-confidence) | 0 |
| S8_vs_k141_193530\|\|full | 2746 | No | NA | 3 | 0 | 2 | Not-determined | Genome-fragment | NA | NA | 0 |
| S8_vs_k141_28737\|\|full | 3445 | No | NA | 3 | 0 | 1 | Not-determined | Genome-fragment | NA | NA | 0 |
| S8_vs_k141_124057\|\|full | 1731 | No | NA | 3 | 0 | 0 | Not-determined | Genome-fragment | NA | NA | 0 |
| S8_vs_k141_80607\|\|full | 1183 | No | NA | 3 | 0 | 0 | Not-determined | Genome-fragment | NA | NA | 0 |
| S2_vs_k141_13462\|\|full | 12645 | No | NA | 22 | 3 | 0 | Low-quality | Genome-fragment | 21.44 | AAI-based (high-confidence) | 0 |
| S8_vs_k141_175850\|\|full | 5473 | No | NA | 4 | 0 | 3 | Not-determined | Genome-fragment | NA | NA | 0 |
| S12_extracted_k141_93276 | 12646 | No | NA | 14 | 4 | 0 | Low-quality | Genome-fragment | 42.41 | HMM-based (lower-bound) | 0 |
| S8_vs_k141_132472\|\|full | 20524 | No | NA | 23 | 0 | 11 | Medium-quality | Genome-fragment | 71.49 | AAI-based (medium-confidence) | 0 |
| S7_vs_4729\|\|full | 12648 | No | NA | 13 | 2 | 0 | Low-quality | Genome-fragment | 8.37 | AAI-based (high-confidence) | 0 |
| S8_vs_k141_167247\|\|full | 2174 | No | NA | 3 | 0 | 0 | Not-determined | Genome-fragment | NA | NA | 0 |
| S16_extracted_54812 | 12650 | No | NA | 15 | 12 | 0 | Low-quality | Genome-fragment | 26.44 | AAI-based (high-confidence) | 0 |
| S8_vs_k141_19752\|\|full | 8429 | No | NA | 14 | 0 | 1 | Not-determined | Genome-fragment | NA | NA | 0 |
| S8_vs_k141_141318\|\|full | 1033 | No | NA | 2 | 0 | 1 | Not-determined | Genome-fragment | NA | NA | 0 |
| S8_vs_k141_167266\|\|full | 3992 | No | NA | 4 | 0 | 1 | Not-determined | Genome-fragment | NA | NA | 0 |
| S1_vs_k141_74343\|\|full | 12653 | No | NA | 5 | 0 | 0 | Low-quality | Genome-fragment | 32.61 | AAI-based (high-confidence) | 0 |
| S8_vs_k141_184122\|\|full | 1275 | No | NA | 3 | 0 | 0 | Not-determined | Genome-fragment | NA | NA | 0 |
| S8_vs_k141_63277\|\|full | 18362 | No | NA | 26 | 15 | 0 | Medium-quality | Genome-fragment | 50.42 | AAI-based (high-confidence) | 0 |
| S8_vs_k141_158265\|\|full | 29476 | No | NA | 34 | 0 | 6 | Not-determined | Genome-fragment | NA | NA | 0 |
| S17_vs_k141_81387\|\|full | 12654 | No | NA | 18 | 8 | 0 | Low-quality | Genome-fragment | 33.65 | AAI-based (high-confidence) | 0 |
| S8_vs_k141_184135\|\|full | 3181 | No | NA | 4 | 0 | 1 | Not-determined | Genome-fragment | NA | NA | 0 |
| S10_vs_87403\|\|0_partial | 12654 | No | NA | 11 | 0 | 8 | Low-quality | Genome-fragment | 3.93 | AAI-based (medium-confidence) | 0 |
| S8_vs_k141_132514\|\|full | 2278 | No | NA | 3 | 0 | 3 | Not-determined | Genome-fragment | NA | NA | 0 |
| S5_vs_k141_120504\|\|full | 12683 | No | NA | 17 | 9 | 0 | Low-quality | Genome-fragment | 28.01 | AAI-based (high-confidence) | 0 |
| S14_vs_k141_47308\|\|0_partial | 12687 | No | NA | 15 | 4 | 2 | Low-quality | Genome-fragment | 44.22 | AAI-based (high-confidence) | 0 |
| S8_vs_k141_63301\|\|full | 1032 | No | NA | 4 | 0 | 0 | Not-determined | Genome-fragment | NA | NA | 0 |
| S8_vs_k141_80687\|\|full | 4069 | No | NA | 4 | 0 | 2 | Not-determined | Genome-fragment | NA | NA | 0 |
| S8_vs_k141_141347\|\|full | 5932 | No | NA | 6 | 0 | 3 | Not-determined | Genome-fragment | NA | NA | 0 |
| S8_vs_k141_149899\|\|full | 2546 | No | NA | 2 | 0 | 0 | Not-determined | Genome-fragment | NA | NA | 0 |
| S14_extracted_k141_20590 | 12708 | No | NA | 16 | 0 | 3 | Low-quality | Genome-fragment | 24.46 | AAI-based (medium-confidence) | 0 |
| S8_vs_k141_19805\|\|full | 2436 | No | NA | 3 | 0 | 0 | Not-determined | Genome-fragment | NA | NA | 0 |
| S14_vs_k141_106224\|\|full | 12710 | No | NA | 15 | 1 | 0 | Low-quality | Genome-fragment | 28.43 | AAI-based (medium-confidence) | 0 |
| S8_vs_k141_80713\|\|full | 3511 | No | NA | 4 | 0 | 0 | Not-determined | Genome-fragment | NA | NA | 0 |
| S3_vs_k141_34600\|\|full | 12717 | No | NA | 15 | 11 | 0 | Low-quality | Genome-fragment | 37.12 | AAI-based (high-confidence) | 0 |
| S8_vs_k141_132569\|\|full | 1197 | No | NA | 3 | 0 | 2 | Not-determined | Genome-fragment | NA | NA | 0 |
| S8_vs_k141_115213\|\|full | 4363 | No | NA | 6 | 0 | 4 | Not-determined | Genome-fragment | NA | NA | 0 |
| S16_extracted_41171 | 12718 | No | NA | 18 | 5 | 0 | Low-quality | Genome-fragment | 20.84 | AAI-based (high-confidence) | 0 |
| S8_vs_k141_89258\|\|full | 1293 | No | NA | 3 | 0 | 0 | Not-determined | Genome-fragment | NA | NA | 0 |
| S12_vs_k141_72635\|\|0_partial | 12726 | No | NA | 16 | 5 | 0 | Low-quality | Genome-fragment | 16.86 | HMM-based (lower-bound) | 0 |
| S8_vs_k141_149927\|\|full | 16615 | No | NA | 13 | 0 | 2 | Not-determined | Genome-fragment | NA | NA | 0 |
| S16_extracted_144673 | 12735 | No | NA | 13 | 3 | 0 | Low-quality | Genome-fragment | 20.04 | AAI-based (high-confidence) | 0 |
| S8_vs_k141_97878\|\|full | 3180 | No | NA | 4 | 0 | 1 | Not-determined | Genome-fragment | NA | NA | 0 |
| S1_vs_k141_123748\|\|full | 12737 | No | NA | 19 | 6 | 1 | Low-quality | Genome-fragment | 29.08 | AAI-based (high-confidence) | 0 |
| S12_extracted_k141_98849 | 12744 | No | NA | 10 | 2 | 0 | Low-quality | Genome-fragment | 25.16 | AAI-based (high-confidence) | 0 |
| S8_vs_k141_63411\|\|full | 1371 | No | NA | 4 | 0 | 0 | Not-determined | Genome-fragment | NA | NA | 0 |
| S7_vs_14581\|\|full | 12745 | No | NA | 28 | 5 | 0 | Low-quality | Genome-fragment | 44.36 | AAI-based (high-confidence) | 0 |
| S10_extracted_50078 | 12749 | No | NA | 22 | 2 | 0 | Low-quality | Genome-fragment | 9.38 | AAI-based (high-confidence) | 0 |
| S11_vs_k141_78031\|\|full | 12755 | No | NA | 12 | 7 | 0 | Low-quality | Genome-fragment | 21.99 | AAI-based (high-confidence) | 0 |
| S8_vs_k141_37441\|\|full | 6914 | No | NA | 8 | 0 | 2 | Not-determined | Genome-fragment | NA | NA | 0 |
| S8_vs_k141_89339\|\|full | 5144 | No | NA | 4 | 0 | 2 | Not-determined | Genome-fragment | NA | NA | 0 |
| S8_vs_k141_80771\|\|full | 1599 | No | NA | 3 | 0 | 1 | Not-determined | Genome-fragment | NA | NA | 0 |
| S11_vs_k141_14780\|\|full | 12765 | No | NA | 14 | 1 | 4 | Low-quality | Genome-fragment | 11.53 | AAI-based (medium-confidence) | 0 |
| S8_vs_k141_25687\|\|full | 12775 | No | NA | 24 | 11 | 0 | Low-quality | Genome-fragment | 35.69 | AAI-based (high-confidence) | 0 |
| S16_vs_36771\|\|full | 12776 | No | NA | 7 | 0 | 3 | Low-quality | Genome-fragment | 13.37 | AAI-based (medium-confidence) | 0 |
| S1_vs_k141_157821\|\|full | 12789 | No | NA | 14 | 11 | 0 | Low-quality | Genome-fragment | 35.43 | AAI-based (high-confidence) | 0 |
| S7_vs_4472\|\|0_partial | 12790 | Yes | 6734 | 16 | 3 | 3 | Low-quality | Genome-fragment | 6.77 | AAI-based (medium-confidence) | 47.35 |
| S8_vs_k141_54718\|\|full | 1385 | No | NA | 3 | 0 | 0 | Not-determined | Genome-fragment | NA | NA | 0 |
| S3_vs_k141_3235\|\|0_partial | 12793 | No | NA | 11 | 1 | 3 | Low-quality | Genome-fragment | 11.55 | AAI-based (medium-confidence) | 0 |
| S8_vs_k141_115311\|\|full | 1088 | No | NA | 3 | 0 | 0 | Not-determined | Genome-fragment | NA | NA | 0 |
| S8_extracted_k141_99414 | 12797 | No | NA | 21 | 1 | 2 | Low-quality | Genome-fragment | 26.83 | AAI-based (medium-confidence) | 0 |
| S11_vs_k141_80473\|\|full | 12800 | No | NA | 7 | 1 | 0 | Low-quality | Genome-fragment | 7.87 | AAI-based (high-confidence) | 0 |
| S8_vs_k141_115318\|\|full | 2892 | No | NA | 4 | 0 | 1 | Not-determined | Genome-fragment | NA | NA | 0 |
| S16_vs_14739\|\|full | 12815 | No | NA | 12 | 1 | 1 | Low-quality | Genome-fragment | 23.25 | AAI-based (high-confidence) | 0 |
| S8_vs_k141_19939\|\|full | 3042 | No | NA | 7 | 0 | 0 | Not-determined | Genome-fragment | NA | NA | 0 |
| S8_vs_k141_124526\|\|full | 12819 | No | NA | 12 | 1 | 1 | Low-quality | Genome-fragment | 7.51 | HMM-based (lower-bound) | 0 |
| S8_vs_k141_80816\|\|full | 2754 | No | NA | 3 | 0 | 3 | Not-determined | Genome-fragment | NA | NA | 0 |
| S5_extracted_k141_54812 | 12820 | No | NA | 17 | 1 | 3 | Low-quality | Genome-fragment | 6.95 | HMM-based (lower-bound) | 0 |
| S8_vs_k141_11037\|\|full | 2100 | No | NA | 2 | 0 | 0 | Not-determined | Genome-fragment | NA | NA | 0 |
| S11_vs_k141_45024\|\|full | 12822 | No | NA | 19 | 14 | 0 | Low-quality | Genome-fragment | 22.49 | AAI-based (high-confidence) | 0 |
| S5_extracted_k141_51328 | 12824 | No | NA | 15 | 3 | 0 | Low-quality | Genome-fragment | 30.85 | AAI-based (high-confidence) | 0 |
| S8_vs_k141_2718\|\|full | 1532 | No | NA | 2 | 0 | 1 | Not-determined | Genome-fragment | NA | NA | 0 |
| S8_vs_k141_201819\|\|full | 5894 | No | NA | 8 | 0 | 3 | Not-determined | Genome-fragment | NA | NA | 0 |
| S8_vs_k141_2730\|\|full | 1610 | No | NA | 2 | 0 | 1 | Not-determined | Genome-fragment | NA | NA | 0 |
| S8_vs_k141_80835\|\|full | 5371 | No | NA | 7 | 0 | 2 | Not-determined | Genome-fragment | NA | NA | 0 |
| S17_vs_k141_29084\|\|full | 12825 | No | NA | 15 | 1 | 1 | Low-quality | Genome-fragment | 7.16 | HMM-based (lower-bound) | 0 |
| S3_extracted_k141_115541 | 12827 | No | NA | 18 | 12 | 0 | Low-quality | Genome-fragment | 30.54 | AAI-based (high-confidence) | 0 |
| S8_vs_k141_115339\|\|full | 6815 | No | NA | 8 | 0 | 2 | Not-determined | Genome-fragment | NA | NA | 0 |
| S16_vs_146364\|\|full | 12831 | No | NA | 15 | 3 | 0 | Low-quality | Genome-fragment | 11.56 | AAI-based (high-confidence) | 0 |
| S13_extracted_33277 | 12837 | No | NA | 8 | 2 | 1 | Low-quality | Genome-fragment | 7.9 | AAI-based (high-confidence) | 0 |
| S8_vs_k141_158459\|\|full | 1927 | No | NA | 3 | 0 | 2 | Not-determined | Genome-fragment | NA | NA | 0 |
| S8_vs_k141_97995\|\|full | 1129 | No | NA | 3 | 0 | 0 | Not-determined | Genome-fragment | NA | NA | 0 |
| S16_vs_1476\|\|full | 12843 | No | NA | 20 | 3 | 0 | Low-quality | Genome-fragment | 36.92 | AAI-based (high-confidence) | 0 |
| S12_vs_k141_94804\|\|full | 12845 | No | NA | 6 | 2 | 0 | Low-quality | Genome-fragment | 21.91 | AAI-based (high-confidence) | 0 |
| S8_vs_k141_184321\|\|full | 2112 | No | NA | 4 | 0 | 4 | Not-determined | Genome-fragment | NA | NA | 0 |
| S8_vs_k141_201897\|\|full | 2577 | No | NA | 4 | 0 | 0 | Not-determined | Genome-fragment | NA | NA | 0 |
| S16_extracted_164572 | 12869 | No | NA | 16 | 4 | 0 | Low-quality | Genome-fragment | 42.69 | HMM-based (lower-bound) | 0 |
| S16_vs_95178\|\|full | 12869 | No | NA | 11 | 1 | 1 | Low-quality | Genome-fragment | 22.9 | HMM-based (lower-bound) | 0 |
| S8_vs_k141_20039\|\|full | 5456 | No | NA | 9 | 0 | 5 | Not-determined | Genome-fragment | NA | NA | 0 |
| S8_vs_k141_37526\|\|full | 7167 | No | NA | 4 | 0 | 2 | Not-determined | Genome-fragment | NA | NA | 0 |
| S8_vs_k141_98046\|\|full | 2195 | No | NA | 5 | 0 | 0 | Not-determined | Genome-fragment | NA | NA | 0 |
| S8_vs_k141_71601\|\|full | 1799 | No | NA | 3 | 0 | 0 | Not-determined | Genome-fragment | NA | NA | 0 |
| S8_vs_k141_132700\|\|full | 1784 | No | NA | 3 | 0 | 2 | Not-determined | Genome-fragment | NA | NA | 0 |
| S16_extracted_33815 | 12878 | No | NA | 16 | 4 | 0 | Low-quality | Genome-fragment | 27.66 | AAI-based (medium-confidence) | 0 |
| S8_vs_k141_20066\|\|full | 1764 | No | NA | 3 | 0 | 2 | Not-determined | Genome-fragment | NA | NA | 0 |
| S8_vs_k141_80937\|\|full | 1179 | No | NA | 3 | 0 | 1 | Not-determined | Genome-fragment | NA | NA | 0 |
| S8_vs_k141_29025\|\|full | 1887 | No | NA | 2 | 0 | 1 | Not-determined | Genome-fragment | NA | NA | 0 |
| S17_extracted_k141_81839 | 12886 | No | NA | 14 | 1 | 1 | Low-quality | Genome-fragment | 17.29 | AAI-based (high-confidence) | 0 |
| S12_vs_k141_3913\|\|full | 12895 | No | NA | 15 | 1 | 0 | Low-quality | Genome-fragment | 15.29 | HMM-based (lower-bound) | 0 |
| S8_vs_k141_98070\|\|full | 2135 | No | NA | 3 | 0 | 0 | Not-determined | Genome-fragment | NA | NA | 0 |
| S8_vs_k141_132724\|\|full | 5333 | No | NA | 4 | 0 | 3 | Not-determined | Genome-fragment | NA | NA | 0 |
| S8_vs_k141_45863\|\|full | 1257 | No | NA | 2 | 0 | 1 | Not-determined | Genome-fragment | NA | NA | 0 |
| S8_vs_k141_184402\|\|full | 2691 | No | NA | 3 | 0 | 3 | Not-determined | Genome-fragment | NA | NA | 0 |
| S17_vs_k141_90767\|\|full | 12907 | No | NA | 19 | 0 | 1 | Low-quality | Genome-fragment | 12.13 | AAI-based (medium-confidence) | 0 |
| S8_vs_k141_132762\|\|full | 1495 | No | NA | 2 | 0 | 0 | Not-determined | Genome-fragment | NA | NA | 0 |
| S8_vs_k141_150125\|\|full | 3284 | No | NA | 5 | 0 | 2 | Not-determined | Genome-fragment | NA | NA | 0 |
| S8_vs_k141_124419\|\|full | 1961 | No | NA | 1 | 0 | 1 | Not-determined | Genome-fragment | NA | NA | 0 |
| S8_vs_k141_80975\|\|full | 2632 | No | NA | 2 | 0 | 2 | Not-determined | Genome-fragment | NA | NA | 0 |
| S8_vs_k141_98141\|\|full | 1842 | No | NA | 3 | 0 | 2 | Not-determined | Genome-fragment | NA | NA | 0 |
| S8_vs_k141_158598\|\|full | 2037 | No | NA | 4 | 0 | 2 | Not-determined | Genome-fragment | NA | NA | 0 |
| S7_vs_112452\|\|full | 12914 | No | NA | 36 | 4 | 0 | Low-quality | Genome-fragment | 31.54 | AAI-based (high-confidence) | 0 |
| S8_vs_k141_124445\|\|full | 5140 | No | NA | 9 | 0 | 1 | Not-determined | Genome-fragment | NA | NA | 0 |
| S12_extracted_k141_6532 | 12951 | No | NA | 24 | 3 | 0 | Low-quality | Genome-fragment | 20.47 | AAI-based (high-confidence) | 0 |
| S8_vs_k141_106955\|\|full | 1278 | No | NA | 3 | 0 | 3 | Not-determined | Genome-fragment | NA | NA | 0 |
| S11_vs_k141_52627\|\|full | 12956 | No | NA | 31 | 6 | 0 | Low-quality | Genome-fragment | 22.4 | AAI-based (high-confidence) | 0 |
| S8_vs_k141_37634\|\|full | 2645 | No | NA | 4 | 0 | 2 | Not-determined | Genome-fragment | NA | NA | 0 |
| S12_extracted_k141_173402 | 12959 | No | NA | 10 | 1 | 0 | Low-quality | Genome-fragment | 7.79 | AAI-based (high-confidence) | 0 |
| S8_vs_k141_106964\|\|full | 6386 | No | NA | 9 | 0 | 5 | Not-determined | Genome-fragment | NA | NA | 0 |

| | | | | | | | | | | | |
|---|---|---|---|---|---|---|---|---|---|---|---|
| S8_vs_k141_89563\|\|full | 2204 | No | NA | 3 | 0 | 3 | Not-determined | Genome-fragment | NA | NA | 0 |
| S16_vs_155121\|\|full | 12961 | No | NA | 17 | 5 | 1 | Low-quality | Genome-fragment | 33.48 | AAI-based (high-confidence) | 0 |
| S8_vs_k141_29036\|\|full | 1496 | No | NA | 3 | 0 | 1 | Not-determined | Genome-fragment | NA | NA | 0 |
| S8_vs_k141_45969\|\|full | 1190 | No | NA | 3 | 0 | 0 | Not-determined | Genome-fragment | NA | NA | 0 |
| S12_extracted_k141_57353 | 12988 | No | NA | 11 | 0 | 3 | Low-quality | Genome-fragment | 15.51 | AAI-based (medium-confidence) | 0 |
| S7_vs_1137\|\|full | 12992 | No | NA | 14 | 9 | 0 | Low-quality | Genome-fragment | 25.55 | AAI-based (medium-confidence) | 0 |
| S8_vs_k141_158661\|\|full | 6096 | No | NA | 4 | 0 | 1 | Not-determined | Genome-fragment | NA | NA | 0 |
| S16_extracted_133720 | 12993 | No | NA | 34 | 3 | 0 | Low-quality | Genome-fragment | 16.11 | AAI-based (high-confidence) | 0 |
| S8_vs_k141_11254\|\|full | 2481 | No | NA | 2 | 0 | 1 | Not-determined | Genome-fragment | NA | NA | 0 |
| S12_extracted_k141_58717 | 12998 | No | NA | 12 | 0 | 3 | Low-quality | Genome-fragment | 16.67 | AAI-based (medium-confidence) | 0 |
| S8_vs_k141_176413\|\|full | 3193 | No | NA | 4 | 0 | 1 | Not-determined | Genome-fragment | NA | NA | 0 |
| S5_vs_k141_90722\|\|full | 13004 | No | NA | 17 | 8 | 0 | Low-quality | Genome-fragment | 21.66 | AAI-based (medium-confidence) | 0 |
| S8_vs_k141_37674\|\|full | 2679 | No | NA | 2 | 0 | 1 | Not-determined | Genome-fragment | NA | NA | 0 |
| S8_vs_k141_202073\|\|full | 1481 | No | NA | 5 | 0 | 4 | Not-determined | Genome-fragment | NA | NA | 0 |
| S8_vs_k141_2857\|\|full | 5447 | No | NA | 4 | 0 | 4 | Not-determined | Genome-fragment | NA | NA | 0 |
| S8_vs_k141_11286\|\|full | 15110 | No | NA | 16 | 0 | 6 | Not-determined | Genome-fragment | NA | NA | 0 |
| S8_vs_k141_63753\|\|full | 1452 | No | NA | 2 | 0 | 0 | Not-determined | Genome-fragment | NA | NA | 0 |
| S12_extracted_k141_183029 | 13006 | No | NA | 17 | 7 | 0 | Low-quality | Genome-fragment | 23.09 | AAI-based (high-confidence) | 0 |
| S12_vs_k141_29646\|\|full | 13015 | No | NA | 30 | 2 | 0 | Low-quality | Genome-fragment | 17.12 | HMM-based (lower-bound) | 0 |
| S16_vs_81787\|\|full | 13016 | Yes | 4503 | 18 | 4 | 4 | Low-quality | Genome-fragment | 8.03 | HMM-based (lower-bound) | 65.4 |
| S8_vs_k141_71727\|\|full | 4603 | No | NA | 6 | 0 | 2 | Not-determined | Genome-fragment | NA | NA | 0 |
| S8_vs_k141_89628\|\|full | 1585 | No | NA | 4 | 0 | 1 | Not-determined | Genome-fragment | NA | NA | 0 |
| S8_vs_k141_29071\|\|full | 4632 | No | NA | 6 | 0 | 0 | Not-determined | Genome-fragment | NA | NA | 0 |
| S13_vs_4814\|\|0_partial | 13019 | Yes | 5257 | 10 | 2 | 5 | Low-quality | Genome-fragment | 5.65 | HMM-based (lower-bound) | 59.62 |
| S8_vs_k141_78262\|\|full | 13026 | Yes | 10857 | 8 | 1 | 1 | Low-quality | Genome-fragment | 7.3 | HMM-based (lower-bound) | 16.65 |
| S8_vs_k141_158704\|\|full | 1212 | No | NA | 2 | 0 | 0 | Not-determined | Genome-fragment | NA | NA | 0 |
| S8_vs_k141_20282\|\|full | 1382 | No | NA | 2 | 0 | 2 | Not-determined | Genome-fragment | NA | NA | 0 |
| S8_vs_k141_89659\|\|full | 1957 | No | NA | 4 | 0 | 2 | Not-determined | Genome-fragment | NA | NA | 0 |
| S16_extracted_96480 | 13044 | No | NA | 13 | 3 | 0 | Low-quality | Genome-fragment | 13.54 | AAI-based (medium-confidence) | 0 |
| S8_vs_k141_202119\|\|full | 1111 | No | NA | 2 | 0 | 0 | Not-determined | Genome-fragment | NA | NA | 0 |
| S16_vs_48371\|\|full | 13046 | No | NA | 14 | 0 | 8 | Low-quality | Genome-fragment | 6.31 | AAI-based (medium-confidence) | 0 |
| S17_extracted_k141_26633 | 13060 | No | NA | 7 | 0 | 6 | Low-quality | Genome-fragment | 11.31 | AAI-based (medium-confidence) | 0 |
| S3_vs_k141_98369\|\|full | 13073 | No | NA | 12 | 0 | 10 | Low-quality | Genome-fragment | 5.95 | AAI-based (medium-confidence) | 0 |
| S12_vs_k141_132289\|\|full | 13086 | No | NA | 17 | 4 | 0 | Low-quality | Genome-fragment | 43.89 | HMM-based (lower-bound) | 0 |
| S8_vs_k141_115671\|\|full | 2561 | No | NA | 3 | 0 | 2 | Not-determined | Genome-fragment | NA | NA | 0 |
| S8_vs_k141_89679\|\|full | 3177 | No | NA | 5 | 0 | 3 | Not-determined | Genome-fragment | NA | NA | 0 |
| S16_vs_26095\|\|full | 13092 | No | NA | 16 | 15 | 0 | Low-quality | Genome-fragment | 40.05 | AAI-based (high-confidence) | 0 |
| S8_vs_k141_37751\|\|full | 2131 | No | NA | 3 | 0 | 1 | Not-determined | Genome-fragment | NA | NA | 0 |
| S16_vs_120907\|\|full | 13097 | No | NA | 23 | 6 | 0 | Low-quality | Genome-fragment | 26.29 | AAI-based (medium-confidence) | 0 |
| S8_vs_k141_153320\|\|0_partial | 13100 | No | NA | 13 | 1 | 3 | Low-quality | Genome-fragment | 4.69 | HMM-based (lower-bound) | 0 |
| S8_vs_k141_89701\|\|full | 1675 | No | NA | 2 | 0 | 0 | Not-determined | Genome-fragment | NA | NA | 0 |
| S3_vs_k141_80384\|\|full | 13106 | No | NA | 16 | 2 | 0 | Low-quality | Genome-fragment | 36.86 | AAI-based (high-confidence) | 0 |
| S8_vs_k141_71802\|\|full | 1049 | No | NA | 2 | 0 | 0 | Not-determined | Genome-fragment | NA | NA | 0 |
| S14_vs_k141_101601\|\|0_partial | 13113 | Yes | 7089 | 16 | 3 | 5 | Low-quality | Genome-fragment | 11.03 | HMM-based (lower-bound) | 45.94 |
| S8_vs_k141_150263\|\|full | 3094 | No | NA | 3 | 0 | 2 | Not-determined | Genome-fragment | NA | NA | 0 |
| S11_vs_k141_59044\|\|full | 13127 | No | NA | 19 | 6 | 0 | Low-quality | Genome-fragment | 22.05 | AAI-based (high-confidence) | 0 |
| S17_vs_k141_35500\|\|full | 13135 | No | NA | 23 | 2 | 0 | Low-quality | Genome-fragment | 16.31 | AAI-based (high-confidence) | 0 |
| S8_vs_k141_55115\|\|full | 3626 | No | NA | 4 | 0 | 1 | Not-determined | Genome-fragment | NA | NA | 0 |
| S12_vs_k141_87401\|\|full | 13139 | No | NA | 16 | 8 | 1 | Low-quality | Genome-fragment | 36.05 | AAI-based (high-confidence) | 0 |
| S8_vs_k141_141796\|\|full | 1349 | No | NA | 3 | 0 | 0 | Not-determined | Genome-fragment | NA | NA | 0 |
| S8_vs_k141_20357\|\|full | 11834 | No | NA | 12 | 0 | 11 | Not-determined | Genome-fragment | NA | NA | 0 |
| S8_vs_k141_141802\|\|full | 2165 | No | NA | 3 | 0 | 1 | Not-determined | Genome-fragment | NA | NA | 0 |
| S12_vs_k141_97523\|\|full | 13139 | No | NA | 20 | 14 | 1 | Low-quality | Genome-fragment | 21.44 | AAI-based (high-confidence) | 0 |
| S8_vs_k141_202171\|\|full | 1127 | No | NA | 3 | 0 | 2 | Not-determined | Genome-fragment | NA | NA | 0 |
| S8_vs_k141_29205\|\|full | 3902 | No | NA | 7 | 0 | 0 | Not-determined | Genome-fragment | NA | NA | 0 |
| S8_vs_k141_184531\|\|full | 13152 | No | NA | 18 | 1 | 1 | Low-quality | Genome-fragment | 33.83 | AAI-based (high-confidence) | 0 |
| S3_extracted_k141_80559 | 13172 | No | NA | 24 | 1 | 1 | Low-quality | Genome-fragment | 27.48 | AAI-based (high-confidence) | 0 |
| S8_vs_k141_71871\|\|full | 1543 | No | NA | 3 | 0 | 1 | Not-determined | Genome-fragment | NA | NA | 0 |
| S8_vs_k141_55149\|\|full | 2821 | No | NA | 3 | 0 | 1 | Not-determined | Genome-fragment | NA | NA | 0 |
| S8_vs_k141_141827\|\|full | 4844 | No | NA | 7 | 0 | 2 | Not-determined | Genome-fragment | NA | NA | 0 |
| S8_vs_k141_98413\|\|full | 3972 | No | NA | 3 | 0 | 3 | Not-determined | Genome-fragment | NA | NA | 0 |
| S8_vs_k141_81205\|\|full | 2629 | No | NA | 3 | 0 | 2 | Not-determined | Genome-fragment | NA | NA | 0 |
| S7_extracted_114246 | 13178 | No | NA | 22 | 0 | 1 | Low-quality | Genome-fragment | 7.89 | AAI-based (medium-confidence) | 0 |
| S15_vs_8977\|\|full | 13178 | No | NA | 14 | 1 | 6 | Low-quality | Genome-fragment | 18.76 | HMM-based (lower-bound) | 0 |
| S8_vs_k141_186207\|\|full | 13180 | No | NA | 14 | 4 | 0 | Low-quality | Genome-fragment | 44.74 | HMM-based (lower-bound) | 0 |
| S12_extracted_k141_39091 | 13181 | No | NA | 17 | 0 | 5 | Low-quality | Genome-fragment | 45.69 | AAI-based (medium-confidence) | 0 |
| S8_vs_k141_89178\|\|full | 13181 | Yes | 7376 | 17 | 3 | 2 | Low-quality | Genome-fragment | 1.09 | AAI-based (medium-confidence) | 44.04 |
| S8_vs_k141_29236\|\|full | 4303 | No | NA | 4 | 0 | 4 | Not-determined | Genome-fragment | NA | NA | 0 |
| S8_vs_k141_107211\|\|full | 1507 | No | NA | 3 | 0 | 1 | Not-determined | Genome-fragment | NA | NA | 0 |
| S3_vs_k141_14542\|\|0_partial | 13184 | No | NA | 13 | 1 | 8 | Low-quality | Genome-fragment | 18.77 | HMM-based (lower-bound) | 0 |
| S8_vs_k141_176602\|\|full | 1629 | No | NA | 3 | 0 | 1 | Not-determined | Genome-fragment | NA | NA | 0 |
| S8_vs_k141_71932\|\|full | 1139 | No | NA | 4 | 0 | 0 | Not-determined | Genome-fragment | NA | NA | 0 |
| S8_vs_k141_98450\|\|full | 3157 | No | NA | 3 | 0 | 1 | Not-determined | Genome-fragment | NA | NA | 0 |
| S5_vs_k141_123589\|\|full | 13188 | No | NA | 13 | 1 | 2 | Low-quality | Genome-fragment | 11.54 | HMM-based (lower-bound) | 0 |
| S14_vs_k141_98593\|\|full | 13196 | No | NA | 19 | 1 | 0 | Low-quality | Genome-fragment | 27.02 | AAI-based (high-confidence) | 0 |
| S8_vs_k141_3059\|\|full | 1030 | No | NA | 2 | 0 | 1 | Not-determined | Genome-fragment | NA | NA | 0 |
| S8_vs_k141_158895\|\|full | 1159 | No | NA | 2 | 0 | 0 | Not-determined | Genome-fragment | NA | NA | 0 |
| S8_vs_k141_176604\|\|full | 36150 | No | NA | 61 | 19 | 1 | High-quality | High-quality | 100 | AAI-based (high-confidence) | 0 |
| S8_vs_k141_46180\|\|full | 1945 | No | NA | 4 | 0 | 0 | Not-determined | Genome-fragment | NA | NA | 0 |
| S12_vs_k141_151619\|\|full | 13205 | No | NA | 9 | 0 | 7 | Low-quality | Genome-fragment | 4.25 | AAI-based (medium-confidence) | 0 |
| S4_vs_VIRSorter_20401-cat_2 | 13209 | No | NA | 17 | 4 | 0 | Low-quality | Genome-fragment | 44.3 | HMM-based (lower-bound) | 0 |
| S8_vs_k141_176613\|\|full | 2909 | No | NA | 4 | 0 | 3 | Not-determined | Genome-fragment | NA | NA | 0 |
| S8_vs_k141_167779\|\|full | 4999 | No | NA | 5 | 0 | 4 | Not-determined | Genome-fragment | NA | NA | 0 |
| S8_vs_k141_107270\|\|full | 7449 | No | NA | 3 | 0 | 1 | Not-determined | Genome-fragment | NA | NA | 0 |
| S11_vs_k141_23661\|\|full | 13210 | No | NA | 27 | 7 | 1 | Low-quality | Genome-fragment | 17.53 | HMM-based (lower-bound) | 0 |
| S3_vs_k141_102625\|\|full | 13215 | Yes | 7705 | 10 | 3 | 2 | Low-quality | Genome-fragment | 14.48 | AAI-based (medium-confidence) | 41.7 |
| S8_vs_k141_98513\|\|full | 1861 | No | NA | 3 | 0 | 1 | Not-determined | Genome-fragment | NA | NA | 0 |
| S8_vs_k141_158910\|\|full | 7297 | No | NA | 8 | 0 | 4 | Not-determined | Genome-fragment | NA | NA | 0 |
| S11_vs_k141_93968\|\|full | 13218 | No | NA | 30 | 0 | 0 | Low-quality | Genome-fragment | 30.77 | AAI-based (medium-confidence) | 0 |
| S5_vs_k141_42741\|\|full | 13226 | No | NA | 20 | 8 | 1 | Low-quality | Genome-fragment | 32.42 | AAI-based (medium-confidence) | 0 |
| S8_vs_k141_115829\|\|full | 1439 | No | NA | 2 | 0 | 0 | Not-determined | Genome-fragment | NA | NA | 0 |
| S8_vs_k141_158913\|\|full | 2263 | No | NA | 4 | 0 | 0 | Not-determined | Genome-fragment | NA | NA | 0 |
| S5_vs_k141_125013\|\|full | 13229 | No | NA | 16 | 9 | 0 | Low-quality | Genome-fragment | 35.42 | AAI-based (high-confidence) | 0 |
| S3_vs_k141_84030\|\|full | 13231 | No | NA | 32 | 4 | 0 | Low-quality | Genome-fragment | 30.05 | AAI-based (high-confidence) | 0 |
| S8_vs_k141_158924\|\|full | 2034 | No | NA | 4 | 0 | 0 | Not-determined | Genome-fragment | NA | NA | 0 |
| S8_vs_k141_55185\|\|full | 1373 | No | NA | 3 | 0 | 0 | Not-determined | Genome-fragment | NA | NA | 0 |
| S3_vs_k141_121685\|\|full | 13237 | No | NA | 15 | 5 | 1 | Low-quality | Genome-fragment | 33.8 | AAI-based (medium-confidence) | 0 |
| S16_extracted_130617 | 13237 | No | NA | 14 | 6 | 0 | Low-quality | Genome-fragment | 44.55 | HMM-based (lower-bound) | 0 |
| S8_vs_k141_71972\|\|full | 2522 | No | NA | 3 | 0 | 0 | Not-determined | Genome-fragment | NA | NA | 0 |
| S8_vs_k141_184663\|\|full | 2560 | No | NA | 4 | 0 | 3 | Not-determined | Genome-fragment | NA | NA | 0 |
| S8_vs_k141_98546\|\|full | 3020 | No | NA | 5 | 0 | 0 | Not-determined | Genome-fragment | NA | NA | 0 |
| S8_vs_k141_29339\|\|full | 1902 | No | NA | 3 | 0 | 1 | Not-determined | Genome-fragment | NA | NA | 0 |
| S16_extracted_112846 | 13239 | No | NA | 25 | 3 | 0 | Low-quality | Genome-fragment | 28.87 | AAI-based (high-confidence) | 0 |
| S8_vs_k141_55217\|\|full | 3847 | No | NA | 3 | 0 | 1 | Not-determined | Genome-fragment | NA | NA | 0 |
| S8_vs_k141_29356\|\|full | 1220 | No | NA | 4 | 0 | 1 | Not-determined | Genome-fragment | NA | NA | 0 |
| S8_vs_k141_107334\|\|full | 3020 | No | NA | 2 | 0 | 2 | Not-determined | Genome-fragment | NA | NA | 0 |
| S8_vs_k141_71981\|\|full | 10449 | No | NA | 11 | 0 | 1 | Not-determined | Genome-fragment | NA | NA | 0 |
| S5_vs_k141_62565\|\|full | 13240 | No | NA | 13 | 3 | 0 | Low-quality | Genome-fragment | 7.85 | HMM-based (lower-bound) | 0 |
| S8_vs_k141_11530\|\|full | 2608 | No | NA | 2 | 0 | 2 | Not-determined | Genome-fragment | NA | NA | 0 |
| S8_vs_k141_133191\|\|full | 1679 | No | NA | 3 | 0 | 0 | Not-determined | Genome-fragment | NA | NA | 0 |
| S8_vs_k141_158951\|\|full | 3330 | No | NA | 6 | 0 | 0 | Not-determined | Genome-fragment | NA | NA | 0 |
| S8_vs_k141_115896\|\|full | 2931 | No | NA | 4 | 0 | 3 | Not-determined | Genome-fragment | NA | NA | 0 |
| S3_vs_k141_6367\|\|full | 13251 | No | NA | 15 | 12 | 0 | Low-quality | Genome-fragment | 25.15 | HMM-based (lower-bound) | 0 |
| S8_vs_k141_37932\|\|full | 1283 | No | NA | 2 | 0 | 2 | Not-determined | Genome-fragment | NA | NA | 0 |
| S8_vs_k141_167854\|\|full | 1518 | No | NA | 4 | 0 | 0 | Not-determined | Genome-fragment | NA | NA | 0 |
| S8_vs_k141_107341\|\|full | 2352 | No | NA | 4 | 0 | 0 | Not-determined | Genome-fragment | NA | NA | 0 |
| S15_vs_35733\|\|full | 13252 | No | NA | 13 | 0 | 1 | Low-quality | Genome-fragment | 12.9 | AAI-based (medium-confidence) | 0 |
| S6_vs_k141_14331\|\|full | 13254 | No | NA | 21 | 2 | 1 | Low-quality | Genome-fragment | 12.87 | HMM-based (lower-bound) | 0 |
| S14_vs_k141_68168\|\|full | 13260 | No | NA | 31 | 3 | 1 | Low-quality | Genome-fragment | 22.29 | AAI-based (medium-confidence) | 0 |
| S8_vs_k141_63988\|\|full | 3697 | No | NA | 3 | 0 | 1 | Not-determined | Genome-fragment | NA | NA | 0 |
| S17_extracted_k141_35476 | 13261 | No | NA | 29 | 4 | 0 | Low-quality | Genome-fragment | 37.05 | AAI-based (high-confidence) | 0 |
| S8_vs_k141_194186\|\|full | 2393 | No | NA | 3 | 0 | 1 | Not-determined | Genome-fragment | NA | NA | 0 |
| S8_vs_k141_150484\|\|full | 1770 | No | NA | 3 | 0 | 1 | Not-determined | Genome-fragment | NA | NA | 0 |
| S17_vs_k141_122103\|\|full | 13261 | No | NA | 26 | 3 | 0 | Low-quality | Genome-fragment | 19.41 | HMM-based (lower-bound) | 0 |
| S8_vs_k141_107373\|\|full | 2693 | No | NA | 3 | 0 | 1 | Not-determined | Genome-fragment | NA | NA | 0 |
| S12_extracted_k141_111822 | 13261 | No | NA | 20 | 6 | 0 | Low-quality | Genome-fragment | 44.63 | HMM-based (lower-bound) | 0 |
| S7_vs_21301\|\|full | 13274 | No | NA | 23 | 4 | 0 | Low-quality | Genome-fragment | 15.58 | HMM-based (lower-bound) | 0 |
| S8_vs_k141_159007\|\|full | 2189 | No | NA | 3 | 0 | 3 | Not-determined | Genome-fragment | NA | NA | 0 |
| S6_vs_k141_43120\|\|0_partial | 13280 | No | NA | 13 | 0 | 4 | Low-quality | Genome-fragment | 15.14 | AAI-based (medium-confidence) | 0 |
| S8_vs_k141_194213\|\|full | 1456 | No | NA | 3 | 0 | 1 | Not-determined | Genome-fragment | NA | NA | 0 |
| S8_vs_k141_115946\|\|full | 3894 | No | NA | 7 | 0 | 2 | Not-determined | Genome-fragment | NA | NA | 0 |
| S7_vs_93022\|\|full | 13281 | No | NA | 18 | 8 | 0 | Low-quality | Genome-fragment | 44.24 | HMM-based (lower-bound) | 0 |
| S8_vs_k141_20567\|\|full | 3156 | No | NA | 3 | 0 | 2 | Not-determined | Genome-fragment | NA | NA | 0 |
| S8_vs_k141_29476\|\|full | 2700 | No | NA | 4 | 0 | 2 | Not-determined | Genome-fragment | NA | NA | 0 |
| S8_vs_k141_176720\|\|full | 23214 | No | NA | 37 | 14 | 0 | Medium-quality | Genome-fragment | 60.95 | AAI-based (high-confidence) | 0 |
| S2_vs_k141_310\|\|full | 13281 | No | NA | 17 | 7 | 0 | Low-quality | Genome-fragment | 41.69 | HMM-based (lower-bound) | 0 |
| S5_extracted_k141_8532 | 13293 | No | NA | 15 | 4 | 0 | Low-quality | Genome-fragment | 44.58 | HMM-based (lower-bound) | 0 |
| S8_vs_k141_3218\|\|full | 5732 | No | NA | 4 | 0 | 3 | Not-determined | Genome-fragment | NA | NA | 0 |
| S8_vs_k141_29492\|\|full | 7794 | No | NA | 10 | 0 | 2 | Not-determined | Genome-fragment | NA | NA | 0 |
| S12_extracted_k141_10968 | 13295 | No | NA | 15 | 0 | 6 | Low-quality | Genome-fragment | 7.43 | AAI-based (high-confidence) | 0 |
| S4_extracted_18555 | 13297 | No | NA | 18 | 15 | 0 | Low-quality | Genome-fragment | 38.47 | AAI-based (medium-confidence) | 0 |
| S3_vs_k141_58941\|\|full | 13304 | Yes | 10999 | 17 | 3 | 2 | Low-quality | Genome-fragment | 14.6 | HMM-based (lower-bound) | 17.33 |
| S8_vs_k141_184810\|\|full | 1802 | No | NA | 3 | 0 | 2 | Not-determined | Genome-fragment | NA | NA | 0 |
| S3_extracted_k141_18959 | 13312 | No | NA | 21 | 4 | 1 | Low-quality | Genome-fragment | 15.38 | AAI-based (high-confidence) | 0 |
| S8_vs_k141_194246\|\|full | 6133 | No | NA | 7 | 0 | 3 | Not-determined | Genome-fragment | NA | NA | 0 |
| S8_vs_k141_38081\|\|full | 2561 | No | NA | 4 | 0 | 0 | Not-determined | Genome-fragment | NA | NA | 0 |
| S8_vs_k141_55354\|\|full | 3546 | No | NA | 6 | 0 | 2 | Not-determined | Genome-fragment | NA | NA | 0 |
| S15_extracted_50579 | 13319 | No | NA | 11 | 1 | 0 | Low-quality | Genome-fragment | 14.5 | AAI-based (high-confidence) | 0 |
| S8_vs_k141_202487\|\|full | 1102 | No | NA | 2 | 0 | 1 | Not-determined | Genome-fragment | NA | NA | 0 |
| S8_vs_k141_187798\|\|full | 13338 | No | NA | 19 | 1 | 1 | Low-quality | Genome-fragment | 22.45 | AAI-based (high-confidence) | 0 |
| S8_vs_k141_142097\|\|full | 15982 | No | NA | 19 | 7 | 0 | Medium-quality | Genome-fragment | 74.41 | AAI-based (medium-confidence) | 0 |
| S14_vs_k141_20364\|\|full | 13343 | No | NA | 16 | 8 | 0 | Low-quality | Genome-fragment | 36.04 | AAI-based (high-confidence) | 0 |
| S8_vs_k141_3224\|\|full | 41293 | No | NA | 65 | 24 | 1 | High-quality | High-quality | 96.53 | AAI-based (medium-confidence) | 0 |
| S8_vs_k141_20642\|\|full | 2076 | No | NA | 3 | 0 | 0 | Not-determined | Genome-fragment | NA | NA | 0 |
| S8_vs_k141_29531\|\|full | 2539 | No | NA | 4 | 0 | 0 | Not-determined | Genome-fragment | NA | NA | 0 |
| S12_extracted_k141_124441 | 13344 | No | NA | 19 | 3 | 0 | Low-quality | Genome-fragment | 44.63 | HMM-based (lower-bound) | 0 |
| S8_vs_k141_194276\|\|full | 2808 | No | NA | 3 | 0 | 2 | Not-determined | Genome-fragment | NA | NA | 0 |
| S3_vs_k141_98778\|\|full | 13352 | No | NA | 22 | 6 | 0 | Low-quality | Genome-fragment | 44.94 | HMM-based (lower-bound) | 0 |
| S8_vs_k141_46317\|\|full | 2369 | No | NA | 3 | 0 | 0 | Not-determined | Genome-fragment | NA | NA | 0 |
| S8_vs_k141_202526\|\|full | 1193 | No | NA | 2 | 0 | 1 | Not-determined | Genome-fragment | NA | NA | 0 |
| S8_vs_k141_168091\|\|full | 1807 | No | NA | 3 | 0 | 1 | Not-determined | Genome-fragment | NA | NA | 0 |
| S8_vs_k141_81403\|\|full | 2682 | No | NA | 4 | 0 | 2 | Not-determined | Genome-fragment | NA | NA | 0 |
| S8_vs_k141_159165\|\|full | 1803 | No | NA | 3 | 0 | 3 | Not-determined | Genome-fragment | NA | NA | 0 |
| S8_vs_k141_20661\|\|full | 1088 | No | NA | 4 | 0 | 3 | Not-determined | Genome-fragment | NA | NA | 0 |
| S13_extracted_153039 | 13360 | No | NA | 22 | 5 | 0 | Low-quality | Genome-fragment | 22.09 | AAI-based (high-confidence) | 0 |
| S9_extracted_k141_70111 | 13363 | No | NA | 17 | 3 | 0 | Low-quality | Genome-fragment | 41.32 | HMM-based (lower-bound) | 0 |
| S8_vs_k141_142143\|\|full | 1164 | No | NA | 3 | 0 | 0 | Not-determined | Genome-fragment | NA | NA | 0 |
| S10_vs_52220\|\|0_partial | 13365 | No | NA | 16 | 1 | 3 | Low-quality | Genome-fragment | 8.98 | HMM-based (lower-bound) | 0 |
| S8_vs_k141_20670\|\|full | 2760 | No | NA | 2 | 0 | 1 | Not-determined | Genome-fragment | NA | NA | 0 |
| S11_extracted_k141_83592 | 13367 | No | NA | 15 | 1 | 0 | Low-quality | Genome-fragment | 11.19 | AAI-based (medium-confidence) | 0 |
| S8_vs_k141_107521\|\|full | 1030 | No | NA | 3 | 0 | 0 | Not-determined | Genome-fragment | NA | NA | 0 |
| S8_vs_k141_194320\|\|full | 1265 | No | NA | 2 | 0 | 0 | Not-determined | Genome-fragment | NA | NA | 0 |
| S8_vs_k141_107535\|\|full | 1363 | No | NA | 2 | 0 | 1 | Not-determined | Genome-fragment | NA | NA | 0 |
| S11_vs_k141_101873\|\|full | 13370 | No | NA | 22 | 2 | 0 | Low-quality | Genome-fragment | 31.2 | AAI-based (high-confidence) | 0 |
| S8_vs_k141_133309\|\|full | 1500 | No | NA | 2 | 0 | 0 | Not-determined | Genome-fragment | NA | NA | 0 |
| S8_vs_k141_159177\|\|full | 6338 | No | NA | 10 | 0 | 1 | Not-determined | Genome-fragment | NA | NA | 0 |
| S8_extracted_k141_36604 | 13372 | No | NA | 19 | 9 | 1 | Low-quality | Genome-fragment | 20.62 | HMM-based (lower-bound) | 0 |
| S8_vs_k141_115958\|\|full | 137050 | Yes | 57290 | 137 | 27 | 50 | High-quality | High-quality | 100 | AAI-based (high-confidence) | 58.2 |
| S8_vs_k141_159191\|\|full | 2737 | No | NA | 4 | 0 | 2 | Not-determined | Genome-fragment | NA | NA | 0 |
| S5_vs_k141_38139\|\|0_partial | 13373 | Yes | 7481 | 7 | 1 | 4 | Low-quality | Genome-fragment | 4.06 | HMM-based (lower-bound) | 44.06 |
| S8_vs_k141_20710\|\|full | 1404 | No | NA | 4 | 0 | 1 | Not-determined | Genome-fragment | NA | NA | 0 |
| S6_vs_k141_14494\|\|full | 13385 | No | NA | 30 | 1 | 2 | Low-quality | Genome-fragment | 35.55 | AAI-based (high-confidence) | 0 |
| S8_vs_k141_29608\|\|full | 1275 | No | NA | 3 | 0 | 1 | Not-determined | Genome-fragment | NA | NA | 0 |
| S8_vs_k141_38676\|\|full | 13385 | No | NA | 17 | 1 | 2 | Low-quality | Genome-fragment | 20.25 | HMM-based (lower-bound) | 0 |
| S8_vs_k141_124988\|\|full | 5229 | No | NA | 3 | 0 | 2 | Not-determined | Genome-fragment | NA | NA | 0 |
| S14_extracted_k141_107568 | 13388 | No | NA | 21 | 6 | 0 | Low-quality | Genome-fragment | 45.06 | HMM-based (lower-bound) | 0 |
| S16_vs_151237\|\|full | 13404 | No | NA | 19 | 3 | 0 | Low-quality | Genome-fragment | 44.84 | HMM-based (lower-bound) | 0 |
| S8_vs_k141_194381\|\|full | 1102 | No | NA | 3 | 0 | 1 | Not-determined | Genome-fragment | NA | NA | 0 |
| S8_vs_k141_72243\|\|full | 1125 | No | NA | 3 | 0 | 2 | Not-determined | Genome-fragment | NA | NA | 0 |
| S8_vs_k141_159237\|\|full | 2934 | No | NA | 6 | 0 | 3 | Not-determined | Genome-fragment | NA | NA | 0 |
| S16_vs_150500\|\|full | 13434 | Yes | 9471 | 22 | 4 | 3 | Low-quality | Genome-fragment | 25.2 | AAI-based (high-confidence) | 29.5 |
| S8_vs_k141_8283\|\|full | 13443 | No | NA | 15 | 6 | 0 | Low-quality | Genome-fragment | 32.32 | AAI-based (medium-confidence) | 0 |
| S12_vs_k141_66579\|\|full | 13453 | No | NA | 17 | 7 | 0 | Low-quality | Genome-fragment | 27.92 | AAI-based (high-confidence) | 0 |
| S8_vs_k141_142192\|\|full | 2290 | No | NA | 3 | 0 | 2 | Not-determined | Genome-fragment | NA | NA | 0 |
| S8_vs_k141_142195\|\|full | 2465 | No | NA | 4 | 0 | 1 | Not-determined | Genome-fragment | NA | NA | 0 |
| S8_vs_k141_159258\|\|full | 6805 | No | NA | 5 | 0 | 3 | Not-determined | Genome-fragment | NA | NA | 0 |
| S8_vs_k141_38244\|\|full | 3053 | No | NA | 5 | 0 | 2 | Not-determined | Genome-fragment | NA | NA | 0 |
| S8_vs_k141_20799\|\|full | 4792 | No | NA | 6 | 0 | 1 | Not-determined | Genome-fragment | NA | NA | 0 |
| S11_vs_k141_8243\|\|full | 13454 | No | NA | 21 | 5 | 0 | Low-quality | Genome-fragment | 19.08 | HMM-based (lower-bound) | 0 |
| S8_vs_k141_133422\|\|full | 1711 | No | NA | 2 | 0 | 0 | Not-determined | Genome-fragment | NA | NA | 0 |
| S3_vs_k141_17754\|\|full | 13474 | No | NA | 29 | 4 | 0 | Low-quality | Genome-fragment | 36.5 | AAI-based (high-confidence) | 0 |
| S8_vs_k141_72319\|\|full | 1191 | No | NA | 3 | 0 | 0 | Not-determined | Genome-fragment | NA | NA | 0 |
| S8_vs_k141_116048\|\|full | 7869 | No | NA | 7 | 0 | 4 | Not-determined | Genome-fragment | NA | NA | 0 |
| S5_extracted_k141_112816 | 13505 | No | NA | 24 | 6 | 0 | Low-quality | Genome-fragment | 27.33 | AAI-based (high-confidence) | 0 |
| S12_vs_k141_59096\|\|full | 13516 | No | NA | 28 | 2 | 0 | Low-quality | Genome-fragment | 27.39 | AAI-based (high-confidence) | 0 |
| S8_vs_k141_55487\|\|full | 3965 | No | NA | 4 | 0 | 2 | Not-determined | Genome-fragment | NA | NA | 0 |
| S7_vs_126180\|\|full | 13522 | Yes | 6080 | 15 | 2 | 3 | Low-quality | Genome-fragment | 13.28 | AAI-based (medium-confidence) | 55.04 |
| S1_vs_k141_135923\|\|full | 13523 | No | NA | 14 | 2 | 0 | Low-quality | Genome-fragment | 32.28 | AAI-based (high-confidence) | 0 |
| S12_vs_k141_92520\|\|0_partial | 13534 | No | NA | 17 | 1 | 6 | Low-quality | Genome-fragment | 17.72 | HMM-based (lower-bound) | 0 |
| S8_vs_k141_90154\|\|full | 2024 | No | NA | 3 | 0 | 1 | Not-determined | Genome-fragment | NA | NA | 0 |
| S7_vs_24188\|\|full | 13539 | No | NA | 30 | 3 | 0 | Low-quality | Genome-fragment | 37.45 | AAI-based (high-confidence) | 0 |
| S4_extracted_1816 | 13564 | No | NA | 11 | 2 | 1 | Low-quality | Genome-fragment | 30.24 | AAI-based (high-confidence) | 0 |
| S8_vs_k141_176986\|\|full | 1097 | No | NA | 4 | 0 | 1 | Not-determined | Genome-fragment | NA | NA | 0 |
| S16_vs_13486\|\|full | 13570 | No | NA | 11 | 1 | 1 | Low-quality | Genome-fragment | 30.83 | AAI-based (medium-confidence) | 0 |
| S8_vs_k141_194483\|\|full | 2697 | No | NA | 7 | 0 | 0 | Not-determined | Genome-fragment | NA | NA | 0 |
| S8_vs_k141_3472\|\|full | 4017 | No | NA | 4 | 0 | 3 | Not-determined | Genome-fragment | NA | NA | 0 |
| S9_extracted_k141_56387 | 13573 | No | NA | 18 | 7 | 1 | Low-quality | Genome-fragment | 21.74 | AAI-based (high-confidence) | 0 |
| S5_extracted_k141_141608 | 13583 | No | NA | 16 | 15 | 0 | Low-quality | Genome-fragment | 35.87 | AAI-based (high-confidence) | 0 |
| S11_vs_k141_26997\|\|full | 13586 | No | NA | 23 | 8 | 0 | Low-quality | Genome-fragment | 35.48 | AAI-based (high-confidence) | 0 |
| S16_extracted_162641 | 13597 | No | NA | 16 | 10 | 0 | Low-quality | Genome-fragment | 34.61 | AAI-based (high-confidence) | 0 |
| S8_vs_k141_11821\|\|full | 1752 | No | NA | 2 | 0 | 1 | Not-determined | Genome-fragment | NA | NA | 0 |
| S8_vs_k141_172919\|\|full | 13611 | No | NA | 20 | 5 | 0 | Low-quality | Genome-fragment | 22.78 | AAI-based (high-confidence) | 0 |
| S9_extracted_k141_17784 | 13617 | No | NA | 11 | 11 | 0 | Low-quality | Genome-fragment | 19.55 | AAI-based (high-confidence) | 0 |
| S8_vs_k141_185041\|\|full | 49943 | No | NA | 61 | 22 | 2 | High-quality | High-quality | 100 | AAI-based (high-confidence) | 0 |
| S12_vs_k141_47409\|\|full | 13626 | No | NA | 12 | 0 | 1 | Low-quality | Genome-fragment | 21.08 | AAI-based (medium-confidence) | 0 |
| S1_extracted_k141_155233 | 13627 | No | NA | 16 | 6 | 0 | Low-quality | Genome-fragment | 16.95 | AAI-based (high-confidence) | 0 |
| S8_vs_k141_177029\|\|full | 1316 | No | NA | 3 | 0 | 0 | Not-determined | Genome-fragment | NA | NA | 0 |
| S8_vs_k141_194532\|\|full | 2998 | No | NA | 4 | 0 | 2 | Not-determined | Genome-fragment | NA | NA | 0 |
| S8_vs_k141_107727\|\|full | 2672 | No | NA | 4 | 0 | 3 | Not-determined | Genome-fragment | NA | NA | 0 |
| S14_vs_k141_59982\|\|full | 13628 | No | NA | 13 | 13 | 0 | Low-quality | Genome-fragment | 33.09 | AAI-based (high-confidence) | 0 |
| S8_vs_k141_194537\|\|full | 1622 | No | NA | 3 | 0 | 0 | Not-determined | Genome-fragment | NA | NA | 0 |
| S3_vs_k141_13143\|\|full | 13648 | No | NA | 12 | 5 | 1 | Low-quality | Genome-fragment | 34.49 | AAI-based (medium-confidence) | 0 |
| S8_vs_k141_81634\|\|full | 3857 | No | NA | 7 | 0 | 2 | Not-determined | Genome-fragment | NA | NA | 0 |
| S10_vs_75439\|\|full | 13657 | No | NA | 12 | 3 | 3 | Low-quality | Genome-fragment | 13.76 | HMM-based (lower-bound) | 0 |
| S8_vs_k141_72393\|\|full | 1281 | No | NA | 3 | 0 | 0 | Not-determined | Genome-fragment | NA | NA | 0 |
| S8_vs_k141_168282\|\|full | 1609 | No | NA | 4 | 0 | 0 | Not-determined | Genome-fragment | NA | NA | 0 |
| S8_vs_k141_202694\|\|full | 3490 | No | NA | 4 | 0 | 3 | Not-determined | Genome-fragment | NA | NA | 0 |
| S8_vs_k141_150733\|\|full | 3276 | No | NA | 3 | 0 | 0 | Not-determined | Genome-fragment | NA | NA | 0 |
| S8_vs_k141_150734\|\|full | 1063 | No | NA | 2 | 0 | 1 | Not-determined | Genome-fragment | NA | NA | 0 |
| S5_vs_k141_117930\|\|full | 13665 | No | NA | 17 | 1 | 5 | Low-quality | Genome-fragment | 8.84 | HMM-based (lower-bound) | 0 |
| S16_extracted_158311 | 13670 | No | NA | 8 | 0 | 2 | Low-quality | Genome-fragment | 8.69 | AAI-based (medium-confidence) | 0 |
| S8_vs_k141_168306\|\|full | 2493 | No | NA | 2 | 0 | 0 | Not-determined | Genome-fragment | NA | NA | 0 |
| S5_vs_k141_144953\|\|full | 13673 | No | NA | 19 | 10 | 0 | Low-quality | Genome-fragment | 23.94 | HMM-based (lower-bound) | 0 |
| S9_vs_k141_3731\|\|full | 13686 | No | NA | 29 | 5 | 1 | Low-quality | Genome-fragment | 29.88 | AAI-based (high-confidence) | 0 |
| S8_vs_k141_150756\|\|full | 1869 | No | NA | 3 | 0 | 0 | Not-determined | Genome-fragment | NA | NA | 0 |
| S16_vs_152902\|\|full | 13687 | No | NA | 11 | 1 | 4 | Low-quality | Genome-fragment | 12.65 | AAI-based (medium-confidence) | 0 |
| S8_vs_k141_194604\|\|full | 1418 | No | NA | 5 | 0 | 1 | Not-determined | Genome-fragment | NA | NA | 0 |
| S8_vs_k141_72428\|\|full | 2278 | No | NA | 3 | 0 | 1 | Not-determined | Genome-fragment | NA | NA | 0 |
| S8_vs_k141_11854\|\|full | 1874 | No | NA | 4 | 0 | 1 | Not-determined | Genome-fragment | NA | NA | 0 |
| S8_vs_k141_150760\|\|full | 1637 | No | NA | 2 | 0 | 1 | Not-determined | Genome-fragment | NA | NA | 0 |
| S11_extracted_k141_99242 | 13690 | No | NA | 18 | 4 | 0 | Low-quality | Genome-fragment | 14.74 | AAI-based (medium-confidence) | 0 |
| S8_vs_k141_90248\|\|full | 1645 | No | NA | 3 | 0 | 0 | Not-determined | Genome-fragment | NA | NA | 0 |
| S16_extracted_11051 | 13704 | No | NA | 18 | 9 | 0 | Low-quality | Genome-fragment | 35.59 | AAI-based (medium-confidence) | 0 |
| S17_vs_k141_64619\|\|0_partial | 13707 | Yes | 2208 | 14 | 2 | 10 | Low-quality | Genome-fragment | 3.31 | HMM-based (lower-bound) | 83.89 |
| S8_vs_k141_29871\|\|full | 2481 | No | NA | 3 | 0 | 1 | Not-determined | Genome-fragment | NA | NA | 0 |
| S8_vs_k141_90264\|\|full | 2585 | No | NA | 3 | 0 | 2 | Not-determined | Genome-fragment | NA | NA | 0 |
| S8_vs_k141_55659\|\|full | 8351 | No | NA | 6 | 0 | 4 | Not-determined | Genome-fragment | NA | NA | 0 |
| S8_vs_k141_150783\|\|full | 1745 | No | NA | 2 | 0 | 2 | Not-determined | Genome-fragment | NA | NA | 0 |
| S8_vs_k141_150788\|\|full | 5042 | No | NA | 3 | 0 | 2 | Not-determined | Genome-fragment | NA | NA | 0 |
| S3_extracted_k141_95533 | 13713 | No | NA | 11 | 2 | 0 | Low-quality | Genome-fragment | 35.27 | AAI-based (medium-confidence) | 0 |
| S8_vs_k141_81697\|\|full | 1847 | No | NA | 3 | 0 | 0 | Not-determined | Genome-fragment | NA | NA | 0 |
| S8_vs_k141_72471\|\|full | 1745 | No | NA | 2 | 0 | 1 | Not-determined | Genome-fragment | NA | NA | 0 |
| S10_extracted_105406 | 13714 | No | NA | 8 | 0 | 3 | Low-quality | Genome-fragment | 13.81 | AAI-based (high-confidence) | 0 |
| S12_extracted_k141_40819 | 13756 | No | NA | 11 | 1 | 1 | Low-quality | Genome-fragment | 7.46 | HMM-based (lower-bound) | 0 |
| S16_vs_163744\|\|full | 13759 | No | NA | 17 | 9 | 0 | Low-quality | Genome-fragment | 22.12 | AAI-based (high-confidence) | 0 |
| S8_vs_k141_99025\|\|full | 1781 | No | NA | 3 | 0 | 0 | Not-determined | Genome-fragment | NA | NA | 0 |
| S8_vs_k141_202785\|\|full | 8354 | No | NA | 10 | 0 | 4 | Not-determined | Genome-fragment | NA | NA | 0 |
| S16_vs_8659\|\|full | 13776 | Yes | 2481 | 20 | 4 | 6 | Low-quality | Genome-fragment | 3.9 | HMM-based (lower-bound) | 81.99 |
| S17_extracted_k141_100949 | 13778 | No | NA | 14 | 4 | 1 | Low-quality | Genome-fragment | 21.2 | AAI-based (high-confidence) | 0 |
| S8_vs_k141_185161\|\|full | 1981 | No | NA | 4 | 0 | 2 | Not-determined | Genome-fragment | NA | NA | 0 |
| S8_vs_k141_194655\|\|full | 1087 | No | NA | 3 | 0 | 0 | Not-determined | Genome-fragment | NA | NA | 0 |
| S8_vs_k141_72508\|\|full | 6480 | No | NA | 6 | 0 | 1 | Not-determined | Genome-fragment | NA | NA | 0 |
| S5_vs_k141_56922\|\|full | 13788 | No | NA | 29 | 4 | 0 | Low-quality | Genome-fragment | 25.32 | AAI-based (high-confidence) | 0 |
| S13_vs_42736\|\|0_partial | 13792 | No | NA | 14 | 1 | 5 | Low-quality | Genome-fragment | 25.76 | HMM-based (lower-bound) | 0 |
| S8_vs_k141_168416\|\|full | 1167 | No | NA | 3 | 0 | 1 | Not-determined | Genome-fragment | NA | NA | 0 |
| S10_extracted_62694 | 13794 | No | NA | 10 | 0 | 9 | Low-quality | Genome-fragment | 5.11 | AAI-based (medium-confidence) | 0 |
| S8_vs_k141_159570\|\|full | 1154 | No | NA | 2 | 0 | 2 | Not-determined | Genome-fragment | NA | NA | 0 |
| S3_vs_k141_22738\|\|full | 13799 | No | NA | 21 | 7 | 0 | Low-quality | Genome-fragment | 30.13 | AAI-based (high-confidence) | 0 |
| S8_vs_k141_29958\|\|full | 1271 | No | NA | 3 | 0 | 0 | Not-determined | Genome-fragment | NA | NA | 0 |
| S11_extracted_k141_102808 | 13802 | No | NA | 12 | 2 | 0 | Low-quality | Genome-fragment | 15.18 | AAI-based (medium-confidence) | 0 |
| S8_vs_k141_159590\|\|full | 3447 | No | NA | 4 | 0 | 3 | Not-determined | Genome-fragment | NA | NA | 0 |
| S8_vs_k141_99090\|\|full | 2329 | No | NA | 3 | 0 | 2 | Not-determined | Genome-fragment | NA | NA | 0 |
| S10_extracted_101856 | 13804 | No | NA | 20 | 17 | 0 | Low-quality | Genome-fragment | 30.49 | AAI-based (high-confidence) | 0 |
| S12_vs_k141_142347\|\|full | 13809 | No | NA | 14 | 3 | 1 | Low-quality | Genome-fragment | 18.84 | HMM-based (lower-bound) | 0 |
| S3_vs_k141_82496\|\|0_partial | 13845 | Yes | 8344 | 13 | 2 | 2 | Low-quality | Genome-fragment | 8.86 | HMM-based (lower-bound) | 39.73 |
| S11_vs_k141_13930\|\|full | 13846 | No | NA | 27 | 4 | 0 | Low-quality | Genome-fragment | 8.49 | AAI-based (high-confidence) | 0 |
| S3_extracted_k141_49333 | 13846 | No | NA | 22 | 8 | 0 | Low-quality | Genome-fragment | 45.06 | HMM-based (lower-bound) | 0 |
| S12_extracted_k141_63253 | 13856 | No | NA | 21 | 2 | 0 | Low-quality | Genome-fragment | 29.22 | AAI-based (high-confidence) | 0 |
| S8_vs_k141_177178\|\|full | 7366 | No | NA | 4 | 0 | 2 | Not-determined | Genome-fragment | NA | NA | 0 |
| S8_vs_k141_168475\|\|full | 3332 | No | NA | 4 | 0 | 0 | Not-determined | Genome-fragment | NA | NA | 0 |
| S8_vs_k141_125276\|\|full | 1295 | No | NA | 3 | 0 | 0 | Not-determined | Genome-fragment | NA | NA | 0 |
| S8_vs_k141_202882\|\|full | 1303 | No | NA | 3 | 0 | 0 | Not-determined | Genome-fragment | NA | NA | 0 |
| S3_vs_k141_99710\|\|full | 13861 | No | NA | 12 | 8 | 0 | Low-quality | Genome-fragment | 34.02 | AAI-based (high-confidence) | 0 |
| S3_vs_k141_412\|\|full | 13871 | No | NA | 9 | 3 | 0 | Low-quality | Genome-fragment | 6.8 | AAI-based (high-confidence) | 0 |
| S8_vs_k141_142547\|\|full | 2035 | No | NA | 3 | 0 | 0 | Not-determined | Genome-fragment | NA | NA | 0 |
| S8_extracted_k141_205123 | 13885 | No | NA | 24 | 5 | 0 | Low-quality | Genome-fragment | 21.92 | AAI-based (high-confidence) | 0 |
| S16_extracted_116545 | 13895 | No | NA | 29 | 4 | 1 | Low-quality | Genome-fragment | 23.2 | AAI-based (high-confidence) | 0 |
| S8_vs_k141_3811\|\|full | 2095 | No | NA | 3 | 0 | 0 | Not-determined | Genome-fragment | NA | NA | 0 |
| S8_vs_k141_99175\|\|full | 7926 | No | NA | 7 | 0 | 5 | Not-determined | Genome-fragment | NA | NA | 0 |
| S11_extracted_k141_96919 | 13897 | No | NA | 25 | 5 | 1 | Low-quality | Genome-fragment | 43.84 | AAI-based (high-confidence) | 0 |
| S7_vs_121493\|\|full | 13900 | No | NA | 17 | 10 | 0 | Low-quality | Genome-fragment | 28.33 | AAI-based (high-confidence) | 0 |
| S8_vs_k141_30117\|\|full | 3058 | No | NA | 3 | 0 | 2 | Not-determined | Genome-fragment | NA | NA | 0 |
| S8_vs_k141_168580\|\|full | 1346 | No | NA | 3 | 0 | 0 | Not-determined | Genome-fragment | NA | NA | 0 |
| S8_vs_k141_108034\|\|full | 1047 | No | NA | 2 | 0 | 1 | Not-determined | Genome-fragment | NA | NA | 0 |
| S8_vs_k141_168581\|\|full | 1299 | No | NA | 3 | 0 | 1 | Not-determined | Genome-fragment | NA | NA | 0 |
| S8_vs_k141_168584\|\|full | 2327 | No | NA | 5 | 0 | 2 | Not-determined | Genome-fragment | NA | NA | 0 |
| S8_vs_k141_21140\|\|full | 6415 | No | NA | 3 | 0 | 2 | Not-determined | Genome-fragment | NA | NA | 0 |
| S12_extracted_k141_185384 | 13910 | No | NA | 18 | 5 | 0 | Low-quality | Genome-fragment | 43.87 | HMM-based (lower-bound) | 0 |
| S5_extracted_k141_119586 | 13919 | No | NA | 17 | 12 | 0 | Low-quality | Genome-fragment | 37.12 | AAI-based (high-confidence) | 0 |
| S8_vs_k141_99243\|\|full | 3619 | No | NA | 3 | 0 | 0 | Not-determined | Genome-fragment | NA | NA | 0 |
| S3_vs_k141_88927\|\|full | 13937 | No | NA | 8 | 0 | 1 | Low-quality | Genome-fragment | 32.54 | AAI-based (high-confidence) | 0 |
| S10_vs_27783\|\|full | 13953 | No | NA | 17 | 15 | 0 | Low-quality | Genome-fragment | 39.66 | AAI-based (medium-confidence) | 0 |
| S8_vs_k141_55927\|\|full | 2252 | No | NA | 4 | 0 | 0 | Not-determined | Genome-fragment | NA | NA | 0 |
| S3_vs_k141_35609\|\|full | 13958 | No | NA | 16 | 4 | 0 | Low-quality | Genome-fragment | 36.55 | AAI-based (high-confidence) | 0 |
| S8_vs_k141_196354\|\|full | 13958 | No | NA | 17 | 7 | 0 | Low-quality | Genome-fragment | 44.57 | HMM-based (lower-bound) | 0 |
| S14_vs_k141_63500\|\|full | 13960 | No | NA | 15 | 3 | 0 | Low-quality | Genome-fragment | 9.06 | HMM-based (lower-bound) | 0 |
| S8_vs_k141_12011\|\|full | 3657 | No | NA | 4 | 0 | 0 | Not-determined | Genome-fragment | NA | NA | 0 |
| S8_vs_k141_133710\|\|full | 4256 | No | NA | 4 | 0 | 3 | Not-determined | Genome-fragment | NA | NA | 0 |
| S8_vs_k141_151041\|\|full | 6997 | No | NA | 6 | 0 | 2 | Not-determined | Genome-fragment | NA | NA | 0 |
| S8_vs_k141_90518\|\|full | 2539 | No | NA | 5 | 0 | 1 | Not-determined | Genome-fragment | NA | NA | 0 |
| S13_extracted_98473 | 13978 | No | NA | 32 | 5 | 0 | Low-quality | Genome-fragment | 12.82 | AAI-based (high-confidence) | 0 |
| S8_vs_k141_110039\|\|full | 13997 | No | NA | 21 | 11 | 0 | Low-quality | Genome-fragment | 23.76 | AAI-based (high-confidence) | 0 |
| S14_extracted_k141_101406 | 14000 | No | NA | 26 | 2 | 0 | Low-quality | Genome-fragment | 24.06 | AAI-based (high-confidence) | 0 |
| S8_vs_k141_90532\|\|full | 1900 | No | NA | 2 | 0 | 1 | Not-determined | Genome-fragment | NA | NA | 0 |
| S11_vs_k141_46646\|\|full | 14012 | No | NA | 33 | 2 | 0 | Low-quality | Genome-fragment | 25.01 | AAI-based (high-confidence) | 0 |
| S8_vs_k141_151063\|\|full | 2473 | No | NA | 3 | 0 | 0 | Not-determined | Genome-fragment | NA | NA | 0 |
| S5_vs_k141_121196\|\|full | 14019 | Yes | 11778 | 18 | 3 | 2 | Low-quality | Genome-fragment | 22.1 | HMM-based (lower-bound) | 15.99 |
| S7_extracted_110206 | 14019 | No | NA | 20 | 2 | 0 | Low-quality | Genome-fragment | 21.81 | AAI-based (medium-confidence) | 0 |
| S8_vs_k141_159697\|\|full | 2299 | No | NA | 3 | 0 | 2 | Not-determined | Genome-fragment | NA | NA | 0 |
| S8_vs_k141_30224\|\|full | 1154 | No | NA | 3 | 0 | 2 | Not-determined | Genome-fragment | NA | NA | 0 |
| S8_vs_k141_185324\|\|full | 1145 | No | NA | 3 | 0 | 0 | Not-determined | Genome-fragment | NA | NA | 0 |
| S8_vs_k141_99337\|\|full | 2305 | No | NA | 3 | 0 | 3 | Not-determined | Genome-fragment | NA | NA | 0 |
| S8_vs_k141_3918\|\|full | 1847 | No | NA | 7 | 0 | 0 | Not-determined | Genome-fragment | NA | NA | 0 |
| S7_vs_61319\|\|full | 14023 | No | NA | 24 | 8 | 0 | Low-quality | Genome-fragment | 29.07 | AAI-based (high-confidence) | 0 |
| S8_vs_k141_12081\|\|full | 5747 | No | NA | 7 | 0 | 2 | Not-determined | Genome-fragment | NA | NA | 0 |
| S8_vs_k141_159720\|\|full | 3068 | No | NA | 3 | 0 | 1 | Not-determined | Genome-fragment | NA | NA | 0 |
| S13_vs_4410\|\|full | 14028 | No | NA | 14 | 1 | 1 | Low-quality | Genome-fragment | 16.01 | HMM-based (lower-bound) | 0 |
| S8_vs_k141_125471\|\|full | 11616 | No | NA | 15 | 0 | 0 | Not-determined | Genome-fragment | NA | NA | 0 |
| S8_vs_k141_151081\|\|full | 16353 | No | NA | 9 | 0 | 5 | Not-determined | Genome-fragment | NA | NA | 0 |
| S15_vs_26392\|\|full | 14035 | No | NA | 7 | 1 | 5 | Low-quality | Genome-fragment | 8.27 | HMM-based (lower-bound) | 0 |
| S1_vs_k141_123626\|\|full | 14037 | No | NA | 14 | 2 | 0 | Low-quality | Genome-fragment | 34.23 | AAI-based (high-confidence) | 0 |
| S8_vs_k141_168691\|\|full | 1787 | No | NA | 2 | 0 | 1 | Not-determined | Genome-fragment | NA | NA | 0 |
| S16_vs_86895\|\|full | 14050 | No | NA | 17 | 4 | 0 | Low-quality | Genome-fragment | 17.47 | AAI-based (high-confidence) | 0 |
| S16_extracted_49599 | 14052 | No | NA | 28 | 6 | 1 | Low-quality | Genome-fragment | 44 | AAI-based (high-confidence) | 0 |
| S8_vs_k141_194871\|\|full | 1794 | No | NA | 5 | 0 | 0 | Not-determined | Genome-fragment | NA | NA | 0 |
| S8_vs_k141_116584\|\|full | 8037 | No | NA | 14 | 0 | 5 | Not-determined | Genome-fragment | NA | NA | 0 |
| S8_vs_k141_56011\|\|full | 4230 | No | NA | 3 | 0 | 3 | Not-determined | Genome-fragment | NA | NA | 0 |
| S8_vs_k141_38524\|\|full | 3607 | No | NA | 4 | 0 | 0 | Not-determined | Genome-fragment | NA | NA | 0 |
| S5_vs_k141_103493\|\|full | 14093 | No | NA | 19 | 11 | 1 | Low-quality | Genome-fragment | 30.28 | AAI-based (high-confidence) | 0 |
| S1_extracted_k141_72164 | 14094 | No | NA | 23 | 6 | 0 | Low-quality | Genome-fragment | 25.55 | AAI-based (high-confidence) | 0 |
| S8_vs_k141_194906\|\|full | 3380 | No | NA | 4 | 0 | 2 | Not-determined | Genome-fragment | NA | NA | 0 |
| S15_extracted_73048 | 14098 | No | NA | 14 | 0 | 0 | Low-quality | Genome-fragment | 15.35 | AAI-based (high-confidence) | 0 |
| S8_vs_k141_3992\|\|full | 1057 | No | NA | 2 | 0 | 1 | Not-determined | Genome-fragment | NA | NA | 0 |
| S8_vs_k141_90659\|\|full | 1623 | No | NA | 3 | 0 | 1 | Not-determined | Genome-fragment | NA | NA | 0 |
| S8_vs_k141_120471\|\|full | 14107 | No | NA | 23 | 0 | 1 | Low-quality | Genome-fragment | 8.69 | AAI-based (high-confidence) | 0 |
| S14_vs_k141_53134\|\|full | 14132 | No | NA | 29 | 2 | 1 | Low-quality | Genome-fragment | 18.72 | AAI-based (high-confidence) | 0 |
| S8_vs_k141_38557\|\|full | 1420 | No | NA | 3 | 0 | 1 | Not-determined | Genome-fragment | NA | NA | 0 |
| S8_vs_k141_168766\|\|full | 1043 | No | NA | 2 | 0 | 0 | Not-determined | Genome-fragment | NA | NA | 0 |
| S3_vs_k141_89464\|\|full | 14133 | No | NA | 19 | 3 | 0 | Low-quality | Genome-fragment | 18.54 | HMM-based (lower-bound) | 0 |
| S14_vs_k141_82295\|\|1_partial | 14134 | No | NA | 20 | 1 | 2 | Low-quality | Genome-fragment | 5.95 | HMM-based (lower-bound) | 0 |
| S8_vs_k141_177468\|\|full | 2855 | No | NA | 3 | 0 | 2 | Not-determined | Genome-fragment | NA | NA | 0 |
| S4_extracted_31738 | 14135 | No | NA | 4 | 1 | 0 | Low-quality | Genome-fragment | 21.01 | AAI-based (high-confidence) | 0 |
| S11_vs_k141_85982\|\|full | 14140 | No | NA | 8 | 0 | 0 | Low-quality | Genome-fragment | 33.94 | AAI-based (medium-confidence) | 0 |
| S8_vs_k141_159819\|\|full | 3730 | No | NA | 7 | 0 | 0 | Not-determined | Genome-fragment | NA | NA | 0 |
| S8_vs_k141_46906\|\|full | 3499 | No | NA | 6 | 0 | 3 | Not-determined | Genome-fragment | NA | NA | 0 |
| S8_vs_k141_99362\|\|full | 45418 | No | NA | 50 | 8 | 1 | High-quality | High-quality | 100 | AAI-based (medium-confidence) | 0 |
| S5_vs_k141_36618\|\|full | 14140 | No | NA | 20 | 9 | 0 | Low-quality | Genome-fragment | 29.89 | AAI-based (high-confidence) | 0 |
| S13_extracted_144988 | 14142 | No | NA | 15 | 8 | 0 | Low-quality | Genome-fragment | 46.08 | HMM-based (lower-bound) | 0 |
| S8_vs_k141_151174\|\|full | 1032 | No | NA | 2 | 0 | 1 | Not-determined | Genome-fragment | NA | NA | 0 |
| S8_vs_k141_90730\|\|full | 3173 | No | NA | 4 | 0 | 1 | Not-determined | Genome-fragment | NA | NA | 0 |
| S8_vs_k141_194986\|\|full | 1798 | No | NA | 3 | 0 | 1 | Not-determined | Genome-fragment | NA | NA | 0 |
| S8_vs_k141_203129\|\|full | 1044 | No | NA | 2 | 0 | 0 | Not-determined | Genome-fragment | NA | NA | 0 |
| S4_vs_VIRSorter_27216-cat_2 | 14160 | No | NA | 18 | 8 | 0 | Low-quality | Genome-fragment | 42.55 | HMM-based (lower-bound) | 0 |
| S8_vs_k141_159826\|\|full | 3276 | No | NA | 5 | 0 | 2 | Not-determined | Genome-fragment | NA | NA | 0 |
| S16_vs_31530\|\|full | 14193 | Yes | 11974 | 21 | 4 | 4 | Low-quality | Genome-fragment | 23.54 | HMM-based (lower-bound) | 15.63 |
| S8_vs_k141_81928\|\|full | 8452 | No | NA | 8 | 0 | 6 | Not-determined | Genome-fragment | NA | NA | 0 |
| S8_vs_k141_203160\|\|full | 3165 | No | NA | 3 | 0 | 1 | Not-determined | Genome-fragment | NA | NA | 0 |
| S8_vs_k141_195023\|\|full | 1049 | No | NA | 2 | 0 | 0 | Not-determined | Genome-fragment | NA | NA | 0 |
| S5_vs_k141_73470\|\|full | 14195 | No | NA | 29 | 3 | 0 | Low-quality | Genome-fragment | 28.39 | AAI-based (medium-confidence) | 0 |
| S11_vs_k141_51567\|\|full | 14214 | Yes | 7639 | 19 | 6 | 6 | Low-quality | Genome-fragment | 42.97 | AAI-based (medium-confidence) | 46.26 |
| S8_vs_k141_108232\|\|full | 1420 | No | NA | 4 | 0 | 0 | Not-determined | Genome-fragment | NA | NA | 0 |
| S8_vs_k141_38652\|\|full | 1989 | No | NA | 3 | 0 | 2 | Not-determined | Genome-fragment | NA | NA | 0 |
| S8_vs_k141_142902\|\|full | 1968 | No | NA | 3 | 0 | 0 | Not-determined | Genome-fragment | NA | NA | 0 |
| S8_vs_k141_21342\|\|full | 4179 | No | NA | 5 | 0 | 3 | Not-determined | Genome-fragment | NA | NA | 0 |
| S10_extracted_73637 | 14223 | No | NA | 22 | 4 | 0 | Low-quality | Genome-fragment | 31.42 | AAI-based (high-confidence) | 0 |
| S3_vs_k141_65537\|\|0_partial | 14233 | No | NA | 17 | 1 | 5 | Low-quality | Genome-fragment | 23.07 | HMM-based (lower-bound) | 0 |
| S13_vs_118409\|\|full | 14240 | No | NA | 25 | 9 | 0 | Low-quality | Genome-fragment | 36.88 | AAI-based (high-confidence) | 0 |
| S11_extracted_k141_96253 | 14249 | No | NA | 27 | 9 | 2 | Low-quality | Genome-fragment | 24.99 | AAI-based (high-confidence) | 0 |
| S14_extracted_k141_85172 | 14270 | No | NA | 23 | 3 | 0 | Low-quality | Genome-fragment | 27.4 | AAI-based (high-confidence) | 0 |
| S12_vs_k141_25107\|\|full | 14295 | No | NA | 27 | 6 | 0 | Low-quality | Genome-fragment | 23.74 | AAI-based (high-confidence) | 0 |
| S8_vs_k141_38678\|\|full | 1738 | No | NA | 5 | 0 | 2 | Not-determined | Genome-fragment | NA | NA | 0 |
| S8_vs_k141_203221\|\|full | 8577 | No | NA | 10 | 0 | 3 | Not-determined | Genome-fragment | NA | NA | 0 |
| S8_vs_k141_38680\|\|full | 3391 | No | NA | 5 | 0 | 0 | Not-determined | Genome-fragment | NA | NA | 0 |
| S8_vs_k141_168828\|\|full | 1643 | No | NA | 2 | 0 | 1 | Not-determined | Genome-fragment | NA | NA | 0 |
| S8_vs_k141_108305\|\|full | 1597 | No | NA | 2 | 0 | 1 | Not-determined | Genome-fragment | NA | NA | 0 |
| S13_vs_34117\|\|full | 14299 | Yes | 11559 | 14 | 4 | 3 | Low-quality | Genome-fragment | 24.43 | AAI-based (medium-confidence) | 19.16 |
| S8_vs_k141_185512\|\|full | 3443 | No | NA | 5 | 0 | 1 | Not-determined | Genome-fragment | NA | NA | 0 |
| S14_vs_k141_1668\|\|full | 14309 | No | NA | 19 | 4 | 1 | Low-quality | Genome-fragment | 18.28 | AAI-based (high-confidence) | 0 |
| S8_vs_k141_195081\|\|full | 1094 | No | NA | 3 | 0 | 0 | Not-determined | Genome-fragment | NA | NA | 0 |
| S8_vs_k141_116674\|\|full | 2460 | No | NA | 3 | 0 | 0 | Not-determined | Genome-fragment | NA | NA | 0 |
| S14_vs_k141_6877\|\|full | 14349 | No | NA | 33 | 3 | 0 | Low-quality | Genome-fragment | 34.89 | AAI-based (medium-confidence) | 0 |
| S17_vs_k141_83317\|\|0_partial | 14386 | No | NA | 14 | 1 | 6 | Low-quality | Genome-fragment | 8.86 | HMM-based (lower-bound) | 0 |
| S8_vs_k141_64523\|\|full | 1494 | No | NA | 3 | 0 | 0 | Not-determined | Genome-fragment | NA | NA | 0 |
| S8_vs_k141_47053\|\|full | 6061 | No | NA | 6 | 0 | 1 | Not-determined | Genome-fragment | NA | NA | 0 |
| S2_vs_k141_1334\|\|full | 14388 | No | NA | 20 | 0 | 2 | Low-quality | Genome-fragment | 31.36 | AAI-based (high-confidence) | 0 |
| S8_extracted_k141_203092 | 14409 | No | NA | 21 | 3 | 2 | Low-quality | Genome-fragment | 8.72 | AAI-based (high-confidence) | 0 |
| S8_vs_k141_203269\|\|full | 1636 | No | NA | 2 | 0 | 0 | Not-determined | Genome-fragment | NA | NA | 0 |
| S4_extracted_29443 | 14414 | No | NA | 18 | 14 | 0 | Low-quality | Genome-fragment | 35.84 | AAI-based (medium-confidence) | 0 |
| S3_vs_k141_92014\|\|full | 14416 | No | NA | 20 | 6 | 0 | Low-quality | Genome-fragment | 32.68 | AAI-based (high-confidence) | 0 |
| S8_vs_k141_125704\|\|full | 1615 | No | NA | 3 | 0 | 3 | Not-determined | Genome-fragment | NA | NA | 0 |
| S8_vs_k141_185536\|\|full | 4392 | No | NA | 4 | 0 | 1 | Not-determined | Genome-fragment | NA | NA | 0 |
| S8_vs_k141_108324\|\|full | 2261 | No | NA | 6 | 0 | 0 | Not-determined | Genome-fragment | NA | NA | 0 |
| S8_vs_k141_168874\|\|full | 4982 | No | NA | 5 | 0 | 4 | Not-determined | Genome-fragment | NA | NA | 0 |

| | | | | | | | | | | | |
|---|---|---|---|---|---|---|---|---|---|---|---|
| S8_vs_k141_185547||full | 2026 | No | NA | 2 | 0 | 2 | Not-determined | Genome-fragment | NA | NA | 0 |
| S8_vs_k141_30542||full | 3409 | No | NA | 3 | 0 | 1 | Not-determined | Genome-fragment | NA | NA | 0 |
| S8_vs_k141_195108||full | 1223 | No | NA | 2 | 0 | 1 | Not-determined | Genome-fragment | NA | NA | 0 |
| S4_extracted_26861 | 14437 | No | NA | 12 | 4 | 1 | Low-quality | Genome-fragment | 35.68 | AAI-based (high-confidence) | 0 |
| S12_vs_k141_104347||0_partial | 14438 | Yes | 10762 | 19 | 2 | 3 | Low-quality | Genome-fragment | 13.07 | HMM-based (lower-bound) | 25.46 |
| S8_vs_k141_64549||full | 6394 | No | NA | 8 | 0 | 0 | Not-determined | Genome-fragment | NA | NA | 0 |
| S9_vs_k141_26819||full | 14462 | No | NA | 20 | 10 | 0 | Low-quality | Genome-fragment | 33.24 | AAI-based (high-confidence) | 0 |
| S8_vs_k141_168908||full | 1296 | No | NA | 3 | 0 | 1 | Not-determined | Genome-fragment | NA | NA | 0 |
| S16_extracted_91410 | 14465 | No | NA | 19 | 1 | 0 | Low-quality | Genome-fragment | 13.05 | AAI-based (high-confidence) | 0 |
| S8_vs_k141_99604||full | 2418 | No | NA | 3 | 0 | 0 | Not-determined | Genome-fragment | NA | NA | 0 |
| S7_vs_24186||full | 14477 | No | NA | 22 | 1 | 0 | Low-quality | Genome-fragment | 32.2 | AAI-based (high-confidence) | 0 |
| S17_vs_k141_80008||full | 14478 | No | NA | 15 | 5 | 1 | Low-quality | Genome-fragment | 18.38 | AAI-based (high-confidence) | 0 |
| S8_vs_k141_56282||full | 1023 | No | NA | 3 | 0 | 1 | Not-determined | Genome-fragment | NA | NA | 0 |
| S8_vs_k141_56283||full | 1085 | No | NA | 3 | 0 | 0 | Not-determined | Genome-fragment | NA | NA | 0 |
| S8_vs_k141_177750||full | 3230 | No | NA | 5 | 0 | 3 | Not-determined | Genome-fragment | NA | NA | 0 |
| S8_vs_k141_12447||full | 2912 | No | NA | 4 | 0 | 0 | Not-determined | Genome-fragment | NA | NA | 0 |
| S8_vs_k141_125717||full | 52956 | Yes | 19532 | 59 | 12 | 16 | Medium-quality | Genome-fragment | 60.76 | AAI-based (high-confidence) | 63.12 |
| S3_vs_k141_112784||full | 14493 | No | NA | 15 | 2 | 1 | Low-quality | Genome-fragment | 34.25 | AAI-based (high-confidence) | 0 |
| S8_vs_k141_108383||full | 2637 | No | NA | 3 | 0 | 0 | Not-determined | Genome-fragment | NA | NA | 0 |
| S14_vs_k141_83953||full | 14495 | No | NA | 21 | 5 | 1 | Low-quality | Genome-fragment | 28.65 | AAI-based (medium-confidence) | 0 |
| S8_vs_k141_21496||full | 1957 | No | NA | 4 | 0 | 1 | Not-determined | Genome-fragment | NA | NA | 0 |
| S8_vs_k141_203303||full | 1384 | No | NA | 3 | 0 | 2 | Not-determined | Genome-fragment | NA | NA | 0 |
| S2_extracted_k141_10026 | 14499 | No | NA | 22 | 1 | 2 | Low-quality | Genome-fragment | 7.86 | HMM-based (lower-bound) | 0 |
| S1_vs_k141_134365||full | 14500 | No | NA | 29 | 5 | 0 | Low-quality | Genome-fragment | 20.66 | HMM-based (lower-bound) | 0 |
| S8_vs_k141_56326||full | 2474 | No | NA | 4 | 0 | 2 | Not-determined | Genome-fragment | NA | NA | 0 |
| S16_extracted_38573 | 14500 | No | NA | 11 | 0 | 8 | Low-quality | Genome-fragment | 8.15 | AAI-based (medium-confidence) | 0 |
| S8_vs_k141_4153||full | 38039 | No | NA | 39 | 1 | 18 | Medium-quality | Genome-fragment | 56.91 | HMM-based (lower-bound) | 0 |
| S10_vs_29922||full | 14501 | No | NA | 21 | 7 | 0 | Low-quality | Genome-fragment | 34.77 | AAI-based (high-confidence) | 0 |
| S5_extracted_k141_27685 | 14510 | No | NA | 20 | 10 | 0 | Low-quality | Genome-fragment | 24.17 | AAI-based (medium-confidence) | 0 |
| S11_extracted_k141_76399 | 14568 | Yes | 11495 | 22 | 6 | 2 | Low-quality | Genome-fragment | 12.21 | AAI-based (high-confidence) | 21.09 |
| S8_vs_k141_108427||full | 1975 | No | NA | 3 | 0 | 0 | Not-determined | Genome-fragment | NA | NA | 0 |
| S7_extracted_118384 | 14591 | No | NA | 25 | 5 | 0 | Low-quality | Genome-fragment | 30.14 | AAI-based (high-confidence) | 0 |
| S16_vs_58126||full | 14603 | No | NA | 18 | 1 | 5 | Low-quality | Genome-fragment | 22.1 | HMM-based (lower-bound) | 0 |
| S8_vs_k141_64627||full | 1777 | No | NA | 2 | 0 | 1 | Not-determined | Genome-fragment | NA | NA | 0 |
| S8_extracted_k141_181943 | 14672 | No | NA | 18 | 14 | 0 | Low-quality | Genome-fragment | 25.7 | AAI-based (high-confidence) | 0 |
| S8_vs_k141_108446||full | 2166 | No | NA | 3 | 0 | 0 | Not-determined | Genome-fragment | NA | NA | 0 |
| S11_vs_k141_82077||0_partial | 14685 | Yes | 10387 | 15 | 3 | 2 | Low-quality | Genome-fragment | 12.11 | HMM-based (lower-bound) | 29.27 |
| S8_vs_k141_177821||full | 1428 | No | NA | 3 | 0 | 0 | Not-determined | Genome-fragment | NA | NA | 0 |
| S8_vs_k141_168991||full | 1482 | No | NA | 3 | 0 | 1 | Not-determined | Genome-fragment | NA | NA | 0 |
| S8_vs_k141_195258||full | 1851 | No | NA | 3 | 0 | 0 | Not-determined | Genome-fragment | NA | NA | 0 |
| S7_extracted_96823 | 14689 | No | NA | 15 | 7 | 0 | Low-quality | Genome-fragment | 30.9 | AAI-based (high-confidence) | 0 |
| S3_vs_k141_67063||full | 14720 | No | NA | 23 | 5 | 0 | Low-quality | Genome-fragment | 42.57 | AAI-based (medium-confidence) | 0 |
| S8_vs_k141_160108||full | 1298 | No | NA | 2 | 0 | 1 | Not-determined | Genome-fragment | NA | NA | 0 |
| S8_vs_k141_99670||full | 1190 | No | NA | 3 | 0 | 0 | Not-determined | Genome-fragment | NA | NA | 0 |
| S8_vs_k141_203364||full | 1808 | No | NA | 5 | 0 | 0 | Not-determined | Genome-fragment | NA | NA | 0 |
| S8_vs_k141_143090||full | 3331 | No | NA | 4 | 0 | 2 | Not-determined | Genome-fragment | NA | NA | 0 |
| S12_vs_k141_176097||full | 14766 | No | NA | 30 | 4 | 0 | Low-quality | Genome-fragment | 34.37 | AAI-based (high-confidence) | 0 |
| S8_vs_k141_91074||full | 2907 | No | NA | 3 | 0 | 0 | Not-determined | Genome-fragment | NA | NA | 0 |
| S11_vs_k141_11756||full | 14768 | No | NA | 23 | 1 | 0 | Low-quality | Genome-fragment | 9.05 | AAI-based (high-confidence) | 0 |
| S8_vs_k141_47220||full | 1566 | No | NA | 4 | 0 | 0 | Not-determined | Genome-fragment | NA | NA | 0 |
| S8_vs_k141_116828||full | 1434 | No | NA | 3 | 0 | 0 | Not-determined | Genome-fragment | NA | NA | 0 |
| S8_vs_k141_143118||full | 1047 | No | NA | 3 | 0 | 1 | Not-determined | Genome-fragment | NA | NA | 0 |
| S8_vs_k141_91092||full | 1115 | No | NA | 2 | 0 | 1 | Not-determined | Genome-fragment | NA | NA | 0 |
| S8_vs_k141_151490||full | 1883 | No | NA | 3 | 0 | 2 | Not-determined | Genome-fragment | NA | NA | 0 |
| S11_vs_k141_19482||full | 14803 | No | NA | 31 | 3 | 2 | Low-quality | Genome-fragment | 28.02 | AAI-based (medium-confidence) | 0 |
| S12_extracted_k141_21393 | 14828 | No | NA | 18 | 6 | 0 | Low-quality | Genome-fragment | 49.91 | HMM-based (lower-bound) | 0 |
| S9_vs_k141_24869||full | 14833 | No | NA | 27 | 6 | 1 | Low-quality | Genome-fragment | 33.85 | AAI-based (high-confidence) | 0 |
| S8_vs_k141_73209||full | 1134 | No | NA | 2 | 0 | 2 | Not-determined | Genome-fragment | NA | NA | 0 |
| S8_vs_k141_82298||full | 1759 | No | NA | 4 | 0 | 4 | Not-determined | Genome-fragment | NA | NA | 0 |
| S8_vs_k141_91122||full | 3257 | No | NA | 3 | 0 | 2 | Not-determined | Genome-fragment | NA | NA | 0 |
| S8_vs_k141_73221||full | 1090 | No | NA | 2 | 0 | 0 | Not-determined | Genome-fragment | NA | NA | 0 |
| S8_vs_k141_185724||full | 2951 | No | NA | 5 | 0 | 2 | Not-determined | Genome-fragment | NA | NA | 0 |
| S11_vs_k141_37489||full | 14863 | No | NA | 20 | 3 | 0 | Low-quality | Genome-fragment | 38.12 | AAI-based (high-confidence) | 0 |
| S16_vs_25188||full | 14886 | No | NA | 35 | 2 | 0 | Low-quality | Genome-fragment | 10.1 | AAI-based (high-confidence) | 0 |
| S12_vs_k141_79065||full | 14889 | No | NA | 14 | 3 | 0 | Low-quality | Genome-fragment | 29.16 | AAI-based (high-confidence) | 0 |
| S8_vs_k141_12612||full | 2184 | No | NA | 4 | 0 | 0 | Not-determined | Genome-fragment | NA | NA | 0 |
| S10_vs_13617||full | 14927 | No | NA | 35 | 5 | 0 | Low-quality | Genome-fragment | 35.99 | AAI-based (high-confidence) | 0 |
| S8_vs_k141_134301||full | 3614 | No | NA | 5 | 0 | 4 | Not-determined | Genome-fragment | NA | NA | 0 |
| S8_vs_k141_56521||full | 1701 | No | NA | 3 | 0 | 1 | Not-determined | Genome-fragment | NA | NA | 0 |
| S8_vs_k141_125944||full | 1261 | No | NA | 2 | 0 | 0 | Not-determined | Genome-fragment | NA | NA | 0 |
| S8_vs_k141_203486||full | 1547 | No | NA | 2 | 0 | 1 | Not-determined | Genome-fragment | NA | NA | 0 |
| S8_vs_k141_56527||full | 3562 | No | NA | 4 | 0 | 2 | Not-determined | Genome-fragment | NA | NA | 0 |
| S7_vs_61353||full | 14957 | No | NA | 17 | 13 | 1 | Low-quality | Genome-fragment | 29.61 | AAI-based (high-confidence) | 0 |
| S17_vs_k141_99801||full | 14982 | No | NA | 14 | 1 | 2 | Low-quality | Genome-fragment | 10.13 | HMM-based (lower-bound) | 0 |
| S9_vs_k141_64213||full | 14989 | Yes | 9733 | 25 | 2 | 3 | Low-quality | Genome-fragment | 18.89 | AAI-based (high-confidence) | 35.07 |
| S14_vs_k141_83601||full | 14992 | No | NA | 17 | 16 | 0 | Low-quality | Genome-fragment | 44.46 | AAI-based (high-confidence) | 0 |
| S8_vs_k141_47265||full | 76710 | Yes | 48801 | 101 | 20 | 20 | Complete | High-quality | 100 | Provirus (high-confidence) | 36.38 |
| S3_vs_k141_74522||full | 15003 | No | NA | 21 | 2 | 0 | Low-quality | Genome-fragment | 9.58 | AAI-based (medium-confidence) | 0 |
| S8_vs_k141_12683||full | 3196 | No | NA | 7 | 0 | 1 | Not-determined | Genome-fragment | NA | NA | 0 |
| S8_vs_k141_177996||full | 2755 | No | NA | 4 | 0 | 4 | Not-determined | Genome-fragment | NA | NA | 0 |
| S8_vs_k141_108631||full | 1780 | No | NA | 3 | 0 | 2 | Not-determined | Genome-fragment | NA | NA | 0 |
| S8_vs_k141_160162||full | 1009 | No | NA | 2 | 0 | 1 | Not-determined | Genome-fragment | NA | NA | 0 |
| S15_extracted_29387 | 15004 | No | NA | 14 | 0 | 12 | Low-quality | Genome-fragment | 5.72 | AAI-based (high-confidence) | 0 |
| S11_vs_k141_77151||full | 15026 | No | NA | 27 | 5 | 0 | Low-quality | Genome-fragment | 40.04 | AAI-based (high-confidence) | 0 |
| S15_vs_26282||full | 15026 | No | NA | 18 | 2 | 0 | Low-quality | Genome-fragment | 9.04 | HMM-based (lower-bound) | 0 |
| S8_vs_k141_143289||full | 9119 | No | NA | 14 | 0 | 4 | Not-determined | Genome-fragment | NA | NA | 0 |
| S8_vs_k141_4447||full | 2350 | No | NA | 3 | 0 | 1 | Not-determined | Genome-fragment | NA | NA | 0 |
| S17_extracted_k141_96091 | 15041 | No | NA | 17 | 2 | 0 | Low-quality | Genome-fragment | 19.09 | AAI-based (high-confidence) | 0 |
| S8_vs_k141_203573||full | 4691 | No | NA | 3 | 0 | 0 | Not-determined | Genome-fragment | NA | NA | 0 |
| S12_vs_k141_88274||full | 15087 | No | NA | 22 | 1 | 5 | Low-quality | Genome-fragment | 16.29 | HMM-based (lower-bound) | 0 |
| S8_vs_k141_73347||full | 2017 | No | NA | 4 | 0 | 2 | Not-determined | Genome-fragment | NA | NA | 0 |
| S5_vs_k141_108799||full | 15096 | No | NA | 17 | 11 | 0 | Low-quality | Genome-fragment | 42.78 | AAI-based (high-confidence) | 0 |
| S8_vs_k141_143305||full | 2839 | No | NA | 3 | 0 | 3 | Not-determined | Genome-fragment | NA | NA | 0 |
| S8_vs_k141_73355||full | 2895 | No | NA | 3 | 0 | 0 | Not-determined | Genome-fragment | NA | NA | 0 |
| S8_vs_k141_64849||full | 6094 | No | NA | 11 | 0 | 11 | Not-determined | Genome-fragment | NA | NA | 0 |
| S16_extracted_57265 | 15096 | No | NA | 15 | 1 | 8 | Low-quality | Genome-fragment | 28.03 | HMM-based (lower-bound) | 0 |
| S12_vs_k141_90309||0_partial | 15096 | No | NA | 15 | 1 | 5 | Low-quality | Genome-fragment | 32.47 | AAI-based (high-confidence) | 0 |
| S16_vs_95632||full | 15113 | No | NA | 16 | 3 | 0 | Low-quality | Genome-fragment | 6.16 | AAI-based (medium-confidence) | 0 |
| S17_extracted_k141_140072 | 15126 | No | NA | 27 | 10 | 0 | Low-quality | Genome-fragment | 33.6 | AAI-based (high-confidence) | 0 |
| S8_vs_k141_203613||full | 1913 | No | NA | 4 | 0 | 2 | Not-determined | Genome-fragment | NA | NA | 0 |
| S14_vs_k141_22206||full | 15156 | No | NA | 18 | 16 | 0 | Low-quality | Genome-fragment | 43.13 | AAI-based (medium-confidence) | 0 |
| S8_vs_k141_47352||full | 2009 | No | NA | 3 | 0 | 0 | Not-determined | Genome-fragment | NA | NA | 0 |
| S12_vs_k141_57081||full | 15175 | No | NA | 19 | 16 | 1 | Low-quality | Genome-fragment | 37.78 | AAI-based (high-confidence) | 0 |
| S8_vs_k141_30757||full | 1501 | No | NA | 2 | 0 | 1 | Not-determined | Genome-fragment | NA | NA | 0 |
| S8_vs_k141_151663||full | 6782 | No | NA | 6 | 0 | 2 | Not-determined | Genome-fragment | NA | NA | 0 |
| S9_extracted_k141_23565 | 15187 | No | NA | 8 | 2 | 0 | Low-quality | Genome-fragment | 9.83 | AAI-based (high-confidence) | 0 |
| S8_vs_k141_56637||full | 1776 | No | NA | 2 | 0 | 0 | Not-determined | Genome-fragment | NA | NA | 0 |
| S8_vs_k141_91157||full | 6689 | No | NA | 7 | 0 | 2 | Not-determined | Genome-fragment | NA | NA | 0 |
| S16_extracted_50006 | 15223 | No | NA | 16 | 1 | 6 | Low-quality | Genome-fragment | 9.44 | HMM-based (lower-bound) | 0 |
| S9_extracted_k141_15732 | 15224 | No | NA | 20 | 9 | 0 | Low-quality | Genome-fragment | 37.29 | AAI-based (high-confidence) | 0 |
| S7_vs_40961||full | 15224 | No | NA | 16 | 0 | 11 | Low-quality | Genome-fragment | 6.49 | AAI-based (medium-confidence) | 0 |
| S8_vs_k141_169298||full | 1073 | No | NA | 2 | 0 | 1 | Not-determined | Genome-fragment | NA | NA | 0 |
| S11_vs_k141_922||full | 15239 | No | NA | 23 | 4 | 0 | Low-quality | Genome-fragment | 33.08 | AAI-based (high-confidence) | 0 |
| S8_vs_k141_39132||full | 2228 | No | NA | 4 | 0 | 2 | Not-determined | Genome-fragment | NA | NA | 0 |
| S8_vs_k141_160298||full | 1296 | No | NA | 2 | 0 | 2 | Not-determined | Genome-fragment | NA | NA | 0 |
| S17_vs_k141_107014||full | 15257 | No | NA | 17 | 1 | 3 | Low-quality | Genome-fragment | 1.23 | HMM-based (lower-bound) | 0 |
| S16_vs_65814||full | 15274 | Yes | 504 | 10 | 1 | 5 | Low-quality | Genome-fragment | 0.3 | HMM-based (lower-bound) | 96.7 |
| S8_vs_k141_73467||full | 2794 | No | NA | 3 | 0 | 2 | Not-determined | Genome-fragment | NA | NA | 0 |
| S11_vs_k141_84820||full | 15279 | No | NA | 26 | 5 | 0 | Low-quality | Genome-fragment | 36.89 | AAI-based (medium-confidence) | 0 |
| S8_vs_k141_185848||full | 2305 | No | NA | 4 | 0 | 2 | Not-determined | Genome-fragment | NA | NA | 0 |
| S16_vs_148745||full | 15280 | No | NA | 16 | 2 | 1 | Low-quality | Genome-fragment | 19.01 | AAI-based (high-confidence) | 0 |
| S8_vs_k141_99915||full | 1840 | No | NA | 3 | 0 | 0 | Not-determined | Genome-fragment | NA | NA | 0 |
| S16_extracted_36644 | 15286 | No | NA | 9 | 3 | 0 | Low-quality | Genome-fragment | 8.53 | HMM-based (lower-bound) | 0 |
| S8_vs_k141_169338||full | 5545 | No | NA | 6 | 0 | 5 | Not-determined | Genome-fragment | NA | NA | 0 |
| S8_vs_k141_178119||full | 5836 | No | NA | 6 | 0 | 3 | Not-determined | Genome-fragment | NA | NA | 0 |
| S3_vs_k141_70294||full | 15297 | No | NA | 13 | 0 | 9 | Low-quality | Genome-fragment | 7.21 | AAI-based (medium-confidence) | 0 |
| S8_vs_k141_108713||full | 2942 | No | NA | 3 | 0 | 3 | Not-determined | Genome-fragment | NA | NA | 0 |
| S8_vs_k141_117112||full | 2939 | No | NA | 3 | 0 | 2 | Not-determined | Genome-fragment | NA | NA | 0 |
| S8_vs_k141_39162||full | 1517 | No | NA | 3 | 0 | 1 | Not-determined | Genome-fragment | NA | NA | 0 |
| S5_vs_k141_121399||full | 15338 | No | NA | 20 | 15 | 0 | Low-quality | Genome-fragment | 35.12 | AAI-based (medium-confidence) | 0 |
| S8_vs_k141_99937||full | 2021 | No | NA | 3 | 0 | 1 | Not-determined | Genome-fragment | NA | NA | 0 |
| S8_vs_k141_21913||full | 4133 | No | NA | 4 | 0 | 3 | Not-determined | Genome-fragment | NA | NA | 0 |
| S8_vs_k141_151793||full | 4124 | No | NA | 4 | 0 | 2 | Not-determined | Genome-fragment | NA | NA | 0 |
| S9_vs_k141_80427||full | 15341 | No | NA | 23 | 16 | 0 | Low-quality | Genome-fragment | 43.57 | AAI-based (high-confidence) | 0 |
| S8_vs_k141_82570||full | 1950 | No | NA | 2 | 0 | 1 | Not-determined | Genome-fragment | NA | NA | 0 |
| S8_vs_k141_143383||full | 1505 | No | NA | 3 | 0 | 2 | Not-determined | Genome-fragment | NA | NA | 0 |
| S8_vs_k141_169390||full | 2862 | No | NA | 6 | 0 | 0 | Not-determined | Genome-fragment | NA | NA | 0 |
| S8_vs_k141_143403||full | 3351 | No | NA | 6 | 0 | 2 | Not-determined | Genome-fragment | NA | NA | 0 |
| S8_vs_k141_203742||full | 1228 | No | NA | 2 | 0 | 0 | Not-determined | Genome-fragment | NA | NA | 0 |
| S9_vs_k141_23181||full | 15353 | No | NA | 37 | 2 | 0 | Low-quality | Genome-fragment | 41.38 | AAI-based (high-confidence) | 0 |
| S8_vs_k141_47492||full | 2546 | No | NA | 4 | 0 | 2 | Not-determined | Genome-fragment | NA | NA | 0 |
| S8_vs_k141_91261||full | 1774 | No | NA | 2 | 0 | 0 | Not-determined | Genome-fragment | NA | NA | 0 |
| S8_vs_k141_65024||full | 2953 | No | NA | 6 | 0 | 0 | Not-determined | Genome-fragment | NA | NA | 0 |
| S11_vs_k141_99934||full | 15370 | No | NA | 25 | 2 | 0 | Low-quality | Genome-fragment | 16.64 | AAI-based (high-confidence) | 0 |
| S7_vs_126377||full | 15381 | No | NA | 10 | 0 | 7 | Low-quality | Genome-fragment | 4.85 | AAI-based (medium-confidence) | 0 |
| S3_extracted_k141_3519 | 15382 | No | NA | 20 | 1 | 4 | Low-quality | Genome-fragment | 7.48 | AAI-based (high-confidence) | 0 |
| S8_vs_k141_12894||full | 1691 | No | NA | 2 | 0 | 1 | Not-determined | Genome-fragment | NA | NA | 0 |
| S8_vs_k141_39249||full | 2145 | No | NA | 4 | 0 | 1 | Not-determined | Genome-fragment | NA | NA | 0 |
| S5_vs_k141_74464||full | 15392 | No | NA | 24 | 11 | 0 | Low-quality | Genome-fragment | 33.63 | AAI-based (high-confidence) | 0 |
| S8_vs_k141_178256||full | 3186 | No | NA | 4 | 0 | 2 | Not-determined | Genome-fragment | NA | NA | 0 |
| S8_extracted_k141_31758 | 15395 | No | NA | 26 | 2 | 1 | Low-quality | Genome-fragment | 20.78 | AAI-based (high-confidence) | 0 |
| S5_vs_k141_23139||full | 15408 | No | NA | 29 | 3 | 1 | Low-quality | Genome-fragment | 11.25 | AAI-based (high-confidence) | 0 |
| S8_vs_k141_108840||full | 3477 | No | NA | 3 | 0 | 2 | Not-determined | Genome-fragment | NA | NA | 0 |
| S16_vs_71618||full | 15429 | No | NA | 18 | 11 | 0 | Low-quality | Genome-fragment | 43.78 | AAI-based (high-confidence) | 0 |
| S8_vs_k141_184630||0_partial | 15449 | Yes | 3639 | 10 | 1 | 5 | Low-quality | Genome-fragment | 2.45 | HMM-based (lower-bound) | 76.45 |
| S8_vs_k141_91315||full | 1048 | No | NA | 3 | 0 | 0 | Not-determined | Genome-fragment | NA | NA | 0 |
| S3_vs_k141_45799||full | 15478 | No | NA | 34 | 5 | 1 | Low-quality | Genome-fragment | 25.67 | AAI-based (high-confidence) | 0 |
| S8_vs_k141_73562||full | 2146 | No | NA | 3 | 0 | 2 | Not-determined | Genome-fragment | NA | NA | 0 |
| S9_extracted_k141_80840 | 15480 | No | NA | 18 | 7 | 0 | Low-quality | Genome-fragment | 48.59 | HMM-based (lower-bound) | 0 |
| S3_vs_k141_66595||0_partial | 15486 | No | NA | 15 | 1 | 7 | Low-quality | Genome-fragment | 36.99 | AAI-based (medium-confidence) | 0 |
| S5_vs_k141_36355||full | 15487 | No | NA | 17 | 13 | 0 | Low-quality | Genome-fragment | 39.76 | AAI-based (high-confidence) | 0 |
| S10_extracted_67295 | 15494 | No | NA | 23 | 2 | 0 | Low-quality | Genome-fragment | 33.63 | AAI-based (high-confidence) | 0 |
| S8_vs_k141_82581||full | 5787 | No | NA | 10 | 0 | 1 | Not-determined | Genome-fragment | NA | NA | 0 |
| S3_vs_k141_45010||full | 15497 | No | NA | 21 | 2 | 0 | Low-quality | Genome-fragment | 20.24 | HMM-based (lower-bound) | 0 |
| S8_vs_k141_82596||full | 1563 | No | NA | 3 | 0 | 1 | Not-determined | Genome-fragment | NA | NA | 0 |
| S8_vs_k141_195677||full | 1474 | No | NA | 3 | 0 | 0 | Not-determined | Genome-fragment | NA | NA | 0 |
| S8_vs_k141_47601||full | 4679 | No | NA | 3 | 0 | 2 | Not-determined | Genome-fragment | NA | NA | 0 |
| S8_vs_k141_186021||full | 1609 | No | NA | 3 | 0 | 1 | Not-determined | Genome-fragment | NA | NA | 0 |
| S8_vs_k141_186029||full | 1778 | No | NA | 2 | 0 | 1 | Not-determined | Genome-fragment | NA | NA | 0 |
| S9_vs_k141_74182||full | 15537 | No | NA | 7 | 1 | 0 | Low-quality | Genome-fragment | 8.55 | AAI-based (medium-confidence) | 0 |
| S8_vs_k141_108870||full | 1207 | No | NA | 4 | 0 | 0 | Not-determined | Genome-fragment | NA | NA | 0 |
| S8_vs_k141_91367||full | 9034 | No | NA | 11 | 0 | 6 | Not-determined | Genome-fragment | NA | NA | 0 |
| S8_vs_k141_203882||full | 4230 | No | NA | 3 | 0 | 0 | Not-determined | Genome-fragment | NA | NA | 0 |
| S8_vs_k141_30967||full | 2693 | No | NA | 3 | 0 | 3 | Not-determined | Genome-fragment | NA | NA | 0 |
| S8_vs_k141_117325||full | 2640 | No | NA | 3 | 0 | 2 | Not-determined | Genome-fragment | NA | NA | 0 |
| S8_vs_k141_12954||full | 2022 | No | NA | 3 | 0 | 1 | Not-determined | Genome-fragment | NA | NA | 0 |
| S11_extracted_k141_45640 | 15563 | No | NA | 18 | 2 | 0 | Low-quality | Genome-fragment | 35.33 | AAI-based (high-confidence) | 0 |
| S8_vs_k141_100101||full | 1255 | No | NA | 2 | 0 | 0 | Not-determined | Genome-fragment | NA | NA | 0 |
| S17_vs_k141_74850||full | 15569 | No | NA | 12 | 1 | 1 | Low-quality | Genome-fragment | 23.43 | AAI-based (high-confidence) | 0 |
| S8_vs_k141_91403||full | 3008 | No | NA | 3 | 0 | 2 | Not-determined | Genome-fragment | NA | NA | 0 |
| S8_vs_k141_140805||full | 15571 | No | NA | 31 | 8 | 0 | Low-quality | Genome-fragment | 23.85 | AAI-based (high-confidence) | 0 |
| S12_extracted_k141_179056 | 15575 | No | NA | 19 | 1 | 1 | Low-quality | Genome-fragment | 9.51 | AAI-based (medium-confidence) | 0 |
| S8_vs_k141_73644||full | 7117 | No | NA | 10 | 0 | 3 | Not-determined | Genome-fragment | NA | NA | 0 |
| S8_vs_k141_134637||full | 2305 | No | NA | 2 | 0 | 1 | Not-determined | Genome-fragment | NA | NA | 0 |
| S8_vs_k141_143605||full | 3136 | No | NA | 5 | 0 | 2 | Not-determined | Genome-fragment | NA | NA | 0 |
| S8_vs_k141_31036||full | 2417 | No | NA | 3 | 0 | 1 | Not-determined | Genome-fragment | NA | NA | 0 |
| S8_vs_k141_31038||full | 1726 | No | NA | 2 | 0 | 1 | Not-determined | Genome-fragment | NA | NA | 0 |
| S9_vs_k141_47341||full | 15586 | No | NA | 26 | 7 | 0 | Low-quality | Genome-fragment | 34.37 | AAI-based (high-confidence) | 0 |
| S17_vs_k141_15281||full | 15590 | No | NA | 22 | 0 | 0 | Low-quality | Genome-fragment | 10.52 | AAI-based (medium-confidence) | 0 |
| S10_vs_35639||full | 15590 | No | NA | 10 | 0 | 5 | Low-quality | Genome-fragment | 5.86 | AAI-based (medium-confidence) | 0 |
| S8_vs_k141_203962||full | 1207 | No | NA | 3 | 0 | 1 | Not-determined | Genome-fragment | NA | NA | 0 |
| S8_vs_k141_134671||full | 1718 | No | NA | 2 | 0 | 1 | Not-determined | Genome-fragment | NA | NA | 0 |
| S5_extracted_k141_7457 | 15601 | No | NA | 13 | 0 | 10 | Low-quality | Genome-fragment | 7.26 | AAI-based (medium-confidence) | 0 |
| S7_vs_38423||full | 15611 | No | NA | 18 | 1 | 1 | Low-quality | Genome-fragment | 11.25 | HMM-based (lower-bound) | 0 |
| S8_vs_k141_143608||full | 18640 | No | NA | 24 | 10 | 0 | Medium-quality | Genome-fragment | 52.33 | AAI-based (high-confidence) | 0 |
| S8_vs_k141_82671||full | 3150 | No | NA | 5 | 0 | 2 | Not-determined | Genome-fragment | NA | NA | 0 |
| S8_vs_k141_169601||full | 4533 | No | NA | 7 | 0 | 1 | Not-determined | Genome-fragment | NA | NA | 0 |
| S8_vs_k141_100214||full | 2021 | No | NA | 3 | 0 | 1 | Not-determined | Genome-fragment | NA | NA | 0 |
| S8_vs_k141_204011||full | 2470 | No | NA | 3 | 0 | 1 | Not-determined | Genome-fragment | NA | NA | 0 |
| S3_extracted_k141_101149 | 15632 | No | NA | 37 | 2 | 0 | Low-quality | Genome-fragment | 9.87 | AAI-based (high-confidence) | 0 |
| S12_extracted_k141_171343 | 15657 | No | NA | 14 | 7 | 0 | Low-quality | Genome-fragment | 14.55 | AAI-based (high-confidence) | 0 |
| S8_vs_k141_143659||full | 2059 | No | NA | 4 | 0 | 2 | Not-determined | Genome-fragment | NA | NA | 0 |
| S8_vs_k141_117453||full | 3202 | No | NA | 5 | 0 | 0 | Not-determined | Genome-fragment | NA | NA | 0 |
| S8_vs_k141_82688||full | 5028 | No | NA | 4 | 0 | 3 | Not-determined | Genome-fragment | NA | NA | 0 |
| S10_vs_46939||full | 15665 | No | NA | 20 | 10 | 1 | Low-quality | Genome-fragment | 37.23 | AAI-based (medium-confidence) | 0 |
| S8_vs_k141_65293||full | 2120 | No | NA | 2 | 0 | 1 | Not-determined | Genome-fragment | NA | NA | 0 |
| S8_vs_k141_143687||full | 3341 | No | NA | 3 | 0 | 2 | Not-determined | Genome-fragment | NA | NA | 0 |
| S14_vs_k141_55896||full | 15678 | No | NA | 23 | 15 | 0 | Low-quality | Genome-fragment | 35.8 | AAI-based (high-confidence) | 0 |
| S8_vs_k141_47765||full | 1122 | No | NA | 3 | 0 | 1 | Not-determined | Genome-fragment | NA | NA | 0 |
| S8_vs_k141_169620||full | 3563 | No | NA | 3 | 0 | 2 | Not-determined | Genome-fragment | NA | NA | 0 |
| S8_vs_k141_169627||full | 2570 | No | NA | 3 | 0 | 1 | Not-determined | Genome-fragment | NA | NA | 0 |
| S8_vs_k141_82717||full | 14349 | No | NA | 11 | 0 | 8 | Not-determined | Genome-fragment | NA | NA | 0 |
| S8_vs_k141_22335||full | 3193 | No | NA | 3 | 0 | 2 | Not-determined | Genome-fragment | NA | NA | 0 |
| S17_vs_k141_64850||full | 15681 | No | NA | 28 | 7 | 1 | Low-quality | Genome-fragment | 28.01 | AAI-based (high-confidence) | 0 |
| S8_vs_k141_143707||full | 2064 | No | NA | 3 | 0 | 1 | Not-determined | Genome-fragment | NA | NA | 0 |
| S8_vs_k141_82722||full | 2550 | No | NA | 4 | 0 | 0 | Not-determined | Genome-fragment | NA | NA | 0 |
| S8_vs_k141_13113||full | 2684 | No | NA | 2 | 0 | 1 | Not-determined | Genome-fragment | NA | NA | 0 |
| S9_vs_k141_30339||full | 15695 | No | NA | 21 | 8 | 0 | Low-quality | Genome-fragment | 37.4 | AAI-based (medium-confidence) | 0 |
| S8_vs_k141_82738||full | 1083 | No | NA | 2 | 0 | 2 | Not-determined | Genome-fragment | NA | NA | 0 |
| S16_vs_148517||full | 15727 | No | NA | 23 | 6 | 0 | Low-quality | Genome-fragment | 39.81 | AAI-based (high-confidence) | 0 |
| S8_vs_k141_65336||full | 1392 | No | NA | 4 | 0 | 2 | Not-determined | Genome-fragment | NA | NA | 0 |
| S5_extracted_k141_93906 | 15739 | Yes | 5274 | 14 | 2 | 4 | Low-quality | Genome-fragment | 2.9 | HMM-based (lower-bound) | 66.49 |
| S8_vs_k141_65343||full | 5652 | No | NA | 5 | 0 | 3 | Not-determined | Genome-fragment | NA | NA | 0 |
| S8_vs_k141_47822||full | 1091 | No | NA | 2 | 0 | 1 | Not-determined | Genome-fragment | NA | NA | 0 |
| S8_vs_k141_117543||full | 1928 | No | NA | 4 | 0 | 0 | Not-determined | Genome-fragment | NA | NA | 0 |
| S8_vs_k141_47839||full | 2075 | No | NA | 4 | 0 | 0 | Not-determined | Genome-fragment | NA | NA | 0 |
| S5_vs_k141_110560||full | 15747 | No | NA | 21 | 14 | 0 | Low-quality | Genome-fragment | 42.86 | AAI-based (medium-confidence) | 0 |
| S8_vs_k141_152162||full | 2231 | No | NA | 7 | 0 | 0 | Not-determined | Genome-fragment | NA | NA | 0 |
| S8_vs_k141_117571||full | 1452 | No | NA | 3 | 0 | 0 | Not-determined | Genome-fragment | NA | NA | 0 |
| S8_vs_k141_73879||full | 2233 | No | NA | 5 | 0 | 2 | Not-determined | Genome-fragment | NA | NA | 0 |
| S11_vs_k141_4007||full | 15749 | No | NA | 17 | 12 | 0 | Low-quality | Genome-fragment | 44.63 | AAI-based (high-confidence) | 0 |
| S8_vs_k141_169700||full | 2301 | No | NA | 4 | 0 | 4 | Not-determined | Genome-fragment | NA | NA | 0 |
| S16_vs_165292||0_partial | 15760 | No | NA | 14 | 1 | 1 | Low-quality | Genome-fragment | 12.82 | AAI-based (medium-confidence) | 0 |
| S3_extracted_k141_81902 | 15761 | No | NA | 16 | 2 | 1 | Low-quality | Genome-fragment | 25.39 | AAI-based (medium-confidence) | 0 |
| S11_vs_k141_62008||full | 15776 | Yes | 8129 | 19 | 5 | 4 | Low-quality | Genome-fragment | 13.29 | AAI-based (medium-confidence) | 48.47 |
| S5_vs_k141_55252||full | 15786 | No | NA | 24 | 4 | 1 | Low-quality | Genome-fragment | 34.21 | AAI-based (medium-confidence) | 0 |
| S9_extracted_k141_78685 | 15801 | No | NA | 18 | 1 | 0 | Low-quality | Genome-fragment | 9.48 | AAI-based (high-confidence) | 0 |
| S8_vs_k141_134805||full | 37347 | No | NA | 51 | 15 | 2 | High-quality | High-quality | 100 | AAI-based (medium-confidence) | 0 |
| S8_vs_k141_160565||full | 2278 | No | NA | 4 | 0 | 2 | Not-determined | Genome-fragment | NA | NA | 0 |
| S8_vs_k141_57108||full | 2128 | No | NA | 2 | 0 | 1 | Not-determined | Genome-fragment | NA | NA | 0 |
| S8_vs_k141_195924||full | 1795 | No | NA | 1 | 0 | 1 | Not-determined | Genome-fragment | NA | NA | 0 |
| S8_vs_k141_160578||full | 2762 | No | NA | 5 | 0 | 2 | Not-determined | Genome-fragment | NA | NA | 0 |
| S17_extracted_k141_155230 | 15803 | No | NA | 25 | 10 | 0 | Low-quality | Genome-fragment | 35.08 | AAI-based (high-confidence) | 0 |
| S8_vs_k141_117618||full | 3910 | No | NA | 4 | 0 | 3 | Not-determined | Genome-fragment | NA | NA | 0 |
| S8_vs_k141_169733||full | 2528 | No | NA | 3 | 0 | 2 | Not-determined | Genome-fragment | NA | NA | 0 |
| S8_vs_k141_117621||full | 3871 | No | NA | 5 | 0 | 0 | Not-determined | Genome-fragment | NA | NA | 0 |
| S8_vs_k141_195935||full | 2371 | No | NA | 4 | 0 | 0 | Not-determined | Genome-fragment | NA | NA | 0 |
| S8_vs_k141_65440||full | 1513 | No | NA | 2 | 0 | 1 | Not-determined | Genome-fragment | NA | NA | 0 |
| S8_vs_k141_126384||full | 4074 | No | NA | 7 | 0 | 3 | Not-determined | Genome-fragment | NA | NA | 0 |
| S8_vs_k141_65451||full | 2588 | No | NA | 2 | 0 | 1 | Not-determined | Genome-fragment | NA | NA | 0 |
| S12_vs_k141_30812||full | 15817 | No | NA | 31 | 7 | 1 | Low-quality | Genome-fragment | 33.34 | AAI-based (high-confidence) | 0 |
| S15_extracted_12335 | 15828 | No | NA | 27 | 5 | 0 | Low-quality | Genome-fragment | 8.78 | HMM-based (lower-bound) | 0 |
| S3_extracted_k141_74064 | 15831 | No | NA | 23 | 4 | 1 | Low-quality | Genome-fragment | 47.39 | HMM-based (lower-bound) | 0 |
| S16_vs_88961||full | 15865 | No | NA | 26 | 6 | 0 | Low-quality | Genome-fragment | 36.23 | AAI-based (high-confidence) | 0 |
| S8_vs_k141_143868||full | 1256 | No | NA | 2 | 0 | 1 | Not-determined | Genome-fragment | NA | NA | 0 |
| S8_vs_k141_169783||full | 1664 | No | NA | 3 | 0 | 1 | Not-determined | Genome-fragment | NA | NA | 0 |
| S8_vs_k141_126428||full | 1652 | No | NA | 3 | 0 | 1 | Not-determined | Genome-fragment | NA | NA | 0 |
| S13_vs_45795||full | 15866 | No | NA | 15 | 2 | 1 | Low-quality | Genome-fragment | 26.95 | AAI-based (high-confidence) | 0 |
| S8_vs_k141_57160||full | 1160 | No | NA | 3 | 0 | 2 | Not-determined | Genome-fragment | NA | NA | 0 |
| S8_vs_k141_143875||full | 3562 | No | NA | 8 | 0 | 2 | Not-determined | Genome-fragment | NA | NA | 0 |
| S10_vs_70704||full | 15895 | No | NA | 21 | 5 | 3 | Low-quality | Genome-fragment | 32.32 | AAI-based (medium-confidence) | 0 |
| S8_vs_k141_82853||full | 4085 | No | NA | 4 | 0 | 2 | Not-determined | Genome-fragment | NA | NA | 0 |
| S2_extracted_k141_11738 | 15920 | No | NA | 32 | 11 | 2 | Low-quality | Genome-fragment | 39.43 | AAI-based (high-confidence) | 0 |
| S12_extracted_k141_129616 | 15926 | No | NA | 23 | 9 | 1 | Low-quality | Genome-fragment | 42.41 | AAI-based (medium-confidence) | 0 |
| S8_vs_k141_73935||full | 3987 | No | NA | 6 | 0 | 1 | Not-determined | Genome-fragment | NA | NA | 0 |
| S13_extracted_138148 | 15930 | No | NA | 16 | 6 | 0 | Low-quality | Genome-fragment | 14.6 | AAI-based (high-confidence) | 0 |
| S8_vs_k141_39660||full | 2086 | No | NA | 2 | 0 | 2 | Not-determined | Genome-fragment | NA | NA | 0 |
| S10_vs_48684||full | 15935 | No | NA | 14 | 0 | 8 | Low-quality | Genome-fragment | 12.22 | AAI-based (medium-confidence) | 0 |
| S8_vs_k141_109170||full | 2660 | No | NA | 4 | 0 | 2 | Not-determined | Genome-fragment | NA | NA | 0 |
| S5_extracted_k141_68532 | 15944 | No | NA | 13 | 0 | 2 | Low-quality | Genome-fragment | 15.69 | AAI-based (medium-confidence) | 0 |
| S8_vs_k141_39678||full | 3452 | No | NA | 4 | 0 | 1 | Not-determined | Genome-fragment | NA | NA | 0 |
| S8_vs_k141_13290||full | 2936 | No | NA | 7 | 0 | 0 | Not-determined | Genome-fragment | NA | NA | 0 |
| S8_vs_k141_178532||full | 4628 | No | NA | 4 | 0 | 3 | Not-determined | Genome-fragment | NA | NA | 0 |
| S8_vs_k141_169870||full | 1454 | No | NA | 3 | 0 | 0 | Not-determined | Genome-fragment | NA | NA | 0 |
| S1_extracted_k141_107267 | 15956 | No | NA | 36 | 4 | 0 | Low-quality | Genome-fragment | 28.35 | AAI-based (high-confidence) | 0 |
| S8_vs_k141_186208||full | 1821 | No | NA | 3 | 0 | 0 | Not-determined | Genome-fragment | NA | NA | 0 |
| S6_vs_k141_20421||0_partial | 15960 | No | NA | 13 | 0 | 3 | Low-quality | Genome-fragment | 30.28 | AAI-based (medium-confidence) | 0 |
| S8_vs_k141_109196||full | 1857 | No | NA | 3 | 0 | 1 | Not-determined | Genome-fragment | NA | NA | 0 |
| S8_vs_k141_204260||full | 35012 | No | NA | 52 | 14 | 0 | Complete | High-quality | 100 | ITR (medium-confidence) | 0 |
| S5_extracted_k141_55167 | 15965 | Yes | 12626 | 23 | 9 | 3 | Low-quality | Genome-fragment | 30.77 | AAI-based (high-confidence) | 20.91 |
| S8_vs_k141_57231||full | 7294 | No | NA | 11 | 0 | 1 | Not-determined | Genome-fragment | NA | NA | 0 |
| S9_vs_k141_32651||full | 15977 | No | NA | 25 | 15 | 0 | Low-quality | Genome-fragment | 41.95 | AAI-based (high-confidence) | 0 |
| S8_vs_k141_4883||full | 27347 | No | NA | 44 | 12 | 0 | Medium-quality | Genome-fragment | 67.76 | AAI-based (medium-confidence) | 0 |
| S8_vs_k141_186219||full | 2977 | No | NA | 6 | 0 | 1 | Not-determined | Genome-fragment | NA | NA | 0 |
| S10_vs_3596||full | 15985 | Yes | 8955 | 24 | 2 | 4 | Low-quality | Genome-fragment | 16.67 | AAI-based (medium-confidence) | 43.98 |
| S9_vs_k141_45763||full | 15997 | No | NA | 21 | 11 | 1 | Low-quality | Genome-fragment | 39.69 | AAI-based (medium-confidence) | 0 |
| S8_vs_k141_109230||full | 1223 | No | NA | 2 | 0 | 0 | Not-determined | Genome-fragment | NA | NA | 0 |
| S8_vs_k141_186230||full | 2765 | No | NA | 2 | 0 | 1 | Not-determined | Genome-fragment | NA | NA | 0 |
| S8_vs_k141_82932||full | 2337 | No | NA | 3 | 0 | 2 | Not-determined | Genome-fragment | NA | NA | 0 |
| S12_extracted_k141_33099 | 16000 | No | NA | 15 | 0 | 4 | Low-quality | Genome-fragment | 14.92 | AAI-based (medium-confidence) | 0 |
| S14_vs_k141_91818||full | 16014 | No | NA | 34 | 14 | 0 | Low-quality | Genome-fragment | 39.05 | AAI-based (high-confidence) | 0 |
| S8_vs_k141_160690||full | 4537 | No | NA | 5 | 0 | 4 | Not-determined | Genome-fragment | NA | NA | 0 |
| S8_vs_k141_109250||full | 8196 | No | NA | 6 | 0 | 5 | Not-determined | Genome-fragment | NA | NA | 0 |
| S9_vs_k141_6787||full | 16042 | No | NA | 19 | 2 | 1 | Low-quality | Genome-fragment | 34.46 | AAI-based (medium-confidence) | 0 |
| S8_vs_k141_186293||full | 1764 | No | NA | 3 | 0 | 1 | Not-determined | Genome-fragment | NA | NA | 0 |
| S12_extracted_k141_88086 | 16054 | No | NA | 33 | 6 | 0 | Low-quality | Genome-fragment | 31.47 | AAI-based (high-confidence) | 0 |
| S11_extracted_k141_16070 | 16059 | No | NA | 13 | 1 | 0 | Low-quality | Genome-fragment | 6.22 | AAI-based (medium-confidence) | 0 |
| S2_extracted_k141_9957 | 16059 | No | NA | 26 | 4 | 0 | Low-quality | Genome-fragment | 25.41 | AAI-based (high-confidence) | 0 |
| S11_vs_k141_43481||0_partial | 16069 | Yes | 9968 | 18 | 3 | 2 | Low-quality | Genome-fragment | 6.13 | AAI-based (medium-confidence) | 37.97 |
| S8_vs_k141_178665||full | 1492 | No | NA | 3 | 0 | 0 | Not-determined | Genome-fragment | NA | NA | 0 |
| S8_vs_k141_83006||full | 2120 | No | NA | 4 | 0 | 3 | Not-determined | Genome-fragment | NA | NA | 0 |
| S14_vs_k141_8525||full | 16072 | No | NA | 26 | 10 | 2 | Low-quality | Genome-fragment | 32.33 | AAI-based (medium-confidence) | 0 |
| S6_extracted_k141_47690 | 16079 | No | NA | 18 | 0 | 3 | Low-quality | Genome-fragment | 23.49 | AAI-based (medium-confidence) | 0 |
| S8_vs_k141_169996||full | 1232 | No | NA | 2 | 0 | 1 | Not-determined | Genome-fragment | NA | NA | 0 |
| S8_vs_k141_65603||full | 1917 | No | NA | 2 | 0 | 1 | Not-determined | Genome-fragment | NA | NA | 0 |
| S8_vs_k141_144085||full | 2248 | No | NA | 2 | 0 | 0 | Not-determined | Genome-fragment | NA | NA | 0 |
| S13_extracted_92590 | 16085 | No | NA | 17 | 1 | 1 | Low-quality | Genome-fragment | 2.97 | HMM-based (lower-bound) | 0 |
| S3_vs_k141_76719||full | 16098 | No | NA | 24 | 12 | 0 | Low-quality | Genome-fragment | 39.34 | AAI-based (high-confidence) | 0 |
| S10_vs_83674||full | 16103 | No | NA | 20 | 15 | 0 | Low-quality | Genome-fragment | 31.53 | AAI-based (high-confidence) | 0 |
| S17_vs_k141_31662||full | 16107 | No | NA | 24 | 5 | 2 | Low-quality | Genome-fragment | 23.26 | AAI-based (high-confidence) | 0 |
| S16_extracted_104509 | 16118 | No | NA | 38 | 3 | 1 | Low-quality | Genome-fragment | 36.3 | AAI-based (high-confidence) | 0 |
| S8_vs_k141_4974||full | 1277 | No | NA | 2 | 0 | 1 | Not-determined | Genome-fragment | NA | NA | 0 |
| S9_extracted_k141_36262 | 16143 | No | NA | 9 | 0 | 7 | Low-quality | Genome-fragment | 6.9 | AAI-based (medium-confidence) | 0 |
| S8_vs_k141_170017||full | 1758 | No | NA | 3 | 0 | 1 | Not-determined | Genome-fragment | NA | NA | 0 |
| S8_vs_k141_57385||full | 6831 | No | NA | 5 | 0 | 3 | Not-determined | Genome-fragment | NA | NA | 0 |
| S13_extracted_54510 | 16153 | No | NA | 12 | 0 | 8 | Low-quality | Genome-fragment | 6.49 | AAI-based (medium-confidence) | 0 |
| S16_vs_136989||full | 16154 | No | NA | 16 | 6 | 0 | Low-quality | Genome-fragment | 6.87 | AAI-based (medium-confidence) | 0 |
| S8_vs_k141_4984||full | 2205 | No | NA | 2 | 0 | 2 | Not-determined | Genome-fragment | NA | NA | 0 |
| S8_vs_k141_196163||full | 2340 | No | NA | 3 | 0 | 2 | Not-determined | Genome-fragment | NA | NA | 0 |
| S12_vs_k141_184313||0_partial | 16163 | No | NA | 15 | 0 | 10 | Low-quality | Genome-fragment | 15.85 | AAI-based (medium-confidence) | 0 |
| S14_vs_k141_77141||full | 16172 | No | NA | 31 | 2 | 0 | Low-quality | Genome-fragment | 43.08 | AAI-based (high-confidence) | 0 |
| S8_vs_k141_109348||full | 1342 | No | NA | 3 | 0 | 0 | Not-determined | Genome-fragment | NA | NA | 0 |
| S8_vs_k141_74082||full | 1505 | No | NA | 3 | 0 | 1 | Not-determined | Genome-fragment | NA | NA | 0 |
| S5_vs_k141_73497||full | 16186 | No | NA | 23 | 3 | 0 | Low-quality | Genome-fragment | 29.9 | AAI-based (medium-confidence) | 0 |
| S8_vs_k141_196174||full | 1118 | No | NA | 3 | 0 | 0 | Not-determined | Genome-fragment | NA | NA | 0 |
| S10_extracted_10743 | 16212 | Yes | 6078 | 10 | 1 | 3 | Low-quality | Genome-fragment | 1.7 | HMM-based (lower-bound) | 62.51 |
| S8_vs_k141_22757||full | 7275 | No | NA | 10 | 0 | 1 | Not-determined | Genome-fragment | NA | NA | 0 |
| S8_vs_k141_39896||full | 1628 | No | NA | 3 | 0 | 0 | Not-determined | Genome-fragment | NA | NA | 0 |
| S16_vs_28728||full | 16221 | No | NA | 19 | 4 | 0 | Low-quality | Genome-fragment | 6.54 | AAI-based (medium-confidence) | 0 |
| S17_vs_k141_108547||full | 16225 | No | NA | 23 | 8 | 0 | Low-quality | Genome-fragment | 45.33 | AAI-based (high-confidence) | 0 |
| S12_extracted_k141_18900 | 16234 | Yes | 7204 | 23 | 2 | 4 | Low-quality | Genome-fragment | 5.37 | AAI-based (high-confidence) | 55.62 |
| S8_vs_k141_22770||full | 1881 | No | NA | 3 | 0 | 0 | Not-determined | Genome-fragment | NA | NA | 0 |
| S8_vs_k141_196192||full | 1428 | No | NA | 2 | 0 | 0 | Not-determined | Genome-fragment | NA | NA | 0 |
| S9_vs_k141_54746||full | 16242 | No | NA | 23 | 9 | 0 | Low-quality | Genome-fragment | 43.77 | AAI-based (medium-confidence) | 0 |
| S8_vs_k141_13544||full | 9084 | No | NA | 4 | 0 | 4 | Not-determined | Genome-fragment | NA | NA | 0 |
| S7_vs_61448||full | 16265 | No | NA | 15 | 2 | 0 | Low-quality | Genome-fragment | 16.71 | AAI-based (medium-confidence) | 0 |
| S8_vs_k141_22803||full | 2374 | No | NA | 4 | 0 | 3 | Not-determined | Genome-fragment | NA | NA | 0 |
| S8_vs_k141_117871||full | 9410 | No | NA | 6 | 0 | 5 | Not-determined | Genome-fragment | NA | NA | 0 |
| S8_vs_k141_5047||full | 1434 | No | NA | 3 | 0 | 1 | Not-determined | Genome-fragment | NA | NA | 0 |
| S14_vs_k141_9168||full | 16280 | No | NA | 10 | 1 | 0 | Low-quality | Genome-fragment | 42.64 | AAI-based (high-confidence) | 0 |
| S8_vs_k141_135041||full | 4110 | No | NA | 6 | 0 | 0 | Not-determined | Genome-fragment | NA | NA | 0 |
| S8_vs_k141_5069||full | 2222 | No | NA | 3 | 0 | 1 | Not-determined | Genome-fragment | NA | NA | 0 |
| S8_vs_k141_186515||full | 1377 | No | NA | 3 | 0 | 2 | Not-determined | Genome-fragment | NA | NA | 0 |
| S8_vs_k141_170149||full | 1982 | No | NA | 3 | 0 | 1 | Not-determined | Genome-fragment | NA | NA | 0 |
| S8_vs_k141_152553||full | 31203 | No | NA | 41 | 15 | 2 | Medium-quality | Genome-fragment | 70.88 | AAI-based (high-confidence) | 0 |
| S8_vs_k141_204432||full | 1470 | No | NA | 5 | 0 | 0 | Not-determined | Genome-fragment | NA | NA | 0 |
| S11_extracted_k141_33350 | 16299 | No | NA | 15 | 8 | 0 | Low-quality | Genome-fragment | 10.75 | AAI-based (medium-confidence) | 0 |
| S14_vs_k141_66216||full | 16315 | No | NA | 20 | 6 | 0 | Low-quality | Genome-fragment | 44.58 | AAI-based (high-confidence) | 0 |
| S8_vs_k141_126582||full | 60732 | No | NA | 64 | 9 | 0 | Medium-quality | Genome-fragment | 74.07 | AAI-based (high-confidence) | 0 |
| S3_extracted_k141_125443 | 16322 | No | NA | 34 | 3 | 0 | Low-quality | Genome-fragment | 9.23 | AAI-based (high-confidence) | 0 |
| S8_vs_k141_152575||full | 2049 | No | NA | 2 | 0 | 0 | Not-determined | Genome-fragment | NA | NA | 0 |
| S8_vs_k141_22847||full | 2982 | No | NA | 4 | 0 | 3 | Not-determined | Genome-fragment | NA | NA | 0 |
| S8_vs_k141_204464||full | 2197 | No | NA | 3 | 0 | 0 | Not-determined | Genome-fragment | NA | NA | 0 |
| S8_vs_k141_196247||full | 14237 | No | NA | 19 | 1 | 0 | Medium-quality | Genome-fragment | 88.12 | AAI-based (medium-confidence) | 0 |
| S8_vs_k141_117929||full | 2615 | No | NA | 2 | 0 | 1 | Not-determined | Genome-fragment | NA | NA | 0 |
| S8_vs_k141_196254||full | 1319 | No | NA | 2 | 0 | 1 | Not-determined | Genome-fragment | NA | NA | 0 |
| S8_vs_k141_170196||full | 1784 | No | NA | 4 | 0 | 0 | Not-determined | Genome-fragment | NA | NA | 0 |
| S8_vs_k141_91951||full | 7286 | No | NA | 9 | 0 | 4 | Not-determined | Genome-fragment | NA | NA | 0 |
| S10_vs_98758||0_partial | 16338 | No | NA | 16 | 2 | 10 | Low-quality | Genome-fragment | 5.85 | HMM-based (lower-bound) | 0 |
| S12_extracted_k141_149688 | 16338 | No | NA | 27 | 5 | 2 | Low-quality | Genome-fragment | 31.77 | AAI-based (high-confidence) | 0 |
| S6_vs_k141_43694||full | 16338 | No | NA | 20 | 0 | 14 | Low-quality | Genome-fragment | 6.73 | AAI-based (medium-confidence) | 0 |
| S8_vs_k141_186572||full | 3208 | No | NA | 3 | 0 | 2 | Not-determined | Genome-fragment | NA | NA | 0 |
| S8_vs_k141_152645||full | 4622 | No | NA | 4 | 0 | 3 | Not-determined | Genome-fragment | NA | NA | 0 |
| S8_vs_k141_109497||full | 1990 | No | NA | 3 | 0 | 0 | Not-determined | Genome-fragment | NA | NA | 0 |
| S17_extracted_k141_140590 | 16351 | No | NA | 19 | 1 | 2 | Low-quality | Genome-fragment | 24.74 | HMM-based (lower-bound) | 0 |
| S8_vs_k141_54518||full | 16372 | No | NA | 23 | 9 | 1 | Low-quality | Genome-fragment | 40.1 | AAI-based (medium-confidence) | 0 |
| S8_vs_k141_92001||full | 2894 | No | NA | 3 | 0 | 3 | Not-determined | Genome-fragment | NA | NA | 0 |
| S3_vs_k141_47385||full | 16380 | No | NA | 27 | 4 | 0 | Low-quality | Genome-fragment | 9 | AAI-based (high-confidence) | 0 |
| S3_vs_k141_117114||full | 16384 | No | NA | 26 | 3 | 1 | Low-quality | Genome-fragment | 38.25 | AAI-based (medium-confidence) | 0 |
| S15_extracted_777 | 16419 | No | NA | 15 | 0 | 15 | Low-quality | Genome-fragment | 7.06 | AAI-based (medium-confidence) | 0 |
| S8_vs_k141_178901||full | 1214 | No | NA | 3 | 0 | 1 | Not-determined | Genome-fragment | NA | NA | 0 |
| S9_vs_k141_24121||full | 16446 | No | NA | 22 | 1 | 0 | Low-quality | Genome-fragment | 14.13 | HMM-based (lower-bound) | 0 |
| S8_vs_k141_109543||full | 2691 | No | NA | 3 | 0 | 0 | Not-determined | Genome-fragment | NA | NA | 0 |
| S8_vs_k141_48241||full | 2216 | No | NA | 2 | 0 | 0 | Not-determined | Genome-fragment | NA | NA | 0 |
| S8_extracted_k141_143081 | 16447 | No | NA | 13 | 0 | 13 | Low-quality | Genome-fragment | 9.75 | AAI-based (high-confidence) | 0 |
| S8_vs_k141_13713||full | 3543 | No | NA | 6 | 0 | 5 | Not-determined | Genome-fragment | NA | NA | 0 |
| S8_vs_k141_178979||full | 2572 | No | NA | 2 | 0 | 2 | Not-determined | Genome-fragment | NA | NA | 0 |
| S8_vs_k141_5211||full | 26189 | No | NA | 37 | 13 | 0 | Medium-quality | Genome-fragment | 76.35 | AAI-based (high-confidence) | 0 |
| S15_vs_64960||full | 16449 | No | NA | 10 | 0 | 8 | Low-quality | Genome-fragment | 6.23 | AAI-based (medium-confidence) | 0 |
| S8_vs_k141_13723||full | 11808 | No | NA | 8 | 0 | 2 | Not-determined | Genome-fragment | NA | NA | 0 |
| S8_vs_k141_152764||full | 1375 | No | NA | 2 | 0 | 0 | Not-determined | Genome-fragment | NA | NA | 0 |
| S3_vs_k141_34796||full | 16453 | Yes | 9653 | 33 | 2 | 3 | Low-quality | Genome-fragment | 11.37 | HMM-based (lower-bound) | 41.33 |
| S8_vs_k141_161091||full | 1524 | No | NA | 4 | 0 | 1 | Not-determined | Genome-fragment | NA | NA | 0 |
| S8_vs_k141_179011||full | 1310 | No | NA | 2 | 0 | 0 | Not-determined | Genome-fragment | NA | NA | 0 |
| S12_extracted_k141_82399 | 16466 | No | NA | 21 | 6 | 0 | Low-quality | Genome-fragment | 25.81 | AAI-based (high-confidence) | 0 |
| S3_vs_k141_96207||full | 16481 | No | NA | 18 | 14 | 0 | Low-quality | Genome-fragment | 27.35 | AAI-based (high-confidence) | 0 |
| S8_vs_k141_161102||full | 1441 | No | NA | 2 | 0 | 0 | Not-determined | Genome-fragment | NA | NA | 0 |
| S8_vs_k141_83419||full | 1324 | No | NA | 3 | 0 | 3 | Not-determined | Genome-fragment | NA | NA | 0 |
| S14_vs_k141_29918||full | 16498 | No | NA | 25 | 1 | 6 | Low-quality | Genome-fragment | 9.75 | HMM-based (lower-bound) | 0 |
| S8_vs_k141_133942||full | 16517 | No | NA | 28 | 3 | 0 | Low-quality | Genome-fragment | 25.96 | AAI-based (high-confidence) | 0 |
| S8_vs_k141_196429||full | 2258 | No | NA | 4 | 0 | 0 | Not-determined | Genome-fragment | NA | NA | 0 |
| S12_extracted_k141_17137 | 16532 | No | NA | 15 | 0 | 12 | Low-quality | Genome-fragment | 10.1 | AAI-based (medium-confidence) | 0 |
| S9_vs_k141_16826||full | 16546 | No | NA | 26 | 1 | 2 | Low-quality | Genome-fragment | 29.71 | AAI-based (high-confidence) | 0 |
| S17_vs_k141_166099||full | 16547 | No | NA | 25 | 14 | 0 | Low-quality | Genome-fragment | 48.52 | AAI-based (high-confidence) | 0 |
| S8_vs_k141_65894||full | 2131 | No | NA | 4 | 0 | 0 | Not-determined | Genome-fragment | NA | NA | 0 |
| S8_vs_k141_152829||full | 1562 | No | NA | 3 | 0 | 1 | Not-determined | Genome-fragment | NA | NA | 0 |
| S9_vs_k141_8552||full | 16588 | No | NA | 16 | 4 | 0 | Low-quality | Genome-fragment | 29.07 | AAI-based (high-confidence) | 0 |
| S10_extracted_16833 | 16629 | No | NA | 21 | 13 | 0 | Low-quality | Genome-fragment | 42.56 | AAI-based (high-confidence) | 0 |
| S8_vs_k141_23104||full | 1650 | No | NA | 2 | 0 | 1 | Not-determined | Genome-fragment | NA | NA | 0 |
| S9_vs_k141_31202||full | 16638 | No | NA | 21 | 10 | 0 | Low-quality | Genome-fragment | 49.71 | AAI-based (high-confidence) | 0 |
| S16_vs_6181||full | 16639 | No | NA | 26 | 5 | 0 | Low-quality | Genome-fragment | 23.59 | HMM-based (lower-bound) | 0 |
| S11_vs_k141_23935||full | 16646 | No | NA | 33 | 6 | 1 | Low-quality | Genome-fragment | 39.71 | AAI-based (medium-confidence) | 0 |
| S8_vs_k141_31715||full | 2557 | No | NA | 2 | 0 | 2 | Not-determined | Genome-fragment | NA | NA | 0 |
| S8_vs_k141_40150||full | 2068 | No | NA | 3 | 0 | 1 | Not-determined | Genome-fragment | NA | NA | 0 |
| S8_vs_k141_40157||full | 1204 | No | NA | 2 | 0 | 0 | Not-determined | Genome-fragment | NA | NA | 0 |
| S1_extracted_k141_20127 | 16657 | No | NA | 18 | 6 | 0 | Low-quality | Genome-fragment | 37.12 | AAI-based (high-confidence) | 0 |
| S8_vs_k141_48449||full | 3067 | No | NA | 5 | 0 | 3 | Not-determined | Genome-fragment | NA | NA | 0 |
| S8_vs_k141_196544||full | 2160 | No | NA | 3 | 0 | 2 | Not-determined | Genome-fragment | NA | NA | 0 |
| S8_vs_k141_161216||full | 1819 | No | NA | 4 | 0 | 2 | Not-determined | Genome-fragment | NA | NA | 0 |
| S8_vs_k141_161222||full | 1179 | No | NA | 3 | 0 | 0 | Not-determined | Genome-fragment | NA | NA | 0 |
| S8_vs_k141_109691||full | 1176 | No | NA | 3 | 0 | 1 | Not-determined | Genome-fragment | NA | NA | 0 |
| S3_extracted_k141_108362 | 16658 | No | NA | 22 | 2 | 1 | Low-quality | Genome-fragment | 21.28 | HMM-based (lower-bound) | 0 |
| S11_vs_k141_731||full | 16666 | No | NA | 22 | 5 | 0 | Low-quality | Genome-fragment | 36.6 | AAI-based (high-confidence) | 0 |

| | | | | | | | | | | | |
|---|---|---|---|---|---|---|---|---|---|---|---|
| S8_vs_k141_170439\|\|full | 3901 | No | NA | 4 | 0 | 1 | Not-determined | Genome-fragment | NA | NA | 0 |
| S12_vs_k141_12137\|\|full | 16710 | Yes | 5077 | 23 | 4 | 8 | Low-quality | Genome-fragment | 12.41 | AAI-based (medium-confidence) | 69.62 |
| S8_vs_k141_100484\|\|full | 1326 | No | NA | 3 | 0 | 2 | Not-determined | Genome-fragment | NA | NA | 0 |
| S4_extracted_27624 | 16720 | No | NA | 19 | 15 | 0 | Low-quality | Genome-fragment | 42.04 | AAI-based (high-confidence) | 0 |
| S8_vs_k141_83520\|\|full | 2011 | No | NA | 2 | 0 | 1 | Not-determined | Genome-fragment | NA | NA | 0 |
| S8_vs_k141_186774\|\|full | 1096 | No | NA | 2 | 0 | 0 | Not-determined | Genome-fragment | NA | NA | 0 |
| S3_extracted_k141_126367 | 16729 | No | NA | 25 | 1 | 0 | Low-quality | Genome-fragment | 19.43 | AAI-based (high-confidence) | 0 |
| S15_extracted_33563 | 16748 | No | NA | 20 | 0 | 4 | Low-quality | Genome-fragment | 18.67 | AAI-based (high-confidence) | 0 |
| S8_vs_k141_196598\|\|full | 4295 | No | NA | 6 | 0 | 2 | Not-determined | Genome-fragment | NA | NA | 0 |
| S8_vs_k141_118275\|\|full | 1407 | No | NA | 3 | 0 | 0 | Not-determined | Genome-fragment | NA | NA | 0 |
| S8_vs_k141_74470\|\|full | 2522 | No | NA | 3 | 0 | 1 | Not-determined | Genome-fragment | NA | NA | 0 |
| S8_vs_k141_204735\|\|full | 1311 | No | NA | 2 | 0 | 1 | Not-determined | Genome-fragment | NA | NA | 0 |
| S6_vs_k141_34170\|\|full | 16776 | No | NA | 40 | 9 | 1 | Low-quality | Genome-fragment | 38.71 | AAI-based (high-confidence) | 0 |
| S8_vs_k141_109713\|\|full | 9226 | No | NA | 7 | 0 | 6 | Not-determined | Genome-fragment | NA | NA | 0 |
| S11_extracted_k141_75122 | 16806 | No | NA | 38 | 4 | 0 | Low-quality | Genome-fragment | 28.32 | AAI-based (high-confidence) | 0 |
| S16_vs_103881\|\|full | 16814 | No | NA | 10 | 2 | 2 | Low-quality | Genome-fragment | 25.46 | AAI-based (medium-confidence) | 0 |
| S8_vs_k141_109718\|\|full | 1554 | No | NA | 3 | 0 | 2 | Not-determined | Genome-fragment | NA | NA | 0 |
| S16_vs_20849\|\|full | 16822 | No | NA | 18 | 0 | 12 | Low-quality | Genome-fragment | 6.97 | AAI-based (medium-confidence) | 0 |
| S8_vs_k141_83575\|\|full | 2547 | No | NA | 4 | 0 | 2 | Not-determined | Genome-fragment | NA | NA | 0 |
| S5_vs_k141_138145\|\|full | 16870 | No | NA | 19 | 15 | 0 | Low-quality | Genome-fragment | 41.49 | AAI-based (high-confidence) | 0 |
| S8_vs_k141_170490\|\|full | 1856 | No | NA | 3 | 0 | 2 | Not-determined | Genome-fragment | NA | NA | 0 |
| S8_vs_k141_13862\|\|full | 2460 | No | NA | 3 | 0 | 2 | Not-determined | Genome-fragment | NA | NA | 0 |
| S7_extracted_79190 | 16872 | No | NA | 19 | 13 | 0 | Low-quality | Genome-fragment | 12.34 | AAI-based (medium-confidence) | 0 |
| S8_vs_k141_135497\|\|full | 3178 | No | NA | 3 | 0 | 1 | Not-determined | Genome-fragment | NA | NA | 0 |
| S8_vs_k141_109752\|\|full | 3924 | No | NA | 5 | 0 | 2 | Not-determined | Genome-fragment | NA | NA | 0 |
| S17_extracted_k141_140349 | 16914 | No | NA | 15 | 0 | 15 | Low-quality | Genome-fragment | 9.3 | AAI-based (medium-confidence) | 0 |
| S7_vs_97822\|\|full | 16914 | No | NA | 24 | 11 | 0 | Low-quality | Genome-fragment | 45.86 | AAI-based (medium-confidence) | 0 |
| S3_extracted_k141_35217 | 16918 | No | NA | 15 | 1 | 3 | Low-quality | Genome-fragment | 1.36 | HMM-based (lower-bound) | 0 |
| S8_vs_k141_100595\|\|full | 2168 | No | NA | 3 | 0 | 0 | Not-determined | Genome-fragment | NA | NA | 0 |
| S16_vs_9234\|\|full | 16919 | No | NA | 13 | 4 | 0 | Low-quality | Genome-fragment | 6.72 | AAI-based (medium-confidence) | 0 |
| S8_vs_k141_153050\|\|full | 1924 | No | NA | 4 | 0 | 2 | Not-determined | Genome-fragment | NA | NA | 0 |
| S11_vs_k141_53039\|\|full | 16922 | No | NA | 39 | 2 | 4 | Low-quality | Genome-fragment | 40.73 | AAI-based (high-confidence) | 0 |
| S11_extracted_k141_12206 | 16938 | No | NA | 34 | 11 | 0 | Low-quality | Genome-fragment | 36.38 | AAI-based (high-confidence) | 0 |
| S8_vs_k141_118383\|\|full | 1789 | No | NA | 3 | 0 | 1 | Not-determined | Genome-fragment | NA | NA | 0 |
| S9_extracted_k141_5906 | 16951 | No | NA | 27 | 5 | 0 | Low-quality | Genome-fragment | 37.88 | AAI-based (high-confidence) | 0 |
| S14_vs_k141_11417\|\|full | 16968 | No | NA | 22 | 3 | 0 | Low-quality | Genome-fragment | 23.62 | AAI-based (high-confidence) | 0 |
| S8_vs_k141_40337\|\|full | 1001 | No | NA | 2 | 0 | 0 | Not-determined | Genome-fragment | NA | NA | 0 |
| S8_vs_k141_109825\|\|full | 5105 | No | NA | 5 | 0 | 3 | Not-determined | Genome-fragment | NA | NA | 0 |
| S13_vs_69774\|\|full | 16969 | No | NA | 15 | 1 | 1 | Low-quality | Genome-fragment | 30.04 | AAI-based (high-confidence) | 0 |
| S4_extracted_12852 | 16993 | No | NA | 32 | 7 | 0 | Low-quality | Genome-fragment | 31.73 | AAI-based (high-confidence) | 0 |
| S8_vs_k141_170603\|\|full | 1409 | No | NA | 3 | 0 | 1 | Not-determined | Genome-fragment | NA | NA | 0 |
| S8_vs_k141_144516\|\|full | 6277 | No | NA | 4 | 0 | 4 | Not-determined | Genome-fragment | NA | NA | 0 |
| S8_vs_k141_40366\|\|full | 1142 | No | NA | 2 | 0 | 1 | Not-determined | Genome-fragment | NA | NA | 0 |
| S11_vs_k141_48039\|\|full | 17028 | No | NA | 17 | 9 | 0 | Low-quality | Genome-fragment | 30.26 | AAI-based (high-confidence) | 0 |
| S8_vs_k141_204806\|\|full | 1498 | No | NA | 3 | 0 | 0 | Not-determined | Genome-fragment | NA | NA | 0 |
| S8_vs_k141_186924\|\|full | 2373 | No | NA | 2 | 0 | 1 | Not-determined | Genome-fragment | NA | NA | 0 |
| S8_vs_k141_179208\|\|full | 109051 | No | NA | 118 | 2 | 40 | Medium-quality | Genome-fragment | 72.26 | HMM-based (lower-bound) | 0 |
| S11_vs_k141_11166\|\|0_partial | 17044 | No | NA | 18 | 1 | 9 | Low-quality | Genome-fragment | 7.32 | HMM-based (lower-bound) | 0 |
| S8_vs_k141_204812\|\|full | 1580 | No | NA | 3 | 0 | 0 | Not-determined | Genome-fragment | NA | NA | 0 |
| S8_vs_k141_13959\|\|full | 1533 | No | NA | 3 | 0 | 1 | Not-determined | Genome-fragment | NA | NA | 0 |
| S8_vs_k141_127016\|\|full | 2455 | No | NA | 4 | 0 | 1 | Not-determined | Genome-fragment | NA | NA | 0 |
| S5_vs_k141_101924\|\|full | 17050 | No | NA | 19 | 14 | 0 | Low-quality | Genome-fragment | 47.3 | AAI-based (medium-confidence) | 0 |
| S5_extracted_k141_89402 | 17066 | No | NA | 16 | 14 | 0 | Low-quality | Genome-fragment | 41.37 | AAI-based (high-confidence) | 0 |
| S8_vs_k141_5582\|\|full | 1200 | No | NA | 2 | 0 | 1 | Not-determined | Genome-fragment | NA | NA | 0 |
| S5_vs_k141_69896\|\|full | 17066 | No | NA | 29 | 12 | 2 | Low-quality | Genome-fragment | 45.04 | AAI-based (medium-confidence) | 0 |
| S8_vs_k141_109882\|\|full | 2721 | No | NA | 6 | 0 | 0 | Not-determined | Genome-fragment | NA | NA | 0 |
| S14_extracted_k141_95950 | 17106 | No | NA | 22 | 5 | 0 | Low-quality | Genome-fragment | 28.02 | AAI-based (high-confidence) | 0 |
| S5_extracted_k141_32774 | 17109 | No | NA | 39 | 4 | 0 | Low-quality | Genome-fragment | 16.62 | HMM-based (lower-bound) | 0 |
| S12_vs_k141_48289\|\|0_partial | 17114 | No | NA | 17 | 1 | 2 | Low-quality | Genome-fragment | 24.36 | HMM-based (lower-bound) | 0 |
| S3_extracted_k141_38325 | 17115 | No | NA | 12 | 0 | 10 | Low-quality | Genome-fragment | 4.61 | AAI-based (medium-confidence) | 0 |
| S8_vs_k141_196763\|\|full | 2374 | No | NA | 2 | 0 | 1 | Not-determined | Genome-fragment | NA | NA | 0 |
| S8_vs_k141_118515\|\|full | 2131 | No | NA | 4 | 0 | 0 | Not-determined | Genome-fragment | NA | NA | 0 |
| S8_vs_k141_66129\|\|full | 1122 | No | NA | 2 | 0 | 1 | Not-determined | Genome-fragment | NA | NA | 0 |
| S10_vs_17793\|\|0_partial | 17124 | No | NA | 17 | 1 | 13 | Low-quality | Genome-fragment | 9.21 | HMM-based (lower-bound) | 0 |
| S16_extracted_57471 | 17144 | No | NA | 20 | 1 | 5 | Low-quality | Genome-fragment | 15.09 | AAI-based (medium-confidence) | 0 |
| S14_vs_k141_114544\|\|full | 17159 | No | NA | 29 | 7 | 0 | Low-quality | Genome-fragment | 43.79 | AAI-based (high-confidence) | 0 |
| S8_vs_k141_31977\|\|full | 5590 | No | NA | 7 | 0 | 7 | Not-determined | Genome-fragment | NA | NA | 0 |
| S8_vs_k141_196789\|\|full | 3810 | No | NA | 5 | 0 | 0 | Not-determined | Genome-fragment | NA | NA | 0 |
| S5_vs_k141_76537\|\|full | 17159 | No | NA | 20 | 11 | 0 | Low-quality | Genome-fragment | 49.07 | AAI-based (high-confidence) | 0 |
| S8_vs_k141_153182\|\|full | 1008 | No | NA | 2 | 0 | 0 | Not-determined | Genome-fragment | NA | NA | 0 |
| S8_vs_k141_66151\|\|full | 5584 | No | NA | 5 | 0 | 4 | Not-determined | Genome-fragment | NA | NA | 0 |
| S8_vs_k141_109919\|\|full | 5097 | No | NA | 7 | 0 | 3 | Not-determined | Genome-fragment | NA | NA | 0 |
| S8_vs_k141_144598\|\|full | 2449 | No | NA | 4 | 0 | 2 | Not-determined | Genome-fragment | NA | NA | 0 |
| S8_vs_k141_144602\|\|full | 2797 | No | NA | 3 | 0 | 0 | Not-determined | Genome-fragment | NA | NA | 0 |
| S17_vs_k141_57689\|\|full | 17160 | No | NA | 40 | 3 | 1 | Low-quality | Genome-fragment | 49.69 | AAI-based (high-confidence) | 0 |
| S8_vs_k141_92532\|\|full | 4088 | No | NA | 6 | 0 | 2 | Not-determined | Genome-fragment | NA | NA | 0 |
| S8_vs_k141_14078\|\|full | 6562 | No | NA | 7 | 0 | 6 | Not-determined | Genome-fragment | NA | NA | 0 |
| S8_vs_k141_118572\|\|full | 3486 | No | NA | 5 | 0 | 1 | Not-determined | Genome-fragment | NA | NA | 0 |
| S8_vs_k141_74711\|\|full | 1173 | No | NA | 3 | 0 | 0 | Not-determined | Genome-fragment | NA | NA | 0 |
| S8_vs_k141_196815\|\|full | 7455 | No | NA | 5 | 0 | 1 | Not-determined | Genome-fragment | NA | NA | 0 |
| S8_vs_k141_32013\|\|full | 2428 | No | NA | 4 | 0 | 2 | Not-determined | Genome-fragment | NA | NA | 0 |
| S8_vs_k141_23333\|\|full | 3095 | No | NA | 3 | 0 | 1 | Not-determined | Genome-fragment | NA | NA | 0 |
| S3_extracted_k141_49949 | 17164 | No | NA | 19 | 15 | 0 | Low-quality | Genome-fragment | 46.43 | AAI-based (high-confidence) | 0 |
| S4_vs_VIRSorter_23805-cat_1 | 17170 | No | NA | 17 | 13 | 0 | Low-quality | Genome-fragment | 28.1 | AAI-based (high-confidence) | 0 |
| S8_vs_k141_207729\|\|full | 17171 | Yes | 10480 | 24 | 4 | 4 | Low-quality | Genome-fragment | 16.98 | AAI-based (high-confidence) | 38.97 |
| S8_vs_k141_170710\|\|full | 4936 | No | NA | 4 | 0 | 3 | Not-determined | Genome-fragment | NA | NA | 0 |
| S8_vs_k141_66219\|\|full | 2338 | No | NA | 6 | 0 | 1 | Not-determined | Genome-fragment | NA | NA | 0 |
| S6_vs_k141_36309\|\|full | 17180 | No | NA | 19 | 1 | 7 | Low-quality | Genome-fragment | 15.51 | AAI-based (high-confidence) | 0 |
| S16_vs_134707\|\|full | 17183 | No | NA | 17 | 0 | 5 | Low-quality | Genome-fragment | 30.32 | AAI-based (medium-confidence) | 0 |
| S8_vs_k141_161570\|\|full | 2585 | No | NA | 3 | 0 | 1 | Not-determined | Genome-fragment | NA | NA | 0 |
| S8_vs_k141_179381\|\|full | 1828 | No | NA | 2 | 0 | 1 | Not-determined | Genome-fragment | NA | NA | 0 |
| S8_vs_k141_179384\|\|full | 2029 | No | NA | 3 | 0 | 2 | Not-determined | Genome-fragment | NA | NA | 0 |
| S8_vs_k141_204980\|\|full | 3918 | No | NA | 3 | 0 | 0 | Not-determined | Genome-fragment | NA | NA | 0 |
| S8_vs_k141_93093\|\|full | 17256 | No | NA | 38 | 6 | 0 | Low-quality | Genome-fragment | 28.3 | AAI-based (high-confidence) | 0 |
| S7_vs_23653\|\|full | 17269 | Yes | 5165 | 19 | 2 | 4 | Low-quality | Genome-fragment | 3.21 | HMM-based (lower-bound) | 70.09 |
| S8_vs_k141_83680\|\|full | 2464 | No | NA | 2 | 0 | 1 | Not-determined | Genome-fragment | NA | NA | 0 |
| S8_vs_k141_58154\|\|full | 1800 | No | NA | 3 | 0 | 1 | Not-determined | Genome-fragment | NA | NA | 0 |
| S3_vs_k141_88141\|\|full | 17275 | Yes | 4966 | 19 | 3 | 7 | Low-quality | Genome-fragment | 6.3 | HMM-based (lower-bound) | 71.25 |
| S8_vs_k141_205008\|\|full | 1547 | No | NA | 4 | 0 | 1 | Not-determined | Genome-fragment | NA | NA | 0 |
| S3_extracted_k141_7573 | 17309 | No | NA | 21 | 8 | 0 | Low-quality | Genome-fragment | 19.43 | AAI-based (high-confidence) | 0 |
| S11_extracted_k141_22731 | 17336 | No | NA | 36 | 5 | 1 | Low-quality | Genome-fragment | 35.82 | AAI-based (high-confidence) | 0 |
| S8_vs_k141_32070\|\|full | 1566 | No | NA | 3 | 0 | 0 | Not-determined | Genome-fragment | NA | NA | 0 |
| S8_vs_k141_70746\|\|full | 17341 | No | NA | 17 | 0 | 0 | Low-quality | Genome-fragment | 28.86 | AAI-based (medium-confidence) | 0 |
| S7_vs_11173\|\|full | 17378 | No | NA | 40 | 6 | 1 | Low-quality | Genome-fragment | 43.73 | AAI-based (medium-confidence) | 0 |
| S14_vs_k141_10416\|\|full | 17380 | Yes | 16379 | 25 | 17 | 2 | Low-quality | Genome-fragment | 34.26 | AAI-based (medium-confidence) | 5.76 |
| S10_extracted_94555 | 17382 | No | NA | 24 | 1 | 0 | Low-quality | Genome-fragment | 10.76 | AAI-based (medium-confidence) | 0 |
| S8_vs_k141_92539\|\|full | 4929 | No | NA | 3 | 0 | 2 | Not-determined | Genome-fragment | NA | NA | 0 |
| S8_vs_k141_144722\|\|full | 1237 | No | NA | 3 | 0 | 0 | Not-determined | Genome-fragment | NA | NA | 0 |
| S8_vs_k141_161638\|\|full | 2525 | No | NA | 4 | 0 | 0 | Not-determined | Genome-fragment | NA | NA | 0 |
| S6_vs_k141_4492\|\|full | 17420 | No | NA | 34 | 4 | 1 | Low-quality | Genome-fragment | 33.48 | AAI-based (high-confidence) | 0 |
| S8_vs_k141_196927\|\|full | 1419 | No | NA | 2 | 0 | 1 | Not-determined | Genome-fragment | NA | NA | 0 |
| S10_vs_21154\|\|full | 17425 | No | NA | 12 | 9 | 0 | Low-quality | Genome-fragment | 27.57 | AAI-based (high-confidence) | 0 |
| S8_vs_k141_161654\|\|full | 1427 | No | NA | 2 | 0 | 1 | Not-determined | Genome-fragment | NA | NA | 0 |
| S14_vs_k141_97007\|\|full | 17454 | No | NA | 20 | 7 | 1 | Low-quality | Genome-fragment | 35.74 | AAI-based (high-confidence) | 0 |
| S8_vs_k141_66307\|\|full | 4727 | No | NA | 6 | 0 | 6 | Not-determined | Genome-fragment | NA | NA | 0 |
| S11_vs_k141_61682\|\|full | 17460 | No | NA | 16 | 1 | 3 | Low-quality | Genome-fragment | 31.36 | AAI-based (high-confidence) | 0 |
| S8_vs_k141_92596\|\|full | 1723 | No | NA | 3 | 0 | 0 | Not-determined | Genome-fragment | NA | NA | 0 |
| S7_vs_118876\|\|full | 17467 | No | NA | 23 | 9 | 1 | Low-quality | Genome-fragment | 37.24 | AAI-based (high-confidence) | 0 |
| S8_vs_k141_58209\|\|full | 1448 | No | NA | 3 | 0 | 1 | Not-determined | Genome-fragment | NA | NA | 0 |
| S8_vs_k141_166769\|\|full | 17467 | No | NA | 27 | 6 | 0 | Low-quality | Genome-fragment | 10.86 | AAI-based (high-confidence) | 0 |
| S5_extracted_k141_86657 | 17493 | No | NA | 17 | 5 | 0 | Low-quality | Genome-fragment | 14.82 | AAI-based (high-confidence) | 0 |
| S16_vs_8144\|\|full | 17541 | No | NA | 15 | 0 | 3 | Low-quality | Genome-fragment | 22.96 | AAI-based (medium-confidence) | 0 |
| S8_vs_k141_161694\|\|full | 1605 | No | NA | 4 | 0 | 3 | Not-determined | Genome-fragment | NA | NA | 0 |
| S8_vs_k141_100924\|\|full | 1000 | No | NA | 2 | 0 | 2 | Not-determined | Genome-fragment | NA | NA | 0 |
| S9_vs_k141_12228\|\|full | 17542 | No | NA | 36 | 4 | 0 | Low-quality | Genome-fragment | 48.91 | AAI-based (high-confidence) | 0 |
| S14_vs_k141_17330\|\|full | 17549 | No | NA | 29 | 0 | 0 | Low-quality | Genome-fragment | 29.41 | AAI-based (high-confidence) | 0 |
| S8_vs_k141_66349\|\|full | 1239 | No | NA | 3 | 0 | 1 | Not-determined | Genome-fragment | NA | NA | 0 |
| S5_vs_k141_33103\|\|0_partial | 17578 | No | NA | 23 | 0 | 5 | Low-quality | Genome-fragment | 25.3 | AAI-based (medium-confidence) | 0 |
| S17_vs_k141_69105\|\|full | 17613 | No | NA | 25 | 9 | 1 | Low-quality | Genome-fragment | 37.49 | AAI-based (medium-confidence) | 0 |
| S9_vs_k141_20608\|\|full | 17628 | No | NA | 19 | 0 | 12 | Low-quality | Genome-fragment | 14.27 | AAI-based (medium-confidence) | 0 |
| S5_vs_k141_79604\|\|full | 17647 | No | NA | 24 | 9 | 0 | Low-quality | Genome-fragment | 43.19 | AAI-based (high-confidence) | 0 |
| S8_vs_k141_118752\|\|full | 6039 | No | NA | 6 | 0 | 5 | Not-determined | Genome-fragment | NA | NA | 0 |
| S8_vs_k141_179505\|\|full | 1505 | No | NA | 2 | 0 | 2 | Not-determined | Genome-fragment | NA | NA | 0 |
| S3_extracted_k141_57987 | 17667 | No | NA | 17 | 4 | 0 | Low-quality | Genome-fragment | 20.39 | AAI-based (high-confidence) | 0 |
| S17_vs_k141_136494\|\|full | 17675 | Yes | 13481 | 17 | 3 | 4 | Low-quality | Genome-fragment | 33.13 | AAI-based (high-confidence) | 23.73 |
| S8_vs_k141_66376\|\|full | 1317 | No | NA | 2 | 0 | 0 | Not-determined | Genome-fragment | NA | NA | 0 |
| S8_vs_k141_179516\|\|full | 1185 | No | NA | 2 | 0 | 0 | Not-determined | Genome-fragment | NA | NA | 0 |
| S3_extracted_k141_117113 | 17708 | No | NA | 18 | 9 | 0 | Low-quality | Genome-fragment | 8.56 | AAI-based (high-confidence) | 0 |
| S8_vs_k141_14246\|\|full | 1765 | No | NA | 4 | 0 | 0 | Not-determined | Genome-fragment | NA | NA | 0 |
| S16_extracted_168736 | 17716 | No | NA | 42 | 5 | 0 | Low-quality | Genome-fragment | 28.6 | AAI-based (high-confidence) | 0 |
| S8_vs_k141_14252\|\|full | 2320 | No | NA | 3 | 0 | 3 | Not-determined | Genome-fragment | NA | NA | 0 |
| S8_vs_k141_187229\|\|full | 1698 | No | NA | 4 | 0 | 0 | Not-determined | Genome-fragment | NA | NA | 0 |
| S10_vs_109342\|\|full | 17725 | No | NA | 20 | 3 | 0 | Low-quality | Genome-fragment | 25.57 | AAI-based (high-confidence) | 0 |
| S8_vs_k141_110187\|\|full | 1956 | No | NA | 2 | 0 | 2 | Not-determined | Genome-fragment | NA | NA | 0 |
| S12_vs_k141_35218\|\|full | 17742 | No | NA | 17 | 13 | 0 | Low-quality | Genome-fragment | 28.63 | AAI-based (high-confidence) | 0 |
| S5_vs_k141_111002\|\|full | 17781 | No | NA | 34 | 12 | 0 | Low-quality | Genome-fragment | 41.44 | AAI-based (high-confidence) | 0 |
| S8_vs_k141_110211\|\|full | 1714 | No | NA | 4 | 0 | 1 | Not-determined | Genome-fragment | NA | NA | 0 |
| S8_vs_k141_58302\|\|full | 1532 | No | NA | 2 | 0 | 2 | Not-determined | Genome-fragment | NA | NA | 0 |
| S8_vs_k141_74992\|\|full | 1093 | No | NA | 2 | 0 | 1 | Not-determined | Genome-fragment | NA | NA | 0 |
| S8_vs_k141_197015\|\|full | 1279 | No | NA | 2 | 0 | 1 | Not-determined | Genome-fragment | NA | NA | 0 |
| S8_vs_k141_144927\|\|full | 1480 | No | NA | 3 | 0 | 0 | Not-determined | Genome-fragment | NA | NA | 0 |
| S8_vs_k141_153356\|\|full | 1727 | No | NA | 2 | 0 | 1 | Not-determined | Genome-fragment | NA | NA | 0 |
| S8_vs_k141_101020\|\|full | 1906 | No | NA | 3 | 0 | 1 | Not-determined | Genome-fragment | NA | NA | 0 |
| S8_vs_k141_40783\|\|full | 3402 | No | NA | 3 | 0 | 2 | Not-determined | Genome-fragment | NA | NA | 0 |
| S8_vs_k141_179529\|\|full | 54041 | No | NA | 81 | 28 | 0 | High-quality | High-quality | 100 | AAI-based (high-confidence) | 0 |
| S9_vs_k141_3111\|\|full | 17795 | No | NA | 37 | 3 | 1 | Low-quality | Genome-fragment | 46.32 | AAI-based (medium-confidence) | 0 |
| S8_vs_k141_58328\|\|full | 6678 | No | NA | 6 | 0 | 6 | Not-determined | Genome-fragment | NA | NA | 0 |
| S14_extracted_k141_81481 | 17796 | No | NA | 32 | 15 | 1 | Low-quality | Genome-fragment | 49.98 | AAI-based (high-confidence) | 0 |
| S8_vs_k141_127350\|\|full | 1557 | No | NA | 3 | 0 | 2 | Not-determined | Genome-fragment | NA | NA | 0 |
| S8_vs_k141_118803\|\|full | 11207 | No | NA | 6 | 0 | 6 | Not-determined | Genome-fragment | NA | NA | 0 |
| S8_vs_k141_101048\|\|full | 1340 | No | NA | 2 | 0 | 1 | Not-determined | Genome-fragment | NA | NA | 0 |
| S8_vs_k141_40800\|\|full | 3755 | No | NA | 5 | 0 | 2 | Not-determined | Genome-fragment | NA | NA | 0 |
| S8_vs_k141_118811\|\|full | 1412 | No | NA | 2 | 0 | 0 | Not-determined | Genome-fragment | NA | NA | 0 |
| S13_vs_93446\|\|full | 17832 | No | NA | 23 | 5 | 4 | Low-quality | Genome-fragment | 12.82 | AAI-based (high-confidence) | 0 |
| S5_vs_k141_87139\|\|full | 17849 | No | NA | 27 | 12 | 0 | Low-quality | Genome-fragment | 40.22 | AAI-based (medium-confidence) | 0 |
| S8_vs_k141_187314\|\|full | 1061 | No | NA | 2 | 0 | 0 | Not-determined | Genome-fragment | NA | NA | 0 |
| S8_vs_k141_179584\|\|full | 3083 | No | NA | 2 | 0 | 2 | Not-determined | Genome-fragment | NA | NA | 0 |
| S8_vs_k141_92741\|\|full | 1762 | No | NA | 2 | 0 | 1 | Not-determined | Genome-fragment | NA | NA | 0 |
| S14_vs_k141_44198\|\|0_partial | 17862 | No | NA | 13 | 1 | 6 | Low-quality | Genome-fragment | 12.01 | HMM-based (lower-bound) | 0 |
| S8_vs_k141_127395\|\|full | 2684 | No | NA | 3 | 0 | 2 | Not-determined | Genome-fragment | NA | NA | 0 |
| S8_vs_k141_66462\|\|full | 1102 | No | NA | 2 | 0 | 0 | Not-determined | Genome-fragment | NA | NA | 0 |
| S5_vs_k141_140805\|\|full | 17866 | Yes | 15942 | 10 | 3 | 1 | Low-quality | Genome-fragment | 19.36 | AAI-based (high-confidence) | 10.77 |
| S8_vs_k141_75063\|\|full | 1363 | No | NA | 2 | 0 | 2 | Not-determined | Genome-fragment | NA | NA | 0 |
| S8_vs_k141_205257\|\|full | 58264 | No | NA | 74 | 7 | 5 | High-quality | High-quality | 100 | AAI-based (high-confidence) | 0 |
| S8_vs_k141_179605\|\|full | 2512 | No | NA | 4 | 0 | 1 | Not-determined | Genome-fragment | NA | NA | 0 |
| S13_vs_12239\|\|full | 17878 | No | NA | 39 | 4 | 0 | Low-quality | Genome-fragment | 45.62 | AAI-based (high-confidence) | 0 |
| S8_vs_k141_23678\|\|full | 2153 | No | NA | 5 | 0 | 0 | Not-determined | Genome-fragment | NA | NA | 0 |
| S8_vs_k141_116675\|\|0_partial | 17882 | No | NA | 18 | 1 | 6 | Low-quality | Genome-fragment | 20.63 | HMM-based (lower-bound) | 0 |
| S8_extracted_k141_192008 | 17917 | No | NA | 26 | 4 | 3 | Low-quality | Genome-fragment | 24.02 | AAI-based (high-confidence) | 0 |
| S4_extracted_26777 | 17931 | No | NA | 19 | 8 | 1 | Low-quality | Genome-fragment | 39.75 | AAI-based (high-confidence) | 0 |
| S8_vs_k141_118881\|\|full | 1864 | No | NA | 2 | 0 | 0 | Not-determined | Genome-fragment | NA | NA | 0 |
| S5_vs_k141_127162\|\|1_partial | 17965 | No | NA | 15 | 1 | 2 | Low-quality | Genome-fragment | 33.56 | HMM-based (lower-bound) | 0 |
| S8_extracted_k141_160477 | 17976 | No | NA | 23 | 12 | 0 | Low-quality | Genome-fragment | 49.79 | AAI-based (high-confidence) | 0 |
| S10_extracted_20935 | 18030 | No | NA | 13 | 1 | 5 | Low-quality | Genome-fragment | 11.3 | HMM-based (lower-bound) | 0 |
| S8_vs_k141_153487\|\|full | 1638 | No | NA | 2 | 0 | 2 | Not-determined | Genome-fragment | NA | NA | 0 |
| S8_vs_k141_110338\|\|full | 1451 | No | NA | 2 | 0 | 2 | Not-determined | Genome-fragment | NA | NA | 0 |
| S8_vs_k141_110339\|\|full | 1588 | No | NA | 3 | 0 | 0 | Not-determined | Genome-fragment | NA | NA | 0 |
| S13_vs_112079\|\|full | 18034 | No | NA | 10 | 0 | 3 | Low-quality | Genome-fragment | 42.11 | AAI-based (medium-confidence) | 0 |
| S8_vs_k141_49038\|\|full | 1121 | No | NA | 1 | 0 | 1 | Not-determined | Genome-fragment | NA | NA | 0 |
| S5_vs_k141_9744\|\|full | 18053 | No | NA | 35 | 1 | 0 | Low-quality | Genome-fragment | 40.22 | AAI-based (high-confidence) | 0 |
| S16_vs_127713\|\|full | 18061 | No | NA | 22 | 16 | 0 | Low-quality | Genome-fragment | 30.2 | AAI-based (high-confidence) | 0 |
| S7_vs_60969\|\|full | 18084 | No | NA | 25 | 17 | 1 | Low-quality | Genome-fragment | 48.39 | AAI-based (medium-confidence) | 0 |
| S8_vs_k141_101130\|\|full | 1220 | No | NA | 3 | 0 | 1 | Not-determined | Genome-fragment | NA | NA | 0 |
| S8_vs_k141_118914\|\|full | 3457 | No | NA | 3 | 0 | 2 | Not-determined | Genome-fragment | NA | NA | 0 |
| S5_extracted_k141_54961 | 18116 | No | NA | 26 | 12 | 1 | Low-quality | Genome-fragment | 44.86 | AAI-based (high-confidence) | 0 |
| S16_extracted_66299 | 18135 | Yes | 13385 | 18 | 3 | 2 | Low-quality | Genome-fragment | 7.76 | AAI-based (high-confidence) | 26.19 |
| S8_vs_k141_83916\|\|full | 4749 | No | NA | 5 | 0 | 3 | Not-determined | Genome-fragment | NA | NA | 0 |
| S8_vs_k141_6107\|\|full | 1718 | No | NA | 2 | 0 | 2 | Not-determined | Genome-fragment | NA | NA | 0 |
| S8_vs_k141_23759\|\|full | 2784 | No | NA | 2 | 0 | 2 | Not-determined | Genome-fragment | NA | NA | 0 |
| S14_extracted_k141_7016 | 18139 | No | NA | 20 | 16 | 0 | Low-quality | Genome-fragment | 46.62 | AAI-based (high-confidence) | 0 |
| S11_extracted_k141_60471 | 18177 | No | NA | 19 | 11 | 0 | Low-quality | Genome-fragment | 46.81 | AAI-based (medium-confidence) | 0 |
| S8_vs_k141_161860\|\|full | 2041 | No | NA | 2 | 0 | 1 | Not-determined | Genome-fragment | NA | NA | 0 |
| S13_extracted_87573 | 18182 | No | NA | 39 | 6 | 0 | Low-quality | Genome-fragment | 29.98 | AAI-based (high-confidence) | 0 |
| S8_vs_k141_179654\|\|full | 2790 | No | NA | 4 | 0 | 1 | Not-determined | Genome-fragment | NA | NA | 0 |
| S8_vs_k141_14445\|\|full | 7651 | No | NA | 8 | 0 | 4 | Not-determined | Genome-fragment | NA | NA | 0 |
| S14_vs_k141_4124\|\|0_partial | 18209 | No | NA | 19 | 1 | 5 | Low-quality | Genome-fragment | 11.52 | HMM-based (lower-bound) | 0 |
| S8_vs_k141_110400\|\|full | 2322 | No | NA | 3 | 0 | 3 | Not-determined | Genome-fragment | NA | NA | 0 |
| S8_vs_k141_75163\|\|full | 3391 | No | NA | 5 | 0 | 2 | Not-determined | Genome-fragment | NA | NA | 0 |
| S8_vs_k141_118957\|\|full | 1743 | No | NA | 2 | 0 | 1 | Not-determined | Genome-fragment | NA | NA | 0 |
| S8_vs_k141_40937\|\|full | 1667 | No | NA | 3 | 0 | 2 | Not-determined | Genome-fragment | NA | NA | 0 |
| S5_vs_k141_41599\|\|full | 18217 | No | NA | 22 | 10 | 0 | Low-quality | Genome-fragment | 27.95 | AAI-based (high-confidence) | 0 |
| S12_vs_k141_139215\|\|full | 18235 | No | NA | 11 | 3 | 0 | Low-quality | Genome-fragment | 42.97 | AAI-based (high-confidence) | 0 |
| S8_vs_k141_110425\|\|full | 2269 | No | NA | 3 | 0 | 3 | Not-determined | Genome-fragment | NA | NA | 0 |
| S8_vs_k141_161891\|\|full | 2621 | No | NA | 4 | 0 | 1 | Not-determined | Genome-fragment | NA | NA | 0 |
| S8_vs_k141_32296\|\|full | 3069 | No | NA | 4 | 0 | 2 | Not-determined | Genome-fragment | NA | NA | 0 |
| S16_vs_144031\|\|full | 18250 | Yes | 11606 | 24 | 3 | 6 | Low-quality | Genome-fragment | 15.3 | HMM-based (lower-bound) | 36.41 |
| S8_vs_k141_153598\|\|full | 1151 | No | NA | 3 | 0 | 1 | Not-determined | Genome-fragment | NA | NA | 0 |
| S8_vs_k141_101210\|\|full | 2322 | No | NA | 3 | 0 | 1 | Not-determined | Genome-fragment | NA | NA | 0 |
| S8_vs_k141_83967\|\|full | 3094 | No | NA | 4 | 0 | 4 | Not-determined | Genome-fragment | NA | NA | 0 |
| S8_vs_k141_23833\|\|full | 1574 | No | NA | 2 | 0 | 1 | Not-determined | Genome-fragment | NA | NA | 0 |
| S5_vs_k141_99996\|\|0_partial | 18312 | No | NA | 24 | 1 | 1 | Low-quality | Genome-fragment | 20.56 | HMM-based (lower-bound) | 0 |
| S16_vs_172367\|\|full | 18318 | No | NA | 11 | 0 | 8 | Low-quality | Genome-fragment | 6.84 | AAI-based (medium-confidence) | 0 |
| S14_vs_k141_76570\|\|full | 18321 | No | NA | 34 | 11 | 1 | Low-quality | Genome-fragment | 41.4 | AAI-based (high-confidence) | 0 |
| S8_extracted_k141_164809 | 18321 | No | NA | 14 | 1 | 0 | Low-quality | Genome-fragment | 18.88 | AAI-based (medium-confidence) | 0 |
| S16_vs_48988\|\|full | 18322 | No | NA | 23 | 10 | 0 | Low-quality | Genome-fragment | 42.56 | AAI-based (high-confidence) | 0 |
| S8_vs_k141_110479\|\|full | 1401 | No | NA | 3 | 0 | 1 | Not-determined | Genome-fragment | NA | NA | 0 |
| S14_vs_k141_28459\|\|0_partial | 18341 | No | NA | 35 | 9 | 1 | Low-quality | Genome-fragment | 46.02 | AAI-based (high-confidence) | 0 |
| S8_vs_k141_145098\|\|full | 3848 | No | NA | 5 | 0 | 2 | Not-determined | Genome-fragment | NA | NA | 0 |
| S8_vs_k141_101268\|\|full | 7745 | No | NA | 8 | 0 | 1 | Not-determined | Genome-fragment | NA | NA | 0 |
| S12_vs_k141_98571\|\|full | 18357 | No | NA | 15 | 1 | 0 | Low-quality | Genome-fragment | 39.37 | AAI-based (medium-confidence) | 0 |
| S16_vs_15773\|\|full | 18375 | No | NA | 13 | 0 | 9 | Low-quality | Genome-fragment | 7.49 | AAI-based (medium-confidence) | 0 |
| S8_vs_k141_41037\|\|full | 2683 | No | NA | 4 | 0 | 2 | Not-determined | Genome-fragment | NA | NA | 0 |
| S8_vs_k141_14551\|\|full | 1073 | No | NA | 2 | 0 | 1 | Not-determined | Genome-fragment | NA | NA | 0 |
| S8_vs_k141_14556\|\|full | 1025 | No | NA | 3 | 0 | 1 | Not-determined | Genome-fragment | NA | NA | 0 |
| S16_vs_170648\|\|full | 18375 | No | NA | 24 | 6 | 1 | Low-quality | Genome-fragment | 30.74 | AAI-based (high-confidence) | 0 |
| S8_vs_k141_179751\|\|full | 1466 | No | NA | 2 | 0 | 1 | Not-determined | Genome-fragment | NA | NA | 0 |
| S8_vs_k141_32340\|\|full | 2862 | No | NA | 3 | 0 | 0 | Not-determined | Genome-fragment | NA | NA | 0 |
| S8_vs_k141_14558\|\|full | 4855 | No | NA | 5 | 0 | 1 | Not-determined | Genome-fragment | NA | NA | 0 |
| S8_vs_k141_110507\|\|full | 2401 | No | NA | 3 | 0 | 2 | Not-determined | Genome-fragment | NA | NA | 0 |
| S8_vs_k141_179765\|\|full | 1373 | No | NA | 3 | 0 | 1 | Not-determined | Genome-fragment | NA | NA | 0 |
| S10_vs_15612\|\|full | 18382 | No | NA | 23 | 1 | 10 | Low-quality | Genome-fragment | 11.9 | HMM-based (lower-bound) | 0 |
| S8_vs_k141_92877\|\|full | 3350 | No | NA | 4 | 0 | 3 | Not-determined | Genome-fragment | NA | NA | 0 |
| S8_vs_k141_6249\|\|full | 3047 | No | NA | 2 | 0 | 2 | Not-determined | Genome-fragment | NA | NA | 0 |
| S8_vs_k141_133535\|\|0_partial | 18438 | Yes | 11472 | 20 | 3 | 6 | Low-quality | Genome-fragment | 21.83 | HMM-based (lower-bound) | 37.78 |
| S8_vs_k141_179801\|\|full | 2076 | No | NA | 4 | 0 | 0 | Not-determined | Genome-fragment | NA | NA | 0 |
| S8_vs_k141_110545\|\|full | 3005 | No | NA | 4 | 0 | 1 | Not-determined | Genome-fragment | NA | NA | 0 |
| S16_vs_105031\|\|0_partial | 18441 | No | NA | 11 | 0 | 10 | Low-quality | Genome-fragment | 25.14 | AAI-based (high-confidence) | 0 |
| S8_vs_k141_23924\|\|full | 1080 | No | NA | 3 | 0 | 0 | Not-determined | Genome-fragment | NA | NA | 0 |
| S11_vs_k141_50916\|\|full | 18448 | No | NA | 36 | 8 | 0 | Low-quality | Genome-fragment | 48.64 | AAI-based (high-confidence) | 0 |
| S3_vs_k141_122179\|\|0_partial | 18477 | No | NA | 17 | 3 | 7 | Low-quality | Genome-fragment | 27.4 | HMM-based (lower-bound) | 0 |
| S8_vs_k141_197305\|\|full | 1241 | No | NA | 4 | 0 | 1 | Not-determined | Genome-fragment | NA | NA | 0 |
| S2_vs_k141_11335\|\|full | 18479 | No | NA | 37 | 3 | 0 | Low-quality | Genome-fragment | 41.17 | AAI-based (high-confidence) | 0 |
| S17_extracted_k141_121637 | 18532 | No | NA | 25 | 4 | 0 | Low-quality | Genome-fragment | 34.02 | AAI-based (high-confidence) | 0 |
| S3_vs_k141_55137\|\|full | 18539 | No | NA | 24 | 15 | 0 | Low-quality | Genome-fragment | 47.85 | AAI-based (high-confidence) | 0 |
| S12_extracted_k141_125394 | 18541 | No | NA | 21 | 17 | 0 | Low-quality | Genome-fragment | 44.31 | AAI-based (high-confidence) | 0 |
| S3_extracted_k141_15330 | 18575 | No | NA | 40 | 3 | 0 | Low-quality | Genome-fragment | 35.59 | AAI-based (high-confidence) | 0 |
| S7_vs_56677\|\|full | 18577 | No | NA | 18 | 1 | 8 | Low-quality | Genome-fragment | 12.49 | HMM-based (lower-bound) | 0 |
| S16_vs_29367\|\|0_partial | 18590 | No | NA | 13 | 1 | 5 | Low-quality | Genome-fragment | 12.85 | HMM-based (lower-bound) | 0 |
| S8_vs_k141_75408\|\|full | 1742 | No | NA | 2 | 0 | 2 | Not-determined | Genome-fragment | NA | NA | 0 |
| S8_vs_k141_32444\|\|full | 3247 | No | NA | 5 | 0 | 2 | Not-determined | Genome-fragment | NA | NA | 0 |
| S8_vs_k141_6349\|\|full | 1973 | No | NA | 2 | 0 | 1 | Not-determined | Genome-fragment | NA | NA | 0 |
| S8_vs_k141_162069\|\|full | 6635 | No | NA | 10 | 0 | 2 | Not-determined | Genome-fragment | NA | NA | 0 |
| S8_vs_k141_119126\|\|full | 1324 | No | NA | 2 | 0 | 1 | Not-determined | Genome-fragment | NA | NA | 0 |
| S8_vs_k141_92990\|\|full | 1314 | No | NA | 2 | 0 | 1 | Not-determined | Genome-fragment | NA | NA | 0 |
| S8_vs_k141_153827\|\|full | 1077 | No | NA | 2 | 0 | 0 | Not-determined | Genome-fragment | NA | NA | 0 |
| S4_extracted_18103 | 18590 | No | NA | 31 | 10 | 0 | Low-quality | Genome-fragment | 33.27 | AAI-based (high-confidence) | 0 |
| S8_vs_k141_14696\|\|full | 9459 | No | NA | 12 | 0 | 5 | Not-determined | Genome-fragment | NA | NA | 0 |
| S8_vs_k141_32478\|\|full | 5365 | No | NA | 6 | 0 | 5 | Not-determined | Genome-fragment | NA | NA | 0 |
| S7_vs_31786\|\|full | 18618 | No | NA | 9 | 2 | 0 | Low-quality | Genome-fragment | 45.08 | AAI-based (high-confidence) | 0 |
| S8_vs_k141_145173\|\|full | 44625 | No | NA | 49 | 7 | 0 | High-quality | High-quality | 100 | AAI-based (high-confidence) | 0 |
| S10_vs_13639\|\|full | 18619 | No | NA | 22 | 2 | 0 | Low-quality | Genome-fragment | 12.11 | AAI-based (high-confidence) | 0 |
| S8_vs_k141_58795\|\|full | 1578 | No | NA | 2 | 0 | 2 | Not-determined | Genome-fragment | NA | NA | 0 |
| S16_vs_47730\|\|full | 18624 | No | NA | 25 | 1 | 1 | Low-quality | Genome-fragment | 11.49 | HMM-based (lower-bound) | 0 |
| S8_vs_k141_187706\|\|full | 3751 | No | NA | 4 | 0 | 1 | Not-determined | Genome-fragment | NA | NA | 0 |
| S8_vs_k141_145196\|\|full | 2047 | No | NA | 3 | 0 | 3 | Not-determined | Genome-fragment | NA | NA | 0 |
| S16_vs_6545\|\|full | 18627 | No | NA | 23 | 3 | 2 | Low-quality | Genome-fragment | 44.73 | AAI-based (medium-confidence) | 0 |
| S10_vs_36174\|\|full | 18631 | No | NA | 21 | 13 | 1 | Low-quality | Genome-fragment | 30.76 | AAI-based (high-confidence) | 0 |
| S8_vs_k141_136212\|\|full | 2852 | No | NA | 2 | 0 | 0 | Not-determined | Genome-fragment | NA | NA | 0 |
| S8_vs_k141_41122\|\|full | 2397 | No | NA | 3 | 0 | 1 | Not-determined | Genome-fragment | NA | NA | 0 |
| S8_vs_k141_171234\|\|full | 1877 | No | NA | 2 | 0 | 1 | Not-determined | Genome-fragment | NA | NA | 0 |
| S16_vs_38663\|\|full | 18638 | No | NA | 29 | 8 | 0 | Low-quality | Genome-fragment | 23.36 | AAI-based (high-confidence) | 0 |
| S8_vs_k141_162160\|\|full | 1159 | No | NA | 3 | 0 | 1 | Not-determined | Genome-fragment | NA | NA | 0 |
| S3_vs_k141_99701\|\|0_partial | 18639 | No | NA | 13 | 1 | 11 | Low-quality | Genome-fragment | 14.28 | HMM-based (lower-bound) | 0 |
| S8_vs_k141_41138\|\|full | 1265 | No | NA | 4 | 0 | 0 | Not-determined | Genome-fragment | NA | NA | 0 |
| S8_vs_k141_119238\|\|full | 1419 | No | NA | 4 | 0 | 1 | Not-determined | Genome-fragment | NA | NA | 0 |
| S8_vs_k141_110692\|\|full | 19724 | No | NA | 31 | 17 | 0 | Medium-quality | Genome-fragment | 69.42 | AAI-based (high-confidence) | 0 |
| S8_vs_k141_205529\|\|full | 3279 | No | NA | 3 | 0 | 2 | Not-determined | Genome-fragment | NA | NA | 0 |
| S8_vs_k141_66816\|\|full | 2175 | No | NA | 4 | 0 | 2 | Not-determined | Genome-fragment | NA | NA | 0 |
| S8_vs_k141_127852\|\|full | 8379 | No | NA | 4 | 0 | 3 | Not-determined | Genome-fragment | NA | NA | 0 |
| S11_vs_k141_1212\|\|full | 18692 | No | NA | 17 | 2 | 2 | Low-quality | Genome-fragment | 43.66 | AAI-based (high-confidence) | 0 |
| S17_vs_k141_117455\|\|full | 18697 | No | NA | 21 | 0 | 7 | Low-quality | Genome-fragment | 17.56 | AAI-based (medium-confidence) | 0 |
| S16_extracted_35820 | 18707 | Yes | 12789 | 27 | 6 | 5 | Low-quality | Genome-fragment | 20.63 | AAI-based (high-confidence) | 31.64 |
| S12_vs_k141_59958\|\|0_partial | 18745 | No | NA | 22 | 2 | 8 | Low-quality | Genome-fragment | 32.4 | AAI-based (medium-confidence) | 0 |
| S7_vs_4208\|\|full | 18792 | No | NA | 14 | 1 | 4 | Low-quality | Genome-fragment | 10.39 | AAI-based (medium-confidence) | 0 |
| S7_vs_17278\|\|full | 18823 | No | NA | 24 | 3 | 2 | Low-quality | Genome-fragment | 16.02 | AAI-based (medium-confidence) | 0 |
| S9_extracted_k141_89614 | 18833 | No | NA | 10 | 1 | 0 | Low-quality | Genome-fragment | 11.82 | AAI-based (high-confidence) | 0 |
| S8_vs_k141_41180\|\|full | 2472 | No | NA | 3 | 0 | 0 | Not-determined | Genome-fragment | NA | NA | 0 |
| S9_vs_k141_2057\|\|full | 18872 | No | NA | 23 | 12 | 0 | Low-quality | Genome-fragment | 43.98 | AAI-based (high-confidence) | 0 |
| S8_vs_k141_145294\|\|full | 2169 | No | NA | 2 | 0 | 1 | Not-determined | Genome-fragment | NA | NA | 0 |
| S8_vs_k141_153950\|\|full | 17117 | No | NA | 19 | 0 | 5 | Not-determined | Genome-fragment | NA | NA | 0 |
| S8_vs_k141_58913\|\|full | 4095 | No | NA | 3 | 0 | 2 | Not-determined | Genome-fragment | NA | NA | 0 |
| S8_vs_k141_41204\|\|full | 2627 | No | NA | 3 | 0 | 0 | Not-determined | Genome-fragment | NA | NA | 0 |
| S13_extracted_141129 | 18877 | No | NA | 17 | 0 | 11 | Low-quality | Genome-fragment | 4.37 | AAI-based (medium-confidence) | 0 |
| S8_vs_k141_197510\|\|full | 2129 | No | NA | 3 | 0 | 0 | Not-determined | Genome-fragment | NA | NA | 0 |
| S5_vs_k141_84840\|\|full | 18888 | No | NA | 34 | 3 | 0 | Low-quality | Genome-fragment | 41.84 | AAI-based (medium-confidence) | 0 |
| S5_vs_k141_85533\|\|full | 18905 | No | NA | 14 | 2 | 0 | Low-quality | Genome-fragment | 45.77 | AAI-based (high-confidence) | 0 |
| S8_vs_k141_32627\|\|full | 1759 | No | NA | 3 | 0 | 1 | Not-determined | Genome-fragment | NA | NA | 0 |
| S8_vs_k141_153967\|\|full | 8985 | No | NA | 9 | 0 | 5 | Not-determined | Genome-fragment | NA | NA | 0 |
| S8_vs_k141_58944\|\|full | 1820 | No | NA | 3 | 0 | 0 | Not-determined | Genome-fragment | NA | NA | 0 |
| S8_vs_k141_205624\|\|full | 1397 | No | NA | 2 | 0 | 0 | Not-determined | Genome-fragment | NA | NA | 0 |
| S8_vs_k141_171391\|\|full | 1369 | No | NA | 2 | 0 | 1 | Not-determined | Genome-fragment | NA | NA | 0 |
| S8_vs_k141_145348\|\|full | 1764 | No | NA | 2 | 0 | 1 | Not-determined | Genome-fragment | NA | NA | 0 |
| S5_vs_k141_44949\|\|full | 18944 | No | NA | 20 | 5 | 0 | Low-quality | Genome-fragment | 45.79 | AAI-based (high-confidence) | 0 |
| S8_vs_k141_14886\|\|full | 1187 | No | NA | 3 | 0 | 1 | Not-determined | Genome-fragment | NA | NA | 0 |
| S4_vs_VIRSorter_8597-cat_2 | 18952 | No | NA | 43 | 4 | 1 | Low-quality | Genome-fragment | 41.88 | AAI-based (medium-confidence) | 0 |
| S17_vs_k141_125155\|\|full | 18961 | Yes | 15066 | 28 | 2 | 2 | Low-quality | Genome-fragment | 20.64 | HMM-based (lower-bound) | 20.54 |
| S8_vs_k141_162206\|\|full | 3273 | No | NA | 3 | 0 | 2 | Not-determined | Genome-fragment | NA | NA | 0 |
| S8_vs_k141_93202\|\|full | 1212 | No | NA | 3 | 0 | 1 | Not-determined | Genome-fragment | NA | NA | 0 |
| S8_vs_k141_75628\|\|full | 1419 | No | NA | 3 | 0 | 1 | Not-determined | Genome-fragment | NA | NA | 0 |
| S8_vs_k141_154026\|\|full | 1371 | No | NA | 2 | 0 | 0 | Not-determined | Genome-fragment | NA | NA | 0 |
| S8_vs_k141_93216\|\|full | 3494 | No | NA | 5 | 0 | 2 | Not-determined | Genome-fragment | NA | NA | 0 |
| S9_vs_k141_66977\|\|full | 18962 | No | NA | 11 | 3 | 2 | Low-quality | Genome-fragment | 27.87 | AAI-based (medium-confidence) | 0 |
| S8_vs_k141_197610\|\|full | 2488 | No | NA | 5 | 0 | 1 | Not-determined | Genome-fragment | NA | NA | 0 |
| S8_vs_k141_162241\|\|full | 1038 | No | NA | 3 | 0 | 0 | Not-determined | Genome-fragment | NA | NA | 0 |
| S16_vs_44268\|\|full | 19047 | No | NA | 25 | 14 | 1 | Low-quality | Genome-fragment | 47.49 | AAI-based (medium-confidence) | 0 |
| S8_vs_k141_101652\|\|full | 3707 | No | NA | 4 | 0 | 2 | Not-determined | Genome-fragment | NA | NA | 0 |
| S16_extracted_165886 | 19086 | No | NA | 21 | 1 | 3 | Low-quality | Genome-fragment | 1.62 | HMM-based (lower-bound) | 0 |
| S13_vs_124618\|\|full | 19105 | Yes | 5749 | 18 | 3 | 9 | Low-quality | Genome-fragment | 13.86 | AAI-based (high-confidence) | 69.91 |
| S5_vs_k141_15478\|\|0_partial | 19121 | No | NA | 19 | 3 | 8 | Low-quality | Genome-fragment | 36.39 | HMM-based (lower-bound) | 0 |
| S8_vs_k141_59021\|\|full | 2132 | No | NA | 3 | 0 | 3 | Not-determined | Genome-fragment | NA | NA | 0 |
| S13_vs_128267\|\|full | 19134 | No | NA | 18 | 12 | 0 | Low-quality | Genome-fragment | 47.89 | AAI-based (medium-confidence) | 0 |
| S8_vs_k141_162254\|\|full | 2478 | No | NA | 4 | 0 | 0 | Not-determined | Genome-fragment | NA | NA | 0 |
| S8_vs_k141_171475\|\|full | 4065 | No | NA | 3 | 0 | 1 | Not-determined | Genome-fragment | NA | NA | 0 |
| S8_vs_k141_119372\|\|full | 1578 | No | NA | 2 | 0 | 1 | Not-determined | Genome-fragment | NA | NA | 0 |
| S8_vs_k141_6653\|\|full | 2816 | No | NA | 3 | 0 | 1 | Not-determined | Genome-fragment | NA | NA | 0 |
| S8_vs_k141_101713\|\|full | 1416 | No | NA | 3 | 0 | 0 | Not-determined | Genome-fragment | NA | NA | 0 |
| S13_extracted_523 | 19157 | No | NA | 14 | 0 | 11 | Low-quality | Genome-fragment | 11.94 | AAI-based (high-confidence) | 0 |
| S8_vs_k141_145459\|\|full | 2057 | No | NA | 3 | 0 | 1 | Not-determined | Genome-fragment | NA | NA | 0 |
| S2_vs_k141_18431\|\|full | 19192 | No | NA | 17 | 3 | 1 | Low-quality | Genome-fragment | 41.78 | AAI-based (high-confidence) | 0 |
| S8_vs_k141_24146\|\|full | 1594 | No | NA | 3 | 0 | 3 | Not-determined | Genome-fragment | NA | NA | 0 |
| S3_vs_k141_19804\|\|full | 19199 | Yes | 2529 | 21 | 3 | 6 | Low-quality | Genome-fragment | 4.39 | AAI-based (high-confidence) | 86.83 |
| S5_vs_k141_9971\|\|full | 19232 | No | NA | 24 | 12 | 0 | Low-quality | Genome-fragment | 43.23 | AAI-based (medium-confidence) | 0 |
| S8_extracted_k141_130106 | 19245 | No | NA | 19 | 0 | 14 | Low-quality | Genome-fragment | 2.6 | AAI-based (medium-confidence) | 0 |
| S8_vs_k141_145498\|\|full | 1630 | No | NA | 2 | 0 | 1 | Not-determined | Genome-fragment | NA | NA | 0 |
| S9_extracted_k141_81703 | 19266 | No | NA | 29 | 2 | 1 | Low-quality | Genome-fragment | 24 | AAI-based (high-confidence) | 0 |
| S4_vs_VIRSorter_24856-cat_2 | 19303 | No | NA | 21 | 11 | 0 | Low-quality | Genome-fragment | 48.17 | AAI-based (high-confidence) | 0 |
| S8_vs_k141_145505\|\|full | 1639 | No | NA | 2 | 0 | 1 | Not-determined | Genome-fragment | NA | NA | 0 |
| S8_vs_k141_93268\|\|full | 1555 | No | NA | 3 | 0 | 0 | Not-determined | Genome-fragment | NA | NA | 0 |
| S16_vs_168934\|\|full | 19416 | No | NA | 35 | 11 | 1 | Low-quality | Genome-fragment | 45.83 | AAI-based (medium-confidence) | 0 |
| S8_vs_k141_101757\|\|full | 1426 | No | NA | 3 | 0 | 0 | Not-determined | Genome-fragment | NA | NA | 0 |
| S8_vs_k141_145507\|\|full | 7962 | No | NA | 7 | 0 | 0 | Not-determined | Genome-fragment | NA | NA | 0 |
| S8_vs_k141_154160\|\|full | 1415 | No | NA | 4 | 0 | 0 | Not-determined | Genome-fragment | NA | NA | 0 |
| S12_vs_k141_57348\|\|full | 19455 | No | NA | 21 | 14 | 0 | Low-quality | Genome-fragment | 28.15 | AAI-based (high-confidence) | 0 |
| S9_vs_k141_22413\|\|full | 19480 | No | NA | 18 | 2 | 3 | Low-quality | Genome-fragment | 41.59 | AAI-based (high-confidence) | 0 |
| S8_vs_k141_171570\|\|full | 1819 | No | NA | 3 | 0 | 2 | Not-determined | Genome-fragment | NA | NA | 0 |
| S17_extracted_k141_1262 | 19499 | No | NA | 30 | 9 | 0 | Low-quality | Genome-fragment | 22.74 | HMM-based (lower-bound) | 0 |
| S8_vs_k141_124782\|\|0_partial | 19499 | Yes | 9536 | 19 | 2 | 3 | Low-quality | Genome-fragment | 18.28 | AAI-based (high-confidence) | 51.09 |
| S8_vs_k141_191594\|\|full | 19567 | No | NA | 38 | 5 | 0 | Low-quality | Genome-fragment | 48.03 | AAI-based (high-confidence) | 0 |
| S8_vs_k141_49370\|\|full | 3734 | No | NA | 3 | 0 | 2 | Not-determined | Genome-fragment | NA | NA | 0 |
| S8_vs_k141_197736\|\|full | 7497 | No | NA | 7 | 0 | 3 | Not-determined | Genome-fragment | NA | NA | 0 |
| S8_extracted_k141_206579 | 19599 | No | NA | 23 | 11 | 0 | Low-quality | Genome-fragment | 31.62 | AAI-based (high-confidence) | 0 |
| S7_vs_75727\|\|full | 19606 | No | NA | 12 | 1 | 1 | Low-quality | Genome-fragment | 17.71 | AAI-based (high-confidence) | 0 |
| S13_extracted_134560 | 19610 | No | NA | 22 | 11 | 0 | Low-quality | Genome-fragment | 29.21 | AAI-based (high-confidence) | 0 |
| S8_vs_k141_32822\|\|full | 1887 | No | NA | 3 | 0 | 0 | Not-determined | Genome-fragment | NA | NA | 0 |
| S8_vs_k141_110895\|\|full | 2834 | No | NA | 2 | 0 | 2 | Not-determined | Genome-fragment | NA | NA | 0 |
| S8_vs_k141_162405\|\|full | 1814 | No | NA | 3 | 0 | 1 | Not-determined | Genome-fragment | NA | NA | 0 |
| S14_vs_k141_8705\|\|full | 19651 | No | NA | 23 | 2 | 0 | Low-quality | Genome-fragment | 43.31 | AAI-based (high-confidence) | 0 |
| S7_vs_38926\|\|full | 19664 | No | NA | 32 | 8 | 0 | Low-quality | Genome-fragment | 33.65 | AAI-based (high-confidence) | 0 |
| S8_vs_k141_6765\|\|full | 1755 | No | NA | 2 | 0 | 0 | Not-determined | Genome-fragment | NA | NA | 0 |
| S10_vs_109744\|\|full | 19691 | No | NA | 30 | 11 | 0 | Low-quality | Genome-fragment | 43.18 | AAI-based (medium-confidence) | 0 |
| S8_vs_k141_101867\|\|full | 1666 | No | NA | 2 | 0 | 1 | Not-determined | Genome-fragment | NA | NA | 0 |
| S8_vs_k141_32847\|\|full | 5480 | No | NA | 7 | 0 | 0 | Not-determined | Genome-fragment | NA | NA | 0 |
| S8_vs_k141_67144\|\|full | 3562 | No | NA | 4 | 0 | 3 | Not-determined | Genome-fragment | NA | NA | 0 |
| S8_vs_k141_49423\|\|full | 1971 | No | NA | 3 | 0 | 1 | Not-determined | Genome-fragment | NA | NA | 0 |
| S8_vs_k141_128171\|\|full | 1620 | No | NA | 3 | 0 | 1 | Not-determined | Genome-fragment | NA | NA | 0 |
| S12_extracted_k141_160747 | 19717 | No | NA | 26 | 11 | 0 | Low-quality | Genome-fragment | 45.89 | AAI-based (high-confidence) | 0 |
| S8_vs_k141_59234\|\|full | 1608 | No | NA | 4 | 0 | 0 | Not-determined | Genome-fragment | NA | NA | 0 |
| S8_vs_k141_32889\|\|full | 1180 | No | NA | 3 | 0 | 2 | Not-determined | Genome-fragment | NA | NA | 0 |
| S8_vs_k141_59235\|\|full | 2600 | No | NA | 3 | 0 | 2 | Not-determined | Genome-fragment | NA | NA | 0 |
| S12_vs_k141_35213\|\|0_partial | 19719 | No | NA | 20 | 1 | 11 | Low-quality | Genome-fragment | 20.07 | HMM-based (lower-bound) | 0 |
| S5_vs_k141_100774\|\|full | 19735 | No | NA | 18 | 0 | 13 | Low-quality | Genome-fragment | 6.25 | AAI-based (medium-confidence) | 0 |
| S8_vs_k141_24317\|\|full | 1631 | No | NA | 3 | 0 | 1 | Not-determined | Genome-fragment | NA | NA | 0 |
| S8_vs_k141_145654\|\|full | 3093 | No | NA | 4 | 0 | 0 | Not-determined | Genome-fragment | NA | NA | 0 |
| S8_vs_k141_154231\|\|full | 33882 | No | NA | 59 | 17 | 0 | High-quality | High-quality | 99.09 | AAI-based (high-confidence) | 0 |
| S12_extracted_k141_170419 | 19740 | No | NA | 22 | 4 | 1 | Low-quality | Genome-fragment | 18.17 | AAI-based (high-confidence) | 0 |
| S4_extracted_16270 | 19762 | No | NA | 38 | 5 | 0 | Low-quality | Genome-fragment | 11.14 | HMM-based (lower-bound) | 0 |
| S8_vs_k141_75902\|\|full | 1315 | No | NA | 3 | 0 | 0 | Not-determined | Genome-fragment | NA | NA | 0 |
| S8_vs_k141_110920\|\|full | 1325 | No | NA | 3 | 0 | 0 | Not-determined | Genome-fragment | NA | NA | 0 |
| S8_vs_k141_15121\|\|full | 1676 | No | NA | 1 | 0 | 1 | Not-determined | Genome-fragment | NA | NA | 0 |
| S8_vs_k141_15123\|\|full | 1105 | No | NA | 2 | 0 | 0 | Not-determined | Genome-fragment | NA | NA | 0 |
| S7_extracted_40712 | 19833 | No | NA | 19 | 6 | 0 | Low-quality | Genome-fragment | 36.41 | AAI-based (high-confidence) | 0 |
| S8_vs_k141_49484\|\|full | 2235 | No | NA | 2 | 0 | 0 | Not-determined | Genome-fragment | NA | NA | 0 |
| S8_vs_k141_180432\|\|full | 2328 | No | NA | 2 | 0 | 0 | Not-determined | Genome-fragment | NA | NA | 0 |
| S8_vs_k141_41375\|\|full | 2882 | No | NA | 3 | 0 | 0 | Not-determined | Genome-fragment | NA | NA | 0 |
| S8_vs_k141_136601\|\|full | 1911 | No | NA | 4 | 0 | 0 | Not-determined | Genome-fragment | NA | NA | 0 |
| S12_vs_k141_181884\|\|full | 19837 | No | NA | 19 | 1 | 5 | Low-quality | Genome-fragment | 17.76 | HMM-based (lower-bound) | 0 |
| S14_vs_k141_61186\|\|0_partial | 19860 | No | NA | 13 | 0 | 10 | Low-quality | Genome-fragment | 24.4 | AAI-based (medium-confidence) | 0 |
| S8_vs_k141_180437\|\|full | 2912 | No | NA | 4 | 0 | 0 | Not-determined | Genome-fragment | NA | NA | 0 |
| S8_vs_k141_162516\|\|full | 3868 | No | NA | 4 | 0 | 4 | Not-determined | Genome-fragment | NA | NA | 0 |
| S8_vs_k141_128239\|\|full | 1067 | No | NA | 3 | 0 | 0 | Not-determined | Genome-fragment | NA | NA | 0 |
| S8_vs_k141_145673\|\|full | 2947 | No | NA | 4 | 0 | 2 | Not-determined | Genome-fragment | NA | NA | 0 |
| S11_extracted_k141_41709 | 19908 | No | NA | 21 | 7 | 0 | Low-quality | Genome-fragment | 14.56 | AAI-based (medium-confidence) | 0 |
| S8_vs_k141_24351\|\|full | 4039 | No | NA | 6 | 0 | 0 | Not-determined | Genome-fragment | NA | NA | 0 |
| S8_vs_k141_6887\|\|full | 7040 | No | NA | 5 | 0 | 2 | Not-determined | Genome-fragment | NA | NA | 0 |
| S10_extracted_101738 | 19920 | No | NA | 33 | 17 | 0 | Low-quality | Genome-fragment | 48.67 | AAI-based (high-confidence) | 0 |
| S8_extracted_k141_17541 | 19925 | No | NA | 29 | 10 | 0 | Low-quality | Genome-fragment | 47.49 | AAI-based (high-confidence) | 0 |
| S8_vs_k141_93424\|\|full | 1751 | No | NA | 3 | 0 | 0 | Not-determined | Genome-fragment | NA | NA | 0 |
| S8_vs_k141_6897\|\|full | 1167 | No | NA | 3 | 0 | 2 | Not-determined | Genome-fragment | NA | NA | 0 |
| S14_vs_k141_27377\|\|full | 19947 | No | NA | 22 | 2 | 0 | Low-quality | Genome-fragment | 45.38 | AAI-based (high-confidence) | 0 |
| S8_vs_k141_6900\|\|full | 1110 | No | NA | 3 | 0 | 0 | Not-determined | Genome-fragment | NA | NA | 0 |
| S8_extracted_k141_51542 | 20023 | No | NA | 34 | 15 | 0 | Low-quality | Genome-fragment | 32.31 | AAI-based (high-confidence) | 0 |
| S5_extracted_k141_133649 | 20024 | No | NA | 29 | 9 | 0 | Low-quality | Genome-fragment | 43.71 | AAI-based (high-confidence) | 0 |
| S3_vs_k141_95123\|\|full | 20055 | Yes | 10603 | 22 | 4 | 7 | Low-quality | Genome-fragment | 17.36 | AAI-based (medium-confidence) | 47.13 |

| | | | | | | | | | | | |
|---|---|---|---|---|---|---|---|---|---|---|---|
| S8_vs_k141_49574\|\|full | 1222 | No | NA | 3 | 0 | 1 | Not-determined | Genome-fragment | NA | NA | 0 |
| S8_vs_k141_32934\|\|full | 2989 | No | NA | 4 | 0 | 1 | Not-determined | Genome-fragment | NA | NA | 0 |
| S5_vs_k141_131082\|\|full | 20059 | No | NA | 29 | 11 | 0 | Low-quality | Genome-fragment | 43.77 | AAI-based (high-confidence) | 0 |
| S8_vs_k141_59391\|\|full | 1169 | No | NA | 2 | 0 | 0 | Not-determined | Genome-fragment | NA | NA | 0 |
| S8_vs_k141_84792\|\|full | 3371 | No | NA | 4 | 0 | 1 | Not-determined | Genome-fragment | NA | NA | 0 |
| S8_vs_k141_145772\|\|full | 3163 | No | NA | 3 | 0 | 2 | Not-determined | Genome-fragment | NA | NA | 0 |
| S8_vs_k141_197925\|\|full | 1385 | No | NA | 2 | 0 | 1 | Not-determined | Genome-fragment | NA | NA | 0 |
| S8_vs_k141_162564\|\|full | 1168 | No | NA | 3 | 0 | 2 | Not-determined | Genome-fragment | NA | NA | 0 |
| S5_vs_k141_66521\|\|0_partial | 20090 | Yes | 14209 | 22 | 2 | 5 | Low-quality | Genome-fragment | 8.97 | HMM-based (lower-bound) | 29.27 |
| S8_vs_k141_32955\|\|full | 2968 | No | NA | 3 | 0 | 0 | Not-determined | Genome-fragment | NA | NA | 0 |
| S8_vs_k141_32959\|\|full | 2276 | No | NA | 2 | 0 | 1 | Not-determined | Genome-fragment | NA | NA | 0 |
| S8_vs_k141_93490\|\|full | 2155 | No | NA | 4 | 0 | 1 | Not-determined | Genome-fragment | NA | NA | 0 |
| S5_vs_k141_20434\|\|full | 20124 | No | NA | 19 | 0 | 8 | Low-quality | Genome-fragment | 46.06 | AAI-based (medium-confidence) | 0 |
| S8_vs_k141_128343\|\|full | 1312 | No | NA | 3 | 0 | 0 | Not-determined | Genome-fragment | NA | NA | 0 |
| S8_vs_k141_93523\|\|full | 2202 | No | NA | 2 | 0 | 1 | Not-determined | Genome-fragment | NA | NA | 0 |
| S8_vs_k141_205967\|\|full | 1499 | No | NA | 2 | 0 | 0 | Not-determined | Genome-fragment | NA | NA | 0 |
| S8_vs_k141_145838\|\|full | 2171 | No | NA | 3 | 0 | 2 | Not-determined | Genome-fragment | NA | NA | 0 |
| S16_vs_89491\|\|full | 20130 | No | NA | 26 | 1 | 10 | Low-quality | Genome-fragment | 12.44 | HMM-based (lower-bound) | 0 |
| S5_vs_k141_111186\|\|0_partial | 20133 | Yes | 14764 | 27 | 7 | 4 | Low-quality | Genome-fragment | 19.89 | HMM-based (lower-bound) | 26.67 |
| S13_extracted_77087 | 20179 | No | NA | 34 | 7 | 0 | Low-quality | Genome-fragment | 18.49 | AAI-based (high-confidence) | 0 |
| S5_vs_k141_91909\|\|full | 20258 | No | NA | 21 | 1 | 4 | Low-quality | Genome-fragment | 44.91 | AAI-based (medium-confidence) | 0 |
| S8_vs_k141_205993\|\|full | 1291 | No | NA | 4 | 0 | 0 | Not-determined | Genome-fragment | NA | NA | 0 |
| S11_vs_k141_64382\|\|full | 20262 | No | NA | 32 | 6 | 0 | Low-quality | Genome-fragment | 37.62 | AAI-based (high-confidence) | 0 |
| S16_extracted_142146 | 20267 | No | NA | 21 | 2 | 1 | Low-quality | Genome-fragment | 8.6 | AAI-based (medium-confidence) | 0 |
| S8_vs_k141_171934\|\|full | 2479 | No | NA | 2 | 0 | 1 | Not-determined | Genome-fragment | NA | NA | 0 |
| S13_extracted_104626 | 20290 | No | NA | 38 | 3 | 1 | Low-quality | Genome-fragment | 37.97 | AAI-based (high-confidence) | 0 |
| S8_vs_k141_119622\|\|full | 1768 | No | NA | 4 | 0 | 2 | Not-determined | Genome-fragment | NA | NA | 0 |
| S5_vs_k141_77499\|\|full | 20296 | No | NA | 14 | 0 | 9 | Low-quality | Genome-fragment | 7.85 | AAI-based (medium-confidence) | 0 |
| S12_vs_k141_177540\|\|full | 20371 | No | NA | 34 | 4 | 0 | Low-quality | Genome-fragment | 43.26 | AAI-based (high-confidence) | 0 |
| S8_vs_k141_162691\|\|full | 2029 | No | NA | 2 | 0 | 0 | Not-determined | Genome-fragment | NA | NA | 0 |
| S8_vs_k141_102137\|\|full | 3269 | No | NA | 3 | 0 | 3 | Not-determined | Genome-fragment | NA | NA | 0 |
| S8_vs_k141_188167\|\|full | 8386 | No | NA | 8 | 0 | 5 | Not-determined | Genome-fragment | NA | NA | 0 |
| S5_extracted_k141_70522 | 20376 | No | NA | 12 | 0 | 2 | Low-quality | Genome-fragment | 20.53 | AAI-based (medium-confidence) | 0 |
| S12_vs_k141_88221\|\|0_partial | 20393 | Yes | 12347 | 24 | 5 | 6 | Low-quality | Genome-fragment | 16.36 | HMM-based (lower-bound) | 39.45 |
| S3_vs_k141_45342\|\|full | 20450 | No | NA | 19 | 0 | 14 | Low-quality | Genome-fragment | 11.52 | AAI-based (medium-confidence) | 0 |
| S8_vs_k141_119660\|\|full | 1666 | No | NA | 2 | 0 | 1 | Not-determined | Genome-fragment | NA | NA | 0 |
| S8_vs_k141_41513\|\|full | 1153 | No | NA | 2 | 0 | 0 | Not-determined | Genome-fragment | NA | NA | 0 |
| S8_vs_k141_6991\|\|full | 3540 | No | NA | 4 | 0 | 1 | Not-determined | Genome-fragment | NA | NA | 0 |
| S8_vs_k141_102191\|\|full | 4518 | No | NA | 6 | 0 | 0 | Not-determined | Genome-fragment | NA | NA | 0 |
| S8_vs_k141_145941\|\|full | 2143 | No | NA | 3 | 0 | 3 | Not-determined | Genome-fragment | NA | NA | 0 |
| S8_vs_k141_162753\|\|full | 2720 | No | NA | 6 | 0 | 1 | Not-determined | Genome-fragment | NA | NA | 0 |
| S8_vs_k141_128388\|\|full | 2274 | No | NA | 3 | 0 | 3 | Not-determined | Genome-fragment | NA | NA | 0 |
| S13_vs_2497\|\|full | 20457 | No | NA | 31 | 8 | 1 | Low-quality | Genome-fragment | 12.26 | AAI-based (high-confidence) | 0 |
| S8_vs_k141_128411\|\|full | 2592 | No | NA | 4 | 0 | 0 | Not-determined | Genome-fragment | NA | NA | 0 |
| S15_extracted_11650 | 20475 | No | NA | 42 | 3 | 0 | Low-quality | Genome-fragment | 21.61 | AAI-based (high-confidence) | 0 |
| S13_vs_34483\|\|full | 20620 | No | NA | 24 | 2 | 1 | Low-quality | Genome-fragment | 31.95 | AAI-based (medium-confidence) | 0 |
| S8_vs_k141_93671\|\|full | 2537 | No | NA | 3 | 0 | 1 | Not-determined | Genome-fragment | NA | NA | 0 |
| S8_vs_k141_198105\|\|full | 1323 | No | NA | 2 | 0 | 1 | Not-determined | Genome-fragment | NA | NA | 0 |
| S3_extracted_k141_111839 | 20636 | No | NA | 23 | 1 | 10 | Low-quality | Genome-fragment | 13.81 | AAI-based (medium-confidence) | 0 |
| S8_vs_k141_7053\|\|full | 1956 | No | NA | 3 | 0 | 2 | Not-determined | Genome-fragment | NA | NA | 0 |
| S8_vs_k141_49803\|\|full | 8704 | No | NA | 8 | 0 | 3 | Not-determined | Genome-fragment | NA | NA | 0 |
| S8_vs_k141_41588\|\|full | 5880 | No | NA | 8 | 0 | 2 | Not-determined | Genome-fragment | NA | NA | 0 |
| S13_extracted_102684 | 20665 | No | NA | 28 | 7 | 1 | Low-quality | Genome-fragment | 34.12 | AAI-based (high-confidence) | 0 |
| S8_vs_k141_102285\|\|full | 2198 | No | NA | 3 | 0 | 0 | Not-determined | Genome-fragment | NA | NA | 0 |
| S8_vs_k141_180734\|\|full | 1138 | No | NA | 2 | 0 | 2 | Not-determined | Genome-fragment | NA | NA | 0 |
| S8_vs_k141_24602\|\|full | 1884 | No | NA | 3 | 0 | 1 | Not-determined | Genome-fragment | NA | NA | 0 |
| S5_extracted_k141_77264 | 20728 | No | NA | 25 | 17 | 0 | Low-quality | Genome-fragment | 49.55 | AAI-based (high-confidence) | 0 |
| S13_vs_53416\|\|0_partial | 20730 | No | NA | 18 | 0 | 8 | Low-quality | Genome-fragment | 9.64 | AAI-based (medium-confidence) | 0 |
| S8_vs_k141_162862\|\|full | 10280 | No | NA | 16 | 0 | 1 | Not-determined | Genome-fragment | NA | NA | 0 |
| S8_vs_k141_206117\|\|full | 2062 | No | NA | 2 | 0 | 0 | Not-determined | Genome-fragment | NA | NA | 0 |
| S8_vs_k141_67451\|\|full | 1231 | No | NA | 2 | 0 | 1 | Not-determined | Genome-fragment | NA | NA | 0 |
| S8_vs_k141_136922\|\|full | 39446 | No | NA | 61 | 18 | 3 | Medium-quality | Genome-fragment | 89.59 | AAI-based (high-confidence) | 0 |
| S2_vs_k141_14070\|\|full | 20777 | No | NA | 41 | 5 | 3 | Low-quality | Genome-fragment | 34.94 | AAI-based (high-confidence) | 0 |
| S7_vs_47104\|\|full | 20799 | No | NA | 20 | 2 | 3 | Low-quality | Genome-fragment | 12.62 | AAI-based (medium-confidence) | 0 |
| S14_vs_k141_82571\|\|full | 20801 | No | NA | 22 | 1 | 0 | Low-quality | Genome-fragment | 48.69 | AAI-based (medium-confidence) | 0 |
| S8_vs_k141_198163\|\|full | 1591 | No | NA | 2 | 0 | 1 | Not-determined | Genome-fragment | NA | NA | 0 |
| S11_vs_k141_20005\|\|full | 20823 | No | NA | 47 | 10 | 0 | Low-quality | Genome-fragment | 25.75 | AAI-based (high-confidence) | 0 |
| S14_vs_k141_82295\|\|0_partial | 20862 | Yes | 18048 | 24 | 2 | 4 | Low-quality | Genome-fragment | 11.39 | HMM-based (lower-bound) | 13.49 |
| S8_vs_k141_24647\|\|full | 1534 | No | NA | 4 | 0 | 0 | Not-determined | Genome-fragment | NA | NA | 0 |
| S8_vs_k141_162903\|\|full | 3269 | No | NA | 4 | 0 | 3 | Not-determined | Genome-fragment | NA | NA | 0 |
| S8_vs_k141_7116\|\|full | 3048 | No | NA | 4 | 0 | 0 | Not-determined | Genome-fragment | NA | NA | 0 |
| S11_vs_k141_20627\|\|full | 20878 | No | NA | 32 | 4 | 0 | Low-quality | Genome-fragment | 48.36 | AAI-based (medium-confidence) | 0 |
| S3_vs_k141_121413\|\|full | 20993 | No | NA | 39 | 6 | 0 | Low-quality | Genome-fragment | 10.3 | AAI-based (high-confidence) | 0 |
| S8_vs_k141_206149\|\|full | 2078 | No | NA | 3 | 0 | 0 | Not-determined | Genome-fragment | NA | NA | 0 |
| S8_vs_k141_162933\|\|full | 1655 | No | NA | 3 | 0 | 0 | Not-determined | Genome-fragment | NA | NA | 0 |
| S8_vs_k141_111369\|\|full | 1674 | No | NA | 3 | 0 | 0 | Not-determined | Genome-fragment | NA | NA | 0 |
| S8_vs_k141_206155\|\|full | 2795 | No | NA | 3 | 0 | 2 | Not-determined | Genome-fragment | NA | NA | 0 |
| S8_vs_k141_7135\|\|full | 4149 | No | NA | 8 | 0 | 5 | Not-determined | Genome-fragment | NA | NA | 0 |
| S9_vs_k141_40765\|\|full | 20994 | No | NA | 40 | 10 | 0 | Low-quality | Genome-fragment | 46.75 | AAI-based (high-confidence) | 0 |
| S3_vs_k141_82389\|\|full | 20996 | No | NA | 27 | 0 | 1 | Low-quality | Genome-fragment | 47.72 | AAI-based (medium-confidence) | 0 |
| S8_vs_k141_172171\|\|full | 4202 | No | NA | 3 | 0 | 1 | Not-determined | Genome-fragment | NA | NA | 0 |
| S8_vs_k141_7147\|\|full | 1774 | No | NA | 2 | 0 | 2 | Not-determined | Genome-fragment | NA | NA | 0 |
| S8_vs_k141_154252\|\|full | 2929 | No | NA | 6 | 0 | 0 | Not-determined | Genome-fragment | NA | NA | 0 |
| S3_vs_k141_6553\|\|full | 21020 | No | NA | 45 | 5 | 4 | Low-quality | Genome-fragment | 25.34 | AAI-based (high-confidence) | 0 |
| S14_vs_k141_22491\|\|full | 21044 | Yes | 14161 | 33 | 7 | 3 | Low-quality | Genome-fragment | 18.96 | AAI-based (high-confidence) | 32.71 |
| S13_vs_139426\|\|0_partial | 21072 | No | NA | 23 | 1 | 0 | Low-quality | Genome-fragment | 37.26 | AAI-based (medium-confidence) | 0 |
| S8_vs_k141_146117\|\|full | 2486 | No | NA | 3 | 0 | 0 | Not-determined | Genome-fragment | NA | NA | 0 |
| S10_extracted_43345 | 21078 | No | NA | 20 | 0 | 5 | Low-quality | Genome-fragment | 12.42 | AAI-based (medium-confidence) | 0 |
| S16_extracted_118692 | 21108 | No | NA | 23 | 1 | 4 | Low-quality | Genome-fragment | 11.45 | HMM-based (lower-bound) | 0 |
| S12_vs_k141_23569\|\|0_partial | 21127 | No | NA | 21 | 1 | 14 | Low-quality | Genome-fragment | 11.31 | HMM-based (lower-bound) | 0 |
| S10_vs_44784\|\|full | 21129 | No | NA | 19 | 2 | 1 | Low-quality | Genome-fragment | 39.74 | AAI-based (high-confidence) | 0 |
| S8_vs_k141_33282\|\|full | 3570 | No | NA | 3 | 0 | 1 | Not-determined | Genome-fragment | NA | NA | 0 |
| S17_extracted_k141_112285 | 21168 | No | NA | 19 | 1 | 5 | Low-quality | Genome-fragment | 3.4 | HMM-based (lower-bound) | 0 |
| S10_extracted_74281 | 21178 | No | NA | 22 | 0 | 14 | Low-quality | Genome-fragment | 3.57 | AAI-based (medium-confidence) | 0 |
| S13_extracted_73717 | 21187 | No | NA | 30 | 6 | 0 | Low-quality | Genome-fragment | 15.78 | AAI-based (high-confidence) | 0 |
| S3_vs_k141_57692\|\|full | 21213 | No | NA | 26 | 13 | 0 | Low-quality | Genome-fragment | 45.87 | AAI-based (high-confidence) | 0 |
| S8_vs_k141_163020\|\|full | 1124 | No | NA | 3 | 0 | 1 | Not-determined | Genome-fragment | NA | NA | 0 |
| S5_vs_k141_132206\|\|full | 21294 | No | NA | 29 | 2 | 6 | Low-quality | Genome-fragment | 25.86 | AAI-based (high-confidence) | 0 |
| S7_extracted_54364 | 21299 | No | NA | 32 | 2 | 2 | Low-quality | Genome-fragment | 47.85 | AAI-based (medium-confidence) | 0 |
| S11_vs_k141_1269\|\|full | 21318 | No | NA | 22 | 6 | 0 | Low-quality | Genome-fragment | 16.24 | AAI-based (medium-confidence) | 0 |
| S8_vs_k141_59700\|\|full | 2901 | No | NA | 3 | 0 | 1 | Not-determined | Genome-fragment | NA | NA | 0 |
| S8_extracted_k141_175014 | 21361 | No | NA | 33 | 9 | 0 | Low-quality | Genome-fragment | 48.36 | AAI-based (high-confidence) | 0 |
| S8_vs_k141_102438\|\|full | 1336 | No | NA | 2 | 0 | 1 | Not-determined | Genome-fragment | NA | NA | 0 |
| S16_extracted_153608 | 21362 | No | NA | 19 | 1 | 12 | Low-quality | Genome-fragment | 12.72 | HMM-based (lower-bound) | 0 |
| S8_vs_k141_24817\|\|full | 1301 | No | NA | 3 | 0 | 2 | Not-determined | Genome-fragment | NA | NA | 0 |
| S13_vs_58880\|\|0_partial | 21425 | No | NA | 26 | 1 | 7 | Low-quality | Genome-fragment | 24.05 | HMM-based (lower-bound) | 0 |
| S8_vs_k141_206332\|\|full | 2121 | No | NA | 4 | 0 | 1 | Not-determined | Genome-fragment | NA | NA | 0 |
| S8_vs_k141_128654\|\|full | 1509 | No | NA | 4 | 0 | 0 | Not-determined | Genome-fragment | NA | NA | 0 |
| S8_vs_k141_198411\|\|full | 3810 | No | NA | 5 | 0 | 2 | Not-determined | Genome-fragment | NA | NA | 0 |
| S6_vs_k141_5186\|\|full | 21428 | No | NA | 26 | 14 | 1 | Low-quality | Genome-fragment | 47.74 | AAI-based (high-confidence) | 0 |
| S17_extracted_k141_38776 | 21476 | Yes | 11474 | 26 | 2 | 2 | Low-quality | Genome-fragment | 20.76 | AAI-based (high-confidence) | 46.57 |
| S8_vs_k141_181010\|\|full | 1235 | No | NA | 2 | 0 | 1 | Not-determined | Genome-fragment | NA | NA | 0 |
| S8_vs_k141_15675\|\|full | 1001 | No | NA | 3 | 0 | 1 | Not-determined | Genome-fragment | NA | NA | 0 |
| S16_vs_79597\|\|full | 21484 | No | NA | 18 | 5 | 0 | Low-quality | Genome-fragment | 12.26 | AAI-based (high-confidence) | 0 |
| S8_vs_k141_50077\|\|full | 1549 | No | NA | 3 | 0 | 1 | Not-determined | Genome-fragment | NA | NA | 0 |
| S8_vs_k141_172378\|\|full | 1558 | No | NA | 2 | 0 | 1 | Not-determined | Genome-fragment | NA | NA | 0 |
| S7_vs_64876\|\|full | 21495 | No | NA | 41 | 8 | 0 | Low-quality | Genome-fragment | 42.06 | AAI-based (high-confidence) | 0 |
| S8_vs_k141_128696\|\|full | 6695 | No | NA | 5 | 0 | 3 | Not-determined | Genome-fragment | NA | NA | 0 |
| S14_extracted_k141_8342 | 21507 | No | NA | 24 | 7 | 2 | Low-quality | Genome-fragment | 34.2 | AAI-based (high-confidence) | 0 |
| S8_vs_k141_120018\|\|full | 4230 | No | NA | 6 | 0 | 5 | Not-determined | Genome-fragment | NA | NA | 0 |
| S8_vs_k141_33485\|\|full | 2033 | No | NA | 3 | 0 | 1 | Not-determined | Genome-fragment | NA | NA | 0 |
| S13_extracted_27468 | 21594 | No | NA | 41 | 6 | 1 | Low-quality | Genome-fragment | 13.57 | AAI-based (high-confidence) | 0 |
| S5_vs_k141_122923\|\|0_partial | 21608 | Yes | 16186 | 21 | 3 | 3 | Low-quality | Genome-fragment | 30.94 | HMM-based (lower-bound) | 25.09 |
| S3_vs_k141_12256\|\|full | 21736 | No | NA | 23 | 1 | 8 | Low-quality | Genome-fragment | 42.45 | AAI-based (high-confidence) | 0 |
| S8_vs_k141_33505\|\|full | 1106 | No | NA | 1 | 0 | 0 | Not-determined | Genome-fragment | NA | NA | 0 |
| S8_vs_k141_172437\|\|full | 2869 | No | NA | 3 | 0 | 0 | Not-determined | Genome-fragment | NA | NA | 0 |
| S8_vs_k141_33514\|\|full | 2201 | No | NA | 2 | 0 | 2 | Not-determined | Genome-fragment | NA | NA | 0 |
| S8_vs_k141_206451\|\|full | 2166 | No | NA | 3 | 0 | 2 | Not-determined | Genome-fragment | NA | NA | 0 |
| S8_vs_k141_181085\|\|full | 8546 | No | NA | 15 | 0 | 5 | Not-determined | Genome-fragment | NA | NA | 0 |
| S8_vs_k141_50180\|\|full | 2872 | No | NA | 3 | 0 | 1 | Not-determined | Genome-fragment | NA | NA | 0 |
| S8_vs_k141_100346\|\|0_partial | 21807 | No | NA | 19 | 1 | 8 | Low-quality | Genome-fragment | 10.19 | AAI-based (medium-confidence) | 0 |
| S8_vs_k141_137288\|\|full | 1096 | No | NA | 3 | 0 | 1 | Not-determined | Genome-fragment | NA | NA | 0 |
| S8_vs_k141_33554\|\|full | 3135 | No | NA | 4 | 0 | 3 | Not-determined | Genome-fragment | NA | NA | 0 |
| S8_extracted_k141_96559 | 21821 | No | NA | 26 | 1 | 4 | Low-quality | Genome-fragment | 26.12 | AAI-based (medium-confidence) | 0 |
| S8_vs_k141_137305\|\|full | 1685 | No | NA | 2 | 0 | 1 | Not-determined | Genome-fragment | NA | NA | 0 |
| S8_vs_k141_33559\|\|full | 5121 | No | NA | 7 | 0 | 3 | Not-determined | Genome-fragment | NA | NA | 0 |
| S8_vs_k141_42045\|\|full | 2421 | No | NA | 4 | 0 | 2 | Not-determined | Genome-fragment | NA | NA | 0 |
| S8_vs_k141_42047\|\|full | 1410 | No | NA | 2 | 0 | 2 | Not-determined | Genome-fragment | NA | NA | 0 |
| S8_vs_k141_120094\|\|full | 2070 | No | NA | 3 | 0 | 0 | Not-determined | Genome-fragment | NA | NA | 0 |
| S9_vs_k141_51168\|\|0_partial | 21831 | No | NA | 16 | 1 | 3 | Low-quality | Genome-fragment | 17.47 | HMM-based (lower-bound) | 0 |
| S8_extracted_k141_207623 | 21832 | No | NA | 41 | 0 | 0 | Low-quality | Genome-fragment | 12.03 | AAI-based (medium-confidence) | 0 |
| S7_vs_72746\|\|0_partial | 21848 | No | NA | 15 | 1 | 5 | Low-quality | Genome-fragment | 10.32 | AAI-based (medium-confidence) | 0 |
| S8_vs_k141_50225\|\|full | 3145 | No | NA | 4 | 0 | 2 | Not-determined | Genome-fragment | NA | NA | 0 |
| S8_vs_k141_206508\|\|full | 8081 | No | NA | 9 | 0 | 2 | Not-determined | Genome-fragment | NA | NA | 0 |
| S6_vs_k141_40014\|\|0_partial | 21937 | Yes | 16032 | 32 | 2 | 6 | Low-quality | Genome-fragment | 23.64 | AAI-based (high-confidence) | 26.92 |
| S8_vs_k141_163226\|\|full | 1378 | No | NA | 3 | 0 | 0 | Not-determined | Genome-fragment | NA | NA | 0 |
| S8_vs_k141_102608\|\|full | 35873 | No | NA | 51 | 10 | 0 | Medium-quality | Genome-fragment | 67.58 | AAI-based (high-confidence) | 0 |
| S1_vs_k141_98843\|\|full | 21966 | No | NA | 31 | 4 | 1 | Low-quality | Genome-fragment | 16.4 | AAI-based (high-confidence) | 0 |
| S8_vs_k141_33591\|\|full | 2611 | No | NA | 3 | 0 | 0 | Not-determined | Genome-fragment | NA | NA | 0 |
| S8_vs_k141_172507\|\|full | 2924 | No | NA | 3 | 0 | 1 | Not-determined | Genome-fragment | NA | NA | 0 |
| S8_vs_k141_154530\|\|full | 1502 | No | NA | 2 | 0 | 1 | Not-determined | Genome-fragment | NA | NA | 0 |
| S8_vs_k141_146282\|\|full | 3087 | No | NA | 4 | 0 | 2 | Not-determined | Genome-fragment | NA | NA | 0 |
| S8_vs_k141_76631\|\|full | 1716 | No | NA | 2 | 0 | 1 | Not-determined | Genome-fragment | NA | NA | 0 |
| S8_vs_k141_172530\|\|full | 2830 | No | NA | 3 | 0 | 2 | Not-determined | Genome-fragment | NA | NA | 0 |
| S9_vs_k141_82414\|\|0_partial | 22007 | Yes | 10735 | 27 | 4 | 6 | Low-quality | Genome-fragment | 13.66 | HMM-based (lower-bound) | 51.22 |
| S7_vs_23790\|\|full | 22019 | No | NA | 20 | 2 | 1 | Low-quality | Genome-fragment | 42.93 | AAI-based (medium-confidence) | 0 |
| S8_vs_k141_67828\|\|full | 1714 | No | NA | 3 | 0 | 3 | Not-determined | Genome-fragment | NA | NA | 0 |
| S5_extracted_k141_85342 | 22065 | No | NA | 37 | 6 | 0 | Low-quality | Genome-fragment | 48.9 | AAI-based (high-confidence) | 0 |
| S8_vs_k141_111784\|\|full | 2288 | No | NA | 2 | 0 | 1 | Not-determined | Genome-fragment | NA | NA | 0 |
| S12_vs_k141_181185\|\|full | 22086 | Yes | 8906 | 30 | 6 | 9 | Low-quality | Genome-fragment | 24.22 | AAI-based (medium-confidence) | 59.68 |
| S8_vs_k141_188693\|\|full | 9854 | No | NA | 10 | 0 | 5 | Not-determined | Genome-fragment | NA | NA | 0 |
| S8_vs_k141_42077\|\|full | 4447 | No | NA | 6 | 0 | 1 | Not-determined | Genome-fragment | NA | NA | 0 |
| S8_vs_k141_163292\|\|full | 1066 | No | NA | 2 | 0 | 0 | Not-determined | Genome-fragment | NA | NA | 0 |
| S8_vs_k141_24927\|\|full | 1519 | No | NA | 4 | 0 | 2 | Not-determined | Genome-fragment | NA | NA | 0 |
| S8_vs_k141_206526\|\|full | 45665 | No | NA | 63 | 15 | 0 | High-quality | High-quality | 98.67 | AAI-based (high-confidence) | 0 |
| S14_vs_k141_5307\|\|full | 22119 | No | NA | 40 | 9 | 0 | Low-quality | Genome-fragment | 42.02 | AAI-based (high-confidence) | 0 |
| S3_vs_k141_115349\|\|full | 22128 | No | NA | 18 | 3 | 0 | Low-quality | Genome-fragment | 19.96 | AAI-based (high-confidence) | 0 |
| S8_vs_k141_181240\|\|full | 2056 | No | NA | 2 | 0 | 2 | Not-determined | Genome-fragment | NA | NA | 0 |
| S8_vs_k141_128915\|\|full | 1961 | No | NA | 3 | 0 | 2 | Not-determined | Genome-fragment | NA | NA | 0 |
| S6_extracted_k141_9716 | 22130 | No | NA | 45 | 4 | 1 | Low-quality | Genome-fragment | 18.57 | AAI-based (high-confidence) | 0 |
| S8_vs_k141_111805\|\|full | 11520 | No | NA | 16 | 0 | 5 | Not-determined | Genome-fragment | NA | NA | 0 |
| S8_vs_k141_146390\|\|full | 1607 | No | NA | 3 | 0 | 0 | Not-determined | Genome-fragment | NA | NA | 0 |
| S8_vs_k141_154629\|\|full | 5561 | No | NA | 4 | 0 | 4 | Not-determined | Genome-fragment | NA | NA | 0 |
| S16_extracted_138733 | 22161 | No | NA | 33 | 2 | 0 | Low-quality | Genome-fragment | 37.05 | AAI-based (high-confidence) | 0 |
| S8_vs_k141_163352\|\|full | 1996 | No | NA | 3 | 0 | 1 | Not-determined | Genome-fragment | NA | NA | 0 |
| S8_vs_k141_76722\|\|full | 1395 | No | NA | 2 | 0 | 1 | Not-determined | Genome-fragment | NA | NA | 0 |
| S8_vs_k141_85455\|\|full | 2017 | No | NA | 3 | 0 | 0 | Not-determined | Genome-fragment | NA | NA | 0 |
| S5_vs_k141_112721\|\|full | 22194 | No | NA | 29 | 15 | 0 | Low-quality | Genome-fragment | 47.51 | AAI-based (medium-confidence) | 0 |
| S8_vs_k141_15778\|\|full | 2040 | No | NA | 2 | 0 | 0 | Not-determined | Genome-fragment | NA | NA | 0 |
| S12_extracted_k141_116416 | 22249 | No | NA | 34 | 4 | 0 | Low-quality | Genome-fragment | 17.68 | AAI-based (high-confidence) | 0 |
| S8_vs_k141_24999\|\|full | 2195 | No | NA | 2 | 0 | 1 | Not-determined | Genome-fragment | NA | NA | 0 |
| S11_extracted_k141_59955 | 22312 | No | NA | 48 | 1 | 0 | Low-quality | Genome-fragment | 13.86 | AAI-based (high-confidence) | 0 |
| S8_vs_k141_206580\|\|full | 2490 | No | NA | 3 | 0 | 2 | Not-determined | Genome-fragment | NA | NA | 0 |
| S8_vs_k141_50376\|\|full | 1707 | No | NA | 2 | 0 | 0 | Not-determined | Genome-fragment | NA | NA | 0 |
| S5_extracted_k141_57448 | 22326 | No | NA | 30 | 16 | 1 | Low-quality | Genome-fragment | 21.27 | HMM-based (lower-bound) | 0 |
| S8_vs_k141_181300\|\|full | 2538 | No | NA | 2 | 0 | 0 | Not-determined | Genome-fragment | NA | NA | 0 |
| S8_vs_k141_76836\|\|full | 2701 | No | NA | 4 | 0 | 0 | Not-determined | Genome-fragment | NA | NA | 0 |
| S8_vs_k141_7640\|\|full | 2777 | No | NA | 2 | 0 | 2 | Not-determined | Genome-fragment | NA | NA | 0 |
| S8_vs_k141_85539\|\|full | 2493 | No | NA | 3 | 0 | 1 | Not-determined | Genome-fragment | NA | NA | 0 |
| S8_vs_k141_33845\|\|full | 1658 | No | NA | 3 | 0 | 0 | Not-determined | Genome-fragment | NA | NA | 0 |
| S1_vs_k141_103749\|\|full | 22329 | No | NA | 27 | 13 | 0 | Low-quality | Genome-fragment | 44.92 | AAI-based (high-confidence) | 0 |
| S8_vs_k141_137504\|\|full | 2480 | No | NA | 4 | 0 | 1 | Not-determined | Genome-fragment | NA | NA | 0 |
| S8_vs_k141_15843\|\|full | 3403 | No | NA | 4 | 0 | 0 | Not-determined | Genome-fragment | NA | NA | 0 |
| S8_vs_k141_15847\|\|full | 1567 | No | NA | 2 | 0 | 0 | Not-determined | Genome-fragment | NA | NA | 0 |
| S8_vs_k141_120372\|\|full | 1913 | No | NA | 3 | 0 | 2 | Not-determined | Genome-fragment | NA | NA | 0 |
| S8_vs_k141_76852\|\|full | 22932 | No | NA | 37 | 15 | 1 | Medium-quality | Genome-fragment | 75.16 | AAI-based (high-confidence) | 0 |
| S8_vs_k141_33869\|\|full | 3586 | No | NA | 3 | 0 | 3 | Not-determined | Genome-fragment | NA | NA | 0 |
| S8_vs_k141_15873\|\|full | 4405 | No | NA | 5 | 0 | 3 | Not-determined | Genome-fragment | NA | NA | 0 |
| S8_vs_k141_50431\|\|full | 26518 | No | NA | 40 | 12 | 0 | Medium-quality | Genome-fragment | 67.77 | AAI-based (high-confidence) | 0 |
| S11_vs_k141_81497\|\|full | 22334 | Yes | 6520 | 20 | 3 | 4 | Low-quality | Genome-fragment | 13.52 | AAI-based (high-confidence) | 70.81 |
| S8_vs_k141_15874\|\|full | 5826 | No | NA | 7 | 0 | 7 | Not-determined | Genome-fragment | NA | NA | 0 |
| S8_vs_k141_50440\|\|full | 1835 | No | NA | 3 | 0 | 0 | Not-determined | Genome-fragment | NA | NA | 0 |
| S9_extracted_k141_80079 | 22340 | No | NA | 24 | 8 | 0 | Low-quality | Genome-fragment | 13.18 | AAI-based (high-confidence) | 0 |
| S12_extracted_k141_35096 | 22381 | No | NA | 48 | 8 | 0 | Low-quality | Genome-fragment | 37.93 | AAI-based (high-confidence) | 0 |
| S3_vs_k141_15975\|\|0_partial | 22384 | Yes | 11513 | 21 | 3 | 9 | Low-quality | Genome-fragment | 27.99 | AAI-based (high-confidence) | 48.57 |
| S3_extracted_k141_3563 | 22406 | No | NA | 34 | 8 | 0 | Low-quality | Genome-fragment | 38.37 | AAI-based (high-confidence) | 0 |
| S15_extracted_17969 | 22468 | No | NA | 43 | 3 | 1 | Low-quality | Genome-fragment | 27.91 | AAI-based (high-confidence) | 0 |
| S8_vs_k141_15914\|\|full | 1328 | No | NA | 3 | 0 | 0 | Not-determined | Genome-fragment | NA | NA | 0 |
| S8_vs_k141_50467\|\|full | 2597 | No | NA | 5 | 0 | 3 | Not-determined | Genome-fragment | NA | NA | 0 |
| S8_vs_k141_137549\|\|full | 9670 | No | NA | 5 | 0 | 5 | Not-determined | Genome-fragment | NA | NA | 0 |
| S16_extracted_10938 | 22473 | No | NA | 26 | 2 | 0 | Low-quality | Genome-fragment | 36.56 | AAI-based (high-confidence) | 0 |
| S8_vs_k141_33910\|\|full | 2195 | No | NA | 2 | 0 | 2 | Not-determined | Genome-fragment | NA | NA | 0 |
| S8_vs_k141_94331\|\|full | 2003 | No | NA | 3 | 0 | 1 | Not-determined | Genome-fragment | NA | NA | 0 |
| S8_vs_k141_188824\|\|full | 10100 | No | NA | 12 | 0 | 8 | Not-determined | Genome-fragment | NA | NA | 0 |
| S12_vs_k141_129444\|\|full | 22516 | No | NA | 38 | 10 | 1 | Low-quality | Genome-fragment | 40.44 | AAI-based (high-confidence) | 0 |
| S11_extracted_k141_34671 | 22526 | No | NA | 46 | 2 | 2 | Low-quality | Genome-fragment | 12.1 | HMM-based (lower-bound) | 0 |
| S14_vs_k141_28350\|\|full | 22548 | No | NA | 32 | 11 | 0 | Low-quality | Genome-fragment | 48.77 | AAI-based (medium-confidence) | 0 |
| S1_extracted_k141_104061 | 22615 | No | NA | 37 | 3 | 2 | Low-quality | Genome-fragment | 28.14 | AAI-based (high-confidence) | 0 |
| S8_vs_k141_129174\|\|full | 2314 | No | NA | 3 | 0 | 2 | Not-determined | Genome-fragment | NA | NA | 0 |
| S8_vs_k141_198909\|\|full | 2393 | No | NA | 3 | 0 | 2 | Not-determined | Genome-fragment | NA | NA | 0 |
| S8_vs_k141_188839\|\|full | 2712 | No | NA | 3 | 0 | 0 | Not-determined | Genome-fragment | NA | NA | 0 |
| S10_vs_5400\|\|full | 22627 | Yes | 14710 | 39 | 4 | 4 | Low-quality | Genome-fragment | 23.86 | AAI-based (high-confidence) | 34.99 |
| S8_vs_k141_181415\|\|full | 1363 | No | NA | 3 | 0 | 2 | Not-determined | Genome-fragment | NA | NA | 0 |
| S17_extracted_k141_176282 | 22641 | No | NA | 26 | 1 | 3 | Low-quality | Genome-fragment | 34.26 | HMM-based (lower-bound) | 0 |
| S8_vs_k141_165854\|\|full | 22682 | No | NA | 20 | 0 | 2 | Low-quality | Genome-fragment | 25.74 | AAI-based (medium-confidence) | 0 |
| S8_vs_k141_198918\|\|full | 3187 | No | NA | 3 | 0 | 2 | Not-determined | Genome-fragment | NA | NA | 0 |
| S14_extracted_k141_1716 | 22730 | No | NA | 40 | 5 | 0 | Low-quality | Genome-fragment | 38.9 | AAI-based (high-confidence) | 0 |
| S8_vs_k141_33975\|\|full | 1337 | No | NA | 2 | 0 | 0 | Not-determined | Genome-fragment | NA | NA | 0 |
| S11_vs_k141_46044\|\|full | 22769 | No | NA | 24 | 1 | 0 | Low-quality | Genome-fragment | 42.1 | AAI-based (high-confidence) | 0 |
| S15_vs_74565\|\|full | 22784 | No | NA | 23 | 0 | 2 | Low-quality | Genome-fragment | 23.58 | AAI-based (medium-confidence) | 0 |
| S8_vs_k141_102882\|\|full | 7547 | No | NA | 4 | 0 | 3 | Not-determined | Genome-fragment | NA | NA | 0 |
| S8_vs_k141_25151\|\|full | 6137 | No | NA | 7 | 0 | 0 | Not-determined | Genome-fragment | NA | NA | 0 |
| S6_extracted_k141_56022 | 22802 | No | NA | 31 | 3 | 1 | Low-quality | Genome-fragment | 19.32 | AAI-based (high-confidence) | 0 |
| S8_vs_k141_102898\|\|full | 2089 | No | NA | 3 | 0 | 0 | Not-determined | Genome-fragment | NA | NA | 0 |
| S12_vs_k141_7527\|\|full | 22948 | No | NA | 39 | 7 | 1 | Low-quality | Genome-fragment | 41.94 | AAI-based (high-confidence) | 0 |
| S8_vs_k141_15971\|\|full | 1191 | No | NA | 2 | 0 | 2 | Not-determined | Genome-fragment | NA | NA | 0 |
| S8_extracted_k141_173273 | 22948 | No | NA | 31 | 5 | 2 | Low-quality | Genome-fragment | 34.05 | AAI-based (high-confidence) | 0 |
| S8_vs_k141_137667\|\|full | 2751 | No | NA | 3 | 0 | 2 | Not-determined | Genome-fragment | NA | NA | 0 |
| S8_vs_k141_25173\|\|full | 3696 | No | NA | 3 | 0 | 2 | Not-determined | Genome-fragment | NA | NA | 0 |
| S14_vs_k141_48104\|\|full | 22989 | No | NA | 24 | 9 | 1 | Low-quality | Genome-fragment | 32.48 | AAI-based (high-confidence) | 0 |
| S11_vs_k141_43841\|\|full | 23001 | Yes | 18722 | 28 | 15 | 2 | Low-quality | Genome-fragment | 45.1 | AAI-based (high-confidence) | 18.6 |
| S8_vs_k141_94431\|\|full | 1045 | No | NA | 3 | 0 | 0 | Not-determined | Genome-fragment | NA | NA | 0 |
| S8_vs_k141_34042\|\|full | 3893 | No | NA | 5 | 0 | 2 | Not-determined | Genome-fragment | NA | NA | 0 |
| S5_vs_k141_104756\|\|full | 23022 | Yes | 14745 | 41 | 6 | 5 | Low-quality | Genome-fragment | 39.71 | AAI-based (medium-confidence) | 35.95 |
| S16_vs_98140\|\|full | 23040 | No | NA | 15 | 1 | 6 | Low-quality | Genome-fragment | 1.85 | HMM-based (lower-bound) | 0 |
| S8_vs_k141_77064\|\|full | 1100 | No | NA | 2 | 0 | 0 | Not-determined | Genome-fragment | NA | NA | 0 |
| S8_vs_k141_68160\|\|full | 3179 | No | NA | 4 | 0 | 1 | Not-determined | Genome-fragment | NA | NA | 0 |
| S8_vs_k141_137703\|\|full | 2659 | No | NA | 3 | 0 | 2 | Not-determined | Genome-fragment | NA | NA | 0 |
| S8_vs_k141_7837\|\|full | 2047 | No | NA | 2 | 0 | 2 | Not-determined | Genome-fragment | NA | NA | 0 |
| S8_vs_k141_199022\|\|full | 2525 | No | NA | 4 | 0 | 2 | Not-determined | Genome-fragment | NA | NA | 0 |
| S8_extracted_k141_64910 | 23073 | No | NA | 38 | 6 | 2 | Low-quality | Genome-fragment | 30.91 | AAI-based (high-confidence) | 0 |
| S10_vs_36954\|\|full | 23088 | No | NA | 28 | 1 | 0 | Low-quality | Genome-fragment | 14.15 | AAI-based (high-confidence) | 0 |
| S8_vs_k141_7871\|\|full | 2404 | No | NA | 2 | 0 | 1 | Not-determined | Genome-fragment | NA | NA | 0 |
| S10_vs_62668\|\|0_partial | 23259 | No | NA | 25 | 2 | 10 | Low-quality | Genome-fragment | 30.79 | HMM-based (lower-bound) | 0 |
| S5_vs_k141_133796\|\|0_partial | 23273 | Yes | 12459 | 26 | 5 | 6 | Low-quality | Genome-fragment | 16.93 | HMM-based (lower-bound) | 46.47 |
| S16_vs_76992\|\|full | 23274 | No | NA | 22 | 16 | 0 | Low-quality | Genome-fragment | 36.44 | AAI-based (high-confidence) | 0 |
| S3_extracted_k141_50486 | 23290 | No | NA | 59 | 2 | 0 | Low-quality | Genome-fragment | 15.92 | AAI-based (high-confidence) | 0 |
| S8_vs_k141_146740\|\|full | 3336 | No | NA | 3 | 0 | 1 | Not-determined | Genome-fragment | NA | NA | 0 |
| S11_extracted_k141_33736 | 23305 | No | NA | 39 | 8 | 1 | Low-quality | Genome-fragment | 40.78 | AAI-based (high-confidence) | 0 |
| S1_vs_k141_43763\|\|full | 23349 | No | NA | 41 | 12 | 1 | Low-quality | Genome-fragment | 17.15 | AAI-based (high-confidence) | 0 |
| S7_vs_101631\|\|full | 23367 | Yes | 14162 | 29 | 5 | 3 | Low-quality | Genome-fragment | 27.71 | AAI-based (medium-confidence) | 39.39 |
| S8_vs_k141_77117\|\|full | 1136 | No | NA | 2 | 0 | 0 | Not-determined | Genome-fragment | NA | NA | 0 |
| S17_vs_k141_37726\|\|full | 23387 | Yes | 7135 | 32 | 3 | 8 | Low-quality | Genome-fragment | 11.04 | AAI-based (high-confidence) | 69.49 |
| S8_vs_k141_146774\|\|full | 4949 | No | NA | 3 | 0 | 1 | Not-determined | Genome-fragment | NA | NA | 0 |
| S8_vs_k141_103052\|\|full | 1370 | No | NA | 3 | 0 | 1 | Not-determined | Genome-fragment | NA | NA | 0 |
| S16_extracted_36832 | 23417 | No | NA | 19 | 0 | 5 | Low-quality | Genome-fragment | 29.67 | AAI-based (medium-confidence) | 0 |
| S8_vs_k141_146783\|\|full | 1801 | No | NA | 2 | 0 | 1 | Not-determined | Genome-fragment | NA | NA | 0 |
| S8_vs_k141_206927\|\|full | 1799 | No | NA | 3 | 0 | 2 | Not-determined | Genome-fragment | NA | NA | 0 |
| S8_vs_k141_7930\|\|full | 2721 | No | NA | 3 | 0 | 3 | Not-determined | Genome-fragment | NA | NA | 0 |
| S8_vs_k141_68225\|\|full | 2120 | No | NA | 2 | 0 | 1 | Not-determined | Genome-fragment | NA | NA | 0 |
| S8_vs_k141_206939\|\|full | 1512 | No | NA | 3 | 0 | 1 | Not-determined | Genome-fragment | NA | NA | 0 |
| S13_vs_43656\|\|0_partial | 23520 | Yes | 2501 | 30 | 4 | 6 | Low-quality | Genome-fragment | 1.97 | HMM-based (lower-bound) | 89.37 |
| S8_vs_k141_154984\|\|full | 4304 | No | NA | 5 | 0 | 3 | Not-determined | Genome-fragment | NA | NA | 0 |
| S12_vs_k141_147259\|\|full | 23558 | No | NA | 37 | 6 | 0 | Low-quality | Genome-fragment | 49.36 | AAI-based (medium-confidence) | 0 |
| S1_vs_k141_151707\|\|full | 23627 | No | NA | 36 | 9 | 0 | Low-quality | Genome-fragment | 40.64 | AAI-based (high-confidence) | 0 |
| S8_vs_k141_206945\|\|full | 3570 | No | NA | 5 | 0 | 2 | Not-determined | Genome-fragment | NA | NA | 0 |
| S3_extracted_k141_118072 | 23659 | No | NA | 18 | 1 | 3 | Low-quality | Genome-fragment | 8.42 | HMM-based (lower-bound) | 0 |
| S8_vs_k141_163595\|\|full | 1994 | No | NA | 3 | 0 | 0 | Not-determined | Genome-fragment | NA | NA | 0 |
| S8_vs_k141_129334\|\|full | 1892 | No | NA | 3 | 0 | 1 | Not-determined | Genome-fragment | NA | NA | 0 |
| S8_vs_k141_42472\|\|full | 1393 | No | NA | 3 | 0 | 2 | Not-determined | Genome-fragment | NA | NA | 0 |
| S8_vs_k141_163630\|\|full | 1809 | No | NA | 2 | 0 | 0 | Not-determined | Genome-fragment | NA | NA | 0 |
| S11_extracted_k141_32021 | 23670 | No | NA | 39 | 9 | 1 | Low-quality | Genome-fragment | 45.97 | AAI-based (medium-confidence) | 0 |
| S15_extracted_57944 | 23710 | No | NA | 5 | 0 | 0 | Low-quality | Genome-fragment | 25.82 | AAI-based (high-confidence) | 0 |
| S8_vs_k141_206967\|\|full | 6235 | No | NA | 7 | 0 | 3 | Not-determined | Genome-fragment | NA | NA | 0 |
| S17_vs_k141_90265\|\|full | 23720 | No | NA | 24 | 1 | 18 | Low-quality | Genome-fragment | 14.6 | HMM-based (lower-bound) | 0 |
| S17_extracted_k141_82072 | 23748 | No | NA | 24 | 8 | 0 | Low-quality | Genome-fragment | 43.6 | AAI-based (high-confidence) | 0 |
| S8_vs_k141_103148\|\|0_partial | 23773 | Yes | 2583 | 27 | 3 | 9 | Low-quality | Genome-fragment | 2.97 | HMM-based (lower-bound) | 89.13 |
| S8_vs_k141_112189\|\|full | 10630 | No | NA | 14 | 0 | 0 | Not-determined | Genome-fragment | NA | NA | 0 |
| S14_vs_k141_54872\|\|full | 23919 | Yes | 19071 | 26 | 12 | 4 | Low-quality | Genome-fragment | 27.54 | AAI-based (high-confidence) | 20.27 |
| S17_extracted_k141_145006 | 23989 | No | NA | 30 | 9 | 1 | Low-quality | Genome-fragment | 22.03 | AAI-based (high-confidence) | 0 |
| S8_vs_k141_199204\|\|full | 2524 | No | NA | 2 | 0 | 1 | Not-determined | Genome-fragment | NA | NA | 0 |
| S16_extracted_145356 | 24080 | No | NA | 50 | 5 | 0 | Low-quality | Genome-fragment | 40.75 | AAI-based (high-confidence) | 0 |
| S6_vs_k141_50185\|\|1_partial | 24099 | Yes | 21450 | 20 | 2 | 3 | Low-quality | Genome-fragment | 27.64 | AAI-based (medium-confidence) | 10.99 |
| S8_vs_k141_25427\|\|full | 1787 | No | NA | 3 | 0 | 1 | Not-determined | Genome-fragment | NA | NA | 0 |
| S12_vs_k141_165925\|\|0_partial | 24115 | Yes | 14768 | 25 | 3 | 8 | Low-quality | Genome-fragment | 20.64 | HMM-based (lower-bound) | 38.76 |
| S8_vs_k141_68328\|\|full | 1366 | No | NA | 3 | 0 | 0 | Not-determined | Genome-fragment | NA | NA | 0 |
| S8_vs_k141_77250\|\|full | 1761 | No | NA | 4 | 0 | 0 | Not-determined | Genome-fragment | NA | NA | 0 |
| S11_extracted_k141_38453 | 24128 | No | NA | 29 | 3 | 0 | Low-quality | Genome-fragment | 25.74 | AAI-based (medium-confidence) | 0 |
| S8_vs_k141_68331\|\|full | 2700 | No | NA | 2 | 0 | 1 | Not-determined | Genome-fragment | NA | NA | 0 |
| S14_extracted_k141_62570 | 24206 | No | NA | 45 | 7 | 1 | Low-quality | Genome-fragment | 39.5 | AAI-based (high-confidence) | 0 |
| S8_vs_k141_181739\|\|full | 1365 | No | NA | 3 | 0 | 1 | Not-determined | Genome-fragment | NA | NA | 0 |
| S8_vs_k141_60357\|\|full | 2354 | No | NA | 3 | 0 | 2 | Not-determined | Genome-fragment | NA | NA | 0 |
| S17_vs_k141_134275\|\|0_partial | 24289 | Yes | 10384 | 28 | 4 | 6 | Low-quality | Genome-fragment | 12.59 | HMM-based (lower-bound) | 57.25 |
| S8_vs_k141_172974\|\|full | 2551 | No | NA | 3 | 0 | 1 | Not-determined | Genome-fragment | NA | NA | 0 |
| S3_vs_k141_52619\|\|full | 24369 | No | NA | 43 | 7 | 1 | Low-quality | Genome-fragment | 11.85 | AAI-based (high-confidence) | 0 |
| S8_vs_k141_60376\|\|full | 1128 | No | NA | 2 | 0 | 0 | Not-determined | Genome-fragment | NA | NA | 0 |
| S11_extracted_k141_89853 | 24373 | No | NA | 7 | 1 | 0 | Low-quality | Genome-fragment | 27.12 | AAI-based (high-confidence) | 0 |
| S9_vs_k141_75356\|\|full | 24387 | No | NA | 31 | 7 | 0 | Low-quality | Genome-fragment | 14.66 | AAI-based (high-confidence) | 0 |
| S8_vs_k141_60397\|\|full | 2702 | No | NA | 3 | 0 | 1 | Not-determined | Genome-fragment | NA | NA | 0 |
| S3_vs_k141_25654\|\|full | 24394 | Yes | 4426 | 24 | 2 | 13 | Low-quality | Genome-fragment | 3.51 | HMM-based (lower-bound) | 81.86 |
| S8_vs_k141_86033\|\|full | 1372 | No | NA | 2 | 0 | 1 | Not-determined | Genome-fragment | NA | NA | 0 |
| S13_vs_13947\|\|full | 24425 | Yes | 16092 | 29 | 5 | 3 | Low-quality | Genome-fragment | 37.57 | AAI-based (high-confidence) | 34.12 |
| S5_vs_k141_141116\|\|0_partial | 24432 | No | NA | 16 | 2 | 9 | Low-quality | Genome-fragment | 41.68 | AAI-based (medium-confidence) | 0 |
| S11_extracted_k141_93111 | 24484 | No | NA | 22 | 0 | 6 | Low-quality | Genome-fragment | 30.24 | AAI-based (medium-confidence) | 0 |
| S8_vs_k141_120717\|\|full | 1204 | No | NA | 3 | 0 | 0 | Not-determined | Genome-fragment | NA | NA | 0 |
| S13_extracted_26348 | 24540 | No | NA | 25 | 6 | 1 | Low-quality | Genome-fragment | 15.43 | AAI-based (high-confidence) | 0 |
| S9_extracted_k141_52162 | 24555 | No | NA | 27 | 3 | 1 | Low-quality | Genome-fragment | 15.9 | AAI-based (high-confidence) | 0 |
| S13_vs_114932\|\|0_partial | 24660 | Yes | 11027 | 31 | 3 | 5 | Low-quality | Genome-fragment | 25.45 | AAI-based (high-confidence) | 55.28 |
| S8_vs_k141_146981\|\|full | 5477 | No | NA | 9 | 0 | 2 | Not-determined | Genome-fragment | NA | NA | 0 |
| S6_vs_k141_49311\|\|full | 24664 | Yes | 18226 | 29 | 3 | 7 | Low-quality | Genome-fragment | 29.59 | HMM-based (lower-bound) | 26.1 |
| S7_vs_87587\|\|full | 24718 | No | NA | 31 | 16 | 0 | Low-quality | Genome-fragment | 40.98 | AAI-based (high-confidence) | 0 |
| S8_vs_k141_60463\|\|full | 2447 | No | NA | 3 | 0 | 1 | Not-determined | Genome-fragment | NA | NA | 0 |
| S8_vs_k141_138019\|\|full | 1169 | No | NA | 2 | 0 | 0 | Not-determined | Genome-fragment | NA | NA | 0 |
| S8_vs_k141_189239\|\|full | 1138 | No | NA | 3 | 0 | 1 | Not-determined | Genome-fragment | NA | NA | 0 |
| S8_vs_k141_16324\|\|full | 1649 | No | NA | 2 | 0 | 0 | Not-determined | Genome-fragment | NA | NA | 0 |
| S8_vs_k141_155163\|\|full | 1085 | No | NA | 2 | 0 | 1 | Not-determined | Genome-fragment | NA | NA | 0 |
| S16_vs_14095\|\|full | 24819 | No | NA | 26 | 3 | 16 | Low-quality | Genome-fragment | 47.23 | HMM-based (lower-bound) | 0 |
| S8_vs_k141_8155\|\|full | 2822 | No | NA | 5 | 0 | 1 | Not-determined | Genome-fragment | NA | NA | 0 |
| S8_vs_k141_207139\|\|full | 1770 | No | NA | 2 | 0 | 1 | Not-determined | Genome-fragment | NA | NA | 0 |
| S8_vs_k141_34345\|\|full | 1602 | No | NA | 3 | 0 | 0 | Not-determined | Genome-fragment | NA | NA | 0 |
| S8_vs_k141_42672\|\|full | 3254 | No | NA | 4 | 0 | 3 | Not-determined | Genome-fragment | NA | NA | 0 |
| S6_vs_k141_19068\|\|full | 24868 | No | NA | 40 | 1 | 1 | Low-quality | Genome-fragment | 39.82 | AAI-based (high-confidence) | 0 |
| S8_vs_k141_8167\|\|full | 1928 | No | NA | 5 | 0 | 1 | Not-determined | Genome-fragment | NA | NA | 0 |
| S10_extracted_19259 | 25058 | Yes | 16419 | 36 | 3 | 3 | Low-quality | Genome-fragment | 40.5 | AAI-based (high-confidence) | 34.48 |
| S1_vs_k141_79563\|\|full | 25118 | No | NA | 30 | 7 | 3 | Low-quality | Genome-fragment | 18.73 | AAI-based (high-confidence) | 0 |
| S8_vs_k141_189320\|\|full | 2101 | No | NA | 3 | 0 | 1 | Not-determined | Genome-fragment | NA | NA | 0 |
| S5_vs_k141_132364\|\|0_partial | 25196 | Yes | 11747 | 29 | 3 | 4 | Low-quality | Genome-fragment | 31.74 | AAI-based (high-confidence) | 53.38 |
| S8_vs_k141_207190\|\|full | 1845 | No | NA | 3 | 0 | 1 | Not-determined | Genome-fragment | NA | NA | 0 |
| S8_vs_k141_189347\|\|full | 3645 | No | NA | 3 | 0 | 0 | Not-determined | Genome-fragment | NA | NA | 0 |
| S8_vs_k141_49120\|\|full | 25438 | No | NA | 34 | 1 | 7 | Low-quality | Genome-fragment | 38.49 | HMM-based (lower-bound) | 0 |
| S8_vs_k141_8208\|\|full | 1420 | No | NA | 2 | 0 | 1 | Not-determined | Genome-fragment | NA | NA | 0 |
| S13_vs_73280\|\|full | 25460 | No | NA | 36 | 12 | 0 | Low-quality | Genome-fragment | 19.01 | AAI-based (high-confidence) | 0 |
| S8_vs_k141_24820\|\|0_partial | 25483 | Yes | 17702 | 25 | 4 | 10 | Low-quality | Genome-fragment | 42.66 | AAI-based (medium-confidence) | 30.53 |
| S8_vs_k141_60565\|\|full | 2477 | No | NA | 4 | 0 | 0 | Not-determined | Genome-fragment | NA | NA | 0 |
| S8_vs_k141_42719\|\|full | 3185 | No | NA | 4 | 0 | 0 | Not-determined | Genome-fragment | NA | NA | 0 |
| S8_vs_k141_103283\|\|full | 2486 | No | NA | 2 | 0 | 2 | Not-determined | Genome-fragment | NA | NA | 0 |
| S12_vs_k141_99010\|\|full | 25520 | No | NA | 38 | 0 | 2 | Low-quality | Genome-fragment | 40.45 | AAI-based (medium-confidence) | 0 |
| S8_vs_k141_163820\|\|full | 2202 | No | NA | 3 | 0 | 2 | Not-determined | Genome-fragment | NA | NA | 0 |
| S3_extracted_k141_100996 | 25573 | No | NA | 25 | 1 | 13 | Low-quality | Genome-fragment | 31.51 | HMM-based (lower-bound) | 0 |
| S11_extracted_k141_79240 | 25610 | Yes | 20298 | 32 | 7 | 3 | Low-quality | Genome-fragment | 14.48 | AAI-based (medium-confidence) | 20.74 |
| S5_vs_k141_60964\|\|full | 25742 | Yes | 18370 | 25 | 2 | 7 | Low-quality | Genome-fragment | 11.39 | HMM-based (lower-bound) | 28.64 |
| S3_extracted_k141_36418 | 25820 | No | NA | 38 | 5 | 2 | Low-quality | Genome-fragment | 24.3 | AAI-based (medium-confidence) | 0 |
| S8_vs_k141_60589\|\|full | 1746 | No | NA | 3 | 0 | 0 | Not-determined | Genome-fragment | NA | NA | 0 |
| S8_vs_k141_138162\|\|full | 1514 | No | NA | 2 | 0 | 1 | Not-determined | Genome-fragment | NA | NA | 0 |
| S8_vs_k141_5092\|\|full | 25858 | No | NA | 43 | 12 | 0 | Low-quality | Genome-fragment | 45.32 | AAI-based (high-confidence) | 0 |
| S8_vs_k141_155312\|\|full | 2449 | No | NA | 3 | 0 | 3 | Not-determined | Genome-fragment | NA | NA | 0 |
| S8_vs_k141_199399\|\|full | 3493 | No | NA | 3 | 0 | 2 | Not-determined | Genome-fragment | NA | NA | 0 |
| S8_vs_k141_77561\|\|full | 1942 | No | NA | 4 | 0 | 2 | Not-determined | Genome-fragment | NA | NA | 0 |
| S17_extracted_k141_110700 | 25859 | No | NA | 43 | 14 | 0 | Low-quality | Genome-fragment | 43.08 | AAI-based (high-confidence) | 0 |
| S8_vs_k141_8266\|\|full | 1345 | No | NA | 2 | 0 | 1 | Not-determined | Genome-fragment | NA | NA | 0 |
| S13_vs_15122\|\|full | 25898 | No | NA | 32 | 2 | 2 | Low-quality | Genome-fragment | 23.33 | AAI-based (medium-confidence) | 0 |
| S8_vs_k141_155325\|\|full | 1175 | No | NA | 3 | 0 | 0 | Not-determined | Genome-fragment | NA | NA | 0 |
| S8_vs_k141_34465\|\|full | 5005 | No | NA | 6 | 0 | 4 | Not-determined | Genome-fragment | NA | NA | 0 |
| S11_extracted_k141_21747 | 25932 | No | NA | 26 | 5 | 2 | Low-quality | Genome-fragment | 42.38 | AAI-based (medium-confidence) | 0 |
| S10_extracted_11488 | 25933 | No | NA | 10 | 0 | 0 | Low-quality | Genome-fragment | 15.91 | AAI-based (high-confidence) | 0 |
| S10_extracted_12211 | 26038 | No | NA | 24 | 1 | 3 | Low-quality | Genome-fragment | 27.16 | HMM-based (lower-bound) | 0 |
| S9_vs_k141_46532\|\|full | 26069 | No | NA | 28 | 17 | 0 | Low-quality | Genome-fragment | 43.96 | AAI-based (high-confidence) | 0 |
| S8_vs_k141_189454\|\|full | 3372 | No | NA | 3 | 0 | 3 | Not-determined | Genome-fragment | NA | NA | 0 |
| S8_vs_k141_103356\|\|full | 1141 | No | NA | 2 | 0 | 2 | Not-determined | Genome-fragment | NA | NA | 0 |
| S16_vs_164461\|\|full | 26091 | No | NA | 32 | 8 | 0 | Low-quality | Genome-fragment | 41.25 | HMM-based (lower-bound) | 0 |
| S5_vs_k141_32370\|\|full | 26101 | Yes | 18181 | 30 | 14 | 4 | Low-quality | Genome-fragment | 47.44 | AAI-based (medium-confidence) | 30.34 |
| S11_extracted_k141_91744 | 26102 | No | NA | 6 | 0 | 0 | Low-quality | Genome-fragment | 28.67 | AAI-based (high-confidence) | 0 |
| S8_vs_k141_155357\|\|full | 5420 | No | NA | 5 | 0 | 1 | Not-determined | Genome-fragment | NA | NA | 0 |
| S8_vs_k141_42789\|\|full | 6149 | No | NA | 9 | 0 | 3 | Not-determined | Genome-fragment | NA | NA | 0 |
| S8_vs_k141_103381\|\|full | 2283 | No | NA | 2 | 0 | 1 | Not-determined | Genome-fragment | NA | NA | 0 |
| S8_vs_k141_50731\|\|full | 6279 | No | NA | 4 | 0 | 1 | Not-determined | Genome-fragment | NA | NA | 0 |
| S8_vs_k141_182043\|\|full | 2139 | No | NA | 2 | 0 | 1 | Not-determined | Genome-fragment | NA | NA | 0 |
| S8_vs_k141_16552\|\|full | 3822 | No | NA | 3 | 0 | 3 | Not-determined | Genome-fragment | NA | NA | 0 |
| S1_vs_k141_38622\|\|full | 26129 | No | NA | 35 | 5 | 1 | Low-quality | Genome-fragment | 19.41 | AAI-based (high-confidence) | 0 |
| S16_extracted_5250 | 26283 | No | NA | 45 | 5 | 0 | Low-quality | Genome-fragment | 17.62 | AAI-based (high-confidence) | 0 |
| S8_vs_k141_163962\|\|full | 1690 | No | NA | 3 | 0 | 0 | Not-determined | Genome-fragment | NA | NA | 0 |
| S9_vs_k141_60649\|\|full | 26335 | No | NA | 29 | 5 | 1 | Low-quality | Genome-fragment | 48.35 | AAI-based (high-confidence) | 0 |
| S8_vs_k141_182093\|\|full | 1155 | No | NA | 3 | 0 | 1 | Not-determined | Genome-fragment | NA | NA | 0 |
| S8_vs_k141_207334\|\|full | 1541 | No | NA | 3 | 0 | 1 | Not-determined | Genome-fragment | NA | NA | 0 |
| S16_extracted_32772 | 26515 | No | NA | 40 | 8 | 2 | Low-quality | Genome-fragment | 32.88 | AAI-based (high-confidence) | 0 |
| S13_vs_148640\|\|0_partial | 26597 | No | NA | 36 | 1 | 12 | Low-quality | Genome-fragment | 39.79 | HMM-based (lower-bound) | 0 |
| S17_extracted_k141_127748 | 26633 | No | NA | 27 | 15 | 0 | Low-quality | Genome-fragment | 40.14 | AAI-based (high-confidence) | 0 |
| S8_vs_k141_68687\|\|full | 6965 | No | NA | 4 | 0 | 2 | Not-determined | Genome-fragment | NA | NA | 0 |
| S16_vs_119992\|\|full | 26634 | No | NA | 29 | 3 | 1 | Low-quality | Genome-fragment | 44.5 | AAI-based (medium-confidence) | 0 |
| S15_vs_40291\|\|full | 26661 | Yes | 13768 | 38 | 7 | 7 | Low-quality | Genome-fragment | 25.77 | AAI-based (high-confidence) | 48.36 |
| S3_vs_k141_85380\|\|full | 26747 | No | NA | 31 | 8 | 0 | Low-quality | Genome-fragment | 20.01 | AAI-based (high-confidence) | 0 |
| S10_vs_13807\|\|full | 26891 | No | NA | 38 | 1 | 12 | Low-quality | Genome-fragment | 15.9 | HMM-based (lower-bound) | 0 |
| S6_vs_k141_30015\|\|full | 26900 | Yes | 8809 | 29 | 3 | 15 | Low-quality | Genome-fragment | 16.76 | HMM-based (lower-bound) | 67.25 |
| S8_vs_k141_34539\|\|full | 2393 | No | NA | 4 | 0 | 2 | Not-determined | Genome-fragment | NA | NA | 0 |
| S16_vs_85316\|\|full | 26931 | No | NA | 40 | 13 | 0 | Low-quality | Genome-fragment | 46.12 | AAI-based (high-confidence) | 0 |
| S8_vs_k141_86292\|\|full | 2304 | No | NA | 3 | 0 | 0 | Not-determined | Genome-fragment | NA | NA | 0 |
| S8_vs_k141_155407\|\|full | 1507 | No | NA | 4 | 0 | 2 | Not-determined | Genome-fragment | NA | NA | 0 |
| S7_extracted_94949 | 26939 | No | NA | 67 | 2 | 0 | Low-quality | Genome-fragment | 16.29 | AAI-based (high-confidence) | 0 |
| S16_vs_102633\|\|0_partial | 26952 | Yes | 13776 | 16 | 2 | 2 | Low-quality | Genome-fragment | 18.24 | HMM-based (lower-bound) | 48.89 |
| S8_vs_k141_77731\|\|full | 1311 | No | NA | 2 | 0 | 0 | Not-determined | Genome-fragment | NA | NA | 0 |
| S8_vs_k141_68710\|\|full | 3480 | No | NA | 4 | 0 | 1 | Not-determined | Genome-fragment | NA | NA | 0 |
| S13_extracted_105454 | 26996 | Yes | 24404 | 31 | 6 | 3 | Low-quality | Genome-fragment | 18.18 | AAI-based (high-confidence) | 9.6 |
| S8_vs_k141_182122\|\|full | 1313 | No | NA | 3 | 0 | 0 | Not-determined | Genome-fragment | NA | NA | 0 |
| S8_vs_k141_155418\|\|full | 2331 | No | NA | 3 | 0 | 1 | Not-determined | Genome-fragment | NA | NA | 0 |
| S8_vs_k141_147178\|\|full | 2961 | No | NA | 3 | 0 | 1 | Not-determined | Genome-fragment | NA | NA | 0 |
| S8_vs_k141_42866\|\|full | 1370 | No | NA | 3 | 0 | 1 | Not-determined | Genome-fragment | NA | NA | 0 |
| S5_vs_k141_135844\|\|full | 27012 | No | NA | 28 | 0 | 12 | Low-quality | Genome-fragment | 9.43 | AAI-based (medium-confidence) | 0 |
| S8_vs_k141_189578\|\|full | 1219 | No | NA | 3 | 0 | 1 | Not-determined | Genome-fragment | NA | NA | 0 |
| S8_vs_k141_182136\|\|full | 1655 | No | NA | 3 | 0 | 0 | Not-determined | Genome-fragment | NA | NA | 0 |
| S3_vs_k141_59142\|\|full | 27048 | No | NA | 50 | 12 | 0 | Low-quality | Genome-fragment | 47.87 | AAI-based (high-confidence) | 0 |
| S10_extracted_58078 | 27111 | No | NA | 32 | 20 | 0 | Low-quality | Genome-fragment | 44.49 | AAI-based (high-confidence) | 0 |
| S16_extracted_31401 | 27215 | No | NA | 23 | 1 | 12 | Low-quality | Genome-fragment | 2.19 | HMM-based (lower-bound) | 0 |
| S8_extracted_k141_58141 | 27297 | No | NA | 50 | 12 | 1 | Low-quality | Genome-fragment | 48.89 | AAI-based (high-confidence) | 0 |

| | | | | | | | | | | | |
|---|---|---|---|---|---|---|---|---|---|---|---|
| S8_vs_k141_68736\|\|full | 1819 | No | NA | 3 | 0 | 2 | Not-determined | Genome-fragment | NA | NA | 0 |
| S8_vs_k141_199576\|\|full | 1444 | No | NA | 2 | 0 | 1 | Not-determined | Genome-fragment | NA | NA | 0 |
| S8_vs_k141_129759\|\|full | 1057 | No | NA | 2 | 0 | 1 | Not-determined | Genome-fragment | NA | NA | 0 |
| S8_vs_k141_16643\|\|full | 1170 | No | NA | 2 | 0 | 1 | Not-determined | Genome-fragment | NA | NA | 0 |
| S8_vs_k141_60671\|\|full | 1401 | No | NA | 2 | 0 | 0 | Not-determined | Genome-fragment | NA | NA | 0 |
| S16_vs_2864\|\|full | 27398 | Yes | 9964 | 27 | 4 | 12 | Low-quality | Genome-fragment | 23.96 | AAI-based (medium-confidence) | 63.63 |
| S10_vs_73341\|\|full | 27472 | No | NA | 27 | 3 | 1 | Low-quality | Genome-fragment | 39.68 | AAI-based (medium-confidence) | 0 |
| S8_vs_k141_94916\|\|full | 3625 | No | NA | 7 | 0 | 3 | Not-determined | Genome-fragment | NA | NA | 0 |
| S8_vs_k141_8407\|\|full | 2176 | No | NA | 4 | 0 | 2 | Not-determined | Genome-fragment | NA | NA | 0 |
| S8_vs_k141_34580\|\|full | 6759 | No | NA | 8 | 0 | 4 | Not-determined | Genome-fragment | NA | NA | 0 |
| S8_vs_k141_16667\|\|full | 3144 | No | NA | 5 | 0 | 3 | Not-determined | Genome-fragment | NA | NA | 0 |
| S14_vs_k141_72262\|\|0_partial | 27532 | No | NA | 27 | 1 | 6 | Low-quality | Genome-fragment | 21.1 | HMM-based (lower-bound) | 0 |
| S8_vs_k141_164035\|\|full | 4625 | No | NA | 5 | 0 | 1 | Not-determined | Genome-fragment | NA | NA | 0 |
| S16_vs_112895\|\|full | 27645 | No | NA | 29 | 20 | 0 | Low-quality | Genome-fragment | 48.8 | AAI-based (high-confidence) | 0 |
| S8_vs_k141_207428\|\|full | 3340 | No | NA | 3 | 0 | 2 | Not-determined | Genome-fragment | NA | NA | 0 |
| S6_vs_k141_35840\|\|full | 27658 | Yes | 14251 | 22 | 5 | 5 | Low-quality | Genome-fragment | 5.2 | HMM-based (lower-bound) | 48.47 |
| S8_vs_k141_16690\|\|full | 2755 | No | NA | 3 | 0 | 1 | Not-determined | Genome-fragment | NA | NA | 0 |
| S7_vs_52264\|\|full | 27693 | No | NA | 25 | 2 | 6 | Low-quality | Genome-fragment | 20.33 | HMM-based (lower-bound) | 0 |
| S8_vs_k141_77827\|\|full | 2491 | No | NA | 3 | 0 | 1 | Not-determined | Genome-fragment | NA | NA | 0 |
| S8_vs_k141_155507\|\|full | 4634 | No | NA | 5 | 0 | 1 | Not-determined | Genome-fragment | NA | NA | 0 |
| S17_vs_k141_145557\|\|full | 27694 | No | NA | 32 | 9 | 1 | Low-quality | Genome-fragment | 42.85 | AAI-based (high-confidence) | 0 |
| S10_vs_112631\|\|full | 27746 | No | NA | 21 | 0 | 15 | Low-quality | Genome-fragment | 14.93 | AAI-based (medium-confidence) | 0 |
| S8_vs_k141_207441\|\|full | 18913 | No | NA | 19 | 0 | 12 | Not-determined | Genome-fragment | NA | NA | 0 |
| S8_extracted_k141_460 | 28054 | No | NA | 60 | 2 | 0 | Low-quality | Genome-fragment | 22.41 | AAI-based (high-confidence) | 0 |
| S11_vs_k141_47316\|\|full | 28074 | No | NA | 33 | 4 | 0 | Low-quality | Genome-fragment | 40.6 | AAI-based (medium-confidence) | 0 |
| S16_extracted_97619 | 28246 | No | NA | 38 | 4 | 0 | Low-quality | Genome-fragment | 23.93 | AAI-based (high-confidence) | 0 |
| S8_vs_k141_182222\|\|full | 5031 | No | NA | 7 | 0 | 5 | Not-determined | Genome-fragment | NA | NA | 0 |
| S7_vs_80749\|\|full | 28481 | No | NA | 32 | 1 | 10 | Low-quality | Genome-fragment | 42.23 | HMM-based (lower-bound) | 0 |
| S7_vs_51416\|\|full | 28533 | No | NA | 38 | 2 | 3 | Low-quality | Genome-fragment | 19.12 | AAI-based (medium-confidence) | 0 |
| S8_vs_k141_8477\|\|full | 2039 | No | NA | 4 | 0 | 0 | Not-determined | Genome-fragment | NA | NA | 0 |
| S8_vs_k141_60736\|\|full | 5296 | No | NA | 3 | 0 | 1 | Not-determined | Genome-fragment | NA | NA | 0 |
| S8_vs_k141_86425\|\|full | 9205 | No | NA | 9 | 0 | 6 | Not-determined | Genome-fragment | NA | NA | 0 |
| S9_extracted_k141_39660 | 28589 | No | NA | 45 | 5 | 1 | Low-quality | Genome-fragment | 29.75 | AAI-based (high-confidence) | 0 |
| S8_vs_k141_207467\|\|full | 1596 | No | NA | 2 | 0 | 0 | Not-determined | Genome-fragment | NA | NA | 0 |
| S5_vs_k141_46047\|\|0_partial | 28633 | Yes | 15186 | 27 | 4 | 6 | Low-quality | Genome-fragment | 36.5 | AAI-based (medium-confidence) | 46.96 |
| S16_extracted_165719 | 28704 | Yes | 23461 | 39 | 14 | 3 | Low-quality | Genome-fragment | 48.28 | AAI-based (high-confidence) | 18.27 |
| S8_vs_k141_95018\|\|full | 2894 | No | NA | 2 | 0 | 2 | Not-determined | Genome-fragment | NA | NA | 0 |
| S6_vs_k141_6271\|\|full | 28736 | Yes | 3986 | 26 | 2 | 8 | Low-quality | Genome-fragment | 3.59 | HMM-based (lower-bound) | 86.13 |
| S11_vs_k141_1692\|\|full | 28752 | No | NA | 41 | 9 | 0 | Low-quality | Genome-fragment | 48.87 | AAI-based (high-confidence) | 0 |
| S5_extracted_k141_128217 | 28783 | No | NA | 27 | 1 | 6 | Low-quality | Genome-fragment | 15.61 | HMM-based (lower-bound) | 0 |
| S16_vs_56685\|\|full | 28882 | Yes | 14158 | 36 | 6 | 13 | Low-quality | Genome-fragment | 29.77 | HMM-based (lower-bound) | 50.98 |
| S15_vs_32352\|\|full | 28927 | Yes | 18379 | 34 | 6 | 4 | Low-quality | Genome-fragment | 38.99 | HMM-based (lower-bound) | 36.46 |
| S12_vs_k141_129490\|\|full | 29011 | Yes | 8609 | 31 | 5 | 10 | Low-quality | Genome-fragment | 16.94 | HMM-based (lower-bound) | 70.33 |
| S16_vs_54071\|\|full | 29049 | No | NA | 51 | 16 | 0 | Low-quality | Genome-fragment | 48.1 | AAI-based (high-confidence) | 0 |
| S8_vs_k141_68916\|\|full | 3249 | No | NA | 4 | 0 | 2 | Not-determined | Genome-fragment | NA | NA | 0 |
| S8_vs_k141_50857\|\|full | 1274 | No | NA | 2 | 0 | 0 | Not-determined | Genome-fragment | NA | NA | 0 |
| S8_vs_k141_121149\|\|full | 1770 | No | NA | 4 | 0 | 0 | Not-determined | Genome-fragment | NA | NA | 0 |
| S3_extracted_k141_44954 | 29143 | No | NA | 20 | 5 | 0 | Low-quality | Genome-fragment | 17.21 | AAI-based (high-confidence) | 0 |
| S16_vs_143298\|\|0_partial | 29157 | No | NA | 28 | 1 | 5 | Low-quality | Genome-fragment | 26.19 | AAI-based (medium-confidence) | 0 |
| S7_extracted_3110 | 29179 | No | NA | 39 | 6 | 1 | Low-quality | Genome-fragment | 19.8 | AAI-based (high-confidence) | 0 |
| S8_vs_k141_138523\|\|full | 2202 | No | NA | 2 | 0 | 2 | Not-determined | Genome-fragment | NA | NA | 0 |
| S8_vs_k141_8607\|\|full | 2952 | No | NA | 4 | 0 | 3 | Not-determined | Genome-fragment | NA | NA | 0 |
| S16_vs_162767\|\|full | 29269 | No | NA | 27 | 1 | 2 | Low-quality | Genome-fragment | 26.41 | AAI-based (high-confidence) | 0 |
| S8_vs_k141_68953\|\|full | 2526 | No | NA | 3 | 0 | 1 | Not-determined | Genome-fragment | NA | NA | 0 |
| S11_extracted_k141_46426 | 29385 | No | NA | 30 | 3 | 1 | Low-quality | Genome-fragment | 37.41 | AAI-based (high-confidence) | 0 |
| S8_vs_k141_164218\|\|full | 1577 | No | NA | 4 | 0 | 0 | Not-determined | Genome-fragment | NA | NA | 0 |
| S5_vs_k141_144748\|\|full | 29463 | No | NA | 24 | 1 | 8 | Low-quality | Genome-fragment | 23.58 | HMM-based (lower-bound) | 0 |
| S8_vs_k141_103748\|\|full | 1627 | No | NA | 3 | 0 | 1 | Not-determined | Genome-fragment | NA | NA | 0 |
| S5_vs_k141_75320\|\|full | 29713 | Yes | 11412 | 35 | 6 | 15 | Low-quality | Genome-fragment | 21.36 | AAI-based (medium-confidence) | 61.59 |
| S16_vs_110480\|\|full | 29779 | Yes | 23062 | 36 | 10 | 3 | Low-quality | Genome-fragment | 37.58 | AAI-based (high-confidence) | 22.56 |
| S14_vs_k141_48351\|\|full | 29857 | Yes | 18264 | 28 | 2 | 6 | Low-quality | Genome-fragment | 23.44 | AAI-based (medium-confidence) | 38.83 |
| S8_vs_k141_121250\|\|full | 1068 | No | NA | 3 | 0 | 0 | Not-determined | Genome-fragment | NA | NA | 0 |
| S8_vs_k141_189811\|\|full | 1646 | No | NA | 3 | 0 | 0 | Not-determined | Genome-fragment | NA | NA | 0 |
| S8_vs_k141_147449\|\|full | 1487 | No | NA | 2 | 0 | 0 | Not-determined | Genome-fragment | NA | NA | 0 |
| S8_vs_k141_95243\|\|full | 2194 | No | NA | 4 | 0 | 1 | Not-determined | Genome-fragment | NA | NA | 0 |
| S8_vs_k141_173430\|\|full | 2140 | No | NA | 5 | 0 | 4 | Not-determined | Genome-fragment | NA | NA | 0 |
| S16_vs_119059\|\|full | 29888 | Yes | 19121 | 29 | 6 | 9 | Low-quality | Genome-fragment | 42.88 | AAI-based (high-confidence) | 36.02 |
| S8_vs_k141_189845\|\|full | 1015 | No | NA | 3 | 0 | 1 | Not-determined | Genome-fragment | NA | NA | 0 |
| S8_vs_k141_155732\|\|full | 10896 | No | NA | 7 | 0 | 5 | Not-determined | Genome-fragment | NA | NA | 0 |
| S16_vs_121030\|\|full | 30080 | No | NA | 39 | 19 | 0 | Low-quality | Genome-fragment | 49.52 | AAI-based (high-confidence) | 0 |
| S8_vs_k141_130113\|\|full | 1507 | No | NA | 3 | 0 | 1 | Not-determined | Genome-fragment | NA | NA | 0 |
| S8_extracted_k141_192408 | 30246 | No | NA | 50 | 1 | 0 | Low-quality | Genome-fragment | 14.62 | HMM-based (lower-bound) | 0 |
| S8_vs_k141_16989\|\|full | 1004 | No | NA | 2 | 0 | 1 | Not-determined | Genome-fragment | NA | NA | 0 |
| S10_extracted_101000 | 30643 | No | NA | 29 | 11 | 0 | Low-quality | Genome-fragment | 20.19 | AAI-based (medium-confidence) | 0 |
| S8_vs_k141_86699\|\|full | 6322 | No | NA | 4 | 0 | 2 | Not-determined | Genome-fragment | NA | NA | 0 |
| S8_vs_k141_121351\|\|full | 1042 | No | NA | 2 | 0 | 0 | Not-determined | Genome-fragment | NA | NA | 0 |
| S8_vs_k141_155805\|\|full | 3344 | No | NA | 2 | 0 | 2 | Not-determined | Genome-fragment | NA | NA | 0 |
| S15_vs_53150\|\|full | 30906 | Yes | 20552 | 39 | 10 | 7 | Low-quality | Genome-fragment | 31.87 | AAI-based (high-confidence) | 33.5 |
| S8_vs_k141_69146\|\|full | 2443 | No | NA | 3 | 0 | 2 | Not-determined | Genome-fragment | NA | NA | 0 |
| S8_vs_k141_51129\|\|full | 1538 | No | NA | 2 | 0 | 0 | Not-determined | Genome-fragment | NA | NA | 0 |
| S8_vs_k141_17007\|\|full | 6423 | No | NA | 6 | 0 | 4 | Not-determined | Genome-fragment | NA | NA | 0 |
| S10_extracted_73144 | 30971 | No | NA | 27 | 10 | 0 | Low-quality | Genome-fragment | 20.13 | AAI-based (medium-confidence) | 0 |
| S3_vs_k141_81867\|\|full | 31029 | Yes | 9475 | 41 | 5 | 13 | Low-quality | Genome-fragment | 17.58 | HMM-based (lower-bound) | 69.46 |
| S7_extracted_89258 | 31464 | No | NA | 53 | 10 | 1 | Low-quality | Genome-fragment | 19.54 | AAI-based (high-confidence) | 0 |
| S17_vs_k141_89792\|\|full | 31489 | No | NA | 46 | 6 | 3 | Low-quality | Genome-fragment | 46.01 | HMM-based (lower-bound) | 0 |
| S8_vs_k141_95392\|\|full | 1217 | No | NA | 2 | 0 | 0 | Not-determined | Genome-fragment | NA | NA | 0 |
| S8_vs_k141_155840\|\|full | 8031 | No | NA | 10 | 0 | 8 | Not-determined | Genome-fragment | NA | NA | 0 |
| S9_vs_k141_37919\|\|full | 31564 | Yes | 17383 | 25 | 6 | 6 | Low-quality | Genome-fragment | 25.15 | AAI-based (high-confidence) | 44.93 |
| S11_vs_k141_63209\|\|0_partial | 31629 | Yes | 10268 | 34 | 4 | 14 | Low-quality | Genome-fragment | 10.77 | HMM-based (lower-bound) | 67.54 |
| S8_vs_k141_69226\|\|full | 1250 | No | NA | 3 | 0 | 0 | Not-determined | Genome-fragment | NA | NA | 0 |
| S7_vs_20717\|\|full | 31778 | No | NA | 29 | 1 | 13 | Low-quality | Genome-fragment | 48.08 | HMM-based (lower-bound) | 0 |
| S8_vs_k141_121472\|\|full | 1053 | No | NA | 2 | 0 | 1 | Not-determined | Genome-fragment | NA | NA | 0 |
| S3_vs_k141_58326\|\|full | 31992 | Yes | 11598 | 42 | 5 | 12 | Low-quality | Genome-fragment | 15.76 | HMM-based (lower-bound) | 63.75 |
| S15_vs_41946\|\|full | 32129 | No | NA | 24 | 1 | 16 | Low-quality | Genome-fragment | 45.52 | HMM-based (lower-bound) | 0 |
| S8_vs_k141_164443\|\|full | 1245 | No | NA | 3 | 0 | 1 | Not-determined | Genome-fragment | NA | NA | 0 |
| S8_vs_k141_43257\|\|full | 7239 | No | NA | 11 | 0 | 2 | Not-determined | Genome-fragment | NA | NA | 0 |
| S13_extracted_8315 | 32520 | No | NA | 28 | 8 | 0 | Low-quality | Genome-fragment | 24.19 | AAI-based (high-confidence) | 0 |
| S16_extracted_7056 | 33331 | No | NA | 55 | 3 | 1 | Low-quality | Genome-fragment | 23.6 | AAI-based (high-confidence) | 0 |
| S6_vs_k141_38776\|\|full | 33463 | No | NA | 33 | 1 | 16 | Low-quality | Genome-fragment | 22.49 | HMM-based (lower-bound) | 0 |
| S7_vs_23557\|\|full | 34197 | No | NA | 46 | 6 | 4 | Low-quality | Genome-fragment | 39.41 | HMM-based (lower-bound) | 0 |
| S17_extracted_k141_163533 | 34298 | No | NA | 65 | 10 | 0 | Low-quality | Genome-fragment | 43.6 | AAI-based (high-confidence) | 0 |
| S8_extracted_k141_183339 | 34387 | No | NA | 49 | 5 | 2 | Low-quality | Genome-fragment | 27.45 | AAI-based (high-confidence) | 0 |
| S8_vs_k141_69370\|\|full | 3369 | No | NA | 4 | 0 | 1 | Not-determined | Genome-fragment | NA | NA | 0 |
| S5_vs_k141_79370\|\|full | 34526 | Yes | 30655 | 68 | 6 | 4 | Low-quality | Genome-fragment | 38.7 | AAI-based (medium-confidence) | 11.21 |
| S8_vs_k141_69387\|\|full | 1075 | No | NA | 3 | 0 | 1 | Not-determined | Genome-fragment | NA | NA | 0 |
| S16_vs_80873\|\|full | 34544 | No | NA | 34 | 5 | 1 | Low-quality | Genome-fragment | 13.97 | AAI-based (medium-confidence) | 0 |
| S8_vs_k141_164598\|\|full | 1942 | No | NA | 3 | 0 | 0 | Not-determined | Genome-fragment | NA | NA | 0 |
| S12_extracted_k141_49398 | 34746 | No | NA | 34 | 17 | 0 | Low-quality | Genome-fragment | 25.43 | AAI-based (medium-confidence) | 0 |
| S7_vs_17785\|\|full | 34788 | No | NA | 45 | 17 | 1 | Low-quality | Genome-fragment | 46.39 | HMM-based (lower-bound) | 0 |
| S9_extracted_k141_45682 | 34829 | No | NA | 28 | 17 | 0 | Low-quality | Genome-fragment | 22.36 | AAI-based (medium-confidence) | 0 |
| S8_vs_k141_156045\|\|full | 2068 | No | NA | 2 | 0 | 2 | Not-determined | Genome-fragment | NA | NA | 0 |
| S8_vs_k141_138835\|\|full | 7065 | No | NA | 12 | 0 | 2 | Not-determined | Genome-fragment | NA | NA | 0 |
| S8_vs_k141_125031\|\|full | 34907 | No | NA | 41 | 1 | 6 | Low-quality | Genome-fragment | 31.45 | AAI-based (medium-confidence) | 0 |
| S16_extracted_118678 | 34996 | No | NA | 35 | 8 | 1 | Low-quality | Genome-fragment | 21.06 | AAI-based (high-confidence) | 0 |
| S16_extracted_162486 | 35495 | No | NA | 27 | 10 | 1 | Low-quality | Genome-fragment | 33.39 | AAI-based (high-confidence) | 0 |
| S8_vs_k141_190302\|\|full | 1230 | No | NA | 3 | 0 | 0 | Not-determined | Genome-fragment | NA | NA | 0 |
| S8_vs_k141_207868\|\|full | 1084 | No | NA | 1 | 0 | 0 | Not-determined | Genome-fragment | NA | NA | 0 |
| S8_vs_k141_104032\|\|full | 3078 | No | NA | 5 | 0 | 0 | Not-determined | Genome-fragment | NA | NA | 0 |
| S8_vs_k141_164700\|\|full | 1537 | No | NA | 3 | 0 | 0 | Not-determined | Genome-fragment | NA | NA | 0 |
| S8_vs_k141_156087\|\|full | 1247 | No | NA | 2 | 0 | 1 | Not-determined | Genome-fragment | NA | NA | 0 |
| S13_extracted_84148 | 35534 | No | NA | 39 | 7 | 2 | Low-quality | Genome-fragment | 26.47 | AAI-based (high-confidence) | 0 |
| S8_vs_k141_190331\|\|full | 1924 | No | NA | 2 | 0 | 2 | Not-determined | Genome-fragment | NA | NA | 0 |
| S17_vs_k141_57702\|\|full | 35897 | Yes | 12631 | 49 | 5 | 18 | Low-quality | Genome-fragment | 33.87 | AAI-based (medium-confidence) | 64.81 |
| S15_extracted_8485 | 35950 | No | NA | 34 | 6 | 1 | Low-quality | Genome-fragment | 37.92 | AAI-based (high-confidence) | 0 |
| S15_extracted_7130 | 36448 | No | NA | 60 | 5 | 1 | Low-quality | Genome-fragment | 38.46 | AAI-based (high-confidence) | 0 |
| S8_vs_k141_156141\|\|full | 1109 | No | NA | 3 | 0 | 0 | Not-determined | Genome-fragment | NA | NA | 0 |
| S8_vs_k141_164754\|\|full | 1175 | No | NA | 2 | 0 | 0 | Not-determined | Genome-fragment | NA | NA | 0 |
| S8_vs_k141_17231\|\|full | 2822 | No | NA | 4 | 0 | 4 | Not-determined | Genome-fragment | NA | NA | 0 |
| S16_vs_53197\|\|full | 36661 | No | NA | 44 | 15 | 1 | Low-quality | Genome-fragment | 22.98 | AAI-based (high-confidence) | 0 |
| S4_vs_VIRSorter_23724-cat_2 | 37015 | No | NA | 55 | 7 | 1 | Low-quality | Genome-fragment | 39.37 | AAI-based (high-confidence) | 0 |
| S8_vs_k141_190519\|\|full | 3071 | No | NA | 4 | 0 | 3 | Not-determined | Genome-fragment | NA | NA | 0 |
| S7_vs_8597\|\|full | 37161 | Yes | 16057 | 43 | 5 | 7 | Low-quality | Genome-fragment | 42.83 | AAI-based (high-confidence) | 56.79 |
| S12_vs_k141_167336\|\|full | 37181 | No | NA | 53 | 0 | 5 | Low-quality | Genome-fragment | 27.53 | AAI-based (medium-confidence) | 0 |
| S8_vs_k141_51566\|\|full | 1542 | No | NA | 2 | 0 | 1 | Not-determined | Genome-fragment | NA | NA | 0 |
| S5_extracted_k141_110500 | 37214 | No | NA | 41 | 1 | 12 | Low-quality | Genome-fragment | 18.25 | HMM-based (lower-bound) | 0 |
| S12_vs_k141_62940\|\|full | 37268 | Yes | 33748 | 55 | 8 | 2 | Low-quality | Genome-fragment | 46.18 | HMM-based (lower-bound) | 9.45 |
| S8_vs_k141_190619\|\|full | 1763 | No | NA | 2 | 0 | 1 | Not-determined | Genome-fragment | NA | NA | 0 |
| S8_vs_k141_51819\|\|full | 1100 | No | NA | 2 | 0 | 1 | Not-determined | Genome-fragment | NA | NA | 0 |
| S11_vs_k141_37914\|\|full | 37606 | Yes | 10744 | 40 | 5 | 19 | Low-quality | Genome-fragment | 30.91 | AAI-based (medium-confidence) | 71.43 |
| S3_vs_k141_126959\|\|full | 37730 | Yes | 23235 | 45 | 10 | 11 | Low-quality | Genome-fragment | 45.46 | HMM-based (lower-bound) | 38.42 |
| S7_vs_19412\|\|full | 37966 | Yes | 30182 | 40 | 3 | 9 | Low-quality | Genome-fragment | 42.92 | AAI-based (medium-confidence) | 20.5 |
| S8_vs_k141_52054\|\|full | 11547 | No | NA | 16 | 0 | 13 | Not-determined | Genome-fragment | NA | NA | 0 |
| S7_extracted_61175 | 38005 | No | NA | 62 | 5 | 4 | Low-quality | Genome-fragment | 22.26 | AAI-based (high-confidence) | 0 |
| S8_vs_k141_35390\|\|0_partial | 30637 | No | NA | 23 | 0 | 18 | Not-determined | Genome-fragment | NA | NA | 0 |
| S13_vs_12830\|\|full | 38028 | No | NA | 62 | 19 | 1 | Low-quality | Genome-fragment | 45.56 | AAI-based (high-confidence) | 0 |
| S13_extracted_12330 | 38274 | No | NA | 26 | 1 | 7 | Low-quality | Genome-fragment | 21.95 | HMM-based (lower-bound) | 0 |
| S8_vs_k141_62364\|\|0_partial | 13084 | No | NA | 14 | 0 | 4 | Not-determined | Genome-fragment | NA | NA | 0 |
| S2_vs_k141_3914\|\|full | 38307 | Yes | 27363 | 46 | 7 | 3 | Low-quality | Genome-fragment | 38.84 | HMM-based (lower-bound) | 28.57 |
| S6_vs_k141_34279\|\|full | 38335 | Yes | 30591 | 45 | 13 | 8 | Low-quality | Genome-fragment | 46.56 | HMM-based (lower-bound) | 20.2 |
| S9_extracted_k141_12293 | 38474 | No | NA | 31 | 0 | 28 | Low-quality | Genome-fragment | 13.55 | AAI-based (medium-confidence) | 0 |
| S8_vs_k141_2668\|\|0_partial | 11651 | No | NA | 11 | 0 | 9 | Not-determined | Genome-fragment | NA | NA | 0 |
| S8_vs_k141_11189\|\|full | 38851 | Yes | 26180 | 45 | 6 | 9 | Low-quality | Genome-fragment | 49.01 | AAI-based (medium-confidence) | 32.61 |
| S8_vs_k141_120627\|\|full | 39189 | No | NA | 35 | 9 | 0 | Low-quality | Genome-fragment | 35.3 | AAI-based (medium-confidence) | 0 |
| S8_vs_k141_29825\|\|0_partial | 10990 | No | NA | 12 | 0 | 9 | Not-determined | Genome-fragment | NA | NA | 0 |
| S8_extracted_k141_64313 | 39803 | No | NA | 41 | 11 | 1 | Low-quality | Genome-fragment | 28.98 | AAI-based (medium-confidence) | 0 |
| S12_vs_k141_124042\|\|full | 39972 | Yes | 13614 | 35 | 4 | 12 | Low-quality | Genome-fragment | 24.93 | HMM-based (lower-bound) | 65.94 |
| S8_vs_k141_185981\|\|0_partial | 13735 | No | NA | 17 | 0 | 10 | Not-determined | Genome-fragment | NA | NA | 0 |
| S5_extracted_k141_137325 | 40094 | No | NA | 48 | 12 | 0 | Low-quality | Genome-fragment | 33.97 | AAI-based (high-confidence) | 0 |
| S4_extracted_20987 | 40120 | No | NA | 38 | 12 | 0 | Low-quality | Genome-fragment | 27.45 | AAI-based (high-confidence) | 0 |
| S8_extracted_k141_122972 | 40510 | No | NA | 55 | 8 | 0 | Low-quality | Genome-fragment | 25.17 | AAI-based (high-confidence) | 0 |
| S11_vs_k141_60428\|\|full | 40617 | No | NA | 64 | 12 | 2 | Low-quality | Genome-fragment | 25.26 | AAI-based (high-confidence) | 0 |
| S12_extracted_k141_28003 | 41890 | No | NA | 69 | 12 | 0 | Low-quality | Genome-fragment | 25.49 | AAI-based (high-confidence) | 0 |
| S16_extracted_41661 | 42002 | No | NA | 34 | 18 | 0 | Low-quality | Genome-fragment | 35.57 | AAI-based (high-confidence) | 0 |
| S12_vs_k141_47540\|\|full | 42209 | Yes | 3614 | 49 | 3 | 20 | Low-quality | Genome-fragment | 5.11 | AAI-based (high-confidence) | 91.44 |
| S14_vs_k141_52308\|\|full | 42531 | Yes | 39448 | 47 | 5 | 4 | Low-quality | Genome-fragment | 48.93 | AAI-based (high-confidence) | 7.25 |
| S8_vs_k141_95659\|\|lt2gene | 1493 | No | NA | 2 | 0 | 0 | Not-determined | Genome-fragment | NA | NA | 0 |
| S8_extracted_k141_357 | 43276 | No | NA | 34 | 3 | 0 | Low-quality | Genome-fragment | 45.47 | AAI-based (high-confidence) | 0 |
| S8_extracted_k141_47914 | 43357 | No | NA | 84 | 12 | 2 | Low-quality | Genome-fragment | 34.69 | AAI-based (high-confidence) | 0 |
| S12_extracted_k141_148065 | 43513 | No | NA | 41 | 19 | 0 | Low-quality | Genome-fragment | 36.57 | AAI-based (high-confidence) | 0 |
| S16_vs_67352\|\|full | 43778 | No | NA | 60 | 6 | 1 | Low-quality | Genome-fragment | 39.43 | AAI-based (medium-confidence) | 0 |
| S13_vs_104879\|\|full | 44008 | No | NA | 51 | 1 | 15 | Low-quality | Genome-fragment | 11.58 | HMM-based (lower-bound) | 0 |
| S14_vs_k141_17325\|\|full | 44871 | Yes | 11148 | 35 | 6 | 17 | Low-quality | Genome-fragment | 14.67 | HMM-based (lower-bound) | 75.16 |
| S3_extracted_k141_97741 | 44923 | No | NA | 83 | 9 | 0 | Low-quality | Genome-fragment | 27.93 | AAI-based (high-confidence) | 0 |
| S8_vs_k141_27234\|\|lt2gene | 1012 | No | NA | 2 | 0 | 1 | Not-determined | Genome-fragment | NA | NA | 0 |
| S8_extracted_k141_99358 | 45147 | No | NA | 46 | 10 | 1 | Low-quality | Genome-fragment | 45.2 | AAI-based (high-confidence) | 0 |
| S10_vs_16005\|\|full | 45426 | No | NA | 66 | 13 | 3 | Low-quality | Genome-fragment | 28.26 | AAI-based (high-confidence) | 0 |
| S8_vs_k141_44578\|\|lt2gene | 2678 | No | NA | 1 | 0 | 1 | Not-determined | Genome-fragment | NA | NA | 0 |
| S7_extracted_81458 | 45921 | No | NA | 42 | 12 | 1 | Low-quality | Genome-fragment | 27.31 | AAI-based (medium-confidence) | 0 |
| S8_extracted_k141_131551 | 46265 | No | NA | 67 | 12 | 0 | Low-quality | Genome-fragment | 46.73 | AAI-based (high-confidence) | 0 |
| S5_vs_k141_19188\|\|full | 46306 | No | NA | 52 | 1 | 8 | Low-quality | Genome-fragment | 18.82 | HMM-based (lower-bound) | 0 |
| S13_extracted_150247 | 47158 | No | NA | 56 | 18 | 0 | Low-quality | Genome-fragment | 43.2 | AAI-based (high-confidence) | 0 |
| S17_extracted_k141_8382 | 47192 | No | NA | 52 | 0 | 48 | Low-quality | Genome-fragment | 23.99 | AAI-based (medium-confidence) | 0 |
| S8_vs_k141_54341\|\|lt2gene | 1385 | No | NA | 2 | 0 | 0 | Not-determined | Genome-fragment | NA | NA | 0 |
| S4_extracted_9769 | 47540 | No | NA | 61 | 14 | 1 | Low-quality | Genome-fragment | 30.22 | AAI-based (medium-confidence) | 0 |
| S8_vs_k141_80357\|\|lt2gene | 2046 | No | NA | 2 | 0 | 0 | Not-determined | Genome-fragment | NA | NA | 0 |
| S8_vs_k141_175750\|\|lt2gene | 1738 | No | NA | 1 | 0 | 0 | Not-determined | Genome-fragment | NA | NA | 0 |
| S8_vs_k141_132393\|\|lt2gene | 2112 | No | NA | 1 | 0 | 0 | Not-determined | Genome-fragment | NA | NA | 0 |
| S8_vs_k141_71340\|\|lt2gene | 1052 | No | NA | 1 | 0 | 0 | Not-determined | Genome-fragment | NA | NA | 0 |
| S14_extracted_k141_37090 | 48080 | No | NA | 51 | 2 | 17 | Low-quality | Genome-fragment | 32.83 | HMM-based (lower-bound) | 0 |
| S14_vs_k141_38235\|\|full | 48094 | No | NA | 49 | 17 | 2 | Low-quality | Genome-fragment | 36.47 | AAI-based (medium-confidence) | 0 |
| S8_vs_k141_97844\|\|lt2gene | 2584 | No | NA | 1 | 0 | 0 | Not-determined | Genome-fragment | NA | NA | 0 |
| S17_vs_k141_176606\|\|full | 48195 | Yes | 17313 | 43 | 7 | 10 | Low-quality | Genome-fragment | 48.37 | AAI-based (medium-confidence) | 64.08 |
| S17_vs_k141_63384\|\|full | 48935 | No | NA | 45 | 1 | 18 | Low-quality | Genome-fragment | 33.82 | HMM-based (lower-bound) | 0 |
| S8_extracted_k141_197122 | 49397 | No | NA | 41 | 5 | 0 | Low-quality | Genome-fragment | 49.77 | AAI-based (high-confidence) | 0 |
| S16_vs_118433\|\|full | 49513 | Yes | 39194 | 53 | 10 | 4 | Low-quality | Genome-fragment | 25.23 | AAI-based (high-confidence) | 20.84 |
| S8_vs_k141_3037\|\|lt2gene | 1332 | No | NA | 1 | 0 | 0 | Not-determined | Genome-fragment | NA | NA | 0 |
| S16_vs_26408\|\|full | 49569 | No | NA | 85 | 27 | 0 | Low-quality | Genome-fragment | 43.53 | AAI-based (high-confidence) | 0 |
| S16_extracted_1715 | 49711 | No | NA | 35 | 8 | 0 | Low-quality | Genome-fragment | 19.93 | AAI-based (high-confidence) | 0 |
| S9_vs_k141_60854\|\|full | 49942 | No | NA | 83 | 7 | 3 | Low-quality | Genome-fragment | 30.02 | AAI-based (high-confidence) | 0 |
| S5_vs_k141_118648\|\|full | 50053 | Yes | 7238 | 45 | 2 | 26 | Low-quality | Genome-fragment | 18.62 | AAI-based (medium-confidence) | 85.54 |
| S9_vs_k141_16635\|\|full | 51022 | Yes | 16375 | 54 | 8 | 9 | Low-quality | Genome-fragment | 38.82 | AAI-based (high-confidence) | 67.91 |
| S16_vs_25976\|\|full | 51282 | Yes | 13215 | 48 | 8 | 19 | Low-quality | Genome-fragment | 24.71 | HMM-based (lower-bound) | 74.23 |
| S14_vs_k141_48012\|\|full | 51580 | No | NA | 56 | 1 | 27 | Low-quality | Genome-fragment | 32.36 | HMM-based (lower-bound) | 0 |
| S8_vs_k141_133435\|\|lt2gene | 4421 | No | NA | 2 | 0 | 1 | Not-determined | Genome-fragment | NA | NA | 0 |
| S17_extracted_k141_145540 | 52253 | No | NA | 42 | 12 | 1 | Low-quality | Genome-fragment | 30.3 | AAI-based (high-confidence) | 0 |
| S8_vs_k141_168250\|\|lt2gene | 1031 | No | NA | 2 | 0 | 0 | Not-determined | Genome-fragment | NA | NA | 0 |
| S3_vs_k141_1357\|\|full | 53897 | Yes | 20373 | 68 | 6 | 13 | Low-quality | Genome-fragment | 25.27 | HMM-based (lower-bound) | 62.2 |
| S8_vs_k141_150726\|\|lt2gene | 1263 | No | NA | 1 | 0 | 0 | Not-determined | Genome-fragment | NA | NA | 0 |
| S8_vs_k141_21251\|\|lt2gene | 1022 | No | NA | 1 | 0 | 0 | Not-determined | Genome-fragment | NA | NA | 0 |
| S8_vs_k141_17608\|\|full | 54731 | Yes | 9678 | 58 | 5 | 25 | Low-quality | Genome-fragment | 20.29 | AAI-based (high-confidence) | 82.32 |
| S14_extracted_k141_45680 | 55322 | No | NA | 55 | 1 | 13 | Low-quality | Genome-fragment | 19.7 | HMM-based (lower-bound) | 0 |
| S3_extracted_k141_1184 | 55979 | No | NA | 70 | 13 | 0 | Low-quality | Genome-fragment | 38.27 | AAI-based (high-confidence) | 0 |
| S8_vs_k141_15240\|\|full | 56536 | Yes | 13482 | 48 | 12 | 19 | Low-quality | Genome-fragment | 31.68 | AAI-based (medium-confidence) | 76.15 |
| S10_vs_33943\|\|full | 58316 | No | NA | 96 | 15 | 0 | Low-quality | Genome-fragment | 35.78 | AAI-based (high-confidence) | 0 |
| S17_vs_k141_2294\|\|full | 58804 | Yes | 16182 | 59 | 14 | 24 | Low-quality | Genome-fragment | 44.41 | AAI-based (medium-confidence) | 72.48 |
| S6_vs_k141_1156\|\|full | 59458 | Yes | 24598 | 61 | 6 | 25 | Low-quality | Genome-fragment | 44.46 | HMM-based (lower-bound) | 58.63 |
| S3_vs_k141_99638\|\|full | 60212 | Yes | 33064 | 64 | 2 | 17 | Low-quality | Genome-fragment | 29.82 | AAI-based (medium-confidence) | 45.09 |
| S6_vs_k141_32125\|\|full | 61741 | No | NA | 63 | 2 | 27 | Low-quality | Genome-fragment | 25.39 | HMM-based (lower-bound) | 0 |
| S6_vs_k141_13728\|\|full | 62293 | No | NA | 74 | 3 | 15 | Low-quality | Genome-fragment | 48.74 | HMM-based (lower-bound) | 0 |
| S11_vs_k141_67166\|\|full | 62328 | Yes | 10781 | 63 | 4 | 38 | Low-quality | Genome-fragment | 14.24 | HMM-based (lower-bound) | 82.7 |
| S17_vs_k141_93952\|\|full | 65549 | Yes | 15238 | 85 | 10 | 19 | Low-quality | Genome-fragment | 30.13 | HMM-based (lower-bound) | 76.75 |
| S8_vs_k141_67230\|\|full | 66029 | No | NA | 67 | 15 | 1 | Low-quality | Genome-fragment | 37.6 | AAI-based (high-confidence) | 0 |
| S5_vs_k141_46296\|\|0_partial | 66760 | Yes | 49232 | 80 | 6 | 18 | Low-quality | Genome-fragment | 25.86 | HMM-based (lower-bound) | 26.26 |
| S6_vs_k141_19225\|\|full | 68262 | Yes | 12650 | 68 | 5 | 30 | Low-quality | Genome-fragment | 19.45 | HMM-based (lower-bound) | 81.47 |
| S8_vs_k141_19188\|\|full | 68392 | Yes | 24565 | 66 | 3 | 30 | Low-quality | Genome-fragment | 18.11 | HMM-based (lower-bound) | 64.08 |
| S9_extracted_k141_23590 | 68734 | No | NA | 94 | 13 | 4 | Low-quality | Genome-fragment | 44.13 | AAI-based (medium-confidence) | 0 |
| S9_vs_k141_59237\|\|full | 70068 | Yes | 32078 | 85 | 14 | 32 | Low-quality | Genome-fragment | 47.52 | HMM-based (lower-bound) | 54.22 |
| S8_vs_k141_83545\|\|lt2gene | 2145 | No | NA | 1 | 0 | 1 | Not-determined | Genome-fragment | NA | NA | 0 |
| S13_vs_44103\|\|full | 71732 | Yes | 4788 | 61 | 3 | 41 | Low-quality | Genome-fragment | 4.06 | HMM-based (lower-bound) | 93.33 |
| S6_vs_k141_14664\|\|full | 73483 | Yes | 39291 | 79 | 12 | 20 | Low-quality | Genome-fragment | 47.18 | HMM-based (lower-bound) | 46.53 |
| S13_extracted_58022 | 74146 | No | NA | 87 | 21 | 2 | Low-quality | Genome-fragment | 46.66 | AAI-based (high-confidence) | 0 |
| S17_vs_k141_176943\|\|full | 74469 | Yes | 31444 | 90 | 7 | 25 | Low-quality | Genome-fragment | 36.17 | HMM-based (lower-bound) | 57.78 |
| S8_vs_k141_144636\|\|lt2gene | 1568 | No | NA | 1 | 0 | 1 | Not-determined | Genome-fragment | NA | NA | 0 |
| S16_extracted_114510 | 75059 | No | NA | 78 | 4 | 0 | Low-quality | Genome-fragment | 18.72 | AAI-based (high-confidence) | 0 |
| S8_extracted_k141_92536 | 77195 | Yes | 3417 | 86 | 2 | 33 | Low-quality | Genome-fragment | 4.39 | HMM-based (lower-bound) | 95.57 |
| S8_vs_k141_58458\|\|lt2gene | 1538 | No | NA | 2 | 0 | 1 | Not-determined | Genome-fragment | NA | NA | 0 |
| S10_extracted_44505 | 77467 | No | NA | 86 | 1 | 10 | Low-quality | Genome-fragment | 35.94 | HMM-based (lower-bound) | 0 |
| S7_vs_94744\|\|full | 78218 | Yes | 24219 | 77 | 3 | 36 | Low-quality | Genome-fragment | 29.26 | HMM-based (lower-bound) | 69.04 |
| S17_vs_k141_106218\|\|full | 79477 | Yes | 31273 | 85 | 13 | 27 | Low-quality | Genome-fragment | 48.42 | HMM-based (lower-bound) | 60.65 |
| S16_vs_49947\|\|full | 79772 | Yes | 15492 | 69 | 5 | 17 | Low-quality | Genome-fragment | 36.17 | AAI-based (high-confidence) | 80.58 |
| S16_vs_98507\|\|full | 81243 | Yes | 26593 | 84 | 9 | 33 | Low-quality | Genome-fragment | 38.43 | HMM-based (lower-bound) | 67.27 |
| S6_vs_k141_1082\|\|full | 81978 | Yes | 18826 | 82 | 8 | 19 | Low-quality | Genome-fragment | 27.63 | HMM-based (lower-bound) | 77.04 |
| S12_vs_k141_72440\|\|full | 82708 | Yes | 29162 | 82 | 13 | 16 | Low-quality | Genome-fragment | 44.66 | HMM-based (lower-bound) | 64.74 |
| S16_vs_76645\|\|full | 84674 | Yes | 26660 | 112 | 17 | 52 | Low-quality | Genome-fragment | 47.15 | HMM-based (lower-bound) | 68.51 |
| S15_vs_41725\|\|full | 84882 | Yes | 11631 | 97 | 8 | 33 | Low-quality | Genome-fragment | 18.32 | AAI-based (high-confidence) | 86.3 |
| S8_vs_k141_157545\|\|full | 84917 | Yes | 32355 | 82 | 5 | 33 | Low-quality | Genome-fragment | 29.98 | HMM-based (lower-bound) | 61.9 |
| S3_vs_k141_125844\|\|full | 85228 | Yes | 51984 | 71 | 8 | 17 | Low-quality | Genome-fragment | 46.73 | AAI-based (medium-confidence) | 39.01 |
| S8_vs_k141_60613\|\|full | 87243 | No | NA | 83 | 2 | 39 | Low-quality | Genome-fragment | 48.46 | HMM-based (lower-bound) | 0 |
| S12_vs_k141_82177\|\|full | 89027 | Yes | 27382 | 85 | 6 | 30 | Low-quality | Genome-fragment | 40.01 | HMM-based (lower-bound) | 69.24 |
| S8_vs_k141_102446\|\|lt2gene | 1315 | No | NA | 2 | 0 | 0 | Not-determined | Genome-fragment | NA | NA | 0 |
| S12_vs_k141_186523\|\|full | 89646 | No | NA | 91 | 1 | 21 | Low-quality | Genome-fragment | 38.89 | HMM-based (lower-bound) | 0 |
| S8_vs_k141_24914\|\|lt2gene | 1265 | No | NA | 1 | 0 | 0 | Not-determined | Genome-fragment | NA | NA | 0 |
| S8_vs_k141_163404\|\|lt2gene | 1689 | No | NA | 1 | 0 | 1 | Not-determined | Genome-fragment | NA | NA | 0 |
| S8_vs_k141_33779\|\|lt2gene | 1017 | No | NA | 1 | 0 | 0 | Not-determined | Genome-fragment | NA | NA | 0 |
| S8_vs_k141_60123\|\|lt2gene | 1306 | No | NA | 1 | 0 | 1 | Not-determined | Genome-fragment | NA | NA | 0 |
| S8_vs_k141_16220\|\|lt2gene | 1865 | No | NA | 1 | 0 | 0 | Not-determined | Genome-fragment | NA | NA | 0 |
| S17_extracted_k141_164870 | 90117 | No | NA | 120 | 8 | 0 | Low-quality | Genome-fragment | 22.47 | AAI-based (high-confidence) | 0 |
| S3_vs_k141_90657\|\|full | 103390 | Yes | 22263 | 96 | 7 | 30 | Low-quality | Genome-fragment | 23.34 | HMM-based (lower-bound) | 78.47 |
| S13_vs_112313\|\|full | 104172 | Yes | 49680 | 115 | 3 | 35 | Low-quality | Genome-fragment | 42.21 | HMM-based (lower-bound) | 52.31 |
| S8_vs_k141_16814\|\|lt2gene | 2148 | No | NA | 2 | 0 | 0 | Not-determined | Genome-fragment | NA | NA | 0 |
| S13_vs_38692\|\|full | 122930 | Yes | 35190 | 130 | 12 | 52 | Low-quality | Genome-fragment | 43.07 | HMM-based (lower-bound) | 71.37 |
| S3_vs_k141_119996\|\|full | 163313 | Yes | 22218 | 157 | 14 | 87 | Low-quality | Genome-fragment | 36.18 | HMM-based (lower-bound) | 86.4 |
| S8_vs_k141_86619\|\|lt2gene | 1020 | No | NA | 2 | 0 | 0 | Not-determined | Genome-fragment | NA | NA | 0 |
| S5_vs_k141_101425\|\|full | 190816 | Yes | 18468 | 194 | 18 | 109 | Low-quality | Genome-fragment | 27.31 | HMM-based (lower-bound) | 90.32 |

| | Count | Mean | Median | SD |
|---|---|---|---|---|
| Complete (100% completeness) | 193 | 62030.8 | 51608 | 40950.2 |
| High quality (90-100% completeness) | 488 | 48150.1 | 41459.5 | 28976.3 |
| Medium quality (50-90% completeness) | 550 | 35293 | 28721.5 | 30273 |
| Low quality (< 50% completeness) | 21851 | 3602.8 | 2005 | 5622.5 |
| Undetermined | 31697 | 2728.2 | 1777 | 2995.7 |
| Total OTUs >= 1kb | 54779 | 30361.0 | 28721.5 | 16623.2 |
| Sum of 50-100% genome completeness | 1231 | 48491.3 | 41459.5 | 6570.9 |
